## Supplementary Tables for "CausalCellInfer: Resolving cell-type-specific disease mechanisms from biobank-scale GWAS"

Table S1 Comparision of differentially expressed genes identified

| Tissue | Cell Type | PPV | scRef Control Count | scRef Condition Count | Overall count |
|---|---|---|---|---|---|
| Pancreas | Acinar Cell | 0.96 | 25437 | 6193 | 31630 |
| | Alpha Cell | 0.831 | 27877 | 13541 | 41418 |
| | Beta Cell | 0.941 | 21866 | 6234 | 28100 |
| | Delta Cell | 0.624 | 2387 | 706 | 3093 |
| | Duct Epithelial Cell | 0.701 | 6148 | 3530 | 9678 |
| | Endothelial Cell | 0.473 | 2064 | 893 | 2957 |
| | Epsilon Cell | 0.036 | 68 | 19 | 87 |
| | Leukocyte | 0.081 | 493 | 258 | 751 |
| | Mesenchymal Cell | 0.627 | 3881 | 1887 | 5768 |
| | PP cell | 0.163 | 814 | 287 | 1101 |
| Frontal cortex | Astrocytes | 0.103 | 172 | 459 | 631 |
| | Excitatory neurons | 0.701 | 10336 | 16368 | 26704 |
| | Inhibitory neurons | 0.47 | 3582 | 5893 | 9475 |
| | Microglia/Macrophage | 0.055 | 248 | 428 | 676 |
| | Oligodendrocytes | 0.183 | 1887 | 1574 | 3461 |
| | Oligodendrocyte progenitor cells | 0.191 | 819 | 942 | 1761 |
| SAT | Adipocyte | 0.906 | 6242 | 6194 | 12436 |
| | Adipose stem and progenitor cells (ASPC) | 0.797 | 5314 | 9417 | 14731 |
| | B cell | 0.074 | 55 | 227 | 282 |
| | Dendritic Cell | 0.229 | 156 | 287 | 443 |
| | Endothelial Cell | 0.707 | 2457 | 4740 | 7197 |
| | Lymphatic endothelial cells (LEC) | 0.198 | 105 | 430 | 535 |
| | Macrophage | 0.88 | 2857 | 4409 | 7266 |
| | Mast Cell | 0.129 | 183 | 572 | 755 |
| | Monocyte | 0.13 | 168 | 629 | 797 |
| | Neutrophil | 0.052 | 52 | 26 | 78 |
| | NK cell | 0.279 | 351 | 855 | 1206 |
| | T cell | 0.214 | 606 | 1766 | 2372 |
| | Smooth muscle cells (SMC) | 0.371 | 398 | 850 | 1248 |
| | Pericyte | 0.204 | 232 | 300 | 532 |
| VAT | Adipocyte | 0.705 | 4425 | 4943 | 9368 |
| | Adipose stem and progenitor cells (ASPC) | 0.621 | 4553 | 6914 | 11467 |
| | B cell | 0.024 | 25 | 42 | 67 |
| | Dendritic Cell | 0.102 | 116 | 300 | 416 |
| | Endometrium | 0.093 | 43 | 262 | 305 |
| | Endothelial Cell | 0.509 | 2081 | 2983 | 5064 |
| | Lymphatic endothelial cells (LEC) | 0.248 | 577 | 1077 | 1654 |
| | Macrophage | 0.478 | 1597 | 5731 | 7328 |
| | Mast Cell | 0.053 | 81 | 127 | 208 |
| | Mesothelium | 0.723 | 5030 | 19658 | 24688 |
| | Monocyte | 0.071 | 90 | 187 | 277 |
| | NK cell | 0.097 | 219 | 276 | 495 |
| | Pericyte | 0.138 | 326 | 281 | 607 |
| | T cell | 0.264 | 686 | 1835 | 2521 |
| | Smooth muscle cells (SMC) | 0.121 | 388 | 692 | 1080 |

Note: SAT is the abbreviation for subcutaneous adipose tissue, VAT is the abbreviation for visceral adipose tissue

Table S2 Estimated associations between estimated cell proprotions and disease

| Disease | celltype | beta | pvalue | qvalue |
|---|---|---|---|---|
| Depression | Astrocyte | 0.349344479 | 0.157297493 | 0.188756991 |
| Depression | Excitory neuron | 0.035768245 | 0.018036671 | 0.027055006 |
| Depression | Inhibitory neuron | 0.077088977 | 0.000498927 | 0.001496782 |
| Depression | Microglia/Macrophage | -0.510795951 | 0.003812823 | 0.007625646 |
| Depression | Oligodendrocyte precursor | 0.056779997 | 0.27037295 | 0.27037295 |
| Depression | Oligodendrocyte | -0.085276671 | 1.45E-07 | 8.69E-07 |
| T2DM | Acinar cell | 0.355020726 | 0.001615201 | 0.003553442 |
| T2DM | Duct epithelial cell | 0.373331282 | 3.81E-06 | 1.40E-05 |
| T2DM | Endothelial cell | 0.095142994 | 0.336647671 | 0.431430617 |
| T2DM | Leukocyte | -0.06040816 | 0.837904613 | 0.913066206 |
| T2DM | Mesenchymal cell | 0.656294611 | 4.58E-08 | 5.04E-07 |
| T2DM | Alpha cell | 0.002653652 | 0.913066206 | 0.913066206 |
| T2DM | Delta cell | 0.188143407 | 0.000636075 | 0.001749206 |
| T2DM | PP cell | 0.215849341 | 0.07091862 | 0.111443546 |
| T2DM | Epsilon cell | -0.881452643 | 0.352988686 | 0.431430617 |
| T2DM | Beta cell | -0.110780929 | 7.76E-07 | 4.27E-06 |
| T2DM | Alpha/beta ratio | 0.017953392 | 0.014327101 | 0.026266352 |
| SAT BMI | Adipose stem and precursor cell | 1.260269017 | 0.076820149 | 0.097771099 |
| SAT BMI | Lymphatic endothelial cell | 1.07717506 | 0.44254914 | 0.476591381 |
| SAT BMI | Smooth muscle cell | 9.57666729 | 0.000186335 | 0.000372671 |
| SAT BMI | Adipocyte | 0.599559889 | 0.622823291 | 0.622823291 |
| SAT BMI | B cell | -29.08081915 | 2.46E-02 | 3.45E-02 |
| SAT BMI | Dendritic cell | -7.40076279 | 0.107269022 | 0.125147193 |
| SAT BMI | Endothelial | -1.510014365 | 4.12E-04 | 7.20E-04 |
| SAT BMI | Macrophage | 4.013504669 | 4.05526E-05 | 9.46227E-05 |
| SAT BMI | Mast cell | 16.66351679 | 1.73983E-05 | 4.87153E-05 |
| SAT BMI | Monocyte | 12.1057829 | 1.76E-03 | 2.73E-03 |
| SAT BMI | Neutrophil | 56.30990704 | 1.94992E-15 | 1.36494E-14 |
| SAT BMI | NK cell | -20.94766299 | 3.28034E-32 | 4.59247E-31 |
| SAT BMI | Pericyte | 18.7857525 | 1.94857E-06 | 6.82001E-06 |
| SAT BMI | T cell | 25.81402129 | 1.41858E-13 | 6.62004E-13 |
| VAT BMI | Adipose stem and precursor cell | -1.062876878 | 1.42E-01 | 1.93E-01 |
| VAT BMI | Lymphatic endothelial cell | -2.255397112 | 0.462572815 | 0.533737863 |

| | | | | |
|---|---|---|---|---|
| VAT BMI | Smooth muscle cell | 45.16195 | 4.57948E-10 | 2.28974E-09 |
| VAT BMI | Adipocyte | 4.153365712 | 0.003342695 | 0.007162919 |
| VAT BMI | B cell | -0.925319782 | 0.931587331 | 0.931587331 |
| VAT BMI | Dendritic cell | 46.03050829 | 1.06245E-09 | 3.98418E-09 |
| VAT BMI | Endometrium | 9.167961312 | 4.16E-01 | 5.20E-01 |
| VAT BMI | Endothelial | -1.605687331 | 0.041210599 | 0.068684332 |
| VAT BMI | Macrophage | 0.33648321 | 8.40E-01 | 9.00E-01 |
| VAT BMI | Mast cell | 37.32441676 | 7.21365E-05 | 0.000180341 |
| VAT BMI | Mesothelium | 4.289093879 | 5.17983E-09 | 1.55395E-08 |
| VAT BMI | Monocyte | -8.539173391 | 1.25E-01 | 1.88E-01 |
| VAT BMI | NK cell | -27.67223717 | 4.56164E-36 | 6.84246E-35 |
| VAT BMI | Pericyte | 28.95595643 | 0.011895107 | 0.022303326 |
| VAT BMI | T cell | -50.07611457 | 2.72392E-20 | 2.04294E-19 |

Note:qvalue indicates the qvaule after multiple testing correction by Benjamini-Hochberg method;Astros is the abbreviation for Astrocytes, Ex is the abbreviation for Excitatory neuron, Inhib is the abbreviation for Inhibitory neuron, MicroMacro is the abbreviation for Microglia/Macrophage, OPCs is the abbreviation for Oligodendrocyte precursor; ASPC is the abbreviation for Adipose stem and precursor cell, LEC is the abbreviation for Lymphatic endothelial cell, SMC is the abbreviation for Smooth muscle cell.

Table S3 Cell type interaction effcts on disorder status

a. Cell type interaction effects on depression in the frontal cortex

| Tissue | Trait | Cell type interaction | Pvalue |
|---|---|---|---|
| Frontal cortex | Depression | Ex:Inhib | 0.9755645 |
| Frontal cortex | Depression | Ex:Micro_Macro | 0.70178907 |
| Frontal cortex | Depression | Ex:OPCs | 0.68071621 |
| Frontal cortex | Depression | Ex:Oligos | **0.01111706** |
| Frontal cortex | Depression | Inhib:Micro_Macro | 0.58126025 |
| Frontal cortex | Depression | Inhib:OPCs | 0.35885833 |
| Frontal cortex | Depression | Inhib:Oligos | ***0.0805781*** |
| Frontal cortex | Depression | Micro_Macro:OPCs | 0.67716352 |
| Frontal cortex | Depression | Micro_Macro:Oligos | 0.81555145 |
| Frontal cortex | Depression | OPCs:Oligos | 0.71161884 |

Note: Astros is the abbreviation for Astrocytes; MicroMacro is the abbreviation for Microglia/Macrophage; OPCs is the abbreviation for Oligodendrocyte precursor

b. Cell type interaction effects on T2DM in the pancreas

| Tissue | Trait | Cell type interaction | Pvalue |
|---|---|---|---|
| Pancreas | T2DM | duct_epithelial_cell:endothelial_cell | ***0.05030018*** |
| Pancreas | T2DM | duct_epithelial_cell:leukocyte | ***0.05670365*** |
| Pancreas | T2DM | duct_epithelial_cell:mesenchymal_cell | **0.00322339** |
| Pancreas | T2DM | duct_epithelial_cell:alpha_cell | 0.9123302 |
| Pancreas | T2DM | duct_epithelial_cell:delta_cell | 3.32E-01 |
| Pancreas | T2DM | duct_epithelial_cell:PP_cell | 0.7080235 |
| Pancreas | T2DM | duct_epithelial_cell:epsilon_cell | 0.34485085 |
| Pancreas | T2DM | duct_epithelial_cell:beta_cell | 0.87473075 |
| Pancreas | T2DM | endothelial_cell:leukocyte | 0.57353405 |
| Pancreas | T2DM | endothelial_cell:mesenchymal_cell | 0.16214993 |
| Pancreas | T2DM | endothelial_cell:alpha_cell | 0.35015599 |
| Pancreas | T2DM | endothelial_cell:delta_cell | 0.73737204 |
| Pancreas | T2DM | endothelial_cell:PP_cell | 1.62E-01 |
| Pancreas | T2DM | endothelial_cell:epsilon_cell | 2.33E-01 |
| Pancreas | T2DM | endothelial_cell:beta_cell | 0.21542553 |
| Pancreas | T2DM | leukocyte:mesenchymal_cell | 0.22532 |
| Pancreas | T2DM | leukocyte:alpha_cell | ***0.0785611*** |
| Pancreas | T2DM | leukocyte:delta_cell | 0.67711279 |
| Pancreas | T2DM | leukocyte:PP_cell | 0.27246319 |
| Pancreas | T2DM | leukocyte:epsilon_cell | 0.19485244 |
| Pancreas | T2DM | leukocyte:beta_cell | **0.00318546** |
| Pancreas | T2DM | mesenchymal_cell:alpha_cell | **3.06E-02** |
| Pancreas | T2DM | mesenchymal_cell:delta_cell | 0.82653135 |
| Pancreas | T2DM | mesenchymal_cell:PP_cell | **0.00565235** |
| Pancreas | T2DM | mesenchymal_cell:epsilon_cell | 0.11614666 |
| Pancreas | T2DM | mesenchymal_cell:beta_cell | ***0.08513141*** |

| | | | |
|---|---|---|---|
| Pancreas | T2DM | alpha_cell:delta_cell | 0.51248043 |
| Pancreas | T2DM | alpha_cell:PP_cell | **0.00327598** |
| Pancreas | T2DM | alpha_cell:epsilon_cell | 0.49339086 |
| Pancreas | T2DM | alpha_cell:beta_cell | 0.43256464 |
| Pancreas | T2DM | delta_cell:PP_cell | 0.80508856 |
| Pancreas | T2DM | delta_cell:epsilon_cell | 7.83E-01 |
| Pancreas | T2DM | delta_cell:beta_cell | ***0.05781763*** |
| Pancreas | T2DM | PP_cell:epsilon_cell | 0.59060423 |
| Pancreas | T2DM | PP_cell:beta_cell | 0.10990806 |
| Pancreas | T2DM | epsilon_cell:beta_cell | 9.57E-01 |

c. Cell type interaction effects on obesity in the subcutaneous adipose

| Tissue | Trait | Cell type interaction | Pvalue |
|---|---|---|---|
| SAT | Obesity | ASPC:LEC | **4.73E-03** |
| SAT | Obesity | ASPC:SMC | 0.24506652 |
| SAT | Obesity | ASPC:adipocyte | 0.57539881 |
| SAT | Obesity | ASPC:b_cell | 0.13532347 |
| SAT | Obesity | ASPC:dendritic_cell | 0.23602351 |
| SAT | Obesity | ASPC:macrophage | 0.85546771 |
| SAT | Obesity | ASPC:mast_cell | **1.78E-03** |
| SAT | Obesity | ASPC:monocyte | 0.6860553 |
| SAT | Obesity | ASPC:neutrophil | 9.40E-01 |
| SAT | Obesity | ASPC:nk_cell | **0.00031683** |
| SAT | Obesity | ASPC:pericyte | **1.43E-03** |
| SAT | Obesity | ASPC:t_cell | **4.13E-07** |
| SAT | Obesity | LEC:SMC | **0.01814628** |
| SAT | Obesity | LEC:adipocyte | **1.88E-02** |
| SAT | Obesity | LEC:b_cell | **0.00025249** |
| SAT | Obesity | LEC:dendritic_cell | **0.00176223** |
| SAT | Obesity | LEC:macrophage | 3.52E-01 |
| SAT | Obesity | LEC:mast_cell | **0.00974052** |
| SAT | Obesity | LEC:monocyte | 0.24483908 |
| SAT | Obesity | LEC:neutrophil | 4.79E-01 |
| SAT | Obesity | LEC:nk_cell | **9.25E-06** |
| SAT | Obesity | LEC:pericyte | **4.09E-02** |
| SAT | Obesity | LEC:t_cell | **4.59E-03** |
| SAT | Obesity | SMC:adipocyte | **4.13E-03** |
| SAT | Obesity | SMC:b_cell | 0.24747253 |
| SAT | Obesity | SMC:dendritic_cell | 2.67E-01 |
| SAT | Obesity | SMC:macrophage | **0.00921008** |
| SAT | Obesity | SMC:mast_cell | **2.89E-02** |
| SAT | Obesity | SMC:monocyte | 0.55301892 |
| SAT | Obesity | SMC:neutrophil | **2.40E-02** |
| SAT | Obesity | SMC:nk_cell | **4.25E-05** |
| SAT | Obesity | SMC:pericyte | **1.31E-06** |
| SAT | Obesity | SMC:t_cell | **0.00346203** |
| SAT | Obesity | adipocyte:b_cell | 0.10736334 |
| SAT | Obesity | adipocyte:dendritic_cell | 0.80851702 |
| SAT | Obesity | adipocyte:macrophage | 1.71E-01 |

| | | | |
|---|---|---|---|
| SAT | Obesity | adipocyte:mast_cell | **0.00424343** |
| SAT | Obesity | adipocyte:monocyte | 4.25E-01 |
| SAT | Obesity | adipocyte:neutrophil | **6.03E-02** |
| SAT | Obesity | adipocyte:nk_cell | 0.31759755 |
| SAT | Obesity | adipocyte:pericyte | 9.39E-01 |
| SAT | Obesity | adipocyte:t_cell | **0.00689157** |
| SAT | Obesity | b_cell:dendritic_cell | 0.95177954 |
| SAT | Obesity | b_cell:macrophage | 4.51E-01 |
| SAT | Obesity | b_cell:mast_cell | **4.21E-03** |
| SAT | Obesity | b_cell:monocyte | 0.23358375 |
| SAT | Obesity | b_cell:neutrophil | **0.03839472** |
| SAT | Obesity | b_cell:nk_cell | **0.01046318** |
| SAT | Obesity | b_cell:pericyte | **7.65E-03** |
| SAT | Obesity | b_cell:t_cell | **1.97E-04** |
| SAT | Obesity | dendritic_cell:macrophage | ***5.17E-02*** |
| SAT | Obesity | dendritic_cell:mast_cell | 3.09E-01 |
| SAT | Obesity | dendritic_cell:monocyte | 0.59036409 |
| SAT | Obesity | dendritic_cell:neutrophil | **0.00238595** |
| SAT | Obesity | dendritic_cell:nk_cell | 0.57412387 |
| SAT | Obesity | dendritic_cell:pericyte | ***0.06021844*** |
| SAT | Obesity | dendritic_cell:t_cell | ***8.19E-02*** |
| SAT | Obesity | macrophage:mast_cell | 0.13139713 |
| SAT | Obesity | macrophage:monocyte | 0.30064477 |
| SAT | Obesity | macrophage:neutrophil | ***0.08357006*** |
| SAT | Obesity | macrophage:nk_cell | **0.00424697** |
| SAT | Obesity | macrophage:pericyte | **1.24E-02** |
| SAT | Obesity | macrophage:t_cell | **2.48E-04** |

| | | | |
|---|---|---|---|
| SAT | Obesity | mast_cell:monocyte | 0.4251265 |
| SAT | Obesity | mast_cell:neutrophil | 4.81E-01 |
| SAT | Obesity | mast_cell:nk_cell | **0.00131752** |
| SAT | Obesity | mast_cell:pericyte | **0.02824009** |
| SAT | Obesity | mast_cell:t_cell | ***9.94E-02*** |
| SAT | Obesity | monocyte:neutrophil | 0.44130355 |
| SAT | Obesity | monocyte:nk_cell | 0.46298214 |
| SAT | Obesity | monocyte:pericyte | 1.19E-01 |
| SAT | Obesity | monocyte:t_cell | 0.46989198 |
| SAT | Obesity | neutrophil:nk_cell | **7.85E-07** |
| SAT | Obesity | neutrophil:pericyte | **1.37E-03** |
| SAT | Obesity | neutrophil:t_cell | 0.36684085 |
| SAT | Obesity | nk_cell:pericyte | **0.02456507** |
| SAT | Obesity | nk_cell:t_cell | ***0.07740871*** |
| SAT | Obesity | pericyte:t_cell | **0.02233631** |

d. Cell type interaction effects on obesity in the visceral adipose tissue

| Tissue | Trait | Cell type interaction | Pvalue |
|---|---|---|---|
| VAT | Obesity | ASPC:LEC | 7.64E-01 |
| VAT | Obesity | ASPC:SMC | ***6.44E-02*** |
| VAT | Obesity | ASPC:adipocyte | **7.95E-03** |
| VAT | Obesity | ASPC:b_cell | **1.36E-02** |
| VAT | Obesity | ASPC:dendritic_cell | 6.01E-01 |
| VAT | Obesity | ASPC:endometrium | **5.41E-03** |
| VAT | Obesity | ASPC:macrophage | 2.57E-01 |
| VAT | Obesity | ASPC:mast_cell | 2.88E-01 |
| VAT | Obesity | ASPC:mesothelium | **3.07E-02** |
| VAT | Obesity | ASPC:monocyte | 8.68E-01 |
| VAT | Obesity | ASPC:nk_cell | 8.36E-01 |
| VAT | Obesity | ASPC:pericyte | 4.18E-01 |
| VAT | Obesity | ASPC:t_cell | 5.92E-01 |
| VAT | Obesity | LEC:SMC | 2.99E-01 |
| VAT | Obesity | LEC:adipocyte | **9.66E-03** |
| VAT | Obesity | LEC:b_cell | ***7.48E-02*** |
| VAT | Obesity | LEC:dendritic_cell | ***8.02E-02*** |
| VAT | Obesity | LEC:endometrium | **5.57E-08** |
| VAT | Obesity | LEC:macrophage | **4.76E-02** |
| VAT | Obesity | LEC:mast_cell | 9.68E-01 |
| VAT | Obesity | LEC:mesothelium | **5.68E-03** |
| VAT | Obesity | LEC:monocyte | 9.46E-01 |
| VAT | Obesity | LEC:nk_cell | **3.75E-03** |
| VAT | Obesity | LEC:pericyte | **1.67E-04** |
| VAT | Obesity | LEC:t_cell | 3.47E-01 |
| VAT | Obesity | SMC:adipocyte | 2.42E-01 |
| VAT | Obesity | SMC:b_cell | 1.13E-01 |
| VAT | Obesity | SMC:dendritic_cell | 6.17E-01 |
| VAT | Obesity | SMC:endometrium | **1.03E-04** |
| VAT | Obesity | SMC:macrophage | 5.82E-01 |
| VAT | Obesity | SMC:mast_cell | 1.49E-01 |
| VAT | Obesity | SMC:mesothelium | 5.21E-01 |
| VAT | Obesity | SMC:monocyte | ***7.26E-02*** |
| VAT | Obesity | SMC:nk_cell | **1.28E-04** |

| | | | |
|---|---|---|---|
| VAT | Obesity | SMC:pericyte | **3.45E-05** |
| VAT | Obesity | SMC:t_cell | 3.92E-01 |
| VAT | Obesity | adipocyte:b_cell | 1.24E-01 |
| VAT | Obesity | adipocyte:dendritic_cell | 2.50E-01 |
| VAT | Obesity | adipocyte:endometrium | **2.78E-09** |
| VAT | Obesity | adipocyte:macrophage | 2.09E-01 |
| VAT | Obesity | adipocyte:mast_cell | 2.81E-01 |
| VAT | Obesity | adipocyte:mesothelium | **2.23E-02** |
| VAT | Obesity | adipocyte:monocyte | 6.39E-01 |
| VAT | Obesity | adipocyte:nk_cell | **3.19E-05** |
| VAT | Obesity | adipocyte:pericyte | **2.73E-05** |
| VAT | Obesity | adipocyte:t_cell | ***6.71E-02*** |
| VAT | Obesity | b_cell:dendritic_cell | 1.21E-01 |
| VAT | Obesity | b_cell:endometrium | **2.91E-08** |
| VAT | Obesity | b_cell:macrophage | 2.86E-01 |
| VAT | Obesity | b_cell:mast_cell | 2.14E-01 |
| VAT | Obesity | b_cell:mesothelium | **1.70E-04** |
| VAT | Obesity | b_cell:monocyte | ***7.49E-02*** |
| VAT | Obesity | b_cell:nk_cell | **1.46E-06** |
| VAT | Obesity | b_cell:pericyte | **1.22E-07** |
| VAT | Obesity | b_cell:t_cell | 1.37E-01 |
| VAT | Obesity | dendritic_cell:endometrium | **1.53E-02** |
| VAT | Obesity | dendritic_cell:macrophage | **4.10E-02** |
| VAT | Obesity | dendritic_cell:mast_cell | 4.94E-01 |
| VAT | Obesity | dendritic_cell:mesothelium | **7.23E-03** |
| VAT | Obesity | dendritic_cell:monocyte | 6.98E-01 |
| VAT | Obesity | dendritic_cell:nk_cell | ***8.91E-02*** |
| VAT | Obesity | dendritic_cell:pericyte | **1.41E-03** |

| | | | |
|---|---|---|---|
| VAT | Obesity | dendritic_cell:t_cell | 3.51E-01 |
| VAT | Obesity | endometrium:macrophage | **5.64E-04** |
| VAT | Obesity | endometrium:mast_cell | 4.12E-01 |
| VAT | Obesity | endometrium:mesothelium | 2.78E-01 |
| VAT | Obesity | endometrium:monocyte | 4.62E-01 |
| VAT | Obesity | endometrium:nk_cell | **2.82E-07** |
| VAT | Obesity | endometrium:pericyte | ***7.23E-02*** |
| VAT | Obesity | endometrium:t_cell | ***5.21E-02*** |
| VAT | Obesity | macrophage:mast_cell | 1.78E-01 |
| VAT | Obesity | macrophage:mesothelium | 8.85E-01 |
| VAT | Obesity | macrophage:monocyte | 1.63E-01 |
| VAT | Obesity | macrophage:nk_cell | 9.19E-01 |
| VAT | Obesity | macrophage:pericyte | **1.90E-02** |
| VAT | Obesity | macrophage:t_cell | 2.10E-01 |
| VAT | Obesity | mast_cell:mesothelium | 1.03E-01 |
| VAT | Obesity | mast_cell:monocyte | **2.42E-02** |
| VAT | Obesity | mast_cell:nk_cell | 5.62E-01 |
| VAT | Obesity | mast_cell:pericyte | 3.16E-01 |
| VAT | Obesity | mast_cell:t_cell | 1.29E-01 |
| VAT | Obesity | mesothelium:monocyte | 4.62E-01 |
| VAT | Obesity | mesothelium:nk_cell | **1.27E-04** |
| VAT | Obesity | mesothelium:pericyte | 5.85E-01 |
| VAT | Obesity | mesothelium:t_cell | 6.89E-01 |
| VAT | Obesity | monocyte:nk_cell | 3.69E-01 |

| | | | |
|---|---|---|---|
| VAT | Obesity | monocyte:pericyte | 3.07E-01 |
| VAT | Obesity | monocyte:t_cell | 5.20E-01 |
| VAT | Obesity | nk_cell:pericyte | **9.47E-04** |
| VAT | Obesity | nk_cell:t_cell | 8.20E-01 |
| VAT | Obesity | pericyte:t_cell | 2.24E-01 |

| Trait | First tissue | Second tissue | Variable | Pvalue |
|---|---|---|---|---|
| Depression | Frontal cortex | Pancrease | Ex | 0.77972662 |
| Depression | Frontal cortex | Pancrease | Inhib | 0.82393658 |
| Depression | Frontal cortex | Pancrease | MicroMacro | 0.91598358 |
| Depression | Frontal cortex | Pancrease | OPCs | 0.73195791 |
| Depression | Frontal cortex | Pancrease | Oligos | 0.74543721 |
| Depression | Frontal cortex | Pancrease | duct_epithelial_cell | 0.78472255 |
| Depression | Frontal cortex | Pancrease | endothelial_cell | 0.63059751 |
| Depression | Frontal cortex | Pancrease | leukocyte | 0.68635775 |
| Depression | Frontal cortex | Pancrease | mesenchymal_cell | 0.48448636 |
| Depression | Frontal cortex | Pancrease | alpha_cell | 0.94239018 |
| Depression | Frontal cortex | Pancrease | delta_cell | 0.90215208 |
| Depression | Frontal cortex | Pancrease | PP_cell | 0.26641881 |
| Depression | Frontal cortex | Pancrease | epsilon_cell | 0.49130186 |
| Depression | Frontal cortex | Pancrease | beta_cell | 0.68523411 |
| Depression | Frontal cortex | Pancrease | Ex:Inhib | 0.99103426 |
| Depression | Frontal cortex | Pancrease | Ex:MicroMacro | 0.71797195 |
| Depression | Frontal cortex | Pancrease | Ex:OPCs | 0.62986962 |
| Depression | Frontal cortex | Pancrease | Ex:Oligos | **0.02913141** |
| Depression | Frontal cortex | Pancrease | Ex:duct_epithelial_cell | 0.70081449 |
| Depression | Frontal cortex | Pancrease | Ex:endothelial_cell | 0.71122987 |
| Depression | Frontal cortex | Pancrease | Ex:leukocyte | 0.68047182 |
| Depression | Frontal cortex | Pancrease | Ex:mesenchymal_cell | 0.4396112 |
| Depression | Frontal cortex | Pancrease | Ex:alpha_cell | 0.91598342 |
| Depression | Frontal cortex | Pancrease | Ex:delta_cell | 0.81748885 |
| Depression | Frontal cortex | Pancrease | Ex:PP_cell | 0.29209388 |
| Depression | Frontal cortex | Pancrease | Ex:epsilon_cell | 0.67799809 |
| Depression | Frontal cortex | Pancrease | Ex:beta_cell | 0.66573396 |
| Depression | Frontal cortex | Pancrease | Inhib:MicroMacro | 0.58517127 |
| Depression | Frontal cortex | Pancrease | Inhib:OPCs | 0.31725008 |
| Depression | Frontal cortex | Pancrease | Inhib:Oligos | 0.18159341 |
| Depression | Frontal cortex | Pancrease | Inhib:duct_epithelial_cell | 0.72814371 |
| Depression | Frontal cortex | Pancrease | Inhib:endothelial_cell | 0.63183261 |
| Depression | Frontal cortex | Pancrease | Inhib:leukocyte | 0.67643079 |
| Depression | Frontal cortex | Pancrease | Inhib:mesenchymal_cell | 0.48385566 |
| Depression | Frontal cortex | Pancrease | Inhib:alpha_cell | 0.97475649 |
| Depression | Frontal cortex | Pancrease | Inhib:delta_cell | 0.87002002 |
| Depression | Frontal cortex | Pancrease | Inhib:PP_cell | 0.27700243 |

| | | | | |
|---|---|---|---|---|
| Depression | Frontal cortex | Pancrease | Inhib:epsilon_cell | 0.54713128 |
| Depression | Frontal cortex | Pancrease | Inhib:beta_cell | 0.70346486 |
| Depression | Frontal cortex | Pancrease | MicroMacro:OPCs | 0.69879793 |
| Depression | Frontal cortex | Pancrease | MicroMacro:Oligos | 0.76581727 |
| Depression | Frontal cortex | Pancrease | MicroMacro:duct_epithelial_cell | 0.52647261 |
| Depression | Frontal cortex | Pancrease | MicroMacro:endothelial_cell | 0.80108911 |
| Depression | Frontal cortex | Pancrease | MicroMacro:leukocyte | 0.82036589 |
| Depression | Frontal cortex | Pancrease | MicroMacro:mesenchymal_cell | 0.21652239 |
| Depression | Frontal cortex | Pancrease | MicroMacro:alpha_cell | 0.92825213 |
| Depression | Frontal cortex | Pancrease | MicroMacro:delta_cell | 0.61997013 |
| Depression | Frontal cortex | Pancrease | MicroMacro:PP_cell | 0.28528554 |
| Depression | Frontal cortex | Pancrease | MicroMacro:epsilon_cell | 0.72819515 |
| Depression | Frontal cortex | Pancrease | MicroMacro:beta_cell | 0.95994792 |
| Depression | Frontal cortex | Pancrease | OPCs:Oligos | 0.59002932 |
| Depression | Frontal cortex | Pancrease | OPCs:duct_epithelial_cell | 0.59200821 |
| Depression | Frontal cortex | Pancrease | OPCs:endothelial_cell | 0.75927277 |
| Depression | Frontal cortex | Pancrease | OPCs:leukocyte | 0.72876039 |
| Depression | Frontal cortex | Pancrease | OPCs:mesenchymal_cell | 0.4803858 |
| Depression | Frontal cortex | Pancrease | OPCs:alpha_cell | 0.89033379 |
| Depression | Frontal cortex | Pancrease | OPCs:delta_cell | 0.78524398 |
| Depression | Frontal cortex | Pancrease | OPCs:PP_cell | 0.25818782 |
| Depression | Frontal cortex | Pancrease | OPCs:epsilon_cell | 0.73411885 |
| Depression | Frontal cortex | Pancrease | OPCs:beta_cell | 0.65957138 |
| Depression | Frontal cortex | Pancrease | Oligos:duct_epithelial_cell | 0.65570306 |
| Depression | Frontal cortex | Pancrease | Oligos:endothelial_cell | 0.70541023 |
| Depression | Frontal cortex | Pancrease | Oligos:leukocyte | 0.63301816 |
| Depression | Frontal cortex | Pancrease | Oligos:mesenchymal_cell | 0.42129957 |
| Depression | Frontal cortex | Pancrease | Oligos:alpha_cell | 0.88629613 |

| | | | | |
|---|---|---|---|---|
| Depression | Frontal cortex | Pancrease | Oligos:delta_cell | 0.7679387 |
| Depression | Frontal cortex | Pancrease | Oligos:PP_cell | 0.28486914 |
| Depression | Frontal cortex | Pancrease | Oligos:epsilon_cell | 0.63662798 |
| Depression | Frontal cortex | Pancrease | Oligos:beta_cell | 0.62069494 |
| Depression | Frontal cortex | Pancrease | duct_epithelial_cell:endothelial_cell | 0.36678455 |
| Depression | Frontal cortex | Pancrease | duct_epithelial_cell:leukocyte | 0.42735462 |
| Depression | Frontal cortex | Pancrease | duct_epithelial_cell:mesenchymal_cell | 0.50826743 |
| Depression | Frontal cortex | Pancrease | duct_epithelial_cell:alpha_cell | 0.48260383 |
| Depression | Frontal cortex | Pancrease | duct_epithelial_cell:delta_cell | 0.38773058 |
| Depression | Frontal cortex | Pancrease | duct_epithelial_cell:PP_cell | 0.94296843 |
| Depression | Frontal cortex | Pancrease | duct_epithelial_cell:epsilon_cell | 0.50261319 |
| Depression | Frontal cortex | Pancrease | duct_epithelial_cell:beta_cell | 0.38625726 |
| Depression | Frontal cortex | Pancrease | endothelial_cell:leukocyte | 0.9634569 |
| Depression | Frontal cortex | Pancrease | endothelial_cell:mesenchymal_cell | 0.54449106 |
| Depression | Frontal cortex | Pancrease | endothelial_cell:alpha_cell | 0.80651466 |
| Depression | Frontal cortex | Pancrease | endothelial_cell:delta_cell | 0.74597747 |
| Depression | Frontal cortex | Pancrease | endothelial_cell:PP_cell | 0.520598 |
| Depression | Frontal cortex | Pancrease | endothelial_cell:epsilon_cell | 0.76118736 |
| Depression | Frontal cortex | Pancrease | endothelial_cell:beta_cell | 0.62273765 |
| Depression | Frontal cortex | Pancrease | leukocyte:mesenchymal_cell | 0.82565192 |
| Depression | Frontal cortex | Pancrease | leukocyte:alpha_cell | 0.79572581 |
| Depression | Frontal cortex | Pancrease | leukocyte:delta_cell | 0.51225656 |
| Depression | Frontal cortex | Pancrease | leukocyte:PP_cell | 0.43518179 |
| Depression | Frontal cortex | Pancrease | leukocyte:epsilon_cell | ***0.05438721*** |

| | | | | |
|---|---|---|---|---|
| Depression | Frontal cortex | Pancrease | leukocyte:beta_cell | 0.88285545 |
| Depression | Frontal cortex | Pancrease | mesenchymal_cell:alpha_cell | 0.90412403 |
| Depression | Frontal cortex | Pancrease | mesenchymal_cell:delta_cell | 0.45813856 |
| Depression | Frontal cortex | Pancrease | mesenchymal_cell:PP_cell | 0.9602634 |
| Depression | Frontal cortex | Pancrease | mesenchymal_cell:epsilon_cell | 0.27522734 |
| Depression | Frontal cortex | Pancrease | mesenchymal_cell:beta_cell | 0.59006751 |
| Depression | Frontal cortex | Pancrease | alpha_cell:delta_cell | 0.23329265 |
| Depression | Frontal cortex | Pancrease | alpha_cell:PP_cell | 0.7843447 |
| Depression | Frontal cortex | Pancrease | alpha_cell:epsilon_cell | 0.65385421 |
| Depression | Frontal cortex | Pancrease | alpha_cell:beta_cell | 0.48237566 |
| Depression | Frontal cortex | Pancrease | delta_cell:PP_cell | 0.33832434 |
| Depression | Frontal cortex | Pancrease | delta_cell:epsilon_cell | 0.82189991 |
| Depression | Frontal cortex | Pancrease | delta_cell:beta_cell | 0.17709732 |
| Depression | Frontal cortex | Pancrease | PP_cell:epsilon_cell | 0.31407615 |
| Depression | Frontal cortex | Pancrease | PP_cell:beta_cell | 0.71154434 |
| Depression | Frontal cortex | Pancrease | epsilon_cell:beta_cell | 0.49666533 |
| Depression | Frontal cortex | SAT | Ex | 0.71514166 |
| Depression | Frontal cortex | SAT | Inhib | 0.75717705 |
| Depression | Frontal cortex | SAT | MicroMacro | 0.91749027 |
| Depression | Frontal cortex | SAT | OPCs | 0.61880318 |
| Depression | Frontal cortex | SAT | Oligos | 0.80515319 |
| Depression | Frontal cortex | SAT | ASPC | 0.5390469 |
| Depression | Frontal cortex | SAT | LEC | 0.79653786 |
| Depression | Frontal cortex | SAT | SMC | ***0.0507791*** |
| Depression | Frontal cortex | SAT | adipocyte | 0.8053499 |
| Depression | Frontal cortex | SAT | b_cell | 0.94401259 |
| Depression | Frontal cortex | SAT | dendritic_cell | 0.81799245 |
| Depression | Frontal cortex | SAT | macrophage | 0.77838479 |
| Depression | Frontal cortex | SAT | mast_cell | 0.94801254 |
| Depression | Frontal cortex | SAT | monocyte | 0.51914066 |
| Depression | Frontal cortex | SAT | neutrophil | 0.89712814 |
| Depression | Frontal cortex | SAT | nk_cell | 0.79409881 |
| Depression | Frontal cortex | SAT | pericyte | 0.75915093 |

| | | | | |
|---|---|---|---|---|
| Depression | Frontal cortex | SAT | t_cell | 0.3570095 |
| Depression | Frontal cortex | SAT | Ex:Inhib | 0.86751388 |
| Depression | Frontal cortex | SAT | Ex:MicroMacro | 0.96455162 |
| Depression | Frontal cortex | SAT | Ex:OPCs | 0.67442311 |
| Depression | Frontal cortex | SAT | Ex:Oligos | 0.47123777 |
| Depression | Frontal cortex | SAT | Ex:ASPC | 0.52898904 |
| Depression | Frontal cortex | SAT | Ex:LEC | 0.81946707 |
| Depression | Frontal cortex | SAT | Ex:SMC | 0.0488227 |
| Depression | Frontal cortex | SAT | Ex:adipocyte | 0.80417435 |
| Depression | Frontal cortex | SAT | Ex:b_cell | 0.85079094 |
| Depression | Frontal cortex | SAT | Ex:dendritic_cell | 0.9524783 |
| Depression | Frontal cortex | SAT | Ex:macrophage | 0.8040401 |
| Depression | Frontal cortex | SAT | Ex:mast_cell | 0.9675978 |
| Depression | Frontal cortex | SAT | Ex:monocyte | 0.56823908 |
| Depression | Frontal cortex | SAT | Ex:neutrophil | 0.73325973 |
| Depression | Frontal cortex | SAT | Ex:nk_cell | 0.62354932 |
| Depression | Frontal cortex | SAT | Ex:pericyte | 0.58921938 |
| Depression | Frontal cortex | SAT | Ex:t_cell | 0.27017588 |
| Depression | Frontal cortex | SAT | Inhib:MicroMacro | 0.89685474 |
| Depression | Frontal cortex | SAT | Inhib:OPCs | 0.34001228 |
| Depression | Frontal cortex | SAT | Inhib:Oligos | 0.37677095 |
| Depression | Frontal cortex | SAT | Inhib:ASPC | 0.58296609 |
| Depression | Frontal cortex | SAT | Inhib:LEC | 0.79647992 |
| Depression | Frontal cortex | SAT | Inhib:SMC | 0.05046435 |
| Depression | Frontal cortex | SAT | Inhib:adipocyte | 0.67412999 |
| Depression | Frontal cortex | SAT | Inhib:b_cell | 0.85700621 |
| Depression | Frontal cortex | SAT | Inhib:dendritic_cell | 0.94825064 |
| Depression | Frontal cortex | SAT | Inhib:macrophage | 0.74060609 |
| Depression | Frontal cortex | SAT | Inhib:mast_cell | 0.94265592 |
| Depression | Frontal cortex | SAT | Inhib:monocyte | 0.49095357 |
| Depression | Frontal cortex | SAT | Inhib:neutrophil | 0.7110502 |
| Depression | Frontal cortex | SAT | Inhib:nk_cell | 0.73966163 |
| Depression | Frontal cortex | SAT | Inhib:pericyte | 0.52410218 |
| Depression | Frontal cortex | SAT | Inhib:t_cell | 0.28994241 |
| Depression | Frontal cortex | SAT | MicroMacro:OPCs | 0.56951091 |
| Depression | Frontal cortex | SAT | MicroMacro:Oligos | 0.9264086 |
| Depression | Frontal cortex | SAT | MicroMacro:ASPC | 0.76231768 |
| Depression | Frontal cortex | SAT | MicroMacro:LEC | 0.68016308 |
| Depression | Frontal cortex | SAT | MicroMacro:SMC | 0.33462081 |

| | | | | |
|---|---|---|---|---|
| Depression | Frontal cortex | SAT | MicroMacro:adipocyte | 0.56271663 |
| Depression | Frontal cortex | SAT | MicroMacro:b_cell | 0.5087087 |
| Depression | Frontal cortex | SAT | MicroMacro:dendritic_cell | 0.80362074 |
| Depression | Frontal cortex | SAT | MicroMacro:macrophage | 0.69061333 |
| Depression | Frontal cortex | SAT | MicroMacro:mast_cell | 0.66844033 |
| Depression | Frontal cortex | SAT | MicroMacro:monocyte | 0.85617912 |
| Depression | Frontal cortex | SAT | MicroMacro:neutrophil | 0.84167034 |
| Depression | Frontal cortex | SAT | MicroMacro:nk_cell | 0.8551138 |
| Depression | Frontal cortex | SAT | MicroMacro:pericyte | 0.23862515 |
| Depression | Frontal cortex | SAT | MicroMacro:t_cell | 0.0928086 |
| Depression | Frontal cortex | SAT | OPCs:Oligos | 0.53173169 |
| Depression | Frontal cortex | SAT | OPCs:ASPC | 0.54779196 |
| Depression | Frontal cortex | SAT | OPCs:LEC | 0.80011699 |
| Depression | Frontal cortex | SAT | OPCs:SMC | 0.05427109 |
| Depression | Frontal cortex | SAT | OPCs:adipocyte | 0.97568576 |
| Depression | Frontal cortex | SAT | OPCs:b_cell | 0.91651241 |
| Depression | Frontal cortex | SAT | OPCs:dendritic_cell | 0.9768748 |
| Depression | Frontal cortex | SAT | OPCs:macrophage | 0.75339458 |
| Depression | Frontal cortex | SAT | OPCs:mast_cell | 0.8467622 |
| Depression | Frontal cortex | SAT | OPCs:monocyte | 0.43576125 |
| Depression | Frontal cortex | SAT | OPCs:neutrophil | 0.52524644 |
| Depression | Frontal cortex | SAT | OPCs:nk_cell | 0.44801769 |
| Depression | Frontal cortex | SAT | OPCs:pericyte | 0.48226167 |
| Depression | Frontal cortex | SAT | OPCs:t_cell | 0.2829173 |
| Depression | Frontal cortex | SAT | Oligos:ASPC | 0.61630409 |
| Depression | Frontal cortex | SAT | Oligos:LEC | 0.7971185 |
| Depression | Frontal cortex | SAT | Oligos:SMC | 0.04564157 |
| Depression | Frontal cortex | SAT | Oligos:adipocyte | 0.71927306 |
| Depression | Frontal cortex | SAT | Oligos:b_cell | 0.77126064 |
| Depression | Frontal cortex | SAT | Oligos:dendritic_cell | 0.89986759 |
| Depression | Frontal cortex | SAT | Oligos:macrophage | 0.82822948 |
| Depression | Frontal cortex | SAT | Oligos:mast_cell | 0.99237962 |
| Depression | Frontal cortex | SAT | Oligos:monocyte | 0.51643498 |

| | | | | |
|---|---|---|---|---|
| Depression | Frontal cortex | SAT | Oligos:neutrophil | 0.64952236 |
| Depression | Frontal cortex | SAT | Oligos:nk_cell | 0.63105383 |
| Depression | Frontal cortex | SAT | Oligos:pericyte | 0.56936286 |
| Depression | Frontal cortex | SAT | Oligos:t_cell | 0.3077139 |
| Depression | Frontal cortex | SAT | ASPC:LEC | 0.96394394 |
| Depression | Frontal cortex | SAT | ASPC:SMC | 0.64504332 |
| Depression | Frontal cortex | SAT | ASPC:adipocyte | 0.78566173 |
| Depression | Frontal cortex | SAT | ASPC:b_cell | 0.48587412 |
| Depression | Frontal cortex | SAT | ASPC:dendritic_cell | 0.46362859 |
| Depression | Frontal cortex | SAT | ASPC:macrophage | 0.88127704 |
| Depression | Frontal cortex | SAT | ASPC:mast_cell | 0.41758334 |
| Depression | Frontal cortex | SAT | ASPC:monocyte | 0.79638099 |
| Depression | Frontal cortex | SAT | ASPC:neutrophil | 0.34772146 |
| Depression | Frontal cortex | SAT | ASPC:nk_cell | 0.24863903 |
| Depression | Frontal cortex | SAT | ASPC:pericyte | 0.23512227 |
| Depression | Frontal cortex | SAT | ASPC:t_cell | 0.40397702 |
| Depression | Frontal cortex | SAT | LEC:SMC | 0.74558144 |
| Depression | Frontal cortex | SAT | LEC:adipocyte | 0.62891682 |
| Depression | Frontal cortex | SAT | LEC:b_cell | 0.05889586 |
| Depression | Frontal cortex | SAT | LEC:dendritic_cell | 0.98197928 |
| Depression | Frontal cortex | SAT | LEC:macrophage | 0.53729488 |
| Depression | Frontal cortex | SAT | LEC:mast_cell | 0.64686872 |
| Depression | Frontal cortex | SAT | LEC:monocyte | 0.76666571 |
| Depression | Frontal cortex | SAT | LEC:neutrophil | 0.25815038 |
| Depression | Frontal cortex | SAT | LEC:nk_cell | 0.49198868 |
| Depression | Frontal cortex | SAT | LEC:pericyte | 0.78628903 |
| Depression | Frontal cortex | SAT | LEC:t_cell | 0.75114937 |
| Depression | Frontal cortex | SAT | SMC:adipocyte | 0.82897551 |
| Depression | Frontal cortex | SAT | SMC:b_cell | 0.54965878 |
| Depression | Frontal cortex | SAT | SMC:dendritic_cell | 0.43040182 |
| Depression | Frontal cortex | SAT | SMC:macrophage | 0.46725396 |
| Depression | Frontal cortex | SAT | SMC:mast_cell | 0.8190997 |
| Depression | Frontal cortex | SAT | SMC:monocyte | 0.17848367 |
| Depression | Frontal cortex | SAT | SMC:neutrophil | 0.68309292 |
| Depression | Frontal cortex | SAT | SMC:nk_cell | 0.68229912 |
| Depression | Frontal cortex | SAT | SMC:pericyte | 0.1573641 |
| Depression | Frontal cortex | SAT | SMC:t_cell | 0.98621158 |
| Depression | Frontal cortex | SAT | adipocyte:b_cell | 0.56040819 |
| Depression | Frontal cortex | SAT | adipocyte:dendritic_cell | 0.38824077 |

| | | | | |
|---|---|---|---|---|
| Depression | Frontal cortex | SAT | adipocyte:macrophage | 0.68366593 |
| Depression | Frontal cortex | SAT | adipocyte:mast_cell | 0.81508571 |
| Depression | Frontal cortex | SAT | adipocyte:monocyte | 0.38478645 |
| Depression | Frontal cortex | SAT | adipocyte:neutrophil | 0.10685972 |
| Depression | Frontal cortex | SAT | adipocyte:nk_cell | 0.33507572 |
| Depression | Frontal cortex | SAT | adipocyte:pericyte | ***0.06975813*** |
| Depression | Frontal cortex | SAT | adipocyte:t_cell | 0.29899958 |
| Depression | Frontal cortex | SAT | b_cell:dendritic_cell | 0.34051921 |
| Depression | Frontal cortex | SAT | b_cell:macrophage | 0.32649462 |
| Depression | Frontal cortex | SAT | b_cell:mast_cell | 0.75051681 |
| Depression | Frontal cortex | SAT | b_cell:monocyte | 0.99152915 |
| Depression | Frontal cortex | SAT | b_cell:neutrophil | 0.21680802 |
| Depression | Frontal cortex | SAT | b_cell:nk_cell | 0.04662689 |
| Depression | Frontal cortex | SAT | b_cell:pericyte | ***0.05719241*** |
| Depression | Frontal cortex | SAT | b_cell:t_cell | 0.20859647 |
| Depression | Frontal cortex | SAT | dendritic_cell:macrophage | 0.47189577 |
| Depression | Frontal cortex | SAT | dendritic_cell:mast_cell | 0.72594595 |
| Depression | Frontal cortex | SAT | dendritic_cell:monocyte | 0.30024762 |
| Depression | Frontal cortex | SAT | dendritic_cell:neutrophil | 0.68882511 |
| Depression | Frontal cortex | SAT | dendritic_cell:nk_cell | 0.65686821 |
| Depression | Frontal cortex | SAT | dendritic_cell:pericyte | 0.59682601 |
| Depression | Frontal cortex | SAT | dendritic_cell:t_cell | 0.89023556 |
| Depression | Frontal cortex | SAT | macrophage:mast_cell | 0.46890482 |
| Depression | Frontal cortex | SAT | macrophage:monocyte | 0.89139344 |
| Depression | Frontal cortex | SAT | macrophage:neutrophil | ***0.08712037*** |
| Depression | Frontal cortex | SAT | macrophage:nk_cell | 0.53692264 |
| Depression | Frontal cortex | SAT | macrophage:pericyte | 0.28249348 |

| | | | | |
|---|---|---|---|---|
| Depression | Frontal cortex | SAT | macrophage:t_cell | 0.80323022 |
| Depression | Frontal cortex | SAT | mast_cell:monocyte | 0.71402465 |
| Depression | Frontal cortex | SAT | mast_cell:neutrophil | 0.35820097 |
| Depression | Frontal cortex | SAT | mast_cell:nk_cell | 0.20320996 |
| Depression | Frontal cortex | SAT | mast_cell:pericyte | 0.16265483 |
| Depression | Frontal cortex | SAT | mast_cell:t_cell | 0.82298204 |
| Depression | Frontal cortex | SAT | monocyte:neutrophil | 0.47728495 |
| Depression | Frontal cortex | SAT | monocyte:nk_cell | 0.43648405 |
| Depression | Frontal cortex | SAT | monocyte:pericyte | 0.13072367 |
| Depression | Frontal cortex | SAT | monocyte:t_cell | 0.28034269 |
| Depression | Frontal cortex | SAT | neutrophil:nk_cell | **0.00397429** |
| Depression | Frontal cortex | SAT | neutrophil:pericyte | 0.22825061 |
| Depression | Frontal cortex | SAT | neutrophil:t_cell | 0.73237156 |
| Depression | Frontal cortex | SAT | nk_cell:pericyte | 0.71568582 |
| Depression | Frontal cortex | SAT | nk_cell:t_cell | 0.93813562 |
| Depression | Frontal cortex | SAT | pericyte:t_cell | 0.65974165 |
| Depression | Frontal cortex | VAT | Ex | 0.71514166 |
| Depression | Frontal cortex | VAT | Inhib | 0.75717705 |
| Depression | Frontal cortex | VAT | MicroMacro | 0.91749027 |
| Depression | Frontal cortex | VAT | OPCs | 0.61880318 |
| Depression | Frontal cortex | VAT | Oligos | 0.80515319 |
| Depression | Frontal cortex | VAT | ASPC | 0.5390469 |
| Depression | Frontal cortex | VAT | LEC | 0.79653786 |
| Depression | Frontal cortex | VAT | SMC | 0.0507791 |
| Depression | Frontal cortex | VAT | adipocyte | 0.8053499 |
| Depression | Frontal cortex | VAT | b_cell | 0.94401259 |
| Depression | Frontal cortex | VAT | dendritic_cell | 0.81799245 |
| Depression | Frontal cortex | VAT | macrophage | 0.77838479 |
| Depression | Frontal cortex | VAT | mast_cell | 0.94801254 |
| Depression | Frontal cortex | VAT | monocyte | 0.51914066 |
| Depression | Frontal cortex | VAT | neutrophil | 0.89712814 |
| Depression | Frontal cortex | VAT | nk_cell | 0.79409881 |
| Depression | Frontal cortex | VAT | pericyte | 0.75915093 |
| Depression | Frontal cortex | VAT | t_cell | 0.3570095 |
| Depression | Frontal cortex | VAT | Ex:Inhib | 0.86751388 |
| Depression | Frontal cortex | VAT | Ex:MicroMacro | 0.96455162 |
| Depression | Frontal cortex | VAT | Ex:OPCs | 0.67442311 |
| Depression | Frontal cortex | VAT | Ex:Oligos | 0.47123777 |

| | | | | |
|---|---|---|---|---|
| Depression | Frontal cortex | VAT | Ex:ASPC | 0.52898904 |
| Depression | Frontal cortex | VAT | Ex:LEC | 0.81946707 |
| Depression | Frontal cortex | VAT | Ex:SMC | **0.0488227** |
| Depression | Frontal cortex | VAT | Ex:adipocyte | 0.80417435 |
| Depression | Frontal cortex | VAT | Ex:b_cell | 0.85079094 |
| Depression | Frontal cortex | VAT | Ex:dendritic_cell | 0.9524783 |
| Depression | Frontal cortex | VAT | Ex:macrophage | 0.8040401 |
| Depression | Frontal cortex | VAT | Ex:mast_cell | 0.9675978 |
| Depression | Frontal cortex | VAT | Ex:monocyte | 0.56823908 |
| Depression | Frontal cortex | VAT | Ex:neutrophil | 0.73325973 |
| Depression | Frontal cortex | VAT | Ex:nk_cell | 0.62354932 |
| Depression | Frontal cortex | VAT | Ex:pericyte | 0.58921938 |
| Depression | Frontal cortex | VAT | Ex:t_cell | 0.27017588 |
| Depression | Frontal cortex | VAT | Inhib:MicroMacro | 0.89685474 |
| Depression | Frontal cortex | VAT | Inhib:OPCs | 0.34001228 |
| Depression | Frontal cortex | VAT | Inhib:Oligos | 0.37677095 |
| Depression | Frontal cortex | VAT | Inhib:ASPC | 0.58296609 |
| Depression | Frontal cortex | VAT | Inhib:LEC | 0.79647992 |
| Depression | Frontal cortex | VAT | Inhib:SMC | 0.05046435 |
| Depression | Frontal cortex | VAT | Inhib:adipocyte | 0.67412999 |
| Depression | Frontal cortex | VAT | Inhib:b_cell | 0.85700621 |
| Depression | Frontal cortex | VAT | Inhib:dendritic_cell | 0.94825064 |
| Depression | Frontal cortex | VAT | Inhib:macrophage | 0.74060609 |
| Depression | Frontal cortex | VAT | Inhib:mast_cell | 0.94265592 |
| Depression | Frontal cortex | VAT | Inhib:monocyte | 0.49095357 |
| Depression | Frontal cortex | VAT | Inhib:neutrophil | 0.7110502 |
| Depression | Frontal cortex | VAT | Inhib:nk_cell | 0.73966163 |
| Depression | Frontal cortex | VAT | Inhib:pericyte | 0.52410218 |
| Depression | Frontal cortex | VAT | Inhib:t_cell | 0.28994241 |
| Depression | Frontal cortex | VAT | MicroMacro:OPCs | 0.56951091 |
| Depression | Frontal cortex | VAT | MicroMacro:Oligos | 0.9264086 |
| Depression | Frontal cortex | VAT | MicroMacro:ASPC | 0.76231768 |
| Depression | Frontal cortex | VAT | MicroMacro:LEC | 0.68016308 |
| Depression | Frontal cortex | VAT | MicroMacro:SMC | 0.33462081 |
| Depression | Frontal cortex | VAT | MicroMacro:adipocyte | 0.56271663 |
| Depression | Frontal cortex | VAT | MicroMacro:b_cell | 0.5087087 |
| Depression | Frontal cortex | VAT | MicroMacro:dendritic_cell | 0.80362074 |

| | | | | |
|---|---|---|---|---|
| Depression | Frontal cortex | VAT | MicroMacro:macrophage | 0.69061333 |
| Depression | Frontal cortex | VAT | MicroMacro:mast_cell | 0.66844033 |
| Depression | Frontal cortex | VAT | MicroMacro:monocyte | 0.85617912 |
| Depression | Frontal cortex | VAT | MicroMacro:neutrophil | 0.84167034 |
| Depression | Frontal cortex | VAT | MicroMacro:nk_cell | 0.8551138 |
| Depression | Frontal cortex | VAT | MicroMacro:pericyte | 0.23862515 |
| Depression | Frontal cortex | VAT | MicroMacro:t_cell | ***0.0928086*** |
| Depression | Frontal cortex | VAT | OPCs:Oligos | 0.53173169 |
| Depression | Frontal cortex | VAT | OPCs:ASPC | 0.54779196 |
| Depression | Frontal cortex | VAT | OPCs:LEC | 0.80011699 |
| Depression | Frontal cortex | VAT | OPCs:SMC | ***0.05427109*** |
| Depression | Frontal cortex | VAT | OPCs:adipocyte | 0.97568576 |
| Depression | Frontal cortex | VAT | OPCs:b_cell | 0.91651241 |
| Depression | Frontal cortex | VAT | OPCs:dendritic_cell | 0.9768748 |
| Depression | Frontal cortex | VAT | OPCs:macrophage | 0.75339458 |
| Depression | Frontal cortex | VAT | OPCs:mast_cell | 0.8467622 |
| Depression | Frontal cortex | VAT | OPCs:monocyte | 0.43576125 |
| Depression | Frontal cortex | VAT | OPCs:neutrophil | 0.52524644 |
| Depression | Frontal cortex | VAT | OPCs:nk_cell | 0.44801769 |
| Depression | Frontal cortex | VAT | OPCs:pericyte | 0.48226167 |
| Depression | Frontal cortex | VAT | OPCs:t_cell | 0.2829173 |
| Depression | Frontal cortex | VAT | Oligos:ASPC | 0.61630409 |
| Depression | Frontal cortex | VAT | Oligos:LEC | 0.7971185 |
| Depression | Frontal cortex | VAT | Oligos:SMC | **0.04564157** |
| Depression | Frontal cortex | VAT | Oligos:adipocyte | 0.71927306 |
| Depression | Frontal cortex | VAT | Oligos:b_cell | 0.77126064 |
| Depression | Frontal cortex | VAT | Oligos:dendritic_cell | 0.89986759 |
| Depression | Frontal cortex | VAT | Oligos:macrophage | 0.82822948 |
| Depression | Frontal cortex | VAT | Oligos:mast_cell | 0.99237962 |
| Depression | Frontal cortex | VAT | Oligos:monocyte | 0.51643498 |
| Depression | Frontal cortex | VAT | Oligos:neutrophil | 0.64952236 |
| Depression | Frontal cortex | VAT | Oligos:nk_cell | 0.63105383 |
| Depression | Frontal cortex | VAT | Oligos:pericyte | 0.56936286 |
| Depression | Frontal cortex | VAT | Oligos:t_cell | 0.3077139 |
| Depression | Frontal cortex | VAT | ASPC:LEC | 0.96394394 |
| Depression | Frontal cortex | VAT | ASPC:SMC | 0.64504332 |

| | | | | |
|---|---|---|---|---|
| Depression | Frontal cortex | VAT | ASPC:adipocyte | 0.78566173 |
| Depression | Frontal cortex | VAT | ASPC:b_cell | 0.48587412 |
| Depression | Frontal cortex | VAT | ASPC:dendritic_cell | 0.46362859 |
| Depression | Frontal cortex | VAT | ASPC:macrophage | 0.88127704 |
| Depression | Frontal cortex | VAT | ASPC:mast_cell | 0.41758334 |
| Depression | Frontal cortex | VAT | ASPC:monocyte | 0.79638099 |
| Depression | Frontal cortex | VAT | ASPC:neutrophil | 0.34772146 |
| Depression | Frontal cortex | VAT | ASPC:nk_cell | 0.24863903 |
| Depression | Frontal cortex | VAT | ASPC:pericyte | 0.23512227 |
| Depression | Frontal cortex | VAT | ASPC:t_cell | 0.40397702 |
| Depression | Frontal cortex | VAT | LEC:SMC | 0.74558144 |
| Depression | Frontal cortex | VAT | LEC:adipocyte | 0.62891682 |
| Depression | Frontal cortex | VAT | LEC:b_cell | 0.05889586 |
| Depression | Frontal cortex | VAT | LEC:dendritic_cell | 0.98197928 |
| Depression | Frontal cortex | VAT | LEC:macrophage | 0.53729488 |
| Depression | Frontal cortex | VAT | LEC:mast_cell | 0.64686872 |
| Depression | Frontal cortex | VAT | LEC:monocyte | 0.76666571 |
| Depression | Frontal cortex | VAT | LEC:neutrophil | 0.25815038 |
| Depression | Frontal cortex | VAT | LEC:nk_cell | 0.49198868 |
| Depression | Frontal cortex | VAT | LEC:pericyte | 0.78628903 |
| Depression | Frontal cortex | VAT | LEC:t_cell | 0.75114937 |
| Depression | Frontal cortex | VAT | SMC:adipocyte | 0.82897551 |
| Depression | Frontal cortex | VAT | SMC:b_cell | 0.54965878 |
| Depression | Frontal cortex | VAT | SMC:dendritic_cell | 0.43040182 |
| Depression | Frontal cortex | VAT | SMC:macrophage | 0.46725396 |
| Depression | Frontal cortex | VAT | SMC:mast_cell | 0.8190997 |
| Depression | Frontal cortex | VAT | SMC:monocyte | 0.17848367 |
| Depression | Frontal cortex | VAT | SMC:neutrophil | 0.68309292 |
| Depression | Frontal cortex | VAT | SMC:nk_cell | 0.68229912 |
| Depression | Frontal cortex | VAT | SMC:pericyte | 0.1573641 |
| Depression | Frontal cortex | VAT | SMC:t_cell | 0.98621158 |
| Depression | Frontal cortex | VAT | adipocyte:b_cell | 0.56040819 |
| Depression | Frontal cortex | VAT | adipocyte:dendritic_cell | 0.38824077 |
| Depression | Frontal cortex | VAT | adipocyte:macrophage | 0.68366593 |
| Depression | Frontal cortex | VAT | adipocyte:mast_cell | 0.81508571 |
| Depression | Frontal cortex | VAT | adipocyte:monocyte | 0.38478645 |
| Depression | Frontal cortex | VAT | adipocyte:neutrophil | 0.10685972 |

| | | | | |
|---|---|---|---|---|
| Depression | Frontal cortex | VAT | adipocyte:nk_cell | 0.33507572 |
| Depression | Frontal cortex | VAT | adipocyte:pericyte | 0.06975813 |
| Depression | Frontal cortex | VAT | adipocyte:t_cell | 0.29899958 |
| Depression | Frontal cortex | VAT | b_cell:dendritic_cell | 0.34051921 |
| Depression | Frontal cortex | VAT | b_cell:macrophage | 0.32649462 |
| Depression | Frontal cortex | VAT | b_cell:mast_cell | 0.75051681 |
| Depression | Frontal cortex | VAT | b_cell:monocyte | 0.99152915 |
| Depression | Frontal cortex | VAT | b_cell:neutrophil | 0.21680802 |
| Depression | Frontal cortex | VAT | b_cell:nk_cell | 0.04662689 |
| Depression | Frontal cortex | VAT | b_cell:pericyte | 0.05719241 |
| Depression | Frontal cortex | VAT | b_cell:t_cell | 0.20859647 |
| Depression | Frontal cortex | VAT | dendritic_cell:macrophage | 0.47189577 |
| Depression | Frontal cortex | VAT | dendritic_cell:mast_cell | 0.72594595 |
| Depression | Frontal cortex | VAT | dendritic_cell:monocyte | 0.30024762 |
| Depression | Frontal cortex | VAT | dendritic_cell:neutrophil | 0.68882511 |
| Depression | Frontal cortex | VAT | dendritic_cell:nk_cell | 0.65686821 |
| Depression | Frontal cortex | VAT | dendritic_cell:pericyte | 0.59682601 |
| Depression | Frontal cortex | VAT | dendritic_cell:t_cell | 0.89023556 |
| Depression | Frontal cortex | VAT | macrophage:mast_cell | 0.46890482 |
| Depression | Frontal cortex | VAT | macrophage:monocyte | 0.89139344 |
| Depression | Frontal cortex | VAT | macrophage:neutrophil | 0.08712037 |
| Depression | Frontal cortex | VAT | macrophage:nk_cell | 0.53692264 |
| Depression | Frontal cortex | VAT | macrophage:pericyte | 0.28249348 |
| Depression | Frontal cortex | VAT | macrophage:t_cell | 0.80323022 |
| Depression | Frontal cortex | VAT | mast_cell:monocyte | 0.71402465 |
| Depression | Frontal cortex | VAT | mast_cell:neutrophil | 0.35820097 |
| Depression | Frontal cortex | VAT | mast_cell:nk_cell | 0.20320996 |

| | | | | |
|---|---|---|---|---|
| Depression | Frontal cortex | VAT | mast_cell:pericyte | 0.16265483 |
| Depression | Frontal cortex | VAT | mast_cell:t_cell | 0.82298204 |
| Depression | Frontal cortex | VAT | monocyte:neutrophil | 0.47728495 |
| Depression | Frontal cortex | VAT | monocyte:nk_cell | 0.43648405 |
| Depression | Frontal cortex | VAT | monocyte:pericyte | 0.13072367 |
| Depression | Frontal cortex | VAT | monocyte:t_cell | 0.28034269 |
| Depression | Frontal cortex | VAT | neutrophil:nk_cell | 0.00397429 |
| Depression | Frontal cortex | VAT | neutrophil:pericyte | 0.22825061 |
| Depression | Frontal cortex | VAT | neutrophil:t_cell | 0.73237156 |
| Depression | Frontal cortex | VAT | nk_cell:pericyte | 0.71568582 |
| Depression | Frontal cortex | VAT | nk_cell:t_cell | 0.93813562 |
| Depression | Frontal cortex | VAT | pericyte:t_cell | 0.65974165 |
| T2DM | Pancreas | SAT | duct_epithelial_cell | 0.96348802 |
| T2DM | Pancreas | SAT | endothelial_cell | 0.000121 |
| T2DM | Pancreas | SAT | leukocyte | 0.68724051 |
| T2DM | Pancreas | SAT | mesenchymal_cell | 0.15992551 |
| T2DM | Pancreas | SAT | alpha_cell | 0.58297637 |
| T2DM | Pancreas | SAT | delta_cell | 0.53356158 |
| T2DM | Pancreas | SAT | PP_cell | 0.27282945 |
| T2DM | Pancreas | SAT | epsilon_cell | 0.12414599 |
| T2DM | Pancreas | SAT | beta_cell | 0.42718069 |
| T2DM | Pancreas | SAT | ASPC | 0.76036518 |
| T2DM | Pancreas | SAT | LEC | 0.76118893 |
| T2DM | Pancreas | SAT | SMC | 0.33267956 |
| T2DM | Pancreas | SAT | adipocyte | 0.00679661 |
| T2DM | Pancreas | SAT | b_cell | 0.17941452 |
| T2DM | Pancreas | SAT | dendritic_cell | 0.36903864 |
| T2DM | Pancreas | SAT | macrophage | 0.99211718 |
| T2DM | Pancreas | SAT | mast_cell | 0.34731047 |
| T2DM | Pancreas | SAT | monocyte | 0.94736611 |
| T2DM | Pancreas | SAT | neutrophil | 0.79777987 |
| T2DM | Pancreas | SAT | nk_cell | 0.20549195 |
| T2DM | Pancreas | SAT | pericyte | 0.47318617 |
| T2DM | Pancreas | SAT | t_cell | 0.48153555 |
| T2DM | Pancreas | SAT | duct_epithelial_cell:endothelial_cell | 0.32997503 |
| T2DM | Pancreas | SAT | duct_epithelial_cell:leukocyte | 0.61614605 |

| | | | | |
|---|---|---|---|---|
| T2DM | Pancreas | SAT | duct_epithelial_cell:mesenchymal_cell | 0.18611637 |
| T2DM | Pancreas | SAT | duct_epithelial_cell:alpha_cell | 0.95796137 |
| T2DM | Pancreas | SAT | duct_epithelial_cell:delta_cell | 0.40195852 |
| T2DM | Pancreas | SAT | duct_epithelial_cell:PP_cell | 0.90363127 |
| T2DM | Pancreas | SAT | duct_epithelial_cell:epsilon_cell | 0.40215133 |
| T2DM | Pancreas | SAT | duct_epithelial_cell:beta_cell | 0.43418157 |
| T2DM | Pancreas | SAT | duct_epithelial_cell:ASPC | 0.83514629 |
| T2DM | Pancreas | SAT | duct_epithelial_cell:LEC | 0.13890262 |
| T2DM | Pancreas | SAT | duct_epithelial_cell:SMC | 0.28105392 |
| T2DM | Pancreas | SAT | duct_epithelial_cell:adipocyte | **0.02318777** |
| T2DM | Pancreas | SAT | duct_epithelial_cell:b_cell | 0.89934261 |
| T2DM | Pancreas | SAT | duct_epithelial_cell:dendritic_cell | 0.85276286 |
| T2DM | Pancreas | SAT | duct_epithelial_cell:macrophage | 0.93470887 |
| T2DM | Pancreas | SAT | duct_epithelial_cell:mast_cell | 0.81536183 |
| T2DM | Pancreas | SAT | duct_epithelial_cell:monocyte | 0.46963148 |
| T2DM | Pancreas | SAT | duct_epithelial_cell:neutrophil | 0.72438364 |
| T2DM | Pancreas | SAT | duct_epithelial_cell:nk_cell | 0.35514209 |
| T2DM | Pancreas | SAT | duct_epithelial_cell:pericyte | 0.31399246 |
| T2DM | Pancreas | SAT | duct_epithelial_cell:t_cell | 0.74338113 |
| T2DM | Pancreas | SAT | endothelial_cell:leukocyte | 0.03573003 |
| T2DM | Pancreas | SAT | endothelial_cell:mesenchymal_cell | 0.11992245 |
| T2DM | Pancreas | SAT | endothelial_cell:alpha_cell | **0.00344697** |
| T2DM | Pancreas | SAT | endothelial_cell:delta_cell | **0.00201649** |

| | | | | |
|---|---|---|---|---|
| T2DM | Pancreas | SAT | endothelial_cell:PP_cell | 0.12120876 |
| T2DM | Pancreas | SAT | endothelial_cell:epsilon_cell | 0.88931359 |
| T2DM | Pancreas | SAT | endothelial_cell:beta_cell | **0.00026039** |
| T2DM | Pancreas | SAT | endothelial_cell:ASPC | **0.0490978** |
| T2DM | Pancreas | SAT | endothelial_cell:LEC | 0.89542088 |
| T2DM | Pancreas | SAT | endothelial_cell:SMC | 0.27077759 |
| T2DM | Pancreas | SAT | endothelial_cell:adipocyte | 0.79067967 |
| T2DM | Pancreas | SAT | endothelial_cell:b_cell | 0.29341569 |
| T2DM | Pancreas | SAT | endothelial_cell:dendritic_cell | 0.32297288 |
| T2DM | Pancreas | SAT | endothelial_cell:macrophage | 0.22577514 |
| T2DM | Pancreas | SAT | endothelial_cell:mast_cell | 0.74824625 |
| T2DM | Pancreas | SAT | endothelial_cell:monocyte | 0.37677253 |
| T2DM | Pancreas | SAT | endothelial_cell:neutrophil | **0.0389968** |
| T2DM | Pancreas | SAT | endothelial_cell:nk_cell | **0.0060122** |
| T2DM | Pancreas | SAT | endothelial_cell:pericyte | **0.02256553** |
| T2DM | Pancreas | SAT | endothelial_cell:t_cell | ***0.08179028*** |
| T2DM | Pancreas | SAT | leukocyte:mesenchymal_cell | 0.83145597 |
| T2DM | Pancreas | SAT | leukocyte:alpha_cell | 0.88451886 |
| T2DM | Pancreas | SAT | leukocyte:delta_cell | 0.68503079 |
| T2DM | Pancreas | SAT | leukocyte:PP_cell | 0.46590921 |
| T2DM | Pancreas | SAT | leukocyte:epsilon_cell | 0.28943154 |
| T2DM | Pancreas | SAT | leukocyte:beta_cell | 0.86672317 |
| T2DM | Pancreas | SAT | leukocyte:ASPC | 0.461492 |
| T2DM | Pancreas | SAT | leukocyte:LEC | 0.98401611 |
| T2DM | Pancreas | SAT | leukocyte:SMC | 0.88432805 |

| | | | | |
|---|---|---|---|---|
| T2DM | Pancreas | SAT | leukocyte:adipocyte | 0.2961349 |
| T2DM | Pancreas | SAT | leukocyte:b_cell | 0.43021527 |
| T2DM | Pancreas | SAT | leukocyte:dendritic_cell | 0.15361512 |
| T2DM | Pancreas | SAT | leukocyte:macrophage | 0.61660868 |
| T2DM | Pancreas | SAT | leukocyte:mast_cell | 0.17559163 |
| T2DM | Pancreas | SAT | leukocyte:monocyte | 0.29776933 |
| T2DM | Pancreas | SAT | leukocyte:neutrophil | 0.51463911 |
| T2DM | Pancreas | SAT | leukocyte:nk_cell | 0.61567035 |
| T2DM | Pancreas | SAT | leukocyte:pericyte | 0.50406664 |
| T2DM | Pancreas | SAT | leukocyte:t_cell | 0.76962909 |
| T2DM | Pancreas | SAT | mesenchymal_cell:alpha_cell | 0.43266364 |
| T2DM | Pancreas | SAT | mesenchymal_cell:delta_cell | 0.77483438 |
| T2DM | Pancreas | SAT | mesenchymal_cell:PP_cell | 0.26462523 |
| T2DM | Pancreas | SAT | mesenchymal_cell:epsilon_cell | 0.52767765 |
| T2DM | Pancreas | SAT | mesenchymal_cell:beta_cell | 0.37973062 |
| T2DM | Pancreas | SAT | mesenchymal_cell:ASPC | 0.44796197 |
| T2DM | Pancreas | SAT | mesenchymal_cell:LEC | 0.12190628 |
| T2DM | Pancreas | SAT | mesenchymal_cell:SMC | 0.58850023 |
| T2DM | Pancreas | SAT | mesenchymal_cell:adipocyte | 0.90278043 |
| T2DM | Pancreas | SAT | mesenchymal_cell:b_cell | 0.42072091 |
| T2DM | Pancreas | SAT | mesenchymal_cell:dendritic_cell | 0.88197174 |
| T2DM | Pancreas | SAT | mesenchymal_cell:macrophage | 0.72586491 |
| T2DM | Pancreas | SAT | mesenchymal_cell:mast_cell | 0.78484502 |
| T2DM | Pancreas | SAT | mesenchymal_cell:monocyte | 0.17974576 |
| T2DM | Pancreas | SAT | mesenchymal_cell:neutrophil | 0.12236134 |

| | | | | |
|---|---|---|---|---|
| T2DM | Pancreas | SAT | mesenchymal_cell:nk_cell | **0.03670891** |
| T2DM | Pancreas | SAT | mesenchymal_cell:pericyte | 0.21107796 |
| T2DM | Pancreas | SAT | mesenchymal_cell:t_cell | 0.39198268 |
| T2DM | Pancreas | SAT | alpha_cell:delta_cell | ***0.09451958*** |
| T2DM | Pancreas | SAT | alpha_cell:PP_cell | 0.60379232 |
| T2DM | Pancreas | SAT | alpha_cell:epsilon_cell | 0.16224869 |
| T2DM | Pancreas | SAT | alpha_cell:beta_cell | 0.16015178 |
| T2DM | Pancreas | SAT | alpha_cell:ASPC | 0.80561448 |
| T2DM | Pancreas | SAT | alpha_cell:LEC | 0.62474367 |
| T2DM | Pancreas | SAT | alpha_cell:SMC | 0.3593575 |
| T2DM | Pancreas | SAT | alpha_cell:adipocyte | **0.01329023** |
| T2DM | Pancreas | SAT | alpha_cell:b_cell | 0.55563113 |
| T2DM | Pancreas | SAT | alpha_cell:dendritic_cell | 0.70830232 |
| T2DM | Pancreas | SAT | alpha_cell:macrophage | 0.65439971 |
| T2DM | Pancreas | SAT | alpha_cell:mast_cell | 0.1485094 |
| T2DM | Pancreas | SAT | alpha_cell:monocyte | 0.52806743 |
| T2DM | Pancreas | SAT | alpha_cell:neutrophil | 0.23760405 |
| T2DM | Pancreas | SAT | alpha_cell:nk_cell | 0.46947705 |
| T2DM | Pancreas | SAT | alpha_cell:pericyte | 0.27026983 |
| T2DM | Pancreas | SAT | alpha_cell:t_cell | 0.85311208 |
| T2DM | Pancreas | SAT | delta_cell:PP_cell | 0.86696666 |
| T2DM | Pancreas | SAT | delta_cell:epsilon_cell | 0.18472022 |
| T2DM | Pancreas | SAT | delta_cell:beta_cell | 0.06963777 |
| T2DM | Pancreas | SAT | delta_cell:ASPC | 0.30627782 |
| T2DM | Pancreas | SAT | delta_cell:LEC | 0.61606291 |
| T2DM | Pancreas | SAT | delta_cell:SMC | 0.16775962 |
| T2DM | Pancreas | SAT | delta_cell:adipocyte | **0.0122907** |
| T2DM | Pancreas | SAT | delta_cell:b_cell | 0.29261235 |

| | | | | |
|---|---|---|---|---|
| T2DM | Pancreas | SAT | delta_cell:dendritic_cell | 0.46455625 |
| T2DM | Pancreas | SAT | delta_cell:macrophage | 0.83272949 |
| T2DM | Pancreas | SAT | delta_cell:mast_cell | **0.03810678** |
| T2DM | Pancreas | SAT | delta_cell:monocyte | 0.90698398 |
| T2DM | Pancreas | SAT | delta_cell:neutrophil | 0.36292943 |
| T2DM | Pancreas | SAT | delta_cell:nk_cell | 0.83339262 |
| T2DM | Pancreas | SAT | delta_cell:pericyte | **0.02716698** |
| T2DM | Pancreas | SAT | delta_cell:t_cell | 0.33280378 |
| T2DM | Pancreas | SAT | PP_cell:epsilon_cell | ***0.09081388*** |
| T2DM | Pancreas | SAT | PP_cell:beta_cell | 0.83677567 |
| T2DM | Pancreas | SAT | PP_cell:ASPC | 0.35688684 |
| T2DM | Pancreas | SAT | PP_cell:LEC | 0.99158559 |
| T2DM | Pancreas | SAT | PP_cell:SMC | 0.1993489 |
| T2DM | Pancreas | SAT | PP_cell:adipocyte | **0.00764883** |
| T2DM | Pancreas | SAT | PP_cell:b_cell | 0.28743724 |
| T2DM | Pancreas | SAT | PP_cell:dendritic_cell | 0.74882022 |
| T2DM | Pancreas | SAT | PP_cell:macrophage | 0.77744561 |
| T2DM | Pancreas | SAT | PP_cell:mast_cell | ***0.0786356*** |
| T2DM | Pancreas | SAT | PP_cell:monocyte | 0.80934329 |
| T2DM | Pancreas | SAT | PP_cell:neutrophil | 0.20753636 |
| T2DM | Pancreas | SAT | PP_cell:nk_cell | 0.8125891 |
| T2DM | Pancreas | SAT | PP_cell:pericyte | 0.05333262 |
| T2DM | Pancreas | SAT | PP_cell:t_cell | 0.88536955 |
| T2DM | Pancreas | SAT | epsilon_cell:beta_cell | 0.15208521 |
| T2DM | Pancreas | SAT | epsilon_cell:ASPC | 0.41633605 |
| T2DM | Pancreas | SAT | epsilon_cell:LEC | ***0.05309682*** |
| T2DM | Pancreas | SAT | epsilon_cell:SMC | 0.76341049 |
| T2DM | Pancreas | SAT | epsilon_cell:adipocyte | 0.25447149 |

| | | | | |
|---|---|---|---|---|
| T2DM | Pancreas | SAT | epsilon_cell:b_cell | 0.19870482 |
| T2DM | Pancreas | SAT | epsilon_cell:dendritic_cell | **0.04753288** |
| T2DM | Pancreas | SAT | epsilon_cell:macrophage | 0.77884542 |
| T2DM | Pancreas | SAT | epsilon_cell:mast_cell | ***0.097459*** |
| T2DM | Pancreas | SAT | epsilon_cell:monocyte | 0.16348134 |
| T2DM | Pancreas | SAT | epsilon_cell:neutrophil | 0.14791614 |
| T2DM | Pancreas | SAT | epsilon_cell:nk_cell | 0.4502471 |
| T2DM | Pancreas | SAT | epsilon_cell:pericyte | 0.15688054 |
| T2DM | Pancreas | SAT | epsilon_cell:t_cell | 0.69701501 |
| T2DM | Pancreas | SAT | beta_cell:ASPC | 0.60216297 |
| T2DM | Pancreas | SAT | beta_cell:LEC | 0.66680427 |
| T2DM | Pancreas | SAT | beta_cell:SMC | 0.32362217 |
| T2DM | Pancreas | SAT | beta_cell:adipocyte | 0.00887285 |
| T2DM | Pancreas | SAT | beta_cell:b_cell | 0.21318757 |
| T2DM | Pancreas | SAT | beta_cell:dendritic_cell | 0.51691788 |
| T2DM | Pancreas | SAT | beta_cell:macrophage | 0.95856785 |
| T2DM | Pancreas | SAT | beta_cell:mast_cell | 0.14583174 |
| T2DM | Pancreas | SAT | beta_cell:monocyte | 0.6378686 |
| T2DM | Pancreas | SAT | beta_cell:neutrophil | 0.22384218 |
| T2DM | Pancreas | SAT | beta_cell:nk_cell | 0.34073849 |
| T2DM | Pancreas | SAT | beta_cell:pericyte | 0.22378215 |
| T2DM | Pancreas | SAT | beta_cell:t_cell | 0.70066687 |
| T2DM | Pancreas | SAT | ASPC:LEC | **0.01828592** |
| T2DM | Pancreas | SAT | ASPC:SMC | 0.25762784 |
| T2DM | Pancreas | SAT | ASPC:adipocyte | 0.69317723 |
| T2DM | Pancreas | SAT | ASPC:b_cell | ***0.07544378*** |
| T2DM | Pancreas | SAT | ASPC:dendritic_cell | 0.23257235 |
| T2DM | Pancreas | SAT | ASPC:macrophage | 0.91222797 |
| T2DM | Pancreas | SAT | ASPC:mast_cell | 0.80415197 |
| T2DM | Pancreas | SAT | ASPC:monocyte | 0.29884771 |

| | | | | |
|---|---|---|---|---|
| T2DM | Pancreas | SAT | ASPC:neutrophil | 0.19439491 |
| T2DM | Pancreas | SAT | ASPC:nk_cell | 0.30017419 |
| T2DM | Pancreas | SAT | ASPC:pericyte | 0.2261871 |
| T2DM | Pancreas | SAT | ASPC:t_cell | 0.27499603 |
| T2DM | Pancreas | SAT | LEC:SMC | 0.63284265 |
| T2DM | Pancreas | SAT | LEC:adipocyte | 0.47695014 |
| T2DM | Pancreas | SAT | LEC:b_cell | **0.00794548** |
| T2DM | Pancreas | SAT | LEC:dendritic_cell | 0.54004226 |
| T2DM | Pancreas | SAT | LEC:macrophage | 0.21195816 |
| T2DM | Pancreas | SAT | LEC:mast_cell | 0.94392561 |
| T2DM | Pancreas | SAT | LEC:monocyte | 0.4759678 |
| T2DM | Pancreas | SAT | LEC:neutrophil | 0.33226424 |
| T2DM | Pancreas | SAT | LEC:nk_cell | **0.00583383** |
| T2DM | Pancreas | SAT | LEC:pericyte | 0.26603199 |
| T2DM | Pancreas | SAT | LEC:t_cell | 0.3719512 |
| T2DM | Pancreas | SAT | SMC:adipocyte | 0.11448421 |
| T2DM | Pancreas | SAT | SMC:b_cell | 0.34808955 |
| T2DM | Pancreas | SAT | SMC:dendritic_cell | 0.25045711 |
| T2DM | Pancreas | SAT | SMC:macrophage | **0.01652624** |
| T2DM | Pancreas | SAT | SMC:mast_cell | 0.41672 |
| T2DM | Pancreas | SAT | SMC:monocyte | 0.36827814 |
| T2DM | Pancreas | SAT | SMC:neutrophil | ***0.05250651*** |
| T2DM | Pancreas | SAT | SMC:nk_cell | 0.13900647 |
| T2DM | Pancreas | SAT | SMC:pericyte | **0.00198425** |
| T2DM | Pancreas | SAT | SMC:t_cell | **0.0303994** |
| T2DM | Pancreas | SAT | adipocyte:b_cell | 0.19834354 |
| T2DM | Pancreas | SAT | adipocyte:dendritic_cell | 0.96614959 |
| T2DM | Pancreas | SAT | adipocyte:macrophage | **0.02831957** |
| T2DM | Pancreas | SAT | adipocyte:mast_cell | 0.10579177 |
| T2DM | Pancreas | SAT | adipocyte:monocyte | 0.15107268 |
| T2DM | Pancreas | SAT | adipocyte:neutrophil | **0.00419385** |
| T2DM | Pancreas | SAT | adipocyte:nk_cell | 0.58328826 |
| T2DM | Pancreas | SAT | adipocyte:pericyte | **0.03746546** |
| T2DM | Pancreas | SAT | adipocyte:t_cell | 0.73179449 |
| T2DM | Pancreas | SAT | b_cell:dendritic_cell | **0.00984223** |

| | | | | |
|---|---|---|---|---|
| T2DM | Pancreas | SAT | b_cell:macrophage | 0.1745086 |
| T2DM | Pancreas | SAT | b_cell:mast_cell | ***0.08808755*** |
| T2DM | Pancreas | SAT | b_cell:monocyte | 0.69580254 |
| T2DM | Pancreas | SAT | b_cell:neutrophil | **7.91E-06** |
| T2DM | Pancreas | SAT | b_cell:nk_cell | 0.34283673 |
| T2DM | Pancreas | SAT | b_cell:pericyte | **0.02558668** |
| T2DM | Pancreas | SAT | b_cell:t_cell | 0.50619276 |
| T2DM | Pancreas | SAT | dendritic_cell:macrophage | 0.12179187 |
| T2DM | Pancreas | SAT | dendritic_cell:mast_cell | 0.16603902 |
| T2DM | Pancreas | SAT | dendritic_cell:monocyte | 0.32733716 |
| T2DM | Pancreas | SAT | dendritic_cell:neutrophil | 0.1586055 |
| T2DM | Pancreas | SAT | dendritic_cell:nk_cell | 0.93197384 |
| T2DM | Pancreas | SAT | dendritic_cell:pericyte | 0.51965566 |
| T2DM | Pancreas | SAT | dendritic_cell:t_cell | 0.69921381 |
| T2DM | Pancreas | SAT | macrophage:mast_cell | 0.39443955 |
| T2DM | Pancreas | SAT | macrophage:monocyte | 0.99869765 |
| T2DM | Pancreas | SAT | macrophage:neutrophil | 0.24111 |
| T2DM | Pancreas | SAT | macrophage:nk_cell | 0.28302848 |
| T2DM | Pancreas | SAT | macrophage:pericyte | 0.91033296 |
| T2DM | Pancreas | SAT | macrophage:t_cell | ***0.07417619*** |
| T2DM | Pancreas | SAT | mast_cell:monocyte | 0.56029284 |
| T2DM | Pancreas | SAT | mast_cell:neutrophil | 0.65921315 |
| T2DM | Pancreas | SAT | mast_cell:nk_cell | 0.9863878 |
| T2DM | Pancreas | SAT | mast_cell:pericyte | 0.77392751 |
| T2DM | Pancreas | SAT | mast_cell:t_cell | 0.27041492 |
| T2DM | Pancreas | SAT | monocyte:neutrophil | 0.39988367 |
| T2DM | Pancreas | SAT | monocyte:nk_cell | 0.20427782 |

| | | | | |
|---|---|---|---|---|
| T2DM | Pancreas | SAT | monocyte:pericyte | 0.29685812 |
| T2DM | Pancreas | SAT | monocyte:t_cell | ***0.0654075*** |
| T2DM | Pancreas | SAT | neutrophil:nk_cell | **0.01071561** |
| T2DM | Pancreas | SAT | neutrophil:pericyte | **0.03955965** |
| T2DM | Pancreas | SAT | neutrophil:t_cell | **0.03977717** |
| T2DM | Pancreas | SAT | nk_cell:pericyte | 0.3729832 |
| T2DM | Pancreas | SAT | nk_cell:t_cell | 0.28358561 |
| T2DM | Pancreas | SAT | pericyte:t_cell | 0.93082578 |
| T2DM | Pancreas | VAT | duct_epithelial_cell | ***0.07809328*** |
| T2DM | Pancreas | VAT | endothelial_cell | **0.04682661** |
| T2DM | Pancreas | VAT | leukocyte | 0.65813829 |
| T2DM | Pancreas | VAT | mesenchymal_cell | 0.652908 |
| T2DM | Pancreas | VAT | alpha_cell | ***0.09755256*** |
| T2DM | Pancreas | VAT | delta_cell | 0.23204584 |
| T2DM | Pancreas | VAT | PP_cell | 0.17239847 |
| T2DM | Pancreas | VAT | epsilon_cell | 0.90540356 |
| T2DM | Pancreas | VAT | beta_cell | 0.13418268 |
| T2DM | Pancreas | VAT | ASPC | 0.13264185 |
| T2DM | Pancreas | VAT | LEC | 0.25880323 |
| T2DM | Pancreas | VAT | SMC | 0.20815095 |
| T2DM | Pancreas | VAT | adipocyte | ***0.07969284*** |
| T2DM | Pancreas | VAT | b_cell | **0.00305949** |
| T2DM | Pancreas | VAT | dendritic_cell | 0.79775727 |
| T2DM | Pancreas | VAT | endometrium | 0.71990995 |
| T2DM | Pancreas | VAT | macrophage | 0.98212345 |
| T2DM | Pancreas | VAT | mast_cell | 0.21342428 |
| T2DM | Pancreas | VAT | mesothelium | 0.11876995 |
| T2DM | Pancreas | VAT | monocyte | 0.7554106 |
| T2DM | Pancreas | VAT | nk_cell | **0.00055607** |
| T2DM | Pancreas | VAT | pericyte | 0.8519925 |
| T2DM | Pancreas | VAT | t_cell | **0.01281705** |
| T2DM | Pancreas | VAT | duct_epithelial_cell:endothelial_cell | 0.28218308 |
| T2DM | Pancreas | VAT | duct_epithelial_cell:leukocyte | 0.94173824 |
| T2DM | Pancreas | VAT | duct_epithelial_cell:mesenchymal_cell | 0.26677191 |
| T2DM | Pancreas | VAT | duct_epithelial_cell:alpha_cell | 0.99602893 |
| T2DM | Pancreas | VAT | duct_epithelial_cell:delta_cell | 0.35984935 |

| | | | | |
|---|---|---|---|---|
| T2DM | Pancreas | VAT | duct_epithelial_cell:PP_cell | 0.95996981 |
| T2DM | Pancreas | VAT | duct_epithelial_cell:epsilon_cell | 0.27706535 |
| T2DM | Pancreas | VAT | duct_epithelial_cell:beta_cell | 0.384384 |
| T2DM | Pancreas | VAT | duct_epithelial_cell:ASPC | **0.01697742** |
| T2DM | Pancreas | VAT | duct_epithelial_cell:LEC | 0.9909473 |
| T2DM | Pancreas | VAT | duct_epithelial_cell:SMC | 0.44163217 |
| T2DM | Pancreas | VAT | duct_epithelial_cell:adipocyte | **0.04657927** |
| T2DM | Pancreas | VAT | duct_epithelial_cell:b_cell | **0.00034608** |
| T2DM | Pancreas | VAT | duct_epithelial_cell:dendritic_cell | 0.70877395 |
| T2DM | Pancreas | VAT | duct_epithelial_cell:endometrium | 0.41377739 |
| T2DM | Pancreas | VAT | duct_epithelial_cell:macrophage | 0.90745635 |
| T2DM | Pancreas | VAT | duct_epithelial_cell:mast_cell | 0.74723944 |
| T2DM | Pancreas | VAT | duct_epithelial_cell:mesothelium | **0.0103271** |
| T2DM | Pancreas | VAT | duct_epithelial_cell:monocyte | 0.922322 |
| T2DM | Pancreas | VAT | duct_epithelial_cell:nk_cell | 2.03E-05 |
| T2DM | Pancreas | VAT | duct_epithelial_cell:pericyte | 0.60051389 |
| T2DM | Pancreas | VAT | duct_epithelial_cell:t_cell | **0.0094299** |
| T2DM | Pancreas | VAT | endothelial_cell:leukocyte | **0.04659236** |
| T2DM | Pancreas | VAT | endothelial_cell:mesenchymal_cell | ***0.05802082*** |
| T2DM | Pancreas | VAT | endothelial_cell:alpha_cell | **0.00136219** |
| T2DM | Pancreas | VAT | endothelial_cell:delta_cell | **0.00052322** |
| T2DM | Pancreas | VAT | endothelial_cell:PP_cell | 0.11548804 |
| T2DM | Pancreas | VAT | endothelial_cell:epsilon_cell | 0.78214809 |

| | | | | |
|---|---|---|---|---|
| T2DM | Pancreas | VAT | endothelial_cell:beta_cell | **6.76E-05** |
| T2DM | Pancreas | VAT | endothelial_cell:ASPC | 0.67080626 |
| T2DM | Pancreas | VAT | endothelial_cell:LEC | **0.0251075** |
| T2DM | Pancreas | VAT | endothelial_cell:SMC | 0.54381246 |
| T2DM | Pancreas | VAT | endothelial_cell:adipocyte | 0.99361037 |
| T2DM | Pancreas | VAT | endothelial_cell:b_cell | 0.32971092 |
| T2DM | Pancreas | VAT | endothelial_cell:dendritic_cell | 0.71760299 |
| T2DM | Pancreas | VAT | endothelial_cell:endometrium | 0.51025662 |
| T2DM | Pancreas | VAT | endothelial_cell:macrophage | 0.34483827 |
| T2DM | Pancreas | VAT | endothelial_cell:mast_cell | 0.48774691 |
| T2DM | Pancreas | VAT | endothelial_cell:mesothelium | 0.93340323 |
| T2DM | Pancreas | VAT | endothelial_cell:monocyte | 0.30853692 |
| T2DM | Pancreas | VAT | endothelial_cell:nk_cell | 0.41193483 |
| T2DM | Pancreas | VAT | endothelial_cell:pericyte | 0.81283548 |
| T2DM | Pancreas | VAT | endothelial_cell:t_cell | 0.26677339 |
| T2DM | Pancreas | VAT | leukocyte:mesenchymal_cell | 0.32481768 |
| T2DM | Pancreas | VAT | leukocyte:alpha_cell | 0.59977003 |
| T2DM | Pancreas | VAT | leukocyte:delta_cell | 0.09580941 |
| T2DM | Pancreas | VAT | leukocyte:PP_cell | 0.6093176 |
| T2DM | Pancreas | VAT | leukocyte:epsilon_cell | 0.3524143 |
| T2DM | Pancreas | VAT | leukocyte:beta_cell | 0.34140521 |
| T2DM | Pancreas | VAT | leukocyte:ASPC | 0.42942941 |
| T2DM | Pancreas | VAT | leukocyte:LEC | 0.20285164 |
| T2DM | Pancreas | VAT | leukocyte:SMC | 0.26344646 |
| T2DM | Pancreas | VAT | leukocyte:adipocyte | 0.53003148 |
| T2DM | Pancreas | VAT | leukocyte:b_cell | 0.35294782 |

| | | | | |
|---|---|---|---|---|
| T2DM | Pancreas | VAT | leukocyte:dendritic_cell | 0.59166957 |
| T2DM | Pancreas | VAT | leukocyte:endometrium | 0.64281266 |
| T2DM | Pancreas | VAT | leukocyte:macrophage | 0.46818609 |
| T2DM | Pancreas | VAT | leukocyte:mast_cell | 0.1913251 |
| T2DM | Pancreas | VAT | leukocyte:mesothelium | 0.62796363 |
| T2DM | Pancreas | VAT | leukocyte:monocyte | 0.54573875 |
| T2DM | Pancreas | VAT | leukocyte:nk_cell | 0.22390733 |
| T2DM | Pancreas | VAT | leukocyte:pericyte | 0.97081939 |
| T2DM | Pancreas | VAT | leukocyte:t_cell | 0.04566505 |
| T2DM | Pancreas | VAT | mesenchymal_cell:alpha_cell | 0.57453048 |
| T2DM | Pancreas | VAT | mesenchymal_cell:delta_cell | 0.88150498 |
| T2DM | Pancreas | VAT | mesenchymal_cell:PP_cell | 0.36522754 |
| T2DM | Pancreas | VAT | mesenchymal_cell:epsilon_cell | 0.75115525 |
| T2DM | Pancreas | VAT | mesenchymal_cell:beta_cell | 0.38366435 |
| T2DM | Pancreas | VAT | mesenchymal_cell:ASPC | 0.30341468 |
| T2DM | Pancreas | VAT | mesenchymal_cell:LEC | 0.17166597 |
| T2DM | Pancreas | VAT | mesenchymal_cell:SMC | 0.31947178 |
| T2DM | Pancreas | VAT | mesenchymal_cell:adipocyte | 0.97324771 |
| T2DM | Pancreas | VAT | mesenchymal_cell:b_cell | 0.12227294 |
| T2DM | Pancreas | VAT | mesenchymal_cell:dendritic_cell | 0.45650506 |
| T2DM | Pancreas | VAT | mesenchymal_cell:endometrium | 0.14842188 |
| T2DM | Pancreas | VAT | mesenchymal_cell:macrophage | 0.66589376 |
| T2DM | Pancreas | VAT | mesenchymal_cell:mast_cell | 0.71744886 |
| T2DM | Pancreas | VAT | mesenchymal_cell:mesothelium | 0.08148733 |

| | | | | |
|---|---|---|---|---|
| T2DM | Pancreas | VAT | mesenchymal_cell:monocyte | 0.71977789 |
| T2DM | Pancreas | VAT | mesenchymal_cell:nk_cell | 0.05127272 |
| T2DM | Pancreas | VAT | mesenchymal_cell:pericyte | 0.32538963 |
| T2DM | Pancreas | VAT | mesenchymal_cell:t_cell | ***0.05655125*** |
| T2DM | Pancreas | VAT | alpha_cell:delta_cell | 0.10075931 |
| T2DM | Pancreas | VAT | alpha_cell:PP_cell | 0.76628939 |
| T2DM | Pancreas | VAT | alpha_cell:epsilon_cell | 0.10167912 |
| T2DM | Pancreas | VAT | alpha_cell:beta_cell | 0.18101797 |
| T2DM | Pancreas | VAT | alpha_cell:ASPC | ***0.06421838*** |
| T2DM | Pancreas | VAT | alpha_cell:LEC | 0.66803107 |
| T2DM | Pancreas | VAT | alpha_cell:SMC | 0.59032758 |
| T2DM | Pancreas | VAT | alpha_cell:adipocyte | 0.11894493 |
| T2DM | Pancreas | VAT | alpha_cell:b_cell | **0.00275569** |
| T2DM | Pancreas | VAT | alpha_cell:dendritic_cell | 0.6517685 |
| T2DM | Pancreas | VAT | alpha_cell:endometrium | 0.95406352 |
| T2DM | Pancreas | VAT | alpha_cell:macrophage | 0.71546863 |
| T2DM | Pancreas | VAT | alpha_cell:mast_cell | 0.52975426 |
| T2DM | Pancreas | VAT | alpha_cell:mesothelium | 0.05607374 |
| T2DM | Pancreas | VAT | alpha_cell:monocyte | 0.85595202 |
| T2DM | Pancreas | VAT | alpha_cell:nk_cell | **0.00118179** |
| T2DM | Pancreas | VAT | alpha_cell:pericyte | 0.46282141 |
| T2DM | Pancreas | VAT | alpha_cell:t_cell | **0.02231581** |
| T2DM | Pancreas | VAT | delta_cell:PP_cell | 0.63141188 |
| T2DM | Pancreas | VAT | delta_cell:epsilon_cell | ***0.07099051*** |
| T2DM | Pancreas | VAT | delta_cell:beta_cell | ***0.07344051*** |
| T2DM | Pancreas | VAT | delta_cell:ASPC | ***0.0724172*** |
| T2DM | Pancreas | VAT | delta_cell:LEC | 0.62687413 |
| T2DM | Pancreas | VAT | delta_cell:SMC | 0.84933548 |

| | | | | |
|---|---|---|---|---|
| T2DM | Pancreas | VAT | delta_cell:adipocyte | 0.24403061 |
| T2DM | Pancreas | VAT | delta_cell:b_cell | **0.02705046** |
| T2DM | Pancreas | VAT | delta_cell:dendritic_cell | 0.52523745 |
| T2DM | Pancreas | VAT | delta_cell:endometrium | 0.63937646 |
| T2DM | Pancreas | VAT | delta_cell:macrophage | 0.92682532 |
| T2DM | Pancreas | VAT | delta_cell:mast_cell | 0.20819836 |
| T2DM | Pancreas | VAT | delta_cell:mesothelium | 0.0990026 |
| T2DM | Pancreas | VAT | delta_cell:monocyte | 0.77730783 |
| T2DM | Pancreas | VAT | delta_cell:nk_cell | 0.00364014 |
| T2DM | Pancreas | VAT | delta_cell:pericyte | 0.24548818 |
| T2DM | Pancreas | VAT | delta_cell:t_cell | **0.04060968** |
| T2DM | Pancreas | VAT | PP_cell:epsilon_cell | ***0.07073076*** |
| T2DM | Pancreas | VAT | PP_cell:beta_cell | 0.61889084 |
| T2DM | Pancreas | VAT | PP_cell:ASPC | 0.13410537 |
| T2DM | Pancreas | VAT | PP_cell:LEC | 0.61567533 |
| T2DM | Pancreas | VAT | PP_cell:SMC | 0.53709744 |
| T2DM | Pancreas | VAT | PP_cell:adipocyte | 0.52950587 |
| T2DM | Pancreas | VAT | PP_cell:b_cell | ***0.07695005*** |
| T2DM | Pancreas | VAT | PP_cell:dendritic_cell | 0.80516253 |
| T2DM | Pancreas | VAT | PP_cell:endometrium | 0.10408507 |
| T2DM | Pancreas | VAT | PP_cell:macrophage | 0.7018505 |
| T2DM | Pancreas | VAT | PP_cell:mast_cell | 0.94114526 |
| T2DM | Pancreas | VAT | PP_cell:mesothelium | 0.23444261 |
| T2DM | Pancreas | VAT | PP_cell:monocyte | 0.96573889 |
| T2DM | Pancreas | VAT | PP_cell:nk_cell | **0.01133093** |
| T2DM | Pancreas | VAT | PP_cell:pericyte | 0.71173355 |
| T2DM | Pancreas | VAT | PP_cell:t_cell | 0.38199992 |
| T2DM | Pancreas | VAT | epsilon_cell:beta_cell | ***0.0848577*** |
| T2DM | Pancreas | VAT | epsilon_cell:ASPC | **0.03696924** |

| | | | | |
|---|---|---|---|---|
| T2DM | Pancreas | VAT | epsilon_cell:LEC | 0.27078956 |
| T2DM | Pancreas | VAT | epsilon_cell:SMC | **0.00499444** |
| T2DM | Pancreas | VAT | epsilon_cell:adipocyte | 0.22210833 |
| T2DM | Pancreas | VAT | epsilon_cell:b_cell | 0.28715126 |
| T2DM | Pancreas | VAT | epsilon_cell:dendritic_cell | 0.682497 |
| T2DM | Pancreas | VAT | epsilon_cell:endometrium | 0.52105331 |
| T2DM | Pancreas | VAT | epsilon_cell:macrophage | 0.51037673 |
| T2DM | Pancreas | VAT | epsilon_cell:mast_cell | 0.93178586 |
| T2DM | Pancreas | VAT | epsilon_cell:mesothelium | ***0.08227178*** |
| T2DM | Pancreas | VAT | epsilon_cell:monocyte | ***0.07749121*** |
| T2DM | Pancreas | VAT | epsilon_cell:nk_cell | 0.2523439 |
| T2DM | Pancreas | VAT | epsilon_cell:pericyte | 0.91224114 |
| T2DM | Pancreas | VAT | epsilon_cell:t_cell | 0.53556408 |
| T2DM | Pancreas | VAT | beta_cell:ASPC | 0.10338746 |
| T2DM | Pancreas | VAT | beta_cell:LEC | 0.4365188 |
| T2DM | Pancreas | VAT | beta_cell:SMC | 0.92410549 |
| T2DM | Pancreas | VAT | beta_cell:adipocyte | 0.19475614 |
| T2DM | Pancreas | VAT | beta_cell:b_cell | **0.00302432** |
| T2DM | Pancreas | VAT | beta_cell:dendritic_cell | 0.7136117 |
| T2DM | Pancreas | VAT | beta_cell:endometrium | 0.87585576 |
| T2DM | Pancreas | VAT | beta_cell:macrophage | 0.96829026 |
| T2DM | Pancreas | VAT | beta_cell:mast_cell | 0.22917949 |
| T2DM | Pancreas | VAT | beta_cell:mesothelium | ***0.07422136*** |
| T2DM | Pancreas | VAT | beta_cell:monocyte | 0.91218422 |
| T2DM | Pancreas | VAT | beta_cell:nk_cell | **0.00346773** |
| T2DM | Pancreas | VAT | beta_cell:pericyte | 0.39138067 |
| T2DM | Pancreas | VAT | beta_cell:t_cell | **0.02274358** |
| T2DM | Pancreas | VAT | ASPC:LEC | 0.47401717 |

| | | | | |
|---|---|---|---|---|
| T2DM | Pancreas | VAT | ASPC:SMC | **0.00790156** |
| T2DM | Pancreas | VAT | ASPC:adipocyte | **0.02285139** |
| T2DM | Pancreas | VAT | ASPC:b_cell | 0.65021814 |
| T2DM | Pancreas | VAT | ASPC:dendritic_cell | 0.35084117 |
| T2DM | Pancreas | VAT | ASPC:endometrium | 0.43185478 |
| T2DM | Pancreas | VAT | ASPC:macrophage | 0.96858651 |
| T2DM | Pancreas | VAT | ASPC:mast_cell | 0.70876975 |
| T2DM | Pancreas | VAT | ASPC:mesothelium | 0.64276565 |
| T2DM | Pancreas | VAT | ASPC:monocyte | 0.48324922 |
| T2DM | Pancreas | VAT | ASPC:nk_cell | 0.12320917 |
| T2DM | Pancreas | VAT | ASPC:pericyte | 0.25369284 |
| T2DM | Pancreas | VAT | ASPC:t_cell | 0.38261655 |
| T2DM | Pancreas | VAT | LEC:SMC | ***0.05291415*** |
| T2DM | Pancreas | VAT | LEC:adipocyte | 0.55187537 |
| T2DM | Pancreas | VAT | LEC:b_cell | 0.64771721 |
| T2DM | Pancreas | VAT | LEC:dendritic_cell | ***0.0503179*** |
| T2DM | Pancreas | VAT | LEC:endometrium | 0.16829462 |
| T2DM | Pancreas | VAT | LEC:macrophage | 0.23342018 |
| T2DM | Pancreas | VAT | LEC:mast_cell | 0.36273323 |
| T2DM | Pancreas | VAT | LEC:mesothelium | 0.11591034 |
| T2DM | Pancreas | VAT | LEC:monocyte | 0.31721287 |
| T2DM | Pancreas | VAT | LEC:nk_cell | 0.24949828 |
| T2DM | Pancreas | VAT | LEC:pericyte | **0.03467235** |
| T2DM | Pancreas | VAT | LEC:t_cell | 0.89829889 |
| T2DM | Pancreas | VAT | SMC:adipocyte | ***0.06749887*** |
| T2DM | Pancreas | VAT | SMC:b_cell | **0.00343835** |
| T2DM | Pancreas | VAT | SMC:dendritic_cell | 0.45376708 |
| T2DM | Pancreas | VAT | SMC:endometrium | 0.16097557 |
| T2DM | Pancreas | VAT | SMC:macrophage | **0.00686134** |
| T2DM | Pancreas | VAT | SMC:mast_cell | 0.21399553 |
| T2DM | Pancreas | VAT | SMC:mesothelium | **0.01328534** |
| T2DM | Pancreas | VAT | SMC:monocyte | **0.04992094** |
| T2DM | Pancreas | VAT | SMC:nk_cell | 0.27662675 |
| T2DM | Pancreas | VAT | SMC:pericyte | 0.96216014 |
| T2DM | Pancreas | VAT | SMC:t_cell | 0.61588723 |
| T2DM | Pancreas | VAT | adipocyte:b_cell | 0.15108652 |

| | | | | |
|---|---|---|---|---|
| T2DM | Pancreas | VAT | adipocyte:dendritic_cell | 0.34425329 |
| T2DM | Pancreas | VAT | adipocyte:endometrium | 0.17031061 |
| T2DM | Pancreas | VAT | adipocyte:macrophage | **0.0093081** |
| T2DM | Pancreas | VAT | adipocyte:mast_cell | ***0.09927229*** |
| T2DM | Pancreas | VAT | adipocyte:mesothelium | 0.85568307 |
| T2DM | Pancreas | VAT | adipocyte:monocyte | **0.02556316** |
| T2DM | Pancreas | VAT | adipocyte:nk_cell | **0.00282693** |
| T2DM | Pancreas | VAT | adipocyte:pericyte | ***0.08490673*** |
| T2DM | Pancreas | VAT | adipocyte:t_cell | 0.36594408 |
| T2DM | Pancreas | VAT | b_cell:dendritic_cell | 0.24945421 |
| T2DM | Pancreas | VAT | b_cell:endometrium | 0.16188815 |
| T2DM | Pancreas | VAT | b_cell:macrophage | **0.00764174** |
| T2DM | Pancreas | VAT | b_cell:mast_cell | 0.10089324 |
| T2DM | Pancreas | VAT | b_cell:mesothelium | 0.53308568 |
| T2DM | Pancreas | VAT | b_cell:monocyte | **0.00190559** |
| T2DM | Pancreas | VAT | b_cell:nk_cell | 0.25922822 |
| T2DM | Pancreas | VAT | b_cell:pericyte | ***0.06839117*** |
| T2DM | Pancreas | VAT | b_cell:t_cell | 0.69971966 |
| T2DM | Pancreas | VAT | dendritic_cell:endometrium | 0.3030885 |
| T2DM | Pancreas | VAT | dendritic_cell:macrophage | 0.3834226 |
| T2DM | Pancreas | VAT | dendritic_cell:mast_cell | 0.2368336 |
| T2DM | Pancreas | VAT | dendritic_cell:mesothelium | 0.16460869 |
| T2DM | Pancreas | VAT | dendritic_cell:monocyte | 0.04524262 |
| T2DM | Pancreas | VAT | dendritic_cell:nk_cell | 0.12784787 |
| T2DM | Pancreas | VAT | dendritic_cell:pericyte | 0.75218023 |
| T2DM | Pancreas | VAT | dendritic_cell:t_cell | 0.80543254 |
| T2DM | Pancreas | VAT | endometrium:macrophage | 0.22618651 |

| | | | | |
|---|---|---|---|---|
| T2DM | Pancreas | VAT | endometrium:mast_cell | 0.32156948 |
| T2DM | Pancreas | VAT | endometrium:mesothelium | 0.21031176 |
| T2DM | Pancreas | VAT | endometrium:monocyte | 0.53933523 |
| T2DM | Pancreas | VAT | endometrium:nk_cell | 0.58024444 |
| T2DM | Pancreas | VAT | endometrium:pericyte | 0.6273574 |
| T2DM | Pancreas | VAT | endometrium:t_cell | 0.78099385 |
| T2DM | Pancreas | VAT | macrophage:mast_cell | 0.41179425 |
| T2DM | Pancreas | VAT | macrophage:mesothelium | 0.14838199 |
| T2DM | Pancreas | VAT | macrophage:monocyte | 0.5109686 |
| T2DM | Pancreas | VAT | macrophage:nk_cell | 0.14050227 |
| T2DM | Pancreas | VAT | macrophage:pericyte | 0.61260784 |
| T2DM | Pancreas | VAT | macrophage:t_cell | 0.14660742 |
| T2DM | Pancreas | VAT | mast_cell:mesothelium | 0.23407552 |
| T2DM | Pancreas | VAT | mast_cell:monocyte | 0.46204858 |
| T2DM | Pancreas | VAT | mast_cell:nk_cell | 0.95480475 |
| T2DM | Pancreas | VAT | mast_cell:pericyte | 0.2127622 |
| T2DM | Pancreas | VAT | mast_cell:t_cell | ***0.05878174*** |
| T2DM | Pancreas | VAT | mesothelium:monocyte | **0.04467139** |
| T2DM | Pancreas | VAT | mesothelium:nk_cell | 0.81910548 |
| T2DM | Pancreas | VAT | mesothelium:pericyte | 0.10063517 |
| T2DM | Pancreas | VAT | mesothelium:t_cell | 0.45671048 |
| T2DM | Pancreas | VAT | monocyte:nk_cell | ***0.09301825*** |
| T2DM | Pancreas | VAT | monocyte:pericyte | 0.65982395 |
| T2DM | Pancreas | VAT | monocyte:t_cell | **0.03169538** |
| T2DM | Pancreas | VAT | nk_cell:pericyte | 0.1732244 |
| T2DM | Pancreas | VAT | nk_cell:t_cell | 0.9134381 |

| | | | | |
|---|---|---|---|---|
| T2DM | Pancreas | VAT | pericyte:t_cell | 0.49692325 |
| Obesity | SAT | VAT | ASPC_SAT:LEC_SAT | 0.139886882 |
| Obesity | SAT | VAT | ASPC_SAT:SMC_SAT | 0.25099973 |
| Obesity | SAT | VAT | ASPC_SAT:adipocyte_SAT | ***0.074404683*** |
| Obesity | SAT | VAT | ASPC_SAT:b_cell_SAT | 0.74005422 |
| Obesity | SAT | VAT | ASPC_SAT:dendritic_cell_SAT | 0.71718851 |
| Obesity | SAT | VAT | ASPC_SAT:macrophage_SAT | 0.523372342 |
| Obesity | SAT | VAT | ASPC_SAT:mast_cell_SAT | **0.007274007** |
| Obesity | SAT | VAT | ASPC_SAT:monocyte_SAT | 0.084956974 |
| Obesity | SAT | VAT | ASPC_SAT:nk_cell_SAT | **0.015089054** |
| Obesity | SAT | VAT | ASPC_SAT:pericyte_SAT | 0.822458654 |
| Obesity | SAT | VAT | ASPC_SAT:t_cell_SAT | **0.000649308** |
| Obesity | SAT | VAT | ASPC_SAT:ASPC_VAT | **0.011339492** |
| Obesity | SAT | VAT | ASPC_SAT:LEC_VAT | **0.024187599** |
| Obesity | SAT | VAT | ASPC_SAT:SMC_VAT | 0.785940583 |
| Obesity | SAT | VAT | ASPC_SAT:adipocyte_VAT | ***0.0833612*** |
| Obesity | SAT | VAT | ASPC_SAT:b_cell_VAT | 0.474037861 |
| Obesity | SAT | VAT | ASPC_SAT:dendritic_cell_VAT | 0.464150051 |
| Obesity | SAT | VAT | ASPC_SAT:macrophage_VAT | 0.641273776 |
| Obesity | SAT | VAT | ASPC_SAT:mast_cell_VAT | 0.515731796 |
| Obesity | SAT | VAT | ASPC_SAT:monocyte_VAT | 0.39663188 |
| Obesity | SAT | VAT | ASPC_SAT:nk_cell_VAT | **0.001656984** |
| Obesity | SAT | VAT | ASPC_SAT:pericyte_VAT | **0.016745403** |
| Obesity | SAT | VAT | ASPC_SAT:t_cell_VAT | 0.687139626 |

| | | | | |
|---|---|---|---|---|
| Obesity | SAT | VAT | LEC_SAT:SMC_SAT | 0.188717305 |
| Obesity | SAT | VAT | LEC_SAT:adipocyte_SAT | 0.159578622 |
| Obesity | SAT | VAT | LEC_SAT:b_cell_SAT | 0.456924067 |
| Obesity | SAT | VAT | LEC_SAT:dendritic_cell_SAT | 0.97158446 |
| Obesity | SAT | VAT | LEC_SAT:macrophage_SAT | 0.557310431 |
| Obesity | SAT | VAT | LEC_SAT:mast_cell_SAT | **0.041021157** |
| Obesity | SAT | VAT | LEC_SAT:monocyte_SAT | 0.30102665 |
| Obesity | SAT | VAT | LEC_SAT:nk_cell_SAT | **0.047380066** |
| Obesity | SAT | VAT | LEC_SAT:pericyte_SAT | 0.111762136 |
| Obesity | SAT | VAT | LEC_SAT:t_cell_SAT | ***0.062695689*** |
| Obesity | SAT | VAT | LEC_SAT:ASPC_VAT | 0.144062083 |
| Obesity | SAT | VAT | LEC_SAT:LEC_VAT | **0.041976758** |
| Obesity | SAT | VAT | LEC_SAT:SMC_VAT | 0.213718846 |
| Obesity | SAT | VAT | LEC_SAT:adipocyte_VAT | **0.010299179** |
| Obesity | SAT | VAT | LEC_SAT:b_cell_VAT | **0.00691476** |
| Obesity | SAT | VAT | LEC_SAT:dendritic_cell_VAT | **0.006699365** |
| Obesity | SAT | VAT | LEC_SAT:macrophage_VAT | 0.736130895 |
| Obesity | SAT | VAT | LEC_SAT:mast_cell_VAT | 0.856837866 |
| Obesity | SAT | VAT | LEC_SAT:monocyte_VAT | 0.780200627 |
| Obesity | SAT | VAT | LEC_SAT:nk_cell_VAT | 0.712713757 |
| Obesity | SAT | VAT | LEC_SAT:pericyte_VAT | 0.838848373 |
| Obesity | SAT | VAT | LEC_SAT:t_cell_VAT | 0.368323341 |
| Obesity | SAT | VAT | SMC_SAT:adipocyte_SAT | ***0.085512836*** |
| Obesity | SAT | VAT | SMC_SAT:b_cell_SAT | 0.611367546 |

| | | | | |
|---|---|---|---|---|
| Obesity | SAT | VAT | SMC_SAT:dendritic_cell_SAT | 0.95782989 |
| Obesity | SAT | VAT | SMC_SAT:macrophage_SAT | ***0.063166895*** |
| Obesity | SAT | VAT | SMC_SAT:mast_cell_SAT | 0.168103691 |
| Obesity | SAT | VAT | SMC_SAT:monocyte_SAT | 0.326959236 |
| Obesity | SAT | VAT | SMC_SAT:nk_cell_SAT | 0.166240582 |
| Obesity | SAT | VAT | SMC_SAT:pericyte_SAT | ***0.076261492*** |
| Obesity | SAT | VAT | SMC_SAT:t_cell_SAT | ***0.050007071*** |
| Obesity | SAT | VAT | SMC_SAT:ASPC_VAT | 0.696128311 |
| Obesity | SAT | VAT | SMC_SAT:LEC_VAT | 0.215970577 |
| Obesity | SAT | VAT | SMC_SAT:SMC_VAT | 0.854794108 |
| Obesity | SAT | VAT | SMC_SAT:adipocyte_VAT | ***0.099048187*** |
| Obesity | SAT | VAT | SMC_SAT:b_cell_VAT | 0.162602859 |
| Obesity | SAT | VAT | SMC_SAT:dendritic_cell_VAT | 0.718715205 |
| Obesity | SAT | VAT | SMC_SAT:macrophage_VAT | 0.828063409 |
| Obesity | SAT | VAT | SMC_SAT:mast_cell_VAT | 0.332014562 |
| Obesity | SAT | VAT | SMC_SAT:monocyte_VAT | 0.880672727 |
| Obesity | SAT | VAT | SMC_SAT:nk_cell_VAT | 0.896403404 |
| Obesity | SAT | VAT | SMC_SAT:pericyte_VAT | 0.429567076 |
| Obesity | SAT | VAT | SMC_SAT:t_cell_VAT | 0.684622679 |
| Obesity | SAT | VAT | adipocyte_SAT:b_cell_SAT | 0.494297399 |
| Obesity | SAT | VAT | adipocyte_SAT:dendritic_cell_SAT | 0.398654351 |
| Obesity | SAT | VAT | adipocyte_SAT:macrophage_SAT | 0.327451836 |
| Obesity | SAT | VAT | adipocyte_SAT:mast_cell_SAT | 0.106293401 |

| | | | | |
|---|---|---|---|---|
| Obesity | SAT | VAT | adipocyte_SAT:monocyte_SAT | ***0.051837511*** |
| Obesity | SAT | VAT | adipocyte_SAT:nk_cell_SAT | 0.108170654 |
| Obesity | SAT | VAT | adipocyte_SAT:pericyte_SAT | 0.103632268 |
| Obesity | SAT | VAT | adipocyte_SAT:t_cell_SAT | 0.610404428 |
| Obesity | SAT | VAT | adipocyte_SAT:ASPC_VAT | 0.73026057 |
| Obesity | SAT | VAT | adipocyte_SAT:LEC_VAT | 0.31193788 |
| Obesity | SAT | VAT | adipocyte_SAT:SMC_VAT | **0.026485201** |
| Obesity | SAT | VAT | adipocyte_SAT:adipocyte_VAT | 0.208417533 |
| Obesity | SAT | VAT | adipocyte_SAT:b_cell_VAT | 0.191119915 |
| Obesity | SAT | VAT | adipocyte_SAT:dendritic_cell_VAT | 0.830946148 |
| Obesity | SAT | VAT | adipocyte_SAT:macrophage_VAT | 0.268924114 |
| Obesity | SAT | VAT | adipocyte_SAT:mast_cell_VAT | 0.112012949 |
| Obesity | SAT | VAT | adipocyte_SAT:monocyte_VAT | 0.712311917 |
| Obesity | SAT | VAT | adipocyte_SAT:nk_cell_VAT | **0.001283656** |
| Obesity | SAT | VAT | adipocyte_SAT:pericyte_VAT | **0.004184319** |
| Obesity | SAT | VAT | adipocyte_SAT:t_cell_VAT | 0.446602771 |
| Obesity | SAT | VAT | b_cell_SAT:dendritic_cell_SAT | 0.375752314 |
| Obesity | SAT | VAT | b_cell_SAT:macrophage_SAT | 0.841839128 |
| Obesity | SAT | VAT | b_cell_SAT:mast_cell_SAT | 0.268884537 |
| Obesity | SAT | VAT | b_cell_SAT:monocyte_SAT | **0.013754236** |
| Obesity | SAT | VAT | b_cell_SAT:nk_cell_SAT | 0.127014476 |
| Obesity | SAT | VAT | b_cell_SAT:pericyte_SAT | 0.427608166 |
| Obesity | SAT | VAT | b_cell_SAT:t_cell_SAT | 0.628019826 |

| | | | | |
|---|---|---|---|---|
| Obesity | SAT | VAT | b_cell_SAT:ASPC_VAT | 0.106146675 |
| Obesity | SAT | VAT | b_cell_SAT:LEC_VAT | 0.312375871 |
| Obesity | SAT | VAT | b_cell_SAT:SMC_VAT | 0.121025397 |
| Obesity | SAT | VAT | b_cell_SAT:adipocyte_VAT | 0.647478307 |
| Obesity | SAT | VAT | b_cell_SAT:b_cell_VAT | 0.051830401 |
| Obesity | SAT | VAT | b_cell_SAT:dendritic_cell_VAT | 0.059983975 |
| Obesity | SAT | VAT | b_cell_SAT:macrophage_VAT | 0.89196523 |
| Obesity | SAT | VAT | b_cell_SAT:mast_cell_VAT | 0.354341204 |
| Obesity | SAT | VAT | b_cell_SAT:monocyte_VAT | 0.256007969 |
| Obesity | SAT | VAT | b_cell_SAT:nk_cell_VAT | **0.000172856** |
| Obesity | SAT | VAT | b_cell_SAT:pericyte_VAT | **0.005405607** |
| Obesity | SAT | VAT | b_cell_SAT:t_cell_VAT | 0.13427837 |
| Obesity | SAT | VAT | dendritic_cell_SAT:macrophage_SAT | 0.9415823 |
| Obesity | SAT | VAT | dendritic_cell_SAT:mast_cell_SAT | 0.548379602 |
| Obesity | SAT | VAT | dendritic_cell_SAT:monocyte_SAT | 0.11227383 |
| Obesity | SAT | VAT | dendritic_cell_SAT:nk_cell_SAT | 0.527962611 |
| Obesity | SAT | VAT | dendritic_cell_SAT:pericyte_SAT | 0.14241599 |
| Obesity | SAT | VAT | dendritic_cell_SAT:t_cell_SAT | 0.887292027 |
| Obesity | SAT | VAT | dendritic_cell_SAT:ASPC_VAT | 0.718006678 |
| Obesity | SAT | VAT | dendritic_cell_SAT:LEC_VAT | 0.481540126 |
| Obesity | SAT | VAT | dendritic_cell_SAT:SMC_VAT | 0.778402798 |

| | | | | |
|---|---|---|---|---|
| Obesity | SAT | VAT | dendritic_cell_SAT:adipocyte_VAT | 0.017304898 |
| Obesity | SAT | VAT | dendritic_cell_SAT:b_cell_VAT | 0.121432717 |
| Obesity | SAT | VAT | dendritic_cell_SAT:dendritic_cell_VAT | ***0.066649316*** |
| Obesity | SAT | VAT | dendritic_cell_SAT:macrophage_VAT | 0.113397125 |
| Obesity | SAT | VAT | dendritic_cell_SAT:mast_cell_VAT | 0.281961623 |
| Obesity | SAT | VAT | dendritic_cell_SAT:monocyte_VAT | 0.96449162 |
| Obesity | SAT | VAT | dendritic_cell_SAT:nk_cell_VAT | 0.552875762 |
| Obesity | SAT | VAT | dendritic_cell_SAT:pericyte_VAT | 0.645446192 |
| Obesity | SAT | VAT | dendritic_cell_SAT:t_cell_VAT | 0.11308887 |
| Obesity | SAT | VAT | macrophage_SAT:mast_cell_SAT | 0.127998641 |
| Obesity | SAT | VAT | macrophage_SAT:monocyte_SAT | 0.161953908 |
| Obesity | SAT | VAT | macrophage_SAT:nk_cell_SAT | **0.000401693** |
| Obesity | SAT | VAT | macrophage_SAT:pericyte_SAT | 0.933379155 |
| Obesity | SAT | VAT | macrophage_SAT:t_cell_SAT | 0.114298162 |
| Obesity | SAT | VAT | macrophage_SAT:ASPC_VAT | 0.275412406 |
| Obesity | SAT | VAT | macrophage_SAT:LEC_VAT | 0.577611367 |
| Obesity | SAT | VAT | macrophage_SAT:SMC_VAT | 0.233827614 |
| Obesity | SAT | VAT | macrophage_SAT:adipocyte_VAT | 0.119273735 |
| Obesity | SAT | VAT | macrophage_SAT:b_cell_VAT | 0.294025613 |

| | | | | |
|---|---|---|---|---|
| Obesity | SAT | VAT | macrophage_SAT:dendritic_cell_VAT | 0.148864891 |
| Obesity | SAT | VAT | macrophage_SAT:macrophage_VAT | 0.304880887 |
| Obesity | SAT | VAT | macrophage_SAT:mast_cell_VAT | 0.38410412 |
| Obesity | SAT | VAT | macrophage_SAT:monocyte_VAT | 0.145783605 |
| Obesity | SAT | VAT | macrophage_SAT:nk_cell_VAT | **7.03E-05** |
| Obesity | SAT | VAT | macrophage_SAT:pericyte_VAT | **0.006387076** |
| Obesity | SAT | VAT | macrophage_SAT:t_cell_VAT | 0.372696992 |
| Obesity | SAT | VAT | mast_cell_SAT:monocyte_SAT | 0.356001563 |
| Obesity | SAT | VAT | mast_cell_SAT:nk_cell_SAT | 0.293662247 |
| Obesity | SAT | VAT | mast_cell_SAT:pericyte_SAT | 0.274887221 |
| Obesity | SAT | VAT | mast_cell_SAT:t_cell_SAT | 0.338656501 |
| Obesity | SAT | VAT | mast_cell_SAT:ASPC_VAT | 0.243959966 |
| Obesity | SAT | VAT | mast_cell_SAT:LEC_VAT | **0.014554844** |
| Obesity | SAT | VAT | mast_cell_SAT:SMC_VAT | 0.75218811 |
| Obesity | SAT | VAT | mast_cell_SAT:adipocyte_VAT | **0.001269535** |
| Obesity | SAT | VAT | mast_cell_SAT:b_cell_VAT | 0.371387579 |
| Obesity | SAT | VAT | mast_cell_SAT:dendritic_cell_VAT | 0.895288729 |
| Obesity | SAT | VAT | mast_cell_SAT:macrophage_VAT | 0.392496545 |
| Obesity | SAT | VAT | mast_cell_SAT:mast_cell_VAT | **0.014604581** |
| Obesity | SAT | VAT | mast_cell_SAT:monocyte_VAT | ***0.07704629*** |
| Obesity | SAT | VAT | mast_cell_SAT:nk_cell_VAT | 0.499588032 |

| | | | | |
|---|---|---|---|---|
| Obesity | SAT | VAT | mast_cell_SAT:pericyte_VAT | **0.005582189** |
| Obesity | SAT | VAT | mast_cell_SAT:t_cell_VAT | **0.017382586** |
| Obesity | SAT | VAT | monocyte_SAT:nk_cell_SAT | 0.196552771 |
| Obesity | SAT | VAT | monocyte_SAT:pericyte_SAT | 0.101710601 |
| Obesity | SAT | VAT | monocyte_SAT:t_cell_SAT | 0.156666624 |
| Obesity | SAT | VAT | monocyte_SAT:ASPC_VAT | 0.401955977 |
| Obesity | SAT | VAT | monocyte_SAT:LEC_VAT | 0.385931313 |
| Obesity | SAT | VAT | monocyte_SAT:SMC_VAT | 0.618987479 |
| Obesity | SAT | VAT | monocyte_SAT:adipocyte_VAT | **0.00149331** |
| Obesity | SAT | VAT | monocyte_SAT:b_cell_VAT | 0.122284369 |
| Obesity | SAT | VAT | monocyte_SAT:dendritic_cell_VAT | 0.038212353 |
| Obesity | SAT | VAT | monocyte_SAT:macrophage_VAT | 0.656918845 |
| Obesity | SAT | VAT | monocyte_SAT:mast_cell_VAT | 0.595456052 |
| Obesity | SAT | VAT | monocyte_SAT:monocyte_VAT | ***0.093954309*** |
| Obesity | SAT | VAT | monocyte_SAT:nk_cell_VAT | **0.028440375** |
| Obesity | SAT | VAT | monocyte_SAT:pericyte_VAT | ***0.099924536*** |
| Obesity | SAT | VAT | monocyte_SAT:t_cell_VAT | 0.200040201 |
| Obesity | SAT | VAT | nk_cell_SAT:pericyte_SAT | 0.165414234 |
| Obesity | SAT | VAT | nk_cell_SAT:t_cell_SAT | 0.315329781 |
| Obesity | SAT | VAT | nk_cell_SAT:ASPC_VAT | **0.00890831** |
| Obesity | SAT | VAT | nk_cell_SAT:LEC_VAT | 0.999667441 |
| Obesity | SAT | VAT | nk_cell_SAT:SMC_VAT | 0.273690258 |
| Obesity | SAT | VAT | nk_cell_SAT:adipocyte_VAT | 0.300103344 |

| | | | | |
|---|---|---|---|---|
| Obesity | SAT | VAT | nk_cell_SAT:b_cell_VAT | 0.554110131 |
| Obesity | SAT | VAT | nk_cell_SAT:dendritic_cell_VAT | 0.863313336 |
| Obesity | SAT | VAT | nk_cell_SAT:macrophage_VAT | 0.951511822 |
| Obesity | SAT | VAT | nk_cell_SAT:mast_cell_VAT | 0.114634409 |
| Obesity | SAT | VAT | nk_cell_SAT:monocyte_VAT | 0.141800371 |
| Obesity | SAT | VAT | nk_cell_SAT:nk_cell_VAT | **0.000769738** |
| Obesity | SAT | VAT | nk_cell_SAT:pericyte_VAT | **0.002933408** |
| Obesity | SAT | VAT | nk_cell_SAT:t_cell_VAT | 0.146690097 |
| Obesity | SAT | VAT | pericyte_SAT:t_cell_SAT | 0.844868205 |
| Obesity | SAT | VAT | pericyte_SAT:ASPC_VAT | 0.833322666 |
| Obesity | SAT | VAT | pericyte_SAT:LEC_VAT | 0.523766211 |
| Obesity | SAT | VAT | pericyte_SAT:SMC_VAT | 0.644500362 |
| Obesity | SAT | VAT | pericyte_SAT:adipocyte_VAT | 0.351519905 |
| Obesity | SAT | VAT | pericyte_SAT:b_cell_VAT | 0.342620982 |
| Obesity | SAT | VAT | pericyte_SAT:dendritic_cell_VAT | 0.279917256 |
| Obesity | SAT | VAT | pericyte_SAT:macrophage_VAT | 0.576528651 |
| Obesity | SAT | VAT | pericyte_SAT:mast_cell_VAT | **0.01078661** |
| Obesity | SAT | VAT | pericyte_SAT:monocyte_VAT | ***0.051482444*** |
| Obesity | SAT | VAT | pericyte_SAT:nk_cell_VAT | 0.114021343 |
| Obesity | SAT | VAT | pericyte_SAT:pericyte_VAT | 0.28873594 |
| Obesity | SAT | VAT | pericyte_SAT:t_cell_VAT | 0.328213881 |
| Obesity | SAT | VAT | t_cell_SAT:ASPC_VAT | 0.258946974 |
| Obesity | SAT | VAT | t_cell_SAT:LEC_VAT | 0.308157919 |

| | | | | |
|---|---|---|---|---|
| Obesity | SAT | VAT | t_cell_SAT:SMC_VAT | 0.352711188 |
| Obesity | SAT | VAT | t_cell_SAT:adipocyte_VAT | **9.78E-06** |
| Obesity | SAT | VAT | t_cell_SAT:b_cell_VAT | 0.362500559 |
| Obesity | SAT | VAT | t_cell_SAT:dendritic_cell_VAT | 0.663371045 |
| Obesity | SAT | VAT | t_cell_SAT:macrophage_VAT | 0.031095081 |
| Obesity | SAT | VAT | t_cell_SAT:mast_cell_VAT | 0.682840301 |
| Obesity | SAT | VAT | t_cell_SAT:monocyte_VAT | 0.667225994 |
| Obesity | SAT | VAT | t_cell_SAT:nk_cell_VAT | **0.028879768** |
| Obesity | SAT | VAT | t_cell_SAT:pericyte_VAT | **0.001900505** |
| Obesity | SAT | VAT | t_cell_SAT:t_cell_VAT | 0.201970288 |
| Obesity | SAT | VAT | ASPC_VAT:LEC_VAT | **0.009320911** |
| Obesity | SAT | VAT | ASPC_VAT:SMC_VAT | **0.030477553** |
| Obesity | SAT | VAT | ASPC_VAT:adipocyte_VAT | **0.000338604** |
| Obesity | SAT | VAT | ASPC_VAT:b_cell_VAT | **1.97E-09** |
| Obesity | SAT | VAT | ASPC_VAT:dendritic_cell_VAT | **0.001131651** |
| Obesity | SAT | VAT | ASPC_VAT:macrophage_VAT | 0.686925993 |
| Obesity | SAT | VAT | ASPC_VAT:mast_cell_VAT | 0.146792576 |
| Obesity | SAT | VAT | ASPC_VAT:monocyte_VAT | 0.600775065 |
| Obesity | SAT | VAT | ASPC_VAT:nk_cell_VAT | 0.286842808 |
| Obesity | SAT | VAT | ASPC_VAT:pericyte_VAT | 0.166510366 |
| Obesity | SAT | VAT | ASPC_VAT:t_cell_VAT | **0.022665254** |
| Obesity | SAT | VAT | LEC_VAT:SMC_VAT | 0.648068081 |
| Obesity | SAT | VAT | LEC_VAT:adipocyte_VAT | 0.296929529 |
| Obesity | SAT | VAT | LEC_VAT:b_cell_VAT | 0.312555809 |

| | | | | |
|---|---|---|---|---|
| Obesity | SAT | VAT | LEC_VAT:dendritic_cell_VAT | 0.213991908 |
| Obesity | SAT | VAT | LEC_VAT:macrophage_VAT | 0.801660969 |
| Obesity | SAT | VAT | LEC_VAT:mast_cell_VAT | 0.24934883 |
| Obesity | SAT | VAT | LEC_VAT:monocyte_VAT | **0.027720115** |
| Obesity | SAT | VAT | LEC_VAT:nk_cell_VAT | **0.007178859** |
| Obesity | SAT | VAT | LEC_VAT:pericyte_VAT | **0.077948698** |
| Obesity | SAT | VAT | LEC_VAT:t_cell_VAT | 0.253330356 |
| Obesity | SAT | VAT | SMC_VAT:adipocyte_VAT | **0.007790049** |
| Obesity | SAT | VAT | SMC_VAT:b_cell_VAT | **0.043087747** |
| Obesity | SAT | VAT | SMC_VAT:dendritic_cell_VAT | 0.814846243 |
| Obesity | SAT | VAT | SMC_VAT:macrophage_VAT | 0.770444202 |
| Obesity | SAT | VAT | SMC_VAT:mast_cell_VAT | 0.121023271 |
| Obesity | SAT | VAT | SMC_VAT:monocyte_VAT | 0.11756166 |
| Obesity | SAT | VAT | SMC_VAT:nk_cell_VAT | 0.433360755 |
| Obesity | SAT | VAT | SMC_VAT:pericyte_VAT | **0.003509705** |
| Obesity | SAT | VAT | SMC_VAT:t_cell_VAT | 0.367739613 |
| Obesity | SAT | VAT | adipocyte_VAT:b_cell_VAT | 0.844825107 |
| Obesity | SAT | VAT | adipocyte_VAT:dendritic_cell_VAT | 0.526634155 |
| Obesity | SAT | VAT | adipocyte_VAT:macrophage_VAT | **0.030842076** |
| Obesity | SAT | VAT | adipocyte_VAT:mast_cell_VAT | 0.301812603 |
| Obesity | SAT | VAT | adipocyte_VAT:monocyte_VAT | 0.241468826 |
| Obesity | SAT | VAT | adipocyte_VAT:nk_cell_VAT | 0.052966861 |
| Obesity | SAT | VAT | adipocyte_VAT:pericyte_VAT | 0.008628862 |

| | | | | |
|---|---|---|---|---|
| Obesity | SAT | VAT | adipocyte_VAT:t_cell_VAT | 0.883291987 |
| Obesity | SAT | VAT | b_cell_VAT:dendritic_cell_VAT | 0.166482247 |
| Obesity | SAT | VAT | b_cell_VAT:macrophage_VAT | 0.130268429 |
| Obesity | SAT | VAT | b_cell_VAT:mast_cell_VAT | 0.889517962 |
| Obesity | SAT | VAT | b_cell_VAT:monocyte_VAT | **0.007997572** |
| Obesity | SAT | VAT | b_cell_VAT:nk_cell_VAT | **0.030289561** |
| Obesity | SAT | VAT | b_cell_VAT:pericyte_VAT | **9.38E-05** |
| Obesity | SAT | VAT | b_cell_VAT:t_cell_VAT | ***0.050626336*** |
| Obesity | SAT | VAT | dendritic_cell_VAT:macrophage_VAT | 0.154146182 |
| Obesity | SAT | VAT | dendritic_cell_VAT:mast_cell_VAT | 0.982544755 |
| Obesity | SAT | VAT | dendritic_cell_VAT:monocyte_VAT | 0.542564088 |
| Obesity | SAT | VAT | dendritic_cell_VAT:nk_cell_VAT | 0.617219184 |
| Obesity | SAT | VAT | dendritic_cell_VAT:pericyte_VAT | 0.677454009 |
| Obesity | SAT | VAT | dendritic_cell_VAT:t_cell_VAT | 0.562413046 |
| Obesity | SAT | VAT | macrophage_VAT:mast_cell_VAT | 0.590375674 |
| Obesity | SAT | VAT | macrophage_VAT:monocyte_VAT | 0.792171094 |
| Obesity | SAT | VAT | macrophage_VAT:nk_cell_VAT | 0.570160431 |
| Obesity | SAT | VAT | macrophage_VAT:pericyte_VAT | 0.126548169 |
| Obesity | SAT | VAT | macrophage_VAT:t_cell_VAT | 0.505378935 |
| Obesity | SAT | VAT | mast_cell_VAT:monocyte_VAT | ***0.053237842*** |

| | | | | |
|---|---|---|---|---|
| Obesity | SAT | VAT | mast_cell_VAT:nk_cell_VAT | **0.041613926** |
| Obesity | SAT | VAT | mast_cell_VAT:pericyte_VAT | 0.650789506 |
| Obesity | SAT | VAT | mast_cell_VAT:t_cell_VAT | 0.723983297 |
| Obesity | SAT | VAT | monocyte_VAT:nk_cell_VAT | 0.583831841 |
| Obesity | SAT | VAT | monocyte_VAT:pericyte_VAT | 0.110081913 |
| Obesity | SAT | VAT | monocyte_VAT:t_cell_VAT | ***0.053467944*** |
| Obesity | SAT | VAT | nk_cell_VAT:pericyte_VAT | 0.713059534 |
| Obesity | SAT | VAT | nk_cell_VAT:t_cell_VAT | **0.041916389** |
| Obesity | SAT | VAT | pericyte_VAT:t_cell_VAT | 0.306876684 |

Table S4 Identified causal genes from inferred CTS expression profiles

a. Identified cell type specific causal genes for depression and related disorders from frontal cortex

| disorder | celltype | gene | assoc | zMin |
|---|---|---|---|---|
| Anxiety | Astros | *ITIH4* | TRUE | 2.214292376 |
| Anxiety | Astros | *KCNN4* | TRUE | 2.249295278 |
| Anxiety | Astros | *CHRNE* | TRUE | 2.477637898 |
| Anxiety | Astros | *SGCA* | TRUE | 2.411862304 |
| Anxiety | Astros | *S1PR4* | TRUE | 2.14671706 |
| Anxiety | Astros | *ZSCAN9* | TRUE | 2.442976446 |
| Anxiety | Astros | *ZNF503* | TRUE | 2.153121139 |
| Anxiety | Astros | *KLK5* | TRUE | 1.998220687 |
| Anxiety | Astros | *FAHD1* | TRUE | 2.573587046 |
| Anxiety | Astros | *CLCNKA* | TRUE | 2.417504283 |
| Anxiety | Astros | *ZKSCAN3* | TRUE | 2.774118275 |
| Anxiety | Astros | *RP11.63A1.2* | TRUE | 2.760243442 |
| Anxiety | Ex | *ITIH4* | TRUE | 2.202417902 |
| Anxiety | Ex | *F7* | TRUE | 2.123384478 |
| Anxiety | Ex | *APBB1IP* | TRUE | 2.114672621 |
| Anxiety | Ex | *SLC5A4* | TRUE | 2.252436482 |
| Anxiety | Ex | *ZC3H14* | TRUE | 2.058084983 |
| Anxiety | Ex | *CD40* | TRUE | 2.192334067 |
| Anxiety | Ex | *KAT8* | TRUE | 2.013787833 |
| Anxiety | Ex | *KCNN4* | TRUE | 2.261680954 |
| Anxiety | Ex | *HAS1* | TRUE | 3.023398156 |
| Anxiety | Ex | *RPL18A* | TRUE | 2.146794906 |
| Anxiety | Ex | *SUSD1* | TRUE | 2.908752771 |
| Anxiety | Ex | *CHRNE* | TRUE | 2.740870039 |
| Anxiety | Ex | *SGCA* | TRUE | 2.794979197 |
| Anxiety | Ex | *RCVRN* | TRUE | 2.378387677 |
| Anxiety | Ex | *GNB3* | TRUE | 2.165877691 |
| Anxiety | Ex | *NME5* | TRUE | 2.904763868 |
| Anxiety | Ex | *CNN3* | TRUE | 2.459891366 |
| Anxiety | Ex | *DIEXF* | TRUE | 1.969520023 |
| Anxiety | Ex | *SOHLH2* | TRUE | 2.00086708 |
| Anxiety | Ex | *POLM* | TRUE | 2.310027669 |
| Anxiety | Ex | *WFDC3* | TRUE | 2.298650999 |
| Anxiety | Ex | *PAK4* | TRUE | 2.138155231 |
| Anxiety | Ex | *ZSCAN9* | TRUE | 2.550143144 |
| Anxiety | Ex | *ETV6* | TRUE | 2.349465916 |
| Anxiety | Ex | *C18orf8* | TRUE | 2.133512974 |
| Anxiety | Ex | *TCTE1* | TRUE | 2.070871703 |
| Anxiety | Ex | *CBR3* | TRUE | 2.294728708 |
| Anxiety | Ex | *ADCK3* | TRUE | 2.839657212 |
| Anxiety | Ex | *TMEM183A* | TRUE | 2.035957136 |
| Anxiety | Ex | *CCDC127* | TRUE | 2.448333275 |
| Anxiety | Ex | *ZNF503* | TRUE | 2.721059895 |
| Anxiety | Ex | *FAM96B* | TRUE | 2.273376372 |
| Anxiety | Ex | *KLK5* | TRUE | 2.222649804 |

| | | | | |
|---|---|---|---|---|
| Anxiety | Ex | *NADSYN1* | TRUE | 2.029578349 |
| Anxiety | Ex | *MAMSTR* | TRUE | 2.169196911 |
| Anxiety | Ex | *C9orf50* | TRUE | 2.363321545 |
| Anxiety | Ex | *C2orf82* | TRUE | 2.137249191 |
| Anxiety | Ex | *NPIPA5* | TRUE | 2.012724922 |
| Anxiety | Ex | *NUDT17* | TRUE | 2.353783772 |
| Anxiety | Ex | *CLCNKA* | TRUE | 3.068949799 |
| Anxiety | Ex | *HYAL3* | TRUE | 2.174818917 |
| Anxiety | Ex | *DHRS4L2* | TRUE | 2.160944944 |
| Anxiety | Ex | *ZFP69* | TRUE | 2.863923067 |
| Anxiety | Ex | *NWD1* | TRUE | 1.991927904 |
| Anxiety | Ex | *JAKMIP3* | TRUE | 2.026530355 |
| Anxiety | Ex | *NBR1* | TRUE | 2.303043882 |
| Anxiety | Ex | *ZKSCAN3* | TRUE | 2.655464339 |
| Anxiety | Ex | *TRAPPC4* | TRUE | 1.96045222 |
| Anxiety | Ex | *TPM2* | TRUE | 2.201206539 |
| Anxiety | Ex | *HLA.DRB5* | TRUE | 2.177416827 |
| Anxiety | Ex | *SLC2A1.AS1* | TRUE | 2.118159959 |
| Anxiety | Ex | *AC093642.3* | TRUE | 2.167348805 |
| Anxiety | Ex | *RP11.108M9.4* | TRUE | 2.455361025 |
| Anxiety | Ex | *RP11.584P21.2* | TRUE | 2.114699488 |
| Anxiety | Ex | *RP13.977J11.2* | TRUE | 3.103136033 |
| Anxiety | Ex | *RP11.63A1.2* | TRUE | 3.101558302 |
| Anxiety | Ex | *RP11.250B2.6* | TRUE | 2.864290769 |
| Anxiety | Ex | *CASC15* | TRUE | 2.094504548 |
| Anxiety | Ex | *AP000350.5* | TRUE | 1.975106351 |
| Anxiety | Ex | *RP11.686O6.2* | TRUE | 2.226459048 |
| Anxiety | Inhib | *ITIH4* | TRUE | 2.205369407 |
| Anxiety | Inhib | *F7* | TRUE | 1.985963481 |
| Anxiety | Inhib | *APBB1IP* | TRUE | 2.275552881 |
| Anxiety | Inhib | *SLC5A4* | TRUE | 2.297307057 |
| Anxiety | Inhib | *ZC3H14* | TRUE | 2.058766978 |
| Anxiety | Inhib | *CD40* | TRUE | 2.085764742 |
| Anxiety | Inhib | *KCNN4* | TRUE | 2.289672591 |
| Anxiety | Inhib | *HAS1* | TRUE | 2.987728376 |
| Anxiety | Inhib | *RPL18A* | TRUE | 2.122287219 |
| Anxiety | Inhib | *SUSD1* | TRUE | 2.773019084 |
| Anxiety | Inhib | *DHRS11* | TRUE | 2.061535132 |
| Anxiety | Inhib | *CHRNE* | TRUE | 2.69787944 |
| Anxiety | Inhib | *SGCA* | TRUE | 2.827457121 |
| Anxiety | Inhib | *RCVRN* | TRUE | 2.521011604 |
| Anxiety | Inhib | *GNB3* | TRUE | 2.042569715 |
| Anxiety | Inhib | *NME5* | TRUE | 2.739897443 |
| Anxiety | Inhib | *CNN3* | TRUE | 2.204814371 |
| Anxiety | Inhib | *SOHLH2* | TRUE | 2.147777906 |
| Anxiety | Inhib | *POLM* | TRUE | 2.234292027 |
| Anxiety | Inhib | *WFDC3* | TRUE | 2.29975841 |
| Anxiety | Inhib | *PAK4* | TRUE | 2.048037267 |

| | | | | |
|---|---|---|---|---|
| Anxiety | Inhib | *ZSCAN9* | TRUE | 2.557136734 |
| Anxiety | Inhib | *ETV6* | TRUE | 2.411365185 |
| Anxiety | Inhib | *C18orf8* | TRUE | 2.036752186 |
| Anxiety | Inhib | *TCTE1* | TRUE | 2.124518805 |
| Anxiety | Inhib | *CBR3* | TRUE | 2.005885414 |
| Anxiety | Inhib | *CCDC64B* | TRUE | 2.091561984 |
| Anxiety | Inhib | *ADCK3* | TRUE | 2.687321106 |
| Anxiety | Inhib | *CCDC127* | TRUE | 2.13249636 |
| Anxiety | Inhib | *ZNF503* | TRUE | 2.513293592 |
| Anxiety | Inhib | *KLK5* | TRUE | 2.156021646 |
| Anxiety | Inhib | *MAMSTR* | TRUE | 2.175189593 |
| Anxiety | Inhib | *C9orf50* | TRUE | 2.202027076 |
| Anxiety | Inhib | *C2orf82* | TRUE | 2.093847539 |
| Anxiety | Inhib | *NPIPA5* | TRUE | 1.961464244 |
| Anxiety | Inhib | *NUDT17* | TRUE | 2.165821821 |
| Anxiety | Inhib | *CLCNKA* | TRUE | 2.748149413 |
| Anxiety | Inhib | *HYAL3* | TRUE | 2.093268208 |
| Anxiety | Inhib | *DHRS4L2* | TRUE | 2.019314041 |
| Anxiety | Inhib | *ZFP69* | TRUE | 2.784097492 |
| Anxiety | Inhib | *NWD1* | TRUE | 1.972463218 |
| Anxiety | Inhib | *NBR1* | TRUE | 1.991412109 |
| Anxiety | Inhib | *ZKSCAN3* | TRUE | 2.662382971 |
| Anxiety | Inhib | *TPM2* | TRUE | 2.140683857 |
| Anxiety | Inhib | *HLA.DRB5* | TRUE | 1.97375404 |
| Anxiety | Inhib | *SLC2A1.AS1* | TRUE | 2.053499737 |
| Anxiety | Inhib | *AC093642.3* | TRUE | 2.086487135 |
| Anxiety | Inhib | *RP11.108M9.4* | TRUE | 2.352713379 |
| Anxiety | Inhib | *GS1.259H13.2* | TRUE | 1.974644829 |
| Anxiety | Inhib | *RP11.584P21.2* | TRUE | 2.070745071 |
| Anxiety | Inhib | *RP13.977J11.2* | TRUE | 2.951074635 |
| Anxiety | Inhib | *RP11.250B2.6* | TRUE | 2.430786209 |
| Anxiety | Inhib | *CASC15* | TRUE | 1.994218117 |
| Anxiety | Micro_Macro | *SGCA* | TRUE | 2.24228395 |
| Anxiety | Micro_Macro | *ZSCAN9* | TRUE | 2.658650527 |
| Anxiety | Micro_Macro | *ZKSCAN3* | TRUE | 2.300699798 |
| Anxiety | Micro_Macro | *RP11.63A1.2* | TRUE | 2.836933203 |
| Anxiety | Oligos | *KCNN4* | TRUE | 2.148672972 |
| Anxiety | Oligos | *HAS1* | TRUE | 1.961602234 |
| Anxiety | Oligos | *SUSD1* | TRUE | 2.023288867 |
| Anxiety | Oligos | *SGCA* | TRUE | 2.839026115 |
| Anxiety | Oligos | *NME5* | TRUE | 2.633878255 |
| Anxiety | Oligos | *S1PR4* | TRUE | 1.972747858 |
| Anxiety | Oligos | *PAK4* | TRUE | 1.98247722 |
| Anxiety | Oligos | *ZSCAN9* | TRUE | 2.594384304 |
| Anxiety | Oligos | *ADCK3* | TRUE | 2.433805788 |
| Anxiety | Oligos | *ZNF503* | TRUE | 2.505925754 |
| Anxiety | Oligos | *FAHD1* | TRUE | 2.204420477 |
| Anxiety | Oligos | *C2orf82* | TRUE | 2.004907253 |
| Anxiety | Oligos | *CLCNKA* | TRUE | 2.775728151 |
| Anxiety | Oligos | *HYAL3* | TRUE | 2.176181195 |

| | | | | |
|---|---|---|---|---|
| Anxiety | Oligos | *ZFP69* | TRUE | 2.08929014 |
| Anxiety | Oligos | *ZKSCAN3* | TRUE | 2.487597856 |
| Anxiety | Oligos | *AC093642.3* | TRUE | 2.114189766 |
| Anxiety | Oligos | *RP13.977J11.2* | TRUE | 2.273672568 |
| Anxiety | Oligos | *RP11.63A1.2* | TRUE | 2.815649417 |
| Anxiety | OPCs | *CHRNE* | TRUE | 2.024351039 |
| Anxiety | OPCs | *GNB3* | TRUE | 1.999178755 |
| Anxiety | OPCs | *NME5* | TRUE | 2.393184107 |
| Anxiety | OPCs | *ZSCAN9* | TRUE | 2.586230891 |
| Anxiety | OPCs | *ADCK3* | TRUE | 2.02099209 |
| Anxiety | OPCs | *ZNF503* | TRUE | 2.375489311 |
| Anxiety | OPCs | *HYAL3* | TRUE | 2.181230302 |
| Anxiety | OPCs | *ZFP69* | TRUE | 2.137494463 |
| Anxiety | OPCs | *ZKSCAN3* | TRUE | 2.770429641 |
| Anxiety | OPCs | *TPM2* | TRUE | 2.221973159 |
| Anxiety | OPCs | *RP11.584P21.2* | TRUE | 2.004428804 |
| Anxiety | OPCs | *RP13.977J11.2* | TRUE | 2.862100691 |
| Anxiety | OPCs | *RP11.63A1.2* | TRUE | 3.066347264 |
| Bipolar | Astros | *PLA2G4C* | TRUE | 2.074189322 |
| Bipolar | Astros | *ZNF623* | TRUE | 2.275630145 |
| Bipolar | Astros | *NUTM2B* | TRUE | 2.424265837 |
| Bipolar | Astros | *AC093159.1* | TRUE | 2.094660726 |
| Bipolar | Astros | *LILRA4* | TRUE | 2.573358251 |
| Bipolar | Astros | *HLA.DOB* | TRUE | 2.442964642 |
| Bipolar | Astros | *CTD.3074O7.2* | TRUE | 2.126353658 |
| Bipolar | Ex | *SCYL3* | TRUE | 2.092474046 |
| Bipolar | Ex | *LIG3* | TRUE | 1.990917801 |
| Bipolar | Ex | *LRRC23* | TRUE | 2.158737466 |
| Bipolar | Ex | *DRD4* | TRUE | 2.311704076 |
| Bipolar | Ex | *IFT74* | TRUE | 2.962816383 |
| Bipolar | Ex | *PCK2* | TRUE | 2.076675051 |
| Bipolar | Ex | *POLI* | TRUE | 2.276661012 |
| Bipolar | Ex | *PLA2G4C* | TRUE | 2.731974226 |
| Bipolar | Ex | *HAS1* | TRUE | 2.136882594 |
| Bipolar | Ex | *TSPAN11* | TRUE | 2.430850887 |
| Bipolar | Ex | *AKAP3* | TRUE | 2.154087921 |
| Bipolar | Ex | *DDX55* | TRUE | 2.053275977 |
| Bipolar | Ex | *TFG* | TRUE | 2.328855245 |
| Bipolar | Ex | *ABCC5* | TRUE | 3.004309798 |
| Bipolar | Ex | *SMYD1* | TRUE | 2.045775065 |
| Bipolar | Ex | *DNAJC16* | TRUE | 1.999946415 |
| Bipolar | Ex | *MORN1* | TRUE | 2.066804557 |
| Bipolar | Ex | *DNAL1* | TRUE | 3.12423148 |
| Bipolar | Ex | *ZNF549* | TRUE | 2.172592997 |
| Bipolar | Ex | *SARDH* | TRUE | 2.265055871 |
| Bipolar | Ex | *PEX6* | TRUE | 2.5144419 |
| Bipolar | Ex | *POT1* | TRUE | 2.081731923 |
| Bipolar | Ex | *NINJ1* | TRUE | 2.495498481 |
| Bipolar | Ex | *ZNF304* | TRUE | 1.984130533 |
| Bipolar | Ex | *RAF1* | TRUE | 2.261890501 |

| | | | | |
|---|---|---|---|---|
| Bipolar | Ex | *CCDC64* | TRUE | 2.872320568 |
| Bipolar | Ex | *DSCC1* | TRUE | 2.173942761 |
| Bipolar | Ex | *TMEM8B* | TRUE | 2.136791671 |
| Bipolar | Ex | *BPHL* | TRUE | 1.976151874 |
| Bipolar | Ex | *POLG* | TRUE | 2.53023666 |
| Bipolar | Ex | *DUS3L* | TRUE | 2.237614276 |
| Bipolar | Ex | *C7orf50* | TRUE | 2.603757595 |
| Bipolar | Ex | *PSPH* | TRUE | 2.11677712 |
| Bipolar | Ex | *REXO4* | TRUE | 1.961563952 |
| Bipolar | Ex | *TTC12* | TRUE | 1.989797782 |
| Bipolar | Ex | *FADS1* | TRUE | 2.198882289 |
| Bipolar | Ex | *ZNF208* | TRUE | 2.614542492 |
| Bipolar | Ex | *ZNF75A* | TRUE | 1.975926066 |
| Bipolar | Ex | *PPP1R21* | TRUE | 2.532621447 |
| Bipolar | Ex | *EOGT* | TRUE | 2.057084903 |
| Bipolar | Ex | *ADORA1* | TRUE | 1.990546757 |
| Bipolar | Ex | *YWHAZ* | TRUE | 3.190620074 |
| Bipolar | Ex | *CCDC171* | TRUE | 2.248805216 |
| Bipolar | Ex | *TRPV6* | TRUE | 2.149790289 |
| Bipolar | Ex | *HIF1AN* | TRUE | 2.205743589 |
| Bipolar | Ex | *THAP5* | TRUE | 2.00695914 |
| Bipolar | Ex | *ZNF623* | TRUE | 3.518583817 |
| Bipolar | Ex | *COA5* | TRUE | 2.102002585 |
| Bipolar | Ex | *ACSM5* | TRUE | 2.378089663 |
| Bipolar | Ex | *CIB1* | TRUE | 2.400579383 |
| Bipolar | Ex | *CDK10* | TRUE | 2.031885555 |
| Bipolar | Ex | *ZKSCAN4* | TRUE | 2.23645745 |
| Bipolar | Ex | *NWD1* | TRUE | 2.63255267 |
| Bipolar | Ex | *NUTM2B* | TRUE | 2.428808051 |
| Bipolar | Ex | *TCEA3* | TRUE | 2.090238757 |
| Bipolar | Ex | *AC093159.1* | TRUE | 2.222305901 |
| Bipolar | Ex | *LRRC37A2* | TRUE | 2.121248063 |
| Bipolar | Ex | *LILRA4* | TRUE | 2.259933064 |
| Bipolar | Ex | *HLA.DOB* | TRUE | 2.543215338 |
| Bipolar | Ex | *RP11.332J15.2* | TRUE | 2.258129293 |
| Bipolar | Ex | *HPR* | TRUE | 2.209439259 |
| Bipolar | Ex | *CTD.2619J13.19* | TRUE | 2.023725571 |
| Bipolar | Ex | *RP11.438L19.1* | TRUE | 2.104467107 |
| Bipolar | Inhib | *TSSC1* | TRUE | 2.941220771 |
| Bipolar | Inhib | *PTGER3* | TRUE | 2.279605657 |
| Bipolar | Inhib | *DRD4* | TRUE | 2.084332915 |
| Bipolar | Inhib | *IFT74* | TRUE | 2.920586315 |
| Bipolar | Inhib | *AKAP3* | TRUE | 2.290416234 |
| Bipolar | Inhib | *DNAJC16* | TRUE | 2.02124449 |
| Bipolar | Inhib | *MORN1* | TRUE | 2.414579038 |
| Bipolar | Inhib | *DNAL1* | TRUE | 2.711042388 |
| Bipolar | Inhib | *ZNF549* | TRUE | 2.427565184 |
| Bipolar | Inhib | *SARDH* | TRUE | 2.461551855 |
| Bipolar | Inhib | *PEX6* | TRUE | 1.97628592 |
| Bipolar | Inhib | *NINJ1* | TRUE | 3.074285764 |

| | | | | |
|---|---|---|---|---|
| Bipolar | Inhib | *ZNF304* | TRUE | 2.321009722 |
| Bipolar | Inhib | *RAF1* | TRUE | 2.641051712 |
| Bipolar | Inhib | *CCDC64* | TRUE | 2.335587481 |
| Bipolar | Inhib | *DSCC1* | TRUE | 2.211405678 |
| Bipolar | Inhib | *TMEM8B* | TRUE | 2.385511842 |
| Bipolar | Inhib | *POLG* | TRUE | 2.71921971 |
| Bipolar | Inhib | *TRIM65* | TRUE | 2.265764297 |
| Bipolar | Inhib | *DUS3L* | TRUE | 2.025266749 |
| Bipolar | Inhib | *PSPH* | TRUE | 2.047234053 |
| Bipolar | Inhib | *FADS1* | TRUE | 2.783974636 |
| Bipolar | Inhib | *MRPL39* | TRUE | 2.711065139 |
| Bipolar | Inhib | *TRMT44* | TRUE | 2.022563786 |
| Bipolar | Inhib | *JMJD8* | TRUE | 2.071809501 |
| Bipolar | Inhib | *TRPV6* | TRUE | 2.277924719 |
| Bipolar | Inhib | *AFAP1L2* | TRUE | 2.050671673 |
| Bipolar | Inhib | *METTL18* | TRUE | 2.110021399 |
| Bipolar | Inhib | *DPY19L1* | TRUE | 2.159401313 |
| Bipolar | Inhib | *THAP5* | TRUE | 2.201051894 |
| Bipolar | Inhib | *STAP2* | TRUE | 2.060319707 |
| Bipolar | Inhib | *ZNF623* | TRUE | 3.077543457 |
| Bipolar | Inhib | *CIB1* | TRUE | 2.359015318 |
| Bipolar | Inhib | *NWD1* | TRUE | 2.330950103 |
| Bipolar | Inhib | *NUTM2B* | TRUE | 2.48614767 |
| Bipolar | Inhib | *ASAH2* | TRUE | 1.991612002 |
| Bipolar | Inhib | *AC093159.1* | TRUE | 2.223648027 |
| Bipolar | Inhib | *LILRA4* | TRUE | 2.710552242 |
| Bipolar | Inhib | *HLA.DOB* | TRUE | 2.464249135 |
| Bipolar | Inhib | *ZNF585B* | TRUE | 2.265875107 |
| Bipolar | Inhib | *HPR* | TRUE | 2.787415371 |
| Bipolar | Inhib | *RP11.438L19.1* | TRUE | 2.280396265 |
| Bipolar | Micro_Macro | *IFT74* | TRUE | 2.277108701 |
| Bipolar | Micro_Macro | *DSCC1* | TRUE | 2.006200992 |
| Bipolar | Micro_Macro | *NUTM2B* | TRUE | 2.348890926 |
| Bipolar | Micro_Macro | *HLA.DOB* | TRUE | 2.143565398 |
| Bipolar | Micro_Macro | *HPR* | TRUE | 1.96468158 |
| Bipolar | Oligos | *IFT74* | TRUE | 2.779262803 |
| Bipolar | Oligos | *CDCA3* | TRUE | 2.14437385 |
| Bipolar | Oligos | *NINJ1* | TRUE | 2.253277335 |
| Bipolar | Oligos | *RAF1* | TRUE | 2.081297355 |
| Bipolar | Oligos | *DSCC1* | TRUE | 2.191501797 |
| Bipolar | Oligos | *POLG* | TRUE | 2.274467895 |
| Bipolar | Oligos | *ZNF623* | TRUE | 2.446017346 |
| Bipolar | Oligos | *CIB1* | TRUE | 2.008476793 |
| Bipolar | Oligos | *NWD1* | TRUE | 1.975034086 |
| Bipolar | Oligos | *NUTM2B* | TRUE | 2.398737599 |
| Bipolar | Oligos | *AC093159.1* | TRUE | 2.007460151 |
| Bipolar | Oligos | *HLA.DOB* | TRUE | 2.593403872 |
| Bipolar | Oligos | *HPR* | TRUE | 2.583861769 |
| Bipolar | OPCs | *IFT74* | TRUE | 2.665099153 |
| Bipolar | OPCs | *PLA2G4C* | TRUE | 2.335260797 |

| | | | | |
|---|---|---|---|---|
| Bipolar | OPCs | *NINJ1* | TRUE | 2.094319254 |
| Bipolar | OPCs | *POLG* | TRUE | 2.092644501 |
| Bipolar | OPCs | *ZNF623* | TRUE | 2.244416968 |
| Bipolar | OPCs | *NUTM2B* | TRUE | 2.445958882 |
| Bipolar | OPCs | *AC093159.1* | TRUE | 1.999833034 |
| Bipolar | OPCs | *HLA.DOB* | TRUE | 2.523254839 |
| Bipolar | OPCs | *CTD.3074O7.2* | TRUE | 2.133668144 |
| Bipolar | OPCs | *HPR* | TRUE | 2.193457768 |
| Depression | Astros | *PPP2R3C* | TRUE | 1.972889809 |
| Depression | Astros | *CLEC4M* | TRUE | 2.453037479 |
| Depression | Astros | *ENPP5* | TRUE | 2.111052117 |
| Depression | Astros | *ZSCAN9* | TRUE | 3.139634959 |
| Depression | Astros | *NBEAL2* | TRUE | 1.971706998 |
| Depression | Astros | *RP9* | TRUE | 1.980994358 |
| Depression | Astros | *ZSCAN21* | TRUE | 2.30180792 |
| Depression | Astros | *EHBP1L1* | TRUE | 2.21087923 |
| Depression | Astros | *FAHD1* | TRUE | 2.042151719 |
| Depression | Astros | *YBEY* | TRUE | 1.993333074 |
| Depression | Astros | *RP3.470B24.5* | TRUE | 2.153885738 |
| Depression | Astros | *CTD.3074O7.2* | TRUE | 2.884027491 |
| Depression | Astros | *CTD.3126B10.4* | TRUE | 2.814599909 |
| Depression | Ex | *ITGA2B* | TRUE | 2.121082813 |
| Depression | Ex | *MED24* | TRUE | 3.129876769 |
| Depression | Ex | *DNAH5* | TRUE | 2.237105889 |
| Depression | Ex | *CDH3* | TRUE | 2.096221084 |
| Depression | Ex | *NFYC* | TRUE | 2.108751089 |
| Depression | Ex | *NUCKS1* | TRUE | 2.836011253 |
| Depression | Ex | *SLC4A11* | TRUE | 2.312291446 |
| Depression | Ex | *MYO15A* | TRUE | 2.463660253 |
| Depression | Ex | *PPP2R3C* | TRUE | 3.003122933 |
| Depression | Ex | *POLI* | TRUE | 2.563500983 |
| Depression | Ex | *KLC3* | TRUE | 2.117875385 |
| Depression | Ex | *CLEC4M* | TRUE | 2.4339663 |
| Depression | Ex | *MTPAP* | TRUE | 2.132898601 |
| Depression | Ex | *ENPP5* | TRUE | 2.769576503 |
| Depression | Ex | *C1orf109* | TRUE | 2.118036974 |
| Depression | Ex | *AVPI1* | TRUE | 2.7312129 |
| Depression | Ex | *PRB2* | TRUE | 2.105208127 |
| Depression | Ex | *MAP1B* | TRUE | 2.301770167 |
| Depression | Ex | *NIP7* | TRUE | 2.890800453 |
| Depression | Ex | *GSTT2B* | TRUE | 2.07877267 |
| Depression | Ex | *SMYD5* | TRUE | 2.68880157 |
| Depression | Ex | *ZSCAN9* | TRUE | 3.365628192 |
| Depression | Ex | *PPA2* | TRUE | 2.421193787 |
| Depression | Ex | *COL6A2* | TRUE | 2.062932022 |
| Depression | Ex | *GIGYF1* | TRUE | 1.970765663 |
| Depression | Ex | *MTG1* | TRUE | 2.022974219 |
| Depression | Ex | *TRPT1* | TRUE | 2.380215459 |
| Depression | Ex | *GUF1* | TRUE | 2.014131365 |
| Depression | Ex | *HEATR3* | TRUE | 2.633793217 |

| | | | | |
|---|---|---|---|---|
| Depression | Ex | *SCUBE1* | TRUE | 2.025980673 |
| Depression | Ex | *SPATC1L* | TRUE | 1.993138303 |
| Depression | Ex | *CCDC64B* | TRUE | 2.717961478 |
| Depression | Ex | *PPP1R21* | TRUE | 2.078350648 |
| Depression | Ex | *ANXA5* | TRUE | 2.407850547 |
| Depression | Ex | *RP9* | TRUE | 3.366126397 |
| Depression | Ex | *SLC18A2* | TRUE | 2.063801972 |
| Depression | Ex | *ZSCAN21* | TRUE | 2.494756757 |
| Depression | Ex | *SAMD14* | TRUE | 2.570833349 |
| Depression | Ex | *FAM21C* | TRUE | 1.967014753 |
| Depression | Ex | *ZNF738* | TRUE | 2.310031767 |
| Depression | Ex | *SLC2A14* | TRUE | 2.233628098 |
| Depression | Ex | *EHBP1L1* | TRUE | 2.53245312 |
| Depression | Ex | *RNF26* | TRUE | 2.140391866 |
| Depression | Ex | *PAAF1* | TRUE | 2.591980204 |
| Depression | Ex | *ZNF619* | TRUE | 2.171166103 |
| Depression | Ex | *PFAS* | TRUE | 2.079263469 |
| Depression | Ex | *PCED1B* | TRUE | 2.427204548 |
| Depression | Ex | *FAHD1* | TRUE | 2.274934164 |
| Depression | Ex | *GLIPR1L2* | TRUE | 2.451457784 |
| Depression | Ex | *FAM89A* | TRUE | 2.60432917 |
| Depression | Ex | *ALG12* | TRUE | 2.446452991 |
| Depression | Ex | *IRS2* | TRUE | 2.517275433 |
| Depression | Ex | *ZNF749* | TRUE | 2.350372563 |
| Depression | Ex | *NUDT17* | TRUE | 1.992718764 |
| Depression | Ex | *GPR179* | TRUE | 2.821801606 |
| Depression | Ex | *FAM180B* | TRUE | 2.667303864 |
| Depression | Ex | *RPL12* | TRUE | 2.304615627 |
| Depression | Ex | *HLA.C* | TRUE | 2.246727887 |
| Depression | Ex | *MALRD1* | TRUE | 2.158272437 |
| Depression | Ex | *LINC00487* | TRUE | 2.168261062 |
| Depression | Ex | *C19orf73* | TRUE | 2.261547652 |
| Depression | Ex | *RP11.197K6.1* | TRUE | 2.466033816 |
| Depression | Ex | *CTA.150C2.13* | TRUE | 2.115531547 |
| Depression | Ex | *KIAA0040* | TRUE | 2.704880449 |
| Depression | Ex | *RP3.470B24.5* | TRUE | 2.508093999 |
| Depression | Ex | *RP1.156L9.1* | TRUE | 2.124834762 |
| Depression | Ex | *LILRA4* | TRUE | 2.440473888 |
| Depression | Ex | *PPIL3* | TRUE | 2.401306509 |
| Depression | Ex | *CTD.3074O7.2* | TRUE | 2.044744105 |
| Depression | Ex | *CTC.498J12.1* | TRUE | 2.296115071 |
| Depression | Ex | *AC133633.2* | TRUE | 2.033341737 |
| Depression | Ex | *RP11.665C16.6* | TRUE | 2.249779506 |
| Depression | Ex | *RP5.1021I20.2* | TRUE | 2.129262194 |
| Depression | Ex | *SMIM6* | TRUE | 2.009539945 |
| Depression | Ex | *CTD.3126B10.4* | TRUE | 2.700701481 |
| Depression | Ex | *CTC.548K16.1* | TRUE | 1.99384064 |
| Depression | Ex | *RP11.678G14.3* | TRUE | 2.531851272 |
| Depression | Ex | *RP11.54O7.17* | TRUE | 3.4616737 |
| Depression | Inhib | *MED24* | TRUE | 2.518394768 |

| | | | | |
|---|---|---|---|---|
| Depression | Inhib | *DNAH5* | TRUE | 2.16576143 |
| Depression | Inhib | *CDH3* | TRUE | 2.257666273 |
| Depression | Inhib | *NFYC* | TRUE | 2.302283615 |
| Depression | Inhib | *NUCKS1* | TRUE | 2.99580921 |
| Depression | Inhib | *NAALAD2* | TRUE | 1.97358534 |
| Depression | Inhib | *SLC4A11* | TRUE | 2.161798269 |
| Depression | Inhib | *MYO15A* | TRUE | 2.341398062 |
| Depression | Inhib | *PPP2R3C* | TRUE | 2.626876841 |
| Depression | Inhib | *OSGEP* | TRUE | 1.985911107 |
| Depression | Inhib | *POLI* | TRUE | 2.635507079 |
| Depression | Inhib | *TAF1C* | TRUE | 2.005967979 |
| Depression | Inhib | *CLEC4M* | TRUE | 2.393077354 |
| Depression | Inhib | *SYT10* | TRUE | 1.994279489 |
| Depression | Inhib | *ENPP5* | TRUE | 2.883216352 |
| Depression | Inhib | *ZNHIT6* | TRUE | 2.025594177 |
| Depression | Inhib | *HEATR1* | TRUE | 2.203339771 |
| Depression | Inhib | *AVPI1* | TRUE | 2.200443405 |
| Depression | Inhib | *AKAP1* | TRUE | 2.063966218 |
| Depression | Inhib | *PRB2* | TRUE | 2.167263006 |
| Depression | Inhib | *RIBC2* | TRUE | 1.99492613 |
| Depression | Inhib | *MAP1B* | TRUE | 2.175927783 |
| Depression | Inhib | *DLG4* | TRUE | 1.993902497 |
| Depression | Inhib | *NIP7* | TRUE | 3.059982239 |
| Depression | Inhib | *SMYD5* | TRUE | 2.869568925 |
| Depression | Inhib | *ZSCAN9* | TRUE | 3.781321087 |
| Depression | Inhib | *PPA2* | TRUE | 2.439878451 |
| Depression | Inhib | *NARF* | TRUE | 2.039984879 |
| Depression | Inhib | *COL6A2* | TRUE | 2.093757845 |
| Depression | Inhib | *MTG1* | TRUE | 2.026431731 |
| Depression | Inhib | *HEATR3* | TRUE | 2.487550656 |
| Depression | Inhib | *SPATC1L* | TRUE | 2.014116609 |
| Depression | Inhib | *NBEAL2* | TRUE | 2.074353284 |
| Depression | Inhib | *CCDC64B* | TRUE | 2.900846865 |
| Depression | Inhib | *PPP1R21* | TRUE | 2.044066663 |
| Depression | Inhib | *ANXA5* | TRUE | 2.133370675 |
| Depression | Inhib | *RP9* | TRUE | 3.585573274 |
| Depression | Inhib | *WBSCR27* | TRUE | 2.138725419 |
| Depression | Inhib | *ZSCAN21* | TRUE | 2.200543927 |
| Depression | Inhib | *SAMD14* | TRUE | 2.339285377 |
| Depression | Inhib | *ZNF738* | TRUE | 2.181269645 |
| Depression | Inhib | *EHBP1L1* | TRUE | 2.437220723 |
| Depression | Inhib | *PAAF1* | TRUE | 2.353194446 |
| Depression | Inhib | *ZNF619* | TRUE | 2.166362249 |
| Depression | Inhib | *PCED1B* | TRUE | 2.457975543 |
| Depression | Inhib | *FAHD1* | TRUE | 2.323599205 |
| Depression | Inhib | *GLIPR1L2* | TRUE | 2.513983099 |
| Depression | Inhib | *ALG12* | TRUE | 2.209730665 |
| Depression | Inhib | *IRS2* | TRUE | 2.532709244 |
| Depression | Inhib | *ZNF749* | TRUE | 2.387616579 |
| Depression | Inhib | *NUDT17* | TRUE | 2.054086168 |

| | | | | |
|---|---|---|---|---|
| Depression | Inhib | *GPR179* | TRUE | 3.027295224 |
| Depression | Inhib | *RPL12* | TRUE | 2.089251465 |
| Depression | Inhib | *HLA.C* | TRUE | 2.499773416 |
| Depression | Inhib | *LINC00487* | TRUE | 2.073132489 |
| Depression | Inhib | *C19orf73* | TRUE | 2.289961705 |
| Depression | Inhib | *RP11.197K6.1* | TRUE | 2.434267788 |
| Depression | Inhib | *CTA.150C2.13* | TRUE | 2.068048402 |
| Depression | Inhib | *KIAA0040* | TRUE | 2.833257423 |
| Depression | Inhib | *RP3.470B24.5* | TRUE | 2.25748251 |
| Depression | Inhib | *RP1.156L9.1* | TRUE | 2.213425945 |
| Depression | Inhib | *LILRA4* | TRUE | 2.098940235 |
| Depression | Inhib | *PPIL3* | TRUE | 2.391453402 |
| Depression | Inhib | *RP11.826N14.2* | TRUE | 1.984165486 |
| Depression | Inhib | *CTC.498J12.1* | TRUE | 2.275481407 |
| Depression | Inhib | *AC133633.2* | TRUE | 1.974789464 |
| Depression | Inhib | *RP5.1021I20.2* | TRUE | 1.993938642 |
| Depression | Inhib | *SMIM6* | TRUE | 2.000619583 |
| Depression | Inhib | *CTD.3126B10.4* | TRUE | 2.832868389 |
| Depression | Inhib | *RP11.107F6.3* | TRUE | 1.96422538 |
| Depression | Inhib | *RP11.678G14.3* | TRUE | 2.578409635 |
| Depression | Inhib | *RP11.54O7.17* | TRUE | 3.174358534 |
| Depression | Micro_Macro | *GSTT2B* | TRUE | 1.975810774 |
| Depression | Micro_Macro | *ZSCAN9* | TRUE | 3.496382076 |
| Depression | Micro_Macro | *NBEAL2* | TRUE | 1.992032249 |
| Depression | Micro_Macro | *CCDC64B* | TRUE | 2.804148048 |
| Depression | Micro_Macro | *ZSCAN21* | TRUE | 2.302935546 |
| Depression | Micro_Macro | *YBEY* | TRUE | 1.975802536 |
| Depression | Micro_Macro | *BTN3A2* | TRUE | 2.280824811 |
| Depression | Micro_Macro | *CTD.3074O7.2* | TRUE | 2.724176897 |
| Depression | Micro_Macro | *CTD.3126B10.4* | TRUE | 2.453527115 |
| Depression | Oligos | *CLEC4M* | TRUE | 2.21730073 |
| Depression | Oligos | *ENPP5* | TRUE | 2.592662501 |
| Depression | Oligos | *NIP7* | TRUE | 2.723463775 |
| Depression | Oligos | *ZSCAN9* | TRUE | 3.660236702 |
| Depression | Oligos | *HEATR3* | TRUE | 2.277434742 |
| Depression | Oligos | *CCDC64B* | TRUE | 2.575787066 |
| Depression | Oligos | *RP9* | TRUE | 3.018742068 |
| Depression | Oligos | *ZSCAN21* | TRUE | 2.782536971 |
| Depression | Oligos | *ZNF738* | TRUE | 2.033151445 |
| Depression | Oligos | *EHBP1L1* | TRUE | 2.435126781 |
| Depression | Oligos | *PCED1B* | TRUE | 1.990756859 |
| Depression | Oligos | *FAHD1* | TRUE | 2.241884518 |
| Depression | Oligos | *ZNF749* | TRUE | 2.156167841 |
| Depression | Oligos | *HLA.C* | TRUE | 2.358467499 |
| Depression | Oligos | *C19orf73* | TRUE | 1.971232342 |
| Depression | Oligos | *CTA.150C2.13* | TRUE | 1.986288775 |
| Depression | Oligos | *KIAA0040* | TRUE | 2.458168324 |
| Depression | Oligos | *RP1.156L9.1* | TRUE | 2.025964453 |
| Depression | Oligos | *PPIL3* | TRUE | 2.048416462 |
| Depression | Oligos | *CTD.3126B10.4* | TRUE | 2.688639672 |

| | | | | |
|---|---|---|---|---|
| Depression | Oligos | *RP11.678G14.3* | TRUE | 2.179811068 |
| Depression | OPCs | *ENPP5* | TRUE | 2.424660161 |
| Depression | OPCs | *GSTT2B* | TRUE | 2.735171095 |
| Depression | OPCs | *ZSCAN9* | TRUE | 3.746482394 |
| Depression | OPCs | *COL6A2* | TRUE | 1.979425345 |
| Depression | OPCs | *SPATC1L* | TRUE | 2.412703051 |
| Depression | OPCs | *CCDC64B* | TRUE | 1.973296649 |
| Depression | OPCs | *RP9* | TRUE | 2.869410294 |
| Depression | OPCs | *ZSCAN21* | TRUE | 2.175805195 |
| Depression | OPCs | *ZNF738* | TRUE | 2.246745443 |
| Depression | OPCs | *EHBP1L1* | TRUE | 1.972020626 |
| Depression | OPCs | *PAAF1* | TRUE | 2.153388342 |
| Depression | OPCs | *FAHD1* | TRUE | 2.29378294 |
| Depression | OPCs | *ZNF749* | TRUE | 2.088208668 |
| Depression | OPCs | *FAM180B* | TRUE | 2.421544208 |
| Depression | OPCs | *CTD.3074O7.2* | TRUE | 2.285637397 |
| Depression | OPCs | *CTD.3126B10.4* | TRUE | 2.859233629 |
| Psychosis | Astros | *GP6* | TRUE | 2.223118573 |
| Psychosis | Astros | *SSH3* | TRUE | 2.408339967 |
| Psychosis | Astros | *CCDC163P* | TRUE | 2.042823605 |
| Psychosis | Astros | *RP11.430H10.1* | TRUE | 3.014324467 |
| Psychosis | Astros | *CTD.3126B10.4* | TRUE | 2.110603529 |
| Psychosis | Astros | *RP13.539J13.1* | TRUE | 1.987879669 |
| Psychosis | Ex | *LIG3* | TRUE | 2.182419421 |
| Psychosis | Ex | *RPS20* | TRUE | 2.385336225 |
| Psychosis | Ex | *ALOX5* | TRUE | 2.756109838 |
| Psychosis | Ex | *RNASET2* | TRUE | 2.078898276 |
| Psychosis | Ex | *ERCC8* | TRUE | 2.472180377 |
| Psychosis | Ex | *GP6* | TRUE | 2.898575377 |
| Psychosis | Ex | *ACR* | TRUE | 2.411333571 |
| Psychosis | Ex | *CLUAP1* | TRUE | 2.364753561 |
| Psychosis | Ex | *TMEM176B* | TRUE | 2.003921377 |
| Psychosis | Ex | *SYT10* | TRUE | 2.585684327 |
| Psychosis | Ex | *BRD8* | TRUE | 1.99225968 |
| Psychosis | Ex | *NID1* | TRUE | 2.362772513 |
| Psychosis | Ex | *DIEXF* | TRUE | 2.254247387 |
| Psychosis | Ex | *PCDHB10* | TRUE | 3.4526547 |
| Psychosis | Ex | *SLC2A8* | TRUE | 2.532524931 |
| Psychosis | Ex | *FRAS1* | TRUE | 1.967518195 |
| Psychosis | Ex | *DUS3L* | TRUE | 2.517329954 |
| Psychosis | Ex | *RSPH10B* | TRUE | 2.070715619 |
| Psychosis | Ex | *SETD9* | TRUE | 2.123908336 |
| Psychosis | Ex | *DRAM2* | TRUE | 2.375114694 |
| Psychosis | Ex | *DRC1* | TRUE | 2.219675108 |
| Psychosis | Ex | *HIST1H4H* | TRUE | 2.268725046 |
| Psychosis | Ex | *C1orf123* | TRUE | 2.38413498 |
| Psychosis | Ex | *RNF149* | TRUE | 2.433279422 |
| Psychosis | Ex | *DUS2* | TRUE | 2.924717314 |
| Psychosis | Ex | *KLK5* | TRUE | 2.07294361 |
| Psychosis | Ex | *ANO5* | TRUE | 2.713812361 |

| | | | | |
|---|---|---|---|---|
| Psychosis | Ex | *SSH3* | TRUE | 2.784028773 |
| Psychosis | Ex | *SLCO4C1* | TRUE | 2.040206336 |
| Psychosis | Ex | *ZNF619* | TRUE | 2.251489579 |
| Psychosis | Ex | *CCDC125* | TRUE | 2.035680543 |
| Psychosis | Ex | *ACSM5* | TRUE | 1.966663401 |
| Psychosis | Ex | *GOLGA6L4* | TRUE | 1.987856262 |
| Psychosis | Ex | *ACOT1* | TRUE | 2.262876504 |
| Psychosis | Ex | *KIF24* | TRUE | 2.293780733 |
| Psychosis | Ex | *RDM1* | TRUE | 2.648970171 |
| Psychosis | Ex | *KBTBD12* | TRUE | 2.81007284 |
| Psychosis | Ex | *S100A6* | TRUE | 2.333317509 |
| Psychosis | Ex | *SH3BGRL2* | TRUE | 1.979650071 |
| Psychosis | Ex | *SH3D21* | TRUE | 2.243880448 |
| Psychosis | Ex | *RP1.60O19.2* | TRUE | 2.342959932 |
| Psychosis | Ex | *LINC00443* | TRUE | 2.14131147 |
| Psychosis | Ex | *CCDC163P* | TRUE | 2.473170475 |
| Psychosis | Ex | *AC079922.3* | TRUE | 2.165520799 |
| Psychosis | Ex | *RP11.430H10.1* | TRUE | 3.540475542 |
| Psychosis | Ex | *RP11.377D9.3* | TRUE | 2.397437194 |
| Psychosis | Ex | *CHURC1* | TRUE | 3.04607523 |
| Psychosis | Ex | *RP11.410E4.1* | TRUE | 2.663023994 |
| Psychosis | Ex | *CTD.3126B10.4* | TRUE | 2.233774257 |
| Psychosis | Ex | *GOLGA8H* | TRUE | 2.194008599 |
| Psychosis | Ex | *CTD.2623N2.11* | TRUE | 2.206742012 |
| Psychosis | Ex | *RP13.539J13.1* | TRUE | 2.144646102 |
| Psychosis | Ex | *AP000350.5* | TRUE | 2.033804716 |
| Psychosis | Inhib | *RPS20* | TRUE | 2.8141125 |
| Psychosis | Inhib | *LARS2* | TRUE | 2.245562637 |
| Psychosis | Inhib | *ALOX5* | TRUE | 2.32677981 |
| Psychosis | Inhib | *RNASET2* | TRUE | 1.976659446 |
| Psychosis | Inhib | *ERCC8* | TRUE | 2.684710229 |
| Psychosis | Inhib | *GP6* | TRUE | 3.068156829 |
| Psychosis | Inhib | *FMO2* | TRUE | 1.965456991 |
| Psychosis | Inhib | *ACR* | TRUE | 2.372223786 |
| Psychosis | Inhib | *CLUAP1* | TRUE | 2.502770386 |
| Psychosis | Inhib | *RIPK2* | TRUE | 2.34239322 |
| Psychosis | Inhib | *TMEM176B* | TRUE | 1.996988758 |
| Psychosis | Inhib | *SYT10* | TRUE | 2.443002942 |
| Psychosis | Inhib | *BRD8* | TRUE | 2.14037127 |
| Psychosis | Inhib | *PCDHB10* | TRUE | 3.181965756 |
| Psychosis | Inhib | *SLC2A8* | TRUE | 2.751757186 |
| Psychosis | Inhib | *CAPN11* | TRUE | 2.194768184 |
| Psychosis | Inhib | *DIAPH3* | TRUE | 2.139495776 |
| Psychosis | Inhib | *GAS8* | TRUE | 2.19980757 |
| Psychosis | Inhib | *DUS3L* | TRUE | 2.451288134 |
| Psychosis | Inhib | *SETD9* | TRUE | 2.249618434 |
| Psychosis | Inhib | *MIER3* | TRUE | 2.258922943 |
| Psychosis | Inhib | *RMND1* | TRUE | 2.0535126 |
| Psychosis | Inhib | *DRC1* | TRUE | 2.141558804 |
| Psychosis | Inhib | *RNF149* | TRUE | 2.747266135 |

| | | | | |
|---|---|---|---|---|
| Psychosis | Inhib | *ELP6* | TRUE | 2.034375669 |
| Psychosis | Inhib | *SCN3B* | TRUE | 2.306581122 |
| Psychosis | Inhib | *AKIP1* | TRUE | 2.257995742 |
| Psychosis | Inhib | *SLC33A1* | TRUE | 2.096305929 |
| Psychosis | Inhib | *FRMPD2* | TRUE | 2.012271688 |
| Psychosis | Inhib | *ANO5* | TRUE | 2.403864302 |
| Psychosis | Inhib | *FIBP* | TRUE | 2.186252779 |
| Psychosis | Inhib | *TMEM134* | TRUE | 2.293507985 |
| Psychosis | Inhib | *SSH3* | TRUE | 2.936458332 |
| Psychosis | Inhib | *SLCO4C1* | TRUE | 2.346077618 |
| Psychosis | Inhib | *TOM1L2* | TRUE | 2.105699622 |
| Psychosis | Inhib | *ZNF619* | TRUE | 2.321181244 |
| Psychosis | Inhib | *PAK2* | TRUE | 2.231172114 |
| Psychosis | Inhib | *FAM87A* | TRUE | 2.355812883 |
| Psychosis | Inhib | *TTC3* | TRUE | 2.239857459 |
| Psychosis | Inhib | *ACOT1* | TRUE | 2.188719054 |
| Psychosis | Inhib | *TOR3A* | TRUE | 2.305876468 |
| Psychosis | Inhib | *RDM1* | TRUE | 2.550814089 |
| Psychosis | Inhib | *KBTBD12* | TRUE | 2.954629593 |
| Psychosis | Inhib | *P2RX2* | TRUE | 2.028218913 |
| Psychosis | Inhib | *S100A6* | TRUE | 2.79115632 |
| Psychosis | Inhib | *C6orf47* | TRUE | 2.258290322 |
| Psychosis | Inhib | *SH3D21* | TRUE | 2.336989 |
| Psychosis | Inhib | *C19orf73* | TRUE | 1.99253968 |
| Psychosis | Inhib | *RP1.60O19.2* | TRUE | 2.477611995 |
| Psychosis | Inhib | *ANKRD65* | TRUE | 2.203088275 |
| Psychosis | Inhib | *CCDC163P* | TRUE | 2.074156907 |
| Psychosis | Inhib | *AC079922.3* | TRUE | 2.284076801 |
| Psychosis | Inhib | *APOBEC3G* | TRUE | 2.103310088 |
| Psychosis | Inhib | *RP11.430H10.1* | TRUE | 3.320403003 |
| Psychosis | Inhib | *RP11.377D9.3* | TRUE | 2.658971013 |
| Psychosis | Inhib | *CHURC1* | TRUE | 2.870699474 |
| Psychosis | Inhib | *RP11.410E4.1* | TRUE | 2.551245343 |
| Psychosis | Inhib | *CTD.3126B10.4* | TRUE | 2.23181891 |
| Psychosis | Inhib | *GOLGA8H* | TRUE | 2.136492641 |
| Psychosis | Inhib | *CTD.2623N2.11* | TRUE | 2.037797311 |
| Psychosis | Inhib | *XXbac.BPGBPG55C20.3* | TRUE | 2.198201773 |
| Psychosis | Inhib | *RP13.539J13.1* | TRUE | 1.962545887 |
| Psychosis | Inhib | *AP000350.5* | TRUE | 2.116199506 |
| Psychosis | Micro_Macro | *RDM1* | TRUE | 2.337963705 |
| Psychosis | Micro_Macro | *CCDC163P* | TRUE | 2.751237169 |
| Psychosis | Micro_Macro | *AC079922.3* | TRUE | 2.016716411 |
| Psychosis | Micro_Macro | *CTD.3074O7.2* | TRUE | 2.146294343 |
| Psychosis | Micro_Macro | *RP11.410E4.1* | TRUE | 2.394342928 |
| Psychosis | Micro_Macro | *RP13.539J13.1* | TRUE | 2.329079819 |
| Psychosis | Oligos | *ALOX5* | TRUE | 2.051135007 |
| Psychosis | Oligos | *ERCC8* | TRUE | 2.193703149 |
| Psychosis | Oligos | *ACR* | TRUE | 2.09469818 |
| Psychosis | Oligos | *PCDHB10* | TRUE | 2.819810567 |
| Psychosis | Oligos | *SLC2A8* | TRUE | 2.129319824 |

| | | | | |
|---|---|---|---|---|
| Psychosis | Oligos | *DUS3L* | TRUE | 2.305893857 |
| Psychosis | Oligos | *ANO5* | TRUE | 2.172426136 |
| Psychosis | Oligos | *SSH3* | TRUE | 2.614607168 |
| Psychosis | Oligos | *ACOT1* | TRUE | 2.065262327 |
| Psychosis | Oligos | *RDM1* | TRUE | 2.117119849 |
| Psychosis | Oligos | *KBTBD12* | TRUE | 2.012339301 |
| Psychosis | Oligos | *C6orf47* | TRUE | 2.062275154 |
| Psychosis | Oligos | *RFPL3S* | TRUE | 2.286518101 |
| Psychosis | Oligos | *ARL2* | TRUE | 2.021997123 |
| Psychosis | Oligos | *CCDC163P* | TRUE | 2.292788821 |
| Psychosis | Oligos | *AC079922.3* | TRUE | 1.979485824 |
| Psychosis | Oligos | *RP11.430H10.1* | TRUE | 3.13052125 |
| Psychosis | Oligos | *CHURC1* | TRUE | 2.812558452 |
| Psychosis | Oligos | *RP11.410E4.1* | TRUE | 2.515768088 |
| Psychosis | Oligos | *CTD.3126B10.4* | TRUE | 2.440320157 |
| Psychosis | Oligos | *RP13.539J13.1* | TRUE | 2.301337952 |
| Psychosis | OPCs | *ERCC8* | TRUE | 2.133233881 |
| Psychosis | OPCs | *SLC2A8* | TRUE | 2.3328283 |
| Psychosis | OPCs | *DUS3L* | TRUE | 2.358269793 |
| Psychosis | OPCs | *ANO5* | TRUE | 2.204222184 |
| Psychosis | OPCs | *CCDC163P* | TRUE | 2.269603691 |
| Psychosis | OPCs | *RP11.430H10.1* | TRUE | 2.66218067 |
| Psychosis | OPCs | *RP11.410E4.1* | TRUE | 2.592622509 |
| Psychosis | OPCs | *CTD.3126B10.4* | TRUE | 2.094242033 |
| Psychosis | OPCs | *RP13.539J13.1* | TRUE | 2.066785655 |
| Depression_admission_freq | Astros | *OBP2A* | TRUE | 2.048776242 |
| Depression_admission_freq | Astros | *AHI1* | TRUE | 2.226903742 |
| Depression_admission_freq | Astros | *EFHB* | TRUE | 2.415744431 |
| Depression_admission_freq | Astros | *RP11.826N14.2* | TRUE | 2.133455818 |
| Depression_admission_freq | Astros | *CTD.3126B10.4* | TRUE | 2.184480486 |
| Depression_admission_freq | Astros | *CTA.246H3.12* | TRUE | 2.206822357 |
| Depression_admission_freq | Ex | *STAG3* | TRUE | 2.174797955 |
| Depression_admission_freq | Ex | *NUCKS1* | TRUE | 2.299737767 |
| Depression_admission_freq | Ex | *FBXW11* | TRUE | 2.320496663 |
| Depression_admission_freq | Ex | *PVR* | TRUE | 2.27100136 |
| Depression_admission_freq | Ex | *ICAM3* | TRUE | 2.076031046 |
| Depression_admission_freq | Ex | *HBQ1* | TRUE | 1.980686971 |
| Depression_admission_freq | Ex | *SPAG7* | TRUE | 2.091440554 |
| Depression_admission_freq | Ex | *ARSB* | TRUE | 2.098487042 |
| Depression_admission_freq | Ex | *ACOT2* | TRUE | 2.132028041 |
| Depression_admission_freq | Ex | *PAX8* | TRUE | 2.389779957 |
| Depression_admission_freq | Ex | *TPH1* | TRUE | 2.108528853 |
| Depression_admission_freq | Ex | *KLK10* | TRUE | 1.962825338 |
| Depression_admission_freq | Ex | *QRSL1* | TRUE | 2.139609896 |
| Depression_admission_freq | Ex | *PSRC1* | TRUE | 3.115073088 |
| Depression_admission_freq | Ex | *DHX34* | TRUE | 2.400152581 |
| Depression_admission_freq | Ex | *AHI1* | TRUE | 2.649882913 |
| Depression_admission_freq | Ex | *CIB2* | TRUE | 2.136337491 |
| Depression_admission_freq | Ex | *TUBGCP4* | TRUE | 1.984683164 |
| Depression_admission_freq | Ex | *CNKSR1* | TRUE | 2.908231754 |

| | | | | |
|---|---|---|---|---|
| Depression_admission_freq | Ex | *FBXL17* | TRUE | 2.043538098 |
| Depression_admission_freq | Ex | *ASAP1* | TRUE | 2.109129748 |
| Depression_admission_freq | Ex | *NRG1* | TRUE | 2.133075717 |
| Depression_admission_freq | Ex | *EYA3* | TRUE | 2.074351702 |
| Depression_admission_freq | Ex | *POGLUT1* | TRUE | 2.224800125 |
| Depression_admission_freq | Ex | *EFHB* | TRUE | 2.442621059 |
| Depression_admission_freq | Ex | *FAM178B* | TRUE | 2.018877574 |
| Depression_admission_freq | Ex | *RNASE2* | TRUE | 2.165712034 |
| Depression_admission_freq | Ex | *PLEKHG5* | TRUE | 1.985388862 |
| Depression_admission_freq | Ex | *TMEM134* | TRUE | 2.512525676 |
| Depression_admission_freq | Ex | *RNF26* | TRUE | 2.076019294 |
| Depression_admission_freq | Ex | *GEN1* | TRUE | 2.406259928 |
| Depression_admission_freq | Ex | *RESP18* | TRUE | 2.121591714 |
| Depression_admission_freq | Ex | *ARHGEF37* | TRUE | 2.175244885 |
| Depression_admission_freq | Ex | *ZNF681* | TRUE | 2.093558734 |
| Depression_admission_freq | Ex | *TNFRSF25* | TRUE | 2.040536019 |
| Depression_admission_freq | Ex | *ZNF737* | TRUE | 2.341087619 |
| Depression_admission_freq | Ex | *HLA.DQA2* | TRUE | 2.051477932 |
| Depression_admission_freq | Ex | *RP11.826N14.2* | TRUE | 2.54530363 |
| Depression_admission_freq | Ex | *RP11.377D9.3* | TRUE | 2.159313924 |
| Depression_admission_freq | Ex | *CTD.3126B10.4* | TRUE | 2.098706008 |
| Depression_admission_freq | Ex | *RP13.539J13.1* | TRUE | 2.045256755 |
| Depression_admission_freq | Ex | *CTA.246H3.12* | TRUE | 2.358782181 |
| Depression_admission_freq | Inhib | *THOC3* | TRUE | 2.203874009 |
| Depression_admission_freq | Inhib | *PVR* | TRUE | 2.002037071 |
| Depression_admission_freq | Inhib | *ICAM3* | TRUE | 2.075543205 |
| Depression_admission_freq | Inhib | *SDR39U1* | TRUE | 1.975398036 |
| Depression_admission_freq | Inhib | *CCDC170* | TRUE | 2.135445961 |
| Depression_admission_freq | Inhib | *ATP5S* | TRUE | 1.965318486 |
| Depression_admission_freq | Inhib | *PAX8* | TRUE | 2.578379052 |
| Depression_admission_freq | Inhib | *AP5S1* | TRUE | 2.321901989 |
| Depression_admission_freq | Inhib | *TPH1* | TRUE | 2.243696451 |
| Depression_admission_freq | Inhib | *CHIT1* | TRUE | 2.238717475 |
| Depression_admission_freq | Inhib | *PSRC1* | TRUE | 2.991714226 |
| Depression_admission_freq | Inhib | *DHX34* | TRUE | 2.096053968 |
| Depression_admission_freq | Inhib | *AHI1* | TRUE | 2.966019404 |
| Depression_admission_freq | Inhib | *CIB2* | TRUE | 1.974831735 |
| Depression_admission_freq | Inhib | *CNKSR1* | TRUE | 3.009047637 |
| Depression_admission_freq | Inhib | *FBXL17* | TRUE | 2.553637144 |
| Depression_admission_freq | Inhib | *TTC12* | TRUE | 2.063161669 |
| Depression_admission_freq | Inhib | *FBXL2* | TRUE | 2.286192127 |
| Depression_admission_freq | Inhib | *VPS11* | TRUE | 2.078659527 |
| Depression_admission_freq | Inhib | *EFHB* | TRUE | 2.564524727 |
| Depression_admission_freq | Inhib | *SLC38A8* | TRUE | 2.902027846 |
| Depression_admission_freq | Inhib | *RNASE2* | TRUE | 2.100582151 |
| Depression_admission_freq | Inhib | *FAM21C* | TRUE | 2.209009274 |
| Depression_admission_freq | Inhib | *TLR1* | TRUE | 2.173774589 |
| Depression_admission_freq | Inhib | *PPP1R42* | TRUE | 2.136052066 |
| Depression_admission_freq | Inhib | *GEN1* | TRUE | 2.037897507 |
| Depression_admission_freq | Inhib | *ATOH7* | TRUE | 2.29057225 |

| | | | | |
|---|---|---|---|---|
| Depression_admission_freq | Inhib | *RESP18* | TRUE | 2.130406989 |
| Depression_admission_freq | Inhib | *ARHGEF37* | TRUE | 2.45256013 |
| Depression_admission_freq | Inhib | *COA5* | TRUE | 2.231708131 |
| Depression_admission_freq | Inhib | *RDM1* | TRUE | 2.064198889 |
| Depression_admission_freq | Inhib | *NWD1* | TRUE | 1.97239086 |
| Depression_admission_freq | Inhib | *ZNF737* | TRUE | 1.982487033 |
| Depression_admission_freq | Inhib | *GS1.259H13.2* | TRUE | 2.049745873 |
| Depression_admission_freq | Inhib | *RP11.826N14.2* | TRUE | 2.355618867 |
| Depression_admission_freq | Inhib | *RP11.377D9.3* | TRUE | 1.982557333 |
| Depression_admission_freq | Inhib | *CTD.3126B10.4* | TRUE | 2.134301556 |
| Depression_admission_freq | Inhib | *XXbac.BPG181B23.7* | TRUE | 1.986659795 |
| Depression_admission_freq | Inhib | *RP13.539J13.1* | TRUE | 2.076930138 |
| Depression_admission_freq | Inhib | *CTA.246H3.12* | TRUE | 2.470126949 |
| Depression_admission_freq | Micro_Macro | *TIPIN* | TRUE | 2.014102675 |
| Depression_admission_freq | Micro_Macro | *AHI1* | TRUE | 1.982940487 |
| Depression_admission_freq | Micro_Macro | *EFHB* | TRUE | 2.225109033 |
| Depression_admission_freq | Oligos | *AP5S1* | TRUE | 2.02531113 |
| Depression_admission_freq | Oligos | *TPH1* | TRUE | 2.00400125 |
| Depression_admission_freq | Oligos | *AHI1* | TRUE | 2.393921578 |
| Depression_admission_freq | Oligos | *CNKSR1* | TRUE | 2.757219636 |
| Depression_admission_freq | Oligos | *VPS11* | TRUE | 2.08283695 |
| Depression_admission_freq | Oligos | *EFHB* | TRUE | 2.398870175 |
| Depression_admission_freq | Oligos | *SLC38A8* | TRUE | 2.051462907 |
| Depression_admission_freq | Oligos | *RNASE2* | TRUE | 1.996661321 |
| Depression_admission_freq | Oligos | *POLE* | TRUE | 2.016746352 |
| Depression_admission_freq | Oligos | *RP11.826N14.2* | TRUE | 2.177054766 |
| Depression_admission_freq | Oligos | *CTD.3126B10.4* | TRUE | 2.153833448 |
| Depression_admission_freq | Oligos | *CTA.246H3.12* | TRUE | 2.058065861 |
| Depression_admission_freq | Oligos | *RBAKDN* | TRUE | 1.974889788 |
| Depression_admission_freq | OPCs | *ICAM3* | TRUE | 2.09623747 |
| Depression_admission_freq | OPCs | *PAX8* | TRUE | 2.440730624 |
| Depression_admission_freq | OPCs | *PSRC1* | TRUE | 2.326777402 |
| Depression_admission_freq | OPCs | *AHI1* | TRUE | 2.788946833 |
| Depression_admission_freq | OPCs | *CNKSR1* | TRUE | 2.862668697 |
| Depression_admission_freq | OPCs | *FBXL17* | TRUE | 2.12682389 |
| Depression_admission_freq | OPCs | *TTC12* | TRUE | 2.011869859 |
| Depression_admission_freq | OPCs | *EFHB* | TRUE | 2.125157358 |
| Depression_admission_freq | OPCs | *RNASE2* | TRUE | 2.018038033 |
| Depression_admission_freq | OPCs | *POLE* | TRUE | 2.067859779 |
| Depression_admission_freq | OPCs | *HLA.DQA2* | TRUE | 2.234249592 |
| Depression_admission_freq | OPCs | *RP11.826N14.2* | TRUE | 2.212079688 |
| Depression_admission_freq | OPCs | *CTD.3126B10.4* | TRUE | 2.022196628 |
| Depression_admission_freq | OPCs | *XXbac.BPG181B23.7* | TRUE | 1.990507432 |
| TRD | Astros | *TNFRSF10C* | TRUE | 1.961284831 |
| TRD | Astros | *ZNF664* | TRUE | 1.974287076 |
| TRD | Astros | *FAHD1* | TRUE | 1.96552785 |
| TRD | Astros | *AC159540.1* | TRUE | 2.043864819 |
| TRD | Astros | *LILRA4* | TRUE | 2.190605943 |
| TRD | Astros | *SMIM22* | TRUE | 2.331348197 |
| TRD | Ex | *SLC7A9* | TRUE | 2.090892779 |

| | | | | |
|---|---|---|---|---|
| TRD | Ex | *VCL* | TRUE | 2.023023341 |
| TRD | Ex | *NUCKS1* | TRUE | 2.418340358 |
| TRD | Ex | *CPSF1* | TRUE | 2.500437462 |
| TRD | Ex | *MRVI1* | TRUE | 2.302924112 |
| TRD | Ex | *SEC31B* | TRUE | 2.201682273 |
| TRD | Ex | *ABCB1* | TRUE | 2.176050035 |
| TRD | Ex | *FCGBP* | TRUE | 2.163079771 |
| TRD | Ex | *CLUAP1* | TRUE | 2.104188136 |
| TRD | Ex | *PYCRL* | TRUE | 2.036507102 |
| TRD | Ex | *CHCHD2* | TRUE | 2.124352769 |
| TRD | Ex | *PNPO* | TRUE | 2.133743081 |
| TRD | Ex | *LRRC42* | TRUE | 2.315225746 |
| TRD | Ex | *DNAL1* | TRUE | 2.244562677 |
| TRD | Ex | *CCDC170* | TRUE | 1.966011328 |
| TRD | Ex | *PCDHB10* | TRUE | 2.616024208 |
| TRD | Ex | *MRS2* | TRUE | 1.96266658 |
| TRD | Ex | *ZNF391* | TRUE | 2.079786172 |
| TRD | Ex | *GPR108* | TRUE | 2.403346321 |
| TRD | Ex | *MASP1* | TRUE | 3.051550944 |
| TRD | Ex | *CCT6B* | TRUE | 2.008083188 |
| TRD | Ex | *SCLY* | TRUE | 1.973266227 |
| TRD | Ex | *TSGA10* | TRUE | 2.691392204 |
| TRD | Ex | *NEDD1* | TRUE | 2.069007849 |
| TRD | Ex | *THNSL2* | TRUE | 1.962174963 |
| TRD | Ex | *EXOG* | TRUE | 2.075684237 |
| TRD | Ex | *TCHH* | TRUE | 1.970731719 |
| TRD | Ex | *PRKCD* | TRUE | 2.115869258 |
| TRD | Ex | *RFT1* | TRUE | 2.023612496 |
| TRD | Ex | *ANXA5* | TRUE | 2.206540471 |
| TRD | Ex | *OR51E2* | TRUE | 2.061918044 |
| TRD | Ex | *CCDC88B* | TRUE | 2.043401828 |
| TRD | Ex | *FGGY* | TRUE | 2.299010553 |
| TRD | Ex | *PARP14* | TRUE | 2.460984316 |
| TRD | Ex | *AHSA2* | TRUE | 2.212207727 |
| TRD | Ex | *TNFRSF10C* | TRUE | 2.326697522 |
| TRD | Ex | *ZNF613* | TRUE | 2.052952123 |
| TRD | Ex | *ZNF664* | TRUE | 2.583641627 |
| TRD | Ex | *C11orf54* | TRUE | 2.033855368 |
| TRD | Ex | *CCDC125* | TRUE | 2.613400648 |
| TRD | Ex | *PCGF3* | TRUE | 2.042184458 |
| TRD | Ex | *KIF24* | TRUE | 2.337819837 |
| TRD | Ex | *FAM26F* | TRUE | 2.245372491 |
| TRD | Ex | *APRT* | TRUE | 2.794235012 |
| TRD | Ex | *ZNF891* | TRUE | 2.224804595 |
| TRD | Ex | *AC004540.5* | TRUE | 2.123516743 |
| TRD | Ex | *LINC00152* | TRUE | 1.969929397 |
| TRD | Ex | *LINC00937* | TRUE | 1.972535294 |
| TRD | Ex | *RP11.15I11.3* | TRUE | 2.131749911 |
| TRD | Ex | *AC097713.3* | TRUE | 2.43995092 |
| TRD | Ex | *LILRA4* | TRUE | 2.031394782 |

| | | | | |
|---|---|---|---|---|
| TRD | Ex | *AF146191.4* | TRUE | 2.159059301 |
| TRD | Ex | *LINC00926* | TRUE | 1.963730898 |
| TRD | Ex | *CTD.2589M5.5* | TRUE | 2.450375092 |
| TRD | Ex | *CTD.2349P21.9* | TRUE | 2.864837982 |
| TRD | Ex | *CTD.2623N2.11* | TRUE | 2.576775484 |
| TRD | Ex | *RP11.147L13.8* | TRUE | 2.167018653 |
| TRD | Inhib | *SLC7A9* | TRUE | 2.066329419 |
| TRD | Inhib | *VCL* | TRUE | 2.126358549 |
| TRD | Inhib | *CASP8* | TRUE | 2.082896977 |
| TRD | Inhib | *NUCKS1* | TRUE | 2.087543289 |
| TRD | Inhib | *CPSF1* | TRUE | 2.114021955 |
| TRD | Inhib | *MRVI1* | TRUE | 2.164092861 |
| TRD | Inhib | *SEC31B* | TRUE | 2.033760244 |
| TRD | Inhib | *FCGBP* | TRUE | 2.16694986 |
| TRD | Inhib | *CDC7* | TRUE | 1.996637742 |
| TRD | Inhib | *CLUAP1* | TRUE | 2.043045294 |
| TRD | Inhib | *PYCRL* | TRUE | 2.225122257 |
| TRD | Inhib | *CHCHD2* | TRUE | 2.083555438 |
| TRD | Inhib | *PNPO* | TRUE | 1.993101446 |
| TRD | Inhib | *LRRC42* | TRUE | 2.424096966 |
| TRD | Inhib | *DNAL1* | TRUE | 2.198032006 |
| TRD | Inhib | *CCDC170* | TRUE | 2.053817875 |
| TRD | Inhib | *PCDHB10* | TRUE | 2.321102506 |
| TRD | Inhib | *ZNF391* | TRUE | 2.030014139 |
| TRD | Inhib | *GPR108* | TRUE | 2.458375514 |
| TRD | Inhib | *MASP1* | TRUE | 3.070999119 |
| TRD | Inhib | *SCLY* | TRUE | 2.156662492 |
| TRD | Inhib | *TSGA10* | TRUE | 2.477714127 |
| TRD | Inhib | *THNSL2* | TRUE | 2.163529496 |
| TRD | Inhib | *RMND1* | TRUE | 1.980831491 |
| TRD | Inhib | *TCHH* | TRUE | 2.015329243 |
| TRD | Inhib | *PRKCD* | TRUE | 2.2069345 |
| TRD | Inhib | *ANXA5* | TRUE | 2.031044439 |
| TRD | Inhib | *OR51E2* | TRUE | 2.362556832 |
| TRD | Inhib | *CCDC88B* | TRUE | 1.970230242 |
| TRD | Inhib | *FGGY* | TRUE | 2.313804297 |
| TRD | Inhib | *PARP14* | TRUE | 2.464459399 |
| TRD | Inhib | *TNFRSF10C* | TRUE | 2.284828796 |
| TRD | Inhib | *ZNF613* | TRUE | 2.222340345 |
| TRD | Inhib | *MUC20* | TRUE | 1.967925032 |
| TRD | Inhib | *ZNF664* | TRUE | 2.646092608 |
| TRD | Inhib | *CCDC125* | TRUE | 2.30330677 |
| TRD | Inhib | *KIF24* | TRUE | 2.32578127 |
| TRD | Inhib | *FAM26F* | TRUE | 2.526880375 |
| TRD | Inhib | *APRT* | TRUE | 2.767036837 |
| TRD | Inhib | *ZNF891* | TRUE | 2.23583854 |
| TRD | Inhib | *RP1.137D17.1* | TRUE | 2.070956203 |
| TRD | Inhib | *RP11.15I11.3* | TRUE | 2.13086465 |
| TRD | Inhib | *AC097713.3* | TRUE | 2.446733928 |
| TRD | Inhib | *LILRA4* | TRUE | 2.229780456 |

| | | | | |
|---|---|---|---|---|
| TRD | Inhib | *CTD.2589M5.5* | TRUE | 2.188907837 |
| TRD | Inhib | *RP11.107F6.3* | TRUE | 2.319068847 |
| TRD | Inhib | *CTD.2349P21.9* | TRUE | 2.497158082 |
| TRD | Inhib | *CTD.2623N2.11* | TRUE | 2.526896337 |
| TRD | Inhib | *RP11.147L13.8* | TRUE | 2.167280916 |
| TRD | Micro_Macro | *MASP1* | TRUE | 2.533299743 |
| TRD | Micro_Macro | *KIF24* | TRUE | 2.044612772 |
| TRD | Oligos | *MRVI1* | TRUE | 2.214566822 |
| TRD | Oligos | *PYCRL* | TRUE | 1.968327893 |
| TRD | Oligos | *CHCHD2* | TRUE | 1.979683587 |
| TRD | Oligos | *GPR108* | TRUE | 2.026774986 |
| TRD | Oligos | *MASP1* | TRUE | 2.497831437 |
| TRD | Oligos | *FGGY* | TRUE | 2.108542107 |
| TRD | Oligos | *PARP14* | TRUE | 2.417444666 |
| TRD | Oligos | *ZNF664* | TRUE | 2.261107021 |
| TRD | Oligos | *APRT* | TRUE | 2.30780318 |
| TRD | Oligos | *ZNF891* | TRUE | 2.003515685 |
| TRD | OPCs | *PYCRL* | TRUE | 2.305825469 |
| TRD | OPCs | *LRRC42* | TRUE | 2.316820291 |
| TRD | OPCs | *SCLY* | TRUE | 2.101094152 |
| TRD | OPCs | *ZNF664* | TRUE | 2.456162801 |
| TRD | OPCs | *FAHD1* | TRUE | 2.006768152 |
| TRD | OPCs | *APRT* | TRUE | 2.301357597 |
| TRD | OPCs | *AC097713.3* | TRUE | 2.187766808 |
| TRD | OPCs | *RP11.384P7.7* | TRUE | 2.047554975 |
| TRD | OPCs | *CTD.2349P21.9* | TRUE | 1.975612165 |

b. Identified cell type specific causal genes for T2DM and related disorders from pancreas

| disorder | celltype | gene | assoc | zMin |
|---|---|---|---|---|
| Prediabetes | acinar_cell | *STPG1* | TRUE | 1.968466597 |
| Prediabetes | acinar_cell | *OTX1* | TRUE | 2.679296292 |
| Prediabetes | acinar_cell | *KIAA0087* | TRUE | 2.320711183 |
| Prediabetes | acinar_cell | *SH2D3A* | TRUE | 2.676365519 |
| Prediabetes | acinar_cell | *A4GALT* | TRUE | 2.388182485 |
| Prediabetes | acinar_cell | *NEK3* | TRUE | 1.967639749 |
| Prediabetes | acinar_cell | *SPTBN5* | TRUE | 1.974824951 |
| Prediabetes | acinar_cell | *RGS3* | TRUE | 2.515002227 |
| Prediabetes | acinar_cell | *CAPN5* | TRUE | 2.071145345 |
| Prediabetes | acinar_cell | *CNNM4* | TRUE | 2.389037647 |
| Prediabetes | acinar_cell | *RPRML* | TRUE | 1.999195573 |
| Prediabetes | acinar_cell | *GPR88* | TRUE | 2.444648854 |
| Prediabetes | acinar_cell | *FAM166A* | TRUE | 2.091930808 |
| Prediabetes | acinar_cell | *ZNF559* | TRUE | 1.985066148 |
| Prediabetes | acinar_cell | *PIWIL2* | TRUE | 2.225069491 |
| Prediabetes | acinar_cell | *NOTCH4* | TRUE | 2.12222411 |
| Prediabetes | alpha_cell | *STPG1* | TRUE | 2.65342763 |
| Prediabetes | alpha_cell | *ST3GAL6* | TRUE | 1.962215815 |
| Prediabetes | alpha_cell | *TESK2* | TRUE | 2.210712172 |
| Prediabetes | alpha_cell | *OTX1* | TRUE | 3.575564914 |
| Prediabetes | alpha_cell | *KIAA0087* | TRUE | 2.514262081 |
| Prediabetes | alpha_cell | *SH2D3A* | TRUE | 3.103958974 |
| Prediabetes | alpha_cell | *A4GALT* | TRUE | 2.56571506 |
| Prediabetes | alpha_cell | *NEK3* | TRUE | 2.140550366 |
| Prediabetes | alpha_cell | *CIB2* | TRUE | 1.99502752 |
| Prediabetes | alpha_cell | *SPTBN5* | TRUE | 2.502567281 |
| Prediabetes | alpha_cell | *RGS3* | TRUE | 2.304939402 |
| Prediabetes | alpha_cell | *ZMYND15* | TRUE | 2.019468352 |
| Prediabetes | alpha_cell | *CAPN5* | TRUE | 2.30737618 |
| Prediabetes | alpha_cell | *PDE3B* | TRUE | 1.973540161 |
| Prediabetes | alpha_cell | *CNNM4* | TRUE | 2.334619736 |
| Prediabetes | alpha_cell | *NBEAL2* | TRUE | 2.028503001 |
| Prediabetes | alpha_cell | *CPSF2* | TRUE | 2.021651792 |
| Prediabetes | alpha_cell | *RAD9A* | TRUE | 2.138245866 |
| Prediabetes | alpha_cell | *HAP1* | TRUE | 2.652267508 |
| Prediabetes | alpha_cell | *RPRML* | TRUE | 2.127133663 |
| Prediabetes | alpha_cell | *GPR88* | TRUE | 2.461672629 |
| Prediabetes | alpha_cell | *SETD4* | TRUE | 2.081376 |
| Prediabetes | alpha_cell | *KBTBD12* | TRUE | 2.590319436 |
| Prediabetes | alpha_cell | *FAM166A* | TRUE | 3.081240949 |
| Prediabetes | alpha_cell | *PIWIL2* | TRUE | 2.730164018 |
| Prediabetes | alpha_cell | *NOTCH4* | TRUE | 2.222706115 |
| Prediabetes | alpha_cell | *PINLYP* | TRUE | 2.129869085 |
| Prediabetes | beta_cell | *STPG1* | TRUE | 3.018156693 |
| Prediabetes | beta_cell | *APBA3* | TRUE | 2.069528056 |
| Prediabetes | beta_cell | *ST3GAL6* | TRUE | 2.324661521 |

| | | | | |
|---|---|---|---|---|
| Prediabetes | beta_cell | *FGF10* | TRUE | 2.042036362 |
| Prediabetes | beta_cell | *TESK2* | TRUE | 2.040689514 |
| Prediabetes | beta_cell | *NDE1* | TRUE | 2.238593079 |
| Prediabetes | beta_cell | *CACNG5* | TRUE | 2.000730554 |
| Prediabetes | beta_cell | *APBB1IP* | TRUE | 2.013320169 |
| Prediabetes | beta_cell | *ZNF500* | TRUE | 2.048778999 |
| Prediabetes | beta_cell | *NOMO3* | TRUE | 2.050889145 |
| Prediabetes | beta_cell | *NAGLU* | TRUE | 2.662969516 |
| Prediabetes | beta_cell | *UFSP2* | TRUE | 2.131499378 |
| Prediabetes | beta_cell | *OTX1* | TRUE | 3.607728808 |
| Prediabetes | beta_cell | *WDR77* | TRUE | 2.247634983 |
| Prediabetes | beta_cell | *FILIP1* | TRUE | 2.10616405 |
| Prediabetes | beta_cell | *ZC2HC1C* | TRUE | 2.228508854 |
| Prediabetes | beta_cell | *SLC17A5* | TRUE | 2.6978888 |
| Prediabetes | beta_cell | *KIAA0087* | TRUE | 2.608001358 |
| Prediabetes | beta_cell | *INSIG2* | TRUE | 1.982283858 |
| Prediabetes | beta_cell | *SH2D3A* | TRUE | 3.11109603 |
| Prediabetes | beta_cell | *A4GALT* | TRUE | 2.573562365 |
| Prediabetes | beta_cell | *MAP3K10* | TRUE | 2.102170261 |
| Prediabetes | beta_cell | *FLNB* | TRUE | 2.099500373 |
| Prediabetes | beta_cell | *NEK3* | TRUE | 2.298526848 |
| Prediabetes | beta_cell | *CIB2* | TRUE | 2.190703649 |
| Prediabetes | beta_cell | *BLK* | TRUE | 1.966236314 |
| Prediabetes | beta_cell | *SPTBN5* | TRUE | 2.481874391 |
| Prediabetes | beta_cell | *RGS3* | TRUE | 2.759294036 |
| Prediabetes | beta_cell | *ZMYND15* | TRUE | 2.181473688 |
| Prediabetes | beta_cell | *IQGAP2* | TRUE | 2.105743178 |
| Prediabetes | beta_cell | *TRIM7* | TRUE | 2.655369286 |
| Prediabetes | beta_cell | *MTG1* | TRUE | 1.996069326 |
| Prediabetes | beta_cell | *CAPN5* | TRUE | 2.358826958 |
| Prediabetes | beta_cell | *DCUN1D2* | TRUE | 1.96152049 |
| Prediabetes | beta_cell | *PDE3B* | TRUE | 2.257430702 |
| Prediabetes | beta_cell | *DDAH1* | TRUE | 2.892198065 |
| Prediabetes | beta_cell | *LRGUK* | TRUE | 2.095378722 |
| Prediabetes | beta_cell | *CNNM4* | TRUE | 2.488239044 |
| Prediabetes | beta_cell | *NBEAL2* | TRUE | 2.178097191 |
| Prediabetes | beta_cell | *SPC24* | TRUE | 1.994893066 |
| Prediabetes | beta_cell | *ERCC3* | TRUE | 1.969730102 |
| Prediabetes | beta_cell | *RYBP* | TRUE | 2.000409044 |
| Prediabetes | beta_cell | *RPL9* | TRUE | 2.003227356 |
| Prediabetes | beta_cell | *CPSF2* | TRUE | 2.052783398 |
| Prediabetes | beta_cell | *KIFC2* | TRUE | 2.167171415 |
| Prediabetes | beta_cell | *HAP1* | TRUE | 2.54687089 |
| Prediabetes | beta_cell | *PFN4* | TRUE | 2.064549681 |
| Prediabetes | beta_cell | *TMEM80* | TRUE | 2.259600496 |
| Prediabetes | beta_cell | *RPRML* | TRUE | 2.109693144 |
| Prediabetes | beta_cell | *GPR88* | TRUE | 2.599469743 |
| Prediabetes | beta_cell | *CALN1* | TRUE | 2.240710456 |

| | | | | |
|---|---|---|---|---|
| Prediabetes | beta_cell | *SH3BGR* | TRUE | 1.987525007 |
| Prediabetes | beta_cell | *SETD4* | TRUE | 2.627092815 |
| Prediabetes | beta_cell | *KBTBD12* | TRUE | 2.822638228 |
| Prediabetes | beta_cell | *FAM166A* | TRUE | 2.336513445 |
| Prediabetes | beta_cell | *ZNF763* | TRUE | 2.225679808 |
| Prediabetes | beta_cell | *PIWIL2* | TRUE | 2.925023622 |
| Prediabetes | beta_cell | *OGDHL* | TRUE | 1.977139651 |
| Prediabetes | beta_cell | *ENPP1* | TRUE | 2.145023107 |
| Prediabetes | beta_cell | *NOTCH4* | TRUE | 2.3162522 |
| Prediabetes | beta_cell | *LINC00865* | TRUE | 2.193767651 |
| Prediabetes | beta_cell | *PINLYP* | TRUE | 2.348198758 |
| Prediabetes | beta_cell | *LRRC37A2* | TRUE | 2.068454661 |
| Prediabetes | delta_cell | *OTX1* | TRUE | 3.150436428 |
| Prediabetes | delta_cell | *SH2D3A* | TRUE | 3.047455399 |
| Prediabetes | delta_cell | *RGS3* | TRUE | 2.543411379 |
| Prediabetes | delta_cell | *PDE3B* | TRUE | 2.00392137 |
| Prediabetes | delta_cell | *CNNM4* | TRUE | 2.142621429 |
| Prediabetes | delta_cell | *NBEAL2* | TRUE | 2.074286704 |
| Prediabetes | delta_cell | *CALN1* | TRUE | 2.134376263 |
| Prediabetes | delta_cell | *FAM166A* | TRUE | 2.061360089 |
| Prediabetes | delta_cell | *PIWIL2* | TRUE | 1.994118079 |
| Prediabetes | delta_cell | *PINLYP* | TRUE | 2.445515608 |
| Prediabetes | duct_epithelial_cell | *STPG1* | TRUE | 1.986533264 |
| Prediabetes | duct_epithelial_cell | *ST3GAL6* | TRUE | 1.983396217 |
| Prediabetes | duct_epithelial_cell | *NAGLU* | TRUE | 2.055169201 |
| Prediabetes | duct_epithelial_cell | *OTX1* | TRUE | 3.170004028 |
| Prediabetes | duct_epithelial_cell | *KIAA0087* | TRUE | 2.309600546 |
| Prediabetes | duct_epithelial_cell | *MAP3K10* | TRUE | 2.115670188 |
| Prediabetes | duct_epithelial_cell | *CIB2* | TRUE | 2.014884646 |
| Prediabetes | duct_epithelial_cell | *RPRML* | TRUE | 2.33776216 |
| Prediabetes | duct_epithelial_cell | *KBTBD12* | TRUE | 2.415083287 |
| Prediabetes | duct_epithelial_cell | *ZNF559* | TRUE | 2.143719424 |
| Prediabetes | duct_epithelial_cell | *PIWIL2* | TRUE | 2.578528929 |
| Prediabetes | duct_epithelial_cell | *BGLAP* | TRUE | 1.974513813 |
| Prediabetes | endothelial_cell | *ST3GAL6* | TRUE | 2.017885735 |
| Prediabetes | endothelial_cell | *OTX1* | TRUE | 3.425651481 |
| Prediabetes | endothelial_cell | *SLC17A5* | TRUE | 1.993181825 |
| Prediabetes | endothelial_cell | *KIAA0087* | TRUE | 2.491446777 |
| Prediabetes | endothelial_cell | *SH2D3A* | TRUE | 3.08875326 |
| Prediabetes | endothelial_cell | *TRIM7* | TRUE | 2.361309123 |
| Prediabetes | endothelial_cell | *CAPN5* | TRUE | 2.19366652 |
| Prediabetes | endothelial_cell | *HAP1* | TRUE | 2.189330856 |
| Prediabetes | endothelial_cell | *RPRML* | TRUE | 2.303360667 |
| Prediabetes | endothelial_cell | *GPR88* | TRUE | 2.393190655 |
| Prediabetes | endothelial_cell | *KBTBD12* | TRUE | 2.175450756 |
| Prediabetes | endothelial_cell | *FAM166A* | TRUE | 2.333497457 |
| Prediabetes | endothelial_cell | *PIWIL2* | TRUE | 2.741115252 |
| Prediabetes | epsilon_cell | *OTX1* | TRUE | 3.231007925 |
| Prediabetes | epsilon_cell | *SPTBN5* | TRUE | 2.359788808 |
| Prediabetes | epsilon_cell | *RPRML* | TRUE | 2.020362268 |

| | | | | |
|---|---|---|---|---|
| Prediabetes | epsilon_cell | *GPR88* | TRUE | 2.171435491 |
| Prediabetes | leukocyte | *OTX1* | TRUE | 3.310923318 |
| Prediabetes | leukocyte | *KIAA0087* | TRUE | 2.12990719 |
| Prediabetes | leukocyte | *SH2D3A* | TRUE | 2.091193932 |
| Prediabetes | leukocyte | *GPR88* | TRUE | 2.293530854 |
| Prediabetes | leukocyte | *FAM166A* | TRUE | 1.972793728 |
| Prediabetes | mesenchymal_cell | *OTX1* | TRUE | 2.467097891 |
| Prediabetes | mesenchymal_cell | *KIAA0087* | TRUE | 2.252648072 |
| Prediabetes | mesenchymal_cell | *SH2D3A* | TRUE | 2.662575857 |
| Prediabetes | mesenchymal_cell | *CIB2* | TRUE | 2.035322709 |
| Prediabetes | mesenchymal_cell | *ZMYND15* | TRUE | 2.035935239 |
| Prediabetes | mesenchymal_cell | *RPRML* | TRUE | 2.282905061 |
| Prediabetes | mesenchymal_cell | *FAM166A* | TRUE | 2.537194537 |
| Prediabetes | mesenchymal_cell | *ZNF559* | TRUE | 2.114099354 |
| Prediabetes | mesenchymal_cell | *PIWIL2* | TRUE | 2.392253974 |
| Prediabetes | mesenchymal_cell | *HLA.DQA2* | TRUE | 2.35416309 |
| Prediabetes | PP_cell | *NAGLU* | TRUE | 1.989406272 |
| Prediabetes | PP_cell | *OTX1* | TRUE | 3.216783482 |
| Prediabetes | PP_cell | *SH2D3A* | TRUE | 2.464207433 |
| Prediabetes | PP_cell | *SPTBN5* | TRUE | 2.292526115 |
| Prediabetes | PP_cell | *RGS3* | TRUE | 2.49350808 |
| Prediabetes | PP_cell | *GPR88* | TRUE | 2.212160255 |
| Prediabetes | PP_cell | *ZNF559* | TRUE | 2.13706378 |
| Prediabetes | PP_cell | *NOTCH4* | TRUE | 2.026833656 |
| T1DM | acinar_cell | *CEP68* | TRUE | 2.039816542 |
| T1DM | acinar_cell | *ARHGAP22* | TRUE | 2.338562117 |
| T1DM | acinar_cell | *ARHGEF19* | TRUE | 2.056157083 |
| T1DM | acinar_cell | *CLEC18A* | TRUE | 2.208844175 |
| T1DM | acinar_cell | *EXTL1* | TRUE | 2.486988106 |
| T1DM | acinar_cell | *RFTN2* | TRUE | 2.280101024 |
| T1DM | acinar_cell | *ZNF589* | TRUE | 2.003447813 |
| T1DM | acinar_cell | *ENGASE* | TRUE | 3.022904695 |
| T1DM | acinar_cell | *MORN4* | TRUE | 1.977746871 |
| T1DM | acinar_cell | *PCP2* | TRUE | 2.130160503 |
| T1DM | acinar_cell | *IRX3* | TRUE | 2.256876722 |
| T1DM | acinar_cell | *RPRML* | TRUE | 2.59619996 |
| T1DM | acinar_cell | *CDK10* | TRUE | 2.09906341 |
| T1DM | acinar_cell | *FAM228A* | TRUE | 2.59118772 |
| T1DM | acinar_cell | *SFTA2* | TRUE | 2.681572037 |
| T1DM | acinar_cell | *CCDC154* | TRUE | 2.219157151 |
| T1DM | acinar_cell | *HSD17B8* | TRUE | 8.330615626 |
| T1DM | acinar_cell | *NOTCH4* | TRUE | 3.472857145 |
| T1DM | acinar_cell | *PSORS1C1* | TRUE | 2.057809738 |
| T1DM | acinar_cell | *HLA.DQB2* | TRUE | 8.115633058 |
| T1DM | acinar_cell | *PSMB9* | TRUE | 2.136022887 |
| T1DM | acinar_cell | *HLA.DOB* | TRUE | 6.198798961 |
| T1DM | acinar_cell | *HLA.DMB* | TRUE | 5.269886796 |
| T1DM | acinar_cell | *GOLGA8K* | TRUE | 2.281748653 |
| T1DM | alpha_cell | *CEP68* | TRUE | 2.414224708 |
| T1DM | alpha_cell | *ZNF213* | TRUE | 2.478459952 |

| | | | | |
|---|---|---|---|---|
| T1DM | alpha_cell | *STX1B* | TRUE | 2.024618568 |
| T1DM | alpha_cell | *KHNYN* | TRUE | 1.976084697 |
| T1DM | alpha_cell | *CTSH* | TRUE | 2.822831157 |
| T1DM | alpha_cell | *FTSJ3* | TRUE | 2.362316392 |
| T1DM | alpha_cell | *GPRC5D* | TRUE | 1.97285872 |
| T1DM | alpha_cell | *VIPR1* | TRUE | 2.357327369 |
| T1DM | alpha_cell | *ARHGAP22* | TRUE | 2.481264638 |
| T1DM | alpha_cell | *EPO* | TRUE | 2.993773722 |
| T1DM | alpha_cell | *PTPN22* | TRUE | 2.466363149 |
| T1DM | alpha_cell | *ULK3* | TRUE | 2.239548473 |
| T1DM | alpha_cell | *ARHGEF19* | TRUE | 1.962610812 |
| T1DM | alpha_cell | *NBPF3* | TRUE | 1.974147033 |
| T1DM | alpha_cell | *CABLES2* | TRUE | 2.838390836 |
| T1DM | alpha_cell | *SLC25A27* | TRUE | 2.086856361 |
| T1DM | alpha_cell | *CDK20* | TRUE | 2.513258955 |
| T1DM | alpha_cell | *SFXN2* | TRUE | 1.971594739 |
| T1DM | alpha_cell | *CLEC18A* | TRUE | 2.580312432 |
| T1DM | alpha_cell | *EXTL1* | TRUE | 2.477253402 |
| T1DM | alpha_cell | *SLC5A11* | TRUE | 2.147199784 |
| T1DM | alpha_cell | *RFTN2* | TRUE | 2.402269383 |
| T1DM | alpha_cell | *GNPDA2* | TRUE | 2.000176054 |
| T1DM | alpha_cell | *SRGAP2* | TRUE | 2.035861656 |
| T1DM | alpha_cell | *LMBRD2* | TRUE | 1.993756954 |
| T1DM | alpha_cell | *ENGASE* | TRUE | 3.097226652 |
| T1DM | alpha_cell | *EVPL* | TRUE | 2.050724425 |
| T1DM | alpha_cell | *MORN4* | TRUE | 2.258744306 |
| T1DM | alpha_cell | *SNTB1* | TRUE | 2.249962881 |
| T1DM | alpha_cell | *PCP2* | TRUE | 2.160643134 |
| T1DM | alpha_cell | *UBE2U* | TRUE | 2.353229262 |
| T1DM | alpha_cell | *IRX3* | TRUE | 2.429888303 |
| T1DM | alpha_cell | *RPRML* | TRUE | 2.804592848 |
| T1DM | alpha_cell | *F2R* | TRUE | 2.085562033 |
| T1DM | alpha_cell | *ANO9* | TRUE | 1.998095872 |
| T1DM | alpha_cell | *CDK10* | TRUE | 2.308407765 |
| T1DM | alpha_cell | *FAM228A* | TRUE | 2.54874806 |
| T1DM | alpha_cell | *ZKSCAN3* | TRUE | 2.000283978 |
| T1DM | alpha_cell | *SFTA2* | TRUE | 2.461808135 |
| T1DM | alpha_cell | *ZNF517* | TRUE | 2.119405728 |
| T1DM | alpha_cell | *CCDC154* | TRUE | 2.322190197 |
| T1DM | alpha_cell | *HSPB9* | TRUE | 2.147530455 |
| T1DM | alpha_cell | *HLA.DRB5* | TRUE | 2.533092923 |
| T1DM | alpha_cell | *HSD17B8* | TRUE | 7.523365461 |
| T1DM | alpha_cell | *NOTCH4* | TRUE | 2.593408811 |
| T1DM | alpha_cell | *HLA.DQB2* | TRUE | 8.938393619 |
| T1DM | alpha_cell | *PSMB9* | TRUE | 2.291112894 |
| T1DM | alpha_cell | *HLA.DOB* | TRUE | 6.269361722 |
| T1DM | alpha_cell | *HLA.DMB* | TRUE | 5.836751043 |
| T1DM | alpha_cell | *GOLGA8K* | TRUE | 2.782592619 |
| T1DM | beta_cell | *USH1C* | TRUE | 2.059272889 |
| T1DM | beta_cell | *BAIAP3* | TRUE | 2.058337341 |

| | | | | |
|---|---|---|---|---|
| T1DM | beta_cell | *CEP68* | TRUE | 3.032492485 |
| T1DM | beta_cell | *ZDHHC6* | TRUE | 2.052419674 |
| T1DM | beta_cell | *RNASET2* | TRUE | 2.371258778 |
| T1DM | beta_cell | *RNF19A* | TRUE | 1.973906797 |
| T1DM | beta_cell | *SPTB* | TRUE | 2.056260109 |
| T1DM | beta_cell | *GPATCH1* | TRUE | 1.96856575 |
| T1DM | beta_cell | *ZNF213* | TRUE | 2.856989636 |
| T1DM | beta_cell | *PPP1R13B* | TRUE | 2.145889441 |
| T1DM | beta_cell | *KIF16B* | TRUE | 2.182137334 |
| T1DM | beta_cell | *STX1B* | TRUE | 2.028384796 |
| T1DM | beta_cell | *KHNYN* | TRUE | 2.375870158 |
| T1DM | beta_cell | *POLI* | TRUE | 2.067977134 |
| T1DM | beta_cell | *CCDC113* | TRUE | 2.18814712 |
| T1DM | beta_cell | *CTSH* | TRUE | 2.837237992 |
| T1DM | beta_cell | *ECH1* | TRUE | 2.204543819 |
| T1DM | beta_cell | *KLC3* | TRUE | 2.116563357 |
| T1DM | beta_cell | *OGN* | TRUE | 1.98516172 |
| T1DM | beta_cell | *BACH2* | TRUE | 2.252146711 |
| T1DM | beta_cell | *DNPH1* | TRUE | 1.968354809 |
| T1DM | beta_cell | *VIPR1* | TRUE | 2.825924131 |
| T1DM | beta_cell | *FANCL* | TRUE | 1.965996158 |
| T1DM | beta_cell | *PYROXD2* | TRUE | 1.963253347 |
| T1DM | beta_cell | *HELLS* | TRUE | 2.050575522 |
| T1DM | beta_cell | *YEATS4* | TRUE | 2.043820396 |
| T1DM | beta_cell | *ARHGAP22* | TRUE | 2.56171327 |
| T1DM | beta_cell | *TPH1* | TRUE | 1.961424769 |
| T1DM | beta_cell | *MAU2* | TRUE | 2.122299491 |
| T1DM | beta_cell | *SYT5* | TRUE | 2.45141832 |
| T1DM | beta_cell | *EPO* | TRUE | 2.774312911 |
| T1DM | beta_cell | *PIAS3* | TRUE | 2.529801733 |
| T1DM | beta_cell | *RHPN2* | TRUE | 2.565120382 |
| T1DM | beta_cell | *EIF5A* | TRUE | 2.21895126 |
| T1DM | beta_cell | *PTPN22* | TRUE | 2.454072041 |
| T1DM | beta_cell | *NOTCH2* | TRUE | 2.020438851 |
| T1DM | beta_cell | *AVIL* | TRUE | 2.306220225 |
| T1DM | beta_cell | *TBRG4* | TRUE | 2.179573793 |
| T1DM | beta_cell | *PLCB2* | TRUE | 1.966678987 |
| T1DM | beta_cell | *SPTBN5* | TRUE | 2.028268733 |
| T1DM | beta_cell | *CALHM2* | TRUE | 2.193108523 |
| T1DM | beta_cell | *ULK3* | TRUE | 2.50514336 |
| T1DM | beta_cell | *ARHGEF19* | TRUE | 2.231805854 |
| T1DM | beta_cell | *NBPF3* | TRUE | 2.021023454 |
| T1DM | beta_cell | *GIGYF1* | TRUE | 2.145361135 |
| T1DM | beta_cell | *EML3* | TRUE | 2.417431081 |
| T1DM | beta_cell | *CABLES2* | TRUE | 3.057641678 |
| T1DM | beta_cell | *AKR1C2* | TRUE | 2.068118499 |
| T1DM | beta_cell | *SPC25* | TRUE | 2.271936221 |
| T1DM | beta_cell | *SLC25A27* | TRUE | 2.260578095 |
| T1DM | beta_cell | *PTPRD* | TRUE | 2.605638962 |
| T1DM | beta_cell | *MRPL39* | TRUE | 2.030552915 |

| | | | | |
|---|---|---|---|---|
| T1DM | beta_cell | *CCT8* | TRUE | 2.058733547 |
| T1DM | beta_cell | *CDK20* | TRUE | 2.699009513 |
| T1DM | beta_cell | *CLEC18A* | TRUE | 2.602509591 |
| T1DM | beta_cell | *EXTL1* | TRUE | 2.513024275 |
| T1DM | beta_cell | *AHCYL2* | TRUE | 2.074207776 |
| T1DM | beta_cell | *SLC5A11* | TRUE | 2.169704279 |
| T1DM | beta_cell | *OMA1* | TRUE | 2.083602976 |
| T1DM | beta_cell | *RFTN2* | TRUE | 3.067113012 |
| T1DM | beta_cell | *GNPDA2* | TRUE | 2.429226593 |
| T1DM | beta_cell | *NIPAL1* | TRUE | 2.271920482 |
| T1DM | beta_cell | *SRGAP2* | TRUE | 2.665570781 |
| T1DM | beta_cell | *ZNF589* | TRUE | 2.037575323 |
| T1DM | beta_cell | *LMBRD2* | TRUE | 2.46346972 |
| T1DM | beta_cell | *ENGASE* | TRUE | 3.241125211 |
| T1DM | beta_cell | *CACNB3* | TRUE | 2.014524062 |
| T1DM | beta_cell | *EVPL* | TRUE | 2.079122765 |
| T1DM | beta_cell | *SLC38A11* | TRUE | 2.264769971 |
| T1DM | beta_cell | *KCNMB3* | TRUE | 1.996778123 |
| T1DM | beta_cell | *MORN4* | TRUE | 2.399524914 |
| T1DM | beta_cell | *ZNF440* | TRUE | 2.260219385 |
| T1DM | beta_cell | *PPID* | TRUE | 2.011925583 |
| T1DM | beta_cell | *CLSTN1* | TRUE | 2.589899802 |
| T1DM | beta_cell | *PTEN* | TRUE | 2.180546234 |
| T1DM | beta_cell | *SNTB1* | TRUE | 2.466644196 |
| T1DM | beta_cell | *B3GALT1* | TRUE | 1.970166439 |
| T1DM | beta_cell | *SMPDL3A* | TRUE | 2.152379726 |
| T1DM | beta_cell | *PCP2* | TRUE | 2.155665704 |
| T1DM | beta_cell | *CHRNA7* | TRUE | 2.199927865 |
| T1DM | beta_cell | *UBE2U* | TRUE | 2.361705473 |
| T1DM | beta_cell | *IRX3* | TRUE | 2.630705317 |
| T1DM | beta_cell | *SAMD12* | TRUE | 1.984957998 |
| T1DM | beta_cell | *IL17RA* | TRUE | 2.586700226 |
| T1DM | beta_cell | *DALRD3* | TRUE | 2.371862453 |
| T1DM | beta_cell | *TMTC2* | TRUE | 2.021975247 |
| T1DM | beta_cell | *RPRML* | TRUE | 2.798134769 |
| T1DM | beta_cell | *ZNF154* | TRUE | 2.247548433 |
| T1DM | beta_cell | *TH* | TRUE | 1.977401528 |
| T1DM | beta_cell | *F2R* | TRUE | 2.461712183 |
| T1DM | beta_cell | *CAMK1D* | TRUE | 2.298335054 |
| T1DM | beta_cell | *ANO9* | TRUE | 2.097187314 |
| T1DM | beta_cell | *RBM11* | TRUE | 2.280463346 |
| T1DM | beta_cell | *CDK10* | TRUE | 2.551725632 |
| T1DM | beta_cell | *FAM228A* | TRUE | 2.610270717 |
| T1DM | beta_cell | *LYPD6* | TRUE | 2.099192019 |
| T1DM | beta_cell | *BCAM* | TRUE | 1.978429637 |
| T1DM | beta_cell | *ZKSCAN3* | TRUE | 1.985122616 |
| T1DM | beta_cell | *SFTA2* | TRUE | 2.701207225 |
| T1DM | beta_cell | *MVB12B* | TRUE | 2.447950535 |
| T1DM | beta_cell | *ZNF517* | TRUE | 2.356242474 |
| T1DM | beta_cell | *CCDC154* | TRUE | 2.259594379 |

| | | | | |
|---|---|---|---|---|
| T1DM | beta_cell | *HSPB9* | TRUE | 1.96651729 |
| T1DM | beta_cell | *RPS26* | TRUE | 3.089749274 |
| T1DM | beta_cell | *C2orf27A* | TRUE | 2.070515242 |
| T1DM | beta_cell | *HLA.DRB5* | TRUE | 2.524555138 |
| T1DM | beta_cell | *HSD17B8* | TRUE | 7.656909119 |
| T1DM | beta_cell | *NOTCH4* | TRUE | 2.584206907 |
| T1DM | beta_cell | *PSORS1C1* | TRUE | 1.982120884 |
| T1DM | beta_cell | *HLA.DQB2* | TRUE | 9.028860491 |
| T1DM | beta_cell | *PSMB9* | TRUE | 2.388460186 |
| T1DM | beta_cell | *HLA.DOB* | TRUE | 6.262548882 |
| T1DM | beta_cell | *HLA.DMB* | TRUE | 5.880130148 |
| T1DM | beta_cell | *GOLGA8K* | TRUE | 3.014647256 |
| T1DM | delta_cell | *ARHGAP22* | TRUE | 2.303790005 |
| T1DM | delta_cell | *ZNF589* | TRUE | 2.842909926 |
| T1DM | delta_cell | *ENGASE* | TRUE | 2.304319419 |
| T1DM | delta_cell | *FAM228A* | TRUE | 2.178024995 |
| T1DM | delta_cell | *SFTA2* | TRUE | 2.977025014 |
| T1DM | delta_cell | *HLA.DRB5* | TRUE | 2.158470401 |
| T1DM | delta_cell | *HSD17B8* | TRUE | 6.917263192 |
| T1DM | delta_cell | *NOTCH4* | TRUE | 4.173463039 |
| T1DM | delta_cell | *HLA.DQB2* | TRUE | 7.910569819 |
| T1DM | delta_cell | *PSMB9* | TRUE | 2.228337488 |
| T1DM | delta_cell | *HLA.DOB* | TRUE | 6.359277496 |
| T1DM | delta_cell | *TAS2R43* | TRUE | 2.055118216 |
| T1DM | duct_epithelial_cell | *ARHGAP22* | TRUE | 2.304773146 |
| T1DM | duct_epithelial_cell | *CDK20* | TRUE | 2.191666092 |
| T1DM | duct_epithelial_cell | *ZNF589* | TRUE | 2.200697662 |
| T1DM | duct_epithelial_cell | *ENGASE* | TRUE | 2.487327545 |
| T1DM | duct_epithelial_cell | *MORN4* | TRUE | 1.97230932 |
| T1DM | duct_epithelial_cell | *RPRML* | TRUE | 1.961464783 |
| T1DM | duct_epithelial_cell | *FAM228A* | TRUE | 1.985759106 |
| T1DM | duct_epithelial_cell | *SFTA2* | TRUE | 2.755481765 |
| T1DM | duct_epithelial_cell | *HLA.DRB5* | TRUE | 2.317664633 |
| T1DM | duct_epithelial_cell | *HSD17B8* | TRUE | 7.044755751 |
| T1DM | duct_epithelial_cell | *NOTCH4* | TRUE | 2.273477623 |
| T1DM | duct_epithelial_cell | *HLA.DQB2* | TRUE | 9.023931516 |
| T1DM | duct_epithelial_cell | *HLA.DOB* | TRUE | 5.521327075 |
| T1DM | duct_epithelial_cell | *HLA.DMB* | TRUE | 5.665865059 |
| T1DM | duct_epithelial_cell | *GOLGA8K* | TRUE | 2.092175859 |
| T1DM | endothelial_cell | *ARHGAP22* | TRUE | 2.017191669 |
| T1DM | endothelial_cell | *VPS11* | TRUE | 2.261124675 |
| T1DM | endothelial_cell | *ENGASE* | TRUE | 2.877059185 |
| T1DM | endothelial_cell | *SNTB1* | TRUE | 2.008521637 |
| T1DM | endothelial_cell | *HSD17B8* | TRUE | 5.798181612 |
| T1DM | endothelial_cell | *ATF6B* | TRUE | 2.80913099 |
| T1DM | endothelial_cell | *HLA.DQB2* | TRUE | 9.604418917 |
| T1DM | endothelial_cell | *HLA.DOB* | TRUE | 7.117175505 |
| T1DM | endothelial_cell | *HLA.DMB* | TRUE | 4.256928155 |
| T1DM | endothelial_cell | *TAS2R43* | TRUE | 2.069265431 |
| T1DM | epsilon_cell | *SFTA2* | TRUE | 3.265879749 |

| | | | | |
|---|---|---|---|---|
| T1DM | epsilon_cell | *HLA.DMA* | TRUE | 3.232776767 |
| T1DM | epsilon_cell | *NOTCH4* | TRUE | 2.743097643 |
| T1DM | epsilon_cell | *PSORS1C1* | TRUE | 2.23303672 |
| T1DM | epsilon_cell | *HLA.DQB2* | TRUE | 2.997978075 |
| T1DM | epsilon_cell | *HLA.DOB* | TRUE | 6.830137518 |
| T1DM | leukocyte | *SFTA2* | TRUE | 2.530126897 |
| T1DM | leukocyte | *PSORS1C1* | TRUE | 2.518054906 |
| T1DM | leukocyte | *HLA.DQB2* | TRUE | 2.128458218 |
| T1DM | leukocyte | *HLA.DOB* | TRUE | 4.128528944 |
| T1DM | leukocyte | *C4A* | TRUE | 4.644546888 |
| T1DM | mesenchymal_cell | *CTSH* | TRUE | 2.19684091 |
| T1DM | mesenchymal_cell | *ENGASE* | TRUE | 3.087012529 |
| T1DM | mesenchymal_cell | *RPRML* | TRUE | 2.133280828 |
| T1DM | mesenchymal_cell | *CDK10* | TRUE | 2.054201903 |
| T1DM | mesenchymal_cell | *LYPD6* | TRUE | 1.997658674 |
| T1DM | mesenchymal_cell | *SFTA2* | TRUE | 3.646186202 |
| T1DM | mesenchymal_cell | *HSD17B8* | TRUE | 4.741012407 |
| T1DM | mesenchymal_cell | *HLA.DMA* | TRUE | 4.958232859 |
| T1DM | mesenchymal_cell | *PSORS1C1* | TRUE | 1.991733606 |
| T1DM | mesenchymal_cell | *HLA.DQB2* | TRUE | 4.687684094 |
| T1DM | mesenchymal_cell | *HLA.DOB* | TRUE | 6.282607679 |
| T1DM | PP_cell | *SFTA2* | TRUE | 2.572871659 |
| T1DM | PP_cell | *HLA.DMA* | TRUE | 3.500176417 |
| T1DM | PP_cell | *NOTCH4* | TRUE | 3.736808286 |
| T1DM | PP_cell | *PSORS1C1* | TRUE | 2.408580169 |
| T1DM | PP_cell | *ATF6B* | TRUE | 2.456807306 |
| T1DM | PP_cell | *HLA.DQB2* | TRUE | 7.451785622 |
| T1DM | PP_cell | *HLA.DOB* | TRUE | 6.777560107 |
| T2DM | acinar_cell | *RBM6* | TRUE | 2.576545433 |
| T2DM | acinar_cell | *MLXIPL* | TRUE | 2.457919973 |
| T2DM | acinar_cell | *QPCTL* | TRUE | 2.083857893 |
| T2DM | acinar_cell | *CEP68* | TRUE | 3.294729016 |
| T2DM | acinar_cell | *BAK1* | TRUE | 2.65831765 |
| T2DM | acinar_cell | *TCF3* | TRUE | 2.699186947 |
| T2DM | acinar_cell | *ST6GAL1* | TRUE | 2.133222151 |
| T2DM | acinar_cell | *CWF19L1* | TRUE | 2.970279289 |
| T2DM | acinar_cell | *CCDC134* | TRUE | 2.494564823 |
| T2DM | acinar_cell | *SLC1A1* | TRUE | 2.182805078 |
| T2DM | acinar_cell | *CNTNAP1* | TRUE | 2.089252718 |
| T2DM | acinar_cell | *BTNL8* | TRUE | 2.13 |
| T2DM | acinar_cell | *DNAH7* | TRUE | 2.228588852 |
| T2DM | acinar_cell | *ZNF211* | TRUE | 2.054908908 |
| T2DM | acinar_cell | *FLT3* | TRUE | 3.260980927 |
| T2DM | acinar_cell | *OPRL1* | TRUE | 3.085659707 |
| T2DM | acinar_cell | *RIBC2* | TRUE | 2.483235675 |
| T2DM | acinar_cell | *NOTCH2* | TRUE | 4.902154906 |
| T2DM | acinar_cell | *LRP4* | TRUE | 2.689001752 |
| T2DM | acinar_cell | *MAN2C1* | TRUE | 3.295470717 |
| T2DM | acinar_cell | *HSD17B12* | TRUE | 2.790330299 |
| T2DM | acinar_cell | *APIP* | TRUE | 2.274434877 |

| | | | | |
|---|---|---|---|---|
| T2DM | acinar_cell | *PRSS53* | TRUE | 2.130611577 |
| T2DM | acinar_cell | *PTPN14* | TRUE | 2.760929446 |
| T2DM | acinar_cell | *POC5* | TRUE | 2.016237569 |
| T2DM | acinar_cell | *JAZF1* | TRUE | 5.412746096 |
| T2DM | acinar_cell | *CLEC18A* | TRUE | 5.32058161 |
| T2DM | acinar_cell | *EXTL1* | TRUE | 2.100875034 |
| T2DM | acinar_cell | *ANKLE1* | TRUE | 2.454977248 |
| T2DM | acinar_cell | *ICA1L* | TRUE | 2.036983589 |
| T2DM | acinar_cell | *PPM1M* | TRUE | 2.142453458 |
| T2DM | acinar_cell | *TMC8* | TRUE | 2.045990126 |
| T2DM | acinar_cell | *IRX3* | TRUE | 9.566213635 |
| T2DM | acinar_cell | *GPR88* | TRUE | 1.962244756 |
| T2DM | acinar_cell | *VMO1* | TRUE | 3.18168685 |
| T2DM | acinar_cell | *ZNF320* | TRUE | 2.56206333 |
| T2DM | acinar_cell | *FAM43B* | TRUE | 3.117137547 |
| T2DM | acinar_cell | *PIWIL2* | TRUE | 2.416840807 |
| T2DM | acinar_cell | *NOTCH4* | TRUE | 3.220598918 |
| T2DM | acinar_cell | *HCG27* | TRUE | 2.659216208 |
| T2DM | acinar_cell | *DNLZ* | TRUE | 3.274865271 |
| T2DM | acinar_cell | *SLC12A8* | TRUE | 2.757676871 |
| T2DM | acinar_cell | *HLA.DQA2* | TRUE | 3.749651891 |
| T2DM | acinar_cell | *HMBS* | TRUE | 2.184343307 |
| T2DM | alpha_cell | *RBM6* | TRUE | 2.976751944 |
| T2DM | alpha_cell | *MLXIPL* | TRUE | 3.058946996 |
| T2DM | alpha_cell | *GIPR* | TRUE | 2.307092364 |
| T2DM | alpha_cell | *CEP68* | TRUE | 4.203392815 |
| T2DM | alpha_cell | *LTF* | TRUE | 2.237029979 |
| T2DM | alpha_cell | *BAK1* | TRUE | 3.434895929 |
| T2DM | alpha_cell | *ITIH4* | TRUE | 2.734774845 |
| T2DM | alpha_cell | *TCF3* | TRUE | 3.133363765 |
| T2DM | alpha_cell | *ST6GAL1* | TRUE | 2.440809565 |
| T2DM | alpha_cell | *MARK3* | TRUE | 2.689954655 |
| T2DM | alpha_cell | *CWF19L1* | TRUE | 3.436667661 |
| T2DM | alpha_cell | *CCDC134* | TRUE | 2.888777255 |
| T2DM | alpha_cell | *SLC1A1* | TRUE | 2.199326536 |
| T2DM | alpha_cell | *CDK2AP1* | TRUE | 2.210137074 |
| T2DM | alpha_cell | *NNT* | TRUE | 2.196550625 |
| T2DM | alpha_cell | *BTNL8* | TRUE | 2.746462089 |
| T2DM | alpha_cell | *DNAH7* | TRUE | 3.311372097 |
| T2DM | alpha_cell | *ZNF211* | TRUE | 2.338087774 |
| T2DM | alpha_cell | *FLT3* | TRUE | 3.148931401 |
| T2DM | alpha_cell | *PTPN12* | TRUE | 2.166191181 |
| T2DM | alpha_cell | *RIBC2* | TRUE | 2.651603696 |
| T2DM | alpha_cell | *MAU2* | TRUE | 2.765915027 |
| T2DM | alpha_cell | *SYT5* | TRUE | 2.17717667 |
| T2DM | alpha_cell | *PPARG* | TRUE | 2.445737115 |
| T2DM | alpha_cell | *NOTCH2* | TRUE | 5.189354821 |
| T2DM | alpha_cell | *LRP4* | TRUE | 2.442637036 |
| T2DM | alpha_cell | *MYPN* | TRUE | 2.585576929 |
| T2DM | alpha_cell | *MAN2C1* | TRUE | 3.88376582 |

| | | | | |
|---|---|---|---|---|
| T2DM | alpha_cell | *ZNF514* | TRUE | 2.167530562 |
| T2DM | alpha_cell | *CDCA7* | TRUE | 2.497253106 |
| T2DM | alpha_cell | *CNTN4* | TRUE | 2.771421753 |
| T2DM | alpha_cell | *RAB19* | TRUE | 2.602875183 |
| T2DM | alpha_cell | *ADHFE1* | TRUE | 2.075832787 |
| T2DM | alpha_cell | *HSD17B12* | TRUE | 3.299178686 |
| T2DM | alpha_cell | *APIP* | TRUE | 2.801287099 |
| T2DM | alpha_cell | *KLHL35* | TRUE | 2.13791723 |
| T2DM | alpha_cell | *CCDC15* | TRUE | 2.102551542 |
| T2DM | alpha_cell | *PRSS53* | TRUE | 2.253966079 |
| T2DM | alpha_cell | *PTPN14* | TRUE | 3.137631812 |
| T2DM | alpha_cell | *POC5* | TRUE | 2.446938423 |
| T2DM | alpha_cell | *JAZF1* | TRUE | 7.106285034 |
| T2DM | alpha_cell | *CC2D1B* | TRUE | 2.567656439 |
| T2DM | alpha_cell | *CLEC18A* | TRUE | 5.307842964 |
| T2DM | alpha_cell | *CIB4* | TRUE | 2.552236291 |
| T2DM | alpha_cell | *EXTL1* | TRUE | 2.613552467 |
| T2DM | alpha_cell | *ANKLE1* | TRUE | 2.396666484 |
| T2DM | alpha_cell | *ICA1L* | TRUE | 2.505810632 |
| T2DM | alpha_cell | *BDH2* | TRUE | 2.150220062 |
| T2DM | alpha_cell | *PPM1M* | TRUE | 2.119514454 |
| T2DM | alpha_cell | *CASP7* | TRUE | 3.295959952 |
| T2DM | alpha_cell | *CYB5R2* | TRUE | 2.145537665 |
| T2DM | alpha_cell | *TMC8* | TRUE | 2.803663337 |
| T2DM | alpha_cell | *INO80E* | TRUE | 3.067875584 |
| T2DM | alpha_cell | *CTSW* | TRUE | 1.97071294 |
| T2DM | alpha_cell | *ZNF738* | TRUE | 2.253214722 |
| T2DM | alpha_cell | *TUBB8* | TRUE | 2.162365059 |
| T2DM | alpha_cell | *PHYHD1* | TRUE | 2.905977225 |
| T2DM | alpha_cell | *DSCAML1* | TRUE | 2.472192286 |
| T2DM | alpha_cell | *IRX3* | TRUE | 9.385655156 |
| T2DM | alpha_cell | *FARSA* | TRUE | 2.159906949 |
| T2DM | alpha_cell | *GPR88* | TRUE | 2.601678955 |
| T2DM | alpha_cell | *CAPN12* | TRUE | 2.117372409 |
| T2DM | alpha_cell | *VMO1* | TRUE | 3.155844566 |
| T2DM | alpha_cell | *ZNF320* | TRUE | 3.308018729 |
| T2DM | alpha_cell | *CAMK1D* | TRUE | 2.202904901 |
| T2DM | alpha_cell | *FAM43B* | TRUE | 2.697097222 |
| T2DM | alpha_cell | *CEACAM19* | TRUE | 2.066055129 |
| T2DM | alpha_cell | *ZNF385C* | TRUE | 1.967267386 |
| T2DM | alpha_cell | *NUTM2B* | TRUE | 3.306569017 |
| T2DM | alpha_cell | *ZDHHC11* | TRUE | 2.022305292 |
| T2DM | alpha_cell | *PIWIL2* | TRUE | 3.076017766 |
| T2DM | alpha_cell | *PCDHB11* | TRUE | 2.146399703 |
| T2DM | alpha_cell | *TBKBP1* | TRUE | 2.111870126 |
| T2DM | alpha_cell | *NOTCH4* | TRUE | 3.416745696 |
| T2DM | alpha_cell | *HCG27* | TRUE | 2.827369438 |
| T2DM | alpha_cell | *DNLZ* | TRUE | 3.416659019 |
| T2DM | alpha_cell | *SLC12A8* | TRUE | 3.712861387 |
| T2DM | alpha_cell | *LINC00957* | TRUE | 2.126418598 |

| | | | | |
|---|---|---|---|---|
| T2DM | alpha_cell | *HLA.DQA2* | TRUE | 3.674970447 |
| T2DM | alpha_cell | *JMJD7* | TRUE | 2.18597019 |
| T2DM | alpha_cell | *OR2A7* | TRUE | 2.006470987 |
| T2DM | beta_cell | *RBM6* | TRUE | 2.441592207 |
| T2DM | beta_cell | *BAIAP3* | TRUE | 2.129697803 |
| T2DM | beta_cell | *MLXIPL* | TRUE | 3.77544523 |
| T2DM | beta_cell | *GIPR* | TRUE | 2.530736973 |
| T2DM | beta_cell | *CEP68* | TRUE | 5.24414491 |
| T2DM | beta_cell | *LTF* | TRUE | 2.646812768 |
| T2DM | beta_cell | *ANGEL1* | TRUE | 2.012180226 |
| T2DM | beta_cell | *BAK1* | TRUE | 3.771321536 |
| T2DM | beta_cell | *ANAPC4* | TRUE | 2.072619006 |
| T2DM | beta_cell | *ITIH4* | TRUE | 3.164791684 |
| T2DM | beta_cell | *MYLK* | TRUE | 1.990882616 |
| T2DM | beta_cell | *SPTB* | TRUE | 2.596246142 |
| T2DM | beta_cell | *TCF3* | TRUE | 3.674320977 |
| T2DM | beta_cell | *NLE1* | TRUE | 2.516969538 |
| T2DM | beta_cell | *ST6GAL1* | TRUE | 3.223649501 |
| T2DM | beta_cell | *MARK3* | TRUE | 3.962145402 |
| T2DM | beta_cell | *SEC31B* | TRUE | 1.986481092 |
| T2DM | beta_cell | *MECOM* | TRUE | 2.024841031 |
| T2DM | beta_cell | *MGST2* | TRUE | 2.40069319 |
| T2DM | beta_cell | *SH3BP2* | TRUE | 2.072719838 |
| T2DM | beta_cell | *CWF19L1* | TRUE | 3.695147595 |
| T2DM | beta_cell | *CCDC134* | TRUE | 2.905795672 |
| T2DM | beta_cell | *AP1B1* | TRUE | 2.067602322 |
| T2DM | beta_cell | *TOM1* | TRUE | 2.309383724 |
| T2DM | beta_cell | *BIK* | TRUE | 2.344073754 |
| T2DM | beta_cell | *DIDO1* | TRUE | 2.029953786 |
| T2DM | beta_cell | *RBL2* | TRUE | 2.732297786 |
| T2DM | beta_cell | *NOMO1* | TRUE | 2.81315716 |
| T2DM | beta_cell | *CTSH* | TRUE | 2.045562937 |
| T2DM | beta_cell | *VIPR2* | TRUE | 2.014607138 |
| T2DM | beta_cell | *SLC1A1* | TRUE | 2.918811709 |
| T2DM | beta_cell | *DHRS11* | TRUE | 2.840751719 |
| T2DM | beta_cell | *SLC2A9* | TRUE | 2.193192156 |
| T2DM | beta_cell | *COQ5* | TRUE | 2.201440098 |
| T2DM | beta_cell | *CDK2AP1* | TRUE | 2.431949565 |
| T2DM | beta_cell | *SEMA5A* | TRUE | 2.23036194 |
| T2DM | beta_cell | *NNT* | TRUE | 2.937351507 |
| T2DM | beta_cell | *TMCO6* | TRUE | 2.459942326 |
| T2DM | beta_cell | *BTNL8* | TRUE | 2.772132391 |
| T2DM | beta_cell | *ANGPTL1* | TRUE | 2.271778762 |
| T2DM | beta_cell | *WDR77* | TRUE | 2.219304979 |
| T2DM | beta_cell | *DNAH7* | TRUE | 3.635122577 |
| T2DM | beta_cell | *BCL11A* | TRUE | 2.360782465 |
| T2DM | beta_cell | *IAPP* | TRUE | 3.203147668 |
| T2DM | beta_cell | *ZNF211* | TRUE | 2.458947401 |
| T2DM | beta_cell | *FLT3* | TRUE | 3.493880844 |
| T2DM | beta_cell | *GRK4* | TRUE | 1.980130144 |

| | | | | |
|---|---|---|---|---|
| T2DM | beta_cell | *OPRL1* | TRUE | 2.046735465 |
| T2DM | beta_cell | *WFIKKN1* | TRUE | 2.17495517 |
| T2DM | beta_cell | *PTPN12* | TRUE | 2.722958139 |
| T2DM | beta_cell | *RIBC2* | TRUE | 2.687890027 |
| T2DM | beta_cell | *MAU2* | TRUE | 2.082740369 |
| T2DM | beta_cell | *SYT5* | TRUE | 3.062069215 |
| T2DM | beta_cell | *GTPBP3* | TRUE | 2.113430679 |
| T2DM | beta_cell | *TRAF3* | TRUE | 2.452724921 |
| T2DM | beta_cell | *PPARG* | TRUE | 3.002579107 |
| T2DM | beta_cell | *INPP5K* | TRUE | 2.092543641 |
| T2DM | beta_cell | *ITGB4* | TRUE | 2.486212013 |
| T2DM | beta_cell | *BHMT2* | TRUE | 1.972308535 |
| T2DM | beta_cell | *NOTCH2* | TRUE | 5.435588748 |
| T2DM | beta_cell | *LRP4* | TRUE | 2.912522301 |
| T2DM | beta_cell | *NHSL1* | TRUE | 2.174443554 |
| T2DM | beta_cell | *MYPN* | TRUE | 2.63823858 |
| T2DM | beta_cell | *NUP54* | TRUE | 2.259209687 |
| T2DM | beta_cell | *C1RL* | TRUE | 2.256722116 |
| T2DM | beta_cell | *VPS53* | TRUE | 2.088386379 |
| T2DM | beta_cell | *SPACA3* | TRUE | 2.13102598 |
| T2DM | beta_cell | *NPC1* | TRUE | 3.016381366 |
| T2DM | beta_cell | *DUS3L* | TRUE | 2.332948287 |
| T2DM | beta_cell | *EVI5L* | TRUE | 2.222016381 |
| T2DM | beta_cell | *POGZ* | TRUE | 3.047356489 |
| T2DM | beta_cell | *ZNF514* | TRUE | 2.606459952 |
| T2DM | beta_cell | *CDCA7* | TRUE | 2.744222896 |
| T2DM | beta_cell | *TAMM41* | TRUE | 2.042140218 |
| T2DM | beta_cell | *CNTN4* | TRUE | 3.230010464 |
| T2DM | beta_cell | *TAGLN3* | TRUE | 2.185912202 |
| T2DM | beta_cell | *GIGYF1* | TRUE | 2.131332064 |
| T2DM | beta_cell | *RAB19* | TRUE | 2.609048847 |
| T2DM | beta_cell | *ADHFE1* | TRUE | 2.358774295 |
| T2DM | beta_cell | *SEC16A* | TRUE | 2.363857685 |
| T2DM | beta_cell | *HSD17B12* | TRUE | 3.603719245 |
| T2DM | beta_cell | *APIP* | TRUE | 2.019585872 |
| T2DM | beta_cell | *KLHL35* | TRUE | 2.519515016 |
| T2DM | beta_cell | *CCDC15* | TRUE | 2.058777425 |
| T2DM | beta_cell | *MAGI1* | TRUE | 2.173883982 |
| T2DM | beta_cell | *PTPN14* | TRUE | 3.381098644 |
| T2DM | beta_cell | *POC5* | TRUE | 2.878412644 |
| T2DM | beta_cell | *NMRAL1* | TRUE | 2.013367593 |
| T2DM | beta_cell | *JAZF1* | TRUE | 7.630344523 |
| T2DM | beta_cell | *CC2D1B* | TRUE | 2.866417137 |
| T2DM | beta_cell | *CLEC18A* | TRUE | 5.524846709 |
| T2DM | beta_cell | *AP3S2* | TRUE | 4.776205894 |
| T2DM | beta_cell | *CIB4* | TRUE | 2.927378729 |
| T2DM | beta_cell | *EXTL1* | TRUE | 2.596282703 |
| T2DM | beta_cell | *ANKLE1* | TRUE | 2.483222887 |
| T2DM | beta_cell | *ZNF208* | TRUE | 2.179766127 |
| T2DM | beta_cell | *NIPAL1* | TRUE | 2.184418214 |

| | | | | |
|---|---|---|---|---|
| T2DM | beta_cell | *SRGAP2* | TRUE | 2.658663811 |
| T2DM | beta_cell | *ICA1L* | TRUE | 2.849605838 |
| T2DM | beta_cell | *BDH2* | TRUE | 2.409353023 |
| T2DM | beta_cell | *PPM1M* | TRUE | 2.136586721 |
| T2DM | beta_cell | *ERAP2* | TRUE | 2.022466624 |
| T2DM | beta_cell | *HCN1* | TRUE | 2.136158142 |
| T2DM | beta_cell | *RELL2* | TRUE | 2.153895392 |
| T2DM | beta_cell | *DLC1* | TRUE | 1.991810871 |
| T2DM | beta_cell | *NOXRED1* | TRUE | 1.999319533 |
| T2DM | beta_cell | *CASP7* | TRUE | 3.62965135 |
| T2DM | beta_cell | *CYB5R2* | TRUE | 2.346922522 |
| T2DM | beta_cell | *RCCD1* | TRUE | 2.055667093 |
| T2DM | beta_cell | *PRR15L* | TRUE | 2.126814769 |
| T2DM | beta_cell | *SLC43A2* | TRUE | 2.130235476 |
| T2DM | beta_cell | *TMC8* | TRUE | 3.026210362 |
| T2DM | beta_cell | *TRANK1* | TRUE | 2.217446916 |
| T2DM | beta_cell | *INO80E* | TRUE | 2.802534281 |
| T2DM | beta_cell | *SMAGP* | TRUE | 2.813176286 |
| T2DM | beta_cell | *ARNT2* | TRUE | 2.186418939 |
| T2DM | beta_cell | *ZNF738* | TRUE | 2.709228418 |
| T2DM | beta_cell | *PPP1R3B* | TRUE | 2.802608535 |
| T2DM | beta_cell | *PEAK1* | TRUE | 2.514474874 |
| T2DM | beta_cell | *TUBB8* | TRUE | 2.312027609 |
| T2DM | beta_cell | *BTC* | TRUE | 2.203884076 |
| T2DM | beta_cell | *PHYHD1* | TRUE | 3.268872941 |
| T2DM | beta_cell | *TCP11L1* | TRUE | 2.081418306 |
| T2DM | beta_cell | *DSCAML1* | TRUE | 2.428576244 |
| T2DM | beta_cell | *IRX3* | TRUE | 10.06812724 |
| T2DM | beta_cell | *IL17RA* | TRUE | 2.061974221 |
| T2DM | beta_cell | *IMP3* | TRUE | 2.227981749 |
| T2DM | beta_cell | *FARSA* | TRUE | 2.178523232 |
| T2DM | beta_cell | *GPR88* | TRUE | 2.617856792 |
| T2DM | beta_cell | *CAPN12* | TRUE | 2.26962379 |
| T2DM | beta_cell | *VMO1* | TRUE | 3.44641396 |
| T2DM | beta_cell | *ZNF320* | TRUE | 3.349355416 |
| T2DM | beta_cell | *CAMK1D* | TRUE | 5.494862163 |
| T2DM | beta_cell | *FAM43B* | TRUE | 3.328959003 |
| T2DM | beta_cell | *ZBTB40* | TRUE | 3.012108404 |
| T2DM | beta_cell | *DMWD* | TRUE | 2.173099771 |
| T2DM | beta_cell | *CEACAM19* | TRUE | 2.388643044 |
| T2DM | beta_cell | *NUTM2B* | TRUE | 3.221623874 |
| T2DM | beta_cell | *NDOR1* | TRUE | 2.32124926 |
| T2DM | beta_cell | *ZDHHC11* | TRUE | 2.284286497 |
| T2DM | beta_cell | *ZNF567* | TRUE | 2.428848 |
| T2DM | beta_cell | *ZNF605* | TRUE | 2.356593992 |
| T2DM | beta_cell | *PIWIL2* | TRUE | 3.142947475 |
| T2DM | beta_cell | *PCDHB11* | TRUE | 2.290515957 |
| T2DM | beta_cell | *ENPP1* | TRUE | 2.054216016 |
| T2DM | beta_cell | *DDX42* | TRUE | 2.320247464 |
| T2DM | beta_cell | *KIAA0753* | TRUE | 2.186058552 |

| | | | | |
|---|---|---|---|---|
| T2DM | beta_cell | *TBKBP1* | TRUE | 2.350272733 |
| T2DM | beta_cell | *NOTCH4* | TRUE | 2.894665577 |
| T2DM | beta_cell | *ZNF425* | TRUE | 2.121965272 |
| T2DM | beta_cell | *HCG27* | TRUE | 2.893580463 |
| T2DM | beta_cell | *DNLZ* | TRUE | 3.31262243 |
| T2DM | beta_cell | *SLC12A8* | TRUE | 4.409199149 |
| T2DM | beta_cell | *EPPK1* | TRUE | 1.994168766 |
| T2DM | beta_cell | *LINC00957* | TRUE | 2.596825954 |
| T2DM | beta_cell | *HLA.DQA2* | TRUE | 3.921472999 |
| T2DM | beta_cell | *JMJD7* | TRUE | 2.465950996 |
| T2DM | beta_cell | *OR2A7* | TRUE | 2.062279896 |
| T2DM | beta_cell | *P2RY11* | TRUE | 2.391698933 |
| T2DM | beta_cell | *BCKDHA* | TRUE | 2.235185854 |
| T2DM | beta_cell | *FPGT* | TRUE | 2.394562033 |
| T2DM | beta_cell | *HMBS* | TRUE | 2.901305206 |
| T2DM | beta_cell | *TAS2R31* | TRUE | 2.026106102 |
| T2DM | delta_cell | *RBM6* | TRUE | 2.713325941 |
| T2DM | delta_cell | *SH2D2A* | TRUE | 2.262813531 |
| T2DM | delta_cell | *CCDC134* | TRUE | 2.438162093 |
| T2DM | delta_cell | *FLT3* | TRUE | 2.267514043 |
| T2DM | delta_cell | *NOTCH2* | TRUE | 3.140123462 |
| T2DM | delta_cell | *MAN2C1* | TRUE | 2.88374485 |
| T2DM | delta_cell | *HSD17B12* | TRUE | 2.085401854 |
| T2DM | delta_cell | *POC5* | TRUE | 1.970862954 |
| T2DM | delta_cell | *JAZF1* | TRUE | 5.318832384 |
| T2DM | delta_cell | *CLEC18A* | TRUE | 2.002918578 |
| T2DM | delta_cell | *CILP2* | TRUE | 2.251151281 |
| T2DM | delta_cell | *CASP7* | TRUE | 2.160497509 |
| T2DM | delta_cell | *TMC8* | TRUE | 2.481810904 |
| T2DM | delta_cell | *CTSW* | TRUE | 2.497908153 |
| T2DM | delta_cell | *IRX3* | TRUE | 6.060713716 |
| T2DM | delta_cell | *FAM43B* | TRUE | 2.724330389 |
| T2DM | delta_cell | *NOTCH4* | TRUE | 2.832601259 |
| T2DM | delta_cell | *HLA.DQA2* | TRUE | 3.444935089 |
| T2DM | delta_cell | *TAS2R20* | TRUE | 2.238574174 |
| T2DM | duct_epithelial_cell | *CEP68* | TRUE | 3.829672282 |
| T2DM | duct_epithelial_cell | *BAK1* | TRUE | 2.203461808 |
| T2DM | duct_epithelial_cell | *CWF19L1* | TRUE | 2.812842676 |
| T2DM | duct_epithelial_cell | *CNTNAP1* | TRUE | 2.080349419 |
| T2DM | duct_epithelial_cell | *BTNL8* | TRUE | 2.280811284 |
| T2DM | duct_epithelial_cell | *ZNF211* | TRUE | 2.275895098 |
| T2DM | duct_epithelial_cell | *RIBC2* | TRUE | 2.535798516 |
| T2DM | duct_epithelial_cell | *NOTCH2* | TRUE | 3.337420104 |
| T2DM | duct_epithelial_cell | *LRP4* | TRUE | 2.030118736 |
| T2DM | duct_epithelial_cell | *MAN2C1* | TRUE | 3.2034066 |
| T2DM | duct_epithelial_cell | *CDCA7* | TRUE | 2.064024502 |
| T2DM | duct_epithelial_cell | *CNTN4* | TRUE | 2.67933297 |
| T2DM | duct_epithelial_cell | *HSD17B12* | TRUE | 2.160866115 |
| T2DM | duct_epithelial_cell | *APIP* | TRUE | 2.62840324 |
| T2DM | duct_epithelial_cell | *PTPN14* | TRUE | 1.968476853 |

| | | | | |
|---|---|---|---|---|
| T2DM | duct_epithelial_cell | *POC5* | TRUE | 2.190607144 |
| T2DM | duct_epithelial_cell | *JAZF1* | TRUE | 4.585462214 |
| T2DM | duct_epithelial_cell | *CC2D1B* | TRUE | 2.261532524 |
| T2DM | duct_epithelial_cell | *CLEC18A* | TRUE | 2.503503956 |
| T2DM | duct_epithelial_cell | *ANKLE1* | TRUE | 2.402139977 |
| T2DM | duct_epithelial_cell | *CTSW* | TRUE | 2.292876031 |
| T2DM | duct_epithelial_cell | *FARSA* | TRUE | 2.007238479 |
| T2DM | duct_epithelial_cell | *VMO1* | TRUE | 2.219127719 |
| T2DM | duct_epithelial_cell | *ZNF320* | TRUE | 3.351384334 |
| T2DM | duct_epithelial_cell | *CAMK1D* | TRUE | 3.161653471 |
| T2DM | duct_epithelial_cell | *PIWIL2* | TRUE | 2.980899584 |
| T2DM | duct_epithelial_cell | *NOTCH4* | TRUE | 3.340958694 |
| T2DM | duct_epithelial_cell | *DNLZ* | TRUE | 3.33287889 |
| T2DM | duct_epithelial_cell | *SLC12A8* | TRUE | 2.070471381 |
| T2DM | duct_epithelial_cell | *HLA.DQA2* | TRUE | 3.741807637 |
| T2DM | duct_epithelial_cell | *HMBS* | TRUE | 2.212612901 |
| T2DM | endothelial_cell | *RBM6* | TRUE | 3.89599485 |
| T2DM | endothelial_cell | *CEP68* | TRUE | 3.760714201 |
| T2DM | endothelial_cell | *BAK1* | TRUE | 2.713624189 |
| T2DM | endothelial_cell | *ITIH4* | TRUE | 2.269227592 |
| T2DM | endothelial_cell | *CWF19L1* | TRUE | 3.425850317 |
| T2DM | endothelial_cell | *ZNF211* | TRUE | 2.318015403 |
| T2DM | endothelial_cell | *MAU2* | TRUE | 2.366976448 |
| T2DM | endothelial_cell | *NOTCH2* | TRUE | 4.796408821 |
| T2DM | endothelial_cell | *MAN2C1* | TRUE | 3.582469158 |
| T2DM | endothelial_cell | *HSD17B12* | TRUE | 2.443312044 |
| T2DM | endothelial_cell | *APIP* | TRUE | 2.830194306 |
| T2DM | endothelial_cell | *JAZF1* | TRUE | 4.20460242 |
| T2DM | endothelial_cell | *CC2D1B* | TRUE | 2.023897167 |
| T2DM | endothelial_cell | *EXTL1* | TRUE | 2.4344008 |
| T2DM | endothelial_cell | *ANKLE1* | TRUE | 2.106224384 |
| T2DM | endothelial_cell | *INO80E* | TRUE | 2.200510148 |
| T2DM | endothelial_cell | *IRX3* | TRUE | 6.580233749 |
| T2DM | endothelial_cell | *GPR88* | TRUE | 2.132412371 |
| T2DM | endothelial_cell | *CAPN12* | TRUE | 2.139109919 |
| T2DM | endothelial_cell | *ZNF320* | TRUE | 3.247204716 |
| T2DM | endothelial_cell | *FAM43B* | TRUE | 3.199212766 |
| T2DM | endothelial_cell | *PIWIL2* | TRUE | 2.946186508 |
| T2DM | endothelial_cell | *PCDHB11* | TRUE | 2.020121776 |
| T2DM | endothelial_cell | *RNF5* | TRUE | 2.865388819 |
| T2DM | endothelial_cell | *DNLZ* | TRUE | 3.156286918 |
| T2DM | endothelial_cell | *HLA.DQA2* | TRUE | 4.668022347 |
| T2DM | endothelial_cell | *JMJD7* | TRUE | 3.200757257 |
| T2DM | epsilon_cell | *BTNL8* | TRUE | 2.166451722 |
| T2DM | epsilon_cell | *CILP2* | TRUE | 2.890407974 |
| T2DM | epsilon_cell | *IRX3* | TRUE | 5.917803869 |
| T2DM | epsilon_cell | *UBA7* | TRUE | 4.270001918 |
| T2DM | epsilon_cell | *NOTCH4* | TRUE | 2.033136859 |
| T2DM | epsilon_cell | *HLA.DQA2* | TRUE | 5.47048731 |
| T2DM | leukocyte | *QPCTL* | TRUE | 2.089579782 |

| | | | | |
|---|---|---|---|---|
| T2DM | leukocyte | *NOTCH2* | TRUE | 4.757076166 |
| T2DM | leukocyte | *CC2D1B* | TRUE | 1.983036503 |
| T2DM | leukocyte | *ART3* | TRUE | 2.05294946 |
| T2DM | leukocyte | *EXTL1* | TRUE | 2.079576681 |
| T2DM | leukocyte | *CILP2* | TRUE | 2.629291141 |
| T2DM | leukocyte | *IRX3* | TRUE | 3.328168402 |
| T2DM | leukocyte | *GPR88* | TRUE | 1.960048263 |
| T2DM | leukocyte | *UBA7* | TRUE | 2.63499007 |
| T2DM | leukocyte | *NUTM2B* | TRUE | 2.343700505 |
| T2DM | leukocyte | *NOTCH4* | TRUE | 2.278348873 |
| T2DM | leukocyte | *DNLZ* | TRUE | 2.256858481 |
| T2DM | mesenchymal_cell | *CEP68* | TRUE | 2.846267998 |
| T2DM | mesenchymal_cell | *ITIH4* | TRUE | 2.114961144 |
| T2DM | mesenchymal_cell | *CWF19L1* | TRUE | 2.625458722 |
| T2DM | mesenchymal_cell | *SLC1A1* | TRUE | 1.995996574 |
| T2DM | mesenchymal_cell | *HSD17B1* | TRUE | 2.917811432 |
| T2DM | mesenchymal_cell | *BTNL8* | TRUE | 2.076689893 |
| T2DM | mesenchymal_cell | *ZNF211* | TRUE | 2.132072481 |
| T2DM | mesenchymal_cell | *RIBC2* | TRUE | 2.169389544 |
| T2DM | mesenchymal_cell | *NOTCH2* | TRUE | 1.962938942 |
| T2DM | mesenchymal_cell | *MAN2C1* | TRUE | 3.537466593 |
| T2DM | mesenchymal_cell | *CDCA7* | TRUE | 2.398782989 |
| T2DM | mesenchymal_cell | *CNTN4* | TRUE | 2.364365324 |
| T2DM | mesenchymal_cell | *CLEC18A* | TRUE | 2.273054733 |
| T2DM | mesenchymal_cell | *PPM1M* | TRUE | 2.287880911 |
| T2DM | mesenchymal_cell | *INO80E* | TRUE | 2.693714751 |
| T2DM | mesenchymal_cell | *CTSW* | TRUE | 2.36046353 |
| T2DM | mesenchymal_cell | *IRX3* | TRUE | 7.59264719 |
| T2DM | mesenchymal_cell | *ZNF320* | TRUE | 3.214740665 |
| T2DM | mesenchymal_cell | *PIWIL2* | TRUE | 2.281711582 |
| T2DM | mesenchymal_cell | *LIME1* | TRUE | 2.106711172 |
| T2DM | mesenchymal_cell | *HCG27* | TRUE | 2.06824214 |
| T2DM | mesenchymal_cell | *SLC12A8* | TRUE | 3.154212449 |
| T2DM | mesenchymal_cell | *JMJD7* | TRUE | 2.464442083 |
| T2DM | PP_cell | *RBM6* | TRUE | 2.409900913 |
| T2DM | PP_cell | *CEP68* | TRUE | 2.486734822 |
| T2DM | PP_cell | *CCDC134* | TRUE | 2.389161863 |
| T2DM | PP_cell | *OPRL1* | TRUE | 3.125195299 |
| T2DM | PP_cell | *RIBC2* | TRUE | 2.593177169 |
| T2DM | PP_cell | *SPTBN5* | TRUE | 2.253293458 |
| T2DM | PP_cell | *MAN2C1* | TRUE | 2.309690263 |
| T2DM | PP_cell | *HSD17B12* | TRUE | 2.377174632 |
| T2DM | PP_cell | *APIP* | TRUE | 2.25847276 |
| T2DM | PP_cell | *PTPN14* | TRUE | 2.319081518 |
| T2DM | PP_cell | *CLEC18A* | TRUE | 2.979866604 |
| T2DM | PP_cell | *CILP2* | TRUE | 2.564722325 |
| T2DM | PP_cell | *TMC8* | TRUE | 2.640148361 |
| T2DM | PP_cell | *IRX3* | TRUE | 2.915446669 |
| T2DM | PP_cell | *VMO1* | TRUE | 2.069487155 |
| T2DM | PP_cell | *ZNF320* | TRUE | 2.715362701 |

| | | | | |
|---|---|---|---|---|
| T2DM | PP_cell | *DNLZ* | TRUE | 2.006827921 |
| T2DM_admission | acinar_cell | *RBM6* | TRUE | 3.050975925 |
| T2DM_admission | acinar_cell | *MLXIPL* | TRUE | 2.22303948 |
| T2DM_admission | acinar_cell | *QPCTL* | TRUE | 2.166003561 |
| T2DM_admission | acinar_cell | *CEP68* | TRUE | 4.46563948 |
| T2DM_admission | acinar_cell | *BAK1* | TRUE | 3.332516336 |
| T2DM_admission | acinar_cell | *TCF3* | TRUE | 2.530763563 |
| T2DM_admission | acinar_cell | *ST6GAL1* | TRUE | 2.416874089 |
| T2DM_admission | acinar_cell | *CWF19L1* | TRUE | 2.321376805 |
| T2DM_admission | acinar_cell | *CCDC134* | TRUE | 2.41736115 |
| T2DM_admission | acinar_cell | *CRHR2* | TRUE | 2.079899981 |
| T2DM_admission | acinar_cell | *SLC1A1* | TRUE | 2.051158548 |
| T2DM_admission | acinar_cell | *CNTNAP1* | TRUE | 2.158522779 |
| T2DM_admission | acinar_cell | *ZNF211* | TRUE | 2.382631787 |
| T2DM_admission | acinar_cell | *FLT3* | TRUE | 3.516854694 |
| T2DM_admission | acinar_cell | *MOCS1* | TRUE | 2.167923285 |
| T2DM_admission | acinar_cell | *OPRL1* | TRUE | 3.420476055 |
| T2DM_admission | acinar_cell | *RIBC2* | TRUE | 2.178797841 |
| T2DM_admission | acinar_cell | *NOTCH2* | TRUE | 5.8768114 |
| T2DM_admission | acinar_cell | *MAN2C1* | TRUE | 3.296907327 |
| T2DM_admission | acinar_cell | *HSD17B12* | TRUE | 3.723518302 |
| T2DM_admission | acinar_cell | *APIP* | TRUE | 2.252838726 |
| T2DM_admission | acinar_cell | *CCDC15* | TRUE | 2.389136349 |
| T2DM_admission | acinar_cell | *PTPN14* | TRUE | 2.733776043 |
| T2DM_admission | acinar_cell | *POC5* | TRUE | 3.096895716 |
| T2DM_admission | acinar_cell | *JAZF1* | TRUE | 6.529028673 |
| T2DM_admission | acinar_cell | *CLEC18A* | TRUE | 6.517472336 |
| T2DM_admission | acinar_cell | *EXTL1* | TRUE | 2.751624592 |
| T2DM_admission | acinar_cell | *ANKLE1* | TRUE | 2.688459536 |
| T2DM_admission | acinar_cell | *SRGAP2* | TRUE | 2.060759663 |
| T2DM_admission | acinar_cell | *ICA1L* | TRUE | 2.317550956 |
| T2DM_admission | acinar_cell | *BDH2* | TRUE | 2.765511805 |
| T2DM_admission | acinar_cell | *TMEM129* | TRUE | 1.984566089 |
| T2DM_admission | acinar_cell | *PEAK1* | TRUE | 2.304625651 |
| T2DM_admission | acinar_cell | *GPR4* | TRUE | 2.106721233 |
| T2DM_admission | acinar_cell | *IRX3* | TRUE | 10.55301929 |
| T2DM_admission | acinar_cell | *FARSA* | TRUE | 2.597134172 |
| T2DM_admission | acinar_cell | *VMO1* | TRUE | 3.047950918 |
| T2DM_admission | acinar_cell | *ZNF320* | TRUE | 2.344073943 |
| T2DM_admission | acinar_cell | *FAM43B* | TRUE | 2.255205803 |
| T2DM_admission | acinar_cell | *PLGLB1* | TRUE | 1.969755154 |
| T2DM_admission | acinar_cell | *LIME1* | TRUE | 3.11265782 |
| T2DM_admission | acinar_cell | *NOTCH4* | TRUE | 3.309785579 |
| T2DM_admission | acinar_cell | *HCG27* | TRUE | 3.000021117 |
| T2DM_admission | acinar_cell | *DNLZ* | TRUE | 2.668558564 |
| T2DM_admission | acinar_cell | *SLC12A8* | TRUE | 2.35007725 |
| T2DM_admission | acinar_cell | *HLA.DQA2* | TRUE | 3.528563443 |
| T2DM_admission | acinar_cell | *CNTF* | TRUE | 2.110730023 |
| T2DM_admission | acinar_cell | *JMJD7* | TRUE | 2.901593377 |
| T2DM_admission | acinar_cell | *OR2A7* | TRUE | 2.164755598 |

| | | | | |
|---|---|---|---|---|
| T2DM_admission | acinar_cell | *P2RY11* | TRUE | 1.964266442 |
| T2DM_admission | acinar_cell | *HMBS* | TRUE | 2.40491079 |
| T2DM_admission | alpha_cell | *RBM6* | TRUE | 3.157918094 |
| T2DM_admission | alpha_cell | *MLXIPL* | TRUE | 2.926750745 |
| T2DM_admission | alpha_cell | *GIPR* | TRUE | 3.476131629 |
| T2DM_admission | alpha_cell | *CEP68* | TRUE | 5.192093176 |
| T2DM_admission | alpha_cell | *BAK1* | TRUE | 3.908055401 |
| T2DM_admission | alpha_cell | *ITIH4* | TRUE | 2.110622284 |
| T2DM_admission | alpha_cell | *TCF3* | TRUE | 3.08980857 |
| T2DM_admission | alpha_cell | *ST6GAL1* | TRUE | 2.896973148 |
| T2DM_admission | alpha_cell | *MYH7B* | TRUE | 2.195841922 |
| T2DM_admission | alpha_cell | *DCT* | TRUE | 1.996379008 |
| T2DM_admission | alpha_cell | *CWF19L1* | TRUE | 2.785163155 |
| T2DM_admission | alpha_cell | *PALMD* | TRUE | 2.149128283 |
| T2DM_admission | alpha_cell | *CCDC134* | TRUE | 3.114514894 |
| T2DM_admission | alpha_cell | *CRHR2* | TRUE | 2.279581276 |
| T2DM_admission | alpha_cell | *SLC1A1* | TRUE | 2.068350606 |
| T2DM_admission | alpha_cell | *CNTNAP1* | TRUE | 2.055681687 |
| T2DM_admission | alpha_cell | *CDK2AP1* | TRUE | 2.497477342 |
| T2DM_admission | alpha_cell | *CD83* | TRUE | 2.138814329 |
| T2DM_admission | alpha_cell | *BTNL8* | TRUE | 2.215709072 |
| T2DM_admission | alpha_cell | *DNAH7* | TRUE | 2.593801556 |
| T2DM_admission | alpha_cell | *ZNF211* | TRUE | 2.650981161 |
| T2DM_admission | alpha_cell | *FLT3* | TRUE | 3.438520401 |
| T2DM_admission | alpha_cell | *MOCS1* | TRUE | 2.428218971 |
| T2DM_admission | alpha_cell | *OPRL1* | TRUE | 2.483461667 |
| T2DM_admission | alpha_cell | *RIBC2* | TRUE | 2.339222061 |
| T2DM_admission | alpha_cell | *PPARG* | TRUE | 2.452525463 |
| T2DM_admission | alpha_cell | *NOTCH2* | TRUE | 6.060722301 |
| T2DM_admission | alpha_cell | *MYPN* | TRUE | 2.685694276 |
| T2DM_admission | alpha_cell | *MAN2C1* | TRUE | 3.607010228 |
| T2DM_admission | alpha_cell | *SPACA3* | TRUE | 2.278468514 |
| T2DM_admission | alpha_cell | *FN3KRP* | TRUE | 2.644073236 |
| T2DM_admission | alpha_cell | *POGZ* | TRUE | 1.987544223 |
| T2DM_admission | alpha_cell | *CDCA7* | TRUE | 2.848268829 |
| T2DM_admission | alpha_cell | *ADHFE1* | TRUE | 2.113374368 |
| T2DM_admission | alpha_cell | *INPP5E* | TRUE | 1.962994936 |
| T2DM_admission | alpha_cell | *HSD17B12* | TRUE | 4.265878884 |
| T2DM_admission | alpha_cell | *APIP* | TRUE | 2.405102563 |
| T2DM_admission | alpha_cell | *KLHL35* | TRUE | 2.106643205 |
| T2DM_admission | alpha_cell | *CCDC15* | TRUE | 2.778526454 |
| T2DM_admission | alpha_cell | *PTPN14* | TRUE | 3.039690922 |
| T2DM_admission | alpha_cell | *POC5* | TRUE | 3.053674361 |
| T2DM_admission | alpha_cell | *JAZF1* | TRUE | 8.155551661 |
| T2DM_admission | alpha_cell | *CC2D1B* | TRUE | 2.153688483 |
| T2DM_admission | alpha_cell | *CLEC18A* | TRUE | 6.430380032 |
| T2DM_admission | alpha_cell | *AP3S2* | TRUE | 2.18180227 |
| T2DM_admission | alpha_cell | *CIB4* | TRUE | 2.466792899 |
| T2DM_admission | alpha_cell | *EXTL1* | TRUE | 3.364259831 |
| T2DM_admission | alpha_cell | *ANKLE1* | TRUE | 2.512900566 |

| | | | | |
|---|---|---|---|---|
| T2DM_admission | alpha_cell | *SRGAP2* | TRUE | 2.608495365 |
| T2DM_admission | alpha_cell | *ICA1L* | TRUE | 2.807559815 |
| T2DM_admission | alpha_cell | *BDH2* | TRUE | 2.874833389 |
| T2DM_admission | alpha_cell | *CASP7* | TRUE | 3.715004099 |
| T2DM_admission | alpha_cell | *EVPL* | TRUE | 2.129867839 |
| T2DM_admission | alpha_cell | *TMC8* | TRUE | 2.620978906 |
| T2DM_admission | alpha_cell | *TMEM129* | TRUE | 2.621308456 |
| T2DM_admission | alpha_cell | *INO80E* | TRUE | 2.743875515 |
| T2DM_admission | alpha_cell | *PEAK1* | TRUE | 2.838792101 |
| T2DM_admission | alpha_cell | *ZNF408* | TRUE | 2.260929568 |
| T2DM_admission | alpha_cell | *FUT2* | TRUE | 2.338818447 |
| T2DM_admission | alpha_cell | *DSCAML1* | TRUE | 2.59151451 |
| T2DM_admission | alpha_cell | *GPR4* | TRUE | 2.165556485 |
| T2DM_admission | alpha_cell | *IRX3* | TRUE | 10.34683147 |
| T2DM_admission | alpha_cell | *TH* | TRUE | 3.190226721 |
| T2DM_admission | alpha_cell | *VMO1* | TRUE | 2.969598698 |
| T2DM_admission | alpha_cell | *ZNF320* | TRUE | 2.961615518 |
| T2DM_admission | alpha_cell | *PLGLB1* | TRUE | 1.979378768 |
| T2DM_admission | alpha_cell | *EIF3C* | TRUE | 2.000405415 |
| T2DM_admission | alpha_cell | *NUTM2B* | TRUE | 3.559829011 |
| T2DM_admission | alpha_cell | *PIWIL2* | TRUE | 2.379400011 |
| T2DM_admission | alpha_cell | *LIME1* | TRUE | 2.715563458 |
| T2DM_admission | alpha_cell | *INPP5B* | TRUE | 1.980471019 |
| T2DM_admission | alpha_cell | *NOTCH4* | TRUE | 3.445004954 |
| T2DM_admission | alpha_cell | *HCG27* | TRUE | 3.258688654 |
| T2DM_admission | alpha_cell | *DNLZ* | TRUE | 2.951411338 |
| T2DM_admission | alpha_cell | *SLC12A8* | TRUE | 3.628170823 |
| T2DM_admission | alpha_cell | *EPPK1* | TRUE | 2.90130265 |
| T2DM_admission | alpha_cell | *HLA.DQA2* | TRUE | 3.426463187 |
| T2DM_admission | alpha_cell | *CNTF* | TRUE | 2.239555208 |
| T2DM_admission | alpha_cell | *JMJD7* | TRUE | 2.932501712 |
| T2DM_admission | alpha_cell | *OR2A7* | TRUE | 2.462014081 |
| T2DM_admission | alpha_cell | *P2RY11* | TRUE | 2.190673107 |
| T2DM_admission | alpha_cell | *HMBS* | TRUE | 2.320933808 |
| T2DM_admission | beta_cell | *STPG1* | TRUE | 2.270062378 |
| T2DM_admission | beta_cell | *RBM6* | TRUE | 2.938137549 |
| T2DM_admission | beta_cell | *ZNF195* | TRUE | 2.117381393 |
| T2DM_admission | beta_cell | *USH1C* | TRUE | 2.196782181 |
| T2DM_admission | beta_cell | *ST7L* | TRUE | 2.354559669 |
| T2DM_admission | beta_cell | *BAIAP3* | TRUE | 2.24630044 |
| T2DM_admission | beta_cell | *MLXIPL* | TRUE | 3.655157808 |
| T2DM_admission | beta_cell | *GIPR* | TRUE | 3.819763309 |
| T2DM_admission | beta_cell | *CEP68* | TRUE | 6.314835567 |
| T2DM_admission | beta_cell | *LTF* | TRUE | 2.377333816 |
| T2DM_admission | beta_cell | *BAK1* | TRUE | 4.63977353 |
| T2DM_admission | beta_cell | *ERCC8* | TRUE | 2.268743876 |
| T2DM_admission | beta_cell | *ANAPC4* | TRUE | 2.256920361 |
| T2DM_admission | beta_cell | *ITIH4* | TRUE | 2.73374573 |
| T2DM_admission | beta_cell | *MYLK* | TRUE | 2.623408425 |
| T2DM_admission | beta_cell | *SPTB* | TRUE | 2.948937666 |

| | | | | |
|---|---|---|---|---|
| T2DM_admission | beta_cell | *GBA2* | TRUE | 2.067759884 |
| T2DM_admission | beta_cell | *TCF3* | TRUE | 3.815196928 |
| T2DM_admission | beta_cell | *SCARB1* | TRUE | 1.975561607 |
| T2DM_admission | beta_cell | *NLE1* | TRUE | 2.396994092 |
| T2DM_admission | beta_cell | *ST6GAL1* | TRUE | 3.652726999 |
| T2DM_admission | beta_cell | *MARK3* | TRUE | 3.928684289 |
| T2DM_admission | beta_cell | *MYH7B* | TRUE | 2.166055767 |
| T2DM_admission | beta_cell | *DCT* | TRUE | 2.160757078 |
| T2DM_admission | beta_cell | *MGST2* | TRUE | 2.494252473 |
| T2DM_admission | beta_cell | *LYZ* | TRUE | 2.180699259 |
| T2DM_admission | beta_cell | *RPGRIP1* | TRUE | 2.096359059 |
| T2DM_admission | beta_cell | *CWF19L1* | TRUE | 3.133611115 |
| T2DM_admission | beta_cell | *PALMD* | TRUE | 2.450057575 |
| T2DM_admission | beta_cell | *CCDC134* | TRUE | 2.354883237 |
| T2DM_admission | beta_cell | *ARSA* | TRUE | 2.256984212 |
| T2DM_admission | beta_cell | *DIDO1* | TRUE | 2.061891228 |
| T2DM_admission | beta_cell | *OLFM4* | TRUE | 2.200266817 |
| T2DM_admission | beta_cell | *RBL2* | TRUE | 2.875156406 |
| T2DM_admission | beta_cell | *ATP8B4* | TRUE | 1.996998064 |
| T2DM_admission | beta_cell | *VIPR2* | TRUE | 2.518140093 |
| T2DM_admission | beta_cell | *CRHR2* | TRUE | 2.331138857 |
| T2DM_admission | beta_cell | *SLC1A1* | TRUE | 2.728405414 |
| T2DM_admission | beta_cell | *DHRS11* | TRUE | 2.752876723 |
| T2DM_admission | beta_cell | *CNTNAP1* | TRUE | 2.247315579 |
| T2DM_admission | beta_cell | *FAM149A* | TRUE | 2.243598058 |
| T2DM_admission | beta_cell | *CDK2AP1* | TRUE | 2.163474784 |
| T2DM_admission | beta_cell | *CD83* | TRUE | 2.348361036 |
| T2DM_admission | beta_cell | *SEMA5A* | TRUE | 1.980734004 |
| T2DM_admission | beta_cell | *NNT* | TRUE | 2.473913647 |
| T2DM_admission | beta_cell | *TMCO6* | TRUE | 2.008003889 |
| T2DM_admission | beta_cell | *BTNL8* | TRUE | 2.305316974 |
| T2DM_admission | beta_cell | *ANGPTL1* | TRUE | 2.226351765 |
| T2DM_admission | beta_cell | *WDR77* | TRUE | 2.06348739 |
| T2DM_admission | beta_cell | *ATP10B* | TRUE | 2.168192502 |
| T2DM_admission | beta_cell | *DNAH7* | TRUE | 3.040766901 |
| T2DM_admission | beta_cell | *BCL11A* | TRUE | 2.240311529 |
| T2DM_admission | beta_cell | *CCDC77* | TRUE | 2.524247099 |
| T2DM_admission | beta_cell | *FAM53C* | TRUE | 2.30319617 |
| T2DM_admission | beta_cell | *IAPP* | TRUE | 2.503418815 |
| T2DM_admission | beta_cell | *ZNF211* | TRUE | 2.731840145 |
| T2DM_admission | beta_cell | *FLT3* | TRUE | 3.758027051 |
| T2DM_admission | beta_cell | *BHLHE41* | TRUE | 2.249515023 |
| T2DM_admission | beta_cell | *MOCS1* | TRUE | 2.651564829 |
| T2DM_admission | beta_cell | *GRK4* | TRUE | 2.12448699 |
| T2DM_admission | beta_cell | *OPRL1* | TRUE | 2.612717375 |
| T2DM_admission | beta_cell | *ABHD8* | TRUE | 2.701952919 |
| T2DM_admission | beta_cell | *RIBC2* | TRUE | 2.370111037 |
| T2DM_admission | beta_cell | *CGNL1* | TRUE | 2.070766385 |
| T2DM_admission | beta_cell | *MAU2* | TRUE | 2.37782754 |
| T2DM_admission | beta_cell | *SYT5* | TRUE | 2.509282491 |

| | | | | |
|---|---|---|---|---|
| T2DM_admission | beta_cell | *GTPBP3* | TRUE | 2.196317747 |
| T2DM_admission | beta_cell | *TULP4* | TRUE | 2.110453981 |
| T2DM_admission | beta_cell | *TRAF3* | TRUE | 2.88661416 |
| T2DM_admission | beta_cell | *PPARG* | TRUE | 3.328506507 |
| T2DM_admission | beta_cell | *BHMT2* | TRUE | 2.595216804 |
| T2DM_admission | beta_cell | *NOTCH2* | TRUE | 6.461922382 |
| T2DM_admission | beta_cell | *NHSL1* | TRUE | 2.321550539 |
| T2DM_admission | beta_cell | *NEK1* | TRUE | 2.123457495 |
| T2DM_admission | beta_cell | *BTBD16* | TRUE | 2.059974836 |
| T2DM_admission | beta_cell | *MYPN* | TRUE | 2.715314056 |
| T2DM_admission | beta_cell | *NUP54* | TRUE | 2.482727877 |
| T2DM_admission | beta_cell | *C1RL* | TRUE | 2.163189185 |
| T2DM_admission | beta_cell | *SPACA3* | TRUE | 2.539108818 |
| T2DM_admission | beta_cell | *NPC1* | TRUE | 3.828866605 |
| T2DM_admission | beta_cell | *FN3KRP* | TRUE | 2.383489236 |
| T2DM_admission | beta_cell | *DUS3L* | TRUE | 2.343535178 |
| T2DM_admission | beta_cell | *EVI5L* | TRUE | 2.95743037 |
| T2DM_admission | beta_cell | *POGZ* | TRUE | 3.375319263 |
| T2DM_admission | beta_cell | *ZNF514* | TRUE | 2.093545233 |
| T2DM_admission | beta_cell | *CDCA7* | TRUE | 3.085745007 |
| T2DM_admission | beta_cell | *IQGAP2* | TRUE | 2.358228707 |
| T2DM_admission | beta_cell | *SCML4* | TRUE | 2.737726652 |
| T2DM_admission | beta_cell | *SLC22A3* | TRUE | 1.961277257 |
| T2DM_admission | beta_cell | *ADHFE1* | TRUE | 2.562151861 |
| T2DM_admission | beta_cell | *INPP5E* | TRUE | 2.019971137 |
| T2DM_admission | beta_cell | *HSD17B12* | TRUE | 4.691160776 |
| T2DM_admission | beta_cell | *KLHL35* | TRUE | 2.50735322 |
| T2DM_admission | beta_cell | *CCDC15* | TRUE | 2.743325562 |
| T2DM_admission | beta_cell | *PTPN14* | TRUE | 3.464755464 |
| T2DM_admission | beta_cell | *POC5* | TRUE | 3.147154136 |
| T2DM_admission | beta_cell | *PTPRD* | TRUE | 2.079974969 |
| T2DM_admission | beta_cell | *ZDHHC7* | TRUE | 2.431381279 |
| T2DM_admission | beta_cell | *JAZF1* | TRUE | 8.931030687 |
| T2DM_admission | beta_cell | *NUS1* | TRUE | 2.122682242 |
| T2DM_admission | beta_cell | *CC2D1B* | TRUE | 2.472508563 |
| T2DM_admission | beta_cell | *CLEC18A* | TRUE | 6.663642948 |
| T2DM_admission | beta_cell | *AP3S2* | TRUE | 5.501512794 |
| T2DM_admission | beta_cell | *CIB4* | TRUE | 2.856934943 |
| T2DM_admission | beta_cell | *EXTL1* | TRUE | 3.307543136 |
| T2DM_admission | beta_cell | *ANKLE1* | TRUE | 2.715422104 |
| T2DM_admission | beta_cell | *LY6K* | TRUE | 2.182349266 |
| T2DM_admission | beta_cell | *CLHC1* | TRUE | 2.191285079 |
| T2DM_admission | beta_cell | *DUSP19* | TRUE | 2.007571306 |
| T2DM_admission | beta_cell | *NIPAL1* | TRUE | 2.539769498 |
| T2DM_admission | beta_cell | *SRGAP2* | TRUE | 3.684447564 |
| T2DM_admission | beta_cell | *ICA1L* | TRUE | 3.289760335 |
| T2DM_admission | beta_cell | *BDH2* | TRUE | 3.116144577 |
| T2DM_admission | beta_cell | *ERAP2* | TRUE | 1.972993946 |
| T2DM_admission | beta_cell | *RELL2* | TRUE | 1.988449784 |
| T2DM_admission | beta_cell | *NOXRED1* | TRUE | 2.328893486 |

| | | | | |
|---|---|---|---|---|
| T2DM_admission | beta_cell | *CASP7* | TRUE | 4.077540696 |
| T2DM_admission | beta_cell | *CEP57* | TRUE | 2.145443643 |
| T2DM_admission | beta_cell | *PROCA1* | TRUE | 2.066227982 |
| T2DM_admission | beta_cell | *TP53I13* | TRUE | 2.368004647 |
| T2DM_admission | beta_cell | *SLC43A2* | TRUE | 2.007085692 |
| T2DM_admission | beta_cell | *EVPL* | TRUE | 2.286218197 |
| T2DM_admission | beta_cell | *TMC8* | TRUE | 2.806402658 |
| T2DM_admission | beta_cell | *TMEM129* | TRUE | 2.407497371 |
| T2DM_admission | beta_cell | *INO80E* | TRUE | 2.809920619 |
| T2DM_admission | beta_cell | *KCNAB3* | TRUE | 2.142322152 |
| T2DM_admission | beta_cell | *SMAGP* | TRUE | 2.301254529 |
| T2DM_admission | beta_cell | *ARNT2* | TRUE | 2.58409781 |
| T2DM_admission | beta_cell | *RNASEH2C* | TRUE | 2.22746217 |
| T2DM_admission | beta_cell | *PPP1R3B* | TRUE | 2.550916569 |
| T2DM_admission | beta_cell | *PEAK1* | TRUE | 3.562995579 |
| T2DM_admission | beta_cell | *ZNF266* | TRUE | 2.274862994 |
| T2DM_admission | beta_cell | *YIF1A* | TRUE | 1.98878481 |
| T2DM_admission | beta_cell | *FUT1* | TRUE | 1.96453563 |
| T2DM_admission | beta_cell | *ZNF408* | TRUE | 2.789657219 |
| T2DM_admission | beta_cell | *PHYHD1* | TRUE | 2.147383424 |
| T2DM_admission | beta_cell | *TCP11L1* | TRUE | 2.197292972 |
| T2DM_admission | beta_cell | *FUT2* | TRUE | 2.308608072 |
| T2DM_admission | beta_cell | *DSCAML1* | TRUE | 2.550250618 |
| T2DM_admission | beta_cell | *GPR4* | TRUE | 2.159236756 |
| T2DM_admission | beta_cell | *IRX3* | TRUE | 10.99429859 |
| T2DM_admission | beta_cell | *COX14* | TRUE | 2.935952566 |
| T2DM_admission | beta_cell | *TRIM73* | TRUE | 2.227040946 |
| T2DM_admission | beta_cell | *RMI1* | TRUE | 2.104690822 |
| T2DM_admission | beta_cell | *FARSA* | TRUE | 2.25443105 |
| T2DM_admission | beta_cell | *TH* | TRUE | 3.248704845 |
| T2DM_admission | beta_cell | *ACTL10* | TRUE | 2.56542579 |
| T2DM_admission | beta_cell | *TTC3* | TRUE | 2.237209565 |
| T2DM_admission | beta_cell | *VMO1* | TRUE | 3.351475086 |
| T2DM_admission | beta_cell | *ZNF320* | TRUE | 3.010949229 |
| T2DM_admission | beta_cell | *CAMK1D* | TRUE | 4.977343257 |
| T2DM_admission | beta_cell | *FAM43B* | TRUE | 2.653848071 |
| T2DM_admission | beta_cell | *EIF3C* | TRUE | 2.079356829 |
| T2DM_admission | beta_cell | *PGP* | TRUE | 2.296974787 |
| T2DM_admission | beta_cell | *ZBTB40* | TRUE | 2.500907914 |
| T2DM_admission | beta_cell | *NDUFA6* | TRUE | 2.055715085 |
| T2DM_admission | beta_cell | *DMWD* | TRUE | 2.495873315 |
| T2DM_admission | beta_cell | *CYP4X1* | TRUE | 2.113784877 |
| T2DM_admission | beta_cell | *NUTM2B* | TRUE | 3.431703162 |
| T2DM_admission | beta_cell | *NDOR1* | TRUE | 2.034856784 |
| T2DM_admission | beta_cell | *ZNF567* | TRUE | 2.533778107 |
| T2DM_admission | beta_cell | *ZNF605* | TRUE | 2.100927209 |
| T2DM_admission | beta_cell | *PIWIL2* | TRUE | 2.524745754 |
| T2DM_admission | beta_cell | *ENPP1* | TRUE | 2.383637419 |
| T2DM_admission | beta_cell | *DPP4* | TRUE | 2.728950956 |
| T2DM_admission | beta_cell | *DDX42* | TRUE | 2.319995914 |

| | | | | |
|---|---|---|---|---|
| T2DM_admission | beta_cell | *KIAA0753* | TRUE | 2.05675933 |
| T2DM_admission | beta_cell | *LIME1* | TRUE | 2.55641902 |
| T2DM_admission | beta_cell | *INPP5B* | TRUE | 2.328401053 |
| T2DM_admission | beta_cell | *AGAP6* | TRUE | 1.966079918 |
| T2DM_admission | beta_cell | *NOTCH4* | TRUE | 3.427135561 |
| T2DM_admission | beta_cell | *NYNRIN* | TRUE | 2.078600015 |
| T2DM_admission | beta_cell | *HCG27* | TRUE | 3.326923897 |
| T2DM_admission | beta_cell | *DNLZ* | TRUE | 2.890356864 |
| T2DM_admission | beta_cell | *SLC12A8* | TRUE | 4.561866859 |
| T2DM_admission | beta_cell | *EPPK1* | TRUE | 3.001394741 |
| T2DM_admission | beta_cell | *PSMG3.AS1* | TRUE | 2.037884028 |
| T2DM_admission | beta_cell | *LINC00957* | TRUE | 2.432470031 |
| T2DM_admission | beta_cell | *HLA.DQA2* | TRUE | 3.632292361 |
| T2DM_admission | beta_cell | *CNTF* | TRUE | 2.099050699 |
| T2DM_admission | beta_cell | *JMJD7* | TRUE | 3.122839795 |
| T2DM_admission | beta_cell | *OR2A7* | TRUE | 2.536860944 |
| T2DM_admission | beta_cell | *P2RY11* | TRUE | 2.642159191 |
| T2DM_admission | beta_cell | *HMBS* | TRUE | 2.589847297 |
| T2DM_admission | beta_cell | *TAS2R31* | TRUE | 2.401313237 |
| T2DM_admission | delta_cell | *RBM6* | TRUE | 2.581248072 |
| T2DM_admission | delta_cell | *CEP68* | TRUE | 2.711119889 |
| T2DM_admission | delta_cell | *SH2D2A* | TRUE | 2.264729431 |
| T2DM_admission | delta_cell | *BAK1* | TRUE | 2.272588017 |
| T2DM_admission | delta_cell | *CCDC134* | TRUE | 2.487485482 |
| T2DM_admission | delta_cell | *FLT3* | TRUE | 2.212432964 |
| T2DM_admission | delta_cell | *NOTCH2* | TRUE | 3.671907689 |
| T2DM_admission | delta_cell | *MAN2C1* | TRUE | 2.803871775 |
| T2DM_admission | delta_cell | *HSD17B12* | TRUE | 2.803749514 |
| T2DM_admission | delta_cell | *POC5* | TRUE | 3.04382622 |
| T2DM_admission | delta_cell | *JAZF1* | TRUE | 6.121770142 |
| T2DM_admission | delta_cell | *CLEC18A* | TRUE | 2.572153153 |
| T2DM_admission | delta_cell | *CILP2* | TRUE | 2.200147185 |
| T2DM_admission | delta_cell | *CASP7* | TRUE | 2.452095648 |
| T2DM_admission | delta_cell | *TMC8* | TRUE | 2.088212961 |
| T2DM_admission | delta_cell | *FUT2* | TRUE | 2.409324895 |
| T2DM_admission | delta_cell | *IRX3* | TRUE | 6.437678318 |
| T2DM_admission | delta_cell | *FAM43B* | TRUE | 2.007260658 |
| T2DM_admission | delta_cell | *EPPK1* | TRUE | 2.096118881 |
| T2DM_admission | delta_cell | *HLA.DQA2* | TRUE | 2.175152578 |
| T2DM_admission | delta_cell | *TAS2R43* | TRUE | 2.317160735 |
| T2DM_admission | delta_cell | *TAS2R20* | TRUE | 2.487124538 |
| T2DM_admission | duct_epithelial_cell | *QPCTL* | TRUE | 2.143002278 |
| T2DM_admission | duct_epithelial_cell | *CEP68* | TRUE | 4.96889593 |
| T2DM_admission | duct_epithelial_cell | *BAK1* | TRUE | 2.601577923 |
| T2DM_admission | duct_epithelial_cell | *CWF19L1* | TRUE | 2.012769764 |
| T2DM_admission | duct_epithelial_cell | *PALMD* | TRUE | 2.000732393 |
| T2DM_admission | duct_epithelial_cell | *CNTNAP1* | TRUE | 2.697028203 |
| T2DM_admission | duct_epithelial_cell | *CDK2AP1* | TRUE | 2.023911184 |
| T2DM_admission | duct_epithelial_cell | *ZNF211* | TRUE | 2.810278826 |
| T2DM_admission | duct_epithelial_cell | *RIBC2* | TRUE | 2.194382164 |

| | | | | |
|---|---|---|---|---|
| T2DM_admission | duct_epithelial_cell | *NOTCH2* | TRUE | 4.500853905 |
| T2DM_admission | duct_epithelial_cell | *MAN2C1* | TRUE | 3.022399638 |
| T2DM_admission | duct_epithelial_cell | *FN3KRP* | TRUE | 2.460691336 |
| T2DM_admission | duct_epithelial_cell | *CDCA7* | TRUE | 2.452502437 |
| T2DM_admission | duct_epithelial_cell | *HSD17B12* | TRUE | 2.82059998 |
| T2DM_admission | duct_epithelial_cell | *CCDC15* | TRUE | 2.471828083 |
| T2DM_admission | duct_epithelial_cell | *PTPN14* | TRUE | 2.045035383 |
| T2DM_admission | duct_epithelial_cell | *POC5* | TRUE | 3.21835657 |
| T2DM_admission | duct_epithelial_cell | *JAZF1* | TRUE | 4.993858523 |
| T2DM_admission | duct_epithelial_cell | *CLEC18A* | TRUE | 3.045318241 |
| T2DM_admission | duct_epithelial_cell | *ANKLE1* | TRUE | 2.575019094 |
| T2DM_admission | duct_epithelial_cell | *TMEM129* | TRUE | 1.969731622 |
| T2DM_admission | duct_epithelial_cell | *CTSW* | TRUE | 2.08454582 |
| T2DM_admission | duct_epithelial_cell | *PEAK1* | TRUE | 2.571794989 |
| T2DM_admission | duct_epithelial_cell | *FUT2* | TRUE | 2.268402666 |
| T2DM_admission | duct_epithelial_cell | *FARSA* | TRUE | 2.912451942 |
| T2DM_admission | duct_epithelial_cell | *ZNF320* | TRUE | 3.017879079 |
| T2DM_admission | duct_epithelial_cell | *CAMK1D* | TRUE | 2.590951858 |
| T2DM_admission | duct_epithelial_cell | *PIWIL2* | TRUE | 2.309896038 |
| T2DM_admission | duct_epithelial_cell | *LIME1* | TRUE | 2.237626297 |
| T2DM_admission | duct_epithelial_cell | *NOTCH4* | TRUE | 3.57527807 |
| T2DM_admission | duct_epithelial_cell | *DNLZ* | TRUE | 2.862182718 |
| T2DM_admission | duct_epithelial_cell | *SLC12A8* | TRUE | 2.132343481 |
| T2DM_admission | duct_epithelial_cell | *HLA.DQA2* | TRUE | 2.865186915 |
| T2DM_admission | duct_epithelial_cell | *HMBS* | TRUE | 2.671370075 |
| T2DM_admission | endothelial_cell | *RBM6* | TRUE | 3.938336456 |
| T2DM_admission | endothelial_cell | *CEP68* | TRUE | 4.549017279 |
| T2DM_admission | endothelial_cell | *BAK1* | TRUE | 3.295943692 |
| T2DM_admission | endothelial_cell | *CWF19L1* | TRUE | 2.774716331 |
| T2DM_admission | endothelial_cell | *CRHR2* | TRUE | 2.006427447 |
| T2DM_admission | endothelial_cell | *ZNF211* | TRUE | 2.712361695 |
| T2DM_admission | endothelial_cell | *NOTCH2* | TRUE | 5.657559486 |
| T2DM_admission | endothelial_cell | *MAN2C1* | TRUE | 3.689987623 |
| T2DM_admission | endothelial_cell | *NPC1* | TRUE | 2.044114061 |
| T2DM_admission | endothelial_cell | *FN3KRP* | TRUE | 1.976488435 |
| T2DM_admission | endothelial_cell | *HSD17B12* | TRUE | 3.163671452 |
| T2DM_admission | endothelial_cell | *APIP* | TRUE | 2.935210798 |
| T2DM_admission | endothelial_cell | *CCDC15* | TRUE | 2.389564971 |
| T2DM_admission | endothelial_cell | *POC5* | TRUE | 2.465142653 |
| T2DM_admission | endothelial_cell | *JAZF1* | TRUE | 4.89160374 |
| T2DM_admission | endothelial_cell | *EXTL1* | TRUE | 2.41460634 |
| T2DM_admission | endothelial_cell | *ANKLE1* | TRUE | 2.248397417 |
| T2DM_admission | endothelial_cell | *TMEM129* | TRUE | 2.136718595 |
| T2DM_admission | endothelial_cell | *FUT2* | TRUE | 2.446079716 |
| T2DM_admission | endothelial_cell | *IRX3* | TRUE | 7.49326076 |
| T2DM_admission | endothelial_cell | *ZNF320* | TRUE | 2.974051218 |
| T2DM_admission | endothelial_cell | *FAM43B* | TRUE | 2.41608965 |
| T2DM_admission | endothelial_cell | *PIWIL2* | TRUE | 2.234692993 |
| T2DM_admission | endothelial_cell | *LIME1* | TRUE | 3.135335432 |
| T2DM_admission | endothelial_cell | *RNF5* | TRUE | 3.250279004 |

| | | | | |
|---|---|---|---|---|
| T2DM_admission | endothelial_cell | *DNLZ* | TRUE | 2.63349165 |
| T2DM_admission | endothelial_cell | *HLA.DQA2* | TRUE | 5.013940934 |
| T2DM_admission | endothelial_cell | *JMJD7* | TRUE | 3.716840544 |
| T2DM_admission | endothelial_cell | *TAS2R43* | TRUE | 2.461439019 |
| T2DM_admission | epsilon_cell | *OPRL1* | TRUE | 2.13782944 |
| T2DM_admission | epsilon_cell | *CILP2* | TRUE | 3.012035436 |
| T2DM_admission | epsilon_cell | *IP6K3* | TRUE | 2.177416845 |
| T2DM_admission | epsilon_cell | *IRX3* | TRUE | 5.895579204 |
| T2DM_admission | epsilon_cell | *UBA7* | TRUE | 4.251084578 |
| T2DM_admission | epsilon_cell | *NOTCH4* | TRUE | 2.227472467 |
| T2DM_admission | epsilon_cell | *TAS2R43* | TRUE | 2.383287683 |
| T2DM_admission | leukocyte | *QPCTL* | TRUE | 2.384226893 |
| T2DM_admission | leukocyte | *NOTCH2* | TRUE | 5.540252879 |
| T2DM_admission | leukocyte | *POC5* | TRUE | 2.580255736 |
| T2DM_admission | leukocyte | *CLEC18A* | TRUE | 2.059824543 |
| T2DM_admission | leukocyte | *CILP2* | TRUE | 2.807600299 |
| T2DM_admission | leukocyte | *IRX3* | TRUE | 3.466935934 |
| T2DM_admission | leukocyte | *UBA7* | TRUE | 2.469102687 |
| T2DM_admission | leukocyte | *PLGLB1* | TRUE | 2.073396928 |
| T2DM_admission | leukocyte | *NUTM2B* | TRUE | 2.163305278 |
| T2DM_admission | leukocyte | *OR2A7* | TRUE | 2.326391462 |
| T2DM_admission | leukocyte | *TAS2R20* | TRUE | 2.133428638 |
| T2DM_admission | mesenchymal_cell | *RBM6* | TRUE | 3.307895251 |
| T2DM_admission | mesenchymal_cell | *CEP68* | TRUE | 3.695609535 |
| T2DM_admission | mesenchymal_cell | *BAK1* | TRUE | 2.158783948 |
| T2DM_admission | mesenchymal_cell | *CWF19L1* | TRUE | 1.991038981 |
| T2DM_admission | mesenchymal_cell | *HSD17B1* | TRUE | 3.310378806 |
| T2DM_admission | mesenchymal_cell | *ZNF211* | TRUE | 2.421576816 |
| T2DM_admission | mesenchymal_cell | *NOTCH2* | TRUE | 2.448952326 |
| T2DM_admission | mesenchymal_cell | *MAN2C1* | TRUE | 3.499004503 |
| T2DM_admission | mesenchymal_cell | *FN3KRP* | TRUE | 2.30472128 |
| T2DM_admission | mesenchymal_cell | *CDCA7* | TRUE | 2.725011987 |
| T2DM_admission | mesenchymal_cell | *HSD17B12* | TRUE | 2.771742704 |
| T2DM_admission | mesenchymal_cell | *CCDC15* | TRUE | 2.568755337 |
| T2DM_admission | mesenchymal_cell | *POC5* | TRUE | 2.285083072 |
| T2DM_admission | mesenchymal_cell | *CLEC18A* | TRUE | 2.964138922 |
| T2DM_admission | mesenchymal_cell | *TMEM129* | TRUE | 2.255389963 |
| T2DM_admission | mesenchymal_cell | *INO80E* | TRUE | 2.302083746 |
| T2DM_admission | mesenchymal_cell | *FUT2* | TRUE | 2.576370037 |
| T2DM_admission | mesenchymal_cell | *IRX3* | TRUE | 8.348023457 |
| T2DM_admission | mesenchymal_cell | *ZNF320* | TRUE | 2.760535735 |
| T2DM_admission | mesenchymal_cell | *LIME1* | TRUE | 3.238258732 |
| T2DM_admission | mesenchymal_cell | *HCG27* | TRUE | 2.397921671 |
| T2DM_admission | mesenchymal_cell | *SLC12A8* | TRUE | 2.8741022 |
| T2DM_admission | mesenchymal_cell | *JMJD7* | TRUE | 2.871151081 |
| T2DM_admission | mesenchymal_cell | *TAS2R43* | TRUE | 2.437445876 |
| T2DM_admission | PP_cell | *RBM6* | TRUE | 2.125509595 |
| T2DM_admission | PP_cell | *CEP68* | TRUE | 2.661860631 |
| T2DM_admission | PP_cell | *CCDC134* | TRUE | 2.270347953 |
| T2DM_admission | PP_cell | *OPRL1* | TRUE | 3.504839914 |

| | | | | |
|---|---|---|---|---|
| T2DM_admission | PP_cell | *RIBC2* | TRUE | 2.258778266 |
| T2DM_admission | PP_cell | *SPTBN5* | TRUE | 2.870454911 |
| T2DM_admission | PP_cell | *MAN2C1* | TRUE | 2.019125429 |
| T2DM_admission | PP_cell | *HSD17B12* | TRUE | 3.152729812 |
| T2DM_admission | PP_cell | *APIP* | TRUE | 2.022742486 |
| T2DM_admission | PP_cell | *CCDC15* | TRUE | 2.049961059 |
| T2DM_admission | PP_cell | *PTPN14* | TRUE | 2.15611758 |
| T2DM_admission | PP_cell | *JAZF1* | TRUE | 2.513048958 |
| T2DM_admission | PP_cell | *CLEC18A* | TRUE | 4.047278555 |
| T2DM_admission | PP_cell | *CILP2* | TRUE | 2.417643417 |
| T2DM_admission | PP_cell | *IP6K3* | TRUE | 2.118816149 |
| T2DM_admission | PP_cell | *TMC8* | TRUE | 2.40700774 |
| T2DM_admission | PP_cell | *IRX3* | TRUE | 3.222702474 |
| T2DM_admission | PP_cell | *ZNF320* | TRUE | 2.349159114 |
| T2DM_insulin | acinar_cell | *CEP68* | TRUE | 2.923357142 |
| T2DM_insulin | acinar_cell | *ALOX5* | TRUE | 2.641552327 |
| T2DM_insulin | acinar_cell | *SPTB* | TRUE | 1.965425146 |
| T2DM_insulin | acinar_cell | *TF* | TRUE | 2.227346976 |
| T2DM_insulin | acinar_cell | *OIP5* | TRUE | 2.022327701 |
| T2DM_insulin | acinar_cell | *BTNL8* | TRUE | 3.492986939 |
| T2DM_insulin | acinar_cell | *ZNF211* | TRUE | 2.217596256 |
| T2DM_insulin | acinar_cell | *PKDREJ* | TRUE | 2.274117011 |
| T2DM_insulin | acinar_cell | *NOTCH2* | TRUE | 2.321346937 |
| T2DM_insulin | acinar_cell | *LRP4* | TRUE | 1.968865811 |
| T2DM_insulin | acinar_cell | *HSPG2* | TRUE | 2.020773685 |
| T2DM_insulin | acinar_cell | *RNF175* | TRUE | 2.570075572 |
| T2DM_insulin | acinar_cell | *TAGLN* | TRUE | 1.967232553 |
| T2DM_insulin | acinar_cell | *CLEC18A* | TRUE | 3.447296132 |
| T2DM_insulin | acinar_cell | *EXTL1* | TRUE | 3.579706928 |
| T2DM_insulin | acinar_cell | *NIPAL1* | TRUE | 2.166237779 |
| T2DM_insulin | acinar_cell | *ZNF589* | TRUE | 2.111142252 |
| T2DM_insulin | acinar_cell | *CXCL11* | TRUE | 2.424840719 |
| T2DM_insulin | acinar_cell | *IRX3* | TRUE | 5.858406783 |
| T2DM_insulin | acinar_cell | *ZNF223* | TRUE | 2.706213999 |
| T2DM_insulin | acinar_cell | *DNAJC22* | TRUE | 2.115995261 |
| T2DM_insulin | acinar_cell | *RMI1* | TRUE | 2.298870016 |
| T2DM_insulin | acinar_cell | *RPRML* | TRUE | 2.146972128 |
| T2DM_insulin | acinar_cell | *ZNF320* | TRUE | 2.125595062 |
| T2DM_insulin | acinar_cell | *EIF3C* | TRUE | 2.508569026 |
| T2DM_insulin | acinar_cell | *LRRC19* | TRUE | 2.499023323 |
| T2DM_insulin | acinar_cell | *HYAL3* | TRUE | 2.53914026 |
| T2DM_insulin | acinar_cell | *FAM47E* | TRUE | 1.968177808 |
| T2DM_insulin | acinar_cell | *PIWIL2* | TRUE | 2.221433182 |
| T2DM_insulin | acinar_cell | *PAX9* | TRUE | 2.528663609 |
| T2DM_insulin | acinar_cell | *NOTCH4* | TRUE | 2.5724545 |
| T2DM_insulin | acinar_cell | *PSORS1C1* | TRUE | 2.228650836 |
| T2DM_insulin | acinar_cell | *LILRB3* | TRUE | 1.999880119 |
| T2DM_insulin | acinar_cell | *HLA.DQB2* | TRUE | 3.141611163 |
| T2DM_insulin | acinar_cell | *HLA.DOB* | TRUE | 2.186789301 |
| T2DM_insulin | acinar_cell | *CDK3* | TRUE | 2.874500562 |

| | | | | |
|---|---|---|---|---|
| T2DM_insulin | acinar_cell | *TAS2R43* | TRUE | 2.284242451 |
| T2DM_insulin | alpha_cell | *CEP68* | TRUE | 2.808518176 |
| T2DM_insulin | alpha_cell | *ALOX5* | TRUE | 2.967492067 |
| T2DM_insulin | alpha_cell | *OSBPL5* | TRUE | 2.30280234 |
| T2DM_insulin | alpha_cell | *ZDHHC6* | TRUE | 2.12770309 |
| T2DM_insulin | alpha_cell | *EPHA3* | TRUE | 2.205257736 |
| T2DM_insulin | alpha_cell | *EPN3* | TRUE | 1.986557149 |
| T2DM_insulin | alpha_cell | *SEC61A2* | TRUE | 2.628137066 |
| T2DM_insulin | alpha_cell | *SPTB* | TRUE | 1.994253775 |
| T2DM_insulin | alpha_cell | *ANO8* | TRUE | 2.548224378 |
| T2DM_insulin | alpha_cell | *TF* | TRUE | 2.315106095 |
| T2DM_insulin | alpha_cell | *MYBL2* | TRUE | 2.277161321 |
| T2DM_insulin | alpha_cell | *SLC17A9* | TRUE | 1.961409409 |
| T2DM_insulin | alpha_cell | *OIP5* | TRUE | 2.20590932 |
| T2DM_insulin | alpha_cell | *BTNL8* | TRUE | 3.348280479 |
| T2DM_insulin | alpha_cell | *PDC* | TRUE | 1.990954685 |
| T2DM_insulin | alpha_cell | *ZNF211* | TRUE | 2.206325293 |
| T2DM_insulin | alpha_cell | *MAU2* | TRUE | 2.028733095 |
| T2DM_insulin | alpha_cell | *PKDREJ* | TRUE | 2.865637717 |
| T2DM_insulin | alpha_cell | *RHPN2* | TRUE | 1.967283745 |
| T2DM_insulin | alpha_cell | *NOTCH2* | TRUE | 2.787415683 |
| T2DM_insulin | alpha_cell | *EVI5L* | TRUE | 1.988247817 |
| T2DM_insulin | alpha_cell | *HSPG2* | TRUE | 2.130134458 |
| T2DM_insulin | alpha_cell | *RNF175* | TRUE | 2.252096891 |
| T2DM_insulin | alpha_cell | *RAB19* | TRUE | 2.329418213 |
| T2DM_insulin | alpha_cell | *CABLES2* | TRUE | 2.139512304 |
| T2DM_insulin | alpha_cell | *SETD9* | TRUE | 2.143109219 |
| T2DM_insulin | alpha_cell | *CLEC18A* | TRUE | 3.421138588 |
| T2DM_insulin | alpha_cell | *EXTL1* | TRUE | 3.605321192 |
| T2DM_insulin | alpha_cell | *NIPAL1* | TRUE | 1.973347302 |
| T2DM_insulin | alpha_cell | *ZNF589* | TRUE | 2.06433603 |
| T2DM_insulin | alpha_cell | *CXCL11* | TRUE | 2.291951351 |
| T2DM_insulin | alpha_cell | *FUT10* | TRUE | 2.268679568 |
| T2DM_insulin | alpha_cell | *IRX3* | TRUE | 5.429803187 |
| T2DM_insulin | alpha_cell | *ZNF223* | TRUE | 2.728335556 |
| T2DM_insulin | alpha_cell | *DNAJC22* | TRUE | 2.32509577 |
| T2DM_insulin | alpha_cell | *RMI1* | TRUE | 2.294825135 |
| T2DM_insulin | alpha_cell | *RPRML* | TRUE | 2.160083782 |
| T2DM_insulin | alpha_cell | *ZNF320* | TRUE | 2.281124886 |
| T2DM_insulin | alpha_cell | *EIF3C* | TRUE | 2.313723561 |
| T2DM_insulin | alpha_cell | *PLA2G6* | TRUE | 2.172005509 |
| T2DM_insulin | alpha_cell | *LRRC19* | TRUE | 2.815960061 |
| T2DM_insulin | alpha_cell | *ZBTB40* | TRUE | 2.212955946 |
| T2DM_insulin | alpha_cell | *HYAL3* | TRUE | 2.403067252 |
| T2DM_insulin | alpha_cell | *FAM47E* | TRUE | 2.167148616 |
| T2DM_insulin | alpha_cell | *ZNF676* | TRUE | 1.98566799 |
| T2DM_insulin | alpha_cell | *PIWIL2* | TRUE | 2.972154473 |
| T2DM_insulin | alpha_cell | *HSPB9* | TRUE | 2.148282375 |
| T2DM_insulin | alpha_cell | *PAX9* | TRUE | 2.966914067 |
| T2DM_insulin | alpha_cell | *L3MBTL3* | TRUE | 2.034432946 |

| | | | | |
|---|---|---|---|---|
| T2DM_insulin | alpha_cell | *NOTCH4* | TRUE | 2.542758172 |
| T2DM_insulin | alpha_cell | *PSORS1C1* | TRUE | 2.265996009 |
| T2DM_insulin | alpha_cell | *LILRB3* | TRUE | 1.982953349 |
| T2DM_insulin | alpha_cell | *GMNC* | TRUE | 2.022459986 |
| T2DM_insulin | alpha_cell | *HLA.DQB2* | TRUE | 3.39185583 |
| T2DM_insulin | alpha_cell | *HLA.DOB* | TRUE | 2.15216655 |
| T2DM_insulin | alpha_cell | *OR2A7* | TRUE | 2.137523128 |
| T2DM_insulin | alpha_cell | *TAS2R43* | TRUE | 2.30722153 |
| T2DM_insulin | beta_cell | *CEP68* | TRUE | 3.61954482 |
| T2DM_insulin | beta_cell | *ALOX5* | TRUE | 2.615268808 |
| T2DM_insulin | beta_cell | *OSBPL5* | TRUE | 2.350707524 |
| T2DM_insulin | beta_cell | *ZDHHC6* | TRUE | 2.609783731 |
| T2DM_insulin | beta_cell | *BAK1* | TRUE | 2.027724088 |
| T2DM_insulin | beta_cell | *EPHA3* | TRUE | 2.178943412 |
| T2DM_insulin | beta_cell | *PTGER3* | TRUE | 2.119054754 |
| T2DM_insulin | beta_cell | *SEC61A2* | TRUE | 2.492805019 |
| T2DM_insulin | beta_cell | *SPTB* | TRUE | 2.118820431 |
| T2DM_insulin | beta_cell | *ANO8* | TRUE | 2.754284299 |
| T2DM_insulin | beta_cell | *ACACB* | TRUE | 2.107232283 |
| T2DM_insulin | beta_cell | *CAD* | TRUE | 1.97975518 |
| T2DM_insulin | beta_cell | *DZANK1* | TRUE | 1.984839527 |
| T2DM_insulin | beta_cell | *TF* | TRUE | 2.321345778 |
| T2DM_insulin | beta_cell | *OCEL1* | TRUE | 2.116429464 |
| T2DM_insulin | beta_cell | *MYBL2* | TRUE | 2.192597437 |
| T2DM_insulin | beta_cell | *MAP1LC3A* | TRUE | 2.174156986 |
| T2DM_insulin | beta_cell | *OIP5* | TRUE | 2.367267723 |
| T2DM_insulin | beta_cell | *ETHE1* | TRUE | 2.118968155 |
| T2DM_insulin | beta_cell | *SLC35F2* | TRUE | 2.002031308 |
| T2DM_insulin | beta_cell | *BACH2* | TRUE | 2.232569706 |
| T2DM_insulin | beta_cell | *SEMA5A* | TRUE | 2.200989008 |
| T2DM_insulin | beta_cell | *BTNL8* | TRUE | 3.694425977 |
| T2DM_insulin | beta_cell | *PDC* | TRUE | 2.003577519 |
| T2DM_insulin | beta_cell | *BMP8B* | TRUE | 2.149884965 |
| T2DM_insulin | beta_cell | *IAPP* | TRUE | 2.486910432 |
| T2DM_insulin | beta_cell | *ZNF211* | TRUE | 2.259005695 |
| T2DM_insulin | beta_cell | *PEPD* | TRUE | 2.133709426 |
| T2DM_insulin | beta_cell | *CLYBL* | TRUE | 2.199168238 |
| T2DM_insulin | beta_cell | *PTPN12* | TRUE | 2.149694053 |
| T2DM_insulin | beta_cell | *MAU2* | TRUE | 2.061447476 |
| T2DM_insulin | beta_cell | *PKDREJ* | TRUE | 2.81475999 |
| T2DM_insulin | beta_cell | *F12* | TRUE | 2.363832664 |
| T2DM_insulin | beta_cell | *TRAF3* | TRUE | 2.547919213 |
| T2DM_insulin | beta_cell | *RHPN2* | TRUE | 2.053310557 |
| T2DM_insulin | beta_cell | *NOTCH2* | TRUE | 2.875405319 |
| T2DM_insulin | beta_cell | *LRP4* | TRUE | 2.137069129 |
| T2DM_insulin | beta_cell | *GRHPR* | TRUE | 2.119013659 |
| T2DM_insulin | beta_cell | *EVI5L* | TRUE | 2.612178545 |
| T2DM_insulin | beta_cell | *HSPG2* | TRUE | 2.156179786 |
| T2DM_insulin | beta_cell | *CNTN4* | TRUE | 2.025968218 |
| T2DM_insulin | beta_cell | *NXPE3* | TRUE | 2.197163824 |

| | | | | |
|---|---|---|---|---|
| T2DM_insulin | beta_cell | *RNF175* | TRUE | 2.544833807 |
| T2DM_insulin | beta_cell | *RAB19* | TRUE | 2.354027017 |
| T2DM_insulin | beta_cell | *TAGLN* | TRUE | 2.294533938 |
| T2DM_insulin | beta_cell | *CABLES2* | TRUE | 2.295079009 |
| T2DM_insulin | beta_cell | *ZNF827* | TRUE | 1.972030719 |
| T2DM_insulin | beta_cell | *AKR1C2* | TRUE | 2.070165099 |
| T2DM_insulin | beta_cell | *KCNE4* | TRUE | 2.145601814 |
| T2DM_insulin | beta_cell | *JAZF1* | TRUE | 2.274886558 |
| T2DM_insulin | beta_cell | *SETD9* | TRUE | 2.34592684 |
| T2DM_insulin | beta_cell | *TMED6* | TRUE | 2.086477511 |
| T2DM_insulin | beta_cell | *CLEC18A* | TRUE | 3.563505599 |
| T2DM_insulin | beta_cell | *EXTL1* | TRUE | 3.57431253 |
| T2DM_insulin | beta_cell | *TONSL* | TRUE | 2.13771782 |
| T2DM_insulin | beta_cell | *NIPAL1* | TRUE | 3.148011784 |
| T2DM_insulin | beta_cell | *SRGAP2* | TRUE | 2.34458633 |
| T2DM_insulin | beta_cell | *CRELD1* | TRUE | 2.61269523 |
| T2DM_insulin | beta_cell | *ZNF589* | TRUE | 2.068102852 |
| T2DM_insulin | beta_cell | *KLKB1* | TRUE | 2.452060537 |
| T2DM_insulin | beta_cell | *LIPC* | TRUE | 2.234870547 |
| T2DM_insulin | beta_cell | *KIAA1586* | TRUE | 2.664952916 |
| T2DM_insulin | beta_cell | *SHOX2* | TRUE | 2.211276872 |
| T2DM_insulin | beta_cell | *CXCL11* | TRUE | 2.419444813 |
| T2DM_insulin | beta_cell | *FUT10* | TRUE | 2.525512256 |
| T2DM_insulin | beta_cell | *ANKLE2* | TRUE | 2.186545816 |
| T2DM_insulin | beta_cell | *IRX3* | TRUE | 6.031667436 |
| T2DM_insulin | beta_cell | *SAMD12* | TRUE | 2.415745929 |
| T2DM_insulin | beta_cell | *ZNF223* | TRUE | 2.692923732 |
| T2DM_insulin | beta_cell | *DNAJC22* | TRUE | 2.684920317 |
| T2DM_insulin | beta_cell | *RMI1* | TRUE | 2.584424094 |
| T2DM_insulin | beta_cell | *RPRML* | TRUE | 2.12933326 |
| T2DM_insulin | beta_cell | *TTC3* | TRUE | 2.083405819 |
| T2DM_insulin | beta_cell | *ZNF320* | TRUE | 2.300132792 |
| T2DM_insulin | beta_cell | *KREMEN1* | TRUE | 2.120424534 |
| T2DM_insulin | beta_cell | *TMEM236* | TRUE | 2.192737172 |
| T2DM_insulin | beta_cell | *EIF3C* | TRUE | 2.974327286 |
| T2DM_insulin | beta_cell | *PLA2G6* | TRUE | 2.441058406 |
| T2DM_insulin | beta_cell | *LRRC19* | TRUE | 2.867440492 |
| T2DM_insulin | beta_cell | *ZBTB40* | TRUE | 3.352137578 |
| T2DM_insulin | beta_cell | *NDUFA6* | TRUE | 2.167404307 |
| T2DM_insulin | beta_cell | *ZNF74* | TRUE | 2.256036796 |
| T2DM_insulin | beta_cell | *HYAL3* | TRUE | 2.586081613 |
| T2DM_insulin | beta_cell | *C17orf97* | TRUE | 2.033838116 |
| T2DM_insulin | beta_cell | *AGAP4* | TRUE | 2.000151779 |
| T2DM_insulin | beta_cell | *FAM47E* | TRUE | 2.370235214 |
| T2DM_insulin | beta_cell | *S100A14* | TRUE | 2.346913695 |
| T2DM_insulin | beta_cell | *ZNF676* | TRUE | 2.049031613 |
| T2DM_insulin | beta_cell | *PIWIL2* | TRUE | 2.931273379 |
| T2DM_insulin | beta_cell | *HSPB9* | TRUE | 2.296087163 |
| T2DM_insulin | beta_cell | *OCLN* | TRUE | 2.46475167 |
| T2DM_insulin | beta_cell | *C2orf27A* | TRUE | 2.034675468 |

| | | | | |
|---|---|---|---|---|
| T2DM_insulin | beta_cell | *PAX9* | TRUE | 3.088941254 |
| T2DM_insulin | beta_cell | *L3MBTL3* | TRUE | 2.137251524 |
| T2DM_insulin | beta_cell | *NOTCH4* | TRUE | 2.288387159 |
| T2DM_insulin | beta_cell | *PSORS1C1* | TRUE | 2.266947455 |
| T2DM_insulin | beta_cell | *LILRB3* | TRUE | 2.149463254 |
| T2DM_insulin | beta_cell | *GMNC* | TRUE | 2.1802163 |
| T2DM_insulin | beta_cell | *C1QTNF9* | TRUE | 1.996616496 |
| T2DM_insulin | beta_cell | *HLA.DMB* | TRUE | 2.288611696 |
| T2DM_insulin | beta_cell | *OR2A7* | TRUE | 2.14609967 |
| T2DM_insulin | beta_cell | *LIMS3* | TRUE | 2.167372994 |
| T2DM_insulin | delta_cell | *ALOX5* | TRUE | 2.701911976 |
| T2DM_insulin | delta_cell | *OSBPL5* | TRUE | 2.033626 |
| T2DM_insulin | delta_cell | *BTNL8* | TRUE | 3.050105957 |
| T2DM_insulin | delta_cell | *PKDREJ* | TRUE | 2.442704138 |
| T2DM_insulin | delta_cell | *CXCL11* | TRUE | 2.556342416 |
| T2DM_insulin | delta_cell | *IRX3* | TRUE | 4.128126599 |
| T2DM_insulin | delta_cell | *DNAJC22* | TRUE | 2.163775298 |
| T2DM_insulin | delta_cell | *PAX9* | TRUE | 2.461609601 |
| T2DM_insulin | delta_cell | *NOTCH4* | TRUE | 2.351335313 |
| T2DM_insulin | delta_cell | *PSORS1C1* | TRUE | 2.144091687 |
| T2DM_insulin | delta_cell | *HLA.DQB2* | TRUE | 3.22824947 |
| T2DM_insulin | delta_cell | *HLA.DOB* | TRUE | 2.212442772 |
| T2DM_insulin | delta_cell | *TAS2R43* | TRUE | 2.407662862 |
| T2DM_insulin | duct_epithelial_cell | *CEP68* | TRUE | 2.937238345 |
| T2DM_insulin | duct_epithelial_cell | *ALOX5* | TRUE | 2.737343295 |
| T2DM_insulin | duct_epithelial_cell | *MYBL2* | TRUE | 2.044566184 |
| T2DM_insulin | duct_epithelial_cell | *BTNL8* | TRUE | 3.540506248 |
| T2DM_insulin | duct_epithelial_cell | *ZNF211* | TRUE | 2.201249803 |
| T2DM_insulin | duct_epithelial_cell | *PKDREJ* | TRUE | 2.630054275 |
| T2DM_insulin | duct_epithelial_cell | *EVI5L* | TRUE | 1.967652593 |
| T2DM_insulin | duct_epithelial_cell | *CLEC18A* | TRUE | 2.520366052 |
| T2DM_insulin | duct_epithelial_cell | *EXTL1* | TRUE | 2.22749084 |
| T2DM_insulin | duct_epithelial_cell | *CXCL11* | TRUE | 3.298171863 |
| T2DM_insulin | duct_epithelial_cell | *ZNF223* | TRUE | 2.476746358 |
| T2DM_insulin | duct_epithelial_cell | *RMI1* | TRUE | 2.068529256 |
| T2DM_insulin | duct_epithelial_cell | *FARSA* | TRUE | 2.034168167 |
| T2DM_insulin | duct_epithelial_cell | *ZNF320* | TRUE | 2.363002805 |
| T2DM_insulin | duct_epithelial_cell | *EIF3C* | TRUE | 2.128590763 |
| T2DM_insulin | duct_epithelial_cell | *LRRC19* | TRUE | 2.394593519 |
| T2DM_insulin | duct_epithelial_cell | *PIWIL2* | TRUE | 2.848488487 |
| T2DM_insulin | duct_epithelial_cell | *PAX9* | TRUE | 2.856088011 |
| T2DM_insulin | duct_epithelial_cell | *NOTCH4* | TRUE | 2.102690855 |
| T2DM_insulin | duct_epithelial_cell | *LILRB3* | TRUE | 2.023325126 |
| T2DM_insulin | duct_epithelial_cell | *HLA.DMB* | TRUE | 2.400203306 |
| T2DM_insulin | duct_epithelial_cell | *CDK3* | TRUE | 2.891470531 |
| T2DM_insulin | duct_epithelial_cell | *TAS2R43* | TRUE | 2.008837879 |
| T2DM_insulin | endothelial_cell | *CEP68* | TRUE | 3.05224816 |
| T2DM_insulin | endothelial_cell | *ALOX5* | TRUE | 2.098203906 |
| T2DM_insulin | endothelial_cell | *ANO8* | TRUE | 2.393460336 |
| T2DM_insulin | endothelial_cell | *ZNF211* | TRUE | 2.745045574 |

| | | | | |
|---|---|---|---|---|
| T2DM_insulin | endothelial_cell | *PKDREJ* | TRUE | 2.55012924 |
| T2DM_insulin | endothelial_cell | *NOTCH2* | TRUE | 2.45656319 |
| T2DM_insulin | endothelial_cell | *EVI5L* | TRUE | 2.11004747 |
| T2DM_insulin | endothelial_cell | *EXTL1* | TRUE | 3.156029862 |
| T2DM_insulin | endothelial_cell | *CXCL11* | TRUE | 2.153051988 |
| T2DM_insulin | endothelial_cell | *IRX3* | TRUE | 3.413372235 |
| T2DM_insulin | endothelial_cell | *ZNF223* | TRUE | 2.687847408 |
| T2DM_insulin | endothelial_cell | *DNAJC22* | TRUE | 2.018739629 |
| T2DM_insulin | endothelial_cell | *RMI1* | TRUE | 2.39998013 |
| T2DM_insulin | endothelial_cell | *ZNF320* | TRUE | 2.263913173 |
| T2DM_insulin | endothelial_cell | *EIF3C* | TRUE | 2.431898352 |
| T2DM_insulin | endothelial_cell | *LRRC19* | TRUE | 2.713412235 |
| T2DM_insulin | endothelial_cell | *PIWIL2* | TRUE | 2.7899545 |
| T2DM_insulin | endothelial_cell | *PAX9* | TRUE | 2.516727995 |
| T2DM_insulin | endothelial_cell | *AS3MT* | TRUE | 2.109962146 |
| T2DM_insulin | endothelial_cell | *HLA.DQA2* | TRUE | 2.046481131 |
| T2DM_insulin | endothelial_cell | *HLA.DMB* | TRUE | 3.153317823 |
| T2DM_insulin | endothelial_cell | *TAS2R43* | TRUE | 2.343103561 |
| T2DM_insulin | epsilon_cell | *BTNL8* | TRUE | 3.144691964 |
| T2DM_insulin | epsilon_cell | *EXTL1* | TRUE | 2.703609234 |
| T2DM_insulin | epsilon_cell | *IRX3* | TRUE | 1.974812986 |
| T2DM_insulin | epsilon_cell | *PAX9* | TRUE | 2.228789101 |
| T2DM_insulin | epsilon_cell | *NOTCH4* | TRUE | 2.349703456 |
| T2DM_insulin | epsilon_cell | *PSORS1C1* | TRUE | 1.963065715 |
| T2DM_insulin | epsilon_cell | *HLA.DQA2* | TRUE | 2.326644284 |
| T2DM_insulin | epsilon_cell | *HLA.DOB* | TRUE | 2.265349732 |
| T2DM_insulin | epsilon_cell | *TAS2R43* | TRUE | 2.072568412 |
| T2DM_insulin | leukocyte | *NOTCH2* | TRUE | 2.49567305 |
| T2DM_insulin | leukocyte | *EXTL1* | TRUE | 2.651736108 |
| T2DM_insulin | leukocyte | *LRRC19* | TRUE | 2.745431409 |
| T2DM_insulin | leukocyte | *PAX9* | TRUE | 2.122054854 |
| T2DM_insulin | leukocyte | *NOTCH4* | TRUE | 2.030662236 |
| T2DM_insulin | leukocyte | *PSORS1C1* | TRUE | 2.280476807 |
| T2DM_insulin | leukocyte | *OR2A7* | TRUE | 2.170735233 |
| T2DM_insulin | mesenchymal_cell | *CEP68* | TRUE | 2.84436493 |
| T2DM_insulin | mesenchymal_cell | *BTNL8* | TRUE | 3.194459796 |
| T2DM_insulin | mesenchymal_cell | *ZNF211* | TRUE | 2.617466005 |
| T2DM_insulin | mesenchymal_cell | *PKDREJ* | TRUE | 2.256831337 |
| T2DM_insulin | mesenchymal_cell | *EXTL1* | TRUE | 2.566662535 |
| T2DM_insulin | mesenchymal_cell | *CXCL11* | TRUE | 2.335972926 |
| T2DM_insulin | mesenchymal_cell | *IRX3* | TRUE | 4.656986298 |
| T2DM_insulin | mesenchymal_cell | *ZNF223* | TRUE | 2.365210155 |
| T2DM_insulin | mesenchymal_cell | *RPRML* | TRUE | 2.054840684 |
| T2DM_insulin | mesenchymal_cell | *ZNF320* | TRUE | 2.164964419 |
| T2DM_insulin | mesenchymal_cell | *LRRC19* | TRUE | 2.54655849 |
| T2DM_insulin | mesenchymal_cell | *PIWIL2* | TRUE | 2.415876105 |
| T2DM_insulin | mesenchymal_cell | *PAX9* | TRUE | 2.839985556 |
| T2DM_insulin | mesenchymal_cell | *PSORS1C1* | TRUE | 2.292280476 |
| T2DM_insulin | mesenchymal_cell | *AS3MT* | TRUE | 1.960978167 |
| T2DM_insulin | mesenchymal_cell | *HLA.DQB2* | TRUE | 2.209343188 |

| | | | | |
|---|---|---|---|---|
| T2DM_insulin | mesenchymal_cell | *CDK3* | TRUE | 2.050967333 |
| T2DM_insulin | PP_cell | *CEP68* | TRUE | 2.847814471 |
| T2DM_insulin | PP_cell | *TF* | TRUE | 1.974419471 |
| T2DM_insulin | PP_cell | *BTNL8* | TRUE | 2.959087583 |
| T2DM_insulin | PP_cell | *CLEC18A* | TRUE | 2.024758692 |
| T2DM_insulin | PP_cell | *CXCL11* | TRUE | 3.151811679 |
| T2DM_insulin | PP_cell | *ZNF223* | TRUE | 2.014153393 |
| T2DM_insulin | PP_cell | *EIF3C* | TRUE | 2.029697028 |
| T2DM_insulin | PP_cell | *PAX9* | TRUE | 2.199184259 |
| T2DM_insulin | PP_cell | *NOTCH4* | TRUE | 2.414044795 |
| T2DM_insulin | PP_cell | *PSORS1C1* | TRUE | 2.081042135 |
| T2DM_insulin | PP_cell | *GMNC* | TRUE | 2.0314107 |
| T2DM_insulin | PP_cell | *CDK3* | TRUE | 2.904075662 |
| T2DM_admission_fre | acinar_cell | *SLC7A9* | TRUE | 2.45726525 |
| T2DM_admission_fre | acinar_cell | *SLC4A11* | TRUE | 2.003881646 |
| T2DM_admission_fre | acinar_cell | *BTNL8* | TRUE | 2.073589224 |
| T2DM_admission_fre | acinar_cell | *CYB561D2* | TRUE | 2.098335825 |
| T2DM_admission_fre | acinar_cell | *VIPR1* | TRUE | 2.206324096 |
| T2DM_admission_fre | acinar_cell | *CXCL6* | TRUE | 2.881969025 |
| T2DM_admission_fre | acinar_cell | *IRF5* | TRUE | 2.119655901 |
| T2DM_admission_fre | acinar_cell | *FAM149B1* | TRUE | 2.040451771 |
| T2DM_admission_fre | acinar_cell | *PIK3C2G* | TRUE | 2.67471584 |
| T2DM_admission_fre | acinar_cell | *PLEKHH2* | TRUE | 2.009837307 |
| T2DM_admission_fre | acinar_cell | *C17orf50* | TRUE | 1.97985277 |
| T2DM_admission_fre | acinar_cell | *STOX1* | TRUE | 2.380706194 |
| T2DM_admission_fre | acinar_cell | *KCNAB3* | TRUE | 2.094918522 |
| T2DM_admission_fre | acinar_cell | *PDE3A* | TRUE | 2.658578325 |
| T2DM_admission_fre | acinar_cell | *TUBB8* | TRUE | 2.509135392 |
| T2DM_admission_fre | acinar_cell | *ZNF404* | TRUE | 1.964319876 |
| T2DM_admission_fre | acinar_cell | *GEN1* | TRUE | 1.970924705 |
| T2DM_admission_fre | acinar_cell | *NPIPA5* | TRUE | 2.018301656 |
| T2DM_admission_fre | acinar_cell | *RHCE* | TRUE | 1.978207019 |
| T2DM_admission_fre | acinar_cell | *ZNF79* | TRUE | 2.251788654 |
| T2DM_admission_fre | acinar_cell | *PAX9* | TRUE | 4.082714005 |
| T2DM_admission_fre | alpha_cell | *SLC7A9* | TRUE | 2.777775469 |
| T2DM_admission_fre | alpha_cell | *MRPS10* | TRUE | 2.029049384 |
| T2DM_admission_fre | alpha_cell | *SLC4A11* | TRUE | 2.289261853 |
| T2DM_admission_fre | alpha_cell | *VIPR2* | TRUE | 2.31420984 |
| T2DM_admission_fre | alpha_cell | *APOA5* | TRUE | 2.02122652 |
| T2DM_admission_fre | alpha_cell | *CHPT1* | TRUE | 2.201601274 |
| T2DM_admission_fre | alpha_cell | *BTNL8* | TRUE | 2.01803416 |
| T2DM_admission_fre | alpha_cell | *CYB561D2* | TRUE | 2.064985137 |
| T2DM_admission_fre | alpha_cell | *VIPR1* | TRUE | 2.594607523 |
| T2DM_admission_fre | alpha_cell | *CXCL6* | TRUE | 2.705824028 |
| T2DM_admission_fre | alpha_cell | *GRK4* | TRUE | 1.98890765 |
| T2DM_admission_fre | alpha_cell | *IRF5* | TRUE | 2.331236014 |
| T2DM_admission_fre | alpha_cell | *STK33* | TRUE | 2.703510346 |
| T2DM_admission_fre | alpha_cell | *CEP85* | TRUE | 2.050474133 |
| T2DM_admission_fre | alpha_cell | *FAM149B1* | TRUE | 2.152282176 |
| T2DM_admission_fre | alpha_cell | *PIK3C2G* | TRUE | 2.481701436 |

| | | | | |
|---|---|---|---|---|
| T2DM_admission_fre | alpha_cell | *SPC25* | TRUE | 2.284883959 |
| T2DM_admission_fre | alpha_cell | *PLEKHH2* | TRUE | 2.037971234 |
| T2DM_admission_fre | alpha_cell | *C17orf50* | TRUE | 1.975492811 |
| T2DM_admission_fre | alpha_cell | *CDK20* | TRUE | 2.308229072 |
| T2DM_admission_fre | alpha_cell | *ICOSLG* | TRUE | 1.99773095 |
| T2DM_admission_fre | alpha_cell | *SIGLEC11* | TRUE | 2.063715709 |
| T2DM_admission_fre | alpha_cell | *STOX1* | TRUE | 2.292258387 |
| T2DM_admission_fre | alpha_cell | *SERPINF2* | TRUE | 2.072294723 |
| T2DM_admission_fre | alpha_cell | *MIS12* | TRUE | 2.01257062 |
| T2DM_admission_fre | alpha_cell | *KCNAB3* | TRUE | 2.177296149 |
| T2DM_admission_fre | alpha_cell | *PAQR8* | TRUE | 1.964562581 |
| T2DM_admission_fre | alpha_cell | *PDE3A* | TRUE | 3.032708503 |
| T2DM_admission_fre | alpha_cell | *TUBB8* | TRUE | 2.545154548 |
| T2DM_admission_fre | alpha_cell | *ZNF404* | TRUE | 2.004337647 |
| T2DM_admission_fre | alpha_cell | *GEN1* | TRUE | 1.991287929 |
| T2DM_admission_fre | alpha_cell | *AQP11* | TRUE | 2.096533452 |
| T2DM_admission_fre | alpha_cell | *NPIPA5* | TRUE | 2.073449058 |
| T2DM_admission_fre | alpha_cell | *NWD1* | TRUE | 1.970485775 |
| T2DM_admission_fre | alpha_cell | *RHCE* | TRUE | 2.067044397 |
| T2DM_admission_fre | alpha_cell | *ZNF79* | TRUE | 2.241173342 |
| T2DM_admission_fre | alpha_cell | *ZNF311* | TRUE | 2.228100101 |
| T2DM_admission_fre | alpha_cell | *ENTPD7* | TRUE | 2.032696835 |
| T2DM_admission_fre | alpha_cell | *PAX9* | TRUE | 4.459744591 |
| T2DM_admission_fre | alpha_cell | *PCDHGC3* | TRUE | 2.082851942 |
| T2DM_admission_fre | alpha_cell | *CFHR1* | TRUE | 2.244854883 |
| T2DM_admission_fre | beta_cell | *SLC7A9* | TRUE | 2.8294167 |
| T2DM_admission_fre | beta_cell | *ADRB1* | TRUE | 2.134680057 |
| T2DM_admission_fre | beta_cell | *LMO3* | TRUE | 2.037886971 |
| T2DM_admission_fre | beta_cell | *MRPS10* | TRUE | 2.058359822 |
| T2DM_admission_fre | beta_cell | *ZNF37A* | TRUE | 2.23352282 |
| T2DM_admission_fre | beta_cell | *TTC39A* | TRUE | 2.176975804 |
| T2DM_admission_fre | beta_cell | *SLC4A11* | TRUE | 2.226561381 |
| T2DM_admission_fre | beta_cell | *VIPR2* | TRUE | 2.444006644 |
| T2DM_admission_fre | beta_cell | *AK1* | TRUE | 2.18949765 |
| T2DM_admission_fre | beta_cell | *APOA5* | TRUE | 1.964173572 |
| T2DM_admission_fre | beta_cell | *CEP164* | TRUE | 1.965117903 |
| T2DM_admission_fre | beta_cell | *CHPT1* | TRUE | 2.66199674 |
| T2DM_admission_fre | beta_cell | *BTNL8* | TRUE | 2.049435445 |
| T2DM_admission_fre | beta_cell | *CYB561D2* | TRUE | 2.145281643 |
| T2DM_admission_fre | beta_cell | *VIPR1* | TRUE | 2.743950906 |
| T2DM_admission_fre | beta_cell | *CXCL6* | TRUE | 2.845349055 |
| T2DM_admission_fre | beta_cell | *GRK4* | TRUE | 2.190303556 |
| T2DM_admission_fre | beta_cell | *IRF5* | TRUE | 2.325556741 |
| T2DM_admission_fre | beta_cell | *STK33* | TRUE | 2.989964863 |
| T2DM_admission_fre | beta_cell | *CEP85* | TRUE | 2.239686926 |
| T2DM_admission_fre | beta_cell | *TSGA10* | TRUE | 2.004670174 |
| T2DM_admission_fre | beta_cell | *MRPS9* | TRUE | 2.21419663 |
| T2DM_admission_fre | beta_cell | *TMEM8B* | TRUE | 2.000671569 |
| T2DM_admission_fre | beta_cell | *DUSP5* | TRUE | 2.038377603 |
| T2DM_admission_fre | beta_cell | *FAM149B1* | TRUE | 2.211650785 |

| | | | | |
|---|---|---|---|---|
| T2DM_admission_fre | beta_cell | *PIK3C2G* | TRUE | 2.718851369 |
| T2DM_admission_fre | beta_cell | *RSU1* | TRUE | 2.459007291 |
| T2DM_admission_fre | beta_cell | *CABLES2* | TRUE | 1.999032709 |
| T2DM_admission_fre | beta_cell | *SPC25* | TRUE | 2.318984354 |
| T2DM_admission_fre | beta_cell | *PLEKHH2* | TRUE | 2.329319862 |
| T2DM_admission_fre | beta_cell | *C17orf50* | TRUE | 2.019019976 |
| T2DM_admission_fre | beta_cell | *CDK20* | TRUE | 2.544178126 |
| T2DM_admission_fre | beta_cell | *CLEC18A* | TRUE | 1.980040305 |
| T2DM_admission_fre | beta_cell | *ICOSLG* | TRUE | 2.081092519 |
| T2DM_admission_fre | beta_cell | *SIGLEC11* | TRUE | 2.088461113 |
| T2DM_admission_fre | beta_cell | *OMA1* | TRUE | 2.036355298 |
| T2DM_admission_fre | beta_cell | *NMNAT3* | TRUE | 2.198718912 |
| T2DM_admission_fre | beta_cell | *C9orf64* | TRUE | 2.051531726 |
| T2DM_admission_fre | beta_cell | *STOX1* | TRUE | 2.347574627 |
| T2DM_admission_fre | beta_cell | *SERPINF2* | TRUE | 2.056384732 |
| T2DM_admission_fre | beta_cell | *MIS12* | TRUE | 2.541556454 |
| T2DM_admission_fre | beta_cell | *PXK* | TRUE | 2.214533541 |
| T2DM_admission_fre | beta_cell | *KCNAB3* | TRUE | 2.1755355 |
| T2DM_admission_fre | beta_cell | *PAQR8* | TRUE | 2.018378332 |
| T2DM_admission_fre | beta_cell | *ATP6V0E2* | TRUE | 2.143477789 |
| T2DM_admission_fre | beta_cell | *SPTLC3* | TRUE | 2.144541807 |
| T2DM_admission_fre | beta_cell | *PDE3A* | TRUE | 3.114569758 |
| T2DM_admission_fre | beta_cell | *TUBB8* | TRUE | 2.552866156 |
| T2DM_admission_fre | beta_cell | *GEN1* | TRUE | 2.019369793 |
| T2DM_admission_fre | beta_cell | *AQP11* | TRUE | 2.08443091 |
| T2DM_admission_fre | beta_cell | *FKRP* | TRUE | 1.973721148 |
| T2DM_admission_fre | beta_cell | *NPIPA5* | TRUE | 2.022654924 |
| T2DM_admission_fre | beta_cell | *SETD4* | TRUE | 2.211854752 |
| T2DM_admission_fre | beta_cell | *AKR1C1* | TRUE | 2.219624366 |
| T2DM_admission_fre | beta_cell | *RHCE* | TRUE | 2.065736649 |
| T2DM_admission_fre | beta_cell | *ZNF311* | TRUE | 2.107444524 |
| T2DM_admission_fre | beta_cell | *ENTPD7* | TRUE | 2.42309866 |
| T2DM_admission_fre | beta_cell | *COLGALT2* | TRUE | 1.979960269 |
| T2DM_admission_fre | beta_cell | *PAX9* | TRUE | 4.455086344 |
| T2DM_admission_fre | beta_cell | *PCDHGC3* | TRUE | 2.161988918 |
| T2DM_admission_fre | beta_cell | *CFHR1* | TRUE | 2.249470706 |
| T2DM_admission_fre | beta_cell | *FMN1* | TRUE | 2.117809481 |
| T2DM_admission_fre | beta_cell | *GTF2H5* | TRUE | 2.051400478 |
| T2DM_admission_fre | delta_cell | *SLC7A9* | TRUE | 2.56807932 |
| T2DM_admission_fre | delta_cell | *CYB561D2* | TRUE | 2.346107328 |
| T2DM_admission_fre | delta_cell | *CXCL6* | TRUE | 2.634444563 |
| T2DM_admission_fre | delta_cell | *FAM149B1* | TRUE | 2.045004105 |
| T2DM_admission_fre | delta_cell | *PIK3C2G* | TRUE | 2.502908273 |
| T2DM_admission_fre | delta_cell | *STOX1* | TRUE | 2.150889806 |
| T2DM_admission_fre | delta_cell | *PDE3A* | TRUE | 2.152709398 |
| T2DM_admission_fre | delta_cell | *AQP11* | TRUE | 2.087845067 |
| T2DM_admission_fre | delta_cell | *ZNF311* | TRUE | 2.116664335 |
| T2DM_admission_fre | delta_cell | *PAX9* | TRUE | 4.469011244 |
| T2DM_admission_fre | delta_cell | *PCDHGC3* | TRUE | 1.991531351 |
| T2DM_admission_fre | duct_epithelial_cell | *SLC7A9* | TRUE | 2.755849187 |

| | | | | |
|---|---|---|---|---|
| T2DM_admission_fre | duct_epithelial_cell | *COQ5* | TRUE | 1.962755566 |
| T2DM_admission_fre | duct_epithelial_cell | *BTNL8* | TRUE | 1.984791336 |
| T2DM_admission_fre | duct_epithelial_cell | *CYB561D2* | TRUE | 2.393233252 |
| T2DM_admission_fre | duct_epithelial_cell | *IRF5* | TRUE | 2.029297856 |
| T2DM_admission_fre | duct_epithelial_cell | *FAM149B1* | TRUE | 2.061606936 |
| T2DM_admission_fre | duct_epithelial_cell | *PIK3C2G* | TRUE | 2.340034213 |
| T2DM_admission_fre | duct_epithelial_cell | *PLEKHH2* | TRUE | 2.227090786 |
| T2DM_admission_fre | duct_epithelial_cell | *CDK20* | TRUE | 2.41229218 |
| T2DM_admission_fre | duct_epithelial_cell | *STOX1* | TRUE | 2.245372169 |
| T2DM_admission_fre | duct_epithelial_cell | *SERPINF2* | TRUE | 2.047469493 |
| T2DM_admission_fre | duct_epithelial_cell | *PDE3A* | TRUE | 2.392028265 |
| T2DM_admission_fre | duct_epithelial_cell | *TUBB8* | TRUE | 2.1377933 |
| T2DM_admission_fre | duct_epithelial_cell | *GEN1* | TRUE | 2.006984073 |
| T2DM_admission_fre | duct_epithelial_cell | *AQP11* | TRUE | 2.074416232 |
| T2DM_admission_fre | duct_epithelial_cell | *NPIPA5* | TRUE | 1.9704183 |
| T2DM_admission_fre | duct_epithelial_cell | *ZNF79* | TRUE | 2.003524169 |
| T2DM_admission_fre | duct_epithelial_cell | *ENTPD7* | TRUE | 2.183037991 |
| T2DM_admission_fre | duct_epithelial_cell | *PAX9* | TRUE | 4.419885742 |
| T2DM_admission_fre | duct_epithelial_cell | *CFHR1* | TRUE | 2.049653266 |
| T2DM_admission_fre | endothelial_cell | *SLC7A9* | TRUE | 2.819155397 |
| T2DM_admission_fre | endothelial_cell | *CYB561D2* | TRUE | 2.20697701 |
| T2DM_admission_fre | endothelial_cell | *IRF5* | TRUE | 1.97477706 |
| T2DM_admission_fre | endothelial_cell | *PIK3C2G* | TRUE | 2.73382645 |
| T2DM_admission_fre | endothelial_cell | *STOX1* | TRUE | 2.222618567 |
| T2DM_admission_fre | endothelial_cell | *SERPINF2* | TRUE | 2.062221026 |
| T2DM_admission_fre | endothelial_cell | *PDE3A* | TRUE | 2.975940356 |
| T2DM_admission_fre | endothelial_cell | *TUBB8* | TRUE | 2.308122446 |
| T2DM_admission_fre | endothelial_cell | *ZNF404* | TRUE | 1.983211423 |
| T2DM_admission_fre | endothelial_cell | *AQP11* | TRUE | 2.097203735 |
| T2DM_admission_fre | endothelial_cell | *ZNF79* | TRUE | 2.253023796 |
| T2DM_admission_fre | endothelial_cell | *ZNF311* | TRUE | 2.185482966 |
| T2DM_admission_fre | endothelial_cell | *PAX9* | TRUE | 4.560888079 |
| T2DM_admission_fre | endothelial_cell | *CFHR1* | TRUE | 2.064700095 |
| T2DM_admission_fre | epsilon_cell | *BTNL8* | TRUE | 1.977112405 |
| T2DM_admission_fre | epsilon_cell | *SERPINF2* | TRUE | 1.992861787 |
| T2DM_admission_fre | epsilon_cell | *ZNF311* | TRUE | 2.03616426 |
| T2DM_admission_fre | epsilon_cell | *PAX9* | TRUE | 4.3611974 |
| T2DM_admission_fre | epsilon_cell | *PSORS1C1* | TRUE | 2.385539001 |
| T2DM_admission_fre | leukocyte | *SLC7A9* | TRUE | 2.716188623 |
| T2DM_admission_fre | leukocyte | *SLC4A11* | TRUE | 2.137269122 |
| T2DM_admission_fre | leukocyte | *STOX1* | TRUE | 2.248319976 |
| T2DM_admission_fre | leukocyte | *PDE3A* | TRUE | 2.039114966 |
| T2DM_admission_fre | leukocyte | *ZNF79* | TRUE | 1.998291858 |
| T2DM_admission_fre | leukocyte | *PAX9* | TRUE | 4.361544712 |
| T2DM_admission_fre | mesenchymal_cell | *SLC7A9* | TRUE | 2.872078227 |
| T2DM_admission_fre | mesenchymal_cell | *BTNL8* | TRUE | 2.038045726 |
| T2DM_admission_fre | mesenchymal_cell | *CYB561D2* | TRUE | 2.521897408 |
| T2DM_admission_fre | mesenchymal_cell | *CXCL6* | TRUE | 2.803631042 |
| T2DM_admission_fre | mesenchymal_cell | *FAM149B1* | TRUE | 2.249834218 |
| T2DM_admission_fre | mesenchymal_cell | *C17orf50* | TRUE | 2.018521126 |

| | | | | |
|---|---|---|---|---|
| T2DM_admission_fre | mesenchymal_cell | *STOX1* | TRUE | 2.21377874 |
| T2DM_admission_fre | mesenchymal_cell | *MIS12* | TRUE | 2.318743514 |
| T2DM_admission_fre | mesenchymal_cell | *ZNF79* | TRUE | 1.995231025 |
| T2DM_admission_fre | mesenchymal_cell | *PAX9* | TRUE | 4.064631936 |
| T2DM_admission_fre | mesenchymal_cell | *PSORS1C1* | TRUE | 2.250775727 |
| T2DM_admission_fre | PP_cell | *SLC7A9* | TRUE | 2.152253392 |
| T2DM_admission_fre | PP_cell | *CYB561D2* | TRUE | 2.38815041 |
| T2DM_admission_fre | PP_cell | *PLEKHH2* | TRUE | 2.255877568 |
| T2DM_admission_fre | PP_cell | *SERPINF2* | TRUE | 2.223770627 |
| T2DM_admission_fre | PP_cell | *TUBB8* | TRUE | 2.099450363 |
| T2DM_admission_fre | PP_cell | *ZNF404* | TRUE | 2.004698334 |
| T2DM_admission_fre | PP_cell | *GEN1* | TRUE | 2.055868907 |
| T2DM_admission_fre | PP_cell | *ZNF311* | TRUE | 2.145312351 |
| T2DM_admission_fre | PP_cell | *PAX9* | TRUE | 3.97832369 |
| T2DM_admission_fre | PP_cell | *HLA.DQB2* | TRUE | 1.972976461 |
| T2DM_mortality | acinar_cell | *KCNG1* | TRUE | 2.336291458 |
| T2DM_mortality | acinar_cell | *BAK1* | TRUE | 1.997587658 |
| T2DM_mortality | acinar_cell | *TCF3* | TRUE | 2.024741563 |
| T2DM_mortality | acinar_cell | *CDHR5* | TRUE | 2.028563438 |
| T2DM_mortality | acinar_cell | *MROH8* | TRUE | 2.492701063 |
| T2DM_mortality | acinar_cell | *MLPH* | TRUE | 2.653231294 |
| T2DM_mortality | acinar_cell | *RIBC2* | TRUE | 2.007532631 |
| T2DM_mortality | acinar_cell | *PLVAP* | TRUE | 2.159202038 |
| T2DM_mortality | acinar_cell | *LPIN3* | TRUE | 1.987898488 |
| T2DM_mortality | acinar_cell | *NOTCH2* | TRUE | 1.981506894 |
| T2DM_mortality | acinar_cell | *MYOF* | TRUE | 1.971366688 |
| T2DM_mortality | acinar_cell | *CYP4B1* | TRUE | 2.812600608 |
| T2DM_mortality | acinar_cell | *TRIM58* | TRUE | 2.260465779 |
| T2DM_mortality | acinar_cell | *HCN1* | TRUE | 2.227737917 |
| T2DM_mortality | acinar_cell | *FUT9* | TRUE | 2.789249632 |
| T2DM_mortality | acinar_cell | *C9orf131* | TRUE | 2.064150522 |
| T2DM_mortality | acinar_cell | *IRX3* | TRUE | 3.332544952 |
| T2DM_mortality | acinar_cell | *GPR88* | TRUE | 2.362701171 |
| T2DM_mortality | acinar_cell | *DDO* | TRUE | 2.150080293 |
| T2DM_mortality | acinar_cell | *C6orf163* | TRUE | 2.288392693 |
| T2DM_mortality | acinar_cell | *SMTNL1* | TRUE | 2.594586696 |
| T2DM_mortality | alpha_cell | *KCNG1* | TRUE | 2.066742539 |
| T2DM_mortality | alpha_cell | *BAK1* | TRUE | 2.172526633 |
| T2DM_mortality | alpha_cell | *ITIH4* | TRUE | 1.980951798 |
| T2DM_mortality | alpha_cell | *MSH4* | TRUE | 1.997251844 |
| T2DM_mortality | alpha_cell | *TCF3* | TRUE | 2.369359859 |
| T2DM_mortality | alpha_cell | *CDHR5* | TRUE | 2.026825261 |
| T2DM_mortality | alpha_cell | *MROH8* | TRUE | 2.620616056 |
| T2DM_mortality | alpha_cell | *CHPT1* | TRUE | 1.996999649 |
| T2DM_mortality | alpha_cell | *MLPH* | TRUE | 2.747906914 |
| T2DM_mortality | alpha_cell | *FLRT1* | TRUE | 1.974355853 |
| T2DM_mortality | alpha_cell | *RIBC2* | TRUE | 2.10550909 |
| T2DM_mortality | alpha_cell | *PLVAP* | TRUE | 2.248880543 |
| T2DM_mortality | alpha_cell | *LPIN3* | TRUE | 2.209954167 |
| T2DM_mortality | alpha_cell | *CASP9* | TRUE | 2.111886693 |

| | | | | |
|---|---|---|---|---|
| T2DM_mortality | alpha_cell | *SFTPD* | TRUE | 2.274666839 |
| T2DM_mortality | alpha_cell | *NOTCH2* | TRUE | 2.213543334 |
| T2DM_mortality | alpha_cell | *NDUFAF1* | TRUE | 2.22277711 |
| T2DM_mortality | alpha_cell | *MYOF* | TRUE | 2.001609096 |
| T2DM_mortality | alpha_cell | *CCDC65* | TRUE | 2.006979488 |
| T2DM_mortality | alpha_cell | *NPC1* | TRUE | 2.573748671 |
| T2DM_mortality | alpha_cell | *TRIM65* | TRUE | 2.189349719 |
| T2DM_mortality | alpha_cell | *CYP4B1* | TRUE | 2.646554701 |
| T2DM_mortality | alpha_cell | *SPATA9* | TRUE | 2.206348255 |
| T2DM_mortality | alpha_cell | *TRIM63* | TRUE | 2.113098934 |
| T2DM_mortality | alpha_cell | *FGF17* | TRUE | 2.069347053 |
| T2DM_mortality | alpha_cell | *TRIM58* | TRUE | 2.162316548 |
| T2DM_mortality | alpha_cell | *TUB* | TRUE | 2.143877173 |
| T2DM_mortality | alpha_cell | *FAM153A* | TRUE | 2.159312206 |
| T2DM_mortality | alpha_cell | *FUT9* | TRUE | 2.836150252 |
| T2DM_mortality | alpha_cell | *MRPL48* | TRUE | 2.114229423 |
| T2DM_mortality | alpha_cell | *IRX3* | TRUE | 3.138244594 |
| T2DM_mortality | alpha_cell | *GPR88* | TRUE | 2.479487998 |
| T2DM_mortality | alpha_cell | *SLC35D3* | TRUE | 2.108271315 |
| T2DM_mortality | alpha_cell | *NUTM2B* | TRUE | 1.965278427 |
| T2DM_mortality | alpha_cell | *AGAP4* | TRUE | 2.381829327 |
| T2DM_mortality | alpha_cell | *EVL* | TRUE | 2.209752311 |
| T2DM_mortality | alpha_cell | *ACSL5* | TRUE | 2.163152345 |
| T2DM_mortality | alpha_cell | *DDO* | TRUE | 2.515933592 |
| T2DM_mortality | alpha_cell | *RBM20* | TRUE | 1.995356372 |
| T2DM_mortality | alpha_cell | *C6orf163* | TRUE | 2.36315963 |
| T2DM_mortality | alpha_cell | *TREX1* | TRUE | 2.080636306 |
| T2DM_mortality | alpha_cell | *SMTNL1* | TRUE | 2.569508868 |
| T2DM_mortality | alpha_cell | *WDR46* | TRUE | 1.960857262 |
| T2DM_mortality | alpha_cell | *OR2A7* | TRUE | 2.013204932 |
| T2DM_mortality | beta_cell | *SYNRG* | TRUE | 2.588671542 |
| T2DM_mortality | beta_cell | *KCNG1* | TRUE | 2.468370806 |
| T2DM_mortality | beta_cell | *BAK1* | TRUE | 2.885198314 |
| T2DM_mortality | beta_cell | *PNPLA6* | TRUE | 2.148847478 |
| T2DM_mortality | beta_cell | *GALC* | TRUE | 2.235984458 |
| T2DM_mortality | beta_cell | *ITIH4* | TRUE | 2.321016213 |
| T2DM_mortality | beta_cell | *MSH4* | TRUE | 2.030234084 |
| T2DM_mortality | beta_cell | *TCF3* | TRUE | 2.442665069 |
| T2DM_mortality | beta_cell | *EXOSC7* | TRUE | 2.074915013 |
| T2DM_mortality | beta_cell | *GPR137B* | TRUE | 2.712059575 |
| T2DM_mortality | beta_cell | *ZNF343* | TRUE | 1.978331328 |
| T2DM_mortality | beta_cell | *CDHR5* | TRUE | 2.327791086 |
| T2DM_mortality | beta_cell | *ARFRP1* | TRUE | 1.964128508 |
| T2DM_mortality | beta_cell | *MROH8* | TRUE | 2.666101709 |
| T2DM_mortality | beta_cell | *DHX35* | TRUE | 2.023158122 |
| T2DM_mortality | beta_cell | *NOMO1* | TRUE | 2.221889609 |
| T2DM_mortality | beta_cell | *LIMK1* | TRUE | 1.970037008 |
| T2DM_mortality | beta_cell | *GLRB* | TRUE | 2.332165859 |
| T2DM_mortality | beta_cell | *CHPT1* | TRUE | 2.151836755 |
| T2DM_mortality | beta_cell | *LAMA4* | TRUE | 2.103512275 |

| | | | | |
|---|---|---|---|---|
| T2DM_mortality | beta_cell | *DNAH6* | TRUE | 2.00271778 |
| T2DM_mortality | beta_cell | *MLPH* | TRUE | 2.513895269 |
| T2DM_mortality | beta_cell | *CEP104* | TRUE | 1.969037573 |
| T2DM_mortality | beta_cell | *CDK2* | TRUE | 2.333274089 |
| T2DM_mortality | beta_cell | *MT1G* | TRUE | 2.228056042 |
| T2DM_mortality | beta_cell | *RIBC2* | TRUE | 2.135000279 |
| T2DM_mortality | beta_cell | *PLVAP* | TRUE | 2.299124606 |
| T2DM_mortality | beta_cell | *LAMA5* | TRUE | 2.068275175 |
| T2DM_mortality | beta_cell | *LPIN3* | TRUE | 2.221740546 |
| T2DM_mortality | beta_cell | *CASP9* | TRUE | 2.434329886 |
| T2DM_mortality | beta_cell | *SFTPD* | TRUE | 2.810405042 |
| T2DM_mortality | beta_cell | *NOTCH2* | TRUE | 2.27013323 |
| T2DM_mortality | beta_cell | *SPIRE1* | TRUE | 2.102954274 |
| T2DM_mortality | beta_cell | *FLNB* | TRUE | 2.133163559 |
| T2DM_mortality | beta_cell | *NDUFAF1* | TRUE | 2.617711897 |
| T2DM_mortality | beta_cell | *PRDM5* | TRUE | 2.232238898 |
| T2DM_mortality | beta_cell | *NUP54* | TRUE | 2.593842178 |
| T2DM_mortality | beta_cell | *CCDC65* | TRUE | 2.008920321 |
| T2DM_mortality | beta_cell | *NPC1* | TRUE | 2.751478569 |
| T2DM_mortality | beta_cell | *TRIM65* | TRUE | 2.215377032 |
| T2DM_mortality | beta_cell | *CELA3A* | TRUE | 2.283238287 |
| T2DM_mortality | beta_cell | *CYP4B1* | TRUE | 2.857589707 |
| T2DM_mortality | beta_cell | *RABL2A* | TRUE | 2.340370424 |
| T2DM_mortality | beta_cell | *SPATA9* | TRUE | 2.42873501 |
| T2DM_mortality | beta_cell | *UGCG* | TRUE | 1.962389682 |
| T2DM_mortality | beta_cell | *POC5* | TRUE | 1.975158814 |
| T2DM_mortality | beta_cell | *CC2D1B* | TRUE | 1.995345237 |
| T2DM_mortality | beta_cell | *CDK20* | TRUE | 1.975757845 |
| T2DM_mortality | beta_cell | *TRIM63* | TRUE | 2.240519795 |
| T2DM_mortality | beta_cell | *FGF17* | TRUE | 2.097408417 |
| T2DM_mortality | beta_cell | *VAV2* | TRUE | 2.063807023 |
| T2DM_mortality | beta_cell | *TRIM58* | TRUE | 2.267772271 |
| T2DM_mortality | beta_cell | *CCDC141* | TRUE | 2.216256295 |
| T2DM_mortality | beta_cell | *CADPS* | TRUE | 2.261775722 |
| T2DM_mortality | beta_cell | *PPM1M* | TRUE | 1.963403918 |
| T2DM_mortality | beta_cell | *HCN1* | TRUE | 2.383120245 |
| T2DM_mortality | beta_cell | *TUB* | TRUE | 2.222325328 |
| T2DM_mortality | beta_cell | *C19orf48* | TRUE | 1.966430129 |
| T2DM_mortality | beta_cell | *FAM153A* | TRUE | 2.702792242 |
| T2DM_mortality | beta_cell | *FOXD4* | TRUE | 2.056880664 |
| T2DM_mortality | beta_cell | *PIK3CD* | TRUE | 2.305194279 |
| T2DM_mortality | beta_cell | *FUT9* | TRUE | 2.9318779 |
| T2DM_mortality | beta_cell | *RAB43* | TRUE | 2.042983438 |
| T2DM_mortality | beta_cell | *C9orf131* | TRUE | 2.314391226 |
| T2DM_mortality | beta_cell | *BTC* | TRUE | 1.977962517 |
| T2DM_mortality | beta_cell | *DENND6A* | TRUE | 2.606806343 |
| T2DM_mortality | beta_cell | *CLCF1* | TRUE | 2.226807571 |
| T2DM_mortality | beta_cell | *MRPL48* | TRUE | 2.188214328 |
| T2DM_mortality | beta_cell | *PHLDB3* | TRUE | 2.014453137 |
| T2DM_mortality | beta_cell | *IRX3* | TRUE | 3.522611759 |

| | | | | |
|---|---|---|---|---|
| T2DM_mortality | beta_cell | *SMCO3* | TRUE | 2.058984288 |
| T2DM_mortality | beta_cell | *GPR88* | TRUE | 2.524244959 |
| T2DM_mortality | beta_cell | *SLC35D3* | TRUE | 2.026600982 |
| T2DM_mortality | beta_cell | *CAMK1D* | TRUE | 2.903759129 |
| T2DM_mortality | beta_cell | *LYPD6* | TRUE | 1.964023491 |
| T2DM_mortality | beta_cell | *AGAP4* | TRUE | 2.215030019 |
| T2DM_mortality | beta_cell | *ADAT2* | TRUE | 2.132601615 |
| T2DM_mortality | beta_cell | *EVL* | TRUE | 2.300475441 |
| T2DM_mortality | beta_cell | *ACSL5* | TRUE | 2.235187583 |
| T2DM_mortality | beta_cell | *DDO* | TRUE | 2.627896265 |
| T2DM_mortality | beta_cell | *RBM20* | TRUE | 2.006518278 |
| T2DM_mortality | beta_cell | *C6orf163* | TRUE | 2.324881059 |
| T2DM_mortality | beta_cell | *TMEM231* | TRUE | 2.73843178 |
| T2DM_mortality | beta_cell | *TREX1* | TRUE | 2.114609014 |
| T2DM_mortality | beta_cell | *SMTNL1* | TRUE | 2.4673685 |
| T2DM_mortality | beta_cell | *LINC00205* | TRUE | 2.209904026 |
| T2DM_mortality | beta_cell | *WDR46* | TRUE | 2.774699526 |
| T2DM_mortality | beta_cell | *OR2A7* | TRUE | 2.031831762 |
| T2DM_mortality | beta_cell | *LIMS3* | TRUE | 2.082918482 |
| T2DM_mortality | delta_cell | *MROH8* | TRUE | 2.065353102 |
| T2DM_mortality | delta_cell | *LPIN3* | TRUE | 2.005739547 |
| T2DM_mortality | delta_cell | *CYP4B1* | TRUE | 2.815051621 |
| T2DM_mortality | delta_cell | *FGF17* | TRUE | 1.973240208 |
| T2DM_mortality | delta_cell | *IRX3* | TRUE | 2.0843938 |
| T2DM_mortality | delta_cell | *DDO* | TRUE | 2.040323006 |
| T2DM_mortality | delta_cell | *SMTNL1* | TRUE | 2.31344799 |
| T2DM_mortality | duct_epithelial_cell | *CDHR5* | TRUE | 2.026949759 |
| T2DM_mortality | duct_epithelial_cell | *MROH8* | TRUE | 2.047737506 |
| T2DM_mortality | duct_epithelial_cell | *RIBC2* | TRUE | 2.079362306 |
| T2DM_mortality | duct_epithelial_cell | *LPIN3* | TRUE | 1.985514364 |
| T2DM_mortality | duct_epithelial_cell | *CASP9* | TRUE | 2.077501675 |
| T2DM_mortality | duct_epithelial_cell | *NDUFAF1* | TRUE | 2.149219213 |
| T2DM_mortality | duct_epithelial_cell | *TRIM58* | TRUE | 2.145309542 |
| T2DM_mortality | duct_epithelial_cell | *FUT9* | TRUE | 2.385110207 |
| T2DM_mortality | duct_epithelial_cell | *DDO* | TRUE | 2.16900015 |
| T2DM_mortality | duct_epithelial_cell | *C6orf163* | TRUE | 2.23446136 |
| T2DM_mortality | duct_epithelial_cell | *TMEM231* | TRUE | 2.075737712 |
| T2DM_mortality | duct_epithelial_cell | *TREX1* | TRUE | 2.012047175 |
| T2DM_mortality | duct_epithelial_cell | *HLA.DQA2* | TRUE | 2.127182565 |
| T2DM_mortality | endothelial_cell | *BAK1* | TRUE | 2.17595251 |
| T2DM_mortality | endothelial_cell | *ITIH4* | TRUE | 2.033995717 |
| T2DM_mortality | endothelial_cell | *MROH8* | TRUE | 2.311272502 |
| T2DM_mortality | endothelial_cell | *MLPH* | TRUE | 2.521244444 |
| T2DM_mortality | endothelial_cell | *CASP9* | TRUE | 2.03531936 |
| T2DM_mortality | endothelial_cell | *NOTCH2* | TRUE | 2.120601659 |
| T2DM_mortality | endothelial_cell | *MYOF* | TRUE | 1.98624805 |
| T2DM_mortality | endothelial_cell | *NPC1* | TRUE | 2.378417506 |
| T2DM_mortality | endothelial_cell | *TRIM65* | TRUE | 2.012838021 |
| T2DM_mortality | endothelial_cell | *FUT9* | TRUE | 2.837503689 |
| T2DM_mortality | endothelial_cell | *GPR88* | TRUE | 2.396895737 |

| | | | | |
|---|---|---|---|---|
| T2DM_mortality | endothelial_cell | *SMTNL1* | TRUE | 2.488469871 |
| T2DM_mortality | epsilon_cell | *CYP4B1* | TRUE | 2.908042065 |
| T2DM_mortality | epsilon_cell | *FUT9* | TRUE | 2.358538248 |
| T2DM_mortality | epsilon_cell | *GPR88* | TRUE | 2.228369147 |
| T2DM_mortality | epsilon_cell | *SMTNL1* | TRUE | 2.324589228 |
| T2DM_mortality | leukocyte | MROH8 | TRUE | 2.121033155 |
| T2DM_mortality | leukocyte | FUT9 | TRUE | 2.184595168 |
| T2DM_mortality | leukocyte | GPR88 | TRUE | 2.324811726 |
| T2DM_mortality | leukocyte | SMTNL1 | TRUE | 2.148790799 |
| T2DM_mortality | mesenchymal_cell | MROH8 | TRUE | 2.409097015 |
| T2DM_mortality | mesenchymal_cell | NDUFAF1 | TRUE | 2.101914614 |
| T2DM_mortality | mesenchymal_cell | TRIM63 | TRUE | 2.182656755 |
| T2DM_mortality | mesenchymal_cell | FUT9 | TRUE | 1.996096189 |
| T2DM_mortality | mesenchymal_cell | IRX3 | TRUE | 2.623270538 |
| T2DM_mortality | mesenchymal_cell | LYPD6 | TRUE | 1.97114281 |
| T2DM_mortality | mesenchymal_cell | DDO | TRUE | 2.290532938 |
| T2DM_mortality | mesenchymal_cell | C6orf163 | TRUE | 2.240441843 |
| T2DM_mortality | mesenchymal_cell | TREX1 | TRUE | 1.975736469 |
| T2DM_mortality | mesenchymal_cell | SMTNL1 | TRUE | 2.181080219 |
| T2DM_mortality | PP_cell | MLPH | TRUE | 2.1446663 |
| T2DM_mortality | PP_cell | RIBC2 | TRUE | 2.303896452 |
| T2DM_mortality | PP_cell | CYP4B1 | TRUE | 2.559864217 |
| T2DM_mortality | PP_cell | GPR88 | TRUE | 2.214974098 |
| T2DM_mortality | PP_cell | SMTNL1 | TRUE | 2.093450854 |

c. Identified cell type specific causal genes for obesity and related disorders from SAT

| disorder | celltype | gene | assoc | zMin |
|---|---|---|---|---|
| CAD | adipocyte | *DLX6* | TRUE | 3.05768278 |
| CAD | adipocyte | *LAMC2* | TRUE | 2.079349762 |
| CAD | adipocyte | *ATP2A3* | TRUE | 2.152364946 |
| CAD | adipocyte | *TIPIN* | TRUE | 3.389545984 |
| CAD | adipocyte | *TXNDC16* | TRUE | 2.035752693 |
| CAD | adipocyte | *RPGRIP1* | TRUE | 2.250177492 |
| CAD | adipocyte | *SUSD2* | TRUE | 2.225892949 |
| CAD | adipocyte | *FOXRED2* | TRUE | 2.214976703 |
| CAD | adipocyte | *ABHD12* | TRUE | 2.269059626 |
| CAD | adipocyte | *MAP1LC3A* | TRUE | 2.938530229 |
| CAD | adipocyte | *CTSH* | TRUE | 2.006163569 |
| CAD | adipocyte | *AMH* | TRUE | 2.045671176 |
| CAD | adipocyte | *MAG* | TRUE | 2.156847402 |
| CAD | adipocyte | *STEAP1B* | TRUE | 2.552130938 |
| CAD | adipocyte | *ICAM2* | TRUE | 2.040423616 |
| CAD | adipocyte | *ENDOU* | TRUE | 2.595457735 |
| CAD | adipocyte | *BVES* | TRUE | 2.002075557 |
| CAD | adipocyte | *APBB3* | TRUE | 2.204270282 |
| CAD | adipocyte | *BNIP1* | TRUE | 2.669579675 |
| CAD | adipocyte | *BCHE* | TRUE | 3.229938448 |
| CAD | adipocyte | *UMPS* | TRUE | 2.197242896 |
| CAD | adipocyte | *PLXNA1* | TRUE | 3.09421374 |
| CAD | adipocyte | *GGCX* | TRUE | 3.628574866 |
| CAD | adipocyte | *TNFSF4* | TRUE | 2.644293724 |
| CAD | adipocyte | *EIF2B2* | TRUE | 2.914922339 |
| CAD | adipocyte | *CKS2* | TRUE | 1.989006238 |
| CAD | adipocyte | *HIF3A* | TRUE | 2.196026373 |
| CAD | adipocyte | *GRK4* | TRUE | 3.316240964 |
| CAD | adipocyte | *REEP2* | TRUE | 1.986100995 |
| CAD | adipocyte | *B4GALNT1* | TRUE | 2.538602116 |
| CAD | adipocyte | *TMBIM1* | TRUE | 2.05694508 |
| CAD | adipocyte | *POLR1E* | TRUE | 2.775350236 |
| CAD | adipocyte | *PIANP* | TRUE | 2.025595455 |
| CAD | adipocyte | *HORMAD1* | TRUE | 2.29651234 |
| CAD | adipocyte | *NKD2* | TRUE | 2.030286464 |
| CAD | adipocyte | *PSD2* | TRUE | 3.576874189 |
| CAD | adipocyte | *SLC18B1* | TRUE | 2.659822255 |
| CAD | adipocyte | *HSD17B12* | TRUE | 2.996461567 |
| CAD | adipocyte | *TTC12* | TRUE | 2.312263904 |
| CAD | adipocyte | *DUSP15* | TRUE | 2.602588602 |
| CAD | adipocyte | *DISP1* | TRUE | 3.221600725 |
| CAD | adipocyte | *MED19* | TRUE | 2.032124753 |
| CAD | adipocyte | *CLEC18A* | TRUE | 2.696215425 |
| CAD | adipocyte | *MX1* | TRUE | 1.982178471 |
| CAD | adipocyte | *U2AF1* | TRUE | 2.066401218 |
| CAD | adipocyte | *NAGS* | TRUE | 2.236997308 |

| | | | | |
|---|---|---|---|---|
| CAD | adipocyte | *JMJD8* | TRUE | 2.095011659 |
| CAD | adipocyte | *LMOD1* | TRUE | 3.135623885 |
| CAD | adipocyte | *ICA1L* | TRUE | 2.457228877 |
| CAD | adipocyte | *APEH* | TRUE | 2.862504721 |
| CAD | adipocyte | *GPER1* | TRUE | 2.509650895 |
| CAD | adipocyte | *OXGR1* | TRUE | 3.439545116 |
| CAD | adipocyte | *KIFC2* | TRUE | 2.202290995 |
| CAD | adipocyte | *ULK4* | TRUE | 2.729962589 |
| CAD | adipocyte | *MLKL* | TRUE | 2.165113775 |
| CAD | adipocyte | *IRS1* | TRUE | 4.667595416 |
| CAD | adipocyte | *PCSK9* | TRUE | 4.051216077 |
| CAD | adipocyte | *CALML6* | TRUE | 1.997065246 |
| CAD | adipocyte | *C3AR1* | TRUE | 2.131530386 |
| CAD | adipocyte | *ZNF738* | TRUE | 1.995108244 |
| CAD | adipocyte | *TLR6* | TRUE | 2.115002253 |
| CAD | adipocyte | *LPL* | TRUE | 4.532917016 |
| CAD | adipocyte | *PAAF1* | TRUE | 2.595644471 |
| CAD | adipocyte | *RIMKLA* | TRUE | 2.419740863 |
| CAD | adipocyte | *PPFIA3* | TRUE | 1.997343838 |
| CAD | adipocyte | *C12orf54* | TRUE | 2.21756941 |
| CAD | adipocyte | *MPI* | TRUE | 2.02998061 |
| CAD | adipocyte | *TIGD5* | TRUE | 2.221339656 |
| CAD | adipocyte | *LYNX1* | TRUE | 2.288578216 |
| CAD | adipocyte | *ZNF443* | TRUE | 2.302078699 |
| CAD | adipocyte | *MRPL14* | TRUE | 2.101945164 |
| CAD | adipocyte | *ADIPOQ* | TRUE | 2.345544328 |
| CAD | adipocyte | *TMEM136* | TRUE | 3.098515473 |
| CAD | adipocyte | *PLCXD3* | TRUE | 2.591928019 |
| CAD | adipocyte | *ACOT1* | TRUE | 2.446667294 |
| CAD | adipocyte | *GDF3* | TRUE | 1.969648528 |
| CAD | adipocyte | *A3GALT2* | TRUE | 2.31935041 |
| CAD | adipocyte | *MSL1* | TRUE | 2.937947718 |
| CAD | adipocyte | *C6orf106* | TRUE | 3.238989676 |
| CAD | adipocyte | *KIAA1211L* | TRUE | 1.965755825 |
| CAD | adipocyte | *WDR5B* | TRUE | 2.063410386 |
| CAD | adipocyte | *RPS26* | TRUE | 2.178447697 |
| CAD | adipocyte | *ZNF71* | TRUE | 2.059794643 |
| CAD | adipocyte | *ZNF334* | TRUE | 2.435719305 |
| CAD | adipocyte | *TMEM116* | TRUE | 3.238650643 |
| CAD | adipocyte | *TBKBP1* | TRUE | 2.341659237 |
| CAD | adipocyte | *CENPW* | TRUE | 3.701389562 |
| CAD | adipocyte | *PRCD* | TRUE | 2.03389422 |
| CAD | adipocyte | *PROB1* | TRUE | 2.188790768 |
| CAD | adipocyte | *AC112229.4* | TRUE | 2.212225006 |
| CAD | adipocyte | *LINC00957* | TRUE | 2.195781915 |
| CAD | adipocyte | *LY6G5B* | TRUE | 2.268335802 |
| CAD | adipocyte | *SHANK3* | TRUE | 2.023691892 |
| CAD | adipocyte | *ARMS2* | TRUE | 2.538403523 |

| | | | | |
|---|---|---|---|---|
| CAD | adipocyte | *ZNF10* | TRUE | 2.506854166 |
| CAD | adipocyte | *HMBS* | TRUE | 3.607661975 |
| CAD | adipocyte | *GALNT4* | TRUE | 2.471979892 |
| CAD | ASPC | *TNFRSF12A* | TRUE | 1.972073829 |
| CAD | ASPC | *DLX6* | TRUE | 3.014625977 |
| CAD | ASPC | *MAPK8IP2* | TRUE | 2.147446193 |
| CAD | ASPC | *MTMR11* | TRUE | 2.345198182 |
| CAD | ASPC | *NPC1L1* | TRUE | 2.967140149 |
| CAD | ASPC | *LAMC2* | TRUE | 2.128126088 |
| CAD | ASPC | *SDK2* | TRUE | 2.001922829 |
| CAD | ASPC | *ASNS* | TRUE | 2.381549242 |
| CAD | ASPC | *ATP2A3* | TRUE | 2.535126438 |
| CAD | ASPC | *TIPIN* | TRUE | 3.683140554 |
| CAD | ASPC | *FBLN1* | TRUE | 2.327983516 |
| CAD | ASPC | *SCGN* | TRUE | 2.096824193 |
| CAD | ASPC | *COL16A1* | TRUE | 2.227620815 |
| CAD | ASPC | *TXNDC16* | TRUE | 2.646310593 |
| CAD | ASPC | *RPGRIP1* | TRUE | 2.513912429 |
| CAD | ASPC | *MARCH2* | TRUE | 2.271872075 |
| CAD | ASPC | *SUSD2* | TRUE | 2.245453618 |
| CAD | ASPC | *SLC25A1* | TRUE | 2.111749214 |
| CAD | ASPC | *FOXRED2* | TRUE | 2.507883921 |
| CAD | ASPC | *TRIM9* | TRUE | 1.985537663 |
| CAD | ASPC | *ABHD12* | TRUE | 3.021598966 |
| CAD | ASPC | *ACTR5* | TRUE | 2.58858499 |
| CAD | ASPC | *MAP1LC3A* | TRUE | 3.62442116 |
| CAD | ASPC | *PIGU* | TRUE | 1.962832113 |
| CAD | ASPC | *DHODH* | TRUE | 2.130101538 |
| CAD | ASPC | *NAGPA* | TRUE | 2.624476957 |
| CAD | ASPC | *CTSH* | TRUE | 2.5298685 |
| CAD | ASPC | *AMH* | TRUE | 2.177995007 |
| CAD | ASPC | *DMPK* | TRUE | 2.566073937 |
| CAD | ASPC | *MAG* | TRUE | 2.215008883 |
| CAD | ASPC | *STEAP1B* | TRUE | 2.947188931 |
| CAD | ASPC | *COA1* | TRUE | 2.483443759 |
| CAD | ASPC | *AK1* | TRUE | 2.220231719 |
| CAD | ASPC | *ICAM2* | TRUE | 3.121948999 |
| CAD | ASPC | *VAT1* | TRUE | 2.077427895 |
| CAD | ASPC | *KLHL5* | TRUE | 2.096934792 |
| CAD | ASPC | *TRPV4* | TRUE | 2.578201767 |
| CAD | ASPC | *MDN1* | TRUE | 2.276191214 |
| CAD | ASPC | *BVES* | TRUE | 2.296545735 |
| CAD | ASPC | *APBB3* | TRUE | 2.363618403 |
| CAD | ASPC | *BNIP1* | TRUE | 2.942649648 |
| CAD | ASPC | *NIT2* | TRUE | 2.104036153 |
| CAD | ASPC | *BCHE* | TRUE | 3.741208038 |
| CAD | ASPC | *UMPS* | TRUE | 3.42475634 |
| CAD | ASPC | *PLXNA1* | TRUE | 3.527471265 |
| CAD | ASPC | *GGCX* | TRUE | 4.091885175 |
| CAD | ASPC | *ZNF142* | TRUE | 2.287200102 |

| | | | | |
|---|---|---|---|---|
| CAD | ASPC | *CFHR3* | TRUE | 2.00038848 |
| CAD | ASPC | *TNFSF4* | TRUE | 2.939128095 |
| CAD | ASPC | *DIEXF* | TRUE | 2.024488832 |
| CAD | ASPC | *EIF2B2* | TRUE | 2.089881485 |
| CAD | ASPC | *CKS2* | TRUE | 2.568516441 |
| CAD | ASPC | *HIF3A* | TRUE | 3.080203608 |
| CAD | ASPC | *GRK4* | TRUE | 3.65185118 |
| CAD | ASPC | *YEATS4* | TRUE | 2.271775546 |
| CAD | ASPC | *SMARCA4* | TRUE | 2.064672999 |
| CAD | ASPC | *EMC4* | TRUE | 2.143774999 |
| CAD | ASPC | *KLHDC10* | TRUE | 3.262597965 |
| CAD | ASPC | *PLVAP* | TRUE | 2.032648114 |
| CAD | ASPC | *REEP2* | TRUE | 2.015461509 |
| CAD | ASPC | *SPIRE1* | TRUE | 2.011600327 |
| CAD | ASPC | *HRK* | TRUE | 2.052233094 |
| CAD | ASPC | *TMBIM1* | TRUE | 1.966820123 |
| CAD | ASPC | *POLR1E* | TRUE | 3.396231439 |
| CAD | ASPC | *RMDN3* | TRUE | 2.116718814 |
| CAD | ASPC | *BTBD16* | TRUE | 1.99433635 |
| CAD | ASPC | *PLCE1* | TRUE | 2.131536219 |
| CAD | ASPC | *NUP54* | TRUE | 2.379116049 |
| CAD | ASPC | *GPR84* | TRUE | 2.047630685 |
| CAD | ASPC | *TICRR* | TRUE | 2.517480026 |
| CAD | ASPC | *TGFB1I1* | TRUE | 2.08075758 |
| CAD | ASPC | *UNC45B* | TRUE | 2.021223791 |
| CAD | ASPC | *TMEM91* | TRUE | 2.099884847 |
| CAD | ASPC | *NME7* | TRUE | 2.88821873 |
| CAD | ASPC | *WNT9A* | TRUE | 2.251808282 |
| CAD | ASPC | *AFF3* | TRUE | 2.123665712 |
| CAD | ASPC | *NKD2* | TRUE | 2.043138525 |
| CAD | ASPC | *PSD2* | TRUE | 3.548732891 |
| CAD | ASPC | *SLC18B1* | TRUE | 3.027091511 |
| CAD | ASPC | *DNAJB12* | TRUE | 2.021157195 |
| CAD | ASPC | *PDCD11* | TRUE | 2.304896774 |
| CAD | ASPC | *HSD17B12* | TRUE | 3.354442977 |
| CAD | ASPC | *TTC12* | TRUE | 2.665661248 |
| CAD | ASPC | *DUSP15* | TRUE | 2.64460057 |
| CAD | ASPC | *TMEM86A* | TRUE | 1.988210006 |
| CAD | ASPC | *ARL14EP* | TRUE | 2.196838525 |
| CAD | ASPC | *PTPRR* | TRUE | 2.154645336 |
| CAD | ASPC | *ADPRHL1* | TRUE | 2.018740912 |
| CAD | ASPC | *MIA3* | TRUE | 2.928588189 |
| CAD | ASPC | *DISP1* | TRUE | 3.688850449 |
| CAD | ASPC | *MED19* | TRUE | 2.361384603 |
| CAD | ASPC | *CLEC18A* | TRUE | 1.997963167 |
| CAD | ASPC | *KCNJ15* | TRUE | 2.122762826 |
| CAD | ASPC | *MX1* | TRUE | 2.313953501 |
| CAD | ASPC | *EPB41* | TRUE | 2.581701397 |
| CAD | ASPC | *U2AF1* | TRUE | 2.37992496 |
| CAD | ASPC | *S100A1* | TRUE | 2.01779496 |

| | | | | |
|---|---|---|---|---|
| CAD | ASPC | *FBXO27* | TRUE | 2.337603361 |
| CAD | ASPC | *ALDH16A1* | TRUE | 2.061203875 |
| CAD | ASPC | *MPP3* | TRUE | 2.341293732 |
| CAD | ASPC | *LARP4* | TRUE | 2.048996596 |
| CAD | ASPC | *JMJD8* | TRUE | 2.52324085 |
| CAD | ASPC | *CGGBP1* | TRUE | 1.965483167 |
| CAD | ASPC | *LMOD1* | TRUE | 3.618345545 |
| CAD | ASPC | *C1orf50* | TRUE | 2.068300736 |
| CAD | ASPC | *APEH* | TRUE | 2.883932128 |
| CAD | ASPC | *GPER1* | TRUE | 2.906907724 |
| CAD | ASPC | *PIGO* | TRUE | 2.155284322 |
| CAD | ASPC | *OXGR1* | TRUE | 3.635140514 |
| CAD | ASPC | *STK32C* | TRUE | 2.190109904 |
| CAD | ASPC | *RIMKLB* | TRUE | 2.030493917 |
| CAD | ASPC | *ANKDD1A* | TRUE | 2.141276754 |
| CAD | ASPC | *SP2* | TRUE | 2.001108553 |
| CAD | ASPC | *KIFC2* | TRUE | 2.146697878 |
| CAD | ASPC | *ULK4* | TRUE | 3.010441786 |
| CAD | ASPC | *MLKL* | TRUE | 2.588371847 |
| CAD | ASPC | *LGALS9* | TRUE | 2.191520583 |
| CAD | ASPC | *IRS1* | TRUE | 4.942158062 |
| CAD | ASPC | *PCSK9* | TRUE | 4.127945484 |
| CAD | ASPC | *CALML6* | TRUE | 2.432296603 |
| CAD | ASPC | *TSNARE1* | TRUE | 2.289729254 |
| CAD | ASPC | *KCNMB3* | TRUE | 2.062404604 |
| CAD | ASPC | *RSL1D1* | TRUE | 2.111140182 |
| CAD | ASPC | *C3AR1* | TRUE | 3.453714936 |
| CAD | ASPC | *TLN2* | TRUE | 2.123077148 |
| CAD | ASPC | *SULT1B1* | TRUE | 2.040567443 |
| CAD | ASPC | *GOLIM4* | TRUE | 2.060382993 |
| CAD | ASPC | *TLR6* | TRUE | 2.282893324 |
| CAD | ASPC | *LPL* | TRUE | 4.734183314 |
| CAD | ASPC | *PAAF1* | TRUE | 2.296333552 |
| CAD | ASPC | *TEX26* | TRUE | 2.059885184 |
| CAD | ASPC | *CHMP6* | TRUE | 2.1057001 |
| CAD | ASPC | *CRLF3* | TRUE | 3.425020419 |
| CAD | ASPC | *RIMKLA* | TRUE | 2.160730746 |
| CAD | ASPC | *FBXO39* | TRUE | 2.010098178 |
| CAD | ASPC | *AC024940.1* | TRUE | 2.064669214 |
| CAD | ASPC | *PPFIA3* | TRUE | 2.195862908 |
| CAD | ASPC | *C12orf54* | TRUE | 2.515493397 |
| CAD | ASPC | *TIGD5* | TRUE | 2.471029452 |
| CAD | ASPC | *TMEM86B* | TRUE | 2.019172576 |
| CAD | ASPC | *LYNX1* | TRUE | 2.503222444 |
| CAD | ASPC | *ZNF443* | TRUE | 2.800832458 |
| CAD | ASPC | *MRPL14* | TRUE | 2.760179848 |
| CAD | ASPC | *ADIPOQ* | TRUE | 2.404821778 |
| CAD | ASPC | *TMEM136* | TRUE | 3.285341473 |
| CAD | ASPC | *FES* | TRUE | 4.516768607 |
| CAD | ASPC | *PLCXD3* | TRUE | 3.041031504 |

| | | | | |
|---|---|---|---|---|
| CAD | ASPC | *ZNF320* | TRUE | 2.083137147 |
| CAD | ASPC | *PP2D1* | TRUE | 2.407688378 |
| CAD | ASPC | *ACOT1* | TRUE | 2.66668853 |
| CAD | ASPC | *GDF3* | TRUE | 2.209181033 |
| CAD | ASPC | *CSF1* | TRUE | 2.439543268 |
| CAD | ASPC | *A3GALT2* | TRUE | 2.366366117 |
| CAD | ASPC | *TCN2* | TRUE | 2.244835734 |
| CAD | ASPC | *SP1* | TRUE | 2.051931919 |
| CAD | ASPC | *ADH1A* | TRUE | 2.078592736 |
| CAD | ASPC | *C2orf66* | TRUE | 2.328907594 |
| CAD | ASPC | *MSL1* | TRUE | 3.37696159 |
| CAD | ASPC | *SULT1A1* | TRUE | 2.054086576 |
| CAD | ASPC | *MYO6* | TRUE | 2.592502931 |
| CAD | ASPC | *C6orf106* | TRUE | 3.272080359 |
| CAD | ASPC | *ZFP28* | TRUE | 2.16552546 |
| CAD | ASPC | *KIAA1211L* | TRUE | 2.1708891 |
| CAD | ASPC | *ZNF100* | TRUE | 2.485095766 |
| CAD | ASPC | *ZNF665* | TRUE | 2.261237783 |
| CAD | ASPC | *MRPL42* | TRUE | 2.195922075 |
| CAD | ASPC | *HIBCH* | TRUE | 2.047621128 |
| CAD | ASPC | *ZNF334* | TRUE | 2.746243527 |
| CAD | ASPC | *TMEM116* | TRUE | 3.484630193 |
| CAD | ASPC | *TBKBP1* | TRUE | 2.027455882 |
| CAD | ASPC | *CENPW* | TRUE | 4.121036616 |
| CAD | ASPC | *FAM216A* | TRUE | 2.058713543 |
| CAD | ASPC | *ZNF425* | TRUE | 2.139864547 |
| CAD | ASPC | *ARHGEF35* | TRUE | 2.105094545 |
| CAD | ASPC | *SIPA1* | TRUE | 2.478695442 |
| CAD | ASPC | *PRCD* | TRUE | 2.710010679 |
| CAD | ASPC | *LINC00863* | TRUE | 2.037602416 |
| CAD | ASPC | *PROB1* | TRUE | 2.079404 |
| CAD | ASPC | *AC112229.4* | TRUE | 2.256054836 |
| CAD | ASPC | *LINC00865* | TRUE | 1.993745885 |
| CAD | ASPC | *GOLGA8N* | TRUE | 2.326215895 |
| CAD | ASPC | *HLA.B* | TRUE | 2.170809172 |
| CAD | ASPC | *LINC00957* | TRUE | 2.375288923 |
| CAD | ASPC | *B3GALT4* | TRUE | 2.038983754 |
| CAD | ASPC | *LY6G5B* | TRUE | 2.157875353 |
| CAD | ASPC | *TDGF1* | TRUE | 2.008638762 |
| CAD | ASPC | *C22orf39* | TRUE | 2.457907485 |
| CAD | ASPC | *BCKDHA* | TRUE | 2.046975367 |
| CAD | ASPC | *ZNF10* | TRUE | 2.767849077 |
| CAD | ASPC | *HMBS* | TRUE | 3.968547686 |
| CAD | ASPC | *GALNT4* | TRUE | 2.491186628 |
| CAD | ASPC | *IQCH.AS1* | TRUE | 2.241557153 |
| CAD | ASPC | *AC144652.1* | TRUE | 2.149410631 |
| CAD | b_cell | *DLX6* | TRUE | 2.023318715 |
| CAD | b_cell | *MAG* | TRUE | 1.975578797 |
| CAD | b_cell | *BNIP1* | TRUE | 2.127887229 |
| CAD | b_cell | *AMT* | TRUE | 1.975563106 |

| | | | | |
|---|---|---|---|---|
| CAD | b_cell | *DISP1* | TRUE | 2.648513381 |
| CAD | b_cell | *CLEC18A* | TRUE | 2.656290534 |
| CAD | b_cell | *OXGR1* | TRUE | 2.823102342 |
| CAD | b_cell | *LY6G5B* | TRUE | 3.182475375 |
| CAD | dendritic_cell | *SUSD2* | TRUE | 2.23171107 |
| CAD | dendritic_cell | *STEAP1B* | TRUE | 2.125954466 |
| CAD | dendritic_cell | *GGCX* | TRUE | 2.29362084 |
| CAD | dendritic_cell | *SNRPC* | TRUE | 2.405908264 |
| CAD | dendritic_cell | *HORMAD1* | TRUE | 2.231064927 |
| CAD | dendritic_cell | *SLC18B1* | TRUE | 2.282181041 |
| CAD | dendritic_cell | *HSD17B12* | TRUE | 2.301192854 |
| CAD | dendritic_cell | *LMOD1* | TRUE | 2.044784769 |
| CAD | dendritic_cell | *IRS1* | TRUE | 2.197510581 |
| CAD | dendritic_cell | *RPS26* | TRUE | 2.126907427 |
| CAD | dendritic_cell | *LY6G5B* | TRUE | 2.562710878 |
| CAD | endothelial | *MAPK8IP2* | TRUE | 2.055647747 |
| CAD | endothelial | *MTMR11* | TRUE | 2.00881158 |
| CAD | endothelial | *TIPIN* | TRUE | 2.090121365 |
| CAD | endothelial | *BCHE* | TRUE | 2.767159137 |
| CAD | endothelial | *SNRPC* | TRUE | 2.031101176 |
| CAD | endothelial | *GRK4* | TRUE | 2.785583095 |
| CAD | endothelial | *REEP2* | TRUE | 2.040257069 |
| CAD | endothelial | *POLR1E* | TRUE | 2.271453174 |
| CAD | endothelial | *AMT* | TRUE | 2.155172436 |
| CAD | endothelial | *IRS1* | TRUE | 3.084076827 |
| CAD | endothelial | *ZNF443* | TRUE | 2.162994259 |
| CAD | endothelial | *PROB1* | TRUE | 2.183659754 |
| CAD | endothelial | *LY6G5B* | TRUE | 2.690644156 |
| CAD | endothelial | *TDGF1* | TRUE | 2.306845259 |
| CAD | LEC | *SEMA3F* | TRUE | 2.350681446 |
| CAD | LEC | *TIPIN* | TRUE | 2.674204855 |
| CAD | LEC | *KCNH4* | TRUE | 2.717109524 |
| CAD | LEC | *SUSD2* | TRUE | 2.245179744 |
| CAD | LEC | *BNIP1* | TRUE | 2.149195351 |
| CAD | LEC | *SNRPC* | TRUE | 2.858267946 |
| CAD | LEC | *GRK4* | TRUE | 2.856727826 |
| CAD | LEC | *POLR1E* | TRUE | 2.410053175 |
| CAD | LEC | *CLEC18A* | TRUE | 2.486157797 |
| CAD | LEC | *IRS1* | TRUE | 2.802293763 |
| CAD | LEC | *ACOT1* | TRUE | 2.219057042 |
| CAD | LEC | *TAS2R14* | TRUE | 2.092158661 |
| CAD | LEC | *LY6G5B* | TRUE | 2.510296679 |
| CAD | macrophage | *DLX6* | TRUE | 2.930276455 |
| CAD | macrophage | *UHRF1BP1* | TRUE | 2.308445902 |
| CAD | macrophage | *TIE1* | TRUE | 2.310768448 |
| CAD | macrophage | *TIPIN* | TRUE | 3.386678977 |
| CAD | macrophage | *SUSD2* | TRUE | 2.248024667 |
| CAD | macrophage | *MAP1LC3A* | TRUE | 3.014619225 |
| CAD | macrophage | *CTSH* | TRUE | 2.09898779 |
| CAD | macrophage | *MAG* | TRUE | 2.175770046 |

| | | | | |
|---|---|---|---|---|
| CAD | macrophage | *STEAP1B* | TRUE | 1.972830136 |
| CAD | macrophage | *ENDOU* | TRUE | 2.464887648 |
| CAD | macrophage | *BVES* | TRUE | 2.122105146 |
| CAD | macrophage | *BNIP1* | TRUE | 2.720277316 |
| CAD | macrophage | *BCHE* | TRUE | 2.962551905 |
| CAD | macrophage | *PLXNA1* | TRUE | 2.942534072 |
| CAD | macrophage | *GGCX* | TRUE | 3.474000563 |
| CAD | macrophage | *EIF2B2* | TRUE | 2.615341747 |
| CAD | macrophage | *GRK4* | TRUE | 3.290424662 |
| CAD | macrophage | *REEP2* | TRUE | 2.014952345 |
| CAD | macrophage | *POLR1E* | TRUE | 3.153632038 |
| CAD | macrophage | *TMEM91* | TRUE | 2.615429246 |
| CAD | macrophage | *HORMAD1* | TRUE | 2.087007064 |
| CAD | macrophage | *NKD2* | TRUE | 1.98893723 |
| CAD | macrophage | *PSD2* | TRUE | 2.95189354 |
| CAD | macrophage | *SLC18B1* | TRUE | 1.993059478 |
| CAD | macrophage | *HSD17B12* | TRUE | 2.785605496 |
| CAD | macrophage | *TTC12* | TRUE | 2.289087677 |
| CAD | macrophage | *DUSP15* | TRUE | 2.594143569 |
| CAD | macrophage | *DISP1* | TRUE | 2.591068098 |
| CAD | macrophage | *CLEC18A* | TRUE | 2.689571402 |
| CAD | macrophage | *MX1* | TRUE | 2.030848638 |
| CAD | macrophage | *LMOD1* | TRUE | 2.241234559 |
| CAD | macrophage | *ICA1L* | TRUE | 3.436455347 |
| CAD | macrophage | *APEH* | TRUE | 2.604573955 |
| CAD | macrophage | *GPER1* | TRUE | 2.483142643 |
| CAD | macrophage | *OXGR1* | TRUE | 3.389402635 |
| CAD | macrophage | *MEI1* | TRUE | 2.005891976 |
| CAD | macrophage | *ZNF641* | TRUE | 2.390036252 |
| CAD | macrophage | *KIFC2* | TRUE | 2.368234425 |
| CAD | macrophage | *ULK4* | TRUE | 2.457117488 |
| CAD | macrophage | *MLKL* | TRUE | 2.009027475 |
| CAD | macrophage | *IRS1* | TRUE | 4.538399626 |
| CAD | macrophage | *PCSK9* | TRUE | 3.477465989 |
| CAD | macrophage | *LPL* | TRUE | 3.969263325 |
| CAD | macrophage | *PAAF1* | TRUE | 2.405030655 |
| CAD | macrophage | *RIMKLA* | TRUE | 2.143152329 |
| CAD | macrophage | *TIGD5* | TRUE | 1.965063697 |
| CAD | macrophage | *ZNF443* | TRUE | 2.613962978 |
| CAD | macrophage | *TMEM136* | TRUE | 2.361721821 |
| CAD | macrophage | *PLCXD3* | TRUE | 2.018455444 |
| CAD | macrophage | *ZNF320* | TRUE | 1.996363798 |
| CAD | macrophage | *BMP8A* | TRUE | 2.026544514 |
| CAD | macrophage | *ACOT1* | TRUE | 2.264665172 |
| CAD | macrophage | *GDF3* | TRUE | 2.01753766 |
| CAD | macrophage | *A3GALT2* | TRUE | 2.129018775 |
| CAD | macrophage | *MSL1* | TRUE | 2.497255846 |
| CAD | macrophage | *C6orf106* | TRUE | 2.680975981 |
| CAD | macrophage | *KIAA1211L* | TRUE | 2.018634659 |
| CAD | macrophage | *WDR5B* | TRUE | 2.760916881 |

| | | | | |
|---|---|---|---|---|
| CAD | macrophage | *ZNF334* | TRUE | 2.081625606 |
| CAD | macrophage | *TMEM116* | TRUE | 2.806013242 |
| CAD | macrophage | *TBKBP1* | TRUE | 2.361449035 |
| CAD | macrophage | *CENPW* | TRUE | 2.615036568 |
| CAD | macrophage | *PROB1* | TRUE | 2.088870011 |
| CAD | macrophage | *AC112229.4* | TRUE | 2.08572987 |
| CAD | macrophage | *GOLGA8N* | TRUE | 1.988686136 |
| CAD | macrophage | *LINC00957* | TRUE | 1.984304024 |
| CAD | macrophage | *LY6G5B* | TRUE | 2.784369296 |
| CAD | macrophage | *ARMS2* | TRUE | 2.140304427 |
| CAD | macrophage | *ZNF10* | TRUE | 2.000533004 |
| CAD | macrophage | *HMBS* | TRUE | 3.629335938 |
| CAD | macrophage | *GALNT4* | TRUE | 2.342183723 |
| CAD | macrophage | *AC144652.1* | TRUE | 1.97647797 |
| CAD | mast_cell | *DLX6* | TRUE | 2.017693815 |
| CAD | mast_cell | *UHRF1BP1* | TRUE | 1.981500504 |
| CAD | mast_cell | *TIPIN* | TRUE | 2.57328573 |
| CAD | mast_cell | *BCHE* | TRUE | 2.549755995 |
| CAD | mast_cell | *GRK4* | TRUE | 3.015009824 |
| CAD | mast_cell | *POLR1E* | TRUE | 2.730811721 |
| CAD | mast_cell | *APEH* | TRUE | 2.058192528 |
| CAD | mast_cell | *OXGR1* | TRUE | 2.70052901 |
| CAD | mast_cell | *IRS1* | TRUE | 4.141678725 |
| CAD | mast_cell | *LPL* | TRUE | 2.694073594 |
| CAD | mast_cell | *ZNF443* | TRUE | 2.156038737 |
| CAD | mast_cell | *C6orf106* | TRUE | 1.988811002 |
| CAD | mast_cell | *RPS26* | TRUE | 1.976798981 |
| CAD | mast_cell | *TMEM116* | TRUE | 1.999004096 |
| CAD | mast_cell | *CENPW* | TRUE | 3.062914363 |
| CAD | mast_cell | *LY6G5B* | TRUE | 2.968903839 |
| CAD | monocyte | *UHRF1BP1* | TRUE | 2.461823054 |
| CAD | monocyte | *MAG* | TRUE | 2.102730688 |
| CAD | monocyte | *GGCX* | TRUE | 2.809831934 |
| CAD | monocyte | *WFDC3* | TRUE | 2.176435791 |
| CAD | monocyte | *PSD2* | TRUE | 3.47306611 |
| CAD | monocyte | *DUSP15* | TRUE | 2.390614082 |
| CAD | monocyte | *DISP1* | TRUE | 2.22382412 |
| CAD | monocyte | *OXGR1* | TRUE | 2.803042257 |
| CAD | monocyte | *RPS26* | TRUE | 2.186482648 |
| CAD | monocyte | *LY6G5B* | TRUE | 2.632236354 |
| CAD | monocyte | *HMBS* | TRUE | 2.83041155 |
| CAD | neutrophil | *TIPIN* | TRUE | 2.161490597 |
| CAD | neutrophil | *CTSH* | TRUE | 1.964151594 |
| CAD | neutrophil | *CLEC18A* | TRUE | 2.230786239 |
| CAD | neutrophil | *APEH* | TRUE | 2.227861318 |
| CAD | neutrophil | *OXGR1* | TRUE | 2.465778307 |
| CAD | neutrophil | *ACOT1* | TRUE | 2.182341051 |
| CAD | neutrophil | *LY6G5B* | TRUE | 2.731785267 |
| CAD | nk_cell | *UHRF1BP1* | TRUE | 2.213446403 |
| CAD | nk_cell | *SUSD2* | TRUE | 2.196145852 |

| | | | | |
|---|---|---|---|---|
| CAD | nk_cell | *BNIP1* | TRUE | 2.215638145 |
| CAD | nk_cell | *GRK4* | TRUE | 3.132143275 |
| CAD | nk_cell | *POLR1E* | TRUE | 2.481015759 |
| CAD | nk_cell | *CLEC18A* | TRUE | 2.69241437 |
| CAD | nk_cell | *OXGR1* | TRUE | 3.195215735 |
| CAD | nk_cell | *KIFC2* | TRUE | 2.027537601 |
| CAD | nk_cell | *IRS1* | TRUE | 2.980911345 |
| CAD | nk_cell | *C8orf46* | TRUE | 1.995719019 |
| CAD | nk_cell | *PPFIA3* | TRUE | 2.169129644 |
| CAD | nk_cell | *BMP8A* | TRUE | 1.985923514 |
| CAD | nk_cell | *TMEM116* | TRUE | 2.408320388 |
| CAD | nk_cell | *LY6G5B* | TRUE | 2.687885556 |
| CAD | nk_cell | *ARMS2* | TRUE | 2.376374411 |
| CAD | nk_cell | *HMBS* | TRUE | 3.437293498 |
| CAD | pericyte | *TIPIN* | TRUE | 2.891701056 |
| CAD | pericyte | *KCNH4* | TRUE | 2.275439972 |
| CAD | pericyte | *MAP1LC3A* | TRUE | 2.305001676 |
| CAD | pericyte | *AMH* | TRUE | 2.178911667 |
| CAD | pericyte | *BNIP1* | TRUE | 2.714138397 |
| CAD | pericyte | *PLXNA1* | TRUE | 2.570512706 |
| CAD | pericyte | *GGCX* | TRUE | 3.284086892 |
| CAD | pericyte | *EIF2B2* | TRUE | 2.535011136 |
| CAD | pericyte | *HIF3A* | TRUE | 2.240865204 |
| CAD | pericyte | *GRK4* | TRUE | 2.323336927 |
| CAD | pericyte | *B4GALNT1* | TRUE | 2.426697937 |
| CAD | pericyte | *HSD17B12* | TRUE | 2.459785247 |
| CAD | pericyte | *TTC12* | TRUE | 2.198577907 |
| CAD | pericyte | *DISP1* | TRUE | 2.839889488 |
| CAD | pericyte | *ICA1L* | TRUE | 4.301414328 |
| CAD | pericyte | *RNF123* | TRUE | 3.062461261 |
| CAD | pericyte | *OXGR1* | TRUE | 3.054938133 |
| CAD | pericyte | *KIFC2* | TRUE | 2.307829131 |
| CAD | pericyte | *IRS1* | TRUE | 2.737092815 |
| CAD | pericyte | *LPL* | TRUE | 2.448487243 |
| CAD | pericyte | *PAAF1* | TRUE | 2.049830411 |
| CAD | pericyte | *PLCXD3* | TRUE | 2.28958668 |
| CAD | pericyte | *A3GALT2* | TRUE | 2.226877844 |
| CAD | pericyte | *C6orf106* | TRUE | 2.376115362 |
| CAD | pericyte | *WDR5B* | TRUE | 2.366441057 |
| CAD | pericyte | *RPS26* | TRUE | 2.102459967 |
| CAD | pericyte | *ZNF717* | TRUE | 2.063464115 |
| CAD | pericyte | *HMBS* | TRUE | 3.094397254 |
| CAD | pericyte | *GALNT4* | TRUE | 1.961010224 |
| CAD | SMC | *DLX6* | TRUE | 2.341780013 |
| CAD | SMC | *TIE1* | TRUE | 2.323356877 |
| CAD | SMC | *TIPIN* | TRUE | 2.677980488 |
| CAD | SMC | *MAP1LC3A* | TRUE | 2.538028985 |
| CAD | SMC | *CTSH* | TRUE | 1.980312181 |
| CAD | SMC | *MAG* | TRUE | 2.064346172 |
| CAD | SMC | *ENDOU* | TRUE | 2.364815093 |

| | | | | |
|---|---|---|---|---|
| CAD | SMC | *BNIP1* | TRUE | 2.084100945 |
| CAD | SMC | *BCHE* | TRUE | 3.046769744 |
| CAD | SMC | *GGCX* | TRUE | 3.007453187 |
| CAD | SMC | *EIF2B2* | TRUE | 2.01165929 |
| CAD | SMC | *GRK4* | TRUE | 3.248969846 |
| CAD | SMC | *POLR1E* | TRUE | 2.926446278 |
| CAD | SMC | *CARF* | TRUE | 1.984887838 |
| CAD | SMC | *PSD2* | TRUE | 3.227535884 |
| CAD | SMC | *SLC18B1* | TRUE | 2.329688168 |
| CAD | SMC | *HSD17B12* | TRUE | 2.668953365 |
| CAD | SMC | *DISP1* | TRUE | 2.827228609 |
| CAD | SMC | *CLEC18A* | TRUE | 2.800506576 |
| CAD | SMC | *GPER1* | TRUE | 2.193387273 |
| CAD | SMC | *OXGR1* | TRUE | 3.19744978 |
| CAD | SMC | *ZNF641* | TRUE | 1.991044827 |
| CAD | SMC | *KIFC2* | TRUE | 2.153501086 |
| CAD | SMC | *ULK4* | TRUE | 2.153978901 |
| CAD | SMC | *IRS1* | TRUE | 4.293967954 |
| CAD | SMC | *LPL* | TRUE | 3.440138753 |
| CAD | SMC | *PAAF1* | TRUE | 2.189294592 |
| CAD | SMC | *PLCXD3* | TRUE | 2.100344035 |
| CAD | SMC | *A3GALT2* | TRUE | 2.164096169 |
| CAD | SMC | *C6orf106* | TRUE | 2.614582032 |
| CAD | SMC | *RPS26* | TRUE | 2.103965675 |
| CAD | SMC | *TMEM116* | TRUE | 2.816241195 |
| CAD | SMC | *LY6G5B* | TRUE | 2.137342533 |
| CAD | SMC | *HMBS* | TRUE | 3.457274449 |
| CAD | SMC | *GALNT4* | TRUE | 2.08190783 |
| CAD | t_cell | *DLX6* | TRUE | 2.288933742 |
| CAD | t_cell | *TIPIN* | TRUE | 2.686978623 |
| CAD | t_cell | *MAG* | TRUE | 2.099578452 |
| CAD | t_cell | *BNIP1* | TRUE | 2.197307988 |
| CAD | t_cell | *GGCX* | TRUE | 2.666313952 |
| CAD | t_cell | *GRK4* | TRUE | 2.821626948 |
| CAD | t_cell | *HORMAD1* | TRUE | 1.996406938 |
| CAD | t_cell | *SLC18B1* | TRUE | 2.301510607 |
| CAD | t_cell | *CLEC18A* | TRUE | 2.142923865 |
| CAD | t_cell | *APEH* | TRUE | 3.459601377 |
| CAD | t_cell | *OXGR1* | TRUE | 2.598095029 |
| CAD | t_cell | *ULK4* | TRUE | 2.08382799 |
| CAD | t_cell | *IRS1* | TRUE | 2.676964775 |
| CAD | t_cell | *BANF1* | TRUE | 2.149693563 |
| CAD | t_cell | *RIMKLA* | TRUE | 2.056546569 |
| CAD | t_cell | *GDF3* | TRUE | 2.092093497 |
| CAD | t_cell | *LY6G5B* | TRUE | 2.484267661 |
| CAD | t_cell | *HMBS* | TRUE | 3.094333325 |
| T2DM | adipocyte | *RBM6* | TRUE | 2.123004519 |
| T2DM | adipocyte | *CEP68* | TRUE | 5.137788013 |
| T2DM | adipocyte | *BAK1* | TRUE | 2.069604233 |
| T2DM | adipocyte | *TUBG2* | TRUE | 2.56955796 |

| | | | | |
|---|---|---|---|---|
| T2DM | adipocyte | *RAB27B* | TRUE | 2.363349114 |
| T2DM | adipocyte | *TSPAN17* | TRUE | 2.065109323 |
| T2DM | adipocyte | *ITIH4* | TRUE | 2.626923602 |
| T2DM | adipocyte | *ASNS* | TRUE | 1.986379847 |
| T2DM | adipocyte | *STXBP2* | TRUE | 2.33279497 |
| T2DM | adipocyte | *LRCH4* | TRUE | 2.631248262 |
| T2DM | adipocyte | *COBLL1* | TRUE | 2.908374924 |
| T2DM | adipocyte | *SNX10* | TRUE | 2.582655729 |
| T2DM | adipocyte | *C3orf18* | TRUE | 1.961481327 |
| T2DM | adipocyte | *CWF19L1* | TRUE | 2.627690609 |
| T2DM | adipocyte | *ISM2* | TRUE | 2.263778199 |
| T2DM | adipocyte | *RBL2* | TRUE | 2.836879663 |
| T2DM | adipocyte | *IGDCC4* | TRUE | 2.057743866 |
| T2DM | adipocyte | *EYA1* | TRUE | 2.549025243 |
| T2DM | adipocyte | *TLE6* | TRUE | 2.02033537 |
| T2DM | adipocyte | *ATP13A1* | TRUE | 4.092632628 |
| T2DM | adipocyte | *CDK2AP1* | TRUE | 2.199772431 |
| T2DM | adipocyte | *ENDOU* | TRUE | 2.732664831 |
| T2DM | adipocyte | *BVES* | TRUE | 2.49930332 |
| T2DM | adipocyte | *NPHP3* | TRUE | 2.361135099 |
| T2DM | adipocyte | *SDC1* | TRUE | 2.266824152 |
| T2DM | adipocyte | *GPX7* | TRUE | 2.947060504 |
| T2DM | adipocyte | *PLA2G2D* | TRUE | 2.19195994 |
| T2DM | adipocyte | *CD3EAP* | TRUE | 2.452388762 |
| T2DM | adipocyte | *ELOVL4* | TRUE | 2.900031604 |
| T2DM | adipocyte | *DNAH7* | TRUE | 3.611225987 |
| T2DM | adipocyte | *ADRA1A* | TRUE | 2.895577027 |
| T2DM | adipocyte | *TEX14* | TRUE | 2.718853519 |
| T2DM | adipocyte | *GJB6* | TRUE | 2.177984587 |
| T2DM | adipocyte | *GRK4* | TRUE | 2.949232707 |
| T2DM | adipocyte | *ZNF436* | TRUE | 2.45691385 |
| T2DM | adipocyte | *PRR12* | TRUE | 2.533385322 |
| T2DM | adipocyte | *RIBC2* | TRUE | 2.755455083 |
| T2DM | adipocyte | *OPN1SW* | TRUE | 2.205358165 |
| T2DM | adipocyte | *PKDREJ* | TRUE | 2.479969347 |
| T2DM | adipocyte | *P2RX4* | TRUE | 2.314869115 |
| T2DM | adipocyte | *NACAD* | TRUE | 2.578596147 |
| T2DM | adipocyte | *MYOF* | TRUE | 2.010753219 |
| T2DM | adipocyte | *LRRC46* | TRUE | 2.674047196 |
| T2DM | adipocyte | *FN3KRP* | TRUE | 2.128025078 |
| T2DM | adipocyte | *DUS3L* | TRUE | 2.290355502 |
| T2DM | adipocyte | *ARHGEF19* | TRUE | 2.232602028 |
| T2DM | adipocyte | *PLA1A* | TRUE | 2.338996578 |
| T2DM | adipocyte | *BHMT* | TRUE | 2.803779577 |
| T2DM | adipocyte | *PSD2* | TRUE | 2.174117218 |
| T2DM | adipocyte | *HSD17B12* | TRUE | 4.716345533 |
| T2DM | adipocyte | *APIP* | TRUE | 3.31384862 |
| T2DM | adipocyte | *DUSP15* | TRUE | 2.279950585 |
| T2DM | adipocyte | *PPP1R1C* | TRUE | 2.042785557 |
| T2DM | adipocyte | *PRSS53* | TRUE | 1.994926147 |

| | | | | |
|---|---|---|---|---|
| T2DM | adipocyte | *TSEN2* | TRUE | 2.939756845 |
| T2DM | adipocyte | *CLEC18A* | TRUE | 2.570465636 |
| T2DM | adipocyte | *KCNJ15* | TRUE | 2.181156136 |
| T2DM | adipocyte | *AP3S2* | TRUE | 5.348464272 |
| T2DM | adipocyte | *C12orf43* | TRUE | 2.02060351 |
| T2DM | adipocyte | *CD1E* | TRUE | 2.773513974 |
| T2DM | adipocyte | *VPS28* | TRUE | 2.158849177 |
| T2DM | adipocyte | *LMX1A* | TRUE | 2.561715303 |
| T2DM | adipocyte | *SPTA1* | TRUE | 2.648783257 |
| T2DM | adipocyte | *BDH2* | TRUE | 2.264412595 |
| T2DM | adipocyte | *TRMT61A* | TRUE | 3.606485223 |
| T2DM | adipocyte | *PCBD1* | TRUE | 3.311256647 |
| T2DM | adipocyte | *LTBP3* | TRUE | 3.145677764 |
| T2DM | adipocyte | *PXK* | TRUE | 2.206381394 |
| T2DM | adipocyte | *IRS1* | TRUE | 9.972414838 |
| T2DM | adipocyte | *C1QTNF4* | TRUE | 2.760470913 |
| T2DM | adipocyte | *FIBP* | TRUE | 2.064705239 |
| T2DM | adipocyte | *VEGFB* | TRUE | 3.087011103 |
| T2DM | adipocyte | *IL20RB* | TRUE | 2.022100075 |
| T2DM | adipocyte | *KLF14* | TRUE | 3.018199544 |
| T2DM | adipocyte | *STX19* | TRUE | 3.056439546 |
| T2DM | adipocyte | *B3GNT3* | TRUE | 3.074123683 |
| T2DM | adipocyte | *ZNF320* | TRUE | 2.96271273 |
| T2DM | adipocyte | *DGCR6* | TRUE | 2.060394564 |
| T2DM | adipocyte | *RP1L1* | TRUE | 2.954753953 |
| T2DM | adipocyte | *MROH7* | TRUE | 2.211693686 |
| T2DM | adipocyte | *NDUFA6* | TRUE | 2.323298882 |
| T2DM | adipocyte | *LINC00839* | TRUE | 3.260223073 |
| T2DM | adipocyte | *RGS7BP* | TRUE | 2.250172338 |
| T2DM | adipocyte | *MCRS1* | TRUE | 2.079953887 |
| T2DM | adipocyte | *NUTM2B* | TRUE | 2.627287164 |
| T2DM | adipocyte | *TMEM201* | TRUE | 2.047988178 |
| T2DM | adipocyte | *ALKBH2* | TRUE | 2.149698453 |
| T2DM | adipocyte | *TMEM116* | TRUE | 2.12283746 |
| T2DM | adipocyte | *HLA.DRB5* | TRUE | 2.23923545 |
| T2DM | adipocyte | *CENPW* | TRUE | 3.81589737 |
| T2DM | adipocyte | *FAM229B* | TRUE | 2.237720207 |
| T2DM | adipocyte | *NELFE* | TRUE | 2.239326743 |
| T2DM | adipocyte | *LINC01123* | TRUE | 2.261323516 |
| T2DM | adipocyte | *AC016586.1* | TRUE | 1.990290456 |
| T2DM | adipocyte | *SERF1B* | TRUE | 2.392719752 |
| T2DM | adipocyte | *LIN52* | TRUE | 2.147077541 |
| T2DM | adipocyte | *DNLZ* | TRUE | 3.627030801 |
| T2DM | adipocyte | *SIPA1* | TRUE | 2.039938586 |
| T2DM | adipocyte | *STARD10* | TRUE | 4.79453349 |
| T2DM | adipocyte | *AC073133.1* | TRUE | 2.096561035 |
| T2DM | adipocyte | *GS1.124K5.4* | TRUE | 2.216529451 |
| T2DM | adipocyte | *HLA.DQA2* | TRUE | 4.734210266 |
| T2DM | adipocyte | *HMBS* | TRUE | 3.694390722 |
| T2DM | adipocyte | *LINC00639* | TRUE | 2.249279336 |

| | | | | |
|---|---|---|---|---|
| T2DM | ASPC | *RBM6* | TRUE | 2.663426375 |
| T2DM | ASPC | *ZNF195* | TRUE | 2.657226648 |
| T2DM | ASPC | *GGNBP2* | TRUE | 2.442958539 |
| T2DM | ASPC | *CEP68* | TRUE | 5.536380814 |
| T2DM | ASPC | *BAK1* | TRUE | 2.032409976 |
| T2DM | ASPC | *TUBG2* | TRUE | 2.83320425 |
| T2DM | ASPC | *RAB27B* | TRUE | 2.526378425 |
| T2DM | ASPC | *MED17* | TRUE | 2.015385772 |
| T2DM | ASPC | *TSPAN17* | TRUE | 2.348158857 |
| T2DM | ASPC | *EPN3* | TRUE | 2.176081041 |
| T2DM | ASPC | *ITIH4* | TRUE | 2.466739413 |
| T2DM | ASPC | *ASNS* | TRUE | 2.145536354 |
| T2DM | ASPC | *SDHA* | TRUE | 2.471446738 |
| T2DM | ASPC | *MARK3* | TRUE | 2.94832361 |
| T2DM | ASPC | *XAB2* | TRUE | 2.630670586 |
| T2DM | ASPC | *STXBP2* | TRUE | 2.415610893 |
| T2DM | ASPC | *CTTNBP2* | TRUE | 3.526144533 |
| T2DM | ASPC | *LRCH4* | TRUE | 3.121305496 |
| T2DM | ASPC | *KCNN2* | TRUE | 2.015510347 |
| T2DM | ASPC | *COBLL1* | TRUE | 4.902917954 |
| T2DM | ASPC | *OVGP1* | TRUE | 2.248177245 |
| T2DM | ASPC | *CPNE3* | TRUE | 2.005357523 |
| T2DM | ASPC | *EPDR1* | TRUE | 2.393225228 |
| T2DM | ASPC | *CWF19L1* | TRUE | 3.359276589 |
| T2DM | ASPC | *PPP6R2* | TRUE | 2.144886535 |
| T2DM | ASPC | *ATP6V1D* | TRUE | 2.089395066 |
| T2DM | ASPC | *ISM2* | TRUE | 2.580599705 |
| T2DM | ASPC | *PSMB5* | TRUE | 2.029037659 |
| T2DM | ASPC | *DHODH* | TRUE | 1.974950378 |
| T2DM | ASPC | *RBL2* | TRUE | 2.530265162 |
| T2DM | ASPC | *IGDCC4* | TRUE | 2.046926698 |
| T2DM | ASPC | *EYA1* | TRUE | 3.805854641 |
| T2DM | ASPC | *DMPK* | TRUE | 2.921438368 |
| T2DM | ASPC | *TLE6* | TRUE | 2.487911002 |
| T2DM | ASPC | *CCDC61* | TRUE | 2.560609701 |
| T2DM | ASPC | *ICAM5* | TRUE | 2.583365958 |
| T2DM | ASPC | *GCDH* | TRUE | 2.347344294 |
| T2DM | ASPC | *ATP13A1* | TRUE | 4.140529164 |
| T2DM | ASPC | *CRHR2* | TRUE | 1.978554951 |
| T2DM | ASPC | *POLD2* | TRUE | 3.613585573 |
| T2DM | ASPC | *SYNGR2* | TRUE | 1.975134284 |
| T2DM | ASPC | *WFS1* | TRUE | 7.14971946 |
| T2DM | ASPC | *C11orf21* | TRUE | 2.51461039 |
| T2DM | ASPC | *CDK2AP1* | TRUE | 2.211904207 |
| T2DM | ASPC | *ENDOU* | TRUE | 3.107038141 |
| T2DM | ASPC | *BVES* | TRUE | 3.198866622 |
| T2DM | ASPC | *NPHP3* | TRUE | 2.597643549 |
| T2DM | ASPC | *UMPS* | TRUE | 2.005694943 |
| T2DM | ASPC | *EFCC1* | TRUE | 2.264244776 |
| T2DM | ASPC | *DNAJC27* | TRUE | 2.837074822 |

| | | | | |
|---|---|---|---|---|
| T2DM | ASPC | *ITGB6* | TRUE | 2.010610616 |
| T2DM | ASPC | *RTN4* | TRUE | 2.022913203 |
| T2DM | ASPC | *CHMP3* | TRUE | 4.006200453 |
| T2DM | ASPC | *SDC1* | TRUE | 2.357808281 |
| T2DM | ASPC | *GPX7* | TRUE | 3.121032696 |
| T2DM | ASPC | *PLA2G2D* | TRUE | 2.620685534 |
| T2DM | ASPC | *TNFSF4* | TRUE | 2.524679669 |
| T2DM | ASPC | *CD3EAP* | TRUE | 3.577499201 |
| T2DM | ASPC | *ELOVL4* | TRUE | 2.778092431 |
| T2DM | ASPC | *DNAH7* | TRUE | 2.991953888 |
| T2DM | ASPC | *HDHD3* | TRUE | 2.321641835 |
| T2DM | ASPC | *EPCAM* | TRUE | 1.966401788 |
| T2DM | ASPC | *MASTL* | TRUE | 2.312245396 |
| T2DM | ASPC | *ADRA1A* | TRUE | 3.678564059 |
| T2DM | ASPC | *TEX14* | TRUE | 2.898684408 |
| T2DM | ASPC | *ECHDC2* | TRUE | 2.122374175 |
| T2DM | ASPC | *SEC22A* | TRUE | 2.151451203 |
| T2DM | ASPC | *GJB6* | TRUE | 2.27572471 |
| T2DM | ASPC | *RASL11A* | TRUE | 2.714050952 |
| T2DM | ASPC | *PMS2* | TRUE | 2.349750432 |
| T2DM | ASPC | *SSPN* | TRUE | 1.998560193 |
| T2DM | ASPC | *EBPL* | TRUE | 2.011894473 |
| T2DM | ASPC | *GLO1* | TRUE | 2.05931237 |
| T2DM | ASPC | *GRK4* | TRUE | 3.385712553 |
| T2DM | ASPC | *ZNF436* | TRUE | 3.315588578 |
| T2DM | ASPC | *PRR12* | TRUE | 2.95062673 |
| T2DM | ASPC | *WFIKKN1* | TRUE | 1.983213474 |
| T2DM | ASPC | *BAIAP2L2* | TRUE | 2.047230424 |
| T2DM | ASPC | *RIBC2* | TRUE | 2.689305391 |
| T2DM | ASPC | *OPN1SW* | TRUE | 2.654722159 |
| T2DM | ASPC | *DPP6* | TRUE | 3.40494568 |
| T2DM | ASPC | *GTPBP3* | TRUE | 2.721038307 |
| T2DM | ASPC | *HBZ* | TRUE | 2.051224474 |
| T2DM | ASPC | *PKDREJ* | TRUE | 2.617817447 |
| T2DM | ASPC | *P2RX4* | TRUE | 2.173129482 |
| T2DM | ASPC | *SMYD5* | TRUE | 2.136792826 |
| T2DM | ASPC | *TMBIM1* | TRUE | 1.971681108 |
| T2DM | ASPC | *AOAH* | TRUE | 3.153689369 |
| T2DM | ASPC | *NACAD* | TRUE | 2.282655989 |
| T2DM | ASPC | *SYTL2* | TRUE | 2.529137029 |
| T2DM | ASPC | *DCUN1D5* | TRUE | 2.566297413 |
| T2DM | ASPC | *MYOF* | TRUE | 2.498686393 |
| T2DM | ASPC | *BTBD16* | TRUE | 2.090996514 |
| T2DM | ASPC | *C1RL* | TRUE | 2.114353595 |
| T2DM | ASPC | *GPR84* | TRUE | 2.618601791 |
| T2DM | ASPC | *HMG20A* | TRUE | 3.27001049 |
| T2DM | ASPC | *SCAMP2* | TRUE | 2.341661367 |
| T2DM | ASPC | *VPS53* | TRUE | 2.316384137 |
| T2DM | ASPC | *SPATA22* | TRUE | 2.012623739 |
| T2DM | ASPC | *LRRC46* | TRUE | 2.182981589 |

| | | | | |
|---|---|---|---|---|
| T2DM | ASPC | *NPC1* | TRUE | 2.481166281 |
| T2DM | ASPC | *FN3KRP* | TRUE | 2.891783543 |
| T2DM | ASPC | *DUS3L* | TRUE | 2.561126727 |
| T2DM | ASPC | *GEMIN7* | TRUE | 2.32554949 |
| T2DM | ASPC | *ARHGEF19* | TRUE | 2.21211627 |
| T2DM | ASPC | *TARS2* | TRUE | 2.307456709 |
| T2DM | ASPC | *WNT9A* | TRUE | 2.051341078 |
| T2DM | ASPC | *THSD7B* | TRUE | 1.982553031 |
| T2DM | ASPC | *PLA1A* | TRUE | 2.997447039 |
| T2DM | ASPC | *PPIP5K2* | TRUE | 2.469512357 |
| T2DM | ASPC | *CCDC25* | TRUE | 2.615269033 |
| T2DM | ASPC | *ZNF7* | TRUE | 2.09656394 |
| T2DM | ASPC | *HSD17B12* | TRUE | 5.459795253 |
| T2DM | ASPC | *APIP* | TRUE | 3.739425888 |
| T2DM | ASPC | *EIF3M* | TRUE | 2.47729353 |
| T2DM | ASPC | *DUSP15* | TRUE | 2.688093884 |
| T2DM | ASPC | *PPP1R1C* | TRUE | 2.065735741 |
| T2DM | ASPC | *ABCB9* | TRUE | 2.615457 |
| T2DM | ASPC | *ARL14EP* | TRUE | 4.330037758 |
| T2DM | ASPC | *POC5* | TRUE | 3.170648278 |
| T2DM | ASPC | *SAR1B* | TRUE | 2.132328025 |
| T2DM | ASPC | *PLA2R1* | TRUE | 2.109145237 |
| T2DM | ASPC | *JAZF1* | TRUE | 7.709060541 |
| T2DM | ASPC | *LONRF1* | TRUE | 2.818210522 |
| T2DM | ASPC | *TSEN2* | TRUE | 3.628872863 |
| T2DM | ASPC | *NDUFAF6* | TRUE | 3.895361262 |
| T2DM | ASPC | *WHAMM* | TRUE | 2.444200085 |
| T2DM | ASPC | *TIMP4* | TRUE | 2.678692835 |
| T2DM | ASPC | *CLEC18A* | TRUE | 2.529193428 |
| T2DM | ASPC | *KCNJ15* | TRUE | 3.350286121 |
| T2DM | ASPC | *AP3S2* | TRUE | 6.199059306 |
| T2DM | ASPC | *C12orf43* | TRUE | 2.410123166 |
| T2DM | ASPC | *CD1E* | TRUE | 2.487014411 |
| T2DM | ASPC | *ZNF276* | TRUE | 2.286960593 |
| T2DM | ASPC | *DIP2A* | TRUE | 2.542576915 |
| T2DM | ASPC | *FDPS* | TRUE | 2.093872772 |
| T2DM | ASPC | *EXTL2* | TRUE | 2.139424753 |
| T2DM | ASPC | *LMX1A* | TRUE | 2.360235506 |
| T2DM | ASPC | *LMOD1* | TRUE | 2.656928622 |
| T2DM | ASPC | *SPTA1* | TRUE | 2.563237143 |
| T2DM | ASPC | *RFT1* | TRUE | 2.681163826 |
| T2DM | ASPC | *BDH2* | TRUE | 3.219077895 |
| T2DM | ASPC | *REEP3* | TRUE | 2.245860274 |
| T2DM | ASPC | *CEP57* | TRUE | 2.030073774 |
| T2DM | ASPC | *PCBD1* | TRUE | 3.762238211 |
| T2DM | ASPC | *SAAL1* | TRUE | 4.878734999 |
| T2DM | ASPC | *VKORC1* | TRUE | 2.019245899 |
| T2DM | ASPC | *PSCA* | TRUE | 2.496223137 |
| T2DM | ASPC | *LTBP3* | TRUE | 3.154510034 |
| T2DM | ASPC | *KIF5C* | TRUE | 2.559351142 |

| | | | | |
|---|---|---|---|---|
| T2DM | ASPC | *PXK* | TRUE | 2.773738183 |
| T2DM | ASPC | *IRS1* | TRUE | 10.6087442 |
| T2DM | ASPC | *LRRC45* | TRUE | 2.283266069 |
| T2DM | ASPC | *EMB* | TRUE | 3.415411977 |
| T2DM | ASPC | *KCNS3* | TRUE | 1.970117057 |
| T2DM | ASPC | *ZNF561* | TRUE | 2.107739844 |
| T2DM | ASPC | *RGS19* | TRUE | 2.693442939 |
| T2DM | ASPC | *LAMB2* | TRUE | 2.329410065 |
| T2DM | ASPC | *C1QTNF4* | TRUE | 2.721007792 |
| T2DM | ASPC | *MYEOV* | TRUE | 2.366480602 |
| T2DM | ASPC | *HPSE2* | TRUE | 2.116301482 |
| T2DM | ASPC | *VEGFB* | TRUE | 4.011853487 |
| T2DM | ASPC | *IL20RB* | TRUE | 2.324865748 |
| T2DM | ASPC | *KLF14* | TRUE | 5.06575773 |
| T2DM | ASPC | *LPL* | TRUE | 2.075509463 |
| T2DM | ASPC | *HORMAD2* | TRUE | 2.242496232 |
| T2DM | ASPC | *C12orf54* | TRUE | 2.089778284 |
| T2DM | ASPC | *GOLGA8Q* | TRUE | 1.960515554 |
| T2DM | ASPC | *STX19* | TRUE | 3.548276829 |
| T2DM | ASPC | *B3GNT3* | TRUE | 2.850446481 |
| T2DM | ASPC | *GRID1* | TRUE | 2.084839716 |
| T2DM | ASPC | *ZNF320* | TRUE | 3.247320772 |
| T2DM | ASPC | *DGCR6* | TRUE | 2.053940504 |
| T2DM | ASPC | *RP1L1* | TRUE | 2.948365911 |
| T2DM | ASPC | *BMP8A* | TRUE | 3.447068112 |
| T2DM | ASPC | *EIF3C* | TRUE | 2.341012062 |
| T2DM | ASPC | *NDUFA6* | TRUE | 2.420088915 |
| T2DM | ASPC | *SH3BGR* | TRUE | 2.019486642 |
| T2DM | ASPC | *RASA3* | TRUE | 2.958923857 |
| T2DM | ASPC | *RGS7BP* | TRUE | 3.228725591 |
| T2DM | ASPC | *CEACAM19* | TRUE | 2.083475121 |
| T2DM | ASPC | *ADH1A* | TRUE | 2.088671441 |
| T2DM | ASPC | *MCRS1* | TRUE | 2.597668916 |
| T2DM | ASPC | *NUTM2B* | TRUE | 2.416830026 |
| T2DM | ASPC | *TMEM201* | TRUE | 2.589753832 |
| T2DM | ASPC | *ALKBH2* | TRUE | 2.560986976 |
| T2DM | ASPC | *GTF2IRD2* | TRUE | 2.611126767 |
| T2DM | ASPC | *ZNF775* | TRUE | 1.976030759 |
| T2DM | ASPC | *MYO6* | TRUE | 2.818077315 |
| T2DM | ASPC | *FAM180B* | TRUE | 2.389941946 |
| T2DM | ASPC | *TRPV1* | TRUE | 2.19110306 |
| T2DM | ASPC | *ZNF33B* | TRUE | 2.272240747 |
| T2DM | ASPC | *C6orf106* | TRUE | 1.985985744 |
| T2DM | ASPC | *TBC1D9B* | TRUE | 2.408509376 |
| T2DM | ASPC | *TTC30A* | TRUE | 2.119830778 |
| T2DM | ASPC | *PRIM1* | TRUE | 3.139404227 |
| T2DM | ASPC | *TMEM116* | TRUE | 2.844059813 |
| T2DM | ASPC | *ZNF568* | TRUE | 2.484986223 |
| T2DM | ASPC | *CENPW* | TRUE | 4.258509016 |
| T2DM | ASPC | *FAM229B* | TRUE | 2.84779676 |

| | | | | |
|---|---|---|---|---|
| T2DM | ASPC | *ASAH2B* | TRUE | 2.044204013 |
| T2DM | ASPC | *AGAP6* | TRUE | 2.116494743 |
| T2DM | ASPC | *OXLD1* | TRUE | 2.698211851 |
| T2DM | ASPC | *NELFE* | TRUE | 2.186369767 |
| T2DM | ASPC | *LINC01123* | TRUE | 3.096652264 |
| T2DM | ASPC | *SERF1B* | TRUE | 2.703285016 |
| T2DM | ASPC | *LIN52* | TRUE | 2.893965293 |
| T2DM | ASPC | *HCG27* | TRUE | 2.603986188 |
| T2DM | ASPC | *FGFR1OP* | TRUE | 2.908418156 |
| T2DM | ASPC | *DNLZ* | TRUE | 3.479743981 |
| T2DM | ASPC | *STARD10* | TRUE | 4.874119353 |
| T2DM | ASPC | *KCNU1* | TRUE | 2.597346709 |
| T2DM | ASPC | *AC062017.1* | TRUE | 2.624284821 |
| T2DM | ASPC | *AC073133.1* | TRUE | 2.241501026 |
| T2DM | ASPC | *GS1.124K5.4* | TRUE | 2.720421854 |
| T2DM | ASPC | *HLA.DQA2* | TRUE | 1.9640459 |
| T2DM | ASPC | *NME1.NME2* | TRUE | 2.481532077 |
| T2DM | ASPC | *JMJD7* | TRUE | 2.064874966 |
| T2DM | ASPC | *CCDC13* | TRUE | 2.183105272 |
| T2DM | ASPC | *ZNF286B* | TRUE | 2.042455404 |
| T2DM | ASPC | *DNAH10OS* | TRUE | 2.207458985 |
| T2DM | ASPC | *HMBS* | TRUE | 4.127560543 |
| T2DM | ASPC | *LINC00639* | TRUE | 2.693230294 |
| T2DM | ASPC | *AC083843.1* | TRUE | 3.224863475 |
| T2DM | ASPC | *SH3RF3.AS1* | TRUE | 2.043556027 |
| T2DM | ASPC | *LINC01140* | TRUE | 2.102249238 |
| T2DM | b_cell | *HSD17B1* | TRUE | 2.781436496 |
| T2DM | b_cell | *ELOVL4* | TRUE | 2.274684984 |
| T2DM | b_cell | *CLEC18A* | TRUE | 4.731854925 |
| T2DM | b_cell | *IRS1* | TRUE | 4.947658968 |
| T2DM | b_cell | *C1QTNF4* | TRUE | 2.027687111 |
| T2DM | b_cell | *HLA.DRB5* | TRUE | 3.573328328 |
| T2DM | b_cell | *HCG27* | TRUE | 2.04413938 |
| T2DM | b_cell | *DNLZ* | TRUE | 3.277140028 |
| T2DM | b_cell | *STARD10* | TRUE | 3.135235559 |
| T2DM | b_cell | *PLA2G4B* | TRUE | 3.033452217 |
| T2DM | dendritic_cell | *TUBG2* | TRUE | 2.154334835 |
| T2DM | dendritic_cell | *ITIH4* | TRUE | 2.243828394 |
| T2DM | dendritic_cell | *BVES* | TRUE | 2.616038881 |
| T2DM | dendritic_cell | *GPX7* | TRUE | 2.96729314 |
| T2DM | dendritic_cell | *ADRA1A* | TRUE | 2.072001007 |
| T2DM | dendritic_cell | *SLC12A5* | TRUE | 2.189298263 |
| T2DM | dendritic_cell | *MAN2C1* | TRUE | 2.710871506 |
| T2DM | dendritic_cell | *HSD17B12* | TRUE | 4.188215128 |
| T2DM | dendritic_cell | *KCNJ15* | TRUE | 2.598510303 |
| T2DM | dendritic_cell | *TRMT61A* | TRUE | 3.191888447 |
| T2DM | dendritic_cell | *IRS1* | TRUE | 7.014624126 |
| T2DM | dendritic_cell | *ZNF320* | TRUE | 2.978021522 |
| T2DM | dendritic_cell | *NOTCH4* | TRUE | 5.01507201 |
| T2DM | dendritic_cell | *DNLZ* | TRUE | 3.319620962 |

| | | | | |
|---|---|---|---|---|
| T2DM | dendritic_cell | *STARD10* | TRUE | 4.456834413 |
| T2DM | dendritic_cell | *PLA2G4B* | TRUE | 2.911348345 |
| T2DM | endothelial | *RBM6* | TRUE | 2.513309128 |
| T2DM | endothelial | *TUBG2* | TRUE | 3.88361392 |
| T2DM | endothelial | *IGDCC4* | TRUE | 2.226054887 |
| T2DM | endothelial | *CDK2AP1* | TRUE | 3.236833785 |
| T2DM | endothelial | *GNB3* | TRUE | 2.238358743 |
| T2DM | endothelial | *GPX7* | TRUE | 2.448051897 |
| T2DM | endothelial | *DNAH7* | TRUE | 2.049098036 |
| T2DM | endothelial | *MAN2C1* | TRUE | 2.407121864 |
| T2DM | endothelial | *LRRC46* | TRUE | 2.533174871 |
| T2DM | endothelial | *BHMT* | TRUE | 2.647080953 |
| T2DM | endothelial | *APIP* | TRUE | 2.369083356 |
| T2DM | endothelial | *CLEC18A* | TRUE | 2.577332424 |
| T2DM | endothelial | *AP3S2* | TRUE | 3.623424637 |
| T2DM | endothelial | *IRS1* | TRUE | 8.936685996 |
| T2DM | endothelial | *RGS19* | TRUE | 2.459362287 |
| T2DM | endothelial | *C1QTNF4* | TRUE | 2.884938911 |
| T2DM | endothelial | *CTSW* | TRUE | 2.486470295 |
| T2DM | endothelial | *ZNF320* | TRUE | 2.105499684 |
| T2DM | endothelial | *DNLZ* | TRUE | 3.027718399 |
| T2DM | endothelial | *HLA.DQA2* | TRUE | 3.496705018 |
| T2DM | endothelial | *JMJD7* | TRUE | 2.421783512 |
| T2DM | LEC | *ITIH4* | TRUE | 2.838004095 |
| T2DM | LEC | *CDK2AP1* | TRUE | 2.761094619 |
| T2DM | LEC | *ADRA1A* | TRUE | 2.367124293 |
| T2DM | LEC | *GRK4* | TRUE | 2.346105552 |
| T2DM | LEC | *BHMT* | TRUE | 2.882790717 |
| T2DM | LEC | *HSD17B12* | TRUE | 2.601725479 |
| T2DM | LEC | *CLEC18A* | TRUE | 4.462760124 |
| T2DM | LEC | *AP3S2* | TRUE | 4.48815426 |
| T2DM | LEC | *IRS1* | TRUE | 5.593120424 |
| T2DM | LEC | *ZNF320* | TRUE | 2.369142412 |
| T2DM | LEC | *HLA.DRB5* | TRUE | 2.481371155 |
| T2DM | LEC | *DNLZ* | TRUE | 2.360591976 |
| T2DM | LEC | *STARD10* | TRUE | 2.157439832 |
| T2DM | LEC | *HLA.DQA2* | TRUE | 5.471976001 |
| T2DM | LEC | *JMJD7* | TRUE | 2.713109768 |
| T2DM | macrophage | *CEP68* | TRUE | 3.937092829 |
| T2DM | macrophage | *BAK1* | TRUE | 1.987434141 |
| T2DM | macrophage | *RAB27B* | TRUE | 2.312886769 |
| T2DM | macrophage | *ITIH4* | TRUE | 2.177687168 |
| T2DM | macrophage | *LRCH4* | TRUE | 2.225328836 |
| T2DM | macrophage | *SNX10* | TRUE | 2.614854829 |
| T2DM | macrophage | *CWF19L1* | TRUE | 2.359613004 |
| T2DM | macrophage | *ISM2* | TRUE | 2.390754878 |
| T2DM | macrophage | *EYA1* | TRUE | 2.787650698 |
| T2DM | macrophage | *TLE6* | TRUE | 2.156751213 |
| T2DM | macrophage | *ICAM5* | TRUE | 2.070108255 |
| T2DM | macrophage | *ATP13A1* | TRUE | 3.450155194 |

| | | | | |
|---|---|---|---|---|
| T2DM | macrophage | *WFS1* | TRUE | 3.504447116 |
| T2DM | macrophage | *C11orf21* | TRUE | 2.162142614 |
| T2DM | macrophage | *COQ5* | TRUE | 2.210126034 |
| T2DM | macrophage | *ENDOU* | TRUE | 2.698423245 |
| T2DM | macrophage | *BVES* | TRUE | 2.894382242 |
| T2DM | macrophage | *NPHP3* | TRUE | 2.034424878 |
| T2DM | macrophage | *CHMP3* | TRUE | 2.361120107 |
| T2DM | macrophage | *GPX7* | TRUE | 2.285750663 |
| T2DM | macrophage | *SERPINC1* | TRUE | 2.15398942 |
| T2DM | macrophage | *ELOVL4* | TRUE | 2.624432831 |
| T2DM | macrophage | *DNAH7* | TRUE | 2.8739288 |
| T2DM | macrophage | *ADRA1A* | TRUE | 3.309982996 |
| T2DM | macrophage | *GRK4* | TRUE | 2.99158606 |
| T2DM | macrophage | *HBZ* | TRUE | 2.039164141 |
| T2DM | macrophage | *PKDREJ* | TRUE | 2.546209489 |
| T2DM | macrophage | *FN3KRP* | TRUE | 2.264648592 |
| T2DM | macrophage | *ARHGEF19* | TRUE | 2.1968391 |
| T2DM | macrophage | *PLA1A* | TRUE | 2.669005774 |
| T2DM | macrophage | *BHMT* | TRUE | 2.518089515 |
| T2DM | macrophage | *HSD17B12* | TRUE | 4.525547696 |
| T2DM | macrophage | *APIP* | TRUE | 3.134255406 |
| T2DM | macrophage | *DUSP15* | TRUE | 2.373699975 |
| T2DM | macrophage | *TSEN2* | TRUE | 2.399427798 |
| T2DM | macrophage | *CLEC18A* | TRUE | 4.796961889 |
| T2DM | macrophage | *KCNJ15* | TRUE | 2.014266129 |
| T2DM | macrophage | *AP3S2* | TRUE | 5.450561353 |
| T2DM | macrophage | *TRIM63* | TRUE | 2.815454981 |
| T2DM | macrophage | *CD1E* | TRUE | 2.452894248 |
| T2DM | macrophage | *TRMT61A* | TRUE | 2.414300942 |
| T2DM | macrophage | *PCBD1* | TRUE | 2.878256692 |
| T2DM | macrophage | *IRS1* | TRUE | 9.813354449 |
| T2DM | macrophage | *C8orf46* | TRUE | 2.505019139 |
| T2DM | macrophage | *RGS19* | TRUE | 3.71956081 |
| T2DM | macrophage | *C1QTNF4* | TRUE | 3.14626282 |
| T2DM | macrophage | *SNX32* | TRUE | 2.168754252 |
| T2DM | macrophage | *VEGFB* | TRUE | 2.846563653 |
| T2DM | macrophage | *HORMAD2* | TRUE | 2.050197163 |
| T2DM | macrophage | *TRIM73* | TRUE | 2.727806471 |
| T2DM | macrophage | *B3GNT3* | TRUE | 2.422038405 |
| T2DM | macrophage | *ZNF320* | TRUE | 2.943651788 |
| T2DM | macrophage | *DGCR6* | TRUE | 1.994846286 |
| T2DM | macrophage | *RP1L1* | TRUE | 2.729501668 |
| T2DM | macrophage | *BMP8A* | TRUE | 5.018950449 |
| T2DM | macrophage | *ZNF703* | TRUE | 2.202140358 |
| T2DM | macrophage | *NDUFA6* | TRUE | 2.090987303 |
| T2DM | macrophage | *LINC00839* | TRUE | 2.720423862 |
| T2DM | macrophage | *NUTM2B* | TRUE | 2.071047795 |
| T2DM | macrophage | *HLA.DRB1* | TRUE | 2.247708682 |
| T2DM | macrophage | *ZNF775* | TRUE | 1.994009761 |
| T2DM | macrophage | *FAM180B* | TRUE | 2.239753395 |

| | | | | |
|---|---|---|---|---|
| T2DM | macrophage | *HCG27* | TRUE | 2.98397739 |
| T2DM | macrophage | *DNLZ* | TRUE | 3.601924991 |
| T2DM | macrophage | *STARD10* | TRUE | 4.647814089 |
| T2DM | macrophage | *GS1.124K5.4* | TRUE | 2.435598898 |
| T2DM | macrophage | *HLA.DQA2* | TRUE | 3.035355499 |
| T2DM | macrophage | *HMBS* | TRUE | 3.835300507 |
| T2DM | mast_cell | *CEP68* | TRUE | 2.698612296 |
| T2DM | mast_cell | *IGDCC4* | TRUE | 2.027708126 |
| T2DM | mast_cell | *ATP13A1* | TRUE | 2.249330403 |
| T2DM | mast_cell | *HSD17B1* | TRUE | 3.560554199 |
| T2DM | mast_cell | *CDK2AP1* | TRUE | 2.139288648 |
| T2DM | mast_cell | *NPHP3* | TRUE | 1.963716205 |
| T2DM | mast_cell | *GPX7* | TRUE | 2.364795805 |
| T2DM | mast_cell | *ELOVL4* | TRUE | 2.257076703 |
| T2DM | mast_cell | *DNAH7* | TRUE | 2.551278599 |
| T2DM | mast_cell | *ADRA1A* | TRUE | 2.74737956 |
| T2DM | mast_cell | *GRK4* | TRUE | 2.935141647 |
| T2DM | mast_cell | *OPRL1* | TRUE | 2.376273539 |
| T2DM | mast_cell | *PKDREJ* | TRUE | 2.480193447 |
| T2DM | mast_cell | *MAN2C1* | TRUE | 2.265964908 |
| T2DM | mast_cell | *HSD17B12* | TRUE | 2.462446757 |
| T2DM | mast_cell | *CLEC18A* | TRUE | 2.079484857 |
| T2DM | mast_cell | *AP3S2* | TRUE | 4.834457856 |
| T2DM | mast_cell | *RNF123* | TRUE | 3.538327002 |
| T2DM | mast_cell | *PCBD1* | TRUE | 2.582360683 |
| T2DM | mast_cell | *IRS1* | TRUE | 9.448609989 |
| T2DM | mast_cell | *C8orf46* | TRUE | 2.405020577 |
| T2DM | mast_cell | *ZNF320* | TRUE | 2.566642403 |
| T2DM | mast_cell | *CENPW* | TRUE | 2.559829737 |
| T2DM | mast_cell | *HCG27* | TRUE | 2.213629564 |
| T2DM | mast_cell | *DNLZ* | TRUE | 3.59552318 |
| T2DM | mast_cell | *HLA.DQA2* | TRUE | 3.371848849 |
| T2DM | mast_cell | *JMJD7* | TRUE | 3.302683265 |
| T2DM | mast_cell | *HMBS* | TRUE | 2.126405114 |
| T2DM | monocyte | *ITIH4* | TRUE | 3.554075564 |
| T2DM | monocyte | *C3orf18* | TRUE | 2.336481097 |
| T2DM | monocyte | *ATP13A1* | TRUE | 3.57573678 |
| T2DM | monocyte | *CLEC18A* | TRUE | 3.004384099 |
| T2DM | monocyte | *AP3S2* | TRUE | 4.015930577 |
| T2DM | monocyte | *IRS1* | TRUE | 2.435763228 |
| T2DM | monocyte | *C1QTNF4* | TRUE | 2.957373351 |
| T2DM | monocyte | *ZNF320* | TRUE | 2.77183161 |
| T2DM | monocyte | *HCG27* | TRUE | 3.073206014 |
| T2DM | monocyte | *DNLZ* | TRUE | 3.468993802 |
| T2DM | monocyte | *HLA.DQB2* | TRUE | 2.27156721 |
| T2DM | monocyte | *JMJD7* | TRUE | 2.466033244 |
| T2DM | monocyte | *HMBS* | TRUE | 3.011010432 |
| T2DM | neutrophil | *CDK2AP1* | TRUE | 2.520547201 |
| T2DM | neutrophil | *CLEC18A* | TRUE | 4.596507401 |
| T2DM | neutrophil | *C1QTNF4* | TRUE | 3.465896503 |

| | | | | |
|---|---|---|---|---|
| T2DM | neutrophil | *HLA.DQB1* | TRUE | 3.323520229 |
| T2DM | neutrophil | *NOTCH4* | TRUE | 3.634106395 |
| T2DM | neutrophil | *DNLZ* | TRUE | 3.669687852 |
| T2DM | neutrophil | *STARD10* | TRUE | 4.102438953 |
| T2DM | neutrophil | *HLA.DQA2* | TRUE | 3.987521252 |
| T2DM | nk_cell | *TUBG2* | TRUE | 2.812356349 |
| T2DM | nk_cell | *ITIH4* | TRUE | 3.469890839 |
| T2DM | nk_cell | *SDC1* | TRUE | 2.139015064 |
| T2DM | nk_cell | *GRK4* | TRUE | 2.447314707 |
| T2DM | nk_cell | *MAN2C1* | TRUE | 2.070244065 |
| T2DM | nk_cell | *HSD17B12* | TRUE | 2.846566039 |
| T2DM | nk_cell | *APIP* | TRUE | 2.076227027 |
| T2DM | nk_cell | *CLEC18A* | TRUE | 4.713698458 |
| T2DM | nk_cell | *AP3S2* | TRUE | 3.362098118 |
| T2DM | nk_cell | *RNF123* | TRUE | 1.997017389 |
| T2DM | nk_cell | *IRS1* | TRUE | 7.961525761 |
| T2DM | nk_cell | *RGS19* | TRUE | 2.057835025 |
| T2DM | nk_cell | *C1QTNF4* | TRUE | 3.020503137 |
| T2DM | nk_cell | *ZNF320* | TRUE | 2.319009137 |
| T2DM | nk_cell | *BMP8A* | TRUE | 2.907438252 |
| T2DM | nk_cell | *EIF3C* | TRUE | 2.160861682 |
| T2DM | nk_cell | *DNLZ* | TRUE | 3.499354622 |
| T2DM | nk_cell | *STARD10* | TRUE | 4.455253775 |
| T2DM | nk_cell | *PLA2G4B* | TRUE | 2.676418365 |
| T2DM | nk_cell | *HMBS* | TRUE | 3.238033788 |
| T2DM | pericyte | *CEP68* | TRUE | 3.507213883 |
| T2DM | pericyte | *ITIH4* | TRUE | 4.190404549 |
| T2DM | pericyte | *ATP13A1* | TRUE | 3.282342938 |
| T2DM | pericyte | *WFS1* | TRUE | 2.529199 |
| T2DM | pericyte | *CDK2AP1* | TRUE | 2.414709139 |
| T2DM | pericyte | *NPHP3* | TRUE | 2.105422064 |
| T2DM | pericyte | *CD3EAP* | TRUE | 2.034968743 |
| T2DM | pericyte | *ELOVL4* | TRUE | 2.03694906 |
| T2DM | pericyte | *ADRA1A* | TRUE | 3.127992843 |
| T2DM | pericyte | *GRK4* | TRUE | 2.233906855 |
| T2DM | pericyte | *MAN2C1* | TRUE | 2.080365396 |
| T2DM | pericyte | *ARHGEF19* | TRUE | 2.198817812 |
| T2DM | pericyte | *PLA1A* | TRUE | 2.541453933 |
| T2DM | pericyte | *HSD17B12* | TRUE | 4.237729372 |
| T2DM | pericyte | *TSEN2* | TRUE | 2.163078625 |
| T2DM | pericyte | *CLEC18A* | TRUE | 4.344083473 |
| T2DM | pericyte | *AP3S2* | TRUE | 3.892042002 |
| T2DM | pericyte | *RNF123* | TRUE | 2.999903613 |
| T2DM | pericyte | *PCBD1* | TRUE | 3.101046772 |
| T2DM | pericyte | *IRS1* | TRUE | 7.021617613 |
| T2DM | pericyte | *C1QTNF4* | TRUE | 1.970626271 |
| T2DM | pericyte | *ZNF320* | TRUE | 2.676670124 |
| T2DM | pericyte | *HLA.DRB5* | TRUE | 2.188527319 |
| T2DM | pericyte | *HCG27* | TRUE | 2.75764131 |
| T2DM | pericyte | *DNLZ* | TRUE | 3.338512508 |

| | | | | |
|---|---|---|---|---|
| T2DM | pericyte | *STARD10* | TRUE | 4.57679239 |
| T2DM | pericyte | *HLA.DQA2* | TRUE | 4.985181301 |
| T2DM | pericyte | *JMJD7* | TRUE | 2.683016334 |
| T2DM | pericyte | *HMBS* | TRUE | 3.319200284 |
| T2DM | SMC | *CEP68* | TRUE | 3.698301756 |
| T2DM | SMC | *TUBG2* | TRUE | 2.033609582 |
| T2DM | SMC | *LRCH4* | TRUE | 2.476100524 |
| T2DM | SMC | *COBLL1* | TRUE | 2.205954558 |
| T2DM | SMC | *IGDCC4* | TRUE | 2.0022171 |
| T2DM | SMC | *ATP13A1* | TRUE | 2.156349473 |
| T2DM | SMC | *CDK2AP1* | TRUE | 2.504036229 |
| T2DM | SMC | *ENDOU* | TRUE | 2.69598799 |
| T2DM | SMC | *GPX7* | TRUE | 2.850440429 |
| T2DM | SMC | *SERPINC1* | TRUE | 2.313291482 |
| T2DM | SMC | *DNAH7* | TRUE | 2.101134048 |
| T2DM | SMC | *ADRA1A* | TRUE | 2.193465531 |
| T2DM | SMC | *GRK4* | TRUE | 2.862898861 |
| T2DM | SMC | *RIBC2* | TRUE | 2.478354563 |
| T2DM | SMC | *OPN1SW* | TRUE | 2.156838946 |
| T2DM | SMC | *PKDREJ* | TRUE | 2.113593795 |
| T2DM | SMC | *MAN2C1* | TRUE | 3.022859229 |
| T2DM | SMC | *FN3KRP* | TRUE | 2.127449338 |
| T2DM | SMC | *PLA1A* | TRUE | 2.799520647 |
| T2DM | SMC | *HSD17B12* | TRUE | 4.59826092 |
| T2DM | SMC | *APIP* | TRUE | 2.434387262 |
| T2DM | SMC | *CLEC18A* | TRUE | 3.985012014 |
| T2DM | SMC | *AP3S2* | TRUE | 5.247681874 |
| T2DM | SMC | *CD1E* | TRUE | 2.110296247 |
| T2DM | SMC | *BDH2* | TRUE | 2.113905121 |
| T2DM | SMC | *RNF123* | TRUE | 1.994011378 |
| T2DM | SMC | *TRMT61A* | TRUE | 3.536623995 |
| T2DM | SMC | *PCBD1* | TRUE | 2.015305998 |
| T2DM | SMC | *IRS1* | TRUE | 9.659558283 |
| T2DM | SMC | *C8orf46* | TRUE | 2.310066014 |
| T2DM | SMC | *RGS19* | TRUE | 2.421213584 |
| T2DM | SMC | *C1QTNF4* | TRUE | 2.848511046 |
| T2DM | SMC | *B3GNT3* | TRUE | 1.960446181 |
| T2DM | SMC | *ZNF320* | TRUE | 2.952353176 |
| T2DM | SMC | *ZNF703* | TRUE | 2.808347007 |
| T2DM | SMC | *NDUFA6* | TRUE | 2.030867869 |
| T2DM | SMC | *TMEM116* | TRUE | 1.995513354 |
| T2DM | SMC | *HCG27* | TRUE | 2.619058766 |
| T2DM | SMC | *DNLZ* | TRUE | 3.597636933 |
| T2DM | SMC | *STARD10* | TRUE | 4.453426907 |
| T2DM | SMC | *GS1.124K5.4* | TRUE | 2.550748167 |
| T2DM | SMC | *NME1.NME2* | TRUE | 2.05323325 |
| T2DM | SMC | *JMJD7* | TRUE | 2.25041293 |
| T2DM | SMC | *HMBS* | TRUE | 3.417269306 |
| T2DM | t_cell | *CEP68* | TRUE | 3.502300635 |
| T2DM | t_cell | *EPN3* | TRUE | 2.051840615 |

| | | | | |
|---|---|---|---|---|
| T2DM | t_cell | *ITIH4* | TRUE | 3.447098804 |
| T2DM | t_cell | *PABPC4* | TRUE | 2.254602911 |
| T2DM | t_cell | *CDK2AP1* | TRUE | 2.052274354 |
| T2DM | t_cell | *GNB3* | TRUE | 2.263373793 |
| T2DM | t_cell | *ADRA1A* | TRUE | 2.627815325 |
| T2DM | t_cell | *GRK4* | TRUE | 2.087751121 |
| T2DM | t_cell | *RIBC2* | TRUE | 2.207018188 |
| T2DM | t_cell | *MAN2C1* | TRUE | 1.983108991 |
| T2DM | t_cell | *CLEC18A* | TRUE | 4.391325352 |
| T2DM | t_cell | *KCNJ15* | TRUE | 2.214816428 |
| T2DM | t_cell | *AP3S2* | TRUE | 5.074153104 |
| T2DM | t_cell | *IRS1* | TRUE | 7.472346283 |
| T2DM | t_cell | *C8orf46* | TRUE | 2.520911718 |
| T2DM | t_cell | *C1QTNF4* | TRUE | 2.013621181 |
| T2DM | t_cell | *KLF14* | TRUE | 3.854777999 |
| T2DM | t_cell | *BANF1* | TRUE | 1.969327413 |
| T2DM | t_cell | *ZNF320* | TRUE | 2.745709155 |
| T2DM | t_cell | *DGCR6* | TRUE | 2.022817088 |
| T2DM | t_cell | *EIF3C* | TRUE | 2.317633868 |
| T2DM | t_cell | *HCG27* | TRUE | 3.277533942 |
| T2DM | t_cell | *DNLZ* | TRUE | 3.517519232 |
| T2DM | t_cell | *STARD10* | TRUE | 4.260740725 |
| T2DM | t_cell | *GS1.124K5.4* | TRUE | 2.190471633 |
| T2DM | t_cell | *HLA.DQA2* | TRUE | 2.598593743 |
| T2DM | t_cell | *HMBS* | TRUE | 3.038366206 |
| HTN | adipocyte | *WNT16* | TRUE | 2.265284722 |
| HTN | adipocyte | *COPZ2* | TRUE | 2.365426091 |
| HTN | adipocyte | *HFE* | TRUE | 3.344896765 |
| HTN | adipocyte | *CEP68* | TRUE | 3.373523315 |
| HTN | adipocyte | *SDCCAG8* | TRUE | 2.015726735 |
| HTN | adipocyte | *PIGV* | TRUE | 3.31109912 |
| HTN | adipocyte | *POLR1A* | TRUE | 2.896716806 |
| HTN | adipocyte | *SCGN* | TRUE | 2.596328683 |
| HTN | adipocyte | *SNX10* | TRUE | 3.883118588 |
| HTN | adipocyte | *PABPC4* | TRUE | 2.721580599 |
| HTN | adipocyte | *SLC7A4* | TRUE | 2.403749691 |
| HTN | adipocyte | *DNTTIP1* | TRUE | 2.407954387 |
| HTN | adipocyte | *POLI* | TRUE | 3.28664977 |
| HTN | adipocyte | *MTHFSD* | TRUE | 2.430511193 |
| HTN | adipocyte | *IGDCC4* | TRUE | 2.187388001 |
| HTN | adipocyte | *EYA1* | TRUE | 2.592630617 |
| HTN | adipocyte | *ICAM5* | TRUE | 2.36266001 |
| HTN | adipocyte | *ATP13A1* | TRUE | 2.944721264 |
| HTN | adipocyte | *HOXA2* | TRUE | 2.492898805 |
| HTN | adipocyte | *MEST* | TRUE | 2.121031865 |
| HTN | adipocyte | *KCTD10* | TRUE | 2.466810314 |
| HTN | adipocyte | *ENDOU* | TRUE | 2.533007733 |
| HTN | adipocyte | *PCDHB6* | TRUE | 2.442652558 |
| HTN | adipocyte | *NPHP3* | TRUE | 2.187273491 |
| HTN | adipocyte | *DNAJC27* | TRUE | 2.261867099 |

| | | | | |
|---|---|---|---|---|
| HTN | adipocyte | *IRF6* | TRUE | 2.430150209 |
| HTN | adipocyte | *TCF21* | TRUE | 3.002125576 |
| HTN | adipocyte | *HDHD3* | TRUE | 2.376835078 |
| HTN | adipocyte | *VSX2* | TRUE | 2.231733603 |
| HTN | adipocyte | *EPHX2* | TRUE | 1.966473974 |
| HTN | adipocyte | *TEX14* | TRUE | 4.170060218 |
| HTN | adipocyte | *GJB6* | TRUE | 2.708246832 |
| HTN | adipocyte | *FAM124B* | TRUE | 3.303191098 |
| HTN | adipocyte | *TOX2* | TRUE | 2.505080683 |
| HTN | adipocyte | *ZNF391* | TRUE | 2.774231165 |
| HTN | adipocyte | *GRK4* | TRUE | 2.874312409 |
| HTN | adipocyte | *TRAP1* | TRUE | 4.252065582 |
| HTN | adipocyte | *APOL5* | TRUE | 2.750041092 |
| HTN | adipocyte | *ATP8B3* | TRUE | 2.894428538 |
| HTN | adipocyte | *HAUS8* | TRUE | 2.952924272 |
| HTN | adipocyte | *REEP2* | TRUE | 2.250202721 |
| HTN | adipocyte | *RSG1* | TRUE | 2.025371417 |
| HTN | adipocyte | *ACP2* | TRUE | 6.831272431 |
| HTN | adipocyte | *AGT* | TRUE | 3.679371814 |
| HTN | adipocyte | *NMT1* | TRUE | 4.603658276 |
| HTN | adipocyte | *FBN2* | TRUE | 3.312575342 |
| HTN | adipocyte | *GPR84* | TRUE | 2.024839957 |
| HTN | adipocyte | *SCRN2* | TRUE | 2.55778411 |
| HTN | adipocyte | *CREB3L4* | TRUE | 2.588145711 |
| HTN | adipocyte | *SLC10A4* | TRUE | 3.53597715 |
| HTN | adipocyte | *SLC18B1* | TRUE | 2.923084441 |
| HTN | adipocyte | *NRBF2* | TRUE | 2.796610467 |
| HTN | adipocyte | *PDCD11* | TRUE | 6.262518634 |
| HTN | adipocyte | *HSD17B12* | TRUE | 4.998584698 |
| HTN | adipocyte | *TTC12* | TRUE | 2.060440994 |
| HTN | adipocyte | *DUSP15* | TRUE | 3.761428869 |
| HTN | adipocyte | *NEIL2* | TRUE | 3.089789194 |
| HTN | adipocyte | *CLEC18A* | TRUE | 2.469651553 |
| HTN | adipocyte | *SLC5A11* | TRUE | 4.46034057 |
| HTN | adipocyte | *S100B* | TRUE | 3.461702476 |
| HTN | adipocyte | *ABCF3* | TRUE | 3.736887129 |
| HTN | adipocyte | *TNFSF13* | TRUE | 4.242560668 |
| HTN | adipocyte | *LMOD1* | TRUE | 4.677496554 |
| HTN | adipocyte | *EXO5* | TRUE | 3.199544965 |
| HTN | adipocyte | *C1orf50* | TRUE | 2.649509237 |
| HTN | adipocyte | *BDH2* | TRUE | 2.080925217 |
| HTN | adipocyte | *RNF123* | TRUE | 2.876272827 |
| HTN | adipocyte | *ERAP1* | TRUE | 3.251981347 |
| HTN | adipocyte | *TRMT61A* | TRUE | 3.484207075 |
| HTN | adipocyte | *XRRA1* | TRUE | 1.974699109 |
| HTN | adipocyte | *TRIM66* | TRUE | 2.865516899 |
| HTN | adipocyte | *RIMKLB* | TRUE | 3.840002888 |
| HTN | adipocyte | *KIR3DL1* | TRUE | 3.829604544 |
| HTN | adipocyte | *SRR* | TRUE | 3.163297762 |
| HTN | adipocyte | *CYP7A1* | TRUE | 3.105603443 |

| | | | | |
|---|---|---|---|---|
| HTN | adipocyte | *HRASLS5* | TRUE | 2.024915327 |
| HTN | adipocyte | *IRS1* | TRUE | 5.046458719 |
| HTN | adipocyte | *INO80E* | TRUE | 2.380230785 |
| HTN | adipocyte | *CALML6* | TRUE | 2.851232063 |
| HTN | adipocyte | *RGS19* | TRUE | 4.443010076 |
| HTN | adipocyte | *HOXC5* | TRUE | 2.026225545 |
| HTN | adipocyte | *VEGFB* | TRUE | 4.5793333 |
| HTN | adipocyte | *NRROS* | TRUE | 1.999833118 |
| HTN | adipocyte | *TLR6* | TRUE | 2.532777381 |
| HTN | adipocyte | *SNX31* | TRUE | 3.533145167 |
| HTN | adipocyte | *GLIS1* | TRUE | 2.028405695 |
| HTN | adipocyte | *RSRC1* | TRUE | 1.974578475 |
| HTN | adipocyte | *NRIP3* | TRUE | 2.357789428 |
| HTN | adipocyte | *PPFIA3* | TRUE | 2.419687781 |
| HTN | adipocyte | *COX14* | TRUE | 2.553212281 |
| HTN | adipocyte | *CPNE7* | TRUE | 1.970459913 |
| HTN | adipocyte | *MPI* | TRUE | 4.786839121 |
| HTN | adipocyte | *HYI* | TRUE | 2.783769731 |
| HTN | adipocyte | *CTC1* | TRUE | 2.141183034 |
| HTN | adipocyte | *DEFB124* | TRUE | 2.749713851 |
| HTN | adipocyte | *ZNF443* | TRUE | 2.341831593 |
| HTN | adipocyte | *CLN8* | TRUE | 2.373046106 |
| HTN | adipocyte | *ZNF320* | TRUE | 3.139293635 |
| HTN | adipocyte | *RP1L1* | TRUE | 3.888295454 |
| HTN | adipocyte | *KREMEN1* | TRUE | 3.080464327 |
| HTN | adipocyte | *EIF3C* | TRUE | 2.634839299 |
| HTN | adipocyte | *A3GALT2* | TRUE | 4.123063891 |
| HTN | adipocyte | *LPAR5* | TRUE | 2.239965449 |
| HTN | adipocyte | *NUTM2A* | TRUE | 2.104189549 |
| HTN | adipocyte | *DDX51* | TRUE | 2.538609038 |
| HTN | adipocyte | *CDK10* | TRUE | 2.074652758 |
| HTN | adipocyte | *TCN2* | TRUE | 1.986531752 |
| HTN | adipocyte | *ARL17A* | TRUE | 2.184865836 |
| HTN | adipocyte | *MCRS1* | TRUE | 3.130674817 |
| HTN | adipocyte | *ZNF559* | TRUE | 2.500650055 |
| HTN | adipocyte | *FBF1* | TRUE | 4.133579171 |
| HTN | adipocyte | *HLA.DRB1* | TRUE | 2.632261933 |
| HTN | adipocyte | *C6orf106* | TRUE | 2.810414712 |
| HTN | adipocyte | *HIST1H3E* | TRUE | 2.038193719 |
| HTN | adipocyte | *ZNF100* | TRUE | 2.361459946 |
| HTN | adipocyte | *RPS26* | TRUE | 2.143893944 |
| HTN | adipocyte | *TMEM116* | TRUE | 5.898335079 |
| HTN | adipocyte | *PNP* | TRUE | 2.622352359 |
| HTN | adipocyte | *AL590822.1* | TRUE | 2.526779598 |
| HTN | adipocyte | *HSD17B8* | TRUE | 3.294593518 |
| HTN | adipocyte | *STK19* | TRUE | 2.991144873 |
| HTN | adipocyte | *MXD3* | TRUE | 2.322728811 |
| HTN | adipocyte | *SIPA1* | TRUE | 3.887884793 |
| HTN | adipocyte | *ATF6B* | TRUE | 3.485932695 |
| HTN | adipocyte | *AC016773.1* | TRUE | 2.187561934 |

| | | | | |
|---|---|---|---|---|
| HTN | adipocyte | *LINC00092* | TRUE | 2.021849892 |
| HTN | adipocyte | *AC017074.2* | TRUE | 2.264387615 |
| HTN | adipocyte | *PROB1* | TRUE | 2.860985231 |
| HTN | adipocyte | *AC067945.3* | TRUE | 2.201906932 |
| HTN | adipocyte | *AC112229.4* | TRUE | 2.124678698 |
| HTN | adipocyte | *PLA2G4B* | TRUE | 1.98346037 |
| HTN | adipocyte | *ZNF10* | TRUE | 3.337434455 |
| HTN | adipocyte | *HMBS* | TRUE | 3.541612296 |
| HTN | adipocyte | *AC144652.1* | TRUE | 2.916058619 |
| HTN | ASPC | *WNT16* | TRUE | 2.55568668 |
| HTN | ASPC | *M6PR* | TRUE | 2.418109157 |
| HTN | ASPC | *ARHGAP33* | TRUE | 2.077504322 |
| HTN | ASPC | *COPZ2* | TRUE | 3.231588878 |
| HTN | ASPC | *HFE* | TRUE | 3.723046191 |
| HTN | ASPC | *CEP68* | TRUE | 5.001044329 |
| HTN | ASPC | *NR1H3* | TRUE | 2.514515712 |
| HTN | ASPC | *SNX1* | TRUE | 2.324111487 |
| HTN | ASPC | *EPN3* | TRUE | 2.371462793 |
| HTN | ASPC | *LIMA1* | TRUE | 2.325317499 |
| HTN | ASPC | *ANAPC4* | TRUE | 2.235832829 |
| HTN | ASPC | *ITIH4* | TRUE | 2.189340332 |
| HTN | ASPC | *PITHD1* | TRUE | 2.145894556 |
| HTN | ASPC | *PIGV* | TRUE | 3.362275374 |
| HTN | ASPC | *HAGH* | TRUE | 2.886853375 |
| HTN | ASPC | *RRP15* | TRUE | 2.129287137 |
| HTN | ASPC | *POLR1A* | TRUE | 2.982924087 |
| HTN | ASPC | *ASNS* | TRUE | 2.534650763 |
| HTN | ASPC | *ATP2A3* | TRUE | 3.642385719 |
| HTN | ASPC | *TIPIN* | TRUE | 2.414704877 |
| HTN | ASPC | *SCGN* | TRUE | 2.949622875 |
| HTN | ASPC | *DDX43* | TRUE | 2.449668865 |
| HTN | ASPC | *CCNT2* | TRUE | 2.777207076 |
| HTN | ASPC | *COL16A1* | TRUE | 2.351237453 |
| HTN | ASPC | *FMO2* | TRUE | 2.202507325 |
| HTN | ASPC | *MARCH2* | TRUE | 4.189501186 |
| HTN | ASPC | *SLC7A4* | TRUE | 3.08792333 |
| HTN | ASPC | *CARD10* | TRUE | 1.967551552 |
| HTN | ASPC | *TXN2* | TRUE | 2.10603708 |
| HTN | ASPC | *FAM118A* | TRUE | 2.144850138 |
| HTN | ASPC | *MTG2* | TRUE | 2.079800615 |
| HTN | ASPC | *MANBAL* | TRUE | 2.089770493 |
| HTN | ASPC | *DNTTIP1* | TRUE | 2.856951311 |
| HTN | ASPC | *POLI* | TRUE | 3.76006783 |
| HTN | ASPC | *VWA8* | TRUE | 2.55001851 |
| HTN | ASPC | *CRISPLD2* | TRUE | 2.571701771 |
| HTN | ASPC | *MTHFSD* | TRUE | 3.184481358 |
| HTN | ASPC | *KAT8* | TRUE | 3.25727258 |
| HTN | ASPC | *ZFAND1* | TRUE | 2.014695372 |
| HTN | ASPC | *EYA1* | TRUE | 3.876907708 |
| HTN | ASPC | *PEX11G* | TRUE | 2.340128949 |

| | | | | |
|---|---|---|---|---|
| HTN | ASPC | *CCDC94* | TRUE | 2.547179541 |
| HTN | ASPC | *ICAM5* | TRUE | 2.838468003 |
| HTN | ASPC | *ATP13A1* | TRUE | 3.705262517 |
| HTN | ASPC | *HOXA2* | TRUE | 4.389903009 |
| HTN | ASPC | *CRHR2* | TRUE | 2.408194558 |
| HTN | ASPC | *EPHB6* | TRUE | 2.758873961 |
| HTN | ASPC | *MEST* | TRUE | 3.197733799 |
| HTN | ASPC | *ACTR3C* | TRUE | 3.186157936 |
| HTN | ASPC | *SLC1A1* | TRUE | 2.110285891 |
| HTN | ASPC | *WNT3* | TRUE | 2.813542441 |
| HTN | ASPC | *ICAM2* | TRUE | 2.780909258 |
| HTN | ASPC | *DDX5* | TRUE | 3.386157021 |
| HTN | ASPC | *CYTH1* | TRUE | 2.385620889 |
| HTN | ASPC | *SLC2A9* | TRUE | 2.158324069 |
| HTN | ASPC | *CAND1* | TRUE | 3.855195994 |
| HTN | ASPC | *BVES* | TRUE | 2.050421153 |
| HTN | ASPC | *GUCA1B* | TRUE | 2.261538559 |
| HTN | ASPC | *CEP72* | TRUE | 2.542475949 |
| HTN | ASPC | *NME5* | TRUE | 1.967147053 |
| HTN | ASPC | *PCDHB6* | TRUE | 2.370074589 |
| HTN | ASPC | *BNIP1* | TRUE | 2.421096851 |
| HTN | ASPC | *NPHP3* | TRUE | 2.765452087 |
| HTN | ASPC | *PCCB* | TRUE | 4.420690315 |
| HTN | ASPC | *TP53I3* | TRUE | 3.00878378 |
| HTN | ASPC | *DNAJC27* | TRUE | 4.189173501 |
| HTN | ASPC | *RTN4* | TRUE | 2.119942191 |
| HTN | ASPC | *ZNF142* | TRUE | 2.571388388 |
| HTN | ASPC | *CD207* | TRUE | 2.441246151 |
| HTN | ASPC | *C1orf21* | TRUE | 2.970209482 |
| HTN | ASPC | *GBP3* | TRUE | 2.053699142 |
| HTN | ASPC | *NSUN4* | TRUE | 2.292551588 |
| HTN | ASPC | *IRF6* | TRUE | 2.840339466 |
| HTN | ASPC | *DDX59* | TRUE | 2.00045948 |
| HTN | ASPC | *FILIP1* | TRUE | 2.212290043 |
| HTN | ASPC | *TCF21* | TRUE | 3.101680656 |
| HTN | ASPC | *TRPM6* | TRUE | 3.059474598 |
| HTN | ASPC | *HDHD3* | TRUE | 2.73348774 |
| HTN | ASPC | *HSDL2* | TRUE | 2.019823522 |
| HTN | ASPC | *VSX2* | TRUE | 2.114376857 |
| HTN | ASPC | *HELLS* | TRUE | 2.086667102 |
| HTN | ASPC | *EPHX2* | TRUE | 3.255297759 |
| HTN | ASPC | *TEX14* | TRUE | 5.194404465 |
| HTN | ASPC | *TRIM6* | TRUE | 2.944483039 |
| HTN | ASPC | *GJB6* | TRUE | 1.990293349 |
| HTN | ASPC | *SLC25A51* | TRUE | 2.148512392 |
| HTN | ASPC | *NDUFAF4* | TRUE | 2.739241623 |
| HTN | ASPC | *ACVR1C* | TRUE | 3.747528467 |
| HTN | ASPC | *FAM124B* | TRUE | 3.4684445 |
| HTN | ASPC | *TOX2* | TRUE | 2.71550888 |
| HTN | ASPC | *PEPD* | TRUE | 2.050534794 |

| | | | | |
|---|---|---|---|---|
| HTN | ASPC | *ZNF391* | TRUE | 2.537109473 |
| HTN | ASPC | *GRK4* | TRUE | 3.439640141 |
| HTN | ASPC | *GPR108* | TRUE | 2.198094911 |
| HTN | ASPC | *TRAP1* | TRUE | 4.688737157 |
| HTN | ASPC | *RHOT1* | TRUE | 2.337540671 |
| HTN | ASPC | *YEATS4* | TRUE | 2.283759082 |
| HTN | ASPC | *STYXL1* | TRUE | 2.136738047 |
| HTN | ASPC | *APOL5* | TRUE | 3.262101764 |
| HTN | ASPC | *KLHDC10* | TRUE | 3.335054907 |
| HTN | ASPC | *RIPK3* | TRUE | 2.03468886 |
| HTN | ASPC | *GALNT8* | TRUE | 2.165051661 |
| HTN | ASPC | *CDC16* | TRUE | 3.530079752 |
| HTN | ASPC | *ATP8B3* | TRUE | 3.241153184 |
| HTN | ASPC | *DDA1* | TRUE | 2.321206649 |
| HTN | ASPC | *PGPEP1* | TRUE | 2.896499576 |
| HTN | ASPC | *CHMP1A* | TRUE | 2.615773776 |
| HTN | ASPC | *HAUS8* | TRUE | 4.659946303 |
| HTN | ASPC | *EXOC4* | TRUE | 2.477264034 |
| HTN | ASPC | *ANO1* | TRUE | 2.650593132 |
| HTN | ASPC | *KDM6B* | TRUE | 2.73145058 |
| HTN | ASPC | *REEP2* | TRUE | 2.157149176 |
| HTN | ASPC | *RSG1* | TRUE | 2.394436334 |
| HTN | ASPC | *WNT2B* | TRUE | 4.374728681 |
| HTN | ASPC | *ACP2* | TRUE | 7.611350447 |
| HTN | ASPC | *APLNR* | TRUE | 2.087696821 |
| HTN | ASPC | *TMEM165* | TRUE | 4.079992979 |
| HTN | ASPC | *TMEM60* | TRUE | 2.166341001 |
| HTN | ASPC | *POM121C* | TRUE | 2.061645901 |
| HTN | ASPC | *BLOC1S1* | TRUE | 2.062901727 |
| HTN | ASPC | *AGT* | TRUE | 3.484660746 |
| HTN | ASPC | *NACAD* | TRUE | 2.29430418 |
| HTN | ASPC | *CIB2* | TRUE | 2.605804217 |
| HTN | ASPC | *NMT1* | TRUE | 5.158630488 |
| HTN | ASPC | *RTP4* | TRUE | 2.468568701 |
| HTN | ASPC | *SLC2A8* | TRUE | 2.449137881 |
| HTN | ASPC | *PAQR5* | TRUE | 2.065213494 |
| HTN | ASPC | *RMDN3* | TRUE | 4.702780601 |
| HTN | ASPC | *IFT172* | TRUE | 3.202488103 |
| HTN | ASPC | *DYNC2LI1* | TRUE | 3.025257334 |
| HTN | ASPC | *TRPC3* | TRUE | 1.986472152 |
| HTN | ASPC | *G3BP2* | TRUE | 2.447169348 |
| HTN | ASPC | *FBN2* | TRUE | 3.591401302 |
| HTN | ASPC | *GLTP* | TRUE | 2.596524683 |
| HTN | ASPC | *GPR84* | TRUE | 2.736024551 |
| HTN | ASPC | *TICRR* | TRUE | 2.877173919 |
| HTN | ASPC | *SCRN2* | TRUE | 2.683005279 |
| HTN | ASPC | *GREB1L* | TRUE | 2.983802166 |
| HTN | ASPC | *RERE* | TRUE | 2.457608596 |
| HTN | ASPC | *SYPL2* | TRUE | 2.423867163 |
| HTN | ASPC | *RGS5* | TRUE | 3.124144393 |

| | | | | |
|---|---|---|---|---|
| HTN | ASPC | *ARNT* | TRUE | 2.145496698 |
| HTN | ASPC | *WNT9A* | TRUE | 2.815500557 |
| HTN | ASPC | *RABL2A* | TRUE | 2.449166864 |
| HTN | ASPC | *GPR17* | TRUE | 2.071800574 |
| HTN | ASPC | *PHOSPHO2* | TRUE | 2.043389085 |
| HTN | ASPC | *SLC10A4* | TRUE | 3.362404823 |
| HTN | ASPC | *TSLP* | TRUE | 2.637817729 |
| HTN | ASPC | *GFOD1* | TRUE | 2.287668736 |
| HTN | ASPC | *PSD2* | TRUE | 2.218369614 |
| HTN | ASPC | *SLC18B1* | TRUE | 3.493754954 |
| HTN | ASPC | *CCDC25* | TRUE | 2.455198364 |
| HTN | ASPC | *SURF1* | TRUE | 2.184845269 |
| HTN | ASPC | *NRBF2* | TRUE | 2.237740372 |
| HTN | ASPC | *PDCD11* | TRUE | 8.595116013 |
| HTN | ASPC | *HSD17B12* | TRUE | 5.602301646 |
| HTN | ASPC | *TTC12* | TRUE | 2.678083867 |
| HTN | ASPC | *DUSP15* | TRUE | 3.895683986 |
| HTN | ASPC | *ARID5B* | TRUE | 2.796809667 |
| HTN | ASPC | *ARL14EP* | TRUE | 2.592540351 |
| HTN | ASPC | *CWF19L2* | TRUE | 4.838018259 |
| HTN | ASPC | *PRDM8* | TRUE | 3.090749735 |
| HTN | ASPC | *HS3ST3A1* | TRUE | 2.156621262 |
| HTN | ASPC | *DISP1* | TRUE | 2.495531277 |
| HTN | ASPC | *NEIL2* | TRUE | 4.301411783 |
| HTN | ASPC | *TSEN2* | TRUE | 2.098779242 |
| HTN | ASPC | *PROM2* | TRUE | 2.592577399 |
| HTN | ASPC | *AK9* | TRUE | 2.437044663 |
| HTN | ASPC | *TTC39B* | TRUE | 2.062859031 |
| HTN | ASPC | *DCK* | TRUE | 2.150429921 |
| HTN | ASPC | *NDUFAF6* | TRUE | 4.23201128 |
| HTN | ASPC | *C8orf37* | TRUE | 2.073968886 |
| HTN | ASPC | *ANKRD9* | TRUE | 2.461846081 |
| HTN | ASPC | *MED19* | TRUE | 2.034161732 |
| HTN | ASPC | *VPS8* | TRUE | 2.732773593 |
| HTN | ASPC | *STEAP2* | TRUE | 1.98174008 |
| HTN | ASPC | *CLEC18A* | TRUE | 2.063080097 |
| HTN | ASPC | *KCNJ15* | TRUE | 2.65050185 |
| HTN | ASPC | *ZNF276* | TRUE | 2.289191353 |
| HTN | ASPC | *SLC5A11* | TRUE | 4.65708226 |
| HTN | ASPC | *S100B* | TRUE | 3.616090345 |
| HTN | ASPC | *FDPS* | TRUE | 2.322412686 |
| HTN | ASPC | *PMF1* | TRUE | 1.973884039 |
| HTN | ASPC | *ABCF3* | TRUE | 4.539128123 |
| HTN | ASPC | *TNFSF13* | TRUE | 4.160443745 |
| HTN | ASPC | *LMOD1* | TRUE | 5.639208269 |
| HTN | ASPC | *FAM208A* | TRUE | 3.641791725 |
| HTN | ASPC | *EXO5* | TRUE | 3.831478401 |
| HTN | ASPC | *C1orf50* | TRUE | 3.55784781 |
| HTN | ASPC | *BDH2* | TRUE | 3.016502594 |
| HTN | ASPC | *RNF123* | TRUE | 2.988048113 |

| | | | | |
|---|---|---|---|---|
| HTN | ASPC | *ERAP1* | TRUE | 4.169878036 |
| HTN | ASPC | *TMEM184A* | TRUE | 2.40279785 |
| HTN | ASPC | *C9orf64* | TRUE | 2.148635446 |
| HTN | ASPC | *ZNF367* | TRUE | 2.784627156 |
| HTN | ASPC | *REEP3* | TRUE | 2.387185089 |
| HTN | ASPC | *HSPA12A* | TRUE | 1.996602389 |
| HTN | ASPC | *XRRA1* | TRUE | 2.849508187 |
| HTN | ASPC | *TRIM66* | TRUE | 3.033042258 |
| HTN | ASPC | *RIMKLB* | TRUE | 4.676888259 |
| HTN | ASPC | *ZNF641* | TRUE | 2.705955167 |
| HTN | ASPC | *KIR3DL1* | TRUE | 3.785066416 |
| HTN | ASPC | *SRR* | TRUE | 3.688221285 |
| HTN | ASPC | *CYP7A1* | TRUE | 2.948998085 |
| HTN | ASPC | *HRASLS5* | TRUE | 3.087956706 |
| HTN | ASPC | *PXK* | TRUE | 2.080460344 |
| HTN | ASPC | *TNIP2* | TRUE | 2.39683721 |
| HTN | ASPC | *IRS1* | TRUE | 5.79324694 |
| HTN | ASPC | *CXCL10* | TRUE | 2.018222694 |
| HTN | ASPC | *STK32A* | TRUE | 1.991175384 |
| HTN | ASPC | *INO80E* | TRUE | 2.564797705 |
| HTN | ASPC | *FAM103A1* | TRUE | 2.042622365 |
| HTN | ASPC | *LIMS1* | TRUE | 2.451008053 |
| HTN | ASPC | *CALML6* | TRUE | 3.810023465 |
| HTN | ASPC | *HOXD4* | TRUE | 2.346505555 |
| HTN | ASPC | *RNF34* | TRUE | 2.094208079 |
| HTN | ASPC | *PAQR8* | TRUE | 4.500278546 |
| HTN | ASPC | *SOX7* | TRUE | 2.240024486 |
| HTN | ASPC | *GIMAP8* | TRUE | 2.220831774 |
| HTN | ASPC | *DLK2* | TRUE | 2.791637225 |
| HTN | ASPC | *RGS19* | TRUE | 5.58276287 |
| HTN | ASPC | *CTSW* | TRUE | 3.050546543 |
| HTN | ASPC | *ZNF738* | TRUE | 2.252273172 |
| HTN | ASPC | *HOXC5* | TRUE | 3.005593503 |
| HTN | ASPC | *MYEOV* | TRUE | 2.839930594 |
| HTN | ASPC | *AHSA2* | TRUE | 2.206162534 |
| HTN | ASPC | *DMRT2* | TRUE | 3.196565893 |
| HTN | ASPC | *VEGFB* | TRUE | 5.360586395 |
| HTN | ASPC | *CCS* | TRUE | 3.035283464 |
| HTN | ASPC | *NRROS* | TRUE | 2.899739466 |
| HTN | ASPC | *TLR6* | TRUE | 4.157076536 |
| HTN | ASPC | *CTU2* | TRUE | 2.238514236 |
| HTN | ASPC | *SNX31* | TRUE | 3.903210748 |
| HTN | ASPC | *GLIS1* | TRUE | 3.344894702 |
| HTN | ASPC | *KLF14* | TRUE | 4.340130657 |
| HTN | ASPC | *RSRC1* | TRUE | 3.989020396 |
| HTN | ASPC | *NRIP3* | TRUE | 2.748841548 |
| HTN | ASPC | *PPP2R2D* | TRUE | 3.089890062 |
| HTN | ASPC | *CRLF3* | TRUE | 2.554957668 |
| HTN | ASPC | *MTHFR* | TRUE | 5.204580584 |
| HTN | ASPC | *TOP3A* | TRUE | 2.488063526 |

| | | | | |
|---|---|---|---|---|
| HTN | ASPC | *PPFIA3* | TRUE | 2.782165405 |
| HTN | ASPC | *GEN1* | TRUE | 1.97257341 |
| HTN | ASPC | *COX14* | TRUE | 5.173036654 |
| HTN | ASPC | *MPI* | TRUE | 3.304614647 |
| HTN | ASPC | *HYI* | TRUE | 2.190368034 |
| HTN | ASPC | *CTC1* | TRUE | 3.313122777 |
| HTN | ASPC | *DEFB124* | TRUE | 3.530822163 |
| HTN | ASPC | *ZNF443* | TRUE | 3.186915268 |
| HTN | ASPC | *TMEM136* | TRUE | 2.433959257 |
| HTN | ASPC | *CLDN7* | TRUE | 3.092270216 |
| HTN | ASPC | *ADO* | TRUE | 2.55061892 |
| HTN | ASPC | *FAM89A* | TRUE | 2.485224655 |
| HTN | ASPC | *CLN8* | TRUE | 4.763915278 |
| HTN | ASPC | *FES* | TRUE | 7.519877567 |
| HTN | ASPC | *MXRA7* | TRUE | 2.167156608 |
| HTN | ASPC | *COL18A1* | TRUE | 2.884896145 |
| HTN | ASPC | *ZNF320* | TRUE | 3.425138589 |
| HTN | ASPC | *FAM43B* | TRUE | 2.424470928 |
| HTN | ASPC | *KLRC4* | TRUE | 2.128494531 |
| HTN | ASPC | *RP1L1* | TRUE | 4.121847666 |
| HTN | ASPC | *BMP8A* | TRUE | 2.040389392 |
| HTN | ASPC | *KREMEN1* | TRUE | 3.529013888 |
| HTN | ASPC | *EIF3C* | TRUE | 2.597690583 |
| HTN | ASPC | *A3GALT2* | TRUE | 4.208984409 |
| HTN | ASPC | *CCR10* | TRUE | 3.087351309 |
| HTN | ASPC | *LPAR5* | TRUE | 2.562318273 |
| HTN | ASPC | *NUTM2A* | TRUE | 2.103004132 |
| HTN | ASPC | *DDX51* | TRUE | 3.213097107 |
| HTN | ASPC | *TCN2* | TRUE | 2.721352961 |
| HTN | ASPC | *LSAMP* | TRUE | 2.248588646 |
| HTN | ASPC | *ARL17A* | TRUE | 3.201383224 |
| HTN | ASPC | *ZNF396* | TRUE | 2.072304673 |
| HTN | ASPC | *FOXI2* | TRUE | 2.063560547 |
| HTN | ASPC | *RHD* | TRUE | 2.014366997 |
| HTN | ASPC | *MCRS1* | TRUE | 4.610890481 |
| HTN | ASPC | *ZNF559* | TRUE | 2.902592215 |
| HTN | ASPC | *ZDHHC11* | TRUE | 2.71882805 |
| HTN | ASPC | *FBF1* | TRUE | 3.614114805 |
| HTN | ASPC | *FAM47E* | TRUE | 2.448349751 |
| HTN | ASPC | *HLA.DRB1* | TRUE | 3.120463955 |
| HTN | ASPC | *ZNF34* | TRUE | 2.07161108 |
| HTN | ASPC | *C6orf106* | TRUE | 3.135159324 |
| HTN | ASPC | *ZFP28* | TRUE | 2.137648279 |
| HTN | ASPC | *KIAA1211L* | TRUE | 2.070744797 |
| HTN | ASPC | *HCAR1* | TRUE | 2.021406747 |
| HTN | ASPC | *HIST1H3E* | TRUE | 2.555262382 |
| HTN | ASPC | *FUT11* | TRUE | 2.411190982 |
| HTN | ASPC | *ZNF100* | TRUE | 3.298676605 |
| HTN | ASPC | *C6orf141* | TRUE | 2.020254735 |
| HTN | ASPC | *SLC22A5* | TRUE | 2.352275608 |

| | | | | |
|---|---|---|---|---|
| HTN | ASPC | *RPS26* | TRUE | 2.195900234 |
| HTN | ASPC | *RPS6KL1* | TRUE | 2.801477461 |
| HTN | ASPC | *TMEM116* | TRUE | 4.717998428 |
| HTN | ASPC | *GFPT1* | TRUE | 3.066504382 |
| HTN | ASPC | *PNP* | TRUE | 2.836270088 |
| HTN | ASPC | *PRMT6* | TRUE | 2.100323302 |
| HTN | ASPC | *TGM2* | TRUE | 2.257067001 |
| HTN | ASPC | *AL590822.1* | TRUE | 3.236953111 |
| HTN | ASPC | *CENPW* | TRUE | 2.673104654 |
| HTN | ASPC | *INPP5B* | TRUE | 2.182807511 |
| HTN | ASPC | *HSD17B8* | TRUE | 3.227207561 |
| HTN | ASPC | *STK19* | TRUE | 3.722350679 |
| HTN | ASPC | *NOTCH2NL* | TRUE | 2.025181653 |
| HTN | ASPC | *MXD3* | TRUE | 2.914093863 |
| HTN | ASPC | *SIPA1* | TRUE | 5.144951364 |
| HTN | ASPC | *ATF6B* | TRUE | 4.010474589 |
| HTN | ASPC | *PPM1N* | TRUE | 3.002853996 |
| HTN | ASPC | *GALT* | TRUE | 2.376941929 |
| HTN | ASPC | *REPIN1* | TRUE | 1.961378665 |
| HTN | ASPC | *WEE2* | TRUE | 2.387519739 |
| HTN | ASPC | *PPME1* | TRUE | 2.149227945 |
| HTN | ASPC | *STARD10* | TRUE | 2.007654624 |
| HTN | ASPC | *SLC12A8* | TRUE | 2.495433014 |
| HTN | ASPC | *LINC00092* | TRUE | 2.155465946 |
| HTN | ASPC | *SLC26A6* | TRUE | 2.179210284 |
| HTN | ASPC | *AC017074.2* | TRUE | 2.225586847 |
| HTN | ASPC | *AC108463.2* | TRUE | 2.35603797 |
| HTN | ASPC | *PROB1* | TRUE | 3.292174645 |
| HTN | ASPC | *AC067945.3* | TRUE | 2.086821214 |
| HTN | ASPC | *AC112229.4* | TRUE | 2.204196332 |
| HTN | ASPC | *GOLGA8N* | TRUE | 2.049143547 |
| HTN | ASPC | *PSG4* | TRUE | 2.980393371 |
| HTN | ASPC | *ZNF10* | TRUE | 2.967919738 |
| HTN | ASPC | *HMBS* | TRUE | 4.22319156 |
| HTN | ASPC | *SH3RF3.AS1* | TRUE | 2.267056629 |
| HTN | ASPC | *LINC00668* | TRUE | 2.217563487 |
| HTN | ASPC | *SRXN1* | TRUE | 2.201647414 |
| HTN | ASPC | *AC144652.1* | TRUE | 3.53088287 |
| HTN | b_cell | *SNX10* | TRUE | 3.149642544 |
| HTN | b_cell | *ACP2* | TRUE | 3.520003386 |
| HTN | b_cell | *HSD17B12* | TRUE | 2.684642805 |
| HTN | b_cell | *CLEC18A* | TRUE | 3.66680268 |
| HTN | b_cell | *ZNF589* | TRUE | 2.366421763 |
| HTN | b_cell | *KIR3DL1* | TRUE | 3.0404597 |
| HTN | b_cell | *RPP25* | TRUE | 3.883862359 |
| HTN | b_cell | *A3GALT2* | TRUE | 2.103218258 |
| HTN | b_cell | *RPS26* | TRUE | 2.266187686 |
| HTN | b_cell | *NOTCH2NL* | TRUE | 2.435476115 |
| HTN | dendritic_cell | *ITIH4* | TRUE | 2.706980525 |
| HTN | dendritic_cell | *SNX10* | TRUE | 4.16335725 |

| | | | | |
|---|---|---|---|---|
| HTN | dendritic_cell | *SNRPC* | TRUE | 2.351101936 |
| HTN | dendritic_cell | *ACP2* | TRUE | 6.019122053 |
| HTN | dendritic_cell | *AGT* | TRUE | 2.386527081 |
| HTN | dendritic_cell | *SLC10A4* | TRUE | 1.991917614 |
| HTN | dendritic_cell | *SLC18B1* | TRUE | 2.548771744 |
| HTN | dendritic_cell | *HSD17B12* | TRUE | 4.117686029 |
| HTN | dendritic_cell | *TNFSF13* | TRUE | 2.467389434 |
| HTN | dendritic_cell | *LMOD1* | TRUE | 3.63310135 |
| HTN | dendritic_cell | *RNF123* | TRUE | 2.60059514 |
| HTN | dendritic_cell | *TRMT61A* | TRUE | 2.878261745 |
| HTN | dendritic_cell | *TRIM66* | TRUE | 2.376966737 |
| HTN | dendritic_cell | *KIR3DL1* | TRUE | 2.789886591 |
| HTN | dendritic_cell | *SNX31* | TRUE | 2.222995727 |
| HTN | dendritic_cell | *RPP25* | TRUE | 2.327653381 |
| HTN | dendritic_cell | *HYI* | TRUE | 3.644615209 |
| HTN | dendritic_cell | *CLDN7* | TRUE | 3.901980528 |
| HTN | dendritic_cell | *ZNF320* | TRUE | 2.866782701 |
| HTN | dendritic_cell | *EIF3C* | TRUE | 2.917819398 |
| HTN | dendritic_cell | *A3GALT2* | TRUE | 2.283244954 |
| HTN | dendritic_cell | *FBF1* | TRUE | 2.220692979 |
| HTN | dendritic_cell | *HIST1H3E* | TRUE | 3.10994969 |
| HTN | dendritic_cell | *RPS26* | TRUE | 3.780348306 |
| HTN | dendritic_cell | *TMEM116* | TRUE | 3.599475562 |
| HTN | dendritic_cell | *HSD17B8* | TRUE | 2.659727563 |
| HTN | dendritic_cell | *LTB4R* | TRUE | 2.483783194 |
| HTN | endothelial | *PCDHA6* | TRUE | 2.034750289 |
| HTN | endothelial | *IGDCC4* | TRUE | 2.105680469 |
| HTN | endothelial | *NDUFAF4* | TRUE | 2.104698292 |
| HTN | endothelial | *ATP8B3* | TRUE | 2.208106237 |
| HTN | endothelial | *PKDREJ* | TRUE | 3.105727211 |
| HTN | endothelial | *RSG1* | TRUE | 2.033473903 |
| HTN | endothelial | *CIB2* | TRUE | 2.172463141 |
| HTN | endothelial | *FBN2* | TRUE | 1.974366342 |
| HTN | endothelial | *TTC12* | TRUE | 1.97213686 |
| HTN | endothelial | *DOC2A* | TRUE | 2.953640799 |
| HTN | endothelial | *NEIL2* | TRUE | 1.989651627 |
| HTN | endothelial | *SLC5A11* | TRUE | 2.593477878 |
| HTN | endothelial | *TNFSF13* | TRUE | 2.906996839 |
| HTN | endothelial | *RGS19* | TRUE | 2.799183536 |
| HTN | endothelial | *RPP25* | TRUE | 3.237285574 |
| HTN | endothelial | *CPNE7* | TRUE | 2.419992243 |
| HTN | endothelial | *TRIM73* | TRUE | 2.318728717 |
| HTN | endothelial | *HYI* | TRUE | 2.073684763 |
| HTN | endothelial | *NPIPA1* | TRUE | 2.013876367 |
| HTN | endothelial | *EIF3C* | TRUE | 2.291125579 |
| HTN | endothelial | *HIST1H3E* | TRUE | 3.360754349 |
| HTN | endothelial | *RPS26* | TRUE | 2.718626643 |
| HTN | endothelial | *HSD17B8* | TRUE | 2.180555017 |
| HTN | endothelial | *AC017074.2* | TRUE | 2.202429675 |
| HTN | endothelial | *PROB1* | TRUE | 2.214900645 |

| | | | | |
|---|---|---|---|---|
| HTN | endothelial | *PSG4* | TRUE | 2.320758883 |
| HTN | LEC | *HFE* | TRUE | 2.409055182 |
| HTN | LEC | *HSD17B12* | TRUE | 2.605661059 |
| HTN | LEC | *DUSP15* | TRUE | 2.076730017 |
| HTN | LEC | *CLEC18A* | TRUE | 2.418108346 |
| HTN | LEC | *SLC5A11* | TRUE | 3.081738621 |
| HTN | LEC | *TNFSF13* | TRUE | 2.489860735 |
| HTN | LEC | *HIST1H3E* | TRUE | 2.535525402 |
| HTN | LEC | *RPS26* | TRUE | 2.860556585 |
| HTN | LEC | *PLA2G4B* | TRUE | 2.053947055 |
| HTN | macrophage | *COPZ2* | TRUE | 2.732645099 |
| HTN | macrophage | *HFE* | TRUE | 3.249489247 |
| HTN | macrophage | *ITIH4* | TRUE | 2.285676677 |
| HTN | macrophage | *SCGN* | TRUE | 2.467615524 |
| HTN | macrophage | *PCDHA6* | TRUE | 2.29635273 |
| HTN | macrophage | *SNX10* | TRUE | 2.935789289 |
| HTN | macrophage | *POLI* | TRUE | 1.999099352 |
| HTN | macrophage | *MTHFSD* | TRUE | 2.967222774 |
| HTN | macrophage | *EYA1* | TRUE | 2.369689628 |
| HTN | macrophage | *ICAM5* | TRUE | 2.022567544 |
| HTN | macrophage | *ENDOU* | TRUE | 2.463217988 |
| HTN | macrophage | *IRF6* | TRUE | 2.184504468 |
| HTN | macrophage | *ALDH8A1* | TRUE | 2.021076415 |
| HTN | macrophage | *TCF21* | TRUE | 2.868906749 |
| HTN | macrophage | *HDHD3* | TRUE | 2.131246833 |
| HTN | macrophage | *VSX2* | TRUE | 2.124607534 |
| HTN | macrophage | *TEX14* | TRUE | 3.102155228 |
| HTN | macrophage | *TRIM6* | TRUE | 2.016147712 |
| HTN | macrophage | *NDUFAF4* | TRUE | 2.68985873 |
| HTN | macrophage | *FAM124B* | TRUE | 2.197620636 |
| HTN | macrophage | *TOX2* | TRUE | 3.583861044 |
| HTN | macrophage | *TCP11* | TRUE | 2.624661847 |
| HTN | macrophage | *TRAP1* | TRUE | 2.563017351 |
| HTN | macrophage | *APOL5* | TRUE | 2.87059002 |
| HTN | macrophage | *AANAT* | TRUE | 2.232539204 |
| HTN | macrophage | *GALNT8* | TRUE | 2.318222455 |
| HTN | macrophage | *ATP8B3* | TRUE | 3.418700427 |
| HTN | macrophage | *PKDREJ* | TRUE | 3.350492389 |
| HTN | macrophage | *RSG1* | TRUE | 2.275416888 |
| HTN | macrophage | *ACP2* | TRUE | 2.531303394 |
| HTN | macrophage | *AGT* | TRUE | 2.406039196 |
| HTN | macrophage | *CIB2* | TRUE | 2.875954697 |
| HTN | macrophage | *NMT1* | TRUE | 3.46432182 |
| HTN | macrophage | *RTP4* | TRUE | 3.479368269 |
| HTN | macrophage | *SLC10A4* | TRUE | 2.098801473 |
| HTN | macrophage | *SLC18B1* | TRUE | 2.011444068 |
| HTN | macrophage | *NRBF2* | TRUE | 2.666943673 |
| HTN | macrophage | *PDCD11* | TRUE | 2.821884557 |
| HTN | macrophage | *HSD17B12* | TRUE | 3.561315562 |
| HTN | macrophage | *TTC12* | TRUE | 3.007221664 |

| | | | | |
|---|---|---|---|---|
| HTN | macrophage | *DUSP15* | TRUE | 3.576450165 |
| HTN | macrophage | *DOC2A* | TRUE | 2.661209686 |
| HTN | macrophage | *NEIL2* | TRUE | 2.542344942 |
| HTN | macrophage | *SLC5A11* | TRUE | 3.368233889 |
| HTN | macrophage | *S100B* | TRUE | 2.632303144 |
| HTN | macrophage | *FCN2* | TRUE | 2.736329306 |
| HTN | macrophage | *PPP1R16A* | TRUE | 2.251263538 |
| HTN | macrophage | *ABCF3* | TRUE | 3.406473178 |
| HTN | macrophage | *TNFSF13* | TRUE | 3.077874986 |
| HTN | macrophage | *LMOD1* | TRUE | 2.186319272 |
| HTN | macrophage | *EXO5* | TRUE | 2.016039341 |
| HTN | macrophage | *RNF123* | TRUE | 2.582904336 |
| HTN | macrophage | *ERAP1* | TRUE | 2.967932215 |
| HTN | macrophage | *KIR3DL1* | TRUE | 2.624622714 |
| HTN | macrophage | *C19orf33* | TRUE | 2.241093741 |
| HTN | macrophage | *CYP7A1* | TRUE | 2.453758721 |
| HTN | macrophage | *HRASLS5* | TRUE | 2.618520063 |
| HTN | macrophage | *RGS19* | TRUE | 3.888510174 |
| HTN | macrophage | *NIPAL4* | TRUE | 2.151168535 |
| HTN | macrophage | *ZNF738* | TRUE | 2.032543373 |
| HTN | macrophage | *VEGFB* | TRUE | 2.212837481 |
| HTN | macrophage | *PPFIA3* | TRUE | 2.729190169 |
| HTN | macrophage | *ZNF223* | TRUE | 2.297810992 |
| HTN | macrophage | *CPNE7* | TRUE | 2.644535441 |
| HTN | macrophage | *MPI* | TRUE | 2.064981462 |
| HTN | macrophage | *TRIM73* | TRUE | 2.753463062 |
| HTN | macrophage | *CTC1* | TRUE | 2.437378895 |
| HTN | macrophage | *NPIPA1* | TRUE | 2.130048653 |
| HTN | macrophage | *RP1L1* | TRUE | 2.723271186 |
| HTN | macrophage | *A3GALT2* | TRUE | 2.488770959 |
| HTN | macrophage | *DDX51* | TRUE | 2.75286854 |
| HTN | macrophage | *MCRS1* | TRUE | 2.513499855 |
| HTN | macrophage | *C6orf106* | TRUE | 2.008947711 |
| HTN | macrophage | *HIST1H3E* | TRUE | 2.033995518 |
| HTN | macrophage | *RPS26* | TRUE | 3.232054613 |
| HTN | macrophage | *TMEM116* | TRUE | 3.4849253 |
| HTN | macrophage | *TGM2* | TRUE | 2.512703418 |
| HTN | macrophage | *HSD17B8* | TRUE | 2.384426411 |
| HTN | macrophage | *STK19* | TRUE | 3.208894585 |
| HTN | macrophage | *LTB4R* | TRUE | 2.182565612 |
| HTN | macrophage | *SLC26A6* | TRUE | 3.142691922 |
| HTN | macrophage | *PROB1* | TRUE | 2.383938049 |
| HTN | macrophage | *PSG4* | TRUE | 2.348055096 |
| HTN | macrophage | *PLA2G4B* | TRUE | 2.629501201 |
| HTN | macrophage | *ZNF10* | TRUE | 2.035694359 |
| HTN | macrophage | *RTEL1* | TRUE | 3.01189 |
| HTN | macrophage | *LINC00668* | TRUE | 2.08764697 |
| HTN | mast_cell | *HFE* | TRUE | 2.168077585 |
| HTN | mast_cell | *MTHFSD* | TRUE | 2.23670788 |
| HTN | mast_cell | *IGDCC4* | TRUE | 2.150324861 |

| | | | | |
|---|---|---|---|---|
| HTN | mast_cell | *HDHD3* | TRUE | 2.192251955 |
| HTN | mast_cell | *TEX14* | TRUE | 2.663033142 |
| HTN | mast_cell | *NDUFAF4* | TRUE | 2.179524173 |
| HTN | mast_cell | *FAM124B* | TRUE | 2.065033932 |
| HTN | mast_cell | *PKDREJ* | TRUE | 3.361616505 |
| HTN | mast_cell | *ACP2* | TRUE | 3.081987356 |
| HTN | mast_cell | *AGT* | TRUE | 1.997218531 |
| HTN | mast_cell | *NMT1* | TRUE | 2.092719681 |
| HTN | mast_cell | *NRBF2* | TRUE | 2.101159012 |
| HTN | mast_cell | *HSD17B12* | TRUE | 2.316693431 |
| HTN | mast_cell | *S100B* | TRUE | 2.476887016 |
| HTN | mast_cell | *ABCF3* | TRUE | 2.642754582 |
| HTN | mast_cell | *RNF123* | TRUE | 2.328539992 |
| HTN | mast_cell | *ERAP1* | TRUE | 2.420444951 |
| HTN | mast_cell | *RGS19* | TRUE | 2.414179743 |
| HTN | mast_cell | *HYI* | TRUE | 2.008415625 |
| HTN | mast_cell | *NPIPA1* | TRUE | 2.341257772 |
| HTN | mast_cell | *DDX51* | TRUE | 2.245873538 |
| HTN | mast_cell | *RPS26* | TRUE | 2.775466064 |
| HTN | mast_cell | *TMEM116* | TRUE | 2.669229506 |
| HTN | mast_cell | *TGM2* | TRUE | 2.645897761 |
| HTN | mast_cell | *PLA2G4B* | TRUE | 2.019821233 |
| HTN | mast_cell | *RTEL1* | TRUE | 2.79828856 |
| HTN | monocyte | *ITIH4* | TRUE | 2.071379152 |
| HTN | monocyte | *YPEL3* | TRUE | 2.120914535 |
| HTN | monocyte | *HOXA2* | TRUE | 2.514193278 |
| HTN | monocyte | *SNX15* | TRUE | 2.022709612 |
| HTN | monocyte | *TEX14* | TRUE | 3.026650403 |
| HTN | monocyte | *PKDREJ* | TRUE | 2.476539369 |
| HTN | monocyte | *ACP2* | TRUE | 4.495321234 |
| HTN | monocyte | *AGT* | TRUE | 2.667112824 |
| HTN | monocyte | *CREB3L4* | TRUE | 2.26090023 |
| HTN | monocyte | *DUSP15* | TRUE | 3.043552274 |
| HTN | monocyte | *ABCF3* | TRUE | 2.588991749 |
| HTN | monocyte | *NME6* | TRUE | 3.924315107 |
| HTN | monocyte | *HYI* | TRUE | 2.053196 |
| HTN | monocyte | *RPS26* | TRUE | 3.248102003 |
| HTN | monocyte | *PLA2G4B* | TRUE | 2.23332595 |
| HTN | monocyte | *RTEL1* | TRUE | 2.493177384 |
| HTN | neutrophil | *ZNF391* | TRUE | 2.610806572 |
| HTN | neutrophil | *ACP2* | TRUE | 4.056735712 |
| HTN | neutrophil | *CIB2* | TRUE | 2.834389962 |
| HTN | neutrophil | *CLEC18A* | TRUE | 2.438975258 |
| HTN | neutrophil | *S100B* | TRUE | 2.198913137 |
| HTN | neutrophil | *CDC25A* | TRUE | 2.46705797 |
| HTN | neutrophil | *CTSW* | TRUE | 2.469343259 |
| HTN | neutrophil | *HSD17B8* | TRUE | 2.521472897 |
| HTN | nk_cell | *ITIH4* | TRUE | 2.376419586 |
| HTN | nk_cell | *SNX10* | TRUE | 3.733070241 |
| HTN | nk_cell | *POLI* | TRUE | 2.33619102 |

| | | | | |
|---|---|---|---|---|
| HTN | nk_cell | *HDHD3* | TRUE | 2.009483531 |
| HTN | nk_cell | *GRK4* | TRUE | 2.130849417 |
| HTN | nk_cell | *ATP8B3* | TRUE | 1.970442618 |
| HTN | nk_cell | *ACP2* | TRUE | 4.775808462 |
| HTN | nk_cell | *AGT* | TRUE | 1.974294023 |
| HTN | nk_cell | *KHK* | TRUE | 2.096494473 |
| HTN | nk_cell | *SCRN2* | TRUE | 2.225118221 |
| HTN | nk_cell | *CREB3L4* | TRUE | 2.634861217 |
| HTN | nk_cell | *SLC10A4* | TRUE | 2.65455238 |
| HTN | nk_cell | *SLC18B1* | TRUE | 2.158453243 |
| HTN | nk_cell | *HSD17B12* | TRUE | 3.529785241 |
| HTN | nk_cell | *NEIL2* | TRUE | 2.688018608 |
| HTN | nk_cell | *CLEC18A* | TRUE | 3.793958103 |
| HTN | nk_cell | *SLC5A11* | TRUE | 3.250437292 |
| HTN | nk_cell | *S100B* | TRUE | 3.28537304 |
| HTN | nk_cell | *TNFSF13* | TRUE | 4.38780516 |
| HTN | nk_cell | *ZNF589* | TRUE | 3.396916449 |
| HTN | nk_cell | *RNF123* | TRUE | 3.321363589 |
| HTN | nk_cell | *CHMP4C* | TRUE | 1.968290381 |
| HTN | nk_cell | *TRIM66* | TRUE | 2.685802982 |
| HTN | nk_cell | *SRR* | TRUE | 2.320901279 |
| HTN | nk_cell | *IRS1* | TRUE | 3.461647426 |
| HTN | nk_cell | *RGS19* | TRUE | 2.197372501 |
| HTN | nk_cell | *VEGFB* | TRUE | 2.864365219 |
| HTN | nk_cell | *PPFIA3* | TRUE | 1.96144941 |
| HTN | nk_cell | *HYI* | TRUE | 2.593434699 |
| HTN | nk_cell | *ZNF320* | TRUE | 2.443652913 |
| HTN | nk_cell | *KREMEN1* | TRUE | 1.980340974 |
| HTN | nk_cell | *EIF3C* | TRUE | 2.544772736 |
| HTN | nk_cell | *ZNF793* | TRUE | 2.004486353 |
| HTN | nk_cell | *FBF1* | TRUE | 2.735828462 |
| HTN | nk_cell | *SLC22A5* | TRUE | 2.786248871 |
| HTN | nk_cell | *TMEM116* | TRUE | 4.474721737 |
| HTN | nk_cell | *PNP* | TRUE | 2.392678796 |
| HTN | nk_cell | *HSD17B8* | TRUE | 2.362600677 |
| HTN | nk_cell | *LTB4R2* | TRUE | 3.186127051 |
| HTN | nk_cell | *AC017074.2* | TRUE | 2.367184171 |
| HTN | nk_cell | *ZNF709* | TRUE | 2.189142487 |
| HTN | nk_cell | *PLA2G4B* | TRUE | 2.707129065 |
| HTN | nk_cell | *HMBS* | TRUE | 2.791731744 |
| HTN | pericyte | *CEP68* | TRUE | 2.278780309 |
| HTN | pericyte | *ITIH4* | TRUE | 3.270761432 |
| HTN | pericyte | *UHRF1BP1* | TRUE | 2.092454688 |
| HTN | pericyte | *TIE1* | TRUE | 2.001150902 |
| HTN | pericyte | *SNX10* | TRUE | 2.903178382 |
| HTN | pericyte | *SLC7A4* | TRUE | 2.09385111 |
| HTN | pericyte | *POLI* | TRUE | 2.858864287 |
| HTN | pericyte | *IGDCC4* | TRUE | 2.067690225 |
| HTN | pericyte | *ATP13A1* | TRUE | 2.165146099 |
| HTN | pericyte | *MEST* | TRUE | 2.015188255 |

| | | | | |
|---|---|---|---|---|
| HTN | pericyte | *KCTD10* | TRUE | 2.041622779 |
| HTN | pericyte | *PCDHB6* | TRUE | 2.306190103 |
| HTN | pericyte | *NPHP3* | TRUE | 1.996482248 |
| HTN | pericyte | *TCF21* | TRUE | 2.073051957 |
| HTN | pericyte | *TRAP1* | TRUE | 3.341127316 |
| HTN | pericyte | *APOL5* | TRUE | 2.578746017 |
| HTN | pericyte | *ATP8B3* | TRUE | 2.648091539 |
| HTN | pericyte | *HAUS8* | TRUE | 2.068780909 |
| HTN | pericyte | *AGT* | TRUE | 3.540472092 |
| HTN | pericyte | *CIB2* | TRUE | 2.287340947 |
| HTN | pericyte | *NMT1* | TRUE | 4.045661036 |
| HTN | pericyte | *FBN2* | TRUE | 3.27629336 |
| HTN | pericyte | *SCRN2* | TRUE | 2.832781337 |
| HTN | pericyte | *SLC10A4* | TRUE | 3.653282611 |
| HTN | pericyte | *NRBF2* | TRUE | 2.58402898 |
| HTN | pericyte | *PDCD11* | TRUE | 4.20399074 |
| HTN | pericyte | *HSD17B12* | TRUE | 4.725927926 |
| HTN | pericyte | *PRDM8* | TRUE | 2.096454411 |
| HTN | pericyte | *NEIL2* | TRUE | 2.876836895 |
| HTN | pericyte | *CLEC18A* | TRUE | 3.10662856 |
| HTN | pericyte | *SLC5A11* | TRUE | 3.179823219 |
| HTN | pericyte | *S100B* | TRUE | 3.24144946 |
| HTN | pericyte | *ABCF3* | TRUE | 2.642407692 |
| HTN | pericyte | *TNFSF13* | TRUE | 3.947533042 |
| HTN | pericyte | *EXO5* | TRUE | 2.758083024 |
| HTN | pericyte | *RNF123* | TRUE | 3.962377687 |
| HTN | pericyte | *ERAP1* | TRUE | 2.148327187 |
| HTN | pericyte | *TRIM66* | TRUE | 2.48019585 |
| HTN | pericyte | *RIMKLB* | TRUE | 2.047373133 |
| HTN | pericyte | *CYP7A1* | TRUE | 2.061004387 |
| HTN | pericyte | *HRASLS5* | TRUE | 2.086174658 |
| HTN | pericyte | *IRS1* | TRUE | 2.235244034 |
| HTN | pericyte | *C8orf46* | TRUE | 2.447865813 |
| HTN | pericyte | *NME6* | TRUE | 2.360222278 |
| HTN | pericyte | *VEGFB* | TRUE | 2.240806792 |
| HTN | pericyte | *ZNF320* | TRUE | 2.593474793 |
| HTN | pericyte | *A3GALT2* | TRUE | 4.052998375 |
| HTN | pericyte | *FBF1* | TRUE | 3.352245017 |
| HTN | pericyte | *HLA.DRB1* | TRUE | 2.976443164 |
| HTN | pericyte | *SULT1A2* | TRUE | 2.289678566 |
| HTN | pericyte | *SLC22A5* | TRUE | 3.569445419 |
| HTN | pericyte | *RPS26* | TRUE | 3.825664461 |
| HTN | pericyte | *TMEM116* | TRUE | 3.674694949 |
| HTN | pericyte | *PNP* | TRUE | 1.999255051 |
| HTN | pericyte | *HSD17B8* | TRUE | 2.884964674 |
| HTN | pericyte | *STK19* | TRUE | 2.618295431 |
| HTN | pericyte | *NOTCH2NL* | TRUE | 2.371080313 |
| HTN | pericyte | *SIPA1* | TRUE | 3.354240298 |
| HTN | pericyte | *ZNF10* | TRUE | 2.977063314 |
| HTN | pericyte | *HMBS* | TRUE | 2.912500833 |

| | | | | |
|---|---|---|---|---|
| HTN | SMC | *HFE* | TRUE | 3.151677291 |
| HTN | SMC | *POLR1A* | TRUE | 2.094201847 |
| HTN | SMC | *SNX10* | TRUE | 2.752741485 |
| HTN | SMC | *AAMDC* | TRUE | 2.486152338 |
| HTN | SMC | *PABPC4* | TRUE | 2.431622664 |
| HTN | SMC | *SLC7A4* | TRUE | 2.141870222 |
| HTN | SMC | *POLI* | TRUE | 3.137645346 |
| HTN | SMC | *IGDCC4* | TRUE | 2.120860482 |
| HTN | SMC | *ENDOU* | TRUE | 2.236826347 |
| HTN | SMC | *IRF6* | TRUE | 2.758401247 |
| HTN | SMC | *HDHD3* | TRUE | 2.333086306 |
| HTN | SMC | *ACOT2* | TRUE | 1.968129569 |
| HTN | SMC | *TOX2* | TRUE | 2.43841944 |
| HTN | SMC | *SNRPC* | TRUE | 3.31738425 |
| HTN | SMC | *GRK4* | TRUE | 2.614233419 |
| HTN | SMC | *TRAP1* | TRUE | 3.041544586 |
| HTN | SMC | *ATP8B3* | TRUE | 2.656473958 |
| HTN | SMC | *ACP2* | TRUE | 5.229379928 |
| HTN | SMC | *AGT* | TRUE | 3.31372937 |
| HTN | SMC | *NMT1* | TRUE | 2.97239367 |
| HTN | SMC | *FBN2* | TRUE | 3.055070514 |
| HTN | SMC | *SCRN2* | TRUE | 2.975756565 |
| HTN | SMC | *SLC10A4* | TRUE | 2.414946477 |
| HTN | SMC | *SLC18B1* | TRUE | 2.739093815 |
| HTN | SMC | *PDCD11* | TRUE | 2.26677731 |
| HTN | SMC | *HSD17B12* | TRUE | 4.866335757 |
| HTN | SMC | *DUSP15* | TRUE | 2.312254545 |
| HTN | SMC | *NEIL2* | TRUE | 2.436047959 |
| HTN | SMC | *CLEC18A* | TRUE | 2.5014285 |
| HTN | SMC | *SLC5A11* | TRUE | 4.453838785 |
| HTN | SMC | *S100B* | TRUE | 2.77577618 |
| HTN | SMC | *ABCF3* | TRUE | 2.784231136 |
| HTN | SMC | *TNFSF13* | TRUE | 3.069670304 |
| HTN | SMC | *C1orf50* | TRUE | 2.683845294 |
| HTN | SMC | *ZNF589* | TRUE | 3.051153509 |
| HTN | SMC | *RNF123* | TRUE | 2.000740745 |
| HTN | SMC | *ERAP1* | TRUE | 2.988662177 |
| HTN | SMC | *TRMT61A* | TRUE | 3.317219958 |
| HTN | SMC | *TRIM66* | TRUE | 2.677468149 |
| HTN | SMC | *RIMKLB* | TRUE | 3.622458652 |
| HTN | SMC | *KIR3DL1* | TRUE | 2.415537429 |
| HTN | SMC | *SRR* | TRUE | 2.566971771 |
| HTN | SMC | *CYP7A1* | TRUE | 2.026543288 |
| HTN | SMC | *IRS1* | TRUE | 4.925208671 |
| HTN | SMC | *INO80E* | TRUE | 3.568516704 |
| HTN | SMC | *CALML6* | TRUE | 2.050048392 |
| HTN | SMC | *PAQR8* | TRUE | 2.034497899 |
| HTN | SMC | *RGS19* | TRUE | 4.909768972 |
| HTN | SMC | *CTSW* | TRUE | 3.000243531 |
| HTN | SMC | *VEGFB* | TRUE | 3.377497093 |

| | | | | |
|---|---|---|---|---|
| HTN | SMC | *NRIP3* | TRUE | 2.187574834 |
| HTN | SMC | *MTHFR* | TRUE | 2.168910976 |
| HTN | SMC | *CPNE7* | TRUE | 1.984258796 |
| HTN | SMC | *HYI* | TRUE | 2.673861499 |
| HTN | SMC | *DEFB124* | TRUE | 2.837765946 |
| HTN | SMC | *ZNF320* | TRUE | 2.937202464 |
| HTN | SMC | *KREMEN1* | TRUE | 2.763502909 |
| HTN | SMC | *EIF3C* | TRUE | 2.340788486 |
| HTN | SMC | *A3GALT2* | TRUE | 3.956647603 |
| HTN | SMC | *CDK10* | TRUE | 2.516476506 |
| HTN | SMC | *ZNF559* | TRUE | 2.33467433 |
| HTN | SMC | *FBF1* | TRUE | 3.251524908 |
| HTN | SMC | *HLA.DRB1* | TRUE | 3.395629523 |
| HTN | SMC | *HIST1H3E* | TRUE | 2.382473294 |
| HTN | SMC | *SLC22A5* | TRUE | 2.489625629 |
| HTN | SMC | *RPS26* | TRUE | 2.013652681 |
| HTN | SMC | *TMEM116* | TRUE | 5.750878345 |
| HTN | SMC | *PNP* | TRUE | 2.486208821 |
| HTN | SMC | *AL590822.1* | TRUE | 2.388436702 |
| HTN | SMC | *HSD17B8* | TRUE | 3.396552752 |
| HTN | SMC | *MXD3* | TRUE | 2.188777713 |
| HTN | SMC | *LTB4R* | TRUE | 2.064703239 |
| HTN | SMC | *AC016773.1* | TRUE | 2.321679561 |
| HTN | SMC | *ZNF10* | TRUE | 2.711843981 |
| HTN | SMC | *HMBS* | TRUE | 3.083476874 |
| HTN | t_cell | *EPN3* | TRUE | 2.189409954 |
| HTN | t_cell | *PCDHA6* | TRUE | 2.499326966 |
| HTN | t_cell | *SNX10* | TRUE | 2.392132214 |
| HTN | t_cell | *YPEL3* | TRUE | 3.069158761 |
| HTN | t_cell | *MTHFSD* | TRUE | 2.63442834 |
| HTN | t_cell | *TEX14* | TRUE | 3.284059199 |
| HTN | t_cell | *NDUFAF4* | TRUE | 2.386558673 |
| HTN | t_cell | *TOX2* | TRUE | 2.834139953 |
| HTN | t_cell | *TCP11* | TRUE | 2.475926521 |
| HTN | t_cell | *APOL5* | TRUE | 2.245699896 |
| HTN | t_cell | *AANAT* | TRUE | 2.256400542 |
| HTN | t_cell | *GALNT8* | TRUE | 1.984266498 |
| HTN | t_cell | *ATP8B3* | TRUE | 2.776061104 |
| HTN | t_cell | *PKDREJ* | TRUE | 2.635820221 |
| HTN | t_cell | *ACP2* | TRUE | 3.118593573 |
| HTN | t_cell | *AGT* | TRUE | 2.448813379 |
| HTN | t_cell | *NMT1* | TRUE | 2.482210364 |
| HTN | t_cell | *SLC18B1* | TRUE | 2.582970126 |
| HTN | t_cell | *PLEKHS1* | TRUE | 2.574344936 |
| HTN | t_cell | *SLC5A11* | TRUE | 2.236513909 |
| HTN | t_cell | *TNFSF13* | TRUE | 4.192692858 |
| HTN | t_cell | *LMOD1* | TRUE | 2.224273396 |
| HTN | t_cell | *ERAP1* | TRUE | 2.492496628 |
| HTN | t_cell | *KIR3DL1* | TRUE | 2.115310759 |
| HTN | t_cell | *C19orf33* | TRUE | 2.121625747 |

| | | | | |
|---|---|---|---|---|
| HTN | t_cell | *NME6* | TRUE | 2.453243186 |
| HTN | t_cell | *NRIP3* | TRUE | 2.192787457 |
| HTN | t_cell | *TRIM73* | TRUE | 2.08544761 |
| HTN | t_cell | *NPIPA1* | TRUE | 2.162389084 |
| HTN | t_cell | *RP1L1* | TRUE | 2.067665643 |
| HTN | t_cell | *NUTM2A* | TRUE | 2.137130892 |
| HTN | t_cell | *HLA.DRB1* | TRUE | 1.999633621 |
| HTN | t_cell | *HIST1H3E* | TRUE | 4.131201165 |
| HTN | t_cell | *RPS26* | TRUE | 3.182888426 |
| HTN | t_cell | *TGM2* | TRUE | 2.341690626 |
| HTN | t_cell | *STK19* | TRUE | 2.176424899 |
| HTN | t_cell | *PLA2G4B* | TRUE | 2.802422209 |
| HTN | t_cell | *ZNF10* | TRUE | 2.052590307 |
| Obesity | adipocyte | *RBM6* | TRUE | 3.17408595 |
| Obesity | adipocyte | *MAPK8IP2* | TRUE | 2.0539476 |
| Obesity | adipocyte | *LSG1* | TRUE | 2.239377791 |
| Obesity | adipocyte | *ITIH4* | TRUE | 4.444239586 |
| Obesity | adipocyte | *UHRF1BP1* | TRUE | 3.802116996 |
| Obesity | adipocyte | *TIPIN* | TRUE | 3.576317138 |
| Obesity | adipocyte | *PCDHGA2* | TRUE | 2.271461094 |
| Obesity | adipocyte | *EPYC* | TRUE | 2.188074689 |
| Obesity | adipocyte | *COL16A1* | TRUE | 2.02633097 |
| Obesity | adipocyte | *PABPC4* | TRUE | 2.966969899 |
| Obesity | adipocyte | *STX1B* | TRUE | 2.28080814 |
| Obesity | adipocyte | *SMARCB1* | TRUE | 2.05929429 |
| Obesity | adipocyte | *RASD2* | TRUE | 2.033685219 |
| Obesity | adipocyte | *DESI1* | TRUE | 2.954839623 |
| Obesity | adipocyte | *ABHD12* | TRUE | 2.405681172 |
| Obesity | adipocyte | *SLC52A3* | TRUE | 2.070004161 |
| Obesity | adipocyte | *ELMO3* | TRUE | 2.169561439 |
| Obesity | adipocyte | *CLUAP1* | TRUE | 2.430465136 |
| Obesity | adipocyte | *KAT8* | TRUE | 3.489762782 |
| Obesity | adipocyte | *MCM4* | TRUE | 2.1098627 |
| Obesity | adipocyte | *PPP1R37* | TRUE | 2.076924502 |
| Obesity | adipocyte | *ICAM5* | TRUE | 1.974244072 |
| Obesity | adipocyte | *ATP13A1* | TRUE | 2.33584097 |
| Obesity | adipocyte | *SLC26A8* | TRUE | 2.113557817 |
| Obesity | adipocyte | *BNIP1* | TRUE | 2.597631098 |
| Obesity | adipocyte | *MFAP2* | TRUE | 2.03574362 |
| Obesity | adipocyte | *SERPINC1* | TRUE | 4.265469368 |
| Obesity | adipocyte | *COLEC11* | TRUE | 2.194717624 |
| Obesity | adipocyte | *CCDC92* | TRUE | 4.897412465 |
| Obesity | adipocyte | *DCAF4* | TRUE | 2.625282398 |
| Obesity | adipocyte | *HOXB3* | TRUE | 2.889940035 |
| Obesity | adipocyte | *ADRA1A* | TRUE | 3.061410095 |
| Obesity | adipocyte | *GJB6* | TRUE | 3.388384178 |
| Obesity | adipocyte | *SMUG1* | TRUE | 2.069420848 |
| Obesity | adipocyte | *GRK4* | TRUE | 3.469494025 |
| Obesity | adipocyte | *PCSK2* | TRUE | 2.463475303 |
| Obesity | adipocyte | *RIBC2* | TRUE | 3.444154759 |

| | | | | |
|---|---|---|---|---|
| Obesity | adipocyte | *ATP8B3* | TRUE | 4.967241449 |
| Obesity | adipocyte | *PKDREJ* | TRUE | 3.088333295 |
| Obesity | adipocyte | *EMILIN2* | TRUE | 3.53723634 |
| Obesity | adipocyte | *LARS* | TRUE | 1.970011063 |
| Obesity | adipocyte | *GSTM5* | TRUE | 2.418126751 |
| Obesity | adipocyte | *P2RX4* | TRUE | 1.980998161 |
| Obesity | adipocyte | *TMEM60* | TRUE | 2.021779112 |
| Obesity | adipocyte | *POM121C* | TRUE | 2.03155076 |
| Obesity | adipocyte | *EPHA7* | TRUE | 2.596926902 |
| Obesity | adipocyte | *DSCC1* | TRUE | 2.264005249 |
| Obesity | adipocyte | *CARF* | TRUE | 2.150496251 |
| Obesity | adipocyte | *UBL7* | TRUE | 1.972696746 |
| Obesity | adipocyte | *KLRG1* | TRUE | 2.186340471 |
| Obesity | adipocyte | *RITA1* | TRUE | 2.429310323 |
| Obesity | adipocyte | *PTGR2* | TRUE | 1.988807172 |
| Obesity | adipocyte | *WDR93* | TRUE | 2.049982802 |
| Obesity | adipocyte | *C18orf8* | TRUE | 3.267288583 |
| Obesity | adipocyte | *TRIM65* | TRUE | 3.164099467 |
| Obesity | adipocyte | *TUFT1* | TRUE | 2.265422209 |
| Obesity | adipocyte | *SLC25A48* | TRUE | 2.938627939 |
| Obesity | adipocyte | *ATP6V0D2* | TRUE | 2.61479925 |
| Obesity | adipocyte | *NRBF2* | TRUE | 2.263013426 |
| Obesity | adipocyte | *HSD17B12* | TRUE | 5.656222854 |
| Obesity | adipocyte | *TTC12* | TRUE | 3.401575632 |
| Obesity | adipocyte | *CCDC15* | TRUE | 3.289691551 |
| Obesity | adipocyte | *DUSP15* | TRUE | 2.190844169 |
| Obesity | adipocyte | *DOC2A* | TRUE | 3.80317559 |
| Obesity | adipocyte | *DPYSL4* | TRUE | 3.032690995 |
| Obesity | adipocyte | *BEND6* | TRUE | 2.120255892 |
| Obesity | adipocyte | *POC5* | TRUE | 2.613164557 |
| Obesity | adipocyte | *TCTEX1D1* | TRUE | 2.309398402 |
| Obesity | adipocyte | *FAM167A* | TRUE | 1.997766422 |
| Obesity | adipocyte | *STEAP2* | TRUE | 2.93634563 |
| Obesity | adipocyte | *CLEC18A* | TRUE | 3.629458568 |
| Obesity | adipocyte | *CD1E* | TRUE | 2.421751958 |
| Obesity | adipocyte | *SLC5A11* | TRUE | 4.892872874 |
| Obesity | adipocyte | *NAGS* | TRUE | 2.025045507 |
| Obesity | adipocyte | *CLHC1* | TRUE | 2.527582701 |
| Obesity | adipocyte | *LMOD1* | TRUE | 6.336067239 |
| Obesity | adipocyte | *DNALI1* | TRUE | 2.97804976 |
| Obesity | adipocyte | *TEX264* | TRUE | 2.038819876 |
| Obesity | adipocyte | *LRFN5* | TRUE | 2.089352204 |
| Obesity | adipocyte | *TRMT61A* | TRUE | 2.362938763 |
| Obesity | adipocyte | *SRR* | TRUE | 3.8985683 |
| Obesity | adipocyte | *BTNL3* | TRUE | 2.04451703 |
| Obesity | adipocyte | *RAB24* | TRUE | 3.0313158 |
| Obesity | adipocyte | *TRMT61B* | TRUE | 3.481564557 |
| Obesity | adipocyte | *C1QTNF4* | TRUE | 5.256647561 |
| Obesity | adipocyte | *FIBP* | TRUE | 3.276496477 |
| Obesity | adipocyte | *FUT10* | TRUE | 2.529809384 |

| | | | | |
|---|---|---|---|---|
| Obesity | adipocyte | *HPSE2* | TRUE | 4.300359329 |
| Obesity | adipocyte | *CADM2* | TRUE | 2.513577382 |
| Obesity | adipocyte | *RUFY1* | TRUE | 2.179427673 |
| Obesity | adipocyte | *PRSS42* | TRUE | 1.993861418 |
| Obesity | adipocyte | *STX19* | TRUE | 3.495972643 |
| Obesity | adipocyte | *B3GNT3* | TRUE | 3.278047913 |
| Obesity | adipocyte | *AMIGO1* | TRUE | 2.124083155 |
| Obesity | adipocyte | *CAV3* | TRUE | 2.52739837 |
| Obesity | adipocyte | *BRICD5* | TRUE | 2.51039822 |
| Obesity | adipocyte | *LYSMD4* | TRUE | 2.169249217 |
| Obesity | adipocyte | *CCBE1* | TRUE | 2.21100448 |
| Obesity | adipocyte | *PSMG1* | TRUE | 3.581270856 |
| Obesity | adipocyte | *RP1L1* | TRUE | 4.480474674 |
| Obesity | adipocyte | *PLA2G6* | TRUE | 2.385416483 |
| Obesity | adipocyte | *KLHL33* | TRUE | 2.226979615 |
| Obesity | adipocyte | *CENPP* | TRUE | 2.345395308 |
| Obesity | adipocyte | *RPS26* | TRUE | 3.023417962 |
| Obesity | adipocyte | *ZNF780A* | TRUE | 2.452367803 |
| Obesity | adipocyte | *ADH4* | TRUE | 3.255308344 |
| Obesity | adipocyte | *ANKRD13B* | TRUE | 2.48764608 |
| Obesity | adipocyte | *CENPW* | TRUE | 2.417274442 |
| Obesity | adipocyte | *CCHCR1* | TRUE | 1.983609688 |
| Obesity | adipocyte | *HLA.E* | TRUE | 3.795603531 |
| Obesity | adipocyte | *EIF3CL* | TRUE | 3.341459278 |
| Obesity | adipocyte | *ASIC3* | TRUE | 3.35060965 |
| Obesity | adipocyte | *NOTCH2NL* | TRUE | 2.066675658 |
| Obesity | adipocyte | *SLC12A8* | TRUE | 2.514072755 |
| Obesity | adipocyte | *PLA2G4B* | TRUE | 2.20488535 |
| Obesity | adipocyte | *PCDHAC1* | TRUE | 2.817779061 |
| Obesity | adipocyte | *MPV17L2* | TRUE | 3.86267166 |
| Obesity | adipocyte | *HMBS* | TRUE | 5.876746714 |
| Obesity | adipocyte | *HP* | TRUE | 3.10039738 |
| Obesity | adipocyte | *DUXA* | TRUE | 2.424032088 |
| Obesity | adipocyte | *SH3RF3.AS1* | TRUE | 3.179465097 |
| Obesity | adipocyte | *LINC00562* | TRUE | 2.855000689 |
| Obesity | adipocyte | *GTF2H5* | TRUE | 2.045034791 |
| Obesity | ASPC | *WNT16* | TRUE | 2.138115464 |
| Obesity | ASPC | *RBM6* | TRUE | 2.888663473 |
| Obesity | ASPC | *SLC22A16* | TRUE | 2.438850062 |
| Obesity | ASPC | *COPZ2* | TRUE | 2.419984198 |
| Obesity | ASPC | *MAPK8IP2* | TRUE | 2.268841382 |
| Obesity | ASPC | *HFE* | TRUE | 2.320205815 |
| Obesity | ASPC | *CENPQ* | TRUE | 2.57071184 |
| Obesity | ASPC | *ZCCHC8* | TRUE | 2.23805265 |
| Obesity | ASPC | *SH3YL1* | TRUE | 3.813700602 |
| Obesity | ASPC | *VCL* | TRUE | 2.167084358 |
| Obesity | ASPC | *TUBG2* | TRUE | 2.11630152 |
| Obesity | ASPC | *LSG1* | TRUE | 2.661735622 |
| Obesity | ASPC | *TSPAN17* | TRUE | 2.094728086 |
| Obesity | ASPC | *UTS2* | TRUE | 2.072791509 |

| | | | | |
|---|---|---|---|---|
| Obesity | ASPC | *RCN1* | TRUE | 2.254413909 |
| Obesity | ASPC | *JKAMP* | TRUE | 2.132954807 |
| Obesity | ASPC | *ITIH4* | TRUE | 2.548863226 |
| Obesity | ASPC | *ATP11B* | TRUE | 2.278554601 |
| Obesity | ASPC | *MBD3* | TRUE | 2.145765723 |
| Obesity | ASPC | *TIPIN* | TRUE | 3.258485278 |
| Obesity | ASPC | *MARK3* | TRUE | 2.025780717 |
| Obesity | ASPC | *XAB2* | TRUE | 2.424517213 |
| Obesity | ASPC | *DYNC1I2* | TRUE | 2.235898558 |
| Obesity | ASPC | *MYH7B* | TRUE | 2.229352802 |
| Obesity | ASPC | *PCDHGA2* | TRUE | 2.441697305 |
| Obesity | ASPC | *CCNT2* | TRUE | 3.459132712 |
| Obesity | ASPC | *COBLL1* | TRUE | 2.977722648 |
| Obesity | ASPC | *DOPEY1* | TRUE | 2.373052067 |
| Obesity | ASPC | *EPYC* | TRUE | 2.502000125 |
| Obesity | ASPC | *COL16A1* | TRUE | 3.89553944 |
| Obesity | ASPC | *SCAMP1* | TRUE | 2.085485921 |
| Obesity | ASPC | *OGFOD1* | TRUE | 4.34744839 |
| Obesity | ASPC | *PABPC4* | TRUE | 3.156307747 |
| Obesity | ASPC | *RGS17* | TRUE | 2.721975404 |
| Obesity | ASPC | *PDE6C* | TRUE | 2.579862316 |
| Obesity | ASPC | *OCEL1* | TRUE | 2.000148638 |
| Obesity | ASPC | *SMARCB1* | TRUE | 2.658411374 |
| Obesity | ASPC | *TFIP11* | TRUE | 2.267158349 |
| Obesity | ASPC | *FBXO7* | TRUE | 1.986890247 |
| Obesity | ASPC | *RASD2* | TRUE | 2.09923308 |
| Obesity | ASPC | *DESI1* | TRUE | 3.352041415 |
| Obesity | ASPC | *DICER1* | TRUE | 2.028688257 |
| Obesity | ASPC | *PSMB5* | TRUE | 2.041435432 |
| Obesity | ASPC | *ABHD12* | TRUE | 3.350052699 |
| Obesity | ASPC | *ANGPT4* | TRUE | 2.216487901 |
| Obesity | ASPC | *CEP192* | TRUE | 2.104700871 |
| Obesity | ASPC | *CORO1A* | TRUE | 3.422172857 |
| Obesity | ASPC | *NAGPA* | TRUE | 2.314638664 |
| Obesity | ASPC | *CLUAP1* | TRUE | 2.855171267 |
| Obesity | ASPC | *KAT8* | TRUE | 4.148402851 |
| Obesity | ASPC | *KNOP1* | TRUE | 3.463667103 |
| Obesity | ASPC | *ZDHHC2* | TRUE | 3.726189874 |
| Obesity | ASPC | *RAB2A* | TRUE | 2.074880336 |
| Obesity | ASPC | *MCM4* | TRUE | 2.509759773 |
| Obesity | ASPC | *SNRNP70* | TRUE | 2.338621407 |
| Obesity | ASPC | *PPP1R37* | TRUE | 2.688662457 |
| Obesity | ASPC | *ICAM5* | TRUE | 3.15272668 |
| Obesity | ASPC | *ZNF175* | TRUE | 2.300292388 |
| Obesity | ASPC | *ELL* | TRUE | 2.072428034 |
| Obesity | ASPC | *ATP13A1* | TRUE | 3.84990908 |
| Obesity | ASPC | *DLX5* | TRUE | 1.99092707 |
| Obesity | ASPC | *HOXA2* | TRUE | 2.40495951 |
| Obesity | ASPC | *MEST* | TRUE | 2.538032244 |
| Obesity | ASPC | *PLGRKT* | TRUE | 2.304770656 |

| | | | | |
|---|---|---|---|---|
| Obesity | ASPC | *ACBD5* | TRUE | 1.974287557 |
| Obesity | ASPC | *RAD51C* | TRUE | 2.578429505 |
| Obesity | ASPC | *FOXRED1* | TRUE | 2.177107939 |
| Obesity | ASPC | *TRPV4* | TRUE | 3.079464187 |
| Obesity | ASPC | *GABRR2* | TRUE | 2.271131931 |
| Obesity | ASPC | *SLC26A8* | TRUE | 2.463141375 |
| Obesity | ASPC | *CAP2* | TRUE | 2.998391302 |
| Obesity | ASPC | *NME5* | TRUE | 3.323869488 |
| Obesity | ASPC | *BNIP1* | TRUE | 3.004209189 |
| Obesity | ASPC | *C3orf14* | TRUE | 2.397804288 |
| Obesity | ASPC | *KLHL24* | TRUE | 2.356218185 |
| Obesity | ASPC | *PLCH1* | TRUE | 2.759479858 |
| Obesity | ASPC | *FAHD2A* | TRUE | 2.025261396 |
| Obesity | ASPC | *ACTR1B* | TRUE | 2.840225605 |
| Obesity | ASPC | *TFCP2L1* | TRUE | 2.006911773 |
| Obesity | ASPC | *ITGB6* | TRUE | 2.107824141 |
| Obesity | ASPC | *CHMP3* | TRUE | 4.660358566 |
| Obesity | ASPC | *CD207* | TRUE | 2.668143604 |
| Obesity | ASPC | *DNAJC16* | TRUE | 2.492995156 |
| Obesity | ASPC | *MFAP2* | TRUE | 2.647078714 |
| Obesity | ASPC | *SERPINC1* | TRUE | 3.216111489 |
| Obesity | ASPC | *CD3EAP* | TRUE | 2.072122994 |
| Obesity | ASPC | *COLEC11* | TRUE | 2.735661588 |
| Obesity | ASPC | *DNAH7* | TRUE | 2.010366013 |
| Obesity | ASPC | *CCDC92* | TRUE | 4.743219261 |
| Obesity | ASPC | *HDHD3* | TRUE | 2.256531134 |
| Obesity | ASPC | *INVS* | TRUE | 1.999482295 |
| Obesity | ASPC | *DCAF4* | TRUE | 3.480410926 |
| Obesity | ASPC | *CNRIP1* | TRUE | 2.14458506 |
| Obesity | ASPC | *CRIPT* | TRUE | 2.525575575 |
| Obesity | ASPC | *HOXB3* | TRUE | 1.962841352 |
| Obesity | ASPC | *SNX19* | TRUE | 3.416831199 |
| Obesity | ASPC | *ADRA1A* | TRUE | 3.883932314 |
| Obesity | ASPC | *SEC22A* | TRUE | 2.091019523 |
| Obesity | ASPC | *GJB6* | TRUE | 3.455318963 |
| Obesity | ASPC | *SLC25A51* | TRUE | 2.302783703 |
| Obesity | ASPC | *ATP7B* | TRUE | 2.184352019 |
| Obesity | ASPC | *ARHGAP9* | TRUE | 2.780088735 |
| Obesity | ASPC | *SMUG1* | TRUE | 2.238251999 |
| Obesity | ASPC | *CHST8* | TRUE | 1.963461399 |
| Obesity | ASPC | *SNRPC* | TRUE | 2.656325963 |
| Obesity | ASPC | *EEF1E1* | TRUE | 2.624254882 |
| Obesity | ASPC | *GRK4* | TRUE | 4.018539352 |
| Obesity | ASPC | *PCSK2* | TRUE | 4.361413845 |
| Obesity | ASPC | *YEATS4* | TRUE | 2.821350608 |
| Obesity | ASPC | *RIBC2* | TRUE | 3.431454916 |
| Obesity | ASPC | *RPAIN* | TRUE | 2.680544597 |
| Obesity | ASPC | *ATP8B3* | TRUE | 5.482420812 |
| Obesity | ASPC | *PGPEP1* | TRUE | 2.358764089 |
| Obesity | ASPC | *CYP2E1* | TRUE | 2.078794283 |

| | | | | |
|---|---|---|---|---|
| Obesity | ASPC | *PKDREJ* | TRUE | 3.17872432 |
| Obesity | ASPC | *EMILIN2* | TRUE | 4.426287586 |
| Obesity | ASPC | *IGHMBP2* | TRUE | 2.827486719 |
| Obesity | ASPC | *PEMT* | TRUE | 2.128372905 |
| Obesity | ASPC | *BTBD2* | TRUE | 2.055498307 |
| Obesity | ASPC | *LARS* | TRUE | 3.396644856 |
| Obesity | ASPC | *GSTM5* | TRUE | 2.385698552 |
| Obesity | ASPC | *P2RX4* | TRUE | 2.561976819 |
| Obesity | ASPC | *TMEM60* | TRUE | 3.89512269 |
| Obesity | ASPC | *POM121C* | TRUE | 5.729142504 |
| Obesity | ASPC | *EPHA7* | TRUE | 2.666001444 |
| Obesity | ASPC | *MRPS9* | TRUE | 2.367403445 |
| Obesity | ASPC | *NMT1* | TRUE | 2.311055661 |
| Obesity | ASPC | *DSCC1* | TRUE | 2.843318083 |
| Obesity | ASPC | *TMEM63B* | TRUE | 2.307217227 |
| Obesity | ASPC | *TTPA* | TRUE | 2.152007737 |
| Obesity | ASPC | *MAP2K5* | TRUE | 2.654707652 |
| Obesity | ASPC | *SLC44A5* | TRUE | 2.081537226 |
| Obesity | ASPC | *IFT172* | TRUE | 3.32540492 |
| Obesity | ASPC | *CARF* | TRUE | 2.544262563 |
| Obesity | ASPC | *UBL7* | TRUE | 2.298851338 |
| Obesity | ASPC | *MMRN1* | TRUE | 2.406494797 |
| Obesity | ASPC | *KLRG1* | TRUE | 2.447491378 |
| Obesity | ASPC | *RITA1* | TRUE | 2.8397358 |
| Obesity | ASPC | *GPR84* | TRUE | 2.07142371 |
| Obesity | ASPC | *PTGR2* | TRUE | 2.350790459 |
| Obesity | ASPC | *CYP11A1* | TRUE | 2.63388696 |
| Obesity | ASPC | *WDR93* | TRUE | 3.1025107 |
| Obesity | ASPC | *SH3GL3* | TRUE | 1.976084397 |
| Obesity | ASPC | *MYO19* | TRUE | 2.423664413 |
| Obesity | ASPC | *C18orf8* | TRUE | 2.319823005 |
| Obesity | ASPC | *TRIM65* | TRUE | 2.671518699 |
| Obesity | ASPC | *CBX8* | TRUE | 2.154136664 |
| Obesity | ASPC | *TUFT1* | TRUE | 2.739799523 |
| Obesity | ASPC | *SEMA6C* | TRUE | 1.96679243 |
| Obesity | ASPC | *S100A8* | TRUE | 2.832202554 |
| Obesity | ASPC | *HAX1* | TRUE | 2.511174431 |
| Obesity | ASPC | *FBLN7* | TRUE | 2.255248186 |
| Obesity | ASPC | *AFF3* | TRUE | 2.799176008 |
| Obesity | ASPC | *THSD7B* | TRUE | 2.30167677 |
| Obesity | ASPC | *PHOSPHO2* | TRUE | 2.028613699 |
| Obesity | ASPC | *EIF2A* | TRUE | 2.162365451 |
| Obesity | ASPC | *MARCH1* | TRUE | 2.731984755 |
| Obesity | ASPC | *SLC25A48* | TRUE | 2.978449337 |
| Obesity | ASPC | *PCYOX1L* | TRUE | 3.445175414 |
| Obesity | ASPC | *RPL7L1* | TRUE | 1.974506404 |
| Obesity | ASPC | *SDK1* | TRUE | 2.278245228 |
| Obesity | ASPC | *ATP6V0D2* | TRUE | 4.110941965 |
| Obesity | ASPC | *NRBF2* | TRUE | 2.248873628 |
| Obesity | ASPC | *HSD17B12* | TRUE | 6.715533498 |

| | | | | |
|---|---|---|---|---|
| Obesity | ASPC | *TTC12* | TRUE | 3.818286609 |
| Obesity | ASPC | *CCDC15* | TRUE | 4.152984727 |
| Obesity | ASPC | *DUSP15* | TRUE | 2.543828314 |
| Obesity | ASPC | *TRPT1* | TRUE | 3.332931294 |
| Obesity | ASPC | *DOC2A* | TRUE | 2.584374109 |
| Obesity | ASPC | *DPYSL4* | TRUE | 3.312771499 |
| Obesity | ASPC | *BEND6* | TRUE | 2.516317249 |
| Obesity | ASPC | *ARL14EP* | TRUE | 3.474093646 |
| Obesity | ASPC | *POC5* | TRUE | 4.962664876 |
| Obesity | ASPC | *TCTEX1D1* | TRUE | 2.498164187 |
| Obesity | ASPC | *JAZF1* | TRUE | 2.153925429 |
| Obesity | ASPC | *CEP112* | TRUE | 2.031800845 |
| Obesity | ASPC | *ZKSCAN2* | TRUE | 2.256164554 |
| Obesity | ASPC | *TIMP4* | TRUE | 2.15521834 |
| Obesity | ASPC | *STEAP2* | TRUE | 3.555743505 |
| Obesity | ASPC | *CLEC18A* | TRUE | 3.506476152 |
| Obesity | ASPC | *HPD* | TRUE | 2.258062834 |
| Obesity | ASPC | *CD1E* | TRUE | 2.264264654 |
| Obesity | ASPC | *COPG2* | TRUE | 2.268373616 |
| Obesity | ASPC | *SLC5A11* | TRUE | 5.12977861 |
| Obesity | ASPC | *SIM2* | TRUE | 2.123500962 |
| Obesity | ASPC | *DFFA* | TRUE | 1.991424177 |
| Obesity | ASPC | *C21orf33* | TRUE | 2.28581157 |
| Obesity | ASPC | *C19orf47* | TRUE | 2.606099086 |
| Obesity | ASPC | *ZBTB7B* | TRUE | 3.981428244 |
| Obesity | ASPC | *FBXO27* | TRUE | 2.01878252 |
| Obesity | ASPC | *FUBP1* | TRUE | 5.926101093 |
| Obesity | ASPC | *ACP6* | TRUE | 3.033256628 |
| Obesity | ASPC | *ANTXR2* | TRUE | 2.957751198 |
| Obesity | ASPC | *SLC15A2* | TRUE | 2.614118118 |
| Obesity | ASPC | *LMOD1* | TRUE | 7.880652908 |
| Obesity | ASPC | *GYG1* | TRUE | 2.624931604 |
| Obesity | ASPC | *DNALI1* | TRUE | 3.697727676 |
| Obesity | ASPC | *RFT1* | TRUE | 2.342808107 |
| Obesity | ASPC | *SLC51A* | TRUE | 3.289061458 |
| Obesity | ASPC | *C1orf50* | TRUE | 2.242355453 |
| Obesity | ASPC | *BDH2* | TRUE | 2.057645225 |
| Obesity | ASPC | *TEX264* | TRUE | 3.02479331 |
| Obesity | ASPC | *ERAP1* | TRUE | 3.489907281 |
| Obesity | ASPC | *TMEM184A* | TRUE | 2.624103816 |
| Obesity | ASPC | *YWHAZ* | TRUE | 3.374695544 |
| Obesity | ASPC | *LRFN5* | TRUE | 2.17206155 |
| Obesity | ASPC | *RIMKLB* | TRUE | 2.584049177 |
| Obesity | ASPC | *SAAL1* | TRUE | 2.112692929 |
| Obesity | ASPC | *MS4A6E* | TRUE | 2.147862434 |
| Obesity | ASPC | *URM1* | TRUE | 2.926511361 |
| Obesity | ASPC | *SP2* | TRUE | 1.980572199 |
| Obesity | ASPC | *VKORC1* | TRUE | 3.553232827 |
| Obesity | ASPC | *PSCA* | TRUE | 3.348691474 |
| Obesity | ASPC | *UBXN6* | TRUE | 3.392692281 |

| | | | | |
|---|---|---|---|---|
| Obesity | ASPC | *SRR* | TRUE | 4.256588212 |
| Obesity | ASPC | *MLST8* | TRUE | 2.165124707 |
| Obesity | ASPC | *VWCE* | TRUE | 2.264108711 |
| Obesity | ASPC | *REEP4* | TRUE | 1.962076019 |
| Obesity | ASPC | *COL4A3* | TRUE | 2.022931493 |
| Obesity | ASPC | *RAB24* | TRUE | 3.646839757 |
| Obesity | ASPC | *CTNND2* | TRUE | 2.014062973 |
| Obesity | ASPC | *HOXD4* | TRUE | 2.110981968 |
| Obesity | ASPC | *ADORA2B* | TRUE | 2.988001669 |
| Obesity | ASPC | *SOX7* | TRUE | 2.471642046 |
| Obesity | ASPC | *CYP4F22* | TRUE | 2.226218688 |
| Obesity | ASPC | *ZNF554* | TRUE | 2.037291606 |
| Obesity | ASPC | *CXCR6* | TRUE | 2.143717238 |
| Obesity | ASPC | *C1QTNF4* | TRUE | 6.20158116 |
| Obesity | ASPC | *CTSW* | TRUE | 2.140405815 |
| Obesity | ASPC | *SNTG2* | TRUE | 3.845385168 |
| Obesity | ASPC | *CCL19* | TRUE | 2.297299872 |
| Obesity | ASPC | *FUT10* | TRUE | 2.077832412 |
| Obesity | ASPC | *HPSE2* | TRUE | 5.369950168 |
| Obesity | ASPC | *KLF14* | TRUE | 2.60297733 |
| Obesity | ASPC | *HRAS* | TRUE | 2.49982106 |
| Obesity | ASPC | *SRP72* | TRUE | 2.291447468 |
| Obesity | ASPC | *RSRC1* | TRUE | 2.344102588 |
| Obesity | ASPC | *CADM2* | TRUE | 3.120429916 |
| Obesity | ASPC | *PSMD2* | TRUE | 2.313677486 |
| Obesity | ASPC | *TMEM9B* | TRUE | 3.16080157 |
| Obesity | ASPC | *CCDC57* | TRUE | 3.014946706 |
| Obesity | ASPC | *SMIM19* | TRUE | 2.093848828 |
| Obesity | ASPC | *RUFY1* | TRUE | 3.210263141 |
| Obesity | ASPC | *TALDO1* | TRUE | 2.494677789 |
| Obesity | ASPC | *AP006621.1* | TRUE | 2.719888842 |
| Obesity | ASPC | *PPFIA3* | TRUE | 2.492404151 |
| Obesity | ASPC | *FAM87B* | TRUE | 2.633256794 |
| Obesity | ASPC | *RPP25* | TRUE | 2.227568579 |
| Obesity | ASPC | *STX19* | TRUE | 4.505674349 |
| Obesity | ASPC | *CTC1* | TRUE | 2.17599314 |
| Obesity | ASPC | *FARSA* | TRUE | 1.992434233 |
| Obesity | ASPC | *GIPC3* | TRUE | 2.049179588 |
| Obesity | ASPC | *B3GNT3* | TRUE | 3.194237247 |
| Obesity | ASPC | *ZNF322* | TRUE | 2.664848741 |
| Obesity | ASPC | *AMIGO1* | TRUE | 2.192811832 |
| Obesity | ASPC | *IZUMO1* | TRUE | 2.087671527 |
| Obesity | ASPC | *FBXL6* | TRUE | 2.473281804 |
| Obesity | ASPC | *CAV3* | TRUE | 3.02893621 |
| Obesity | ASPC | *BRICD5* | TRUE | 2.796553601 |
| Obesity | ASPC | *GRID1* | TRUE | 3.401966582 |
| Obesity | ASPC | *LYSMD4* | TRUE | 2.680207931 |
| Obesity | ASPC | *CCBE1* | TRUE | 2.217888448 |
| Obesity | ASPC | *LRIT3* | TRUE | 2.177594331 |
| Obesity | ASPC | *PSMG1* | TRUE | 3.689081254 |

| | | | | |
|---|---|---|---|---|
| Obesity | ASPC | *RP1L1* | TRUE | 3.182829825 |
| Obesity | ASPC | *MAATS1* | TRUE | 2.09273425 |
| Obesity | ASPC | *PLA2G6* | TRUE | 3.889255146 |
| Obesity | ASPC | *SNN* | TRUE | 2.285715337 |
| Obesity | ASPC | *NOC4L* | TRUE | 2.45699001 |
| Obesity | ASPC | *MAFF* | TRUE | 2.569349987 |
| Obesity | ASPC | *GP1BA* | TRUE | 2.371845324 |
| Obesity | ASPC | *KLHL33* | TRUE | 2.249823677 |
| Obesity | ASPC | *SRL* | TRUE | 2.035531008 |
| Obesity | ASPC | *TRABD2A* | TRUE | 2.415871605 |
| Obesity | ASPC | *ADH1A* | TRUE | 2.199512951 |
| Obesity | ASPC | *HYKK* | TRUE | 2.416302063 |
| Obesity | ASPC | *CENPP* | TRUE | 2.763200558 |
| Obesity | ASPC | *JAKMIP3* | TRUE | 2.023487376 |
| Obesity | ASPC | *FAM47E* | TRUE | 3.200529899 |
| Obesity | ASPC | *HCAR1* | TRUE | 2.490158631 |
| Obesity | ASPC | *SLC25A29* | TRUE | 2.051862508 |
| Obesity | ASPC | *SULT1A2* | TRUE | 2.3401438 |
| Obesity | ASPC | *PCDHB11* | TRUE | 2.540594848 |
| Obesity | ASPC | *C14orf178* | TRUE | 1.994137818 |
| Obesity | ASPC | *ZNF780A* | TRUE | 3.132926575 |
| Obesity | ASPC | *LEKR1* | TRUE | 2.130638947 |
| Obesity | ASPC | *MRPL42* | TRUE | 1.9873508 |
| Obesity | ASPC | *ZNF649* | TRUE | 2.42353615 |
| Obesity | ASPC | *ADH4* | TRUE | 3.383313222 |
| Obesity | ASPC | *ZKSCAN8* | TRUE | 2.068860265 |
| Obesity | ASPC | *ANKRD13B* | TRUE | 2.410880022 |
| Obesity | ASPC | *TYW1* | TRUE | 2.169715293 |
| Obesity | ASPC | *PRMT6* | TRUE | 3.096065722 |
| Obesity | ASPC | *CENPW* | TRUE | 2.764297281 |
| Obesity | ASPC | *SYT15* | TRUE | 2.029616309 |
| Obesity | ASPC | *NOTCH4* | TRUE | 2.046417823 |
| Obesity | ASPC | *SKIV2L* | TRUE | 2.624415977 |
| Obesity | ASPC | *HLA.E* | TRUE | 4.08217347 |
| Obesity | ASPC | *VGLL3* | TRUE | 1.987498615 |
| Obesity | ASPC | *COLQ* | TRUE | 1.997175851 |
| Obesity | ASPC | *ASIC3* | TRUE | 3.784679251 |
| Obesity | ASPC | *NOTCH2NL* | TRUE | 2.63323079 |
| Obesity | ASPC | *CPNE1* | TRUE | 2.338352048 |
| Obesity | ASPC | *C17orf67* | TRUE | 1.968700268 |
| Obesity | ASPC | *ATAD3C* | TRUE | 2.531089409 |
| Obesity | ASPC | *SLC12A8* | TRUE | 3.463751061 |
| Obesity | ASPC | *EXOSC6* | TRUE | 2.827645598 |
| Obesity | ASPC | *AC069213.1* | TRUE | 2.011471384 |
| Obesity | ASPC | *LINC00240* | TRUE | 2.181921467 |
| Obesity | ASPC | *RPL41* | TRUE | 2.117195571 |
| Obesity | ASPC | *RPS18* | TRUE | 2.036754072 |
| Obesity | ASPC | *LINC00484* | TRUE | 2.176324954 |
| Obesity | ASPC | *LINC00545* | TRUE | 2.283571967 |
| Obesity | ASPC | *TXNDC5* | TRUE | 1.98448542 |

| | | | | |
|---|---|---|---|---|
| Obesity | ASPC | *CCDC169* | TRUE | 2.170804705 |
| Obesity | ASPC | *PLA2G4B* | TRUE | 2.266760926 |
| Obesity | ASPC | *SRP14.AS1* | TRUE | 2.180315224 |
| Obesity | ASPC | *SNHG18* | TRUE | 2.657449426 |
| Obesity | ASPC | *PCDHGA6* | TRUE | 2.005830032 |
| Obesity | ASPC | *MPV17L2* | TRUE | 4.185548729 |
| Obesity | ASPC | *HMBS* | TRUE | 6.258539697 |
| Obesity | ASPC | *AC093668.2* | TRUE | 2.511956641 |
| Obesity | ASPC | *PGA5* | TRUE | 2.031211731 |
| Obesity | ASPC | *HP* | TRUE | 2.687319925 |
| Obesity | ASPC | *DUXA* | TRUE | 2.504506423 |
| Obesity | ASPC | *UBE2Q2L* | TRUE | 2.217134023 |
| Obesity | ASPC | *AC083843.1* | TRUE | 2.700745198 |
| Obesity | ASPC | *SH3RF3.AS1* | TRUE | 3.674419062 |
| Obesity | ASPC | *LINC00562* | TRUE | 3.184523157 |
| Obesity | ASPC | *LINC01140* | TRUE | 2.620250287 |
| Obesity | ASPC | *GTF2H5* | TRUE | 2.748847804 |
| Obesity | b_cell | *ITIH4* | TRUE | 2.862143514 |
| Obesity | b_cell | *CHRNE* | TRUE | 1.969396231 |
| Obesity | b_cell | *SNRPC* | TRUE | 4.389287651 |
| Obesity | b_cell | *PKDREJ* | TRUE | 2.409191935 |
| Obesity | b_cell | *PRSS53* | TRUE | 2.192760586 |
| Obesity | b_cell | *DPYSL4* | TRUE | 2.339348316 |
| Obesity | b_cell | *CLEC18A* | TRUE | 7.527964202 |
| Obesity | b_cell | *RNF123* | TRUE | 3.703288117 |
| Obesity | b_cell | *C1QTNF4* | TRUE | 6.353463238 |
| Obesity | b_cell | *CTSW* | TRUE | 2.2651154 |
| Obesity | b_cell | *TRIM73* | TRUE | 4.951161845 |
| Obesity | b_cell | *SULT1A2* | TRUE | 4.104755643 |
| Obesity | b_cell | *POU5F1* | TRUE | 2.816018375 |
| Obesity | b_cell | *NOTCH2NL* | TRUE | 3.080096999 |
| Obesity | dendritic_cell | *ITIH4* | TRUE | 5.923736035 |
| Obesity | dendritic_cell | *CHRNE* | TRUE | 2.364528414 |
| Obesity | dendritic_cell | *SERPINC1* | TRUE | 5.367454686 |
| Obesity | dendritic_cell | *ADRA1A* | TRUE | 2.35937321 |
| Obesity | dendritic_cell | *PKDREJ* | TRUE | 2.174270524 |
| Obesity | dendritic_cell | *IGHMBP2* | TRUE | 2.536241535 |
| Obesity | dendritic_cell | *DSCC1* | TRUE | 1.968621701 |
| Obesity | dendritic_cell | *HSD17B12* | TRUE | 5.077962654 |
| Obesity | dendritic_cell | *TTC12* | TRUE | 2.39691368 |
| Obesity | dendritic_cell | *DPYSL4* | TRUE | 3.092604396 |
| Obesity | dendritic_cell | *LMOD1* | TRUE | 4.582431836 |
| Obesity | dendritic_cell | *RNF123* | TRUE | 2.080452253 |
| Obesity | dendritic_cell | *TRMT61A* | TRUE | 2.142219066 |
| Obesity | dendritic_cell | *C8orf46* | TRUE | 2.424513171 |
| Obesity | dendritic_cell | *C1QTNF4* | TRUE | 5.379203865 |
| Obesity | dendritic_cell | *TRIM73* | TRUE | 4.023350055 |
| Obesity | dendritic_cell | *EIF3C* | TRUE | 4.096527092 |
| Obesity | dendritic_cell | *BTN3A2* | TRUE | 2.821839546 |
| Obesity | dendritic_cell | *RPS26* | TRUE | 3.133241542 |

| | | | | |
|---|---|---|---|---|
| Obesity | dendritic_cell | *ZNF780A* | TRUE | 3.26235296 |
| Obesity | dendritic_cell | *NOTCH4* | TRUE | 2.438208846 |
| Obesity | dendritic_cell | *SKIV2L* | TRUE | 2.997162837 |
| Obesity | endothelial | *RBM6* | TRUE | 3.115452345 |
| Obesity | endothelial | *ITIH4* | TRUE | 3.447589415 |
| Obesity | endothelial | *CHRNE* | TRUE | 2.336186325 |
| Obesity | endothelial | *MFAP2* | TRUE | 2.612754373 |
| Obesity | endothelial | *SERPINC1* | TRUE | 2.552047395 |
| Obesity | endothelial | *HIST1H2BJ* | TRUE | 2.324827522 |
| Obesity | endothelial | *ATP8B3* | TRUE | 3.899754769 |
| Obesity | endothelial | *PKDREJ* | TRUE | 2.456682375 |
| Obesity | endothelial | *DOC2A* | TRUE | 3.52563015 |
| Obesity | endothelial | *DPYSL4* | TRUE | 2.348900733 |
| Obesity | endothelial | *SLC5A11* | TRUE | 3.878947677 |
| Obesity | endothelial | *LMOD1* | TRUE | 1.969703798 |
| Obesity | endothelial | *ADORA2B* | TRUE | 3.448810034 |
| Obesity | endothelial | *C1QTNF4* | TRUE | 7.150885439 |
| Obesity | endothelial | *CTSW* | TRUE | 4.253129836 |
| Obesity | endothelial | *TRIM73* | TRUE | 4.815546663 |
| Obesity | endothelial | *PSMG1* | TRUE | 2.081799308 |
| Obesity | endothelial | *CCL4L2* | TRUE | 1.976491793 |
| Obesity | endothelial | *ZNF780A* | TRUE | 2.91879801 |
| Obesity | endothelial | *ADH4* | TRUE | 2.14332473 |
| Obesity | endothelial | *MRPL38* | TRUE | 1.999684249 |
| Obesity | endothelial | *SKIV2L* | TRUE | 2.211457739 |
| Obesity | endothelial | *ZFP57* | TRUE | 2.098240821 |
| Obesity | endothelial | *HP* | TRUE | 2.330981273 |
| Obesity | endothelial | *DUXA* | TRUE | 2.045677855 |
| Obesity | LEC | *ITIH4* | TRUE | 4.253499999 |
| Obesity | LEC | *TIPIN* | TRUE | 2.227676722 |
| Obesity | LEC | *SERPINC1* | TRUE | 2.498888174 |
| Obesity | LEC | *ADRA1A* | TRUE | 2.304673154 |
| Obesity | LEC | *GRK4* | TRUE | 2.905162028 |
| Obesity | LEC | *HSD17B12* | TRUE | 2.724131222 |
| Obesity | LEC | *DPYSL4* | TRUE | 3.257270845 |
| Obesity | LEC | *CLEC18A* | TRUE | 7.543023477 |
| Obesity | LEC | *SLC5A11* | TRUE | 3.487644911 |
| Obesity | LEC | *C1QTNF4* | TRUE | 3.854333081 |
| Obesity | LEC | *BTN3A2* | TRUE | 2.848294317 |
| Obesity | LEC | *RPS26* | TRUE | 2.382566948 |
| Obesity | LEC | *ZNF780A* | TRUE | 2.076364321 |
| Obesity | LEC | *SKIV2L* | TRUE | 2.328456539 |
| Obesity | LEC | *C17orf107* | TRUE | 2.015390582 |
| Obesity | LEC | *NOTCH2NL* | TRUE | 2.785030839 |
| Obesity | macrophage | *WNT16* | TRUE | 2.346449499 |
| Obesity | macrophage | *RBM6* | TRUE | 3.201714157 |
| Obesity | macrophage | *ITIH4* | TRUE | 4.75746538 |
| Obesity | macrophage | *TIPIN* | TRUE | 3.580041723 |
| Obesity | macrophage | *DESI1* | TRUE | 3.18314756 |
| Obesity | macrophage | *CORO1A* | TRUE | 2.148748791 |

| | | | | |
|---|---|---|---|---|
| Obesity | macrophage | *ELMO3* | TRUE | 1.962452553 |
| Obesity | macrophage | *CLUAP1* | TRUE | 2.332283872 |
| Obesity | macrophage | *ICAM5* | TRUE | 2.393108438 |
| Obesity | macrophage | *ATP13A1* | TRUE | 2.391810374 |
| Obesity | macrophage | *DLX5* | TRUE | 2.022761766 |
| Obesity | macrophage | *BNIP1* | TRUE | 2.259663879 |
| Obesity | macrophage | *CHMP3* | TRUE | 2.435342454 |
| Obesity | macrophage | *SERPINC1* | TRUE | 5.679609483 |
| Obesity | macrophage | *CCDC92* | TRUE | 3.164863258 |
| Obesity | macrophage | *DCAF4* | TRUE | 2.403107566 |
| Obesity | macrophage | *ADRA1A* | TRUE | 3.529216681 |
| Obesity | macrophage | *GJB6* | TRUE | 2.788493285 |
| Obesity | macrophage | *SMUG1* | TRUE | 2.114380287 |
| Obesity | macrophage | *GRK4* | TRUE | 3.575481917 |
| Obesity | macrophage | *OPRL1* | TRUE | 2.406675487 |
| Obesity | macrophage | *YEATS4* | TRUE | 2.110982386 |
| Obesity | macrophage | *ATP8B3* | TRUE | 5.122799078 |
| Obesity | macrophage | *PKDREJ* | TRUE | 3.11662732 |
| Obesity | macrophage | *TMEM60* | TRUE | 2.04852031 |
| Obesity | macrophage | *EPHA7* | TRUE | 2.470458901 |
| Obesity | macrophage | *DSCC1* | TRUE | 2.5512707 |
| Obesity | macrophage | *CARF* | TRUE | 2.62276932 |
| Obesity | macrophage | *C18orf8* | TRUE | 2.910776983 |
| Obesity | macrophage | *TRIM65* | TRUE | 2.629369835 |
| Obesity | macrophage | *SLC25A48* | TRUE | 2.650843508 |
| Obesity | macrophage | *PCYOX1L* | TRUE | 2.354358962 |
| Obesity | macrophage | *MTG1* | TRUE | 2.565510125 |
| Obesity | macrophage | *HSD17B12* | TRUE | 5.479955641 |
| Obesity | macrophage | *TTC12* | TRUE | 3.303428689 |
| Obesity | macrophage | *CCDC15* | TRUE | 2.986732967 |
| Obesity | macrophage | *DUSP15* | TRUE | 2.239154346 |
| Obesity | macrophage | *DOC2A* | TRUE | 2.52209808 |
| Obesity | macrophage | *DPYSL4* | TRUE | 3.73156787 |
| Obesity | macrophage | *POC5* | TRUE | 2.390230822 |
| Obesity | macrophage | *TCTEX1D1* | TRUE | 2.290992068 |
| Obesity | macrophage | *NEIL2* | TRUE | 2.257305482 |
| Obesity | macrophage | *CLEC18A* | TRUE | 5.741623713 |
| Obesity | macrophage | *HPD* | TRUE | 3.424298527 |
| Obesity | macrophage | *CD1E* | TRUE | 2.122406707 |
| Obesity | macrophage | *SLC5A11* | TRUE | 4.655745782 |
| Obesity | macrophage | *CLHC1* | TRUE | 2.422393962 |
| Obesity | macrophage | *LMOD1* | TRUE | 4.838537359 |
| Obesity | macrophage | *ZNF646* | TRUE | 2.940108208 |
| Obesity | macrophage | *VKORC1* | TRUE | 2.754559905 |
| Obesity | macrophage | *PSCA* | TRUE | 2.619081208 |
| Obesity | macrophage | *SRR* | TRUE | 3.961862717 |
| Obesity | macrophage | *C8orf46* | TRUE | 3.306456141 |
| Obesity | macrophage | *RAB24* | TRUE | 2.661831851 |
| Obesity | macrophage | *FAM103A1* | TRUE | 2.589662013 |
| Obesity | macrophage | *TRMT61B* | TRUE | 3.264702748 |

| | | | | |
|---|---|---|---|---|
| Obesity | macrophage | *C1QTNF4* | TRUE | 6.842600008 |
| Obesity | macrophage | *EFEMP2* | TRUE | 3.163270442 |
| Obesity | macrophage | *FUT10* | TRUE | 1.999976557 |
| Obesity | macrophage | *HPSE2* | TRUE | 3.452402368 |
| Obesity | macrophage | *STX19* | TRUE | 2.641910635 |
| Obesity | macrophage | *TRIM73* | TRUE | 4.56067287 |
| Obesity | macrophage | *B3GNT3* | TRUE | 2.632710121 |
| Obesity | macrophage | *ZNF322* | TRUE | 2.273195407 |
| Obesity | macrophage | *ZFP41* | TRUE | 1.963685884 |
| Obesity | macrophage | *BRICD5* | TRUE | 2.299049251 |
| Obesity | macrophage | *PSMG1* | TRUE | 3.664990547 |
| Obesity | macrophage | *RP1L1* | TRUE | 4.198414012 |
| Obesity | macrophage | *PLA2G6* | TRUE | 3.675619925 |
| Obesity | macrophage | *CENPP* | TRUE | 1.963648533 |
| Obesity | macrophage | *FAM47E* | TRUE | 2.657990403 |
| Obesity | macrophage | *HCAR1* | TRUE | 2.297421714 |
| Obesity | macrophage | *CCL4L2* | TRUE | 2.236482575 |
| Obesity | macrophage | *RPS26* | TRUE | 3.446396085 |
| Obesity | macrophage | *ZNF780A* | TRUE | 2.569967686 |
| Obesity | macrophage | *ADH4* | TRUE | 2.854052539 |
| Obesity | macrophage | *CCHCR1* | TRUE | 2.100785767 |
| Obesity | macrophage | *HLA.E* | TRUE | 3.620654394 |
| Obesity | macrophage | *EIF3CL* | TRUE | 3.292862465 |
| Obesity | macrophage | *ASIC3* | TRUE | 3.691357908 |
| Obesity | macrophage | *NOTCH2NL* | TRUE | 2.114733021 |
| Obesity | macrophage | *LINC00545* | TRUE | 2.044931392 |
| Obesity | macrophage | *PLA2G4B* | TRUE | 2.142953941 |
| Obesity | macrophage | *PCDHAC1* | TRUE | 2.490681411 |
| Obesity | macrophage | *MPV17L2* | TRUE | 3.742098851 |
| Obesity | macrophage | *HMBS* | TRUE | 6.04572154 |
| Obesity | macrophage | *DUXA* | TRUE | 2.503250547 |
| Obesity | macrophage | *SH3RF3.AS1* | TRUE | 2.716810578 |
| Obesity | mast_cell | *ITIH4* | TRUE | 2.979242143 |
| Obesity | mast_cell | *UHRF1BP1* | TRUE | 7.824117756 |
| Obesity | mast_cell | *TIPIN* | TRUE | 2.458324998 |
| Obesity | mast_cell | *DESI1* | TRUE | 2.111638523 |
| Obesity | mast_cell | *CORO1A* | TRUE | 3.095193997 |
| Obesity | mast_cell | *DLX5* | TRUE | 1.973047628 |
| Obesity | mast_cell | *SERPINC1* | TRUE | 3.730264663 |
| Obesity | mast_cell | *ADRA1A* | TRUE | 1.960144202 |
| Obesity | mast_cell | *GRK4* | TRUE | 3.271221288 |
| Obesity | mast_cell | *PKDREJ* | TRUE | 3.137193706 |
| Obesity | mast_cell | *IGHMBP2* | TRUE | 2.250257829 |
| Obesity | mast_cell | *DSCC1* | TRUE | 2.283725649 |
| Obesity | mast_cell | *HSD17B12* | TRUE | 2.811009012 |
| Obesity | mast_cell | *DPYSL4* | TRUE | 3.340432809 |
| Obesity | mast_cell | *CLEC18A* | TRUE | 2.241636101 |
| Obesity | mast_cell | *HPD* | TRUE | 3.278803862 |
| Obesity | mast_cell | *ZNF646* | TRUE | 2.45526027 |
| Obesity | mast_cell | *PSCA* | TRUE | 2.729501709 |

| | | | | |
|---|---|---|---|---|
| Obesity | mast_cell | *SRR* | TRUE | 3.082450719 |
| Obesity | mast_cell | *C8orf46* | TRUE | 2.927291977 |
| Obesity | mast_cell | *TRMT61B* | TRUE | 2.122835821 |
| Obesity | mast_cell | *C1QTNF4* | TRUE | 5.687877558 |
| Obesity | mast_cell | *TRIM73* | TRUE | 2.960252459 |
| Obesity | mast_cell | *PSMG1* | TRUE | 3.368247659 |
| Obesity | mast_cell | *BTN3A2* | TRUE | 2.194768637 |
| Obesity | mast_cell | *SULT1A2* | TRUE | 3.153972542 |
| Obesity | mast_cell | *RPS26* | TRUE | 2.117454817 |
| Obesity | mast_cell | *HMBS* | TRUE | 2.27667498 |
| Obesity | monocyte | *RBM6* | TRUE | 2.90977825 |
| Obesity | monocyte | *CENPQ* | TRUE | 2.337001379 |
| Obesity | monocyte | *ITIH4* | TRUE | 3.007340976 |
| Obesity | monocyte | *UHRF1BP1* | TRUE | 3.806003657 |
| Obesity | monocyte | *DESI1* | TRUE | 1.978035625 |
| Obesity | monocyte | *DLX5* | TRUE | 2.486748751 |
| Obesity | monocyte | *PRSS16* | TRUE | 3.340050671 |
| Obesity | monocyte | *GSTM1* | TRUE | 2.005036147 |
| Obesity | monocyte | *SLC25A48* | TRUE | 2.171924161 |
| Obesity | monocyte | *CLEC18A* | TRUE | 2.501562948 |
| Obesity | monocyte | *C1QTNF4* | TRUE | 4.803922712 |
| Obesity | monocyte | *TRIM73* | TRUE | 2.872122002 |
| Obesity | monocyte | *EIF3C* | TRUE | 3.028591506 |
| Obesity | monocyte | *TRABD2A* | TRUE | 2.852850954 |
| Obesity | monocyte | *SP6* | TRUE | 2.841357392 |
| Obesity | monocyte | *POU5F1* | TRUE | 2.216417625 |
| Obesity | neutrophil | *TIPIN* | TRUE | 2.771740374 |
| Obesity | neutrophil | *SERPINC1* | TRUE | 1.967708414 |
| Obesity | neutrophil | *CLEC18A* | TRUE | 7.61942465 |
| Obesity | neutrophil | *C1QTNF4* | TRUE | 7.198257012 |
| Obesity | neutrophil | *CTSW* | TRUE | 3.031529387 |
| Obesity | neutrophil | *TRIM73* | TRUE | 2.723150093 |
| Obesity | neutrophil | *EIF3C* | TRUE | 4.918312713 |
| Obesity | neutrophil | *HYAL3* | TRUE | 2.909261002 |
| Obesity | neutrophil | *NOTCH4* | TRUE | 2.354542619 |
| Obesity | neutrophil | *SKIV2L* | TRUE | 2.592395946 |
| Obesity | neutrophil | *AC084018.1* | TRUE | 2.282049134 |
| Obesity | nk_cell | *ITIH4* | TRUE | 5.508391392 |
| Obesity | nk_cell | *UHRF1BP1* | TRUE | 2.940670732 |
| Obesity | nk_cell | *PRSS16* | TRUE | 2.361485905 |
| Obesity | nk_cell | *MFAP2* | TRUE | 2.221757499 |
| Obesity | nk_cell | *GRK4* | TRUE | 2.775357337 |
| Obesity | nk_cell | *ATP8B3* | TRUE | 4.113992947 |
| Obesity | nk_cell | *PKDREJ* | TRUE | 2.218126151 |
| Obesity | nk_cell | *HSD17B12* | TRUE | 3.136835257 |
| Obesity | nk_cell | *DPYSL4* | TRUE | 2.908459632 |
| Obesity | nk_cell | *CLEC18A* | TRUE | 7.772949403 |
| Obesity | nk_cell | *HPD* | TRUE | 2.824402375 |
| Obesity | nk_cell | *SLC5A11* | TRUE | 2.799347276 |
| Obesity | nk_cell | *SRR* | TRUE | 3.325849765 |

| | | | | |
|---|---|---|---|---|
| Obesity | nk_cell | *FAM103A1* | TRUE | 1.968588751 |
| Obesity | nk_cell | *ADORA2B* | TRUE | 3.948645586 |
| Obesity | nk_cell | *C1QTNF4* | TRUE | 5.21358165 |
| Obesity | nk_cell | *TRIM73* | TRUE | 4.365831149 |
| Obesity | nk_cell | *PSMG1* | TRUE | 2.601459789 |
| Obesity | nk_cell | *BMP8A* | TRUE | 3.247260783 |
| Obesity | nk_cell | *HCAR1* | TRUE | 2.2168667 |
| Obesity | nk_cell | *ZNF780A* | TRUE | 2.641471911 |
| Obesity | nk_cell | *NOTCH4* | TRUE | 2.67107901 |
| Obesity | nk_cell | *SKIV2L* | TRUE | 3.158113287 |
| Obesity | nk_cell | *POU5F1* | TRUE | 2.433227678 |
| Obesity | nk_cell | *C17orf107* | TRUE | 2.164286114 |
| Obesity | nk_cell | *HMBS* | TRUE | 5.190198766 |
| Obesity | pericyte | *ITIH4* | TRUE | 5.961262843 |
| Obesity | pericyte | *UHRF1BP1* | TRUE | 6.908488353 |
| Obesity | pericyte | *TIPIN* | TRUE | 2.942201601 |
| Obesity | pericyte | *YPEL3* | TRUE | 4.024346239 |
| Obesity | pericyte | *STX1B* | TRUE | 2.128645178 |
| Obesity | pericyte | *KAT8* | TRUE | 1.973066402 |
| Obesity | pericyte | *ATP13A1* | TRUE | 2.086091589 |
| Obesity | pericyte | *BNIP1* | TRUE | 2.155086309 |
| Obesity | pericyte | *SERPINC1* | TRUE | 4.40357426 |
| Obesity | pericyte | *CCDC92* | TRUE | 2.465484247 |
| Obesity | pericyte | *ADRA1A* | TRUE | 2.998754292 |
| Obesity | pericyte | *SMUG1* | TRUE | 2.003780283 |
| Obesity | pericyte | *GRK4* | TRUE | 2.330673676 |
| Obesity | pericyte | *RIBC2* | TRUE | 2.14628658 |
| Obesity | pericyte | *ATP8B3* | TRUE | 4.878092177 |
| Obesity | pericyte | *PKDREJ* | TRUE | 2.234804785 |
| Obesity | pericyte | *IGHMBP2* | TRUE | 2.8703246 |
| Obesity | pericyte | *EPHA7* | TRUE | 2.192368651 |
| Obesity | pericyte | *HSD17B12* | TRUE | 5.64989039 |
| Obesity | pericyte | *TTC12* | TRUE | 3.175097653 |
| Obesity | pericyte | *DPYSL4* | TRUE | 3.461168664 |
| Obesity | pericyte | *TCTEX1D1* | TRUE | 2.259658753 |
| Obesity | pericyte | *CLEC18A* | TRUE | 6.702127013 |
| Obesity | pericyte | *SLC5A11* | TRUE | 2.863298179 |
| Obesity | pericyte | *SRR* | TRUE | 2.430056557 |
| Obesity | pericyte | *C8orf46* | TRUE | 2.158418261 |
| Obesity | pericyte | *TRMT61B* | TRUE | 2.670261311 |
| Obesity | pericyte | *C1QTNF4* | TRUE | 5.103130038 |
| Obesity | pericyte | *TRIM73* | TRUE | 4.364505389 |
| Obesity | pericyte | *CAV3* | TRUE | 2.188595302 |
| Obesity | pericyte | *CENPP* | TRUE | 2.498955639 |
| Obesity | pericyte | *SULT1A2* | TRUE | 2.846313131 |
| Obesity | pericyte | *RPS26* | TRUE | 2.873072597 |
| Obesity | pericyte | *HLA.E* | TRUE | 2.990419439 |
| Obesity | pericyte | *ASIC3* | TRUE | 2.516258668 |
| Obesity | pericyte | *NOTCH2NL* | TRUE | 2.555377338 |
| Obesity | pericyte | *LINC00240* | TRUE | 2.164128663 |

| | | | | |
|---|---|---|---|---|
| Obesity | pericyte | *DPP3* | TRUE | 2.26014111 |
| Obesity | pericyte | *HMBS* | TRUE | 5.342325351 |
| Obesity | pericyte | *SH3RF3.AS1* | TRUE | 2.343096775 |
| Obesity | pericyte | *LINC00562* | TRUE | 2.048248241 |
| Obesity | SMC | *WNT16* | TRUE | 2.30892939 |
| Obesity | SMC | *ITIH4* | TRUE | 2.397866031 |
| Obesity | SMC | *TIPIN* | TRUE | 2.702208822 |
| Obesity | SMC | *COL16A1* | TRUE | 2.198319152 |
| Obesity | SMC | *PABPC4* | TRUE | 3.303666026 |
| Obesity | SMC | *DESI1* | TRUE | 2.708401852 |
| Obesity | SMC | *ELMO3* | TRUE | 2.416173534 |
| Obesity | SMC | *MFAP2* | TRUE | 2.289294238 |
| Obesity | SMC | *SERPINC1* | TRUE | 5.448194497 |
| Obesity | SMC | *CCDC92* | TRUE | 3.442064773 |
| Obesity | SMC | *GJB6* | TRUE | 2.34465217 |
| Obesity | SMC | *GRK4* | TRUE | 3.484720993 |
| Obesity | SMC | *RIBC2* | TRUE | 3.166005972 |
| Obesity | SMC | *ATP8B3* | TRUE | 4.708860496 |
| Obesity | SMC | *PKDREJ* | TRUE | 2.570510285 |
| Obesity | SMC | *IGHMBP2* | TRUE | 2.553674518 |
| Obesity | SMC | *EPHA7* | TRUE | 2.064455335 |
| Obesity | SMC | *DSCC1* | TRUE | 2.499177495 |
| Obesity | SMC | *HSD17B12* | TRUE | 5.578113859 |
| Obesity | SMC | *TTC12* | TRUE | 2.752180395 |
| Obesity | SMC | *DPYSL4* | TRUE | 3.448581369 |
| Obesity | SMC | *CLEC18A* | TRUE | 6.382257294 |
| Obesity | SMC | *HPD* | TRUE | 2.796478687 |
| Obesity | SMC | *SLC5A11* | TRUE | 4.807086797 |
| Obesity | SMC | *CLHC1* | TRUE | 1.968045087 |
| Obesity | SMC | *RNF123* | TRUE | 2.998270867 |
| Obesity | SMC | *TRMT61A* | TRUE | 2.654926412 |
| Obesity | SMC | *SRR* | TRUE | 3.643160751 |
| Obesity | SMC | *C8orf46* | TRUE | 3.125770396 |
| Obesity | SMC | *TRMT61B* | TRUE | 2.193269518 |
| Obesity | SMC | *C1QTNF4* | TRUE | 6.515763481 |
| Obesity | SMC | *CAV3* | TRUE | 2.056619971 |
| Obesity | SMC | *PSMG1* | TRUE | 3.562162464 |
| Obesity | SMC | *RP1L1* | TRUE | 2.442202622 |
| Obesity | SMC | *PLA2G6* | TRUE | 2.28914915 |
| Obesity | SMC | *GP1BA* | TRUE | 1.964190316 |
| Obesity | SMC | *BTN3A2* | TRUE | 2.182163729 |
| Obesity | SMC | *CENPP* | TRUE | 2.874190329 |
| Obesity | SMC | *CCL4L2* | TRUE | 2.830560877 |
| Obesity | SMC | *PCDHB11* | TRUE | 1.983107056 |
| Obesity | SMC | *RPS26* | TRUE | 3.095146866 |
| Obesity | SMC | *ZNF780A* | TRUE | 3.014186564 |
| Obesity | SMC | *ADH4* | TRUE | 3.326919178 |
| Obesity | SMC | *SKIV2L* | TRUE | 2.491631773 |
| Obesity | SMC | *HLA.E* | TRUE | 2.670864895 |
| Obesity | SMC | *ASIC3* | TRUE | 2.338992941 |

| | | | | |
|---|---|---|---|---|
| Obesity | SMC | *NOTCH2NL* | TRUE | 2.557855287 |
| Obesity | SMC | *PCDHAC1* | TRUE | 2.633528207 |
| Obesity | SMC | *MPV17L2* | TRUE | 3.008788922 |
| Obesity | SMC | *HMBS* | TRUE | 5.647514339 |
| Obesity | SMC | *DUXA* | TRUE | 2.361024527 |
| Obesity | t_cell | *RBM6* | TRUE | 3.060033121 |
| Obesity | t_cell | *ITIH4* | TRUE | 3.074054585 |
| Obesity | t_cell | *TIPIN* | TRUE | 2.808990312 |
| Obesity | t_cell | *ELMO3* | TRUE | 2.024358978 |
| Obesity | t_cell | *SERPINC1* | TRUE | 4.23059354 |
| Obesity | t_cell | *CCDC92* | TRUE | 2.745316682 |
| Obesity | t_cell | *ADRA1A* | TRUE | 3.018932337 |
| Obesity | t_cell | *GRK4* | TRUE | 2.907611074 |
| Obesity | t_cell | *PCSK2* | TRUE | 2.297374774 |
| Obesity | t_cell | *RIBC2* | TRUE | 3.059302428 |
| Obesity | t_cell | *ATP8B3* | TRUE | 3.735872742 |
| Obesity | t_cell | *SH3GL3* | TRUE | 2.015447949 |
| Obesity | t_cell | *HSD17B12* | TRUE | 2.12324404 |
| Obesity | t_cell | *DPYSL4* | TRUE | 3.691188801 |
| Obesity | t_cell | *CLEC18A* | TRUE | 6.258795416 |
| Obesity | t_cell | *HPD* | TRUE | 2.041686793 |
| Obesity | t_cell | *SLC5A11* | TRUE | 2.489103147 |
| Obesity | t_cell | *LMOD1* | TRUE | 3.823614165 |
| Obesity | t_cell | *PSCA* | TRUE | 2.115312729 |
| Obesity | t_cell | *SRR* | TRUE | 3.567663736 |
| Obesity | t_cell | *C8orf46* | TRUE | 3.468893243 |
| Obesity | t_cell | *ADORA2B* | TRUE | 3.916407456 |
| Obesity | t_cell | *TRMT61B* | TRUE | 2.524077436 |
| Obesity | t_cell | *C1QTNF4* | TRUE | 5.115189612 |
| Obesity | t_cell | *TRIM73* | TRUE | 4.659679169 |
| Obesity | t_cell | *PSMG1* | TRUE | 3.464047622 |
| Obesity | t_cell | *RP1L1* | TRUE | 3.926154931 |
| Obesity | t_cell | *PLA2G6* | TRUE | 2.264651661 |
| Obesity | t_cell | *CENPP* | TRUE | 2.578296415 |
| Obesity | t_cell | *SULT1A2* | TRUE | 3.950010917 |
| Obesity | t_cell | *RPS26* | TRUE | 3.206728536 |
| Obesity | t_cell | *ASIC3* | TRUE | 2.711412812 |
| Obesity | t_cell | *HMBS* | TRUE | 5.444626684 |
| Stroke | adipocyte | *CENPQ* | TRUE | 3.090126877 |
| Stroke | adipocyte | *CYP2D6* | TRUE | 2.820884266 |
| Stroke | adipocyte | *MGRN1* | TRUE | 2.049953032 |
| Stroke | adipocyte | *IGDCC4* | TRUE | 2.091191773 |
| Stroke | adipocyte | *C17orf75* | TRUE | 2.730624137 |
| Stroke | adipocyte | *BVES* | TRUE | 2.319668743 |
| Stroke | adipocyte | *PCDHB6* | TRUE | 2.414111173 |
| Stroke | adipocyte | *MFAP2* | TRUE | 2.14261014 |
| Stroke | adipocyte | *EPCAM* | TRUE | 2.553048338 |
| Stroke | adipocyte | *STIL* | TRUE | 2.309127539 |
| Stroke | adipocyte | *BTN2A2* | TRUE | 2.212874994 |
| Stroke | adipocyte | *TRAP1* | TRUE | 2.365871642 |

| | | | | |
|---|---|---|---|---|
| Stroke | adipocyte | *LARS* | TRUE | 1.984747441 |
| Stroke | adipocyte | *ITGA7* | TRUE | 2.015906974 |
| Stroke | adipocyte | *LYG1* | TRUE | 2.455783419 |
| Stroke | adipocyte | *NBEAL1* | TRUE | 1.984102884 |
| Stroke | adipocyte | *ILDR1* | TRUE | 2.082802691 |
| Stroke | adipocyte | *C1orf50* | TRUE | 2.478533073 |
| Stroke | adipocyte | *TMEM184A* | TRUE | 2.145897345 |
| Stroke | adipocyte | *KIR3DL1* | TRUE | 2.017483632 |
| Stroke | adipocyte | *CHAF1A* | TRUE | 2.085252021 |
| Stroke | adipocyte | *FAM57A* | TRUE | 2.485336003 |
| Stroke | adipocyte | *ULK4* | TRUE | 2.359389018 |
| Stroke | adipocyte | *KCNAB3* | TRUE | 2.427912562 |
| Stroke | adipocyte | *GEN1* | TRUE | 2.00433866 |
| Stroke | adipocyte | *FUCA1* | TRUE | 2.059965239 |
| Stroke | adipocyte | *FJX1* | TRUE | 2.041168552 |
| Stroke | adipocyte | *TMEM86B* | TRUE | 2.103404809 |
| Stroke | adipocyte | *CLDN7* | TRUE | 2.864433202 |
| Stroke | adipocyte | *RP1L1* | TRUE | 2.324008629 |
| Stroke | adipocyte | *PDE6G* | TRUE | 2.104661623 |
| Stroke | adipocyte | *FFAR4* | TRUE | 1.982886066 |
| Stroke | adipocyte | *ZNF749* | TRUE | 2.896579015 |
| Stroke | adipocyte | *GPAT2* | TRUE | 2.15015192 |
| Stroke | adipocyte | *TMEM116* | TRUE | 2.556173173 |
| Stroke | adipocyte | *ANKRD13B* | TRUE | 2.227647787 |
| Stroke | adipocyte | *COL15A1* | TRUE | 2.633510247 |
| Stroke | adipocyte | *DNLZ* | TRUE | 2.027312298 |
| Stroke | adipocyte | *NUTM2D* | TRUE | 2.040389815 |
| Stroke | adipocyte | *LILRA2* | TRUE | 2.22551278 |
| Stroke | adipocyte | *TDGF1* | TRUE | 3.247932047 |
| Stroke | adipocyte | *LINC00942* | TRUE | 2.085812264 |
| Stroke | adipocyte | *SHANK3* | TRUE | 2.024870114 |
| Stroke | adipocyte | *ARMS2* | TRUE | 2.059704776 |
| Stroke | ASPC | *ZDHHC6* | TRUE | 1.964118155 |
| Stroke | ASPC | *NR1H3* | TRUE | 2.079218967 |
| Stroke | ASPC | *CENPQ* | TRUE | 3.471412158 |
| Stroke | ASPC | *FSCN1* | TRUE | 2.102640051 |
| Stroke | ASPC | *GPR137B* | TRUE | 2.240283502 |
| Stroke | ASPC | *EPDR1* | TRUE | 2.078730168 |
| Stroke | ASPC | *NECAP1* | TRUE | 2.359791266 |
| Stroke | ASPC | *SPTLC1* | TRUE | 2.305127014 |
| Stroke | ASPC | *TPSD1* | TRUE | 2.306317577 |
| Stroke | ASPC | *MARCH2* | TRUE | 2.973924125 |
| Stroke | ASPC | *CYP2D6* | TRUE | 3.019762085 |
| Stroke | ASPC | *CNIH1* | TRUE | 2.410647749 |
| Stroke | ASPC | *MGRN1* | TRUE | 2.13044261 |
| Stroke | ASPC | *IGDCC4* | TRUE | 2.073073238 |
| Stroke | ASPC | *RAB2A* | TRUE | 2.324263853 |
| Stroke | ASPC | *CAV1* | TRUE | 2.270158566 |
| Stroke | ASPC | *TBL2* | TRUE | 2.015409524 |
| Stroke | ASPC | *C17orf75* | TRUE | 3.86761454 |

| | | | | |
|---|---|---|---|---|
| Stroke | ASPC | *TMEM104* | TRUE | 1.96203411 |
| Stroke | ASPC | *BVES* | TRUE | 2.648322587 |
| Stroke | ASPC | *CEP72* | TRUE | 2.015808052 |
| Stroke | ASPC | *PCDHB6* | TRUE | 2.444180242 |
| Stroke | ASPC | *MRPL37* | TRUE | 2.273854047 |
| Stroke | ASPC | *MFAP2* | TRUE | 2.191013329 |
| Stroke | ASPC | *GPR75* | TRUE | 2.108782443 |
| Stroke | ASPC | *EPCAM* | TRUE | 2.850179364 |
| Stroke | ASPC | *MRC1* | TRUE | 2.403313149 |
| Stroke | ASPC | *FMOD* | TRUE | 1.989634939 |
| Stroke | ASPC | *FAM213A* | TRUE | 2.288538645 |
| Stroke | ASPC | *CCZ1* | TRUE | 2.101766869 |
| Stroke | ASPC | *SLC25A16* | TRUE | 1.970823653 |
| Stroke | ASPC | *STIL* | TRUE | 3.068072574 |
| Stroke | ASPC | *BTN2A2* | TRUE | 2.014193654 |
| Stroke | ASPC | *TRAP1* | TRUE | 2.561104502 |
| Stroke | ASPC | *RHOT1* | TRUE | 2.010213442 |
| Stroke | ASPC | *SHC2* | TRUE | 2.134615326 |
| Stroke | ASPC | *QRSL1* | TRUE | 2.246955575 |
| Stroke | ASPC | *EMILIN2* | TRUE | 2.45918648 |
| Stroke | ASPC | *LARS* | TRUE | 2.527710193 |
| Stroke | ASPC | *CNTN6* | TRUE | 2.109478792 |
| Stroke | ASPC | *WNT2B* | TRUE | 1.967672646 |
| Stroke | ASPC | *PRR5L* | TRUE | 2.094593523 |
| Stroke | ASPC | *ITGA7* | TRUE | 2.289007097 |
| Stroke | ASPC | *KLHL36* | TRUE | 2.662175242 |
| Stroke | ASPC | *RNASEL* | TRUE | 2.355326526 |
| Stroke | ASPC | *SPAG8* | TRUE | 2.176653139 |
| Stroke | ASPC | *RMDN3* | TRUE | 2.457196543 |
| Stroke | ASPC | *SLC35A5* | TRUE | 2.115934323 |
| Stroke | ASPC | *C14orf79* | TRUE | 2.212099043 |
| Stroke | ASPC | *RPRD1A* | TRUE | 2.245481899 |
| Stroke | ASPC | *FKBP10* | TRUE | 2.818494335 |
| Stroke | ASPC | *CELSR2* | TRUE | 2.068357297 |
| Stroke | ASPC | *ARNT* | TRUE | 2.189517627 |
| Stroke | ASPC | *LYG1* | TRUE | 2.743053505 |
| Stroke | ASPC | *PLA1A* | TRUE | 2.126466939 |
| Stroke | ASPC | *ILDR1* | TRUE | 2.107817093 |
| Stroke | ASPC | *KLHL3* | TRUE | 2.659979465 |
| Stroke | ASPC | *ATP6V0D2* | TRUE | 2.324042049 |
| Stroke | ASPC | *NTMT1* | TRUE | 2.145330525 |
| Stroke | ASPC | *GSTO1* | TRUE | 4.696306051 |
| Stroke | ASPC | *GLYAT* | TRUE | 1.997406691 |
| Stroke | ASPC | *NMT2* | TRUE | 2.162979501 |
| Stroke | ASPC | *WDYHV1* | TRUE | 2.269350316 |
| Stroke | ASPC | *SLC6A1* | TRUE | 2.142091263 |
| Stroke | ASPC | *CLEC18A* | TRUE | 1.982921349 |
| Stroke | ASPC | *B4GALT3* | TRUE | 2.42322188 |
| Stroke | ASPC | *CBR3* | TRUE | 2.551681497 |
| Stroke | ASPC | *DIP2A* | TRUE | 2.066718029 |

| | | | | |
|---|---|---|---|---|
| Stroke | ASPC | *ZBTB7B* | TRUE | 2.116619128 |
| Stroke | ASPC | *FDPS* | TRUE | 2.005066769 |
| Stroke | ASPC | *RFC4* | TRUE | 2.548941949 |
| Stroke | ASPC | *C1orf50* | TRUE | 2.693059833 |
| Stroke | ASPC | *TMEM184A* | TRUE | 2.838755876 |
| Stroke | ASPC | *YWHAZ* | TRUE | 2.540420321 |
| Stroke | ASPC | *HSPA12A* | TRUE | 2.524716461 |
| Stroke | ASPC | *ZFYVE19* | TRUE | 1.987690166 |
| Stroke | ASPC | *KIR3DL1* | TRUE | 1.982073075 |
| Stroke | ASPC | *CHAF1A* | TRUE | 2.393479352 |
| Stroke | ASPC | *FAM57A* | TRUE | 2.615454914 |
| Stroke | ASPC | *SRR* | TRUE | 2.012287765 |
| Stroke | ASPC | *ULK4* | TRUE | 2.635779605 |
| Stroke | ASPC | *RFWD3* | TRUE | 2.008267081 |
| Stroke | ASPC | *IRS1* | TRUE | 2.008167111 |
| Stroke | ASPC | *UPF3A* | TRUE | 2.564131811 |
| Stroke | ASPC | *SH3TC2* | TRUE | 2.024337884 |
| Stroke | ASPC | *GAA* | TRUE | 2.125785314 |
| Stroke | ASPC | *MRPL52* | TRUE | 1.983275636 |
| Stroke | ASPC | *FAM192A* | TRUE | 1.984350821 |
| Stroke | ASPC | *GMPPB* | TRUE | 1.964890361 |
| Stroke | ASPC | *DDX23* | TRUE | 2.254921073 |
| Stroke | ASPC | *CTDNEP1* | TRUE | 1.977606876 |
| Stroke | ASPC | *CRLF3* | TRUE | 3.211616774 |
| Stroke | ASPC | *ACSF3* | TRUE | 1.985692214 |
| Stroke | ASPC | *IRX3* | TRUE | 2.158747962 |
| Stroke | ASPC | *GEN1* | TRUE | 2.372130221 |
| Stroke | ASPC | *FUCA1* | TRUE | 2.596042974 |
| Stroke | ASPC | *FJX1* | TRUE | 2.104821591 |
| Stroke | ASPC | *TMEM86B* | TRUE | 2.232626785 |
| Stroke | ASPC | *SPNS3* | TRUE | 2.246238666 |
| Stroke | ASPC | *GRID1* | TRUE | 2.082495277 |
| Stroke | ASPC | *PLGLB1* | TRUE | 1.976384999 |
| Stroke | ASPC | *RP1L1* | TRUE | 2.246625364 |
| Stroke | ASPC | *PDE6G* | TRUE | 2.197205898 |
| Stroke | ASPC | *FFAR4* | TRUE | 2.965187587 |
| Stroke | ASPC | *ZNF749* | TRUE | 3.009114693 |
| Stroke | ASPC | *ADH1A* | TRUE | 2.281154399 |
| Stroke | ASPC | *BEND4* | TRUE | 2.004667876 |
| Stroke | ASPC | *ZFP28* | TRUE | 2.179479998 |
| Stroke | ASPC | *PIWIL2* | TRUE | 2.107190299 |
| Stroke | ASPC | *TMEM116* | TRUE | 2.303809973 |
| Stroke | ASPC | *SH3BGRL2* | TRUE | 2.277304477 |
| Stroke | ASPC | *COL15A1* | TRUE | 3.529724364 |
| Stroke | ASPC | *LRRC10B* | TRUE | 2.471656095 |
| Stroke | ASPC | *DNLZ* | TRUE | 1.993223497 |
| Stroke | ASPC | *NUTM2D* | TRUE | 2.155379533 |
| Stroke | ASPC | *PPP3R1* | TRUE | 2.120146823 |
| Stroke | ASPC | *CDKN2AIPNL* | TRUE | 1.987232819 |
| Stroke | ASPC | *IRGM* | TRUE | 1.998495858 |

| | | | | |
|---|---|---|---|---|
| Stroke | ASPC | *LILRA2* | TRUE | 2.195145889 |
| Stroke | ASPC | *TDGF1* | TRUE | 2.898546993 |
| Stroke | ASPC | *MRPL20* | TRUE | 2.228287531 |
| Stroke | ASPC | *NME1.NME2* | TRUE | 1.96297386 |
| Stroke | ASPC | *LINC00942* | TRUE | 2.259336658 |
| Stroke | ASPC | *SHANK3* | TRUE | 2.424833018 |
| Stroke | ASPC | *TMEM200B* | TRUE | 2.417649185 |
| Stroke | ASPC | *ARMS2* | TRUE | 2.325858471 |
| Stroke | ASPC | *GOLGA8H* | TRUE | 2.016858338 |
| Stroke | b_cell | *IGDCC4* | TRUE | 2.005338368 |
| Stroke | b_cell | *FAM57A* | TRUE | 2.182553552 |
| Stroke | b_cell | *LILRA2* | TRUE | 2.119379776 |
| Stroke | b_cell | *TDGF1* | TRUE | 2.933708497 |
| Stroke | dendritic_cell | *BVES* | TRUE | 2.587831683 |
| Stroke | dendritic_cell | *HORMAD1* | TRUE | 1.965764724 |
| Stroke | dendritic_cell | *FJX1* | TRUE | 1.960904124 |
| Stroke | dendritic_cell | *CLDN7* | TRUE | 2.777175877 |
| Stroke | dendritic_cell | *PIGP* | TRUE | 2.002291571 |
| Stroke | dendritic_cell | *ZNF749* | TRUE | 2.643628506 |
| Stroke | dendritic_cell | *TDGF1* | TRUE | 2.975802058 |
| Stroke | endothelial | *CYP2D6* | TRUE | 2.04591217 |
| Stroke | endothelial | *IGDCC4* | TRUE | 2.062800179 |
| Stroke | endothelial | *PCDHB6* | TRUE | 2.097757101 |
| Stroke | endothelial | *EPCAM* | TRUE | 1.972900962 |
| Stroke | endothelial | *BTN2A2* | TRUE | 2.004354151 |
| Stroke | endothelial | *HBZ* | TRUE | 1.995744808 |
| Stroke | endothelial | *FKBP10* | TRUE | 2.223981072 |
| Stroke | endothelial | *GPAT2* | TRUE | 2.049170273 |
| Stroke | endothelial | *HLA.DQB2* | TRUE | 2.05288553 |
| Stroke | endothelial | *TDGF1* | TRUE | 3.218915041 |
| Stroke | LEC | *CENPQ* | TRUE | 2.643385581 |
| Stroke | LEC | *MFAP2* | TRUE | 2.109118075 |
| Stroke | LEC | *FKBP10* | TRUE | 3.335414018 |
| Stroke | LEC | *ILDR1* | TRUE | 2.060704258 |
| Stroke | LEC | *CLEC18A* | TRUE | 2.033996229 |
| Stroke | LEC | *FAM57A* | TRUE | 1.96623733 |
| Stroke | LEC | *NUTM2D* | TRUE | 2.149951579 |
| Stroke | LEC | *TDGF1* | TRUE | 2.024559132 |
| Stroke | macrophage | *CENPQ* | TRUE | 2.989593953 |
| Stroke | macrophage | *CYP2D6* | TRUE | 2.956790449 |
| Stroke | macrophage | *IGDCC4* | TRUE | 2.047070092 |
| Stroke | macrophage | *DLX5* | TRUE | 2.005950829 |
| Stroke | macrophage | *BVES* | TRUE | 2.438633809 |
| Stroke | macrophage | *PCDHB6* | TRUE | 2.126024832 |
| Stroke | macrophage | *MFAP2* | TRUE | 2.109137501 |
| Stroke | macrophage | *EPCAM* | TRUE | 2.62534582 |
| Stroke | macrophage | *TRAP1* | TRUE | 2.235034721 |
| Stroke | macrophage | *FKBP10* | TRUE | 2.548818316 |
| Stroke | macrophage | *LYG1* | TRUE | 2.176287847 |
| Stroke | macrophage | *NBEAL1* | TRUE | 2.288888738 |

| | | | | |
|---|---|---|---|---|
| Stroke | macrophage | *PLA1A* | TRUE | 2.04588254 |
| Stroke | macrophage | *GSTO1* | TRUE | 2.157192047 |
| Stroke | macrophage | *C1orf50* | TRUE | 2.297858957 |
| Stroke | macrophage | *TMEM184A* | TRUE | 2.110759212 |
| Stroke | macrophage | *CHAF1A* | TRUE | 2.195040733 |
| Stroke | macrophage | *FAM57A* | TRUE | 2.366994262 |
| Stroke | macrophage | *ULK4* | TRUE | 2.209281787 |
| Stroke | macrophage | *KCNAB3* | TRUE | 2.306413658 |
| Stroke | macrophage | *RP1L1* | TRUE | 2.023696005 |
| Stroke | macrophage | *FFAR4* | TRUE | 2.700951755 |
| Stroke | macrophage | *ZNF749* | TRUE | 2.762856386 |
| Stroke | macrophage | *GPAT2* | TRUE | 2.287747925 |
| Stroke | macrophage | *TMEM116* | TRUE | 2.341927844 |
| Stroke | macrophage | *AC084018.1* | TRUE | 2.106044374 |
| Stroke | macrophage | *DNLZ* | TRUE | 2.006160776 |
| Stroke | macrophage | *NUTM2D* | TRUE | 2.086276145 |
| Stroke | macrophage | *TDGF1* | TRUE | 3.277241326 |
| Stroke | macrophage | *LINC00942* | TRUE | 2.087975532 |
| Stroke | macrophage | *ARMS2* | TRUE | 2.100336122 |
| Stroke | mast_cell | *IGDCC4* | TRUE | 2.176126754 |
| Stroke | mast_cell | *DLX5* | TRUE | 1.99216303 |
| Stroke | mast_cell | *FKBP10* | TRUE | 2.725217782 |
| Stroke | mast_cell | *ULK4* | TRUE | 2.150862146 |
| Stroke | mast_cell | *TMEM116* | TRUE | 1.995528707 |
| Stroke | mast_cell | *TDGF1* | TRUE | 2.29433021 |
| Stroke | monocyte | *CENPQ* | TRUE | 3.169078912 |
| Stroke | monocyte | *IGDCC4* | TRUE | 1.99096254 |
| Stroke | monocyte | *ILDR1* | TRUE | 2.105999427 |
| Stroke | monocyte | *CLDN7* | TRUE | 2.187004435 |
| Stroke | monocyte | *HLA.DQB2* | TRUE | 2.57931495 |
| Stroke | monocyte | *TDGF1* | TRUE | 2.354665124 |
| Stroke | neutrophil | *ILDR1* | TRUE | 1.984321431 |
| Stroke | neutrophil | *HLA.DQB1* | TRUE | 2.048778893 |
| Stroke | neutrophil | *TDGF1* | TRUE | 2.421380753 |
| Stroke | nk_cell | *CENPQ* | TRUE | 2.456694505 |
| Stroke | nk_cell | *DLX5* | TRUE | 2.009003425 |
| Stroke | nk_cell | *BVES* | TRUE | 2.086089847 |
| Stroke | nk_cell | *BTN2A2* | TRUE | 2.128629958 |
| Stroke | nk_cell | *FKBP10* | TRUE | 2.116152718 |
| Stroke | nk_cell | *ILDR1* | TRUE | 2.079434397 |
| Stroke | nk_cell | *CHAF1A* | TRUE | 1.965442752 |
| Stroke | nk_cell | *FAM57A* | TRUE | 2.302483351 |
| Stroke | nk_cell | *TMEM116* | TRUE | 2.121293412 |
| Stroke | nk_cell | *TDGF1* | TRUE | 2.681727465 |
| Stroke | pericyte | *CENPQ* | TRUE | 1.997981936 |
| Stroke | pericyte | *CYP2D6* | TRUE | 2.960426939 |
| Stroke | pericyte | *IGDCC4* | TRUE | 1.998189945 |
| Stroke | pericyte | *PCDHB6* | TRUE | 2.115777935 |
| Stroke | pericyte | *FKBP10* | TRUE | 2.444341501 |
| Stroke | pericyte | *FFAR4* | TRUE | 2.593083978 |

| | | | | |
|---|---|---|---|---|
| Stroke | pericyte | *COL15A1* | TRUE | 2.469609871 |
| Stroke | pericyte | *NUTM2D* | TRUE | 2.084051255 |
| Stroke | pericyte | *TDGF1* | TRUE | 3.077453391 |
| Stroke | pericyte | *TMEM200B* | TRUE | 2.086188066 |
| Stroke | SMC | *CENPQ* | TRUE | 2.208000864 |
| Stroke | SMC | *CYP2D6* | TRUE | 2.476125279 |
| Stroke | SMC | *FKBP10* | TRUE | 2.463491233 |
| Stroke | SMC | *LYG1* | TRUE | 2.226549435 |
| Stroke | SMC | *PLA1A* | TRUE | 2.111719285 |
| Stroke | SMC | *C1orf50* | TRUE | 2.264934601 |
| Stroke | SMC | *ULK4* | TRUE | 2.176226171 |
| Stroke | SMC | *HLA.DQB1* | TRUE | 2.163332804 |
| Stroke | SMC | *CLDN7* | TRUE | 2.813012916 |
| Stroke | SMC | *YBEY* | TRUE | 2.094117005 |
| Stroke | SMC | *PIGP* | TRUE | 2.01049075 |
| Stroke | SMC | *ZNF749* | TRUE | 2.812407475 |
| Stroke | SMC | *TMEM116* | TRUE | 2.36322714 |
| Stroke | SMC | *NUTM2D* | TRUE | 1.967703905 |
| Stroke | SMC | *TDGF1* | TRUE | 2.770674506 |
| Stroke | t_cell | *CD79B* | TRUE | 2.109913838 |
| Stroke | t_cell | *CENPQ* | TRUE | 2.578620214 |
| Stroke | t_cell | *MFAP2* | TRUE | 1.977363704 |
| Stroke | t_cell | *EPCAM* | TRUE | 2.163846527 |
| Stroke | t_cell | *TRAP1* | TRUE | 2.180167487 |
| Stroke | t_cell | *FKBP10* | TRUE | 2.93074714 |
| Stroke | t_cell | *CHAF1A* | TRUE | 2.190116637 |
| Stroke | t_cell | *FAM57A* | TRUE | 2.029555247 |
| Stroke | t_cell | *ULK4* | TRUE | 2.076064436 |
| Stroke | t_cell | *RP1L1* | TRUE | 2.106079489 |
| Stroke | t_cell | *GPAT2* | TRUE | 3.029271396 |
| Stroke | t_cell | *LRRC10B* | TRUE | 2.401628848 |
| Stroke | t_cell | *DNLZ* | TRUE | 2.126795894 |
| Stroke | t_cell | *LILRA2* | TRUE | 2.153649675 |
| Stroke | t_cell | *TDGF1* | TRUE | 2.16824111 |
| Heart_Failure | adipocyte | *MAP3K9* | TRUE | 1.981314771 |
| Heart_Failure | adipocyte | *SLC7A9* | TRUE | 2.550724617 |
| Heart_Failure | adipocyte | *LAMC2* | TRUE | 2.701560396 |
| Heart_Failure | adipocyte | *TIPIN* | TRUE | 2.031170154 |
| Heart_Failure | adipocyte | *OXCT1* | TRUE | 2.034362405 |
| Heart_Failure | adipocyte | *COL16A1* | TRUE | 3.282580561 |
| Heart_Failure | adipocyte | *BIRC7* | TRUE | 2.759953212 |
| Heart_Failure | adipocyte | *NDUFAF5* | TRUE | 2.126263197 |
| Heart_Failure | adipocyte | *CORO1A* | TRUE | 1.968980698 |
| Heart_Failure | adipocyte | *CTSH* | TRUE | 2.203848311 |
| Heart_Failure | adipocyte | *TIMM44* | TRUE | 2.261165791 |
| Heart_Failure | adipocyte | *CDK2AP1* | TRUE | 3.247369391 |
| Heart_Failure | adipocyte | *PCDHB6* | TRUE | 2.900218304 |
| Heart_Failure | adipocyte | *BNIP1* | TRUE | 2.294912428 |
| Heart_Failure | adipocyte | *HEMK1* | TRUE | 2.735344285 |
| Heart_Failure | adipocyte | *GRIN3B* | TRUE | 2.145949348 |

| | | | | |
|---|---|---|---|---|
| Heart_Failure | adipocyte | *CFHR3* | TRUE | 2.944966137 |
| Heart_Failure | adipocyte | *ALDH8A1* | TRUE | 1.959996484 |
| Heart_Failure | adipocyte | *CTNNAL1* | TRUE | 2.363846508 |
| Heart_Failure | adipocyte | *LDB3* | TRUE | 2.007132051 |
| Heart_Failure | adipocyte | *TBCC* | TRUE | 2.000847912 |
| Heart_Failure | adipocyte | *GRK4* | TRUE | 1.991281074 |
| Heart_Failure | adipocyte | *UCK1* | TRUE | 2.344368753 |
| Heart_Failure | adipocyte | *CHMP1A* | TRUE | 2.568479226 |
| Heart_Failure | adipocyte | *MRPL35* | TRUE | 2.263687088 |
| Heart_Failure | adipocyte | *REEP2* | TRUE | 3.583390573 |
| Heart_Failure | adipocyte | *ADAM19* | TRUE | 2.036492714 |
| Heart_Failure | adipocyte | *POM121C* | TRUE | 2.531975602 |
| Heart_Failure | adipocyte | *LGR5* | TRUE | 2.462444605 |
| Heart_Failure | adipocyte | *FLG* | TRUE | 2.495768755 |
| Heart_Failure | adipocyte | *CISD2* | TRUE | 2.005609206 |
| Heart_Failure | adipocyte | *BHMT* | TRUE | 2.898113139 |
| Heart_Failure | adipocyte | *TSLP* | TRUE | 3.477019667 |
| Heart_Failure | adipocyte | *SLC18B1* | TRUE | 1.967903094 |
| Heart_Failure | adipocyte | *DUSP15* | TRUE | 3.22275141 |
| Heart_Failure | adipocyte | *DPYSL4* | TRUE | 2.187716721 |
| Heart_Failure | adipocyte | *GAL3ST2* | TRUE | 2.903948753 |
| Heart_Failure | adipocyte | *SLFN13* | TRUE | 1.967599034 |
| Heart_Failure | adipocyte | *WHAMM* | TRUE | 1.993811394 |
| Heart_Failure | adipocyte | *FCRL3* | TRUE | 2.481975674 |
| Heart_Failure | adipocyte | *EFHB* | TRUE | 2.779848421 |
| Heart_Failure | adipocyte | *DCLK3* | TRUE | 2.743499044 |
| Heart_Failure | adipocyte | *HDHD2* | TRUE | 2.608844042 |
| Heart_Failure | adipocyte | *GSTM4* | TRUE | 1.960227786 |
| Heart_Failure | adipocyte | *RGS14* | TRUE | 2.69456646 |
| Heart_Failure | adipocyte | *ORMDL3* | TRUE | 5.562462294 |
| Heart_Failure | adipocyte | *DCAKD* | TRUE | 2.000262566 |
| Heart_Failure | adipocyte | *NRROS* | TRUE | 2.603283175 |
| Heart_Failure | adipocyte | *CHST1* | TRUE | 2.479480804 |
| Heart_Failure | adipocyte | *PAAF1* | TRUE | 2.235091347 |
| Heart_Failure | adipocyte | *FBXO39* | TRUE | 2.3575031 |
| Heart_Failure | adipocyte | *NIM1K* | TRUE | 2.33286384 |
| Heart_Failure | adipocyte | *ZNF454* | TRUE | 2.85841248 |
| Heart_Failure | adipocyte | *KLHDC7A* | TRUE | 2.498555058 |
| Heart_Failure | adipocyte | *HLA.DQB1* | TRUE | 3.866291603 |
| Heart_Failure | adipocyte | *FKRP* | TRUE | 2.379261108 |
| Heart_Failure | adipocyte | *TDRKH* | TRUE | 2.450670864 |
| Heart_Failure | adipocyte | *ACOT1* | TRUE | 2.048876453 |
| Heart_Failure | adipocyte | *A3GALT2* | TRUE | 2.225591158 |
| Heart_Failure | adipocyte | *MUC1* | TRUE | 3.68142814 |
| Heart_Failure | adipocyte | *MYBL1* | TRUE | 2.04076106 |
| Heart_Failure | adipocyte | *MCRS1* | TRUE | 3.163280416 |
| Heart_Failure | adipocyte | *SLC22A5* | TRUE | 2.251906415 |
| Heart_Failure | adipocyte | *KLHL14* | TRUE | 2.262738338 |
| Heart_Failure | adipocyte | *RPS26* | TRUE | 2.748888673 |
| Heart_Failure | adipocyte | *MICA* | TRUE | 2.962858574 |

| | | | | |
|---|---|---|---|---|
| Heart_Failure | adipocyte | *FAM153C* | TRUE | 2.35126502 |
| Heart_Failure | adipocyte | *ATF6B* | TRUE | 2.065005048 |
| Heart_Failure | adipocyte | *CPNE1* | TRUE | 2.733121011 |
| Heart_Failure | adipocyte | *TBC1D26* | TRUE | 2.031188866 |
| Heart_Failure | adipocyte | *PROB1* | TRUE | 2.022620854 |
| Heart_Failure | adipocyte | *ZBTB22* | TRUE | 2.187794809 |
| Heart_Failure | adipocyte | *HLA.DQA2* | TRUE | 3.064180109 |
| Heart_Failure | adipocyte | *UPK3B* | TRUE | 2.243119541 |
| Heart_Failure | adipocyte | *TTC4* | TRUE | 2.772428659 |
| Heart_Failure | adipocyte | *MPV17L2* | TRUE | 2.111458007 |
| Heart_Failure | ASPC | *RBM6* | TRUE | 2.236295116 |
| Heart_Failure | ASPC | *MAPK8IP2* | TRUE | 2.005099058 |
| Heart_Failure | ASPC | *MED24* | TRUE | 2.44739495 |
| Heart_Failure | ASPC | *SLC7A9* | TRUE | 2.597639749 |
| Heart_Failure | ASPC | *PHLPP2* | TRUE | 2.567756367 |
| Heart_Failure | ASPC | *RRP12* | TRUE | 2.180787945 |
| Heart_Failure | ASPC | *LAMC2* | TRUE | 2.939809786 |
| Heart_Failure | ASPC | *TAF2* | TRUE | 2.762505488 |
| Heart_Failure | ASPC | *LIMS2* | TRUE | 2.70024634 |
| Heart_Failure | ASPC | *OXCT1* | TRUE | 3.6192932 |
| Heart_Failure | ASPC | *COL16A1* | TRUE | 4.246267782 |
| Heart_Failure | ASPC | *KCNH4* | TRUE | 2.416414229 |
| Heart_Failure | ASPC | *PABPC4* | TRUE | 2.193371589 |
| Heart_Failure | ASPC | *ISM2* | TRUE | 2.075369451 |
| Heart_Failure | ASPC | *PRPF6* | TRUE | 2.279403354 |
| Heart_Failure | ASPC | *SLC17A9* | TRUE | 2.900459339 |
| Heart_Failure | ASPC | *BIRC7* | TRUE | 2.854060636 |
| Heart_Failure | ASPC | *NDUFAF5* | TRUE | 2.472924962 |
| Heart_Failure | ASPC | *CORO1A* | TRUE | 3.044136506 |
| Heart_Failure | ASPC | *DHODH* | TRUE | 2.118669936 |
| Heart_Failure | ASPC | *CTSH* | TRUE | 2.787063467 |
| Heart_Failure | ASPC | *TIMM44* | TRUE | 2.948978676 |
| Heart_Failure | ASPC | *GRWD1* | TRUE | 2.211330408 |
| Heart_Failure | ASPC | *HOXA6* | TRUE | 2.509410544 |
| Heart_Failure | ASPC | *RGP1* | TRUE | 2.083271618 |
| Heart_Failure | ASPC | *PEX12* | TRUE | 2.108721218 |
| Heart_Failure | ASPC | *CTSC* | TRUE | 1.974269317 |
| Heart_Failure | ASPC | *CDK2AP1* | TRUE | 3.108943611 |
| Heart_Failure | ASPC | *CHPT1* | TRUE | 2.397129766 |
| Heart_Failure | ASPC | *PCDHB6* | TRUE | 2.489617634 |
| Heart_Failure | ASPC | *BNIP1* | TRUE | 2.421823793 |
| Heart_Failure | ASPC | *HEMK1* | TRUE | 2.458040591 |
| Heart_Failure | ASPC | *INO80B* | TRUE | 2.427047345 |
| Heart_Failure | ASPC | *CD207* | TRUE | 2.020511147 |
| Heart_Failure | ASPC | *CFHR3* | TRUE | 3.117077055 |
| Heart_Failure | ASPC | *OSCP1* | TRUE | 2.293226501 |
| Heart_Failure | ASPC | *CTNNAL1* | TRUE | 2.650576646 |
| Heart_Failure | ASPC | *ARHGAP9* | TRUE | 3.004130138 |
| Heart_Failure | ASPC | *IKZF4* | TRUE | 1.971272297 |
| Heart_Failure | ASPC | *TBCC* | TRUE | 2.589108559 |

| | | | | |
|---|---|---|---|---|
| Heart_Failure | ASPC | *GRK4* | TRUE | 2.156450541 |
| Heart_Failure | ASPC | *KRI1* | TRUE | 2.110030821 |
| Heart_Failure | ASPC | *TOMM40* | TRUE | 2.828831188 |
| Heart_Failure | ASPC | *UCK1* | TRUE | 2.526646373 |
| Heart_Failure | ASPC | *CHMP1A* | TRUE | 2.502670826 |
| Heart_Failure | ASPC | *MRPL35* | TRUE | 1.984178344 |
| Heart_Failure | ASPC | *REEP2* | TRUE | 4.001663926 |
| Heart_Failure | ASPC | *PCNA* | TRUE | 2.444868352 |
| Heart_Failure | ASPC | *BTBD2* | TRUE | 2.451310071 |
| Heart_Failure | ASPC | *SPIRE1* | TRUE | 2.275591802 |
| Heart_Failure | ASPC | *GOLM1* | TRUE | 2.593089275 |
| Heart_Failure | ASPC | *ADAM19* | TRUE | 2.374179868 |
| Heart_Failure | ASPC | *POM121C* | TRUE | 4.003968692 |
| Heart_Failure | ASPC | *AHI1* | TRUE | 2.17840331 |
| Heart_Failure | ASPC | *ZDHHC4* | TRUE | 2.003657214 |
| Heart_Failure | ASPC | *RMDN3* | TRUE | 2.785107795 |
| Heart_Failure | ASPC | *SLC35A5* | TRUE | 2.022985643 |
| Heart_Failure | ASPC | *GSTCD* | TRUE | 2.483338837 |
| Heart_Failure | ASPC | *LGR5* | TRUE | 3.180237872 |
| Heart_Failure | ASPC | *MFGE8* | TRUE | 2.037215762 |
| Heart_Failure | ASPC | *FN3KRP* | TRUE | 1.97560936 |
| Heart_Failure | ASPC | *DUS3L* | TRUE | 2.050658705 |
| Heart_Failure | ASPC | *NME7* | TRUE | 2.238489592 |
| Heart_Failure | ASPC | *GPA33* | TRUE | 2.251056442 |
| Heart_Failure | ASPC | *ADCY10* | TRUE | 2.310493911 |
| Heart_Failure | ASPC | *FLG* | TRUE | 2.541971739 |
| Heart_Failure | ASPC | *CISD2* | TRUE | 2.667779141 |
| Heart_Failure | ASPC | *TSLP* | TRUE | 4.279650486 |
| Heart_Failure | ASPC | *SLC18B1* | TRUE | 2.393096764 |
| Heart_Failure | ASPC | *EPHA1* | TRUE | 2.139525613 |
| Heart_Failure | ASPC | *HSD17B12* | TRUE | 2.358662182 |
| Heart_Failure | ASPC | *DUSP15* | TRUE | 3.364525767 |
| Heart_Failure | ASPC | *DPYSL4* | TRUE | 2.287873944 |
| Heart_Failure | ASPC | *POC5* | TRUE | 2.139757288 |
| Heart_Failure | ASPC | *GAL3ST2* | TRUE | 2.879916039 |
| Heart_Failure | ASPC | *TTC39B* | TRUE | 2.530128276 |
| Heart_Failure | ASPC | *NDUFAF6* | TRUE | 2.992517786 |
| Heart_Failure | ASPC | *WHAMM* | TRUE | 2.739030641 |
| Heart_Failure | ASPC | *EPB41* | TRUE | 2.202549362 |
| Heart_Failure | ASPC | *TEPP* | TRUE | 2.158623123 |
| Heart_Failure | ASPC | *C21orf33* | TRUE | 2.810993459 |
| Heart_Failure | ASPC | *FCRL3* | TRUE | 2.752433519 |
| Heart_Failure | ASPC | *FUBP1* | TRUE | 2.765536398 |
| Heart_Failure | ASPC | *EFHB* | TRUE | 2.793993102 |
| Heart_Failure | ASPC | *DCLK3* | TRUE | 2.765882076 |
| Heart_Failure | ASPC | *DNALI1* | TRUE | 2.791857839 |
| Heart_Failure | ASPC | *SAP30L* | TRUE | 2.557739333 |
| Heart_Failure | ASPC | *AKR1E2* | TRUE | 2.233510002 |
| Heart_Failure | ASPC | *HSPA12A* | TRUE | 2.118155664 |
| Heart_Failure | ASPC | *CHRFAM7A* | TRUE | 2.055339449 |

| | | | | |
|---|---|---|---|---|
| Heart_Failure | ASPC | *HDHD2* | TRUE | 2.727433696 |
| Heart_Failure | ASPC | *UBXN6* | TRUE | 1.962487923 |
| Heart_Failure | ASPC | *HRASLS5* | TRUE | 2.179285585 |
| Heart_Failure | ASPC | *GSTM4* | TRUE | 2.226213757 |
| Heart_Failure | ASPC | *RGS14* | TRUE | 2.886121879 |
| Heart_Failure | ASPC | *CALML6* | TRUE | 2.340088775 |
| Heart_Failure | ASPC | *RPS9* | TRUE | 2.181740895 |
| Heart_Failure | ASPC | *LRRC34* | TRUE | 2.462765181 |
| Heart_Failure | ASPC | *ORMDL3* | TRUE | 5.932102175 |
| Heart_Failure | ASPC | *DCAKD* | TRUE | 2.390525026 |
| Heart_Failure | ASPC | *PARP15* | TRUE | 1.986090306 |
| Heart_Failure | ASPC | *GOLIM4* | TRUE | 2.580683144 |
| Heart_Failure | ASPC | *NRROS* | TRUE | 3.069431899 |
| Heart_Failure | ASPC | *CHST1* | TRUE | 2.851653912 |
| Heart_Failure | ASPC | *PAAF1* | TRUE | 2.207764166 |
| Heart_Failure | ASPC | *ACSF3* | TRUE | 2.1977151 |
| Heart_Failure | ASPC | *FBXO39* | TRUE | 2.442455086 |
| Heart_Failure | ASPC | *NIM1K* | TRUE | 2.67975606 |
| Heart_Failure | ASPC | *ZNF454* | TRUE | 2.912444461 |
| Heart_Failure | ASPC | *KLHDC7A* | TRUE | 2.6149323 |
| Heart_Failure | ASPC | *HLA.DQB1* | TRUE | 3.202219159 |
| Heart_Failure | ASPC | *HIGD1A* | TRUE | 2.446551315 |
| Heart_Failure | ASPC | *TDRKH* | TRUE | 2.785805396 |
| Heart_Failure | ASPC | *NPLOC4* | TRUE | 2.090469891 |
| Heart_Failure | ASPC | *GLRX5* | TRUE | 2.102524671 |
| Heart_Failure | ASPC | *GRID1* | TRUE | 1.97387918 |
| Heart_Failure | ASPC | *ZNF623* | TRUE | 2.444321008 |
| Heart_Failure | ASPC | *ACOT1* | TRUE | 2.663506708 |
| Heart_Failure | ASPC | *CSF1* | TRUE | 2.221223797 |
| Heart_Failure | ASPC | *A3GALT2* | TRUE | 2.308732071 |
| Heart_Failure | ASPC | *FAM227A* | TRUE | 2.043365551 |
| Heart_Failure | ASPC | *MUC1* | TRUE | 3.317743361 |
| Heart_Failure | ASPC | *MYBL1* | TRUE | 2.529173247 |
| Heart_Failure | ASPC | *MRPS21* | TRUE | 2.549116446 |
| Heart_Failure | ASPC | *ADH1A* | TRUE | 2.471911283 |
| Heart_Failure | ASPC | *MCRS1* | TRUE | 3.497494 |
| Heart_Failure | ASPC | *ZNF626* | TRUE | 1.972898588 |
| Heart_Failure | ASPC | *MSL1* | TRUE | 2.197089479 |
| Heart_Failure | ASPC | *UTS2B* | TRUE | 2.407307059 |
| Heart_Failure | ASPC | *FAM47E* | TRUE | 1.977089896 |
| Heart_Failure | ASPC | *FAM180B* | TRUE | 2.156053748 |
| Heart_Failure | ASPC | *SLC22A5* | TRUE | 2.979126149 |
| Heart_Failure | ASPC | *KLHL14* | TRUE | 2.710865024 |
| Heart_Failure | ASPC | *RPS26* | TRUE | 3.108544454 |
| Heart_Failure | ASPC | *MICB* | TRUE | 2.072523596 |
| Heart_Failure | ASPC | *MICA* | TRUE | 2.168219489 |
| Heart_Failure | ASPC | *FAM153C* | TRUE | 2.979843797 |
| Heart_Failure | ASPC | *CLLU1OS* | TRUE | 2.158081673 |
| Heart_Failure | ASPC | *FAM205A* | TRUE | 2.360260514 |
| Heart_Failure | ASPC | *SERF1B* | TRUE | 2.157372685 |

| | | | | |
|---|---|---|---|---|
| Heart_Failure | ASPC | *ATF6B* | TRUE | 2.153131437 |
| Heart_Failure | ASPC | *CPNE1* | TRUE | 3.502565491 |
| Heart_Failure | ASPC | *TBC1D26* | TRUE | 2.138724279 |
| Heart_Failure | ASPC | *CYB5RL* | TRUE | 2.21999671 |
| Heart_Failure | ASPC | *PROB1* | TRUE | 2.086292763 |
| Heart_Failure | ASPC | *ZNF736* | TRUE | 2.198990538 |
| Heart_Failure | ASPC | *ZBTB22* | TRUE | 2.191884393 |
| Heart_Failure | ASPC | *HLA.DQA2* | TRUE | 2.882010287 |
| Heart_Failure | ASPC | *EIF6* | TRUE | 2.109781547 |
| Heart_Failure | ASPC | *UPK3B* | TRUE | 2.245671092 |
| Heart_Failure | ASPC | *TTC4* | TRUE | 2.955862477 |
| Heart_Failure | ASPC | *ARMS2* | TRUE | 2.053528263 |
| Heart_Failure | ASPC | *MPV17L2* | TRUE | 2.176418079 |
| Heart_Failure | ASPC | *GTF2H5* | TRUE | 2.06019633 |
| Heart_Failure | b_cell | *PDLIM4* | TRUE | 2.986175029 |
| Heart_Failure | b_cell | *REEP2* | TRUE | 2.275937724 |
| Heart_Failure | b_cell | *HLA.DQB1* | TRUE | 2.060475793 |
| Heart_Failure | b_cell | *RPS26* | TRUE | 2.34099437 |
| Heart_Failure | b_cell | *FAM205A* | TRUE | 2.549119659 |
| Heart_Failure | b_cell | *UPK3B* | TRUE | 2.193125993 |
| Heart_Failure | dendritic_cell | *SEMA3F* | TRUE | 3.010085151 |
| Heart_Failure | dendritic_cell | *DBNDD1* | TRUE | 2.175274426 |
| Heart_Failure | dendritic_cell | *BIRC7* | TRUE | 2.636806843 |
| Heart_Failure | dendritic_cell | *CDK2AP1* | TRUE | 2.350337051 |
| Heart_Failure | dendritic_cell | *IL4* | TRUE | 2.286034605 |
| Heart_Failure | dendritic_cell | *CFHR3* | TRUE | 2.138173493 |
| Heart_Failure | dendritic_cell | *REEP2* | TRUE | 3.301403548 |
| Heart_Failure | dendritic_cell | *CYP2C9* | TRUE | 2.215453219 |
| Heart_Failure | dendritic_cell | *FLG* | TRUE | 2.447901685 |
| Heart_Failure | dendritic_cell | *DUSP15* | TRUE | 2.007189457 |
| Heart_Failure | dendritic_cell | *KLHDC9* | TRUE | 1.974839467 |
| Heart_Failure | dendritic_cell | *DCLK3* | TRUE | 2.333656854 |
| Heart_Failure | dendritic_cell | *ORMDL3* | TRUE | 5.587455059 |
| Heart_Failure | dendritic_cell | *DCAKD* | TRUE | 2.582325493 |
| Heart_Failure | dendritic_cell | *CPNE7* | TRUE | 1.973130685 |
| Heart_Failure | dendritic_cell | *KLHDC7A* | TRUE | 2.508857343 |
| Heart_Failure | dendritic_cell | *SH3BGR* | TRUE | 2.047083943 |
| Heart_Failure | dendritic_cell | *RPS26* | TRUE | 3.367973886 |
| Heart_Failure | dendritic_cell | *MICB* | TRUE | 3.268231994 |
| Heart_Failure | dendritic_cell | *FAM153C* | TRUE | 2.313058472 |
| Heart_Failure | dendritic_cell | *FAM205A* | TRUE | 2.886753831 |
| Heart_Failure | dendritic_cell | *CPNE1* | TRUE | 2.303635231 |
| Heart_Failure | dendritic_cell | *ZBTB22* | TRUE | 2.621439862 |
| Heart_Failure | dendritic_cell | *UPK3B* | TRUE | 2.448546682 |
| Heart_Failure | endothelial | *SLC7A9* | TRUE | 2.021259839 |
| Heart_Failure | endothelial | *CDK2AP1* | TRUE | 2.765000599 |
| Heart_Failure | endothelial | *PCDHB6* | TRUE | 2.538659714 |
| Heart_Failure | endothelial | *CFHR3* | TRUE | 2.244421932 |
| Heart_Failure | endothelial | *REEP2* | TRUE | 3.459851261 |
| Heart_Failure | endothelial | *BHMT* | TRUE | 2.237699873 |

| | | | | |
|---|---|---|---|---|
| Heart_Failure | endothelial | *TSLP* | TRUE | 3.124376247 |
| Heart_Failure | endothelial | *GAL3ST2* | TRUE | 2.686389526 |
| Heart_Failure | endothelial | *ORMDL3* | TRUE | 2.627435758 |
| Heart_Failure | endothelial | *CPNE7* | TRUE | 1.961821011 |
| Heart_Failure | endothelial | *TRIM73* | TRUE | 2.192504606 |
| Heart_Failure | endothelial | *HLA.DQB1* | TRUE | 2.918494983 |
| Heart_Failure | endothelial | *SH3BGR* | TRUE | 2.068769137 |
| Heart_Failure | endothelial | *MUC1* | TRUE | 2.152675959 |
| Heart_Failure | endothelial | *SLC22A5* | TRUE | 2.120727035 |
| Heart_Failure | endothelial | *RPS26* | TRUE | 3.202063403 |
| Heart_Failure | endothelial | *FAM205A* | TRUE | 2.28380071 |
| Heart_Failure | endothelial | *HLA.DQA2* | TRUE | 2.860994578 |
| Heart_Failure | LEC | *KCNH4* | TRUE | 2.156905613 |
| Heart_Failure | LEC | *CDK2AP1* | TRUE | 1.970493012 |
| Heart_Failure | LEC | *FLG* | TRUE | 2.479572537 |
| Heart_Failure | LEC | *BHMT* | TRUE | 2.344148772 |
| Heart_Failure | LEC | *ROM1* | TRUE | 2.606208765 |
| Heart_Failure | LEC | *DUSP15* | TRUE | 2.051829 |
| Heart_Failure | LEC | *HLA.DQB1* | TRUE | 3.888114689 |
| Heart_Failure | LEC | *RPS26* | TRUE | 3.365954627 |
| Heart_Failure | LEC | *MICA* | TRUE | 2.603708998 |
| Heart_Failure | LEC | *FAM205A* | TRUE | 2.38875791 |
| Heart_Failure | LEC | *CPNE1* | TRUE | 2.046257391 |
| Heart_Failure | LEC | *HLA.DQA2* | TRUE | 3.381435866 |
| Heart_Failure | macrophage | *SLC7A9* | TRUE | 2.494481847 |
| Heart_Failure | macrophage | *LAMC2* | TRUE | 2.454837365 |
| Heart_Failure | macrophage | *COL16A1* | TRUE | 2.31080587 |
| Heart_Failure | macrophage | *BIRC7* | TRUE | 2.7752976 |
| Heart_Failure | macrophage | *NDUFAF5* | TRUE | 2.006716643 |
| Heart_Failure | macrophage | *CORO1A* | TRUE | 2.018781463 |
| Heart_Failure | macrophage | *CTSH* | TRUE | 1.998098922 |
| Heart_Failure | macrophage | *CDK2AP1* | TRUE | 3.189940243 |
| Heart_Failure | macrophage | *PCDHB6* | TRUE | 2.339548336 |
| Heart_Failure | macrophage | *BNIP1* | TRUE | 2.243671778 |
| Heart_Failure | macrophage | *HEMK1* | TRUE | 2.635601526 |
| Heart_Failure | macrophage | *GRIN3B* | TRUE | 2.052824232 |
| Heart_Failure | macrophage | *CFHR3* | TRUE | 2.785850513 |
| Heart_Failure | macrophage | *ALDH8A1* | TRUE | 2.050890699 |
| Heart_Failure | macrophage | *UCK1* | TRUE | 2.216088503 |
| Heart_Failure | macrophage | *CHMP1A* | TRUE | 2.005961632 |
| Heart_Failure | macrophage | *MRPL35* | TRUE | 2.325496128 |
| Heart_Failure | macrophage | *REEP2* | TRUE | 3.756347658 |
| Heart_Failure | macrophage | *FLG* | TRUE | 2.819276828 |
| Heart_Failure | macrophage | *BHMT* | TRUE | 2.758642784 |
| Heart_Failure | macrophage | *TSLP* | TRUE | 4.29321349 |
| Heart_Failure | macrophage | *DUSP15* | TRUE | 3.064357801 |
| Heart_Failure | macrophage | *DPYSL4* | TRUE | 2.335503783 |
| Heart_Failure | macrophage | *GAL3ST2* | TRUE | 2.956099579 |
| Heart_Failure | macrophage | *C21orf33* | TRUE | 2.334087619 |
| Heart_Failure | macrophage | *FCRL3* | TRUE | 2.344691136 |

| | | | | |
|---|---|---|---|---|
| Heart_Failure | macrophage | *EFHB* | TRUE | 2.669601215 |
| Heart_Failure | macrophage | *DCLK3* | TRUE | 2.046773515 |
| Heart_Failure | macrophage | *CHRFAM7A* | TRUE | 1.992419909 |
| Heart_Failure | macrophage | *HDHD2* | TRUE | 2.522176763 |
| Heart_Failure | macrophage | *HRASLS5* | TRUE | 2.04316415 |
| Heart_Failure | macrophage | *RGS14* | TRUE | 2.411414162 |
| Heart_Failure | macrophage | *ORMDL3* | TRUE | 5.752058705 |
| Heart_Failure | macrophage | *DCAKD* | TRUE | 2.186849931 |
| Heart_Failure | macrophage | *CHST1* | TRUE | 2.201910609 |
| Heart_Failure | macrophage | *PAAF1* | TRUE | 2.277544257 |
| Heart_Failure | macrophage | *FBXO39* | TRUE | 2.259175909 |
| Heart_Failure | macrophage | *NIM1K* | TRUE | 2.010776843 |
| Heart_Failure | macrophage | *KLHDC7A* | TRUE | 2.492558585 |
| Heart_Failure | macrophage | *HLA.DQB1* | TRUE | 3.691619788 |
| Heart_Failure | macrophage | *FKRP* | TRUE | 2.366514329 |
| Heart_Failure | macrophage | *ACOT1* | TRUE | 2.273378709 |
| Heart_Failure | macrophage | *A3GALT2* | TRUE | 2.142675487 |
| Heart_Failure | macrophage | *MUC1* | TRUE | 3.295025883 |
| Heart_Failure | macrophage | *MCRS1* | TRUE | 2.760447928 |
| Heart_Failure | macrophage | *SLC22A5* | TRUE | 2.710539987 |
| Heart_Failure | macrophage | *RPS26* | TRUE | 2.835549651 |
| Heart_Failure | macrophage | *MICB* | TRUE | 2.183537102 |
| Heart_Failure | macrophage | *MICA* | TRUE | 2.308540824 |
| Heart_Failure | macrophage | *FAM205A* | TRUE | 2.716728954 |
| Heart_Failure | macrophage | *CPNE1* | TRUE | 2.890533834 |
| Heart_Failure | macrophage | *ZBTB22* | TRUE | 2.105449712 |
| Heart_Failure | macrophage | *HLA.DQA2* | TRUE | 2.747223844 |
| Heart_Failure | macrophage | *UPK3B* | TRUE | 2.218091169 |
| Heart_Failure | macrophage | *TTC4* | TRUE | 2.665898152 |
| Heart_Failure | macrophage | *ARMS2* | TRUE | 2.074308474 |
| Heart_Failure | mast_cell | *SLC7A9* | TRUE | 2.219555544 |
| Heart_Failure | mast_cell | *LAMC2* | TRUE | 2.376630421 |
| Heart_Failure | mast_cell | *REEP2* | TRUE | 2.420062785 |
| Heart_Failure | mast_cell | *FLG* | TRUE | 2.683797252 |
| Heart_Failure | mast_cell | *GAL3ST2* | TRUE | 1.989659747 |
| Heart_Failure | mast_cell | *ORMDL3* | TRUE | 5.589273831 |
| Heart_Failure | mast_cell | *KLHDC7A* | TRUE | 2.116679677 |
| Heart_Failure | mast_cell | *HLA.DQB1* | TRUE | 2.177428596 |
| Heart_Failure | mast_cell | *FKRP* | TRUE | 2.255880438 |
| Heart_Failure | mast_cell | *MUC1* | TRUE | 3.02839433 |
| Heart_Failure | mast_cell | *SLC22A5* | TRUE | 3.012986483 |
| Heart_Failure | mast_cell | *RPS26* | TRUE | 3.188573592 |
| Heart_Failure | mast_cell | *MICB* | TRUE | 2.324885624 |
| Heart_Failure | mast_cell | *FAM205A* | TRUE | 2.735828357 |
| Heart_Failure | mast_cell | *ZBTB22* | TRUE | 2.334915743 |
| Heart_Failure | mast_cell | *HLA.DQA2* | TRUE | 3.133462918 |
| Heart_Failure | mast_cell | *TNFSF12* | TRUE | 1.99938631 |
| Heart_Failure | monocyte | *BIRC7* | TRUE | 2.248910996 |
| Heart_Failure | monocyte | *DHODH* | TRUE | 2.076510716 |
| Heart_Failure | monocyte | *UCK1* | TRUE | 1.989988026 |

| | | | | |
|---|---|---|---|---|
| Heart_Failure | monocyte | *DUSP15* | TRUE | 2.572324922 |
| Heart_Failure | monocyte | *RPS26* | TRUE | 3.510164098 |
| Heart_Failure | monocyte | *MICA* | TRUE | 2.803020529 |
| Heart_Failure | monocyte | *FAM205A* | TRUE | 2.293260845 |
| Heart_Failure | monocyte | *ATF6B* | TRUE | 2.982123602 |
| Heart_Failure | monocyte | *HLA.DQA2* | TRUE | 1.999220437 |
| Heart_Failure | neutrophil | *NRROS* | TRUE | 2.528332176 |
| Heart_Failure | neutrophil | *HLA.DQB1* | TRUE | 8.350750182 |
| Heart_Failure | neutrophil | *ACOT1* | TRUE | 1.968139932 |
| Heart_Failure | neutrophil | *FAM205A* | TRUE | 2.607496799 |
| Heart_Failure | nk_cell | *MRPL35* | TRUE | 2.177644571 |
| Heart_Failure | nk_cell | *REEP2* | TRUE | 2.098362051 |
| Heart_Failure | nk_cell | *RNF123* | TRUE | 2.554327282 |
| Heart_Failure | nk_cell | *HDHD2* | TRUE | 2.482721519 |
| Heart_Failure | nk_cell | *ORMDL3* | TRUE | 3.812469253 |
| Heart_Failure | nk_cell | *HLA.DQB1* | TRUE | 3.217759652 |
| Heart_Failure | nk_cell | *BMP8A* | TRUE | 2.117855194 |
| Heart_Failure | nk_cell | *SLC22A5* | TRUE | 3.625326882 |
| Heart_Failure | nk_cell | *RPS26* | TRUE | 2.623848851 |
| Heart_Failure | nk_cell | *HLA.DQA2* | TRUE | 2.840685623 |
| Heart_Failure | pericyte | *DBNDD1* | TRUE | 1.979540965 |
| Heart_Failure | pericyte | *KCNH4* | TRUE | 2.102500056 |
| Heart_Failure | pericyte | *CDK2AP1* | TRUE | 2.611727674 |
| Heart_Failure | pericyte | *PCDHB6* | TRUE | 2.502092151 |
| Heart_Failure | pericyte | *BNIP1* | TRUE | 2.329373963 |
| Heart_Failure | pericyte | *HEMK1* | TRUE | 2.681153047 |
| Heart_Failure | pericyte | *UCK1* | TRUE | 2.108857379 |
| Heart_Failure | pericyte | *REEP2* | TRUE | 2.965010338 |
| Heart_Failure | pericyte | *ADAM19* | TRUE | 2.04656636 |
| Heart_Failure | pericyte | *NMT1* | TRUE | 2.556369797 |
| Heart_Failure | pericyte | *ROM1* | TRUE | 2.595303328 |
| Heart_Failure | pericyte | *DPYSL4* | TRUE | 2.25445109 |
| Heart_Failure | pericyte | *TTC39B* | TRUE | 1.965987579 |
| Heart_Failure | pericyte | *NIT1* | TRUE | 2.034776849 |
| Heart_Failure | pericyte | *EFHB* | TRUE | 2.3247683 |
| Heart_Failure | pericyte | *RNF123* | TRUE | 3.227493097 |
| Heart_Failure | pericyte | *SAP30L* | TRUE | 2.09084574 |
| Heart_Failure | pericyte | *HDHD2* | TRUE | 2.106080113 |
| Heart_Failure | pericyte | *RGS14* | TRUE | 2.122915998 |
| Heart_Failure | pericyte | *ORMDL3* | TRUE | 2.967658992 |
| Heart_Failure | pericyte | *KLHDC7A* | TRUE | 2.30156134 |
| Heart_Failure | pericyte | *HLA.DQB1* | TRUE | 2.698171415 |
| Heart_Failure | pericyte | *FKRP* | TRUE | 2.296610733 |
| Heart_Failure | pericyte | *A3GALT2* | TRUE | 2.124258044 |
| Heart_Failure | pericyte | *SLC22A5* | TRUE | 5.097878071 |
| Heart_Failure | pericyte | *RPS26* | TRUE | 3.487318421 |
| Heart_Failure | pericyte | *MICB* | TRUE | 2.460621653 |
| Heart_Failure | pericyte | *FAM205A* | TRUE | 2.84339239 |
| Heart_Failure | pericyte | *CPNE1* | TRUE | 1.999971671 |
| Heart_Failure | pericyte | *HLA.DQA2* | TRUE | 3.273883476 |

| | | | | |
|---|---|---|---|---|
| Heart_Failure | pericyte | *TMEM200B* | TRUE | 2.380135909 |
| Heart_Failure | SMC | *COL16A1* | TRUE | 2.840955693 |
| Heart_Failure | SMC | *BIRC7* | TRUE | 2.889716705 |
| Heart_Failure | SMC | *CTSH* | TRUE | 2.057044922 |
| Heart_Failure | SMC | *CDK2AP1* | TRUE | 2.615560462 |
| Heart_Failure | SMC | *HEMK1* | TRUE | 2.135927822 |
| Heart_Failure | SMC | *GRIN3B* | TRUE | 2.03292185 |
| Heart_Failure | SMC | *CFHR3* | TRUE | 2.748028331 |
| Heart_Failure | SMC | *ALDH8A1* | TRUE | 2.108529053 |
| Heart_Failure | SMC | *CTNNAL1* | TRUE | 2.233562298 |
| Heart_Failure | SMC | *UCK1* | TRUE | 2.183917206 |
| Heart_Failure | SMC | *MRPL35* | TRUE | 2.118973 |
| Heart_Failure | SMC | *REEP2* | TRUE | 3.274675517 |
| Heart_Failure | SMC | *ADAM19* | TRUE | 2.070352649 |
| Heart_Failure | SMC | *ROM1* | TRUE | 2.05615311 |
| Heart_Failure | SMC | *DUSP15* | TRUE | 2.214233617 |
| Heart_Failure | SMC | *DPYSL4* | TRUE | 2.120337661 |
| Heart_Failure | SMC | *GAL3ST2* | TRUE | 2.313300602 |
| Heart_Failure | SMC | *EFHB* | TRUE | 2.543868763 |
| Heart_Failure | SMC | *DCLK3* | TRUE | 2.593922823 |
| Heart_Failure | SMC | *CHRFAM7A* | TRUE | 1.966790979 |
| Heart_Failure | SMC | *ORMDL3* | TRUE | 5.624567621 |
| Heart_Failure | SMC | *DCAKD* | TRUE | 2.399910861 |
| Heart_Failure | SMC | *CHST1* | TRUE | 2.082750794 |
| Heart_Failure | SMC | *PAAF1* | TRUE | 2.108200413 |
| Heart_Failure | SMC | *FBXO39* | TRUE | 2.253431609 |
| Heart_Failure | SMC | *ZNF454* | TRUE | 2.510937706 |
| Heart_Failure | SMC | *KLHDC7A* | TRUE | 2.324244361 |
| Heart_Failure | SMC | *HLA.DQB1* | TRUE | 3.83211278 |
| Heart_Failure | SMC | *FKRP* | TRUE | 2.123975736 |
| Heart_Failure | SMC | *TDRKH* | TRUE | 2.303973209 |
| Heart_Failure | SMC | *A3GALT2* | TRUE | 2.108127763 |
| Heart_Failure | SMC | *MUC1* | TRUE | 3.183947436 |
| Heart_Failure | SMC | *MCRS1* | TRUE | 2.013181621 |
| Heart_Failure | SMC | *SLC22A5* | TRUE | 3.551192329 |
| Heart_Failure | SMC | *RPS26* | TRUE | 2.868309124 |
| Heart_Failure | SMC | *MICA* | TRUE | 2.489056297 |
| Heart_Failure | SMC | *FAM205A* | TRUE | 2.923976629 |
| Heart_Failure | SMC | *CPNE1* | TRUE | 2.660625619 |
| Heart_Failure | SMC | *HLA.DQA2* | TRUE | 2.441840001 |
| Heart_Failure | SMC | *TTC4* | TRUE | 2.438696157 |
| Heart_Failure | t_cell | *SLC7A9* | TRUE | 2.21887474 |
| Heart_Failure | t_cell | *UCK1* | TRUE | 1.976584542 |
| Heart_Failure | t_cell | *MRPL35* | TRUE | 2.099063555 |
| Heart_Failure | t_cell | *REEP2* | TRUE | 3.029638434 |
| Heart_Failure | t_cell | *DPYSL4* | TRUE | 2.342065051 |
| Heart_Failure | t_cell | *ORMDL3* | TRUE | 3.768566614 |
| Heart_Failure | t_cell | *CHST1* | TRUE | 2.712045958 |
| Heart_Failure | t_cell | *KLHDC7A* | TRUE | 1.96543216 |
| Heart_Failure | t_cell | *HLA.DQB1* | TRUE | 3.951443346 |

| | | | | |
|---|---|---|---|---|
| Heart_Failure | t_cell | *FKRP* | TRUE | 2.004536115 |
| Heart_Failure | t_cell | *MUC1* | TRUE | 2.551222509 |
| Heart_Failure | t_cell | *SLC22A5* | TRUE | 2.927958151 |
| Heart_Failure | t_cell | *RPS26* | TRUE | 3.219138437 |
| Heart_Failure | t_cell | *MICB* | TRUE | 2.223862981 |
| Heart_Failure | t_cell | *FAM205A* | TRUE | 2.735326238 |
| Heart_Failure | t_cell | *ATF6B* | TRUE | 2.310607017 |
| Heart_Failure | t_cell | *CPNE1* | TRUE | 2.164168444 |
| Heart_Failure | t_cell | *ZBTB22* | TRUE | 2.355309645 |
| CAD_admission_freq | adipocyte | *F7* | TRUE | 2.90847062 |
| CAD_admission_freq | adipocyte | *LAMC2* | TRUE | 2.01741734 |
| CAD_admission_freq | adipocyte | *CLTCL1* | TRUE | 2.42525931 |
| CAD_admission_freq | adipocyte | *RPGRIP1* | TRUE | 2.28268388 |
| CAD_admission_freq | adipocyte | *IGDCC4* | TRUE | 2.0306909 |
| CAD_admission_freq | adipocyte | *CCDC61* | TRUE | 2.22724831 |
| CAD_admission_freq | adipocyte | *CAPS* | TRUE | 2.19675446 |
| CAD_admission_freq | adipocyte | *CYP27B1* | TRUE | 2.03384363 |
| CAD_admission_freq | adipocyte | *TOX2* | TRUE | 2.55184934 |
| CAD_admission_freq | adipocyte | *NRM* | TRUE | 2.36444441 |
| CAD_admission_freq | adipocyte | *KHK* | TRUE | 2.38746746 |
| CAD_admission_freq | adipocyte | *DMTN* | TRUE | 2.26400921 |
| CAD_admission_freq | adipocyte | *GSTM4* | TRUE | 1.97650472 |
| CAD_admission_freq | adipocyte | *RAB24* | TRUE | 1.99666243 |
| CAD_admission_freq | adipocyte | *HOXD4* | TRUE | 2.13716925 |
| CAD_admission_freq | adipocyte | *CSDC2* | TRUE | 1.98479415 |
| CAD_admission_freq | adipocyte | *SLC22A1* | TRUE | 3.12157785 |
| CAD_admission_freq | adipocyte | *PAAF1* | TRUE | 3.12881035 |
| CAD_admission_freq | adipocyte | *GOLGA8Q* | TRUE | 2.67057785 |
| CAD_admission_freq | adipocyte | *ACTL10* | TRUE | 2.28384808 |
| CAD_admission_freq | adipocyte | *PLA2G6* | TRUE | 2.29691344 |
| CAD_admission_freq | adipocyte | *SHISA4* | TRUE | 2.01908593 |
| CAD_admission_freq | adipocyte | *PSORS1C1* | TRUE | 2.0652492 |
| CAD_admission_freq | adipocyte | *AC084018.1* | TRUE | 2.05943039 |
| CAD_admission_freq | adipocyte | *GS1.124K5.4* | TRUE | 2.26677472 |
| CAD_admission_freq | adipocyte | *ZNF286B* | TRUE | 2.20606858 |
| CAD_admission_freq | adipocyte | *HMBS* | TRUE | 2.11348006 |
| CAD_admission_freq | adipocyte | *SRXN1* | TRUE | 2.46994373 |
| CAD_admission_freq | ASPC | *TYMP* | TRUE | 2.1130286 |
| CAD_admission_freq | ASPC | *MED17* | TRUE | 2.10998282 |
| CAD_admission_freq | ASPC | *F7* | TRUE | 2.40314068 |
| CAD_admission_freq | ASPC | *LAMC2* | TRUE | 2.04879382 |
| CAD_admission_freq | ASPC | *CLTCL1* | TRUE | 2.56584873 |
| CAD_admission_freq | ASPC | *TPD52* | TRUE | 2.27580725 |
| CAD_admission_freq | ASPC | *STX7* | TRUE | 2.05525908 |
| CAD_admission_freq | ASPC | *GEMIN5* | TRUE | 2.10560566 |
| CAD_admission_freq | ASPC | *TXNDC16* | TRUE | 2.29400936 |
| CAD_admission_freq | ASPC | *RPGRIP1* | TRUE | 2.44847445 |
| CAD_admission_freq | ASPC | *TMED8* | TRUE | 2.02513224 |
| CAD_admission_freq | ASPC | *MON1B* | TRUE | 2.592485 |
| CAD_admission_freq | ASPC | *TRIP4* | TRUE | 2.20697284 |

| | | | | |
|---|---|---|---|---|
| CAD_admission_freq | ASPC | *CCDC61* | TRUE | 2.6146719 |
| CAD_admission_freq | ASPC | *CAPS* | TRUE | 2.54239619 |
| CAD_admission_freq | ASPC | *ELAVL2* | TRUE | 2.00238693 |
| CAD_admission_freq | ASPC | *AKAP10* | TRUE | 2.10382961 |
| CAD_admission_freq | ASPC | *CYP27B1* | TRUE | 2.23112368 |
| CAD_admission_freq | ASPC | *CHPT1* | TRUE | 2.03963585 |
| CAD_admission_freq | ASPC | *EFCC1* | TRUE | 2.33570152 |
| CAD_admission_freq | ASPC | *SDC1* | TRUE | 1.98856316 |
| CAD_admission_freq | ASPC | *DNAJC6* | TRUE | 2.05772062 |
| CAD_admission_freq | ASPC | *MED28* | TRUE | 2.24058252 |
| CAD_admission_freq | ASPC | *C4BPB* | TRUE | 2.09242299 |
| CAD_admission_freq | ASPC | *TOX2* | TRUE | 2.60676534 |
| CAD_admission_freq | ASPC | *CLPP* | TRUE | 1.960804 |
| CAD_admission_freq | ASPC | *CCL4* | TRUE | 2.21214409 |
| CAD_admission_freq | ASPC | *MRPL34* | TRUE | 2.01157261 |
| CAD_admission_freq | ASPC | *DHX34* | TRUE | 1.99326027 |
| CAD_admission_freq | ASPC | *TOR1B* | TRUE | 1.99691793 |
| CAD_admission_freq | ASPC | *NRM* | TRUE | 2.36575854 |
| CAD_admission_freq | ASPC | *KHK* | TRUE | 2.3672599 |
| CAD_admission_freq | ASPC | *LHCGR* | TRUE | 2.1527545 |
| CAD_admission_freq | ASPC | *ABCG5* | TRUE | 2.08189511 |
| CAD_admission_freq | ASPC | *ARL3* | TRUE | 2.03562157 |
| CAD_admission_freq | ASPC | *PAPSS1* | TRUE | 2.33912158 |
| CAD_admission_freq | ASPC | *PPFIA2* | TRUE | 2.1782352 |
| CAD_admission_freq | ASPC | *LYSMD2* | TRUE | 2.29094351 |
| CAD_admission_freq | ASPC | *KIFC3* | TRUE | 2.3017993 |
| CAD_admission_freq | ASPC | *MAP3K6* | TRUE | 2.36217672 |
| CAD_admission_freq | ASPC | *SCCPDH* | TRUE | 2.69632409 |
| CAD_admission_freq | ASPC | *SNAP47* | TRUE | 2.20899469 |
| CAD_admission_freq | ASPC | *ESPNL* | TRUE | 2.60472891 |
| CAD_admission_freq | ASPC | *TSLP* | TRUE | 2.02324337 |
| CAD_admission_freq | ASPC | *ATP6V0D2* | TRUE | 2.10439258 |
| CAD_admission_freq | ASPC | *VIPAS39* | TRUE | 2.08735755 |
| CAD_admission_freq | ASPC | *FAM213B* | TRUE | 1.98624333 |
| CAD_admission_freq | ASPC | *DMTN* | TRUE | 3.0004541 |
| CAD_admission_freq | ASPC | *CCDC127* | TRUE | 1.96594995 |
| CAD_admission_freq | ASPC | *FAM227B* | TRUE | 2.02818923 |
| CAD_admission_freq | ASPC | *GPRC5B* | TRUE | 2.08967627 |
| CAD_admission_freq | ASPC | *CX3CR1* | TRUE | 1.96288317 |
| CAD_admission_freq | ASPC | *GSTM4* | TRUE | 1.97091518 |
| CAD_admission_freq | ASPC | *RAB24* | TRUE | 2.36072078 |
| CAD_admission_freq | ASPC | *HOXD4* | TRUE | 2.29847007 |
| CAD_admission_freq | ASPC | *GRIK1* | TRUE | 1.97561259 |
| CAD_admission_freq | ASPC | *CSDC2* | TRUE | 1.99787743 |
| CAD_admission_freq | ASPC | *DMRT2* | TRUE | 2.31166917 |
| CAD_admission_freq | ASPC | *SLC22A1* | TRUE | 3.83873534 |
| CAD_admission_freq | ASPC | *ZNF25* | TRUE | 2.02887206 |
| CAD_admission_freq | ASPC | *PAAF1* | TRUE | 2.92883611 |
| CAD_admission_freq | ASPC | *TMEM80* | TRUE | 2.07636027 |
| CAD_admission_freq | ASPC | *CD163L1* | TRUE | 1.98557466 |

| | | | | |
|---|---|---|---|---|
| CAD_admission_freq | ASPC | *GOLGA8Q* | TRUE | 2.71655971 |
| CAD_admission_freq | ASPC | *TMPRSS9* | TRUE | 2.53995487 |
| CAD_admission_freq | ASPC | *HYI* | TRUE | 2.08947594 |
| CAD_admission_freq | ASPC | *CLEC4G* | TRUE | 2.41245845 |
| CAD_admission_freq | ASPC | *ACTL10* | TRUE | 2.33725374 |
| CAD_admission_freq | ASPC | *PLA2G6* | TRUE | 3.00825682 |
| CAD_admission_freq | ASPC | *RASA3* | TRUE | 2.18733612 |
| CAD_admission_freq | ASPC | *GSAP* | TRUE | 2.27536363 |
| CAD_admission_freq | ASPC | *ISG15* | TRUE | 1.97305194 |
| CAD_admission_freq | ASPC | *SEMA4A* | TRUE | 2.27515916 |
| CAD_admission_freq | ASPC | *GM2A* | TRUE | 1.97158516 |
| CAD_admission_freq | ASPC | *SHISA4* | TRUE | 2.05894671 |
| CAD_admission_freq | ASPC | *PSORS1C1* | TRUE | 2.08082945 |
| CAD_admission_freq | ASPC | *PCDHA5* | TRUE | 2.38634751 |
| CAD_admission_freq | ASPC | *GIMAP1* | TRUE | 2.09420083 |
| CAD_admission_freq | ASPC | *GS1.124K5.4* | TRUE | 1.96211214 |
| CAD_admission_freq | ASPC | *ZNF286B* | TRUE | 2.39016173 |
| CAD_admission_freq | ASPC | *HMBS* | TRUE | 2.2639158 |
| CAD_admission_freq | ASPC | *ZNF578* | TRUE | 1.98149039 |
| CAD_admission_freq | ASPC | *HOXB7* | TRUE | 1.97725432 |
| CAD_admission_freq | ASPC | *SRXN1* | TRUE | 2.73738404 |
| CAD_admission_freq | b_cell | *F7* | TRUE | 2.63818105 |
| CAD_admission_freq | b_cell | *MICB* | TRUE | 2.66677283 |
| CAD_admission_freq | b_cell | *SRXN1* | TRUE | 2.22217696 |
| CAD_admission_freq | dendritic_cell | *SNX15* | TRUE | 1.99434912 |
| CAD_admission_freq | dendritic_cell | *KHK* | TRUE | 2.15827213 |
| CAD_admission_freq | dendritic_cell | *NUDT13* | TRUE | 2.10907034 |
| CAD_admission_freq | dendritic_cell | *PAAF1* | TRUE | 2.42736327 |
| CAD_admission_freq | dendritic_cell | *GOLGA8Q* | TRUE | 2.32177767 |
| CAD_admission_freq | dendritic_cell | *SMDT1* | TRUE | 2.15880403 |
| CAD_admission_freq | dendritic_cell | *FAM228A* | TRUE | 1.997153 |
| CAD_admission_freq | dendritic_cell | *MICB* | TRUE | 2.34241892 |
| CAD_admission_freq | dendritic_cell | *PCDHA5* | TRUE | 2.03585185 |
| CAD_admission_freq | dendritic_cell | *GS1.124K5.4* | TRUE | 2.13329679 |
| CAD_admission_freq | dendritic_cell | *SRXN1* | TRUE | 2.7141939 |
| CAD_admission_freq | endothelial | *F7* | TRUE | 2.33139437 |
| CAD_admission_freq | endothelial | *LAMC2* | TRUE | 2.04412644 |
| CAD_admission_freq | endothelial | *KHK* | TRUE | 2.27095979 |
| CAD_admission_freq | endothelial | *PAAF1* | TRUE | 2.91410124 |
| CAD_admission_freq | endothelial | *GOLGA8Q* | TRUE | 2.21989649 |
| CAD_admission_freq | endothelial | *MICB* | TRUE | 2.70358901 |
| CAD_admission_freq | endothelial | *PSORS1C1* | TRUE | 2.15150417 |
| CAD_admission_freq | endothelial | *AC084018.1* | TRUE | 1.98884019 |
| CAD_admission_freq | endothelial | *GS1.124K5.4* | TRUE | 1.96854138 |
| CAD_admission_freq | endothelial | *SRXN1* | TRUE | 2.06615117 |
| CAD_admission_freq | LEC | *KHK* | TRUE | 2.28797042 |
| CAD_admission_freq | LEC | *PAAF1* | TRUE | 2.48924406 |
| CAD_admission_freq | LEC | *GOLGA8Q* | TRUE | 2.26711375 |
| CAD_admission_freq | LEC | *SMDT1* | TRUE | 2.14717705 |
| CAD_admission_freq | LEC | *HYAL3* | TRUE | 1.98907162 |

| | | | | |
|---|---|---|---|---|
| CAD_admission_freq | LEC | *GS1.124K5.4* | TRUE | 2.03817197 |
| CAD_admission_freq | LEC | *SRXN1* | TRUE | 2.45124589 |
| CAD_admission_freq | macrophage | *F7* | TRUE | 2.16543393 |
| CAD_admission_freq | macrophage | *CLTCL1* | TRUE | 2.57509824 |
| CAD_admission_freq | macrophage | *IGDCC4* | TRUE | 1.96978336 |
| CAD_admission_freq | macrophage | *CCDC61* | TRUE | 2.05529177 |
| CAD_admission_freq | macrophage | *CAPS* | TRUE | 1.99170551 |
| CAD_admission_freq | macrophage | *TOX2* | TRUE | 2.47426058 |
| CAD_admission_freq | macrophage | *NRM* | TRUE | 2.29425265 |
| CAD_admission_freq | macrophage | *KHK* | TRUE | 2.36818135 |
| CAD_admission_freq | macrophage | *DMTN* | TRUE | 2.72590747 |
| CAD_admission_freq | macrophage | *RAB24* | TRUE | 2.04389008 |
| CAD_admission_freq | macrophage | *HOXD4* | TRUE | 2.21631976 |
| CAD_admission_freq | macrophage | *SLC22A1* | TRUE | 3.34171705 |
| CAD_admission_freq | macrophage | *PAAF1* | TRUE | 3.12178205 |
| CAD_admission_freq | macrophage | *GOLGA8Q* | TRUE | 2.89346058 |
| CAD_admission_freq | macrophage | *HYI* | TRUE | 1.9622752 |
| CAD_admission_freq | macrophage | *ACTL10* | TRUE | 1.9687866 |
| CAD_admission_freq | macrophage | *SMDT1* | TRUE | 1.96503618 |
| CAD_admission_freq | macrophage | *PLA2G6* | TRUE | 2.41249947 |
| CAD_admission_freq | macrophage | *FAM228A* | TRUE | 1.99066354 |
| CAD_admission_freq | macrophage | *PSORS1C1* | TRUE | 2.19042399 |
| CAD_admission_freq | macrophage | *AC084018.1* | TRUE | 2.05425055 |
| CAD_admission_freq | macrophage | *GS1.124K5.4* | TRUE | 2.30708577 |
| CAD_admission_freq | macrophage | *ZNF286B* | TRUE | 2.1245186 |
| CAD_admission_freq | macrophage | *HMBS* | TRUE | 2.19530084 |
| CAD_admission_freq | macrophage | *SRXN1* | TRUE | 2.68107173 |
| CAD_admission_freq | mast_cell | *F7* | TRUE | 2.86525807 |
| CAD_admission_freq | mast_cell | *IGDCC4* | TRUE | 2.08581928 |
| CAD_admission_freq | mast_cell | *NRM* | TRUE | 2.33143309 |
| CAD_admission_freq | mast_cell | *SLC22A1* | TRUE | 2.55842953 |
| CAD_admission_freq | mast_cell | *PAAF1* | TRUE | 2.51584241 |
| CAD_admission_freq | mast_cell | *GOLGA8Q* | TRUE | 2.3976914 |
| CAD_admission_freq | mast_cell | *HYI* | TRUE | 1.97328163 |
| CAD_admission_freq | mast_cell | *SMN2* | TRUE | 2.04786338 |
| CAD_admission_freq | mast_cell | *SRXN1* | TRUE | 2.50524045 |
| CAD_admission_freq | monocyte | *SLC22A1* | TRUE | 2.13439621 |
| CAD_admission_freq | monocyte | *GOLGA8Q* | TRUE | 2.38951796 |
| CAD_admission_freq | monocyte | *SMDT1* | TRUE | 1.96776148 |
| CAD_admission_freq | monocyte | *MICB* | TRUE | 2.1024977 |
| CAD_admission_freq | monocyte | *SRXN1* | TRUE | 2.06227881 |
| CAD_admission_freq | neutrophil | *SRXN1* | TRUE | 2.15799376 |
| CAD_admission_freq | nk_cell | *F7* | TRUE | 2.42492538 |
| CAD_admission_freq | nk_cell | *SDC1* | TRUE | 1.99475271 |
| CAD_admission_freq | nk_cell | *TOX2* | TRUE | 2.1970605 |
| CAD_admission_freq | nk_cell | *NRM* | TRUE | 3.04958673 |
| CAD_admission_freq | nk_cell | *KHK* | TRUE | 2.25441241 |
| CAD_admission_freq | nk_cell | *SLC22A1* | TRUE | 2.630457 |
| CAD_admission_freq | nk_cell | *PAAF1* | TRUE | 2.75622112 |
| CAD_admission_freq | nk_cell | *GOLGA8Q* | TRUE | 2.37439554 |

| | | | | |
|---|---|---|---|---|
| CAD_admission_freq | pericyte | *F7* | TRUE | 2.67747432 |
| CAD_admission_freq | pericyte | *IGDCC4* | TRUE | 2.04297819 |
| CAD_admission_freq | pericyte | *TOX2* | TRUE | 1.9945387 |
| CAD_admission_freq | pericyte | *KHK* | TRUE | 2.13887286 |
| CAD_admission_freq | pericyte | *PAAF1* | TRUE | 3.13542164 |
| CAD_admission_freq | pericyte | *GOLGA8Q* | TRUE | 2.6145165 |
| CAD_admission_freq | pericyte | *SMDT1* | TRUE | 2.11679727 |
| CAD_admission_freq | pericyte | *HYAL3* | TRUE | 2.02613444 |
| CAD_admission_freq | pericyte | *MICB* | TRUE | 2.20562597 |
| CAD_admission_freq | pericyte | *PSORS1C1* | TRUE | 2.09847078 |
| CAD_admission_freq | pericyte | *GS1.124K5.4* | TRUE | 2.1914403 |
| CAD_admission_freq | pericyte | *SRXN1* | TRUE | 2.57002766 |
| CAD_admission_freq | SMC | *F7* | TRUE | 2.4348322 |
| CAD_admission_freq | SMC | *IGDCC4* | TRUE | 2.1162553 |
| CAD_admission_freq | SMC | *TOX2* | TRUE | 2.48295478 |
| CAD_admission_freq | SMC | *NRM* | TRUE | 2.79868634 |
| CAD_admission_freq | SMC | *KHK* | TRUE | 2.07061469 |
| CAD_admission_freq | SMC | *DMTN* | TRUE | 2.72350333 |
| CAD_admission_freq | SMC | *HOXD4* | TRUE | 2.33837521 |
| CAD_admission_freq | SMC | *PAAF1* | TRUE | 2.91363234 |
| CAD_admission_freq | SMC | *GOLGA8Q* | TRUE | 2.73790959 |
| CAD_admission_freq | SMC | *GPANK1* | TRUE | 2.24949577 |
| CAD_admission_freq | SMC | *GSTM2* | TRUE | 2.08838341 |
| CAD_admission_freq | SMC | *GS1.124K5.4* | TRUE | 2.34965988 |
| CAD_admission_freq | SMC | *HMBS* | TRUE | 2.09296632 |
| CAD_admission_freq | SMC | *SRXN1* | TRUE | 2.51100181 |
| CAD_admission_freq | t_cell | *F7* | TRUE | 2.59222378 |
| CAD_admission_freq | t_cell | *TOX2* | TRUE | 1.99843811 |
| CAD_admission_freq | t_cell | *KHK* | TRUE | 2.19318604 |
| CAD_admission_freq | t_cell | *DMTN* | TRUE | 2.26794839 |
| CAD_admission_freq | t_cell | *SLC22A1* | TRUE | 2.76500155 |
| CAD_admission_freq | t_cell | *PAAF1* | TRUE | 2.81136504 |
| CAD_admission_freq | t_cell | *GOLGA8Q* | TRUE | 2.61634109 |
| CAD_admission_freq | t_cell | *MICB* | TRUE | 2.10788285 |
| CAD_admission_freq | t_cell | *GS1.124K5.4* | TRUE | 2.03997287 |
| CAD_admission_freq | t_cell | *HMBS* | TRUE | 1.99987137 |
| CAD_admission_freq | t_cell | *SRXN1* | TRUE | 2.70145589 |
| CAD_mortality | adipocyte | *ICAM3* | TRUE | 2.38686283 |
| CAD_mortality | adipocyte | *LYRM2* | TRUE | 1.98825774 |
| CAD_mortality | adipocyte | *MANBAL* | TRUE | 2.15814287 |
| CAD_mortality | adipocyte | *TNFRSF10A* | TRUE | 2.41101905 |
| CAD_mortality | adipocyte | *IRF6* | TRUE | 2.50156458 |
| CAD_mortality | adipocyte | *EPCAM* | TRUE | 2.09732392 |
| CAD_mortality | adipocyte | *TOX2* | TRUE | 1.96521034 |
| CAD_mortality | adipocyte | *TOR1B* | TRUE | 2.65914709 |
| CAD_mortality | adipocyte | *KANSL2* | TRUE | 2.48081336 |
| CAD_mortality | adipocyte | *POMZP3* | TRUE | 2.48272946 |
| CAD_mortality | adipocyte | *MPP7* | TRUE | 2.09367765 |
| CAD_mortality | adipocyte | *RCAN1* | TRUE | 2.83542178 |
| CAD_mortality | adipocyte | *PLCD3* | TRUE | 2.25448698 |

| | | | | |
|---|---|---|---|---|
| CAD_mortality | adipocyte | *PDZK1IP1* | TRUE | 2.59125367 |
| CAD_mortality | adipocyte | *TRIM66* | TRUE | 2.45257163 |
| CAD_mortality | adipocyte | *PCSK9* | TRUE | 2.20768142 |
| CAD_mortality | adipocyte | *STK32A* | TRUE | 2.20764376 |
| CAD_mortality | adipocyte | *TSNARE1* | TRUE | 2.89198711 |
| CAD_mortality | adipocyte | *COL24A1* | TRUE | 2.2546817 |
| CAD_mortality | adipocyte | *HAP1* | TRUE | 2.15912654 |
| CAD_mortality | adipocyte | *TMEM81* | TRUE | 2.20140719 |
| CAD_mortality | adipocyte | *BANF1* | TRUE | 1.96005109 |
| CAD_mortality | adipocyte | *TACSTD2* | TRUE | 2.03221492 |
| CAD_mortality | adipocyte | *GDF3* | TRUE | 2.60805853 |
| CAD_mortality | adipocyte | *TOP1MT* | TRUE | 1.98368198 |
| CAD_mortality | adipocyte | *PDE6G* | TRUE | 2.15519134 |
| CAD_mortality | adipocyte | *FCAR* | TRUE | 3.47437567 |
| CAD_mortality | adipocyte | *JAKMIP3* | TRUE | 2.92290728 |
| CAD_mortality | adipocyte | *XPNPEP3* | TRUE | 2.22754278 |
| CAD_mortality | adipocyte | *KIAA1211L* | TRUE | 2.97394114 |
| CAD_mortality | adipocyte | *ZNF100* | TRUE | 2.09456147 |
| CAD_mortality | adipocyte | *TBKBP1* | TRUE | 2.00671856 |
| CAD_mortality | adipocyte | *NELFE* | TRUE | 2.08129317 |
| CAD_mortality | adipocyte | *AC112229.4* | TRUE | 2.35558507 |
| CAD_mortality | adipocyte | *PCDHGA11* | TRUE | 2.0991968 |
| CAD_mortality | ASPC | *RHOBTB2* | TRUE | 2.06181802 |
| CAD_mortality | ASPC | *ATP2A3* | TRUE | 1.97293076 |
| CAD_mortality | ASPC | *ICAM3* | TRUE | 2.36420512 |
| CAD_mortality | ASPC | *NMRK2* | TRUE | 1.97157952 |
| CAD_mortality | ASPC | *LYRM2* | TRUE | 2.13010642 |
| CAD_mortality | ASPC | *MRPS33* | TRUE | 2.64975844 |
| CAD_mortality | ASPC | *TPD52L2* | TRUE | 2.50810737 |
| CAD_mortality | ASPC | *MANBAL* | TRUE | 2.6438733 |
| CAD_mortality | ASPC | *TNFRSF10A* | TRUE | 2.44798754 |
| CAD_mortality | ASPC | *ALDH3A1* | TRUE | 1.96804877 |
| CAD_mortality | ASPC | *CAND1* | TRUE | 2.33355444 |
| CAD_mortality | ASPC | *CHMP3* | TRUE | 2.3890642 |
| CAD_mortality | ASPC | *CD207* | TRUE | 1.97049017 |
| CAD_mortality | ASPC | *IRF6* | TRUE | 2.61378524 |
| CAD_mortality | ASPC | *EPCAM* | TRUE | 2.47117283 |
| CAD_mortality | ASPC | *RWDD3* | TRUE | 2.09232987 |
| CAD_mortality | ASPC | *EBPL* | TRUE | 2.44079904 |
| CAD_mortality | ASPC | *TOX2* | TRUE | 2.11196893 |
| CAD_mortality | ASPC | *CHCHD5* | TRUE | 2.07691164 |
| CAD_mortality | ASPC | *LRRC8A* | TRUE | 2.19014469 |
| CAD_mortality | ASPC | *TOR1B* | TRUE | 2.75663058 |
| CAD_mortality | ASPC | *PRPF4* | TRUE | 2.10155733 |
| CAD_mortality | ASPC | *ERP27* | TRUE | 1.99200003 |
| CAD_mortality | ASPC | *KANSL2* | TRUE | 2.45123919 |
| CAD_mortality | ASPC | *TRIM65* | TRUE | 2.0336088 |
| CAD_mortality | ASPC | *PYCR2* | TRUE | 2.18696301 |
| CAD_mortality | ASPC | *LYG1* | TRUE | 1.98070851 |
| CAD_mortality | ASPC | *ZNF660* | TRUE | 1.99256649 |

| | | | | |
|---|---|---|---|---|
| CAD_mortality | ASPC | *POMZP3* | TRUE | 2.62384003 |
| CAD_mortality | ASPC | *GRIK4* | TRUE | 2.24469793 |
| CAD_mortality | ASPC | *MPP7* | TRUE | 2.09537727 |
| CAD_mortality | ASPC | *ATG10* | TRUE | 2.12834298 |
| CAD_mortality | ASPC | *RAD17* | TRUE | 2.00488195 |
| CAD_mortality | ASPC | *WDR66* | TRUE | 2.21398629 |
| CAD_mortality | ASPC | *RCAN1* | TRUE | 2.86659765 |
| CAD_mortality | ASPC | *TEPP* | TRUE | 2.41533783 |
| CAD_mortality | ASPC | *MUM1* | TRUE | 2.96192264 |
| CAD_mortality | ASPC | *PLCD3* | TRUE | 2.86216756 |
| CAD_mortality | ASPC | *PDZK1IP1* | TRUE | 2.79887205 |
| CAD_mortality | ASPC | *COA7* | TRUE | 2.20347605 |
| CAD_mortality | ASPC | *TSSK3* | TRUE | 2.30405347 |
| CAD_mortality | ASPC | *ARPC5* | TRUE | 2.03107708 |
| CAD_mortality | ASPC | *LRRC2* | TRUE | 1.99976539 |
| CAD_mortality | ASPC | *AKR1E2* | TRUE | 2.05432235 |
| CAD_mortality | ASPC | *TRUB1* | TRUE | 2.09304711 |
| CAD_mortality | ASPC | *TRIM66* | TRUE | 2.35183095 |
| CAD_mortality | ASPC | *VKORC1* | TRUE | 2.00887414 |
| CAD_mortality | ASPC | *LRP1B* | TRUE | 2.03067093 |
| CAD_mortality | ASPC | *LGALS9* | TRUE | 2.13141231 |
| CAD_mortality | ASPC | *PCSK9* | TRUE | 2.18200078 |
| CAD_mortality | ASPC | *STK32A* | TRUE | 2.31070272 |
| CAD_mortality | ASPC | *TSNARE1* | TRUE | 2.92156149 |
| CAD_mortality | ASPC | *HOPX* | TRUE | 2.80672679 |
| CAD_mortality | ASPC | *COL24A1* | TRUE | 2.6426626 |
| CAD_mortality | ASPC | *RAB43* | TRUE | 2.86563135 |
| CAD_mortality | ASPC | *RHOD* | TRUE | 1.99272751 |
| CAD_mortality | ASPC | *HAP1* | TRUE | 2.25802397 |
| CAD_mortality | ASPC | *LPL* | TRUE | 2.36695186 |
| CAD_mortality | ASPC | *ETV4* | TRUE | 2.32499626 |
| CAD_mortality | ASPC | *TMEM39A* | TRUE | 3.51055536 |
| CAD_mortality | ASPC | *CTC1* | TRUE | 2.26366277 |
| CAD_mortality | ASPC | *SF3A3* | TRUE | 2.4331797 |
| CAD_mortality | ASPC | *B3GALT5* | TRUE | 2.21290885 |
| CAD_mortality | ASPC | *TACSTD2* | TRUE | 2.06680745 |
| CAD_mortality | ASPC | *GDF3* | TRUE | 2.62033415 |
| CAD_mortality | ASPC | *TMED9* | TRUE | 2.10421253 |
| CAD_mortality | ASPC | *PDE6G* | TRUE | 2.10940809 |
| CAD_mortality | ASPC | *FCAR* | TRUE | 3.61277606 |
| CAD_mortality | ASPC | *HYKK* | TRUE | 2.02922502 |
| CAD_mortality | ASPC | *JAKMIP3* | TRUE | 3.63983718 |
| CAD_mortality | ASPC | *SEMA4A* | TRUE | 1.96683636 |
| CAD_mortality | ASPC | *XPNPEP3* | TRUE | 2.57501492 |
| CAD_mortality | ASPC | *MYO18A* | TRUE | 2.05093368 |
| CAD_mortality | ASPC | *KIAA1211L* | TRUE | 2.63569726 |
| CAD_mortality | ASPC | *ZNF429* | TRUE | 2.14610789 |
| CAD_mortality | ASPC | *ADH4* | TRUE | 2.07746507 |
| CAD_mortality | ASPC | *RPS6KL1* | TRUE | 2.14190211 |
| CAD_mortality | ASPC | *ITPRIPL1* | TRUE | 2.47594656 |

| | | | | |
|---|---|---|---|---|
| CAD_mortality | ASPC | *TBKBP1* | TRUE | 2.29986047 |
| CAD_mortality | ASPC | *ZNF525* | TRUE | 2.05189707 |
| CAD_mortality | ASPC | *ASAH2B* | TRUE | 2.11440185 |
| CAD_mortality | ASPC | *NELFE* | TRUE | 2.09380032 |
| CAD_mortality | ASPC | *AC112229.4* | TRUE | 2.34635985 |
| CAD_mortality | ASPC | *LINC00484* | TRUE | 2.20569626 |
| CAD_mortality | b_cell | *IRF6* | TRUE | 2.05574148 |
| CAD_mortality | b_cell | *RAB43* | TRUE | 3.13063779 |
| CAD_mortality | b_cell | *HAP1* | TRUE | 2.36661602 |
| CAD_mortality | b_cell | *GDF3* | TRUE | 2.12819624 |
| CAD_mortality | dendritic_cell | *ICAM3* | TRUE | 2.00934735 |
| CAD_mortality | dendritic_cell | *IRF6* | TRUE | 2.01101247 |
| CAD_mortality | dendritic_cell | *TRIM66* | TRUE | 2.11715466 |
| CAD_mortality | dendritic_cell | *TSNARE1* | TRUE | 2.22492902 |
| CAD_mortality | dendritic_cell | *FCAR* | TRUE | 2.90318768 |
| CAD_mortality | endothelial | *EPCAM* | TRUE | 2.11010774 |
| CAD_mortality | endothelial | *PLCD3* | TRUE | 2.03154125 |
| CAD_mortality | endothelial | *PDZK1IP1* | TRUE | 2.1623807 |
| CAD_mortality | endothelial | *TRIM66* | TRUE | 2.20216535 |
| CAD_mortality | endothelial | *COL24A1* | TRUE | 1.96873613 |
| CAD_mortality | endothelial | *HAP1* | TRUE | 2.02172463 |
| CAD_mortality | LEC | *OPN3* | TRUE | 1.99645181 |
| CAD_mortality | LEC | *RAB43* | TRUE | 2.83476278 |
| CAD_mortality | LEC | *KIAA1211L* | TRUE | 2.46615361 |
| CAD_mortality | LEC | *AC112229.4* | TRUE | 2.27115807 |
| CAD_mortality | LEC | *HLA.DQA2* | TRUE | 2.0327369 |
| CAD_mortality | macrophage | *F7* | TRUE | 2.02992231 |
| CAD_mortality | macrophage | *ICAM3* | TRUE | 2.1047245 |
| CAD_mortality | macrophage | *MANBAL* | TRUE | 2.26553938 |
| CAD_mortality | macrophage | *TNFRSF10A* | TRUE | 2.22305739 |
| CAD_mortality | macrophage | *ALDH3A1* | TRUE | 1.98563688 |
| CAD_mortality | macrophage | *IRF6* | TRUE | 2.43236659 |
| CAD_mortality | macrophage | *EPCAM* | TRUE | 2.16746153 |
| CAD_mortality | macrophage | *TOX2* | TRUE | 1.96399852 |
| CAD_mortality | macrophage | *TOR1B* | TRUE | 2.37891929 |
| CAD_mortality | macrophage | *KANSL2* | TRUE | 2.1490865 |
| CAD_mortality | macrophage | *ZNF660* | TRUE | 1.98222945 |
| CAD_mortality | macrophage | *POMZP3* | TRUE | 2.27973218 |
| CAD_mortality | macrophage | *ANKLE1* | TRUE | 1.98193553 |
| CAD_mortality | macrophage | *PDZK1IP1* | TRUE | 2.23494239 |
| CAD_mortality | macrophage | *TRIM66* | TRUE | 2.34688936 |
| CAD_mortality | macrophage | *PCSK9* | TRUE | 2.04049594 |
| CAD_mortality | macrophage | *STK32A* | TRUE | 2.01727665 |
| CAD_mortality | macrophage | *TSNARE1* | TRUE | 2.54943386 |
| CAD_mortality | macrophage | *COL24A1* | TRUE | 2.31876469 |
| CAD_mortality | macrophage | *HAP1* | TRUE | 2.22975207 |
| CAD_mortality | macrophage | *GOLGA8Q* | TRUE | 2.08025615 |
| CAD_mortality | macrophage | *B3GALT5* | TRUE | 2.07347327 |
| CAD_mortality | macrophage | *TACSTD2* | TRUE | 1.98983978 |
| CAD_mortality | macrophage | *GDF3* | TRUE | 2.61144305 |

| | | | | |
|---|---|---|---|---|
| CAD_mortality | macrophage | *FCAR* | TRUE | 3.32929212 |
| CAD_mortality | macrophage | *JAKMIP3* | TRUE | 3.42581578 |
| CAD_mortality | macrophage | *XPNPEP3* | TRUE | 2.11113921 |
| CAD_mortality | macrophage | *KIAA1211L* | TRUE | 2.83984492 |
| CAD_mortality | macrophage | *RPS6KL1* | TRUE | 2.05112964 |
| CAD_mortality | macrophage | *TBKBP1* | TRUE | 2.2423836 |
| CAD_mortality | macrophage | *AC112229.4* | TRUE | 2.3562552 |
| CAD_mortality | macrophage | *LINC00484* | TRUE | 2.14021043 |
| CAD_mortality | macrophage | *PCDHGA11* | TRUE | 1.99926997 |
| CAD_mortality | macrophage | *LINC00668* | TRUE | 1.98096096 |
| CAD_mortality | mast_cell | *F7* | TRUE | 2.00280275 |
| CAD_mortality | mast_cell | *TNFRSF10A* | TRUE | 2.18515677 |
| CAD_mortality | mast_cell | *TOR1B* | TRUE | 2.19964186 |
| CAD_mortality | mast_cell | *POMZP3* | TRUE | 1.9898792 |
| CAD_mortality | mast_cell | *RCAN1* | TRUE | 2.17419711 |
| CAD_mortality | mast_cell | *HAP1* | TRUE | 2.30016012 |
| CAD_mortality | mast_cell | *KIAA1211L* | TRUE | 2.42915273 |
| CAD_mortality | mast_cell | *HLA.DQA2* | TRUE | 2.18632774 |
| CAD_mortality | monocyte | *TNFRSF10A* | TRUE | 2.19120755 |
| CAD_mortality | monocyte | *RAB43* | TRUE | 2.92017959 |
| CAD_mortality | monocyte | *GDF3* | TRUE | 2.04864477 |
| CAD_mortality | monocyte | *HLA.DQB2* | TRUE | 2.07679835 |
| CAD_mortality | neutrophil | *JAKMIP3* | TRUE | 3.05102547 |
| CAD_mortality | nk_cell | *TSNARE1* | TRUE | 2.1074391 |
| CAD_mortality | nk_cell | *HAP1* | TRUE | 2.1325662 |
| CAD_mortality | nk_cell | *FCAR* | TRUE | 2.45141521 |
| CAD_mortality | nk_cell | *HLA.DQA2* | TRUE | 2.13593032 |
| CAD_mortality | pericyte | *IRF6* | TRUE | 2.11371466 |
| CAD_mortality | pericyte | *RCAN1* | TRUE | 2.72010697 |
| CAD_mortality | pericyte | *TRIM66* | TRUE | 2.08829377 |
| CAD_mortality | pericyte | *TSNARE1* | TRUE | 2.33544115 |
| CAD_mortality | pericyte | *RAB43* | TRUE | 2.94302636 |
| CAD_mortality | pericyte | *HAP1* | TRUE | 2.2616866 |
| CAD_mortality | pericyte | *TMEM80* | TRUE | 2.05195802 |
| CAD_mortality | pericyte | *GDF3* | TRUE | 2.11894492 |
| CAD_mortality | pericyte | *FCAR* | TRUE | 3.14988467 |
| CAD_mortality | pericyte | *JAKMIP3* | TRUE | 2.08004644 |
| CAD_mortality | pericyte | *KIAA1211L* | TRUE | 2.4851677 |
| CAD_mortality | pericyte | *CHML* | TRUE | 2.12625192 |
| CAD_mortality | pericyte | *AC112229.4* | TRUE | 2.13136821 |
| CAD_mortality | pericyte | *LINC00484* | TRUE | 2.25289355 |
| CAD_mortality | pericyte | *HLA.DQA2* | TRUE | 2.1133294 |
| CAD_mortality | SMC | *IRF6* | TRUE | 2.48492436 |
| CAD_mortality | SMC | *TOR1B* | TRUE | 2.03480841 |
| CAD_mortality | SMC | *KANSL2* | TRUE | 2.03907808 |
| CAD_mortality | SMC | *POMZP3* | TRUE | 2.13191864 |
| CAD_mortality | SMC | *PDZK1IP1* | TRUE | 1.97693434 |
| CAD_mortality | SMC | *TRIM66* | TRUE | 2.16496946 |
| CAD_mortality | SMC | *TSNARE1* | TRUE | 2.51231531 |
| CAD_mortality | SMC | *COL24A1* | TRUE | 2.04632399 |

| | | | | |
|---|---|---|---|---|
| CAD_mortality | SMC | *RAB43* | TRUE | 2.74601319 |
| CAD_mortality | SMC | *B3GALT5* | TRUE | 2.12744742 |
| CAD_mortality | SMC | *GDF3* | TRUE | 2.00301893 |
| CAD_mortality | SMC | *JAKMIP3* | TRUE | 3.23377229 |
| CAD_mortality | SMC | *NELFE* | TRUE | 2.02911988 |
| CAD_mortality | SMC | *LINC00484* | TRUE | 2.10667612 |
| CAD_mortality | SMC | *HLA.DQA2* | TRUE | 2.14389011 |
| CAD_mortality | t_cell | *ALDH3A1* | TRUE | 2.0372145 |
| CAD_mortality | t_cell | *TOR1B* | TRUE | 1.98272868 |
| CAD_mortality | t_cell | *TRIM66* | TRUE | 2.0636015 |
| CAD_mortality | t_cell | *TSNARE1* | TRUE | 2.2341198 |
| CAD_mortality | t_cell | *RAB43* | TRUE | 2.66885942 |
| CAD_mortality | t_cell | *HAP1* | TRUE | 1.96541843 |
| CAD_mortality | t_cell | *BANF1* | TRUE | 2.07805983 |
| CAD_mortality | t_cell | *GDF3* | TRUE | 2.65008727 |
| CAD_mortality | t_cell | *FCAR* | TRUE | 3.15387315 |
| CAD_mortality | t_cell | *KIAA1211L* | TRUE | 2.45217158 |
| CAD_mortality | t_cell | *CHML* | TRUE | 2.00939497 |

d. Identified cell type specific causal genes for obesity and related disorders from VAT

| disorder | celltype | gene | assoc | zMin |
|---|---|---|---|---|
| CAD | adipocyte | *UHRF1BP1* | TRUE | 3.393733818 |
| CAD | adipocyte | *ZMYND12* | TRUE | 1.996840264 |
| CAD | adipocyte | *TIPIN* | TRUE | 3.070426959 |
| CAD | adipocyte | *CACNG4* | TRUE | 2.055818263 |
| CAD | adipocyte | *FMO4* | TRUE | 3.579666418 |
| CAD | adipocyte | *ITPR3* | TRUE | 2.682334956 |
| CAD | adipocyte | *MTAP* | TRUE | 2.20116992 |
| CAD | adipocyte | *SUSD2* | TRUE | 2.658618336 |
| CAD | adipocyte | *LIPA* | TRUE | 2.338472599 |
| CAD | adipocyte | *RAB5C* | TRUE | 1.994469361 |
| CAD | adipocyte | *BVES* | TRUE | 2.159419095 |
| CAD | adipocyte | *BNIP1* | TRUE | 2.995144556 |
| CAD | adipocyte | *PCCB* | TRUE | 2.277385404 |
| CAD | adipocyte | *GGCX* | TRUE | 2.742268414 |
| CAD | adipocyte | *CFHR3* | TRUE | 2.217021664 |
| CAD | adipocyte | *EIF2B2* | TRUE | 1.999905089 |
| CAD | adipocyte | *PTGIS* | TRUE | 2.378844758 |
| CAD | adipocyte | *TCP11* | TRUE | 2.859153123 |
| CAD | adipocyte | *GRK4* | TRUE | 3.536755833 |
| CAD | adipocyte | *FAM98C* | TRUE | 1.962006333 |
| CAD | adipocyte | *DDC* | TRUE | 2.491223789 |
| CAD | adipocyte | *B4GALNT1* | TRUE | 2.562923995 |
| CAD | adipocyte | *POLR1E* | TRUE | 2.567974698 |
| CAD | adipocyte | *TMEM91* | TRUE | 3.157018071 |
| CAD | adipocyte | *EVA1B* | TRUE | 2.3390181 |
| CAD | adipocyte | *WDPCP* | TRUE | 2.416576767 |
| CAD | adipocyte | *FBXO10* | TRUE | 3.265674117 |
| CAD | adipocyte | *HSD17B12* | TRUE | 2.981459042 |
| CAD | adipocyte | *TTC12* | TRUE | 2.22049331 |
| CAD | adipocyte | *MMP16* | TRUE | 2.083691884 |
| CAD | adipocyte | *CLEC18A* | TRUE | 2.144680577 |
| CAD | adipocyte | *S100A1* | TRUE | 2.015175436 |
| CAD | adipocyte | *CPSF4* | TRUE | 2.124093999 |
| CAD | adipocyte | *ALDH16A1* | TRUE | 1.995363272 |
| CAD | adipocyte | *DAPL1* | TRUE | 2.061587535 |
| CAD | adipocyte | *RNF123* | TRUE | 2.533791488 |
| CAD | adipocyte | *PIGO* | TRUE | 2.040986574 |
| CAD | adipocyte | *OXGR1* | TRUE | 1.992245247 |
| CAD | adipocyte | *C16orf46* | TRUE | 2.8653684 |
| CAD | adipocyte | *ENGASE* | TRUE | 2.200127193 |
| CAD | adipocyte | *LGALS9* | TRUE | 2.210893193 |
| CAD | adipocyte | *NPIPB3* | TRUE | 2.141111863 |
| CAD | adipocyte | *MYO7B* | TRUE | 2.15234102 |
| CAD | adipocyte | *TTLL6* | TRUE | 2.099445201 |
| CAD | adipocyte | *RGS19* | TRUE | 2.213111459 |
| CAD | adipocyte | *NXNL1* | TRUE | 2.227692043 |

| | | | | |
|---|---|---|---|---|
| CAD | adipocyte | *C3AR1* | TRUE | 2.264236812 |
| CAD | adipocyte | *NME6* | TRUE | 2.071016274 |
| CAD | adipocyte | *ZNF738* | TRUE | 2.051063992 |
| CAD | adipocyte | *CCNE2* | TRUE | 2.419117342 |
| CAD | adipocyte | *PAAF1* | TRUE | 2.231659426 |
| CAD | adipocyte | *SLC25A22* | TRUE | 2.380243806 |
| CAD | adipocyte | *LYNX1* | TRUE | 2.501051025 |
| CAD | adipocyte | *ADIPOQ* | TRUE | 2.536176622 |
| CAD | adipocyte | *TMEM136* | TRUE | 2.021535376 |
| CAD | adipocyte | *FES* | TRUE | 2.61765554 |
| CAD | adipocyte | *VSIG10L* | TRUE | 2.298535218 |
| CAD | adipocyte | *COLCA1* | TRUE | 2.087290561 |
| CAD | adipocyte | *RPS26* | TRUE | 2.003267326 |
| CAD | adipocyte | *MRPL42* | TRUE | 2.092995621 |
| CAD | adipocyte | *TMEM116* | TRUE | 3.505752212 |
| CAD | adipocyte | *TBKBP1* | TRUE | 2.298571707 |
| CAD | adipocyte | *AP001468.1* | TRUE | 2.124479614 |
| CAD | adipocyte | *LINC00707* | TRUE | 1.990800831 |
| CAD | adipocyte | *LY6G5B* | TRUE | 1.973817124 |
| CAD | adipocyte | *GOLGA8K* | TRUE | 2.048830393 |
| CAD | ASPC | *UHRF1BP1* | TRUE | 2.410792779 |
| CAD | ASPC | *TIE1* | TRUE | 2.140619418 |
| CAD | ASPC | *ZMYND12* | TRUE | 2.011425463 |
| CAD | ASPC | *TIPIN* | TRUE | 3.304212238 |
| CAD | ASPC | *ADD2* | TRUE | 2.140456604 |
| CAD | ASPC | *CACNG4* | TRUE | 2.057018036 |
| CAD | ASPC | *FMO4* | TRUE | 3.507972285 |
| CAD | ASPC | *RPH3A* | TRUE | 2.392641836 |
| CAD | ASPC | *PABPC4* | TRUE | 1.997777681 |
| CAD | ASPC | *ITPR3* | TRUE | 2.860495615 |
| CAD | ASPC | *SCD* | TRUE | 2.361646608 |
| CAD | ASPC | *MTAP* | TRUE | 2.423961832 |
| CAD | ASPC | *SUSD2* | TRUE | 2.710782133 |
| CAD | ASPC | *THOC5* | TRUE | 2.034597367 |
| CAD | ASPC | *RABAC1* | TRUE | 1.99545928 |
| CAD | ASPC | *LIPA* | TRUE | 2.667275143 |
| CAD | ASPC | *RAB5C* | TRUE | 2.583051865 |
| CAD | ASPC | *BVES* | TRUE | 2.337598439 |
| CAD | ASPC | *BNIP1* | TRUE | 3.054808929 |
| CAD | ASPC | *PCCB* | TRUE | 2.76894673 |
| CAD | ASPC | *C3orf14* | TRUE | 2.223439407 |
| CAD | ASPC | *STEAP3* | TRUE | 2.024068334 |
| CAD | ASPC | *GGCX* | TRUE | 3.015213233 |
| CAD | ASPC | *CFHR3* | TRUE | 2.725334726 |
| CAD | ASPC | *C2orf40* | TRUE | 2.006476151 |
| CAD | ASPC | *POPDC2* | TRUE | 2.312062161 |
| CAD | ASPC | *PTGIS* | TRUE | 2.8054053 |
| CAD | ASPC | *TCP11* | TRUE | 2.741814583 |

| | | | | |
|---|---|---|---|---|
| CAD | ASPC | *GRK4* | TRUE | 3.702971649 |
| CAD | ASPC | *TRAP1* | TRUE | 2.067358767 |
| CAD | ASPC | *ABHD8* | TRUE | 2.091624942 |
| CAD | ASPC | *OPN1SW* | TRUE | 2.060516429 |
| CAD | ASPC | *SERGEF* | TRUE | 2.193348255 |
| CAD | ASPC | *FAM98C* | TRUE | 2.065514957 |
| CAD | ASPC | *PLVAP* | TRUE | 2.130578178 |
| CAD | ASPC | *CCT6B* | TRUE | 2.157350845 |
| CAD | ASPC | *TIMM10B* | TRUE | 2.484736857 |
| CAD | ASPC | *DDC* | TRUE | 2.526290221 |
| CAD | ASPC | *B4GALNT1* | TRUE | 2.739199777 |
| CAD | ASPC | *POLR1E* | TRUE | 2.642651422 |
| CAD | ASPC | *TCF19* | TRUE | 2.691819765 |
| CAD | ASPC | *DIAPH3* | TRUE | 2.165372943 |
| CAD | ASPC | *TMEM91* | TRUE | 3.258414387 |
| CAD | ASPC | *SIRT3* | TRUE | 1.990221875 |
| CAD | ASPC | *EVA1B* | TRUE | 2.521251891 |
| CAD | ASPC | *ARNT* | TRUE | 2.490805465 |
| CAD | ASPC | *TBC1D7* | TRUE | 2.184010355 |
| CAD | ASPC | *IRAK1BP1* | TRUE | 2.23702871 |
| CAD | ASPC | *CSGALNACT1* | TRUE | 2.207945461 |
| CAD | ASPC | *FBXO10* | TRUE | 3.328130187 |
| CAD | ASPC | *LRRC27* | TRUE | 2.049315044 |
| CAD | ASPC | *HSD17B12* | TRUE | 3.121418942 |
| CAD | ASPC | *TTC12* | TRUE | 2.348583155 |
| CAD | ASPC | *ROBO3* | TRUE | 2.523151368 |
| CAD | ASPC | *MMP16* | TRUE | 2.472879986 |
| CAD | ASPC | *CLEC18A* | TRUE | 2.394790801 |
| CAD | ASPC | *S100A1* | TRUE | 2.328768608 |
| CAD | ASPC | *CPSF4* | TRUE | 2.697098824 |
| CAD | ASPC | *ABCF3* | TRUE | 2.101593164 |
| CAD | ASPC | *ALDH16A1* | TRUE | 2.043037328 |
| CAD | ASPC | *DAPL1* | TRUE | 2.581503886 |
| CAD | ASPC | *RNF123* | TRUE | 2.333062222 |
| CAD | ASPC | *DAGLB* | TRUE | 2.009004335 |
| CAD | ASPC | *PIGO* | TRUE | 2.366805363 |
| CAD | ASPC | *OXGR1* | TRUE | 2.105484964 |
| CAD | ASPC | *C16orf46* | TRUE | 2.880321801 |
| CAD | ASPC | *CCDC68* | TRUE | 2.111514638 |
| CAD | ASPC | *ENGASE* | TRUE | 2.451353743 |
| CAD | ASPC | *ATOH8* | TRUE | 2.359642143 |
| CAD | ASPC | *LGALS9* | TRUE | 2.524091298 |
| CAD | ASPC | *IRS1* | TRUE | 2.043354338 |
| CAD | ASPC | *NPIPB3* | TRUE | 2.244854193 |
| CAD | ASPC | *MYO7B* | TRUE | 2.227566 |
| CAD | ASPC | *DNAJC18* | TRUE | 2.36717926 |
| CAD | ASPC | *TTLL6* | TRUE | 1.99641588 |
| CAD | ASPC | *RGS19* | TRUE | 2.506306464 |
| CAD | ASPC | *NXNL1* | TRUE | 2.535974271 |
| CAD | ASPC | *C3AR1* | TRUE | 2.744694311 |

| | | | | |
|---|---|---|---|---|
| CAD | ASPC | *NME6* | TRUE | 2.129988226 |
| CAD | ASPC | *ZNF738* | TRUE | 2.149164898 |
| CAD | ASPC | *MAP3K11* | TRUE | 1.978040483 |
| CAD | ASPC | *CCNE2* | TRUE | 2.545255959 |
| CAD | ASPC | *PAAF1* | TRUE | 2.288921408 |
| CAD | ASPC | *SPRYD4* | TRUE | 2.112902306 |
| CAD | ASPC | *SLC25A22* | TRUE | 2.462152342 |
| CAD | ASPC | *LYNX1* | TRUE | 2.576182568 |
| CAD | ASPC | *DEFB119* | TRUE | 2.900666365 |
| CAD | ASPC | *MRPL14* | TRUE | 2.179490916 |
| CAD | ASPC | *ADIPOQ* | TRUE | 2.494678497 |
| CAD | ASPC | *TMEM136* | TRUE | 2.886163578 |
| CAD | ASPC | *CLN8* | TRUE | 1.966162526 |
| CAD | ASPC | *FES* | TRUE | 3.144389387 |
| CAD | ASPC | *A3GALT2* | TRUE | 2.057044082 |
| CAD | ASPC | *SEPT5* | TRUE | 2.036087275 |
| CAD | ASPC | *NBPF12* | TRUE | 2.113185461 |
| CAD | ASPC | *VSIG10L* | TRUE | 2.426965315 |
| CAD | ASPC | *COLCA1* | TRUE | 2.099422729 |
| CAD | ASPC | *SEMA4A* | TRUE | 1.968724698 |
| CAD | ASPC | *ZNF605* | TRUE | 2.173625021 |
| CAD | ASPC | *WDR5B* | TRUE | 1.978151695 |
| CAD | ASPC | *RPS26* | TRUE | 2.106354066 |
| CAD | ASPC | *MRPL42* | TRUE | 2.281399655 |
| CAD | ASPC | *TMEM116* | TRUE | 3.459136594 |
| CAD | ASPC | *TBKBP1* | TRUE | 2.395636399 |
| CAD | ASPC | *STK19* | TRUE | 1.98809866 |
| CAD | ASPC | *BAG6* | TRUE | 1.985300054 |
| CAD | ASPC | *LILRB3* | TRUE | 2.722265493 |
| CAD | ASPC | *LINC00484* | TRUE | 2.019441813 |
| CAD | ASPC | *AP001468.1* | TRUE | 2.205786069 |
| CAD | ASPC | *LINC00707* | TRUE | 2.455809833 |
| CAD | ASPC | *SRP14.AS1* | TRUE | 2.417361575 |
| CAD | ASPC | *GOLGA8K* | TRUE | 2.580828882 |
| CAD | ASPC | *HMBS* | TRUE | 2.293670369 |
| CAD | ASPC | *GRIN2B* | TRUE | 2.610355257 |
| CAD | b_cell | *TCP11* | TRUE | 3.25351524 |
| CAD | b_cell | *CLEC18A* | TRUE | 2.344965584 |
| CAD | b_cell | *NLRP6* | TRUE | 2.212029786 |
| CAD | b_cell | *HYAL3* | TRUE | 1.99281422 |
| CAD | b_cell | *LY6G5B* | TRUE | 2.14962493 |
| CAD | b_cell | *LINC00960* | TRUE | 2.495317062 |
| CAD | dendritic_cell | *SUSD2* | TRUE | 2.30585158 |
| CAD | dendritic_cell | *CENPM* | TRUE | 2.053607737 |
| CAD | dendritic_cell | *TCP11* | TRUE | 2.755376598 |
| CAD | dendritic_cell | *GRK4* | TRUE | 3.045944862 |
| CAD | dendritic_cell | *TMEM91* | TRUE | 3.021310057 |
| CAD | dendritic_cell | *FBXO10* | TRUE | 3.266754364 |
| CAD | dendritic_cell | *CLEC18A* | TRUE | 2.311915623 |
| CAD | dendritic_cell | *ENGASE* | TRUE | 2.055253466 |

| | | | | |
|---|---|---|---|---|
| CAD | dendritic_cell | *LGALS9* | TRUE | 2.117923127 |
| CAD | dendritic_cell | *NXNL1* | TRUE | 2.600241435 |
| CAD | dendritic_cell | *CCNE2* | TRUE | 2.446750699 |
| CAD | dendritic_cell | *TMEM116* | TRUE | 2.133996328 |
| CAD | dendritic_cell | *TBKBP1* | TRUE | 2.034757962 |
| CAD | dendritic_cell | *LINC00960* | TRUE | 2.691754616 |
| CAD | dendritic_cell | *GOLGA8K* | TRUE | 1.987483877 |
| CAD | dendritic_cell | *TAS2R20* | TRUE | 2.646892121 |
| CAD | endometrium | *TIPIN* | TRUE | 2.323194231 |
| CAD | endometrium | *SUSD2* | TRUE | 2.303242901 |
| CAD | endometrium | *TCP11* | TRUE | 3.245432244 |
| CAD | endometrium | *TCF19* | TRUE | 1.990697005 |
| CAD | endometrium | *TMEM91* | TRUE | 2.127358395 |
| CAD | endometrium | *TMEM116* | TRUE | 2.65330628 |
| CAD | endometrium | *LINC00960* | TRUE | 1.991233887 |
| CAD | endothelial | *SUSD2* | TRUE | 2.124490566 |
| CAD | endothelial | *CENPM* | TRUE | 2.04587885 |
| CAD | endothelial | *WFDC3* | TRUE | 2.055221206 |
| CAD | endothelial | *TCP11* | TRUE | 2.639379715 |
| CAD | endothelial | *GRK4* | TRUE | 2.974882003 |
| CAD | endothelial | *B4GALNT1* | TRUE | 1.998678296 |
| CAD | endothelial | *TCF19* | TRUE | 1.961969151 |
| CAD | endothelial | *SLC37A4* | TRUE | 2.179367156 |
| CAD | endothelial | *TMEM91* | TRUE | 2.420130087 |
| CAD | endothelial | *AMT* | TRUE | 2.150444458 |
| CAD | endothelial | *FBXO10* | TRUE | 2.69000207 |
| CAD | endothelial | *HSD17B12* | TRUE | 2.138457968 |
| CAD | endothelial | *CLEC18A* | TRUE | 2.082602339 |
| CAD | endothelial | *RGS19* | TRUE | 1.966047494 |
| CAD | endothelial | *NXNL1* | TRUE | 2.54363595 |
| CAD | endothelial | *KCNK7* | TRUE | 2.432331824 |
| CAD | endothelial | *LYNX1* | TRUE | 2.05394471 |
| CAD | endothelial | *TMEM116* | TRUE | 2.361174248 |
| CAD | endothelial | *LY6G5B* | TRUE | 2.054102624 |
| CAD | endothelial | *LINC00960* | TRUE | 2.599558125 |
| CAD | endothelial | *TAS2R20* | TRUE | 2.637160725 |
| CAD | LEC | *TIPIN* | TRUE | 2.730015303 |
| CAD | LEC | *ITPR3* | TRUE | 3.24790752 |
| CAD | LEC | *SUSD2* | TRUE | 2.488634172 |
| CAD | LEC | *BNIP1* | TRUE | 2.155806515 |
| CAD | LEC | *C3orf14* | TRUE | 2.158881348 |
| CAD | LEC | *EIF2B2* | TRUE | 2.477770758 |
| CAD | LEC | *TCP11* | TRUE | 2.663643531 |
| CAD | LEC | *GRK4* | TRUE | 3.297236057 |
| CAD | LEC | *POLR1E* | TRUE | 1.989397532 |
| CAD | LEC | *TCF19* | TRUE | 2.762799171 |
| CAD | LEC | *SLC37A4* | TRUE | 2.643763016 |
| CAD | LEC | *TMEM91* | TRUE | 2.70763168 |
| CAD | LEC | *HSD17B12* | TRUE | 2.65988103 |
| CAD | LEC | *RNF123* | TRUE | 2.20300982 |

| | | | | |
|---|---|---|---|---|
| CAD | LEC | *RGS19* | TRUE | 2.204871271 |
| CAD | LEC | *NXNL1* | TRUE | 2.526556519 |
| CAD | LEC | *ZNF738* | TRUE | 2.017432809 |
| CAD | LEC | *NLRP6* | TRUE | 2.429436074 |
| CAD | LEC | *RPS26* | TRUE | 2.005193842 |
| CAD | LEC | *TMEM116* | TRUE | 2.775350324 |
| CAD | LEC | *TBKBP1* | TRUE | 2.155998817 |
| CAD | LEC | *LY6G5B* | TRUE | 2.100550252 |
| CAD | LEC | *GOLGA8K* | TRUE | 2.230285986 |
| CAD | macrophage | *UHRF1BP1* | TRUE | 3.023247317 |
| CAD | macrophage | *ZMYND12* | TRUE | 2.041704316 |
| CAD | macrophage | *TIPIN* | TRUE | 2.728041884 |
| CAD | macrophage | *FMO4* | TRUE | 2.926906502 |
| CAD | macrophage | *ITPR3* | TRUE | 2.298787511 |
| CAD | macrophage | *SUSD2* | TRUE | 2.670269092 |
| CAD | macrophage | *CENPM* | TRUE | 2.009909618 |
| CAD | macrophage | *BVES* | TRUE | 2.290861759 |
| CAD | macrophage | *BNIP1* | TRUE | 2.815033442 |
| CAD | macrophage | *PCCB* | TRUE | 2.195062788 |
| CAD | macrophage | *GGCX* | TRUE | 2.052896966 |
| CAD | macrophage | *CFHR3* | TRUE | 2.248454586 |
| CAD | macrophage | *EIF2B2* | TRUE | 2.192688561 |
| CAD | macrophage | *POPDC2* | TRUE | 2.12821945 |
| CAD | macrophage | *TCP11* | TRUE | 2.665605177 |
| CAD | macrophage | *GRK4* | TRUE | 3.442537195 |
| CAD | macrophage | *DDC* | TRUE | 2.491653301 |
| CAD | macrophage | *POLR1E* | TRUE | 2.483546397 |
| CAD | macrophage | *TCF19* | TRUE | 2.318537017 |
| CAD | macrophage | *TMEM91* | TRUE | 3.162314946 |
| CAD | macrophage | *FBXO10* | TRUE | 2.691010456 |
| CAD | macrophage | *HSD17B12* | TRUE | 2.920455926 |
| CAD | macrophage | *CLEC18A* | TRUE | 2.36001278 |
| CAD | macrophage | *RNF123* | TRUE | 2.475510207 |
| CAD | macrophage | *C16orf46* | TRUE | 2.700225524 |
| CAD | macrophage | *ATOH8* | TRUE | 2.090515913 |
| CAD | macrophage | *NXNL1* | TRUE | 2.558705796 |
| CAD | macrophage | *KCNK7* | TRUE | 2.080696539 |
| CAD | macrophage | *PAAF1* | TRUE | 1.993847308 |
| CAD | macrophage | *SPRYD4* | TRUE | 1.968771963 |
| CAD | macrophage | *TMEM116* | TRUE | 2.95414163 |
| CAD | macrophage | *TBKBP1* | TRUE | 2.228664881 |
| CAD | macrophage | *GOLGA8K* | TRUE | 2.330492968 |
| CAD | macrophage | *HMBS* | TRUE | 3.120412193 |
| CAD | mast_cell | *ITPR3* | TRUE | 2.153893386 |
| CAD | mast_cell | *SUSD2* | TRUE | 2.536932538 |
| CAD | mast_cell | *BVES* | TRUE | 2.169784022 |
| CAD | mast_cell | *BNIP1* | TRUE | 2.4983257 |
| CAD | mast_cell | *GRK4* | TRUE | 2.07475856 |
| CAD | mast_cell | *EVA1B* | TRUE | 2.310089199 |
| CAD | mast_cell | *AMT* | TRUE | 2.676966336 |

| | | | | |
|---|---|---|---|---|
| CAD | mast_cell | *SLC39A4* | TRUE | 2.082652092 |
| CAD | mast_cell | *CLEC18A* | TRUE | 2.281561045 |
| CAD | mast_cell | *RGS19* | TRUE | 2.095937497 |
| CAD | mast_cell | *NXNL1* | TRUE | 2.561265907 |
| CAD | mast_cell | *SEPT5* | TRUE | 1.986664009 |
| CAD | mast_cell | *AP001468.1* | TRUE | 2.024996493 |
| CAD | mast_cell | *LINC00960* | TRUE | 2.1006242 |
| CAD | mast_cell | *TAS2R20* | TRUE | 2.148723453 |
| CAD | mesothelium | *PAX6* | TRUE | 2.235399868 |
| CAD | mesothelium | *CYB561* | TRUE | 2.060803492 |
| CAD | mesothelium | *TUBG2* | TRUE | 1.965385074 |
| CAD | mesothelium | *LAMC2* | TRUE | 1.99101651 |
| CAD | mesothelium | *TIE1* | TRUE | 3.001128639 |
| CAD | mesothelium | *ZMYND12* | TRUE | 2.081282256 |
| CAD | mesothelium | *TIPIN* | TRUE | 2.993314081 |
| CAD | mesothelium | *FMO4* | TRUE | 2.844061939 |
| CAD | mesothelium | *TXNDC16* | TRUE | 2.365445571 |
| CAD | mesothelium | *PABPC4* | TRUE | 2.606394978 |
| CAD | mesothelium | *ITPR3* | TRUE | 2.861735714 |
| CAD | mesothelium | *SCD* | TRUE | 2.902522971 |
| CAD | mesothelium | *SUSD2* | TRUE | 2.730404356 |
| CAD | mesothelium | *THOC5* | TRUE | 2.40747584 |
| CAD | mesothelium | *ZNF500* | TRUE | 2.056367053 |
| CAD | mesothelium | *CTSH* | TRUE | 2.053290568 |
| CAD | mesothelium | *NFKBIB* | TRUE | 2.308108567 |
| CAD | mesothelium | *LIPA* | TRUE | 2.033111408 |
| CAD | mesothelium | *ST3GAL4* | TRUE | 2.234961622 |
| CAD | mesothelium | *KCTD10* | TRUE | 2.254820784 |
| CAD | mesothelium | *BVES* | TRUE | 2.46016649 |
| CAD | mesothelium | *BNIP1* | TRUE | 2.825575902 |
| CAD | mesothelium | *PCCB* | TRUE | 2.208113397 |
| CAD | mesothelium | *C3orf14* | TRUE | 2.050335429 |
| CAD | mesothelium | *GGCX* | TRUE | 2.276033951 |
| CAD | mesothelium | *PASK* | TRUE | 2.618028183 |
| CAD | mesothelium | *CFHR3* | TRUE | 2.818654711 |
| CAD | mesothelium | *HSDL2* | TRUE | 2.100786175 |
| CAD | mesothelium | *EIF2B2* | TRUE | 2.270538932 |
| CAD | mesothelium | *POPDC2* | TRUE | 2.479157392 |
| CAD | mesothelium | *PTGIS* | TRUE | 2.755746562 |
| CAD | mesothelium | *SNRPC* | TRUE | 3.370312878 |
| CAD | mesothelium | *TCP11* | TRUE | 2.434374015 |
| CAD | mesothelium | *GRK4* | TRUE | 3.473175295 |
| CAD | mesothelium | *ABHD8* | TRUE | 2.54251339 |
| CAD | mesothelium | *OPN1SW* | TRUE | 2.197459305 |
| CAD | mesothelium | *CCT6B* | TRUE | 2.422250825 |
| CAD | mesothelium | *B4GALNT1* | TRUE | 2.135251525 |
| CAD | mesothelium | *RCBTB1* | TRUE | 1.996430622 |
| CAD | mesothelium | *POLR1E* | TRUE | 2.447144751 |
| CAD | mesothelium | *TCF19* | TRUE | 2.235374159 |
| CAD | mesothelium | *POU2F3* | TRUE | 1.97746396 |

| | | | | |
|---|---|---|---|---|
| CAD | mesothelium | *ASCC1* | TRUE | 2.528912262 |
| CAD | mesothelium | *GFOD2* | TRUE | 2.179133656 |
| CAD | mesothelium | *TMEM91* | TRUE | 3.102626517 |
| CAD | mesothelium | *EVA1B* | TRUE | 2.532910297 |
| CAD | mesothelium | *NME7* | TRUE | 2.78774796 |
| CAD | mesothelium | *ARNT* | TRUE | 3.471845434 |
| CAD | mesothelium | *NBEAL1* | TRUE | 2.853948892 |
| CAD | mesothelium | *AMT* | TRUE | 2.337855021 |
| CAD | mesothelium | *FBXO10* | TRUE | 3.177792784 |
| CAD | mesothelium | *HSD17B12* | TRUE | 3.347100753 |
| CAD | mesothelium | *MIA3* | TRUE | 2.467846696 |
| CAD | mesothelium | *MMP16* | TRUE | 2.710735031 |
| CAD | mesothelium | *WDYHV1* | TRUE | 2.061916062 |
| CAD | mesothelium | *MX1* | TRUE | 2.104836323 |
| CAD | mesothelium | *S100A1* | TRUE | 2.40233796 |
| CAD | mesothelium | *DAPL1* | TRUE | 2.195846541 |
| CAD | mesothelium | *DAGLB* | TRUE | 2.228773717 |
| CAD | mesothelium | *OXGR1* | TRUE | 2.232732384 |
| CAD | mesothelium | *C16orf46* | TRUE | 2.858191864 |
| CAD | mesothelium | *CCDC68* | TRUE | 2.551061083 |
| CAD | mesothelium | *MS4A7* | TRUE | 1.974938267 |
| CAD | mesothelium | *ENGASE* | TRUE | 1.993604224 |
| CAD | mesothelium | *ULK4* | TRUE | 2.354150678 |
| CAD | mesothelium | *ATOH8* | TRUE | 2.121147968 |
| CAD | mesothelium | *LGALS9* | TRUE | 2.717313012 |
| CAD | mesothelium | *IRS1* | TRUE | 2.237081032 |
| CAD | mesothelium | *NPIPB3* | TRUE | 2.418179875 |
| CAD | mesothelium | *DNAJC18* | TRUE | 2.768179674 |
| CAD | mesothelium | *TTLL6* | TRUE | 2.250260672 |
| CAD | mesothelium | *RGS19* | TRUE | 2.676880555 |
| CAD | mesothelium | *NXNL1* | TRUE | 2.465367893 |
| CAD | mesothelium | *C3AR1* | TRUE | 3.076809468 |
| CAD | mesothelium | *C11orf80* | TRUE | 2.981187712 |
| CAD | mesothelium | *PAAF1* | TRUE | 2.351510987 |
| CAD | mesothelium | *MYPOP* | TRUE | 2.469939641 |
| CAD | mesothelium | *SPRYD4* | TRUE | 2.331341817 |
| CAD | mesothelium | *SLC25A22* | TRUE | 2.560266708 |
| CAD | mesothelium | *MPI* | TRUE | 2.018149106 |
| CAD | mesothelium | *LINC00174* | TRUE | 2.166365476 |
| CAD | mesothelium | *LYNX1* | TRUE | 2.857126963 |
| CAD | mesothelium | *DEFB119* | TRUE | 2.254126763 |
| CAD | mesothelium | *ADIPOQ* | TRUE | 2.706969115 |
| CAD | mesothelium | *TMEM136* | TRUE | 2.390590056 |
| CAD | mesothelium | *FES* | TRUE | 3.638613238 |
| CAD | mesothelium | *A3GALT2* | TRUE | 2.11332493 |
| CAD | mesothelium | *SEPT5* | TRUE | 2.599726982 |
| CAD | mesothelium | *PSMD13* | TRUE | 2.254614426 |
| CAD | mesothelium | *VSIG10L* | TRUE | 2.588095488 |
| CAD | mesothelium | *MSL1* | TRUE | 2.323497871 |
| CAD | mesothelium | *COLCA1* | TRUE | 2.751737307 |

| | | | | |
|---|---|---|---|---|
| CAD | mesothelium | *MYO6* | TRUE | 2.107919764 |
| CAD | mesothelium | *ZNF100* | TRUE | 2.223420652 |
| CAD | mesothelium | *ZNF334* | TRUE | 2.161692574 |
| CAD | mesothelium | *TMEM116* | TRUE | 3.451695607 |
| CAD | mesothelium | *SHISA4* | TRUE | 2.252456409 |
| CAD | mesothelium | *TBKBP1* | TRUE | 2.52186229 |
| CAD | mesothelium | *C6orf163* | TRUE | 2.513399126 |
| CAD | mesothelium | *STK19* | TRUE | 2.339463809 |
| CAD | mesothelium | *LINC00484* | TRUE | 2.088556655 |
| CAD | mesothelium | *LINC00707* | TRUE | 2.356137965 |
| CAD | mesothelium | *LINC00960* | TRUE | 2.285998106 |
| CAD | mesothelium | *GOLGA8K* | TRUE | 2.578261987 |
| CAD | mesothelium | *HMBS* | TRUE | 2.004027779 |
| CAD | mesothelium | *GRIN2B* | TRUE | 2.143718842 |
| CAD | monocyte | *SUSD2* | TRUE | 2.631443608 |
| CAD | monocyte | *BNIP1* | TRUE | 2.38120635 |
| CAD | monocyte | *DDC* | TRUE | 2.133255471 |
| CAD | monocyte | *TMEM91* | TRUE | 2.732560903 |
| CAD | monocyte | *CLEC18A* | TRUE | 2.083805981 |
| CAD | monocyte | *NXNL1* | TRUE | 2.378024921 |
| CAD | monocyte | *HYAL3* | TRUE | 1.970637507 |
| CAD | monocyte | *TMEM116* | TRUE | 2.301442065 |
| CAD | monocyte | *LY6G5B* | TRUE | 2.214645298 |
| CAD | monocyte | *TAS2R20* | TRUE | 2.318215274 |
| CAD | nk_cell | *TIPIN* | TRUE | 2.273623791 |
| CAD | nk_cell | *EIF2B2* | TRUE | 2.038682079 |
| CAD | nk_cell | *TCP11* | TRUE | 3.033180395 |
| CAD | nk_cell | *GRK4* | TRUE | 1.980298227 |
| CAD | nk_cell | *POLR1E* | TRUE | 2.577604288 |
| CAD | nk_cell | *CLEC18A* | TRUE | 2.112658436 |
| CAD | nk_cell | *NXNL1* | TRUE | 2.625422548 |
| CAD | nk_cell | *LINC00960* | TRUE | 2.193111563 |
| CAD | nk_cell | *TAS2R20* | TRUE | 2.317742548 |
| CAD | pericyte | *PAX6* | TRUE | 2.055373349 |
| CAD | pericyte | *ZMYND12* | TRUE | 2.174454846 |
| CAD | pericyte | *TIPIN* | TRUE | 2.095938378 |
| CAD | pericyte | *CENPM* | TRUE | 2.361538856 |
| CAD | pericyte | *GGCX* | TRUE | 2.176373711 |
| CAD | pericyte | *CFHR3* | TRUE | 2.363659098 |
| CAD | pericyte | *EIF2B2* | TRUE | 2.188532903 |
| CAD | pericyte | *TCP11* | TRUE | 2.542357892 |
| CAD | pericyte | *GRK4* | TRUE | 3.286907144 |
| CAD | pericyte | *DDC* | TRUE | 2.154642674 |
| CAD | pericyte | *POLR1E* | TRUE | 2.459111539 |
| CAD | pericyte | *TMEM91* | TRUE | 3.115810328 |
| CAD | pericyte | *SLC39A4* | TRUE | 2.223688509 |
| CAD | pericyte | *HSD17B12* | TRUE | 2.909797032 |
| CAD | pericyte | *MMP16* | TRUE | 2.200684798 |
| CAD | pericyte | *CLEC18A* | TRUE | 2.086827312 |
| CAD | pericyte | *C16orf46* | TRUE | 2.305531422 |

| | | | | |
|---|---|---|---|---|
| CAD | pericyte | *ENGASE* | TRUE | 1.969352602 |
| CAD | pericyte | *LGALS9* | TRUE | 2.288208061 |
| CAD | pericyte | *RGS19* | TRUE | 2.08734614 |
| CAD | pericyte | *NXNL1* | TRUE | 2.185583381 |
| CAD | pericyte | *C3AR1* | TRUE | 2.093826062 |
| CAD | pericyte | *CCNE2* | TRUE | 2.060631518 |
| CAD | pericyte | *LYNX1* | TRUE | 2.097122881 |
| CAD | pericyte | *ADIPOQ* | TRUE | 2.164177328 |
| CAD | pericyte | *FES* | TRUE | 2.035567804 |
| CAD | pericyte | *TMEM116* | TRUE | 3.125909749 |
| CAD | pericyte | *TBKBP1* | TRUE | 2.174421731 |
| CAD | pericyte | *C6orf163* | TRUE | 2.23363571 |
| CAD | pericyte | *LINC00960* | TRUE | 2.041086901 |
| CAD | pericyte | *SRP14.AS1* | TRUE | 2.014954483 |
| CAD | pericyte | *GOLGA8K* | TRUE | 2.193024449 |
| CAD | pericyte | *TAS2R20* | TRUE | 2.173256843 |
| CAD | SMC | *CD79B* | TRUE | 2.000524187 |
| CAD | SMC | *ZMYND12* | TRUE | 2.103738427 |
| CAD | SMC | *TIPIN* | TRUE | 2.51173508 |
| CAD | SMC | *BNIP1* | TRUE | 2.437485508 |
| CAD | SMC | *SNRPC* | TRUE | 2.65851186 |
| CAD | SMC | *TCP11* | TRUE | 2.514877675 |
| CAD | SMC | *GRK4* | TRUE | 3.423735611 |
| CAD | SMC | *POLR1E* | TRUE | 2.716693115 |
| CAD | SMC | *TMEM91* | TRUE | 2.718340013 |
| CAD | SMC | *HSD17B12* | TRUE | 3.006230149 |
| CAD | SMC | *MMP16* | TRUE | 2.369209601 |
| CAD | SMC | *NXNL1* | TRUE | 2.435898276 |
| CAD | SMC | *FES* | TRUE | 2.138328139 |
| CAD | SMC | *HYAL3* | TRUE | 2.04362283 |
| CAD | SMC | *TMEM116* | TRUE | 3.302983084 |
| CAD | SMC | *TBKBP1* | TRUE | 2.050075008 |
| CAD | SMC | *LY6G5B* | TRUE | 2.167210613 |
| CAD | SMC | *LINC00960* | TRUE | 2.216913402 |
| CAD | SMC | *GOLGA8K* | TRUE | 2.26720405 |
| CAD | t_cell | *UHRF1BP1* | TRUE | 3.169648187 |
| CAD | t_cell | *ZMYND12* | TRUE | 2.075230686 |
| CAD | t_cell | *TIPIN* | TRUE | 2.749645177 |
| CAD | t_cell | *ITPR3* | TRUE | 3.113609876 |
| CAD | t_cell | *SUSD2* | TRUE | 2.151334229 |
| CAD | t_cell | *EIF2B2* | TRUE | 2.318163426 |
| CAD | t_cell | *TCP11* | TRUE | 2.741738324 |
| CAD | t_cell | *GRK4* | TRUE | 3.393135747 |
| CAD | t_cell | *DDC* | TRUE | 2.371912757 |
| CAD | t_cell | *POLR1E* | TRUE | 2.595880163 |
| CAD | t_cell | *TMEM91* | TRUE | 2.037674131 |
| CAD | t_cell | *EVA1B* | TRUE | 2.494499641 |
| CAD | t_cell | *HSD17B12* | TRUE | 2.541350102 |
| CAD | t_cell | *RNF123* | TRUE | 2.861079256 |
| CAD | t_cell | *MS4A14* | TRUE | 2.059113024 |

| | | | | |
|---|---|---|---|---|
| CAD | t_cell | *LGALS9* | TRUE | 2.211502162 |
| CAD | t_cell | *NXNL1* | TRUE | 2.463490152 |
| CAD | t_cell | *C3AR1* | TRUE | 2.046330377 |
| CAD | t_cell | *LYNX1* | TRUE | 2.218861296 |
| CAD | t_cell | *HMBS* | TRUE | 2.058633731 |
| T2DM | adipocyte | *SNX11* | TRUE | 2.078921042 |
| T2DM | adipocyte | *CEP68* | TRUE | 4.194511934 |
| T2DM | adipocyte | *CD5L* | TRUE | 2.023003415 |
| T2DM | adipocyte | *TACR2* | TRUE | 2.653055369 |
| T2DM | adipocyte | *OVGP1* | TRUE | 2.232586597 |
| T2DM | adipocyte | *PABPC4* | TRUE | 4.398579411 |
| T2DM | adipocyte | *CWF19L1* | TRUE | 2.709550751 |
| T2DM | adipocyte | *NOMO3* | TRUE | 2.547306824 |
| T2DM | adipocyte | *TLE6* | TRUE | 2.588178512 |
| T2DM | adipocyte | *CHN2* | TRUE | 3.407812546 |
| T2DM | adipocyte | *CNTNAP1* | TRUE | 3.124377412 |
| T2DM | adipocyte | *GNRHR* | TRUE | 1.982818949 |
| T2DM | adipocyte | *C11orf21* | TRUE | 2.40816879 |
| T2DM | adipocyte | *CDK2AP1* | TRUE | 2.180630591 |
| T2DM | adipocyte | *BVES* | TRUE | 2.943122791 |
| T2DM | adipocyte | *NPHP3* | TRUE | 2.027705234 |
| T2DM | adipocyte | *GPX7* | TRUE | 2.882133565 |
| T2DM | adipocyte | *APOA1* | TRUE | 2.277946862 |
| T2DM | adipocyte | *ELOVL4* | TRUE | 2.555063777 |
| T2DM | adipocyte | *ADRA1A* | TRUE | 3.707836732 |
| T2DM | adipocyte | *CD80* | TRUE | 2.047474244 |
| T2DM | adipocyte | *GRK4* | TRUE | 2.952493239 |
| T2DM | adipocyte | *RIBC2* | TRUE | 2.119836241 |
| T2DM | adipocyte | *OPN1SW* | TRUE | 2.401356537 |
| T2DM | adipocyte | *PKDREJ* | TRUE | 2.200399385 |
| T2DM | adipocyte | *IRF4* | TRUE | 3.06457889 |
| T2DM | adipocyte | *TCF19* | TRUE | 1.976117337 |
| T2DM | adipocyte | *SLC37A4* | TRUE | 2.904047725 |
| T2DM | adipocyte | *MAN2C1* | TRUE | 3.190679243 |
| T2DM | adipocyte | *RPL3L* | TRUE | 2.045784852 |
| T2DM | adipocyte | *FN3KRP* | TRUE | 2.01155088 |
| T2DM | adipocyte | *DUS3L* | TRUE | 2.9067452 |
| T2DM | adipocyte | *ARHGEF19* | TRUE | 2.446417123 |
| T2DM | adipocyte | *HSD17B12* | TRUE | 4.726625119 |
| T2DM | adipocyte | *APIP* | TRUE | 3.001354123 |
| T2DM | adipocyte | *JAZF1* | TRUE | 4.602493767 |
| T2DM | adipocyte | *PCDH1* | TRUE | 1.970651107 |
| T2DM | adipocyte | *CLEC18A* | TRUE | 5.170874308 |
| T2DM | adipocyte | *KRTCAP3* | TRUE | 3.971695008 |
| T2DM | adipocyte | *FTCD* | TRUE | 1.96261682 |
| T2DM | adipocyte | *SPTA1* | TRUE | 2.974753598 |
| T2DM | adipocyte | *RFT1* | TRUE | 2.776138782 |
| T2DM | adipocyte | *ANPEP* | TRUE | 2.384444048 |
| T2DM | adipocyte | *ZSWIM1* | TRUE | 2.576274528 |
| T2DM | adipocyte | *IRS1* | TRUE | 3.468094074 |

| | | | | |
|---|---|---|---|---|
| T2DM | adipocyte | *TTLL6* | TRUE | 4.658522723 |
| T2DM | adipocyte | *RGS19* | TRUE | 3.287370809 |
| T2DM | adipocyte | *RNASEH2C* | TRUE | 2.06962675 |
| T2DM | adipocyte | *CCNE2* | TRUE | 3.573494825 |
| T2DM | adipocyte | *SLC25A22* | TRUE | 2.166301624 |
| T2DM | adipocyte | *ZNF223* | TRUE | 2.025086187 |
| T2DM | adipocyte | *STX19* | TRUE | 2.005718484 |
| T2DM | adipocyte | *TRIM73* | TRUE | 2.49373911 |
| T2DM | adipocyte | *RPRML* | TRUE | 2.726597408 |
| T2DM | adipocyte | *TTC3* | TRUE | 2.211870169 |
| T2DM | adipocyte | *ZNF320* | TRUE | 2.671600392 |
| T2DM | adipocyte | *NDUFA6* | TRUE | 2.460519615 |
| T2DM | adipocyte | *GP1BA* | TRUE | 2.295664705 |
| T2DM | adipocyte | *ZKSCAN7* | TRUE | 2.428907523 |
| T2DM | adipocyte | *CCDC154* | TRUE | 2.05351616 |
| T2DM | adipocyte | *ZNF780A* | TRUE | 1.987123247 |
| T2DM | adipocyte | *TMEM116* | TRUE | 2.035475547 |
| T2DM | adipocyte | *SYCE1L* | TRUE | 2.031050147 |
| T2DM | adipocyte | *LIN52* | TRUE | 2.445861194 |
| T2DM | adipocyte | *HCG27* | TRUE | 2.583277709 |
| T2DM | adipocyte | *ASIC3* | TRUE | 1.979490588 |
| T2DM | adipocyte | *DNLZ* | TRUE | 3.49712892 |
| T2DM | adipocyte | *STARD10* | TRUE | 5.280456696 |
| T2DM | adipocyte | *AC016773.1* | TRUE | 2.01793066 |
| T2DM | adipocyte | *GS1.124K5.4* | TRUE | 2.27495518 |
| T2DM | adipocyte | *HLA.DQA2* | TRUE | 3.647546555 |
| T2DM | adipocyte | *NME1.NME2* | TRUE | 2.477292423 |
| T2DM | adipocyte | *SRP14.AS1* | TRUE | 2.064689588 |
| T2DM | adipocyte | *AP001172.2* | TRUE | 2.649471837 |
| T2DM | ASPC | *RBM6* | TRUE | 2.023745927 |
| T2DM | ASPC | *TRAF3IP3* | TRUE | 2.002399204 |
| T2DM | ASPC | *CEP68* | TRUE | 4.298846306 |
| T2DM | ASPC | *SEC61A2* | TRUE | 2.24104952 |
| T2DM | ASPC | *CD5L* | TRUE | 2.072630807 |
| T2DM | ASPC | *TACR2* | TRUE | 2.624954626 |
| T2DM | ASPC | *OVGP1* | TRUE | 2.327461213 |
| T2DM | ASPC | *PABPC4* | TRUE | 5.156154884 |
| T2DM | ASPC | *CWF19L1* | TRUE | 3.121510562 |
| T2DM | ASPC | *NOMO3* | TRUE | 2.81597165 |
| T2DM | ASPC | *TLE6* | TRUE | 2.674707473 |
| T2DM | ASPC | *ICAM5* | TRUE | 2.489018655 |
| T2DM | ASPC | *GCDH* | TRUE | 2.365403311 |
| T2DM | ASPC | *CHN2* | TRUE | 2.306132354 |
| T2DM | ASPC | *GNRHR* | TRUE | 1.97139657 |
| T2DM | ASPC | *C11orf21* | TRUE | 2.705304218 |
| T2DM | ASPC | *CDK2AP1* | TRUE | 2.389480218 |
| T2DM | ASPC | *BVES* | TRUE | 3.320155808 |
| T2DM | ASPC | *NPHP3* | TRUE | 1.963953778 |
| T2DM | ASPC | *GPX7* | TRUE | 3.149166965 |
| T2DM | ASPC | *APOA1* | TRUE | 2.289862563 |

| | | | | |
|---|---|---|---|---|
| T2DM | ASPC | *ELOVL4* | TRUE | 2.162749124 |
| T2DM | ASPC | *TECTB* | TRUE | 2.0871882 |
| T2DM | ASPC | *ADRA1A* | TRUE | 3.764035988 |
| T2DM | ASPC | *PDZRN3* | TRUE | 2.211243588 |
| T2DM | ASPC | *CD80* | TRUE | 2.257209975 |
| T2DM | ASPC | *KCNK17* | TRUE | 3.029197996 |
| T2DM | ASPC | *GRK4* | TRUE | 2.975580668 |
| T2DM | ASPC | *ABHD8* | TRUE | 2.221150296 |
| T2DM | ASPC | *RIBC2* | TRUE | 2.096576658 |
| T2DM | ASPC | *OPN1SW* | TRUE | 2.798359824 |
| T2DM | ASPC | *STK33* | TRUE | 2.273044581 |
| T2DM | ASPC | *PKDREJ* | TRUE | 2.67145531 |
| T2DM | ASPC | *MICAL1* | TRUE | 2.078890721 |
| T2DM | ASPC | *IRF4* | TRUE | 3.187630086 |
| T2DM | ASPC | *TCF19* | TRUE | 3.065099628 |
| T2DM | ASPC | *SLC37A4* | TRUE | 3.014873765 |
| T2DM | ASPC | *TRPC3* | TRUE | 2.377680568 |
| T2DM | ASPC | *MAN2C1* | TRUE | 2.665081199 |
| T2DM | ASPC | *RPL3L* | TRUE | 2.264931607 |
| T2DM | ASPC | *C18orf8* | TRUE | 2.585376891 |
| T2DM | ASPC | *FN3KRP* | TRUE | 1.966391232 |
| T2DM | ASPC | *DUS3L* | TRUE | 2.920879996 |
| T2DM | ASPC | *ARHGEF19* | TRUE | 2.497836553 |
| T2DM | ASPC | *BHMT* | TRUE | 2.155087632 |
| T2DM | ASPC | *HSD17B12* | TRUE | 5.01780832 |
| T2DM | ASPC | *APIP* | TRUE | 3.167036009 |
| T2DM | ASPC | *POC5* | TRUE | 2.570536013 |
| T2DM | ASPC | *JAZF1* | TRUE | 4.705151748 |
| T2DM | ASPC | *PCDH1* | TRUE | 1.973617318 |
| T2DM | ASPC | *CLEC18A* | TRUE | 5.653811432 |
| T2DM | ASPC | *KRTCAP3* | TRUE | 4.04861657 |
| T2DM | ASPC | *FTCD* | TRUE | 2.074710272 |
| T2DM | ASPC | *SPTA1* | TRUE | 3.107351642 |
| T2DM | ASPC | *RFT1* | TRUE | 3.168236119 |
| T2DM | ASPC | *ZSWIM1* | TRUE | 2.709565602 |
| T2DM | ASPC | *IRS1* | TRUE | 4.668822625 |
| T2DM | ASPC | *C8orf46* | TRUE | 2.556957422 |
| T2DM | ASPC | *INO80E* | TRUE | 1.98049075 |
| T2DM | ASPC | *SEMA3E* | TRUE | 2.194471312 |
| T2DM | ASPC | *TTLL6* | TRUE | 4.40148998 |
| T2DM | ASPC | *RGS19* | TRUE | 3.600653971 |
| T2DM | ASPC | *RNASEH2C* | TRUE | 2.525464805 |
| T2DM | ASPC | *CCNE2* | TRUE | 3.970532781 |
| T2DM | ASPC | *SPRYD4* | TRUE | 2.221966993 |
| T2DM | ASPC | *SLC25A22* | TRUE | 2.236311221 |
| T2DM | ASPC | *ZNF223* | TRUE | 2.148602439 |
| T2DM | ASPC | *STX19* | TRUE | 2.383148323 |
| T2DM | ASPC | *TRIM73* | TRUE | 2.893599541 |
| T2DM | ASPC | *RPRML* | TRUE | 2.856062284 |
| T2DM | ASPC | *ZNF320* | TRUE | 2.616044865 |

| | | | | |
|---|---|---|---|---|
| T2DM | ASPC | *EMILIN3* | TRUE | 2.019741628 |
| T2DM | ASPC | *ACOT1* | TRUE | 2.04798351 |
| T2DM | ASPC | *NDUFA6* | TRUE | 2.485482393 |
| T2DM | ASPC | *GP1BA* | TRUE | 2.78024748 |
| T2DM | ASPC | *EVI2B* | TRUE | 2.106235006 |
| T2DM | ASPC | *FFAR4* | TRUE | 5.420390861 |
| T2DM | ASPC | *ZKSCAN7* | TRUE | 2.286722341 |
| T2DM | ASPC | *FAM110D* | TRUE | 2.136119373 |
| T2DM | ASPC | *CCDC154* | TRUE | 2.005132683 |
| T2DM | ASPC | *ZNF780A* | TRUE | 2.02365776 |
| T2DM | ASPC | *TMEM116* | TRUE | 2.318494297 |
| T2DM | ASPC | *NOTCH4* | TRUE | 2.352627232 |
| T2DM | ASPC | *FAM153C* | TRUE | 2.215380332 |
| T2DM | ASPC | *LIN52* | TRUE | 2.242021795 |
| T2DM | ASPC | *HCG27* | TRUE | 2.520013746 |
| T2DM | ASPC | *ASIC3* | TRUE | 1.96049014 |
| T2DM | ASPC | *DNLZ* | TRUE | 3.487046752 |
| T2DM | ASPC | *ARHGAP19* | TRUE | 3.043443865 |
| T2DM | ASPC | *STARD10* | TRUE | 5.346118964 |
| T2DM | ASPC | *EVPLL* | TRUE | 2.097412255 |
| T2DM | ASPC | *AC016773.1* | TRUE | 2.347828977 |
| T2DM | ASPC | *POLR2J2* | TRUE | 2.20339205 |
| T2DM | ASPC | *LINC00941* | TRUE | 2.759029716 |
| T2DM | ASPC | *GS1.124K5.4* | TRUE | 2.233202863 |
| T2DM | ASPC | *HLA.DQA2* | TRUE | 3.904232775 |
| T2DM | ASPC | *NME1.NME2* | TRUE | 2.409127103 |
| T2DM | ASPC | *PLA2G4B* | TRUE | 2.084380109 |
| T2DM | ASPC | *SRP14.AS1* | TRUE | 2.191556879 |
| T2DM | ASPC | *MC1R* | TRUE | 2.28554455 |
| T2DM | ASPC | *AP001172.2* | TRUE | 2.500504563 |
| T2DM | ASPC | *YTHDF3.AS1* | TRUE | 2.666587873 |
| T2DM | b_cell | *ITIH4* | TRUE | 2.478392217 |
| T2DM | b_cell | *PKDREJ* | TRUE | 2.092639669 |
| T2DM | b_cell | *CLEC18A* | TRUE | 5.484473331 |
| T2DM | b_cell | *CTSW* | TRUE | 3.48575657 |
| T2DM | b_cell | *TRIM73* | TRUE | 2.313255375 |
| T2DM | b_cell | *HLA.DRB5* | TRUE | 2.807325517 |
| T2DM | b_cell | *POLR2J2* | TRUE | 2.167102953 |
| T2DM | b_cell | *HLA.DQA2* | TRUE | 5.565624227 |
| T2DM | dendritic_cell | *RBM6* | TRUE | 2.369591725 |
| T2DM | dendritic_cell | *PABPC4* | TRUE | 2.548553545 |
| T2DM | dendritic_cell | *CWF19L1* | TRUE | 2.134464871 |
| T2DM | dendritic_cell | *OPN1SW* | TRUE | 2.02871465 |
| T2DM | dendritic_cell | *PKDREJ* | TRUE | 2.064807699 |
| T2DM | dendritic_cell | *SPTBN5* | TRUE | 2.864409607 |
| T2DM | dendritic_cell | *APIP* | TRUE | 2.176878568 |
| T2DM | dendritic_cell | *CLEC18A* | TRUE | 5.222258316 |
| T2DM | dendritic_cell | *SPTA1* | TRUE | 3.020374555 |
| T2DM | dendritic_cell | *C1QTNF4* | TRUE | 2.44399065 |
| T2DM | dendritic_cell | *CTSW* | TRUE | 1.967524954 |

| | | | | |
|---|---|---|---|---|
| T2DM | dendritic_cell | *CCNE2* | TRUE | 2.8514478 |
| T2DM | dendritic_cell | *TRIM73* | TRUE | 1.960697998 |
| T2DM | dendritic_cell | *NOTCH4* | TRUE | 2.785077148 |
| T2DM | dendritic_cell | *DNLZ* | TRUE | 3.12314523 |
| T2DM | dendritic_cell | *TMEM110* | TRUE | 2.187720316 |
| T2DM | dendritic_cell | *STARD10* | TRUE | 3.344349492 |
| T2DM | dendritic_cell | *POLR2J2* | TRUE | 2.211143648 |
| T2DM | dendritic_cell | *LINC00941* | TRUE | 2.343783792 |
| T2DM | dendritic_cell | *NME1.NME2* | TRUE | 2.623632573 |
| T2DM | dendritic_cell | *TAS2R20* | TRUE | 2.312874616 |
| T2DM | endometrium | *ADRA1A* | TRUE | 2.015704569 |
| T2DM | endometrium | *TCF19* | TRUE | 2.682302651 |
| T2DM | endometrium | *MAN2C1* | TRUE | 2.318173086 |
| T2DM | endometrium | *KRTCAP3* | TRUE | 3.576869811 |
| T2DM | endometrium | *ZSWIM1* | TRUE | 2.26158804 |
| T2DM | endometrium | *GP1BA* | TRUE | 2.057841979 |
| T2DM | endometrium | *HLA.DRB5* | TRUE | 3.074164763 |
| T2DM | endometrium | *DNLZ* | TRUE | 3.316193336 |
| T2DM | endometrium | *STARD10* | TRUE | 2.435837607 |
| T2DM | endometrium | *POLR2J2* | TRUE | 2.266633218 |
| T2DM | endometrium | *HLA.DQA2* | TRUE | 5.74557069 |
| T2DM | endometrium | *PLA2G4B* | TRUE | 3.486212388 |
| T2DM | endometrium | *AP001172.2* | TRUE | 2.285222336 |
| T2DM | endothelial | *PABPC4* | TRUE | 2.240831542 |
| T2DM | endothelial | *BVES* | TRUE | 2.51882362 |
| T2DM | endothelial | *GPX7* | TRUE | 2.500919175 |
| T2DM | endothelial | *PKDREJ* | TRUE | 2.330883737 |
| T2DM | endothelial | *TCF19* | TRUE | 2.649444744 |
| T2DM | endothelial | *MAN2C1* | TRUE | 2.575921981 |
| T2DM | endothelial | *HSD17B12* | TRUE | 2.684787212 |
| T2DM | endothelial | *APIP* | TRUE | 2.163764338 |
| T2DM | endothelial | *CLEC18A* | TRUE | 5.354913073 |
| T2DM | endothelial | *KRTCAP3* | TRUE | 2.620734309 |
| T2DM | endothelial | *SPTA1* | TRUE | 3.08866963 |
| T2DM | endothelial | *IRS1* | TRUE | 2.845254122 |
| T2DM | endothelial | *TTLL6* | TRUE | 3.944206291 |
| T2DM | endothelial | *RGS19* | TRUE | 2.766168895 |
| T2DM | endothelial | *CTSW* | TRUE | 3.927879441 |
| T2DM | endothelial | *CCNE2* | TRUE | 2.145642845 |
| T2DM | endothelial | *DNLZ* | TRUE | 2.377159339 |
| T2DM | endothelial | *STARD10* | TRUE | 3.029250062 |
| T2DM | endothelial | *HLA.DQA2* | TRUE | 3.677523866 |
| T2DM | endothelial | *TDGF1* | TRUE | 2.015755802 |
| T2DM | endothelial | *PLA2G4B* | TRUE | 4.617331245 |
| T2DM | endothelial | *TAS2R20* | TRUE | 2.800157315 |
| T2DM | LEC | *PABPC4* | TRUE | 2.929777279 |
| T2DM | LEC | *TLE6* | TRUE | 2.451383669 |
| T2DM | LEC | *GPX7* | TRUE | 2.244997743 |
| T2DM | LEC | *ADRA1A* | TRUE | 2.570164223 |
| T2DM | LEC | *GRK4* | TRUE | 2.548484734 |

| | | | | |
|---|---|---|---|---|
| T2DM | LEC | *OPN1SW* | TRUE | 1.995026364 |
| T2DM | LEC | *PKDREJ* | TRUE | 1.993111892 |
| T2DM | LEC | *TCF19* | TRUE | 3.368132877 |
| T2DM | LEC | *SLC37A4* | TRUE | 2.400841238 |
| T2DM | LEC | *MAN2C1* | TRUE | 3.163551052 |
| T2DM | LEC | *ARHGEF19* | TRUE | 2.420069891 |
| T2DM | LEC | *BHMT* | TRUE | 2.010447498 |
| T2DM | LEC | *HSD17B12* | TRUE | 4.243875272 |
| T2DM | LEC | *APIP* | TRUE | 2.306640986 |
| T2DM | LEC | *CLEC18A* | TRUE | 4.765170054 |
| T2DM | LEC | *KRTCAP3* | TRUE | 3.486629855 |
| T2DM | LEC | *RFT1* | TRUE | 2.262430402 |
| T2DM | LEC | *MST1R* | TRUE | 2.266864243 |
| T2DM | LEC | *ZSWIM1* | TRUE | 2.557022615 |
| T2DM | LEC | *IRS1* | TRUE | 2.638379647 |
| T2DM | LEC | *C8orf46* | TRUE | 2.306919484 |
| T2DM | LEC | *INO80E* | TRUE | 1.979747812 |
| T2DM | LEC | *TTLL6* | TRUE | 3.461426221 |
| T2DM | LEC | *RGS19* | TRUE | 3.131613544 |
| T2DM | LEC | *TRIM73* | TRUE | 2.211597103 |
| T2DM | LEC | *RPRML* | TRUE | 2.130527406 |
| T2DM | LEC | *ZNF320* | TRUE | 2.286762657 |
| T2DM | LEC | *HLA.DRB5* | TRUE | 2.348013112 |
| T2DM | LEC | *NOTCH4* | TRUE | 2.078772017 |
| T2DM | LEC | *DNLZ* | TRUE | 3.333233956 |
| T2DM | LEC | *STARD10* | TRUE | 5.089798388 |
| T2DM | LEC | *POLR2J2* | TRUE | 2.224678833 |
| T2DM | LEC | *GS1.124K5.4* | TRUE | 2.355815185 |
| T2DM | LEC | *HLA.DQA2* | TRUE | 2.714659563 |
| T2DM | macrophage | *RBM6* | TRUE | 2.138304909 |
| T2DM | macrophage | *CEP68* | TRUE | 3.250942003 |
| T2DM | macrophage | *OVGP1* | TRUE | 2.039950802 |
| T2DM | macrophage | *PABPC4* | TRUE | 2.606257897 |
| T2DM | macrophage | *CWF19L1* | TRUE | 2.423013966 |
| T2DM | macrophage | *NOMO3* | TRUE | 2.322638464 |
| T2DM | macrophage | *TLE6* | TRUE | 2.625897081 |
| T2DM | macrophage | *CDK2AP1* | TRUE | 3.044051429 |
| T2DM | macrophage | *BVES* | TRUE | 3.058791922 |
| T2DM | macrophage | *GPX7* | TRUE | 2.184317018 |
| T2DM | macrophage | *ADRA1A* | TRUE | 2.818472965 |
| T2DM | macrophage | *GRK4* | TRUE | 2.658805108 |
| T2DM | macrophage | *PKDREJ* | TRUE | 2.392327324 |
| T2DM | macrophage | *TCF19* | TRUE | 2.391820863 |
| T2DM | macrophage | *MAN2C1* | TRUE | 3.104566308 |
| T2DM | macrophage | *RPL3L* | TRUE | 2.130697124 |
| T2DM | macrophage | *FN3KRP* | TRUE | 2.139552186 |
| T2DM | macrophage | *DUS3L* | TRUE | 2.739603107 |
| T2DM | macrophage | *ARHGEF19* | TRUE | 2.045580238 |
| T2DM | macrophage | *HSD17B12* | TRUE | 4.629983575 |
| T2DM | macrophage | *APIP* | TRUE | 2.629440908 |

| | | | | |
|---|---|---|---|---|
| T2DM | macrophage | *JAZF1* | TRUE | 4.423416328 |
| T2DM | macrophage | *CLEC18A* | TRUE | 5.58401526 |
| T2DM | macrophage | *KRTCAP3* | TRUE | 3.856090564 |
| T2DM | macrophage | *PAQR6* | TRUE | 2.085666349 |
| T2DM | macrophage | *SPTA1* | TRUE | 3.49079786 |
| T2DM | macrophage | *RFT1* | TRUE | 2.78637183 |
| T2DM | macrophage | *RAPSN* | TRUE | 2.216516085 |
| T2DM | macrophage | *ANPEP* | TRUE | 2.399509306 |
| T2DM | macrophage | *ZSWIM1* | TRUE | 2.973887301 |
| T2DM | macrophage | *IRS1* | TRUE | 2.99157252 |
| T2DM | macrophage | *C8orf46* | TRUE | 1.98731983 |
| T2DM | macrophage | *TTLL6* | TRUE | 3.648546527 |
| T2DM | macrophage | *CCNE2* | TRUE | 2.82438912 |
| T2DM | macrophage | *TRIM73* | TRUE | 2.56857574 |
| T2DM | macrophage | *RPRML* | TRUE | 2.22874237 |
| T2DM | macrophage | *ZNF320* | TRUE | 2.533788303 |
| T2DM | macrophage | *NDUFA6* | TRUE | 2.280057715 |
| T2DM | macrophage | *GP1BA* | TRUE | 2.140223909 |
| T2DM | macrophage | *NOTCH4* | TRUE | 2.290863347 |
| T2DM | macrophage | *SYCE1L* | TRUE | 2.00598743 |
| T2DM | macrophage | *HCG27* | TRUE | 2.153407309 |
| T2DM | macrophage | *DNLZ* | TRUE | 3.559817109 |
| T2DM | macrophage | *STARD10* | TRUE | 5.064403829 |
| T2DM | macrophage | *AC016773.1* | TRUE | 1.985209804 |
| T2DM | macrophage | *POLR2J2* | TRUE | 2.19516844 |
| T2DM | macrophage | *GS1.124K5.4* | TRUE | 2.126753514 |
| T2DM | macrophage | *HLA.DQA2* | TRUE | 4.419101108 |
| T2DM | macrophage | *NME1.NME2* | TRUE | 2.686825973 |
| T2DM | macrophage | *PLA2G4B* | TRUE | 2.085607672 |
| T2DM | macrophage | *AP001172.2* | TRUE | 2.105449616 |
| T2DM | macrophage | *YTHDF3.AS1* | TRUE | 2.178346911 |
| T2DM | mast_cell | *GNB3* | TRUE | 2.354845523 |
| T2DM | mast_cell | *BVES* | TRUE | 2.482596398 |
| T2DM | mast_cell | *GPX7* | TRUE | 2.623828349 |
| T2DM | mast_cell | *ADRA1A* | TRUE | 3.391373627 |
| T2DM | mast_cell | *PKDREJ* | TRUE | 2.170016172 |
| T2DM | mast_cell | *IRF4* | TRUE | 2.112658322 |
| T2DM | mast_cell | *CLEC18A* | TRUE | 5.304514374 |
| T2DM | mast_cell | *SPTA1* | TRUE | 3.011750288 |
| T2DM | mast_cell | *RAPSN* | TRUE | 2.931151028 |
| T2DM | mast_cell | *ANPEP* | TRUE | 2.662455801 |
| T2DM | mast_cell | *ZSWIM1* | TRUE | 2.452089611 |
| T2DM | mast_cell | *RGS19* | TRUE | 3.469603905 |
| T2DM | mast_cell | *RPRML* | TRUE | 2.383208263 |
| T2DM | mast_cell | *ACOT1* | TRUE | 2.189195496 |
| T2DM | mast_cell | *HLA.DRB5* | TRUE | 2.305585046 |
| T2DM | mast_cell | *DNLZ* | TRUE | 3.418652813 |
| T2DM | mast_cell | *TMEM110* | TRUE | 2.029520239 |
| T2DM | mast_cell | *STARD10* | TRUE | 3.935899718 |
| T2DM | mast_cell | *HLA.DQA2* | TRUE | 6.409162694 |

| | | | | |
|---|---|---|---|---|
| T2DM | mast_cell | *NME1.NME2* | TRUE | 2.092377388 |
| T2DM | mast_cell | *PLA2G4B* | TRUE | 2.801461669 |
| T2DM | mesothelium | *RBM6* | TRUE | 4.141768141 |
| T2DM | mesothelium | *CEP68* | TRUE | 3.978079428 |
| T2DM | mesothelium | *TUBG2* | TRUE | 2.97710571 |
| T2DM | mesothelium | *PIGV* | TRUE | 2.176447961 |
| T2DM | mesothelium | *SEC61A2* | TRUE | 3.402223655 |
| T2DM | mesothelium | *CD5L* | TRUE | 1.999271437 |
| T2DM | mesothelium | *TACR2* | TRUE | 2.571210332 |
| T2DM | mesothelium | *DCT* | TRUE | 2.185020761 |
| T2DM | mesothelium | *OVGP1* | TRUE | 2.533139996 |
| T2DM | mesothelium | *SNX10* | TRUE | 2.258958901 |
| T2DM | mesothelium | *PABPC4* | TRUE | 6.069441518 |
| T2DM | mesothelium | *CWF19L1* | TRUE | 3.555958842 |
| T2DM | mesothelium | *SCD* | TRUE | 2.017603581 |
| T2DM | mesothelium | *NOMO3* | TRUE | 3.121441946 |
| T2DM | mesothelium | *TLE6* | TRUE | 2.546865843 |
| T2DM | mesothelium | *ICAM5* | TRUE | 2.477401328 |
| T2DM | mesothelium | *GCDH* | TRUE | 2.350297837 |
| T2DM | mesothelium | *RNF32* | TRUE | 2.308086383 |
| T2DM | mesothelium | *C11orf21* | TRUE | 3.056023308 |
| T2DM | mesothelium | *CDK2AP1* | TRUE | 3.565920676 |
| T2DM | mesothelium | *BVES* | TRUE | 3.288002723 |
| T2DM | mesothelium | *CHMP3* | TRUE | 2.092661869 |
| T2DM | mesothelium | *GPX7* | TRUE | 3.03059546 |
| T2DM | mesothelium | *APOA1* | TRUE | 2.169314158 |
| T2DM | mesothelium | *ELOVL4* | TRUE | 3.026444685 |
| T2DM | mesothelium | *TECTB* | TRUE | 2.135349981 |
| T2DM | mesothelium | *ADRA1A* | TRUE | 3.728027566 |
| T2DM | mesothelium | *TEX14* | TRUE | 2.071248598 |
| T2DM | mesothelium | *PDZRN3* | TRUE | 2.53163324 |
| T2DM | mesothelium | *CD80* | TRUE | 2.478932965 |
| T2DM | mesothelium | *RPS10* | TRUE | 2.062304121 |
| T2DM | mesothelium | *KCNK17* | TRUE | 3.074819818 |
| T2DM | mesothelium | *GRK4* | TRUE | 2.969377267 |
| T2DM | mesothelium | *XRCC3* | TRUE | 2.099084039 |
| T2DM | mesothelium | *ABHD8* | TRUE | 2.842729624 |
| T2DM | mesothelium | *RIBC2* | TRUE | 1.973966094 |
| T2DM | mesothelium | *OPN1SW* | TRUE | 2.742268944 |
| T2DM | mesothelium | *PKDREJ* | TRUE | 2.67703097 |
| T2DM | mesothelium | *MICAL1* | TRUE | 2.012715808 |
| T2DM | mesothelium | *RCBTB1* | TRUE | 2.070213613 |
| T2DM | mesothelium | *IRF4* | TRUE | 3.104074284 |
| T2DM | mesothelium | *TCF19* | TRUE | 2.708464659 |
| T2DM | mesothelium | *SLC37A4* | TRUE | 2.915989903 |
| T2DM | mesothelium | *TRPC3* | TRUE | 2.37381824 |
| T2DM | mesothelium | *GCSH* | TRUE | 2.065246239 |
| T2DM | mesothelium | *RPL3L* | TRUE | 2.041590634 |
| T2DM | mesothelium | *VPS53* | TRUE | 2.208007397 |
| T2DM | mesothelium | *C18orf8* | TRUE | 3.629226267 |

| | | | | |
|---|---|---|---|---|
| T2DM | mesothelium | *FN3KRP* | TRUE | 2.756685508 |
| T2DM | mesothelium | *DUS3L* | TRUE | 2.85686726 |
| T2DM | mesothelium | *SAE1* | TRUE | 2.412894438 |
| T2DM | mesothelium | *GEMIN7* | TRUE | 2.0097632 |
| T2DM | mesothelium | *ARHGEF19* | TRUE | 2.535522224 |
| T2DM | mesothelium | *PLEKHA6* | TRUE | 2.308247597 |
| T2DM | mesothelium | *BHMT* | TRUE | 2.130356098 |
| T2DM | mesothelium | *HSD17B12* | TRUE | 5.350269618 |
| T2DM | mesothelium | *APIP* | TRUE | 3.424179838 |
| T2DM | mesothelium | *CCDC15* | TRUE | 1.965518656 |
| T2DM | mesothelium | *POC5* | TRUE | 2.939734911 |
| T2DM | mesothelium | *JAZF1* | TRUE | 5.350626432 |
| T2DM | mesothelium | *PCDH1* | TRUE | 2.140259572 |
| T2DM | mesothelium | *CLEC18A* | TRUE | 5.196138473 |
| T2DM | mesothelium | *KRTCAP3* | TRUE | 3.72489215 |
| T2DM | mesothelium | *SPATA2L* | TRUE | 2.174413722 |
| T2DM | mesothelium | *FTCD* | TRUE | 2.212845761 |
| T2DM | mesothelium | *PAQR6* | TRUE | 2.237631825 |
| T2DM | mesothelium | *SPTA1* | TRUE | 3.129158939 |
| T2DM | mesothelium | *RFT1* | TRUE | 3.731624745 |
| T2DM | mesothelium | *TMEM63C* | TRUE | 2.003925419 |
| T2DM | mesothelium | *RAPSN* | TRUE | 2.522754398 |
| T2DM | mesothelium | *ANPEP* | TRUE | 2.810389461 |
| T2DM | mesothelium | *SERINC2* | TRUE | 2.043263744 |
| T2DM | mesothelium | *ZSWIM1* | TRUE | 2.623254512 |
| T2DM | mesothelium | *IRS1* | TRUE | 4.710583862 |
| T2DM | mesothelium | *INO80E* | TRUE | 3.118572994 |
| T2DM | mesothelium | *LRRC45* | TRUE | 2.133015532 |
| T2DM | mesothelium | *SEMA3E* | TRUE | 1.984065241 |
| T2DM | mesothelium | *EMB* | TRUE | 3.085509476 |
| T2DM | mesothelium | *TTLL6* | TRUE | 5.075730352 |
| T2DM | mesothelium | *RGS19* | TRUE | 3.829221971 |
| T2DM | mesothelium | *CTSW* | TRUE | 2.386787842 |
| T2DM | mesothelium | *RNASEH2C* | TRUE | 2.612160665 |
| T2DM | mesothelium | *HOXB2* | TRUE | 2.290383734 |
| T2DM | mesothelium | *SPRYD4* | TRUE | 2.26698801 |
| T2DM | mesothelium | *SLC25A22* | TRUE | 2.756393594 |
| T2DM | mesothelium | *STX19* | TRUE | 2.213569362 |
| T2DM | mesothelium | *ZNF852* | TRUE | 2.344545448 |
| T2DM | mesothelium | *RPRML* | TRUE | 2.769557182 |
| T2DM | mesothelium | *ZNF320* | TRUE | 2.566996872 |
| T2DM | mesothelium | *ACOT1* | TRUE | 2.063347136 |
| T2DM | mesothelium | *NDUFA6* | TRUE | 2.149922776 |
| T2DM | mesothelium | *GP1BA* | TRUE | 2.531137178 |
| T2DM | mesothelium | *DMWD* | TRUE | 2.588503412 |
| T2DM | mesothelium | *EVI2B* | TRUE | 2.053880575 |
| T2DM | mesothelium | *RASA3* | TRUE | 2.510081953 |
| T2DM | mesothelium | *FFAR4* | TRUE | 4.13460683 |
| T2DM | mesothelium | *COLCA1* | TRUE | 2.022914213 |
| T2DM | mesothelium | *ZNF780A* | TRUE | 2.153574154 |

| | | | | |
|---|---|---|---|---|
| T2DM | mesothelium | *TMEM116* | TRUE | 2.521258446 |
| T2DM | mesothelium | *HLA.DRB5* | TRUE | 2.653547224 |
| T2DM | mesothelium | *SHISA4* | TRUE | 2.486425884 |
| T2DM | mesothelium | *LIN52* | TRUE | 1.997623218 |
| T2DM | mesothelium | *HCG27* | TRUE | 2.378137758 |
| T2DM | mesothelium | *DNLZ* | TRUE | 3.624306497 |
| T2DM | mesothelium | *ARHGAP19* | TRUE | 3.366030913 |
| T2DM | mesothelium | *STARD10* | TRUE | 5.2731063 |
| T2DM | mesothelium | *AC016773.1* | TRUE | 2.426065758 |
| T2DM | mesothelium | *AC108463.2* | TRUE | 2.797641394 |
| T2DM | mesothelium | *POLR2J2* | TRUE | 2.190514536 |
| T2DM | mesothelium | *LINC00332* | TRUE | 1.997187955 |
| T2DM | mesothelium | *LINC00957* | TRUE | 2.294355418 |
| T2DM | mesothelium | *LINC00941* | TRUE | 2.561988407 |
| T2DM | mesothelium | *GS1.124K5.4* | TRUE | 2.49558881 |
| T2DM | mesothelium | *HLA.DQA2* | TRUE | 3.530644534 |
| T2DM | mesothelium | *NME1.NME2* | TRUE | 2.713494861 |
| T2DM | mesothelium | *SMIM6* | TRUE | 1.974591594 |
| T2DM | mesothelium | *LINC00675* | TRUE | 2.269833366 |
| T2DM | mesothelium | *AP001172.2* | TRUE | 2.593210422 |
| T2DM | mesothelium | *YTHDF3.AS1* | TRUE | 2.567172367 |
| T2DM | monocyte | *ITIH4* | TRUE | 3.273797569 |
| T2DM | monocyte | *NOMO3* | TRUE | 2.244670157 |
| T2DM | monocyte | *TLE6* | TRUE | 2.301988594 |
| T2DM | monocyte | *IRF4* | TRUE | 2.396271329 |
| T2DM | monocyte | *CLEC18A* | TRUE | 5.054702533 |
| T2DM | monocyte | *SPTA1* | TRUE | 3.42986924 |
| T2DM | monocyte | *C1QTNF4* | TRUE | 2.057875725 |
| T2DM | monocyte | *HLA.DRB5* | TRUE | 2.075650421 |
| T2DM | monocyte | *NOTCH4* | TRUE | 2.467129398 |
| T2DM | monocyte | *DNLZ* | TRUE | 2.991949016 |
| T2DM | monocyte | *STARD10* | TRUE | 4.62310333 |
| T2DM | monocyte | *HLA.DQA2* | TRUE | 3.477285722 |
| T2DM | monocyte | *TAS2R20* | TRUE | 2.154656928 |
| T2DM | nk_cell | *RIBC2* | TRUE | 1.963101166 |
| T2DM | nk_cell | *IRF4* | TRUE | 2.881432962 |
| T2DM | nk_cell | *SPTBN5* | TRUE | 2.502915333 |
| T2DM | nk_cell | *ARHGEF19* | TRUE | 2.075541683 |
| T2DM | nk_cell | *HSD17B12* | TRUE | 2.759275946 |
| T2DM | nk_cell | *APIP* | TRUE | 2.012390725 |
| T2DM | nk_cell | *CLEC18A* | TRUE | 4.635218185 |
| T2DM | nk_cell | *SPTA1* | TRUE | 2.444894547 |
| T2DM | nk_cell | *IRS1* | TRUE | 2.656902717 |
| T2DM | nk_cell | *HLA.DQA1* | TRUE | 2.998391541 |
| T2DM | nk_cell | *NOTCH4* | TRUE | 2.256742135 |
| T2DM | pericyte | *RBM6* | TRUE | 3.333646094 |
| T2DM | pericyte | *OVGP1* | TRUE | 2.097630949 |
| T2DM | pericyte | *PABPC4* | TRUE | 3.178466948 |
| T2DM | pericyte | *CHN2* | TRUE | 2.110381863 |
| T2DM | pericyte | *CDK2AP1* | TRUE | 2.073647373 |

| | | | | |
|---|---|---|---|---|
| T2DM | pericyte | *BVES* | TRUE | 2.563512935 |
| T2DM | pericyte | *GPX7* | TRUE | 2.70180145 |
| T2DM | pericyte | *ADRA1A* | TRUE | 2.290473199 |
| T2DM | pericyte | *GRK4* | TRUE | 2.430863734 |
| T2DM | pericyte | *PKDREJ* | TRUE | 2.308956589 |
| T2DM | pericyte | *IRF4* | TRUE | 2.667861955 |
| T2DM | pericyte | *TCF19* | TRUE | 2.188710889 |
| T2DM | pericyte | *MAN2C1* | TRUE | 2.608083955 |
| T2DM | pericyte | *DUS3L* | TRUE | 2.521267993 |
| T2DM | pericyte | *ARHGEF19* | TRUE | 1.992386061 |
| T2DM | pericyte | *HSD17B12* | TRUE | 4.392532143 |
| T2DM | pericyte | *APIP* | TRUE | 2.602917265 |
| T2DM | pericyte | *JAZF1* | TRUE | 3.019632301 |
| T2DM | pericyte | *CLEC18A* | TRUE | 5.235651894 |
| T2DM | pericyte | *KRTCAP3* | TRUE | 3.01532231 |
| T2DM | pericyte | *SPTA1* | TRUE | 2.928377562 |
| T2DM | pericyte | *RFT1* | TRUE | 2.458000224 |
| T2DM | pericyte | *RAPSN* | TRUE | 2.180885858 |
| T2DM | pericyte | *ZSWIM1* | TRUE | 2.422130305 |
| T2DM | pericyte | *IRS1* | TRUE | 2.233034833 |
| T2DM | pericyte | *RGS19* | TRUE | 2.524118573 |
| T2DM | pericyte | *CTSW* | TRUE | 2.236886419 |
| T2DM | pericyte | *CCNE2* | TRUE | 2.319348749 |
| T2DM | pericyte | *TRIM73* | TRUE | 2.344287717 |
| T2DM | pericyte | *RPRML* | TRUE | 2.356103964 |
| T2DM | pericyte | *ZNF320* | TRUE | 2.320847958 |
| T2DM | pericyte | *NOTCH4* | TRUE | 2.260839262 |
| T2DM | pericyte | *DNLZ* | TRUE | 3.402514008 |
| T2DM | pericyte | *STARD10* | TRUE | 4.957298202 |
| T2DM | pericyte | *POLR2J2* | TRUE | 2.2062748 |
| T2DM | pericyte | *GS1.124K5.4* | TRUE | 2.092955554 |
| T2DM | pericyte | *HLA.DQA2* | TRUE | 3.082852897 |
| T2DM | pericyte | *NME1.NME2* | TRUE | 2.008966787 |
| T2DM | pericyte | *JMJD7* | TRUE | 1.983864382 |
| T2DM | SMC | *RBM6* | TRUE | 3.324282676 |
| T2DM | SMC | *PABPC4* | TRUE | 3.414110872 |
| T2DM | SMC | *TLE6* | TRUE | 2.167987154 |
| T2DM | SMC | *GPX7* | TRUE | 2.507171468 |
| T2DM | SMC | *ADRA1A* | TRUE | 3.014844642 |
| T2DM | SMC | *GRK4* | TRUE | 2.738470141 |
| T2DM | SMC | *TCF19* | TRUE | 2.094974707 |
| T2DM | SMC | *MAN2C1* | TRUE | 3.183459746 |
| T2DM | SMC | *ARHGEF19* | TRUE | 2.316599896 |
| T2DM | SMC | *HSD17B12* | TRUE | 4.594166729 |
| T2DM | SMC | *APIP* | TRUE | 2.02381446 |
| T2DM | SMC | *JAZF1* | TRUE | 3.061958504 |
| T2DM | SMC | *SPTA1* | TRUE | 3.26666004 |
| T2DM | SMC | *ANPEP* | TRUE | 2.420988439 |
| T2DM | SMC | *ZSWIM1* | TRUE | 2.348507993 |
| T2DM | SMC | *IRS1* | TRUE | 2.481853146 |

| | | | | |
|---|---|---|---|---|
| T2DM | SMC | *TTLL6* | TRUE | 4.139676013 |
| T2DM | SMC | *RPRML* | TRUE | 2.034523689 |
| T2DM | SMC | *PIGP* | TRUE | 2.233470248 |
| T2DM | SMC | *HLA.DRB5* | TRUE | 2.578939892 |
| T2DM | SMC | *FAM153C* | TRUE | 2.005794598 |
| T2DM | SMC | *DNLZ* | TRUE | 3.427427628 |
| T2DM | SMC | *STARD10* | TRUE | 3.850756309 |
| T2DM | SMC | *AC108463.2* | TRUE | 2.125506491 |
| T2DM | SMC | *LINC00941* | TRUE | 2.097066558 |
| T2DM | SMC | *HLA.DQA2* | TRUE | 2.824716737 |
| T2DM | t_cell | *PABPC4* | TRUE | 3.725922852 |
| T2DM | t_cell | *CWF19L1* | TRUE | 2.282378626 |
| T2DM | t_cell | *NOMO3* | TRUE | 2.374105364 |
| T2DM | t_cell | *TLE6* | TRUE | 2.340801189 |
| T2DM | t_cell | *CHN2* | TRUE | 3.734175578 |
| T2DM | t_cell | *GNB3* | TRUE | 2.442193779 |
| T2DM | t_cell | *BVES* | TRUE | 2.483380206 |
| T2DM | t_cell | *ADRA1A* | TRUE | 2.075030165 |
| T2DM | t_cell | *GRK4* | TRUE | 2.135511374 |
| T2DM | t_cell | *OPN1SW* | TRUE | 2.148293968 |
| T2DM | t_cell | *PKDREJ* | TRUE | 2.532716516 |
| T2DM | t_cell | *IRF4* | TRUE | 2.232430929 |
| T2DM | t_cell | *FN3KRP* | TRUE | 2.086380454 |
| T2DM | t_cell | *ARHGEF19* | TRUE | 2.402035481 |
| T2DM | t_cell | *HSD17B12* | TRUE | 4.52819912 |
| T2DM | t_cell | *APIP* | TRUE | 2.619054451 |
| T2DM | t_cell | *PCDH1* | TRUE | 2.031951365 |
| T2DM | t_cell | *CLEC18A* | TRUE | 4.166157791 |
| T2DM | t_cell | *KRTCAP3* | TRUE | 3.403905298 |
| T2DM | t_cell | *PAQR6* | TRUE | 1.988300666 |
| T2DM | t_cell | *SPTA1* | TRUE | 3.214796173 |
| T2DM | t_cell | *ANPEP* | TRUE | 2.141011457 |
| T2DM | t_cell | *ZSWIM1* | TRUE | 2.928800443 |
| T2DM | t_cell | *IRS1* | TRUE | 3.88418071 |
| T2DM | t_cell | *TTLL6* | TRUE | 3.800914212 |
| T2DM | t_cell | *CCNE2* | TRUE | 3.051492164 |
| T2DM | t_cell | *TRIM73* | TRUE | 2.297609891 |
| T2DM | t_cell | *ACOT1* | TRUE | 2.179657613 |
| T2DM | t_cell | *NDUFA6* | TRUE | 2.102368412 |
| T2DM | t_cell | *NOTCH4* | TRUE | 2.470051652 |
| T2DM | t_cell | *DNLZ* | TRUE | 3.391589639 |
| T2DM | t_cell | *STARD10* | TRUE | 4.886955038 |
| T2DM | t_cell | *GS1.124K5.4* | TRUE | 2.238174483 |
| T2DM | t_cell | *HLA.DQA2* | TRUE | 2.380565196 |
| HTN | adipocyte | *CD79B* | TRUE | 2.435092866 |
| HTN | adipocyte | *CEP68* | TRUE | 2.76916073 |
| HTN | adipocyte | *SDCCAG8* | TRUE | 3.38780082 |
| HTN | adipocyte | *PIGV* | TRUE | 3.795722243 |
| HTN | adipocyte | *UHRF1BP1* | TRUE | 3.197984687 |
| HTN | adipocyte | *POLR1A* | TRUE | 2.117674676 |

| | | | | |
|---|---|---|---|---|
| HTN | adipocyte | *DUSP13* | TRUE | 2.078074017 |
| HTN | adipocyte | *SNX10* | TRUE | 3.687943819 |
| HTN | adipocyte | *AAMDC* | TRUE | 2.069794283 |
| HTN | adipocyte | *MUL1* | TRUE | 2.257011178 |
| HTN | adipocyte | *PABPC4* | TRUE | 2.668444721 |
| HTN | adipocyte | *SCD* | TRUE | 2.369149381 |
| HTN | adipocyte | *MARCH2* | TRUE | 3.477110574 |
| HTN | adipocyte | *BIRC7* | TRUE | 2.872976852 |
| HTN | adipocyte | *POLI* | TRUE | 3.053940802 |
| HTN | adipocyte | *MTHFSD* | TRUE | 3.048813068 |
| HTN | adipocyte | *ICAM5* | TRUE | 2.276591 |
| HTN | adipocyte | *SGCA* | TRUE | 2.650832229 |
| HTN | adipocyte | *CCND1* | TRUE | 2.357709158 |
| HTN | adipocyte | *BNIP1* | TRUE | 1.970892959 |
| HTN | adipocyte | *NPHP3* | TRUE | 1.966213487 |
| HTN | adipocyte | *PCCB* | TRUE | 3.865370109 |
| HTN | adipocyte | *IRF6* | TRUE | 2.630044212 |
| HTN | adipocyte | *ACOT2* | TRUE | 2.465798248 |
| HTN | adipocyte | *ZFP30* | TRUE | 2.091720716 |
| HTN | adipocyte | *ADRA1A* | TRUE | 1.997146288 |
| HTN | adipocyte | *TBX2* | TRUE | 5.040321368 |
| HTN | adipocyte | *TEX14* | TRUE | 2.474015244 |
| HTN | adipocyte | *BTN2A2* | TRUE | 3.213263832 |
| HTN | adipocyte | *ZNF391* | TRUE | 2.339470288 |
| HTN | adipocyte | *TCP11* | TRUE | 1.987801258 |
| HTN | adipocyte | *GRK4* | TRUE | 2.710977402 |
| HTN | adipocyte | *TSKS* | TRUE | 2.347658368 |
| HTN | adipocyte | *TRAP1* | TRUE | 4.072804472 |
| HTN | adipocyte | *ABHD8* | TRUE | 2.538008495 |
| HTN | adipocyte | *ADCY4* | TRUE | 2.880117164 |
| HTN | adipocyte | *ATP8B3* | TRUE | 2.955910493 |
| HTN | adipocyte | *PGPEP1* | TRUE | 2.360267283 |
| HTN | adipocyte | *HAUS8* | TRUE | 4.374981136 |
| HTN | adipocyte | *RSG1* | TRUE | 2.35208348 |
| HTN | adipocyte | *ACP2* | TRUE | 4.284909786 |
| HTN | adipocyte | *AGT* | TRUE | 3.007240763 |
| HTN | adipocyte | *NMT1* | TRUE | 3.896399033 |
| HTN | adipocyte | *SLC37A4* | TRUE | 2.789949245 |
| HTN | adipocyte | *NUSAP1* | TRUE | 3.022577511 |
| HTN | adipocyte | *FBN2* | TRUE | 3.811396437 |
| HTN | adipocyte | *FITM1* | TRUE | 2.537215287 |
| HTN | adipocyte | *RPL3L* | TRUE | 2.157454702 |
| HTN | adipocyte | *DEF8* | TRUE | 2.440502614 |
| HTN | adipocyte | *SCRN2* | TRUE | 2.275412295 |
| HTN | adipocyte | *C18orf8* | TRUE | 2.169186584 |
| HTN | adipocyte | *MMEL1* | TRUE | 2.210119178 |
| HTN | adipocyte | *CREB3L4* | TRUE | 3.4330296 |
| HTN | adipocyte | *PLEKHA6* | TRUE | 2.053864052 |
| HTN | adipocyte | *IRAK1BP1* | TRUE | 2.157212283 |
| HTN | adipocyte | *ZNF215* | TRUE | 2.238505387 |

| | | | | |
|---|---|---|---|---|
| HTN | adipocyte | *HSD17B12* | TRUE | 5.355486765 |
| HTN | adipocyte | *TNKS1BP1* | TRUE | 4.352803049 |
| HTN | adipocyte | *NEIL2* | TRUE | 3.543290603 |
| HTN | adipocyte | *ART3* | TRUE | 2.615897567 |
| HTN | adipocyte | *SFXN2* | TRUE | 4.918352001 |
| HTN | adipocyte | *CLEC18A* | TRUE | 2.794213121 |
| HTN | adipocyte | *SLC5A11* | TRUE | 4.60305368 |
| HTN | adipocyte | *S100B* | TRUE | 3.50860745 |
| HTN | adipocyte | *ABCF3* | TRUE | 4.746788753 |
| HTN | adipocyte | *MRPL55* | TRUE | 5.6538278 |
| HTN | adipocyte | *LYSMD1* | TRUE | 3.387814337 |
| HTN | adipocyte | *RFT1* | TRUE | 2.383548685 |
| HTN | adipocyte | *RNF123* | TRUE | 4.649806432 |
| HTN | adipocyte | *ERAP1* | TRUE | 1.963188832 |
| HTN | adipocyte | *CYB5R2* | TRUE | 2.664788959 |
| HTN | adipocyte | *TRIM66* | TRUE | 3.565939864 |
| HTN | adipocyte | *RIMKLB* | TRUE | 3.486477515 |
| HTN | adipocyte | *SRR* | TRUE | 3.412489298 |
| HTN | adipocyte | *SOAT2* | TRUE | 2.551907865 |
| HTN | adipocyte | *SERINC2* | TRUE | 2.453963585 |
| HTN | adipocyte | *CPT1C* | TRUE | 3.479782686 |
| HTN | adipocyte | *INO80E* | TRUE | 4.383188723 |
| HTN | adipocyte | *RGPD8* | TRUE | 2.923416626 |
| HTN | adipocyte | *MUC3A* | TRUE | 2.010132333 |
| HTN | adipocyte | *MYO7B* | TRUE | 3.523402652 |
| HTN | adipocyte | *TTLL6* | TRUE | 5.448183147 |
| HTN | adipocyte | *PAQR8* | TRUE | 2.028002237 |
| HTN | adipocyte | *MRPL36* | TRUE | 2.720585085 |
| HTN | adipocyte | *RGS19* | TRUE | 4.848111934 |
| HTN | adipocyte | *NME6* | TRUE | 2.49253504 |
| HTN | adipocyte | *CTSW* | TRUE | 2.738687899 |
| HTN | adipocyte | *MST1* | TRUE | 3.143805034 |
| HTN | adipocyte | *C11orf80* | TRUE | 3.046223075 |
| HTN | adipocyte | *CTU2* | TRUE | 2.209061382 |
| HTN | adipocyte | *SNX31* | TRUE | 4.299490352 |
| HTN | adipocyte | *CCNE2* | TRUE | 4.403567401 |
| HTN | adipocyte | *ORAI3* | TRUE | 3.97471976 |
| HTN | adipocyte | *ODF3B* | TRUE | 2.246363037 |
| HTN | adipocyte | *GEN1* | TRUE | 2.475295645 |
| HTN | adipocyte | *MPI* | TRUE | 6.17800662 |
| HTN | adipocyte | *TRIM73* | TRUE | 2.388117597 |
| HTN | adipocyte | *HYI* | TRUE | 3.581730225 |
| HTN | adipocyte | *CLDN7* | TRUE | 2.264310892 |
| HTN | adipocyte | *ADO* | TRUE | 5.473750758 |
| HTN | adipocyte | *CLN8* | TRUE | 3.519391604 |
| HTN | adipocyte | *FES* | TRUE | 6.868552594 |
| HTN | adipocyte | *ZNF320* | TRUE | 3.46879707 |
| HTN | adipocyte | *A3GALT2* | TRUE | 3.75863654 |
| HTN | adipocyte | *NUTM2A* | TRUE | 3.581737117 |
| HTN | adipocyte | *FOXI2* | TRUE | 2.20791163 |

| | | | | |
|---|---|---|---|---|
| HTN | adipocyte | *KLHL17* | TRUE | 2.173838145 |
| HTN | adipocyte | *ZNF559* | TRUE | 2.530353649 |
| HTN | adipocyte | *FBF1* | TRUE | 3.081039701 |
| HTN | adipocyte | *COLCA1* | TRUE | 2.541918762 |
| HTN | adipocyte | *HRCT1* | TRUE | 2.496053821 |
| HTN | adipocyte | *HIST1H3E* | TRUE | 4.990549733 |
| HTN | adipocyte | *WDR5B* | TRUE | 2.542163695 |
| HTN | adipocyte | *ZNF100* | TRUE | 2.163465088 |
| HTN | adipocyte | *SULT1A2* | TRUE | 3.320129162 |
| HTN | adipocyte | *C6orf141* | TRUE | 1.967762752 |
| HTN | adipocyte | *RPS26* | TRUE | 3.875630571 |
| HTN | adipocyte | *ZNF311* | TRUE | 2.316166789 |
| HTN | adipocyte | *AGAP9* | TRUE | 2.082262357 |
| HTN | adipocyte | *TMEM116* | TRUE | 6.680691784 |
| HTN | adipocyte | *PNP* | TRUE | 2.761130468 |
| HTN | adipocyte | *SHISA4* | TRUE | 4.072941023 |
| HTN | adipocyte | *HSD17B8* | TRUE | 3.030170478 |
| HTN | adipocyte | *HLA.C* | TRUE | 2.294475863 |
| HTN | adipocyte | *ATF6B* | TRUE | 3.362110165 |
| HTN | adipocyte | *PPM1N* | TRUE | 2.425504633 |
| HTN | adipocyte | *AC016773.1* | TRUE | 2.419413981 |
| HTN | adipocyte | *SLC12A8* | TRUE | 2.625957542 |
| HTN | adipocyte | *TIAF1* | TRUE | 2.827603431 |
| HTN | adipocyte | *OSER1.AS1* | TRUE | 2.042850127 |
| HTN | adipocyte | *HLA.DQA2* | TRUE | 2.404768676 |
| HTN | adipocyte | *TNFSF12* | TRUE | 3.065930791 |
| HTN | adipocyte | *ZNF709* | TRUE | 2.278011027 |
| HTN | adipocyte | *LINC00638* | TRUE | 2.876215969 |
| HTN | ASPC | *ARHGAP33* | TRUE | 2.003063637 |
| HTN | ASPC | *CD79B* | TRUE | 2.242524 |
| HTN | ASPC | *CEP68* | TRUE | 3.88742929 |
| HTN | ASPC | *CHDH* | TRUE | 2.924319778 |
| HTN | ASPC | *RNASET2* | TRUE | 2.062312632 |
| HTN | ASPC | *ANAPC4* | TRUE | 2.185407242 |
| HTN | ASPC | *SDCCAG8* | TRUE | 3.971006044 |
| HTN | ASPC | *PLEKHH1* | TRUE | 2.138705105 |
| HTN | ASPC | *PIGV* | TRUE | 3.867925667 |
| HTN | ASPC | *POLR1A* | TRUE | 2.313341261 |
| HTN | ASPC | *DUSP13* | TRUE | 2.158903529 |
| HTN | ASPC | *SNX10* | TRUE | 3.23253013 |
| HTN | ASPC | *AAMDC* | TRUE | 2.540652533 |
| HTN | ASPC | *TBX5* | TRUE | 1.971722261 |
| HTN | ASPC | *MUL1* | TRUE | 1.98366033 |
| HTN | ASPC | *PABPC4* | TRUE | 2.883441265 |
| HTN | ASPC | *RGS17* | TRUE | 2.715097992 |
| HTN | ASPC | *SCD* | TRUE | 3.006486127 |
| HTN | ASPC | *SETD1A* | TRUE | 2.297780012 |
| HTN | ASPC | *MARCH2* | TRUE | 4.229499534 |
| HTN | ASPC | *BIRC7* | TRUE | 2.802382501 |
| HTN | ASPC | *POLI* | TRUE | 3.346142375 |

| | | | | |
|---|---|---|---|---|
| HTN | ASPC | *MTHFSD* | TRUE | 3.28822488 |
| HTN | ASPC | *SNX16* | TRUE | 2.826944373 |
| HTN | ASPC | *ICAM5* | TRUE | 2.837813145 |
| HTN | ASPC | *MEST* | TRUE | 1.984623823 |
| HTN | ASPC | *SLC1A1* | TRUE | 2.261975913 |
| HTN | ASPC | *NAGLU* | TRUE | 2.022590844 |
| HTN | ASPC | *SGCA* | TRUE | 2.637253485 |
| HTN | ASPC | *CCND1* | TRUE | 3.501579281 |
| HTN | ASPC | *BVES* | TRUE | 2.077614259 |
| HTN | ASPC | *NME5* | TRUE | 2.688649369 |
| HTN | ASPC | *BNIP1* | TRUE | 2.17927391 |
| HTN | ASPC | *CLDN16* | TRUE | 2.00801914 |
| HTN | ASPC | *NPHP3* | TRUE | 2.06952678 |
| HTN | ASPC | *PCCB* | TRUE | 4.64802231 |
| HTN | ASPC | *KIAA1324* | TRUE | 1.990372228 |
| HTN | ASPC | *IRF6* | TRUE | 2.566142212 |
| HTN | ASPC | *ACOT2* | TRUE | 2.979134039 |
| HTN | ASPC | *ZFP30* | TRUE | 2.774895381 |
| HTN | ASPC | *ADRA1A* | TRUE | 2.173577963 |
| HTN | ASPC | *TBX2* | TRUE | 5.386652229 |
| HTN | ASPC | *TEX14* | TRUE | 2.430603883 |
| HTN | ASPC | *TOX2* | TRUE | 1.970543283 |
| HTN | ASPC | *BTN2A2* | TRUE | 3.475253825 |
| HTN | ASPC | *ZNF391* | TRUE | 2.088656247 |
| HTN | ASPC | *GRK4* | TRUE | 2.772909101 |
| HTN | ASPC | *TSKS* | TRUE | 2.89924096 |
| HTN | ASPC | *TRAP1* | TRUE | 4.551716359 |
| HTN | ASPC | *ABHD8* | TRUE | 3.325656133 |
| HTN | ASPC | *GFER* | TRUE | 2.062963176 |
| HTN | ASPC | *GAD1* | TRUE | 2.272947768 |
| HTN | ASPC | *ADCY4* | TRUE | 4.33054174 |
| HTN | ASPC | *CDC16* | TRUE | 2.134970213 |
| HTN | ASPC | *ATP8B3* | TRUE | 3.073507351 |
| HTN | ASPC | *PGPEP1* | TRUE | 2.58991653 |
| HTN | ASPC | *HAUS8* | TRUE | 4.576343145 |
| HTN | ASPC | *GCH1* | TRUE | 2.4717296 |
| HTN | ASPC | *RSG1* | TRUE | 2.081442288 |
| HTN | ASPC | *SLC2A11* | TRUE | 2.241338702 |
| HTN | ASPC | *ACP2* | TRUE | 5.374876581 |
| HTN | ASPC | *CLDN10* | TRUE | 2.361448106 |
| HTN | ASPC | *AGT* | TRUE | 3.388903173 |
| HTN | ASPC | *NMT1* | TRUE | 4.344448427 |
| HTN | ASPC | *SLC37A4* | TRUE | 2.955342656 |
| HTN | ASPC | *PPP2R1B* | TRUE | 2.306759065 |
| HTN | ASPC | *NUSAP1* | TRUE | 2.743251032 |
| HTN | ASPC | *G3BP2* | TRUE | 2.063538941 |
| HTN | ASPC | *ENPEP* | TRUE | 2.219765032 |
| HTN | ASPC | *FBN2* | TRUE | 4.00025587 |
| HTN | ASPC | *FITM1* | TRUE | 2.739186453 |
| HTN | ASPC | *RTN1* | TRUE | 2.984496353 |

| | | | | |
|---|---|---|---|---|
| HTN | ASPC | *RPL3L* | TRUE | 2.40113064 |
| HTN | ASPC | *DEF8* | TRUE | 3.927666187 |
| HTN | ASPC | *LRRC46* | TRUE | 1.963220952 |
| HTN | ASPC | *SCRN2* | TRUE | 2.4596661 |
| HTN | ASPC | *C18orf8* | TRUE | 2.383746039 |
| HTN | ASPC | *MMEL1* | TRUE | 2.199667277 |
| HTN | ASPC | *CREB3L4* | TRUE | 3.21851299 |
| HTN | ASPC | *CISD2* | TRUE | 2.005465318 |
| HTN | ASPC | *TSLP* | TRUE | 2.154473051 |
| HTN | ASPC | *IRAK1BP1* | TRUE | 3.651353415 |
| HTN | ASPC | *ZNF215* | TRUE | 2.301190514 |
| HTN | ASPC | *HSD17B12* | TRUE | 5.398453172 |
| HTN | ASPC | *TNKS1BP1* | TRUE | 4.553516485 |
| HTN | ASPC | *TTC12* | TRUE | 2.265241577 |
| HTN | ASPC | *PRDM8* | TRUE | 2.929587621 |
| HTN | ASPC | *MED21* | TRUE | 1.965777734 |
| HTN | ASPC | *NEIL2* | TRUE | 2.062730005 |
| HTN | ASPC | *MMP16* | TRUE | 2.380364357 |
| HTN | ASPC | *ART3* | TRUE | 2.615377469 |
| HTN | ASPC | *SFXN2* | TRUE | 5.096879542 |
| HTN | ASPC | *CLEC18A* | TRUE | 2.835321149 |
| HTN | ASPC | *ETS2* | TRUE | 2.023237169 |
| HTN | ASPC | *SLC5A11* | TRUE | 4.50148411 |
| HTN | ASPC | *LRRC36* | TRUE | 2.180937448 |
| HTN | ASPC | *ATAD3B* | TRUE | 1.998769551 |
| HTN | ASPC | *S100B* | TRUE | 3.717466598 |
| HTN | ASPC | *CCR5* | TRUE | 2.030703412 |
| HTN | ASPC | *ABCF3* | TRUE | 5.05303829 |
| HTN | ASPC | *AQP5* | TRUE | 1.984045397 |
| HTN | ASPC | *MRPL55* | TRUE | 6.38707822 |
| HTN | ASPC | *LYSMD1* | TRUE | 3.558209958 |
| HTN | ASPC | *DNALI1* | TRUE | 2.103639922 |
| HTN | ASPC | *RFT1* | TRUE | 2.245868828 |
| HTN | ASPC | *RNF123* | TRUE | 4.807521271 |
| HTN | ASPC | *ERAP1* | TRUE | 2.852447473 |
| HTN | ASPC | *SOX17* | TRUE | 2.232612747 |
| HTN | ASPC | *DCSTAMP* | TRUE | 3.013615469 |
| HTN | ASPC | *AQP7* | TRUE | 2.175888833 |
| HTN | ASPC | *STOX1* | TRUE | 2.107868095 |
| HTN | ASPC | *CYB5R2* | TRUE | 2.894303686 |
| HTN | ASPC | *TRIM66* | TRUE | 3.523247457 |
| HTN | ASPC | *GALR1* | TRUE | 2.110262053 |
| HTN | ASPC | *CHAF1A* | TRUE | 2.003913013 |
| HTN | ASPC | *SRR* | TRUE | 3.546288759 |
| HTN | ASPC | *C8orf46* | TRUE | 2.441338261 |
| HTN | ASPC | *CPT1C* | TRUE | 4.56197495 |
| HTN | ASPC | *INO80E* | TRUE | 3.292211657 |
| HTN | ASPC | *RGPD8* | TRUE | 3.135707744 |
| HTN | ASPC | *MYO7B* | TRUE | 3.830710136 |
| HTN | ASPC | *DNAJC18* | TRUE | 2.800649513 |

| | | | | |
|---|---|---|---|---|
| HTN | ASPC | *TTLL6* | TRUE | 4.739370465 |
| HTN | ASPC | *PAQR8* | TRUE | 2.477022264 |
| HTN | ASPC | *GIMAP8* | TRUE | 1.968855277 |
| HTN | ASPC | *MRPL36* | TRUE | 2.872347073 |
| HTN | ASPC | *RGS19* | TRUE | 4.822157804 |
| HTN | ASPC | *NME6* | TRUE | 2.599712961 |
| HTN | ASPC | *ZNF738* | TRUE | 1.989495463 |
| HTN | ASPC | *PPP1R14B* | TRUE | 2.211246029 |
| HTN | ASPC | *MST1* | TRUE | 2.585974174 |
| HTN | ASPC | *C11orf80* | TRUE | 2.987250528 |
| HTN | ASPC | *GPR160* | TRUE | 2.48729336 |
| HTN | ASPC | *CTU2* | TRUE | 2.445883562 |
| HTN | ASPC | *SNX31* | TRUE | 4.195645949 |
| HTN | ASPC | *CCNE2* | TRUE | 4.863893347 |
| HTN | ASPC | *TMEM9B* | TRUE | 2.088599656 |
| HTN | ASPC | *CTDNEP1* | TRUE | 2.059974193 |
| HTN | ASPC | *ORAI3* | TRUE | 2.890542405 |
| HTN | ASPC | *ODF3B* | TRUE | 2.671182563 |
| HTN | ASPC | *GEN1* | TRUE | 2.291595294 |
| HTN | ASPC | *MPI* | TRUE | 2.407426235 |
| HTN | ASPC | *TRIM73* | TRUE | 2.71195608 |
| HTN | ASPC | *HYI* | TRUE | 3.91420354 |
| HTN | ASPC | *CTC1* | TRUE | 2.679816678 |
| HTN | ASPC | *ZFP41* | TRUE | 1.978204395 |
| HTN | ASPC | *ADO* | TRUE | 5.725346825 |
| HTN | ASPC | *CLN8* | TRUE | 4.157622145 |
| HTN | ASPC | *FES* | TRUE | 7.326148597 |
| HTN | ASPC | *ZNF320* | TRUE | 3.488457809 |
| HTN | ASPC | *KREMEN1* | TRUE | 2.131720054 |
| HTN | ASPC | *ALDH1A3* | TRUE | 2.761609486 |
| HTN | ASPC | *A3GALT2* | TRUE | 3.889054933 |
| HTN | ASPC | *NUTM2A* | TRUE | 2.76596477 |
| HTN | ASPC | *FFAR4* | TRUE | 2.489218242 |
| HTN | ASPC | *FOXI2* | TRUE | 2.382723749 |
| HTN | ASPC | *KLHL17* | TRUE | 3.054816921 |
| HTN | ASPC | *ZNF559* | TRUE | 2.662794272 |
| HTN | ASPC | *FZD9* | TRUE | 2.581995064 |
| HTN | ASPC | *FAM26F* | TRUE | 1.968495273 |
| HTN | ASPC | *FBF1* | TRUE | 3.338645358 |
| HTN | ASPC | *FAM47E* | TRUE | 3.01099499 |
| HTN | ASPC | *COLCA1* | TRUE | 3.234005836 |
| HTN | ASPC | *HRCT1* | TRUE | 2.091075404 |
| HTN | ASPC | *ZNF605* | TRUE | 2.016307062 |
| HTN | ASPC | *HIST1H3E* | TRUE | 5.149133872 |
| HTN | ASPC | *WDR5B* | TRUE | 3.317775563 |
| HTN | ASPC | *ZNF100* | TRUE | 2.343476888 |
| HTN | ASPC | *SULT1A2* | TRUE | 3.282477932 |
| HTN | ASPC | *CCL4L2* | TRUE | 2.18670714 |
| HTN | ASPC | *MFAP5* | TRUE | 2.90608183 |
| HTN | ASPC | *RPS26* | TRUE | 3.831808852 |

| | | | | |
|---|---|---|---|---|
| HTN | ASPC | *ZNF311* | TRUE | 2.625379818 |
| HTN | ASPC | *AGAP9* | TRUE | 2.2598391 |
| HTN | ASPC | *RPS6KL1* | TRUE | 2.780186058 |
| HTN | ASPC | *TMEM116* | TRUE | 6.722541838 |
| HTN | ASPC | *ZNF442* | TRUE | 2.022896238 |
| HTN | ASPC | *PNP* | TRUE | 2.642982416 |
| HTN | ASPC | *SHISA4* | TRUE | 4.066847069 |
| HTN | ASPC | *MFAP3L* | TRUE | 2.045073341 |
| HTN | ASPC | *TGM2* | TRUE | 2.35113833 |
| HTN | ASPC | *HSD17B8* | TRUE | 2.786625858 |
| HTN | ASPC | *HLA.C* | TRUE | 2.446631325 |
| HTN | ASPC | *TMEM170B* | TRUE | 2.528542498 |
| HTN | ASPC | *MXD3* | TRUE | 2.005155024 |
| HTN | ASPC | *ATF6B* | TRUE | 3.562731444 |
| HTN | ASPC | *PPM1N* | TRUE | 2.981636131 |
| HTN | ASPC | *GALT* | TRUE | 2.062038692 |
| HTN | ASPC | *UBXN2B* | TRUE | 2.402985785 |
| HTN | ASPC | *SLC12A8* | TRUE | 3.120167895 |
| HTN | ASPC | *TIAF1* | TRUE | 4.015108613 |
| HTN | ASPC | *OSER1.AS1* | TRUE | 2.224902996 |
| HTN | ASPC | *AC108463.2* | TRUE | 2.334457184 |
| HTN | ASPC | *HLA.DQA2* | TRUE | 2.625141519 |
| HTN | ASPC | *TNFSF12* | TRUE | 3.639806675 |
| HTN | ASPC | *PNMA2* | TRUE | 2.061772174 |
| HTN | ASPC | *PLA2G4B* | TRUE | 2.136763069 |
| HTN | ASPC | *LINC00638* | TRUE | 3.066042024 |
| HTN | ASPC | *ZNF285* | TRUE | 1.97377213 |
| HTN | b_cell | *ITIH4* | TRUE | 2.337731564 |
| HTN | b_cell | *BIRC7* | TRUE | 2.721211191 |
| HTN | b_cell | *CLEC18A* | TRUE | 3.979641447 |
| HTN | b_cell | *CTSW* | TRUE | 2.961350462 |
| HTN | b_cell | *SNX31* | TRUE | 3.01334377 |
| HTN | b_cell | *RPP25* | TRUE | 2.334647301 |
| HTN | b_cell | *TRIM73* | TRUE | 2.350750298 |
| HTN | b_cell | *ADO* | TRUE | 3.073902526 |
| HTN | b_cell | *HIST1H3E* | TRUE | 3.788638347 |
| HTN | b_cell | *HLA.DRB5* | TRUE | 2.181184461 |
| HTN | b_cell | *POU5F1* | TRUE | 2.042488774 |
| HTN | b_cell | *HLA.DQA2* | TRUE | 2.378828057 |
| HTN | b_cell | *WFDC6* | TRUE | 2.727738113 |
| HTN | dendritic_cell | *CD79B* | TRUE | 2.038968464 |
| HTN | dendritic_cell | *MARCH2* | TRUE | 2.272451943 |
| HTN | dendritic_cell | *BIRC7* | TRUE | 2.647868331 |
| HTN | dendritic_cell | *IRF6* | TRUE | 2.588257825 |
| HTN | dendritic_cell | *ACOT2* | TRUE | 2.008847351 |
| HTN | dendritic_cell | *TBX2* | TRUE | 2.087508422 |
| HTN | dendritic_cell | *TCP11* | TRUE | 2.111491818 |
| HTN | dendritic_cell | *RIPK3* | TRUE | 2.073288999 |
| HTN | dendritic_cell | *ATP8B3* | TRUE | 2.004154768 |
| HTN | dendritic_cell | *HAUS8* | TRUE | 3.097272008 |

| | | | | |
|---|---|---|---|---|
| HTN | dendritic_cell | *ART3* | TRUE | 2.258863779 |
| HTN | dendritic_cell | *CLEC18A* | TRUE | 4.272709235 |
| HTN | dendritic_cell | *S100B* | TRUE | 2.774752896 |
| HTN | dendritic_cell | *SRR* | TRUE | 2.343183585 |
| HTN | dendritic_cell | *RGPD8* | TRUE | 2.852558715 |
| HTN | dendritic_cell | *RGS19* | TRUE | 3.271962106 |
| HTN | dendritic_cell | *NME6* | TRUE | 4.303552563 |
| HTN | dendritic_cell | *CTSW* | TRUE | 2.807312867 |
| HTN | dendritic_cell | *SNX31* | TRUE | 2.283252563 |
| HTN | dendritic_cell | *CCNE2* | TRUE | 3.468665761 |
| HTN | dendritic_cell | *TRIM73* | TRUE | 2.724952587 |
| HTN | dendritic_cell | *CLDN7* | TRUE | 2.929402979 |
| HTN | dendritic_cell | *ADO* | TRUE | 4.821504762 |
| HTN | dendritic_cell | *ZNF320* | TRUE | 2.53629936 |
| HTN | dendritic_cell | *A3GALT2* | TRUE | 2.097573369 |
| HTN | dendritic_cell | *NUTM2A* | TRUE | 2.338858648 |
| HTN | dendritic_cell | *FOXI2* | TRUE | 2.072242822 |
| HTN | dendritic_cell | *RPS26* | TRUE | 3.378467226 |
| HTN | dendritic_cell | *TMEM116* | TRUE | 3.04478332 |
| HTN | dendritic_cell | *TMEM110* | TRUE | 2.082355853 |
| HTN | endometrium | *BIRC7* | TRUE | 2.911644373 |
| HTN | endometrium | *SGCA* | TRUE | 2.215087499 |
| HTN | endometrium | *TCP11* | TRUE | 2.313901133 |
| HTN | endometrium | *TNKS1BP1* | TRUE | 2.605026664 |
| HTN | endometrium | *SLC5A11* | TRUE | 2.848249799 |
| HTN | endometrium | *LYSMD1* | TRUE | 2.585950251 |
| HTN | endometrium | *RNF123* | TRUE | 2.12927781 |
| HTN | endometrium | *TRIM66* | TRUE | 2.203638872 |
| HTN | endometrium | *MST1* | TRUE | 1.986628308 |
| HTN | endometrium | *RPP25* | TRUE | 2.783645052 |
| HTN | endometrium | *TRIM73* | TRUE | 2.485229901 |
| HTN | endometrium | *NUTM2A* | TRUE | 2.970117728 |
| HTN | endometrium | *HIST1H3E* | TRUE | 3.982078319 |
| HTN | endometrium | *TMEM116* | TRUE | 5.535716198 |
| HTN | endometrium | *PSORS1C2* | TRUE | 2.549470777 |
| HTN | endometrium | *WFDC6* | TRUE | 2.830936326 |
| HTN | endometrium | *PLA2G4B* | TRUE | 2.203686355 |
| HTN | endothelial | *BIRC7* | TRUE | 2.420947371 |
| HTN | endothelial | *SGCA* | TRUE | 2.480957392 |
| HTN | endothelial | *ACOT2* | TRUE | 2.096649876 |
| HTN | endothelial | *TBX2* | TRUE | 3.457755059 |
| HTN | endothelial | *BTN2A2* | TRUE | 1.971083338 |
| HTN | endothelial | *TCP11* | TRUE | 2.337068479 |
| HTN | endothelial | *AGT* | TRUE | 2.10319135 |
| HTN | endothelial | *FBN2* | TRUE | 3.09952747 |
| HTN | endothelial | *HSD17B12* | TRUE | 3.230942993 |
| HTN | endothelial | *NEIL2* | TRUE | 2.957606198 |
| HTN | endothelial | *ART3* | TRUE | 2.067022194 |
| HTN | endothelial | *SFXN2* | TRUE | 2.292553073 |
| HTN | endothelial | *CLEC18A* | TRUE | 3.873919891 |

| | | | | |
|---|---|---|---|---|
| HTN | endothelial | *SLC5A11* | TRUE | 2.606118974 |
| HTN | endothelial | *S100B* | TRUE | 2.478893813 |
| HTN | endothelial | *LYSMD1* | TRUE | 2.210840508 |
| HTN | endothelial | *RNF123* | TRUE | 2.340620671 |
| HTN | endothelial | *NYAP1* | TRUE | 2.030209941 |
| HTN | endothelial | *SRR* | TRUE | 2.511976184 |
| HTN | endothelial | *CPT1C* | TRUE | 2.054424293 |
| HTN | endothelial | *INO80E* | TRUE | 2.18583022 |
| HTN | endothelial | *RGPD8* | TRUE | 2.603448053 |
| HTN | endothelial | *TTLL6* | TRUE | 4.031628652 |
| HTN | endothelial | *RGS19* | TRUE | 3.826551118 |
| HTN | endothelial | *NME6* | TRUE | 2.682821071 |
| HTN | endothelial | *CTSW* | TRUE | 3.637866512 |
| HTN | endothelial | *MST1* | TRUE | 2.576882222 |
| HTN | endothelial | *SNX31* | TRUE | 3.560378166 |
| HTN | endothelial | *TRIM73* | TRUE | 2.001007844 |
| HTN | endothelial | *HYI* | TRUE | 2.067287729 |
| HTN | endothelial | *ADO* | TRUE | 3.955479914 |
| HTN | endothelial | *CLN8* | TRUE | 2.333484378 |
| HTN | endothelial | *ZNF320* | TRUE | 2.478275386 |
| HTN | endothelial | *A3GALT2* | TRUE | 2.106961196 |
| HTN | endothelial | *NUTM2A* | TRUE | 3.532025975 |
| HTN | endothelial | *HIST1H3E* | TRUE | 4.701108485 |
| HTN | endothelial | *RPS26* | TRUE | 2.964321273 |
| HTN | endothelial | *TMEM116* | TRUE | 4.1157047 |
| HTN | endothelial | *PRRT1* | TRUE | 3.262299271 |
| HTN | endothelial | *AC016773.1* | TRUE | 1.986506711 |
| HTN | endothelial | *HLA.DQA2* | TRUE | 2.279608058 |
| HTN | endothelial | *PLA2G4B* | TRUE | 2.305004428 |
| HTN | LEC | *CD79B* | TRUE | 2.080200043 |
| HTN | LEC | *PIGV* | TRUE | 1.974051569 |
| HTN | LEC | *MARCH2* | TRUE | 2.688593991 |
| HTN | LEC | *BIRC7* | TRUE | 2.438204851 |
| HTN | LEC | *IRF6* | TRUE | 2.341086258 |
| HTN | LEC | *ZNF391* | TRUE | 2.866849744 |
| HTN | LEC | *RSG1* | TRUE | 2.09256159 |
| HTN | LEC | *AGT* | TRUE | 3.129571125 |
| HTN | LEC | *SLC37A4* | TRUE | 2.608320586 |
| HTN | LEC | *FBN2* | TRUE | 2.781802631 |
| HTN | LEC | *SCRN2* | TRUE | 2.032052422 |
| HTN | LEC | *HSD17B12* | TRUE | 4.477639126 |
| HTN | LEC | *NEIL2* | TRUE | 3.342979615 |
| HTN | LEC | *PROM2* | TRUE | 2.257522235 |
| HTN | LEC | *SFXN2* | TRUE | 4.505455698 |
| HTN | LEC | *CLEC18A* | TRUE | 2.142800493 |
| HTN | LEC | *SLC5A11* | TRUE | 3.200786124 |
| HTN | LEC | *S100B* | TRUE | 2.767443346 |
| HTN | LEC | *ABCF3* | TRUE | 2.634212031 |
| HTN | LEC | *LYSMD1* | TRUE | 3.362868657 |
| HTN | LEC | *TRIM66* | TRUE | 2.116265539 |

| | | | | |
|---|---|---|---|---|
| HTN | LEC | *SRR* | TRUE | 2.826534542 |
| HTN | LEC | *CPT1C* | TRUE | 4.261980169 |
| HTN | LEC | *INO80E* | TRUE | 4.154307605 |
| HTN | LEC | *RGPD8* | TRUE | 2.931196736 |
| HTN | LEC | *MYO7B* | TRUE | 2.829383353 |
| HTN | LEC | *ADORA2B* | TRUE | 1.981358505 |
| HTN | LEC | *TTLL6* | TRUE | 3.312274512 |
| HTN | LEC | *MRPL36* | TRUE | 2.300258918 |
| HTN | LEC | *RGS19* | TRUE | 4.51696302 |
| HTN | LEC | *NME6* | TRUE | 2.142975104 |
| HTN | LEC | *CTSW* | TRUE | 2.621487881 |
| HTN | LEC | *MST1* | TRUE | 2.636756456 |
| HTN | LEC | *CTU2* | TRUE | 2.366371241 |
| HTN | LEC | *SNX31* | TRUE | 2.858046028 |
| HTN | LEC | *CCNE2* | TRUE | 2.728835333 |
| HTN | LEC | *ORAI3* | TRUE | 3.428765229 |
| HTN | LEC | *MPI* | TRUE | 3.692143859 |
| HTN | LEC | *TRIM73* | TRUE | 2.305710866 |
| HTN | LEC | *HYI* | TRUE | 2.21242126 |
| HTN | LEC | *ADO* | TRUE | 2.00406746 |
| HTN | LEC | *CLN8* | TRUE | 3.386705831 |
| HTN | LEC | *FES* | TRUE | 4.576868508 |
| HTN | LEC | *ZNF320* | TRUE | 2.649463575 |
| HTN | LEC | *NPIPA1* | TRUE | 1.988556935 |
| HTN | LEC | *A3GALT2* | TRUE | 2.663506074 |
| HTN | LEC | *NUTM2A* | TRUE | 3.240745568 |
| HTN | LEC | *HRCT1* | TRUE | 2.737732512 |
| HTN | LEC | *HIST1H3E* | TRUE | 3.546277477 |
| HTN | LEC | *RPS26* | TRUE | 3.643225299 |
| HTN | LEC | *TMEM116* | TRUE | 5.232096279 |
| HTN | LEC | *HLA.DRB5* | TRUE | 2.695689624 |
| HTN | LEC | *PNP* | TRUE | 2.428348078 |
| HTN | LEC | *HLA.C* | TRUE | 2.174518335 |
| HTN | LEC | *TIAF1* | TRUE | 3.278877787 |
| HTN | LEC | *TNFSF12* | TRUE | 2.946806843 |
| HTN | macrophage | *SDCCAG8* | TRUE | 2.125033662 |
| HTN | macrophage | *PIGV* | TRUE | 3.690352898 |
| HTN | macrophage | *UHRF1BP1* | TRUE | 3.17700991 |
| HTN | macrophage | *MUL1* | TRUE | 2.02341487 |
| HTN | macrophage | *SCD* | TRUE | 2.409761406 |
| HTN | macrophage | *MARCH2* | TRUE | 3.089468026 |
| HTN | macrophage | *BIRC7* | TRUE | 3.00803116 |
| HTN | macrophage | *POLI* | TRUE | 3.025046681 |
| HTN | macrophage | *MTHFSD* | TRUE | 2.527488935 |
| HTN | macrophage | *ICAM5* | TRUE | 2.273076076 |
| HTN | macrophage | *SGCA* | TRUE | 2.738929947 |
| HTN | macrophage | *PCCB* | TRUE | 3.270492755 |
| HTN | macrophage | *IRF6* | TRUE | 2.231491999 |
| HTN | macrophage | *ACOT2* | TRUE | 2.478366166 |
| HTN | macrophage | *TBX2* | TRUE | 4.731890892 |

| | | | | |
|---|---|---|---|---|
| HTN | macrophage | *BTN2A2* | TRUE | 3.008632919 |
| HTN | macrophage | *ZNF391* | TRUE | 1.982117966 |
| HTN | macrophage | *GRK4* | TRUE | 2.185842901 |
| HTN | macrophage | *TSKS* | TRUE | 2.023923825 |
| HTN | macrophage | *TRAP1* | TRUE | 2.658803742 |
| HTN | macrophage | *ABHD8* | TRUE | 2.352271473 |
| HTN | macrophage | *ATP8B3* | TRUE | 2.856808181 |
| HTN | macrophage | *PGPEP1* | TRUE | 2.087707422 |
| HTN | macrophage | *HAUS8* | TRUE | 3.692602002 |
| HTN | macrophage | *NMT1* | TRUE | 2.675295333 |
| HTN | macrophage | *SLC37A4* | TRUE | 2.452969994 |
| HTN | macrophage | *FBN2* | TRUE | 2.607295654 |
| HTN | macrophage | *RPL3L* | TRUE | 1.9831937 |
| HTN | macrophage | *SCRN2* | TRUE | 2.350563491 |
| HTN | macrophage | *MMEL1* | TRUE | 2.023657695 |
| HTN | macrophage | *CREB3L4* | TRUE | 3.252057361 |
| HTN | macrophage | *TSLP* | TRUE | 2.191329544 |
| HTN | macrophage | *ZNF215* | TRUE | 2.042782127 |
| HTN | macrophage | *HSD17B12* | TRUE | 5.252932834 |
| HTN | macrophage | *NEIL2* | TRUE | 4.973662718 |
| HTN | macrophage | *ART3* | TRUE | 2.747915843 |
| HTN | macrophage | *SFXN2* | TRUE | 3.96744111 |
| HTN | macrophage | *CLEC18A* | TRUE | 3.03443237 |
| HTN | macrophage | *SLC5A11* | TRUE | 4.467514943 |
| HTN | macrophage | *S100B* | TRUE | 3.547471445 |
| HTN | macrophage | *ABCF3* | TRUE | 4.60867629 |
| HTN | macrophage | *MRPL55* | TRUE | 4.565656013 |
| HTN | macrophage | *LYSMD1* | TRUE | 2.903942403 |
| HTN | macrophage | *RFT1* | TRUE | 2.247536501 |
| HTN | macrophage | *RNF123* | TRUE | 4.468003947 |
| HTN | macrophage | *RPP25L* | TRUE | 2.017080706 |
| HTN | macrophage | *CYB5R2* | TRUE | 2.0516688 |
| HTN | macrophage | *TRIM66* | TRUE | 3.313059354 |
| HTN | macrophage | *RIMKLB* | TRUE | 2.914676305 |
| HTN | macrophage | *SRR* | TRUE | 3.347950416 |
| HTN | macrophage | *INO80E* | TRUE | 4.73221386 |
| HTN | macrophage | *RGPD8* | TRUE | 3.073000684 |
| HTN | macrophage | *MUC3A* | TRUE | 2.170610946 |
| HTN | macrophage | *TTLL6* | TRUE | 4.269999224 |
| HTN | macrophage | *MRPL36* | TRUE | 2.482288698 |
| HTN | macrophage | *CTSW* | TRUE | 2.660826873 |
| HTN | macrophage | *MST1* | TRUE | 2.994033367 |
| HTN | macrophage | *CTU2* | TRUE | 2.378634967 |
| HTN | macrophage | *SNX31* | TRUE | 4.298224057 |
| HTN | macrophage | *CCNE2* | TRUE | 4.267682815 |
| HTN | macrophage | *ORAI3* | TRUE | 3.748747995 |
| HTN | macrophage | *MPI* | TRUE | 6.14290418 |
| HTN | macrophage | *TRIM73* | TRUE | 2.438889537 |
| HTN | macrophage | *HYI* | TRUE | 3.535858986 |
| HTN | macrophage | *ADO* | TRUE | 5.240328578 |

| | | | | |
|---|---|---|---|---|
| HTN | macrophage | *ZNF320* | TRUE | 3.296769715 |
| HTN | macrophage | *A3GALT2* | TRUE | 3.440268603 |
| HTN | macrophage | *NUTM2A* | TRUE | 3.326293316 |
| HTN | macrophage | *FOXI2* | TRUE | 2.041594452 |
| HTN | macrophage | *KLHL17* | TRUE | 2.492852279 |
| HTN | macrophage | *ZNF559* | TRUE | 2.440255937 |
| HTN | macrophage | *HIST1H3E* | TRUE | 4.955654048 |
| HTN | macrophage | *WDR5B* | TRUE | 2.101679217 |
| HTN | macrophage | *SULT1A2* | TRUE | 2.665587352 |
| HTN | macrophage | *C6orf141* | TRUE | 2.226302529 |
| HTN | macrophage | *RPS26* | TRUE | 3.528943225 |
| HTN | macrophage | *ZNF311* | TRUE | 2.01545452 |
| HTN | macrophage | *TMEM116* | TRUE | 6.257741832 |
| HTN | macrophage | *SHISA4* | TRUE | 4.23325889 |
| HTN | macrophage | *HSD17B8* | TRUE | 2.874842862 |
| HTN | macrophage | *HLA.C* | TRUE | 2.871395337 |
| HTN | macrophage | *MXD3* | TRUE | 2.052007359 |
| HTN | macrophage | *ATF6B* | TRUE | 2.787946033 |
| HTN | macrophage | *AC016773.1* | TRUE | 2.395263267 |
| HTN | macrophage | *TIAF1* | TRUE | 3.238551484 |
| HTN | macrophage | *LINC00092* | TRUE | 1.981753568 |
| HTN | macrophage | *HLA.DQA2* | TRUE | 2.537300245 |
| HTN | macrophage | *TNFSF12* | TRUE | 3.648512839 |
| HTN | macrophage | *LINC00638* | TRUE | 2.089196995 |
| HTN | mast_cell | *SGCA* | TRUE | 2.137589653 |
| HTN | mast_cell | *TEX14* | TRUE | 2.999314022 |
| HTN | mast_cell | *BTN2A2* | TRUE | 3.466737603 |
| HTN | mast_cell | *RIPK3* | TRUE | 1.974092161 |
| HTN | mast_cell | *HAUS8* | TRUE | 3.329138521 |
| HTN | mast_cell | *FBN2* | TRUE | 3.151726252 |
| HTN | mast_cell | *NEIL2* | TRUE | 2.266136228 |
| HTN | mast_cell | *CLEC18A* | TRUE | 4.33944633 |
| HTN | mast_cell | *SLC5A11* | TRUE | 4.397678502 |
| HTN | mast_cell | *S100B* | TRUE | 2.902600954 |
| HTN | mast_cell | *SRR* | TRUE | 2.572133529 |
| HTN | mast_cell | *RGS19* | TRUE | 4.510679984 |
| HTN | mast_cell | *CTSW* | TRUE | 3.014250972 |
| HTN | mast_cell | *SNX31* | TRUE | 2.362241674 |
| HTN | mast_cell | *SEZ6L2* | TRUE | 1.999520193 |
| HTN | mast_cell | *ORAI3* | TRUE | 2.302779354 |
| HTN | mast_cell | *RPP25* | TRUE | 2.829928162 |
| HTN | mast_cell | *ZNF320* | TRUE | 1.987669414 |
| HTN | mast_cell | *ACOT1* | TRUE | 2.11354592 |
| HTN | mast_cell | *A3GALT2* | TRUE | 2.146520614 |
| HTN | mast_cell | *NUTM2A* | TRUE | 3.274808122 |
| HTN | mast_cell | *HIST1H3E* | TRUE | 3.99715597 |
| HTN | mast_cell | *PCDHB11* | TRUE | 1.966532706 |
| HTN | mast_cell | *TMEM116* | TRUE | 3.324055673 |
| HTN | mast_cell | *PPM1N* | TRUE | 2.129165037 |
| HTN | mast_cell | *HLA.DQA2* | TRUE | 2.197077505 |

| | | | | |
|---|---|---|---|---|
| HTN | mast_cell | *TNFSF12* | TRUE | 3.165886478 |
| HTN | mesothelium | *M6PR* | TRUE | 2.265014751 |
| HTN | mesothelium | *ARHGAP33* | TRUE | 2.144287037 |
| HTN | mesothelium | *CD79B* | TRUE | 2.372758825 |
| HTN | mesothelium | *CEP68* | TRUE | 3.300829473 |
| HTN | mesothelium | *RNASET2* | TRUE | 2.819208402 |
| HTN | mesothelium | *CENPQ* | TRUE | 2.112719443 |
| HTN | mesothelium | *SDCCAG8* | TRUE | 3.891994141 |
| HTN | mesothelium | *PIGV* | TRUE | 4.226725255 |
| HTN | mesothelium | *POLR1A* | TRUE | 3.067920596 |
| HTN | mesothelium | *SNX10* | TRUE | 4.346132808 |
| HTN | mesothelium | *AAMDC* | TRUE | 3.257907752 |
| HTN | mesothelium | *MUL1* | TRUE | 2.762118826 |
| HTN | mesothelium | *PABPC4* | TRUE | 3.956733537 |
| HTN | mesothelium | *RGS17* | TRUE | 2.111850996 |
| HTN | mesothelium | *ITPR3* | TRUE | 2.126780502 |
| HTN | mesothelium | *SCD* | TRUE | 3.980970734 |
| HTN | mesothelium | *MARCH2* | TRUE | 3.554366062 |
| HTN | mesothelium | *THOC5* | TRUE | 2.345655541 |
| HTN | mesothelium | *FAM118A* | TRUE | 1.961280969 |
| HTN | mesothelium | *BIRC7* | TRUE | 2.920585028 |
| HTN | mesothelium | *SIRPB1* | TRUE | 2.244215523 |
| HTN | mesothelium | *POLI* | TRUE | 3.59386901 |
| HTN | mesothelium | *MTHFSD* | TRUE | 3.187962983 |
| HTN | mesothelium | *LIN7B* | TRUE | 2.61281731 |
| HTN | mesothelium | *PEX11G* | TRUE | 2.023086148 |
| HTN | mesothelium | *ICAM5* | TRUE | 2.869019542 |
| HTN | mesothelium | *MEST* | TRUE | 3.019700606 |
| HTN | mesothelium | *SLC1A1* | TRUE | 1.993114069 |
| HTN | mesothelium | *SGCA* | TRUE | 2.66764968 |
| HTN | mesothelium | *CCND1* | TRUE | 4.257719269 |
| HTN | mesothelium | *C11orf21* | TRUE | 2.052950368 |
| HTN | mesothelium | *BVES* | TRUE | 2.037778699 |
| HTN | mesothelium | *NME5* | TRUE | 2.389406963 |
| HTN | mesothelium | *BNIP1* | TRUE | 2.073267871 |
| HTN | mesothelium | *NPHP3* | TRUE | 2.169113153 |
| HTN | mesothelium | *PCCB* | TRUE | 4.284053651 |
| HTN | mesothelium | *IRF6* | TRUE | 2.752773345 |
| HTN | mesothelium | *ELOVL4* | TRUE | 2.046448684 |
| HTN | mesothelium | *ACOT2* | TRUE | 2.909619278 |
| HTN | mesothelium | *ZFP30* | TRUE | 2.15909632 |
| HTN | mesothelium | *ADRA1A* | TRUE | 2.227150551 |
| HTN | mesothelium | *TBX2* | TRUE | 5.273790542 |
| HTN | mesothelium | *TEX14* | TRUE | 3.711703078 |
| HTN | mesothelium | *TOX2* | TRUE | 2.284230523 |
| HTN | mesothelium | *BTN2A2* | TRUE | 3.247350346 |
| HTN | mesothelium | *ZNF391* | TRUE | 2.402738471 |
| HTN | mesothelium | *GRK4* | TRUE | 2.872207393 |
| HTN | mesothelium | *UROD* | TRUE | 2.113083779 |
| HTN | mesothelium | *TSKS* | TRUE | 2.590018496 |

| | | | | |
|---|---|---|---|---|
| HTN | mesothelium | *TRAP1* | TRUE | 4.257279353 |
| HTN | mesothelium | *ABHD8* | TRUE | 3.587489268 |
| HTN | mesothelium | *GFER* | TRUE | 2.009637668 |
| HTN | mesothelium | *STYXL1* | TRUE | 2.067873585 |
| HTN | mesothelium | *GAD1* | TRUE | 2.324817519 |
| HTN | mesothelium | *EIF2AK4* | TRUE | 2.147480875 |
| HTN | mesothelium | *ATP8B3* | TRUE | 3.225429908 |
| HTN | mesothelium | *PGPEP1* | TRUE | 2.539348241 |
| HTN | mesothelium | *HAUS8* | TRUE | 5.150969613 |
| HTN | mesothelium | *RSG1* | TRUE | 2.138982719 |
| HTN | mesothelium | *ACP2* | TRUE | 4.209128271 |
| HTN | mesothelium | *AGT* | TRUE | 3.208536548 |
| HTN | mesothelium | *SLC37A4* | TRUE | 2.582526565 |
| HTN | mesothelium | *PPP2R1B* | TRUE | 2.937058548 |
| HTN | mesothelium | *NUSAP1* | TRUE | 2.850573539 |
| HTN | mesothelium | *ENPEP* | TRUE | 2.224083796 |
| HTN | mesothelium | *FBN2* | TRUE | 3.872248945 |
| HTN | mesothelium | *FITM1* | TRUE | 2.320644678 |
| HTN | mesothelium | *RTN1* | TRUE | 2.175010545 |
| HTN | mesothelium | *HAPLN3* | TRUE | 2.415673463 |
| HTN | mesothelium | *RPL3L* | TRUE | 2.265278574 |
| HTN | mesothelium | *DEF8* | TRUE | 2.475615461 |
| HTN | mesothelium | *VPS53* | TRUE | 2.050403341 |
| HTN | mesothelium | *LRRC46* | TRUE | 2.360758277 |
| HTN | mesothelium | *SCRN2* | TRUE | 2.657303623 |
| HTN | mesothelium | *C18orf8* | TRUE | 3.173706524 |
| HTN | mesothelium | *CREB3L4* | TRUE | 2.591243039 |
| HTN | mesothelium | *PLEKHA6* | TRUE | 2.833566862 |
| HTN | mesothelium | *TSLP* | TRUE | 2.450540339 |
| HTN | mesothelium | *IRAK1BP1* | TRUE | 3.251435664 |
| HTN | mesothelium | *SLC18B1* | TRUE | 2.596004277 |
| HTN | mesothelium | *ZNF215* | TRUE | 2.239531843 |
| HTN | mesothelium | *HSD17B12* | TRUE | 5.831481044 |
| HTN | mesothelium | *TNKS1BP1* | TRUE | 4.592866128 |
| HTN | mesothelium | *TTC12* | TRUE | 2.201297683 |
| HTN | mesothelium | *DUSP15* | TRUE | 2.022543175 |
| HTN | mesothelium | *TRPT1* | TRUE | 2.01140731 |
| HTN | mesothelium | *PRDM8* | TRUE | 2.708981459 |
| HTN | mesothelium | *NEIL2* | TRUE | 3.67279926 |
| HTN | mesothelium | *PROM2* | TRUE | 2.310777291 |
| HTN | mesothelium | *MMP16* | TRUE | 2.395653427 |
| HTN | mesothelium | *ART3* | TRUE | 2.681255732 |
| HTN | mesothelium | *SFXN2* | TRUE | 5.04246131 |
| HTN | mesothelium | *SLC5A11* | TRUE | 4.753307468 |
| HTN | mesothelium | *HSF2BP* | TRUE | 1.99111381 |
| HTN | mesothelium | *S100B* | TRUE | 3.746622007 |
| HTN | mesothelium | *ZNF208* | TRUE | 2.160863206 |
| HTN | mesothelium | *ABCF3* | TRUE | 4.89676831 |
| HTN | mesothelium | *MRPL55* | TRUE | 5.970439209 |
| HTN | mesothelium | *LYSMD1* | TRUE | 3.036686752 |

| | | | | |
|---|---|---|---|---|
| HTN | mesothelium | *DCAF16* | TRUE | 3.463302357 |
| HTN | mesothelium | *RFT1* | TRUE | 2.91326618 |
| HTN | mesothelium | *ZNF589* | TRUE | 2.355012266 |
| HTN | mesothelium | *RNF123* | TRUE | 3.821670113 |
| HTN | mesothelium | *ERAP1* | TRUE | 4.181951695 |
| HTN | mesothelium | *RPP25L* | TRUE | 2.766904829 |
| HTN | mesothelium | *OXGR1* | TRUE | 2.159258012 |
| HTN | mesothelium | *CASP7* | TRUE | 2.081899399 |
| HTN | mesothelium | *ARL5B* | TRUE | 3.096952794 |
| HTN | mesothelium | *CYB5R2* | TRUE | 2.685304652 |
| HTN | mesothelium | *TRIM66* | TRUE | 3.977733185 |
| HTN | mesothelium | *RIMKLB* | TRUE | 4.772893138 |
| HTN | mesothelium | *GALR1* | TRUE | 2.071509847 |
| HTN | mesothelium | *ZNF23* | TRUE | 2.150566708 |
| HTN | mesothelium | *CHAF1A* | TRUE | 2.221654445 |
| HTN | mesothelium | *SRR* | TRUE | 3.400853518 |
| HTN | mesothelium | *SOAT2* | TRUE | 2.771917559 |
| HTN | mesothelium | *ZNF598* | TRUE | 2.519151086 |
| HTN | mesothelium | *SERINC2* | TRUE | 2.623954594 |
| HTN | mesothelium | *TCTN2* | TRUE | 1.998122515 |
| HTN | mesothelium | *CPT1C* | TRUE | 3.361721952 |
| HTN | mesothelium | *RAB24* | TRUE | 2.85770103 |
| HTN | mesothelium | *INO80E* | TRUE | 4.904382954 |
| HTN | mesothelium | *RGPD8* | TRUE | 3.305098168 |
| HTN | mesothelium | *MUC3A* | TRUE | 2.043563003 |
| HTN | mesothelium | *MYO7B* | TRUE | 3.304810865 |
| HTN | mesothelium | *DNAJC18* | TRUE | 3.299218843 |
| HTN | mesothelium | *SMAGP* | TRUE | 1.970706015 |
| HTN | mesothelium | *TTLL6* | TRUE | 5.144097006 |
| HTN | mesothelium | *PAQR8* | TRUE | 2.71065598 |
| HTN | mesothelium | *MRPL36* | TRUE | 2.7907448 |
| HTN | mesothelium | *RGS19* | TRUE | 5.317990145 |
| HTN | mesothelium | *CTSW* | TRUE | 3.056707339 |
| HTN | mesothelium | *MST1* | TRUE | 2.733658889 |
| HTN | mesothelium | *C11orf80* | TRUE | 4.170399444 |
| HTN | mesothelium | *CTU2* | TRUE | 2.279777808 |
| HTN | mesothelium | *SNX31* | TRUE | 4.453537491 |
| HTN | mesothelium | *ORAI3* | TRUE | 4.240134955 |
| HTN | mesothelium | *MTHFR* | TRUE | 3.57555457 |
| HTN | mesothelium | *ODF3B* | TRUE | 2.457935974 |
| HTN | mesothelium | *GEN1* | TRUE | 2.866119181 |
| HTN | mesothelium | *MPI* | TRUE | 7.049378809 |
| HTN | mesothelium | *HYI* | TRUE | 3.438494912 |
| HTN | mesothelium | *CTC1* | TRUE | 2.190241116 |
| HTN | mesothelium | *CLDN7* | TRUE | 2.871481279 |
| HTN | mesothelium | *ADO* | TRUE | 5.636459714 |
| HTN | mesothelium | *CLN8* | TRUE | 3.820611085 |
| HTN | mesothelium | *FES* | TRUE | 8.126442456 |
| HTN | mesothelium | *ZNF320* | TRUE | 3.513871467 |
| HTN | mesothelium | *NPIPA1* | TRUE | 2.10732923 |

| | | | | |
|---|---|---|---|---|
| HTN | mesothelium | *ACOT1* | TRUE | 2.429985604 |
| HTN | mesothelium | *ALDH1A3* | TRUE | 2.048312091 |
| HTN | mesothelium | *A3GALT2* | TRUE | 3.917055524 |
| HTN | mesothelium | *NUTM2A* | TRUE | 3.069988445 |
| HTN | mesothelium | *FOXI2* | TRUE | 2.371788932 |
| HTN | mesothelium | *KLHL17* | TRUE | 1.986023316 |
| HTN | mesothelium | *ZNF559* | TRUE | 2.772896236 |
| HTN | mesothelium | *FAM26F* | TRUE | 2.551136536 |
| HTN | mesothelium | *FBF1* | TRUE | 3.90600605 |
| HTN | mesothelium | *MSL1* | TRUE | 1.980770889 |
| HTN | mesothelium | *COLCA1* | TRUE | 3.231820363 |
| HTN | mesothelium | *HRCT1* | TRUE | 3.010000524 |
| HTN | mesothelium | *HIST1H3E* | TRUE | 5.006394582 |
| HTN | mesothelium | *WDR5B* | TRUE | 2.863339783 |
| HTN | mesothelium | *ZNF100* | TRUE | 3.143261928 |
| HTN | mesothelium | *SULT1A2* | TRUE | 3.35693663 |
| HTN | mesothelium | *C6orf141* | TRUE | 2.418830318 |
| HTN | mesothelium | *PCDHB11* | TRUE | 1.985192912 |
| HTN | mesothelium | *RPS26* | TRUE | 3.875741755 |
| HTN | mesothelium | *ZNF311* | TRUE | 2.34911183 |
| HTN | mesothelium | *AGAP9* | TRUE | 2.472549298 |
| HTN | mesothelium | *TMEM116* | TRUE | 6.599013798 |
| HTN | mesothelium | *PNP* | TRUE | 2.910954223 |
| HTN | mesothelium | *SHISA4* | TRUE | 5.249397533 |
| HTN | mesothelium | *AL590822.1* | TRUE | 2.48249547 |
| HTN | mesothelium | *HSD17B8* | TRUE | 3.015218973 |
| HTN | mesothelium | *HLA.C* | TRUE | 3.180796091 |
| HTN | mesothelium | *TMEM170B* | TRUE | 2.02296282 |
| HTN | mesothelium | *ATF6B* | TRUE | 3.317193777 |
| HTN | mesothelium | *PPM1N* | TRUE | 2.617416632 |
| HTN | mesothelium | *LTB4R* | TRUE | 2.572503815 |
| HTN | mesothelium | *AC016773.1* | TRUE | 1.982013011 |
| HTN | mesothelium | *SLC12A8* | TRUE | 2.532098871 |
| HTN | mesothelium | *TIAF1* | TRUE | 3.651650085 |
| HTN | mesothelium | *OSER1.AS1* | TRUE | 2.160095723 |
| HTN | mesothelium | *AL049840.1* | TRUE | 2.711612665 |
| HTN | mesothelium | *PLEKHM1* | TRUE | 2.1002566 |
| HTN | mesothelium | *LINC00092* | TRUE | 2.015537973 |
| HTN | mesothelium | *AC108463.2* | TRUE | 2.317732837 |
| HTN | mesothelium | *HLA.DQA2* | TRUE | 2.149108734 |
| HTN | mesothelium | *LINC00211* | TRUE | 2.178782343 |
| HTN | mesothelium | *TNFSF12* | TRUE | 3.222864054 |
| HTN | mesothelium | *ZNF709* | TRUE | 3.238936469 |
| HTN | mesothelium | *LINC00638* | TRUE | 2.818521138 |
| HTN | monocyte | *ITIH4* | TRUE | 2.076290437 |
| HTN | monocyte | *PIGV* | TRUE | 2.870135536 |
| HTN | monocyte | *BIRC7* | TRUE | 2.211461404 |
| HTN | monocyte | *MTHFSD* | TRUE | 1.981564632 |
| HTN | monocyte | *SGCA* | TRUE | 2.396120266 |
| HTN | monocyte | *WFDC3* | TRUE | 2.093977117 |

| | | | | |
|---|---|---|---|---|
| HTN | monocyte | *BTN2A2* | TRUE | 2.367654335 |
| HTN | monocyte | *ZNF391* | TRUE | 2.205224463 |
| HTN | monocyte | *AGT* | TRUE | 2.947221424 |
| HTN | monocyte | *SFXN2* | TRUE | 4.03803994 |
| HTN | monocyte | *CLEC18A* | TRUE | 3.382491918 |
| HTN | monocyte | *CDC25A* | TRUE | 3.760130428 |
| HTN | monocyte | *TRIM66* | TRUE | 1.996957122 |
| HTN | monocyte | *TTLL6* | TRUE | 2.027189089 |
| HTN | monocyte | *MST1* | TRUE | 2.611964611 |
| HTN | monocyte | *ORAI3* | TRUE | 2.380487509 |
| HTN | monocyte | *MPI* | TRUE | 2.099034953 |
| HTN | monocyte | *HYI* | TRUE | 2.86603087 |
| HTN | monocyte | *CLN8* | TRUE | 2.910777192 |
| HTN | monocyte | *ZNF320* | TRUE | 2.209155187 |
| HTN | monocyte | *NUTM2A* | TRUE | 3.172552487 |
| HTN | monocyte | *HIST1H3E* | TRUE | 3.124326084 |
| HTN | monocyte | *TMEM116* | TRUE | 4.094161259 |
| HTN | monocyte | *HLA.DQA2* | TRUE | 2.171903058 |
| HTN | nk_cell | *BTN2A2* | TRUE | 1.970700341 |
| HTN | nk_cell | *HSD17B12* | TRUE | 2.064445544 |
| HTN | nk_cell | *PROM2* | TRUE | 2.245570601 |
| HTN | nk_cell | *CLEC18A* | TRUE | 3.227513839 |
| HTN | nk_cell | *ABCF3* | TRUE | 3.442639232 |
| HTN | nk_cell | *INO80E* | TRUE | 2.444106294 |
| HTN | nk_cell | *RGPD8* | TRUE | 2.678321476 |
| HTN | nk_cell | *SNX31* | TRUE | 3.771776745 |
| HTN | nk_cell | *MPI* | TRUE | 3.54179157 |
| HTN | nk_cell | *TRIM73* | TRUE | 2.04783346 |
| HTN | nk_cell | *ADO* | TRUE | 4.117028702 |
| HTN | nk_cell | *TMEM116* | TRUE | 3.933150263 |
| HTN | nk_cell | *PRRT1* | TRUE | 2.115499025 |
| HTN | nk_cell | *TIAF1* | TRUE | 2.550622039 |
| HTN | pericyte | *PIGV* | TRUE | 2.62694283 |
| HTN | pericyte | *BIRC7* | TRUE | 2.863172675 |
| HTN | pericyte | *POLI* | TRUE | 2.916434869 |
| HTN | pericyte | *IRF6* | TRUE | 2.289746756 |
| HTN | pericyte | *BTN2A2* | TRUE | 2.769105244 |
| HTN | pericyte | *TRAP1* | TRUE | 2.741459131 |
| HTN | pericyte | *ATP8B3* | TRUE | 2.442505336 |
| HTN | pericyte | *HAUS8* | TRUE | 2.941553722 |
| HTN | pericyte | *ACP2* | TRUE | 2.687864004 |
| HTN | pericyte | *AGT* | TRUE | 3.079003464 |
| HTN | pericyte | *FBN2* | TRUE | 3.286760654 |
| HTN | pericyte | *SCRN2* | TRUE | 2.253706413 |
| HTN | pericyte | *CREB3L4* | TRUE | 2.750866756 |
| HTN | pericyte | *TSLP* | TRUE | 2.188873181 |
| HTN | pericyte | *HSD17B12* | TRUE | 4.990231152 |
| HTN | pericyte | *TNKS1BP1* | TRUE | 2.406338707 |
| HTN | pericyte | *ART3* | TRUE | 2.527941033 |
| HTN | pericyte | *SFXN2* | TRUE | 4.160144999 |

| | | | | |
|---|---|---|---|---|
| HTN | pericyte | *SLC5A11* | TRUE | 3.52126529 |
| HTN | pericyte | *S100B* | TRUE | 3.43717788 |
| HTN | pericyte | *ABCF3* | TRUE | 4.154620577 |
| HTN | pericyte | *MRPL55* | TRUE | 2.589384938 |
| HTN | pericyte | *LYSMD1* | TRUE | 2.760634358 |
| HTN | pericyte | *TRIM66* | TRUE | 2.943141171 |
| HTN | pericyte | *RIMKLB* | TRUE | 2.48088179 |
| HTN | pericyte | *SRR* | TRUE | 2.540858189 |
| HTN | pericyte | *CPT1C* | TRUE | 3.420395051 |
| HTN | pericyte | *INO80E* | TRUE | 3.399426563 |
| HTN | pericyte | *RGPD8* | TRUE | 2.857518001 |
| HTN | pericyte | *TTLL6* | TRUE | 2.796788506 |
| HTN | pericyte | *PAQR8* | TRUE | 2.206501093 |
| HTN | pericyte | *MRPL36* | TRUE | 1.96296958 |
| HTN | pericyte | *RGS19* | TRUE | 4.383390943 |
| HTN | pericyte | *CTSW* | TRUE | 3.035770685 |
| HTN | pericyte | *MST1* | TRUE | 3.76470951 |
| HTN | pericyte | *CCNE2* | TRUE | 3.875491386 |
| HTN | pericyte | *ORAI3* | TRUE | 3.275103142 |
| HTN | pericyte | *GEN1* | TRUE | 2.081561107 |
| HTN | pericyte | *MPI* | TRUE | 5.965724376 |
| HTN | pericyte | *ADO* | TRUE | 3.939246675 |
| HTN | pericyte | *CLN8* | TRUE | 2.55265505 |
| HTN | pericyte | *FES* | TRUE | 4.786941114 |
| HTN | pericyte | *ZNF320* | TRUE | 3.125634288 |
| HTN | pericyte | *A3GALT2* | TRUE | 3.101456706 |
| HTN | pericyte | *NUTM2A* | TRUE | 3.60377747 |
| HTN | pericyte | *KLHL17* | TRUE | 2.179213432 |
| HTN | pericyte | *ZNF559* | TRUE | 2.110899784 |
| HTN | pericyte | *FAM26F* | TRUE | 2.165826888 |
| HTN | pericyte | *HRCT1* | TRUE | 2.552921827 |
| HTN | pericyte | *HIST1H3E* | TRUE | 5.024049823 |
| HTN | pericyte | *C6orf141* | TRUE | 2.207564642 |
| HTN | pericyte | *RPS26* | TRUE | 3.703787658 |
| HTN | pericyte | *TMEM116* | TRUE | 5.887087301 |
| HTN | pericyte | *PNP* | TRUE | 2.48071754 |
| HTN | pericyte | *HSD17B8* | TRUE | 2.671991996 |
| HTN | pericyte | *HLA.C* | TRUE | 2.509483937 |
| HTN | pericyte | *ATF6B* | TRUE | 2.036166664 |
| HTN | pericyte | *AC016773.1* | TRUE | 2.767947295 |
| HTN | pericyte | *TIAF1* | TRUE | 2.932268141 |
| HTN | pericyte | *LINC00960* | TRUE | 2.170227653 |
| HTN | SMC | *CD79B* | TRUE | 2.393903716 |
| HTN | SMC | *POLI* | TRUE | 2.864734473 |
| HTN | SMC | *ACOT2* | TRUE | 2.288525816 |
| HTN | SMC | *ZNF391* | TRUE | 2.850517536 |
| HTN | SMC | *GRK4* | TRUE | 2.31512947 |
| HTN | SMC | *ATP8B3* | TRUE | 2.707846648 |
| HTN | SMC | *HAUS8* | TRUE | 2.340055123 |
| HTN | SMC | *AGT* | TRUE | 3.047170908 |

| | | | | |
|---|---|---|---|---|
| HTN | SMC | *LRRC46* | TRUE | 2.168406963 |
| HTN | SMC | *MMEL1* | TRUE | 2.179011124 |
| HTN | SMC | *CREB3L4* | TRUE | 2.44613212 |
| HTN | SMC | *HSD17B12* | TRUE | 5.069684963 |
| HTN | SMC | *TNKS1BP1* | TRUE | 2.075589383 |
| HTN | SMC | *NEIL2* | TRUE | 3.904919661 |
| HTN | SMC | *SFXN2* | TRUE | 3.058294937 |
| HTN | SMC | *SLC5A11* | TRUE | 3.746520686 |
| HTN | SMC | *S100B* | TRUE | 3.06807955 |
| HTN | SMC | *ABCF3* | TRUE | 3.619118777 |
| HTN | SMC | *ZNF589* | TRUE | 3.125423388 |
| HTN | SMC | *TRIM66* | TRUE | 2.786000358 |
| HTN | SMC | *SRR* | TRUE | 2.653228625 |
| HTN | SMC | *INO80E* | TRUE | 2.67864931 |
| HTN | SMC | *RGPD8* | TRUE | 2.938679843 |
| HTN | SMC | *TTLL6* | TRUE | 4.651732067 |
| HTN | SMC | *RGS19* | TRUE | 3.271131807 |
| HTN | SMC | *CTSW* | TRUE | 2.312283187 |
| HTN | SMC | *MST1* | TRUE | 2.411468799 |
| HTN | SMC | *SNX31* | TRUE | 2.704548375 |
| HTN | SMC | *CCNE2* | TRUE | 2.447543306 |
| HTN | SMC | *MPI* | TRUE | 3.691834227 |
| HTN | SMC | *HYI* | TRUE | 3.521057719 |
| HTN | SMC | *FES* | TRUE | 5.490366263 |
| HTN | SMC | *A3GALT2* | TRUE | 2.452549273 |
| HTN | SMC | *NUTM2A* | TRUE | 3.330076026 |
| HTN | SMC | *CDK10* | TRUE | 2.700499509 |
| HTN | SMC | *GLDN* | TRUE | 2.218114887 |
| HTN | SMC | *HIST1H3E* | TRUE | 3.302897741 |
| HTN | SMC | *TMEM116* | TRUE | 6.310479446 |
| HTN | SMC | *HLA.DRB5* | TRUE | 2.271412979 |
| HTN | SMC | *HLA.C* | TRUE | 3.089690102 |
| HTN | SMC | *TNFSF12* | TRUE | 2.989565026 |
| HTN | SMC | *LINC00960* | TRUE | 2.541200641 |
| HTN | t_cell | *CD79B* | TRUE | 2.291898988 |
| HTN | t_cell | *SNX10* | TRUE | 2.089697113 |
| HTN | t_cell | *BIRC7* | TRUE | 2.048853456 |
| HTN | t_cell | *POLI* | TRUE | 2.430620125 |
| HTN | t_cell | *MTHFSD* | TRUE | 2.182403438 |
| HTN | t_cell | *SGCA* | TRUE | 2.633968686 |
| HTN | t_cell | *IRF6* | TRUE | 2.110620785 |
| HTN | t_cell | *ACOT2* | TRUE | 2.719533499 |
| HTN | t_cell | *TBX2* | TRUE | 3.163538512 |
| HTN | t_cell | *TEX14* | TRUE | 2.477880551 |
| HTN | t_cell | *WFDC3* | TRUE | 2.145726159 |
| HTN | t_cell | *BTN2A2* | TRUE | 3.10004195 |
| HTN | t_cell | *GRK4* | TRUE | 2.079890052 |
| HTN | t_cell | *TRAP1* | TRUE | 2.453196698 |
| HTN | t_cell | *ATP8B3* | TRUE | 2.601334389 |
| HTN | t_cell | *HAUS8* | TRUE | 2.045820372 |

| | | | | |
|---|---|---|---|---|
| HTN | t_cell | *ACP2* | TRUE | 3.523655288 |
| HTN | t_cell | *AGT* | TRUE | 2.698265642 |
| HTN | t_cell | *NMT1* | TRUE | 2.945546775 |
| HTN | t_cell | *FBN2* | TRUE | 2.502722349 |
| HTN | t_cell | *FITM1* | TRUE | 2.360046399 |
| HTN | t_cell | *SCRN2* | TRUE | 2.153708281 |
| HTN | t_cell | *HSD17B12* | TRUE | 4.141415511 |
| HTN | t_cell | *NEIL2* | TRUE | 2.361677516 |
| HTN | t_cell | *ART3* | TRUE | 2.346479933 |
| HTN | t_cell | *SFXN2* | TRUE | 4.298197733 |
| HTN | t_cell | *CLEC18A* | TRUE | 3.202464767 |
| HTN | t_cell | *SLC5A11* | TRUE | 4.057225693 |
| HTN | t_cell | *S100B* | TRUE | 3.153383096 |
| HTN | t_cell | *ABCF3* | TRUE | 3.645802883 |
| HTN | t_cell | *MRPL55* | TRUE | 3.607566949 |
| HTN | t_cell | *LYSMD1* | TRUE | 2.025804774 |
| HTN | t_cell | *RNF123* | TRUE | 4.462405975 |
| HTN | t_cell | *TRIM66* | TRUE | 2.778203088 |
| HTN | t_cell | *SRR* | TRUE | 2.977505453 |
| HTN | t_cell | *RGPD8* | TRUE | 2.483917984 |
| HTN | t_cell | *TTLL6* | TRUE | 3.332673876 |
| HTN | t_cell | *MRPL36* | TRUE | 2.508288957 |
| HTN | t_cell | *RGS19* | TRUE | 2.963722989 |
| HTN | t_cell | *CTSW* | TRUE | 2.159636197 |
| HTN | t_cell | *ZNF738* | TRUE | 2.03283858 |
| HTN | t_cell | *MST1* | TRUE | 3.059341676 |
| HTN | t_cell | *SNX31* | TRUE | 3.046882003 |
| HTN | t_cell | *CCNE2* | TRUE | 2.799331528 |
| HTN | t_cell | *ORAI3* | TRUE | 2.920243449 |
| HTN | t_cell | *MPI* | TRUE | 5.254785307 |
| HTN | t_cell | *TRIM73* | TRUE | 2.797769472 |
| HTN | t_cell | *HYI* | TRUE | 3.186738339 |
| HTN | t_cell | *ADO* | TRUE | 4.775944117 |
| HTN | t_cell | *CLN8* | TRUE | 2.581138619 |
| HTN | t_cell | *FES* | TRUE | 3.575089992 |
| HTN | t_cell | *ZNF320* | TRUE | 2.912225181 |
| HTN | t_cell | *ACOT1* | TRUE | 2.433393711 |
| HTN | t_cell | *A3GALT2* | TRUE | 2.514633134 |
| HTN | t_cell | *NUTM2A* | TRUE | 3.115762736 |
| HTN | t_cell | *CDK10* | TRUE | 1.965285607 |
| HTN | t_cell | *HIST1H3E* | TRUE | 4.92856209 |
| HTN | t_cell | *C6orf141* | TRUE | 2.145391374 |
| HTN | t_cell | *RPS26* | TRUE | 3.327948987 |
| HTN | t_cell | *TMEM116* | TRUE | 3.850873264 |
| HTN | t_cell | *HLA.DRB5* | TRUE | 2.173674639 |
| HTN | t_cell | *HSD17B8* | TRUE | 2.767825328 |
| HTN | t_cell | *TMEM110* | TRUE | 2.273769763 |
| HTN | t_cell | *ATF6B* | TRUE | 2.315774399 |
| HTN | t_cell | *AC016773.1* | TRUE | 2.711983075 |
| HTN | t_cell | *TIAF1* | TRUE | 3.301714122 |

| | | | | |
|---|---|---|---|---|
| HTN | t_cell | *TNFSF12* | TRUE | 3.317957145 |
| Obesity | adipocyte | *CENPQ* | TRUE | 2.176942112 |
| Obesity | adipocyte | *LSG1* | TRUE | 2.267938048 |
| Obesity | adipocyte | *CD5L* | TRUE | 2.637644991 |
| Obesity | adipocyte | *TIPIN* | TRUE | 2.261014229 |
| Obesity | adipocyte | *PABPC4* | TRUE | 3.658270538 |
| Obesity | adipocyte | *RGS17* | TRUE | 2.109532667 |
| Obesity | adipocyte | *TBX15* | TRUE | 3.258994568 |
| Obesity | adipocyte | *ITPR3* | TRUE | 2.367324261 |
| Obesity | adipocyte | *ANKRD54* | TRUE | 2.495216458 |
| Obesity | adipocyte | *CENPM* | TRUE | 3.630221322 |
| Obesity | adipocyte | *ELMO3* | TRUE | 2.3318332 |
| Obesity | adipocyte | *CLUAP1* | TRUE | 2.400924933 |
| Obesity | adipocyte | *PYCARD* | TRUE | 2.197951225 |
| Obesity | adipocyte | *SLC22A18* | TRUE | 2.491545181 |
| Obesity | adipocyte | *BTN3A3* | TRUE | 2.005599252 |
| Obesity | adipocyte | *GABRR2* | TRUE | 2.17909303 |
| Obesity | adipocyte | *GLP1R* | TRUE | 2.035829984 |
| Obesity | adipocyte | *NME5* | TRUE | 1.988185328 |
| Obesity | adipocyte | *BNIP1* | TRUE | 2.099090311 |
| Obesity | adipocyte | *PCCB* | TRUE | 2.667449365 |
| Obesity | adipocyte | *CHMP3* | TRUE | 2.074033113 |
| Obesity | adipocyte | *SERPINC1* | TRUE | 4.698451302 |
| Obesity | adipocyte | *INVS* | TRUE | 1.988106944 |
| Obesity | adipocyte | *SNX19* | TRUE | 2.187374258 |
| Obesity | adipocyte | *ADRA1A* | TRUE | 2.295289843 |
| Obesity | adipocyte | *OPN4* | TRUE | 3.264768856 |
| Obesity | adipocyte | *SLC25A51* | TRUE | 2.112110436 |
| Obesity | adipocyte | *BTN2A2* | TRUE | 2.612576549 |
| Obesity | adipocyte | *GRK4* | TRUE | 3.065989573 |
| Obesity | adipocyte | *RIBC2* | TRUE | 3.233830514 |
| Obesity | adipocyte | *AP1M2* | TRUE | 2.772965389 |
| Obesity | adipocyte | *ATP8B3* | TRUE | 5.838034815 |
| Obesity | adipocyte | *PKDREJ* | TRUE | 2.653526926 |
| Obesity | adipocyte | *NDUFA2* | TRUE | 2.135948064 |
| Obesity | adipocyte | *MYBBP1A* | TRUE | 2.193726252 |
| Obesity | adipocyte | *GSTM5* | TRUE | 2.206286835 |
| Obesity | adipocyte | *MICAL1* | TRUE | 2.018070326 |
| Obesity | adipocyte | *DSCC1* | TRUE | 2.213073359 |
| Obesity | adipocyte | *SLC37A4* | TRUE | 4.065268679 |
| Obesity | adipocyte | *NDUFAF1* | TRUE | 2.04399428 |
| Obesity | adipocyte | *CENPO* | TRUE | 6.452184182 |
| Obesity | adipocyte | *CYP2C8* | TRUE | 2.221996099 |
| Obesity | adipocyte | *VPS33A* | TRUE | 2.903236525 |
| Obesity | adipocyte | *DIAPH3* | TRUE | 2.680071175 |
| Obesity | adipocyte | *FITM1* | TRUE | 2.349872066 |
| Obesity | adipocyte | *CYP11A1* | TRUE | 2.442033457 |
| Obesity | adipocyte | *MYO19* | TRUE | 2.565784479 |
| Obesity | adipocyte | *C18orf8* | TRUE | 4.655108456 |
| Obesity | adipocyte | *CBX8* | TRUE | 2.537168251 |

| | | | | |
|---|---|---|---|---|
| Obesity | adipocyte | *COL6A1* | TRUE | 2.221701375 |
| Obesity | adipocyte | *MMEL1* | TRUE | 2.131955705 |
| Obesity | adipocyte | *FAHD2B* | TRUE | 2.058922708 |
| Obesity | adipocyte | *ADHFE1* | TRUE | 2.203343188 |
| Obesity | adipocyte | *RGR* | TRUE | 2.355362247 |
| Obesity | adipocyte | *HSD17B12* | TRUE | 5.695887748 |
| Obesity | adipocyte | *TNKS1BP1* | TRUE | 3.028530796 |
| Obesity | adipocyte | *TTC12* | TRUE | 2.704924932 |
| Obesity | adipocyte | *RGS18* | TRUE | 2.729461732 |
| Obesity | adipocyte | *DPYSL4* | TRUE | 2.696966043 |
| Obesity | adipocyte | *POC5* | TRUE | 2.759325171 |
| Obesity | adipocyte | *MMP16* | TRUE | 1.982026923 |
| Obesity | adipocyte | *CLEC18A* | TRUE | 5.861585408 |
| Obesity | adipocyte | *ETS2* | TRUE | 2.395624755 |
| Obesity | adipocyte | *SLC5A11* | TRUE | 4.773016064 |
| Obesity | adipocyte | *CILP2* | TRUE | 2.28810914 |
| Obesity | adipocyte | *ABCF3* | TRUE | 2.461063483 |
| Obesity | adipocyte | *HS3ST6* | TRUE | 2.114773592 |
| Obesity | adipocyte | *CAMK2N1* | TRUE | 2.554219517 |
| Obesity | adipocyte | *CLHC1* | TRUE | 2.340462223 |
| Obesity | adipocyte | *DNALI1* | TRUE | 3.460386383 |
| Obesity | adipocyte | *RFT1* | TRUE | 2.51808696 |
| Obesity | adipocyte | *MST1R* | TRUE | 2.822537433 |
| Obesity | adipocyte | *RAPSN* | TRUE | 2.669484807 |
| Obesity | adipocyte | *SMG8* | TRUE | 2.246586575 |
| Obesity | adipocyte | *SRR* | TRUE | 3.781611545 |
| Obesity | adipocyte | *FNTA* | TRUE | 2.117762734 |
| Obesity | adipocyte | *NPNT* | TRUE | 1.983152146 |
| Obesity | adipocyte | *GPRIN1* | TRUE | 3.005486488 |
| Obesity | adipocyte | *INO80E* | TRUE | 3.629925055 |
| Obesity | adipocyte | *RGPD8* | TRUE | 2.261941735 |
| Obesity | adipocyte | *MYO7B* | TRUE | 2.248049827 |
| Obesity | adipocyte | *ADORA2B* | TRUE | 4.785461654 |
| Obesity | adipocyte | *TTLL6* | TRUE | 6.66805566 |
| Obesity | adipocyte | *TSNARE1* | TRUE | 2.58163944 |
| Obesity | adipocyte | *RGS19* | TRUE | 2.43216149 |
| Obesity | adipocyte | *FUT10* | TRUE | 2.528392662 |
| Obesity | adipocyte | *MRGPRF* | TRUE | 2.055719064 |
| Obesity | adipocyte | *CADM2* | TRUE | 4.233592355 |
| Obesity | adipocyte | *ORAI3* | TRUE | 5.057489954 |
| Obesity | adipocyte | *SPRYD4* | TRUE | 2.114852431 |
| Obesity | adipocyte | *STX19* | TRUE | 3.462054952 |
| Obesity | adipocyte | *MPI* | TRUE | 1.98062859 |
| Obesity | adipocyte | *ZNF852* | TRUE | 2.526178986 |
| Obesity | adipocyte | *C3orf38* | TRUE | 3.93457845 |
| Obesity | adipocyte | *RPRML* | TRUE | 3.974131918 |
| Obesity | adipocyte | *PPP1R27* | TRUE | 3.423830151 |
| Obesity | adipocyte | *PSMG1* | TRUE | 2.44632929 |
| Obesity | adipocyte | *NPIPA5* | TRUE | 2.312484069 |
| Obesity | adipocyte | *EMILIN3* | TRUE | 3.221647589 |

| | | | | |
|---|---|---|---|---|
| Obesity | adipocyte | *ACOT1* | TRUE | 2.131821737 |
| Obesity | adipocyte | *NUTM2A* | TRUE | 3.067894416 |
| Obesity | adipocyte | *GP1BA* | TRUE | 2.71070471 |
| Obesity | adipocyte | *CENPP* | TRUE | 2.941225374 |
| Obesity | adipocyte | *SKOR1* | TRUE | 2.650370713 |
| Obesity | adipocyte | *ZNF605* | TRUE | 2.072387875 |
| Obesity | adipocyte | *SULT1A2* | TRUE | 9.393845978 |
| Obesity | adipocyte | *DNAH10* | TRUE | 4.067429145 |
| Obesity | adipocyte | *RPS26* | TRUE | 2.948047025 |
| Obesity | adipocyte | *ZNF780A* | TRUE | 3.735740575 |
| Obesity | adipocyte | *SHISA4* | TRUE | 4.667379691 |
| Obesity | adipocyte | *C6orf163* | TRUE | 2.085232855 |
| Obesity | adipocyte | *GPANK1* | TRUE | 3.387634764 |
| Obesity | adipocyte | *C15orf59* | TRUE | 2.287450891 |
| Obesity | adipocyte | *SMN2* | TRUE | 2.213046378 |
| Obesity | adipocyte | *AC084018.1* | TRUE | 3.655705184 |
| Obesity | adipocyte | *ASIC3* | TRUE | 2.662468263 |
| Obesity | adipocyte | *SLC23A3* | TRUE | 2.349933524 |
| Obesity | adipocyte | *LINC00960* | TRUE | 2.313040568 |
| Obesity | adipocyte | *PLA2G4B* | TRUE | 2.084926142 |
| Obesity | adipocyte | *PDF* | TRUE | 2.132444627 |
| Obesity | adipocyte | *AP001172.2* | TRUE | 3.084424453 |
| Obesity | ASPC | *RBM6* | TRUE | 3.189373737 |
| Obesity | ASPC | *CENPQ* | TRUE | 2.35307605 |
| Obesity | ASPC | *FLT4* | TRUE | 2.353188643 |
| Obesity | ASPC | *LSG1* | TRUE | 2.502374353 |
| Obesity | ASPC | *CD5L* | TRUE | 2.64391232 |
| Obesity | ASPC | *TIPIN* | TRUE | 2.777862254 |
| Obesity | ASPC | *FDFT1* | TRUE | 3.837924004 |
| Obesity | ASPC | *GSTP1* | TRUE | 2.382957394 |
| Obesity | ASPC | *CD82* | TRUE | 2.06666315 |
| Obesity | ASPC | *PABPC4* | TRUE | 4.543790978 |
| Obesity | ASPC | *RGS17* | TRUE | 4.1413682 |
| Obesity | ASPC | *TBX15* | TRUE | 3.724511575 |
| Obesity | ASPC | *ITPR3* | TRUE | 2.368990631 |
| Obesity | ASPC | *IL12RB1* | TRUE | 2.01029235 |
| Obesity | ASPC | *CENPM* | TRUE | 3.24586488 |
| Obesity | ASPC | *ABHD12* | TRUE | 2.563467539 |
| Obesity | ASPC | *ELMO3* | TRUE | 2.478749786 |
| Obesity | ASPC | *CLCN7* | TRUE | 2.098585859 |
| Obesity | ASPC | *CLUAP1* | TRUE | 2.605472152 |
| Obesity | ASPC | *PYCARD* | TRUE | 3.015305188 |
| Obesity | ASPC | *MEST* | TRUE | 2.198512354 |
| Obesity | ASPC | *SLC22A18* | TRUE | 2.547732166 |
| Obesity | ASPC | *BTN3A3* | TRUE | 2.694183573 |
| Obesity | ASPC | *GABRR2* | TRUE | 2.823235326 |
| Obesity | ASPC | *GLP1R* | TRUE | 2.109875108 |
| Obesity | ASPC | *NME5* | TRUE | 3.294257749 |
| Obesity | ASPC | *BNIP1* | TRUE | 2.284417871 |
| Obesity | ASPC | *PCCB* | TRUE | 3.754221242 |

| | | | | |
|---|---|---|---|---|
| Obesity | ASPC | *CHMP3* | TRUE | 2.933860205 |
| Obesity | ASPC | *SERPINC1* | TRUE | 6.15171884 |
| Obesity | ASPC | *INVS* | TRUE | 2.53489522 |
| Obesity | ASPC | *SNX19* | TRUE | 2.707156404 |
| Obesity | ASPC | *ADRA1A* | TRUE | 2.805645841 |
| Obesity | ASPC | *TEX14* | TRUE | 2.637380594 |
| Obesity | ASPC | *OPN4* | TRUE | 3.184267399 |
| Obesity | ASPC | *SLC25A51* | TRUE | 2.778751868 |
| Obesity | ASPC | *NEURL2* | TRUE | 2.262991439 |
| Obesity | ASPC | *BTN2A2* | TRUE | 2.420524225 |
| Obesity | ASPC | *MED20* | TRUE | 2.085561075 |
| Obesity | ASPC | *TREM1* | TRUE | 2.330402217 |
| Obesity | ASPC | *KCNK17* | TRUE | 2.313483386 |
| Obesity | ASPC | *GRK4* | TRUE | 3.449731917 |
| Obesity | ASPC | *RIBC2* | TRUE | 3.313711055 |
| Obesity | ASPC | *AP1M2* | TRUE | 2.906842713 |
| Obesity | ASPC | *ATP8B3* | TRUE | 6.239212258 |
| Obesity | ASPC | *STK33* | TRUE | 3.147716502 |
| Obesity | ASPC | *PGPEP1* | TRUE | 2.334200116 |
| Obesity | ASPC | *PKDREJ* | TRUE | 2.974302576 |
| Obesity | ASPC | *MYBBP1A* | TRUE | 2.414340219 |
| Obesity | ASPC | *LARS* | TRUE | 2.136772526 |
| Obesity | ASPC | *MICAL1* | TRUE | 2.385151731 |
| Obesity | ASPC | *SCN2A* | TRUE | 3.016797482 |
| Obesity | ASPC | *DSCC1* | TRUE | 2.254536059 |
| Obesity | ASPC | *IRF4* | TRUE | 2.004124262 |
| Obesity | ASPC | *SLC37A4* | TRUE | 3.462570399 |
| Obesity | ASPC | *NDUFAF1* | TRUE | 3.102610683 |
| Obesity | ASPC | *CENPO* | TRUE | 6.724166973 |
| Obesity | ASPC | *CYP2C8* | TRUE | 2.320830506 |
| Obesity | ASPC | *KLRG1* | TRUE | 2.683939018 |
| Obesity | ASPC | *VPS33A* | TRUE | 3.33244857 |
| Obesity | ASPC | *DIAPH3* | TRUE | 3.009884773 |
| Obesity | ASPC | *FITM1* | TRUE | 2.501984161 |
| Obesity | ASPC | *DUOXA1* | TRUE | 1.973724576 |
| Obesity | ASPC | *CYP11A1* | TRUE | 2.268695355 |
| Obesity | ASPC | *MYO19* | TRUE | 3.585807729 |
| Obesity | ASPC | *C18orf8* | TRUE | 5.595124917 |
| Obesity | ASPC | *CBX8* | TRUE | 2.937125635 |
| Obesity | ASPC | *COL6A1* | TRUE | 2.162868867 |
| Obesity | ASPC | *SAE1* | TRUE | 3.629055667 |
| Obesity | ASPC | *MMEL1* | TRUE | 2.539515536 |
| Obesity | ASPC | *SEMA6C* | TRUE | 2.58615713 |
| Obesity | ASPC | *FAHD2B* | TRUE | 2.499039782 |
| Obesity | ASPC | *RGR* | TRUE | 2.942774653 |
| Obesity | ASPC | *MTG1* | TRUE | 2.075946549 |
| Obesity | ASPC | *HSD17B12* | TRUE | 5.987696349 |
| Obesity | ASPC | *TNKS1BP1* | TRUE | 4.026746913 |
| Obesity | ASPC | *TTC12* | TRUE | 3.027160677 |
| Obesity | ASPC | *RGS18* | TRUE | 3.183914507 |

| | | | | |
|---|---|---|---|---|
| Obesity | ASPC | *DPYSL4* | TRUE | 2.70288147 |
| Obesity | ASPC | *POC5* | TRUE | 3.488323826 |
| Obesity | ASPC | *JAZF1* | TRUE | 2.294818502 |
| Obesity | ASPC | *MMP16* | TRUE | 2.827705124 |
| Obesity | ASPC | *RWDD2B* | TRUE | 2.261588864 |
| Obesity | ASPC | *STEAP2* | TRUE | 2.296693324 |
| Obesity | ASPC | *TMED6* | TRUE | 2.821300965 |
| Obesity | ASPC | *CLEC18A* | TRUE | 4.333813028 |
| Obesity | ASPC | *ETS2* | TRUE | 2.783843908 |
| Obesity | ASPC | *SLC5A11* | TRUE | 4.783947221 |
| Obesity | ASPC | *CILP2* | TRUE | 3.308370538 |
| Obesity | ASPC | *ABCF3* | TRUE | 2.715011877 |
| Obesity | ASPC | *HS3ST6* | TRUE | 2.270568291 |
| Obesity | ASPC | *CLHC1* | TRUE | 2.52194847 |
| Obesity | ASPC | *DCST2* | TRUE | 2.269925153 |
| Obesity | ASPC | *DCLK3* | TRUE | 2.045278967 |
| Obesity | ASPC | *DNALI1* | TRUE | 3.963446778 |
| Obesity | ASPC | *RFT1* | TRUE | 2.655427934 |
| Obesity | ASPC | *RASEF* | TRUE | 2.087579804 |
| Obesity | ASPC | *CCDC68* | TRUE | 2.665061875 |
| Obesity | ASPC | *SPDYE6* | TRUE | 2.332937214 |
| Obesity | ASPC | *PIP4K2C* | TRUE | 2.20552413 |
| Obesity | ASPC | *PIP5KL1* | TRUE | 2.058209204 |
| Obesity | ASPC | *SRR* | TRUE | 3.964121743 |
| Obesity | ASPC | *FNTA* | TRUE | 2.592675984 |
| Obesity | ASPC | *NPNT* | TRUE | 1.980368514 |
| Obesity | ASPC | *CPLX1* | TRUE | 2.302186992 |
| Obesity | ASPC | *IRS1* | TRUE | 2.292531351 |
| Obesity | ASPC | *GPRIN1* | TRUE | 2.513332248 |
| Obesity | ASPC | *INO80E* | TRUE | 3.23872436 |
| Obesity | ASPC | *RGPD8* | TRUE | 2.550875836 |
| Obesity | ASPC | *MYO7B* | TRUE | 2.501209941 |
| Obesity | ASPC | *CNGB3* | TRUE | 2.011029306 |
| Obesity | ASPC | *ADORA2B* | TRUE | 4.86155004 |
| Obesity | ASPC | *TTLL6* | TRUE | 5.804848569 |
| Obesity | ASPC | *TSNARE1* | TRUE | 3.041767576 |
| Obesity | ASPC | *TRMT61B* | TRUE | 2.564089484 |
| Obesity | ASPC | *RGS19* | TRUE | 2.777850281 |
| Obesity | ASPC | *CTSW* | TRUE | 2.724528288 |
| Obesity | ASPC | *FUT10* | TRUE | 2.81470052 |
| Obesity | ASPC | *HOXB2* | TRUE | 1.964715078 |
| Obesity | ASPC | *CADM2* | TRUE | 4.965731005 |
| Obesity | ASPC | *ORAI3* | TRUE | 5.010343317 |
| Obesity | ASPC | *SPRYD4* | TRUE | 2.240535029 |
| Obesity | ASPC | *RUFY1* | TRUE | 2.459213166 |
| Obesity | ASPC | *AP006621.1* | TRUE | 3.408866513 |
| Obesity | ASPC | *STX19* | TRUE | 3.897228319 |
| Obesity | ASPC | *MPI* | TRUE | 2.031599434 |
| Obesity | ASPC | *ZNF852* | TRUE | 3.014013143 |
| Obesity | ASPC | *C3orf38* | TRUE | 4.411246395 |

| | | | | |
|---|---|---|---|---|
| Obesity | ASPC | *RPRML* | TRUE | 3.949357778 |
| Obesity | ASPC | *FKRP* | TRUE | 2.0437254 |
| Obesity | ASPC | *ZFP41* | TRUE | 2.532570551 |
| Obesity | ASPC | *CLN8* | TRUE | 2.082683605 |
| Obesity | ASPC | *PPP1R27* | TRUE | 3.086581733 |
| Obesity | ASPC | *PSMG1* | TRUE | 2.884338087 |
| Obesity | ASPC | *UPP1* | TRUE | 2.119465566 |
| Obesity | ASPC | *NPIPA5* | TRUE | 2.648706539 |
| Obesity | ASPC | *EMILIN3* | TRUE | 3.299931701 |
| Obesity | ASPC | *ACOT1* | TRUE | 3.366049141 |
| Obesity | ASPC | *NUTM2A* | TRUE | 2.011178678 |
| Obesity | ASPC | *MAFF* | TRUE | 2.118074749 |
| Obesity | ASPC | *GP1BA* | TRUE | 3.507731043 |
| Obesity | ASPC | *TRABD2A* | TRUE | 2.149206283 |
| Obesity | ASPC | *CENPP* | TRUE | 3.353263269 |
| Obesity | ASPC | *OSTN* | TRUE | 2.462015046 |
| Obesity | ASPC | *SKOR1* | TRUE | 2.2245205 |
| Obesity | ASPC | *FBF1* | TRUE | 2.362354849 |
| Obesity | ASPC | *CPNE4* | TRUE | 3.328900232 |
| Obesity | ASPC | *ZNF605* | TRUE | 2.750242199 |
| Obesity | ASPC | *SULT1A2* | TRUE | 8.892268688 |
| Obesity | ASPC | *MFAP5* | TRUE | 2.197489969 |
| Obesity | ASPC | *DNAH10* | TRUE | 4.463094555 |
| Obesity | ASPC | *RPS26* | TRUE | 3.027004761 |
| Obesity | ASPC | *ZNF780A* | TRUE | 3.764948765 |
| Obesity | ASPC | *ZNF560* | TRUE | 1.984894313 |
| Obesity | ASPC | *ADH4* | TRUE | 2.071837519 |
| Obesity | ASPC | *SHISA4* | TRUE | 5.056061272 |
| Obesity | ASPC | *C6orf163* | TRUE | 2.341244714 |
| Obesity | ASPC | *GPANK1* | TRUE | 3.405344706 |
| Obesity | ASPC | *C15orf59* | TRUE | 2.322847078 |
| Obesity | ASPC | *SMN2* | TRUE | 2.459004913 |
| Obesity | ASPC | *AC084018.1* | TRUE | 3.444338373 |
| Obesity | ASPC | *ASIC3* | TRUE | 2.653668928 |
| Obesity | ASPC | *SLC23A3* | TRUE | 2.149307351 |
| Obesity | ASPC | *GALT* | TRUE | 2.240901119 |
| Obesity | ASPC | *RNU6ATAC35P* | TRUE | 2.057784142 |
| Obesity | ASPC | *GS1.124K5.4* | TRUE | 2.094832122 |
| Obesity | ASPC | *ISY1* | TRUE | 2.549545934 |
| Obesity | ASPC | *LINC00960* | TRUE | 2.352385616 |
| Obesity | ASPC | *PLA2G4B* | TRUE | 1.99750839 |
| Obesity | ASPC | *BRK1* | TRUE | 2.226169167 |
| Obesity | ASPC | *AP001172.2* | TRUE | 3.009606817 |
| Obesity | ASPC | *GTF2H5* | TRUE | 2.128285333 |
| Obesity | b_cell | *ITIH4* | TRUE | 5.279223765 |
| Obesity | b_cell | *MFAP2* | TRUE | 2.004646682 |
| Obesity | b_cell | *ATP8B3* | TRUE | 3.320941987 |
| Obesity | b_cell | *DPYSL4* | TRUE | 2.410297212 |
| Obesity | b_cell | *CLEC18A* | TRUE | 7.864561823 |
| Obesity | b_cell | *RNF123* | TRUE | 3.296715155 |

| | | | | |
|---|---|---|---|---|
| Obesity | b_cell | *RAPSN* | TRUE | 2.050231336 |
| Obesity | b_cell | *CTSW* | TRUE | 6.306802679 |
| Obesity | b_cell | *TRIM73* | TRUE | 5.98076843 |
| Obesity | b_cell | *RPRML* | TRUE | 2.414309775 |
| Obesity | b_cell | *ZFP57* | TRUE | 2.646839299 |
| Obesity | b_cell | *LINC00664* | TRUE | 2.278340764 |
| Obesity | dendritic_cell | *RBM6* | TRUE | 3.652415122 |
| Obesity | dendritic_cell | *UHRF1BP1* | TRUE | 2.823350055 |
| Obesity | dendritic_cell | *PABPC4* | TRUE | 2.863415527 |
| Obesity | dendritic_cell | *RIBC2* | TRUE | 3.059004571 |
| Obesity | dendritic_cell | *ATP8B3* | TRUE | 4.82741285 |
| Obesity | dendritic_cell | *TBX6* | TRUE | 4.467539959 |
| Obesity | dendritic_cell | *DPYSL4* | TRUE | 2.919583327 |
| Obesity | dendritic_cell | *CLEC18A* | TRUE | 7.928725366 |
| Obesity | dendritic_cell | *SRR* | TRUE | 2.035721958 |
| Obesity | dendritic_cell | *RGPD8* | TRUE | 2.059055077 |
| Obesity | dendritic_cell | *CTSW* | TRUE | 3.771209606 |
| Obesity | dendritic_cell | *TRIM73* | TRUE | 6.236058019 |
| Obesity | dendritic_cell | *RPRML* | TRUE | 2.655886934 |
| Obesity | dendritic_cell | *NUTM2A* | TRUE | 2.272509657 |
| Obesity | dendritic_cell | *GP1BA* | TRUE | 2.750525266 |
| Obesity | dendritic_cell | *SULT1A2* | TRUE | 6.832152515 |
| Obesity | dendritic_cell | *RPS26* | TRUE | 2.764802587 |
| Obesity | dendritic_cell | *NOTCH4* | TRUE | 2.079328741 |
| Obesity | dendritic_cell | *TMEM110* | TRUE | 2.713817671 |
| Obesity | dendritic_cell | *LINC00664* | TRUE | 2.363376144 |
| Obesity | endometrium | *CENPM* | TRUE | 2.310906006 |
| Obesity | endometrium | *SNRPC* | TRUE | 1.990312292 |
| Obesity | endometrium | *TBX6* | TRUE | 3.62807079 |
| Obesity | endometrium | *DPYSL4* | TRUE | 2.788635387 |
| Obesity | endometrium | *RNF123* | TRUE | 3.667925188 |
| Obesity | endometrium | *ADORA2B* | TRUE | 3.076478125 |
| Obesity | endometrium | *C1QTNF4* | TRUE | 3.642877735 |
| Obesity | endometrium | *CTSW* | TRUE | 2.110772 |
| Obesity | endometrium | *ORAI3* | TRUE | 3.18249664 |
| Obesity | endometrium | *TRIM73* | TRUE | 5.363718462 |
| Obesity | endometrium | *RPRML* | TRUE | 2.794633059 |
| Obesity | endometrium | *EMILIN3* | TRUE | 2.654691051 |
| Obesity | endometrium | *NUTM2A* | TRUE | 2.762594193 |
| Obesity | endometrium | *GP1BA* | TRUE | 2.867860952 |
| Obesity | endometrium | *SULT1A2* | TRUE | 3.425971339 |
| Obesity | endometrium | *DNAH10* | TRUE | 3.053257615 |
| Obesity | endometrium | *ZFP57* | TRUE | 2.756972498 |
| Obesity | endometrium | *LINC00664* | TRUE | 2.336520203 |
| Obesity | endometrium | *AP001172.2* | TRUE | 2.547276039 |
| Obesity | endothelial | *SNRPC* | TRUE | 4.587986162 |
| Obesity | endothelial | *AP1M2* | TRUE | 2.156876408 |
| Obesity | endothelial | *ATP8B3* | TRUE | 4.748823602 |
| Obesity | endothelial | *PKDREJ* | TRUE | 2.475092685 |
| Obesity | endothelial | *CBX8* | TRUE | 2.141068725 |

| | | | | |
|---|---|---|---|---|
| Obesity | endothelial | *HSD17B12* | TRUE | 3.062129726 |
| Obesity | endothelial | *DPYSL4* | TRUE | 2.392804969 |
| Obesity | endothelial | *CLEC18A* | TRUE | 4.812351196 |
| Obesity | endothelial | *SLC5A11* | TRUE | 3.199567434 |
| Obesity | endothelial | *CILP2* | TRUE | 3.285862073 |
| Obesity | endothelial | *RAPSN* | TRUE | 2.593842596 |
| Obesity | endothelial | *SRR* | TRUE | 2.224644777 |
| Obesity | endothelial | *ADORA2B* | TRUE | 3.789445286 |
| Obesity | endothelial | *TTLL6* | TRUE | 5.263222447 |
| Obesity | endothelial | *CTSW* | TRUE | 5.965572309 |
| Obesity | endothelial | *RPP25* | TRUE | 2.564452033 |
| Obesity | endothelial | *TRIM73* | TRUE | 4.473860447 |
| Obesity | endothelial | *RPRML* | TRUE | 2.297071716 |
| Obesity | endothelial | *NUTM2A* | TRUE | 3.351475578 |
| Obesity | endothelial | *BTN3A2* | TRUE | 2.211654693 |
| Obesity | endothelial | *ZNF780A* | TRUE | 3.250551014 |
| Obesity | endothelial | *AC084018.1* | TRUE | 2.884729101 |
| Obesity | LEC | *ITIH4* | TRUE | 3.286197227 |
| Obesity | LEC | *ITPR3* | TRUE | 2.263583378 |
| Obesity | LEC | *CENPM* | TRUE | 2.042137275 |
| Obesity | LEC | *BTN3A3* | TRUE | 2.008089938 |
| Obesity | LEC | *SERPINC1* | TRUE | 2.211407672 |
| Obesity | LEC | *OPN4* | TRUE | 2.607939696 |
| Obesity | LEC | *SLC25A51* | TRUE | 2.448066608 |
| Obesity | LEC | *GRK4* | TRUE | 2.49022637 |
| Obesity | LEC | *AP1M2* | TRUE | 2.359877755 |
| Obesity | LEC | *ATP8B3* | TRUE | 3.26818699 |
| Obesity | LEC | *PKDREJ* | TRUE | 2.361979823 |
| Obesity | LEC | *IGHMBP2* | TRUE | 2.37269295 |
| Obesity | LEC | *SLC37A4* | TRUE | 3.80188218 |
| Obesity | LEC | *CYP2C8* | TRUE | 2.007380156 |
| Obesity | LEC | *CBX8* | TRUE | 2.719209182 |
| Obesity | LEC | *MTG1* | TRUE | 2.228726978 |
| Obesity | LEC | *HSD17B12* | TRUE | 4.918814109 |
| Obesity | LEC | *DPYSL4* | TRUE | 2.235895011 |
| Obesity | LEC | *CLEC18A* | TRUE | 6.354389826 |
| Obesity | LEC | *ETS2* | TRUE | 2.397690607 |
| Obesity | LEC | *SLC5A11* | TRUE | 3.191916858 |
| Obesity | LEC | *CILP2* | TRUE | 2.158823739 |
| Obesity | LEC | *DNALI1* | TRUE | 2.325214884 |
| Obesity | LEC | *MST1R* | TRUE | 3.758459283 |
| Obesity | LEC | *SRR* | TRUE | 3.338419902 |
| Obesity | LEC | *C8orf46* | TRUE | 2.080789669 |
| Obesity | LEC | *GPRIN1* | TRUE | 2.714306246 |
| Obesity | LEC | *INO80E* | TRUE | 2.614508884 |
| Obesity | LEC | *RGPD8* | TRUE | 2.140296632 |
| Obesity | LEC | *ADORA2B* | TRUE | 4.676641059 |
| Obesity | LEC | *TTLL6* | TRUE | 4.005793107 |
| Obesity | LEC | *C1QTNF4* | TRUE | 2.965596748 |
| Obesity | LEC | *CTSW* | TRUE | 3.440903713 |

| | | | | |
|---|---|---|---|---|
| Obesity | LEC | *ORAI3* | TRUE | 4.952153865 |
| Obesity | LEC | *STX19* | TRUE | 1.988177241 |
| Obesity | LEC | *TRIM73* | TRUE | 5.061358114 |
| Obesity | LEC | *RPRML* | TRUE | 2.794574982 |
| Obesity | LEC | *PPP1R27* | TRUE | 3.325008303 |
| Obesity | LEC | *ACOT1* | TRUE | 2.043192657 |
| Obesity | LEC | *NUTM2A* | TRUE | 3.015279083 |
| Obesity | LEC | *SULT1A2* | TRUE | 5.680361108 |
| Obesity | LEC | *DNAH10* | TRUE | 4.985772722 |
| Obesity | LEC | *RPS26* | TRUE | 2.788658106 |
| Obesity | LEC | *ZNF780A* | TRUE | 3.496466868 |
| Obesity | LEC | *GPANK1* | TRUE | 2.294777189 |
| Obesity | LEC | *C15orf59* | TRUE | 1.976057102 |
| Obesity | LEC | *SMN2* | TRUE | 2.047407162 |
| Obesity | LEC | *AC084018.1* | TRUE | 2.76370559 |
| Obesity | LEC | *ASIC3* | TRUE | 2.58187143 |
| Obesity | LEC | *GS1.124K5.4* | TRUE | 2.197820867 |
| Obesity | LEC | *DPP3* | TRUE | 2.372758189 |
| Obesity | LEC | *LINC00664* | TRUE | 2.360878503 |
| Obesity | macrophage | *RBM6* | TRUE | 3.116525178 |
| Obesity | macrophage | *CENPQ* | TRUE | 2.010492387 |
| Obesity | macrophage | *TBX15* | TRUE | 3.164117177 |
| Obesity | macrophage | *CENPM* | TRUE | 2.976265003 |
| Obesity | macrophage | *ELMO3* | TRUE | 2.485664186 |
| Obesity | macrophage | *BTN3A3* | TRUE | 2.448459362 |
| Obesity | macrophage | *PCCB* | TRUE | 2.361819802 |
| Obesity | macrophage | *MFAP2* | TRUE | 2.147511358 |
| Obesity | macrophage | *SERPINC1* | TRUE | 5.145205232 |
| Obesity | macrophage | *SLC25A51* | TRUE | 2.447260423 |
| Obesity | macrophage | *GRK4* | TRUE | 2.764854231 |
| Obesity | macrophage | *RIBC2* | TRUE | 2.232812273 |
| Obesity | macrophage | *AP1M2* | TRUE | 2.332809671 |
| Obesity | macrophage | *ATP8B3* | TRUE | 5.889859484 |
| Obesity | macrophage | *PKDREJ* | TRUE | 2.797387663 |
| Obesity | macrophage | *IGHMBP2* | TRUE | 2.044268285 |
| Obesity | macrophage | *MICAL1* | TRUE | 2.069370344 |
| Obesity | macrophage | *CENPO* | TRUE | 2.16150987 |
| Obesity | macrophage | *VPS33A* | TRUE | 2.433386104 |
| Obesity | macrophage | *DIAPH3* | TRUE | 2.249538705 |
| Obesity | macrophage | *C18orf8* | TRUE | 3.590905756 |
| Obesity | macrophage | *CBX8* | TRUE | 2.783635706 |
| Obesity | macrophage | *MMEL1* | TRUE | 2.000545327 |
| Obesity | macrophage | *HSD17B12* | TRUE | 5.468788781 |
| Obesity | macrophage | *DPYSL4* | TRUE | 2.736493708 |
| Obesity | macrophage | *POC5* | TRUE | 2.489933778 |
| Obesity | macrophage | *NEIL2* | TRUE | 2.43585996 |
| Obesity | macrophage | *CLEC18A* | TRUE | 5.75746055 |
| Obesity | macrophage | *SLC5A11* | TRUE | 4.45413658 |
| Obesity | macrophage | *CILP2* | TRUE | 3.156787502 |
| Obesity | macrophage | *ABCF3* | TRUE | 2.266300327 |

| | | | | |
|---|---|---|---|---|
| Obesity | macrophage | *DNALI1* | TRUE | 2.239754553 |
| Obesity | macrophage | *RFT1* | TRUE | 2.685815894 |
| Obesity | macrophage | *SRR* | TRUE | 3.664891293 |
| Obesity | macrophage | *GPRIN1* | TRUE | 2.58630778 |
| Obesity | macrophage | *INO80E* | TRUE | 3.203424422 |
| Obesity | macrophage | *RGPD8* | TRUE | 2.389764283 |
| Obesity | macrophage | *ADORA2B* | TRUE | 3.668018165 |
| Obesity | macrophage | *TTLL6* | TRUE | 4.913203571 |
| Obesity | macrophage | *C1QTNF4* | TRUE | 2.983417274 |
| Obesity | macrophage | *CTSW* | TRUE | 3.511992446 |
| Obesity | macrophage | *CADM2* | TRUE | 2.039008588 |
| Obesity | macrophage | *ORAI3* | TRUE | 5.119400024 |
| Obesity | macrophage | *STX19* | TRUE | 1.969418914 |
| Obesity | macrophage | *C3orf38* | TRUE | 3.462061318 |
| Obesity | macrophage | *RPRML* | TRUE | 3.473468562 |
| Obesity | macrophage | *ZFP41* | TRUE | 2.490320327 |
| Obesity | macrophage | *NPIPA5* | TRUE | 2.599689085 |
| Obesity | macrophage | *ACOT1* | TRUE | 2.879853135 |
| Obesity | macrophage | *NUTM2A* | TRUE | 2.995458833 |
| Obesity | macrophage | *GP1BA* | TRUE | 2.982026938 |
| Obesity | macrophage | *CENPP* | TRUE | 2.397611347 |
| Obesity | macrophage | *SKOR1* | TRUE | 2.407599962 |
| Obesity | macrophage | *CPNE4* | TRUE | 2.529430609 |
| Obesity | macrophage | *SULT1A2* | TRUE | 8.776585569 |
| Obesity | macrophage | *DNAH10* | TRUE | 3.382555137 |
| Obesity | macrophage | *RPS26* | TRUE | 2.825369595 |
| Obesity | macrophage | *ZNF780A* | TRUE | 3.640109712 |
| Obesity | macrophage | *SHISA4* | TRUE | 5.190388275 |
| Obesity | macrophage | *C6orf163* | TRUE | 2.026274672 |
| Obesity | macrophage | *GPANK1* | TRUE | 2.725299928 |
| Obesity | macrophage | *C15orf59* | TRUE | 2.134822302 |
| Obesity | macrophage | *SMN2* | TRUE | 2.104714797 |
| Obesity | macrophage | *AC084018.1* | TRUE | 3.30510914 |
| Obesity | macrophage | *ASIC3* | TRUE | 2.34547195 |
| Obesity | macrophage | *GS1.124K5.4* | TRUE | 2.077824566 |
| Obesity | macrophage | *ISY1* | TRUE | 2.130697781 |
| Obesity | macrophage | *LINC00664* | TRUE | 2.276661311 |
| Obesity | macrophage | *AP001172.2* | TRUE | 2.518040369 |
| Obesity | mast_cell | *RBM6* | TRUE | 1.979941287 |
| Obesity | mast_cell | *CENPM* | TRUE | 2.400026998 |
| Obesity | mast_cell | *SERPINC1* | TRUE | 3.098673746 |
| Obesity | mast_cell | *TEX14* | TRUE | 2.171176379 |
| Obesity | mast_cell | *BTN2A2* | TRUE | 3.187502863 |
| Obesity | mast_cell | *RIBC2* | TRUE | 2.146528353 |
| Obesity | mast_cell | *PKDREJ* | TRUE | 2.504732875 |
| Obesity | mast_cell | *TBX6* | TRUE | 4.301816865 |
| Obesity | mast_cell | *DPYSL4* | TRUE | 2.686247318 |
| Obesity | mast_cell | *CLEC18A* | TRUE | 8.175526768 |
| Obesity | mast_cell | *SLC5A11* | TRUE | 4.41747633 |
| Obesity | mast_cell | *SPDYE6* | TRUE | 2.123616678 |

| | | | | |
|---|---|---|---|---|
| Obesity | mast_cell | *SRR* | TRUE | 2.984523643 |
| Obesity | mast_cell | *ADORA2B* | TRUE | 2.550161545 |
| Obesity | mast_cell | *CTSW* | TRUE | 2.887517867 |
| Obesity | mast_cell | *ORAI3* | TRUE | 3.788170914 |
| Obesity | mast_cell | *TRIM73* | TRUE | 4.791383115 |
| Obesity | mast_cell | *RPRML* | TRUE | 2.914753548 |
| Obesity | mast_cell | *ACOT1* | TRUE | 3.423248556 |
| Obesity | mast_cell | *NUTM2A* | TRUE | 3.251352239 |
| Obesity | mast_cell | *BTN3A2* | TRUE | 3.533217888 |
| Obesity | mast_cell | *SULT1A2* | TRUE | 6.145792127 |
| Obesity | mast_cell | *C17orf107* | TRUE | 2.373775121 |
| Obesity | mast_cell | *LINC00664* | TRUE | 2.338418835 |
| Obesity | mesothelium | *RBM6* | TRUE | 3.470363239 |
| Obesity | mesothelium | *FMO3* | TRUE | 2.067526957 |
| Obesity | mesothelium | *CENPQ* | TRUE | 2.473119334 |
| Obesity | mesothelium | *FLT4* | TRUE | 2.341943698 |
| Obesity | mesothelium | *LSG1* | TRUE | 2.539776222 |
| Obesity | mesothelium | *CD5L* | TRUE | 2.630677601 |
| Obesity | mesothelium | *TIPIN* | TRUE | 2.746314282 |
| Obesity | mesothelium | *NT5C2* | TRUE | 2.144084574 |
| Obesity | mesothelium | *FDFT1* | TRUE | 3.834854008 |
| Obesity | mesothelium | *GSTP1* | TRUE | 2.544487634 |
| Obesity | mesothelium | *CD82* | TRUE | 1.971277889 |
| Obesity | mesothelium | *OGFOD1* | TRUE | 2.433582933 |
| Obesity | mesothelium | *PABPC4* | TRUE | 5.048926233 |
| Obesity | mesothelium | *RGS17* | TRUE | 4.464359758 |
| Obesity | mesothelium | *TBX15* | TRUE | 3.417853561 |
| Obesity | mesothelium | *ITPR3* | TRUE | 2.911399666 |
| Obesity | mesothelium | *IL12RB1* | TRUE | 2.213892687 |
| Obesity | mesothelium | *CENPM* | TRUE | 3.289251072 |
| Obesity | mesothelium | *ABHD12* | TRUE | 2.544643989 |
| Obesity | mesothelium | *ELMO3* | TRUE | 2.561792788 |
| Obesity | mesothelium | *CLCN7* | TRUE | 2.082140474 |
| Obesity | mesothelium | *CLUAP1* | TRUE | 2.56857971 |
| Obesity | mesothelium | *PYCARD* | TRUE | 3.111933449 |
| Obesity | mesothelium | *MEST* | TRUE | 2.578940812 |
| Obesity | mesothelium | *SLC22A18* | TRUE | 2.361909133 |
| Obesity | mesothelium | *BTN3A3* | TRUE | 2.435185332 |
| Obesity | mesothelium | *GABRR2* | TRUE | 2.790850916 |
| Obesity | mesothelium | *GLP1R* | TRUE | 2.100707297 |
| Obesity | mesothelium | *PRPH2* | TRUE | 1.978926736 |
| Obesity | mesothelium | *NME5* | TRUE | 3.029942188 |
| Obesity | mesothelium | *BNIP1* | TRUE | 2.437499441 |
| Obesity | mesothelium | *PCCB* | TRUE | 3.858242908 |
| Obesity | mesothelium | *CHMP3* | TRUE | 3.469916163 |
| Obesity | mesothelium | *SERPINC1* | TRUE | 6.187865208 |
| Obesity | mesothelium | *INVS* | TRUE | 2.867727726 |
| Obesity | mesothelium | *SNX19* | TRUE | 3.15052845 |
| Obesity | mesothelium | *ADRA1A* | TRUE | 2.832305264 |
| Obesity | mesothelium | *TEX14* | TRUE | 2.525343599 |

| | | | | |
|---|---|---|---|---|
| Obesity | mesothelium | *OPN4* | TRUE | 3.220910828 |
| Obesity | mesothelium | *SLC25A51* | TRUE | 2.765335236 |
| Obesity | mesothelium | *NEURL2* | TRUE | 2.285851988 |
| Obesity | mesothelium | *BTN2A2* | TRUE | 2.460845832 |
| Obesity | mesothelium | *MED20* | TRUE | 2.033916017 |
| Obesity | mesothelium | *TREM1* | TRUE | 2.150979309 |
| Obesity | mesothelium | *KCNK17* | TRUE | 2.224451012 |
| Obesity | mesothelium | *GRK4* | TRUE | 3.543376948 |
| Obesity | mesothelium | *RIBC2* | TRUE | 3.175080841 |
| Obesity | mesothelium | *AP1M2* | TRUE | 2.733985008 |
| Obesity | mesothelium | *ATP8B3* | TRUE | 6.380437238 |
| Obesity | mesothelium | *PGPEP1* | TRUE | 2.503814021 |
| Obesity | mesothelium | *PKDREJ* | TRUE | 2.942102106 |
| Obesity | mesothelium | *MYBBP1A* | TRUE | 2.286028052 |
| Obesity | mesothelium | *LARS* | TRUE | 2.471696848 |
| Obesity | mesothelium | *GSTM5* | TRUE | 2.564082349 |
| Obesity | mesothelium | *POM121C* | TRUE | 2.581661629 |
| Obesity | mesothelium | *MICAL1* | TRUE | 2.550458108 |
| Obesity | mesothelium | *SCN2A* | TRUE | 2.494431685 |
| Obesity | mesothelium | *DSCC1* | TRUE | 2.357539107 |
| Obesity | mesothelium | *SLC37A4* | TRUE | 3.465661438 |
| Obesity | mesothelium | *NDUFAF1* | TRUE | 2.842981927 |
| Obesity | mesothelium | *CENPO* | TRUE | 7.388959186 |
| Obesity | mesothelium | *CYP2C8* | TRUE | 2.167493999 |
| Obesity | mesothelium | *KLRG1* | TRUE | 2.64337465 |
| Obesity | mesothelium | *VPS33A* | TRUE | 3.347491675 |
| Obesity | mesothelium | *NOVA1* | TRUE | 2.195408751 |
| Obesity | mesothelium | *FITM1* | TRUE | 2.543514038 |
| Obesity | mesothelium | *CYP11A1* | TRUE | 2.28561875 |
| Obesity | mesothelium | *MYO19* | TRUE | 3.878455311 |
| Obesity | mesothelium | *C18orf8* | TRUE | 5.935076043 |
| Obesity | mesothelium | *CBX8* | TRUE | 2.734493577 |
| Obesity | mesothelium | *COL6A1* | TRUE | 2.531106162 |
| Obesity | mesothelium | *SAE1* | TRUE | 3.684745221 |
| Obesity | mesothelium | *MMEL1* | TRUE | 2.506346606 |
| Obesity | mesothelium | *SEMA6C* | TRUE | 2.871792496 |
| Obesity | mesothelium | *FAHD2B* | TRUE | 2.645491352 |
| Obesity | mesothelium | *RGR* | TRUE | 3.106567205 |
| Obesity | mesothelium | *HSD17B12* | TRUE | 6.155975119 |
| Obesity | mesothelium | *TNKS1BP1* | TRUE | 3.669533738 |
| Obesity | mesothelium | *TTC12* | TRUE | 2.880551678 |
| Obesity | mesothelium | *RGS18* | TRUE | 3.045013346 |
| Obesity | mesothelium | *DPYSL4* | TRUE | 2.672582903 |
| Obesity | mesothelium | *POC5* | TRUE | 3.584634758 |
| Obesity | mesothelium | *PLA2R1* | TRUE | 2.218854061 |
| Obesity | mesothelium | *JAZF1* | TRUE | 2.472015139 |
| Obesity | mesothelium | *MMP16* | TRUE | 2.868959193 |
| Obesity | mesothelium | *RWDD2B* | TRUE | 2.280239157 |
| Obesity | mesothelium | *STEAP2* | TRUE | 2.604104137 |
| Obesity | mesothelium | *TMED6* | TRUE | 2.673632011 |

| | | | | |
|---|---|---|---|---|
| Obesity | mesothelium | *CLEC18A* | TRUE | 4.471797046 |
| Obesity | mesothelium | *ETS2* | TRUE | 2.680621612 |
| Obesity | mesothelium | *SLC5A11* | TRUE | 4.920302856 |
| Obesity | mesothelium | *CILP2* | TRUE | 3.234984993 |
| Obesity | mesothelium | *GPSM1* | TRUE | 2.362898019 |
| Obesity | mesothelium | *ABCF3* | TRUE | 2.679541272 |
| Obesity | mesothelium | *CAMK2N1* | TRUE | 3.116515432 |
| Obesity | mesothelium | *CLHC1* | TRUE | 2.494812145 |
| Obesity | mesothelium | *ANTXR2* | TRUE | 2.140066939 |
| Obesity | mesothelium | *DCST2* | TRUE | 2.334242791 |
| Obesity | mesothelium | *DCLK3* | TRUE | 1.965770567 |
| Obesity | mesothelium | *DNALI1* | TRUE | 3.72275255 |
| Obesity | mesothelium | *RFT1* | TRUE | 2.910841964 |
| Obesity | mesothelium | *TEX264* | TRUE | 2.378722331 |
| Obesity | mesothelium | *CCDC68* | TRUE | 2.444741311 |
| Obesity | mesothelium | *CHRFAM7A* | TRUE | 2.009835265 |
| Obesity | mesothelium | *SPDYE6* | TRUE | 2.2995722 |
| Obesity | mesothelium | *SRR* | TRUE | 3.927296002 |
| Obesity | mesothelium | *FNTA* | TRUE | 2.734296001 |
| Obesity | mesothelium | *IRS1* | TRUE | 2.436370134 |
| Obesity | mesothelium | *GPRIN1* | TRUE | 2.522672248 |
| Obesity | mesothelium | *INO80E* | TRUE | 4.251230597 |
| Obesity | mesothelium | *RGPD8* | TRUE | 2.584465307 |
| Obesity | mesothelium | *CALML6* | TRUE | 2.049878888 |
| Obesity | mesothelium | *MYO7B* | TRUE | 2.303135222 |
| Obesity | mesothelium | *ADORA2B* | TRUE | 4.912083298 |
| Obesity | mesothelium | *TTLL6* | TRUE | 5.808223398 |
| Obesity | mesothelium | *TSNARE1* | TRUE | 3.130918547 |
| Obesity | mesothelium | *RGS19* | TRUE | 2.751961088 |
| Obesity | mesothelium | *CTSW* | TRUE | 3.631631535 |
| Obesity | mesothelium | *FUT10* | TRUE | 2.920092632 |
| Obesity | mesothelium | *HOXB2* | TRUE | 2.170983653 |
| Obesity | mesothelium | *CADM2* | TRUE | 4.739622895 |
| Obesity | mesothelium | *ORAI3* | TRUE | 5.089886703 |
| Obesity | mesothelium | *SPRYD4* | TRUE | 2.492311138 |
| Obesity | mesothelium | *RUFY1* | TRUE | 2.443742928 |
| Obesity | mesothelium | *AP006621.1* | TRUE | 3.34518941 |
| Obesity | mesothelium | *STX19* | TRUE | 3.728932908 |
| Obesity | mesothelium | *ZNF852* | TRUE | 3.1482199 |
| Obesity | mesothelium | *C3orf38* | TRUE | 4.489466093 |
| Obesity | mesothelium | *RPRML* | TRUE | 4.074701633 |
| Obesity | mesothelium | *FKRP* | TRUE | 2.035558824 |
| Obesity | mesothelium | *ZFP41* | TRUE | 2.212473442 |
| Obesity | mesothelium | *CLDN7* | TRUE | 2.420807213 |
| Obesity | mesothelium | *CLN8* | TRUE | 2.063347143 |
| Obesity | mesothelium | *PPP1R27* | TRUE | 2.968407832 |
| Obesity | mesothelium | *PSMG1* | TRUE | 2.738638021 |
| Obesity | mesothelium | *NPIPA5* | TRUE | 2.636604697 |
| Obesity | mesothelium | *EMILIN3* | TRUE | 2.993578301 |
| Obesity | mesothelium | *ACOT1* | TRUE | 3.726208185 |

| | | | | |
|---|---|---|---|---|
| Obesity | mesothelium | *OAF* | TRUE | 2.114249316 |
| Obesity | mesothelium | *NUTM2A* | TRUE | 2.408177577 |
| Obesity | mesothelium | *GP1BA* | TRUE | 3.619518691 |
| Obesity | mesothelium | *TRABD2A* | TRUE | 2.094601282 |
| Obesity | mesothelium | *CENPP* | TRUE | 2.901280745 |
| Obesity | mesothelium | *JAKMIP3* | TRUE | 1.986678228 |
| Obesity | mesothelium | *OSTN* | TRUE | 2.334119579 |
| Obesity | mesothelium | *SKOR1* | TRUE | 2.296491458 |
| Obesity | mesothelium | *FBF1* | TRUE | 2.070215725 |
| Obesity | mesothelium | *CPNE4* | TRUE | 3.317823007 |
| Obesity | mesothelium | *ZNF605* | TRUE | 2.727959579 |
| Obesity | mesothelium | *SULT1A2* | TRUE | 8.476164186 |
| Obesity | mesothelium | *MFAP5* | TRUE | 2.446849655 |
| Obesity | mesothelium | *DNAH10* | TRUE | 4.482720193 |
| Obesity | mesothelium | *RPS26* | TRUE | 3.157749808 |
| Obesity | mesothelium | *ZNF780A* | TRUE | 3.859022904 |
| Obesity | mesothelium | *ADH4* | TRUE | 2.264974466 |
| Obesity | mesothelium | *SH3BGRL2* | TRUE | 1.968569149 |
| Obesity | mesothelium | *SHISA4* | TRUE | 5.504562917 |
| Obesity | mesothelium | *C6orf163* | TRUE | 2.339239171 |
| Obesity | mesothelium | *GPANK1* | TRUE | 3.72705833 |
| Obesity | mesothelium | *C15orf59* | TRUE | 2.062019686 |
| Obesity | mesothelium | *SMN2* | TRUE | 2.471806811 |
| Obesity | mesothelium | *AC084018.1* | TRUE | 3.625620799 |
| Obesity | mesothelium | *ASIC3* | TRUE | 2.46196668 |
| Obesity | mesothelium | *SLC23A3* | TRUE | 2.087762164 |
| Obesity | mesothelium | *CPNE1* | TRUE | 1.963107686 |
| Obesity | mesothelium | *AL049840.1* | TRUE | 1.986953956 |
| Obesity | mesothelium | *GS1.124K5.4* | TRUE | 2.086207169 |
| Obesity | mesothelium | *ISY1* | TRUE | 2.655217912 |
| Obesity | mesothelium | *LINC00960* | TRUE | 2.328554384 |
| Obesity | mesothelium | *PLA2G4B* | TRUE | 2.117493309 |
| Obesity | mesothelium | *BRK1* | TRUE | 2.580774185 |
| Obesity | mesothelium | *AP001172.2* | TRUE | 3.177271403 |
| Obesity | mesothelium | *GTF2H5* | TRUE | 2.037820937 |
| Obesity | monocyte | *RBM6* | TRUE | 3.47065819 |
| Obesity | monocyte | *ITIH4* | TRUE | 4.469042551 |
| Obesity | monocyte | *MFAP2* | TRUE | 2.099209111 |
| Obesity | monocyte | *ATP8B3* | TRUE | 2.950605883 |
| Obesity | monocyte | *DPYSL4* | TRUE | 2.070777806 |
| Obesity | monocyte | *CLEC18A* | TRUE | 7.372504141 |
| Obesity | monocyte | *RNF123* | TRUE | 2.177177512 |
| Obesity | monocyte | *TTLL6* | TRUE | 2.214241091 |
| Obesity | monocyte | *C1QTNF4* | TRUE | 4.90451996 |
| Obesity | monocyte | *ORAI3* | TRUE | 3.665439882 |
| Obesity | monocyte | *NUTM2A* | TRUE | 3.440048479 |
| Obesity | monocyte | *BTN3A2* | TRUE | 3.147276531 |
| Obesity | monocyte | *NOTCH4* | TRUE | 2.181698155 |
| Obesity | monocyte | *AC084018.1* | TRUE | 3.087258658 |
| Obesity | monocyte | *LINC00664* | TRUE | 2.495978227 |

| | | | | |
|---|---|---|---|---|
| Obesity | nk_cell | *RBM6* | TRUE | 5.713580655 |
| Obesity | nk_cell | *ITIH4* | TRUE | 3.330537017 |
| Obesity | nk_cell | *BTN3A3* | TRUE | 2.000511744 |
| Obesity | nk_cell | *MFAP2* | TRUE | 2.109178044 |
| Obesity | nk_cell | *RIBC2* | TRUE | 3.092226424 |
| Obesity | nk_cell | *HSD17B12* | TRUE | 2.061994654 |
| Obesity | nk_cell | *CLEC18A* | TRUE | 6.772615042 |
| Obesity | nk_cell | *GPRIN1* | TRUE | 2.318803905 |
| Obesity | nk_cell | *INO80E* | TRUE | 3.261369134 |
| Obesity | nk_cell | *ADORA2B* | TRUE | 2.265095034 |
| Obesity | nk_cell | *TRIM73* | TRUE | 4.841060096 |
| Obesity | nk_cell | *SULT1A2* | TRUE | 6.946175093 |
| Obesity | nk_cell | *ZNF780A* | TRUE | 2.342826832 |
| Obesity | nk_cell | *ZFP57* | TRUE | 2.204394541 |
| Obesity | pericyte | *RBM6* | TRUE | 3.198840644 |
| Obesity | pericyte | *ITIH4* | TRUE | 2.968237967 |
| Obesity | pericyte | *PABPC4* | TRUE | 2.033359221 |
| Obesity | pericyte | *CENPM* | TRUE | 2.91577482 |
| Obesity | pericyte | *SERPINC1* | TRUE | 3.286651943 |
| Obesity | pericyte | *GRK4* | TRUE | 2.468690187 |
| Obesity | pericyte | *RIBC2* | TRUE | 2.422237556 |
| Obesity | pericyte | *ATP8B3* | TRUE | 5.233210711 |
| Obesity | pericyte | *PKDREJ* | TRUE | 2.650460567 |
| Obesity | pericyte | *CBX8* | TRUE | 2.302611618 |
| Obesity | pericyte | *HSD17B12* | TRUE | 5.213581751 |
| Obesity | pericyte | *DPYSL4* | TRUE | 2.708135747 |
| Obesity | pericyte | *POC5* | TRUE | 2.012108956 |
| Obesity | pericyte | *CLEC18A* | TRUE | 3.840103701 |
| Obesity | pericyte | *ETS2* | TRUE | 2.052983809 |
| Obesity | pericyte | *SLC5A11* | TRUE | 3.25386019 |
| Obesity | pericyte | *CILP2* | TRUE | 2.79651672 |
| Obesity | pericyte | *HS3ST6* | TRUE | 2.036136852 |
| Obesity | pericyte | *RAPSN* | TRUE | 4.000848786 |
| Obesity | pericyte | *SRR* | TRUE | 2.937407721 |
| Obesity | pericyte | *GPRIN1* | TRUE | 2.125503257 |
| Obesity | pericyte | *ADORA2B* | TRUE | 4.02271791 |
| Obesity | pericyte | *TTLL6* | TRUE | 2.755989899 |
| Obesity | pericyte | *CTSW* | TRUE | 4.553706212 |
| Obesity | pericyte | *CADM2* | TRUE | 2.042228361 |
| Obesity | pericyte | *ORAI3* | TRUE | 5.019397922 |
| Obesity | pericyte | *RPP25* | TRUE | 2.174432005 |
| Obesity | pericyte | *TRIM73* | TRUE | 2.134256096 |
| Obesity | pericyte | *C3orf38* | TRUE | 2.387609363 |
| Obesity | pericyte | *RPRML* | TRUE | 3.199979558 |
| Obesity | pericyte | *ZFP41* | TRUE | 2.065070682 |
| Obesity | pericyte | *NUTM2A* | TRUE | 3.957462897 |
| Obesity | pericyte | *SULT1A2* | TRUE | 6.858564297 |
| Obesity | pericyte | *DNAH10* | TRUE | 4.323641795 |
| Obesity | pericyte | *RPS26* | TRUE | 2.778904717 |
| Obesity | pericyte | *ZNF780A* | TRUE | 3.150381417 |

| | | | | |
|---|---|---|---|---|
| Obesity | pericyte | *GPANK1* | TRUE | 2.274047347 |
| Obesity | pericyte | *C17orf107* | TRUE | 2.047175849 |
| Obesity | pericyte | *SLC23A3* | TRUE | 2.119190305 |
| Obesity | pericyte | *GS1.124K5.4* | TRUE | 2.014149574 |
| Obesity | pericyte | *LINC00960* | TRUE | 2.02668045 |
| Obesity | pericyte | *LINC00664* | TRUE | 2.32685937 |
| Obesity | SMC | *CD79B* | TRUE | 2.062686203 |
| Obesity | SMC | *CENPM* | TRUE | 2.130556552 |
| Obesity | SMC | *SLC22A18* | TRUE | 2.220370982 |
| Obesity | SMC | *SERPINC1* | TRUE | 2.170842969 |
| Obesity | SMC | *GRK4* | TRUE | 2.977234901 |
| Obesity | SMC | *RIBC2* | TRUE | 2.878985629 |
| Obesity | SMC | *ATP8B3* | TRUE | 5.73994882 |
| Obesity | SMC | *PKDREJ* | TRUE | 2.416837524 |
| Obesity | SMC | *SLC37A4* | TRUE | 2.561681696 |
| Obesity | SMC | *C18orf8* | TRUE | 2.231556983 |
| Obesity | SMC | *MMEL1* | TRUE | 2.319958472 |
| Obesity | SMC | *HSD17B12* | TRUE | 5.31995192 |
| Obesity | SMC | *DPYSL4* | TRUE | 2.96260229 |
| Obesity | SMC | *MMP16* | TRUE | 2.152371438 |
| Obesity | SMC | *SLC5A11* | TRUE | 3.91373291 |
| Obesity | SMC | *SRR* | TRUE | 3.018483985 |
| Obesity | SMC | *INO80E* | TRUE | 2.600772818 |
| Obesity | SMC | *RGPD8* | TRUE | 2.25886015 |
| Obesity | SMC | *ADORA2B* | TRUE | 3.841590414 |
| Obesity | SMC | *TTLL6* | TRUE | 5.812309868 |
| Obesity | SMC | *CTSW* | TRUE | 3.837113795 |
| Obesity | SMC | *RPRML* | TRUE | 2.813834845 |
| Obesity | SMC | *PPP1R27* | TRUE | 2.730555767 |
| Obesity | SMC | *EMILIN3* | TRUE | 2.829115671 |
| Obesity | SMC | *ACOT1* | TRUE | 2.645551626 |
| Obesity | SMC | *NUTM2A* | TRUE | 3.792541638 |
| Obesity | SMC | *GP1BA* | TRUE | 2.336892224 |
| Obesity | SMC | *BTN3A2* | TRUE | 2.977553063 |
| Obesity | SMC | *SULT1A2* | TRUE | 5.701033895 |
| Obesity | SMC | *DNAH10* | TRUE | 4.034161381 |
| Obesity | SMC | *ZNF780A* | TRUE | 3.145522079 |
| Obesity | SMC | *GPANK1* | TRUE | 2.388530318 |
| Obesity | SMC | *C15orf59* | TRUE | 2.077389606 |
| Obesity | SMC | *AC084018.1* | TRUE | 1.965495112 |
| Obesity | SMC | *TMEM110* | TRUE | 2.354456935 |
| Obesity | SMC | *LINC00960* | TRUE | 2.271910601 |
| Obesity | t_cell | *RBM6* | TRUE | 2.78566623 |
| Obesity | t_cell | *PABPC4* | TRUE | 2.309091304 |
| Obesity | t_cell | *BTN3A3* | TRUE | 3.134817571 |
| Obesity | t_cell | *SERPINC1* | TRUE | 1.976790546 |
| Obesity | t_cell | *GRK4* | TRUE | 2.22783984 |
| Obesity | t_cell | *AP1M2* | TRUE | 2.573651146 |
| Obesity | t_cell | *ATP8B3* | TRUE | 5.961838724 |
| Obesity | t_cell | *CYP2E1* | TRUE | 2.053949744 |

| | | | | |
|---|---|---|---|---|
| Obesity | t_cell | *PKDREJ* | TRUE | 2.733683636 |
| Obesity | t_cell | *FITM1* | TRUE | 2.310243011 |
| Obesity | t_cell | *HSD17B12* | TRUE | 5.227429744 |
| Obesity | t_cell | *DPYSL4* | TRUE | 2.610996406 |
| Obesity | t_cell | *CLEC18A* | TRUE | 6.294185899 |
| Obesity | t_cell | *SLC5A11* | TRUE | 4.130911147 |
| Obesity | t_cell | *CILP2* | TRUE | 3.229613369 |
| Obesity | t_cell | *SRR* | TRUE | 3.102639602 |
| Obesity | t_cell | *INO80E* | TRUE | 2.510046781 |
| Obesity | t_cell | *RGPD8* | TRUE | 2.06926909 |
| Obesity | t_cell | *ADORA2B* | TRUE | 4.332597046 |
| Obesity | t_cell | *TTLL6* | TRUE | 5.066265337 |
| Obesity | t_cell | *C1QTNF4* | TRUE | 3.805577175 |
| Obesity | t_cell | *ORAI3* | TRUE | 3.401162834 |
| Obesity | t_cell | *RPP25* | TRUE | 2.475366053 |
| Obesity | t_cell | *C3orf38* | TRUE | 2.469920005 |
| Obesity | t_cell | *RPRML* | TRUE | 2.173089864 |
| Obesity | t_cell | *PPP1R27* | TRUE | 2.151296092 |
| Obesity | t_cell | *ACOT1* | TRUE | 3.499853422 |
| Obesity | t_cell | *NUTM2A* | TRUE | 2.658004672 |
| Obesity | t_cell | *GP1BA* | TRUE | 3.604849723 |
| Obesity | t_cell | *SULT1A2* | TRUE | 5.221392215 |
| Obesity | t_cell | *MRPL21* | TRUE | 2.482669448 |
| Obesity | t_cell | *RPS26* | TRUE | 2.552709744 |
| Obesity | t_cell | *ZNF780A* | TRUE | 2.562994878 |
| Obesity | t_cell | *GPANK1* | TRUE | 2.766575661 |
| Obesity | t_cell | *AC084018.1* | TRUE | 1.982904992 |
| Obesity | t_cell | *TMEM110* | TRUE | 3.907825656 |
| Obesity | t_cell | *GS1.124K5.4* | TRUE | 2.216484613 |
| Obesity | t_cell | *DNAH10OS* | TRUE | 2.929107254 |
| Obesity | t_cell | *PDF* | TRUE | 2.320133176 |
| Stroke | adipocyte | *SNX11* | TRUE | 2.012468032 |
| Stroke | adipocyte | *CD79B* | TRUE | 1.999049511 |
| Stroke | adipocyte | *CENPQ* | TRUE | 3.017709695 |
| Stroke | adipocyte | *TBX5* | TRUE | 2.778310385 |
| Stroke | adipocyte | *CHRD* | TRUE | 2.326567185 |
| Stroke | adipocyte | *MARCH2* | TRUE | 2.102573247 |
| Stroke | adipocyte | *OSM* | TRUE | 2.092672975 |
| Stroke | adipocyte | *MLC1* | TRUE | 2.399614835 |
| Stroke | adipocyte | *C17orf75* | TRUE | 2.184675179 |
| Stroke | adipocyte | *BVES* | TRUE | 2.509157244 |
| Stroke | adipocyte | *DR1* | TRUE | 2.480430579 |
| Stroke | adipocyte | *EPCAM* | TRUE | 2.271837387 |
| Stroke | adipocyte | *BTN2A2* | TRUE | 2.003715348 |
| Stroke | adipocyte | *TRAP1* | TRUE | 2.190876566 |
| Stroke | adipocyte | *SLC2A11* | TRUE | 2.162209434 |
| Stroke | adipocyte | *LARS* | TRUE | 2.157534722 |
| Stroke | adipocyte | *TPMT* | TRUE | 2.179238556 |
| Stroke | adipocyte | *NOVA1* | TRUE | 2.000243365 |
| Stroke | adipocyte | *DOPEY2* | TRUE | 2.473945209 |

| | | | | |
|---|---|---|---|---|
| Stroke | adipocyte | *LYG1* | TRUE | 2.139365905 |
| Stroke | adipocyte | *CLIC6* | TRUE | 2.083482936 |
| Stroke | adipocyte | *ICOSLG* | TRUE | 2.470569425 |
| Stroke | adipocyte | *ALDH16A1* | TRUE | 2.517941215 |
| Stroke | adipocyte | *DNASE1L3* | TRUE | 2.232315187 |
| Stroke | adipocyte | *CHRFAM7A* | TRUE | 3.025986026 |
| Stroke | adipocyte | *FAM57A* | TRUE | 2.562223636 |
| Stroke | adipocyte | *SCARA5* | TRUE | 2.066956606 |
| Stroke | adipocyte | *GPRIN1* | TRUE | 2.186420536 |
| Stroke | adipocyte | *GPR156* | TRUE | 2.333041222 |
| Stroke | adipocyte | *MYPOP* | TRUE | 2.337999359 |
| Stroke | adipocyte | *ZNF404* | TRUE | 2.060679726 |
| Stroke | adipocyte | *GEN1* | TRUE | 2.450795607 |
| Stroke | adipocyte | *YBEY* | TRUE | 2.119328751 |
| Stroke | adipocyte | *CLN8* | TRUE | 2.114405798 |
| Stroke | adipocyte | *EMILIN3* | TRUE | 2.045474253 |
| Stroke | adipocyte | *ZNF749* | TRUE | 2.95382722 |
| Stroke | adipocyte | *FAM86B1* | TRUE | 1.98138865 |
| Stroke | adipocyte | *QRFPR* | TRUE | 2.051439423 |
| Stroke | adipocyte | *JAKMIP3* | TRUE | 2.277674462 |
| Stroke | adipocyte | *WDR5B* | TRUE | 2.386801174 |
| Stroke | adipocyte | *CCDC154* | TRUE | 2.275273611 |
| Stroke | adipocyte | *TMEM116* | TRUE | 2.828800698 |
| Stroke | adipocyte | *AC084018.1* | TRUE | 2.079999382 |
| Stroke | adipocyte | *DNLZ* | TRUE | 2.237046908 |
| Stroke | adipocyte | *LINC01166* | TRUE | 2.251292122 |
| Stroke | adipocyte | *LILRA2* | TRUE | 2.092771545 |
| Stroke | adipocyte | *PNMA2* | TRUE | 2.243035499 |
| Stroke | adipocyte | *SPRR2F* | TRUE | 2.06475897 |
| Stroke | adipocyte | *ARMS2* | TRUE | 1.977043317 |
| Stroke | adipocyte | *LIN37* | TRUE | 2.017534439 |
| Stroke | adipocyte | *AC026740.1* | TRUE | 2.15058548 |
| Stroke | ASPC | *AOC1* | TRUE | 2.259683176 |
| Stroke | ASPC | *CD79B* | TRUE | 2.005201507 |
| Stroke | ASPC | *CENPQ* | TRUE | 3.32602858 |
| Stroke | ASPC | *TIPIN* | TRUE | 2.148041987 |
| Stroke | ASPC | *TBX5* | TRUE | 2.544368641 |
| Stroke | ASPC | *MARCH2* | TRUE | 2.641228205 |
| Stroke | ASPC | *OSM* | TRUE | 2.111934527 |
| Stroke | ASPC | *MLC1* | TRUE | 2.615924258 |
| Stroke | ASPC | *PYCARD* | TRUE | 2.195463919 |
| Stroke | ASPC | *TBL2* | TRUE | 2.093470893 |
| Stroke | ASPC | *ENO3* | TRUE | 2.328516652 |
| Stroke | ASPC | *BVES* | TRUE | 2.441454143 |
| Stroke | ASPC | *RASGRF2* | TRUE | 2.16808069 |
| Stroke | ASPC | *DR1* | TRUE | 2.118289798 |
| Stroke | ASPC | *EPCAM* | TRUE | 2.599682933 |
| Stroke | ASPC | *TRAP1* | TRUE | 2.60110013 |
| Stroke | ASPC | *QRSL1* | TRUE | 2.193388546 |
| Stroke | ASPC | *RGS22* | TRUE | 2.00888748 |

| | | | | |
|---|---|---|---|---|
| Stroke | ASPC | *SLC2A11* | TRUE | 2.797347204 |
| Stroke | ASPC | *KLHL36* | TRUE | 2.674719246 |
| Stroke | ASPC | *CAPN9* | TRUE | 2.475079471 |
| Stroke | ASPC | *TPMT* | TRUE | 2.100401922 |
| Stroke | ASPC | *NOVA1* | TRUE | 2.337631164 |
| Stroke | ASPC | *DOPEY2* | TRUE | 3.118876778 |
| Stroke | ASPC | *LYG1* | TRUE | 1.964144704 |
| Stroke | ASPC | *NBEAL1* | TRUE | 2.59974047 |
| Stroke | ASPC | *PLA2R1* | TRUE | 2.223551744 |
| Stroke | ASPC | *NEIL2* | TRUE | 2.05291467 |
| Stroke | ASPC | *CLIC6* | TRUE | 2.256599136 |
| Stroke | ASPC | *ICOSLG* | TRUE | 2.54371399 |
| Stroke | ASPC | *CCR5* | TRUE | 2.008453055 |
| Stroke | ASPC | *ZNF577* | TRUE | 2.287043439 |
| Stroke | ASPC | *ALDH16A1* | TRUE | 2.361746046 |
| Stroke | ASPC | *DNASE1L3* | TRUE | 2.584181133 |
| Stroke | ASPC | *HSPA12A* | TRUE | 2.221778398 |
| Stroke | ASPC | *CHRFAM7A* | TRUE | 2.433777933 |
| Stroke | ASPC | *CNPY4* | TRUE | 2.410375198 |
| Stroke | ASPC | *PROCA1* | TRUE | 2.016533825 |
| Stroke | ASPC | *CHAF1A* | TRUE | 2.07099775 |
| Stroke | ASPC | *FAM57A* | TRUE | 2.827203792 |
| Stroke | ASPC | *GPRIN1* | TRUE | 2.149772632 |
| Stroke | ASPC | *GPR156* | TRUE | 2.078787175 |
| Stroke | ASPC | *CTDNEP1* | TRUE | 2.603344447 |
| Stroke | ASPC | *ZNF404* | TRUE | 1.991497447 |
| Stroke | ASPC | *GEN1* | TRUE | 2.316788405 |
| Stroke | ASPC | *MRPL14* | TRUE | 2.040242322 |
| Stroke | ASPC | *CLN8* | TRUE | 2.445025212 |
| Stroke | ASPC | *EMILIN3* | TRUE | 1.971713249 |
| Stroke | ASPC | *TMEM106A* | TRUE | 2.131545626 |
| Stroke | ASPC | *ZNF749* | TRUE | 2.755363625 |
| Stroke | ASPC | *FAM86B1* | TRUE | 2.121486032 |
| Stroke | ASPC | *QRFPR* | TRUE | 2.200673525 |
| Stroke | ASPC | *JAKMIP3* | TRUE | 2.19634468 |
| Stroke | ASPC | *STK31* | TRUE | 2.150292776 |
| Stroke | ASPC | *WDR5B* | TRUE | 3.040565067 |
| Stroke | ASPC | *CCDC154* | TRUE | 2.147982166 |
| Stroke | ASPC | *ZNF181* | TRUE | 2.271354663 |
| Stroke | ASPC | *TMEM116* | TRUE | 3.158273127 |
| Stroke | ASPC | *C20orf203* | TRUE | 2.169186504 |
| Stroke | ASPC | *COL15A1* | TRUE | 2.005173686 |
| Stroke | ASPC | *AC084018.1* | TRUE | 2.04337204 |
| Stroke | ASPC | *TCTEX1D2* | TRUE | 1.986279229 |
| Stroke | ASPC | *DNLZ* | TRUE | 2.215252903 |
| Stroke | ASPC | *AC009495.3* | TRUE | 2.194857787 |
| Stroke | ASPC | *LINC01166* | TRUE | 2.573571577 |
| Stroke | ASPC | *PSMB9* | TRUE | 2.131351418 |
| Stroke | ASPC | *PNMA2* | TRUE | 2.593107159 |
| Stroke | ASPC | *SPRR2F* | TRUE | 2.192194464 |

| | | | | |
|---|---|---|---|---|
| Stroke | ASPC | *ARMS2* | TRUE | 2.101184919 |
| Stroke | ASPC | *LIN37* | TRUE | 2.346222825 |
| Stroke | b_cell | *CHRFAM7A* | TRUE | 2.784975996 |
| Stroke | dendritic_cell | *CENPQ* | TRUE | 2.711239831 |
| Stroke | dendritic_cell | *MLC1* | TRUE | 2.057967631 |
| Stroke | dendritic_cell | *DOPEY2* | TRUE | 2.257406619 |
| Stroke | dendritic_cell | *CHRFAM7A* | TRUE | 2.800368867 |
| Stroke | dendritic_cell | *ZNF404* | TRUE | 1.973760712 |
| Stroke | dendritic_cell | *YBEY* | TRUE | 2.423911712 |
| Stroke | dendritic_cell | *DNLZ* | TRUE | 2.013551633 |
| Stroke | endometrium | *MLC1* | TRUE | 2.004405514 |
| Stroke | endometrium | *ICOSLG* | TRUE | 2.332899967 |
| Stroke | endometrium | *CHRFAM7A* | TRUE | 3.072563104 |
| Stroke | endometrium | *CCDC154* | TRUE | 1.99293583 |
| Stroke | endometrium | *TMEM116* | TRUE | 2.698889565 |
| Stroke | endometrium | *DNLZ* | TRUE | 2.117780284 |
| Stroke | endothelial | *CENPQ* | TRUE | 2.585174257 |
| Stroke | endothelial | *OSM* | TRUE | 2.062636396 |
| Stroke | endothelial | *BVES* | TRUE | 2.120169524 |
| Stroke | endothelial | *ICOSLG* | TRUE | 2.254266888 |
| Stroke | endothelial | *CHRFAM7A* | TRUE | 2.543229275 |
| Stroke | endothelial | *YBEY* | TRUE | 1.960418426 |
| Stroke | endothelial | *CLN8* | TRUE | 2.216821536 |
| Stroke | endothelial | *ZNF749* | TRUE | 2.359671666 |
| Stroke | endothelial | *TMEM116* | TRUE | 2.35209193 |
| Stroke | endothelial | *HLA.DQB2* | TRUE | 2.075279634 |
| Stroke | endothelial | *LINC01166* | TRUE | 2.031174736 |
| Stroke | endothelial | *LIN37* | TRUE | 2.167406849 |
| Stroke | LEC | *CENPQ* | TRUE | 2.521038418 |
| Stroke | LEC | *CHRD* | TRUE | 1.980823252 |
| Stroke | LEC | *OSM* | TRUE | 2.285327918 |
| Stroke | LEC | *BVES* | TRUE | 2.150087601 |
| Stroke | LEC | *TPMT* | TRUE | 2.280658424 |
| Stroke | LEC | *RASL10B* | TRUE | 2.155218929 |
| Stroke | LEC | *DOPEY2* | TRUE | 1.985644176 |
| Stroke | LEC | *ICOSLG* | TRUE | 2.335058276 |
| Stroke | LEC | *CHRFAM7A* | TRUE | 2.90675144 |
| Stroke | LEC | *FAM57A* | TRUE | 2.182511416 |
| Stroke | LEC | *GPRIN1* | TRUE | 2.244151257 |
| Stroke | LEC | *GPR156* | TRUE | 2.27539799 |
| Stroke | LEC | *GEN1* | TRUE | 2.384216766 |
| Stroke | LEC | *YBEY* | TRUE | 2.085051774 |
| Stroke | LEC | *ZNF749* | TRUE | 2.704636829 |
| Stroke | LEC | *CCDC154* | TRUE | 2.103641797 |
| Stroke | LEC | *TMEM116* | TRUE | 2.486336194 |
| Stroke | LEC | *DNLZ* | TRUE | 2.091820814 |
| Stroke | LEC | *LINC01166* | TRUE | 2.385913849 |
| Stroke | macrophage | *CENPQ* | TRUE | 3.377216357 |
| Stroke | macrophage | *CHRD* | TRUE | 2.1632069 |
| Stroke | macrophage | *MARCH2* | TRUE | 2.018044299 |

| | | | | |
|---|---|---|---|---|
| Stroke | macrophage | *MLC1* | TRUE | 2.411228005 |
| Stroke | macrophage | *BVES* | TRUE | 2.429495733 |
| Stroke | macrophage | *SLC2A11* | TRUE | 2.092302736 |
| Stroke | macrophage | *CLIC6* | TRUE | 1.98297975 |
| Stroke | macrophage | *ICOSLG* | TRUE | 2.213730266 |
| Stroke | macrophage | *DNASE1L3* | TRUE | 2.261137959 |
| Stroke | macrophage | *CHRFAM7A* | TRUE | 2.9563358 |
| Stroke | macrophage | *CHAF1A* | TRUE | 1.988259842 |
| Stroke | macrophage | *FAM57A* | TRUE | 2.158172956 |
| Stroke | macrophage | *ZNF404* | TRUE | 2.038202607 |
| Stroke | macrophage | *GEN1* | TRUE | 2.085108792 |
| Stroke | macrophage | *ZNF749* | TRUE | 2.932017904 |
| Stroke | macrophage | *QRFPR* | TRUE | 2.226052668 |
| Stroke | macrophage | *WDR5B* | TRUE | 2.582463838 |
| Stroke | macrophage | *TMEM116* | TRUE | 2.977607066 |
| Stroke | macrophage | *AC084018.1* | TRUE | 2.027060802 |
| Stroke | macrophage | *DNLZ* | TRUE | 2.160286302 |
| Stroke | macrophage | *HLA.DQB2* | TRUE | 2.192756444 |
| Stroke | macrophage | *SPRR2F* | TRUE | 2.141169069 |
| Stroke | macrophage | *AC026740.1* | TRUE | 1.981173343 |
| Stroke | mast_cell | *CENPQ* | TRUE | 3.273189823 |
| Stroke | mast_cell | *MLC1* | TRUE | 1.988890932 |
| Stroke | mast_cell | *BVES* | TRUE | 2.291070513 |
| Stroke | mast_cell | *BTN2A2* | TRUE | 2.01134538 |
| Stroke | mast_cell | *ICOSLG* | TRUE | 2.392710827 |
| Stroke | mast_cell | *ALDH16A1* | TRUE | 2.512854714 |
| Stroke | mast_cell | *CHRFAM7A* | TRUE | 2.995220559 |
| Stroke | mast_cell | *CHAF1A* | TRUE | 1.987196366 |
| Stroke | mast_cell | *YBEY* | TRUE | 2.555691884 |
| Stroke | mast_cell | *DNLZ* | TRUE | 2.1399111 |
| Stroke | mast_cell | *AC026740.1* | TRUE | 1.961185954 |
| Stroke | mesothelium | *AOC1* | TRUE | 2.142053683 |
| Stroke | mesothelium | *SNX11* | TRUE | 2.039317235 |
| Stroke | mesothelium | *CENPQ* | TRUE | 2.878815569 |
| Stroke | mesothelium | *ALDH18A1* | TRUE | 2.010159414 |
| Stroke | mesothelium | *TIPIN* | TRUE | 1.961059866 |
| Stroke | mesothelium | *TBX5* | TRUE | 2.735083835 |
| Stroke | mesothelium | *SPTLC1* | TRUE | 2.06974791 |
| Stroke | mesothelium | *CHRD* | TRUE | 2.360072604 |
| Stroke | mesothelium | *MARCH2* | TRUE | 2.359462408 |
| Stroke | mesothelium | *OSM* | TRUE | 2.135487746 |
| Stroke | mesothelium | *SLC35E4* | TRUE | 2.009641693 |
| Stroke | mesothelium | *MLC1* | TRUE | 2.391362987 |
| Stroke | mesothelium | *ZNF500* | TRUE | 2.047877128 |
| Stroke | mesothelium | *ZDHHC2* | TRUE | 2.044636441 |
| Stroke | mesothelium | *ENO3* | TRUE | 2.178912394 |
| Stroke | mesothelium | *C17orf75* | TRUE | 2.28498408 |
| Stroke | mesothelium | *BVES* | TRUE | 2.539647687 |
| Stroke | mesothelium | *SLC41A3* | TRUE | 2.101896449 |
| Stroke | mesothelium | *DR1* | TRUE | 2.724036761 |

| | | | | |
|---|---|---|---|---|
| Stroke | mesothelium | *EPCAM* | TRUE | 2.440237435 |
| Stroke | mesothelium | *BTN2A2* | TRUE | 2.087722743 |
| Stroke | mesothelium | *TRAP1* | TRUE | 2.350443866 |
| Stroke | mesothelium | *SLC2A11* | TRUE | 2.52552104 |
| Stroke | mesothelium | *LARS* | TRUE | 2.467921259 |
| Stroke | mesothelium | *CAPN9* | TRUE | 2.35774878 |
| Stroke | mesothelium | *TPMT* | TRUE | 2.416626753 |
| Stroke | mesothelium | *ASCC1* | TRUE | 2.012722472 |
| Stroke | mesothelium | *NOVA1* | TRUE | 2.246524597 |
| Stroke | mesothelium | *RASL10B* | TRUE | 2.400060173 |
| Stroke | mesothelium | *SCRN2* | TRUE | 2.13012386 |
| Stroke | mesothelium | *MIEN1* | TRUE | 2.199612399 |
| Stroke | mesothelium | *DOPEY2* | TRUE | 2.837448599 |
| Stroke | mesothelium | *ARNT* | TRUE | 2.320968838 |
| Stroke | mesothelium | *LYG1* | TRUE | 2.093493211 |
| Stroke | mesothelium | *RGR* | TRUE | 2.06761107 |
| Stroke | mesothelium | *NMT2* | TRUE | 2.442846514 |
| Stroke | mesothelium | *ATP5G3* | TRUE | 1.993340903 |
| Stroke | mesothelium | *CLIC6* | TRUE | 2.168311173 |
| Stroke | mesothelium | *ICOSLG* | TRUE | 2.460131185 |
| Stroke | mesothelium | *CCR5* | TRUE | 2.104510343 |
| Stroke | mesothelium | *ALDH16A1* | TRUE | 2.560495859 |
| Stroke | mesothelium | *DNASE1L3* | TRUE | 2.438144699 |
| Stroke | mesothelium | *CHRFAM7A* | TRUE | 3.08070772 |
| Stroke | mesothelium | *CNPY4* | TRUE | 2.160012211 |
| Stroke | mesothelium | *CHAF1A* | TRUE | 2.075066753 |
| Stroke | mesothelium | *FAM57A* | TRUE | 2.690725854 |
| Stroke | mesothelium | *ULK4* | TRUE | 2.314035844 |
| Stroke | mesothelium | *SCARA5* | TRUE | 2.423983391 |
| Stroke | mesothelium | *GPRIN1* | TRUE | 2.229287881 |
| Stroke | mesothelium | *USP38* | TRUE | 2.192043121 |
| Stroke | mesothelium | *GAA* | TRUE | 2.064359627 |
| Stroke | mesothelium | *RGS19* | TRUE | 2.001797278 |
| Stroke | mesothelium | *GPR156* | TRUE | 2.571512575 |
| Stroke | mesothelium | *MYPOP* | TRUE | 2.914287565 |
| Stroke | mesothelium | *GEN1* | TRUE | 2.576841615 |
| Stroke | mesothelium | *ZFPM1* | TRUE | 2.076068694 |
| Stroke | mesothelium | *CHST13* | TRUE | 2.020374007 |
| Stroke | mesothelium | *YBEY* | TRUE | 2.122809641 |
| Stroke | mesothelium | *CLN8* | TRUE | 2.305221825 |
| Stroke | mesothelium | *EMILIN3* | TRUE | 2.061643826 |
| Stroke | mesothelium | *PIGP* | TRUE | 2.004826998 |
| Stroke | mesothelium | *ZNF749* | TRUE | 2.995868542 |
| Stroke | mesothelium | *CLCNKA* | TRUE | 2.00381894 |
| Stroke | mesothelium | *JAKMIP3* | TRUE | 2.316148759 |
| Stroke | mesothelium | *FAM26F* | TRUE | 2.113190963 |
| Stroke | mesothelium | *WDR5B* | TRUE | 2.753928134 |
| Stroke | mesothelium | *PIWIL2* | TRUE | 2.127236229 |
| Stroke | mesothelium | *CCDC154* | TRUE | 2.162345097 |
| Stroke | mesothelium | *ZNF181* | TRUE | 1.998793061 |

| | | | | |
|---|---|---|---|---|
| Stroke | mesothelium | *TMEM116* | TRUE | 2.619317763 |
| Stroke | mesothelium | *COL15A1* | TRUE | 2.070614276 |
| Stroke | mesothelium | *AC084018.1* | TRUE | 2.104571973 |
| Stroke | mesothelium | *DNLZ* | TRUE | 2.007352723 |
| Stroke | mesothelium | *LTB4R* | TRUE | 1.990394763 |
| Stroke | mesothelium | *NBPF1* | TRUE | 2.256499192 |
| Stroke | mesothelium | *AC009495.3* | TRUE | 2.252402334 |
| Stroke | mesothelium | *LINC01166* | TRUE | 2.462895803 |
| Stroke | mesothelium | *LINC00342* | TRUE | 1.962976103 |
| Stroke | mesothelium | *CDKN2AIPNL* | TRUE | 2.077594688 |
| Stroke | mesothelium | *LILRA2* | TRUE | 2.151204672 |
| Stroke | mesothelium | *PNMA2* | TRUE | 2.133470616 |
| Stroke | mesothelium | *SPRR2F* | TRUE | 2.22307037 |
| Stroke | mesothelium | *ARMS2* | TRUE | 2.050572199 |
| Stroke | mesothelium | *LIN37* | TRUE | 2.266804835 |
| Stroke | mesothelium | *AC026740.1* | TRUE | 2.047656151 |
| Stroke | monocyte | *ICOSLG* | TRUE | 2.442432255 |
| Stroke | monocyte | *HLA.DQB1* | TRUE | 1.978442934 |
| Stroke | monocyte | *ZNF749* | TRUE | 2.931938618 |
| Stroke | monocyte | *TMEM116* | TRUE | 2.28389238 |
| Stroke | monocyte | *DNLZ* | TRUE | 2.130602669 |
| Stroke | nk_cell | *CENPQ* | TRUE | 3.136545856 |
| Stroke | nk_cell | *ICOSLG* | TRUE | 2.029325904 |
| Stroke | nk_cell | *CHRFAM7A* | TRUE | 2.836707382 |
| Stroke | nk_cell | *CHAF1A* | TRUE | 2.012822691 |
| Stroke | nk_cell | *GPRIN1* | TRUE | 2.065784252 |
| Stroke | nk_cell | *YBEY* | TRUE | 2.043144141 |
| Stroke | nk_cell | *ZNF749* | TRUE | 2.184687172 |
| Stroke | nk_cell | *LINC01166* | TRUE | 2.194841329 |
| Stroke | pericyte | *CENPQ* | TRUE | 3.057503769 |
| Stroke | pericyte | *OSM* | TRUE | 1.976294466 |
| Stroke | pericyte | *MLC1* | TRUE | 2.071735415 |
| Stroke | pericyte | *BVES* | TRUE | 2.334980432 |
| Stroke | pericyte | *EPCAM* | TRUE | 2.070681988 |
| Stroke | pericyte | *BTN2A2* | TRUE | 2.005538793 |
| Stroke | pericyte | *TRAP1* | TRUE | 2.096083059 |
| Stroke | pericyte | *TPMT* | TRUE | 1.986029058 |
| Stroke | pericyte | *NOVA1* | TRUE | 1.996088636 |
| Stroke | pericyte | *DOPEY2* | TRUE | 2.469757963 |
| Stroke | pericyte | *CLIC6* | TRUE | 2.036503865 |
| Stroke | pericyte | *ICOSLG* | TRUE | 2.228773092 |
| Stroke | pericyte | *ALDH16A1* | TRUE | 2.292841168 |
| Stroke | pericyte | *CHRFAM7A* | TRUE | 2.998998369 |
| Stroke | pericyte | *GEN1* | TRUE | 2.072686241 |
| Stroke | pericyte | *ZNF749* | TRUE | 2.84019226 |
| Stroke | pericyte | *TMEM116* | TRUE | 2.647540303 |
| Stroke | pericyte | *DNLZ* | TRUE | 2.168058165 |
| Stroke | pericyte | *LINC01166* | TRUE | 2.415858981 |
| Stroke | pericyte | *PNMA2* | TRUE | 2.245530092 |
| Stroke | pericyte | *SPRR2F* | TRUE | 2.197149465 |

| | | | | |
|---|---|---|---|---|
| Stroke | SMC | *CENPQ* | TRUE | 2.98468532 |
| Stroke | SMC | *C17orf75* | TRUE | 2.042551716 |
| Stroke | SMC | *DR1* | TRUE | 2.372960992 |
| Stroke | SMC | *TPMT* | TRUE | 2.033528384 |
| Stroke | SMC | *LYG1* | TRUE | 2.022892242 |
| Stroke | SMC | *ALDH16A1* | TRUE | 2.42314239 |
| Stroke | SMC | *CHRFAM7A* | TRUE | 2.625824307 |
| Stroke | SMC | *ZNF404* | TRUE | 1.974108196 |
| Stroke | SMC | *GEN1* | TRUE | 2.255810729 |
| Stroke | SMC | *YBEY* | TRUE | 2.607224659 |
| Stroke | SMC | *ZNF749* | TRUE | 2.962934123 |
| Stroke | SMC | *TMEM116* | TRUE | 3.105257644 |
| Stroke | SMC | *CR1* | TRUE | 2.079201394 |
| Stroke | SMC | *DNLZ* | TRUE | 2.109806782 |
| Stroke | t_cell | *CENPQ* | TRUE | 2.786970162 |
| Stroke | t_cell | *MLC1* | TRUE | 2.103903081 |
| Stroke | t_cell | *BTN2A2* | TRUE | 1.999667841 |
| Stroke | t_cell | *SLC2A11* | TRUE | 2.001696587 |
| Stroke | t_cell | *SCRN2* | TRUE | 1.963655516 |
| Stroke | t_cell | *DOPEY2* | TRUE | 2.54692767 |
| Stroke | t_cell | *ICOSLG* | TRUE | 2.152815318 |
| Stroke | t_cell | *CHRFAM7A* | TRUE | 2.828556413 |
| Stroke | t_cell | *GEN1* | TRUE | 2.13135521 |
| Stroke | t_cell | *ZNF749* | TRUE | 2.561807651 |
| Stroke | t_cell | *CCDC154* | TRUE | 2.054078681 |
| Stroke | t_cell | *TMEM116* | TRUE | 2.404924816 |
| Stroke | t_cell | *DNLZ* | TRUE | 2.223558505 |
| Heart_Failure | adipocyte | *SLC7A9* | TRUE | 2.476123053 |
| Heart_Failure | adipocyte | *LAMC2* | TRUE | 2.515965089 |
| Heart_Failure | adipocyte | *CROCC* | TRUE | 2.024056745 |
| Heart_Failure | adipocyte | *MAST2* | TRUE | 2.608176231 |
| Heart_Failure | adipocyte | *SETD1A* | TRUE | 2.78116359 |
| Heart_Failure | adipocyte | *SMARCB1* | TRUE | 2.59315539 |
| Heart_Failure | adipocyte | *TOX3* | TRUE | 2.600949122 |
| Heart_Failure | adipocyte | *CTSH* | TRUE | 2.224099378 |
| Heart_Failure | adipocyte | *TIMM44* | TRUE | 2.256766455 |
| Heart_Failure | adipocyte | *CD33* | TRUE | 1.996417998 |
| Heart_Failure | adipocyte | *RAB5C* | TRUE | 2.009944357 |
| Heart_Failure | adipocyte | *KIF20A* | TRUE | 3.697263857 |
| Heart_Failure | adipocyte | *BNIP1* | TRUE | 2.491604059 |
| Heart_Failure | adipocyte | *IL18R1* | TRUE | 3.847624129 |
| Heart_Failure | adipocyte | *CTNNAL1* | TRUE | 2.84153713 |
| Heart_Failure | adipocyte | *GRK4* | TRUE | 2.082833785 |
| Heart_Failure | adipocyte | *ANGPTL6* | TRUE | 2.437436854 |
| Heart_Failure | adipocyte | *NMT1* | TRUE | 2.954100037 |
| Heart_Failure | adipocyte | *ALG10* | TRUE | 1.96533639 |
| Heart_Failure | adipocyte | *PNMT* | TRUE | 2.90810301 |
| Heart_Failure | adipocyte | *EVA1B* | TRUE | 2.199510271 |
| Heart_Failure | adipocyte | *FLG* | TRUE | 2.821792245 |
| Heart_Failure | adipocyte | *TSLP* | TRUE | 4.27586407 |

| | | | | |
|---|---|---|---|---|
| Heart_Failure | adipocyte | *ROM1* | TRUE | 3.103881609 |
| Heart_Failure | adipocyte | *DPYSL4* | TRUE | 2.130959346 |
| Heart_Failure | adipocyte | *FZD7* | TRUE | 2.13354253 |
| Heart_Failure | adipocyte | *FCRL3* | TRUE | 2.597895847 |
| Heart_Failure | adipocyte | *FGFR4* | TRUE | 2.669693778 |
| Heart_Failure | adipocyte | *KLHDC9* | TRUE | 2.687052015 |
| Heart_Failure | adipocyte | *EFHB* | TRUE | 2.7583911 |
| Heart_Failure | adipocyte | *MST1R* | TRUE | 2.463217477 |
| Heart_Failure | adipocyte | *GPER1* | TRUE | 2.515929689 |
| Heart_Failure | adipocyte | *HSPA12A* | TRUE | 2.203905443 |
| Heart_Failure | adipocyte | *HDHD2* | TRUE | 2.714087395 |
| Heart_Failure | adipocyte | *ZNF23* | TRUE | 2.95421401 |
| Heart_Failure | adipocyte | *TRPV3* | TRUE | 2.286519824 |
| Heart_Failure | adipocyte | *GSDMA* | TRUE | 2.143603203 |
| Heart_Failure | adipocyte | *CHRNB1* | TRUE | 2.527115784 |
| Heart_Failure | adipocyte | *TMEM133* | TRUE | 2.306843484 |
| Heart_Failure | adipocyte | *PAAF1* | TRUE | 2.025768319 |
| Heart_Failure | adipocyte | *RPS6KB2* | TRUE | 2.712724808 |
| Heart_Failure | adipocyte | *OR4N2* | TRUE | 2.003350542 |
| Heart_Failure | adipocyte | *DPY19L2* | TRUE | 2.207415407 |
| Heart_Failure | adipocyte | *ZNF454* | TRUE | 2.708512492 |
| Heart_Failure | adipocyte | *LINC00314* | TRUE | 2.32635356 |
| Heart_Failure | adipocyte | *TRIM73* | TRUE | 2.113618122 |
| Heart_Failure | adipocyte | *HLA.DQB1* | TRUE | 4.161773076 |
| Heart_Failure | adipocyte | *D2HGDH* | TRUE | 2.710643939 |
| Heart_Failure | adipocyte | *FKRP* | TRUE | 2.051652489 |
| Heart_Failure | adipocyte | *A3GALT2* | TRUE | 2.277809488 |
| Heart_Failure | adipocyte | *QRFPR* | TRUE | 2.517079897 |
| Heart_Failure | adipocyte | *TMEM232* | TRUE | 2.489641262 |
| Heart_Failure | adipocyte | *WDR5B* | TRUE | 2.182178509 |
| Heart_Failure | adipocyte | *PDCD1LG2* | TRUE | 2.826753591 |
| Heart_Failure | adipocyte | *RPS26* | TRUE | 2.990053597 |
| Heart_Failure | adipocyte | *MICA* | TRUE | 3.566591027 |
| Heart_Failure | adipocyte | *ATF6B* | TRUE | 2.728278958 |
| Heart_Failure | adipocyte | *CPNE1* | TRUE | 2.310527217 |
| Heart_Failure | adipocyte | *OR2A1* | TRUE | 2.88559975 |
| Heart_Failure | adipocyte | *ZNF736* | TRUE | 2.495496521 |
| Heart_Failure | adipocyte | *ZBTB22* | TRUE | 2.721540426 |
| Heart_Failure | adipocyte | *TTC4* | TRUE | 2.702398086 |
| Heart_Failure | adipocyte | *LILRA6* | TRUE | 2.366427753 |
| Heart_Failure | adipocyte | *MC1R* | TRUE | 2.101035743 |
| Heart_Failure | adipocyte | *GTF2H5* | TRUE | 2.382966672 |
| Heart_Failure | ASPC | *CLCA4* | TRUE | 2.054357424 |
| Heart_Failure | ASPC | *SLC7A9* | TRUE | 2.536497227 |
| Heart_Failure | ASPC | *LAMC2* | TRUE | 2.633414899 |
| Heart_Failure | ASPC | *CROCC* | TRUE | 2.128700626 |
| Heart_Failure | ASPC | *BPIFB2* | TRUE | 2.019187336 |
| Heart_Failure | ASPC | *MAST2* | TRUE | 2.820241975 |
| Heart_Failure | ASPC | *FXYD5* | TRUE | 2.006474279 |
| Heart_Failure | ASPC | *PABPC4* | TRUE | 2.52088209 |

| | | | | |
|---|---|---|---|---|
| Heart_Failure | ASPC | *SETD1A* | TRUE | 2.812154019 |
| Heart_Failure | ASPC | *SMARCB1* | TRUE | 2.62897569 |
| Heart_Failure | ASPC | *TOX3* | TRUE | 2.566661704 |
| Heart_Failure | ASPC | *CTSH* | TRUE | 2.399828084 |
| Heart_Failure | ASPC | *TIMM44* | TRUE | 2.167476311 |
| Heart_Failure | ASPC | *CD33* | TRUE | 2.139314139 |
| Heart_Failure | ASPC | *RAB5C* | TRUE | 2.234126084 |
| Heart_Failure | ASPC | *PRDM4* | TRUE | 2.272160443 |
| Heart_Failure | ASPC | *VNN2* | TRUE | 2.074966297 |
| Heart_Failure | ASPC | *KIF20A* | TRUE | 3.701543596 |
| Heart_Failure | ASPC | *BNIP1* | TRUE | 2.517869745 |
| Heart_Failure | ASPC | *IL18R1* | TRUE | 4.085580936 |
| Heart_Failure | ASPC | *ALDH8A1* | TRUE | 2.204299082 |
| Heart_Failure | ASPC | *CTNNAL1* | TRUE | 2.864217402 |
| Heart_Failure | ASPC | *ITIH5* | TRUE | 2.525075905 |
| Heart_Failure | ASPC | *GRK4* | TRUE | 2.173976 |
| Heart_Failure | ASPC | *PROZ* | TRUE | 2.035669691 |
| Heart_Failure | ASPC | *ANGPTL6* | TRUE | 2.486540067 |
| Heart_Failure | ASPC | *ADAM19* | TRUE | 2.313603307 |
| Heart_Failure | ASPC | *NMT1* | TRUE | 2.889192497 |
| Heart_Failure | ASPC | *ALG10* | TRUE | 2.508591273 |
| Heart_Failure | ASPC | *PNMT* | TRUE | 3.348022671 |
| Heart_Failure | ASPC | *EVA1B* | TRUE | 2.832306462 |
| Heart_Failure | ASPC | *FLG* | TRUE | 3.106300913 |
| Heart_Failure | ASPC | *GPR17* | TRUE | 2.156620592 |
| Heart_Failure | ASPC | *BHMT* | TRUE | 2.039880947 |
| Heart_Failure | ASPC | *TSLP* | TRUE | 4.667439173 |
| Heart_Failure | ASPC | *ROM1* | TRUE | 3.042609004 |
| Heart_Failure | ASPC | *DPYSL4* | TRUE | 2.132430361 |
| Heart_Failure | ASPC | *FZD7* | TRUE | 2.720827023 |
| Heart_Failure | ASPC | *SIM2* | TRUE | 2.165211706 |
| Heart_Failure | ASPC | *FCRL3* | TRUE | 2.698886332 |
| Heart_Failure | ASPC | *FGFR4* | TRUE | 2.803452938 |
| Heart_Failure | ASPC | *KLHDC9* | TRUE | 2.960151358 |
| Heart_Failure | ASPC | *NOSTRIN* | TRUE | 2.170529993 |
| Heart_Failure | ASPC | *EFHB* | TRUE | 2.759092833 |
| Heart_Failure | ASPC | *DNALI1* | TRUE | 2.179172155 |
| Heart_Failure | ASPC | *MST1R* | TRUE | 2.824465019 |
| Heart_Failure | ASPC | *SAP30L* | TRUE | 2.011812374 |
| Heart_Failure | ASPC | *GPER1* | TRUE | 2.559817273 |
| Heart_Failure | ASPC | *HSPA12A* | TRUE | 2.084940293 |
| Heart_Failure | ASPC | *PIP4K2C* | TRUE | 2.366313779 |
| Heart_Failure | ASPC | *HDHD2* | TRUE | 2.126866229 |
| Heart_Failure | ASPC | *ZNF23* | TRUE | 2.925406088 |
| Heart_Failure | ASPC | *TRPV3* | TRUE | 2.12775905 |
| Heart_Failure | ASPC | *GSDMA* | TRUE | 2.22104226 |
| Heart_Failure | ASPC | *ZSWIM1* | TRUE | 2.041632888 |
| Heart_Failure | ASPC | *CHRNB1* | TRUE | 2.607453057 |
| Heart_Failure | ASPC | *TMEM133* | TRUE | 2.29278841 |
| Heart_Failure | ASPC | *LGALS4* | TRUE | 1.985356257 |

| | | | | |
|---|---|---|---|---|
| Heart_Failure | ASPC | *PARP15* | TRUE | 2.096117526 |
| Heart_Failure | ASPC | *PAAF1* | TRUE | 2.600860957 |
| Heart_Failure | ASPC | *RPS6KB2* | TRUE | 2.581868252 |
| Heart_Failure | ASPC | *OR4N2* | TRUE | 2.225691556 |
| Heart_Failure | ASPC | *DPY19L2* | TRUE | 2.779047125 |
| Heart_Failure | ASPC | *ZNF454* | TRUE | 2.802565899 |
| Heart_Failure | ASPC | *LINC00314* | TRUE | 2.282086249 |
| Heart_Failure | ASPC | *HLA.DQB1* | TRUE | 4.110523959 |
| Heart_Failure | ASPC | *LBX2* | TRUE | 2.120252474 |
| Heart_Failure | ASPC | *D2HGDH* | TRUE | 2.733138195 |
| Heart_Failure | ASPC | *FKRP* | TRUE | 2.236523966 |
| Heart_Failure | ASPC | *PSMG1* | TRUE | 2.006433151 |
| Heart_Failure | ASPC | *A3GALT2* | TRUE | 2.391020385 |
| Heart_Failure | ASPC | *SIGIRR* | TRUE | 2.292293316 |
| Heart_Failure | ASPC | *C14orf80* | TRUE | 1.97619241 |
| Heart_Failure | ASPC | *GOLGA8R* | TRUE | 2.064828075 |
| Heart_Failure | ASPC | *QRFPR* | TRUE | 2.881672444 |
| Heart_Failure | ASPC | *TMEM232* | TRUE | 2.948374329 |
| Heart_Failure | ASPC | *MRPS21* | TRUE | 2.485022553 |
| Heart_Failure | ASPC | *SLC15A5* | TRUE | 2.014609183 |
| Heart_Failure | ASPC | *WDR5B* | TRUE | 2.232682818 |
| Heart_Failure | ASPC | *PDCD1LG2* | TRUE | 2.873469204 |
| Heart_Failure | ASPC | *RPS26* | TRUE | 2.902225703 |
| Heart_Failure | ASPC | *MICA* | TRUE | 3.332231963 |
| Heart_Failure | ASPC | *ATF6B* | TRUE | 2.390904079 |
| Heart_Failure | ASPC | *ZNF891* | TRUE | 2.10667481 |
| Heart_Failure | ASPC | *CPNE1* | TRUE | 2.540193077 |
| Heart_Failure | ASPC | *OR2A1* | TRUE | 2.850038581 |
| Heart_Failure | ASPC | *ZNF736* | TRUE | 2.712221046 |
| Heart_Failure | ASPC | *ZBTB22* | TRUE | 2.394869611 |
| Heart_Failure | ASPC | *TTC4* | TRUE | 3.082561614 |
| Heart_Failure | ASPC | *LILRA6* | TRUE | 2.36844137 |
| Heart_Failure | ASPC | *MEX3A* | TRUE | 2.013334352 |
| Heart_Failure | ASPC | *MC1R* | TRUE | 2.146435084 |
| Heart_Failure | ASPC | *PWRN1* | TRUE | 2.260170694 |
| Heart_Failure | ASPC | *GTF2H5* | TRUE | 2.674366584 |
| Heart_Failure | b_cell | *TSLP* | TRUE | 3.898578905 |
| Heart_Failure | b_cell | *GSDMA* | TRUE | 2.270986701 |
| Heart_Failure | b_cell | *HLA.DQA2* | TRUE | 6.850664572 |
| Heart_Failure | dendritic_cell | *RBM6* | TRUE | 2.862298577 |
| Heart_Failure | dendritic_cell | *SLC7A9* | TRUE | 2.000039966 |
| Heart_Failure | dendritic_cell | *IL18R1* | TRUE | 2.046401711 |
| Heart_Failure | dendritic_cell | *FLG* | TRUE | 2.825255203 |
| Heart_Failure | dendritic_cell | *TSLP* | TRUE | 2.852377133 |
| Heart_Failure | dendritic_cell | *ROM1* | TRUE | 2.188731939 |
| Heart_Failure | dendritic_cell | *KLHDC9* | TRUE | 2.21724931 |
| Heart_Failure | dendritic_cell | *GSDMA* | TRUE | 5.308672637 |
| Heart_Failure | dendritic_cell | *ZNF454* | TRUE | 2.30794781 |
| Heart_Failure | dendritic_cell | *ZNF385C* | TRUE | 1.984048852 |
| Heart_Failure | dendritic_cell | *RPS26* | TRUE | 2.76495863 |

| | | | | |
|---|---|---|---|---|
| Heart_Failure | dendritic_cell | *PSMB9* | TRUE | 2.906506457 |
| Heart_Failure | endometrium | *TSLP* | TRUE | 4.159514581 |
| Heart_Failure | endometrium | *ROM1* | TRUE | 2.654664394 |
| Heart_Failure | endometrium | *HDHD2* | TRUE | 2.516169456 |
| Heart_Failure | endometrium | *GSDMA* | TRUE | 2.653378162 |
| Heart_Failure | endometrium | *LINC00314* | TRUE | 2.130818436 |
| Heart_Failure | endometrium | *HLA.DQB1* | TRUE | 3.587299127 |
| Heart_Failure | endometrium | *HLA.DQB2* | TRUE | 3.41265777 |
| Heart_Failure | endometrium | *ZNF736* | TRUE | 2.668998611 |
| Heart_Failure | endothelial | *SLC7A9* | TRUE | 2.260449439 |
| Heart_Failure | endothelial | *CDK2AP1* | TRUE | 2.395548332 |
| Heart_Failure | endothelial | *TSLP* | TRUE | 3.148163269 |
| Heart_Failure | endothelial | *FCRL3* | TRUE | 2.389640702 |
| Heart_Failure | endothelial | *MST1R* | TRUE | 3.469881499 |
| Heart_Failure | endothelial | *GSDMA* | TRUE | 4.810977516 |
| Heart_Failure | endothelial | *CHRNB1* | TRUE | 2.52291724 |
| Heart_Failure | endothelial | *HLA.DQB1* | TRUE | 3.350352835 |
| Heart_Failure | endothelial | *RPS26* | TRUE | 2.611286754 |
| Heart_Failure | endothelial | *LILRA6* | TRUE | 2.00724219 |
| Heart_Failure | LEC | *SLC7A9* | TRUE | 2.409006223 |
| Heart_Failure | LEC | *LAMC2* | TRUE | 2.321946986 |
| Heart_Failure | LEC | *PNMT* | TRUE | 2.19839101 |
| Heart_Failure | LEC | *FLG* | TRUE | 2.430723407 |
| Heart_Failure | LEC | *TSLP* | TRUE | 3.992547053 |
| Heart_Failure | LEC | *FGFR4* | TRUE | 2.46739806 |
| Heart_Failure | LEC | *KLHDC9* | TRUE | 2.290575804 |
| Heart_Failure | LEC | *MST1R* | TRUE | 3.004428547 |
| Heart_Failure | LEC | *GSDMA* | TRUE | 2.777852508 |
| Heart_Failure | LEC | *CHRNB1* | TRUE | 2.453757665 |
| Heart_Failure | LEC | *DPY19L2* | TRUE | 2.163916079 |
| Heart_Failure | LEC | *ZNF454* | TRUE | 2.729620981 |
| Heart_Failure | LEC | *LINC00314* | TRUE | 2.139572666 |
| Heart_Failure | LEC | *HLA.DQB1* | TRUE | 4.403319376 |
| Heart_Failure | LEC | *RPS26* | TRUE | 3.162160176 |
| Heart_Failure | LEC | *MICA* | TRUE | 2.675637223 |
| Heart_Failure | macrophage | *SLC7A9* | TRUE | 2.532419314 |
| Heart_Failure | macrophage | *MAST2* | TRUE | 2.048207897 |
| Heart_Failure | macrophage | *SETD1A* | TRUE | 2.21104569 |
| Heart_Failure | macrophage | *SMARCB1* | TRUE | 2.340669692 |
| Heart_Failure | macrophage | *KIF20A* | TRUE | 2.493122154 |
| Heart_Failure | macrophage | *BNIP1* | TRUE | 2.33921991 |
| Heart_Failure | macrophage | *ALDH8A1* | TRUE | 2.077396016 |
| Heart_Failure | macrophage | *ANGPTL6* | TRUE | 2.15543009 |
| Heart_Failure | macrophage | *NMT1* | TRUE | 2.326999813 |
| Heart_Failure | macrophage | *PNMT* | TRUE | 5.31643103 |
| Heart_Failure | macrophage | *EVA1B* | TRUE | 2.933684511 |
| Heart_Failure | macrophage | *FLG* | TRUE | 2.683023821 |
| Heart_Failure | macrophage | *TSLP* | TRUE | 4.518360384 |
| Heart_Failure | macrophage | *ROM1* | TRUE | 2.256484906 |
| Heart_Failure | macrophage | *DPYSL4* | TRUE | 2.124075162 |

| | | | | |
|---|---|---|---|---|
| Heart_Failure | macrophage | *SIM2* | TRUE | 1.960583222 |
| Heart_Failure | macrophage | *FCRL3* | TRUE | 2.203986793 |
| Heart_Failure | macrophage | *FGFR4* | TRUE | 1.985776101 |
| Heart_Failure | macrophage | *KLHDC9* | TRUE | 2.535304969 |
| Heart_Failure | macrophage | *EFHB* | TRUE | 2.679817183 |
| Heart_Failure | macrophage | *HDHD2* | TRUE | 2.591653403 |
| Heart_Failure | macrophage | *CHRNB1* | TRUE | 2.509517184 |
| Heart_Failure | macrophage | *PAAF1* | TRUE | 2.182367415 |
| Heart_Failure | macrophage | *RPS6KB2* | TRUE | 2.192475484 |
| Heart_Failure | macrophage | *LINC00314* | TRUE | 1.984079969 |
| Heart_Failure | macrophage | *TRIM73* | TRUE | 2.119701697 |
| Heart_Failure | macrophage | *FKRP* | TRUE | 2.084606155 |
| Heart_Failure | macrophage | *GOLGA8R* | TRUE | 2.012784394 |
| Heart_Failure | macrophage | *QRFPR* | TRUE | 2.808453054 |
| Heart_Failure | macrophage | *WDR5B* | TRUE | 1.984677592 |
| Heart_Failure | macrophage | *PDCD1LG2* | TRUE | 2.587381506 |
| Heart_Failure | macrophage | *RPS26* | TRUE | 2.942637938 |
| Heart_Failure | macrophage | *HSD17B8* | TRUE | 1.965651283 |
| Heart_Failure | macrophage | *MICA* | TRUE | 2.940135048 |
| Heart_Failure | macrophage | *ATF6B* | TRUE | 1.989648224 |
| Heart_Failure | macrophage | *CPNE1* | TRUE | 2.477075264 |
| Heart_Failure | macrophage | *ZNF736* | TRUE | 2.361354104 |
| Heart_Failure | macrophage | *ZBTB22* | TRUE | 2.442405017 |
| Heart_Failure | macrophage | *HLA.DQA2* | TRUE | 3.624910918 |
| Heart_Failure | macrophage | *TTC4* | TRUE | 2.481236699 |
| Heart_Failure | mast_cell | *PNMT* | TRUE | 2.576738464 |
| Heart_Failure | mast_cell | *EVA1B* | TRUE | 3.395010061 |
| Heart_Failure | mast_cell | *TSLP* | TRUE | 3.090753323 |
| Heart_Failure | mast_cell | *EFHB* | TRUE | 2.527361842 |
| Heart_Failure | mast_cell | *GSDMA* | TRUE | 2.731641155 |
| Heart_Failure | mast_cell | *HLA.DQB1* | TRUE | 5.516632707 |
| Heart_Failure | mesothelium | *RBM6* | TRUE | 3.221842511 |
| Heart_Failure | mesothelium | *CYB561* | TRUE | 2.106496628 |
| Heart_Failure | mesothelium | *CLCA4* | TRUE | 2.216150054 |
| Heart_Failure | mesothelium | *SLC7A9* | TRUE | 2.490195568 |
| Heart_Failure | mesothelium | *LAMC2* | TRUE | 2.950672218 |
| Heart_Failure | mesothelium | *AHRR* | TRUE | 2.099067051 |
| Heart_Failure | mesothelium | *ADD2* | TRUE | 2.109115959 |
| Heart_Failure | mesothelium | *BPIFB2* | TRUE | 2.144605656 |
| Heart_Failure | mesothelium | *OXCT1* | TRUE | 2.346141509 |
| Heart_Failure | mesothelium | *MLF2* | TRUE | 2.049961417 |
| Heart_Failure | mesothelium | *PABPC4* | TRUE | 3.510457096 |
| Heart_Failure | mesothelium | *SMARCB1* | TRUE | 2.177333623 |
| Heart_Failure | mesothelium | *NDUFAF5* | TRUE | 2.499124859 |
| Heart_Failure | mesothelium | *TOX3* | TRUE | 2.545307172 |
| Heart_Failure | mesothelium | *CTSH* | TRUE | 2.643896179 |
| Heart_Failure | mesothelium | *CD33* | TRUE | 2.236931996 |
| Heart_Failure | mesothelium | *RGP1* | TRUE | 1.972895879 |
| Heart_Failure | mesothelium | *KCTD10* | TRUE | 2.755500694 |
| Heart_Failure | mesothelium | *CDK2AP1* | TRUE | 2.532644691 |

| | | | | |
|---|---|---|---|---|
| Heart_Failure | mesothelium | *KIF20A* | TRUE | 3.756807541 |
| Heart_Failure | mesothelium | *BNIP1* | TRUE | 2.215963631 |
| Heart_Failure | mesothelium | *IL18R1* | TRUE | 3.444695559 |
| Heart_Failure | mesothelium | *CTNNAL1* | TRUE | 2.229612447 |
| Heart_Failure | mesothelium | *ITIH5* | TRUE | 3.06343408 |
| Heart_Failure | mesothelium | *PROZ* | TRUE | 2.031682824 |
| Heart_Failure | mesothelium | *ZNF384* | TRUE | 2.037814329 |
| Heart_Failure | mesothelium | *ANGPTL6* | TRUE | 2.445740404 |
| Heart_Failure | mesothelium | *LRRC41* | TRUE | 2.00770317 |
| Heart_Failure | mesothelium | *FIGNL1* | TRUE | 2.441570811 |
| Heart_Failure | mesothelium | *ADAM19* | TRUE | 2.047810148 |
| Heart_Failure | mesothelium | *POM121C* | TRUE | 2.994134495 |
| Heart_Failure | mesothelium | *TSGA10* | TRUE | 2.010372865 |
| Heart_Failure | mesothelium | *ALG10* | TRUE | 3.020772782 |
| Heart_Failure | mesothelium | *FN3KRP* | TRUE | 2.46574147 |
| Heart_Failure | mesothelium | *PNMT* | TRUE | 3.020118672 |
| Heart_Failure | mesothelium | *EVA1B* | TRUE | 2.789405229 |
| Heart_Failure | mesothelium | *NME7* | TRUE | 2.084331452 |
| Heart_Failure | mesothelium | *FLG* | TRUE | 3.060191 |
| Heart_Failure | mesothelium | *GPR17* | TRUE | 2.193000608 |
| Heart_Failure | mesothelium | *TSLP* | TRUE | 4.159175006 |
| Heart_Failure | mesothelium | *SLC18B1* | TRUE | 2.369303517 |
| Heart_Failure | mesothelium | *HSD17B12* | TRUE | 2.250341947 |
| Heart_Failure | mesothelium | *ROM1* | TRUE | 2.803250529 |
| Heart_Failure | mesothelium | *DPYSL4* | TRUE | 2.133437549 |
| Heart_Failure | mesothelium | *POC5* | TRUE | 2.083309081 |
| Heart_Failure | mesothelium | *FZD7* | TRUE | 2.031364247 |
| Heart_Failure | mesothelium | *WHAMM* | TRUE | 2.288061432 |
| Heart_Failure | mesothelium | *DHRS1* | TRUE | 2.033249626 |
| Heart_Failure | mesothelium | *RNF166* | TRUE | 2.231852237 |
| Heart_Failure | mesothelium | *CDC42SE2* | TRUE | 2.003590286 |
| Heart_Failure | mesothelium | *SIM2* | TRUE | 1.984631292 |
| Heart_Failure | mesothelium | *C21orf33* | TRUE | 2.102958283 |
| Heart_Failure | mesothelium | *FCRL3* | TRUE | 2.754065281 |
| Heart_Failure | mesothelium | *FGFR4* | TRUE | 2.981270374 |
| Heart_Failure | mesothelium | *KLHDC9* | TRUE | 2.551125782 |
| Heart_Failure | mesothelium | *EFHB* | TRUE | 2.603771575 |
| Heart_Failure | mesothelium | *RCHY1* | TRUE | 2.133801665 |
| Heart_Failure | mesothelium | *RAD54L2* | TRUE | 2.03948695 |
| Heart_Failure | mesothelium | *ZFYVE19* | TRUE | 2.045027004 |
| Heart_Failure | mesothelium | *C16orf46* | TRUE | 1.968214417 |
| Heart_Failure | mesothelium | *TMEM170A* | TRUE | 2.869026914 |
| Heart_Failure | mesothelium | *KATNAL2* | TRUE | 2.467046357 |
| Heart_Failure | mesothelium | *ZNF23* | TRUE | 2.536311997 |
| Heart_Failure | mesothelium | *TRPV3* | TRUE | 2.082517567 |
| Heart_Failure | mesothelium | *NUDT8* | TRUE | 1.978206962 |
| Heart_Failure | mesothelium | *GSDMA* | TRUE | 2.429416425 |
| Heart_Failure | mesothelium | *DTWD2* | TRUE | 2.053963664 |
| Heart_Failure | mesothelium | *CHRNB1* | TRUE | 2.74223888 |
| Heart_Failure | mesothelium | *DNAJC18* | TRUE | 2.217775573 |

| | | | | |
|---|---|---|---|---|
| Heart_Failure | mesothelium | *TMEM133* | TRUE | 2.381391753 |
| Heart_Failure | mesothelium | *C11orf80* | TRUE | 3.385734662 |
| Heart_Failure | mesothelium | *GPR160* | TRUE | 2.175036302 |
| Heart_Failure | mesothelium | *PAAF1* | TRUE | 2.710554976 |
| Heart_Failure | mesothelium | *SPRYD4* | TRUE | 2.501400636 |
| Heart_Failure | mesothelium | *DPY19L2* | TRUE | 2.388176277 |
| Heart_Failure | mesothelium | *ZNF454* | TRUE | 2.578855826 |
| Heart_Failure | mesothelium | *LINC00314* | TRUE | 2.223323257 |
| Heart_Failure | mesothelium | *HLA.DQB1* | TRUE | 3.580698544 |
| Heart_Failure | mesothelium | *FKRP* | TRUE | 1.994339864 |
| Heart_Failure | mesothelium | *PSMG1* | TRUE | 3.114963353 |
| Heart_Failure | mesothelium | *FAM19A1* | TRUE | 2.048417493 |
| Heart_Failure | mesothelium | *ZNF703* | TRUE | 2.309193604 |
| Heart_Failure | mesothelium | *A3GALT2* | TRUE | 2.41783828 |
| Heart_Failure | mesothelium | *CDK10* | TRUE | 2.30163432 |
| Heart_Failure | mesothelium | *GOLGA8R* | TRUE | 2.073843291 |
| Heart_Failure | mesothelium | *TMEM232* | TRUE | 2.209330414 |
| Heart_Failure | mesothelium | *MRPS21* | TRUE | 2.175337788 |
| Heart_Failure | mesothelium | *ZNF385C* | TRUE | 2.200412843 |
| Heart_Failure | mesothelium | *ZP3* | TRUE | 2.028287048 |
| Heart_Failure | mesothelium | *PDCD1LG2* | TRUE | 2.609657488 |
| Heart_Failure | mesothelium | *RPS26* | TRUE | 3.158214627 |
| Heart_Failure | mesothelium | *TATDN3* | TRUE | 2.158084832 |
| Heart_Failure | mesothelium | *MICA* | TRUE | 2.887538962 |
| Heart_Failure | mesothelium | *ZNF155* | TRUE | 2.141033315 |
| Heart_Failure | mesothelium | *SLFN12L* | TRUE | 2.192551366 |
| Heart_Failure | mesothelium | *CPNE1* | TRUE | 2.99412135 |
| Heart_Failure | mesothelium | *OR2A1* | TRUE | 2.537173299 |
| Heart_Failure | mesothelium | *ZNF736* | TRUE | 2.137211159 |
| Heart_Failure | mesothelium | *ZBTB22* | TRUE | 2.122415117 |
| Heart_Failure | mesothelium | *TTC4* | TRUE | 3.511751741 |
| Heart_Failure | mesothelium | *LILRA6* | TRUE | 2.689710888 |
| Heart_Failure | mesothelium | *MEX3A* | TRUE | 2.162229542 |
| Heart_Failure | mesothelium | *GTF2H5* | TRUE | 2.177880622 |
| Heart_Failure | monocyte | *GSDMA* | TRUE | 2.016396854 |
| Heart_Failure | monocyte | *HLA.DQB1* | TRUE | 4.962505119 |
| Heart_Failure | monocyte | *HLA.DQA2* | TRUE | 2.454520512 |
| Heart_Failure | nk_cell | *EVA1B* | TRUE | 2.876060026 |
| Heart_Failure | nk_cell | *TSLP* | TRUE | 2.260108876 |
| Heart_Failure | nk_cell | *KLHDC9* | TRUE | 2.103994415 |
| Heart_Failure | nk_cell | *GSDMA* | TRUE | 2.53309544 |
| Heart_Failure | nk_cell | *GPRIN1* | TRUE | 2.114271457 |
| Heart_Failure | nk_cell | *CHRNB1* | TRUE | 2.803839469 |
| Heart_Failure | nk_cell | *ZNF454* | TRUE | 2.582543778 |
| Heart_Failure | nk_cell | *HLA.DQB1* | TRUE | 5.01288932 |
| Heart_Failure | nk_cell | *MICB* | TRUE | 2.137545957 |
| Heart_Failure | pericyte | *RBM6* | TRUE | 2.452426973 |
| Heart_Failure | pericyte | *PNMT* | TRUE | 2.770873292 |
| Heart_Failure | pericyte | *FLG* | TRUE | 2.821562717 |
| Heart_Failure | pericyte | *TSLP* | TRUE | 4.334520143 |

| | | | | |
|---|---|---|---|---|
| Heart_Failure | pericyte | *DPYSL4* | TRUE | 2.040610402 |
| Heart_Failure | pericyte | *SIM2* | TRUE | 2.061909918 |
| Heart_Failure | pericyte | *FCRL3* | TRUE | 2.085886021 |
| Heart_Failure | pericyte | *FGFR4* | TRUE | 1.979706133 |
| Heart_Failure | pericyte | *EFHB* | TRUE | 2.395052992 |
| Heart_Failure | pericyte | *HDHD2* | TRUE | 2.095980804 |
| Heart_Failure | pericyte | *GSDMA* | TRUE | 3.15602552 |
| Heart_Failure | pericyte | *CHRNB1* | TRUE | 2.765917606 |
| Heart_Failure | pericyte | *ZNF454* | TRUE | 2.205288204 |
| Heart_Failure | pericyte | *HLA.DQB1* | TRUE | 4.261080535 |
| Heart_Failure | pericyte | *LBX2* | TRUE | 2.186225188 |
| Heart_Failure | pericyte | *PDCD1LG2* | TRUE | 1.967806679 |
| Heart_Failure | pericyte | *RPS26* | TRUE | 3.03730821 |
| Heart_Failure | pericyte | *CPNE1* | TRUE | 2.442457543 |
| Heart_Failure | pericyte | *ZBTB22* | TRUE | 2.483699118 |
| Heart_Failure | pericyte | *LILRA6* | TRUE | 2.064809456 |
| Heart_Failure | SMC | *RBM6* | TRUE | 3.5425593 |
| Heart_Failure | SMC | *ADAM11* | TRUE | 2.230573437 |
| Heart_Failure | SMC | *INO80B* | TRUE | 1.96463558 |
| Heart_Failure | SMC | *ALG10* | TRUE | 1.965156291 |
| Heart_Failure | SMC | *FLG* | TRUE | 1.999751448 |
| Heart_Failure | SMC | *TSLP* | TRUE | 2.296157806 |
| Heart_Failure | SMC | *ROM1* | TRUE | 2.264983105 |
| Heart_Failure | SMC | *DPYSL4* | TRUE | 2.082322165 |
| Heart_Failure | SMC | *FCRL3* | TRUE | 2.307129699 |
| Heart_Failure | SMC | *KLHDC9* | TRUE | 2.792228903 |
| Heart_Failure | SMC | *GSDMA* | TRUE | 3.515322819 |
| Heart_Failure | SMC | *PAAF1* | TRUE | 2.083185541 |
| Heart_Failure | SMC | *HLA.DQB1* | TRUE | 4.560790967 |
| Heart_Failure | SMC | *PDCD1LG2* | TRUE | 2.065741041 |
| Heart_Failure | SMC | *MICA* | TRUE | 2.811040122 |
| Heart_Failure | SMC | *CPNE1* | TRUE | 2.631082159 |
| Heart_Failure | SMC | *ZBTB22* | TRUE | 2.563814188 |
| Heart_Failure | t_cell | *SLC7A9* | TRUE | 2.415374168 |
| Heart_Failure | t_cell | *KIF20A* | TRUE | 2.855633402 |
| Heart_Failure | t_cell | *ALDH8A1* | TRUE | 1.995055764 |
| Heart_Failure | t_cell | *NMT1* | TRUE | 2.353751294 |
| Heart_Failure | t_cell | *EVA1B* | TRUE | 3.355637471 |
| Heart_Failure | t_cell | *FLG* | TRUE | 2.385178999 |
| Heart_Failure | t_cell | *TSLP* | TRUE | 2.454209 |
| Heart_Failure | t_cell | *DPYSL4* | TRUE | 1.971371077 |
| Heart_Failure | t_cell | *SIM2* | TRUE | 2.025480735 |
| Heart_Failure | t_cell | *EFHB* | TRUE | 2.245699667 |
| Heart_Failure | t_cell | *RNF123* | TRUE | 1.993468197 |
| Heart_Failure | t_cell | *HDHD2* | TRUE | 2.567370427 |
| Heart_Failure | t_cell | *ZNF23* | TRUE | 2.416410173 |
| Heart_Failure | t_cell | *GSDMA* | TRUE | 2.875359446 |
| Heart_Failure | t_cell | *CHRNB1* | TRUE | 2.416645608 |
| Heart_Failure | t_cell | *PAAF1* | TRUE | 1.996805213 |
| Heart_Failure | t_cell | *HLA.DQB1* | TRUE | 4.435107578 |

| | | | | |
|---|---|---|---|---|
| Heart_Failure | t_cell | *LBX2* | TRUE | 2.076232588 |
| Heart_Failure | t_cell | *RPS26* | TRUE | 3.247469366 |
| Heart_Failure | t_cell | *MICA* | TRUE | 2.236690086 |
| Heart_Failure | t_cell | *CPNE1* | TRUE | 2.122053855 |
| CAD_admission_freq | adipocyte | *ZNF200* | TRUE | 3.0341879 |
| CAD_admission_freq | adipocyte | *LAMC2* | TRUE | 2.046419116 |
| CAD_admission_freq | adipocyte | *GNA15* | TRUE | 4.123063719 |
| CAD_admission_freq | adipocyte | *NAALAD2* | TRUE | 2.055434266 |
| CAD_admission_freq | adipocyte | *SLC7A5* | TRUE | 2.061024632 |
| CAD_admission_freq | adipocyte | *EPHX3* | TRUE | 2.231185907 |
| CAD_admission_freq | adipocyte | *CCL24* | TRUE | 2.185277474 |
| CAD_admission_freq | adipocyte | *DNAJC12* | TRUE | 2.029139222 |
| CAD_admission_freq | adipocyte | *PEX12* | TRUE | 2.111799899 |
| CAD_admission_freq | adipocyte | *TAS2R10* | TRUE | 2.898793682 |
| CAD_admission_freq | adipocyte | *TOX2* | TRUE | 2.144678676 |
| CAD_admission_freq | adipocyte | *WDFY4* | TRUE | 2.40725209 |
| CAD_admission_freq | adipocyte | *ENPEP* | TRUE | 2.596832402 |
| CAD_admission_freq | adipocyte | *ADPRH* | TRUE | 2.487361989 |
| CAD_admission_freq | adipocyte | *TMEM236* | TRUE | 2.629599972 |
| CAD_admission_freq | adipocyte | *PROM2* | TRUE | 2.081436564 |
| CAD_admission_freq | adipocyte | *POLR2J3* | TRUE | 2.857090954 |
| CAD_admission_freq | adipocyte | *TTLL6* | TRUE | 2.182592713 |
| CAD_admission_freq | adipocyte | *SMPDL3A* | TRUE | 2.204385488 |
| CAD_admission_freq | adipocyte | *PAAF1* | TRUE | 2.948931768 |
| CAD_admission_freq | adipocyte | *B3GNT3* | TRUE | 2.094658537 |
| CAD_admission_freq | adipocyte | *PPP1R27* | TRUE | 3.082048292 |
| CAD_admission_freq | adipocyte | *AGAP9* | TRUE | 2.486530794 |
| CAD_admission_freq | adipocyte | *MICB* | TRUE | 2.39887364 |
| CAD_admission_freq | adipocyte | *AC084018.1* | TRUE | 2.013355713 |
| CAD_admission_freq | adipocyte | *ARHGAP19* | TRUE | 2.003418702 |
| CAD_admission_freq | adipocyte | *ACSM4* | TRUE | 2.114812267 |
| CAD_admission_freq | adipocyte | *ZNF883* | TRUE | 2.566464576 |
| CAD_admission_freq | adipocyte | *JMJD7* | TRUE | 2.040271433 |
| CAD_admission_freq | adipocyte | *RNASE4* | TRUE | 2.129617423 |
| CAD_admission_freq | adipocyte | *AC004466.1* | TRUE | 2.905662374 |
| CAD_admission_freq | ASPC | *ZNF200* | TRUE | 3.051659672 |
| CAD_admission_freq | ASPC | *RNASET2* | TRUE | 1.963994531 |
| CAD_admission_freq | ASPC | *LAMC2* | TRUE | 2.15339074 |
| CAD_admission_freq | ASPC | *GNA15* | TRUE | 3.417502461 |
| CAD_admission_freq | ASPC | *NAALAD2* | TRUE | 2.123690359 |
| CAD_admission_freq | ASPC | *PPP1R37* | TRUE | 1.961337949 |
| CAD_admission_freq | ASPC | *EPHX3* | TRUE | 2.286274869 |
| CAD_admission_freq | ASPC | *CCL24* | TRUE | 2.106481513 |
| CAD_admission_freq | ASPC | *DNAJC12* | TRUE | 2.214099809 |
| CAD_admission_freq | ASPC | *PEX12* | TRUE | 2.27958199 |
| CAD_admission_freq | ASPC | *TAS2R10* | TRUE | 2.675164099 |
| CAD_admission_freq | ASPC | *TOX2* | TRUE | 2.341583818 |
| CAD_admission_freq | ASPC | *WDFY4* | TRUE | 2.809519033 |
| CAD_admission_freq | ASPC | *ENPEP* | TRUE | 2.701059206 |
| CAD_admission_freq | ASPC | *ADPRH* | TRUE | 2.43174877 |

| | | | | |
|---|---|---|---|---|
| CAD_admission_freq | ASPC | *TMEM236* | TRUE | 2.658569278 |
| CAD_admission_freq | ASPC | *PROM2* | TRUE | 1.979142324 |
| CAD_admission_freq | ASPC | *CBR1* | TRUE | 1.97530937 |
| CAD_admission_freq | ASPC | *GPER1* | TRUE | 2.015114744 |
| CAD_admission_freq | ASPC | *NUDT13* | TRUE | 2.134823933 |
| CAD_admission_freq | ASPC | *DCTN5* | TRUE | 1.972682819 |
| CAD_admission_freq | ASPC | *POLR2J3* | TRUE | 2.809109235 |
| CAD_admission_freq | ASPC | *PRNP* | TRUE | 1.984491343 |
| CAD_admission_freq | ASPC | *SMPDL3A* | TRUE | 2.332347845 |
| CAD_admission_freq | ASPC | *SEZ6L2* | TRUE | 2.233505743 |
| CAD_admission_freq | ASPC | *PAAF1* | TRUE | 3.059907834 |
| CAD_admission_freq | ASPC | *B3GNT3* | TRUE | 2.139446142 |
| CAD_admission_freq | ASPC | *CLEC4G* | TRUE | 2.141742203 |
| CAD_admission_freq | ASPC | *PPP1R27* | TRUE | 3.185415804 |
| CAD_admission_freq | ASPC | *ZFP62* | TRUE | 2.263735202 |
| CAD_admission_freq | ASPC | *AGAP9* | TRUE | 2.645260556 |
| CAD_admission_freq | ASPC | *INPP5B* | TRUE | 2.016935321 |
| CAD_admission_freq | ASPC | *MICB* | TRUE | 2.383427775 |
| CAD_admission_freq | ASPC | *SERF1B* | TRUE | 2.092635909 |
| CAD_admission_freq | ASPC | *AC084018.1* | TRUE | 1.994400221 |
| CAD_admission_freq | ASPC | *ARHGAP19* | TRUE | 2.32540185 |
| CAD_admission_freq | ASPC | *ACSM4* | TRUE | 2.024979282 |
| CAD_admission_freq | ASPC | *TMEM242* | TRUE | 1.972823948 |
| CAD_admission_freq | ASPC | *ZNF883* | TRUE | 2.554363266 |
| CAD_admission_freq | ASPC | *JMJD7* | TRUE | 2.15434909 |
| CAD_admission_freq | ASPC | *RNASE4* | TRUE | 1.991580471 |
| CAD_admission_freq | ASPC | *AC004466.1* | TRUE | 2.813313115 |
| CAD_admission_freq | ASPC | *DOC2B* | TRUE | 2.101616256 |
| CAD_admission_freq | dendritic_cell | *ZNF200* | TRUE | 2.476083176 |
| CAD_admission_freq | dendritic_cell | *EPHX3* | TRUE | 1.992856387 |
| CAD_admission_freq | dendritic_cell | *TAS2R10* | TRUE | 2.740876545 |
| CAD_admission_freq | dendritic_cell | *SPTBN5* | TRUE | 2.069112773 |
| CAD_admission_freq | dendritic_cell | *DPYSL4* | TRUE | 1.984005563 |
| CAD_admission_freq | dendritic_cell | *POLR2J3* | TRUE | 2.665742903 |
| CAD_admission_freq | dendritic_cell | *PAAF1* | TRUE | 2.321236442 |
| CAD_admission_freq | dendritic_cell | *ZNF575* | TRUE | 2.011328502 |
| CAD_admission_freq | dendritic_cell | *AC004466.1* | TRUE | 2.459597271 |
| CAD_admission_freq | endometrium | *TAS2R10* | TRUE | 2.870032441 |
| CAD_admission_freq | endometrium | *PPP1R27* | TRUE | 1.983305317 |
| CAD_admission_freq | endothelial | *GNA15* | TRUE | 3.329026416 |
| CAD_admission_freq | endothelial | *EPHX3* | TRUE | 2.027179645 |
| CAD_admission_freq | endothelial | *TAS2R10* | TRUE | 2.394104387 |
| CAD_admission_freq | endothelial | *PROM2* | TRUE | 2.13203202 |
| CAD_admission_freq | endothelial | *PAAF1* | TRUE | 2.105956561 |
| CAD_admission_freq | endothelial | *MICB* | TRUE | 2.420769143 |
| CAD_admission_freq | LEC | *ZNF200* | TRUE | 2.950100381 |
| CAD_admission_freq | LEC | *GNA15* | TRUE | 3.185129233 |
| CAD_admission_freq | LEC | *TAS2R10* | TRUE | 3.033096482 |
| CAD_admission_freq | LEC | *WDFY4* | TRUE | 2.288244172 |
| CAD_admission_freq | LEC | *ADPRH* | TRUE | 2.06897996 |

| | | | | |
|---|---|---|---|---|
| CAD_admission_freq | LEC | *PROM2* | TRUE | 1.977278325 |
| CAD_admission_freq | LEC | *POLR2J3* | TRUE | 2.717901908 |
| CAD_admission_freq | LEC | *PAAF1* | TRUE | 2.565307037 |
| CAD_admission_freq | LEC | *B3GNT3* | TRUE | 2.049838014 |
| CAD_admission_freq | LEC | *PPP1R27* | TRUE | 3.125898965 |
| CAD_admission_freq | LEC | *MICB* | TRUE | 2.307368738 |
| CAD_admission_freq | LEC | *ZNF883* | TRUE | 1.985075271 |
| CAD_admission_freq | LEC | *AC004466.1* | TRUE | 2.344926724 |
| CAD_admission_freq | macrophage | *ZNF200* | TRUE | 2.932291875 |
| CAD_admission_freq | macrophage | *EPHX3* | TRUE | 2.139949192 |
| CAD_admission_freq | macrophage | *TAS2R10* | TRUE | 2.713767401 |
| CAD_admission_freq | macrophage | *TOX2* | TRUE | 2.042203808 |
| CAD_admission_freq | macrophage | *ENPEP* | TRUE | 2.019859816 |
| CAD_admission_freq | macrophage | *POLR2J3* | TRUE | 2.706983106 |
| CAD_admission_freq | macrophage | *PAAF1* | TRUE | 2.99756031 |
| CAD_admission_freq | macrophage | *PPP1R27* | TRUE | 2.317819328 |
| CAD_admission_freq | macrophage | *AGAP9* | TRUE | 2.347993258 |
| CAD_admission_freq | macrophage | *MICB* | TRUE | 2.469960826 |
| CAD_admission_freq | macrophage | *SERF1B* | TRUE | 2.046385161 |
| CAD_admission_freq | macrophage | *ARHGAP19* | TRUE | 2.103454294 |
| CAD_admission_freq | macrophage | *JMJD7* | TRUE | 2.02521718 |
| CAD_admission_freq | macrophage | *AC004466.1* | TRUE | 2.806581167 |
| CAD_admission_freq | macrophage | *DOC2B* | TRUE | 1.990986747 |
| CAD_admission_freq | mast_cell | *ZNF200* | TRUE | 2.465132261 |
| CAD_admission_freq | mast_cell | *GNA15* | TRUE | 3.725660203 |
| CAD_admission_freq | mast_cell | *EPHX3* | TRUE | 1.974788486 |
| CAD_admission_freq | mast_cell | *TAS2R10* | TRUE | 2.770589085 |
| CAD_admission_freq | mast_cell | *TMEM236* | TRUE | 2.386512017 |
| CAD_admission_freq | mast_cell | *POLR2J3* | TRUE | 2.665457576 |
| CAD_admission_freq | mast_cell | *ZNF575* | TRUE | 2.158847643 |
| CAD_admission_freq | mast_cell | *GPAT2* | TRUE | 2.06016334 |
| CAD_admission_freq | mast_cell | *MICB* | TRUE | 2.320382089 |
| CAD_admission_freq | mast_cell | *JMJD7* | TRUE | 2.081331181 |
| CAD_admission_freq | mast_cell | *AC004466.1* | TRUE | 2.364962609 |
| CAD_admission_freq | mesothelium | *SPATA20* | TRUE | 2.04357797 |
| CAD_admission_freq | mesothelium | *ZNF200* | TRUE | 3.076645065 |
| CAD_admission_freq | mesothelium | *RNASET2* | TRUE | 2.03419978 |
| CAD_admission_freq | mesothelium | *CHPF2* | TRUE | 1.9621745 |
| CAD_admission_freq | mesothelium | *PNKP* | TRUE | 2.07407616 |
| CAD_admission_freq | mesothelium | *LAMC2* | TRUE | 2.06011015 |
| CAD_admission_freq | mesothelium | *GNA15* | TRUE | 3.47205811 |
| CAD_admission_freq | mesothelium | *NAALAD2* | TRUE | 2.360221345 |
| CAD_admission_freq | mesothelium | *SLC7A5* | TRUE | 2.030413663 |
| CAD_admission_freq | mesothelium | *PPP1R37* | TRUE | 2.044422151 |
| CAD_admission_freq | mesothelium | *EPHX3* | TRUE | 2.242727824 |
| CAD_admission_freq | mesothelium | *CCL24* | TRUE | 2.222763144 |
| CAD_admission_freq | mesothelium | *LIPA* | TRUE | 1.980804271 |
| CAD_admission_freq | mesothelium | *DNAJC12* | TRUE | 1.979663044 |
| CAD_admission_freq | mesothelium | *PEX12* | TRUE | 2.351946497 |
| CAD_admission_freq | mesothelium | *TAS2R10* | TRUE | 2.64678314 |

| | | | | |
|---|---|---|---|---|
| CAD_admission_freq | mesothelium | *TOX2* | TRUE | 2.300669233 |
| CAD_admission_freq | mesothelium | *L3HYPDH* | TRUE | 1.989991171 |
| CAD_admission_freq | mesothelium | *WDFY4* | TRUE | 2.898415915 |
| CAD_admission_freq | mesothelium | *ENPEP* | TRUE | 2.617601792 |
| CAD_admission_freq | mesothelium | *ADPRH* | TRUE | 2.51712726 |
| CAD_admission_freq | mesothelium | *TMEM236* | TRUE | 2.676406134 |
| CAD_admission_freq | mesothelium | *PROM2* | TRUE | 2.003801397 |
| CAD_admission_freq | mesothelium | *GPER1* | TRUE | 2.111148922 |
| CAD_admission_freq | mesothelium | *NUDT13* | TRUE | 2.165109418 |
| CAD_admission_freq | mesothelium | *DCTN5* | TRUE | 2.0998912 |
| CAD_admission_freq | mesothelium | *POLR2J3* | TRUE | 2.84341621 |
| CAD_admission_freq | mesothelium | *TTLL6* | TRUE | 2.131662046 |
| CAD_admission_freq | mesothelium | *SMPDL3A* | TRUE | 2.266349303 |
| CAD_admission_freq | mesothelium | *PAAF1* | TRUE | 3.07063268 |
| CAD_admission_freq | mesothelium | *B3GNT3* | TRUE | 2.121005811 |
| CAD_admission_freq | mesothelium | *CLEC4G* | TRUE | 2.274765078 |
| CAD_admission_freq | mesothelium | *PPP1R27* | TRUE | 3.052773039 |
| CAD_admission_freq | mesothelium | *RNPC3* | TRUE | 2.196597121 |
| CAD_admission_freq | mesothelium | *ZFP62* | TRUE | 2.609967784 |
| CAD_admission_freq | mesothelium | *AGAP9* | TRUE | 2.551358774 |
| CAD_admission_freq | mesothelium | *INPP5B* | TRUE | 1.977096103 |
| CAD_admission_freq | mesothelium | *MICB* | TRUE | 2.391650268 |
| CAD_admission_freq | mesothelium | *SERF1B* | TRUE | 2.033445181 |
| CAD_admission_freq | mesothelium | *AC084018.1* | TRUE | 2.090845338 |
| CAD_admission_freq | mesothelium | *ARHGAP19* | TRUE | 2.242926842 |
| CAD_admission_freq | mesothelium | *ACSM4* | TRUE | 2.200594232 |
| CAD_admission_freq | mesothelium | *TMEM242* | TRUE | 1.985129414 |
| CAD_admission_freq | mesothelium | *ZNF883* | TRUE | 2.42382764 |
| CAD_admission_freq | mesothelium | *JMJD7* | TRUE | 2.001092734 |
| CAD_admission_freq | mesothelium | *RNASE4* | TRUE | 2.317689413 |
| CAD_admission_freq | mesothelium | *AC004466.1* | TRUE | 2.848915962 |
| CAD_admission_freq | mesothelium | *DOC2B* | TRUE | 2.143250368 |
| CAD_admission_freq | monocyte | *TAS2R10* | TRUE | 2.414446834 |
| CAD_admission_freq | monocyte | *ENPEP* | TRUE | 2.420856402 |
| CAD_admission_freq | monocyte | *PPP1R27* | TRUE | 2.605043582 |
| CAD_admission_freq | monocyte | *MICB* | TRUE | 2.323064512 |
| CAD_admission_freq | nk_cell | *ZNF200* | TRUE | 2.694214175 |
| CAD_admission_freq | nk_cell | *TAS2R10* | TRUE | 2.657182242 |
| CAD_admission_freq | nk_cell | *WDFY4* | TRUE | 2.070364369 |
| CAD_admission_freq | nk_cell | *PROM2* | TRUE | 2.604276067 |
| CAD_admission_freq | nk_cell | *POLR2J3* | TRUE | 2.2727187 |
| CAD_admission_freq | nk_cell | *PAAF1* | TRUE | 2.670278013 |
| CAD_admission_freq | nk_cell | *MICB* | TRUE | 2.658667525 |
| CAD_admission_freq | nk_cell | *JMJD7* | TRUE | 1.98914701 |
| CAD_admission_freq | nk_cell | *AC004466.1* | TRUE | 2.783936474 |
| CAD_admission_freq | pericyte | *ZNF200* | TRUE | 2.772551076 |
| CAD_admission_freq | pericyte | *GNA15* | TRUE | 3.571452235 |
| CAD_admission_freq | pericyte | *EPHX3* | TRUE | 2.18183592 |
| CAD_admission_freq | pericyte | *TAS2R10* | TRUE | 2.577283194 |
| CAD_admission_freq | pericyte | *TOX2* | TRUE | 1.993780897 |

| | | | | |
|---|---|---|---|---|
| CAD_admission_freq | pericyte | *WDFY4* | TRUE | 2.493543795 |
| CAD_admission_freq | pericyte | *ADPRH* | TRUE | 2.155678371 |
| CAD_admission_freq | pericyte | *POLR2J3* | TRUE | 2.285700264 |
| CAD_admission_freq | pericyte | *SMPDL3A* | TRUE | 1.991440669 |
| CAD_admission_freq | pericyte | *PAAF1* | TRUE | 2.707982503 |
| CAD_admission_freq | pericyte | *PPP1R27* | TRUE | 2.315324943 |
| CAD_admission_freq | pericyte | *AGAP9* | TRUE | 2.184833724 |
| CAD_admission_freq | pericyte | *MICB* | TRUE | 2.281238136 |
| CAD_admission_freq | pericyte | *SERF1B* | TRUE | 2.001103237 |
| CAD_admission_freq | pericyte | *JMJD7* | TRUE | 2.067190152 |
| CAD_admission_freq | pericyte | *AC004466.1* | TRUE | 2.593534481 |
| CAD_admission_freq | SMC | *ZNF200* | TRUE | 2.846113867 |
| CAD_admission_freq | SMC | *GNA15* | TRUE | 3.56779177 |
| CAD_admission_freq | SMC | *TAS2R10* | TRUE | 2.701998442 |
| CAD_admission_freq | SMC | *WDFY4* | TRUE | 1.960613948 |
| CAD_admission_freq | SMC | *ADPRH* | TRUE | 2.103553378 |
| CAD_admission_freq | SMC | *TMEM236* | TRUE | 2.462447073 |
| CAD_admission_freq | SMC | *POLR2J3* | TRUE | 2.40404501 |
| CAD_admission_freq | SMC | *PAAF1* | TRUE | 2.88267979 |
| CAD_admission_freq | SMC | *ZNF575* | TRUE | 2.030394057 |
| CAD_admission_freq | SMC | *PPP1R27* | TRUE | 2.515041341 |
| CAD_admission_freq | SMC | *MICB* | TRUE | 2.45771392 |
| CAD_admission_freq | SMC | *JMJD7* | TRUE | 2.070535676 |
| CAD_admission_freq | SMC | *DOC2B* | TRUE | 2.09920863 |
| CAD_admission_freq | t_cell | *ZNF200* | TRUE | 2.519115673 |
| CAD_admission_freq | t_cell | *GNA15* | TRUE | 3.528552556 |
| CAD_admission_freq | t_cell | *EPHX3* | TRUE | 2.095666384 |
| CAD_admission_freq | t_cell | *CCL24* | TRUE | 2.044613792 |
| CAD_admission_freq | t_cell | *TAS2R10* | TRUE | 2.107355427 |
| CAD_admission_freq | t_cell | *WDFY4* | TRUE | 2.33647352 |
| CAD_admission_freq | t_cell | *ENPEP* | TRUE | 1.989464246 |
| CAD_admission_freq | t_cell | *TMEM236* | TRUE | 2.091928419 |
| CAD_admission_freq | t_cell | *POLR2J3* | TRUE | 2.439315603 |
| CAD_admission_freq | t_cell | *SEZ6L2* | TRUE | 1.969105818 |
| CAD_admission_freq | t_cell | *PAAF1* | TRUE | 2.823779947 |
| CAD_admission_freq | t_cell | *ZNF575* | TRUE | 2.1769168 |
| CAD_admission_freq | t_cell | *B3GNT3* | TRUE | 2.123251177 |
| CAD_admission_freq | t_cell | *CLEC4G* | TRUE | 2.120181965 |
| CAD_admission_freq | t_cell | *MICB* | TRUE | 2.553762985 |
| CAD_admission_freq | t_cell | *ACSM4* | TRUE | 2.074910122 |
| CAD_admission_freq | t_cell | *JMJD7* | TRUE | 2.03011916 |
| CAD_admission_freq | t_cell | *AC004466.1* | TRUE | 2.513903623 |
| CAD_mortality | adipocyte | *BAK1* | TRUE | 1.983031575 |
| CAD_mortality | adipocyte | *ARHGEF5* | TRUE | 2.062229679 |
| CAD_mortality | adipocyte | *OPN3* | TRUE | 2.203990173 |
| CAD_mortality | adipocyte | *CDC6* | TRUE | 2.103946287 |
| CAD_mortality | adipocyte | *CYB5R3* | TRUE | 2.015080003 |
| CAD_mortality | adipocyte | *PPP1R37* | TRUE | 1.990922829 |
| CAD_mortality | adipocyte | *MTRF1L* | TRUE | 2.446584132 |
| CAD_mortality | adipocyte | *IRF6* | TRUE | 2.283838458 |

| | | | | |
|---|---|---|---|---|
| CAD_mortality | adipocyte | *OPN4* | TRUE | 2.164757524 |
| CAD_mortality | adipocyte | *LYG1* | TRUE | 2.318197259 |
| CAD_mortality | adipocyte | *ZNF660* | TRUE | 1.985063464 |
| CAD_mortality | adipocyte | *POMZP3* | TRUE | 2.531018167 |
| CAD_mortality | adipocyte | *POLE3* | TRUE | 2.004067178 |
| CAD_mortality | adipocyte | *ROM1* | TRUE | 2.414586322 |
| CAD_mortality | adipocyte | *UVSSA* | TRUE | 2.049204498 |
| CAD_mortality | adipocyte | *TRIM66* | TRUE | 2.328409132 |
| CAD_mortality | adipocyte | *ENDOG* | TRUE | 2.037018499 |
| CAD_mortality | adipocyte | *RAB43* | TRUE | 2.521850047 |
| CAD_mortality | adipocyte | *GOLGA6L4* | TRUE | 2.716687086 |
| CAD_mortality | adipocyte | *C11orf87* | TRUE | 2.324297454 |
| CAD_mortality | adipocyte | *TBKBP1* | TRUE | 1.98216485 |
| CAD_mortality | adipocyte | *GOLGA8B* | TRUE | 2.000876628 |
| CAD_mortality | adipocyte | *LINC00937* | TRUE | 2.046479827 |
| CAD_mortality | adipocyte | *TDGF1* | TRUE | 2.314582255 |
| CAD_mortality | adipocyte | *LEFTY1* | TRUE | 2.247858496 |
| CAD_mortality | ASPC | *BAK1* | TRUE | 2.030363319 |
| CAD_mortality | ASPC | *ARHGEF5* | TRUE | 2.157653046 |
| CAD_mortality | ASPC | *OPN3* | TRUE | 2.310398336 |
| CAD_mortality | ASPC | *SYNJ2* | TRUE | 2.144529111 |
| CAD_mortality | ASPC | *CDC6* | TRUE | 2.200653082 |
| CAD_mortality | ASPC | *CYB5R3* | TRUE | 2.094959475 |
| CAD_mortality | ASPC | *TNFRSF10A* | TRUE | 2.131618004 |
| CAD_mortality | ASPC | *PPP1R37* | TRUE | 2.098471052 |
| CAD_mortality | ASPC | *MTRF1L* | TRUE | 2.439147159 |
| CAD_mortality | ASPC | *IRF6* | TRUE | 2.299290499 |
| CAD_mortality | ASPC | *OPN4* | TRUE | 2.090224715 |
| CAD_mortality | ASPC | *GTF2F1* | TRUE | 2.14952767 |
| CAD_mortality | ASPC | *NAAA* | TRUE | 2.11509565 |
| CAD_mortality | ASPC | *CIAO1* | TRUE | 2.023354142 |
| CAD_mortality | ASPC | *LYG1* | TRUE | 2.339137072 |
| CAD_mortality | ASPC | *POMZP3* | TRUE | 2.842935111 |
| CAD_mortality | ASPC | *POLE3* | TRUE | 2.183692843 |
| CAD_mortality | ASPC | *ROM1* | TRUE | 2.503408112 |
| CAD_mortality | ASPC | *GBP5* | TRUE | 1.971820936 |
| CAD_mortality | ASPC | *STEAP2* | TRUE | 1.989075738 |
| CAD_mortality | ASPC | *LRRC56* | TRUE | 1.960666127 |
| CAD_mortality | ASPC | *DCD* | TRUE | 2.029376029 |
| CAD_mortality | ASPC | *UVSSA* | TRUE | 2.33275295 |
| CAD_mortality | ASPC | *TRIM66* | TRUE | 2.240976548 |
| CAD_mortality | ASPC | *ACSM1* | TRUE | 2.027633279 |
| CAD_mortality | ASPC | *ENDOG* | TRUE | 2.165811513 |
| CAD_mortality | ASPC | *IRS1* | TRUE | 2.025871251 |
| CAD_mortality | ASPC | *ZNF703* | TRUE | 2.072612044 |
| CAD_mortality | ASPC | *GOLGA6L4* | TRUE | 2.777578164 |
| CAD_mortality | ASPC | *C11orf87* | TRUE | 2.319336636 |
| CAD_mortality | ASPC | *ZNF100* | TRUE | 2.118761892 |
| CAD_mortality | ASPC | *RPS6KL1* | TRUE | 2.051461541 |
| CAD_mortality | ASPC | *GOLGA8B* | TRUE | 2.152170999 |

| | | | | |
|---|---|---|---|---|
| CAD_mortality | ASPC | *LINC00937* | TRUE | 1.995372664 |
| CAD_mortality | ASPC | *TDGF1* | TRUE | 2.362459731 |
| CAD_mortality | ASPC | *LEFTY1* | TRUE | 2.514879715 |
| CAD_mortality | ASPC | *TMEM158* | TRUE | 2.027394727 |
| CAD_mortality | b_cell | *IRF6* | TRUE | 1.990434098 |
| CAD_mortality | b_cell | *RAB43* | TRUE | 3.23392267 |
| CAD_mortality | dendritic_cell | *TNFRSF10A* | TRUE | 2.004646502 |
| CAD_mortality | dendritic_cell | *IRF6* | TRUE | 2.126269477 |
| CAD_mortality | dendritic_cell | *ROM1* | TRUE | 2.336407524 |
| CAD_mortality | dendritic_cell | *PROM2* | TRUE | 2.271652051 |
| CAD_mortality | dendritic_cell | *CYP4A22* | TRUE | 1.979793383 |
| CAD_mortality | dendritic_cell | *ENDOG* | TRUE | 2.063326084 |
| CAD_mortality | dendritic_cell | *RAB43* | TRUE | 2.839976187 |
| CAD_mortality | dendritic_cell | *GOLGA6L4* | TRUE | 2.044138174 |
| CAD_mortality | dendritic_cell | *TDGF1* | TRUE | 2.608573668 |
| CAD_mortality | dendritic_cell | *ITGB3* | TRUE | 2.040612615 |
| CAD_mortality | endometrium | *OPN3* | TRUE | 2.093892514 |
| CAD_mortality | endometrium | *ROM1* | TRUE | 2.458566295 |
| CAD_mortality | endometrium | *C11orf87* | TRUE | 2.091915478 |
| CAD_mortality | endometrium | *TDGF1* | TRUE | 2.194413658 |
| CAD_mortality | endothelial | *MTRF1L* | TRUE | 2.309143219 |
| CAD_mortality | endothelial | *PROM2* | TRUE | 2.339488806 |
| CAD_mortality | endothelial | *TRIM66* | TRUE | 2.045514404 |
| CAD_mortality | endothelial | *RAB43* | TRUE | 2.888742201 |
| CAD_mortality | endothelial | *TOP1MT* | TRUE | 1.981348651 |
| CAD_mortality | endothelial | *TDGF1* | TRUE | 2.580937133 |
| CAD_mortality | LEC | *OPN3* | TRUE | 2.022355789 |
| CAD_mortality | LEC | *IRF6* | TRUE | 2.152262517 |
| CAD_mortality | LEC | *ZNF660* | TRUE | 2.075547595 |
| CAD_mortality | LEC | *POMZP3* | TRUE | 2.373706582 |
| CAD_mortality | LEC | *POLE3* | TRUE | 2.02833565 |
| CAD_mortality | LEC | *UVSSA* | TRUE | 2.280225611 |
| CAD_mortality | LEC | *GOLGA6L4* | TRUE | 2.505662087 |
| CAD_mortality | LEC | *TBKBP1* | TRUE | 2.052518169 |
| CAD_mortality | LEC | *TDGF1* | TRUE | 2.452358372 |
| CAD_mortality | macrophage | *ARHGEF5* | TRUE | 1.962751101 |
| CAD_mortality | macrophage | *TNFRSF10A* | TRUE | 2.062722377 |
| CAD_mortality | macrophage | *PPP1R37* | TRUE | 2.095541095 |
| CAD_mortality | macrophage | *MTRF1L* | TRUE | 2.063838585 |
| CAD_mortality | macrophage | *IRF6* | TRUE | 2.182605886 |
| CAD_mortality | macrophage | *POMZP3* | TRUE | 2.233122934 |
| CAD_mortality | macrophage | *POLE3* | TRUE | 2.038765277 |
| CAD_mortality | macrophage | *ROM1* | TRUE | 2.201174152 |
| CAD_mortality | macrophage | *CYP4A22* | TRUE | 2.242527222 |
| CAD_mortality | macrophage | *UVSSA* | TRUE | 2.03539153 |
| CAD_mortality | macrophage | *TRIM66* | TRUE | 2.178060005 |
| CAD_mortality | macrophage | *ENDOG* | TRUE | 1.992420832 |
| CAD_mortality | macrophage | *GOLGA6L4* | TRUE | 2.325755801 |
| CAD_mortality | macrophage | *C11orf87* | TRUE | 2.221740958 |
| CAD_mortality | macrophage | *HLA.DQA2* | TRUE | 1.981703029 |

| | | | | |
|---|---|---|---|---|
| CAD_mortality | macrophage | *LEFTY1* | TRUE | 2.472790388 |
| CAD_mortality | mast_cell | *POMZP3* | TRUE | 2.37025147 |
| CAD_mortality | mast_cell | *CYP4A22* | TRUE | 2.123607136 |
| CAD_mortality | mast_cell | *HLA.DQB1* | TRUE | 2.005568104 |
| CAD_mortality | mast_cell | *GOLGA6L4* | TRUE | 2.11212547 |
| CAD_mortality | mast_cell | *C11orf87* | TRUE | 2.194528577 |
| CAD_mortality | mast_cell | *HCG20* | TRUE | 2.1100468 |
| CAD_mortality | mast_cell | *TDGF1* | TRUE | 2.412649935 |
| CAD_mortality | mesothelium | *OPN3* | TRUE | 2.272437896 |
| CAD_mortality | mesothelium | *SYNJ2* | TRUE | 2.412913439 |
| CAD_mortality | mesothelium | *MRPS33* | TRUE | 1.9632844 |
| CAD_mortality | mesothelium | *CDC6* | TRUE | 2.12653001 |
| CAD_mortality | mesothelium | *TOP3B* | TRUE | 2.136669063 |
| CAD_mortality | mesothelium | *CYB5R3* | TRUE | 2.465297827 |
| CAD_mortality | mesothelium | *MANBAL* | TRUE | 2.185613675 |
| CAD_mortality | mesothelium | *ACP5* | TRUE | 2.020617855 |
| CAD_mortality | mesothelium | *TNFRSF10A* | TRUE | 2.117393346 |
| CAD_mortality | mesothelium | *PPP1R37* | TRUE | 1.981361662 |
| CAD_mortality | mesothelium | *UST* | TRUE | 2.079981395 |
| CAD_mortality | mesothelium | *MTRF1L* | TRUE | 2.73448391 |
| CAD_mortality | mesothelium | *IRF6* | TRUE | 2.320792431 |
| CAD_mortality | mesothelium | *EPCAM* | TRUE | 2.043569782 |
| CAD_mortality | mesothelium | *OPN4* | TRUE | 2.019557717 |
| CAD_mortality | mesothelium | *EBPL* | TRUE | 2.103115222 |
| CAD_mortality | mesothelium | *PRPF4* | TRUE | 2.039103286 |
| CAD_mortality | mesothelium | *CENPO* | TRUE | 2.329793392 |
| CAD_mortality | mesothelium | *NAAA* | TRUE | 1.999792501 |
| CAD_mortality | mesothelium | *CCDC40* | TRUE | 2.024610646 |
| CAD_mortality | mesothelium | *LYG1* | TRUE | 2.520788347 |
| CAD_mortality | mesothelium | *ZNF660* | TRUE | 2.287945405 |
| CAD_mortality | mesothelium | *POMZP3* | TRUE | 2.642432478 |
| CAD_mortality | mesothelium | *POLE3* | TRUE | 2.138484628 |
| CAD_mortality | mesothelium | *TMEM236* | TRUE | 2.038484041 |
| CAD_mortality | mesothelium | *GBP5* | TRUE | 1.977826892 |
| CAD_mortality | mesothelium | *STEAP2* | TRUE | 2.362713223 |
| CAD_mortality | mesothelium | *DIP2A* | TRUE | 2.127767326 |
| CAD_mortality | mesothelium | *LRRC56* | TRUE | 2.110142931 |
| CAD_mortality | mesothelium | *DCD* | TRUE | 1.964549357 |
| CAD_mortality | mesothelium | *CYP4A22* | TRUE | 2.313674122 |
| CAD_mortality | mesothelium | *UVSSA* | TRUE | 2.238372791 |
| CAD_mortality | mesothelium | *TRIM66* | TRUE | 2.008737735 |
| CAD_mortality | mesothelium | *ENDOG* | TRUE | 2.096953889 |
| CAD_mortality | mesothelium | *IRS1* | TRUE | 2.048034203 |
| CAD_mortality | mesothelium | *TTLL6* | TRUE | 1.977211414 |
| CAD_mortality | mesothelium | *SCAND1* | TRUE | 1.96715089 |
| CAD_mortality | mesothelium | *RAB43* | TRUE | 2.556132216 |
| CAD_mortality | mesothelium | *CHRNA7* | TRUE | 2.085413285 |
| CAD_mortality | mesothelium | *RBM44* | TRUE | 2.095556818 |
| CAD_mortality | mesothelium | *ODF3B* | TRUE | 2.128324538 |
| CAD_mortality | mesothelium | *CDH4* | TRUE | 2.213689009 |

| | | | | |
|---|---|---|---|---|
| CAD_mortality | mesothelium | *GOLGA6L4* | TRUE | 2.824743691 |
| CAD_mortality | mesothelium | *C11orf87* | TRUE | 2.323610597 |
| CAD_mortality | mesothelium | *XPNPEP3* | TRUE | 2.090564677 |
| CAD_mortality | mesothelium | *ZNF100* | TRUE | 1.980555372 |
| CAD_mortality | mesothelium | *RPS6KL1* | TRUE | 1.965685388 |
| CAD_mortality | mesothelium | *TBKBP1* | TRUE | 2.021865366 |
| CAD_mortality | mesothelium | *LINC00937* | TRUE | 2.216300988 |
| CAD_mortality | mesothelium | *TDGF1* | TRUE | 2.168210337 |
| CAD_mortality | mesothelium | *LEFTY1* | TRUE | 2.348746991 |
| CAD_mortality | monocyte | *CYP4A22* | TRUE | 2.016130002 |
| CAD_mortality | monocyte | *TRIM66* | TRUE | 2.04930788 |
| CAD_mortality | monocyte | *HLA.DQB1* | TRUE | 2.195058476 |
| CAD_mortality | monocyte | *C11orf87* | TRUE | 2.29342431 |
| CAD_mortality | monocyte | *TBKBP1* | TRUE | 2.255019217 |
| CAD_mortality | nk_cell | *LYG1* | TRUE | 2.11337816 |
| CAD_mortality | nk_cell | *PROM2* | TRUE | 2.26477583 |
| CAD_mortality | nk_cell | *CYP4A22* | TRUE | 2.062160124 |
| CAD_mortality | nk_cell | *RAB43* | TRUE | 3.381391989 |
| CAD_mortality | nk_cell | *HLA.DQB1* | TRUE | 2.042916423 |
| CAD_mortality | nk_cell | *C11orf87* | TRUE | 2.023011807 |
| CAD_mortality | pericyte | *MTRF1L* | TRUE | 2.020710477 |
| CAD_mortality | pericyte | *IRF6* | TRUE | 2.229029494 |
| CAD_mortality | pericyte | *ROM1* | TRUE | 2.331266958 |
| CAD_mortality | pericyte | *CYP4A22* | TRUE | 2.188833332 |
| CAD_mortality | pericyte | *GOLGA6L4* | TRUE | 2.565498853 |
| CAD_mortality | pericyte | *C11orf87* | TRUE | 2.120299468 |
| CAD_mortality | pericyte | *TDGF1* | TRUE | 2.359245024 |
| CAD_mortality | pericyte | *LEFTY1* | TRUE | 1.960510453 |
| CAD_mortality | SMC | *ARHGEF5* | TRUE | 1.971590998 |
| CAD_mortality | SMC | *IRF6* | TRUE | 2.031926644 |
| CAD_mortality | SMC | *LYG1* | TRUE | 2.396458894 |
| CAD_mortality | SMC | *ZNF660* | TRUE | 2.038445265 |
| CAD_mortality | SMC | *ROM1* | TRUE | 2.265194053 |
| CAD_mortality | SMC | *CYP4A22* | TRUE | 2.069950649 |
| CAD_mortality | SMC | *TRIM66* | TRUE | 2.236989221 |
| CAD_mortality | SMC | *RAB43* | TRUE | 2.192207838 |
| CAD_mortality | SMC | *C11orf87* | TRUE | 2.084557937 |
| CAD_mortality | SMC | *TBKBP1* | TRUE | 2.09242149 |
| CAD_mortality | SMC | *TDGF1* | TRUE | 2.381968003 |
| CAD_mortality | t_cell | *CDC6* | TRUE | 1.998990462 |
| CAD_mortality | t_cell | *MTRF1L* | TRUE | 2.138817861 |
| CAD_mortality | t_cell | *IRF6* | TRUE | 2.237830807 |
| CAD_mortality | t_cell | *LYG1* | TRUE | 2.043063417 |
| CAD_mortality | t_cell | *DCD* | TRUE | 2.033533308 |
| CAD_mortality | t_cell | *CYP4A22* | TRUE | 2.135861497 |
| CAD_mortality | t_cell | *TRIM66* | TRUE | 2.209307281 |
| CAD_mortality | t_cell | *RAB43* | TRUE | 2.871455049 |
| CAD_mortality | t_cell | *TOP1MT* | TRUE | 2.112937424 |
| CAD_mortality | t_cell | *TBKBP1* | TRUE | 2.175042622 |
| CAD_mortality | t_cell | *TDGF1* | TRUE | 2.314855544 |

Anxiety

| pathway name | set size | candidates contained | p-value | q-value | pathway source |
|---|---|---|---|---|---|
| Viral myocarditis - Homo sapiens (human) | 60 | 3 (5.1%) | 0.000331 | 0.17 | KEGG |
| Asthma - Homo sapiens (human) | 31 | 2 (6.7%) | 0.00215 | 0.222 | KEGG |
| Allograft rejection - Homo sapiens (human) | 38 | 2 (5.4%) | 0.00326 | 0.222 | KEGG |
| Intestinal immune network for IgA production - Homo sapiens (human) | 49 | 2 (4.3%) | 0.00522 | 0.222 | KEGG |
| Autoimmune thyroid disease - Homo sapiens (human) | 53 | 2 (3.8%) | 0.00636 | 0.222 | KEGG |
| Signaling by membrane-tethered fusions of PDGFRA or PDGFRB | 5 | 1 (20.0%) | 0.0115 | 0.222 | Reactome |
| Extrinsic Pathway of Fibrin Clot Formation | 5 | 1 (20.0%) | 0.0115 | 0.222 | Reactome |
| NAD biosynthesis from 2-amino-3-carboxymuconate semialdehyde | 5 | 1 (20.0%) | 0.0115 | 0.222 | HumanCyc |
| Highly sodium permeable postsynaptic acetylcholine nicotinic receptors | 7 | 1 (14.3%) | 0.016 | 0.222 | Reactome |
| CS/DS degradation | 7 | 1 (14.3%) | 0.016 | 0.222 | Reactome |
| Hypertrophic cardiomyopathy - Homo sapiens (human) | 90 | 2 (2.2%) | 0.0182 | 0.222 | KEGG |
| NAD Biosynthesis II (from tryptophan) | 8 | 1 (12.5%) | 0.0183 | 0.222 | Wikipathways |
| Allograft Rejection | 93 | 2 (2.2%) | 0.0194 | 0.222 | Wikipathways |
| Dilated cardiomyopathy - Homo sapiens (human) | 96 | 2 (2.1%) | 0.0201 | 0.222 | KEGG |
| cd40l signaling pathway | 9 | 1 (11.1%) | 0.0206 | 0.222 | BioCarta |
| Ca2+ activated K+ channels | 9 | 1 (11.1%) | 0.0206 | 0.222 | Reactome |
| Platelet activation, signaling and aggregation | 263 | 3 (1.1%) | 0.0223 | 0.222 | Reactome |
| NLR Proteins | 10 | 1 (10.0%) | 0.0228 | 0.222 | Wikipathways |
| Gamma-carboxylation of protein precursors | 10 | 1 (10.0%) | 0.0228 | 0.222 | Reactome |
| Removal of aminoterminal propeptides from gamma-carboxylated proteins | 10 | 1 (10.0%) | 0.0228 | 0.222 | Reactome |
| Pexophagy | 10 | 1 (10.0%) | 0.0228 | 0.222 | Reactome |
| Non-homologous end joining | 10 | 1 (10.0%) | 0.0228 | 0.222 | Wikipathways |
| Potassium Channels | 103 | 2 (1.9%) | 0.0234 | 0.222 | Reactome |
| Gamma-carboxylation, transport, and amino-terminal cleavage of proteins | 11 | 1 (9.1%) | 0.0251 | 0.222 | Reactome |
| CMP phosphorylation | 11 | 1 (9.1%) | 0.0251 | 0.222 | HumanCyc |
| Presynaptic nicotinic acetylcholine receptors | 12 | 1 (8.3%) | 0.0274 | 0.222 | Reactome |
| pyrimidine deoxyribonucleotide phosphorylation | 12 | 1 (8.3%) | 0.0274 | 0.222 | HumanCyc |
| guanosine deoxyribonucleotides de novo biosynthesis | 12 | 1 (8.3%) | 0.0274 | 0.222 | HumanCyc |
| adenosine deoxyribonucleotides de novo biosynthesis | 12 | 1 (8.3%) | 0.0274 | 0.222 | HumanCyc |
| NAD de novo biosynthesis | 12 | 1 (8.3%) | 0.0274 | 0.222 | HumanCyc |
| Toxoplasmosis - Homo sapiens (human) | 112 | 2 (1.8%) | 0.0274 | 0.222 | KEGG |
| extrinsic prothrombin activation pathway | 13 | 1 (7.7%) | 0.0296 | 0.222 | BioCarta |
| BMAL1:CLOCK,NPAS2 activates circadian gene expression | 13 | 1 (7.7%) | 0.0296 | 0.222 | Reactome |
| visual signal transduction | 13 | 1 (7.7%) | 0.0296 | 0.222 | BioCarta |
| purine deoxyribonucleosides salvage | 13 | 1 (7.7%) | 0.0296 | 0.222 | HumanCyc |
| guanosine ribonucleotides de novo biosynthesis | 13 | 1 (7.7%) | 0.0296 | 0.222 | HumanCyc |
| UTP and CTP de novo biosynthesis | 13 | 1 (7.7%) | 0.0296 | 0.222 | HumanCyc |
| Activation of RAC1 | 13 | 1 (7.7%) | 0.0296 | 0.222 | Reactome |
| Non-homologous end-joining - Homo sapiens (human) | 13 | 1 (7.7%) | 0.0296 | 0.222 | KEGG |
| DNA-PK pathway in nonhomologous end joining | 13 | 1 (7.7%) | 0.0296 | 0.222 | PID |
| Postsynaptic nicotinic acetylcholine receptors | 14 | 1 (7.1%) | 0.0318 | 0.222 | Reactome |
| Acetylcholine binding and downstream events | 14 | 1 (7.1%) | 0.0318 | 0.222 | Reactome |
| S1P4 pathway | 14 | 1 (7.1%) | 0.0318 | 0.222 | PID |
| Lysosphingolipid and LPA receptors | 14 | 1 (7.1%) | 0.0318 | 0.222 | Reactome |
| Nicotinate and Nicotinamide Metabolism | 14 | 1 (7.1%) | 0.0318 | 0.222 | SMPDB |
| superpathway of pyrimidine ribonucleotides de novo biosynthesis | 15 | 1 (6.7%) | 0.0341 | 0.222 | HumanCyc |
| p130Cas linkage to MAPK signaling for integrins | 15 | 1 (6.7%) | 0.0341 | 0.222 | Reactome |
| GRB2:SOS provides linkage to MAPK signaling for Integrins | 15 | 1 (6.7%) | 0.0341 | 0.222 | Reactome |
| pyrimidine deoxyribonucleotides biosynthesis from CTP | 16 | 1 (6.2%) | 0.0363 | 0.222 | HumanCyc |
| guanosine nucleotides de novo biosynthesis | 16 | 1 (6.2%) | 0.0363 | 0.222 | HumanCyc |
| pyrimidine deoxyribonucleotides de novo biosynthesis | 16 | 1 (6.2%) | 0.0363 | 0.222 | HumanCyc |
| Systemic lupus erythematosus - Homo sapiens (human) | 136 | 2 (1.5%) | 0.038 | 0.222 | KEGG |
| Doxorubicin Metabolism Pathway | 17 | 1 (5.9%) | 0.0385 | 0.222 | SMPDB |
| TNF receptor superfamily (TNFSF) members mediating non-canonical NF-kB pathway | 17 | 1 (5.9%) | 0.0385 | 0.222 | Reactome |
| superpathway of pyrimidine deoxyribonucleoside salvage | 17 | 1 (5.9%) | 0.0385 | 0.222 | HumanCyc |
| Signaling by FLT3 fusion proteins | 18 | 1 (5.6%) | 0.0408 | 0.222 | Reactome |
| G beta:gamma signalling through BTK | 18 | 1 (5.6%) | 0.0408 | 0.222 | Reactome |
| NRF2 pathway | 143 | 2 (1.4%) | 0.0428 | 0.222 | Wikipathways |
| Hematopoietic Stem Cell Gene Regulation by GABP alpha-beta Complex | 19 | 1 (5.3%) | 0.043 | 0.222 | Wikipathways |
| Prostacyclin signalling through prostacyclin receptor | 19 | 1 (5.3%) | 0.043 | 0.222 | Reactome |
| Glycosaminoglycan degradation - Homo sapiens (human) | 19 | 1 (5.3%) | 0.043 | 0.222 | KEGG |
| Small Ligand GPCRs | 19 | 1 (5.3%) | 0.043 | 0.222 | Wikipathways |
| Signaling by PDGFR in disease | 20 | 1 (5.0%) | 0.0452 | 0.222 | Reactome |
| Bivalirudin Action Pathway | 20 | 1 (5.0%) | 0.0452 | 0.222 | SMPDB |
| Argatroban Action Pathway | 20 | 1 (5.0%) | 0.0452 | 0.222 | SMPDB |
| Ximelagatran Action Pathway | 20 | 1 (5.0%) | 0.0452 | 0.222 | SMPDB |
| Lepirudin Action Pathway | 20 | 1 (5.0%) | 0.0452 | 0.222 | SMPDB |

| | | | | | |
|---|---|---|---|---|---|
| Coagulation | 20 | 1 (5.0%) | 0.0452 | 0.222 | SMPDB |
| Aminocaproic Acid Action Pathway | 20 | 1 (5.0%) | 0.0452 | 0.222 | SMPDB |
| Tranexamic Acid Action Pathway | 20 | 1 (5.0%) | 0.0452 | 0.222 | SMPDB |
| Urokinase Action Pathway | 20 | 1 (5.0%) | 0.0452 | 0.222 | SMPDB |
| Reteplase Action Pathway | 20 | 1 (5.0%) | 0.0452 | 0.222 | SMPDB |
| Streptokinase Action Pathway | 20 | 1 (5.0%) | 0.0452 | 0.222 | SMPDB |
| Tenecteplase Action Pathway | 20 | 1 (5.0%) | 0.0452 | 0.222 | SMPDB |
| Alteplase Action Pathway | 20 | 1 (5.0%) | 0.0452 | 0.222 | SMPDB |
| Anistreplase Action Pathway | 20 | 1 (5.0%) | 0.0452 | 0.222 | SMPDB |
| Aprotinin Action Pathway | 20 | 1 (5.0%) | 0.0452 | 0.222 | SMPDB |
| Phenindione Action Pathway | 20 | 1 (5.0%) | 0.0452 | 0.222 | SMPDB |
| Dicoumarol Action Pathway | 20 | 1 (5.0%) | 0.0452 | 0.222 | SMPDB |
| Warfarin Action Pathway | 20 | 1 (5.0%) | 0.0452 | 0.222 | SMPDB |
| Acenocoumarol Action Pathway | 20 | 1 (5.0%) | 0.0452 | 0.222 | SMPDB |
| Phenprocoumon Action Pathway | 20 | 1 (5.0%) | 0.0452 | 0.222 | SMPDB |
| Dicumarol Action Pathway | 20 | 1 (5.0%) | 0.0452 | 0.222 | SMPDB |
| G beta:gamma signalling through PLC beta | 20 | 1 (5.0%) | 0.0452 | 0.222 | Reactome |
| G beta:gamma signalling through CDC42 | 20 | 1 (5.0%) | 0.0452 | 0.222 | Reactome |
| Cellular hexose transport | 20 | 1 (5.0%) | 0.0452 | 0.222 | Reactome |
| Syndecan interactions | 20 | 1 (5.0%) | 0.0452 | 0.222 | Reactome |
| Cell adhesion molecules - Homo sapiens (human) | 149 | 2 (1.4%) | 0.0455 | 0.222 | KEGG |
| FLT3 signaling in disease | 21 | 1 (4.8%) | 0.0474 | 0.222 | Reactome |
| Ardeparin Action Pathway | 21 | 1 (4.8%) | 0.0474 | 0.222 | SMPDB |
| Heparin Action Pathway | 21 | 1 (4.8%) | 0.0474 | 0.222 | SMPDB |
| Fondaparinux Action Pathway | 21 | 1 (4.8%) | 0.0474 | 0.222 | SMPDB |
| Enoxaparin Action Pathway | 21 | 1 (4.8%) | 0.0474 | 0.222 | SMPDB |
| Presynaptic function of Kainate receptors | 21 | 1 (4.8%) | 0.0474 | 0.222 | Reactome |
| Sphingosine 1-phosphate (S1P) pathway | 21 | 1 (4.8%) | 0.0474 | 0.222 | PID |
| ADP signalling through P2Y purinoceptor 12 | 22 | 1 (4.5%) | 0.0496 | 0.222 | Reactome |
| superpathway of pyrimidine deoxyribonucleotides de novo biosynthesis | 22 | 1 (4.5%) | 0.0496 | 0.222 | HumanCyc |
| NAD+ biosynthetic pathways | 22 | 1 (4.5%) | 0.0496 | 0.222 | Wikipathways |
| Citric acid cycle (TCA cycle) | 22 | 1 (4.5%) | 0.0496 | 0.222 | Reactome |

Bipolar

| pathway name | set size | candidates contained | p-value | q-value | pathway source |
|---|---|---|---|---|---|
| GP1b-IX-V activation signalling | 10 | 2 (20.0%) | 0.000464 | 0.162 | Reactome |
| eicosapentaenoate biosynthesis | 13 | 2 (15.4%) | 0.000799 | 0.162 | HumanCyc |
| Rap1 signalling | 16 | 2 (12.5%) | 0.00122 | 0.162 | Reactome |
| p38 signaling mediated by MAPKAP kinases | 21 | 2 (9.5%) | 0.00212 | 0.162 | PID |
| 3-Phosphoglycerate dehydrogenase deficiency | 27 | 2 (7.4%) | 0.00349 | 0.162 | SMPDB |
| Non Ketotic Hyperglycinemia | 27 | 2 (7.4%) | 0.00349 | 0.162 | SMPDB |
| Glycine and Serine Metabolism | 27 | 2 (7.4%) | 0.00349 | 0.162 | SMPDB |
| Dimethylglycine Dehydrogenase Deficiency | 27 | 2 (7.4%) | 0.00349 | 0.162 | SMPDB |
| Hyperglycinemia, non-ketotic | 27 | 2 (7.4%) | 0.00349 | 0.162 | SMPDB |
| Dimethylglycine Dehydrogenase Deficiency | 27 | 2 (7.4%) | 0.00349 | 0.162 | SMPDB |
| Sarcosinemia | 27 | 2 (7.4%) | 0.00349 | 0.162 | SMPDB |
| Dihydropyrimidine Dehydrogenase Deficiency (DHPD) | 27 | 2 (7.4%) | 0.00349 | 0.162 | SMPDB |
| Polymerase switching on the C-strand of the telomere | 27 | 2 (7.4%) | 0.00349 | 0.162 | Reactome |
| EGF | 28 | 2 (7.1%) | 0.00375 | 0.162 | INOH |
| IGF1 pathway | 30 | 2 (6.7%) | 0.0043 | 0.173 | PID |
| Herpes simplex virus 1 infection - Homo sapiens (human) | 498 | 6 (1.2%) | 0.00537 | 0.195 | KEGG |
| Class I PI3K signaling events mediated by Akt | 35 | 2 (5.7%) | 0.00582 | 0.195 | PID |
| Telomere C-strand (Lagging Strand) Synthesis | 35 | 2 (5.7%) | 0.00582 | 0.195 | Reactome |
| GMCSF-mediated signaling events | 37 | 2 (5.4%) | 0.00649 | 0.197 | PID |
| Hyaluronan biosynthesis and export | 2 | 1 (50.0%) | 0.00654 | 0.197 | Reactome |
| Sphingolipid signaling pathway - Homo sapiens (human) | 119 | 3 (2.5%) | 0.00689 | 0.198 | KEGG |
| Glycine, serine and threonine metabolism - Homo sapiens (human) | 40 | 2 (5.0%) | 0.00756 | 0.207 | KEGG |
| Common Pathways Underlying Drug Addiction | 41 | 2 (4.9%) | 0.00793 | 0.208 | Wikipathways |
| Hydrolysis of LPE | 3 | 1 (33.3%) | 0.00979 | 0.246 | Reactome |
| IL5 | 48 | 2 (4.2%) | 0.0107 | 0.254 | NetPath |
| IL3 | 51 | 2 (3.9%) | 0.0121 | 0.254 | NetPath |
| Extension of Telomeres | 51 | 2 (3.9%) | 0.0121 | 0.254 | Reactome |
| Resolution of AP sites via the single-nucleotide replacement pathway | 4 | 1 (25.0%) | 0.013 | 0.254 | Reactome |
| Linoleic acid metabolism known to be affected by coronavirus infection | 4 | 1 (25.0%) | 0.013 | 0.254 | Wikipathways |
| serine biosynthesis (phosphorylated route) | 4 | 1 (25.0%) | 0.013 | 0.254 | HumanCyc |
| PTK6 Activates STAT3 | 4 | 1 (25.0%) | 0.013 | 0.254 | Reactome |
| Regulation of lipolysis in adipocytes - Homo sapiens (human) | 57 | 2 (3.6%) | 0.0139 | 0.263 | KEGG |
| VEGF signaling pathway - Homo sapiens (human) | 59 | 2 (3.4%) | 0.0159 | 0.268 | KEGG |
| Adenosine P1 receptors | 5 | 1 (20.0%) | 0.0163 | 0.268 | Reactome |
| glycine betaine degradation | 5 | 1 (20.0%) | 0.0163 | 0.268 | HumanCyc |
| Dopamine receptors | 5 | 1 (20.0%) | 0.0163 | 0.268 | Reactome |
| Long-term depression - Homo sapiens (human) | 60 | 2 (3.3%) | 0.0165 | 0.268 | KEGG |
| mTOR signaling pathway | 64 | 2 (3.1%) | 0.0186 | 0.279 | PID |
| Choline catabolism | 6 | 1 (16.7%) | 0.0195 | 0.279 | Reactome |
| Alpha Linolenic Acid and Linoleic Acid Metabolism | 6 | 1 (16.7%) | 0.0195 | 0.279 | SMPDB |
| Pilocytic astrocytoma | 6 | 1 (16.7%) | 0.0195 | 0.279 | Wikipathways |
| Fc epsilon RI signaling pathway - Homo sapiens (human) | 68 | 2 (3.0%) | 0.0203 | 0.279 | KEGG |
| FGF | 67 | 2 (3.0%) | 0.0203 | 0.279 | INOH |
| Prolactin | 70 | 2 (2.9%) | 0.022 | 0.279 | NetPath |
| superpathway of choline degradation to L-serine | 7 | 1 (14.3%) | 0.0227 | 0.279 | HumanCyc |
| APEX1-Independent Resolution of AP Sites via the Single Nucleotide Replacement Pathway | 7 | 1 (14.3%) | 0.0227 | 0.279 | Reactome |
| serine and glycine biosynthesis | 7 | 1 (14.3%) | 0.0227 | 0.279 | HumanCyc |
| Negative feedback regulation of MAPK pathway | 7 | 1 (14.3%) | 0.0227 | 0.279 | Reactome |
| MicroRNA for Targeting Cancer Growth and Vascularization in Glioblastoma | 7 | 1 (14.3%) | 0.0227 | 0.279 | Wikipathways |
| Linoleate metabolism | 72 | 2 (2.8%) | 0.0232 | 0.279 | EHMN |
| Prolactin Signaling Pathway | 76 | 2 (2.6%) | 0.0256 | 0.279 | Wikipathways |
| oleate biosynthesis | 8 | 1 (12.5%) | 0.0259 | 0.279 | HumanCyc |
| Linoleic acid (LA) metabolism | 8 | 1 (12.5%) | 0.0259 | 0.279 | Reactome |
| MAPK and NFkB signaling pathways inhibited by Yersinia YopJ | 8 | 1 (12.5%) | 0.0259 | 0.279 | Wikipathways |
| Conjugation of salicylate with glycine | 8 | 1 (12.5%) | 0.0259 | 0.279 | Reactome |
| B cell receptor signaling pathway - Homo sapiens (human) | 82 | 2 (2.5%) | 0.0282 | 0.279 | KEGG |
| Serine biosynthesis | 9 | 1 (11.1%) | 0.0291 | 0.279 | Reactome |
| Prostanoid ligand receptors | 9 | 1 (11.1%) | 0.0291 | 0.279 | Reactome |
| Signaling by MRAS-complex mutants | 9 | 1 (11.1%) | 0.0291 | 0.279 | Reactome |
| MAP kinase cascade | 9 | 1 (11.1%) | 0.0291 | 0.279 | HumanCyc |
| Conjugation of carboxylic acids | 9 | 1 (11.1%) | 0.0291 | 0.279 | Reactome |
| Amino Acid conjugation | 9 | 1 (11.1%) | 0.0291 | 0.279 | Reactome |
| sphingosine and sphingosine-1-phosphate metabolism | 9 | 1 (11.1%) | 0.0291 | 0.279 | HumanCyc |
| HDR through MMEJ (alt-NHEJ) | 10 | 1 (10.0%) | 0.0323 | 0.293 | Reactome |
| regulation of splicing through sam68 | 10 | 1 (10.0%) | 0.0323 | 0.293 | BioCarta |

| | | | | | |
|---|---|---|---|---|---|
| cAMP signaling pathway - Homo sapiens (human) | 216 | 3 (1.4%) | 0.0335 | 0.293 | KEGG |
| Nucleotide GPCRs | 11 | 1 (9.1%) | 0.0355 | 0.293 | Wikipathways |
| Hydrolysis of LPC | 11 | 1 (9.1%) | 0.0355 | 0.293 | Reactome |
| Purine metabolism | 222 | 3 (1.4%) | 0.0359 | 0.293 | EHMN |
| GnRH signaling pathway - Homo sapiens (human) | 93 | 2 (2.2%) | 0.0372 | 0.293 | KEGG |
| Base Excision Repair | 94 | 2 (2.2%) | 0.0372 | 0.293 | Reactome |
| Stimuli-sensing channels | 94 | 2 (2.1%) | 0.0379 | 0.293 | Reactome |
| Scavenging of heme from plasma | 12 | 1 (8.3%) | 0.0386 | 0.293 | Reactome |
| Omega-9 FA synthesis | 12 | 1 (8.3%) | 0.0386 | 0.293 | Wikipathways |
| Negative regulation of NOTCH4 signaling | 12 | 1 (8.3%) | 0.0386 | 0.293 | Reactome |
| Regulation of localization of FOXO transcription factors | 12 | 1 (8.3%) | 0.0386 | 0.293 | Reactome |
| Hyaluronan metabolism | 12 | 1 (8.3%) | 0.0386 | 0.293 | Reactome |
| Fc gamma R-mediated phagocytosis - Homo sapiens (human) | 97 | 2 (2.1%) | 0.0394 | 0.293 | KEGG |
| Choline metabolism in cancer - Homo sapiens (human) | 98 | 2 (2.0%) | 0.0409 | 0.293 | KEGG |
| sarcosine oncometabolite pathway | 13 | 1 (7.7%) | 0.0418 | 0.293 | SMPDB |
| alpha-linolenic acid (ALA) metabolism | 13 | 1 (7.7%) | 0.0418 | 0.293 | Reactome |
| alpha-linolenic (omega3) and linoleic (omega6) acid metabolism | 13 | 1 (7.7%) | 0.0418 | 0.293 | Reactome |
| melanocyte development and pigmentation pathway | 13 | 1 (7.7%) | 0.0418 | 0.293 | BioCarta |
| multiple antiapoptotic pathways from igf-1r signaling lead to bad phosphorylation | 13 | 1 (7.7%) | 0.0418 | 0.293 | BioCarta |
| Chk1/Chk2(Cds1) mediated inactivation of Cyclin B:Cdk1 complex | 13 | 1 (7.7%) | 0.0418 | 0.293 | Reactome |
| Telomere C-strand synthesis initiation | 13 | 1 (7.7%) | 0.0418 | 0.293 | Reactome |
| Omega-3-Omega-6 FA synthesis | 14 | 1 (7.1%) | 0.0449 | 0.295 | Wikipathways |
| Ras signaling in the CD4+ TCR pathway | 14 | 1 (7.1%) | 0.0449 | 0.295 | PID |
| MAPK pathway in congenital thyroid cancer | 14 | 1 (7.1%) | 0.0449 | 0.295 | Wikipathways |
| phosphorylation of mek1 by cdk5/p35 down regulates the map kinase pathway | 14 | 1 (7.1%) | 0.0449 | 0.295 | BioCarta |
| γ-linolenate biosynthesis | 14 | 1 (7.1%) | 0.0449 | 0.295 | HumanCyc |
| Telomere Maintenance | 105 | 2 (1.9%) | 0.0455 | 0.295 | Reactome |
| ErbB1 downstream signaling | 106 | 2 (1.9%) | 0.0471 | 0.295 | PID |
| Eicosanoid ligand-binding receptors | 15 | 1 (6.7%) | 0.048 | 0.295 | Reactome |
| ERK Pathway in Huntington,s Disease | 15 | 1 (6.7%) | 0.048 | 0.295 | Wikipathways |
| Activation of BAD and translocation to mitochondria | 15 | 1 (6.7%) | 0.048 | 0.295 | Reactome |
| Mercaptopurine Metabolism Pathway | 15 | 1 (6.7%) | 0.048 | 0.295 | SMPDB |

Depression

| pathway name | set size | candidates contained | p-value | q-value | pathway source |
|---|---|---|---|---|---|
| Eptifibatide Action Pathway | 2 | 1 (50.0%) | 0.00624 | 0.307 | SMPDB |
| Tirofiban Action Pathway | 2 | 1 (50.0%) | 0.00624 | 0.307 | SMPDB |
| Abciximab Action Pathway | 2 | 1 (50.0%) | 0.00624 | 0.307 | SMPDB |
| Pyrophosphate hydrolysis | 2 | 1 (50.0%) | 0.00624 | 0.307 | Reactome |
| 5-aminoimidazole ribonucleotide biosynthesis | 3 | 1 (33.3%) | 0.00935 | 0.307 | HumanCyc |
| Cocaine addiction - Homo sapiens (human) | 49 | 2 (4.1%) | 0.0102 | 0.307 | KEGG |
| IRS activation | 4 | 1 (25.0%) | 0.0124 | 0.307 | Reactome |
| Neurexins and neuroligins | 57 | 2 (3.5%) | 0.0137 | 0.307 | Reactome |
| Focal Adhesion-PI3K-Akt-mTOR-signaling pathway | 302 | 4 (1.3%) | 0.0143 | 0.307 | Wikipathways |
| tRNA splicing | 5 | 1 (20.0%) | 0.0155 | 0.307 | HumanCyc |
| Human papillomavirus infection - Homo sapiens (human) | 331 | 4 (1.2%) | 0.0194 | 0.307 | KEGG |
| NrCAM interactions | 7 | 1 (14.3%) | 0.0217 | 0.307 | Reactome |
| overview of telomerase rna component gene hterc transcriptional regulation | 7 | 1 (14.3%) | 0.0217 | 0.307 | BioCarta |
| SOS-mediated signalling | 7 | 1 (14.3%) | 0.0217 | 0.307 | Reactome |
| Erythropoietin activates Phospholipase C gamma (PLCG) | 7 | 1 (14.3%) | 0.0217 | 0.307 | Reactome |
| Erythropoietin activates STAT5 | 7 | 1 (14.3%) | 0.0217 | 0.307 | Reactome |
| Axon guidance | 356 | 4 (1.1%) | 0.0246 | 0.307 | Reactome |
| Antigen processing and presentation - Homo sapiens (human) | 78 | 2 (2.6%) | 0.0247 | 0.307 | KEGG |
| PI3K/AKT activation | 9 | 1 (11.1%) | 0.0278 | 0.307 | Reactome |
| Nervous system development | 379 | 4 (1.1%) | 0.03 | 0.307 | Reactome |
| Protein-protein interactions at synapses | 88 | 2 (2.3%) | 0.0308 | 0.307 | Reactome |
| ECM-receptor interaction - Homo sapiens (human) | 88 | 2 (2.3%) | 0.0308 | 0.307 | KEGG |
| IL-13 signaling | 10 | 1 (10.0%) | 0.0308 | 0.307 | INOH |
| Signal attenuation | 10 | 1 (10.0%) | 0.0308 | 0.307 | Reactome |
| Signaling by Leptin | 11 | 1 (9.1%) | 0.0339 | 0.307 | Reactome |
| insulin signaling pathway | 11 | 1 (9.1%) | 0.0339 | 0.307 | BioCarta |
| RHO GTPases activate KTN1 | 11 | 1 (9.1%) | 0.0339 | 0.307 | Reactome |
| Complement system | 93 | 2 (2.2%) | 0.0341 | 0.307 | Wikipathways |
| RAF/MAP kinase cascade | 235 | 3 (1.3%) | 0.0368 | 0.307 | Reactome |
| Activation of Ca-permeable Kainate Receptor | 12 | 1 (8.3%) | 0.0369 | 0.307 | Reactome |
| Ionotropic activity of kainate receptors | 12 | 1 (8.3%) | 0.0369 | 0.307 | Reactome |
| Purine ribonucleoside monophosphate biosynthesis | 12 | 1 (8.3%) | 0.0369 | 0.307 | Reactome |
| Erythropoietin activates Phosphoinositide-3-kinase (PI3K) | 12 | 1 (8.3%) | 0.0369 | 0.307 | Reactome |
| Aspartate and asparagine metabolism | 12 | 1 (8.3%) | 0.0369 | 0.307 | Reactome |
| Butyrophilin (BTN) family interactions | 12 | 1 (8.3%) | 0.0369 | 0.307 | Reactome |
| MAPK1/MAPK3 signaling | 236 | 3 (1.3%) | 0.0372 | 0.307 | Reactome |
| Endosomal/Vacuolar pathway | 13 | 1 (7.7%) | 0.0399 | 0.307 | Reactome |
| L1CAM interactions | 102 | 2 (2.0%) | 0.0404 | 0.307 | Reactome |
| LGI-ADAM interactions | 14 | 1 (7.1%) | 0.0429 | 0.307 | Reactome |
| Erythropoietin activates RAS | 14 | 1 (7.1%) | 0.0429 | 0.307 | Reactome |
| il-7 signal transduction | 15 | 1 (6.7%) | 0.0459 | 0.307 | BioCarta |
| p130Cas linkage to MAPK signaling for integrins | 15 | 1 (6.7%) | 0.0459 | 0.307 | Reactome |
| GRB2:SOS provides linkage to MAPK signaling for Integrins | 15 | 1 (6.7%) | 0.0459 | 0.307 | Reactome |
| FOXO-mediated transcription of cell death genes | 15 | 1 (6.7%) | 0.0459 | 0.307 | Reactome |
| Nucleobase biosynthesis | 15 | 1 (6.7%) | 0.0459 | 0.307 | Reactome |
| Leucine Stimulation on Insulin Signaling | 15 | 1 (6.7%) | 0.0459 | 0.307 | SMPDB |
| RHO GTPases activate CIT | 16 | 1 (6.2%) | 0.0489 | 0.307 | Reactome |
| FOXA2 pathway | 16 | 1 (6.2%) | 0.0489 | 0.307 | Wikipathways |

Psychosis

| pathway name | set size | candidates contained | p-value | q-value | pathway source |
|---|---|---|---|---|---|
| Transport of vitamins, nucleosides, and related molecules | 44 | 3 (6.8%) | 0.00032 | 0.13 | Reactome |
| eicosapentaenoate biosynthesis | 13 | 2 (15.4%) | 0.000694 | 0.141 | HumanCyc |
| fas signaling pathway (cd95) | 20 | 2 (10.0%) | 0.00167 | 0.181 | BioCarta |
| Biosynthesis of DPAn-3-derived 13-series resolvins | 1 | 1 (100.0%) | 0.00305 | 0.181 | Reactome |
| Biosynthesis of DPAn-3-derived maresins | 2 | 1 (50.0%) | 0.00609 | 0.181 | Reactome |
| Biosynthesis of electrophilic ?-3 PUFA oxo-derivatives | 2 | 1 (50.0%) | 0.00609 | 0.181 | Reactome |
| aspirin triggered resolvin E biosynthesis | 2 | 1 (50.0%) | 0.00609 | 0.181 | HumanCyc |
| aspirin triggered resolvin D biosynthesis | 2 | 1 (50.0%) | 0.00609 | 0.181 | HumanCyc |
| aspirin-triggered lipoxin biosynthesis | 2 | 1 (50.0%) | 0.00609 | 0.181 | HumanCyc |
| Biosynthesis of DPAn-3-derived protectins and resolvins | 2 | 1 (50.0%) | 0.00609 | 0.181 | Reactome |
| resolvin D biosynthesis | 2 | 1 (50.0%) | 0.00609 | 0.181 | HumanCyc |
| Stimulation of the cell death response by PAK-2p34 | 2 | 1 (50.0%) | 0.00609 | 0.181 | Reactome |
| Regulation of PAK-2p34 activity by PS-GAP/RHG10 | 2 | 1 (50.0%) | 0.00609 | 0.181 | Reactome |
| SLC-mediated transmembrane transport | 243 | 4 (1.6%) | 0.00626 | 0.181 | Reactome |
| Fas ligand pathway and stress induction of heat shock proteins | 43 | 2 (4.7%) | 0.00758 | 0.183 | Wikipathways |
| Biosynthesis of aspirin-triggered D-series resolvins | 3 | 1 (33.3%) | 0.00913 | 0.183 | Reactome |
| lipoxin biosynthesis | 3 | 1 (33.3%) | 0.00913 | 0.183 | HumanCyc |
| FMO oxidises nucleophiles | 3 | 1 (33.3%) | 0.00913 | 0.183 | Reactome |
| Ovarian steroidogenesis - Homo sapiens (human) | 51 | 2 (3.9%) | 0.0105 | 0.183 | KEGG |
| Regulation of Actin Cytoskeleton | 151 | 3 (2.0%) | 0.0109 | 0.183 | Wikipathways |
| Apoptosis-related network due to altered Notch3 in ovarian cancer | 53 | 2 (3.8%) | 0.0114 | 0.183 | Wikipathways |
| Biosynthesis of E-series 18(R)-resolvins | 4 | 1 (25.0%) | 0.0122 | 0.183 | Reactome |
| Biosynthesis of DPAn-3 SPMs | 4 | 1 (25.0%) | 0.0122 | 0.183 | Reactome |
| Biosynthesis of DPA-derived SPMs | 4 | 1 (25.0%) | 0.0122 | 0.183 | Reactome |
| Biosynthesis of D-series resolvins | 4 | 1 (25.0%) | 0.0122 | 0.183 | Reactome |
| Resolution of AP sites via the single-nucleotide replacement pathway | 4 | 1 (25.0%) | 0.0122 | 0.183 | Reactome |
| Regulation of Apoptosis | 4 | 1 (25.0%) | 0.0122 | 0.183 | Reactome |
| RHOJ GTPase cycle | 56 | 2 (3.6%) | 0.0126 | 0.183 | Reactome |
| RHOQ GTPase cycle | 60 | 2 (3.3%) | 0.0144 | 0.192 | Reactome |
| Biosynthesis of E-series 18(S)-resolvins | 5 | 1 (20.0%) | 0.0152 | 0.192 | Reactome |
| Catalytic cycle of mammalian Flavin-containing MonoOxygenases (FMOs) | 5 | 1 (20.0%) | 0.0152 | 0.192 | Wikipathways |
| hiv-1 defeats host-mediated resistance by cem15 | 5 | 1 (20.0%) | 0.0152 | 0.192 | BioCarta |
| Gap-filling DNA repair synthesis and ligation in TC-NER | 66 | 2 (3.0%) | 0.0172 | 0.192 | Reactome |
| Ciliopathies | 183 | 3 (1.6%) | 0.0181 | 0.192 | Wikipathways |
| Acrosome Reaction and Sperm:Oocyte Membrane Binding | 6 | 1 (16.7%) | 0.0182 | 0.192 | Reactome |
| Synthesis of Lipoxins (LX) | 6 | 1 (16.7%) | 0.0182 | 0.192 | Reactome |
| Biosynthesis of EPA-derived SPMs | 6 | 1 (16.7%) | 0.0182 | 0.192 | Reactome |
| leukotriene biosynthesis | 6 | 1 (16.7%) | 0.0182 | 0.192 | HumanCyc |
| CDC42 signaling events | 71 | 2 (2.8%) | 0.0198 | 0.192 | PID |
| TCR signaling | 73 | 2 (2.7%) | 0.0208 | 0.192 | Reactome |
| APEX1-Independent Resolution of AP Sites via the Single Nucleotide Replacement Pathway | 7 | 1 (14.3%) | 0.0212 | 0.192 | Reactome |
| APOBEC3G mediated resistance to HIV-1 infection | 7 | 1 (14.3%) | 0.0212 | 0.192 | Reactome |
| RHOG GTPase cycle | 74 | 2 (2.7%) | 0.0214 | 0.192 | Reactome |
| Fibroblast growth factor-1 | 74 | 2 (2.7%) | 0.0214 | 0.192 | NetPath |
| Transcription-Coupled Nucleotide Excision Repair (TC-NER) | 79 | 2 (2.5%) | 0.0241 | 0.192 | Reactome |
| Biosynthesis of maresins | 8 | 1 (12.5%) | 0.0242 | 0.192 | Reactome |
| Nef and signal transduction | 8 | 1 (12.5%) | 0.0242 | 0.192 | Reactome |
| Conjugation of salicylate with glycine | 8 | 1 (12.5%) | 0.0242 | 0.192 | Reactome |
| Dual hijack model of Vif in HIV infection | 8 | 1 (12.5%) | 0.0242 | 0.192 | Wikipathways |
| oleate biosynthesis | 8 | 1 (12.5%) | 0.0242 | 0.192 | HumanCyc |
| palmitate biosynthesis | 9 | 1 (12.5%) | 0.0242 | 0.192 | HumanCyc |
| Synthesis of 5-eicosatetraenoic acids | 9 | 1 (11.1%) | 0.0271 | 0.2 | Reactome |
| p75NTR recruits signalling complexes | 9 | 1 (11.1%) | 0.0271 | 0.2 | Reactome |
| Conjugation of carboxylic acids | 9 | 1 (11.1%) | 0.0271 | 0.2 | Reactome |
| Amino Acid conjugation | 9 | 1 (11.1%) | 0.0271 | 0.2 | Reactome |
| RAC2 GTPase cycle | 85 | 2 (2.4%) | 0.0277 | 0.2 | Reactome |
| Regulation of actin cytoskeleton - Homo sapiens (human) | 218 | 3 (1.4%) | 0.0286 | 0.2 | KEGG |
| ECM-receptor interaction - Homo sapiens (human) | 88 | 2 (2.3%) | 0.0295 | 0.2 | KEGG |
| Host Interactions of HIV factors | 89 | 2 (2.2%) | 0.0301 | 0.2 | Reactome |
| HDR through MMEJ (alt-NHEJ) | 10 | 1 (10.0%) | 0.0301 | 0.2 | Reactome |
| Vif-mediated degradation of APOBEC3G | 10 | 1 (10.0%) | 0.0301 | 0.2 | Reactome |
| 3-oxo-10R-octadecatrienoate beta-oxidation | 11 | 1 (9.1%) | 0.0331 | 0.215 | EHMN |
| RAC3 GTPase cycle | 94 | 2 (2.1%) | 0.0333 | 0.215 | Reactome |
| Nucleotide Excision Repair | 97 | 2 (2.1%) | 0.0353 | 0.216 | Reactome |
| Transport of nucleosides and free purine and pyrimidine bases across the plasma membrane | 12 | 1 (8.3%) | 0.036 | 0.216 | Reactome |
| CD28 dependent Vav1 pathway | 12 | 1 (8.3%) | 0.036 | 0.216 | Reactome |
| Transport of organic anions | 12 | 1 (8.3%) | 0.036 | 0.216 | Reactome |
| Integrin-mediated Cell Adhesion | 101 | 2 (2.0%) | 0.038 | 0.216 | Wikipathways |
| Leukotriene metabolism | 102 | 2 (2.0%) | 0.0386 | 0.216 | EHMN |
| Activation of RAC1 | 13 | 1 (7.7%) | 0.039 | 0.216 | Reactome |

| | | | | | |
|---|---|---|---|---|---|
| Ganglio Sphingolipid Metabolism | 13 | 1 (7.7%) | 0.039 | 0.216 | **Wikipathways** |
| Platelet Adhesion to exposed collagen | 13 | 1 (7.7%) | 0.039 | 0.216 | **Reactome** |
| stearate biosynthesis | 13 | 1 (7.7%) | 0.039 | 0.216 | **HumanCyc** |
| Degradation of the extracellular matrix | 103 | 2 (1.9%) | 0.0393 | 0.216 | **Reactome** |
| γ-linolenate biosynthesis | 14 | 1 (7.1%) | 0.0419 | 0.227 | **HumanCyc** |
| Transport of small molecules | 641 | 5 (0.8%) | 0.0442 | 0.231 | **Reactome** |
| Glycosphingolipid biosynthesis - ganglio series - Homo sapiens (human) | 15 | 1 (6.7%) | 0.0448 | 0.231 | **KEGG** |
| Sema3A PAK dependent Axon repulsion | 16 | 1 (6.2%) | 0.0478 | 0.231 | **Reactome** |
| p75NTR signals via NF-kB | 16 | 1 (6.2%) | 0.0478 | 0.231 | **Reactome** |
| Elevation of cytosolic Ca2+ levels | 16 | 1 (6.2%) | 0.0478 | 0.231 | **Reactome** |

Prediabetes

| pathway name | set size | candidates contained | p-value | q-value | pathway source |
|---|---|---|---|---|---|
| Pre-NOTCH Processing in Golgi | 12 | 2 (16.7%) | 0.000773 | 0.355 | Reactome |
| Ephrin B reverse signaling | 24 | 2 (8.3%) | 0.00315 | 0.355 | PID |
| Glycosphingolipid biosynthesis - lacto and neolacto series - Homo sapiens (human) | 27 | 2 (7.4%) | 0.00398 | 0.355 | KEGG |
| Sialic acid metabolism | 33 | 2 (6.1%) | 0.0059 | 0.355 | Reactome |
| PDE3B signalling | 2 | 1 (50.0%) | 0.00699 | 0.355 | Reactome |
| PKB-mediated events | 2 | 1 (50.0%) | 0.00699 | 0.355 | Reactome |
| PI3K Cascade | 44 | 2 (4.5%) | 0.0103 | 0.355 | Reactome |
| DNA IR-double strand breaks and cellular response via ATM | 3 | 1 (33.3%) | 0.0105 | 0.355 | Wikipathways |
| IRS-mediated signalling | 48 | 2 (4.2%) | 0.0122 | 0.355 | Reactome |
| IRS-related events triggered by IGF1R | 52 | 2 (3.8%) | 0.0142 | 0.355 | Reactome |
| IGF1R signaling cascade | 53 | 2 (3.8%) | 0.0147 | 0.355 | Reactome |
| Signaling by Type 1 Insulin-like Growth Factor 1 Receptor (IGF1R) | 53 | 2 (3.8%) | 0.0147 | 0.355 | Reactome |
| Insulin receptor signalling cascade | 54 | 2 (3.7%) | 0.0153 | 0.355 | Reactome |
| Class IB PI3K non-lipid kinase events | 5 | 1 (20.0%) | 0.0174 | 0.355 | PID |
| Vitamin B2 (riboflavin) metabolism | 5 | 1 (20.0%) | 0.0174 | 0.355 | EHMN |
| Riboflavin Metabolism | 5 | 1 (20.0%) | 0.0174 | 0.355 | SMPDB |
| Synthesis of substrates in N-glycan biosythesis | 63 | 2 (3.2%) | 0.0204 | 0.355 | Reactome |
| RUNX1 regulates transcription of genes involved in BCR signaling | 6 | 1 (16.7%) | 0.0208 | 0.355 | Reactome |
| FGFR1b ligand binding and activation | 6 | 1 (16.7%) | 0.0208 | 0.355 | Reactome |
| Pre-NOTCH Processing in the Endoplasmic Reticulum | 6 | 1 (16.7%) | 0.0208 | 0.355 | Reactome |
| Pantothenate and CoA Biosynthesis | 6 | 1 (16.7%) | 0.0208 | 0.355 | SMPDB |
| Vitamin B2 (riboflavin) metabolism | 7 | 1 (14.3%) | 0.0242 | 0.355 | Reactome |
| Gene expression (Transcription) | 1439 | 10 (0.7%) | 0.0248 | 0.355 | Reactome |
| NOTCH4 Intracellular Domain Regulates Transcription | 8 | 1 (12.5%) | 0.0277 | 0.355 | Reactome |
| Riboflavin metabolism - Homo sapiens (human) | 8 | 1 (12.5%) | 0.0277 | 0.355 | KEGG |
| Biosynthesis of the N-glycan precursor (dolichol lipid-linked oligosaccharide, LLO) and transfer to a nascent protein | 77 | 2 (2.6%) | 0.0297 | 0.355 | Reactome |
| Signaling by Insulin receptor | 78 | 2 (2.6%) | 0.0304 | 0.355 | Reactome |
| NOTCH4 Activation and Transmission of Signal to the Nucleus | 9 | 1 (11.1%) | 0.0311 | 0.355 | Reactome |
| ATR Signaling | 9 | 1 (11.1%) | 0.0311 | 0.355 | Wikipathways |
| polyadenylation of mrna | 9 | 1 (11.1%) | 0.0311 | 0.355 | BioCarta |
| Focal Adhesion | 198 | 3 (1.5%) | 0.0318 | 0.355 | Wikipathways |
| Gamma-carboxylation of protein precursors | 10 | 1 (10.0%) | 0.0345 | 0.355 | Reactome |
| Removal of aminoterminal propeptides from gamma-carboxylated proteins | 10 | 1 (10.0%) | 0.0345 | 0.355 | Reactome |
| FGFR2b ligand binding and activation | 10 | 1 (10.0%) | 0.0345 | 0.355 | Reactome |
| Organic anion transporters | 10 | 1 (10.0%) | 0.0345 | 0.355 | Reactome |
| RNA Polymerase II Transcription | 1307 | 9 (0.7%) | 0.035 | 0.355 | Reactome |
| Rap1 signaling pathway - Homo sapiens (human) | 210 | 3 (1.4%) | 0.0369 | 0.355 | KEGG |
| Gamma-carboxylation, transport, and amino-terminal cleavage of proteins | 11 | 1 (9.1%) | 0.0378 | 0.355 | Reactome |
| PTF1A related regulatory pathway | 11 | 1 (9.1%) | 0.0378 | 0.355 | Wikipathways |
| regulators of bone mineralization | 11 | 1 (9.1%) | 0.0378 | 0.355 | BioCarta |
| Regulation of actin cytoskeleton - Homo sapiens (human) | 218 | 3 (1.4%) | 0.0405 | 0.355 | KEGG |
| Negative regulation of NOTCH4 signaling | 12 | 1 (8.3%) | 0.0412 | 0.355 | Reactome |
| Transport of Mature mRNA Derived from an Intronless Transcript | 12 | 1 (8.3%) | 0.0412 | 0.355 | Reactome |
| Amplification of signal from unattached kinetochores via a MAD2 inhibitory signal | 96 | 2 (2.1%) | 0.0444 | 0.355 | Reactome |
| Amplification of signal from the kinetochores | 96 | 2 (2.1%) | 0.0444 | 0.355 | Reactome |
| FGFRL1 modulation of FGFR1 signaling | 13 | 1 (7.7%) | 0.0446 | 0.355 | Reactome |
| RHO GTPases activate IQGAPs | 13 | 1 (7.7%) | 0.0446 | 0.355 | Reactome |
| eNOS activation | 13 | 1 (7.7%) | 0.0446 | 0.355 | Reactome |
| NO metabolism in cystic fibrosis | 13 | 1 (7.7%) | 0.0446 | 0.355 | Wikipathways |
| Transport of Mature mRNAs Derived from Intronless Transcripts | 13 | 1 (7.7%) | 0.0446 | 0.355 | Reactome |
| EML4 and NUDC in mitotic spindle formation | 100 | 2 (2.0%) | 0.0478 | 0.355 | Reactome |
| Osteoblast Signaling | 14 | 1 (7.1%) | 0.0479 | 0.355 | Wikipathways |
| Nicotinate and Nicotinamide Metabolism | 14 | 1 (7.1%) | 0.0479 | 0.355 | SMPDB |
| PIWI-interacting RNA (piRNA) biogenesis | 14 | 1 (7.1%) | 0.0479 | 0.355 | Reactome |
| RAF/MAP kinase cascade | 235 | 3 (1.3%) | 0.0488 | 0.355 | Reactome |
| MAPK1/MAPK3 signaling | 236 | 3 (1.3%) | 0.0493 | 0.355 | Reactome |
| Generic Transcription Pathway | 1178 | 8 (0.7%) | 0.0497 | 0.355 | |

T1DM

| pathway name | set size | candidates contained | p-value | q-value | pathway source |
|---|---|---|---|---|---|
| MHC class II antigen presentation | 60 | 6 (10.0%) | 1.91E-06 | 0.00148 | Reactome |
| Asthma - Homo sapiens (human) | 31 | 4 (13.3%) | 3.44E-05 | 0.0134 | KEGG |
| Allograft rejection - Homo sapiens (human) | 38 | 4 (10.8%) | 8.03E-05 | 0.0208 | KEGG |
| Graft-versus-host disease - Homo sapiens (human) | 42 | 4 (9.5%) | 0.000133 | 0.0208 | KEGG |
| Type I diabetes mellitus - Homo sapiens (human) | 43 | 4 (9.3%) | 0.000146 | 0.0208 | KEGG |
| Ebola Virus Pathway on Host | 130 | 6 (4.6%) | 0.00016 | 0.0208 | Wikipathways |
| Intestinal immune network for IgA production - Homo sapiens (human) | 49 | 4 (8.5%) | 0.000207 | 0.0217 | KEGG |
| Th1 and Th2 cell differentiation - Homo sapiens (human) | 92 | 5 (5.4%) | 0.000272 | 0.0217 | KEGG |
| Allograft Rejection | 93 | 5 (5.4%) | 0.000286 | 0.0217 | Wikipathways |
| Staphylococcus aureus infection - Homo sapiens (human) | 96 | 5 (5.3%) | 0.000301 | 0.0217 | KEGG |
| Autoimmune thyroid disease - Homo sapiens (human) | 53 | 4 (7.7%) | 0.000306 | 0.0217 | KEGG |
| Hematopoietic cell lineage - Homo sapiens (human) | 99 | 5 (5.1%) | 0.000365 | 0.0237 | KEGG |
| Viral myocarditis - Homo sapiens (human) | 60 | 4 (6.8%) | 0.000498 | 0.0298 | KEGG |
| Pre-NOTCH Processing in the Endoplasmic Reticulum | 6 | 2 (33.3%) | 0.00057 | 0.0317 | Reactome |
| Translocation of ZAP-70 to Immunological synapse | 28 | 3 (10.7%) | 0.00069 | 0.035 | Reactome |
| Inflammatory bowel disease - Homo sapiens (human) | 65 | 4 (6.2%) | 0.000721 | 0.035 | KEGG |
| Phosphorylation of CD3 and TCR zeta chains | 31 | 3 (9.7%) | 0.000934 | 0.0428 | Reactome |
| TCR signaling | 73 | 4 (5.5%) | 0.00112 | 0.0482 | Reactome |
| Leishmaniasis - Homo sapiens (human) | 77 | 4 (5.3%) | 0.0013 | 0.0531 | KEGG |
| Antigen processing and presentation - Homo sapiens (human) | 78 | 4 (5.1%) | 0.00143 | 0.0555 | KEGG |
| Systemic lupus erythematosus - Homo sapiens (human) | 136 | 5 (3.7%) | 0.00151 | 0.0558 | KEGG |
| Pre-NOTCH Processing in Golgi | 12 | 2 (16.7%) | 0.00245 | 0.0815 | Reactome |
| Human T-cell leukemia virus 1 infection - Homo sapiens (human) | 219 | 6 (2.7%) | 0.00248 | 0.0815 | KEGG |
| Renin-angiotensin-aldosterone system (RAAS) | 44 | 3 (6.8%) | 0.0026 | 0.0815 | Wikipathways |
| Rheumatoid arthritis - Homo sapiens (human) | 93 | 4 (4.3%) | 0.00262 | 0.0815 | KEGG |
| Sudden Infant Death Syndrome (SIDS) Susceptibility Pathways | 159 | 5 (3.1%) | 0.00318 | 0.095 | Wikipathways |
| Biogenic Amine Synthesis | 15 | 2 (13.3%) | 0.00385 | 0.111 | Wikipathways |
| Downstream TCR signaling | 52 | 3 (5.8%) | 0.00418 | 0.116 | Reactome |
| Metabolism of amine-derived hormones | 16 | 2 (12.5%) | 0.00438 | 0.117 | Reactome |
| Th17 cell differentiation - Homo sapiens (human) | 107 | 4 (3.7%) | 0.0045 | 0.117 | KEGG |
| Toxoplasmosis - Homo sapiens (human) | 112 | 4 (3.6%) | 0.0053 | 0.133 | KEGG |
| NCAM signaling for neurite out-growth | 59 | 3 (5.1%) | 0.00596 | 0.145 | Reactome |
| Folate biosynthesis - Homo sapiens (human) | 26 | 2 (7.7%) | 0.0114 | 0.26 | KEGG |
| Canonical and non-canonical Notch signaling | 27 | 2 (7.4%) | 0.0123 | 0.26 | Wikipathways |
| Herpes simplex virus 1 infection - Homo sapiens (human) | 498 | 8 (1.6%) | 0.0125 | 0.26 | KEGG |
| Notch | 78 | 3 (3.8%) | 0.0128 | 0.26 | INOH |
| Regulatory circuits of the STAT3 signaling pathway | 78 | 3 (3.8%) | 0.0128 | 0.26 | Wikipathways |
| Notch-HLH transcription pathway | 28 | 2 (7.1%) | 0.0131 | 0.26 | Reactome |
| Cell adhesion molecules - Homo sapiens (human) | 149 | 4 (2.7%) | 0.0138 | 0.26 | KEGG |
| Adaptive Immune System | 719 | 10 (1.4%) | 0.014 | 0.26 | Reactome |
| Synthesis of IP3 and IP4 in the cytosol | 29 | 2 (6.9%) | 0.0141 | 0.26 | Reactome |
| Interaction between L1 and Ankyrins | 29 | 2 (6.9%) | 0.0141 | 0.26 | Reactome |
| Phagosome - Homo sapiens (human) | 152 | 4 (2.6%) | 0.0148 | 0.267 | KEGG |
| C-MYB transcription factor network | 85 | 3 (3.5%) | 0.0161 | 0.281 | PID |
| Insulin secretion - Homo sapiens (human) | 86 | 3 (3.5%) | 0.0166 | 0.281 | KEGG |
| PD-1 signaling | 32 | 2 (6.2%) | 0.017 | 0.281 | Reactome |
| Alpha-synuclein signaling | 32 | 2 (6.2%) | 0.017 | 0.281 | PID |
| Role of Osx and miRNAs in tooth development | 33 | 2 (6.1%) | 0.018 | 0.284 | Wikipathways |
| Toxicity of botulinum toxin type C (botC) | 3 | 1 (33.3%) | 0.0186 | 0.284 | Reactome |
| Inositol transporters | 3 | 1 (33.3%) | 0.0186 | 0.284 | Reactome |
| *N-acetylglucosamine degradation I* | 3 | 1 (33.3%) | 0.0186 | 0.284 | HumanCyc |
| Influenza A - Homo sapiens (human) | 172 | 4 (2.3%) | 0.0227 | 0.306 | KEGG |
| Aldosterone synthesis and secretion - Homo sapiens (human) | 98 | 3 (3.1%) | 0.0234 | 0.306 | KEGG |
| eIF5A regulation in response to inhibition of the nuclear export system | 4 | 1 (25.0%) | 0.0248 | 0.306 | Wikipathways |
| Hypusine synthesis from eIF5A-lysine | 4 | 1 (25.0%) | 0.0248 | 0.306 | Reactome |
| Fosphenytoin (Antiarrhythmic) Metabolism Pathway | 4 | 1 (25.0%) | 0.0248 | 0.306 | SMPDB |
| Catecholamine biosynthesis | 4 | 1 (25.0%) | 0.0248 | 0.306 | Reactome |
| catecholamine biosynthesis | 4 | 1 (25.0%) | 0.0248 | 0.306 | HumanCyc |
| Tyrosine hydroxylase deficiency | 4 | 1 (25.0%) | 0.0248 | 0.306 | SMPDB |
| Catecholamine Biosynthesis | 4 | 1 (25.0%) | 0.0248 | 0.306 | SMPDB |
| Aromatic L-Aminoacid Decarboxylase Deficiency | 4 | 1 (25.0%) | 0.0248 | 0.306 | SMPDB |

| | | | | | |
|---|---|---|---|---|---|
| Regulation of Apoptosis | 4 | 1 (25.0%) | 0.0248 | 0.306 | **Reactome** |
| *N-acetylglucosamine degradation II* | 4 | 1 (25.0%) | 0.0248 | 0.306 | **HumanCyc** |
| Tuberculosis - Homo sapiens (human) | 180 | 4 (2.2%) | 0.0258 | 0.313 | **KEGG** |
| Inositol phosphate metabolism | 41 | 2 (4.9%) | 0.0271 | 0.324 | **INOH** |
| Generation of second messenger molecules | 42 | 2 (4.8%) | 0.0283 | 0.334 | **Reactome** |
| PAR1-mediated thrombin signaling events | 43 | 2 (4.7%) | 0.0296 | 0.334 | **PID** |
| Kinesins | 43 | 2 (4.7%) | 0.0296 | 0.334 | **Reactome** |
| Serotonin and melatonin biosynthesis | 5 | 1 (20.0%) | 0.0309 | 0.334 | **Reactome** |
| serotonin and melatonin biosynthesis | 5 | 1 (20.0%) | 0.0309 | 0.334 | **HumanCyc** |
| The fatty acid cycling model | 5 | 1 (20.0%) | 0.0309 | 0.334 | **Reactome** |
| The proton buffering model | 5 | 1 (20.0%) | 0.0309 | 0.334 | **Reactome** |
| Notch Signaling | 45 | 2 (4.4%) | 0.0322 | 0.343 | **Wikipathways** |
| Activation of C3 and C5 | 6 | 1 (16.7%) | 0.0369 | 0.366 | **Reactome** |
| allopregnanolone biosynthesis | 6 | 1 (16.7%) | 0.0369 | 0.366 | **HumanCyc** |
| Somatroph axis (GH) and its relationship to dietary restriction and aging | 6 | 1 (16.7%) | 0.0369 | 0.366 | **Wikipathways** |
| Mitochondrial Uncoupling | 6 | 1 (16.7%) | 0.0369 | 0.366 | **Reactome** |
| methionine degradation | 6 | 1 (16.7%) | 0.0369 | 0.366 | **HumanCyc** |
| Epstein-Barr virus infection - Homo sapiens (human) | 202 | 4 (2.0%) | 0.0372 | 0.366 | **KEGG** |
| Cocaine addiction - Homo sapiens (human) | 49 | 2 (4.1%) | 0.0376 | 0.366 | **KEGG** |
| Thyroid hormone signaling pathway - Homo sapiens (human) | 121 | 3 (2.5%) | 0.0401 | 0.384 | **KEGG** |
| Inositol phosphate metabolism | 51 | 2 (3.9%) | 0.0404 | 0.384 | **Reactome** |
| Signaling events mediated by Stem cell factor receptor (c-Kit) | 52 | 2 (3.8%) | 0.0419 | 0.388 | **PID** |
| Erythropoietin activates Phospholipase C gamma (PLCG) | 7 | 1 (14.3%) | 0.043 | 0.388 | **Reactome** |
| Erythropoietin activates STAT5 | 7 | 1 (14.3%) | 0.043 | 0.388 | **Reactome** |
| Apoptosis-related network due to altered Notch3 in ovarian cancer | 53 | 2 (3.8%) | 0.0433 | 0.388 | **Wikipathways** |
| Notch signaling pathway | 53 | 2 (3.8%) | 0.0433 | 0.388 | **PID** |
| Fanconi anemia pathway - Homo sapiens (human) | 54 | 2 (3.7%) | 0.0448 | 0.396 | **KEGG** |
| The alternative pathway of fetal androgen synthesis | 8 | 1 (12.5%) | 0.049 | 0.401 | **Wikipathways** |
| NOTCH4 Intracellular Domain Regulates Transcription | 8 | 1 (12.5%) | 0.049 | 0.401 | **Reactome** |
| Fatty Acids bound to GPR40 (FFAR1) regulate insulin secretion | 8 | 1 (12.5%) | 0.049 | 0.401 | **Reactome** |
| Regulation of PTEN localization | 8 | 1 (12.5%) | 0.049 | 0.401 | **Reactome** |
| Inactivation of CDC42 and RAC1 | 8 | 1 (12.5%) | 0.049 | 0.401 | **Reactome** |
| cysteine biosynthesis | 8 | 1 (12.5%) | 0.049 | 0.401 | **HumanCyc** |
| FTO Obesity Variant Mechanism | 8 | 1 (12.5%) | 0.049 | 0.401 | **Wikipathways** |
| Dopaminergic synapse - Homo sapiens (human) | 132 | 3 (2.3%) | 0.0497 | 0.403 | **KEGG** |

T2DM

| pathway name | set size | candidates contained | p-value | q-value | pathway source |
|---|---|---|---|---|---|
| Vitamin B3 (nicotinate and nicotinamide) metabolism | 47 | 4 (8.5%) | 0.000803 | 0.544 | EHMN |
| Pre-NOTCH Processing in the Endoplasmic Reticulum | 6 | 2 (33.3%) | 0.00116 | 0.544 | Reactome |
| Cytochrome c-mediated apoptotic response | 12 | 2 (16.7%) | 0.00493 | 0.544 | Reactome |
| Pre-NOTCH Processing in Golgi | 12 | 2 (16.7%) | 0.00493 | 0.544 | Reactome |
| Lysosome - Homo sapiens (human) | 128 | 5 (3.9%) | 0.00578 | 0.544 | KEGG |
| role of mitochondria in apoptotic signaling | 13 | 2 (15.4%) | 0.00579 | 0.544 | BioCarta |
| Nicotinate and Nicotinamide Metabolism | 14 | 2 (14.3%) | 0.00671 | 0.544 | SMPDB |
| Transcriptional misregulation in cancer - Homo sapiens (human) | 192 | 6 (3.1%) | 0.00732 | 0.544 | KEGG |
| Notch Signaling | 45 | 3 (6.7%) | 0.00751 | 0.544 | Wikipathways |
| Apoptotic factor-mediated response | 16 | 2 (12.5%) | 0.00875 | 0.544 | Reactome |
| FLT3 mutants bind TKIs | 1 | 1 (100.0%) | 0.00893 | 0.544 | Reactome |
| Mtb iron assimilation by chelation | 1 | 1 (100.0%) | 0.00893 | 0.544 | Reactome |
| Latent infection - Other responses of Mtb to phagocytosis | 1 | 1 (100.0%) | 0.00893 | 0.544 | Reactome |
| Apoptosis Modulation and Signaling | 92 | 4 (4.3%) | 0.00925 | 0.544 | Wikipathways |
| Intrinsic Pathway for Apoptosis | 52 | 3 (5.8%) | 0.0112 | 0.544 | Reactome |
| Nanomaterial induced apoptosis | 20 | 2 (10.0%) | 0.0135 | 0.544 | Wikipathways |
| Herpes simplex virus 1 infection - Homo sapiens (human) | 498 | 10 (2.0%) | 0.0137 | 0.544 | KEGG |
| Regulation of apoptosis by parathyroid hormone-related protein | 21 | 2 (9.5%) | 0.0149 | 0.544 | Wikipathways |
| Notch | 58 | 3 (5.2%) | 0.015 | 0.544 | NetPath |
| SHC1 events in ERBB2 signaling | 22 | 2 (9.1%) | 0.0163 | 0.544 | Reactome |
| Wnt-beta-catenin signaling pathway in leukemia | 22 | 2 (9.1%) | 0.0163 | 0.544 | Wikipathways |
| MHC class II antigen presentation | 60 | 3 (5.0%) | 0.0164 | 0.544 | Reactome |
| Steroid hormone biosynthesis - Homo sapiens (human) | 61 | 3 (4.9%) | 0.0172 | 0.544 | KEGG |
| Negative regulators of DDX58/IFIH1 signaling | 23 | 2 (8.7%) | 0.0177 | 0.544 | Reactome |
| GPCRs, Class B Secretin-like | 23 | 2 (8.7%) | 0.0177 | 0.544 | Wikipathways |
| Activation and oligomerization of BAK protein | 2 | 1 (50.0%) | 0.0178 | 0.544 | Reactome |
| GDP-L-fucose biosynthesis II (from L-fucose) | 2 | 1 (50.0%) | 0.0178 | 0.544 | HumanCyc |
| NAD phosphorylation and dephosphorylation | 2 | 1 (50.0%) | 0.0178 | 0.544 | HumanCyc |
| Cytosolic iron-sulfur cluster assembly | 2 | 1 (50.0%) | 0.0178 | 0.544 | Reactome |
| ChREBP activates metabolic gene expression | 2 | 1 (50.0%) | 0.0178 | 0.544 | Reactome |
| Pathways in cancer - Homo sapiens (human) | 531 | 10 (1.9%) | 0.0205 | 0.544 | KEGG |
| ID | 26 | 2 (7.7%) | 0.0224 | 0.544 | NetPath |
| Canonical and non-canonical Notch signaling | 27 | 2 (7.4%) | 0.024 | 0.544 | Wikipathways |
| Sulfur amino acid metabolism | 27 | 2 (7.4%) | 0.024 | 0.544 | Reactome |
| Notch-HLH transcription pathway | 28 | 2 (7.1%) | 0.0257 | 0.544 | Reactome |
| methionine salvage | 3 | 1 (33.3%) | 0.0266 | 0.544 | HumanCyc |
| Interconversion of 2-oxoglutarate and 2-hydroxyglutarate | 3 | 1 (33.3%) | 0.0266 | 0.544 | Reactome |
| Downregulation of ERBB2 signaling | 29 | 2 (6.9%) | 0.0275 | 0.544 | Reactome |
| Interaction between L1 and Ankyrins | 29 | 2 (6.9%) | 0.0275 | 0.544 | Reactome |
| Human papillomavirus infection - Homo sapiens (human) | 331 | 7 (2.1%) | 0.0289 | 0.544 | KEGG |
| EGFR downregulation | 30 | 2 (6.7%) | 0.0292 | 0.544 | Reactome |
| ADORA2B mediated anti-inflammatory cytokines production | 132 | 4 (3.0%) | 0.0306 | 0.544 | Reactome |
| Vitamin B5 - CoA biosynthesis from pantothenate | 31 | 2 (6.5%) | 0.0311 | 0.544 | EHMN |
| Nuclear Envelope (NE) Reassembly | 78 | 3 (3.8%) | 0.0326 | 0.544 | Reactome |
| Apoptosis - multiple species - Homo sapiens (human) | 32 | 2 (6.2%) | 0.033 | 0.544 | KEGG |
| White fat cell differentiation | 32 | 2 (6.2%) | 0.033 | 0.544 | Wikipathways |
| IL17 signaling pathway | 32 | 2 (6.2%) | 0.033 | 0.544 | Wikipathways |
| Apoptosis - Homo sapiens (human) | 136 | 4 (2.9%) | 0.0336 | 0.544 | KEGG |
| Glucagon-type ligand receptors | 33 | 2 (6.1%) | 0.0349 | 0.544 | Reactome |
| Role of Osx and miRNAs in tooth development | 33 | 2 (6.1%) | 0.0349 | 0.544 | Wikipathways |
| Sealing of the nuclear envelope (NE) by ESCRT-III | 33 | 2 (6.1%) | 0.0349 | 0.544 | Reactome |
| 2-oxoisovalerate decarboxylation to isobutanoyl-CoA | 4 | 1 (25.0%) | 0.0353 | 0.544 | HumanCyc |
| Lysosomal oligosaccharide catabolism | 4 | 1 (25.0%) | 0.0353 | 0.544 | Reactome |
| phospholipase c delta in phospholipid associated cell signaling | 4 | 1 (25.0%) | 0.0353 | 0.544 | BioCarta |
| *S-methyl-5-thio-α-D-ribose 1-phosphate degradation* | 4 | 1 (25.0%) | 0.0353 | 0.544 | HumanCyc |
| HCN channels | 4 | 1 (25.0%) | 0.0353 | 0.544 | Reactome |
| RHOU GTPase cycle | 34 | 2 (5.9%) | 0.0369 | 0.544 | Reactome |
| Nicotinate and nicotinamide metabolism - Homo sapiens (human) | 35 | 2 (5.7%) | 0.0389 | 0.544 | KEGG |
| G alpha (s) signalling events | 143 | 4 (2.8%) | 0.0392 | 0.544 | Reactome |
| Asparagine N-linked glycosylation | 283 | 6 (2.1%) | 0.0414 | 0.544 | Reactome |
| Apoptosis | 87 | 3 (3.4%) | 0.0429 | 0.544 | Wikipathways |
| Thyroid cancer - Homo sapiens (human) | 37 | 2 (5.4%) | 0.043 | 0.544 | KEGG |
| Activation of caspases through apoptosome-mediated cleavage | 5 | 1 (20.0%) | 0.0439 | 0.544 | Reactome |
| FLT3 signaling through SRC family kinases | 5 | 1 (20.0%) | 0.0439 | 0.544 | Reactome |
| tetrapyrrole biosynthesis | 5 | 1 (20.0%) | 0.0439 | 0.544 | HumanCyc |
| Vitamin B2 (riboflavin) metabolism | 5 | 1 (20.0%) | 0.0439 | 0.544 | EHMN |
| Riboflavin Metabolism | 5 | 1 (20.0%) | 0.0439 | 0.544 | SMPDB |
| MECP2 regulates transcription factors | 5 | 1 (20.0%) | 0.0439 | 0.544 | Reactome |
| basic mechanism of action of ppara pparb(d) and pparg and effects on gene expression | 5 | 1 (20.0%) | 0.0439 | 0.544 | BioCarta |
| role of ppar-gamma coactivators in obesity and thermogenesis | 5 | 1 (20.0%) | 0.0439 | 0.544 | BioCarta |

| | | | | | |
|---|---|---|---|---|---|
| PKA-mediated phosphorylation of key metabolic factors | 5 | 1 (20.0%) | 0.0439 | 0.544 | Reactome |
| ketolysis | 5 | 1 (20.0%) | 0.0439 | 0.544 | HumanCyc |
| Ubiquinone Biosynthesis | 5 | 1 (20.0%) | 0.0439 | 0.544 | SMPDB |
| Class B/2 (Secretin family receptors) | 91 | 3 (3.3%) | 0.0479 | 0.544 | Reactome |
| Oxidative Damage | 40 | 2 (5.0%) | 0.0495 | 0.544 | Wikipathways |
| Class C/3 (Metabotropic glutamate/pheromone receptors) | 40 | 2 (5.0%) | 0.0495 | 0.544 | Reactome |

T2DM_related_admission

| pathway name | set size | candidates contained | p-value | q-value | pathway source |
|---|---|---|---|---|---|
| ABO blood group biosynthesis | 3 | 2 (66.7%) | 0.000299 | 0.327 | Reactome |
| Globo Sphingolipid Metabolism | 19 | 3 (15.8%) | 0.000855 | 0.415 | Wikipathways |
| Pre-NOTCH Processing in the Endoplasmic Reticulum | 6 | 2 (33.3%) | 0.00146 | 0.415 | Reactome |
| GPCRs, Class B Secretin-like | 23 | 3 (13.0%) | 0.00152 | 0.415 | Wikipathways |
| Glycosphingolipid biosynthesis - lactoseries | 7 | 2 (28.6%) | 0.00204 | 0.446 | EHMN |
| Macrophage markers | 9 | 2 (22.2%) | 0.00345 | 0.594 | Wikipathways |
| Cytochrome c-mediated apoptotic response | 12 | 2 (16.7%) | 0.00619 | 0.594 | Reactome |
| Pre-NOTCH Processing in Golgi | 12 | 2 (16.7%) | 0.00619 | 0.594 | Reactome |
| Class C/3 (Metabotropic glutamate/pheromone receptors) | 40 | 3 (7.5%) | 0.00747 | 0.594 | Reactome |
| Nicotinate and Nicotinamide Metabolism | 14 | 2 (14.3%) | 0.00843 | 0.594 | SMPDB |
| Glycosphingolipid biosynthesis - globo and isoglobo series - Homo sapiens (human) | 15 | 2 (13.3%) | 0.00966 | 0.594 | KEGG |
| FLT3 mutants bind TKIs | 1 | 1 (100.0%) | 0.01 | 0.594 | Reactome |
| Mtb iron assimilation by chelation | 1 | 1 (100.0%) | 0.01 | 0.594 | Reactome |
| Latent infection - Other responses of Mtb to phagocytosis | 1 | 1 (100.0%) | 0.01 | 0.594 | Reactome |
| Molybdenum cofactor (Moco) biosynthesis | 1 | 1 (100.0%) | 0.01 | 0.594 | Wikipathways |
| ADORA2B mediated anti-inflammatory cytokines production | 132 | 5 (3.8%) | 0.0107 | 0.594 | Reactome |
| Apoptotic factor-mediated response | 16 | 2 (12.5%) | 0.011 | 0.594 | Reactome |
| Glycosphingolipid biosynthesis - neolactoseries | 16 | 2 (12.5%) | 0.011 | 0.594 | EHMN |
| Vitamin B3 (nicotinate and nicotinamide) metabolism | 47 | 3 (6.4%) | 0.0117 | 0.594 | EHMN |
| Transcriptional misregulation in cancer - Homo sapiens (human) | 192 | 6 (3.1%) | 0.0127 | 0.594 | KEGG |
| Class B/2 (Secretin family receptors) | 91 | 4 (4.4%) | 0.0133 | 0.594 | Reactome |
| Glycosphingolipid biosynthesis - globoseries | 18 | 2 (11.1%) | 0.0138 | 0.594 | EHMN |
| Other glycan degradation - Homo sapiens (human) | 18 | 2 (11.1%) | 0.0138 | 0.594 | KEGG |
| G alpha (s) signalling events | 143 | 5 (3.5%) | 0.0147 | 0.594 | Reactome |
| Intrinsic Pathway for Apoptosis | 52 | 3 (5.8%) | 0.0153 | 0.594 | Reactome |
| Nanomaterial induced apoptosis | 20 | 2 (10.0%) | 0.0169 | 0.594 | Wikipathways |
| Blood group systems biosynthesis | 20 | 2 (10.0%) | 0.0169 | 0.594 | Reactome |
| 3-phosphoinositide degradation | 20 | 2 (10.0%) | 0.0169 | 0.594 | HumanCyc |
| p38 signaling mediated by MAPKAP kinases | 21 | 2 (9.5%) | 0.0186 | 0.594 | PID |
| Leukotriene metabolism | 102 | 4 (3.9%) | 0.0194 | 0.594 | EHMN |
| C21-steroid hormone biosynthesis and metabolism | 57 | 3 (5.3%) | 0.0196 | 0.594 | EHMN |
| Activation and oligomerization of BAK protein | 2 | 1 (50.0%) | 0.02 | 0.594 | Reactome |
| NAD phosphorylation and dephosphorylation | 2 | 1 (50.0%) | 0.02 | 0.594 | HumanCyc |
| Cytosolic iron-sulfur cluster assembly | 2 | 1 (50.0%) | 0.02 | 0.594 | Reactome |
| ChREBP activates metabolic gene expression | 2 | 1 (50.0%) | 0.02 | 0.594 | Reactome |
| Wnt-beta-catenin signaling pathway in leukemia | 22 | 2 (9.1%) | 0.0203 | 0.594 | Wikipathways |
| Notch | 58 | 3 (5.2%) | 0.0205 | 0.594 | NetPath |
| GPCR ligand binding | 476 | 10 (2.1%) | 0.0218 | 0.594 | Reactome |
| Negative regulators of DDX58/IFIH1 signaling | 23 | 2 (8.7%) | 0.0221 | 0.594 | Reactome |
| Steroid hormone biosynthesis - Homo sapiens (human) | 61 | 3 (4.9%) | 0.0234 | 0.594 | KEGG |
| ID | 26 | 2 (7.7%) | 0.0278 | 0.594 | NetPath |
| Folate biosynthesis - Homo sapiens (human) | 26 | 2 (7.7%) | 0.0278 | 0.594 | KEGG |
| SARS-CoV-2 and COVID-19 Pathway | 3 | 1 (33.3%) | 0.0298 | 0.594 | Wikipathways |
| Melanin biosynthesis | 3 | 1 (33.3%) | 0.0298 | 0.594 | Reactome |
| methionine salvage | 3 | 1 (33.3%) | 0.0298 | 0.594 | HumanCyc |
| ARL13B-mediated ciliary trafficking of INPP5E | 3 | 1 (33.3%) | 0.0298 | 0.594 | Reactome |
| Interconversion of 2-oxoglutarate and 2-hydroxyglutarate | 3 | 1 (33.3%) | 0.0298 | 0.594 | Reactome |
| Glycosphingolipid biosynthesis - lacto and neolacto series - Homo sapiens (human) | 27 | 2 (7.4%) | 0.0299 | 0.594 | KEGG |
| Canonical and non-canonical Notch signaling | 27 | 2 (7.4%) | 0.0299 | 0.594 | Wikipathways |
| Sulfur amino acid metabolism | 27 | 2 (7.4%) | 0.0299 | 0.594 | Reactome |
| Notch-HLH transcription pathway | 28 | 2 (7.1%) | 0.032 | 0.594 | Reactome |
| NOTCH-Ncore | 28 | 2 (7.1%) | 0.032 | 0.594 | Signalink |
| Plasma lipoprotein clearance | 29 | 2 (6.9%) | 0.0341 | 0.594 | Reactome |
| Interaction between L1 and Ankyrins | 29 | 2 (6.9%) | 0.0341 | 0.594 | Reactome |
| Parkinson,s disease pathway | 71 | 3 (4.2%) | 0.0346 | 0.594 | Wikipathways |
| Vitamin B5 - CoA biosynthesis from pantothenate | 31 | 2 (6.5%) | 0.0385 | 0.594 | EHMN |
| eumelanin biosynthesis | 4 | 1 (25.0%) | 0.0396 | 0.594 | HumanCyc |
| Lysosomal oligosaccharide catabolism | 4 | 1 (25.0%) | 0.0396 | 0.594 | Reactome |
| Catecholamine biosynthesis | 4 | 1 (25.0%) | 0.0396 | 0.594 | Reactome |
| catecholamine biosynthesis | 4 | 1 (25.0%) | 0.0396 | 0.594 | HumanCyc |
| Tyrosine hydroxylase deficiency | 4 | 1 (25.0%) | 0.0396 | 0.594 | SMPDB |
| Catecholamine Biosynthesis | 4 | 1 (25.0%) | 0.0396 | 0.594 | SMPDB |
| Aromatic L-Aminoacid Decarboxylase Deficiency | 4 | 1 (25.0%) | 0.0396 | 0.594 | SMPDB |
| *S-methyl-5-thio-α-D-ribose 1-phosphate degradation* | 4 | 1 (25.0%) | 0.0396 | 0.594 | HumanCyc |

| | | | | | |
|---|---|---|---|---|---|
| Lysosome - Homo sapiens (human) | 128 | 4 (3.1%) | 0.0401 | 0.594 | KEGG |
| Apoptosis - multiple species - Homo sapiens (human) | 32 | 2 (6.2%) | 0.0409 | 0.594 | KEGG |
| Monoamine Transport | 32 | 2 (6.2%) | 0.0409 | 0.594 | Wikipathways |
| Glucagon-type ligand receptors | 33 | 2 (6.1%) | 0.0432 | 0.594 | Reactome |
| Role of Osx and miRNAs in tooth development | 33 | 2 (6.1%) | 0.0432 | 0.594 | Wikipathways |
| Tyrosine metabolism | 33 | 2 (6.1%) | 0.0432 | 0.594 | INOH |
| RHOU GTPase cycle | 34 | 2 (5.9%) | 0.0456 | 0.594 | Reactome |
| Nicotinate and nicotinamide metabolism - Homo sapiens (human) | 35 | 2 (5.7%) | 0.0481 | 0.594 | KEGG |
| Histidine metabolism | 35 | 2 (5.7%) | 0.0481 | 0.594 | EHMN |
| Degradation pathway of sphingolipids, including diseases | 5 | 1 (20.0%) | 0.0493 | 0.594 | Wikipathways |
| Activation of caspases through apoptosome-mediated cleavage | 5 | 1 (20.0%) | 0.0493 | 0.594 | Reactome |
| HDL clearance | 5 | 1 (20.0%) | 0.0493 | 0.594 | Reactome |
| FLT3 signaling through SRC family kinases | 5 | 1 (20.0%) | 0.0493 | 0.594 | Reactome |
| tetrapyrrole biosynthesis | 5 | 1 (20.0%) | 0.0493 | 0.594 | HumanCyc |
| Vitamin B2 (riboflavin) metabolism | 5 | 1 (20.0%) | 0.0493 | 0.594 | EHMN |
| Riboflavin Metabolism | 5 | 1 (20.0%) | 0.0493 | 0.594 | SMPDB |
| MECP2 regulates transcription factors | 5 | 1 (20.0%) | 0.0493 | 0.594 | Reactome |
| basic mechanism of action of ppara pparb(d) and pparg and effects on gene expression | 5 | 1 (20.0%) | 0.0493 | 0.594 | BioCarta |
| role of ppar-gamma coactivators in obesity and thermogenesis | 5 | 1 (20.0%) | 0.0493 | 0.594 | BioCarta |
| Abacavir transmembrane transport | 5 | 1 (20.0%) | 0.0493 | 0.594 | Reactome |
| PKA-mediated phosphorylation of key metabolic factors | 5 | 1 (20.0%) | 0.0493 | 0.594 | Reactome |
| ketolysis | 5 | 1 (20.0%) | 0.0493 | 0.594 | HumanCyc |
| bupropion degradation | 5 | 1 (20.0%) | 0.0493 | 0.594 | HumanCyc |
| Metabolism | 1954 | 27 (1.4%) | 0.0499 | 0.594 | Reactome |

T2DM_requires_insulin

| pathway name | set size | candidates contained | p-value | q-value | pathway source |
|---|---|---|---|---|---|
| Defective factor XII causes hereditary angioedema | 2 | 2 (100.0%) | 2.99E-05 | 0.0214 | Reactome |
| Herpes simplex virus 1 infection - Homo sapiens (human) | 498 | 11 (2.2%) | 8.21E-05 | 0.0294 | KEGG |
| Defects of contact activation system (CAS) and kallikrein/kinin system (KKS) | 5 | 2 (40.0%) | 0.000296 | 0.0469 | Reactome |
| Diseases of hemostasis | 5 | 2 (40.0%) | 0.000296 | 0.0469 | Reactome |
| MHC class II antigen presentation | 60 | 4 (6.7%) | 0.000328 | 0.0469 | Reactome |
| Pre-NOTCH Processing in the Endoplasmic Reticulum | 6 | 2 (33.3%) | 0.000443 | 0.0528 | Reactome |
| Asthma - Homo sapiens (human) | 31 | 3 (10.0%) | 0.000585 | 0.0599 | KEGG |
| Allograft rejection - Homo sapiens (human) | 38 | 3 (8.1%) | 0.00109 | 0.0895 | KEGG |
| Graft-versus-host disease - Homo sapiens (human) | 42 | 3 (7.1%) | 0.00158 | 0.0895 | KEGG |
| Th1 and Th2 cell differentiation - Homo sapiens (human) | 92 | 4 (4.3%) | 0.00164 | 0.0895 | KEGG |
| Type I diabetes mellitus - Homo sapiens (human) | 43 | 3 (7.0%) | 0.00169 | 0.0895 | KEGG |
| Allograft Rejection | 93 | 4 (4.3%) | 0.00171 | 0.0895 | Wikipathways |
| Pre-NOTCH Processing in Golgi | 12 | 2 (16.7%) | 0.00191 | 0.0895 | Reactome |
| Intestinal immune network for IgA production - Homo sapiens (human) | 49 | 3 (6.4%) | 0.00219 | 0.0895 | KEGG |
| Influenza A - Homo sapiens (human) | 172 | 5 (2.9%) | 0.00256 | 0.0895 | KEGG |
| Autoimmune thyroid disease - Homo sapiens (human) | 53 | 3 (5.8%) | 0.00292 | 0.0895 | KEGG |
| Toxoplasmosis - Homo sapiens (human) | 112 | 4 (3.6%) | 0.00336 | 0.0895 | KEGG |
| Intrinsic Pathway of Fibrin Clot Formation | 17 | 2 (11.8%) | 0.00386 | 0.0895 | Reactome |
| Viral myocarditis - Homo sapiens (human) | 60 | 3 (5.1%) | 0.00418 | 0.0895 | KEGG |
| Epstein-Barr virus infection - Homo sapiens (human) | 202 | 5 (2.5%) | 0.00499 | 0.0895 | KEGG |
| Bivalirudin Action Pathway | 20 | 2 (10.0%) | 0.00533 | 0.0895 | SMPDB |
| Argatroban Action Pathway | 20 | 2 (10.0%) | 0.00533 | 0.0895 | SMPDB |
| Ximelagatran Action Pathway | 20 | 2 (10.0%) | 0.00533 | 0.0895 | SMPDB |
| Lepirudin Action Pathway | 20 | 2 (10.0%) | 0.00533 | 0.0895 | SMPDB |
| Coagulation | 20 | 2 (10.0%) | 0.00533 | 0.0895 | SMPDB |
| Aminocaproic Acid Action Pathway | 20 | 2 (10.0%) | 0.00533 | 0.0895 | SMPDB |
| Tranexamic Acid Action Pathway | 20 | 2 (10.0%) | 0.00533 | 0.0895 | SMPDB |
| Urokinase Action Pathway | 20 | 2 (10.0%) | 0.00533 | 0.0895 | SMPDB |
| Reteplase Action Pathway | 20 | 2 (10.0%) | 0.00533 | 0.0895 | SMPDB |
| Streptokinase Action Pathway | 20 | 2 (10.0%) | 0.00533 | 0.0895 | SMPDB |
| Tenecteplase Action Pathway | 20 | 2 (10.0%) | 0.00533 | 0.0895 | SMPDB |
| Alteplase Action Pathway | 20 | 2 (10.0%) | 0.00533 | 0.0895 | SMPDB |
| Anistreplase Action Pathway | 20 | 2 (10.0%) | 0.00533 | 0.0895 | SMPDB |
| Aprotinin Action Pathway | 20 | 2 (10.0%) | 0.00533 | 0.0895 | SMPDB |
| Phenindione Action Pathway | 20 | 2 (10.0%) | 0.00533 | 0.0895 | SMPDB |
| Dicoumarol Action Pathway | 20 | 2 (10.0%) | 0.00533 | 0.0895 | SMPDB |
| Warfarin Action Pathway | 20 | 2 (10.0%) | 0.00533 | 0.0895 | SMPDB |
| Acenocoumarol Action Pathway | 20 | 2 (10.0%) | 0.00533 | 0.0895 | SMPDB |
| Phenprocoumon Action Pathway | 20 | 2 (10.0%) | 0.00533 | 0.0895 | SMPDB |
| Dicumarol Action Pathway | 20 | 2 (10.0%) | 0.00533 | 0.0895 | SMPDB |
| Inflammatory bowel disease - Homo sapiens (human) | 65 | 3 (4.6%) | 0.00549 | 0.0895 | KEGG |
| Biosynthesis of DPAn-3-derived 13-series resolvins | 1 | 1 (100.0%) | 0.00551 | 0.0895 | Reactome |
| Ebola Virus Pathway on Host | 130 | 4 (3.1%) | 0.00571 | 0.0895 | Wikipathways |
| Ardeparin Action Pathway | 21 | 2 (9.5%) | 0.00587 | 0.0895 | SMPDB |
| Heparin Action Pathway | 21 | 2 (9.5%) | 0.00587 | 0.0895 | SMPDB |
| Fondaparinux Action Pathway | 21 | 2 (9.5%) | 0.00587 | 0.0895 | SMPDB |
| Enoxaparin Action Pathway | 21 | 2 (9.5%) | 0.00587 | 0.0895 | SMPDB |
| eicosanoid metabolism | 23 | 2 (8.7%) | 0.00703 | 0.0987 | BioCarta |
| intrinsic prothrombin activation pathway | 23 | 2 (8.7%) | 0.00703 | 0.0987 | BioCarta |
| Blood Clotting Cascade | 23 | 2 (8.7%) | 0.00703 | 0.0987 | Wikipathways |
| Eicosanoid Synthesis | 23 | 2 (8.7%) | 0.00703 | 0.0987 | Wikipathways |
| Leishmaniasis - Homo sapiens (human) | 77 | 3 (3.9%) | 0.00846 | 0.116 | KEGG |
| Cell adhesion molecules - Homo sapiens (human) | 149 | 4 (2.7%) | 0.00897 | 0.116 | KEGG |
| Antigen processing and presentation - Homo sapiens (human) | 78 | 3 (3.8%) | 0.00908 | 0.116 | KEGG |
| Phagosome - Homo sapiens (human) | 152 | 4 (2.6%) | 0.0096 | 0.116 | KEGG |
| Canonical and non-canonical Notch signaling | 27 | 2 (7.4%) | 0.00961 | 0.116 | Wikipathways |
| Translocation of ZAP-70 to Immunological synapse | 28 | 2 (7.1%) | 0.0103 | 0.116 | Reactome |
| RAS and bradykinin pathways in COVID-19 | 28 | 2 (7.1%) | 0.0103 | 0.116 | Wikipathways |
| Notch-HLH transcription pathway | 28 | 2 (7.1%) | 0.0103 | 0.116 | Reactome |
| Biosynthesis of DPAn-3-derived maresins | 2 | 1 (50.0%) | 0.011 | 0.116 | Reactome |
| Biosynthesis of electrophilic ?-3 PUFA oxo-derivatives | 2 | 1 (50.0%) | 0.011 | 0.116 | Reactome |
| aspirin triggered resolvin E biosynthesis | 2 | 1 (50.0%) | 0.011 | 0.116 | HumanCyc |
| aspirin triggered resolvin D biosynthesis | 2 | 1 (50.0%) | 0.011 | 0.116 | HumanCyc |
| aspirin-triggered lipoxin biosynthesis | 2 | 1 (50.0%) | 0.011 | 0.116 | HumanCyc |
| Biosynthesis of DPAn-3-derived protectins and resolvins | 2 | 1 (50.0%) | 0.011 | 0.116 | Reactome |
| resolvin D biosynthesis | 2 | 1 (50.0%) | 0.011 | 0.116 | HumanCyc |
| Activation and oligomerization of BAK protein | 2 | 1 (50.0%) | 0.011 | 0.116 | Reactome |

| | | | | | |
|---|---|---|---|---|---|
| UMP biosynthesis | 2 | 1 (50.0%) | 0.011 | 0.116 | HumanCyc |
| Phosphorylation of CD3 and TCR zeta chains | 31 | 2 (6.5%) | 0.0126 | 0.13 | Reactome |
| PD-1 signaling | 32 | 2 (6.2%) | 0.0133 | 0.136 | Reactome |
| Role of Osx and miRNAs in tooth development | 33 | 2 (6.1%) | 0.0142 | 0.141 | Wikipathways |
| Rheumatoid arthritis - Homo sapiens (human) | 93 | 3 (3.3%) | 0.0142 | 0.141 | KEGG |
| Staphylococcus aureus infection - Homo sapiens (human) | 96 | 3 (3.2%) | 0.015 | 0.147 | KEGG |
| biotin-carboxyl carrier protein assembly | 3 | 1 (33.3%) | 0.0164 | 0.153 | HumanCyc |
| Biosynthesis of aspirin-triggered D-series resolvins | 3 | 1 (33.3%) | 0.0164 | 0.153 | Reactome |
| lipoxin biosynthesis | 3 | 1 (33.3%) | 0.0164 | 0.153 | HumanCyc |
| Pyrimidine biosynthesis | 3 | 1 (33.3%) | 0.0164 | 0.153 | Reactome |
| Hematopoietic cell lineage - Homo sapiens (human) | 99 | 3 (3.1%) | 0.0168 | 0.154 | KEGG |
| Formation of Fibrin Clot (Clotting Cascade) | 37 | 2 (5.4%) | 0.0176 | 0.16 | Reactome |
| Oxidative Damage | 40 | 2 (5.0%) | 0.0204 | 0.176 | Wikipathways |
| Ferroptosis | 40 | 2 (5.0%) | 0.0204 | 0.176 | Wikipathways |
| Th17 cell differentiation - Homo sapiens (human) | 107 | 3 (2.8%) | 0.0212 | 0.176 | KEGG |
| Ferroptosis - Homo sapiens (human) | 41 | 2 (4.9%) | 0.0214 | 0.176 | KEGG |
| fatty acid biosynthesis initiation | 4 | 1 (25.0%) | 0.0219 | 0.176 | HumanCyc |
| Biosynthesis of E-series 18(R)-resolvins | 4 | 1 (25.0%) | 0.0219 | 0.176 | Reactome |
| Biosynthesis of DPAn-3 SPMs | 4 | 1 (25.0%) | 0.0219 | 0.176 | Reactome |
| Biosynthesis of DPA-derived SPMs | 4 | 1 (25.0%) | 0.0219 | 0.176 | Reactome |
| Biosynthesis of D-series resolvins | 4 | 1 (25.0%) | 0.0219 | 0.176 | Reactome |
| arsenate detoxification I (glutaredoxin) | 4 | 1 (25.0%) | 0.0219 | 0.176 | HumanCyc |
| Generation of second messenger molecules | 42 | 2 (4.8%) | 0.0224 | 0.178 | Reactome |
| Prostaglandin Synthesis and Regulation | 44 | 2 (4.5%) | 0.0244 | 0.188 | Wikipathways |
| CD4 T cell receptor signaling-ERK cascade | 44 | 2 (4.5%) | 0.0244 | 0.188 | INOH |
| Neurodegeneration with brain iron accumulation (NBIA) subtypes pathway | 44 | 2 (4.5%) | 0.0244 | 0.188 | Wikipathways |
| Notch Signaling | 45 | 2 (4.4%) | 0.0254 | 0.194 | Wikipathways |
| Generic Transcription Pathway | 1178 | 12 (1.0%) | 0.0265 | 0.199 | Reactome |
| Biosynthesis of E-series 18(S)-resolvins | 5 | 1 (20.0%) | 0.0272 | 0.199 | Reactome |
| Sulfide oxidation to sulfate | 5 | 1 (20.0%) | 0.0272 | 0.199 | Reactome |
| RUNX1 regulates expression of components of tight junctions | 5 | 1 (20.0%) | 0.0272 | 0.199 | Reactome |
| Pyruvate metabolism - Homo sapiens (human) | 47 | 2 (4.3%) | 0.0276 | 0.199 | KEGG |
| Chondroitin sulfate/dermatan sulfate metabolism | 50 | 2 (4.0%) | 0.0309 | 0.219 | Reactome |
| Cholesterol metabolism - Homo sapiens (human) | 50 | 2 (4.0%) | 0.0309 | 0.219 | KEGG |
| Synthesis of Lipoxins (LX) | 6 | 1 (16.7%) | 0.0326 | 0.22 | Reactome |
| Biosynthesis of EPA-derived SPMs | 6 | 1 (16.7%) | 0.0326 | 0.22 | Reactome |
| leukotriene biosynthesis | 6 | 1 (16.7%) | 0.0326 | 0.22 | HumanCyc |
| allopregnanolone biosynthesis | 6 | 1 (16.7%) | 0.0326 | 0.22 | HumanCyc |
| Acyl chain remodeling of CL | 6 | 1 (16.7%) | 0.0326 | 0.22 | Reactome |
| Downstream TCR signaling | 52 | 2 (3.8%) | 0.0332 | 0.222 | Reactome |
| Notch signaling pathway | 53 | 2 (3.8%) | 0.0344 | 0.228 | PID |
| Systemic lupus erythematosus - Homo sapiens (human) | 136 | 3 (2.2%) | 0.0377 | 0.241 | KEGG |
| TFAP2A acts as a transcriptional repressor during retinoic acid induced cell differentiation | 7 | 1 (14.3%) | 0.0379 | 0.241 | Reactome |
| Activation of NLRP3 Inflammasome by SARS-CoV-2 | 7 | 1 (14.3%) | 0.0379 | 0.241 | Wikipathways |
| CS/DS degradation | 7 | 1 (14.3%) | 0.0379 | 0.241 | Reactome |
| ECM proteoglycans | 56 | 2 (3.6%) | 0.0381 | 0.241 | Reactome |
| CD4 T cell receptor signaling-JNK cascade | 57 | 2 (3.5%) | 0.0393 | 0.246 | INOH |
| Notch | 58 | 2 (3.4%) | 0.0406 | 0.246 | NetPath |
| Complement and Coagulation Cascades | 59 | 2 (3.4%) | 0.0419 | 0.246 | Wikipathways |
| Notch signaling pathway - Homo sapiens (human) | 59 | 2 (3.4%) | 0.0419 | 0.246 | KEGG |
| SARS-CoV-2 innate immunity evasion and cell-specific immune response | 60 | 2 (3.4%) | 0.0419 | 0.246 | Wikipathways |
| Biosynthesis of maresins | 8 | 1 (12.5%) | 0.0432 | 0.246 | Reactome |
| The alternative pathway of fetal androgen synthesis | 8 | 1 (12.5%) | 0.0432 | 0.246 | Wikipathways |
| NOTCH4 Intracellular Domain Regulates Transcription | 8 | 1 (12.5%) | 0.0432 | 0.246 | Reactome |
| TICAM1-dependent activation of IRF3/IRF7 | 8 | 1 (12.5%) | 0.0432 | 0.246 | Reactome |
| yaci and bcma stimulation of b cell immune responses | 8 | 1 (12.5%) | 0.0432 | 0.246 | BioCarta |
| Inactivation of CDC42 and RAC1 | 8 | 1 (12.5%) | 0.0432 | 0.246 | Reactome |
| Phase 3 - rapid repolarisation | 8 | 1 (12.5%) | 0.0432 | 0.246 | Reactome |
| FTO Obesity Variant Mechanism | 8 | 1 (12.5%) | 0.0432 | 0.246 | Wikipathways |
| Notch Signaling Pathway Netpath | 61 | 2 (3.3%) | 0.0445 | 0.249 | Wikipathways |
| Arachidonic acid metabolism - Homo sapiens (human) | 61 | 2 (3.3%) | 0.0445 | 0.249 | KEGG |
| Breast cancer - Homo sapiens (human) | 147 | 3 (2.0%) | 0.0474 | 0.257 | KEGG |
| Synthesis of 5-eicosatetraenoic acids | 9 | 1 (11.1%) | 0.0485 | 0.257 | Reactome |
| Benzo(a)pyrene metabolism | 9 | 1 (11.1%) | 0.0485 | 0.257 | Wikipathways |
| NOTCH4 Activation and Transmission of Signal to the Nucleus | 9 | 1 (11.1%) | 0.0485 | 0.257 | Reactome |
| Prostanoid ligand receptors | 9 | 1 (11.1%) | 0.0485 | 0.257 | Reactome |
| cd40l signaling pathway | 9 | 1 (11.1%) | 0.0485 | 0.257 | BioCarta |
| RHO GTPases Activate Rhotekin and Rhophilins | 9 | 1 (11.1%) | 0.0485 | 0.257 | Reactome |
| Gene expression (Transcription) | 1439 | 13 (0.9%) | 0.0492 | 0.259 | Reactome |

CAD_SAT

| pathway name | set size | candidates contained | p-value | q-value | pathway source |
|---|---|---|---|---|---|
| Ganglio Sphingolipid Metabolism | 13 | 3 (23.1%) | 0.000359 | 0.21 | Wikipathways |
| Pyrimidine biosynthesis | 3 | 2 (66.7%) | 0.000369 | 0.21 | Reactome |
| Alanine, aspartate and glutamate metabolism - Homo sapiens (human) | 37 | 4 (10.8%) | 0.00074 | 0.281 | KEGG |
| Vitamin A and carotenoid metabolism | 43 | 4 (9.3%) | 0.00131 | 0.373 | Wikipathways |
| Metabolic pathway of LDL, HDL and TG, including diseases | 6 | 2 (33.3%) | 0.0018 | 0.41 | Wikipathways |
| visceral fat deposits and the metabolic syndrome | 8 | 2 (25.0%) | 0.00332 | 0.629 | BioCarta |
| spliceosomal assembly | 9 | 2 (22.2%) | 0.00423 | 0.632 | BioCarta |
| Factors and pathways affecting insulin-like growth factor (IGF1)-Akt signaling | 36 | 3 (8.3%) | 0.00744 | 0.632 | Wikipathways |
| Ammonia Recycling | 12 | 2 (16.7%) | 0.0076 | 0.632 | SMPDB |
| valine degradation | 13 | 2 (15.4%) | 0.00891 | 0.632 | HumanCyc |
| Herpes simplex virus 1 infection - Homo sapiens (human) | 498 | 12 (2.4%) | 0.00996 | 0.632 | KEGG |
| SEMA3A-Plexin repulsion signaling by inhibiting Integrin adhesion | 14 | 2 (14.3%) | 0.0103 | 0.632 | Reactome |
| serotonin degradation | 14 | 2 (14.3%) | 0.0103 | 0.632 | HumanCyc |
| D-mannose degradation | 1 | 1 (100.0%) | 0.0112 | 0.632 | HumanCyc |
| Intestinal lipid absorption | 1 | 1 (100.0%) | 0.0112 | 0.632 | Reactome |
| Nucleobase biosynthesis | 15 | 2 (13.3%) | 0.0118 | 0.632 | Reactome |
| superpathway of pyrimidine ribonucleotides de novo biosynthesis | 15 | 2 (13.3%) | 0.0118 | 0.632 | HumanCyc |
| Glycosphingolipid biosynthesis - ganglio series - Homo sapiens (human) | 15 | 2 (13.3%) | 0.0118 | 0.632 | KEGG |
| Glutamate and glutamine metabolism | 15 | 2 (13.3%) | 0.0118 | 0.632 | Reactome |
| Sema3A PAK dependent Axon repulsion | 16 | 2 (12.5%) | 0.0134 | 0.632 | Reactome |
| CRMPs in Sema3A signaling | 16 | 2 (12.5%) | 0.0134 | 0.632 | Reactome |
| Regulation of TLR by endogenous ligand | 16 | 2 (12.5%) | 0.0134 | 0.632 | Reactome |
| Pyrimidine metabolism | 84 | 4 (4.8%) | 0.0145 | 0.632 | Wikipathways |
| Sulfation Biotransformation Reaction | 17 | 2 (11.8%) | 0.0151 | 0.632 | Wikipathways |
| Rett syndrome causing genes | 48 | 3 (6.2%) | 0.0164 | 0.632 | Wikipathways |
| Pyrimidine nucleotides nucleosides metabolism | 51 | 3 (5.9%) | 0.0192 | 0.632 | INOH |
| UMP biosynthesis | 2 | 1 (50.0%) | 0.0222 | 0.632 | HumanCyc |
| SDK interactions | 2 | 1 (50.0%) | 0.0222 | 0.632 | Reactome |
| Proprotein convertase subtilisin-kexin type 9 (PCSK9) mediated LDL receptor degradation | 2 | 1 (50.0%) | 0.0222 | 0.632 | Wikipathways |
| Evolocumab Mechanism | 2 | 1 (50.0%) | 0.0222 | 0.632 | Wikipathways |
| Glycosphingolipid biosynthesis - ganglioseries | 21 | 2 (9.5%) | 0.0226 | 0.632 | EHMN |
| Branched-chain amino acid catabolism | 21 | 2 (9.5%) | 0.0226 | 0.632 | Reactome |
| Transcription factor regulation in adipogenesis | 22 | 2 (9.1%) | 0.0247 | 0.632 | Wikipathways |
| Type II diabetes mellitus | 22 | 2 (9.1%) | 0.0247 | 0.632 | Wikipathways |
| superpathway of pyrimidine deoxyribonucleotides de novo biosynthesis | 22 | 2 (9.1%) | 0.0247 | 0.632 | HumanCyc |
| Pyrimidine metabolism - Homo sapiens (human) | 57 | 3 (5.3%) | 0.0257 | 0.632 | KEGG |
| Estrogen signaling pathway | 23 | 2 (8.7%) | 0.0269 | 0.632 | Wikipathways |
| Cytosolic sulfonation of small molecules | 25 | 2 (8.0%) | 0.0314 | 0.632 | Reactome |
| glycine cleavage | 3 | 1 (33.3%) | 0.0331 | 0.632 | HumanCyc |
| glutamine degradation/glutamate biosynthesis | 3 | 1 (33.3%) | 0.0331 | 0.632 | HumanCyc |
| asparagine biosynthesis | 3 | 1 (33.3%) | 0.0331 | 0.632 | HumanCyc |
| Heroin metabolism | 3 | 1 (33.3%) | 0.0331 | 0.632 | Wikipathways |
| Integration of viral DNA into host genomic DNA | 3 | 1 (33.3%) | 0.0331 | 0.632 | Reactome |
| Autointegration results in viral DNA circles | 3 | 1 (33.3%) | 0.0331 | 0.632 | Reactome |
| EGR2 and SOX10-mediated initiation of Schwann cell myelination | 26 | 2 (7.7%) | 0.0338 | 0.632 | Reactome |
| Glycosylphosphatidylinositol (GPI)-anchor biosynthesis - Homo sapiens (human) | 26 | 2 (7.7%) | 0.0338 | 0.632 | KEGG |
| IL4-mediated signaling events | 65 | 3 (4.6%) | 0.036 | 0.632 | PID |
| Fatty acid elongation - Homo sapiens (human) | 27 | 2 (7.4%) | 0.0362 | 0.632 | KEGG |
| Biosynthesis of unsaturated fatty acids - Homo sapiens (human) | 27 | 2 (7.4%) | 0.0362 | 0.632 | KEGG |
| Metabolism of amino acids and derivatives | 339 | 8 (2.4%) | 0.0364 | 0.632 | Reactome |
| One-carbon metabolism | 28 | 2 (7.1%) | 0.0387 | 0.632 | Wikipathways |
| Felbamate Metabolism Pathway | 4 | 1 (25.0%) | 0.0439 | 0.632 | SMPDB |
| Glycine degradation | 4 | 1 (25.0%) | 0.0439 | 0.632 | Reactome |
| Heroin Metabolism Pathway | 4 | 1 (25.0%) | 0.0439 | 0.632 | SMPDB |
| 2-oxoisovalerate decarboxylation to isobutanoyl-CoA | 4 | 1 (25.0%) | 0.0439 | 0.632 | HumanCyc |
| Vitamin K Metabolism | 4 | 1 (25.0%) | 0.0439 | 0.632 | SMPDB |
| IRS activation | 4 | 1 (25.0%) | 0.0439 | 0.632 | Reactome |
| Composition of Lipid Particles | 4 | 1 (25.0%) | 0.0439 | 0.632 | Wikipathways |
| Intestinal absorption | 4 | 1 (25.0%) | 0.0439 | 0.632 | Reactome |
| Regulation of gene expression by Hypoxia-inducible Factor | 4 | 1 (25.0%) | 0.0439 | 0.632 | Reactome |
| Microbial modulation of RIPK1-mediated regulated necrosis | 4 | 1 (25.0%) | 0.0439 | 0.632 | Reactome |
| 3-Methylglutaconic Aciduria Type I | 30 | 2 (6.7%) | 0.0439 | 0.632 | SMPDB |
| Valine, Leucine and Isoleucine Degradation | 30 | 2 (6.7%) | 0.0439 | 0.632 | SMPDB |
| 2-Methyl-3-Hydroxybutryl CoA Dehydrogenase Deficiency | 30 | 2 (6.7%) | 0.0439 | 0.632 | SMPDB |
| Isovaleric Aciduria | 30 | 2 (6.7%) | 0.0439 | 0.632 | SMPDB |
| 3-Methylcrotonyl Coa Carboxylase Deficiency Type I | 30 | 2 (6.7%) | 0.0439 | 0.632 | SMPDB |
| Propionic Acidemia | 30 | 2 (6.7%) | 0.0439 | 0.632 | SMPDB |

| | | | | | |
|---|---|---|---|---|---|
| Maple Syrup Urine Disease | 30 | 2 (6.7%) | 0.0439 | 0.632 | **SMPDB** |
| 3-Hydroxy-3-Methylglutaryl-CoA Lyase Deficiency | 30 | 2 (6.7%) | 0.0439 | 0.632 | **SMPDB** |
| Isobutyryl-coa dehydrogenase deficiency | 30 | 2 (6.7%) | 0.0439 | 0.632 | **SMPDB** |
| 3-hydroxyisobutyric aciduria | 30 | 2 (6.7%) | 0.0439 | 0.632 | **SMPDB** |
| 3-hydroxyisobutyric acid dehydrogenase deficiency | 30 | 2 (6.7%) | 0.0439 | 0.632 | **SMPDB** |
| Isovaleric acidemia | 30 | 2 (6.7%) | 0.0439 | 0.632 | **SMPDB** |
| Methylmalonate Semialdehyde Dehydrogenase Deficiency | 30 | 2 (6.7%) | 0.0439 | 0.632 | **SMPDB** |
| Methylmalonic Aciduria | 30 | 2 (6.7%) | 0.0439 | 0.632 | **SMPDB** |
| 3-Methylglutaconic Aciduria Type IV | 30 | 2 (6.7%) | 0.0439 | 0.632 | **SMPDB** |
| 3-Methylglutaconic Aciduria Type III | 30 | 2 (6.7%) | 0.0439 | 0.632 | **SMPDB** |
| Beta-Ketothiolase Deficiency | 30 | 2 (6.7%) | 0.0439 | 0.632 | **SMPDB** |
| Folate metabolism | 31 | 2 (6.5%) | 0.0466 | 0.632 | **INOH** |
| MECP2 and Associated Rett Syndrome | 73 | 3 (4.1%) | 0.0482 | 0.632 | **Wikipathways** |
| Glyoxylate metabolism and glycine degradation | 32 | 2 (6.2%) | 0.0494 | 0.632 | **Reactome** |
| Constitutive Androstane Receptor Pathway | 32 | 2 (6.2%) | 0.0494 | 0.632 | **Wikipathways** |

T2DM_SAT

| pathway name | set size | candidates contained | p-value | q-value | pathway source |
|---|---|---|---|---|---|
| Translocation of ZAP-70 to Immunological synapse | 28 | 5 (17.9%) | 2.18E-05 | 0.0191 | Reactome |
| Phosphorylation of CD3 and TCR zeta chains | 31 | 5 (16.1%) | 3.66E-05 | 0.0191 | Reactome |
| PD-1 signaling | 32 | 5 (15.6%) | 4.29E-05 | 0.0191 | Reactome |
| MHC class II antigen presentation | 60 | 6 (10.0%) | 9.69E-05 | 0.0323 | Reactome |
| Generation of second messenger molecules | 42 | 5 (11.9%) | 0.000164 | 0.0437 | Reactome |
| Downstream TCR signaling | 52 | 5 (9.6%) | 0.000453 | 0.0817 | Reactome |
| Pyrimidine biosynthesis | 3 | 2 (66.7%) | 0.000457 | 0.0817 | Reactome |
| Asthma - Homo sapiens (human) | 31 | 4 (13.3%) | 0.000491 | 0.0817 | KEGG |
| Allograft rejection - Homo sapiens (human) | 38 | 4 (10.8%) | 0.0011 | 0.164 | KEGG |
| Graft-versus-host disease - Homo sapiens (human) | 42 | 4 (9.5%) | 0.00178 | 0.212 | KEGG |
| Type I diabetes mellitus - Homo sapiens (human) | 43 | 4 (9.3%) | 0.00195 | 0.212 | KEGG |
| CD4 T cell receptor signaling-ERK cascade | 44 | 4 (9.1%) | 0.00212 | 0.212 | INOH |
| TCR signaling | 73 | 5 (6.8%) | 0.00213 | 0.212 | Reactome |
| Alpha Linolenic Acid and Linoleic Acid Metabolism | 6 | 2 (33.3%) | 0.00223 | 0.212 | SMPDB |
| Acyl chain remodelling of PS | 22 | 3 (13.6%) | 0.00244 | 0.213 | Reactome |
| Costimulation by the CD28 family | 77 | 5 (6.5%) | 0.00269 | 0.213 | Reactome |
| Intestinal immune network for IgA production - Homo sapiens (human) | 49 | 4 (8.5%) | 0.00271 | 0.213 | KEGG |
| Pyrimidine metabolism | 84 | 5 (6.0%) | 0.00392 | 0.275 | Wikipathways |
| Autoimmune thyroid disease - Homo sapiens (human) | 53 | 4 (7.7%) | 0.00392 | 0.275 | KEGG |
| CD4 T cell receptor signaling-JNK cascade | 57 | 4 (7.0%) | 0.00545 | 0.349 | INOH |
| Ebola Virus Pathway on Host | 130 | 6 (4.6%) | 0.00567 | 0.349 | Wikipathways |
| Rheumatoid arthritis - Homo sapiens (human) | 93 | 5 (5.4%) | 0.00576 | 0.349 | KEGG |
| Viral myocarditis - Homo sapiens (human) | 60 | 4 (6.8%) | 0.00616 | 0.357 | KEGG |
| Interferon gamma signaling | 96 | 5 (5.2%) | 0.00689 | 0.382 | Reactome |
| Hematopoietic cell lineage - Homo sapiens (human) | 99 | 5 (5.1%) | 0.0075 | 0.387 | KEGG |
| CD4 T cell receptor signaling-NFkB cascade | 99 | 5 (5.1%) | 0.00782 | 0.387 | INOH |
| Ubiquinone and other terpenoid-quinone biosynthesis - Homo sapiens (human) | 11 | 2 (18.2%) | 0.00785 | 0.387 | KEGG |
| Inflammatory bowel disease - Homo sapiens (human) | 65 | 4 (6.2%) | 0.00866 | 0.412 | KEGG |
| Visual phototransduction | 103 | 5 (4.9%) | 0.00921 | 0.423 | Reactome |
| antigen processing and presentation | 13 | 2 (15.4%) | 0.011 | 0.448 | BioCarta |
| Phagosome - Homo sapiens (human) | 152 | 6 (4.0%) | 0.0114 | 0.448 | KEGG |
| ALKBH2 mediated reversal of alkylation damage | 1 | 1 (100.0%) | 0.0124 | 0.448 | Reactome |
| Mismatch repair (MMR) directed by MSH2:MSH3 (MutSbeta) | 14 | 2 (14.3%) | 0.0127 | 0.448 | Reactome |
| Mismatch repair (MMR) directed by MSH2:MSH6 (MutSalpha) | 14 | 2 (14.3%) | 0.0127 | 0.448 | Reactome |
| Removal of the Flap Intermediate | 14 | 2 (14.3%) | 0.0127 | 0.448 | Reactome |
| Polymerase switching | 14 | 2 (14.3%) | 0.0127 | 0.448 | Reactome |
| Leading Strand Synthesis | 14 | 2 (14.3%) | 0.0127 | 0.448 | Reactome |
| Toxoplasmosis - Homo sapiens (human) | 112 | 5 (4.5%) | 0.0129 | 0.448 | KEGG |
| Endocytosis - Homo sapiens (human) | 252 | 8 (3.2%) | 0.0135 | 0.448 | KEGG |
| Mismatch Repair | 15 | 2 (13.3%) | 0.0145 | 0.448 | Reactome |
| Processive synthesis on the lagging strand | 15 | 2 (13.3%) | 0.0145 | 0.448 | Reactome |
| Nucleobase biosynthesis | 15 | 2 (13.3%) | 0.0145 | 0.448 | Reactome |
| superpathway of pyrimidine ribonucleotides de novo biosynthesis | 15 | 2 (13.3%) | 0.0145 | 0.448 | HumanCyc |
| Leishmaniasis - Homo sapiens (human) | 77 | 4 (5.3%) | 0.0148 | 0.448 | KEGG |
| Antigen processing and presentation - Homo sapiens (human) | 78 | 4 (5.1%) | 0.0161 | 0.466 | KEGG |
| Deregulation of Rab and Rab Effector Genes in Bladder Cancer | 16 | 2 (12.5%) | 0.0164 | 0.466 | Wikipathways |
| Transcription of E2F targets under negative control by DREAM complex | 16 | 2 (12.5%) | 0.0164 | 0.466 | Reactome |
| Beta5 beta6 beta7 and beta8 integrin cell surface interactions | 17 | 2 (11.8%) | 0.0185 | 0.503 | PID |
| Serotonin and anxiety | 17 | 2 (11.8%) | 0.0185 | 0.503 | Wikipathways |
| Influenza A - Homo sapiens (human) | 172 | 6 (3.5%) | 0.0205 | 0.539 | KEGG |
| Acyl chain remodelling of PG | 18 | 2 (11.1%) | 0.0206 | 0.539 | Reactome |
| Herpes simplex virus 1 infection - Homo sapiens (human) | 498 | 12 (2.4%) | 0.0217 | 0.542 | KEGG |
| Metabolism of fat-soluble vitamins | 49 | 3 (6.1%) | 0.0229 | 0.542 | Reactome |
| Glycerophospholipid biosynthesis | 130 | 5 (3.8%) | 0.0231 | 0.542 | Reactome |
| Platelet degranulation | 131 | 5 (3.8%) | 0.0238 | 0.542 | Reactome |
| CD4 T cell receptor signaling | 132 | 5 (3.8%) | 0.0245 | 0.542 | INOH |
| Activation and oligomerization of BAK protein | 2 | 1 (50.0%) | 0.0247 | 0.542 | Reactome |
| UMP biosynthesis | 2 | 1 (50.0%) | 0.0247 | 0.542 | HumanCyc |

| | | | | | |
|---|---|---|---|---|---|
| Lagging Strand Synthesis | 20 | 2 (10.0%) | 0.0252 | 0.542 | Reactome |
| Pyrimidine nucleotides nucleosides metabolism | 51 | 3 (5.9%) | 0.0254 | 0.542 | INOH |
| Response to elevated platelet cytosolic Ca2+ | 136 | 5 (3.7%) | 0.0274 | 0.542 | Reactome |
| Ardeparin Action Pathway | 21 | 2 (9.5%) | 0.0276 | 0.542 | SMPDB |
| Heparin Action Pathway | 21 | 2 (9.5%) | 0.0276 | 0.542 | SMPDB |
| Fondaparinux Action Pathway | 21 | 2 (9.5%) | 0.0276 | 0.542 | SMPDB |
| Enoxaparin Action Pathway | 21 | 2 (9.5%) | 0.0276 | 0.542 | SMPDB |
| HS-GAG degradation | 21 | 2 (9.5%) | 0.0276 | 0.542 | Reactome |
| Methionine De Novo and Salvage Pathway | 21 | 2 (9.5%) | 0.0276 | 0.542 | Wikipathways |
| Th1 and Th2 cell differentiation - Homo sapiens (human) | 92 | 4 (4.3%) | 0.0277 | 0.542 | KEGG |
| Allograft Rejection | 93 | 4 (4.3%) | 0.0286 | 0.553 | Wikipathways |
| Staphylococcus aureus infection - Homo sapiens (human) | 96 | 4 (4.3%) | 0.0296 | 0.564 | KEGG |
| superpathway of pyrimidine deoxyribonucleotides de novo biosynthesis | 22 | 2 (9.1%) | 0.0302 | 0.564 | HumanCyc |
| Mismatch repair - Homo sapiens (human) | 23 | 2 (8.7%) | 0.0328 | 0.564 | KEGG |
| DNA Mismatch Repair | 23 | 2 (8.7%) | 0.0328 | 0.564 | Wikipathways |
| Butanoate metabolism | 23 | 2 (8.7%) | 0.0328 | 0.564 | EHMN |
| Pyrimidine metabolism - Homo sapiens (human) | 57 | 3 (5.3%) | 0.0339 | 0.564 | KEGG |
| Synthesis of very long-chain fatty acyl-CoAs | 24 | 2 (8.3%) | 0.0354 | 0.564 | Reactome |
| glutamine degradation/glutamate biosynthesis | 3 | 1 (33.3%) | 0.0368 | 0.564 | HumanCyc |
| asparagine biosynthesis | 3 | 1 (33.3%) | 0.0368 | 0.564 | HumanCyc |
| methionine salvage | 3 | 1 (33.3%) | 0.0368 | 0.564 | HumanCyc |
| methylglyoxal degradation I | 3 | 1 (33.3%) | 0.0368 | 0.564 | HumanCyc |
| Pyruvaldehyde Degradation | 3 | 1 (33.3%) | 0.0368 | 0.564 | SMPDB |
| Integration of viral DNA into host genomic DNA | 3 | 1 (33.3%) | 0.0368 | 0.564 | Reactome |
| Autointegration results in viral DNA circles | 3 | 1 (33.3%) | 0.0368 | 0.564 | Reactome |
| Metabolism of vitamin K | 3 | 1 (33.3%) | 0.0368 | 0.564 | Reactome |
| Direct Reversal Repair | 3 | 1 (33.3%) | 0.0368 | 0.564 | Wikipathways |
| Cell adhesion molecules - Homo sapiens (human) | 149 | 5 (3.4%) | 0.0374 | 0.564 | KEGG |
| G0 and Early G1 | 25 | 2 (8.0%) | 0.0382 | 0.564 | Reactome |
| alpha-Linolenic acid metabolism - Homo sapiens (human) | 25 | 2 (8.0%) | 0.0382 | 0.564 | KEGG |
| Potassium Channels | 103 | 4 (3.9%) | 0.0395 | 0.564 | Reactome |
| Arachidonic acid metabolism - Homo sapiens (human) | 61 | 3 (4.9%) | 0.0402 | 0.564 | KEGG |
| Calcium Regulation in the Cardiac Cell | 151 | 5 (3.3%) | 0.0403 | 0.564 | Wikipathways |
| Fertilization | 26 | 2 (7.7%) | 0.0411 | 0.564 | Reactome |
| Polymerase switching on the C-strand of the telomere | 27 | 2 (7.4%) | 0.044 | 0.564 | Reactome |
| Fatty acid elongation - Homo sapiens (human) | 27 | 2 (7.4%) | 0.044 | 0.564 | KEGG |
| Biosynthesis of unsaturated fatty acids - Homo sapiens (human) | 27 | 2 (7.4%) | 0.044 | 0.564 | KEGG |
| Acyl chain remodelling of PC | 27 | 2 (7.4%) | 0.044 | 0.564 | Reactome |
| Sulfur amino acid metabolism | 27 | 2 (7.4%) | 0.044 | 0.564 | Reactome |
| Th17 cell differentiation - Homo sapiens (human) | 107 | 4 (3.7%) | 0.0444 | 0.564 | KEGG |
| Phospholipid Biosynthesis | 28 | 2 (7.1%) | 0.047 | 0.564 | SMPDB |
| Felbamate Metabolism Pathway | 4 | 1 (25.0%) | 0.0488 | 0.564 | SMPDB |
| *trans, trans-farnesyl diphosphate biosynthesis* | 4 | 1 (25.0%) | 0.0488 | 0.564 | HumanCyc |
| glutaryl-CoA degradation | 4 | 1 (25.0%) | 0.0488 | 0.564 | HumanCyc |
| IRS activation | 4 | 1 (25.0%) | 0.0488 | 0.564 | Reactome |
| Composition of Lipid Particles | 4 | 1 (25.0%) | 0.0488 | 0.564 | Wikipathways |
| Lysosomal oligosaccharide catabolism | 4 | 1 (25.0%) | 0.0488 | 0.564 | Reactome |
| phenylalanine degradation/tyrosine biosynthesis | 4 | 1 (25.0%) | 0.0488 | 0.564 | HumanCyc |
| Defective pyroptosis | 4 | 1 (25.0%) | 0.0488 | 0.564 | Reactome |
| Proteoglycan syndecan-mediated signaling events | 4 | 1 (25.0%) | 0.0488 | 0.564 | PID |
| *S-methyl-5-thio-α-D-ribose 1-phosphate degradation* | 4 | 1 (25.0%) | 0.0488 | 0.564 | HumanCyc |
| Vitamin K Metabolism | 4 | 1 (25.0%) | 0.0488 | 0.564 | SMPDB |

HTN_SAT

| pathway name | set size | candidates contained | p-value | q-value | pathway source |
|---|---|---|---|---|---|
| Felbamate Metabolism Pathway | 4 | 2 (50.0%) | 0.00162 | 0.588 | SMPDB |
| Fatty Acid Omega Oxidation | 15 | 3 (20.0%) | 0.00179 | 0.588 | Wikipathways |
| Chemical carcinogenesis - Homo sapiens (human) | 83 | 6 (7.2%) | 0.00261 | 0.588 | KEGG |
| Benzene metabolism | 6 | 2 (33.3%) | 0.00397 | 0.588 | Wikipathways |
| Drug metabolism - cytochrome P450 - Homo sapiens (human) | 72 | 5 (7.1%) | 0.0062 | 0.588 | KEGG |
| tRNA processing | 103 | 6 (5.8%) | 0.00751 | 0.588 | Reactome |
| Ion transport by P-type ATPases | 47 | 4 (8.5%) | 0.00766 | 0.588 | Reactome |
| Disulfiram Action Pathway | 25 | 3 (12.0%) | 0.00803 | 0.588 | SMPDB |
| Metabolism of xenobiotics by cytochrome P450 - Homo sapiens (human) | 78 | 5 (6.6%) | 0.00873 | 0.588 | KEGG |
| Surfactant metabolism | 26 | 3 (11.5%) | 0.00896 | 0.588 | Reactome |
| spliceosomal assembly | 9 | 2 (22.2%) | 0.00922 | 0.588 | BioCarta |
| Ethanol metabolism resulting in production of ROS by CYP2E1 | 9 | 2 (22.2%) | 0.00922 | 0.588 | Wikipathways |
| acetone degradation I (to methylglyoxal) | 10 | 2 (20.0%) | 0.0114 | 0.588 | HumanCyc |
| GP1b-IX-V activation signalling | 10 | 2 (20.0%) | 0.0114 | 0.588 | Reactome |
| Linoleic acid metabolism - Homo sapiens (human) | 29 | 3 (10.3%) | 0.0121 | 0.588 | KEGG |
| Molecules associated with elastic fibres | 30 | 3 (10.0%) | 0.0133 | 0.588 | Reactome |
| Eicosanoid metabolism via cyclooxygenases (COX) | 30 | 3 (10.0%) | 0.0133 | 0.588 | Wikipathways |
| C-MYB transcription factor network | 85 | 5 (5.9%) | 0.0138 | 0.588 | PID |
| Phenylalanine and Tyrosine Metabolism | 11 | 2 (18.2%) | 0.0138 | 0.588 | SMPDB |
| Phenylketonuria | 11 | 2 (18.2%) | 0.0138 | 0.588 | SMPDB |
| Tyrosinemia Type 3 (TYRO3) | 11 | 2 (18.2%) | 0.0138 | 0.588 | SMPDB |
| Tyrosinemia Type 2 (or Richner-Hanhart syndrome) | 11 | 2 (18.2%) | 0.0138 | 0.588 | SMPDB |
| Ubiquinone and other terpenoid-quinone biosynthesis - Homo sapiens (human) | 11 | 2 (18.2%) | 0.0138 | 0.588 | KEGG |
| Translation | 307 | 11 (3.6%) | 0.0141 | 0.588 | Reactome |
| Transferrin endocytosis and recycling | 31 | 3 (9.7%) | 0.0146 | 0.588 | Reactome |
| C21-steroid hormone biosynthesis and metabolism | 57 | 4 (7.0%) | 0.0149 | 0.588 | EHMN |
| Ethanol oxidation | 12 | 2 (16.7%) | 0.0164 | 0.588 | Reactome |
| Negative regulation of NOTCH4 signaling | 12 | 2 (16.7%) | 0.0164 | 0.588 | Reactome |
| Metabolism of Dichloroethylene by CYP450 | 1 | 1 (100.0%) | 0.0167 | 0.588 | Wikipathways |
| Acrylamide Biotransformation and Exposure Biomarkers | 1 | 1 (100.0%) | 0.0167 | 0.588 | Wikipathways |
| pregnenolone biosynthesis | 1 | 1 (100.0%) | 0.0167 | 0.588 | HumanCyc |
| Vitamin E | 1 | 1 (100.0%) | 0.0167 | 0.588 | Reactome |
| dicer pathway | 1 | 1 (100.0%) | 0.0167 | 0.588 | BioCarta |
| Formation of the ternary complex, and subsequently, the 43S complex | 59 | 4 (6.8%) | 0.0168 | 0.588 | Reactome |
| Tyrosine metabolism | 33 | 3 (9.1%) | 0.0173 | 0.588 | INOH |
| SNARE interactions in vesicular transport - Homo sapiens (human) | 33 | 3 (9.1%) | 0.0173 | 0.588 | KEGG |
| noradrenaline and adrenaline degradation | 13 | 2 (15.4%) | 0.0191 | 0.588 | HumanCyc |
| Elastic fibre formation | 35 | 3 (8.6%) | 0.0202 | 0.588 | Reactome |
| Ion channel transport | 165 | 7 (4.2%) | 0.0207 | 0.588 | Reactome |
| Glycerophospholipid biosynthesis | 130 | 6 (4.6%) | 0.0217 | 0.588 | Reactome |
| Celecoxib Action Pathway | 36 | 3 (8.3%) | 0.0218 | 0.588 | SMPDB |
| Tyrosine metabolism - Homo sapiens (human) | 36 | 3 (8.3%) | 0.0218 | 0.588 | KEGG |
| serotonin degradation | 14 | 2 (14.3%) | 0.0221 | 0.588 | HumanCyc |
| Omega-3-Omega-6 FA synthesis | 14 | 2 (14.3%) | 0.0221 | 0.588 | Wikipathways |
| Platelet degranulation | 131 | 6 (4.6%) | 0.0225 | 0.588 | Reactome |
| Integrin cell surface interactions | 66 | 4 (6.1%) | 0.0243 | 0.588 | Reactome |
| Ribosomal scanning and start codon recognition | 66 | 4 (6.1%) | 0.0243 | 0.588 | Reactome |
| Translation initiation complex formation | 66 | 4 (6.1%) | 0.0243 | 0.588 | Reactome |
| MAP2K and MAPK activation | 38 | 3 (7.9%) | 0.0251 | 0.588 | Reactome |
| Activation of the mRNA upon binding of the cap-binding complex and eIFs, and subsequent binding to 43S | 67 | 4 (6.0%) | 0.0255 | 0.588 | Reactome |
| Response to elevated platelet cytosolic Ca2+ | 136 | 6 (4.4%) | 0.0264 | 0.588 | Reactome |
| Integrin-mediated Cell Adhesion | 101 | 5 (5.0%) | 0.0269 | 0.588 | Wikipathways |
| Phase I - Functionalization of compounds | 101 | 5 (5.0%) | 0.0269 | 0.588 | Reactome |
| Smooth Muscle Contraction | 39 | 3 (7.7%) | 0.0269 | 0.588 | Reactome |
| mRNA decay by 3, to 5, exoribonuclease | 16 | 2 (12.5%) | 0.0285 | 0.588 | Reactome |
| Biological oxidations | 219 | 8 (3.7%) | 0.0306 | 0.588 | Reactome |
| tRNA modification in the nucleus and cytosol | 41 | 3 (7.3%) | 0.0306 | 0.588 | Reactome |
| phospholipases | 41 | 3 (7.3%) | 0.0306 | 0.588 | HumanCyc |
| Late Phase of HIV Life Cycle | 141 | 6 (4.3%) | 0.0308 | 0.588 | Reactome |
| Platelet activation, signaling and aggregation | 263 | 9 (3.4%) | 0.0324 | 0.588 | Reactome |
| Signaling downstream of RAS mutants | 42 | 3 (7.1%) | 0.0326 | 0.588 | Reactome |
| Signaling by RAS mutants | 42 | 3 (7.1%) | 0.0326 | 0.588 | Reactome |
| Paradoxical activation of RAF signaling by kinase inactive BRAF | 42 | 3 (7.1%) | 0.0326 | 0.588 | Reactome |
| Signaling by moderate kinase activity BRAF mutants | 42 | 3 (7.1%) | 0.0326 | 0.588 | Reactome |
| Signaling by RAF1 mutants | 42 | 3 (7.1%) | 0.0326 | 0.588 | Reactome |
| Felbamate Metabolism | 2 | 1 (50.0%) | 0.0331 | 0.588 | Wikipathways |
| Signalling to ERK5 | 2 | 1 (50.0%) | 0.0331 | 0.588 | Reactome |
| Insulin effects increased synthesis of Xylulose-5-Phosphate | 2 | 1 (50.0%) | 0.0331 | 0.588 | Reactome |
| Hypoxia-mediated EMT and Stemness | 2 | 1 (50.0%) | 0.0331 | 0.588 | Wikipathways |
| 7-(3-amino-3-carboxypropyl)-wyosine biosynthesis | 2 | 1 (50.0%) | 0.0331 | 0.588 | HumanCyc |
| SDK interactions | 2 | 1 (50.0%) | 0.0331 | 0.588 | Reactome |
| Neutrophil degranulation | 487 | 14 (2.9%) | 0.0333 | 0.588 | Reactome |
| Thermogenesis | 108 | 5 (4.6%) | 0.0345 | 0.588 | Wikipathways |

| | | | | | |
|---|---|---|---|---|---|
| Vitamin A and carotenoid metabolism | 43 | 3 (7.0%) | 0.0346 | 0.588 | Wikipathways |
| Deposition of new CENPA-containing nucleosomes at the centromere | 43 | 3 (7.0%) | 0.0346 | 0.588 | Reactome |
| Nucleosome assembly | 43 | 3 (7.0%) | 0.0346 | 0.588 | Reactome |
| Metabolism of RNA | 584 | 16 (2.7%) | 0.035 | 0.588 | Reactome |
| Tyrosinemia, transient, of the newborn | 18 | 2 (11.1%) | 0.0355 | 0.588 | SMPDB |
| Dopamine beta-hydroxylase deficiency | 18 | 2 (11.1%) | 0.0355 | 0.588 | SMPDB |
| Tyrosine Metabolism | 18 | 2 (11.1%) | 0.0355 | 0.588 | SMPDB |
| Alkaptonuria | 18 | 2 (11.1%) | 0.0355 | 0.588 | SMPDB |
| Monoamine oxidase-a deficiency (MAO-A) | 18 | 2 (11.1%) | 0.0355 | 0.588 | SMPDB |
| Hawkinsinuria | 18 | 2 (11.1%) | 0.0355 | 0.588 | SMPDB |
| Tyrosinemia Type I | 18 | 2 (11.1%) | 0.0355 | 0.588 | SMPDB |
| Tie2 Signaling | 18 | 2 (11.1%) | 0.0355 | 0.588 | Reactome |
| Metapathway biotransformation Phase I and II | 186 | 7 (3.8%) | 0.0364 | 0.588 | Wikipathways |
| Glycosaminoglycan degradation - Homo sapiens (human) | 19 | 2 (10.5%) | 0.0393 | 0.588 | KEGG |
| Formation of a pool of free 40S subunits | 113 | 5 (4.4%) | 0.0407 | 0.588 | Reactome |
| HIV Life Cycle | 151 | 6 (4.0%) | 0.0409 | 0.588 | Reactome |
| Golgi-to-ER retrograde transport | 116 | 5 (4.3%) | 0.0447 | 0.588 | Reactome |
| Ardeparin Action Pathway | 21 | 2 (9.5%) | 0.0472 | 0.588 | SMPDB |
| Heparin Action Pathway | 21 | 2 (9.5%) | 0.0472 | 0.588 | SMPDB |
| Fondaparinux Action Pathway | 21 | 2 (9.5%) | 0.0472 | 0.588 | SMPDB |
| Enoxaparin Action Pathway | 21 | 2 (9.5%) | 0.0472 | 0.588 | SMPDB |
| Formation of HIV elongation complex in the absence of HIV Tat | 49 | 3 (6.1%) | 0.0481 | 0.588 | Reactome |
| methylglyoxal degradation III | 3 | 1 (33.3%) | 0.0492 | 0.588 | HumanCyc |
| Hydroxycarboxylic acid-binding receptors | 3 | 1 (33.3%) | 0.0492 | 0.588 | Reactome |
| wybutosine biosynthesis | 3 | 1 (33.3%) | 0.0492 | 0.588 | HumanCyc |
| Metabolism of vitamin K | 3 | 1 (33.3%) | 0.0492 | 0.588 | Reactome |
| Toxicity of botulinum toxin type C (botC) | 3 | 1 (33.3%) | 0.0492 | 0.588 | Reactome |
| Inositol transporters | 3 | 1 (33.3%) | 0.0492 | 0.588 | Reactome |

Obesity_SAT

| pathway name | set size | candidates contained | p-value | q-value | pathway source |
|---|---|---|---|---|---|
| Felbamate Metabolism Pathway | 4 | 2 (50.0%) | 0.00162 | 0.588 | SMPDB |
| Fatty Acid Omega Oxidation | 15 | 3 (20.0%) | 0.00179 | 0.588 | Wikipathways |
| Chemical carcinogenesis - Homo sapiens (human) | 83 | 6 (7.2%) | 0.00261 | 0.588 | KEGG |
| Benzene metabolism | 6 | 2 (33.3%) | 0.00397 | 0.588 | Wikipathways |
| Drug metabolism - cytochrome P450 - Homo sapiens (human) | 72 | 5 (7.1%) | 0.0062 | 0.588 | KEGG |
| tRNA processing | 103 | 6 (5.8%) | 0.00751 | 0.588 | Reactome |
| Ion transport by P-type ATPases | 47 | 4 (8.5%) | 0.00766 | 0.588 | Reactome |
| Disulfiram Action Pathway | 25 | 3 (12.0%) | 0.00803 | 0.588 | SMPDB |
| Metabolism of xenobiotics by cytochrome P450 - Homo sapiens (human) | 78 | 5 (6.6%) | 0.00873 | 0.588 | KEGG |
| Surfactant metabolism | 26 | 3 (11.5%) | 0.00896 | 0.588 | Reactome |
| spliceosomal assembly | 9 | 2 (22.2%) | 0.00922 | 0.588 | BioCarta |
| Ethanol metabolism resulting in production of ROS by CYP2E1 | 9 | 2 (22.2%) | 0.00922 | 0.588 | Wikipathways |
| acetone degradation I (to methylglyoxal) | 10 | 2 (20.0%) | 0.0114 | 0.588 | HumanCyc |
| GP1b-IX-V activation signalling | 10 | 2 (20.0%) | 0.0114 | 0.588 | Reactome |
| Linoleic acid metabolism - Homo sapiens (human) | 29 | 3 (10.3%) | 0.0121 | 0.588 | KEGG |
| Molecules associated with elastic fibres | 30 | 3 (10.0%) | 0.0133 | 0.588 | Reactome |
| Eicosanoid metabolism via cyclooxygenases (COX) | 30 | 3 (10.0%) | 0.0133 | 0.588 | Wikipathways |
| C-MYB transcription factor network | 85 | 5 (5.9%) | 0.0138 | 0.588 | PID |
| Phenylalanine and Tyrosine Metabolism | 11 | 2 (18.2%) | 0.0138 | 0.588 | SMPDB |
| Phenylketonuria | 11 | 2 (18.2%) | 0.0138 | 0.588 | SMPDB |
| Tyrosinemia Type 3 (TYRO3) | 11 | 2 (18.2%) | 0.0138 | 0.588 | SMPDB |
| Tyrosinemia Type 2 (or Richner-Hanhart syndrome) | 11 | 2 (18.2%) | 0.0138 | 0.588 | SMPDB |
| Ubiquinone and other terpenoid-quinone biosynthesis - Homo sapiens (human) | 11 | 2 (18.2%) | 0.0138 | 0.588 | KEGG |
| Translation | 307 | 11 (3.6%) | 0.0141 | 0.588 | Reactome |
| Transferrin endocytosis and recycling | 31 | 3 (9.7%) | 0.0146 | 0.588 | Reactome |
| C21-steroid hormone biosynthesis and metabolism | 57 | 4 (7.0%) | 0.0149 | 0.588 | EHMN |
| Ethanol oxidation | 12 | 2 (16.7%) | 0.0164 | 0.588 | Reactome |
| Negative regulation of NOTCH4 signaling | 12 | 2 (16.7%) | 0.0164 | 0.588 | Reactome |
| Metabolism of Dichloroethylene by CYP450 | 1 | 1 (100.0%) | 0.0167 | 0.588 | Wikipathways |
| Acrylamide Biotransformation and Exposure Biomarkers | 1 | 1 (100.0%) | 0.0167 | 0.588 | Wikipathways |
| pregnenolone biosynthesis | 1 | 1 (100.0%) | 0.0167 | 0.588 | HumanCyc |
| Vitamin E | 1 | 1 (100.0%) | 0.0167 | 0.588 | Reactome |
| dicer pathway | 1 | 1 (100.0%) | 0.0167 | 0.588 | BioCarta |
| Formation of the ternary complex, and subsequently, the 43S complex | 59 | 4 (6.8%) | 0.0168 | 0.588 | Reactome |
| Tyrosine metabolism | 33 | 3 (9.1%) | 0.0173 | 0.588 | INOH |
| SNARE interactions in vesicular transport - Homo sapiens (human) | 33 | 3 (9.1%) | 0.0173 | 0.588 | KEGG |
| noradrenaline and adrenaline degradation | 13 | 2 (15.4%) | 0.0191 | 0.588 | HumanCyc |
| Elastic fibre formation | 35 | 3 (8.6%) | 0.0202 | 0.588 | Reactome |
| Ion channel transport | 165 | 7 (4.2%) | 0.0207 | 0.588 | Reactome |
| Glycerophospholipid biosynthesis | 130 | 6 (4.6%) | 0.0217 | 0.588 | Reactome |
| Celecoxib Action Pathway | 36 | 3 (8.3%) | 0.0218 | 0.588 | SMPDB |
| Tyrosine metabolism - Homo sapiens (human) | 36 | 3 (8.3%) | 0.0218 | 0.588 | KEGG |
| serotonin degradation | 14 | 2 (14.3%) | 0.0221 | 0.588 | HumanCyc |
| Omega-3-Omega-6 FA synthesis | 14 | 2 (14.3%) | 0.0221 | 0.588 | Wikipathways |
| Platelet degranulation | 131 | 6 (4.6%) | 0.0225 | 0.588 | Reactome |
| Integrin cell surface interactions | 66 | 4 (6.1%) | 0.0243 | 0.588 | Reactome |
| Ribosomal scanning and start codon recognition | 66 | 4 (6.1%) | 0.0243 | 0.588 | Reactome |
| Translation initiation complex formation | 66 | 4 (6.1%) | 0.0243 | 0.588 | Reactome |
| MAP2K and MAPK activation | 38 | 3 (7.9%) | 0.0251 | 0.588 | Reactome |
| Activation of the mRNA upon binding of the cap-binding complex and eIFs, and subsequent binding to 43S | 67 | 4 (6.0%) | 0.0255 | 0.588 | Reactome |
| Response to elevated platelet cytosolic Ca2+ | 136 | 6 (4.4%) | 0.0264 | 0.588 | Reactome |
| Integrin-mediated Cell Adhesion | 101 | 5 (5.0%) | 0.0269 | 0.588 | Wikipathways |
| Phase I - Functionalization of compounds | 101 | 5 (5.0%) | 0.0269 | 0.588 | Reactome |
| Smooth Muscle Contraction | 39 | 3 (7.7%) | 0.0269 | 0.588 | Reactome |
| mRNA decay by 3, to 5, exoribonuclease | 16 | 2 (12.5%) | 0.0285 | 0.588 | Reactome |
| Biological oxidations | 219 | 8 (3.7%) | 0.0306 | 0.588 | Reactome |
| tRNA modification in the nucleus and cytosol | 41 | 3 (7.3%) | 0.0306 | 0.588 | Reactome |
| phospholipases | 41 | 3 (7.3%) | 0.0306 | 0.588 | HumanCyc |
| Late Phase of HIV Life Cycle | 141 | 6 (4.3%) | 0.0308 | 0.588 | Reactome |
| Platelet activation, signaling and aggregation | 263 | 9 (3.4%) | 0.0324 | 0.588 | Reactome |
| Signaling downstream of RAS mutants | 42 | 3 (7.1%) | 0.0326 | 0.588 | Reactome |
| Signaling by RAS mutants | 42 | 3 (7.1%) | 0.0326 | 0.588 | Reactome |
| Paradoxical activation of RAF signaling by kinase inactive BRAF | 42 | 3 (7.1%) | 0.0326 | 0.588 | Reactome |
| Signaling by moderate kinase activity BRAF mutants | 42 | 3 (7.1%) | 0.0326 | 0.588 | Reactome |
| Signaling by RAF1 mutants | 42 | 3 (7.1%) | 0.0326 | 0.588 | Reactome |
| Felbamate Metabolism | 2 | 1 (50.0%) | 0.0331 | 0.588 | Wikipathways |
| Signalling to ERK5 | 2 | 1 (50.0%) | 0.0331 | 0.588 | Reactome |
| Insulin effects increased synthesis of Xylulose-5-Phosphate | 2 | 1 (50.0%) | 0.0331 | 0.588 | Reactome |
| Hypoxia-mediated EMT and Stemness | 2 | 1 (50.0%) | 0.0331 | 0.588 | Wikipathways |
| 7-(3-amino-3-carboxypropyl)-wyosine biosynthesis | 2 | 1 (50.0%) | 0.0331 | 0.588 | HumanCyc |
| SDK interactions | 2 | 1 (50.0%) | 0.0331 | 0.588 | Reactome |
| Neutrophil degranulation | 487 | 14 (2.9%) | 0.0333 | 0.588 | Reactome |

| | | | | | |
|---|---|---|---|---|---|
| Thermogenesis | 108 | 5 (4.6%) | 0.0345 | 0.588 | **Wikipathways** |
| Vitamin A and carotenoid metabolism | 43 | 3 (7.0%) | 0.0346 | 0.588 | **Wikipathways** |
| Deposition of new CENPA-containing nucleosomes at the centromere | 43 | 3 (7.0%) | 0.0346 | 0.588 | **Reactome** |
| Nucleosome assembly | 43 | 3 (7.0%) | 0.0346 | 0.588 | **Reactome** |
| Metabolism of RNA | 584 | 16 (2.7%) | 0.035 | 0.588 | **Reactome** |
| Tyrosinemia, transient, of the newborn | 18 | 2 (11.1%) | 0.0355 | 0.588 | **SMPDB** |
| Dopamine beta-hydroxylase deficiency | 18 | 2 (11.1%) | 0.0355 | 0.588 | **SMPDB** |
| Tyrosine Metabolism | 18 | 2 (11.1%) | 0.0355 | 0.588 | **SMPDB** |
| Alkaptonuria | 18 | 2 (11.1%) | 0.0355 | 0.588 | **SMPDB** |
| Monoamine oxidase-a deficiency (MAO-A) | 18 | 2 (11.1%) | 0.0355 | 0.588 | **SMPDB** |
| Hawkinsinuria | 18 | 2 (11.1%) | 0.0355 | 0.588 | **SMPDB** |
| Tyrosinemia Type I | 18 | 2 (11.1%) | 0.0355 | 0.588 | **SMPDB** |
| Tie2 Signaling | 18 | 2 (11.1%) | 0.0355 | 0.588 | **Reactome** |
| Metapathway biotransformation Phase I and II | 186 | 7 (3.8%) | 0.0364 | 0.588 | **Wikipathways** |
| Glycosaminoglycan degradation - Homo sapiens (human) | 19 | 2 (10.5%) | 0.0393 | 0.588 | **KEGG** |
| Formation of a pool of free 40S subunits | 113 | 5 (4.4%) | 0.0407 | 0.588 | **Reactome** |
| HIV Life Cycle | 151 | 6 (4.0%) | 0.0409 | 0.588 | **Reactome** |
| Golgi-to-ER retrograde transport | 116 | 5 (4.3%) | 0.0447 | 0.588 | **Reactome** |
| Ardeparin Action Pathway | 21 | 2 (9.5%) | 0.0472 | 0.588 | **SMPDB** |
| Heparin Action Pathway | 21 | 2 (9.5%) | 0.0472 | 0.588 | **SMPDB** |
| Fondaparinux Action Pathway | 21 | 2 (9.5%) | 0.0472 | 0.588 | **SMPDB** |
| Enoxaparin Action Pathway | 21 | 2 (9.5%) | 0.0472 | 0.588 | **SMPDB** |
| Formation of HIV elongation complex in the absence of HIV Tat | 49 | 3 (6.1%) | 0.0481 | 0.588 | **Reactome** |
| methylglyoxal degradation III | 3 | 1 (33.3%) | 0.0492 | 0.588 | **HumanCyc** |
| Hydroxycarboxylic acid-binding receptors | 3 | 1 (33.3%) | 0.0492 | 0.588 | **Reactome** |
| wybutosine biosynthesis | 3 | 1 (33.3%) | 0.0492 | 0.588 | **HumanCyc** |
| Metabolism of vitamin K | 3 | 1 (33.3%) | 0.0492 | 0.588 | **Reactome** |
| Toxicity of botulinum toxin type C (botC) | 3 | 1 (33.3%) | 0.0492 | 0.588 | **Reactome** |
| Inositol transporters | 3 | 1 (33.3%) | 0.0492 | 0.588 | **Reactome** |

Stroke_SAT

| pathway name | set size | candidates contained | p-value | q-value | pathway source |
|---|---|---|---|---|---|
| Celecoxib Metabolism Pathway | 8 | 2 (25.0%) | 0.00132 | 0.441 | SMPDB |
| Metabolism of xenobiotics by cytochrome P450 - Homo sapiens (human) | 78 | 4 (5.3%) | 0.00196 | 0.441 | KEGG |
| Biological oxidations | 219 | 6 (2.7%) | 0.00434 | 0.441 | Reactome |
| Fatty Acid Omega Oxidation | 15 | 2 (13.3%) | 0.00479 | 0.441 | Wikipathways |
| Activation of BAD and translocation to mitochondria | 15 | 2 (13.3%) | 0.00479 | 0.441 | Reactome |
| Phase I - Functionalization of compounds | 101 | 4 (4.0%) | 0.00547 | 0.441 | Reactome |
| CD4 T cell receptor signaling-JNK cascade | 57 | 3 (5.3%) | 0.00739 | 0.441 | INOH |
| Integrin | 124 | 4 (3.2%) | 0.0111 | 0.441 | INOH |
| Adaptive Immune System | 719 | 11 (1.5%) | 0.0115 | 0.441 | Reactome |
| Drug metabolism - cytochrome P450 - Homo sapiens (human) | 72 | 3 (4.3%) | 0.013 | 0.441 | KEGG |
| Aripiprazole Metabolic Pathway | 2 | 1 (50.0%) | 0.0139 | 0.441 | Wikipathways |
| ascorbate recycling (cytosolic) | 2 | 1 (50.0%) | 0.0139 | 0.441 | HumanCyc |
| L-glutamine tRNA biosynthesis | 2 | 1 (50.0%) | 0.0139 | 0.441 | HumanCyc |
| N-glycan antennae elongation in the medial/trans-Golgi | 26 | 2 (7.7%) | 0.0141 | 0.441 | Reactome |
| Canonical and non-canonical Notch signaling | 27 | 2 (7.4%) | 0.0152 | 0.441 | Wikipathways |
| Translocation of ZAP-70 to Immunological synapse | 28 | 2 (7.1%) | 0.0163 | 0.441 | Reactome |
| EGF | 28 | 2 (7.1%) | 0.0163 | 0.441 | INOH |
| Keratan sulfate biosynthesis | 28 | 2 (7.1%) | 0.0163 | 0.441 | Reactome |
| insulin | 78 | 3 (3.8%) | 0.0173 | 0.441 | INOH |
| VEGFR1 specific signals | 29 | 2 (6.9%) | 0.0174 | 0.441 | PID |
| B cell receptor signaling pathway - Homo sapiens (human) | 82 | 3 (3.8%) | 0.0185 | 0.441 | KEGG |
| vegf hypoxia and angiogenesis | 30 | 2 (6.7%) | 0.0186 | 0.441 | BioCarta |
| Activation of BH3-only proteins | 30 | 2 (6.7%) | 0.0186 | 0.441 | Reactome |
| IGF1 pathway | 30 | 2 (6.7%) | 0.0186 | 0.441 | PID |
| insulin Mam | 82 | 3 (3.7%) | 0.0197 | 0.441 | INOH |
| Phosphorylation of CD3 and TCR zeta chains | 31 | 2 (6.5%) | 0.0197 | 0.441 | Reactome |
| Chemical carcinogenesis - Homo sapiens (human) | 83 | 3 (3.6%) | 0.0204 | 0.441 | KEGG |
| Desipramine Metabolism Pathway | 3 | 1 (33.3%) | 0.0208 | 0.441 | SMPDB |
| Phenylacetate Metabolism | 3 | 1 (33.3%) | 0.0208 | 0.441 | SMPDB |
| PD-1 signaling | 32 | 2 (6.2%) | 0.021 | 0.441 | Reactome |
| 15q13.3 copy number variation syndrome | 32 | 2 (6.2%) | 0.021 | 0.441 | Wikipathways |
| Keratan sulfate/keratin metabolism | 33 | 2 (6.1%) | 0.0222 | 0.441 | Reactome |
| Mitochondrial translation termination | 88 | 3 (3.4%) | 0.0237 | 0.441 | Reactome |
| Mitochondrial translation elongation | 88 | 3 (3.4%) | 0.0237 | 0.441 | Reactome |
| Mitochondrial translation initiation | 88 | 3 (3.4%) | 0.0237 | 0.441 | Reactome |
| Hepatitis C - Homo sapiens (human) | 157 | 4 (2.5%) | 0.0243 | 0.441 | KEGG |
| Celecoxib Action Pathway | 36 | 2 (5.6%) | 0.0262 | 0.441 | SMPDB |
| IGF signaling | 36 | 2 (5.6%) | 0.0262 | 0.441 | INOH |
| Inactivation, recovery and regulation of the phototransduction cascade | 36 | 2 (5.6%) | 0.0262 | 0.441 | Reactome |
| ATR signaling pathway | 37 | 2 (5.4%) | 0.0275 | 0.441 | PID |
| The phototransduction cascade | 37 | 2 (5.4%) | 0.0275 | 0.441 | Reactome |
| Felbamate Metabolism Pathway | 4 | 1 (25.0%) | 0.0277 | 0.441 | SMPDB |
| ahr signal transduction pathway | 4 | 1 (25.0%) | 0.0277 | 0.441 | BioCarta |
| Regulation of gene expression by Hypoxia-inducible Factor | 4 | 1 (25.0%) | 0.0277 | 0.441 | Reactome |
| Clomipramine Metabolism Pathway | 4 | 1 (25.0%) | 0.0277 | 0.441 | SMPDB |
| Codeine Metabolism Pathway | 4 | 1 (25.0%) | 0.0277 | 0.441 | SMPDB |
| *trans, trans-farnesyl diphosphate biosynthesis* | 4 | 1 (25.0%) | 0.0277 | 0.441 | HumanCyc |
| IRS activation | 4 | 1 (25.0%) | 0.0277 | 0.441 | Reactome |
| Reuptake of GABA | 4 | 1 (25.0%) | 0.0277 | 0.441 | Reactome |
| arsenate detoxification I (glutaredoxin) | 4 | 1 (25.0%) | 0.0277 | 0.441 | HumanCyc |
| Amino acid conjugation of benzoic acid | 4 | 1 (25.0%) | 0.0277 | 0.441 | Wikipathways |
| Mitochondrial translation | 94 | 3 (3.2%) | 0.0281 | 0.441 | Reactome |
| Signaling events regulated by Ret tyrosine kinase | 39 | 2 (5.1%) | 0.0304 | 0.441 | PID |
| Influenza A - Homo sapiens (human) | 172 | 4 (2.3%) | 0.0325 | 0.441 | KEGG |
| Abacavir metabolism | 5 | 1 (20.0%) | 0.0345 | 0.441 | Reactome |
| Doxepin Metabolism Pathway | 5 | 1 (20.0%) | 0.0345 | 0.441 | SMPDB |
| Cross-presentation of soluble exogenous antigens (endosomes) | 5 | 1 (20.0%) | 0.0345 | 0.441 | Reactome |
| NR1H2 & NR1H3 regulate gene expression linked to triglyceride lipolysis in adipose | 5 | 1 (20.0%) | 0.0345 | 0.441 | Reactome |
| NR1H2 & NR1H3 regulate gene expression linked to gluconeogenesis | 5 | 1 (20.0%) | 0.0345 | 0.441 | Reactome |
| NR1H2 & NR1H3 regulate gene expression to limit cholesterol uptake | 5 | 1 (20.0%) | 0.0345 | 0.441 | Reactome |
| Synthesis of GDP-mannose | 5 | 1 (20.0%) | 0.0345 | 0.441 | Reactome |
| RHOT1 GTPase cycle | 5 | 1 (20.0%) | 0.0345 | 0.441 | Reactome |

| | | | | | |
|---|---|---|---|---|---|
| Free fatty acid receptors | 5 | 1 (20.0%) | 0.0345 | 0.441 | Reactome |
| Adefovir Dipivoxil Metabolism Pathway | 5 | 1 (20.0%) | 0.0345 | 0.441 | SMPDB |
| Tenofovir Metabolism Pathway | 5 | 1 (20.0%) | 0.0345 | 0.441 | SMPDB |
| Graft-versus-host disease - Homo sapiens (human) | 42 | 2 (4.8%) | 0.0348 | 0.441 | KEGG |
| Generation of second messenger molecules | 42 | 2 (4.8%) | 0.0348 | 0.441 | Reactome |
| Tuberculosis - Homo sapiens (human) | 180 | 4 (2.2%) | 0.0368 | 0.441 | KEGG |
| Galactose metabolism | 44 | 2 (4.5%) | 0.0379 | 0.441 | EHMN |
| CD4 T cell receptor signaling-ERK cascade | 44 | 2 (4.5%) | 0.0379 | 0.441 | INOH |
| Insulin Pathway | 46 | 2 (4.3%) | 0.0411 | 0.441 | PID |
| Cholesterol metabolism (includes both Bloch and Kandutsch-Russell pathways) | 46 | 2 (4.3%) | 0.0411 | 0.441 | Wikipathways |
| NOSTRIN mediated eNOS trafficking | 6 | 1 (16.7%) | 0.0413 | 0.441 | Reactome |
| Biosynthesis of maresin-like SPMs | 6 | 1 (16.7%) | 0.0413 | 0.441 | Reactome |
| Venlafaxine Metabolism Pathway | 6 | 1 (16.7%) | 0.0413 | 0.441 | SMPDB |
| Imipramine Metabolism Pathway | 6 | 1 (16.7%) | 0.0413 | 0.441 | SMPDB |
| Fluoxetine Metabolism Pathway | 6 | 1 (16.7%) | 0.0413 | 0.441 | SMPDB |
| Activated NTRK3 signals through PI3K | 6 | 1 (16.7%) | 0.0413 | 0.441 | Reactome |
| Conjugation of benzoate with glycine | 6 | 1 (16.7%) | 0.0413 | 0.441 | Reactome |
| GDP-mannose biosynthesis | 6 | 1 (16.7%) | 0.0413 | 0.441 | HumanCyc |
| Metapathway biotransformation Phase I and II | 186 | 4 (2.2%) | 0.0415 | 0.441 | Wikipathways |
| Notch-mediated HES/HEY network | 48 | 2 (4.2%) | 0.0444 | 0.441 | PID |
| Aryl hydrocarbon receptor signalling | 7 | 1 (14.3%) | 0.048 | 0.441 | Reactome |
| Tramadol Metabolism Pathway | 7 | 1 (14.3%) | 0.048 | 0.441 | SMPDB |
| SOS-mediated signalling | 7 | 1 (14.3%) | 0.048 | 0.441 | Reactome |
| Modulation by Mtb of host immune system | 7 | 1 (14.3%) | 0.048 | 0.441 | Reactome |
| ceramide de novo biosynthesis | 7 | 1 (14.3%) | 0.048 | 0.441 | HumanCyc |
| serine and glycine biosynthesis | 7 | 1 (14.3%) | 0.048 | 0.441 | |

Heart_failure_SAT

| pathway name | set size | candidates contained | p-value | q-value | pathway source |
|---|---|---|---|---|---|
| MHC class II antigen presentation | 60 | 4 (6.7%) | 0.00172 | 0.497 | Reactome |
| Celecoxib Metabolism Pathway | 8 | 2 (25.0%) | 0.00197 | 0.497 | SMPDB |
| Asthma - Homo sapiens (human) | 31 | 3 (10.0%) | 0.0021 | 0.497 | KEGG |
| Allograft rejection - Homo sapiens (human) | 38 | 3 (8.1%) | 0.00384 | 0.497 | KEGG |
| Intestinal immune network for IgA production - Homo sapiens (human) | 49 | 3 (6.4%) | 0.00754 | 0.497 | KEGG |
| Allograft Rejection | 93 | 4 (4.3%) | 0.00829 | 0.497 | Wikipathways |
| Metabolism of Tetrahydrocannabinol (THC) | 1 | 1 (100.0%) | 0.00856 | 0.497 | Wikipathways |
| Biosynthesis of DHA-derived SPMs | 17 | 2 (11.8%) | 0.00908 | 0.497 | Reactome |
| Pyrimidine nucleotides nucleosides metabolism | 51 | 3 (5.9%) | 0.00944 | 0.497 | INOH |
| Hematopoietic cell lineage - Homo sapiens (human) | 99 | 4 (4.1%) | 0.00993 | 0.497 | KEGG |
| Autoimmune thyroid disease - Homo sapiens (human) | 53 | 3 (5.8%) | 0.00996 | 0.497 | KEGG |
| Biosynthesis of specialized proresolving mediators (SPMs) | 19 | 2 (10.5%) | 0.0113 | 0.497 | Reactome |
| Translesion Synthesis by POLH | 19 | 2 (10.5%) | 0.0113 | 0.497 | Reactome |
| Pyrimidine metabolism - Homo sapiens (human) | 57 | 3 (5.3%) | 0.0128 | 0.497 | KEGG |
| RA biosynthesis pathway | 22 | 2 (9.1%) | 0.015 | 0.497 | Reactome |
| Toxoplasmosis - Homo sapiens (human) | 112 | 4 (3.6%) | 0.0156 | 0.497 | KEGG |
| Biosynthesis of maresin conjugates in tissue regeneration (MCTR) | 2 | 1 (50.0%) | 0.017 | 0.497 | Reactome |
| Biosynthesis of DHA-derived sulfido conjugates | 2 | 1 (50.0%) | 0.017 | 0.497 | Reactome |
| Fatty acid metabolism | 177 | 5 (2.8%) | 0.0176 | 0.497 | Reactome |
| Synthesis of very long-chain fatty acyl-CoAs | 24 | 2 (8.3%) | 0.0177 | 0.497 | Reactome |
| Inflammatory bowel disease - Homo sapiens (human) | 65 | 3 (4.6%) | 0.0182 | 0.497 | KEGG |
| Amphetamine addiction - Homo sapiens (human) | 69 | 3 (4.3%) | 0.0213 | 0.497 | KEGG |
| Metapathway biotransformation Phase I and II | 186 | 5 (2.7%) | 0.0218 | 0.497 | Wikipathways |
| Drug metabolism - cytochrome P450 - Homo sapiens (human) | 72 | 3 (4.3%) | 0.0221 | 0.497 | KEGG |
| Fatty acid elongation - Homo sapiens (human) | 27 | 2 (7.4%) | 0.0222 | 0.497 | KEGG |
| Biosynthesis of unsaturated fatty acids - Homo sapiens (human) | 27 | 2 (7.4%) | 0.0222 | 0.497 | KEGG |
| Cytokines and Inflammatory Response | 29 | 2 (7.1%) | 0.0238 | 0.497 | Wikipathways |
| Translocation of ZAP-70 to Immunological synapse | 28 | 2 (7.1%) | 0.0238 | 0.497 | Reactome |
| Ebola Virus Pathway on Host | 130 | 4 (3.1%) | 0.0254 | 0.497 | Wikipathways |
| methionine salvage | 3 | 1 (33.3%) | 0.0255 | 0.497 | HumanCyc |
| Rosiglitazone Metabolism Pathway | 3 | 1 (33.3%) | 0.0255 | 0.497 | SMPDB |
| Pyrimidine biosynthesis | 3 | 1 (33.3%) | 0.0255 | 0.497 | Reactome |
| Metabolism of xenobiotics by cytochrome P450 - Homo sapiens (human) | 78 | 3 (3.9%) | 0.0273 | 0.497 | KEGG |
| Leishmaniasis - Homo sapiens (human) | 77 | 3 (3.9%) | 0.0273 | 0.497 | KEGG |
| Phosphorylation of CD3 and TCR zeta chains | 31 | 2 (6.5%) | 0.0287 | 0.497 | Reactome |
| PD-1 signaling | 32 | 2 (6.2%) | 0.0305 | 0.497 | Reactome |
| Development and heterogeneity of the ILC family | 32 | 2 (6.2%) | 0.0305 | 0.497 | Wikipathways |
| Drug metabolism - other enzymes - Homo sapiens (human) | 80 | 3 (3.8%) | 0.0312 | 0.497 | KEGG |
| Inflammatory Response Pathway | 33 | 2 (6.1%) | 0.0323 | 0.497 | Wikipathways |
| Late Phase of HIV Life Cycle | 141 | 4 (2.8%) | 0.0329 | 0.497 | Reactome |
| Felbamate Metabolism Pathway | 4 | 1 (25.0%) | 0.0338 | 0.497 | SMPDB |
| Activation, myristolyation of BID and translocation to mitochondria | 4 | 1 (25.0%) | 0.0338 | 0.497 | Reactome |
| Utilization of Ketone Bodies | 4 | 1 (25.0%) | 0.0338 | 0.497 | Reactome |
| HCMV Late Events | 4 | 1 (25.0%) | 0.0338 | 0.497 | Reactome |
| Invadopodia formation | 4 | 1 (25.0%) | 0.0338 | 0.497 | Reactome |
| pyrimidine ribonucleosides salvage I | 4 | 1 (25.0%) | 0.0338 | 0.497 | HumanCyc |
| Chemical carcinogenesis - Homo sapiens (human) | 83 | 3 (3.6%) | 0.0343 | 0.497 | KEGG |
| Pyrimidine metabolism | 84 | 3 (3.6%) | 0.0353 | 0.497 | Wikipathways |
| Celecoxib Action Pathway | 36 | 2 (5.6%) | 0.0379 | 0.497 | SMPDB |
| Inactivation, recovery and regulation of the phototransduction cascade | 36 | 2 (5.6%) | 0.0379 | 0.497 | Reactome |
| The phototransduction cascade | 37 | 2 (5.4%) | 0.0398 | 0.497 | Reactome |
| Fatty acyl-CoA biosynthesis | 37 | 2 (5.4%) | 0.0398 | 0.497 | Reactome |
| HIV Life Cycle | 151 | 4 (2.6%) | 0.0407 | 0.497 | Reactome |
| Translesion synthesis by Y family DNA polymerases bypasses lesions on DNA template | 38 | 2 (5.3%) | 0.0418 | 0.497 | Reactome |
| Abacavir metabolism | 5 | 1 (20.0%) | 0.0421 | 0.497 | Reactome |
| glycine betaine degradation | 5 | 1 (20.0%) | 0.0421 | 0.497 | HumanCyc |
| Doxepin Metabolism Pathway | 5 | 1 (20.0%) | 0.0421 | 0.497 | SMPDB |
| Succinyl CoA: 3-ketoacid CoA transferase deficiency | 5 | 1 (20.0%) | 0.0421 | 0.497 | SMPDB |
| Ketone Body Metabolism | 5 | 1 (20.0%) | 0.0421 | 0.497 | SMPDB |
| ketolysis | 5 | 1 (20.0%) | 0.0421 | 0.497 | HumanCyc |
| Synthesis and Degradation of Ketone Bodies | 5 | 1 (20.0%) | 0.0421 | 0.497 | Wikipathways |
| Cross-presentation of soluble exogenous antigens (endosomes) | 5 | 1 (20.0%) | 0.0421 | 0.497 | Reactome |
| SUMOylation of DNA replication proteins | 39 | 2 (5.1%) | 0.0438 | 0.497 | Reactome |
| Rheumatoid arthritis - Homo sapiens (human) | 93 | 3 (3.3%) | 0.0443 | 0.497 | KEGG |
| Th1 and Th2 cell differentiation - Homo sapiens (human) | 92 | 3 (3.3%) | 0.0443 | 0.497 | KEGG |
| Glycine Serine metabolism | 40 | 2 (5.0%) | 0.0459 | 0.497 | INOH |
| Ribosome - Homo sapiens (human) | 158 | 4 (2.5%) | 0.0467 | 0.497 | KEGG |

| | | | | | |
|---|---|---|---|---|---|
| Dual Incision in GG-NER | 41 | 2 (4.9%) | 0.048 | 0.497 | **Reactome** |
| Metabolism of lipids | 646 | 10 (1.6%) | 0.0497 | 0.497 | **Reactome** |

CAD_VAT

| pathway name | set size | candidates contained | p-value | q-value | pathway source |
|---|---|---|---|---|---|
| Folate metabolism | 31 | 3 (9.7%) | 0.00168 | 0.426 | INOH |
| CMP phosphorylation | 11 | 2 (18.2%) | 0.00306 | 0.426 | HumanCyc |
| pyrimidine deoxyribonucleotide phosphorylation | 12 | 2 (16.7%) | 0.00365 | 0.426 | HumanCyc |
| guanosine deoxyribonucleotides de novo biosynthesis | 12 | 2 (16.7%) | 0.00365 | 0.426 | HumanCyc |
| adenosine deoxyribonucleotides de novo biosynthesis | 12 | 2 (16.7%) | 0.00365 | 0.426 | HumanCyc |
| Ganglio Sphingolipid Metabolism | 13 | 2 (15.4%) | 0.0043 | 0.426 | Wikipathways |
| UTP and CTP de novo biosynthesis | 13 | 2 (15.4%) | 0.0043 | 0.426 | HumanCyc |
| purine deoxyribonucleosides salvage | 13 | 2 (15.4%) | 0.0043 | 0.426 | HumanCyc |
| guanosine ribonucleotides de novo biosynthesis | 13 | 2 (15.4%) | 0.0043 | 0.426 | HumanCyc |
| superpathway of pyrimidine ribonucleotides de novo biosynthesis | 15 | 2 (13.3%) | 0.00573 | 0.426 | HumanCyc |
| pyrimidine deoxyribonucleotides biosynthesis from CTP | 16 | 2 (12.5%) | 0.00651 | 0.426 | HumanCyc |
| pyrimidine deoxyribonucleotides de novo biosynthesis | 16 | 2 (12.5%) | 0.00651 | 0.426 | HumanCyc |
| guanosine nucleotides de novo biosynthesis | 16 | 2 (12.5%) | 0.00651 | 0.426 | HumanCyc |
| superpathway of pyrimidine deoxyribonucleoside salvage | 17 | 2 (11.8%) | 0.00735 | 0.426 | HumanCyc |
| D-mannose degradation | 1 | 1 (100.0%) | 0.00767 | 0.426 | HumanCyc |
| *S-methyl-5,-thioadenosine degradation* | 1 | 1 (100.0%) | 0.00767 | 0.426 | HumanCyc |
| TNFalpha | 234 | 6 (2.6%) | 0.00921 | 0.426 | NetPath |
| Glycosphingolipid biosynthesis - ganglioseries | 21 | 2 (9.5%) | 0.0111 | 0.426 | EHMN |
| Transcription factor regulation in adipogenesis | 22 | 2 (9.1%) | 0.0122 | 0.426 | Wikipathways |
| Type II diabetes mellitus | 22 | 2 (9.1%) | 0.0122 | 0.426 | Wikipathways |
| superpathway of pyrimidine deoxyribonucleotides de novo biosynthesis | 22 | 2 (9.1%) | 0.0122 | 0.426 | HumanCyc |
| Sterols are 12-hydroxylated by CYP8B1 | 2 | 1 (50.0%) | 0.0153 | 0.426 | Reactome |
| Inhibition of Host mRNA Processing and RNA Silencing | 2 | 1 (50.0%) | 0.0153 | 0.426 | Reactome |
| Adipocytokine signaling pathway - Homo sapiens (human) | 69 | 3 (4.3%) | 0.0159 | 0.426 | KEGG |
| Vitamin B9 (folate) metabolism | 26 | 2 (7.7%) | 0.0168 | 0.426 | EHMN |
| CDC42 signaling events | 71 | 3 (4.2%) | 0.0172 | 0.426 | PID |
| Biosynthesis of unsaturated fatty acids - Homo sapiens (human) | 27 | 2 (7.4%) | 0.018 | 0.426 | KEGG |
| Effects of PIP2 hydrolysis | 27 | 2 (7.4%) | 0.018 | 0.426 | Reactome |
| Adipogenesis | 131 | 4 (3.1%) | 0.0182 | 0.426 | Wikipathways |
| Reelin signaling pathway | 28 | 2 (7.1%) | 0.0193 | 0.426 | PID |
| B cell receptor signaling | 134 | 4 (3.0%) | 0.0196 | 0.426 | INOH |
| Glyoxylate and dicarboxylate metabolism - Homo sapiens (human) | 30 | 2 (6.7%) | 0.022 | 0.426 | KEGG |
| Trafficking of AMPA receptors | 30 | 2 (6.7%) | 0.022 | 0.426 | Reactome |
| Glutamate binding, activation of AMPA receptors and synaptic plasticity | 30 | 2 (6.7%) | 0.022 | 0.426 | Reactome |
| glycine cleavage | 3 | 1 (33.3%) | 0.0228 | 0.426 | HumanCyc |
| Tandem of pore domain in a weak inwardly rectifying K+ channels (TWIK) | 3 | 1 (33.3%) | 0.0228 | 0.426 | Reactome |
| B cell receptor signaling pathway - Homo sapiens (human) | 82 | 3 (3.8%) | 0.0235 | 0.426 | KEGG |
| Disruption of postsynaptic signaling by CNV | 33 | 2 (6.1%) | 0.0263 | 0.426 | Wikipathways |
| Alpha 6 Beta 4 signaling pathway | 33 | 2 (6.1%) | 0.0263 | 0.426 | Wikipathways |
| Pyrimidine metabolism | 84 | 3 (3.6%) | 0.0267 | 0.426 | Wikipathways |
| Vitamin D-sensitive calcium signaling in depression | 35 | 2 (5.7%) | 0.0294 | 0.426 | Wikipathways |
| Glycine degradation | 4 | 1 (25.0%) | 0.0303 | 0.426 | Reactome |
| ahr signal transduction pathway | 4 | 1 (25.0%) | 0.0303 | 0.426 | BioCarta |
| Regulation of gene expression by Hypoxia-inducible Factor | 4 | 1 (25.0%) | 0.0303 | 0.426 | Reactome |
| tryptophan degradation X (mammalian, via tryptamine) | 4 | 1 (25.0%) | 0.0303 | 0.426 | HumanCyc |
| Catecholamine biosynthesis | 4 | 1 (25.0%) | 0.0303 | 0.426 | Reactome |
| catecholamine biosynthesis | 4 | 1 (25.0%) | 0.0303 | 0.426 | HumanCyc |
| Tyrosine hydroxylase deficiency | 4 | 1 (25.0%) | 0.0303 | 0.426 | SMPDB |
| Catecholamine Biosynthesis | 4 | 1 (25.0%) | 0.0303 | 0.426 | SMPDB |
| Aromatic L-Aminoacid Decarboxylase Deficiency | 4 | 1 (25.0%) | 0.0303 | 0.426 | SMPDB |
| Vitamin K Metabolism | 4 | 1 (25.0%) | 0.0303 | 0.426 | SMPDB |
| IRS activation | 4 | 1 (25.0%) | 0.0303 | 0.426 | Reactome |
| propionyl-CoA degradation | 4 | 1 (25.0%) | 0.0303 | 0.426 | HumanCyc |
| Phosphorylation of proteins involved in G1/S transition by active Cyclin E:Cdk2 complexes | 4 | 1 (25.0%) | 0.0303 | 0.426 | Reactome |
| ALKBH3 mediated reversal of alkylation damage | 4 | 1 (25.0%) | 0.0303 | 0.426 | Reactome |
| Factors and pathways affecting insulin-like growth factor (IGF1)-Akt signaling | 36 | 2 (5.6%) | 0.031 | 0.426 | Wikipathways |
| Cushing syndrome - Homo sapiens (human) | 155 | 4 (2.6%) | 0.0313 | 0.426 | KEGG |
| Fatty acyl-CoA biosynthesis | 37 | 2 (5.4%) | 0.0326 | 0.426 | Reactome |
| TBC/RABGAPs | 38 | 2 (5.3%) | 0.0342 | 0.426 | Reactome |
| G alpha (i) signalling events | 316 | 6 (1.9%) | 0.0346 | 0.426 | Reactome |
| Association of TriC/CCT with target proteins during biosynthesis | 39 | 2 (5.1%) | 0.0359 | 0.426 | Reactome |
| Small cell lung cancer | 96 | 3 (3.1%) | 0.0375 | 0.426 | Wikipathways |
| Serotonin and melatonin biosynthesis | 5 | 1 (20.0%) | 0.0378 | 0.426 | Reactome |
| serotonin and melatonin biosynthesis | 5 | 1 (20.0%) | 0.0378 | 0.426 | HumanCyc |
| Catalytic cycle of mammalian Flavin-containing MonoOxygenases (FMOs) | 5 | 1 (20.0%) | 0.0378 | 0.426 | Wikipathways |
| tetrapyrrole biosynthesis | 5 | 1 (20.0%) | 0.0378 | 0.426 | HumanCyc |
| Synthesis of GDP-mannose | 5 | 1 (20.0%) | 0.0378 | 0.426 | Reactome |

| | | | | | |
|---|---|---|---|---|---|
| Propionyl-CoA catabolism | 5 | 1 (20.0%) | 0.0378 | 0.426 | Reactome |
| Ultraconserved region 339 modulation of tumor suppressor microRNAs in cancer | 5 | 1 (20.0%) | 0.0378 | 0.426 | Wikipathways |
| Arachidonate production from DAG | 5 | 1 (20.0%) | 0.0378 | 0.426 | Reactome |
| CD4 T cell receptor signaling-NFkB cascade | 99 | 3 (3.0%) | 0.0405 | 0.426 | INOH |
| RHOF GTPase cycle | 42 | 2 (4.8%) | 0.0411 | 0.426 | Reactome |
| Activated NTRK3 signals through PI3K | 6 | 1 (16.7%) | 0.0451 | 0.426 | Reactome |
| GDP-mannose biosynthesis | 6 | 1 (16.7%) | 0.0451 | 0.426 | HumanCyc |
| Methionine salvage pathway | 6 | 1 (16.7%) | 0.0451 | 0.426 | Reactome |
| Mitochondrial fatty acid synthesis pathway | 6 | 1 (16.7%) | 0.0451 | 0.426 | Wikipathways |
| chondroitin and dermatan biosynthesis | 6 | 1 (16.7%) | 0.0451 | 0.426 | HumanCyc |
| Fatty acid metabolism | 177 | 4 (2.3%) | 0.0464 | 0.426 | Reactome |
| Cardiac conduction | 105 | 3 (2.9%) | 0.0468 | 0.426 | Reactome |
| Type II diabetes mellitus - Homo sapiens (human) | 46 | 2 (4.3%) | 0.0484 | 0.426 | KEGG |

T2DM_VAT

| pathway name | set size | candidates contained | p-value | q-value | pathway source |
|---|---|---|---|---|---|
| Biosynthesis of unsaturated fatty acids - Homo sapiens (human) | 27 | 4 (14.8%) | 2.94E-05 | 0.0285 | KEGG |
| Allograft rejection - Homo sapiens (human) | 38 | 4 (10.8%) | 0.000105 | 0.0462 | KEGG |
| Graft-versus-host disease - Homo sapiens (human) | 42 | 4 (9.5%) | 0.000174 | 0.0462 | KEGG |
| Type I diabetes mellitus - Homo sapiens (human) | 43 | 4 (9.3%) | 0.00019 | 0.0462 | KEGG |
| Intestinal immune network for IgA production - Homo sapiens (human) | 49 | 4 (8.5%) | 0.00027 | 0.0524 | KEGG |
| Autoimmune thyroid disease - Homo sapiens (human) | 53 | 4 (7.7%) | 0.000399 | 0.0606 | KEGG |
| Metabolism of amino acids and derivatives | 339 | 9 (2.7%) | 0.000437 | 0.0606 | Reactome |
| Hematopoietic cell lineage - Homo sapiens (human) | 99 | 5 (5.1%) | 0.000502 | 0.0609 | KEGG |
| Viral myocarditis - Homo sapiens (human) | 60 | 4 (6.8%) | 0.000647 | 0.0613 | KEGG |
| Alpha Linolenic Acid and Linoleic Acid Metabolism | 6 | 2 (33.3%) | 0.000654 | 0.0613 | SMPDB |
| G alpha (q) signalling events | 221 | 7 (3.2%) | 0.000697 | 0.0613 | Reactome |
| Fatty acid elongation - Homo sapiens (human) | 27 | 3 (11.1%) | 0.000757 | 0.0613 | KEGG |
| Translocation of ZAP-70 to Immunological synapse | 28 | 3 (10.7%) | 0.000844 | 0.063 | Reactome |
| Asthma - Homo sapiens (human) | 31 | 3 (10.0%) | 0.00104 | 0.0716 | KEGG |
| Phosphorylation of CD3 and TCR zeta chains | 31 | 3 (9.7%) | 0.00114 | 0.0716 | Reactome |
| oleate biosynthesis | 8 | 2 (25.0%) | 0.00121 | 0.0716 | HumanCyc |
| PD-1 signaling | 32 | 3 (9.4%) | 0.00125 | 0.0716 | Reactome |
| Costimulation by the CD28 family | 77 | 4 (5.2%) | 0.00176 | 0.0947 | Reactome |
| Fatty acyl-CoA biosynthesis | 37 | 3 (8.1%) | 0.00192 | 0.0979 | Reactome |
| Generation of second messenger molecules | 42 | 3 (7.1%) | 0.00276 | 0.134 | Reactome |
| Cell adhesion molecules - Homo sapiens (human) | 149 | 5 (3.4%) | 0.00315 | 0.141 | KEGG |
| GPCR downstream signalling | 633 | 11 (1.7%) | 0.00329 | 0.141 | Reactome |
| Rheumatoid arthritis - Homo sapiens (human) | 93 | 4 (4.3%) | 0.00336 | 0.141 | KEGG |
| Allograft Rejection | 93 | 4 (4.3%) | 0.00349 | 0.141 | Wikipathways |
| Interferon gamma signaling | 96 | 4 (4.2%) | 0.00391 | 0.152 | Reactome |
| Downstream TCR signaling | 52 | 3 (5.8%) | 0.00507 | 0.189 | Reactome |
| Th17 cell differentiation - Homo sapiens (human) | 107 | 4 (3.7%) | 0.00575 | 0.207 | KEGG |
| er associated degradation (erad) pathway | 18 | 2 (11.1%) | 0.00633 | 0.212 | BioCarta |
| Synthesis of glycosylphosphatidylinositol (GPI) | 18 | 2 (11.1%) | 0.00633 | 0.212 | Reactome |
| CD4 T cell receptor signaling-JNK cascade | 57 | 3 (5.3%) | 0.00655 | 0.212 | INOH |
| Signaling by GPCR | 706 | 11 (1.6%) | 0.00739 | 0.229 | Reactome |
| MHC class II antigen presentation | 60 | 3 (5.0%) | 0.00756 | 0.229 | Reactome |
| Platelet activation, signaling and aggregation | 263 | 6 (2.3%) | 0.00841 | 0.238 | Reactome |
| Methionine De Novo and Salvage Pathway | 21 | 2 (9.5%) | 0.00858 | 0.238 | Wikipathways |
| Metabolism | 1954 | 22 (1.1%) | 0.00858 | 0.238 | Reactome |
| Inflammatory bowel disease - Homo sapiens (human) | 65 | 3 (4.6%) | 0.00942 | 0.254 | KEGG |
| Synthesis of very long-chain fatty acyl-CoAs | 24 | 2 (8.3%) | 0.0111 | 0.292 | Reactome |
| G0 and Early G1 | 25 | 2 (8.0%) | 0.012 | 0.297 | Reactome |
| Systemic lupus erythematosus - Homo sapiens (human) | 136 | 4 (3.0%) | 0.0125 | 0.297 | KEGG |
| TCR signaling | 73 | 3 (4.1%) | 0.0129 | 0.297 | Reactome |
| Glycosylphosphatidylinositol (GPI)-anchor biosynthesis - Homo sapiens (human) | 26 | 2 (7.7%) | 0.013 | 0.297 | KEGG |
| Vitamin B9 (folate) metabolism | 26 | 2 (7.7%) | 0.013 | 0.297 | EHMN |
| TWIK-related alkaline pH activated K+ channel (TALK) | 2 | 1 (50.0%) | 0.0134 | 0.297 | Reactome |
| Peptide GPCRs | 75 | 3 (4.0%) | 0.0139 | 0.297 | Wikipathways |
| Sulfur amino acid metabolism | 27 | 2 (7.4%) | 0.014 | 0.297 | Reactome |
| GPCR ligand binding | 476 | 8 (1.7%) | 0.0144 | 0.297 | Reactome |
| Leishmaniasis - Homo sapiens (human) | 77 | 3 (3.9%) | 0.0144 | 0.297 | KEGG |
| One-carbon metabolism | 28 | 2 (7.1%) | 0.015 | 0.303 | Wikipathways |
| Antigen processing and presentation - Homo sapiens (human) | 78 | 3 (3.8%) | 0.0154 | 0.305 | KEGG |
| Interaction between L1 and Ankyrins | 29 | 2 (6.9%) | 0.016 | 0.311 | Reactome |
| G alpha (12/13) signalling events | 80 | 3 (3.8%) | 0.0165 | 0.314 | Reactome |
| Folate metabolism | 31 | 2 (6.5%) | 0.0182 | 0.334 | INOH |
| Phagosome - Homo sapiens (human) | 152 | 4 (2.6%) | 0.0186 | 0.334 | KEGG |
| Calcium Regulation in the Cardiac Cell | 151 | 4 (2.6%) | 0.0186 | 0.334 | Wikipathways |
| methionine salvage | 3 | 1 (33.3%) | 0.02 | 0.34 | HumanCyc |
| Melanin biosynthesis | 3 | 1 (33.3%) | 0.02 | 0.34 | Reactome |
| Signaling by MST1 | 3 | 1 (33.3%) | 0.02 | 0.34 | Reactome |
| Interferon Signaling | 162 | 4 (2.5%) | 0.0234 | 0.357 | Reactome |
| Th1 and Th2 cell differentiation - Homo sapiens (human) | 92 | 3 (3.3%) | 0.0238 | 0.357 | KEGG |
| Staphylococcus aureus infection - Homo sapiens (human) | 96 | 3 (3.2%) | 0.0252 | 0.357 | KEGG |
| Post-translational modification: synthesis of GPI-anchored proteins | 94 | 3 (3.2%) | 0.0252 | 0.357 | Reactome |
| eumelanin biosynthesis | 4 | 1 (25.0%) | 0.0265 | 0.357 | HumanCyc |
| glutaryl-CoA degradation | 4 | 1 (25.0%) | 0.0265 | 0.357 | HumanCyc |
| Glycine degradation | 4 | 1 (25.0%) | 0.0265 | 0.357 | Reactome |
| IRS activation | 4 | 1 (25.0%) | 0.0265 | 0.357 | Reactome |
| Lysosomal oligosaccharide catabolism | 4 | 1 (25.0%) | 0.0265 | 0.357 | Reactome |

| | | | | | |
|---|---|---|---|---|---|
| Phosphorylation of proteins involved in G1/S transition by active Cyclin E:Cdk2 complexes | 4 | 1 (25.0%) | 0.0265 | 0.357 | Reactome |
| Amino Acid Metabolism Pathway Excerpt (Histidine catabolism extension) | 4 | 1 (25.0%) | 0.0265 | 0.357 | Wikipathways |
| *S-methyl-5-thio-α-D-ribose 1-phosphate degradation* | 4 | 1 (25.0%) | 0.0265 | 0.357 | HumanCyc |
| Vitamin B6-dependent and responsive disorders | 4 | 1 (25.0%) | 0.0265 | 0.357 | Wikipathways |
| Class A/1 (Rhodopsin-like receptors) | 346 | 6 (1.7%) | 0.0286 | 0.357 | Reactome |
| CD4 T cell receptor signaling-NFkB cascade | 99 | 3 (3.0%) | 0.0288 | 0.357 | INOH |
| Glycine Serine metabolism | 40 | 2 (5.0%) | 0.0294 | 0.357 | INOH |
| Glycine, serine and threonine metabolism - Homo sapiens (human) | 40 | 2 (5.0%) | 0.0294 | 0.357 | KEGG |
| Fatty acid metabolism | 177 | 4 (2.3%) | 0.0305 | 0.357 | Reactome |
| L1CAM interactions | 102 | 3 (2.9%) | 0.031 | 0.357 | Reactome |
| Visual phototransduction | 103 | 3 (2.9%) | 0.0318 | 0.357 | Reactome |
| HDL clearance | 5 | 1 (20.0%) | 0.0331 | 0.357 | Reactome |
| glycine betaine degradation | 5 | 1 (20.0%) | 0.0331 | 0.357 | HumanCyc |
| Tachykinin receptors bind tachykinins | 5 | 1 (20.0%) | 0.0331 | 0.357 | Reactome |
| Ultraconserved region 339 modulation of tumor suppressor microRNAs in cancer | 5 | 1 (20.0%) | 0.0331 | 0.357 | Wikipathways |
| SUMO is conjugated to E1 (UBA2:SAE1) | 5 | 1 (20.0%) | 0.0331 | 0.357 | Reactome |
| Trafficking of myristoylated proteins to the cilium | 5 | 1 (20.0%) | 0.0331 | 0.357 | Reactome |
| Free fatty acid receptors | 5 | 1 (20.0%) | 0.0331 | 0.357 | Reactome |
| Adefovir Dipivoxil Metabolism Pathway | 5 | 1 (20.0%) | 0.0331 | 0.357 | SMPDB |
| Tenofovir Metabolism Pathway | 5 | 1 (20.0%) | 0.0331 | 0.357 | SMPDB |
| De novo fatty acid biosynthesis | 43 | 2 (4.7%) | 0.0336 | 0.357 | EHMN |
| CD4 T cell receptor signaling-ERK cascade | 44 | 2 (4.5%) | 0.035 | 0.357 | INOH |
| Cholesterol metabolism (includes both Bloch and Kandutsch-Russell pathways) | 46 | 2 (4.3%) | 0.038 | 0.357 | Wikipathways |
| Toxoplasmosis - Homo sapiens (human) | 112 | 3 (2.7%) | 0.0393 | 0.357 | KEGG |
| Choline catabolism | 6 | 1 (16.7%) | 0.0395 | 0.357 | Reactome |
| Activated NTRK3 signals through PI3K | 6 | 1 (16.7%) | 0.0395 | 0.357 | Reactome |
| Pre-NOTCH Processing in the Endoplasmic Reticulum | 6 | 1 (16.7%) | 0.0395 | 0.357 | Reactome |
| SUMO is transferred from E1 to E2 (UBE2I, UBC9) | 6 | 1 (16.7%) | 0.0395 | 0.357 | Reactome |
| basic mechanisms of sumoylation | 6 | 1 (16.7%) | 0.0395 | 0.357 | BioCarta |
| histidine degradation | 6 | 1 (16.7%) | 0.0395 | 0.357 | HumanCyc |
| Methionine salvage pathway | 6 | 1 (16.7%) | 0.0395 | 0.357 | Reactome |
| Mitochondrial fatty acid synthesis pathway | 6 | 1 (16.7%) | 0.0395 | 0.357 | Wikipathways |
| Proton-coupled monocarboxylate transport | 6 | 1 (16.7%) | 0.0395 | 0.357 | Reactome |
| Cysteine and methionine metabolism - Homo sapiens (human) | 50 | 2 (4.0%) | 0.0442 | 0.357 | KEGG |
| Ovarian steroidogenesis - Homo sapiens (human) | 51 | 2 (3.9%) | 0.0458 | 0.357 | KEGG |
| Scavenging by Class B Receptors | 7 | 1 (14.3%) | 0.046 | 0.357 | Reactome |
| HDL assembly | 7 | 1 (14.3%) | 0.046 | 0.357 | Reactome |
| The retinoid cycle in cones (daylight vision) | 7 | 1 (14.3%) | 0.046 | 0.357 | Reactome |
| superpathway of choline degradation to L-serine | 7 | 1 (14.3%) | 0.046 | 0.357 | HumanCyc |
| SOS-mediated signalling | 7 | 1 (14.3%) | 0.046 | 0.357 | Reactome |
| Neurofascin interactions | 7 | 1 (14.3%) | 0.046 | 0.357 | Reactome |
| Epstein-Barr virus infection - Homo sapiens (human) | 202 | 4 (2.0%) | 0.046 | 0.357 | KEGG |
| AMPK signaling pathway - Homo sapiens (human) | 120 | 3 (2.5%) | 0.0466 | 0.357 | KEGG |
| One-carbon metabolism and related pathways | 52 | 2 (3.8%) | 0.0474 | 0.357 | Wikipathways |
| SRP-dependent cotranslational protein targeting to membrane | 123 | 3 (2.4%) | 0.0495 | 0.357 | Reactome |

HTN_VAT

| pathway name | set size | candidates contained | p-value | q-value | pathway source |
|---|---|---|---|---|---|
| Biosynthesis of unsaturated fatty acids - Homo sapiens (human) | 27 | 5 (18.5%) | 2.27E-05 | 0.0333 | KEGG |
| oleate biosynthesis | 8 | 3 (37.5%) | 0.000116 | 0.068 | HumanCyc |
| Human T-cell leukemia virus 1 infection - Homo sapiens (human) | 219 | 11 (5.0%) | 0.000139 | 0.068 | KEGG |
| Cushing syndrome - Homo sapiens (human) | 155 | 9 (5.8%) | 0.000194 | 0.0711 | KEGG |
| Fatty acid elongation - Homo sapiens (human) | 27 | 4 (14.8%) | 0.000386 | 0.113 | KEGG |
| Folate metabolism | 31 | 4 (12.9%) | 0.000665 | 0.163 | INOH |
| Herpes simplex virus 1 infection - Homo sapiens (human) | 498 | 16 (3.2%) | 0.000802 | 0.168 | KEGG |
| Viral myocarditis - Homo sapiens (human) | 60 | 5 (8.5%) | 0.001 | 0.184 | KEGG |
| Fatty acyl-CoA biosynthesis | 37 | 4 (10.8%) | 0.00131 | 0.201 | Reactome |
| G1 to S cell cycle control | 64 | 5 (7.8%) | 0.00145 | 0.201 | Wikipathways |
| Cortisol synthesis and secretion - Homo sapiens (human) | 65 | 5 (7.7%) | 0.00156 | 0.201 | KEGG |
| Taurine and Hypotaurine Metabolism | 5 | 2 (40.0%) | 0.00164 | 0.201 | SMPDB |
| Type I diabetes mellitus - Homo sapiens (human) | 43 | 4 (9.3%) | 0.00231 | 0.246 | KEGG |
| Alpha Linolenic Acid and Linoleic Acid Metabolism | 6 | 2 (33.3%) | 0.00245 | 0.246 | SMPDB |
| Renin-angiotensin-aldosterone system (RAAS) | 44 | 4 (9.1%) | 0.00252 | 0.246 | Wikipathways |
| Cholesterol metabolism (includes both Bloch and Kandutsch-Russell pathways) | 46 | 4 (8.7%) | 0.00296 | 0.272 | Wikipathways |
| Myometrial relaxation and contraction pathways | 155 | 7 (4.5%) | 0.00418 | 0.324 | Wikipathways |
| Ovarian steroidogenesis - Homo sapiens (human) | 51 | 4 (7.8%) | 0.00432 | 0.324 | KEGG |
| palmitate biosynthesis | 9 | 2 (25.0%) | 0.00449 | 0.324 | HumanCyc |
| AMPK signaling pathway - Homo sapiens (human) | 120 | 6 (5.0%) | 0.00482 | 0.324 | KEGG |
| btg family proteins and cell cycle regulation | 9 | 2 (22.2%) | 0.00572 | 0.324 | BioCarta |
| Serine biosynthesis | 9 | 2 (22.2%) | 0.00572 | 0.324 | Reactome |
| Cyclin E associated events during G1/S transition | 29 | 3 (10.3%) | 0.00619 | 0.324 | Reactome |
| Cyclin A:Cdk2-associated events at S phase entry | 29 | 3 (10.3%) | 0.00619 | 0.324 | Reactome |
| CDK-mediated phosphorylation and removal of Cdc6 | 29 | 3 (10.3%) | 0.00619 | 0.324 | Reactome |
| -arrestins in gpcr desensitization | 30 | 3 (10.0%) | 0.00681 | 0.324 | BioCarta |
| Oocyte meiosis - Homo sapiens (human) | 129 | 6 (4.7%) | 0.00682 | 0.324 | KEGG |
| Tight junction interactions | 31 | 3 (9.7%) | 0.00747 | 0.324 | Reactome |
| activation of camp-dependent protein kinase pka | 31 | 3 (9.7%) | 0.00747 | 0.324 | BioCarta |
| Vascular smooth muscle contraction - Homo sapiens (human) | 133 | 6 (4.5%) | 0.00788 | 0.324 | KEGG |
| role of -arrestins in the activation and targeting of map kinases | 32 | 3 (9.4%) | 0.00817 | 0.324 | BioCarta |
| Fatty acid metabolism | 177 | 7 (4.0%) | 0.00825 | 0.324 | Reactome |
| G alpha (i) signalling events | 316 | 10 (3.2%) | 0.00847 | 0.324 | Reactome |
| Nucleotide GPCRs | 11 | 2 (18.2%) | 0.00859 | 0.324 | Wikipathways |
| Taurine and hypotaurine metabolism - Homo sapiens (human) | 11 | 2 (18.2%) | 0.00859 | 0.324 | KEGG |
| CMP phosphorylation | 11 | 2 (18.2%) | 0.00859 | 0.324 | HumanCyc |
| pertussis toxin-insensitive ccr5 signaling in macrophage | 11 | 2 (18.2%) | 0.00859 | 0.324 | BioCarta |
| Autosomal recessive Osteopetrosis pathways | 11 | 2 (18.2%) | 0.00859 | 0.324 | Wikipathways |
| Aldosterone synthesis and secretion - Homo sapiens (human) | 98 | 5 (5.1%) | 0.00908 | 0.324 | KEGG |
| Human cytomegalovirus infection - Homo sapiens (human) | 225 | 8 (3.6%) | 0.00932 | 0.324 | KEGG |
| pyrimidine deoxyribonucleotide phosphorylation | 12 | 2 (16.7%) | 0.0102 | 0.324 | HumanCyc |
| guanosine deoxyribonucleotides de novo biosynthesis | 12 | 2 (16.7%) | 0.0102 | 0.324 | HumanCyc |
| adenosine deoxyribonucleotides de novo biosynthesis | 12 | 2 (16.7%) | 0.0102 | 0.324 | HumanCyc |
| g-protein signaling through tubby proteins | 12 | 2 (16.7%) | 0.0102 | 0.324 | BioCarta |
| roles of arrestin dependent recruitment of src kinases in gpcr signaling | 36 | 3 (8.3%) | 0.0113 | 0.324 | BioCarta |
| Mitochondrial Fatty Acid Beta-Oxidation | 37 | 3 (8.3%) | 0.0113 | 0.324 | Reactome |
| S Phase | 104 | 5 (4.8%) | 0.0116 | 0.324 | Reactome |
| Passive transport by Aquaporins | 13 | 2 (15.4%) | 0.012 | 0.324 | Reactome |
| MFAP5-mediated ovarian cancer cell motility and invasiveness | 13 | 2 (15.4%) | 0.012 | 0.324 | Wikipathways |
| purine deoxyribonucleosides salvage | 13 | 2 (15.4%) | 0.012 | 0.324 | HumanCyc |
| guanosine ribonucleotides de novo biosynthesis | 13 | 2 (15.4%) | 0.012 | 0.324 | HumanCyc |
| UTP and CTP de novo biosynthesis | 13 | 2 (15.4%) | 0.012 | 0.324 | HumanCyc |
| activation of pkc through g-protein coupled receptors | 13 | 2 (15.4%) | 0.012 | 0.324 | BioCarta |
| stearate biosynthesis | 13 | 2 (15.4%) | 0.012 | 0.324 | HumanCyc |
| Allograft rejection - Homo sapiens (human) | 38 | 3 (8.1%) | 0.0122 | 0.324 | KEGG |
| Parathyroid hormone synthesis, secretion and action - Homo sapiens (human) | 106 | 5 (4.7%) | 0.0125 | 0.324 | KEGG |
| Cell adhesion molecules - Homo sapiens (human) | 149 | 6 (4.1%) | 0.0129 | 0.324 | KEGG |
| Purine nucleotides nucleosides metabolism | 107 | 5 (4.7%) | 0.013 | 0.324 | INOH |
| D-mannose degradation | 1 | 1 (100.0%) | 0.013 | 0.324 | HumanCyc |
| Adrenergic signaling in cardiomyocytes - Homo sapiens (human) | 150 | 6 (4.0%) | 0.0137 | 0.333 | KEGG |
| Omega-3-Omega-6 FA synthesis | 14 | 2 (14.3%) | 0.0139 | 0.333 | Wikipathways |
| Calcium Regulation in the Cardiac Cell | 151 | 6 (4.0%) | 0.0142 | 0.335 | Wikipathways |
| superpathway of pyrimidine ribonucleotides de novo biosynthesis | 15 | 2 (13.3%) | 0.0158 | 0.369 | HumanCyc |
| Thyroid hormone synthesis - Homo sapiens (human) | 75 | 4 (5.3%) | 0.0165 | 0.371 | KEGG |
| Hepatitis C - Homo sapiens (human) | 157 | 6 (3.8%) | 0.0169 | 0.371 | KEGG |
| Graft-versus-host disease - Homo sapiens (human) | 42 | 3 (7.1%) | 0.0172 | 0.371 | KEGG |
| chrebp regulation by carbohydrates and camp | 42 | 3 (7.1%) | 0.0172 | 0.371 | BioCarta |
| G1/S Transition | 76 | 4 (5.3%) | 0.0173 | 0.371 | Reactome |
| pyrimidine deoxyribonucleotides biosynthesis from CTP | 16 | 2 (12.5%) | 0.018 | 0.371 | HumanCyc |
| guanosine nucleotides de novo biosynthesis | 16 | 2 (12.5%) | 0.018 | 0.371 | HumanCyc |
| pyrimidine deoxyribonucleotides de novo biosynthesis | 16 | 2 (12.5%) | 0.018 | 0.371 | HumanCyc |
| De novo fatty acid biosynthesis | 43 | 3 (7.0%) | 0.0183 | 0.373 | EHMN |
| Acute viral myocarditis | 78 | 4 (5.1%) | 0.0188 | 0.378 | Wikipathways |
| superpathway of pyrimidine deoxyribonucleoside salvage | 17 | 2 (11.8%) | 0.0202 | 0.398 | HumanCyc |
| Cell cycle | 120 | 5 (4.2%) | 0.0203 | 0.398 | Wikipathways |
| activation of csk by camp-dependent protein kinase inhibits signaling through the t cell receptor | 46 | 3 (6.5%) | 0.0219 | 0.403 | BioCarta |
| Metabolism of Angiotensinogen to Angiotensins | 18 | 2 (11.1%) | 0.0225 | 0.403 | Reactome |
| SCF(Skp2)-mediated degradation of p27/p21 | 18 | 2 (11.1%) | 0.0225 | 0.403 | Reactome |
| Cell cycle - Homo sapiens (human) | 124 | 5 (4.0%) | 0.023 | 0.403 | KEGG |
| Switching of origins to a post-replicative state | 47 | 3 (6.4%) | 0.0232 | 0.403 | Reactome |
| ion channels and their functional role in vascular endothelium | 47 | 3 (6.4%) | 0.0232 | 0.403 | BioCarta |
| Tight junction - Homo sapiens (human) | 169 | 6 (3.6%) | 0.0233 | 0.403 | KEGG |
| Amino sugar and nucleotide sugar metabolism - Homo sapiens (human) | 48 | 3 (6.2%) | 0.0245 | 0.403 | KEGG |

| | | | | | |
|---|---|---|---|---|---|
| G alpha (q) signalling events | 221 | 7 (3.2%) | 0.0257 | 0.403 | Reactome |
| Aquaporin-mediated transport | 49 | 3 (6.1%) | 0.0258 | 0.403 | Reactome |
| Insulin secretion - Homo sapiens (human) | 86 | 4 (4.7%) | 0.0259 | 0.403 | KEGG |
| Transport of glycerol from adipocytes to the liver by Aquaporins | 2 | 1 (50.0%) | 0.0259 | 0.403 | Reactome |
| GABA synthesis | 2 | 1 (50.0%) | 0.0259 | 0.403 | Reactome |
| purine deoxyribonucleosides degradation | 2 | 1 (50.0%) | 0.0259 | 0.403 | HumanCyc |
| guanine and guanosine salvage | 2 | 1 (50.0%) | 0.0259 | 0.403 | HumanCyc |
| choline degradation | 2 | 1 (50.0%) | 0.0259 | 0.403 | HumanCyc |
| Neurotransmitter release cycle | 50 | 3 (6.0%) | 0.0272 | 0.403 | Reactome |
| S-Adenosylhomocysteine (SAH) Hydrolase Deficiency | 20 | 2 (10.0%) | 0.0275 | 0.403 | SMPDB |
| Methionine Metabolism | 20 | 2 (10.0%) | 0.0275 | 0.403 | SMPDB |
| Methionine Adenosyltransferase Deficiency | 20 | 2 (10.0%) | 0.0275 | 0.403 | SMPDB |
| Glycine N-methyltransferase Deficiency | 20 | 2 (10.0%) | 0.0275 | 0.403 | SMPDB |
| Hypermethioninemia | 20 | 2 (10.0%) | 0.0275 | 0.403 | SMPDB |
| Methylenetetrahydrofolate Reductase Deficiency (MTHFRD) | 20 | 2 (10.0%) | 0.0275 | 0.403 | SMPDB |
| Homocystinuria-megaloblastic anemia due to defect in cobalamin metabolism, cblG complementation type | 20 | 2 (10.0%) | 0.0275 | 0.403 | SMPDB |
| Cystathionine Beta-Synthase Deficiency | 20 | 2 (10.0%) | 0.0275 | 0.403 | SMPDB |
| ncRNAs involved in Wnt signaling in hepatocellular carcinoma | 90 | 4 (4.4%) | 0.0299 | 0.405 | Wikipathways |
| Phosphorylation of the APC/C | 21 | 2 (9.5%) | 0.0301 | 0.405 | Reactome |
| G1/S DNA Damage Checkpoints | 21 | 2 (9.5%) | 0.0301 | 0.405 | Reactome |
| Conversion from APC/C:Cdc20 to APC/C:Cdh1 in late anaphase | 21 | 2 (9.5%) | 0.0301 | 0.405 | Reactome |
| Autoimmune thyroid disease - Homo sapiens (human) | 53 | 3 (5.8%) | 0.0301 | 0.405 | KEGG |
| One-carbon metabolism and related pathways | 52 | 3 (5.8%) | 0.0301 | 0.405 | Wikipathways |
| Netrin-UNC5B signaling pathway | 52 | 3 (5.8%) | 0.0301 | 0.405 | Wikipathways |
| Cardiac Progenitor Differentiation | 53 | 3 (5.7%) | 0.0317 | 0.405 | Wikipathways |
| Bile acid biosynthesis | 53 | 3 (5.7%) | 0.0317 | 0.405 | EHMN |
| superpathway of pyrimidine deoxyribonucleotides de novo biosynthesis | 22 | 2 (9.1%) | 0.0329 | 0.405 | HumanCyc |
| cystic fibrosis transmembrane conductance regulator (cftr) and beta 2 adrenergic receptor (b2ar) pathway | 22 | 2 (9.1%) | 0.0329 | 0.405 | BioCarta |
| Inactivation of APC/C via direct inhibition of the APC/C complex | 22 | 2 (9.1%) | 0.0329 | 0.405 | Reactome |
| Inhibition of the proteolytic activity of APC/C required for the onset of anaphase by mitotic spindle checkpoint components | 22 | 2 (9.1%) | 0.0329 | 0.405 | Reactome |
| Allograft Rejection | 93 | 4 (4.3%) | 0.0332 | 0.405 | Wikipathways |
| Salivary secretion - Homo sapiens (human) | 93 | 4 (4.3%) | 0.0332 | 0.405 | KEGG |
| Renin-angiotensin system - Homo sapiens (human) | 23 | 2 (8.7%) | 0.0357 | 0.405 | KEGG |
| cyclins and cell cycle regulation | 23 | 2 (8.7%) | 0.0357 | 0.405 | BioCarta |
| induction of apoptosis through dr3 and dr4/5 death receptors | 23 | 2 (8.7%) | 0.0357 | 0.405 | BioCarta |
| Autodegradation of Cdh1 by Cdh1:APC/C | 23 | 2 (8.7%) | 0.0357 | 0.405 | Reactome |
| Interferon gamma signaling | 96 | 4 (4.2%) | 0.0367 | 0.405 | Reactome |
| GPCR downstream signalling | 633 | 14 (2.2%) | 0.0371 | 0.405 | Reactome |
| Class A/1 (Rhodopsin-like receptors) | 346 | 9 (2.6%) | 0.0375 | 0.405 | Reactome |
| LncRNA involvement in canonical Wnt signaling and colorectal cancer | 97 | 4 (4.1%) | 0.0379 | 0.405 | Wikipathways |
| Anchoring of the basal body to the plasma membrane | 97 | 4 (4.1%) | 0.0379 | 0.405 | Reactome |
| glutamate dependent acid resistance | 3 | 1 (33.3%) | 0.0386 | 0.405 | HumanCyc |
| MECP2 regulates transcription of genes involved in GABA signaling | 3 | 1 (33.3%) | 0.0386 | 0.405 | Reactome |
| tetrahydrobiopterin de novo biosynthesis | 3 | 1 (33.3%) | 0.0386 | 0.405 | HumanCyc |
| Signaling by MST1 | 3 | 1 (33.3%) | 0.0386 | 0.405 | Reactome |
| Glycine Metabolism | 3 | 1 (33.3%) | 0.0386 | 0.405 | Wikipathways |
| adenine and adenosine salvage III | 3 | 1 (33.3%) | 0.0386 | 0.405 | HumanCyc |
| purine ribonucleosides degradation to ribose-1-phosphate | 3 | 1 (33.3%) | 0.0386 | 0.405 | HumanCyc |
| taurine biosynthesis | 3 | 1 (33.3%) | 0.0386 | 0.405 | HumanCyc |
| Inositol transporters | 3 | 1 (33.3%) | 0.0386 | 0.405 | Reactome |
| Cyclin A/B1/B2 associated events during G2/M transition | 24 | 2 (8.3%) | 0.0386 | 0.405 | Reactome |
| tumor suppressor arf inhibits ribosomal biogenesis | 24 | 2 (8.3%) | 0.0386 | 0.405 | BioCarta |
| APC/C:Cdc20 mediated degradation of Securin | 24 | 2 (8.3%) | 0.0386 | 0.405 | Reactome |
| Synthesis of very long-chain fatty acyl-CoAs | 24 | 2 (8.3%) | 0.0386 | 0.405 | Reactome |
| superpathway of purine nucleotide salvage | 59 | 3 (5.1%) | 0.0415 | 0.405 | HumanCyc |
| cell cycle: g1/s check point | 25 | 2 (8.0%) | 0.0416 | 0.405 | BioCarta |
| G0 and Early G1 | 25 | 2 (8.0%) | 0.0416 | 0.405 | Reactome |
| APC/C:Cdc20 mediated degradation of Cyclin B | 25 | 2 (8.0%) | 0.0416 | 0.405 | Reactome |
| Progesterone-mediated oocyte maturation - Homo sapiens (human) | 100 | 4 (4.0%) | 0.0416 | 0.405 | KEGG |
| Signaling by GPCR | 706 | 15 (2.1%) | 0.0425 | 0.405 | Reactome |
| Wnt signaling pathway and pluripotency | 101 | 4 (4.0%) | 0.0429 | 0.405 | Wikipathways |
| Long-term depression - Homo sapiens (human) | 60 | 3 (5.0%) | 0.0433 | 0.405 | KEGG |
| Mitotic G1 phase and G1/S transition | 104 | 4 (3.8%) | 0.0469 | 0.405 | Reactome |
| corticosteroids and cardioprotection | 27 | 2 (7.4%) | 0.0479 | 0.405 | BioCarta |
| APC-Cdc20 mediated degradation of Nek2A | 27 | 2 (7.4%) | 0.0479 | 0.405 | Reactome |
| Viral carcinogenesis - Homo sapiens (human) | 204 | 6 (3.0%) | 0.0498 | 0.405 | KEGG |

Heart_failure_VAT

| pathway name | set size | candidates contained | p-value | q-value | pathway source |
|---|---|---|---|---|---|
| PD-1 signaling | 32 | 4 (12.5%) | 6.96E-05 | 0.0533 | Reactome |
| Interleukin-37 signaling | 5 | 2 (40.0%) | 0.000478 | 0.183 | Reactome |
| MHC class II antigen presentation | 60 | 4 (6.7%) | 0.000812 | 0.183 | Reactome |
| Translocation of ZAP-70 to Immunological synapse | 28 | 3 (10.7%) | 0.000957 | 0.183 | Reactome |
| Phosphorylation of CD3 and TCR zeta chains | 31 | 3 (9.7%) | 0.00129 | 0.198 | Reactome |
| Costimulation by the CD28 family | 77 | 4 (5.2%) | 0.00206 | 0.263 | Reactome |
| Generation of second messenger molecules | 42 | 3 (7.1%) | 0.00313 | 0.342 | Reactome |
| CD4 T cell receptor signaling-ERK cascade | 44 | 3 (6.8%) | 0.00357 | 0.342 | INOH |
| Adaptive Immune System | 719 | 12 (1.7%) | 0.00427 | 0.363 | Reactome |
| CD4 T cell receptor signaling-NFkB cascade | 99 | 4 (4.0%) | 0.0051 | 0.391 | INOH |
| Downstream TCR signaling | 52 | 3 (5.8%) | 0.00572 | 0.399 | Reactome |
| FGFR4 mutant receptor activation | 1 | 1 (100.0%) | 0.007 | 0.435 | Reactome |
| CD4 T cell receptor signaling-JNK cascade | 57 | 3 (5.3%) | 0.00739 | 0.435 | INOH |
| Inflammatory bowel disease - Homo sapiens (human) | 65 | 3 (4.6%) | 0.0106 | 0.507 | KEGG |
| CD4 T cell receptor signaling | 132 | 4 (3.0%) | 0.0138 | 0.507 | INOH |
| TCR signaling | 73 | 3 (4.1%) | 0.0145 | 0.507 | Reactome |
| Asthma - Homo sapiens (human) | 31 | 2 (6.7%) | 0.0186 | 0.507 | KEGG |
| methionine salvage | 3 | 1 (33.3%) | 0.0208 | 0.507 | HumanCyc |
| betaKlotho-mediated ligand binding | 3 | 1 (33.3%) | 0.0208 | 0.507 | Reactome |
| Signaling by MST1 | 3 | 1 (33.3%) | 0.0208 | 0.507 | Reactome |
| Interconversion of 2-oxoglutarate and 2-hydroxyglutarate | 3 | 1 (33.3%) | 0.0208 | 0.507 | Reactome |
| Inactivation, recovery and regulation of the phototransduction cascade | 36 | 2 (5.6%) | 0.0262 | 0.507 | Reactome |
| Allograft Rejection | 93 | 3 (3.2%) | 0.0274 | 0.507 | Wikipathways |
| The phototransduction cascade | 37 | 2 (5.4%) | 0.0275 | 0.507 | Reactome |
| Allograft rejection - Homo sapiens (human) | 38 | 2 (5.4%) | 0.0275 | 0.507 | KEGG |
| Fatty acyl-CoA biosynthesis | 37 | 2 (5.4%) | 0.0275 | 0.507 | Reactome |
| Activation, myristolyation of BID and translocation to mitochondria | 4 | 1 (25.0%) | 0.0277 | 0.507 | Reactome |
| Utilization of Ketone Bodies | 4 | 1 (25.0%) | 0.0277 | 0.507 | Reactome |
| Catecholamine biosynthesis | 4 | 1 (25.0%) | 0.0277 | 0.507 | Reactome |
| catecholamine biosynthesis | 4 | 1 (25.0%) | 0.0277 | 0.507 | HumanCyc |
| Tyrosine hydroxylase deficiency | 4 | 1 (25.0%) | 0.0277 | 0.507 | SMPDB |
| Catecholamine Biosynthesis | 4 | 1 (25.0%) | 0.0277 | 0.507 | SMPDB |
| Aromatic L-Aminoacid Decarboxylase Deficiency | 4 | 1 (25.0%) | 0.0277 | 0.507 | SMPDB |
| Invadopodia formation | 4 | 1 (25.0%) | 0.0277 | 0.507 | Reactome |
| Interleukin-18 signaling | 4 | 1 (25.0%) | 0.0277 | 0.507 | Reactome |
| Synthesis of PIPs in the nucleus | 4 | 1 (25.0%) | 0.0277 | 0.507 | Reactome |
| Interferon gamma signaling | 96 | 3 (3.1%) | 0.0297 | 0.507 | Reactome |
| Hematopoietic cell lineage - Homo sapiens (human) | 99 | 3 (3.1%) | 0.0313 | 0.507 | KEGG |
| Glycine Serine metabolism | 40 | 2 (5.0%) | 0.0318 | 0.507 | INOH |
| Gamma carboxylation, hypusine formation and arylsulfatase activation | 41 | 2 (4.9%) | 0.0333 | 0.507 | Reactome |
| glycine betaine degradation | 5 | 1 (20.0%) | 0.0345 | 0.507 | HumanCyc |
| Leukotriene receptors | 5 | 1 (20.0%) | 0.0345 | 0.507 | Reactome |
| Succinyl CoA: 3-ketoacid CoA transferase deficiency | 5 | 1 (20.0%) | 0.0345 | 0.507 | SMPDB |
| Ketone Body Metabolism | 5 | 1 (20.0%) | 0.0345 | 0.507 | SMPDB |
| ketolysis | 5 | 1 (20.0%) | 0.0345 | 0.507 | HumanCyc |
| Synthesis and Degradation of Ketone Bodies | 5 | 1 (20.0%) | 0.0345 | 0.507 | Wikipathways |
| Graft-versus-host disease - Homo sapiens (human) | 42 | 2 (4.8%) | 0.0348 | 0.507 | KEGG |
| Type I diabetes mellitus - Homo sapiens (human) | 43 | 2 (4.7%) | 0.0363 | 0.507 | KEGG |
| a6b1 and a6b4 Integrin signaling | 45 | 2 (4.4%) | 0.0395 | 0.507 | PID |
| Choline catabolism | 6 | 1 (16.7%) | 0.0413 | 0.507 | Reactome |
| p75NTR negatively regulates cell cycle via SC1 | 6 | 1 (16.7%) | 0.0413 | 0.507 | Reactome |
| Mitochondrial fatty acid synthesis pathway | 6 | 1 (16.7%) | 0.0413 | 0.507 | Wikipathways |
| NOSTRIN mediated eNOS trafficking | 6 | 1 (16.7%) | 0.0413 | 0.507 | Reactome |
| Intestinal immune network for IgA production - Homo sapiens (human) | 49 | 2 (4.3%) | 0.0427 | 0.507 | KEGG |
| Toxoplasmosis - Homo sapiens (human) | 112 | 3 (2.7%) | 0.0438 | 0.507 | KEGG |
| superpathway of choline degradation to L-serine | 7 | 1 (14.3%) | 0.048 | 0.507 | HumanCyc |

| | | | | | |
|---|---|---|---|---|---|
| Anchoring fibril formation | 7 | 1 (14.3%) | 0.048 | 0.507 | Reactome |
| Aryl hydrocarbon receptor signalling | 7 | 1 (14.3%) | 0.048 | 0.507 | Reactome |
| RHOD GTPase cycle | 51 | 2 (3.9%) | 0.0495 | 0.507 | Reactome |

Obesity_VAT

| pathway name | set size | candidates contained | p-value | q-value | pathway source |
|---|---|---|---|---|---|
| Drug metabolism - cytochrome P450 - Homo sapiens (human) | 72 | 6 (8.6%) | 9.44E-05 | 0.0879 | KEGG |
| Butyrophilin (BTN) family interactions | 12 | 3 (25.0%) | 0.000237 | 0.0879 | Reactome |
| Chemical carcinogenesis - Homo sapiens (human) | 83 | 6 (7.2%) | 0.000243 | 0.0879 | KEGG |
| Metapathway biotransformation Phase I and II | 186 | 8 (4.3%) | 0.000804 | 0.0879 | Wikipathways |
| Deposition of new CENPA-containing nucleosomes at the centromere | 43 | 4 (9.3%) | 0.00107 | 0.0879 | Reactome |
| Nucleosome assembly | 43 | 4 (9.3%) | 0.00107 | 0.0879 | Reactome |
| Myometrial relaxation and contraction pathways | 155 | 7 (4.5%) | 0.00129 | 0.0879 | Wikipathways |
| Tamoxifen metabolism | 21 | 3 (14.3%) | 0.00134 | 0.0879 | Wikipathways |
| Biosynthesis of maresin-like SPMs | 6 | 2 (33.3%) | 0.00162 | 0.0879 | Reactome |
| Arachidonate Epoxygenase - Epoxide Hydrolase | 7 | 2 (28.6%) | 0.00225 | 0.0879 | Wikipathways |
| Caffeine Metabolism | 7 | 2 (28.6%) | 0.00225 | 0.0879 | SMPDB |
| Biological oxidations | 219 | 8 (3.7%) | 0.00227 | 0.0879 | Reactome |
| Biosynthesis of maresins | 8 | 2 (25.0%) | 0.00298 | 0.0879 | Reactome |
| NOTCH4 Intracellular Domain Regulates Transcription | 8 | 2 (25.0%) | 0.00298 | 0.0879 | Reactome |
| C21-steroid hormone biosynthesis and metabolism | 57 | 4 (7.0%) | 0.00306 | 0.0879 | EHMN |
| Linoleic acid metabolism - Homo sapiens (human) | 29 | 3 (10.3%) | 0.00345 | 0.0879 | KEGG |
| Ethanol metabolism resulting in production of ROS by CYP2E1 | 9 | 2 (22.2%) | 0.0038 | 0.0879 | Wikipathways |
| Ketoprofen Action Pathway | 30 | 3 (10.0%) | 0.00381 | 0.0879 | SMPDB |
| Acetylsalicylic Acid Action Pathway | 30 | 3 (10.0%) | 0.00381 | 0.0879 | SMPDB |
| Diflunisal Action Pathway | 30 | 3 (10.0%) | 0.00381 | 0.0879 | SMPDB |
| Acetaminophen Action Pathway | 30 | 3 (10.0%) | 0.00381 | 0.0879 | SMPDB |
| Sulindac Action Pathway | 30 | 3 (10.0%) | 0.00381 | 0.0879 | SMPDB |
| Ketorolac Action Pathway | 30 | 3 (10.0%) | 0.00381 | 0.0879 | SMPDB |
| Naproxen Action Pathway | 30 | 3 (10.0%) | 0.00381 | 0.0879 | SMPDB |
| Flurbiprofen Action Pathway | 30 | 3 (10.0%) | 0.00381 | 0.0879 | SMPDB |
| Antrafenine Action Pathway | 30 | 3 (10.0%) | 0.00381 | 0.0879 | SMPDB |
| Trisalicylate-choline Action Pathway | 30 | 3 (10.0%) | 0.00381 | 0.0879 | SMPDB |
| Nepafenac Action Pathway | 30 | 3 (10.0%) | 0.00381 | 0.0879 | SMPDB |
| Phenylbutazone Action Pathway | 30 | 3 (10.0%) | 0.00381 | 0.0879 | SMPDB |
| Lornoxicam Action Pathway | 30 | 3 (10.0%) | 0.00381 | 0.0879 | SMPDB |
| Salsalate Action Pathway | 30 | 3 (10.0%) | 0.00381 | 0.0879 | SMPDB |
| Salicylic Acid Action Pathway | 30 | 3 (10.0%) | 0.00381 | 0.0879 | SMPDB |
| Salicylate-sodium Action Pathway | 30 | 3 (10.0%) | 0.00381 | 0.0879 | SMPDB |
| Oxaprozin Action Pathway | 30 | 3 (10.0%) | 0.00381 | 0.0879 | SMPDB |
| Nabumetone Action Pathway | 30 | 3 (10.0%) | 0.00381 | 0.0879 | SMPDB |
| Bromfenac Action Pathway | 30 | 3 (10.0%) | 0.00381 | 0.0879 | SMPDB |
| Mefenamic Acid Action Pathway | 30 | 3 (10.0%) | 0.00381 | 0.0879 | SMPDB |
| Piroxicam Action Pathway | 30 | 3 (10.0%) | 0.00381 | 0.0879 | SMPDB |
| Carprofen Action Pathway | 30 | 3 (10.0%) | 0.00381 | 0.0879 | SMPDB |
| Fenoprofen Action Pathway | 30 | 3 (10.0%) | 0.00381 | 0.0879 | SMPDB |
| Antipyrine Action Pathway | 30 | 3 (10.0%) | 0.00381 | 0.0879 | SMPDB |
| Magnesium salicylate Action Pathway | 30 | 3 (10.0%) | 0.00381 | 0.0879 | SMPDB |
| Tenoxicam Action Pathway | 30 | 3 (10.0%) | 0.00381 | 0.0879 | SMPDB |
| Tiaprofenic Acid Action Pathway | 30 | 3 (10.0%) | 0.00381 | 0.0879 | SMPDB |
| Tolmetin Action Pathway | 30 | 3 (10.0%) | 0.00381 | 0.0879 | SMPDB |
| Suprofen Action Pathway | 30 | 3 (10.0%) | 0.00381 | 0.0879 | SMPDB |
| Etodolac Action Pathway | 30 | 3 (10.0%) | 0.00381 | 0.0879 | SMPDB |
| Rofecoxib Action Pathway | 30 | 3 (10.0%) | 0.00381 | 0.0879 | SMPDB |
| Diclofenac Action Pathway | 30 | 3 (10.0%) | 0.00381 | 0.0879 | SMPDB |
| Etoricoxib Action Pathway | 30 | 3 (10.0%) | 0.00381 | 0.0879 | SMPDB |
| Lumiracoxib Action Pathway | 30 | 3 (10.0%) | 0.00381 | 0.0879 | SMPDB |
| Valdecoxib Action Pathway | 30 | 3 (10.0%) | 0.00381 | 0.0879 | SMPDB |
| Meloxicam Action Pathway | 30 | 3 (10.0%) | 0.00381 | 0.0879 | SMPDB |
| Leukotriene C4 Synthesis Deficiency | 30 | 3 (10.0%) | 0.00381 | 0.0879 | SMPDB |
| Arachidonic Acid Metabolism | 30 | 3 (10.0%) | 0.00381 | 0.0879 | SMPDB |
| Indomethacin Action Pathway | 31 | 3 (9.7%) | 0.00419 | 0.0948 | SMPDB |
| Phase I - Functionalization of compounds | 101 | 5 (5.0%) | 0.00433 | 0.0963 | Reactome |
| Opsins | 10 | 2 (20.0%) | 0.00472 | 0.103 | Reactome |
| Calcium Regulation in the Cardiac Cell | 151 | 6 (4.0%) | 0.00534 | 0.115 | Wikipathways |
| CYP2E1 reactions | 11 | 2 (18.2%) | 0.00573 | 0.121 | Reactome |
| Celecoxib Action Pathway | 36 | 3 (8.3%) | 0.0064 | 0.133 | SMPDB |
| MFAP5-mediated ovarian cancer cell motility and invasiveness | 13 | 2 (15.4%) | 0.00802 | 0.164 | Wikipathways |
| Metabolism of xenobiotics by cytochrome P450 - Homo sapiens (human) | 78 | 4 (5.3%) | 0.00853 | 0.172 | KEGG |

| | | | | | |
|---|---|---|---|---|---|
| G alpha (q) signalling events | 221 | 7 (3.2%) | 0.00907 | 0.18 | Reactome |
| RHO GTPases Activate Formins | 123 | 5 (4.1%) | 0.00982 | 0.19 | Reactome |
| Drug metabolism - other enzymes - Homo sapiens (human) | 80 | 4 (5.0%) | 0.0102 | 0.19 | KEGG |
| Metabolism of Dichloroethylene by CYP450 | 1 | 1 (100.0%) | 0.0106 | 0.19 | Wikipathways |
| Acrylamide Biotransformation and Exposure Biomarkers | 1 | 1 (100.0%) | 0.0106 | 0.19 | Wikipathways |
| pregnenolone biosynthesis | 1 | 1 (100.0%) | 0.0106 | 0.19 | HumanCyc |
| D-mannose degradation | 1 | 1 (100.0%) | 0.0106 | 0.19 | HumanCyc |
| Fatty Acid Omega Oxidation | 15 | 2 (13.3%) | 0.0106 | 0.19 | Wikipathways |
| Renin-angiotensin-aldosterone system (RAAS) | 44 | 3 (6.8%) | 0.0112 | 0.197 | Wikipathways |
| Metabolism | 1954 | 31 (1.6%) | 0.0119 | 0.208 | Reactome |
| Chromosome Maintenance | 134 | 5 (3.8%) | 0.0134 | 0.224 | Reactome |
| Vascular smooth muscle contraction - Homo sapiens (human) | 133 | 5 (3.8%) | 0.0134 | 0.224 | KEGG |
| Calcium signaling pathway - Homo sapiens (human) | 240 | 7 (2.9%) | 0.0135 | 0.224 | KEGG |
| Biosynthesis of DHA-derived SPMs | 17 | 2 (11.8%) | 0.0136 | 0.224 | Reactome |
| Ibuprofen Action Pathway | 50 | 3 (6.0%) | 0.0158 | 0.257 | SMPDB |
| Ovarian steroidogenesis - Homo sapiens (human) | 51 | 3 (5.9%) | 0.0166 | 0.262 | KEGG |
| Pyrimidine nucleotides nucleosides metabolism | 51 | 3 (5.9%) | 0.0166 | 0.262 | INOH |
| Biosynthesis of specialized proresolving mediators (SPMs) | 19 | 2 (10.5%) | 0.0169 | 0.262 | Reactome |
| Phosphatidylinositol phosphate metabolism | 93 | 4 (4.3%) | 0.0169 | 0.262 | EHMN |
| Mitotic Anaphase | 195 | 6 (3.1%) | 0.0174 | 0.267 | Reactome |
| Mitotic Metaphase and Anaphase | 196 | 6 (3.1%) | 0.0178 | 0.269 | Reactome |
| Notch signaling pathway | 53 | 3 (5.7%) | 0.0184 | 0.274 | PID |
| Amplification of signal from unattached kinetochores via a MAD2 inhibitory signal | 96 | 4 (4.2%) | 0.0188 | 0.274 | Reactome |
| Amplification of signal from the kinetochores | 96 | 4 (4.2%) | 0.0188 | 0.274 | Reactome |
| Inflammatory mediator regulation of TRP channels - Homo sapiens (human) | 98 | 4 (4.1%) | 0.0201 | 0.29 | KEGG |
| Xenobiotics | 21 | 2 (9.5%) | 0.0204 | 0.29 | Reactome |
| Felbamate Metabolism | 2 | 1 (50.0%) | 0.021 | 0.29 | Wikipathways |
| epoxysqualene biosynthesis | 2 | 1 (50.0%) | 0.021 | 0.29 | HumanCyc |
| TWIK-related alkaline pH activated K+ channel (TALK) | 2 | 1 (50.0%) | 0.021 | 0.29 | Reactome |
| EML4 and NUDC in mitotic spindle formation | 100 | 4 (4.0%) | 0.0215 | 0.293 | Reactome |
| glutathione-mediated detoxification | 22 | 2 (9.1%) | 0.0223 | 0.298 | HumanCyc |
| Cytochrome P450 - arranged by substrate type | 57 | 3 (5.3%) | 0.0223 | 0.298 | Reactome |
| Leukotriene metabolism | 102 | 4 (3.9%) | 0.0229 | 0.303 | EHMN |
| Arachidonic acid metabolism - Homo sapiens (human) | 61 | 3 (4.9%) | 0.0267 | 0.345 | KEGG |
| Steroid hormone biosynthesis - Homo sapiens (human) | 61 | 3 (4.9%) | 0.0267 | 0.345 | KEGG |
| Oxidation by Cytochrome P450 | 62 | 3 (4.8%) | 0.0278 | 0.356 | Wikipathways |
| Resolution of Sister Chromatid Cohesion | 109 | 4 (3.7%) | 0.0284 | 0.36 | Reactome |
| Rosiglitazone Metabolism Pathway | 3 | 1 (33.3%) | 0.0314 | 0.362 | SMPDB |
| methylglyoxal degradation III | 3 | 1 (33.3%) | 0.0314 | 0.362 | HumanCyc |
| FMO oxidises nucleophiles | 3 | 1 (33.3%) | 0.0314 | 0.362 | Reactome |
| Signaling by MST1 | 3 | 1 (33.3%) | 0.0314 | 0.362 | Reactome |
| pyrimidine ribonucleosides degradation | 3 | 1 (33.3%) | 0.0314 | 0.362 | HumanCyc |
| The AIM2 inflammasome | 3 | 1 (33.3%) | 0.0314 | 0.362 | Reactome |
| Inositol transporters | 3 | 1 (33.3%) | 0.0314 | 0.362 | Reactome |
| Interconversion of 2-oxoglutarate and 2-hydroxyglutarate | 3 | 1 (33.3%) | 0.0314 | 0.362 | Reactome |
| Mitotic Spindle Checkpoint | 114 | 4 (3.5%) | 0.0327 | 0.362 | Reactome |
| Signaling by NOTCH4 | 27 | 2 (7.4%) | 0.0328 | 0.362 | Reactome |
| Canonical and non-canonical Notch signaling | 27 | 2 (7.4%) | 0.0328 | 0.362 | Wikipathways |
| Effects of PIP2 hydrolysis | 27 | 2 (7.4%) | 0.0328 | 0.362 | Reactome |
| Fatty acid elongation - Homo sapiens (human) | 27 | 2 (7.4%) | 0.0328 | 0.362 | KEGG |
| Biosynthesis of unsaturated fatty acids - Homo sapiens (human) | 27 | 2 (7.4%) | 0.0328 | 0.362 | KEGG |
| D-myo-inositol (1,4,5)-trisphosphate biosynthesis | 27 | 2 (7.4%) | 0.0328 | 0.362 | HumanCyc |
| Acetaminophen Metabolism Pathway | 30 | 2 (6.7%) | 0.0398 | 0.416 | SMPDB |
| Molecules associated with elastic fibres | 30 | 2 (6.7%) | 0.0398 | 0.416 | Reactome |
| Metabolism of lipids | 646 | 12 (1.9%) | 0.0406 | 0.416 | Reactome |
| Linoleate metabolism | 72 | 3 (4.2%) | 0.0406 | 0.416 | EHMN |
| Felbamate Metabolism Pathway | 4 | 1 (25.0%) | 0.0416 | 0.416 | SMPDB |
| IRS activation | 4 | 1 (25.0%) | 0.0416 | 0.416 | Reactome |
| propionyl-CoA degradation | 4 | 1 (25.0%) | 0.0416 | 0.416 | HumanCyc |
| pyrimidine deoxyribonucleosides degradation | 4 | 1 (25.0%) | 0.0416 | 0.416 | HumanCyc |
| guanosine nucleotides degradation | 4 | 1 (25.0%) | 0.0416 | 0.416 | HumanCyc |
| Nanomaterial-induced inflammasome activation | 4 | 1 (25.0%) | 0.0416 | 0.416 | Wikipathways |
| CLEC7A/inflammasome pathway | 4 | 1 (25.0%) | 0.0416 | 0.416 | Reactome |
| Synthesis of PIPs in the nucleus | 4 | 1 (25.0%) | 0.0416 | 0.416 | Reactome |
| Metabolism of steroid hormones | 31 | 2 (6.5%) | 0.0422 | 0.419 | Reactome |
| Tyrosine metabolism | 33 | 2 (6.1%) | 0.0473 | 0.462 | INOH |
| SNARE interactions in vesicular transport - Homo sapiens (human) | 33 | 2 (6.1%) | 0.0473 | 0.462 | KEGG |

| | | | | | |
|---|---|---|---|---|---|
| Separation of Sister Chromatids | 130 | 4 (3.1%) | 0.049 | 0.464 | **Reactome** |

Stroke_VAT

| pathway name | set size | candidates contained | p-value | q-value | pathway source |
|---|---|---|---|---|---|
| Costimulation by the CD28 family | 77 | 3 (3.9%) | 0.00639 | 0.392 | Reactome |
| Translocation of ZAP-70 to Immunological synapse | 28 | 2 (7.1%) | 0.00828 | 0.392 | Reactome |
| histamine degradation | 2 | 1 (50.0%) | 0.0098 | 0.392 | HumanCyc |
| L-glutamine tRNA biosynthesis | 2 | 1 (50.0%) | 0.0098 | 0.392 | HumanCyc |
| Phosphorylation of CD3 and TCR zeta chains | 31 | 2 (6.5%) | 0.0101 | 0.392 | Reactome |
| PD-1 signaling | 32 | 2 (6.2%) | 0.0107 | 0.392 | Reactome |
| CD4 T cell receptor signaling-NFkB cascade | 99 | 3 (3.0%) | 0.0127 | 0.392 | INOH |
| The AIM2 inflammasome | 3 | 1 (33.3%) | 0.0147 | 0.392 | Reactome |
| Generation of second messenger molecules | 42 | 2 (4.8%) | 0.018 | 0.392 | Reactome |
| ahr signal transduction pathway | 4 | 1 (25.0%) | 0.0195 | 0.392 | BioCarta |
| Regulation of gene expression by Hypoxia-inducible Factor | 4 | 1 (25.0%) | 0.0195 | 0.392 | Reactome |
| proline biosynthesis | 4 | 1 (25.0%) | 0.0195 | 0.392 | HumanCyc |
| Nanomaterial-induced inflammasome activation | 4 | 1 (25.0%) | 0.0195 | 0.392 | Wikipathways |
| CLEC7A/inflammasome pathway | 4 | 1 (25.0%) | 0.0195 | 0.392 | Reactome |
| ALKBH3 mediated reversal of alkylation damage | 4 | 1 (25.0%) | 0.0195 | 0.392 | Reactome |
| CD4 T cell receptor signaling-ERK cascade | 44 | 2 (4.5%) | 0.0197 | 0.392 | INOH |
| Galactose metabolism | 44 | 2 (4.5%) | 0.0197 | 0.392 | EHMN |
| Intestinal immune network for IgA production - Homo sapiens (human) | 49 | 2 (4.3%) | 0.0223 | 0.392 | KEGG |
| Notch-mediated HES/HEY network | 48 | 2 (4.2%) | 0.0232 | 0.392 | PID |
| Binding and entry of HIV virion | 5 | 1 (20.0%) | 0.0243 | 0.392 | Reactome |
| Leukotriene receptors | 5 | 1 (20.0%) | 0.0243 | 0.392 | Reactome |
| ornithine de novo biosynthesis | 5 | 1 (20.0%) | 0.0243 | 0.392 | HumanCyc |
| Arginine and proline metabolism - Homo sapiens (human) | 51 | 2 (4.0%) | 0.025 | 0.392 | KEGG |
| Downstream TCR signaling | 52 | 2 (3.8%) | 0.0269 | 0.392 | Reactome |
| CD4 T cell receptor signaling | 132 | 3 (2.3%) | 0.0271 | 0.392 | INOH |
| CD4 T cell receptor signaling-JNK cascade | 57 | 2 (3.5%) | 0.0319 | 0.392 | INOH |
| Legionellosis - Homo sapiens (human) | 57 | 2 (3.5%) | 0.0319 | 0.392 | KEGG |
| Aryl hydrocarbon receptor signalling | 7 | 1 (14.3%) | 0.0339 | 0.392 | Reactome |
| ceramide de novo biosynthesis | 7 | 1 (14.3%) | 0.0339 | 0.392 | HumanCyc |
| Activation of NLRP3 Inflammasome by SARS-CoV-2 | 7 | 1 (14.3%) | 0.0339 | 0.392 | Wikipathways |
| Reversal of alkylation damage by DNA dioxygenases | 7 | 1 (14.3%) | 0.0339 | 0.392 | Reactome |
| APEX1-Independent Resolution of AP Sites via the Single Nucleotide Replacement Pathway | 7 | 1 (14.3%) | 0.0339 | 0.392 | Reactome |
| MHC class II antigen presentation | 60 | 2 (3.3%) | 0.0351 | 0.392 | Reactome |
| TYROBP causal network in microglia | 60 | 2 (3.3%) | 0.0351 | 0.392 | Wikipathways |
| DNA Damage Reversal | 8 | 1 (12.5%) | 0.0386 | 0.392 | Reactome |
| Orexin and neuropeptides FF and QRFP bind to their respective receptors | 8 | 1 (12.5%) | 0.0386 | 0.392 | Reactome |
| YAP1- and WWTR1 (TAZ)-stimulated gene expression | 9 | 1 (11.1%) | 0.0434 | 0.392 | Reactome |
| Methylation Pathways | 9 | 1 (11.1%) | 0.0434 | 0.392 | Wikipathways |
| btg family proteins and cell cycle regulation | 9 | 1 (11.1%) | 0.0434 | 0.392 | BioCarta |
| RUNX1 and FOXP3 control the development of regulatory T lymphocytes (Tregs) | 10 | 1 (10.0%) | 0.0481 | 0.392 | Reactome |
| Opsins | 10 | 1 (10.0%) | 0.0481 | 0.392 | Reactome |
| The NLRP3 inflammasome | 10 | 1 (10.0%) | 0.0481 | 0.392 | Reactome |
| TCR signaling | 73 | 2 (2.7%) | 0.05 | 0.392 | Reactome |

Depression-related admission

| pathway name | set size | candidates contained | p-value | q-value | pathway source |
|---|---|---|---|---|---|
| DNA Double-Strand Break Repair | 169 | 4 (2.4%) | 0.000817 | 0.248 | Reactome |
| Ebola Virus Pathway on Host | 130 | 3 (2.3%) | 0.00426 | 0.248 | Wikipathways |
| HDR through Homologous Recombination (HRR) or Single Strand Annealing (SSA) | 134 | 3 (2.3%) | 0.00454 | 0.248 | Reactome |
| L-glutamine tRNA biosynthesis | 2 | 1 (50.0%) | 0.00506 | 0.248 | HumanCyc |
| Homology Directed Repair | 140 | 3 (2.2%) | 0.00513 | 0.248 | Reactome |
| Cell adhesion molecules - Homo sapiens (human) | 149 | 3 (2.0%) | 0.00611 | 0.248 | KEGG |
| TNFR2 non-canonical NF-kB pathway | 50 | 2 (4.0%) | 0.00706 | 0.248 | Reactome |
| DNA Repair | 323 | 4 (1.2%) | 0.00852 | 0.248 | Reactome |
| NRAGE signals death through JNK | 59 | 2 (3.4%) | 0.00972 | 0.248 | Reactome |
| Linoleic acid metabolism known to be affected by coronavirus infection | 4 | 1 (25.0%) | 0.0101 | 0.248 | Wikipathways |
| GRB7 events in ERBB2 signaling | 5 | 1 (20.0%) | 0.0126 | 0.248 | Reactome |
| Serotonin and melatonin biosynthesis | 5 | 1 (20.0%) | 0.0126 | 0.248 | Reactome |
| serotonin and melatonin biosynthesis | 5 | 1 (20.0%) | 0.0126 | 0.248 | HumanCyc |
| HDR through Homologous Recombination (HRR) | 68 | 2 (2.9%) | 0.0128 | 0.248 | Reactome |
| neuroregulin receptor degredation protein-1 controls erbb3 receptor recycling | 6 | 1 (16.7%) | 0.0151 | 0.248 | BioCarta |
| Pre-NOTCH Processing in the Endoplasmic Reticulum | 6 | 1 (16.7%) | 0.0151 | 0.248 | Reactome |
| Cell death signalling via NRAGE, NRIF and NADE | 76 | 2 (2.6%) | 0.0158 | 0.248 | Reactome |
| G alpha (12/13) signalling events | 80 | 2 (2.5%) | 0.0174 | 0.248 | Reactome |
| chondroitin sulfate degradation (metazoa) | 7 | 1 (14.3%) | 0.0176 | 0.248 | HumanCyc |
| CS/DS degradation | 7 | 1 (14.3%) | 0.0176 | 0.248 | Reactome |
| Nectin/Necl trans heterodimerization | 7 | 1 (14.3%) | 0.0176 | 0.248 | Reactome |
| C-type lectin receptors (CLRs) | 84 | 2 (2.4%) | 0.019 | 0.248 | Reactome |
| DNA replication initiation | 8 | 1 (12.5%) | 0.0201 | 0.248 | Reactome |
| oleate biosynthesis | 8 | 1 (12.5%) | 0.0201 | 0.248 | HumanCyc |
| palmitate biosynthesis | 9 | 1 (12.5%) | 0.0201 | 0.248 | HumanCyc |
| Dectin-1 mediated noncanonical NF-kB signaling | 8 | 1 (12.5%) | 0.0201 | 0.248 | Reactome |
| NIK-->noncanonical NF-kB signaling | 8 | 1 (12.5%) | 0.0201 | 0.248 | Reactome |
| MyD88:MAL(TIRAP) cascade initiated on plasma membrane | 91 | 2 (2.2%) | 0.0221 | 0.248 | Reactome |
| Toll Like Receptor TLR1:TLR2 Cascade | 91 | 2 (2.2%) | 0.0221 | 0.248 | Reactome |
| Toll Like Receptor TLR6:TLR2 Cascade | 91 | 2 (2.2%) | 0.0221 | 0.248 | Reactome |
| Toll Like Receptor 2 (TLR2) Cascade | 91 | 2 (2.2%) | 0.0221 | 0.248 | Reactome |
| p75 NTR receptor-mediated signalling | 96 | 2 (2.1%) | 0.0244 | 0.248 | Reactome |
| Digestion of dietary carbohydrate | 10 | 1 (10.0%) | 0.025 | 0.248 | Reactome |
| PI3K events in ERBB4 signaling | 10 | 1 (10.0%) | 0.025 | 0.248 | Reactome |
| 15q11.2 copy number variation syndrome | 10 | 1 (10.0%) | 0.025 | 0.248 | Wikipathways |
| Nucleotide Excision Repair | 11 | 1 (9.1%) | 0.0275 | 0.248 | SMPDB |
| Regulation of BACH1 activity | 11 | 1 (9.1%) | 0.0275 | 0.248 | Reactome |
| Omega-9 FA synthesis | 12 | 1 (8.3%) | 0.03 | 0.248 | Wikipathways |
| The activation of arylsulfatases | 13 | 1 (7.7%) | 0.0324 | 0.248 | Reactome |
| ERBB2 Activates PTK6 Signaling | 13 | 1 (7.7%) | 0.0324 | 0.248 | Reactome |
| Downregulation of ERBB2:ERBB3 signaling | 13 | 1 (7.7%) | 0.0324 | 0.248 | Reactome |
| stearate biosynthesis | 13 | 1 (7.7%) | 0.0324 | 0.248 | HumanCyc |
| Toll Like Receptor 4 (TLR4) Cascade | 112 | 2 (1.8%) | 0.0325 | 0.248 | Reactome |
| Immune System | 1828 | 9 (0.5%) | 0.034 | 0.248 | Reactome |
| SHC1 events in ERBB4 signaling | 14 | 1 (7.1%) | 0.0349 | 0.248 | Reactome |
| Omega-3-Omega-6 FA synthesis | 14 | 1 (7.1%) | 0.0349 | 0.248 | Wikipathways |
| ERBB2 Regulates Cell Motility | 15 | 1 (6.7%) | 0.0373 | 0.248 | Reactome |
| ErbB receptor signaling network | 15 | 1 (6.7%) | 0.0373 | 0.248 | PID |
| Biogenic Amine Synthesis | 15 | 1 (6.7%) | 0.0373 | 0.248 | Wikipathways |
| PI3K events in ERBB2 signaling | 16 | 1 (6.2%) | 0.0398 | 0.248 | Reactome |
| GRB2 events in ERBB2 signaling | 16 | 1 (6.2%) | 0.0398 | 0.248 | Reactome |
| Regulation of TLR by endogenous ligand | 16 | 1 (6.2%) | 0.0398 | 0.248 | Reactome |
| Metabolism of amine-derived hormones | 16 | 1 (6.2%) | 0.0398 | 0.248 | Reactome |
| ID signaling pathway | 16 | 1 (6.2%) | 0.0398 | 0.248 | Wikipathways |
| MAP3K8 (TPL2)-dependent MAPK1/3 activation | 16 | 1 (6.2%) | 0.0398 | 0.248 | Reactome |
| Oocyte meiosis - Homo sapiens (human) | 129 | 2 (1.6%) | 0.042 | 0.248 | KEGG |
| Glycosaminoglycan degradation | 17 | 1 (5.9%) | 0.0422 | 0.248 | Wikipathways |
| Sialuria or French Type Sialuria | 17 | 1 (5.9%) | 0.0422 | 0.248 | SMPDB |
| Sialuria or French Type Sialuria | 17 | 1 (5.9%) | 0.0422 | 0.248 | SMPDB |
| Amino Sugar Metabolism | 17 | 1 (5.9%) | 0.0422 | 0.248 | SMPDB |
| G(M2)-Gangliosidosis: Variant B, Tay-sachs disease | 17 | 1 (5.9%) | 0.0422 | 0.248 | SMPDB |
| Tay-Sachs Disease | 17 | 1 (5.9%) | 0.0422 | 0.248 | SMPDB |
| Salla Disease/Infantile Sialic Acid Storage Disease | 17 | 1 (5.9%) | 0.0422 | 0.248 | SMPDB |
| Simplified Depiction of MYD88 Distinct Input-Output Pathway | 17 | 1 (5.9%) | 0.0422 | 0.248 | Wikipathways |

| | | | | | |
|---|---|---|---|---|---|
| Regulation of HMOX1 expression and activity | 17 | 1 (5.9%) | 0.0422 | 0.248 | **Reactome** |
| Toll-like Receptor Cascades | 130 | 2 (1.5%) | 0.0426 | 0.248 | **Reactome** |
| Activation of NF-kappaB in B cells | 18 | 1 (5.6%) | 0.0446 | 0.248 | **Reactome** |
| Death Receptor Signalling | 134 | 2 (1.5%) | 0.045 | 0.248 | **Reactome** |
| Glycosaminoglycan degradation - Homo sapiens (human) | 19 | 1 (5.3%) | 0.047 | 0.248 | **KEGG** |
| CD209 (DC-SIGN) signaling | 19 | 1 (5.3%) | 0.047 | 0.248 | **Reactome** |
| Tryptophan Metabolism | 19 | 1 (5.3%) | 0.047 | 0.248 | **SMPDB** |
| G2/M Transition | 138 | 2 (1.4%) | 0.0475 | 0.248 | **Reactome** |
| Innate Immune System | 1065 | 6 (0.6%) | 0.0484 | 0.248 | **Reactome** |
| Mitotic G2-G2/M phases | 140 | 2 (1.4%) | 0.0487 | 0.248 | **Reactome** |
| Arf1 pathway | 20 | 1 (5.0%) | 0.0495 | 0.248 | **PID** |

TRD

| pathway name | set size | candidates contained | p-value | q-value | pathway source |
|---|---|---|---|---|---|
| C-type lectin receptors (CLRs) | 84 | 3 (3.6%) | 0.00103 | 0.207 | Reactome |
| TRAIL signaling pathway | 25 | 2 (8.0%) | 0.00159 | 0.207 | PID |
| adenine and adenosine salvage I | 1 | 1 (100.0%) | 0.00238 | 0.207 | HumanCyc |
| Apoptotic cleavage of cellular proteins | 38 | 2 (5.3%) | 0.00366 | 0.207 | Reactome |
| CLEC7A (Dectin-1) signaling | 41 | 2 (4.9%) | 0.00425 | 0.207 | Reactome |
| RAGE | 45 | 2 (4.4%) | 0.00511 | 0.207 | NetPath |
| Ceramide signaling pathway | 48 | 2 (4.2%) | 0.00579 | 0.207 | PID |
| Apoptotic execution phase | 52 | 2 (3.8%) | 0.00677 | 0.207 | Reactome |
| hiv-1 nef: negative effector of fas and tnf | 53 | 2 (3.8%) | 0.00702 | 0.207 | BioCarta |
| Erlotinib Action Pathway | 3 | 1 (33.3%) | 0.00713 | 0.207 | SMPDB |
| Vitamins B6 activation to pyridoxal phosphate | 3 | 1 (33.3%) | 0.00713 | 0.207 | Reactome |
| Proximal tubule transport | 57 | 2 (3.5%) | 0.00809 | 0.207 | Wikipathways |
| Cilium Assembly | 178 | 3 (1.7%) | 0.00856 | 0.207 | Reactome |
| Activation, myristolyation of BID and translocation to mitochondria | 4 | 1 (25.0%) | 0.0095 | 0.207 | Reactome |
| CLEC7A/inflammasome pathway | 4 | 1 (25.0%) | 0.0095 | 0.207 | Reactome |
| Microbial modulation of RIPK1-mediated regulated necrosis | 4 | 1 (25.0%) | 0.0095 | 0.207 | Reactome |
| Vitamin B6 metabolism | 4 | 1 (25.0%) | 0.0095 | 0.207 | INOH |
| Vitamin B6-dependent and responsive disorders | 4 | 1 (25.0%) | 0.0095 | 0.207 | Wikipathways |
| Integrins in angiogenesis | 63 | 2 (3.2%) | 0.00981 | 0.207 | PID |
| AGE-RAGE pathway | 66 | 2 (3.0%) | 0.0107 | 0.207 | Wikipathways |
| Signaling events mediated by VEGFR1 and VEGFR2 | 68 | 2 (2.9%) | 0.0114 | 0.207 | PID |
| FasL/ CD95L signaling | 5 | 1 (20.0%) | 0.0119 | 0.207 | Reactome |
| Abacavir transmembrane transport | 5 | 1 (20.0%) | 0.0119 | 0.207 | Reactome |
| Ficolins bind to repetitive carbohydrate structures on the target cell surface | 5 | 1 (20.0%) | 0.0119 | 0.207 | Reactome |
| Hypophosphatasia | 5 | 1 (20.0%) | 0.0119 | 0.207 | SMPDB |
| Vitamin B6 Metabolism | 5 | 1 (20.0%) | 0.0119 | 0.207 | SMPDB |
| pyridoxal 5,-phosphate salvage | 5 | 1 (20.0%) | 0.0119 | 0.207 | HumanCyc |
| TLR3-mediated TICAM1-dependent programmed cell death | 6 | 1 (16.7%) | 0.0142 | 0.207 | Reactome |
| multi-drug resistance factors | 6 | 1 (16.7%) | 0.0142 | 0.207 | BioCarta |
| Venlafaxine Metabolism Pathway | 6 | 1 (16.7%) | 0.0142 | 0.207 | SMPDB |
| Vitamin B6 metabolism - Homo sapiens (human) | 6 | 1 (16.7%) | 0.0142 | 0.207 | KEGG |
| 16p11.2 proximal deletion syndrome | 79 | 2 (2.5%) | 0.0151 | 0.207 | Wikipathways |
| TNFR1-induced proapoptotic signaling | 7 | 1 (14.3%) | 0.0166 | 0.207 | Reactome |
| TRAIL signaling | 7 | 1 (14.3%) | 0.0166 | 0.207 | Reactome |
| Paclitaxel Action Pathway | 7 | 1 (14.3%) | 0.0166 | 0.207 | SMPDB |
| Docetaxel Action Pathway | 7 | 1 (14.3%) | 0.0166 | 0.207 | SMPDB |
| Organelle biogenesis and maintenance | 231 | 3 (1.3%) | 0.0172 | 0.207 | Reactome |
| HuR (ELAVL1) binds and stabilizes mRNA | 8 | 1 (12.5%) | 0.0189 | 0.207 | Reactome |
| Lectin pathway of complement activation | 8 | 1 (12.5%) | 0.0189 | 0.207 | Reactome |
| Apoptosis Modulation and Signaling | 92 | 2 (2.2%) | 0.0201 | 0.207 | Wikipathways |
| Allograft Rejection | 93 | 2 (2.2%) | 0.0206 | 0.207 | Wikipathways |
| TRIF-mediated programmed cell death | 9 | 1 (11.1%) | 0.0212 | 0.207 | Reactome |
| Clopidogrel Metabolism Pathway | 9 | 1 (11.1%) | 0.0212 | 0.207 | SMPDB |
| Clopidogrel Action Pathway | 9 | 1 (11.1%) | 0.0212 | 0.207 | SMPDB |
| ROS sensing by NFE2L2 | 9 | 1 (11.1%) | 0.0212 | 0.207 | Reactome |
| polyadenylation of mrna | 9 | 1 (11.1%) | 0.0212 | 0.207 | BioCarta |
| Metabolism of ingested SeMet, Sec, MeSec into H2Se | 9 | 1 (11.1%) | 0.0212 | 0.207 | Reactome |
| Anchoring of the basal body to the plasma membrane | 97 | 2 (2.1%) | 0.0222 | 0.207 | Reactome |
| Dimerization of procaspase-8 | 10 | 1 (10.0%) | 0.0236 | 0.207 | Reactome |
| Nanoparticle triggered regulated necrosis | 10 | 1 (10.0%) | 0.0236 | 0.207 | Wikipathways |
| Abacavir transport and metabolism | 10 | 1 (10.0%) | 0.0236 | 0.207 | Reactome |
| Vindesine Action Pathway | 10 | 1 (10.0%) | 0.0236 | 0.207 | SMPDB |
| Vincristine Action Pathway | 10 | 1 (10.0%) | 0.0236 | 0.207 | SMPDB |
| Folding of actin by CCT/TriC | 10 | 1 (10.0%) | 0.0236 | 0.207 | Reactome |
| Maturation of nucleoprotein | 10 | 1 (10.0%) | 0.0236 | 0.207 | Reactome |
| Maturation of nucleoprotein | 10 | 1 (10.0%) | 0.0236 | 0.207 | Reactome |
| Platelet activation, signaling and aggregation | 263 | 3 (1.1%) | 0.0242 | 0.207 | Reactome |
| C-type lectin receptor signaling pathway - Homo sapiens (human) | 104 | 2 (1.9%) | 0.0253 | 0.207 | KEGG |
| Regulation by c-FLIP | 11 | 1 (9.1%) | 0.0259 | 0.207 | Reactome |
| CASP8 activity is inhibited | 11 | 1 (9.1%) | 0.0259 | 0.207 | Reactome |
| Vinblastine Action Pathway | 11 | 1 (9.1%) | 0.0259 | 0.207 | SMPDB |
| Vinorelbine Action Pathway | 11 | 1 (9.1%) | 0.0259 | 0.207 | SMPDB |
| MET activates RAS signaling | 11 | 1 (9.1%) | 0.0259 | 0.207 | Reactome |
| Caspase-mediated cleavage of cytoskeletal proteins | 12 | 1 (8.3%) | 0.0282 | 0.207 | Reactome |
| NF-kB activation through FADD/RIP-1 pathway mediated by caspase-8 and -10 | 12 | 1 (8.3%) | 0.0282 | 0.207 | Reactome |
| LncRNA-mediated mechanisms of therapeutic resistance | 12 | 1 (8.3%) | 0.0282 | 0.207 | Wikipathways |
| lectin induced complement pathway | 12 | 1 (8.3%) | 0.0282 | 0.207 | BioCarta |
| TP53 Regulates Transcription of Death Receptors and Ligands | 12 | 1 (8.3%) | 0.0282 | 0.207 | Reactome |

| | | | | | |
|---|---|---|---|---|---|
| Transport of Mature mRNA Derived from an Intronless Transcript | 12 | 1 (8.3%) | 0.0282 | 0.207 | Reactome |
| Selenoamino Acid Metabolism | 12 | 1 (8.3%) | 0.0282 | 0.207 | SMPDB |
| Purine salvage | 13 | 1 (7.7%) | 0.0305 | 0.207 | Reactome |
| Transport of Mature mRNAs Derived from Intronless Transcripts | 13 | 1 (7.7%) | 0.0305 | 0.207 | Reactome |
| Apoptosis | 127 | 2 (1.6%) | 0.0366 | 0.207 | Reactome |
| Innate Immune System | 1065 | 6 (0.6%) | 0.0372 | 0.207 | Reactome |
| Caspase activation via Death Receptors in the presence of ligand | 16 | 1 (6.2%) | 0.0375 | 0.207 | Reactome |
| Mycophenolic Acid Metabolism Pathway | 16 | 1 (6.2%) | 0.0375 | 0.207 | SMPDB |
| Codeine and Morphine Metabolism | 16 | 1 (6.2%) | 0.0375 | 0.207 | Wikipathways |
| Platelet degranulation | 131 | 2 (1.5%) | 0.0387 | 0.207 | Reactome |
| tnfr1 signaling pathway | 17 | 1 (5.9%) | 0.0398 | 0.207 | BioCarta |
| Drug Induction of Bile Acid Pathway | 17 | 1 (5.9%) | 0.0398 | 0.207 | Wikipathways |
| Doxorubicin Metabolism Pathway | 17 | 1 (5.9%) | 0.0398 | 0.207 | SMPDB |
| Regulation of HMOX1 expression and activity | 17 | 1 (5.9%) | 0.0398 | 0.207 | Reactome |
| Selenocompound metabolism - Homo sapiens (human) | 17 | 1 (5.9%) | 0.0398 | 0.207 | KEGG |
| Response to elevated platelet cytosolic Ca2+ | 136 | 2 (1.5%) | 0.0414 | 0.207 | Reactome |
| Simplified Interaction Map Between LOXL4 and Oxidative Stress Pathway | 18 | 1 (5.6%) | 0.042 | 0.207 | Wikipathways |
| internal ribosome entry pathway | 18 | 1 (5.6%) | 0.042 | 0.207 | BioCarta |
| Lamivudine Metabolism Pathway | 18 | 1 (5.6%) | 0.042 | 0.207 | SMPDB |
| MFAP5 effect on permeability and motility of endothelial cells via cytoskeleton rearrangement | 18 | 1 (5.6%) | 0.042 | 0.207 | Wikipathways |
| Apoptosis Modulation by HSP70 | 19 | 1 (5.3%) | 0.0443 | 0.207 | Wikipathways |
| Etoposide Action Pathway | 19 | 1 (5.3%) | 0.0443 | 0.207 | SMPDB |
| Etoposide Metabolism Pathway | 19 | 1 (5.3%) | 0.0443 | 0.207 | SMPDB |
| Scavenging by Class A Receptors | 19 | 1 (5.3%) | 0.0443 | 0.207 | Reactome |
| Processing of Intronless Pre-mRNAs | 19 | 1 (5.3%) | 0.0443 | 0.207 | Reactome |
| Programmed Cell Death | 142 | 2 (1.4%) | 0.0448 | 0.207 | Reactome |
| Regucalcin in proximal tubule epithelial kidney cells | 20 | 1 (5.0%) | 0.0466 | 0.207 | Wikipathways |
| Nanomaterial induced apoptosis | 20 | 1 (5.0%) | 0.0466 | 0.207 | Wikipathways |
| fas signaling pathway (cd95) | 20 | 1 (5.0%) | 0.0466 | 0.207 | BioCarta |
| VEGFR2 mediated cell proliferation | 20 | 1 (5.0%) | 0.0466 | 0.207 | Reactome |
| Host-pathogen interaction of human coronaviruses - apoptosis | 21 | 1 (4.8%) | 0.0489 | 0.207 | Wikipathways |
| Imatinib and Chronic Myeloid Leukemia | 21 | 1 (4.8%) | 0.0489 | 0.207 | Wikipathways |
| NGF | 21 | 1 (4.8%) | 0.0489 | 0.207 | INOH |
| Galanin receptor pathway | 21 | 1 (4.8%) | 0.0489 | 0.207 | Wikipathways |

T2DM_admission_freq

| pathway name | set size | candidates contained | p-value | q-value | pathway source |
|---|---|---|---|---|---|
| G alpha (s) signalling events | 143 | 4 (2.8%) | 0.00167 | 0.313 | Reactome |
| GPCRs, Class B Secretin-like | 23 | 2 (8.7%) | 0.00301 | 0.313 | Wikipathways |
| Glucagon-type ligand receptors | 33 | 2 (6.1%) | 0.00615 | 0.313 | Reactome |
| ADORA2B mediated anti-inflammatory cytokines production | 132 | 3 (2.3%) | 0.0116 | 0.313 | Reactome |
| Regulation of Apoptosis | 4 | 1 (25.0%) | 0.0142 | 0.313 | Reactome |
| NAD salvage | 4 | 1 (25.0%) | 0.0142 | 0.313 | HumanCyc |
| Adefovir Dipivoxil Metabolism Pathway | 5 | 1 (20.0%) | 0.0177 | 0.313 | SMPDB |
| Tenofovir Metabolism Pathway | 5 | 1 (20.0%) | 0.0177 | 0.313 | SMPDB |
| Ubiquinone Biosynthesis | 5 | 1 (20.0%) | 0.0177 | 0.313 | SMPDB |
| NAD biosynthesis from 2-amino-3-carboxymuconate semialdehyde | 5 | 1 (20.0%) | 0.0177 | 0.313 | HumanCyc |
| allopregnanolone biosynthesis | 6 | 1 (16.7%) | 0.0213 | 0.313 | HumanCyc |
| Phosphonate and phosphinate metabolism - Homo sapiens (human) | 6 | 1 (16.7%) | 0.0213 | 0.313 | KEGG |
| ceramide de novo biosynthesis | 7 | 1 (14.3%) | 0.0248 | 0.313 | HumanCyc |
| phosphatidylcholine biosynthesis | 7 | 1 (14.3%) | 0.0248 | 0.313 | HumanCyc |
| ubiquinol-10 biosynthesis | 7 | 1 (14.3%) | 0.0248 | 0.313 | HumanCyc |
| Renin secretion - Homo sapiens (human) | 69 | 2 (2.9%) | 0.0252 | 0.313 | KEGG |
| Metabolism of vitamins and cofactors | 180 | 3 (1.7%) | 0.0263 | 0.313 | Reactome |
| Endoderm differentiation | 71 | 2 (2.8%) | 0.0266 | 0.313 | Wikipathways |
| Regulation of cytoskeletal remodeling and cell spreading by IPP complex components | 8 | 1 (12.5%) | 0.0282 | 0.313 | Reactome |
| Phosphate bond hydrolysis by NTPDase proteins | 8 | 1 (12.5%) | 0.0282 | 0.313 | Reactome |
| Ubiquinol biosynthesis | 8 | 1 (12.5%) | 0.0282 | 0.313 | Reactome |
| Costimulation by the CD28 family | 77 | 2 (2.6%) | 0.0309 | 0.313 | Reactome |
| Mitotic Prometaphase | 192 | 3 (1.6%) | 0.0311 | 0.313 | Reactome |
| Adrenoceptors | 9 | 1 (11.1%) | 0.0317 | 0.313 | Reactome |
| Benzo(a)pyrene metabolism | 9 | 1 (11.1%) | 0.0317 | 0.313 | Wikipathways |
| Mitotic Anaphase | 195 | 3 (1.5%) | 0.0323 | 0.313 | Reactome |
| Mitotic Metaphase and Anaphase | 196 | 3 (1.5%) | 0.0327 | 0.313 | Reactome |
| Complement and coagulation cascades - Homo sapiens (human) | 85 | 2 (2.4%) | 0.037 | 0.313 | KEGG |
| Synthesis of bile acids and bile salts via 27-hydroxycholesterol | 11 | 1 (9.1%) | 0.0386 | 0.313 | Reactome |
| cxcr4 signaling pathway | 11 | 1 (9.1%) | 0.0386 | 0.313 | BioCarta |
| Dissolution of Fibrin Clot | 11 | 1 (9.1%) | 0.0386 | 0.313 | Reactome |
| Ubiquinone and other terpenoid-quinone biosynthesis - Homo sapiens (human) | 11 | 1 (9.1%) | 0.0386 | 0.313 | KEGG |
| Mitochondrial translation termination | 88 | 2 (2.3%) | 0.0394 | 0.313 | Reactome |
| Mitochondrial translation elongation | 88 | 2 (2.3%) | 0.0394 | 0.313 | Reactome |
| Mitochondrial translation initiation | 88 | 2 (2.3%) | 0.0394 | 0.313 | Reactome |
| Gap junction - Homo sapiens (human) | 88 | 2 (2.3%) | 0.0394 | 0.313 | KEGG |
| cAMP signaling pathway - Homo sapiens (human) | 216 | 3 (1.4%) | 0.0418 | 0.313 | KEGG |
| Morphine addiction - Homo sapiens (human) | 91 | 2 (2.2%) | 0.0419 | 0.313 | KEGG |
| Class B/2 (Secretin family receptors) | 91 | 2 (2.2%) | 0.0419 | 0.313 | Reactome |
| Butyrophilin (BTN) family interactions | 12 | 1 (8.3%) | 0.0421 | 0.313 | Reactome |
| NAD de novo biosynthesis | 12 | 1 (8.3%) | 0.0421 | 0.313 | HumanCyc |
| Rheumatoid arthritis - Homo sapiens (human) | 93 | 2 (2.2%) | 0.0427 | 0.313 | KEGG |
| Mitochondrial translation | 94 | 2 (2.1%) | 0.0444 | 0.313 | Reactome |
| attenuation of gpcr signaling | 13 | 1 (7.7%) | 0.0455 | 0.313 | BioCarta |
| Passive transport by Aquaporins | 13 | 1 (7.7%) | 0.0455 | 0.313 | Reactome |
| Interferon gamma signaling | 96 | 2 (2.1%) | 0.0461 | 0.313 | Reactome |
| Amplification of signal from unattached kinetochores via a MAD2 inhibitory signal | 96 | 2 (2.1%) | 0.0461 | 0.313 | Reactome |
| Amplification of signal from the kinetochores | 96 | 2 (2.1%) | 0.0461 | 0.313 | Reactome |
| M Phase | 383 | 4 (1.0%) | 0.047 | 0.313 | Reactome |
| Synthesis of bile acids and bile salts via 24-hydroxycholesterol | 14 | 1 (7.1%) | 0.0489 | 0.313 | Reactome |
| Kennedy pathway from sphingolipids | 14 | 1 (7.1%) | 0.0489 | 0.313 | Wikipathways |
| EML4 and NUDC in mitotic spindle formation | 100 | 2 (2.0%) | 0.0496 | 0.313 | Reactome |

CAD_admission_freq_SAT

| pathway name | set size | candidates contained | p-value | q-value | pathway source |
|---|---|---|---|---|---|
| Lysosome - Homo sapiens (human) | 128 | 4 (3.1%) | 0.00254 | 0.258 | KEGG |
| Norepinephrine Neurotransmitter Release Cycle | 18 | 2 (11.1%) | 0.00287 | 0.258 | Reactome |
| Lamivudine Metabolism Pathway | 18 | 2 (11.1%) | 0.00287 | 0.258 | SMPDB |
| glutathione-mediated detoxification | 22 | 2 (9.1%) | 0.00428 | 0.258 | HumanCyc |
| snRNP Assembly | 24 | 2 (8.3%) | 0.00508 | 0.258 | Reactome |
| Metabolism of non-coding RNA | 24 | 2 (8.3%) | 0.00508 | 0.258 | Reactome |
| Drug metabolism - other enzymes - Homo sapiens (human) | 80 | 3 (3.8%) | 0.00545 | 0.258 | KEGG |
| Activation of Na-permeable kainate receptors | 2 | 1 (50.0%) | 0.00891 | 0.258 | Reactome |
| Biosynthesis of maresin conjugates in tissue regeneration (MCTR) | 2 | 1 (50.0%) | 0.00891 | 0.258 | Reactome |
| Biosynthesis of DHA-derived sulfido conjugates | 2 | 1 (50.0%) | 0.00891 | 0.258 | Reactome |
| sulfate activation for sulfonation | 2 | 1 (50.0%) | 0.00891 | 0.258 | HumanCyc |
| Nuclear Receptors in Lipid Metabolism and Toxicity | 35 | 2 (5.7%) | 0.0106 | 0.258 | Wikipathways |
| Glutathione conjugation | 37 | 2 (5.4%) | 0.0118 | 0.258 | Reactome |
| 1,25-dihydroxyvitamin D3 biosynthesis | 3 | 1 (33.3%) | 0.0133 | 0.258 | HumanCyc |
| Phase II - Conjugation of compounds | 111 | 3 (2.7%) | 0.0134 | 0.258 | Reactome |
| Non-integrin membrane-ECM interactions | 42 | 2 (4.8%) | 0.0151 | 0.258 | Reactome |
| Glycosaminoglycan metabolism | 119 | 3 (2.5%) | 0.0161 | 0.258 | Reactome |
| Biological oxidations | 219 | 4 (1.8%) | 0.0165 | 0.258 | Reactome |
| pyrimidine deoxyribonucleosides degradation | 4 | 1 (25.0%) | 0.0177 | 0.258 | HumanCyc |
| Polyol Pathway | 4 | 1 (25.0%) | 0.0177 | 0.258 | Wikipathways |
| Proteoglycan syndecan-mediated signaling events | 4 | 1 (25.0%) | 0.0177 | 0.258 | PID |
| Metabolism of ingested H2SeO4 and H2SeO3 into H2Se | 4 | 1 (25.0%) | 0.0177 | 0.258 | Reactome |
| Transcriptional regulation of white adipocyte differentiation | 47 | 2 (4.3%) | 0.0186 | 0.258 | Reactome |
| Ether lipid metabolism - Homo sapiens (human) | 49 | 2 (4.1%) | 0.0202 | 0.258 | KEGG |
| Chondroitin sulfate/dermatan sulfate metabolism | 50 | 2 (4.0%) | 0.0209 | 0.258 | Reactome |
| Neurotransmitter release cycle | 50 | 2 (4.0%) | 0.0209 | 0.258 | Reactome |
| Trafficking of myristoylated proteins to the cilium | 5 | 1 (20.0%) | 0.0221 | 0.258 | Reactome |
| Capecitabine Action Pathway | 5 | 1 (20.0%) | 0.0221 | 0.258 | SMPDB |
| Capecitabine Metabolism Pathway | 5 | 1 (20.0%) | 0.0221 | 0.258 | SMPDB |
| Extrinsic Pathway of Fibrin Clot Formation | 5 | 1 (20.0%) | 0.0221 | 0.258 | Reactome |
| Degradation pathway of sphingolipids, including diseases | 5 | 1 (20.0%) | 0.0221 | 0.258 | Wikipathways |
| tetrapyrrole biosynthesis | 5 | 1 (20.0%) | 0.0221 | 0.258 | HumanCyc |
| Fructose catabolism | 5 | 1 (20.0%) | 0.0221 | 0.258 | Reactome |
| Abacavir transmembrane transport | 5 | 1 (20.0%) | 0.0221 | 0.258 | Reactome |
| Fluid shear stress and atherosclerosis - Homo sapiens (human) | 139 | 3 (2.2%) | 0.0242 | 0.258 | KEGG |
| Clathrin-mediated endocytosis | 142 | 3 (2.1%) | 0.0255 | 0.258 | Reactome |
| Golgi Associated Vesicle Biogenesis | 56 | 2 (3.6%) | 0.0259 | 0.258 | Reactome |
| NRF2 pathway | 143 | 3 (2.1%) | 0.026 | 0.258 | Wikipathways |
| Vitamins | 6 | 1 (16.7%) | 0.0265 | 0.258 | Reactome |
| Acyl chain remodeling of CL | 6 | 1 (16.7%) | 0.0265 | 0.258 | Reactome |
| Transport and synthesis of PAPS | 6 | 1 (16.7%) | 0.0265 | 0.258 | Reactome |
| Phosphonate and phosphinate metabolism - Homo sapiens (human) | 6 | 1 (16.7%) | 0.0265 | 0.258 | KEGG |
| Glutathione metabolism - Homo sapiens (human) | 57 | 2 (3.5%) | 0.0268 | 0.258 | KEGG |
| Porphyrin metabolism | 7 | 1 (14.3%) | 0.0309 | 0.258 | INOH |
| Fructose metabolism | 7 | 1 (14.3%) | 0.0309 | 0.258 | Reactome |
| Anchoring fibril formation | 7 | 1 (14.3%) | 0.0309 | 0.258 | Reactome |
| Tramadol Metabolism Pathway | 7 | 1 (14.3%) | 0.0309 | 0.258 | SMPDB |
| CS/DS degradation | 7 | 1 (14.3%) | 0.0309 | 0.258 | Reactome |
| phosphatidylcholine biosynthesis | 7 | 1 (14.3%) | 0.0309 | 0.258 | HumanCyc |
| Chemokine receptors bind chemokines | 62 | 2 (3.2%) | 0.0313 | 0.258 | Reactome |
| sucrose degradation | 8 | 1 (12.5%) | 0.0352 | 0.258 | HumanCyc |
| Organic cation transport | 8 | 1 (12.5%) | 0.0352 | 0.258 | Reactome |
| Non-genomic actions of 1,25 dihydroxyvitamin D3 | 68 | 2 (2.9%) | 0.037 | 0.258 | Wikipathways |
| Metabolism of carbohydrates | 285 | 4 (1.4%) | 0.0379 | 0.258 | Reactome |
| Drug metabolism - cytochrome P450 - Homo sapiens (human) | 72 | 2 (2.9%) | 0.039 | 0.258 | KEGG |
| Vitamin D (calciferol) metabolism | 9 | 1 (11.1%) | 0.0395 | 0.258 | Reactome |
| heme biosynthesis | 9 | 1 (11.1%) | 0.0395 | 0.258 | HumanCyc |
| Heme Biosynthesis | 9 | 1 (11.1%) | 0.0395 | 0.258 | Wikipathways |
| Type I hemidesmosome assembly | 9 | 1 (11.1%) | 0.0395 | 0.258 | Reactome |
| Formation of annular gap junctions | 9 | 1 (11.1%) | 0.0395 | 0.258 | Reactome |
| Bile acids synthesis and enterohepatic circulation | 9 | 1 (11.1%) | 0.0395 | 0.258 | Wikipathways |
| trans-Golgi Network Vesicle Budding | 72 | 2 (2.8%) | 0.0411 | 0.258 | Reactome |
| Cytosolic DNA-sensing pathway | 73 | 2 (2.7%) | 0.0421 | 0.258 | Wikipathways |
| Membrane Trafficking | 578 | 6 (1.0%) | 0.0435 | 0.258 | Reactome |
| Vitamin D Metabolism | 10 | 1 (10.0%) | 0.0438 | 0.258 | Wikipathways |
| Gamma-carboxylation of protein precursors | 10 | 1 (10.0%) | 0.0438 | 0.258 | Reactome |
| Removal of aminoterminal propeptides from gamma-carboxylated proteins | 10 | 1 (10.0%) | 0.0438 | 0.258 | Reactome |
| Intracellular Signalling Through LHCGR Receptor and Luteinizing Hormone/Choriogonadotropin | 10 | 1 (10.0%) | 0.0438 | 0.258 | SMPDB |
| Neurotransmitter clearance | 10 | 1 (10.0%) | 0.0438 | 0.258 | Reactome |
| Abacavir transport and metabolism | 10 | 1 (10.0%) | 0.0438 | 0.258 | Reactome |

| | | | | | |
|---|---|---|---|---|---|
| Gap junction degradation | 10 | 1 (10.0%) | 0.0438 | 0.258 | **Reactome** |
| Sulfur metabolism - Homo sapiens (human) | 10 | 1 (10.0%) | 0.0438 | 0.258 | **KEGG** |
| Liver X receptor pathway | 10 | 1 (10.0%) | 0.0438 | 0.258 | **Wikipathways** |
| Peptide GPCRs | 75 | 2 (2.7%) | 0.0442 | 0.258 | **Wikipathways** |
| Metabolism of xenobiotics by cytochrome P450 - Homo sapiens (human) | 78 | 2 (2.6%) | 0.0453 | 0.258 | **KEGG** |
| Metabolism | 1954 | 14 (0.7%) | 0.0458 | 0.258 | **Reactome** |
| Synaptic vesicle cycle - Homo sapiens (human) | 78 | 2 (2.6%) | 0.0474 | 0.258 | **KEGG** |
| Pyrimidine salvage | 11 | 1 (9.1%) | 0.0481 | 0.258 | **Reactome** |
| Gamma-carboxylation, transport, and amino-terminal cleavage of proteins | 11 | 1 (9.1%) | 0.0481 | 0.258 | **Reactome** |
| Alpha6 beta4 integrin-ligand interactions | 11 | 1 (9.1%) | 0.0481 | 0.258 | **PID** |
| pertussis toxin-insensitive ccr5 signaling in macrophage | 11 | 1 (9.1%) | 0.0481 | 0.258 | **BioCarta** |

CAD_mortality_SAT

| pathway name | set size | candidates contained | p-value | q-value | pathway source |
|---|---|---|---|---|---|
| Metabolic pathway of LDL, HDL and TG, including diseases | 6 | 2 (33.3%) | 0.000311 | 0.078 | Wikipathways |
| Familial lipoprotein lipase deficiency | 13 | 2 (15.4%) | 0.00158 | 0.078 | SMPDB |
| Glycerolipid Metabolism | 13 | 2 (15.4%) | 0.00158 | 0.078 | SMPDB |
| Glycerol Kinase Deficiency | 13 | 2 (15.4%) | 0.00158 | 0.078 | SMPDB |
| D-glyceric acidura | 13 | 2 (15.4%) | 0.00158 | 0.078 | SMPDB |
| Bivalirudin Action Pathway | 20 | 2 (10.0%) | 0.00377 | 0.078 | SMPDB |
| Argatroban Action Pathway | 20 | 2 (10.0%) | 0.00377 | 0.078 | SMPDB |
| Ximelagatran Action Pathway | 20 | 2 (10.0%) | 0.00377 | 0.078 | SMPDB |
| Lepirudin Action Pathway | 20 | 2 (10.0%) | 0.00377 | 0.078 | SMPDB |
| Coagulation | 20 | 2 (10.0%) | 0.00377 | 0.078 | SMPDB |
| Aminocaproic Acid Action Pathway | 20 | 2 (10.0%) | 0.00377 | 0.078 | SMPDB |
| Tranexamic Acid Action Pathway | 20 | 2 (10.0%) | 0.00377 | 0.078 | SMPDB |
| Urokinase Action Pathway | 20 | 2 (10.0%) | 0.00377 | 0.078 | SMPDB |
| Reteplase Action Pathway | 20 | 2 (10.0%) | 0.00377 | 0.078 | SMPDB |
| Streptokinase Action Pathway | 20 | 2 (10.0%) | 0.00377 | 0.078 | SMPDB |
| Tenecteplase Action Pathway | 20 | 2 (10.0%) | 0.00377 | 0.078 | SMPDB |
| Alteplase Action Pathway | 20 | 2 (10.0%) | 0.00377 | 0.078 | SMPDB |
| Anistreplase Action Pathway | 20 | 2 (10.0%) | 0.00377 | 0.078 | SMPDB |
| Aprotinin Action Pathway | 20 | 2 (10.0%) | 0.00377 | 0.078 | SMPDB |
| Phenindione Action Pathway | 20 | 2 (10.0%) | 0.00377 | 0.078 | SMPDB |
| Dicoumarol Action Pathway | 20 | 2 (10.0%) | 0.00377 | 0.078 | SMPDB |
| Warfarin Action Pathway | 20 | 2 (10.0%) | 0.00377 | 0.078 | SMPDB |
| Acenocoumarol Action Pathway | 20 | 2 (10.0%) | 0.00377 | 0.078 | SMPDB |
| Phenprocoumon Action Pathway | 20 | 2 (10.0%) | 0.00377 | 0.078 | SMPDB |
| Dicumarol Action Pathway | 20 | 2 (10.0%) | 0.00377 | 0.078 | SMPDB |
| Ardeparin Action Pathway | 21 | 2 (9.5%) | 0.00416 | 0.078 | SMPDB |
| Heparin Action Pathway | 21 | 2 (9.5%) | 0.00416 | 0.078 | SMPDB |
| Fondaparinux Action Pathway | 21 | 2 (9.5%) | 0.00416 | 0.078 | SMPDB |
| Enoxaparin Action Pathway | 21 | 2 (9.5%) | 0.00416 | 0.078 | SMPDB |
| Translocation of ZAP-70 to Immunological synapse | 28 | 2 (7.1%) | 0.00733 | 0.133 | Reactome |
| Phosphorylation of CD3 and TCR zeta chains | 31 | 2 (6.5%) | 0.00894 | 0.152 | Reactome |
| Proprotein convertase subtilisin-kexin type 9 (PCSK9) mediated LDL receptor degradation | 2 | 1 (50.0%) | 0.00921 | 0.152 | Wikipathways |
| Evolocumab Mechanism | 2 | 1 (50.0%) | 0.00921 | 0.152 | Wikipathways |
| PD-1 signaling | 32 | 2 (6.2%) | 0.00951 | 0.152 | Reactome |
| Interferon gamma signaling | 96 | 3 (3.1%) | 0.00986 | 0.153 | Reactome |
| Tyrosine metabolism | 33 | 2 (6.1%) | 0.0101 | 0.153 | INOH |
| Tyrosine metabolism - Homo sapiens (human) | 36 | 2 (5.6%) | 0.0119 | 0.175 | KEGG |
| Integration of viral DNA into host genomic DNA | 3 | 1 (33.3%) | 0.0138 | 0.187 | Reactome |
| Autointegration results in viral DNA circles | 3 | 1 (33.3%) | 0.0138 | 0.187 | Reactome |
| Metabolism of vitamin K | 3 | 1 (33.3%) | 0.0138 | 0.187 | Reactome |
| Generation of second messenger molecules | 42 | 2 (4.8%) | 0.016 | 0.204 | Reactome |
| Vitamin A and carotenoid metabolism | 43 | 2 (4.7%) | 0.0168 | 0.204 | Wikipathways |
| CD4 T cell receptor signaling-ERK cascade | 44 | 2 (4.5%) | 0.0175 | 0.204 | INOH |
| Thyroid hormone signaling pathway - Homo sapiens (human) | 121 | 3 (2.5%) | 0.0183 | 0.204 | KEGG |
| Felbamate Metabolism Pathway | 4 | 1 (25.0%) | 0.0183 | 0.204 | SMPDB |
| Composition of Lipid Particles | 4 | 1 (25.0%) | 0.0183 | 0.204 | Wikipathways |
| proline biosynthesis | 4 | 1 (25.0%) | 0.0183 | 0.204 | HumanCyc |
| Vitamin K Metabolism | 4 | 1 (25.0%) | 0.0183 | 0.204 | SMPDB |
| hop pathway in cardiac development | 4 | 1 (25.0%) | 0.0183 | 0.204 | BioCarta |
| Rett syndrome causing genes | 48 | 2 (4.2%) | 0.0206 | 0.222 | Wikipathways |
| Metabolism of fat-soluble vitamins | 49 | 2 (4.1%) | 0.0214 | 0.222 | Reactome |
| Ebola Virus Pathway on Host | 130 | 3 (2.3%) | 0.0221 | 0.222 | Wikipathways |
| Cholesterol metabolism - Homo sapiens (human) | 50 | 2 (4.0%) | 0.0223 | 0.222 | KEGG |
| Extrinsic Pathway of Fibrin Clot Formation | 5 | 1 (20.0%) | 0.0229 | 0.222 | Reactome |
| role of ppar-gamma coactivators in obesity and thermogenesis | 5 | 1 (20.0%) | 0.0229 | 0.222 | BioCarta |
| Cross-presentation of soluble exogenous antigens (endosomes) | 5 | 1 (20.0%) | 0.0229 | 0.222 | Reactome |
| Downstream TCR signaling | 52 | 2 (3.8%) | 0.024 | 0.229 | Reactome |
| 2,-deoxy-α-D-ribose 1-phosphate degradation | 6 | 1 (16.7%) | 0.0274 | 0.257 | HumanCyc |
| CD4 T cell receptor signaling-JNK cascade | 57 | 2 (3.5%) | 0.0284 | 0.262 | INOH |
| MHC class II antigen presentation | 60 | 2 (3.3%) | 0.0313 | 0.271 | Reactome |
| Plasma lipoprotein assembly, remodeling, and clearance | 60 | 2 (3.3%) | 0.0313 | 0.271 | Reactome |
| TRAIL signaling | 7 | 1 (14.3%) | 0.0319 | 0.271 | Reactome |
| 2-LTR circle formation | 7 | 1 (14.3%) | 0.0319 | 0.271 | Reactome |
| APOBEC3G mediated resistance to HIV-1 infection | 7 | 1 (14.3%) | 0.0319 | 0.271 | Reactome |
| HIV Life Cycle | 151 | 3 (2.0%) | 0.0325 | 0.272 | Reactome |
| RAB geranylgeranylation | 65 | 2 (3.1%) | 0.0362 | 0.282 | Reactome |
| Ifosfamide Action Pathway | 8 | 1 (12.5%) | 0.0363 | 0.282 | SMPDB |
| Ifosfamide Metabolism Pathway | 8 | 1 (12.5%) | 0.0363 | 0.282 | SMPDB |

| | | | | | |
|---|---|---|---|---|---|
| Hypothetical Craniofacial Development Pathway | 8 | 1 (12.5%) | 0.0363 | 0.282 | Wikipathways |
| visceral fat deposits and the metabolic syndrome | 8 | 1 (12.5%) | 0.0363 | 0.282 | BioCarta |
| Glycolysis / Gluconeogenesis - Homo sapiens (human) | 67 | 2 (3.0%) | 0.0382 | 0.283 | KEGG |
| Interferon Signaling | 162 | 3 (1.9%) | 0.0388 | 0.283 | Reactome |
| Cyclophosphamide Action Pathway | 9 | 1 (11.1%) | 0.0408 | 0.283 | SMPDB |
| Cyclophosphamide Metabolism Pathway | 9 | 1 (11.1%) | 0.0408 | 0.283 | SMPDB |
| Chylomicron remodeling | 9 | 1 (11.1%) | 0.0408 | 0.283 | Reactome |
| Integration of provirus | 9 | 1 (11.1%) | 0.0408 | 0.283 | Reactome |
| Drug metabolism - cytochrome P450 - Homo sapiens (human) | 72 | 2 (2.9%) | 0.0414 | 0.283 | KEGG |
| Endoderm differentiation | 71 | 2 (2.8%) | 0.0425 | 0.283 | Wikipathways |
| TCR signaling | 73 | 2 (2.7%) | 0.0447 | 0.283 | Reactome |
| putrescine degradation III | 10 | 1 (10.0%) | 0.0452 | 0.283 | HumanCyc |
| Reduction of cytosolic Ca++ levels | 10 | 1 (10.0%) | 0.0452 | 0.283 | Reactome |
| Gamma-carboxylation of protein precursors | 10 | 1 (10.0%) | 0.0452 | 0.283 | Reactome |
| Removal of aminoterminal propeptides from gamma-carboxylated proteins | 10 | 1 (10.0%) | 0.0452 | 0.283 | Reactome |
| Activation of the phototransduction cascade | 10 | 1 (10.0%) | 0.0452 | 0.283 | Reactome |
| Dimerization of procaspase-8 | 10 | 1 (10.0%) | 0.0452 | 0.283 | Reactome |
| Opsins | 10 | 1 (10.0%) | 0.0452 | 0.283 | Reactome |
| VLDLR internalisation and degradation | 10 | 1 (10.0%) | 0.0452 | 0.283 | Reactome |
| Metabolism of xenobiotics by cytochrome P450 - Homo sapiens (human) | 78 | 2 (2.6%) | 0.048 | 0.285 | KEGG |
| Costimulation by the CD28 family | 77 | 2 (2.6%) | 0.0492 | 0.285 | Reactome |
| Gamma-carboxylation, transport, and amino-terminal cleavage of proteins | 11 | 1 (9.1%) | 0.0496 | 0.285 | Reactome |
| Regulation by c-FLIP | 11 | 1 (9.1%) | 0.0496 | 0.285 | Reactome |
| CASP8 activity is inhibited | 11 | 1 (9.1%) | 0.0496 | 0.285 | Reactome |
| Ubiquinone and other terpenoid-quinone biosynthesis - Homo sapiens (human) | 11 | 1 (9.1%) | 0.0496 | 0.285 | KEGG |
| Membrane Trafficking | 578 | 6 (1.0%) | 0.0498 | 0.285 | Reactome |

T2DM_mortality

| pathway name | set size | candidates contained | p-value | q-value | pathway source |
|---|---|---|---|---|---|
| Small cell lung cancer - Homo sapiens (human) | 92 | 6 (6.5%) | 4.77E-06 | 0.00272 | KEGG |
| Small cell lung cancer | 96 | 6 (6.2%) | 6.11E-06 | 0.00272 | Wikipathways |
| Ebola Virus Pathway on Host | 130 | 5 (3.8%) | 0.000379 | 0.112 | Wikipathways |
| Genes related to primary cilium development (based on CRISPR | 103 | 4 (3.9%) | 0.00146 | 0.133 | Wikipathways |
| PI3K/AKT Signaling in Cancer | 106 | 4 (3.8%) | 0.00162 | 0.133 | Reactome |
| role of mitochondria in apoptotic signaling | 13 | 2 (15.4%) | 0.00168 | 0.133 | BioCarta |
| Ciliopathies | 183 | 5 (2.7%) | 0.00177 | 0.133 | Wikipathways |
| Toxoplasmosis - Homo sapiens (human) | 112 | 4 (3.6%) | 0.00198 | 0.133 | KEGG |
| Hepatitis C and Hepatocellular Carcinoma | 56 | 3 (5.4%) | 0.00239 | 0.133 | Wikipathways |
| Focal Adhesion | 198 | 5 (2.5%) | 0.00249 | 0.133 | Wikipathways |
| Endometrial cancer - Homo sapiens (human) | 58 | 3 (5.2%) | 0.00264 | 0.133 | KEGG |
| Epstein-Barr virus infection - Homo sapiens (human) | 202 | 5 (2.5%) | 0.00266 | 0.133 | KEGG |
| Focal adhesion - Homo sapiens (human) | 201 | 5 (2.5%) | 0.00266 | 0.133 | KEGG |
| regulation of cell cycle progression by plk3 | 18 | 2 (11.1%) | 0.00325 | 0.133 | BioCarta |
| Endometrial cancer | 63 | 3 (4.8%) | 0.00334 | 0.133 | Wikipathways |
| Pathways in cancer - Homo sapiens (human) | 531 | 8 (1.5%) | 0.00357 | 0.133 | KEGG |
| prion pathway | 19 | 2 (10.5%) | 0.00363 | 0.133 | BioCarta |
| Regucalcin in proximal tubule epithelial kidney cells | 20 | 2 (10.0%) | 0.00402 | 0.133 | Wikipathways |
| Nanomaterial induced apoptosis | 20 | 2 (10.0%) | 0.00402 | 0.133 | Wikipathways |
| Measles - Homo sapiens (human) | 139 | 4 (2.9%) | 0.00431 | 0.133 | KEGG |
| Glycosphingolipid metabolism | 69 | 3 (4.3%) | 0.00432 | 0.133 | EHMN |
| Regulation by TREX1 | 1 | 1 (100.0%) | 0.00476 | 0.133 | Reactome |
| Human papillomavirus infection - Homo sapiens (human) | 331 | 6 (1.8%) | 0.00483 | 0.133 | KEGG |
| Sphingolipid Metabolism | 22 | 2 (9.1%) | 0.00485 | 0.133 | SMPDB |
| Gaucher Disease | 22 | 2 (9.1%) | 0.00485 | 0.133 | SMPDB |
| Globoid Cell Leukodystrophy | 22 | 2 (9.1%) | 0.00485 | 0.133 | SMPDB |
| Metachromatic Leukodystrophy (MLD) | 22 | 2 (9.1%) | 0.00485 | 0.133 | SMPDB |
| Fabry disease | 22 | 2 (9.1%) | 0.00485 | 0.133 | SMPDB |
| Krabbe disease | 22 | 2 (9.1%) | 0.00485 | 0.133 | SMPDB |
| Non-small cell lung cancer | 72 | 3 (4.2%) | 0.00487 | 0.133 | Wikipathways |
| Non-small cell lung cancer - Homo sapiens (human) | 72 | 3 (4.2%) | 0.00487 | 0.133 | KEGG |
| Melanoma - Homo sapiens (human) | 72 | 3 (4.2%) | 0.00487 | 0.133 | KEGG |
| Breast cancer - Homo sapiens (human) | 147 | 4 (2.7%) | 0.00526 | 0.133 | KEGG |
| Laminin interactions | 23 | 2 (8.7%) | 0.0053 | 0.133 | Reactome |
| PI3K-Akt signaling pathway | 340 | 6 (1.8%) | 0.00541 | 0.133 | Wikipathways |
| Glioma - Homo sapiens (human) | 75 | 3 (4.0%) | 0.00546 | 0.133 | KEGG |
| Gastric cancer - Homo sapiens (human) | 149 | 4 (2.7%) | 0.00552 | 0.133 | KEGG |
| Pancreatic cancer - Homo sapiens (human) | 76 | 3 (3.9%) | 0.00566 | 0.133 | KEGG |
| Breast cancer pathway | 154 | 4 (2.6%) | 0.00619 | 0.14 | Wikipathways |
| Constitutive Signaling by Aberrant PI3K in Cancer | 79 | 3 (3.8%) | 0.0063 | 0.14 | Reactome |
| PI3K-Akt signaling pathway - Homo sapiens (human) | 354 | 6 (1.7%) | 0.00657 | 0.14 | KEGG |
| Hepatitis C - Homo sapiens (human) | 157 | 4 (2.5%) | 0.00662 | 0.14 | KEGG |
| ID | 26 | 2 (7.7%) | 0.00675 | 0.14 | NetPath |
| Colorectal cancer - Homo sapiens (human) | 86 | 3 (3.5%) | 0.00797 | 0.161 | KEGG |
| Synthesis of PC | 29 | 2 (6.9%) | 0.00835 | 0.165 | Reactome |
| Pancreatic adenocarcinoma pathway | 89 | 3 (3.4%) | 0.00875 | 0.169 | Wikipathways |
| Influenza A - Homo sapiens (human) | 172 | 4 (2.3%) | 0.00908 | 0.172 | KEGG |
| Downstream signaling of activated FGFR1 | 31 | 2 (6.5%) | 0.00951 | 0.173 | Reactome |
| Activation and oligomerization of BAK protein | 2 | 1 (50.0%) | 0.00951 | 0.173 | Reactome |
| Apoptosis - multiple species - Homo sapiens (human) | 32 | 2 (6.2%) | 0.0101 | 0.18 | KEGG |
| Fc gamma R-mediated phagocytosis - Homo sapiens (human) | 97 | 3 (3.1%) | 0.0108 | 0.187 | KEGG |
| Prostate cancer - Homo sapiens (human) | 97 | 3 (3.1%) | 0.0111 | 0.187 | KEGG |
| Metabolism of lipids | 646 | 8 (1.2%) | 0.0112 | 0.187 | Reactome |
| Glycerophospholipid metabolism - Homo sapiens (human) | 98 | 3 (3.1%) | 0.0114 | 0.187 | KEGG |
| PI5P, PP2A and IER3 Regulate PI3K/AKT Signaling | 100 | 3 (3.0%) | 0.012 | 0.194 | Reactome |
| Amoebiasis - Homo sapiens (human) | 102 | 3 (3.0%) | 0.0123 | 0.196 | KEGG |
| Leukotriene metabolism | 102 | 3 (2.9%) | 0.0127 | 0.198 | EHMN |
| G13 Signaling Pathway | 38 | 2 (5.3%) | 0.0141 | 0.214 | Wikipathways |
| Focal Adhesion-PI3K-Akt-mTOR-signaling pathway | 302 | 5 (1.7%) | 0.0144 | 0.214 | Wikipathways |
| Negative regulation of the PI3K/AKT network | 107 | 3 (2.8%) | 0.0144 | 0.214 | Reactome |

| | | | | | |
|---|---|---|---|---|---|
| Oxidative Damage | 40 | 2 (5.0%) | 0.0155 | 0.223 | Wikipathways |
| Microglia Pathogen Phagocytosis Pathway | 40 | 2 (5.0%) | 0.0155 | 0.223 | Wikipathways |
| Generation of second messenger molecules | 42 | 2 (4.8%) | 0.017 | 0.233 | Reactome |
| FOXM1 transcription factor network | 42 | 2 (4.8%) | 0.017 | 0.233 | PID |
| Non-integrin membrane-ECM interactions | 42 | 2 (4.8%) | 0.017 | 0.233 | Reactome |
| Rap1 signaling pathway - Homo sapiens (human) | 210 | 4 (1.9%) | 0.0178 | 0.239 | KEGG |
| Human immunodeficiency virus 1 infection - Homo sapiens (hu | 212 | 4 (1.9%) | 0.0183 | 0.239 | KEGG |
| Phospholipid metabolism | 213 | 4 (1.9%) | 0.0186 | 0.239 | Reactome |
| Phosphorylation of proteins involved in G1/S transition by activ | 4 | 1 (25.0%) | 0.0189 | 0.239 | Reactome |
| HCN channels | 4 | 1 (25.0%) | 0.0189 | 0.239 | Reactome |
| a6b1 and a6b4 Integrin signaling | 45 | 2 (4.4%) | 0.0194 | 0.239 | PID |
| Thyroid hormone signaling pathway - Homo sapiens (human) | 121 | 3 (2.5%) | 0.0199 | 0.239 | KEGG |
| PIP3 activates AKT signaling | 218 | 4 (1.8%) | 0.0201 | 0.239 | Reactome |
| Regulation of actin cytoskeleton - Homo sapiens (human) | 218 | 4 (1.8%) | 0.0201 | 0.239 | KEGG |
| Glycosphingolipid metabolism | 46 | 2 (4.3%) | 0.0202 | 0.239 | Reactome |
| Human T-cell leukemia virus 1 infection - Homo sapiens (huma | 219 | 4 (1.8%) | 0.0204 | 0.239 | KEGG |
| agrin in postsynaptic differentiation | 47 | 2 (4.3%) | 0.021 | 0.243 | BioCarta |
| Sphingolipid metabolism - Homo sapiens (human) | 49 | 2 (4.1%) | 0.0228 | 0.256 | KEGG |
| Retrograde transport at the Trans-Golgi-Network | 49 | 2 (4.1%) | 0.0228 | 0.256 | Reactome |
| BCR | 129 | 3 (2.3%) | 0.0236 | 0.256 | NetPath |
| Activation of caspases through apoptosome-mediated cleavage | 5 | 1 (20.0%) | 0.0236 | 0.256 | Reactome |
| G2 Phase | 5 | 1 (20.0%) | 0.0236 | 0.256 | Reactome |
| Signaling by FGFR1 | 51 | 2 (3.9%) | 0.0245 | 0.263 | Reactome |
| Intrinsic Pathway for Apoptosis | 52 | 2 (3.8%) | 0.0254 | 0.269 | Reactome |
| Nuclear Envelope Breakdown | 53 | 2 (3.8%) | 0.0263 | 0.27 | Reactome |
| Apoptosis - Homo sapiens (human) | 136 | 3 (2.2%) | 0.027 | 0.27 | KEGG |
| Yersinia infection - Homo sapiens (human) | 137 | 3 (2.2%) | 0.027 | 0.27 | KEGG |
| Glycerophospholipid metabolism | 136 | 3 (2.2%) | 0.027 | 0.27 | EHMN |
| Signaling by PTK6 | 54 | 2 (3.7%) | 0.0273 | 0.27 | Reactome |
| Signaling by Non-Receptor Tyrosine Kinases | 54 | 2 (3.7%) | 0.0273 | 0.27 | Reactome |
| PTK6 Regulates Cell Cycle | 6 | 1 (16.7%) | 0.0283 | 0.27 | Reactome |
| Pre-NOTCH Processing in the Endoplasmic Reticulum | 6 | 1 (16.7%) | 0.0283 | 0.27 | Reactome |
| Phosphonate and phosphinate metabolism - Homo sapiens (hum | 6 | 1 (16.7%) | 0.0283 | 0.27 | KEGG |
| TCR | 244 | 4 (1.6%) | 0.0285 | 0.27 | NetPath |
| ECM proteoglycans | 56 | 2 (3.6%) | 0.0292 | 0.273 | Reactome |
| Caspase Cascade in Apoptosis | 57 | 2 (3.5%) | 0.0301 | 0.277 | PID |
| C21-steroid hormone biosynthesis and metabolism | 57 | 2 (3.5%) | 0.0301 | 0.277 | EHMN |
| Notch | 58 | 2 (3.4%) | 0.0311 | 0.279 | NetPath |
| VEGF signaling pathway - Homo sapiens (human) | 59 | 2 (3.4%) | 0.0321 | 0.279 | KEGG |
| Viral myocarditis - Homo sapiens (human) | 60 | 2 (3.4%) | 0.0321 | 0.279 | KEGG |
| SMAC (DIABLO) binds to IAPs | 7 | 1 (14.3%) | 0.0329 | 0.279 | Reactome |
| SMAC(DIABLO)-mediated dissociation of IAP:caspase comple | 7 | 1 (14.3%) | 0.0329 | 0.279 | Reactome |
| FGFR3b ligand binding and activation | 7 | 1 (14.3%) | 0.0329 | 0.279 | Reactome |
| Glycerophospholipid catabolism | 7 | 1 (14.3%) | 0.0329 | 0.279 | Reactome |
| phosphatidylcholine biosynthesis | 7 | 1 (14.3%) | 0.0329 | 0.279 | HumanCyc |
| Intracellular signaling by second messengers | 259 | 4 (1.5%) | 0.0348 | 0.288 | Reactome |
| Regulation of Actin Cytoskeleton | 151 | 3 (2.0%) | 0.0352 | 0.288 | Wikipathways |
| Regulation of retinoblastoma protein | 64 | 2 (3.1%) | 0.0373 | 0.288 | PID |
| GnRH secretion - Homo sapiens (human) | 64 | 2 (3.1%) | 0.0373 | 0.288 | KEGG |
| Inhibition of Signaling by Overexpressed EGFR | 8 | 1 (12.5%) | 0.0375 | 0.288 | Reactome |
| Signaling by Overexpressed Wild-Type EGFR in Cancer | 8 | 1 (12.5%) | 0.0375 | 0.288 | Reactome |
| west nile virus | 8 | 1 (12.5%) | 0.0375 | 0.288 | BioCarta |
| SMAC, XIAP-regulated apoptotic response | 8 | 1 (12.5%) | 0.0375 | 0.288 | Reactome |
| EPH-ephrin mediated repulsion of cells | 8 | 1 (12.5%) | 0.0375 | 0.288 | Reactome |
| fatty acid activation | 8 | 1 (12.5%) | 0.0375 | 0.288 | HumanCyc |
| FTO Obesity Variant Mechanism | 8 | 1 (12.5%) | 0.0375 | 0.288 | Wikipathways |
| Beta1 integrin cell surface interactions | 66 | 2 (3.0%) | 0.0394 | 0.289 | PID |
| Fc epsilon RI signaling pathway - Homo sapiens (human) | 68 | 2 (3.0%) | 0.0405 | 0.289 | KEGG |
| FGF | 67 | 2 (3.0%) | 0.0405 | 0.289 | INOH |
| Diseases of signal transduction by growth factor receptors and se | 274 | 4 (1.5%) | 0.0415 | 0.289 | Reactome |
| DNA damage response | 68 | 2 (2.9%) | 0.0416 | 0.289 | Wikipathways |
| Melanoma | 68 | 2 (2.9%) | 0.0416 | 0.289 | Wikipathways |
| Hepatitis B - Homo sapiens (human) | 162 | 3 (1.9%) | 0.042 | 0.289 | KEGG |

| | | | | | |
|---|---|---|---|---|---|
| EGFR interacts with phospholipase C-gamma | 9 | 1 (11.1%) | 0.0421 | 0.289 | **Reactome** |
| cyclin e destruction pathway | 9 | 1 (11.1%) | 0.0421 | 0.289 | **BioCarta** |
| Alpha6Beta4Integrin | 71 | 2 (2.8%) | 0.045 | 0.289 | **NetPath** |
| CDC42 signaling events | 71 | 2 (2.8%) | 0.045 | 0.289 | **PID** |
| Joubert Syndrome | 72 | 2 (2.8%) | 0.0461 | 0.289 | **Wikipathways** |
| PI3K events in ERBB4 signaling | 10 | 1 (10.0%) | 0.0466 | 0.289 | **Reactome** |
| MTHFR deficiency | 10 | 1 (10.0%) | 0.0466 | 0.289 | **Wikipathways** |
| Regulation of the apoptosome activity | 10 | 1 (10.0%) | 0.0466 | 0.289 | **Reactome** |
| Formation of apoptosome | 10 | 1 (10.0%) | 0.0466 | 0.289 | **Reactome** |
| Caspase activation via Dependence Receptors in the absence of | 10 | 1 (10.0%) | 0.0466 | 0.289 | **Reactome** |
| e2f1 destruction pathway | 10 | 1 (10.0%) | 0.0466 | 0.289 | **BioCarta** |
| Chromosomal and microsatellite instability in colorectal cancer | 73 | 2 (2.7%) | 0.0473 | 0.289 | **Wikipathways** |
| p53 signaling pathway - Homo sapiens (human) | 73 | 2 (2.7%) | 0.0473 | 0.289 | **KEGG** |
| TCR signaling | 73 | 2 (2.7%) | 0.0473 | 0.289 | **Reactome** |
| EPH-Ephrin signaling | 74 | 2 (2.7%) | 0.0485 | 0.289 | **Reactome** |

CAD_admission_freq_VAT

| pathway name | set size | candidates contained | p-value | q-value | pathway source |
|---|---|---|---|---|---|
| prion pathway | 19 | 2 (10.5%) | 0.00103 | 0.148 | BioCarta |
| NCAM signaling for neurite out-growth | 59 | 2 (3.4%) | 0.00972 | 0.148 | Reactome |
| chondroitin and dermatan biosynthesis | 6 | 1 (16.7%) | 0.0151 | 0.148 | HumanCyc |
| Anchoring fibril formation | 7 | 1 (14.3%) | 0.0176 | 0.148 | Reactome |
| APEX1-Independent Resolution of AP Sites via the Single Nucleotide Replacement Pathway | 7 | 1 (14.3%) | 0.0176 | 0.148 | Reactome |
| COPI-mediated anterograde transport | 83 | 2 (2.4%) | 0.0186 | 0.148 | Reactome |
| Metabolism | 1954 | 10 (0.5%) | 0.0194 | 0.148 | Reactome |
| Fatty Acids bound to GPR40 (FFAR1) regulate insulin secretion | 8 | 1 (12.5%) | 0.0201 | 0.148 | Reactome |
| Phase I biotransformations, non P450 | 8 | 1 (12.5%) | 0.0201 | 0.148 | Wikipathways |
| Conjugation of salicylate with glycine | 8 | 1 (12.5%) | 0.0201 | 0.148 | Reactome |
| Sepiapterin reductase deficiency | 9 | 1 (11.1%) | 0.0226 | 0.148 | SMPDB |
| Segawa syndrome | 9 | 1 (11.1%) | 0.0226 | 0.148 | SMPDB |
| Pterine Biosynthesis | 9 | 1 (11.1%) | 0.0226 | 0.148 | SMPDB |
| Dopa-responsive dystonia | 9 | 1 (11.1%) | 0.0226 | 0.148 | SMPDB |
| Hyperphenylalaniemia due to guanosine triphosphate cyclohydrolase deficiency | 9 | 1 (11.1%) | 0.0226 | 0.148 | SMPDB |
| Hyperphenylalaninemia due to 6-pyruvoyltetrahydropterin synthase deficiency (ptps) | 9 | 1 (11.1%) | 0.0226 | 0.148 | SMPDB |
| Hyperphenylalaninemia due to dhpr-deficiency | 9 | 1 (11.1%) | 0.0226 | 0.148 | SMPDB |
| Type I hemidesmosome assembly | 9 | 1 (11.1%) | 0.0226 | 0.148 | Reactome |
| Conjugation of carboxylic acids | 9 | 1 (11.1%) | 0.0226 | 0.148 | Reactome |
| Amino Acid conjugation | 9 | 1 (11.1%) | 0.0226 | 0.148 | Reactome |
| Complement system | 93 | 2 (2.2%) | 0.023 | 0.148 | Wikipathways |
| Acetylcholine regulates insulin secretion | 10 | 1 (10.0%) | 0.025 | 0.148 | Reactome |
| Amoebiasis - Homo sapiens (human) | 102 | 2 (2.0%) | 0.0269 | 0.148 | KEGG |
| Free fatty acids regulate insulin secretion | 11 | 1 (9.1%) | 0.0275 | 0.148 | Reactome |
| Alpha6 beta4 integrin-ligand interactions | 11 | 1 (9.1%) | 0.0275 | 0.148 | PID |
| Aspartate and asparagine metabolism | 12 | 1 (8.3%) | 0.03 | 0.148 | Reactome |
| D-myo-inositol (1,4,5)-trisphosphate degradation | 13 | 1 (7.7%) | 0.0324 | 0.148 | HumanCyc |
| DNA-PK pathway in nonhomologous end joining | 13 | 1 (7.7%) | 0.0324 | 0.148 | PID |
| eicosapentaenoate biosynthesis | 13 | 1 (7.7%) | 0.0324 | 0.148 | HumanCyc |
| Synthesis of IP2, IP, and Ins in the cytosol | 14 | 1 (7.1%) | 0.0349 | 0.148 | Reactome |
| Tryptophan catabolism | 14 | 1 (7.1%) | 0.0349 | 0.148 | Reactome |
| Triglyceride biosynthesis | 14 | 1 (7.1%) | 0.0349 | 0.148 | Reactome |
| γ-linolenate biosynthesis | 14 | 1 (7.1%) | 0.0349 | 0.148 | HumanCyc |
| Glycosaminoglycan metabolism | 119 | 2 (1.7%) | 0.0363 | 0.148 | Reactome |
| PAR4-mediated thrombin signaling events | 15 | 1 (6.7%) | 0.0373 | 0.148 | PID |
| 1D-myo-inositol hexakisphosphate biosynthesis II (mammalian) | 15 | 1 (6.7%) | 0.0373 | 0.148 | HumanCyc |
| D-myo-inositol (1,3,4)-trisphosphate biosynthesis | 15 | 1 (6.7%) | 0.0373 | 0.148 | HumanCyc |
| Leucine Stimulation on Insulin Signaling | 15 | 1 (6.7%) | 0.0373 | 0.148 | SMPDB |
| Synthesis of Prostaglandins (PG) and Thromboxanes (TX) | 16 | 1 (6.2%) | 0.0398 | 0.148 | Reactome |
| CRMPs in Sema3A signaling | 16 | 1 (6.2%) | 0.0398 | 0.148 | Reactome |
| Doxorubicin Metabolism Pathway | 17 | 1 (5.9%) | 0.0422 | 0.148 | SMPDB |
| Bile Acid Biosynthesis | 17 | 1 (5.9%) | 0.0422 | 0.148 | SMPDB |
| 27-Hydroxylase Deficiency | 17 | 1 (5.9%) | 0.0422 | 0.148 | SMPDB |
| Congenital Bile Acid Synthesis Defect Type II | 17 | 1 (5.9%) | 0.0422 | 0.148 | SMPDB |
| Cerebrotendinous Xanthomatosis (CTX) | 17 | 1 (5.9%) | 0.0422 | 0.148 | SMPDB |
| Zellweger Syndrome | 17 | 1 (5.9%) | 0.0422 | 0.148 | SMPDB |
| Familial Hypercholanemia (FHCA) | 17 | 1 (5.9%) | 0.0422 | 0.148 | SMPDB |
| Congenital Bile Acid Synthesis Defect Type III | 17 | 1 (5.9%) | 0.0422 | 0.148 | SMPDB |
| dermatan sulfate biosynthesis | 17 | 1 (5.9%) | 0.0422 | 0.148 | HumanCyc |
| Metabolism of Angiotensinogen to Angiotensins | 18 | 1 (5.6%) | 0.0446 | 0.148 | Reactome |
| LDL clearance | 18 | 1 (5.6%) | 0.0446 | 0.148 | Reactome |
| G alpha (i) signalling events | 316 | 3 (0.9%) | 0.0451 | 0.148 | Reactome |
| ER to Golgi Anterograde Transport | 136 | 2 (1.5%) | 0.0462 | 0.148 | Reactome |
| superpathway of D-myo-inositol (1,4,5)-trisphosphate metabolism | 20 | 1 (5.0%) | 0.0495 | 0.148 | HumanCyc |
| 3-phosphoinositide degradation | 20 | 1 (5.0%) | 0.0495 | 0.148 | HumanCyc |
| Steroid biosynthesis - Homo sapiens (human) | 20 | 1 (5.0%) | 0.0495 | 0.148 | KEGG |
| Class I peroxisomal membrane protein import | 20 | 1 (5.0%) | 0.0495 | 0.148 | Reactome |
| Glycosaminoglycan biosynthesis - chondroitin sulfate / dermatan sulfate - Homo sapiens (human) | 20 | 1 (5.0%) | 0.0495 | 0.148 | KEGG |
| Chondroitin sulfate biosynthesis | 20 | 1 (5.0%) | 0.0495 | 0.148 | Reactome |

CAD_mortality_VAT

| pathway name | set size | candidates contained | p-value | q-value | pathway source |
|---|---|---|---|---|---|
| Opsins | 10 | 2 (20.0%) | 0.000443 | 0.189 | Reactome |
| Regulation of signaling by NODAL | 12 | 2 (16.7%) | 0.000647 | 0.189 | Reactome |
| Osteoclast Signaling | 16 | 2 (12.5%) | 0.00117 | 0.214 | Wikipathways |
| Nanomaterial induced apoptosis | 20 | 2 (10.0%) | 0.00183 | 0.214 | Wikipathways |
| Signaling by NODAL | 20 | 2 (10.0%) | 0.00183 | 0.214 | Reactome |
| Apoptosis Modulation and Signaling | 92 | 3 (3.3%) | 0.00315 | 0.249 | Wikipathways |
| Interferon gamma signaling | 96 | 3 (3.1%) | 0.00355 | 0.249 | Reactome |
| the information processing pathway at the ifn beta enhancer | 29 | 2 (6.9%) | 0.00385 | 0.249 | BioCarta |
| Activation of the pre-replicative complex | 33 | 2 (6.1%) | 0.00496 | 0.249 | Reactome |
| DNA Replication Pre-Initiation | 37 | 2 (5.4%) | 0.00621 | 0.249 | Reactome |
| Activation and oligomerization of BAK protein | 2 | 1 (50.0%) | 0.00639 | 0.249 | Reactome |
| Eptifibatide Action Pathway | 2 | 1 (50.0%) | 0.00639 | 0.249 | SMPDB |
| Tirofiban Action Pathway | 2 | 1 (50.0%) | 0.00639 | 0.249 | SMPDB |
| Abciximab Action Pathway | 2 | 1 (50.0%) | 0.00639 | 0.249 | SMPDB |
| Conjugation of phenylacetate with glutamine | 2 | 1 (50.0%) | 0.00639 | 0.249 | Reactome |
| Apoptosis - Homo sapiens (human) | 136 | 3 (2.2%) | 0.00932 | 0.278 | KEGG |
| Phenylacetate Metabolism | 3 | 1 (33.3%) | 0.00957 | 0.278 | SMPDB |
| Signaling events mediated by PTP1B | 52 | 2 (3.8%) | 0.012 | 0.278 | PID |
| IRS activation | 4 | 1 (25.0%) | 0.0127 | 0.278 | Reactome |
| dermatan sulfate biosynthesis (late stages) | 4 | 1 (25.0%) | 0.0127 | 0.278 | HumanCyc |
| Lipoic acid metabolism - Homo sapiens (human) | 4 | 1 (25.0%) | 0.0127 | 0.278 | KEGG |
| Interferon Signaling | 162 | 3 (1.9%) | 0.0149 | 0.278 | Reactome |
| Oxidation by Cytochrome P450 | 62 | 2 (3.2%) | 0.0168 | 0.278 | Wikipathways |
| Integrins in angiogenesis | 63 | 2 (3.2%) | 0.0173 | 0.278 | PID |
| Influenza A - Homo sapiens (human) | 172 | 3 (1.7%) | 0.0175 | 0.278 | KEGG |
| IL4-mediated signaling events | 65 | 2 (3.1%) | 0.0183 | 0.278 | PID |
| Gap-filling DNA repair synthesis and ligation in TC-NER | 66 | 2 (3.0%) | 0.0189 | 0.278 | Reactome |
| Activated NTRK3 signals through PI3K | 6 | 1 (16.7%) | 0.0191 | 0.278 | Reactome |
| Conjugation of benzoate with glycine | 6 | 1 (16.7%) | 0.0191 | 0.278 | Reactome |
| Dual incision in TC-NER | 67 | 2 (3.0%) | 0.0194 | 0.278 | Reactome |
| Vitamin B2 (riboflavin) metabolism | 7 | 1 (14.3%) | 0.0222 | 0.278 | Reactome |
| SOS-mediated signalling | 7 | 1 (14.3%) | 0.0222 | 0.278 | Reactome |
| TRAIL signaling | 7 | 1 (14.3%) | 0.0222 | 0.278 | Reactome |
| Primary focal segmental glomerulosclerosis (FSGS) | 73 | 2 (2.7%) | 0.0228 | 0.278 | Wikipathways |
| Synthesis of DNA | 76 | 2 (2.6%) | 0.0246 | 0.278 | Reactome |
| G1/S Transition | 76 | 2 (2.6%) | 0.0246 | 0.278 | Reactome |
| Riboflavin metabolism - Homo sapiens (human) | 8 | 1 (12.5%) | 0.0253 | 0.278 | KEGG |
| CDC6 association with the ORC:origin complex | 8 | 1 (12.5%) | 0.0253 | 0.278 | Reactome |
| Vitamin C (ascorbate) metabolism | 8 | 1 (12.5%) | 0.0253 | 0.278 | Reactome |
| Hypothetical Craniofacial Development Pathway | 8 | 1 (12.5%) | 0.0253 | 0.278 | Wikipathways |
| DNA replication initiation | 8 | 1 (12.5%) | 0.0253 | 0.278 | Reactome |
| Mitochondrial Beta-Oxidation of Medium Chain Saturated Fatty Acids | 8 | 1 (12.5%) | 0.0253 | 0.278 | SMPDB |
| Butyrate Metabolism | 8 | 1 (12.5%) | 0.0253 | 0.278 | SMPDB |
| Eicosanoid metabolism via cytochrome P450 monooxygenases (CYP) pathway | 8 | 1 (12.5%) | 0.0253 | 0.278 | Wikipathways |
| Transcription-Coupled Nucleotide Excision Repair (TC-NER) | 79 | 2 (2.5%) | 0.0264 | 0.278 | Reactome |
| DNA Replication | 81 | 2 (2.5%) | 0.0276 | 0.278 | Reactome |
| Signaling by NTRK3 (TRKC) | 9 | 1 (11.1%) | 0.0284 | 0.278 | Reactome |
| PI3K/AKT activation | 9 | 1 (11.1%) | 0.0284 | 0.278 | Reactome |
| Conjugation of carboxylic acids | 9 | 1 (11.1%) | 0.0284 | 0.278 | Reactome |
| Amino Acid conjugation | 9 | 1 (11.1%) | 0.0284 | 0.278 | Reactome |
| Cell surface interactions at the vascular wall | 215 | 3 (1.4%) | 0.0312 | 0.278 | Reactome |
| Apoptosis | 87 | 2 (2.3%) | 0.0315 | 0.278 | Wikipathways |
| IL-13 signaling | 10 | 1 (10.0%) | 0.0316 | 0.278 | INOH |
| Signal attenuation | 10 | 1 (10.0%) | 0.0316 | 0.278 | Reactome |
| Dimerization of procaspase-8 | 10 | 1 (10.0%) | 0.0316 | 0.278 | Reactome |
| Mitochondrial translation termination | 88 | 2 (2.3%) | 0.0322 | 0.278 | Reactome |
| Biological oxidations | 219 | 3 (1.4%) | 0.0327 | 0.278 | Reactome |
| Highly calcium permeable postsynaptic nicotinic acetylcholine receptors | 11 | 1 (9.1%) | 0.0347 | 0.278 | Reactome |
| apoptotic dna-fragmentation and tissue homeostasis | 11 | 1 (9.1%) | 0.0347 | 0.278 | BioCarta |
| Signaling by Leptin | 11 | 1 (9.1%) | 0.0347 | 0.278 | Reactome |
| insulin signaling pathway | 11 | 1 (9.1%) | 0.0347 | 0.278 | BioCarta |
| Serotonin Transporter Activity | 11 | 1 (9.1%) | 0.0347 | 0.278 | Wikipathways |

| | | | | | |
|---|---|---|---|---|---|
| Regulation by c-FLIP | 11 | 1 (9.1%) | 0.0347 | 0.278 | Reactome |
| CASP8 activity is inhibited | 11 | 1 (9.1%) | 0.0347 | 0.278 | Reactome |
| Dermatan sulfate biosynthesis | 11 | 1 (9.1%) | 0.0347 | 0.278 | Reactome |
| Valproic Acid Metabolism Pathway | 11 | 1 (9.1%) | 0.0347 | 0.278 | SMPDB |
| Rheumatoid arthritis - Homo sapiens (human) | 93 | 2 (2.2%) | 0.0349 | 0.278 | KEGG |
| Mitochondrial translation | 94 | 2 (2.1%) | 0.0363 | 0.278 | Reactome |
| carm1 and regulation of the estrogen receptor | 12 | 1 (8.3%) | 0.0378 | 0.278 | BioCarta |
| PECAM1 interactions | 12 | 1 (8.3%) | 0.0378 | 0.278 | Reactome |
| POU5F1 (OCT4), SOX2, NANOG activate genes related to proliferation | 12 | 1 (8.3%) | 0.0378 | 0.278 | Reactome |
| TP53 Regulates Transcription of Death Receptors and Ligands | 12 | 1 (8.3%) | 0.0378 | 0.278 | Reactome |
| Eicosanoids | 12 | 1 (8.3%) | 0.0378 | 0.278 | Reactome |
| Miscellaneous substrates | 12 | 1 (8.3%) | 0.0378 | 0.278 | Reactome |
| Nucleotide Excision Repair | 97 | 2 (2.1%) | 0.0385 | 0.278 | Reactome |
| Hematopoietic cell lineage - Homo sapiens (human) | 99 | 2 (2.0%) | 0.0392 | 0.278 | KEGG |
| role of mitochondria in apoptotic signaling | 13 | 1 (7.7%) | 0.0408 | 0.278 | BioCarta |
| multiple antiapoptotic pathways from igf-1r signaling lead to bad phosphorylation | 13 | 1 (7.7%) | 0.0408 | 0.278 | BioCarta |
| Osteopontin Signaling | 13 | 1 (7.7%) | 0.0408 | 0.278 | Wikipathways |
| MFAP5-mediated ovarian cancer cell motility and invasiveness | 13 | 1 (7.7%) | 0.0408 | 0.278 | Wikipathways |
| D-myo-inositol (1,4,5)-trisphosphate degradation | 13 | 1 (7.7%) | 0.0408 | 0.278 | HumanCyc |
| chondroitin sulfate biosynthesis (late stages) | 13 | 1 (7.7%) | 0.0408 | 0.278 | HumanCyc |
| Valproic acid pathway | 13 | 1 (7.7%) | 0.0408 | 0.278 | Wikipathways |
| eicosapentaenoate biosynthesis | 13 | 1 (7.7%) | 0.0408 | 0.278 | HumanCyc |
| Phase I - Functionalization of compounds | 101 | 2 (2.0%) | 0.0414 | 0.278 | Reactome |
| S Phase | 104 | 2 (1.9%) | 0.0436 | 0.278 | Reactome |
| Mitotic G1 phase and G1/S transition | 104 | 2 (1.9%) | 0.0436 | 0.278 | Reactome |
| CDT1 association with the CDC6:ORC:origin complex | 14 | 1 (7.1%) | 0.0439 | 0.278 | Reactome |
| Postsynaptic nicotinic acetylcholine receptors | 14 | 1 (7.1%) | 0.0439 | 0.278 | Reactome |
| Acetylcholine binding and downstream events | 14 | 1 (7.1%) | 0.0439 | 0.278 | Reactome |
| Osteoblast Signaling | 14 | 1 (7.1%) | 0.0439 | 0.278 | Wikipathways |
| γ-linolenate biosynthesis | 14 | 1 (7.1%) | 0.0439 | 0.278 | HumanCyc |
| Leucine Stimulation on Insulin Signaling | 15 | 1 (6.7%) | 0.047 | 0.278 | SMPDB |
| p130Cas linkage to MAPK signaling for integrins | 15 | 1 (6.7%) | 0.047 | 0.278 | Reactome |
| GRB2:SOS provides linkage to MAPK signaling for Integrins | 15 | 1 (6.7%) | 0.047 | 0.278 | Reactome |
| 1D-myo-inositol hexakisphosphate biosynthesis II (mammalian) | 15 | 1 (6.7%) | 0.047 | 0.278 | HumanCyc |
| D-myo-inositol (1,3,4)-trisphosphate biosynthesis | 15 | 1 (6.7%) | 0.047 | 0.278 | HumanCyc |
| Fatty acids | 15 | 1 (6.7%) | 0.047 | 0.278 | Reactome |
| DNA damage response (only ATM dependent) | 110 | 2 (1.8%) | 0.0483 | 0.278 | Wikipathways |
| Metabolism of water-soluble vitamins and cofactors | 111 | 2 (1.8%) | 0.0491 | 0.278 | Reactome |

| Table S6 Genes identified from TWAS analysis with adjustment for estimated cell proportions | | | | | | | |
|---|---|---|---|---|---|---|---|
| tissue | disorder | Genes | Estimates_cell prop | Pvalues_cell prop | Pvalue_Bonferroni_cell prop | Pvalue_BH_cell prop | gene_name |
| Frontal Cortex | Depression | ENSG00000137185.7 | -0.007446263 | 1.27E-07 | 0.000255456 | 0.000255456 | ZSCAN9 |
| Frontal Cortex | Depression | ENSG00000186470.9 | 0.006024303 | 6.26E-06 | 0.012601627 | 0.006300813 | BTN3A2 |
| Frontal Cortex | Depression | ENSG00000244731.3 | 0.007779012 | 6.25E-05 | 0.125838035 | 0.041946012 | C4A |
| Pancreas | T1DM | ENSG00000011523.9 | 0.001320601 | 0.000993831 | 1 | 0.04640285 | CEP68 |
| Pancreas | T1DM | ENSG00000076650.2 | 0.003483687 | 0.001078733 | 1 | 0.049467622 | GPATCH1 |
| Pancreas | T1DM | ENSG00000099785.6 | 0.001726473 | 0.000873976 | 1 | 0.042346594 | MARCH2 |
| Pancreas | T1DM | ENSG00000124508.12 | -0.002359585 | 0.000255627 | 0.656451124 | 0.014587803 | BTN2A2 |
| Pancreas | T1DM | ENSG00000137288.5 | 0.002643162 | 0.000293663 | 0.754127073 | 0.016394067 | UQCC2 |
| Pancreas | T1DM | ENSG00000167280.12 | 0.001354907 | 0.000775642 | 1 | 0.03830479 | ENGASE |
| Pancreas | T1DM | ENSG00000179344.12 | -0.005476016 | 2.96E-21 | 7.61E-18 | 6.34E-19 | HLA-DQB1 |
| Pancreas | T1DM | ENSG00000182477.5 | 0.001240491 | 5.71E-05 | 0.146623614 | 0.003491038 | OR2B8P |
| Pancreas | T1DM | ENSG00000186470.9 | -0.000958064 | 1.28E-05 | 0.032914354 | 0.000843958 | BTN3A2 |
| Pancreas | T1DM | ENSG00000189298.9 | -0.004961783 | 1.41E-09 | 3.62E-06 | 1.58E-07 | ZKSCAN3 |
| Pancreas | T1DM | ENSG00000196260.3 | -0.003202614 | 3.94E-11 | 1.01E-07 | 5.06E-09 | SFTA2 |
| Pancreas | T1DM | ENSG00000197062.7 | -0.002154522 | 2.03E-08 | 5.22E-05 | 2.01E-06 | RP5-874C20.3 |
| Pancreas | T1DM | ENSG00000197728.5 | 0.00118086 | 0.000466793 | 1 | 0.024973406 | RPS26 |
| Pancreas | T1DM | ENSG00000198502.5 | -0.003063224 | 1.81E-39 | 4.64E-36 | 1.16E-36 | HLA-DRB5 |
| Pancreas | T1DM | ENSG00000204228.3 | 0.0047664 | 3.40E-28 | 8.73E-25 | 1.25E-25 | HSD17B8 |
| Pancreas | T1DM | ENSG00000204257.10 | 0.0031622 | 1.01E-29 | 2.58E-26 | 5.17E-27 | HLA-DMA |
| Pancreas | T1DM | ENSG00000204301.5 | -0.006629097 | 9.03E-26 | 2.32E-22 | 2.32E-23 | NOTCH4 |
| Pancreas | T1DM | ENSG00000204305.9 | 0.003713229 | 0.000235442 | 0.604614121 | 0.01374123 | AGER |
| Pancreas | T1DM | ENSG00000204308.6 | -0.002142507 | 1.90E-14 | 4.88E-11 | 2.87E-12 | RNF5 |
| Pancreas | T1DM | ENSG00000204338.4 | -0.003504122 | 4.50E-29 | 1.16E-25 | 1.93E-26 | CYP21A1P |
| Pancreas | T1DM | ENSG00000204351.7 | -0.002830754 | 2.10E-23 | 5.40E-20 | 4.91E-21 | SKIV2L |
| Pancreas | T1DM | ENSG00000204371.7 | 0.004453456 | 2.63E-05 | 0.067635975 | 0.001690899 | EHMT2 |
| Pancreas | T1DM | ENSG00000204385.6 | -0.001364878 | 0.000328449 | 0.843457414 | 0.017945902 | SLC44A4 |
| Pancreas | T1DM | ENSG00000204520.8 | 0.001432358 | 3.86E-07 | 0.000990548 | 3.20E-05 | MICA |
| Pancreas | T1DM | ENSG00000204525.10 | 0.000970859 | 3.03E-05 | 0.077936275 | 0.001900885 | HLA-C |
| Pancreas | T1DM | ENSG00000204536.9 | 0.003534407 | 5.25E-09 | 1.35E-05 | 5.40E-07 | CCHCR1 |
| Pancreas | T1DM | ENSG00000204540.6 | -0.002118881 | 1.52E-11 | 3.91E-08 | 2.06E-09 | PSORS1C1 |
| Pancreas | T1DM | ENSG00000204616.6 | -0.002149228 | 1.71E-07 | 0.000438408 | 1.51E-05 | TRIM31 |
| Pancreas | T1DM | ENSG00000204618.4 | 0.004892466 | 6.37E-16 | 1.64E-12 | 1.02E-13 | RNF39 |
| Pancreas | T1DM | ENSG00000204644.5 | 0.001914015 | 1.84E-07 | 0.000472293 | 1.57E-05 | ZFP57 |
| Pancreas | T1DM | ENSG00000206344.6 | 0.002092894 | 3.72E-06 | 0.009546279 | 0.000260615 | HCG27 |
| Pancreas | T1DM | ENSG00000213676.6 | 0.002074921 | 4.48E-09 | 1.15E-05 | 4.80E-07 | ATF6B |
| Pancreas | T1DM | ENSG00000213760.6 | -0.002161277 | 0.000760322 | 1 | 0.038284453 | ATP6V1G2 |
| Pancreas | T1DM | ENSG00000219392.1 | -0.002945179 | 3.84E-06 | 0.009864654 | 0.000260615 | RP1-265C24.5 |
| Pancreas | T1DM | ENSG00000221988.8 | 0.00623609 | 8.79E-11 | 2.26E-07 | 1.03E-08 | PPT2 |
| Pancreas | T1DM | ENSG00000224389.4 | 0.005486048 | 3.86E-06 | 0.009903386 | 0.000260615 | C4B |
| Pancreas | T1DM | ENSG00000224796.1 | 0.005930985 | 1.33E-17 | 3.42E-14 | 2.28E-15 | RPL32P1 |
| Pancreas | T1DM | ENSG00000229391.3 | 0.005878019 | 1.51E-53 | 3.88E-50 | 3.88E-50 | HLA-DRB6 |
| Pancreas | T1DM | ENSG00000229759.1 | -0.000990963 | 0.00096253 | 1 | 0.045773668 | MRPS18AP1 |
| Pancreas | T1DM | ENSG00000229836.1 | 0.002601908 | 2.55E-14 | 6.56E-11 | 3.64E-12 | XXbac-BPG248L24.10 |
| Pancreas | T1DM | ENSG00000231402.1 | 0.001939407 | 6.79E-11 | 1.74E-07 | 8.30E-09 | WASF5P |
| Pancreas | T1DM | ENSG00000232629.4 | 0.004904399 | 1.57E-47 | 4.02E-44 | 2.01E-44 | HLA-DQB2 |
| Pancreas | T1DM | ENSG00000234127.4 | -0.001865492 | 0.000676758 | 1 | 0.034758299 | TRIM26 |
| Pancreas | T1DM | ENSG00000235109.3 | 0.001098206 | 1.96E-06 | 0.005031361 | 0.000143753 | ZSCAN31 |
| Pancreas | T1DM | ENSG00000235863.2 | -0.001878393 | 0.000528441 | 1 | 0.027694627 | B3GALT4 |
| Pancreas | T1DM | ENSG00000236529.1 | 0.005179134 | 9.10E-05 | 0.233721676 | 0.005435388 | RP13-254B10.1 |
| Pancreas | T1DM | ENSG00000237541.3 | 0.005887442 | 6.44E-27 | 1.65E-23 | 1.84E-24 | HLA-DQA2 |
| Pancreas | T1DM | ENSG00000237669.1 | 0.002010631 | 7.94E-07 | 0.002039598 | 6.37E-05 | HCG4P3 |
| Pancreas | T1DM | ENSG00000240065.3 | 0.008117937 | 6.51E-20 | 1.67E-16 | 1.29E-17 | PSMB9 |
| Pancreas | T1DM | ENSG00000241106.2 | 0.002981786 | 1.15E-19 | 2.94E-16 | 2.10E-17 | HLA-DOB |
| Pancreas | T1DM | ENSG00000244731.3 | -0.005878779 | 4.52E-46 | 1.16E-42 | 3.87E-43 | C4A |
| Pancreas | T1DM | ENSG00000256166.1 | 0.004916826 | 6.19E-27 | 1.59E-23 | 1.84E-24 | XXbac-BPG248L24.13 |
| Pancreas | T1DM | ENSG00000260302.1 | 0.003374929 | 1.17E-06 | 0.00301007 | 9.12E-05 | RP11-973H7.1 |
| Pancreas | T1DM | ENSG00000271821.1 | 0.003872823 | 1.56E-06 | 0.004004901 | 0.000117791 | XXbac-BPG299F13.14 |
| Pancreas | T1DM | ENSG00000272221.1 | 0.001179828 | 9.10E-08 | 0.000233599 | 8.34E-06 | XXbac-BPG181B23.7 |
| Pancreas | T1DM | ENSG00000273340.1 | -0.001647747 | 2.43E-08 | 6.24E-05 | 2.31E-06 | MICE |
| Pancreas | T2DM | ENSG00000004534.10 | -0.003393908 | 2.04E-07 | 0.000523424 | 5.23E-05 | RBM6 |
| Pancreas | T2DM | ENSG00000009950.11 | 0.005485409 | 7.27E-05 | 0.186710079 | 0.006438279 | MLXIPL |
| Pancreas | T2DM | ENSG00000010310.4 | -0.00764268 | 0.00119303 | 1 | 0.040849362 | GIPR |
| Pancreas | T2DM | ENSG00000011523.9 | 0.005797831 | 2.76E-08 | 7.08E-05 | 8.85E-06 | CEP68 |
| Pancreas | T2DM | ENSG00000030110.8 | 0.011017341 | 2.02E-06 | 0.00519481 | 0.000431985 | BAK1 |
| Pancreas | T2DM | ENSG00000066294.10 | 0.005912261 | 0.000436359 | 1 | 0.020225759 | CD84 |
| Pancreas | T2DM | ENSG00000071564.10 | -0.010878643 | 3.16E-05 | 0.081240829 | 0.003692765 | TCF3 |
| Pancreas | T2DM | ENSG00000073849.10 | 0.008251104 | 0.000914691 | 1 | 0.035306133 | ST6GAL1 |
| Pancreas | T2DM | ENSG00000075413.13 | -0.015487526 | 1.45E-08 | 3.73E-05 | 6.22E-06 | MARK3 |
| Pancreas | T2DM | ENSG00000095485.12 | -0.002744305 | 9.36E-05 | 0.240323682 | 0.007510115 | CWF19L1 |
| Pancreas | T2DM | ENSG00000103479.10 | 0.005922199 | 0.00093469 | 1 | 0.035306133 | RBL2 |
| Pancreas | T2DM | ENSG00000103512.10 | 0.008459255 | 0.001261266 | 1 | 0.041428975 | NOMO1 |
| Pancreas | T2DM | ENSG00000108272.9 | 0.008331831 | 0.000663435 | 1 | 0.028395025 | DHRS11 |
| Pancreas | T2DM | ENSG00000108786.6 | -0.004851247 | 0.00044106 | 1 | 0.020225759 | HSD17B1 |
| Pancreas | T2DM | ENSG00000108797.7 | 0.007219314 | 7.79E-05 | 0.199995047 | 0.006666502 | CNTNAP1 |
| Pancreas | T2DM | ENSG00000111215.7 | 0.005412917 | 0.001254073 | 1 | 0.041428975 | PRR4 |

| | | | | | | | |
|---|---|---|---|---|---|---|---|
| Pancreas | T2DM | ENSG00000111328.2 | 0.005531994 | 0.000100877 | 0.259052616 | 0.007669642 | CDK2AP1 |
| Pancreas | T2DM | ENSG00000112992.12 | -0.005734172 | 0.000396943 | 1 | 0.01997046 | NNT |
| Pancreas | T2DM | ENSG00000118997.9 | -0.005724881 | 7.19E-05 | 0.184518162 | 0.006438279 | DNAH7 |
| Pancreas | T2DM | ENSG00000122025.10 | 0.010222286 | 0.000313939 | 0.806195284 | 0.017915451 | FLT3 |
| Pancreas | T2DM | ENSG00000125510.11 | -0.006830479 | 0.000348317 | 0.894477061 | 0.018634939 | OPRL1 |
| Pancreas | T2DM | ENSG00000127947.11 | -0.006256751 | 0.001469256 | 1 | 0.045626317 | PTPN12 |
| Pancreas | T2DM | ENSG00000127957.12 | -0.009593219 | 0.000181782 | 0.466817424 | 0.011385791 | PMS2P3 |
| Pancreas | T2DM | ENSG00000129933.16 | -0.008815974 | 7.45E-06 | 0.019127487 | 0.001195468 | MAU2 |
| Pancreas | T2DM | ENSG00000129990.10 | 0.006880814 | 0.000934898 | 1 | 0.035306133 | SYT5 |
| Pancreas | T2DM | ENSG00000134250.13 | -0.007991324 | 2.28E-08 | 5.85E-05 | 8.36E-06 | NOTCH2 |
| Pancreas | T2DM | ENSG00000137731.9 | -0.009595427 | 0.000810902 | 1 | 0.03305389 | FXYD2 |
| Pancreas | T2DM | ENSG00000140400.10 | -0.002869621 | 1.43E-05 | 0.036608117 | 0.002096587 | MAN2C1 |
| Pancreas | T2DM | ENSG00000141699.6 | 0.007479622 | 0.001499514 | 1 | 0.045626317 | FAM134C |
| Pancreas | T2DM | ENSG00000143442.17 | 0.006702144 | 0.000420847 | 1 | 0.020225759 | POGZ |
| Pancreas | T2DM | ENSG00000144619.10 | -0.005594357 | 0.000623919 | 1 | 0.027624536 | CNTN4 |
| Pancreas | T2DM | ENSG00000146830.9 | -0.010270288 | 0.000309471 | 0.794721893 | 0.017915451 | GIGYF1 |
| Pancreas | T2DM | ENSG00000148396.14 | 0.005482624 | 0.000902131 | 1 | 0.035306133 | SEC16A |
| Pancreas | T2DM | ENSG00000149084.7 | -0.002294707 | 0.000124611 | 0.320000702 | 0.009142877 | HSD17B12 |
| Pancreas | T2DM | ENSG00000149089.8 | -0.002348758 | 0.000404386 | 1 | 0.01997046 | APIP |
| Pancreas | T2DM | ENSG00000152104.7 | -0.0067859 | 0.000325618 | 0.836187396 | 0.018177987 | PTPN14 |
| Pancreas | T2DM | ENSG00000153814.7 | -0.014963784 | 1.35E-16 | 3.46E-13 | 1.73E-13 | JAZF1 |
| Pancreas | T2DM | ENSG00000154222.10 | -0.003491239 | 0.001510217 | 1 | 0.045626317 | CC2D1B |
| Pancreas | T2DM | ENSG00000158008.5 | -0.007988942 | 0.001004224 | 1 | 0.036840659 | EXTL1 |
| Pancreas | T2DM | ENSG00000160161.5 | -0.00635158 | 9.02E-05 | 0.231587465 | 0.007470563 | CILP2 |
| Pancreas | T2DM | ENSG00000165806.15 | 0.004285223 | 0.00016621 | 0.426826694 | 0.010998318 | CASP7 |
| Pancreas | T2DM | ENSG00000166165.8 | -0.006736253 | 0.000782146 | 1 | 0.032927049 | CKB |
| Pancreas | T2DM | ENSG00000166228.4 | -0.005793195 | 0.000914678 | 1 | 0.035306133 | PCBD1 |
| Pancreas | T2DM | ENSG00000169592.10 | -0.006715716 | 3.58E-05 | 0.091878487 | 0.003994717 | INO80E |
| Pancreas | T2DM | ENSG00000171282.9 | 0.009477228 | 0.001166528 | 1 | 0.040481675 | RP11-1055B8.7 |
| Pancreas | T2DM | ENSG00000172543.3 | -0.002823507 | 0.001482795 | 1 | 0.045626317 | CTSW |
| Pancreas | T2DM | ENSG00000175287.14 | 0.005441456 | 0.000639173 | 1 | 0.027820271 | PHYHD1 |
| Pancreas | T2DM | ENSG00000177508.11 | 0.034536061 | 1.01E-24 | 2.59E-21 | 2.59E-21 | IRX3 |
| Pancreas | T2DM | ENSG00000179115.6 | -0.009705202 | 6.68E-05 | 0.171549729 | 0.006353694 | FARSA |
| Pancreas | T2DM | ENSG00000179344.12 | -0.005587074 | 0.000205921 | 0.528805957 | 0.012297813 | HLA-DQB1 |
| Pancreas | T2DM | ENSG00000182179.6 | -0.006125673 | 3.55E-06 | 0.00911166 | 0.000607444 | UBA7 |
| Pancreas | T2DM | ENSG00000182196.9 | 0.006299 | 0.001046325 | 1 | 0.037844535 | ARL6IP4 |
| Pancreas | T2DM | ENSG00000182853.7 | 0.009116853 | 0.000428728 | 1 | 0.020225759 | VMO1 |
| Pancreas | T2DM | ENSG00000182986.8 | 0.002505139 | 0.00049976 | 1 | 0.022515516 | ZNF320 |
| Pancreas | T2DM | ENSG00000196126.6 | -0.007545411 | 2.19E-06 | 0.005615803 | 0.000431985 | HLA-DRB1 |
| Pancreas | T2DM | ENSG00000197181.7 | -0.006174536 | 0.000981695 | 1 | 0.036536121 | PIWIL2 |
| Pancreas | T2DM | ENSG00000198231.8 | 0.005185353 | 0.001290622 | 1 | 0.041428975 | DDX42 |
| Pancreas | T2DM | ENSG00000204301.5 | -0.008497019 | 2.33E-07 | 0.00059716 | 5.43E-05 | NOTCH4 |
| Pancreas | T2DM | ENSG00000204308.6 | -0.003071697 | 2.44E-05 | 0.06277343 | 0.003239188 | RNF5 |
| Pancreas | T2DM | ENSG00000204618.4 | 0.00587238 | 0.000192334 | 0.493914241 | 0.011759863 | RNF39 |
| Pancreas | T2DM | ENSG00000206344.6 | 0.004772884 | 5.00E-05 | 0.128287656 | 0.005131506 | HCG27 |
| Pancreas | T2DM | ENSG00000212127.5 | 0.004598987 | 0.001135688 | 1 | 0.040083532 | TAS2R14 |
| Pancreas | T2DM | ENSG00000213221.4 | 0.003469077 | 0.000156025 | 0.400672926 | 0.010998318 | DNLZ |
| Pancreas | T2DM | ENSG00000213676.6 | 0.003302169 | 0.000333084 | 0.855358434 | 0.018199116 | ATF6B |
| Pancreas | T2DM | ENSG00000221988.8 | 0.010536438 | 2.52E-05 | 0.064783756 | 0.003239188 | PPT2 |
| Pancreas | T2DM | ENSG00000226232.4 | -0.005851814 | 0.000172863 | 0.443911521 | 0.011097788 | RP11-419C5.2 |
| Pancreas | T2DM | ENSG00000229358.3 | 0.008032456 | 0.001283993 | 1 | 0.041428975 | DPY19L1P1 |
| Pancreas | T2DM | ENSG00000229391.3 | 0.005808094 | 4.89E-09 | 1.26E-05 | 3.14E-06 | HLA-DRB6 |
| Pancreas | T2DM | ENSG00000232629.4 | 0.003697468 | 2.71E-05 | 0.069568765 | 0.003312798 | HLA-DQB2 |
| Pancreas | T2DM | ENSG00000232709.1 | 0.01625128 | 1.64E-07 | 0.000420512 | 4.67E-05 | MARK2P9 |
| Pancreas | T2DM | ENSG00000233609.3 | 0.006472533 | 0.001447058 | 1 | 0.045626317 | RP11-62H7.2 |
| Pancreas | T2DM | ENSG00000234745.5 | 0.003641763 | 0.00016404 | 0.421253663 | 0.010998318 | HLA-B |
| Pancreas | T2DM | ENSG00000237541.3 | 0.008534871 | 2.15E-09 | 5.52E-06 | 1.84E-06 | HLA-DQA2 |
| Pancreas | T2DM | ENSG00000242142.1 | 0.005630956 | 1.47E-05 | 0.037738565 | 0.002096587 | SERBP1P3 |
| Pancreas | T2DM | ENSG00000243753.1 | 0.003968251 | 0.000167031 | 0.428934408 | 0.010998318 | HLA-L |
| Pancreas | T2DM | ENSG00000243789.6 | 0.010325273 | 3.85E-05 | 0.098896375 | 0.004120682 | JMJD7 |
| Pancreas | T2DM | ENSG00000244731.3 | -0.003821207 | 0.000370817 | 0.952258785 | 0.019433853 | C4A |
| Pancreas | T2DM | ENSG00000247157.2 | 0.002652802 | 0.001267528 | 1 | 0.041428975 | RP11-434C1.1 |
| Pancreas | T2DM | ENSG00000248278.1 | 0.010742648 | 7.85E-09 | 2.02E-05 | 4.03E-06 | SUMO2P17 |
| Pancreas | T2DM | ENSG00000251417.1 | -0.011279226 | 0.000101545 | 0.260767811 | 0.007669642 | RP11-1348G14.4 |
| Pancreas | T2DM | ENSG00000255837.1 | 0.005339102 | 0.000390917 | 1 | 0.01997046 | TAS2R20 |
| Pancreas | T2DM | ENSG00000260274.1 | -0.00863994 | 3.34E-06 | 0.008587928 | 0.000607444 | RP11-817O13.8 |
| Pancreas | T2DM | ENSG00000260814.2 | -0.009833664 | 6.40E-05 | 0.164258423 | 0.006317632 | RP11-107F6.3 |
| Pancreas | T2DM | ENSG00000260911.1 | 0.003020388 | 0.001139446 | 1 | 0.040083532 | RP11-196G11.2 |
| Pancreas | T2DM | ENSG00000268225.1 | 0.005456772 | 0.00080574 | 1 | 0.03305389 | CTD-2331H12.5 |
| Pancreas | T2DM_admission | ENSG00000004534.10 | -0.004083911 | 5.37E-08 | 0.000137948 | 1.25E-05 | RBM6 |
| Pancreas | T2DM_admission | ENSG00000009950.11 | 0.006173441 | 0.000102825 | 0.26405371 | 0.005063095 | MLXIPL |
| Pancreas | T2DM_admission | ENSG00000010310.4 | -0.012730988 | 2.66E-06 | 0.00682726 | 0.00031033 | GIPR |
| Pancreas | T2DM_admission | ENSG00000011523.9 | 0.00800987 | 2.44E-11 | 6.25E-08 | 1.56E-08 | CEP68 |
| Pancreas | T2DM_admission | ENSG00000030110.8 | 0.015031688 | 1.71E-08 | 4.40E-05 | 5.50E-06 | BAK1 |
| Pancreas | T2DM_admission | ENSG00000066294.10 | 0.007160392 | 0.000211199 | 0.542359703 | 0.00904011 | CD84 |
| Pancreas | T2DM_admission | ENSG00000070182.13 | 0.007574605 | 0.001475735 | 1 | 0.033836506 | SPTB |
| Pancreas | T2DM_admission | ENSG00000071564.10 | -0.012807298 | 2.03E-05 | 0.052127595 | 0.001468422 | TCF3 |
| Pancreas | T2DM_admission | ENSG00000073849.10 | 0.010792528 | 0.000161688 | 0.415215244 | 0.007414558 | ST6GAL1 |
| Pancreas | T2DM_admission | ENSG00000075413.13 | -0.01756302 | 2.27E-08 | 5.83E-05 | 6.47E-06 | MARK3 |
| Pancreas | T2DM_admission | ENSG00000095485.12 | -0.002692489 | 0.000855789 | 1 | 0.022656349 | CWF19L1 |
| Pancreas | T2DM_admission | ENSG00000103479.10 | 0.007222671 | 0.000446474 | 1 | 0.014331807 | RBL2 |

| | | | | | | | |
|---|---|---|---|---|---|---|---|
| Pancreas | T2DM_admission | ENSG00000106460.14 | 0.014501798 | 0.000110616 | 0.284062315 | 0.005260413 | TMEM106B |
| Pancreas | T2DM_admission | ENSG00000108272.9 | 0.009844182 | 0.000467426 | 1 | 0.014819145 | DHRS11 |
| Pancreas | T2DM_admission | ENSG00000108786.6 | -0.006265681 | 7.88E-05 | 0.20223339 | 0.004302838 | HSD17B1 |
| Pancreas | T2DM_admission | ENSG00000108797.7 | 0.009343987 | 8.66E-06 | 0.022243711 | 0.000794418 | CNTNAP1 |
| Pancreas | T2DM_admission | ENSG00000109332.15 | 0.008572443 | 0.001747972 | 1 | 0.038040607 | UBE2D3 |
| Pancreas | T2DM_admission | ENSG00000110435.7 | 0.005635664 | 0.000406141 | 1 | 0.013545063 | PDHX |
| Pancreas | T2DM_admission | ENSG00000111328.2 | 0.00700922 | 1.82E-05 | 0.046783846 | 0.001443011 | CDK2AP1 |
| Pancreas | T2DM_admission | ENSG00000112992.12 | -0.005681812 | 0.00226552 | 1 | 0.046778499 | NNT |
| Pancreas | T2DM_admission | ENSG00000118997.9 | -0.005559873 | 0.000796877 | 1 | 0.021449685 | DNAH7 |
| Pancreas | T2DM_admission | ENSG00000120662.11 | 0.007685578 | 0.001543586 | 1 | 0.034468939 | MTRF1 |
| Pancreas | T2DM_admission | ENSG00000120709.6 | -0.009370029 | 0.000391155 | 1 | 0.013393155 | FAM53C |
| Pancreas | T2DM_admission | ENSG00000121417.9 | 0.004836372 | 0.000284686 | 0.731073748 | 0.010911548 | ZNF211 |
| Pancreas | T2DM_admission | ENSG00000122008.11 | -0.01264184 | 7.06E-05 | 0.181426286 | 0.004031695 | POLK |
| Pancreas | T2DM_admission | ENSG00000122025.10 | 0.012704947 | 9.78E-05 | 0.251273667 | 0.004964677 | FLT3 |
| Pancreas | T2DM_admission | ENSG00000125510.11 | -0.008957477 | 4.51E-05 | 0.1157578 | 0.002756138 | OPRL1 |
| Pancreas | T2DM_admission | ENSG00000127220.5 | 0.011263469 | 0.000969452 | 1 | 0.02417042 | ABHD8 |
| Pancreas | T2DM_admission | ENSG00000127957.12 | -0.011578497 | 8.50E-05 | 0.218336501 | 0.004548677 | PMS2P3 |
| Pancreas | T2DM_admission | ENSG00000129933.16 | -0.01085965 | 1.58E-06 | 0.004056744 | 0.000213513 | MAU2 |
| Pancreas | T2DM_admission | ENSG00000130338.8 | 0.006364416 | 0.002335635 | 1 | 0.047333554 | TULP4 |
| Pancreas | T2DM_admission | ENSG00000131323.10 | 0.010107775 | 0.000377244 | 0.968763585 | 0.013188885 | TRAF3 |
| Pancreas | T2DM_admission | ENSG00000134250.13 | -0.01084365 | 4.18E-11 | 1.07E-07 | 2.15E-08 | NOTCH2 |
| Pancreas | T2DM_admission | ENSG00000137731.9 | -0.011652882 | 0.000403358 | 1 | 0.013545063 | FXYD2 |
| Pancreas | T2DM_admission | ENSG00000137877.8 | 0.006688412 | 0.000603246 | 1 | 0.017806153 | SPTBN5 |
| Pancreas | T2DM_admission | ENSG00000140105.13 | -0.00801426 | 0.002276991 | 1 | 0.046778499 | WARS |
| Pancreas | T2DM_admission | ENSG00000140400.10 | -0.003381778 | 8.63E-06 | 0.022165888 | 0.000794418 | MAN2C1 |
| Pancreas | T2DM_admission | ENSG00000141560.10 | -0.002701629 | 0.002461443 | 1 | 0.048622958 | FN3KRP |
| Pancreas | T2DM_admission | ENSG00000141699.6 | 0.009972005 | 0.000231525 | 0.59455591 | 0.009437395 | FAM134C |
| Pancreas | T2DM_admission | ENSG00000142459.4 | 0.011132138 | 0.000980269 | 1 | 0.024205102 | EVI5L |
| Pancreas | T2DM_admission | ENSG00000143442.17 | 0.008559146 | 8.94E-05 | 0.229564541 | 0.004684991 | POGZ |
| Pancreas | T2DM_admission | ENSG00000144354.9 | 0.008407045 | 0.00152402 | 1 | 0.03433055 | CDCA7 |
| Pancreas | T2DM_admission | ENSG00000146830.9 | -0.009931678 | 0.00241051 | 1 | 0.048360847 | GIGYF1 |
| Pancreas | T2DM_admission | ENSG00000148384.11 | 0.011727967 | 0.000228947 | 0.587935065 | 0.009437395 | INPP5E |
| Pancreas | T2DM_admission | ENSG00000148396.14 | 0.007144747 | 0.000168028 | 0.43149599 | 0.007570105 | SEC16A |
| Pancreas | T2DM_admission | ENSG00000149084.7 | -0.003366714 | 9.74E-07 | 0.002501751 | 0.000156359 | HSD17B12 |
| Pancreas | T2DM_admission | ENSG00000149089.8 | -0.002765679 | 0.000291134 | 0.74763117 | 0.010994576 | APIP |
| Pancreas | T2DM_admission | ENSG00000152104.7 | -0.007931525 | 0.000258075 | 0.662737678 | 0.01004148 | PTPN14 |
| Pancreas | T2DM_admission | ENSG00000152359.10 | -0.004003392 | 1.58E-05 | 0.040614291 | 0.001310138 | POC5 |
| Pancreas | T2DM_admission | ENSG00000153814.7 | -0.019917769 | 1.02E-21 | 2.62E-18 | 1.31E-18 | JAZF1 |
| Pancreas | T2DM_admission | ENSG00000158008.5 | -0.009607447 | 0.000579648 | 1 | 0.017308569 | EXTL1 |
| Pancreas | T2DM_admission | ENSG00000160161.5 | -0.007941133 | 2.06E-05 | 0.05286319 | 0.001468422 | CILP2 |
| Pancreas | T2DM_admission | ENSG00000163486.8 | 0.011089539 | 5.68E-05 | 0.145772969 | 0.003390069 | SRGAP2 |
| Pancreas | T2DM_admission | ENSG00000163596.12 | -0.003561478 | 0.000375795 | 0.965040757 | 0.013188885 | ICA1L |
| Pancreas | T2DM_admission | ENSG00000164039.10 | -0.012681047 | 0.000116967 | 0.300371847 | 0.005461306 | BDH2 |
| Pancreas | T2DM_admission | ENSG00000165806.15 | 0.005547722 | 2.23E-05 | 0.057269899 | 0.001547835 | CASP7 |
| Pancreas | T2DM_admission | ENSG00000166037.6 | -0.006004083 | 0.001510123 | 1 | 0.034318558 | CEP57 |
| Pancreas | T2DM_admission | ENSG00000166165.8 | -0.00819247 | 0.000380054 | 0.975977497 | 0.013188885 | CKB |
| Pancreas | T2DM_admission | ENSG00000166224.12 | -0.006799718 | 0.000638376 | 1 | 0.018419672 | SGPL1 |
| Pancreas | T2DM_admission | ENSG00000166228.4 | -0.008258884 | 3.93E-05 | 0.100905808 | 0.002522645 | PCBD1 |
| Pancreas | T2DM_admission | ENSG00000167543.11 | -0.007448719 | 0.002050509 | 1 | 0.043161544 | TP53I13 |
| Pancreas | T2DM_admission | ENSG00000168936.6 | -0.004770644 | 0.000318695 | 0.818407761 | 0.011691539 | TMEM129 |
| Pancreas | T2DM_admission | ENSG00000169085.7 | 0.002628433 | 0.001820281 | 1 | 0.039281359 | C8orf46 |
| Pancreas | T2DM_admission | ENSG00000169592.10 | -0.007456002 | 6.56E-05 | 0.168518426 | 0.003829964 | INO80E |
| Pancreas | T2DM_admission | ENSG00000171282.9 | 0.011583739 | 0.000556064 | 1 | 0.016799675 | RP11-1055B8.7 |
| Pancreas | T2DM_admission | ENSG00000171700.9 | -0.012516904 | 0.00096127 | 1 | 0.02417042 | RGS19 |
| Pancreas | T2DM_admission | ENSG00000171962.13 | 0.006632691 | 0.002493368 | 1 | 0.048877622 | LRRC48 |
| Pancreas | T2DM_admission | ENSG00000172379.14 | 0.010953569 | 0.002178824 | 1 | 0.045489587 | ARNT2 |
| Pancreas | T2DM_admission | ENSG00000172543.3 | -0.0034789 | 0.000658865 | 1 | 0.018799603 | CTSW |
| Pancreas | T2DM_admission | ENSG00000172766.14 | 0.007166531 | 0.00023918 | 0.614214725 | 0.009597105 | NAA16 |
| Pancreas | T2DM_admission | ENSG00000172803.13 | 0.006369343 | 0.00053312 | 1 | 0.016386194 | SNX32 |
| Pancreas | T2DM_admission | ENSG00000172922.4 | -0.007626802 | 0.000535997 | 1 | 0.016386194 | RNASEH2C |
| Pancreas | T2DM_admission | ENSG00000173517.6 | 0.009606885 | 9.86E-05 | 0.253198519 | 0.004964677 | PEAK1 |
| Pancreas | T2DM_admission | ENSG00000174851.10 | -0.004649186 | 0.000710318 | 1 | 0.019613942 | YIF1A |
| Pancreas | T2DM_admission | ENSG00000175213.2 | -0.008626582 | 0.001712257 | 1 | 0.037581849 | ZNF408 |
| Pancreas | T2DM_admission | ENSG00000176920.10 | 0.002119068 | 0.001226974 | 1 | 0.028907053 | FUT2 |
| Pancreas | T2DM_admission | ENSG00000177103.9 | -0.01109344 | 0.001188154 | 1 | 0.028251653 | DSCAML1 |
| Pancreas | T2DM_admission | ENSG00000177508.11 | 0.043414047 | 3.06E-29 | 7.87E-26 | 7.87E-26 | IRX3 |
| Pancreas | T2DM_admission | ENSG00000178449.4 | 0.013288302 | 3.78E-05 | 0.097093342 | 0.002489573 | COX14 |
| Pancreas | T2DM_admission | ENSG00000179115.6 | -0.014549899 | 1.99E-07 | 0.000511908 | 3.59E-05 | FARSA |
| Pancreas | T2DM_admission | ENSG00000179218.9 | 0.006814805 | 0.002437495 | 1 | 0.048523163 | CALR |
| Pancreas | T2DM_admission | ENSG00000179344.12 | -0.006158764 | 0.000372283 | 0.956022431 | 0.013188885 | HLA-DQB1 |
| Pancreas | T2DM_admission | ENSG00000182179.6 | -0.006853497 | 6.42E-06 | 0.016486244 | 0.000634086 | UBA7 |
| Pancreas | T2DM_admission | ENSG00000182196.9 | 0.009413479 | 2.04E-05 | 0.052265626 | 0.001468422 | ARL6IP4 |
| Pancreas | T2DM_admission | ENSG00000182853.7 | 0.010182242 | 0.000622931 | 1 | 0.018178262 | VMO1 |
| Pancreas | T2DM_admission | ENSG00000182986.8 | 0.002605872 | 0.001634655 | 1 | 0.036187889 | ZNF320 |
| Pancreas | T2DM_admission | ENSG00000184110.10 | 0.006701508 | 0.001251815 | 1 | 0.029224201 | EIF3C |
| Pancreas | T2DM_admission | ENSG00000184207.8 | -0.00888625 | 0.000748824 | 1 | 0.020457225 | PGP |
| Pancreas | T2DM_admission | ENSG00000185800.7 | -0.008368071 | 0.000419679 | 1 | 0.013642227 | DMWD |
| Pancreas | T2DM_admission | ENSG00000189398.5 | -0.009125156 | 0.001107595 | 1 | 0.02658228 | OR7E12P |
| Pancreas | T2DM_admission | ENSG00000196126.6 | -0.009983638 | 5.03E-08 | 0.000129215 | 1.25E-05 | HLA-DRB1 |
| Pancreas | T2DM_admission | ENSG00000197062.7 | -0.003698379 | 0.001283949 | 1 | 0.029704334 | RP5-874C20.3 |
| Pancreas | T2DM_admission | ENSG00000198231.8 | 0.006915491 | 0.000188997 | 0.48534387 | 0.008367998 | DDX42 |

| | | | | | | | |
|---|---|---|---|---|---|---|---|
| Pancreas | T2DM_admission | ENSG00000198502.5 | -0.003050984 | 1.20E-05 | 0.030809414 | 0.00102698 | HLA-DRB5 |
| Pancreas | T2DM_admission | ENSG00000203896.5 | -0.005821256 | 0.000417944 | 1 | 0.013642227 | LIME1 |
| Pancreas | T2DM_admission | ENSG00000204301.5 | -0.010163301 | 7.43E-08 | 0.000190837 | 1.59E-05 | NOTCH4 |
| Pancreas | T2DM_admission | ENSG00000204305.9 | 0.010503577 | 0.000504287 | 1 | 0.015792794 | AGER |
| Pancreas | T2DM_admission | ENSG00000204308.6 | -0.003832033 | 4.66E-06 | 0.011978467 | 0.000479139 | RNF5 |
| Pancreas | T2DM_admission | ENSG00000204536.9 | 0.005460227 | 0.0025594 | 1 | 0.049791969 | CCHCR1 |
| Pancreas | T2DM_admission | ENSG00000204618.4 | 0.007550476 | 3.04E-05 | 0.078039375 | 0.002053668 | RNF39 |
| Pancreas | T2DM_admission | ENSG00000206344.6 | 0.006293701 | 3.28E-06 | 0.008433577 | 0.000351399 | HCG27 |
| Pancreas | T2DM_admission | ENSG00000213221.4 | 0.003583132 | 0.000680445 | 1 | 0.019202006 | DNLZ |
| Pancreas | T2DM_admission | ENSG00000213676.6 | 0.004333867 | 4.20E-05 | 0.107791619 | 0.002629064 | ATF6B |
| Pancreas | T2DM_admission | ENSG00000221988.8 | 0.011155718 | 0.000104495 | 0.268344037 | 0.005063095 | PPT2 |
| Pancreas | T2DM_admission | ENSG00000224997.1 | -0.010115069 | 0.002340873 | 1 | 0.047333554 | AL049840.1 |
| Pancreas | T2DM_admission | ENSG00000226232.4 | -0.00933826 | 1.85E-07 | 0.000476149 | 3.59E-05 | RP11-419C5.2 |
| Pancreas | T2DM_admission | ENSG00000228536.1 | -0.011339387 | 0.000801857 | 1 | 0.021449685 | RP11-392O17.1 |
| Pancreas | T2DM_admission | ENSG00000228782.3 | 0.010024914 | 0.00202592 | 1 | 0.042996391 | MRPL45P2 |
| Pancreas | T2DM_admission | ENSG00000229358.3 | 0.010599914 | 0.000219259 | 0.563056426 | 0.009230433 | DPY19L1P1 |
| Pancreas | T2DM_admission | ENSG00000229391.3 | 0.008072664 | 1.51E-12 | 3.87E-09 | 1.29E-09 | HLA-DRB6 |
| Pancreas | T2DM_admission | ENSG00000232629.4 | 0.004006725 | 7.63E-05 | 0.195854535 | 0.004257707 | HLA-DQB2 |
| Pancreas | T2DM_admission | ENSG00000232709.1 | 0.018519455 | 2.10E-07 | 0.000538064 | 3.59E-05 | MARK2P9 |
| Pancreas | T2DM_admission | ENSG00000233609.3 | 0.007748022 | 0.00091124 | 1 | 0.023400641 | RP11-62H7.2 |
| Pancreas | T2DM_admission | ENSG00000234745.5 | 0.004756387 | 1.85E-05 | 0.047619377 | 0.001443011 | HLA-B |
| Pancreas | T2DM_admission | ENSG00000235728.1 | 0.01061569 | 0.000900044 | 1 | 0.023400641 | AC007349.5 |
| Pancreas | T2DM_admission | ENSG00000237541.3 | 0.010257914 | 3.89E-10 | 1.00E-06 | 1.67E-07 | HLA-DQA2 |
| Pancreas | T2DM_admission | ENSG00000242142.1 | 0.006560262 | 1.13E-05 | 0.028925455 | 0.000997429 | SERBP1P3 |
| Pancreas | T2DM_admission | ENSG00000243753.1 | 0.005638929 | 3.27E-06 | 0.008400931 | 0.000351399 | HLA-L |
| Pancreas | T2DM_admission | ENSG00000243789.6 | 0.013769293 | 1.80E-06 | 0.004617304 | 0.000230865 | JMJD7 |
| Pancreas | T2DM_admission | ENSG00000244731.3 | -0.004520624 | 0.000248734 | 0.638747882 | 0.00982689 | C4A |
| Pancreas | T2DM_admission | ENSG00000248278.1 | 0.012578111 | 4.14E-09 | 1.06E-05 | 1.52E-06 | SUMO2P17 |
| Pancreas | T2DM_admission | ENSG00000251417.1 | -0.016108067 | 1.38E-06 | 0.003531518 | 0.000196195 | RP11-1348G14.4 |
| Pancreas | T2DM_admission | ENSG00000255837.1 | 0.005865702 | 0.000701718 | 1 | 0.019587077 | TAS2R20 |
| Pancreas | T2DM_admission | ENSG00000256166.1 | 0.005070308 | 0.000211218 | 0.542406597 | 0.00904011 | XXbac-BPG248L24.13 |
| Pancreas | T2DM_admission | ENSG00000256269.2 | 0.003973338 | 0.000305163 | 0.783659739 | 0.011357388 | HMBS |
| Pancreas | T2DM_admission | ENSG00000259015.1 | 0.010991632 | 0.00090317 | 1 | 0.023400641 | RP11-109N23.6 |
| Pancreas | T2DM_admission | ENSG00000259982.1 | 0.00376819 | 0.001090039 | 1 | 0.02640773 | CDC37P1 |
| Pancreas | T2DM_admission | ENSG00000260274.1 | -0.010065856 | 2.47E-06 | 0.006334161 | 0.000301627 | RP11-817O13.8 |
| Pancreas | T2DM_admission | ENSG00000260814.2 | -0.013790315 | 1.08E-06 | 0.002770896 | 0.000162994 | RP11-107F6.3 |
| Pancreas | T2DM_admission | ENSG00000260911.1 | 0.00351099 | 0.001002146 | 1 | 0.024509633 | RP11-196G11.2 |
| Pancreas | T2DM_admission | ENSG00000268225.1 | 0.00579192 | 0.001976261 | 1 | 0.042291985 | CTD-2331H12.5 |
| Pancreas | T2DM_admission | ENSG00000271751.1 | -0.006650697 | 0.000955898 | 1 | 0.02417042 | RP11-110I1.14 |
| Pancreas | T2DM_insulin | ENSG00000011523.9 | 0.001833919 | 0.000118228 | 0.303610495 | 0.0202407 | CEP68 |
| Pancreas | T2DM_insulin | ENSG00000156219.12 | -0.001326335 | 3.08E-05 | 0.07912119 | 0.006593433 | ART3 |
| Pancreas | T2DM_insulin | ENSG00000158008.5 | -0.004053963 | 0.000255866 | 0.657064958 | 0.03865088 | EXTL1 |
| Pancreas | T2DM_insulin | ENSG00000169248.8 | 0.001353192 | 2.35E-05 | 0.060260938 | 0.005478267 | CXCL11 |
| Pancreas | T2DM_insulin | ENSG00000177508.11 | 0.009364345 | 1.08E-09 | 2.78E-06 | 5.55E-07 | IRX3 |
| Pancreas | T2DM_insulin | ENSG00000179344.12 | -0.003073255 | 7.74E-06 | 0.019877522 | 0.002208614 | HLA-DQB1 |
| Pancreas | T2DM_insulin | ENSG00000198502.5 | -0.001748696 | 2.64E-10 | 6.79E-07 | 1.70E-07 | HLA-DRB5 |
| Pancreas | T2DM_insulin | ENSG00000204257.10 | 0.001495763 | 6.49E-06 | 0.016667122 | 0.00208339 | HLA-DMA |
| Pancreas | T2DM_insulin | ENSG00000204301.5 | -0.004171586 | 2.68E-08 | 6.88E-05 | 1.15E-05 | NOTCH4 |
| Pancreas | T2DM_insulin | ENSG00000204351.7 | -0.001446573 | 1.79E-05 | 0.046073901 | 0.00460739 | SKIV2L |
| Pancreas | T2DM_insulin | ENSG00000204540.6 | -0.001441721 | 0.000111166 | 0.285475097 | 0.0202407 | PSORS1C1 |
| Pancreas | T2DM_insulin | ENSG00000204618.4 | 0.002611841 | 0.000280247 | 0.719673768 | 0.039981876 | RNF39 |
| Pancreas | T2DM_insulin | ENSG00000229391.3 | 0.003267528 | 5.59E-13 | 1.43E-09 | 1.43E-09 | HLA-DRB6 |
| Pancreas | T2DM_insulin | ENSG00000232629.4 | 0.002794741 | 3.69E-12 | 9.48E-09 | 4.74E-09 | HLA-DQB2 |
| Pancreas | T2DM_insulin | ENSG00000237541.3 | 0.004150398 | 1.81E-10 | 4.66E-07 | 1.55E-07 | HLA-DQA2 |
| Pancreas | T2DM_insulin | ENSG00000242142.1 | 0.002117592 | 0.000357618 | 0.918363457 | 0.048334919 | SERBP1P3 |
| Pancreas | T2DM_insulin | ENSG00000244731.3 | -0.002611987 | 9.79E-08 | 0.000251466 | 3.59E-05 | C4A |
| Pancreas | T2DM_insulin | ENSG00000251417.1 | -0.004905942 | 0.00021283 | 0.546547838 | 0.03415924 | RP11-1348G14.4 |
| Pancreas | T2DM_insulin | ENSG00000256166.1 | 0.002186259 | 5.74E-05 | 0.147433792 | 0.011341061 | XXbac-BPG248L24.13 |
| SAT | CAD | ENSG00000001617.7 | -0.014757532 | 5.22E-05 | 0.171185343 | 0.009510297 | SEMA3F |
| SAT | CAD | ENSG00000004534.10 | -0.002869433 | 0.000780112 | 1 | 0.049222079 | RBM6 |
| SAT | CAD | ENSG00000065060.12 | 0.007031212 | 8.23E-06 | 0.026991917 | 0.002453811 | UHRF1BP1 |
| SAT | CAD | ENSG00000075131.5 | 0.005546056 | 0.000164166 | 0.538627495 | 0.020332943 | TIPIN |
| SAT | CAD | ENSG00000087301.4 | -0.01651292 | 0.000196849 | 0.645862115 | 0.021008952 | TXNDC16 |
| SAT | CAD | ENSG00000089558.4 | 0.01354567 | 0.0006759 | 1 | 0.044352545 | KCNH4 |
| SAT | CAD | ENSG00000090621.9 | -0.007792787 | 0.000618717 | 1 | 0.041428814 | PABPC4 |
| SAT | CAD | ENSG00000100997.14 | 0.014054407 | 0.000209189 | 0.686349333 | 0.021008952 | ABHD12 |
| SAT | CAD | ENSG00000101460.8 | 0.02217467 | 6.33E-06 | 0.020772867 | 0.002077287 | MAP1LC3A |
| SAT | CAD | ENSG00000101464.6 | 0.011067404 | 0.000518914 | 1 | 0.037834601 | PIGU |
| SAT | CAD | ENSG00000114023.11 | -0.017405772 | 0.000401638 | 1 | 0.031912061 | FAM162A |
| SAT | CAD | ENSG00000114200.5 | -0.018311052 | 5.12E-05 | 0.168023676 | 0.009510297 | BCHE |
| SAT | CAD | ENSG00000114491.9 | 0.018732502 | 0.000101289 | 0.332328498 | 0.015825167 | UMPS |
| SAT | CAD | ENSG00000114554.7 | 0.012283305 | 0.000201054 | 0.659657844 | 0.021008952 | PLXNA1 |
| SAT | CAD | ENSG00000115486.7 | 0.009401774 | 1.23E-05 | 0.040309209 | 0.003359101 | GGCX |
| SAT | CAD | ENSG00000119718.6 | 0.006710809 | 0.000398018 | 1 | 0.031912061 | EIF2B2 |
| SAT | CAD | ENSG00000124562.5 | -0.008949368 | 5.83E-06 | 0.019112485 | 0.002077287 | SNRPC |
| SAT | CAD | ENSG00000125388.15 | 0.004794537 | 0.00022184 | 0.727855892 | 0.021008952 | GRK4 |
| SAT | CAD | ENSG00000137054.11 | 0.008517123 | 0.000577705 | 1 | 0.040328707 | POLR1E |
| SAT | CAD | ENSG00000138380.13 | -0.016780187 | 1.74E-08 | 5.72E-05 | 4.20E-05 | CARF |
| SAT | CAD | ENSG00000142046.10 | 0.011165307 | 0.000146644 | 0.481139752 | 0.019375329 | TMEM91 |
| SAT | CAD | ENSG00000143156.9 | -0.022267702 | 5.16E-06 | 0.016939967 | 0.002077287 | NME7 |
| SAT | CAD | ENSG00000144426.14 | -0.013521673 | 2.56E-08 | 8.40E-05 | 4.20E-05 | NBEAL1 |

| | | | | | | | |
|---|---|---|---|---|---|---|---|
| SAT | CAD | ENSG00000145020.10 | -0.004954599 | 7.48E-05 | 0.245401176 | 0.012270059 | AMT |
| SAT | CAD | ENSG00000145029.7 | -0.008297714 | 7.13E-05 | 0.234070148 | 0.012270059 | NICN1 |
| SAT | CAD | ENSG00000149084.7 | -0.003361131 | 0.00039596 | 1 | 0.031912061 | HSD17B12 |
| SAT | CAD | ENSG00000154309.7 | 0.010130475 | 0.000147633 | 0.484383225 | 0.019375329 | DISP1 |
| SAT | CAD | ENSG00000163431.11 | -0.017129977 | 4.82E-05 | 0.158283524 | 0.009510297 | LMOD1 |
| SAT | CAD | ENSG00000163596.12 | 0.016834253 | 5.00E-08 | 0.000164174 | 5.47E-05 | ICA1L |
| SAT | CAD | ENSG00000164062.8 | -0.013190522 | 1.51E-06 | 0.00495548 | 0.000707926 | APEH |
| SAT | CAD | ENSG00000164068.11 | 0.005461084 | 0.000221335 | 0.726200621 | 0.021008952 | RNF123 |
| SAT | CAD | ENSG00000169047.5 | -0.013297668 | 5.00E-07 | 0.001639411 | 0.000327882 | IRS1 |
| SAT | CAD | ENSG00000175445.10 | -0.020132155 | 8.84E-08 | 0.00028992 | 7.25E-05 | LPL |
| SAT | CAD | ENSG00000176390.10 | 0.020520044 | 2.40E-05 | 0.078716095 | 0.006055084 | CRLF3 |
| SAT | CAD | ENSG00000181264.4 | 0.01218491 | 0.000266654 | 0.874891114 | 0.023645706 | TMEM136 |
| SAT | CAD | ENSG00000188895.7 | -0.007167999 | 0.000229173 | 0.751916536 | 0.021008952 | MSL1 |
| SAT | CAD | ENSG00000196821.5 | 0.012407021 | 1.14E-06 | 0.003747179 | 0.00062453 | C6orf106 |
| SAT | CAD | ENSG00000196981.2 | 0.017052005 | 0.000408506 | 1 | 0.031912061 | WDR5B |
| SAT | CAD | ENSG00000198270.8 | -0.006554776 | 0.000230516 | 0.756322289 | 0.021008952 | TMEM116 |
| SAT | CAD | ENSG00000204978.2 | -0.011016248 | 0.000134903 | 0.442615565 | 0.019244155 | C19orf69 |
| SAT | CAD | ENSG00000213277.3 | -0.01436543 | 0.000219656 | 0.720691506 | 0.021008952 | MARCKSL1P1 |
| SAT | CAD | ENSG00000213689.5 | 0.007456711 | 0.000448786 | 1 | 0.033465169 | TREX1 |
| SAT | CAD | ENSG00000215452.3 | -0.019187442 | 0.00033043 | 1 | 0.028529979 | ZNF663P |
| SAT | CAD | ENSG00000249685.1 | -0.011005016 | 0.000609622 | 1 | 0.041428814 | RP11-360F5.3 |
| SAT | CAD | ENSG00000256269.2 | 0.010887665 | 4.81E-05 | 0.157810392 | 0.009510297 | HMBS |
| SAT | CAD | ENSG00000261143.1 | -0.011409273 | 0.000128153 | 0.420471326 | 0.019112333 | ADAMTS7P3 |
| SAT | CAD | ENSG00000264538.2 | 0.005846046 | 0.000441234 | 1 | 0.033465169 | SUZ12P |
| SAT | CAD | ENSG00000269001.1 | -0.012157662 | 0.00021491 | 0.70512093 | 0.021008952 | ZNF818P |
| SAT | CAD | ENSG00000269051.1 | -0.019823445 | 0.000167324 | 0.548989452 | 0.020332943 | CTD-2245F17.3 |
| SAT | CAD | ENSG00000269202.1 | -0.01010399 | 0.000775577 | 1 | 0.049222079 | RP4-614O4.12 |
| SAT | CAD | ENSG00000270018.1 | 0.014404178 | 0.000569487 | 1 | 0.040328707 | RP3-462E2.5 |
| SAT | CAD | ENSG00000271751.1 | -0.018429148 | 4.29E-05 | 0.140719533 | 0.009510297 | RP11-110I1.14 |
| SAT | T2DM | ENSG00000001617.7 | -0.01381935 | 7.46E-07 | 0.002447204 | 0.0001064 | SEMA3F |
| SAT | T2DM | ENSG00000004534.10 | -0.00357068 | 4.72E-08 | 0.000154812 | 1.19E-05 | RBM6 |
| SAT | T2DM | ENSG00000005955.8 | -0.012528307 | 1.22E-06 | 0.003992826 | 0.000137684 | GGNBP2 |
| SAT | T2DM | ENSG00000011523.9 | 0.009709795 | 1.18E-09 | 3.87E-06 | 6.46E-07 | CEP68 |
| SAT | T2DM | ENSG00000037042.8 | -0.008467234 | 7.67E-09 | 2.52E-05 | 2.80E-06 | TUBG2 |
| SAT | T2DM | ENSG00000055955.11 | 0.005059899 | 8.41E-06 | 0.027589044 | 0.000689726 | ITIH4 |
| SAT | T2DM | ENSG00000073578.12 | 0.007899305 | 0.001738269 | 1 | 0.044907563 | SDHA |
| SAT | T2DM | ENSG00000077454.11 | -0.00967636 | 0.000930098 | 1 | 0.027246889 | LRCH4 |
| SAT | T2DM | ENSG00000090621.9 | -0.010218366 | 4.52E-09 | 1.48E-05 | 1.85E-06 | PABPC4 |
| SAT | T2DM | ENSG00000091542.8 | -0.006521455 | 0.002000284 | 1 | 0.049345351 | ALKBH5 |
| SAT | T2DM | ENSG00000095485.12 | -0.00414476 | 0.000116993 | 0.383854135 | 0.006618175 | CWF19L1 |
| SAT | T2DM | ENSG00000103479.10 | 0.005953016 | 0.000107304 | 0.352063407 | 0.006213443 | RBL2 |
| SAT | T2DM | ENSG00000104313.13 | 0.012667694 | 5.61E-05 | 0.184031207 | 0.003472287 | EYA1 |
| SAT | T2DM | ENSG00000105726.12 | 0.010044259 | 3.60E-07 | 0.001181578 | 6.22E-05 | ATP13A1 |
| SAT | T2DM | ENSG00000108272.9 | -0.009583119 | 0.001686433 | 1 | 0.043914172 | DHRS11 |
| SAT | T2DM | ENSG00000108786.6 | -0.006297286 | 4.16E-07 | 0.001363469 | 6.82E-05 | HSD17B1 |
| SAT | T2DM | ENSG00000111328.2 | 0.00583938 | 0.000140401 | 0.460654189 | 0.007718167 | CDK2AP1 |
| SAT | T2DM | ENSG00000111405.4 | -0.010784648 | 0.001251031 | 1 | 0.034785013 | ENDOU |
| SAT | T2DM | ENSG00000112276.9 | -0.007489759 | 0.000818288 | 1 | 0.024700393 | BVES |
| SAT | T2DM | ENSG00000115216.9 | -0.012635055 | 8.70E-07 | 0.002854673 | 0.000109795 | NRBP1 |
| SAT | T2DM | ENSG00000115241.9 | -0.017497646 | 4.51E-07 | 0.001479264 | 7.04E-05 | PPM1G |
| SAT | T2DM | ENSG00000115561.10 | -0.007094507 | 9.17E-06 | 0.030097647 | 0.000734089 | CHMP3 |
| SAT | T2DM | ENSG00000116157.5 | -0.004077206 | 0.000543105 | 1 | 0.019368778 | GPX7 |
| SAT | T2DM | ENSG00000117280.8 | -0.006528854 | 0.000784418 | 1 | 0.024351274 | RAB7L1 |
| SAT | T2DM | ENSG00000117877.6 | -0.011219767 | 3.59E-05 | 0.117693459 | 0.002401907 | CD3EAP |
| SAT | T2DM | ENSG00000118402.5 | 0.008916566 | 0.001651815 | 1 | 0.04335685 | ELOVL4 |
| SAT | T2DM | ENSG00000118997.9 | -0.006637526 | 1.68E-05 | 0.055028986 | 0.001250659 | DNAH7 |
| SAT | T2DM | ENSG00000119242.4 | -0.008684463 | 0.001120496 | 1 | 0.03169265 | CCDC92 |
| SAT | T2DM | ENSG00000120093.7 | -0.010823248 | 4.23E-05 | 0.138943375 | 0.002749283 | HOXB3 |
| SAT | T2DM | ENSG00000120907.13 | 0.005156029 | 0.000181567 | 0.595721453 | 0.008583862 | ADRA1A |
| SAT | T2DM | ENSG00000121101.11 | 0.01516347 | 0.000144956 | 0.475600177 | 0.00776917 | TEX14 |
| SAT | T2DM | ENSG00000125388.15 | 0.003432358 | 0.000553525 | 1 | 0.019528141 | GRK4 |
| SAT | T2DM | ENSG00000125510.11 | -0.014209361 | 7.89E-06 | 0.025878571 | 0.000663553 | OPRL1 |
| SAT | T2DM | ENSG00000126464.9 | 0.006654648 | 0.000602382 | 1 | 0.02037543 | PRR12 |
| SAT | T2DM | ENSG00000127957.12 | -0.007799487 | 9.84E-06 | 0.032271237 | 0.000768363 | PMS2P3 |
| SAT | T2DM | ENSG00000130299.12 | -0.010153632 | 0.000748955 | 1 | 0.023857503 | GTPBP3 |
| SAT | T2DM | ENSG00000130921.3 | 0.0141348 | 0.000271393 | 0.890441137 | 0.011415912 | C12orf65 |
| SAT | T2DM | ENSG00000133059.12 | -0.010468085 | 0.000791863 | 1 | 0.024351274 | DSTYK |
| SAT | T2DM | ENSG00000135124.10 | 0.009671505 | 0.00186461 | 1 | 0.04742469 | P2RX4 |
| SAT | T2DM | ENSG00000135926.8 | -0.008560775 | 0.001183505 | 1 | 0.033188714 | TMBIM1 |
| SAT | T2DM | ENSG00000137877.8 | 0.006913974 | 4.57E-05 | 0.149831054 | 0.002881366 | SPTBN5 |
| SAT | T2DM | ENSG00000138380.13 | 0.007705973 | 0.00072242 | 1 | 0.023237829 | CARF |
| SAT | T2DM | ENSG00000140400.10 | -0.00267677 | 2.84E-05 | 0.093085174 | 0.001980536 | MAN2C1 |
| SAT | T2DM | ENSG00000141252.15 | -0.008527486 | 0.001477396 | 1 | 0.039732269 | VPS53 |
| SAT | T2DM | ENSG00000141452.5 | -0.010261089 | 0.000141143 | 0.463090023 | 0.007718167 | C18orf8 |
| SAT | T2DM | ENSG00000141458.8 | -0.016433383 | 7.27E-06 | 0.023857506 | 0.000627829 | NPC1 |
| SAT | T2DM | ENSG00000142082.10 | 0.011993739 | 0.000590798 | 1 | 0.020191744 | SIRT3 |
| SAT | T2DM | ENSG00000144426.14 | 0.006406391 | 0.000567287 | 1 | 0.019800723 | NBEAL1 |
| SAT | T2DM | ENSG00000144837.4 | -0.010389887 | 0.002088159 | 1 | 0.049971446 | PLA1A |
| SAT | T2DM | ENSG00000145692.10 | -0.006711055 | 0.000153347 | 0.503132281 | 0.007957713 | BHMT |
| SAT | T2DM | ENSG00000145725.15 | -0.006594666 | 0.000103874 | 0.340811119 | 0.006196566 | PPIP5K2 |
| SAT | T2DM | ENSG00000147419.12 | 0.012799654 | 0.000517662 | 1 | 0.018677439 | CCDC25 |

| | | | | | | | |
|---|---|---|---|---|---|---|---|
| SAT | T2DM | ENSG00000149084.7 | -0.004111705 | 1.50E-08 | 4.92E-05 | 4.10E-06 | HSD17B12 |
| SAT | T2DM | ENSG00000149089.8 | -0.002683842 | 0.000165478 | 0.542934157 | 0.00820853 | APIP |
| SAT | T2DM | ENSG00000149100.8 | -0.010746473 | 0.000518027 | 1 | 0.018677439 | EIF3M |
| SAT | T2DM | ENSG00000152219.4 | -0.017520121 | 2.40E-07 | 0.000785911 | 4.37E-05 | ARL14EP |
| SAT | T2DM | ENSG00000152359.10 | -0.006039661 | 0.000224734 | 0.737353088 | 0.009831375 | POC5 |
| SAT | T2DM | ENSG00000153814.7 | -0.030438495 | 8.69E-21 | 2.85E-17 | 1.42E-17 | JAZF1 |
| SAT | T2DM | ENSG00000154743.13 | 0.017860588 | 1.01E-06 | 0.003326096 | 0.000120866 | TSEN2 |
| SAT | T2DM | ENSG00000156170.8 | -0.016574211 | 1.79E-06 | 0.005860807 | 0.000184017 | NDUFAF6 |
| SAT | T2DM | ENSG00000157322.12 | -0.008678203 | 1.17E-08 | 3.84E-05 | 3.84E-06 | CLEC18A |
| SAT | T2DM | ENSG00000157823.12 | 0.010120838 | 1.74E-10 | 5.71E-07 | 1.14E-07 | AP3S2 |
| SAT | T2DM | ENSG00000157895.7 | 0.008349819 | 0.002100165 | 1 | 0.049971446 | C12orf43 |
| SAT | T2DM | ENSG00000158022.6 | 0.008912887 | 0.001285858 | 1 | 0.03545294 | TRIM63 |
| SAT | T2DM | ENSG00000158805.7 | -0.012003329 | 0.000633986 | 1 | 0.021225603 | ZNF276 |
| SAT | T2DM | ENSG00000160305.13 | -0.003829623 | 0.000895113 | 1 | 0.02645824 | DIP2A |
| SAT | T2DM | ENSG00000160752.10 | 0.008317422 | 0.001369688 | 1 | 0.03714006 | FDPS |
| SAT | T2DM | ENSG00000163554.7 | -0.004697565 | 0.002098839 | 1 | 0.049971446 | SPTA1 |
| SAT | T2DM | ENSG00000163596.12 | -0.008032222 | 0.000679613 | 1 | 0.022298088 | ICA1L |
| SAT | T2DM | ENSG00000163933.5 | -0.013872735 | 3.10E-06 | 0.010154793 | 0.00029867 | RFT1 |
| SAT | T2DM | ENSG00000164039.10 | -0.007877137 | 0.000229539 | 0.753118457 | 0.009909453 | BDH2 |
| SAT | T2DM | ENSG00000164068.11 | 0.005567757 | 8.70E-07 | 0.002854234 | 0.000109795 | RNF123 |
| SAT | T2DM | ENSG00000165178.8 | 0.008500072 | 0.0003751 | 1 | 0.014946268 | NCF1C |
| SAT | T2DM | ENSG00000165476.8 | -0.009984603 | 0.001043225 | 1 | 0.029763665 | REEP3 |
| SAT | T2DM | ENSG00000166166.8 | -0.019052313 | 1.03E-06 | 0.003384238 | 0.000120866 | TRMT61A |
| SAT | T2DM | ENSG00000166228.4 | -0.005479857 | 0.000107945 | 0.354166225 | 0.006213443 | PCBD1 |
| SAT | T2DM | ENSG00000166278.10 | 0.01966209 | 2.08E-06 | 0.006826591 | 0.000206866 | C2 |
| SAT | T2DM | ENSG00000167397.10 | -0.010582631 | 0.000697092 | 1 | 0.022645139 | VKORC1 |
| SAT | T2DM | ENSG00000167528.8 | 0.004292774 | 0.001998061 | 1 | 0.049345351 | ZNF641 |
| SAT | T2DM | ENSG00000167930.11 | 0.010005045 | 0.00149276 | 1 | 0.039819067 | ITFG3 |
| SAT | T2DM | ENSG00000168056.10 | 0.011064621 | 0.000208443 | 0.68390041 | 0.009368499 | LTBP3 |
| SAT | T2DM | ENSG00000168297.11 | 0.011804575 | 0.000155225 | 0.509293604 | 0.007957713 | PXK |
| SAT | T2DM | ENSG00000168394.9 | -0.021736455 | 1.78E-07 | 0.000582522 | 3.64E-05 | TAP1 |
| SAT | T2DM | ENSG00000169047.5 | -0.021698235 | 8.68E-27 | 2.85E-23 | 2.85E-23 | IRS1 |
| SAT | T2DM | ENSG00000169592.10 | -0.008776437 | 0.000642758 | 1 | 0.021301893 | INO80E |
| SAT | T2DM | ENSG00000170092.10 | 0.009154091 | 0.001007324 | 1 | 0.029248051 | AC018720.10 |
| SAT | T2DM | ENSG00000170571.7 | -0.011055356 | 0.000260389 | 0.854336845 | 0.011095284 | EMB |
| SAT | T2DM | ENSG00000171700.9 | -0.010305651 | 1.91E-07 | 0.000626829 | 3.69E-05 | RGS19 |
| SAT | T2DM | ENSG00000171723.11 | -0.008564429 | 0.001354533 | 1 | 0.037035189 | GPHN |
| SAT | T2DM | ENSG00000172037.9 | -0.014680252 | 1.74E-05 | 0.057116562 | 0.001269257 | LAMB2 |
| SAT | T2DM | ENSG00000172247.3 | -0.00357062 | 0.000186796 | 0.612876884 | 0.008632069 | C1QTNF4 |
| SAT | T2DM | ENSG00000173511.5 | -0.010267639 | 6.96E-06 | 0.022826296 | 0.000616927 | VEGFB |
| SAT | T2DM | ENSG00000173917.9 | -0.010213979 | 3.31E-05 | 0.10844693 | 0.002259311 | HOXB2 |
| SAT | T2DM | ENSG00000175334.3 | 0.007415485 | 0.000496071 | 1 | 0.018287726 | BANF1 |
| SAT | T2DM | ENSG00000178750.2 | 0.013686705 | 4.27E-05 | 0.140213418 | 0.002749283 | STX19 |
| SAT | T2DM | ENSG00000179115.6 | -0.018846323 | 1.58E-06 | 0.005183293 | 0.000172776 | FARSA |
| SAT | T2DM | ENSG00000179218.9 | 0.016085737 | 2.62E-05 | 0.085805061 | 0.001865327 | CALR |
| SAT | T2DM | ENSG00000182986.8 | 0.002406435 | 0.000887987 | 1 | 0.02645824 | ZNF320 |
| SAT | T2DM | ENSG00000183779.5 | -0.013951765 | 6.77E-05 | 0.22220762 | 0.004114956 | ZNF703 |
| SAT | T2DM | ENSG00000184110.10 | 0.007364308 | 0.000483352 | 1 | 0.018228475 | EIF3C |
| SAT | T2DM | ENSG00000185904.7 | 0.013950684 | 0.000416109 | 1 | 0.016061818 | LINC00839 |
| SAT | T2DM | ENSG00000187166.1 | 0.005316135 | 0.000791175 | 1 | 0.024351274 | H1FNT |
| SAT | T2DM | ENSG00000187778.9 | 0.010189324 | 0.001547911 | 1 | 0.040957227 | MCRS1 |
| SAT | T2DM | ENSG00000189398.5 | -0.011963901 | 0.002117047 | 1 | 0.049971446 | OR7E12P |
| SAT | T2DM | ENSG00000196126.6 | -0.016018717 | 1.11E-07 | 0.000365797 | 2.44E-05 | HLA-DRB1 |
| SAT | T2DM | ENSG00000196693.10 | -0.005335639 | 0.000326552 | 1 | 0.013392728 | ZNF33B |
| SAT | T2DM | ENSG00000198056.9 | -0.011652568 | 0.000183136 | 0.600870316 | 0.008583862 | PRIM1 |
| SAT | T2DM | ENSG00000198453.8 | 0.011664832 | 0.000575495 | 1 | 0.019875797 | ZNF568 |
| SAT | T2DM | ENSG00000198502.5 | -0.002858768 | 0.00033098 | 1 | 0.013406744 | HLA-DRB5 |
| SAT | T2DM | ENSG00000203896.5 | -0.008794209 | 0.000405167 | 1 | 0.015825616 | LIME1 |
| SAT | T2DM | ENSG00000204237.4 | 0.011762554 | 0.001893709 | 1 | 0.047650044 | OXLD1 |
| SAT | T2DM | ENSG00000204287.9 | 0.014789878 | 2.87E-09 | 9.41E-06 | 1.34E-06 | HLA-DRA |
| SAT | T2DM | ENSG00000204622.6 | 0.004299143 | 0.002105722 | 1 | 0.049971446 | HLA-J |
| SAT | T2DM | ENSG00000205583.9 | -0.004911484 | 0.000215074 | 0.705657617 | 0.009535914 | STAG3L1 |
| SAT | T2DM | ENSG00000205659.6 | 0.010944158 | 0.000146812 | 0.48168855 | 0.00776917 | LIN52 |
| SAT | T2DM | ENSG00000206344.6 | 0.004371075 | 6.79E-07 | 0.002227894 | 0.000101268 | HCG27 |
| SAT | T2DM | ENSG00000213221.4 | 0.00343402 | 0.000193347 | 0.63436993 | 0.008810693 | DNLZ |
| SAT | T2DM | ENSG00000214530.3 | 0.005513693 | 8.07E-07 | 0.002647908 | 0.000109795 | STARD10 |
| SAT | T2DM | ENSG00000223756.2 | -0.009024297 | 0.000453644 | 1 | 0.017307053 | TSSC2 |
| SAT | T2DM | ENSG00000226232.4 | -0.005889907 | 1.79E-06 | 0.00588853 | 0.000184017 | RP11-419C5.2 |
| SAT | T2DM | ENSG00000229391.3 | 0.007004399 | 1.89E-11 | 6.22E-08 | 2.07E-08 | HLA-DRB6 |
| SAT | T2DM | ENSG00000230953.2 | -0.012322511 | 0.000820586 | 1 | 0.024700393 | RP4-631H13.6 |
| SAT | T2DM | ENSG00000231852.2 | 0.006404531 | 4.68E-06 | 0.015356883 | 0.000430718 | CYP21A2 |
| SAT | T2DM | ENSG00000235081.1 | 0.013909923 | 0.000167623 | 0.549971522 | 0.00820853 | AC010492.2 |
| SAT | T2DM | ENSG00000237541.3 | 0.014020545 | 3.25E-11 | 1.07E-07 | 2.66E-08 | HLA-DQA2 |
| SAT | T2DM | ENSG00000237984.2 | -0.01208302 | 0.001902516 | 1 | 0.047650044 | PTENP1 |
| SAT | T2DM | ENSG00000239557.1 | 0.013394316 | 0.001026754 | 1 | 0.029550704 | RP11-168J18.6 |
| SAT | T2DM | ENSG00000242600.2 | 0.00749259 | 0.000378098 | 1 | 0.014946268 | MBL1P |
| SAT | T2DM | ENSG00000243708.4 | 0.007847131 | 6.40E-08 | 0.000210073 | 1.50E-05 | PLA2G4B |
| SAT | T2DM | ENSG00000243789.6 | 0.012285133 | 1.50E-08 | 4.91E-05 | 4.10E-06 | JMJD7 |
| SAT | T2DM | ENSG00000246523.3 | -0.009302668 | 0.000282444 | 0.926699671 | 0.011730376 | RP11-736K20.6 |
| SAT | T2DM | ENSG00000253813.1 | 0.009791969 | 0.000167324 | 0.548990588 | 0.00820853 | COX6B1P4 |
| SAT | T2DM | ENSG00000253816.2 | -0.008605434 | 0.001801112 | 1 | 0.046167565 | RP11-1415C14.3 |

| | | | | | | | |
|---|---|---|---|---|---|---|---|
| SAT | T2DM | ENSG00000256269.2 | 0.009006153 | 1.12E-05 | 0.036810281 | 0.000856053 | HMBS |
| SAT | T2DM | ENSG00000258602.1 | 0.008314567 | 0.002044449 | 1 | 0.049971446 | RP11-7F17.7 |
| SAT | T2DM | ENSG00000260274.1 | -0.004337331 | 0.000794144 | 1 | 0.024351274 | RP11-817O13.8 |
| SAT | T2DM | ENSG00000269963.1 | -0.010815618 | 0.000493927 | 1 | 0.018287726 | RP11-73M18.9 |
| SAT | T2DM | ENSG00000271751.1 | -0.012907761 | 0.00018156 | 0.595698457 | 0.008583862 | RP11-110I1.14 |
| SAT | T2DM | ENSG00000272221.1 | -0.003233973 | 4.73E-06 | 0.01550583 | 0.000430718 | XXbac-BPG181B23.7 |
| SAT | Obesity | ENSG00000001617.7 | -0.042278276 | 7.59E-24 | 2.49E-20 | 3.11E-21 | SEMA3F |
| SAT | Obesity | ENSG00000004534.10 | -0.010277366 | 1.41E-25 | 4.63E-22 | 6.62E-23 | RBM6 |
| SAT | Obesity | ENSG00000005955.8 | -0.025349744 | 6.61E-11 | 2.17E-07 | 6.57E-09 | GGNBP2 |
| SAT | Obesity | ENSG00000010704.14 | -0.016780294 | 1.51E-07 | 0.000495766 | 8.70E-06 | HFE |
| SAT | Obesity | ENSG00000011295.11 | 0.020785063 | 3.82E-05 | 0.125325841 | 0.001046683 | TTC19 |
| SAT | Obesity | ENSG00000013275.3 | 0.012990825 | 0.000773095 | 1 | 0.012176893 | PSMC4 |
| SAT | Obesity | ENSG00000026950.12 | -0.006436415 | 0.001248371 | 1 | 0.017886049 | BTN3A1 |
| SAT | Obesity | ENSG00000029725.12 | 0.013705185 | 1.77E-05 | 0.058046705 | 0.00054761 | RABEP1 |
| SAT | Obesity | ENSG00000035115.17 | -0.01384339 | 1.37E-05 | 0.044874705 | 0.000439948 | SH3YL1 |
| SAT | Obesity | ENSG00000041802.6 | 0.003202284 | 0.002899481 | 1 | 0.035691396 | LSG1 |
| SAT | Obesity | ENSG00000049449.4 | -0.017530283 | 0.001091641 | 1 | 0.016106882 | RCN1 |
| SAT | Obesity | ENSG00000050130.13 | 0.010533917 | 0.003623772 | 1 | 0.042161685 | JKAMP |
| SAT | Obesity | ENSG00000053900.6 | 0.016325778 | 0.000596746 | 1 | 0.009888499 | ANAPC4 |
| SAT | Obesity | ENSG00000054282.11 | 0.010568712 | 0.000253019 | 0.830156695 | 0.004854718 | SDCCAG8 |
| SAT | Obesity | ENSG00000055955.11 | 0.010930379 | 1.57E-10 | 5.15E-07 | 1.47E-08 | ITIH4 |
| SAT | Obesity | ENSG00000058063.11 | 0.008410033 | 0.002324537 | 1 | 0.02956126 | ATP11B |
| SAT | Obesity | ENSG00000064995.12 | -0.008993872 | 0.001731465 | 1 | 0.022999738 | TAF11 |
| SAT | Obesity | ENSG00000065060.12 | 0.017877663 | 7.04E-23 | 2.31E-19 | 2.31E-20 | UHRF1BP1 |
| SAT | Obesity | ENSG00000071794.11 | -0.011929378 | 0.004136335 | 1 | 0.046318486 | HLTF |
| SAT | Obesity | ENSG00000075131.5 | 0.007142579 | 2.48E-05 | 0.081257563 | 0.00073805 | TIPIN |
| SAT | Obesity | ENSG00000076685.14 | 0.027327284 | 3.57E-06 | 0.011724467 | 0.000160609 | NT5C2 |
| SAT | Obesity | ENSG00000077380.11 | 0.024794871 | 4.63E-06 | 0.015187396 | 0.000192246 | DYNC1I2 |
| SAT | Obesity | ENSG00000082258.8 | -0.009146466 | 6.69E-05 | 0.219488925 | 0.001602715 | CCNT2 |
| SAT | Obesity | ENSG00000083097.10 | 0.005179793 | 0.00429089 | 1 | 0.047531447 | DOPEY1 |
| SAT | Obesity | ENSG00000085365.13 | 0.017474778 | 0.000244244 | 0.80136467 | 0.00471391 | SCAMP1 |
| SAT | Obesity | ENSG00000087263.12 | -0.020899752 | 5.74E-07 | 0.001884118 | 2.98E-05 | OGFOD1 |
| SAT | Obesity | ENSG00000090238.7 | 0.006676745 | 8.90E-09 | 2.92E-05 | 6.64E-07 | YPEL3 |
| SAT | Obesity | ENSG00000090621.9 | -0.016852852 | 1.26E-10 | 4.15E-07 | 1.22E-08 | PABPC4 |
| SAT | Obesity | ENSG00000099956.13 | 0.00767829 | 0.001282303 | 1 | 0.018134645 | SMARCB1 |
| SAT | Obesity | ENSG00000100124.8 | -0.012590642 | 8.06E-05 | 0.264521688 | 0.001862829 | ANKRD54 |
| SAT | Obesity | ENSG00000100225.13 | 0.013094019 | 0.00056832 | 1 | 0.009661437 | FBXO7 |
| SAT | Obesity | ENSG00000100418.7 | 0.009406871 | 2.48E-05 | 0.081304379 | 0.00073805 | DESI1 |
| SAT | Obesity | ENSG00000100697.10 | 0.009578081 | 0.000337454 | 1 | 0.006185402 | DICER1 |
| SAT | Obesity | ENSG00000100997.14 | 0.018638295 | 1.95E-05 | 0.063843665 | 0.00059667 | ABHD12 |
| SAT | Obesity | ENSG00000102595.14 | -0.019278175 | 0.000893975 | 1 | 0.013579312 | UGGT2 |
| SAT | Obesity | ENSG00000102890.10 | 0.007148976 | 0.002061819 | 1 | 0.026738446 | ELMO3 |
| SAT | Obesity | ENSG00000103051.14 | -0.020206062 | 1.48E-06 | 0.004853661 | 6.93E-05 | COG4 |
| SAT | Obesity | ENSG00000103351.8 | 0.009005263 | 0.0003845 | 1 | 0.00693156 | CLUAP1 |
| SAT | Obesity | ENSG00000103550.9 | -0.017974194 | 4.37E-06 | 0.014325206 | 0.000186042 | KNOP1 |
| SAT | Obesity | ENSG00000104219.8 | -0.011961943 | 8.71E-06 | 0.028562241 | 0.000299495 | ZDHHC2 |
| SAT | Obesity | ENSG00000104738.12 | 0.014983574 | 0.002942468 | 1 | 0.036023276 | MCM4 |
| SAT | Obesity | ENSG00000104852.10 | -0.024042722 | 4.89E-05 | 0.16030374 | 0.001292772 | SNRNP70 |
| SAT | Obesity | ENSG00000104866.6 | 0.020159468 | 0.000490047 | 1 | 0.008507117 | PPP1R37 |
| SAT | Obesity | ENSG00000105656.8 | -0.020595188 | 0.000712265 | 1 | 0.01139972 | ELL |
| SAT | Obesity | ENSG00000105726.12 | -0.013128225 | 9.77E-06 | 0.032061565 | 0.000323854 | ATP13A1 |
| SAT | Obesity | ENSG00000108384.10 | -0.005753469 | 0.000421317 | 1 | 0.007477175 | RAD51C |
| SAT | Obesity | ENSG00000108556.7 | 0.00785795 | 0.001099647 | 1 | 0.016106882 | CHRNE |
| SAT | Obesity | ENSG00000110906.8 | -0.007453852 | 0.001712482 | 1 | 0.022842502 | KCTD10 |
| SAT | Obesity | ENSG00000111364.11 | 0.010011036 | 9.08E-05 | 0.297799924 | 0.00202585 | DDX55 |
| SAT | Obesity | ENSG00000112053.9 | -0.008512259 | 0.002371971 | 1 | 0.030048015 | SLC26A8 |
| SAT | Obesity | ENSG00000112812.11 | 0.017307355 | 3.78E-06 | 0.012414489 | 0.000167763 | PRSS16 |
| SAT | Obesity | ENSG00000113141.11 | -0.011799974 | 0.000228853 | 0.750867837 | 0.004550714 | IK |
| SAT | Obesity | ENSG00000113734.13 | -0.005273578 | 0.001810209 | 1 | 0.023757177 | BNIP1 |
| SAT | Obesity | ENSG00000114302.11 | 0.013108288 | 0.003297945 | 1 | 0.039204918 | PRKAR2A |
| SAT | Obesity | ENSG00000114378.12 | -0.008814288 | 0.000409199 | 1 | 0.007336518 | HYAL1 |
| SAT | Obesity | ENSG00000114796.11 | 0.018763661 | 0.000639387 | 1 | 0.010446545 | KLHL24 |
| SAT | Obesity | ENSG00000115310.13 | -0.014383428 | 0.003917062 | 1 | 0.044436554 | RTN4 |
| SAT | Obesity | ENSG00000115561.10 | -0.012536593 | 1.88E-07 | 0.000616035 | 1.06E-05 | CHMP3 |
| SAT | Obesity | ENSG00000116138.8 | -0.015460011 | 0.00083258 | 1 | 0.012764933 | DNAJC16 |
| SAT | Obesity | ENSG00000117122.9 | -0.008629217 | 0.000775669 | 1 | 0.012176893 | MFAP2 |
| SAT | Obesity | ENSG00000117523.11 | 0.018647624 | 0.00059242 | 1 | 0.009866658 | PRRC2C |
| SAT | Obesity | ENSG00000117601.9 | 0.021715825 | 5.63E-10 | 1.85E-06 | 5.00E-08 | SERPINC1 |
| SAT | Obesity | ENSG00000118004.13 | -0.017575243 | 0.00372367 | 1 | 0.043018874 | COLEC11 |
| SAT | Obesity | ENSG00000119242.4 | 0.027390299 | 8.21E-12 | 2.69E-08 | 8.69E-10 | CCDC92 |
| SAT | Obesity | ENSG00000119599.12 | 0.016618221 | 8.71E-05 | 0.285612737 | 0.001973489 | DCAF4 |
| SAT | Obesity | ENSG00000119865.4 | -0.013705844 | 0.001403582 | 1 | 0.019431029 | CNRIP1 |
| SAT | Obesity | ENSG00000119878.5 | -0.020584036 | 0.000723481 | 1 | 0.011523022 | CRIPT |
| SAT | Obesity | ENSG00000120093.7 | 0.017886924 | 6.84E-06 | 0.02243978 | 0.000249331 | HOXB3 |
| SAT | Obesity | ENSG00000120314.14 | -0.021424112 | 0.000173122 | 0.568012556 | 0.003641106 | WDR55 |
| SAT | Obesity | ENSG00000120451.6 | -0.017572975 | 3.82E-05 | 0.12529416 | 0.001046683 | SNX19 |
| SAT | Obesity | ENSG00000120907.13 | 0.00823833 | 6.98E-05 | 0.229108206 | 0.001660204 | ADRA1A |
| SAT | Obesity | ENSG00000121101.11 | 0.019050009 | 0.001503146 | 1 | 0.020513962 | TEX14 |
| SAT | Obesity | ENSG00000121957.8 | 0.017593969 | 0.003691149 | 1 | 0.042793854 | GPSM2 |
| SAT | Obesity | ENSG00000124562.5 | -0.02280671 | 1.09E-23 | 3.57E-20 | 3.96E-21 | SNRPC |
| SAT | Obesity | ENSG00000124635.7 | 0.027951843 | 6.71E-07 | 0.002202038 | 3.34E-05 | HIST1H2BJ |

| | | | | | | | |
|---|---|---|---|---|---|---|---|
| SAT | Obesity | ENSG00000125388.15 | 0.006154441 | 3.83E-05 | 0.12560192 | 0.001046683 | GRK4 |
| SAT | Obesity | ENSG00000125510.11 | 0.015497886 | 0.00119393 | 1 | 0.017256766 | OPRL1 |
| SAT | Obesity | ENSG00000127337.2 | 0.006090088 | 0.0017743 | 1 | 0.023473705 | YEATS4 |
| SAT | Obesity | ENSG00000127957.12 | -0.024926563 | 5.78E-21 | 1.90E-17 | 1.46E-18 | PMS2P3 |
| SAT | Obesity | ENSG00000128408.7 | 0.013371095 | 0.000305384 | 1 | 0.005692976 | RIBC2 |
| SAT | Obesity | ENSG00000129197.10 | -0.032204003 | 3.65E-08 | 0.000119856 | 2.45E-06 | RPAIN |
| SAT | Obesity | ENSG00000130270.12 | -0.01400295 | 1.47E-08 | 4.83E-05 | 1.03E-06 | ATP8B3 |
| SAT | Obesity | ENSG00000130517.9 | 0.009807601 | 0.001624761 | 1 | 0.021937612 | PGPEP1 |
| SAT | Obesity | ENSG00000130943.5 | -0.008874988 | 0.001784837 | 1 | 0.02351828 | PKDREJ |
| SAT | Obesity | ENSG00000132205.6 | 0.026821135 | 5.01E-07 | 0.001642551 | 2.65E-05 | EMILIN2 |
| SAT | Obesity | ENSG00000132740.4 | 0.008664672 | 1.17E-05 | 0.038497372 | 0.000381162 | IGHMBP2 |
| SAT | Obesity | ENSG00000133243.4 | -0.015967862 | 0.002319207 | 1 | 0.02956126 | BTBD2 |
| SAT | Obesity | ENSG00000133706.13 | -0.00508441 | 0.00022355 | 0.733468235 | 0.004472367 | LARS |
| SAT | Obesity | ENSG00000134201.6 | 0.013463108 | 0.004302603 | 1 | 0.047531447 | GSTM5 |
| SAT | Obesity | ENSG00000134575.5 | 0.017574904 | 5.48E-05 | 0.17982338 | 0.001415932 | ACP2 |
| SAT | Obesity | ENSG00000135124.10 | 0.014759029 | 0.001597735 | 1 | 0.021661855 | P2RX4 |
| SAT | Obesity | ENSG00000135211.5 | -0.01668482 | 1.50E-05 | 0.049339487 | 0.000479024 | TMEM60 |
| SAT | Obesity | ENSG00000135213.8 | -0.046739279 | 1.65E-19 | 5.41E-16 | 3.61E-17 | POM121C |
| SAT | Obesity | ENSG00000135913.6 | 0.01635167 | 7.54E-05 | 0.247256916 | 0.001778827 | USP37 |
| SAT | Obesity | ENSG00000135972.4 | 0.006534016 | 0.004170656 | 1 | 0.046543954 | MRPS9 |
| SAT | Obesity | ENSG00000136982.5 | 0.003482088 | 0.003226864 | 1 | 0.038781468 | DSCC1 |
| SAT | Obesity | ENSG00000137185.7 | 0.009808909 | 8.76E-05 | 0.287531101 | 0.001973489 | ZSCAN9 |
| SAT | Obesity | ENSG00000137216.14 | -0.012363997 | 0.003313348 | 1 | 0.039245833 | TMEM63B |
| SAT | Obesity | ENSG00000137404.10 | 0.015205567 | 0.002904481 | 1 | 0.035691396 | NRM |
| SAT | Obesity | ENSG00000138380.13 | 0.015750905 | 4.30E-06 | 0.014119163 | 0.000185778 | CARF |
| SAT | Obesity | ENSG00000139187.5 | 0.006733262 | 0.003389699 | 1 | 0.040005764 | KLRG1 |
| SAT | Obesity | ENSG00000139405.11 | -0.007247464 | 0.002727848 | 1 | 0.034160566 | RITA1 |
| SAT | Obesity | ENSG00000140600.12 | -0.018049471 | 0.000203285 | 0.666978035 | 0.004142721 | SH3GL3 |
| SAT | Obesity | ENSG00000141076.13 | 0.008823096 | 0.000643158 | 1 | 0.010446545 | CIRH1A |
| SAT | Obesity | ENSG00000141452.5 | -0.029765758 | 2.11E-13 | 6.94E-10 | 2.78E-11 | C18orf8 |
| SAT | Obesity | ENSG00000141458.8 | -0.040230733 | 2.84E-13 | 9.32E-10 | 3.58E-11 | NPC1 |
| SAT | Obesity | ENSG00000141569.6 | 0.013012468 | 1.76E-05 | 0.057832235 | 0.00054761 | TRIM65 |
| SAT | Obesity | ENSG00000142534.2 | 0.013970509 | 0.002499151 | 1 | 0.031537368 | RPS11 |
| SAT | Obesity | ENSG00000143367.11 | -0.01534896 | 0.000815486 | 1 | 0.012561551 | TUFT1 |
| SAT | Obesity | ENSG00000143575.10 | 0.01914416 | 0.000573508 | 1 | 0.009667596 | HAX1 |
| SAT | Obesity | ENSG00000143702.11 | 0.01722168 | 0.000360136 | 1 | 0.00654777 | CEP170 |
| SAT | Obesity | ENSG00000144218.14 | -0.020145986 | 0.000809544 | 1 | 0.012532499 | AFF3 |
| SAT | Obesity | ENSG00000144426.14 | 0.011405036 | 4.48E-05 | 0.147047397 | 0.001205307 | NBEAL1 |
| SAT | Obesity | ENSG00000145020.10 | -0.004643925 | 0.00126081 | 1 | 0.017985728 | AMT |
| SAT | Obesity | ENSG00000145029.7 | -0.007688364 | 0.001389118 | 1 | 0.019312276 | NICN1 |
| SAT | Obesity | ENSG00000145241.6 | -0.014759239 | 8.36E-06 | 0.027435231 | 0.00029764 | CENPC |
| SAT | Obesity | ENSG00000145416.9 | -0.012797829 | 0.001277164 | 1 | 0.018134645 | MARCH1 |
| SAT | Obesity | ENSG00000145832.8 | -0.013198781 | 0.000751391 | 1 | 0.011909726 | SLC25A48 |
| SAT | Obesity | ENSG00000145882.6 | -0.012551354 | 0.000215879 | 0.70830057 | 0.004372226 | PCYOX1L |
| SAT | Obesity | ENSG00000146109.3 | 0.012643625 | 0.000270544 | 0.887656266 | 0.005101473 | ABT1 |
| SAT | Obesity | ENSG00000148572.10 | -0.010926754 | 0.000361215 | 1 | 0.00654777 | NRBF2 |
| SAT | Obesity | ENSG00000148824.14 | 0.006880983 | 0.000952249 | 1 | 0.014397835 | MTG1 |
| SAT | Obesity | ENSG00000149084.7 | -0.007552758 | 4.68E-12 | 1.54E-08 | 5.12E-10 | HSD17B12 |
| SAT | Obesity | ENSG00000149292.12 | 0.005345496 | 6.25E-05 | 0.204985657 | 0.001541246 | TTC12 |
| SAT | Obesity | ENSG00000149548.10 | -0.012312527 | 9.13E-06 | 0.029954828 | 0.000305662 | CCDC15 |
| SAT | Obesity | ENSG00000149922.6 | 0.01776672 | 5.99E-08 | 0.000196444 | 3.71E-06 | TBX6 |
| SAT | Obesity | ENSG00000151006.7 | -0.023778637 | 1.16E-06 | 0.003818547 | 5.62E-05 | PRSS53 |
| SAT | Obesity | ENSG00000151640.8 | 0.005947517 | 0.000153636 | 0.504078468 | 0.003273237 | DPYSL4 |
| SAT | Obesity | ENSG00000151917.13 | -0.016022363 | 0.001481122 | 1 | 0.020332893 | BEND6 |
| SAT | Obesity | ENSG00000152061.17 | 0.00993509 | 0.001712666 | 1 | 0.022842502 | RABGAP1L |
| SAT | Obesity | ENSG00000152219.4 | -0.022440645 | 1.08E-05 | 0.035524439 | 0.000355244 | ARL14EP |
| SAT | Obesity | ENSG00000152359.10 | -0.013535617 | 3.82E-08 | 0.000125486 | 2.51E-06 | POC5 |
| SAT | Obesity | ENSG00000153814.7 | 0.020498389 | 2.82E-05 | 0.092484676 | 0.000818448 | JAZF1 |
| SAT | Obesity | ENSG00000154240.12 | 0.015441913 | 0.003927644 | 1 | 0.044436554 | CEP112 |
| SAT | Obesity | ENSG00000154328.11 | 0.015725558 | 0.000238861 | 0.783703498 | 0.004690985 | NEIL2 |
| SAT | Obesity | ENSG00000154710.11 | -0.004344335 | 0.003169214 | 1 | 0.03822864 | RABGEF1 |
| SAT | Obesity | ENSG00000157214.9 | -0.011628705 | 0.000197743 | 0.648793464 | 0.004054959 | STEAP2 |
| SAT | Obesity | ENSG00000157322.12 | -0.019597129 | 1.07E-17 | 3.51E-14 | 2.06E-15 | CLEC18A |
| SAT | Obesity | ENSG00000158104.7 | 0.027885679 | 3.90E-06 | 0.012798118 | 0.000170642 | HPD |
| SAT | Obesity | ENSG00000158623.10 | -0.005595692 | 0.001306911 | 1 | 0.018403323 | COPG2 |
| SAT | Obesity | ENSG00000158865.8 | -0.009120169 | 1.43E-07 | 0.000469438 | 8.38E-06 | SLC5A11 |
| SAT | Obesity | ENSG00000160049.7 | 0.012898403 | 0.00435504 | 1 | 0.047949285 | DFFA |
| SAT | Obesity | ENSG00000160200.13 | -0.010800855 | 0.002094583 | 1 | 0.027056402 | CBS |
| SAT | Obesity | ENSG00000162341.11 | 0.016427474 | 0.004189299 | 1 | 0.046593526 | TPCN2 |
| SAT | Obesity | ENSG00000162994.11 | 0.002986389 | 0.003855174 | 1 | 0.043920336 | CLHC1 |
| SAT | Obesity | ENSG00000163297.12 | 0.024608233 | 3.11E-05 | 0.102058163 | 0.000895247 | ANTXR2 |
| SAT | Obesity | ENSG00000163406.6 | -0.012292034 | 0.002569742 | 1 | 0.032303922 | SLC15A2 |
| SAT | Obesity | ENSG00000163431.11 | -0.038194812 | 3.52E-15 | 1.16E-11 | 5.50E-13 | LMOD1 |
| SAT | Obesity | ENSG00000163596.12 | -0.014307922 | 5.69E-05 | 0.186685093 | 0.001429286 | ICA1L |
| SAT | Obesity | ENSG00000163933.5 | -0.022945619 | 2.91E-07 | 0.000955264 | 1.59E-05 | RFT1 |
| SAT | Obesity | ENSG00000164062.8 | -0.022516413 | 9.88E-13 | 3.24E-09 | 1.20E-10 | APEH |
| SAT | Obesity | ENSG00000164068.11 | 0.015964241 | 6.72E-21 | 2.20E-17 | 1.57E-18 | RNF123 |
| SAT | Obesity | ENSG00000164078.8 | 0.021221784 | 5.81E-07 | 0.001905393 | 2.98E-05 | MST1R |
| SAT | Obesity | ENSG00000164081.8 | -0.00872573 | 0.000308979 | 1 | 0.005727464 | TEX264 |
| SAT | Obesity | ENSG00000164088.13 | -0.01339176 | 4.99E-05 | 0.163870549 | 0.00130056 | PPM1M |
| SAT | Obesity | ENSG00000164307.8 | -0.012260691 | 5.64E-05 | 0.185177429 | 0.001429286 | ERAP1 |

| | | | | | | | |
|---|---|---|---|---|---|---|---|
| SAT | Obesity | ENSG00000164924.13 | -0.010821797 | 0.000121501 | 0.398643746 | 0.002622656 | YWHAZ |
| SAT | Obesity | ENSG00000165178.8 | 0.016283427 | 5.89E-06 | 0.019317811 | 0.000225872 | NCF1C |
| SAT | Obesity | ENSG00000165476.8 | -0.01650235 | 0.000313681 | 1 | 0.005781949 | REEP3 |
| SAT | Obesity | ENSG00000165916.4 | 0.009940708 | 0.003985623 | 1 | 0.044937557 | PSMC3 |
| SAT | Obesity | ENSG00000166166.8 | -0.027505078 | 2.74E-06 | 0.008979114 | 0.00012471 | TRMT61A |
| SAT | Obesity | ENSG00000166529.10 | -0.006685048 | 0.002858769 | 1 | 0.035394795 | ZSCAN21 |
| SAT | Obesity | ENSG00000166532.11 | -0.004663568 | 0.003842084 | 1 | 0.043920336 | RIMKLB |
| SAT | Obesity | ENSG00000166913.8 | -0.014732008 | 0.002302946 | 1 | 0.029515494 | YWHAB |
| SAT | Obesity | ENSG00000167118.6 | -0.018423772 | 0.000240469 | 0.788979083 | 0.004690985 | URM1 |
| SAT | Obesity | ENSG00000167395.10 | 0.036937091 | 2.40E-09 | 7.87E-06 | 1.92E-07 | ZNF646 |
| SAT | Obesity | ENSG00000167397.10 | -0.028526597 | 1.22E-09 | 3.99E-06 | 1.05E-07 | VKORC1 |
| SAT | Obesity | ENSG00000167671.7 | -0.021454305 | 6.41E-05 | 0.210389038 | 0.001558437 | UBXN6 |
| SAT | Obesity | ENSG00000167720.8 | 0.005885123 | 8.76E-06 | 0.028751567 | 0.000299495 | SRR |
| SAT | Obesity | ENSG00000168090.5 | -0.016659455 | 0.001031198 | 1 | 0.015449135 | COPS6 |
| SAT | Obesity | ENSG00000168268.6 | -0.017166669 | 0.003602275 | 1 | 0.042161685 | NT5DC2 |
| SAT | Obesity | ENSG00000168405.10 | 0.023295629 | 4.43E-05 | 0.145510061 | 0.001202562 | CMAHP |
| SAT | Obesity | ENSG00000169085.7 | 0.003778689 | 0.004081271 | 1 | 0.04585839 | C8orf46 |
| SAT | Obesity | ENSG00000169203.12 | 0.016424187 | 1.57E-13 | 5.14E-10 | 2.14E-11 | RP11-231C14.4 |
| SAT | Obesity | ENSG00000169592.10 | -0.030717024 | 1.97E-15 | 6.47E-12 | 3.60E-13 | INO80E |
| SAT | Obesity | ENSG00000169612.3 | -0.011326705 | 0.000147915 | 0.485307476 | 0.003171944 | FAM103A1 |
| SAT | Obesity | ENSG00000170092.10 | 0.035993839 | 8.11E-18 | 2.66E-14 | 1.66E-15 | AC018720.10 |
| SAT | Obesity | ENSG00000170425.3 | -0.01180501 | 9.26E-07 | 0.003038024 | 4.53E-05 | ADORA2B |
| SAT | Obesity | ENSG00000171103.6 | 0.006621874 | 3.14E-05 | 0.103008686 | 0.000895728 | TRMT61B |
| SAT | Obesity | ENSG00000172247.3 | -0.011342034 | 2.98E-15 | 9.78E-12 | 4.89E-13 | C1QTNF4 |
| SAT | Obesity | ENSG00000172728.11 | -0.008454642 | 5.71E-05 | 0.187236523 | 0.001429286 | FUT10 |
| SAT | Obesity | ENSG00000172780.12 | 0.01313391 | 0.000526879 | 1 | 0.009009799 | RAB43 |
| SAT | Obesity | ENSG00000173226.12 | -0.006389418 | 0.000861686 | 1 | 0.013149728 | IQCB1 |
| SAT | Obesity | ENSG00000173531.11 | -0.013773754 | 8.53E-06 | 0.027978132 | 0.00029764 | MST1 |
| SAT | Obesity | ENSG00000173540.8 | 0.031979962 | 8.01E-08 | 0.000262732 | 4.78E-06 | GMPPB |
| SAT | Obesity | ENSG00000173917.9 | 0.010888814 | 0.003255678 | 1 | 0.038984968 | HOXB2 |
| SAT | Obesity | ENSG00000174775.12 | 0.018442261 | 0.002056266 | 1 | 0.026738446 | HRAS |
| SAT | Obesity | ENSG00000174891.8 | -0.007745753 | 0.001706672 | 1 | 0.022842502 | RSRC1 |
| SAT | Obesity | ENSG00000176095.7 | 0.013326345 | 0.000291662 | 0.956943251 | 0.005468247 | IP6K1 |
| SAT | Obesity | ENSG00000176155.14 | -0.011091601 | 0.000221105 | 0.725444264 | 0.004450578 | CCDC57 |
| SAT | Obesity | ENSG00000176783.10 | 0.012548425 | 4.67E-05 | 0.153264522 | 0.001246053 | RUFY1 |
| SAT | Obesity | ENSG00000177156.6 | -0.017488416 | 0.000999924 | 1 | 0.015049313 | TALDO1 |
| SAT | Obesity | ENSG00000177380.9 | -0.021047357 | 0.000179414 | 0.588655802 | 0.0037494 | PPFIA3 |
| SAT | Obesity | ENSG00000177595.13 | 0.01415513 | 6.69E-06 | 0.021963429 | 0.00024678 | PIDD |
| SAT | Obesity | ENSG00000177697.13 | 0.015511684 | 0.001156523 | 1 | 0.016790048 | CD151 |
| SAT | Obesity | ENSG00000177700.5 | -0.017361272 | 0.001419596 | 1 | 0.019570148 | POLR2L |
| SAT | Obesity | ENSG00000178188.10 | -0.036534084 | 1.96E-31 | 6.44E-28 | 3.22E-28 | SH2B1 |
| SAT | Obesity | ENSG00000178467.13 | -0.024224317 | 4.97E-05 | 0.162925248 | 0.00130056 | P4HTM |
| SAT | Obesity | ENSG00000178718.5 | -0.012410505 | 0.000467919 | 1 | 0.008253984 | RPP25 |
| SAT | Obesity | ENSG00000178750.2 | 0.025963157 | 2.44E-07 | 0.000799682 | 1.36E-05 | STX19 |
| SAT | Obesity | ENSG00000178809.7 | -0.037929965 | 1.80E-12 | 5.91E-09 | 2.11E-10 | TRIM73 |
| SAT | Obesity | ENSG00000178952.4 | 0.02279601 | 8.03E-23 | 2.63E-19 | 2.39E-20 | TUFM |
| SAT | Obesity | ENSG00000179115.6 | -0.019354201 | 0.001044378 | 1 | 0.015575476 | FARSA |
| SAT | Obesity | ENSG00000179673.3 | 0.008953975 | 0.000481406 | 1 | 0.008401553 | RPRML |
| SAT | Obesity | ENSG00000180385.4 | 0.015022513 | 2.39E-05 | 0.078356152 | 0.00072552 | EMC3-AS1 |
| SAT | Obesity | ENSG00000181315.6 | -0.01832876 | 4.55E-08 | 0.000149321 | 2.93E-06 | ZNF322 |
| SAT | Obesity | ENSG00000182533.6 | -0.00964656 | 0.001506816 | 1 | 0.020513962 | CAV3 |
| SAT | Obesity | ENSG00000182685.3 | -0.00977826 | 0.002277223 | 1 | 0.029300269 | BRICD5 |
| SAT | Obesity | ENSG00000182841.8 | 0.026747317 | 9.08E-06 | 0.029782437 | 0.000305662 | RRP7B |
| SAT | Obesity | ENSG00000183060.11 | 0.006281201 | 0.003160748 | 1 | 0.03822864 | LYSMD4 |
| SAT | Obesity | ENSG00000183527.7 | 0.006590148 | 3.40E-05 | 0.111687929 | 0.000954598 | PSMG1 |
| SAT | Obesity | ENSG00000183604.10 | -0.027129336 | 1.74E-14 | 5.72E-11 | 2.60E-12 | RP11-347C12.2 |
| SAT | Obesity | ENSG00000184110.10 | 0.034581887 | 1.16E-27 | 3.81E-24 | 6.35E-25 | EIF3C |
| SAT | Obesity | ENSG00000184381.14 | 0.028566128 | 6.66E-07 | 0.002184204 | 3.34E-05 | PLA2G6 |
| SAT | Obesity | ENSG00000185245.6 | 0.024790052 | 7.54E-06 | 0.02472934 | 0.000271751 | GP1BA |
| SAT | Obesity | ENSG00000186470.9 | -0.005600123 | 5.80E-08 | 0.000190203 | 3.66E-06 | BTN3A2 |
| SAT | Obesity | ENSG00000186792.11 | -0.008932908 | 5.69E-06 | 0.018657521 | 0.000222113 | HYAL3 |
| SAT | Obesity | ENSG00000188312.9 | 0.00826765 | 1.20E-06 | 0.003948386 | 5.72E-05 | CENPP |
| SAT | Obesity | ENSG00000188878.12 | 0.006909775 | 0.000527242 | 1 | 0.009009799 | FBF1 |
| SAT | Obesity | ENSG00000189157.9 | 0.020440046 | 0.000473803 | 1 | 0.008313089 | FAM47E |
| SAT | Obesity | ENSG00000196502.7 | -0.02228088 | 7.37E-08 | 0.000241689 | 4.48E-06 | SULT1A1 |
| SAT | Obesity | ENSG00000196821.5 | 0.016928598 | 8.12E-09 | 2.66E-05 | 6.20E-07 | C6orf106 |
| SAT | Obesity | ENSG00000196968.6 | 0.017456293 | 0.000501354 | 1 | 0.008657585 | FUT11 |
| SAT | Obesity | ENSG00000197062.7 | -0.007421931 | 0.000112433 | 0.368891926 | 0.002475785 | RP5-874C20.3 |
| SAT | Obesity | ENSG00000197119.8 | 0.015802193 | 0.003617774 | 1 | 0.042161685 | SLC25A29 |
| SAT | Obesity | ENSG00000197165.6 | 0.025056674 | 4.73E-29 | 1.55E-25 | 3.88E-26 | SULT1A2 |
| SAT | Obesity | ENSG00000197262.8 | -0.022032746 | 5.92E-06 | 0.019424959 | 0.000225872 | CCL4L2 |
| SAT | Obesity | ENSG00000197279.3 | -0.013316878 | 0.003458675 | 1 | 0.040673517 | ZNF165 |
| SAT | Obesity | ENSG00000197345.8 | -0.00459975 | 0.000260086 | 0.853342504 | 0.004934546 | MRPL21 |
| SAT | Obesity | ENSG00000197728.5 | -0.004951639 | 7.76E-05 | 0.254532644 | 0.00181809 | RPS26 |
| SAT | Obesity | ENSG00000197782.10 | -0.005772295 | 6.58E-06 | 0.021573699 | 0.000245156 | ZNF780A |
| SAT | Obesity | ENSG00000198093.6 | -0.011036791 | 0.004524819 | 1 | 0.049651945 | ZNF649 |
| SAT | Obesity | ENSG00000198099.4 | -0.012664397 | 6.69E-05 | 0.219571995 | 0.001602715 | ADH4 |
| SAT | Obesity | ENSG00000198315.6 | 0.024473063 | 4.94E-06 | 0.016193189 | 0.000199916 | ZKSCAN8 |
| SAT | Obesity | ENSG00000204316.8 | -0.016294726 | 0.00064131 | 1 | 0.010446545 | MRPL38 |
| SAT | Obesity | ENSG00000204351.7 | -0.004266343 | 6.31E-05 | 0.207024255 | 0.001544957 | SKIV2L |
| SAT | Obesity | ENSG00000204356.7 | 0.008249118 | 0.000181971 | 0.597045615 | 0.00377877 | NELFE |

| | | | | | | | |
|---|---|---|---|---|---|---|---|
| SAT | Obesity | ENSG00000204531.11 | 0.004670615 | 0.000629435 | 1 | 0.010377772 | POU5F1 |
| SAT | Obesity | ENSG00000204536.9 | 0.004687221 | 0.000241626 | 0.792776436 | 0.004690985 | CCHCR1 |
| SAT | Obesity | ENSG00000204574.8 | 0.010231213 | 0.003855244 | 1 | 0.043920336 | ABCF1 |
| SAT | Obesity | ENSG00000204592.5 | 0.022714046 | 1.39E-09 | 4.56E-06 | 1.17E-07 | HLA-E |
| SAT | Obesity | ENSG00000204644.5 | 0.004463343 | 0.000109397 | 0.358933116 | 0.002425224 | ZFP57 |
| SAT | Obesity | ENSG00000204789.3 | 0.012733339 | 4.93E-06 | 0.016168856 | 0.000199916 | ZNF204P |
| SAT | Obesity | ENSG00000205571.8 | -0.006592112 | 0.003294321 | 1 | 0.039204918 | SMN2 |
| SAT | Obesity | ENSG00000205583.9 | -0.018817342 | 4.17E-21 | 1.37E-17 | 1.14E-18 | STAG3L1 |
| SAT | Obesity | ENSG00000205710.3 | 0.007777583 | 0.000231813 | 0.760578827 | 0.0045818 | C17orf107 |
| SAT | Obesity | ENSG00000212694.4 | -0.010187849 | 0.000187365 | 0.614743135 | 0.003866309 | AC084018.1 |
| SAT | Obesity | ENSG00000213199.3 | 0.014354611 | 5.79E-05 | 0.190112536 | 0.001440246 | ASIC3 |
| SAT | Obesity | ENSG00000213240.7 | 0.01155662 | 0.000795788 | 1 | 0.012433244 | NOTCH2NL |
| SAT | Obesity | ENSG00000213277.3 | 0.017674233 | 7.88E-05 | 0.258687005 | 0.00183466 | MARCKSL1P1 |
| SAT | Obesity | ENSG00000213639.5 | -0.020123905 | 2.50E-05 | 0.081923556 | 0.00073805 | PPP1CB |
| SAT | Obesity | ENSG00000214279.8 | -0.013128371 | 0.00193562 | 1 | 0.025301871 | RP11-108K14.4 |
| SAT | Obesity | ENSG00000214659.4 | 0.02253179 | 0.000115136 | 0.377760746 | 0.002501727 | KRT8P26 |
| SAT | Obesity | ENSG00000223496.1 | -0.033906615 | 2.24E-15 | 7.35E-12 | 3.87E-13 | EXOSC6 |
| SAT | Obesity | ENSG00000224389.4 | 0.011035607 | 0.000578937 | 1 | 0.009691288 | C4B |
| SAT | Obesity | ENSG00000224843.2 | 0.027332415 | 3.56E-07 | 0.001166758 | 1.91E-05 | LINC00240 |
| SAT | Obesity | ENSG00000224997.1 | -0.01067946 | 8.47E-06 | 0.027775013 | 0.00029764 | AL049840.1 |
| SAT | Obesity | ENSG00000225726.1 | -0.014695957 | 0.002756575 | 1 | 0.034389064 | AC007000.10 |
| SAT | Obesity | ENSG00000226232.4 | -0.010804259 | 5.71E-09 | 1.87E-05 | 4.46E-07 | RP11-419C5.2 |
| SAT | Obesity | ENSG00000227370.1 | -0.019186444 | 0.001106192 | 1 | 0.016130738 | RP4-669P10.19 |
| SAT | Obesity | ENSG00000230521.1 | 0.004482198 | 0.003829703 | 1 | 0.043920336 | HCG4P7 |
| SAT | Obesity | ENSG00000231500.2 | 0.008023035 | 0.000260188 | 0.853676482 | 0.004934546 | RPS18 |
| SAT | Obesity | ENSG00000231852.2 | 0.008473329 | 5.63E-05 | 0.184626332 | 0.001429286 | CYP21A2 |
| SAT | Obesity | ENSG00000232561.2 | -0.030197708 | 3.72E-12 | 1.22E-08 | 4.21E-10 | GTF2IP1 |
| SAT | Obesity | ENSG00000233137.2 | 0.025925009 | 2.52E-05 | 0.082800682 | 0.000739292 | RP11-220I1.1 |
| SAT | Obesity | ENSG00000233609.3 | 0.017348264 | 0.000809781 | 1 | 0.012532499 | RP11-62H7.2 |
| SAT | Obesity | ENSG00000235109.3 | 0.00630703 | 0.002836556 | 1 | 0.035252799 | ZSCAN31 |
| SAT | Obesity | ENSG00000235821.1 | -0.006112398 | 0.003086069 | 1 | 0.037501449 | IFITM4P |
| SAT | Obesity | ENSG00000239557.1 | 0.03522512 | 9.39E-09 | 3.08E-05 | 6.85E-07 | RP11-168J18.6 |
| SAT | Obesity | ENSG00000245958.2 | -0.007466345 | 0.001369965 | 1 | 0.019127047 | RP11-33B1.1 |
| SAT | Obesity | ENSG00000248290.1 | -0.006363871 | 3.32E-05 | 0.109084671 | 0.000940385 | TNXA |
| SAT | Obesity | ENSG00000248408.1 | -0.026588763 | 5.52E-06 | 0.01812343 | 0.000218355 | RP11-452C8.1 |
| SAT | Obesity | ENSG00000250412.1 | 0.015571498 | 0.000574575 | 1 | 0.009667596 | KLHL2P1 |
| SAT | Obesity | ENSG00000251417.1 | -0.022797794 | 4.83E-30 | 1.59E-26 | 5.29E-27 | RP11-1348G14.4 |
| SAT | Obesity | ENSG00000255298.2 | -0.013003033 | 0.00122897 | 1 | 0.017685316 | OR8G5 |
| SAT | Obesity | ENSG00000256269.2 | 0.020655418 | 2.11E-11 | 6.91E-08 | 2.16E-09 | HMBS |
| SAT | Obesity | ENSG00000256604.1 | -0.012777939 | 0.003015237 | 1 | 0.036776926 | AC093668.2 |
| SAT | Obesity | ENSG00000259863.1 | -0.012834804 | 0.000157677 | 0.517338 | 0.003337665 | SH3RF3-AS1 |
| SAT | Obesity | ENSG00000259982.1 | 0.030223046 | 3.88E-33 | 1.27E-29 | 1.27E-29 | CDC37P1 |
| SAT | Obesity | ENSG00000260388.1 | -0.011396776 | 0.00066081 | 1 | 0.010628026 | LINC00562 |
| SAT | Obesity | ENSG00000260911.1 | 0.016932484 | 2.39E-08 | 7.84E-05 | 1.63E-06 | RP11-196G11.2 |
| SAT | Obesity | ENSG00000260947.1 | -0.00530445 | 8.78E-05 | 0.288129407 | 0.001973489 | RP11-384P7.7 |
| SAT | Obesity | ENSG00000261556.4 | -0.026703012 | 1.48E-13 | 4.86E-10 | 2.11E-11 | RP11-296I10.6 |
| SAT | Obesity | ENSG00000265055.1 | 0.016273156 | 4.51E-06 | 0.014813626 | 0.000189918 | AC145343.2 |
| SAT | Obesity | ENSG00000265798.2 | -0.01151471 | 0.001096313 | 1 | 0.016106882 | RP11-271K11.5 |
| SAT | Obesity | ENSG00000266173.1 | -0.016871158 | 0.000113659 | 0.372914451 | 0.002486096 | STRADA |
| SAT | Obesity | ENSG00000267352.1 | 0.02820439 | 5.11E-06 | 0.016753551 | 0.000204312 | SH3GL1P3 |
| SAT | Obesity | ENSG00000269928.1 | 0.013509007 | 0.000421602 | 1 | 0.007477175 | RP11-849F2.8 |
| SAT | Obesity | ENSG00000269963.1 | -0.026754741 | 1.00E-08 | 3.28E-05 | 7.14E-07 | RP11-73M18.9 |
| SAT | Obesity | ENSG00000269990.1 | 0.023598735 | 6.52E-06 | 0.021379575 | 0.000245156 | CTD-3074O7.12 |
| SAT | Obesity | ENSG00000270692.1 | 0.020520586 | 1.76E-05 | 0.057669744 | 0.00054761 | RP11-89F17.5 |
| SAT | Obesity | ENSG00000271623.1 | -0.051129969 | 2.18E-28 | 7.15E-25 | 1.43E-25 | RP11-435I10.5 |
| SAT | Obesity | ENSG00000271751.1 | -0.031308692 | 1.56E-09 | 5.13E-06 | 1.28E-07 | RP11-110I1.14 |
| SAT | Obesity | ENSG00000271780.1 | -0.008895472 | 0.001061782 | 1 | 0.015763375 | RP11-1017G21.5 |
| SAT | Obesity | ENSG00000272047.1 | 0.013983929 | 0.000653312 | 1 | 0.010559198 | GTF2H5 |
| SAT | Obesity | ENSG00000272462.2 | 0.006008114 | 3.73E-10 | 1.22E-06 | 3.40E-08 | U91328.19 |
| SAT | Obesity | ENSG00000272810.1 | 0.012607958 | 1.58E-06 | 0.005186625 | 7.31E-05 | U91328.22 |
| SAT | Obesity | ENSG00000272849.1 | -0.013163843 | 8.61E-05 | 0.282504754 | 0.001973489 | RP11-347I19.8 |
| SAT | Obesity | ENSG00000272983.1 | 0.006323255 | 0.001352385 | 1 | 0.01896229 | RP11-508N22.12 |
| SAT | HTN | ENSG00000001617.7 | -0.024725353 | 2.55E-07 | 0.000836316 | 1.49E-05 | SEMA3F |
| SAT | HTN | ENSG00000003056.3 | -0.014748332 | 0.000175393 | 0.575464512 | 0.003585193 | M6PR |
| SAT | HTN | ENSG00000004534.10 | -0.005129629 | 4.95E-06 | 0.016234673 | 0.000186605 | RBM6 |
| SAT | HTN | ENSG00000005243.5 | 0.017019524 | 6.62E-05 | 0.21727313 | 0.001621441 | COPZ2 |
| SAT | HTN | ENSG00000005955.8 | -0.015963459 | 0.000319207 | 1 | 0.00557084 | GGNBP2 |
| SAT | HTN | ENSG00000007312.8 | -0.020285484 | 0.001894128 | 1 | 0.0223548 | CD79B |
| SAT | HTN | ENSG00000007341.14 | -0.005382131 | 0.004320172 | 1 | 0.040731275 | ST7L |
| SAT | HTN | ENSG00000010704.14 | -0.019455954 | 9.86E-08 | 0.000323437 | 6.88E-06 | HFE |
| SAT | HTN | ENSG00000011523.9 | 0.014929688 | 5.21E-08 | 0.000170892 | 4.07E-06 | CEP68 |
| SAT | HTN | ENSG00000028528.10 | 0.013614597 | 0.000313311 | 1 | 0.005497191 | SNX1 |
| SAT | HTN | ENSG00000050405.9 | 0.015528929 | 0.000112615 | 0.369488452 | 0.002548196 | LIMA1 |
| SAT | HTN | ENSG00000053900.6 | 0.020535732 | 0.000156636 | 0.513923815 | 0.003315638 | ANAPC4 |
| SAT | HTN | ENSG00000054282.11 | 0.020938725 | 2.20E-10 | 7.23E-07 | 4.82E-08 | SDCCAG8 |
| SAT | HTN | ENSG00000055955.11 | 0.007016938 | 0.000323567 | 1 | 0.005617053 | ITIH4 |
| SAT | HTN | ENSG00000060642.6 | -0.03512346 | 5.42E-07 | 0.001779741 | 2.78E-05 | PIGV |
| SAT | HTN | ENSG00000063854.8 | 0.010984352 | 0.001279202 | 1 | 0.016552068 | HAGH |
| SAT | HTN | ENSG00000065060.12 | 0.010163766 | 9.52E-07 | 0.003123699 | 4.53E-05 | UHRF1BP1 |
| SAT | HTN | ENSG00000068654.11 | -0.006997956 | 0.000100095 | 0.328412008 | 0.002296587 | POLR1A |
| SAT | HTN | ENSG00000073584.14 | 0.017514597 | 0.004242158 | 1 | 0.040343539 | SMARCE1 |

| | | | | | | | |
|---|---|---|---|---|---|---|---|
| SAT | HTN | ENSG00000074201.4 | 0.010995411 | 1.49E-05 | 0.048847232 | 0.000460823 | CLNS1A |
| SAT | HTN | ENSG00000076685.14 | 0.019531652 | 0.003737122 | 1 | 0.037382609 | NT5C2 |
| SAT | HTN | ENSG00000082258.8 | -0.009231772 | 0.0004251 | 1 | 0.0068707 | CCNT2 |
| SAT | HTN | ENSG00000085365.13 | 0.015609337 | 0.00412717 | 1 | 0.039725318 | SCAMP1 |
| SAT | HTN | ENSG00000086015.16 | 0.018917631 | 0.001649091 | 1 | 0.019892164 | MAST2 |
| SAT | HTN | ENSG00000086300.11 | -0.007553247 | 4.93E-07 | 0.001618651 | 2.57E-05 | SNX10 |
| SAT | HTN | ENSG00000087884.10 | -0.011124874 | 2.35E-05 | 0.077161004 | 0.000682841 | AAMDC |
| SAT | HTN | ENSG00000090238.7 | 0.005156261 | 0.000101286 | 0.332320906 | 0.002307784 | YPEL3 |
| SAT | HTN | ENSG00000090621.9 | -0.013853296 | 3.70E-06 | 0.012151461 | 0.000150018 | PABPC4 |
| SAT | HTN | ENSG00000091542.8 | 0.010360647 | 0.004268478 | 1 | 0.040476523 | ALKBH5 |
| SAT | HTN | ENSG00000092295.7 | 0.019496423 | 0.000180944 | 0.593676249 | 0.003619977 | TGM1 |
| SAT | HTN | ENSG00000095787.17 | 0.025705085 | 3.20E-05 | 0.105016618 | 0.000867906 | WAC |
| SAT | HTN | ENSG00000100106.15 | -0.008560302 | 0.004547328 | 1 | 0.042219581 | TRIOBP |
| SAT | HTN | ENSG00000100116.12 | 0.019493943 | 0.001997123 | 1 | 0.023153921 | GCAT |
| SAT | HTN | ENSG00000100731.11 | 0.008406302 | 0.000203554 | 0.667861435 | 0.003975366 | PCNX |
| SAT | HTN | ENSG00000101457.8 | -0.009135436 | 0.000131996 | 0.433079389 | 0.002873849 | DNTTIP1 |
| SAT | HTN | ENSG00000101751.6 | -0.006201173 | 9.63E-05 | 0.315865668 | 0.002234893 | POLI |
| SAT | HTN | ENSG00000102858.8 | 0.012405398 | 0.000439709 | 1 | 0.007003334 | MGRN1 |
| SAT | HTN | ENSG00000103196.7 | 0.021845417 | 0.000180824 | 0.593283879 | 0.003619977 | CRISPLD2 |
| SAT | HTN | ENSG00000103199.9 | -0.010081731 | 0.004172775 | 1 | 0.040031798 | ZNF500 |
| SAT | HTN | ENSG00000103248.13 | 0.007554851 | 0.000728042 | 1 | 0.010472448 | MTHFSD |
| SAT | HTN | ENSG00000103671.5 | 0.008892772 | 0.005381657 | 1 | 0.048776843 | TRIP4 |
| SAT | HTN | ENSG00000104231.6 | 0.012707267 | 0.001309759 | 1 | 0.016591972 | ZFAND1 |
| SAT | HTN | ENSG00000104313.13 | 0.022268515 | 3.75E-05 | 0.123109758 | 0.000992821 | EYA1 |
| SAT | HTN | ENSG00000105726.12 | 0.014676056 | 1.50E-05 | 0.049362984 | 0.000461336 | ATP13A1 |
| SAT | HTN | ENSG00000106290.10 | -0.020268034 | 0.001091175 | 1 | 0.014494516 | TAF6 |
| SAT | HTN | ENSG00000107960.6 | -0.03814924 | 9.77E-08 | 0.000320627 | 6.88E-06 | OBFC1 |
| SAT | HTN | ENSG00000108384.10 | -0.007310663 | 8.77E-05 | 0.287714717 | 0.002055105 | RAD51C |
| SAT | HTN | ENSG00000108639.3 | -0.015873155 | 0.000755404 | 1 | 0.010637259 | SYNGR2 |
| SAT | HTN | ENSG00000108654.7 | 0.016376652 | 2.67E-05 | 0.087626082 | 0.000742594 | DDX5 |
| SAT | HTN | ENSG00000108669.12 | 0.022036613 | 0.000702286 | 1 | 0.010195574 | CYTH1 |
| SAT | HTN | ENSG00000110906.8 | -0.010308762 | 0.000146464 | 0.480547383 | 0.003120438 | KCTD10 |
| SAT | HTN | ENSG00000111405.4 | -0.017124075 | 0.002859541 | 1 | 0.030167694 | ENDOU |
| SAT | HTN | ENSG00000111530.8 | -0.017413605 | 4.90E-07 | 0.001608484 | 2.57E-05 | CAND1 |
| SAT | HTN | ENSG00000112877.6 | 0.015779019 | 0.000695415 | 1 | 0.010185969 | CEP72 |
| SAT | HTN | ENSG00000112992.12 | -0.012979067 | 0.000789709 | 1 | 0.011072796 | NNT |
| SAT | HTN | ENSG00000113971.14 | 0.00372186 | 0.003569222 | 1 | 0.036032666 | NPHP3 |
| SAT | HTN | ENSG00000114023.11 | -0.021046116 | 0.001138794 | 1 | 0.014945536 | FAM162A |
| SAT | HTN | ENSG00000114378.12 | -0.008549243 | 0.00269345 | 1 | 0.028974455 | HYAL1 |
| SAT | HTN | ENSG00000115084.8 | -0.011234598 | 0.003132345 | 1 | 0.032116323 | SLC35F5 |
| SAT | HTN | ENSG00000115310.13 | -0.020337191 | 0.000356452 | 1 | 0.006123144 | RTN4 |
| SAT | HTN | ENSG00000115568.11 | -0.018198099 | 0.000334844 | 1 | 0.005782235 | ZNF142 |
| SAT | HTN | ENSG00000116489.8 | -0.020197665 | 0.001291475 | 1 | 0.016552068 | CAPZA1 |
| SAT | HTN | ENSG00000116667.8 | -0.016096114 | 0.000479798 | 1 | 0.007532147 | C1orf21 |
| SAT | HTN | ENSG00000117450.9 | 0.017664502 | 0.003010834 | 1 | 0.031261227 | PRDX1 |
| SAT | HTN | ENSG00000117481.6 | 0.018062531 | 0.002538167 | 1 | 0.027986366 | NSUN4 |
| SAT | HTN | ENSG00000117595.6 | -0.012764197 | 0.002669851 | 1 | 0.028815066 | IRF6 |
| SAT | HTN | ENSG00000117877.6 | -0.013060575 | 0.005108722 | 1 | 0.046690022 | CD3EAP |
| SAT | HTN | ENSG00000118197.9 | -0.009474786 | 0.002704779 | 1 | 0.028984413 | DDX59 |
| SAT | HTN | ENSG00000118407.10 | 0.016554398 | 0.005413566 | 1 | 0.048930885 | FILIP1 |
| SAT | HTN | ENSG00000119431.5 | -0.006120676 | 0.003954199 | 1 | 0.038497707 | HDHD3 |
| SAT | HTN | ENSG00000119969.10 | -0.015576292 | 0.003821897 | 1 | 0.037884123 | HELLS |
| SAT | HTN | ENSG00000120784.11 | 0.008265877 | 0.000755278 | 1 | 0.010637259 | ZFP30 |
| SAT | HTN | ENSG00000121101.11 | 0.042853366 | 4.10E-10 | 1.34E-06 | 8.40E-08 | TEX14 |
| SAT | HTN | ENSG00000122565.14 | -0.026013402 | 4.84E-07 | 0.00158637 | 2.57E-05 | CBX3 |
| SAT | HTN | ENSG00000123545.5 | 0.007468869 | 0.004128721 | 1 | 0.039725318 | NDUFAF4 |
| SAT | HTN | ENSG00000124116.14 | -0.004629516 | 0.004466909 | 1 | 0.041729241 | WFDC3 |
| SAT | HTN | ENSG00000124191.13 | -0.006294108 | 0.004024871 | 1 | 0.039069831 | TOX2 |
| SAT | HTN | ENSG00000124508.12 | 0.007426189 | 0.001391382 | 1 | 0.017448468 | BTN2A2 |
| SAT | HTN | ENSG00000124562.5 | -0.013184049 | 3.83E-07 | 0.00125572 | 2.13E-05 | SNRPC |
| SAT | HTN | ENSG00000124575.5 | 0.029707316 | 2.31E-07 | 0.000758423 | 1.38E-05 | HIST1H1D |
| SAT | HTN | ENSG00000124613.4 | -0.010722599 | 6.15E-06 | 0.020162759 | 0.000224031 | ZNF391 |
| SAT | HTN | ENSG00000125388.15 | 0.006005631 | 0.000436206 | 1 | 0.006981431 | GRK4 |
| SAT | HTN | ENSG00000125510.11 | -0.029883644 | 4.51E-08 | 0.00014783 | 3.61E-06 | OPRL1 |
| SAT | HTN | ENSG00000126602.6 | 0.005779148 | 7.34E-07 | 0.002407481 | 3.63E-05 | TRAP1 |
| SAT | HTN | ENSG00000126858.12 | 0.017778398 | 0.002360345 | 1 | 0.026466726 | RHOT1 |
| SAT | HTN | ENSG00000127957.12 | -0.019512502 | 1.21E-10 | 3.98E-07 | 2.84E-08 | PMS2P3 |
| SAT | HTN | ENSG00000128829.7 | -0.007856602 | 0.003362488 | 1 | 0.034155797 | EIF2AK4 |
| SAT | HTN | ENSG00000130177.10 | 0.013551523 | 3.37E-05 | 0.110524543 | 0.000905939 | CDC16 |
| SAT | HTN | ENSG00000130270.12 | -0.009512404 | 0.000755286 | 1 | 0.010637259 | ATP8B3 |
| SAT | HTN | ENSG00000130517.9 | 0.01263956 | 0.000377823 | 1 | 0.006324679 | PGPEP1 |
| SAT | HTN | ENSG00000131165.10 | 0.021509591 | 1.27E-06 | 0.004169694 | 5.79E-05 | CHMP1A |
| SAT | HTN | ENSG00000131435.8 | 0.035750399 | 1.22E-07 | 0.000401808 | 8.04E-06 | PDLIM4 |
| SAT | HTN | ENSG00000133731.5 | 0.00571594 | 0.001539978 | 1 | 0.018853233 | IMPA1 |
| SAT | HTN | ENSG00000134245.13 | -0.026443864 | 3.36E-08 | 0.000110153 | 2.90E-06 | WNT2B |
| SAT | HTN | ENSG00000134574.7 | 0.045911632 | 1.09E-11 | 3.58E-08 | 4.04E-09 | DDB2 |
| SAT | HTN | ENSG00000134575.5 | 0.049714348 | 1.69E-23 | 5.54E-20 | 5.54E-20 | ACP2 |
| SAT | HTN | ENSG00000134851.8 | 0.033051563 | 1.04E-07 | 0.00033991 | 7.08E-06 | TMEM165 |
| SAT | HTN | ENSG00000135213.8 | -0.024117316 | 4.50E-05 | 0.147797145 | 0.001172993 | POM121C |
| SAT | HTN | ENSG00000135744.7 | 0.007112811 | 0.000134662 | 0.441825993 | 0.00290675 | AGT |
| SAT | HTN | ENSG00000135913.6 | 0.015258363 | 0.001222842 | 1 | 0.015921202 | USP37 |

| | | | | | | | |
|---|---|---|---|---|---|---|---|
| SAT | HTN | ENSG00000135968.15 | 0.010006925 | 0.002174492 | 1 | 0.024858915 | GCC2 |
| SAT | HTN | ENSG00000136425.8 | -0.0173279 | 0.002794756 | 1 | 0.029675062 | CIB2 |
| SAT | HTN | ENSG00000136448.7 | 0.015703223 | 6.52E-10 | 2.14E-06 | 1.07E-07 | NMT1 |
| SAT | HTN | ENSG00000136856.13 | 0.015911073 | 0.00274977 | 1 | 0.029292189 | SLC2A8 |
| SAT | HTN | ENSG00000137411.12 | 0.00372299 | 0.002599218 | 1 | 0.028332336 | VARS2 |
| SAT | HTN | ENSG00000137944.12 | -0.024764378 | 0.000386707 | 1 | 0.006408012 | CCBL2 |
| SAT | HTN | ENSG00000138036.14 | 0.017231602 | 0.000381063 | 1 | 0.006346531 | DYNC2LI1 |
| SAT | HTN | ENSG00000138175.8 | 0.021719004 | 7.91E-07 | 0.002595531 | 3.82E-05 | ARL3 |
| SAT | HTN | ENSG00000138757.10 | 0.013615797 | 0.000569228 | 1 | 0.008727282 | G3BP2 |
| SAT | HTN | ENSG00000138829.6 | 0.005630305 | 0.000230312 | 0.755652406 | 0.004297476 | FBN2 |
| SAT | HTN | ENSG00000139636.11 | -0.017951079 | 0.00197106 | 1 | 0.02301441 | LMBR1L |
| SAT | HTN | ENSG00000140995.12 | 0.017160074 | 3.86E-05 | 0.126664381 | 0.001013315 | DEF8 |
| SAT | HTN | ENSG00000141076.13 | 0.012713997 | 1.68E-05 | 0.054958636 | 0.000508876 | CIRH1A |
| SAT | HTN | ENSG00000141252.15 | -0.014723285 | 0.001398643 | 1 | 0.017448468 | VPS53 |
| SAT | HTN | ENSG00000141295.9 | -0.009163468 | 0.000227008 | 0.74481305 | 0.004297476 | SCRN2 |
| SAT | HTN | ENSG00000141452.5 | -0.020245865 | 1.24E-05 | 0.040532123 | 0.000401308 | C18orf8 |
| SAT | HTN | ENSG00000141458.8 | -0.02993074 | 1.98E-06 | 0.006498719 | 8.33E-05 | NPC1 |
| SAT | HTN | ENSG00000141569.6 | -0.012649129 | 0.000257874 | 0.846086136 | 0.004726738 | TRIM65 |
| SAT | HTN | ENSG00000142599.13 | 0.024103573 | 8.38E-05 | 0.274874119 | 0.001977512 | RERE |
| SAT | HTN | ENSG00000143028.7 | 0.01619207 | 0.002541889 | 1 | 0.027986366 | SYPL2 |
| SAT | HTN | ENSG00000143156.9 | -0.021484027 | 0.000826253 | 1 | 0.011487014 | NME7 |
| SAT | HTN | ENSG00000143248.8 | -0.029395488 | 2.47E-05 | 0.081163813 | 0.000693708 | RGS5 |
| SAT | HTN | ENSG00000143437.16 | -0.021409627 | 0.000265265 | 0.870335062 | 0.004782061 | ARNT |
| SAT | HTN | ENSG00000143578.11 | -0.017464012 | 2.39E-05 | 0.07854582 | 0.000683007 | CREB3L4 |
| SAT | HTN | ENSG00000143702.11 | 0.036247951 | 4.91E-11 | 1.61E-07 | 1.34E-08 | CEP170 |
| SAT | HTN | ENSG00000144677.10 | -0.021331105 | 0.001343634 | 1 | 0.016955623 | CTDSPL |
| SAT | HTN | ENSG00000145020.10 | -0.005006805 | 0.002340835 | 1 | 0.026392712 | AMT |
| SAT | HTN | ENSG00000145029.7 | -0.008353365 | 0.002363533 | 1 | 0.026466726 | NICN1 |
| SAT | HTN | ENSG00000145241.6 | -0.011371229 | 0.002660035 | 1 | 0.028803875 | CENPC |
| SAT | HTN | ENSG00000145777.10 | -0.016608027 | 0.003852015 | 1 | 0.038067659 | TSLP |
| SAT | HTN | ENSG00000145990.6 | 0.01182034 | 0.005283333 | 1 | 0.048151705 | GFOD1 |
| SAT | HTN | ENSG00000146109.3 | 0.014008239 | 0.000411778 | 1 | 0.006721602 | ABT1 |
| SAT | HTN | ENSG00000146409.6 | -0.008010662 | 0.000192284 | 0.63088224 | 0.003777738 | SLC18B1 |
| SAT | HTN | ENSG00000146556.10 | -0.028580901 | 9.32E-06 | 0.030588524 | 0.000321984 | WASH2P |
| SAT | HTN | ENSG00000147419.12 | 0.021396087 | 0.000730933 | 1 | 0.010472448 | CCDC25 |
| SAT | HTN | ENSG00000148572.10 | -0.016184001 | 3.77E-06 | 0.012367156 | 0.000150819 | NRBF2 |
| SAT | HTN | ENSG00000148843.9 | 0.065679101 | 1.66E-22 | 5.45E-19 | 2.72E-19 | PDCD11 |
| SAT | HTN | ENSG00000149084.7 | -0.007351138 | 3.82E-09 | 1.25E-05 | 4.32E-07 | HSD17B12 |
| SAT | HTN | ENSG00000149100.8 | -0.016022636 | 0.002590911 | 1 | 0.028332336 | EIF3M |
| SAT | HTN | ENSG00000149179.9 | -0.021343736 | 0.001412147 | 1 | 0.017483974 | C11orf49 |
| SAT | HTN | ENSG00000149292.12 | 0.004365936 | 0.004206087 | 1 | 0.040173476 | TTC12 |
| SAT | HTN | ENSG00000152219.4 | -0.023451201 | 5.71E-05 | 0.1871961 | 0.001428978 | ARL14EP |
| SAT | HTN | ENSG00000152784.11 | -0.023839423 | 0.000699009 | 1 | 0.010193108 | PRDM8 |
| SAT | HTN | ENSG00000153113.19 | -0.010020763 | 0.000844023 | 1 | 0.011635455 | CAST |
| SAT | HTN | ENSG00000153814.7 | -0.018301822 | 0.001065844 | 1 | 0.014273607 | JAZF1 |
| SAT | HTN | ENSG00000154328.11 | 0.027123923 | 2.90E-08 | 9.51E-05 | 2.72E-06 | NEIL2 |
| SAT | HTN | ENSG00000155085.11 | 0.021832095 | 0.001073954 | 1 | 0.014323756 | AK9 |
| SAT | HTN | ENSG00000156170.8 | -0.035283255 | 3.25E-09 | 1.07E-05 | 3.83E-07 | NDUFAF6 |
| SAT | HTN | ENSG00000156398.8 | 0.027534525 | 1.82E-05 | 0.059592552 | 0.000546721 | SFXN2 |
| SAT | HTN | ENSG00000156931.11 | 0.013108317 | 0.000220024 | 0.721897342 | 0.004271582 | VPS8 |
| SAT | HTN | ENSG00000157322.12 | -0.011707892 | 7.49E-06 | 0.024585509 | 0.000261923 | CLEC18A |
| SAT | HTN | ENSG00000158022.6 | 0.017157923 | 0.00030963 | 1 | 0.005461812 | TRIM63 |
| SAT | HTN | ENSG00000158805.7 | 0.026253204 | 1.36E-05 | 0.044732652 | 0.000430122 | ZNF276 |
| SAT | HTN | ENSG00000158865.8 | -0.009692173 | 9.86E-07 | 0.0032355 | 4.62E-05 | SLC5A11 |
| SAT | HTN | ENSG00000159479.12 | 0.0257854 | 4.09E-06 | 0.013410438 | 0.000159648 | MED8 |
| SAT | HTN | ENSG00000160307.5 | -0.009359372 | 5.90E-05 | 0.193638209 | 0.001466956 | S100B |
| SAT | HTN | ENSG00000160752.10 | 0.014940592 | 0.000817375 | 1 | 0.011411947 | FDPS |
| SAT | HTN | ENSG00000160783.15 | 0.016540556 | 0.000899106 | 1 | 0.012139779 | PMF1 |
| SAT | HTN | ENSG00000161204.7 | 0.014170824 | 1.89E-06 | 0.006187277 | 8.04E-05 | ABCF3 |
| SAT | HTN | ENSG00000161955.12 | 0.034166373 | 1.93E-07 | 0.000634652 | 1.18E-05 | TNFSF13 |
| SAT | HTN | ENSG00000163431.11 | -0.034253391 | 6.44E-10 | 2.11E-06 | 1.07E-07 | LMOD1 |
| SAT | HTN | ENSG00000163933.5 | -0.019082181 | 0.000188064 | 0.617038859 | 0.003717102 | RFT1 |
| SAT | HTN | ENSG00000163946.9 | 0.016315152 | 4.21E-06 | 0.013819791 | 0.000162586 | FAM208A |
| SAT | HTN | ENSG00000164002.7 | -0.020486811 | 2.11E-05 | 0.069345539 | 0.000619157 | EXO5 |
| SAT | HTN | ENSG00000164039.10 | -0.0122778 | 0.000830657 | 1 | 0.011499513 | BDH2 |
| SAT | HTN | ENSG00000164045.7 | -0.027088268 | 8.41E-11 | 2.76E-07 | 2.12E-08 | CDC25A |
| SAT | HTN | ENSG00000164048.9 | -0.010724119 | 1.32E-11 | 4.33E-08 | 4.33E-09 | ZNF589 |
| SAT | HTN | ENSG00000164062.8 | -0.021557081 | 2.27E-09 | 7.45E-06 | 3.10E-07 | APEH |
| SAT | HTN | ENSG00000164068.11 | 0.013241839 | 9.78E-12 | 3.21E-08 | 4.04E-09 | RNF123 |
| SAT | HTN | ENSG00000164307.8 | -0.018727963 | 7.22E-08 | 0.000236849 | 5.38E-06 | ERAP1 |
| SAT | HTN | ENSG00000164308.12 | 0.002794442 | 0.001682424 | 1 | 0.020219897 | ERAP2 |
| SAT | HTN | ENSG00000164587.7 | -0.020819257 | 0.001787916 | 1 | 0.021361157 | RPS14 |
| SAT | HTN | ENSG00000164695.4 | -0.00982677 | 0.004212032 | 1 | 0.040173476 | CHMP4C |
| SAT | HTN | ENSG00000165118.10 | 0.015284365 | 0.005509073 | 1 | 0.049385977 | C9orf64 |
| SAT | HTN | ENSG00000165178.8 | 0.014610602 | 0.000372526 | 1 | 0.006267988 | NCF1C |
| SAT | HTN | ENSG00000165244.6 | 0.020100828 | 0.002498579 | 1 | 0.027695394 | ZNF367 |
| SAT | HTN | ENSG00000165476.8 | -0.028113071 | 7.73E-08 | 0.000253571 | 5.63E-06 | REEP3 |
| SAT | HTN | ENSG00000165490.8 | -0.017045303 | 0.005556178 | 1 | 0.049672537 | C11orf82 |
| SAT | HTN | ENSG00000166037.6 | -0.014508221 | 0.000503241 | 1 | 0.007862535 | CEP57 |
| SAT | HTN | ENSG00000166166.8 | -0.032214089 | 1.52E-06 | 0.004995927 | 6.75E-05 | TRMT61A |
| SAT | HTN | ENSG00000166435.11 | -0.007249154 | 0.000605426 | 1 | 0.009196305 | XRRA1 |

| | | | | | | | |
|---|---|---|---|---|---|---|---|
| SAT | HTN | ENSG00000166436.11 | -0.004834316 | 3.46E-05 | 0.113639107 | 0.000923895 | TRIM66 |
| SAT | HTN | ENSG00000166532.11 | -0.009600905 | 1.89E-07 | 0.000619154 | 1.17E-05 | RIMKLB |
| SAT | HTN | ENSG00000167395.10 | 0.024064028 | 0.00066502 | 1 | 0.009917863 | ZNF646 |
| SAT | HTN | ENSG00000167397.10 | -0.015480994 | 0.003887291 | 1 | 0.038072244 | VKORC1 |
| SAT | HTN | ENSG00000167528.8 | 0.007155712 | 0.002712043 | 1 | 0.028984413 | ZNF641 |
| SAT | HTN | ENSG00000167633.12 | 0.026542571 | 0.000120062 | 0.393923321 | 0.002698105 | KIR3DL1 |
| SAT | HTN | ENSG00000167720.8 | 0.005780621 | 0.000132262 | 0.433951171 | 0.002873849 | SRR |
| SAT | HTN | ENSG00000167910.3 | 0.023350792 | 0.000421602 | 1 | 0.006847904 | CYP7A1 |
| SAT | HTN | ENSG00000168004.5 | 0.01246669 | 0.000578545 | 1 | 0.008828861 | HRASLS5 |
| SAT | HTN | ENSG00000168297.11 | -0.017016637 | 0.001504431 | 1 | 0.018487036 | PXK |
| SAT | HTN | ENSG00000168394.9 | -0.021694434 | 0.002418657 | 1 | 0.026900389 | TAP1 |
| SAT | HTN | ENSG00000168702.12 | -0.022536887 | 0.000402906 | 1 | 0.00660968 | LRP1B |
| SAT | HTN | ENSG00000168884.10 | -0.008691035 | 0.003444834 | 1 | 0.034884257 | TNIP2 |
| SAT | HTN | ENSG00000168936.6 | 0.010356987 | 0.003928514 | 1 | 0.038361473 | TMEM129 |
| SAT | HTN | ENSG00000169047.5 | -0.020587958 | 3.27E-09 | 1.07E-05 | 3.83E-07 | IRS1 |
| SAT | HTN | ENSG00000169203.12 | 0.01143981 | 6.80E-06 | 0.02231238 | 0.000245191 | RP11-231C14.4 |
| SAT | HTN | ENSG00000169592.10 | -0.026623454 | 1.68E-09 | 5.50E-06 | 2.39E-07 | INO80E |
| SAT | HTN | ENSG00000169629.7 | -0.004582065 | 0.004459906 | 1 | 0.041729241 | RGPD8 |
| SAT | HTN | ENSG00000170049.5 | -0.015140591 | 0.002917253 | 1 | 0.030579894 | KCNAB3 |
| SAT | HTN | ENSG00000170092.10 | 0.021828856 | 5.01E-06 | 0.016432151 | 0.000186729 | AC018720.10 |
| SAT | HTN | ENSG00000170633.12 | -0.017256325 | 0.000231836 | 0.760653289 | 0.004297476 | RNF34 |
| SAT | HTN | ENSG00000170955.9 | 0.01733567 | 7.30E-05 | 0.239584078 | 0.001774697 | PRKCDBP |
| SAT | HTN | ENSG00000171421.8 | 0.020897637 | 0.002376693 | 1 | 0.026523566 | MRPL36 |
| SAT | HTN | ENSG00000171700.9 | -0.025908773 | 2.52E-14 | 8.26E-11 | 1.38E-11 | RGS19 |
| SAT | HTN | ENSG00000172113.4 | 0.017943362 | 5.92E-10 | 1.94E-06 | 1.07E-07 | NME6 |
| SAT | HTN | ENSG00000172687.9 | 0.005910219 | 0.001405167 | 1 | 0.017463464 | ZNF738 |
| SAT | HTN | ENSG00000172789.3 | 0.0219391 | 0.001285255 | 1 | 0.016552068 | HOXC5 |
| SAT | HTN | ENSG00000172927.3 | -0.016180647 | 0.002043549 | 1 | 0.023608743 | MYEOV |
| SAT | HTN | ENSG00000172992.7 | -0.008149312 | 0.000370583 | 1 | 0.006267431 | DCAKD |
| SAT | HTN | ENSG00000173209.18 | 0.014235339 | 0.002987563 | 1 | 0.031118074 | AHSA2 |
| SAT | HTN | ENSG00000173511.5 | -0.0233707 | 2.59E-09 | 8.51E-06 | 3.41E-07 | VEGFB |
| SAT | HTN | ENSG00000173531.11 | -0.016085402 | 5.37E-06 | 0.017622722 | 0.000198008 | MST1 |
| SAT | HTN | ENSG00000174004.5 | 0.015382697 | 0.001963171 | 1 | 0.023004153 | NRROS |
| SAT | HTN | ENSG00000174177.8 | -0.021717258 | 0.001641157 | 1 | 0.019869511 | CTU2 |
| SAT | HTN | ENSG00000174226.4 | -0.026903752 | 3.09E-05 | 0.101233222 | 0.00084361 | SNX31 |
| SAT | HTN | ENSG00000174891.8 | -0.01476271 | 1.66E-07 | 0.000545032 | 1.07E-05 | RSRC1 |
| SAT | HTN | ENSG00000175220.7 | 0.01778731 | 0.000170162 | 0.558302901 | 0.003511339 | ARHGAP1 |
| SAT | HTN | ENSG00000175334.3 | 0.010312241 | 0.004814434 | 1 | 0.044371228 | BANF1 |
| SAT | HTN | ENSG00000175470.14 | -0.02015348 | 1.42E-05 | 0.046684881 | 0.000444618 | PPP2R2D |
| SAT | HTN | ENSG00000175489.9 | 0.009821118 | 0.003224548 | 1 | 0.032958701 | LRRC25 |
| SAT | HTN | ENSG00000176390.10 | 0.023940506 | 0.000178765 | 0.586527662 | 0.003619977 | CRLF3 |
| SAT | HTN | ENSG00000177000.6 | -0.054668639 | 3.95E-17 | 1.29E-13 | 3.24E-14 | MTHFR |
| SAT | HTN | ENSG00000177311.6 | 0.019990895 | 0.000514385 | 1 | 0.007998565 | ZBTB38 |
| SAT | HTN | ENSG00000177380.9 | -0.02259175 | 0.000430774 | 1 | 0.00692828 | PPFIA3 |
| SAT | HTN | ENSG00000177697.13 | 0.016151435 | 0.003055603 | 1 | 0.03162597 | CD151 |
| SAT | HTN | ENSG00000177700.5 | -0.018292018 | 0.003251771 | 1 | 0.033133733 | POLR2L |
| SAT | HTN | ENSG00000177839.4 | 0.014908018 | 0.001288495 | 1 | 0.016552068 | PCDHB9 |
| SAT | HTN | ENSG00000178188.10 | -0.013482943 | 0.000164981 | 0.541304063 | 0.003447797 | SH2B1 |
| SAT | HTN | ENSG00000178467.13 | -0.020786655 | 0.002309686 | 1 | 0.026131312 | P4HTM |
| SAT | HTN | ENSG00000178718.5 | 0.024589834 | 1.30E-09 | 4.27E-06 | 1.94E-07 | RPP25 |
| SAT | HTN | ENSG00000178809.7 | -0.021310413 | 0.000525392 | 1 | 0.008131178 | TRIM73 |
| SAT | HTN | ENSG00000178922.12 | -0.015000191 | 2.84E-07 | 0.000932114 | 1.64E-05 | HYI |
| SAT | HTN | ENSG00000178971.9 | 0.010463225 | 0.000261805 | 0.858981653 | 0.004769464 | CTC1 |
| SAT | HTN | ENSG00000180855.11 | 0.01984662 | 0.000139471 | 0.457603234 | 0.002990871 | ZNF443 |
| SAT | HTN | ENSG00000181885.14 | -0.039664138 | 6.52E-10 | 2.14E-06 | 1.07E-07 | CLDN7 |
| SAT | HTN | ENSG00000182372.6 | 0.029691644 | 1.06E-07 | 0.000347083 | 7.08E-06 | CLN8 |
| SAT | HTN | ENSG00000182871.10 | 0.010930603 | 0.001548288 | 1 | 0.018884508 | COL18A1 |
| SAT | HTN | ENSG00000182986.8 | 0.004501667 | 0.000295536 | 0.969653396 | 0.005269855 | ZNF320 |
| SAT | HTN | ENSG00000183604.10 | -0.013430969 | 0.000890578 | 1 | 0.012122569 | RP11-347C12.2 |
| SAT | HTN | ENSG00000183762.8 | -0.008017406 | 0.000224702 | 0.737248672 | 0.004297476 | KREMEN1 |
| SAT | HTN | ENSG00000184110.10 | 0.017946603 | 7.41E-07 | 0.002431272 | 3.63E-05 | EIF3C |
| SAT | HTN | ENSG00000184385.2 | -0.009934233 | 0.000388661 | 1 | 0.006408027 | C21orf128 |
| SAT | HTN | ENSG00000184389.8 | -0.014988451 | 1.33E-05 | 0.043766819 | 0.000424921 | A3GALT2 |
| SAT | HTN | ENSG00000184574.5 | -0.017681058 | 0.000875585 | 1 | 0.011992756 | LPAR5 |
| SAT | HTN | ENSG00000184923.8 | -0.018817736 | 0.004719878 | 1 | 0.043622307 | NUTM2A |
| SAT | HTN | ENSG00000185163.5 | 0.009052041 | 0.00069333 | 1 | 0.010185969 | DDX51 |
| SAT | HTN | ENSG00000185324.17 | -0.009445977 | 1.64E-06 | 0.005374204 | 7.07E-05 | CDK10 |
| SAT | HTN | ENSG00000185339.4 | -0.007823014 | 0.001790405 | 1 | 0.021361157 | TCN2 |
| SAT | HTN | ENSG00000185829.11 | -0.015682496 | 4.55E-05 | 0.14941435 | 0.001176491 | ARL17A |
| SAT | HTN | ENSG00000186792.11 | -0.006543891 | 0.00361868 | 1 | 0.036308532 | HYAL3 |
| SAT | HTN | ENSG00000187583.6 | 0.02089978 | 0.000686367 | 1 | 0.01015143 | PLEKHN1 |
| SAT | HTN | ENSG00000187778.9 | 0.03047864 | 3.56E-08 | 0.000116657 | 2.92E-06 | MCRS1 |
| SAT | HTN | ENSG00000188227.8 | -0.008217238 | 0.000653723 | 1 | 0.009793908 | ZNF793 |
| SAT | HTN | ENSG00000188321.9 | -0.003648849 | 0.002087143 | 1 | 0.024027772 | ZNF559 |
| SAT | HTN | ENSG00000188383.7 | -0.016129133 | 0.00110814 | 1 | 0.014637359 | AC018892.9 |
| SAT | HTN | ENSG00000188629.7 | -0.006637237 | 0.001882042 | 1 | 0.022292345 | ZNF177 |
| SAT | HTN | ENSG00000188878.12 | -0.011550304 | 3.93E-07 | 0.001288415 | 2.15E-05 | FBF1 |
| SAT | HTN | ENSG00000188976.6 | -0.006951196 | 0.001303591 | 1 | 0.016590986 | NOC2L |
| SAT | HTN | ENSG00000196126.6 | -0.031825735 | 8.40E-10 | 2.76E-06 | 1.31E-07 | HLA-DRB1 |
| SAT | HTN | ENSG00000196502.7 | -0.017733121 | 0.000175927 | 0.577216082 | 0.003585193 | SULT1A1 |
| SAT | HTN | ENSG00000196821.5 | 0.016134623 | 1.50E-06 | 0.004925645 | 6.75E-05 | C6orf106 |

| | | | | | | | |
|---|---|---|---|---|---|---|---|
| SAT | HTN | ENSG00000196961.8 | -0.013587551 | 0.004406645 | 1 | 0.041427517 | AP2A1 |
| SAT | HTN | ENSG00000196966.3 | 0.024088968 | 1.06E-08 | 3.49E-05 | 1.09E-06 | HIST1H3E |
| SAT | HTN | ENSG00000196968.6 | -0.021102971 | 0.000230554 | 0.756447477 | 0.004297476 | FUT11 |
| SAT | HTN | ENSG00000196981.2 | 0.02111026 | 0.000877251 | 1 | 0.011992756 | WDR5B |
| SAT | HTN | ENSG00000197020.6 | -0.008765391 | 1.19E-05 | 0.039040158 | 0.000394345 | ZNF100 |
| SAT | HTN | ENSG00000197062.7 | 0.008927332 | 4.77E-05 | 0.156577668 | 0.001223263 | RP5-874C20.3 |
| SAT | HTN | ENSG00000197165.6 | 0.010937042 | 1.92E-05 | 0.063035388 | 0.000572975 | SULT1A2 |
| SAT | HTN | ENSG00000197279.3 | 0.017284393 | 0.000894136 | 1 | 0.012122569 | ZNF165 |
| SAT | HTN | ENSG00000197375.8 | 0.02341602 | 1.33E-08 | 4.35E-05 | 1.32E-06 | SLC22A5 |
| SAT | HTN | ENSG00000197535.10 | -0.017775822 | 0.005488053 | 1 | 0.049332333 | MYO5A |
| SAT | HTN | ENSG00000197728.5 | -0.006573244 | 4.39E-06 | 0.014402644 | 0.000167473 | RPS26 |
| SAT | HTN | ENSG00000198208.7 | 0.02037609 | 0.002570519 | 1 | 0.028206929 | RPS6KL1 |
| SAT | HTN | ENSG00000198270.8 | -0.015802767 | 1.46E-11 | 4.79E-08 | 4.36E-09 | TMEM116 |
| SAT | HTN | ENSG00000198646.9 | -0.017593379 | 0.003613698 | 1 | 0.036308532 | NCOA6 |
| SAT | HTN | ENSG00000198805.7 | -0.005441425 | 0.002625827 | 1 | 0.028527611 | PNP |
| SAT | HTN | ENSG00000198837.5 | -0.037618236 | 6.43E-08 | 0.000211112 | 4.91E-06 | DENND4B |
| SAT | HTN | ENSG00000203301.2 | -0.01907133 | 0.000369931 | 1 | 0.006267431 | AL590822.1 |
| SAT | HTN | ENSG00000204228.3 | -0.022113084 | 2.60E-06 | 0.008531918 | 0.000107999 | HSD17B8 |
| SAT | HTN | ENSG00000204252.8 | -0.012935674 | 0.003793237 | 1 | 0.037713967 | HLA-DOA |
| SAT | HTN | ENSG00000204287.9 | 0.012161025 | 0.004476895 | 1 | 0.041729241 | HLA-DRA |
| SAT | HTN | ENSG00000204314.6 | 0.015864263 | 0.000231365 | 0.75910892 | 0.004297476 | PRRT1 |
| SAT | HTN | ENSG00000204316.8 | 0.017387823 | 0.001428342 | 1 | 0.017618005 | MRPL38 |
| SAT | HTN | ENSG00000204338.4 | 0.020866772 | 8.98E-15 | 2.95E-11 | 5.90E-12 | CYP21A1P |
| SAT | HTN | ENSG00000204344.10 | 0.014356281 | 1.55E-06 | 0.005088711 | 6.78E-05 | STK19 |
| SAT | HTN | ENSG00000204387.8 | 0.014596686 | 2.88E-05 | 0.094393682 | 0.000793224 | C6orf48 |
| SAT | HTN | ENSG00000204438.6 | 0.018295014 | 9.89E-06 | 0.032435179 | 0.000334383 | GPANK1 |
| SAT | HTN | ENSG00000204463.8 | 0.011548621 | 1.09E-05 | 0.035620101 | 0.00036347 | BAG6 |
| SAT | HTN | ENSG00000204516.5 | -0.004195103 | 0.004120911 | 1 | 0.039725318 | MICB |
| SAT | HTN | ENSG00000204525.10 | -0.008179527 | 0.000228991 | 0.751320646 | 0.004297476 | HLA-C |
| SAT | HTN | ENSG00000204531.11 | -0.004663311 | 0.002803926 | 1 | 0.029676388 | POU5F1 |
| SAT | HTN | ENSG00000204536.9 | -0.005751687 | 8.04E-05 | 0.263683982 | 0.001918893 | CCHCR1 |
| SAT | HTN | ENSG00000204789.3 | 0.01346018 | 2.37E-05 | 0.077854105 | 0.000682931 | ZNF204P |
| SAT | HTN | ENSG00000205583.9 | -0.013540687 | 2.88E-09 | 9.43E-06 | 3.63E-07 | STAG3L1 |
| SAT | HTN | ENSG00000206077.6 | 0.016987818 | 0.003114828 | 1 | 0.032036838 | ZDHHC11B |
| SAT | HTN | ENSG00000213240.7 | 0.012207423 | 0.001915064 | 1 | 0.02252088 | NOTCH2NL |
| SAT | HTN | ENSG00000213246.2 | 0.023350234 | 0.000160524 | 0.526677971 | 0.003376141 | SUPT4H1 |
| SAT | HTN | ENSG00000213277.3 | -0.046311774 | 1.32E-19 | 4.33E-16 | 1.44E-16 | MARCKSL1P1 |
| SAT | HTN | ENSG00000213347.6 | -0.011579372 | 0.001397672 | 1 | 0.017448468 | MXD3 |
| SAT | HTN | ENSG00000213445.4 | 0.027571394 | 2.75E-08 | 9.02E-05 | 2.65E-06 | SIPA1 |
| SAT | HTN | ENSG00000213676.6 | 0.013173741 | 6.91E-09 | 2.27E-05 | 7.31E-07 | ATF6B |
| SAT | HTN | ENSG00000213719.4 | 0.04350156 | 3.54E-08 | 0.000116308 | 2.92E-06 | CLIC1 |
| SAT | HTN | ENSG00000213903.4 | -0.013586873 | 9.71E-06 | 0.031868696 | 0.000331966 | LTB4R |
| SAT | HTN | ENSG00000214331.4 | -0.014354367 | 0.003779971 | 1 | 0.037696311 | RP11-252A24.2 |
| SAT | HTN | ENSG00000214517.4 | -0.009065405 | 0.002938107 | 1 | 0.030700413 | PPME1 |
| SAT | HTN | ENSG00000216901.1 | -0.005755527 | 0.001304625 | 1 | 0.016590986 | AL022393.7 |
| SAT | HTN | ENSG00000218422.2 | 0.009341112 | 0.000470573 | 1 | 0.00742284 | AC016773.1 |
| SAT | HTN | ENSG00000219392.1 | 0.006301656 | 0.000737134 | 1 | 0.010515378 | RP1-265C24.5 |
| SAT | HTN | ENSG00000221838.5 | -0.004671752 | 0.003877625 | 1 | 0.038072244 | AP4M1 |
| SAT | HTN | ENSG00000223496.1 | -0.020875224 | 1.94E-05 | 0.063600177 | 0.000572975 | EXOSC6 |
| SAT | HTN | ENSG00000223718.2 | 0.017611431 | 0.004316934 | 1 | 0.040731275 | AC093107.7 |
| SAT | HTN | ENSG00000223865.6 | 0.014349701 | 0.001640129 | 1 | 0.019869511 | HLA-DPB1 |
| SAT | HTN | ENSG00000224389.4 | -0.010576109 | 0.003886077 | 1 | 0.038072244 | C4B |
| SAT | HTN | ENSG00000224997.1 | -0.011093593 | 5.13E-05 | 0.168425256 | 0.001305622 | AL049840.1 |
| SAT | HTN | ENSG00000225398.2 | 0.013847765 | 8.07E-05 | 0.26480729 | 0.001918893 | RP11-395L14.5 |
| SAT | HTN | ENSG00000225851.1 | 0.009155689 | 0.000364728 | 1 | 0.006232678 | HLA-S |
| SAT | HTN | ENSG00000226232.4 | -0.007238731 | 0.00063537 | 1 | 0.009606677 | RP11-419C5.2 |
| SAT | HTN | ENSG00000228638.1 | 0.014252047 | 2.84E-06 | 0.009310949 | 0.000116387 | FCF1P2 |
| SAT | HTN | ENSG00000229391.3 | 0.006871832 | 0.000126107 | 0.413756274 | 0.002814669 | HLA-DRB6 |
| SAT | HTN | ENSG00000229759.1 | -0.014069908 | 1.11E-11 | 3.64E-08 | 4.04E-09 | MRPS18AP1 |
| SAT | HTN | ENSG00000231130.1 | 0.006676282 | 0.005039457 | 1 | 0.046258571 | HLA-T |
| SAT | HTN | ENSG00000232561.2 | -0.015235432 | 0.002154753 | 1 | 0.024719383 | GTF2IP1 |
| SAT | HTN | ENSG00000233137.2 | 0.021873393 | 0.001859759 | 1 | 0.022108226 | RP11-220I1.1 |
| SAT | HTN | ENSG00000236397.2 | 0.010275443 | 7.50E-06 | 0.024620795 | 0.000261923 | DDX11L2 |
| SAT | HTN | ENSG00000237541.3 | 0.018590358 | 3.04E-07 | 0.00099886 | 1.72E-05 | HLA-DQA2 |
| SAT | HTN | ENSG00000238024.1 | -0.024601528 | 0.000234573 | 0.769635591 | 0.004323795 | DDX39BP2 |
| SAT | HTN | ENSG00000239557.1 | 0.027326064 | 9.67E-05 | 0.317354766 | 0.002234893 | RP11-168J18.6 |
| SAT | HTN | ENSG00000240399.1 | -0.025047249 | 7.43E-05 | 0.243622707 | 0.001791343 | RP1-228P16.1 |
| SAT | HTN | ENSG00000241170.2 | 0.011680517 | 0.002219304 | 1 | 0.025283117 | RP11-147I3.1 |
| SAT | HTN | ENSG00000242852.2 | -0.01621878 | 0.004555237 | 1 | 0.042219581 | ZNF709 |
| SAT | HTN | ENSG00000243697.1 | -0.02281377 | 0.001163228 | 1 | 0.015205383 | RP11-463O9.1 |
| SAT | HTN | ENSG00000243708.4 | 0.009320828 | 0.000185195 | 0.607623764 | 0.003682568 | PLA2G4B |
| SAT | HTN | ENSG00000243789.6 | 0.012603156 | 0.000722235 | 1 | 0.010439001 | JMJD7 |
| SAT | HTN | ENSG00000248290.1 | -0.006705231 | 0.000128516 | 0.421662093 | 0.002849068 | TNXA |
| SAT | HTN | ENSG00000249740.1 | 0.018112597 | 0.003099877 | 1 | 0.031983321 | CTD-2127H9.1 |
| SAT | HTN | ENSG00000251417.1 | -0.008446506 | 0.000222022 | 0.728452935 | 0.004285017 | RP11-1348G14.4 |
| SAT | HTN | ENSG00000251600.1 | -0.013531364 | 0.002894746 | 1 | 0.030441221 | RP11-673E1.1 |
| SAT | HTN | ENSG00000253203.2 | -0.013051401 | 0.001110851 | 1 | 0.014637359 | GUSBP3 |
| SAT | HTN | ENSG00000253320.1 | 0.013503547 | 0.000268763 | 0.881812692 | 0.004818649 | KB-1507C5.2 |
| SAT | HTN | ENSG00000253669.3 | 0.025048887 | 3.09E-08 | 0.000101338 | 2.81E-06 | KB-1732A1.1 |
| SAT | HTN | ENSG00000254851.1 | 0.009851543 | 1.78E-07 | 0.000585025 | 1.13E-05 | RP11-109L13.1 |
| SAT | HTN | ENSG00000256223.1 | -0.017832451 | 3.91E-06 | 0.01281652 | 0.000154416 | ZNF10 |

| | | | | | | | |
|---|---|---|---|---|---|---|---|
| SAT | HTN | ENSG00000256269.2 | 0.015386624 | 1.25E-05 | 0.04117433 | 0.00040367 | HMBS |
| SAT | HTN | ENSG00000256885.1 | -0.008673306 | 0.000536282 | 1 | 0.00826075 | AP001877.1 |
| SAT | HTN | ENSG00000259295.2 | 0.01513138 | 0.000949996 | 1 | 0.012774334 | CSPG4P12 |
| SAT | HTN | ENSG00000259982.1 | 0.012156686 | 2.42E-05 | 0.079480759 | 0.000685179 | CDC37P1 |
| SAT | HTN | ENSG00000260075.1 | 0.011145285 | 0.005301518 | 1 | 0.0481836 | NSFP1 |
| SAT | HTN | ENSG00000260077.1 | -0.013121778 | 0.001992879 | 1 | 0.023153921 | RP11-254F7.2 |
| SAT | HTN | ENSG00000260877.1 | -0.021502211 | 1.17E-06 | 0.003847323 | 5.42E-05 | RP11-211G23.2 |
| SAT | HTN | ENSG00000261556.4 | -0.016697628 | 5.25E-05 | 0.172303871 | 0.001325414 | RP11-296I10.6 |
| SAT | HTN | ENSG00000264538.2 | 0.008750856 | 6.34E-05 | 0.208043723 | 0.001564239 | SUZ12P |
| SAT | HTN | ENSG00000266497.1 | 0.015301626 | 0.005440137 | 1 | 0.04903596 | RP11-995C19.2 |
| SAT | HTN | ENSG00000267592.1 | 0.01762604 | 0.005047415 | 1 | 0.046258571 | CTC-507E2.2 |
| SAT | HTN | ENSG00000268117.1 | -0.010878449 | 0.000299234 | 0.981787206 | 0.005306958 | VN1R84P |
| SAT | HTN | ENSG00000268225.1 | 0.01241561 | 0.000447546 | 1 | 0.00709371 | CTD-2331H12.5 |
| SAT | HTN | ENSG00000268535.1 | -0.018618942 | 0.000646199 | 1 | 0.009725584 | RP11-420K14.3 |
| SAT | HTN | ENSG00000268555.1 | -0.014587446 | 0.000130331 | 0.42761721 | 0.002869914 | RP11-678G14.3 |
| SAT | HTN | ENSG00000269963.1 | -0.026641592 | 5.87E-07 | 0.001926583 | 2.96E-05 | RP11-73M18.9 |
| SAT | HTN | ENSG00000270018.1 | 0.030384429 | 3.26E-08 | 0.000106961 | 2.89E-06 | RP3-462E2.5 |
| SAT | HTN | ENSG00000270020.1 | -0.008318399 | 0.000167471 | 0.54947322 | 0.003477679 | RP11-463O9.9 |
| SAT | HTN | ENSG00000270091.1 | -0.016506116 | 0.000686869 | 1 | 0.01015143 | RP11-78O7.2 |
| SAT | HTN | ENSG00000271581.1 | 0.01143513 | 4.68E-09 | 1.54E-05 | 5.12E-07 | XXbac-BPG248L24.12 |
| SAT | HTN | ENSG00000271584.1 | 0.009297776 | 0.002297031 | 1 | 0.02607806 | RP11-89C3.4 |
| SAT | HTN | ENSG00000271623.1 | -0.023676658 | 7.50E-06 | 0.024609466 | 0.000261923 | RP11-435I10.5 |
| SAT | HTN | ENSG00000271751.1 | -0.025899965 | 1.23E-05 | 0.040377418 | 0.000401308 | RP11-110I1.14 |
| SAT | HTN | ENSG00000273117.1 | 0.014651574 | 0.000263113 | 0.863273011 | 0.004769464 | AC144652.1 |
| SAT | Heart_Failure | ENSG00000001617.7 | -0.013173139 | 1.05E-06 | 0.003449752 | 0.000581811 | SEMA3F |
| SAT | Heart_Failure | ENSG00000004534.10 | -0.003315477 | 1.54E-07 | 0.000505253 | 0.000168418 | RBM6 |
| SAT | Heart_Failure | ENSG00000008838.13 | -0.010686635 | 0.000182938 | 0.600219466 | 0.025009144 | MED24 |
| SAT | Heart_Failure | ENSG00000083720.8 | -0.014315518 | 0.000100061 | 0.328301019 | 0.015633382 | OXCT1 |
| SAT | Heart_Failure | ENSG00000090621.9 | -0.006657589 | 7.70E-05 | 0.252746332 | 0.012637317 | PABPC4 |
| SAT | Heart_Failure | ENSG00000114735.5 | 0.011309655 | 0.000265749 | 0.871922754 | 0.027247586 | HEMK1 |
| SAT | Heart_Failure | ENSG00000116883.8 | -0.006508555 | 4.34E-05 | 0.142549737 | 0.009503316 | RP11-268J15.5 |
| SAT | Heart_Failure | ENSG00000127957.12 | -0.007254743 | 2.09E-05 | 0.068565762 | 0.006233251 | PMS2P3 |
| SAT | Heart_Failure | ENSG00000131165.10 | 0.008755673 | 0.00045558 | 1 | 0.039533553 | CHMP1A |
| SAT | Heart_Failure | ENSG00000131435.8 | 0.017314273 | 5.25E-06 | 0.017229775 | 0.002461396 | PDLIM4 |
| SAT | Heart_Failure | ENSG00000132563.11 | -0.007308444 | 6.30E-05 | 0.206637533 | 0.011955478 | REEP2 |
| SAT | Heart_Failure | ENSG00000135213.8 | -0.016235475 | 1.05E-06 | 0.003430974 | 0.000581811 | POM121C |
| SAT | Heart_Failure | ENSG00000137312.10 | 0.009188357 | 6.79E-05 | 0.222937851 | 0.011955478 | FLOT1 |
| SAT | Heart_Failure | ENSG00000145692.10 | -0.005991572 | 0.00046992 | 1 | 0.039533553 | BHMT |
| SAT | Heart_Failure | ENSG00000145777.10 | 0.014444087 | 7.88E-06 | 0.025849598 | 0.00258496 | TSLP |
| SAT | Heart_Failure | ENSG00000156170.8 | -0.012287641 | 0.000248167 | 0.814234976 | 0.027247586 | NDUFAF6 |
| SAT | Heart_Failure | ENSG00000164068.11 | 0.004947231 | 6.10E-06 | 0.020018014 | 0.002502252 | RNF123 |
| SAT | Heart_Failure | ENSG00000170092.10 | 0.009495741 | 0.000415699 | 1 | 0.038968793 | AC018720.10 |
| SAT | Heart_Failure | ENSG00000172057.5 | -0.019872989 | 7.34E-12 | 2.41E-08 | 2.41E-08 | ORMDL3 |
| SAT | Heart_Failure | ENSG00000172992.7 | -0.005162731 | 6.08E-05 | 0.1994629 | 0.011955478 | DCAKD |
| SAT | Heart_Failure | ENSG00000173540.8 | 0.013994653 | 0.00025701 | 0.843250484 | 0.027247586 | GMPPB |
| SAT | Heart_Failure | ENSG00000173905.4 | 0.011184706 | 0.000337862 | 1 | 0.033591688 | GOLIM4 |
| SAT | Heart_Failure | ENSG00000174004.5 | -0.009783529 | 0.000464676 | 1 | 0.039533553 | NRROS |
| SAT | Heart_Failure | ENSG00000176715.11 | -0.008858515 | 0.000210541 | 0.690786369 | 0.027247586 | ACSF3 |
| SAT | Heart_Failure | ENSG00000179344.12 | -0.010040039 | 0.00025911 | 0.850138983 | 0.027247586 | HLA-DQB1 |
| SAT | Heart_Failure | ENSG00000182134.11 | -0.010459075 | 0.000572017 | 1 | 0.046919683 | TDRKH |
| SAT | Heart_Failure | ENSG00000183604.10 | -0.007986835 | 0.000443093 | 1 | 0.039533553 | RP11-347C12.2 |
| SAT | Heart_Failure | ENSG00000185499.12 | 0.011829975 | 3.77E-05 | 0.123763084 | 0.009269033 | MUC1 |
| SAT | Heart_Failure | ENSG00000187778.9 | 0.012376657 | 6.92E-05 | 0.227154091 | 0.011955478 | MCRS1 |
| SAT | Heart_Failure | ENSG00000188895.7 | -0.005474804 | 0.000142301 | 0.46688838 | 0.020801401 | MSL1 |
| SAT | Heart_Failure | ENSG00000197375.8 | 0.012305926 | 1.10E-07 | 0.000360941 | 0.000168418 | SLC22A5 |
| SAT | Heart_Failure | ENSG00000197728.5 | 0.00305817 | 0.000145819 | 0.478432218 | 0.020801401 | RPS26 |
| SAT | Heart_Failure | ENSG00000204438.6 | -0.009692416 | 3.15E-05 | 0.103222284 | 0.008601857 | GPANK1 |
| SAT | Heart_Failure | ENSG00000204516.5 | 0.00338079 | 3.96E-05 | 0.129766464 | 0.009269033 | MICB |
| SAT | Heart_Failure | ENSG00000205583.9 | -0.006258194 | 1.06E-06 | 0.003490867 | 0.000581811 | STAG3L1 |
| SAT | Heart_Failure | ENSG00000214078.7 | -0.002434439 | 0.000412456 | 1 | 0.038968793 | CPNE1 |
| SAT | Heart_Failure | ENSG00000225851.1 | -0.005296477 | 0.000246642 | 0.809233072 | 0.027247586 | HLA-S |
| SAT | Heart_Failure | ENSG00000232491.1 | 0.0133915 | 7.78E-06 | 0.025520246 | 0.00258496 | RP4-756H11.1 |
| SAT | Heart_Failure | ENSG00000243753.1 | -0.002967725 | 0.000261247 | 0.857152676 | 0.027247586 | HLA-L |
| SAT | Heart_Failure | ENSG00000267369.1 | -0.013093376 | 0.00023909 | 0.784454911 | 0.027247586 | RP11-1094M14.8 |
| VAT | CAD | ENSG00000065060.12 | 0.009846128 | 1.82E-05 | 0.050297922 | 0.00628724 | UHRF1BP1 |
| VAT | CAD | ENSG00000075131.5 | 0.006277787 | 0.000441759 | 1 | 0.043413258 | TIPIN |
| VAT | CAD | ENSG00000076258.5 | 0.025806204 | 9.11E-07 | 0.002514751 | 0.001257375 | FMO4 |
| VAT | CAD | ENSG00000096433.6 | 0.020058784 | 4.21E-05 | 0.116207024 | 0.0124879 | ITPR3 |
| VAT | CAD | ENSG00000099194.5 | -0.018167814 | 0.000317871 | 0.877323535 | 0.038787669 | SCD |
| VAT | CAD | ENSG00000115486.7 | 0.012781827 | 0.000297342 | 0.820663296 | 0.038787669 | GGCX |
| VAT | CAD | ENSG00000124562.5 | -0.014130006 | 4.71E-06 | 0.013007468 | 0.004335823 | SNRPC |
| VAT | CAD | ENSG00000124678.13 | 0.009730688 | 0.00020691 | 0.571071418 | 0.032700425 | TCP11 |
| VAT | CAD | ENSG00000125388.15 | 0.004473991 | 0.000110826 | 0.305880846 | 0.025490071 | GRK4 |
| VAT | CAD | ENSG00000137700.12 | 0.01458388 | 0.000216117 | 0.596482349 | 0.032700425 | SLC37A4 |
| VAT | CAD | ENSG00000138303.13 | -0.012234845 | 0.000425295 | 1 | 0.043413258 | ASCC1 |
| VAT | CAD | ENSG00000142046.10 | 0.006917361 | 0.000471883 | 1 | 0.043413258 | TMEM91 |
| VAT | CAD | ENSG00000143156.9 | -0.01984382 | 1.10E-05 | 0.030430146 | 0.006086029 | NME7 |
| VAT | CAD | ENSG00000143437.16 | -0.02122307 | 6.05E-07 | 0.001668724 | 0.001257375 | ARNT |
| VAT | CAD | ENSG00000143951.11 | 0.01971114 | 0.000142036 | 0.392019033 | 0.03015531 | WDPCP |
| VAT | CAD | ENSG00000145020.10 | -0.004068616 | 0.000457551 | 1 | 0.043413258 | AMT |
| VAT | CAD | ENSG00000145029.7 | -0.010245957 | 1.01E-05 | 0.027853713 | 0.006086029 | NICN1 |

| | | | | | | | |
|---|---|---|---|---|---|---|---|
| VAT | CAD | ENSG00000147912.8 | 0.018442494 | 0.000198155 | 0.546908391 | 0.032700425 | FBXO10 |
| VAT | CAD | ENSG00000149084.7 | -0.003180787 | 0.000323231 | 0.892116391 | 0.038787669 | HSD17B12 |
| VAT | CAD | ENSG00000164068.11 | 0.007463329 | 7.65E-05 | 0.211276889 | 0.01920699 | RNF123 |
| VAT | CAD | ENSG00000168038.6 | -0.009214595 | 0.000458819 | 1 | 0.043413258 | ULK4 |
| VAT | CAD | ENSG00000172171.6 | 0.015175036 | 0.000410809 | 1 | 0.043413258 | TEFM |
| VAT | CAD | ENSG00000176390.10 | 0.011282356 | 0.000219483 | 0.605772978 | 0.032700425 | CRLF3 |
| VAT | CAD | ENSG00000181092.5 | -0.011737578 | 0.000338054 | 0.933027817 | 0.038876159 | ADIPOQ |
| VAT | CAD | ENSG00000182511.7 | -0.020917584 | 1.66E-05 | 0.045721642 | 0.00628724 | FES |
| VAT | CAD | ENSG00000197081.8 | -0.016782083 | 0.000225112 | 0.621308073 | 0.032700425 | IGF2R |
| VAT | CAD | ENSG00000198270.8 | -0.006122141 | 4.52E-05 | 0.124879004 | 0.0124879 | TMEM116 |
| VAT | CAD | ENSG00000213689.5 | 0.005765878 | 0.000157238 | 0.433977799 | 0.030998414 | TREX1 |
| VAT | CAD | ENSG00000264538.2 | 0.008847486 | 0.000280086 | 0.773038338 | 0.038651917 | SUZ12P |
| VAT | CAD | ENSG00000271581.1 | 0.025678587 | 1.54E-05 | 0.042601867 | 0.00628724 | XXbac-BPG248L24.12 |
| VAT | T2DM | ENSG00000004534.10 | -0.004332966 | 8.35E-09 | 2.31E-05 | 2.88E-06 | RBM6 |
| VAT | T2DM | ENSG00000011105.7 | -0.007855696 | 0.001608371 | 1 | 0.045352953 | TSPAN9 |
| VAT | T2DM | ENSG00000011523.9 | 0.017150923 | 3.96E-11 | 1.09E-07 | 2.18E-08 | CEP68 |
| VAT | T2DM | ENSG00000037042.8 | -0.018185417 | 1.75E-07 | 0.000483299 | 3.72E-05 | TUBG2 |
| VAT | T2DM | ENSG00000055955.11 | 0.004708028 | 7.06E-06 | 0.019493733 | 0.000749759 | ITIH4 |
| VAT | T2DM | ENSG00000065665.16 | -0.015980757 | 2.44E-06 | 0.006744149 | 0.000293224 | SEC61A2 |
| VAT | T2DM | ENSG00000078618.15 | 0.011965055 | 0.000268002 | 0.739685085 | 0.012455159 | NRD1 |
| VAT | T2DM | ENSG00000082258.8 | 0.01172735 | 0.000873292 | 1 | 0.028693864 | CCNT2 |
| VAT | T2DM | ENSG00000086619.9 | 0.010205169 | 4.33E-06 | 0.011941327 | 0.000497555 | ERO1LB |
| VAT | T2DM | ENSG00000090621.9 | -0.021186856 | 1.68E-12 | 4.64E-09 | 1.16E-09 | PABPC4 |
| VAT | T2DM | ENSG00000091542.8 | -0.013200608 | 8.26E-05 | 0.228005177 | 0.005066782 | ALKBH5 |
| VAT | T2DM | ENSG00000095485.12 | -0.003234458 | 6.19E-05 | 0.170837118 | 0.004163595 | CWF19L1 |
| VAT | T2DM | ENSG00000099194.5 | -0.012184002 | 0.001610358 | 1 | 0.045352953 | SCD |
| VAT | T2DM | ENSG00000103226.13 | -0.009507402 | 0.000629956 | 1 | 0.022877354 | NOMO3 |
| VAT | T2DM | ENSG00000103479.10 | 0.013294817 | 9.75E-05 | 0.269144509 | 0.005726479 | RBL2 |
| VAT | T2DM | ENSG00000108785.7 | -0.016199637 | 4.07E-05 | 0.112285703 | 0.003034749 | HSD17B1P1 |
| VAT | T2DM | ENSG00000111215.7 | 0.005219074 | 0.00139763 | 1 | 0.041478058 | PRR4 |
| VAT | T2DM | ENSG00000112276.9 | -0.006415103 | 0.000417895 | 1 | 0.017475613 | BVES |
| VAT | T2DM | ENSG00000116157.5 | -0.003528954 | 0.000819006 | 1 | 0.027234428 | GPX7 |
| VAT | T2DM | ENSG00000119242.4 | -0.011382156 | 0.000475094 | 1 | 0.018940823 | CCDC92 |
| VAT | T2DM | ENSG00000120907.13 | 0.005038235 | 3.90E-05 | 0.10772573 | 0.002992381 | ADRA1A |
| VAT | T2DM | ENSG00000124767.6 | 0.013392092 | 0.000985229 | 1 | 0.030900358 | GLO1 |
| VAT | T2DM | ENSG00000125388.15 | 0.00292367 | 0.000967516 | 1 | 0.030693597 | GRK4 |
| VAT | T2DM | ENSG00000126464.9 | 0.009820216 | 0.001381229 | 1 | 0.041436863 | PRR12 |
| VAT | T2DM | ENSG00000127220.5 | -0.008076621 | 0.000594626 | 1 | 0.022177952 | ABHD8 |
| VAT | T2DM | ENSG00000127957.12 | -0.012346798 | 2.65E-05 | 0.073082294 | 0.002149479 | PMS2P3 |
| VAT | T2DM | ENSG00000130921.3 | 0.009891253 | 0.000270764 | 0.747309516 | 0.012455159 | C12orf65 |
| VAT | T2DM | ENSG00000137265.10 | 0.006881096 | 0.000947059 | 1 | 0.030693597 | IRF4 |
| VAT | T2DM | ENSG00000137310.7 | 0.015733453 | 4.31E-05 | 0.118967945 | 0.003109431 | TCF19 |
| VAT | T2DM | ENSG00000137501.12 | -0.012988865 | 0.000541869 | 1 | 0.020487111 | SYTL2 |
| VAT | T2DM | ENSG00000137700.12 | 0.01110075 | 0.000234761 | 0.647941435 | 0.011907029 | SLC37A4 |
| VAT | T2DM | ENSG00000137877.8 | 0.005685689 | 6.61E-05 | 0.182401528 | 0.004241896 | SPTBN5 |
| VAT | T2DM | ENSG00000140398.9 | -0.0116514 | 2.93E-05 | 0.081000887 | 0.002314311 | NEIL1 |
| VAT | T2DM | ENSG00000140400.10 | -0.002826645 | 1.04E-05 | 0.028831068 | 0.001029681 | MAN2C1 |
| VAT | T2DM | ENSG00000141140.12 | 0.016661083 | 1.11E-06 | 0.003071798 | 0.00015359 | MYO19 |
| VAT | T2DM | ENSG00000141252.15 | -0.006793446 | 0.000230029 | 0.634880333 | 0.011907029 | VPS53 |
| VAT | T2DM | ENSG00000141452.5 | -0.015369868 | 2.94E-07 | 0.000812085 | 5.41E-05 | C18orf8 |
| VAT | T2DM | ENSG00000141994.11 | 0.004915812 | 0.001111163 | 1 | 0.034458528 | DUS3L |
| VAT | T2DM | ENSG00000149084.7 | -0.003847924 | 1.33E-08 | 3.66E-05 | 4.07E-06 | HSD17B12 |
| VAT | T2DM | ENSG00000149089.8 | -0.003930868 | 0.000170065 | 0.46937914 | 0.009203513 | APIP |
| VAT | T2DM | ENSG00000152359.10 | -0.004289249 | 0.000230042 | 0.634916231 | 0.011907029 | POC5 |
| VAT | T2DM | ENSG00000153814.7 | -0.018729946 | 4.56E-21 | 1.26E-17 | 1.26E-17 | JAZF1 |
| VAT | T2DM | ENSG00000156931.11 | 0.013325172 | 0.001246879 | 1 | 0.038237629 | VPS8 |
| VAT | T2DM | ENSG00000157322.12 | -0.010306535 | 3.56E-08 | 9.82E-05 | 8.92E-06 | CLEC18A |
| VAT | T2DM | ENSG00000157992.8 | 0.009026748 | 1.14E-05 | 0.031349587 | 0.001044986 | KRTCAP3 |
| VAT | T2DM | ENSG00000163554.7 | -0.006730558 | 0.000277531 | 0.765985439 | 0.012557138 | SPTA1 |
| VAT | T2DM | ENSG00000163933.5 | -0.008955202 | 5.77E-07 | 0.001591183 | 9.94E-05 | RFT1 |
| VAT | T2DM | ENSG00000164068.11 | 0.006429384 | 8.56E-06 | 0.023631569 | 0.000875243 | RNF123 |
| VAT | T2DM | ENSG00000164078.8 | 0.018391917 | 7.57E-07 | 0.002088443 | 0.000121943 | MST1R |
| VAT | T2DM | ENSG00000165917.5 | -0.014135316 | 2.11E-05 | 0.058128376 | 0.001816512 | RAPSN |
| VAT | T2DM | ENSG00000166825.9 | 0.006954768 | 0.000655704 | 1 | 0.023226937 | ANPEP |
| VAT | T2DM | ENSG00000168612.4 | 0.004580254 | 0.000500891 | 1 | 0.019471269 | ZSWIM1 |
| VAT | T2DM | ENSG00000169047.5 | -0.012800604 | 2.62E-07 | 0.000722378 | 5.16E-05 | IRS1 |
| VAT | T2DM | ENSG00000169592.10 | -0.005985636 | 0.000247041 | 0.681832834 | 0.011907029 | INO80E |
| VAT | T2DM | ENSG00000170092.10 | 0.010439489 | 0.000757196 | 1 | 0.025800768 | AC018720.10 |
| VAT | T2DM | ENSG00000170571.7 | -0.013382621 | 0.000136639 | 0.377124602 | 0.007667187 | EMB |
| VAT | T2DM | ENSG00000171700.9 | -0.008129381 | 2.61E-05 | 0.072044667 | 0.002149479 | RGS19 |
| VAT | T2DM | ENSG00000172247.3 | -0.00611146 | 0.000138898 | 0.383359364 | 0.007667187 | C1QTNF4 |
| VAT | T2DM | ENSG00000173917.9 | -0.008600179 | 0.000808705 | 1 | 0.027219831 | HOXB2 |
| VAT | T2DM | ENSG00000175305.12 | -0.01287363 | 7.95E-07 | 0.002194966 | 0.000121943 | CCNE2 |
| VAT | T2DM | ENSG00000178750.2 | 0.009638938 | 0.000656413 | 1 | 0.023226937 | STX19 |
| VAT | T2DM | ENSG00000178917.10 | -0.010530923 | 0.00138109 | 1 | 0.041436863 | ZNF852 |
| VAT | T2DM | ENSG00000179344.12 | -0.010906808 | 5.89E-09 | 1.63E-05 | 2.32E-06 | HLA-DQB1 |
| VAT | T2DM | ENSG00000182670.9 | 0.002819548 | 0.001485644 | 1 | 0.043621024 | TTC3 |
| VAT | T2DM | ENSG00000185245.6 | 0.011856977 | 0.001534186 | 1 | 0.044404773 | GP1BA |
| VAT | T2DM | ENSG00000185800.7 | -0.01187574 | 0.000101738 | 0.280797392 | 0.005849946 | DMWD |
| VAT | T2DM | ENSG00000185989.9 | -0.012188646 | 7.09E-05 | 0.195566885 | 0.004444702 | RASA3 |
| VAT | T2DM | ENSG00000196126.6 | -0.014329638 | 3.23E-10 | 8.92E-07 | 1.49E-07 | HLA-DRB1 |

| | | | | | | | |
|---|---|---|---|---|---|---|---|
| VAT | T2DM | ENSG00000196301.3 | 0.00636131 | 2.05E-05 | 0.056686469 | 0.001816512 | HLA-DRB9 |
| VAT | T2DM | ENSG00000196735.7 | -0.016587164 | 9.85E-07 | 0.002717469 | 0.000143025 | HLA-DQA1 |
| VAT | T2DM | ENSG00000198502.5 | -0.003934515 | 4.39E-05 | 0.12126781 | 0.003109431 | HLA-DRB5 |
| VAT | T2DM | ENSG00000198909.3 | -0.014350704 | 1.13E-05 | 0.031065747 | 0.001044986 | MAP3K3 |
| VAT | T2DM | ENSG00000204344.10 | -0.010911555 | 0.00025022 | 0.690607666 | 0.011907029 | STK19 |
| VAT | T2DM | ENSG00000204525.10 | -0.00392544 | 0.000707124 | 1 | 0.024704568 | HLA-C |
| VAT | T2DM | ENSG00000204560.5 | 0.010150218 | 0.000474877 | 1 | 0.018940823 | DHX16 |
| VAT | T2DM | ENSG00000204622.6 | 0.003817344 | 0.000729002 | 1 | 0.025150583 | HLA-J |
| VAT | T2DM | ENSG00000205583.9 | -0.004657484 | 0.000615596 | 1 | 0.022653918 | STAG3L1 |
| VAT | T2DM | ENSG00000205659.6 | 0.013875109 | 6.34E-05 | 0.174870981 | 0.004163595 | LIN52 |
| VAT | T2DM | ENSG00000206344.6 | 0.006194087 | 1.60E-06 | 0.004405667 | 0.000200258 | HCG27 |
| VAT | T2DM | ENSG00000212127.5 | 0.006917457 | 0.000468892 | 1 | 0.018940823 | TAS2R14 |
| VAT | T2DM | ENSG00000213221.4 | 0.003317361 | 0.000249452 | 0.688488117 | 0.011907029 | DNLZ |
| VAT | T2DM | ENSG00000214530.3 | 0.008081462 | 2.72E-08 | 7.52E-05 | 7.52E-06 | STARD10 |
| VAT | T2DM | ENSG00000214706.6 | -0.007967616 | 0.001544514 | 1 | 0.044404773 | IFRD2 |
| VAT | T2DM | ENSG00000226232.4 | -0.011696758 | 1.57E-06 | 0.00433226 | 0.000200258 | RP11-419C5.2 |
| VAT | T2DM | ENSG00000229391.3 | 0.009970144 | 8.93E-16 | 2.46E-12 | 1.23E-12 | HLA-DRB6 |
| VAT | T2DM | ENSG00000230224.1 | -0.008905757 | 8.59E-05 | 0.237220907 | 0.005156976 | PHBP9 |
| VAT | T2DM | ENSG00000231742.1 | -0.010218315 | 0.000408506 | 1 | 0.017475613 | RP11-112L6.4 |
| VAT | T2DM | ENSG00000232063.1 | 0.003214853 | 0.000966094 | 1 | 0.030693597 | RP11-307E17.8 |
| VAT | T2DM | ENSG00000233569.1 | -0.014553418 | 0.000364413 | 1 | 0.015964759 | RP11-500B12.1 |
| VAT | T2DM | ENSG00000233609.3 | 0.010851893 | 0.000523609 | 1 | 0.020071668 | RP11-62H7.2 |
| VAT | T2DM | ENSG00000237541.3 | 0.009016539 | 3.46E-15 | 9.55E-12 | 3.18E-12 | HLA-DQA2 |
| VAT | T2DM | ENSG00000237854.3 | 0.009228254 | 0.000480383 | 1 | 0.018940823 | LINC00674 |
| VAT | T2DM | ENSG00000243789.6 | 0.00914322 | 1.21E-07 | 0.00033434 | 2.79E-05 | JMJD7 |
| VAT | T2DM | ENSG00000253813.1 | 0.007287578 | 5.29E-05 | 0.145903188 | 0.00364758 | COX6B1P4 |
| VAT | T2DM | ENSG00000261760.2 | 0.011217579 | 0.000245298 | 0.677023394 | 0.011907029 | RP11-1223D19.1 |
| VAT | T2DM | ENSG00000268879.1 | -0.007729438 | 0.001767767 | 1 | 0.0492832 | IGFL1P1 |
| VAT | T2DM | ENSG00000269963.1 | -0.014579698 | 0.00041611 | 1 | 0.017475613 | RP11-73M18.9 |
| VAT | T2DM | ENSG00000271581.1 | 0.020804375 | 4.77E-06 | 0.013171662 | 0.000526866 | XXbac-BPG248L24.12 |
| VAT | T2DM | ENSG00000272221.1 | -0.002981503 | 0.00028465 | 0.785633404 | 0.012671507 | XXbac-BPG181B23.7 |
| VAT | Obesity | ENSG00000004534.10 | -0.012003509 | 2.58E-26 | 7.13E-23 | 2.38E-23 | RBM6 |
| VAT | Obesity | ENSG00000011021.17 | 0.013500691 | 0.000885796 | 1 | 0.013658085 | CLCN6 |
| VAT | Obesity | ENSG00000015676.13 | 0.007905229 | 0.003327451 | 1 | 0.03573449 | NUDCD3 |
| VAT | Obesity | ENSG00000031691.6 | -0.00386726 | 0.004294372 | 1 | 0.043575249 | CENPQ |
| VAT | Obesity | ENSG00000033800.9 | 0.033988019 | 1.05E-07 | 0.000290608 | 6.46E-06 | PIAS1 |
| VAT | Obesity | ENSG00000035115.17 | -0.026225758 | 2.47E-06 | 0.006811713 | 0.000111667 | SH3YL1 |
| VAT | Obesity | ENSG00000037280.11 | -0.018298065 | 0.000481932 | 1 | 0.008365619 | FLT4 |
| VAT | Obesity | ENSG00000041802.6 | 0.00351383 | 0.002751449 | 1 | 0.031393572 | LSG1 |
| VAT | Obesity | ENSG00000049449.4 | -0.016994951 | 0.000684696 | 1 | 0.010986495 | RCN1 |
| VAT | Obesity | ENSG00000053900.6 | 0.01925026 | 0.000688646 | 1 | 0.010986495 | ANAPC4 |
| VAT | Obesity | ENSG00000054282.11 | 0.007832158 | 0.0012334 | 1 | 0.017587781 | SDCCAG8 |
| VAT | Obesity | ENSG00000055955.11 | 0.010291055 | 6.65E-11 | 1.84E-07 | 5.74E-09 | ITIH4 |
| VAT | Obesity | ENSG00000058063.11 | 0.0176696 | 0.001732351 | 1 | 0.022553246 | ATP11B |
| VAT | Obesity | ENSG00000065060.12 | 0.026202429 | 3.92E-23 | 1.08E-19 | 1.80E-20 | UHRF1BP1 |
| VAT | Obesity | ENSG00000066855.11 | -0.0158627 | 0.002044103 | 1 | 0.02463635 | MTFR1 |
| VAT | Obesity | ENSG00000070540.8 | -0.012133575 | 0.00298535 | 1 | 0.033090625 | WIPI1 |
| VAT | Obesity | ENSG00000073754.5 | 0.017280654 | 0.005173729 | 1 | 0.048735468 | CD5L |
| VAT | Obesity | ENSG00000075131.5 | 0.006857447 | 0.000853405 | 1 | 0.013232578 | TIPIN |
| VAT | Obesity | ENSG00000076685.14 | 0.009746773 | 0.000149162 | 0.41168648 | 0.003293492 | NT5C2 |
| VAT | Obesity | ENSG00000082258.8 | -0.015736066 | 0.002982103 | 1 | 0.033090625 | CCNT2 |
| VAT | Obesity | ENSG00000083097.10 | 0.006290028 | 0.004378496 | 1 | 0.0439491 | DOPEY1 |
| VAT | Obesity | ENSG00000084207.11 | 0.013535305 | 2.61E-05 | 0.071910214 | 0.00080798 | GSTP1 |
| VAT | Obesity | ENSG00000085832.12 | -0.011717602 | 0.000469143 | 1 | 0.00819516 | EPS15 |
| VAT | Obesity | ENSG00000086015.16 | 0.014842102 | 0.001143441 | 1 | 0.016436964 | MAST2 |
| VAT | Obesity | ENSG00000087263.12 | -0.020889599 | 6.35E-06 | 0.017512614 | 0.000261382 | OGFOD1 |
| VAT | Obesity | ENSG00000088833.13 | 0.012381224 | 0.004378799 | 1 | 0.0439491 | NSFL1C |
| VAT | Obesity | ENSG00000090238.7 | 0.014557413 | 2.79E-09 | 7.70E-06 | 2.08E-07 | YPEL3 |
| VAT | Obesity | ENSG00000090621.9 | -0.029430877 | 6.96E-11 | 1.92E-07 | 5.82E-09 | PABPC4 |
| VAT | Obesity | ENSG00000092020.6 | 0.016177942 | 0.000134442 | 0.371058792 | 0.003016738 | PPP2R3C |
| VAT | Obesity | ENSG00000096433.6 | 0.026003821 | 4.00E-06 | 0.011048227 | 0.000172629 | ITPR3 |
| VAT | Obesity | ENSG00000100124.8 | -0.013321151 | 0.000161725 | 0.446360912 | 0.003514653 | ANKRD54 |
| VAT | Obesity | ENSG00000100162.10 | 0.018974267 | 4.14E-05 | 0.114208827 | 0.001165396 | CENPM |
| VAT | Obesity | ENSG00000100997.14 | 0.023030399 | 1.48E-05 | 0.040738325 | 0.000515675 | ABHD12 |
| VAT | Obesity | ENSG00000102595.14 | -0.017286049 | 0.001626071 | 1 | 0.021473469 | UGGT2 |
| VAT | Obesity | ENSG00000102890.10 | 0.009346793 | 0.003406491 | 1 | 0.036161216 | ELMO3 |
| VAT | Obesity | ENSG00000103051.14 | -0.011816616 | 0.000818241 | 1 | 0.012759014 | COG4 |
| VAT | Obesity | ENSG00000103351.8 | 0.005819632 | 0.001246349 | 1 | 0.01764063 | CLUAP1 |
| VAT | Obesity | ENSG00000103472.5 | -0.020124905 | 0.000567481 | 1 | 0.009378732 | RRN3P2 |
| VAT | Obesity | ENSG00000103932.7 | -0.01914295 | 0.001489429 | 1 | 0.020451858 | RPAP1 |
| VAT | Obesity | ENSG00000104219.8 | -0.019595967 | 0.000505744 | 1 | 0.008563512 | ZDHHC2 |
| VAT | Obesity | ENSG00000104714.9 | -0.019871922 | 0.000337953 | 0.932749424 | 0.006218329 | ERICH1 |
| VAT | Obesity | ENSG00000106290.10 | -0.008079442 | 0.004432314 | 1 | 0.044318348 | TAF6 |
| VAT | Obesity | ENSG00000106638.11 | 0.012038339 | 0.001948608 | 1 | 0.023938382 | TBL2 |
| VAT | Obesity | ENSG00000107521.14 | -0.018214315 | 0.002335128 | 1 | 0.027779968 | HPS1 |
| VAT | Obesity | ENSG00000108384.10 | -0.011455808 | 5.43E-05 | 0.149901575 | 0.001441361 | RAD51C |
| VAT | Obesity | ENSG00000108556.7 | 0.005360431 | 0.000502248 | 1 | 0.008563512 | CHRNE |
| VAT | Obesity | ENSG00000110906.8 | -0.015803936 | 0.001068617 | 1 | 0.015746258 | KCTD10 |
| VAT | Obesity | ENSG00000111801.11 | 0.016884885 | 6.22E-08 | 0.000171758 | 3.90E-06 | BTN3A3 |
| VAT | Obesity | ENSG00000112041.8 | -0.016852345 | 0.000368778 | 1 | 0.006696239 | TULP1 |
| VAT | Obesity | ENSG00000112514.11 | -0.01119687 | 7.77E-05 | 0.214367329 | 0.001981293 | CUTA |

| | | | | | | | |
|---|---|---|---|---|---|---|---|
| VAT | Obesity | ENSG00000112981.3 | 0.015932553 | 0.000190646 | 0.526181988 | 0.004057373 | NME5 |
| VAT | Obesity | ENSG00000113119.8 | -0.017969029 | 0.000285796 | 0.788795859 | 0.005533516 | TMCO6 |
| VAT | Obesity | ENSG00000113141.11 | -0.010831715 | 0.000327891 | 0.904978408 | 0.006073681 | IK |
| VAT | Obesity | ENSG00000113734.13 | -0.00636403 | 0.00446395 | 1 | 0.044318348 | BNIP1 |
| VAT | Obesity | ENSG00000114054.9 | 0.02095936 | 1.65E-06 | 0.004551112 | 7.59E-05 | PCCB |
| VAT | Obesity | ENSG00000114904.8 | -0.012848176 | 0.004768645 | 1 | 0.046019087 | NEK4 |
| VAT | Obesity | ENSG00000115561.10 | -0.015518132 | 9.93E-07 | 0.002739306 | 4.72E-05 | CHMP3 |
| VAT | Obesity | ENSG00000117481.6 | -0.018949557 | 3.82E-05 | 0.10542661 | 0.001109754 | NSUN4 |
| VAT | Obesity | ENSG00000117523.11 | 0.01663217 | 0.000519791 | 1 | 0.0087477 | PRRC2C |
| VAT | Obesity | ENSG00000119242.4 | 0.022350386 | 5.05E-06 | 0.013927662 | 0.000214272 | CCDC92 |
| VAT | Obesity | ENSG00000119509.8 | 0.011394059 | 5.94E-07 | 0.001640224 | 2.98E-05 | INVS |
| VAT | Obesity | ENSG00000119865.4 | -0.014601175 | 0.001236243 | 1 | 0.017587781 | CNRIP1 |
| VAT | Obesity | ENSG00000120451.6 | -0.019453561 | 4.02E-07 | 0.001110843 | 2.10E-05 | SNX19 |
| VAT | Obesity | ENSG00000120907.13 | 0.005940957 | 0.001259209 | 1 | 0.017731721 | ADRA1A |
| VAT | Obesity | ENSG00000121101.11 | 0.02223548 | 3.06E-05 | 0.084422637 | 0.000927721 | TEX14 |
| VAT | Obesity | ENSG00000122696.8 | 0.007546505 | 0.002925596 | 1 | 0.032690866 | SLC25A51 |
| VAT | Obesity | ENSG00000123243.10 | 0.013920304 | 0.004620509 | 1 | 0.044966202 | ITIH5 |
| VAT | Obesity | ENSG00000124508.12 | 0.010322219 | 4.46E-05 | 0.123161055 | 0.001207461 | BTN2A2 |
| VAT | Obesity | ENSG00000124562.5 | -0.036021943 | 3.86E-24 | 1.07E-20 | 2.13E-21 | SNRPC |
| VAT | Obesity | ENSG00000124678.13 | 0.008563181 | 0.004562759 | 1 | 0.044966202 | TCP11 |
| VAT | Obesity | ENSG00000125388.15 | 0.005089653 | 0.000133419 | 0.368235569 | 0.003016738 | GRK4 |
| VAT | Obesity | ENSG00000127337.2 | 0.006435319 | 0.002997626 | 1 | 0.033093789 | YEATS4 |
| VAT | Obesity | ENSG00000127957.12 | -0.042314049 | 1.02E-21 | 2.80E-18 | 3.51E-19 | PMS2P3 |
| VAT | Obesity | ENSG00000128408.7 | 0.014354065 | 0.000584391 | 1 | 0.009600702 | RIBC2 |
| VAT | Obesity | ENSG00000128829.7 | 0.018408673 | 0.001863703 | 1 | 0.023559873 | EIF2AK4 |
| VAT | Obesity | ENSG00000129197.10 | -0.020264101 | 1.82E-05 | 0.050340929 | 0.000606517 | RPAIN |
| VAT | Obesity | ENSG00000129354.7 | -0.017183279 | 0.000709502 | 1 | 0.01125417 | AP1M2 |
| VAT | Obesity | ENSG00000130270.12 | -0.018745558 | 3.93E-11 | 1.08E-07 | 3.50E-09 | ATP8B3 |
| VAT | Obesity | ENSG00000130517.9 | 0.008814609 | 0.001112362 | 1 | 0.016244016 | PGPEP1 |
| VAT | Obesity | ENSG00000130649.5 | -0.011128478 | 0.001770611 | 1 | 0.02256353 | CYP2E1 |
| VAT | Obesity | ENSG00000130921.3 | 0.013453053 | 0.000990195 | 1 | 0.01501615 | C12orf65 |
| VAT | Obesity | ENSG00000130943.5 | -0.009607804 | 0.003403407 | 1 | 0.036161216 | PKDREJ |
| VAT | Obesity | ENSG00000131495.4 | 0.013137927 | 0.000127808 | 0.352748922 | 0.00291528 | NDUFA2 |
| VAT | Obesity | ENSG00000131558.10 | 0.019289287 | 0.000101199 | 0.279309184 | 0.002428776 | EXOC4 |
| VAT | Obesity | ENSG00000132207.13 | -0.01004113 | 0.00025522 | 0.704408389 | 0.004995804 | SLX1A |
| VAT | Obesity | ENSG00000132382.10 | 0.011151368 | 0.001702489 | 1 | 0.02237557 | MYBBP1A |
| VAT | Obesity | ENSG00000132740.4 | 0.007165858 | 4.02E-05 | 0.110868798 | 0.001142977 | IGHMBP2 |
| VAT | Obesity | ENSG00000132781.13 | 0.013527145 | 0.003114806 | 1 | 0.034114543 | MUTYH |
| VAT | Obesity | ENSG00000133706.13 | -0.005728604 | 0.0002489 | 0.686965306 | 0.004942196 | LARS |
| VAT | Obesity | ENSG00000134201.6 | 0.016020355 | 0.001869425 | 1 | 0.023559873 | GSTM5 |
| VAT | Obesity | ENSG00000135213.8 | -0.03665955 | 9.92E-18 | 2.74E-14 | 2.11E-15 | POM121C |
| VAT | Obesity | ENSG00000135596.13 | 0.011002853 | 0.00278482 | 1 | 0.031500426 | MICAL1 |
| VAT | Obesity | ENSG00000135913.6 | 0.011651197 | 0.00073972 | 1 | 0.011600152 | USP37 |
| VAT | Obesity | ENSG00000137700.12 | 0.024583661 | 6.05E-08 | 0.000166866 | 3.88E-06 | SLC37A4 |
| VAT | Obesity | ENSG00000137806.4 | 0.021405284 | 2.25E-05 | 0.062222594 | 0.000715202 | NDUFAF1 |
| VAT | Obesity | ENSG00000137876.5 | -0.021077465 | 0.000117355 | 0.32389977 | 0.002744913 | RSL24D1 |
| VAT | Obesity | ENSG00000139187.5 | 0.00559315 | 0.003635112 | 1 | 0.037860038 | KLRG1 |
| VAT | Obesity | ENSG00000139719.5 | 0.020459415 | 8.39E-06 | 0.023154862 | 0.000340513 | VPS33A |
| VAT | Obesity | ENSG00000139910.15 | 0.011640891 | 0.005013493 | 1 | 0.047714623 | NOVA1 |
| VAT | Obesity | ENSG00000141076.13 | 0.010079368 | 0.001936628 | 1 | 0.023938382 | CIRH1A |
| VAT | Obesity | ENSG00000141140.12 | 0.034991488 | 1.03E-11 | 2.84E-08 | 1.06E-09 | MYO19 |
| VAT | Obesity | ENSG00000141452.5 | -0.037094171 | 1.91E-16 | 5.27E-13 | 3.76E-14 | C18orf8 |
| VAT | Obesity | ENSG00000141570.6 | 0.013444641 | 0.00268575 | 1 | 0.031015357 | CBX8 |
| VAT | Obesity | ENSG00000142156.10 | 0.008914991 | 0.000524442 | 1 | 0.008772485 | COL6A1 |
| VAT | Obesity | ENSG00000144199.7 | -0.015170688 | 0.001740586 | 1 | 0.022554068 | FAHD2B |
| VAT | Obesity | ENSG00000144791.5 | 0.018998099 | 0.000169904 | 0.468935712 | 0.00366356 | LIMD1 |
| VAT | Obesity | ENSG00000145020.10 | -0.003929861 | 0.003274455 | 1 | 0.035441163 | AMT |
| VAT | Obesity | ENSG00000145029.7 | -0.010973657 | 4.00E-05 | 0.110283331 | 0.001142977 | NICN1 |
| VAT | Obesity | ENSG00000146001.4 | 0.008686514 | 0.005038134 | 1 | 0.047784366 | PCDHB18 |
| VAT | Obesity | ENSG00000146109.3 | 0.009540872 | 0.001138869 | 1 | 0.016436964 | ABT1 |
| VAT | Obesity | ENSG00000146555.14 | -0.011270494 | 0.003849018 | 1 | 0.03963914 | SDK1 |
| VAT | Obesity | ENSG00000147576.11 | -0.010850738 | 0.00099774 | 1 | 0.015047886 | ADHFE1 |
| VAT | Obesity | ENSG00000148824.14 | 0.007243889 | 0.000466278 | 1 | 0.00819516 | MTG1 |
| VAT | Obesity | ENSG00000148843.9 | 0.016097028 | 0.004821483 | 1 | 0.046205883 | PDCD11 |
| VAT | Obesity | ENSG00000149084.7 | -0.006785865 | 2.67E-11 | 7.38E-08 | 2.54E-09 | HSD17B12 |
| VAT | Obesity | ENSG00000149115.9 | 0.015175782 | 3.37E-07 | 0.000929397 | 1.79E-05 | TNKS1BP1 |
| VAT | Obesity | ENSG00000149292.12 | 0.012034421 | 4.82E-05 | 0.132935468 | 0.001290636 | TTC12 |
| VAT | Obesity | ENSG00000149922.6 | 0.013463047 | 2.53E-11 | 6.98E-08 | 2.49E-09 | TBX6 |
| VAT | Obesity | ENSG00000151640.8 | 0.004053569 | 0.002018203 | 1 | 0.024430883 | DPYSL4 |
| VAT | Obesity | ENSG00000152359.10 | -0.008208809 | 2.75E-06 | 0.00759498 | 0.0001225 | POC5 |
| VAT | Obesity | ENSG00000152404.11 | 0.015522862 | 0.000898503 | 1 | 0.01370093 | CWF19L2 |
| VAT | Obesity | ENSG00000153246.7 | 0.008905953 | 0.002358135 | 1 | 0.027933277 | PLA2R1 |
| VAT | Obesity | ENSG00000153814.7 | 0.014463193 | 1.32E-06 | 0.00364871 | 6.18E-05 | JAZF1 |
| VAT | Obesity | ENSG00000154328.11 | 0.015737866 | 9.14E-05 | 0.252314893 | 0.002232875 | NEIL2 |
| VAT | Obesity | ENSG00000156103.11 | -0.011650417 | 0.000314157 | 0.867072481 | 0.005858598 | MMP16 |
| VAT | Obesity | ENSG00000156253.2 | -0.010235138 | 0.001951499 | 1 | 0.023938382 | RWDD2B |
| VAT | Obesity | ENSG00000157214.9 | -0.014480599 | 0.000390016 | 1 | 0.007035575 | STEAP2 |
| VAT | Obesity | ENSG00000157322.12 | -0.024588954 | 2.22E-18 | 6.14E-15 | 5.11E-16 | CLEC18A |
| VAT | Obesity | ENSG00000157557.7 | 0.009213634 | 0.001716884 | 1 | 0.022457821 | ETS2 |
| VAT | Obesity | ENSG00000158865.8 | -0.011617253 | 2.33E-07 | 0.000643665 | 1.31E-05 | SLC5A11 |
| VAT | Obesity | ENSG00000160294.6 | -0.00694216 | 0.001524038 | 1 | 0.020753408 | MCM3AP |

| | | | | | | | |
|---|---|---|---|---|---|---|---|
| VAT | Obesity | ENSG00000160360.7 | 0.017604068 | 0.003300264 | 1 | 0.035580971 | GPSM1 |
| VAT | Obesity | ENSG00000161036.6 | -0.012490064 | 0.002480624 | 1 | 0.029010686 | LRWD1 |
| VAT | Obesity | ENSG00000161204.7 | 0.010987081 | 0.001477941 | 1 | 0.020395589 | ABCF3 |
| VAT | Obesity | ENSG00000162994.11 | 0.00306279 | 0.003575483 | 1 | 0.037380051 | CLHC1 |
| VAT | Obesity | ENSG00000163297.12 | 0.021509946 | 1.30E-05 | 0.035924051 | 0.000466546 | ANTXR2 |
| VAT | Obesity | ENSG00000163728.6 | -0.014450928 | 0.003169532 | 1 | 0.034440579 | TTC14 |
| VAT | Obesity | ENSG00000163933.5 | -0.014200955 | 1.36E-07 | 0.000375284 | 7.88E-06 | RFT1 |
| VAT | Obesity | ENSG00000164068.11 | 0.019480678 | 3.06E-19 | 8.43E-16 | 8.43E-17 | RNF123 |
| VAT | Obesity | ENSG00000164078.8 | 0.054426081 | 2.18E-22 | 6.00E-19 | 8.58E-20 | MST1R |
| VAT | Obesity | ENSG00000164081.8 | -0.012301614 | 0.000291542 | 0.804656486 | 0.005549355 | TEX264 |
| VAT | Obesity | ENSG00000164144.10 | 0.017306391 | 0.003531265 | 1 | 0.037199588 | ARFIP1 |
| VAT | Obesity | ENSG00000164307.8 | -0.018613439 | 5.22E-06 | 0.014406061 | 0.000218274 | ERAP1 |
| VAT | Obesity | ENSG00000164574.11 | -0.025235181 | 1.52E-05 | 0.041869671 | 0.000523371 | GALNT10 |
| VAT | Obesity | ENSG00000164733.16 | -0.009357891 | 0.005172656 | 1 | 0.048735468 | CTSB |
| VAT | Obesity | ENSG00000164924.13 | -0.015663421 | 7.89E-05 | 0.217643783 | 0.001981293 | YWHAZ |
| VAT | Obesity | ENSG00000164967.5 | -0.019022999 | 0.000408121 | 1 | 0.007220609 | RPP25L |
| VAT | Obesity | ENSG00000165138.12 | 0.023991911 | 6.28E-05 | 0.173410635 | 0.00162066 | ANKS6 |
| VAT | Obesity | ENSG00000165917.5 | -0.040591581 | 4.58E-16 | 1.26E-12 | 7.90E-14 | RAPSN |
| VAT | Obesity | ENSG00000166529.10 | -0.00753569 | 0.001980414 | 1 | 0.024079037 | ZSCAN21 |
| VAT | Obesity | ENSG00000166667.8 | 0.009434995 | 0.004492196 | 1 | 0.044438931 | SPDYE6 |
| VAT | Obesity | ENSG00000167720.8 | 0.006216213 | 1.92E-05 | 0.052932954 | 0.000626206 | SRR |
| VAT | Obesity | ENSG00000168116.9 | -0.012692844 | 0.00106302 | 1 | 0.015746258 | KIAA1586 |
| VAT | Obesity | ENSG00000168268.6 | -0.0205891 | 3.50E-06 | 0.009673111 | 0.000153541 | NT5DC2 |
| VAT | Obesity | ENSG00000168405.10 | 0.010253168 | 0.001290601 | 1 | 0.017990194 | CMAHP |
| VAT | Obesity | ENSG00000168522.8 | 0.012501451 | 6.24E-05 | 0.172290581 | 0.00162066 | FNTA |
| VAT | Obesity | ENSG00000168653.6 | 0.014903049 | 0.00481777 | 1 | 0.046205883 | NDUFS5 |
| VAT | Obesity | ENSG00000168743.8 | 0.018508377 | 0.001310122 | 1 | 0.018170536 | NPNT |
| VAT | Obesity | ENSG00000169085.7 | 0.006408825 | 0.001604268 | 1 | 0.021287397 | C8orf46 |
| VAT | Obesity | ENSG00000169203.12 | 0.03701837 | 5.21E-14 | 1.44E-10 | 7.19E-12 | RP11-231C14.4 |
| VAT | Obesity | ENSG00000169592.10 | -0.019555013 | 1.68E-15 | 4.63E-12 | 2.72E-13 | INO80E |
| VAT | Obesity | ENSG00000170092.10 | 0.041701703 | 3.66E-19 | 1.01E-15 | 9.19E-17 | AC018720.10 |
| VAT | Obesity | ENSG00000170289.8 | 0.019824786 | 0.000683629 | 1 | 0.010986495 | CNGB3 |
| VAT | Obesity | ENSG00000170425.3 | -0.011051696 | 6.17E-07 | 0.001703607 | 3.04E-05 | ADORA2B |
| VAT | Obesity | ENSG00000171045.10 | 0.01441411 | 1.13E-05 | 0.031313061 | 0.000417507 | TSNARE1 |
| VAT | Obesity | ENSG00000171103.6 | 0.016348216 | 2.55E-05 | 0.070366984 | 0.000799625 | TRMT61B |
| VAT | Obesity | ENSG00000171700.9 | 0.008799469 | 0.002473166 | 1 | 0.029010686 | RGS19 |
| VAT | Obesity | ENSG00000171735.14 | 0.017679161 | 0.001774017 | 1 | 0.02256353 | CAMTA1 |
| VAT | Obesity | ENSG00000172247.3 | -0.019118297 | 2.27E-15 | 6.25E-12 | 3.47E-13 | C1QTNF4 |
| VAT | Obesity | ENSG00000172650.8 | -0.013083155 | 0.001526428 | 1 | 0.020753408 | AGAP5 |
| VAT | Obesity | ENSG00000172671.15 | -0.012765018 | 0.002583273 | 1 | 0.030083688 | ZFAND4 |
| VAT | Obesity | ENSG00000172728.11 | -0.004941714 | 0.000246046 | 0.67908709 | 0.004920921 | FUT10 |
| VAT | Obesity | ENSG00000172780.12 | 0.017862012 | 0.000161043 | 0.444477774 | 0.003514653 | RAB43 |
| VAT | Obesity | ENSG00000172935.8 | 0.020836133 | 3.32E-05 | 0.091525667 | 0.000984147 | MRGPRF |
| VAT | Obesity | ENSG00000173226.12 | -0.010749192 | 0.002321423 | 1 | 0.027736481 | IQCB1 |
| VAT | Obesity | ENSG00000173531.11 | -0.01511439 | 0.000208654 | 0.575885908 | 0.004290423 | MST1 |
| VAT | Obesity | ENSG00000173917.9 | 0.013230121 | 0.00061136 | 1 | 0.009984341 | HOXB2 |
| VAT | Obesity | ENSG00000174780.11 | 0.005328571 | 0.004639881 | 1 | 0.044966202 | SRP72 |
| VAT | Obesity | ENSG00000174943.5 | 0.017536839 | 2.85E-07 | 0.000785888 | 1.54E-05 | KCTD13 |
| VAT | Obesity | ENSG00000175115.7 | -0.015369721 | 0.000120833 | 0.333499937 | 0.00280252 | PACS1 |
| VAT | Obesity | ENSG00000175279.17 | 0.00686758 | 0.003530537 | 1 | 0.037199588 | APITD1 |
| VAT | Obesity | ENSG00000175938.6 | 0.028725649 | 1.04E-11 | 2.87E-08 | 1.06E-09 | ORAI3 |
| VAT | Obesity | ENSG00000176155.14 | -0.009931938 | 0.001125781 | 1 | 0.016353444 | CCDC57 |
| VAT | Obesity | ENSG00000176422.10 | -0.010975319 | 0.001757854 | 1 | 0.02256353 | SPRYD4 |
| VAT | Obesity | ENSG00000176623.7 | 0.007339807 | 0.002385276 | 1 | 0.028134026 | RMDN1 |
| VAT | Obesity | ENSG00000176783.10 | 0.013696833 | 1.36E-05 | 0.037411072 | 0.000479629 | RUFY1 |
| VAT | Obesity | ENSG00000177697.13 | 0.013533477 | 0.0029159 | 1 | 0.032690866 | CD151 |
| VAT | Obesity | ENSG00000178718.5 | -0.01375462 | 0.000125888 | 0.347451702 | 0.002895431 | RPP25 |
| VAT | Obesity | ENSG00000178750.2 | 0.021002931 | 7.90E-07 | 0.002181146 | 3.83E-05 | STX19 |
| VAT | Obesity | ENSG00000178802.13 | 0.016566589 | 7.90E-05 | 0.217942244 | 0.001981293 | MPI |
| VAT | Obesity | ENSG00000178809.7 | -0.04038699 | 1.58E-13 | 4.36E-10 | 1.98E-11 | TRIM73 |
| VAT | Obesity | ENSG00000178917.10 | -0.021790144 | 1.08E-05 | 0.029835669 | 0.00040653 | ZNF852 |
| VAT | Obesity | ENSG00000178952.4 | 0.021672909 | 3.26E-11 | 9.01E-08 | 3.00E-09 | TUFM |
| VAT | Obesity | ENSG00000179021.5 | 0.0153587 | 3.90E-08 | 0.000107762 | 2.63E-06 | C3orf38 |
| VAT | Obesity | ENSG00000181019.8 | -0.013665818 | 0.003714534 | 1 | 0.038541785 | NQO1 |
| VAT | Obesity | ENSG00000181315.6 | -0.012943813 | 0.000294877 | 0.813859235 | 0.005574378 | ZNF322 |
| VAT | Obesity | ENSG00000181625.13 | 0.009859835 | 0.000191108 | 0.527458497 | 0.004057373 | SLX1B |
| VAT | Obesity | ENSG00000182871.10 | 0.020748871 | 0.000209858 | 0.57920706 | 0.004290423 | COL18A1 |
| VAT | Obesity | ENSG00000183060.11 | 0.01255758 | 0.002882733 | 1 | 0.032474874 | LYSMD4 |
| VAT | Obesity | ENSG00000183250.7 | -0.012185771 | 0.00460266 | 1 | 0.044966202 | C21orf67 |
| VAT | Obesity | ENSG00000183281.10 | 0.01237416 | 0.001033511 | 1 | 0.015502666 | PLGLB1 |
| VAT | Obesity | ENSG00000183336.7 | -0.014480323 | 2.47E-12 | 6.82E-09 | 2.73E-10 | BOLA2 |
| VAT | Obesity | ENSG00000183506.12 | -0.008746361 | 0.001060625 | 1 | 0.015746258 | PI4KAP2 |
| VAT | Obesity | ENSG00000183527.7 | 0.016017532 | 0.000310513 | 0.85701644 | 0.005830044 | PSMG1 |
| VAT | Obesity | ENSG00000184227.3 | 0.014981728 | 0.000116174 | 0.32064074 | 0.002740519 | ACOT1 |
| VAT | Obesity | ENSG00000184923.8 | -0.014593362 | 1.04E-05 | 0.028754851 | 0.000403836 | NUTM2A |
| VAT | Obesity | ENSG00000185022.7 | -0.014628714 | 0.000255125 | 0.704144699 | 0.004995804 | MAFF |
| VAT | Obesity | ENSG00000185245.6 | 0.028348483 | 4.72E-07 | 0.001303903 | 2.41E-05 | GP1BA |
| VAT | Obesity | ENSG00000185344.9 | -0.023173714 | 5.96E-05 | 0.164384688 | 0.001565568 | ATP6V0A2 |
| VAT | Obesity | ENSG00000186470.9 | -0.004828131 | 2.76E-07 | 0.000760503 | 1.52E-05 | BTN3A2 |
| VAT | Obesity | ENSG00000186715.6 | 0.015239998 | 0.000738525 | 1 | 0.011600152 | MST1L |
| VAT | Obesity | ENSG00000188312.9 | 0.009515507 | 1.03E-05 | 0.02850231 | 0.000403836 | CENPP |

| | | | | | | | |
|---|---|---|---|---|---|---|---|
| VAT | Obesity | ENSG00000188878.12 | 0.015175032 | 0.000505567 | 1 | 0.008563512 | FBF1 |
| VAT | Obesity | ENSG00000189298.9 | -0.015262222 | 1.03E-05 | 0.02831071 | 0.000403836 | ZKSCAN3 |
| VAT | Obesity | ENSG00000196458.6 | 0.013017935 | 0.000207675 | 0.57318292 | 0.004290423 | ZNF605 |
| VAT | Obesity | ENSG00000197062.7 | -0.007990855 | 4.26E-05 | 0.117648929 | 0.001170502 | RP5-874C20.3 |
| VAT | Obesity | ENSG00000197165.6 | 0.062849037 | 7.74E-29 | 2.14E-25 | 1.07E-25 | SULT1A2 |
| VAT | Obesity | ENSG00000197345.8 | -0.005043928 | 0.000289787 | 0.799810892 | 0.005549355 | MRPL21 |
| VAT | Obesity | ENSG00000197653.10 | 0.038775569 | 8.50E-13 | 2.35E-09 | 9.77E-11 | DNAH10 |
| VAT | Obesity | ENSG00000197728.5 | -0.007396076 | 0.000346933 | 0.957535616 | 0.006341295 | RPS26 |
| VAT | Obesity | ENSG00000197782.10 | -0.004433373 | 4.27E-05 | 0.117906757 | 0.001170502 | ZNF780A |
| VAT | Obesity | ENSG00000198099.4 | -0.012934222 | 0.004378986 | 1 | 0.0439491 | ADH4 |
| VAT | Obesity | ENSG00000198792.8 | -0.014408282 | 0.00022285 | 0.61506588 | 0.004522543 | TMEM184B |
| VAT | Obesity | ENSG00000198909.3 | -0.026013503 | 1.20E-07 | 0.000332085 | 7.22E-06 | MAP3K3 |
| VAT | Obesity | ENSG00000204351.7 | -0.006943576 | 1.75E-05 | 0.048301174 | 0.000589039 | SKIV2L |
| VAT | Obesity | ENSG00000204438.6 | -0.019733244 | 1.37E-07 | 0.00037817 | 7.88E-06 | GPANK1 |
| VAT | Obesity | ENSG00000204618.4 | -0.018908854 | 0.001756312 | 1 | 0.02256353 | RNF39 |
| VAT | Obesity | ENSG00000204644.5 | 0.004078199 | 3.77E-05 | 0.103922864 | 0.001105562 | ZFP57 |
| VAT | Obesity | ENSG00000204709.4 | 0.011580548 | 2.70E-05 | 0.074453131 | 0.000827257 | C6orf100 |
| VAT | Obesity | ENSG00000204713.6 | -0.010341312 | 0.001891382 | 1 | 0.023620883 | TRIM27 |
| VAT | Obesity | ENSG00000205583.9 | -0.018842512 | 3.15E-20 | 8.68E-17 | 9.65E-18 | STAG3L1 |
| VAT | Obesity | ENSG00000205710.3 | 0.011234371 | 0.0002867 | 0.791292807 | 0.005533516 | C17orf107 |
| VAT | Obesity | ENSG00000212694.4 | -0.011770043 | 8.19E-05 | 0.226036704 | 0.002018185 | AC084018.1 |
| VAT | Obesity | ENSG00000213199.3 | 0.014590844 | 0.003387617 | 1 | 0.036161216 | ASIC3 |
| VAT | Obesity | ENSG00000213402.2 | 0.007284576 | 0.00155021 | 1 | 0.020868421 | PTPRCAP |
| VAT | Obesity | ENSG00000213639.5 | -0.015846028 | 1.54E-05 | 0.042523325 | 0.000524979 | PPP1CB |
| VAT | Obesity | ENSG00000213689.5 | 0.005209813 | 0.00301453 | 1 | 0.033147822 | TREX1 |
| VAT | Obesity | ENSG00000214063.6 | 0.014773845 | 0.004032406 | 1 | 0.041068046 | TSPAN4 |
| VAT | Obesity | ENSG00000214659.4 | 0.014177506 | 0.001072571 | 1 | 0.015746258 | KRT8P26 |
| VAT | Obesity | ENSG00000216901.1 | 0.004908492 | 0.00023558 | 0.650200703 | 0.004745991 | AL022393.7 |
| VAT | Obesity | ENSG00000223496.1 | -0.039467672 | 7.61E-15 | 2.10E-11 | 1.11E-12 | EXOSC6 |
| VAT | Obesity | ENSG00000223861.1 | -0.022070677 | 0.000141769 | 0.39128379 | 0.003155514 | RP11-74C1.2 |
| VAT | Obesity | ENSG00000224843.2 | 0.006041627 | 0.003900377 | 1 | 0.040018734 | LINC00240 |
| VAT | Obesity | ENSG00000224997.1 | -0.028787917 | 4.70E-08 | 0.000129621 | 3.09E-06 | AL049840.1 |
| VAT | Obesity | ENSG00000225498.1 | -0.019430182 | 0.001593722 | 1 | 0.021249624 | AC002064.5 |
| VAT | Obesity | ENSG00000226232.4 | -0.021343988 | 5.63E-09 | 1.55E-05 | 3.98E-07 | RP11-419C5.2 |
| VAT | Obesity | ENSG00000228022.1 | -0.013910012 | 0.003132945 | 1 | 0.03417758 | HCG20 |
| VAT | Obesity | ENSG00000228078.1 | 0.005851993 | 0.002134658 | 1 | 0.025615891 | HLA-U |
| VAT | Obesity | ENSG00000230772.1 | -0.013784895 | 0.001541298 | 1 | 0.020852861 | VN1R108P |
| VAT | Obesity | ENSG00000231043.2 | 0.01697534 | 0.001976643 | 1 | 0.024079037 | AC007238.1 |
| VAT | Obesity | ENSG00000231925.7 | 0.013618858 | 0.000396691 | 1 | 0.007109524 | TAPBP |
| VAT | Obesity | ENSG00000232075.1 | 0.018270716 | 0.000488716 | 1 | 0.008430344 | MRPL35P2 |
| VAT | Obesity | ENSG00000232112.3 | 0.011467787 | 0.004643249 | 1 | 0.044966202 | TMA7 |
| VAT | Obesity | ENSG00000232561.2 | -0.01750575 | 3.88E-08 | 0.000107003 | 2.63E-06 | GTF2IP1 |
| VAT | Obesity | ENSG00000233569.1 | -0.025122472 | 4.28E-05 | 0.118220734 | 0.001170502 | RP11-500B12.1 |
| VAT | Obesity | ENSG00000233609.3 | 0.016265015 | 0.000548708 | 1 | 0.009123097 | RP11-62H7.2 |
| VAT | Obesity | ENSG00000233633.1 | 0.047956137 | 2.62E-16 | 7.24E-13 | 4.83E-14 | AC093326.3 |
| VAT | Obesity | ENSG00000235109.3 | 0.006747713 | 1.93E-05 | 0.053308014 | 0.000626206 | ZSCAN31 |
| VAT | Obesity | ENSG00000235374.1 | -0.012762803 | 0.000617721 | 1 | 0.01002889 | SSR4P1 |
| VAT | Obesity | ENSG00000237854.3 | 0.016517246 | 3.25E-05 | 0.089627377 | 0.000974211 | LINC00674 |
| VAT | Obesity | ENSG00000240065.3 | 0.005532217 | 0.00190308 | 1 | 0.023659912 | PSMB9 |
| VAT | Obesity | ENSG00000240682.5 | -0.012017661 | 0.000400438 | 1 | 0.007130384 | ISY1 |
| VAT | Obesity | ENSG00000242193.5 | -0.016776811 | 0.00010237 | 0.282541352 | 0.002435701 | RP11-568K15.1 |
| VAT | Obesity | ENSG00000244219.2 | 0.020286129 | 1.95E-05 | 0.053853684 | 0.000626206 | GS1-259H13.2 |
| VAT | Obesity | ENSG00000245958.2 | -0.009238838 | 0.001884532 | 1 | 0.023620883 | RP11-33B1.1 |
| VAT | Obesity | ENSG00000251417.1 | -0.035332435 | 3.46E-32 | 9.54E-29 | 9.54E-29 | RP11-1348G14.4 |
| VAT | Obesity | ENSG00000253704.1 | -0.022984484 | 5.38E-09 | 1.49E-05 | 3.91E-07 | RP11-267M23.4 |
| VAT | Obesity | ENSG00000253813.1 | 0.008117002 | 0.002752625 | 1 | 0.031393572 | COX6B1P4 |
| VAT | Obesity | ENSG00000253981.1 | 0.037653507 | 1.85E-13 | 5.11E-10 | 2.22E-11 | ALG1L13P |
| VAT | Obesity | ENSG00000254634.3 | -0.021242474 | 3.14E-10 | 8.66E-07 | 2.47E-08 | RP11-231C14.3 |
| VAT | Obesity | ENSG00000254986.3 | 0.006169819 | 0.004452802 | 1 | 0.044318348 | DPP3 |
| VAT | Obesity | ENSG00000254999.2 | -0.009747345 | 0.00010041 | 0.277131732 | 0.002428776 | BRK1 |
| VAT | Obesity | ENSG00000257595.2 | 0.014405593 | 0.001282352 | 1 | 0.017965945 | RP3-473L9.4 |
| VAT | Obesity | ENSG00000258512.1 | 0.012873274 | 0.004879288 | 1 | 0.046598047 | RP11-796G6.2 |
| VAT | Obesity | ENSG00000259982.1 | 0.027019652 | 1.35E-24 | 3.72E-21 | 9.29E-22 | CDC37P1 |
| VAT | Obesity | ENSG00000260911.1 | 0.012569447 | 3.77E-10 | 1.04E-06 | 2.89E-08 | RP11-196G11.2 |
| VAT | Obesity | ENSG00000260947.1 | -0.005234709 | 1.16E-05 | 0.031990896 | 0.000420933 | RP11-384P7.7 |
| VAT | Obesity | ENSG00000261556.4 | -0.017054478 | 7.31E-14 | 2.02E-10 | 9.61E-12 | RP11-296I10.6 |
| VAT | Obesity | ENSG00000261706.1 | 0.009476289 | 0.003780966 | 1 | 0.03908414 | AP001505.9 |
| VAT | Obesity | ENSG00000265055.1 | 0.0196158 | 1.09E-05 | 0.030083252 | 0.00040653 | AC145343.2 |
| VAT | Obesity | ENSG00000266173.1 | -0.012274283 | 0.003573702 | 1 | 0.037380051 | STRADA |
| VAT | Obesity | ENSG00000267731.1 | 0.008832746 | 0.002764996 | 1 | 0.031404892 | RP11-147L13.8 |
| VAT | Obesity | ENSG00000268171.1 | 0.014386346 | 0.002620104 | 1 | 0.030384397 | AC068620.1 |
| VAT | Obesity | ENSG00000269963.1 | -0.027372986 | 1.05E-05 | 0.029076197 | 0.000403836 | RP11-73M18.9 |
| VAT | Obesity | ENSG00000270071.1 | 0.008374599 | 0.000897283 | 1 | 0.01370093 | AP001172.2 |
| VAT | Obesity | ENSG00000271751.1 | -0.019079697 | 0.000195191 | 0.538727649 | 0.004112425 | RP11-110I1.14 |
| VAT | Obesity | ENSG00000272047.1 | 0.006412849 | 0.003997369 | 1 | 0.040861995 | GTF2H5 |
| VAT | Obesity | ENSG00000272275.1 | -0.008568137 | 0.00462921 | 1 | 0.044966202 | RP11-791G15.2 |
| VAT | Obesity | ENSG00000272338.1 | -0.020686037 | 0.000208345 | 0.575030975 | 0.004290423 | RP11-722E23.2 |
| VAT | Obesity | ENSG00000272462.2 | 0.00669793 | 1.25E-10 | 3.44E-07 | 1.01E-08 | U91328.19 |
| VAT | Obesity | ENSG00000272505.1 | 0.018175902 | 0.002725409 | 1 | 0.031342199 | RP11-981G7.6 |
| VAT | Obesity | ENSG00000272849.1 | -0.01652107 | 8.09E-05 | 0.223329467 | 0.002011977 | RP11-347I19.8 |
| VAT | Obesity | ENSG00000272983.1 | 0.010873305 | 0.001557571 | 1 | 0.020868421 | RP11-508N22.12 |

| | | | | | | | |
|---|---|---|---|---|---|---|---|
| VAT | HTN | ENSG00000003056.3 | -0.014919899 | 0.000411253 | 1 | 0.00822505 | M6PR |
| VAT | HTN | ENSG00000004534.10 | -0.005959478 | 3.98E-06 | 0.010983982 | 0.000183066 | RBM6 |
| VAT | HTN | ENSG00000007341.14 | -0.006490431 | 0.003495248 | 1 | 0.038281288 | ST7L |
| VAT | HTN | ENSG00000009950.11 | -0.019846191 | 0.00263143 | 1 | 0.030644506 | MLXIPL |
| VAT | HTN | ENSG00000011021.17 | -0.015481276 | 0.000843934 | 1 | 0.013542191 | CLCN6 |
| VAT | HTN | ENSG00000011523.9 | 0.026018476 | 5.44E-09 | 1.50E-05 | 6.52E-07 | CEP68 |
| VAT | HTN | ENSG00000026297.11 | -0.015855893 | 0.000614532 | 1 | 0.010600675 | RNASET2 |
| VAT | HTN | ENSG00000035115.17 | -0.018036684 | 0.004561216 | 1 | 0.046861116 | SH3YL1 |
| VAT | HTN | ENSG00000053900.6 | 0.030408377 | 2.69E-06 | 0.007423149 | 0.000125816 | ANAPC4 |
| VAT | HTN | ENSG00000054282.11 | 0.016840477 | 1.19E-09 | 3.28E-06 | 1.93E-07 | SDCCAG8 |
| VAT | HTN | ENSG00000054523.12 | 0.013449302 | 0.001486974 | 1 | 0.020939026 | KIF1B |
| VAT | HTN | ENSG00000055955.11 | 0.006195101 | 0.00058058 | 1 | 0.010205917 | ITIH4 |
| VAT | HTN | ENSG00000060642.6 | -0.013759292 | 7.55E-06 | 0.020832632 | 0.000306731 | PIGV |
| VAT | HTN | ENSG00000063854.8 | 0.012754888 | 0.004448854 | 1 | 0.046161039 | HAGH |
| VAT | HTN | ENSG00000064703.7 | 0.015665492 | 0.003231204 | 1 | 0.036252527 | DDX20 |
| VAT | HTN | ENSG00000065060.12 | 0.015011965 | 6.75E-07 | 0.001862089 | 3.88E-05 | UHRF1BP1 |
| VAT | HTN | ENSG00000068654.11 | -0.010460665 | 0.000140691 | 0.388308383 | 0.003463845 | POLR1A |
| VAT | HTN | ENSG00000085788.9 | 0.023991964 | 0.00037489 | 1 | 0.007608054 | DDHD2 |
| VAT | HTN | ENSG00000086300.11 | -0.025971325 | 4.88E-07 | 0.001346797 | 3.28E-05 | SNX10 |
| VAT | HTN | ENSG00000087884.10 | -0.012506741 | 1.63E-05 | 0.044983217 | 0.000576708 | AAMDC |
| VAT | HTN | ENSG00000090238.7 | 0.011233713 | 5.95E-05 | 0.164102622 | 0.001676064 | YPEL3 |
| VAT | HTN | ENSG00000090432.5 | -0.011921128 | 0.001592175 | 1 | 0.022193958 | MUL1 |
| VAT | HTN | ENSG00000090621.9 | -0.025704807 | 6.19E-07 | 0.001709597 | 3.64E-05 | PABPC4 |
| VAT | HTN | ENSG00000096433.6 | 0.021988685 | 0.000640344 | 1 | 0.01096636 | ITPR3 |
| VAT | HTN | ENSG00000099194.5 | -0.032533335 | 9.47E-07 | 0.00261343 | 5.03E-05 | SCD |
| VAT | HTN | ENSG00000099785.6 | 0.018580431 | 2.45E-06 | 0.006761129 | 0.000118124 | MARCH2 |
| VAT | HTN | ENSG00000099810.14 | -0.021152367 | 0.00125882 | 1 | 0.018581684 | MTAP |
| VAT | HTN | ENSG00000100296.9 | -0.009097575 | 0.003172839 | 1 | 0.03594872 | THOC5 |
| VAT | HTN | ENSG00000100731.11 | 0.014266314 | 0.001945963 | 1 | 0.025575518 | PCNX |
| VAT | HTN | ENSG00000101197.8 | -0.008692573 | 0.000743136 | 1 | 0.012305555 | BIRC7 |
| VAT | HTN | ENSG00000101363.8 | 0.011510974 | 0.001292639 | 1 | 0.018581684 | MANBAL |
| VAT | HTN | ENSG00000101457.8 | -0.007879223 | 0.000549212 | 1 | 0.010098967 | DNTTIP1 |
| VAT | HTN | ENSG00000101751.6 | -0.007145307 | 0.000114777 | 0.316783726 | 0.002960596 | POLI |
| VAT | HTN | ENSG00000102858.8 | 0.028526491 | 1.05E-05 | 0.029020646 | 0.000397543 | MGRN1 |
| VAT | HTN | ENSG00000103248.13 | 0.007122091 | 0.000237708 | 0.656073133 | 0.005248585 | MTHFSD |
| VAT | HTN | ENSG00000105376.4 | -0.018148762 | 0.00228798 | 1 | 0.028307557 | ICAM5 |
| VAT | HTN | ENSG00000105404.6 | 0.019912277 | 0.000401054 | 1 | 0.008079637 | RABAC1 |
| VAT | HTN | ENSG00000106125.14 | -0.015579308 | 0.000369529 | 1 | 0.007554817 | FAM188B |
| VAT | HTN | ENSG00000106133.13 | -0.007756566 | 0.00315018 | 1 | 0.035927668 | NSUN5P2 |
| VAT | HTN | ENSG00000106290.10 | -0.011183109 | 0.000564757 | 1 | 0.010098967 | TAF6 |
| VAT | HTN | ENSG00000108384.10 | -0.009884795 | 0.002297425 | 1 | 0.028307557 | RAD51C |
| VAT | HTN | ENSG00000108823.11 | 0.009940583 | 0.00474155 | 1 | 0.048112786 | SGCA |
| VAT | HTN | ENSG00000110851.7 | 0.01889672 | 0.002177712 | 1 | 0.027474564 | PRDM4 |
| VAT | HTN | ENSG00000111801.11 | -0.010838049 | 0.002357226 | 1 | 0.02884247 | BTN3A3 |
| VAT | HTN | ENSG00000112981.3 | 0.014165448 | 0.003695866 | 1 | 0.040002317 | NME5 |
| VAT | HTN | ENSG00000113269.9 | 0.021451891 | 0.000556264 | 1 | 0.010098967 | RNF130 |
| VAT | HTN | ENSG00000113971.14 | 0.004635239 | 0.003954922 | 1 | 0.042308471 | NPHP3 |
| VAT | HTN | ENSG00000114054.9 | 0.027607456 | 3.29E-08 | 9.08E-05 | 3.24E-06 | PCCB |
| VAT | HTN | ENSG00000114904.8 | -0.015327133 | 0.003199378 | 1 | 0.036041975 | NEK4 |
| VAT | HTN | ENSG00000116151.9 | -0.014851418 | 0.003178075 | 1 | 0.03594872 | MORN1 |
| VAT | HTN | ENSG00000117595.6 | -0.010551096 | 0.004434898 | 1 | 0.046161039 | IRF6 |
| VAT | HTN | ENSG00000119673.10 | 0.007104652 | 0.001268536 | 1 | 0.018581684 | ACOT2 |
| VAT | HTN | ENSG00000120784.11 | 0.013377832 | 4.84E-05 | 0.133575816 | 0.001451911 | ZFP30 |
| VAT | HTN | ENSG00000121068.9 | -0.03001438 | 3.52E-07 | 0.000972808 | 2.55E-05 | TBX2 |
| VAT | HTN | ENSG00000121101.11 | 0.03300021 | 6.10E-08 | 0.000168359 | 5.71E-06 | TEX14 |
| VAT | HTN | ENSG00000123374.6 | -0.019071512 | 0.00367685 | 1 | 0.039953174 | CDK2 |
| VAT | HTN | ENSG00000124116.14 | -0.006972357 | 0.000329 | 0.908040755 | 0.006869451 | WFDC3 |
| VAT | HTN | ENSG00000124508.12 | 0.012935719 | 7.56E-06 | 0.020857722 | 0.000306731 | BTN2A2 |
| VAT | HTN | ENSG00000124562.5 | -0.017368984 | 1.88E-05 | 0.051980452 | 0.000641734 | SNRPC |
| VAT | HTN | ENSG00000124613.4 | -0.007007258 | 2.71E-05 | 0.074933901 | 0.000902818 | ZNF391 |
| VAT | HTN | ENSG00000125388.15 | 0.004801917 | 0.001606016 | 1 | 0.02227439 | GRK4 |
| VAT | HTN | ENSG00000125734.9 | 0.011865387 | 0.002751575 | 1 | 0.031775514 | GPR108 |
| VAT | HTN | ENSG00000126088.8 | -0.017037331 | 0.0024555 | 1 | 0.02959467 | UROD |
| VAT | HTN | ENSG00000126602.6 | 0.007726321 | 7.10E-07 | 0.001958312 | 3.92E-05 | TRAP1 |
| VAT | HTN | ENSG00000127220.5 | -0.018191378 | 6.72E-06 | 0.018544774 | 0.000280981 | ABHD8 |
| VAT | HTN | ENSG00000127957.12 | -0.031705797 | 3.37E-10 | 9.29E-07 | 9.29E-08 | PMS2P3 |
| VAT | HTN | ENSG00000128829.7 | -0.033522651 | 7.06E-07 | 0.001948855 | 3.92E-05 | EIF2AK4 |
| VAT | HTN | ENSG00000130177.10 | 0.009688663 | 0.000213751 | 0.589951873 | 0.004835671 | CDC16 |
| VAT | HTN | ENSG00000130270.12 | -0.011239855 | 0.000524744 | 1 | 0.009785759 | ATP8B3 |
| VAT | HTN | ENSG00000130517.9 | 0.010688188 | 0.000538294 | 1 | 0.009971089 | PGPEP1 |
| VAT | HTN | ENSG00000130529.11 | -0.023118435 | 0.000149318 | 0.412116568 | 0.003552729 | TRPM4 |
| VAT | HTN | ENSG00000131351.10 | -0.030883699 | 9.33E-09 | 2.58E-05 | 1.03E-06 | HAUS8 |
| VAT | HTN | ENSG00000131558.10 | 0.024846442 | 1.17E-05 | 0.032209103 | 0.000435258 | EXOC4 |
| VAT | HTN | ENSG00000132781.13 | 0.01526582 | 0.003494113 | 1 | 0.038281288 | MUTYH |
| VAT | HTN | ENSG00000133731.5 | 0.007317355 | 0.000889237 | 1 | 0.014105132 | IMPA1 |
| VAT | HTN | ENSG00000134490.9 | -0.014812412 | 0.003532448 | 1 | 0.038535799 | TMEM241 |
| VAT | HTN | ENSG00000135213.8 | -0.015189366 | 0.001873194 | 1 | 0.02509716 | POM121C |
| VAT | HTN | ENSG00000135744.7 | 0.014794462 | 0.000428479 | 1 | 0.008447166 | AGT |
| VAT | HTN | ENSG00000135968.15 | 0.015347173 | 0.002027909 | 1 | 0.026401085 | GCC2 |
| VAT | HTN | ENSG00000136448.7 | 0.012507012 | 1.03E-07 | 0.000282914 | 8.57E-06 | NMT1 |
| VAT | HTN | ENSG00000137700.12 | 0.021209868 | 4.29E-05 | 0.118512663 | 0.001331603 | SLC37A4 |

| | | | | | | | |
|---|---|---|---|---|---|---|---|
| VAT | HTN | ENSG00000137944.12 | -0.013915527 | 5.26E-05 | 0.145214526 | 0.001530313 | CCBL2 |
| VAT | HTN | ENSG00000138036.14 | 0.01678463 | 0.00093551 | 1 | 0.014754326 | DYNC2LI1 |
| VAT | HTN | ENSG00000138111.10 | -0.004453934 | 0.004155056 | 1 | 0.04410752 | TMEM180 |
| VAT | HTN | ENSG00000138175.8 | 0.009614262 | 0.002361738 | 1 | 0.02884247 | ARL3 |
| VAT | HTN | ENSG00000138600.5 | -0.019039935 | 5.27E-05 | 0.145379761 | 0.001530313 | SPPL2A |
| VAT | HTN | ENSG00000138757.10 | 0.011789096 | 0.000869199 | 1 | 0.013866987 | G3BP2 |
| VAT | HTN | ENSG00000138829.6 | 0.009324679 | 4.81E-05 | 0.132857361 | 0.001451911 | FBN2 |
| VAT | HTN | ENSG00000139970.12 | 0.022602858 | 0.000298124 | 0.822821044 | 0.006329393 | RTN1 |
| VAT | HTN | ENSG00000140995.12 | 0.025940009 | 2.48E-06 | 0.006851187 | 0.000118124 | DEF8 |
| VAT | HTN | ENSG00000141002.14 | 0.020059327 | 0.000514019 | 1 | 0.009771271 | TCF25 |
| VAT | HTN | ENSG00000141076.13 | 0.01350772 | 0.000277123 | 0.764859141 | 0.005975462 | CIRH1A |
| VAT | HTN | ENSG00000141140.12 | 0.021897048 | 0.000194458 | 0.536703269 | 0.004472527 | MYO19 |
| VAT | HTN | ENSG00000141252.15 | -0.010920708 | 0.000567152 | 1 | 0.010098967 | VPS53 |
| VAT | HTN | ENSG00000141295.9 | -0.009612055 | 0.000585838 | 1 | 0.010205917 | SCRN2 |
| VAT | HTN | ENSG00000141452.5 | -0.02660741 | 2.38E-07 | 0.000658012 | 1.88E-05 | C18orf8 |
| VAT | HTN | ENSG00000143156.9 | -0.020820691 | 0.000452738 | 1 | 0.008738169 | NME7 |
| VAT | HTN | ENSG00000143376.8 | 0.023751049 | 5.24E-05 | 0.144656132 | 0.001530313 | SNX27 |
| VAT | HTN | ENSG00000143437.16 | -0.019035908 | 0.000666141 | 1 | 0.011210665 | ARNT |
| VAT | HTN | ENSG00000143578.11 | -0.033300218 | 5.73E-07 | 0.001580135 | 3.51E-05 | CREB3L4 |
| VAT | HTN | ENSG00000144026.7 | -0.010839827 | 0.004002226 | 1 | 0.042649204 | ZNF514 |
| VAT | HTN | ENSG00000144134.14 | -0.012308273 | 0.003073313 | 1 | 0.03519645 | RABL2A |
| VAT | HTN | ENSG00000145029.7 | -0.010775132 | 0.000414484 | 1 | 0.008230051 | NICN1 |
| VAT | HTN | ENSG00000146109.3 | 0.010266399 | 0.002180047 | 1 | 0.027474564 | ABT1 |
| VAT | HTN | ENSG00000146243.9 | 0.040514616 | 3.20E-09 | 8.82E-06 | 4.45E-07 | IRAK1BP1 |
| VAT | HTN | ENSG00000146409.6 | -0.016244236 | 0.000517647 | 1 | 0.009771271 | SLC18B1 |
| VAT | HTN | ENSG00000146556.10 | -0.019096743 | 5.94E-06 | 0.016393759 | 0.000252212 | WASH2P |
| VAT | HTN | ENSG00000147576.11 | -0.01213279 | 0.001274589 | 1 | 0.018581684 | ADHFE1 |
| VAT | HTN | ENSG00000149084.7 | -0.007132402 | 8.71E-10 | 2.40E-06 | 1.60E-07 | HSD17B12 |
| VAT | HTN | ENSG00000149115.9 | 0.022034126 | 8.91E-11 | 2.46E-07 | 3.66E-08 | TNKS1BP1 |
| VAT | HTN | ENSG00000149292.12 | 0.010890964 | 0.001282493 | 1 | 0.018581684 | TTC12 |
| VAT | HTN | ENSG00000149743.9 | -0.014109572 | 0.001800158 | 1 | 0.024236277 | TRPT1 |
| VAT | HTN | ENSG00000149922.6 | 0.007556028 | 0.001046632 | 1 | 0.016048355 | TBX6 |
| VAT | HTN | ENSG00000152404.11 | 0.026630832 | 6.13E-07 | 0.001691057 | 3.64E-05 | CWF19L2 |
| VAT | HTN | ENSG00000153113.19 | -0.00999079 | 0.000744575 | 1 | 0.012305555 | CAST |
| VAT | HTN | ENSG00000154328.11 | 0.026998523 | 4.21E-09 | 1.16E-05 | 5.29E-07 | NEIL2 |
| VAT | HTN | ENSG00000156103.11 | -0.010395134 | 0.004885205 | 1 | 0.049388889 | MMP16 |
| VAT | HTN | ENSG00000156219.12 | -0.015385284 | 0.002466677 | 1 | 0.029600124 | ART3 |
| VAT | HTN | ENSG00000156398.8 | 0.013865929 | 2.46E-08 | 6.79E-05 | 2.52E-06 | SFXN2 |
| VAT | HTN | ENSG00000156603.10 | 0.012466744 | 0.00209083 | 1 | 0.026978653 | MED19 |
| VAT | HTN | ENSG00000156931.11 | 0.025349329 | 0.000350872 | 0.968405953 | 0.00722691 | VPS8 |
| VAT | HTN | ENSG00000157322.12 | -0.015574217 | 1.25E-06 | 0.003441552 | 6.37E-05 | CLEC18A |
| VAT | HTN | ENSG00000158865.8 | -0.012855734 | 5.47E-07 | 0.001510664 | 3.51E-05 | SLC5A11 |
| VAT | HTN | ENSG00000160307.5 | -0.007833076 | 3.54E-05 | 0.097632917 | 0.001135266 | S100B |
| VAT | HTN | ENSG00000160321.10 | -0.026928126 | 5.98E-05 | 0.165182137 | 0.001676064 | ZNF208 |
| VAT | HTN | ENSG00000161204.7 | 0.021370919 | 6.21E-08 | 0.000171308 | 5.71E-06 | ABCF3 |
| VAT | HTN | ENSG00000162910.14 | 0.036422041 | 2.02E-12 | 5.58E-09 | 1.40E-09 | MRPL55 |
| VAT | HTN | ENSG00000163155.7 | -0.01359099 | 3.45E-05 | 0.095173587 | 0.001119689 | LYSMD1 |
| VAT | HTN | ENSG00000163933.5 | -0.01179084 | 0.000127485 | 0.351859582 | 0.00322807 | RFT1 |
| VAT | HTN | ENSG00000164048.9 | -0.018465782 | 8.69E-10 | 2.40E-06 | 1.60E-07 | ZNF589 |
| VAT | HTN | ENSG00000164068.11 | 0.015964479 | 1.25E-10 | 3.45E-07 | 4.32E-08 | RNF123 |
| VAT | HTN | ENSG00000164078.8 | 0.026040659 | 4.57E-05 | 0.126141889 | 0.001401577 | MST1R |
| VAT | HTN | ENSG00000164307.8 | -0.028869176 | 6.11E-10 | 1.69E-06 | 1.30E-07 | ERAP1 |
| VAT | HTN | ENSG00000164308.12 | 0.002988645 | 0.001575253 | 1 | 0.022069535 | ERAP2 |
| VAT | HTN | ENSG00000164967.5 | -0.021529835 | 0.000461586 | 1 | 0.008847074 | RPP25L |
| VAT | HTN | ENSG00000166037.6 | -0.017208976 | 0.000449318 | 1 | 0.008733218 | CEP57 |
| VAT | HTN | ENSG00000166275.11 | -0.004383795 | 0.003431034 | 1 | 0.037878616 | C10orf32 |
| VAT | HTN | ENSG00000166394.10 | 0.01541718 | 0.002154223 | 1 | 0.027474564 | CYB5R2 |
| VAT | HTN | ENSG00000166435.11 | -0.011762891 | 0.002446103 | 1 | 0.02959467 | XRRA1 |
| VAT | HTN | ENSG00000166436.11 | -0.007144876 | 2.21E-05 | 0.061025527 | 0.000744214 | TRIM66 |
| VAT | HTN | ENSG00000166532.11 | -0.018036485 | 9.29E-11 | 2.57E-07 | 3.66E-08 | RIMKLB |
| VAT | HTN | ENSG00000166924.4 | 0.012405556 | 0.00419028 | 1 | 0.044311011 | NYAP1 |
| VAT | HTN | ENSG00000166997.3 | 0.017815928 | 0.002615291 | 1 | 0.030644506 | CNPY4 |
| VAT | HTN | ENSG00000167720.8 | 0.006317744 | 0.000143279 | 0.395449219 | 0.003468853 | SRR |
| VAT | HTN | ENSG00000168268.6 | -0.015564998 | 0.002142838 | 1 | 0.027474564 | NT5DC2 |
| VAT | HTN | ENSG00000169062.10 | 0.013600007 | 0.001290836 | 1 | 0.018581684 | UPF3A |
| VAT | HTN | ENSG00000169085.7 | 0.007202816 | 0.001907316 | 1 | 0.025341822 | C8orf46 |
| VAT | HTN | ENSG00000169169.10 | 0.033613689 | 1.60E-07 | 0.000442463 | 1.30E-05 | CPT1C |
| VAT | HTN | ENSG00000169203.12 | 0.016495278 | 0.003328411 | 1 | 0.037041994 | RP11-231C14.4 |
| VAT | HTN | ENSG00000169592.10 | -0.017507831 | 4.35E-10 | 1.20E-06 | 1.00E-07 | INO80E |
| VAT | HTN | ENSG00000169629.7 | -0.004698666 | 0.000775424 | 1 | 0.012663725 | RGPD8 |
| VAT | HTN | ENSG00000170092.10 | 0.023544548 | 9.80E-06 | 0.027037163 | 0.000386245 | AC018720.10 |
| VAT | HTN | ENSG00000170464.5 | -0.023156457 | 6.81E-05 | 0.187992018 | 0.001843059 | DNAJC18 |
| VAT | HTN | ENSG00000170915.8 | -0.021779112 | 0.000831422 | 1 | 0.013419446 | PAQR8 |
| VAT | HTN | ENSG00000171421.8 | 0.007907723 | 0.001671174 | 1 | 0.022833865 | MRPL36 |
| VAT | HTN | ENSG00000171700.9 | -0.019110622 | 8.70E-09 | 2.40E-05 | 1.00E-06 | RGS19 |
| VAT | HTN | ENSG00000172113.4 | 0.034405149 | 4.16E-10 | 1.15E-06 | 1.00E-07 | NME6 |
| VAT | HTN | ENSG00000172687.9 | 0.006137524 | 0.00199575 | 1 | 0.026105551 | ZNF738 |
| VAT | HTN | ENSG00000172746.6 | -0.018830743 | 0.002585544 | 1 | 0.030627042 | RP11-344H11.4 |
| VAT | HTN | ENSG00000173531.11 | -0.021166612 | 5.48E-06 | 0.015114334 | 0.000236161 | MST1 |
| VAT | HTN | ENSG00000174177.8 | -0.013211707 | 0.002406173 | 1 | 0.029255675 | CTU2 |
| VAT | HTN | ENSG00000174226.4 | -0.01448089 | 5.12E-06 | 0.014132487 | 0.000224325 | SNX31 |

| | | | | | | | |
|---|---|---|---|---|---|---|---|
| VAT | HTN | ENSG00000175305.12 | -0.026414887 | 3.72E-09 | 1.03E-05 | 4.89E-07 | CCNE2 |
| VAT | HTN | ENSG00000175470.14 | -0.018926511 | 0.000133152 | 0.367500698 | 0.003310817 | PPP2R2D |
| VAT | HTN | ENSG00000175772.10 | 0.006596755 | 0.001919 | 1 | 0.025341822 | LINC01106 |
| VAT | HTN | ENSG00000175826.7 | 0.023690121 | 8.00E-05 | 0.220732361 | 0.002143033 | CTDNEP1 |
| VAT | HTN | ENSG00000175873.3 | -0.023150149 | 0.00031646 | 0.873428867 | 0.006667396 | AC004840.9 |
| VAT | HTN | ENSG00000175938.6 | 0.024739225 | 2.93E-07 | 0.000809814 | 2.19E-05 | ORAI3 |
| VAT | HTN | ENSG00000176390.10 | 0.013701373 | 0.000643678 | 1 | 0.01096636 | CRLF3 |
| VAT | HTN | ENSG00000177989.9 | 0.010021384 | 0.002164701 | 1 | 0.027474564 | ODF3B |
| VAT | HTN | ENSG00000178295.10 | -0.006371754 | 0.001623851 | 1 | 0.022409137 | GEN1 |
| VAT | HTN | ENSG00000178718.5 | 0.019526007 | 1.90E-06 | 0.005245327 | 9.37E-05 | RPP25 |
| VAT | HTN | ENSG00000178802.13 | -0.039096741 | 3.51E-16 | 9.69E-13 | 4.85E-13 | MPI |
| VAT | HTN | ENSG00000178809.7 | -0.02155279 | 0.00056573 | 1 | 0.010098967 | TRIM73 |
| VAT | HTN | ENSG00000178922.12 | -0.015185959 | 2.77E-07 | 0.000763296 | 2.12E-05 | HYI |
| VAT | HTN | ENSG00000178971.9 | 0.020207273 | 0.000130681 | 0.360678915 | 0.003278899 | CTC1 |
| VAT | HTN | ENSG00000181915.3 | 0.026755312 | 1.02E-09 | 2.82E-06 | 1.76E-07 | ADO |
| VAT | HTN | ENSG00000182372.6 | 0.015439995 | 1.04E-05 | 0.028610072 | 0.000397362 | CLN8 |
| VAT | HTN | ENSG00000182511.7 | -0.057726927 | 1.60E-19 | 4.40E-16 | 4.40E-16 | FES |
| VAT | HTN | ENSG00000182986.8 | 0.00485425 | 0.000185348 | 0.511559856 | 0.004298822 | ZNF320 |
| VAT | HTN | ENSG00000183336.7 | -0.007210534 | 0.002263462 | 1 | 0.028267668 | BOLA2 |
| VAT | HTN | ENSG00000184254.12 | -0.018789251 | 0.001755846 | 1 | 0.023755566 | ALDH1A3 |
| VAT | HTN | ENSG00000184385.2 | -0.00734495 | 0.000141817 | 0.391414429 | 0.003463845 | C21orf128 |
| VAT | HTN | ENSG00000184389.8 | -0.014317631 | 6.24E-05 | 0.17231603 | 0.00172316 | A3GALT2 |
| VAT | HTN | ENSG00000184923.8 | -0.014551063 | 0.000118929 | 0.328245316 | 0.003039308 | NUTM2A |
| VAT | HTN | ENSG00000185324.17 | -0.006119511 | 9.37E-05 | 0.25854422 | 0.002439096 | CDK10 |
| VAT | HTN | ENSG00000186409.10 | 0.012520681 | 0.001653737 | 1 | 0.022708031 | CCDC30 |
| VAT | HTN | ENSG00000186417.9 | 0.011377274 | 0.000998015 | 1 | 0.015650693 | GLDN |
| VAT | HTN | ENSG00000187961.9 | 0.018659186 | 0.001291011 | 1 | 0.018581684 | KLHL17 |
| VAT | HTN | ENSG00000188321.9 | -0.003730897 | 0.002653153 | 1 | 0.030767652 | ZNF559 |
| VAT | HTN | ENSG00000188629.7 | -0.007576082 | 0.002603289 | 1 | 0.030644506 | ZNF177 |
| VAT | HTN | ENSG00000188820.8 | -0.012959504 | 0.004214612 | 1 | 0.044398198 | FAM26F |
| VAT | HTN | ENSG00000188878.12 | -0.024923496 | 5.73E-07 | 0.001580927 | 3.51E-05 | FBF1 |
| VAT | HTN | ENSG00000188895.7 | -0.008566505 | 0.002933341 | 1 | 0.033733423 | MSL1 |
| VAT | HTN | ENSG00000196126.6 | -0.013586704 | 0.000520426 | 1 | 0.009771271 | HLA-DRB1 |
| VAT | HTN | ENSG00000196196.2 | 0.016311348 | 0.000689577 | 1 | 0.011534741 | HRCT1 |
| VAT | HTN | ENSG00000196966.3 | 0.017547845 | 9.62E-08 | 0.000265476 | 8.30E-06 | HIST1H3E |
| VAT | HTN | ENSG00000197020.6 | -0.013644606 | 1.03E-05 | 0.028399626 | 0.000397362 | ZNF100 |
| VAT | HTN | ENSG00000197165.6 | 0.027713692 | 1.70E-05 | 0.046990952 | 0.000594822 | SULT1A2 |
| VAT | HTN | ENSG00000197448.9 | -0.013008176 | 0.002243486 | 1 | 0.028145547 | GSTK1 |
| VAT | HTN | ENSG00000197479.4 | -0.008101643 | 0.004605358 | 1 | 0.047053283 | PCDHB11 |
| VAT | HTN | ENSG00000197728.5 | -0.010540702 | 8.08E-06 | 0.022291892 | 0.000323071 | RPS26 |
| VAT | HTN | ENSG00000197935.6 | 0.012240733 | 0.004620087 | 1 | 0.047053283 | ZNF311 |
| VAT | HTN | ENSG00000198270.8 | -0.014726784 | 8.58E-14 | 2.37E-10 | 7.90E-11 | TMEM116 |
| VAT | HTN | ENSG00000198502.5 | -0.005686134 | 0.00058795 | 1 | 0.010205917 | HLA-DRB5 |
| VAT | HTN | ENSG00000198646.9 | -0.010396593 | 0.003259267 | 1 | 0.036419338 | NCOA6 |
| VAT | HTN | ENSG00000198805.7 | -0.005642425 | 0.002623473 | 1 | 0.030644506 | PNP |
| VAT | HTN | ENSG00000203896.5 | 0.018836878 | 0.002529076 | 1 | 0.030217531 | LIME1 |
| VAT | HTN | ENSG00000204228.3 | -0.019993031 | 4.14E-05 | 0.114384039 | 0.001299819 | HSD17B8 |
| VAT | HTN | ENSG00000204314.6 | 0.017060208 | 4.63E-06 | 0.012768436 | 0.000209319 | PRRT1 |
| VAT | HTN | ENSG00000204338.4 | 0.009463215 | 1.19E-05 | 0.032787313 | 0.000437164 | CYP21A1P |
| VAT | HTN | ENSG00000204525.10 | -0.01185149 | 2.65E-09 | 7.30E-06 | 4.06E-07 | HLA-C |
| VAT | HTN | ENSG00000204531.11 | -0.00413257 | 0.000224101 | 0.618518387 | 0.005028605 | POU5F1 |
| VAT | HTN | ENSG00000205269.4 | 0.017507615 | 0.004500833 | 1 | 0.046525467 | TMEM170B |
| VAT | HTN | ENSG00000205583.9 | -0.011889096 | 3.60E-07 | 0.000994519 | 2.55E-05 | STAG3L1 |
| VAT | HTN | ENSG00000213347.6 | -0.014064225 | 0.001084872 | 1 | 0.016542793 | MXD3 |
| VAT | HTN | ENSG00000213676.6 | 0.02114734 | 3.23E-09 | 8.90E-06 | 4.45E-07 | ATF6B |
| VAT | HTN | ENSG00000213889.6 | 0.016855429 | 0.002281503 | 1 | 0.028307557 | PPM1N |
| VAT | HTN | ENSG00000214435.3 | -0.00450098 | 0.001112069 | 1 | 0.016772188 | AS3MT |
| VAT | HTN | ENSG00000214659.4 | 0.018023923 | 0.000272832 | 0.753016283 | 0.005929262 | KRT8P26 |
| VAT | HTN | ENSG00000215114.3 | 0.016932557 | 0.002091823 | 1 | 0.026978653 | UBXN2B |
| VAT | HTN | ENSG00000215464.4 | -0.021754908 | 0.000813093 | 1 | 0.013200804 | AP000354.2 |
| VAT | HTN | ENSG00000218422.2 | 0.017156351 | 0.000291116 | 0.803478988 | 0.006228519 | AC016773.1 |
| VAT | HTN | ENSG00000221838.5 | -0.009193481 | 0.00043354 | 1 | 0.008486318 | AP4M1 |
| VAT | HTN | ENSG00000221955.6 | 0.021069758 | 0.000663512 | 1 | 0.011210665 | SLC12A8 |
| VAT | HTN | ENSG00000223496.1 | -0.025385273 | 1.21E-05 | 0.03327451 | 0.000437822 | EXOSC6 |
| VAT | HTN | ENSG00000223891.1 | 0.010124805 | 0.003348943 | 1 | 0.037120812 | OSER1-AS1 |
| VAT | HTN | ENSG00000224997.1 | -0.029228153 | 1.20E-06 | 0.003320986 | 6.27E-05 | AL049840.1 |
| VAT | HTN | ENSG00000225190.4 | -0.025351329 | 0.000172703 | 0.476659174 | 0.004039485 | PLEKHM1 |
| VAT | HTN | ENSG00000225851.1 | 0.005595613 | 0.003883949 | 1 | 0.04171089 | HLA-S |
| VAT | HTN | ENSG00000226232.4 | -0.014415003 | 0.000571181 | 1 | 0.010105514 | RP11-419C5.2 |
| VAT | HTN | ENSG00000228998.3 | -0.0219941 | 0.001043829 | 1 | 0.016048355 | RP11-697E2.7 |
| VAT | HTN | ENSG00000229391.3 | 0.008547019 | 6.01E-05 | 0.165930289 | 0.001676064 | HLA-DRB6 |
| VAT | HTN | ENSG00000229759.1 | -0.020544178 | 1.81E-11 | 4.99E-08 | 9.99E-09 | MRPS18AP1 |
| VAT | HTN | ENSG00000231130.1 | 0.007436088 | 0.004396269 | 1 | 0.045960991 | HLA-T |
| VAT | HTN | ENSG00000232075.1 | 0.027344422 | 4.92E-06 | 0.01358799 | 0.000219161 | MRPL35P2 |
| VAT | HTN | ENSG00000232112.3 | 0.017188242 | 0.000203697 | 0.562203004 | 0.004646306 | TMA7 |
| VAT | HTN | ENSG00000232561.2 | -0.01195486 | 0.001018213 | 1 | 0.015810108 | GTF2IP1 |
| VAT | HTN | ENSG00000236397.2 | 0.008955716 | 6.50E-05 | 0.179533751 | 0.001777562 | DDX11L2 |
| VAT | HTN | ENSG00000237541.3 | 0.008486144 | 1.61E-05 | 0.044397706 | 0.000576594 | HLA-DQA2 |
| VAT | HTN | ENSG00000239697.6 | 0.030712299 | 6.93E-08 | 0.000191319 | 6.17E-06 | TNFSF12 |
| VAT | HTN | ENSG00000240399.1 | -0.012143521 | 0.000331028 | 0.91363702 | 0.006869451 | RP1-228P16.1 |
| VAT | HTN | ENSG00000243789.6 | 0.00968732 | 0.001099957 | 1 | 0.016680674 | JMJD7 |

| | | | | | | | |
|---|---|---|---|---|---|---|---|
| VAT | HTN | ENSG00000244219.2 | 0.018730011 | 0.00055621 | 1 | 0.010098967 | GS1-259H13.2 |
| VAT | HTN | ENSG00000244291.1 | -0.022701594 | 0.001019637 | 1 | 0.015810108 | C7orf13 |
| VAT | HTN | ENSG00000251417.1 | -0.01466861 | 1.78E-05 | 0.04901568 | 0.000612696 | RP11-1348G14.4 |
| VAT | HTN | ENSG00000253981.1 | 0.029243817 | 5.65E-07 | 0.001559224 | 3.51E-05 | ALG1L13P |
| VAT | HTN | ENSG00000254634.3 | -0.010939008 | 0.004567261 | 1 | 0.046861116 | RP11-231C14.3 |
| VAT | HTN | ENSG00000254851.1 | 0.007205896 | 4.67E-07 | 0.001288313 | 3.22E-05 | RP11-109L13.1 |
| VAT | HTN | ENSG00000256885.1 | -0.011771235 | 0.00126024 | 1 | 0.018581684 | AP001877.1 |
| VAT | HTN | ENSG00000257595.2 | -0.03269804 | 1.59E-10 | 4.38E-07 | 4.87E-08 | RP3-473L9.4 |
| VAT | HTN | ENSG00000257877.1 | -0.023318786 | 0.000262543 | 0.724618892 | 0.005750944 | RP3-462E2.3 |
| VAT | HTN | ENSG00000258408.1 | 0.014940924 | 0.001313977 | 1 | 0.018790547 | NT5CP2 |
| VAT | HTN | ENSG00000258512.1 | 0.016735002 | 0.001361483 | 1 | 0.019369546 | RP11-796G6.2 |
| VAT | HTN | ENSG00000258819.1 | -0.021289961 | 0.001381589 | 1 | 0.019554794 | RP11-7F17.3 |
| VAT | HTN | ENSG00000258908.1 | -0.018880547 | 0.002563092 | 1 | 0.030491956 | RP11-203M5.8 |
| VAT | HTN | ENSG00000259982.1 | 0.011369371 | 0.000162907 | 0.449623483 | 0.003842936 | CDC37P1 |
| VAT | HTN | ENSG00000260077.1 | -0.019889877 | 0.001179756 | 1 | 0.017696346 | RP11-254F7.2 |
| VAT | HTN | ENSG00000260265.1 | -0.016919347 | 0.003752784 | 1 | 0.040459698 | RP11-44F21.5 |
| VAT | HTN | ENSG00000260911.1 | 0.008962444 | 9.25E-05 | 0.255413501 | 0.00243251 | RP11-196G11.2 |
| VAT | HTN | ENSG00000261556.4 | -0.010232066 | 8.53E-05 | 0.235331206 | 0.0022628 | RP11-296I10.6 |
| VAT | HTN | ENSG00000264538.2 | 0.013204862 | 3.74E-05 | 0.103156097 | 0.001185702 | SUZ12P |
| VAT | HTN | ENSG00000266912.1 | -0.011378551 | 0.00191062 | 1 | 0.025341822 | CTC-325H20.2 |
| VAT | HTN | ENSG00000266970.1 | 0.01543606 | 0.004347509 | 1 | 0.045624044 | RP11-806H10.4 |
| VAT | HTN | ENSG00000266992.1 | -0.017329672 | 0.000751499 | 1 | 0.012346048 | DHX40P1 |
| VAT | HTN | ENSG00000268225.1 | 0.014209688 | 0.000148836 | 0.410786292 | 0.003552729 | CTD-2331H12.5 |
| VAT | HTN | ENSG00000268555.1 | -0.01913086 | 0.001689173 | 1 | 0.02296609 | RP11-678G14.3 |
| VAT | HTN | ENSG00000269963.1 | -0.034195482 | 1.44E-06 | 0.003982953 | 7.24E-05 | RP11-73M18.9 |
| VAT | HTN | ENSG00000270018.1 | 0.028315386 | 9.06E-07 | 0.002499754 | 4.90E-05 | RP3-462E2.5 |
| VAT | HTN | ENSG00000270020.1 | -0.011914584 | 5.95E-05 | 0.164321217 | 0.001676064 | RP11-463O9.9 |
| VAT | HTN | ENSG00000271581.1 | 0.044446831 | 1.28E-08 | 3.53E-05 | 1.36E-06 | XXbac-BPG248L24.12 |
| VAT | HTN | ENSG00000271751.1 | -0.024355655 | 3.15E-05 | 0.086968195 | 0.001035336 | RP11-110I1.14 |
| VAT | HTN | ENSG00000272505.1 | 0.025490349 | 0.000232852 | 0.642670646 | 0.005182828 | RP11-981G7.6 |
| VAT | Heart_Failure | ENSG00000004534.10 | -0.00396061 | 5.06E-08 | 0.000139782 | 1.75E-05 | RBM6 |
| VAT | Heart_Failure | ENSG00000086015.16 | 0.011575077 | 7.90E-05 | 0.217930737 | 0.011470039 | MAST2 |
| VAT | Heart_Failure | ENSG00000088340.11 | -0.00956845 | 0.000127609 | 0.352200569 | 0.016009117 | FER1L4 |
| VAT | Heart_Failure | ENSG00000090621.9 | -0.011305629 | 9.72E-05 | 0.268293058 | 0.01277586 | PABPC4 |
| VAT | Heart_Failure | ENSG00000116883.8 | -0.006517515 | 3.77E-05 | 0.104013135 | 0.006798613 | RP11-268J15.5 |
| VAT | Heart_Failure | ENSG00000123243.10 | -0.011163945 | 0.000408276 | 1 | 0.037561413 | ITIH5 |
| VAT | Heart_Failure | ENSG00000127957.12 | -0.012652278 | 8.38E-06 | 0.023115823 | 0.00165113 | PMS2P3 |
| VAT | Heart_Failure | ENSG00000135213.8 | -0.00968636 | 0.000423427 | 1 | 0.037698636 | POM121C |
| VAT | Heart_Failure | ENSG00000142694.6 | 0.011048087 | 5.87E-05 | 0.162053448 | 0.009002969 | EVA1B |
| VAT | Heart_Failure | ENSG00000143631.10 | -0.005208116 | 0.000548707 | 1 | 0.04577203 | FLG |
| VAT | Heart_Failure | ENSG00000145777.10 | 0.008131126 | 2.23E-06 | 0.00615126 | 0.000559205 | TSLP |
| VAT | Heart_Failure | ENSG00000164068.11 | 0.006439656 | 3.98E-06 | 0.010975386 | 0.000914615 | RNF123 |
| VAT | Heart_Failure | ENSG00000164078.8 | 0.019788337 | 3.66E-08 | 0.000100932 | 1.44E-05 | MST1R |
| VAT | Heart_Failure | ENSG00000167914.6 | -0.018985077 | 4.61E-11 | 1.27E-07 | 2.12E-08 | GSDMA |
| VAT | Heart_Failure | ENSG00000170092.10 | 0.010967044 | 0.000251127 | 0.693110117 | 0.030135222 | AC018720.10 |
| VAT | Heart_Failure | ENSG00000179344.12 | -0.017737781 | 1.19E-22 | 3.30E-19 | 3.30E-19 | HLA-DQB1 |
| VAT | Heart_Failure | ENSG00000179528.11 | 0.009003784 | 0.000607779 | 1 | 0.046596405 | LBX2 |
| VAT | Heart_Failure | ENSG00000183336.7 | -0.004722502 | 0.000378371 | 1 | 0.036010503 | BOLA2 |
| VAT | Heart_Failure | ENSG00000187145.10 | -0.004703193 | 0.000360208 | 0.994173902 | 0.036010503 | MRPS21 |
| VAT | Heart_Failure | ENSG00000188895.7 | -0.005811648 | 0.000334166 | 0.922297947 | 0.036010503 | MSL1 |
| VAT | Heart_Failure | ENSG00000196301.3 | 0.00989992 | 6.96E-12 | 1.92E-08 | 3.84E-09 | HLA-DRB9 |
| VAT | Heart_Failure | ENSG00000196735.7 | -0.029201905 | 4.80E-19 | 1.32E-15 | 6.62E-16 | HLA-DQA1 |
| VAT | Heart_Failure | ENSG00000197728.5 | 0.004571214 | 0.000580442 | 1 | 0.04577203 | RPS26 |
| VAT | Heart_Failure | ENSG00000204516.5 | 0.002793396 | 0.000577761 | 1 | 0.04577203 | MICB |
| VAT | Heart_Failure | ENSG00000204520.8 | 0.006116252 | 6.79E-06 | 0.018742897 | 0.001441761 | MICA |
| VAT | Heart_Failure | ENSG00000205583.9 | -0.006316305 | 1.54E-06 | 0.004255017 | 0.00047278 | STAG3L1 |
| VAT | Heart_Failure | ENSG00000213676.6 | 0.007146307 | 0.000376644 | 1 | 0.036010503 | ATF6B |
| VAT | Heart_Failure | ENSG00000214078.7 | -0.002215688 | 0.000529886 | 1 | 0.045702697 | CPNE1 |
| VAT | Heart_Failure | ENSG00000214194.4 | 0.014245906 | 4.96E-05 | 0.137026576 | 0.008060387 | LINC00998 |
| VAT | Heart_Failure | ENSG00000229391.3 | 0.0049263 | 3.94E-05 | 0.108777811 | 0.006798613 | HLA-DRB6 |
| VAT | Heart_Failure | ENSG00000232629.4 | 0.008056206 | 2.35E-14 | 6.49E-11 | 2.16E-11 | HLA-DQB2 |
| VAT | Heart_Failure | ENSG00000237541.3 | 0.007866205 | 1.18E-12 | 3.27E-09 | 8.17E-10 | HLA-DQA2 |
| VAT | Heart_Failure | ENSG00000240065.3 | 0.00545849 | 1.87E-06 | 0.005156801 | 0.00051568 | PSMB9 |
| VAT | Heart_Failure | ENSG00000243725.2 | -0.009904762 | 0.000267003 | 0.736928354 | 0.030705348 | TTC4 |
| VAT | Heart_Failure | ENSG00000248290.1 | -0.0045749 | 0.000346877 | 0.957379669 | 0.036010503 | TNXA |
| VAT | Heart_Failure | ENSG00000272221.1 | 0.003118532 | 8.58E-05 | 0.236867022 | 0.011843351 | XXbac-BPG181B23.7 |

Table S7 Number of genes with significant coefficient changes after adjusting for estimated cell proportions

| Tissue | Disorder | No. genes with significant coefficient change in the original bulk level TWAS analysis | No. significant genes from original bulk level TWAS analysis | No. significant genes(original TWAS analysis) with significant coefficient change | No. significant genes from adjusted(by estimated cell proportions) bulk level TWAS analysis | No. common significant genes | Proprotion of significant genes with significant change |
|---|---|---|---|---|---|---|---|
| Frontal_Cortex | Anxiety | 174 | 0 | 0 | 0 | 0 | |
| Frontal_Cortex | Bipolar | 0 | 0 | 0 | 0 | 0 | |
| Frontal_Cortex | Depression | 6 | 3 | 0 | 3 | 3 | 0 |
| Frontal_Cortex | Psychosis | 109 | 0 | 0 | 0 | 0 | |
| Pancreas | Prediabetes | 24 | 0 | 0 | 0 | 0 | |
| Pancreas | T1DM | 683 | 57 | 34 | 56 | 53 | 0.596491228 |
| Pancreas | T2DM_admission | 368 | 135 | 23 | 132 | 130 | 0.17037037 |
| Pancreas | T2DM_insulin | 151 | 20 | 4 | 19 | 17 | 0.2 |
| Pancreas | T2DM | 333 | 88 | 18 | 85 | 84 | 0.204545455 |
| SAT | CAD | 99 | 55 | 5 | 52 | 52 | 0.090909091 |
| SAT | Heart_Failure | 1280 | 52 | 31 | 40 | 36 | 0.596153846 |
| SAT | HTN | 572 | 364 | 81 | 367 | 350 | 0.222527473 |
| SAT | Obesity | 1031 | 331 | 127 | 299 | 297 | 0.383685801 |
| SAT | Stroke | 28 | 0 | 0 | 0 | 0 | |
| SAT | T2DM | 1189 | 176 | 68 | 139 | 138 | 0.386363636 |
| VAT | CAD | 17 | 30 | 1 | 30 | 28 | 0.033333333 |
| VAT | Heart_Failure | 392 | 36 | 7 | 36 | 34 | 0.194444444 |
| VAT | HTN | 696 | 275 | 68 | 273 | 267 | 0.247272727 |
| VAT | Obesity | 966 | 326 | 152 | 293 | 291 | 0.466257669 |
| VAT | Stroke | 1 | 0 | 0 | 0 | 0 | |
| VAT | T2DM | 679 | 124 | 42 | 99 | 98 | 0.338709677 |

Note: No. genes with significant coefficient change in the original bulk level TWAS analysis indicates genes with significant coefficient changes, some genes with significant coefficient changes are not associated with the target trait based on the TWAS analysis results, here significant genes indicated those that have a adjusted pvalue(FDR) <=0.05

| Tissue | Disorder | Genes | Estimates | Pvalues (standard TWAS) | SD (standard TWAS) | Residual variance (standard TWAS) | Estimates (conditional TWAS) | Pvalues (conditional TWAS) | SD (conditional TWAS) | Residual variance (conditional TWAS) | coefficient change | pvalue of coefficient change | Pvalue Bonferroni (standard TWAS) | Pvalue BH (standard TWAS) | Pvalue Bonferroni (conditional TWAS) | Pvalue BH (conditional TWAS) | gene_id | version | gene_name |
|---|---|---|---|---|---|---|---|---|---|---|---|---|---|---|---|---|---|---|---|
| Frontal Cortex | Anxiety | ENSG0000000 | 0.001670908 | 0.293181019 | 0.001589569 | 0.022786724 | 0.00165484 | 0.297843647 | 0.001589556 | 0.022785788 | 1.61E-05 | 0.046159201 | 1 | 0.947573237 | 1 | 0.953162532 | ENSG00000000 | 9 | SCYL3 |
| Frontal Cortex | Anxiety | ENSG0000001 | 0.000357985 | 0.744130591 | 0.001096805 | 0.022786797 | 0.000370957 | 0.735199999 | 0.001096802 | 0.022785859 | -1.30E-05 | 0.047354966 | 1 | 0.978113594 | 1 | 0.977376329 | ENSG00000000 | 9 | SCYL3 |
| Frontal Cortex | Anxiety | ENSG0000005 | -0.00033361 | 0.688372135 | 0.000831802 | 0.022786793 | -0.00034543 | 0.67793979 | 0.000831801 | 0.022785855 | 1.18E-05 | 0.02246455 | 1 | 0.975684776 | 1 | 0.977376329 | ENSG00000000 | 13 | STPG1 |
| Frontal Cortex | Anxiety | ENSG0000006 | 0.00225482 | 0.216490814 | 0.001824412 | 0.022786693 | 0.002285599 | 0.21028472 | 0.001824414 | 0.022785753 | -3.08E-05 | 0.0096653 | 1 | 0.903356609 | 1 | 0.892631846 | ENSG00000000 | 12 | NIPAL3 |
| Frontal Cortex | Anxiety | ENSG0000006 | -0.0005206 | 0.79521483 | 0.002005802 | 0.0227868 | -0.00054826 | 0.784594886 | 0.00200581 | 0.022785862 | 2.77E-05 | 0.048438773 | 1 | 0.982406987 | 1 | 0.985862728 | ENSG00000000 | 6 | ENPP4 |
| Frontal Cortex | Anxiety | ENSG0000006 | -0.000867 | 0.718632083 | 0.002406415 | 0.022786795 | -0.00083473 | 0.728684151 | 0.002406409 | 0.022785859 | -3.23E-05 | 0.024923512 | 1 | 0.975684776 | 1 | 0.977376329 | ENSG00000000 | 6 | ENPP4 |
| Frontal Cortex | Anxiety | ENSG0000006 | -0.00265599 | 0.05467061 | 0.001382255 | 0.022786535 | -0.00267159 | 0.053263151 | 0.001382244 | 0.022785595 | 1.56E-05 | 0.027187821 | 1 | 0.732084207 | 1 | 0.754532031 | ENSG00000000 | 6 | ENPP4 |
| Frontal Cortex | Anxiety | ENSG0000007 | 0.000528103 | 0.801321271 | 0.002098666 | 0.0227868 | 0.000652836 | 0.755782947 | 0.002098993 | 0.02278586 | -0.00012473 | 0.001548793 | 1 | 0.982406987 | 1 | 0.982577711 | ENSG00000000 | 6 | ENPP4 |
| Frontal Cortex | Anxiety | ENSG0000007 | 0.00435098 | 0.072659814 | 0.002423988 | 0.022786569 | 0.004305207 | 0.075722032 | 0.002424005 | 0.022785637 | 4.58E-05 | 0.010907812 | 1 | 0.791500747 | 1 | 0.810468204 | ENSG00000000 | 6 | ENPP4 |
| Frontal Cortex | Anxiety | ENSG0000007 | -0.00037844 | 0.692798559 | 0.00095792 | 0.022786793 | -0.00181543 | 0.101316289 | 0.001107975 | 0.022785672 | 0.001436992 | 0.009859026 | 1 | 0.975684776 | 1 | 0.827410035 | ENSG00000000 | 6 | ENPP4 |
| Frontal Cortex | Anxiety | ENSG0000008 | 0.001442453 | 0.344241812 | 0.001525084 | 0.022786739 | 0.001424273 | 0.350355336 | 0.001525078 | 0.022785804 | 1.82E-05 | 0.038163628 | 1 | 0.954139853 | 1 | 0.960761585 | ENSG00000000 | 7 | SEMA3F |
| Frontal Cortex | Anxiety | ENSG0000008 | -0.00136753 | 0.437017341 | 0.001759468 | 0.02278676 | -0.00139044 | 0.429374678 | 0.001759465 | 0.022785822 | 2.29E-05 | 0.032809492 | 1 | 0.974419575 | 1 | 0.976777864 | ENSG00000000 | 10 | SNX11 |
| Frontal Cortex | Anxiety | ENSG0000008 | -0.00082041 | 0.68950032 | 0.002053419 | 0.022786793 | -0.00078581 | 0.701953581 | 0.002053419 | 0.022785857 | -3.46E-05 | 0.00849836 | 1 | 0.975684776 | 1 | 0.977376329 | ENSG00000000 | 10 | SNX11 |
| Frontal Cortex | Anxiety | ENSG0000009 | -0.00149849 | 0.487347481 | 0.002157549 | 0.022786769 | -0.0014642 | 0.497369098 | 0.002157556 | 0.022785834 | -3.43E-05 | 0.021514182 | 1 | 0.975684776 | 1 | 0.977376329 | ENSG00000000 | 10 | SNX11 |
| Frontal Cortex | Anxiety | ENSG0000009 | -6.57E-05 | 0.967990253 | 0.001636309 | 0.022786804 | -8.64E-05 | 0.957898773 | 0.001636307 | 0.022785867 | 2.07E-05 | 0.042418125 | 1 | 0.996527112 | 1 | 0.995692427 | ENSG00000000 | 10 | SNX11 |
| Frontal Cortex | Anxiety | ENSG0000009 | 0.000691669 | 0.775918026 | 0.002429954 | 0.022786799 | 0.000723757 | 0.765819462 | 0.002429955 | 0.022785861 | -3.21E-05 | 0.042215464 | 1 | 0.982406987 | 1 | 0.982577711 | ENSG00000000 | 3 | M6PR |
| Frontal Cortex | Anxiety | ENSG0000010 | -0.00038145 | 0.66449982 | 0.000879514 | 0.022786791 | -0.00039426 | 0.653960221 | 0.000879519 | 0.022785853 | 1.28E-05 | 0.04860299 | 1 | 0.975684776 | 1 | 0.977376329 | ENSG00000000 | 3 | M6PR |
| Frontal Cortex | Anxiety | ENSG0000010 | -0.00072516 | 0.514516461 | 0.001112515 | 0.022786773 | -0.00070728 | 0.524939745 | 0.001112516 | 0.022785838 | -1.79E-05 | 0.013376137 | 1 | 0.975684776 | 1 | 0.977376329 | ENSG00000000 | 9 | DBNDD1 |
| Frontal Cortex | Anxiety | ENSG0000010 | -0.00071313 | 0.210210963 | 0.000569143 | 0.02278669 | -0.00070638 | 0.214553742 | 0.00056914 | 0.022785755 | -6.74E-06 | 0.035910329 | 1 | 0.89263614 | 1 | 0.89933777 | ENSG00000000 | 9 | DBNDD1 |
| Frontal Cortex | Anxiety | ENSG0000010 | -0.00436475 | 0.054107698 | 0.002266246 | 0.022786534 | -0.00438838 | 0.052817103 | 0.002266231 | 0.022785594 | 2.36E-05 | 0.049658898 | 1 | 0.732084207 | 1 | 0.754532031 | ENSG00000000 | 9 | SLC22A16 |
| Frontal Cortex | Anxiety | ENSG0000010 | 0.002834983 | 0.180848517 | 0.002118584 | 0.022786674 | 0.002861863 | 0.176747911 | 0.002118581 | 0.022785734 | -2.69E-05 | 0.041766883 | 1 | 0.879106816 | 1 | 0.887019637 | ENSG00000000 | 9 | SLC22A16 |
| Frontal Cortex | Anxiety | ENSG0000010 | 0.002928076 | 0.026014652 | 0.001315396 | 0.022786443 | 0.003251814 | 0.013783719 | 0.001320338 | 0.022785425 | -0.00032374 | 0.004683021 | 1 | 0.732084207 | 1 | 0.746228219 | ENSG00000000 | 9 | SLC22A16 |
| Frontal Cortex | Anxiety | ENSG0000010 | 0.001001653 | 0.512517041 | 0.001529418 | 0.022786773 | 0.001022114 | 0.503939409 | 0.001529413 | 0.022785835 | -2.05E-05 | 0.023128845 | 1 | 0.975684776 | 1 | 0.977376329 | ENSG00000000 | 9 | SLC22A16 |
| Frontal Cortex | Anxiety | ENSG0000010 | -0.00017872 | 0.872043671 | 0.00110961 | 0.022786803 | -0.00019023 | 0.86388138 | 0.001109603 | 0.022785865 | 1.15E-05 | 0.048358759 | 1 | 0.983816039 | 1 | 0.988452375 | ENSG00000000 | 7 | CDC27 |
| Frontal Cortex | Anxiety | ENSG0000011 | 2.45E-05 | 0.983930396 | 0.001215311 | 0.022786804 | 0.000452608 | 0.71159749 | 0.001224219 | 0.022785858 | -0.00042813 | 0.003728897 | 1 | 0.999327591 | 1 | 0.977376329 | ENSG00000000 | 8 | MEOX1 |
| Frontal Cortex | Anxiety | ENSG0000011 | 7.21E-05 | 0.968626824 | 0.001832007 | 0.022786804 | -5.34E-05 | 0.976768403 | 0.001832494 | 0.022785867 | 0.000125417 | 0.004248055 | 1 | 0.996527112 | 1 | 0.998606721 | ENSG00000000 | 8 | MEOX1 |
| Frontal Cortex | Anxiety | ENSG0000011 | 0.001097273 | 0.496720683 | 0.001614446 | 0.022786771 | 0.001126142 | 0.485467256 | 0.001614454 | 0.022785832 | -2.89E-05 | 0.01261057 | 1 | 0.975684776 | 1 | 0.977376329 | ENSG00000000 | 8 | MEOX1 |
| Frontal Cortex | Anxiety | ENSG0000011 | 0.000204424 | 0.772325094 | 0.000706531 | 0.022786798 | 0.000214018 | 0.7619557 | 0.000706532 | 0.022785861 | -9.59E-06 | 0.038159926 | 1 | 0.982406987 | 1 | 0.982577711 | ENSG00000000 | 7 | LIG3 |
| Frontal Cortex | Anxiety | ENSG0000011 | -0.00177489 | 0.502747227 | 0.002648408 | 0.022786772 | -0.00173596 | 0.512163269 | 0.002648411 | 0.022785836 | -3.89E-05 | 0.025462979 | 1 | 0.975684776 | 1 | 0.977376329 | ENSG00000000 | 7 | LIG3 |
| Frontal Cortex | Anxiety | ENSG0000012 | -0.00334056 | 0.038914418 | 0.001617625 | 0.022786493 | -0.00336755 | 0.037363565 | 0.001617631 | 0.022785551 | 2.70E-05 | 0.016981543 | 1 | 0.732084207 | 1 | 0.754532031 | ENSG00000000 | 7 | LIG3 |
| Frontal Cortex | Anxiety | ENSG0000012 | -0.00304088 | 0.049411809 | 0.001547499 | 0.022786523 | -0.00305734 | 0.048191649 | 0.001547488 | 0.022785583 | 1.65E-05 | 0.043340381 | 1 | 0.732084207 | 1 | 0.754532031 | ENSG00000000 | 7 | LIG3 |
| Frontal Cortex | Anxiety | ENSG0000012 | -0.00053172 | 0.693261838 | 0.001348072 | 0.022786793 | -0.00062817 | 0.64133107 | 0.001348481 | 0.022785852 | 9.65E-05 | 0.00495967 | 1 | 0.975684776 | 1 | 0.977376329 | ENSG00000000 | 15 | AC004381.6 |
| Frontal Cortex | Anxiety | ENSG0000012 | 0.000871764 | 0.558439749 | 0.001489785 | 0.02278678 | 0.000931727 | 0.531735562 | 0.001489907 | 0.022785839 | -6.00E-05 | 0.004970063 | 1 | 0.975684776 | 1 | 0.977376329 | ENSG00000000 | 5 | COPZ2 |
| Frontal Cortex | Anxiety | ENSG0000012 | 0.000635472 | 0.51970682 | 0.000987068 | 0.022786774 | 0.000617798 | 0.531388529 | 0.000987074 | 0.022785839 | 1.77E-05 | 0.014185771 | 1 | 0.975684776 | 1 | 0.977376329 | ENSG00000000 | 5 | COPZ2 |
| Frontal Cortex | Anxiety | ENSG0000013 | -0.00018541 | 0.935961379 | 0.002307654 | 0.022786804 | -0.00014642 | 0.949407412 | 0.00230766 | 0.022785867 | -3.90E-05 | 0.013355873 | 1 | 0.99194522 | 1 | 0.995681012 | ENSG00000000 | 5 | COPZ2 |
| Frontal Cortex | Anxiety | ENSG0000013 | 0.002486058 | 0.07917927 | 0.001416175 | 0.02278658 | 0.00250436 | 0.076995099 | 0.001416173 | 0.022785639 | -1.83E-05 | 0.038592778 | 1 | 0.791500747 | 1 | 0.812107305 | ENSG00000000 | 5 | COPZ2 |
| Frontal Cortex | Anxiety | ENSG0000013 | 0.001163959 | 0.530923193 | 0.001857585 | 0.022786776 | 0.001183831 | 0.523930239 | 0.001857574 | 0.022785838 | -1.99E-05 | 0.046306871 | 1 | 0.975684776 | 1 | 0.977376329 | ENSG00000000 | 9 | SOX8 |
| Frontal Cortex | Anxiety | ENSG0000013 | -0.00233842 | 0.189874995 | 0.00178376 | 0.022786679 | -0.00230622 | 0.196047218 | 0.001783763 | 0.022785745 | -3.22E-05 | 0.006760069 | 1 | 0.879106816 | 1 | 0.888056607 | ENSG00000000 | 9 | SOX8 |
| Frontal Cortex | Anxiety | ENSG0000013 | 0.000600203 | 0.811856198 | 0.002521519 | 0.0227868 | 0.00062575 | 0.804006256 | 0.002521501 | 0.022785863 | -2.55E-05 | 0.04902638 | 1 | 0.983816039 | 1 | 0.986980224 | ENSG00000000 | 9 | SOX8 |
| Frontal Cortex | Anxiety | ENSG0000013 | 0.00217071 | 0.120154012 | 0.001396735 | 0.022786628 | 0.002189716 | 0.116942773 | 0.001396734 | 0.022785688 | -1.90E-05 | 0.030704626 | 1 | 0.831449405 | 1 | 0.837078333 | ENSG00000000 | 12 | ZNF195 |
| Frontal Cortex | Anxiety | ENSG0000013 | -0.00284743 | 0.000132231 | 0.000744948 | 0.022785738 | -0.00277742 | 0.000194437 | 0.00074538 | 0.022784854 | -7.00E-05 | 0.006601883 | 0.26604846 | 0.13302423 | 0.391207122 | 0.195603561 | ENSG00000000 | 12 | ZNF195 |
| Frontal Cortex | Anxiety | ENSG0000013 | -0.00108238 | 0.330391782 | 0.001112041 | 0.022786735 | -0.00089444 | 0.422054534 | 0.001114062 | 0.02278582 | -0.00018794 | 0.00532259 | 1 | 0.954139853 | 1 | 0.97340047 | ENSG00000000 | 12 | ZNF195 |
| Frontal Cortex | Anxiety | ENSG0000013 | 0.000375595 | 0.467153429 | 0.000516552 | 0.022786766 | 0.000351038 | 0.49684182 | 0.000516636 | 0.022785834 | 2.46E-05 | 0.01293654 | 1 | 0.975684776 | 1 | 0.977376329 | ENSG00000000 | 8 | GGNBP2 |
| Frontal Cortex | Anxiety | ENSG0000013 | -0.00314551 | 0.120259715 | 0.00202454 | 0.022786628 | -0.00317003 | 0.117394284 | 0.002024532 | 0.022785689 | 2.45E-05 | 0.034440889 | 1 | 0.831449405 | 1 | 0.837078333 | ENSG00000000 | 8 | GGNBP2 |
| Frontal Cortex | Anxiety | ENSG0000013 | 0.000416857 | 0.706586332 | 0.001107351 | 0.022786794 | 0.000435941 | 0.693821785 | 0.001107364 | 0.022785856 | -1.91E-05 | 0.033255067 | 1 | 0.975684776 | 1 | 0.977376329 | ENSG00000000 | 8 | ASB4 |
| Frontal Cortex | Anxiety | ENSG0000014 | -0.0023746 | 0.268009497 | 0.002143802 | 0.022786715 | -0.00240027 | 0.262869828 | 0.002143794 | 0.022785776 | 2.57E-05 | 0.03978872 | 1 | 0.942322745 | 1 | 0.948868815 | ENSG00000000 | 8 | ASB4 |
| Frontal Cortex | Anxiety | ENSG0000014 | -0.00324302 | 0.15067775 | 0.002256569 | 0.022786654 | -0.00327751 | 0.146382343 | 0.002256566 | 0.022785714 | 3.45E-05 | 0.01377728 | 1 | 0.870476788 | 1 | 0.875203335 | ENSG00000000 | 8 | ASB4 |
| Frontal Cortex | Anxiety | ENSG0000014 | -1.52E-06 | 0.999428767 | 0.00211829 | 0.022786805 | -3.78E-05 | 0.985761882 | 0.002118292 | 0.022785868 | 3.63E-05 | 0.008689849 | 1 | 0.999973246 | 1 | 0.999485958 | ENSG00000000 | 6 | CRLF1 |
| Frontal Cortex | Anxiety | ENSG0000014 | 0.000828372 | 0.591403684 | 0.001543157 | 0.022786783 | 0.000853641 | 0.580145449 | 0.001543172 | 0.022785845 | -2.53E-05 | 0.03557899 | 1 | 0.975684776 | 1 | 0.977376329 | ENSG00000000 | 7 | ABCC8 |
| Frontal Cortex | Anxiety | ENSG0000014 | -0.00071674 | 0.383845638 | 0.000823051 | 0.022786749 | -0.00070751 | 0.389996246 | 0.000823048 | 0.022785814 | -9.23E-06 | 0.049936376 | 1 | 0.962964367 | 1 | 0.970014921 | ENSG00000000 | 7 | ABCC8 |
| Frontal Cortex | Anxiety | ENSG0000014 | 0.004031796 | 0.124805105 | 0.002626718 | 0.022786632 | 0.002867818 | 0.27948781 | 0.002651771 | 0.022785782 | 0.001163978 | 0.001385308 | 1 | 0.839825657 | 1 | 0.953162532 | ENSG00000000 | 15 | SPATA20 |
| Frontal Cortex | Anxiety | ENSG0000015 | 0.001410842 | 0.569170342 | 0.002478317 | 0.022786781 | 0.001449465 | 0.558644522 | 0.002478324 | 0.022785843 | -3.86E-05 | 0.023441118 | 1 | 0.975684776 | 1 | 0.977376329 | ENSG00000000 | 11 | MAP3K9 |
| Frontal Cortex | Anxiety | ENSG0000015 | 0.001047792 | 0.486090508 | 0.001504279 | 0.022786769 | 0.001066999 | 0.478131093 | 0.001504272 | 0.022785831 | -1.92E-05 | 0.025016133 | 1 | 0.975684776 | 1 | 0.977376329 | ENSG00000000 | 11 | AGK |
| Frontal Cortex | Anxiety | ENSG0000015 | 0.00023493 | 0.599455956 | 0.00044733 | 0.022786784 | 0.000241167 | 0.589800643 | 0.000447329 | 0.022785846 | -6.24E-06 | 0.024207941 | 1 | 0.975684776 | 1 | 0.977376329 | ENSG00000000 | 6 | GTF2IRD1 |
| Frontal Cortex | Anxiety | ENSG0000016 | 0.001161767 | 0.573592421 | 0.002064375 | 0.022786781 | 0.001187918 | 0.564995571 | 0.002064372 | 0.022785843 | -2.62E-05 | 0.039975349 | 1 | 0.975684776 | 1 | 0.977376329 | ENSG00000000 | 11 | VPS41 |
| Frontal Cortex | Anxiety | ENSG0000016 | 0.001714547 | 0.306470754 | 0.001676561 | 0.022786728 | 0.001695639 | 0.31183291 | 0.001676551 | 0.022785793 | 1.89E-05 | 0.035994682 | 1 | 0.947573237 | 1 | 0.953162532 | ENSG00000000 | 11 | VPS41 |
| Frontal Cortex | Anxiety | ENSG0000016 | -0.00057351 | 0.817348444 | 0.002483193 | 0.022786801 | -0.0006011 | 0.808726285 | 0.00248318 | 0.022785863 | 2.76E-05 | 0.044759122 | 1 | 0.983816039 | 1 | 0.988452375 | ENSG00000000 | 11 | VPS41 |
| Frontal Cortex | Anxiety | ENSG0000016 | -0.00408461 | 0.040303854 | 0.00199188 | 0.022786497 | -0.00405905 | 0.041570739 | 0.001991876 | 0.022785564 | -2.56E-05 | 0.03429987 | 1 | 0.732084207 | 1 | 0.754532031 | ENSG00000000 | 14 | ELAC2 |
| Frontal Cortex | Anxiety | ENSG0000016 | 0.001773418 | 0.349490198 | 0.001895528 | 0.022786741 | 0.00174283 | 0.357867205 | 0.001895538 | 0.022785806 | 3.06E-05 | 0.024957939 | 1 | 0.954139853 | 1 | 0.960761585 | ENSG00000000 | 14 | ELAC2 |
| Frontal Cortex | Anxiety | ENSG0000016 | 0.001746212 | 0.293650091 | 0.001662822 | 0.022786724 | 0.001723015 | 0.300108581 | 0.001662817 | 0.022785789 | 2.32E-05 | 0.018736452 | 1 | 0.947573237 | 1 | 0.953162532 | ENSG00000000 | 14 | ST7L |
| Frontal Cortex | Anxiety | ENSG0000016 | -0.00063773 | 0.440630125 | 0.000827005 | 0.022786761 | -0.00062805 | 0.447598475 | 0.000827002 | 0.022785825 | -9.68E-06 | 0.042591235 | 1 | 0.974419575 | 1 | 0.977376329 | ENSG00000000 | 14 | ST7L |
| Frontal Cortex | Anxiety | ENSG0000016 | -0.00141706 | 0.346249071 | 0.001504484 | 0.02278674 | -0.00143404 | 0.340500181 | 0.001504477 | 0.022785801 | 1.70E-05 | 0.045078042 | 1 | 0.954139853 | 1 | 0.960761585 | ENSG00000000 | 16 | PAX6 |
| Frontal Cortex | Anxiety | ENSG0000016 | 0.002421912 | 0.200781702 | 0.001893109 | 0.022786685 | 0.002443168 | 0.19685549 | 0.0018931 | 0.022785746 | -2.13E-05 | 0.044139354 | 1 | 0.88859291 | 1 | 0.888056607 | ENSG00000000 | 16 | PAX6 |
| Frontal Cortex | Anxiety | ENSG0000016 | -0.00331862 | 0.063362458 | 0.001787428 | 0.022786553 | -0.00334337 | 0.061415623 | 0.001787421 | 0.022785612 | 2.47E-05 | 0.01732634 | 1 | 0.749913332 | 1 | 0.767504561 | ENSG00000000 | 10 | PIGQ |
| Frontal Cortex | Anxiety | ENSG0000017 | -0.00187678 | 0.132140734 | 0.001246435 | 0.022786639 | -0.00189552 | 0.128321583 | 0.001246434 | 0.022785699 | 1.87E-05 | 0.016103928 | 1 | 0.846710687 | 1 | 0.856073301 | ENSG00000000 | 10 | PIGQ |
| Frontal Cortex | Anxiety | ENSG0000017 | -0.00131684 | 0.572767819 | 0.002334911 | 0.022786781 | -0.00126791 | 0.587119674 | 0.002334936 | 0.022785846 | -4.89E-05 | 0.008031266 | 1 | 0.975684776 | 1 | 0.977376329 | ENSG00000000 | 6 | SS18L2 |
| Frontal Cortex | Anxiety | ENSG0000017 | -0.0032428 | 0.017578072 | 0.001365733 | 0.022786393 | -0.0032215 | 0.018334392 | 0.001365731 | 0.022785461 | -2.13E-05 | 0.011318474 | 1 | 0.732084207 | 1 | 0.754532031 | ENSG00000000 | 6 | SS18L2 |
| Frontal Cortex | Anxiety | ENSG0000017 | -2.37E-05 | 0.978709941 | 0.000888236 | 0.022786804 | -3.55E-05 | 0.968146776 | 0.000888237 | 0.022785867 | 1.18E-05 | 0.043360118 | 1 | 0.998018934 | 1 | 0.995959201 | ENSG00000000 | 8 | MGST1 |
| Frontal Cortex | Anxiety | ENSG0000017 | -0.00268009 | 0.159434529 | 0.001904855 | 0.02278666 | -0.00264671 | 0.164699105 | 0.001904874 | 0.022785727 | -3.34E-05 | 0.024813069 | 1 | 0.875855548 | 1 | 0.887019637 | ENSG00000000 | 8 | MGST1 |
| Frontal Cortex | Anxiety | ENSG0000017 | 0.000354801 | 0.756192016 | 0.00114273 | 0.022786797 | 0.000333022 | 0.770725948 | 0.001142735 | 0.022785861 | 2.18E-05 | 0.006778774 | 1 | 0.982406987 | 1 | 0.982577711 | ENSG00000000 | 8 | MGST1 |
| Frontal Cortex | Anxiety | ENSG0000017 | 0.000210836 | 0.775182647 | 0.000738213 | 0.022786799 | 9.63E-05 | 0.896338798 | 0.000739433 | 0.022785866 | 0.000114497 | 0.007363793 | 1 | 0.982406987 | 1 | 0.990584607 | ENSG00000000 | 8 | MGST1 |
| Frontal Cortex | Anxiety | ENSG0000017 | -0.00052977 | 0.696122232 | 0.001356422 | 0.022786793 | -0.00049772 | 0.713671781 | 0.001356442 | 0.022785858 | -3.20E-05 | 0.004749935 | 1 | 0.975684776 | 1 | 0.977376329 | ENSG00000000 | 12 | NFIX |
| Frontal Cortex | Anxiety | ENSG0000017 | 0.001681103 | 0.394381892 | 0.001973824 | 0.022786752 | 0.001714741 | 0.384989555 | 0.001973823 | 0.022785812 | -3.36E-05 | 0.007712329 | 1 | 0.965610146 | 1 | 0.969930908 | ENSG00000000 | 13 | PKD1 |
| Frontal Cortex | Anxiety | ENSG0000017 | 0.001888505 | 0.224272876 | 0.001554009 | 0.022786697 | 0.000552845 | 0.735580495 | 0.001637031 | 0.022785859 | 0.00133566 | 0.009471699 | 1 | 0.903356609 | 1 | 0.977376329 | ENSG00000000 | 12 | RHOBTB2 |
| Frontal Cortex | Anxiety | ENSG0000017 | 0.002477184 | 0.321957849 | 0.002501084 | 0.022786733 | 0.002510055 | 0.315578995 | 0.002501086 | 0.022785794 | -3.29E-05 | 0.044572755 | 1 | 0.954139853 | 1 | 0.954804417 | ENSG00000000 | 12 | RHOBTB2 |
| Frontal Cortex | Anxiety | ENSG0000018 | -0.00065356 | 0.443622431 | 0.000853112 | 0.022786762 | -0.00051863 | 0.543866942 | 0.000854446 | 0.022785841 | -0.00013493 | 0.004980815 | 1 | 0.974419575 | 1 | 0.977376329 | ENSG00000000 | 12 | RHOBTB2 |
| Frontal Cortex | Anxiety | ENSG0000018 | 0.000886678 | 0.73776822 | 0.002648297 | 0.022786796 | 0.001045561 | 0.693101781 | 0.002649341 | 0.022785856 | -0.00015888 | 0.037223651 | 1 | 0.975684776 | 1 | 0.977376329 | ENSG00000000 | 12 | RHOBTB2 |
| Frontal Cortex | Anxiety | ENSG0000018 | 0.000650041 | 0.718139232 | 0.00180093 | 0.022786795 | 0.000627224 | 0.727631744 | 0.001800931 | 0.022785859 | 2.28E-05 | 0.049949756 | 1 | 0.975684776 | 1 | 0.977376329 | ENSG00000000 | 12 | RHOBTB2 |
| Frontal Cortex | Anxiety | ENSG0000018 | -0.00078327 | 0.409062379 | 0.000948791 | 0.022786755 | -0.00076737 | 0.418640956 | 0.000948791 | 0.02278582 | -1.59E-05 | 0.008919793 | 1 | 0.96914464 | 1 | 0.97340047 | ENSG00000000 | 11 | REV3L |
| Frontal Cortex | Anxiety | ENSG0000018 | 0.002524252 | 0.161156428 | 0.001801498 | 0.022786661 | 0.002556162 | 0.155929551 | 0.001801511 | 0.022785721 | -3.19E-05 | 0.017951906 | 1 | 0.875855548 | 1 | 0.880880974 | ENSG00000000 | 11 | REV3L |
| Frontal Cortex | Anxiety | ENSG0000018 | -0.00024581 | 0.667574136 | 0.000572339 | 0.022786791 | -0.00025473 | 0.656269935 | 0.000572338 | 0.022785853 | 8.92E-06 | 0.014179861 | 1 | 0.975684776 | 1 | 0.977376329 | ENSG00000000 | 11 | REV3L |
| Frontal Cortex | Anxiety | ENSG0000018 | 0.001349495 | 0.055491434 | 0.000704691 | 0.022786537 | 0.001271631 | 0.071387995 | 0.000705283 | 0.02278563 | 7.79E-05 | 0.007725625 | 1 | 0.732084207 | 1 | 0.810468204 | ENSG00000000 | 7 | POMT2 |
| Frontal Cortex | Anxiety | ENSG0000018 | 0.002165015 | 0.051606185 | 0.001112311 | 0.022786528 | 0.002177732 | 0.050247901 | 0.001112305 | 0.022785588 | -1.27E-05 | 0.038402606 | 1 | 0.732084207 | 1 | 0.754532031 | ENSG00000000 | 7 | POMT2 |
| Frontal Cortex | Anxiety | ENSG0000018 | -0.00472453 | 0.030397065 | 0.002182346 | 0.022786462 | -0.00445133 | 0.041598579 | 0.00218468 | 0.022785564 | -0.0002732 | 0.007324394 | 1 | 0.732084207 | 1 | 0.754532031 | ENSG00000000 | 11 | MLXIPL |
| Frontal Cortex | Anxiety | ENSG0000018 | -0.00213808 | 0.112435166 | 0.001346951 | 0.022786621 | -0.00200504 | 0.1368258 | 0.00134773 | 0.022785706 | -0.00013303 | 0.004292134 | 1 | 0.831449405 | 1 | 0.874752649 | ENSG00000000 | 11 | MLXIPL |
| Frontal Cortex | Anxiety | ENSG0000018 | -0.00273998 | 0.265988262 | 0.002463236 | 0.022786714 | -0.00277339 | 0.260201912 | 0.002463226 | 0.022785775 | 3.34E-05 | 0.018319142 | 1 | 0.942197859 | 1 | 0.948868815 | ENSG00000000 | 11 | MLXIPL |
| Frontal Cortex | Anxiety | ENSG0000018 | 0.003311552 | 0.000100365 | 0.000851352 | 0.0227857 | 0.003246697 | 0.000137873 | 0.000851704 | 0.022784807 | 6.49E-05 | 0.009728474 | 0.20193404 | 0.13302423 | 0.277401391 | 0.195603561 | ENSG00000000 | 11 | MLXIPL |
| Frontal Cortex | Anxiety | ENSG0000019 | 0.002247675 | 0.040671388 | 0.001098102 | 0.022786499 | 0.000434765 | 0.759593584 | 0.001420731 | 0.022785861 | 0.00181291 | 0.044323519 | 1 | 0.732084207 | 1 | 0.982577711 | ENSG0000001 | 9 | DYRK4 |
| Frontal Cortex | Anxiety | ENSG0000019 | 0.001593673 | 0.429305232 | 0.002016332 | 0.022786759 | 0.001617345 | 0.42248109 | 0.002016325 | 0.022785821 | -2.37E-05 | 0.045513751 | 1 | 0.973801722 | 1 | 0.97340047 | ENSG0000001 | 9 | DYRK4 |
| Frontal Cortex | Anxiety | ENSG0000019 | 0.000472465 | 0.732886055 | 0.001384355 | 0.022786796 | 0.00041087 | 0.766645486 | 0.001384493 | 0.022785861 | 6.16E-05 | 0.004178642 | 1 | 0.975684776 | 1 | 0.982577711 | ENSG0000001 | 8 | NCAPD2 |
| Frontal Cortex | Anxiety | ENSG0000019 | 0.002278325 | 0.037518784 | 0.001095305 | 0.022786489 | 0.002120589 | 0.05317977 | 0.001096781 | 0.022785595 | 0.000157736 | 0.00590922 | 1 | 0.732084207 | 1 | 0.754532031 | ENSG0000001 | 8 | NCAPD2 |
| Frontal Cortex | Anxiety | ENSG0000019 | 7.22E-05 | 0.974621887 | 0.00227025 | 0.022786804 | 0.000107422 | 0.962260481 | 0.002270262 | 0.022785867 | -3.52E-05 | 0.030837662 | 1 | 0.997933454 | 1 | 0.995959201 | ENSG0000001 | 7 | ZNF200 |
| Frontal Cortex | Anxiety | ENSG0000019 | 0.004932974 | 0.009286334 | 0.001896316 | 0.02278631 | 0.004575881 | 0.01608039 | 0.001900994 | 0.022785444 | 0.000357092 | 0.007607114 | 1 | 0.584946729 | 1 | 0.754532031 | ENSG0000001 | 14 | HFE |
| Frontal Cortex | Anxiety | ENSG0000019 | -0.00123722 | 0.229013282 | 0.001028528 | 0.022786699 | -0.00124765 | 0.225110908 | 0.00102852 | 0.02278576 | 1.04E-05 | 0.048879873 | 1 | 0.903356609 | 1 | 0.908010471 | ENSG0000001 | 14 | HFE |
| Frontal Cortex | Anxiety | ENSG0000020 | 0.002489794 | 0.022231302 | 0.001088956 | 0.022786423 | 0.002502286 | 0.021568844 | 0.00108895 | 0.022785482 | -1.25E-05 | 0.036887442 | 1 | 0.732084207 | 1 | 0.754532031 | ENSG0000001 | 12 | SCMH1 |
| Frontal Cortex | Anxiety | ENSG0000020 | 0.001711118 | 0.398459247 | 0.002026485 | 0.022786752 | 0.001668783 | 0.410236488 | 0.002026504 | 0.022785818 | 4.23E-05 | 0.007084574 | 1 | 0.968693242 | 1 | 0.97340047 | ENSG0000001 | 12 | SCMH1 |
| Frontal Cortex | Anxiety | ENSG0000020 | -0.00050947 | 0.439808925 | 0.000659499 | 0.022786761 | -0.00050182 | 0.446705349 | 0.000659496 | 0.022785825 | -7.65E-06 | 0.038345267 | 1 | 0.974419575 | 1 | 0.977376329 | ENSG0000001 | 12 | SCMH1 |
| Frontal Cortex | Anxiety | ENSG0000020 | 5.32E-05 | 0.950263977 | 0.000852436 | 0.022786804 | -0.00024017 | 0.779698467 | 0.000858628 | 0.022785862 | 0.00029334 | 0.004428932 | 1 | 0.99542388 | 1 | 0.982928143 | ENSG0000001 | 7 | TSPAN9 |
| Frontal Cortex | Anxiety | ENSG0000020 | -0.00167947 | 0.021309575 | 0.000729422 | 0.022786417 | -0.00146681 | 0.045649139 | 0.00073391 | 0.022785576 | -0.00021265 | 0.008789578 | 1 | 0.732084207 | 1 | 0.754532031 | ENSG0000001 | 7 | TSPAN9 |
| Frontal Cortex | Anxiety | ENSG0000020 | -0.00217883 | 0.01432337 | 0.000889658 | 0.022786367 | -0.00209374 | 0.018675188 | 0.000890211 | 0.022785464 | -8.51E-05 | 0.007550402 | 1 | 0.719280595 | 1 | 0.754532031 | ENSG0000001 | 10 | BTBD7 |
| Frontal Cortex | Anxiety | ENSG0000020 | -0.00084907 | 0.419337293 | 0.001051389 | 0.022786757 | -0.00069634 | 0.508308766 | 0.001052715 | 0.022785836 | -0.00015273 | 0.004118809 | 1 | 0.96914464 | 1 | 0.977376329 | ENSG0000001 | 10 | BTBD7 |
| Frontal Cortex | Anxiety | ENSG0000020 | 0.000472855 | 0.580440733 | 0.000855472 | 0.022786782 | 6.08E-06 | 0.994438127 | 0.000871617 | 0.022785868 | 0.000466779 | 0.005207341 | 1 | 0.975684776 | 1 | 0.999485958 | ENSG0000001 | 5 | LARS2 |
| Frontal Cortex | Anxiety | ENSG0000020 | -0.00011992 | 0.854617841 | 0.000654485 | 0.022786802 | 5.74E-05 | 0.930467013 | 0.000657427 | 0.022785867 | -0.00017729 | 0.004411915 | 1 | 0.983816039 | 1 | 0.99421287 | ENSG0000001 | 5 | LARS2 |
| Frontal Cortex | Anxiety | ENSG0000020 | -0.00144326 | 0.290659578 | 0.00136585 | 0.022786723 | -0.00062169 | 0.656842386 | 0.001399338 | 0.022785853 | -0.00082157 | 0.006958894 | 1 | 0.947573237 | 1 | 0.977376329 | ENSG0000001 | 10 | PPP5C |
| Frontal Cortex | Anxiety | ENSG0000020 | 0.000368463 | 0.568084525 | 0.000645436 | 0.022786781 | 0.000490929 | 0.447842046 | 0.000646795 | 0.022785825 | -0.00012247 | 0.003638555 | 1 | 0.975684776 | 1 | 0.977376329 | ENSG0000001 | 10 | PPP5C |
| Frontal Cortex | Anxiety | ENSG0000020 | -0.00055727 | 0.443302484 | 0.000726915 | 0.022786762 | -0.00048378 | 0.506089914 | 0.000727549 | 0.022785835 | -7.35E-05 | 0.016736784 | 1 | 0.974419575 | 1 | 0.977376329 | ENSG0000001 | 9 | CEP68 |
| Frontal Cortex | Anxiety | ENSG0000020 | -0.00069286 | 0.161823252 | 0.000495266 | 0.022786662 | -0.0007675 | 0.121698353 | 0.000495904 | 0.022785693 | 7.46E-05 | 0.003240462 | 1 | 0.875855548 | 1 | 0.837078333 | ENSG0000001 | 9 | CEP68 |
| Frontal Cortex | Anxiety | ENSG0000020 | -0.00177941 | 0.462120803 | 0.002419778 | 0.022786765 | -0.00163015 | 0.500609303 | 0.002420295 | 0.022785834 | -0.00014926 | 0.004355564 | 1 | 0.975684776 | 1 | 0.977376329 | ENSG0000001 | 9 | CEP68 |
| Frontal Cortex | Anxiety | ENSG0000020 | 0.000642715 | 0.611855146 | 0.001266614 | 0.022786786 | 0.000392543 | 0.757247243 | 0.001269964 | 0.022785861 | 0.000250172 | 0.006858578 | 1 | 0.975684776 | 1 | 0.982577711 | ENSG0000001 | 9 | CEP68 |
| Frontal Cortex | Anxiety | ENSG0000020 | -0.00090556 | 0.228544805 | 0.000752051 | 0.022786699 | -0.000888 | 0.237710677 | 0.000752079 | 0.022785766 | -1.76E-05 | 0.029498361 | 1 | 0.903356609 | 1 | 0.914522914 | ENSG0000001 | 9 | CEP68 |
| Frontal Cortex | Anxiety | ENSG0000020 | 0.000650262 | 0.493830512 | 0.000950355 | 0.02278677 | 0.00063913 | 0.50125306 | 0.00095035 | 0.022785835 | 1.11E-05 | 0.03271001 | 1 | 0.975684776 | 1 | 0.977376329 | ENSG0000001 | 9 | CEP68 |
| Frontal Cortex | Anxiety | ENSG0000020 | -0.00024334 | 0.876445632 | 0.001565136 | 0.022786803 | -0.00026764 | 0.864224858 | 0.001565152 | 0.022785865 | 2.43E-05 | 0.047845025 | 1 | 0.983816039 | 1 | 0.988452375 | ENSG0000001 | 9 | CEP68 |
| Frontal Cortex | Anxiety | ENSG0000020 | 0.000254506 | 0.798944713 | 0.000999186 | 0.0227868 | 0.000380148 | 0.703896682 | 0.001000216 | 0.022785857 | -0.00012564 | 0.006128383 | 1 | 0.982406987 | 1 | 0.977376329 | ENSG0000001 | 9 | CEP68 |
| Frontal Cortex | Anxiety | ENSG0000020 | 0.000132302 | 0.799002092 | 0.000519567 | 0.0227868 | 0.000194687 | 0.708124991 | 0.000520031 | 0.022785857 | -6.24E-05 | 0.005002832 | 1 | 0.982406987 | 1 | 0.977376329 | ENSG0000001 | 9 | ZBTB32 |
| Frontal Cortex | Anxiety | ENSG0000020 | 0.000814736 | 0.599973227 | 0.001553537 | 0.022786784 | 0.000635488 | 0.682748424 | 0.001554841 | 0.022785855 | 0.000179248 | 0.005405565 | 1 | 0.975684776 | 1 | 0.977376329 | ENSG0000001 | 11 | ERCC1 |
| Frontal Cortex | Anxiety | ENSG0000020 | -0.00086795 | 0.348900723 | 0.000926581 | 0.02278674 | -0.00085424 | 0.356569648 | 0.000926582 | 0.022785805 | -1.37E-05 | 0.026220436 | 1 | 0.954139853 | 1 | 0.960761585 | ENSG0000001 | 11 | ERCC1 |
| Frontal Cortex | Anxiety | ENSG0000021 | 0.001110979 | 0.559983023 | 0.001906059 | 0.02278678 | 0.001138983 | 0.5501339 | 0.001906057 | 0.022785841 | -2.80E-05 | 0.018402999 | 1 | 0.975684776 | 1 | 0.977376329 | ENSG0000001 | 11 | ERCC1 |
| Frontal Cortex | Anxiety | ENSG0000021 | 0.00167847 | 0.265053192 | 0.001505991 | 0.022786714 | 0.001660794 | 0.2701153 | 0.001505981 | 0.022785779 | 1.77E-05 | 0.025779033 | 1 | 0.942197859 | 1 | 0.953162532 | ENSG0000001 | 4 | EXTL3 |
| Frontal Cortex | Anxiety | ENSG0000021 | -0.00046817 | 0.69199774 | 0.001181802 | 0.022786793 | -0.00044954 | 0.70366218 | 0.001181807 | 0.022785857 | -1.86E-05 | 0.024335182 | 1 | 0.975684776 | 1 | 0.977376329 | ENSG0000001 | 4 | EXTL3 |
| Frontal Cortex | Anxiety | ENSG0000021 | -0.00177734 | 0.282788088 | 0.001654755 | 0.02278672 | -0.00175815 | 0.288016478 | 0.001654749 | 0.022785785 | -1.92E-05 | 0.045089374 | 1 | 0.947573237 | 1 | 0.953162532 | ENSG0000001 | 4 | EXTL3 |
| Frontal Cortex | Anxiety | ENSG0000021 | -0.00116763 | 0.00919111 | 0.000448246 | 0.022786309 | -0.00110432 | 0.013882058 | 0.000448853 | 0.022785426 | -6.33E-05 | 0.007039041 | 1 | 0.584946729 | 1 | 0.746228219 | ENSG0000001 | 4 | EXTL3 |
| Frontal Cortex | Anxiety | ENSG0000021 | 0.002847555 | 0.00062163 | 0.000832124 | 0.02278595 | 0.00274994 | 0.00096139 | 0.000832906 | 0.022785072 | 9.76E-05 | 0.007411476 | 1 | 0.250143712 | 1 | 0.386863274 | ENSG0000001 | 9 | UBR7 |

| | | | | | | | | | | | | | | | | | | | |
|---|---|---|---|---|---|---|---|---|---|---|---|---|---|---|---|---|---|---|---|
| Frontal Cortex | Anxiety | ENSG0000022 | 0.00340446 | 0.106440562 | 0.002108801 | 0.022786614 | 0.004098277 | 0.053546468 | 0.002122915 | 0.022785595 | -0.00069382 | 0.004593122 | 1 | 0.821912463 | 1 | 0.754532031 | ENSG0000001 | 9 | UBR7 |
| Frontal Cortex | Anxiety | ENSG0000022 | 0.000411405 | 0.693448637 | 0.001043698 | 0.022786793 | 0.000433926 | 0.677590002 | 0.001043707 | 0.022785855 | -2.25E-05 | 0.00503671 | 1 | 0.975684776 | 1 | 0.977376329 | ENSG0000001 | 9 | UBR7 |
| Frontal Cortex | Anxiety | ENSG0000022 | 0.000708882 | 0.174053646 | 0.000521506 | 0.02278667 | 0.0007354 | 0.158774508 | 0.000521598 | 0.022785723 | -2.62E-05 | 0.011368776 | 1 | 0.879106816 | 1 | 0.884914984 | ENSG0000001 | 11 | SLC25A39 |
| Frontal Cortex | Anxiety | ENSG0000022 | -0.00069432 | 0.575802763 | 0.00124091 | 0.022786782 | -0.00072881 | 0.55700379 | 0.001240953 | 0.022785842 | 3.45E-05 | 0.008024142 | 1 | 0.975684776 | 1 | 0.977376329 | ENSG0000001 | 11 | SLC25A39 |
| Frontal Cortex | Anxiety | ENSG0000022 | 0.003143329 | 0.011914227 | 0.001249987 | 0.022786343 | 0.003126268 | 0.012383679 | 0.001249991 | 0.022785411 | 1.71E-05 | 0.045625721 | 1 | 0.665872926 | 1 | 0.746228219 | ENSG0000001 | 5 | ANGEL1 |
| Frontal Cortex | Anxiety | ENSG0000022 | -0.00138976 | 0.00168778 | 0.000442554 | 0.022786084 | -0.00135659 | 0.002183157 | 0.00044273 | 0.022785182 | -3.32E-05 | 0.009510701 | 1 | 0.40325044 | 1 | 0.403493216 | ENSG0000001 | 12 | DDX11 |
| Frontal Cortex | Anxiety | ENSG0000022 | 0.000983473 | 0.365420892 | 0.001086608 | 0.022786745 | 0.000954338 | 0.379810403 | 0.00108664 | 0.022785811 | 2.91E-05 | 0.007025277 | 1 | 0.956084178 | 1 | 0.96652945 | ENSG0000001 | 14 | TACC3 |
| Frontal Cortex | Anxiety | ENSG0000022 | 0.002528038 | 0.072136506 | 0.001405823 | 0.022786568 | 0.002511692 | 0.073996017 | 0.001405816 | 0.022785634 | 1.63E-05 | 0.034692946 | 1 | 0.791500747 | 1 | 0.810468204 | ENSG0000001 | 14 | TACC3 |
| Frontal Cortex | Anxiety | ENSG0000022 | 0.00108296 | 0.077756942 | 0.000613972 | 0.022786577 | 0.001042709 | 0.089568037 | 0.000614194 | 0.022785657 | 4.03E-05 | 0.01764599 | 1 | 0.791500747 | 1 | 0.825400182 | ENSG0000001 | 6 | ZC3H3 |
| Frontal Cortex | Anxiety | ENSG0000022 | 0.000178353 | 0.905736818 | 0.001506132 | 0.022786803 | 0.000148752 | 0.921327479 | 0.001506167 | 0.022785867 | 2.96E-05 | 0.036089497 | 1 | 0.990619017 | 1 | 0.993905606 | ENSG0000001 | 6 | ZC3H3 |
| Frontal Cortex | Anxiety | ENSG0000022 | 0.000627512 | 0.731457563 | 0.001828481 | 0.022786796 | 0.000650612 | 0.72197405 | 0.001828475 | 0.022785858 | -2.31E-05 | 0.030370357 | 1 | 0.975684776 | 1 | 0.977376329 | ENSG0000001 | 15 | MTMR11 |
| Frontal Cortex | Anxiety | ENSG0000022 | 0.000279118 | 0.785939666 | 0.001027728 | 0.022786799 | 0.000445909 | 0.664936246 | 0.001029556 | 0.022785854 | -0.00016679 | 0.006843497 | 1 | 0.982406987 | 1 | 0.977376329 | ENSG0000001 | 15 | MTMR11 |
| Frontal Cortex | Anxiety | ENSG0000022 | -0.00178635 | 0.045690762 | 0.000893956 | 0.022786513 | -0.00092375 | 0.343455307 | 0.000975071 | 0.022785802 | -0.0008626 | 0.026745374 | 1 | 0.732084207 | 1 | 0.960761585 | ENSG0000001 | 14 | CCDC88C |
| Frontal Cortex | Anxiety | ENSG0000023 | 0.000233975 | 0.733090794 | 0.000686108 | 0.022786796 | 0.000174883 | 0.798909256 | 0.000686465 | 0.022785863 | 5.91E-05 | 0.00882665 | 1 | 0.975684776 | 1 | 0.98622766 | ENSG0000001 | 13 | NUDCD3 |
| Frontal Cortex | Anxiety | ENSG0000023 | 0.003838973 | 0.082661641 | 0.002212087 | 0.022786585 | 0.003522945 | 0.111718496 | 0.002214963 | 0.022785683 | 0.000316028 | 0.00543926 | 1 | 0.800053168 | 1 | 0.837078333 | ENSG0000001 | 12 | GLT8D1 |
| Frontal Cortex | Anxiety | ENSG0000023 | -0.00112304 | 0.570938501 | 0.001981821 | 0.022786781 | -0.00116551 | 0.556471406 | 0.001981846 | 0.022785842 | 4.25E-05 | 0.008639752 | 1 | 0.975684776 | 1 | 0.977376329 | ENSG0000001 | 12 | GLT8D1 |
| Frontal Cortex | Anxiety | ENSG0000023 | -0.00092114 | 0.597997287 | 0.001746948 | 0.022786784 | -0.00094577 | 0.588243436 | 0.001746956 | 0.022785846 | 2.46E-05 | 0.045896343 | 1 | 0.975684776 | 1 | 0.977376329 | ENSG0000001 | 12 | GLT8D1 |
| Frontal Cortex | Anxiety | ENSG0000023 | -0.00053993 | 0.277256272 | 0.00049694 | 0.022786718 | -0.00021355 | 0.676272504 | 0.000511424 | 0.022785855 | -0.00032638 | 0.006938025 | 1 | 0.947278828 | 1 | 0.977376329 | ENSG0000001 | 12 | GLT8D1 |
| Frontal Cortex | Anxiety | ENSG0000023 | 0.000526402 | 0.459986999 | 0.000712443 | 0.022786765 | 0.00106049 | 0.14790449 | 0.000732901 | 0.022785715 | -0.00053409 | 0.001904388 | 1 | 0.975684776 | 1 | 0.875203335 | ENSG0000001 | 8 | RUFY3 |
| Frontal Cortex | Anxiety | ENSG0000023 | -0.00170154 | 0.003400861 | 0.00058093 | 0.022786178 | -0.00162303 | 0.005263069 | 0.00058163 | 0.022785299 | -7.85E-05 | 0.006298994 | 1 | 0.40325044 | 1 | 0.423571825 | ENSG0000001 | 8 | RUFY3 |
| Frontal Cortex | Anxiety | ENSG0000023 | 0.001683966 | 0.016351332 | 0.000701363 | 0.022786384 | 0.001613505 | 0.021510853 | 0.000701857 | 0.022785482 | 7.05E-05 | 0.00833203 | 1 | 0.728293434 | 1 | 0.754532031 | ENSG0000001 | 12 | SLC11A1 |
| Frontal Cortex | Anxiety | ENSG0000023 | 0.007050282 | 0.002804551 | 0.002359208 | 0.022786152 | 0.007010924 | 0.002961505 | 0.002359207 | 0.022785223 | 3.94E-05 | 0.007990495 | 1 | 0.40325044 | 1 | 0.403493216 | ENSG0000001 | 12 | SLC11A1 |
| Frontal Cortex | Anxiety | ENSG0000023 | 0.003738441 | 0.100369137 | 0.002275276 | 0.022786607 | 0.003708292 | 0.103139824 | 0.002275272 | 0.022785674 | 3.01E-05 | 0.030326105 | 1 | 0.812485947 | 1 | 0.827410035 | ENSG0000001 | 8 | AGPS |
| Frontal Cortex | Anxiety | ENSG0000023 | 0.001755171 | 0.154781445 | 0.001233563 | 0.022786657 | 0.001602871 | 0.194280494 | 0.001234856 | 0.022785745 | 0.0001523 | 0.007586534 | 1 | 0.870476788 | 1 | 0.888056607 | ENSG0000001 | 8 | AGPS |
| Frontal Cortex | Anxiety | ENSG0000023 | 0.001789929 | 0.4096897 | 0.002171082 | 0.022786755 | 0.001765746 | 0.416042545 | 0.002171069 | 0.022785819 | 2.42E-05 | 0.03867666 | 1 | 0.96914464 | 1 | 0.97340047 | ENSG0000001 | 8 | AGPS |
| Frontal Cortex | Anxiety | ENSG0000023 | -0.00039518 | 0.576062316 | 0.00070676 | 0.022786782 | -0.00038788 | 0.583131703 | 0.000706755 | 0.022785846 | -7.30E-06 | 0.041324788 | 1 | 0.975684776 | 1 | 0.977376329 | ENSG0000001 | 7 | TTC27 |
| Frontal Cortex | Anxiety | ENSG0000023 | -0.00574087 | 0.005226492 | 0.002055633 | 0.022786235 | -0.0062324 | 0.002507776 | 0.002062052 | 0.022785201 | 0.000491525 | 0.002590716 | 1 | 0.420628052 | 1 | 0.403493216 | ENSG0000001 | 7 | TTC27 |
| Frontal Cortex | Anxiety | ENSG0000023 | 0.000131543 | 0.799809709 | 0.000518716 | 0.0227868 | 0.000107697 | 0.835536434 | 0.000518758 | 0.022785864 | 2.38E-05 | 0.001214006 | 1 | 0.982406987 | 1 | 0.988452375 | ENSG0000001 | 9 | MARCO |
| Frontal Cortex | Anxiety | ENSG0000023 | -0.00105112 | 0.250054349 | 0.000913845 | 0.022786708 | 0.000145744 | 0.886486041 | 0.00102096 | 0.022785866 | -0.00119687 | 0.008566956 | 1 | 0.928246033 | 1 | 0.988543414 | ENSG0000002 | 8 | MRE11A |
| Frontal Cortex | Anxiety | ENSG0000023 | -0.00057248 | 0.327496095 | 0.000584659 | 0.022786735 | -0.00052809 | 0.36661055 | 0.000584917 | 0.022785808 | -4.44E-05 | 0.012555932 | 1 | 0.954139853 | 1 | 0.96212728 | ENSG0000002 | 8 | MRE11A |
| Frontal Cortex | Anxiety | ENSG0000023 | 0.002646373 | 0.333968256 | 0.002739082 | 0.022786736 | 0.002607978 | 0.341028189 | 0.002739079 | 0.022785801 | 3.84E-05 | 0.023483132 | 1 | 0.954139853 | 1 | 0.960761585 | ENSG0000002 | 8 | MRE11A |
| Frontal Cortex | Anxiety | ENSG0000024 | 0.001172059 | 0.034360162 | 0.000553948 | 0.022786478 | 0.00101794 | 0.067654485 | 0.000557071 | 0.022785624 | 0.000154119 | 0.008998844 | 1 | 0.732084207 | 1 | 0.79602821 | ENSG0000002 | 8 | MRE11A |
| Frontal Cortex | Anxiety | ENSG0000024 | 0.000338957 | 0.449775401 | 0.00044848 | 0.022786763 | 0.000429495 | 0.339362953 | 0.000449533 | 0.022785801 | -9.05E-05 | 0.00337355 | 1 | 0.974440587 | 1 | 0.960761585 | ENSG0000002 | 8 | MRE11A |
| Frontal Cortex | Anxiety | ENSG0000024 | -0.00047223 | 0.671107713 | 0.001112109 | 0.022786791 | -0.00048659 | 0.661722535 | 0.001112104 | 0.022785854 | 1.44E-05 | 0.021246046 | 1 | 0.975684776 | 1 | 0.977376329 | ENSG0000002 | 11 | NLRP2 |
| Frontal Cortex | Anxiety | ENSG0000024 | -0.00030411 | 0.798897065 | 0.001193629 | 0.0227868 | 0.000908729 | 0.496739946 | 0.001337096 | 0.022785834 | -0.00121284 | 0.044152274 | 1 | 0.982406987 | 1 | 0.977376329 | ENSG0000002 | 11 | NLRP2 |
| Frontal Cortex | Anxiety | ENSG0000024 | 0.00164555 | 0.427513769 | 0.002073906 | 0.022786759 | 0.001622586 | 0.433988725 | 0.002073896 | 0.022785823 | 2.30E-05 | 0.047371012 | 1 | 0.971929609 | 1 | 0.976777864 | ENSG0000002 | 11 | NLRP2 |
| Frontal Cortex | Anxiety | ENSG0000024 | 0.001430028 | 0.161030923 | 0.001020271 | 0.022786661 | 0.001116198 | 0.277349501 | 0.001027528 | 0.022785781 | 0.00031383 | 0.010143294 | 1 | 0.875855548 | 1 | 0.953162532 | ENSG0000002 | 11 | NLRP2 |
| Frontal Cortex | Anxiety | ENSG0000024 | -0.0004797 | 0.767759709 | 0.001624417 | 0.022786798 | -0.00045675 | 0.778573436 | 0.001624413 | 0.022785862 | -2.30E-05 | 0.019579855 | 1 | 0.982406987 | 1 | 0.982741376 | ENSG0000002 | 11 | NLRP2 |
| Frontal Cortex | Anxiety | ENSG0000025 | -0.00099198 | 0.28305256 | 0.000924069 | 0.02278672 | -0.00100716 | 0.275749065 | 0.000924071 | 0.022785781 | 1.52E-05 | 0.014590584 | 1 | 0.947573237 | 1 | 0.953162532 | ENSG0000002 | 7 | ZDHHC6 |
| Frontal Cortex | Anxiety | ENSG0000025 | 0.00175418 | 0.506469444 | 0.002640462 | 0.022786772 | 0.001796139 | 0.496355638 | 0.002640465 | 0.022785834 | -4.20E-05 | 0.015844074 | 1 | 0.975684776 | 1 | 0.977376329 | ENSG0000002 | 7 | ZDHHC6 |
| Frontal Cortex | Anxiety | ENSG0000025 | 0.001228883 | 0.590747472 | 0.002285215 | 0.022786783 | 0.00119707 | 0.60039329 | 0.002285204 | 0.022785847 | 3.18E-05 | 0.013497013 | 1 | 0.975684776 | 1 | 0.977376329 | ENSG0000002 | 7 | ZDHHC6 |
| Frontal Cortex | Anxiety | ENSG0000025 | 0.000222541 | 0.858289945 | 0.001246371 | 0.022786802 | 0.000239192 | 0.847811562 | 0.001246366 | 0.022785865 | -1.67E-05 | 0.022643683 | 1 | 0.983816039 | 1 | 0.988452375 | ENSG0000002 | 12 | RNH1 |
| Frontal Cortex | Anxiety | ENSG0000026 | 3.56E-05 | 0.989910452 | 0.002817794 | 0.022786804 | -4.92E-07 | 0.999860687 | 0.002817793 | 0.022785868 | 3.61E-05 | 0.044321121 | 1 | 0.999849312 | 1 | 0.999860687 | ENSG0000002 | 12 | RNH1 |
| Frontal Cortex | Anxiety | ENSG0000026 | -0.00184394 | 0.04152817 | 0.00090468 | 0.022786501 | -0.0017996 | 0.046709808 | 0.000904808 | 0.022785579 | -4.43E-05 | 0.006415781 | 1 | 0.732084207 | 1 | 0.754532031 | ENSG0000002 | 4 | GLRX2 |
| Frontal Cortex | Anxiety | ENSG0000026 | -7.28E-06 | 0.994909617 | 0.001141739 | 0.022786805 | -8.65E-05 | 0.939638677 | 0.001142044 | 0.022785867 | 7.92E-05 | 0.003792643 | 1 | 0.999973246 | 1 | 0.99421287 | ENSG0000002 | 4 | GLRX2 |
| Frontal Cortex | Anxiety | ENSG0000026 | 0.001810401 | 0.417404523 | 0.00223249 | 0.022786756 | 0.001844367 | 0.408723414 | 0.002232503 | 0.022785818 | -3.40E-05 | 0.03652099 | 1 | 0.96914464 | 1 | 0.97340047 | ENSG0000002 | 8 | HSF2 |
| Frontal Cortex | Anxiety | ENSG0000026 | 0.00277256 | 0.174732002 | 0.002042916 | 0.02278667 | 0.002817854 | 0.167805398 | 0.00204297 | 0.022785729 | -4.53E-05 | 0.021955597 | 1 | 0.879106816 | 1 | 0.887019637 | ENSG0000002 | 8 | TYMP |
| Frontal Cortex | Anxiety | ENSG0000027 | 0.003144131 | 0.026563494 | 0.001417627 | 0.022786445 | 0.002594357 | 0.070549172 | 0.001434634 | 0.022785629 | 0.000549775 | 0.012621141 | 1 | 0.732084207 | 1 | 0.810468204 | ENSG0000002 | 9 | KPNA6 |
| Frontal Cortex | Anxiety | ENSG0000027 | 0.000457827 | 0.642796061 | 0.000987135 | 0.022786789 | 0.000495467 | 0.615752297 | 0.000987222 | 0.022785849 | -3.76E-05 | 0.009889497 | 1 | 0.975684776 | 1 | 0.977376329 | ENSG0000002 | 9 | KPNA6 |
| Frontal Cortex | Anxiety | ENSG0000027 | -0.00013675 | 0.875561581 | 0.000873226 | 0.022786803 | -0.00017462 | 0.841514651 | 0.00087331 | 0.022785865 | 3.79E-05 | 0.004522349 | 1 | 0.983816039 | 1 | 0.988452375 | ENSG0000002 | 11 | RNASET2 |
| Frontal Cortex | Anxiety | ENSG0000027 | -0.00135395 | 0.032065978 | 0.000631625 | 0.022786469 | -0.0012987 | 0.039888434 | 0.000631998 | 0.022785559 | -5.53E-05 | 0.012296222 | 1 | 0.732084207 | 1 | 0.754532031 | ENSG0000002 | 11 | RNASET2 |
| Frontal Cortex | Anxiety | ENSG0000027 | 0.000300676 | 0.648754158 | 0.00066011 | 0.022786789 | 0.000294313 | 0.655700365 | 0.000660105 | 0.022785853 | 6.36E-06 | 0.046797677 | 1 | 0.975684776 | 1 | 0.977376329 | ENSG0000002 | 12 | BTN3A1 |
| Frontal Cortex | Depression | ENSG0000007 | 0.000849714 | 0.830405725 | 0.003967261 | 0.082019693 | 0.001003516 | 0.800339573 | 0.003967911 | 0.082018088 | -0.0001538 | 0.037428979 | 1 | 0.996212728 | 1 | 0.989691378 | ENSG0000002 | 12 | BTN3A1 |
| Frontal Cortex | Depression | ENSG0000010 | -0.00012119 | 0.639867634 | 0.00218258 | 0.082019648 | -0.00094096 | 0.6664265 | 0.002182912 | 0.082018057 | -8.02E-05 | 0.04101176 | 1 | 0.965289072 | 1 | 0.966991696 | ENSG0000002 | 12 | BTN3A1 |
| Frontal Cortex | Depression | ENSG0000011 | 0.00411902 | 0.365432429 | 0.004551084 | 0.082019491 | 0.004775356 | 0.295289028 | 0.004562772 | 0.082017819 | -0.00065634 | 0.044748914 | 1 | 0.948993265 | 1 | 0.950067467 | ENSG0000002 | 12 | BTN3A1 |
| Frontal Cortex | Depression | ENSG0000014 | 0.000558198 | 0.910493102 | 0.004965439 | 0.082019702 | -0.00083409 | 0.867846695 | 0.005012747 | 0.082018098 | 0.001392291 | 0.042822757 | 1 | 0.996212728 | 1 | 0.997190658 | ENSG0000002 | 12 | BTN3A1 |
| Frontal Cortex | Depression | ENSG0000017 | -0.00090294 | 0.837987644 | 0.004416042 | 0.082019694 | -0.00083151 | 0.850650558 | 0.004416136 | 0.082018096 | -7.14E-05 | 0.039515495 | 1 | 0.996212728 | 1 | 0.997190658 | ENSG0000002 | 12 | BTN3A1 |
| Frontal Cortex | Depression | ENSG0000023 | 8.45E-05 | 0.931365821 | 0.000980814 | 0.082019703 | 5.50E-05 | 0.955299205 | 0.000980901 | 0.082018104 | 2.95E-05 | 0.031775457 | 1 | 0.996212728 | 1 | 0.997190658 | ENSG0000002 | 12 | RABEP1 |
| Frontal Cortex | Psychosis | ENSG0000000 | -0.00029054 | 0.597567361 | 0.000550367 | 0.003450919 | -0.00029959 | 0.586207109 | 0.000550369 | 0.003450787 | 9.05E-06 | 0.011670804 | 1 | 0.996311095 | 1 | 0.996713278 | ENSG0000003 | 8 | BAK1 |
| Frontal Cortex | Psychosis | ENSG0000004 | 0.001035876 | 0.087529149 | 0.000606278 | 0.00345089 | 0.001042842 | 0.085419436 | 0.000606277 | 0.003450758 | -6.97E-06 | 0.049673707 | 1 | 0.836091108 | 1 | 0.838360512 | ENSG0000003 | 8 | BAK1 |
| Frontal Cortex | Psychosis | ENSG0000004 | -0.00053133 | 0.320930092 | 0.000535314 | 0.003450911 | -0.00052229 | 0.329233129 | 0.000535318 | 0.00345078 | -9.04E-06 | 0.022327112 | 1 | 0.950491103 | 1 | 0.951230466 | ENSG0000003 | 8 | BAK1 |
| Frontal Cortex | Psychosis | ENSG0000006 | -0.00035189 | 0.686368618 | 0.000871462 | 0.00345092 | -0.0003615 | 0.67827475 | 0.000871459 | 0.003450789 | 9.61E-06 | 0.045564666 | 1 | 0.996561629 | 1 | 0.996713278 | ENSG0000003 | 8 | BAK1 |
| Frontal Cortex | Psychosis | ENSG0000007 | -2.10E-05 | 0.983010244 | 0.000987196 | 0.003450922 | 0.000140332 | 0.887115744 | 0.000988568 | 0.003450791 | -0.00016135 | 0.002082002 | 1 | 0.998166036 | 1 | 0.997262509 | ENSG0000003 | 8 | BAK1 |
| Frontal Cortex | Psychosis | ENSG0000008 | 0.00016595 | 0.707970535 | 0.000443026 | 0.00345092 | 4.68E-05 | 0.916517051 | 0.000446575 | 0.00345079 | 0.000119139 | 0.03418111 | 1 | 0.996561629 | 1 | 0.998402312 | ENSG0000003 | 8 | BAK1 |
| Frontal Cortex | Psychosis | ENSG0000008 | 0.001545284 | 0.069031419 | 0.000849888 | 0.003450885 | 0.001558187 | 0.066743452 | 0.000849888 | 0.003450753 | -1.29E-05 | 0.012227998 | 1 | 0.808945891 | 1 | 0.822157607 | ENSG0000003 | 8 | BAK1 |
| Frontal Cortex | Psychosis | ENSG0000009 | -0.00039103 | 0.452293367 | 0.000520261 | 0.003450916 | -0.00038177 | 0.46306844 | 0.000520263 | 0.003450785 | -9.26E-06 | 0.008269305 | 1 | 0.977814099 | 1 | 0.981288252 | ENSG0000003 | 8 | BAK1 |
| Frontal Cortex | Psychosis | ENSG0000010 | -0.00089973 | 0.289165822 | 0.000848834 | 0.003450909 | -0.00088927 | 0.294806301 | 0.000848831 | 0.003450778 | -1.05E-05 | 0.025318208 | 1 | 0.945359859 | 1 | 0.950813143 | ENSG0000003 | 8 | BAK1 |
| Frontal Cortex | Psychosis | ENSG0000010 | 9.93E-05 | 0.82532789 | 0.000449771 | 0.003450921 | 7.63E-05 | 0.865340488 | 0.000449834 | 0.00345079 | 2.30E-05 | 0.004263594 | 1 | 0.998166036 | 1 | 0.997262509 | ENSG0000003 | 6 | CENPQ |
| Frontal Cortex | Psychosis | ENSG0000010 | 0.000711906 | 0.186236767 | 0.000538589 | 0.003450902 | 0.001014368 | 0.066828881 | 0.000553444 | 0.003450753 | -0.00030246 | 0.017601872 | 1 | 0.927900596 | 1 | 0.822157607 | ENSG0000003 | 6 | CENPQ |
| Frontal Cortex | Psychosis | ENSG0000011 | -0.00042363 | 0.346154772 | 0.000449677 | 0.003450912 | -0.00042993 | 0.339033358 | 0.000449679 | 0.003450781 | 6.30E-06 | 0.036118326 | 1 | 0.955368176 | 1 | 0.951230466 | ENSG0000003 | 11 | PNPLA6 |
| Frontal Cortex | Psychosis | ENSG0000011 | 0.000428378 | 0.330307713 | 0.000440041 | 0.003450911 | 0.00043469 | 0.323233534 | 0.000440042 | 0.00345078 | -6.31E-06 | 0.035542609 | 1 | 0.950491103 | 1 | 0.951230466 | ENSG0000003 | 13 | IFT88 |
| Frontal Cortex | Psychosis | ENSG0000011 | -0.00069075 | 0.440465293 | 0.000895437 | 0.003450915 | -0.00067908 | 0.448228899 | 0.000895439 | 0.003450784 | -1.17E-05 | 0.046916702 | 1 | 0.977814099 | 1 | 0.981288252 | ENSG0000003 | 13 | IFT88 |
| Frontal Cortex | Psychosis | ENSG0000011 | -0.00084028 | 0.354374363 | 0.00090729 | 0.003450912 | -0.00082831 | 0.36127113 | 0.000907287 | 0.003450781 | -1.20E-05 | 0.022029854 | 1 | 0.958388154 | 1 | 0.959429486 | ENSG0000003 | 10 | CHPF2 |
| Frontal Cortex | Psychosis | ENSG0000011 | 0.000834638 | 0.373577898 | 0.000938014 | 0.003450913 | 0.000647806 | 0.490916699 | 0.000940414 | 0.003450785 | 0.000186832 | 0.005559264 | 1 | 0.966142504 | 1 | 0.981288252 | ENSG0000003 | 8 | UBA6 |
| Frontal Cortex | Psychosis | ENSG0000011 | 0.000160692 | 0.852689003 | 0.000865383 | 0.003450921 | 0.000173016 | 0.841536493 | 0.000865387 | 0.00345079 | -1.23E-05 | 0.040209742 | 1 | 0.998166036 | 1 | 0.997262509 | ENSG0000003 | 8 | UBA6 |
| Frontal Cortex | Psychosis | ENSG0000011 | 1.58E-05 | 0.971230903 | 0.000439227 | 0.003450922 | 9.26E-06 | 0.983182819 | 0.000439229 | 0.003450791 | 6.58E-06 | 0.023795209 | 1 | 0.998166036 | 1 | 0.998725145 | ENSG0000003 | 8 | UBA6 |
| Frontal Cortex | Psychosis | ENSG0000012 | -2.12E-05 | 0.966989939 | 0.000513474 | 0.003450922 | -1.53E-05 | 0.976274916 | 0.000513472 | 0.003450791 | -5.98E-06 | 0.040335864 | 1 | 0.998166036 | 1 | 0.998725145 | ENSG0000003 | 8 | UBA6 |
| Frontal Cortex | Psychosis | ENSG0000012 | -0.00012325 | 0.887755902 | 0.000873237 | 0.003450922 | -0.0001336 | 0.878402432 | 0.000873236 | 0.00345079 | 1.03E-05 | 0.049252857 | 1 | 0.998166036 | 1 | 0.997262509 | ENSG0000003 | 9 | PIAS1 |
| Frontal Cortex | Psychosis | ENSG0000012 | -0.00076831 | 0.163241576 | 0.000551055 | 0.0034509 | -0.00076171 | 0.166888609 | 0.000551053 | 0.003450769 | -6.61E-06 | 0.029184867 | 1 | 0.923358565 | 1 | 0.921921371 | ENSG0000003 | 7 | ASTE1 |
| Frontal Cortex | Psychosis | ENSG0000012 | -2.90E-06 | 0.994160924 | 0.000395666 | 0.003450922 | 2.53E-06 | 0.994893984 | 0.000395667 | 0.003450791 | -5.43E-06 | 0.034186907 | 1 | 0.998166036 | 1 | 0.998725145 | ENSG0000003 | 7 | ASTE1 |
| Frontal Cortex | Psychosis | ENSG0000012 | -0.00052707 | 0.408771881 | 0.00063806 | 0.003450914 | -0.00053581 | 0.401053378 | 0.000638063 | 0.003450783 | 8.73E-06 | 0.043713025 | 1 | 0.972923465 | 1 | 0.980415059 | ENSG0000003 | 7 | ASTE1 |
| Frontal Cortex | Psychosis | ENSG0000012 | -5.06E-05 | 0.842601867 | 0.000254893 | 0.003450921 | -4.69E-05 | 0.854070945 | 0.000254893 | 0.003450791 | -3.73E-06 | 0.021403313 | 1 | 0.998166036 | 1 | 0.997262509 | ENSG0000003 | 7 | ASTE1 |
| Frontal Cortex | Psychosis | ENSG0000012 | -6.23E-05 | 0.75235166 | 0.000197402 | 0.003450921 | -6.56E-05 | 0.739831071 | 0.000197403 | 0.003450789 | 3.26E-06 | 0.021830657 | 1 | 0.998166036 | 1 | 0.997262509 | ENSG0000003 | 17 | SH3YL1 |
| Frontal Cortex | Psychosis | ENSG0000013 | -2.08E-05 | 0.979456915 | 0.000806072 | 0.003450922 | 0.000136006 | 0.86633608 | 0.000808049 | 0.003450791 | -0.00015676 | 0.005701566 | 1 | 0.998166036 | 1 | 0.997262509 | ENSG0000003 | 17 | SH3YL1 |
| Frontal Cortex | Psychosis | ENSG0000013 | 0.000355428 | 0.583877953 | 0.000648911 | 0.003450919 | 0.000345614 | 0.594308328 | 0.000648914 | 0.003450787 | 9.81E-06 | 0.026455123 | 1 | 0.996311095 | 1 | 0.996713278 | ENSG0000003 | 17 | SH3YL1 |
| Frontal Cortex | Psychosis | ENSG0000013 | -0.00021706 | 0.827240435 | 0.000994593 | 0.003450921 | -0.00020298 | 0.838290504 | 0.000994594 | 0.00345079 | -1.41E-05 | 0.027734961 | 1 | 0.998166036 | 1 | 0.997262509 | ENSG0000003 | 17 | SH3YL1 |
| Frontal Cortex | Psychosis | ENSG0000014 | -0.00113548 | 0.068888237 | 0.00062418 | 0.003450885 | -0.00111279 | 0.070760158 | 0.000624179 | 0.003450754 | -7.58E-06 | 0.037940997 | 1 | 0.808945891 | 1 | 0.822157607 | ENSG0000003 | 17 | SH3YL1 |
| Frontal Cortex | Psychosis | ENSG0000014 | -0.00012272 | 0.657345575 | 0.000276657 | 0.00345092 | -0.0001259 | 0.649053219 | 0.000276656 | 0.003450788 | 3.18E-06 | 0.043627053 | 1 | 0.996561629 | 1 | 0.996713278 | ENSG0000003 | 8 | TUBG2 |
| Frontal Cortex | Psychosis | ENSG0000014 | 0.00098384 | 0.009553684 | 0.000379627 | 0.003450847 | 0.000991076 | 0.009038044 | 0.000379631 | 0.003450715 | -7.24E-06 | 0.013316193 | 1 | 0.665039569 | 1 | 0.717109479 | ENSG0000003 | 10 | NSUN2 |
| Frontal Cortex | Psychosis | ENSG0000014 | -0.00048458 | 0.22776842 | 0.000401764 | 0.003450906 | -0.00125836 | 0.016336415 | 0.000524027 | 0.003450727 | 0.00077378 | 0.02145739 | 1 | 0.927900596 | 1 | 0.777758283 | ENSG0000003 | 10 | NSUN2 |
| Frontal Cortex | Psychosis | ENSG0000014 | 0.00018513 | 0.831334611 | 0.000869196 | 0.003450921 | 0.000196153 | 0.821456325 | 0.000869194 | 0.00345079 | -1.10E-05 | 0.029551218 | 1 | 0.998166036 | 1 | 0.997262509 | ENSG0000003 | 10 | NSUN2 |
| Frontal Cortex | Psychosis | ENSG0000014 | -0.00034506 | 0.524252747 | 0.000541858 | 0.003450917 | -0.00033696 | 0.534030447 | 0.000541862 | 0.003450786 | -8.09E-06 | 0.044551059 | 1 | 0.984848837 | 1 | 0.981288252 | ENSG0000003 | 7 | MFAP3 |
| Frontal Cortex | Psychosis | ENSG0000014 | -0.00042785 | 0.455280786 | 0.000573034 | 0.003450916 | -0.00041937 | 0.464265575 | 0.000573035 | 0.003450785 | -8.48E-06 | 0.021227359 | 1 | 0.977814099 | 1 | 0.981288252 | ENSG0000003 | 7 | MFAP3 |
| Frontal Cortex | Psychosis | ENSG0000014 | 0.000216129 | 0.737403522 | 0.000644596 | 0.003450921 | 0.00022437 | 0.727781265 | 0.000644596 | 0.003450789 | -8.24E-06 | 0.035169529 | 1 | 0.998166036 | 1 | 0.997262509 | ENSG0000003 | 9 | MRI1 |
| Frontal Cortex | Psychosis | ENSG0000015 | 8.44E-05 | 0.843680341 | 0.000428069 | 0.003450921 | 8.98E-05 | 0.83378565 | 0.00042807 | 0.00345079 | -5.42E-06 | 0.048392084 | 1 | 0.998166036 | 1 | 0.997262509 | ENSG0000003 | 9 | MRI1 |
| Frontal Cortex | Psychosis | ENSG0000015 | -0.00112013 | 0.031082671 | 0.000519534 | 0.00345087 | -0.00112711 | 0.030047933 | 0.000519534 | 0.003450738 | 6.99E-06 | 0.031193657 | 1 | 0.808945891 | 1 | 0.822157607 | ENSG0000003 | 9 | MRI1 |
| Frontal Cortex | Psychosis | ENSG0000015 | -0.00115132 | 0.118977756 | 0.00073846 | 0.003450895 | -0.00115998 | 0.116225509 | 0.000738458 | 0.003450763 | 8.66E-06 | 0.036591804 | 1 | 0.863116848 | 1 | 0.867850871 | ENSG0000003 | 9 | MRI1 |
| Frontal Cortex | Psychosis | ENSG0000015 | 0.000553047 | 0.600051249 | 0.001054775 | 0.003450919 | 0.000565985 | 0.591547706 | 0.001054772 | 0.003450787 | -1.29E-05 | 0.03194102 | 1 | 0.996311095 | 1 | 0.996713278 | ENSG0000003 | 9 | MRI1 |
| Frontal Cortex | Psychosis | ENSG0000015 | -0.00027937 | 0.706409982 | 0.000741672 | 0.00345092 | -0.00026768 | 0.718167106 | 0.000741673 | 0.003450789 | -1.17E-05 | 0.013786431 | 1 | 0.996561629 | 1 | 0.997262509 | ENSG0000003 | 9 | MRI1 |
| Frontal Cortex | Psychosis | ENSG0000016 | -0.0004378 | 0.538778079 | 0.000712259 | 0.003450918 | -0.00044791 | 0.529442849 | 0.000712259 | 0.003450786 | 1.01E-05 | 0.020800896 | 1 | 0.984848837 | 1 | 0.981288252 | ENSG0000003 | 8 | BOD1L1 |
| Frontal Cortex | Psychosis | ENSG0000016 | 2.95E-05 | 0.976089332 | 0.000984129 | 0.003450922 | 4.31E-05 | 0.965048216 | 0.000984132 | 0.003450791 | -1.36E-05 | 0.03397524 | 1 | 0.998166036 | 1 | 0.998725145 | ENSG0000003 | 8 | BOD1L1 |
| Frontal Cortex | Psychosis | ENSG0000016 | 0.00056532 | 0.395460948 | 0.000665272 | 0.003450914 | 0.00057465 | 0.387708389 | 0.000665272 | 0.003450782 | -9.33E-06 | 0.021319699 | 1 | 0.972923465 | 1 | 0.980415059 | ENSG0000003 | 8 | BOD1L1 |
| Frontal Cortex | Psychosis | ENSG0000016 | 0.000285992 | 0.779100132 | 0.001019606 | 0.003450921 | 0.000252102 | 0.804728582 | 0.001019697 | 0.00345079 | 3.39E-05 | 0.023669728 | 1 | 0.998166036 | 1 | 0.997262509 | ENSG0000003 | 8 | BOD1L1 |
| Frontal Cortex | Psychosis | ENSG0000016 | -0.00083975 | 0.175057161 | 0.000619225 | 0.003450901 | -0.00083263 | 0.178744497 | 0.000619223 | 0.003450771 | -7.13E-06 | 0.035132202 | 1 | 0.926881598 | 1 | 0.926773198 | ENSG0000004 | 14 | PHLPP2 |
| Frontal Cortex | Psychosis | ENSG0000016 | 0.000108597 | 0.711306382 | 0.000293425 | 0.00345092 | 0.000104543 | 0.721627233 | 0.000293426 | 0.003450789 | 4.05E-06 | 0.045309882 | 1 | 0.996561629 | 1 | 0.997262509 | ENSG0000004 | 14 | PHLPP2 |
| Frontal Cortex | Psychosis | ENSG0000016 | 0.000128955 | 0.847543829 | 0.000670758 | 0.003450921 | 0.000137997 | 0.836999982 | 0.000670758 | 0.00345079 | -9.04E-06 | 0.032823364 | 1 | 0.998166036 | 1 | 0.997262509 | ENSG0000004 | 10 | CTNS |
| Frontal Cortex | Psychosis | ENSG0000016 | -0.00066245 | 0.385280745 | 0.000763013 | 0.003450914 | -0.00067644 | 0.375333451 | 0.000763016 | 0.003450782 | 1.40E-05 | 0.007581829 | 1 | 0.972923465 | 1 | 0.967521421 | ENSG0000004 | 10 | CTNS |
| Frontal Cortex | Psychosis | ENSG0000016 | 0.000534482 | 0.584033371 | 0.000976218 | 0.003450919 | 0.000606983 | 0.5342356 | 0.000976556 | 0.003450786 | -7.25E-05 | 0.006073889 | 1 | 0.996311095 | 1 | 0.981288252 | ENSG0000004 | 10 | CTNS |
| Frontal Cortex | Psychosis | ENSG0000016 | -0.00025432 | 0.372978877 | 0.000285463 | 0.003450913 | -0.00026031 | 0.36184469 | 0.000285468 | 0.003450781 | 5.99E-06 | 0.017697628 | 1 | 0.966142504 | 1 | 0.959429486 | ENSG0000004 | 10 | CTNS |
| Frontal Cortex | Psychosis | ENSG0000017 | -0.0011044 | 0.063521229 | 0.000595193 | 0.003450884 | -0.00111438 | 0.061169784 | 0.0005952 | 0.003450752 | 9.98E-06 | 0.032712187 | 1 | 0.808945891 | 1 | 0.822157607 | ENSG0000004 | 11 | INPP4A |
| Frontal Cortex | Psychosis | ENSG0000017 | -0.00052228 | 0.54818745 | 0.000869769 | 0.003450918 | -0.00051158 | 0.556414375 | 0.000869765 | 0.003450787 | -1.07E-05 | 0.026071471 | 1 | 0.984848837 | 1 | 0.988534574 | ENSG0000004 | 5 | RAB27B |
| Frontal Cortex | Psychosis | ENSG0000017 | -0.00147371 | 0.070717963 | 0.000815425 | 0.003450886 | -0.00146357 | 0.072676223 | 0.000815422 | 0.003450755 | -1.01E-05 | 0.028973026 | 1 | 0.808945891 | 1 | 0.822157607 | ENSG0000004 | 5 | RAB27B |
| Frontal Cortex | Psychosis | ENSG0000017 | 0.000990441 | 0.32654215 | 0.001009516 | 0.003450911 | 0.000974448 | 0.334414779 | 0.001009518 | 0.00345078 | 1.60E-05 | 0.013551308 | 1 | 0.950491103 | 1 | 0.951230466 | ENSG0000004 | 5 | RAB27B |
| Frontal Cortex | Psychosis | ENSG0000017 | 0.001700046 | 0.105879471 | 0.001051359 | 0.003450893 | 0.001717054 | 0.102435269 | 0.001051366 | 0.003450761 | -1.70E-05 | 0.024974624 | 1 | 0.858989904 | 1 | 0.857620674 | ENSG0000004 | 5 | RAB27B |
| Frontal Cortex | Psychosis | ENSG0000017 | 0.000108965 | 0.835463963 | 0.000524631 | 0.003450921 | -8.34E-05 | 0.875215488 | 0.000530813 | 0.00345079 | 0.000192323 | 0.017357496 | 1 | 0.998166036 | 1 | 0.997262509 | ENSG0000004 | 5 | RAB27B |
| Frontal Cortex | Psychosis | ENSG0000017 | -0.00062449 | 0.031359569 | 0.000290127 | 0.003450871 | -0.00062026 | 0.032526596 | 0.000290127 | 0.00345074 | -4.24E-06 | 0.020736229 | 1 | 0.808945891 | 1 | 0.822157607 | ENSG0000004 | 6 | LSG1 |
| Frontal Cortex | Psychosis | ENSG0000017 | -0.00040664 | 0.507188738 | 0.000613127 | 0.003450917 | -0.0003974 | 0.516885589 | 0.000613128 | 0.003450786 | -9.24E-06 | 0.019928216 | 1 | 0.984848837 | 1 | 0.981288252 | ENSG0000004 | 6 | LSG1 |
| Frontal Cortex | Psychosis | ENSG0000017 | 0.000164122 | 0.679197971 | 0.000396854 | 0.00345092 | 0.000172274 | 0.664218832 | 0.000396858 | 0.003450788 | -8.15E-06 | 0.00507065 | 1 | 0.996561629 | 1 | 0.996713278 | ENSG0000004 | 6 | LSG1 |
| Frontal Cortex | Psychosis | ENSG0000017 | -0.00031729 | 0.525581744 | 0.000499861 | 0.003450917 | -0.00031162 | 0.533016047 | 0.00049986 | 0.003450786 | -5.68E-06 | 0.041685869 | 1 | 0.984848837 | 1 | 0.981288252 | ENSG0000004 | 6 | MED17 |
| Frontal Cortex | Psychosis | ENSG0000018 | -0.00015847 | 0.820245433 | 0.000697404 | 0.003450921 | -0.00016891 | 0.80862818 | 0.000697408 | 0.00345079 | 1.04E-05 | 0.032146343 | 1 | 0.998166036 | 1 | 0.997262509 | ENSG0000004 | 6 | MED17 |
| Frontal Cortex | Psychosis | ENSG0000018 | -0.00015131 | 0.765051191 | 0.000506306 | 0.003450921 | -0.00015741 | 0.75588239 | 0.000506305 | 0.00345079 | 6.09E-06 | 0.035682112 | 1 | 0.998166036 | 1 | 0.997262509 | ENSG0000004 | 11 | CAPG |
| Frontal Cortex | Psychosis | ENSG0000018 | -0.00171916 | 0.01873771 | 0.000731332 | 0.003450861 | -0.00173106 | 0.017934329 | 0.000731335 | 0.003450728 | 1.19E-05 | 0.016270967 | 1 | 0.808945891 | 1 | 0.801863787 | ENSG0000004 | 11 | CAPG |
| Frontal Cortex | Psychosis | ENSG0000018 | 1.48E-05 | 0.959913324 | 0.000293653 | 0.003450922 | 1.86E-05 | 0.949375677 | 0.000293654 | 0.003450791 | -3.88E-06 | 0.049171365 | 1 | 0.998166036 | 1 | 0.998725145 | ENSG0000004 | 11 | TRIT1 |
| Frontal Cortex | Psychosis | ENSG0000018 | -0.00087902 | 0.14503557 | 0.000603184 | 0.003450898 | -0.00086791 | 0.150187319 | 0.000603187 | 0.003450768 | -1.11E-05 | 0.007063024 | 1 | 0.896136687 | 1 | 0.910171345 | ENSG0000004 | 11 | TRIT1 |
| Frontal Cortex | Psychosis | ENSG0000019 | 0.001604639 | 0.032605294 | 0.000750912 | 0.003450871 | 0.001613304 | 0.031677371 | 0.000750909 | 0.003450739 | -8.67E-06 | 0.035522111 | 1 | 0.808945891 | 1 | 0.822157607 | ENSG0000004 | 4 | CUL7 |
| Frontal Cortex | Psychosis | ENSG0000020 | 0.000979452 | 0.215887855 | 0.000791452 | 0.003450905 | 0.000963183 | 0.2236172 | 0.000791461 | 0.003450774 | 1.63E-05 | 0.009479759 | 1 | 0.927900596 | 1 | 0.932819306 | ENSG0000004 | 4 | CUL7 |
| Frontal Cortex | Psychosis | ENSG0000020 | -6.42E-05 | 0.936245952 | 0.000803081 | 0.003450922 | -5.34E-05 | 0.946947546 | 0.00080308 | 0.003450791 | -1.08E-05 | 0.020873077 | 1 | 0.998166036 | 1 | 0.998725145 | ENSG0000004 | 4 | CUL7 |

| | | | | | | | | | | | | | | | | | | | |
|---|---|---|---|---|---|---|---|---|---|---|---|---|---|---|---|---|---|---|---|
| Frontal Cortex | Psychosis | ENSG0000020 | 0.00060096 | 0.093681918 | 0.000358505 | 0.003450891 | 0.000595532 | 0.096687433 | 0.000358509 | 0.00345076 | 5.43E-06 | 0.04655432 | 1 | 0.83985898 | 1 | 0.853224191 | ENSG0000004 | 6 | ROS1 |
| Frontal Cortex | Psychosis | ENSG0000021 | 0.001360824 | 0.011970313 | 0.000541507 | 0.003450852 | 0.001353645 | 0.012427606 | 0.000541506 | 0.003450721 | 7.18E-06 | 0.025829149 | 1 | 0.752633419 | 1 | 0.777758283 | ENSG0000004 | 13 | TSPAN17 |
| Frontal Cortex | Psychosis | ENSG0000021 | 0.000310686 | 0.72382603 | 0.000879257 | 0.00345092 | 0.000296297 | 0.736129127 | 0.000879261 | 0.003450789 | 1.44E-05 | 0.017234691 | 1 | 0.997218736 | 1 | 0.997262509 | ENSG0000004 | 13 | TSPAN17 |
| Frontal Cortex | Psychosis | ENSG0000021 | -0.00038275 | 0.547651034 | 0.000636546 | 0.003450918 | -0.00037518 | 0.555596598 | 0.000636544 | 0.003450787 | -7.57E-06 | 0.03274261 | 1 | 0.984848837 | 1 | 0.988534574 | ENSG0000004 | 13 | TSPAN17 |
| Frontal Cortex | Psychosis | ENSG0000021 | -0.00104374 | 0.164916784 | 0.00075158 | 0.0034509 | -0.00103059 | 0.17030746 | 0.000751584 | 0.00345077 | -1.32E-05 | 0.011447255 | 1 | 0.923358565 | 1 | 0.921921371 | ENSG0000004 | 10 | LMO3 |
| Frontal Cortex | Psychosis | ENSG0000022 | -0.00119908 | 0.255482632 | 0.001054478 | 0.003450908 | -0.00116524 | 0.269163009 | 0.001054523 | 0.003450777 | -3.38E-05 | 0.003648478 | 1 | 0.936726396 | 1 | 0.940460229 | ENSG0000004 | 5 | MRPS10 |
| Frontal Cortex | Psychosis | ENSG0000022 | 8.37E-05 | 0.897969922 | 0.000652825 | 0.003450922 | 9.08E-05 | 0.889326007 | 0.000652822 | 0.00345079 | -7.14E-06 | 0.041834778 | 1 | 0.998166036 | 1 | 0.997262509 | ENSG0000004 | 5 | MRPS10 |
| Frontal Cortex | Psychosis | ENSG0000022 | -0.00042755 | 0.627330112 | 0.000880658 | 0.003450919 | -0.00045355 | 0.60655815 | 0.000880695 | 0.003450788 | 2.60E-05 | 0.007726708 | 1 | 0.996311095 | 1 | 0.996713278 | ENSG0000004 | 5 | MRPS10 |
| Frontal Cortex | Psychosis | ENSG0000022 | 0.000117075 | 0.856056059 | 0.00064541 | 0.003450922 | 0.000123662 | 0.848052678 | 0.000645406 | 0.00345079 | -6.59E-06 | 0.047904991 | 1 | 0.998166036 | 1 | 0.997262509 | ENSG0000004 | 5 | MRPS10 |
| Frontal Cortex | Psychosis | ENSG0000023 | 0.0001241 | 0.85257802 | 0.000667813 | 0.003450921 | 0.000113082 | 0.865537085 | 0.000667819 | 0.00345079 | 1.10E-05 | 0.028633498 | 1 | 0.998166036 | 1 | 0.997262509 | ENSG0000004 | 9 | GUCA1A |
| Frontal Cortex | Psychosis | ENSG0000023 | 4.99E-05 | 0.910441045 | 0.000443364 | 0.003450922 | 5.48E-05 | 0.901715732 | 0.000443363 | 0.00345079 | -4.88E-06 | 0.049461462 | 1 | 0.998166036 | 1 | 0.997262509 | ENSG0000004 | 9 | COL9A2 |
| Frontal Cortex | Psychosis | ENSG0000023 | 0.001414568 | 0.169366355 | 0.001029341 | 0.003450901 | 0.001402259 | 0.173106353 | 0.001029337 | 0.00345077 | 1.23E-05 | 0.027700677 | 1 | 0.923358565 | 1 | 0.923878279 | ENSG0000004 | 10 | ADAMTS6 |
| Frontal Cortex | Psychosis | ENSG0000023 | -4.44E-05 | 0.922690436 | 0.000457362 | 0.003450922 | -5.13E-05 | 0.910633486 | 0.000457367 | 0.00345079 | 6.95E-06 | 0.049682364 | 1 | 0.998166036 | 1 | 0.997262509 | ENSG0000004 | 9 | UTS2 |
| Frontal Cortex | Psychosis | ENSG0000023 | 0.000256357 | 0.644716668 | 0.000555951 | 0.00345092 | 0.000249654 | 0.653389051 | 0.00055595 | 0.003450788 | 6.70E-06 | 0.032691909 | 1 | 0.996561629 | 1 | 0.996713278 | ENSG0000004 | 4 | RCN1 |
| Frontal Cortex | Psychosis | ENSG0000024 | -0.00085765 | 0.210635215 | 0.000685121 | 0.003450904 | -0.0008674 | 0.205498017 | 0.000685122 | 0.003450773 | 9.74E-06 | 0.027634975 | 1 | 0.927900596 | 1 | 0.930914275 | ENSG0000004 | 4 | RCN1 |
| Frontal Cortex | Psychosis | ENSG0000024 | 0.000374972 | 0.420128162 | 0.00046511 | 0.003450915 | -0.00020123 | 0.699329419 | 0.00052101 | 0.003450789 | 0.000576199 | 0.014128249 | 1 | 0.972923465 | 1 | 0.996713278 | ENSG0000004 | 4 | RCN1 |
| Frontal Cortex | Psychosis | ENSG0000024 | -4.55E-05 | 0.901217195 | 0.000366298 | 0.003450922 | -5.08E-05 | 0.88968202 | 0.00036663 | 0.00345079 | 5.34E-06 | 0.030211179 | 1 | 0.998166036 | 1 | 0.997262509 | ENSG0000004 | 4 | RCN1 |
| Frontal Cortex | Psychosis | ENSG0000024 | 0.000510793 | 0.451007291 | 0.00067768 | 0.003450916 | 0.000500805 | 0.459907476 | 0.000677679 | 0.003450785 | 9.99E-06 | 0.015114468 | 1 | 0.977814099 | 1 | 0.981288252 | ENSG0000005 | 13 | JKAMP |
| Frontal Cortex | Psychosis | ENSG0000024 | 0.000463222 | 0.322405601 | 0.000468124 | 0.003450911 | 0.000468134 | 0.317298046 | 0.000468121 | 0.003450779 | -4.91E-06 | 0.045412107 | 1 | 0.950491103 | 1 | 0.951230466 | ENSG0000005 | 13 | JKAMP |
| Frontal Cortex | Psychosis | ENSG0000024 | -0.0015644 | 0.131867887 | 0.001038238 | 0.003450897 | -0.00157549 | 0.129148493 | 0.001038233 | 0.003450765 | 1.11E-05 | 0.041118086 | 1 | 0.88836141 | 1 | 0.880701241 | ENSG0000005 | 13 | JKAMP |
| Frontal Cortex | Psychosis | ENSG0000025 | -0.00032155 | 0.718343741 | 0.000891534 | 0.00345092 | -0.00030753 | 0.730136343 | 0.000891543 | 0.003450789 | -1.40E-05 | 0.039716551 | 1 | 0.997197795 | 1 | 0.997262509 | ENSG0000005 | 13 | JKAMP |
| Frontal Cortex | Psychosis | ENSG0000025 | -0.00038865 | 0.622257224 | 0.000788888 | 0.003450919 | -0.0003796 | 0.630380932 | 0.000788885 | 0.003450788 | -9.04E-06 | 0.034079705 | 1 | 0.996311095 | 1 | 0.996713278 | ENSG0000005 | 13 | JKAMP |
| Frontal Cortex | Psychosis | ENSG0000025 | 0.000185358 | 0.828704354 | 0.000856698 | 0.003450921 | 0.000202759 | 0.812910727 | 0.000856704 | 0.00345079 | -1.74E-05 | 0.005654761 | 1 | 0.998166036 | 1 | 0.997262509 | ENSG0000005 | 7 | MCUR1 |
| Frontal Cortex | Psychosis | ENSG0000025 | -0.00025807 | 0.50970444 | 0.00039143 | 0.003450917 | -0.00025299 | 0.518065277 | 0.00039143 | 0.003450786 | -5.08E-06 | 0.038987733 | 1 | 0.984848837 | 1 | 0.981288252 | ENSG0000005 | 7 | MCUR1 |
| Frontal Cortex | Psychosis | ENSG0000025 | -0.00022772 | 0.804239059 | 0.000918725 | 0.003450921 | -0.00021808 | 0.812365513 | 0.00091872 | 0.00345079 | -9.64E-06 | 0.043556868 | 1 | 0.998166036 | 1 | 0.997262509 | ENSG0000005 | 7 | MCUR1 |
| Frontal Cortex | Psychosis | ENSG0000025 | 0.001410317 | 0.095992939 | 0.000847239 | 0.003450891 | 0.00142379 | 0.092861634 | 0.000847243 | 0.003450759 | -1.35E-05 | 0.022218146 | 1 | 0.83985898 | 1 | 0.841610847 | ENSG0000005 | 9 | LIMA1 |
| Frontal Cortex | Psychosis | ENSG0000025 | -7.63E-05 | 0.811319956 | 0.000319526 | 0.003450921 | -7.09E-05 | 0.824424848 | 0.000319527 | 0.00345079 | -5.39E-06 | 0.013632246 | 1 | 0.998166036 | 1 | 0.997262509 | ENSG0000005 | 9 | LIMA1 |
| Frontal Cortex | Psychosis | ENSG0000026 | 5.56E-05 | 0.767826235 | 0.000188287 | 0.003450921 | 5.86E-05 | 0.755734729 | 0.000188288 | 0.00345079 | -2.99E-06 | 0.017086706 | 1 | 0.998166036 | 1 | 0.997262509 | ENSG0000005 | 13 | LAMC3 |
| Frontal Cortex | Psychosis | ENSG0000026 | -6.59E-05 | 0.882395797 | 0.000445388 | 0.003450922 | -7.20E-05 | 0.871626393 | 0.00044539 | 0.00345079 | 6.08E-06 | 0.043123136 | 1 | 0.998166036 | 1 | 0.997262509 | ENSG0000005 | 13 | LAMC3 |
| Frontal Cortex | Psychosis | ENSG0000026 | 0.000686475 | 0.291776158 | 0.00065116 | 0.00345091 | 0.000693384 | 0.286944636 | 0.000651157 | 0.003450778 | -6.91E-06 | 0.047269745 | 1 | 0.945359859 | 1 | 0.950813143 | ENSG0000005 | 13 | LAMC3 |
| Frontal Cortex | Psychosis | ENSG0000026 | 0.000258311 | 0.565565045 | 0.000449553 | 0.003450918 | 0.000250119 | 0.577959404 | 0.000449556 | 0.003450787 | 8.19E-06 | 0.012048449 | 1 | 0.994682579 | 1 | 0.996713278 | ENSG0000005 | 13 | LAMC3 |
| Frontal Cortex | Psychosis | ENSG0000026 | -0.00031324 | 0.756990259 | 0.001012288 | 0.003450921 | -0.00030028 | 0.766742952 | 0.001012286 | 0.00345079 | -1.30E-05 | 0.031016199 | 1 | 0.998166036 | 1 | 0.997262509 | ENSG0000005 | 4 | PIK3CB |
| Frontal Cortex | Psychosis | ENSG0000026 | 0.000170849 | 0.857576499 | 0.000952017 | 0.003450922 | 0.000182185 | 0.848238186 | 0.000952016 | 0.00345079 | -1.13E-05 | 0.047583617 | 1 | 0.998166036 | 1 | 0.997262509 | ENSG0000005 | 6 | HEBP2 |
| Frontal Cortex | Psychosis | ENSG0000026 | 0.000229686 | 0.49679294 | 0.000337999 | 0.003450917 | 0.000234713 | 0.487423901 | 0.000338002 | 0.003450785 | -5.03E-06 | 0.042953134 | 1 | 0.984848837 | 1 | 0.981288252 | ENSG0000005 | 4 | MRTO4 |
| Frontal Cortex | Psychosis | ENSG0000027 | 0.000445757 | 0.437403132 | 0.000573995 | 0.003450915 | 0.00045534 | 0.427614816 | 0.000573996 | 0.003450784 | -9.58E-06 | 0.011705634 | 1 | 0.977814099 | 1 | 0.981288252 | ENSG0000005 | 6 | ANAPC4 |
| Frontal Cortex | Psychosis | ENSG0000027 | 0.000258752 | 0.7606504 | 0.000849405 | 0.003450921 | 0.000269958 | 0.750620945 | 0.000849404 | 0.00345079 | -1.12E-05 | 0.026887819 | 1 | 0.998166036 | 1 | 0.997262509 | ENSG0000005 | 6 | ANAPC4 |
| Frontal Cortex | Psychosis | ENSG0000027 | 0.000472711 | 0.514160911 | 0.000724601 | 0.003450917 | 0.000482024 | 0.505906575 | 0.000724602 | 0.003450786 | -9.31E-06 | 0.03986657 | 1 | 0.984848837 | 1 | 0.981288252 | ENSG0000005 | 6 | ANAPC4 |
| Frontal Cortex | Psychosis | ENSG0000027 | 0.000113999 | 0.894385147 | 0.000858699 | 0.003450922 | 0.00010065 | 0.906692469 | 0.00085874 | 0.00345079 | 1.33E-05 | 0.027936389 | 1 | 0.998166036 | 1 | 0.997262509 | ENSG0000005 | 8 | OPN3 |
| Frontal Cortex | Psychosis | ENSG0000027 | -0.00034584 | 0.637003034 | 0.000732887 | 0.003450919 | -0.00035839 | 0.624835416 | 0.0007329 | 0.003450788 | 1.26E-05 | 0.045204485 | 1 | 0.996311095 | 1 | 0.996713278 | ENSG0000005 | 8 | OPN3 |
| Frontal Cortex | Psychosis | ENSG0000027 | 0.001012984 | 0.255577722 | 0.000890999 | 0.003450908 | 0.000998235 | 0.26256727 | 0.000891006 | 0.003450777 | 1.47E-05 | 0.025312849 | 1 | 0.936726396 | 1 | 0.940460229 | ENSG0000005 | 11 | SDCCAG8 |
| Pancreas | Prediabetes | ENSG0000000 | -0.0002299 | 0.720258092 | 0.000641988 | 0.002176049 | 0.00104959 | 0.194553328 | 0.000809102 | 0.002175991 | -0.00127949 | 0.009370972 | 1 | 0.980154834 | 1 | 0.933545778 | ENSG0000005 | 12 | KIF1B |
| Pancreas | Prediabetes | ENSG0000000 | 0.000251681 | 0.361082334 | 0.000275571 | 0.002176046 | 0.000644747 | 0.052702938 | 0.000332798 | 0.002175982 | -0.00039307 | 0.035158106 | 1 | 0.931097229 | 1 | 0.931226496 | ENSG0000005 | 11 | CHRDL2 |
| Pancreas | Prediabetes | ENSG0000000 | -0.00015918 | 0.684791813 | 0.000399033 | 0.002176049 | -0.00015616 | 0.695538391 | 0.000399039 | 0.002175998 | -5.82E-06 | 0.043940017 | 1 | 0.975612224 | 1 | 0.975423656 | ENSG0000005 | 11 | CHRDL2 |
| Pancreas | Prediabetes | ENSG0000001 | 0.000700088 | 0.205361186 | 0.000552806 | 0.002176042 | 0.000719322 | 0.193245348 | 0.000552882 | 0.002175991 | -1.92E-05 | 0.043481357 | 1 | 0.931097229 | 1 | 0.933545778 | ENSG0000005 | 11 | CHRDL2 |
| Pancreas | Prediabetes | ENSG0000001 | 0.000183833 | 0.622093117 | 0.000372977 | 0.002176049 | 0.00018917 | 0.612022559 | 0.000372977 | 0.002175998 | -5.34E-06 | 0.04028706 | 1 | 0.968763492 | 1 | 0.964723844 | ENSG0000005 | 12 | GALC |
| Pancreas | Prediabetes | ENSG0000001 | -0.00047917 | 0.022136681 | 0.000209425 | 0.002176026 | -0.00047512 | 0.023291382 | 0.000209432 | 0.002175976 | -4.04E-06 | 0.044910734 | 1 | 0.926755475 | 1 | 0.931226496 | ENSG0000005 | 13 | FAM114A2 |
| Pancreas | Prediabetes | ENSG0000001 | -6.67E-05 | 0.801680766 | 0.000265705 | 0.002176049 | -7.03E-05 | 0.791296448 | 0.000265707 | 0.002175999 | 3.58E-06 | 0.030832539 | 1 | 0.982693856 | 1 | 0.981881037 | ENSG0000005 | 13 | FAM114A2 |
| Pancreas | Prediabetes | ENSG0000001 | 0.000143018 | 0.751837849 | 0.000452278 | 0.002176049 | 0.000149306 | 0.741311999 | 0.000452282 | 0.002175998 | -6.29E-06 | 0.034124062 | 1 | 0.980154834 | 1 | 0.981881037 | ENSG0000005 | 8 | GINM1 |
| Pancreas | Prediabetes | ENSG0000001 | 0.000196678 | 0.671133419 | 0.000463216 | 0.002176049 | 0.000204634 | 0.658663576 | 0.000463225 | 0.002175998 | -7.96E-06 | 0.028904513 | 1 | 0.975612224 | 1 | 0.973329377 | ENSG0000005 | 8 | GINM1 |
| Pancreas | Prediabetes | ENSG0000001 | 0.000106021 | 0.65426481 | 0.000236736 | 0.002176049 | 0.000109305 | 0.644286216 | 0.000236738 | 0.002175998 | -3.28E-06 | 0.029105158 | 1 | 0.974245837 | 1 | 0.9720762 | ENSG0000005 | 12 | MRPL43 |
| Pancreas | Prediabetes | ENSG0000001 | -0.00074998 | 0.009188245 | 0.000287903 | 0.002176019 | -0.00075525 | 0.00871063 | 0.00028791 | 0.002175968 | 5.27E-06 | 0.029768736 | 1 | 0.926755475 | 1 | 0.931226496 | ENSG0000005 | 11 | ITIH4 |
| Pancreas | Prediabetes | ENSG0000001 | 0.000491648 | 0.364190175 | 0.000541814 | 0.002176046 | 0.000502118 | 0.354080923 | 0.000541832 | 0.002175995 | -1.05E-05 | 0.041562892 | 1 | 0.931097229 | 1 | 0.933545778 | ENSG0000005 | 11 | ITIH4 |
| Pancreas | Prediabetes | ENSG0000001 | 5.67E-05 | 0.564056024 | 9.83E-05 | 0.002176048 | 5.51E-05 | 0.575333679 | 9.83E-05 | 0.002175998 | 1.63E-06 | 0.01940657 | 1 | 0.96510431 | 1 | 0.964589659 | ENSG0000005 | 9 | PCGF2 |
| Pancreas | Prediabetes | ENSG0000001 | 0.000101763 | 0.762559955 | 0.000336829 | 0.002176049 | 0.000106101 | 0.752764959 | 0.000336832 | 0.002175999 | -4.34E-06 | 0.037350225 | 1 | 0.980154834 | 1 | 0.981881037 | ENSG0000005 | 9 | PCGF2 |
| Pancreas | Prediabetes | ENSG0000001 | 0.001123634 | 0.063871487 | 0.00060636 | 0.002176034 | 0.001110739 | 0.066989037 | 0.000606381 | 0.002175984 | 1.29E-05 | 0.02644547 | 1 | 0.926755475 | 1 | 0.931226496 | ENSG0000005 | 6 | MSH4 |
| Pancreas | Prediabetes | ENSG0000001 | 5.49E-05 | 0.87658077 | 0.000353541 | 0.00217605 | 4.89E-05 | 0.889891332 | 0.000353547 | 0.002175999 | 5.96E-06 | 0.025331249 | 1 | 0.983178195 | 1 | 0.985769389 | ENSG0000005 | 6 | MSH4 |
| Pancreas | Prediabetes | ENSG0000001 | -9.25E-05 | 0.759761759 | 0.000302382 | 0.002176049 | -8.86E-05 | 0.769589899 | 0.000302385 | 0.002175999 | -3.89E-06 | 0.048953617 | 1 | 0.980154834 | 1 | 0.981881037 | ENSG0000005 | 6 | MSH4 |
| Pancreas | Prediabetes | ENSG0000001 | 0.000260628 | 0.595786573 | 0.000491316 | 0.002176048 | 0.00026756 | 0.586047585 | 0.000491321 | 0.002175998 | -6.93E-06 | 0.033399173 | 1 | 0.96510431 | 1 | 0.964589659 | ENSG0000005 | 12 | GDI2 |
| Pancreas | Prediabetes | ENSG0000001 | 0.000221881 | 0.689936815 | 0.000556174 | 0.002176049 | 0.000231345 | 0.67744824 | 0.000556188 | 0.002175998 | -9.46E-06 | 0.044486578 | 1 | 0.975612224 | 1 | 0.974993264 | ENSG0000005 | 12 | GDI2 |
| Pancreas | Prediabetes | ENSG0000002 | -0.00020818 | 0.523627913 | 0.000326423 | 0.002176048 | -0.00020381 | 0.532380394 | 0.000326426 | 0.002175997 | -4.37E-06 | 0.035770065 | 1 | 0.96510431 | 1 | 0.964589659 | ENSG0000005 | 12 | GDI2 |
| Pancreas | Prediabetes | ENSG0000002 | -0.00039818 | 0.392803968 | 0.000465958 | 0.002176046 | -0.00031378 | 0.502326974 | 0.000467745 | 0.002175997 | -8.44E-05 | 0.039076195 | 1 | 0.941475052 | 1 | 0.964589659 | ENSG0000005 | 11 | ATP11B |
| Pancreas | Prediabetes | ENSG0000002 | -0.00069208 | 0.164150561 | 0.000497451 | 0.002176041 | -0.00068468 | 0.16870958 | 0.000497458 | 0.002175991 | -7.40E-06 | 0.033100399 | 1 | 0.931097229 | 1 | 0.933545778 | ENSG0000005 | 10 | LAMC2 |
| Pancreas | Prediabetes | ENSG0000002 | -0.00067395 | 0.202237395 | 0.000531624 | 0.002176042 | -0.00068599 | 0.196930007 | 0.000531633 | 0.002175992 | 8.06E-06 | 0.043931424 | 1 | 0.931097229 | 1 | 0.933545778 | ENSG0000005 | 10 | LAMC2 |
| Pancreas | Prediabetes | ENSG0000002 | 0.00032754 | 0.526639067 | 0.000517322 | 0.002176048 | 0.000320858 | 0.535110767 | 0.000517327 | 0.002175997 | 6.68E-06 | 0.046094748 | 1 | 0.96510431 | 1 | 0.964589659 | ENSG0000005 | 10 | LAMC2 |
| Pancreas | T1DM | ENSG0000000 | -8.70E-05 | 0.926232476 | 0.000940201 | 0.008204775 | -0.00011864 | 0.899589056 | 0.000940242 | 0.00820389 | 3.16E-05 | 0.015772547 | 1 | 0.997898334 | 1 | 0.994038165 | ENSG0000005 | 10 | LAMC2 |
| Pancreas | T1DM | ENSG0000000 | 0.000302992 | 0.736780576 | 0.00090144 | 0.008204774 | 0.000289813 | 0.747823577 | 0.000901411 | 0.008203888 | 1.32E-05 | 0.024418123 | 1 | 0.975415042 | 1 | 0.968082788 | ENSG0000005 | 12 | CROCC |
| Pancreas | T1DM | ENSG0000000 | -0.00018233 | 0.806555627 | 0.000744584 | 0.008204775 | -0.00019493 | 0.793477084 | 0.000744564 | 0.008203889 | 1.26E-05 | 0.019756276 | 1 | 0.98201188 | 1 | 0.978923745 | ENSG0000005 | 12 | CROCC |
| Pancreas | T1DM | ENSG0000000 | 0.000710182 | 0.528101569 | 0.001125653 | 0.008204769 | 0.000686589 | 0.541882355 | 0.001125612 | 0.008203884 | 2.36E-05 | 0.000460206 | 1 | 0.919664009 | 1 | 0.928420568 | ENSG0000005 | 12 | CROCC |
| Pancreas | T1DM | ENSG0000000 | -0.00037573 | 0.620822424 | 0.000759542 | 0.008204771 | -0.00039313 | 0.604733261 | 0.00075952 | 0.008203885 | 1.74E-05 | 0.000979919 | 1 | 0.95220407 | 1 | 0.939938439 | ENSG0000005 | 12 | CROCC |
| Pancreas | T1DM | ENSG0000000 | 1.99E-05 | 0.987496499 | 0.001271343 | 0.008204776 | 5.04E-05 | 0.968365903 | 0.001271307 | 0.00820389 | -3.05E-05 | 0.000724632 | 1 | 0.997898334 | 1 | 0.99966149 | ENSG0000005 | 12 | CROCC |
| Pancreas | T1DM | ENSG0000001 | -0.0003732 | 0.6385104 | 0.000794411 | 0.008204772 | -0.00034637 | 0.662824744 | 0.000794394 | 0.008203887 | -2.68E-05 | 2.85E-05 | 1 | 0.960781248 | 1 | 0.961035047 | ENSG0000005 | 4 | ALDH18A1 |
| Pancreas | T1DM | ENSG0000001 | 6.58E-06 | 0.992607705 | 0.000709783 | 0.008204776 | 2.18E-05 | 0.975527042 | 0.00070976 | 0.00820389 | -1.52E-05 | 0.000990099 | 1 | 0.998452063 | 1 | 0.99966149 | ENSG0000005 | 4 | ALDH18A1 |
| Pancreas | T1DM | ENSG0000001 | 0.000293677 | 0.703781363 | 0.000772385 | 0.008204773 | 0.000279614 | 0.717333545 | 0.00077236 | 0.008203888 | 1.41E-05 | 0.00605114 | 1 | 0.974645207 | 1 | 0.968082788 | ENSG0000006 | 3 | GNA15 |
| Pancreas | T1DM | ENSG0000002 | -1.79E-05 | 0.932704322 | 0.000212393 | 0.008204775 | -1.16E-05 | 0.956320427 | 0.000212396 | 0.00820389 | -6.30E-06 | 0.011717587 | 1 | 0.997898334 | 1 | 0.999388266 | ENSG0000006 | 3 | GNA15 |
| Pancreas | T1DM | ENSG0000002 | -0.0014297 | 0.1234426 | 0.000928064 | 0.008204736 | -0.00144703 | 0.11894591 | 0.000928052 | 0.008203849 | 1.73E-05 | 0.003045661 | 1 | 0.680843282 | 1 | 0.666556371 | ENSG0000006 | 3 | PIGV |
| Pancreas | T1DM | ENSG0000002 | -0.00043532 | 0.501730748 | 0.00064798 | 0.008204768 | -0.00042612 | 0.510777039 | 0.000647958 | 0.008203883 | -9.20E-06 | 0.025670187 | 1 | 0.914783379 | 1 | 0.915335266 | ENSG0000006 | 10 | PRDM6 |
| Pancreas | T1DM | ENSG0000003 | -0.00114116 | 0.198170146 | 0.000886828 | 0.008204748 | -0.00173262 | 0.051929547 | 0.000891395 | 0.008203826 | 0.000591468 | 6.67E-11 | 1 | 0.761827748 | 1 | 0.509067603 | ENSG0000006 | 10 | PRDM6 |
| Pancreas | T1DM | ENSG0000003 | -0.00155681 | 0.157545632 | 0.001101484 | 0.008204742 | -0.00154125 | 0.161725368 | 0.001101451 | 0.008203857 | -1.56E-05 | 0.038723559 | 1 | 0.719888227 | 1 | 0.723580189 | ENSG0000006 | 10 | PRDM6 |
| Pancreas | T1DM | ENSG0000003 | -0.00046209 | 0.516669377 | 0.000712564 | 0.008204768 | -0.00049216 | 0.489753937 | 0.000712548 | 0.008203882 | 3.01E-05 | 7.65E-08 | 1 | 0.914783379 | 1 | 0.914417995 | ENSG0000006 | 12 | TNK2 |
| Pancreas | T1DM | ENSG0000003 | 0.000223751 | 0.586991106 | 0.000411911 | 0.008204771 | 0.000230561 | 0.575649402 | 0.000411899 | 0.008203885 | -6.81E-06 | 0.017118442 | 1 | 0.937588291 | 1 | 0.936315417 | ENSG0000006 | 9 | CDH3 |
| Pancreas | T1DM | ENSG0000004 | -0.00113146 | 0.12882902 | 0.000744997 | 0.008204737 | -0.00194302 | 0.015975836 | 0.000806407 | 0.008203792 | 0.000811561 | 0.008578067 | 1 | 0.68791014 | 1 | 0.301309092 | ENSG0000006 | 12 | ZNF112 |
| Pancreas | T1DM | ENSG0000005 | 0.001195078 | 0.329492963 | 0.00122555 | 0.00820476 | 0.001217714 | 0.320396737 | 0.001225504 | 0.008203874 | -2.26E-05 | 0.001366205 | 1 | 0.83462449 | 1 | 0.830335577 | ENSG0000006 | 13 | EIF4B |
| Pancreas | T1DM | ENSG0000005 | -0.00083977 | 0.39406325 | 0.000985328 | 0.008204763 | -0.0008522 | 0.387081753 | 0.000985291 | 0.008203877 | 1.24E-05 | 0.029048621 | 1 | 0.873903579 | 1 | 0.867022428 | ENSG0000006 | 13 | EIF4B |
| Pancreas | T1DM | ENSG0000005 | 0.000620171 | 0.528071803 | 0.000982912 | 0.008204769 | 0.000598498 | 0.542580632 | 0.000982891 | 0.008203884 | 2.17E-05 | 0.005797114 | 1 | 0.919664009 | 1 | 0.928420568 | ENSG0000006 | 13 | EIF4B |
| Pancreas | T1DM | ENSG0000006 | 0.000967032 | 0.293110817 | 0.000919824 | 0.008204757 | 0.000980812 | 0.286271963 | 0.000919795 | 0.008203871 | -1.38E-05 | 0.025688523 | 1 | 0.810235282 | 1 | 0.801351316 | ENSG0000006 | 13 | EIF4B |
| Pancreas | T1DM | ENSG0000006 | -0.00100074 | 0.13328891 | 0.000666599 | 0.008204738 | -0.00099159 | 0.136860411 | 0.000666574 | 0.008203853 | -9.15E-06 | 0.014457551 | 1 | 0.692886478 | 1 | 0.692461116 | ENSG0000006 | 13 | EIF4B |
| Pancreas | T1DM | ENSG0000006 | 0.00014468 | 0.742814915 | 0.000440925 | 0.008204774 | 0.000152587 | 0.729288574 | 0.000440911 | 0.008203888 | -7.91E-06 | 0.008941009 | 1 | 0.975415042 | 1 | 0.968082788 | ENSG0000006 | 13 | EIF4B |
| Pancreas | T1DM | ENSG0000006 | -0.00060175 | 0.60318421 | 0.001157591 | 0.008204771 | -0.00056265 | 0.626923488 | 0.001157566 | 0.008203886 | -3.91E-05 | 0.0028224 | 1 | 0.941627387 | 1 | 0.953755638 | ENSG0000006 | 8 | HAGH |
| Pancreas | T1DM | ENSG0000006 | -0.00071548 | 0.380989588 | 0.000816684 | 0.008204763 | -0.00070112 | 0.390605698 | 0.000816661 | 0.008203878 | -1.44E-05 | 0.012544694 | 1 | 0.862440233 | 1 | 0.868463578 | ENSG0000006 | 8 | HAGH |
| Pancreas | T1DM | ENSG0000006 | 0.001148324 | 0.059885745 | 0.000610279 | 0.008204716 | 0.001134589 | 0.063003539 | 0.000610266 | 0.008203832 | 1.37E-05 | 0.005521953 | 1 | 0.527104393 | 1 | 0.528735579 | ENSG0000006 | 8 | HAGH |
| Pancreas | T1DM | ENSG0000006 | 0.000112087 | 0.834081759 | 0.000535098 | 0.008204775 | 0.002053123 | 0.001486705 | 0.000646188 | 0.00820372 | -0.00194104 | 8.44E-08 | 1 | 0.984098328 | 1 | 0.064162591 | ENSG0000006 | 8 | HAGH |
| Pancreas | T1DM | ENSG0000006 | 0.000722708 | 0.491656298 | 0.00105094 | 0.008204768 | 0.000739287 | 0.481758812 | 0.001050904 | 0.008203882 | -1.66E-05 | 0.0115784 | 1 | 0.914693561 | 1 | 0.913030722 | ENSG0000006 | 8 | ST3GAL6 |
| Pancreas | T1DM | ENSG0000007 | 0.001007627 | 0.214369406 | 0.000811528 | 0.00820475 | 0.001018056 | 0.209646791 | 0.000811498 | 0.008203863 | -1.04E-05 | 0.029531004 | 1 | 0.772090651 | 1 | 0.769778216 | ENSG0000006 | 8 | ATP2C2 |
| Pancreas | T1DM | ENSG0000007 | 0.001765808 | 0.026653535 | 0.000796643 | 0.008204693 | 0.00177863 | 0.025567974 | 0.000796621 | 0.008203806 | -1.28E-05 | 0.026538311 | 1 | 0.395643231 | 1 | 0.374802485 | ENSG0000006 | 8 | ATP2C2 |
| Pancreas | T1DM | ENSG0000007 | -0.00105111 | 0.319437124 | 0.001055736 | 0.008204759 | -0.00103967 | 0.324712616 | 0.001055694 | 0.008203874 | -1.14E-05 | 0.045627896 | 1 | 0.829307456 | 1 | 0.830335577 | ENSG0000006 | 10 | CDON |
| Pancreas | T1DM | ENSG0000007 | 0.0004284 | 0.469847572 | 0.000592754 | 0.008204767 | 0.000420042 | 0.478538821 | 0.000592731 | 0.008203882 | 8.36E-06 | 0.010606694 | 1 | 0.908712989 | 1 | 0.911454131 | ENSG0000006 | 10 | CDON |
| Pancreas | T1DM | ENSG0000007 | 0.000562288 | 0.556698674 | 0.000956674 | 0.00820477 | 0.00054701 | 0.567454509 | 0.000956638 | 0.008203884 | 1.53E-05 | 0.00539663 | 1 | 0.93270646 | 1 | 0.936315417 | ENSG0000006 | 7 | TAF2 |
| Pancreas | T1DM | ENSG0000007 | 9.42E-05 | 0.928092728 | 0.001043453 | 0.008204775 | 0.000108948 | 0.916840058 | 0.001043413 | 0.00820389 | -1.48E-05 | 0.011617259 | 1 | 0.997898334 | 1 | 0.997092545 | ENSG0000006 | 7 | TAF2 |
| Pancreas | T1DM | ENSG0000007 | 0.001074193 | 0.08186078 | 0.000617354 | 0.008204725 | 0.001065012 | 0.084496085 | 0.000617334 | 0.00820384 | 9.18E-06 | 0.023220308 | 1 | 0.587627507 | 1 | 0.580557814 | ENSG0000006 | 12 | SUGP2 |
| Pancreas | T1DM | ENSG0000007 | -0.00133131 | 0.12252765 | 0.000862109 | 0.008204735 | -0.0013156 | 0.126994963 | 0.00086209 | 0.008203851 | -1.57E-05 | 0.022083737 | 1 | 0.680843282 | 1 | 0.673807988 | ENSG0000006 | 12 | SUGP2 |
| Pancreas | T1DM | ENSG0000007 | 0.001843035 | 0.075831502 | 0.001038092 | 0.008204722 | 0.001829484 | 0.077998867 | 0.001038052 | 0.008203838 | 1.36E-05 | 0.016802929 | 1 | 0.57107125 | 1 | 0.562287494 | ENSG0000006 | 8 | ABCA7 |
| Pancreas | T1DM | ENSG0000007 | 0.000403097 | 0.701074828 | 0.001050088 | 0.008204773 | 0.000419348 | 0.689628887 | 0.001050052 | 0.008203887 | -1.63E-05 | 0.014498348 | 1 | 0.974645207 | 1 | 0.966685034 | ENSG0000006 | 8 | ABCA7 |
| Pancreas | T1DM | ENSG0000007 | 0.000353366 | 0.601635494 | 0.000676881 | 0.008204771 | 0.000362659 | 0.592096038 | 0.000676854 | 0.008203885 | -9.29E-06 | 0.012230851 | 1 | 0.940352982 | 1 | 0.937577789 | ENSG0000006 | 8 | ABCA7 |
| Pancreas | T1DM | ENSG0000007 | 0.000448987 | 0.670236671 | 0.001054398 | 0.008204773 | 0.000431789 | 0.682154297 | 0.001054363 | 0.008203887 | 1.72E-05 | 0.011804409 | 1 | 0.963860935 | 1 | 0.966338775 | ENSG0000006 | 7 | DDX20 |
| Pancreas | T1DM | ENSG0000007 | -0.00153653 | 0.031763821 | 0.000715538 | 0.008204698 | -0.00152361 | 0.033221326 | 0.000715511 | 0.008203814 | -1.29E-05 | 0.001459671 | 1 | 0.427517623 | 1 | 0.43087053 | ENSG0000006 | 12 | TAF11 |
| Pancreas | T1DM | ENSG0000007 | 0.001201371 | 0.203369713 | 0.000944466 | 0.008204748 | 0.001068324 | 0.258093619 | 0.000944659 | 0.008203868 | 0.000133047 | 5.67E-10 | 1 | 0.766891956 | 1 | 0.788955599 | ENSG0000006 | 12 | TAF11 |
| Pancreas | T1DM | ENSG0000008 | -2.65E-05 | 0.950202489 | 0.000424894 | 0.008204775 | -3.36E-05 | 0.936934667 | 0.00042488 | 0.00820389 | 7.08E-06 | 0.011296863 | 1 | 0.997898334 | 1 | 0.999388266 | ENSG0000006 | 3 | NTHL1 |
| Pancreas | T1DM | ENSG0000008 | -0.00035377 | 0.655764546 | 0.00079361 | 0.008204772 | -0.00036772 | 0.643098834 | 0.000793581 | 0.008203886 | 1.40E-05 | 0.003149444 | 1 | 0.962467202 | 1 | 0.960161515 | ENSG0000006 | 12 | UHRF1BP1 |
| Pancreas | T1DM | ENSG0000008 | 0.001649236 | 0.084292709 | 0.000955354 | 0.008204725 | 0.001636802 | 0.08664718 | 0.000955317 | 0.008203841 | 1.24E-05 | 0.018207069 | 1 | 0.589819284 | 1 | 0.580557814 | ENSG0000006 | 12 | UHRF1BP1 |
| Pancreas | T1DM | ENSG0000008 | -0.00111839 | 0.148649427 | 0.000774337 | 0.00820474 | -0.0011062 | 0.153111989 | 0.000774313 | 0.008203856 | -1.22E-05 | 0.018459746 | 1 | 0.712186059 | 1 | 0.711161998 | ENSG0000006 | 12 | UHRF1BP1 |
| Pancreas | T1DM | ENSG0000008 | 0.001013868 | 0.375422313 | 0.001143852 | 0.008204762 | 0.001033721 | 0.366126393 | 0.001143808 | 0.008203876 | -1.99E-05 | 0.002095696 | 1 | 0.860999343 | 1 | 0.850635855 | ENSG0000006 | 12 | UHRF1BP1 |
| Pancreas | T1DM | ENSG0000008 | -0.00064891 | 0.353446015 | 0.000699312 | 0.008204761 | -0.00065855 | 0.346326083 | 0.000699286 | 0.008203875 | 9.64E-06 | 0.015591862 | 1 | 0.854061442 | 1 | 0.847091708 | ENSG0000006 | 12 | UHRF1BP1 |
| Pancreas | T1DM | ENSG0000008 | 0.000701934 | 0.26754324 | 0.000633092 | 0.008204755 | 0.000693001 | 0.273664071 | 0.000633072 | 0.00820387 | 8.93E-06 | 0.03202559 | 1 | 0.798582374 | 1 | 0.79588826 | ENSG0000006 | 12 | UHRF1BP1 |
| Pancreas | T1DM | ENSG0000008 | -0.00146571 | 0.035977312 | 0.000698894 | 0.008204702 | -0.00147694 | 0.034573842 | 0.000698871 | 0.008203815 | 1.12E-05 | 0.013228217 | 1 | 0.443514005 | 1 | 0.43733359 | ENSG0000006 | 12 | UHRF1BP1 |
| Pancreas | T1DM | ENSG0000008 | 0.000424607 | 0.736582748 | 0.001262276 | 0.008204774 | 0.000440479 | 0.727112906 | 0.001262231 | 0.008203888 | -1.59E-05 | 0.037009391 | 1 | 0.975415042 | 1 | 0.968082788 | ENSG0000006 | 6 | WDR18 |
| Pancreas | T1DM | ENSG0000008 | -0.00037867 | 0.455087924 | 0.000506947 | 0.008204766 | -0.00045552 | 0.370088718 | 0.00050822 | 0.008203879 | 7.69E-05 | 0.034376052 | 1 | 0.899832468 | 1 | 0.851044876 | ENSG0000006 | 6 | WDR18 |
| Pancreas | T1DM | ENSG0000008 | 0.000378668 | 0.636827613 | 0.000802025 | 0.008204772 | 0.000386585 | 0.629784195 | 0.000801991 | 0.008203886 | -7.92E-06 | 0.040576672 | 1 | 0.960748332 | 1 | 0.955974304 | ENSG0000006 | 6 | ADAT1 |
| Pancreas | T1DM | ENSG0000008 | 0.001753765 | 0.012702629 | 0.000703755 | 0.008204671 | 0.001766354 | 0.012074454 | 0.000703735 | 0.008203784 | -1.26E-05 | 0.011189782 | 1 | 0.276443652 | 1 | 0.258393321 | ENSG0000006 | 16 | SEC61A2 |
| Pancreas | T1DM | ENSG0000009 | -0.00013033 | 0.805787362 | 0.000530083 | 0.008204775 | -0.00012179 | 0.818278115 | 0.000530067 | 0.008203889 | -8.54E-06 | 0.020564672 | 1 | 0.98201188 | 1 | 0.981810269 | ENSG0000006 | 16 | SEC61A2 |
| Pancreas | T1DM | ENSG0000009 | -0.0001614 | 0.728995875 | 0.000465856 | 0.008204774 | -0.00015142 | 0.745151443 | 0.000465843 | 0.008203888 | -9.98E-06 | 0.002210113 | 1 | 0.975415042 | 1 | 0.968082788 | ENSG0000006 | 16 | SEC61A2 |
| Pancreas | T1DM | ENSG0000009 | -0.00013016 | 0.788575996 | 0.000485391 | 0.008204774 | -0.00014952 | 0.758057145 | 0.000485403 | 0.008203888 | 1.94E-05 | 0.001523767 | 1 | 0.981920629 | 1 | 0.968082788 | ENSG0000006 | 8 | ZMYND12 |
| Pancreas | T1DM | ENSG0000009 | 0.001042317 | 0.396750105 | 0.001229959 | 0.008204763 | 0.001028551 | 0.402996653 | 0.001229909 | 0.008203878 | 1.38E-05 | 0.027292914 | 1 | 0.874941428 | 1 | 0.877030004 | ENSG0000006 | 8 | ZMYND12 |
| Pancreas | T1DM | ENSG0000009 | 0.001516205 | 0.184921318 | 0.001143653 | 0.008204746 | 0.001498476 | 0.190097269 | 0.001143619 | 0.008203861 | 1.77E-05 | 0.024544301 | 1 | 0.746326771 | 1 | 0.756852382 | ENSG0000006 | 8 | ZMYND12 |
| Pancreas | T1DM | ENSG0000009 | -0.00040083 | 0.6343312 | 0.000842712 | 0.008204772 | -0.00038358 | 0.648972828 | 0.000842684 | 0.008203886 | -1.72E-05 | 0.001462247 | 1 | 0.959806249 | 1 | 0.961035047 | ENSG0000006 | 8 | ZMYND12 |
| Pancreas | T1DM | ENSG0000009 | 0.000207337 | 0.8179642 | 0.000900821 | 0.008204775 | 0.000191113 | 0.831980961 | 0.000900787 | 0.008203889 | 1.62E-05 | 0.001775148 | 1 | 0.982079052 | 1 | 0.984766677 | ENSG0000006 | 6 | SLC9A3 |
| Pancreas | T1DM | ENSG0000009 | -0.00043026 | 0.626189752 | 0.000883313 | 0.008204772 | -0.00041692 | 0.636917011 | 0.000883279 | 0.008203886 | -1.33E-05 | 0.007962407 | 1 | 0.953769445 | 1 | 0.955974304 | ENSG0000006 | 10 | CD84 |
| Pancreas | T1DM | ENSG0000009 | -0.00017924 | 0.889366855 | 0.001288525 | 0.008204775 | -0.00016303 | 0.89931587 | 0.001288474 | 0.00820389 | -1.62E-05 | 0.019452199 | 1 | 0.99200447 | 1 | 0.994038165 | ENSG0000006 | 10 | CD84 |

| | | | | | | | | | | | | | | | | | | | |
|---|---|---|---|---|---|---|---|---|---|---|---|---|---|---|---|---|---|---|---|
| Pancreas | T1DM | ENSG000001( | -0.00025034 | 0.804257193 | 0.001010093 | 0.008204775 | -0.0002223 | 0.825806446 | 0.001010061 | 0.008203889 | -2.80E-05 | 3.45E-05 | 1 | 0.98201188 | 1 | 0.984037376 | ENSG0000006 | 10 | CD84 |
| Pancreas | T1DM | ENSG000001( | 0.000519648 | 0.520087124 | 0.000807895 | 0.008204769 | 0.000508535 | 0.529034903 | 0.000807865 | 0.008203883 | 1.11E-05 | 0.016305309 | 1 | 0.914783379 | 1 | 0.920313283 | ENSG0000006 | 10 | CD84 |
| Pancreas | T1DM | ENSG000001( | 0.000239645 | 0.784468381 | 0.000876212 | 0.008204774 | 0.000250683 | 0.774795403 | 0.000876182 | 0.008203889 | -1.10E-05 | 0.042809469 | 1 | 0.981920629 | 1 | 0.972640681 | ENSG0000006 | 10 | CD84 |
| Pancreas | T1DM | ENSG000001( | -0.00051296 | 0.23001115 | 0.000427349 | 0.008204751 | -0.00051885 | 0.2246907 | 0.000427333 | 0.008203865 | 5.89E-06 | 0.013549121 | 1 | 0.785512902 | 1 | 0.780702981 | ENSG0000006 | 10 | ZNRD1 |
| Pancreas | T1DM | ENSG000001( | 0.000490968 | 0.639127102 | 0.001047018 | 0.008204772 | 0.000502201 | 0.631464097 | 0.001046977 | 0.008203886 | -1.12E-05 | 0.048275164 | 1 | 0.96093583 | 1 | 0.955974304 | ENSG0000006 | 10 | ZNRD1 |
| Pancreas | T1DM | ENSG000001( | 2.90E-05 | 0.923045773 | 0.000300001 | 0.008204775 | 3.34E-05 | 0.911230922 | 0.000299993 | 0.00820389 | -4.47E-06 | 0.038578721 | 1 | 0.997898334 | 1 | 0.996366272 | ENSG0000006 | 10 | ZNRD1 |
| Pancreas | T1DM | ENSG000001( | 0.000283034 | 0.604494056 | 0.000546452 | 0.008204771 | 0.000293492 | 0.591196336 | 0.000546435 | 0.008203885 | -1.05E-05 | 0.004167682 | 1 | 0.941811128 | 1 | 0.937577789 | ENSG0000006 | 10 | ZNRD1 |
| Pancreas | T1DM | ENSG000001( | 0.000541241 | 0.580763707 | 0.000980031 | 0.00820477 | 0.000521842 | 0.594383427 | 0.000979994 | 0.008203885 | 1.94E-05 | 0.000665394 | 1 | 0.937588291 | 1 | 0.937577789 | ENSG0000006 | 10 | ZNRD1 |
| Pancreas | T1DM | ENSG000001( | -1.01E-06 | 0.998309284 | 0.000477462 | 0.008204776 | -1.19E-05 | 0.980143645 | 0.000477465 | 0.00820389 | 1.09E-05 | 0.0370828 | 1 | 0.999322433 | 1 | 0.99966149 | ENSG0000006 | 10 | ZNRD1 |
| Pancreas | T1DM | ENSG000001( | 0.00085473 | 0.511357497 | 0.001301502 | 0.008204768 | 0.000878553 | 0.49964071 | 0.001301453 | 0.008203882 | -2.38E-05 | 0.001456798 | 1 | 0.914783379 | 1 | 0.914417995 | ENSG0000006 | 17 | ATXN3 |
| Pancreas | T1DM | ENSG000001( | 0.000226303 | 0.820109949 | 0.00099516 | 0.008204775 | 0.000246042 | 0.804717648 | 0.000995127 | 0.008203889 | -1.97E-05 | 0.002183227 | 1 | 0.982562562 | 1 | 0.979350575 | ENSG0000006 | 11 | MTFR1 |
| Pancreas | T1DM | ENSG000001( | 0.000289433 | 0.589223333 | 0.000536025 | 0.008204771 | 0.000904981 | 0.134316297 | 0.000604409 | 0.008203852 | -0.00061555 | 0.027543344 | 1 | 0.937588291 | 1 | 0.685736609 | ENSG0000006 | 6 | FECH |
| Pancreas | T1DM | ENSG000001( | -0.00061619 | 0.529715674 | 0.00098051 | 0.008204769 | -0.00063214 | 0.519103609 | 0.000980478 | 0.008203883 | 1.59E-05 | 0.012889284 | 1 | 0.920284891 | 1 | 0.915978195 | ENSG0000006 | 6 | FECH |
| Pancreas | T1DM | ENSG000001( | 0.000259482 | 0.808987651 | 0.001073427 | 0.008204775 | 0.000314462 | 0.769580563 | 0.001073529 | 0.008203889 | -5.50E-05 | 0.002953965 | 1 | 0.98201188 | 1 | 0.972640681 | ENSG0000006 | 11 | MYO9A |
| Pancreas | T1DM | ENSG000001( | -0.00133561 | 0.268768943 | 0.001207706 | 0.008204755 | -0.0013604 | 0.259967614 | 0.001207669 | 0.008203869 | 2.48E-05 | 0.002366221 | 1 | 0.798582374 | 1 | 0.788955599 | ENSG0000006 | 11 | MYO9A |
| Pancreas | T1DM | ENSG000001( | -0.00106567 | 0.238425863 | 0.000903925 | 0.008204752 | -0.00030343 | 0.744170269 | 0.000929803 | 0.008203888 | -0.00076224 | 0.000472389 | 1 | 0.785512902 | 1 | 0.968082788 | ENSG0000006 | 12 | NEO1 |
| Pancreas | T1DM | ENSG000001( | -0.00044046 | 0.563862825 | 0.000763211 | 0.00820477 | -0.00042107 | 0.581140364 | 0.000763191 | 0.008203885 | -1.94E-05 | 0.000599084 | 1 | 0.936396473 | 1 | 0.936315417 | ENSG0000006 | 3 | TRAM1 |
| Pancreas | T1DM | ENSG000001( | 0.000601874 | 0.299082193 | 0.000579614 | 0.008204757 | 0.000611452 | 0.291441258 | 0.000579594 | 0.008203871 | -9.58E-06 | 0.008259867 | 1 | 0.817067098 | 1 | 0.805533316 | ENSG0000006 | 10 | METTL22 |
| Pancreas | T1DM | ENSG000001( | -0.00029614 | 0.666833282 | 0.0006879 | 0.008204772 | -0.00028456 | 0.679114314 | 0.000687879 | 0.008203887 | -1.16E-05 | 0.01398634 | 1 | 0.963860935 | 1 | 0.964582721 | ENSG0000006 | 5 | RRP15 |
| Pancreas | T1DM | ENSG000001( | -0.0003946 | 0.662061063 | 0.000902829 | 0.008204772 | -0.00038313 | 0.671290077 | 0.000902795 | 0.008203887 | -1.15E-05 | 0.025538653 | 1 | 0.962467202 | 1 | 0.961035047 | ENSG0000006 | 5 | RRP15 |
| Pancreas | T1DM | ENSG000001( | -0.00040823 | 0.212209593 | 0.000327238 | 0.008204749 | -0.00041228 | 0.207691587 | 0.000327226 | 0.008203863 | 4.05E-06 | 0.025485076 | 1 | 0.772090651 | 1 | 0.769778216 | ENSG0000006 | 5 | RRP15 |
| Pancreas | T1DM | ENSG000001( | -0.00030316 | 0.683138454 | 0.000742699 | 0.008204773 | -0.00025801 | 0.728319874 | 0.000742777 | 0.008203888 | -4.51E-05 | 0.000657588 | 1 | 0.967740456 | 1 | 0.968082788 | ENSG0000006 | 6 | DHX8 |
| Pancreas | T1DM | ENSG000001( | -0.00217021 | 0.011570811 | 0.000859497 | 0.008204668 | -0.00215168 | 0.012299543 | 0.00085949 | 0.008203784 | -1.85E-05 | 0.024847952 | 1 | 0.260075765 | 1 | 0.258895305 | ENSG0000006 | 9 | SYT1 |
| Pancreas | T1DM | ENSG000001( | 0.000572447 | 0.435276555 | 0.000733724 | 0.008204765 | 0.000559483 | 0.445733859 | 0.000733702 | 0.00820388 | 1.30E-05 | 0.011833693 | 1 | 0.89813429 | 1 | 0.896838446 | ENSG0000006 | 13 | RASSF1 |
| Pancreas | T1DM | ENSG000001( | -0.00061712 | 0.571305568 | 0.001090069 | 0.00820477 | -0.00063355 | 0.561089944 | 0.001090027 | 0.008203884 | 1.64E-05 | 0.005365192 | 1 | 0.937588291 | 1 | 0.936315417 | ENSG0000006 | 13 | RASSF1 |
| Pancreas | T1DM | ENSG000001( | 6.45E-05 | 0.867438209 | 0.000386536 | 0.008204775 | 7.52E-05 | 0.845770454 | 0.000386552 | 0.008203889 | -1.07E-05 | 0.045498743 | 1 | 0.99183698 | 1 | 0.986098414 | ENSG0000006 | 13 | FGFR3 |
| Pancreas | T1DM | ENSG000001( | -0.00102921 | 0.15418469 | 0.000722299 | 0.008204741 | -0.00133623 | 0.069745778 | 0.000736798 | 0.008203835 | 0.000307019 | 0.035029549 | 1 | 0.717060869 | 1 | 0.538522335 | ENSG0000006 | 13 | FGFR3 |
| Pancreas | T1DM | ENSG000001( | 2.45E-05 | 0.952157927 | 0.000407634 | 0.008204776 | 3.06E-05 | 0.940132263 | 0.00040762 | 0.00820389 | -6.16E-06 | 0.020728218 | 1 | 0.997898334 | 1 | 0.999388266 | ENSG0000006 | 3 | IFI35 |
| Pancreas | T1DM | ENSG000001( | 0.001918235 | 0.126193516 | 0.001254331 | 0.008204736 | 0.001934908 | 0.122919237 | 0.001254285 | 0.00820385 | -1.67E-05 | 0.022689332 | 1 | 0.684395521 | 1 | 0.671609788 | ENSG0000006 | 11 | POLR1A |
| Pancreas | T1DM | ENSG000001( | 0.000719123 | 0.393648214 | 0.000843031 | 0.008204763 | 0.000705312 | 0.402779535 | 0.000843001 | 0.008203878 | 1.38E-05 | 0.006512192 | 1 | 0.873903579 | 1 | 0.877030004 | ENSG0000006 | 11 | POLR1A |
| Pancreas | T1DM | ENSG000001( | -7.02E-05 | 0.934008135 | 0.000848299 | 0.008204775 | -5.90E-05 | 0.944527428 | 0.000848268 | 0.00820389 | -1.12E-05 | 0.023887522 | 1 | 0.997898334 | 1 | 0.999388266 | ENSG0000006 | 12 | NUCKS1 |
| Pancreas | T1DM | ENSG000001( | 0.000775186 | 0.143244232 | 0.000529564 | 0.008204739 | 0.000782211 | 0.139640701 | 0.000529547 | 0.008203853 | -7.03E-06 | 0.041091121 | 1 | 0.706048346 | 1 | 0.693219864 | ENSG0000006 | 10 | CLEC2D |
| Pancreas | T1DM | ENSG000001( | -0.00068689 | 0.521059655 | 0.001070406 | 0.008204769 | -0.00069859 | 0.513970627 | 0.001070363 | 0.008203883 | 1.17E-05 | 0.03322963 | 1 | 0.914876612 | 1 | 0.915381179 | ENSG0000006 | 11 | NEDD4 |
| Pancreas | T1DM | ENSG000001( | 2.24E-05 | 0.962916549 | 0.000481059 | 0.008204776 | 3.09E-05 | 0.948780881 | 0.000481043 | 0.00820389 | -8.53E-06 | 0.005855287 | 1 | 0.997898334 | 1 | 0.999388266 | ENSG0000006 | 11 | NEDD4 |
| Pancreas | T1DM | ENSG000001( | -0.00010125 | 0.795266989 | 0.000390197 | 0.008204774 | -9.31E-05 | 0.811444492 | 0.000390184 | 0.008203889 | -8.16E-06 | 0.000976117 | 1 | 0.981920629 | 1 | 0.981619417 | ENSG0000006 | 11 | NEDD4 |
| Pancreas | T1DM | ENSG000001( | -0.00041056 | 0.634570345 | 0.000863779 | 0.008204772 | -0.0004212 | 0.625801973 | 0.000863747 | 0.008203886 | 1.06E-05 | 0.034090308 | 1 | 0.959806249 | 1 | 0.953755638 | ENSG0000006 | 14 | GNB5 |
| Pancreas | T1DM | ENSG000001( | -0.00041426 | 0.556639169 | 0.000704715 | 0.00820477 | -0.00043953 | 0.532842486 | 0.000704751 | 0.008203883 | 2.53E-05 | 0.013230416 | 1 | 0.93270646 | 1 | 0.920352808 | ENSG0000006 | 14 | GNB5 |
| Pancreas | T1DM | ENSG000001( | -0.00029321 | 0.787729514 | 0.001088963 | 0.008204774 | -0.00027785 | 0.798597207 | 0.001088923 | 0.008203889 | -1.54E-05 | 0.01730699 | 1 | 0.981920629 | 1 | 0.978923745 | ENSG0000006 | 14 | GNB5 |
| Pancreas | T1DM | ENSG000001( | 0.002417565 | 0.008296655 | 0.000915826 | 0.008204658 | 0.002429157 | 0.007989422 | 0.00091579 | 0.008203772 | -1.16E-05 | 0.016819757 | 1 | 0.221935509 | 1 | 0.204622746 | ENSG0000006 | 14 | GNB5 |
| Pancreas | T1DM | ENSG000001( | 2.43E-05 | 0.977670847 | 0.000866786 | 0.008204776 | 9.93E-06 | 0.990859777 | 0.000866752 | 0.00820389 | 1.43E-05 | 0.00258485 | 1 | 0.997898334 | 1 | 0.99966149 | ENSG0000006 | 14 | GNB5 |
| Pancreas | T1DM | ENSG0000011 | 0.000332239 | 0.727119388 | 0.000952084 | 0.008204774 | 0.000316563 | 0.739508932 | 0.000952058 | 0.008203888 | 1.57E-05 | 0.025504112 | 1 | 0.975415042 | 1 | 0.968082788 | ENSG0000006 | 15 | RAB27A |
| Pancreas | T1DM | ENSG0000011 | 0.00110406 | 0.28504016 | 0.001032733 | 0.008204756 | 0.001091361 | 0.290599572 | 0.001032695 | 0.008203871 | 1.27E-05 | 0.03802037 | 1 | 0.804377068 | 1 | 0.805533316 | ENSG0000006 | 8 | CECR5 |
| Pancreas | T1DM | ENSG0000011 | 0.000422144 | 0.406683003 | 0.000508763 | 0.008204764 | 0.000415506 | 0.414083829 | 0.000508744 | 0.008203879 | 6.64E-06 | 0.027770893 | 1 | 0.88063625 | 1 | 0.883839257 | ENSG0000006 | 8 | CECR5 |
| Pancreas | T1DM | ENSG0000011 | 0.000788115 | 0.190513909 | 0.000602046 | 0.008204747 | 0.00077799 | 0.196255431 | 0.000602022 | 0.008203862 | 1.01E-05 | 0.002043977 | 1 | 0.752840936 | 1 | 0.762792436 | ENSG0000007 | 13 | SPTB |
| Pancreas | T1DM | ENSG0000011 | 0.000345058 | 0.477016176 | 0.000485239 | 0.008204767 | 0.000354533 | 0.464991214 | 0.000485227 | 0.008203881 | -9.47E-06 | 0.012076623 | 1 | 0.911441622 | 1 | 0.903250709 | ENSG0000007 | 13 | SPTB |
| Pancreas | T1DM | ENSG0000011 | 0.000869393 | 0.177666192 | 0.000644961 | 0.008204745 | 0.000861357 | 0.181690406 | 0.000644935 | 0.00820386 | 8.04E-06 | 0.018609907 | 1 | 0.732338331 | 1 | 0.739805892 | ENSG0000007 | 13 | SPTB |
| Pancreas | T1DM | ENSG0000011 | -0.00066663 | 0.505706486 | 0.001001642 | 0.008204768 | -0.00067923 | 0.497679454 | 0.001001604 | 0.008203882 | 1.26E-05 | 0.026801779 | 1 | 0.914783379 | 1 | 0.914417995 | ENSG0000007 | 8 | DAPP1 |
| Pancreas | T1DM | ENSG0000011 | -0.00046243 | 0.519900555 | 0.000718613 | 0.008204769 | -0.00047108 | 0.512105968 | 0.000718584 | 0.008203883 | 8.65E-06 | 0.017499881 | 1 | 0.914783379 | 1 | 0.915381179 | ENSG0000007 | 8 | DAPP1 |
| Pancreas | T1DM | ENSG0000011 | -8.52E-05 | 0.901833543 | 0.000691019 | 0.008204775 | -7.45E-05 | 0.914185041 | 0.000690995 | 0.00820389 | -1.08E-05 | 0.011693526 | 1 | 0.99317359 | 1 | 0.996869293 | ENSG0000007 | 14 | DGCR2 |
| Pancreas | T1DM | ENSG0000011 | -0.00085685 | 0.385882533 | 0.000988171 | 0.008204763 | -0.0008723 | 0.377356214 | 0.000988132 | 0.008203877 | 1.54E-05 | 0.003776681 | 1 | 0.865581925 | 1 | 0.858326624 | ENSG0000007 | 7 | POLB |
| Pancreas | T1DM | ENSG0000011 | 0.00020495 | 0.807628394 | 0.000841728 | 0.008204775 | 0.000191844 | 0.819704523 | 0.000841699 | 0.008203889 | 1.31E-05 | 0.014004428 | 1 | 0.98201188 | 1 | 0.981810269 | ENSG0000007 | 8 | WIPI1 |
| Pancreas | T1DM | ENSG0000011 | -0.00048126 | 0.323799239 | 0.000487756 | 0.008204759 | -0.0004752 | 0.329913228 | 0.000487739 | 0.008203874 | -6.06E-06 | 0.042205625 | 1 | 0.832533491 | 1 | 0.832236906 | ENSG0000007 | 8 | WIPI1 |
| Pancreas | T1DM | ENSG0000011 | 3.08E-05 | 0.959716235 | 0.00060972 | 0.008204776 | 0.001073224 | 0.089125591 | 0.000631297 | 0.008203841 | -0.00104243 | 1.95E-10 | 1 | 0.997898334 | 1 | 0.585431045 | ENSG0000007 | 12 | EIF2B3 |
| Pancreas | T1DM | ENSG0000011 | -0.00068402 | 0.537131049 | 0.00110834 | 0.008204769 | -0.0007171 | 0.51762459 | 0.001108323 | 0.008203883 | 3.31E-05 | 0.000671713 | 1 | 0.924272254 | 1 | 0.915978195 | ENSG0000007 | 12 | EIF2B3 |
| Pancreas | T1DM | ENSG0000011 | -0.00062624 | 0.392891204 | 0.000732968 | 0.008204763 | -0.00059826 | 0.414385695 | 0.000732983 | 0.008203879 | -2.80E-05 | 0.0017201 | 1 | 0.873903579 | 1 | 0.883839257 | ENSG0000007 | 7 | WBSCR22 |
| Pancreas | T1DM | ENSG0000011 | 0.001460508 | 0.01995432 | 0.000627578 | 0.008204684 | 0.001450942 | 0.020775731 | 0.000627559 | 0.0082038 | 9.57E-06 | 0.024948685 | 1 | 0.346717973 | 1 | 0.339822154 | ENSG0000007 | 7 | WBSCR22 |
| Pancreas | T1DM | ENSG0000011 | 0.000340808 | 0.757725907 | 0.00110484 | 0.008204774 | 0.000360915 | 0.74391147 | 0.001104803 | 0.008203888 | -2.01E-05 | 0.005014531 | 1 | 0.976337245 | 1 | 0.968082788 | ENSG0000007 | 7 | WBSCR22 |
| Pancreas | T1DM | ENSG0000011 | 0.000310031 | 0.554783971 | 0.000524936 | 0.00820477 | 0.000317564 | 0.54519401 | 0.000524918 | 0.008203884 | -7.53E-06 | 0.025302306 | 1 | 0.93270646 | 1 | 0.928420568 | ENSG0000007 | 7 | TRIB2 |
| Pancreas | T1DM | ENSG0000011 | 0.001153519 | 0.243075991 | 0.000988165 | 0.008204753 | 0.001137831 | 0.249530419 | 0.000988136 | 0.008203868 | 1.57E-05 | 0.023466731 | 1 | 0.787779958 | 1 | 0.788955599 | ENSG0000007 | 7 | TRIB2 |
| Pancreas | T1DM | ENSG0000011 | 0.001746102 | 0.098272024 | 0.001056137 | 0.00820473 | 0.001764309 | 0.094802231 | 0.001056096 | 0.008203843 | -1.82E-05 | 0.00206278 | 1 | 0.633704355 | 1 | 0.605602312 | ENSG0000007 | 7 | TRIB2 |
| Pancreas | T1DM | ENSG0000011 | 9.57E-05 | 0.802184984 | 0.000382072 | 0.008204775 | 9.03E-05 | 0.813189899 | 0.000382058 | 0.008203889 | 5.43E-06 | 0.021187459 | 1 | 0.98201188 | 1 | 0.981619417 | ENSG0000007 | 7 | CYBRD1 |
| Pancreas | T1DM | ENSG0000011 | -0.00095553 | 0.412767414 | 0.001166653 | 0.008204764 | -0.00094192 | 0.41943545 | 0.001166604 | 0.008203879 | -1.36E-05 | 0.018873995 | 1 | 0.883403823 | 1 | 0.888704815 | ENSG0000007 | 7 | CYBRD1 |
| Pancreas | T1DM | ENSG0000011 | -0.0002489 | 0.438275518 | 0.000321111 | 0.008204765 | -0.00025336 | 0.4300849 | 0.000321098 | 0.00820388 | 4.47E-06 | 0.007335331 | 1 | 0.89813429 | 1 | 0.890691955 | ENSG0000007 | 11 | EPN2 |
| Pancreas | T1DM | ENSG0000011 | 0.001080707 | 0.359951899 | 0.00118051 | 0.008204761 | 0.001099768 | 0.351522208 | 0.001180463 | 0.008203875 | -1.91E-05 | 0.001886732 | 1 | 0.854209198 | 1 | 0.847508521 | ENSG0000007 | 8 | AFF4 |
| Pancreas | T1DM | ENSG0000011 | -0.00024766 | 0.783954475 | 0.00090332 | 0.008204774 | -0.0002358 | 0.794055066 | 0.00090329 | 0.008203889 | -1.19E-05 | 0.039497143 | 1 | 0.981920629 | 1 | 0.978923745 | ENSG0000007 | 14 | NFATC3 |
| Pancreas | T1DM | ENSG0000011 | -0.0006523 | 0.43630758 | 0.000837956 | 0.008204765 | -0.00066932 | 0.424419212 | 0.000837933 | 0.008203879 | 1.70E-05 | 0.005507918 | 1 | 0.89813429 | 1 | 0.889250668 | ENSG0000007 | 6 | SIDT1 |
| Pancreas | T1DM | ENSG0000011 | 0.001291097 | 0.187886178 | 0.000980436 | 0.008204746 | 0.002063326 | 0.042046131 | 0.001014883 | 0.00820382 | -0.00077223 | 0.00324806 | 1 | 0.750693896 | 1 | 0.471816773 | ENSG0000007 | 8 | NDE1 |
| Pancreas | T1DM | ENSG0000011 | -0.0005188 | 0.087853656 | 0.000303956 | 0.008204727 | -0.00051204 | 0.09205993 | 0.000303946 | 0.008203842 | -6.76E-06 | 0.000455637 | 1 | 0.595273618 | 1 | 0.598506077 | ENSG0000007 | 8 | NDE1 |
| Pancreas | T1DM | ENSG0000011 | 0.000251617 | 0.711901226 | 0.000681328 | 0.008204773 | 0.000244674 | 0.719499838 | 0.0006813 | 0.008203888 | 6.94E-06 | 0.048754124 | 1 | 0.975415042 | 1 | 0.968082788 | ENSG0000007 | 8 | NDE1 |
| Pancreas | T1DM | ENSG0000011 | 7.78E-05 | 0.890378559 | 0.000564128 | 0.008204775 | 8.46E-05 | 0.880723325 | 0.000564107 | 0.008203889 | -6.89E-06 | 0.036458887 | 1 | 0.99200447 | 1 | 0.993128823 | ENSG0000007 | 4 | AP1M1 |
| Pancreas | T1DM | ENSG0000011 | 0.000214867 | 0.659534917 | 0.000487717 | 0.008204772 | 0.000203266 | 0.676836651 | 0.0004877 | 0.008203887 | 1.16E-05 | 8.22E-05 | 1 | 0.962467202 | 1 | 0.96294544 | ENSG0000007 | 10 | PVR |
| Pancreas | T1DM | ENSG0000011 | 0.000567751 | 0.654498281 | 0.001268655 | 0.008204772 | 0.000546712 | 0.666501259 | 0.001268607 | 0.008203887 | 2.10E-05 | 0.003158774 | 1 | 0.962467202 | 1 | 0.961035047 | ENSG0000007 | 11 | MOV10L1 |
| Pancreas | T1DM | ENSG0000011 | -0.00048923 | 0.509452928 | 0.000741609 | 0.008204768 | -0.00050086 | 0.49942125 | 0.00074158 | 0.008203882 | 1.16E-05 | 0.004308947 | 1 | 0.914783379 | 1 | 0.914417995 | ENSG0000007 | 13 | ALPK1 |
| Pancreas | T1DM | ENSG0000011 | 0.000302579 | 0.743554177 | 0.000924893 | 0.008204774 | 0.000288698 | 0.754923885 | 0.000924857 | 0.008203888 | 1.39E-05 | 0.007803943 | 1 | 0.975415042 | 1 | 0.968082788 | ENSG0000007 | 9 | LLGL2 |
| Pancreas | T1DM | ENSG0000011 | 0.000198596 | 0.720149952 | 0.000554336 | 0.008204773 | 0.000206748 | 0.709165629 | 0.000554318 | 0.008203888 | -8.15E-06 | 0.023436003 | 1 | 0.975415042 | 1 | 0.968082788 | ENSG0000007 | 10 | PDE8A |
| Pancreas | T1DM | ENSG0000011 | -0.00088637 | 0.494125494 | 0.001296313 | 0.008204768 | -0.00086793 | 0.503139341 | 0.001296271 | 0.008203882 | -1.84E-05 | 0.030019971 | 1 | 0.914693561 | 1 | 0.914417995 | ENSG0000007 | 12 | SDHA |
| Pancreas | T1DM | ENSG0000011 | -0.00075507 | 0.231989404 | 0.000631728 | 0.008204752 | -0.00076282 | 0.227217968 | 0.000631705 | 0.008203865 | 7.75E-06 | 0.041496248 | 1 | 0.785512902 | 1 | 0.780702981 | ENSG0000007 | 14 | SMARCE1 |
| Pancreas | T1DM | ENSG0000011 | -0.00070644 | 0.241155839 | 0.000602709 | 0.008204752 | -0.0007156 | 0.23509371 | 0.00060269 | 0.008203866 | 9.16E-06 | 0.021526584 | 1 | 0.786897324 | 1 | 0.780702981 | ENSG0000007 | 9 | ADAM11 |
| Pancreas | T1DM | ENSG0000011 | 0.000787222 | 0.38263922 | 0.000901698 | 0.008204763 | 0.000775063 | 0.390017596 | 0.00090167 | 0.008203878 | 1.22E-05 | 0.047526292 | 1 | 0.864072637 | 1 | 0.867930487 | ENSG0000007 | 10 | ST6GAL1 |
| Pancreas | T1DM | ENSG0000012 | -0.00028308 | 0.395292088 | 0.000333006 | 0.008204763 | 0.000809872 | 0.164926047 | 0.000583189 | 0.008203858 | -0.00109295 | 0.022443223 | 1 | 0.873903579 | 1 | 0.726605951 | ENSG0000007 | 10 | ST6GAL1 |
| Pancreas | T1DM | ENSG0000012 | 0.000101554 | 0.747409786 | 0.000315328 | 0.008204774 | 0.000105403 | 0.738170077 | 0.000315317 | 0.008203888 | -3.85E-06 | 0.04162637 | 1 | 0.975415042 | 1 | 0.968082788 | ENSG0000007 | 10 | ST6GAL1 |
| Pancreas | T1DM | ENSG0000012 | 2.17E-05 | 0.985749482 | 0.001217373 | 0.008204776 | 9.10E-06 | 0.994036843 | 0.001217323 | 0.00820389 | 1.26E-05 | 0.038679977 | 1 | 0.997898334 | 1 | 0.999770584 | ENSG0000007 | 13 | CLASP1 |
| Pancreas | T1DM | ENSG0000012 | 2.76E-05 | 0.971031999 | 0.000761063 | 0.008204776 | 1.22E-05 | 0.987262157 | 0.000761036 | 0.00820389 | 1.55E-05 | 0.001010872 | 1 | 0.997898334 | 1 | 0.99966149 | ENSG0000007 | 9 | IPCEF1 |
| Pancreas | T1DM | ENSG0000012 | -0.00068452 | 0.485206211 | 0.000980747 | 0.008204767 | -0.00066259 | 0.499280119 | 0.000980713 | 0.008203882 | -2.19E-05 | 0.000321745 | 1 | 0.914693561 | 1 | 0.914417995 | ENSG0000007 | 9 | IPCEF1 |
| Pancreas | T1DM | ENSG0000012 | 2.61E-05 | 0.981443232 | 0.001120712 | 0.008204776 | 4.04E-06 | 0.997121781 | 0.001120677 | 0.00820389 | 2.20E-05 | 0.003305501 | 1 | 0.997898334 | 1 | 0.999770584 | ENSG0000007 | 9 | IPCEF1 |
| Pancreas | T1DM | ENSG0000012 | 0.000350257 | 0.719103506 | 0.000973864 | 0.008204773 | 0.000366201 | 0.706886317 | 0.000973832 | 0.008203888 | -1.59E-05 | 0.011776901 | 1 | 0.975415042 | 1 | 0.968082788 | ENSG0000007 | 15 | EIF4G3 |
| Pancreas | T1DM | ENSG0000012 | -0.00100632 | 0.292035992 | 0.000955069 | 0.008204757 | -0.0009944 | 0.297772862 | 0.000955036 | 0.008203872 | -1.19E-05 | 0.045818474 | 1 | 0.810235282 | 1 | 0.810652485 | ENSG0000007 | 15 | EIF4G3 |
| Pancreas | T1DM | ENSG0000012 | -0.00058211 | 0.489861774 | 0.000842988 | 0.008204768 | -0.00056913 | 0.499575994 | 0.000842963 | 0.008203882 | -1.30E-05 | 0.026653187 | 1 | 0.914693561 | 1 | 0.914417995 | ENSG0000007 | 5 | CACNG4 |
| Pancreas | T1DM | ENSG0000012 | 3.72E-05 | 0.974729086 | 0.001174306 | 0.008204776 | 5.12E-05 | 0.965216377 | 0.001174262 | 0.00820389 | -1.40E-05 | 0.035757709 | 1 | 0.997898334 | 1 | 0.999388266 | ENSG0000007 | 5 | CACNG4 |
| Pancreas | T1DM | ENSG0000012 | -0.00043791 | 0.331588398 | 0.000451024 | 0.00820476 | -0.00043097 | 0.339288772 | 0.000451011 | 0.008203875 | -6.93E-06 | 0.029735527 | 1 | 0.83462449 | 1 | 0.839770819 | ENSG0000007 | 5 | CACNG4 |
| Pancreas | T1DM | ENSG0000012 | -0.00101579 | 0.317868148 | 0.001016961 | 0.008204759 | -0.0009946 | 0.32805234 | 0.001016931 | 0.008203874 | -2.12E-05 | 0.00291871 | 1 | 0.829307456 | 1 | 0.830795913 | ENSG0000007 | 5 | MOCOS |
| Pancreas | T1DM | ENSG0000012 | -0.00014193 | 0.871988052 | 0.000880845 | 0.008204775 | -0.00012675 | 0.885578936 | 0.000880813 | 0.00820389 | -1.52E-05 | 0.002990139 | 1 | 0.99200447 | 1 | 0.993128823 | ENSG0000007 | 5 | MOCOS |
| Pancreas | T1DM | ENSG0000012 | -0.00071084 | 0.50825913 | 0.001074511 | 0.008204768 | -0.00068731 | 0.522386766 | 0.001074473 | 0.008203883 | -2.35E-05 | 0.000389448 | 1 | 0.914783379 | 1 | 0.917005275 | ENSG0000007 | 7 | ATP12A |
| Pancreas | T1DM | ENSG0000012 | 0.000620067 | 0.465297849 | 0.00084923 | 0.008204767 | 0.000604496 | 0.476562242 | 0.000849198 | 0.008203881 | 1.56E-05 | 0.001091923 | 1 | 0.908712989 | 1 | 0.911454131 | ENSG0000007 | 12 | SEC31B |
| Pancreas | T1DM | ENSG0000012 | 3.29E-06 | 0.996896842 | 0.000844799 | 0.008204776 | -1.37E-05 | 0.987095813 | 0.000844767 | 0.00820389 | 1.69E-05 | 0.000502356 | 1 | 0.998452063 | 1 | 0.99966149 | ENSG0000007 | 12 | SEC31B |
| Pancreas | T1DM | ENSG0000012 | -9.33E-05 | 0.859348072 | 0.00052671 | 0.008204775 | -8.44E-05 | 0.872665253 | 0.000526691 | 0.00820389 | -8.92E-06 | 0.00380453 | 1 | 0.990317162 | 1 | 0.993128823 | ENSG0000007 | 11 | MKRN2 |
| Pancreas | T1DM | ENSG0000012 | 0.000429069 | 0.657816713 | 0.000968706 | 0.008204772 | 0.00041438 | 0.668811971 | 0.000968677 | 0.008203887 | 1.47E-05 | 0.028742242 | 1 | 0.962467202 | 1 | 0.961035047 | ENSG0000007 | 10 | MLH1 |
| Pancreas | T1DM | ENSG0000012 | 0.000277292 | 0.572921297 | 0.000491866 | 0.00820477 | 0.000296059 | 0.547237113 | 0.000491869 | 0.008203884 | -1.88E-05 | 0.000477088 | 1 | 0.937588291 | 1 | 0.929364427 | ENSG0000007 | 10 | MLH1 |
| Pancreas | T1DM | ENSG0000012 | -0.00016807 | 0.858928668 | 0.000945615 | 0.008204775 | -0.00014917 | 0.874653525 | 0.000945599 | 0.00820389 | -1.89E-05 | 0.01987623 | 1 | 0.990317162 | 1 | 0.993128823 | ENSG0000007 | 10 | MLH1 |
| Pancreas | T1DM | ENSG0000012 | -0.00176495 | 0.132018264 | 0.001171794 | 0.008204737 | -0.00179102 | 0.126390095 | 0.001171756 | 0.008203851 | 2.61E-05 | 0.000645639 | 1 | 0.690950163 | 1 | 0.673807988 | ENSG0000007 | 5 | FMO4 |
| Pancreas | T1DM | ENSG0000012 | -0.00026372 | 0.590431913 | 0.000490002 | 0.008204771 | -0.00027465 | 0.575128008 | 0.000489987 | 0.008203885 | 1.09E-05 | 0.000890531 | 1 | 0.937680367 | 1 | 0.936315417 | ENSG0000007 | 11 | RGS11 |
| Pancreas | T1DM | ENSG0000012 | -0.00089967 | 0.005618459 | 0.000324877 | 0.008204646 | -0.00093985 | 0.003843221 | 0.000325122 | 0.008203749 | 4.02E-05 | 0.002114768 | 1 | 0.179978441 | 1 | 0.129860443 | ENSG0000007 | 11 | RGS11 |
| Pancreas | T1DM | ENSG0000012 | 3.57E-05 | 0.959043238 | 0.000694757 | 0.008204776 | 5.17E-05 | 0.940673647 | 0.000694747 | 0.00820389 | -1.60E-05 | 0.009687191 | 1 | 0.997898334 | 1 | 0.999388266 | ENSG0000007 | 11 | RGS11 |
| Pancreas | T1DM | ENSG0000012 | 0.000539347 | 0.173776254 | 0.000396529 | 0.008204744 | 0.000472014 | 0.234081206 | 0.000396679 | 0.008203866 | 6.73E-05 | 7.91E-09 | 1 | 0.729268002 | 1 | 0.780702981 | ENSG0000007 | 11 | TPD52 |
| Pancreas | T1DM | ENSG0000012 | -0.00153786 | 0.220873746 | 0.001256203 | 0.00820475 | -0.00150513 | 0.230848308 | 0.001256186 | 0.008203866 | -3.27E-05 | 0.003842161 | 1 | 0.780835021 | 1 | 0.780702981 | ENSG0000007 | 11 | TPD52 |
| Pancreas | T1DM | ENSG0000012 | -5.19E-05 | 0.943649503 | 0.000734806 | 0.008204775 | -4.00E-05 | 0.956614744 | 0.000734782 | 0.00820389 | -1.20E-05 | 0.012187931 | 1 | 0.997898334 | 1 | 0.999388266 | ENSG0000007 | 5 | ICAM3 |
| Pancreas | T1DM | ENSG0000012 | -9.95E-05 | 0.847268785 | 0.000516462 | 0.008204775 | -8.78E-05 | 0.864977574 | 0.000516447 | 0.008203889 | -1.17E-05 | 0.001012161 | 1 | 0.989913489 | 1 | 0.990577333 | ENSG0000007 | 5 | ICAM3 |
| Pancreas | T1DM | ENSG0000012 | 0.00148391 | 0.083504357 | 0.0008574 | 0.008204725 | 0.001470063 | 0.086415878 | 0.00085737 | 0.00820384 | 1.38E-05 | 0.008718723 | 1 | 0.589819284 | 1 | 0.580557814 | ENSG0000007 | 5 | ICAM3 |
| Pancreas | T1DM | ENSG0000012 | -0.00150498 | 0.137413457 | 0.001013115 | 0.008204738 | -0.00149103 | 0.141080043 | 0.00101308 | 0.008203854 | -1.39E-05 | 0.027783784 | 1 | 0.69738343 | 1 | 0.693219864 | ENSG0000007 | 5 | ICAM3 |
| Pancreas | T1DM | ENSG0000012 | -0.0003322 | 0.649284994 | 0.000730499 | 0.008204772 | -0.0003147 | 0.666609506 | 0.000730484 | 0.008203887 | -1.75E-05 | 0.003414596 | 1 | 0.962467202 | 1 | 0.961035047 | ENSG0000007 | 14 | NT5C2 |
| Pancreas | T1DM | ENSG0000012 | -0.00020231 | 0.758125883 | 0.000656957 | 0.008204774 | -0.00019167 | 0.770464977 | 0.000656937 | 0.008203889 | -1.06E-05 | 0.016941434 | 1 | 0.976362722 | 1 | 0.972640681 | ENSG0000007 | 14 | NT5C2 |
| Pancreas | T1DM | ENSG0000012 | 0.000464378 | 0.467497101 | 0.000639149 | 0.008204767 | 0.000453094 | 0.478368463 | 0.000639123 | 0.008203882 | 1.13E-05 | 0.001061706 | 1 | 0.908712989 | 1 | 0.911454131 | ENSG0000007 | 14 | NT5C2 |
| Pancreas | T1DM | ENSG0000012 | 0.000202242 | 0.645686481 | 0.000439883 | 0.008204772 | 0.000194261 | 0.65875862 | 0.000439875 | 0.008203887 | 7.98E-06 | 0.032873626 | 1 | 0.961788216 | 1 | 0.961035047 | ENSG0000007 | 14 | NT5C2 |
| Pancreas | T1DM | ENSG0000012 | -0.00040439 | 0.743822652 | 0.001237453 | 0.008204774 | -0.00038769 | 0.754406006 | 0.001237406 | 0.008203888 | -1.67E-05 | 0.017545243 | 1 | 0.975415042 | 1 | 0.968082788 | ENSG0000007 | 5 | ACTL6B |
| Pancreas | T1DM | ENSG0000012 | 0.000581286 | 0.380069575 | 0.000662231 | 0.008204763 | 0.000597283 | 0.367084995 | 0.000662215 | 0.008203876 | -1.60E-05 | 0.001477302 | 1 | 0.862207304 | 1 | 0.850635855 | ENSG0000007 | 10 | TM9SF3 |
| Pancreas | T1DM | ENSG0000012 | 0.000590576 | 0.481039923 | 0.000838135 | 0.008204767 | 0.00061565 | 0.462602216 | 0.0008381 | 0.008203881 | -2.51E-05 | 1.65E-05 | 1 | 0.913691214 | 1 | 0.901757398 | ENSG0000007 | 9 | IL4R |
| Pancreas | T1DM | ENSG0000012 | -0.00185465 | 0.014209628 | 0.000756404 | 0.008204674 | -0.00186449 | 0.013700416 | 0.000756374 | 0.008203788 | 9.84E-06 | 0.013940132 | 1 | 0.2919226 | 1 | 0.277028884 | ENSG0000007 | 11 | APBB1IP |
| Pancreas | T1DM | ENSG0000013 | -0.00025268 | 0.699630708 | 0.000654908 | 0.008204773 | -0.00026479 | 0.685976223 | 0.000654889 | 0.008203887 | 1.21E-05 | 0.010159595 | 1 | 0.974470668 | 1 | 0.966338775 | ENSG0000007 | 9 | GPR137B |
| Pancreas | T1DM | ENSG0000013 | -0.00053442 | 0.230617073 | 0.00044581 | 0.008204751 | -0.00055716 | 0.211506337 | 0.000445927 | 0.008203864 | 2.27E-05 | 0.043199695 | 1 | 0.785512902 | 1 | 0.769778216 | ENSG0000007 | 9 | GPR137B |
| Pancreas | T1DM | ENSG0000013 | 0.000388518 | 0.423325991 | 0.000485244 | 0.008204765 | 0.000396336 | 0.414042308 | 0.000485229 | 0.008203879 | -7.82E-06 | 0.017599201 | 1 | 0.889410424 | 1 | 0.883839257 | ENSG0000007 | 9 | GPR137B |
| Pancreas | T1DM | ENSG0000013 | 0.001652188 | 0.181521923 | 0.001236588 | 0.008204746 | 0.001626113 | 0.188494278 | 0.001236542 | 0.008203861 | 2.61E-05 | 0.000241839 | 1 | 0.737576421 | 1 | 0.753977114 | ENSG0000007 | 9 | GPR137B |
| Pancreas | T1DM | ENSG0000013 | -0.00045801 | 0.284836181 | 0.000428242 | 0.008204756 | -0.00045258 | 0.290567788 | 0.000428226 | 0.008203871 | -5.43E-06 | 0.031441908 | 1 | 0.804377068 | 1 | 0.805533316 | ENSG0000007 | 6 | NAALAD2 |
| Pancreas | T1DM | ENSG0000013 | -0.00054704 | 0.137312684 | 0.000368161 | 0.008204738 | -0.00054197 | 0.140976267 | 0.000368148 | 0.008203853 | -5.07E-06 | 0.022425979 | 1 | 0.69738343 | 1 | 0.693219864 | ENSG0000007 | 6 | NAALAD2 |
| Pancreas | T1DM | ENSG0000013 | 0.000257372 | 0.796553225 | 0.000998298 | 0.008204774 | 0.000272725 | 0.784702469 | 0.000998274 | 0.008203889 | -1.54E-05 | 0.046289227 | 1 | 0.981920629 | 1 | 0.976789113 | ENSG0000007 | 15 | MAP2 |
| Pancreas | T1DM | ENSG0000013 | -0.00080095 | 0.304082205 | 0.000779345 | 0.008204758 | -0.0008133 | 0.296671121 | 0.000779322 | 0.008203872 | 1.24E-05 | 0.021667642 | 1 | 0.82025536 | 1 | 0.809618957 | ENSG0000007 | 7 | MCCC1 |
| Pancreas | T1DM | ENSG0000013 | 0.000665742 | 0.469833389 | 0.000921122 | 0.008204767 | 0.000648384 | 0.481477474 | 0.000921094 | 0.008203882 | 1.74E-05 | 0.005313078 | 1 | 0.908712989 | 1 | 0.913030722 | ENSG0000007 | 7 | MCCC1 |
| Pancreas | T1DM | ENSG0000013 | -0.00265613 | 0.004651242 | 0.000938475 | 0.008204641 | -0.00267396 | 0.004381181 | 0.000938447 | 0.008203753 | 1.78E-05 | 0.006578463 | 1 | 0.151194791 | 1 | 0.138899658 | ENSG0000007 | 9 | SYNJ2 |

| | | | | | | | | | | | | | | | | | | | |
|---|---|---|---|---|---|---|---|---|---|---|---|---|---|---|---|---|---|---|---|
| Pancreas | T1DM | ENSG0000013 | -7.41E-05 | 0.935168321 | 0.000911021 | 0.008204775 | -6.26E-05 | 0.945226393 | 0.00091099 | 0.00820389 | -1.15E-05 | 0.041364769 | 1 | 0.997898334 | 1 | 0.999388266 | ENSG0000007 | 11 | ADCY2 |
| Pancreas | T1DM | ENSG0000013 | 0.000537225 | 0.375990118 | 0.000606821 | 0.008204762 | 0.000530457 | 0.382014746 | 0.000606798 | 0.008203877 | 6.77E-06 | 0.044973072 | 1 | 0.860999343 | 1 | 0.860591675 | ENSG0000007 | 15 | NRD1 |
| Pancreas | T1DM | ENSG0000013 | 0.002642411 | 0.038488242 | 0.00127676 | 0.008204703 | 0.002624793 | 0.039793175 | 0.001276715 | 0.008203819 | 1.76E-05 | 0.026170232 | 1 | 0.459710727 | 1 | 0.458621792 | ENSG0000007 | 13 | FKBP7 |
| Pancreas | T1DM | ENSG0000013 | 9.90E-05 | 0.85550284 | 0.000543386 | 0.008204775 | 8.85E-05 | 0.870594175 | 0.000543366 | 0.00820389 | 1.04E-05 | 0.000566081 | 1 | 0.990317162 | 1 | 0.992315064 | ENSG0000007 | 13 | FKBP7 |
| Pancreas | T1DM | ENSG0000013 | -0.0001448 | 0.831176476 | 0.000679179 | 0.008204775 | -0.00013726 | 0.83983486 | 0.000679152 | 0.008203889 | -7.54E-06 | 0.033142885 | 1 | 0.984098328 | 1 | 0.98524254 | ENSG0000007 | 13 | FKBP7 |
| Pancreas | T1DM | ENSG0000013 | -0.00091489 | 0.358676047 | 0.000996735 | 0.008204761 | -0.00090067 | 0.36617861 | 0.000996696 | 0.008203876 | -1.42E-05 | 0.008775806 | 1 | 0.854209198 | 1 | 0.850635855 | ENSG0000007 | 13 | FKBP7 |
| Pancreas | T1DM | ENSG0000013 | 0.000173029 | 0.694594293 | 0.000440695 | 0.008204773 | 0.000166348 | 0.705818867 | 0.000440682 | 0.008203888 | 6.68E-06 | 0.028125872 | 1 | 0.972276902 | 1 | 0.968082788 | ENSG0000007 | 13 | FKBP7 |
| Pancreas | T1DM | ENSG0000013 | -0.00105241 | 0.410447484 | 0.001278582 | 0.008204764 | -0.00103334 | 0.418966357 | 0.001278538 | 0.008203879 | -1.91E-05 | 0.017473264 | 1 | 0.88063625 | 1 | 0.888443934 | ENSG0000007 | 3 | LXN |
| Pancreas | T1DM | ENSG0000013 | -0.00034675 | 0.658215865 | 0.000783839 | 0.008204772 | -0.00035697 | 0.648799138 | 0.000783811 | 0.008203886 | 1.02E-05 | 0.030434049 | 1 | 0.962467202 | 1 | 0.961035047 | ENSG0000007 | 3 | LXN |
| Pancreas | T1DM | ENSG0000013 | -0.00018969 | 0.857695787 | 0.001057904 | 0.008204775 | -0.00020449 | 0.84671824 | 0.001057863 | 0.008203889 | 1.48E-05 | 0.011702719 | 1 | 0.990317162 | 1 | 0.986098414 | ENSG0000007 | 3 | LXN |
| Pancreas | T1DM | ENSG0000013 | -0.0003558 | 0.511138224 | 0.000541494 | 0.008204768 | -0.00034243 | 0.527128364 | 0.000541475 | 0.008203883 | -1.34E-05 | 9.33E-05 | 1 | 0.914783379 | 1 | 0.920313283 | ENSG0000007 | 3 | LXN |
| Pancreas | T1DM | ENSG0000013 | -0.00074923 | 0.257149133 | 0.000661191 | 0.008204754 | -0.00075643 | 0.252588631 | 0.000661165 | 0.008203868 | 7.20E-06 | 0.045412353 | 1 | 0.79720158 | 1 | 0.788955599 | ENSG0000007 | 3 | LXN |
| Pancreas | T1DM | ENSG0000013 | 0.000868505 | 0.436495516 | 0.001116151 | 0.008204765 | 0.000685602 | 0.54035752 | 0.001119765 | 0.008203884 | 0.000182903 | 0.043606063 | 1 | 0.89813429 | 1 | 0.928420568 | ENSG0000007 | 14 | SP140 |
| Pancreas | T1DM | ENSG0000013 | -0.00012218 | 0.910567467 | 0.001087791 | 0.008204775 | -0.00013752 | 0.899397851 | 0.001087757 | 0.00820389 | 1.53E-05 | 0.034698803 | 1 | 0.995698473 | 1 | 0.994038165 | ENSG0000007 | 14 | SP140 |
| Pancreas | T1DM | ENSG0000013 | 0.000380564 | 0.232048938 | 0.000318437 | 0.008204752 | 0.000374626 | 0.239399323 | 0.000318427 | 0.008203867 | 5.94E-06 | 0.003832229 | 1 | 0.785512902 | 1 | 0.780702981 | ENSG0000007 | 14 | SP140 |
| Pancreas | T1DM | ENSG0000013 | 0.000157112 | 0.750042153 | 0.000493157 | 0.008204774 | 0.000149764 | 0.761361999 | 0.000493143 | 0.008203888 | 7.35E-06 | 0.036706758 | 1 | 0.975415042 | 1 | 0.968355942 | ENSG0000007 | 13 | CDC14A |
| Pancreas | T1DM | ENSG0000013 | 0.001507355 | 0.032821152 | 0.000706263 | 0.008204699 | 0.001520877 | 0.031281635 | 0.000706242 | 0.008203812 | -1.35E-05 | 0.006950098 | 1 | 0.434431116 | 1 | 0.417001664 | ENSG0000007 | 13 | CDC14A |
| Pancreas | T1DM | ENSG0000013 | 0.000189971 | 0.849446058 | 0.001000768 | 0.008204775 | 0.000177023 | 0.859591988 | 0.001000733 | 0.008203889 | 1.29E-05 | 0.033560969 | 1 | 0.989913489 | 1 | 0.990577333 | ENSG0000007 | 13 | CDC14A |
| Pancreas | T1DM | ENSG0000013 | 0.00069815 | 0.512875512 | 0.001066909 | 0.008204768 | 0.000673495 | 0.52786791 | 0.001066899 | 0.008203883 | 2.47E-05 | 0.014336587 | 1 | 0.914783379 | 1 | 0.920313283 | ENSG0000007 | 8 | FDFT1 |
| Pancreas | T1DM | ENSG0000013 | 0.000991492 | 0.385938981 | 0.001143581 | 0.008204763 | 0.001007229 | 0.378427546 | 0.001143541 | 0.008203877 | -1.57E-05 | 0.028256572 | 1 | 0.865581925 | 1 | 0.858482278 | ENSG0000007 | 9 | SCGN |
| Pancreas | T1DM | ENSG0000013 | 0.000125828 | 0.898778743 | 0.00098918 | 0.008204775 | 0.000112721 | 0.909270813 | 0.000989144 | 0.00820389 | 1.31E-05 | 0.02501854 | 1 | 0.99281063 | 1 | 0.996366272 | ENSG0000007 | 9 | SCGN |
| Pancreas | T1DM | ENSG0000013 | 0.000664581 | 0.106568907 | 0.000411808 | 0.008204732 | 0.000654562 | 0.11195248 | 0.000411808 | 0.008203847 | 1.00E-05 | 0.019106463 | 1 | 0.661051879 | 1 | 0.666335944 | ENSG0000007 | 9 | SCGN |
| Pancreas | T1DM | ENSG0000013 | 0.003197771 | 8.88E-06 | 0.000719771 | 0.008204443 | 0.002643162 | 0.000293663 | 0.000729983 | 0.008203669 | 0.000554609 | 5.35E-06 | 0.022811571 | 0.000543133 | 0.754127073 | 0.016394067 | ENSG0000007 | 9 | SCGN |
| Pancreas | T1DM | ENSG0000013 | 0.00079295 | 0.10340664 | 0.000486902 | 0.008204731 | 0.000743768 | 0.126838879 | 0.000487177 | 0.008203851 | 4.92E-05 | 0.004052413 | 1 | 0.654059731 | 1 | 0.673807988 | ENSG0000007 | 9 | STX7 |
| Pancreas | T1DM | ENSG0000013 | 0.000939376 | 0.361242675 | 0.001028889 | 0.008204762 | 0.000954637 | 0.353476656 | 0.001028851 | 0.008203875 | -1.53E-05 | 0.01128258 | 1 | 0.854209198 | 1 | 0.847990418 | ENSG0000007 | 9 | STX7 |
| Pancreas | T1DM | ENSG0000013 | 0.000727922 | 0.216223252 | 0.000588631 | 0.00820475 | 0.000747939 | 0.20384305 | 0.000588613 | 0.008203863 | -2.00E-05 | 1.01E-06 | 1 | 0.77457828 | 1 | 0.76794547 | ENSG0000008 | 13 | RIF1 |
| Pancreas | T1DM | ENSG0000013 | 0.000629403 | 0.23809428 | 0.000533499 | 0.008204752 | 0.000621229 | 0.244233062 | 0.000533485 | 0.008203867 | 8.17E-06 | 0.041236596 | 1 | 0.785512902 | 1 | 0.78207287 | ENSG0000008 | 14 | PSEN1 |
| Pancreas | T1DM | ENSG0000013 | 0.000440018 | 0.732007083 | 0.001284886 | 0.008204774 | 0.00046394 | 0.718034933 | 0.001284844 | 0.008203888 | -2.39E-05 | 0.004693324 | 1 | 0.975415042 | 1 | 0.968082788 | ENSG0000008 | 9 | ZNF506 |
| Pancreas | T1DM | ENSG0000013 | 0.000193062 | 0.826963659 | 0.000883184 | 0.008204775 | -0.00033173 | 0.717535271 | 0.000917001 | 0.008203888 | 0.000524792 | 0.033546645 | 1 | 0.984098328 | 1 | 0.968082788 | ENSG0000008 | 4 | KIAA0141 |
| Pancreas | T1DM | ENSG0000013 | 0.000317371 | 0.741545578 | 0.000962293 | 0.008204774 | 0.000357701 | 0.710130918 | 0.000962388 | 0.008203888 | -4.03E-05 | 0.016703516 | 1 | 0.975415042 | 1 | 0.968082788 | ENSG0000008 | 9 | PHLPP1 |
| Pancreas | T1DM | ENSG0000013 | 0.001063149 | 0.363590485 | 0.001170166 | 0.008204762 | 0.001022088 | 0.382401784 | 0.001170132 | 0.008203877 | 4.11E-05 | 7.11E-07 | 1 | 0.854831234 | 1 | 0.860591675 | ENSG0000008 | 8 | GEMIN5 |
| Pancreas | T1DM | ENSG0000013 | 0.001146333 | 0.352874629 | 0.001233905 | 0.008204761 | 0.001167713 | 0.343948716 | 0.001233856 | 0.008203875 | -2.14E-05 | 0.00130294 | 1 | 0.854061442 | 1 | 0.846034773 | ENSG0000008 | 8 | GEMIN5 |
| Pancreas | T1DM | ENSG0000013 | -0.00022419 | 0.637985445 | 0.000476469 | 0.008204772 | -0.00021756 | 0.647940829 | 0.000476449 | 0.008203886 | -6.63E-06 | 0.004739601 | 1 | 0.960781248 | 1 | 0.961035047 | ENSG0000008 | 8 | GEMIN5 |
| Pancreas | T1DM | ENSG0000013 | 0.000129832 | 0.871328984 | 0.000801579 | 0.008204775 | 0.000112047 | 0.888828209 | 0.000801553 | 0.00820389 | 1.78E-05 | 0.000611504 | 1 | 0.99200447 | 1 | 0.993128823 | ENSG0000008 | 8 | GEMIN5 |
| Pancreas | T1DM | ENSG0000013 | 0.000289981 | 0.738044526 | 0.000867054 | 0.008204774 | 0.000300708 | 0.728720539 | 0.000867022 | 0.008203888 | -1.07E-05 | 0.036637615 | 1 | 0.975415042 | 1 | 0.968082788 | ENSG0000008 | 10 | SEMA5B |
| Pancreas | T1DM | ENSG0000013 | 0.000262747 | 0.578214206 | 0.00047257 | 0.00820477 | 0.000252532 | 0.593069474 | 0.000472559 | 0.008203885 | 1.02E-05 | 0.005023471 | 1 | 0.937588291 | 1 | 0.937577789 | ENSG0000008 | 10 | DOPEY1 |
| Pancreas | T1DM | ENSG0000013 | -0.00065959 | 0.201100006 | 0.000515939 | 0.008204748 | -0.00066615 | 0.19663886 | 0.000515921 | 0.008203862 | 6.56E-06 | 0.041270833 | 1 | 0.7650738 | 1 | 0.762792436 | ENSG0000008 | 10 | DOPEY1 |
| Pancreas | T1DM | ENSG0000013 | -0.0002414 | 0.652927041 | 0.000536794 | 0.008204772 | -0.00024729 | 0.645017611 | 0.000536774 | 0.008203886 | 5.89E-06 | 0.046169391 | 1 | 0.962467202 | 1 | 0.961035047 | ENSG0000008 | 10 | DOPEY1 |
| Pancreas | T1DM | ENSG0000013 | 0.000646852 | 0.60983979 | 0.001267584 | 0.008204771 | 0.000679452 | 0.591934376 | 0.001267551 | 0.008203885 | -3.26E-05 | 0.000580792 | 1 | 0.943530179 | 1 | 0.937577789 | ENSG0000008 | 10 | DOPEY1 |
| Pancreas | T1DM | ENSG0000013 | -9.19E-06 | 0.994258331 | 0.001277017 | 0.008204776 | -6.34E-05 | 0.960391043 | 0.001277188 | 0.00820389 | 5.42E-05 | 0.028590631 | 1 | 0.998452063 | 1 | 0.999388266 | ENSG0000008 | 6 | LYRM2 |
| Pancreas | T1DM | ENSG0000013 | -0.00067923 | 0.245625802 | 0.000585016 | 0.008204753 | -0.0006889 | 0.23894801 | 0.000584995 | 0.008203867 | 9.68E-06 | 0.007039465 | 1 | 0.788479116 | 1 | 0.780702981 | ENSG0000008 | 6 | LYRM2 |
| Pancreas | T1DM | ENSG0000013 | -0.00023402 | 0.816802659 | 0.001010171 | 0.008204775 | -0.00021311 | 0.832910133 | 0.001010132 | 0.008203889 | -2.09E-05 | 0.000210485 | 1 | 0.98201188 | 1 | 0.984766677 | ENSG0000008 | 6 | LYRM2 |
| Pancreas | T1DM | ENSG0000013 | 0.000844118 | 0.454952367 | 0.001129731 | 0.008204766 | 0.000875226 | 0.438506312 | 0.001129734 | 0.00820388 | -3.11E-05 | 0.009706188 | 1 | 0.899832468 | 1 | 0.896263433 | ENSG0000008 | 15 | ULK2 |
| Pancreas | T1DM | ENSG0000013 | -0.00025999 | 0.307636281 | 0.000254844 | 0.008204758 | -0.00025622 | 0.31468764 | 0.000254837 | 0.008203873 | -3.77E-06 | 0.036744247 | 1 | 0.824645062 | 1 | 0.826271144 | ENSG0000008 | 8 | OXCT1 |
| Pancreas | T1DM | ENSG0000013 | -0.00048609 | 0.678755564 | 0.001173664 | 0.008204773 | -0.0004658 | 0.691452774 | 0.001173633 | 0.008203887 | -2.03E-05 | 0.020006521 | 1 | 0.966210802 | 1 | 0.966923215 | ENSG0000008 | 11 | ZNF586 |
| Pancreas | T1DM | ENSG0000013 | 0.000221191 | 0.792415945 | 0.000840473 | 0.008204774 | 0.000208139 | 0.804402604 | 0.000840444 | 0.008203889 | 1.31E-05 | 0.012489161 | 1 | 0.981920629 | 1 | 0.979350575 | ENSG0000008 | 11 | ZNF586 |
| Pancreas | T1DM | ENSG0000014 | 0.000380714 | 0.651803184 | 0.000843672 | 0.008204772 | 0.000363644 | 0.66644021 | 0.000843644 | 0.008203887 | 1.71E-05 | 0.001811708 | 1 | 0.962467202 | 1 | 0.961035047 | ENSG0000008 | 4 | RPS5 |
| Pancreas | T1DM | ENSG0000014 | -0.00024496 | 0.7859343 | 0.000901926 | 0.008204774 | -0.00023301 | 0.796131263 | 0.000901897 | 0.008203889 | -1.19E-05 | 0.042685041 | 1 | 0.981920629 | 1 | 0.978923745 | ENSG0000008 | 4 | CHMP2B |
| Pancreas | T1DM | ENSG0000014 | 3.28E-05 | 0.975419844 | 0.001063216 | 0.008204776 | 5.39E-05 | 0.959560501 | 0.001063186 | 0.00820389 | -2.11E-05 | 0.005487693 | 1 | 0.997898334 | 1 | 0.999388266 | ENSG0000008 | 12 | PPIE |
| Pancreas | T1DM | ENSG0000014 | 0.000260877 | 0.669618667 | 0.000611423 | 0.008204772 | 0.000249442 | 0.683286193 | 0.000611403 | 0.008203887 | 1.14E-05 | 0.00328313 | 1 | 0.963860935 | 1 | 0.966338775 | ENSG0000008 | 12 | PPIE |
| Pancreas | T1DM | ENSG0000014 | 0.000420798 | 0.675756583 | 0.001006066 | 0.008204773 | 0.00043355 | 0.666503155 | 0.001006028 | 0.008203887 | -1.28E-05 | 0.029236868 | 1 | 0.963860935 | 1 | 0.961035047 | ENSG0000008 | 6 | NOA1 |
| Pancreas | T1DM | ENSG0000014 | -0.00115849 | 0.359040063 | 0.001263075 | 0.008204761 | -0.00118203 | 0.349341584 | 0.001263026 | 0.008203875 | 2.35E-05 | 0.000749911 | 1 | 0.854209198 | 1 | 0.847508521 | ENSG0000008 | 6 | NOA1 |
| Pancreas | T1DM | ENSG0000014 | -3.81E-05 | 0.966245054 | 0.000899461 | 0.008204776 | -1.88E-05 | 0.983298747 | 0.00089944 | 0.00820389 | -1.92E-05 | 0.006517581 | 1 | 0.997898334 | 1 | 0.99966149 | ENSG0000008 | 6 | NOA1 |
| Pancreas | T1DM | ENSG0000014 | 0.000112732 | 0.893408749 | 0.000841335 | 0.008204775 | 0.000123823 | 0.882989833 | 0.000841303 | 0.00820389 | -1.11E-05 | 0.016723735 | 1 | 0.99200447 | 1 | 0.993128823 | ENSG0000008 | 11 | GSTP1 |
| Pancreas | T1DM | ENSG0000014 | -0.00031985 | 0.283685347 | 0.000298344 | 0.008204756 | -0.00031186 | 0.295869959 | 0.000298335 | 0.008203872 | -7.99E-06 | 7.86E-05 | 1 | 0.804088269 | 1 | 0.809618957 | ENSG0000008 | 11 | GSTP1 |
| Pancreas | T1DM | ENSG0000014 | -0.00021513 | 0.755594163 | 0.000691131 | 0.008204774 | -0.00020573 | 0.765946913 | 0.000691108 | 0.008203888 | -9.40E-06 | 0.031697947 | 1 | 0.975415042 | 1 | 0.971814067 | ENSG0000008 | 11 | GSTP1 |
| Pancreas | T1DM | ENSG0000014 | 0.000164903 | 0.813110788 | 0.000697514 | 0.008204775 | 0.000177324 | 0.799318201 | 0.000697497 | 0.008203889 | -1.24E-05 | 0.021281045 | 1 | 0.98201188 | 1 | 0.978923745 | ENSG0000008 | 11 | AGBL5 |
| Pancreas | T1DM | ENSG0000014 | 0.000580643 | 0.596859712 | 0.001097786 | 0.008204771 | 0.000597293 | 0.586369575 | 0.001097754 | 0.008203885 | -1.66E-05 | 0.029378614 | 1 | 0.938170574 | 1 | 0.936945142 | ENSG0000008 | 9 | CAD |
| Pancreas | T1DM | ENSG0000014 | -0.00019112 | 0.849803406 | 0.001009254 | 0.008204775 | -0.0001554 | 0.877640567 | 0.001009356 | 0.00820389 | -3.57E-05 | 0.044462721 | 1 | 0.989913489 | 1 | 0.993128823 | ENSG0000008 | 10 | CD59 |
| Pancreas | T1DM | ENSG0000014 | -0.00071481 | 0.248643879 | 0.000619609 | 0.008204753 | -0.00072472 | 0.242127131 | 0.000619585 | 0.008203867 | 9.91E-06 | 0.003175046 | 1 | 0.789267592 | 1 | 0.780845092 | ENSG0000008 | 7 | CD82 |
| Pancreas | T1DM | ENSG0000014 | 0.000229469 | 0.444100538 | 0.000299847 | 0.008204766 | 0.000224916 | 0.453177522 | 0.000299836 | 0.008203881 | 4.55E-06 | 0.014126067 | 1 | 0.898699907 | 1 | 0.899350755 | ENSG0000008 | 7 | CD82 |
| Pancreas | T1DM | ENSG0000014 | 0.0006362 | 0.596170359 | 0.001200568 | 0.008204771 | 0.000620887 | 0.605030467 | 0.001200524 | 0.008203885 | 1.53E-05 | 0.029058385 | 1 | 0.938170574 | 1 | 0.939938439 | ENSG0000008 | 7 | CD82 |
| Pancreas | T1DM | ENSG0000014 | -0.00013007 | 0.697546843 | 0.00033468 | 0.008204773 | -0.00013496 | 0.686747767 | 0.000334668 | 0.008203887 | 4.89E-06 | 0.018010521 | 1 | 0.974062149 | 1 | 0.966338775 | ENSG0000008 | 7 | CD82 |
| Pancreas | T1DM | ENSG0000014 | 0.000211638 | 0.807484963 | 0.000868537 | 0.008204775 | 0.000229027 | 0.792008652 | 0.000868503 | 0.008203889 | -1.74E-05 | 0.000283038 | 1 | 0.98201188 | 1 | 0.978923745 | ENSG0000008 | 7 | CD82 |
| Pancreas | T1DM | ENSG0000014 | -0.00057262 | 0.584540797 | 0.001047295 | 0.008204771 | -0.00052919 | 0.613377385 | 0.001047363 | 0.008203886 | -4.34E-05 | 0.007066927 | 1 | 0.937588291 | 1 | 0.946218353 | ENSG0000008 | 7 | CD82 |
| Pancreas | T1DM | ENSG0000014 | 0.000154728 | 0.809427442 | 0.000641586 | 0.008204775 | 0.000165314 | 0.796660334 | 0.000641569 | 0.008203889 | -1.06E-05 | 0.024977669 | 1 | 0.98201188 | 1 | 0.978923745 | ENSG0000008 | 7 | CD82 |
| Pancreas | T1DM | ENSG0000014 | 0.000517063 | 0.308184157 | 0.000507401 | 0.008204758 | 0.000525392 | 0.300440446 | 0.000507385 | 0.008203872 | -8.33E-06 | 0.011268513 | 1 | 0.825252257 | 1 | 0.812137964 | ENSG0000008 | 7 | CD82 |
| Pancreas | T1DM | ENSG0000014 | -0.00254689 | 0.048833978 | 0.001292809 | 0.00820471 | -0.0025749 | 0.046395761 | 0.001292764 | 0.008203823 | 2.80E-05 | 0.000572885 | 1 | 0.507019677 | 1 | 0.490305822 | ENSG0000008 | 7 | CD82 |
| Pancreas | T1DM | ENSG0000014 | 0.000227931 | 0.693255435 | 0.000577858 | 0.008204773 | 0.000219622 | 0.703890808 | 0.000577839 | 0.008203888 | 8.31E-06 | 0.028136221 | 1 | 0.971848816 | 1 | 0.968082788 | ENSG0000008 | 7 | CD82 |
| Pancreas | T1DM | ENSG0000014 | 0.000684064 | 0.4395478 | 0.000884996 | 0.008204766 | 0.000700369 | 0.428706199 | 0.000884965 | 0.008203879 | -1.63E-05 | 0.003132904 | 1 | 0.89813429 | 1 | 0.890691955 | ENSG0000008 | 7 | CD82 |
| Pancreas | T1DM | ENSG0000014 | 0.000151516 | 0.870104515 | 0.000926558 | 0.008204775 | 0.000163816 | 0.859659597 | 0.000926521 | 0.008203889 | -1.23E-05 | 0.01388643 | 1 | 0.99200447 | 1 | 0.990577333 | ENSG0000008 | 13 | SCAMP1 |
| Pancreas | T1DM | ENSG0000014 | 0.000341518 | 0.768302505 | 0.001159276 | 0.008204774 | 0.000358003 | 0.757452879 | 0.001159233 | 0.008203888 | -1.65E-05 | 0.014619908 | 1 | 0.977622335 | 1 | 0.968082788 | ENSG0000008 | 13 | SCAMP1 |
| Pancreas | T1DM | ENSG0000014 | 0.000268515 | 0.826252941 | 0.001223247 | 0.008204775 | 0.000248767 | 0.838844776 | 0.001223213 | 0.008203889 | 1.97E-05 | 0.025750463 | 1 | 0.984098328 | 1 | 0.98524254 | ENSG0000008 | 13 | SCAMP1 |
| Pancreas | T1DM | ENSG0000014 | -0.00060842 | 0.207980663 | 0.000483201 | 0.008204749 | -0.00060209 | 0.212733713 | 0.000483182 | 0.008203864 | -6.33E-06 | 0.018037498 | 1 | 0.770698909 | 1 | 0.769778216 | ENSG0000008 | 11 | SLC25A24 |
| Pancreas | T1DM | ENSG0000014 | -0.0001331 | 0.894978465 | 0.001008281 | 0.008204775 | -0.0001119 | 0.911626391 | 0.001008263 | 0.00820389 | -2.12E-05 | 0.01335978 | 1 | 0.99200447 | 1 | 0.996366272 | ENSG0000008 | 11 | PILRA |
| Pancreas | T1DM | ENSG0000014 | -0.00032772 | 0.507641835 | 0.000494662 | 0.008204768 | -0.0003355 | 0.497598835 | 0.000494645 | 0.008203882 | 7.78E-06 | 0.01391215 | 1 | 0.914783379 | 1 | 0.914417995 | ENSG0000008 | 9 | ZNF213 |
| Pancreas | T1DM | ENSG0000014 | -0.00047527 | 0.395487266 | 0.000559337 | 0.008204763 | -0.00048345 | 0.387391774 | 0.000559316 | 0.008203877 | 8.18E-06 | 0.009062018 | 1 | 0.873903579 | 1 | 0.867022428 | ENSG0000008 | 9 | ZNF213 |
| Pancreas | T1DM | ENSG0000014 | -0.00026415 | 0.245862036 | 0.000227623 | 0.008204753 | -0.00026008 | 0.253195105 | 0.000227617 | 0.008203868 | -4.07E-06 | 0.017795751 | 1 | 0.788479116 | 1 | 0.788955599 | ENSG0000008 | 11 | CTTN |
| Pancreas | T1DM | ENSG0000014 | -0.0008127 | 0.22959434 | 0.000676457 | 0.008204751 | -0.00083837 | 0.215203895 | 0.000676443 | 0.008203864 | 2.57E-05 | 4.45E-06 | 1 | 0.785512902 | 1 | 0.77077211 | ENSG0000008 | 8 | WNT11 |
| Pancreas | T1DM | ENSG0000014 | 0.001287456 | 0.117092668 | 0.000821554 | 0.008204734 | 0.001275105 | 0.120636555 | 0.00082153 | 0.008203849 | 1.24E-05 | 0.030321699 | 1 | 0.67493056 | 1 | 0.669849249 | ENSG0000008 | 9 | USP40 |
| Pancreas | T1DM | ENSG0000014 | 0.000189094 | 0.691919513 | 0.000477206 | 0.008204773 | 0.000177902 | 0.709290132 | 0.000477193 | 0.008203888 | 1.12E-05 | 0.001549042 | 1 | 0.971486774 | 1 | 0.968082788 | ENSG0000008 | 16 | MAST2 |
| Pancreas | T1DM | ENSG0000014 | 0.000799552 | 0.336130467 | 0.000831273 | 0.00820476 | 0.000783435 | 0.34595488 | 0.000831261 | 0.008203875 | 1.61E-05 | 0.029448828 | 1 | 0.838041785 | 1 | 0.847091708 | ENSG0000008 | 16 | MAST2 |
| Pancreas | T1DM | ENSG0000014 | -0.00127083 | 0.135737347 | 0.000851843 | 0.008204738 | -0.00123931 | 0.145700252 | 0.000851822 | 0.008203854 | -3.15E-05 | 1.07E-06 | 1 | 0.694369535 | 1 | 0.700862587 | ENSG0000008 | 12 | FOLH1 |
| Pancreas | T1DM | ENSG0000014 | -0.00153333 | 0.091915565 | 0.000909779 | 0.008204728 | -0.00150255 | 0.098615629 | 0.000909753 | 0.008203844 | -3.08E-05 | 2.17E-06 | 1 | 0.607172104 | 1 | 0.623755999 | ENSG0000008 | 8 | EIF2AK1 |
| Pancreas | T1DM | ENSG0000015 | 0.000286707 | 0.635138566 | 0.000604221 | 0.008204772 | 0.000292597 | 0.628190694 | 0.000604196 | 0.008203886 | -5.89E-06 | 0.042959912 | 1 | 0.95999755 | 1 | 0.954552486 | ENSG0000008 | 11 | SNX10 |
| Pancreas | T1DM | ENSG0000015 | -0.00045544 | 0.55876046 | 0.000778949 | 0.00820477 | -0.00044023 | 0.571956367 | 0.00077893 | 0.008203885 | -1.52E-05 | 0.011253118 | 1 | 0.93270646 | 1 | 0.936315417 | ENSG0000008 | 11 | SNX10 |
| Pancreas | T1DM | ENSG0000015 | -0.00101034 | 0.312447924 | 0.001000239 | 0.008204758 | -0.00099376 | 0.320440483 | 0.001000209 | 0.008203873 | -1.66E-05 | 0.017422637 | 1 | 0.828095461 | 1 | 0.830335577 | ENSG0000008 | 10 | SEPHS1 |
| Pancreas | T1DM | ENSG0000015 | -0.00110066 | 0.180908058 | 0.000822637 | 0.008204745 | -0.00111331 | 0.175930573 | 0.000822604 | 0.008203859 | 1.26E-05 | 0.004516832 | 1 | 0.736247058 | 1 | 0.729595175 | ENSG0000008 | 10 | SEPHS1 |
| Pancreas | T1DM | ENSG0000015 | 1.28E-05 | 0.963435335 | 0.000278144 | 0.008204776 | 1.82E-05 | 0.94770575 | 0.000278137 | 0.00820389 | -5.49E-06 | 0.012856295 | 1 | 0.997898334 | 1 | 0.999388266 | ENSG0000008 | 2 | ITPKC |
| Pancreas | T1DM | ENSG0000015 | 0.000624005 | 0.166686146 | 0.000451218 | 0.008204743 | 0.000614239 | 0.173406981 | 0.000451202 | 0.008203859 | 9.77E-06 | 0.000290078 | 1 | 0.729268002 | 1 | 0.728578649 | ENSG0000008 | 6 | TMED2 |
| Pancreas | T1DM | ENSG0000015 | 6.21E-06 | 0.989748066 | 0.000483671 | 0.008204776 | -1.10E-06 | 0.998190543 | 0.000483651 | 0.00820389 | 7.31E-06 | 0.003364686 | 1 | 0.997898334 | 1 | 0.999770584 | ENSG0000008 | 6 | TMED2 |
| Pancreas | T1DM | ENSG0000015 | -0.00013367 | 0.832899087 | 0.000633537 | 0.008204775 | -0.00014462 | 0.819432767 | 0.000633524 | 0.008203889 | 1.10E-05 | 0.03795823 | 1 | 0.984098328 | 1 | 0.981810269 | ENSG0000008 | 9 | ERO1LB |
| Pancreas | T1DM | ENSG0000015 | 0.001058792 | 0.262027171 | 0.000943989 | 0.008204754 | 0.001118938 | 0.236104431 | 0.000944426 | 0.008203866 | -6.01E-05 | 0.047399824 | 1 | 0.79720158 | 1 | 0.780702981 | ENSG0000008 | 11 | ACOX3 |
| Pancreas | T1DM | ENSG0000015 | 0.000905107 | 0.402162926 | 0.001080382 | 0.008204764 | 0.000887026 | 0.411612664 | 0.001080343 | 0.008203879 | 1.81E-05 | 0.005256356 | 1 | 0.87893991 | 1 | 0.88342201 | ENSG0000008 | 8 | PSMC5 |
| Pancreas | T1DM | ENSG0000015 | -0.00020391 | 0.704675541 | 0.000537985 | 0.008204773 | -0.00021213 | 0.693346836 | 0.000537964 | 0.008203887 | 8.22E-06 | 0.006819661 | 1 | 0.974645207 | 1 | 0.968082788 | ENSG0000008 | 8 | PSMC5 |
| Pancreas | T1DM | ENSG0000015 | 0.00111653 | 0.27252271 | 0.001017551 | 0.008204755 | 0.00108985 | 0.284129865 | 0.001017516 | 0.008203871 | 2.67E-05 | 3.33E-05 | 1 | 0.798582374 | 1 | 0.799151365 | ENSG0000008 | 12 | UIMC1 |
| Pancreas | T1DM | ENSG0000015 | -0.00066397 | 0.174016315 | 0.000488423 | 0.008204744 | -0.00065739 | 0.178305054 | 0.000488403 | 0.008203859 | -6.58E-06 | 0.00850491 | 1 | 0.729268002 | 1 | 0.731367165 | ENSG0000008 | 12 | OGFOD1 |
| Pancreas | T1DM | ENSG0000015 | 0.001408802 | 0.016870034 | 0.000589571 | 0.008204679 | 0.001419845 | 0.016026311 | 0.000589557 | 0.008203792 | -1.10E-05 | 0.01569707 | 1 | 0.328198842 | 1 | 0.301309092 | ENSG0000008 | 12 | OGFOD1 |
| Pancreas | T1DM | ENSG0000015 | 0.000115978 | 0.886244361 | 0.000810706 | 0.008204775 | 0.000106357 | 0.895620498 | 0.000810674 | 0.00820389 | 9.62E-06 | 0.031857545 | 1 | 0.99200447 | 1 | 0.993128823 | ENSG0000008 | 11 | SH3BP2 |
| Pancreas | T1DM | ENSG0000015 | 0.001155225 | 0.290503988 | 0.00109291 | 0.008204757 | 0.001169641 | 0.284507916 | 0.001092868 | 0.008203871 | -1.44E-05 | 0.02062253 | 1 | 0.809076913 | 1 | 0.799151365 | ENSG0000008 | 4 | TXNDC16 |
| Pancreas | T1DM | ENSG0000015 | -0.00073457 | 0.125059281 | 0.000478898 | 0.008204736 | -0.00074678 | 0.118895554 | 0.00047888 | 0.008203849 | 1.22E-05 | 1.04E-05 | 1 | 0.680843282 | 1 | 0.666556371 | ENSG0000008 | 4 | TXNDC16 |
| Pancreas | T1DM | ENSG0000015 | 0.000356509 | 0.604952365 | 0.000689182 | 0.008204771 | 0.000371777 | 0.589568601 | 0.000689163 | 0.008203885 | -1.53E-05 | 0.002223719 | 1 | 0.941811128 | 1 | 0.937577789 | ENSG0000008 | 4 | TXNDC16 |
| Pancreas | T1DM | ENSG0000015 | -0.00048344 | 0.071251678 | 0.000268001 | 0.008204721 | -0.0004881 | 0.068557118 | 0.000267992 | 0.008203834 | 4.66E-06 | 0.003538467 | 1 | 0.557848504 | 1 | 0.536516198 | ENSG0000008 | 4 | TXNDC16 |
| Pancreas | T1DM | ENSG0000015 | -0.00072327 | 0.270442045 | 0.000656294 | 0.008204755 | -0.00071632 | 0.275047698 | 0.000656266 | 0.00820387 | -6.94E-06 | 0.029170566 | 1 | 0.798582374 | 1 | 0.795892573 | ENSG0000008 | 10 | SF3B2 |
| Pancreas | T1DM | ENSG0000015 | -0.00060613 | 0.239309457 | 0.000515099 | 0.008204752 | -0.00058916 | 0.252707205 | 0.000515089 | 0.008203868 | -1.70E-05 | 5.83E-05 | 1 | 0.785512902 | 1 | 0.788955599 | ENSG0000008 | 10 | SF3B2 |
| Pancreas | T1DM | ENSG0000015 | -0.00025593 | 0.827329744 | 0.001173316 | 0.008204775 | -0.00028226 | 0.809884941 | 0.001173275 | 0.008203889 | 2.63E-05 | 0.000279881 | 1 | 0.984098328 | 1 | 0.981245796 | ENSG0000008 | 10 | SF3B2 |
| Pancreas | T1DM | ENSG0000015 | 0.000561516 | 0.315177973 | 0.000559046 | 0.008204759 | 0.000549538 | 0.325602775 | 0.000559036 | 0.008203874 | 1.20E-05 | 0.010947983 | 1 | 0.829307456 | 1 | 0.830335577 | ENSG0000008 | 10 | SF3B2 |
| Pancreas | T1DM | ENSG0000015 | 0.000451488 | 0.503748544 | 0.00067527 | 0.008204768 | 0.000334062 | 0.621060782 | 0.000675763 | 0.008203886 | 0.000117427 | 1.11E-05 | 1 | 0.914783379 | 1 | 0.950611338 | ENSG0000008 | 5 | KLHL42 |
| Pancreas | T1DM | ENSG0000015 | 0.000627802 | 0.257804987 | 0.000554794 | 0.008204754 | 0.00063785 | 0.250252604 | 0.000554779 | 0.008203868 | -1.00E-05 | 0.013918958 | 1 | 0.79720158 | 1 | 0.788955599 | ENSG0000008 | 5 | KLHL42 |
| Pancreas | T1DM | ENSG0000015 | 0.000224123 | 0.729049985 | 0.000647025 | 0.008204774 | 0.000211835 | 0.743359778 | 0.000647006 | 0.008203888 | 1.23E-05 | 0.006674424 | 1 | 0.975415042 | 1 | 0.968082788 | ENSG0000008 | 5 | KLHL42 |
| Pancreas | T1DM | ENSG0000015 | 0.000592436 | 0.295996602 | 0.000566893 | 0.008204757 | 0.000602363 | 0.287963816 | 0.000566876 | 0.008203871 | -9.93E-06 | 0.013611829 | 1 | 0.81296179 | 1 | 0.801351316 | ENSG0000008 | 12 | PHACTR3 |
| Pancreas | T1DM | ENSG0000016 | -0.00045148 | 0.678698709 | 0.001089906 | 0.008204773 | -0.00043546 | 0.689485781 | 0.001089866 | 0.008203887 | -1.60E-05 | 0.011113436 | 1 | 0.966210802 | 1 | 0.966685034 | ENSG0000008 | 10 | AAMDC |
| Pancreas | T1DM | ENSG0000016 | 0.000433878 | 0.277321354 | 0.000399388 | 0.008204756 | 0.000441039 | 0.269452293 | 0.000399374 | 0.008203869 | -7.16E-06 | 0.004137334 | 1 | 0.798582374 | 1 | 0.795306713 | ENSG0000008 | 7 | GP6 |
| Pancreas | T1DM | ENSG0000016 | -0.00200695 | 0.002087299 | 0.000652129 | 0.008204616 | -0.00202686 | 0.001883754 | 0.000652143 | 0.008203727 | 1.99E-05 | 0.013225722 | 1 | 0.083752862 | 1 | 0.078023894 | ENSG0000008 | 8 | DDX18 |
| Pancreas | T1DM | ENSG0000016 | -6.33E-06 | 0.990277623 | 0.000519667 | 0.008204776 | 5.45E-06 | 0.991638436 | 0.000519652 | 0.00820389 | -1.18E-05 | 0.001585304 | 1 | 0.997898334 | 1 | 0.99966149 | ENSG0000008 | 8 | EDEM2 |
| Pancreas | T1DM | ENSG0000016 | -0.00032488 | 0.681028528 | 0.000790338 | 0.008204773 | -0.00031607 | 0.689210269 | 0.000790308 | 0.008203887 | -8.81E-06 | 0.046773609 | 1 | 0.967301581 | 1 | 0.966685034 | ENSG0000008 | 8 | EDEM2 |
| Pancreas | T1DM | ENSG0000016 | -0.00030873 | 0.725886516 | 0.000880574 | 0.008204773 | -0.00029618 | 0.736597993 | 0.000880546 | 0.008203888 | -1.25E-05 | 0.030518822 | 1 | 0.975415042 | 1 | 0.968082788 | ENSG0000008 | 8 | EDEM2 |
| Pancreas | T1DM | ENSG0000016 | 0.000567922 | 0.609053924 | 0.001110471 | 0.008204771 | 0.000587461 | 0.596777225 | 0.001110426 | 0.008203885 | -1.95E-05 | 0.00062791 | 1 | 0.943530179 | 1 | 0.939046515 | ENSG0000008 | 11 | TPX2 |
| Pancreas | T1DM | ENSG0000016 | 0.000686446 | 0.262130713 | 0.000612149 | 0.008204754 | 0.000675654 | 0.269691003 | 0.00061213 | 0.008203869 | 1.08E-05 | 0.009058337 | 1 | 0.79720158 | 1 | 0.795306713 | ENSG0000008 | 11 | TPX2 |
| Pancreas | T1DM | ENSG0000016 | -0.00192853 | 0.090710892 | 0.001140021 | 0.008204727 | -0.00191348 | 0.093247168 | 0.001139984 | 0.008203843 | -1.51E-05 | 0.044638421 | 1 | 0.603485936 | 1 | 0.60170043 | ENSG0000008 | 11 | TPX2 |
| Pancreas | T1DM | ENSG0000016 | 7.74E-05 | 0.868392087 | 0.000466888 | 0.008204775 | 0.000174795 | 0.708506581 | 0.000467537 | 0.008203888 | -9.74E-05 | 0.00010356 | 1 | 0.99183698 | 1 | 0.968082788 | ENSG0000008 | 11 | TPX2 |
| Pancreas | T1DM | ENSG0000016 | -0.00094246 | 0.122357382 | 0.000610023 | 0.008204735 | -0.00112891 | 0.064617972 | 0.000610914 | 0.008203832 | 0.000186446 | 2.85E-08 | 1 | 0.680843282 | 1 | 0.536001791 | ENSG0000008 | 11 | TPX2 |
| Pancreas | T1DM | ENSG0000016 | -0.00151384 | 0.204788307 | 0.001193853 | 0.008204748 | -0.00153085 | 0.19973074 | 0.001193816 | 0.008203862 | 1.70E-05 | 0.035868839 | 1 | 0.769258416 | 1 | 0.764823318 | ENSG0000008 | 11 | FER1L4 |
| Pancreas | T1DM | ENSG0000016 | 0.001340425 | 0.172342305 | 0.000982199 | 0.008204744 | 0.00135201 | 0.168647359 | 0.000982162 | 0.008203858 | -1.16E-05 | 0.035069561 | 1 | 0.729268002 | 1 | 0.726605951 | ENSG0000008 | 6 | TGDS |
| Pancreas | T1DM | ENSG0000016 | 0.000601536 | 0.587803167 | 0.001109798 | 0.008204771 | 0.000615305 | 0.579270618 | 0.001109756 | 0.008203885 | -1.38E-05 | 0.027853268 | 1 | 0.937588291 | 1 | 0.936315417 | ENSG0000008 | 6 | TGDS |
| Pancreas | T1DM | ENSG0000016 | 0.000846231 | 0.384659941 | 0.000973414 | 0.008204763 | 0.000862979 | 0.375305344 | 0.000973379 | 0.008203877 | -1.67E-05 | 0.004160125 | 1 | 0.865581925 | 1 | 0.855936166 | ENSG0000008 | 6 | TGDS |
| Pancreas | T1DM | ENSG0000016 | 1.44E-06 | 0.998965386 | 0.001109845 | 0.008204776 | -5.78E-05 | 0.9584421 | 0.001109953 | 0.00820389 | 5.93E-05 | 0.002194308 | 1 | 0.999322433 | 1 | 0.999388266 | ENSG0000008 | 12 | DOCK3 |

| | | | | | | | | | | | | | | | | | | | |
|---|---|---|---|---|---|---|---|---|---|---|---|---|---|---|---|---|---|---|---|
| Pancreas | T1DM | ENSG000001( | -0.00011154 | 0.740801815 | 0.000337188 | 0.008204774 | -0.00010648 | 0.752149959 | 0.000337176 | 0.008203888 | -5.06E-06 | 0.009439015 | 1 | 0.975415042 | 1 | 0.968082788 | ENSG0000008 | 12 | DOCK3 |
| Pancreas | T1DM | ENSG000001( | -0.00063406 | 0.466244697 | 0.000870242 | 0.008204767 | -0.00064727 | 0.456994725 | 0.000870215 | 0.008203881 | 1.32E-05 | 0.023551967 | 1 | 0.908712989 | 1 | 0.899776609 | ENSG0000008 | 13 | NSFL1C |
| Pancreas | T1DM | ENSG000001( | -0.00019548 | 0.795180983 | 0.000753025 | 0.008204774 | -0.00017697 | 0.814193675 | 0.000752998 | 0.008203889 | -1.85E-05 | 6.46E-05 | 1 | 0.981920629 | 1 | 0.981619417 | ENSG0000008 | 13 | NSFL1C |
| Pancreas | T1DM | ENSG000001( | 0.00163715 | 0.194061357 | 0.001260646 | 0.008204747 | 0.001721084 | 0.172325425 | 0.001261078 | 0.008203859 | -8.39E-05 | 0.018212792 | 1 | 0.757378621 | 1 | 0.728578649 | ENSG0000008 | 13 | NSFL1C |
| Pancreas | T1DM | ENSG000001( | 0.000912249 | 0.009407721 | 0.000351286 | 0.008204662 | 0.000918052 | 0.008962488 | 0.000351274 | 0.008203775 | -5.80E-06 | 0.006837679 | 1 | 0.232298348 | 1 | 0.208629512 | ENSG0000008 | 7 | ZNF343 |
| Pancreas | T1DM | ENSG000001( | -0.00041299 | 0.249529387 | 0.00035866 | 0.008204753 | -0.00041931 | 0.242341721 | 0.000358646 | 0.008203867 | 6.32E-06 | 0.0015197 | 1 | 0.789394255 | 1 | 0.780845092 | ENSG0000008 | 7 | ZNF343 |
| Pancreas | T1DM | ENSG000001( | -0.0006428 | 0.547822722 | 0.001069502 | 0.008204769 | -0.00062337 | 0.559972751 | 0.001069466 | 0.008203884 | -1.94E-05 | 0.004767004 | 1 | 0.930855143 | 1 | 0.936315417 | ENSG0000008 | 13 | MAVS |
| Pancreas | T1DM | ENSG000001( | 0.002632351 | 0.004265555 | 0.000921098 | 0.008204638 | 0.002653552 | 0.003964987 | 0.000921069 | 0.00820375 | -2.12E-05 | 0.000575145 | 1 | 0.142259026 | 1 | 0.132234879 | ENSG0000008 | 10 | ESF1 |
| Pancreas | T1DM | ENSG000001( | -0.00058254 | 0.49383333 | 0.000851382 | 0.008204768 | -0.00056903 | 0.503888726 | 0.000851359 | 0.008203882 | -1.35E-05 | 0.030430452 | 1 | 0.914693561 | 1 | 0.914417995 | ENSG0000008 | 10 | RBBP9 |
| Pancreas | T1DM | ENSG000001( | 9.78E-05 | 0.914516698 | 0.000910794 | 0.008204775 | 7.63E-05 | 0.933208433 | 0.000910763 | 0.00820389 | 2.14E-05 | 0.000169795 | 1 | 0.995870671 | 1 | 0.999388266 | ENSG0000008 | 7 | SLC8B1 |
| Pancreas | T1DM | ENSG000001( | -6.15E-05 | 0.852155439 | 0.000330002 | 0.008204775 | -6.72E-05 | 0.838630339 | 0.000329991 | 0.008203889 | 5.70E-06 | 0.005129694 | 1 | 0.989913489 | 1 | 0.98524254 | ENSG0000008 | 10 | TMEM230 |
| Pancreas | T1DM | ENSG000001( | -2.94E-05 | 0.930068504 | 0.000334859 | 0.008204775 | -2.51E-05 | 0.94033212 | 0.000334847 | 0.00820389 | -4.32E-06 | 0.027457839 | 1 | 0.997898334 | 1 | 0.999388266 | ENSG0000008 | 10 | TMEM230 |
| Pancreas | T1DM | ENSG000001( | 0.000658787 | 0.405194308 | 0.000791449 | 0.008204764 | 0.000524684 | 0.508840855 | 0.0007942 | 0.008203883 | 0.000134104 | 0.043891191 | 1 | 0.88063625 | 1 | 0.914417995 | ENSG0000008 | 13 | KIF16B |
| Pancreas | T1DM | ENSG000001( | 0.000611827 | 0.630653935 | 0.0012725 | 0.008204772 | 0.000627533 | 0.621895268 | 0.001272456 | 0.008203886 | -1.57E-05 | 0.047483836 | 1 | 0.957728744 | 1 | 0.950611338 | ENSG0000008 | 16 | ZNF302 |
| Pancreas | T1DM | ENSG000001( | 0.000447766 | 0.459966216 | 0.000605988 | 0.008204766 | 0.000455174 | 0.452557105 | 0.000605962 | 0.00820388 | -7.41E-06 | 0.012154003 | 1 | 0.903743873 | 1 | 0.899052624 | ENSG0000008 | 16 | ZNF302 |
| Pancreas | T1DM | ENSG000001( | 0.000496484 | 0.377215773 | 0.000562245 | 0.008204762 | 0.000484612 | 0.388720549 | 0.000562234 | 0.008203877 | 1.19E-05 | 0.009835411 | 1 | 0.860999343 | 1 | 0.867275734 | ENSG0000008 | 4 | KCNH4 |
| Pancreas | T1DM | ENSG000001( | 0.000820901 | 0.492460024 | 0.001195948 | 0.008204768 | 0.00083546 | 0.484801458 | 0.001195904 | 0.008203882 | -1.46E-05 | 0.040444243 | 1 | 0.914693561 | 1 | 0.914417995 | ENSG0000008 | 6 | MLF2 |
| Pancreas | T1DM | ENSG000001( | 0.00221566 | 0.050996993 | 0.001135357 | 0.008204711 | 0.002256071 | 0.046915178 | 0.001135376 | 0.008203824 | -4.04E-05 | 0.002815994 | 1 | 0.511511249 | 1 | 0.491747656 | ENSG0000008 | 13 | GPATCH2L |
| Pancreas | T1DM | ENSG000001( | 0.000278422 | 0.777107285 | 0.000983506 | 0.008204774 | 0.000305463 | 0.756107257 | 0.000983472 | 0.008203888 | -2.70E-05 | 1.12E-05 | 1 | 0.979738629 | 1 | 0.968082788 | ENSG0000009 | 9 | SPTLC1 |
| Pancreas | T1DM | ENSG000001( | -0.00041017 | 0.474811031 | 0.000573921 | 0.008204767 | -0.00049675 | 0.387931764 | 0.000575361 | 0.008203877 | 8.66E-05 | 0.035243232 | 1 | 0.911384054 | 1 | 0.867022428 | ENSG0000009 | 9 | SPTLC1 |
| Pancreas | T1DM | ENSG000001( | 0.000322685 | 0.687195499 | 0.000801375 | 0.008204773 | 0.00033525 | 0.675685307 | 0.000801348 | 0.008203887 | -1.26E-05 | 0.013052696 | 1 | 0.968560944 | 1 | 0.962373748 | ENSG0000009 | 9 | SPTLC1 |
| Pancreas | T1DM | ENSG000001( | -0.00033256 | 0.644907297 | 0.000721627 | 0.008204772 | -0.0003426 | 0.634944702 | 0.000721602 | 0.008203886 | 1.00E-05 | 0.025363362 | 1 | 0.961788216 | 1 | 0.955974304 | ENSG0000009 | 9 | SPTLC1 |
| Pancreas | T1DM | ENSG000001( | 0.000383579 | 0.287260481 | 0.000360455 | 0.008204756 | 0.000388976 | 0.280514496 | 0.000360442 | 0.00820387 | -5.40E-06 | 0.010995407 | 1 | 0.809076913 | 1 | 0.797009688 | ENSG0000009 | 9 | SPTLC1 |
| Pancreas | T1DM | ENSG000001( | 0.00059405 | 0.48292956 | 0.000846708 | 0.008204767 | 0.00058425 | 0.490162159 | 0.000846676 | 0.00820382 | 9.80E-06 | 0.038989193 | 1 | 0.914693561 | 1 | 0.914417995 | ENSG0000009 | 9 | SPTLC1 |
| Pancreas | T1DM | ENSG000001( | 0.001025173 | 0.182530763 | 0.000769069 | 0.008204746 | 0.001100974 | 0.152513499 | 0.000769529 | 0.008203856 | -7.58E-05 | 0.00635907 | 1 | 0.739746982 | 1 | 0.711161998 | ENSG0000009 | 9 | SPTLC1 |
| Pancreas | T1DM | ENSG000001( | 0.000764531 | 0.13612757 | 0.00051298 | 0.008204738 | 0.000757028 | 0.139999674 | 0.000512963 | 0.008203853 | 7.50E-06 | 0.020475641 | 1 | 0.694981311 | 1 | 0.693219864 | ENSG0000009 | 9 | SPTLC1 |
| Pancreas | T1DM | ENSG000001( | 0.000612594 | 0.15851381 | 0.000434436 | 0.008204742 | 0.000605825 | 0.163150014 | 0.000434421 | 0.008203857 | 6.77E-06 | 0.011258676 | 1 | 0.720466306 | 1 | 0.726116528 | ENSG0000009 | 7 | YPEL3 |
| Pancreas | T1DM | ENSG000001( | 3.59E-05 | 0.85448368 | 0.000195868 | 0.008204775 | 2.44E-05 | 0.900891362 | 0.000195898 | 0.00820389 | 1.15E-05 | 0.003781724 | 1 | 0.990317162 | 1 | 0.99419382 | ENSG0000009 | 7 | YPEL3 |
| Pancreas | T1DM | ENSG000001( | 0.000532002 | 0.117600025 | 0.000339953 | 0.008204734 | 0.000536306 | 0.114647643 | 0.00033994 | 0.008203848 | -4.30E-06 | 0.036267371 | 1 | 0.67493056 | 1 | 0.666335944 | ENSG0000009 | 7 | YPEL3 |
| Pancreas | T1DM | ENSG000001( | 0.000504526 | 0.162584862 | 0.000361296 | 0.008204743 | 0.000512766 | 0.155817231 | 0.000361285 | 0.008203856 | -8.24E-06 | 0.000750543 | 1 | 0.727383146 | 1 | 0.713429173 | ENSG0000009 | 7 | YPEL3 |
| Pancreas | T1DM | ENSG000001( | 0.00216611 | 0.060101595 | 0.001152154 | 0.008204716 | 0.00217992 | 0.05847622 | 0.001152108 | 0.00820383 | -1.38E-05 | 0.021304385 | 1 | 0.527104393 | 1 | 0.510771882 | ENSG0000009 | 7 | YPEL3 |
| Pancreas | T1DM | ENSG000001( | -0.00010956 | 0.862551844 | 0.000632808 | 0.008204775 | -0.00011782 | 0.8522892 | 0.000632782 | 0.008203889 | 8.27E-06 | 0.010235023 | 1 | 0.990623048 | 1 | 0.989215428 | ENSG0000009 | 7 | YPEL3 |
| Pancreas | T1DM | ENSG000001( | -0.00091894 | 0.171278678 | 0.000671688 | 0.008204744 | -0.0009064 | 0.177181809 | 0.000671665 | 0.008203859 | -1.25E-05 | 0.002837131 | 1 | 0.729268002 | 1 | 0.730341709 | ENSG0000009 | 11 | MRPS33 |
| Pancreas | T1DM | ENSG000001( | -0.00048216 | 0.140429794 | 0.000327064 | 0.008204739 | -0.00048633 | 0.13701259 | 0.000327052 | 0.008203853 | 4.17E-06 | 0.028108415 | 1 | 0.703643077 | 1 | 0.692461116 | ENSG0000009 | 11 | MRPS33 |
| Pancreas | T1DM | ENSG000001( | 0.000953102 | 0.145050189 | 0.000654045 | 0.00820474 | 0.000962018 | 0.141312254 | 0.000654023 | 0.008203854 | -8.92E-06 | 0.027535434 | 1 | 0.70950264 | 1 | 0.693219864 | ENSG0000009 | 11 | MAEA |
| Pancreas | T1DM | ENSG000001( | -0.00062646 | 0.546135343 | 0.001037944 | 0.008204769 | -0.00060569 | 0.559514396 | 0.001037912 | 0.008203884 | -2.08E-05 | 0.003158168 | 1 | 0.930023581 | 1 | 0.936315417 | ENSG0000009 | 11 | MAEA |
| Pancreas | T1DM | ENSG000001( | -0.00038794 | 0.329667898 | 0.000397979 | 0.00820476 | -0.00039266 | 0.323799548 | 0.000397964 | 0.008203874 | 4.72E-06 | 0.045007647 | 1 | 0.83462449 | 1 | 0.830335577 | ENSG0000009 | 11 | MAEA |
| Pancreas | T1DM | ENSG000001( | -0.00037084 | 0.663230346 | 0.000851617 | 0.008204772 | -0.00035911 | 0.673251037 | 0.000851584 | 0.008203887 | -1.17E-05 | 0.011059851 | 1 | 0.962467202 | 1 | 0.961035047 | ENSG0000009 | 11 | MAEA |
| Pancreas | T1DM | ENSG000001( | -0.0019638 | 0.096211863 | 0.001180516 | 0.008204729 | -0.00197899 | 0.093653029 | 0.001180475 | 0.008203843 | 1.52E-05 | 0.037228724 | 1 | 0.625289471 | 1 | 0.60170043 | ENSG0000009 | 5 | MUL1 |
| Pancreas | T1DM | ENSG000001( | 0.000243846 | 0.681858419 | 0.000594851 | 0.008204773 | 0.000253973 | 0.669403861 | 0.000594832 | 0.008203887 | -1.01E-05 | 0.010224441 | 1 | 0.967410176 | 1 | 0.961035047 | ENSG0000009 | 8 | GOLGA3 |
| Pancreas | T1DM | ENSG000001( | -0.0017529 | 0.088930859 | 0.001030469 | 0.008204727 | -0.00193697 | 0.060998625 | 0.001033873 | 0.008203833 | 0.000184075 | 0.029425106 | 1 | 0.5974078 | 1 | 0.519575945 | ENSG0000009 | 9 | PABPC4 |
| Pancreas | T1DM | ENSG000001( | 0.001807925 | 0.092536626 | 0.001074758 | 0.008204728 | 0.001825703 | 0.089364864 | 0.001074726 | 0.008203841 | -1.78E-05 | 0.017761895 | 1 | 0.60775973 | 1 | 0.585431045 | ENSG0000009 | 9 | PABPC4 |
| Pancreas | T1DM | ENSG000001( | 0.000973964 | 0.343719963 | 0.001028645 | 0.00820476 | 0.000957635 | 0.35185692 | 0.001028615 | 0.008203875 | 1.63E-05 | 0.023416314 | 1 | 0.85026773 | 1 | 0.847508521 | ENSG0000009 | 9 | PABPC4 |
| Pancreas | T1DM | ENSG000001( | 0.000201608 | 0.738828455 | 0.000604695 | 0.008204774 | 0.000212636 | 0.725097684 | 0.000604674 | 0.008203888 | -1.10E-05 | 0.003997827 | 1 | 0.975415042 | 1 | 0.968082788 | ENSG0000009 | 11 | USP48 |
| Pancreas | T1DM | ENSG000001( | 0.00043785 | 0.320151138 | 0.000440427 | 0.008204759 | 0.000447889 | 0.3091686 | 0.000440415 | 0.008203873 | -1.00E-05 | 0.001579728 | 1 | 0.829307456 | 1 | 0.82064652 | ENSG0000009 | 11 | USP48 |
| Pancreas | T1DM | ENSG000001( | -1.04E-05 | 0.987931589 | 0.00068613 | 0.008204776 | -1.91E-05 | 0.977763416 | 0.000686102 | 0.00820389 | 8.75E-06 | 0.014512794 | 1 | 0.997898334 | 1 | 0.99966149 | ENSG0000009 | 11 | USP48 |
| Pancreas | T1DM | ENSG000001( | -0.00026359 | 0.645471614 | 0.000572945 | 0.008204772 | -0.00027258 | 0.634241031 | 0.000572925 | 0.008203886 | 8.99E-06 | 0.01082818 | 1 | 0.961788216 | 1 | 0.955974304 | ENSG0000009 | 11 | USP48 |
| Pancreas | T1DM | ENSG000001( | 4.15E-05 | 0.890947859 | 0.000302387 | 0.008204775 | 3.34E-05 | 0.912151318 | 0.000302377 | 0.00820389 | 8.10E-06 | 1.99E-05 | 1 | 0.99200447 | 1 | 0.996366272 | ENSG0000009 | 8 | ALKBH5 |
| Pancreas | T1DM | ENSG000001( | 0.000743951 | 0.271888403 | 0.000677104 | 0.008204755 | 0.000770941 | 0.254942899 | 0.000677199 | 0.008203868 | -2.70E-05 | 0.04285227 | 1 | 0.798582374 | 1 | 0.788955599 | ENSG0000009 | 3 | SPAG7 |
| Pancreas | T1DM | ENSG000001( | -0.00147886 | 0.045855096 | 0.000740643 | 0.008204708 | -0.00149333 | 0.043765882 | 0.000740616 | 0.008203822 | 1.45E-05 | 0.001003939 | 1 | 0.494772636 | 1 | 0.47422272 | ENSG0000009 | 3 | SPAG7 |
| Pancreas | T1DM | ENSG000001( | -0.00138501 | 0.239472026 | 0.001177417 | 0.008204752 | -0.00142111 | 0.227437988 | 0.001177408 | 0.008203865 | 3.61E-05 | 0.001369074 | 1 | 0.785512902 | 1 | 0.780702981 | ENSG0000009 | 3 | SPAG7 |
| Pancreas | T1DM | ENSG000001( | -0.00029379 | 0.357756882 | 0.000319461 | 0.008204761 | -0.00028777 | 0.367681386 | 0.000319448 | 0.008203876 | -6.02E-06 | 0.000705482 | 1 | 0.854209198 | 1 | 0.850635855 | ENSG0000009 | 3 | SPAG7 |
| Pancreas | T1DM | ENSG000001( | 0.00088117 | 0.278520788 | 0.00081315 | 0.008204756 | 0.000891076 | 0.273134211 | 0.000813119 | 0.00820387 | -9.91E-06 | 0.029203669 | 1 | 0.798679956 | 1 | 0.79588826 | ENSG0000009 | 6 | OSGEP |
| Pancreas | T1DM | ENSG000001( | -0.00167969 | 0.030776231 | 0.000777649 | 0.008204697 | -0.00167081 | 0.031664846 | 0.000777618 | 0.008203812 | -8.88E-06 | 0.030082419 | 1 | 0.424384765 | 1 | 0.417001664 | ENSG0000009 | 16 | SCFD1 |
| Pancreas | T1DM | ENSG000001( | -0.00016652 | 0.793470027 | 0.000636042 | 0.008204774 | -0.00018388 | 0.772499162 | 0.000636031 | 0.008203889 | 1.74E-05 | 0.001345382 | 1 | 0.981920629 | 1 | 0.972640681 | ENSG0000009 | 16 | SCFD1 |
| Pancreas | T1DM | ENSG000001( | 0.000528292 | 0.049914739 | 0.000269441 | 0.008204711 | 0.000533166 | 0.047832092 | 0.000269431 | 0.008203824 | -4.87E-06 | 0.001928895 | 1 | 0.507019677 | 1 | 0.498304494 | ENSG0000009 | 8 | RPGRIP1 |
| Pancreas | T1DM | ENSG000001( | -0.0017228 | 0.14908391 | 0.001194086 | 0.008204741 | -0.00173966 | 0.145131044 | 0.001194044 | 0.008203854 | 1.69E-05 | 0.019365588 | 1 | 0.71293758 | 1 | 0.700862587 | ENSG0000009 | 7 | TGM1 |
| Pancreas | T1DM | ENSG000001( | -0.00041663 | 0.67385491 | 0.000989944 | 0.008204773 | -0.0004887 | 0.621633627 | 0.000990196 | 0.008203886 | 7.21E-05 | 0.003392611 | 1 | 0.963860935 | 1 | 0.950611338 | ENSG0000009 | 7 | TGM1 |
| Pancreas | T1DM | ENSG000001( | 9.76E-06 | 0.993007118 | 0.001113674 | 0.008204776 | -5.25E-06 | 0.99623902 | 0.00111363 | 0.00820389 | 1.50E-05 | 0.014767842 | 1 | 0.998452063 | 1 | 0.999770584 | ENSG0000009 | 7 | TGM1 |
| Pancreas | T1DM | ENSG000001 | -0.00010654 | 0.918361716 | 0.001039442 | 0.008204775 | -8.24E-05 | 0.936786816 | 0.001039419 | 0.00820389 | -2.41E-05 | 0.003391812 | 1 | 0.997611204 | 1 | 0.999388266 | ENSG0000009 | 7 | MFSD11 |
| Pancreas | T1DM | ENSG000001 | 0.000460427 | 0.62219259 | 0.000934411 | 0.008204771 | 0.00047311 | 0.612620349 | 0.000934378 | 0.008203886 | -1.27E-05 | 0.027066896 | 1 | 0.95220407 | 1 | 0.946218353 | ENSG0000009 | 7 | TGFB2 |
| Pancreas | T1DM | ENSG000001 | -0.00142709 | 0.180575127 | 0.001065795 | 0.008204745 | -0.00140963 | 0.185949147 | 0.001065757 | 0.008203861 | -1.75E-05 | 0.007544871 | 1 | 0.736247058 | 1 | 0.748740319 | ENSG0000009 | 14 | NUP50 |
| Pancreas | T1DM | ENSG000001 | -0.00162048 | 0.105544336 | 0.001001192 | 0.008204731 | -0.00155682 | 0.120042473 | 0.00100143 | 0.008203849 | -6.37E-05 | 0.008484607 | 1 | 0.661051879 | 1 | 0.669849249 | ENSG0000009 | 14 | NUP50 |
| Pancreas | T1DM | ENSG000001 | -0.00022845 | 0.735452043 | 0.000676127 | 0.008204774 | -0.0002435 | 0.718743755 | 0.000676125 | 0.008203888 | 1.50E-05 | 0.026643793 | 1 | 0.975415042 | 1 | 0.968082788 | ENSG0000009 | 14 | NUP50 |
| Pancreas | T1DM | ENSG000001 | -0.00089102 | 0.202802801 | 0.000699604 | 0.008204748 | -0.00090579 | 0.195402681 | 0.000699583 | 0.008203862 | 1.48E-05 | 0.002464903 | 1 | 0.766675365 | 1 | 0.762604991 | ENSG0000009 | 14 | NUP50 |
| Pancreas | T1DM | ENSG000001 | -0.00178598 | 0.009279422 | 0.000686493 | 0.008204662 | -0.00179758 | 0.008830262 | 0.000686476 | 0.008203775 | 1.16E-05 | 0.024943996 | 1 | 0.231681817 | 1 | 0.208037741 | ENSG0000009 | 14 | NUP50 |
| Pancreas | T1DM | ENSG000001 | 0.001069455 | 0.293989643 | 0.0010191 | 0.008204757 | 0.001050848 | 0.302455516 | 0.00101907 | 0.008203872 | 1.86E-05 | 0.009010543 | 1 | 0.810918801 | 1 | 0.81428005 | ENSG0000009 | 11 | CECR1 |
| Pancreas | T1DM | ENSG000001 | -9.09E-05 | 0.88965131 | 0.000655131 | 0.008204775 | -7.21E-05 | 0.912358757 | 0.000655115 | 0.00820389 | -1.88E-05 | 0.000195484 | 1 | 0.99200447 | 1 | 0.996366272 | ENSG0000009 | 11 | CECR1 |
| Pancreas | T1DM | ENSG000001 | -0.00055132 | 0.558501572 | 0.00094231 | 0.00820477 | -0.0005359 | 0.569543779 | 0.000942286 | 0.008203885 | -1.54E-05 | 0.029134732 | 1 | 0.93270646 | 1 | 0.936315417 | ENSG0000009 | 9 | VNN3 |
| Pancreas | T1DM | ENSG000001 | -0.00029689 | 0.74039579 | 0.000896064 | 0.008204774 | -0.00028264 | 0.752430956 | 0.00089632 | 0.008203888 | -1.43E-05 | 0.008068262 | 1 | 0.975415042 | 1 | 0.968082788 | ENSG0000009 | 14 | ECHDC1 |
| Pancreas | T1DM | ENSG000001 | 0.001859945 | 0.116442163 | 0.001184764 | 0.008204734 | 0.001875903 | 0.113329915 | 0.001184727 | 0.008203848 | -1.60E-05 | 0.046832112 | 1 | 0.67493056 | 1 | 0.666335944 | ENSG0000009 | 9 | CBX5 |
| Pancreas | T1DM | ENSG000001 | 0.001177947 | 0.160789029 | 0.000839936 | 0.008204742 | 0.001191958 | 0.15585427 | 0.000839906 | 0.008203856 | -1.40E-05 | 0.006073817 | 1 | 0.724477654 | 1 | 0.713429173 | ENSG0000009 | 9 | CBX5 |
| Pancreas | T1DM | ENSG000001 | 9.02E-05 | 0.900865391 | 0.000723935 | 0.008204775 | 0.00010682 | 0.882690399 | 0.00072391 | 0.00820389 | -1.66E-05 | 0.000219746 | 1 | 0.99317359 | 1 | 0.993128823 | ENSG0000009 | 9 | CBX5 |
| Pancreas | T1DM | ENSG000001 | -7.39E-05 | 0.910880258 | 0.000660168 | 0.008204775 | -5.36E-05 | 0.935271268 | 0.000660163 | 0.00820389 | -2.03E-05 | 0.001462201 | 1 | 0.995698473 | 1 | 0.999388266 | ENSG0000009 | 9 | FMO2 |
| Pancreas | T1DM | ENSG000001 | -0.00062621 | 0.341875517 | 0.000658843 | 0.00820476 | -0.00063551 | 0.334734896 | 0.00065882 | 0.008203874 | 9.30E-06 | 0.026023101 | 1 | 0.847428888 | 1 | 0.835993972 | ENSG0000009 | 9 | FMO2 |
| Pancreas | T1DM | ENSG000001 | -7.98E-05 | 0.914880878 | 0.000746562 | 0.008204775 | -6.07E-05 | 0.935214061 | 0.000746537 | 0.00820389 | -1.91E-05 | 5.46E-05 | 1 | 0.995870671 | 1 | 0.999388266 | ENSG0000009 | 9 | FMO2 |
| Pancreas | T1DM | ENSG000001 | -0.00016059 | 0.518378511 | 0.000248655 | 0.008204769 | -0.00015776 | 0.525770583 | 0.000248645 | 0.008203883 | -2.83E-06 | 0.034795991 | 1 | 0.914783379 | 1 | 0.920313283 | ENSG0000009 | 9 | FMO2 |
| Pancreas | T1DM | ENSG000001 | 6.47E-05 | 0.957013232 | 0.001199805 | 0.008204776 | 8.13E-05 | 0.945978319 | 0.00119976 | 0.00820389 | -1.66E-05 | 0.014731287 | 1 | 0.997898334 | 1 | 0.999388266 | ENSG0000009 | 7 | TMEM38B |
| Pancreas | T1DM | ENSG000001 | -0.00053769 | 0.657959033 | 0.001214486 | 0.008204772 | -0.0005136 | 0.67233216 | 0.00121444 | 0.008203887 | -2.40E-05 | 0.000536237 | 1 | 0.962467202 | 1 | 0.961035047 | ENSG0000009 | 12 | CWF19L1 |
| Pancreas | T1DM | ENSG000001 | 0.00032101 | 0.76385843 | 0.001068544 | 0.008204774 | 0.000337197 | 0.752324224 | 0.00106851 | 0.008203888 | -1.62E-05 | 0.023155512 | 1 | 0.977622335 | 1 | 0.968082788 | ENSG0000009 | 12 | CWF19L1 |
| Pancreas | T1DM | ENSG000001 | -0.00028127 | 0.484037443 | 0.000401913 | 0.008204767 | -0.00028758 | 0.474266061 | 0.000401902 | 0.008203881 | 6.32E-06 | 0.033834337 | 1 | 0.914693561 | 1 | 0.911454131 | ENSG0000009 | 12 | CWF19L1 |
| Pancreas | T1DM | ENSG000001 | -0.00047468 | 0.55405127 | 0.000802223 | 0.00820477 | -0.00048602 | 0.544609825 | 0.000802198 | 0.008203884 | 1.13E-05 | 0.033418728 | 1 | 0.93270646 | 1 | 0.928420568 | ENSG0000009 | 12 | CWF19L1 |
| Pancreas | T1DM | ENSG000001 | 0.001055961 | 0.242754944 | 0.000903975 | 0.008204753 | 0.00106719 | 0.237764579 | 0.000903943 | 0.008203867 | -1.12E-05 | 0.039700253 | 1 | 0.787779958 | 1 | 0.780702981 | ENSG0000009 | 12 | CWF19L1 |
| Pancreas | T1DM | ENSG000001 | -0.00190324 | 0.026201675 | 0.000856077 | 0.008204692 | -0.00191505 | 0.025281783 | 0.000856049 | 0.008203806 | 1.18E-05 | 0.034285528 | 1 | 0.393484808 | 1 | 0.374802485 | ENSG0000009 | 9 | CRTAC1 |
| Pancreas | T1DM | ENSG000001 | -0.00060525 | 0.108307621 | 0.000376906 | 0.008204732 | -0.00059846 | 0.112316225 | 0.000376892 | 0.008203848 | -6.80E-06 | 0.003043652 | 1 | 0.661051879 | 1 | 0.666335944 | ENSG0000009 | 9 | CRTAC1 |
| Pancreas | T1DM | ENSG000001 | -0.00031998 | 0.642378106 | 0.000689062 | 0.008204772 | -0.00030519 | 0.657827287 | 0.00068904 | 0.008203887 | -1.48E-05 | 0.000945306 | 1 | 0.961788216 | 1 | 0.961035047 | ENSG0000009 | 17 | WAC |
| Pancreas | T1DM | ENSG000001 | -0.00020702 | 0.808763027 | 0.000855386 | 0.008204775 | -0.00021628 | 0.800380043 | 0.000855353 | 0.008203889 | 9.26E-06 | 0.046121637 | 1 | 0.98201188 | 1 | 0.978923745 | ENSG0000009 | 9 | TPSD1 |
| Pancreas | T1DM | ENSG000001 | 0.0008166 | 0.187698865 | 0.000619848 | 0.008204746 | 0.000801504 | 0.195974598 | 0.000619828 | 0.008203862 | 1.51E-05 | 0.000175097 | 1 | 0.750693896 | 1 | 0.762792436 | ENSG0000009 | 6 | ITPR3 |
| Pancreas | T1DM | ENSG000001 | -0.00088949 | 0.378245735 | 0.00100948 | 0.008204762 | -0.00090538 | 0.369768429 | 0.001009442 | 0.008203876 | 1.59E-05 | 0.006630479 | 1 | 0.860999343 | 1 | 0.851044876 | ENSG0000009 | 6 | ITPR3 |
| Pancreas | T1DM | ENSG000001 | 0.002097007 | 0.008476586 | 0.000796589 | 0.008204659 | 0.002106853 | 0.008170587 | 0.000796557 | 0.008203772 | -9.85E-06 | 0.020848601 | 1 | 0.222121144 | 1 | 0.204622746 | ENSG0000009 | 6 | ITPR3 |
| Pancreas | T1DM | ENSG000001 | -0.00074411 | 0.342959718 | 0.000784648 | 0.00820476 | -0.00072013 | 0.358724414 | 0.000784627 | 0.008203876 | -2.40E-05 | 3.46E-05 | 1 | 0.849296581 | 1 | 0.850635855 | ENSG0000009 | 6 | ITPR3 |
| Pancreas | T1DM | ENSG000001 | 0.000117723 | 0.815751215 | 0.000505222 | 0.008204775 | 0.000130628 | 0.795971221 | 0.000505205 | 0.008203889 | -1.29E-05 | 0.000111815 | 1 | 0.98201188 | 1 | 0.978923745 | ENSG0000009 | 6 | ITPR3 |
| Pancreas | T1DM | ENSG000001 | 0.000428283 | 0.494202545 | 0.000626474 | 0.008204768 | 0.000439665 | 0.482787224 | 0.000626457 | 0.008203882 | -1.14E-05 | 0.011681638 | 1 | 0.914693561 | 1 | 0.91349461 | ENSG0000009 | 11 | IFT74 |
| Pancreas | T1DM | ENSG000001 | -0.00126017 | 0.076280067 | 0.000710881 | 0.008204723 | -0.00126951 | 0.074118962 | 0.000710858 | 0.008203836 | 9.33E-06 | 0.044638175 | 1 | 0.571784974 | 1 | 0.550108364 | ENSG0000009 | 11 | IL12RB1 |
| Pancreas | T1DM | ENSG000001 | -1.07E-05 | 0.98928725 | 0.000797421 | 0.008204776 | 4.39E-07 | 0.999560723 | 0.000797394 | 0.00820389 | -1.11E-05 | 0.028123673 | 1 | 0.997898334 | 1 | 0.999800859 | ENSG0000009 | 11 | IL12RB1 |
| Pancreas | T1DM | ENSG000001 | 0.000109496 | 0.871676829 | 0.000677871 | 0.008204775 | 0.000121155 | 0.858145099 | 0.000677845 | 0.008203889 | -1.17E-05 | 0.002659519 | 1 | 0.99200447 | 1 | 0.990577333 | ENSG0000009 | 8 | CDC7 |
| Pancreas | T1DM | ENSG000001 | 0.000562484 | 0.555465334 | 0.000954024 | 0.00820477 | 0.000577566 | 0.544901146 | 0.000953994 | 0.008203884 | -1.51E-05 | 0.017142863 | 1 | 0.93270646 | 1 | 0.928420568 | ENSG0000009 | 8 | CDC7 |
| Pancreas | T1DM | ENSG000001 | 0.000150717 | 0.839457375 | 0.00074396 | 0.008204775 | 0.000124442 | 0.867154116 | 0.000743941 | 0.00820389 | 2.63E-05 | 2.97E-06 | 1 | 0.986602535 | 1 | 0.990577333 | ENSG0000009 | 8 | CDC7 |
| Pancreas | T1DM | ENSG000001 | -0.00093734 | 0.12739937 | 0.000614877 | 0.008204736 | -0.0009304 | 0.130227277 | 0.000614851 | 0.008203851 | -6.94E-06 | 0.024411753 | 1 | 0.684395521 | 1 | 0.676232745 | ENSG0000009 | 8 | CDC7 |
| Pancreas | T1DM | ENSG000001 | 7.68E-06 | 0.986366474 | 0.000449446 | 0.008204776 | 1.12E-07 | 0.999800859 | 0.000449429 | 0.00820389 | 7.57E-06 | 0.003592288 | 1 | 0.997898334 | 1 | 0.999800859 | ENSG0000009 | 12 | RAB18 |
| Pancreas | T1DM | ENSG000001 | 0.001585797 | 0.189734618 | 0.001209271 | 0.008204747 | 0.001605644 | 0.184236463 | 0.001209229 | 0.00820386 | -1.98E-05 | 0.007840426 | 1 | 0.752840936 | 1 | 0.747990804 | ENSG0000009 | 14 | PRTFDC1 |
| Pancreas | T1DM | ENSG000001 | -0.00041522 | 0.127657887 | 0.000272563 | 0.008204736 | -0.00040781 | 0.13458382 | 0.000272554 | 0.008203852 | -7.41E-06 | 1.83E-05 | 1 | 0.684395521 | 1 | 0.685736609 | ENSG0000009 | 14 | PRTFDC1 |
| Pancreas | T1DM | ENSG000001 | 0.000831021 | 0.442593726 | 0.00108231 | 0.008204766 | 0.000849374 | 0.432566876 | 0.001082271 | 0.00820388 | -1.84E-05 | 0.004475112 | 1 | 0.89813429 | 1 | 0.890803318 | ENSG0000009 | 5 | TSPAN15 |
| Pancreas | T1DM | ENSG000001 | -3.91E-05 | 0.97131832 | 0.001088328 | 0.008204776 | -2.43E-05 | 0.982153738 | 0.00108829 | 0.00820389 | -1.48E-05 | 0.02948026 | 1 | 0.997898334 | 1 | 0.99966149 | ENSG0000009 | 4 | OCEL1 |
| Pancreas | T1DM | ENSG000001 | -0.00176627 | 0.043812324 | 0.000876174 | 0.008204707 | -0.00175649 | 0.044984316 | 0.000876138 | 0.008203822 | -9.78E-06 | 0.032746282 | 1 | 0.48081217 | 1 | 0.485049395 | ENSG0000009 | 4 | OCEL1 |
| Pancreas | T1DM | ENSG000001 | 0.000460439 | 0.116028188 | 0.000292962 | 0.008204734 | 0.00036119 | 0.218909892 | 0.000293786 | 0.008203865 | 9.92E-05 | 7.75E-06 | 1 | 0.67493056 | 1 | 0.775393934 | ENSG0000009 | 4 | OCEL1 |
| Pancreas | T1DM | ENSG000001 | -5.10E-05 | 0.961900202 | 0.001066622 | 0.008204776 | -3.69E-05 | 0.972393551 | 0.0010658 | 0.00820389 | -1.40E-05 | 0.01510388 | 1 | 0.997898334 | 1 | 0.99966149 | ENSG0000009 | 9 | MYO9B |
| Pancreas | T1DM | ENSG000001 | -0.00111359 | 0.318031431 | 0.00111525 | 0.008204759 | -0.00109933 | 0.324250458 | 0.00111521 | 0.008203874 | -1.43E-05 | 0.033087004 | 1 | 0.829307456 | 1 | 0.830335577 | ENSG0000009 | 18 | CATSPERG |
| Pancreas | T1DM | ENSG000001 | 0.000396197 | 0.275845275 | 0.000363583 | 0.008204756 | 0.000400805 | 0.270283113 | 0.000363571 | 0.00820387 | -4.61E-06 | 0.045385601 | 1 | 0.798582374 | 1 | 0.795306713 | ENSG0000009 | 7 | HNRNPM |
| Pancreas | T1DM | ENSG000001 | -0.00084167 | 0.21021777 | 0.000671741 | 0.008204749 | -0.00085797 | 0.201507202 | 0.000671723 | 0.008203863 | 1.63E-05 | 0.000913357 | 1 | 0.771198903 | 1 | 0.766509931 | ENSG0000009 | 14 | MTAP |
| Pancreas | T1DM | ENSG000001 | -0.00053981 | 0.177551098 | 0.000400352 | 0.008204745 | -0.00048848 | 0.223099797 | 0.000400945 | 0.008203865 | -5.13E-05 | 0.020662898 | 1 | 0.732338331 | 1 | 0.78054534 | ENSG0000009 | 14 | MTAP |
| Pancreas | T1DM | ENSG000001 | -0.00012985 | 0.814310538 | 0.000552841 | 0.008204775 | -0.00020466 | 0.711374154 | 0.00055312 | 0.008203888 | 7.48E-05 | 5.15E-05 | 1 | 0.98201188 | 1 | 0.968082788 | ENSG0000009 | 11 | CEP170B |
| Pancreas | T1DM | ENSG000001 | 0.000275603 | 0.424503786 | 0.000345094 | 0.008204765 | 0.000271297 | 0.431760397 | 0.000345081 | 0.00820388 | 4.31E-06 | 0.025987288 | 1 | 0.889410424 | 1 | 0.890803318 | ENSG0000009 | 7 | SNAP29 |
| Pancreas | T1DM | ENSG000001 | 0.000904178 | 0.129458917 | 0.000596329 | 0.008204737 | 0.000911169 | 0.126507322 | 0.000596306 | 0.008203851 | -6.99E-06 | 0.038011709 | 1 | 0.688303313 | 1 | 0.673807988 | ENSG0000009 | 7 | SNAP29 |
| Pancreas | T1DM | ENSG000001 | -0.00235087 | 0.025384996 | 0.00105161 | 0.008204691 | -0.0023650 | 0.024510004 | 0.001051574 | 0.008203805 | 1.42E-05 | 0.029617873 | 1 | 0.392702831 | 1 | 0.374802485 | ENSG0000009 | 14 | LZTR1 |
| Pancreas | T1DM | ENSG000001 | -0.00027922 | 0.625799389 | 0.000572588 | 0.008204772 | -0.00027053 | 0.636575286 | 0.000572567 | 0.008203886 | -8.69E-06 | 0.010159859 | 1 | 0.953740553 | 1 | 0.955974304 | ENSG0000009 | 14 | LZTR1 |
| Pancreas | T1DM | ENSG000001 | 0.001868999 | 0.092016623 | 0.001109292 | 0.008204728 | 0.001890853 | 0.088267987 | 0.001109259 | 0.008203841 | -2.19E-05 | 0.00509926 | 1 | 0.607172104 | 1 | 0.582754369 | ENSG0000009 | 14 | LZTR1 |
| Pancreas | T1DM | ENSG000001 | -0.00057785 | 0.202730791 | 0.000453642 | 0.008204748 | -0.00057189 | 0.207414584 | 0.000453624 | 0.008203863 | -5.97E-06 | 0.015385079 | 1 | 0.766675365 | 1 | 0.769778216 | ENSG0000009 | 14 | LZTR1 |
| Pancreas | T1DM | ENSG000001 | -0.00051201 | 0.164982197 | 0.000368746 | 0.008204743 | -0.0005176 | 0.160402911 | 0.000368733 | 0.008203857 | 5.59E-06 | 0.011445451 | 1 | 0.729268002 | 1 | 0.723297686 | ENSG0000009 | 13 | SMARCB1 |
| Pancreas | T1DM | ENSG000001 | 2.77E-05 | 0.968139831 | 0.000694471 | 0.008204776 | 1.74E-05 | 0.979981967 | 0.00069445 | 0.00820389 | 1.03E-05 | 0.031045716 | 1 | 0.997898334 | 1 | 0.99966149 | ENSG0000009 | 13 | SMARCB1 |
| Pancreas | T1DM | ENSG000001 | -0.00077298 | 0.333006782 | 0.000798472 | 0.00820476 | -0.00079725 | 0.31804026 | 0.000798451 | 0.008203873 | 2.43E-05 | 4.83E-05 | 1 | 0.83462449 | 1 | 0.830006722 | ENSG0000009 | 13 | SMARCB1 |
| Pancreas | T1DM | ENSG000001 | -0.00095409 | 0.174946229 | 0.000703354 | 0.008204745 | -0.00097047 | 0.16742257 | 0.00070333 | 0.008203858 | 1.64E-05 | 0.000221843 | 1 | 0.729321291 | 1 | 0.726605951 | ENSG0000009 | 13 | SMARCB1 |
| Pancreas | T1DM | ENSG000001 | -0.00051821 | 0.469384763 | 0.000716274 | 0.008204767 | -0.00052918 | 0.460015922 | 0.000716251 | 0.008203881 | 1.10E-05 | 0.018418075 | 1 | 0.908712989 | 1 | 0.901310005 | ENSG0000009 | 7 | DDTL |
| Pancreas | T1DM | ENSG000001 | 0.000998341 | 0.310081085 | 0.000983536 | 0.008204758 | 0.000974796 | 0.321618907 | 0.000983512 | 0.008203873 | 2.35E-05 | 0.002095 | 1 | 0.826025131 | 1 | 0.830335577 | ENSG0000009 | 7 | DDTL |
| Pancreas | T1DM | ENSG000001 | -3.15E-05 | 0.939421726 | 0.000414207 | 0.008204775 | -3.96E-05 | 0.923889554 | 0.000414193 | 0.00820389 | 8.09E-06 | 0.002368383 | 1 | 0.997898334 | 1 | 0.997889334 | ENSG0000009 | 7 | DDTL |
| Pancreas | T1DM | ENSG000001 | -0.00107922 | 0.24762865 | 0.000933476 | 0.008204753 | -0.00109618 | 0.240257474 | 0.000933441 | 0.008203867 | 1.70E-05 | 0.001389004 | 1 | 0.788479116 | 1 | 0.780702981 | ENSG0000009 | 9 | DDT |
| Pancreas | T1DM | ENSG000001 | 0.000576768 | 0.064620654 | 0.000312125 | 0.008204718 | 0.000567658 | 0.068951997 | 0.000312116 | 0.008203834 | 9.11E-06 | 8.26E-05 | 1 | 0.544685729 | 1 | 0.536516198 | ENSG0000009 | 6 | GSTT2 |

| | | | | | | | | | | | | | | | | | | |
|---|---|---|---|---|---|---|---|---|---|---|---|---|---|---|---|---|---|---|
| Pancreas | T1DM | ENSG0000018 | 0.001812203 | 0.023112301 | 0.000797772 | 0.008204689 | 0.001833324 | 0.021560333 | 0.00079778 | 0.008203801 | -2.11E-05 | 0.019264292 | 1 | 0.382918644 | 1 | 0.349355274 | ENSG0000009 | 6 | GSTT2 |
| Pancreas | T1DM | ENSG0000018 | -0.00050013 | 0.625182767 | 0.00102377 | 0.008204772 | -0.00048532 | 0.635451462 | 0.00102373 | 0.008203886 | -1.48E-05 | 0.008882936 | 1 | 0.953366595 | 1 | 0.955974304 | ENSG0000009 | 3 | OSM |
| Pancreas | T1DM | ENSG0000018 | -0.00013166 | 0.739461467 | 0.000395902 | 0.008204774 | -0.00013998 | 0.723646224 | 0.000395888 | 0.008203888 | 8.32E-06 | 0.000608356 | 1 | 0.975415042 | 1 | 0.968082788 | ENSG0000009 | 12 | CABIN1 |
| Pancreas | T1DM | ENSG0000018 | -5.19E-05 | 0.966958691 | 0.001253907 | 0.008204776 | -3.65E-05 | 0.976773582 | 0.001253861 | 0.00820389 | -1.54E-05 | 0.037836986 | 1 | 0.997898334 | 1 | 0.99966149 | ENSG0000009 | 12 | CABIN1 |
| Pancreas | T1DM | ENSG0000018 | -0.00134606 | 0.289720349 | 0.001271386 | 0.008204757 | -0.00136154 | 0.284194934 | 0.001271342 | 0.008203871 | 1.55E-05 | 0.049223431 | 1 | 0.809076913 | 1 | 0.799151365 | ENSG0000009 | 10 | SUSD2 |
| Pancreas | T1DM | ENSG0000018 | -0.00027878 | 0.651627359 | 0.000617453 | 0.008204772 | -0.00028807 | 0.640819547 | 0.000617434 | 0.008203886 | 9.28E-06 | 0.029139688 | 1 | 0.962467202 | 1 | 0.957315064 | ENSG0000010 | 15 | SPECC1L |
| Pancreas | T1DM | ENSG0000018 | 0.000824096 | 0.296719187 | 0.000789745 | 0.008204757 | 0.000812425 | 0.303596014 | 0.000789718 | 0.008203872 | 1.17E-05 | 0.016445221 | 1 | 0.813206908 | 1 | 0.815517326 | ENSG0000010 | 15 | SPECC1L |
| Pancreas | T1DM | ENSG0000018 | -0.00081732 | 0.140838217 | 0.000554988 | 0.008204739 | -0.00083214 | 0.13376351 | 0.000554971 | 0.008203852 | 1.48E-05 | 0.000114749 | 1 | 0.703643077 | 1 | 0.685736609 | ENSG0000010 | 15 | SPECC1L |
| Pancreas | T1DM | ENSG0000018 | 0.002211815 | 0.082937297 | 0.001275634 | 0.008204725 | 0.002240766 | 0.078982925 | 0.001275611 | 0.008203838 | -2.90E-05 | 0.007709429 | 1 | 0.589329992 | 1 | 0.56498092 | ENSG0000010 | 15 | SPECC1L |
| Pancreas | T1DM | ENSG0000018 | 0.002660471 | 0.031797456 | 0.001239184 | 0.008204698 | 0.002371499 | 0.055945612 | 0.001240672 | 0.008203828 | 0.000288972 | 3.23E-06 | 1 | 0.427517623 | 1 | 0.510771882 | ENSG0000010 | 15 | SPECC1L |
| Pancreas | T1DM | ENSG0000018 | -0.00012068 | 0.650258237 | 0.00026616 | 0.008204772 | -0.00011567 | 0.663840497 | 0.000266149 | 0.008203887 | -5.00E-06 | 0.000901143 | 1 | 0.962467202 | 1 | 0.961035047 | ENSG0000010 | 13 | PPIL2 |
| Pancreas | T1DM | ENSG0000018 | -0.00028406 | 0.514211778 | 0.000435473 | 0.008204768 | -0.00027683 | 0.524961686 | 0.000435456 | 0.008203883 | -7.23E-06 | 0.001826405 | 1 | 0.914783379 | 1 | 0.919578178 | ENSG0000010 | 13 | PPIL2 |
| Pancreas | T1DM | ENSG0000018 | -0.00022455 | 0.59218393 | 0.000419184 | 0.008204771 | -0.00023141 | 0.580900363 | 0.00041917 | 0.008203885 | 6.87E-06 | 0.011884471 | 1 | 0.938170574 | 1 | 0.936315417 | ENSG0000010 | 10 | UPB1 |
| Pancreas | T1DM | ENSG0000018 | 0.000791113 | 0.522056258 | 0.001235768 | 0.008204769 | 0.000807946 | 0.513225483 | 0.001235724 | 0.008203883 | -1.68E-05 | 0.024577729 | 1 | 0.915738026 | 1 | 0.915381179 | ENSG0000010 | 10 | UPB1 |
| Pancreas | T1DM | ENSG0000018 | 0.00026527 | 0.766743839 | 0.000894257 | 0.008204774 | 0.000253285 | 0.776988985 | 0.000894226 | 0.008203889 | 1.20E-05 | 0.031019011 | 1 | 0.977622335 | 1 | 0.972830721 | ENSG0000010 | 12 | PRODH |
| Pancreas | T1DM | ENSG0000018 | 0.001170642 | 0.172351293 | 0.000857809 | 0.008204744 | 0.001184555 | 0.167296042 | 0.000857785 | 0.008203858 | -1.39E-05 | 0.025177236 | 1 | 0.729268002 | 1 | 0.726605951 | ENSG0000010 | 12 | PRODH |
| Pancreas | T1DM | ENSG0000018 | 0.000325328 | 0.710590798 | 0.000876747 | 0.008204773 | 0.000369879 | 0.673137876 | 0.000876811 | 0.008203887 | -4.46E-05 | 0.001482998 | 1 | 0.975415042 | 1 | 0.961035047 | ENSG0000010 | 7 | LRP5L |
| Pancreas | T1DM | ENSG0000018 | 0.001276739 | 0.075810066 | 0.000719072 | 0.008204722 | 0.001285493 | 0.07381264 | 0.000719046 | 0.008203836 | -8.75E-06 | 0.035203766 | 1 | 0.57107125 | 1 | 0.550108364 | ENSG0000010 | 7 | LRP5L |
| Pancreas | T1DM | ENSG0000018 | -0.00508613 | 4.91E-10 | 0.000817428 | 0.008204124 | -0.00496178 | 1.41E-09 | 0.000819543 | 0.008203273 | -0.00012435 | 0.036397457 | 1.26E-06 | 5.48E-08 | 3.62E-06 | 1.58E-07 | ENSG0000010 | 5 | LGALS2 |
| Pancreas | T1DM | ENSG0000018 | -8.93E-05 | 0.922299043 | 0.000915685 | 0.008204775 | -0.00010988 | 0.904483553 | 0.000915653 | 0.00820389 | 2.06E-05 | 0.000285034 | 1 | 0.997898334 | 1 | 0.995067586 | ENSG0000010 | 5 | LGALS2 |
| Pancreas | T1DM | ENSG0000019 | -0.00264574 | 1.22E-09 | 0.000435334 | 0.008204154 | -0.00161516 | 0.008366584 | 0.000612519 | 0.008203773 | -0.00103058 | 0.016770568 | 3.14E-06 | 1.31E-07 | 1 | 0.204622746 | ENSG0000010 | 14 | GGA1 |
| Pancreas | T1DM | ENSG0000019 | -0.00080216 | 0.221662278 | 0.000656368 | 0.00820475 | -0.00082697 | 0.207717244 | 0.000656396 | 0.008203863 | 2.48E-05 | 0.00649261 | 1 | 0.780835021 | 1 | 0.769778216 | ENSG0000010 | 16 | HPS4 |
| Pancreas | T1DM | ENSG0000019 | -0.00288651 | 2.21E-09 | 0.000482535 | 0.008204173 | -0.00320261 | 3.94E-11 | 0.00048477 | 0.008203155 | 0.000316103 | 1.44E-11 | 5.67E-06 | 2.18E-07 | 1.01E-07 | 5.06E-09 | ENSG0000010 | 16 | HPS4 |
| Pancreas | T1DM | ENSG0000019 | 0.00139306 | 0.110469699 | 0.000872796 | 0.008204733 | 0.001405427 | 0.107328676 | 0.000872763 | 0.008203846 | -1.24E-05 | 0.011375218 | 1 | 0.665930015 | 1 | 0.652153441 | ENSG0000010 | 16 | HPS4 |
| Pancreas | T1DM | ENSG0000019 | -0.0004399 | 0.353577714 | 0.000474193 | 0.008204761 | 0.001595421 | 0.004219253 | 0.000557586 | 0.008203752 | -0.00203532 | 3.99E-12 | 1 | 0.854061442 | 1 | 0.135438011 | ENSG0000010 | 16 | HPS4 |
| Pancreas | T1DM | ENSG0000019 | -0.00088855 | 0.394508381 | 0.00104472 | 0.008204763 | -0.00068961 | 0.510137705 | 0.001047051 | 0.008203883 | -0.00019994 | 0.004666178 | 1 | 0.873903579 | 1 | 0.915335266 | ENSG0000010 | 16 | HPS4 |
| Pancreas | T1DM | ENSG0000019 | 0.000190855 | 0.51969723 | 0.000296445 | 0.008204769 | 0.000202192 | 0.495208521 | 0.000296447 | 0.008203882 | -1.13E-05 | 0.000653004 | 1 | 0.914783379 | 1 | 0.914417995 | ENSG0000010 | 15 | TRIOBP |
| Pancreas | T1DM | ENSG0000019 | -0.00110731 | 0.241061519 | 0.000944534 | 0.008204752 | -0.00112192 | 0.234895638 | 0.000944502 | 0.008203866 | 1.46E-05 | 0.014858715 | 1 | 0.786897324 | 1 | 0.780702981 | ENSG0000010 | 12 | TFIP11 |
| Pancreas | T1DM | ENSG0000019 | 0.000553593 | 0.237461431 | 0.000468608 | 0.008204752 | 0.0005462 | 0.24376667 | 0.00046859 | 0.008203867 | 7.39E-06 | 0.005482304 | 1 | 0.785512902 | 1 | 0.78207287 | ENSG0000010 | 12 | TFIP11 |
| Pancreas | T1DM | ENSG0000019 | 0.000375297 | 0.625123904 | 0.000768104 | 0.008204772 | 0.000387543 | 0.613868598 | 0.000768082 | 0.008203886 | -1.22E-05 | 0.026185163 | 1 | 0.953366595 | 1 | 0.946218353 | ENSG0000010 | 13 | EIF3L |
| Pancreas | T1DM | ENSG0000019 | -0.00039142 | 0.710639177 | 0.001055052 | 0.008204773 | -0.00045585 | 0.665709469 | 0.001055098 | 0.008203887 | 6.44E-05 | 1.23E-05 | 1 | 0.975415042 | 1 | 0.961035047 | ENSG0000010 | 13 | EIF3L |
| Pancreas | T1DM | ENSG0000019 | -0.00223642 | 5.09E-09 | 0.000382665 | 0.008204201 | -0.00215452 | 2.03E-08 | 0.0003841 | 0.00820336 | -8.19E-05 | 0.014202592 | 1.31E-05 | 4.84E-07 | 5.22E-05 | 2.01E-06 | ENSG0000010 | 13 | EIF3L |
| Pancreas | T1DM | ENSG0000019 | -0.00021673 | 0.778734578 | 0.000771377 | 0.008204774 | -0.00023041 | 0.765162 | 0.000771348 | 0.008203888 | 1.37E-05 | 0.001852027 | 1 | 0.979738629 | 1 | 0.971298081 | ENSG0000010 | 9 | CCDC134 |
| Pancreas | T1DM | ENSG0000019 | -2.01E-05 | 0.980575869 | 0.000824887 | 0.008204776 | -3.11E-06 | 0.996992451 | 0.000824859 | 0.00820389 | -1.70E-05 | 0.001309443 | 1 | 0.997898334 | 1 | 0.999770584 | ENSG0000010 | 11 | PICK1 |
| Pancreas | T1DM | ENSG0000019 | -0.0028832 | 0.014048883 | 0.001173931 | 0.008204674 | -0.0028671 | 0.014590495 | 0.00117389 | 0.00820379 | -1.61E-05 | 0.025383337 | 1 | 0.2919226 | 1 | 0.288218403 | ENSG0000010 | 10 | CENPM |
| Pancreas | T1DM | ENSG0000019 | -0.00027777 | 0.80603252 | 0.001131232 | 0.008204775 | -0.00025108 | 0.824341644 | 0.001131194 | 0.008203889 | -2.67E-05 | 0.000221223 | 1 | 0.98201188 | 1 | 0.984037376 | ENSG0000010 | 17 | TPTEP1 |
| Pancreas | T1DM | ENSG0000019 | 0.001664289 | 0.051459012 | 0.000854517 | 0.008204712 | 0.001652289 | 0.053155451 | 0.000854487 | 0.008203827 | 1.20E-05 | 0.023711032 | 1 | 0.512196676 | 1 | 0.510771882 | ENSG0000010 | 17 | TPTEP1 |
| Pancreas | T1DM | ENSG0000019 | -0.0006187 | 0.385450618 | 0.000712872 | 0.008204763 | -0.00060741 | 0.394169938 | 0.00071285 | 0.008203878 | -1.13E-05 | 0.01877078 | 1 | 0.865581925 | 1 | 0.871859088 | ENSG0000010 | 6 | KDELR3 |
| Pancreas | T1DM | ENSG0000019 | -0.00073687 | 0.239813908 | 0.000626883 | 0.008204752 | -0.00074532 | 0.234447989 | 0.000626859 | 0.008203866 | 8.45E-06 | 0.019192647 | 1 | 0.785512902 | 1 | 0.780702981 | ENSG0000010 | 6 | KDELR3 |
| Pancreas | T1DM | ENSG0000019 | 0.001184472 | 0.000448646 | 0.000337485 | 0.008204568 | 0.00118086 | 0.000466793 | 0.000337471 | 0.008203684 | 3.61E-06 | 0.033916602 | 1 | 0.022156213 | 1 | 0.024973406 | ENSG0000010 | 16 | CYP2D6 |
| Pancreas | T1DM | ENSG0000019 | -0.00034106 | 0.705674775 | 0.000903071 | 0.008204773 | -0.00032739 | 0.716944107 | 0.000903043 | 0.008203888 | -1.37E-05 | 0.025967888 | 1 | 0.974645207 | 1 | 0.968082788 | ENSG0000010 | 16 | CYP2D6 |
| Pancreas | T1DM | ENSG0000019 | -0.00072145 | 0.330748605 | 0.000741772 | 0.00820476 | -0.00073227 | 0.323534892 | 0.000741748 | 0.008203874 | 1.08E-05 | 0.025433747 | 1 | 0.83462449 | 1 | 0.830335577 | ENSG0000010 | 14 | DDX17 |
| Pancreas | T1DM | ENSG0000019 | 0.000670958 | 0.288555714 | 0.000632206 | 0.008204757 | 0.000773936 | 0.221187809 | 0.000632622 | 0.008203865 | -0.00010298 | 1.62E-05 | 1 | 0.809076913 | 1 | 0.776261656 | ENSG0000010 | 14 | DDX17 |
| Pancreas | T1DM | ENSG0000019 | -0.00094338 | 0.073871611 | 0.000527792 | 0.008204722 | -0.00121218 | 0.022096416 | 0.000529633 | 0.008203802 | 0.000268796 | 1.50E-09 | 1 | 0.563394551 | 1 | 0.350269115 | ENSG0000010 | 14 | DDX17 |
| Pancreas | T1DM | ENSG0000019 | 0.000993824 | 0.30537611 | 0.00096961 | 0.008204758 | 0.000979342 | 0.312459938 | 0.000969574 | 0.008203873 | 1.45E-05 | 0.009906343 | 1 | 0.822018771 | 1 | 0.825219355 | ENSG0000010 | 5 | HSCB |
| Pancreas | T1DM | ENSG0000019 | -0.00068829 | 0.35955525 | 0.00075123 | 0.008204761 | -0.00067867 | 0.36629375 | 0.000751203 | 0.008203876 | -9.62E-06 | 0.029181835 | 1 | 0.854209198 | 1 | 0.850635855 | ENSG0000010 | 6 | CBY1 |
| Pancreas | T1DM | ENSG0000019 | 0.001988148 | 0.117435393 | 0.001269869 | 0.008204734 | 0.002007162 | 0.11395463 | 0.001269819 | 0.008203848 | -1.90E-05 | 0.005257596 | 1 | 0.67493056 | 1 | 0.666335944 | ENSG0000010 | 6 | CBY1 |
| Pancreas | T1DM | ENSG0000019 | 0.000571097 | 0.156941239 | 0.000403478 | 0.008204742 | 0.000565776 | 0.160826705 | 0.000403464 | 0.008203857 | 5.32E-06 | 0.028706533 | 1 | 0.719687683 | 1 | 0.723297686 | ENSG0000010 | 6 | CBY1 |
| Pancreas | T1DM | ENSG0000019 | -0.00039985 | 0.584220079 | 0.000721546 | 0.008204771 | -0.00027361 | 0.705010869 | 0.000722752 | 0.008203888 | -0.00012124 | 0.00424502 | 1 | 0.937588291 | 1 | 0.968082788 | ENSG0000010 | 6 | CBY1 |
| Pancreas | T1DM | ENSG0000019 | 0.000427397 | 0.544015019 | 0.0007044 | 0.008204769 | 0.000404777 | 0.565526582 | 0.000704385 | 0.008203884 | 2.26E-05 | 6.56E-05 | 1 | 0.929438478 | 1 | 0.936315417 | ENSG0000010 | 7 | RTCB |
| Pancreas | T1DM | ENSG0000019 | 4.37E-05 | 0.943373185 | 0.000615468 | 0.008204775 | 3.56E-05 | 0.953901599 | 0.000615446 | 0.00820389 | 8.14E-06 | 0.027789416 | 1 | 0.997898334 | 1 | 0.999388266 | ENSG0000010 | 13 | FBXO7 |
| Pancreas | T1DM | ENSG0000019 | -0.00023559 | 0.617825082 | 0.000472183 | 0.008204771 | -0.00024275 | 0.607173693 | 0.000472167 | 0.008203886 | 7.16E-06 | 0.016839082 | 1 | 0.950614027 | 1 | 0.940990975 | ENSG0000010 | 13 | FBXO7 |
| Pancreas | T1DM | ENSG0000019 | 0.000299301 | 0.655186433 | 0.000670222 | 0.008204772 | 0.000307431 | 0.646438505 | 0.000670199 | 0.008203886 | -8.13E-06 | 0.047784506 | 1 | 0.962467202 | 1 | 0.961035047 | ENSG0000010 | 13 | FBXO7 |
| Pancreas | T1DM | ENSG0000020 | 0.000514339 | 0.50055935 | 0.000763555 | 0.008204768 | 0.000541237 | 0.478415042 | 0.000763538 | 0.008203882 | -2.69E-05 | 6.76E-06 | 1 | 0.914783379 | 1 | 0.911454131 | ENSG0000010 | 16 | CYB5R3 |
| Pancreas | T1DM | ENSG0000020 | 0.004910809 | 4.88E-30 | 0.000431254 | 0.008202593 | 0.0047664 | 3.40E-28 | 0.000432867 | 0.008201849 | 0.000144409 | 0.000121084 | 1.25E-26 | 1.79E-27 | 8.73E-25 | 1.25E-25 | ENSG0000010 | 13 | LMF2 |
| Pancreas | T1DM | ENSG0000020 | -0.00702496 | 1.13E-29 | 0.0006209 | 0.008202621 | -0.0066291 | 9.03E-26 | 0.00063156 | 0.008202036 | -0.00039586 | 0.000620636 | 2.89E-26 | 3.62E-27 | 2.32E-22 | 2.32E-23 | ENSG0000010 | 13 | LMF2 |
| Pancreas | T1DM | ENSG0000020 | 0.004690598 | 2.39E-06 | 0.000994321 | 0.008204401 | 0.003713229 | 0.000235442 | 0.001009681 | 0.008203662 | 0.000977369 | 2.66E-08 | 0.006136418 | 0.00015341 | 0.604614121 | 0.01374123 | ENSG0000010 | 13 | LMF2 |
| Pancreas | T1DM | ENSG0000020 | -0.00018094 | 0.864220972 | 0.001058107 | 0.008204775 | -0.00016265 | 0.87782677 | 0.001058081 | 0.00820389 | -1.83E-05 | 0.025051467 | 1 | 0.990858518 | 1 | 0.993128823 | ENSG0000010 | 16 | TOM1 |
| Pancreas | T1DM | ENSG0000020 | -0.0033546 | 1.85E-27 | 0.000308967 | 0.008202791 | -0.00350412 | 4.50E-29 | 0.000313095 | 0.008201782 | 0.00014952 | 0.003239236 | 4.75E-24 | 4.75E-25 | 1.16E-25 | 1.93E-26 | ENSG0000010 | 16 | TOM1 |
| Pancreas | T1DM | ENSG0000020 | -0.00203178 | 4.26E-14 | 0.000268999 | 0.008203815 | -0.00283075 | 2.10E-23 | 0.000283966 | 0.008202217 | 0.000798974 | 1.71E-18 | 1.09E-10 | 6.83E-12 | 5.40E-20 | 4.91E-21 | ENSG0000010 | 16 | TOM1 |
| Pancreas | T1DM | ENSG0000020 | 0.004136711 | 9.16E-05 | 0.001057448 | 0.008204518 | 0.004453456 | 2.63E-05 | 0.001059575 | 0.008203593 | -0.00031674 | 3.24E-06 | 0.235115938 | 0.005002467 | 0.067635975 | 0.001690899 | ENSG0000010 | 2 | BIK |
| Pancreas | T1DM | ENSG0000020 | 0.000708123 | 0.033326686 | 0.000332744 | 0.008204699 | 0.000608904 | 0.068135351 | 0.000333808 | 0.008203834 | 9.92E-05 | 0.000219671 | 1 | 0.434431116 | 1 | 0.536516198 | ENSG0000010 | 9 | THOC5 |
| Pancreas | T1DM | ENSG0000020 | 0.000710321 | 0.001797712 | 0.000227537 | 0.008204611 | 0.000970859 | 3.03E-05 | 0.000232771 | 0.008203597 | -0.00026054 | 1.15E-07 | 1 | 0.075680736 | 0.077936275 | 0.001900885 | ENSG0000010 | 9 | THOC5 |
| Pancreas | T1DM | ENSG0000020 | 0.001882674 | 0.000308418 | 0.000521784 | 0.008204556 | 0.00168047 | 0.001308986 | 0.000522856 | 0.008203716 | 0.000202204 | 2.41E-09 | 0.792016331 | 0.015529732 | 1 | 0.05795648 | ENSG0000010 | 9 | THOC5 |
| Pancreas | T1DM | ENSG0000020 | 0.002509043 | 0.026825859 | 0.001133237 | 0.008204693 | 0.002789045 | 0.014012056 | 0.00113516 | 0.008203788 | -0.00028 | 3.01E-05 | 1 | 0.395912673 | 1 | 0.281029074 | ENSG0000010 | 13 | ARSA |
| Pancreas | T1DM | ENSG0000020 | -0.00224511 | 4.32E-08 | 0.000409903 | 0.00820427 | -0.00214923 | 1.71E-07 | 0.00041104 | 0.008220343 | -9.59E-05 | 0.001864791 | 0.000111035 | 3.58E-06 | 0.000438408 | 1.51E-05 | ENSG0000010 | 13 | TSPO |
| Pancreas | T1DM | ENSG0000020 | 0.004451946 | 8.74E-14 | 0.000596871 | 0.008203839 | 0.004892466 | 6.37E-16 | 0.000605336 | 0.00820279 | -0.00044052 | 1.32E-05 | 2.25E-10 | 1.25E-11 | 1.64E-12 | 1.02E-13 | ENSG0000010 | 13 | TSPO |
| Pancreas | T1DM | ENSG0000020 | 0.00101445 | 0.190886824 | 0.000775596 | 0.008204747 | 0.000839992 | 0.279864139 | 0.00077732 | 0.00820387 | 0.000174458 | 0.000862237 | 1 | 0.752840936 | 1 | 0.797009688 | ENSG0000010 | 13 | TSPO |
| Pancreas | T1DM | ENSG0000020 | 0.001775607 | 0.010242653 | 0.00069156 | 0.008204665 | 0.00119589 | 0.086467829 | 0.000697582 | 0.008203841 | 0.000579717 | 2.61E-10 | 1 | 0.24354753 | 1 | 0.580557814 | ENSG0000010 | 13 | TSPO |
| Pancreas | T1DM | ENSG0000020 | 0.000738628 | 0.151725073 | 0.000515274 | 0.008204741 | 0.00077949 | 0.130394119 | 0.000515348 | 0.008203851 | -4.09E-05 | 6.69E-05 | 1 | 0.717060869 | 1 | 0.676232745 | ENSG0000010 | 6 | RASD2 |
| Pancreas | T1DM | ENSG0000020 | -0.00090207 | 0.003996583 | 0.000313388 | 0.008204636 | -0.00077713 | 0.013392186 | 0.000314222 | 0.008203787 | -0.00012494 | 6.33E-08 | 1 | 0.135042429 | 1 | 0.275576512 | ENSG0000010 | 6 | RASD2 |
| Pancreas | T1DM | ENSG0000020 | 0.000497807 | 0.111746007 | 0.000313008 | 0.008204733 | 0.000434657 | 0.165434031 | 0.000313373 | 0.008203858 | 6.31E-05 | 4.46E-05 | 1 | 0.670476041 | 1 | 0.726605951 | ENSG0000010 | 11 | MTMR3 |
| Pancreas | T1DM | ENSG0000020 | -0.00012187 | 0.914029623 | 0.001128869 | 0.008204775 | -0.00010703 | 0.924461722 | 0.001128826 | 0.00820389 | -1.48E-05 | 0.019329492 | 1 | 0.995870671 | 1 | 0.999388266 | ENSG0000010 | 13 | APOL4 |
| Pancreas | T1DM | ENSG0000020 | -0.00130779 | 0.085008097 | 0.000759307 | 0.008204726 | -0.00131775 | 0.082646932 | 0.000759277 | 0.008203839 | 9.96E-06 | 0.01475093 | 1 | 0.590002147 | 1 | 0.578303331 | ENSG0000010 | 10 | FOXRED2 |
| Pancreas | T1DM | ENSG0000020 | -0.00021683 | 0.699736726 | 0.000562209 | 0.008204773 | -0.00023077 | 0.681455731 | 0.000562193 | 0.008203887 | 1.39E-05 | 0.000625978 | 1 | 0.974470668 | 1 | 0.966338775 | ENSG0000010 | 10 | FOXRED2 |
| Pancreas | T1DM | ENSG0000020 | 0.000313907 | 0.607924635 | 0.000611859 | 0.008204771 | 0.000326414 | 0.593692158 | 0.000611842 | 0.008203885 | -1.25E-05 | 0.003924735 | 1 | 0.943530179 | 1 | 0.937577789 | ENSG0000010 | 10 | SLC25A17 |
| Pancreas | T1DM | ENSG0000020 | -0.00019161 | 0.842723932 | 0.000965738 | 0.008204775 | -0.0002029 | 0.833583566 | 0.0009657 | 0.008203889 | 1.13E-05 | 0.027527212 | 1 | 0.987729373 | 1 | 0.985109341 | ENSG0000010 | 7 | FAM118A |
| Pancreas | T1DM | ENSG0000020 | 0.000604592 | 0.417846521 | 0.000746259 | 0.008204765 | 0.000567672 | 0.446871616 | 0.000746307 | 0.00820388 | 3.69E-05 | 0.001263198 | 1 | 0.886967947 | 1 | 0.896838446 | ENSG0000010 | 10 | L3MBTL2 |
| Pancreas | T1DM | ENSG0000020 | -0.00072503 | 0.249913368 | 0.000630155 | 0.008204753 | -0.00073918 | 0.240777555 | 0.000630136 | 0.008203867 | 1.41E-05 | 0.001172792 | 1 | 0.789394255 | 1 | 0.780702981 | ENSG0000010 | 12 | POLR3H |
| Pancreas | T1DM | ENSG0000020 | 0.00068591 | 0.275246055 | 0.000566962 | 0.008204756 | 0.000628807 | 0.267381899 | 0.000566945 | 0.008203869 | -1.02E-05 | 0.009929491 | 1 | 0.798582374 | 1 | 0.795306713 | ENSG0000010 | 13 | HDAC10 |
| Pancreas | T1DM | ENSG0000020 | -0.00058784 | 0.353702022 | 0.000633839 | 0.008204761 | -0.00057416 | 0.36500293 | 0.000633818 | 0.008203876 | -1.37E-05 | 0.000705081 | 1 | 0.854061442 | 1 | 0.850635855 | ENSG0000010 | 13 | HDAC10 |
| Pancreas | T1DM | ENSG0000020 | 0.001025825 | 0.292738186 | 0.000974994 | 0.008204757 | 0.001056298 | 0.27866902 | 0.00097506 | 0.00820387 | -3.05E-05 | 0.045870968 | 1 | 0.810235282 | 1 | 0.796020071 | ENSG0000010 | 13 | HDAC10 |
| Pancreas | T1DM | ENSG0000020 | 3.08E-05 | 0.97035886 | 0.000827759 | 0.008204776 | 1.12E-05 | 0.989197603 | 0.000827756 | 0.00820389 | 1.96E-05 | 0.018496236 | 1 | 0.997898334 | 1 | 0.99966149 | ENSG0000010 | 5 | KHNYN |
| Pancreas | T1DM | ENSG0000020 | 0.00093338 | 0.030303433 | 0.000430902 | 0.008204697 | 0.000731396 | 0.090589145 | 0.00043219 | 0.008203842 | 0.000201984 | 1.91E-09 | 1 | 0.424384765 | 1 | 0.590438896 | ENSG0000010 | 5 | KHNYN |
| Pancreas | T1DM | ENSG0000020 | 0.00255688 | 8.16E-09 | 0.000443502 | 0.008204216 | 0.002092894 | 3.72E-06 | 0.000452357 | 0.00820353 | 0.000463986 | 1.95E-07 | 2.10E-05 | 7.23E-07 | 0.009546279 | 0.000260615 | ENSG0000010 | 12 | SDR39U1 |
| Pancreas | T1DM | ENSG0000020 | 0.000615738 | 0.120676213 | 0.000396753 | 0.008204735 | 0.001102708 | 0.006356598 | 0.000404099 | 0.008203765 | -0.00048697 | 2.31E-10 | 1 | 0.677664852 | 1 | 0.186420941 | ENSG0000010 | 12 | SDR39U1 |
| Pancreas | T1DM | ENSG0000021 | 0.000804727 | 0.138499046 | 0.000543219 | 0.008204739 | 0.000796172 | 0.14272718 | 0.000543198 | 0.008203853 | 8.56E-06 | 0.004938267 | 1 | 0.69738343 | 1 | 0.696812542 | ENSG0000010 | 12 | SDR39U1 |
| Pancreas | T1DM | ENSG0000021 | 0.000449854 | 0.202122142 | 0.00035268 | 0.008204748 | 0.000445742 | 0.206260144 | 0.000352667 | 0.008203863 | 4.11E-06 | 0.04138566 | 1 | 0.766675365 | 1 | 0.769778216 | ENSG0000010 | 12 | SDR39U1 |
| Pancreas | T1DM | ENSG0000021 | 0.00194083 | 0.015571065 | 0.000802385 | 0.008204677 | 0.002120752 | 0.008315004 | 0.000803615 | 0.008203773 | -0.00017992 | 6.95E-05 | 1 | 0.309972834 | 1 | 0.204622746 | ENSG0000010 | 13 | RBM23 |
| Pancreas | T1DM | ENSG0000021 | 0.002281016 | 7.87E-11 | 0.00035075 | 0.008204064 | 0.002074921 | 4.48E-09 | 0.000353759 | 0.008203311 | 0.000206095 | 8.03E-06 | 2.02E-07 | 9.62E-09 | 1.15E-05 | 4.80E-07 | ENSG0000010 | 13 | RBM23 |
| Pancreas | T1DM | ENSG0000021 | -0.00106508 | 0.085719432 | 0.000619797 | 0.008204726 | -0.00216128 | 0.000760322 | 0.000641924 | 0.008203699 | 0.0010962 | 5.56E-11 | 1 | 0.592504613 | 1 | 0.038284453 | ENSG0000010 | 13 | RBM23 |
| Pancreas | T1DM | ENSG0000021 | 0.000796424 | 0.416439292 | 0.000980075 | 0.008204764 | 0.000814264 | 0.406060451 | 0.000980041 | 0.008203878 | -1.78E-05 | 0.002900864 | 1 | 0.886746354 | 1 | 0.882202402 | ENSG0000010 | 10 | AP4S1 |
| Pancreas | T1DM | ENSG0000021 | 0.002015448 | 0.109052243 | 0.001257713 | 0.008204732 | 0.002040559 | 0.104698621 | 0.00125767 | 0.008203846 | -2.51E-05 | 0.001361345 | 1 | 0.661051879 | 1 | 0.646312642 | ENSG0000010 | 10 | AP4S1 |
| Pancreas | T1DM | ENSG0000021 | -0.00088984 | 0.266660499 | 0.000801093 | 0.008204755 | -0.00213488 | 0.01509487 | 0.000878515 | 0.008203791 | 0.001245039 | 0.00055727 | 1 | 0.797304864 | 1 | 0.293663832 | ENSG0000010 | 9 | VCPKMT |
| Pancreas | T1DM | ENSG0000021 | 0.001098734 | 0.257961223 | 0.000971279 | 0.008204754 | 0.001113174 | 0.25174084 | 0.000971244 | 0.008203868 | -1.44E-05 | 0.013578644 | 1 | 0.79720158 | 1 | 0.788955599 | ENSG0000010 | 9 | TRIM9 |
| Pancreas | T1DM | ENSG0000021 | 3.88E-05 | 0.932631508 | 0.000459251 | 0.008204775 | 2.95E-05 | 0.948795638 | 0.000459233 | 0.00820389 | 9.33E-06 | 0.000444993 | 1 | 0.997898334 | 1 | 0.999388266 | ENSG0000010 | 9 | TRIM9 |
| Pancreas | T1DM | ENSG0000022 | -0.00014384 | 0.860542188 | 0.000818728 | 0.008204775 | -0.00013593 | 0.868133962 | 0.000818694 | 0.00820389 | -7.91E-06 | 0.049128286 | 1 | 0.990511029 | 1 | 0.990577333 | ENSG0000010 | 7 | CNIH1 |
| Pancreas | T1DM | ENSG0000022 | 0.007009714 | 1.09E-13 | 0.000943467 | 0.008203846 | 0.00623609 | 8.79E-11 | 0.000961389 | 0.008203182 | 0.000773625 | 2.88E-05 | 2.80E-10 | 1.47E-11 | 2.26E-07 | 1.03E-08 | ENSG0000010 | 7 | ATP6V1D |
| Pancreas | T1DM | ENSG0000022 | -0.00038683 | 0.659896957 | 0.000879055 | 0.008204772 | -0.00040653 | 0.643736964 | 0.0008703 | 0.008203886 | 1.97E-05 | 0.001820355 | 1 | 0.962467202 | 1 | 0.960555796 | ENSG0000010 | 4 | PLEK2 |
| Pancreas | T1DM | ENSG0000022 | -0.00037624 | 0.555709166 | 0.000638534 | 0.0082047 | -0.00038782 | 0.543598121 | 0.000638515 | 0.008203884 | 1.16E-05 | 0.009866343 | 1 | 0.93270646 | 1 | 0.928420568 | ENSG0000010 | 4 | PIGH |
| Pancreas | T1DM | ENSG0000022 | -0.00106613 | 0.008764201 | 0.000406743 | 0.00820466 | -0.00047502 | 0.299093973 | 0.000457466 | 0.008203872 | -0.0005911 | 0.004761386 | 1 | 0.225064674 | 1 | 0.810652485 | ENSG0000010 | 4 | PIGH |
| Pancreas | T1DM | ENSG0000022 | 0.00589385 | 5.07E-07 | 0.001173243 | 0.008204351 | 0.005486048 | 3.86E-06 | 0.001187707 | 0.008203531 | 0.000407802 | 0.027640292 | 0.001303041 | 3.62E-05 | 0.009903386 | 0.000260615 | ENSG0000010 | 7 | TMED8 |
| Pancreas | T1DM | ENSG0000022 | -0.00134991 | 0.135707628 | 0.000904784 | 0.008204738 | -0.0011578 | 0.202377487 | 0.000908216 | 0.008203863 | -0.00019211 | 0.015573678 | 1 | 0.694369535 | 1 | 0.766509931 | ENSG0000010 | 7 | TMED8 |
| Pancreas | T1DM | ENSG0000022 | 0.005772603 | 8.88E-17 | 0.000693893 | 0.00820361 | 0.005930985 | 1.33E-17 | 0.000694387 | 0.008202662 | -0.00015838 | 6.09E-09 | 2.28E-13 | 1.63E-14 | 3.42E-14 | 2.28E-15 | ENSG0000010 | 7 | TMED8 |
| Pancreas | T1DM | ENSG0000022 | 0.000152366 | 0.895962183 | 0.001165196 | 0.008204775 | 0.000180394 | 0.876961921 | 0.001165173 | 0.00820389 | -2.80E-05 | 0.0035103 | 1 | 0.99200447 | 1 | 0.993128823 | ENSG0000010 | 7 | TMED8 |
| Pancreas | T1DM | ENSG0000022 | 0.001354953 | 0.145431883 | 0.000930689 | 0.00820474 | 0.001344232 | 0.148626776 | 0.00093065 | 0.008203855 | 1.07E-05 | 0.019052584 | 1 | 0.709552898 | 1 | 0.706802888 | ENSG0000010 | 13 | PPM1A |
| Pancreas | T1DM | ENSG0000022 | -0.00042123 | 0.417793848 | 0.000519875 | 0.008204765 | -0.00041031 | 0.429950332 | 0.000519859 | 0.008203879 | -1.09E-05 | 0.001824704 | 1 | 0.886967947 | 1 | 0.890691955 | ENSG0000010 | 13 | PPM1A |
| Pancreas | T1DM | ENSG0000022 | 0.000163312 | 0.842230267 | 0.00082049 | 0.008204775 | 0.000175448 | 0.830672449 | 0.000820465 | 0.008203889 | -1.21E-05 | 0.028446894 | 1 | 0.987601519 | 1 | 0.984297508 | ENSG0000010 | 10 | TELO2 |
| Pancreas | T1DM | ENSG0000022 | 0.001877649 | 0.001899047 | 0.000604601 | 0.008204613 | 0.001797701 | 0.002968927 | 0.000604857 | 0.008203741 | 8.06E-05 | 1.54E-05 | 1 | 0.078657282 | 1 | 0.112120641 | ENSG0000010 | 11 | PCNX |
| Pancreas | T1DM | ENSG0000022 | -0.00048038 | 0.622487952 | 0.000975739 | 0.008204771 | -0.00046688 | 0.632289226 | 0.000975705 | 0.008203886 | -1.35E-05 | 0.027559675 | 1 | 0.95220407 | 1 | 0.955974304 | ENSG0000010 | 11 | PCNX |
| Pancreas | T1DM | ENSG0000022 | 0.000188293 | 0.794754071 | 0.000723806 | 0.008204774 | 0.000207573 | 0.774273507 | 0.000723782 | 0.008203889 | -1.93E-05 | 3.07E-05 | 1 | 0.981920629 | 1 | 0.972640681 | ENSG0000010 | 11 | PCNX |
| Pancreas | T1DM | ENSG0000022 | -0.00078449 | 0.422723236 | 0.000978529 | 0.008204765 | -0.00077076 | 0.430871009 | 0.000978493 | 0.00820388 | -1.37E-05 | 0.016765051 | 1 | 0.889410424 | 1 | 0.890803318 | ENSG0000010 | 11 | PCNX |
| Pancreas | T1DM | ENSG0000022 | -0.00095492 | 0.369781727 | 0.001064713 | 0.008204762 | -0.0009706 | 0.361961702 | 0.001064676 | 0.008203876 | 1.57E-05 | 0.018924232 | 1 | 0.856244109 | 1 | 0.850635855 | ENSG0000010 | 11 | PCNX |
| Pancreas | T1DM | ENSG0000022 | -0.00213321 | 0.045101875 | 0.00106463 | 0.008204708 | -0.00215073 | 0.043360108 | 0.001064594 | 0.008203821 | 1.75E-05 | 0.009838765 | 1 | 0.490769555 | 1 | 0.471816773 | ENSG0000010 | 11 | PCNX |
| Pancreas | T1DM | ENSG0000022 | 0.000777921 | 0.473839874 | 0.001086108 | 0.008204767 | 0.000756758 | 0.485947707 | 0.001086096 | 0.008203881 | 2.12E-05 | 0.03395431 | 1 | 0.911384054 | 1 | 0.914417995 | ENSG0000010 | 11 | PCNX |
| Pancreas | T1DM | ENSG0000022 | 0.001352507 | 0.271834962 | 0.001230841 | 0.008204755 | 0.001331006 | 0.279515023 | 0.001230806 | 0.00820387 | 2.15E-05 | 0.013724392 | 1 | 0.798582374 | 1 | 0.797009688 | ENSG0000010 | 9 | EMC9 |
| Pancreas | T1DM | ENSG0000022 | -0.00038111 | 0.645376379 | 0.000828154 | 0.008204772 | -0.00037143 | 0.653775972 | 0.000828118 | 0.008203887 | -9.68E-06 | 0.01347867 | 1 | 0.961788216 | 1 | 0.961035047 | ENSG0000010 | 9 | EMC9 |
| Pancreas | T1DM | ENSG0000022 | -0.000888 | 0.467500002 | 0.001222215 | 0.008204767 | -0.00091322 | 0.454938685 | 0.001222173 | 0.008203881 | 2.52E-05 | 0.001068549 | 1 | 0.908712989 | 1 | 0.899776609 | ENSG0000010 | 9 | EMC9 |
| Pancreas | T1DM | ENSG0000022 | -0.00170926 | 0.011503191 | 0.00067639 | 0.008204668 | -0.00196755 | 0.003697345 | 0.000677788 | 0.008203748 | 0.00025829 | 4.69E-09 | 1 | 0.260075765 | 1 | 0.128265071 | ENSG0000010 | 9 | EMC9 |
| Pancreas | T1DM | ENSG0000022 | 0.005122867 | 5.98E-59 | 0.000316388 | 0.008200364 | 0.005878019 | 1.51E-53 | 0.000381513 | 0.008199895 | -0.00075515 | 0.000397251 | 1.54E-55 | 1.54E-55 | 3.88E-50 | 3.88E-50 | ENSG0000010 | 10 | PLTP |
| Pancreas | T1DM | ENSG0000022 | -0.00098562 | 0.001025772 | 0.000300186 | 0.008204594 | -0.00099096 | 0.00096253 | 0.000300176 | 0.008203707 | 5.34E-06 | 0.005733977 | 1 | 0.047038972 | 1 | 0.045773668 | ENSG0000010 | 7 | PYGB |
| Pancreas | T1DM | ENSG0000023 | 0.000244935 | 0.748053599 | 0.000762546 | 0.008204774 | 0.00022557 | 0.767373932 | 0.000762544 | 0.008203889 | 1.94E-05 | 0.012498543 | 1 | 0.975415042 | 1 | 0.972640681 | ENSG0000010 | 7 | PYGB |
| Pancreas | T1DM | ENSG0000023 | -5.74E-05 | 0.911560401 | 0.000516375 | 0.008204775 | -6.76E-05 | 0.895780752 | 0.000516367 | 0.00820389 | 1.03E-05 | 0.021923025 | 1 | 0.995698473 | 1 | 0.993128823 | ENSG0000010 | 14 | ABHD12 |
| Pancreas | T1DM | ENSG0000023 | -0.00054326 | 0.405464882 | 0.000653029 | 0.008204764 | -0.00055343 | 0.396709297 | 0.000653004 | 0.008203878 | 1.02E-05 | 0.004211125 | 1 | 0.88063625 | 1 | 0.871863634 | ENSG0000010 | 4 | PROCR |
| Pancreas | T1DM | ENSG0000023 | -2.18E-05 | 0.938743813 | 0.000283945 | 0.008204775 | -0.00011427 | 0.688377092 | 0.000284918 | 0.008203887 | 9.24E-05 | 9.59E-05 | 1 | 0.997898334 | 1 | 0.966685034 | ENSG0000010 | 4 | PROCR |
| Pancreas | T1DM | ENSG0000023 | -0.00112602 | 0.026107633 | 0.000506164 | 0.008204692 | -0.00111606 | 0.027453998 | 0.000506149 | 0.008203808 | -9.96E-06 | 0.004092989 | 1 | 0.393484808 | 1 | 0.393865178 | ENSG0000010 | 4 | PROCR |
| Pancreas | T1DM | ENSG0000023 | -0.00027787 | 0.714872875 | 0.000760632 | 0.008204773 | -0.00025735 | 0.735100102 | 0.000760609 | 0.008203888 | -2.05E-05 | 7.97E-05 | 1 | 0.975415042 | 1 | 0.968082788 | ENSG0000010 | 17 | UQCC1 |

| | | | | | | | | | | | | | | | | | | | |
|---|---|---|---|---|---|---|---|---|---|---|---|---|---|---|---|---|---|---|---|
| Pancreas | T1DM | ENSG0000023 | 0.001752144 | 2.00E-09 | 0.000292109 | 0.00820417 | 0.001939407 | 6.79E-11 | 0.000297211 | 0.008203173 | -0.00018726 | 0.000651418 | 5.13E-06 | 2.05E-07 | 1.74E-07 | 8.30E-09 | ENSG000001( | 8 | IFT52 |
| Pancreas | T1DM | ENSG0000023 | 0.001189497 | 0.14208444 | 0.000810242 | 0.008204739 | 0.001202657 | 0.137711216 | 0.000810213 | 0.008203853 | -1.32E-05 | 0.006697566 | 1 | 0.705750175 | 1 | 0.692461116 | ENSG000001( | 8 | IFT52 |
| Pancreas | T1DM | ENSG0000023 | -0.00024471 | 0.733599279 | 0.000719023 | 0.008204774 | -0.00058409 | 0.418267373 | 0.00072161 | 0.008203879 | 0.000339381 | 3.44E-08 | 1 | 0.975415042 | 1 | 0.88842896 | ENSG000001( | 8 | IFT52 |
| Pancreas | T1DM | ENSG0000023 | 0.000521515 | 0.361884422 | 0.000571974 | 0.008204762 | 0.000336832 | 0.556430399 | 0.000572695 | 0.008203884 | 0.000184683 | 3.44E-08 | 1 | 0.854831234 | 1 | 0.93515266 | ENSG000001( | 8 | IFT52 |
| Pancreas | T1DM | ENSG0000023 | -4.86E-05 | 0.926609199 | 0.000527781 | 0.008204775 | -3.75E-05 | 0.943317993 | 0.000527777 | 0.00820389 | -1.11E-05 | 0.029081205 | 1 | 0.997898334 | 1 | 0.999388266 | ENSG000001( | 13 | MTG2 |
| Pancreas | T1DM | ENSG0000023 | -0.0007243 | 0.564996623 | 0.001258699 | 0.00820477 | -0.00069198 | 0.582473849 | 0.001258654 | 0.008203883 | -3.23E-05 | 1 | | 0.936676131 | 1 | 0.936315417 | ENSG000001( | 13 | MTG2 |
| Pancreas | T1DM | ENSG0000023 | 4.93E-05 | 0.899244942 | 0.000389653 | 0.008204775 | 4.28E-05 | 0.912559763 | 0.000389639 | 0.00820389 | 6.55E-06 | 0.003897409 | 1 | 0.99281063 | 1 | 0.996366272 | ENSG000001( | 8 | TCFL5 |
| Pancreas | T1DM | ENSG0000023 | 2.16E-05 | 0.977933636 | 0.000781152 | 0.008204776 | -4.45E-06 | 0.995451665 | 0.000781161 | 0.00820389 | 2.61E-05 | 0.003509354 | 1 | 0.997898334 | 1 | 0.999770584 | ENSG000001( | 12 | DIDO1 |
| Pancreas | T1DM | ENSG0000023 | 0.000379619 | 0.752885572 | 0.001205762 | 0.008204774 | 0.000395957 | 0.742609483 | 0.001205715 | 0.008203889 | -1.63E-05 | 0.013675767 | 1 | 0.975415042 | 1 | 0.968082788 | ENSG000001( | 8 | BIRC7 |
| Pancreas | T1DM | ENSG0000023 | -0.00123938 | 0.021921401 | 0.000540805 | 0.008204687 | -0.00186549 | 0.000676758 | 0.00054886 | 0.008203695 | 0.000626111 | 2.57E-11 | 1 | 0.366243427 | 1 | 0.034758299 | ENSG000001( | 8 | BIRC7 |
| Pancreas | T1DM | ENSG0000023 | 0.00146043 | 4.65E-05 | 0.000358581 | 0.008204496 | 0.000962971 | 0.009528653 | 0.000371446 | 0.008203777 | 0.000497458 | 2.90E-07 | 0.11930418 | 0.002651204 | 1 | 0.216544969 | ENSG000001( | 10 | NKAIN4 |
| Pancreas | T1DM | ENSG0000023 | -0.00080963 | 0.456221979 | 0.001086634 | 0.008204766 | -0.00082779 | 0.446170952 | 0.001086599 | 0.00820388 | 1.82E-05 | 0.011583075 | 1 | 0.899832468 | 1 | 0.896838446 | ENSG000001( | 13 | CDC25B |
| Pancreas | T1DM | ENSG0000023 | 0.001183503 | 2.76E-07 | 0.000230302 | 0.008204331 | 0.001098206 | 1.96E-06 | 0.00023083 | 0.008203509 | 8.53E-05 | 6.28E-08 | 0.00070991 | 2.03E-05 | 0.005031361 | 0.000143753 | ENSG000001( | 13 | CDC25B |
| Pancreas | T1DM | ENSG0000023 | -9.53E-05 | 0.813415736 | 0.000403627 | 0.008204775 | -8.80E-05 | 0.827462334 | 0.000403615 | 0.008203889 | -7.29E-06 | 0.010609469 | 1 | 0.98201188 | 1 | 0.984037376 | ENSG000001( | 13 | CDC25B |
| Pancreas | T1DM | ENSG0000023 | 0.001153289 | 0.04770598 | 0.000582474 | 0.00820471 | 0.001372884 | 0.018656361 | 0.000583626 | 0.008203797 | -0.0002196 | 3.45E-09 | 1 | 0.50640868 | 1 | 0.329178568 | ENSG000001( | 5 | ISM1 |
| Pancreas | T1DM | ENSG0000023 | -0.00251849 | 0.00347586 | 0.000861846 | 0.008204632 | -0.00255261 | 0.00306095 | 0.000861914 | 0.008203742 | 3.41E-05 | 0.014893725 | 1 | 0.123976875 | 1 | 0.112293137 | ENSG000001( | 4 | RSPO4 |
| Pancreas | T1DM | ENSG0000023 | 0.00085416 | 0.240744466 | 0.000728104 | 0.008204752 | 0.000844635 | 0.246011427 | 0.000728075 | 0.008203867 | 9.53E-06 | 0.015234452 | 1 | 0.786897324 | 1 | 0.784493531 | ENSG000001( | 11 | SIRPB1 |
| Pancreas | T1DM | ENSG0000023 | 3.95E-05 | 0.968924943 | 0.001012731 | 0.008204776 | 0.000364097 | 0.719708278 | 0.001014625 | 0.008203888 | -0.00032464 | 2.40E-07 | 1 | 0.997898334 | 1 | 0.968082788 | ENSG000001( | 10 | SEC23B |
| Pancreas | T1DM | ENSG0000023 | -0.00272534 | 5.93E-08 | 0.000502731 | 0.008204281 | -0.00187839 | 0.000528441 | 0.000541959 | 0.008203688 | -0.00084694 | 2.88E-05 | 0.000152211 | 4.76E-06 | 1 | 0.027694627 | ENSG000001( | 10 | SEC23B |
| Pancreas | T1DM | ENSG0000023 | -0.00111696 | 0.257718642 | 0.000986887 | 0.008204754 | -0.00113008 | 0.252156268 | 0.000986854 | 0.008203868 | 1.31E-05 | 0.0362861 | 1 | 0.79720158 | 1 | 0.788955599 | ENSG000001( | 10 | SEC23B |
| Pancreas | T1DM | ENSG0000023 | 0.000919359 | 0.098707844 | 0.000556798 | 0.00820473 | 0.000910377 | 0.102032855 | 0.000556778 | 0.008203845 | 8.98E-06 | 0.008153658 | 1 | 0.633704355 | 1 | 0.637519152 | ENSG000001( | 10 | SEC23B |
| Pancreas | T1DM | ENSG0000023 | 0.001089748 | 0.076371591 | 0.000614933 | 0.008204723 | 0.001021151 | 0.097117144 | 0.000615523 | 0.008203844 | 6.86E-05 | 0.013276791 | 1 | 0.571784974 | 1 | 0.617318874 | ENSG000001( | 12 | PLCB4 |
| Pancreas | T1DM | ENSG0000023 | 0.00075968 | 0.357385442 | 0.000825416 | 0.008204761 | 0.000773341 | 0.348791615 | 0.000825393 | 0.008203875 | -1.37E-05 | 0.02047818 | 1 | 0.854209198 | 1 | 0.847508521 | ENSG000001( | 12 | PLCB4 |
| Pancreas | T1DM | ENSG0000023 | 0.001805601 | 8.42E-06 | 0.000405371 | 0.008204442 | 0.002010631 | 7.94E-07 | 0.000407271 | 0.00820348 | -0.00020503 | 2.15E-07 | 0.021629374 | 0.000527546 | 0.002039598 | 6.37E-05 | ENSG000001( | 5 | MYL9 |
| Pancreas | T1DM | ENSG0000023 | 0.000253395 | 0.722127215 | 0.000712549 | 0.008204773 | 0.000263553 | 0.711468379 | 0.000712528 | 0.008203888 | -1.02E-05 | 0.039811249 | 1 | 0.975415042 | 1 | 0.968082788 | ENSG000001( | 5 | MYL9 |
| Pancreas | T1DM | ENSG0000023 | 0.000886374 | 0.322283326 | 0.000895529 | 0.008204759 | 0.000910082 | 0.309496296 | 0.000895503 | 0.008203873 | -2.37E-05 | 0.000176319 | 1 | 0.832533491 | 1 | 0.82064652 | ENSG000001( | 10 | CRNKL1 |
| Pancreas | T1DM | ENSG0000023 | 0.000982921 | 0.258194566 | 0.000869326 | 0.008204754 | 0.000998596 | 0.250663323 | 0.000869296 | 0.008203868 | -1.57E-05 | 0.00353551 | 1 | 0.79720158 | 1 | 0.788955599 | ENSG000001( | 10 | MROH8 |
| Pancreas | T1DM | ENSG0000023 | -5.93E-05 | 0.890871112 | 0.000432216 | 0.008204775 | -0.00023673 | 0.584753415 | 0.000433216 | 0.008203885 | 0.000177432 | 2.48E-09 | 1 | 0.99200447 | 1 | 0.936772783 | ENSG000001( | 10 | NOP56 |
| Pancreas | T1DM | ENSG0000023 | -0.00018744 | 0.779973857 | 0.000670985 | 0.008204774 | -0.00017847 | 0.790242753 | 0.000670962 | 0.008203889 | -8.97E-06 | 0.034641258 | 1 | 0.980404234 | 1 | 0.97879227 | ENSG000001( | 10 | NOP56 |
| Pancreas | T1DM | ENSG0000024 | -0.00030215 | 0.493962824 | 0.000441724 | 0.008204768 | -4.71E-05 | 0.916140551 | 0.000447489 | 0.00820389 | -0.00025503 | 0.000378443 | 1 | 0.914693561 | 1 | 0.997092545 | ENSG000001( | 8 | MANBAL |
| Pancreas | T1DM | ENSG0000024 | 0.008406519 | 2.42E-21 | 0.000886254 | 0.008203261 | 0.008117937 | 6.51E-20 | 0.000888578 | 0.008202485 | 0.000288582 | 8.46E-06 | 6.22E-18 | 5.19E-19 | 1.67E-16 | 1.29E-17 | ENSG000001( | 8 | MANBAL |
| Pancreas | T1DM | ENSG0000024 | 0.000196572 | 0.518951653 | 0.00030478 | 0.008204769 | 0.000191691 | 0.529368633 | 0.00030477 | 0.008203883 | 4.88E-06 | 0.014442056 | 1 | 0.914783379 | 1 | 0.920313283 | ENSG000001( | 8 | MANBAL |
| Pancreas | T1DM | ENSG0000024 | 3.82E-05 | 0.889587231 | 0.00027522 | 0.008204775 | 2.53E-05 | 0.926903285 | 0.000275246 | 0.00820389 | 1.30E-05 | 0.006361831 | 1 | 0.99200447 | 1 | 0.999388266 | ENSG000001( | 7 | PXMP4 |
| Pancreas | T1DM | ENSG0000024 | -0.00025112 | 0.740992035 | 0.000759718 | 0.008204774 | -0.00023556 | 0.756504685 | 0.000759698 | 0.008203888 | -1.56E-05 | 0.005922047 | 1 | 0.975415042 | 1 | 0.968082788 | ENSG000001( | 8 | BPI |
| Pancreas | T1DM | ENSG0000024 | 0.000791347 | 0.329413134 | 0.0008139 | 0.00820476 | 0.000775192 | 0.339365834 | 0.000811363 | 0.008203875 | 1.62E-05 | 0.001753246 | 1 | 0.83462449 | 1 | 0.839770819 | ENSG000001( | 10 | DHX35 |
| Pancreas | T1DM | ENSG0000024 | -0.00170226 | 0.082540202 | 0.000980487 | 0.008204725 | -0.00168889 | 0.084969397 | 0.000980455 | 0.00820384 | -1.34E-05 | 0.032385119 | 1 | 0.589329992 | 1 | 0.580557814 | ENSG000001( | 10 | DHX35 |
| Pancreas | T1DM | ENSG0000024 | 0.00148345 | 0.000240012 | 0.000403911 | 0.008204548 | 0.001306587 | 0.001262876 | 0.000405231 | 0.008203715 | 0.000176863 | 7.84E-08 | 0.616352091 | 0.012840669 | 1 | 0.056895894 | ENSG000001( | 10 | DHX35 |
| Pancreas | T1DM | ENSG0000024 | -0.00108071 | 0.263218586 | 0.000965939 | 0.008204754 | -0.00106132 | 0.271864514 | 0.00096591 | 0.008203869 | -1.94E-05 | 0.004019711 | 1 | 0.79720158 | 1 | 0.79588826 | ENSG000001( | 8 | DNTTIP1 |
| Pancreas | T1DM | ENSG0000024 | 0.000468557 | 0.71388598 | 0.001277969 | 0.008204773 | 0.000491517 | 0.700519152 | 0.001277929 | 0.008203887 | -2.30E-05 | 0.008417052 | 1 | 0.975415042 | 1 | 0.968082788 | ENSG000001( | 8 | DNTTIP1 |
| Pancreas | T1DM | ENSG0000024 | 0.000294935 | 0.351234257 | 0.000316386 | 0.008204761 | 0.000289365 | 0.360378484 | 0.000316375 | 0.008203876 | 5.57E-06 | 0.002724456 | 1 | 0.854061442 | 1 | 0.850635855 | ENSG000001( | 8 | DNTTIP1 |
| Pancreas | T1DM | ENSG0000024 | 0.001145022 | 0.118050197 | 0.000732574 | 0.008204734 | 0.001137052 | 0.120615832 | 0.000732544 | 0.008203849 | 7.97E-06 | 0.032643502 | 1 | 0.67493056 | 1 | 0.669849249 | ENSG000001( | 8 | DNTTIP1 |
| Pancreas | T1DM | ENSG0000024 | -0.00067923 | 0.469926343 | 0.000939979 | 0.008204767 | -0.00069395 | 0.460337815 | 0.000939942 | 0.00820381 | 1.47E-05 | 0.003982042 | 1 | 0.908712989 | 1 | 0.901310005 | ENSG000001( | 8 | MAP1LC3A |
| Pancreas | T1DM | ENSG0000024 | -2.54E-05 | 0.976570716 | 0.000864626 | 0.008204776 | -1.16E-05 | 0.989323816 | 0.000864601 | 0.00820389 | -1.38E-05 | 0.022632414 | 1 | 0.997898334 | 1 | 0.99966149 | ENSG000001( | 6 | PIGU |
| Pancreas | T1DM | ENSG0000024 | -0.00077092 | 0.455182853 | 0.001032296 | 0.008204766 | -0.00080408 | 0.436012726 | 0.00103227 | 0.00820388 | 3.32E-05 | 1.98E-05 | 1 | 0.899832468 | 1 | 0.894313643 | ENSG000001( | 8 | RBFA |
| Pancreas | T1DM | ENSG0000024 | -0.00011318 | 0.893446593 | 0.000845014 | 0.008204775 | -9.18E-05 | 0.913475053 | 0.000844986 | 0.00820389 | -2.14E-05 | 6.91E-05 | 1 | 0.99200447 | 1 | 0.996818865 | ENSG000001( | 14 | CEP192 |
| Pancreas | T1DM | ENSG0000024 | 0.000137355 | 0.873912995 | 0.000865555 | 0.008204775 | 0.000154373 | 0.858443052 | 0.00086553 | 0.008203889 | -1.70E-05 | 0.005275693 | 1 | 0.99200447 | 1 | 0.990577333 | ENSG000001( | 6 | POLI |
| Pancreas | T1DM | ENSG0000025 | -1.45E-05 | 0.988763035 | 0.001026609 | 0.008204776 | -2.91E-05 | 0.977363819 | 0.001026571 | 0.00820389 | 1.47E-05 | 0.012165248 | 1 | 0.997898334 | 1 | 0.99966149 | ENSG000001( | 6 | POLI |
| Pancreas | T1DM | ENSG0000025 | -0.00053228 | 0.197693578 | 0.000413209 | 0.008204748 | -0.00053834 | 0.192622862 | 0.000413195 | 0.008203861 | 6.06E-06 | 0.021052791 | 1 | 0.761135096 | 1 | 0.761003345 | ENSG000001( | 6 | POLI |
| Pancreas | T1DM | ENSG0000025 | 0.000856703 | 0.026075995 | 0.00038502 | 0.008204692 | 0.000800378 | 0.037826469 | 0.000385402 | 0.008203817 | 5.63E-05 | 0.001372155 | 1 | 0.393484808 | 1 | 0.449714688 | ENSG000001( | 6 | ACP5 |
| Pancreas | T1DM | ENSG0000025 | 0.001058632 | 0.405036864 | 0.001271385 | 0.008204764 | 0.001079724 | 0.395725782 | 0.00127134 | 0.008203878 | -2.11E-05 | 0.006501137 | 1 | 0.88063625 | 1 | 0.871863634 | ENSG000001( | 6 | ACP5 |
| Pancreas | T1DM | ENSG0000025 | -0.00056111 | 0.435906661 | 0.000720179 | 0.008204765 | -0.00057339 | 0.425910794 | 0.000720153 | 0.008203879 | 1.23E-05 | 0.00497619 | 1 | 0.89813429 | 1 | 0.889527368 | ENSG000001( | 5 | PARP4 |
| Pancreas | T1DM | ENSG0000025 | -0.00016986 | 0.877046613 | 0.001097905 | 0.008204775 | -0.0001864 | 0.865177656 | 0.001097862 | 0.008203889 | 1.65E-05 | 0.005802518 | 1 | 0.99200447 | 1 | 0.990577333 | ENSG000001( | 12 | RGCC |
| Pancreas | T1DM | ENSG0000025 | -0.0007509 | 0.49658219 | 0.001104458 | 0.008204768 | -0.0007316 | 0.507699366 | 0.001104426 | 0.008203883 | -1.93E-05 | 0.0145484 | 1 | 0.914783379 | 1 | 0.914417995 | ENSG000001( | 12 | DGKH |
| Pancreas | T1DM | ENSG0000025 | -0.00108584 | 0.330363558 | 0.00111553 | 0.00820476 | -0.00106394 | 0.340193967 | 0.001115498 | 0.008203875 | -2.19E-05 | 0.005450065 | 1 | 0.83462449 | 1 | 0.840825897 | ENSG000001( | 12 | DGKH |
| Pancreas | T1DM | ENSG0000025 | -0.00164187 | 0.188823597 | 0.001249452 | 0.008204746 | -0.00165846 | 0.184376238 | 0.001249404 | 0.00820386 | 1.66E-05 | 0.017888367 | 1 | 0.751239289 | 1 | 0.747990804 | ENSG000001( | 6 | OLFM4 |
| Pancreas | T1DM | ENSG0000025 | 0.000458175 | 0.662372888 | 0.00104932 | 0.008204772 | 0.000442137 | 0.67348364 | 0.00104928 | 0.008203887 | 1.60E-05 | 0.005686269 | 1 | 0.962467202 | 1 | 0.961035047 | ENSG000001( | 6 | OLFM4 |
| Pancreas | T1DM | ENSG0000025 | -0.00086276 | 0.450438053 | 0.001143198 | 0.008204766 | -0.00084824 | 0.458076952 | 0.00114316 | 0.008203881 | -1.45E-05 | 0.046014923 | 1 | 0.899061289 | 1 | 0.900031838 | ENSG000001( | 6 | OLFM4 |
| Pancreas | T1DM | ENSG0000025 | 0.000763963 | 0.306140065 | 0.000746527 | 0.008204758 | 0.00077668 | 0.298144052 | 0.000746504 | 0.008203872 | -1.27E-05 | 0.012668172 | 1 | 0.822290036 | 1 | 0.810652485 | ENSG000001( | 8 | MGRN1 |
| Pancreas | T1DM | ENSG0000025 | 0.000245061 | 0.810247562 | 0.001020631 | 0.008204775 | 0.000258455 | 0.80008376 | 0.001020599 | 0.008203889 | -1.34E-05 | 0.048938385 | 1 | 0.98201188 | 1 | 0.978923745 | ENSG000001( | 7 | DHODH |
| Pancreas | T1DM | ENSG0000025 | 0.00021151 | 0.850898636 | 0.001125219 | 0.008204775 | 0.000195722 | 0.8619698 | 0.001125181 | 0.008203889 | 1.58E-05 | 0.024674025 | 1 | 0.989913489 | 1 | 0.990577333 | ENSG000001( | 9 | ACD |
| Pancreas | T1DM | ENSG0000025 | 0.0006701 | 0.071875172 | 0.000372296 | 0.008204721 | 0.000659704 | 0.07638796 | 0.000372284 | 0.008203837 | 1.04E-05 | 2.80E-05 | 1 | 0.560516232 | 1 | 0.562075305 | ENSG000001( | 4 | MMP15 |
| Pancreas | T1DM | ENSG0000025 | 0.002688338 | 0.001797366 | 0.000861139 | 0.008204611 | 0.00266918 | 0.001937575 | 0.000861122 | 0.008203728 | 1.92E-05 | 0.006411044 | 1 | 0.075680736 | 1 | 0.078979241 | ENSG000001( | 4 | MMP15 |
| Pancreas | T1DM | ENSG0000025 | -0.00036987 | 0.235190515 | 0.000311572 | 0.008204752 | -0.00037545 | 0.228185223 | 0.000311562 | 0.008203866 | 5.58E-06 | 0.00743254 | 1 | 0.785512902 | 1 | 0.780702981 | ENSG000001( | 11 | NPRL3 |
| Pancreas | T1DM | ENSG0000025 | 7.65E-05 | 0.932663148 | 0.000905817 | 0.008204775 | 6.21E-05 | 0.945376623 | 0.000905786 | 0.00820389 | 1.45E-05 | 0.009245703 | 1 | 0.997898334 | 1 | 0.999388266 | ENSG000001( | 11 | NPRL3 |
| Pancreas | T1DM | ENSG0000025 | 0.000546917 | 0.662338242 | 0.001252421 | 0.008204772 | 0.000597204 | 0.63347999 | 0.001252435 | 0.008203886 | -5.03E-05 | 0.000422047 | 1 | 0.962467202 | 1 | 0.955974304 | ENSG000001( | 11 | NPRL3 |
| Pancreas | T1DM | ENSG0000025 | -0.00114045 | 0.209748111 | 0.000909263 | 0.008204749 | -0.00113059 | 0.213696023 | 0.000909227 | 0.008203864 | -9.86E-06 | 0.045631431 | 1 | 0.771198903 | 1 | 0.769778216 | ENSG000001( | 12 | TAF1C |
| Pancreas | T1DM | ENSG0000025 | 0.000259217 | 0.585559814 | 0.000475381 | 0.008204771 | 0.000246415 | 0.604227101 | 0.0004754 | 0.008203885 | 1.28E-05 | 0.049187396 | 1 | 0.937588291 | 1 | 0.939938439 | ENSG000001( | 12 | TAF1C |
| Pancreas | T1DM | ENSG0000025 | 0.000920482 | 0.260904897 | 0.000818748 | 0.008204754 | 0.000934136 | 0.253882864 | 0.00081872 | 0.008203868 | -1.37E-05 | 0.009064687 | 1 | 0.79720158 | 1 | 0.788955599 | ENSG000001( | 12 | TAF1C |
| Pancreas | T1DM | ENSG0000025 | 0.000887597 | 0.12513942 | 0.000578783 | 0.008204736 | 0.000879716 | 0.128513203 | 0.000578763 | 0.008203851 | 7.88E-06 | 0.023504386 | 1 | 0.680843282 | 1 | 0.676232745 | ENSG000001( | 7 | NAGPA |
| Pancreas | T1DM | ENSG0000025 | -9.89E-05 | 0.83254263 | 0.000467733 | 0.008204775 | -9.07E-05 | 0.846244202 | 0.000467715 | 0.008203889 | -8.20E-06 | 0.002501772 | 1 | 0.984098328 | 1 | 0.986098414 | ENSG000001( | 7 | NAGPA |
| Pancreas | T1DM | ENSG0000025 | 0.00471818 | 1.50E-25 | 0.000451575 | 0.008202938 | 0.004916826 | 6.19E-27 | 0.000457522 | 0.008201946 | -0.00019865 | 0.007027382 | 3.86E-22 | 3.51E-23 | 1.59E-23 | 1.84E-24 | ENSG000001( | 7 | CRISPLD2 |
| Pancreas | T1DM | ENSG0000025 | -0.00015256 | 0.855132148 | 0.000835606 | 0.008204775 | -0.00016165 | 0.846600938 | 0.000835573 | 0.008203889 | 9.09E-06 | 0.046734146 | 1 | 0.990317162 | 1 | 0.986098414 | ENSG000001( | 7 | CRISPLD2 |
| Pancreas | T1DM | ENSG0000025 | -0.00015017 | 0.832045851 | 0.000708072 | 0.008204775 | -0.00016085 | 0.820292245 | 0.000708043 | 0.008203889 | 1.07E-05 | 0.003412607 | 1 | 0.984098328 | 1 | 0.98205617 | ENSG000001( | 9 | ZNF500 |
| Pancreas | T1DM | ENSG0000025 | -0.00143472 | 0.222724202 | 0.00117666 | 0.00820475 | -0.00140805 | 0.23142748 | 0.001176617 | 0.008203866 | -2.67E-05 | 0.000139973 | 1 | 0.783501029 | 1 | 0.780702981 | ENSG000001( | 13 | NOMO3 |
| Pancreas | T1DM | ENSG0000025 | -0.00211421 | 0.087504839 | 0.001237312 | 0.008204726 | -0.00213337 | 0.084661727 | 0.00123727 | 0.00820384 | 1.92E-05 | 0.015945015 | 1 | 0.595273618 | 1 | 0.580557814 | ENSG000001( | 13 | MTHFSD |
| Pancreas | T1DM | ENSG0000025 | -0.00140325 | 0.067100106 | 0.00076638 | 0.008204719 | -0.00141219 | 0.065367531 | 0.000766351 | 0.008203833 | 8.94E-06 | 0.036058565 | 1 | 0.548630246 | 1 | 0.536001791 | ENSG000001( | 9 | RHBDL1 |
| Pancreas | T1DM | ENSG0000025 | -0.00034483 | 0.582688343 | 0.000627586 | 0.00820477 | -0.00035539 | 0.57118371 | 0.000627563 | 0.008203885 | 1.06E-05 | 0.004201821 | 1 | 0.937588291 | 1 | 0.936315417 | ENSG000001( | 9 | RHBDL1 |
| Pancreas | T1DM | ENSG0000025 | -0.00016831 | 0.671320216 | 0.000396641 | 0.008204773 | -0.0001733 | 0.662161812 | 0.000396625 | 0.008203887 | 4.99E-06 | 0.01686453 | 1 | 0.963860935 | 1 | 0.961035047 | ENSG000001( | 9 | RHBDL1 |
| Pancreas | T1DM | ENSG0000025 | -0.00137684 | 0.135567666 | 0.000922504 | 0.008204738 | -0.00139499 | 0.130478003 | 0.000922475 | 0.008203851 | 1.81E-05 | 0.003508336 | 1 | 0.694369535 | 1 | 0.676232745 | ENSG000001( | 14 | UBE2I |
| Pancreas | T1DM | ENSG0000025 | 0.000857829 | 0.033891261 | 0.000404375 | 0.0082047 | 0.000851696 | 0.035180896 | 0.000404361 | 0.008203815 | 6.13E-06 | 0.014279517 | 1 | 0.436429495 | 1 | 0.43733359 | ENSG000001( | 8 | CLUAP1 |
| Pancreas | T1DM | ENSG0000025 | -0.00013209 | 0.891877232 | 0.000971765 | 0.008204775 | -0.00014566 | 0.880849148 | 0.000971733 | 0.00820389 | 1.36E-05 | 0.030787093 | 1 | 0.99200447 | 1 | 0.993128823 | ENSG000001( | 8 | CLUAP1 |
| Pancreas | T1DM | ENSG0000025 | 7.19E-06 | 0.985126293 | 0.000385784 | 0.008204776 | 1.45E-06 | 0.997008559 | 0.000385773 | 0.00820389 | 5.75E-06 | 0.035348063 | 1 | 0.997898334 | 1 | 0.999770584 | ENSG000001( | 11 | EARS2 |
| Pancreas | T1DM | ENSG0000026 | 0.00336409 | 1.27E-06 | 0.000694403 | 0.00820438 | 0.003374929 | 1.17E-06 | 0.00069438 | 0.008203492 | -1.08E-05 | 0.016274433 | 0.003259257 | 8.63E-05 | 0.00301007 | 9.12E-05 | ENSG000001( | 11 | EARS2 |
| Pancreas | T1DM | ENSG0000026 | 0.000681607 | 0.588847034 | 0.001261049 | 0.008204771 | 0.000656893 | 0.602415463 | 0.001261 | 0.008203885 | 2.47E-05 | 0.000335155 | 1 | 0.937588291 | 1 | 0.939938439 | ENSG000001( | 11 | EARS2 |
| Pancreas | T1DM | ENSG0000026 | 0.000916801 | 0.246840422 | 0.00079167 | 0.008204753 | 0.000942388 | 0.233886249 | 0.00079165 | 0.008203866 | -2.56E-05 | 2.14E-05 | 1 | 0.788479116 | 1 | 0.780702981 | ENSG000001( | 11 | EARS2 |
| Pancreas | T1DM | ENSG0000026 | -0.00093986 | 0.020750815 | 0.000406426 | 0.008204686 | -0.00090181 | 0.026515993 | 0.00040648 | 0.008203807 | -3.81E-05 | 1.14E-06 | 1 | 0.35290128 | 1 | 0.382545334 | ENSG000001( | 11 | EARS2 |
| Pancreas | T1DM | ENSG0000026 | -0.00013804 | 0.915599996 | 0.00130256 | 0.008204775 | -0.00016506 | 0.899159874 | 0.001302534 | 0.00820389 | 2.70E-05 | 0.012230987 | 1 | 0.995870671 | 1 | 0.994038165 | ENSG000001( | 7 | HMOX2 |
| Pancreas | T1DM | ENSG0000026 | 0.000582968 | 0.370105514 | 0.000650434 | 0.008204762 | 0.000595102 | 0.360213432 | 0.000650413 | 0.008203876 | -1.21E-05 | 0.00576012 | 1 | 0.856244109 | 1 | 0.850635855 | ENSG000001( | 7 | HMOX2 |
| Pancreas | T1DM | ENSG0000026 | -0.00034093 | 0.675252624 | 0.000813774 | 0.008204773 | -0.00033016 | 0.684939951 | 0.000813748 | 0.008203887 | -1.08E-05 | 0.044769729 | 1 | 0.963860935 | 1 | 0.966338775 | ENSG000001( | 7 | HMOX2 |
| Pancreas | T1DM | ENSG0000026 | -0.00152321 | 0.17436548 | 0.001121401 | 0.008204744 | -0.00155091 | 0.166649978 | 0.001121367 | 0.008203858 | 2.77E-05 | 0.000343969 | 1 | 0.729268002 | 1 | 0.726605951 | ENSG000001( | 5 | RRN3P2 |
| Pancreas | T1DM | ENSG0000026 | 0.000552632 | 0.36317565 | 0.000607734 | 0.008204762 | 0.000537563 | 0.376409473 | 0.000607738 | 0.008203877 | 1.51E-05 | 0.02354872 | 1 | 0.854831234 | 1 | 0.857692571 | ENSG000001( | 5 | RRN3P2 |
| Pancreas | T1DM | ENSG0000026 | 0.001108064 | 0.15981502 | 0.000788267 | 0.008204742 | 0.001088511 | 0.167297849 | 0.000788239 | 0.008203858 | 1.96E-05 | 3.37E-05 | 1 | 0.724477654 | 1 | 0.726605951 | ENSG000001( | 5 | RRN3P2 |
| Pancreas | T1DM | ENSG0000026 | 0.000529321 | 0.403865143 | 0.000634115 | 0.008204764 | 0.000539576 | 0.39480465 | 0.000634096 | 0.008203878 | -1.03E-05 | 0.01685294 | 1 | 0.879665554 | 1 | 0.871863634 | ENSG000001( | 10 | RBL2 |
| Pancreas | T1DM | ENSG0000026 | -0.00010164 | 0.850808891 | 0.000540407 | 0.008204775 | -9.24E-05 | 0.864183854 | 0.000540386 | 0.008203889 | -9.21E-06 | 0.002280723 | 1 | 0.989913489 | 1 | 0.990577333 | ENSG000001( | 10 | RBL2 |
| Pancreas | T1DM | ENSG0000026 | 0.00126068 | 0.148544256 | 0.000872627 | 0.00820474 | 0.001279922 | 0.142431914 | 0.000872596 | 0.008203854 | -1.92E-05 | 0.000344558 | 1 | 0.712186059 | 1 | 0.696695532 | ENSG000001( | 9 | KNOP1 |
| Pancreas | T1DM | ENSG0000026 | -2.38E-05 | 0.961353034 | 0.000491947 | 0.008204776 | -1.64E-05 | 0.973367387 | 0.000491932 | 0.00820389 | -7.41E-06 | 0.022655139 | 1 | 0.997898334 | 1 | 0.99966149 | ENSG000001( | 9 | KNOP1 |
| Pancreas | T1DM | ENSG0000026 | -0.0001182 | 0.868178372 | 0.000712171 | 0.008204775 | -0.00013198 | 0.852972529 | 0.000712145 | 0.008203889 | 1.38E-05 | 0.000910303 | 1 | 0.99183698 | 1 | 0.989215428 | ENSG000001( | 9 | KNOP1 |
| Pancreas | T1DM | ENSG0000026 | -0.00138849 | 0.200339641 | 0.001084262 | 0.008204748 | -0.00140403 | 0.195333024 | 0.001084223 | 0.008203862 | 1.55E-05 | 0.017613739 | 1 | 0.7650738 | 1 | 0.762604991 | ENSG000001( | 5 | TRIP4 |
| Pancreas | T1DM | ENSG0000026 | 0.000515611 | 0.618276427 | 0.00103475 | 0.008204771 | 0.000502061 | 0.627522382 | 0.001034713 | 0.008203886 | 1.36E-05 | 0.028408873 | 1 | 0.95073884 | 1 | 0.954101526 | ENSG000001( | 7 | IGDCC4 |
| Pancreas | T1DM | ENSG0000027 | 0.000122945 | 0.860240302 | 0.000698281 | 0.008204775 | 0.000230429 | 0.741808038 | 0.000699412 | 0.008203888 | -0.00010748 | 0.007842431 | 1 | 0.990511029 | 1 | 0.968082788 | ENSG000001( | 7 | IGDCC4 |
| Pancreas | T1DM | ENSG0000027 | -0.00010189 | 0.752661778 | 0.000290372 | 0.008204773 | -0.00010635 | 0.714156914 | 0.000290361 | 0.008203888 | 4.46E-06 | 0.004703015 | 1 | 0.975415042 | 1 | 0.968082788 | ENSG000001( | 11 | CTSH |
| Pancreas | T1DM | ENSG0000027 | 0.000177476 | 0.879664065 | 0.001172267 | 0.008204775 | 0.000199315 | 0.864986506 | 0.001172236 | 0.008203889 | -2.18E-05 | 0.011896548 | 1 | 0.99200447 | 1 | 0.990577333 | ENSG000001( | 11 | CTSH |
| Pancreas | T1DM | ENSG0000027 | -0.00040991 | 0.66179306 | 0.000937069 | 0.008204772 | -0.00042394 | 0.650956909 | 0.000937031 | 0.008203887 | 1.40E-05 | 0.004605483 | 1 | 0.962467202 | 1 | 0.961035047 | ENSG000001( | 11 | CTSH |
| Pancreas | T1DM | ENSG0000027 | 0.004981549 | 1.08E-10 | 0.000771607 | 0.008204074 | 0.003872823 | 1.56E-06 | 0.000806248 | 0.008203502 | 0.001108726 | 2.13E-06 | 2.76E-07 | 1.26E-08 | 0.004004901 | 0.000117791 | ENSG000001( | 11 | CTSH |
| Pancreas | T1DM | ENSG0000027 | 0.000937929 | 0.221310826 | 0.000766876 | 0.00820475 | 0.000926409 | 0.227020271 | 0.00076685 | 0.008203865 | 1.15E-05 | 0.015102417 | 1 | 0.780835021 | 1 | 0.780702981 | ENSG000001( | 11 | CTSH |
| Pancreas | T1DM | ENSG0000027 | 0.001379181 | 0.174067392 | 0.001014662 | 0.008204744 | 0.001367173 | 0.177830207 | 0.001014623 | 0.008203865 | 1.20E-05 | 0.038162405 | 1 | 0.729268002 | 1 | 0.731367165 | ENSG000001( | 7 | RPAP1 |
| Pancreas | T1DM | ENSG0000027 | 0.001054874 | 1.58E-06 | 0.000219696 | 0.008204387 | 0.001179828 | 9.10E-08 | 0.000220777 | 0.008203409 | -0.00012495 | 1.24E-08 | 0.004045142 | 0.000103722 | 0.000233599 | 8.34E-06 | ENSG000001( | 7 | RPAP1 |
| Pancreas | T1DM | ENSG0000027 | 0.001031465 | 0.215199232 | 0.000832232 | 0.00820475 | 0.00102124 | 0.219765285 | 0.000832202 | 0.008203865 | 1.02E-05 | 0.044234835 | 1 | 0.773993876 | 1 | 0.776261656 | ENSG000001( | 11 | OCA2 |
| Pancreas | T1DM | ENSG0000027 | 0.000326959 | 0.581277709 | 0.000592834 | 0.00820477 | 0.000333959 | 0.573196934 | 0.000592809 | 0.008203885 | -7.00E-06 | 0.021631784 | 1 | 0.937588291 | 1 | 0.936315417 | ENSG000001( | 6 | RHOV |
| Pancreas | T1DM | ENSG0000027 | 2.51E-05 | 0.965067058 | 0.000573188 | 0.008204776 | 3.24E-05 | 0.954871115 | 0.000573167 | 0.00820389 | -7.33E-06 | 0.028397922 | 1 | 0.997898334 | 1 | 0.999388266 | ENSG000001( | 8 | ZDHHC2 |
| Pancreas | T1DM | ENSG0000027 | -0.000492 | 0.630120898 | 0.001021693 | 0.008204772 | -0.00047704 | 0.640556048 | 0.001021652 | 0.008203886 | -1.50E-05 | 0.006368559 | 1 | 0.95754589 | 1 | 0.957315064 | ENSG000001( | 8 | ZDHHC2 |
| Pancreas | T1DM | ENSG0000027 | -0.0017083 | 6.75E-09 | 0.000294678 | 0.00820421 | -0.00164775 | 2.43E-08 | 0.000295384 | 0.008203366 | -6.06E-05 | 0.003328182 | 1.73E-05 | 6.19E-07 | 6.24E-05 | 2.31E-06 | ENSG000001( | 6 | ZFAND1 |
| Pancreas | T2DM admiss | ENSG0000000 | 0.001249152 | 0.33262339 | 0.001289322 | 0.073378338 | 0.001227799 | 0.340952034 | 0.001289317 | 0.07337387 | 2.14E-05 | 0.022647671 | 1 | 0.708860469 | 1 | 0.715170664 | ENSG000001( | 11 | CPQ |
| Pancreas | T2DM admiss | ENSG0000000 | 0.005294099 | 0.059717802 | 0.002811709 | 0.073377945 | 0.005187869 | 0.065042229 | 0.002811896 | 0.073373494 | 0.000106229 | 0.006637847 | 1 | 0.322853295 | 1 | 0.338114259 | ENSG000001( | 4 | IMPAD1 |
| Pancreas | T2DM admiss | ENSG0000000 | 0.004103606 | 0.127953577 | 0.002695796 | 0.07337813 | 0.004061112 | 0.131945914 | 0.002695771 | 0.073373665 | 4.25E-05 | 0.015248037 | 1 | 0.483297266 | 1 | 0.490357609 | ENSG000001( | 10 | RAB2A |
| Pancreas | T2DM admiss | ENSG0000000 | 0.003074695 | 0.361048536 | 0.00336632 | 0.073378354 | 0.003022825 | 0.369200562 | 0.003366277 | 0.073373885 | 5.19E-05 | 0.010021531 | 1 | 0.723010568 | 1 | 0.727627319 | ENSG000001( | 10 | RAB2A |
| Pancreas | T2DM admiss | ENSG0000000 | -0.00129455 | 0.733487233 | 0.003802015 | 0.073378462 | -0.00120564 | 0.751162698 | 0.003801994 | 0.073373991 | -8.89E-05 | 0.000982518 | 1 | 0.919728783 | 1 | 0.925863522 | ENSG000001( | 5 | ARMC1 |
| Pancreas | T2DM admiss | ENSG0000001 | 0.00352401 | 0.331658926 | 0.003630085 | 0.073378337 | 0.003599353 | 0.321425729 | 0.003630088 | 0.073373859 | -7.53E-05 | 0.008676559 | 1 | 0.707897253 | 1 | 0.706943978 | ENSG000001( | 5 | ARMC1 |
| Pancreas | T2DM admiss | ENSG0000001 | -0.00184032 | 0.438556732 | 0.002375726 | 0.073378389 | -0.00180212 | 0.448120769 | 0.002375731 | 0.07337392 | -3.82E-05 | 0.046322742 | 1 | 0.774562372 | 1 | 0.779994504 | ENSG000001( | 5 | ARMC1 |
| Pancreas | T2DM admiss | ENSG0000001 | 0.004244308 | 0.045549621 | 0.002122634 | 0.073377877 | 0.004282629 | 0.043631314 | 0.002122614 | 0.073373394 | -3.83E-05 | 0.005489301 | 1 | 0.285670854 | 1 | 0.28152064 | ENSG000001( | 5 | ARMC1 |
| Pancreas | T2DM admiss | ENSG0000002 | 0.000256711 | 0.926306962 | 0.002775488 | 0.073378478 | 0.000217506 | 0.937535644 | 0.002775458 | 0.073374006 | 3.92E-05 | 0.025028505 | 1 | 0.979054639 | 1 | 0.98183872 | ENSG000001( | 5 | ARMC1 |
| Pancreas | T2DM admiss | ENSG0000003 | 0.016024865 | 1.52E-09 | 0.002652009 | 0.073372982 | 0.015031688 | 1.71E-08 | 0.002665748 | 0.073369219 | 0.000993178 | 0.000246949 | 3.90E-06 | 4.87E-07 | 4.40E-05 | 5.50E-06 | ENSG000001( | 5 | ARMC1 |
| Pancreas | T2DM admiss | ENSG0000003 | -0.00187726 | 0.378347296 | 0.002130957 | 0.073378362 | -0.0019512 | 0.359853134 | 0.002130958 | 0.0733788 | 7.39E-05 | 9.87E-06 | 1 | 0.733271857 | 1 | 0.72195535 | ENSG000001( | 8 | SPAG1 |
| Pancreas | T2DM admiss | ENSG0000003 | 0.000782718 | 0.525164807 | 0.001231841 | 0.073378418 | 0.00080257 | 0.514707885 | 0.001231833 | 0.073373943 | -1.99E-05 | 0.02012302 | 1 | 0.827884115 | 1 | 0.81978735 | ENSG000001( | 6 | GSDMD |
| Pancreas | T2DM admiss | ENSG0000004 | -0.00032774 | 0.90086644 | 0.002631006 | 0.073378477 | -0.0003757 | 0.886450704 | 0.002631035 | 0.073374003 | 4.80E-05 | 0.044855867 | 1 | 0.972507979 | 1 | 0.972260921 | ENSG000001( | 11 | TSTA3 |
| Pancreas | T2DM admiss | ENSG0000000 | 0.005622553 | 0.040954942 | 0.002750772 | 0.07337785 | 0.005670307 | 0.03926847 | 0.00275075 | 0.073373367 | -4.78E-05 | 0.009730729 | 1 | 0.274649 | 1 | 0.27460182 | ENSG000001( | 6 | R3HCC1 |
| Pancreas | T2DM admiss | ENSG0000000 | 0.000709474 | 0.590544244 | 0.001318607 | 0.073378436 | 0.000739876 | 0.574724173 | 0.001318598 | 0.073373959 | -3.04E-05 | 0.000776641 | 1 | 0.858891717 | 1 | 0.84967857 | ENSG000001( | 5 | TNFRSF10A |

| | | | | | | | | | | | | | | | | | | | | |
|---|---|---|---|---|---|---|---|---|---|---|---|---|---|---|---|---|---|---|---|---|
| Pancreas | T2DM | admiss | ENSG0000006 | -0.01021832 | 0.003160101 | 0.003461804 | 0.073377167 | -0.01013471 | 0.00341634 | 0.003461811 | 0.073372716 | -8.36E-05 | 0.002660116 | 1 | 0.057554185 | 1 | 0.062665442 | ENSG000001 | 5 | TNFRSF10A |
| Pancreas | T2DM | admiss | ENSG0000006 | 0.000302326 | 0.889633692 | 0.002178649 | 0.073378476 | 0.000256225 | 0.90637876 | 0.002178644 | 0.073374004 | 4.61E-05 | 0.004812447 | 1 | 0.972507979 | 1 | 0.972260921 | ENSG000001 | 5 | TNFRSF10A |
| Pancreas | T2DM | admiss | ENSG0000006 | -0.00103804 | 0.670822654 | 0.002442337 | 0.073378452 | -0.00099379 | 0.684078142 | 0.002442323 | 0.073373982 | -4.42E-05 | 0.010092476 | 1 | 0.894197754 | 1 | 0.899034119 | ENSG000001 | 9 | ERICH1 |
| Pancreas | T2DM | admiss | ENSG0000006 | 0.002385396 | 0.136052744 | 0.001600234 | 0.073378145 | 0.007160392 | 0.000211199 | 0.001932495 | 0.073371939 | -0.004775 | 1.05E-05 | 1 | 0.496989255 | 0.542359703 | 0.00904011 | ENSG000001 | 9 | KLHDC4 |
| Pancreas | T2DM | admiss | ENSG0000006 | 0.005094226 | 0.105044432 | 0.003142881 | 0.073378084 | 0.005142243 | 0.101803815 | 0.003142846 | 0.073373603 | -4.80E-05 | 0.014483039 | 1 | 0.438624554 | 1 | 0.434949328 | ENSG000001 | 9 | KLHDC4 |
| Pancreas | T2DM | admiss | ENSG0000007 | 0.007528394 | 0.001577677 | 0.002382388 | 0.073376976 | 0.007574605 | 0.001475735 | 0.002382377 | 0.073372484 | -4.62E-05 | 0.007511067 | 1 | 0.03523021 | 1 | 0.033836506 | ENSG000001 | 8 | MAN2B1 |
| Pancreas | T2DM | admiss | ENSG0000007 | 0.01083172 | 0.000153067 | 0.00286094 | 0.073376321 | 0.010792528 | 0.000161688 | 0.0028609 | 0.073371864 | 3.92E-05 | 0.017000187 | 0.393075034 | 0.00680233 | 0.415215244 | 0.007414558 | ENSG000001 | 8 | MAN2B1 |
| Pancreas | T2DM | admiss | ENSG0000007 | -0.00543189 | 0.081733365 | 0.003120488 | 0.073378023 | -0.00538514 | 0.084390828 | 0.003120442 | 0.073373558 | -4.68E-05 | 0.007608974 | 1 | 0.383490365 | 1 | 0.395012569 | ENSG000001 | 8 | MAN2B1 |
| Pancreas | T2DM | admiss | ENSG0000007 | -0.00226732 | 0.379171014 | 0.002578183 | 0.073378363 | -0.00222083 | 0.389022041 | 0.002578186 | 0.073373895 | -4.65E-05 | 0.023537009 | 1 | 0.733271857 | 1 | 0.743863441 | ENSG000001 | 8 | MAN2B1 |
| Pancreas | T2DM | admiss | ENSG0000007 | -0.00145186 | 0.560180005 | 0.002492137 | 0.073378428 | -0.00149315 | 0.549073526 | 0.002492117 | 0.073373952 | 4.13E-05 | 0.01281163 | 1 | 0.847939375 | 1 | 0.84635103 | ENSG000001 | 8 | MAN2B1 |
| Pancreas | T2DM | admiss | ENSG0000007 | 0.000419846 | 0.881833452 | 0.002824476 | 0.073378476 | 0.000173814 | 0.950941506 | 0.00282512 | 0.073374006 | 0.000246031 | 0.000126464 | 1 | 0.972507979 | 1 | 0.98349488 | ENSG000001 | 8 | MAN2B1 |
| Pancreas | T2DM | admiss | ENSG0000008 | -5.36E-05 | 0.981532143 | 0.002315699 | 0.073378479 | -1.81E-05 | 0.993759643 | 0.00231568 | 0.073374007 | -3.55E-05 | 0.021755768 | 1 | 0.994701872 | 1 | 0.999041616 | ENSG000001 | 4 | ECH1 |
| Pancreas | T2DM | admiss | ENSG0000008 | -0.00021806 | 0.885630306 | 0.001516052 | 0.073378476 | -0.00055169 | 0.716618783 | 0.001519893 | 0.073373987 | 0.000333631 | 0.002132901 | 1 | 0.972507979 | 1 | 0.913596366 | ENSG000001 | 4 | ECH1 |
| Pancreas | T2DM | admiss | ENSG0000009 | -0.00043332 | 0.765312446 | 0.001451584 | 0.073378466 | -0.0004925 | 0.734407568 | 0.001451655 | 0.073373989 | 5.92E-05 | 0.001191238 | 1 | 0.927632086 | 1 | 0.922046709 | ENSG000001 | 4 | ECH1 |
| Pancreas | T2DM | admiss | ENSG0000009 | -0.00269989 | 0.284029375 | 0.00252017 | 0.073378306 | -0.00264731 | 0.293508085 | 0.002520145 | 0.07337384 | -5.26E-05 | 0.001177961 | 1 | 0.679218679 | 1 | 0.689809485 | ENSG000001 | 10 | SARS2 |
| Pancreas | T2DM | admiss | ENSG000001 | 0.002654587 | 0.379515913 | 0.003020732 | 0.073378363 | 0.002710927 | 0.369480539 | 0.003020707 | 0.073373885 | -5.63E-05 | 0.005400968 | 1 | 0.733271857 | 1 | 0.727627319 | ENSG000001 | 10 | SARS2 |
| Pancreas | T2DM | admiss | ENSG000001 | 0.001625625 | 0.203373976 | 0.001278008 | 0.073378236 | 0.001608935 | 0.208045941 | 0.001277989 | 0.073373768 | 1.67E-05 | 0.019225593 | 1 | 0.584188335 | 1 | 0.592308178 | ENSG000001 | 10 | SARS2 |
| Pancreas | T2DM | admiss | ENSG000001 | 0.002646962 | 0.428921254 | 0.003346181 | 0.073378385 | 0.002610656 | 0.435272362 | 0.003346128 | 0.073373915 | 3.63E-05 | 0.045207829 | 1 | 0.767595622 | 1 | 0.771947117 | ENSG000001 | 10 | SNRNP70 |
| Pancreas | T2DM | admiss | ENSG000001 | 0.002457134 | 0.401820595 | 0.002930829 | 0.073378373 | 0.002407513 | 0.411387311 | 0.002930789 | 0.073373905 | 4.96E-05 | 0.003603246 | 1 | 0.754055489 | 1 | 0.758117931 | ENSG000001 | 6 | PPP1R37 |
| Pancreas | T2DM | admiss | ENSG000001 | 0.001271809 | 0.373089351 | 0.001427872 | 0.07337836 | 0.001237148 | 0.38627012 | 0.001427914 | 0.073373894 | 3.47E-05 | 0.026243464 | 1 | 0.730660228 | 1 | 0.740807819 | ENSG000001 | 6 | PPP1R37 |
| Pancreas | T2DM | admiss | ENSG000001 | -0.0019006 | 0.598727088 | 0.003611709 | 0.073378437 | -0.00195367 | 0.588554933 | 0.003611682 | 0.073373962 | 5.31E-05 | 0.029565634 | 1 | 0.862328189 | 1 | 0.856807861 | ENSG000001 | 6 | PPP1R37 |
| Pancreas | T2DM | admiss | ENSG000001 | -0.00296648 | 0.272475719 | 0.002703233 | 0.073378298 | -0.00162667 | 0.55855496 | 0.002780687 | 0.073373955 | -0.0013398 | 0.039905731 | 1 | 0.670227632 | 1 | 0.847111521 | ENSG000001 | 6 | PPP1R37 |
| Pancreas | T2DM | admiss | ENSG000001 | 0.005563028 | 0.014795657 | 0.002282407 | 0.073377585 | 0.005624561 | 0.013727733 | 0.0022824 | 0.073373092 | -6.15E-05 | 0.000271043 | 1 | 0.163069734 | 1 | 0.152590546 | ENSG000001 | 6 | PPP1R37 |
| Pancreas | T2DM | admiss | ENSG000001 | 0.001675917 | 0.614394862 | 0.003326484 | 0.073378441 | 0.001579471 | 0.634939519 | 0.003326699 | 0.073373973 | 9.64E-05 | 0.035578484 | 1 | 0.871212593 | 1 | 0.880942324 | ENSG000001 | 8 | FCGRT |
| Pancreas | T2DM | admiss | ENSG000001 | -0.00045818 | 0.836566141 | 0.002221078 | 0.073378473 | -0.00036839 | 0.86828296 | 0.002221364 | 0.073374002 | -8.98E-05 | 0.023476416 | 1 | 0.958120026 | 1 | 0.967511091 | ENSG000001 | 4 | AMH |
| Pancreas | T2DM | admiss | ENSG000001 | 0.00026145 | 0.918981898 | 0.002570385 | 0.073378478 | 0.000313636 | 0.9028856 | 0.002570426 | 0.073374004 | -5.22E-05 | 0.034596773 | 1 | 0.977203112 | 1 | 0.972260921 | ENSG000001 | 2 | DKKL1 |
| Pancreas | T2DM | admiss | ENSG000001 | 0.000549574 | 0.876974473 | 0.00355009 | 0.073378476 | 0.000602454 | 0.865244772 | 0.003550077 | 0.073374002 | -5.29E-05 | 0.042657618 | 1 | 0.970211524 | 1 | 0.965225271 | ENSG000001 | 2 | DKKL1 |
| Pancreas | T2DM | admiss | ENSG000001 | 0.000178438 | 0.93073588 | 0.002052917 | 0.073378478 | 0.000150069 | 0.941725829 | 0.002052895 | 0.073374006 | 2.84E-05 | 0.027742581 | 1 | 0.979119408 | 1 | 0.981898834 | ENSG000001 | 2 | DKKL1 |
| Pancreas | T2DM | admiss | ENSG000001 | 0.000790186 | 0.49830216 | 0.001166904 | 0.07337841 | 0.000810033 | 0.487569755 | 0.001166892 | 0.073373934 | -1.98E-05 | 0.007352523 | 1 | 0.808874808 | 1 | 0.804739043 | ENSG000001 | 14 | TLE6 |
| Pancreas | T2DM | admiss | ENSG000001 | -0.00198787 | 0.523249639 | 0.003114094 | 0.073378418 | -0.00194596 | 0.532040938 | 0.00311407 | 0.073373948 | -4.19E-05 | 0.045460393 | 1 | 0.825853281 | 1 | 0.834115463 | ENSG000001 | 14 | TLE6 |
| Pancreas | T2DM | admiss | ENSG000001 | 0.000482456 | 0.865086029 | 0.002839586 | 0.073378475 | 0.000525346 | 0.85322109 | 0.002839553 | 0.073374001 | -4.29E-05 | 0.014060785 | 1 | 0.966374076 | 1 | 0.961211992 | ENSG000001 | 14 | TLE6 |
| Pancreas | T2DM | admiss | ENSG000001 | 0.006696785 | 0.009780967 | 0.002592148 | 0.073377474 | 0.006663016 | 0.010155464 | 0.002592109 | 0.073373012 | 3.38E-05 | 0.017589675 | 1 | 0.120179531 | 1 | 0.123598259 | ENSG000001 | 14 | TLE6 |
| Pancreas | T2DM | admiss | ENSG0000011 | -0.00141289 | 0.711173994 | 0.00381574 | 0.073378459 | -0.00118604 | 0.756022104 | 0.003817194 | 0.073373992 | -0.00022686 | 0.038221367 | 1 | 0.912258576 | 1 | 0.926713491 | ENSG000001 | 14 | TLE6 |
| Pancreas | T2DM | admiss | ENSG0000011 | 0.002276359 | 0.206112245 | 0.001800447 | 0.073378238 | 0.002253559 | 0.210685099 | 0.001800419 | 0.073373771 | 2.28E-05 | 0.02022533 | 1 | 0.590494782 | 1 | 0.597756217 | ENSG000001 | 6 | NOVA2 |
| Pancreas | T2DM | admiss | ENSG0000011 | -0.0004437 | 0.861637279 | 0.002545769 | 0.073378475 | -0.00040865 | 0.872469006 | 0.002545754 | 0.073374003 | -3.51E-05 | 0.049546427 | 1 | 0.965890282 | 1 | 0.967818751 | ENSG000001 | 6 | NOVA2 |
| Pancreas | T2DM | admiss | ENSG0000011 | -0.00466804 | 0.010464694 | 0.001823387 | 0.073377492 | -0.00246868 | 0.191015653 | 0.001887971 | 0.073373749 | -0.00219936 | 7.10E-06 | 1 | 0.125576324 | 1 | 0.570204108 | ENSG000001 | 4 | C19orf53 |
| Pancreas | T2DM | admiss | ENSG0000011 | 0.002762611 | 0.404573797 | 0.003314545 | 0.073378375 | 0.002684894 | 0.417924779 | 0.003314572 | 0.073373908 | 7.77E-05 | 0.007543522 | 1 | 0.755550584 | 1 | 0.763867723 | ENSG000001 | 4 | CCDC61 |
| Pancreas | T2DM | admiss | ENSG0000011 | 0.000551183 | 0.801463074 | 0.00219198 | 0.07337847 | 0.000614829 | 0.77911102 | 0.002192076 | 0.073373995 | -6.36E-05 | 0.017098792 | 1 | 0.939717987 | 1 | 0.936079501 | ENSG000001 | 4 | C19orf44 |
| Pancreas | T2DM | admiss | ENSG0000011 | 0.001685977 | 0.609861451 | 0.003304077 | 0.07337844 | 0.001731792 | 0.600179377 | 0.003304046 | 0.073373965 | -4.58E-05 | 0.032514065 | 1 | 0.868723317 | 1 | 0.863271188 | ENSG000001 | 4 | OLFM2 |
| Pancreas | T2DM | admiss | ENSG0000011 | 0.001122667 | 0.474520155 | 0.001569846 | 0.073378402 | 0.001146254 | 0.465282436 | 0.001569831 | 0.073373926 | -2.36E-05 | 0.019187727 | 1 | 0.795049308 | 1 | 0.789197685 | ENSG000001 | 4 | OLFM2 |
| Pancreas | T2DM | admiss | ENSG0000011 | 0.000455187 | 0.690352963 | 0.001142604 | 0.073378455 | 0.0004367 | 0.702311884 | 0.001142591 | 0.073373985 | 1.85E-05 | 0.008719955 | 1 | 0.903120941 | 1 | 0.908125337 | ENSG000001 | 4 | OLFM2 |
| Pancreas | T2DM | admiss | ENSG0000011 | 0.006374746 | 0.070967417 | 0.003530366 | 0.073377988 | 0.006415495 | 0.069177432 | 0.003530306 | 0.073373509 | -4.07E-05 | 0.026324015 | 1 | 0.35594595 | 1 | 0.35006171 | ENSG000001 | 4 | OLFM2 |
| Pancreas | T2DM | admiss | ENSG0000011 | 0.001469613 | 0.665697215 | 0.003401402 | 0.073378451 | 0.00142526 | 0.675197028 | 0.00340136 | 0.07337398 | 4.44E-05 | 0.030456347 | 1 | 0.891995883 | 1 | 0.894675443 | ENSG000001 | 4 | OLFM2 |
| Pancreas | T2DM | admiss | ENSG0000011 | 0.004785605 | 0.102643048 | 0.00293204 | 0.073378078 | 0.006331542 | 0.036971202 | 0.003035127 | 0.073373351 | -0.00154594 | 0.048816228 | 1 | 0.431403186 | 1 | 0.266843516 | ENSG000001 | 4 | OLFM2 |
| Pancreas | T2DM | admiss | ENSG0000011 | -0.00203442 | 0.025214933 | 0.000908994 | 0.073377725 | -0.00201968 | 0.026289944 | 0.000908984 | 0.073373263 | -1.47E-05 | 0.010648467 | 1 | 0.207485894 | 1 | 0.213574989 | ENSG000001 | 4 | OLFM2 |
| Pancreas | T2DM | admiss | ENSG0000011 | -0.00259908 | 0.12341253 | 0.001687048 | 0.073378122 | -0.00257761 | 0.126537642 | 0.001687026 | 0.073373655 | -2.15E-05 | 0.02937284 | 1 | 0.477599265 | 1 | 0.479275315 | ENSG000001 | 15 | ZNF419 |
| Pancreas | T2DM | admiss | ENSG0000011 | 0.002007397 | 0.168727722 | 0.001458541 | 0.073378194 | 0.001988294 | 0.172812485 | 0.001458523 | 0.073373727 | 1.91E-05 | 0.030133153 | 1 | 0.548084313 | 1 | 0.553785383 | ENSG000001 | 8 | AURKC |
| Pancreas | T2DM | admiss | ENSG0000011 | 0.000911192 | 0.814174982 | 0.003876691 | 0.073378471 | 0.000966942 | 0.803030733 | 0.003876656 | 0.073373997 | -5.58E-05 | 0.02826986 | 1 | 0.946063961 | 1 | 0.939682535 | ENSG000001 | 8 | AURKC |
| Pancreas | T2DM | admiss | ENSG0000011 | 0.001360666 | 0.450306323 | 0.001802434 | 0.073378393 | 0.001337127 | 0.458177523 | 0.001802418 | 0.073373924 | 2.35E-05 | 0.048261028 | 1 | 0.781199104 | 1 | 0.786497245 | ENSG000001 | 8 | AURKC |
| Pancreas | T2DM | admiss | ENSG0000011 | -0.00207141 | 0.44239024 | 0.002696572 | 0.07337839 | -0.00211071 | 0.433777355 | 0.002696552 | 0.073373914 | 3.93E-05 | 0.032211939 | 1 | 0.776526409 | 1 | 0.771947117 | ENSG000001 | 7 | PDCD5 |
| Pancreas | T2DM | admiss | ENSG0000012 | -0.00097029 | 0.307457301 | 0.000950728 | 0.073378322 | -0.00098597 | 0.299701693 | 0.000950722 | 0.073373845 | 1.57E-05 | 0.017409646 | 1 | 0.702883653 | 1 | 0.696408377 | ENSG000001 | 7 | PDCD5 |
| Pancreas | T2DM | admiss | ENSG0000012 | -0.00152907 | 0.674483413 | 0.003640614 | 0.073378453 | -0.00157384 | 0.665517502 | 0.003640549 | 0.073373978 | 4.48E-05 | 0.01438008 | 1 | 0.895591213 | 1 | 0.88982611 | ENSG000001 | 4 | RPS16 |
| Pancreas | T2DM | admiss | ENSG0000012 | -0.00145578 | 0.52241675 | 0.002275994 | 0.073378418 | -0.00149009 | 0.512657084 | 0.002275968 | 0.073373942 | 3.43E-05 | 0.01488395 | 1 | 0.825071473 | 1 | 0.81978735 | ENSG000001 | 4 | RPS16 |
| Pancreas | T2DM | admiss | ENSG0000012 | 0.00141158 | 0.630318105 | 0.002932973 | 0.073378444 | 0.001447602 | 0.621612512 | 0.002932941 | 0.07337397 | -3.60E-05 | 0.048125555 | 1 | 0.879226992 | 1 | 0.875160807 | ENSG000001 | 4 | POLR2I |
| Pancreas | T2DM | admiss | ENSG0000012 | 0.001418535 | 0.60406492 | 0.002735506 | 0.073378439 | 0.001383466 | 0.613032513 | 0.002735476 | 0.073373968 | 3.51E-05 | 0.040063624 | 1 | 0.865912508 | 1 | 0.870241843 | ENSG000001 | 4 | POLR2I |
| Pancreas | T2DM | admiss | ENSG0000012 | 0.001685034 | 0.503878605 | 0.002520997 | 0.073378412 | 0.001736183 | 0.491016553 | 0.002520981 | 0.073373935 | -5.11E-05 | 0.003480925 | 1 | 0.813942548 | 1 | 0.805191895 | ENSG000001 | 4 | POLR2I |
| Pancreas | T2DM | admiss | ENSG0000012 | 0.001208938 | 0.646279279 | 0.002634212 | 0.073378447 | 0.001246726 | 0.636007425 | 0.002634176 | 0.073373973 | -3.78E-05 | 0.013497982 | 1 | 0.886308052 | 1 | 0.880942324 | ENSG000001 | 4 | POLR2I |
| Pancreas | T2DM | admiss | ENSG0000012 | -6.43E-06 | 0.99803215 | 0.002606562 | 0.073378479 | -4.27E-05 | 0.986920843 | 0.002606525 | 0.073374007 | 3.63E-05 | 0.015161072 | 1 | 0.99984869 | 1 | 0.997626211 | ENSG000001 | 4 | POLR2I |
| Pancreas | T2DM | admiss | ENSG0000012 | 0.002580997 | 0.421857351 | 0.003213377 | 0.073378382 | 0.002631553 | 0.412817415 | 0.00321334 | 0.073373906 | -5.06E-05 | 0.010835947 | 1 | 0.764941163 | 1 | 0.758851196 | ENSG000001 | 10 | CLIP3 |
| Pancreas | T2DM | admiss | ENSG0000012 | 0.002370647 | 0.348067043 | 0.00252641 | 0.073378347 | 0.002339724 | 0.354384461 | 0.002526375 | 0.073373877 | 3.09E-05 | 0.033777204 | 1 | 0.714946894 | 1 | 0.720077577 | ENSG000001 | 10 | CLIP3 |
| Pancreas | T2DM | admiss | ENSG0000012 | -6.96E-05 | 0.980826051 | 0.002896963 | 0.073378479 | -0.00012608 | 0.965284416 | 0.002896944 | 0.073374006 | 5.65E-05 | 0.004939641 | 1 | 0.994701872 | 1 | 0.988657083 | ENSG000001 | 8 | PRKD2 |
| Pancreas | T2DM | admiss | ENSG0000012 | -7.03E-05 | 0.96190825 | 0.001470952 | 0.073378479 | -2.68E-05 | 0.985463436 | 0.001470994 | 0.073374007 | -4.34E-05 | 0.006842684 | 1 | 0.987677084 | 1 | 0.997569125 | ENSG000001 | 7 | APLP1 |
| Pancreas | T2DM | admiss | ENSG0000012 | 0.006667655 | 0.018383103 | 0.002827892 | 0.073377642 | 0.006719201 | 0.017500401 | 0.00282791 | 0.073373156 | -5.15E-05 | 0.033731521 | 1 | 0.180872825 | 1 | 0.174867821 | ENSG000001 | 5 | TGFB1 |
| Pancreas | T2DM | admiss | ENSG0000012 | -0.00112174 | 0.748890966 | 0.003504317 | 0.073378464 | -0.0011731 | 0.737804638 | 0.003504285 | 0.07337399 | 5.14E-05 | 0.024636358 | 1 | 0.923704131 | 1 | 0.922650112 | ENSG000001 | 4 | PLIN3 |
| Pancreas | T2DM | admiss | ENSG0000012 | -0.00062874 | 0.667875396 | 0.001465376 | 0.073378451 | -0.00066006 | 0.652392485 | 0.001465364 | 0.073373976 | 3.13E-05 | 0.001440568 | 1 | 0.893283342 | 1 | 0.887364354 | ENSG000001 | 9 | MRPL4 |
| Pancreas | T2DM | admiss | ENSG0000012 | 0.004155986 | 0.111286615 | 0.002609823 | 0.073378097 | 0.004097732 | 0.11640028 | 0.00260991 | 0.073373635 | 5.83E-05 | 0.047743183 | 1 | 0.454346625 | 1 | 0.469231995 | ENSG000001 | 9 | MRPL4 |
| Pancreas | T2DM | admiss | ENSG0000012 | 0.000151216 | 0.898530582 | 0.001185841 | 0.073378477 | 2.66E-05 | 0.98209306 | 0.001186318 | 0.073374006 | 0.000124589 | 0.000356797 | 1 | 0.972507979 | 1 | 0.996056468 | ENSG000001 | 10 | CD33 |
| Pancreas | T2DM | admiss | ENSG0000012 | -0.00024559 | 0.94787761 | 0.003756742 | 0.073378479 | -0.0001385 | 0.970590331 | 0.00375678 | 0.073374006 | -0.00010708 | 0.001559461 | 1 | 0.983097618 | 1 | 0.991832858 | ENSG000001 | 8 | GRWD1 |
| Pancreas | T2DM | admiss | ENSG0000012 | 0.000847568 | 0.769228882 | 0.00288894 | 0.073378466 | 0.000882115 | 0.76010237 | 0.002888898 | 0.073373993 | -3.45E-05 | 0.034712296 | 1 | 0.93002814 | 1 | 0.927749352 | ENSG000001 | 8 | GRWD1 |
| Pancreas | T2DM | admiss | ENSG0000012 | -0.00260558 | 0.184765493 | 0.001964661 | 0.073378214 | -0.00257532 | 0.189915961 | 0.001964646 | 0.073373748 | -3.03E-05 | 0.023026457 | 1 | 0.565800753 | 1 | 0.570204108 | ENSG000001 | 8 | GRWD1 |
| Pancreas | T2DM | admiss | ENSG0000012 | 0.002163275 | 0.257730581 | 0.001911404 | 0.073378286 | 0.002141951 | 0.262444052 | 0.001911373 | 0.073373817 | 2.13E-05 | 0.038603424 | 1 | 0.654004082 | 1 | 0.658289049 | ENSG000001 | 11 | CCDC114 |
| Pancreas | T2DM | admiss | ENSG0000012 | 0.000982777 | 0.619723367 | 0.001980437 | 0.073378442 | 0.001013087 | 0.608967739 | 0.001980434 | 0.073373967 | -3.03E-05 | 0.044001569 | 1 | 0.874422861 | 1 | 0.867518185 | ENSG000001 | 11 | CCDC114 |
| Pancreas | T2DM | admiss | ENSG0000012 | -0.00152903 | 0.541843319 | 0.002506485 | 0.073378423 | -0.00148216 | 0.554297795 | 0.002506469 | 0.073373954 | -4.69E-05 | 0.007089086 | 1 | 0.838486276 | 1 | 0.846590114 | ENSG000001 | 11 | CCDC114 |
| Pancreas | T2DM | admiss | ENSG0000013 | -0.0015888 | 0.679533418 | 0.003846033 | 0.073378453 | 0.002495191 | 0.567025339 | 0.004358883 | 0.073373957 | -0.00408399 | 0.046514008 | 1 | 0.899069186 | 1 | 0.849476828 | ENSG000001 | 11 | CCDC114 |
| Pancreas | T2DM | admiss | ENSG0000013 | 0.002011804 | 0.165638589 | 0.001451145 | 0.07337819 | 0.002032045 | 0.161419517 | 0.001451134 | 0.073373711 | -2.02E-05 | 0.039862354 | 1 | 0.54463495 | 1 | 0.53974651 | ENSG000001 | 3 | ZNF175 |
| Pancreas | T2DM | admiss | ENSG0000013 | 0.004037785 | 0.274895385 | 0.003698081 | 0.0733783 | 0.003981757 | 0.281604085 | 0.003698029 | 0.073373832 | 5.60E-05 | 0.008355734 | 1 | 0.670309634 | 1 | 0.677115441 | ENSG000001 | 7 | SIGLEC5 |
| Pancreas | T2DM | admiss | ENSG0000013 | 0.001717064 | 0.651245684 | 0.003798541 | 0.073378448 | 0.001843891 | 0.627395636 | 0.003798734 | 0.073373971 | -0.00012683 | 0.008873568 | 1 | 0.887189213 | 1 | 0.877200348 | ENSG000001 | 8 | CAPS |
| Pancreas | T2DM | admiss | ENSG0000013 | -0.00191394 | 0.521465446 | 0.002985458 | 0.073378417 | -0.00184743 | 0.536042575 | 0.002985456 | 0.073373949 | -6.65E-05 | 0.003890841 | 1 | 0.824583291 | 1 | 0.836849651 | ENSG000001 | 9 | PRPF31 |
| Pancreas | T2DM | admiss | ENSG0000013 | 0.004163402 | 0.074042003 | 0.002330666 | 0.073377999 | 0.004126058 | 0.076670057 | 0.00233065 | 0.073373535 | 3.73E-05 | 0.020269514 | 1 | 0.360189008 | 1 | 0.370788522 | ENSG000001 | 8 | RPL18A |
| Pancreas | T2DM | admiss | ENSG0000013 | -0.00370241 | 0.187106178 | 0.002806575 | 0.073378217 | -0.00375197 | 0.181269807 | 0.002806557 | 0.073373737 | 4.96E-05 | 0.011541346 | 1 | 0.567450741 | 1 | 0.561218928 | ENSG000001 | 8 | RPL18A |
| Pancreas | T2DM | admiss | ENSG0000013 | -0.00219951 | 0.175888105 | 0.001625021 | 0.073378203 | -0.00222077 | 0.171742034 | 0.001624997 | 0.073373725 | 2.13E-05 | 0.018853248 | 1 | 0.557630438 | 1 | 0.552673613 | ENSG000001 | 8 | RPL18A |
| Pancreas | T2DM | admiss | ENSG0000013 | -0.00079715 | 0.810509989 | 0.003324657 | 0.073378471 | -0.00084515 | 0.799333262 | 0.00332462 | 0.073373997 | 4.80E-05 | 0.020328766 | 1 | 0.943940886 | 1 | 0.939682535 | ENSG000001 | 8 | ELL |
| Pancreas | T2DM | admiss | ENSG0000013 | -0.00136449 | 0.374719439 | 0.001537162 | 0.073378361 | -0.00134359 | 0.382070897 | 0.001537139 | 0.073373892 | -2.09E-05 | 0.015314062 | 1 | 0.730660228 | 1 | 0.736052561 | ENSG000001 | 8 | ELL |
| Pancreas | T2DM | admiss | ENSG0000013 | 0.002088081 | 0.452352684 | 0.002778554 | 0.073378394 | 0.002043545 | 0.462051386 | 0.002778542 | 0.073373925 | 4.45E-05 | 0.026974461 | 1 | 0.781199104 | 1 | 0.787880451 | ENSG000001 | 10 | MAG |
| Pancreas | T2DM | admiss | ENSG0000013 | -0.00505114 | 0.186494034 | 0.003823658 | 0.073378216 | -0.00499786 | 0.191178246 | 0.003823617 | 0.073373749 | -5.33E-05 | 0.026430389 | 1 | 0.567450741 | 1 | 0.570204108 | ENSG000001 | 12 | LSR |
| Pancreas | T2DM | admiss | ENSG0000013 | -0.00027396 | 0.865654791 | 0.001619365 | 0.073378475 | -0.00024269 | 0.880866982 | 0.001619348 | 0.073374003 | -3.13E-05 | 0.002244091 | 1 | 0.966374076 | 1 | 0.971621772 | ENSG000001 | 12 | LSR |
| Pancreas | T2DM | admiss | ENSG0000013 | 0.003995998 | 0.245296079 | 0.003439334 | 0.073378276 | 0.001838448 | 0.601458778 | 0.00351988 | 0.073373965 | 0.00215755 | 0.003976933 | 1 | 0.636926523 | 1 | 0.86327687 | ENSG000001 | 12 | LSR |
| Pancreas | T2DM | admiss | ENSG0000013 | 0.000170424 | 0.959279996 | 0.003337906 | 0.073378479 | -0.00043237 | 0.897269061 | 0.003348794 | 0.073374004 | 0.000602794 | 0.026168489 | 1 | 0.986823205 | 1 | 0.972260921 | ENSG000001 | 10 | ZNF85 |
| Pancreas | T2DM | admiss | ENSG0000013 | 0.000395881 | 0.908129261 | 0.00343055 | 0.073378477 | 0.000348894 | 0.918992511 | 0.003430519 | 0.073374005 | 4.70E-05 | 0.036901788 | 1 | 0.972507979 | 1 | 0.978433155 | ENSG000001 | 3 | ETHE1 |
| Pancreas | T2DM | admiss | ENSG0000013 | 0.001206148 | 0.669932435 | 0.002829732 | 0.073378452 | 0.001244084 | 0.660189837 | 0.002829705 | 0.073373977 | -3.79E-05 | 0.038178951 | 1 | 0.89370727 | 1 | 0.888091933 | ENSG000001 | 3 | ETHE1 |
| Pancreas | T2DM | admiss | ENSG0000013 | 0.002582743 | 0.221398105 | 0.00211212 | 0.073378254 | 0.002614671 | 0.215737484 | 0.002112108 | 0.073373776 | -3.19E-05 | 0.033071554 | 1 | 0.610032546 | 1 | 0.602188978 | ENSG000001 | 9 | SMG9 |
| Pancreas | T2DM | admiss | ENSG0000013 | 0.001859037 | 0.387784307 | 0.002152552 | 0.073378367 | 0.000988814 | 0.650596627 | 0.002183131 | 0.073373976 | 0.000870223 | 0.016965779 | 1 | 0.740290513 | 1 | 0.885860095 | ENSG000001 | 4 | DLX5 |
| Pancreas | T2DM | admiss | ENSG0000013 | 0.001275846 | 0.380920243 | 0.001456108 | 0.073378364 | 0.001095883 | 0.451949371 | 0.001456963 | 0.073373921 | 0.000179962 | 0.000437344 | 1 | 0.733835847 | 1 | 0.783134943 | ENSG000001 | 4 | DLX5 |
| Pancreas | T2DM | admiss | ENSG0000013 | 0.001764029 | 0.316295412 | 0.001760331 | 0.073378328 | 0.0018033 | 0.305639404 | 0.00176032 | 0.073373849 | -3.93E-05 | 0.001338836 | 1 | 0.705815487 | 1 | 0.703146938 | ENSG000001 | 10 | STEAP1B |
| Pancreas | T2DM | admiss | ENSG0000013 | -0.00535946 | 0.163082456 | 0.003842507 | 0.073378186 | -0.00528587 | 0.168933145 | 0.003842474 | 0.073373722 | -7.36E-05 | 0.003633766 | 1 | 0.541071967 | 1 | 0.54913964 | ENSG000001 | 9 | DFNA5 |
| Pancreas | T2DM | admiss | ENSG0000013 | 0.002652436 | 0.315257131 | 0.002641203 | 0.073378327 | 0.000588408 | 0.830111133 | 0.002742403 | 0.073374 | 0.002064028 | 0.005185582 | 1 | 0.705815487 | 1 | 0.955502191 | ENSG000001 | 9 | DFNA5 |
| Pancreas | T2DM | admiss | ENSG0000013 | -0.00069636 | 0.842268845 | 0.003499443 | 0.073378473 | -0.00076913 | 0.826035826 | 0.003499424 | 0.073373999 | 7.28E-05 | 0.003300647 | 1 | 0.959017468 | 1 | 0.954095567 | ENSG000001 | 14 | H2AFV |
| Pancreas | T2DM | admiss | ENSG0000013 | 0.000264028 | 0.942959195 | 0.003690057 | 0.073378478 | 0.000303326 | 0.934485965 | 0.003689998 | 0.073374006 | -3.93E-05 | 0.04812715 | 1 | 0.982923376 | 1 | 0.98183872 | ENSG000001 | 14 | H2AFV |
| Pancreas | T2DM | admiss | ENSG0000013 | -0.0023838 | 0.094335927 | 0.001424901 | 0.073378058 | -0.00236978 | 0.096283215 | 0.001424874 | 0.07337359 | -1.40E-05 | 0.045749087 | 1 | 0.41411053 | 1 | 0.419076774 | ENSG000001 | 14 | H2AFV |
| Pancreas | T2DM | admiss | ENSG0000013 | 0.002332804 | 0.330477722 | 0.002397161 | 0.073378337 | 0.002282176 | 0.341076211 | 0.002397138 | 0.07337387 | 5.06E-05 | 0.001108407 | 1 | 0.707406446 | 1 | 0.715170664 | ENSG000001 | 14 | H2AFV |
| Pancreas | T2DM | admiss | ENSG0000013 | 0.002011575 | 0.154627593 | 0.001413242 | 0.073378174 | 0.001982574 | 0.160660266 | 0.001413241 | 0.07337371 | 2.90E-05 | 0.00773254 | 1 | 0.531571161 | 1 | 0.538610395 | ENSG000001 | 12 | RNF32 |
| Pancreas | T2DM | admiss | ENSG0000013 | 0.000532973 | 0.888188124 | 0.003790775 | 0.073378476 | 0.000625154 | 0.869011008 | 0.003790765 | 0.073374002 | -9.22E-05 | 0.001142543 | 1 | 0.972507979 | 1 | 0.967645366 | ENSG000001 | 12 | RNF32 |
| Pancreas | T2DM | admiss | ENSG0000013 | 0.002182284 | 0.54204713 | 0.003579154 | 0.073378423 | 0.002116681 | 0.55425968 | 0.003579165 | 0.073373954 | 6.56E-05 | 0.025553076 | 1 | 0.838486276 | 1 | 0.846590114 | ENSG000001 | 12 | RNF32 |
| Pancreas | T2DM | admiss | ENSG0000013 | -0.00470128 | 0.218311102 | 0.003818975 | 0.073378251 | -0.00485896 | 0.20333158 | 0.003819577 | 0.073373763 | 0.000157671 | 0.033350007 | 1 | 0.607038146 | 1 | 0.585376119 | ENSG000001 | 15 | LMBR1 |
| Pancreas | T2DM | admiss | ENSG0000013 | -0.00104923 | 0.756135054 | 0.003378518 | 0.073378465 | -0.0009549 | 0.777459145 | 0.003378607 | 0.073373995 | -9.43E-05 | 0.008735663 | 1 | 0.926431379 | 1 | 0.935113451 | ENSG000001 | 11 | BRAT1 |
| Pancreas | T2DM | admiss | ENSG0000013 | 0.001254718 | 0.720732833 | 0.0035099 | 0.07337846 | 0.001315347 | 0.707843206 | 0.00350989 | 0.073373985 | -6.06E-05 | 0.020112564 | 1 | 0.915352085 | 1 | 0.91023603 | ENSG000001 | 11 | BRAT1 |
| Pancreas | T2DM | admiss | ENSG0000014 | -0.00041105 | 0.822133185 | 0.001828492 | 0.073378472 | -0.00043571 | 0.811654041 | 0.001828474 | 0.073373998 | 2.47E-05 | 0.033987046 | 1 | 0.951436692 | 1 | 0.945273278 | ENSG000001 | 11 | BRAT1 |
| Pancreas | T2DM | admiss | ENSG0000014 | -0.00311191 | 0.410026926 | 0.00377729 | 0.073378377 | -0.00315285 | 0.403887182 | 0.003777232 | 0.073373902 | 4.09E-05 | 0.049918138 | 1 | 0.756489657 | 1 | 0.755027733 | ENSG000001 | 4 | HIBADH |
| Pancreas | T2DM | admiss | ENSG0000014 | 0.001303482 | 0.452372732 | 0.001734586 | 0.073378394 | 0.001283598 | 0.459292705 | 0.001734562 | 0.073373924 | 1.99E-05 | 0.047910795 | 1 | 0.781199104 | 1 | 0.78659086 | ENSG000001 | 14 | ABHD11 |
| Pancreas | T2DM | admiss | ENSG0000014 | -0.00271494 | 0.002342802 | 0.000892205 | 0.073377085 | -0.00270163 | 0.002461443 | 0.000892199 | 0.073372626 | -1.33E-05 | 0.027813426 | 1 | 0.04685311 | 1 | 0.048622958 | ENSG000001 | 14 | ABHD11 |
| Pancreas | T2DM | admiss | ENSG0000014 | -0.00172744 | 0.407595524 | 0.002085948 | 0.073378376 | -0.00169355 | 0.41685788 | 0.002085947 | 0.073373907 | -3.39E-05 | 0.035629088 | 1 | 0.755743904 | 1 | 0.763545674 | ENSG000001 | 14 | ABHD11 |
| Pancreas | T2DM | admiss | ENSG0000014 | -0.00572859 | 0.02741833 | 0.00259739 | 0.073377747 | -0.00569487 | 0.028338845 | 0.00259735 | 0.073373283 | -3.37E-05 | 0.018595329 | 1 | 0.216737649 | 1 | 0.223585926 | ENSG000001 | 14 | CRHR2 |
| Pancreas | T2DM | admiss | ENSG0000014 | 0.003366925 | 0.224330079 | 0.002770913 | 0.073378257 | 0.003401534 | 0.219595522 | 0.002770868 | 0.07337378 | -3.46E-05 | 0.02062606 | 1 | 0.614157401 | 1 | 0.606366989 | ENSG000001 | 7 | EPHB6 |
| Pancreas | T2DM | admiss | ENSG0000014 | -0.00107248 | 0.457977234 | 0.001445037 | 0.073378396 | -0.00105661 | 0.464649561 | 0.001445015 | 0.073373926 | -1.59E-05 | 0.047491355 | 1 | 0.783534669 | 1 | 0.788724539 | ENSG000001 | 14 | FAM188B |
| Pancreas | T2DM | admiss | ENSG0000014 | -0.0009891 | 0.146216622 | 0.000680716 | 0.073378161 | -0.00097685 | 0.151279224 | 0.000680714 | 0.073373696 | -1.23E-05 | 0.016947998 | 1 | 0.516484575 | 1 | 0.529993244 | ENSG000001 | 14 | FAM188B |
| Pancreas | T2DM | admiss | ENSG0000014 | -0.00359938 | 0.075199114 | 0.002022973 | 0.073378002 | -0.00363258 | 0.072549746 | 0.002022981 | 0.073373521 | 3.32E-05 | 0.04720457 | 1 | 0.362991211 | 1 | 0.361061527 | ENSG000001 | 13 | NSUN5P2 |
| Pancreas | T2DM | admiss | ENSG0000014 | 0.000899689 | 0.717421526 | 0.002485965 | 0.073378459 | 0.000841302 | 0.73504873 | 0.002485988 | 0.073373989 | 5.84E-05 | 0.00834794 | 1 | 0.915352085 | 1 | 0.922046709 | ENSG000001 | 13 | NSUN5P2 |
| Pancreas | T2DM | admiss | ENSG0000014 | -0.0059033 | 0.02048685 | 0.00254747 | 0.073377671 | -0.00584344 | 0.021801076 | 0.002547466 | 0.073373214 | -5.99E-05 | 0.0019461 | 1 | 0.192167217 | 1 | 0.19439293 | ENSG000001 | 13 | NSUN5P2 |
| Pancreas | T2DM | admiss | ENSG0000014 | -0.0072806 | 0.007451443 | 0.002720728 | 0.073377401 | -0.00722307 | 0.007934832 | 0.002720715 | 0.073372945 | -5.75E-05 | 0.003061914 | 1 | 0.103016032 | 1 | 0.109202996 | ENSG000001 | 13 | NSUN5P2 |
| Pancreas | T2DM | admiss | ENSG0000015 | 0.003887492 | 0.285384025 | 0.003638938 | 0.073378307 | 0.003926512 | 0.280568557 | 0.003638879 | 0.073373831 | -3.90E-05 | 0.044961316 | 1 | 0.679838752 | 1 | 0.675257783 | ENSG000001 | 12 | CHCHD2 |
| Pancreas | T2DM | admiss | ENSG0000015 | 0.002757142 | 0.041027798 | 0.001349388 | 0.073377851 | 0.00273315 | 0.042816647 | 0.001349371 | 0.073373389 | 2.40E-05 | 0.002908069 | 1 | 0.274649 | 1 | 0.28152064 | ENSG000001 | 12 | CHCHD2 |
| Pancreas | T2DM | admiss | ENSG0000015 | 6.55E-05 | 0.981480237 | 0.00282305 | 0.073378479 | 0.000326978 | 0.907836415 | 0.002824422 | 0.073374005 | -0.00026145 | 0.00395359 | 1 | 0.994701872 | 1 | 0.972600715 | ENSG000001 | 2 | CCL24 |
| Pancreas | T2DM | admiss | ENSG0000015 | 0.001059855 | 0.510051818 | 0.001608869 | 0.073378414 | 0.001040726 | 0.517711683 | 0.001608846 | 0.073373944 | 1.91E-05 | 0.03534523 | 1 | 0.815555259 | 1 | 0.821175789 | ENSG000001 | 2 | CCL24 |
| Pancreas | T2DM | admiss | ENSG0000015 | -0.00162768 | 0.592728925 | 0.003043038 | 0.073378436 | -0.00168164 | 0.580521658 | 0.003043006 | 0.073373961 | 5.40E-05 | 0.005011995 | 1 | 0.858891717 | 1 | 0.853336931 | ENSG000001 | 3 | NPTX2 |
| Pancreas | T2DM | admiss | ENSG0000015 | -0.00155668 | 0.286541335 | 0.001460654 | 0.073378308 | -0.00153989 | 0.291761561 | 0.001460629 | 0.073373839 | -1.68E-05 | 0.024807941 | 1 | 0.680701339 | 1 | 0.68737953 | ENSG000001 | 13 | EIF3B |
| Pancreas | T2DM | admiss | ENSG0000015 | 0.00260911 | 0.424706658 | 0.003268402 | 0.073378383 | 0.00265855 | 0.415976847 | 0.003268355 | 0.073373907 | -4.94E-05 | 0.007940662 | 1 | 0.765366105 | 1 | 0.762475763 | ENSG000001 | 13 | EIF3B |
| Pancreas | T2DM | admiss | ENSG0000015 | -0.00274723 | 0.05508167 | 0.001432167 | 0.073377925 | -0.00277412 | 0.052741457 | 0.001432147 | 0.073373442 | 2.69E-05 | 0.001168429 | 1 | 0.312941876 | 1 | 0.309931493 | ENSG000001 | 13 | EIF3B |

| | | | | | | | | | | | | | | | | | | | | |
|---|---|---|---|---|---|---|---|---|---|---|---|---|---|---|---|---|---|---|---|---|
| Pancreas | T2DM | admiss | ENSG0000015 | -0.00096397 | 0.578099217 | 0.001733244 | 0.073378433 | -0.00093454 | 0.589761663 | 0.001733253 | 0.073373963 | -2.94E-05 | 0.044802337 | 1 | 0.85368533 | 1 | 0.857158565 | ENSG000001 | 13 | EIF3B |
| Pancreas | T2DM | admiss | ENSG0000015 | -0.00136751 | 0.087962277 | 0.000801472 | 0.073378041 | -0.00137702 | 0.085771951 | 0.000801462 | 0.073373562 | 9.51E-06 | 0.046571646 | 1 | 0.395599172 | 1 | 0.396836678 | ENSG000001 | 10 | TAF6 |
| Pancreas | T2DM | admiss | ENSG0000015 | 0.000460345 | 0.765061131 | 0.001540431 | 0.073378466 | 0.000501954 | 0.744536436 | 0.001540436 | 0.073373991 | -4.16E-05 | 0.000980508 | 1 | 0.927632086 | 1 | 0.92339769 | ENSG000001 | 7 | WASL |
| Pancreas | T2DM | admiss | ENSG0000015 | 0.00356573 | 0.345937467 | 0.00378327 | 0.073378345 | 0.003637579 | 0.336300951 | 0.00378323 | 0.073373867 | -7.18E-05 | 0.002749894 | 1 | 0.714946894 | 1 | 0.712394628 | ENSG000001 | 7 | MOSPD3 |
| Pancreas | T2DM | admiss | ENSG0000015 | 0.005184465 | 0.139531911 | 0.003508852 | 0.07337815 | 0.00514092 | 0.142881846 | 0.003508812 | 0.073373683 | 4.35E-05 | 0.044499516 | 1 | 0.500672845 | 1 | 0.511031449 | ENSG000001 | 4 | RBM28 |
| Pancreas | T2DM | admiss | ENSG0000015 | 0.002254707 | 0.264205429 | 0.002019428 | 0.073378291 | 0.002028152 | 0.315588813 | 0.002020948 | 0.073373855 | 0.000226554 | 0.004585629 | 1 | 0.661240615 | 1 | 0.706943978 | ENSG000001 | 4 | RBM28 |
| Pancreas | T2DM | admiss | ENSG0000015 | -0.0021191 | 0.201521882 | 0.001659137 | 0.073378234 | -0.00209334 | 0.20705358 | 0.001659132 | 0.073373767 | -2.58E-05 | 0.035039109 | 1 | 0.581469879 | 1 | 0.591085559 | ENSG000001 | 12 | IMPDH1 |
| Pancreas | T2DM | admiss | ENSG0000015 | -0.00266247 | 0.168827032 | 0.001934955 | 0.073378194 | -0.0026956 | 0.163586182 | 0.001934944 | 0.073373714 | 3.31E-05 | 0.014473537 | 1 | 0.548084313 | 1 | 0.54345319 | ENSG000001 | 14 | TMEM106B |
| Pancreas | T2DM | admiss | ENSG0000016 | -0.00093662 | 0.773839103 | 0.003259419 | 0.073378467 | -0.00089161 | 0.7844295 | 0.003259374 | 0.073373995 | -4.50E-05 | 0.017079588 | 1 | 0.930781647 | 1 | 0.936079501 | ENSG000001 | 14 | CEP41 |
| Pancreas | T2DM | admiss | ENSG0000016 | 0.000289294 | 0.644943321 | 0.00062781 | 0.073378447 | 0.000298047 | 0.634969339 | 0.000627804 | 0.073373973 | -8.75E-06 | 0.031539085 | 1 | 0.885748011 | 1 | 0.880942324 | ENSG000001 | 5 | RARRES2 |
| Pancreas | T2DM | admiss | ENSG0000016 | -0.00520125 | 0.049455556 | 0.002647419 | 0.073377898 | -0.00517013 | 0.05082933 | 0.002647383 | 0.073373432 | -3.11E-05 | 0.042994484 | 1 | 0.296041653 | 1 | 0.304975981 | ENSG000001 | 13 | TMEM176B |
| Pancreas | T2DM | admiss | ENSG0000016 | 0.002137906 | 0.557069346 | 0.003640846 | 0.073378427 | 0.002080491 | 0.567704669 | 0.003640814 | 0.073373957 | 5.74E-05 | 0.016977483 | 1 | 0.847939375 | 1 | 0.849476828 | ENSG000001 | 3 | MRPL32 |
| Pancreas | T2DM | admiss | ENSG0000016 | -0.00245124 | 0.114730433 | 0.001554086 | 0.073378105 | -0.0024218 | 0.119150871 | 0.001554079 | 0.073373641 | -2.94E-05 | 0.008295776 | 1 | 0.463251181 | 1 | 0.472809902 | ENSG000001 | 13 | COA1 |
| Pancreas | T2DM | admiss | ENSG0000016 | 0.002592097 | 0.435075567 | 0.003320917 | 0.073378387 | 0.002630466 | 0.428300448 | 0.00332086 | 0.073373912 | -3.84E-05 | 0.024751773 | 1 | 0.771915116 | 1 | 0.769192559 | ENSG000001 | 13 | COA1 |
| Pancreas | T2DM | admiss | ENSG0000016 | -0.00332141 | 0.169285472 | 0.002416438 | 0.073378195 | -0.00329231 | 0.173045879 | 0.002416404 | 0.073373727 | -2.91E-05 | 0.034924093 | 1 | 0.548084313 | 1 | 0.553785383 | ENSG000001 | 13 | COA1 |
| Pancreas | T2DM | admiss | ENSG0000016 | 0.003206127 | 0.021662048 | 0.001396245 | 0.073377685 | 0.003495952 | 0.012409419 | 0.001398217 | 0.073373065 | -0.00028982 | 0.000112732 | 1 | 0.193802603 | 1 | 0.143546788 | ENSG000001 | 10 | STAG3L4 |
| Pancreas | T2DM | admiss | ENSG0000016 | 0.00127816 | 0.483536182 | 0.001824309 | 0.073378405 | 0.000980336 | 0.591559793 | 0.001827017 | 0.073373963 | 0.000297824 | 0.003030032 | 1 | 0.799048209 | 1 | 0.857294328 | ENSG000001 | 10 | STAG3L4 |
| Pancreas | T2DM | admiss | ENSG0000016 | 0.003377017 | 0.246020085 | 0.002911041 | 0.073378277 | 0.003414497 | 0.240811788 | 0.002911005 | 0.073373799 | -3.75E-05 | 0.031997955 | 1 | 0.637517234 | 1 | 0.627263575 | ENSG000001 | 10 | STAG3L4 |
| Pancreas | T2DM | admiss | ENSG0000016 | -0.00110547 | 0.739082246 | 0.003319045 | 0.073378462 | -0.00122948 | 0.711094989 | 0.003319448 | 0.073373986 | 0.000124006 | 0.032168678 | 1 | 0.920031049 | 1 | 0.912283108 | ENSG000001 | 3 | BCL7B |
| Pancreas | T2DM | admiss | ENSG0000016 | 0.000981304 | 0.330478644 | 0.001008378 | 0.073378337 | 0.000994577 | 0.323972861 | 0.001008364 | 0.07337386 | -1.33E-05 | 0.022721109 | 1 | 0.707406446 | 1 | 0.706943978 | ENSG000001 | 3 | BCL7B |
| Pancreas | T2DM | admiss | ENSG0000016 | -0.00139798 | 0.534741113 | 0.002251957 | 0.073378421 | -0.00136779 | 0.543596396 | 0.002251931 | 0.073373951 | -3.02E-05 | 0.029287233 | 1 | 0.832756324 | 1 | 0.842968325 | ENSG000001 | 3 | BCL7B |
| Pancreas | T2DM | admiss | ENSG0000016 | 0.001237718 | 0.238730624 | 0.001050545 | 0.07337827 | 0.001252828 | 0.233040684 | 0.001050533 | 0.073373792 | -1.51E-05 | 0.018523318 | 1 | 0.627977804 | 1 | 0.619840368 | ENSG000001 | 11 | TBL2 |
| Pancreas | T2DM | admiss | ENSG0000016 | -0.00105837 | 0.323769614 | 0.00107259 | 0.073378333 | -0.0010711 | 0.317977804 | 0.001072574 | 0.073373856 | 1.27E-05 | 0.032686258 | 1 | 0.707406446 | 1 | 0.706943978 | ENSG000001 | 11 | TBL2 |
| Pancreas | T2DM | admiss | ENSG0000016 | 0.000922521 | 0.734841676 | 0.002723774 | 0.073378462 | 0.000886708 | 0.744767021 | 0.002723744 | 0.073373991 | 3.58E-05 | 0.035686477 | 1 | 0.919728783 | 1 | 0.92339769 | ENSG000001 | 11 | TBL2 |
| Pancreas | T2DM | admiss | ENSG0000016 | -0.00268434 | 0.480568665 | 0.003805474 | 0.073378404 | -0.00262161 | 0.490879264 | 0.003805432 | 0.073373935 | -6.27E-05 | 0.00811841 | 1 | 0.797318048 | 1 | 0.805191895 | ENSG000001 | 11 | TBL2 |
| Pancreas | T2DM | admiss | ENSG0000016 | 0.002965644 | 0.077770541 | 0.001681419 | 0.073378011 | 0.002928397 | 0.081576027 | 0.001681424 | 0.07337355 | 3.72E-05 | 0.006765159 | 1 | 0.371908286 | 1 | 0.386507818 | ENSG000001 | 7 | SLC1A1 |
| Pancreas | T2DM | admiss | ENSG0000016 | -0.00357966 | 0.223579335 | 0.002941219 | 0.073378256 | -0.00353972 | 0.228782833 | 0.002941187 | 0.073373788 | -3.99E-05 | 0.030040634 | 1 | 0.613012193 | 1 | 0.615924595 | ENSG000001 | 7 | SLC1A1 |
| Pancreas | T2DM | admiss | ENSG0000016 | -0.00065124 | 0.704366106 | 0.001716339 | 0.073378458 | -0.00091453 | 0.595076682 | 0.001720687 | 0.073373964 | 0.000263298 | 0.032279195 | 1 | 0.909407823 | 1 | 0.859705602 | ENSG000001 | 7 | SLC1A1 |
| Pancreas | T2DM | admiss | ENSG0000016 | -0.00059557 | 0.795675398 | 0.002299942 | 0.073378469 | -0.00037318 | 0.871182276 | 0.002301372 | 0.073374003 | -0.00022238 | 0.007433288 | 1 | 0.938428627 | 1 | 0.967645366 | ENSG000001 | 16 | NMRK1 |
| Pancreas | T2DM | admiss | ENSG0000016 | 0.001902998 | 0.214802078 | 0.001534094 | 0.073378247 | 0.001883267 | 0.219589771 | 0.001534078 | 0.07337378 | 1.97E-05 | 0.041550444 | 1 | 0.60288425 | 1 | 0.606366989 | ENSG000001 | 16 | NMRK1 |
| Pancreas | T2DM | admiss | ENSG0000016 | 0.002029868 | 0.118195297 | 0.001299202 | 0.073378112 | 0.002012798 | 0.12131616 | 0.001299187 | 0.073373645 | 1.71E-05 | 0.032582848 | 1 | 0.469127545 | 1 | 0.473464893 | ENSG000001 | 16 | NMRK1 |
| Pancreas | T2DM | admiss | ENSG0000016 | -0.00052441 | 0.37064313 | 0.000585754 | 0.073378359 | -0.00055639 | 0.342267986 | 0.000585857 | 0.073373871 | 3.20E-05 | 0.00722327 | 1 | 0.728887861 | 1 | 0.715170664 | ENSG000001 | 16 | NMRK1 |
| Pancreas | T2DM | admiss | ENSG0000016 | 0.002743189 | 0.412648325 | 0.003348445 | 0.073378378 | 0.002791211 | 0.404510872 | 0.00334841 | 0.073373902 | -4.80E-05 | 0.023942388 | 1 | 0.757001665 | 1 | 0.755027733 | ENSG000001 | 16 | NMRK1 |
| Pancreas | T2DM | admiss | ENSG0000016 | 0.000619819 | 0.566202782 | 0.001080476 | 0.07337843 | 0.000637018 | 0.555474853 | 0.001080468 | 0.073373954 | -1.72E-05 | 0.018644925 | 1 | 0.850297512 | 1 | 0.846590114 | ENSG000001 | 12 | PTGR1 |
| Pancreas | T2DM | admiss | ENSG0000016 | -0.00598505 | 0.001563578 | 0.001892425 | 0.073376973 | -0.00600408 | 0.001510123 | 0.001892391 | 0.073372491 | 1.90E-05 | 0.048048565 | 1 | 0.035221657 | 1 | 0.034318558 | ENSG000001 | 12 | PTGR1 |
| Pancreas | T2DM | admiss | ENSG0000016 | -0.00682917 | 0.000604667 | 0.001991283 | 0.073376708 | -0.00679972 | 0.000638376 | 0.001991258 | 0.073372251 | -2.95E-05 | 0.014071952 | 1 | 0.017666591 | 1 | 0.018419672 | ENSG000001 | 12 | PTGR1 |
| Pancreas | T2DM | admiss | ENSG0000016 | -0.00829836 | 3.61E-05 | 0.002008684 | 0.073375909 | -0.00825888 | 3.93E-05 | 0.002008663 | 0.073371461 | -3.95E-05 | 0.001676631 | 0.092670575 | 0.00215513 | 0.100905808 | 0.002522645 | ENSG000001 | 12 | SUSD1 |
| Pancreas | T2DM | admiss | ENSG0000016 | -0.00027934 | 0.775189557 | 0.000978103 | 0.073378467 | -0.00029261 | 0.764812916 | 0.000978089 | 0.073373993 | 1.33E-05 | 0.019553844 | 1 | 0.931097653 | 1 | 0.928829506 | ENSG000001 | 13 | AK1 |
| Pancreas | T2DM | admiss | ENSG0000016 | 0.002176182 | 0.084509962 | 0.001261481 | 0.073378031 | 0.002193145 | 0.082112486 | 0.00126147 | 0.073373551 | -1.70E-05 | 0.04224197 | 1 | 0.391029878 | 1 | 0.386704232 | ENSG000001 | 13 | AK1 |
| Pancreas | T2DM | admiss | ENSG0000016 | 0.001317355 | 0.458977333 | 0.00177893 | 0.073378397 | 0.001353834 | 0.446630439 | 0.001778915 | 0.073373919 | -3.65E-05 | 0.001979107 | 1 | 0.784014077 | 1 | 0.779994504 | ENSG000001 | 13 | AK1 |
| Pancreas | T2DM | admiss | ENSG0000016 | 0.000128099 | 0.968208685 | 0.003214127 | 0.073378479 | 0.000185731 | 0.953918934 | 0.003214107 | 0.073374006 | -5.76E-05 | 0.010180364 | 1 | 0.989399086 | 1 | 0.984195992 | ENSG000001 | 5 | PLGRKT |
| Pancreas | T2DM | admiss | ENSG0000016 | -0.00079359 | 0.715671082 | 0.002178696 | 0.073378459 | -0.0008206 | 0.706433509 | 0.002178673 | 0.073373985 | 2.70E-05 | 0.049601426 | 1 | 0.914349919 | 1 | 0.909378635 | ENSG000001 | 5 | PLGRKT |
| Pancreas | T2DM | admiss | ENSG0000016 | 0.004515197 | 0.115250751 | 0.002866742 | 0.073378106 | 0.004550069 | 0.112463965 | 0.002866695 | 0.073373627 | -3.49E-05 | 0.021993411 | 1 | 0.46462155 | 1 | 0.461815776 | ENSG000001 | 11 | DOCK8 |
| Pancreas | T2DM | admiss | ENSG0000016 | -0.00491959 | 0.055955181 | 0.002573828 | 0.073377929 | -0.00499931 | 0.052104401 | 0.002573948 | 0.073373439 | 7.97E-05 | 0.012582656 | 1 | 0.313835611 | 1 | 0.308304381 | ENSG000001 | 11 | DOCK8 |
| Pancreas | T2DM | admiss | ENSG0000016 | 0.000658535 | 0.466477061 | 0.000904303 | 0.073378399 | 0.000644091 | 0.47630448 | 0.000904293 | 0.07337393 | 1.44E-05 | 0.010963424 | 1 | 0.790180141 | 1 | 0.79570852 | ENSG000001 | 11 | DOCK8 |
| Pancreas | T2DM | admiss | ENSG0000016 | 0.000581145 | 0.668733887 | 0.001358175 | 0.073378452 | 0.00060196 | 0.657610078 | 0.001358167 | 0.073373977 | -2.08E-05 | 0.027909811 | 1 | 0.893500844 | 1 | 0.887614942 | ENSG000001 | 11 | DOCK8 |
| Pancreas | T2DM | admiss | ENSG0000016 | 0.000494701 | 0.604587666 | 0.000955363 | 0.073378439 | 0.000508357 | 0.594645554 | 0.000955349 | 0.073373964 | -1.37E-05 | 0.010241433 | 1 | 0.865912508 | 1 | 0.859705602 | ENSG000001 | 11 | DOCK8 |
| Pancreas | T2DM | admiss | ENSG0000016 | 0.00359699 | 0.058617706 | 0.001902108 | 0.073377941 | 0.003540358 | 0.062706681 | 0.00190212 | 0.073373485 | 5.66E-05 | 0.000470263 | 1 | 0.320228732 | 1 | 0.334088709 | ENSG000001 | 11 | DOCK8 |
| Pancreas | T2DM | admiss | ENSG0000016 | -0.00102851 | 0.52724445 | 0.00162682 | 0.073378419 | -0.00101144 | 0.534114066 | 0.001626791 | 0.073373948 | -1.71E-05 | 0.038621828 | 1 | 0.82963465 | 1 | 0.835325774 | ENSG000001 | 11 | DOCK8 |
| Pancreas | T2DM | admiss | ENSG0000017 | -0.0013894 | 0.654899157 | 0.003108502 | 0.073378449 | -0.00133558 | 0.66744782 | 0.003108505 | 0.073373979 | -5.38E-05 | 0.028727333 | 1 | 0.889360674 | 1 | 0.88982611 | ENSG000001 | 6 | NCS1 |
| Pancreas | T2DM | admiss | ENSG0000017 | 0.002483643 | 0.374114993 | 0.002794401 | 0.07337836 | 0.002524796 | 0.366245055 | 0.002794368 | 0.073373884 | -4.12E-05 | 0.016469794 | 1 | 0.730660228 | 1 | 0.725397099 | ENSG000001 | 6 | NCS1 |
| Pancreas | T2DM | admiss | ENSG0000017 | 0.002374418 | 0.427762543 | 0.00299412 | 0.073378384 | 0.002516349 | 0.400790631 | 0.002994902 | 0.0733739 | -0.00014193 | 0.049729275 | 1 | 0.767595622 | 1 | 0.753462913 | ENSG000001 | 6 | NCS1 |
| Pancreas | T2DM | admiss | ENSG0000017 | 0.002961061 | 0.156984956 | 0.002092198 | 0.073378178 | 0.002925563 | 0.162015032 | 0.002092185 | 0.073373712 | 3.55E-05 | 0.014986687 | 1 | 0.53381119 | 1 | 0.540033938 | ENSG000001 | 15 | FUBP3 |
| Pancreas | T2DM | admiss | ENSG0000017 | 0.001300009 | 0.627585987 | 0.002679725 | 0.073378444 | 0.001339169 | 0.617253143 | 0.002679691 | 0.073373969 | -3.92E-05 | 0.01493197 | 1 | 0.879226992 | 1 | 0.872396545 | ENSG000001 | 13 | PIP5K1B |
| Pancreas | T2DM | admiss | ENSG0000017 | -0.00186413 | 0.389212935 | 0.002164963 | 0.073378368 | -0.00182546 | 0.399120671 | 0.002164939 | 0.073373899 | -3.87E-05 | 0.004086693 | 1 | 0.741467965 | 1 | 0.752527081 | ENSG000001 | 11 | CCNJ |
| Pancreas | T2DM | admiss | ENSG0000017 | -0.00203128 | 0.592576137 | 0.003796026 | 0.073378436 | -0.00209134 | 0.581680705 | 0.003795997 | 0.073373961 | 6.01E-05 | 0.019178876 | 1 | 0.858891717 | 1 | 0.854282157 | ENSG000001 | 11 | CCNJ |
| Pancreas | T2DM | admiss | ENSG0000017 | -0.00243658 | 0.275119297 | 0.002232629 | 0.0733783 | -0.00240061 | 0.282263125 | 0.002232606 | 0.073373832 | -3.60E-05 | 0.01110782 | 1 | 0.670309634 | 1 | 0.678065206 | ENSG000001 | 11 | CCNJ |
| Pancreas | T2DM | admiss | ENSG0000017 | 0.006767385 | 0.062423088 | 0.003631969 | 0.073377956 | 0.006825661 | 0.060196657 | 0.003631917 | 0.073373475 | -5.83E-05 | 0.005023546 | 1 | 0.32978603 | 1 | 0.32708278 | ENSG000001 | 11 | CCNJ |
| Pancreas | T2DM | admiss | ENSG0000017 | -0.00109718 | 0.36132712 | 0.00120194 | 0.073378354 | -0.00111593 | 0.35317945 | 0.001201936 | 0.073373877 | 1.87E-05 | 0.035160971 | 1 | 0.723010568 | 1 | 0.71888913 | ENSG000001 | 9 | PHYH |
| Pancreas | T2DM | admiss | ENSG0000017 | -0.00154342 | 0.449620448 | 0.002041427 | 0.073378393 | -0.00156962 | 0.441958628 | 0.002041406 | 0.073373918 | 2.62E-05 | 0.04189656 | 1 | 0.781199104 | 1 | 0.777362848 | ENSG000001 | 9 | PHYH |
| Pancreas | T2DM | admiss | ENSG0000017 | 0.002383078 | 0.247495099 | 0.002060673 | 0.073378278 | 0.002422877 | 0.239683037 | 0.002060653 | 0.073373798 | -3.98E-05 | 0.002946183 | 1 | 0.638493224 | 1 | 0.626788227 | ENSG000001 | 11 | DNMBP |
| Pancreas | T2DM | admiss | ENSG0000017 | 0.000677222 | 0.781837422 | 0.002445503 | 0.073378468 | 0.000708904 | 0.771904632 | 0.002445471 | 0.073373994 | -3.17E-05 | 0.02800114 | 1 | 0.934710661 | 1 | 0.931343937 | ENSG000001 | 11 | DNMBP |
| Pancreas | T2DM | admiss | ENSG0000017 | 0.001523294 | 0.516229728 | 0.002346531 | 0.073378416 | 0.001575774 | 0.501879781 | 0.002346523 | 0.073373939 | -5.25E-05 | 0.002444013 | 1 | 0.821323036 | 1 | 0.814167579 | ENSG000001 | 15 | PKD2L1 |
| Pancreas | T2DM | admiss | ENSG0000017 | 0.000882975 | 0.558946795 | 0.00151089 | 0.073378428 | 0.000910234 | 0.546872215 | 0.001510877 | 0.073373952 | -2.73E-05 | 0.006357042 | 1 | 0.847939375 | 1 | 0.845484919 | ENSG000001 | 15 | PKD2L1 |
| Pancreas | T2DM | admiss | ENSG0000017 | -0.00049044 | 0.808835345 | 0.002027206 | 0.07337847 | -0.00046513 | 0.81852071 | 0.002027178 | 0.073373999 | -2.53E-05 | 0.029207054 | 1 | 0.942917767 | 1 | 0.949824303 | ENSG000001 | 10 | NSMCE4A |
| Pancreas | T2DM | admiss | ENSG0000017 | 0.00198957 | 0.371188331 | 0.002224846 | 0.073378359 | 0.001939781 | 0.383277736 | 0.002224842 | 0.073373892 | 4.98E-05 | 0.003067562 | 1 | 0.729283118 | 1 | 0.737196508 | ENSG000001 | 8 | SPOCK2 |
| Pancreas | T2DM | admiss | ENSG0000017 | -0.00222443 | 0.226392923 | 0.001838821 | 0.073378259 | -0.00220578 | 0.230302201 | 0.001838788 | 0.07337379 | -1.87E-05 | 0.04325359 | 1 | 0.617173065 | 1 | 0.617989606 | ENSG000001 | 8 | SPOCK2 |
| Pancreas | T2DM | admiss | ENSG0000018 | -0.00030182 | 0.71117655 | 0.000815115 | 0.073378459 | -0.00028507 | 0.726534781 | 0.000815106 | 0.073373988 | -1.67E-05 | 0.001207851 | 1 | 0.912258576 | 1 | 0.917883561 | ENSG000001 | 7 | BMPR1A |
| Pancreas | T2DM | admiss | ENSG0000018 | 0.000217298 | 0.946473898 | 0.003236704 | 0.073378479 | 0.000256646 | 0.936799298 | 0.003236663 | 0.073374006 | -3.93E-05 | 0.041571421 | 1 | 0.982923376 | 1 | 0.98183872 | ENSG000001 | 7 | BMPR1A |
| Pancreas | T2DM | admiss | ENSG0000018 | 0.006946571 | 0.042011293 | 0.003416211 | 0.073377857 | 0.007001218 | 0.040420661 | 0.003416179 | 0.073373374 | -5.46E-05 | 0.013795004 | 1 | 0.277250207 | 1 | 0.277540798 | ENSG000001 | 13 | LIPA |
| Pancreas | T2DM | admiss | ENSG0000018 | 0.000579311 | 0.508468893 | 0.000876119 | 0.073378413 | 0.000361662 | 0.680608427 | 0.000878603 | 0.073373981 | 0.000217649 | 0.00104118 | 1 | 0.815555259 | 1 | 0.898151305 | ENSG000001 | 13 | LIPA |
| Pancreas | T2DM | admiss | ENSG0000018 | 0.003292359 | 0.101232952 | 0.002008873 | 0.073378075 | 0.00326257 | 0.104358141 | 0.002008865 | 0.073373609 | 2.98E-05 | 0.042750172 | 1 | 0.429184223 | 1 | 0.440775833 | ENSG000001 | 12 | ANKRD26 |
| Pancreas | T2DM | admiss | ENSG0000018 | 0.000746686 | 0.651532695 | 0.001653298 | 0.073378448 | 0.000601446 | 0.71616171 | 0.001654171 | 0.073373987 | 0.00014524 | 0.008584411 | 1 | 0.887189213 | 1 | 0.913596366 | ENSG000001 | 12 | ANKRD26 |
| Pancreas | T2DM | admiss | ENSG0000018 | 0.001477071 | 0.152360734 | 0.001032018 | 0.073378171 | 0.001465104 | 0.155704421 | 0.001032003 | 0.073373703 | 1.20E-05 | 0.038574629 | 1 | 0.52858693 | 1 | 0.534225295 | ENSG000001 | 12 | ANKRD26 |
| Pancreas | T2DM | admiss | ENSG0000018 | -0.00132592 | 0.673311704 | 0.003144908 | 0.073378452 | -0.00136945 | 0.663233467 | 0.003144873 | 0.073373978 | 4.35E-05 | 0.025391432 | 1 | 0.895424369 | 1 | 0.889390884 | ENSG000001 | 12 | ANKRD26 |
| Pancreas | T2DM | admiss | ENSG0000018 | 0.006731407 | 0.00119037 | 0.002076828 | 0.073376897 | 0.006701508 | 0.001251815 | 0.002076814 | 0.073372439 | 2.99E-05 | 0.036578966 | 1 | 0.027789735 | 1 | 0.029224201 | ENSG000001 | 6 | NEURL1 |
| Pancreas | T2DM | admiss | ENSG0000018 | 0.003390942 | 0.155587705 | 0.002387868 | 0.073378176 | 0.003346342 | 0.161096156 | 0.002387862 | 0.073373711 | 4.46E-05 | 0.012509163 | 1 | 0.532022937 | 1 | 0.539367574 | ENSG000001 | 8 | RPL28 |
| Pancreas | T2DM | admiss | ENSG0000018 | -0.00378113 | 0.072238268 | 0.002103412 | 0.073377993 | -0.00381224 | 0.069921066 | 0.00210339 | 0.073373512 | 3.11E-05 | 0.019039753 | 1 | 0.357418609 | 1 | 0.352073131 | ENSG000001 | 8 | RPL28 |
| Pancreas | T2DM | admiss | ENSG0000018 | 0.002992835 | 0.308907987 | 0.002941312 | 0.073378323 | 0.002925157 | 0.319976168 | 0.002941311 | 0.073373858 | 6.77E-05 | 0.003107361 | 1 | 0.702883653 | 1 | 0.706943978 | ENSG000001 | 11 | PBLD |
| Pancreas | T2DM | admiss | ENSG0000018 | -0.00220835 | 0.314885236 | 0.002197306 | 0.073378327 | -0.00217026 | 0.32330217 | 0.002197292 | 0.07337386 | -3.81E-05 | 0.012905913 | 1 | 0.705815487 | 1 | 0.706943978 | ENSG000001 | 11 | PBLD |
| Pancreas | T2DM | admiss | ENSG0000018 | 0.001024086 | 0.408385447 | 0.001238705 | 0.073378376 | 0.001000592 | 0.419217325 | 0.001238693 | 0.073373908 | 2.35E-05 | 0.003167182 | 1 | 0.756116674 | 1 | 0.763867723 | ENSG000001 | 12 | KRT23 |
| Pancreas | T2DM | admiss | ENSG0000018 | 0.002233557 | 0.016717531 | 0.000933422 | 0.073377617 | 0.002218527 | 0.017465127 | 0.000933419 | 0.073373156 | 1.50E-05 | 0.02986839 | 1 | 0.173120979 | 1 | 0.174867821 | ENSG000001 | 12 | TADA2A |
| Pancreas | T2DM | admiss | ENSG0000018 | -0.00130034 | 0.585730137 | 0.002385788 | 0.073378434 | -0.00124216 | 0.602624448 | 0.002385869 | 0.073373966 | -5.82E-05 | 0.031109608 | 1 | 0.856580292 | 1 | 0.863582357 | ENSG000001 | 12 | TADA2A |
| Pancreas | T2DM | admiss | ENSG0000018 | -0.00102689 | 0.737318968 | 0.003061635 | 0.073378462 | -0.00099284 | 0.745718206 | 0.003061589 | 0.073373991 | -3.40E-05 | 0.044328756 | 1 | 0.919728783 | 1 | 0.923784058 | ENSG000001 | 12 | TADA2A |
| Pancreas | T2DM | admiss | ENSG0000018 | -0.00222633 | 0.426848947 | 0.00280184 | 0.073378384 | -0.00219439 | 0.43350663 | 0.002801802 | 0.073373914 | -3.19E-05 | 0.048745908 | 1 | 0.767595622 | 1 | 0.771947117 | ENSG000001 | 12 | TADA2A |
| Pancreas | T2DM | admiss | ENSG0000018 | 0.002530758 | 0.170514119 | 0.00184652 | 0.073378196 | 0.00250374 | 0.17512115 | 0.001846508 | 0.07337373 | 2.70E-05 | 0.033767886 | 1 | 0.548084313 | 1 | 0.556573159 | ENSG000001 | 12 | TADA2A |
| Pancreas | T2DM | admiss | ENSG0000018 | -0.0001608 | 0.922820617 | 0.001659723 | 0.073378478 | -0.00019907 | 0.904530597 | 0.001659712 | 0.073374004 | 3.83E-05 | 0.000866841 | 1 | 0.978091879 | 1 | 0.972260921 | ENSG000001 | 12 | TADA2A |
| Pancreas | T2DM | admiss | ENSG0000018 | 0.005033356 | 0.174395079 | 0.003705852 | 0.073378201 | 0.004483213 | 0.226937327 | 0.003710387 | 0.073373787 | 0.000550143 | 0.003038581 | 1 | 0.553580425 | 1 | 0.614988604 | ENSG000001 | 9 | DHRS11 |
| Pancreas | T2DM | admiss | ENSG0000018 | 0.000591044 | 0.457753478 | 0.000795963 | 0.073378396 | 0.000600542 | 0.450552029 | 0.000795951 | 0.073373921 | -9.50E-06 | 0.035140285 | 1 | 0.783534669 | 1 | 0.78249589 | ENSG000001 | 9 | DHRS11 |
| Pancreas | T2DM | admiss | ENSG0000018 | 0.004659215 | 0.060464892 | 0.002481748 | 0.073377948 | -0.0010024 | 0.758367642 | 0.003258502 | 0.073373992 | 0.005661612 | 0.007337545 | 1 | 0.324840676 | 1 | 0.92717566 | ENSG000001 | 9 | DHRS11 |
| Pancreas | T2DM | admiss | ENSG0000018 | 0.003410811 | 0.231438201 | 0.002850267 | 0.073378264 | 0.003366567 | 0.237547153 | 0.002850266 | 0.073373796 | 4.42E-05 | 0.045573929 | 1 | 0.619742754 | 1 | 0.623743445 | ENSG000001 | 5 | RPL19 |
| Pancreas | T2DM | admiss | ENSG0000018 | -0.00675406 | 0.029895706 | 0.003110353 | 0.073377769 | -0.00669937 | 0.03124919 | 0.003110354 | 0.073373308 | -5.47E-05 | 0.025032839 | 1 | 0.229023436 | 1 | 0.238833099 | ENSG000001 | 10 | RAD51C |
| Pancreas | T2DM | admiss | ENSG0000018 | 0.000995723 | 0.444518817 | 0.001302304 | 0.073378391 | 0.0010098 | 0.438099391 | 0.001302283 | 0.073373916 | -1.41E-05 | 0.042428101 | 1 | 0.777745586 | 1 | 0.774133008 | ENSG000001 | 10 | RAD51C |
| Pancreas | T2DM | admiss | ENSG0000018 | 0.000382656 | 0.886222716 | 0.00267432 | 0.073378476 | 0.000349951 | 0.895888006 | 0.00267429 | 0.073374004 | 3.27E-05 | 0.049052533 | 1 | 0.972507979 | 1 | 0.972260921 | ENSG000001 | 10 | RAD51C |
| Pancreas | T2DM | admiss | ENSG0000018 | 0.001659556 | 0.526768803 | 0.002621953 | 0.073378419 | 0.001777593 | 0.497835785 | 0.002622208 | 0.073373937 | -0.00011804 | 0.004870276 | 1 | 0.829394412 | 1 | 0.809993529 | ENSG000001 | 10 | RAD51C |
| Pancreas | T2DM | admiss | ENSG0000019 | 0.000358391 | 0.855126195 | 0.001962904 | 0.073378474 | 0.000297651 | 0.879480236 | 0.001963033 | 0.073374003 | 6.07E-05 | 0.025879969 | 1 | 0.965328541 | 1 | 0.971621772 | ENSG000001 | 10 | SEPT4 |
| Pancreas | T2DM | admiss | ENSG0000019 | 0.001031803 | 0.474614715 | 0.001443098 | 0.073378402 | 0.000437064 | 0.763064398 | 0.001449828 | 0.073373993 | 0.000594739 | 2.15E-05 | 1 | 0.795049308 | 1 | 0.928625715 | ENSG000001 | 5 | PNPO |
| Pancreas | T2DM | admiss | ENSG0000019 | -0.00185703 | 0.240447462 | 0.001581973 | 0.073378272 | -0.00197132 | 0.213009266 | 0.001582961 | 0.073373773 | 0.000114285 | 0.045959099 | 1 | 0.629908248 | 1 | 0.601107468 | ENSG000001 | 5 | PNPO |
| Pancreas | T2DM | admiss | ENSG0000019 | 0.000959125 | 0.549017417 | 0.001600587 | 0.073378425 | 0.000942038 | 0.556151683 | 0.00160056 | 0.073373954 | 1.71E-05 | 0.041900534 | 1 | 0.841622385 | 1 | 0.846590114 | ENSG000001 | 10 | CDK5RAP3 |
| Pancreas | T2DM | admiss | ENSG0000019 | -0.00407232 | 0.004082908 | 0.001418088 | 0.073377237 | -0.00129242 | 0.438313627 | 0.00166754 | 0.073373916 | -0.0027799 | 0.001532861 | 1 | 0.068540999 | 1 | 0.774133008 | ENSG000001 | 10 | CDK5RAP3 |
| Pancreas | T2DM | admiss | ENSG0000019 | 0.001391234 | 0.656105889 | 0.003124289 | 0.073378449 | 0.002043153 | 0.514087596 | 0.003131333 | 0.073373942 | -0.00065192 | 0.002037763 | 1 | 0.890058068 | 1 | 0.81978735 | ENSG000001 | 10 | CBX1 |
| Pancreas | T2DM | admiss | ENSG0000019 | 0.000994091 | 0.665183629 | 0.002297054 | 0.073378451 | 0.001028966 | 0.654187676 | 0.002297043 | 0.073373976 | -3.49E-05 | 0.034234378 | 1 | 0.891995883 | 1 | 0.887614942 | ENSG000001 | 10 | RECQL5 |
| Pancreas | T2DM | admiss | ENSG0000019 | -0.00167443 | 0.595632887 | 0.003155185 | 0.073378437 | -0.00179332 | 0.569809217 | 0.003155397 | 0.073373958 | 0.000118886 | 0.006989081 | 1 | 0.859482295 | 1 | 0.849574534 | ENSG000001 | 16 | CAMTA2 |
| Pancreas | T2DM | admiss | ENSG0000019 | 0.000505656 | 0.83758849 | 0.002466865 | 0.073378473 | 0.000540025 | 0.826717578 | 0.002466841 | 0.073373999 | -3.44E-05 | 0.029561521 | 1 | 0.958120026 | 1 | 0.954095567 | ENSG000001 | 16 | CAMTA2 |
| Pancreas | T2DM | admiss | ENSG0000019 | -0.00203257 | 0.547961988 | 0.003383003 | 0.073378425 | -0.00198267 | 0.557826136 | 0.003382969 | 0.073373955 | -4.99E-05 | 0.020934737 | 1 | 0.841622385 | 1 | 0.847111521 | ENSG000001 | 7 | CHRNE |
| Pancreas | T2DM | admiss | ENSG0000019 | -0.0017525 | 0.492852859 | 0.002555484 | 0.073378408 | -0.00178499 | 0.484864798 | 0.002555456 | 0.073373933 | 3.25E-05 | 0.040603296 | 1 | 0.805630899 | 1 | 0.803311485 | ENSG000001 | 7 | CHRNE |
| Pancreas | T2DM | admiss | ENSG0000019 | -0.0010128 | 0.634733048 | 0.00213188 | 0.073378445 | -0.00097444 | 0.647612238 | 0.002131863 | 0.073373975 | -3.84E-05 | 0.00759989 | 1 | 0.881724325 | 1 | 0.884341453 | ENSG000001 | 7 | CHRNE |
| Pancreas | T2DM | admiss | ENSG0000019 | 0.001272656 | 0.637472543 | 0.002700677 | 0.073378446 | 0.001312714 | 0.62691626 | 0.002700657 | 0.073373971 | -4.01E-05 | 0.029114354 | 1 | 0.8820202 | 1 | 0.877200348 | ENSG000001 | 7 | CHRNE |
| Pancreas | T2DM | admiss | ENSG0000019 | 0.002742997 | 0.146827949 | 0.00189064 | 0.073378162 | 0.00296845 | 0.116647311 | 0.001891931 | 0.073373636 | -0.00022545 | 0.001595194 | 1 | 0.517931557 | 1 | 0.469231995 | ENSG000001 | 7 | CHRNE |
| Pancreas | T2DM | admiss | ENSG0000019 | 0.000852029 | 0.306421206 | 0.000833067 | 0.073378322 | 0.000837567 | 0.314707286 | 0.000833073 | 0.073373854 | 1.45E-05 | 0.043797303 | 1 | 0.702883653 | 1 | 0.706943978 | ENSG000001 | 12 | FTSJ3 |
| Pancreas | T2DM | admiss | ENSG0000019 | -0.00136799 | 0.3861076 | 0.001578393 | 0.073378366 | -0.00190875 | 0.228176841 | 0.001583936 | 0.073373788 | 0.000540763 | 4.77E-05 | 1 | 0.737741307 | 1 | 0.615685781 | ENSG000001 | 13 | ALDH3A1 |
| Pancreas | T2DM | admiss | ENSG0000019 | 0.000916581 | 0.671003449 | 0.00215782 | 0.073378452 | 0.001302128 | 0.546891501 | 0.002161477 | 0.073373952 | -0.00038555 | 0.002362922 | 1 | 0.894197754 | 1 | 0.845484919 | ENSG000001 | 13 | ALDH3A1 |
| Pancreas | T2DM | admiss | ENSG0000019 | -0.00080593 | 0.702028259 | 0.002106545 | 0.073378457 | -0.00086284 | 0.682099615 | 0.002106549 | 0.073373981 | 5.69E-05 | 0.000784608 | 1 | 0.908467044 | 1 | 0.898858571 | ENSG000001 | 3 | SYNGR2 |
| Pancreas | T2DM | admiss | ENSG0000020 | -0.0023891 | 0.295437642 | 0.002283449 | 0.073378314 | -0.00233134 | 0.30726755 | 0.002283449 | 0.07337385 | -5.78E-05 | 0.001229837 | 1 | 0.689712604 | 1 | 0.703146938 | ENSG000001 | 3 | SYNGR2 |
| Pancreas | T2DM | admiss | ENSG0000020 | 0.004242144 | 0.035177935 | 0.002014017 | 0.073377811 | 0.004199828 | 0.037042898 | 0.00201402 | 0.073373352 | 4.23E-05 | 0.008453073 | 1 | 0.260336995 | 1 | 0.266843516 | ENSG000001 | 3 | SYNGR2 |
| Pancreas | T2DM | admiss | ENSG0000020 | 0.004385182 | 0.006614914 | 0.001614785 | 0.073377369 | 0.004411657 | 0.00629459 | 0.001614785 | 0.073372883 | -2.65E-05 | 0.037203192 | 1 | 0.094172779 | 1 | 0.092368612 | ENSG000001 | 7 | DDX5 |
| Pancreas | T2DM | admiss | ENSG0000020 | 0.000532127 | 0.67993856 | 0.001289857 | 0.073378454 | 0.00024968 | 0.847078568 | 0.001294703 | 0.073374001 | 0.000282447 | 0.011949475 | 1 | 0.899069186 | 1 | 0.961211992 | ENSG000001 | 7 | DDX5 |
| Pancreas | T2DM | admiss | ENSG0000020 | -0.0110728 | 2.48E-09 | 0.001857009 | 0.073373126 | -0.0101633 | 7.43E-08 | 0.001888914 | 0.073369648 | -0.0009095 | 0.008562442 | 6.38E-06 | 7.08E-07 | 0.000190837 | 1.59E-05 | ENSG000001 | 5 | C17orf75 |
| Pancreas | T2DM | admiss | ENSG0000020 | 0.012376063 | 3.16E-05 | 0.002973581 | 0.073375871 | 0.010503577 | 0.000504287 | 0.003019578 | 0.073372185 | 0.001872486 | 0.000365934 | 0.0810249 | 0.001929164 | 1 | 0.015792794 | ENSG000001 | 5 | C17orf75 |
| Pancreas | T2DM | admiss | ENSG0000020 | -0.0005438 | 0.499071758 | 0.0008045 | 0.07337841 | -0.00168716 | 0.046979143 | 0.000849317 | 0.073373412 | 0.00114336 | 2.69E-05 | 1 | 0.809101183 | 1 | 0.292821452 | ENSG000001 | 12 | CYTH1 |
| Pancreas | T2DM | admiss | ENSG0000020 | -0.0041664 | 0.187677349 | 0.003162394 | 0.073378218 | -0.00364194 | 0.25043345 | 0.00316884 | 0.073373808 | -0.00052446 | 0.009960072 | 1 | 0.567450741 | 1 | 0.641749029 | ENSG000001 | 12 | CYTH1 |
| Pancreas | T2DM | admiss | ENSG0000020 | 0.00073224 | 0.461820734 | 0.000995091 | 0.073378398 | 0.000520711 | 0.601949236 | 0.000998296 | 0.073373966 | 0.00021153 | 0.008432103 | 1 | 0.785921567 | 1 | 0.86327687 | ENSG000001 | 5 | PEX12 |
| Pancreas | T2DM | admiss | ENSG0000020 | -0.0020878 | 0.002153586 | 0.000680461 | 0.073377062 | -0.00173166 | 0.012864226 | 0.000696139 | 0.073373075 | -0.00035614 | 0.015399722 | 1 | 0.044962667 | 1 | 0.145219373 | ENSG000001 | 8 | DHX58 |
| Pancreas | T2DM | admiss | ENSG0000020 | 0.004537075 | 0.003642473 | 0.001560427 | 0.073377206 | 0.004170265 | 0.007653994 | 0.001563669 | 0.073372936 | 0.000366809 | 0.000295025 | 1 | 0.064286245 | 1 | 0.107944917 | ENSG000001 | 10 | RAB5C |
| Pancreas | T2DM | admiss | ENSG0000020 | 0.002817196 | 0.405820216 | 0.003389017 | 0.073378375 | 0.003303269 | 0.330541954 | 0.003394849 | 0.073373864 | -0.00048607 | 0.015425547 | 1 | 0.75562027 | 1 | 0.707173382 | ENSG000001 | 10 | RAB5C |
| Pancreas | T2DM | admiss | ENSG0000020 | -0.00264315 | 0.031072721 | 0.001225869 | 0.073377779 | -0.00237958 | 0.052901089 | 0.001229293 | 0.073373442 | -0.00026357 | 0.004245147 | 1 | 0.234690436 | 1 | 0.310159807 | ENSG000001 | 6 | HSD17B1 |
| Pancreas | T2DM | admiss | ENSG0000020 | 0.006737938 | 0.000160248 | 0.001785047 | 0.073376334 | 0.007550476 | 3.04E-05 | 0.001810418 | 0.073371388 | -0.00081254 | 0.007207645 | 0.411516019 | 0.006974848 | 0.078039375 | 0.002053668 | ENSG000001 | 11 | SGCA |

| | | | | | | | | | | | | | | | | | | | | |
|---|---|---|---|---|---|---|---|---|---|---|---|---|---|---|---|---|---|---|---|---|
| Pancreas | T2DM | admiss | ENSG000002( | 0.000298897 | 0.885087476 | 0.002068159 | 0.073378476 | -0.00097117 | 0.641560578 | 0.002086209 | 0.073373974 | 0.001270065 | 3.66E-06 | 1 | 0.972507979 | 1 | 0.883436051 | ENSG000001( | 11 | SGCA |
| Pancreas | T2DM | admiss | ENSG000002( | -0.00014606 | 0.876154576 | 0.00093721 | 0.073378476 | 0.000133859 | 0.886728139 | 0.000939723 | 0.073374003 | -0.00027992 | 5.27E-05 | 1 | 0.970211524 | 1 | 0.972260921 | ENSG000001( | 11 | ABCC3 |
| Pancreas | T2DM | admiss | ENSG000002( | -0.00035743 | 0.702576148 | 0.000936068 | 0.073378457 | -0.00050979 | 0.586470357 | 0.000937182 | 0.073373962 | 0.000152355 | 0.000990901 | 1 | 0.908467044 | 1 | 0.855368726 | ENSG000001( | 11 | ABCC3 |
| Pancreas | T2DM | admiss | ENSG000002( | -0.00024166 | 0.885713049 | 0.001681314 | 0.073378476 | -0.00027853 | 0.86842409 | 0.001681307 | 0.073374002 | 3.69E-05 | 0.002483861 | 1 | 0.972507979 | 1 | 0.967511091 | ENSG000001( | 11 | ABCC3 |
| Pancreas | T2DM | admiss | ENSG000002( | -0.0007482 | 0.737430515 | 0.002231727 | 0.073378462 | -0.00082202 | 0.712648875 | 0.002231921 | 0.073373986 | 7.38E-05 | 0.031125456 | 1 | 0.919728783 | 1 | 0.912759257 | ENSG000001( | 11 | ABCC3 |
| Pancreas | T2DM | admiss | ENSG000002( | -0.00013395 | 0.943332844 | 0.001884512 | 0.073378478 | -0.00016564 | 0.929960694 | 0.0018845 | 0.073374005 | 3.17E-05 | 0.01508656 | 1 | 0.982923376 | 1 | 0.981537798 | ENSG000001( | 9 | HLF |
| Pancreas | T2DM | admiss | ENSG000002( | 0.001824245 | 0.436675823 | 0.002345332 | 0.073378388 | 0.001872159 | 0.424737537 | 0.002345388 | 0.073373911 | -4.79E-05 | 0.04980182 | 1 | 0.77336794 | 1 | 0.768116897 | ENSG000001( | 9 | HLF |
| Pancreas | T2DM | admiss | ENSG000002( | 0.00652243 | 0.025289363 | 0.002915755 | 0.073377726 | 0.006633862 | 0.022908236 | 0.002916023 | 0.073373227 | -0.00011143 | 0.01463471 | 1 | 0.207485894 | 1 | 0.20026655 | ENSG000001( | 9 | HLF |
| Pancreas | T2DM | admiss | ENSG000002( | 0.002478199 | 0.054465726 | 0.001288635 | 0.073377922 | 0.002067117 | 0.109755553 | 0.001292517 | 0.073373621 | 0.000411082 | 4.37E-05 | 1 | 0.311854896 | 1 | 0.455334832 | ENSG000001( | 9 | HLF |
| Pancreas | T2DM | admiss | ENSG000002( | 0.007140656 | 7.30E-08 | 0.001326319 | 0.073374115 | 0.006293701 | 3.28E-06 | 0.001352828 | 0.073370748 | 0.000846955 | 0.00149171 | 0.000187355 | 1.17E-05 | 0.008433577 | 0.000351399 | ENSG000001( | 10 | MYH3 |
| Pancreas | T2DM | admiss | ENSG000002( | 0.000248047 | 0.83440465 | 0.001186512 | 0.073378473 | 0.001369545 | 0.257110768 | 0.001208513 | 0.073373813 | -0.0011215 | 1.05E-06 | 1 | 0.958120026 | 1 | 0.651827256 | ENSG000001( | 9 | TMEM97 |
| Pancreas | T2DM | admiss | ENSG0000021 | 0.004665016 | 0.004083634 | 0.001624512 | 0.07337237 | 0.004644 | 0.004253357 | 0.001624488 | 0.073372776 | 2.10E-05 | 0.02094745 | 1 | 0.068540999 | 1 | 0.073306173 | ENSG000001( | 10 | SLAIN2 |
| Pancreas | T2DM | admiss | ENSG0000021 | 0.003597034 | 0.000648504 | 0.001054696 | 0.073376728 | 0.003583132 | 0.000680445 | 0.001054681 | 0.073372269 | 1.39E-05 | 0.021130899 | 1 | 0.018300632 | 1 | 0.019202006 | ENSG000001( | 8 | KIAA1211 |
| Pancreas | T2DM | admiss | ENSG0000021 | 7.77E-05 | 0.974178753 | 0.002399588 | 0.073378479 | 0.000573046 | 0.811540648 | 0.002403324 | 0.073373998 | -0.00049538 | 0.000250056 | 1 | 0.991496562 | 1 | 0.945273278 | ENSG000001( | 8 | KIAA1211 |
| Pancreas | T2DM | admiss | ENSG0000021 | 0.004774112 | 5.33E-06 | 0.001048959 | 0.07337536 | 0.004333867 | 4.20E-05 | 0.001057978 | 0.07337148 | 0.000440245 | 0.001428732 | 0.013695565 | 0.000456519 | 0.107791619 | 0.002629064 | ENSG000001( | 8 | KIAA1211 |
| Pancreas | T2DM | admiss | ENSG0000021 | 0.003712728 | 0.045172001 | 0.001853533 | 0.073377875 | 0.001999338 | 0.297669492 | 0.001919772 | 0.073373843 | 0.00171339 | 0.000612779 | 1 | 0.285670854 | 1 | 0.694291785 | ENSG000001( | 6 | AREG |
| Pancreas | T2DM | admiss | ENSG0000021 | 0.003622807 | 0.335452838 | 0.00376126 | 0.073378339 | 0.003679286 | 0.327967749 | 0.003761218 | 0.073373862 | -5.65E-05 | 0.016009956 | 1 | 0.71074304 | 1 | 0.706943978 | ENSG000001( | 6 | AREG |
| Pancreas | T2DM | admiss | ENSG0000021 | -0.00114302 | 0.633282967 | 0.002395711 | 0.073378445 | -0.00349638 | 0.183262466 | 0.002627316 | 0.07337374 | 0.002353355 | 0.029143891 | 1 | 0.881247101 | 1 | 0.565681828 | ENSG000001( | 15 | UBE2D3 |
| Pancreas | T2DM | admiss | ENSG0000021 | 0.000110261 | 0.936012659 | 0.001373411 | 0.073378478 | 8.53E-05 | 0.95047603 | 0.001373393 | 0.073374006 | 2.50E-05 | 0.001681946 | 1 | 0.980909228 | 1 | 0.983433635 | ENSG000001( | 9 | CPE |
| Pancreas | T2DM | admiss | ENSG0000022 | 0.01276871 | 6.03E-06 | 0.002821584 | 0.073375396 | 0.011155718 | 0.000104495 | 0.002875228 | 0.07337174 | 0.001612993 | 0.003546609 | 0.015486148 | 0.000499553 | 0.268344037 | 0.005063095 | ENSG000001( | 6 | ANXA10 |
| Pancreas | T2DM | admiss | ENSG0000022 | 0.001187277 | 0.651534008 | 0.002628857 | 0.073378448 | 0.001134444 | 0.666078159 | 0.002628845 | 0.073373979 | 5.28E-05 | 0.005173609 | 1 | 0.887189213 | 1 | 0.88982611 | ENSG000001( | 6 | ANXA10 |
| Pancreas | T2DM | admiss | ENSG0000022 | 0.000619143 | 0.765440962 | 0.002075268 | 0.073378466 | 0.00094586 | 0.648792561 | 0.002076803 | 0.073373975 | -0.00032672 | 6.07E-05 | 1 | 0.927632086 | 1 | 0.884341453 | ENSG000001( | 11 | LRP2BP |
| Pancreas | T2DM | admiss | ENSG0000022 | 0.002285495 | 0.134399433 | 0.001526738 | 0.073378142 | 0.001864976 | 0.22510162 | 0.001537395 | 0.073373785 | 0.000420519 | 0.020220019 | 1 | 0.495887562 | 1 | 0.614988604 | ENSG000001( | 6 | UFSP2 |
| Pancreas | T2DM | admiss | ENSG0000022 | 0.000857858 | 0.635178724 | 0.001808107 | 0.073378445 | 0.000661097 | 0.714763334 | 0.001808913 | 0.073373986 | 0.000196761 | 0.000420779 | 1 | 0.881724325 | 1 | 0.913596366 | ENSG000001( | 9 | FAM149A |
| Pancreas | T2DM | admiss | ENSG0000022 | -0.00302392 | 0.323354576 | 0.003061922 | 0.073378332 | -0.0030627 | 0.317182574 | 0.003061887 | 0.073373856 | 3.88E-05 | 0.03942211 | 1 | 0.707406446 | 1 | 0.706943978 | ENSG000001( | 9 | FAM149A |
| Pancreas | T2DM | admiss | ENSG0000022 | 0.001801145 | 0.405352681 | 0.002164577 | 0.073378375 | 0.00183394 | 0.39685092 | 0.002164555 | 0.073373898 | -3.28E-05 | 0.0177652 | 1 | 0.755550584 | 1 | 0.751558379 | ENSG000001( | 9 | HTATIP2 |
| Pancreas | T2DM | admiss | ENSG0000022 | 0.002761997 | 0.385701212 | 0.003184077 | 0.073378366 | 0.002718837 | 0.393163783 | 0.003184043 | 0.073373897 | 4.32E-05 | 0.030678623 | 1 | 0.737741307 | 1 | 0.748994506 | ENSG000001( | 9 | HTATIP2 |
| Pancreas | T2DM | admiss | ENSG0000022 | -0.00030711 | 0.923154932 | 0.003183843 | 0.073378478 | -0.00035883 | 0.91026364 | 0.00318381 | 0.073374005 | 5.17E-05 | 0.010815264 | 1 | 0.978091879 | 1 | 0.97369587 | ENSG000001( | 9 | HTATIP2 |
| Pancreas | T2DM | admiss | ENSG0000022 | 0.008236264 | 0.02524878 | 0.003680877 | 0.073377725 | 0.008182086 | 0.026224706 | 0.003680858 | 0.073373263 | 5.42E-05 | 0.037850709 | 1 | 0.207485894 | 1 | 0.213574989 | ENSG000001( | 11 | CTSC |
| Pancreas | T2DM | admiss | ENSG0000022 | -0.00509553 | 0.1632908 | 0.003655088 | 0.073378187 | -0.0051749 | 0.156828245 | 0.00365505 | 0.073373705 | 7.94E-05 | 0.000572724 | 1 | 0.541071967 | 1 | 0.534414035 | ENSG000001( | 6 | ZNF259 |
| Pancreas | T2DM | admiss | ENSG0000022 | -0.00141475 | 0.484298592 | 0.002022789 | 0.073378406 | -0.00186034 | 0.358739915 | 0.002027021 | 0.07337388 | 0.000445587 | 0.000727207 | 1 | 0.799279425 | 1 | 0.721412765 | ENSG0000011 | 9 | DNAJC4 |
| Pancreas | T2DM | admiss | ENSG0000023 | -0.00120797 | 0.536214322 | 0.001952917 | 0.073378422 | -0.00123403 | 0.527453138 | 0.001952886 | 0.073373946 | 2.61E-05 | 0.014256611 | 1 | 0.833893716 | 1 | 0.829727017 | ENSG0000011 | 9 | DNAJC4 |
| Pancreas | T2DM | admiss | ENSG0000023 | -0.00015012 | 0.859672685 | 0.000849153 | 0.073378474 | -0.00036018 | 0.672505194 | 0.00085208 | 0.07337398 | 0.000210062 | 0.003039582 | 1 | 0.965890282 | 1 | 0.893214295 | ENSG0000011 | 9 | DNAJC4 |
| Pancreas | T2DM | admiss | ENSG0000023 | 0.000982269 | 0.665870759 | 0.002274707 | 0.073378451 | 0.001019303 | 0.654076866 | 0.002274691 | 0.073373976 | -3.70E-05 | 0.017281355 | 1 | 0.891995883 | 1 | 0.887614942 | ENSG0000011 | 8 | SNX15 |
| Pancreas | T2DM | admiss | ENSG0000023 | 0.007558535 | 0.029965683 | 0.003482313 | 0.07337777 | 0.00750734 | 0.031094687 | 0.003482287 | 0.073373307 | 5.12E-05 | 0.029588519 | 1 | 0.229023436 | 1 | 0.238361663 | ENSG0000011 | 8 | SNX15 |
| Pancreas | T2DM | admiss | ENSG0000023 | 0.004849205 | 0.024123182 | 0.002150263 | 0.073377713 | 0.004153519 | 0.054272724 | 0.002158055 | 0.073373449 | 0.000695687 | 0.00015575 | 1 | 0.203396248 | 1 | 0.314289811 | ENSG0000011 | 8 | SNX15 |
| Pancreas | T2DM | admiss | ENSG0000023 | -0.00059999 | 0.72576652 | 0.001710517 | 0.073378461 | -0.00100873 | 0.555883942 | 0.001712712 | 0.073373954 | 0.000408745 | 3.17E-06 | 1 | 0.915856719 | 1 | 0.846590114 | ENSG0000011 | 8 | SNX15 |
| Pancreas | T2DM | admiss | ENSG0000023 | -0.00321257 | 0.39340759 | 0.0037642 | 0.07337837 | -0.00314362 | 0.403635017 | 0.003764153 | 0.073373902 | -6.90E-05 | 0.002210346 | 1 | 0.746137882 | 1 | 0.755027733 | ENSG0000011 | 8 | SNX15 |
| Pancreas | T2DM | admiss | ENSG0000023 | 0.002584962 | 0.200640549 | 0.002019927 | 0.073378233 | 0.002553805 | 0.206115434 | 0.002019902 | 0.073373766 | 3.12E-05 | 0.010648358 | 1 | 0.580230776 | 1 | 0.590082981 | ENSG0000011 | 8 | SNX15 |
| Pancreas | T2DM | admiss | ENSG0000023 | 0.007807652 | 0.000831084 | 0.002336047 | 0.073376797 | 0.007748022 | 0.00091124 | 0.002336128 | 0.07337235 | 5.96E-05 | 0.025509518 | 1 | 0.021777795 | 1 | 0.023400641 | ENSG0000011 | 3 | DTX4 |
| Pancreas | T2DM | admiss | ENSG0000023 | -0.00284169 | 0.078909509 | 0.001617307 | 0.07337801 | -0.0040936 | 0.012634859 | 0.001641441 | 0.07337307 | 0.00125191 | 8.23E-06 | 1 | 0.375066777 | 1 | 0.144849635 | ENSG0000011 | 4 | PUS3 |
| Pancreas | T2DM | admiss | ENSG0000023 | 0.005618334 | 1.61E-07 | 0.001072342 | 0.073374346 | 0.004756387 | 1.85E-05 | 0.001110842 | 0.073371246 | 0.000861947 | 0.002956379 | 0.000414074 | 2.44E-05 | 0.047619377 | 0.001443011 | ENSG0000011 | 6 | FOXRED1 |
| Pancreas | T2DM | admiss | ENSG0000023 | -0.00285079 | 0.380341944 | 0.003249631 | 0.073378363 | -0.00289595 | 0.372837916 | 0.003249603 | 0.073373887 | 4.52E-05 | 0.035752333 | 1 | 0.733271857 | 1 | 0.730903468 | ENSG0000011 | 6 | FOXRED1 |
| Pancreas | T2DM | admiss | ENSG0000023 | 0.00157975 | 0.02181034 | 0.000688746 | 0.073377687 | 0.001397517 | 0.04292998 | 0.000690338 | 0.073373389 | 0.000182234 | 0.000111023 | 1 | 0.193802603 | 1 | 0.28152064 | ENSG0000011 | 6 | FOXRED1 |
| Pancreas | T2DM | admiss | ENSG0000023 | 0.001402766 | 0.42064851 | 0.001741923 | 0.073378382 | 0.001969061 | 0.259263181 | 0.001745411 | 0.073373815 | -0.00056629 | 3.49E-07 | 1 | 0.764941163 | 1 | 0.654717006 | ENSG0000011 | 6 | HPX |
| Pancreas | T2DM | admiss | ENSG0000023 | -0.00261277 | 0.388306589 | 0.003028619 | 0.073378367 | -0.00192018 | 0.526856329 | 0.003034356 | 0.073373946 | -0.00069259 | 0.00022912 | 1 | 0.740290513 | 1 | 0.829556081 | ENSG0000011 | 6 | HPX |
| Pancreas | T2DM | admiss | ENSG0000023 | -0.00328404 | 0.028940954 | 0.00150348 | 0.073377761 | -0.00151617 | 0.349563965 | 0.001620811 | 0.073373875 | -0.00176788 | 0.00350673 | 1 | 0.224532842 | 1 | 0.717365997 | ENSG0000011 | 4 | FOLR3 |
| Pancreas | T2DM | admiss | ENSG0000023 | 0.001587137 | 0.590737164 | 0.002951339 | 0.073378436 | 0.00154316 | 0.60106265 | 0.002951308 | 0.073373965 | 4.40E-05 | 0.018899362 | 1 | 0.858891717 | 1 | 0.863271188 | ENSG0000011 | 7 | SLC1A2 |
| Pancreas | T2DM | admiss | ENSG0000023 | -0.00116548 | 0.483969799 | 0.001665136 | 0.073378405 | -0.00118563 | 0.476439186 | 0.001665116 | 0.07337393 | 2.01E-05 | 0.047188342 | 1 | 0.799250446 | 1 | 0.79570852 | ENSG0000011 | 9 | ACCS |
| Pancreas | T2DM | admiss | ENSG0000023 | 0.001051561 | 0.38571983 | 0.001212305 | 0.073378366 | 0.001535191 | 0.207526096 | 0.00121802 | 0.073373767 | -0.00048363 | 4.31E-05 | 1 | 0.737741307 | 1 | 0.591483923 | ENSG0000011 | 9 | ACCS |
| Pancreas | T2DM | admiss | ENSG0000023 | 0.001199136 | 0.573616619 | 0.002130913 | 0.073378432 | 0.001237935 | 0.56127787 | 0.002130899 | 0.073373956 | -3.88E-05 | 0.008655094 | 1 | 0.851343964 | 1 | 0.848858403 | ENSG0000011 | 9 | ACCS |
| Pancreas | T2DM | admiss | ENSG0000023 | 0.001416926 | 0.272984895 | 0.001292562 | 0.073378298 | 0.00101676 | 0.432577123 | 0.001295583 | 0.073373914 | 0.000400166 | 6.90E-06 | 1 | 0.670309634 | 1 | 0.771947117 | ENSG0000011 | 9 | ACCS |
| Pancreas | T2DM | admiss | ENSG0000024 | -0.00144304 | 0.274664401 | 0.001320996 | 0.073378 | -0.00099079 | 0.459087121 | 0.001338269 | 0.073373924 | -0.00045225 | 0.035049813 | 1 | 0.670309634 | 1 | 0.78659086 | ENSG0000011 | 9 | SLC22A18 |
| Pancreas | T2DM | admiss | ENSG0000024 | 0.003433994 | 0.194362162 | 0.002650626 | 0.073378226 | 0.002718742 | 0.306309483 | 0.002657623 | 0.073373849 | 0.000721197 | 0.000198375 | 1 | 0.572793457 | 1 | 0.703146938 | ENSG0000011 | 9 | SLC22A18 |
| Pancreas | T2DM | admiss | ENSG0000024 | 0.003681853 | 0.288530856 | 0.003469025 | 0.07337831 | 0.003630344 | 0.295324612 | 0.00346899 | 0.073373842 | 5.15E-05 | 0.019931289 | 1 | 0.682535343 | 1 | 0.690198968 | ENSG0000011 | 10 | SLC35F2 |
| Pancreas | T2DM | admiss | ENSG0000024 | -0.00282547 | 0.262564234 | 0.002521952 | 0.07337829 | -0.00279065 | 0.26848617 | 0.00252192 | 0.073373822 | -3.48E-05 | 0.020203822 | 1 | 0.660396625 | 1 | 0.663592381 | ENSG0000011 | 7 | C11orf21 |
| Pancreas | T2DM | admiss | ENSG0000024 | 0.001364017 | 0.395037134 | 0.001603748 | 0.07337837 | 0.001384045 | 0.388127538 | 0.001603725 | 0.073373894 | -2.00E-05 | 0.030689306 | 1 | 0.747572115 | 1 | 0.742706048 | ENSG0000011 | 2 | RPS13 |
| Pancreas | T2DM | admiss | ENSG0000024 | -0.00044217 | 0.591109809 | 0.000823058 | 0.073378436 | -0.00047371 | 0.56496383 | 0.000823156 | 0.073373957 | 3.15E-05 | 0.02633781 | 1 | 0.858891717 | 1 | 0.849476828 | ENSG0000011 | 2 | RPS13 |
| Pancreas | T2DM | admiss | ENSG0000024 | 0.001414357 | 0.559975355 | 0.002426504 | 0.073378428 | 0.001379469 | 0.569691413 | 0.002426479 | 0.073373958 | 3.49E-05 | 0.023842904 | 1 | 0.847939375 | 1 | 0.849574534 | ENSG0000011 | 2 | RPS13 |
| Pancreas | T2DM | admiss | ENSG0000024 | -0.00490974 | 0.038377557 | 0.002370935 | 0.07337833 | -0.00487528 | 0.03975451 | 0.002370903 | 0.0737337 | -3.45E-05 | 0.012725844 | 1 | 0.265643038 | 1 | 0.275174077 | ENSG0000011 | 5 | AIP |
| Pancreas | T2DM | admiss | ENSG0000024 | -0.00483314 | 0.099291852 | 0.002932197 | 0.0733787 | -0.00478039 | 0.103033072 | 0.002932168 | 0.073373606 | -5.27E-05 | 0.004771574 | 1 | 0.426555097 | 1 | 0.436615395 | ENSG0000011 | 5 | AIP |
| Pancreas | T2DM | admiss | ENSG0000024 | 0.005980814 | 7.37E-07 | 0.001207902 | 0.073374788 | 0.005638929 | 3.27E-06 | 0.001211877 | 0.073370747 | 0.000341884 | 0.000517412 | 0.001892709 | 8.60E-05 | 0.008400931 | 0.000351399 | ENSG0000011 | 5 | AIP |
| Pancreas | T2DM | admiss | ENSG0000024 | 0.004908529 | 0.003499632 | 0.00168096 | 0.073377195 | 0.004942627 | 0.003278413 | 0.001680957 | 0.073372705 | -3.41E-05 | 0.007593206 | 1 | 0.062610489 | 1 | 0.061006986 | ENSG0000011 | 7 | GTF2H1 |
| Pancreas | T2DM | admiss | ENSG0000024 | -0.00254148 | 0.506056265 | 0.003821827 | 0.073378413 | -0.00248223 | 0.516019944 | 0.003821799 | 0.073373943 | -5.92E-05 | 0.022989616 | 1 | 0.814511821 | 1 | 0.820393199 | ENSG0000011 | 7 | GTF2H1 |
| Pancreas | T2DM | admiss | ENSG0000024 | 0.002228509 | 0.018507697 | 0.000946164 | 0.073377644 | 0.002215334 | 0.019211204 | 0.000946152 | 0.073373181 | 1.32E-05 | 0.017753188 | 1 | 0.181403685 | 1 | 0.183399155 | ENSG0000011 | 7 | GTF2H1 |
| Pancreas | T2DM | admiss | ENSG0000024 | -0.00234346 | 0.284763599 | 0.002190801 | 0.073378307 | -0.00236553 | 0.280242719 | 0.002190762 | 0.073373831 | 2.21E-05 | 0.047871865 | 1 | 0.679621676 | 1 | 0.675106288 | ENSG0000011 | 7 | GTF2H1 |
| Pancreas | T2DM | admiss | ENSG0000024 | -0.00017775 | 0.949581547 | 0.002811054 | 0.073378479 | -0.000211 | 0.940164101 | 0.00281101 | 0.073374006 | 3.33E-05 | 0.029637988 | 1 | 0.984069981 | 1 | 0.98183872 | ENSG0000011 | 13 | CAPRIN2 |
| Pancreas | T2DM | admiss | ENSG0000024 | 0.001586434 | 0.530148662 | 0.002527056 | 0.07337842 | 0.001643509 | 0.515452482 | 0.00252703 | 0.073373943 | -5.71E-05 | 0.000380367 | 1 | 0.831148818 | 1 | 0.820125138 | ENSG0000011 | 13 | CAPRIN2 |
| Pancreas | T2DM | admiss | ENSG0000025 | 0.005405326 | 0.078302408 | 0.003070116 | 0.073378012 | 0.00536552 | 0.08051921 | 0.003070073 | 0.073373547 | 3.98E-05 | 0.022905949 | 1 | 0.373062306 | 1 | 0.383122792 | ENSG0000011 | 13 | CAPRIN2 |
| Pancreas | T2DM | admiss | ENSG0000025 | 0.001703719 | 0.138964611 | 0.001151426 | 0.07337815 | 0.001560457 | 0.175779794 | 0.001152593 | 0.073373731 | 0.000143262 | 0.006490227 | 1 | 0.500672845 | 1 | 0.556849121 | ENSG0000011 | 13 | CAPRIN2 |
| Pancreas | T2DM | admiss | ENSG0000025 | 0.008654302 | 0.022835912 | 0.003802123 | 0.073377699 | 0.008708424 | 0.021996359 | 0.003802078 | 0.073373217 | -5.41E-05 | 0.019545775 | 1 | 0.198567397 | 1 | 0.195455536 | ENSG0000011 | 8 | KCTD10 |
| Pancreas | T2DM | admiss | ENSG0000025 | 0.001251422 | 0.56120821 | 0.002153731 | 0.073378428 | 0.001223865 | 0.569858343 | 0.002153705 | 0.073373958 | 2.76E-05 | 0.035138289 | 1 | 0.847939375 | 1 | 0.849574534 | ENSG0000011 | 4 | IL23A |
| Pancreas | T2DM | admiss | ENSG0000025 | 0.009501762 | 0.003804359 | 0.00328331 | 0.073377218 | 0.009461067 | 0.003956686 | 0.003283259 | 0.073372756 | 4.07E-05 | 0.023239385 | 1 | 0.065967883 | 1 | 0.069941797 | ENSG0000011 | 3 | ACSS3 |
| Pancreas | T2DM | admiss | ENSG0000025 | -0.01616367 | 1.26E-06 | 0.003335971 | 0.073374944 | -0.01610807 | 1.38E-06 | 0.003335953 | 0.073370496 | -5.56E-05 | 0.01827569 | 0.00324801 | 0.00012992 | 0.003531518 | 0.000196195 | ENSG0000011 | 3 | ACSS3 |
| Pancreas | T2DM | admiss | ENSG0000025 | 0.002963488 | 0.007774102 | 0.001113366 | 0.07337412 | 0.00294359 | 0.008196261 | 0.001113357 | 0.073372954 | 1.99E-05 | 0.00734396 | 1 | 0.105629064 | 1 | 0.109202996 | ENSG0000011 | 9 | METAP2 |
| Pancreas | T2DM | admiss | ENSG0000025 | 0.005365271 | 0.152004775 | 0.00374542 | 0.07337817 | 0.005497829 | 0.142151007 | 0.003745549 | 0.073373682 | -0.00013256 | 0.001884796 | 1 | 0.528211449 | 1 | 0.50983769 | ENSG0000011 | 5 | LTA4H |
| Pancreas | T2DM | admiss | ENSG0000025 | 0.003370121 | 0.168698597 | 0.002448505 | 0.073378194 | 0.003404631 | 0.164375237 | 0.00244848 | 0.073373715 | -3.45E-05 | 0.027409372 | 1 | 0.548084313 | 1 | 0.544665301 | ENSG0000011 | 5 | LTA4H |
| Pancreas | T2DM | admiss | ENSG0000025 | 0.005887506 | 0.000670265 | 0.001730864 | 0.073376737 | 0.005865702 | 0.000701718 | 0.001730843 | 0.073372277 | 2.18E-05 | 0.036143935 | 1 | 0.018709131 | 1 | 0.019587077 | ENSG0000011 | 7 | PRR4 |
| Pancreas | T2DM | admiss | ENSG0000025 | -0.00629598 | 0.073580993 | 0.003518853 | 0.073377997 | -0.00624822 | 0.075789099 | 0.003518809 | 0.073373532 | -4.78E-05 | 0.022601575 | 1 | 0.360189008 | 1 | 0.369310068 | ENSG0000011 | 7 | PRR4 |
| Pancreas | T2DM | admiss | ENSG0000025 | -0.00914132 | 0.013493795 | 0.00370023 | 0.07337756 | -0.00919555 | 0.012949858 | 0.003700193 | 0.073373077 | 5.42E-05 | 0.022568635 | 1 | 0.151982746 | 1 | 0.145219373 | ENSG0000011 | 3 | AKAP3 |
| Pancreas | T2DM | admiss | ENSG0000025 | -0.00080796 | 0.495778682 | 0.001186173 | 0.073378409 | -0.00082233 | 0.488137766 | 0.001186153 | 0.073373934 | 1.44E-05 | 0.021330542 | 1 | 0.807330155 | 1 | 0.804739043 | ENSG0000011 | 6 | LTBR |
| Pancreas | T2DM | admiss | ENSG0000025 | -0.00229936 | 0.404582407 | 0.002758798 | 0.073378375 | -0.00234232 | 0.395858541 | 0.002758777 | 0.073373898 | 4.30E-05 | 0.020857451 | 1 | 0.755550584 | 1 | 0.751558379 | ENSG0000011 | 6 | LTBR |
| Pancreas | T2DM | admiss | ENSG0000025 | 0.004306323 | 0.138388984 | 0.002906105 | 0.073378149 | 0.004260332 | 0.142646874 | 0.002906078 | 0.073373683 | 4.60E-05 | 0.014338475 | 1 | 0.500672845 | 1 | 0.510902611 | ENSG0000011 | 5 | MGP |
| Pancreas | T2DM | admiss | ENSG0000025 | 0.003784703 | 0.001036275 | 0.001153694 | 0.073376859 | 0.00376819 | 0.001090039 | 0.001153688 | 0.0733724 | 1.65E-05 | 0.043144875 | 1 | 0.025105219 | 1 | 0.02640773 | ENSG0000011 | 5 | MGP |
| Pancreas | T2DM | admiss | ENSG0000026 | 0.005951037 | 0.004161691 | 0.002076678 | 0.073377243 | 0.005980856 | 0.003976442 | 0.002076659 | 0.073372758 | -2.98E-05 | 0.027088373 | 1 | 0.069397554 | 1 | 0.069941797 | ENSG0000011 | 5 | MGP |
| Pancreas | T2DM | admiss | ENSG0000026 | -0.00237108 | 0.52952777 | 0.003771229 | 0.07337842 | -0.00241424 | 0.522053642 | 0.00377117 | 0.073373945 | 4.32E-05 | 0.036204643 | 1 | 0.830895322 | 1 | 0.825005386 | ENSG0000011 | 11 | DDX55 |
| Pancreas | T2DM | admiss | ENSG0000026 | 0.0025675 | 0.278159768 | 0.00236753 | 0.073378302 | 0.002626444 | 0.267274443 | 0.002367526 | 0.073373821 | -5.89E-05 | 0.001060649 | 1 | 0.671348012 | 1 | 0.663456699 | ENSG0000011 | 9 | STX2 |
| Pancreas | T2DM | admiss | ENSG0000026 | 0.000834519 | 0.675658697 | 0.001994574 | 0.073378453 | 0.000807321 | 0.68565198 | 0.001994552 | 0.073373982 | 2.72E-05 | 0.027921227 | 1 | 0.896564018 | 1 | 0.900181127 | ENSG0000011 | 9 | STX2 |
| Pancreas | T2DM | admiss | ENSG0000026 | -0.00093403 | 0.442208679 | 0.001215444 | 0.07337839 | -0.00084471 | 0.487136558 | 0.001215632 | 0.073373934 | -8.93E-05 | 0.000133821 | 1 | 0.776526409 | 1 | 0.804739043 | ENSG0000011 | 9 | STX2 |
| Pancreas | T2DM | admiss | ENSG0000026 | -0.00029996 | 0.928729401 | 0.003353612 | 0.073378478 | -0.00034601 | 0.917823956 | 0.00335359 | 0.073374005 | 4.60E-05 | 0.046572996 | 1 | 0.979054639 | 1 | 0.978402623 | ENSG0000011 | 8 | CAND1 |
| Pancreas | T2DM | admiss | ENSG0000026 | -0.00262032 | 0.149372924 | 0.001817456 | 0.073378166 | -0.00267678 | 0.140812229 | 0.001817509 | 0.0737368 | 5.65E-05 | 0.004550966 | 1 | 0.521890706 | 1 | 0.506450707 | ENSG0000011 | 8 | CAND1 |
| Pancreas | T2DM | admiss | ENSG0000026 | 0.006264612 | 0.007872722 | 0.002357341 | 0.073377416 | 0.006219012 | 0.008335485 | 0.002357311 | 0.073372959 | 4.56E-05 | 0.001222627 | 1 | 0.105848958 | 1 | 0.109202996 | ENSG0000011 | 8 | KRR1 |
| Pancreas | T2DM | admiss | ENSG0000026 | -0.0017689 | 0.273717428 | 0.001616112 | 0.073378299 | -0.00174665 | 0.279792741 | 0.001616088 | 0.073373831 | -2.23E-05 | 0.013676918 | 1 | 0.670309634 | 1 | 0.674655172 | ENSG0000011 | 8 | KRR1 |
| Pancreas | T2DM | admiss | ENSG0000026 | 0.000616758 | 0.772131875 | 0.002129782 | 0.073378467 | 0.000577944 | 0.786109031 | 0.002129753 | 0.073373995 | 3.88E-05 | 0.001782414 | 1 | 0.930781647 | 1 | 0.936079501 | ENSG0000011 | 3 | MRPL51 |
| Pancreas | T2DM | admiss | ENSG0000026 | -0.00056159 | 0.517818092 | 0.000868371 | 0.073378416 | -0.00057403 | 0.508576716 | 0.000868358 | 0.073373941 | 1.24E-05 | 0.00836034 | 1 | 0.821852201 | 1 | 0.817366177 | ENSG0000011 | 3 | MRPL51 |
| Pancreas | T2DM | admiss | ENSG0000027 | 0.00855494 | 0.000209505 | 0.002307594 | 0.07337641 | 0.006221137 | 0.00987817 | 0.002411223 | 0.073373004 | 0.002333803 | 0.000848257 | 0.538010056 | 0.008674644 | 1 | 0.122546576 | ENSG0000011 | 6 | GNB3 |
| Pancreas | T2DM | admiss | ENSG0000027 | -0.00195492 | 0.002925999 | 0.000657022 | 0.073377146 | -0.00174323 | 0.008286746 | 0.000660314 | 0.073372957 | -0.00021168 | 0.001258796 | 1 | 0.054846454 | 1 | 0.109202996 | ENSG0000011 | 6 | CHPT1 |
| Pancreas | T2DM | admiss | ENSG0000027 | -0.00192463 | 0.261526622 | 0.001714144 | 0.073378289 | -0.00189683 | 0.268472279 | 0.001714121 | 0.073373822 | -2.78E-05 | 0.005462602 | 1 | 0.659077885 | 1 | 0.663592381 | ENSG0000011 | 6 | CHPT1 |
| Pancreas | T2DM | admiss | ENSG0000027 | -0.00090555 | 0.304166799 | 0.000881278 | 0.07337832 | -0.00078027 | 0.377100893 | 0.000883409 | 0.073373889 | -0.00012528 | 0.042288775 | 1 | 0.701687643 | 1 | 0.733034226 | ENSG0000011 | 6 | CHPT1 |
| Pancreas | T2DM | insulin | ENSG0000001 | 0.000677951 | 0.472376495 | 0.000943409 | 0.011571118 | 0.000692136 | 0.461792539 | 0.000943421 | 0.011570676 | -1.63E-05 | 0.032153147 | 1 | 0.890734922 | 1 | 0.886973253 | ENSG0000011 | 10 | ATN1 |
| Pancreas | T2DM | insulin | ENSG0000003 | 0.003374198 | 0.001355836 | 0.00105315 | 0.011570886 | 0.003063946 | 0.00380025 | 0.001058615 | 0.01157049 | 0.000310252 | 0.003935307 | 1 | 0.112315715 | 1 | 0.17977037 | ENSG0000011 | 7 | NT5DC3 |
| Pancreas | T2DM | insulin | ENSG0000003 | -0.00012045 | 0.837862935 | 0.000588621 | 0.011571129 | -0.0001109 | 0.850560643 | 0.000588623 | 0.011570688 | -9.55E-06 | 0.014793264 | 1 | 0.97568398 | 1 | 0.979057591 | ENSG0000011 | 6 | PRKAB1 |
| Pancreas | T2DM | insulin | ENSG0000003 | 0.000853033 | 0.313424833 | 0.00084621 | 0.011571106 | 0.000839097 | 0.321401734 | 0.00084622 | 0.011570666 | 1.39E-05 | 0.035926996 | 1 | 0.835397773 | 1 | 0.835647835 | ENSG0000011 | 6 | PRKAB1 |
| Pancreas | T2DM | insulin | ENSG0000004 | 0.000319651 | 0.607041406 | 0.000621524 | 0.011571123 | -0.00222718 | 0.086607146 | 0.001299724 | 0.011570619 | 0.002546827 | 0.025672092 | 1 | 0.927884944 | 1 | 0.596265823 | ENSG0000011 | 6 | PRKAB1 |
| Pancreas | T2DM | insulin | ENSG0000006 | 0.000890588 | 0.161068483 | 0.000635458 | 0.011571083 | 0.002385046 | 0.001884275 | 0.000767412 | 0.01157046 | -0.00149446 | 0.000514159 | 1 | 0.6998417 | 1 | 0.15121308 | ENSG0000011 | 4 | SRSF9 |
| Pancreas | T2DM | insulin | ENSG0000007 | -0.00193749 | 0.058432517 | 0.001023802 | 0.011571045 | -0.00191928 | 0.060843298 | 0.001023815 | 0.011570606 | -1.82E-05 | 0.025526452 | 1 | 0.530577607 | 1 | 0.538777894 | ENSG0000011 | 9 | RP11-22B23.1 |
| Pancreas | T2DM | insulin | ENSG0000007 | 0.000359666 | 0.748460766 | 0.001121609 | 0.011571127 | 0.000297058 | 0.791173844 | 0.001121877 | 0.011570687 | 6.26E-05 | 0.014034239 | 1 | 0.965153689 | 1 | 0.967426237 | ENSG0000011 | 11 | BTN3A3 |
| Pancreas | T2DM | insulin | ENSG0000008 | -0.00090316 | 0.326022556 | 0.000919571 | 0.011571107 | -0.00088865 | 0.33385835 | 0.000919574 | 0.011570667 | -1.45E-05 | 0.018139914 | 1 | 0.835554813 | 1 | 0.837432686 | ENSG0000011 | 11 | BTN3A3 |
| Pancreas | T2DM | insulin | ENSG0000009 | -0.00062069 | 0.549231371 | 0.00103636 | 0.011571121 | -0.0006331 | 0.541272577 | 0.001036359 | 0.01157068 | 1.24E-05 | 0.04913899 | 1 | 0.907039349 | 1 | 0.902870944 | ENSG0000011 | 11 | BTN3A3 |
| Pancreas | T2DM | insulin | ENSG000001( | -0.00146472 | 0.222060991 | 0.001199542 | 0.011571094 | -0.00144665 | 0.227818602 | 0.001199546 | 0.011570654 | -1.81E-05 | 0.024631637 | 1 | 0.763006274 | 1 | 0.769631524 | ENSG0000011 | 11 | BTN3A3 |
| Pancreas | T2DM | insulin | ENSG000001( | -5.10E-05 | 0.956094245 | 0.000927008 | 0.01157113 | -3.69E-05 | 0.968222456 | 0.000927012 | 0.011570689 | -1.41E-05 | 0.027327601 | 1 | 0.989356544 | 1 | 0.988759842 | ENSG0000011 | 11 | BTN3A3 |
| Pancreas | T2DM | insulin | ENSG000001( | 0.000687813 | 0.554531102 | 0.001163842 | 0.011571121 | 0.000674273 | 0.562351858 | 0.00116384 | 0.011570681 | 1.35E-05 | 0.045479932 | 1 | 0.910322362 | 1 | 0.908714845 | ENSG0000011 | 11 | BTN3A3 |
| Pancreas | T2DM | insulin | ENSG000001( | -0.00241737 | 0.073708268 | 0.001351676 | 0.011571054 | -0.00239237 | 0.076740818 | 0.001351683 | 0.011570615 | -2.50E-05 | 0.00775162 | 1 | 0.582072897 | 1 | 0.587816164 | ENSG0000011 | 6 | SMIM8 |
| Pancreas | T2DM | insulin | ENSG000001( | -5.31E-05 | 0.942970551 | 0.000742259 | 0.01157113 | -6.18E-05 | 0.933595466 | 0.000742257 | 0.011570689 | 8.75E-06 | 0.039572284 | 1 | 0.989356544 | 1 | 0.988759842 | ENSG0000011 | 6 | SMIM8 |
| Pancreas | T2DM | insulin | ENSG000001( | -0.00065402 | 0.422403375 | 0.00081522 | 0.011571114 | -0.00066452 | 0.414515403 | 0.00081522 | 0.011570673 | 1.12E-05 | 0.028935205 | 1 | 0.870194075 | 1 | 0.86606442 | ENSG0000011 | 14 | FAM184A |
| Pancreas | T2DM | insulin | ENSG000001( | 0.000720445 | 0.577459361 | 0.001293205 | 0.011571122 | 0.000735661 | 0.569446292 | 0.001293203 | 0.011570681 | -1.52E-05 | 0.04711356 | 1 | 0.917471363 | 1 | 0.911776571 | ENSG0000011 | 14 | FAM184A |
| Pancreas | T2DM | insulin | ENSG0000001 | 8.53E-05 | 0.893664944 | 0.000638207 | 0.011571129 | 7.67E-05 | 0.904284137 | 0.000638209 | 0.011570689 | 8.56E-06 | 0.045145999 | 1 | 0.988687703 | 1 | 0.988759842 | ENSG0000011 | 14 | FAM184A |
| Pancreas | T2DM | insulin | ENSG0000001 | -0.00014576 | 0.840464294 | 0.000724078 | 0.011571129 | 0.000562059 | 0.45344639 | 0.000749729 | 0.011570689 | -0.00070782 | 0.000273279 | 1 | 0.97568398 | 1 | 0.880237961 | ENSG0000011 | 9 | GABRR2 |
| Pancreas | T2DM | insulin | ENSG0000001 | 8.42E-05 | 0.949012333 | 0.001316218 | 0.01157113 | 5.85E-05 | 0.964526293 | 0.001316244 | 0.011570689 | 2.56E-05 | 0.026498293 | 1 | 0.989356544 | 1 | 0.988759842 | ENSG0000011 | 11 | MTRF1L |
| Pancreas | T2DM | insulin | ENSG0000001 | 0.000460789 | 0.459806838 | 0.000623391 | 0.011571117 | 0.000468749 | 0.452092016 | 0.000623392 | 0.011570676 | -7.96E-06 | 0.046586257 | 1 | 0.886474445 | 1 | 0.880222754 | ENSG0000011 | 11 | MTRF1L |
| Pancreas | T2DM | insulin | ENSG000001: | -0.00106655 | 0.189633519 | 0.000813127 | 0.011571089 | -0.00105675 | 0.193733067 | 0.000813126 | 0.011570649 | -9.80E-06 | 0.039219247 | 1 | 0.728954236 | 1 | 0.731489476 | ENSG0000011 | 11 | MTRF1L |
| Pancreas | T2DM | insulin | ENSG000001: | -0.0011419 | 0.206432723 | 0.000903804 | 0.011571092 | -0.00115469 | 0.201395516 | 0.000903804 | 0.01157065 | 1.28E-05 | 0.022256801 | 1 | 0.751302486 | 1 | 0.743080008 | ENSG0000011 | 11 | MTRF1L |
| Pancreas | T2DM | insulin | ENSG000001: | -0.00197009 | 0.102831746 | 0.001207697 | 0.011571067 | -0.00195334 | 0.105793116 | 0.001207704 | 0.011570627 | -1.68E-05 | 0.047479641 | 1 | 0.616990479 | 1 | 0.614110331 | ENSG0000011 | 8 | TULP1 |
| Pancreas | T2DM | insulin | ENSG000001: | -9.45E-05 | 0.871457312 | 0.00058412 | 0.011571129 | -7.52E-05 | 0.897576686 | 0.000584143 | 0.011570689 | -1.93E-05 | 0.002457475 | 1 | 0.984420735 | 1 | 0.988759842 | ENSG0000011 | 8 | TULP1 |
| Pancreas | T2DM | insulin | ENSG000001: | -0.00045355 | 0.435728843 | 0.000581905 | 0.011571115 | -0.00046251 | 0.426721697 | 0.000581907 | 0.011570674 | 8.96E-06 | 0.021752904 | 1 | 0.875147029 | 1 | 0.870390246 | ENSG0000011 | 9 | SLC26A8 |
| Pancreas | T2DM | insulin | ENSG000001: | -0.00079546 | 0.039229923 | 0.000385811 | 0.011571029 | -0.00083396 | 0.030782473 | 0.000386116 | 0.011570578 | 3.85E-05 | 0.013122436 | 1 | 0.489907645 | 1 | 0.440318545 | ENSG0000011 | 9 | SLC26A8 |
| Pancreas | T2DM | insulin | ENSG000001: | 0.000481531 | 0.306509772 | 0.000470901 | 0.011571105 | 0.000440285 | 0.349996527 | 0.000471096 | 0.011570668 | 4.12E-05 | 0.002917185 | 1 | 0.835397773 | 1 | 0.842353403 | ENSG0000011 | 9 | SLC26A8 |

| | | | | | | | | | | | | | | | | | | | | |
|---|---|---|---|---|---|---|---|---|---|---|---|---|---|---|---|---|---|---|---|---|
| Pancreas | T2DM | insulin | ENSG0000012 | 0.000398035 | 0.769763499 | 0.001359947 | 0.011571128 | 0.000374569 | 0.782989929 | 0.001359966 | 0.011570687 | 2.35E-05 | 0.034821821 | 1 | 0.965153689 | 1 | 0.965819649 | ENSG0000011 | 9 | SLC26A8 |
| Pancreas | T2DM | insulin | ENSG0000012 | -7.03E-05 | 0.953470156 | 0.001205159 | 0.01157113 | -5.32E-05 | 0.964801822 | 0.001205161 | 0.011570689 | -1.71E-05 | 0.026347094 | 1 | 0.989356544 | 1 | 0.988759842 | ENSG0000011 | 9 | SLC26A8 |
| Pancreas | T2DM | insulin | ENSG0000013 | -0.0027324 | 0.06173312 | 0.001462574 | 0.011571047 | -0.00271071 | 0.063830074 | 0.001462586 | 0.011570607 | -2.17E-05 | 0.04391999 | 1 | 0.546657422 | 1 | 0.551424497 | ENSG0000011 | 9 | SLC26A8 |
| Pancreas | T2DM | insulin | ENSG0000013 | 0.000927343 | 0.232406889 | 0.00077655 | 0.011571096 | 0.000915733 | 0.238309974 | 0.000776557 | 0.011570656 | 1.16E-05 | 0.04379621 | 1 | 0.7689997 | 1 | 0.772107424 | ENSG0000011 | 5 | MRPL18 |
| Pancreas | T2DM | insulin | ENSG0000013 | -0.00047414 | 0.754841312 | 0.00151839 | 0.011571127 | -0.00045374 | 0.765067848 | 0.001518391 | 0.011570687 | -2.04E-05 | 0.032408532 | 1 | 0.965153689 | 1 | 0.965703051 | ENSG0000011 | 5 | MRPL18 |
| Pancreas | T2DM | insulin | ENSG0000013 | 4.41E-05 | 0.959939999 | 0.000877127 | 0.01157113 | 5.65E-05 | 0.9486546 | 0.00087713 | 0.011570689 | -1.24E-05 | 0.03272454 | 1 | 0.989356544 | 1 | 0.988759842 | ENSG0000011 | 5 | MRPL18 |
| Pancreas | T2DM | insulin | ENSG0000013 | 0.000774585 | 0.180378962 | 0.000578225 | 0.011571087 | 0.000726038 | 0.209522079 | 0.000578571 | 0.011570652 | 4.85E-05 | 0.016905538 | 1 | 0.724903247 | 1 | 0.751903832 | ENSG0000011 | 12 | RNF8 |
| Pancreas | T2DM | insulin | ENSG0000013 | 0.001506417 | 0.217619212 | 0.001221863 | 0.011571094 | 0.001522156 | 0.212849766 | 0.001221861 | 0.011570652 | -1.57E-05 | 0.02777792 | 1 | 0.762155249 | 1 | 0.756014105 | ENSG0000011 | 12 | RNF8 |
| Pancreas | T2DM | insulin | ENSG0000013 | 0.001367293 | 0.050467852 | 0.000699031 | 0.011571039 | 0.001376961 | 0.048861496 | 0.000699035 | 0.011570597 | -9.67E-06 | 0.046741574 | 1 | 0.5082124 | 1 | 0.5082859 | ENSG0000011 | 12 | RNF8 |
| Pancreas | T2DM | insulin | ENSG0000013 | 0.000170579 | 0.90230481 | 0.00138964 | 0.011571129 | 0.000144982 | 0.91690734 | 0.001389649 | 0.011570689 | 2.56E-05 | 0.009253046 | 1 | 0.988687703 | 1 | 0.988759842 | ENSG0000011 | 5 | CD83 |
| Pancreas | T2DM | insulin | ENSG0000013 | 0.001427332 | 0.330023899 | 0.001465332 | 0.011571107 | 0.00144328 | 0.324646724 | 0.001465326 | 0.011570666 | -1.59E-05 | 0.043422461 | 1 | 0.836625244 | 1 | 0.835647835 | ENSG0000011 | 5 | CD83 |
| Pancreas | T2DM | insulin | ENSG0000013 | 0.001064439 | 0.467696755 | 0.0014657 | 0.011571117 | 0.001043381 | 0.476550317 | 0.001465706 | 0.011570677 | 2.11E-05 | 0.03433609 | 1 | 0.890322658 | 1 | 0.890022702 | ENSG0000011 | 5 | CD83 |
| Pancreas | T2DM | insulin | ENSG0000013 | 0.000253472 | 0.866281934 | 0.001505329 | 0.011571129 | 0.000276873 | 0.854070934 | 0.001505343 | 0.011570688 | -2.34E-05 | 0.037577549 | 1 | 0.982534385 | 1 | 0.980006326 | ENSG0000011 | 5 | CD83 |
| Pancreas | T2DM | insulin | ENSG0000014 | 9.40E-05 | 0.930093101 | 0.001071089 | 0.01157113 | 0.000109095 | 0.918872859 | 0.001071091 | 0.011570689 | -1.51E-05 | 0.030655343 | 1 | 0.988687703 | 1 | 0.988759842 | ENSG0000011 | 7 | MDN1 |
| Pancreas | T2DM | insulin | ENSG0000014 | -0.00156662 | 0.022897871 | 0.00068858 | 0.011571007 | -0.00157938 | 0.021810216 | 0.000688585 | 0.011570564 | 1.28E-05 | 0.011236409 | 1 | 0.407644283 | 1 | 0.397935172 | ENSG0000011 | 7 | GPR63 |
| Pancreas | T2DM | insulin | ENSG0000014 | -0.00129922 | 0.248947073 | 0.001126903 | 0.011571098 | -0.00128062 | 0.255792428 | 0.001126915 | 0.011570658 | -1.86E-05 | 0.030326606 | 1 | 0.785376024 | 1 | 0.792479149 | ENSG0000011 | 8 | WASF1 |
| Pancreas | T2DM | insulin | ENSG0000014 | -0.00227725 | 0.116967903 | 0.001452672 | 0.011571071 | -0.00230266 | 0.112942433 | 0.001452682 | 0.011570629 | 2.54E-05 | 0.01571211 | 1 | 0.637653295 | 1 | 0.627975408 | ENSG0000011 | 8 | WASF1 |
| Pancreas | T2DM | insulin | ENSG0000014 | -0.0003883 | 0.628842586 | 0.000803332 | 0.011571124 | -0.00040302 | 0.615893821 | 0.000803344 | 0.011570683 | 1.47E-05 | 0.026668353 | 1 | 0.931641109 | 1 | 0.926645564 | ENSG0000011 | 10 | GPLD1 |
| Pancreas | T2DM | insulin | ENSG0000014 | -0.00035456 | 0.721317064 | 0.000994009 | 0.011571127 | -0.00034369 | 0.729518141 | 0.000994004 | 0.011570686 | -1.09E-05 | 0.039477282 | 1 | 0.957777777 | 1 | 0.960804329 | ENSG0000011 | 10 | GPLD1 |
| Pancreas | T2DM | insulin | ENSG0000014 | -0.00013281 | 0.925304539 | 0.001416631 | 0.01157113 | -0.0002923 | 0.836745186 | 0.001418539 | 0.011570688 | 0.000159487 | 0.031282863 | 1 | 0.988687703 | 1 | 0.974053326 | ENSG0000011 | 8 | ALDH5A1 |
| Pancreas | T2DM | insulin | ENSG0000014 | -0.00035473 | 0.702922111 | 0.00093013 | 0.011571126 | -0.00034075 | 0.714105763 | 0.000930139 | 0.011570686 | -1.40E-05 | 0.046121147 | 1 | 0.951524626 | 1 | 0.953885959 | ENSG0000011 | 8 | ALDH5A1 |
| Pancreas | T2DM | insulin | ENSG0000014 | -8.59E-05 | 0.788995992 | 0.000321106 | 0.011571128 | -9.02E-05 | 0.778706494 | 0.000321105 | 0.011570687 | 4.30E-06 | 0.021881376 | 1 | 0.967242173 | 1 | 0.965819649 | ENSG0000011 | 8 | ALDH5A1 |
| Pancreas | T2DM | insulin | ENSG0000014 | -0.00208725 | 0.039085368 | 0.00101161 | 0.011571029 | -0.00207193 | 0.040547413 | 0.00101162 | 0.011570589 | -1.53E-05 | 0.045924702 | 1 | 0.489907645 | 1 | 0.486639905 | ENSG0000011 | 8 | ALDH5A1 |
| Pancreas | T2DM | insulin | ENSG0000015 | 0.000741091 | 0.423051088 | 0.000925046 | 0.011571115 | 0.000755656 | 0.413999366 | 0.000925055 | 0.011570673 | -1.46E-05 | 0.040951315 | 1 | 0.870194075 | 1 | 0.86606442 | ENSG0000011 | 8 | ALDH5A1 |
| Pancreas | T2DM | insulin | ENSG0000015 | 9.75E-05 | 0.767868664 | 0.000330311 | 0.011571128 | 0.000104315 | 0.752150292 | 0.000330316 | 0.011570687 | -6.82E-06 | 0.009288335 | 1 | 0.965153689 | 1 | 0.965703051 | ENSG0000011 | 7 | VNN1 |
| Pancreas | T2DM | insulin | ENSG0000015 | -0.00041243 | 0.583565062 | 0.000752361 | 0.011571123 | -0.00042778 | 0.569641993 | 0.000752372 | 0.011570681 | 1.54E-05 | 0.014304318 | 1 | 0.920512947 | 1 | 0.911776571 | ENSG0000011 | 8 | C6orf62 |
| Pancreas | T2DM | insulin | ENSG0000015 | -0.00148732 | 0.008917387 | 0.000568716 | 0.011570967 | -0.00149377 | 0.008625334 | 0.000568715 | 0.011570525 | 6.45E-06 | 0.049751873 | 1 | 0.260225561 | 1 | 0.255611981 | ENSG0000011 | 8 | C6orf62 |
| Pancreas | T2DM | insulin | ENSG0000015 | -0.00070218 | 0.614305099 | 0.001393379 | 0.011571124 | -0.00072164 | 0.604523927 | 0.001393379 | 0.011570683 | 1.95E-05 | 0.023719848 | 1 | 0.927999789 | 1 | 0.925226219 | ENSG0000011 | 8 | C6orf62 |
| Pancreas | T2DM | insulin | ENSG0000015 | 0.000394973 | 0.622343253 | 0.000801922 | 0.011571124 | 0.000328408 | 0.682383531 | 0.000802535 | 0.011570685 | 6.66E-05 | 0.035956519 | 1 | 0.928033725 | 1 | 0.945452933 | ENSG0000011 | 8 | C6orf62 |
| Pancreas | T2DM | insulin | ENSG0000015 | -0.00013303 | 0.839989096 | 0.00065885 | 0.011571129 | -0.00012252 | 0.852481609 | 0.000658855 | 0.011570688 | -1.05E-05 | 0.030292028 | 1 | 0.97568398 | 1 | 0.979057591 | ENSG0000011 | 10 | HBS1L |
| Pancreas | T2DM | insulin | ENSG0000016 | -0.00079331 | 0.145739472 | 0.000545327 | 0.01157108 | -0.00078706 | 0.148943172 | 0.000545326 | 0.011570639 | -6.26E-06 | 0.043472696 | 1 | 0.676779319 | 1 | 0.685443348 | ENSG0000011 | 10 | HBS1L |
| Pancreas | T2DM | insulin | ENSG0000016 | 6.05E-05 | 0.913130036 | 0.000554457 | 0.011571129 | 0.000123725 | 0.82366918 | 0.000555247 | 0.011570688 | -6.32E-05 | 0.033864467 | 1 | 0.988687703 | 1 | 0.971625322 | ENSG0000011 | 10 | HBS1L |
| Pancreas | T2DM | insulin | ENSG0000016 | -0.00057724 | 0.544149306 | 0.000951678 | 0.011571121 | -0.00056269 | 0.554348946 | 0.000951679 | 0.011570681 | -1.46E-05 | 0.015472344 | 1 | 0.906527682 | 1 | 0.905957993 | ENSG0000011 | 10 | HBS1L |
| Pancreas | T2DM | insulin | ENSG0000016 | -0.00058271 | 0.615950915 | 0.001161702 | 0.011571124 | -0.000604 | 0.603117613 | 0.001161715 | 0.011570683 | 2.13E-05 | 0.017880102 | 1 | 0.928033725 | 1 | 0.925226219 | ENSG0000011 | 8 | TRIM38 |
| Pancreas | T2DM | insulin | ENSG0000016 | -0.00015832 | 0.659301985 | 0.000359102 | 0.011571125 | -0.00016321 | 0.649464568 | 0.000359102 | 0.011570684 | 4.89E-06 | 0.029950115 | 1 | 0.936442199 | 1 | 0.931745816 | ENSG0000011 | 8 | TRIM38 |
| Pancreas | T2DM | insulin | ENSG0000016 | 0.000875019 | 0.276508514 | 0.0008041 | 0.011571102 | 0.000909836 | 0.257929782 | 0.00080424 | 0.011570659 | -3.48E-05 | 0.027822037 | 1 | 0.821426426 | 1 | 0.795356813 | ENSG0000011 | 8 | TRIM38 |
| Pancreas | T2DM | insulin | ENSG0000016 | 0.000396219 | 0.296304591 | 0.000379378 | 0.011571104 | 0.000401358 | 0.290082959 | 0.000379376 | 0.011570662 | -5.14E-06 | 0.014971436 | 1 | 0.8274401 | 1 | 0.823130431 | ENSG0000011 | 8 | PEX7 |
| Pancreas | T2DM | insulin | ENSG0000016 | -0.00069945 | 0.604085267 | 0.001348895 | 0.011571123 | -0.00071599 | 0.595559729 | 0.001348892 | 0.011570682 | 1.65E-05 | 0.036414576 | 1 | 0.927884944 | 1 | 0.922435093 | ENSG0000011 | 8 | PEX7 |
| Pancreas | T2DM | insulin | ENSG0000017 | -0.00070008 | 0.602995546 | 0.001346063 | 0.011571123 | -0.00071584 | 0.594861051 | 0.001346062 | 0.011570682 | 1.58E-05 | 0.049112677 | 1 | 0.927884944 | 1 | 0.921947305 | ENSG0000011 | 8 | PEX7 |
| Pancreas | T2DM | insulin | ENSG0000017 | 0.000244625 | 0.77599549 | 0.000859715 | 0.011571128 | 0.000256237 | 0.765665939 | 0.000859715 | 0.011570687 | -1.16E-05 | 0.029906896 | 1 | 0.965153689 | 1 | 0.965703051 | ENSG0000011 | 11 | PERP |
| Pancreas | T2DM | insulin | ENSG0000017 | -0.00245092 | 0.103968873 | 0.001507411 | 0.011571067 | -0.00247183 | 0.101052093 | 0.001507417 | 0.011570625 | 2.09E-05 | 0.040035591 | 1 | 0.618037191 | 1 | 0.609159095 | ENSG0000011 | 11 | PERP |
| Pancreas | T2DM | insulin | ENSG0000017 | 7.92E-05 | 0.914975886 | 0.000742021 | 0.011571129 | 9.28E-05 | 0.900483976 | 0.000742024 | 0.011570689 | -1.36E-05 | 0.008020866 | 1 | 0.988687703 | 1 | 0.988759842 | ENSG0000011 | 11 | PERP |
| Pancreas | T2DM | insulin | ENSG0000017 | -6.82E-05 | 0.900490579 | 0.00054532 | 0.011571129 | -5.87E-05 | 0.914246573 | 0.000545326 | 0.011570689 | -9.46E-06 | 0.02618616 | 1 | 0.988687703 | 1 | 0.988759842 | ENSG0000011 | 11 | PERP |
| Pancreas | T2DM | insulin | ENSG0000017 | 0.000447259 | 0.631236839 | 0.000931815 | 0.011571124 | 0.000461325 | 0.620544985 | 0.000931823 | 0.011570683 | -1.41E-05 | 0.040829703 | 1 | 0.931641109 | 1 | 0.926645564 | ENSG0000011 | 12 | SLC39A7 |
| Pancreas | T2DM | insulin | ENSG0000017 | -0.00074561 | 0.39870946 | 0.000883494 | 0.011571113 | -0.00075946 | 0.390010489 | 0.000883502 | 0.011570671 | 1.39E-05 | 0.038046536 | 1 | 0.863490577 | 1 | 0.863051851 | ENSG0000011 | 11 | CUTA |
| Pancreas | T2DM | insulin | ENSG0000018 | -0.00019934 | 0.80267484 | 0.000797732 | 0.011571128 | -0.00021346 | 0.789021254 | 0.000797738 | 0.011570687 | 1.41E-05 | 0.015604522 | 1 | 0.969074255 | 1 | 0.96624062 | ENSG0000011 | 11 | CUTA |
| Pancreas | T2DM | insulin | ENSG0000018 | 0.000508763 | 0.699347272 | 0.001317351 | 0.011571126 | 0.000533293 | 0.685610235 | 0.001317358 | 0.011570685 | -2.45E-05 | 0.008123349 | 1 | 0.949787479 | 1 | 0.94546011 | ENSG0000011 | 9 | FAM120B |
| Pancreas | T2DM | insulin | ENSG0000018 | 0.000139281 | 0.883222766 | 0.000948232 | 0.011571129 | 0.000118184 | 0.900812968 | 0.000948241 | 0.011570689 | 2.11E-05 | 0.002929816 | 1 | 0.985280653 | 1 | 0.988759842 | ENSG0000011 | 8 | GUCA1B |
| Pancreas | T2DM | insulin | ENSG0000018 | 0.000108041 | 0.926300475 | 0.001168006 | 0.01157113 | 8.84E-05 | 0.939694888 | 0.001168019 | 0.011570689 | 1.97E-05 | 0.030398367 | 1 | 0.988687703 | 1 | 0.988759842 | ENSG0000011 | 6 | PRPH2 |
| Pancreas | T2DM | insulin | ENSG0000018 | 0.000512549 | 0.673333068 | 0.001215785 | 0.011571126 | 0.000526241 | 0.665129567 | 0.001215781 | 0.011570684 | -1.37E-05 | 0.041688065 | 1 | 0.941892795 | 1 | 0.937945164 | ENSG0000011 | 6 | PRPH2 |
| Pancreas | T2DM | insulin | ENSG0000018 | 0.001901175 | 0.197992459 | 0.001476877 | 0.01157109 | 0.001880989 | 0.202794812 | 0.001476871 | 0.01157065 | 2.02E-05 | 0.012806424 | 1 | 0.740790404 | 1 | 0.743967252 | ENSG0000011 | 6 | PRPH2 |
| Pancreas | T2DM | insulin | ENSG0000018 | 0.000588463 | 0.610530177 | 0.001155395 | 0.011571124 | 0.000572951 | 0.619970754 | 0.001155395 | 0.011570683 | 1.55E-05 | 0.030124608 | 1 | 0.927884944 | 1 | 0.926645564 | ENSG0000011 | 6 | PRPH2 |
| Pancreas | T2DM | insulin | ENSG0000018 | 3.82E-05 | 0.952543848 | 0.000642555 | 0.01157113 | 4.65E-05 | 0.94231791 | 0.000642554 | 0.011570689 | -8.25E-06 | 0.027275009 | 1 | 0.989356544 | 1 | 0.988759842 | ENSG0000011 | 8 | DNPH1 |
| Pancreas | T2DM | insulin | ENSG0000018 | 0.000365099 | 0.218151951 | 0.000296477 | 0.011571094 | 0.000369444 | 0.212726664 | 0.000296479 | 0.011570652 | -4.34E-06 | 0.041544105 | 1 | 0.762155249 | 1 | 0.756014105 | ENSG0000011 | 8 | DNPH1 |
| Pancreas | T2DM | insulin | ENSG0000019 | -0.00043702 | 0.296561293 | 0.00041867 | 0.011571104 | -0.00044319 | 0.289797491 | 0.000418672 | 0.011570662 | 6.17E-06 | 0.032867775 | 1 | 0.8274401 | 1 | 0.823130431 | ENSG0000011 | 8 | DNPH1 |
| Pancreas | T2DM | insulin | ENSG0000019 | -0.00133815 | 0.019538166 | 0.000573056 | 0.011571 | -0.00152437 | 0.008104227 | 0.000575734 | 0.011570522 | 0.000186227 | 0.000807245 | 1 | 0.377248201 | 1 | 0.255611981 | ENSG0000011 | 8 | DNPH1 |
| Pancreas | T2DM | insulin | ENSG0000019 | 0.000756031 | 0.154831302 | 0.000531416 | 0.011571082 | 0.000764406 | 0.150313497 | 0.000531418 | 0.01157064 | -8.38E-06 | 0.022086703 | 1 | 0.692694745 | 1 | 0.686843532 | ENSG0000011 | 8 | DNPH1 |
| Pancreas | T2DM | insulin | ENSG0000019 | -0.00076725 | 0.173048873 | 0.000563131 | 0.011571086 | 0.000183425 | 0.781783889 | 0.000662194 | 0.011570687 | -0.00095068 | 0.006361222 | 1 | 0.716757267 | 1 | 0.965819649 | ENSG0000011 | 11 | TMEM30A |
| Pancreas | T2DM | insulin | ENSG0000019 | -0.00086479 | 0.485777831 | 0.001240665 | 0.011571118 | -0.0006349 | 0.609641132 | 0.001243477 | 0.011570683 | -0.00022989 | 0.006158154 | 1 | 0.89524472 | 1 | 0.925226219 | ENSG0000011 | 11 | BTN2A1 |
| Pancreas | T2DM | insulin | ENSG0000019 | -0.00284319 | 0.041408159 | 0.001394113 | 0.011571031 | -0.00282632 | 0.042629909 | 0.001394113 | 0.011570591 | -1.69E-05 | 0.048460841 | 1 | 0.490028351 | 1 | 0.494154798 | ENSG0000011 | 11 | BTN2A1 |
| Pancreas | T2DM | insulin | ENSG0000019 | 0.000991668 | 0.186551954 | 0.00075078 | 0.011571088 | 0.001080122 | 0.150528517 | 0.000751301 | 0.01157064 | -8.85E-05 | 0.001815275 | 1 | 0.726940181 | 1 | 0.686843532 | ENSG0000011 | 11 | BTN2A1 |
| Pancreas | T2DM | insulin | ENSG0000019 | -0.00106868 | 0.088191241 | 0.000626784 | 0.011571061 | -0.00125261 | 0.046431271 | 0.000628992 | 0.011570595 | 0.000183931 | 0.000495907 | 1 | 0.597559646 | 1 | 0.504240075 | ENSG0000011 | 11 | BTN2A1 |
| Pancreas | T2DM | insulin | ENSG0000019 | -0.00054368 | 0.525758776 | 0.000856877 | 0.01157112 | -0.00040669 | 0.635631637 | 0.00085834 | 0.011570684 | -0.00013699 | 0.00652133 | 1 | 0.904925293 | 1 | 0.928352537 | ENSG0000011 | 11 | PRSS16 |
| Pancreas | T2DM | insulin | ENSG0000019 | 0.000901025 | 0.281428591 | 0.000836515 | 0.011571102 | 0.000885408 | 0.289858325 | 0.000836526 | 0.011570662 | 1.56E-05 | 0.020310864 | 1 | 0.82273396 | 1 | 0.823130431 | ENSG0000011 | 11 | PRSS16 |
| Pancreas | T2DM | insulin | ENSG0000020 | 0.00223207 | 0.118651269 | 0.00143039 | 0.011571072 | 0.00220401 | 0.123357857 | 0.0014304 | 0.011570633 | 2.81E-05 | 0.006580572 | 1 | 0.637653295 | 1 | 0.639285058 | ENSG0000011 | 11 | PRSS16 |
| Pancreas | T2DM | insulin | ENSG0000020 | 0.000580077 | 0.522354161 | 0.000906766 | 0.01157112 | 0.000598389 | 0.509313394 | 0.000906776 | 0.011570679 | -1.83E-05 | 0.009876355 | 1 | 0.902196499 | 1 | 0.895220396 | ENSG0000011 | 4 | PCDHB2 |
| Pancreas | T2DM | insulin | ENSG0000020 | -0.00138922 | 0.224895834 | 0.001144696 | 0.011571095 | -0.00137479 | 0.229747079 | 0.001144697 | 0.011570655 | -1.44E-05 | 0.04345084 | 1 | 0.763931881 | 1 | 0.769631524 | ENSG0000011 | 5 | NUDT12 |
| Pancreas | T2DM | insulin | ENSG0000020 | 0.001389163 | 2.21E-05 | 0.000327449 | 0.011570702 | 0.001495763 | 6.49E-06 | 0.000331669 | 0.011570206 | -0.0001066 | 0.043441293 | 0.056804435 | 0.005539789 | 0.016667122 | 0.00208339 | ENSG0000011 | 5 | NUDT12 |
| Pancreas | T2DM | insulin | ENSG0000020 | 0.003702295 | 0.001716662 | 0.001180828 | 0.011570896 | 0.003278703 | 0.00625163 | 0.001199104 | 0.011570511 | 0.000423592 | 0.042356411 | 1 | 0.131838761 | 1 | 0.231056694 | ENSG0000011 | 5 | NUDT12 |
| Pancreas | T2DM | insulin | ENSG0000020 | -0.00112008 | 0.002270769 | 0.000366957 | 0.011570909 | -0.00127568 | 0.000602734 | 0.000371874 | 0.01157041 | 0.0001556 | 0.009893663 | 1 | 0.138841298 | 1 | 0.064492585 | ENSG0000011 | 9 | GHR |
| Pancreas | T2DM | insulin | ENSG0000020 | -0.00095894 | 0.002684916 | 0.000319467 | 0.011570916 | -0.00144657 | 1.79E-05 | 0.000337266 | 0.011570252 | 0.00048763 | 6.51E-06 | 1 | 0.155835303 | 0.046073901 | 0.00460739 | ENSG0000011 | 9 | GHR |
| Pancreas | T2DM | insulin | ENSG0000020 | -0.00101639 | 0.418309398 | 0.001255798 | 0.011571114 | -0.00077004 | 0.540583231 | 0.001258372 | 0.01157068 | -0.00024635 | 0.002299716 | 1 | 0.870194075 | 1 | 0.902870944 | ENSG0000011 | 11 | DAP |
| Pancreas | T2DM | insulin | ENSG0000020 | -0.00015433 | 0.567919361 | 0.000270216 | 0.011571122 | 3.08E-05 | 0.911361341 | 0.000276444 | 0.011570689 | -0.0001851 | 0.001519103 | 1 | 0.915166981 | 1 | 0.988759842 | ENSG0000011 | 11 | DAP |
| Pancreas | T2DM | insulin | ENSG0000020 | 0.001536777 | 0.01313623 | 0.000619652 | 0.011570984 | 0.001419019 | 0.02229845 | 0.000620946 | 0.011570565 | 0.000117758 | 0.003431372 | 1 | 0.318243773 | 1 | 0.400436509 | ENSG0000011 | 11 | DAP |
| Pancreas | T2DM | insulin | ENSG0000020 | 0.001438313 | 0.042972564 | 0.000710635 | 0.011571032 | 0.001659498 | 0.020973953 | 0.000718877 | 0.011570562 | -0.00022119 | 0.041746035 | 1 | 0.494856299 | 1 | 0.393901236 | ENSG0000011 | 11 | DAP |
| Pancreas | T2DM | insulin | ENSG0000020 | -0.00137784 | 0.00021222 | 0.000371983 | 0.011570804 | -0.00144172 | 0.000111166 | 0.000373033 | 0.011570334 | 6.39E-05 | 0.022853586 | 0.544979856 | 0.034061241 | 0.285475097 | 0.0202407 | ENSG0000011 | 11 | DAP |
| Pancreas | T2DM | insulin | ENSG0000020 | 0.001057272 | 0.432093627 | 0.00134579 | 0.011571115 | 0.001304215 | 0.333328858 | 0.001348121 | 0.011570667 | -0.00024694 | 0.001942688 | 1 | 0.874402233 | 1 | 0.837432686 | ENSG0000011 | 11 | DAP |
| Pancreas | T2DM | insulin | ENSG0000020 | 0.002218398 | 0.001750739 | 0.000708851 | 0.011570897 | 0.002611841 | 0.000280247 | 0.000718934 | 0.011570376 | -0.00039344 | 0.001051029 | 1 | 0.131838761 | 0.719673768 | 0.039981876 | ENSG0000011 | 11 | DAP |
| Pancreas | T2DM | insulin | ENSG0000020 | 0.00061444 | 0.504711077 | 0.000921066 | 0.011571119 | 0.000442399 | 0.631775017 | 0.000923145 | 0.011570683 | 0.00017204 | 0.005661337 | 1 | 0.895546057 | 1 | 0.928352537 | ENSG0000011 | 11 | DAP |
| Pancreas | T2DM | insulin | ENSG0000020 | -4.63E-05 | 0.955032728 | 0.000821273 | 0.01157113 | -0.00040815 | 0.622251399 | 0.00082845 | 0.011570683 | 0.000361836 | 0.000895046 | 1 | 0.989356544 | 1 | 0.926830112 | ENSG0000011 | 12 | NNT |
| Pancreas | T2DM | insulin | ENSG0000020 | -0.00049215 | 0.1860399 | 0.000372168 | 0.011571088 | -0.00040168 | 0.281745746 | 0.000373171 | 0.011570661 | -9.05E-05 | 0.00097398 | 1 | 0.726940181 | 1 | 0.822541329 | ENSG0000011 | 5 | PFDN1 |
| Pancreas | T2DM | insulin | ENSG0000020 | -0.00100058 | 0.258881352 | 0.000886225 | 0.011571099 | -0.00102742 | 0.246373347 | 0.000886312 | 0.011570657 | 2.68E-05 | 0.048473349 | 1 | 0.796330558 | 1 | 0.782060264 | ENSG0000011 | 8 | TMCO6 |
| Pancreas | T2DM | insulin | ENSG0000020 | 0.000435658 | 0.39457232 | 0.000511723 | 0.011571113 | 0.000313597 | 0.541213485 | 0.00051327 | 0.01157068 | 0.000122061 | 0.00224343 | 1 | 0.863490577 | 1 | 0.902870944 | ENSG0000011 | 8 | TMCO6 |
| Pancreas | T2DM | insulin | ENSG0000020 | 0.0016944 | 0.001295303 | 0.000526696 | 0.011570884 | 0.001399168 | 0.009203064 | 0.000537227 | 0.011570528 | 0.000295232 | 0.005301854 | 1 | 0.110877909 | 1 | 0.262793788 | ENSG0000011 | 11 | IK |
| Pancreas | T2DM | insulin | ENSG0000020 | 0.000512365 | 0.276843782 | 0.000471167 | 0.011571102 | 0.000834925 | 0.081902358 | 0.000479909 | 0.011570617 | -0.00032256 | 0.000406871 | 1 | 0.821426426 | 1 | 0.587816164 | ENSG0000011 | 11 | IK |
| Pancreas | T2DM | insulin | ENSG0000021 | 0.00138007 | 0.000922741 | 0.000416549 | 0.011570869 | 0.001215498 | 0.0038146 | 0.000420135 | 0.01157049 | 0.000164572 | 0.002684418 | 1 | 0.087762956 | 1 | 0.17977037 | ENSG0000011 | 11 | IK |
| Pancreas | T2DM | insulin | ENSG0000021 | -0.00032802 | 0.655854337 | 0.000736047 | 0.011571125 | -0.00103398 | 0.175006705 | 0.000762356 | 0.011570645 | 0.000705963 | 0.000378548 | 1 | 0.936254796 | 1 | 0.717520656 | ENSG0000011 | 3 | PCDHB15 |
| Pancreas | T2DM | insulin | ENSG0000021 | 0.001322319 | 0.099627323 | 0.000803028 | 0.011571065 | 0.001306792 | 0.103671185 | 0.000803036 | 0.011570626 | 1.55E-05 | 0.011649081 | 1 | 0.61011476 | 1 | 0.612137828 | ENSG0000011 | 3 | PCDHB15 |
| Pancreas | T2DM | insulin | ENSG0000022 | -0.00024468 | 0.854145383 | 0.001330988 | 0.011571129 | -0.00026026 | 0.844969341 | 0.001330986 | 0.011570688 | 1.56E-05 | 0.046408055 | 1 | 0.982534385 | 1 | 0.975833338 | ENSG0000011 | 8 | HAVCR1 |
| Pancreas | T2DM | insulin | ENSG0000022 | -0.00072898 | 0.396391311 | 0.000859561 | 0.011571113 | -0.00071679 | 0.404338902 | 0.000859562 | 0.011570672 | -1.22E-05 | 0.026456425 | 1 | 0.863490577 | 1 | 0.86606442 | ENSG0000011 | 8 | HAVCR1 |
| Pancreas | T2DM | insulin | ENSG0000022 | -0.00374075 | 0.009958877 | 0.001451441 | 0.011570972 | -0.00376185 | 0.009547734 | 0.001451442 | 0.011570529 | 2.11E-05 | 0.021101903 | 1 | 0.272068047 | 1 | 0.262793788 | ENSG0000011 | 11 | ARSB |
| Pancreas | T2DM | insulin | ENSG0000022 | -0.0005802 | 0.470102043 | 0.000803256 | 0.011571117 | -0.00073333 | 0.362277231 | 0.000804946 | 0.011570669 | 0.000153131 | 0.00345209 | 1 | 0.890734922 | 1 | 0.846603325 | ENSG0000011 | 9 | MSH3 |
| Pancreas | T2DM | insulin | ENSG0000022 | 0.001081786 | 0.00728235 | 0.000403101 | 0.011570959 | 0.001172173 | 0.003850222 | 0.000405569 | 0.011570491 | -9.04E-05 | 0.043400934 | 1 | 0.242871117 | 1 | 0.17977037 | ENSG0000011 | 14 | CCNG1 |
| Pancreas | T2DM | insulin | ENSG0000023 | -0.00011133 | 0.741269118 | 0.000337201 | 0.011571127 | -0.00019508 | 0.564261259 | 0.000338368 | 0.011570681 | 8.37E-05 | 0.002927554 | 1 | 0.965153689 | 1 | 0.908714845 | ENSG0000011 | 12 | DROSHA |
| Pancreas | T2DM | insulin | ENSG0000023 | 7.65E-05 | 0.932474808 | 0.000903294 | 0.01157113 | 9.16E-05 | 0.919232738 | 0.000903298 | 0.011570689 | -1.51E-05 | 0.014790819 | 1 | 0.988687703 | 1 | 0.988759842 | ENSG0000011 | 8 | CDH6 |
| Pancreas | T2DM | insulin | ENSG0000023 | 0.000412182 | 0.234772494 | 0.000346909 | 0.011571096 | 0.000590833 | 0.094164101 | 0.000352982 | 0.011570622 | -0.00017865 | 0.006170415 | 1 | 0.7689997 | 1 | 0.601947903 | ENSG0000011 | 6 | IL4 |
| Pancreas | T2DM | insulin | ENSG0000023 | -9.29E-05 | 0.913363938 | 0.000853881 | 0.011571129 | -0.00033476 | 0.696072953 | 0.000856984 | 0.011570685 | 0.000241862 | 0.000930295 | 1 | 0.988687703 | 1 | 0.948517485 | ENSG0000011 | 7 | PPWD1 |
| Pancreas | T2DM | insulin | ENSG0000023 | -0.00037589 | 0.579994158 | 0.000679252 | 0.011571122 | -0.00048371 | 0.476962447 | 0.000680131 | 0.011570677 | 0.000107814 | 0.001967259 | 1 | 0.918264486 | 1 | 0.890145032 | ENSG0000011 | 7 | PPWD1 |
| Pancreas | T2DM | insulin | ENSG0000023 | -0.00098273 | 0.125976607 | 0.000642239 | 0.011571074 | -0.00138559 | 0.033528843 | 0.00065183 | 0.011570582 | 0.000402864 | 0.000302131 | 1 | 0.650322205 | 1 | 0.451379305 | ENSG0000011 | 7 | PPWD1 |
| Pancreas | T2DM | insulin | ENSG0000023 | 0.000511484 | 0.459644001 | 0.000691724 | 0.011571117 | 0.000632842 | 0.361223148 | 0.000693117 | 0.011570669 | -0.00012136 | 0.005966695 | 1 | 0.886474445 | 1 | 0.846567342 | ENSG0000011 | 7 | PPWD1 |
| Pancreas | T2DM | insulin | ENSG0000023 | 0.000410877 | 0.534348941 | 0.000661231 | 0.011571121 | 0.000402015 | 0.543201095 | 0.000661231 | 0.01157068 | 8.86E-06 | 0.027953086 | 1 | 0.906527682 | 1 | 0.902870944 | ENSG0000011 | 13 | TRAPPC13 |
| Pancreas | T2DM | insulin | ENSG0000023 | 0.000881945 | 0.066950875 | 0.000481409 | 0.01157105 | 0.001037087 | 0.032021871 | 0.000483684 | 0.01157058 | -0.00015514 | 0.000952699 | 1 | 0.558070981 | 1 | 0.446913945 | ENSG0000011 | 13 | TRAPPC13 |
| Pancreas | T2DM | insulin | ENSG0000023 | 0.000500906 | 0.553882302 | 0.000846192 | 0.011571121 | 0.000517249 | 0.541025791 | 0.000846197 | 0.01157068 | -1.63E-05 | 0.005357183 | 1 | 0.910322362 | 1 | 0.902870944 | ENSG0000011 | 13 | TRAPPC13 |
| Pancreas | T2DM | insulin | ENSG0000024 | -0.0006259 | 0.232803829 | 0.000524572 | 0.011571096 | -0.00038764 | 0.465744497 | 0.000531437 | 0.011570676 | -0.00023826 | 0.005167983 | 1 | 0.7689997 | 1 | 0.887263998 | ENSG0000011 | 13 | TRAPPC13 |
| Pancreas | T2DM | insulin | ENSG0000024 | 0.00073748 | 0.124180246 | 0.000479673 | 0.011571074 | 0.000642816 | 0.181645838 | 0.000481256 | 0.011570647 | 9.47E-05 | 0.015497207 | 1 | 0.646084789 | 1 | 0.720968335 | ENSG0000011 | 13 | BNIP1 |
| Pancreas | T2DM | insulin | ENSG0000024 | 0.000732781 | 0.272306863 | 0.00066752 | 0.011571101 | 0.000745346 | 0.264174748 | 0.000667526 | 0.011570659 | -1.26E-05 | 0.013238708 | 1 | 0.817876053 | 1 | 0.801805531 | ENSG0000011 | 13 | BNIP1 |
| Pancreas | T2DM | insulin | ENSG0000024 | 0.001047328 | 0.079782746 | 0.000597806 | 0.011571057 | 0.001038124 | 0.082469665 | 0.000597813 | 0.011570617 | 9.20E-06 | 0.046400708 | 1 | 0.592408703 | 1 | 0.587816164 | ENSG0000011 | 13 | BNIP1 |
| Pancreas | T2DM | insulin | ENSG0000024 | -0.00240558 | 5.28E-07 | 0.000479573 | 0.011570532 | -0.00261199 | 9.79E-08 | 0.000489997 | 0.011570014 | 0.000206407 | 0.040158114 | 0.001354727 | 0.000169341 | 0.000251466 | 3.59E-05 | ENSG0000011 | 13 | BNIP1 |
| Pancreas | T2DM | insulin | ENSG0000024 | 0.001923681 | 0.157214993 | 0.00135997 | 0.011571082 | 0.001941872 | 0.153328016 | 0.001359972 | 0.011570641 | -1.82E-05 | 0.037674289 | 1 | 0.696168424 | 1 | 0.688839634 | ENSG0000011 | 13 | BNIP1 |
| Pancreas | T2DM | insulin | ENSG0000025 | 0.000708597 | 0.121203792 | 0.000457235 | 0.011571073 | 0.000649578 | 0.155838316 | 0.000457704 | 0.011570641 | 5.90E-05 | 0.004746466 | 1 | 0.63982072 | 1 | 0.689930464 | ENSG0000011 | 14 | NPHP3 |
| Pancreas | T2DM | insulin | ENSG0000025 | -0.00093116 | 0.276264737 | 0.000855253 | 0.011571102 | -0.00094227 | 0.270572115 | 0.000855252 | 0.01157066 | 1.11E-05 | 0.03235337 | 1 | 0.821426426 | 1 | 0.815055965 | ENSG0000011 | 14 | NPHP3 |
| Pancreas | T2DM | insulin | ENSG0000025 | -0.00148242 | 0.274865253 | 0.001357612 | 0.011571101 | -0.00146015 | 0.282138242 | 0.001357614 | 0.011570661 | -2.23E-05 | 0.00995234 | 1 | 0.821426426 | 1 | 0.822541329 | ENSG0000011 | 14 | NPHP3 |
| Pancreas | T2DM | insulin | ENSG0000025 | 0.00196412 | 0.000250097 | 0.000536324 | 0.011570811 | 0.002186259 | 5.74E-05 | 0.000543408 | 0.011570305 | -0.00022214 | 0.011151126 | 0.642248041 | 0.037403048 | 0.147433792 | 0.011341061 | ENSG0000011 | 14 | NPHP3 |
| Pancreas | T2DM | insulin | ENSG0000025 | 0.00010261 | 0.89962266 | 0.000813471 | 0.011571129 | 9.36E-05 | 0.908368668 | 0.000813468 | 0.011570689 | 8.98E-06 | 0.04834875 | 1 | 0.988687703 | 1 | 0.988759842 | ENSG0000011 | 7 | NIT2 |
| Pancreas | T2DM | insulin | ENSG0000025 | -0.000431 | 0.360181542 | 0.000471033 | 0.01157111 | -0.00043594 | 0.354703278 | 0.000471031 | 0.011570669 | 4.94E-06 | 0.046428851 | 1 | 0.848639286 | 1 | 0.843405573 | ENSG0000011 | 11 | FAM162A |
| Pancreas | T2DM | insulin | ENSG0000026 | 8.61E-05 | 0.93123212 | 0.00099809 | 0.01157113 | 7.04E-05 | 0.943778525 | 0.000998102 | 0.011570689 | 1.57E-05 | 0.043916997 | 1 | 0.988687703 | 1 | 0.988759842 | ENSG0000011 | 9 | PCCB |
| Pancreas | T2DM | insulin | ENSG0000026 | 0.002151593 | 0.009078976 | 0.000824657 | 0.011570968 | 0.002163587 | 0.00870046 | 0.000824659 | 0.011570526 | -1.20E-05 | 0.025170488 | 1 | 0.261507236 | 1 | 0.255611981 | ENSG0000011 | 9 | RNF7 |
| Pancreas | T2DM | insulin | ENSG0000026 | -0.00049065 | 0.309363617 | 0.000482656 | 0.011571105 | -0.00045961 | 0.341049502 | 0.000482737 | 0.011570667 | -3.10E-05 | 0.000831891 | 1 | 0.835397773 | 1 | 0.838317574 | ENSG0000011 | 9 | RNF7 |
| Pancreas | T2DM | insulin | ENSG0000026 | -0.00077521 | 0.282771742 | 0.000721718 | 0.011571102 | -0.0007935 | 0.271586519 | 0.000721748 | 0.01157066 | 1.83E-05 | 0.020618769 | 1 | 0.82273396 | 1 | 0.815055965 | ENSG0000011 | 5 | BCHE |
| Pancreas | T2DM | insulin | ENSG0000027 | -0.00027722 | 0.421435409 | 0.000344833 | 0.011571114 | -0.00028136 | 0.414535183 | 0.000344832 | 0.011570673 | 4.14E-06 | 0.027226752 | 1 | 0.870194075 | 1 | 0.86606442 | ENSG0000011 | 5 | BCHE |
| Pancreas | T2DM | insulin | ENSG0000027 | 0.002589936 | 0.004708597 | 0.000916359 | 0.01157094 | 0.001899157 | 0.047321596 | 0.000957518 | 0.011570596 | 0.00069078 | 0.012885163 | 1 | 0.191931378 | 1 | 0.504240075 | ENSG0000011 | 11 | PRKAR2A |
| Pancreas | T2DM | insulin | ENSG0000027 | 0.000345472 | 0.185464751 | 0.000260908 | 0.011571088 | 0.000435648 | 0.096612783 | 0.000262201 | 0.011570623 | -9.02E-05 | 0.000540601 | 1 | 0.726940181 | 1 | 0.605592158 | ENSG0000011 | 11 | PRKAR2A |
| Pancreas | T2DM | insulin | ENSG0000027 | -0.00028864 | 0.671541876 | 0.000680693 | 0.011571125 | -0.00028057 | 0.680203127 | 0.000680692 | 0.011570685 | -8.06E-06 | 0.042382127 | 1 | 0.941892795 | 1 | 0.945452933 | ENSG0000011 | 12 | HYAL1 |
| Pancreas | T2DM | | ENSG0000000 | 0.001101838 | 0.325884304 | 0.001121533 | 0.055522567 | 0.001085424 | 0.333142444 | 0.001121532 | 0.055519517 | 1.64E-05 | 0.043987851 | 1 | 0.723936757 | 1 | 0.735008377 | ENSG0000011 | 5 | TUSC2 |
| Pancreas | T2DM | | ENSG0000000 | 0.003638725 | 0.136819772 | 0.002445805 | 0.055522425 | 0.003542856 | 0.147493749 | 0.002445975 | 0.055519385 | 9.59E-05 | 0.004859027 | 1 | 0.542179906 | 1 | 0.55791981 | ENSG0000011 | 5 | TUSC2 |
| Pancreas | T2DM | | ENSG0000000 | 0.002074875 | 0.376255929 | 0.002344977 | 0.055522588 | 0.002039064 | 0.384546254 | 0.002344962 | 0.055519538 | 3.58E-05 | 0.018737599 | 1 | 0.763882089 | 1 | 0.772390956 | ENSG0000011 | 6 | CYB561D2 |
| Pancreas | T2DM | | ENSG0000000 | 0.004271553 | 0.144635202 | 0.002928233 | 0.055522435 | 0.004232103 | 0.148377268 | 0.002928205 | 0.055519386 | 3.94E-05 | 0.024354402 | 1 | 0.554532985 | 1 | 0.55791981 | ENSG0000011 | 6 | C3orf14 |

| | | | | | | | | | | | | | | | | | | | |
|---|---|---|---|---|---|---|---|---|---|---|---|---|---|---|---|---|---|---|---|
| Pancreas | T2DM | ENSG0000000 | -0.00474471 | 0.151387299 | 0.003307225 | 0.055522443 | -0.00467951 | 0.157087291 | 0.003307218 | 0.055519396 | -6.52E-05 | 0.005463617 | 1 | 0.556171292 | 1 | 0.561838668 | ENSG0000011 | 6 | C3orf14 |
| Pancreas | T2DM | ENSG0000001 | 0.000579837 | 0.854305831 | 0.00315768 | 0.055522673 | 0.000632558 | 0.841228248 | 0.003157692 | 0.05551962 | -5.27E-05 | 0.034751053 | 1 | 0.97245451 | 1 | 0.968298585 | ENSG0000011 | 4 | IFT57 |
| Pancreas | T2DM | ENSG0000001 | -0.00027472 | 0.894242365 | 0.002066557 | 0.055522675 | -0.00023665 | 0.908829388 | 0.002066568 | 0.055519623 | -3.81E-05 | 0.022445366 | 1 | 0.981247774 | 1 | 0.985211345 | ENSG0000011 | 4 | IFT57 |
| Pancreas | T2DM | ENSG0000001 | 0.003720054 | 0.04393032 | 0.0018464 | 0.055522215 | 0.003746137 | 0.042469028 | 0.001846388 | 0.055519155 | -2.61E-05 | 0.02979463 | 1 | 0.311795766 | 1 | 0.31228686 | ENSG0000011 | 4 | IFT57 |
| Pancreas | T2DM | ENSG0000002 | -0.00147309 | 0.541760674 | 0.002414293 | 0.055522635 | -0.0015058 | 0.532820918 | 0.002414274 | 0.05551958 | 3.27E-05 | 0.031604649 | 1 | 0.860225195 | 1 | 0.854992848 | ENSG0000011 | 6 | NCBP2 |
| Pancreas | T2DM | ENSG0000003 | 0.011934087 | 2.30E-07 | 0.002306908 | 0.055519628 | 0.011017341 | 2.02E-06 | 0.002318864 | 0.055517052 | 0.000916745 | 0.000100475 | 0.000591188 | 3.94E-05 | 0.00519481 | 0.000431985 | ENSG0000011 | 6 | NCBP2 |
| Pancreas | T2DM | ENSG0000003 | -0.00114392 | 0.537154134 | 0.001853642 | 0.055522634 | -0.0012025 | 0.516517418 | 0.001853648 | 0.055519576 | 5.86E-05 | 5.68E-05 | 1 | 0.858495882 | 1 | 0.848610147 | ENSG0000011 | 6 | NCBP2 |
| Pancreas | T2DM | ENSG0000004 | 0.000720066 | 0.75304297 | 0.002288615 | 0.055522666 | 0.000662953 | 0.772068676 | 0.002288646 | 0.055519615 | 5.71E-05 | 0.006035528 | 1 | 0.935675707 | 1 | 0.943354401 | ENSG0000011 | 11 | SLC41A3 |
| Pancreas | T2DM | ENSG0000005 | 0.002797242 | 0.231149613 | 0.002336088 | 0.055522514 | 0.002766521 | 0.236309751 | 0.002336071 | 0.055519464 | 3.07E-05 | 0.038633283 | 1 | 0.648026426 | 1 | 0.655938795 | ENSG0000011 | 11 | SLC41A3 |
| Pancreas | T2DM | ENSG0000006 | 0.004740686 | 0.047565649 | 0.002392795 | 0.05552223 | 0.00478506 | 0.045523627 | 0.002392783 | 0.055519168 | -4.44E-05 | 0.00574995 | 1 | 0.325729564 | 1 | 0.321166687 | ENSG0000011 | 5 | ROPN1B |
| Pancreas | T2DM | ENSG0000006 | 1.74E-05 | 0.987925094 | 0.001147008 | 0.055522677 | 3.92E-05 | 0.972740473 | 0.001147003 | 0.055519624 | -2.18E-05 | 0.005518622 | 1 | 0.997207295 | 1 | 0.996011776 | ENSG0000011 | 3 | UPK1B |
| Pancreas | T2DM | ENSG0000006 | -0.00717816 | 0.017138136 | 0.003011305 | 0.05552203 | -0.00711164 | 0.018194969 | 0.003011319 | 0.055518989 | -6.65E-05 | 0.005994954 | 1 | 0.200048793 | 1 | 0.212384905 | ENSG0000011 | 3 | UPK1B |
| Pancreas | T2DM | ENSG0000006 | 0.000363645 | 0.847832843 | 0.001895126 | 0.055522673 | 0.000326178 | 0.863347979 | 0.001895128 | 0.055519621 | 3.75E-05 | 0.008436691 | 1 | 0.970153451 | 1 | 0.974539608 | ENSG0000011 | 5 | HEMK1 |
| Pancreas | T2DM | ENSG0000006 | -0.00159765 | 0.452044464 | 0.002124497 | 0.055522613 | -0.00156496 | 0.461348848 | 0.002124492 | 0.055519562 | -3.27E-05 | 0.028908463 | 1 | 0.807173989 | 1 | 0.814816947 | ENSG0000011 | 9 | WDR48 |
| Pancreas | T2DM | ENSG0000006 | 0.002183167 | 0.116791917 | 0.001391984 | 0.055522397 | 0.005912261 | 0.000436359 | 0.001681013 | 0.055518215 | -0.00372909 | 7.60E-05 | 1 | 0.510742285 | 1 | 0.020225759 | ENSG0000011 | 9 | WDR48 |
| Pancreas | T2DM | ENSG0000006 | 0.003384382 | 0.21573838 | 0.002733879 | 0.055522503 | 0.003425983 | 0.210144975 | 0.002733857 | 0.055519445 | -4.16E-05 | 0.014879011 | 1 | 0.621094348 | 1 | 0.620720671 | ENSG0000011 | 9 | WDR48 |
| Pancreas | T2DM | ENSG0000007 | 0.006092444 | 0.003283685 | 0.002072354 | 0.055521692 | 0.006123793 | 0.00312686 | 0.002072351 | 0.055518629 | -3.13E-05 | 0.037087816 | 1 | 0.074942369 | 1 | 0.072830925 | ENSG0000011 | 9 | AADAC |
| Pancreas | T2DM | ENSG0000007 | 0.008285735 | 0.0008703 | 0.002488634 | 0.055521414 | 0.008251104 | 0.000914691 | 0.002488607 | 0.055518372 | 3.46E-05 | 0.015329531 | 1 | 0.034383558 | 1 | 0.035306133 | ENSG0000011 | 11 | KLHL24 |
| Pancreas | T2DM | ENSG0000007 | -0.00365134 | 0.178570121 | 0.0027144 | 0.055522471 | -0.00361982 | 0.182343082 | 0.002714369 | 0.055519422 | -3.15E-05 | 0.03860933 | 1 | 0.581719513 | 1 | 0.586401239 | ENSG0000011 | 11 | KLHL24 |
| Pancreas | T2DM | ENSG0000007 | -0.0024369 | 0.147878898 | 0.001684026 | 0.055522439 | -0.00246433 | 0.14336935 | 0.001684022 | 0.05551938 | 2.74E-05 | 0.021232617 | 1 | 0.556171292 | 1 | 0.552027703 | ENSG0000011 | 11 | KLHL24 |
| Pancreas | T2DM | ENSG0000007 | 0.000367509 | 0.869832593 | 0.002242668 | 0.055522674 | 0.000404549 | 0.856849185 | 0.002242677 | 0.05551962 | -3.70E-05 | 0.038053255 | 1 | 0.976734957 | 1 | 0.973048371 | ENSG0000011 | 13 | DNAH1 |
| Pancreas | T2DM | ENSG0000007 | 0.000569526 | 0.792767817 | 0.002167819 | 0.055522669 | 0.000538639 | 0.803769486 | 0.002167807 | 0.055519617 | 3.09E-05 | 0.032341692 | 1 | 0.949920498 | 1 | 0.951905176 | ENSG0000011 | 7 | LMAN2L |
| Pancreas | T2DM | ENSG0000007 | -0.00167937 | 0.494272858 | 0.002456906 | 0.055522624 | -0.00188258 | 0.443637874 | 0.002457473 | 0.055519557 | 0.000203217 | 0.000272786 | 1 | 0.838924455 | 1 | 0.801344566 | ENSG0000011 | 7 | LMAN2L |
| Pancreas | T2DM | ENSG0000008 | -0.00211966 | 0.2926682 | 0.002014338 | 0.055522551 | -0.00209301 | 0.298777934 | 0.002014328 | 0.055519501 | -2.67E-05 | 0.047560462 | 1 | 0.703060747 | 1 | 0.709510872 | ENSG0000011 | 8 | SLC35F5 |
| Pancreas | T2DM | ENSG0000008 | 0.00031139 | 0.813336616 | 0.001318758 | 0.055522671 | 8.12E-05 | 0.951022028 | 0.001322103 | 0.055519624 | 0.000230182 | 0.014858487 | 1 | 0.957217428 | 1 | 0.991564989 | ENSG0000011 | 8 | SLC35F5 |
| Pancreas | T2DM | ENSG0000009 | 0.000504038 | 0.689759533 | 0.00126268 | 0.055522659 | 0.00044965 | 0.721773882 | 0.001262745 | 0.05551961 | 5.44E-05 | 0.000617366 | 1 | 0.91152931 | 1 | 0.921688379 | ENSG0000011 | 8 | SLC35F5 |
| Pancreas | T2DM | ENSG0000009 | -0.00303608 | 0.16606901 | 0.0021922 | 0.055522459 | -0.00299559 | 0.171786712 | 0.002192186 | 0.055519411 | -4.05E-05 | 0.00408166 | 1 | 0.570140665 | 1 | 0.578934746 | ENSG0000011 | 8 | SLC35F5 |
| Pancreas | T2DM | ENSG0000009 | -0.00123305 | 0.514123424 | 0.001889935 | 0.055522629 | -0.00126156 | 0.504441196 | 0.00188993 | 0.055519573 | 2.85E-05 | 0.033724602 | 1 | 0.848731838 | 1 | 0.844462185 | ENSG0000011 | 10 | CENPA |
| Pancreas | T2DM | ENSG0000009 | -0.0004885 | 0.839340424 | 0.00240954 | 0.055522672 | -0.0005252 | 0.827454992 | 0.002409529 | 0.055519619 | 3.67E-05 | 0.024086725 | 1 | 0.966558838 | 1 | 0.961060344 | ENSG0000011 | 6 | ITGB6 |
| Pancreas | T2DM | ENSG0000010 | 0.002501381 | 0.341119799 | 0.002627622 | 0.055522574 | 0.002548429 | 0.332113636 | 0.002627609 | 0.055519517 | -4.70E-05 | 0.007565637 | 1 | 0.738900364 | 1 | 0.735008377 | ENSG0000011 | 6 | ITGB6 |
| Pancreas | T2DM | ENSG0000010 | -0.00213279 | 0.293575051 | 0.002030627 | 0.055522551 | -0.00210331 | 0.300297843 | 0.002030619 | 0.055519502 | -2.95E-05 | 0.035230964 | 1 | 0.703265608 | 1 | 0.709510872 | ENSG0000011 | 6 | ITGB6 |
| Pancreas | T2DM | ENSG0000010 | 0.001290977 | 0.245531849 | 0.001111692 | 0.055522523 | 0.001278459 | 0.250134127 | 0.001111679 | 0.055519473 | 1.25E-05 | 0.043523717 | 1 | 0.658561546 | 1 | 0.666460318 | ENSG0000011 | 6 | REEP6 |
| Pancreas | T2DM | ENSG0000010 | 0.001798388 | 0.480555104 | 0.00254942 | 0.05552262 | 0.001754126 | 0.491417684 | 0.002549393 | 0.05551957 | 4.43E-05 | 0.002835339 | 1 | 0.827121653 | 1 | 0.834196451 | ENSG0000011 | 10 | INO80B |
| Pancreas | T2DM | ENSG0000010 | -0.00024854 | 0.858532513 | 0.001394398 | 0.055522674 | 0.001246123 | 0.428051983 | 0.001572336 | 0.055519553 | -0.00149466 | 0.039689926 | 1 | 0.975540847 | 1 | 0.793088295 | ENSG0000011 | 10 | INO80B |
| Pancreas | T2DM | ENSG0000010 | 0.004371888 | 0.086525315 | 0.00255066 | 0.055522342 | 0.004337597 | 0.089020967 | 0.002550645 | 0.055519295 | 3.43E-05 | 0.03985555 | 1 | 0.45439061 | 1 | 0.455804017 | ENSG0000011 | 7 | MOGS |
| Pancreas | T2DM | ENSG0000010 | -0.00209577 | 0.452935248 | 0.002792378 | 0.055522613 | -0.00199948 | 0.474014441 | 0.002792716 | 0.055519566 | -9.63E-05 | 0.045385456 | 1 | 0.807173989 | 1 | 0.821923758 | ENSG0000011 | 7 | MOGS |
| Pancreas | T2DM | ENSG0000010 | -0.00111016 | 0.723815559 | 0.003141693 | 0.055522663 | -0.00115443 | 0.713279003 | 0.003141678 | 0.055519609 | 4.43E-05 | 0.036955237 | 1 | 0.921476762 | 1 | 0.920472548 | ENSG0000011 | 13 | RTN4 |
| Pancreas | T2DM | ENSG0000010 | 0.003542648 | 0.074365606 | 0.001985387 | 0.055522314 | 0.003592705 | 0.070362454 | 0.001985387 | 0.055519251 | -5.01E-05 | 0.000660411 | 1 | 0.424379724 | 1 | 0.410660871 | ENSG0000011 | 13 | RTN4 |
| Pancreas | T2DM | ENSG0000010 | 0.002291923 | 0.428319757 | 0.002893584 | 0.055522606 | 0.002208041 | 0.445445821 | 0.00289378 | 0.055519558 | 8.39E-05 | 0.035606221 | 1 | 0.792249323 | 1 | 0.8027301 | ENSG0000011 | 14 | DNAH6 |
| Pancreas | T2DM | ENSG0000010 | -0.00025594 | 0.908865637 | 0.002235883 | 0.055522676 | -0.00020505 | 0.92693007 | 0.002235925 | 0.055519623 | -5.09E-05 | 0.017846869 | 1 | 0.98382859 | 1 | 0.987209256 | ENSG0000011 | 14 | DNAH6 |
| Pancreas | T2DM | ENSG0000010 | -0.00256367 | 0.406437008 | 0.003088089 | 0.055522599 | -0.00251189 | 0.415982638 | 0.003088088 | 0.055519549 | -5.18E-05 | 0.022482225 | 1 | 0.784169975 | 1 | 0.788638662 | ENSG0000011 | 14 | DNAH6 |
| Pancreas | T2DM | ENSG0000010 | -2.77E-05 | 0.988853073 | 0.001981754 | 0.055522677 | -0.00145649 | 0.487333672 | 0.00209701 | 0.055519569 | 0.001428805 | 0.037213806 | 1 | 0.997207295 | 1 | 0.831484518 | ENSG0000011 | 14 | DNAH6 |
| Pancreas | T2DM | ENSG0000010 | -0.00073281 | 0.681537355 | 0.001785756 | 0.055522658 | -0.0007563 | 0.671914043 | 0.001785742 | 0.055519604 | 2.35E-05 | 0.036183755 | 1 | 0.91019995 | 1 | 0.907798476 | ENSG0000011 | 5 | IGFBP2 |
| Pancreas | T2DM | ENSG0000010 | 0.000634783 | 0.531726335 | 0.001015047 | 0.055522633 | 0.000651256 | 0.521128583 | 0.001015039 | 0.055519577 | -1.65E-05 | 0.010539985 | 1 | 0.857709315 | 1 | 0.848610147 | ENSG0000011 | 7 | GGCX |
| Pancreas | T2DM | ENSG0000010 | -0.00233841 | 0.41062199 | 0.002842008 | 0.0555226 | -0.00238075 | 0.402197501 | 0.002841995 | 0.055519544 | 4.23E-05 | 0.028050425 | 1 | 0.785561726 | 1 | 0.782159733 | ENSG0000011 | 7 | GGCX |
| Pancreas | T2DM | ENSG0000010 | -0.00031003 | 0.900115927 | 0.002470051 | 0.055522675 | -0.00027515 | 0.911304174 | 0.002470029 | 0.055519623 | -3.49E-05 | 0.021676186 | 1 | 0.981944648 | 1 | 0.985399068 | ENSG0000011 | 7 | GGCX |
| Pancreas | T2DM | ENSG0000010 | 0.001182018 | 0.511039106 | 0.001798509 | 0.055522628 | 0.001156792 | 0.520097041 | 0.001798505 | 0.055519577 | 2.52E-05 | 0.048582176 | 1 | 0.848731838 | 1 | 0.848610147 | ENSG0000011 | 7 | GGCX |
| Pancreas | T2DM | ENSG0000011 | 0.001799618 | 0.502941229 | 0.00268652 | 0.055522626 | 0.001766614 | 0.510801342 | 0.002686493 | 0.055519575 | 3.30E-05 | 0.038206024 | 1 | 0.846716275 | 1 | 0.847548645 | ENSG0000011 | 7 | GGCX |
| Pancreas | T2DM | ENSG0000011 | 0.002202102 | 0.159704464 | 0.001566142 | 0.055522452 | 0.002183666 | 0.163223508 | 0.001566122 | 0.055519403 | 1.84E-05 | 0.030880478 | 1 | 0.563353108 | 1 | 0.568944307 | ENSG0000011 | 10 | CHMP3 |
| Pancreas | T2DM | ENSG0000011 | -0.00337496 | 0.033351076 | 0.0015861 | 0.055522161 | -0.00157581 | 0.337294929 | 0.001642282 | 0.055519519 | -0.00179915 | 2.41E-05 | 1 | 0.280805126 | 1 | 0.739686916 | ENSG0000011 | 10 | CHMP3 |
| Pancreas | T2DM | ENSG0000011 | 0.001324029 | 0.646075032 | 0.002883202 | 0.055522653 | 0.001242131 | 0.666605403 | 0.002883233 | 0.055519603 | 8.19E-05 | 0.001208681 | 1 | 0.898449905 | 1 | 0.905736865 | ENSG0000011 | 10 | CHMP3 |
| Pancreas | T2DM | ENSG0000011 | 0.001299631 | 0.49548915 | 0.001906721 | 0.055522624 | 0.001347771 | 0.479678814 | 0.00190681 | 0.055519567 | -4.81E-05 | 0.038128082 | 1 | 0.84043338 | 1 | 0.827056253 | ENSG0000011 | 11 | ZNF142 |
| Pancreas | T2DM | ENSG0000011 | 0.001597958 | 0.578220365 | 0.002874094 | 0.055522642 | 0.00164436 | 0.567230622 | 0.002874076 | 0.055519587 | -4.64E-05 | 0.012796978 | 1 | 0.878208132 | 1 | 0.868486338 | ENSG0000011 | 11 | ZNF142 |
| Pancreas | T2DM | ENSG0000011 | 0.002862478 | 0.036063657 | 0.001365546 | 0.055522176 | 0.002882141 | 0.034804681 | 0.001365536 | 0.055519117 | -1.97E-05 | 0.024806301 | 1 | 0.285894549 | 1 | 0.288681949 | ENSG0000011 | 11 | ZNF142 |
| Pancreas | T2DM | ENSG0000011 | -5.17E-05 | 0.958551378 | 0.000993909 | 0.055522677 | -6.92E-05 | 0.94449752 | 0.000993901 | 0.055519624 | 1.75E-05 | 0.004227304 | 1 | 0.990168921 | 1 | 0.987209256 | ENSG0000011 | 9 | MLPH |
| Pancreas | T2DM | ENSG0000011 | -0.0018853 | 0.534463556 | 0.003034895 | 0.055522633 | -0.00185482 | 0.54108446 | 0.003034849 | 0.055519582 | -3.05E-05 | 0.043263362 | 1 | 0.858495882 | 1 | 0.858355187 | ENSG0000011 | 12 | HDLBP |
| Pancreas | T2DM | ENSG0000011 | 0.008286376 | 0.006968969 | 0.003070922 | 0.055521848 | 0.008319014 | 0.006748846 | 0.003070879 | 0.055518788 | -3.26E-05 | 0.040812505 | 1 | 0.122577481 | 1 | 0.121196067 | ENSG0000011 | 12 | HDLBP |
| Pancreas | T2DM | ENSG0000011 | 0.0007897 | 0.789544016 | 0.002958754 | 0.055522669 | 0.000750994 | 0.799632034 | 0.002958727 | 0.055519617 | 3.87E-05 | 0.029926447 | 1 | 0.949920498 | 1 | 0.95067364 | ENSG0000011 | 9 | PASK |
| Pancreas | T2DM | ENSG0000011 | -0.00182223 | 0.021190394 | 0.0007907 | 0.055522072 | -0.00180877 | 0.022162883 | 0.000790694 | 0.055519028 | -1.35E-05 | 0.007320419 | 1 | 0.223020208 | 1 | 0.237142843 | ENSG0000011 | 9 | PASK |
| Pancreas | T2DM | ENSG0000011 | -0.00233522 | 0.111544738 | 0.0014675 | 0.055522389 | -0.00231706 | 0.114351929 | 0.001467485 | 0.05551934 | -1.82E-05 | 0.034194657 | 1 | 0.503138201 | 1 | 0.506303024 | ENSG0000011 | 9 | PASK |
| Pancreas | T2DM | ENSG0000011 | 0.002309158 | 0.068750929 | 0.001268729 | 0.0555223 | 0.00229223 | 0.070804894 | 0.001268717 | 0.055519252 | 1.69E-05 | 0.027180809 | 1 | 0.406108759 | 1 | 0.412306048 | ENSG0000011 | 13 | PROC |
| Pancreas | T2DM | ENSG0000011 | 0.002251965 | 0.504258358 | 0.003372189 | 0.055522626 | 0.002304524 | 0.494357524 | 0.003372169 | 0.055519571 | -5.26E-05 | 0.017439511 | 1 | 0.846916588 | 1 | 0.835942127 | ENSG0000011 | 13 | PROC |
| Pancreas | T2DM | ENSG0000011 | 0.001117164 | 0.549982127 | 0.001868831 | 0.055522636 | 0.001139777 | 0.54193237 | 0.001868812 | 0.055519582 | -2.26E-05 | 0.039690157 | 1 | 0.865271405 | 1 | 0.859063164 | ENSG0000011 | 12 | TAF1B |
| Pancreas | T2DM | ENSG0000011 | -0.00134517 | 0.566324069 | 0.002345649 | 0.05552264 | -0.0013797 | 0.55639991 | 0.002345638 | 0.055519585 | 3.45E-05 | 0.030526272 | 1 | 0.872395717 | 1 | 0.864388971 | ENSG0000011 | 12 | TAF1B |
| Pancreas | T2DM | ENSG0000012 | -0.00085923 | 0.298817273 | 0.000827003 | 0.055522554 | -0.00087194 | 0.291729434 | 0.000827 | 0.055519497 | 1.27E-05 | 0.026818582 | 1 | 0.705945498 | 1 | 0.702726916 | ENSG0000011 | 5 | PRKD3 |
| Pancreas | T2DM | ENSG0000012 | -0.00697232 | 0.027688514 | 0.00316682 | 0.055522125 | -0.00700586 | 0.026946259 | 0.003166773 | 0.055519067 | 3.35E-05 | 0.035036903 | 1 | 0.254853419 | 1 | 0.254404388 | ENSG0000011 | 9 | SLC25A12 |
| Pancreas | T2DM | ENSG0000012 | -0.00205252 | 0.299862016 | 0.001979802 | 0.055522555 | -0.00208166 | 0.293050186 | 0.001979785 | 0.055519498 | 2.91E-05 | 0.017439786 | 1 | 0.707112634 | 1 | 0.702726916 | ENSG0000011 | 9 | SLC25A12 |
| Pancreas | T2DM | ENSG0000012 | -0.00080892 | 0.712220448 | 0.002192923 | 0.055522662 | -0.00076576 | 0.726941896 | 0.002192915 | 0.05551961 | -4.32E-05 | 0.004595705 | 1 | 0.921421644 | 1 | 0.922488007 | ENSG0000011 | 9 | ORC4 |
| Pancreas | T2DM | ENSG0000012 | -5.07E-05 | 0.982357678 | 0.002291404 | 0.055522677 | -2.41E-05 | 0.991597337 | 0.002291379 | 0.055519624 | -2.65E-05 | 0.04609251 | 1 | 0.99701908 | 1 | 0.998538835 | ENSG0000011 | 9 | ORC4 |
| Pancreas | T2DM | ENSG0000012 | -0.00162937 | 0.472373677 | 0.00226735 | 0.055522618 | -0.0016567 | 0.464971955 | 0.002267325 | 0.055519563 | 2.73E-05 | 0.035554634 | 1 | 0.817750308 | 1 | 0.815865042 | ENSG0000011 | 9 | ORC4 |
| Pancreas | T2DM | ENSG0000012 | 0.002201835 | 0.430860463 | 0.002795198 | 0.055522606 | 0.002240944 | 0.422716068 | 0.002795174 | 0.055519551 | -3.91E-05 | 0.023461829 | 1 | 0.792249323 | 1 | 0.791410966 | ENSG0000011 | 11 | NFE2L2 |
| Pancreas | T2DM | ENSG0000012 | -0.00034774 | 0.890244431 | 0.002519961 | 0.055522675 | -0.00038899 | 0.877323977 | 0.002519952 | 0.055519621 | 4.12E-05 | 0.018247596 | 1 | 0.981247774 | 1 | 0.980763999 | ENSG0000011 | 13 | ALMS1 |
| Pancreas | T2DM | ENSG0000012 | -0.00034974 | 0.784594961 | 0.001279526 | 0.055522669 | -0.00032101 | 0.80191232 | 0.001279567 | 0.055519617 | -2.87E-05 | 0.039796935 | 1 | 0.949163351 | 1 | 0.951905176 | ENSG0000011 | 8 | DNAJC16 |
| Pancreas | T2DM | ENSG0000012 | 0.004038571 | 0.100637972 | 0.002459886 | 0.05552237 | 0.004091512 | 0.096257483 | 0.002459909 | 0.055519309 | -5.29E-05 | 0.012177264 | 1 | 0.483403189 | 1 | 0.472157259 | ENSG0000011 | 9 | MORN1 |
| Pancreas | T2DM | ENSG0000012 | 0.002946157 | 0.333794347 | 0.003048273 | 0.055522571 | 0.00289869 | 0.3416381 | 0.003048254 | 0.055519521 | 4.75E-05 | 0.016972015 | 1 | 0.733262517 | 1 | 0.741897611 | ENSG0000011 | 9 | MORN1 |
| Pancreas | T2DM | ENSG0000012 | -0.00026968 | 0.832445343 | 0.001274676 | 0.055522672 | -0.00029538 | 0.81674546 | 0.00127467 | 0.055519618 | 2.57E-05 | 0.002642429 | 1 | 0.964161038 | 1 | 0.956844134 | ENSG0000011 | 9 | MORN1 |
| Pancreas | T2DM | ENSG0000012 | 0.001413157 | 0.53362466 | 0.002270193 | 0.055522633 | 0.001358278 | 0.549648132 | 0.002270275 | 0.055519583 | 5.49E-05 | 0.032038127 | 1 | 0.858495882 | 1 | 0.863653712 | ENSG0000011 | 5 | GPX7 |
| Pancreas | T2DM | ENSG0000012 | -0.00048338 | 0.567349818 | 0.000845131 | 0.05552264 | -0.00055814 | 0.509316692 | 0.000845792 | 0.055519575 | 7.48E-05 | 0.027928291 | 1 | 0.872395717 | 1 | 0.847548645 | ENSG0000011 | 5 | GPX7 |
| Pancreas | T2DM | ENSG0000012 | -0.0002135 | 0.836026358 | 0.001031519 | 0.055522672 | -0.00033157 | 0.747976278 | 0.001031937 | 0.055519612 | 0.000118068 | 0.000100483 | 1 | 0.964161038 | 1 | 0.933447328 | ENSG0000011 | 5 | GPX7 |
| Pancreas | T2DM | ENSG0000012 | 0.000811149 | 0.803963657 | 0.003267851 | 0.05552267 | 0.000897749 | 0.783533184 | 0.003267894 | 0.055519616 | -8.66E-05 | 0.0032712 | 1 | 0.952792712 | 1 | 0.945477902 | ENSG0000011 | 5 | GPX7 |
| Pancreas | T2DM | ENSG0000012 | 0.002178316 | 0.386037813 | 0.00251298 | 0.055522592 | 0.002206507 | 0.379914862 | 0.002512951 | 0.055519536 | -2.82E-05 | 0.047595347 | 1 | 0.770733172 | 1 | 0.768582324 | ENSG0000011 | 5 | GPX7 |
| Pancreas | T2DM | ENSG0000012 | -0.00133049 | 0.39598565 | 0.001567467 | 0.055522595 | -0.00135172 | 0.388485243 | 0.001567455 | 0.055519539 | 2.12E-05 | 0.034172266 | 1 | 0.776325713 | 1 | 0.772390956 | ENSG0000011 | 5 | GPX7 |
| Pancreas | T2DM | ENSG0000012 | -0.00299094 | 0.080096308 | 0.001708983 | 0.055522328 | -0.0029638 | 0.082873541 | 0.001708976 | 0.055519281 | -2.71E-05 | 0.019102068 | 1 | 0.438568358 | 1 | 0.449543964 | ENSG0000011 | 5 | GPX7 |
| Pancreas | T2DM | ENSG0000012 | 0.000177895 | 0.917752885 | 0.001722709 | 0.055522676 | 0.000205665 | 0.904970781 | 0.001722712 | 0.055519623 | -2.78E-05 | 0.033886907 | 1 | 0.9849439 | 1 | 0.984551279 | ENSG0000011 | 5 | GPX7 |
| Pancreas | T2DM | ENSG0000012 | -0.00286999 | 0.188064177 | 0.002180295 | 0.05552248 | -0.0028393 | 0.192828657 | 0.002180288 | 0.055519431 | -3.07E-05 | 0.042687924 | 1 | 0.591234665 | 1 | 0.602109922 | ENSG0000011 | 5 | GPX7 |
| Pancreas | T2DM | ENSG0000013 | -0.00278514 | 0.016324644 | 0.00115971 | 0.05552202 | -0.00284314 | 0.014251028 | 0.001160047 | 0.05551894 | 5.80E-05 | 0.047399015 | 1 | 0.197743801 | 1 | 0.18767507 | ENSG0000011 | 8 | CEP104 |
| Pancreas | T2DM | ENSG0000013 | -0.00095281 | 0.775796393 | 0.003345522 | 0.055522668 | 0.002978494 | 0.432136113 | 0.003791641 | 0.055519554 | -0.0039313 | 0.027597918 | 1 | 0.948363389 | 1 | 0.793088295 | ENSG0000011 | 8 | CEP104 |
| Pancreas | T2DM | ENSG0000013 | 0.001703888 | 0.59633415 | 0.003216827 | 0.055522645 | 0.001652563 | 0.60744075 | 0.003216791 | 0.055519594 | 5.13E-05 | 0.005479361 | 1 | 0.88286336 | 1 | 0.882490377 | ENSG0000011 | 11 | MRPL37 |
| Pancreas | T2DM | ENSG0000013 | 0.00026467 | 0.936157211 | 0.003304211 | 0.055522676 | 0.00036174 | 0.912827629 | 0.003304389 | 0.055519623 | -9.71E-05 | 0.021303271 | 1 | 0.987512886 | 1 | 0.985865795 | ENSG0000011 | 11 | MRPL37 |
| Pancreas | T2DM | ENSG0000013 | -0.00132999 | 0.608555747 | 0.002596939 | 0.055522647 | -0.0012886 | 0.619753997 | 0.002596945 | 0.055519596 | -4.14E-05 | 0.038937383 | 1 | 0.884921381 | 1 | 0.888839757 | ENSG0000011 | 11 | MRPL37 |
| Pancreas | T2DM | ENSG0000013 | -0.00066539 | 0.742757171 | 0.002027366 | 0.055522665 | -0.00069487 | 0.731791048 | 0.002027359 | 0.055519611 | 2.95E-05 | 0.035180373 | 1 | 0.930893321 | 1 | 0.924390297 | ENSG0000011 | 6 | ATP5F1 |
| Pancreas | T2DM | ENSG0000013 | 0.004598824 | 0.054954662 | 0.00239617 | 0.055522257 | 0.004562306 | 0.056909771 | 0.002396159 | 0.055519211 | 3.65E-05 | 0.024178057 | 1 | 0.360929851 | 1 | 0.375107923 | ENSG0000011 | 8 | CAPZA1 |
| Pancreas | T2DM | ENSG0000013 | -0.00066525 | 0.785241397 | 0.002441339 | 0.055522669 | -0.00071629 | 0.769215021 | 0.002441332 | 0.055519614 | 5.10E-05 | 0.002787463 | 1 | 0.949163351 | 1 | 0.943143698 | ENSG0000011 | 6 | ASH1L |
| Pancreas | T2DM | ENSG0000013 | -0.00143705 | 0.309330259 | 0.001413547 | 0.055522559 | -0.00145493 | 0.303344837 | 0.00141353 | 0.055519503 | 1.79E-05 | 0.023202542 | 1 | 0.715658284 | 1 | 0.712707723 | ENSG0000011 | 6 | ASH1L |
| Pancreas | T2DM | ENSG0000013 | -0.00192071 | 0.150873002 | 0.001337119 | 0.055522442 | -0.00190525 | 0.154183433 | 0.001337103 | 0.055519393 | -1.55E-05 | 0.039163757 | 1 | 0.556171292 | 1 | 0.561621356 | ENSG0000011 | 6 | ASH1L |
| Pancreas | T2DM | ENSG0000013 | 0.000338782 | 0.888526987 | 0.002416963 | 0.055522675 | 0.000297808 | 0.90193614 | 0.00241696 | 0.055519622 | 4.10E-05 | 0.019314443 | 1 | 0.981247774 | 1 | 0.984551279 | ENSG0000011 | 9 | FBXO2 |
| Pancreas | T2DM | ENSG0000013 | 0.000576017 | 0.682597725 | 0.001408625 | 0.055522658 | 0.000598177 | 0.671087542 | 0.001408615 | 0.055519604 | -2.22E-05 | 0.012796324 | 1 | 0.91019995 | 1 | 0.907798476 | ENSG0000011 | 6 | FBXO6 |
| Pancreas | T2DM | ENSG0000013 | 0.0024912 | 0.405020318 | 0.002991752 | 0.055522598 | 0.001006608 | 0.742336582 | 0.003061823 | 0.055519612 | 0.001484592 | 0.022716971 | 1 | 0.783229955 | 1 | 0.930820479 | ENSG0000011 | 8 | C1orf21 |
| Pancreas | T2DM | ENSG0000013 | -0.00017128 | 0.944524616 | 0.00246148 | 0.055522677 | -0.00013325 | 0.956828687 | 0.002461464 | 0.055519624 | -3.80E-05 | 0.016889969 | 1 | 0.989794375 | 1 | 0.991898954 | ENSG0000011 | 11 | DNAJC6 |
| Pancreas | T2DM | ENSG0000013 | 0.002923348 | 0.111576113 | 0.001837253 | 0.055522389 | 0.002950182 | 0.108327069 | 0.001837248 | 0.05551933 | -2.68E-05 | 0.03947978 | 1 | 0.503138201 | 1 | 0.498212091 | ENSG0000011 | 11 | DNAJC6 |
| Pancreas | T2DM | ENSG0000013 | 0.001509949 | 0.420004709 | 0.001872425 | 0.055522603 | 0.000753056 | 0.691701326 | 0.001899031 | 0.055519606 | 0.000756894 | 0.016977941 | 1 | 0.7882025 | 1 | 0.913082753 | ENSG0000011 | 11 | IVNS1ABP |
| Pancreas | T2DM | ENSG0000013 | 0.000669411 | 0.597149677 | 0.001266615 | 0.055522645 | 0.000520354 | 0.681380338 | 0.001267362 | 0.055519605 | 0.000149057 | 0.000813098 | 1 | 0.88286336 | 1 | 0.910080116 | ENSG0000011 | 12 | MFN2 |
| Pancreas | T2DM | ENSG0000013 | 0.001178799 | 0.441401635 | 0.001531248 | 0.05552261 | 0.001215055 | 0.427481588 | 0.001531242 | 0.055519552 | -3.63E-05 | 0.00066327 | 1 | 0.797847021 | 1 | 0.793088295 | ENSG0000011 | 10 | NCF2 |
| Pancreas | T2DM | ENSG0000013 | -0.0038582 | 0.248377402 | 0.003342457 | 0.055522525 | -0.00380787 | 0.254599951 | 0.003342438 | 0.055519476 | -5.03E-05 | 0.022207437 | 1 | 0.659767299 | 1 | 0.670900145 | ENSG0000011 | 5 | AGMAT |
| Pancreas | T2DM | ENSG0000013 | -0.00197924 | 0.515560671 | 0.003044035 | 0.055522629 | -0.00203247 | 0.504331556 | 0.003044027 | 0.055519573 | 5.32E-05 | 0.013493164 | 1 | 0.848731838 | 1 | 0.844462185 | ENSG0000011 | 5 | AGMAT |
| Pancreas | T2DM | ENSG0000013 | 0.00158431 | 0.447382023 | 0.002085202 | 0.055522611 | 0.001548004 | 0.457856888 | 0.002085188 | 0.055519561 | 3.63E-05 | 0.007172973 | 1 | 0.803972733 | 1 | 0.812561499 | ENSG0000011 | 5 | AGMAT |
| Pancreas | T2DM | ENSG0000013 | 0.001753376 | 0.153785336 | 0.001229326 | 0.055522445 | 0.001731276 | 0.159040166 | 0.001229329 | 0.055519398 | 2.21E-05 | 0.019629813 | 1 | 0.558586622 | 1 | 0.567025251 | ENSG0000011 | 9 | CFHR3 |
| Pancreas | T2DM | ENSG0000013 | -3.46E-05 | 0.991637599 | 0.003297455 | 0.055522677 | 2.25E-05 | 0.99455717 | 0.003297456 | 0.055519624 | -5.71E-05 | 0.020639733 | 1 | 0.997347815 | 1 | 0.998538835 | ENSG0000011 | 9 | CRYZ |
| Pancreas | T2DM | ENSG0000013 | -0.00274949 | 0.34949573 | 0.002938846 | 0.055522577 | -0.00266696 | 0.364164976 | 0.002938931 | 0.05551953 | -8.25E-05 | 0.008351208 | 1 | 0.743483984 | 1 | 0.758455523 | ENSG0000011 | 12 | TMEM9 |
| Pancreas | T2DM | ENSG0000014 | -0.00179055 | 0.585790464 | 0.003285726 | 0.055522643 | -0.00182792 | 0.57798708 | 0.003285685 | 0.055519589 | 3.74E-05 | 0.03965952 | 1 | 0.87968741 | 1 | 0.873565759 | ENSG0000011 | 12 | TMEM9 |
| Pancreas | T2DM | ENSG0000014 | -0.00133583 | 0.40143666 | 0.001592056 | 0.055522597 | -0.00130309 | 0.413084234 | 0.001592085 | 0.055519548 | -3.27E-05 | 0.031283656 | 1 | 0.781568873 | 1 | 0.78811316 | ENSG0000011 | 12 | TMEM9 |
| Pancreas | T2DM | ENSG0000014 | -0.00070271 | 0.365239011 | 0.000776103 | 0.055522584 | -0.00068845 | 0.37504512 | 0.0007761 | 0.055519534 | -1.43E-05 | 0.006759231 | 1 | 0.75545002 | 1 | 0.766201963 | ENSG0000011 | 12 | TMEM9 |
| Pancreas | T2DM | ENSG0000014 | -0.00295043 | 0.191600903 | 0.00225938 | 0.055522483 | -0.00292538 | 0.1953946 | 0.002259353 | 0.055519433 | -2.51E-05 | 0.044354056 | 1 | 0.59495903 | 1 | 0.60527543 | ENSG0000011 | 7 | WARS2 |
| Pancreas | T2DM | ENSG0000014 | -0.00142676 | 0.430062665 | 0.001808114 | 0.055522606 | -0.00145535 | 0.420878677 | 0.001808121 | 0.05551955 | 2.86E-05 | 0.044772804 | 1 | 0.792249323 | 1 | 0.790339598 | ENSG0000011 | 7 | WARS2 |
| Pancreas | T2DM | ENSG0000014 | 0.001704002 | 0.380760326 | 0.001944105 | 0.05552259 | 0.001682979 | 0.38665839 | 0.001944081 | 0.055519539 | 2.10E-05 | 0.049988352 | 1 | 0.767612694 | 1 | 0.772390956 | ENSG0000011 | 7 | WARS2 |
| Pancreas | T2DM | ENSG0000014 | -6.28E-05 | 0.978029635 | 0.002279259 | 0.055522677 | -9.03E-05 | 0.968411273 | 0.002279235 | 0.055519624 | 2.75E-05 | 0.037613349 | 1 | 0.99701908 | 1 | 0.995018895 | ENSG0000011 | 7 | WARS2 |
| Pancreas | T2DM | ENSG0000014 | -0.00116676 | 0.048786406 | 0.000592128 | 0.055522235 | -0.00115605 | 0.050895111 | 0.000592129 | 0.05551919 | -1.07E-05 | 0.016419394 | 1 | 0.329693399 | 1 | 0.344851303 | ENSG0000011 | 8 | RP11-268J15.5 |
| Pancreas | T2DM | ENSG0000014 | -0.00030475 | 0.796523839 | 0.001181899 | 0.05552267 | -0.00040412 | 0.732595311 | 0.001182767 | 0.055519611 | 9.94E-05 | 0.031293991 | 1 | 0.949920498 | 1 | 0.924390297 | ENSG0000011 | 7 | GNPAT |
| Pancreas | T2DM | ENSG0000014 | 0.002413461 | 0.264388945 | 0.002162447 | 0.055522535 | 0.002370501 | 0.272992127 | 0.002162473 | 0.055519487 | 4.30E-05 | 0.025676651 | 1 | 0.67759562 | 1 | 0.68377529 | ENSG0000011 | 14 | LGALS8 |
| Pancreas | T2DM | ENSG0000014 | -0.00286791 | 0.195594632 | 0.002215958 | 0.055522486 | -0.00282453 | 0.20244098 | 0.002215961 | 0.055519439 | -4.34E-05 | 0.009848045 | 1 | 0.599883083 | 1 | 0.615373581 | ENSG0000011 | 8 | PADI2 |
| Pancreas | T2DM | ENSG0000014 | -0.00392164 | 0.097514565 | 0.002366671 | 0.055522364 | -0.00388225 | 0.10092541 | 0.002366667 | 0.055519318 | -3.94E-05 | 0.019767899 | 1 | 0.478030203 | 1 | 0.486257122 | ENSG0000011 | 8 | PADI2 |
| Pancreas | T2DM | ENSG0000015 | 0.000874452 | 0.456281656 | 0.001173787 | 0.055522614 | 0.000855547 | 0.466072436 | 0.001173775 | 0.055519564 | 1.89E-05 | 0.006997541 | 1 | 0.809765924 | 1 | 0.815865042 | ENSG0000011 | 8 | PADI2 |
| Pancreas | T2DM | ENSG0000015 | 0.000673771 | 0.682667405 | 0.001648062 | 0.055522658 | 0.000642671 | 0.696571664 | 0.001648074 | 0.055519607 | 3.11E-05 | 0.023477347 | 1 | 0.91019995 | 1 | 0.9151214 | ENSG0000011 | 8 | PADI2 |
| Pancreas | T2DM | ENSG0000015 | -0.00245406 | 0.317627765 | 0.002455664 | 0.055522563 | -0.0022745 | 0.35456511 | 0.002456865 | 0.055519526 | -0.00017956 | 0.022884385 | 1 | 0.722957701 | 1 | 0.750019112 | ENSG0000011 | 9 | MFAP2 |
| Pancreas | T2DM | ENSG0000015 | 0.003710133 | 0.117262883 | 0.00236862 | 0.055522398 | 0.003820657 | 0.10681976 | 0.002369169 | 0.055519328 | -0.00011052 | 0.040615557 | 1 | 0.510742285 | 1 | 0.494470247 | ENSG0000011 | 9 | MFAP2 |
| Pancreas | T2DM | ENSG0000015 | 0.001014651 | 0.701484649 | 0.002647027 | 0.05552266 | 0.00096797 | 0.714600498 | 0.002647007 | 0.055519609 | 4.67E-05 | 0.005253769 | 1 | 0.917424714 | 1 | 0.920472548 | ENSG0000011 | 9 | MFAP2 |
| Pancreas | T2DM | ENSG0000015 | -0.00385111 | 0.040759395 | 0.001882286 | 0.0555222 | -0.0038173 | 0.042562461 | 0.001882313 | 0.055519156 | -3.38E-05 | 0.048760426 | 1 | 0.304588118 | 1 | 0.31228686 | ENSG0000011 | 9 | MFAP2 |

| | | | | | | | | | | | | | | | | | | | |
|---|---|---|---|---|---|---|---|---|---|---|---|---|---|---|---|---|---|---|---|
| Pancreas | T2DM | ENSG0000015 | -0.00149466 | 0.239448386 | 0.001270569 | 0.055522519 | -0.00148016 | 0.244028082 | 0.00127055 | 0.05551947 | -1.45E-05 | 0.025921755 | 1 | 0.651353501 | 1 | 0.662902761 | ENSG0000011 | 9 | MFAP2 |
| Pancreas | T2DM | ENSG0000015 | -0.00043118 | 0.879454329 | 0.002843064 | 0.055522675 | -0.00039482 | 0.889551716 | 0.002843032 | 0.055519622 | -3.64E-05 | 0.024788893 | 1 | 0.980887799 | 1 | 0.981479231 | ENSG0000011 | 9 | MFAP2 |
| Pancreas | T2DM | ENSG0000015 | -0.00209385 | 0.092813543 | 0.00124579 | 0.055522355 | -0.00211554 | 0.089476068 | 0.001245776 | 0.055519296 | 2.17E-05 | 0.002606514 | 1 | 0.467725388 | 1 | 0.455804017 | ENSG0000011 | 11 | KLHL12 |
| Pancreas | T2DM | ENSG0000015 | 0.001513342 | 0.258733165 | 0.001339962 | 0.055522532 | 0.001546294 | 0.248510625 | 0.00133997 | 0.055519472 | -3.30E-05 | 0.002693661 | 1 | 0.675917364 | 1 | 0.666460318 | ENSG0000011 | 6 | ZNHIT6 |
| Pancreas | T2DM | ENSG0000015 | -0.00123727 | 0.706942894 | 0.003290929 | 0.055522661 | -0.00118057 | 0.719790817 | 0.003290905 | 0.055519609 | -5.67E-05 | 0.006596104 | 1 | 0.918274836 | 1 | 0.92146093 | ENSG0000011 | 6 | ZNHIT6 |
| Pancreas | T2DM | ENSG0000015 | 0.007455189 | 0.014584025 | 0.003052208 | 0.055521997 | 0.007417863 | 0.015084743 | 0.003052183 | 0.055518951 | 3.73E-05 | 0.047688254 | 1 | 0.186327245 | 1 | 0.192724475 | ENSG0000011 | 6 | ZNHIT6 |
| Pancreas | T2DM | ENSG0000015 | 0.002682015 | 0.126811108 | 0.001756624 | 0.055522412 | 0.002516057 | 0.152360964 | 0.001757951 | 0.055519391 | 0.000165958 | 0.01697553 | 1 | 0.527160606 | 1 | 0.558149724 | ENSG0000011 | 6 | ZNHIT6 |
| Pancreas | T2DM | ENSG0000015 | -0.00228541 | 0.113297309 | 0.001443221 | 0.055522391 | -0.00226217 | 0.117010448 | 0.00144322 | 0.055519344 | -2.32E-05 | 0.028803921 | 1 | 0.505335944 | 1 | 0.512769334 | ENSG0000011 | 9 | RBBP5 |
| Pancreas | T2DM | ENSG0000015 | -0.00130472 | 0.438240327 | 0.001683148 | 0.055522609 | -0.00133974 | 0.426046624 | 0.001683143 | 0.055519552 | 3.50E-05 | 0.002963591 | 1 | 0.796485955 | 1 | 0.793088295 | ENSG0000011 | 9 | RBBP5 |
| Pancreas | T2DM | ENSG0000016 | -0.00194499 | 0.492711753 | 0.002835247 | 0.055522624 | -0.00190831 | 0.500901318 | 0.002835216 | 0.055519573 | -3.67E-05 | 0.025460689 | 1 | 0.837381722 | 1 | 0.842380212 | ENSG0000011 | 7 | GBP3 |
| Pancreas | T2DM | ENSG0000016 | -0.00641888 | 0.005314758 | 0.002302882 | 0.055521792 | -0.00638885 | 0.00553204 | 0.002302858 | 0.055518747 | -3.00E-05 | 0.024750028 | 1 | 0.10498691 | 1 | 0.10972011 | ENSG0000011 | 7 | GBP3 |
| Pancreas | T2DM | ENSG0000016 | 0.005239212 | 0.098067498 | 0.003167029 | 0.055522365 | 0.005192248 | 0.101114215 | 0.003167011 | 0.055519318 | 4.70E-05 | 0.024783184 | 1 | 0.479690164 | 1 | 0.486257122 | ENSG0000011 | 9 | PRDX1 |
| Pancreas | T2DM | ENSG0000016 | -0.00193592 | 0.152126634 | 0.001351842 | 0.055522443 | -0.00191281 | 0.157078281 | 0.00135184 | 0.055519396 | -2.31E-05 | 0.017199488 | 1 | 0.557291293 | 1 | 0.561838668 | ENSG0000011 | 9 | PRDX1 |
| Pancreas | T2DM | ENSG0000016 | 0.003856836 | 0.026847217 | 0.001742227 | 0.055522119 | 0.003836508 | 0.027659091 | 0.001742209 | 0.055519072 | 2.03E-05 | 0.044207889 | 1 | 0.251619173 | 1 | 0.256314825 | ENSG0000011 | 9 | PRDX1 |
| Pancreas | T2DM | ENSG0000016 | 0.002407565 | 0.404602557 | 0.002888743 | 0.055522598 | 0.002440804 | 0.398139902 | 0.002888701 | 0.055519543 | -3.32E-05 | 0.025349592 | 1 | 0.783229955 | 1 | 0.778692512 | ENSG0000011 | 9 | PRDX1 |
| Pancreas | T2DM | ENSG0000016 | -0.00425684 | 0.042850488 | 0.002101965 | 0.05552221 | -0.00423081 | 0.044135076 | 0.002101942 | 0.055519163 | -2.60E-05 | 0.030093723 | 1 | 0.311721277 | 1 | 0.31658904 | ENSG0000011 | 8 | CCDC181 |
| Pancreas | T2DM | ENSG0000016 | -0.00296509 | 0.3173979 | 0.002965621 | 0.055522563 | -0.0029216 | 0.324545116 | 0.002965603 | 0.055519514 | -4.35E-05 | 0.025716613 | 1 | 0.722957701 | 1 | 0.729254023 | ENSG0000011 | 11 | FAAH |
| Pancreas | T2DM | ENSG0000016 | 0.001429515 | 0.239198112 | 0.001214547 | 0.055522519 | 0.001683829 | 0.166229098 | 0.001216266 | 0.055519406 | -0.00025431 | 9.81E-05 | 1 | 0.651353501 | 1 | 0.572425504 | ENSG0000011 | 11 | FAAH |
| Pancreas | T2DM | ENSG0000016 | 0.000104001 | 0.94774624 | 0.001586899 | 0.055522677 | -0.00013594 | 0.931836441 | 0.00158926 | 0.055519623 | 0.000239938 | 0.006036893 | 1 | 0.989794375 | 1 | 0.987209256 | ENSG0000011 | 11 | FAAH |
| Pancreas | T2DM | ENSG0000016 | 0.004166166 | 0.099914135 | 0.002532203 | 0.055522369 | 0.004200803 | 0.097123008 | 0.002532179 | 0.055519311 | -3.46E-05 | 0.022711767 | 1 | 0.483200563 | 1 | 0.472157259 | ENSG0000011 | 6 | NSUN4 |
| Pancreas | T2DM | ENSG0000016 | 0.000863958 | 0.324644401 | 0.000877151 | 0.055522567 | 0.000875285 | 0.318336368 | 0.000877141 | 0.055519511 | -1.13E-05 | 0.025426465 | 1 | 0.723686476 | 1 | 0.729012597 | ENSG0000011 | 6 | NSUN4 |
| Pancreas | T2DM | ENSG0000016 | 0.000157237 | 0.863387353 | 0.000913832 | 0.055522674 | 0.000171711 | 0.850952102 | 0.000913824 | 0.05551962 | -1.45E-05 | 0.009507463 | 1 | 0.976495788 | 1 | 0.97099298 | ENSG0000011 | 8 | DR1 |
| Pancreas | T2DM | ENSG0000016 | 0.001365568 | 0.623547857 | 0.002782168 | 0.05552265 | 0.001406806 | 0.613099447 | 0.002782149 | 0.055519595 | -4.12E-05 | 0.021269443 | 1 | 0.89259216 | 1 | 0.885013704 | ENSG0000011 | 11 | PRRC2C |
| Pancreas | T2DM | ENSG0000016 | 0.000924 | 0.696546296 | 0.00236931 | 0.05552266 | 0.00088917 | 0.707445523 | 0.002369291 | 0.055519608 | 3.48E-05 | 0.018852179 | 1 | 0.916358037 | 1 | 0.919392765 | ENSG0000011 | 11 | PRRC2C |
| Pancreas | T2DM | ENSG0000016 | 0.000363282 | 0.672164277 | 0.000858457 | 0.055522657 | 0.000350442 | 0.683108006 | 0.00085845 | 0.055519605 | 1.28E-05 | 0.015352232 | 1 | 0.909524436 | 1 | 0.910080116 | ENSG0000011 | 10 | VAMP4 |
| Pancreas | T2DM | ENSG0000016 | -0.00274273 | 0.001640399 | 0.000871083 | 0.055521548 | -0.00273256 | 0.001706974 | 0.000871074 | 0.055518503 | -1.02E-05 | 0.046084922 | 1 | 0.048420063 | 1 | 0.050971036 | ENSG0000011 | 10 | VAMP4 |
| Pancreas | T2DM | ENSG0000016 | -0.00193033 | 0.559800702 | 0.003310242 | 0.055522638 | -0.00188601 | 0.568845024 | 0.003310215 | 0.055519587 | -4.43E-05 | 0.031559102 | 1 | 0.86976803 | 1 | 0.868486338 | ENSG0000011 | 14 | PTBP2 |
| Pancreas | T2DM | ENSG0000016 | -0.00196099 | 0.44339617 | 0.002558459 | 0.05552261 | -0.00192545 | 0.451697181 | 0.002558439 | 0.05551956 | -3.55E-05 | 0.026482341 | 1 | 0.799538512 | 1 | 0.805155685 | ENSG0000011 | 14 | PTBP2 |
| Pancreas | T2DM | ENSG0000016 | -0.001228 | 0.539342952 | 0.002000634 | 0.055522634 | -0.00102268 | 0.609449787 | 0.002001884 | 0.055519594 | -0.00020532 | 0.004494747 | 1 | 0.858495882 | 1 | 0.883228336 | ENSG0000011 | 6 | IRF6 |
| Pancreas | T2DM | ENSG0000016 | 0.00136915 | 0.304891526 | 0.001334452 | 0.055522557 | 0.001351987 | 0.310990245 | 0.001334442 | 0.055519507 | 1.72E-05 | 0.04156034 | 1 | 0.711360218 | 1 | 0.721874686 | ENSG0000011 | 6 | IRF6 |
| Pancreas | T2DM | ENSG0000016 | 0.001855024 | 0.100708998 | 0.001130127 | 0.05552237 | 0.001839461 | 0.103595009 | 0.001130118 | 0.055519322 | 1.56E-05 | 0.025092576 | 1 | 0.483403189 | 1 | 0.486323959 | ENSG0000011 | 6 | IRF6 |
| Pancreas | T2DM | ENSG0000016 | -0.00039335 | 0.440121803 | 0.000509526 | 0.055522609 | -0.00042034 | 0.409479302 | 0.000509617 | 0.055519547 | 2.70E-05 | 0.009144689 | 1 | 0.797847021 | 1 | 0.785871608 | ENSG0000011 | 6 | IRF6 |
| Pancreas | T2DM | ENSG0000016 | 0.001842036 | 0.527114205 | 0.002912689 | 0.055522632 | 0.001890247 | 0.516355206 | 0.002912667 | 0.055519576 | -4.82E-05 | 0.009158739 | 1 | 0.856215034 | 1 | 0.848610147 | ENSG0000011 | 12 | DHDDS |
| Pancreas | T2DM | ENSG0000016 | -8.65E-05 | 0.926668997 | 0.000939866 | 0.055522676 | -7.15E-05 | 0.939346365 | 0.000939862 | 0.055519623 | -1.50E-05 | 0.018449059 | 1 | 0.986516064 | 1 | 0.987209256 | ENSG0000011 | 10 | NSL1 |
| Pancreas | T2DM | ENSG0000016 | -0.00499386 | 0.003938683 | 0.001732149 | 0.05552173 | -0.00496924 | 0.004119756 | 0.001732133 | 0.055518686 | -2.46E-05 | 0.01829558 | 1 | 0.083591224 | 1 | 0.088903632 | ENSG0000011 | 10 | NSL1 |
| Pancreas | T2DM | ENSG0000016 | -0.00582401 | 0.000858711 | 0.001747291 | 0.055521411 | -0.0057932 | 0.000914678 | 0.001747277 | 0.055518372 | -3.08E-05 | 0.004817152 | 1 | 0.034383558 | 1 | 0.035306133 | ENSG0000011 | 5 | RPA2 |
| Pancreas | T2DM | ENSG0000016 | 0.001560702 | 0.313176433 | 0.001547424 | 0.055522561 | 0.001590308 | 0.304082799 | 0.001547416 | 0.055519504 | -2.96E-05 | 0.003900563 | 1 | 0.719997385 | 1 | 0.713788508 | ENSG0000011 | 11 | ESYT2 |
| Pancreas | T2DM | ENSG0000016 | 0.00097048 | 0.728505361 | 0.002795849 | 0.055522663 | 0.001021502 | 0.714839193 | 0.002795841 | 0.055519609 | -5.10E-05 | 0.008916102 | 1 | 0.923852724 | 1 | 0.920472548 | ENSG0000011 | 3 | A4GNT |
| Pancreas | T2DM | ENSG0000016 | 0.002798512 | 0.261759309 | 0.002493676 | 0.055522534 | 0.002826693 | 0.256979661 | 0.002493642 | 0.055519478 | -2.82E-05 | 0.033343233 | 1 | 0.67693646 | 1 | 0.672705168 | ENSG0000011 | 3 | A4GNT |
| Pancreas | T2DM | ENSG0000016 | -0.00287585 | 0.198967556 | 0.002238883 | 0.055522489 | -0.00293641 | 0.189693663 | 0.002238993 | 0.055519428 | 6.06E-05 | 0.029311659 | 1 | 0.606827415 | 1 | 0.59849 | ENSG0000011 | 3 | IFT46 |
| Pancreas | T2DM | ENSG0000016 | -4.45E-05 | 0.954911395 | 0.00078662 | 0.055522677 | -5.49E-05 | 0.944362042 | 0.000786614 | 0.055519624 | 1.04E-05 | 0.034861373 | 1 | 0.989794375 | 1 | 0.987209256 | ENSG0000011 | 9 | DDX59 |
| Pancreas | T2DM | ENSG0000016 | 0.000158611 | 0.893201882 | 0.001181426 | 0.055522675 | 0.000178158 | 0.880133642 | 0.001181423 | 0.055519622 | -1.95E-05 | 0.017622106 | 1 | 0.981247774 | 1 | 0.980763999 | ENSG0000011 | 9 | DDX59 |
| Pancreas | T2DM | ENSG0000016 | 0.000164271 | 0.843303037 | 0.000831036 | 0.055522673 | 0.000175258 | 0.832970695 | 0.000831026 | 0.055519619 | -1.10E-05 | 0.017560588 | 1 | 0.96894953 | 1 | 0.962244149 | ENSG0000011 | 9 | DDX59 |
| Pancreas | T2DM | ENSG0000016 | 0.003605659 | 0.029316569 | 0.001654572 | 0.055522136 | 0.003560783 | 0.031392454 | 0.001654586 | 0.055519096 | 4.49E-05 | 0.001443884 | 1 | 0.264157714 | 1 | 0.275139321 | ENSG0000011 | 9 | DDX59 |
| Pancreas | T2DM | ENSG0000016 | -9.18E-05 | 0.948281103 | 0.001415111 | 0.055522677 | -7.61E-05 | 0.957136141 | 0.00141509 | 0.055519624 | -1.57E-05 | 0.028383699 | 1 | 0.989794375 | 1 | 0.991898954 | ENSG0000011 | 9 | DDX59 |
| Pancreas | T2DM | ENSG0000017 | 0.003665992 | 0.13150953 | 0.002430742 | 0.055522418 | 0.003706374 | 0.127308543 | 0.002430721 | 0.055519359 | -4.04E-05 | 0.006821596 | 1 | 0.531854333 | 1 | 0.53156386 | ENSG0000011 | 10 | CAMSAP2 |
| Pancreas | T2DM | ENSG0000017 | -0.00253735 | 0.126250557 | 0.001659423 | 0.055522411 | -0.00255775 | 0.123227863 | 0.001659403 | 0.055519353 | 2.04E-05 | 0.028297417 | 1 | 0.526317257 | 1 | 0.521333034 | ENSG0000011 | 14 | CASC1 |
| Pancreas | T2DM | ENSG0000017 | 0.000517256 | 0.776243511 | 0.001819929 | 0.055522668 | 0.000488673 | 0.7883046 | 0.001819924 | 0.055519616 | 2.86E-05 | 0.024331323 | 1 | 0.948363389 | 1 | 0.947789023 | ENSG0000011 | 14 | CASC1 |
| Pancreas | T2DM | ENSG0000017 | 0.001841811 | 0.429446326 | 0.002330992 | 0.055522606 | 0.001880405 | 0.419836827 | 0.00233097 | 0.05551955 | -3.86E-05 | 0.005819781 | 1 | 0.792249323 | 1 | 0.789846866 | ENSG0000011 | 14 | CASC1 |
| Pancreas | T2DM | ENSG0000017 | -0.00302416 | 0.108308546 | 0.001883218 | 0.055522383 | -0.00299301 | 0.111989224 | 0.001883202 | 0.055519336 | -3.11E-05 | 0.007841299 | 1 | 0.49934712 | 1 | 0.500338797 | ENSG0000011 | 14 | CASC1 |
| Pancreas | T2DM | ENSG0000017 | -0.00035997 | 0.913189746 | 0.003302024 | 0.055522676 | -0.00041725 | 0.899446086 | 0.003302008 | 0.055519622 | 5.73E-05 | 0.010239892 | 1 | 0.98441119 | 1 | 0.984551279 | ENSG0000011 | 10 | LRMP |
| Pancreas | T2DM | ENSG0000017 | -0.00069355 | 0.721004169 | 0.001942084 | 0.055522663 | -0.0006694 | 0.730333953 | 0.001942069 | 0.055519611 | -2.42E-05 | 0.049946004 | 1 | 0.921476762 | 1 | 0.924345782 | ENSG0000011 | 5 | ELOVL4 |
| Pancreas | T2DM | ENSG0000017 | 0.007456977 | 0.018259595 | 0.003159308 | 0.055522042 | 0.00750216 | 0.017566094 | 0.003159273 | 0.055518982 | -4.52E-05 | 0.01239789 | 1 | 0.206641906 | 1 | 0.209456125 | ENSG0000011 | 10 | FILIP1 |
| Pancreas | T2DM | ENSG0000017 | -0.00444049 | 0.06348675 | 0.002392802 | 0.055522285 | -0.00447514 | 0.061448663 | 0.002392792 | 0.055519226 | 3.46E-05 | 0.033703455 | 1 | 0.386336431 | 1 | 0.38581948 | ENSG0000011 | 10 | FILIP1 |
| Pancreas | T2DM | ENSG0000017 | -0.00015423 | 0.882722626 | 0.001045524 | 0.055522675 | -0.00017184 | 0.869445822 | 0.001045524 | 0.055519621 | 1.76E-05 | 0.022920516 | 1 | 0.980887799 | 1 | 0.976273228 | ENSG0000011 | 11 | AKAP7 |
| Pancreas | T2DM | ENSG0000017 | -0.00134096 | 0.450161379 | 0.001775762 | 0.055522612 | -0.00136308 | 0.442718852 | 0.001775749 | 0.055519557 | 2.21E-05 | 0.048268883 | 1 | 0.806146738 | 1 | 0.800635219 | ENSG0000011 | 11 | AKAP7 |
| Pancreas | T2DM | ENSG0000017 | 0.000832856 | 0.317063627 | 0.000832431 | 0.055522563 | 0.000845223 | 0.309931166 | 0.000832431 | 0.055519507 | -1.24E-05 | 0.044775264 | 1 | 0.722957701 | 1 | 0.720926844 | ENSG0000011 | 11 | AKAP7 |
| Pancreas | T2DM | ENSG0000017 | 0.002046541 | 0.253570041 | 0.001792503 | 0.055522529 | 0.002074224 | 0.247202061 | 0.001792492 | 0.055519472 | -2.77E-05 | 0.017427548 | 1 | 0.66855017 | 1 | 0.664032315 | ENSG0000011 | 11 | AKAP7 |
| Pancreas | T2DM | ENSG0000017 | -0.00058545 | 0.823580321 | 0.00262603 | 0.055522671 | -0.00061581 | 0.814592253 | 0.002626002 | 0.055519618 | 3.04E-05 | 0.046161783 | 1 | 0.960390921 | 1 | 0.956268122 | ENSG0000011 | 9 | ALDH8A1 |
| Pancreas | T2DM | ENSG0000017 | 0.002316489 | 0.256422276 | 0.002041159 | 0.05552253 | 0.002363015 | 0.246993682 | 0.002041158 | 0.055519471 | -4.65E-05 | 0.002012639 | 1 | 0.67153808 | 1 | 0.664032315 | ENSG0000011 | 9 | ALDH8A1 |
| Pancreas | T2DM | ENSG0000017 | 0.001068366 | 0.416276367 | 0.001314267 | 0.055522602 | 0.001089385 | 0.407163308 | 0.001314259 | 0.055519546 | -2.10E-05 | 0.015568664 | 1 | 0.786828099 | 1 | 0.784981512 | ENSG0000011 | 9 | ALDH8A1 |
| Pancreas | T2DM | ENSG0000017 | -0.00018229 | 0.924957769 | 0.001935313 | 0.055522676 | -0.00022559 | 0.907205736 | 0.001935315 | 0.055519623 | 4.33E-05 | 0.003074394 | 1 | 0.986312425 | 1 | 0.985211345 | ENSG0000011 | 9 | ALDH8A1 |
| Pancreas | T2DM | ENSG0000018 | 1.71E-05 | 0.980760835 | 0.000709038 | 0.055522677 | 2.95E-05 | 0.966763674 | 0.000709033 | 0.055519624 | -1.24E-05 | 0.005675189 | 1 | 0.99701908 | 1 | 0.99385473 | ENSG0000011 | 7 | MED28 |
| Pancreas | T2DM | ENSG0000018 | 0.003769288 | 0.10283518 | 0.002310654 | 0.055522374 | 0.003734178 | 0.106077679 | 0.002310639 | 0.055519327 | 3.51E-05 | 0.018870118 | 1 | 0.487300933 | 1 | 0.493491813 | ENSG0000011 | 7 | MED28 |
| Pancreas | T2DM | ENSG0000018 | 0.000358488 | 0.63807414 | 0.000762103 | 0.055522652 | 0.000197356 | 0.796229722 | 0.000764267 | 0.055519617 | 0.000161132 | 0.00525703 | 1 | 0.898449905 | 1 | 0.948825024 | ENSG0000011 | 7 | MED28 |
| Pancreas | T2DM | ENSG0000018 | 0.001241699 | 0.387915184 | 0.001438142 | 0.055522592 | 0.001142467 | 0.427205479 | 0.001438906 | 0.055519552 | 9.92E-05 | 0.03899189 | 1 | 0.770733172 | 1 | 0.793088295 | ENSG0000011 | 7 | TMEM5 |
| Pancreas | T2DM | ENSG0000018 | 0.000879311 | 0.327334419 | 0.000897716 | 0.055522568 | 0.000868918 | 0.333078405 | 0.000897705 | 0.055519517 | 1.04E-05 | 0.038894491 | 1 | 0.725275917 | 1 | 0.735008377 | ENSG0000011 | 6 | VAMP8 |
| Pancreas | T2DM | ENSG0000018 | 0.002420148 | 0.243960325 | 0.002077119 | 0.055522522 | 0.002386184 | 0.250641336 | 0.00207712 | 0.055519474 | 3.40E-05 | 0.028787765 | 1 | 0.656811468 | 1 | 0.666460318 | ENSG0000011 | 6 | VAMP8 |
| Pancreas | T2DM | ENSG0000018 | -0.00165518 | 0.365663698 | 0.001829685 | 0.055522584 | -0.00168234 | 0.357846203 | 0.001829671 | 0.055519528 | 2.72E-05 | 0.018597619 | 1 | 0.75545002 | 1 | 0.752004132 | ENSG0000011 | 8 | ARMC2 |
| Pancreas | T2DM | ENSG0000018 | 0.003089306 | 0.227259353 | 0.002558537 | 0.055522511 | 0.00303355 | 0.235759192 | 0.002558544 | 0.055519464 | 5.58E-05 | 0.005102935 | 1 | 0.6413209 | 1 | 0.655938795 | ENSG0000011 | 10 | UBN1 |
| Pancreas | T2DM | ENSG0000018 | -0.00088752 | 0.642403363 | 0.001911356 | 0.055522653 | -0.00085623 | 0.654174515 | 0.00191135 | 0.055519601 | -3.13E-05 | 0.018865622 | 1 | 0.898449905 | 1 | 0.900271628 | ENSG0000011 | 10 | UBN1 |
| Pancreas | T2DM | ENSG0000018 | 0.000459079 | 0.670065828 | 0.001077504 | 0.055522656 | 0.000441535 | 0.681968828 | 0.001077497 | 0.055519605 | 1.75E-05 | 0.011294982 | 1 | 0.908515863 | 1 | 0.910080116 | ENSG0000011 | 10 | UBN1 |
| Pancreas | T2DM | ENSG0000018 | 0.001336172 | 0.099840528 | 0.000811951 | 0.055522369 | 0.00132395 | 0.102980174 | 0.000811951 | 0.055519321 | 1.22E-05 | 0.042326108 | 1 | 0.483200563 | 1 | 0.486323959 | ENSG0000011 | 10 | UBN1 |
| Pancreas | T2DM | ENSG0000018 | -0.0016446 | 0.428091362 | 0.002075308 | 0.055522606 | -0.00157842 | 0.446931408 | 0.002075384 | 0.055519558 | -6.62E-05 | 0.004813564 | 1 | 0.792249323 | 1 | 0.8027301 | ENSG0000011 | 10 | C2orf43 |
| Pancreas | T2DM | ENSG0000018 | -0.00232741 | 0.018227452 | 0.000985783 | 0.055522042 | -0.00251901 | 0.010968884 | 0.000990293 | 0.055518887 | 0.0001916 | 0.043043385 | 1 | 0.206641906 | 1 | 0.16247674 | ENSG0000011 | 10 | C2orf43 |
| Pancreas | T2DM | ENSG0000018 | -0.00105224 | 0.466103126 | 0.001443731 | 0.055522617 | -0.00108394 | 0.452777824 | 0.001443726 | 0.05551956 | 3.17E-05 | 0.001517944 | 1 | 0.815461018 | 1 | 0.806333878 | ENSG0000011 | 10 | C2orf43 |
| Pancreas | T2DM | ENSG0000018 | 0.004043275 | 0.209741102 | 0.003223584 | 0.055522498 | 0.003617152 | 0.262409508 | 0.003227538 | 0.055519481 | 0.000426123 | 0.008312403 | 1 | 0.61697039 | 1 | 0.677451547 | ENSG0000011 | 10 | C2orf43 |
| Pancreas | T2DM | ENSG0000018 | -1.81E-05 | 0.979094739 | 0.000692379 | 0.055522677 | -9.28E-06 | 0.989306689 | 0.000692371 | 0.055519624 | -8.86E-06 | 0.023816934 | 1 | 0.99701908 | 1 | 0.998538835 | ENSG0000011 | 10 | C2orf43 |
| Pancreas | T2DM | ENSG0000018 | 0.00234185 | 0.278010939 | 0.002158786 | 0.055522543 | -0.00167703 | 0.554078908 | 0.002834458 | 0.055519584 | 0.004018884 | 0.028675925 | 1 | 0.688459105 | 1 | 0.864388971 | ENSG0000011 | 10 | C2orf43 |
| Pancreas | T2DM | ENSG0000018 | -1.72E-05 | 0.98790529 | 0.001132827 | 0.055522677 | -4.56E-06 | 0.996789086 | 0.001132812 | 0.055519624 | -1.26E-05 | 0.036580676 | 1 | 0.997207295 | 1 | 0.998538835 | ENSG0000011 | 9 | DNAH7 |
| Pancreas | T2DM | ENSG0000018 | -0.0004934 | 0.795714155 | 0.001905767 | 0.05552267 | -0.00104991 | 0.585605189 | 0.001925672 | 0.05551959 | 0.000556508 | 0.044165898 | 1 | 0.949920498 | 1 | 0.876360212 | ENSG0000011 | 4 | NDUFB3 |
| Pancreas | T2DM | ENSG0000018 | 0.001177058 | 0.605794638 | 0.00228074 | 0.055522647 | 0.001267165 | 0.578527088 | 0.002280969 | 0.055519589 | -9.01E-05 | 0.013480526 | 1 | 0.883541492 | 1 | 0.873565759 | ENSG0000011 | 4 | NDUFB3 |
| Pancreas | T2DM | ENSG0000019 | -9.78E-05 | 0.954336415 | 0.001707457 | 0.055522677 | -0.00014757 | 0.931132541 | 0.001707575 | 0.055519623 | 4.98E-05 | 0.03574147 | 1 | 0.989794375 | 1 | 0.987209256 | ENSG0000011 | 4 | NDUFB3 |
| Pancreas | T2DM | ENSG0000019 | 0.001082671 | 0.388423093 | 0.001255297 | 0.055522592 | 0.000564483 | 0.654447642 | 0.001261155 | 0.055519601 | 0.000518187 | 2.08E-05 | 1 | 0.770733172 | 1 | 0.900271628 | ENSG0000011 | 9 | CPSF3 |
| Pancreas | T2DM | ENSG0000019 | -0.00163155 | 0.235768192 | 0.0013761 | 0.055522517 | -0.00173535 | 0.20757235 | 0.001376963 | 0.055519443 | 0.000103798 | 0.037177142 | 1 | 0.65041537 | 1 | 0.617666041 | ENSG0000011 | 9 | CPSF3 |
| Pancreas | T2DM | ENSG0000019 | -0.00434364 | 0.000429489 | 0.001233537 | 0.055521264 | -0.00209523 | 0.148611492 | 0.001450534 | 0.055519386 | -0.00224841 | 0.003219071 | 1 | 0.020102425 | 1 | 0.55791981 | ENSG0000011 | 9 | CPSF3 |
| Pancreas | T2DM | ENSG0000019 | -0.00113667 | 0.675768156 | 0.002717703 | 0.055522657 | -0.00073258 | 0.787968695 | 0.00272384 | 0.055519616 | -0.00040409 | 0.027943931 | 1 | 0.909524436 | 1 | 0.947789023 | ENSG0000011 | 9 | CPSF3 |
| Pancreas | T2DM | ENSG0000019 | 0.000794212 | 0.691014214 | 0.001998122 | 0.055522659 | 0.000826573 | 0.679111933 | 0.001998119 | 0.055519605 | -3.24E-05 | 0.023910968 | 1 | 0.91152931 | 1 | 0.909733669 | ENSG0000011 | 3 | PIGZ |
| Pancreas | T2DM | ENSG0000019 | -0.00333837 | 0.223851339 | 0.002744575 | 0.055522509 | -0.00341897 | 0.212900485 | 0.002744767 | 0.055519447 | 8.06E-05 | 0.035534977 | 1 | 0.636627966 | 1 | 0.621677611 | ENSG0000011 | 3 | PIGZ |
| Pancreas | T2DM | ENSG0000019 | 0.001166176 | 0.586813067 | 0.002145834 | 0.055522643 | 0.001202746 | 0.575134073 | 0.002145819 | 0.055519588 | -3.66E-05 | 0.007775168 | 1 | 0.87968741 | 1 | 0.87300225 | ENSG0000011 | 3 | PIGZ |
| Pancreas | T2DM | ENSG0000019 | 0.001676391 | 0.475477901 | 0.002349218 | 0.055522619 | 0.001713469 | 0.465768671 | 0.002349208 | 0.055519564 | -3.71E-05 | 0.020246698 | 1 | 0.821134667 | 1 | 0.815865042 | ENSG0000011 | 7 | WDR34 |
| Pancreas | T2DM | ENSG0000019 | 0.003547413 | 0.031005031 | 0.001644594 | 0.055522147 | 0.003767599 | 0.022060578 | 0.001645721 | 0.055519027 | -0.00022019 | 0.000393676 | 1 | 0.269901423 | 1 | 0.237035833 | ENSG0000011 | 7 | WDR34 |
| Pancreas | T2DM | ENSG0000019 | -0.00111673 | 0.416012338 | 0.001372986 | 0.055522602 | -0.00162138 | 0.239285256 | 0.001377812 | 0.055519466 | 0.000504645 | 1.29E-05 | 1 | 0.786828099 | 1 | 0.655938795 | ENSG0000011 | 10 | BSPRY |
| Pancreas | T2DM | ENSG0000019 | -0.00255461 | 2.85E-06 | 0.000545723 | 0.055520181 | -0.00189174 | 0.001806254 | 0.000606251 | 0.055518515 | -0.00066287 | 0.012067198 | 0.007328071 | 0.000407115 | 1 | 0.05167457 | ENSG0000011 | 5 | HDHD3 |
| Pancreas | T2DM | ENSG0000019 | -0.00024434 | 0.896426807 | 0.001877008 | 0.055522675 | -4.35E-06 | 0.998152931 | 0.001880195 | 0.055519624 | -0.00023999 | 0.029581708 | 1 | 0.981247774 | 1 | 0.998541772 | ENSG0000011 | 5 | HDHD3 |
| Pancreas | T2DM | ENSG0000019 | -0.00094447 | 0.606255082 | 0.001832405 | 0.055522647 | -0.00099983 | 0.58531571 | 0.001832414 | 0.05551959 | 5.54E-05 | 0.000173014 | 1 | 0.883541492 | 1 | 0.876360212 | ENSG0000011 | 5 | HDHD3 |
| Pancreas | T2DM | ENSG0000019 | 0.000221983 | 0.889728874 | 0.001601058 | 0.055522675 | 0.000202141 | 0.899529706 | 0.001601043 | 0.055519622 | 1.98E-05 | 0.039229125 | 1 | 0.981247774 | 1 | 0.984551279 | ENSG0000011 | 5 | HDHD3 |
| Pancreas | T2DM | ENSG0000020 | -0.00208118 | 0.294743789 | 0.001986287 | 0.055522552 | -0.00203362 | 0.305918516 | 0.001986294 | 0.055519505 | -4.76E-05 | 0.002218077 | 1 | 0.703918761 | 1 | 0.714830528 | ENSG0000011 | 5 | HDHD3 |
| Pancreas | T2DM | ENSG0000020 | 0.004545899 | 0.009464388 | 0.001751914 | 0.05552191 | 0.004515255 | 0.00995741 | 0.001751922 | 0.055518867 | 3.06E-05 | 0.028353185 | 1 | 0.144669926 | 1 | 0.154039932 | ENSG0000011 | 10 | HSDL2 |
| Pancreas | T2DM | ENSG0000020 | 0.001144713 | 0.307612528 | 0.001121997 | 0.055522559 | 0.000934856 | 0.406490719 | 0.001126217 | 0.055519546 | 0.000209857 | 0.031791658 | 1 | 0.715533489 | 1 | 0.784273604 | ENSG0000011 | 10 | HSDL2 |
| Pancreas | T2DM | ENSG0000020 | -0.00922753 | 1.11E-08 | 0.001615348 | 0.055518959 | -0.00849702 | 2.33E-07 | 0.001643105 | 0.055516577 | -0.00073051 | 0.015200963 | 2.86E-05 | 3.58E-06 | 0.00059716 | 5.43E-05 | ENSG0000011 | 10 | HSDL2 |
| Pancreas | T2DM | ENSG0000020 | 0.006540707 | 0.011450431 | 0.002586637 | 0.055521949 | 0.004839405 | 0.065413667 | 0.00262665 | 0.055519237 | 0.001701302 | 0.000197596 | 1 | 0.161564328 | 1 | 0.398062316 | ENSG0000011 | 10 | HSDL2 |
| Pancreas | T2DM | ENSG0000020 | -0.00051166 | 0.464694221 | 0.000699805 | 0.055522616 | -0.00158128 | 0.032326248 | 0.000738792 | 0.055519102 | 0.001069624 | 6.32E-06 | 1 | 0.815461018 | 1 | 0.280452045 | ENSG0000011 | 8 | INVS |
| Pancreas | T2DM | ENSG0000020 | -0.00290924 | 0.290248886 | 0.002750851 | 0.05552255 | -0.00237103 | 0.389696311 | 0.002756466 | 0.05551954 | -0.00053821 | 0.002362183 | 1 | 0.700376625 | 1 | 0.772390956 | ENSG0000011 | 8 | INVS |
| Pancreas | T2DM | ENSG0000020 | 0.001381367 | 0.110519425 | 0.000865591 | 0.055522387 | 0.001233648 | 0.155425201 | 0.000868382 | 0.055519394 | 0.000147719 | 0.034445774 | 1 | 0.503138201 | 1 | 0.561765877 | ENSG0000011 | 12 | DCAF4 |
| Pancreas | T2DM | ENSG0000020 | -0.00148041 | 0.012381934 | 0.00059191 | 0.055521964 | -0.00110209 | 0.068762241 | 0.000605549 | 0.055519247 | -0.00037832 | 0.003088531 | 1 | 0.169131954 | 1 | 0.405934333 | ENSG0000011 | 12 | DCAF4 |
| Pancreas | T2DM | ENSG0000020 | 0.00335196 | 0.013531726 | 0.001357361 | 0.055521982 | 0.003014369 | 0.026681917 | 0.001360185 | 0.055519065 | 0.000337591 | 0.000128313 | 1 | 0.175502385 | 1 | 0.252838238 | ENSG0000011 | 2 | VSX2 |
| Pancreas | T2DM | ENSG0000020 | 0.001267427 | 0.667246568 | 0.002947982 | 0.055522656 | 0.001775548 | 0.547669661 | 0.002953064 | 0.055519583 | -0.00050812 | 0.003603745 | 1 | 0.90774704 | 1 | 0.863361381 | ENSG0000011 | 10 | ACOT2 |
| Pancreas | T2DM | ENSG0000020 | -0.00236213 | 0.026748192 | 0.001066337 | 0.055522118 | -0.00218376 | 0.041132549 | 0.001069319 | 0.055519149 | -0.00017837 | 0.026113185 | 1 | 0.251609367 | 1 | 0.31158816 | ENSG0000011 | 10 | ACOT2 |
| Pancreas | T2DM | ENSG0000020 | 0.005046817 | 0.001153156 | 0.001552752 | 0.055521474 | 0.00587238 | 0.000192334 | 0.001574826 | 0.05551804 | -0.00082556 | 0.001697569 | 1 | 0.039484046 | 0.493914241 | 0.011759863 | ENSG0000011 | 8 | ALDH6A1 |
| Pancreas | T2DM | ENSG0000020 | -0.00029195 | 0.884949226 | 0.002017615 | 0.055522675 | -0.00064562 | 0.749520401 | 0.002022152 | 0.055519613 | 0.000353671 | 0.009416 | 1 | 0.981184043 | 1 | 0.933447328 | ENSG0000011 | 10 | ESRRB |
| Pancreas | T2DM | ENSG0000020 | 0.001000043 | 0.57829048 | 0.001799014 | 0.055522642 | -0.00010516 | 0.953790212 | 0.001814722 | 0.055519624 | 0.001105202 | 3.63E-06 | 1 | 0.878208132 | 1 | 0.991898954 | ENSG0000011 | 10 | ESRRB |
| Pancreas | T2DM | ENSG0000020 | -0.00026346 | 0.746570664 | 0.000815244 | 0.055522665 | -3.65E-06 | 0.996434562 | 0.000817433 | 0.055519624 | -0.00025981 | 1.53E-05 | 1 | 0.932263247 | 1 | 0.998538835 | ENSG0000011 | 10 | ESRRB |
| Pancreas | T2DM | ENSG0000020 | -0.00026937 | 0.740777553 | 0.000814251 | 0.055522665 | -0.00038163 | 0.639692745 | 0.000815223 | 0.055519599 | 0.000112255 | 0.005281361 | 1 | 0.930574024 | 1 | 0.895174073 | ENSG0000011 | 10 | ESRRB |
| Pancreas | T2DM | ENSG0000020 | -0.00051811 | 0.72314398 | 0.001462513 | 0.055522663 | -0.00054726 | 0.708262224 | 0.001462511 | 0.055519608 | 2.91E-05 | 0.005967389 | 1 | 0.921476762 | 1 | 0.919473235 | ENSG0000011 | 10 | ESRRB |
| Pancreas | T2DM | ENSG0000020 | 0.004477673 | 0.077492724 | 0.002536312 | 0.055522322 | 0.004569888 | 0.071606902 | 0.002536553 | 0.055519254 | -9.22E-05 | 0.020206063 | 1 | 0.429808455 | 1 | 0.414158837 | ENSG0000011 | 10 | ESRRB |
| Pancreas | T2DM | ENSG0000020 | 0.001319648 | 0.239088329 | 0.001120939 | 0.055522519 | 0.00094315 | 0.401546012 | 0.001124318 | 0.055519544 | 0.000376497 | 1.68E-05 | 1 | 0.651353501 | 1 | 0.782159733 | ENSG0000011 | 6 | EIF2B2 |
| Pancreas | T2DM | ENSG0000020 | 0.005575874 | 1.35E-06 | 0.001153723 | 0.055520016 | 0.004772884 | 5.00E-05 | 0.001176785 | 0.05551775 | 0.00080299 | 0.000536353 | 0.00345579 | 0.000215987 | 0.128287656 | 0.005131506 | ENSG0000011 | 6 | EIF2B2 |
| Pancreas | T2DM | ENSG0000020 | -0.0002805 | 0.785794892 | 0.001032103 | 0.055522669 | 0.000671714 | 0.522842579 | 0.001051245 | 0.055519578 | -0.00095221 | 1.89E-06 | 1 | 0.949163351 | 1 | 0.850322826 | ENSG0000011 | 6 | EIF2B2 |
| Pancreas | T2DM | ENSG0000021 | 0.004614728 | 0.001092097 | 0.001413099 | 0.055521462 | 0.004598987 | 0.001135688 | 0.001413083 | 0.055518417 | 1.57E-05 | 0.046800895 | 1 | 0.038951466 | 1 | 0.040083532 | ENSG0000011 | 6 | EIF2B2 |
| Pancreas | T2DM | ENSG0000021 | -0.00498589 | 0.023665375 | 0.002203682 | 0.055522094 | -0.00495691 | 0.024487496 | 0.002203659 | 0.055519048 | -2.90E-05 | 0.024822354 | 1 | 0.236493759 | 1 | 0.245759311 | ENSG0000011 | 12 | DNMT3A |
| Pancreas | T2DM | ENSG0000021 | 0.003482854 | 0.0001469 | 0.000917438 | 0.055521035 | 0.003469077 | 0.000156025 | 0.000917428 | 0.055517995 | 1.38E-05 | 0.008620228 | 0.377238052 | 0.009200928 | 0.400672926 | 0.010998318 | ENSG0000011 | 10 | AFTPH |
| Pancreas | T2DM | ENSG0000021 | 0.001828533 | 0.248135127 | 0.001583293 | 0.055522525 | 0.001804442 | 0.254420299 | 0.001583288 | 0.055519476 | 2.41E-05 | 0.029374411 | 1 | 0.659767299 | 1 | 0.670900145 | ENSG0000011 | 4 | CNRIP1 |
| Pancreas | T2DM | ENSG0000021 | 0.000899478 | 0.666522098 | 0.002087312 | 0.055522656 | 0.001315047 | 0.529324292 | 0.002090568 | 0.055519579 | -0.00041557 | 0.000412715 | 1 | 0.90774704 | 1 | 0.854617121 | ENSG0000011 | 4 | CNRIP1 |
| Pancreas | T2DM | ENSG0000021 | 0.003729919 | 4.36E-05 | 0.000912454 | 0.055520773 | 0.003302169 | 0.000333084 | 0.000920302 | 0.055518157 | 0.00042775 | 0.000368358 | 0.111855854 | 0.003857098 | 0.855358434 | 0.018199116 | ENSG0000011 | 5 | CRIPT |
| Pancreas | T2DM | ENSG0000021 | 0.003463129 | 0.031720729 | 0.001612319 | 0.055522151 | 0.001854023 | 0.266899987 | 0.001669943 | 0.055519484 | 0.001609106 | 0.000216698 | 1 | 0.272437568 | 1 | 0.682076264 | ENSG0000011 | 5 | CRIPT |
| Pancreas | T2DM | ENSG0000021 | 0.002549955 | 0.435757431 | 0.003271782 | 0.055522608 | 0.002591779 | 0.428263187 | 0.003271755 | 0.055519553 | -4.18E-05 | 0.040310094 | 1 | 0.795009083 | 1 | 0.793088295 | ENSG0000011 | 11 | SLC17A5 |

| | | | | | | | | | | | | | | | | | | | |
|---|---|---|---|---|---|---|---|---|---|---|---|---|---|---|---|---|---|---|---|
| Pancreas | T2DM | ENSG0000021 | -0.00127361 | 0.541096642 | 0.00208394 | 0.055522635 | -0.00357199 | 0.118063999 | 0.00228541 | 0.055519346 | 0.00229838 | 0.014312843 | 1 | 0.860225195 | 1 | 0.514751016 | ENSG0000011 | 7 | OGFRL1 |
| Pancreas | T2DM | ENSG0000021 | -0.00030233 | 0.778206039 | 0.001073378 | 0.055522668 | -0.00031879 | 0.766471927 | 0.001073377 | 0.055519614 | 1.65E-05 | 0.033807803 | 1 | 0.948363389 | 1 | 0.941828222 | ENSG0000011 | 7 | OGFRL1 |
| Pancreas | T2DM | ENSG0000021 | 0.000451033 | 0.685675043 | 0.001114401 | 0.055522658 | 0.000466004 | 0.675824583 | 0.001114396 | 0.055519604 | -1.50E-05 | 0.046716905 | 1 | 0.91019995 | 1 | 0.907798476 | ENSG0000011 | 6 | PYROXD2 |
| Pancreas | T2DM | ENSG0000021 | -2.82E-05 | 0.98119396 | 0.00119468 | 0.055522677 | -4.69E-05 | 0.968671042 | 0.001194667 | 0.055519624 | 1.88E-05 | 0.006645653 | 1 | 0.99701908 | 1 | 0.995018895 | ENSG0000011 | 6 | PYROXD2 |
| Pancreas | T2DM | ENSG0000022 | 0.011817598 | 1.47E-06 | 0.002454385 | 0.055520036 | 0.010536438 | 2.52E-05 | 0.002501055 | 0.055517602 | 0.001281161 | 0.007755739 | 0.003783263 | 0.000222545 | 0.064783756 | 0.003239188 | ENSG0000011 | 16 | TCTN3 |
| Pancreas | T2DM | ENSG0000022 | -0.00184333 | 0.420190507 | 0.002286745 | 0.055522603 | -0.00189167 | 0.408105857 | 0.002286741 | 0.055519546 | 4.83E-05 | 0.003271893 | 1 | 0.7882025 | 1 | 0.785840676 | ENSG0000011 | 16 | TCTN3 |
| Pancreas | T2DM | ENSG0000022 | 0.002824547 | 0.117659184 | 0.001805194 | 0.055522398 | 0.003099772 | 0.086187059 | 0.001806534 | 0.055519289 | -0.00027523 | 0.000102921 | 1 | 0.510920333 | 1 | 0.452744404 | ENSG0000011 | 16 | TCTN3 |
| Pancreas | T2DM | ENSG0000022 | 0.000152481 | 0.91445956 | 0.001419547 | 0.055522676 | 0.000171056 | 0.904086048 | 0.001419539 | 0.055519622 | -1.86E-05 | 0.046651785 | 1 | 0.9849439 | 1 | 0.984551279 | ENSG0000012 | 5 | C10orf95 |
| Pancreas | T2DM | ENSG0000022 | 0.001923811 | 0.147451496 | 0.001328053 | 0.055522438 | 0.001539772 | 0.249576276 | 0.001337327 | 0.055519473 | 0.000384039 | 0.014766456 | 1 | 0.556171292 | 1 | 0.666460318 | ENSG0000012 | 5 | C10orf95 |
| Pancreas | T2DM | ENSG0000022 | -0.00020769 | 0.89494441 | 0.001572806 | 0.055522675 | -0.00035422 | 0.821888563 | 0.001573511 | 0.055519618 | 0.000146534 | 0.00253305 | 1 | 0.981247774 | 1 | 0.958932226 | ENSG0000012 | 8 | KANSL1 |
| Pancreas | T2DM | ENSG0000022 | -0.00232541 | 0.38261847 | 0.002663453 | 0.05552259 | -0.00235923 | 0.375733218 | 0.002663431 | 0.055519535 | 3.38E-05 | 0.038945534 | 1 | 0.76943166 | 1 | 0.766997539 | ENSG0000012 | 8 | KANSL1 |
| Pancreas | T2DM | ENSG0000022 | 0.000969629 | 0.60657463 | 0.001882886 | 0.055522647 | 0.000996759 | 0.596540829 | 0.001882873 | 0.055519592 | -2.71E-05 | 0.024171163 | 1 | 0.883541492 | 1 | 0.879499402 | ENSG0000012 | 7 | HOXB3 |
| Pancreas | T2DM | ENSG0000022 | 0.000310775 | 0.910654423 | 0.002769507 | 0.055522676 | 0.000273515 | 0.921328641 | 0.002769487 | 0.055519623 | 3.73E-05 | 0.034794463 | 1 | 0.98382859 | 1 | 0.987209256 | ENSG0000012 | 7 | CAAP1 |
| Pancreas | T2DM | ENSG0000022 | -0.00641928 | 0.043486746 | 0.003179419 | 0.055522213 | -0.0064886 | 0.041268409 | 0.003179395 | 0.05551915 | 6.93E-05 | 0.000544225 | 1 | 0.311795766 | 1 | 0.311697862 | ENSG0000012 | 14 | WDR55 |
| Pancreas | T2DM | ENSG0000022 | -0.00231289 | 0.188686001 | 0.001759547 | 0.05552248 | -0.00272478 | 0.12226623 | 0.001763233 | 0.055519352 | 0.000411896 | 0.000329503 | 1 | 0.591234665 | 1 | 0.519878374 | ENSG0000012 | 14 | WDR55 |
| Pancreas | T2DM | ENSG0000023 | -0.00190956 | 0.260977059 | 0.001698768 | 0.055522533 | -0.00193109 | 0.255633037 | 0.001698747 | 0.055519477 | 2.15E-05 | 0.019899359 | 1 | 0.676275568 | 1 | 0.671232759 | ENSG0000012 | 2 | PCDHB8 |
| Pancreas | T2DM | ENSG0000023 | -4.18E-06 | 0.997366516 | 0.001267095 | 0.055522677 | 1.37E-05 | 0.99135995 | 0.001267089 | 0.055519624 | -1.79E-05 | 0.038481598 | 1 | 0.99892247 | 1 | 0.998538835 | ENSG0000012 | 4 | PCDHB12 |
| Pancreas | T2DM | ENSG0000023 | -0.0003433 | 0.64210185 | 0.000738647 | 0.055522653 | -0.00054965 | 0.458346028 | 0.000741195 | 0.055519561 | 0.000206355 | 0.000817023 | 1 | 0.898449905 | 1 | 0.812867817 | ENSG0000012 | 4 | PCDHB12 |
| Pancreas | T2DM | ENSG0000023 | -0.00161154 | 0.415387994 | 0.001978682 | 0.055522602 | -0.00157995 | 0.424586607 | 0.001978674 | 0.055519551 | -3.16E-05 | 0.019586911 | 1 | 0.786828099 | 1 | 0.792397097 | ENSG0000012 | 4 | MRPS14 |
| Pancreas | T2DM | ENSG0000023 | 0.000668723 | 0.378859102 | 0.000759912 | 0.055522589 | 0.000973168 | 0.208172278 | 0.000773209 | 0.055519444 | -0.00030444 | 0.033129145 | 1 | 0.766674685 | 1 | 0.61801897 | ENSG0000012 | 7 | ACAT2 |
| Pancreas | T2DM | ENSG0000023 | 0.003992597 | 0.032795653 | 0.001870435 | 0.055522158 | 0.003340479 | 0.075161278 | 0.001877219 | 0.055519263 | 0.000652118 | 4.60E-05 | 1 | 0.277951279 | 1 | 0.426361119 | ENSG0000012 | 7 | ACAT2 |
| Pancreas | T2DM | ENSG0000023 | -8.02E-05 | 0.957027614 | 0.001487916 | 0.055522677 | -0.00043065 | 0.772536723 | 0.00148983 | 0.055519615 | 0.000350473 | 4.36E-06 | 1 | 0.990168921 | 1 | 0.943354401 | ENSG0000012 | 7 | ACAT2 |
| Pancreas | T2DM | ENSG0000023 | -0.00031031 | 0.924497029 | 0.003274341 | 0.055522676 | -0.00025569 | 0.937756561 | 0.00327431 | 0.055519623 | -5.46E-05 | 0.005319075 | 1 | 0.986312425 | 1 | 0.987209256 | ENSG0000012 | 6 | SNX19 |
| Pancreas | T2DM | ENSG0000023 | 0.001845625 | 0.293532544 | 0.001757061 | 0.055522551 | 0.001824242 | 0.29915671 | 0.001757045 | 0.055519501 | 2.14E-05 | 0.043898777 | 1 | 0.703265608 | 1 | 0.709510872 | ENSG0000012 | 6 | SNX19 |
| Pancreas | T2DM | ENSG0000023 | 0.00651977 | 0.001334464 | 0.002032042 | 0.055521504 | 0.006472533 | 0.001447058 | 0.002032119 | 0.055518468 | 4.72E-05 | 0.041940999 | 1 | 0.041791518 | 1 | 0.045626317 | ENSG0000012 | 7 | KCNJ5 |
| Pancreas | T2DM | ENSG0000023 | -0.00221844 | 0.114819361 | 0.001406836 | 0.055522394 | -0.00335832 | 0.018671419 | 0.001427834 | 0.055518994 | 0.001139876 | 3.05E-06 | 1 | 0.50643167 | 1 | 0.215982899 | ENSG0000012 | 10 | MASTL |
| Pancreas | T2DM | ENSG0000023 | 0.004353564 | 3.05E-06 | 0.000932796 | 0.055520195 | 0.003641763 | 0.00016404 | 0.000966287 | 0.055518006 | 0.000711801 | 0.004777311 | 0.007842296 | 0.000412752 | 0.421253663 | 0.010998318 | ENSG0000012 | 10 | MASTL |
| Pancreas | T2DM | ENSG0000023 | -0.00075722 | 0.763266448 | 0.002514058 | 0.055522667 | -0.00072749 | 0.772296143 | 0.002514025 | 0.055519615 | -2.97E-05 | 0.027742077 | 1 | 0.94281578 | 1 | 0.943354401 | ENSG0000012 | 10 | MASTL |
| Pancreas | T2DM | ENSG0000023 | 0.00080907 | 0.176876133 | 0.000599117 | 0.055522469 | 0.00066029 | 0.271523717 | 0.000600503 | 0.055519486 | 0.00014878 | 0.000285979 | 1 | 0.581719513 | 1 | 0.68377529 | ENSG0000012 | 9 | SEPT7P9 |
| Pancreas | T2DM | ENSG0000023 | 0.000739563 | 0.625490042 | 0.001515235 | 0.05552265 | 0.001194509 | 0.431426136 | 0.001518274 | 0.055519554 | -0.00045495 | 2.53E-06 | 1 | 0.89259216 | 1 | 0.793088295 | ENSG0000012 | 9 | SEPT7P9 |
| Pancreas | T2DM | ENSG0000023 | -0.00215538 | 0.413276437 | 0.002634484 | 0.055522601 | -0.00162822 | 0.537320545 | 0.002639481 | 0.055519581 | -0.00052716 | 0.001263973 | 1 | 0.785561726 | 1 | 0.856222273 | ENSG0000012 | 9 | SEPT7P9 |
| Pancreas | T2DM | ENSG0000023 | -0.00383849 | 0.003335209 | 0.001307816 | 0.055521696 | -0.00229943 | 0.102905415 | 0.001409885 | 0.055519321 | -0.00153906 | 0.003480112 | 1 | 0.075129978 | 1 | 0.486323959 | ENSG0000012 | 11 | SOHLH2 |
| Pancreas | T2DM | ENSG0000023 | 0.000142979 | 0.892150309 | 0.00105454 | 0.055522675 | 0.000562922 | 0.59521026 | 0.001059515 | 0.055519592 | -0.00041994 | 4.45E-05 | 1 | 0.981247774 | 1 | 0.879499402 | ENSG0000012 | 4 | DNAJC15 |
| Pancreas | T2DM | ENSG0000023 | -6.84E-05 | 0.970552382 | 0.001853603 | 0.055522677 | -3.62E-05 | 0.984417492 | 0.001853596 | 0.055519624 | -3.22E-05 | 0.01218646 | 1 | 0.994696741 | 1 | 0.997232395 | ENSG0000012 | 4 | DNAJC15 |
| Pancreas | T2DM | ENSG0000023 | 0.001048472 | 0.351072458 | 0.001124352 | 0.055522578 | 0.000735215 | 0.514160122 | 0.001126983 | 0.055519576 | 0.000313257 | 5.19E-05 | 1 | 0.743483984 | 1 | 0.847609136 | ENSG0000012 | 4 | DNAJC15 |
| Pancreas | T2DM | ENSG0000024 | -0.00041498 | 0.717995925 | 0.001149087 | 0.055522662 | 8.75E-05 | 0.9400928 | 0.001164115 | 0.055519623 | -0.00050247 | 0.007098585 | 1 | 0.921476762 | 1 | 0.987209256 | ENSG0000012 | 8 | SIL1 |
| Pancreas | T2DM | ENSG0000024 | 0.003113355 | 0.176921162 | 0.002305681 | 0.055522469 | 0.00254367 | 0.271197407 | 0.002311774 | 0.055519486 | 0.000569685 | 0.000727309 | 1 | 0.581719513 | 1 | 0.68377529 | ENSG0000012 | 8 | SIL1 |
| Pancreas | T2DM | ENSG0000024 | 0.000112959 | 0.94819611 | 0.001738576 | 0.055522677 | 0.000138939 | 0.936304618 | 0.001738577 | 0.055519623 | -2.60E-05 | 0.045946079 | 1 | 0.989794375 | 1 | 0.987209256 | ENSG0000012 | 8 | SIL1 |
| Pancreas | T2DM | ENSG0000024 | -0.0028465 | 0.194442464 | 0.002193752 | 0.055522485 | -0.00281757 | 0.199011166 | 0.00219373 | 0.055519436 | -2.89E-05 | 0.026562777 | 1 | 0.599023 | 1 | 0.610586229 | ENSG0000012 | 8 | SIL1 |
| Pancreas | T2DM | ENSG0000024 | -0.00036415 | 0.794066543 | 0.001395042 | 0.055522669 | -0.00034776 | 0.803138179 | 0.001395027 | 0.055519617 | -1.64E-05 | 0.042043105 | 1 | 0.949920498 | 1 | 0.951905176 | ENSG0000012 | 8 | PLS1 |
| Pancreas | T2DM | ENSG0000024 | -0.00457988 | 0.026373801 | 0.002062387 | 0.055522115 | -0.00455322 | 0.027260976 | 0.002062366 | 0.055519069 | -2.67E-05 | 0.026740815 | 1 | 0.251609367 | 1 | 0.255898044 | ENSG0000012 | 8 | PLS1 |
| Pancreas | T2DM | ENSG0000024 | -0.00260996 | 0.306182319 | 0.002550614 | 0.055522558 | -0.00256706 | 0.314197664 | 0.002550596 | 0.055519509 | -4.29E-05 | 0.008325224 | 1 | 0.712852398 | 1 | 0.725505793 | ENSG0000012 | 11 | ZFP30 |
| Pancreas | T2DM | ENSG0000024 | 0.00425696 | 5.09E-05 | 0.001050718 | 0.055520807 | 0.003968251 | 0.000167031 | 0.001054178 | 0.05551801 | 0.000288709 | 0.000751196 | 0.130719261 | 0.004357309 | 0.428934408 | 0.010998318 | ENSG0000012 | 11 | ZFP30 |
| Pancreas | T2DM | ENSG0000024 | 0.003847184 | 0.008511672 | 0.001462207 | 0.055521888 | 0.003873344 | 0.008074064 | 0.001462209 | 0.055518825 | -2.62E-05 | 0.018541609 | 1 | 0.13747153 | 1 | 0.13385914 | ENSG0000012 | 11 | ZFP30 |
| Pancreas | T2DM | ENSG0000024 | 3.06E-05 | 0.992649205 | 0.003324467 | 0.055522677 | 8.43E-05 | 0.979762646 | 0.003324453 | 0.055519624 | -5.37E-05 | 0.017836597 | 1 | 0.997347815 | 1 | 0.9961668 | ENSG0000012 | 11 | ZFP30 |
| Pancreas | T2DM | ENSG0000024 | -0.00046832 | 0.831288381 | 0.002198193 | 0.055522672 | -0.00042753 | 0.845790894 | 0.002198177 | 0.05551962 | -4.08E-05 | 0.003501522 | 1 | 0.963990872 | 1 | 0.969100518 | ENSG0000012 | 4 | UTP20 |
| Pancreas | T2DM | ENSG0000025 | 0.004603244 | 0.084765066 | 0.002670581 | 0.055522339 | 0.004568333 | 0.087149521 | 0.002670551 | 0.055519291 | 3.49E-05 | 0.021800108 | 1 | 0.449552839 | 1 | 0.453955316 | ENSG0000012 | 9 | ARL1 |
| Pancreas | T2DM | ENSG0000025 | 0.001432449 | 0.152664447 | 0.001001583 | 0.055522444 | 0.001285087 | 0.199930076 | 0.001002602 | 0.055519437 | 0.000147362 | 0.001287769 | 1 | 0.558464815 | 1 | 0.611164195 | ENSG0000012 | 9 | ARL1 |
| Pancreas | T2DM | ENSG0000025 | 0.004838428 | 0.143484995 | 0.003307337 | 0.055522433 | 0.004890675 | 0.139208172 | 0.003307307 | 0.055519375 | -5.22E-05 | 0.009562346 | 1 | 0.553257458 | 1 | 0.548928759 | ENSG0000012 | 9 | ARL1 |
| Pancreas | T2DM | ENSG0000025 | -0.0113211 | 9.57E-05 | 0.002901862 | 0.055520943 | -0.01127923 | 0.000101545 | 0.002901855 | 0.055517903 | -4.19E-05 | 0.041045994 | 0.245710472 | 0.006466065 | 0.260767811 | 0.007669642 | ENSG0000012 | 9 | ARL1 |
| Pancreas | T2DM | ENSG0000025 | 0.001667709 | 0.085072689 | 0.00096848 | 0.055522339 | 0.001654991 | 0.087476944 | 0.000968475 | 0.055519291 | 1.27E-05 | 0.048862714 | 1 | 0.449552839 | 1 | 0.454738448 | ENSG0000012 | 6 | CCDC53 |
| Pancreas | T2DM | ENSG0000025 | 0.004474255 | 0.045793424 | 0.002240154 | 0.055522223 | 0.004430565 | 0.047953755 | 0.002240167 | 0.055519178 | 4.37E-05 | 0.01686092 | 1 | 0.319558457 | 1 | 0.331035598 | ENSG0000012 | 6 | CCDC53 |
| Pancreas | T2DM | ENSG0000025 | 0.002343607 | 0.471933306 | 0.003258008 | 0.055522618 | 0.002457635 | 0.450664295 | 0.003258129 | 0.055519559 | -0.00011403 | 0.002117119 | 1 | 0.817750308 | 1 | 0.805155685 | ENSG0000012 | 9 | APAF1 |
| Pancreas | T2DM | ENSG0000025 | 0.003500784 | 0.100245619 | 0.002129862 | 0.055522369 | 0.003535594 | 0.096910504 | 0.002129847 | 0.05551931 | -3.48E-05 | 0.010539645 | 1 | 0.483403189 | 1 | 0.472157259 | ENSG0000012 | 13 | ADRA1A |
| Pancreas | T2DM | ENSG0000025 | 9.14E-05 | 0.937133826 | 0.001159103 | 0.055522676 | 0.000111711 | 0.923220792 | 0.0011591 | 0.055519623 | -2.03E-05 | 0.012929221 | 1 | 0.987914477 | 1 | 0.987209256 | ENSG0000012 | 13 | ADRA1A |
| Pancreas | T2DM | ENSG0000025 | -0.0034017 | 0.290579328 | 0.003218709 | 0.05552255 | -0.00344519 | 0.28445294 | 0.003218687 | 0.055519494 | 4.35E-05 | 0.035526366 | 1 | 0.700376625 | 1 | 0.694923899 | ENSG0000012 | 9 | EPHX2 |
| Pancreas | T2DM | ENSG0000025 | 0.000239882 | 0.816160042 | 0.001031808 | 0.055522671 | 0.000229167 | 0.824231868 | 0.001031794 | 0.055519619 | 1.07E-05 | 0.048480965 | 1 | 0.958017935 | 1 | 0.959648084 | ENSG0000012 | 9 | EPHX2 |
| Pancreas | T2DM | ENSG0000025 | 0.001473705 | 0.539149022 | 0.002399777 | 0.055522634 | 0.00143658 | 0.549417582 | 0.002399766 | 0.055519583 | 3.71E-05 | 0.021685092 | 1 | 0.858495882 | 1 | 0.863653712 | ENSG0000012 | 9 | EPHX2 |
| Pancreas | T2DM | ENSG0000025 | 0.004014248 | 0.112293871 | 0.002527913 | 0.05552239 | 0.00397987 | 0.115400176 | 0.002527896 | 0.055519342 | 3.44E-05 | 0.035362881 | 1 | 0.50414451 | 1 | 0.508315014 | ENSG0000012 | 7 | ZNF706 |
| Pancreas | T2DM | ENSG0000026 | 0.0035403 | 0.085601507 | 0.002059423 | 0.05552234 | 0.00357907 | 0.082229505 | 0.002059425 | 0.05551928 | -3.88E-05 | 0.013302319 | 1 | 0.45138536 | 1 | 0.448334114 | ENSG0000012 | 7 | ZNF706 |
| Pancreas | T2DM | ENSG0000026 | -0.00088104 | 0.404666218 | 0.001057269 | 0.055522598 | -0.00079678 | 0.451147877 | 0.001057436 | 0.055519559 | -8.43E-05 | 3.45E-05 | 1 | 0.783229955 | 1 | 0.805155685 | ENSG0000012 | 8 | SCPEP1 |
| Pancreas | T2DM | ENSG0000026 | 0.001063148 | 0.529787118 | 0.001692018 | 0.055522632 | 0.001086892 | 0.520635066 | 0.00169201 | 0.055519577 | -2.37E-05 | 0.037799308 | 1 | 0.856215034 | 1 | 0.848610147 | ENSG0000012 | 8 | SCPEP1 |
| Pancreas | T2DM | ENSG0000026 | -0.00139666 | 0.632101804 | 0.002917183 | 0.055522651 | -0.00144147 | 0.621213578 | 0.002917172 | 0.055519596 | 4.48E-05 | 0.026008571 | 1 | 0.89608366 | 1 | 0.888839757 | ENSG0000012 | 9 | TBX2 |
| Pancreas | T2DM | ENSG0000026 | -0.00199861 | 0.206162648 | 0.001580938 | 0.055522495 | -0.00204783 | 0.195223208 | 0.00158099 | 0.055519433 | 4.92E-05 | 0.004461983 | 1 | 0.612052808 | 1 | 0.60527543 | ENSG0000012 | 11 | TEX14 |
| Pancreas | T2DM | ENSG0000026 | 0.006449132 | 0.001660611 | 0.002050557 | 0.05552155 | 0.006414145 | 0.001759923 | 0.002050538 | 0.055518509 | 3.50E-05 | 0.004342366 | 1 | 0.048459653 | 1 | 0.05167457 | ENSG0000012 | 11 | TEX14 |
| Pancreas | T2DM | ENSG0000026 | 0.000154951 | 0.912232593 | 0.001405797 | 0.055522676 | 0.000172122 | 0.902551521 | 0.001405781 | 0.055519622 | -1.72E-05 | 0.028768399 | 1 | 0.984291302 | 1 | 0.984551279 | ENSG0000012 | 13 | CEP89 |
| Pancreas | T2DM | ENSG0000026 | -0.00274955 | 0.158144051 | 0.00194818 | 0.05552245 | -0.00277816 | 0.153858884 | 0.001948173 | 0.055519392 | 2.86E-05 | 0.034552435 | 1 | 0.562662048 | 1 | 0.561235248 | ENSG0000012 | 11 | PYROXD1 |
| Pancreas | T2DM | ENSG0000026 | 0.000110101 | 0.952609495 | 0.001852619 | 0.055522677 | 8.14E-05 | 0.964947883 | 0.001852599 | 0.055519624 | 2.87E-05 | 0.007938611 | 1 | 0.989794375 | 1 | 0.99332874 | ENSG0000012 | 11 | PYROXD1 |
| Pancreas | T2DM | ENSG0000027 | 0.00018337 | 0.808193453 | 0.000755364 | 0.05552267 | 0.000174154 | 0.81765708 | 0.000755355 | 0.055519618 | 9.22E-06 | 0.024775884 | 1 | 0.955103906 | 1 | 0.957038916 | ENSG0000012 | 11 | PYROXD1 |
| Pancreas | T2DM | ENSG0000027 | 0.006188043 | 0.002050948 | 0.002007299 | 0.055521594 | 0.003960008 | 0.059024717 | 0.002097447 | 0.055519218 | 0.002228036 | 0.00025039 | 1 | 0.055440362 | 1 | 0.378938685 | ENSG0000012 | 11 | PYROXD1 |
| Pancreas | T2DM | ENSG0000027 | -0.00149257 | 0.009012675 | 0.000571521 | 0.0555219 | -0.00128473 | 0.025297221 | 0.000574351 | 0.055519054 | -0.00020784 | 0.000272196 | 1 | 0.142867588 | 1 | 0.248901391 | ENSG0000012 | 4 | ZNF549 |
| Pancreas | T2DM | ENSG0000027 | -0.00107628 | 0.554539678 | 0.001821209 | 0.055522637 | -0.00110202 | 0.545110606 | 0.001821202 | 0.055519582 | 2.57E-05 | 0.039125712 | 1 | 0.867798838 | 1 | 0.861442484 | ENSG0000012 | 8 | ZSCAN18 |
| Pancreas | T2DM | ENSG0000027 | -0.00143265 | 0.33664279 | 0.001491071 | 0.055522572 | -0.00141133 | 0.343879004 | 0.001491056 | 0.055519522 | -2.13E-05 | 0.014282842 | 1 | 0.734309489 | 1 | 0.743948203 | ENSG0000012 | 9 | ZNF211 |
| Pancreas | T2DM | ENSG0000027 | -0.00356148 | 0.080096791 | 0.002034983 | 0.055522328 | -0.00353324 | 0.082518278 | 0.002034967 | 0.055519281 | -2.82E-05 | 0.027681045 | 1 | 0.438568358 | 1 | 0.448955377 | ENSG0000012 | 4 | LHX4 |
| Pancreas | T2DM | ENSG0000027 | 0.000621682 | 0.555543313 | 0.001054638 | 0.055522638 | 0.00063614 | 0.546384952 | 0.001054633 | 0.055519583 | -1.45E-05 | 0.042549105 | 1 | 0.868305521 | 1 | 0.862394934 | ENSG0000012 | 3 | CSTA |
| SAT | CAD | ENSG0000000 | -0.00283586 | 0.506616824 | 0.004270132 | 0.138947771 | -0.00274797 | 0.519889427 | 0.004270242 | 0.138942021 | -8.79E-05 | 0.032850908 | 1 | 0.832236789 | 1 | 0.837215552 | ENSG0000012 | 3 | CSTA |
| SAT | CAD | ENSG0000001 | 0.000236297 | 0.961302676 | 0.004870198 | 0.138947896 | 0.00031916 | 0.947750197 | 0.004870274 | 0.138942138 | -8.29E-05 | 0.04586508 | 1 | 0.991640191 | 1 | 0.985387556 | ENSG0000012 | 3 | CSTA |
| SAT | CAD | ENSG0000002 | 0.001908181 | 0.270567 | 0.00173194 | 0.13894755 | 0.001993502 | 0.249800948 | 0.001732223 | 0.138941762 | -8.53E-05 | 0.010322541 | 1 | 0.669985153 | 1 | 0.656684558 | ENSG0000012 | 3 | CSTA |
| SAT | CAD | ENSG0000004 | -0.00932613 | 0.067351198 | 0.005098119 | 0.138946942 | -0.01978308 | 0.001009338 | 0.006016879 | 0.138939057 | 0.010456951 | 0.001067759 | 1 | 0.415234417 | 1 | 0.058098932 | ENSG0000012 | 3 | CSTA |
| SAT | CAD | ENSG0000005 | 0.00240232 | 0.429097847 | 0.003038077 | 0.138947718 | 0.0015643 | 0.608938597 | 0.003057725 | 0.138942065 | 0.00083802 | 0.015618152 | 1 | 0.794212752 | 1 | 0.869331532 | ENSG0000012 | 9 | POPDC2 |
| SAT | CAD | ENSG0000008 | -0.00362702 | 0.398629372 | 0.004297053 | 0.138947693 | -0.00369358 | 0.390034217 | 0.004297083 | 0.138941929 | 6.66E-05 | 0.038003758 | 1 | 0.776664471 | 1 | 0.772767069 | ENSG0000012 | 7 | CD80 |
| SAT | CAD | ENSG0000008 | 0.001560433 | 0.57793247 | 0.002804478 | 0.138947808 | 0.001610976 | 0.565683115 | 0.00280452 | 0.138942045 | -5.05E-05 | 0.032529203 | 1 | 0.864959129 | 1 | 0.853336231 | ENSG0000012 | 4 | CAT |
| SAT | CAD | ENSG0000010 | -0.00163153 | 0.625284124 | 0.003340733 | 0.138947828 | -0.00170808 | 0.609156508 | 0.003340803 | 0.138942065 | 7.65E-05 | 0.012278148 | 1 | 0.878643356 | 1 | 0.869331532 | ENSG0000012 | 4 | CAT |
| SAT | CAD | ENSG0000010 | -0.00535543 | 0.039402292 | 0.002599767 | 0.138946687 | -0.00530569 | 0.041271653 | 0.002599816 | 0.138940952 | -4.97E-05 | 0.030794032 | 1 | 0.338426494 | 1 | 0.349902564 | ENSG0000012 | 14 | PILRB |
| SAT | CAD | ENSG0000010 | 0.000978885 | 0.766918646 | 0.003302494 | 0.138947871 | 0.001049275 | 0.750700004 | 0.00330255 | 0.13894211 | -7.04E-05 | 0.014108188 | 1 | 0.930280564 | 1 | 0.927351925 | ENSG0000012 | 8 | CPXM2 |
| SAT | CAD | ENSG0000010 | -0.00656785 | 0.207621473 | 0.005212024 | 0.138947444 | -0.00646948 | 0.214518204 | 0.005212113 | 0.1389417 | -9.84E-05 | 0.029574031 | 1 | 0.623552391 | 1 | 0.633044396 | ENSG0000012 | 8 | CPXM2 |
| SAT | CAD | ENSG0000010 | 0.007412605 | 0.169650382 | 0.005397541 | 0.138947359 | 0.007501787 | 0.164579615 | 0.00539761 | 0.138941589 | -8.92E-05 | 0.043975304 | 1 | 0.579212177 | 1 | 0.576272733 | ENSG0000012 | 8 | CPXM2 |
| SAT | CAD | ENSG0000010 | 0.006951097 | 0.038261192 | 0.003354694 | 0.138946672 | 0.007024234 | 0.036277485 | 0.003354763 | 0.138940889 | -7.31E-05 | 0.01675441 | 1 | 0.337586455 | 1 | 0.340087662 | ENSG0000012 | 8 | GPSM2 |
| SAT | CAD | ENSG0000011 | 0.001105245 | 0.632312909 | 0.002309936 | 0.138947831 | 0.00113926 | 0.621874183 | 0.002309952 | 0.13894207 | -3.40E-05 | 0.04899668 | 1 | 0.880256991 | 1 | 0.872418743 | ENSG0000012 | 12 | MTERFD2 |
| SAT | CAD | ENSG0000011 | 0.006674678 | 0.000427677 | 0.001894917 | 0.138944359 | 0.006710809 | 0.000398018 | 0.001894949 | 0.138938563 | -3.61E-05 | 0.028752325 | 1 | 0.031133309 | 1 | 0.031912061 | ENSG0000012 | 12 | MTERFD2 |
| SAT | CAD | ENSG0000011 | -0.0029734 | 0.407502866 | 0.003589777 | 0.138947701 | -0.00303004 | 0.398631308 | 0.003589806 | 0.138941936 | 5.66E-05 | 0.038477077 | 1 | 0.78509507 | 1 | 0.776668242 | ENSG0000012 | 12 | MTERFD2 |
| SAT | CAD | ENSG0000012 | -1.82E-06 | 0.999706633 | 0.004953546 | 0.138947896 | -7.76E-05 | 0.987498905 | 0.004953587 | 0.138942139 | 7.58E-05 | 0.044506289 | 1 | 0.999715369 | 1 | 0.997619483 | ENSG0000012 | 10 | FMOD |
| SAT | CAD | ENSG0000012 | 0.005517646 | 0.107734173 | 0.003430381 | 0.138947159 | 0.005567798 | 0.104573885 | 0.0034304 | 0.138941388 | -5.02E-05 | 0.043025115 | 1 | 0.501109823 | 1 | 0.506174025 | ENSG0000012 | 10 | COPA |
| SAT | CAD | ENSG0000012 | 0.00623342 | 0.222468023 | 0.005109389 | 0.138947472 | 0.006309748 | 0.216860095 | 0.00510943 | 0.138941704 | -7.63E-05 | 0.049004501 | 1 | 0.633338537 | 1 | 0.633679313 | ENSG0000012 | 15 | LDB3 |
| SAT | CAD | ENSG0000012 | 0.004818505 | 0.000206239 | 0.001298342 | 0.138943969 | 0.004794537 | 0.00022184 | 0.001298354 | 0.138938251 | 2.40E-05 | 0.017641472 | 0.676671469 | 0.020521943 | 0.727855892 | 0.021008952 | ENSG0000012 | 7 | FAM35A |
| SAT | CAD | ENSG0000012 | -0.00415546 | 0.206255376 | 0.003287724 | 0.138947441 | -0.00421623 | 0.199702125 | 0.003287758 | 0.13894167 | 6.08E-05 | 0.019152623 | 1 | 0.623552391 | 1 | 0.619657947 | ENSG0000012 | 7 | FAM35A |
| SAT | CAD | ENSG0000013 | 0.00357279 | 0.383073996 | 0.004096075 | 0.13894768 | 0.003654292 | 0.372327332 | 0.004096174 | 0.138941912 | -8.15E-05 | 0.035732128 | 1 | 0.7654481 | 1 | 0.756881026 | ENSG0000012 | 7 | FAM35A |
| SAT | CAD | ENSG0000013 | 0.002254101 | 0.63035977 | 0.00468413 | 0.13894783 | 0.002175085 | 0.642399885 | 0.004684188 | 0.138942078 | 7.90E-05 | 0.038261571 | 1 | 0.879715187 | 1 | 0.881519876 | ENSG0000012 | 7 | FAM35A |
| SAT | CAD | ENSG0000013 | 0.00624555 | 0.158170507 | 0.004425533 | 0.138947329 | 0.006906668 | 0.119499015 | 0.004436216 | 0.138941448 | -0.00066112 | 0.032407465 | 1 | 0.568778433 | 1 | 0.518715438 | ENSG0000012 | 11 | ODF2L |
| SAT | CAD | ENSG0000013 | 0.007161061 | 0.064779481 | 0.003877591 | 0.138946924 | 0.007109954 | 0.066714046 | 0.003877593 | 0.138941181 | 5.11E-05 | 0.043818279 | 1 | 0.412224502 | 1 | 0.422893011 | ENSG0000012 | 11 | ODF2L |
| SAT | CAD | ENSG0000013 | -0.00555945 | 0.188124041 | 0.004224024 | 0.138947403 | -0.00568115 | 0.17866578 | 0.004224294 | 0.138941624 | 0.000121701 | 0.026904105 | 1 | 0.599839629 | 1 | 0.593087199 | ENSG0000012 | 11 | ODF2L |
| SAT | CAD | ENSG0000014 | 0.000216712 | 0.930559356 | 0.002486904 | 0.138947894 | 0.001211376 | 0.631519737 | 0.002525862 | 0.138942074 | -0.00099466 | 0.024491357 | 1 | 0.980888267 | 1 | 0.877853708 | ENSG0000012 | 8 | LRRC39 |
| SAT | CAD | ENSG0000014 | 0.004037462 | 0.330193472 | 0.004146408 | 0.138947626 | 0.004279782 | 0.30213165 | 0.004147582 | 0.138941836 | -0.00024232 | 0.017775945 | 1 | 0.730754359 | 1 | 0.705646132 | ENSG0000012 | 8 | LRRC39 |
| SAT | CAD | ENSG0000014 | -0.00097619 | 0.684903217 | 0.002405708 | 0.13894785 | -0.00087716 | 0.715440927 | 0.002406099 | 0.138942101 | -9.90E-05 | 0.031446406 | 1 | 0.903579074 | 1 | 0.911950925 | ENSG0000012 | 8 | LRRC39 |
| SAT | CAD | ENSG0000015 | -0.00559975 | 0.153035795 | 0.003918947 | 0.138947314 | -0.00553479 | 0.157862689 | 0.003918992 | 0.138941571 | -6.50E-05 | 0.038270596 | 1 | 0.56761721 | 1 | 0.575497204 | ENSG0000012 | 8 | LRRC39 |
| SAT | CAD | ENSG0000015 | 0.005341101 | 0.25037092 | 0.004646657 | 0.13894752 | 0.004928371 | 0.289344346 | 0.004651328 | 0.138941819 | 0.00041273 | 0.049918068 | 1 | 0.652182137 | 1 | 0.691433941 | ENSG0000012 | 12 | RWDD3 |
| SAT | CAD | ENSG0000015 | 0.004305926 | 0.012499823 | 0.001723943 | 0.138946118 | 0.004464766 | 0.009680011 | 0.001725803 | 0.138940231 | -0.00015884 | 0.049601477 | 1 | 0.219747192 | 1 | 0.194847333 | ENSG0000012 | 12 | RWDD3 |
| SAT | CAD | ENSG0000015 | 0.004626933 | 0.42064541 | 0.005745583 | 0.138947712 | 0.004388879 | 0.445016493 | 0.005746485 | 0.138941973 | 0.000238055 | 0.027939679 | 1 | 0.791817322 | 1 | 0.801408187 | ENSG0000012 | 13 | CCDC18 |
| SAT | CAD | ENSG0000016 | 0.010042488 | 0.013180057 | 0.004051227 | 0.138946144 | 0.009414926 | 0.020412167 | 0.004060447 | 0.138940606 | 0.000627562 | 0.022329621 | 1 | 0.222647204 | 1 | 0.275231745 | ENSG0000012 | 13 | CCDC18 |
| SAT | CAD | ENSG0000016 | 0.000131727 | 0.952528287 | 0.002212697 | 0.138947895 | 0.000167933 | 0.939503451 | 0.002212724 | 0.138942138 | -3.62E-05 | 0.043991116 | 1 | 0.988062414 | 1 | 0.984697834 | ENSG0000012 | 13 | CCDC18 |
| SAT | CAD | ENSG0000016 | 0.001842795 | 0.211676676 | 0.001475452 | 0.138947452 | 0.001868489 | 0.205383253 | 0.001475475 | 0.138941682 | -2.57E-05 | 0.040657014 | 1 | 0.625685743 | 1 | 0.623946715 | ENSG0000012 | 16 | BBS9 |
| SAT | CAD | ENSG0000016 | -0.00105853 | 0.75513997 | 0.003394144 | 0.138947869 | -0.0004336 | 0.898722554 | 0.003406823 | 0.138942135 | -0.00062493 | 0.033811448 | 1 | 0.930016551 | 1 | 0.971567941 | ENSG0000012 | 14 | CBX3 |
| SAT | CAD | ENSG0000016 | -0.00512712 | 0.304037104 | 0.004988367 | 0.138947595 | -0.0050464 | 0.311719463 | 0.004988429 | 0.138941847 | -8.07E-05 | 0.046702112 | 1 | 0.706076325 | 1 | 0.711630554 | ENSG0000012 | 8 | CCZ1 |
| SAT | CAD | ENSG0000016 | -0.00221267 | 0.216981788 | 0.00179222 | 0.138947462 | -0.00194886 | 0.278199397 | 0.001797225 | 0.138941804 | -0.0002638 | 0.049868097 | 1 | 0.627239866 | 1 | 0.684502895 | ENSG0000012 | 10 | POLM |
| SAT | CAD | ENSG0000016 | -0.00277949 | 0.122561953 | 0.001800058 | 0.138947217 | -0.00266629 | 0.138715736 | 0.001800823 | 0.138941514 | -0.0001132 | 0.035161326 | 1 | 0.520011049 | 1 | 0.544767853 | ENSG0000012 | 10 | POLM |
| SAT | CAD | ENSG0000016 | 0.003590249 | 0.443227085 | 0.004682389 | 0.138947729 | 0.001051925 | 0.828060931 | 0.004843357 | 0.138942126 | 0.002538324 | 0.040434387 | 1 | 0.800421792 | 1 | 0.953956431 | ENSG0000012 | 7 | SMU1 |
| SAT | CAD | ENSG0000016 | -0.00256181 | 0.456289306 | 0.003438804 | 0.138947738 | -0.00251257 | 0.464992856 | 0.00343881 | 0.138941987 | -4.92E-05 | 0.032552587 | 1 | 0.807488986 | 1 | 0.810911439 | ENSG0000012 | 7 | SMU1 |
| SAT | CAD | ENSG0000017 | 0.001108136 | 0.625738753 | 0.002272011 | 0.138947829 | 0.001068259 | 0.638228535 | 0.002272034 | 0.138942076 | 3.99E-05 | 0.02502442 | 1 | 0.878873651 | 1 | 0.87989599 | ENSG0000012 | 8 | SLC25A51 |
| SAT | CAD | ENSG0000017 | -0.00087285 | 0.759078911 | 0.002846013 | 0.13894787 | -0.00208239 | 0.473562357 | 0.002905537 | 0.138941993 | 0.00120954 | 0.038813191 | 1 | 0.930057215 | 1 | 0.811850954 | ENSG0000012 | 8 | SLC25A51 |
| SAT | CAD | ENSG0000017 | -0.01830284 | 7.19E-07 | 0.003692923 | 0.138940893 | -0.02013215 | 8.84E-08 | 0.003763553 | 0.138933981 | 0.001829319 | 0.011765093 | 0.002359439 | 0.000471888 | 0.00028992 | 7.25E-05 | ENSG0000012 | 8 | SLC25A51 |
| SAT | CAD | ENSG0000017 | 0.000424441 | 0.905501244 | 0.003575296 | 0.138947892 | 0.000879786 | 0.80580301 | 0.003578635 | 0.138942122 | -0.00045535 | 0.003566669 | 1 | 0.972806019 | 1 | 0.945895506 | ENSG0000012 | 8 | SLC25A51 |
| SAT | CAD | ENSG0000018 | -0.00748511 | 0.012191397 | 0.002986202 | 0.138946105 | -0.00967918 | 0.002071165 | 0.003142744 | 0.138939435 | 0.002194071 | 0.025125208 | 1 | 0.217391164 | 1 | 0.090606557 | ENSG0000012 | 8 | SLC25A51 |
| SAT | CAD | ENSG0000018 | 0.003113916 | 0.086078126 | 0.001814148 | 0.138947056 | 0.002627906 | 0.14996908 | 0.001825387 | 0.138941548 | 0.00048601 | 0.016432398 | 1 | 0.456355415 | 1 | 0.570769052 | ENSG0000012 | 8 | SLC25A51 |
| SAT | CAD | ENSG0000018 | -0.00317685 | 0.244159462 | 0.002727718 | 0.13894751 | -0.00432382 | 0.116485216 | 0.002754552 | 0.138941437 | 0.001146976 | 0.002816364 | 1 | 0.651832164 | 1 | 0.516412287 | ENSG0000012 | 11 | KIAA1045 |
| SAT | CAD | ENSG0000018 | -0.00278409 | 0.129559691 | 0.001836662 | 0.138947241 | -0.00275561 | 0.133530299 | 0.001836673 | 0.138941497 | -2.85E-05 | 0.03472134 | 1 | 0.530136906 | 1 | 0.539205888 | ENSG0000012 | 11 | KIAA1045 |
| SAT | CAD | ENSG0000018 | -0.0021783 | 0.627477281 | 0.004488736 | 0.138947829 | -0.00130015 | 0.773110271 | 0.00450958 | 0.138942116 | -0.00087814 | 0.043055588 | 1 | 0.879075282 | 1 | 0.931084711 | ENSG0000012 | 12 | C7orf49 |
| SAT | CAD | ENSG0000018 | -0.00926804 | 0.021784878 | 0.004039937 | 0.138946396 | -0.0091782 | 0.023099809 | 0.00404008 | 0.138940668 | -8.98E-05 | 0.035577533 | 1 | 0.272809861 | 1 | 0.288176704 | ENSG0000012 | 12 | C7orf49 |

| | | | | | | | | | | | | | | | | | | | |
|---|---|---|---|---|---|---|---|---|---|---|---|---|---|---|---|---|---|---|---|
| SAT | CAD | ENSG0000018 | 0.000778616 | 0.85884548 | 0.004378081 | 0.138947887 | 0.000847965 | 0.846424494 | 0.004378119 | 0.138942129 | -6.93E-05 | 0.038843169 | 1 | 0.965404111 | 1 | 0.958221905 | ENSG0000012 | 12 | C7orf49 |
| SAT | CAD | ENSG0000018 | 0.00790488 | 0.041641646 | 0.003880478 | 0.138946713 | 0.007990161 | 0.039490991 | 0.003880528 | 0.13894093 | -8.53E-05 | 0.007496864 | 1 | 0.345015756 | 1 | 0.346299459 | ENSG0000012 | 12 | C7orf49 |
| SAT | CAD | ENSG0000018 | -0.00072364 | 0.596179577 | 0.001365612 | 0.138947816 | -0.00074604 | 0.584860305 | 0.001365619 | 0.138942054 | 2.24E-05 | 0.021662353 | 1 | 0.871671099 | 1 | 0.86205151 | ENSG0000012 | 12 | C7orf49 |
| SAT | CAD | ENSG0000018 | -0.00255747 | 0.353057425 | 0.002753887 | 0.138947651 | -0.00251485 | 0.361143377 | 0.002753915 | 0.138941902 | -4.26E-05 | 0.049120674 | 1 | 0.747968053 | 1 | 0.750418885 | ENSG0000012 | 5 | CHST3 |
| SAT | CAD | ENSG0000019 | 0.00250371 | 0.152876548 | 0.001751524 | 0.138947314 | 9.85E-06 | 0.996111975 | 0.002021113 | 0.138942139 | 0.002493861 | 0.013409004 | 1 | 0.56761721 | 1 | 0.999893579 | ENSG0000012 | 7 | CISD1 |
| SAT | CAD | ENSG0000019 | 0.012308291 | 1.38E-06 | 0.002549715 | 0.138941252 | 0.012407021 | 1.14E-06 | 0.002550004 | 0.13893539 | -9.87E-05 | 0.018183008 | 0.004541695 | 0.000674511 | 0.003747179 | 0.00062453 | ENSG0000012 | 7 | CISD1 |
| SAT | CAD | ENSG0000019 | 0.002555329 | 0.557512574 | 0.004356612 | 0.138947798 | 0.002684534 | 0.53779347 | 0.004356923 | 0.138942031 | -0.0001292 | 0.029070513 | 1 | 0.856342959 | 1 | 0.846069608 | ENSG0000012 | 10 | SLC25A16 |
| SAT | CAD | ENSG0000019 | -0.00372103 | 0.367639824 | 0.004130329 | 0.138947665 | -0.00380994 | 0.35631993 | 0.004130455 | 0.138941897 | 8.89E-05 | 0.033496495 | 1 | 0.751266179 | 1 | 0.748429484 | ENSG0000012 | 10 | SLC25A16 |
| SAT | CAD | ENSG0000019 | -0.00359056 | 0.440266645 | 0.004652533 | 0.138947727 | -0.00393723 | 0.397561139 | 0.004654016 | 0.138941935 | 0.000346668 | 0.004249313 | 1 | 0.799399481 | 1 | 0.776418397 | ENSG0000012 | 10 | SLC25A16 |
| SAT | CAD | ENSG0000020 | 0.000695599 | 0.734744051 | 0.002052994 | 0.138947864 | 0.00066168 | 0.747228359 | 0.002053017 | 0.13894211 | 3.39E-05 | 0.037666001 | 1 | 0.921273782 | 1 | 0.925296786 | ENSG0000012 | 10 | SLC25A16 |
| SAT | CAD | ENSG0000020 | -0.00174127 | 0.683013118 | 0.004264098 | 0.138947849 | -0.00267233 | 0.532514511 | 0.004281392 | 0.138942028 | 0.000931061 | 0.015703825 | 1 | 0.903565193 | 1 | 0.844456312 | ENSG0000012 | 10 | SLC25A16 |
| SAT | CAD | ENSG0000020 | -0.0004663 | 0.895546698 | 0.00355174 | 0.138947892 | -0.00153392 | 0.668088882 | 0.003577473 | 0.138942087 | 0.001067618 | 0.012809164 | 1 | 0.970661333 | 1 | 0.897319103 | ENSG0000012 | 12 | ZWINT |
| SAT | CAD | ENSG0000020 | 0.007507119 | 0.018389926 | 0.003184113 | 0.138946312 | 0.005850467 | 0.074134146 | 0.003276126 | 0.13894123 | 0.001656652 | 0.031708684 | 1 | 0.25766667 | 1 | 0.439050783 | ENSG0000012 | 3 | NME2P1 |
| SAT | CAD | ENSG0000020 | -0.00219152 | 0.332103073 | 0.002259561 | 0.138947628 | -0.00161733 | 0.476569624 | 0.002272075 | 0.138941995 | -0.00057419 | 0.016093629 | 1 | 0.731123848 | 1 | 0.811850954 | ENSG0000012 | 7 | SSPN |
| SAT | CAD | ENSG0000020 | 0.007410801 | 0.044338383 | 0.003685347 | 0.138946743 | 0.005405201 | 0.156560517 | 0.003815247 | 0.138941567 | 0.002005599 | 0.042222422 | 1 | 0.353926215 | 1 | 0.574768189 | ENSG0000012 | 7 | SSPN |
| SAT | CAD | ENSG0000020 | -0.0071422 | 0.013304448 | 0.002885128 | 0.138946149 | -0.00844293 | 0.003792174 | 0.002916417 | 0.13893975 | 0.001300724 | 0.002290483 | 1 | 0.222713752 | 1 | 0.123298635 | ENSG0000012 | 7 | SSPN |
| SAT | CAD | ENSG0000020 | 0.006127681 | 0.00143178 | 0.001921997 | 0.138944998 | 0.004956171 | 0.0130285 | 0.001996043 | 0.138940381 | 0.00117151 | 0.029664113 | 1 | 0.071330396 | 1 | 0.233587473 | ENSG0000012 | 8 | PKN1 |
| SAT | CAD | ENSG0000020 | 0.000433978 | 0.714118468 | 0.001184662 | 0.13894785 | 7.68E-05 | 0.948613161 | 0.001191463 | 0.138942138 | 0.00035719 | 0.005035063 | 1 | 0.912127964 | 1 | 0.985387556 | ENSG0000012 | 8 | PKN1 |
| SAT | CAD | ENSG0000020 | 0.001241723 | 0.13531404 | 0.000831433 | 0.13894726 | 0.000971295 | 0.248810348 | 0.000842228 | 0.13894176 | 0.000270428 | 0.044388103 | 1 | 0.535543265 | 1 | 0.656525015 | ENSG0000012 | 9 | EBPL |
| SAT | CAD | ENSG0000020 | -0.00071545 | 0.691459944 | 0.001802697 | 0.138947852 | -0.00145565 | 0.425459086 | 0.001826443 | 0.138941958 | 0.000740205 | 0.01175202 | 1 | 0.904577383 | 1 | 0.794497018 | ENSG0000012 | 9 | EBPL |
| SAT | CAD | ENSG0000020 | 0.001985428 | 0.360005967 | 0.002169026 | 0.138947658 | 0.001397851 | 0.521998587 | 0.002183227 | 0.138942022 | 0.000587577 | 0.018284524 | 1 | 0.750654992 | 1 | 0.838314911 | ENSG0000012 | 9 | EBPL |
| SAT | CAD | ENSG0000020 | 0.00291739 | 0.009696312 | 0.001127936 | 0.138945989 | 0.002422701 | 0.035072893 | 0.00114955 | 0.138940873 | 0.000494689 | 0.025840009 | 1 | 0.189366663 | 1 | 0.340042469 | ENSG0000012 | 9 | ATP7B |
| SAT | CAD | ENSG0000021 | 0.00466321 | 0.005455483 | 0.001678112 | 0.138945695 | 0.003812417 | 0.027468109 | 0.001729139 | 0.138940753 | 0.000850793 | 0.041366394 | 1 | 0.146128675 | 1 | 0.310609304 | ENSG0000012 | 9 | ATP7B |
| SAT | CAD | ENSG0000021 | 0.002110286 | 0.212760974 | 0.001693634 | 0.138947454 | 0.001231389 | 0.48073529 | 0.001746352 | 0.138941998 | 0.000878897 | 0.039092398 | 1 | 0.626499199 | 1 | 0.814411031 | ENSG0000012 | 9 | ATP7B |
| SAT | CAD | ENSG0000021 | 0.001333989 | 0.118111318 | 0.000853615 | 0.1389472 | 0.001348444 | 0.114183884 | 0.000853626 | 0.138941428 | -1.45E-05 | 0.038909404 | 1 | 0.511544451 | 1 | 0.516412287 | ENSG0000012 | 18 | NLN |
| SAT | CAD | ENSG0000022 | 0.001270849 | 0.709641368 | 0.00341316 | 0.138947857 | 0.002502868 | 0.470136528 | 0.003465341 | 0.138941991 | -0.00123202 | 0.039876809 | 1 | 0.911825726 | 1 | 0.811850954 | ENSG0000012 | 18 | NLN |
| SAT | CAD | ENSG0000022 | -0.00163236 | 0.114322303 | 0.001033749 | 0.138947186 | -0.00108564 | 0.308717331 | 0.001066525 | 0.138941844 | -0.00054672 | 0.037238618 | 1 | 0.511544451 | 1 | 0.709397581 | ENSG0000012 | 18 | NLN |
| SAT | CAD | ENSG0000022 | -0.00250604 | 0.262287905 | 0.00223554 | 0.138947538 | -0.00316513 | 0.159323675 | 0.002248995 | 0.138941575 | 0.000659085 | 0.007398291 | 1 | 0.659183967 | 1 | 0.575903653 | ENSG0000012 | 10 | ITIH5 |
| SAT | CAD | ENSG0000022 | 1.49E-05 | 0.995993452 | 0.002971627 | 0.138947896 | -0.00062362 | 0.834265159 | 0.002980464 | 0.138942127 | 0.000638538 | 0.005527651 | 1 | 0.999715369 | 1 | 0.954845447 | ENSG0000012 | 6 | CDK2 |
| SAT | CAD | ENSG0000023 | 0.002810558 | 0.036733164 | 0.001345587 | 0.138946652 | 0.002914524 | 0.030384141 | 0.001346168 | 0.138940803 | -0.00010397 | 0.010259593 | 1 | 0.337152794 | 1 | 0.323670021 | ENSG0000012 | 6 | CDK2 |
| SAT | CAD | ENSG0000023 | 0.0003086 | 0.894658258 | 0.002330596 | 0.138947891 | 0.000857748 | 0.713695548 | 0.00233784 | 0.138942101 | -0.00054915 | 0.002918219 | 1 | 0.970661333 | 1 | 0.911950925 | ENSG0000012 | 10 | SMUG1 |
| SAT | CAD | ENSG0000023 | 0.001341371 | 0.359539199 | 0.001463988 | 0.138947657 | 0.001030439 | 0.482892619 | 0.001468577 | 0.138941999 | 0.000310932 | 0.007546212 | 1 | 0.750414105 | 1 | 0.814411031 | ENSG0000012 | 10 | SMUG1 |
| SAT | CAD | ENSG0000023 | -0.00785316 | 0.056422538 | 0.004116425 | 0.138946859 | -0.00873404 | 0.034407299 | 0.004129038 | 0.138940864 | 0.000880877 | 0.006489808 | 1 | 0.382484189 | 1 | 0.336986116 | ENSG0000012 | 10 | SMUG1 |
| SAT | CAD | ENSG0000023 | -0.00487866 | 0.037885516 | 0.002349918 | 0.138946667 | -0.0053805 | 0.022597717 | 0.002359689 | 0.138940657 | 0.000501847 | 0.019627 | 1 | 0.337586455 | 1 | 0.287074915 | ENSG0000012 | 11 | METTL21B |
| SAT | CAD | ENSG0000023 | -0.00445109 | 0.118046047 | 0.00284773 | 0.1389472 | -0.0055525 | 0.053620597 | 0.002877101 | 0.138941077 | 0.001101408 | 0.007292043 | 1 | 0.511544451 | 1 | 0.38135058 | ENSG0000012 | 11 | METTL21B |
| SAT | CAD | ENSG0000023 | 0.007457169 | 0.00035946 | 0.00208989 | 0.138944266 | 0.006646833 | 0.001725514 | 0.002120994 | 0.138939339 | 0.000810336 | 0.025237075 | 1 | 0.028765591 | 1 | 0.085131734 | ENSG0000012 | 12 | SARDH |
| SAT | CAD | ENSG0000023 | 0.002442282 | 0.600758502 | 0.004666985 | 0.138947818 | 0.002373189 | 0.611101501 | 0.00466702 | 0.138942066 | 6.91E-05 | 0.04804071 | 1 | 0.871671099 | 1 | 0.869331532 | ENSG0000012 | 12 | SARDH |
| SAT | CAD | ENSG0000023 | 0.00157825 | 0.756205953 | 0.00508347 | 0.138947869 | 0.001659558 | 0.744077114 | 0.005083509 | 0.138942109 | -8.13E-05 | 0.033456109 | 1 | 0.930016551 | 1 | 0.924790073 | ENSG0000012 | 12 | SARDH |
| SAT | CAD | ENSG0000024 | -0.00158454 | 0.172000104 | 0.001160146 | 0.138947365 | -0.00185295 | 0.11244666 | 0.001167365 | 0.138941421 | 0.000268417 | 0.038720186 | 1 | 0.580279266 | 1 | 0.516412287 | ENSG0000012 | 6 | DBH |
| SAT | CAD | ENSG0000024 | -0.00056708 | 0.685188886 | 0.001398836 | 0.13894785 | -0.00049048 | 0.725933462 | 0.001399226 | 0.138942104 | -7.66E-05 | 0.025295129 | 1 | 0.903579074 | 1 | 0.917446451 | ENSG0000012 | 6 | DBH |
| SAT | CAD | ENSG0000025 | 0.006763865 | 0.141291099 | 0.004598133 | 0.138947279 | 0.00640042 | 0.164163833 | 0.004600629 | 0.138941587 | 0.000363446 | 0.018545035 | 1 | 0.545530242 | 1 | 0.576272733 | ENSG0000012 | 6 | DBH |
| SAT | CAD | ENSG0000025 | 0.003645696 | 0.351338003 | 0.0039117 | 0.138947649 | 0.003707894 | 0.343187102 | 0.003911745 | 0.138941883 | -6.22E-05 | 0.047657794 | 1 | 0.745627419 | 1 | 0.742742007 | ENSG0000012 | 5 | NDUFAF4 |
| SAT | CAD | ENSG0000026 | 0.000245779 | 0.834224795 | 0.001174365 | 0.138947884 | 0.000266217 | 0.82066642 | 0.001174376 | 0.138942125 | -2.04E-05 | 0.025294694 | 1 | 0.956013789 | 1 | 0.951114986 | ENSG0000012 | 5 | NDUFAF4 |
| SAT | CAD | ENSG0000026 | 0.00115196 | 0.73593201 | 0.00341579 | 0.138947864 | 0.00132846 | 0.697403069 | 0.00341657 | 0.138942096 | -0.0001765 | 0.020672756 | 1 | 0.921952243 | 1 | 0.905849293 | ENSG0000012 | 5 | NDUFAF4 |
| SAT | CAD | ENSG0000026 | 0.004853072 | 0.015368844 | 0.002002442 | 0.138946222 | 0.004824808 | 0.015976944 | 0.002002452 | 0.138940484 | 2.83E-05 | 0.04878891 | 1 | 0.241268794 | 1 | 0.26048278 | ENSG0000012 | 5 | NDUFAF4 |
| SAT | CAD | ENSG0000027 | -0.00067817 | 0.89581302 | 0.005178732 | 0.138947892 | -0.0007635 | 0.882794232 | 0.00517877 | 0.138942133 | 8.53E-05 | 0.027938342 | 1 | 0.970661333 | 1 | 0.966771654 | ENSG0000012 | 8 | SLC36A1 |
| SAT | CAD | ENSG0000027 | 0.003424596 | 0.133049956 | 0.002279743 | 0.138947253 | 0.003192389 | 0.161751011 | 0.002281567 | 0.138941581 | 0.000232207 | 0.011941262 | 1 | 0.533294064 | 1 | 0.576272733 | ENSG0000012 | 8 | SLC36A1 |
| SAT | Heart_Failure | ENSG0000000 | 0.004567263 | 0.134131938 | 0.003048892 | 0.076055143 | 0.004494403 | 0.140408654 | 0.003048555 | 0.076035001 | 7.29E-05 | 0.000291797 | 1 | 0.659800431 | 1 | 0.655068096 | ENSG0000012 | 8 | SLC36A1 |
| SAT | Heart_Failure | ENSG0000000 | -0.00305141 | 0.062748954 | 0.001639688 | 0.076054953 | -0.00308568 | 0.059826321 | 0.001639507 | 0.076034787 | 3.43E-05 | 0.001818478 | 1 | 0.515988262 | 1 | 0.504627254 | ENSG0000012 | 8 | SLC36A1 |
| SAT | Heart_Failure | ENSG0000000 | -0.01323296 | 9.42E-07 | 0.00269868 | 0.076051741 | -0.01317314 | 1.05E-06 | 0.002698406 | 0.076031622 | -5.98E-05 | 0.004825078 | 0.003089965 | 0.000233648 | 0.003449752 | 0.000581811 | ENSG0000012 | 8 | SLC36A1 |
| SAT | Heart_Failure | ENSG0000000 | 0.003058205 | 0.409107228 | 0.00370482 | 0.076055387 | 0.002974887 | 0.421948702 | 0.003704505 | 0.07603524 | 8.33E-05 | 0.021078539 | 1 | 0.870429337 | 1 | 0.873390936 | ENSG0000012 | 7 | ADCK4 |
| SAT | Heart_Failure | ENSG0000000 | 0.001149478 | 0.367916714 | 0.001276654 | 0.076055366 | 0.00116673 | 0.36071871 | 0.001276514 | 0.07603521 | -1.73E-05 | 0.043817774 | 1 | 0.853098755 | 1 | 0.848283315 | ENSG0000012 | 7 | ADCK4 |
| SAT | Heart_Failure | ENSG0000000 | 0.003076803 | 0.199831098 | 0.002399937 | 0.076055236 | 0.003029419 | 0.206797629 | 0.002399688 | 0.076035091 | 4.74E-05 | 0.009114902 | 1 | 0.734105989 | 1 | 0.73263258 | ENSG0000012 | 7 | ADCK4 |
| SAT | Heart_Failure | ENSG0000000 | -0.00057022 | 0.785017296 | 0.002090369 | 0.076055481 | -0.00062379 | 0.765366292 | 0.002090145 | 0.076035326 | 5.36E-05 | 0.000320848 | 1 | 0.972730696 | 1 | 0.965963214 | ENSG0000012 | 7 | ADCK4 |
| SAT | Heart_Failure | ENSG0000000 | -0.00454512 | 0.068513208 | 0.002495096 | 0.076054975 | -0.00459054 | 0.065767075 | 0.002494841 | 0.076034812 | 4.54E-05 | 0.019419391 | 1 | 0.537536555 | 1 | 0.515872519 | ENSG0000012 | 7 | ADCK4 |
| SAT | Heart_Failure | ENSG0000000 | 0.002448139 | 0.47029459 | 0.003390772 | 0.076055412 | 0.002541269 | 0.453534066 | 0.003390453 | 0.076035252 | -9.31E-05 | 0.001688897 | 1 | 0.893630105 | 1 | 0.886796943 | ENSG0000012 | 7 | ADCK4 |
| SAT | Heart_Failure | ENSG0000000 | 0.002718464 | 0.365926188 | 0.003006713 | 0.076055365 | 0.002654612 | 0.377242412 | 0.003006392 | 0.076035218 | 6.39E-05 | 0.002992921 | 1 | 0.85158824 | 1 | 0.854967426 | ENSG0000012 | 9 | ZNF137P |
| SAT | Heart_Failure | ENSG0000000 | -0.00279235 | 0.008490783 | 0.001060956 | 0.076054412 | -0.00281035 | 0.008069339 | 0.001060843 | 0.076034245 | 1.80E-05 | 0.01884834 | 1 | 0.207897455 | 1 | 0.209225934 | ENSG0000012 | 9 | ZNF137P |
| SAT | Heart_Failure | ENSG0000000 | -0.00218354 | 0.555004099 | 0.003699162 | 0.076055439 | -0.0022353 | 0.54561867 | 0.003698749 | 0.076035283 | 5.18E-05 | 0.030262966 | 1 | 0.921542737 | 1 | 0.912597963 | ENSG0000012 | 9 | CHPF |
| SAT | Heart_Failure | ENSG0000000 | 0.001629777 | 0.549598545 | 0.00272373 | 0.076055437 | 0.001586787 | 0.560135246 | 0.002723439 | 0.076035287 | 4.30E-05 | 0.027529248 | 1 | 0.918172602 | 1 | 0.917983887 | ENSG0000012 | 9 | CHPF |
| SAT | Heart_Failure | ENSG0000000 | 0.001419368 | 0.653232032 | 0.003159226 | 0.076055462 | 0.001481845 | 0.639003084 | 0.003158954 | 0.076035306 | -6.25E-05 | 0.040312447 | 1 | 0.962760695 | 1 | 0.949533115 | ENSG0000012 | 9 | FAM210B |
| SAT | Heart_Failure | ENSG0000001 | -0.0018203 | 0.63650251 | 0.003851711 | 0.076055458 | -0.0017644 | 0.646859034 | 0.003851301 | 0.076035307 | -5.59E-05 | 0.043803552 | 1 | 0.959428651 | 1 | 0.95317119 | ENSG0000012 | 14 | SNX21 |
| SAT | Heart_Failure | ENSG0000001 | -0.00284042 | 0.430516063 | 0.003603177 | 0.076055396 | -0.00268398 | 0.456293075 | 0.003602831 | 0.076035254 | -0.00015644 | 3.48E-07 | 1 | 0.878825835 | 1 | 0.887431878 | ENSG0000012 | 14 | SNX21 |
| SAT | Heart_Failure | ENSG0000001 | 0.002606592 | 0.490517508 | 0.003780483 | 0.076055419 | 0.002483206 | 0.511233943 | 0.003780085 | 0.076035273 | 0.000123386 | 9.00E-06 | 1 | 0.898909993 | 1 | 0.902750226 | ENSG0000012 | 14 | SNX21 |
| SAT | Heart_Failure | ENSG0000001 | 0.00072956 | 0.825143407 | 0.003302153 | 0.076055485 | 0.000780319 | 0.813175575 | 0.003301799 | 0.076035331 | -5.08E-05 | 0.030860685 | 1 | 0.976669362 | 1 | 0.972791643 | ENSG0000012 | 14 | WFDC3 |
| SAT | Heart_Failure | ENSG0000001 | -0.00039781 | 0.796519124 | 0.001542776 | 0.076055483 | -0.00035469 | 0.818144683 | 0.001542608 | 0.076035332 | -4.31E-05 | 4.99E-05 | 1 | 0.976456935 | 1 | 0.972791643 | ENSG0000012 | 14 | WFDC3 |
| SAT | Heart_Failure | ENSG0000001 | 0.002185777 | 0.566557965 | 0.003813764 | 0.076055442 | 0.00206008 | 0.589042367 | 0.003813377 | 0.076035295 | 0.000125696 | 2.90E-05 | 1 | 0.927165893 | 1 | 0.934989637 | ENSG0000012 | 14 | WFDC3 |
| SAT | Heart_Failure | ENSG0000001 | 0.004270483 | 0.052093403 | 0.002198599 | 0.076054904 | 0.004321713 | 0.049314796 | 0.002198378 | 0.076034737 | -5.12E-05 | 0.003677801 | 1 | 0.491119403 | 1 | 0.480049081 | ENSG0000012 | 14 | WFDC3 |
| SAT | Heart_Failure | ENSG0000001 | 0.000258642 | 0.853060529 | 0.001396444 | 0.076055488 | 0.000162302 | 0.907490302 | 0.001396689 | 0.076035338 | 9.63E-05 | 0.005474731 | 1 | 0.976669362 | 1 | 0.982067422 | ENSG0000012 | 14 | WFDC3 |
| SAT | Heart_Failure | ENSG0000001 | 0.001157644 | 0.49518791 | 0.001697224 | 0.07605542 | 0.001184371 | 0.485237873 | 0.001697041 | 0.076035264 | -2.67E-05 | 0.026199616 | 1 | 0.898909993 | 1 | 0.894608312 | ENSG0000012 | 14 | WFDC3 |
| SAT | Heart_Failure | ENSG0000001 | -0.0080157 | 0.004399542 | 0.002814491 | 0.076054227 | -0.00810357 | 0.0039829 | 0.002814202 | 0.076034046 | 8.79E-05 | 5.27E-05 | 1 | 0.154647076 | 1 | 0.161355485 | ENSG0000012 | 14 | WFDC3 |
| SAT | Heart_Failure | ENSG0000001 | 0.001047339 | 0.63370775 | 0.002197913 | 0.076055458 | 0.001090325 | 0.619804511 | 0.00219767 | 0.076035302 | -4.30E-05 | 0.003133932 | 1 | 0.959428651 | 1 | 0.94798204 | ENSG0000012 | 8 | SLC12A5 |
| SAT | Heart_Failure | ENSG0000001 | -0.00462318 | 0.122556062 | 0.002994026 | 0.076055121 | -0.00454478 | 0.128987669 | 0.002993711 | 0.076034981 | -7.84E-05 | 0.00038047 | 1 | 0.646585619 | 1 | 0.649215182 | ENSG0000012 | 11 | VAPB |
| SAT | Heart_Failure | ENSG0000002 | 0.00230165 | 0.495615855 | 0.003377802 | 0.076055421 | 0.002245519 | 0.506144477 | 0.003377463 | 0.076035271 | 5.61E-05 | 0.038137846 | 1 | 0.898909993 | 1 | 0.900571724 | ENSG0000012 | 11 | VAPB |
| SAT | Heart_Failure | ENSG0000002 | 0.000584135 | 0.854847251 | 0.003193088 | 0.076055488 | 0.000633935 | 0.842611116 | 0.003192745 | 0.076035334 | -4.98E-05 | 0.026881631 | 1 | 0.976669362 | 1 | 0.972791643 | ENSG0000012 | 13 | TOX2 |
| SAT | Heart_Failure | ENSG0000002 | 0.000150691 | 0.937251425 | 0.001914147 | 0.076055492 | 0.000194228 | 0.91916868 | 0.001913939 | 0.076035339 | -4.35E-05 | 0.000993173 | 1 | 0.990058572 | 1 | 0.984094962 | ENSG0000012 | 13 | TOX2 |
| SAT | Heart_Failure | ENSG0000002 | 0.00142793 | 0.5596253 | 0.002447606 | 0.07605544 | 0.001334579 | 0.585544082 | 0.002447399 | 0.076035294 | 9.34E-05 | 9.74E-05 | 1 | 0.923606947 | 1 | 0.933194299 | ENSG0000012 | 13 | TOX2 |
| SAT | Heart_Failure | ENSG0000002 | -0.00525649 | 0.005587046 | 0.001896902 | 0.076054295 | -0.00520562 | 0.006061813 | 0.001896794 | 0.076034165 | -5.09E-05 | 0.028889585 | 1 | 0.164958288 | 1 | 0.182466145 | ENSG0000012 | 12 | CSE1L |
| SAT | Heart_Failure | ENSG0000002 | 0.001901419 | 0.589751841 | 0.003526396 | 0.076055448 | 0.001959859 | 0.578329146 | 0.003526022 | 0.076035292 | -5.84E-05 | 0.022571097 | 1 | 0.941136084 | 1 | 0.929235028 | ENSG0000012 | 7 | RNF114 |
| SAT | Heart_Failure | ENSG0000002 | 0.000387168 | 0.762535351 | 0.001281364 | 0.076055479 | 0.000627829 | 0.624173842 | 0.001281431 | 0.076035303 | -0.00024066 | 1.38E-22 | 1 | 0.969898847 | 1 | 0.94798204 | ENSG0000012 | 7 | RNF114 |
| SAT | Heart_Failure | ENSG0000003 | 0.000707921 | 0.617342382 | 0.001416917 | 0.076055454 | 0.000683372 | 0.629559775 | 0.001416764 | 0.076035304 | 2.45E-05 | 0.013034655 | 1 | 0.954216727 | 1 | 0.94798204 | ENSG0000012 | 6 | NEURL2 |
| SAT | Heart_Failure | ENSG0000004 | 0.000187699 | 0.883181091 | 0.001277404 | 0.07605549 | 0.000157532 | 0.901841312 | 0.001277264 | 0.076035338 | 3.02E-05 | 0.000471844 | 1 | 0.979223495 | 1 | 0.980373283 | ENSG0000012 | 12 | ATP8A1 |
| SAT | Heart_Failure | ENSG0000004 | 0.001435264 | 0.325018336 | 0.001458307 | 0.076055342 | 0.001457967 | 0.317373628 | 0.001458156 | 0.076035184 | -2.27E-05 | 0.039932277 | 1 | 0.832463044 | 1 | 0.823487262 | ENSG0000012 | 7 | USP22 |
| SAT | Heart_Failure | ENSG0000004 | 0.001201978 | 0.749974257 | 0.003771818 | 0.076055477 | -0.01343008 | 0.002550661 | 0.004451053 | 0.07603392 | 0.014632058 | 6.05E-10 | 1 | 0.966658386 | 1 | 0.122791633 | ENSG0000012 | 7 | USP22 |
| SAT | Heart_Failure | ENSG0000004 | -0.00016633 | 0.958035525 | 0.003160998 | 0.076055493 | -5.89E-05 | 0.985133672 | 0.003160764 | 0.07603534 | -0.00010743 | 0.001695771 | 1 | 0.994671121 | 1 | 0.999768899 | ENSG0000012 | 12 | BTN2A2 |
| SAT | Heart_Failure | ENSG0000004 | 0.000337694 | 0.646780814 | 0.000736939 | 0.07605546 | 0.000350107 | 0.63469302 | 0.000736862 | 0.076035305 | -1.24E-05 | 0.023859732 | 1 | 0.961525986 | 1 | 0.94798204 | ENSG0000012 | 12 | BTN2A2 |
| SAT | Heart_Failure | ENSG0000004 | -0.00556155 | 0.106856364 | 0.003449051 | 0.076055087 | -0.00563007 | 0.102570088 | 0.003448692 | 0.076034924 | 6.85E-05 | 0.008562879 | 1 | 0.638670167 | 1 | 0.614882715 | ENSG0000012 | 12 | BTN2A2 |
| SAT | Heart_Failure | ENSG0000005 | 0.00202791 | 0.383435299 | 0.002326692 | 0.076055374 | 0.001710271 | 0.462351835 | 0.002326962 | 0.076035256 | 0.000317638 | 9.09E-10 | 1 | 0.85759885 | 1 | 0.892844363 | ENSG0000012 | 12 | BTN2A2 |
| SAT | Heart_Failure | ENSG0000005 | 1.39E-05 | 0.995066858 | 0.0022477 | 0.076055493 | -0.00072439 | 0.748782841 | 0.002261982 | 0.076035324 | 0.000738285 | 0.003985786 | 1 | 0.999479422 | 1 | 0.962415493 | ENSG0000012 | 12 | BTN2A2 |
| SAT | Heart_Failure | ENSG0000005 | 0.000973634 | 0.750031649 | 0.003055997 | 0.076055477 | 0.001086299 | 0.722212967 | 0.003055667 | 0.07603532 | -0.00011266 | 1.61E-07 | 1 | 0.966658386 | 1 | 0.958028875 | ENSG0000012 | 10 | MRS2 |
| SAT | Heart_Failure | ENSG0000005 | 0.005935203 | 0.001385088 | 0.001856051 | 0.076053897 | 0.005971097 | 0.001293564 | 0.001855863 | 0.076033725 | -3.59E-05 | 0.015194877 | 1 | 0.077908515 | 1 | 0.081932833 | ENSG0000012 | 10 | MRS2 |
| SAT | Heart_Failure | ENSG0000005 | 0.004040528 | 0.169446678 | 0.002940734 | 0.076055198 | 0.003995679 | 0.1741869 | 0.002940432 | 0.076035052 | 4.48E-05 | 0.04795248 | 1 | 0.699678071 | 1 | 0.700325828 | ENSG0000012 | 10 | MRS2 |
| SAT | Heart_Failure | ENSG0000005 | -0.00441185 | 0.053664244 | 0.002286476 | 0.076054912 | -0.00432394 | 0.058589405 | 0.002286269 | 0.076034782 | -8.79E-05 | 2.66E-05 | 1 | 0.497196222 | 1 | 0.503576399 | ENSG0000012 | 10 | MRS2 |
| SAT | Heart_Failure | ENSG0000005 | 0.000420186 | 0.701909992 | 0.001097826 | 0.07605547 | 0.000376923 | 0.731320671 | 0.001097719 | 0.076035322 | 4.33E-05 | 2.68E-06 | 1 | 0.966658386 | 1 | 0.958028875 | ENSG0000012 | 10 | MRS2 |
| SAT | Heart_Failure | ENSG0000005 | 0.001706105 | 0.196057155 | 0.001319628 | 0.076055232 | 0.00168372 | 0.201943284 | 0.001319492 | 0.076035086 | 2.24E-05 | 0.028313791 | 1 | 0.726878652 | 1 | 0.728906397 | ENSG0000012 | 10 | MRS2 |
| SAT | Heart_Failure | ENSG0000005 | -0.00151086 | 0.353812797 | 0.001629455 | 0.076055359 | -0.00147225 | 0.366204152 | 0.001629305 | 0.076035213 | -3.86E-05 | 0.008382042 | 1 | 0.848363544 | 1 | 0.849290727 | ENSG0000012 | 10 | BTN2A3P |
| SAT | Heart_Failure | ENSG0000006 | -0.00313768 | 0.242639543 | 0.002685417 | 0.07605528 | -0.00307747 | 0.251749911 | 0.002685145 | 0.076035135 | -6.02E-05 | 0.00436838 | 1 | 0.759865642 | 1 | 0.765423527 | ENSG0000012 | 10 | BTN2A3P |
| SAT | Heart_Failure | ENSG0000006 | -0.00867937 | 0.000962078 | 0.00262899 | 0.076053792 | -0.00873844 | 0.000886737 | 0.00262872 | 0.076033616 | 5.91E-05 | 0.003606746 | 1 | 0.05739234 | 1 | 0.06612236 | ENSG0000012 | 10 | BTN2A3P |
| SAT | Heart_Failure | ENSG0000006 | -0.00237329 | 0.436790142 | 0.003051972 | 0.076055399 | -0.0024403 | 0.423907683 | 0.003051666 | 0.07603524 | 6.70E-05 | 0.006293415 | 1 | 0.880339889 | 1 | 0.873390936 | ENSG0000012 | 10 | BTN2A3P |
| SAT | Heart_Failure | ENSG0000006 | -0.0030697 | 0.120831458 | 0.001978795 | 0.076055117 | -0.00301598 | 0.127435123 | 0.001978601 | 0.076034978 | -5.37E-05 | 0.001086062 | 1 | 0.646585619 | 1 | 0.647874406 | ENSG0000012 | 10 | BTN2A3P |
| SAT | Heart_Failure | ENSG0000006 | -0.00068155 | 0.627927674 | 0.001406278 | 0.076055456 | -0.00065495 | 0.641370097 | 0.001406131 | 0.076035306 | -2.66E-05 | 0.011239999 | 1 | 0.958246837 | 1 | 0.951372499 | ENSG0000012 | 10 | BTN2A3P |
| SAT | Heart_Failure | ENSG0000006 | 0.004230695 | 0.097373613 | 0.002552109 | 0.076055064 | 0.004168724 | 0.102339947 | 0.002551837 | 0.076034924 | 6.20E-05 | 0.000727828 | 1 | 0.61676221 | 1 | 0.614882715 | ENSG0000012 | 5 | SNRPC |
| SAT | Heart_Failure | ENSG0000006 | 0.000929224 | 0.583257981 | 0.001693709 | 0.076055446 | 0.000895207 | 0.597081501 | 0.001693536 | 0.076035297 | 3.40E-05 | 0.01013479 | 1 | 0.937265914 | 1 | 0.938230079 | ENSG0000012 | 5 | SNRPC |
| SAT | Heart_Failure | ENSG0000006 | -0.0024846 | 0.216675469 | 0.002011138 | 0.076055255 | -0.00390822 | 0.057104361 | 0.002054238 | 0.076034775 | 0.001423624 | 0.00069772 | 1 | 0.745333464 | 1 | 0.503576399 | ENSG0000012 | 5 | SNRPC |
| SAT | Heart_Failure | ENSG0000006 | -0.00146868 | 0.202085643 | 0.001151334 | 0.076055239 | -0.00150553 | 0.190949892 | 0.00115121 | 0.076035073 | 3.68E-05 | 7.47E-06 | 1 | 0.735350411 | 1 | 0.720122525 | ENSG0000012 | 5 | SNRPC |
| SAT | Heart_Failure | ENSG0000006 | -0.00392189 | 0.10820947 | 0.002441575 | 0.07605509 | -0.00388122 | 0.111880356 | 0.002441327 | 0.076034946 | -4.07E-05 | 0.033962061 | 1 | 0.640599412 | 1 | 0.640177219 | ENSG0000012 | 5 | SNRPC |
| SAT | Heart_Failure | ENSG0000006 | -0.00255137 | 0.450299291 | 0.003379666 | 0.076055404 | -0.00261333 | 0.439326924 | 0.003379311 | 0.076035247 | 6.20E-05 | 0.013405248 | 1 | 0.887019168 | 1 | 0.879941679 | ENSG0000012 | 5 | SNRPC |
| SAT | Heart_Failure | ENSG0000006 | 0.00587648 | 0.050483208 | 0.003004563 | 0.076054896 | 0.005825159 | 0.052506045 | 0.003004261 | 0.076034754 | 5.13E-05 | 0.031715848 | 1 | 0.48962926 | 1 | 0.490345005 | ENSG0000012 | 5 | SNRPC |
| SAT | Heart_Failure | ENSG0000006 | 0.001110418 | 0.568985561 | 0.001949653 | 0.076055442 | 0.001143518 | 0.557480813 | 0.001949441 | 0.076035286 | -3.31E-05 | 0.013543608 | 1 | 0.92942386 | 1 | 0.915531921 | ENSG0000012 | 13 | SERPINB6 |
| SAT | Heart_Failure | ENSG0000006 | -0.00010169 | 0.977014127 | 0.003529454 | 0.076055493 | -0.00015874 | 0.964121721 | 0.003529066 | 0.07603534 | 5.71E-05 | 0.016425336 | 1 | 0.998920999 | 1 | 0.996055789 | ENSG0000012 | 5 | HIST1H1D |
| SAT | Heart_Failure | ENSG0000006 | -0.00013322 | 0.955045017 | 0.002363234 | 0.076055493 | -8.74E-05 | 0.97049246 | 0.002363005 | 0.07603534 | -4.58E-05 | 0.02144464 | 1 | 0.994131568 | 1 | 0.99755193 | ENSG0000012 | 5 | HIST1H1D |
| SAT | Heart_Failure | ENSG0000006 | 0.003060918 | 0.194086633 | 0.002357116 | 0.07605523 | 0.003024686 | 0.199369878 | 0.002356866 | 0.076035083 | 3.62E-05 | 0.035198196 | 1 | 0.726878652 | 1 | 0.726007291 | ENSG0000012 | 5 | HIST1H1D |
| SAT | Heart_Failure | ENSG0000007 | 0.004183342 | 0.189357396 | 0.003187346 | 0.076055224 | 0.00423513 | 0.183893279 | 0.003187029 | 0.076035065 | -5.18E-05 | 0.045490116 | 1 | 0.722420486 | 1 | 0.715224819 | ENSG0000012 | 9 | PEX6 |
| SAT | Heart_Failure | ENSG0000007 | -0.00075726 | 0.798170561 | 0.002961345 | 0.076055483 | -0.00068769 | 0.816346676 | 0.002961027 | 0.076035332 | -6.96E-05 | 0.000885754 | 1 | 0.976669362 | 1 | 0.972791643 | ENSG0000012 | 9 | PEX6 |
| SAT | Heart_Failure | ENSG0000007 | -0.00136 | 0.697524081 | 0.003499161 | 0.076055469 | -0.00126666 | 0.717334847 | 0.003498845 | 0.07603532 | -9.33E-05 | 0.003709865 | 1 | 0.966658386 | 1 | 0.958028875 | ENSG0000012 | 9 | PEX6 |
| SAT | Heart_Failure | ENSG0000007 | 0.002988902 | 0.23385553 | 0.002510655 | 0.076055272 | 0.002938816 | 0.241734409 | 0.002510383 | 0.076035126 | 5.01E-05 | 0.004342788 | 1 | 0.754244409 | 1 | 0.753454838 | ENSG0000012 | 4 | ZNF391 |
| SAT | Heart_Failure | ENSG0000007 | 0.003694631 | 0.234480127 | 0.003107608 | 0.076055272 | 0.003519413 | 0.257513483 | 0.003108228 | 0.07603514 | 0.000175218 | 0.028677901 | 1 | 0.754244409 | 1 | 0.770781077 | ENSG0000012 | 4 | ZNF391 |
| SAT | Heart_Failure | ENSG0000007 | 0.001585613 | 0.64538377 | 0.003445601 | 0.07605546 | 0.001693222 | 0.623097415 | 0.00344525 | 0.076035302 | -0.00010761 | 6.41E-05 | 1 | 0.960776086 | 1 | 0.94798204 | ENSG0000012 | 4 | ZNF391 |
| SAT | Heart_Failure | ENSG0000007 | 6.32E-05 | 0.985498562 | 0.003475668 | 0.076055493 | 0.000140844 | 0.967672862 | 0.00347531 | 0.07603534 | -7.77E-05 | 0.003598079 | 1 | 0.998924529 | 1 | 0.99755193 | ENSG0000012 | 4 | ZNF391 |
| SAT | Heart_Failure | ENSG0000007 | -0.0007748 | 0.768509587 | 0.002632472 | 0.076055479 | -0.00069101 | 0.792917402 | 0.002632193 | 0.076035329 | -8.38E-05 | 1.21E-05 | 1 | 0.971467105 | 1 | 0.970635778 | ENSG0000012 | 4 | ZNF391 |
| SAT | Heart_Failure | ENSG0000007 | 0.002419669 | 0.523126846 | 0.003789417 | 0.076055429 | 0.002521902 | 0.505683141 | 0.003789059 | 0.076035271 | -0.00010223 | 0.001933522 | 1 | 0.906724424 | 1 | 0.900571724 | ENSG0000012 | 4 | ZNF391 |
| SAT | Heart_Failure | ENSG0000007 | -0.00453578 | 0.148679718 | 0.00314066 | 0.076055167 | -0.00459007 | 0.143834844 | 0.003140309 | 0.076035007 | 5.43E-05 | 0.007463831 | 1 | 0.674894985 | 1 | 0.659101398 | ENSG0000012 | 4 | ZNF391 |
| SAT | Heart_Failure | ENSG0000007 | 0.001573637 | 0.690014464 | 0.003945577 | 0.076055468 | 0.001470552 | 0.709336339 | 0.003945165 | 0.076035318 | 0.000103085 | 0.000497306 | 1 | 0.966658386 | 1 | 0.958028875 | ENSG0000012 | 4 | ZNF391 |
| SAT | Heart_Failure | ENSG0000007 | -0.00149339 | 0.36782241 | 0.001658289 | 0.076055366 | -0.00145533 | 0.380103231 | 0.001658108 | 0.07603522 | -3.81E-05 | 0.000768441 | 1 | 0.853098755 | 1 | 0.854967426 | ENSG0000012 | 4 | ZNF391 |
| SAT | Heart_Failure | ENSG0000007 | 0.002337701 | 0.528763572 | 0.003711255 | 0.076055431 | 0.002223654 | 0.549021327 | 0.003710866 | 0.076035284 | 0.000114047 | 3.63E-05 | 1 | 0.907158447 | 1 | 0.91411374 | ENSG0000012 | 4 | ZNF391 |
| SAT | Heart_Failure | ENSG0000007 | -0.00297905 | 0.203100623 | 0.002340603 | 0.07605524 | -0.00255569 | 0.27569769 | 0.002344591 | 0.076035155 | -0.00042336 | 0.002845235 | 1 | 0.735350411 | 1 | 0.785211912 | ENSG0000012 | 7 | HIST1H2BJ |
| SAT | Heart_Failure | ENSG0000007 | 0.001093729 | 0.704041704 | 0.002879215 | 0.07605547 | 0.000965363 | 0.737389792 | 0.002878998 | 0.076035323 | 0.000128366 | 3.14E-05 | 1 | 0.966658386 | 1 | 0.960499534 | ENSG0000012 | 7 | HIST1H2BJ |

| | | | | | | | | | | | | | | | | | | | |
|---|---|---|---|---|---|---|---|---|---|---|---|---|---|---|---|---|---|---|---|
| SAT | Heart_Failure | ENSG0000008 | 0.000953504 | 0.552319529 | 0.001604446 | 0.076055438 | 0.000998148 | 0.533823697 | 0.001604275 | 0.07603528 | -4.46E-05 | 0.000109971 | 1 | 0.920018143 | 1 | 0.908441675 | ENSG0000012 | 10 | MED20 |
| SAT | Heart_Failure | ENSG0000008 | 0.000140407 | 0.962414372 | 0.00297951 | 0.076055493 | 0.000186815 | 0.95000018 | 0.002979199 | 0.07603534 | -4.64E-05 | 0.037418439 | 1 | 0.995788523 | 1 | 0.990755659 | ENSG0000012 | 10 | MED20 |
| SAT | Heart_Failure | ENSG0000008 | -0.00114152 | 0.46385963 | 0.00155837 | 0.076055409 | -0.00116385 | 0.455110774 | 0.001558199 | 0.076035253 | 2.23E-05 | 0.032614847 | 1 | 0.890605014 | 1 | 0.886989673 | ENSG0000012 | 10 | MED20 |
| SAT | Heart_Failure | ENSG0000008 | -0.0009939 | 0.751230219 | 0.003135149 | 0.076055477 | -0.00104388 | 0.739136494 | 0.003134794 | 0.076035323 | 5.00E-05 | 0.010265387 | 1 | 0.966658386 | 1 | 0.960499534 | ENSG0000012 | 5 | TBCC |
| SAT | Heart_Failure | ENSG0000008 | -0.0074714 | 0.016610468 | 0.003119288 | 0.076054598 | -0.00755946 | 0.015363648 | 0.003118974 | 0.076034424 | 8.81E-05 | 0.0004094 | 1 | 0.287798638 | 1 | 0.276579386 | ENSG0000012 | 5 | TBCC |
| SAT | Heart_Failure | ENSG0000008 | 0.001668641 | 0.522324317 | 0.0026082 | 0.076055429 | 0.001701301 | 0.51416663 | 0.002607902 | 0.076035274 | -3.27E-05 | 0.037177761 | 1 | 0.906724424 | 1 | 0.903395669 | ENSG0000012 | 13 | TCP11 |
| SAT | Heart_Failure | ENSG0000008 | 0.00427832 | 0.149252189 | 0.002966564 | 0.076055168 | 0.004211528 | 0.155661307 | 0.002966245 | 0.076035026 | 6.68E-05 | 0.001466662 | 1 | 0.675317996 | 1 | 0.669469928 | ENSG0000012 | 13 | TCP11 |
| SAT | Heart_Failure | ENSG0000008 | -8.20E-06 | 0.99225804 | 0.00084495 | 0.076055493 | -3.37E-05 | 0.968228696 | 0.000844865 | 0.07603534 | 2.55E-05 | 0.000170132 | 1 | 0.999479422 | 1 | 0.99755193 | ENSG0000012 | 13 | TCP11 |
| SAT | Heart_Failure | ENSG0000008 | 0.002544857 | 0.533896297 | 0.004090952 | 0.076055433 | 0.002443768 | 0.550228113 | 0.004090545 | 0.076035284 | 0.000101089 | 0.002376456 | 1 | 0.908781401 | 1 | 0.914465015 | ENSG0000012 | 13 | TCP11 |
| SAT | Heart_Failure | ENSG0000008 | 0.009661948 | 0.010763432 | 0.003788558 | 0.076054478 | 0.009742199 | 0.010118984 | 0.003788164 | 0.076034308 | -8.03E-05 | 0.005051281 | 1 | 0.232334342 | 1 | 0.230558242 | ENSG0000012 | 13 | TCP11 |
| SAT | Heart_Failure | ENSG0000008 | 0.004330566 | 0.037216579 | 0.002078619 | 0.076054816 | 0.004260946 | 0.040369073 | 0.002078553 | 0.076034684 | 6.96E-05 | 0.01800951 | 1 | 0.436153525 | 1 | 0.452859815 | ENSG0000012 | 5 | GNMT |
| SAT | Heart_Failure | ENSG0000008 | -0.00102068 | 0.748169388 | 0.003179144 | 0.076055477 | -0.00114306 | 0.719155699 | 0.003178812 | 0.07603532 | 0.000122383 | 2.51E-07 | 1 | 0.966658386 | 1 | 0.958028875 | ENSG0000012 | 5 | GNMT |
| SAT | Heart_Failure | ENSG0000008 | -0.00233431 | 0.186661883 | 0.001767717 | 0.076055221 | -0.00223724 | 0.20561039 | 0.001767548 | 0.07603509 | -9.71E-05 | 1.77E-10 | 1 | 0.720656885 | 1 | 0.731678624 | ENSG0000012 | 5 | GNMT |
| SAT | Heart_Failure | ENSG0000008 | -0.00051634 | 0.803474373 | 0.002074873 | 0.076055483 | -0.00042486 | 0.837741823 | 0.002074672 | 0.076035334 | -9.15E-05 | 1.69E-07 | 1 | 0.976669362 | 1 | 0.972791643 | ENSG0000012 | 5 | GNMT |
| SAT | Heart_Failure | ENSG0000008 | -0.00654494 | 0.074664892 | 0.003671731 | 0.076054997 | -0.00650206 | 0.076553372 | 0.003671308 | 0.076034851 | -4.29E-05 | 0.048246619 | 1 | 0.564459699 | 1 | 0.560403102 | ENSG0000012 | 5 | GNMT |
| SAT | Heart_Failure | ENSG0000008 | -0.00658894 | 0.057551806 | 0.003469504 | 0.07605493 | -0.00649876 | 0.061029189 | 0.003469174 | 0.076034793 | -9.02E-05 | 0.002665849 | 1 | 0.503931718 | 1 | 0.505632969 | ENSG0000012 | 5 | GNMT |
| SAT | Heart_Failure | ENSG0000008 | 0.008326065 | 0.004741784 | 0.002948235 | 0.076054248 | 0.00841109 | 0.004328019 | 0.002947927 | 0.07603407 | -8.50E-05 | 0.00011856 | 1 | 0.157149427 | 1 | 0.161355485 | ENSG0000012 | 3 | MEA1 |
| SAT | Heart_Failure | ENSG0000009 | 0.000300058 | 0.908104878 | 0.002599498 | 0.076055491 | 0.00042242 | 0.870900036 | 0.002599263 | 0.076035336 | -0.00012236 | 2.83E-07 | 1 | 0.983702811 | 1 | 0.974665278 | ENSG0000012 | 3 | MEA1 |
| SAT | Heart_Failure | ENSG0000009 | 0.000179575 | 0.809811908 | 0.00074615 | 0.076055484 | 0.000160278 | 0.82990419 | 0.000746086 | 0.076035333 | 1.93E-05 | 0.006709967 | 1 | 0.976669362 | 1 | 0.972791643 | ENSG0000012 | 3 | MEA1 |
| SAT | Heart_Failure | ENSG0000009 | 0.000203719 | 0.888535289 | 0.001453494 | 0.07605549 | 0.000156692 | 0.914143099 | 0.001453355 | 0.076035338 | 4.70E-05 | 0.000157615 | 1 | 0.979270459 | 1 | 0.98402346 | ENSG0000012 | 6 | GLO1 |
| SAT | Heart_Failure | ENSG0000009 | 0.000169506 | 0.959617324 | 0.003347682 | 0.076055493 | 0.000224091 | 0.946624345 | 0.003347319 | 0.076035339 | -5.46E-05 | 0.019011894 | 1 | 0.994780111 | 1 | 0.990755659 | ENSG0000012 | 6 | GLO1 |
| SAT | Heart_Failure | ENSG0000009 | -0.00663377 | 8.19E-05 | 0.001684171 | 0.076053072 | -0.00665759 | 7.70E-05 | 0.001683982 | 0.076032901 | 2.38E-05 | 0.026661498 | 0.268610782 | 0.008953693 | 0.252746332 | 0.012637317 | ENSG0000012 | 4 | RIOK1 |
| SAT | Heart_Failure | ENSG0000009 | 0.005406533 | 0.024378385 | 0.002401708 | 0.076054702 | 0.0053625 | 0.025548509 | 0.002401463 | 0.076034562 | 4.40E-05 | 0.019008513 | 1 | 0.336073452 | 1 | 0.354978312 | ENSG0000012 | 9 | RPP40 |
| SAT | Heart_Failure | ENSG0000009 | 0.000903307 | 0.53081311 | 0.001441219 | 0.076055432 | 0.00094547 | 0.511769406 | 0.001441078 | 0.076035273 | -4.22E-05 | 0.000416513 | 1 | 0.908320477 | 1 | 0.902750226 | ENSG0000012 | 9 | RPP40 |
| SAT | Heart_Failure | ENSG0000009 | 0.000880535 | 0.753744894 | 0.002806885 | 0.076055478 | 0.000776034 | 0.782161749 | 0.002806602 | 0.076035328 | 0.000104501 | 2.83E-06 | 1 | 0.966658386 | 1 | 0.968039494 | ENSG0000012 | 9 | RPP40 |
| SAT | Heart_Failure | ENSG0000009 | 0.002116719 | 0.520497137 | 0.003294096 | 0.076055429 | 0.002033029 | 0.537081006 | 0.003293776 | 0.076035281 | 8.37E-05 | 0.002551023 | 1 | 0.906724424 | 1 | 0.909319572 | ENSG0000012 | 9 | MYRF |
| SAT | Heart_Failure | ENSG0000009 | -0.00257807 | 0.225867583 | 0.002128745 | 0.076055264 | -0.00265278 | 0.212683029 | 0.002128658 | 0.076035098 | 7.47E-05 | 0.00969501 | 1 | 0.751217604 | 1 | 0.733958243 | ENSG0000012 | 9 | MYRF |
| SAT | Heart_Failure | ENSG0000009 | 1.65E-05 | 0.996321599 | 0.003583551 | 0.076055493 | -6.68E-05 | 0.985124474 | 0.003583171 | 0.07603534 | 8.33E-05 | 0.001429001 | 1 | 0.999479422 | 1 | 0.999768899 | ENSG0000012 | 9 | MYRF |
| SAT | Heart_Failure | ENSG0000009 | -0.00269622 | 0.333742915 | 0.002789375 | 0.076055347 | -0.00277169 | 0.320342519 | 0.002789106 | 0.076035186 | 7.55E-05 | 0.001463509 | 1 | 0.841293164 | 1 | 0.825446748 | ENSG0000012 | 9 | MYRF |
| SAT | Heart_Failure | ENSG0000009 | -0.00095236 | 0.346505888 | 0.001011655 | 0.076055355 | -0.00097165 | 0.336776312 | 0.001011548 | 0.076035196 | 1.93E-05 | 0.009885677 | 1 | 0.847011103 | 1 | 0.837075007 | ENSG0000012 | 4 | SCGB2A1 |
| SAT | Heart_Failure | ENSG0000009 | 0.003452816 | 0.024249803 | 0.001532436 | 0.076054701 | 0.003406536 | 0.026203778 | 0.001532277 | 0.076034569 | 4.63E-05 | 7.28E-05 | 1 | 0.336073452 | 1 | 0.356741057 | ENSG0000012 | 4 | SCGB2A1 |
| SAT | Heart_Failure | ENSG000001( | 0.001432894 | 0.457160175 | 0.001927146 | 0.076055407 | 0.001473977 | 0.444312662 | 0.00192694 | 0.076035249 | -4.11E-05 | 0.002961436 | 1 | 0.88796436 | 1 | 0.882462767 | ENSG0000012 | 4 | SCGB2A1 |
| SAT | Heart_Failure | ENSG000001( | -0.0019439 | 0.383163075 | 0.002229026 | 0.076055374 | -0.00198927 | 0.372107336 | 0.002228784 | 0.076035216 | 4.54E-05 | 0.003459644 | 1 | 0.85759885 | 1 | 0.850197891 | ENSG0000012 | 4 | SCGB2A1 |
| SAT | Heart_Failure | ENSG000001( | 0.002444504 | 0.143276823 | 0.001670086 | 0.076055159 | 0.002404281 | 0.149932972 | 0.001669905 | 0.076035017 | 4.02E-05 | 0.00053503 | 1 | 0.669181587 | 1 | 0.665121824 | ENSG0000012 | 12 | CNOT1 |
| SAT | Heart_Failure | ENSG000001( | 0.001725534 | 0.485090006 | 0.002471616 | 0.076055417 | 0.001649247 | 0.504558356 | 0.002471392 | 0.076035271 | 7.63E-05 | 0.000742443 | 1 | 0.897349195 | 1 | 0.900571724 | ENSG0000012 | 12 | CNOT1 |
| SAT | Heart_Failure | ENSG000001( | 0.003539522 | 0.147147879 | 0.002441585 | 0.076055165 | 0.003579519 | 0.142590886 | 0.002441336 | 0.076035005 | -4.00E-05 | 0.036168814 | 1 | 0.674290771 | 1 | 0.658003792 | ENSG0000012 | 7 | BBS2 |
| SAT | Heart_Failure | ENSG000001( | -0.00428038 | 0.126286254 | 0.002799617 | 0.076055128 | -0.0042442 | 0.129478331 | 0.002799297 | 0.076034981 | -3.62E-05 | 0.031407238 | 1 | 0.64967372 | 1 | 0.649215182 | ENSG0000012 | 7 | BBS2 |
| SAT | Heart_Failure | ENSG000001( | 0.003428568 | 0.192116498 | 0.002628574 | 0.076055227 | 0.003481491 | 0.185297411 | 0.002628292 | 0.076035066 | -5.29E-05 | 0.004482631 | 1 | 0.722912434 | 1 | 0.715224819 | ENSG0000012 | 9 | ABCC4 |
| SAT | Heart_Failure | ENSG000001( | -7.61E-05 | 0.976817088 | 0.002617239 | 0.076055493 | -3.65E-05 | 0.988885572 | 0.002616954 | 0.07603534 | -3.96E-05 | 0.027053191 | 1 | 0.998920999 | 1 | 0.999768899 | ENSG0000012 | 9 | ABCC4 |
| SAT | Heart_Failure | ENSG000001( | -0.00093307 | 0.663526143 | 0.002144752 | 0.076055463 | -0.00097425 | 0.649614946 | 0.002144518 | 0.076035308 | 4.12E-05 | 0.005145747 | 1 | 0.962994646 | 1 | 0.953700363 | ENSG0000012 | 9 | ABCC4 |
| SAT | Heart_Failure | ENSG000001( | 0.001083406 | 0.710257927 | 0.002916223 | 0.076055471 | 0.00101493 | 0.727790661 | 0.002915913 | 0.076035321 | 6.85E-05 | 0.001123171 | 1 | 0.966658386 | 1 | 0.958028875 | ENSG0000012 | 15 | GRK4 |
| SAT | Heart_Failure | ENSG000001( | -0.00032556 | 0.815963569 | 0.00139881 | 0.076055485 | -0.00030201 | 0.829042755 | 0.001398667 | 0.076035333 | -2.35E-05 | 0.030688715 | 1 | 0.976669362 | 1 | 0.972791643 | ENSG0000012 | 15 | GRK4 |
| SAT | Heart_Failure | ENSG000001( | -0.00124024 | 0.556779861 | 0.002110566 | 0.076055439 | -0.00119478 | 0.571289569 | 0.002110352 | 0.07603529 | -4.55E-05 | 0.006547972 | 1 | 0.922001323 | 1 | 0.924404562 | ENSG0000012 | 15 | GRK4 |
| SAT | Heart_Failure | ENSG000001( | 0.000388561 | 0.890950883 | 0.002834118 | 0.07605549 | 0.000428402 | 0.879837112 | 0.002833798 | 0.076035337 | -3.98E-05 | 0.024539839 | 1 | 0.979270459 | 1 | 0.974665278 | ENSG0000012 | 15 | GRK4 |
| SAT | Heart_Failure | ENSG000001( | 0.000338601 | 0.889578838 | 0.002438837 | 0.07605549 | 0.000284965 | 0.906973562 | 0.002438578 | 0.076035338 | 5.36E-05 | 0.002436846 | 1 | 0.979270459 | 1 | 0.982067422 | ENSG0000012 | 15 | GRK4 |
| SAT | Heart_Failure | ENSG000001( | -0.0006083 | 0.818538691 | 0.002651413 | 0.076055485 | -0.0006519 | 0.805762491 | 0.002651112 | 0.076035331 | 4.36E-05 | 0.007383747 | 1 | 0.976669362 | 1 | 0.972791643 | ENSG0000012 | 8 | TEKT3 |
| SAT | Heart_Failure | ENSG000001( | 0.000132374 | 0.96492559 | 0.003010306 | 0.076055493 | 4.35E-05 | 0.988481822 | 0.003010057 | 0.07603534 | 8.89E-05 | 0.002993081 | 1 | 0.996432082 | 1 | 0.999768899 | ENSG0000012 | 8 | TEKT3 |
| SAT | Heart_Failure | ENSG000001( | 0.000166091 | 0.896202158 | 0.001273115 | 0.07605549 | 0.000129159 | 0.919183394 | 0.001272975 | 0.076035339 | 3.69E-05 | 1.26E-05 | 1 | 0.979542669 | 1 | 0.984094962 | ENSG0000012 | 6 | NUP85 |
| SAT | Heart_Failure | ENSG000001( | -0.00150182 | 0.434918737 | 0.001923426 | 0.076055398 | -0.00139783 | 0.467343634 | 0.001923246 | 0.076035258 | -0.00010398 | 1.04E-09 | 1 | 0.880339889 | 1 | 0.892844363 | ENSG0000012 | 6 | NUP85 |
| SAT | Heart_Failure | ENSG000001( | -0.00296725 | 0.365967515 | 0.003282164 | 0.076055365 | -0.00305868 | 0.35133189 | 0.003281814 | 0.076035205 | 9.14E-05 | 0.000106385 | 1 | 0.85158824 | 1 | 0.843366096 | ENSG0000012 | 6 | NUP85 |
| SAT | Heart_Failure | ENSG000001( | 0.003351185 | 0.396016066 | 0.003948337 | 0.076055381 | 0.003245583 | 0.411020538 | 0.003947919 | 0.076035235 | 0.000105602 | 0.000231577 | 1 | 0.866797005 | 1 | 0.867451005 | ENSG0000012 | 6 | NUP85 |
| SAT | Heart_Failure | ENSG000001( | -0.00101291 | 0.654194293 | 0.002261253 | 0.076055462 | -0.00104575 | 0.643711542 | 0.002261009 | 0.076035307 | 3.28E-05 | 0.038364962 | 1 | 0.962948172 | 1 | 0.953076521 | ENSG0000012 | 6 | NUP85 |
| SAT | Heart_Failure | ENSG000001( | -0.00119847 | 0.71179645 | 0.00324397 | 0.076055472 | -0.00130216 | 0.688092085 | 0.003243665 | 0.076035315 | 0.000103688 | 0.000268415 | 1 | 0.966658386 | 1 | 0.956878334 | ENSG0000012 | 13 | DDX31 |
| SAT | Heart_Failure | ENSG000001( | 0.001874303 | 0.285878671 | 0.001756268 | 0.076055315 | 0.001845037 | 0.293419794 | 0.001756091 | 0.076035168 | 2.93E-05 | 0.035625371 | 1 | 0.800157 | 1 | 0.807642907 | ENSG0000012 | 15 | KIR2DL1 |
| SAT | Heart_Failure | ENSG000001( | 0.009287858 | 0.018526472 | 0.003944002 | 0.076054627 | 0.008376678 | 0.034099769 | 0.003953322 | 0.07603464 | 0.00091118 | 0.001079115 | 1 | 0.296513921 | 1 | 0.413317418 | ENSG0000012 | 11 | OPRL1 |
| SAT | Heart_Failure | ENSG000001( | 0.001074972 | 0.3364147 | 0.001118278 | 0.076055349 | 0.00102776 | 0.358034928 | 0.001118204 | 0.076035208 | 4.72E-05 | 0.000234008 | 1 | 0.841293164 | 1 | 0.846989344 | ENSG0000012 | 11 | OPRL1 |
| SAT | Heart_Failure | ENSG000001( | -0.00089671 | 0.504775498 | 0.001344398 | 0.076055424 | -0.00091607 | 0.49557269 | 0.001344254 | 0.076035268 | 1.94E-05 | 0.043217076 | 1 | 0.9005699 | 1 | 0.894608312 | ENSG0000012 | 5 | PPDPF |
| SAT | Heart_Failure | ENSG000001( | -0.00111841 | 0.758234119 | 0.003633557 | 0.076055478 | -0.00124349 | 0.732158618 | 0.003633232 | 0.076035322 | 0.000125083 | 0.000201883 | 1 | 0.967625883 | 1 | 0.958028875 | ENSG0000012 | 5 | PPDPF |
| SAT | Heart_Failure | ENSG000001( | 0.001374359 | 0.124428845 | 0.000894503 | 0.076055125 | 0.001319839 | 0.1400527 | 0.000894444 | 0.076035 | 5.45E-05 | 1.23E-07 | 1 | 0.647388543 | 1 | 0.654576795 | ENSG0000012 | 5 | PPDPF |
| SAT | Heart_Failure | ENSG000001( | 0.001804026 | 0.363540766 | 0.001985414 | 0.076055364 | 0.001773513 | 0.371660591 | 0.001985199 | 0.076035216 | 3.05E-05 | 0.026301994 | 1 | 0.85040303 | 1 | 0.849768918 | ENSG0000012 | 11 | CHCHD5 |
| SAT | Heart_Failure | ENSG000001( | 0.001165275 | 0.686202917 | 0.002884247 | 0.076055468 | 0.001098782 | 0.703204828 | 0.002883957 | 0.076035317 | 6.65E-05 | 0.004078305 | 1 | 0.966658386 | 1 | 0.956953563 | ENSG0000012 | 11 | CHCHD5 |
| SAT | Heart_Failure | ENSG000001( | 0.001752389 | 0.426881 | 0.002205536 | 0.076055394 | 0.001789049 | 0.417221859 | 0.002205296 | 0.076035237 | -3.67E-05 | 0.014934883 | 1 | 0.878482333 | 1 | 0.867451005 | ENSG0000012 | 11 | CHCHD5 |
| SAT | Heart_Failure | ENSG000001( | -0.00013412 | 0.882973066 | 0.00091112 | 0.07605549 | -0.00017144 | 0.850745129 | 0.000911076 | 0.076035335 | 3.73E-05 | 0.001585424 | 1 | 0.979223495 | 1 | 0.972791643 | ENSG0000012 | 11 | CHCHD5 |
| SAT | Heart_Failure | ENSG000001( | -0.00010137 | 0.94839158 | 0.001566077 | 0.076055492 | -0.00012477 | 0.936492668 | 0.001565903 | 0.076035339 | 2.34E-05 | 0.021560918 | 1 | 0.992559098 | 1 | 0.988983601 | ENSG0000012 | 11 | POLR1B |
| SAT | Heart_Failure | ENSG000001( | 0.000105721 | 0.954415991 | 0.001849486 | 0.076055493 | 0.000177962 | 0.923336305 | 0.001849304 | 0.076035339 | -7.22E-05 | 2.15E-06 | 1 | 0.994131568 | 1 | 0.985235111 | ENSG0000012 | 10 | SLC25A23 |
| SAT | Heart_Failure | ENSG000001( | 0.002105623 | 0.216783008 | 0.00170478 | 0.076055255 | 0.002038334 | 0.231781323 | 0.0017046 | 0.076035117 | 6.73E-05 | 7.42E-08 | 1 | 0.745333464 | 1 | 0.750242469 | ENSG0000012 | 10 | SLC25A23 |
| SAT | Heart_Failure | ENSG000001( | 0.002492426 | 0.093350448 | 0.001485368 | 0.076055054 | 0.002451641 | 0.098797711 | 0.0014852 | 0.076034915 | 4.08E-05 | 1.35E-05 | 1 | 0.604759516 | 1 | 0.610461938 | ENSG0000012 | 4 | CLPP |
| SAT | Heart_Failure | ENSG000001( | 0.00148573 | 0.554609543 | 0.002514489 | 0.076055439 | 0.001423241 | 0.571347248 | 0.002514254 | 0.07603529 | 6.25E-05 | 0.004861519 | 1 | 0.921542737 | 1 | 0.924404562 | ENSG0000012 | 4 | CLPP |
| SAT | Heart_Failure | ENSG000001( | 6.99E-05 | 0.954109142 | 0.001214814 | 0.076055493 | 9.10E-05 | 0.940263976 | 0.001214685 | 0.076035339 | -2.11E-05 | 0.016915938 | 1 | 0.994131568 | 1 | 0.988983601 | ENSG0000012 | 9 | GPR108 |
| SAT | Heart_Failure | ENSG000001( | -0.00217667 | 0.004033832 | 0.000756965 | 0.076054203 | -0.00218939 | 0.003820581 | 0.000756887 | 0.076034035 | 1.27E-05 | 0.02879651 | 1 | 0.1521008 | 1 | 0.159899049 | ENSG0000012 | 9 | GPR108 |
| SAT | Heart_Failure | ENSG000001( | -0.00314576 | 0.068793061 | 0.001728648 | 0.076054976 | -0.00317951 | 0.065844215 | 0.001728476 | 0.076034812 | 3.37E-05 | 0.01641193 | 1 | 0.537536555 | 1 | 0.515872519 | ENSG0000012 | 6 | TNFSF14 |
| SAT | Heart_Failure | ENSG000001( | 0.003892794 | 0.058704224 | 0.002059237 | 0.076054935 | 0.00392358 | 0.056708618 | 0.002059022 | 0.076034774 | -3.08E-05 | 0.047734751 | 1 | 0.506864626 | 1 | 0.501512062 | ENSG0000012 | 13 | PSMF1 |
| SAT | Heart_Failure | ENSG000001( | -0.0019637 | 0.476109325 | 0.002755786 | 0.076055414 | -0.00205749 | 0.455254681 | 0.002755502 | 0.076035253 | 9.38E-05 | 9.60E-06 | 1 | 0.895715287 | 1 | 0.886989673 | ENSG0000012 | 13 | PSMF1 |
| SAT | Heart_Failure | ENSG000001( | 0.000891568 | 0.531305864 | 0.001424198 | 0.076055432 | 0.000972228 | 0.49489939 | 0.001424432 | 0.076035267 | -8.07E-05 | 0.020047615 | 1 | 0.908320477 | 1 | 0.894608312 | ENSG0000012 | 7 | DTD1 |
| SAT | Heart_Failure | ENSG000001( | 0.001969853 | 0.362817194 | 0.002164648 | 0.076055364 | 0.001942609 | 0.369439223 | 0.002164406 | 0.076035214 | 2.72E-05 | 0.049799178 | 1 | 0.85040303 | 1 | 0.849425432 | ENSG0000012 | 7 | DTD1 |
| SAT | Heart_Failure | ENSG000001( | -0.00083254 | 0.645890186 | 0.00181192 | 0.07605546 | -0.00080463 | 0.656955073 | 0.001811731 | 0.076035309 | -2.79E-05 | 0.040042641 | 1 | 0.960776086 | 1 | 0.956286421 | ENSG0000012 | 7 | DTD1 |
| SAT | Heart_Failure | ENSG000001( | 0.004383119 | 0.047064154 | 0.002207311 | 0.076054878 | 0.004434764 | 0.044503219 | 0.002207091 | 0.07603471 | -5.16E-05 | 0.003894669 | 1 | 0.476597187 | 1 | 0.456297071 | ENSG0000012 | 7 | DTD1 |
| SAT | Heart_Failure | ENSG000001( | 0.002506325 | 0.372465812 | 0.002810207 | 0.076055369 | 0.002607415 | 0.353443248 | 0.00280992 | 0.076035206 | -0.00010109 | 4.29E-06 | 1 | 0.857586195 | 1 | 0.843366096 | ENSG0000012 | 15 | RBCK1 |
| SAT | Heart_Failure | ENSG000001( | 0.004534335 | 0.222350561 | 0.003715745 | 0.076055261 | 0.004445656 | 0.231481324 | 0.003715385 | 0.076035117 | 8.87E-05 | 0.004726086 | 1 | 0.751090303 | 1 | 0.750242469 | ENSG0000012 | 11 | RRBP1 |
| SAT | Heart_Failure | ENSG000001( | 0.003827495 | 0.252335462 | 0.003343676 | 0.076055289 | 0.003767736 | 0.259766708 | 0.003343321 | 0.076035142 | 5.98E-05 | 0.013624691 | 1 | 0.767444733 | 1 | 0.773899606 | ENSG0000012 | 13 | MKKS |
| SAT | Heart_Failure | ENSG000001( | -0.00064862 | 0.858921785 | 0.003649113 | 0.076055488 | -0.0005889 | 0.871780005 | 0.003648718 | 0.076035336 | -5.97E-05 | 0.018457962 | 1 | 0.976669362 | 1 | 0.974665278 | ENSG0000012 | 13 | MKKS |
| SAT | Heart_Failure | ENSG000001( | 0.001034 | 0.513960367 | 0.001584228 | 0.076055427 | 0.001079511 | 0.49556656 | 0.001584062 | 0.076035268 | -4.55E-05 | 0.000115604 | 1 | 0.905155107 | 1 | 0.894608312 | ENSG0000012 | 9 | MCM8 |
| SAT | Heart_Failure | ENSG000001( | -5.88E-05 | 0.980804696 | 0.002443325 | 0.076055493 | 7.08E-07 | 0.999768899 | 0.002443094 | 0.07603534 | -5.95E-05 | 0.0050412 | 1 | 0.998920999 | 1 | 0.999768899 | ENSG0000012 | 5 | MRPS26 |
| SAT | Heart_Failure | ENSG000001( | 0.000129045 | 0.923499766 | 0.001343849 | 0.076055492 | 0.000154495 | 0.908464125 | 0.001343712 | 0.076035338 | -2.55E-05 | 0.015742469 | 1 | 0.988739396 | 1 | 0.982067422 | ENSG0000012 | 5 | MRPS26 |
| SAT | Heart_Failure | ENSG000001( | -0.00473326 | 0.016833564 | 0.001980171 | 0.076054601 | -0.00469137 | 0.01781714 | 0.00197998 | 0.076034464 | -4.19E-05 | 0.01253838 | 1 | 0.287798638 | 1 | 0.299784809 | ENSG0000012 | 6 | NCLN |
| SAT | Heart_Failure | ENSG000001( | -0.00303027 | 0.179332255 | 0.002256654 | 0.076055212 | -0.00298567 | 0.185773693 | 0.002256424 | 0.076035067 | -4.46E-05 | 0.011800675 | 1 | 0.714462461 | 1 | 0.715224819 | ENSG0000012 | 4 | GDF5 |
| SAT | Heart_Failure | ENSG000001( | 0.000210961 | 0.935372952 | 0.002601672 | 0.076055492 | 0.000265838 | 0.918605442 | 0.002601394 | 0.076035339 | -5.49E-05 | 0.003330486 | 1 | 0.989966131 | 1 | 0.984094962 | ENSG0000012 | 4 | GDF5 |
| SAT | Heart_Failure | ENSG000001( | -0.00474965 | 0.210527136 | 0.003793287 | 0.076055248 | -0.00465011 | 0.220197887 | 0.0037929 | 0.076035106 | -9.95E-05 | 0.000777573 | 1 | 0.739879495 | 1 | 0.742028061 | ENSG0000012 | 8 | MMP24 |
| SAT | Heart_Failure | ENSG000001( | 0.000907537 | 0.779883314 | 0.003247334 | 0.076055481 | 0.000818832 | 0.800900341 | 0.003246988 | 0.07603533 | 8.87E-05 | 0.000140556 | 1 | 0.972664863 | 1 | 0.972791643 | ENSG0000012 | 8 | MMP24 |
| SAT | Heart_Failure | ENSG000001( | -0.00022373 | 0.924326631 | 0.002355377 | 0.076055492 | -0.00018867 | 0.936148573 | 0.002355123 | 0.076035339 | -3.51E-05 | 0.034698518 | 1 | 0.988739396 | 1 | 0.988983601 | ENSG0000012 | 8 | MMP24 |
| SAT | Heart_Failure | ENSG000001( | 0.005116452 | 0.06762312 | 0.002799678 | 0.076054972 | 0.005074424 | 0.069878912 | 0.002799375 | 0.076034827 | 4.20E-05 | 0.030496779 | 1 | 0.535424809 | 1 | 0.529994954 | ENSG0000012 | 14 | ERGIC3 |
| SAT | Heart_Failure | ENSG000001( | 0.003365882 | 0.222680368 | 0.002760202 | 0.076055261 | 0.003418739 | 0.215454982 | 0.002759928 | 0.076035101 | -5.29E-05 | 0.018795185 | 1 | 0.751090303 | 1 | 0.736362287 | ENSG0000012 | 15 | ST3GAL3 |
| SAT | Heart_Failure | ENSG000001( | -0.00093025 | 0.787960414 | 0.003458691 | 0.076055482 | -0.00084327 | 0.80735891 | 0.003458365 | 0.076035331 | -8.70E-05 | 0.004143498 | 1 | 0.974113835 | 1 | 0.972791643 | ENSG0000012 | 15 | ST3GAL3 |
| SAT | Heart_Failure | ENSG000001( | -0.0003751 | 0.878490714 | 0.002453524 | 0.076055489 | -0.00041524 | 0.865591092 | 0.002453264 | 0.076035336 | 4.01E-05 | 0.02457632 | 1 | 0.979223495 | 1 | 0.974123167 | ENSG0000012 | 9 | XRCC3 |
| SAT | Heart_Failure | ENSG000001( | -0.00273072 | 0.32152572 | 0.002754604 | 0.07605534 | -0.00266458 | 0.333335326 | 0.002754318 | 0.076035194 | -6.61E-05 | 0.001487356 | 1 | 0.828692762 | 1 | 0.836656731 | ENSG0000012 | 17 | PCID2 |
| SAT | Heart_Failure | ENSG000001( | -0.00835782 | 0.008727173 | 0.003186881 | 0.07605442 | -0.00843148 | 0.00814622 | 0.00318655 | 0.076034248 | 7.37E-05 | 0.00224533 | 1 | 0.210020691 | 1 | 0.209225934 | ENSG0000012 | 17 | PCID2 |
| SAT | Heart_Failure | ENSG000001( | 0.002511517 | 0.322614189 | 0.002539191 | 0.07605534 | 0.002567829 | 0.31183051 | 0.002538912 | 0.076035181 | -5.63E-05 | 0.000909982 | 1 | 0.829398936 | 1 | 0.821155716 | ENSG0000012 | 5 | IGFLR1 |
| SAT | Heart_Failure | ENSG000001( | 0.00018575 | 0.909048133 | 0.001625975 | 0.076055491 | 0.000155806 | 0.923653427 | 0.00162581 | 0.076035339 | 2.99E-05 | 0.019566576 | 1 | 0.983702811 | 1 | 0.985235111 | ENSG0000012 | 6 | CAPNS1 |
| SAT | Heart_Failure | ENSG000001( | 0.001561353 | 0.335654385 | 0.0016217 | 0.076055348 | 0.001539035 | 0.342554456 | 0.00162152 | 0.0760352 | 2.23E-05 | 0.036077389 | 1 | 0.841293164 | 1 | 0.839311387 | ENSG0000012 | 6 | CAPNS1 |
| SAT | Heart_Failure | ENSG000001( | -0.00061258 | 0.811290521 | 0.002565644 | 0.076055484 | -0.00019318 | 0.940123762 | 0.00257184 | 0.076035339 | -0.0004194 | 0.022095151 | 1 | 0.976669362 | 1 | 0.988983601 | ENSG0000012 | 6 | CAPNS1 |
| SAT | Heart_Failure | ENSG000001( | 0.000798817 | 0.841451324 | 0.003993338 | 0.076055487 | 0.000901937 | 0.821292597 | 0.003992943 | 0.076035332 | -0.00010312 | 0.001641194 | 1 | 0.976669362 | 1 | 0.972791643 | ENSG0000012 | 5 | GPR42 |
| SAT | Heart_Failure | ENSG000001( | -0.00023944 | 0.856226221 | 0.001321554 | 0.076055488 | -0.00026214 | 0.842746253 | 0.001321411 | 0.076035334 | 2.27E-05 | 0.014712199 | 1 | 0.976669362 | 1 | 0.972791643 | ENSG0000012 | 5 | GPR42 |
| SAT | Heart_Failure | ENSG000001( | 0.001249516 | 0.590797115 | 0.002323895 | 0.076055448 | 0.000457499 | 0.845938795 | 0.002354568 | 0.076035334 | 0.000792016 | 0.037487328 | 1 | 0.941376309 | 1 | 0.972791643 | ENSG0000012 | 5 | GPR42 |
| SAT | Heart_Failure | ENSG000001( | 0.00229679 | 0.354760486 | 0.002481954 | 0.076055359 | 0.002421758 | 0.329146541 | 0.002481728 | 0.076035192 | -0.00012497 | 3.29E-08 | 1 | 0.848363544 | 1 | 0.833382337 | ENSG0000012 | 16 | PRMT1 |
| SAT | Heart_Failure | ENSG000001( | -0.00105954 | 0.681236812 | 0.002579359 | 0.076055467 | -0.0009977 | 0.698876123 | 0.002579107 | 0.076035317 | -6.18E-05 | 0.00393868 | 1 | 0.966658386 | 1 | 0.956878334 | ENSG0000012 | 9 | PRR12 |
| SAT | Heart_Failure | ENSG000001( | -5.60E-05 | 0.969269069 | 0.00145379 | 0.076055493 | -9.03E-05 | 0.95044991 | 0.001453635 | 0.07603534 | 3.43E-05 | 0.000943044 | 1 | 0.996609664 | 1 | 0.990755659 | ENSG0000012 | 3 | FLRT1 |
| SAT | Heart_Failure | ENSG000001( | -0.00149614 | 0.0732649 | 0.000835284 | 0.076054992 | -0.00145776 | 0.080916706 | 0.000835205 | 0.076034865 | -3.84E-05 | 1.32E-07 | 1 | 0.559028222 | 1 | 0.580120808 | ENSG0000012 | 3 | FLRT1 |
| SAT | Heart_Failure | ENSG000001( | -0.00476414 | 0.214587204 | 0.003838793 | 0.076055253 | -0.00483293 | 0.207993948 | 0.003838394 | 0.076035093 | 6.88E-05 | 0.01757662 | 1 | 0.745086527 | 1 | 0.733958243 | ENSG0000012 | 12 | WNK4 |
| SAT | Heart_Failure | ENSG000001( | -0.00439267 | 0.148469107 | 0.003039989 | 0.076055167 | -0.00434087 | 0.153268619 | 0.00303965 | 0.076035022 | -5.18E-05 | 0.008257833 | 1 | 0.674894985 | 1 | 0.667541522 | ENSG0000012 | 6 | TRAP1 |
| SAT | Heart_Failure | ENSG000001( | -0.00178436 | 0.248845115 | 0.001547365 | 0.076055285 | -0.00181838 | 0.239887525 | 0.001547204 | 0.076035125 | 3.40E-05 | 0.003367988 | 1 | 0.765246076 | 1 | 0.752162449 | ENSG0000012 | 6 | TRAP1 |
| SAT | Heart_Failure | ENSG000001( | -0.00507541 | 0.055530924 | 0.002650758 | 0.076054921 | -0.00498638 | 0.059932977 | 0.00265051 | 0.076034788 | -8.90E-05 | 0.000138316 | 1 | 0.503306524 | 1 | 0.504627254 | ENSG0000012 | 6 | DNAJC8 |
| SAT | Heart_Failure | ENSG000001( | 0.000784352 | 0.720456384 | 0.002191851 | 0.076055473 | 0.000504776 | 0.817985863 | 0.002193376 | 0.076035332 | 0.000279576 | 0.00172296 | 1 | 0.966658386 | 1 | 0.972791643 | ENSG0000012 | 6 | DNAJC8 |
| SAT | Heart_Failure | ENSG000001( | 0.000213613 | 0.95343757 | 0.003658343 | 0.076055492 | 0.000129726 | 0.971709479 | 0.003657933 | 0.07603534 | 8.39E-05 | 0.000330917 | 1 | 0.994131568 | 1 | 0.998061182 | ENSG0000012 | 13 | ZNF384 |
| SAT | Heart_Failure | ENSG000001( | 0.007134404 | 0.015696989 | 0.002953111 | 0.076054582 | 0.007208057 | 0.014643221 | 0.002952799 | 0.07603441 | -7.37E-05 | 0.000655978 | 1 | 0.276891502 | 1 | 0.272970533 | ENSG0000012 | 7 | L3HYPDH |
| SAT | Heart_Failure | ENSG000001( | -0.00147509 | 0.219903846 | 0.001202404 | 0.076055258 | -0.00149945 | 0.212335984 | 0.001202285 | 0.076035097 | 2.44E-05 | 0.013220137 | 1 | 0.749908476 | 1 | 0.733958243 | ENSG0000012 | 7 | L3HYPDH |
| SAT | Heart_Failure | ENSG000001( | 0.000691694 | 0.719859374 | 0.00192862 | 0.076055473 | 0.000740575 | 0.700955226 | 0.001928421 | 0.076035317 | -4.89E-05 | 0.000970307 | 1 | 0.966658386 | 1 | 0.956878334 | ENSG0000012 | 7 | L3HYPDH |
| SAT | Heart_Failure | ENSG000001( | 0.00229048 | 0.468148731 | 0.003157127 | 0.076055411 | 0.002027322 | 0.520786954 | 0.003157167 | 0.076035276 | 0.000263157 | 1.01E-06 | 1 | 0.893397133 | 1 | 0.904369708 | ENSG0000012 | 12 | RHOT1 |
| SAT | Heart_Failure | ENSG0000011 | 0.000996398 | 0.699888602 | 0.002584884 | 0.07605547 | 0.000947588 | 0.713901862 | 0.002584654 | 0.076035319 | 4.88E-05 | 0.042560714 | 1 | 0.966658386 | 1 | 0.958028875 | ENSG0000012 | 11 | WDR60 |
| SAT | Heart_Failure | ENSG0000011 | -4.07E-05 | 0.977902662 | 0.001470843 | 0.076055493 | -6.55E-05 | 0.964483159 | 0.001470698 | 0.07603534 | 2.47E-05 | 0.041592746 | 1 | 0.998920999 | 1 | 0.996055789 | ENSG0000012 | 11 | WDR60 |
| SAT | Heart_Failure | ENSG0000011 | 0.000219216 | 0.844714689 | 0.00111921 | 0.076055487 | 0.000246849 | 0.825418767 | 0.001119087 | 0.076035333 | -2.76E-05 | 0.000253659 | 1 | 0.976669362 | 1 | 0.972791643 | ENSG0000012 | 11 | WDR60 |
| SAT | Heart_Failure | ENSG0000011 | 0.005890389 | 0.120696539 | 0.003795698 | 0.076055117 | 0.005826805 | 0.124717113 | 0.003795286 | 0.076034972 | 6.36E-05 | 0.01545436 | 1 | 0.646585619 | 1 | 0.647874406 | ENSG0000012 | 5 | ABHD8 |
| SAT | Heart_Failure | ENSG0000011 | -0.00021486 | 0.927719494 | 0.002368507 | 0.076055492 | -0.0004194 | 0.859491786 | 0.002369183 | 0.076035335 | 0.000204538 | 0.002814382 | 1 | 0.988739396 | 1 | 0.972791643 | ENSG0000012 | 5 | ABHD8 |
| SAT | Heart_Failure | ENSG0000011 | 0.001080612 | 0.7764573 | 0.003805789 | 0.07605548 | 0.001018427 | 0.788985747 | 0.00380538 | 0.076035329 | 6.22E-05 | 0.021432309 | 1 | 0.972664863 | 1 | 0.970635778 | ENSG0000012 | 2 | YEATS4 |
| SAT | Heart_Failure | ENSG0000011 | -0.00146844 | 0.450847407 | 0.001947525 | 0.076055404 | -0.00151144 | 0.437652643 | 0.001947321 | 0.076035246 | 4.30E-05 | 0.003127505 | 1 | 0.887019168 | 1 | 0.879864169 | ENSG0000012 | 2 | YEATS4 |
| SAT | Heart_Failure | ENSG0000011 | 0.002376216 | 0.374897606 | 0.002677917 | 0.07605537 | 0.002421126 | 0.365888725 | 0.002677639 | 0.076035213 | -4.49E-05 | 0.026607922 | 1 | 0.85759885 | 1 | 0.849290727 | ENSG0000012 | 2 | YEATS4 |
| SAT | Heart_Failure | ENSG0000011 | -0.0006283 | 0.723308856 | 0.001774646 | 0.076055473 | -0.00059787 | 0.736167209 | 0.001774454 | 0.076035322 | -3.04E-05 | 0.013994629 | 1 | 0.966658386 | 1 | 0.960001834 | ENSG0000012 | 2 | YEATS4 |
| SAT | Heart_Failure | ENSG0000011 | -0.00038991 | 0.662389156 | 0.000893014 | 0.076055463 | -0.00027389 | 0.759397845 | 0.00089427 | 0.076035326 | -0.00011602 | 0.019206567 | 1 | 0.962994646 | 1 | 0.963228667 | ENSG0000012 | 2 | YEATS4 |

| | | | | | | | | | | | | | | | | | | | |
|---|---|---|---|---|---|---|---|---|---|---|---|---|---|---|---|---|---|---|---|
| SAT | Heart_Failure | ENSG0000011 | 0.001787476 | 0.320582507 | 0.001799601 | 0.076055339 | 0.001912447 | 0.287894768 | 0.001799523 | 0.076035164 | -0.00012497 | 2.03E-07 | 1 | 0.828692762 | 1 | 0.799139368 | ENSG0000012 | 2 | YEATS4 |
| SAT | Heart_Failure | ENSG0000011 | -0.00143511 | 0.540656792 | 0.002345634 | 0.076055435 | -0.00149352 | 0.524264305 | 0.002345395 | 0.076035277 | 5.84E-05 | 0.001388127 | 1 | 0.913035829 | 1 | 0.904369708 | ENSG0000012 | 2 | YEATS4 |
| SAT | Heart_Failure | ENSG0000011 | -0.00255829 | 0.4297361 | 0.003239804 | 0.076055396 | -0.00247304 | 0.445222035 | 0.003239483 | 0.076035249 | -8.53E-05 | 0.001332178 | 1 | 0.878482333 | 1 | 0.882462767 | ENSG0000012 | 2 | YEATS4 |
| SAT | Heart_Failure | ENSG0000011 | -0.00079362 | 0.7073418 | 0.002113895 | 0.076055471 | -0.00085468 | 0.685952835 | 0.002113693 | 0.076035315 | 6.11E-05 | 0.000777755 | 1 | 0.966658386 | 1 | 0.956878334 | ENSG0000012 | 10 | FGFRL1 |
| SAT | Heart_Failure | ENSG0000011 | 0.008798877 | 0.014917226 | 0.003614406 | 0.076054568 | 0.009008611 | 0.012682119 | 0.003614168 | 0.076034371 | -0.00020973 | 5.30E-07 | 1 | 0.270426545 | 1 | 0.253719716 | ENSG0000012 | 10 | FGFRL1 |
| SAT | Heart_Failure | ENSG0000011 | 0.000668441 | 0.8373327 | 0.003255818 | 0.076055486 | 0.000619732 | 0.849021013 | 0.003255446 | 0.076035334 | 4.87E-05 | 0.013668614 | 1 | 0.976669362 | 1 | 0.972791643 | ENSG0000012 | 10 | FGFRL1 |
| SAT | Heart_Failure | ENSG0000011 | 0.001327405 | 0.732704508 | 0.003886637 | 0.076055475 | 0.001707756 | 0.660547021 | 0.003888702 | 0.07603531 | -0.00038035 | 0.00723645 | 1 | 0.966658386 | 1 | 0.956878334 | ENSG0000012 | 12 | TMEM175 |
| SAT | Heart_Failure | ENSG0000011 | 0.003995445 | 0.016992797 | 0.001673925 | 0.076054604 | 0.003974558 | 0.017565109 | 0.001673732 | 0.07603446 | 2.09E-05 | 0.03142404 | 1 | 0.288877546 | 1 | 0.297067641 | ENSG0000012 | 9 | EMC1 |
| SAT | Heart_Failure | ENSG0000011 | -0.00507736 | 0.114462046 | 0.003216665 | 0.076055104 | -0.00501395 | 0.119017983 | 0.003216315 | 0.076034961 | -6.34E-05 | 0.004102832 | 1 | 0.641850091 | 1 | 0.64481576 | ENSG0000012 | 10 | UBR4 |
| SAT | Heart_Failure | ENSG0000011 | -0.00147104 | 0.389365358 | 0.001708988 | 0.076055377 | -0.00141535 | 0.407520461 | 0.001708809 | 0.076035233 | -5.57E-05 | 1.32E-05 | 1 | 0.861555536 | 1 | 0.867451005 | ENSG0000012 | 13 | EMR2 |
| SAT | Heart_Failure | ENSG0000011 | 0.006141858 | 0.062441642 | 0.00329649 | 0.076054951 | 0.006069202 | 0.065575744 | 0.00329612 | 0.076034811 | 7.27E-05 | 0.000540444 | 1 | 0.515554877 | 1 | 0.515872519 | ENSG0000012 | 13 | SMARCA4 |
| SAT | Heart_Failure | ENSG0000011 | 0.00179013 | 0.44588518 | 0.002348348 | 0.076055402 | 0.002918865 | 0.225043309 | 0.002405867 | 0.07603511 | -0.00112873 | 0.031343446 | 1 | 0.88490557 | 1 | 0.747837725 | ENSG0000012 | 13 | SMARCA4 |
| SAT | Heart_Failure | ENSG0000011 | -0.00035193 | 0.802798561 | 0.001409278 | 0.076055483 | -0.00032628 | 0.816890023 | 0.001409118 | 0.076035332 | -2.57E-05 | 0.00360793 | 1 | 0.976669362 | 1 | 0.972791643 | ENSG0000012 | 13 | SMARCA4 |
| SAT | Heart_Failure | ENSG0000011 | -0.00327728 | 0.269614749 | 0.002968683 | 0.076055303 | -0.0033426 | 0.260133425 | 0.002968362 | 0.076035142 | 6.53E-05 | 0.001540121 | 1 | 0.782624692 | 1 | 0.773899606 | ENSG0000012 | 13 | SMARCA4 |
| SAT | Heart_Failure | ENSG0000011 | 0.001133508 | 0.665121384 | 0.002618688 | 0.076055464 | 0.001062331 | 0.684951478 | 0.002618409 | 0.076035314 | 7.12E-05 | 0.000159963 | 1 | 0.962994646 | 1 | 0.956878334 | ENSG0000012 | 12 | ZNF835 |
| SAT | Heart_Failure | ENSG0000011 | -0.00044361 | 0.764451343 | 0.001480489 | 0.076055479 | -0.0032742 | 0.100375617 | 0.00199277 | 0.076034919 | 0.002830583 | 0.033864254 | 1 | 0.970642331 | 1 | 0.614666997 | ENSG0000012 | 12 | ZNF835 |
| SAT | Heart_Failure | ENSG0000011 | -0.00991988 | 0.009832191 | 0.003842402 | 0.076054453 | -0.00998808 | 0.009330764 | 0.003842012 | 0.076034286 | 6.82E-05 | 0.024163472 | 1 | 0.227178996 | 1 | 0.221316258 | ENSG0000012 | 12 | ZNF835 |
| SAT | Heart_Failure | ENSG0000011 | 0.000635244 | 0.606501129 | 0.001233306 | 0.076055452 | 0.0006644 | 0.590046824 | 0.001233183 | 0.076035295 | -2.92E-05 | 0.003424126 | 1 | 0.948489135 | 1 | 0.934989637 | ENSG0000012 | 12 | ZNF835 |
| SAT | Heart_Failure | ENSG0000011 | -0.00169242 | 0.627833187 | 0.003491113 | 0.076055456 | -0.00162296 | 0.641980711 | 0.003490756 | 0.076035306 | -6.95E-05 | 0.010709222 | 1 | 0.958246837 | 1 | 0.951372499 | ENSG0000012 | 12 | STYXL1 |
| SAT | Heart_Failure | ENSG0000011 | 0.002756468 | 0.011167134 | 0.001086321 | 0.076054488 | 0.00273229 | 0.011888625 | 0.001086206 | 0.076034353 | 2.42E-05 | 0.00248451 | 1 | 0.236383009 | 1 | 0.246278972 | ENSG0000012 | 12 | STYXL1 |
| SAT | Heart_Failure | ENSG0000011 | -0.00230651 | 0.526164158 | 0.003638766 | 0.07605543 | -0.00224218 | 0.537721216 | 0.003638356 | 0.076035281 | -6.43E-05 | 0.005165045 | 1 | 0.906724424 | 1 | 0.909319572 | ENSG0000012 | 12 | PMS2P3 |
| SAT | Heart_Failure | ENSG0000011 | 0.000895738 | 0.585775372 | 0.00164365 | 0.076055447 | 0.00092101 | 0.575205693 | 0.001643482 | 0.076035291 | -2.53E-05 | 0.047868528 | 1 | 0.939818579 | 1 | 0.927160988 | ENSG0000012 | 6 | SRD5A3 |
| SAT | Heart_Failure | ENSG0000011 | -0.0071372 | 0.032865066 | 0.003344936 | 0.076054782 | -0.00706434 | 0.034674318 | 0.003344624 | 0.076034644 | -7.29E-05 | 0.013720054 | 1 | 0.408448042 | 1 | 0.418258961 | ENSG0000012 | 6 | SRD5A3 |
| SAT | Heart_Failure | ENSG0000011 | 0.003802962 | 0.184702265 | 0.002867094 | 0.076055218 | 0.003720911 | 0.19431157 | 0.002866801 | 0.076035077 | 8.21E-05 | 0.000235101 | 1 | 0.720580417 | 1 | 0.723506187 | ENSG0000012 | 5 | DGCR6L |
| SAT | Heart_Failure | ENSG0000011 | 0.001325611 | 0.408193663 | 0.001602765 | 0.076055386 | 0.001350044 | 0.399558257 | 0.001602596 | 0.076035229 | -2.44E-05 | 0.036786529 | 1 | 0.870429337 | 1 | 0.865885497 | ENSG0000012 | 5 | DGCR6L |
| SAT | Heart_Failure | ENSG0000011 | 0.005730545 | 0.104660968 | 0.003531557 | 0.076055082 | 0.005786551 | 0.10127729 | 0.003531193 | 0.076034921 | -5.60E-05 | 0.039457949 | 1 | 0.632398959 | 1 | 0.614882715 | ENSG0000012 | 5 | DGCR6L |
| SAT | Heart_Failure | ENSG0000011 | -0.00033537 | 0.875421539 | 0.002139194 | 0.076055489 | -0.00028302 | 0.894733086 | 0.002138964 | 0.076035337 | -5.24E-05 | 0.000566838 | 1 | 0.979223495 | 1 | 0.97886604 | ENSG0000012 | 7 | VPREB3 |
| SAT | Heart_Failure | ENSG0000011 | 0.000645507 | 0.840239813 | 0.003202129 | 0.076055487 | 0.000897493 | 0.779391054 | 0.00320404 | 0.076035328 | -0.00025199 | 0.039375141 | 1 | 0.976669362 | 1 | 0.966957854 | ENSG0000012 | 7 | VPREB3 |
| SAT | Heart_Failure | ENSG0000011 | 0.00208492 | 0.062219493 | 0.00111808 | 0.07605495 | 0.002559245 | 0.024083042 | 0.001134513 | 0.076034546 | -0.00047432 | 0.014118617 | 1 | 0.515510499 | 1 | 0.342062594 | ENSG0000012 | 4 | POM121L9P |
| SAT | Heart_Failure | ENSG0000011 | 0.001396578 | 0.379982472 | 0.001590764 | 0.076055373 | 0.001354654 | 0.394421446 | 0.001590665 | 0.076035227 | 4.19E-05 | 0.026487371 | 1 | 0.85759885 | 1 | 0.862334842 | ENSG0000012 | 4 | POM121L9P |
| SAT | Heart_Failure | ENSG0000011 | -0.00223754 | 0.147751175 | 0.001545769 | 0.076055166 | -0.0022091 | 0.152925734 | 0.001545605 | 0.076035021 | -2.84E-05 | 0.011611826 | 1 | 0.674894985 | 1 | 0.667541522 | ENSG0000012 | 4 | POM121L9P |
| SAT | Heart_Failure | ENSG0000011 | -0.00121816 | 0.669533185 | 0.002854253 | 0.076055465 | -0.00116632 | 0.682784129 | 0.002853968 | 0.076035314 | -5.18E-05 | 0.024683852 | 1 | 0.96567764 | 1 | 0.956878334 | ENSG0000012 | 15 | ADORA2A |
| SAT | Heart_Failure | ENSG0000011 | 0.000923791 | 0.791314545 | 0.00349122 | 0.076055482 | 0.000987604 | 0.777245621 | 0.003490872 | 0.076035328 | -6.38E-05 | 0.023966644 | 1 | 0.9760395 | 1 | 0.965963214 | ENSG0000012 | 11 | A4GALT |
| SAT | Heart_Failure | ENSG0000011 | -0.00140883 | 0.658679925 | 0.003189297 | 0.076055463 | -0.00135424 | 0.671079441 | 0.00318894 | 0.076035312 | -5.46E-05 | 0.007592877 | 1 | 0.962994646 | 1 | 0.956878334 | ENSG0000012 | 7 | RIBC2 |
| SAT | Heart_Failure | ENSG0000011 | 0.001739741 | 0.616332454 | 0.003472155 | 0.076055454 | 0.001792315 | 0.605679909 | 0.003471796 | 0.076035299 | -5.26E-05 | 0.047645876 | 1 | 0.954216727 | 1 | 0.942350873 | ENSG0000012 | 7 | RIBC2 |
| SAT | Heart_Failure | ENSG0000011 | 0.000645771 | 0.570858587 | 0.001139352 | 0.076055443 | 0.000681248 | 0.549851192 | 0.001139243 | 0.076035284 | -3.55E-05 | 0.000261762 | 1 | 0.930445615 | 1 | 0.914465015 | ENSG0000012 | 8 | EMC4 |
| SAT | Heart_Failure | ENSG0000011 | -0.00900808 | 0.010737512 | 0.003531006 | 0.076054477 | -0.00906673 | 0.010228369 | 0.003530628 | 0.076034311 | 5.87E-05 | 0.02034722 | 1 | 0.232334342 | 1 | 0.230669232 | ENSG0000012 | 8 | EMC4 |
| SAT | Heart_Failure | ENSG0000011 | -0.00070413 | 0.716542818 | 0.001939314 | 0.076055472 | -0.00066447 | 0.731849992 | 0.001939115 | 0.076035322 | -3.97E-05 | 0.00782894 | 1 | 0.966658386 | 1 | 0.958028875 | ENSG0000012 | 9 | PRKRIP1 |
| SAT | Heart_Failure | ENSG0000011 | -0.00312437 | 0.155887394 | 0.002201749 | 0.076055179 | -0.00316312 | 0.150777359 | 0.002201513 | 0.076035018 | 3.87E-05 | 0.013855751 | 1 | 0.677064618 | 1 | 0.665121824 | ENSG0000012 | 11 | FLNC |
| SAT | Heart_Failure | ENSG0000011 | -0.00653563 | 4.04E-05 | 0.001592128 | 0.076052863 | -0.00650856 | 4.34E-05 | 0.001591965 | 0.076032732 | -2.71E-05 | 0.027729187 | 0.132696315 | 0.005529013 | 0.142549737 | 0.009503316 | ENSG0000012 | 14 | IRF5 |
| SAT | Heart_Failure | ENSG0000011 | -0.00234142 | 0.183932447 | 0.001762132 | 0.076055217 | -0.00237771 | 0.177184945 | 0.00176195 | 0.076035056 | 3.63E-05 | 0.007214842 | 1 | 0.720580417 | 1 | 0.704659157 | ENSG0000012 | 14 | IRF5 |
| SAT | Heart_Failure | ENSG0000011 | -0.00043054 | 0.505856252 | 0.000647132 | 0.076055424 | -0.00041809 | 0.518192822 | 0.000647067 | 0.076035275 | -1.24E-05 | 0.015647082 | 1 | 0.9005699 | 1 | 0.903395669 | ENSG0000012 | 14 | IRF5 |
| SAT | Heart_Failure | ENSG0000011 | 0.003414699 | 0.308011945 | 0.003349703 | 0.076055331 | 0.003355581 | 0.316410055 | 0.003349339 | 0.076035184 | 5.91E-05 | 0.010680513 | 1 | 0.816225947 | 1 | 0.823487262 | ENSG0000012 | 14 | IRF5 |
| SAT | Heart_Failure | ENSG0000011 | 0.001170411 | 0.467010211 | 0.001609138 | 0.07605541 | 0.00113816 | 0.479326382 | 0.001608963 | 0.076035262 | 3.23E-05 | 0.003913948 | 1 | 0.892925701 | 1 | 0.894608312 | ENSG0000012 | 14 | IRF5 |
| SAT | Heart_Failure | ENSG0000011 | -0.00397496 | 0.159504169 | 0.002825648 | 0.076055184 | -0.00390346 | 0.167101768 | 0.002825362 | 0.076035042 | -7.15E-05 | 0.001380042 | 1 | 0.678956178 | 1 | 0.684786616 | ENSG0000012 | 14 | IRF5 |
| SAT | Heart_Failure | ENSG0000011 | 0.002019603 | 0.28554793 | 0.00189112 | 0.076055315 | 0.001971494 | 0.297131725 | 0.001890931 | 0.076035171 | 4.81E-05 | 0.001659627 | 1 | 0.800157 | 1 | 0.810520426 | ENSG0000012 | 14 | IRF5 |
| SAT | Heart_Failure | ENSG0000011 | 0.001506092 | 0.528727415 | 0.002390811 | 0.076055431 | 0.001404831 | 0.5567649 | 0.002390575 | 0.076035286 | 0.000101261 | 2.41E-07 | 1 | 0.907158447 | 1 | 0.915531921 | ENSG0000012 | 2 | OPN1SW |
| SAT | Heart_Failure | ENSG0000011 | 0.006922058 | 0.016841615 | 0.00289607 | 0.076054601 | 0.006991073 | 0.015768503 | 0.002895767 | 0.076034431 | -6.90E-05 | 0.00147358 | 1 | 0.287798638 | 1 | 0.278152999 | ENSG0000012 | 11 | HOXD1 |
| SAT | Heart_Failure | ENSG0000011 | 0.000356268 | 0.832817386 | 0.001687756 | 0.076055486 | 0.000394385 | 0.815219027 | 0.001687578 | 0.076035332 | -3.81E-05 | 0.002030459 | 1 | 0.976669362 | 1 | 0.972791643 | ENSG0000012 | 7 | HOXD3 |
| SAT | Heart_Failure | ENSG0000011 | -0.00120241 | 0.49863478 | 0.001777036 | 0.076055422 | -0.00116585 | 0.511738952 | 0.00177685 | 0.076035273 | -3.66E-05 | 0.006015935 | 1 | 0.899263643 | 1 | 0.902750226 | ENSG0000012 | 12 | PDE11A |
| SAT | Heart_Failure | ENSG0000011 | 0.001226358 | 0.736302428 | 0.003641701 | 0.076055475 | 0.001144554 | 0.753275333 | 0.003641322 | 0.076035325 | 8.18E-05 | 0.002902714 | 1 | 0.966658386 | 1 | 0.962420704 | ENSG0000012 | 12 | PDE11A |
| SAT | Heart_Failure | ENSG0000011 | -0.00080343 | 0.778165817 | 0.002851971 | 0.076055481 | -0.00084905 | 0.765904029 | 0.002851683 | 0.076035326 | 4.56E-05 | 0.043956713 | 1 | 0.972664863 | 1 | 0.965963214 | ENSG0000012 | 12 | PDE11A |
| SAT | Heart_Failure | ENSG0000011 | 0.003916127 | 0.242172279 | 0.003348331 | 0.07605528 | 0.003859984 | 0.248937625 | 0.003347963 | 0.076035133 | 5.61E-05 | 0.012501016 | 1 | 0.759865642 | 1 | 0.764045228 | ENSG0000012 | 9 | ORMDL1 |
| SAT | Heart_Failure | ENSG0000011 | -0.00164703 | 0.44702993 | 0.002166073 | 0.076055403 | -0.0015871 | 0.463706587 | 0.002165934 | 0.076035256 | -5.99E-05 | 0.017974283 | 1 | 0.884975994 | 1 | 0.892844363 | ENSG0000012 | 9 | ORMDL1 |
| SAT | Heart_Failure | ENSG0000011 | 0.000580718 | 0.87810257 | 0.003786259 | 0.076055489 | 0.000521365 | 0.890466414 | 0.00378585 | 0.076035337 | 5.94E-05 | 0.025313285 | 1 | 0.979223495 | 1 | 0.977805317 | ENSG0000012 | 10 | HOXD9 |
| SAT | Heart_Failure | ENSG0000011 | -0.00081111 | 0.605662714 | 0.001571078 | 0.076055451 | -0.00085793 | 0.584971387 | 0.001570911 | 0.076035294 | 4.68E-05 | 3.63E-05 | 1 | 0.947817475 | 1 | 0.933194299 | ENSG0000012 | 16 | PSMG2 |
| SAT | Heart_Failure | ENSG0000011 | 6.00E-05 | 0.982758128 | 0.002778228 | 0.076055493 | 1.03E-05 | 0.997055904 | 0.002777946 | 0.07603534 | 4.98E-05 | 0.023201163 | 1 | 0.998920999 | 1 | 0.999768899 | ENSG0000012 | 13 | WDFY4 |
| SAT | Heart_Failure | ENSG0000011 | 0.000439854 | 0.712776319 | 0.001194833 | 0.076055472 | 0.000464427 | 0.697468355 | 0.001194698 | 0.076035317 | -2.46E-05 | 0.001045332 | 1 | 0.966658386 | 1 | 0.956878334 | ENSG0000012 | 13 | WDFY4 |
| SAT | Heart_Failure | ENSG0000011 | 0.002802209 | 0.18677419 | 0.00212259 | 0.076055221 | 0.002764905 | 0.192658657 | 0.00212235 | 0.076035075 | 3.73E-05 | 0.004966407 | 1 | 0.720656885 | 1 | 0.72324148 | ENSG0000012 | 7 | EIF2AK4 |
| SAT | Heart_Failure | ENSG0000011 | 0.001143491 | 0.318599671 | 0.00114654 | 0.076055338 | 0.00111977 | 0.328697597 | 0.001146434 | 0.076035191 | 2.37E-05 | 0.02182673 | 1 | 0.827258183 | 1 | 0.833382337 | ENSG0000012 | 7 | EIF2AK4 |
| SAT | Heart_Failure | ENSG0000011 | 0.001623556 | 0.246837957 | 0.001401956 | 0.076055284 | 0.001690563 | 0.227827919 | 0.001401823 | 0.076035113 | -6.70E-05 | 4.19E-08 | 1 | 0.761161032 | 1 | 0.747837725 | ENSG0000012 | 7 | EIF2AK4 |
| SAT | Heart_Failure | ENSG0000011 | 0.002881191 | 0.296132483 | 0.002757743 | 0.076055323 | 0.002795703 | 0.310645991 | 0.002757461 | 0.07603518 | 8.55E-05 | 6.57E-05 | 1 | 0.813063328 | 1 | 0.821155716 | ENSG0000012 | 7 | ELL3 |
| SAT | Heart_Failure | ENSG0000011 | -0.00039 | 0.883255102 | 0.002655871 | 0.07605549 | -0.000467 | 0.860408074 | 0.002655596 | 0.076035335 | 7.70E-05 | 0.000142763 | 1 | 0.979223495 | 1 | 0.972791643 | ENSG0000012 | 11 | C15orf57 |
| SAT | Heart_Failure | ENSG0000011 | -0.00179959 | 0.635472923 | 0.003796291 | 0.076055458 | -0.00188146 | 0.620136018 | 0.003795884 | 0.076035302 | 8.19E-05 | 0.002401192 | 1 | 0.959428651 | 1 | 0.94798204 | ENSG0000012 | 11 | C15orf57 |
| SAT | Heart_Failure | ENSG0000012 | 0.001156747 | 0.652927437 | 0.002572267 | 0.076055461 | 0.001248903 | 0.627267837 | 0.002572002 | 0.076035303 | -9.22E-05 | 2.91E-06 | 1 | 0.962760695 | 1 | 0.94798204 | ENSG0000012 | 4 | IVD |
| SAT | Heart_Failure | ENSG0000012 | 0.002056706 | 0.420811835 | 0.002554869 | 0.076055392 | 0.00212637 | 0.405206044 | 0.002554628 | 0.076035232 | -6.97E-05 | 0.001770846 | 1 | 0.876068293 | 1 | 0.867451005 | ENSG0000012 | 9 | KNSTRN |
| SAT | Heart_Failure | ENSG0000012 | -7.96E-05 | 0.962984756 | 0.001714509 | 0.076055493 | -5.14E-05 | 0.976064283 | 0.001714324 | 0.07603534 | -2.81E-05 | 0.01947219 | 1 | 0.995788523 | 1 | 0.999521508 | ENSG0000012 | 7 | CHAC1 |
| SAT | Heart_Failure | ENSG0000012 | 0.001270785 | 0.357250697 | 0.001380362 | 0.076055361 | 0.001246962 | 0.366288988 | 0.001380227 | 0.076035213 | 2.38E-05 | 0.036887428 | 1 | 0.848363544 | 1 | 0.849290727 | ENSG0000012 | 11 | VPS13C |
| SAT | Heart_Failure | ENSG0000012 | 0.002137015 | 0.432620672 | 0.002723299 | 0.076055397 | 0.002100968 | 0.440373129 | 0.002722999 | 0.076035247 | 3.60E-05 | 0.047959076 | 1 | 0.879986926 | 1 | 0.879941679 | ENSG0000012 | 11 | VPS13C |
| SAT | Heart_Failure | ENSG0000012 | 0.002308345 | 0.460818657 | 0.003129958 | 0.076055408 | 0.002407521 | 0.441732941 | 0.003129615 | 0.076035248 | -9.92E-05 | 2.60E-06 | 1 | 0.890157253 | 1 | 0.880611614 | ENSG0000012 | 11 | VPS13C |
| SAT | Heart_Failure | ENSG0000012 | -0.00072659 | 0.842841383 | 0.003664844 | 0.076055487 | -0.00088763 | 0.808603858 | 0.003664464 | 0.076035331 | 0.000161041 | 7.88E-09 | 1 | 0.976669362 | 1 | 0.972791643 | ENSG0000012 | 8 | ANAPC13 |
| SAT | Heart_Failure | ENSG0000012 | 0.002359802 | 0.352471091 | 0.002537947 | 0.076055358 | 0.002444555 | 0.335395233 | 0.002537676 | 0.076035195 | -8.48E-05 | 3.79E-06 | 1 | 0.848363544 | 1 | 0.836830236 | ENSG0000012 | 8 | ANAPC13 |
| SAT | Heart_Failure | ENSG0000012 | 0.000608074 | 0.72766858 | 0.001746192 | 0.076055474 | 0.000653745 | 0.708093445 | 0.001746028 | 0.076035318 | -4.57E-05 | 0.002984851 | 1 | 0.966658386 | 1 | 0.958028875 | ENSG0000012 | 8 | ANAPC13 |
| SAT | Heart_Failure | ENSG0000012 | 0.000379184 | 0.896247209 | 0.002907777 | 0.07605549 | 0.000293444 | 0.919608398 | 0.002907484 | 0.076035339 | 8.57E-05 | 0.00021578 | 1 | 0.979542669 | 1 | 0.984094962 | ENSG0000012 | 8 | ANAPC13 |
| SAT | Heart_Failure | ENSG0000012 | 0.005584592 | 0.139582376 | 0.003780141 | 0.076055152 | 0.005675587 | 0.133206777 | 0.003779748 | 0.076034988 | -9.10E-05 | 0.001511832 | 1 | 0.666622671 | 1 | 0.649215182 | ENSG0000012 | 8 | ANAPC13 |
| SAT | Heart_Failure | ENSG0000012 | 0.001465075 | 0.63682594 | 0.003103041 | 0.076055458 | 0.001408129 | 0.649946362 | 0.003102719 | 0.076035308 | 5.69E-05 | 0.015702586 | 1 | 0.959428651 | 1 | 0.953700363 | ENSG0000012 | 10 | DCTD |
| SAT | Heart_Failure | ENSG0000012 | 0.000800479 | 0.808450868 | 0.003301966 | 0.076055484 | 0.000923004 | 0.779814228 | 0.003301615 | 0.076035328 | -0.00012252 | 2.99E-07 | 1 | 0.976669362 | 1 | 0.966957854 | ENSG0000012 | 10 | DCTD |
| SAT | Heart_Failure | ENSG0000012 | 0.000204013 | 0.88062377 | 0.001358471 | 0.076055489 | 0.000221593 | 0.870409865 | 0.001358318 | 0.076035336 | -1.76E-05 | 0.039752222 | 1 | 0.979223495 | 1 | 0.974665278 | ENSG0000012 | 10 | RPAIN |
| SAT | Heart_Failure | ENSG0000012 | -0.00013013 | 0.894324342 | 0.000979637 | 0.07605549 | -9.98E-05 | 0.918816407 | 0.000979537 | 0.076035339 | -3.03E-05 | 6.49E-05 | 1 | 0.979270459 | 1 | 0.984094962 | ENSG0000012 | 12 | USP6 |
| SAT | Heart_Failure | ENSG0000012 | -0.00218007 | 0.345413983 | 0.002310565 | 0.076055354 | -0.00222215 | 0.336133288 | 0.00231032 | 0.076035196 | 4.21E-05 | 0.012933578 | 1 | 0.847011103 | 1 | 0.837075007 | ENSG0000012 | 12 | USP6 |
| SAT | Heart_Failure | ENSG0000012 | 0.001521746 | 0.298907576 | 0.001464936 | 0.076055325 | 0.001496574 | 0.306921532 | 0.001464782 | 0.076035177 | 2.52E-05 | 0.020672632 | 1 | 0.814547972 | 1 | 0.815392345 | ENSG0000012 | 12 | USP6 |
| SAT | Heart_Failure | ENSG0000012 | -0.00265865 | 0.384002152 | 0.003054006 | 0.076055375 | -0.0026057 | 0.393493388 | 0.003053669 | 0.076035227 | -5.30E-05 | 0.009043186 | 1 | 0.858250041 | 1 | 0.862334842 | ENSG0000012 | 12 | USP6 |
| SAT | Heart_Failure | ENSG0000012 | -0.00135721 | 0.621434236 | 0.002748402 | 0.076055455 | -0.00131824 | 0.631447604 | 0.002748103 | 0.076035304 | -3.90E-05 | 0.039687528 | 1 | 0.954216727 | 1 | 0.94798204 | ENSG0000012 | 12 | USP6 |
| SAT | Heart_Failure | ENSG0000012 | -0.00495606 | 0.122785197 | 0.00321157 | 0.076055121 | -0.00489182 | 0.127671792 | 0.003211225 | 0.076034978 | -6.42E-05 | 0.004620035 | 1 | 0.646585619 | 1 | 0.647874406 | ENSG0000012 | 12 | USP6 |
| SAT | Heart_Failure | ENSG0000012 | -0.00241724 | 0.138340591 | 0.001631068 | 0.07605515 | -0.00238437 | 0.143739999 | 0.001630887 | 0.076035007 | -3.29E-05 | 0.00239269 | 1 | 0.664561462 | 1 | 0.659101398 | ENSG0000012 | 9 | PLD2 |
| SAT | Heart_Failure | ENSG0000012 | 0.00297941 | 0.427351873 | 0.003753673 | 0.076055395 | 0.003063643 | 0.414350235 | 0.003753255 | 0.076035236 | -8.42E-05 | 0.000567152 | 1 | 0.878482333 | 1 | 0.867451005 | ENSG0000012 | 9 | PLD2 |
| SAT | Heart_Failure | ENSG0000012 | 0.001126527 | 0.731410552 | 0.003281942 | 0.076055475 | 0.001175911 | 0.720092695 | 0.003281595 | 0.07603532 | -4.94E-05 | 0.039448202 | 1 | 0.966658386 | 1 | 0.958028875 | ENSG0000012 | 10 | PUS7L |
| SAT | Heart_Failure | ENSG0000012 | 0.000984096 | 0.450670349 | 0.00130465 | 0.076055404 | 0.001142358 | 0.38204321 | 0.001306839 | 0.076035221 | -0.00015826 | 0.043874246 | 1 | 0.887019168 | 1 | 0.854967426 | ENSG0000012 | 10 | PUS7L |
| SAT | Heart_Failure | ENSG0000012 | 0.005799644 | 0.04488301 | 0.002891503 | 0.076054865 | 0.005867448 | 0.04241647 | 0.0028912 | 0.076034697 | -6.78E-05 | 0.001697117 | 1 | 0.469267596 | 1 | 0.45628996 | ENSG0000012 | 10 | PUS7L |
| SAT | Heart_Failure | ENSG0000012 | 0.002143471 | 0.142264994 | 0.001460717 | 0.076055157 | 0.002204589 | 0.131208206 | 0.001460614 | 0.076034985 | -6.11E-05 | 0.000186596 | 1 | 0.669181587 | 1 | 0.649215182 | ENSG0000012 | 15 | KRI1 |
| SAT | Heart_Failure | ENSG0000012 | -0.00015854 | 0.884848236 | 0.001094676 | 0.07605549 | -0.00017735 | 0.871282528 | 0.001094556 | 0.076035336 | 1.88E-05 | 0.011991013 | 1 | 0.979270459 | 1 | 0.974665278 | ENSG0000012 | 7 | AP1M2 |
| SAT | Heart_Failure | ENSG0000012 | -0.00798692 | 0.011923296 | 0.003176451 | 0.076054506 | -0.00793697 | 0.01245656 | 0.003176122 | 0.076034366 | -5.00E-05 | 0.038394622 | 1 | 0.24440108 | 1 | 0.252283786 | ENSG0000012 | 6 | CDKN2D |
| SAT | Heart_Failure | ENSG0000012 | -0.00721549 | 0.005092145 | 0.002575896 | 0.076054268 | -0.00714095 | 0.005562795 | 0.002575626 | 0.076034141 | -7.45E-05 | 9.86E-05 | 1 | 0.162207064 | 1 | 0.179428249 | ENSG0000012 | 6 | CDKN2D |
| SAT | Heart_Failure | ENSG0000012 | 0.004374063 | 0.119717384 | 0.00281116 | 0.076055115 | 0.004306998 | 0.125463353 | 0.002810912 | 0.076034974 | 6.71E-05 | 0.011200334 | 1 | 0.646585619 | 1 | 0.647874406 | ENSG0000012 | 6 | CDKN2D |
| SAT | Heart_Failure | ENSG0000012 | -0.0017338 | 0.290793614 | 0.001641261 | 0.076055319 | -0.0016983 | 0.300731128 | 0.001641082 | 0.076035173 | -3.55E-05 | 0.001623881 | 1 | 0.803109301 | 1 | 0.810520426 | ENSG0000012 | 7 | KLK10 |
| SAT | Heart_Failure | ENSG0000012 | 0.000414242 | 0.901782594 | 0.003356633 | 0.076055491 | 0.000479885 | 0.886305444 | 0.003356294 | 0.076035337 | -6.56E-05 | 0.013748156 | 1 | 0.981342849 | 1 | 0.975781755 | ENSG0000012 | 11 | RIPK3 |
| SAT | Heart_Failure | ENSG0000012 | -0.00334724 | 0.220661455 | 0.002732951 | 0.076055259 | -0.00323653 | 0.236290526 | 0.002732835 | 0.076035121 | -0.00011072 | 0.002532339 | 1 | 0.750422029 | 1 | 0.752162449 | ENSG0000012 | 5 | BCL2L2 |
| SAT | Heart_Failure | ENSG0000012 | -0.00275003 | 0.30242988 | 0.002666723 | 0.076055327 | -0.00254061 | 0.340929815 | 0.002667786 | 0.076035199 | -0.00020941 | 0.015977274 | 1 | 0.816013516 | 1 | 0.83914552 | ENSG0000012 | 5 | BCL2L2 |
| SAT | Heart_Failure | ENSG0000012 | -0.00077646 | 0.391614339 | 0.00090635 | 0.076055378 | -0.00079945 | 0.377696022 | 0.000906254 | 0.076035219 | 2.30E-05 | 0.00049987 | 1 | 0.862642401 | 1 | 0.854967426 | ENSG0000012 | 8 | DTD2 |
| SAT | Heart_Failure | ENSG0000012 | 0.002181496 | 0.023145757 | 0.000960577 | 0.076054688 | 0.002148146 | 0.025317073 | 0.000960479 | 0.07603456 | 3.34E-05 | 8.06E-06 | 1 | 0.327199936 | 1 | 0.353469426 | ENSG0000012 | 8 | DTD2 |
| SAT | Heart_Failure | ENSG0000012 | -0.00271424 | 0.377175701 | 0.003073494 | 0.076055371 | -0.00278648 | 0.36456234 | 0.003073184 | 0.076035212 | 7.22E-05 | 0.003209219 | 1 | 0.85759885 | 1 | 0.849290727 | ENSG0000012 | 8 | DTD2 |
| SAT | Heart_Failure | ENSG0000012 | -0.00477395 | 0.067035217 | 0.00260666 | 0.07605497 | -0.00471502 | 0.070445736 | 0.002606369 | 0.076034829 | -5.89E-05 | 0.000452113 | 1 | 0.534977442 | 1 | 0.532563273 | ENSG0000012 | 9 | PARP2 |
| SAT | Heart_Failure | ENSG0000012 | -0.00043588 | 0.885574476 | 0.003028927 | 0.07605549 | -0.00054067 | 0.858316299 | 0.003028638 | 0.076035335 | 0.000104784 | 5.70E-05 | 1 | 0.979270459 | 1 | 0.972791643 | ENSG0000012 | 9 | PARP2 |
| SAT | Heart_Failure | ENSG0000012 | 0.000868176 | 0.695891463 | 0.002221121 | 0.076055469 | 0.000927572 | 0.676198325 | 0.002220896 | 0.076035313 | -5.94E-05 | 0.000698871 | 1 | 0.966658386 | 1 | 0.956878334 | ENSG0000012 | 9 | PARP2 |
| SAT | Heart_Failure | ENSG0000012 | -0.00210129 | 0.484306755 | 0.003004448 | 0.076055417 | -0.00204136 | 0.496818445 | 0.003004185 | 0.076035268 | -5.99E-05 | 0.034901056 | 1 | 0.897349195 | 1 | 0.894608312 | ENSG0000012 | 6 | DAD1 |
| SAT | Heart_Failure | ENSG0000012 | -0.00624544 | 0.10103496 | 0.003808519 | 0.076055073 | -0.00617365 | 0.104976747 | 0.00380809 | 0.07603493 | -7.18E-05 | 0.002619981 | 1 | 0.622638992 | 1 | 0.620855876 | ENSG0000012 | 6 | DAD1 |
| SAT | Heart_Failure | ENSG0000012 | 0.008645909 | 0.00542871 | 0.003109545 | 0.076054286 | 0.008545103 | 0.005990657 | 0.003109227 | 0.076034162 | 0.000100806 | 2.85E-05 | 1 | 0.164958288 | 1 | 0.182466145 | ENSG0000012 | 6 | DAD1 |
| SAT | Heart_Failure | ENSG0000012 | 0.000965522 | 0.803310753 | 0.00387659 | 0.076055483 | 0.001034296 | 0.789595343 | 0.003876146 | 0.076035329 | -6.88E-05 | 0.003121355 | 1 | 0.976669362 | 1 | 0.970635778 | ENSG0000012 | 6 | DAD1 |
| SAT | Heart_Failure | ENSG0000012 | 0.000267015 | 0.880223469 | 0.001771995 | 0.076055489 | 0.000235326 | 0.894338869 | 0.001771815 | 0.076035337 | 3.17E-05 | 0.022190237 | 1 | 0.979223495 | 1 | 0.97886604 | ENSG0000012 | 10 | SEC14L1 |
| SAT | Heart_Failure | ENSG0000012 | 3.82E-05 | 0.983763997 | 0.001874809 | 0.076055493 | -3.31E-06 | 0.998589554 | 0.001874604 | 0.07603534 | 4.15E-05 | 0.001172563 | 1 | 0.998924529 | 1 | 0.999768899 | ENSG0000012 | 10 | SEC14L1 |
| SAT | Heart_Failure | ENSG0000012 | -0.00041211 | 0.530177749 | 0.000656506 | 0.076055432 | -0.00040147 | 0.540802131 | 0.000656432 | 0.076035282 | -1.06E-05 | 0.010616822 | 1 | 0.908320477 | 1 | 0.911336308 | ENSG0000012 | 10 | SEC14L1 |
| SAT | Heart_Failure | ENSG0000012 | 0.002022392 | 0.500833518 | 0.003004239 | 0.076055422 | 0.001865029 | 0.534691024 | 0.003003937 | 0.07603528 | 0.000157363 | 4.88E-11 | 1 | 0.900549813 | 1 | 0.90897474 | ENSG0000012 | 8 | RHBDF2 |
| SAT | Heart_Failure | ENSG0000012 | -0.00165198 | 0.416717123 | 0.00203413 | 0.07605539 | -0.00142029 | 0.48507373 | 0.002034321 | 0.076035264 | -0.00023168 | 8.74E-08 | 1 | 0.87398914 | 1 | 0.894608312 | ENSG0000012 | 8 | RHBDF2 |
| SAT | Heart_Failure | ENSG0000012 | -0.00310267 | 0.308561659 | 0.003047072 | 0.076055331 | -0.00319021 | 0.295059337 | 0.003046745 | 0.076035169 | 8.75E-05 | 4.74E-05 | 1 | 0.816225947 | 1 | 0.808088218 | ENSG0000012 | 5 | AANAT |
| SAT | Heart_Failure | ENSG0000012 | -0.00081019 | 0.517607563 | 0.001252155 | 0.076055428 | -0.00079205 | 0.526982304 | 0.001252023 | 0.076035278 | -1.81E-05 | 0.048264416 | 1 | 0.90671138 | 1 | 0.906199653 | ENSG0000012 | 16 | MAU2 |
| SAT | Heart_Failure | ENSG0000012 | -0.00108884 | 0.718336917 | 0.003018839 | 0.076055473 | -0.00115726 | 0.701433421 | 0.00301853 | 0.076035317 | 6.84E-05 | 0.003350335 | 1 | 0.966658386 | 1 | 0.956878334 | ENSG0000013 | 2 | GALNT8 |
| SAT | Heart_Failure | ENSG0000012 | 0.00109189 | 0.68441654 | 0.002686446 | 0.076055467 | 0.00104237 | 0.697977878 | 0.002686163 | 0.076035317 | 4.95E-05 | 0.012305895 | 1 | 0.966658386 | 1 | 0.956878334 | ENSG0000013 | 2 | GALNT8 |
| SAT | Heart_Failure | ENSG0000012 | 9.67E-05 | 0.944671817 | 0.001392983 | 0.076055492 | 5.42E-05 | 0.968986608 | 0.001392834 | 0.07603534 | 4.25E-05 | 1.76E-05 | 1 | 0.992414898 | 1 | 0.99755193 | ENSG0000013 | 11 | SH3BP4 |
| SAT | Heart_Failure | ENSG0000012 | -0.00335154 | 0.272099346 | 0.003051735 | 0.076055305 | -0.00325099 | 0.286701073 | 0.003051463 | 0.076035163 | -0.00010055 | 0.000397085 | 1 | 0.785878482 | 1 | 0.799139368 | ENSG0000013 | 9 | DOCK6 |
| SAT | Heart_Failure | ENSG0000012 | -0.00141587 | 0.650144893 | 0.003121675 | 0.076055461 | -0.00147129 | 0.637381402 | 0.003121352 | 0.076035305 | 5.54E-05 | 0.020017125 | 1 | 0.962760695 | 1 | 0.94798204 | ENSG0000013 | 10 | CDC16 |
| SAT | Heart_Failure | ENSG0000012 | -0.00196633 | 0.556313863 | 0.003342245 | 0.076055439 | -0.00201676 | 0.546190011 | 0.003341893 | 0.076035283 | 5.04E-05 | 0.040215289 | 1 | 0.922001323 | 1 | 0.912597963 | ENSG0000013 | 10 | CDC16 |
| SAT | Heart_Failure | ENSG0000012 | 0.004224543 | 0.075844495 | 0.002379585 | 0.076055001 | 0.004098684 | 0.084977024 | 0.002379477 | 0.076034877 | 0.000125859 | 6.03E-05 | 1 | 0.57074722 | 1 | 0.58987835 | ENSG0000013 | 7 | THEM6 |
| SAT | Heart_Failure | ENSG0000012 | 0.000288972 | 0.923381345 | 0.003004637 | 0.076055492 | 0.00035832 | 0.905066891 | 0.003004444 | 0.076035338 | -6.93E-05 | 0.048246331 | 1 | 0.988739396 | 1 | 0.981985605 | ENSG0000013 | 8 | FAM98C |
| SAT | Heart_Failure | ENSG0000012 | 0.003182036 | 0.413265109 | 0.003889249 | 0.076055389 | 0.003096891 | 0.425826414 | 0.003888835 | 0.076035241 | 8.51E-05 | 0.002389652 | 1 | 0.872873333 | 1 | 0.873465455 | ENSG0000013 | 8 | FAM98C |
| SAT | Heart_Failure | ENSG0000012 | -0.00047465 | 0.856832006 | 0.002630998 | 0.076055488 | -0.00055109 | 0.834072985 | 0.002630725 | 0.076035333 | 7.64E-05 | 0.000131912 | 1 | 0.976669362 | 1 | 0.972791643 | ENSG0000013 | 12 | ATP8B3 |
| SAT | Heart_Failure | ENSG0000012 | -0.00035819 | 0.840180592 | 0.001776207 | 0.076055487 | -0.00039724 | 0.823014246 | 0.00177602 | 0.076035332 | 3.90E-05 | 0.002898333 | 1 | 0.976669362 | 1 | 0.972791643 | ENSG0000013 | 12 | ATP8B3 |
| SAT | Heart_Failure | ENSG0000012 | -0.00128802 | 0.392778424 | 0.001507183 | 0.076055379 | -0.00131518 | 0.382826877 | 0.001507018 | 0.076035221 | 2.72E-05 | 0.007879255 | 1 | 0.863144683 | 1 | 0.854967426 | ENSG0000013 | 12 | GTPBP3 |

| | | | | | | | | | | | | | | | | | | | |
|---|---|---|---|---|---|---|---|---|---|---|---|---|---|---|---|---|---|---|---|
| SAT | Heart_Failure | ENSG0000012 | -0.00159845 | 0.504399805 | 0.002394383 | 0.076055423 | -0.00153168 | 0.522325402 | 0.002394129 | 0.076035276 | -6.68E-05 | 0.000128801 | 1 | 0.9005699 | 1 | 0.904369708 | ENSG0000013 | 12 | GTPBP3 |
| SAT | Heart_Failure | ENSG0000012 | -0.00068936 | 0.623388053 | 0.001403831 | 0.076055455 | -0.00066801 | 0.634144493 | 0.001403676 | 0.076035305 | -2.13E-05 | 0.023286202 | 1 | 0.955683491 | 1 | 0.94798204 | ENSG0000013 | 4 | PLVAP |
| SAT | Heart_Failure | ENSG0000012 | -0.00315121 | 0.223992168 | 0.00259149 | 0.076055262 | -0.00308662 | 0.23359362 | 0.002591288 | 0.076035119 | -6.46E-05 | 0.016762258 | 1 | 0.751217604 | 1 | 0.750248244 | ENSG0000013 | 4 | PLVAP |
| SAT | Heart_Failure | ENSG0000012 | -0.00248886 | 0.306208748 | 0.0024324 | 0.07605533 | -0.00253131 | 0.297982914 | 0.002432153 | 0.076035171 | 4.25E-05 | 0.02695793 | 1 | 0.816225947 | 1 | 0.810520426 | ENSG0000013 | 4 | PLVAP |
| SAT | Heart_Failure | ENSG0000012 | 0.000108043 | 0.975051031 | 0.00345471 | 0.076055493 | 0.000164029 | 0.962127019 | 0.00345437 | 0.07603534 | -5.60E-05 | 0.049544659 | 1 | 0.998920999 | 1 | 0.99592587 | ENSG0000013 | 4 | PLVAP |
| SAT | Heart_Failure | ENSG0000012 | 0.000627649 | 0.814100981 | 0.002669269 | 0.076055484 | 0.000675419 | 0.800221989 | 0.002669006 | 0.07603533 | -4.78E-05 | 0.029878265 | 1 | 0.976669362 | 1 | 0.972791643 | ENSG0000013 | 2 | MRPL34 |
| SAT | Heart_Failure | ENSG0000012 | 0.001647223 | 0.520974097 | 0.002566387 | 0.076055429 | 0.00171915 | 0.502893418 | 0.002566108 | 0.07603527 | -7.19E-05 | 4.45E-05 | 1 | 0.906724424 | 1 | 0.900571724 | ENSG0000013 | 2 | MRPL34 |
| SAT | Heart_Failure | ENSG0000013 | -0.00030979 | 0.866177825 | 0.001838326 | 0.076055489 | -0.00033761 | 0.854273643 | 0.001838136 | 0.076035335 | 2.78E-05 | 0.047549223 | 1 | 0.978259692 | 1 | 0.972791643 | ENSG0000013 | 10 | LSM7 |
| SAT | Heart_Failure | ENSG0000013 | 0.002415952 | 0.346741047 | 0.002567621 | 0.076055355 | 0.002348948 | 0.360227777 | 0.002567347 | 0.07603521 | 6.70E-05 | 0.000264666 | 1 | 0.847011103 | 1 | 0.848283315 | ENSG0000013 | 10 | LSM7 |
| SAT | Heart_Failure | ENSG0000013 | -0.00035289 | 0.824196712 | 0.001588536 | 0.076055485 | -0.00032376 | 0.838484186 | 0.001588373 | 0.076035334 | -2.91E-05 | 0.017227737 | 1 | 0.976669362 | 1 | 0.972791643 | ENSG0000013 | 10 | LSM7 |
| SAT | Heart_Failure | ENSG0000013 | -0.00078222 | 0.576403103 | 0.001400213 | 0.076055444 | -0.00081107 | 0.5623794 | 0.001400058 | 0.076035288 | 2.88E-05 | 0.001754647 | 1 | 0.932993874 | 1 | 0.918907775 | ENSG0000013 | 7 | QRSL1 |
| SAT | Heart_Failure | ENSG0000013 | 0.003381002 | 0.238994485 | 0.002871327 | 0.076055277 | 0.003293511 | 0.251318301 | 0.002871027 | 0.076035135 | 8.75E-05 | 4.45E-05 | 1 | 0.758532873 | 1 | 0.765423527 | ENSG0000013 | 7 | QRSL1 |
| SAT | Heart_Failure | ENSG0000013 | 0.00155464 | 0.3567559 | 0.001686956 | 0.07605536 | 0.00157717 | 0.349774474 | 0.001686763 | 0.076035204 | -2.25E-05 | 0.026969866 | 1 | 0.848363544 | 1 | 0.842583404 | ENSG0000013 | 7 | RSPH3 |
| SAT | Heart_Failure | ENSG0000013 | 0.001361145 | 0.467717321 | 0.001874343 | 0.076055411 | 0.001335137 | 0.476215374 | 0.001874131 | 0.076035261 | 2.60E-05 | 0.026802329 | 1 | 0.893397133 | 1 | 0.894608312 | ENSG0000013 | 7 | RSPH3 |
| SAT | Heart_Failure | ENSG0000013 | -0.00228231 | 0.53820593 | 0.003707885 | 0.076055434 | -0.00234615 | 0.526856463 | 0.003707492 | 0.076035278 | 6.38E-05 | 0.017926831 | 1 | 0.912011429 | 1 | 0.906199653 | ENSG0000013 | 7 | RSPH3 |
| SAT | Heart_Failure | ENSG0000013 | 0.001124111 | 0.513540538 | 0.001720573 | 0.076055426 | 0.00103596 | 0.547067711 | 0.001720406 | 0.076035284 | 8.82E-05 | 1.16E-09 | 1 | 0.905155107 | 1 | 0.912597963 | ENSG0000013 | 7 | RSPH3 |
| SAT | Heart_Failure | ENSG0000013 | -0.00277356 | 0.48225139 | 0.003947079 | 0.076055416 | -0.00271667 | 0.491234686 | 0.00394666 | 0.076035266 | -5.69E-05 | 0.046315378 | 1 | 0.897349195 | 1 | 0.894608312 | ENSG0000013 | 7 | RSPH3 |
| SAT | Heart_Failure | ENSG0000013 | 0.008828029 | 0.000408947 | 0.00249784 | 0.076053544 | 0.008755673 | 0.00045558 | 0.002497608 | 0.076033423 | 7.24E-05 | 0.001125138 | 1 | 0.031946583 | 1 | 0.039533553 | ENSG0000013 | 11 | STK33 |
| SAT | Heart_Failure | ENSG0000013 | -0.00215357 | 0.582850897 | 0.003921111 | 0.076055446 | -0.00223549 | 0.568561057 | 0.003920721 | 0.076035289 | 8.19E-05 | 0.010412594 | 1 | 0.937265914 | 1 | 0.923489519 | ENSG0000013 | 7 | NDUFA10 |
| SAT | Heart_Failure | ENSG0000013 | 0.002765978 | 0.302816808 | 0.002684339 | 0.076055327 | 0.002685456 | 0.317063785 | 0.002684084 | 0.076035184 | 8.05E-05 | 0.000518628 | 1 | 0.816145643 | 1 | 0.823487262 | ENSG0000013 | 7 | NDUFA10 |
| SAT | Heart_Failure | ENSG0000013 | 0.000710704 | 0.849992928 | 0.003757801 | 0.076055487 | 0.000785113 | 0.834485909 | 0.003757388 | 0.076035333 | -7.44E-05 | 0.003187419 | 1 | 0.976669362 | 1 | 0.972791643 | ENSG0000013 | 11 | SSBP4 |
| SAT | Heart_Failure | ENSG0000013 | 0.002843965 | 0.041740042 | 0.001396765 | 0.076054846 | 0.002818433 | 0.043586063 | 0.001396608 | 0.076034705 | 2.55E-05 | 0.003913555 | 1 | 0.461108001 | 1 | 0.456297071 | ENSG0000013 | 11 | SSBP4 |
| SAT | Heart_Failure | ENSG0000013 | 0.001485599 | 0.652557495 | 0.003299773 | 0.076055461 | 0.001373738 | 0.677149007 | 0.003299424 | 0.076035313 | 0.000111861 | 3.30E-06 | 1 | 0.962760695 | 1 | 0.956878334 | ENSG0000013 | 9 | PGPEP1 |
| SAT | Heart_Failure | ENSG0000013 | 0.001161394 | 0.748987039 | 0.003629633 | 0.076055477 | 0.001071041 | 0.767909462 | 0.003629284 | 0.076035327 | 9.04E-05 | 0.003565737 | 1 | 0.966658386 | 1 | 0.965963214 | ENSG0000013 | 9 | PGPEP1 |
| SAT | Heart_Failure | ENSG0000013 | -0.00349278 | 0.162389667 | 0.002500055 | 0.076055188 | -0.00344907 | 0.167665669 | 0.00249979 | 0.076035043 | -4.37E-05 | 0.016121091 | 1 | 0.686431329 | 1 | 0.685924016 | ENSG0000013 | 12 | KIAA1683 |
| SAT | Heart_Failure | ENSG0000013 | -0.00754021 | 0.053795995 | 0.003909926 | 0.076054913 | -0.00759206 | 0.052143043 | 0.00390949 | 0.076034752 | 5.18E-05 | 0.041954669 | 1 | 0.497196222 | 1 | 0.49020437 | ENSG0000013 | 12 | KIAA1683 |
| SAT | Heart_Failure | ENSG0000013 | -0.00226192 | 0.091748126 | 0.001341389 | 0.076055049 | -0.00230126 | 0.086209234 | 0.00134126 | 0.076034881 | 3.93E-05 | 0.000597605 | 1 | 0.60325772 | 1 | 0.59547894 | ENSG0000013 | 6 | LSM4 |
| SAT | Heart_Failure | ENSG0000013 | 0.001813232 | 0.635387939 | 0.003824115 | 0.076055458 | 0.001751075 | 0.646985508 | 0.00382369 | 0.076035307 | 6.22E-05 | 0.012728173 | 1 | 0.959428651 | 1 | 0.95317119 | ENSG0000013 | 6 | LSM4 |
| SAT | Heart_Failure | ENSG0000013 | 0.004728319 | 0.167935308 | 0.003429125 | 0.076055196 | 0.004830895 | 0.158857216 | 0.003428774 | 0.07603503 | -0.00010258 | 0.00012753 | 1 | 0.694824396 | 1 | 0.670798618 | ENSG0000013 | 6 | LSM4 |
| SAT | Heart_Failure | ENSG0000013 | 0.00408498 | 0.00519247 | 0.001461606 | 0.076054274 | 0.004024597 | 0.005898504 | 0.001461692 | 0.076034157 | 6.04E-05 | 0.034468392 | 1 | 0.163002345 | 1 | 0.182466145 | ENSG0000013 | 6 | LSM4 |
| SAT | Heart_Failure | ENSG0000013 | 0.002399849 | 0.193547966 | 0.001845815 | 0.076055229 | 0.002437759 | 0.186565299 | 0.001845655 | 0.076035068 | -3.79E-05 | 0.031443416 | 1 | 0.726250291 | 1 | 0.715930698 | ENSG0000013 | 11 | TRPM4 |
| SAT | Heart_Failure | ENSG0000013 | -0.00733275 | 5.96E-05 | 0.001826686 | 0.076052978 | -0.00730844 | 6.30E-05 | 0.001826475 | 0.076032842 | -2.43E-05 | 0.023079532 | 0.195701696 | 0.007106166 | 0.206637533 | 0.011955478 | ENSG0000013 | 9 | TUBGCP2 |
| SAT | Heart_Failure | ENSG0000013 | -0.00297638 | 0.087687507 | 0.001742889 | 0.076055038 | -0.00294773 | 0.090746004 | 0.001742695 | 0.076034894 | -2.86E-05 | 0.010983555 | 1 | 0.592308253 | 1 | 0.598355674 | ENSG0000013 | 4 | HBZ |
| SAT | Heart_Failure | ENSG0000013 | 0.000770718 | 0.578138961 | 0.00138592 | 0.076055445 | 0.000798801 | 0.564323887 | 0.001385769 | 0.076035288 | -2.81E-05 | 0.003240736 | 1 | 0.933960577 | 1 | 0.92032393 | ENSG0000013 | 9 | ZNF337 |
| SAT | Heart_Failure | ENSG0000013 | 0.003017268 | 0.049316966 | 0.001534845 | 0.07605489 | 0.002981797 | 0.052025435 | 0.001534693 | 0.076034751 | 3.55E-05 | 0.004659076 | 1 | 0.483011842 | 1 | 0.49020437 | ENSG0000013 | 9 | CEP85 |
| SAT | Heart_Failure | ENSG0000013 | -0.00993272 | 0.003186408 | 0.003367969 | 0.076054136 | -0.00997971 | 0.003042215 | 0.003367593 | 0.07603397 | 4.70E-05 | 0.031586796 | 1 | 0.127495168 | 1 | 0.134046431 | ENSG0000013 | 11 | POMT1 |
| SAT | Heart_Failure | ENSG0000013 | -0.00050033 | 0.671857286 | 0.001181131 | 0.076055465 | -0.00046663 | 0.692763743 | 0.001181013 | 0.076035316 | -3.37E-05 | 0.000353043 | 1 | 0.965985276 | 1 | 0.956878334 | ENSG0000013 | 11 | POMT1 |
| SAT | Heart_Failure | ENSG0000013 | -0.00037894 | 0.708982672 | 0.001015321 | 0.076055471 | -0.00040459 | 0.690237499 | 0.001015209 | 0.076035315 | 2.57E-05 | 0.000176994 | 1 | 0.966658386 | 1 | 0.956878334 | ENSG0000013 | 4 | CHMP2A |
| SAT | Heart_Failure | ENSG0000013 | -0.00161732 | 0.454695127 | 0.002163317 | 0.076055406 | -0.0015734 | 0.466993719 | 0.0021631 | 0.076035258 | -4.39E-05 | 0.011381298 | 1 | 0.887019168 | 1 | 0.892844363 | ENSG0000013 | 6 | YIPF2 |
| SAT | Heart_Failure | ENSG0000013 | 0.001989139 | 0.347004688 | 0.00211517 | 0.076055355 | 0.001935033 | 0.360225393 | 0.002114937 | 0.07603521 | 5.41E-05 | 0.000137583 | 1 | 0.847011103 | 1 | 0.848283315 | ENSG0000013 | 6 | YIPF2 |
| SAT | Heart_Failure | ENSG0000013 | -0.00167677 | 0.54092453 | 0.002742439 | 0.076055435 | -0.00162117 | 0.554382003 | 0.002742138 | 0.076035286 | -5.56E-05 | 0.002542718 | 1 | 0.913035829 | 1 | 0.915531921 | ENSG0000013 | 6 | YIPF2 |
| SAT | Heart_Failure | ENSG0000013 | -0.00207965 | 0.495732181 | 0.003052821 | 0.076055421 | -0.00202306 | 0.507484957 | 0.003052479 | 0.076035272 | -5.66E-05 | 0.003733818 | 1 | 0.898909993 | 1 | 0.901005489 | ENSG0000013 | 3 | MAP3K10 |
| SAT | Heart_Failure | ENSG0000013 | -0.00412557 | 0.169534918 | 0.003003248 | 0.076055198 | -0.00419087 | 0.162841218 | 0.003002956 | 0.076035036 | 6.53E-05 | 0.009519355 | 1 | 0.699678071 | 1 | 0.679583582 | ENSG0000013 | 3 | MAP3K10 |
| SAT | Heart_Failure | ENSG0000013 | 0.001593424 | 0.228497481 | 0.001323178 | 0.076055267 | 0.001653385 | 0.21141561 | 0.001323042 | 0.076035096 | -6.00E-05 | 4.79E-09 | 1 | 0.751217604 | 1 | 0.733958243 | ENSG0000013 | 5 | LRRC47 |
| SAT | Heart_Failure | ENSG0000013 | 0.002447187 | 0.419360327 | 0.003030451 | 0.076055391 | 0.00261904 | 0.387414196 | 0.003030185 | 0.076035224 | -0.00017185 | 2.16E-09 | 1 | 0.874438427 | 1 | 0.858921514 | ENSG0000013 | 13 | ATPIF1 |
| SAT | Heart_Failure | ENSG0000013 | 0.001078777 | 0.387963612 | 0.001249572 | 0.076055377 | 0.001118818 | 0.370543123 | 0.001249438 | 0.076035215 | -4.00E-05 | 6.59E-06 | 1 | 0.86096131 | 1 | 0.8495821 | ENSG0000013 | 13 | ATPIF1 |
| SAT | Heart_Failure | ENSG0000013 | 0.000109171 | 0.966527937 | 0.002601581 | 0.076055493 | 0.000148096 | 0.954599569 | 0.002601296 | 0.07603534 | -3.89E-05 | 0.027591481 | 1 | 0.996432082 | 1 | 0.991911131 | ENSG0000013 | 10 | ZNF317 |
| SAT | Heart_Failure | ENSG0000013 | -0.00115332 | 0.668589884 | 0.002694153 | 0.076055464 | -0.00121314 | 0.652470913 | 0.00269388 | 0.076035308 | 5.98E-05 | 0.004937111 | 1 | 0.965492279 | 1 | 0.954583686 | ENSG0000013 | 11 | ZNF236 |
| SAT | Heart_Failure | ENSG0000013 | -0.00163978 | 0.652111232 | 0.00363724 | 0.076055461 | -0.00156638 | 0.666693447 | 0.003636893 | 0.076035311 | -7.34E-05 | 0.019452609 | 1 | 0.962760695 | 1 | 0.956878334 | ENSG0000013 | 3 | C12orf65 |
| SAT | Heart_Failure | ENSG0000013 | 0.001153017 | 0.683340085 | 0.002826648 | 0.076055467 | 0.00108891 | 0.700039387 | 0.002826373 | 0.076035317 | 6.41E-05 | 0.007015741 | 1 | 0.966658386 | 1 | 0.956878334 | ENSG0000013 | 5 | PKDREJ |
| SAT | Heart_Failure | ENSG0000013 | 0.000161862 | 0.954328818 | 0.002826218 | 0.076055493 | 0.000212787 | 0.939977298 | 0.002825921 | 0.076035339 | -5.09E-05 | 0.015034793 | 1 | 0.994131568 | 1 | 0.988983601 | ENSG0000013 | 5 | PKDREJ |
| SAT | Heart_Failure | ENSG0000013 | 0.006542464 | 0.085243214 | 0.003801448 | 0.076055031 | 0.006658865 | 0.079814159 | 0.003801223 | 0.076034865 | -0.0001164 | 0.011564839 | 1 | 0.592308253 | 1 | 0.576806732 | ENSG0000013 | 8 | ATP6V1E1 |
| SAT | Heart_Failure | ENSG0000013 | 0.001461047 | 0.66266135 | 0.003349167 | 0.076055463 | 0.001376678 | 0.681003371 | 0.00334881 | 0.076035314 | 8.44E-05 | 0.000475899 | 1 | 0.962994646 | 1 | 0.956878334 | ENSG0000013 | 10 | CHMP1A |
| SAT | Heart_Failure | ENSG0000013 | 0.00593309 | 0.069573452 | 0.003269493 | 0.076054979 | 0.006114801 | 0.061489654 | 0.003270013 | 0.076034805 | -0.00018171 | 0.021384541 | 1 | 0.537536555 | 1 | 0.505632969 | ENSG0000013 | 10 | CHMP1A |
| SAT | Heart_Failure | ENSG0000013 | -0.00593778 | 0.045081795 | 0.002963119 | 0.076054866 | -0.00584861 | 0.048395436 | 0.002962995 | 0.076034732 | -8.92E-05 | 0.02535976 | 1 | 0.469267596 | 1 | 0.476833109 | ENSG0000013 | 10 | HAUS8 |
| SAT | Heart_Failure | ENSG0000013 | -0.00090065 | 0.698023336 | 0.002321317 | 0.07605547 | -0.00094346 | 0.68439325 | 0.002321071 | 0.076035314 | 4.28E-05 | 0.011242844 | 1 | 0.966658386 | 1 | 0.956878334 | ENSG0000013 | 7 | NAPSB |
| SAT | Heart_Failure | ENSG0000013 | -0.01629368 | 9.59E-07 | 0.003325287 | 0.076051746 | -0.01623548 | 1.05E-06 | 0.003324967 | 0.07603162 | -5.82E-05 | 0.039327579 | 0.00314652 | 0.000233648 | 0.003430974 | 0.000581811 | ENSG0000013 | 7 | NAPSB |
| SAT | Heart_Failure | ENSG0000013 | -0.00097781 | 0.683854792 | 0.002401246 | 0.076055467 | -0.00090703 | 0.705599284 | 0.002400995 | 0.076035318 | -7.08E-05 | 8.49E-05 | 1 | 0.966658386 | 1 | 0.958028875 | ENSG0000013 | 7 | NAPSB |
| SAT | Heart_Failure | ENSG0000013 | 0.001128549 | 0.7528201 | 0.003583569 | 0.076055478 | 0.001228246 | 0.731765887 | 0.003583214 | 0.076035322 | -9.97E-05 | 0.000677784 | 1 | 0.966658386 | 1 | 0.958028875 | ENSG0000013 | 11 | KIF3A |
| SAT | Heart_Failure | ENSG0000013 | -0.0021562 | 0.023023197 | 0.000948592 | 0.076054687 | -0.00213566 | 0.024344785 | 0.000948436 | 0.076034549 | -2.05E-05 | 0.000665949 | 1 | 0.327199936 | 1 | 0.342903392 | ENSG0000013 | 11 | KIF3A |
| SAT | Heart_Failure | ENSG0000013 | 0.003534599 | 0.123744235 | 0.002296314 | 0.076055123 | 0.003616593 | 0.115230578 | 0.002296082 | 0.076034953 | -8.20E-05 | 6.37E-06 | 1 | 0.647388543 | 1 | 0.64481576 | ENSG0000013 | 11 | KIF3A |
| SAT | Heart_Failure | ENSG0000013 | 9.60E-05 | 0.976305485 | 0.00323306 | 0.076055493 | 1.24E-05 | 0.996934651 | 0.003232709 | 0.07603534 | 8.36E-05 | 0.000189244 | 1 | 0.998920999 | 1 | 0.999768899 | ENSG0000013 | 11 | KIF3A |
| SAT | Heart_Failure | ENSG0000013 | 0.001506142 | 0.45658091 | 0.002023054 | 0.076055407 | 0.001472034 | 0.466793544 | 0.00202284 | 0.076035258 | 3.41E-05 | 0.021606854 | 1 | 0.88796436 | 1 | 0.892844363 | ENSG0000013 | 12 | ACLY |
| SAT | Heart_Failure | ENSG0000013 | -0.00012607 | 0.927464445 | 0.001384878 | 0.076055492 | -9.36E-05 | 0.946114995 | 0.001384733 | 0.076035339 | -3.25E-05 | 0.00156375 | 1 | 0.988739396 | 1 | 0.990755659 | ENSG0000013 | 12 | ACLY |
| SAT | Heart_Failure | ENSG0000013 | 0.001853046 | 0.630573135 | 0.003853122 | 0.076055457 | 0.001763231 | 0.647197843 | 0.00385272 | 0.076035307 | 8.98E-05 | 0.001917047 | 1 | 0.959161083 | 1 | 0.95317119 | ENSG0000013 | 4 | NDUFA2 |
| SAT | Heart_Failure | ENSG0000013 | -0.00125532 | 0.534023329 | 0.002018608 | 0.076055433 | -0.00121933 | 0.545769155 | 0.00201839 | 0.076035283 | -3.60E-05 | 0.011184579 | 1 | 0.908781401 | 1 | 0.912597963 | ENSG0000013 | 4 | NDUFA2 |
| SAT | Heart_Failure | ENSG0000013 | -0.00179907 | 0.547520067 | 0.002991068 | 0.076055437 | -0.00191429 | 0.522126792 | 0.002990752 | 0.076035276 | 0.000115223 | 1.40E-07 | 1 | 0.917005278 | 1 | 0.904369708 | ENSG0000013 | 4 | NDUFA2 |
| SAT | Heart_Failure | ENSG0000013 | 0.006097639 | 0.021618152 | 0.002654589 | 0.07605467 | 0.006047499 | 0.022706038 | 0.00265433 | 0.07603453 | 5.01E-05 | 0.023870538 | 1 | 0.317837587 | 1 | 0.334074035 | ENSG0000013 | 10 | EXOC4 |
| SAT | Heart_Failure | ENSG0000013 | -0.00348099 | 0.17246318 | 0.00255142 | 0.076055203 | -0.00344554 | 0.176828437 | 0.002551146 | 0.076035056 | -3.54E-05 | 0.048663933 | 1 | 0.704583343 | 1 | 0.704659157 | ENSG0000013 | 10 | EXOC4 |
| SAT | Heart_Failure | ENSG0000013 | 0.001308006 | 0.372171101 | 0.001465693 | 0.076055369 | 0.001338752 | 0.360985391 | 0.001465535 | 0.07603521 | -3.07E-05 | 0.003187 | 1 | 0.857509398 | 1 | 0.848283315 | ENSG0000013 | 11 | PPFIA1 |
| SAT | Heart_Failure | ENSG0000013 | 0.002621343 | 0.347532062 | 0.00279048 | 0.076055355 | 0.00258085 | 0.354979756 | 0.002790186 | 0.076035207 | 4.05E-05 | 0.048922143 | 1 | 0.847141677 | 1 | 0.843366096 | ENSG0000013 | 11 | PPFIA1 |
| SAT | Heart_Failure | ENSG0000013 | -0.00058847 | 0.561262052 | 0.001012909 | 0.07605544 | -0.00056603 | 0.576249935 | 0.001012809 | 0.076035291 | -2.24E-05 | 0.006394324 | 1 | 0.924448189 | 1 | 0.9277115 | ENSG0000013 | 8 | TRAF7 |
| SAT | Heart_Failure | ENSG0000013 | -0.00182458 | 0.409808599 | 0.002213671 | 0.076055387 | -0.0017453 | 0.430405435 | 0.002213446 | 0.076035243 | -7.93E-05 | 5.66E-06 | 1 | 0.870429337 | 1 | 0.876213234 | ENSG0000013 | 8 | TRAF7 |
| SAT | Heart_Failure | ENSG0000013 | -0.00203029 | 0.427439125 | 0.002558392 | 0.076055395 | -0.00198147 | 0.43858837 | 0.002558119 | 0.076035247 | -4.88E-05 | 0.008004552 | 1 | 0.878482333 | 1 | 0.879941679 | ENSG0000013 | 13 | NPHP4 |
| SAT | Heart_Failure | ENSG0000013 | -0.00143664 | 0.560391754 | 0.002467376 | 0.07605544 | -0.00151957 | 0.537942483 | 0.002467123 | 0.076035281 | 8.29E-05 | 1.39E-05 | 1 | 0.923681045 | 1 | 0.909319572 | ENSG0000013 | 10 | MAP1B |
| SAT | Heart_Failure | ENSG0000013 | -0.00288508 | 0.36286915 | 0.003170722 | 0.076055364 | -0.00282258 | 0.373308472 | 0.003170391 | 0.076035216 | -6.25E-05 | 0.008621983 | 1 | 0.85040303 | 1 | 0.851164069 | ENSG0000013 | 10 | MAP1B |
| SAT | Heart_Failure | ENSG0000013 | 0.008929145 | 0.02110007 | 0.003871806 | 0.076054663 | 0.008839826 | 0.022409097 | 0.003871397 | 0.076034527 | 8.93E-05 | 0.001608556 | 1 | 0.315614662 | 1 | 0.332367548 | ENSG0000013 | 10 | MAP1B |
| SAT | Heart_Failure | ENSG0000013 | 0.003901635 | 0.077009462 | 0.002206419 | 0.076055005 | 0.003938211 | 0.074247498 | 0.00220617 | 0.076034843 | -3.66E-05 | 0.008887224 | 1 | 0.574245555 | 1 | 0.55673323 | ENSG0000013 | 8 | CLUHP3 |
| SAT | Heart_Failure | ENSG0000013 | 0.000763499 | 0.599224519 | 0.001452855 | 0.07605545 | 0.000744682 | 0.608215161 | 0.001452689 | 0.076035299 | 1.88E-05 | 0.033568863 | 1 | 0.944941904 | 1 | 0.943970645 | ENSG0000013 | 10 | ZNF304 |
| SAT | Heart_Failure | ENSG0000013 | -0.00172406 | 0.59699245 | 0.003260751 | 0.076055449 | -0.00179589 | 0.581762793 | 0.003260434 | 0.076035293 | 7.18E-05 | 0.008612201 | 1 | 0.944127995 | 1 | 0.931104255 | ENSG0000013 | 10 | ZNF304 |
| SAT | Heart_Failure | ENSG0000013 | 0.004892308 | 0.000623825 | 0.001430056 | 0.076053666 | 0.004854534 | 0.000686288 | 0.001429898 | 0.076033542 | 3.78E-05 | 6.76E-05 | 1 | 0.04264103 | 1 | 0.054919768 | ENSG0000013 | 10 | ZNF304 |
| SAT | Heart_Failure | ENSG0000013 | 0.000322683 | 0.928355779 | 0.003588804 | 0.076055492 | 0.000418844 | 0.907082101 | 0.003588452 | 0.076035338 | -9.62E-05 | 0.001205885 | 1 | 0.988739396 | 1 | 0.982067422 | ENSG0000013 | 10 | ZNF132 |
| SAT | Heart_Failure | ENSG0000013 | -0.00094887 | 0.795891609 | 0.003668305 | 0.076055483 | -0.00083368 | 0.820198082 | 0.003667902 | 0.076035332 | -0.00011519 | 2.80E-06 | 1 | 0.976456935 | 1 | 0.972791643 | ENSG0000013 | 10 | ZNF132 |
| SAT | Heart_Failure | ENSG0000013 | 0.000996441 | 0.738874315 | 0.002989223 | 0.076055476 | 0.000953978 | 0.749594204 | 0.00298889 | 0.076035324 | 4.25E-05 | 0.02831516 | 1 | 0.966658386 | 1 | 0.962415493 | ENSG0000013 | 10 | ZNF132 |
| SAT | Heart_Failure | ENSG0000013 | -0.00098318 | 0.195628208 | 0.000759733 | 0.076055232 | -0.00099587 | 0.189872484 | 0.000759654 | 0.076035072 | 1.27E-05 | 0.024043555 | 1 | 0.726878652 | 1 | 0.720122525 | ENSG0000013 | 10 | ZNF132 |
| SAT | Heart_Failure | ENSG0000013 | 0.000121969 | 0.946883031 | 0.001830779 | 0.076055492 | 8.28E-05 | 0.963933143 | 0.001830592 | 0.07603534 | 3.92E-05 | 0.005603878 | 1 | 0.992414898 | 1 | 0.996055789 | ENSG0000013 | 3 | RHPN2 |
| SAT | Heart_Failure | ENSG0000013 | 0.002092657 | 0.285268881 | 0.001958392 | 0.076055315 | 0.002147171 | 0.272855335 | 0.001958183 | 0.076035153 | -5.45E-05 | 0.000122165 | 1 | 0.800157 | 1 | 0.783093703 | ENSG0000013 | 13 | C19orf12 |
| SAT | Heart_Failure | ENSG0000013 | 0.000319409 | 0.840205495 | 0.001584127 | 0.076055487 | 0.001122452 | 0.484846162 | 0.001606877 | 0.076035264 | -0.00080304 | 0.003008225 | 1 | 0.976669362 | 1 | 0.894608312 | ENSG0000013 | 13 | C19orf12 |
| SAT | Heart_Failure | ENSG0000013 | 0.003227854 | 0.232761422 | 0.002705033 | 0.076055271 | 0.003275541 | 0.225880554 | 0.002704733 | 0.076035111 | -4.77E-05 | 0.007655861 | 1 | 0.753146179 | 1 | 0.747837725 | ENSG0000013 | 5 | C19orf40 |
| SAT | Heart_Failure | ENSG0000013 | -0.00161826 | 0.526928641 | 0.002557702 | 0.076055431 | -0.0016951 | 0.507453146 | 0.002557459 | 0.076035272 | 7.68E-05 | 0.000551649 | 1 | 0.906724424 | 1 | 0.901005489 | ENSG0000013 | 5 | C19orf40 |
| SAT | Heart_Failure | ENSG0000013 | -0.00827591 | 0.017783664 | 0.003491799 | 0.076054616 | -0.01193291 | 0.001553435 | 0.003770832 | 0.076033778 | 0.003656996 | 0.010264159 | 1 | 0.290924976 | 1 | 0.09221864 | ENSG0000013 | 3 | DNAJB1 |
| SAT | Heart_Failure | ENSG0000013 | 0.006788396 | 0.002646539 | 0.002258223 | 0.076054083 | 0.009188357 | 6.79E-05 | 0.002306666 | 0.076032864 | -0.00239996 | 3.63E-07 | 1 | 0.113128813 | 0.222937851 | 0.011955478 | ENSG0000013 | 8 | GAS2L2 |
| SAT | Heart_Failure | ENSG0000013 | -0.00220135 | 0.501372287 | 0.003274203 | 0.076055422 | -0.00027891 | 0.932271156 | 0.003281743 | 0.076035339 | -0.00192245 | 4.12E-17 | 1 | 0.900549813 | 1 | 0.988742522 | ENSG0000013 | 8 | GAS2L2 |
| SAT | Heart_Failure | ENSG0000013 | -0.00049669 | 0.463089465 | 0.000676898 | 0.076055409 | -0.00116094 | 0.095017936 | 0.000695381 | 0.076034905 | 0.000664255 | 3.17E-05 | 1 | 0.890529906 | 1 | 0.60300551 | ENSG0000013 | 8 | GAS2L2 |
| SAT | Heart_Failure | ENSG0000013 | -0.00013957 | 0.961197655 | 0.002868817 | 0.076055493 | -0.00020896 | 0.941929284 | 0.002868498 | 0.076035339 | 6.94E-05 | 0.000215938 | 1 | 0.995482799 | 1 | 0.988983601 | ENSG0000013 | 9 | CCT6B |
| SAT | Heart_Failure | ENSG0000013 | -0.0008651 | 0.679616997 | 0.002094747 | 0.076055466 | -0.00089767 | 0.668230669 | 0.00209452 | 0.076035311 | 3.26E-05 | 0.025643925 | 1 | 0.966658386 | 1 | 0.956878334 | ENSG0000013 | 9 | CCT6B |
| SAT | Heart_Failure | ENSG0000013 | -0.0016693 | 0.159460518 | 0.001186517 | 0.076055184 | -0.00169418 | 0.153291078 | 0.001186394 | 0.076035022 | 2.49E-05 | 0.00592776 | 1 | 0.678956178 | 1 | 0.667541522 | ENSG0000013 | 9 | CCT6B |
| SAT | Heart_Failure | ENSG0000013 | 0.004298671 | 0.204911676 | 0.003390966 | 0.076055242 | 0.004359535 | 0.19852749 | 0.003390629 | 0.076035082 | -6.09E-05 | 0.027428964 | 1 | 0.735350411 | 1 | 0.725655076 | ENSG0000013 | 9 | CCT6B |
| SAT | Heart_Failure | ENSG0000013 | -0.00159981 | 0.353943882 | 0.001725855 | 0.076055359 | -0.00156664 | 0.363959481 | 0.001725666 | 0.076035212 | -3.32E-05 | 0.004789246 | 1 | 0.848363544 | 1 | 0.849290727 | ENSG0000013 | 14 | ENOSF1 |
| SAT | Heart_Failure | ENSG0000013 | -0.00250708 | 0.141401379 | 0.001704806 | 0.076055155 | -0.00253264 | 0.13734576 | 0.001704623 | 0.076034996 | 2.56E-05 | 0.033909524 | 1 | 0.669181587 | 1 | 0.649215182 | ENSG0000013 | 6 | EMILIN2 |
| SAT | Heart_Failure | ENSG0000013 | 0.002487681 | 0.361896327 | 0.002728443 | 0.076055363 | 0.002444701 | 0.37019983 | 0.002728156 | 0.076035215 | 4.30E-05 | 0.033300014 | 1 | 0.85040303 | 1 | 0.84956238 | ENSG0000013 | 6 | EMILIN2 |
| SAT | Heart_Failure | ENSG0000013 | 0.004847633 | 0.21444437 | 0.003904851 | 0.076055252 | 0.004915429 | 0.208057932 | 0.003904464 | 0.076035093 | -6.78E-05 | 0.034006927 | 1 | 0.745086527 | 1 | 0.733958243 | ENSG0000013 | 6 | EMILIN2 |
| SAT | Heart_Failure | ENSG0000013 | 0.003491213 | 0.15829016 | 0.00247455 | 0.076055182 | 0.003443714 | 0.163985335 | 0.002474298 | 0.076035038 | 4.75E-05 | 0.014532669 | 1 | 0.67888891 | 1 | 0.679583582 | ENSG0000013 | 6 | EMILIN2 |
| SAT | Heart_Failure | ENSG0000013 | 8.31E-05 | 0.973020798 | 0.002457826 | 0.076055493 | 1.14E-06 | 0.999628555 | 0.002457813 | 0.07603534 | 8.20E-05 | 0.036399614 | 1 | 0.99766005 | 1 | 0.999768899 | ENSG0000013 | 11 | TRIM22 |
| SAT | Heart_Failure | ENSG0000013 | 0.003731137 | 0.09030693 | 0.002202841 | 0.076055045 | 0.003691977 | 0.09370287 | 0.002202609 | 0.076034902 | 3.92E-05 | 0.015989281 | 1 | 0.598579873 | 1 | 0.600153846 | ENSG0000013 | 11 | TRIM22 |
| SAT | Heart_Failure | ENSG0000013 | 0.002456812 | 0.020440504 | 0.001059807 | 0.076054654 | 0.002434757 | 0.021584809 | 0.001059694 | 0.076034516 | 2.21E-05 | 0.004025303 | 1 | 0.312755508 | 1 | 0.326358332 | ENSG0000013 | 11 | TRIM22 |
| SAT | Heart_Failure | ENSG0000013 | 0.001168676 | 0.648114392 | 0.002560732 | 0.076055461 | 0.001232486 | 0.630264984 | 0.002560455 | 0.076035304 | -6.38E-05 | 0.000360717 | 1 | 0.962760695 | 1 | 0.94798204 | ENSG0000013 | 7 | TIMM10B |
| SAT | Heart_Failure | ENSG0000013 | -0.0015073 | 0.13753994 | 0.001015006 | 0.076055149 | -0.00152539 | 0.132839397 | 0.001014895 | 0.076034988 | 1.81E-05 | 0.008657954 | 1 | 0.663104757 | 1 | 0.649215182 | ENSG0000013 | 7 | TIMM10B |
| SAT | Heart_Failure | ENSG0000013 | 0.000835873 | 0.693735191 | 0.002122628 | 0.076055469 | 0.000880582 | 0.678217621 | 0.002122411 | 0.076035313 | -4.47E-05 | 0.006844058 | 1 | 0.966658386 | 1 | 0.956878334 | ENSG0000013 | 16 | IMMT |
| SAT | Heart_Failure | ENSG0000013 | 0.005088956 | 0.022053355 | 0.00222278 | 0.076054675 | 0.004957127 | 0.025727198 | 0.002222616 | 0.076034564 | 0.000131829 | 3.91E-08 | 1 | 0.321586925 | 1 | 0.354978312 | ENSG0000013 | 16 | IMMT |
| SAT | Heart_Failure | ENSG0000013 | -0.00041318 | 0.856955419 | 0.002292238 | 0.076055488 | -0.000271 | 0.905893137 | 0.002292363 | 0.076035338 | -0.00014217 | 0.001356933 | 1 | 0.976669362 | 1 | 0.982067422 | ENSG0000013 | 16 | IMMT |
| SAT | Heart_Failure | ENSG0000013 | 0.003555405 | 0.177836375 | 0.00263862 | 0.07605521 | 0.00363043 | 0.168818529 | 0.002638371 | 0.076035045 | -7.50E-05 | 0.001176145 | 1 | 0.713302136 | 1 | 0.688922379 | ENSG0000013 | 16 | IMMT |
| SAT | Heart_Failure | ENSG0000013 | 0.000782378 | 0.759713899 | 0.002557988 | 0.076055478 | 0.000733065 | 0.774411635 | 0.002557715 | 0.076035327 | 4.93E-05 | 0.007018546 | 1 | 0.969137365 | 1 | 0.965963214 | ENSG0000013 | 16 | IMMT |
| SAT | Heart_Failure | ENSG0000013 | -0.00156001 | 0.290673673 | 0.001476379 | 0.076055319 | -0.00149103 | 0.312595991 | 0.00147657 | 0.076035181 | -6.90E-05 | 0.041110345 | 1 | 0.803109301 | 1 | 0.821155716 | ENSG0000013 | 16 | IMMT |
| SAT | Heart_Failure | ENSG0000013 | 0.000212177 | 0.951449395 | 0.003484771 | 0.076055492 | 0.000520337 | 0.881392418 | 0.003487409 | 0.076035337 | -0.00030816 | 0.036050016 | 1 | 0.994131568 | 1 | 0.974665278 | ENSG0000013 | 10 | MRPL35 |
| SAT | Heart_Failure | ENSG0000013 | 0.000872849 | 0.624500276 | 0.001783207 | 0.076055456 | 0.000814418 | 0.647842656 | 0.001783026 | 0.076035308 | 5.84E-05 | 3.16E-05 | 1 | 0.955683491 | 1 | 0.95317119 | ENSG0000013 | 10 | MRPL35 |
| SAT | Heart_Failure | ENSG0000013 | -0.00154233 | 0.424065006 | 0.00192938 | 0.076055393 | -0.00157214 | 0.415113448 | 0.001929177 | 0.076035237 | 2.98E-05 | 0.037095119 | 1 | 0.876565321 | 1 | 0.867451005 | ENSG0000013 | 10 | MRPL35 |
| SAT | Heart_Failure | ENSG0000013 | -0.00256924 | 0.424256547 | 0.00321533 | 0.076055393 | -0.00261765 | 0.415528865 | 0.003214987 | 0.076035237 | 4.84E-05 | 0.035868219 | 1 | 0.876565321 | 1 | 0.867451005 | ENSG0000013 | 12 | SCLY |
| SAT | Heart_Failure | ENSG0000013 | -0.00077 | 0.739078226 | 0.002311785 | 0.076055476 | -0.00070769 | 0.759485694 | 0.002311547 | 0.076035326 | -6.23E-05 | 0.000464305 | 1 | 0.966658386 | 1 | 0.963228667 | ENSG0000013 | 12 | SCLY |
| SAT | Heart_Failure | ENSG0000013 | 0.000241298 | 0.893211092 | 0.001797481 | 0.07605549 | 0.000275295 | 0.878263317 | 0.001797304 | 0.076035336 | -3.40E-05 | 0.022315337 | 1 | 0.979270459 | 1 | 0.974665278 | ENSG0000013 | 10 | MYBBP1A |
| SAT | Heart_Failure | ENSG0000013 | -0.00548845 | 0.07904742 | 0.003125108 | 0.076055012 | -0.00561357 | 0.072436925 | 0.003124959 | 0.076034837 | 0.000125118 | 0.002102349 | 1 | 0.580653917 | 1 | 0.545104471 | ENSG0000013 | 10 | MYBBP1A |
| SAT | Heart_Failure | ENSG0000013 | -0.00091827 | 0.703538405 | 0.002413024 | 0.07605547 | -0.000866 | 0.719648869 | 0.00241275 | 0.07603532 | -5.23E-05 | 0.000415436 | 1 | 0.966658386 | 1 | 0.958028875 | ENSG0000013 | 7 | RPA1 |
| SAT | Heart_Failure | ENSG0000014 | 0.002230588 | 0.441821696 | 0.002900177 | 0.076055401 | 0.002278753 | 0.431976688 | 0.002899863 | 0.076035244 | -4.82E-05 | 0.016898785 | 1 | 0.882529682 | 1 | 0.876305924 | ENSG0000013 | 7 | TMEM128 |
| SAT | Heart_Failure | ENSG0000014 | -0.00024586 | 0.911269801 | 0.002206309 | 0.076055491 | -0.00027443 | 0.901000911 | 0.002206062 | 0.076035338 | 2.86E-05 | 0.041693186 | 1 | 0.984491733 | 1 | 0.980373283 | ENSG0000013 | 9 | SEC61G |
| SAT | Heart_Failure | ENSG0000014 | 0.001255656 | 0.518480199 | 0.001944672 | 0.076055428 | 0.001284892 | 0.508744692 | 0.001944466 | 0.076035272 | -2.92E-05 | 0.039625393 | 1 | 0.906724424 | 1 | 0.902291622 | ENSG0000013 | 9 | SEC61G |
| SAT | Heart_Failure | ENSG0000014 | -0.00315954 | 0.022563671 | 0.001385309 | 0.076054681 | -0.00319699 | 0.020998974 | 0.001385172 | 0.076034509 | 3.74E-05 | 0.000958115 | 1 | 0.324699143 | 1 | 0.320454116 | ENSG0000013 | 9 | SEC61G |
| SAT | Heart_Failure | ENSG0000014 | 0.004021842 | 0.086409569 | 0.002345573 | 0.076055034 | 0.003967805 | 0.090684173 | 0.002345312 | 0.076034894 | 5.40E-05 | 0.000394643 | 1 | 0.592308253 | 1 | 0.598355674 | ENSG0000013 | 13 | EIF5A |

| | | | | | | | | | | | | | | | | | | | |
|---|---|---|---|---|---|---|---|---|---|---|---|---|---|---|---|---|---|---|---|
| SAT | Heart_Failure | ENSG0000014 | -0.00368711 | 0.285887906 | 0.003454979 | 0.076055315 | -0.00379611 | 0.271835039 | 0.003454629 | 0.076035152 | 0.000109002 | 6.37E-05 | 1 | 0.800157 | 1 | 0.783093703 | ENSG0000013 | 13 | EIF5A |
| SAT | Heart_Failure | ENSG0000014 | 0.001326292 | 0.265779784 | 0.001191813 | 0.0760553 | 0.001301686 | 0.274697147 | 0.001191681 | 0.076035154 | 2.46E-05 | 0.001783254 | 1 | 0.777897832 | 1 | 0.784122993 | ENSG0000013 | 13 | EIF5A |
| SAT | Heart_Failure | ENSG0000014 | 0.003432469 | 0.038860594 | 0.001661677 | 0.076054827 | 0.00347367 | 0.036556936 | 0.001661499 | 0.076034658 | -4.12E-05 | 0.000564825 | 1 | 0.445276442 | 1 | 0.42836895 | ENSG0000013 | 11 | REEP2 |
| SAT | Heart_Failure | ENSG0000014 | -0.00136764 | 0.618167976 | 0.002743801 | 0.076055454 | -0.00131189 | 0.632520856 | 0.002743948 | 0.076035304 | -5.58E-05 | 0.002138568 | 1 | 0.954216727 | 1 | 0.94798204 | ENSG0000013 | 11 | REEP2 |
| SAT | Heart_Failure | ENSG0000014 | -0.00013697 | 0.947633875 | 0.002085458 | 0.076055492 | -8.06E-05 | 0.969173689 | 0.002085259 | 0.07603534 | -5.64E-05 | 0.001729699 | 1 | 0.992414898 | 1 | 0.99755193 | ENSG0000013 | 12 | PRMT7 |
| SAT | Heart_Failure | ENSG0000014 | -0.00123046 | 0.633886662 | 0.002583569 | 0.076055458 | -0.00130057 | 0.614645914 | 0.002583292 | 0.076035301 | 7.01E-05 | 0.001735733 | 1 | 0.959428651 | 1 | 0.945897394 | ENSG0000013 | 12 | PRMT7 |
| SAT | Heart_Failure | ENSG0000014 | -0.00012933 | 0.926300306 | 0.001398191 | 0.076055492 | -0.00010179 | 0.941960912 | 0.001398047 | 0.076035339 | -2.75E-05 | 0.010540886 | 1 | 0.988739396 | 1 | 0.988983601 | ENSG0000013 | 10 | MTSS1L |
| SAT | Heart_Failure | ENSG0000014 | 0.000153751 | 0.933741656 | 0.001849336 | 0.076055492 | 0.000200558 | 0.91363004 | 0.001849127 | 0.076035338 | -4.68E-05 | 5.25E-05 | 1 | 0.989642217 | 1 | 0.98402346 | ENSG0000013 | 12 | RBM38 |
| SAT | Heart_Failure | ENSG0000014 | -0.0032748 | 0.208805887 | 0.00260555 | 0.076055246 | -0.00319426 | 0.220172197 | 0.002605275 | 0.076035106 | -8.05E-05 | 2.64E-05 | 1 | 0.739840299 | 1 | 0.742028061 | ENSG0000013 | 9 | SERINC3 |
| SAT | Heart_Failure | ENSG0000014 | -0.00324045 | 0.360102267 | 0.003540811 | 0.076055362 | -0.00314747 | 0.374000047 | 0.003540432 | 0.076035217 | -9.30E-05 | 0.000240778 | 1 | 0.85040303 | 1 | 0.852148719 | ENSG0000013 | 9 | SERINC3 |
| SAT | Heart_Failure | ENSG0000014 | 0.001787314 | 0.617673204 | 0.003580708 | 0.076055454 | 0.001876281 | 0.600241298 | 0.00358032 | 0.076035297 | -8.90E-05 | 0.000363081 | 1 | 0.954216727 | 1 | 0.938338302 | ENSG0000013 | 9 | SERINC3 |
| SAT | Heart_Failure | ENSG0000014 | 0.003733222 | 0.055213631 | 0.00194723 | 0.076054919 | 0.003641023 | 0.06148105 | 0.001947046 | 0.076034794 | 9.22E-05 | 5.09E-08 | 1 | 0.503210899 | 1 | 0.505632969 | ENSG0000013 | 10 | DMGDH |
| SAT | Heart_Failure | ENSG0000014 | -0.00085924 | 0.707735949 | 0.002291932 | 0.07605471 | -0.0009265 | 0.686005973 | 0.002291709 | 0.076035315 | 6.73E-05 | 0.000473176 | 1 | 0.966658386 | 1 | 0.956878334 | ENSG0000013 | 10 | DMGDH |
| SAT | Heart_Failure | ENSG0000014 | -0.00058428 | 0.876344981 | 0.003754893 | 0.076055489 | -0.00065452 | 0.861606984 | 0.003754488 | 0.076035335 | 7.02E-05 | 0.00783878 | 1 | 0.979223495 | 1 | 0.972791643 | ENSG0000013 | 10 | DMGDH |
| SAT | Heart_Failure | ENSG0000014 | -0.00313336 | 0.220104195 | 0.002555228 | 0.076055258 | -0.00319102 | 0.211686363 | 0.002554974 | 0.076035097 | 5.77E-05 | 0.005390615 | 1 | 0.749908476 | 1 | 0.733958243 | ENSG0000013 | 10 | DMGDH |
| SAT | Heart_Failure | ENSG0000014 | -0.00550293 | 0.110301778 | 0.003446134 | 0.076055095 | -0.00544209 | 0.114254001 | 0.003445753 | 0.07603495 | -6.08E-05 | 0.007544093 | 1 | 0.640599412 | 1 | 0.64481576 | ENSG0000013 | 5 | BHMT2 |
| SAT | Heart_Failure | ENSG0000014 | -0.00111087 | 0.402132614 | 0.001325901 | 0.076055383 | -0.00106405 | 0.422210187 | 0.001325761 | 0.07603524 | -4.68E-05 | 1.56E-06 | 1 | 0.870429337 | 1 | 0.873390936 | ENSG0000013 | 5 | BHMT2 |
| SAT | Heart_Failure | ENSG0000014 | -0.00091676 | 0.543176713 | 0.001507787 | 0.076055435 | -0.00088783 | 0.555937474 | 0.001507647 | 0.076035286 | -2.89E-05 | 0.030702589 | 1 | 0.913779466 | 1 | 0.915531921 | ENSG0000013 | 5 | BHMT2 |
| SAT | Heart_Failure | ENSG0000014 | 0.001564207 | 0.42833476 | 0.001974896 | 0.076055395 | 0.00160946 | 0.415050065 | 0.001974701 | 0.076035236 | -4.53E-05 | 0.005256542 | 1 | 0.878482333 | 1 | 0.867451005 | ENSG0000013 | 7 | RSG1 |
| SAT | Heart_Failure | ENSG0000014 | -0.00341284 | 0.031401426 | 0.001585926 | 0.07605477 | -0.0033773 | 0.033189886 | 0.001585747 | 0.076034632 | -3.55E-05 | 0.000364248 | 1 | 0.396261842 | 1 | 0.407850241 | ENSG0000013 | 13 | CASP9 |
| SAT | Heart_Failure | ENSG0000014 | -0.00061927 | 0.806110058 | 0.002523033 | 0.076055484 | -0.00050704 | 0.840710817 | 0.002522777 | 0.076035334 | -0.00011223 | 1.38E-08 | 1 | 0.976669362 | 1 | 0.972791643 | ENSG0000013 | 13 | CASP9 |
| SAT | Heart_Failure | ENSG0000014 | 0.000380754 | 0.708100411 | 0.001016949 | 0.076055471 | 0.000398895 | 0.694843431 | 0.001016834 | 0.076035316 | -1.81E-05 | 0.004689622 | 1 | 0.966658386 | 1 | 0.956878334 | ENSG0000013 | 13 | CASP9 |
| SAT | Heart_Failure | ENSG0000014 | 0.003219621 | 0.400499077 | 0.00382955 | 0.076055383 | 0.00329899 | 0.388936754 | 0.003829142 | 0.076035224 | -7.94E-05 | 0.004148974 | 1 | 0.870429337 | 1 | 0.858921514 | ENSG0000013 | 13 | CASP9 |
| SAT | Heart_Failure | ENSG0000014 | 0.00108751 | 0.730021116 | 0.00315131 | 0.076055474 | 0.001142402 | 0.716935623 | 0.003150954 | 0.07603532 | -5.49E-05 | 0.005651272 | 1 | 0.966658386 | 1 | 0.958028875 | ENSG0000013 | 6 | SCO1 |
| SAT | Heart_Failure | ENSG0000014 | -0.00192251 | 0.100553412 | 0.001170704 | 0.076055072 | -0.00251902 | 0.031805324 | 0.001173352 | 0.076034621 | 0.000596509 | 1.89E-13 | 1 | 0.622495351 | 1 | 0.401948008 | ENSG0000013 | 8 | CHI3L1 |
| SAT | Heart_Failure | ENSG0000014 | -3.29E-06 | 0.998951098 | 0.002502299 | 0.076055493 | -4.59E-05 | 0.985375884 | 0.002502027 | 0.07603534 | 4.26E-05 | 0.013615371 | 1 | 0.999875583 | 1 | 0.999768899 | ENSG0000013 | 8 | CHI3L1 |
| SAT | Heart_Failure | ENSG0000014 | 0.006561862 | 0.029681029 | 0.003017881 | 0.076054755 | 0.006497797 | 0.031294237 | 0.003017575 | 0.076034617 | 6.41E-05 | 0.007044907 | 1 | 0.380908976 | 1 | 0.397970509 | ENSG0000013 | 12 | DSTYK |
| SAT | Heart_Failure | ENSG0000014 | 0.005066722 | 0.024680375 | 0.002255509 | 0.076054705 | 0.005146585 | 0.022488752 | 0.002255279 | 0.076034528 | -7.99E-05 | 5.78E-06 | 1 | 0.336323411 | 1 | 0.332367548 | ENSG0000013 | 12 | DSTYK |
| SAT | Heart_Failure | ENSG0000014 | -0.00096251 | 0.743576439 | 0.002942359 | 0.076055476 | -0.0005509 | 0.851680513 | 0.00294639 | 0.076035335 | -0.00041161 | 0.01074267 | 1 | 0.966658386 | 1 | 0.972791643 | ENSG0000013 | 10 | TMCC2 |
| SAT | Heart_Failure | ENSG0000014 | 0.002210548 | 0.481804685 | 0.003142646 | 0.076055416 | 0.00308132 | 0.330265997 | 0.00316494 | 0.076035192 | -0.00087077 | 0.02140778 | 1 | 0.897349195 | 1 | 0.834182246 | ENSG0000013 | 10 | TMCC2 |
| SAT | Heart_Failure | ENSG0000014 | -0.00732463 | 0.025883812 | 0.003287598 | 0.076054718 | -0.00665302 | 0.043926481 | 0.003302081 | 0.076034707 | -0.00067161 | 0.03217013 | 1 | 0.348052412 | 1 | 0.456297071 | ENSG0000013 | 4 | BTBD2 |
| SAT | Heart_Failure | ENSG0000014 | 0.000509131 | 0.823620489 | 0.002284214 | 0.076055485 | 0.000594315 | 0.794703903 | 0.002284003 | 0.07603533 | -8.52E-05 | 3.11E-05 | 1 | 0.976669362 | 1 | 0.970635778 | ENSG0000013 | 11 | WDR74 |
| SAT | Heart_Failure | ENSG0000014 | 0.000211928 | 0.826820377 | 0.000968675 | 0.076055486 | 0.000539943 | 0.580793851 | 0.000977758 | 0.076035293 | -0.00032801 | 0.014293121 | 1 | 0.976669362 | 1 | 0.930810832 | ENSG0000013 | 11 | WDR74 |
| SAT | Heart_Failure | ENSG0000014 | -0.00236398 | 0.35694446 | 0.002566183 | 0.076055361 | -0.00230545 | 0.368930818 | 0.002565943 | 0.076035214 | -5.85E-05 | 0.009653062 | 1 | 0.848363544 | 1 | 0.849425432 | ENSG0000013 | 11 | WDR74 |
| SAT | Heart_Failure | ENSG0000014 | -0.00534241 | 0.00066766 | 0.001570121 | 0.076053686 | -0.00529124 | 0.000751165 | 0.001570003 | 0.076033568 | -5.12E-05 | 0.002341759 | 1 | 0.04470599 | 1 | 0.05868032 | ENSG0000013 | 6 | GSTT2B |
| SAT | Heart_Failure | ENSG0000014 | 0.009909371 | 0.001400976 | 0.003102041 | 0.0760539 | 0.009976148 | 0.001298539 | 0.003101726 | 0.076033726 | -6.68E-05 | 0.006467385 | 1 | 0.077908515 | 1 | 0.081932833 | ENSG0000013 | 15 | SLC2A11 |
| SAT | Heart_Failure | ENSG0000014 | -0.00028595 | 0.93474588 | 0.003492562 | 0.076055492 | -0.00021506 | 0.950895938 | 0.003492237 | 0.07603534 | -7.09E-05 | 0.022228178 | 1 | 0.989642217 | 1 | 0.990755659 | ENSG0000013 | 8 | ZDHHC8P1 |
| SAT | Heart_Failure | ENSG0000014 | -0.00257443 | 0.205073339 | 0.00203154 | 0.076055242 | -0.00263687 | 0.194258107 | 0.002031349 | 0.076035077 | 6.24E-05 | 0.000444163 | 1 | 0.735350411 | 1 | 0.723506187 | ENSG0000013 | 5 | ADCK2 |
| SAT | Heart_Failure | ENSG0000014 | -0.00313489 | 0.046362711 | 0.001573677 | 0.076054874 | -0.00316222 | 0.044468723 | 0.001573516 | 0.07603471 | 2.73E-05 | 0.025304228 | 1 | 0.475002578 | 1 | 0.456297071 | ENSG0000013 | 5 | ADCK2 |
| SAT | Heart_Failure | ENSG0000014 | -0.00702789 | 0.05494654 | 0.003661686 | 0.076054918 | -0.00692416 | 0.058600476 | 0.003661284 | 0.076034782 | -0.00010373 | 2.50E-05 | 1 | 0.502171577 | 1 | 0.503576399 | ENSG0000013 | 11 | SFTPD |
| SAT | Heart_Failure | ENSG0000014 | 0.003073219 | 0.087115418 | 0.001796345 | 0.076055036 | 0.003040811 | 0.090464736 | 0.001796156 | 0.076034893 | 3.24E-05 | 0.014685212 | 1 | 0.592308253 | 1 | 0.598355674 | ENSG0000013 | 11 | SFTPD |
| SAT | Heart_Failure | ENSG0000014 | -0.00191888 | 0.608122007 | 0.003742285 | 0.076055452 | -0.00179161 | 0.632093003 | 0.003742009 | 0.076035304 | -0.00012727 | 0.001660364 | 1 | 0.949666018 | 1 | 0.94798204 | ENSG0000013 | 11 | SFTPD |
| SAT | Heart_Failure | ENSG0000014 | -0.00124551 | 0.476113216 | 0.001747912 | 0.076055414 | -0.00129287 | 0.459463423 | 0.00174775 | 0.076035255 | 4.74E-05 | 0.00242847 | 1 | 0.895715287 | 1 | 0.890430886 | ENSG0000013 | 11 | SFTPD |
| SAT | Heart_Failure | ENSG0000014 | -0.00174827 | 0.058904451 | 0.000925542 | 0.076054936 | -0.00171901 | 0.063241996 | 0.000925447 | 0.076034802 | -2.93E-05 | 4.31E-05 | 1 | 0.507258544 | 1 | 0.511069956 | ENSG0000013 | 13 | LARS |
| SAT | Heart_Failure | ENSG0000014 | -0.00294933 | 0.056373051 | 0.001545652 | 0.076054925 | -0.00290154 | 0.060460731 | 0.001545493 | 0.07603479 | -4.78E-05 | 6.02E-05 | 1 | 0.503931718 | 1 | 0.50513473 | ENSG0000013 | 13 | LARS |
| SAT | Heart_Failure | ENSG0000014 | -0.00255717 | 0.298404716 | 0.002459143 | 0.076055324 | -0.00242131 | 0.324772311 | 0.002458931 | 0.076035189 | -0.00013586 | 9.07E-09 | 1 | 0.814547972 | 1 | 0.828598718 | ENSG0000013 | 13 | LARS |
| SAT | Heart_Failure | ENSG0000014 | 0.002416407 | 0.128590746 | 0.00159007 | 0.076055133 | 0.002442792 | 0.12442996 | 0.001589899 | 0.076034972 | -2.64E-05 | 0.018471127 | 1 | 0.649888404 | 1 | 0.647874406 | ENSG0000013 | 13 | LARS |
| SAT | Heart_Failure | ENSG0000014 | 0.000364108 | 0.864212822 | 0.002129111 | 0.076055488 | 0.000408033 | 0.848005373 | 0.002128896 | 0.076035334 | -4.39E-05 | 0.009250901 | 1 | 0.978183841 | 1 | 0.972791643 | ENSG0000013 | 5 | IMPA1 |
| SAT | Heart_Failure | ENSG0000014 | -0.00596089 | 0.000503146 | 0.001713344 | 0.076053604 | -0.00599157 | 0.00046992 | 0.001713167 | 0.076033432 | 3.07E-05 | 0.02024726 | 1 | 0.03682027 | 1 | 0.039533553 | ENSG0000013 | 5 | IMPA1 |
| SAT | Heart_Failure | ENSG0000014 | -0.00037578 | 0.904348198 | 0.00312707 | 0.076055491 | -0.00046128 | 0.882715936 | 0.003126738 | 0.076035337 | 8.55E-05 | 0.000162134 | 1 | 0.981517198 | 1 | 0.974665278 | ENSG0000013 | 11 | LRRCC1 |
| SAT | Heart_Failure | ENSG0000014 | 0.014512329 | 7.15E-06 | 0.00323277 | 0.076052348 | 0.014444087 | 7.88E-06 | 0.00323242 | 0.076032225 | 6.82E-05 | 0.002414828 | 0.023469109 | 0.001235216 | 0.025849598 | 0.00258496 | ENSG0000013 | 11 | LRRCC1 |
| SAT | Heart_Failure | ENSG0000014 | 0.001032381 | 0.685598419 | 0.002550123 | 0.076055467 | 0.001125304 | 0.658986048 | 0.002549897 | 0.07603531 | -9.29E-05 | 9.84E-05 | 1 | 0.966658386 | 1 | 0.956791995 | ENSG0000013 | 9 | MICAL2 |
| SAT | Heart_Failure | ENSG0000014 | 0.000317127 | 0.893425721 | 0.002367132 | 0.07605549 | -0.00054682 | 0.818564372 | 0.002383783 | 0.076035332 | 0.000863947 | 0.0023402 | 1 | 0.979270459 | 1 | 0.972791643 | ENSG0000013 | 9 | MICAL2 |
| SAT | Heart_Failure | ENSG0000014 | 0.001126478 | 0.524869196 | 0.001771588 | 0.07605543 | 0.001179692 | 0.505433333 | 0.001771401 | 0.07603527 | -5.32E-05 | 4.60E-05 | 1 | 0.906724424 | 1 | 0.900571724 | ENSG0000013 | 9 | MICAL2 |
| SAT | Heart_Failure | ENSG0000014 | -0.00401682 | 0.271057678 | 0.003649557 | 0.076055304 | -0.00391444 | 0.283412172 | 0.003649188 | 0.076035161 | -0.00010238 | 0.00039441 | 1 | 0.784141183 | 1 | 0.795445114 | ENSG0000013 | 9 | MICAL2 |
| SAT | Heart_Failure | ENSG0000014 | -0.00327988 | 0.291326864 | 0.003108256 | 0.076055319 | -0.0032203 | 0.300126341 | 0.003107908 | 0.076035173 | -5.96E-05 | 0.002776039 | 1 | 0.803905332 | 1 | 0.810520426 | ENSG0000013 | 5 | DPF2 |
| SAT | Heart_Failure | ENSG0000014 | -0.00093704 | 0.802395962 | 0.00374445 | 0.076055483 | -0.00087989 | 0.814201444 | 0.00374405 | 0.076035332 | -5.72E-05 | 0.033361558 | 1 | 0.976669362 | 1 | 0.972791643 | ENSG0000013 | 5 | DPF2 |
| SAT | Heart_Failure | ENSG0000014 | -0.00117462 | 0.696001111 | 0.003006249 | 0.076055469 | -0.0012257 | 0.683450414 | 0.003005942 | 0.076035314 | 5.11E-05 | 0.029291942 | 1 | 0.966658386 | 1 | 0.956878334 | ENSG0000013 | 5 | DPF2 |
| SAT | Heart_Failure | ENSG0000014 | -0.00161967 | 0.378699903 | 0.001839916 | 0.076055372 | -0.00059528 | 0.750044625 | 0.001868531 | 0.076035324 | -0.00102439 | 0.001739174 | 1 | 0.85759885 | 1 | 0.962415493 | ENSG0000013 | 10 | COX16 |
| SAT | Heart_Failure | ENSG0000014 | 0.000190906 | 0.93206895 | 0.002239576 | 0.076055492 | 0.000136061 | 0.951550731 | 0.002239336 | 0.07603534 | 5.48E-05 | 0.000591048 | 1 | 0.989362092 | 1 | 0.991047336 | ENSG0000013 | 10 | COX16 |
| SAT | Heart_Failure | ENSG0000014 | 0.002411406 | 0.347220339 | 0.002565338 | 0.076055355 | 0.002456581 | 0.338214359 | 0.002565083 | 0.076035197 | -4.52E-05 | 0.030977718 | 1 | 0.847011103 | 1 | 0.837495329 | ENSG0000013 | 10 | COX16 |
| SAT | Heart_Failure | ENSG0000014 | 0.003154281 | 0.303843443 | 0.003067689 | 0.076055328 | 0.003320093 | 0.279211802 | 0.003068214 | 0.076035157 | -0.00016581 | 0.028347154 | 1 | 0.816145643 | 1 | 0.791037893 | ENSG0000013 | 10 | COX16 |
| SAT | Heart_Failure | ENSG0000014 | 0.005946682 | 0.039706894 | 0.002891243 | 0.076054833 | 0.006007387 | 0.037708169 | 0.00289092 | 0.076034666 | -6.07E-05 | 0.001221114 | 1 | 0.448303633 | 1 | 0.434107027 | ENSG0000013 | 7 | MED6 |
| SAT | Heart_Failure | ENSG0000014 | -0.00267513 | 0.162630011 | 0.001915893 | 0.076055189 | -0.00150373 | 0.443870247 | 0.001963928 | 0.076035234 | -0.0011714 | 0.006800434 | 1 | 0.686431329 | 1 | 0.882462767 | ENSG0000013 | 7 | MED6 |
| SAT | Heart_Failure | ENSG0000014 | 0.00075044 | 0.659685154 | 0.001704198 | 0.076055463 | 0.000773628 | 0.649825491 | 0.001704007 | 0.076035308 | -2.32E-05 | 0.034158486 | 1 | 0.962994646 | 1 | 0.953700363 | ENSG0000013 | 7 | MED6 |
| SAT | Heart_Failure | ENSG0000014 | 0.001795481 | 0.484039949 | 0.002565632 | 0.076055417 | 0.001848224 | 0.471246625 | 0.002565365 | 0.076035259 | -5.27E-05 | 0.006473559 | 1 | 0.897349195 | 1 | 0.894608312 | ENSG0000013 | 7 | MED6 |
| SAT | Heart_Failure | ENSG0000014 | 0.003948134 | 0.281693336 | 0.003667482 | 0.076055312 | 0.003895048 | 0.28815952 | 0.003667071 | 0.076035164 | 5.31E-05 | 0.022776072 | 1 | 0.796074522 | 1 | 0.799198129 | ENSG0000013 | 7 | MRPS36 |
| SAT | Heart_Failure | ENSG0000014 | 0.001355897 | 0.720212539 | 0.003785571 | 0.076055473 | 0.001296491 | 0.7319612 | 0.003785175 | 0.076035322 | 5.94E-05 | 0.035251633 | 1 | 0.966658386 | 1 | 0.958028875 | ENSG0000013 | 11 | THUMPD3 |
| SAT | Heart_Failure | ENSG0000014 | -0.00145826 | 0.307999901 | 0.001430468 | 0.076055331 | -0.00142618 | 0.318714202 | 0.001430326 | 0.076035185 | -3.21E-05 | 0.00543291 | 1 | 0.816225947 | 1 | 0.823562856 | ENSG0000013 | 9 | TRPM1 |
| SAT | Heart_Failure | ENSG0000014 | -0.00078215 | 0.79377586 | 0.002992045 | 0.076055482 | -0.00083365 | 0.780514027 | 0.002991755 | 0.076035328 | 5.15E-05 | 0.041222228 | 1 | 0.976267425 | 1 | 0.967288536 | ENSG0000013 | 8 | GSTM1 |
| SAT | Heart_Failure | ENSG0000014 | 0.000889546 | 0.711492645 | 0.002405137 | 0.076055472 | 0.00094646 | 0.693908789 | 0.002404891 | 0.076035316 | -5.69E-05 | 0.002295738 | 1 | 0.966658386 | 1 | 0.956878334 | ENSG0000013 | 8 | GSTM1 |
| SAT | Heart_Failure | ENSG0000014 | 0.001723866 | 0.576351391 | 0.00308537 | 0.076055444 | 0.001825138 | 0.554113858 | 0.00308505 | 0.076035286 | -0.00010127 | 1.60E-05 | 1 | 0.932993874 | 1 | 0.915531921 | ENSG0000013 | 6 | GSTM5 |
| SAT | Heart_Failure | ENSG0000014 | -0.00230047 | 0.442643359 | 0.002996424 | 0.076055401 | -0.00217283 | 0.468328299 | 0.002996173 | 0.076035258 | -0.00012764 | 1.58E-05 | 1 | 0.882758633 | 1 | 0.892844363 | ENSG0000013 | 6 | GSTM5 |
| SAT | Heart_Failure | ENSG0000014 | 0.005053643 | 0.004520101 | 0.001779831 | 0.076054235 | 0.005163903 | 0.003717536 | 0.00177992 | 0.076034027 | -0.00011026 | 0.001203233 | 1 | 0.154647076 | 1 | 0.158405666 | ENSG0000013 | 6 | GSTM5 |
| SAT | Heart_Failure | ENSG0000014 | -0.00425188 | 0.111431009 | 0.002671118 | 0.076055098 | -0.00419711 | 0.11607574 | 0.00267083 | 0.076034955 | -5.48E-05 | 0.003453369 | 1 | 0.640599412 | 1 | 0.64481576 | ENSG0000013 | 6 | GSTM5 |
| SAT | Heart_Failure | ENSG0000015 | 0.001190522 | 0.686119719 | 0.002945913 | 0.076055468 | 0.001260378 | 0.668736471 | 0.002945612 | 0.076035312 | -6.99E-05 | 0.002364437 | 1 | 0.966658386 | 1 | 0.956878334 | ENSG0000013 | 6 | GSTM5 |
| SAT | Heart_Failure | ENSG0000015 | -0.00197183 | 0.359318295 | 0.002151089 | 0.076055362 | -0.00204038 | 0.342809682 | 0.002150877 | 0.0760352 | 6.86E-05 | 0.000111875 | 1 | 0.85040303 | 1 | 0.839311387 | ENSG0000013 | 6 | GSTM5 |
| SAT | Heart_Failure | ENSG0000015 | 0.003311742 | 0.142711223 | 0.002259385 | 0.076055158 | 0.003372121 | 0.135529915 | 0.002259153 | 0.076034992 | -6.04E-05 | 0.000553984 | 1 | 0.669181587 | 1 | 0.649215182 | ENSG0000013 | 6 | GSTM3 |
| SAT | Heart_Failure | ENSG0000015 | -0.00165986 | 0.563942085 | 0.002876723 | 0.076055441 | -0.00170387 | 0.553612296 | 0.002876421 | 0.076035285 | 4.40E-05 | 0.039184911 | 1 | 0.925856651 | 1 | 0.915531921 | ENSG0000013 | 6 | GSTM3 |
| SAT | Heart_Failure | ENSG0000015 | -0.00458997 | 0.1705896 | 0.003349575 | 0.0760552 | -0.00453895 | 0.175346366 | 0.003349218 | 0.076035054 | -5.10E-05 | 0.034232224 | 1 | 0.702264088 | 1 | 0.700744733 | ENSG0000013 | 14 | CHIA |
| SAT | Heart_Failure | ENSG0000015 | -0.00085519 | 0.546771833 | 0.001419151 | 0.076055436 | -0.00089216 | 0.529526178 | 0.001418994 | 0.076035278 | 3.70E-05 | 7.34E-05 | 1 | 0.916219808 | 1 | 0.908441675 | ENSG0000013 | 12 | PSRC1 |
| SAT | Heart_Failure | ENSG0000015 | 0.002360872 | 0.019381434 | 0.001009731 | 0.07605464 | 0.002378772 | 0.018468683 | 0.001009626 | 0.076034474 | -1.79E-05 | 0.018596772 | 1 | 0.302151987 | 1 | 0.304501243 | ENSG0000013 | 12 | PSRC1 |
| SAT | Heart_Failure | ENSG0000015 | 0.000717711 | 0.745038464 | 0.002207062 | 0.076055477 | 0.000753954 | 0.732618852 | 0.002206839 | 0.076035322 | -3.62E-05 | 0.039126419 | 1 | 0.966658386 | 1 | 0.958028875 | ENSG0000013 | 12 | PSRC1 |
| SAT | Heart_Failure | ENSG0000015 | 0.003192672 | 0.326755553 | 0.0032556 | 0.076055343 | 0.003140626 | 0.334655486 | 0.003255277 | 0.076035195 | 5.20E-05 | 0.049914993 | 1 | 0.833010855 | 1 | 0.836830236 | ENSG0000013 | 12 | PSRC1 |
| SAT | Heart_Failure | ENSG0000015 | -0.0028404 | 0.316328051 | 0.002834637 | 0.076055336 | -0.00292622 | 0.301881107 | 0.002834362 | 0.076035174 | 8.58E-05 | 0.000312604 | 1 | 0.824393135 | 1 | 0.810625291 | ENSG0000013 | 13 | WNT2B |
| SAT | Heart_Failure | ENSG0000015 | -0.00585136 | 0.004046292 | 0.002035577 | 0.076054203 | -0.00581936 | 0.004248255 | 0.002035363 | 0.076034065 | -3.20E-05 | 0.033277363 | 1 | 0.1521008 | 1 | 0.161355485 | ENSG0000013 | 8 | AP4B1 |
| SAT | Heart_Failure | ENSG0000015 | -0.00389058 | 0.060305666 | 0.00207105 | 0.076054942 | -0.00353711 | 0.088575875 | 0.002077035 | 0.076034888 | -0.00035347 | 0.028241466 | 1 | 0.508644962 | 1 | 0.596493272 | ENSG0000013 | 8 | AP4B1 |
| SAT | Heart_Failure | ENSG0000015 | -0.00479647 | 0.124505386 | 0.003122424 | 0.076055125 | -0.00451373 | 0.148395841 | 0.003123205 | 0.076035014 | -0.00028274 | 0.001058349 | 1 | 0.647388543 | 1 | 0.665121824 | ENSG0000013 | 13 | PPHLN1 |
| SAT | Heart_Failure | ENSG0000015 | -1.37E-05 | 0.984838343 | 0.000723093 | 0.076055493 | 3.94E-06 | 0.995647559 | 0.000723018 | 0.07603534 | -1.77E-05 | 0.001224757 | 1 | 0.998924529 | 1 | 0.999768899 | ENSG0000013 | 13 | PPHLN1 |
| SAT | Heart_Failure | ENSG0000015 | -0.00374832 | 0.017828758 | 0.00158213 | 0.076054617 | -0.00377154 | 0.017121416 | 0.001581958 | 0.076034543 | 2.32E-05 | 0.034270233 | 1 | 0.290924976 | 1 | 0.294111855 | ENSG0000013 | 6 | FKBP11 |
| SAT | Heart_Failure | ENSG0000015 | -0.00342144 | 0.382339864 | 0.003916513 | 0.076055374 | -0.00335486 | 0.391620926 | 0.003916108 | 0.076035226 | -6.66E-05 | 0.025483942 | 1 | 0.85759885 | 1 | 0.861776162 | ENSG0000013 | 6 | FKBP11 |
| SAT | Heart_Failure | ENSG0000015 | -0.0027602 | 0.079107681 | 0.001572015 | 0.076055012 | -0.00280071 | 0.074784768 | 0.001571858 | 0.076034845 | 4.04E-05 | 0.001595074 | 1 | 0.580653917 | 1 | 0.557368838 | ENSG0000013 | 6 | FKBP11 |
| SAT | Heart_Failure | ENSG0000015 | -0.00392829 | 0.077405674 | 0.002224467 | 0.076055006 | -0.00246653 | 0.292983482 | 0.0023455 | 0.076035168 | -0.00146176 | 0.049624527 | 1 | 0.575515143 | 1 | 0.807119064 | ENSG0000013 | 12 | LPIN1 |
| SAT | Heart_Failure | ENSG0000015 | -0.00617107 | 0.108154027 | 0.003841196 | 0.07605509 | -0.00627777 | 0.102157981 | 0.003840827 | 0.076034923 | 0.000106701 | 0.001180443 | 1 | 0.640599412 | 1 | 0.614882715 | ENSG0000013 | 7 | NARS |
| SAT | Heart_Failure | ENSG0000015 | -0.00313613 | 0.427921902 | 0.003955993 | 0.076055395 | -0.00303976 | 0.442218936 | 0.003955689 | 0.076035248 | -9.64E-05 | 0.020766656 | 1 | 0.878482333 | 1 | 0.880611614 | ENSG0000013 | 7 | NARS |
| SAT | Heart_Failure | ENSG0000015 | -0.00152121 | 0.556966359 | 0.002589943 | 0.076055439 | -0.00146331 | 0.572036262 | 0.002589672 | 0.07603529 | -5.79E-05 | 0.002613197 | 1 | 0.922001323 | 1 | 0.924557132 | ENSG0000013 | 7 | NARS |
| SAT | Heart_Failure | ENSG0000015 | 0.000256933 | 0.822829685 | 0.0011475 | 0.076055485 | 0.000303774 | 0.791200117 | 0.001147389 | 0.076035329 | -4.68E-05 | 1.39E-06 | 1 | 0.976669362 | 1 | 0.970635778 | ENSG0000013 | 7 | NARS |
| SAT | Heart_Failure | ENSG0000015 | 0.003138357 | 0.295865195 | 0.003002229 | 0.076055322 | 0.003081302 | 0.304679555 | 0.003001899 | 0.076035176 | 5.71E-05 | 0.004737433 | 1 | 0.813063328 | 1 | 0.812640695 | ENSG0000013 | 11 | IL2RA |
| SAT | Heart_Failure | ENSG0000015 | 0.000980868 | 0.741109154 | 0.002968861 | 0.076055476 | 0.001065339 | 0.719687157 | 0.002968534 | 0.07603532 | -8.45E-05 | 2.02E-05 | 1 | 0.966658386 | 1 | 0.958028875 | ENSG0000013 | 11 | IL2RA |
| SAT | Heart_Failure | ENSG0000015 | -0.00462834 | 0.110420922 | 0.002899402 | 0.076055095 | -0.00455052 | 0.116502865 | 0.002899111 | 0.076034956 | -7.78E-05 | 0.000794273 | 1 | 0.640599412 | 1 | 0.64481576 | ENSG0000013 | 11 | IL2RA |
| SAT | Heart_Failure | ENSG0000015 | -0.00537115 | 0.08772947 | 0.003145617 | 0.076055038 | -0.00558148 | 0.076018731 | 0.003145788 | 0.076034849 | 0.000210339 | 0.000548104 | 1 | 0.592308253 | 1 | 0.560403102 | ENSG0000013 | 10 | ECHDC3 |
| SAT | Heart_Failure | ENSG0000015 | -0.00428817 | 0.223319425 | 0.00352139 | 0.076055262 | -0.00420713 | 0.232140838 | 0.003521016 | 0.076035117 | -8.10E-05 | 0.001525278 | 1 | 0.751217604 | 1 | 0.750242469 | ENSG0000013 | 9 | TMEM241 |
| SAT | Heart_Failure | ENSG0000015 | -0.00175092 | 0.245372675 | 0.001507254 | 0.076055282 | -0.00178709 | 0.235706849 | 0.001507087 | 0.076035121 | 3.62E-05 | 0.000290636 | 1 | 0.760933598 | 1 | 0.752162449 | ENSG0000013 | 5 | EMP1 |
| SAT | Heart_Failure | ENSG0000015 | -0.00111324 | 0.746072191 | 0.003437799 | 0.076055477 | -0.00164216 | 0.633182204 | 0.003440869 | 0.076035305 | 0.000528926 | 0.000682349 | 1 | 0.966658386 | 1 | 0.94798204 | ENSG0000013 | 5 | EMP1 |
| SAT | Heart_Failure | ENSG0000015 | 0.003231227 | 0.226633017 | 0.002672469 | 0.076055265 | 0.003137627 | 0.240337954 | 0.002672268 | 0.076035125 | 9.36E-05 | 0.001023627 | 1 | 0.751217604 | 1 | 0.752432087 | ENSG0000013 | 7 | DDB2 |
| SAT | Heart_Failure | ENSG0000015 | 0.000729983 | 0.543365967 | 0.00120116 | 0.076055435 | 0.000749068 | 0.532832081 | 0.001201028 | 0.076035279 | -1.91E-05 | 0.017561849 | 1 | 0.913779466 | 1 | 0.908441675 | ENSG0000013 | 10 | AGO4 |
| SAT | Heart_Failure | ENSG0000015 | -0.00825853 | 0.019335798 | 0.003530795 | 0.076054639 | -0.00817416 | 0.020594474 | 0.003530427 | 0.076034504 | -8.44E-05 | 0.001421426 | 1 | 0.302151987 | 1 | 0.320239196 | ENSG0000013 | 9 | ZCCHC11 |
| SAT | Heart_Failure | ENSG0000015 | 0.004574271 | 0.156420293 | 0.003227645 | 0.07605518 | 0.00462639 | 0.151710999 | 0.003227302 | 0.076035019 | -5.21E-05 | 0.026141739 | 1 | 0.677064618 | 1 | 0.666350453 | ENSG0000013 | 11 | FHOD3 |
| SAT | Heart_Failure | ENSG0000015 | 0.002690552 | 0.450066153 | 0.003562204 | 0.076055404 | 0.002558977 | 0.472483843 | 0.003561832 | 0.07603526 | 0.000131575 | 7.42E-07 | 1 | 0.887019168 | 1 | 0.894608312 | ENSG0000013 | 4 | TIMM10 |
| SAT | Heart_Failure | ENSG0000015 | 0.00059632 | 0.869049534 | 0.003616996 | 0.076055489 | 0.00066882 | 0.853285768 | 0.003616655 | 0.076035335 | -7.25E-05 | 0.021999624 | 1 | 0.979223495 | 1 | 0.972791643 | ENSG0000013 | 4 | TIMM10 |
| SAT | Heart_Failure | ENSG0000015 | 0.001702278 | 0.118278055 | 0.001089775 | 0.076055112 | 0.001679687 | 0.123202144 | 0.001089663 | 0.076034969 | 2.26E-05 | 0.0065894 | 1 | 0.646149939 | 1 | 0.647747917 | ENSG0000013 | 4 | TIMM10 |
| SAT | Heart_Failure | ENSG0000015 | 0.00654105 | 0.085092165 | 0.003798787 | 0.07605503 | 0.006484368 | 0.087795368 | 0.003798365 | 0.076034885 | 5.67E-05 | 0.02243421 | 1 | 0.592308253 | 1 | 0.596493272 | ENSG0000013 | 4 | TIMM10 |
| SAT | Heart_Failure | ENSG0000015 | -0.00740686 | 0.051037247 | 0.003796108 | 0.076054899 | -0.0073386 | 0.05318985 | 0.003795734 | 0.076034757 | -6.83E-05 | 0.029157004 | 1 | 0.48962926 | 1 | 0.490345005 | ENSG0000013 | 14 | DHX34 |
| SAT | Heart_Failure | ENSG0000015 | 0.002251153 | 0.410141199 | 0.002733156 | 0.076055387 | 0.002320826 | 0.39576005 | 0.002732896 | 0.076035228 | -6.97E-05 | 0.003198065 | 1 | 0.870429337 | 1 | 0.862334842 | ENSG0000013 | 14 | DHX34 |
| SAT | Heart_Failure | ENSG0000015 | -0.00496576 | 0.002527922 | 0.001644293 | 0.07605407 | -0.00502377 | 0.002246522 | 0.001644141 | 0.076033883 | 5.80E-05 | 9.15E-05 | 1 | 0.113128813 | 1 | 0.11516936 | ENSG0000013 | 9 | TMEM258 |
| SAT | Heart_Failure | ENSG0000015 | 0.001571214 | 0.410445689 | 0.001908874 | 0.076055387 | 0.00154759 | 0.417465753 | 0.001908657 | 0.076035238 | 2.36E-05 | 0.042799507 | 1 | 0.870429337 | 1 | 0.867451005 | ENSG0000013 | 9 | TMEM258 |
| SAT | Heart_Failure | ENSG0000015 | 0.002876138 | 0.255145061 | 0.00252749 | 0.076055291 | 0.002832098 | 0.262441631 | 0.002527215 | 0.076035144 | 4.40E-05 | 0.011118066 | 1 | 0.770621765 | 1 | 0.774729353 | ENSG0000013 | 8 | TMEM165 |
| SAT | Heart_Failure | ENSG0000015 | -5.76E-05 | 0.979197978 | 0.002207639 | 0.076055493 | -8.86E-05 | 0.967965702 | 0.002207388 | 0.07603534 | 3.11E-05 | 0.022038686 | 1 | 0.998920999 | 1 | 0.99755193 | ENSG0000013 | 8 | TMEM165 |
| SAT | Heart_Failure | ENSG0000015 | -0.00200024 | 0.173729937 | 0.001470421 | 0.076055204 | -0.00195128 | 0.18446479 | 0.001470294 | 0.076035065 | -4.90E-05 | 0.000525271 | 1 | 0.704583343 | 1 | 0.715224819 | ENSG0000013 | 10 | CLOCK |
| SAT | Heart_Failure | ENSG0000015 | -0.00127811 | 0.204207677 | 0.001006658 | 0.076055241 | -0.00129368 | 0.198704387 | 0.001006553 | 0.076035082 | 1.56E-05 | 0.038516854 | 1 | 0.735350411 | 1 | 0.725655076 | ENSG0000013 | 10 | CLOCK |
| SAT | Heart_Failure | ENSG0000015 | 0.002407688 | 0.157703081 | 0.001704146 | 0.076055181 | 0.002472123 | 0.146835239 | 0.001703968 | 0.076035012 | -6.44E-05 | 4.14E-07 | 1 | 0.678274948 | 1 | 0.664505404 | ENSG0000013 | 10 | CLOCK |
| SAT | Heart_Failure | ENSG0000015 | 0.006665018 | 0.017896017 | 0.002814892 | 0.076054618 | 0.006620389 | 0.018664057 | 0.002814574 | 0.076034477 | 4.46E-05 | 0.011113078 | 1 | 0.290924976 | 1 | 0.304660546 | ENSG0000013 | 5 | CLDN10 |
| SAT | Heart_Failure | ENSG0000015 | -0.00115692 | 0.705624684 | 0.003062752 | 0.076055471 | -0.0012007 | 0.695002046 | 0.003062415 | 0.076035316 | 4.38E-05 | 0.033107161 | 1 | 0.966658386 | 1 | 0.956878334 | ENSG0000013 | 5 | CLDN10 |
| SAT | Heart_Failure | ENSG0000015 | 0.005474168 | 0.036944658 | 0.002623762 | 0.076054814 | 0.005517556 | 0.03545422 | 0.002623488 | 0.07603465 | -4.34E-05 | 0.0266 | 1 | 0.436153525 | 1 | 0.423805321 | ENSG0000013 | 13 | DZIP1 |
| SAT | Heart_Failure | ENSG0000015 | 0.001512081 | 0.520830099 | 0.002355021 | 0.076055429 | 0.001445866 | 0.539205903 | 0.002354774 | 0.076035281 | 6.62E-05 | 0.000149159 | 1 | 0.906724424 | 1 | 0.91051702 | ENSG0000013 | 13 | DZIP1 |
| SAT | Heart_Failure | ENSG0000015 | 0.007323192 | 0.059242007 | 0.003882104 | 0.076054938 | 0.007269461 | 0.061102147 | 0.003881684 | 0.076034793 | 5.37E-05 | 0.045975296 | 1 | 0.507375885 | 1 | 0.505632969 | ENSG0000013 | 12 | CARS2 |
| SAT | Heart_Failure | ENSG0000015 | -0.00325607 | 0.157868007 | 0.00230554 | 0.076055182 | -0.00319324 | 0.166002098 | 0.002305311 | 0.076035041 | -6.28E-05 | 0.000794357 | 1 | 0.678274948 | 1 | 0.68372624 | ENSG0000013 | 12 | CARS2 |

| | | | | | | | | | | | | | | | | | | | |
|---|---|---|---|---|---|---|---|---|---|---|---|---|---|---|---|---|---|---|---|
| SAT | Heart_Failure | ENSG0000015 | 0.000615883 | 0.87080857 | 0.003786984 | 0.076055489 | 0.000677323 | 0.858036456 | 0.003786585 | 0.076035335 | -6.14E-05 | 0.028022665 | 1 | 0.979223495 | 1 | 0.972791643 | ENSG0000013 | 12 | CARS2 |
| SAT | Heart_Failure | ENSG0000015 | 0.005954646 | 0.037731959 | 0.002865897 | 0.076054819 | 0.006004522 | 0.036138335 | 0.00286561 | 0.076034655 | -4.99E-05 | 0.030885499 | 1 | 0.439253432 | 1 | 0.428050103 | ENSG0000013 | 12 | CARS2 |
| SAT | Heart_Failure | ENSG0000015 | 0.003085431 | 0.179291717 | 0.002297518 | 0.076055212 | 0.003158504 | 0.169171479 | 0.002297306 | 0.076035045 | -7.31E-05 | 0.000399369 | 1 | 0.714462461 | 1 | 0.689505122 | ENSG0000013 | 12 | GOLM1 |
| SAT | Heart_Failure | ENSG0000015 | -0.00105974 | 0.616346832 | 0.002115099 | 0.076055454 | -0.00111276 | 0.598782386 | 0.002114894 | 0.076035297 | 5.30E-05 | 0.002949812 | 1 | 0.954216727 | 1 | 0.938318026 | ENSG0000013 | 12 | GOLM1 |
| SAT | Heart_Failure | ENSG0000015 | -0.00048546 | 0.664242416 | 0.001118411 | 0.076055464 | -0.0005109 | 0.647772182 | 0.00111829 | 0.076035308 | 2.54E-05 | 0.001152392 | 1 | 0.962994646 | 1 | 0.95317119 | ENSG0000013 | 11 | ADAM19 |
| SAT | Heart_Failure | ENSG0000015 | -0.00150257 | 0.334557886 | 0.001557108 | 0.076055348 | -0.00146624 | 0.346322024 | 0.001556934 | 0.076035202 | -3.63E-05 | 0.000310397 | 1 | 0.841293164 | 1 | 0.840027851 | ENSG0000013 | 11 | ADAM19 |
| SAT | Heart_Failure | ENSG0000015 | 0.000137399 | 0.956763348 | 0.002534307 | 0.076055493 | 0.000202782 | 0.93621899 | 0.002534046 | 0.076035339 | -6.54E-05 | 0.000736936 | 1 | 0.994657967 | 1 | 0.988983601 | ENSG0000013 | 11 | ADAM19 |
| SAT | Heart_Failure | ENSG0000015 | 0.001399503 | 0.698356733 | 0.003611248 | 0.07605547 | 0.001520072 | 0.67377712 | 0.003610878 | 0.076035312 | -0.00012057 | 1.65E-05 | 1 | 0.966658386 | 1 | 0.956878334 | ENSG0000013 | 8 | USP30 |
| SAT | Heart_Failure | ENSG0000015 | 0.006369146 | 0.087523179 | 0.003727669 | 0.076055037 | 0.006546592 | 0.079023333 | 0.003727311 | 0.076034859 | -0.00017745 | 2.53E-08 | 1 | 0.592308253 | 1 | 0.572352216 | ENSG0000013 | 14 | DMTF1 |
| SAT | Heart_Failure | ENSG0000015 | 0.000873734 | 0.718123026 | 0.002420526 | 0.076055473 | 0.000829583 | 0.731774095 | 0.002420256 | 0.076035322 | 4.42E-05 | 0.004788778 | 1 | 0.966658386 | 1 | 0.958028875 | ENSG0000013 | 5 | TMEM60 |
| SAT | Heart_Failure | ENSG0000015 | 0.00091718 | 0.783948768 | 0.003345212 | 0.076055481 | 0.000965804 | 0.772777935 | 0.003344853 | 0.076035327 | -4.86E-05 | 0.041032724 | 1 | 0.972730696 | 1 | 0.965963214 | ENSG0000013 | 8 | POM121C |
| SAT | Heart_Failure | ENSG0000016 | 0.001016405 | 0.72658749 | 0.002906767 | 0.076055474 | 0.001109576 | 0.702641144 | 0.002906491 | 0.076035317 | -9.32E-05 | 0.000211011 | 1 | 0.966658386 | 1 | 0.956878334 | ENSG0000013 | 8 | POM121C |
| SAT | Heart_Failure | ENSG0000016 | -0.00052815 | 0.836481737 | 0.002558909 | 0.076055486 | -0.00049032 | 0.848027424 | 0.002558619 | 0.076035334 | -3.78E-05 | 0.017455808 | 1 | 0.976669362 | 1 | 0.972791643 | ENSG0000013 | 8 | POM121C |
| SAT | Heart_Failure | ENSG0000016 | 0.000327244 | 0.676161228 | 0.000783428 | 0.076055466 | 0.000349865 | 0.655142644 | 0.000783344 | 0.076035309 | -2.26E-05 | 3.69E-05 | 1 | 0.966658386 | 1 | 0.95563262 | ENSG0000013 | 13 | TES |
| SAT | Heart_Failure | ENSG0000016 | -0.00332812 | 0.14009862 | 0.002255698 | 0.076055153 | -0.00339845 | 0.131873258 | 0.002255471 | 0.076034986 | 7.03E-05 | 9.85E-05 | 1 | 0.668115657 | 1 | 0.649215182 | ENSG0000013 | 13 | TES |
| SAT | Heart_Failure | ENSG0000016 | -0.00383176 | 0.191073047 | 0.002930792 | 0.076055226 | -0.00375274 | 0.200338785 | 0.002930478 | 0.076035084 | -7.90E-05 | 0.000150641 | 1 | 0.722912434 | 1 | 0.727919771 | ENSG0000013 | 13 | TES |
| SAT | Heart_Failure | ENSG0000016 | -0.00182992 | 0.409397399 | 0.00221821 | 0.076055387 | -0.00179451 | 0.418469704 | 0.002217965 | 0.076035238 | -3.54E-05 | 0.015753855 | 1 | 0.870429337 | 1 | 0.868986772 | ENSG0000013 | 13 | TES |
| SAT | Heart_Failure | ENSG0000016 | -0.00147585 | 0.630169375 | 0.003065181 | 0.076055457 | -0.00155577 | 0.611722158 | 0.003064853 | 0.0760353 | 7.99E-05 | 0.000264078 | 1 | 0.959161083 | 1 | 0.945324871 | ENSG0000013 | 11 | MTO1 |
| SAT | Heart_Failure | ENSG0000016 | -0.00494857 | 0.188605323 | 0.00376398 | 0.076055223 | -0.00480005 | 0.202191973 | 0.003763759 | 0.076035086 | -0.00014853 | 0.001146653 | 1 | 0.722191019 | 1 | 0.729002049 | ENSG0000013 | 8 | KHDC1 |
| SAT | Heart_Failure | ENSG0000016 | -0.0016987 | 0.25223347 | 0.001483655 | 0.076055288 | -0.00166891 | 0.260600831 | 0.001483513 | 0.076035143 | -2.98E-05 | 0.018221804 | 1 | 0.767444733 | 1 | 0.773899606 | ENSG0000013 | 7 | NT5E |
| SAT | Heart_Failure | ENSG0000016 | -0.00211068 | 0.591807206 | 0.003936224 | 0.076055448 | -0.00220672 | 0.575023208 | 0.003935861 | 0.076035291 | 9.60E-05 | 0.006653347 | 1 | 0.942124911 | 1 | 0.927160988 | ENSG0000013 | 11 | ITGA7 |
| SAT | Heart_Failure | ENSG0000016 | 0.000759587 | 0.820264905 | 0.003343188 | 0.076055485 | 0.000652145 | 0.845324238 | 0.003342828 | 0.076035334 | 0.000107442 | 5.27E-06 | 1 | 0.976669362 | 1 | 0.972791643 | ENSG0000013 | 10 | KRT7 |
| SAT | Heart_Failure | ENSG0000016 | 0.000256626 | 0.914861543 | 0.002400429 | 0.076055491 | 0.000534309 | 0.824089138 | 0.002403669 | 0.076035332 | -0.00027768 | 0.033674188 | 1 | 0.985971321 | 1 | 0.972791643 | ENSG0000013 | 10 | KRT7 |
| SAT | Heart_Failure | ENSG0000016 | -0.00834293 | 0.002468433 | 0.00275599 | 0.076054063 | -0.00846457 | 0.002128898 | 0.002755707 | 0.076033868 | 0.000121639 | 1.37E-08 | 1 | 0.113128813 | 1 | 0.111957631 | ENSG0000013 | 10 | KRT7 |
| SAT | Heart_Failure | ENSG0000016 | -0.00377704 | 0.328977484 | 0.003869217 | 0.076055344 | -0.00386332 | 0.31800011 | 0.003868825 | 0.076035185 | 8.63E-05 | 0.004698424 | 1 | 0.836086663 | 1 | 0.823487262 | ENSG0000013 | 10 | KRT7 |
| SAT | Heart_Failure | ENSG0000016 | -0.00216759 | 0.260286205 | 0.001925519 | 0.076055295 | -0.00220362 | 0.252401554 | 0.001925338 | 0.076035136 | 3.60E-05 | 0.03215501 | 1 | 0.774632399 | 1 | 0.765423527 | ENSG0000013 | 14 | MAP7 |
| SAT | Heart_Failure | ENSG0000016 | -0.0012556 | 0.73114904 | 0.003654263 | 0.076055475 | -0.00131918 | 0.718076472 | 0.003653913 | 0.07603532 | 6.36E-05 | 0.042679646 | 1 | 0.966658386 | 1 | 0.958028875 | ENSG0000013 | 16 | AHI1 |
| SAT | Heart_Failure | ENSG0000016 | -0.00046735 | 0.779969904 | 0.001672925 | 0.076055481 | -0.00043433 | 0.795132165 | 0.001672746 | 0.07603533 | -3.30E-05 | 0.005971931 | 1 | 0.972664863 | 1 | 0.970635778 | ENSG0000013 | 16 | AHI1 |
| SAT | Heart_Failure | ENSG0000016 | -0.00106915 | 0.719301655 | 0.002974887 | 0.076055473 | -0.00101316 | 0.733399365 | 0.002974574 | 0.076035322 | -5.60E-05 | 0.010824131 | 1 | 0.966658386 | 1 | 0.958028875 | ENSG0000013 | 4 | SMPD2 |
| SAT | Heart_Failure | ENSG0000016 | -0.00283399 | 0.30713794 | 0.002775028 | 0.07605533 | -0.00287667 | 0.299859468 | 0.002774741 | 0.076035172 | 4.27E-05 | 0.043416816 | 1 | 0.816225947 | 1 | 0.810520426 | ENSG0000013 | 4 | SMPD2 |
| SAT | Heart_Failure | ENSG0000016 | 0.000845485 | 0.669585713 | 0.001981384 | 0.076055465 | 0.000888938 | 0.653658366 | 0.001981202 | 0.076035309 | -4.35E-05 | 0.015094149 | 1 | 0.96567764 | 1 | 0.954876713 | ENSG0000013 | 13 | MICAL1 |
| SAT | Heart_Failure | ENSG0000016 | -0.0028505 | 0.352410042 | 0.003065299 | 0.076055358 | -0.0029045 | 0.343309671 | 0.00306496 | 0.0760352 | 5.40E-05 | 0.007934368 | 1 | 0.848363544 | 1 | 0.839131387 | ENSG0000013 | 9 | DYSF |
| SAT | Heart_Failure | ENSG0000016 | 0.005117755 | 0.087735998 | 0.00299728 | 0.076055038 | 0.004575904 | 0.127661967 | 0.003003765 | 0.076034978 | 0.000541851 | 0.00766156 | 1 | 0.592308253 | 1 | 0.647874406 | ENSG0000013 | 7 | ZNF670 |
| SAT | Heart_Failure | ENSG0000016 | 0.004110574 | 0.116131856 | 0.002616164 | 0.076055108 | 0.003627036 | 0.165773758 | 0.002617072 | 0.07603504 | 0.000483537 | 2.33E-09 | 1 | 0.642397685 | 1 | 0.68372624 | ENSG0000013 | 7 | ZNF670 |
| SAT | Heart_Failure | ENSG0000016 | 0.003713802 | 0.194587457 | 0.002863094 | 0.07605523 | 0.003637991 | 0.203805451 | 0.002862789 | 0.076035088 | 7.58E-05 | 0.000217543 | 1 | 0.726878652 | 1 | 0.729840368 | ENSG0000013 | 7 | ZNF670 |
| SAT | Heart_Failure | ENSG0000016 | 0.001512361 | 0.619164303 | 0.003042762 | 0.076055454 | 0.001565854 | 0.606783884 | 0.003042442 | 0.076035299 | -5.35E-05 | 0.017153525 | 1 | 0.954216727 | 1 | 0.943534562 | ENSG0000013 | 10 | KCNK1 |
| SAT | Heart_Failure | ENSG0000016 | 0.004155511 | 0.071897599 | 0.002308912 | 0.076054987 | 0.004109448 | 0.075075985 | 0.002308674 | 0.076034846 | 4.61E-05 | 0.009066596 | 1 | 0.549874178 | 1 | 0.557368838 | ENSG0000013 | 7 | NTPCR |
| SAT | Heart_Failure | ENSG0000016 | -0.00129932 | 0.454966379 | 0.001739011 | 0.076055406 | -0.00123942 | 0.47597548 | 0.001738834 | 0.076035261 | -5.99E-05 | 1.02E-05 | 1 | 0.887019168 | 1 | 0.894608312 | ENSG0000013 | 7 | NTPCR |
| SAT | Heart_Failure | ENSG0000016 | 7.35E-05 | 0.980151418 | 0.002954458 | 0.076055493 | 0.000196203 | 0.947048146 | 0.002954231 | 0.076035339 | -0.0001227 | 8.29E-05 | 1 | 0.998920999 | 1 | 0.990755659 | ENSG0000013 | 7 | RNASEL |
| SAT | Heart_Failure | ENSG0000016 | -0.00273138 | 0.457744583 | 0.003678292 | 0.076055407 | -0.00278595 | 0.448761908 | 0.003677908 | 0.076035251 | 5.46E-05 | 0.048524427 | 1 | 0.888149011 | 1 | 0.882462767 | ENSG0000013 | 7 | RNASEL |
| SAT | Heart_Failure | ENSG0000016 | -0.00124294 | 0.565841744 | 0.002164696 | 0.076055442 | -0.00131869 | 0.542363261 | 0.002164471 | 0.076035282 | 7.58E-05 | 3.81E-06 | 1 | 0.927165893 | 1 | 0.912245255 | ENSG0000013 | 7 | RNASEL |
| SAT | Heart_Failure | ENSG0000016 | -0.00418207 | 0.196286036 | 0.003236377 | 0.076055232 | -0.00458161 | 0.157161804 | 0.003238609 | 0.076035028 | 0.000399539 | 0.002338284 | 1 | 0.726878652 | 1 | 0.669469928 | ENSG0000013 | 5 | PIGC |
| SAT | Heart_Failure | ENSG0000016 | 0.001611598 | 0.301683493 | 0.00156037 | 0.076055327 | 0.002491462 | 0.118006922 | 0.001593825 | 0.076034959 | -0.00087986 | 0.006928956 | 1 | 0.815859931 | 1 | 0.64481576 | ENSG0000013 | 5 | PIGC |
| SAT | Heart_Failure | ENSG0000016 | -0.00345307 | 0.07979031 | 0.001971037 | 0.076055014 | -0.0034102 | 0.08357008 | 0.001970825 | 0.076034873 | -4.29E-05 | 0.002125315 | 1 | 0.582544121 | 1 | 0.586427958 | ENSG0000013 | 6 | USP37 |
| SAT | Heart_Failure | ENSG0000016 | 0.000398014 | 0.715399977 | 0.001091604 | 0.076055472 | 0.000437015 | 0.688876026 | 0.001091499 | 0.076035315 | -3.90E-05 | 2.66E-05 | 1 | 0.966658386 | 1 | 0.956878334 | ENSG0000013 | 6 | USP37 |
| SAT | Heart_Failure | ENSG0000016 | 0.005375726 | 0.155723648 | 0.003786778 | 0.076055178 | 0.006154267 | 0.105021338 | 0.00379662 | 0.07603493 | -0.00077854 | 0.005441729 | 1 | 0.677064618 | 1 | 0.620855876 | ENSG0000013 | 9 | SLC19A3 |
| SAT | Heart_Failure | ENSG0000016 | -0.00038663 | 0.641853687 | 0.00083126 | 0.076055459 | -0.00033176 | 0.689822283 | 0.000831281 | 0.076035315 | -5.49E-05 | 0.000194988 | 1 | 0.960292726 | 1 | 0.956878334 | ENSG0000013 | 9 | SLC19A3 |
| SAT | Heart_Failure | ENSG0000016 | -0.00475236 | 0.119466806 | 0.00305222 | 0.076055115 | -0.00637331 | 0.040960114 | 0.003118152 | 0.076034688 | 0.00162095 | 0.011290399 | 1 | 0.646585619 | 1 | 0.453733817 | ENSG0000013 | 8 | TMBIM1 |
| SAT | Heart_Failure | ENSG0000016 | -0.0020577 | 0.446565913 | 0.002703398 | 0.076055403 | -0.0021086 | 0.435351649 | 0.002703104 | 0.076035245 | 5.09E-05 | 0.006251396 | 1 | 0.88490557 | 1 | 0.877927942 | ENSG0000013 | 8 | TMBIM1 |
| SAT | Heart_Failure | ENSG0000016 | 0.001754853 | 0.004112231 | 0.000611567 | 0.076054208 | 0.001733569 | 0.004584132 | 0.000611509 | 0.076034086 | 2.13E-05 | 4.77E-05 | 1 | 0.1521008 | 1 | 0.163212403 | ENSG0000013 | 10 | TSGA10 |
| SAT | Heart_Failure | ENSG0000016 | -0.00392933 | 0.085472227 | 0.002284783 | 0.076055031 | -0.00388562 | 0.088975804 | 0.002284544 | 0.076034889 | -4.37E-05 | 0.010585864 | 1 | 0.592308253 | 1 | 0.596493272 | ENSG0000013 | 10 | TSGA10 |
| SAT | Heart_Failure | ENSG0000016 | -0.00665119 | 0.009592291 | 0.00256783 | 0.076054446 | -0.00685227 | 0.007619783 | 0.002567856 | 0.076034229 | 0.000201079 | 3.70E-06 | 1 | 0.224802197 | 1 | 0.202724069 | ENSG0000013 | 15 | GCC2 |
| SAT | Heart_Failure | ENSG0000016 | 0.000217144 | 0.858970213 | 0.001222069 | 0.076055488 | 0.000240139 | 0.844201058 | 0.001221939 | 0.076035334 | -2.30E-05 | 0.009235996 | 1 | 0.976669362 | 1 | 0.972791643 | ENSG0000013 | 15 | GCC2 |
| SAT | Heart_Failure | ENSG0000016 | -0.00381456 | 0.308537528 | 0.00374602 | 0.076055331 | -0.00374361 | 0.317569402 | 0.00374561 | 0.076035184 | -7.09E-05 | 0.005269769 | 1 | 0.816225947 | 1 | 0.823487262 | ENSG0000013 | 4 | MRPS9 |
| SAT | Heart_Failure | ENSG0000016 | -0.00509743 | 0.01199286 | 0.002028938 | 0.076054508 | -0.00502796 | 0.013198096 | 0.002028722 | 0.076034382 | -6.95E-05 | 1.79E-06 | 1 | 0.24440108 | 1 | 0.258578925 | ENSG0000013 | 4 | MRPS9 |
| SAT | Heart_Failure | ENSG0000016 | 0.004967505 | 5.60E-06 | 0.001093929 | 0.076052275 | 0.004947231 | 6.10E-06 | 0.001093818 | 0.076032148 | 2.03E-05 | 0.018865977 | 0.018376036 | 0.001020891 | 0.020018014 | 0.002502252 | ENSG0000013 | 7 | USP44 |
| SAT | Heart_Failure | ENSG0000016 | 0.003885372 | 0.172705616 | 0.002849425 | 0.076055203 | 0.003791137 | 0.183317641 | 0.00284917 | 0.076035064 | 9.42E-05 | 0.000361398 | 1 | 0.704583343 | 1 | 0.715224819 | ENSG0000013 | 7 | USP44 |
| SAT | Heart_Failure | ENSG0000016 | 0.005791965 | 0.033747542 | 0.002728094 | 0.07605479 | 0.005747848 | 0.03510573 | 0.002727793 | 0.076034647 | 4.41E-05 | 0.014932703 | 1 | 0.411793489 | 1 | 0.42191172 | ENSG0000013 | 9 | DRAM1 |
| SAT | Heart_Failure | ENSG0000016 | 0.00492383 | 0.110704368 | 0.003086967 | 0.076055096 | 0.004839678 | 0.116896937 | 0.003086657 | 0.076034957 | 8.42E-05 | 0.000661877 | 1 | 0.640599412 | 1 | 0.64481576 | ENSG0000013 | 9 | DRAM1 |
| SAT | Heart_Failure | ENSG0000016 | -0.0229856 | 0.10599293 | 0.001421962 | 0.076055085 | -0.00231972 | 0.102779553 | 0.001421805 | 0.076034925 | 2.12E-05 | 0.026807108 | 1 | 0.636928212 | 1 | 0.614882715 | ENSG0000013 | 12 | NEK3 |
| SAT | Heart_Failure | ENSG0000016 | 0.001616345 | 0.608904774 | 0.003159158 | 0.076055452 | 0.00167626 | 0.595652357 | 0.003158802 | 0.076035296 | -5.99E-05 | 0.002553746 | 1 | 0.9499841 | 1 | 0.937611754 | ENSG0000013 | 12 | NEK3 |
| SAT | Heart_Failure | ENSG0000016 | -0.00485745 | 0.142001422 | 0.003308031 | 0.076055157 | -0.00500479 | 0.130272549 | 0.003307795 | 0.076034983 | 0.000147345 | 5.77E-05 | 1 | 0.669181587 | 1 | 0.649215182 | ENSG0000013 | 8 | VPS36 |
| SAT | Heart_Failure | ENSG0000016 | -0.00054102 | 0.416897143 | 0.000666433 | 0.07605539 | -0.0005282 | 0.427973376 | 0.00066636 | 0.076035242 | -1.28E-05 | 0.004850972 | 1 | 0.87398914 | 1 | 0.875416354 | ENSG0000013 | 14 | RNASEH2B |
| SAT | Heart_Failure | ENSG0000016 | -0.00119126 | 0.613064446 | 0.00235565 | 0.076055453 | -0.00124738 | 0.596399086 | 0.002355392 | 0.076035296 | 5.61E-05 | 0.000406998 | 1 | 0.954205146 | 1 | 0.937611754 | ENSG0000013 | 14 | RNASEH2B |
| SAT | Heart_Failure | ENSG0000016 | -0.00236726 | 0.210369036 | 0.001889945 | 0.076055248 | -0.00245064 | 0.194713796 | 0.001889813 | 0.076035078 | 8.34E-05 | 7.74E-05 | 1 | 0.739879495 | 1 | 0.723506187 | ENSG0000013 | 14 | RNASEH2B |
| SAT | Heart_Failure | ENSG0000016 | 0.001427273 | 0.352482288 | 0.001535055 | 0.076055358 | 0.001382154 | 0.367860131 | 0.001534892 | 0.076035214 | 4.51E-05 | 4.84E-05 | 1 | 0.848363544 | 1 | 0.849290727 | ENSG0000013 | 9 | SUCLA2 |
| SAT | Heart_Failure | ENSG0000016 | -0.00036729 | 0.915924905 | 0.003479127 | 0.076055491 | -0.00027472 | 0.937057719 | 0.003478793 | 0.076035339 | -9.26E-05 | 0.00183301 | 1 | 0.986559779 | 1 | 0.988983601 | ENSG0000013 | 9 | SUCLA2 |
| SAT | Heart_Failure | ENSG0000016 | -0.00299836 | 0.390361793 | 0.003557782 | 0.076055382 | -0.00289259 | 0.416153378 | 0.003557427 | 0.076035237 | -0.00010577 | 0.000247349 | 1 | 0.870429337 | 1 | 0.867451005 | ENSG0000013 | 7 | RCBTB1 |
| SAT | Heart_Failure | ENSG0000016 | -0.00207325 | 0.503318252 | 0.003097734 | 0.076055423 | -0.00201496 | 0.515353449 | 0.003097442 | 0.076035274 | -5.83E-05 | 0.031068603 | 1 | 0.900549813 | 1 | 0.903395669 | ENSG0000013 | 10 | MED4 |
| SAT | Heart_Failure | ENSG0000016 | 0.001641808 | 0.439424783 | 0.002123488 | 0.0760554 | 0.001597138 | 0.451928173 | 0.002123275 | 0.076035252 | 4.47E-05 | 0.00860693 | 1 | 0.880728596 | 1 | 0.88576842 | ENSG0000013 | 10 | MED4 |
| SAT | Heart_Failure | ENSG0000016 | -0.00084395 | 0.761171333 | 0.002776676 | 0.076055479 | -0.00089137 | 0.748169852 | 0.002776394 | 0.076035324 | 4.74E-05 | 0.029521199 | 1 | 0.969898847 | 1 | 0.962415493 | ENSG0000013 | 10 | MED4 |
| SAT | Heart_Failure | ENSG0000016 | -0.00422376 | 0.229893527 | 0.003517939 | 0.076055268 | -0.00428254 | 0.223427561 | 0.003517588 | 0.076035109 | 5.88E-05 | 0.03850854 | 1 | 0.751389909 | 1 | 0.747263842 | ENSG0000013 | 12 | SETDB2 |
| SAT | Heart_Failure | ENSG0000016 | 0.001096765 | 0.475847553 | 0.001538244 | 0.076055414 | 0.000142778 | 0.926926173 | 0.00155679 | 0.076035339 | 0.000953987 | 7.48E-05 | 1 | 0.895715287 | 1 | 0.987053153 | ENSG0000013 | 12 | SETDB2 |
| SAT | Heart_Failure | ENSG0000016 | -0.00058577 | 0.688726418 | 0.001462281 | 0.076055468 | -0.00055306 | 0.70524062 | 0.001462132 | 0.076035318 | -3.27E-05 | 0.004382976 | 1 | 0.966658386 | 1 | 0.958028875 | ENSG0000013 | 8 | C7orf25 |
| SAT | Heart_Failure | ENSG0000016 | 0.002067949 | 0.378242663 | 0.002346902 | 0.076055372 | 0.002025705 | 0.388011238 | 0.002346656 | 0.076035224 | 4.22E-05 | 0.015583336 | 1 | 0.85759885 | 1 | 0.858921514 | ENSG0000013 | 7 | CHST12 |
| SAT | Heart_Failure | ENSG0000016 | 1.48E-05 | 0.991115203 | 0.001326177 | 0.076055493 | -1.09E-05 | 0.993445012 | 0.001326034 | 0.07603534 | 2.57E-05 | 0.006454395 | 1 | 0.999479422 | 1 | 0.999768899 | ENSG0000013 | 7 | CHST12 |
| SAT | Heart_Failure | ENSG0000016 | -0.00206615 | 0.351784797 | 0.00221896 | 0.076055358 | -0.0021332 | 0.336331362 | 0.002218752 | 0.076035196 | 6.70E-05 | 0.000592111 | 1 | 0.848363544 | 1 | 0.837075007 | ENSG0000013 | 7 | CHST12 |
| SAT | Heart_Failure | ENSG0000016 | -0.00175529 | 0.454335328 | 0.002345992 | 0.076055406 | -0.00178732 | 0.446093233 | 0.002345733 | 0.07603525 | 3.20E-05 | 0.039777743 | 1 | 0.887019168 | 1 | 0.882462767 | ENSG0000013 | 7 | CHST12 |
| SAT | Heart_Failure | ENSG0000016 | -0.00640261 | 0.089368643 | 0.003769031 | 0.076055043 | -0.00655686 | 0.081885552 | 0.00376863 | 0.076034868 | 0.000154254 | 1.74E-08 | 1 | 0.597756036 | 1 | 0.581529209 | ENSG0000013 | 10 | ZDHHC4 |
| SAT | Heart_Failure | ENSG0000016 | 0.00165004 | 0.498487502 | 0.002437743 | 0.076055422 | 0.00168448 | 0.489518366 | 0.002437475 | 0.076035266 | -3.44E-05 | 0.035602284 | 1 | 0.899263643 | 1 | 0.894608312 | ENSG0000013 | 10 | ZDHHC4 |
| SAT | Heart_Failure | ENSG0000016 | 0.002023932 | 0.489261265 | 0.002926937 | 0.076055418 | 0.002104421 | 0.472102938 | 0.00292662 | 0.076035259 | -8.05E-05 | 7.59E-05 | 1 | 0.898909993 | 1 | 0.894608312 | ENSG0000013 | 10 | ZDHHC4 |
| SAT | Heart_Failure | ENSG0000016 | -0.00023822 | 0.777641855 | 0.000843588 | 0.076055481 | -0.00026095 | 0.757044784 | 0.000843499 | 0.076035325 | 2.27E-05 | 0.000243871 | 1 | 0.972664863 | 1 | 0.962699472 | ENSG0000013 | 10 | ZDHHC4 |
| SAT | Heart_Failure | ENSG0000016 | 0.015750908 | 1.69E-05 | 0.003661154 | 0.076052604 | 0.000675242 | 0.866081304 | 0.004004806 | 0.076035336 | 0.015075666 | 1.50E-20 | 0.055503379 | 0.002522881 | 1 | 0.974123167 | ENSG0000013 | 14 | DBNL |
| SAT | Heart_Failure | ENSG0000016 | 0.001495694 | 0.679234924 | 0.003617009 | 0.076055466 | 0.001579084 | 0.662393529 | 0.003616682 | 0.07603531 | -8.34E-05 | 0.000242421 | 1 | 0.966658386 | 1 | 0.956878334 | ENSG0000013 | 14 | DBNL |
| SAT | Heart_Failure | ENSG0000016 | 0.00055497 | 0.730807882 | 0.001613043 | 0.076055475 | 0.00052706 | 0.74383334 | 0.001612884 | 0.076035323 | 2.79E-05 | 0.034610167 | 1 | 0.966658386 | 1 | 0.962349048 | ENSG0000013 | 14 | DBNL |
| SAT | Heart_Failure | ENSG0000016 | 0.004632491 | 0.209402371 | 0.003690605 | 0.076055247 | 0.004748672 | 0.198157614 | 0.003690237 | 0.076035082 | -0.00011618 | 0.00010887 | 1 | 0.739879495 | 1 | 0.725655076 | ENSG0000013 | 14 | DBNL |
| SAT | Heart_Failure | ENSG0000016 | -0.00210836 | 0.417660964 | 0.002601351 | 0.07605539 | -0.0020561 | 0.429245412 | 0.002601064 | 0.076035243 | -5.23E-05 | 0.002575295 | 1 | 0.87398914 | 1 | 0.875416354 | ENSG0000013 | 7 | TTC5 |
| SAT | Heart_Failure | ENSG0000016 | 0.000615914 | 0.82308105 | 0.002754731 | 0.076055485 | 0.000552503 | 0.841023742 | 0.002754468 | 0.076035334 | 6.34E-05 | 0.007452301 | 1 | 0.976669362 | 1 | 0.972791643 | ENSG0000013 | 5 | MTHFS |
| SAT | Heart_Failure | ENSG0000016 | 0.001937632 | 0.407817909 | 0.002340869 | 0.076055386 | 0.001899302 | 0.417108248 | 0.00234063 | 0.076035237 | 3.83E-05 | 0.035374335 | 1 | 0.870429337 | 1 | 0.867451005 | ENSG0000013 | 5 | MTHFS |
| SAT | Heart_Failure | ENSG0000016 | 0.005563327 | 0.154041327 | 0.003902985 | 0.076055176 | 0.005513761 | 0.157696835 | 0.003902547 | 0.076035029 | 4.96E-05 | 0.046082225 | 1 | 0.677064618 | 1 | 0.669469928 | ENSG0000013 | 5 | MTHFS |
| SAT | Heart_Failure | ENSG0000016 | -0.00087284 | 0.569422201 | 0.001534243 | 0.076055443 | -0.00089714 | 0.558678381 | 0.001534074 | 0.076035287 | 2.43E-05 | 0.018844012 | 1 | 0.92942386 | 1 | 0.916248766 | ENSG0000013 | 5 | MTHFS |
| SAT | Heart_Failure | ENSG0000016 | 6.04E-05 | 0.980104003 | 0.002420775 | 0.076055493 | 1.89E-05 | 0.993777617 | 0.002420513 | 0.07603534 | 4.15E-05 | 0.013979582 | 1 | 0.998920999 | 1 | 0.999768899 | ENSG0000013 | 5 | MTHFS |
| SAT | Heart_Failure | ENSG0000016 | 0.0038122 | 0.137633142 | 0.002567717 | 0.076055149 | 0.003850736 | 0.133656711 | 0.002567432 | 0.076034989 | -3.85E-05 | 0.023432524 | 1 | 0.663104757 | 1 | 0.649215182 | ENSG0000013 | 5 | MTHFS |
| SAT | Heart_Failure | ENSG0000016 | -0.00114257 | 0.66545827 | 0.002642443 | 0.076055464 | -0.00105968 | 0.688371891 | 0.002642169 | 0.076035315 | -8.29E-05 | 3.86E-05 | 1 | 0.962994646 | 1 | 0.956878334 | ENSG0000013 | 8 | CIB2 |
| SAT | Heart_Failure | ENSG0000016 | -0.00062681 | 0.651332252 | 0.001387024 | 0.076055461 | -0.00066493 | 0.631621896 | 0.001386874 | 0.076035304 | 3.81E-05 | 8.11E-05 | 1 | 0.962760695 | 1 | 0.94798204 | ENSG0000013 | 8 | CIB2 |
| SAT | Heart_Failure | ENSG0000016 | 0.004510811 | 0.21582402 | 0.00364448 | 0.076055254 | 0.004613795 | 0.205477894 | 0.0036441 | 0.07603509 | -0.00010298 | 0.000180316 | 1 | 0.745333464 | 1 | 0.731678624 | ENSG0000013 | 7 | NMT1 |
| SAT | Heart_Failure | ENSG0000016 | 0.000170144 | 0.899144254 | 0.001342438 | 0.076055491 | 0.000197953 | 0.882759677 | 0.00134231 | 0.076035337 | -2.78E-05 | 0.016855028 | 1 | 0.980097109 | 1 | 0.974665278 | ENSG0000013 | 7 | NMT1 |
| SAT | Heart_Failure | ENSG0000016 | -0.00189122 | 0.252384294 | 0.001652329 | 0.076055289 | -0.00193318 | 0.241961113 | 0.001652147 | 0.076035127 | 4.20E-05 | 0.000160896 | 1 | 0.767444733 | 1 | 0.753454838 | ENSG0000013 | 7 | NMT1 |
| SAT | Heart_Failure | ENSG0000016 | -0.00746091 | 0.030552651 | 0.003449564 | 0.076054763 | -0.00737184 | 0.032579386 | 0.00344924 | 0.076034627 | -8.91E-05 | 0.003209399 | 1 | 0.390051553 | 1 | 0.406437132 | ENSG0000013 | 9 | MYCBPAP |
| SAT | Heart_Failure | ENSG0000016 | 0.003939019 | 0.088958341 | 0.002315814 | 0.076055041 | 0.003980014 | 0.085650525 | 0.00231557 | 0.076034879 | -4.10E-05 | 0.016412735 | 1 | 0.597756036 | 1 | 0.592867877 | ENSG0000013 | 9 | MYCBPAP |
| SAT | Heart_Failure | ENSG0000016 | -0.00016409 | 0.959360038 | 0.003220228 | 0.076055493 | -0.00010775 | 0.973304191 | 0.003219923 | 0.07603534 | -5.63E-05 | 0.044347974 | 1 | 0.994780111 | 1 | 0.998213938 | ENSG0000013 | 4 | LIMD2 |
| SAT | Heart_Failure | ENSG0000016 | -0.00036804 | 0.738669208 | 0.001103194 | 0.076055476 | -0.00034676 | 0.753246011 | 0.001103073 | 0.076035325 | -2.13E-05 | 0.004086698 | 1 | 0.966658386 | 1 | 0.962420704 | ENSG0000013 | 4 | LIMD2 |
| SAT | Heart_Failure | ENSG0000016 | -0.0027899 | 0.164630237 | 0.002007601 | 0.076055192 | -0.00275402 | 0.170080015 | 0.002007379 | 0.076035046 | -3.59E-05 | 0.007235616 | 1 | 0.689582559 | 1 | 0.692348051 | ENSG0000013 | 4 | LIMD2 |
| SAT | Heart_Failure | ENSG0000016 | 0.000215369 | 0.94360509 | 0.003044529 | 0.076055492 | 0.000172051 | 0.954929448 | 0.0030442 | 0.07603534 | 4.33E-05 | 0.041030332 | 1 | 0.992414898 | 1 | 0.991911131 | ENSG0000013 | 4 | BRIP1 |
| SAT | Heart_Failure | ENSG0000016 | -0.0014364 | 0.660271946 | 0.003267976 | 0.076055463 | -0.00149702 | 0.646855027 | 0.003267634 | 0.076035307 | 6.06E-05 | 0.013342604 | 1 | 0.962994646 | 1 | 0.95317119 | ENSG0000013 | 5 | CCDC115 |
| SAT | Heart_Failure | ENSG0000016 | -0.00050123 | 0.706644021 | 0.001331763 | 0.076055471 | -0.00040094 | 0.763443672 | 0.00133218 | 0.076035326 | -0.0001003 | 0.011640823 | 1 | 0.966658386 | 1 | 0.965635578 | ENSG0000013 | 13 | SAP130 |
| SAT | Heart_Failure | ENSG0000016 | -0.00376969 | 0.308516012 | 0.003701787 | 0.076055331 | -0.00387025 | 0.295739226 | 0.003701413 | 0.07603517 | 0.000100569 | 0.000610101 | 1 | 0.816225947 | 1 | 0.809274729 | ENSG0000013 | 13 | SAP130 |
| SAT | Heart_Failure | ENSG0000016 | 0.000873872 | 0.822062279 | 0.003885715 | 0.076055485 | 0.000806748 | 0.835510385 | 0.003885329 | 0.076035333 | 6.71E-05 | 0.033308583 | 1 | 0.976669362 | 1 | 0.972791643 | ENSG0000013 | 13 | SAP130 |
| SAT | Heart_Failure | ENSG0000016 | 0.00177486 | 0.532745526 | 0.002845144 | 0.076055432 | 0.001844944 | 0.516651358 | 0.002844869 | 0.076035275 | -7.01E-05 | 0.003573493 | 1 | 0.908491721 | 1 | 0.903395669 | ENSG0000013 | 13 | SAP130 |
| SAT | Heart_Failure | ENSG0000016 | -0.00013364 | 0.970148438 | 0.00357114 | 0.076055493 | -7.56E-05 | 0.983110455 | 0.003570753 | 0.07603534 | -5.80E-05 | 0.019856771 | 1 | 0.996886008 | 1 | 0.999768899 | ENSG0000013 | 10 | BIN1 |
| SAT | Heart_Failure | ENSG0000016 | -0.00191071 | 0.526417229 | 0.003016187 | 0.07605543 | -0.00196267 | 0.515187186 | 0.003015866 | 0.076035274 | 5.20E-05 | 0.017085598 | 1 | 0.906724424 | 1 | 0.903395669 | ENSG0000013 | 9 | NIPSNAP3A |
| SAT | Heart_Failure | ENSG0000016 | -0.00329456 | 0.397302777 | 0.003892208 | 0.076055381 | 0.003971338 | 0.323564233 | 0.00402299 | 0.076035188 | -0.00726589 | 1.02E-12 | 1 | 0.86729901 | 1 | 0.828598718 | ENSG0000013 | 9 | NIPSNAP3A |
| SAT | Heart_Failure | ENSG0000016 | 0.000256309 | 0.836712579 | 0.00124361 | 0.076055486 | 0.000303714 | 0.807042244 | 0.001243487 | 0.076035331 | -4.74E-05 | 2.90E-06 | 1 | 0.976669362 | 1 | 0.972791643 | ENSG0000013 | 11 | TOR1B |
| SAT | Heart_Failure | ENSG0000016 | -0.0061314 | 0.128270962 | 0.004031272 | 0.076055132 | -0.00598943 | 0.137345326 | 0.004031246 | 0.076034996 | -0.00014196 | 0.026600812 | 1 | 0.649888404 | 1 | 0.649215182 | ENSG0000013 | 11 | TOR1B |
| SAT | Heart_Failure | ENSG0000016 | -0.0043463 | 0.172354377 | 0.003184849 | 0.076055202 | -0.00427053 | 0.179911188 | 0.003184511 | 0.07603506 | -7.58E-05 | 0.001055382 | 1 | 0.704583343 | 1 | 0.709481499 | ENSG0000013 | 11 | TOR1B |
| SAT | Heart_Failure | ENSG0000016 | 0.001468092 | 0.577125479 | 0.002632929 | 0.076055444 | 0.001546371 | 0.556952171 | 0.002632681 | 0.076035286 | -7.83E-05 | 0.000685768 | 1 | 0.933702513 | 1 | 0.915531921 | ENSG0000013 | 13 | SLC2A8 |
| SAT | Heart_Failure | ENSG0000016 | -0.00019959 | 0.954056874 | 0.003464233 | 0.076055493 | -0.00406511 | 0.256550817 | 0.003582917 | 0.076035139 | 0.00386553 | 2.46E-05 | 1 | 0.994131568 | 1 | 0.770741077 | ENSG0000013 | 13 | SLC2A8 |
| SAT | Heart_Failure | ENSG0000016 | 0.00471074 | 0.018162268 | 0.001994132 | 0.076054622 | 0.004789972 | 0.016293457 | 0.00199325 | 0.076034444 | -7.92E-05 | 1.68E-07 | 1 | 0.292109804 | 1 | 0.285876102 | ENSG0000013 | 13 | SLC2A8 |
| SAT | Heart_Failure | ENSG0000016 | 0.00123112 | 0.152856034 | 0.000861214 | 0.076055174 | 0.001245127 | 0.148193986 | 0.000861118 | 0.076035014 | -1.40E-05 | 0.012651031 | 1 | 0.677064618 | 1 | 0.665121824 | ENSG0000013 | 13 | SLC2A8 |
| SAT | Heart_Failure | ENSG0000016 | -3.99E-05 | 0.967328137 | 0.000975134 | 0.076055493 | -2.41E-05 | 0.980291619 | 0.000975033 | 0.07603534 | -1.59E-05 | 0.031605969 | 1 | 0.996432082 | 1 | 0.999768899 | ENSG0000013 | 9 | ZFP37 |
| SAT | Heart_Failure | ENSG0000016 | -0.0003886 | 0.816627914 | 0.001676262 | 0.076055485 | -0.00034122 | 0.838679602 | 0.001676089 | 0.076035334 | -4.74E-05 | 0.000209029 | 1 | 0.976669362 | 1 | 0.972791643 | ENSG0000013 | 6 | STX17 |
| SAT | Heart_Failure | ENSG0000016 | 0.001341731 | 0.103534697 | 0.000824181 | 0.076055079 | 0.000839346 | 0.320818322 | 0.000845451 | 0.076035186 | 0.000502385 | 0.007832121 | 1 | 0.629069151 | 1 | 0.825572481 | ENSG0000013 | 8 | PRPF4 |
| SAT | Heart_Failure | ENSG0000016 | -0.00567469 | 0.145635351 | 0.003899798 | 0.076055163 | -0.00201193 | 0.612077737 | 0.003967448 | 0.0760353 | -0.00366276 | 5.67E-07 | 1 | 0.672999418 | 1 | 0.945324871 | ENSG0000013 | 8 | PRPF4 |
| SAT | Heart_Failure | ENSG0000016 | -0.00465153 | 0.002220424 | 0.001520573 | 0.076054033 | -0.00462226 | 0.00236481 | 0.00152041 | 0.076033898 | -2.93E-05 | 0.006728188 | 1 | 0.10407443 | 1 | 0.119368322 | ENSG0000013 | 8 | PRPF4 |
| SAT | Heart_Failure | ENSG0000016 | -0.00259718 | 0.401692333 | 0.003097032 | 0.076055383 | -0.0026953 | 0.384096917 | 0.003096714 | 0.076035222 | 9.81E-05 | 4.05E-05 | 1 | 0.870429337 | 1 | 0.854976426 | ENSG0000013 | 6 | ATP6V1G1 |
| SAT | Heart_Failure | ENSG0000016 | -0.00620257 | 0.085342643 | 0.003605102 | 0.076055031 | -0.00613857 | 0.088586797 | 0.003604765 | 0.076034888 | -6.40E-05 | 0.044040404 | 1 | 0.592308253 | 1 | 0.596493272 | ENSG0000013 | 9 | C9orf156 |
| SAT | Heart_Failure | ENSG0000016 | -0.00132504 | 0.602498074 | 0.002544174 | 0.076055451 | -0.00126581 | 0.618775845 | 0.002543894 | 0.076035302 | -5.92E-05 | 0.000509753 | 1 | 0.945538861 | 1 | 0.94798204 | ENSG0000013 | 9 | C9orf156 |

| | | | | | | | | | | | | | | | | | | |
|---|---|---|---|---|---|---|---|---|---|---|---|---|---|---|---|---|---|---|
| SAT | Heart_Failure | ENSG0000016 | -0.00012614 | 0.942899443 | 0.001761105 | 0.076055492 | -8.60E-06 | 0.996104796 | 0.001761319 | 0.07603534 | -0.00011754 | 0.003047083 | 1 | 0.992414898 | 1 | 0.999768899 | ENSG0000013 | 9 C9orf156 |
| SAT | Heart_Failure | ENSG0000016 | 0.000726405 | 0.704846718 | 0.001917717 | 0.076055471 | 0.000787663 | 0.681256582 | 0.001917625 | 0.076035314 | -6.13E-05 | 0.014067874 | 1 | 0.966658386 | 1 | 0.956878334 | ENSG0000013 | 9 ARPC5L |
| SAT | Heart_Failure | ENSG0000016 | -0.00130023 | 0.502622498 | 0.001939572 | 0.076055423 | -0.00133471 | 0.491312539 | 0.001939361 | 0.076035266 | 3.45E-05 | 0.010281988 | 1 | 0.900549813 | 1 | 0.894608312 | ENSG0000013 | 5 DSCC1 |
| SAT | Heart_Failure | ENSG0000016 | -0.00043316 | 0.772665634 | 0.001499392 | 0.07605548 | -0.00048467 | 0.746482332 | 0.001499231 | 0.076035324 | 5.15E-05 | 1.48E-06 | 1 | 0.971681082 | 1 | 0.962415493 | ENSG0000013 | 5 DSCC1 |
| SAT | Heart_Failure | ENSG0000017 | -0.00033676 | 0.860811403 | 0.001920578 | 0.076055488 | -0.00029381 | 0.878400144 | 0.001920374 | 0.076035336 | -4.29E-05 | 0.002018423 | 1 | 0.976669362 | 1 | 0.974665278 | ENSG0000013 | 11 POLR1E |
| SAT | Heart_Failure | ENSG0000017 | -0.00322115 | 0.114216366 | 0.002039315 | 0.076055104 | -0.00317305 | 0.119686326 | 0.002039113 | 0.076034962 | -4.81E-05 | 0.00379405 | 1 | 0.641850091 | 1 | 0.64481576 | ENSG0000013 | 11 POLR1E |
| SAT | Heart_Failure | ENSG0000017 | -0.00053155 | 0.839809571 | 0.002629645 | 0.076055487 | -0.00058339 | 0.824411598 | 0.002629376 | 0.076035332 | 5.18E-05 | 0.011180186 | 1 | 0.976669362 | 1 | 0.972791643 | ENSG0000013 | 13 IL11RA |
| SAT | Heart_Failure | ENSG0000017 | -0.00234675 | 0.525750311 | 0.003698546 | 0.07605543 | -0.00220684 | 0.550689037 | 0.003698227 | 0.076035285 | -0.00013992 | 8.53E-05 | 1 | 0.906724424 | 1 | 0.914465015 | ENSG0000013 | 13 IL11RA |
| SAT | Heart_Failure | ENSG0000017 | -0.00123991 | 0.59803221 | 0.002351731 | 0.07605545 | -0.00130523 | 0.578848818 | 0.002351479 | 0.076035292 | 6.53E-05 | 0.000103102 | 1 | 0.944700858 | 1 | 0.929614769 | ENSG0000013 | 13 IL11RA |
| SAT | Heart_Failure | ENSG0000017 | 0.002141156 | 0.41678579 | 0.002636857 | 0.07605539 | 0.002206326 | 0.402698905 | 0.002636586 | 0.076035231 | -6.52E-05 | 0.00136262 | 1 | 0.87398914 | 1 | 0.867451005 | ENSG0000013 | 13 IL11RA |
| SAT | Heart_Failure | ENSG0000017 | -0.00604718 | 0.069537359 | 0.003331935 | 0.076054979 | -0.00611691 | 0.066353692 | 0.003331595 | 0.076034814 | 6.97E-05 | 0.007414671 | 1 | 0.537536555 | 1 | 0.516964783 | ENSG0000013 | 13 GRHPR |
| SAT | Heart_Failure | ENSG0000017 | 0.004093533 | 0.266619509 | 0.003684938 | 0.0760553 | 0.004288786 | 0.244472941 | 0.003684902 | 0.076035129 | -0.00019525 | 0.000720093 | 1 | 0.779024202 | 1 | 0.757427498 | ENSG0000013 | 13 GRHPR |
| SAT | Heart_Failure | ENSG0000017 | -0.00254546 | 0.447747684 | 0.003352926 | 0.076055403 | -0.00262231 | 0.434113248 | 0.003352592 | 0.076035245 | 7.68E-05 | 0.004869993 | 1 | 0.884975994 | 1 | 0.877927942 | ENSG0000013 | 7 ZSCAN9 |
| SAT | Heart_Failure | ENSG0000017 | -0.0008074 | 0.753202436 | 0.002567911 | 0.076055478 | -0.00076126 | 0.766862449 | 0.002567638 | 0.076035326 | -4.61E-05 | 0.013416189 | 1 | 0.966658386 | 1 | 0.965963214 | ENSG0000013 | 7 ZSCAN9 |
| SAT | Heart_Failure | ENSG0000017 | 0.002383044 | 0.332458016 | 0.002458835 | 0.076055346 | 0.002333224 | 0.342617035 | 0.002458593 | 0.0760352 | 4.98E-05 | 0.013689098 | 1 | 0.840365755 | 1 | 0.839311387 | ENSG0000013 | 7 ZSCAN9 |
| SAT | Heart_Failure | ENSG0000017 | -0.00019057 | 0.891623486 | 0.001398675 | 0.07605549 | -0.00016906 | 0.903785062 | 0.001398531 | 0.076035338 | -2.15E-05 | 0.046027889 | 1 | 0.979270459 | 1 | 0.981243808 | ENSG0000013 | 7 ZSCAN9 |
| SAT | Heart_Failure | ENSG0000017 | 0.001230049 | 0.505867213 | 0.0018489 | 0.076055424 | 0.001183368 | 0.522100924 | 0.001848693 | 0.076035276 | 4.67E-05 | 7.61E-05 | 1 | 0.9005699 | 1 | 0.904369708 | ENSG0000013 | 14 TMEM63B |
| SAT | Heart_Failure | ENSG0000017 | -0.00138785 | 0.198903996 | 0.00108031 | 0.076055235 | -0.00137054 | 0.204518093 | 0.001080198 | 0.076035089 | -1.73E-05 | 0.033860838 | 1 | 0.732439966 | 1 | 0.729840368 | ENSG0000013 | 14 TMEM63B |
| SAT | Heart_Failure | ENSG0000017 | 0.000228041 | 0.942263236 | 0.00314862 | 0.076055492 | 0.000313111 | 0.920777465 | 0.003148279 | 0.076035339 | -8.51E-05 | 0.000107209 | 1 | 0.992414898 | 1 | 0.984731983 | ENSG0000013 | 8 CAPN11 |
| SAT | Heart_Failure | ENSG0000017 | -0.00044047 | 0.650120447 | 0.00097107 | 0.076055461 | -0.00042026 | 0.665140219 | 0.000970971 | 0.076035311 | -2.02E-05 | 0.00800492 | 1 | 0.962760695 | 1 | 0.956878334 | ENSG0000013 | 8 CAPN11 |
| SAT | Heart_Failure | ENSG0000017 | -0.00494533 | 0.201168743 | 0.003868891 | 0.076055238 | -0.00502408 | 0.194041941 | 0.0038685 | 0.076035077 | 7.87E-05 | 0.010307431 | 1 | 0.735350411 | 1 | 0.723506187 | ENSG0000013 | 8 CAPN11 |
| SAT | Heart_Failure | ENSG0000017 | -0.00193127 | 0.250585486 | 0.001680929 | 0.076055287 | -0.0020014 | 0.233744341 | 0.001680757 | 0.076035119 | 7.01E-05 | 9.96E-08 | 1 | 0.766457153 | 1 | 0.750248244 | ENSG0000013 | 10 IRF4 |
| SAT | Heart_Failure | ENSG0000017 | 0.004502565 | 0.107168078 | 0.002794791 | 0.076055088 | 0.004576889 | 0.101460738 | 0.002794508 | 0.076034922 | -7.43E-05 | 0.000779706 | 1 | 0.639306298 | 1 | 0.614882715 | ENSG0000013 | 8 BPHL |
| SAT | Heart_Failure | ENSG0000017 | 0.000547144 | 0.744416795 | 0.001678303 | 0.076055476 | 0.000486541 | 0.771868902 | 0.001678126 | 0.076035327 | 6.06E-05 | 8.80E-07 | 1 | 0.966658386 | 1 | 0.965963214 | ENSG0000013 | 9 RIPK1 |
| SAT | Heart_Failure | ENSG0000017 | -0.00286243 | 0.2678371 | 0.002583281 | 0.076055301 | -0.00300018 | 0.245440071 | 0.002583028 | 0.07603513 | 0.000137755 | 1.57E-10 | 1 | 0.780722709 | 1 | 0.758328312 | ENSG0000013 | 9 TUBB2B |
| SAT | Heart_Failure | ENSG0000017 | -0.00114058 | 0.494796317 | 0.001670688 | 0.07605542 | -0.00109518 | 0.512084398 | 0.001670517 | 0.076035273 | -4.54E-05 | 0.000491767 | 1 | 0.898909993 | 1 | 0.902820479 | ENSG0000013 | 9 TUBB2B |
| SAT | Heart_Failure | ENSG0000017 | 0.002024638 | 0.627667406 | 0.004174401 | 0.076055456 | 0.002132378 | 0.609438857 | 0.004173973 | 0.076035299 | -0.00010774 | 0.000864486 | 1 | 0.958246837 | 1 | 0.944452141 | ENSG0000013 | 9 TUBB2B |
| SAT | Heart_Failure | ENSG0000017 | -0.0019812 | 0.382789707 | 0.002270018 | 0.076055374 | -0.00203363 | 0.370275588 | 0.002269777 | 0.076035215 | 5.24E-05 | 0.001498832 | 1 | 0.85759885 | 1 | 0.84956238 | ENSG0000013 | 9 TUBB2B |
| SAT | Heart_Failure | ENSG0000017 | -0.00111257 | 0.228376071 | 0.00092364 | 0.076055267 | -0.00109675 | 0.235011013 | 0.000923541 | 0.07603512 | -1.58E-05 | 0.016511451 | 1 | 0.751217604 | 1 | 0.751870773 | ENSG0000013 | 5 UQCC2 |
| SAT | Heart_Failure | ENSG0000017 | -0.00118059 | 0.62894249 | 0.002443183 | 0.076055457 | -0.00109933 | 0.652707093 | 0.002442932 | 0.076035309 | -8.13E-05 | 1.62E-05 | 1 | 0.95890349 | 1 | 0.954583686 | ENSG0000013 | 5 UQCC2 |
| SAT | Heart_Failure | ENSG0000017 | 0.001860554 | 0.638188758 | 0.003956664 | 0.076055459 | 0.001911985 | 0.628892845 | 0.003956219 | 0.076035304 | -5.14E-05 | 0.039855628 | 1 | 0.959428651 | 1 | 0.94798204 | ENSG0000013 | 5 UQCC2 |
| SAT | Heart_Failure | ENSG0000017 | -0.00028738 | 0.782497683 | 0.001040971 | 0.076055481 | -0.0003307 | 0.7507264 | 0.00104099 | 0.076035324 | 4.33E-05 | 0.016516239 | 1 | 0.972730696 | 1 | 0.962420704 | ENSG0000013 | 5 UQCC2 |
| SAT | Heart_Failure | ENSG0000017 | 0.001481551 | 0.272485077 | 0.001350106 | 0.076055305 | 0.001501649 | 0.2659807 | 0.001349957 | 0.076035147 | -2.01E-05 | 0.025329175 | 1 | 0.786300384 | 1 | 0.774729353 | ENSG0000013 | 7 TCF19 |
| SAT | Heart_Failure | ENSG0000017 | -0.00141728 | 0.711622531 | 0.003833815 | 0.076055472 | -0.00129671 | 0.735163772 | 0.003833414 | 0.076035322 | -0.00012057 | 2.59E-05 | 1 | 0.966658386 | 1 | 0.959837778 | ENSG0000013 | 7 TCF19 |
| SAT | Heart_Failure | ENSG0000017 | -0.00286226 | 0.228847183 | 0.002378602 | 0.076055267 | -0.0125109 | 0.007661623 | 0.004691633 | 0.076034231 | 0.009648641 | 0.017041087 | 1 | 0.751217604 | 1 | 0.202724069 | ENSG0000013 | 7 TCF19 |
| SAT | Heart_Failure | ENSG0000017 | -0.00086478 | 0.724722748 | 0.002455675 | 0.076055474 | -0.00081938 | 0.738602655 | 0.002455409 | 0.076035323 | -4.54E-05 | 0.007812786 | 1 | 0.966658386 | 1 | 0.960499534 | ENSG0000013 | 7 TCF19 |
| SAT | Heart_Failure | ENSG0000017 | 0.000320611 | 0.851756763 | 0.001715623 | 0.076055488 | 0.000264839 | 0.877306212 | 0.001715446 | 0.076035336 | 5.58E-05 | 2.25E-05 | 1 | 0.976669362 | 1 | 0.974665278 | ENSG0000013 | 7 TCF19 |
| SAT | Heart_Failure | ENSG0000017 | -0.00190557 | 0.338014843 | 0.001988915 | 0.07605535 | -0.00187816 | 0.344957335 | 0.001988698 | 0.076035201 | -2.74E-05 | 0.043505754 | 1 | 0.842725455 | 1 | 0.83966685 | ENSG0000013 | 7 TCF19 |
| SAT | Heart_Failure | ENSG0000017 | 0.000185952 | 0.89771036 | 0.001446487 | 0.07605549 | 0.000154832 | 0.914748162 | 0.001446333 | 0.076035338 | 3.11E-05 | 0.002723224 | 1 | 0.979990566 | 1 | 0.984094962 | ENSG0000013 | 7 TCF19 |
| SAT | Heart_Failure | ENSG0000017 | 0.011245539 | 0.000313956 | 0.003120701 | 0.076053466 | 0.011184706 | 0.000337862 | 0.003120368 | 0.076033335 | 6.08E-05 | 0.006514629 | 1 | 0.02641253 | 1 | 0.033591688 | ENSG0000013 | 7 TCF19 |
| SAT | Heart_Failure | ENSG0000017 | 0.000509957 | 0.83019191 | 0.002377913 | 0.076055486 | 0.000553958 | 0.81577535 | 0.002377686 | 0.076035332 | -4.40E-05 | 0.031645761 | 1 | 0.976669362 | 1 | 0.972791643 | ENSG0000013 | 7 TCF19 |
| SAT | Heart_Failure | ENSG0000017 | -0.00972244 | 0.000505002 | 0.002795321 | 0.076053605 | -0.00978353 | 0.000464676 | 0.002795009 | 0.076033428 | 6.11E-05 | 0.000717112 | 1 | 0.03682027 | 1 | 0.039533553 | ENSG0000013 | 7 TCF19 |
| SAT | Heart_Failure | ENSG0000017 | -0.0050446 | 0.03856122 | 0.002438366 | 0.076054825 | -0.00507573 | 0.037357205 | 0.002438093 | 0.076034664 | 3.11E-05 | 0.046719167 | 1 | 0.44392759 | 1 | 0.431580953 | ENSG0000013 | 10 FLOT1 |
| SAT | Heart_Failure | ENSG0000017 | 5.62E-05 | 0.987648174 | 0.003632013 | 0.076055493 | 0.000122436 | 0.973105627 | 0.003631665 | 0.07603534 | -6.62E-05 | 0.033272057 | 1 | 0.999440735 | 1 | 0.998213938 | ENSG0000013 | 10 FLOT1 |
| SAT | Heart_Failure | ENSG0000017 | -0.00348386 | 0.028127314 | 0.001586809 | 0.076054741 | -0.00345814 | 0.029291988 | 0.001586638 | 0.076034599 | -2.57E-05 | 0.021148119 | 1 | 0.369142867 | 1 | 0.383605048 | ENSG0000013 | 10 FLOT1 |
| SAT | Heart_Failure | ENSG0000017 | 4.94E-05 | 0.984512806 | 0.002543949 | 0.076055493 | 8.88E-05 | 0.972145224 | 0.002543685 | 0.07603534 | -3.94E-05 | 0.040911243 | 1 | 0.998924529 | 1 | 0.998061182 | ENSG0000013 | 10 FLOT1 |
| SAT | Heart_Failure | ENSG0000017 | 0.002322202 | 0.261173089 | 0.002066707 | 0.076055296 | 0.002387134 | 0.248026446 | 0.002066501 | 0.076035132 | -6.49E-05 | 9.63E-05 | 1 | 0.774632399 | 1 | 0.763390965 | ENSG0000013 | 10 FLOT1 |
| SAT | Heart_Failure | ENSG0000017 | -0.00110206 | 0.405340869 | 0.001324403 | 0.076055385 | -0.00113725 | 0.390459724 | 0.00132426 | 0.076035225 | 3.52E-05 | 0.000133353 | 1 | 0.870429337 | 1 | 0.859797554 | ENSG0000013 | 10 FLOT1 |
| SAT | Heart_Failure | ENSG0000017 | -0.00296507 | 0.167790508 | 0.002149629 | 0.076055196 | -0.00288949 | 0.178845226 | 0.002149407 | 0.076035058 | -7.56E-05 | 4.33E-06 | 1 | 0.694824396 | 1 | 0.706126577 | ENSG0000013 | 5 CLPS |
| SAT | Heart_Failure | ENSG0000017 | -0.00430676 | 0.068663215 | 0.002365529 | 0.076054976 | -0.00435033 | 0.065879484 | 0.002365276 | 0.076034812 | 4.36E-05 | 0.009992041 | 1 | 0.537536555 | 1 | 0.515872519 | ENSG0000013 | 5 CLPS |
| SAT | Heart_Failure | ENSG0000017 | -0.00306641 | 0.057336998 | 0.001613272 | 0.076054929 | -0.00310095 | 0.054561595 | 0.001613099 | 0.076034764 | 3.45E-05 | 0.002593303 | 1 | 0.503931718 | 1 | 0.492997393 | ENSG0000013 | 10 NRM |
| SAT | Heart_Failure | ENSG0000017 | 0.00085785 | 0.813971364 | 0.003645681 | 0.076055484 | 0.000785337 | 0.829424467 | 0.003645273 | 0.076035333 | 7.25E-05 | 0.001986153 | 1 | 0.976669362 | 1 | 0.972791643 | ENSG0000013 | 10 NRM |
| SAT | Heart_Failure | ENSG0000017 | -0.0018356 | 0.520791523 | 0.002858629 | 0.076055429 | -0.00173451 | 0.543969315 | 0.002858356 | 0.076035283 | -0.00010109 | 4.23E-05 | 1 | 0.906724424 | 1 | 0.912455686 | ENSG0000013 | 10 NRM |
| SAT | Heart_Failure | ENSG0000017 | -0.00277148 | 0.388225025 | 0.003212038 | 0.076055377 | -0.00283145 | 0.377999628 | 0.003211753 | 0.076035219 | 6.00E-05 | 0.046029912 | 1 | 0.86096131 | 1 | 0.854967426 | ENSG0000013 | 12 VARS2 |
| SAT | Heart_Failure | ENSG0000017 | 0.001837062 | 0.225473291 | 0.001515598 | 0.076055264 | 0.001813771 | 0.231360137 | 0.001515436 | 0.076035117 | 2.33E-05 | 0.03080691 | 1 | 0.751217604 | 1 | 0.750242469 | ENSG0000013 | 12 VARS2 |
| SAT | Heart_Failure | ENSG0000017 | -0.00017827 | 0.919714922 | 0.001768685 | 0.076055491 | -0.00020591 | 0.90731105 | 0.001768489 | 0.07603538 | 2.76E-05 | 0.018524825 | 1 | 0.988399823 | 1 | 0.982067422 | ENSG0000013 | 12 VARS2 |
| SAT | Heart_Failure | ENSG0000017 | 0.006727515 | 0.061228955 | 0.00359406 | 0.076054946 | 0.006800128 | 0.058458333 | 0.003593674 | 0.076034781 | -7.26E-05 | 0.004469819 | 1 | 0.510326031 | 1 | 0.503576399 | ENSG0000013 | 12 VARS2 |
| SAT | Heart_Failure | ENSG0000017 | 0.003124141 | 0.216944059 | 0.002530291 | 0.076055255 | 0.003051545 | 0.227781818 | 0.002530105 | 0.076035113 | 7.26E-05 | 0.008277737 | 1 | 0.745333464 | 1 | 0.747837725 | ENSG0000013 | 12 VARS2 |
| SAT | Heart_Failure | ENSG0000017 | -0.0003531 | 0.694375645 | 0.000898648 | 0.076055469 | -0.00033813 | 0.706688338 | 0.000898555 | 0.076035318 | -1.50E-05 | 0.02814697 | 1 | 0.966658386 | 1 | 0.958028875 | ENSG0000013 | 12 VARS2 |
| SAT | Heart_Failure | ENSG0000017 | 0.00066982 | 0.77069273 | 0.002298082 | 0.07605548 | -0.00122283 | 0.605735444 | 0.00236905 | 0.076035299 | 0.001892654 | 0.00103191 | 1 | 0.971467105 | 1 | 0.942350873 | ENSG0000013 | 12 VARS2 |
| SAT | Heart_Failure | ENSG0000017 | -0.00235649 | 0.338437181 | 0.002461706 | 0.07605535 | -0.00230153 | 0.349772354 | 0.002461447 | 0.076035204 | -5.50E-05 | 0.002517527 | 1 | 0.84294423 | 1 | 0.842583404 | ENSG0000013 | 15 MYO7A |
| SAT | Heart_Failure | ENSG0000017 | 0.000472908 | 0.811281845 | 0.001980577 | 0.076055484 | 0.000526679 | 0.790277739 | 0.001980366 | 0.076035329 | -5.38E-05 | 0.000160606 | 1 | 0.976669362 | 1 | 0.970635778 | ENSG0000013 | 10 FCHSD2 |
| SAT | Heart_Failure | ENSG0000017 | -0.0032738 | 0.215842823 | 0.002645152 | 0.076055254 | -0.00208181 | 0.431645133 | 0.002647329 | 0.076035244 | -0.00119199 | 6.20E-25 | 1 | 0.745333464 | 1 | 0.876305924 | ENSG0000013 | 10 FCHSD2 |
| SAT | Heart_Failure | ENSG0000017 | 0.005632262 | 0.147947819 | 0.003892854 | 0.076055166 | 0.005719111 | 0.141752748 | 0.003892412 | 0.076035003 | -8.68E-05 | 0.000286957 | 1 | 0.674894985 | 1 | 0.657837012 | ENSG0000013 | 10 FCHSD2 |
| SAT | Heart_Failure | ENSG0000017 | 0.001756222 | 0.574404032 | 0.003127299 | 0.076055444 | 0.001832508 | 0.557857764 | 0.003127003 | 0.076035287 | -7.63E-05 | 0.004973038 | 1 | 0.932980015 | 1 | 0.915623473 | ENSG0000013 | 10 FCHSD2 |
| SAT | Heart_Failure | ENSG0000017 | 9.02E-05 | 0.965170726 | 0.00206585 | 0.076055493 | 0.000122661 | 0.952647874 | 0.002065621 | 0.07603534 | -3.25E-05 | 0.016865458 | 1 | 0.996432082 | 1 | 0.991228627 | ENSG0000013 | 12 SYTL2 |
| SAT | Heart_Failure | ENSG0000017 | 0.004915945 | 0.150132105 | 0.003416063 | 0.07605517 | 0.004829644 | 0.157377016 | 0.003415709 | 0.076035028 | 8.63E-05 | 0.000864378 | 1 | 0.6766256 | 1 | 0.669469928 | ENSG0000013 | 6 PRCP |
| SAT | Heart_Failure | ENSG0000017 | 0.004707783 | 0.009808579 | 0.001822944 | 0.076054452 | 0.00474939 | 0.009171066 | 0.001822748 | 0.076034281 | -4.16E-05 | 0.001172801 | 1 | 0.227178996 | 1 | 0.221316258 | ENSG0000013 | 6 PRCP |
| SAT | Heart_Failure | ENSG0000017 | 0.009385883 | 0.003747479 | 0.00323798 | 0.076054182 | 0.009433806 | 0.003570734 | 0.003237628 | 0.076034015 | -4.79E-05 | 0.033059851 | 1 | 0.146649141 | 1 | 0.154152365 | ENSG0000013 | 6 PRCP |
| SAT | Heart_Failure | ENSG0000017 | -0.00134171 | 0.270018638 | 0.0012164 | 0.076055303 | -0.00136632 | 0.261281269 | 0.001216273 | 0.076035143 | 2.46E-05 | 0.006986757 | 1 | 0.782624692 | 1 | 0.774402751 | ENSG0000013 | 6 PRCP |
| SAT | Heart_Failure | ENSG0000017 | -0.00840938 | 0.008083753 | 0.003175073 | 0.076054398 | -0.00830222 | 0.008925632 | 0.003174966 | 0.076034273 | -0.00010716 | 0.016263516 | 1 | 0.203936797 | 1 | 0.218544776 | ENSG0000013 | 6 PRCP |
| SAT | Heart_Failure | ENSG0000017 | 0.00070752 | 0.814558277 | 0.003016501 | 0.076055484 | 0.000777244 | 0.796646579 | 0.003016202 | 0.07603533 | -6.97E-05 | 0.00463922 | 1 | 0.976669362 | 1 | 0.971094631 | ENSG0000013 | 4 TTPA |
| SAT | Heart_Failure | ENSG0000017 | -0.0025676 | 0.202204507 | 0.002013332 | 0.076055239 | -0.00250941 | 0.212571325 | 0.002013119 | 0.076035098 | -5.82E-05 | 6.49E-05 | 1 | 0.735350411 | 1 | 0.733958243 | ENSG0000013 | 4 TTPA |
| SAT | Heart_Failure | ENSG0000017 | -0.0004855 | 0.640381753 | 0.001039249 | 0.076055459 | -0.00045571 | 0.660990878 | 0.001039138 | 0.07603531 | -2.98E-05 | 5.97E-05 | 1 | 0.959428651 | 1 | 0.956878334 | ENSG0000013 | 7 GGH |
| SAT | Heart_Failure | ENSG0000017 | -0.00504824 | 0.188341419 | 0.003837494 | 0.076055223 | -0.00494487 | 0.197501429 | 0.003837088 | 0.076035081 | -0.00010336 | 0.00022447 | 1 | 0.722191019 | 1 | 0.725655076 | ENSG0000013 | 7 GGH |
| SAT | Heart_Failure | ENSG0000017 | -0.00058055 | 0.872177747 | 0.003608278 | 0.076055489 | -0.00051904 | 0.885610815 | 0.003607912 | 0.076035337 | -6.15E-05 | 0.02992715 | 1 | 0.979223495 | 1 | 0.975390764 | ENSG0000013 | 7 GGH |
| SAT | Heart_Failure | ENSG0000017 | 0.000892331 | 0.77796173 | 0.003164559 | 0.076055481 | 0.001001805 | 0.751545879 | 0.003164243 | 0.076035324 | -0.00010947 | 1.95E-05 | 1 | 0.972664863 | 1 | 0.962420704 | ENSG0000013 | 6 TGS1 |
| SAT | Heart_Failure | ENSG0000017 | 0.000460032 | 0.853188767 | 0.002485966 | 0.076055488 | 0.00054679 | 0.825891272 | 0.002485701 | 0.076035333 | -8.68E-05 | 1.16E-06 | 1 | 0.976669362 | 1 | 0.972791643 | ENSG0000013 | 6 TGS1 |
| SAT | Heart_Failure | ENSG0000017 | 0.0017645 | 0.344369945 | 0.001866076 | 0.076055353 | 0.001799017 | 0.334963024 | 0.001865878 | 0.076035195 | -3.45E-05 | 0.011470987 | 1 | 0.847011103 | 1 | 0.836830236 | ENSG0000013 | 8 TRPC6 |
| SAT | Heart_Failure | ENSG0000017 | -0.00144762 | 0.378492248 | 0.001643751 | 0.076055372 | -0.0014191 | 0.38790579 | 0.001643581 | 0.076035224 | -2.85E-05 | 0.022946978 | 1 | 0.85759885 | 1 | 0.858921514 | ENSG0000013 | 8 TRPC6 |
| SAT | Heart_Failure | ENSG0000017 | 0.000786845 | 0.59738181 | 0.001489757 | 0.076055449 | 0.000743622 | 0.617632655 | 0.001489603 | 0.076035301 | 4.32E-05 | 0.000129358 | 1 | 0.944127995 | 1 | 0.947942988 | ENSG0000013 | 8 TRPC6 |
| SAT | Heart_Failure | ENSG0000017 | 0.002773691 | 0.277440172 | 0.002553834 | 0.076055309 | 0.002714569 | 0.287759003 | 0.002553563 | 0.076035164 | 5.91E-05 | 0.001370295 | 1 | 0.793619184 | 1 | 0.799139368 | ENSG0000013 | 4 MMP7 |
| SAT | Heart_Failure | ENSG0000017 | 0.001600121 | 0.321029092 | 0.001612462 | 0.076055339 | 0.00157559 | 0.328453128 | 0.001612293 | 0.076035191 | 2.45E-05 | 0.041555428 | 1 | 0.828692762 | 1 | 0.833382337 | ENSG0000013 | 4 MMP7 |
| SAT | Heart_Failure | ENSG0000017 | -0.00282219 | 0.007048596 | 0.001047369 | 0.07605436 | -0.00284326 | 0.006628857 | 0.001047261 | 0.07603419 | 2.11E-05 | 0.009528222 | 1 | 0.182097987 | 1 | 0.191383111 | ENSG0000013 | 4 MMP27 |
| SAT | Heart_Failure | ENSG0000017 | -0.0020621 | 0.516252138 | 0.0031767 | 0.076055427 | -0.00198286 | 0.532459439 | 0.003176353 | 0.076035279 | -7.92E-05 | 0.000246504 | 1 | 0.906069117 | 1 | 0.908441675 | ENSG0000013 | 7 DCUN1D5 |
| SAT | Heart_Failure | ENSG0000017 | 0.000736321 | 0.691961081 | 0.00185848 | 0.076055469 | 0.000654551 | 0.724670221 | 0.001858335 | 0.076035321 | 8.18E-05 | 2.61E-05 | 1 | 0.966658386 | 1 | 0.958028875 | ENSG0000013 | 7 DCUN1D5 |
| SAT | Heart_Failure | ENSG0000017 | 0.003073596 | 0.411591754 | 0.003743283 | 0.076055388 | 0.003175859 | 0.396158315 | 0.003742897 | 0.076035228 | -0.00010226 | 0.000355764 | 1 | 0.870429337 | 1 | 0.862334842 | ENSG0000013 | 12 SLC37A4 |
| SAT | Heart_Failure | ENSG0000017 | -0.00068106 | 0.781802275 | 0.002458941 | 0.076055481 | -0.00073623 | 0.764602668 | 0.002458684 | 0.076035326 | 5.52E-05 | 0.002787344 | 1 | 0.972730696 | 1 | 0.965679451 | ENSG0000013 | 5 POU2F3 |
| SAT | Heart_Failure | ENSG0000017 | -0.00070902 | 0.521268165 | 0.001105445 | 0.076055429 | -0.00069124 | 0.531728017 | 0.001105334 | 0.076035279 | -1.78E-05 | 0.046874549 | 1 | 0.906724424 | 1 | 0.908441675 | ENSG0000013 | 5 POU2F3 |
| SAT | Heart_Failure | ENSG0000018 | -0.00106059 | 0.784719133 | 0.003882477 | 0.076055481 | -0.00089718 | 0.817234747 | 0.003882181 | 0.076035332 | -0.00016341 | 7.22E-05 | 1 | 0.972730696 | 1 | 0.972791643 | ENSG0000013 | 5 POU2F3 |
| SAT | Heart_Failure | ENSG0000018 | 0.000914193 | 0.704807868 | 0.002413145 | 0.076055471 | 0.00095937 | 0.690922391 | 0.002412879 | 0.076035315 | -4.52E-05 | 0.005117811 | 1 | 0.966658386 | 1 | 0.956878334 | ENSG0000013 | 5 POU2F3 |
| SAT | Heart_Failure | ENSG0000018 | -0.00030479 | 0.730789708 | 0.000885832 | 0.076055475 | -0.00027467 | 0.756484747 | 0.000885741 | 0.076035325 | -3.01E-05 | 9.58E-06 | 1 | 0.966658386 | 1 | 0.962699472 | ENSG0000013 | 5 POU2F3 |
| SAT | Heart_Failure | ENSG0000018 | 0.003210185 | 0.228387856 | 0.002665107 | 0.076055267 | 0.003141224 | 0.238489565 | 0.002664827 | 0.076035123 | 6.90E-05 | 0.000471798 | 1 | 0.751217604 | 1 | 0.752162449 | ENSG0000013 | 18 CASP1 |
| SAT | Heart_Failure | ENSG0000018 | 0.00445417 | 0.004373679 | 0.001562929 | 0.076054225 | 0.004497801 | 0.004001389 | 0.001562786 | 0.076034048 | -4.36E-05 | 0.002035712 | 1 | 0.154647076 | 1 | 0.161355485 | ENSG0000013 | 18 CASP1 |
| SAT | Heart_Failure | ENSG0000018 | -0.00010708 | 0.936411782 | 0.001342188 | 0.076055492 | -0.00164534 | 0.223050262 | 0.001350349 | 0.076035108 | 0.001538264 | 1.01E-24 | 1 | 0.989966131 | 1 | 0.746763174 | ENSG0000013 | 18 CASP1 |
| SAT | Heart_Failure | ENSG0000018 | 0.00154404 | 0.694826159 | 0.003935722 | 0.076055469 | 0.001676567 | 0.670087961 | 0.00393535 | 0.076035312 | -0.00013253 | 0.000111421 | 1 | 0.966658386 | 1 | 0.956878334 | ENSG0000013 | 18 CASP1 |
| SAT | Heart_Failure | ENSG0000018 | -0.00559271 | 0.130094679 | 0.003694652 | 0.076055135 | -0.0055115 | 0.135726417 | 0.003694289 | 0.076034993 | -8.12E-05 | 0.007731935 | 1 | 0.649888404 | 1 | 0.649215182 | ENSG0000013 | 9 SQRDL |
| SAT | Heart_Failure | ENSG0000018 | -0.00053204 | 0.804658265 | 0.002151196 | 0.076055483 | -0.00079199 | 0.713098882 | 0.002153909 | 0.076035319 | 0.000259947 | 0.022134043 | 1 | 0.976669362 | 1 | 0.958028875 | ENSG0000013 | 4 NDUFAF1 |
| SAT | Heart_Failure | ENSG0000018 | 0.003131546 | 0.383356371 | 0.003592336 | 0.076055374 | 0.002477043 | 0.490644509 | 0.003593643 | 0.076035266 | 0.000654503 | 7.29E-09 | 1 | 0.85759885 | 1 | 0.894608312 | ENSG0000013 | 4 NDUFAF1 |
| SAT | Heart_Failure | ENSG0000018 | 0.000803008 | 0.655878041 | 0.001802037 | 0.076055462 | 0.000856779 | 0.634432789 | 0.001801858 | 0.076035305 | -5.38E-05 | 0.00026348 | 1 | 0.962994646 | 1 | 0.94798204 | ENSG0000013 | 4 NDUFAF1 |
| SAT | Heart_Failure | ENSG0000018 | 0.001268941 | 0.434937661 | 0.001625242 | 0.076055398 | 0.001230591 | 0.448896787 | 0.001625065 | 0.076035251 | 3.84E-05 | 0.000639586 | 1 | 0.880339889 | 1 | 0.882462767 | ENSG0000013 | 10 UACA |
| SAT | Heart_Failure | ENSG0000018 | 0.003794411 | 0.293141158 | 0.003609405 | 0.076055321 | 0.003693045 | 0.30617457 | 0.003609016 | 0.076035177 | 0.000101366 | 6.93E-05 | 1 | 0.806875957 | 1 | 0.814926763 | ENSG0000013 | 10 UACA |
| SAT | Heart_Failure | ENSG0000018 | -0.00135757 | 0.196162078 | 0.001050288 | 0.076055232 | -0.00138426 | 0.187476735 | 0.00105021 | 0.076035069 | 2.67E-05 | 0.018698171 | 1 | 0.726878652 | 1 | 0.7171788 | ENSG0000013 | 10 UACA |
| SAT | Heart_Failure | ENSG0000018 | 0.001182276 | 0.726809196 | 0.003383989 | 0.076055474 | 0.001092311 | 0.746832411 | 0.003383663 | 0.076035324 | 9.00E-05 | 0.001743797 | 1 | 0.966658386 | 1 | 0.962415493 | ENSG0000013 | 7 PLCB2 |
| SAT | Heart_Failure | ENSG0000018 | 0.000252759 | 0.938535936 | 0.003277893 | 0.076055492 | 0.000379566 | 0.907805695 | 0.00327758 | 0.076035338 | -0.00012681 | 6.69E-06 | 1 | 0.990777479 | 1 | 0.982067422 | ENSG0000013 | 7 PLCB2 |
| SAT | Heart_Failure | ENSG0000018 | 0.000531892 | 0.792116934 | 0.002018084 | 0.076055482 | -0.00130594 | 0.521597695 | 0.002037711 | 0.076035276 | 0.001837831 | 9.80E-11 | 1 | 0.9760395 | 1 | 0.904369708 | ENSG0000013 | 7 PAK6 |
| SAT | Heart_Failure | ENSG0000018 | 0.00078984 | 0.30059309 | 0.00076301 | 0.076055326 | 0.000766669 | 0.314949327 | 0.000762936 | 0.076035183 | 2.32E-05 | 0.000319838 | 1 | 0.814702237 | 1 | 0.823487262 | ENSG0000013 | 7 PAK6 |
| SAT | Heart_Failure | ENSG0000018 | -0.00133402 | 0.679525702 | 0.003229213 | 0.076055466 | -0.0014065 | 0.663125667 | 0.003228864 | 0.076035311 | 7.25E-05 | 0.001317019 | 1 | 0.966658386 | 1 | 0.956878334 | ENSG0000013 | 13 DUOX1 |
| SAT | Heart_Failure | ENSG0000018 | -0.00200062 | 0.140941234 | 0.001358841 | 0.076055155 | -0.00195422 | 0.150348542 | 0.001358698 | 0.076035017 | -4.64E-05 | 3.31E-06 | 1 | 0.669181587 | 1 | 0.665121824 | ENSG0000013 | 13 DUOX1 |
| SAT | Heart_Failure | ENSG0000018 | 0.000676098 | 0.861397619 | 0.003872371 | 0.076055488 | 0.000762301 | 0.843923512 | 0.003871956 | 0.076035334 | -8.62E-05 | 0.0017731 | 1 | 0.97692554 | 1 | 0.972791643 | ENSG0000013 | 13 DUOX1 |
| SAT | Heart_Failure | ENSG0000018 | 0.000646647 | 0.400640795 | 0.00076938 | 0.076055383 | 0.000625905 | 0.415872423 | 0.000769299 | 0.076035237 | 2.07E-05 | 0.000249326 | 1 | 0.870429337 | 1 | 0.867451005 | ENSG0000013 | 5 RSL24D1 |
| SAT | Heart_Failure | ENSG0000018 | -0.01070211 | 0.004641689 | 0.003780441 | 0.076054242 | -0.01085344 | 0.004089179 | 0.003780088 | 0.076034054 | 0.000151327 | 6.27E-06 | 1 | 0.15540184 | 1 | 0.161355485 | ENSG0000013 | 5 RSL24D1 |
| SAT | Heart_Failure | ENSG0000018 | 0.00087987 | 0.326551707 | 0.000896834 | 0.076055343 | 0.000895717 | 0.317861773 | 0.000896737 | 0.076035184 | -1.58E-05 | 0.012198395 | 1 | 0.833010855 | 1 | 0.823487262 | ENSG0000013 | 5 RSL24D1 |
| SAT | Heart_Failure | ENSG0000018 | -0.00238399 | 0.370156036 | 0.002660163 | 0.076055368 | -0.00233422 | 0.380183472 | 0.002659897 | 0.07603522 | -4.98E-05 | 0.020481651 | 1 | 0.855871709 | 1 | 0.854967426 | ENSG0000013 | 5 RSL24D1 |
| SAT | Heart_Failure | ENSG0000018 | 0.000866262 | 0.794210609 | 0.003320958 | 0.076055482 | 0.001979561 | 0.552918627 | 0.003336006 | 0.076035285 | -0.0011133 | 0.000525775 | 1 | 0.976267425 | 1 | 0.915531921 | ENSG0000013 | 8 SPTBN5 |
| SAT | Heart_Failure | ENSG0000018 | 0.000466594 | 0.736527815 | 0.001386794 | 0.076055475 | 0.000432196 | 0.755279089 | 0.001386639 | 0.076035325 | 3.44E-05 | 0.000115084 | 1 | 0.966658386 | 1 | 0.962699472 | ENSG0000013 | 10 DBT |
| SAT | Heart_Failure | ENSG0000018 | 0.000479371 | 0.744285145 | 0.001469633 | 0.076055476 | 0.000511091 | 0.727986772 | 0.001469476 | 0.076035321 | -3.17E-05 | 0.002546544 | 1 | 0.966658386 | 1 | 0.958028875 | ENSG0000013 | 10 DBT |
| SAT | Heart_Failure | ENSG0000018 | 0.000654319 | 0.545176268 | 0.001081512 | 0.076055436 | 0.00068194 | 0.528296141 | 0.001081398 | 0.076035278 | -2.76E-05 | 0.000577075 | 1 | 0.91464723 | 1 | 0.907507664 | ENSG0000013 | 8 RTCA |
| SAT | Heart_Failure | ENSG0000018 | -0.00082589 | 0.499300783 | 0.001222473 | 0.076055422 | -0.00080517 | 0.510077876 | 0.001222334 | 0.076035272 | -2.07E-05 | 0.005226462 | 1 | 0.899263643 | 1 | 0.902678269 | ENSG0000013 | 8 RTCA |
| SAT | Heart_Failure | ENSG0000018 | 0.000136711 | 0.961642647 | 0.002842677 | 0.076055493 | 5.76E-05 | 0.983838691 | 0.0028424 | 0.07603534 | 7.91E-05 | 0.000874163 | 1 | 0.995629386 | 1 | 0.999768899 | ENSG0000013 | 8 RTCA |
| SAT | Heart_Failure | ENSG0000018 | -0.00140149 | 0.185855018 | 0.001059377 | 0.07605522 | -0.00149399 | 0.158486805 | 0.00105943 | 0.07603503 | 9.25E-05 | 5.16E-06 | 1 | 0.720656885 | 1 | 0.670798618 | ENSG0000013 | 8 KHK |
| SAT | Heart_Failure | ENSG0000018 | -4.01E-05 | 0.989707951 | 0.003108655 | 0.076055493 | 4.68E-05 | 0.987990567 | 0.003108313 | 0.07603534 | -8.69E-05 | 2.96E-05 | 1 | 0.999440735 | 1 | 0.999768899 | ENSG0000013 | 8 KHK |
| SAT | Heart_Failure | ENSG0000018 | 0.001785149 | 0.381414616 | 0.002039486 | 0.076055373 | 0.001714806 | 0.400410099 | 0.002039276 | 0.07603523 | 7.03E-05 | 7.45E-06 | 1 | 0.85759885 | 1 | 0.867158768 | ENSG0000013 | 8 KHK |
| SAT | Heart_Failure | ENSG0000018 | -0.00149434 | 0.638993128 | 0.003185507 | 0.076055459 | -0.00154972 | 0.626581274 | 0.003185164 | 0.076035303 | 5.54E-05 | 0.013545513 | 1 | 0.959428651 | 1 | 0.94798204 | ENSG0000013 | 14 DYNC2LI1 |
| SAT | Heart_Failure | ENSG0000018 | -0.00355596 | 0.203531134 | 0.002796543 | 0.076055241 | -0.00347022 | 0.214596172 | 0.002796245 | 0.0760351 | -8.57E-05 | 1.99E-05 | 1 | 0.735350411 | 1 | 0.736362287 | ENSG0000013 | 14 DYNC2LI1 |
| SAT | Heart_Failure | ENSG0000018 | -0.00105679 | 0.731129708 | 0.003075422 | 0.076055475 | -0.00100064 | 0.744876132 | 0.003075092 | 0.076035324 | -5.61E-05 | 0.010443708 | 1 | 0.966658386 | 1 | 0.962415493 | ENSG0000013 | 12 ATRAID |
| SAT | Heart_Failure | ENSG0000018 | -0.00122322 | 0.47694513 | 0.001719874 | 0.076055414 | -0.00114443 | 0.5057434 | 0.001719701 | 0.076035271 | -7.88E-05 | 9.37E-09 | 1 | 0.895739537 | 1 | 0.900571724 | ENSG0000013 | 12 ATRAID |

| | | | | | | | | | | | | | | | | | | | |
|---|---|---|---|---|---|---|---|---|---|---|---|---|---|---|---|---|---|---|---|
| SAT | Heart_Failure | ENSG0000018 | 0.006081321 | 0.007817399 | 0.002286325 | 0.076054389 | 0.003972451 | 0.087643342 | 0.002325838 | 0.076034885 | 0.00210887 | 8.57E-07 | 1 | 0.198828569 | 1 | 0.596493272 | ENSG0000013 | 10 | TMEM180 |
| SAT | Heart_Failure | ENSG0000018 | 0.004000209 | 0.278706988 | 0.003692853 | 0.07605531 | 0.003927953 | 0.28742991 | 0.003692456 | 0.076035164 | 7.23E-05 | 0.005609717 | 1 | 0.793905695 | 1 | 0.799139368 | ENSG0000013 | 12 | MYOF |
| SAT | Heart_Failure | ENSG0000018 | -0.00056889 | 0.718001642 | 0.00157529 | 0.076055473 | -0.0005454 | 0.729150417 | 0.001575121 | 0.076035321 | -2.35E-05 | 0.037029039 | 1 | 0.966658386 | 1 | 0.958028875 | ENSG0000013 | 12 | MYOF |
| SAT | Heart_Failure | ENSG0000018 | -0.00081252 | 0.837382753 | 0.003958812 | 0.076055486 | -0.00074293 | 0.851123077 | 0.003958384 | 0.076035335 | -6.96E-05 | 0.01167313 | 1 | 0.976669362 | 1 | 0.972791643 | ENSG0000013 | 12 | MYOF |
| SAT | Heart_Failure | ENSG0000018 | -0.00190193 | 0.524565691 | 0.002988932 | 0.07605543 | -0.00181006 | 0.544758113 | 0.002988703 | 0.076035283 | -9.19E-05 | 0.003669816 | 1 | 0.906724424 | 1 | 0.912597963 | ENSG0000013 | 12 | MYOF |
| SAT | Heart_Failure | ENSG0000018 | 0.001533581 | 0.363891712 | 0.001689011 | 0.076055364 | 0.001495009 | 0.376033812 | 0.001688839 | 0.076035218 | 3.86E-05 | 0.003437753 | 1 | 0.85040303 | 1 | 0.854967426 | ENSG0000013 | 12 | MYOF |
| SAT | Heart_Failure | ENSG0000018 | -0.00195042 | 0.489323992 | 0.002821035 | 0.076055418 | -0.00159938 | 0.571016281 | 0.00282298 | 0.07603529 | -0.00035105 | 0.002145794 | 1 | 0.898909993 | 1 | 0.924404562 | ENSG0000013 | 5 | CH25H |
| SAT | Heart_Failure | ENSG0000018 | 0.000860458 | 0.765699499 | 0.002887395 | 0.076055479 | 0.000965892 | 0.737962553 | 0.002887112 | 0.076035323 | -0.00010543 | 1.12E-05 | 1 | 0.971109414 | 1 | 0.960499534 | ENSG0000013 | 5 | CH25H |
| SAT | Heart_Failure | ENSG0000018 | 0.000526317 | 0.477822995 | 0.000741491 | 0.076055414 | 0.000514466 | 0.487744949 | 0.000741412 | 0.076035265 | 1.19E-05 | 0.025458428 | 1 | 0.896874856 | 1 | 0.894608312 | ENSG0000013 | 5 | CH25H |
| SAT | Heart_Failure | ENSG0000018 | -0.00113256 | 0.491266728 | 0.001645455 | 0.076055419 | -0.00117036 | 0.476872076 | 0.001645276 | 0.076035261 | 3.78E-05 | 0.000845891 | 1 | 0.898909993 | 1 | 0.894608312 | ENSG0000013 | 6 | CALHM2 |
| SAT | Heart_Failure | ENSG0000018 | 0.00012424 | 0.934040817 | 0.001501178 | 0.076055492 | 0.000148888 | 0.920986474 | 0.001501021 | 0.076035339 | -2.46E-05 | 0.029230958 | 1 | 0.989642217 | 1 | 0.984731983 | ENSG0000013 | 8 | ARL3 |
| SAT | Heart_Failure | ENSG0000018 | -0.00074542 | 0.834036866 | 0.00355764 | 0.076055486 | -0.00079419 | 0.823332688 | 0.003557244 | 0.076035332 | 4.88E-05 | 0.035064042 | 1 | 0.976669362 | 1 | 0.972791643 | ENSG0000013 | 8 | ARL3 |
| SAT | Heart_Failure | ENSG0000018 | 0.002213795 | 0.494315228 | 0.003239089 | 0.07605542 | 0.002376278 | 0.463131227 | 0.003238755 | 0.076035256 | -0.00016248 | 6.03E-11 | 1 | 0.898909993 | 1 | 0.892844363 | ENSG0000013 | 8 | ARL3 |
| SAT | Heart_Failure | ENSG0000018 | 0.002094883 | 0.55218713 | 0.003523854 | 0.076055438 | 0.002164082 | 0.539094798 | 0.003523515 | 0.076035281 | -6.92E-05 | 0.021097086 | 1 | 0.920018143 | 1 | 0.91051702 | ENSG0000013 | 8 | ARL3 |
| SAT | Heart_Failure | ENSG0000018 | 0.011733361 | 4.37E-05 | 0.002870906 | 0.076052886 | 0.011829975 | 3.77E-05 | 0.002870621 | 0.07603269 | -9.66E-05 | 4.22E-05 | 0.143404981 | 0.005736199 | 0.123763084 | 0.009269033 | ENSG0000013 | 8 | ARL3 |
| SAT | Heart_Failure | ENSG0000018 | 0.000613807 | 0.831718164 | 0.002888523 | 0.076055486 | 0.000534774 | 0.853105804 | 0.002888216 | 0.076035335 | 7.90E-05 | 0.000164471 | 1 | 0.976669362 | 1 | 0.972791643 | ENSG0000013 | 10 | PLCE1 |
| SAT | Heart_Failure | ENSG0000018 | 0.0004591 | 0.649539159 | 0.001010338 | 0.076055461 | 0.000426856 | 0.672635659 | 0.00101023 | 0.076035312 | 3.22E-05 | 7.85E-06 | 1 | 0.962760695 | 1 | 0.956878334 | ENSG0000013 | 10 | PLCE1 |
| SAT | Heart_Failure | ENSG0000018 | -0.00798187 | 0.004218584 | 0.002789548 | 0.076054215 | -0.00588494 | 0.046441211 | 0.00295523 | 0.076034721 | -0.00209693 | 0.031784313 | 1 | 0.1521008 | 1 | 0.467403724 | ENSG0000013 | 10 | FAM149B1 |
| SAT | Heart_Failure | ENSG0000018 | -0.00635655 | 0.089105862 | 0.003738849 | 0.076055042 | -0.00627725 | 0.0931347 | 0.003738477 | 0.0760349 | -7.93E-05 | 0.00890211 | 1 | 0.597756036 | 1 | 0.599166569 | ENSG0000013 | 10 | FAM149B1 |
| SAT | Heart_Failure | ENSG0000018 | 0.004188563 | 0.190280741 | 0.003197984 | 0.076055225 | 0.004109615 | 0.198723648 | 0.003197651 | 0.076035082 | 7.89E-05 | 0.000993664 | 1 | 0.722912434 | 1 | 0.725655076 | ENSG0000013 | 10 | FAM149B1 |
| SAT | Heart_Failure | ENSG0000018 | -0.00041695 | 0.881400541 | 0.002794669 | 0.07605549 | -0.00035146 | 0.899911707 | 0.002794367 | 0.076035338 | -6.55E-05 | 0.00079266 | 1 | 0.979223495 | 1 | 0.980373283 | ENSG0000013 | 13 | ASCC1 |
| SAT | Heart_Failure | ENSG0000018 | -0.00121376 | 0.750679388 | 0.003819933 | 0.076055477 | -0.00128854 | 0.735849223 | 0.003819538 | 0.076035322 | 7.48E-05 | 0.010181683 | 1 | 0.966658386 | 1 | 0.960001834 | ENSG0000013 | 13 | ASCC1 |
| SAT | Heart_Failure | ENSG0000018 | -0.0063548 | 0.090688303 | 0.003756279 | 0.076055046 | -0.0062621 | 0.095458783 | 0.003755871 | 0.076034906 | -9.27E-05 | 0.00034927 | 1 | 0.599895813 | 1 | 0.603495 | ENSG0000013 | 14 | RPS24 |
| SAT | Heart_Failure | ENSG0000018 | 0.001857708 | 0.410436665 | 0.002256893 | 0.076055387 | 0.001905063 | 0.398562274 | 0.002256669 | 0.076035229 | -4.74E-05 | 0.009718617 | 1 | 0.870429337 | 1 | 0.864869591 | ENSG0000013 | 10 | DNA2 |
| SAT | Heart_Failure | ENSG0000018 | -0.00150515 | 0.498062092 | 0.002221484 | 0.076055421 | -0.0015525 | 0.484596785 | 0.002221253 | 0.076035264 | 4.73E-05 | 0.004782481 | 1 | 0.899263643 | 1 | 0.894608312 | ENSG0000013 | 10 | DNA2 |
| SAT | Heart_Failure | ENSG0000018 | 0.000950182 | 0.595301105 | 0.001788847 | 0.076055449 | 0.000916086 | 0.608534678 | 0.00178865 | 0.076035299 | 3.41E-05 | 0.003923896 | 1 | 0.943566631 | 1 | 0.94401999 | ENSG0000013 | 10 | DNA2 |
| SAT | Heart_Failure | ENSG0000018 | -0.0043442 | 0.131369218 | 0.002879382 | 0.076055138 | -0.00443679 | 0.123308503 | 0.002879091 | 0.07603497 | 9.26E-05 | 5.36E-05 | 1 | 0.653539914 | 1 | 0.647747917 | ENSG0000013 | 10 | ATIC |
| SAT | Heart_Failure | ENSG0000018 | -0.00020376 | 0.946352399 | 0.00302824 | 0.076055492 | -0.00014458 | 0.961915406 | 0.003027932 | 0.07603534 | -5.92E-05 | 0.012921456 | 1 | 0.992414898 | 1 | 0.99592587 | ENSG0000013 | 10 | ATIC |
| SAT | Heart_Failure | ENSG0000018 | 0.002404121 | 0.129765309 | 0.001586847 | 0.076055135 | 0.002428699 | 0.125847944 | 0.001586674 | 0.076034975 | -2.46E-05 | 0.025105036 | 1 | 0.649888404 | 1 | 0.647874406 | ENSG0000013 | 13 | CARF |
| SAT | Heart_Failure | ENSG0000018 | -0.00354602 | 0.081784158 | 0.002037438 | 0.07605502 | -0.00346799 | 0.088698952 | 0.002037232 | 0.076034888 | -7.80E-05 | 1.12E-06 | 1 | 0.592308253 | 1 | 0.596493272 | ENSG0000013 | 13 | CARF |
| SAT | Heart_Failure | ENSG0000018 | -0.00093142 | 0.461621734 | 0.00126521 | 0.076055408 | -0.00090981 | 0.472036719 | 0.001265089 | 0.076035259 | -2.16E-05 | 0.044669969 | 1 | 0.890157253 | 1 | 0.894608312 | ENSG0000013 | 11 | SSB |
| SAT | Heart_Failure | ENSG0000018 | 0.000211954 | 0.86914922 | 0.001286602 | 0.076055489 | 0.002233514 | 0.133211329 | 0.001487462 | 0.076034988 | -0.00202156 | 0.006786929 | 1 | 0.979223495 | 1 | 0.649215182 | ENSG0000013 | 11 | SSB |
| SAT | Heart_Failure | ENSG0000018 | 0.0006911 | 0.860280253 | 0.003926316 | 0.076055488 | 0.000610625 | 0.876396976 | 0.00392588 | 0.076035336 | 8.05E-05 | 0.001774418 | 1 | 0.976669362 | 1 | 0.974665278 | ENSG0000013 | 7 | ITGAV |
| SAT | Heart_Failure | ENSG0000018 | 0.000771859 | 0.840462297 | 0.003834327 | 0.076055487 | 0.000678389 | 0.859552303 | 0.003833916 | 0.076035335 | 9.35E-05 | 0.000592994 | 1 | 0.976669362 | 1 | 0.972791643 | ENSG0000013 | 4 | SLC35A5 |
| SAT | Heart_Failure | ENSG0000018 | 0.000143482 | 0.949033225 | 0.002244676 | 0.076055492 | 0.000188512 | 0.933064253 | 0.002244453 | 0.076035339 | -4.50E-05 | 0.013442038 | 1 | 0.992913907 | 1 | 0.988742522 | ENSG0000013 | 4 | SLC35A5 |
| SAT | Heart_Failure | ENSG0000018 | -0.01096345 | 0.001747416 | 0.003502559 | 0.076053964 | -0.0110699 | 0.001574271 | 0.003502414 | 0.076033781 | 0.000106453 | 0.024439328 | 1 | 0.088204203 | 1 | 0.09221864 | ENSG0000013 | 11 | SENP7 |
| SAT | Heart_Failure | ENSG0000018 | -0.00100834 | 0.510152677 | 0.001531029 | 0.076055425 | -0.00098539 | 0.519783077 | 0.00153087 | 0.076035275 | -2.29E-05 | 0.04647717 | 1 | 0.903297859 | 1 | 0.904369708 | ENSG0000013 | 11 | SENP7 |
| SAT | Heart_Failure | ENSG0000018 | -0.00015556 | 0.901030583 | 0.001250896 | 0.076055491 | -0.00013084 | 0.916685577 | 0.001250762 | 0.076035338 | -2.47E-05 | 0.005132397 | 1 | 0.980849815 | 1 | 0.984094962 | ENSG0000013 | 11 | SENP7 |
| SAT | Heart_Failure | ENSG0000018 | -0.0011434 | 0.602825162 | 0.0021974 | 0.076055451 | -0.00123804 | 0.573118353 | 0.002197181 | 0.076035291 | 9.46E-05 | 1.08E-07 | 1 | 0.945538861 | 1 | 0.925849984 | ENSG0000013 | 6 | GUCA1C |
| SAT | Heart_Failure | ENSG0000018 | 0.012475282 | 6.06E-05 | 0.00311079 | 0.076052983 | 0.012376657 | 6.92E-05 | 0.003110541 | 0.07603287 | 9.86E-05 | 0.001938011 | 0.198972651 | 0.007106166 | 0.227154091 | 0.011955478 | ENSG0000013 | 9 | USP8 |
| SAT | Heart_Failure | ENSG0000018 | 6.48E-05 | 0.98021267 | 0.002614234 | 0.076055493 | 2.82E-05 | 0.991383098 | 0.002613941 | 0.07603534 | 3.66E-05 | 0.028799397 | 1 | 0.998920999 | 1 | 0.999768899 | ENSG0000013 | 9 | USP8 |
| SAT | Heart_Failure | ENSG0000018 | -0.00398954 | 0.086460333 | 0.002327107 | 0.076055034 | -0.00516872 | 0.026683045 | 0.002332317 | 0.076034574 | 0.00117918 | 1.96E-13 | 1 | 0.592308253 | 1 | 0.360276017 | ENSG0000013 | 9 | USP8 |
| SAT | Heart_Failure | ENSG0000018 | 0.003205736 | 0.23612871 | 0.002705901 | 0.076055274 | 0.003267192 | 0.227223792 | 0.002705653 | 0.076035113 | -6.15E-05 | 0.011997447 | 1 | 0.755348658 | 1 | 0.747837725 | ENSG0000013 | 5 | SPPL2A |
| SAT | Heart_Failure | ENSG0000018 | 0.000500102 | 0.594014395 | 0.000938228 | 0.076055449 | 0.000755674 | 0.424794361 | 0.000946804 | 0.076035241 | -0.00025557 | 0.045958659 | 1 | 0.943350063 | 1 | 0.873390936 | ENSG0000013 | 9 | APH1B |
| SAT | Heart_Failure | ENSG0000018 | 0.001344601 | 0.639528217 | 0.002870878 | 0.076055459 | 0.001419985 | 0.620834922 | 0.002870583 | 0.076035302 | -7.54E-05 | 0.000647904 | 1 | 0.959428651 | 1 | 0.94798204 | ENSG0000013 | 9 | APH1B |
| SAT | Heart_Failure | ENSG0000018 | 0.0020941 | 0.418217272 | 0.002586846 | 0.076055391 | 0.002137721 | 0.408541756 | 0.002586589 | 0.076035234 | -4.36E-05 | 0.038366175 | 1 | 0.873994184 | 1 | 0.867451005 | ENSG0000013 | 9 | APH1B |
| SAT | Heart_Failure | ENSG0000018 | -0.0015534 | 0.155791 | 0.001094428 | 0.076055179 | -0.00152422 | 0.163661749 | 0.001094308 | 0.076035037 | -2.92E-05 | 8.64E-05 | 1 | 0.677064618 | 1 | 0.679583582 | ENSG0000013 | 9 | APH1B |
| SAT | Heart_Failure | ENSG0000018 | -0.00554722 | 0.019014528 | 0.002365295 | 0.076054635 | -0.00559339 | 0.018028918 | 0.00236504 | 0.076034467 | 4.62E-05 | 0.005651411 | 1 | 0.302151987 | 1 | 0.300690097 | ENSG0000013 | 7 | PPCDC |
| SAT | Heart_Failure | ENSG0000018 | 0.004253215 | 0.126303432 | 0.002781978 | 0.076055128 | 0.004167849 | 0.134054035 | 0.002781705 | 0.07603499 | 8.54E-05 | 0.000200945 | 1 | 0.64967372 | 1 | 0.649215182 | ENSG0000013 | 7 | PPCDC |
| SAT | Heart_Failure | ENSG0000018 | -0.00082512 | 0.296117743 | 0.000789746 | 0.076055323 | -0.00081156 | 0.304074334 | 0.000789657 | 0.076035175 | -1.36E-05 | 0.007468886 | 1 | 0.813063328 | 1 | 0.811772083 | ENSG0000013 | 7 | PPCDC |
| SAT | Heart_Failure | ENSG0000018 | -0.00172215 | 0.56658568 | 0.003005041 | 0.076055442 | -0.00178024 | 0.553526613 | 0.00300471 | 0.076035285 | 5.81E-05 | 0.004004391 | 1 | 0.927165893 | 1 | 0.915531921 | ENSG0000013 | 7 | PPCDC |
| SAT | Heart_Failure | ENSG0000018 | -0.00239773 | 0.133522697 | 0.001598109 | 0.076055142 | -0.00236244 | 0.13930418 | 0.001597979 | 0.076034999 | -3.53E-05 | 0.029025043 | 1 | 0.659800431 | 1 | 0.653383835 | ENSG0000013 | 7 | PPCDC |
| SAT | Heart_Failure | ENSG0000018 | 0.000407095 | 0.908950658 | 0.003559696 | 0.076055491 | 0.00035032 | 0.921596122 | 0.003559312 | 0.076035339 | 5.68E-05 | 0.023164572 | 1 | 0.983702811 | 1 | 0.984731983 | ENSG0000013 | 7 | AP1AR |
| SAT | Heart_Failure | ENSG0000018 | 0.001322509 | 0.301875634 | 0.001280979 | 0.076055327 | 0.001288985 | 0.314247788 | 0.001280852 | 0.076035182 | 3.35E-05 | 0.00137229 | 1 | 0.815859931 | 1 | 0.823487262 | ENSG0000013 | 5 | PRKG2 |
| SAT | Heart_Failure | ENSG0000018 | 0.011078993 | 0.00193532 | 0.003573869 | 0.076053993 | 0.011142765 | 0.001819776 | 0.003573475 | 0.076033823 | -6.38E-05 | 0.007456515 | 1 | 0.09524259 | 1 | 0.101198063 | ENSG0000013 | 5 | PRKG2 |
| SAT | Heart_Failure | ENSG0000018 | -0.00131401 | 0.690798368 | 0.003303423 | 0.076055468 | -0.00137399 | 0.677432623 | 0.003303097 | 0.076035313 | 6.00E-05 | 0.027498637 | 1 | 0.966658386 | 1 | 0.956878334 | ENSG0000013 | 5 | BBS7 |
| SAT | Heart_Failure | ENSG0000018 | 0.009109544 | 0.015354992 | 0.00375821 | 0.076054576 | 0.009451556 | 0.01190603 | 0.003758183 | 0.076034353 | -0.00034201 | 9.51E-09 | 1 | 0.273107546 | 1 | 0.246278972 | ENSG0000013 | 5 | BBS7 |
| SAT | Heart_Failure | ENSG0000018 | 0.001518382 | 0.613386423 | 0.003005234 | 0.076055453 | 0.001593752 | 0.595849964 | 0.003004935 | 0.076035296 | -7.54E-05 | 0.00203074 | 1 | 0.954216727 | 1 | 0.937611754 | ENSG0000013 | 10 | NAAA |
| SAT | Heart_Failure | ENSG0000018 | -5.90E-05 | 0.975757198 | 0.001940261 | 0.076055493 | -2.15E-05 | 0.991153364 | 0.001940048 | 0.07603534 | -3.75E-05 | 0.004073164 | 1 | 0.998920999 | 1 | 0.999768899 | ENSG0000013 | 10 | NAAA |
| SAT | Heart_Failure | ENSG0000018 | -0.00081757 | 0.531289087 | 0.001305939 | 0.076055432 | -0.0008507 | 0.5147437 | 0.00130582 | 0.076035274 | 3.31E-05 | 0.005325995 | 1 | 0.908320477 | 1 | 0.903395669 | ENSG0000013 | 10 | NUP54 |
| SAT | Heart_Failure | ENSG0000018 | -0.00016394 | 0.947223436 | 0.002476627 | 0.076055492 | -0.00022945 | 0.926175774 | 0.002476379 | 0.076035339 | 6.55E-05 | 0.00099989 | 1 | 0.992414898 | 1 | 0.986938199 | ENSG0000013 | 13 | BMP2K |
| SAT | Heart_Failure | ENSG0000018 | -0.00103058 | 0.784147484 | 0.003762362 | 0.076055481 | -0.00112651 | 0.764597826 | 0.003761959 | 0.076035326 | 9.59E-05 | 0.000340385 | 1 | 0.972730696 | 1 | 0.965679451 | ENSG0000013 | 13 | BMP2K |
| SAT | Heart_Failure | ENSG0000019 | -0.01441811 | 1.33E-11 | 0.002131234 | 0.076048351 | 0.000333289 | 0.909033454 | 0.002916992 | 0.076035338 | -0.0147514 | 1.30E-13 | 4.37E-08 | 6.25E-09 | 1 | 0.982067422 | ENSG0000013 | 13 | BMP2K |
| SAT | Heart_Failure | ENSG0000019 | -0.0024362 | 0.228959953 | 0.002025027 | 0.076055267 | -0.00250938 | 0.215227492 | 0.002024807 | 0.0760351 | 7.32E-05 | 1.69E-07 | 1 | 0.751217604 | 1 | 0.736362287 | ENSG0000013 | 10 | G3BP2 |
| SAT | Heart_Failure | ENSG0000019 | 0.00254431 | 0.496430559 | 0.003740987 | 0.076055421 | 0.002456596 | 0.511350951 | 0.003740613 | 0.076035273 | 8.77E-05 | 0.003679383 | 1 | 0.898994301 | 1 | 0.902750226 | ENSG0000013 | 10 | G3BP2 |
| SAT | Heart_Failure | ENSG0000019 | -0.000371 | 0.858673716 | 0.002083529 | 0.076055488 | -0.00033346 | 0.872832748 | 0.002083338 | 0.076035336 | -3.75E-05 | 0.045676794 | 1 | 0.976669362 | 1 | 0.974665278 | ENSG0000013 | 10 | G3BP2 |
| SAT | Heart_Failure | ENSG0000019 | 0.007891862 | 1.13E-09 | 0.001295805 | 0.076049704 | 0.002414639 | 0.106311676 | 0.001495134 | 0.076034933 | 0.005477222 | 2.12E-13 | 3.70E-06 | 4.63E-07 | 1 | 0.625105036 | ENSG0000013 | 4 | SCARB2 |
| SAT | Heart_Failure | ENSG0000019 | 0.001024819 | 0.472897628 | 0.001427776 | 0.076055413 | 0.000999585 | 0.483818296 | 0.001427621 | 0.076035264 | 2.52E-05 | 0.010536019 | 1 | 0.895715287 | 1 | 0.894608312 | ENSG0000013 | 8 | CNOT6L |
| SAT | Heart_Failure | ENSG0000019 | 0.002482917 | 0.364213885 | 0.002736402 | 0.076055365 | 0.002520957 | 0.356859508 | 0.002736106 | 0.076035208 | -3.80E-05 | 0.047172658 | 1 | 0.85040303 | 1 | 0.845383425 | ENSG0000013 | 8 | CNOT6L |
| SAT | Heart_Failure | ENSG0000019 | -0.00205139 | 0.406070124 | 0.002469082 | 0.076055385 | -0.00201275 | 0.414918116 | 0.00246881 | 0.076035236 | -3.86E-05 | 0.01859136 | 1 | 0.870429337 | 1 | 0.867451005 | ENSG0000013 | 8 | CNOT6L |
| SAT | Heart_Failure | ENSG0000019 | 0.000106814 | 0.968846304 | 0.002734931 | 0.076055493 | 1.89E-05 | 0.994477058 | 0.002734632 | 0.07603534 | 8.79E-05 | 2.28E-06 | 1 | 0.996609664 | 1 | 0.999768899 | ENSG0000013 | 15 | PPA2 |
| SAT | Heart_Failure | ENSG0000019 | -0.00171715 | 0.23149588 | 0.001435127 | 0.07605527 | -0.00169118 | 0.23857895 | 0.00143497 | 0.076035123 | -2.60E-05 | 0.008519662 | 1 | 0.752763116 | 1 | 0.752162449 | ENSG0000013 | 15 | PPA2 |
| SAT | Heart_Failure | ENSG0000019 | 0.002314383 | 0.456357993 | 0.003107147 | 0.076055406 | 0.002262072 | 0.466551549 | 0.003106806 | 0.076035257 | 5.23E-05 | 0.012766572 | 1 | 0.88796436 | 1 | 0.892844363 | ENSG0000013 | 15 | PPA2 |
| SAT | Heart_Failure | ENSG0000019 | -0.02143179 | 8.15E-21 | 0.002290178 | 0.076041827 | -0.00426991 | 0.323756806 | 0.004327162 | 0.076035188 | -0.01716188 | 2.95E-06 | 2.67E-17 | 8.91E-18 | 1 | 0.828598718 | ENSG0000013 | 15 | PPA2 |
| SAT | Heart_Failure | ENSG0000019 | 0.003859521 | 0.146462917 | 0.002657813 | 0.076055164 | 0.003819361 | 0.15066716 | 0.002657536 | 0.076035018 | 4.02E-05 | 0.044402583 | 1 | 0.673975917 | 1 | 0.665121824 | ENSG0000013 | 5 | ENPEP |
| SAT | Heart_Failure | ENSG0000019 | 0.000872641 | 0.643659938 | 0.001886432 | 0.07605546 | 0.001136073 | 0.547019706 | 0.001886435 | 0.076035284 | -0.00026343 | 1.61E-17 | 1 | 0.960732494 | 1 | 0.912597963 | ENSG0000013 | 4 | PAPSS1 |
| SAT | Heart_Failure | ENSG0000019 | 0.003814364 | 0.114784786 | 0.002418671 | 0.076055105 | 0.00375333 | 0.120669575 | 0.002418427 | 0.076034964 | 6.10E-05 | 0.001442809 | 1 | 0.641850091 | 1 | 0.64481576 | ENSG0000013 | 4 | PAPSS1 |
| SAT | Heart_Failure | ENSG0000019 | 0.000899381 | 0.683920847 | 0.002209135 | 0.076055467 | 0.000939466 | 0.670610552 | 0.002208894 | 0.076035312 | -4.01E-05 | 0.007655858 | 1 | 0.966658386 | 1 | 0.956878334 | ENSG0000013 | 8 | SLC39A8 |
| SAT | Heart_Failure | ENSG0000019 | 9.43E-05 | 0.971968758 | 0.002684371 | 0.076055493 | 3.71E-05 | 0.988969908 | 0.002684117 | 0.07603534 | 5.72E-05 | 0.014305874 | 1 | 0.99766005 | 1 | 0.999768899 | ENSG0000013 | 8 | SLC39A8 |
| SAT | Heart_Failure | ENSG0000019 | 0.00177879 | 0.45258363 | 0.002368198 | 0.076055405 | 0.001375054 | 0.561576735 | 0.002368735 | 0.076035288 | 0.000403736 | 1.98E-10 | 1 | 0.887019168 | 1 | 0.918876404 | ENSG0000013 | 8 | SLC39A8 |
| SAT | Heart_Failure | ENSG0000019 | 0.001551332 | 0.663831569 | 0.003569321 | 0.076055464 | 0.00147581 | 0.679228218 | 0.003568925 | 0.076035313 | 7.55E-05 | 0.001220155 | 1 | 0.962994646 | 1 | 0.956878334 | ENSG0000013 | 6 | FBN2 |
| SAT | Heart_Failure | ENSG0000019 | -0.00227841 | 0.500914817 | 0.003385193 | 0.076055422 | -0.00239005 | 0.480145633 | 0.003385007 | 0.076035262 | 0.000111639 | 0.008145724 | 1 | 0.900549813 | 1 | 0.894608312 | ENSG0000013 | 8 | MAPK8IP3 |
| SAT | Heart_Failure | ENSG0000019 | 0.001304845 | 0.364734698 | 0.001439621 | 0.076055365 | 0.001266484 | 0.378954435 | 0.001439473 | 0.076035219 | 3.84E-05 | 0.000551843 | 1 | 0.85040303 | 1 | 0.854967426 | ENSG0000013 | 8 | MAPK8IP3 |
| SAT | Heart_Failure | ENSG0000019 | -0.00125297 | 0.748820936 | 0.003913148 | 0.076055477 | -0.00114252 | 0.770288948 | 0.003912773 | 0.076035327 | -0.00011045 | 0.000987517 | 1 | 0.966658386 | 1 | 0.965963214 | ENSG0000013 | 8 | MAPK8IP3 |
| SAT | Heart_Failure | ENSG0000019 | -0.00228824 | 0.453965588 | 0.003055794 | 0.076055406 | -0.00243285 | 0.425911331 | 0.003055546 | 0.076035241 | 0.000144609 | 2.94E-06 | 1 | 0.887019168 | 1 | 0.873465455 | ENSG0000013 | 2 | ALG10 |
| SAT | Heart_Failure | ENSG0000019 | -0.00040601 | 0.776477632 | 0.001430048 | 0.07605548 | -0.00036715 | 0.797357188 | 0.001429901 | 0.07603533 | -3.89E-05 | 0.000413142 | 1 | 0.972664863 | 1 | 0.971094631 | ENSG0000013 | 2 | ALG10 |
| SAT | Heart_Failure | ENSG0000019 | 0.00135215 | 0.653574709 | 0.003012799 | 0.076055462 | 0.001263766 | 0.674844413 | 0.003012489 | 0.076035313 | 8.84E-05 | 0.000137238 | 1 | 0.962760695 | 1 | 0.956878334 | ENSG0000013 | 3 | ZCRB1 |
| SAT | Heart_Failure | ENSG0000019 | -0.0029563 | 0.320777154 | 0.002977546 | 0.076055339 | -0.00286978 | 0.335091784 | 0.00297723 | 0.076035195 | -8.65E-05 | 6.08E-05 | 1 | 0.828692762 | 1 | 0.836830236 | ENSG0000013 | 6 | C1RL |
| SAT | Heart_Failure | ENSG0000019 | 0.001550145 | 0.187302244 | 0.001175596 | 0.076055222 | 0.001512225 | 0.198279409 | 0.001175483 | 0.076035082 | 3.79E-05 | 0.000167338 | 1 | 0.72128951 | 1 | 0.725655076 | ENSG0000013 | 6 | C1RL |
| SAT | Heart_Failure | ENSG0000019 | 0.003167305 | 0.120857266 | 0.002041857 | 0.076055117 | 0.003499025 | 0.087050883 | 0.002044818 | 0.076034883 | -0.00033172 | 0.003896035 | 1 | 0.646585619 | 1 | 0.596493272 | ENSG0000013 | 5 | KLRG1 |
| SAT | Heart_Failure | ENSG0000019 | -0.00048176 | 0.819576081 | 0.002112128 | 0.076055485 | -0.00052324 | 0.804323615 | 0.002111933 | 0.076035331 | 4.15E-05 | 0.02866064 | 1 | 0.976669362 | 1 | 0.972791643 | ENSG0000013 | 12 | VAMP1 |
| SAT | Heart_Failure | ENSG0000019 | -0.00083651 | 0.309275568 | 0.000822732 | 0.076055332 | -0.00082015 | 0.318782331 | 0.000822644 | 0.076035185 | -1.64E-05 | 0.004821405 | 1 | 0.81701541 | 1 | 0.823562856 | ENSG0000013 | 12 | VAMP1 |
| SAT | Heart_Failure | ENSG0000019 | 0.001905185 | 0.024704036 | 0.000848254 | 0.076054706 | 0.001889665 | 0.02588594 | 0.000848172 | 0.076034566 | 1.55E-05 | 0.030663166 | 1 | 0.336323411 | 1 | 0.355059412 | ENSG0000013 | 7 | TAPBPL |
| SAT | Heart_Failure | ENSG0000019 | -0.00069671 | 0.772147697 | 0.002406036 | 0.07605548 | -0.00038525 | 0.872952622 | 0.002409193 | 0.076035336 | -0.00031146 | 0.016047446 | 1 | 0.971467105 | 1 | 0.974665278 | ENSG0000013 | 7 | TAPBPL |
| SAT | Heart_Failure | ENSG0000019 | 0.00123626 | 0.403632258 | 0.00148028 | 0.076055384 | 0.001274447 | 0.389216452 | 0.001480125 | 0.076035224 | -3.82E-05 | 0.000556052 | 1 | 0.870429337 | 1 | 0.858921514 | ENSG0000013 | 6 | PEX5 |
| SAT | Heart_Failure | ENSG0000019 | -0.00370702 | 0.131442007 | 0.002457513 | 0.076055138 | -0.00365542 | 0.136855092 | 0.00245725 | 0.076034995 | -5.16E-05 | 0.003257495 | 1 | 0.653539914 | 1 | 0.649215182 | ENSG0000013 | 9 | PIANP |
| SAT | Heart_Failure | ENSG0000019 | -0.00218691 | 0.283976406 | 0.002041112 | 0.076055314 | -0.00221959 | 0.2767927 | 0.002040897 | 0.076035156 | 3.27E-05 | 0.029330371 | 1 | 0.798394677 | 1 | 0.786282985 | ENSG0000013 | 9 | PIANP |
| SAT | Heart_Failure | ENSG0000019 | -0.0047536 | 0.153245994 | 0.003328474 | 0.076055175 | -0.00486448 | 0.143842742 | 0.003328113 | 0.076035007 | 0.000110878 | 1.71E-06 | 1 | 0.677064618 | 1 | 0.659101398 | ENSG0000013 | 12 | PPFIA2 |
| SAT | Heart_Failure | ENSG0000019 | 0.003188614 | 0.393997621 | 0.003740788 | 0.07605538 | 0.002991653 | 0.423817356 | 0.003740423 | 0.07603524 | 0.00019696 | 2.82E-10 | 1 | 0.864419838 | 1 | 0.873390936 | ENSG0000013 | 6 | SNRPF |
| SAT | Heart_Failure | ENSG0000019 | 0.002334196 | 0.413895518 | 0.002856829 | 0.076055389 | 0.002274135 | 0.425960409 | 0.002856509 | 0.076035241 | 6.01E-05 | 0.00098504 | 1 | 0.873306234 | 1 | 0.873465455 | ENSG0000013 | 6 | SNRPF |
| SAT | Heart_Failure | ENSG0000019 | 0.005005094 | 0.188027484 | 0.003801995 | 0.076055223 | 0.005064336 | 0.182808787 | 0.003801603 | 0.076035063 | -5.92E-05 | 0.041510195 | 1 | 0.722191019 | 1 | 0.715224819 | ENSG0000013 | 6 | SNRPF |
| SAT | Heart_Failure | ENSG0000019 | 0.002199043 | 0.17347188 | 0.001615596 | 0.076055204 | 0.002151213 | 0.182970536 | 0.00161543 | 0.076035063 | 4.78E-05 | 0.000117807 | 1 | 0.704583343 | 1 | 0.715224819 | ENSG0000013 | 6 | SNRPF |
| SAT | Heart_Failure | ENSG0000019 | 0.001955953 | 0.569874403 | 0.003442143 | 0.076055443 | 0.001106429 | 0.747930471 | 0.003442856 | 0.076035324 | 0.000849523 | 2.78E-21 | 1 | 0.92942386 | 1 | 0.962415493 | ENSG0000013 | 3 | AMDHD1 |
| SAT | Heart_Failure | ENSG0000019 | 0.008480051 | 0.048286051 | 0.004294029 | 0.076054884 | 0.008553597 | 0.046351601 | 0.004293581 | 0.076034721 | -7.35E-05 | 0.022804887 | 1 | 0.478630006 | 1 | 0.467403724 | ENSG0000013 | 3 | AMDHD1 |
| SAT | Heart_Failure | ENSG0000019 | -0.00137127 | 0.03722127 | 0.000658211 | 0.076054816 | 0.000778606 | 0.311675473 | 0.000769591 | 0.07603518 | -0.00214988 | 7.08E-08 | 1 | 0.436153525 | 1 | 0.821155716 | ENSG0000013 | 3 | AMDHD1 |
| SAT | Heart_Failure | ENSG0000019 | -0.00122504 | 0.718691839 | 0.003400922 | 0.076055473 | -0.00103872 | 0.760029341 | 0.003400714 | 0.076035326 | -0.00018632 | 4.43E-06 | 1 | 0.966658386 | 1 | 0.963545699 | ENSG0000013 | 3 | AMDHD1 |
| SAT | Heart_Failure | ENSG0000019 | 0.00167842 | 0.305041048 | 0.001636391 | 0.076055329 | 0.001730961 | 0.290097803 | 0.001636211 | 0.076035166 | -5.25E-05 | 1.73E-06 | 1 | 0.816225947 | 1 | 0.803578953 | ENSG0000013 | 11 | RITA1 |
| SAT | Heart_Failure | ENSG0000019 | 0.000438977 | 0.811259894 | 0.001838255 | 0.076055484 | 0.000405253 | 0.825498479 | 0.001838059 | 0.076035333 | 3.37E-05 | 0.011190389 | 1 | 0.976669362 | 1 | 0.972791643 | ENSG0000013 | 11 | RITA1 |
| SAT | Heart_Failure | ENSG0000020 | 0.0007781 | 0.796224898 | 0.003013134 | 0.076055483 | 0.00085523 | 0.77651512 | 0.003012822 | 0.076035328 | -7.71E-05 | 0.000772631 | 1 | 0.976456935 | 1 | 0.965963214 | ENSG0000013 | 10 | MTMR6 |
| SAT | Heart_Failure | ENSG0000020 | -0.00026499 | 0.897851765 | 0.00206419 | 0.07605549 | -0.0002304 | 0.911117363 | 0.002063983 | 0.076035338 | -3.46E-05 | 0.036567096 | 1 | 0.979990566 | 1 | 0.982887891 | ENSG0000013 | 10 | SLC46A3 |
| SAT | Heart_Failure | ENSG0000020 | -0.00225662 | 0.137358901 | 0.001518889 | 0.076055149 | -0.00230208 | 0.129573636 | 0.001518736 | 0.076034982 | 4.55E-05 | 0.000165816 | 1 | 0.663104757 | 1 | 0.649215182 | ENSG0000013 | 8 | SUOX |
| SAT | Heart_Failure | ENSG0000020 | 0.006946922 | 0.064423318 | 0.00375663 | 0.076054959 | 0.00699848 | 0.062438539 | 0.003756216 | 0.076034798 | -5.16E-05 | 0.040389695 | 1 | 0.523200263 | 1 | 0.509604095 | ENSG0000013 | 6 | CCDC65 |
| SAT | Heart_Failure | ENSG0000020 | 0.003728359 | 0.202741932 | 0.002927004 | 0.07605524 | 0.003819298 | 0.191897432 | 0.002926691 | 0.076035074 | -9.09E-05 | 1.55E-05 | 1 | 0.735350411 | 1 | 0.721677191 | ENSG0000013 | 6 | CCDC65 |
| SAT | Heart_Failure | ENSG0000020 | 0.001364554 | 0.67472288 | 0.003251452 | 0.076055466 | 0.00141371 | 0.663677371 | 0.003251089 | 0.076035311 | -4.92E-05 | 0.019456375 | 1 | 0.966658386 | 1 | 0.956878334 | ENSG0000013 | 8 | KANSL2 |
| SAT | Heart_Failure | ENSG0000020 | 0.006723585 | 0.05953664 | 0.003568377 | 0.076054939 | 0.006623641 | 0.063396974 | 0.003568 | 0.076034802 | 9.99E-05 | 0.000134133 | 1 | 0.507375885 | 1 | 0.511069956 | ENSG0000013 | 8 | KANSL2 |
| SAT | Heart_Failure | ENSG0000020 | 0.001666853 | 0.489186368 | 0.002410126 | 0.076055418 | 0.001731705 | 0.472395209 | 0.002409872 | 0.07603526 | -6.49E-05 | 0.000245057 | 1 | 0.898909993 | 1 | 0.894608312 | ENSG0000013 | 8 | KANSL2 |
| SAT | Heart_Failure | ENSG0000020 | -0.00047282 | 0.880862637 | 0.003154761 | 0.07605549 | -0.00327489 | 0.301135141 | 0.003167199 | 0.076035173 | 0.002802072 | 8.54E-23 | 1 | 0.979223495 | 1 | 0.810520426 | ENSG0000013 | 8 | CERS5 |
| SAT | Heart_Failure | ENSG0000020 | 0.002952715 | 0.261150543 | 0.002627725 | 0.076055296 | -0.00011293 | 0.965964435 | 0.002646471 | 0.07603534 | 0.00306564 | 4.39E-22 | 1 | 0.774632399 | 1 | 0.996957946 | ENSG0000013 | 8 | CERS5 |
| SAT | Heart_Failure | ENSG0000020 | 0.001488676 | 0.684166508 | 0.003659619 | 0.076055467 | 0.001426236 | 0.696710515 | 0.003659221 | 0.076035316 | 6.24E-05 | 0.012882517 | 1 | 0.966658386 | 1 | 0.956878334 | ENSG0000013 | 8 | CERS5 |
| SAT | Heart_Failure | ENSG0000020 | -0.00525209 | 0.035283043 | 0.002494935 | 0.076054801 | -0.00424088 | 0.091559494 | 0.002513513 | 0.076034896 | -0.00101121 | 0.001014793 | 1 | 0.425601709 | 1 | 0.599166569 | ENSG0000013 | 8 | CERS5 |
| SAT | Heart_Failure | ENSG0000020 | -0.00123815 | 0.266639494 | 0.001114613 | 0.0760553 | -9.02E-05 | 0.936097232 | 0.001124678 | 0.076035339 | -0.00114798 | 3.15E-14 | 1 | 0.779024202 | 1 | 0.988983601 | ENSG0000013 | 11 | LMBR1L |
| SAT | Heart_Failure | ENSG0000020 | 0.010722186 | 8.85E-07 | 0.002181173 | 0.076051722 | 0.003055095 | 0.204270454 | 0.002406572 | 0.076035089 | 0.007667091 | 4.85E-14 | 0.002902194 | 0.000233648 | 1 | 0.729840368 | ENSG0000013 | 9 | ZNF740 |
| SAT | Heart_Failure | ENSG0000020 | 0.002784295 | 0.288140657 | 0.002621228 | 0.076055317 | -0.0001774 | 0.952860436 | 0.003000967 | 0.07603534 | 0.002961698 | 0.04275421 | 1 | 0.801524014 | 1 | 0.991228627 | ENSG0000013 | 9 | ZNF740 |
| SAT | Heart_Failure | ENSG0000020 | 0.007967833 | 0.000718812 | 0.002355728 | 0.076053708 | 0.002764885 | 0.253936918 | 0.002423551 | 0.076035137 | 0.005202948 | 7.54E-20 | 1 | 0.046243566 | 1 | 0.766534574 | ENSG0000013 | 9 | GRTP1 |
| SAT | Heart_Failure | ENSG0000020 | -0.00450796 | 0.141663802 | 0.003067422 | 0.076055156 | -0.00436004 | 0.155159238 | 0.003067118 | 0.076035025 | -0.00014792 | 3.37E-09 | 1 | 0.669181587 | 1 | 0.669469928 | ENSG0000013 | 6 | CBLN3 |
| SAT | Heart_Failure | ENSG0000020 | -0.00043759 | 0.739338878 | 0.001315167 | 0.076055476 | -0.00329825 | 0.0293683 | 0.001513993 | 0.0760346 | 0.00286066 | 0.000137528 | 1 | 0.966658386 | 1 | 0.383605048 | ENSG0000013 | 6 | CBLN3 |
| SAT | Heart_Failure | ENSG0000020 | -0.00027198 | 0.870759425 | 0.001671721 | 0.076055489 | 0.001517976 | 0.366455833 | 0.001680789 | 0.076035213 | -0.00178995 | 3.59E-24 | 1 | 0.979223495 | 1 | 0.849290727 | ENSG0000013 | 6 | CBLN3 |

| | | | | | | | | | | | | | | | | | | | |
|---|---|---|---|---|---|---|---|---|---|---|---|---|---|---|---|---|---|---|---|
| SAT | Heart_Failure | ENSG000002( | 0.00314953 | 0.08630531 | 0.001836224 | 0.076055034 | 0.001941983 | 0.298755713 | 0.001868897 | 0.076035172 | 0.001207547 | 0.00054406 | 1 | 0.592308253 | 1 | 0.810520426 | ENSG0000013 | 15 | NOVA1 |
| SAT | Heart_Failure | ENSG000002( | -0.00063386 | 0.338616426 | 0.000662413 | 0.07605535 | -0.00165883 | 0.015469487 | 0.000685126 | 0.076034425 | 0.001024965 | 5.00E-09 | 1 | 0.84294423 | 1 | 0.276579386 | ENSG0000013 | 12 | RTN1 |
| SAT | Heart_Failure | ENSG000002( | -0.0006344 | 0.745057479 | 0.001951032 | 0.076055477 | -0.0018515 | 0.345545122 | 0.001962865 | 0.076035201 | 0.0012171 | 2.21E-08 | 1 | 0.966658386 | 1 | 0.839802625 | ENSG0000013 | 12 | RTN1 |
| SAT | Heart_Failure | ENSG000002( | -0.0017338 | 0.524850069 | 0.002726587 | 0.07605543 | -0.00911745 | 0.001236007 | 0.002822342 | 0.076033712 | 0.007383652 | 5.03E-24 | 1 | 0.906724424 | 1 | 0.081106807 | ENSG0000013 | 11 | C14orf37 |
| SAT | Heart_Failure | ENSG000002( | 0.001314377 | 0.538051848 | 0.002134553 | 0.076055434 | -0.00151288 | 0.483158352 | 0.002157466 | 0.076035263 | 0.002827262 | 3.22E-19 | 1 | 0.912011429 | 1 | 0.894608312 | ENSG0000013 | 10 | RAB15 |
| SAT | Heart_Failure | ENSG000002( | 0.001714331 | 0.122193054 | 0.001109143 | 0.07605512 | 0.002374532 | 0.033834975 | 0.001118986 | 0.076034638 | -0.0006602 | 9.66E-06 | 1 | 0.646585619 | 1 | 0.412686066 | ENSG0000014 | 14 | ESR2 |
| SAT | Heart_Failure | ENSG000002( | -0.00508376 | 0.020866482 | 0.00220038 | 0.07605466 | -0.00969242 | 3.15E-05 | 0.002328415 | 0.076032636 | 0.004608659 | 1.49E-09 | 1 | 0.31549736 | 0.103222284 | 0.008601857 | ENSG0000014 | 11 | EFCAB11 |
| SAT | Heart_Failure | ENSG000002( | 0.006679661 | 2.63E-06 | 0.001421958 | 0.076052049 | 0.003001342 | 0.042092563 | 0.001476595 | 0.076034695 | 0.003678319 | 2.72E-20 | 0.00864292 | 0.000576195 | 1 | 0.45628996 | ENSG0000014 | 11 | EFCAB11 |
| SAT | Heart_Failure | ENSG000002( | 0.002748059 | 0.000682295 | 0.000809057 | 0.076053692 | 0.00338079 | 3.96E-05 | 0.000822552 | 0.076032704 | -0.00063273 | 2.17E-05 | 1 | 0.044772199 | 0.129766464 | 0.009269033 | ENSG0000014 | 7 | PTGR2 |
| SAT | Heart_Failure | ENSG000002( | 0.00380571 | 1.41E-05 | 0.000876446 | 0.07605255 | 0.002861829 | 0.001166519 | 0.000881387 | 0.076033695 | 0.000943881 | 1.26E-23 | 0.046290748 | 0.002204321 | 1 | 0.078109189 | ENSG0000014 | 13 | WARS |
| SAT | Heart_Failure | ENSG000002( | -0.00357957 | 0.002615442 | 0.001189356 | 0.076054079 | -0.0018402 | 0.140557129 | 0.001248677 | 0.076035001 | -0.00173937 | 4.92E-06 | 1 | 0.113128813 | 1 | 0.655068096 | ENSG0000014 | 8 | DUOXA1 |
| SAT | Heart_Failure | ENSG000002( | 0.000971806 | 0.223992142 | 0.000799194 | 0.076055262 | -0.0004777 | 0.564098611 | 0.000828236 | 0.076035288 | 0.001449503 | 2.82E-11 | 1 | 0.751217604 | 1 | 0.92032393 | ENSG0000014 | 15 | SERF2 |
| SAT | Heart_Failure | ENSG000002( | -0.00023715 | 0.699850986 | 0.000615131 | 0.07605547 | -0.00100645 | 0.106231975 | 0.000623046 | 0.076034933 | 0.0007693 | 1.07E-14 | 1 | 0.966658386 | 1 | 0.625105036 | ENSG0000014 | 15 | SERF2 |
| SAT | Heart_Failure | ENSG000002( | -0.00117873 | 0.376808261 | 0.001333711 | 0.076055371 | -0.0033129 | 0.014208298 | 0.001351122 | 0.076034402 | 0.002134174 | 9.25E-23 | 1 | 0.85759885 | 1 | 0.266385282 | ENSG0000014 | 8 | ZSCAN29 |
| SAT | Heart_Failure | ENSG000002( | -0.00283369 | 0.165583598 | 0.002043719 | 0.076055193 | -0.00484355 | 0.02042942 | 0.002089062 | 0.076034501 | 0.002009861 | 3.68E-06 | 1 | 0.689582559 | 1 | 0.319052908 | ENSG0000014 | 6 | SRP14 |
| SAT | Heart_Failure | ENSG000002( | 0.001465097 | 0.671142924 | 0.003450695 | 0.076055465 | 0.001540833 | 0.6551806 | 0.003450318 | 0.076035309 | -7.57E-05 | 0.00124337 | 1 | 0.965985276 | 1 | 0.95563262 | ENSG0000014 | 6 | SRP14 |
| SAT | Heart_Failure | ENSG000002( | 0.001427852 | 0.257714747 | 0.001261564 | 0.076055293 | 0.001081693 | 0.395108353 | 0.001271995 | 0.076035227 | 0.000346159 | 0.034631131 | 1 | 0.773173321 | 1 | 0.862334842 | ENSG0000014 | 6 | SRP14 |
| SAT | Heart_Failure | ENSG000002( | 3.05E-05 | 0.986225857 | 0.001764607 | 0.076055493 | -0.00076488 | 0.666214748 | 0.001773217 | 0.076035311 | 0.00079534 | 6.91E-06 | 1 | 0.999322741 | 1 | 0.956878334 | ENSG0000014 | 4 | DISP2 |
| SAT | Heart_Failure | ENSG000002( | -0.00261815 | 0.276290445 | 0.002404855 | 0.076055308 | -0.00267024 | 0.266794425 | 0.002404592 | 0.076035148 | 5.21E-05 | 0.001536023 | 1 | 0.792402928 | 1 | 0.775062994 | ENSG0000014 | 4 | DISP2 |
| SAT | Heart_Failure | ENSG000002( | -0.00057939 | 0.878333672 | 0.003784852 | 0.076055489 | -0.00051622 | 0.891500975 | 0.00378445 | 0.076035337 | -6.32E-05 | 0.021330606 | 1 | 0.979223495 | 1 | 0.977805317 | ENSG0000014 | 4 | DISP2 |
| SAT | Heart_Failure | ENSG000002( | -0.00076681 | 0.742192199 | 0.002331069 | 0.076055476 | -0.00080951 | 0.728363302 | 0.002330834 | 0.076035321 | 4.27E-05 | 0.02153504 | 1 | 0.966658386 | 1 | 0.958028875 | ENSG0000014 | 4 | DISP2 |
| SAT | Heart_Failure | ENSG000002( | -0.00033005 | 0.898888146 | 0.002597451 | 0.07605549 | -0.00036487 | 0.888272763 | 0.002597156 | 0.076035337 | 3.48E-05 | 0.028938033 | 1 | 0.980097109 | 1 | 0.977226037 | ENSG0000014 | 8 | SCAPER |
| SAT | Heart_Failure | ENSG000002( | -0.0007887 | 0.77986375 | 0.002821875 | 0.076055481 | -0.00083107 | 0.768345001 | 0.002821568 | 0.076035327 | 4.24E-05 | 0.029479429 | 1 | 0.972664863 | 1 | 0.965963214 | ENSG0000014 | 8 | SCAPER |
| SAT | Heart_Failure | ENSG000002( | -0.0013257 | 0.586407553 | 0.002436722 | 0.076055447 | -0.00137909 | 0.571378376 | 0.002436458 | 0.07603529 | 5.34E-05 | 0.001527552 | 1 | 0.940373012 | 1 | 0.924404562 | ENSG0000014 | 10 | MAN2C1 |
| SAT | Heart_Failure | ENSG000002( | 0.003188289 | 0.225062278 | 0.002628047 | 0.076055263 | 0.003227593 | 0.219346125 | 0.002627754 | 0.07603105 | -3.93E-05 | 0.021990524 | 1 | 0.751217604 | 1 | 0.742028061 | ENSG0000014 | 9 | BBS4 |
| SAT | Heart_Failure | ENSG000002( | -0.00021453 | 0.89111965 | 0.001567179 | 0.07605549 | -0.00026825 | 0.864077138 | 0.001567019 | 0.076035336 | 5.37E-05 | 9.52E-06 | 1 | 0.979270459 | 1 | 0.973904875 | ENSG0000014 | 9 | ADAMTS17 |
| SAT | Heart_Failure | ENSG000002( | -0.00029032 | 0.880207503 | 0.001926424 | 0.076055489 | -0.00034595 | 0.857466394 | 0.001926223 | 0.076035335 | 5.56E-05 | 0.000110422 | 1 | 0.979223495 | 1 | 0.972791643 | ENSG0000014 | 12 | LINS |
| SAT | Heart_Failure | ENSG000002( | 0.009263663 | 0.019523383 | 0.003966642 | 0.076054642 | 0.009203849 | 0.020310231 | 0.003966194 | 0.0760345 | 5.98E-05 | 0.015746482 | 1 | 0.302151987 | 1 | 0.318841471 | ENSG0000014 | 12 | LINS |
| SAT | Heart_Failure | ENSG000002( | -0.00075401 | 0.775178405 | 0.002438663 | 0.076055478 | -0.00070664 | 0.771972119 | 0.00243841 | 0.076035327 | -4.74E-05 | 0.01023946 | 1 | 0.967625883 | 1 | 0.965963214 | ENSG0000014 | 12 | LINS |
| SAT | Heart_Failure | ENSG000002( | 0.001643578 | 0.492165111 | 0.002392852 | 0.076055419 | 0.001596178 | 0.504689311 | 0.002392604 | 0.076035271 | 4.74E-05 | 0.008848735 | 1 | 0.898909993 | 1 | 0.900571724 | ENSG0000014 | 12 | LINS |
| SAT | Heart_Failure | ENSG000002( | -0.01054756 | 0.006090788 | 0.003845453 | 0.076054319 | -0.01062066 | 0.005742141 | 0.003845046 | 0.07603415 | 7.31E-05 | 0.009638519 | 1 | 0.172274799 | 1 | 0.179428249 | ENSG0000014 | 12 | LINS |
| SAT | Heart_Failure | ENSG000002( | -0.0017235 | 0.928293812 | 0.001915123 | 0.076055492 | -0.00013303 | 0.944616183 | 0.001914936 | 0.076035339 | -3.93E-05 | 0.014018973 | 1 | 0.988739396 | 1 | 0.990503578 | ENSG0000014 | 12 | LINS |
| SAT | Heart_Failure | ENSG000002( | 0.002237617 | 0.364872074 | 0.002469446 | 0.076055365 | 0.002159538 | 0.381799335 | 0.00246921 | 0.076035221 | 7.81E-05 | 0.000237233 | 1 | 0.85040303 | 1 | 0.854967426 | ENSG0000014 | 12 | SCAMP2 |
| SAT | Heart_Failure | ENSG000002( | -0.00284385 | 0.460683296 | 0.003854901 | 0.076055408 | -0.00279529 | 0.468324691 | 0.003854468 | 0.076035258 | -4.86E-05 | 0.047071459 | 1 | 0.890157253 | 1 | 0.892844363 | ENSG0000014 | 12 | UNC45A |
| SAT | Heart_Failure | ENSG000002( | -0.0032746 | 0.011841547 | 0.001301075 | 0.076054504 | -0.00330717 | 0.011018062 | 0.001300941 | 0.076034332 | 3.26E-05 | 0.001095707 | 1 | 0.24440108 | 1 | 0.242619208 | ENSG0000014 | 8 | IQGAP1 |
| SAT | Heart_Failure | ENSG000002( | -0.00079612 | 0.580828425 | 0.001441784 | 0.076055445 | -0.00081805 | 0.570407555 | 0.001441629 | 0.07603529 | 2.19E-05 | 0.032253863 | 1 | 0.936460965 | 1 | 0.924404562 | ENSG0000014 | 8 | IQGAP1 |
| SAT | Heart_Failure | ENSG000002( | -0.00192468 | 0.156334748 | 0.001357788 | 0.076055179 | -0.00196142 | 0.148535253 | 0.001357641 | 0.076035014 | 3.67E-05 | 0.000102687 | 1 | 0.677064618 | 1 | 0.665121824 | ENSG0000014 | 8 | IQGAP1 |
| SAT | Heart_Failure | ENSG000002( | 0.001016114 | 0.719051225 | 0.00282468 | 0.076055473 | 0.001060269 | 0.707362643 | 0.002824367 | 0.076035318 | -4.42E-05 | 0.017650502 | 1 | 0.966658386 | 1 | 0.958028875 | ENSG0000014 | 8 | IQGAP1 |
| SAT | Heart_Failure | ENSG000002( | 0.001785735 | 0.580680515 | 0.003232736 | 0.076055445 | 0.001851869 | 0.56671061 | 0.003232431 | 0.076035289 | -6.61E-05 | 0.019014614 | 1 | 0.936460965 | 1 | 0.922318901 | ENSG0000014 | 11 | CRTC3 |
| SAT | Heart_Failure | ENSG000002( | -0.0016296 | 0.309870775 | 0.001604738 | 0.076055332 | -0.00306395 | 0.057813604 | 0.001615059 | 0.076034779 | 0.001434347 | 6.86E-15 | 1 | 0.817271716 | 1 | 0.503576399 | ENSG0000014 | 7 | CLEC18B |
| SAT | Heart_Failure | ENSG000002( | 0.000191185 | 0.818790488 | 0.000834501 | 0.076055485 | -0.00119265 | 0.160775029 | 0.000850393 | 0.076035033 | 0.001383837 | 3.53E-17 | 1 | 0.976066362 | 1 | 0.676259259 | ENSG0000014 | 7 | NUDT7 |
| SAT | Heart_Failure | ENSG000002( | -0.00019748 | 0.956528545 | 0.003622735 | 0.076055493 | -0.0002708 | 0.9403914 | 0.003622331 | 0.076035339 | 7.34E-05 | 0.001799542 | 1 | 0.994657967 | 1 | 0.988983601 | ENSG0000014 | 7 | NUDT7 |
| SAT | Heart_Failure | ENSG0000021 | 0.00128717 | 0.564092486 | 0.002231671 | 0.076055441 | 0.001358259 | 0.542731709 | 0.002231454 | 0.076035282 | -7.11E-05 | 0.000142196 | 1 | 0.925856651 | 1 | 0.912245255 | ENSG0000014 | 7 | NUDT7 |
| SAT | Heart_Failure | ENSG0000021 | 0.001917388 | 0.62396644 | 0.003911145 | 0.076055456 | 0.001974248 | 0.613678481 | 0.003910719 | 0.0760353 | -5.69E-05 | 0.034266995 | 1 | 0.955683491 | 1 | 0.945647463 | ENSG0000014 | 7 | NUDT7 |
| SAT | Heart_Failure | ENSG0000021 | 0.00178015 | 0.604195714 | 0.003434085 | 0.076055451 | 0.001862717 | 0.587488066 | 0.003433705 | 0.076035294 | -8.26E-05 | 0.000279083 | 1 | 0.9466887 | 1 | 0.933194299 | ENSG0000014 | 6 | ADAD2 |
| SAT | Heart_Failure | ENSG0000021 | 0.001335556 | 0.698625352 | 0.003449472 | 0.07605547 | 0.001428701 | 0.678713959 | 0.003449144 | 0.076035313 | -9.31E-05 | 0.001857448 | 1 | 0.966658386 | 1 | 0.956878334 | ENSG0000014 | 6 | ADAD2 |
| SAT | Heart_Failure | ENSG0000021 | -0.00286792 | 0.318699891 | 0.00287616 | 0.076055338 | -0.00280707 | 0.329023117 | 0.002875848 | 0.076035191 | -6.08E-05 | 0.00238916 | 1 | 0.827258183 | 1 | 0.833382337 | ENSG0000014 | 6 | ADAD2 |
| SAT | Heart_Failure | ENSG0000021 | 0.001288708 | 0.650979786 | 0.002848591 | 0.076055461 | 0.001428366 | 0.616036138 | 0.002848321 | 0.076035301 | -0.00013966 | 1.65E-08 | 1 | 0.962760695 | 1 | 0.94759239 | ENSG0000014 | 6 | ADAD2 |
| SAT | Heart_Failure | ENSG0000021 | -0.00038523 | 0.750751689 | 0.001212757 | 0.076055477 | -0.00041717 | 0.730834279 | 0.00121264 | 0.076035322 | 3.19E-05 | 0.001910807 | 1 | 0.966658386 | 1 | 0.958028875 | ENSG0000014 | 6 | TIGD7 |
| SAT | Heart_Failure | ENSG0000021 | 0.002371463 | 0.156957982 | 0.001675496 | 0.07605518 | 0.002406053 | 0.150951669 | 0.001675314 | 0.076035018 | -3.46E-05 | 0.002637134 | 1 | 0.678274948 | 1 | 0.665121824 | ENSG0000014 | 12 | DEF8 |
| SAT | Heart_Failure | ENSG0000021 | -0.00084046 | 0.701215687 | 0.002190535 | 0.07605547 | -0.00087336 | 0.690081874 | 0.002190286 | 0.076035315 | 3.29E-05 | 0.014191811 | 1 | 0.966658386 | 1 | 0.956878334 | ENSG0000014 | 12 | DEF8 |
| SAT | Heart_Failure | ENSG0000021 | 0.005698643 | 4.43E-06 | 0.001241521 | 0.076052205 | 0.003446098 | 0.007058878 | 0.001279144 | 0.076034208 | 0.002252544 | 2.87E-13 | 0.014543183 | 0.000908949 | 1 | 0.194766089 | ENSG0000014 | 14 | TCF25 |
| SAT | Heart_Failure | ENSG0000021 | 0.00028345 | 0.856899097 | 0.001571901 | 0.076055488 | 0.000244434 | 0.876413825 | 0.00157175 | 0.076035336 | 3.90E-05 | 0.003538191 | 1 | 0.976669362 | 1 | 0.974665278 | ENSG0000014 | 14 | TCF25 |
| SAT | Heart_Failure | ENSG0000021 | -0.00729877 | 0.047733228 | 0.004439213 | 0.076054881 | -0.01448584 | 0.001101869 | 0.004439213 | 0.076033679 | 0.007187068 | 0.003665782 | 1 | 0.478075483 | 1 | 0.076874944 | ENSG0000014 | 8 | GALNS |
| SAT | Heart_Failure | ENSG0000021 | 0.002843771 | 0.023230082 | 0.001253018 | 0.076054689 | -3.11E-05 | 0.980808833 | 0.001291882 | 0.07603534 | 0.002874847 | 7.32E-20 | 1 | 0.327199936 | 1 | 0.999768899 | ENSG0000014 | 8 | GALNS |
| SAT | Heart_Failure | ENSG0000021 | 0.000618425 | 0.795355936 | 0.002384406 | 0.076055483 | 0.00058797 | 0.80520408 | 0.002384132 | 0.07603531 | 3.05E-05 | 0.031379335 | 1 | 0.976267425 | 1 | 0.972791643 | ENSG0000014 | 10 | GAS8 |
| SAT | Heart_Failure | ENSG0000021 | -0.00244825 | 0.000382831 | 0.000689325 | 0.076053524 | -0.00243444 | 0.000412456 | 0.000689251 | 0.076033394 | -1.38E-05 | 0.004421867 | 1 | 0.031401733 | 1 | 0.038968793 | ENSG0000014 | 10 | GAS8 |
| SAT | Heart_Failure | ENSG0000021 | -0.00012341 | 0.845066762 | 0.000631542 | 0.076055487 | -0.00010058 | 0.873450793 | 0.00063148 | 0.076035336 | -2.28E-05 | 1.03E-05 | 1 | 0.976669362 | 1 | 0.974665278 | ENSG0000014 | 10 | GAS8 |
| SAT | Heart_Failure | ENSG0000021 | -0.00359871 | 0.186005599 | 0.002721161 | 0.07605522 | -0.00365897 | 0.178696708 | 0.002720865 | 0.076035058 | 6.03E-05 | 0.001335254 | 1 | 0.720656885 | 1 | 0.706126577 | ENSG0000014 | 13 | CIRH1A |
| SAT | Heart_Failure | ENSG0000021 | -0.00194962 | 0.428670553 | 0.002463291 | 0.076055395 | -0.00189293 | 0.442170734 | 0.002463047 | 0.076035248 | -5.67E-05 | 0.004947752 | 1 | 0.878482333 | 1 | 0.880611614 | ENSG0000014 | 6 | RANBP10 |
| SAT | Heart_Failure | ENSG0000021 | -0.00036634 | 0.808696062 | 0.001513137 | 0.076055484 | -0.00039948 | 0.791749095 | 0.0015129 | 0.076035329 | 3.31E-05 | 0.001156489 | 1 | 0.976066362 | 1 | 0.970635778 | ENSG0000014 | 6 | GFOD2 |
| SAT | Heart_Failure | ENSG0000021 | 1.52E-05 | 0.981791685 | 0.000664834 | 0.076055493 | -5.26E-06 | 0.993684723 | 0.000664762 | 0.07603534 | 2.04E-05 | 1.07E-05 | 1 | 0.998920999 | 1 | 0.999768899 | ENSG0000014 | 10 | PRPSAP2 |
| SAT | Heart_Failure | ENSG0000021 | -0.00113704 | 0.507255883 | 0.001714696 | 0.076055424 | -0.00110308 | 0.519984846 | 0.001714538 | 0.076035276 | -3.40E-05 | 0.027489558 | 1 | 0.901211406 | 1 | 0.904369708 | ENSG0000014 | 10 | PRPSAP2 |
| SAT | Heart_Failure | ENSG0000021 | -0.00331546 | 0.085509834 | 0.001928067 | 0.076055032 | -0.00326018 | 0.090820215 | 0.001927858 | 0.076034894 | -5.53E-05 | 3.50E-05 | 1 | 0.592308253 | 1 | 0.598355674 | ENSG0000014 | 10 | PRPSAP2 |
| SAT | Heart_Failure | ENSG0000021 | -0.00109888 | 0.499377738 | 0.001626836 | 0.076055422 | -0.00115839 | 0.476390135 | 0.001626677 | 0.076035261 | 5.95E-05 | 1.10E-05 | 1 | 0.899263643 | 1 | 0.894608312 | ENSG0000014 | 12 | MYO19 |
| SAT | Heart_Failure | ENSG0000021 | -0.00116846 | 0.767602713 | 0.003954031 | 0.076055479 | -0.0012211 | 0.757428892 | 0.003953596 | 0.076035325 | 5.26E-05 | 0.047066373 | 1 | 0.971467105 | 1 | 0.962699472 | ENSG0000014 | 9 | TOM1L1 |
| SAT | Heart_Failure | ENSG0000021 | 0.000525796 | 0.844142487 | 0.002674476 | 0.076055487 | 0.000577726 | 0.828958917 | 0.002674198 | 0.076035333 | -5.19E-05 | 0.010533366 | 1 | 0.976669362 | 1 | 0.972791643 | ENSG0000014 | 11 | C17orf80 |
| SAT | Heart_Failure | ENSG0000021 | 0.000451657 | 0.119918506 | 0.002862584 | 0.076055116 | 0.004366346 | 0.127144099 | 0.002862307 | 0.076034977 | 8.53E-05 | 0.000411745 | 1 | 0.646585619 | 1 | 0.647874406 | ENSG0000014 | 15 | VPS53 |
| SAT | Heart_Failure | ENSG0000021 | -0.00068136 | 0.773432712 | 0.00236675 | 0.07605548 | -0.00062612 | 0.791339565 | 0.002366525 | 0.076035329 | -5.52E-05 | 0.006854216 | 1 | 0.972273076 | 1 | 0.970635778 | ENSG0000014 | 15 | VPS53 |
| SAT | Heart_Failure | ENSG0000021 | 0.001018321 | 0.400053874 | 0.001210089 | 0.076055382 | 0.000999738 | 0.408658056 | 0.001209958 | 0.076035234 | 1.86E-05 | 0.026540417 | 1 | 0.870429337 | 1 | 0.867451005 | ENSG0000014 | 8 | SGSM2 |
| SAT | Heart_Failure | ENSG0000022 | 0.002066788 | 0.563131542 | 0.003574541 | 0.076055441 | 0.002132551 | 0.550741503 | 0.003574209 | 0.076035285 | -6.58E-05 | 0.038956588 | 1 | 0.925856651 | 1 | 0.914465015 | ENSG0000014 | 8 | SGSM2 |
| SAT | Heart_Failure | ENSG0000022 | -0.00107265 | 0.232316468 | 0.000898055 | 0.07605527 | -0.00105558 | 0.239783054 | 0.000897961 | 0.076035125 | -1.71E-05 | 0.011861417 | 1 | 0.752934502 | 1 | 0.752162449 | ENSG0000014 | 9 | SCRN2 |
| SAT | Heart_Failure | ENSG0000022 | 0.002940692 | 0.417340453 | 0.003625808 | 0.07605539 | 0.003006714 | 0.406911144 | 0.00362542 | 0.076035233 | -6.60E-05 | 0.01062055 | 1 | 0.87398914 | 1 | 0.867451005 | ENSG0000014 | 9 | SCRN2 |
| SAT | Heart_Failure | ENSG0000022 | 0.001581721 | 0.305022311 | 0.001542054 | 0.076055329 | 0.001538644 | 0.318329213 | 0.001541884 | 0.076035185 | 4.31E-05 | 3.17E-05 | 1 | 0.816225947 | 1 | 0.823562856 | ENSG0000014 | 8 | RHBDL3 |
| SAT | Heart_Failure | ENSG0000022 | -0.00123033 | 0.724264049 | 0.00348768 | 0.076055474 | -0.00130743 | 0.707730269 | 0.003487363 | 0.076035318 | 7.71E-05 | 0.01518971 | 1 | 0.966658386 | 1 | 0.958028875 | ENSG0000014 | 9 | SLMO1 |
| SAT | Heart_Failure | ENSG0000022 | 0.000902527 | 0.74267882 | 0.002749024 | 0.076055476 | 0.000969492 | 0.724312552 | 0.002748762 | 0.076035321 | -6.70E-05 | 0.004756913 | 1 | 0.966658386 | 1 | 0.958028875 | ENSG0000014 | 9 | SLMO1 |
| SAT | Heart_Failure | ENSG0000022 | 0.000295404 | 0.904546655 | 0.002487167 | 0.076055491 | 0.000227267 | 0.927186168 | 0.002486897 | 0.076035339 | 6.81E-05 | 6.74E-05 | 1 | 0.981697482 | 1 | 0.987053153 | ENSG0000014 | 9 | SLMO1 |
| SAT | Heart_Failure | ENSG0000022 | -0.00224479 | 0.374026784 | 0.002525199 | 0.07605537 | 0.001652948 | 0.519059604 | 0.00256352 | 0.076035275 | -0.00389774 | 1.51E-18 | 1 | 0.85759885 | 1 | 0.903946158 | ENSG0000014 | 9 | SLMO1 |
| SAT | Heart_Failure | ENSG0000022 | -0.00234828 | 0.092660964 | 0.001396517 | 0.076055052 | -0.00231528 | 0.097302744 | 0.001396365 | 0.076034911 | -3.30E-05 | 0.000551558 | 1 | 0.604414756 | 1 | 0.60854036 | ENSG0000014 | 13 | RPRD1A |
| SAT | Heart_Failure | ENSG0000022 | 0.000484243 | 0.784708774 | 0.001772564 | 0.076055481 | 0.004639166 | 0.024351262 | 0.002060433 | 0.076034549 | -0.00415492 | 7.69E-05 | 1 | 0.972730696 | 1 | 0.342903392 | ENSG0000014 | 13 | RPRD1A |
| SAT | Heart_Failure | ENSG0000022 | -0.00444457 | 0.242712342 | 0.003804514 | 0.07605528 | -0.00454363 | 0.23232135 | 0.003804101 | 0.076035118 | 9.91E-05 | 0.000167756 | 1 | 0.759865642 | 1 | 0.750242469 | ENSG0000014 | 5 | C18orf8 |
| SAT | Heart_Failure | ENSG0000022 | 0.002086699 | 0.008142554 | 0.000788591 | 0.0760544 | 0.002067097 | 0.00875462 | 0.000788517 | 0.076034268 | 1.96E-05 | 0.004400447 | 1 | 0.203936797 | 1 | 0.215969227 | ENSG0000014 | 5 | C18orf8 |
| SAT | Heart_Failure | ENSG0000022 | -0.00119591 | 0.070302237 | 0.000660739 | 0.076054982 | -0.00177635 | 0.007637995 | 0.00066588 | 0.07603423 | 0.000580441 | 3.21E-12 | 1 | 0.540191194 | 1 | 0.202724069 | ENSG0000014 | 5 | C18orf8 |
| SAT | Heart_Failure | ENSG0000022 | -0.0007463 | 0.639105904 | 0.001591425 | 0.076055459 | -0.00070721 | 0.656728515 | 0.001591265 | 0.076035309 | -3.91E-05 | 0.002020015 | 1 | 0.959428651 | 1 | 0.956286421 | ENSG0000014 | 5 | C18orf8 |
| SAT | Heart_Failure | ENSG0000022 | 0.000559977 | 0.464062472 | 0.000764813 | 0.076055409 | 0.002254325 | 0.004272628 | 0.000788967 | 0.076034066 | -0.00169435 | 2.61E-18 | 1 | 0.890605014 | 1 | 0.161355485 | ENSG0000014 | 8 | NPC1 |
| SAT | Heart_Failure | ENSG0000022 | 0.000531839 | 0.857690548 | 0.00296595 | 0.076055488 | 0.000461038 | 0.876460048 | 0.002965667 | 0.076035336 | 7.08E-05 | 0.005371629 | 1 | 0.976066362 | 1 | 0.974665278 | ENSG0000014 | 8 | NPC1 |
| SAT | Heart_Failure | ENSG0000022 | -0.00259281 | 0.092491251 | 0.001541136 | 0.076055051 | -0.00263078 | 0.087782804 | 0.001540976 | 0.076034885 | 3.80E-05 | 0.001149212 | 1 | 0.604414756 | 1 | 0.596493272 | ENSG0000014 | 10 | CCDC40 |
| SAT | Heart_Failure | ENSG0000022 | -0.00500886 | 0.111484994 | 0.003147147 | 0.076055098 | -0.00511414 | 0.104129008 | 0.003146851 | 0.076034928 | 0.000105275 | 0.000138083 | 1 | 0.640599412 | 1 | 0.620049499 | ENSG0000014 | 10 | CCDC40 |
| SAT | Heart_Failure | ENSG0000022 | 0.000255685 | 0.923649901 | 0.002667912 | 0.076055492 | 0.000212093 | 0.936630304 | 0.002667639 | 0.076035339 | 4.36E-05 | 0.034606809 | 1 | 0.988739396 | 1 | 0.988983601 | ENSG0000014 | 10 | CCDC40 |
| SAT | Heart_Failure | ENSG0000022 | 0.004095641 | 0.12960752 | 0.002702227 | 0.076055134 | 0.004031371 | 0.135692797 | 0.002701943 | 0.076034993 | 6.43E-05 | 0.001306846 | 1 | 0.649888404 | 1 | 0.649215182 | ENSG0000014 | 16 | TBCD |
| SAT | Heart_Failure | ENSG0000022 | -0.00047274 | 0.880822188 | 0.003153156 | 0.07605549 | -0.00052118 | 0.868702806 | 0.003152822 | 0.076035336 | 4.84E-05 | 0.035303499 | 1 | 0.979223495 | 1 | 0.974432105 | ENSG0000014 | 16 | TBCD |
| SAT | Heart_Failure | ENSG0000022 | -0.0024181 | 0.082302347 | 0.001391726 | 0.076055022 | -0.00529648 | 0.000246642 | 0.001444856 | 0.076033243 | 0.002878376 | 1.35E-13 | 1 | 0.592308253 | 0.809233072 | 0.027247586 | ENSG0000014 | 16 | TBCD |
| SAT | Heart_Failure | ENSG0000022 | -0.00064727 | 0.523034893 | 0.001013452 | 0.076055429 | -0.00061741 | 0.542341255 | 0.001013341 | 0.076035282 | -2.99E-05 | 1.24E-05 | 1 | 0.906724424 | 1 | 0.912245255 | ENSG0000014 | 10 | FN3KRP |
| SAT | Heart_Failure | ENSG0000022 | -0.00034929 | 0.875294627 | 0.002225642 | 0.076055489 | -0.0004002 | 0.857286088 | 0.002225413 | 0.076035335 | 5.09E-05 | 0.003024539 | 1 | 0.979223495 | 1 | 0.972791643 | ENSG0000014 | 10 | FN3KRP |
| SAT | Heart_Failure | ENSG0000022 | -0.00180495 | 0.129992816 | 0.001192066 | 0.076055135 | -0.0017693 | 0.137711302 | 0.001191954 | 0.076034996 | -3.56E-05 | 0.000722645 | 1 | 0.649888404 | 1 | 0.649215182 | ENSG0000014 | 10 | FN3KRP |
| SAT | Heart_Failure | ENSG0000022 | -0.00013647 | 0.969272002 | 0.003542797 | 0.076055493 | -0.00023765 | 0.946512299 | 0.003542405 | 0.076035339 | 0.000101177 | 1.55E-05 | 1 | 0.996609664 | 1 | 0.990755659 | ENSG0000014 | 10 | FN3KRP |
| SAT | Heart_Failure | ENSG0000022 | 0.000710364 | 0.466497224 | 0.000975519 | 0.07605541 | 0.000256246 | 0.793799834 | 0.000980358 | 0.076035329 | 0.000454118 | 4.09E-06 | 1 | 0.892464951 | 1 | 0.970635778 | ENSG0000014 | 10 | FN3KRP |
| SAT | Heart_Failure | ENSG0000022 | -0.00537399 | 0.077530537 | 0.003044405 | 0.076055007 | -0.00542863 | 0.07453292 | 0.003044096 | 0.076034844 | 5.46E-05 | 0.023099124 | 1 | 0.575515143 | 1 | 0.557044443 | ENSG0000014 | 13 | NARF |
| SAT | Heart_Failure | ENSG0000022 | -0.00045761 | 0.681441925 | 0.001114783 | 0.076055467 | -0.00042643 | 0.702043939 | 0.001114661 | 0.076035317 | -3.12E-05 | 3.76E-05 | 1 | 0.966658386 | 1 | 0.956878334 | ENSG0000014 | 6 | TRIM65 |
| SAT | Heart_Failure | ENSG0000022 | -0.00195669 | 0.236793365 | 0.001653947 | 0.076055275 | -0.00380795 | 0.02208954 | 0.001663711 | 0.076034523 | 0.001851266 | 2.71E-24 | 1 | 0.755348658 | 1 | 0.330737054 | ENSG0000014 | 6 | TRIM65 |
| SAT | Heart_Failure | ENSG0000022 | -0.00149931 | 0.096002804 | 0.000900728 | 0.076055061 | -0.00152589 | 0.090222706 | 0.000900639 | 0.076034892 | 2.66E-05 | 0.000296785 | 1 | 0.612811673 | 1 | 0.598355674 | ENSG0000014 | 6 | TRIM65 |
| SAT | Heart_Failure | ENSG0000022 | 0.001105749 | 0.774809728 | 0.003865046 | 0.07605548 | 0.001044791 | 0.786893763 | 0.003864637 | 0.076035329 | 6.10E-05 | 0.030347635 | 1 | 0.972664863 | 1 | 0.969773699 | ENSG0000014 | 6 | TRIM65 |
| SAT | Heart_Failure | ENSG0000022 | -0.00134899 | 0.421552261 | 0.001678406 | 0.076055392 | -0.0013803 | 0.410807901 | 0.001678233 | 0.076035235 | 3.13E-05 | 0.014855744 | 1 | 0.876497446 | 1 | 0.867451005 | ENSG0000014 | 6 | TRIM65 |
| SAT | Heart_Failure | ENSG0000022 | -0.00101818 | 0.751899748 | 0.00322071 | 0.076055477 | -0.00092591 | 0.773716961 | 0.003220385 | 0.076035327 | -9.23E-05 | 0.000315959 | 1 | 0.966658386 | 1 | 0.965963214 | ENSG0000014 | 6 | TRIM65 |
| SAT | Heart_Failure | ENSG0000022 | 0.001130886 | 0.433422393 | 0.001443655 | 0.076055397 | 0.001095592 | 0.447862933 | 0.0014435 | 0.07603525 | 3.53E-05 | 0.000580868 | 1 | 0.879986926 | 1 | 0.882462767 | ENSG0000014 | 3 | SECTM1 |
| SAT | Heart_Failure | ENSG0000022 | -0.0022656 | 0.516752632 | 0.00349435 | 0.076055427 | -0.00218272 | 0.532161065 | 0.00349397 | 0.076035279 | -8.29E-05 | 0.000562269 | 1 | 0.906069117 | 1 | 0.908441675 | ENSG0000014 | 3 | SECTM1 |
| SAT | Heart_Failure | ENSG0000022 | 0.000107234 | 0.904860973 | 0.000897179 | 0.076055491 | 0.000130576 | 0.884272417 | 0.000897087 | 0.076035337 | -2.33E-05 | 0.000659075 | 1 | 0.981517198 | 1 | 0.974895455 | ENSG0000014 | 10 | RNF157 |
| SAT | Heart_Failure | ENSG0000022 | 0.00168951 | 0.640306585 | 0.0036157 | 0.076055459 | 0.001842397 | 0.610372347 | 0.003615787 | 0.0760353 | -0.00015289 | 0.016934694 | 1 | 0.959428651 | 1 | 0.945083374 | ENSG0000014 | 10 | RNF157 |
| SAT | Heart_Failure | ENSG0000022 | 0.002543975 | 0.21814866 | 0.002065808 | 0.076055256 | 0.002573908 | 0.212729411 | 0.002065576 | 0.076035098 | -2.99E-05 | 0.022313753 | 1 | 0.747815981 | 1 | 0.733958243 | ENSG0000014 | 9 | AZI1 |
| SAT | Heart_Failure | ENSG0000022 | -0.00128807 | 0.557957597 | 0.002198535 | 0.076055439 | -0.00292811 | 0.184162374 | 0.002204823 | 0.076035065 | 0.001640041 | 5.79E-22 | 1 | 0.922459771 | 1 | 0.715224819 | ENSG0000014 | 9 | AZI1 |
| SAT | Heart_Failure | ENSG0000022 | 0.005667797 | 5.57E-12 | 0.000822568 | 0.076048084 | 0.00130563 | 0.19531984 | 0.001008206 | 0.076035078 | 0.004362167 | 7.36E-14 | 1.83E-08 | 3.66E-09 | 1 | 0.7243687 | ENSG0000014 | 11 | WDR45B |
| SAT | Heart_Failure | ENSG0000022 | 4.90E-05 | 0.98907484 | 0.003579347 | 0.076055493 | -3.93E-05 | 0.99123883 | 0.003578973 | 0.07603534 | 8.83E-05 | 0.000939567 | 1 | 0.999440735 | 1 | 0.999768899 | ENSG0000014 | 11 | WDR45B |
| SAT | Heart_Failure | ENSG0000023 | 0.000672631 | 0.499260573 | 0.000995526 | 0.076055422 | 0.000942332 | 0.344014283 | 0.000995845 | 0.0760352 | -0.0002697 | 2.23E-19 | 1 | 0.899263643 | 1 | 0.839547455 | ENSG0000014 | 11 | WDR45B |
| SAT | Heart_Failure | ENSG0000023 | -0.00326426 | 0.056670074 | 0.001712755 | 0.076054926 | -0.00322261 | 0.05987415 | 0.001712581 | 0.076034788 | -4.17E-05 | 0.001950781 | 1 | 0.503931718 | 1 | 0.504627254 | ENSG0000014 | 11 | WDR45B |
| SAT | Heart_Failure | ENSG0000023 | -0.00042939 | 0.6426711 | 0.000925464 | 0.076055459 | -0.0017965 | 0.057594068 | 0.000946136 | 0.076034778 | 0.001367118 | 4.22E-12 | 1 | 0.960639581 | 1 | 0.503576399 | ENSG0000014 | 11 | WDR45B |
| SAT | Heart_Failure | ENSG0000023 | -0.00314721 | 0.376612085 | 0.003559564 | 0.076055371 | -0.00320821 | 0.367382339 | 0.003559192 | 0.076035213 | 6.10E-05 | 0.021824184 | 1 | 0.85759885 | 1 | 0.849290727 | ENSG0000014 | 7 | MAPK4 |
| SAT | Heart_Failure | ENSG0000023 | -0.00053548 | 0.508894055 | 0.000810651 | 0.076055425 | -0.00056521 | 0.485613857 | 0.000810572 | 0.076035264 | 2.97E-05 | 1.08E-05 | 1 | 0.901555828 | 1 | 0.894608312 | ENSG0000014 | 7 | MAPK4 |
| SAT | Heart_Failure | ENSG0000023 | -5.20E-05 | 0.967983923 | 0.001295241 | 0.076055493 | -0.00198125 | 0.138985129 | 0.001339058 | 0.076034999 | 0.001929261 | 1.45E-08 | 1 | 0.996534437 | 1 | 0.653383835 | ENSG0000014 | 5 | ZCCHC2 |
| SAT | Heart_Failure | ENSG0000023 | 0.002188631 | 0.311292039 | 0.002161577 | 0.076055333 | 0.002129914 | 0.324399018 | 0.002161342 | 0.076035189 | 5.87E-05 | 7.06E-05 | 1 | 0.819702392 | 1 | 0.828598718 | ENSG0000014 | 12 | NT5C3B |
| SAT | Heart_Failure | ENSG0000023 | 0.00359843 | 0.010188326 | 0.001400505 | 0.076054463 | 0.002118763 | 0.133368214 | 0.001411612 | 0.076034989 | 0.001479667 | 9.91E-17 | 1 | 0.230796464 | 1 | 0.649215182 | ENSG0000014 | 5 | IGFBP4 |
| SAT | Heart_Failure | ENSG0000023 | 0.002475531 | 0.356790895 | 0.00268642 | 0.07605536 | 0.002428082 | 0.366029168 | 0.002686119 | 0.076035213 | 4.74E-05 | 0.005357011 | 1 | 0.848363544 | 1 | 0.849290727 | ENSG0000014 | 5 | IGFBP4 |
| SAT | Heart_Failure | ENSG0000023 | -0.00201624 | 0.242272518 | 0.001724272 | 0.07605528 | -0.00072268 | 0.676043041 | 0.00172944 | 0.076035313 | -0.00129356 | 2.62E-21 | 1 | 0.759865642 | 1 | 0.956878334 | ENSG0000014 | 14 | FKBP10 |
| SAT | Heart_Failure | ENSG0000023 | 0.013354322 | 8.26E-06 | 0.002995372 | 0.076052391 | 0.0133915 | 7.78E-06 | 0.002995025 | 0.076032221 | -3.72E-05 | 0.032835772 | 0.027108694 | 0.001355435 | 0.025520246 | 0.00258496 | ENSG0000014 | 10 | TXNL4A |
| SAT | Heart_Failure | ENSG0000023 | 0.006390158 | 2.01E-14 | 0.000835259 | 0.076046359 | 0.002273927 | 0.05387884 | 0.001179537 | 0.07603476 | 0.004116231 | 7.74E-07 | 6.58E-11 | 1.64E-11 | 1 | 0.491574717 | ENSG0000014 | 10 | TXNL4A |
| SAT | Heart_Failure | ENSG0000023 | -0.0017013 | 0.581814385 | 0.003089134 | 0.076055446 | -0.00176518 | 0.567681831 | 0.003088842 | 0.076035289 | 6.39E-05 | 0.017918811 | 1 | 0.937265914 | 1 | 0.923432865 | ENSG0000014 | 10 | TXNL4A |
| SAT | Heart_Failure | ENSG0000023 | 0.000412518 | 0.840919285 | 0.002055215 | 0.076055487 | 0.00037722 | 0.854357064 | 0.002055001 | 0.076035335 | 3.53E-05 | 0.021700992 | 1 | 0.976669362 | 1 | 0.972791643 | ENSG0000014 | 8 | TPGS1 |
| SAT | Heart_Failure | ENSG0000023 | 0.000482088 | 0.856545194 | 0.002666801 | 0.076055488 | 0.000429795 | 0.871949747 | 0.00266651 | 0.076035336 | 5.23E-05 | 0.004217541 | 1 | 0.976669362 | 1 | 0.974665278 | ENSG0000014 | 8 | TPGS1 |

| | | | | | | | | | | | | | | | | | | | |
|---|---|---|---|---|---|---|---|---|---|---|---|---|---|---|---|---|---|---|---|
| SAT | Heart_Failure | ENSG0000023 | 0.002587547 | 0.20985088 | 0.002063472 | 0.076055248 | 0.002519148 | 0.222102886 | 0.002063257 | 0.076035108 | 6.84E-05 | 9.02E-06 | 1 | 0.739879495 | 1 | 0.746638902 | ENSG0000014 | 5 | PPAP2C |
| SAT | Heart_Failure | ENSG0000023 | -0.00074705 | 0.490369839 | 0.001083121 | 0.076055419 | -0.00163106 | 0.133263574 | 0.001086392 | 0.076034988 | 0.000884012 | 9.80E-25 | 1 | 0.898909993 | 1 | 0.649215182 | ENSG0000014 | 9 | PRDM15 |
| SAT | Heart_Failure | ENSG0000023 | -0.00657095 | 0.04009057 | 0.003200937 | 0.076054835 | -0.0065119 | 0.041892548 | 0.003200594 | 0.076034694 | -5.90E-05 | 0.009443684 | 1 | 0.450042677 | 1 | 0.45628996 | ENSG0000014 | 9 | PRDM15 |
| SAT | Heart_Failure | ENSG0000023 | -0.00067649 | 0.741351993 | 0.002049577 | 0.076055476 | -0.00074641 | 0.715694934 | 0.002049357 | 0.076035319 | 6.99E-05 | 1.60E-06 | 1 | 0.966658386 | 1 | 0.958028875 | ENSG0000014 | 12 | PFKL |
| SAT | Heart_Failure | ENSG0000023 | -0.00657297 | 0.086416688 | 0.003833501 | 0.076055034 | -0.00643783 | 0.093049089 | 0.003833111 | 0.0760349 | -0.00013514 | 6.93E-06 | 1 | 0.592308253 | 1 | 0.599166569 | ENSG0000014 | 12 | PFKL |
| SAT | Heart_Failure | ENSG0000023 | -0.00581938 | 0.05602968 | 0.003045506 | 0.076054923 | -0.00819107 | 0.007326211 | 0.003054488 | 0.076034218 | 0.002371691 | 3.89E-23 | 1 | 0.503931718 | 1 | 0.198655363 | ENSG0000014 | 8 | MVB12A |
| SAT | Heart_Failure | ENSG0000023 | -0.00265478 | 0.126764363 | 0.001738573 | 0.076055129 | -0.00364376 | 0.03685313 | 0.001745604 | 0.07603466 | 0.000988981 | 5.10E-10 | 1 | 0.649888404 | 1 | 0.429320811 | ENSG0000014 | 8 | MVB12A |
| SAT | Heart_Failure | ENSG0000023 | 0.000852403 | 0.685785453 | 0.002106877 | 0.076055467 | -0.00149376 | 0.482783709 | 0.00212837 | 0.076035263 | 0.002346164 | 1.11E-14 | 1 | 0.966658386 | 1 | 0.894608312 | ENSG0000014 | 8 | MVB12A |
| SAT | Heart_Failure | ENSG0000023 | 0.003198455 | 0.008769532 | 0.001220355 | 0.076054421 | 0.003158134 | 0.009649597 | 0.001220229 | 0.076034295 | 4.03E-05 | 1.15E-05 | 1 | 0.210020691 | 1 | 0.22454133 | ENSG0000014 | 11 | DUS3L |
| SAT | Heart_Failure | ENSG0000023 | -0.0004747 | 0.609616097 | 0.000929659 | 0.076055452 | -0.00044867 | 0.629330277 | 0.000929556 | 0.076035304 | -2.60E-05 | 2.61E-05 | 1 | 0.950190221 | 1 | 0.94798204 | ENSG0000014 | 11 | DUS3L |
| SAT | Heart_Failure | ENSG0000023 | -0.00163885 | 0.289182061 | 0.001546208 | 0.076055318 | -0.00415875 | 0.008037085 | 0.001569031 | 0.076034244 | 0.002519891 | 5.12E-21 | 1 | 0.802034101 | 1 | 0.209225934 | ENSG0000014 | 11 | DUS3L |
| SAT | Heart_Failure | ENSG0000023 | 0.002743383 | 0.136597263 | 0.001842947 | 0.076055147 | 0.002703607 | 0.142332352 | 0.001842745 | 0.076035004 | 3.98E-05 | 0.001422678 | 1 | 0.663104757 | 1 | 0.658003792 | ENSG0000014 | 11 | DUS3L |
| SAT | Heart_Failure | ENSG0000023 | -0.00274613 | 0.06778555 | 0.001503547 | 0.076054972 | -0.00272497 | 0.06990072 | 0.001503384 | 0.076034828 | -2.12E-05 | 0.042228672 | 1 | 0.535424809 | 1 | 0.529994954 | ENSG0000014 | 11 | DUS3L |
| SAT | Heart_Failure | ENSG0000023 | -0.00063344 | 0.719559529 | 0.001764224 | 0.076055473 | -0.00057709 | 0.743558578 | 0.001764034 | 0.076035323 | -5.63E-05 | 5.85E-06 | 1 | 0.966658386 | 1 | 0.962349048 | ENSG0000014 | 12 | DPP9 |
| SAT | Heart_Failure | ENSG0000023 | -0.00031768 | 0.842377353 | 0.001597575 | 0.076055487 | -0.0002703 | 0.865630009 | 0.001597408 | 0.076035336 | -4.74E-05 | 6.98E-05 | 1 | 0.976669362 | 1 | 0.974123167 | ENSG0000014 | 12 | DPP9 |
| SAT | Heart_Failure | ENSG0000023 | -0.00042201 | 0.739157117 | 0.001267399 | 0.076055476 | -0.0004505 | 0.722223775 | 0.001267264 | 0.07603532 | 2.85E-05 | 0.001720275 | 1 | 0.966658386 | 1 | 0.958028875 | ENSG0000014 | 10 | TMEM91 |
| SAT | Heart_Failure | ENSG0000023 | -0.00076234 | 0.554738186 | 0.001290622 | 0.076055439 | -0.00164329 | 0.215730942 | 0.001327414 | 0.076035101 | 0.000880948 | 0.004626416 | 1 | 0.921542737 | 1 | 0.736538211 | ENSG0000014 | 10 | TMEM91 |
| SAT | Heart_Failure | ENSG0000023 | 0.000620568 | 0.8751668 | 0.003950171 | 0.076055489 | 0.000553925 | 0.888468537 | 0.003949766 | 0.076035337 | 6.66E-05 | 0.029133215 | 1 | 0.979223495 | 1 | 0.977226037 | ENSG0000014 | 11 | ATHL1 |
| SAT | Heart_Failure | ENSG0000023 | 0.002411603 | 0.246261163 | 0.0020799 | 0.076055283 | 0.00078426 | 0.713089455 | 0.002132822 | 0.076035319 | 0.001627342 | 0.000586502 | 1 | 0.761161032 | 1 | 0.958028875 | ENSG0000014 | 11 | ATHL1 |
| SAT | Heart_Failure | ENSG0000023 | 0.008639788 | 0.000533674 | 0.002494682 | 0.076053621 | 0.007932823 | 0.001500173 | 0.002498788 | 0.076033768 | 0.000706964 | 2.01E-06 | 1 | 0.037254953 | 1 | 0.09221864 | ENSG0000014 | 11 | ATHL1 |
| SAT | Heart_Failure | ENSG0000023 | 0.002077288 | 0.38344941 | 0.002383417 | 0.076055374 | 0.002161953 | 0.364312171 | 0.002383158 | 0.076035212 | -8.47E-05 | 3.14E-07 | 1 | 0.85759885 | 1 | 0.849290727 | ENSG0000014 | 10 | COL6A1 |
| SAT | Heart_Failure | ENSG0000023 | -0.00164577 | 0.447531991 | 0.002166805 | 0.076055403 | -0.0016044 | 0.45898486 | 0.002166591 | 0.076035255 | -4.14E-05 | 0.019462792 | 1 | 0.884975994 | 1 | 0.890430886 | ENSG0000014 | 10 | COL6A1 |
| SAT | Heart_Failure | ENSG0000023 | 0.006820391 | 0.048233597 | 0.003452822 | 0.076054884 | 0.006765758 | 0.050032457 | 0.003452461 | 0.076034741 | 5.46E-05 | 0.034581524 | 1 | 0.478630006 | 1 | 0.480888199 | ENSG0000014 | 8 | IFNAR1 |
| SAT | Heart_Failure | ENSG0000023 | 0.002027555 | 0.553983645 | 0.003426071 | 0.076055438 | 0.001939102 | 0.571365329 | 0.00342572 | 0.07603529 | 8.85E-05 | 0.000870858 | 1 | 0.921261889 | 1 | 0.924404562 | ENSG0000014 | 7 | SIK1 |
| SAT | Heart_Failure | ENSG0000023 | 0.000356035 | 0.71420151 | 0.000972192 | 0.076055472 | 0.000397326 | 0.682736958 | 0.000972096 | 0.076035314 | -4.13E-05 | 3.06E-07 | 1 | 0.966658386 | 1 | 0.956878334 | ENSG0000014 | 12 | TRPM2 |
| SAT | Heart_Failure | ENSG0000023 | 0.013315141 | 1.47E-22 | 0.001362376 | 0.076040588 | 0.006147784 | 0.002608409 | 0.002042122 | 0.076033926 | 0.007167356 | 2.46E-06 | 4.82E-19 | 2.41E-19 | 1 | 0.122791633 | ENSG0000014 | 12 | TRPM2 |
| SAT | Heart_Failure | ENSG0000023 | -0.00067221 | 0.41866741 | 0.00083119 | 0.076055391 | -6.34E-05 | 0.941802048 | 0.000868797 | 0.076035339 | -0.00060879 | 0.016203516 | 1 | 0.874377957 | 1 | 0.988983601 | ENSG0000014 | 12 | TRPM2 |
| SAT | Heart_Failure | ENSG0000023 | -0.00178502 | 0.635060687 | 0.003760967 | 0.076055458 | -0.00169985 | 0.651255599 | 0.003760575 | 0.076035308 | -8.52E-05 | 0.00260346 | 1 | 0.959428651 | 1 | 0.954583686 | ENSG0000014 | 12 | TRPM2 |
| SAT | Heart_Failure | ENSG0000023 | 0.004957063 | 0.115587213 | 0.003150199 | 0.076055107 | 0.005043021 | 0.109372262 | 0.003149872 | 0.07603494 | -8.60E-05 | 0.000335835 | 1 | 0.641850091 | 1 | 0.631382451 | ENSG0000014 | 12 | TMEM50B |
| SAT | Heart_Failure | ENSG0000023 | 0.000471379 | 0.819253307 | 0.002062878 | 0.076055485 | 0.000414908 | 0.840580331 | 0.002062668 | 0.076035334 | 5.65E-05 | 0.000463357 | 1 | 0.976669362 | 1 | 0.972791643 | ENSG0000014 | 8 | DOPEY2 |
| SAT | Heart_Failure | ENSG0000023 | 0.001593208 | 0.527150532 | 0.002519455 | 0.076055431 | 0.001634349 | 0.516490329 | 0.002519167 | 0.076035274 | -4.11E-05 | 0.007153046 | 1 | 0.906724424 | 1 | 0.903395669 | ENSG0000014 | 5 | URB1 |
| SAT | Heart_Failure | ENSG0000023 | 0.003039576 | 0.082766693 | 0.001752061 | 0.076055023 | 0.003064793 | 0.080215072 | 0.001751868 | 0.076034863 | -2.52E-05 | 0.031303946 | 1 | 0.592308253 | 1 | 0.57843 | ENSG0000014 | 5 | URB1 |
| SAT | Heart_Failure | ENSG0000023 | 0.000192079 | 0.928518406 | 0.002141127 | 0.076055492 | 0.000144424 | 0.946216031 | 0.002140907 | 0.076035339 | 4.77E-05 | 0.003811016 | 1 | 0.988739396 | 1 | 0.990755659 | ENSG0000014 | 7 | SAE1 |
| SAT | Heart_Failure | ENSG0000024 | -0.00101593 | 0.222127812 | 0.000832121 | 0.07605526 | 0.000255046 | 0.763314327 | 0.00084696 | 0.076035326 | -0.00127098 | 1.03E-15 | 1 | 0.751090303 | 1 | 0.965635578 | ENSG0000014 | 6 | CBLC |
| SAT | Heart_Failure | ENSG0000024 | 0.006211607 | 0.00160865 | 0.001969218 | 0.07605394 | 0.004737196 | 0.020034034 | 0.002036875 | 0.076034496 | 0.001474411 | 0.004700152 | 1 | 0.083777454 | 1 | 0.318841471 | ENSG0000014 | 6 | CBLC |
| SAT | Heart_Failure | ENSG0000024 | 0.001443798 | 0.702452453 | 0.003779455 | 0.07605547 | 0.00153072 | 0.685439921 | 0.003779077 | 0.076035315 | -8.69E-05 | 0.004392338 | 1 | 0.966658386 | 1 | 0.956878334 | ENSG0000014 | 6 | CBLC |
| SAT | Heart_Failure | ENSG0000024 | 0.005472601 | 0.13212713 | 0.003634423 | 0.076055139 | 0.005537476 | 0.127562978 | 0.003634026 | 0.076034978 | -6.49E-05 | 0.00904416 | 1 | 0.655838294 | 1 | 0.647874406 | ENSG0000014 | 14 | SLC6A3 |
| SAT | Heart_Failure | ENSG0000024 | 0.001252847 | 0.144389321 | 0.000858325 | 0.07605516 | 0.000490458 | 0.570075094 | 0.000863572 | 0.07603529 | 0.000762389 | 2.07E-15 | 1 | 0.669181587 | 1 | 0.924404562 | ENSG0000014 | 15 | CAPN10 |
| SAT | Heart_Failure | ENSG0000024 | -0.00429973 | 0.055439352 | 0.002244796 | 0.07605492 | -0.00437792 | 0.051120996 | 0.002244551 | 0.076034747 | 7.82E-05 | 4.53E-07 | 1 | 0.503306524 | 1 | 0.485739162 | ENSG0000014 | 15 | CAPN10 |
| SAT | Heart_Failure | ENSG0000024 | 0.000228038 | 0.82560358 | 0.001034918 | 0.076055485 | 0.000457141 | 0.658747494 | 0.001035091 | 0.07603531 | -0.0002291 | 1.50E-19 | 1 | 0.976669362 | 1 | 0.956791995 | ENSG0000014 | 13 | RERE |
| SAT | Heart_Failure | ENSG0000024 | -0.00133815 | 0.644421664 | 0.002899405 | 0.07605546 | -0.00140284 | 0.628465665 | 0.002899095 | 0.076035304 | 6.47E-05 | 0.001837052 | 1 | 0.960776086 | 1 | 0.94798204 | ENSG0000014 | 15 | FHAD1 |
| SAT | Heart_Failure | ENSG0000024 | 0.002529966 | 0.381479531 | 0.002890815 | 0.076055373 | 0.002585077 | 0.371144848 | 0.002890512 | 0.076035215 | -5.51E-05 | 0.01071886 | 1 | 0.85759885 | 1 | 0.849768918 | ENSG0000014 | 12 | ARHGEF19 |
| SAT | Heart_Failure | ENSG0000024 | -0.00862629 | 0.004980029 | 0.003071672 | 0.076054262 | -0.00868524 | 0.004687418 | 0.0030714 | 0.076034092 | 5.90E-05 | 0.041041627 | 1 | 0.160190922 | 1 | 0.163610845 | ENSG0000014 | 12 | ARHGEF19 |
| SAT | Heart_Failure | ENSG0000024 | 0.000210186 | 0.93441342 | 0.002554099 | 0.076055492 | 0.000112737 | 0.964789573 | 0.002553847 | 0.07603534 | 9.74E-05 | 3.36E-06 | 1 | 0.989642217 | 1 | 0.996058713 | ENSG0000014 | 8 | PEX14 |
| SAT | Heart_Failure | ENSG0000024 | 4.39E-05 | 0.973273321 | 0.001310454 | 0.076055493 | 2.39E-05 | 0.985442107 | 0.001310305 | 0.07603534 | 2.00E-05 | 0.013168694 | 1 | 0.99766005 | 1 | 0.999768899 | ENSG0000014 | 8 | PEX14 |
| SAT | Heart_Failure | ENSG0000024 | -0.00079482 | 0.501109352 | 0.001181449 | 0.076055422 | -0.0008208 | 0.487174146 | 0.001181326 | 0.076035265 | 2.60E-05 | 0.003480215 | 1 | 0.900549813 | 1 | 0.894608312 | ENSG0000014 | 13 | CNKSR1 |
| SAT | Heart_Failure | ENSG0000024 | 0.001216367 | 0.709239382 | 0.003262105 | 0.076055471 | 0.001349122 | 0.679156705 | 0.003261786 | 0.076035313 | -0.00013275 | 1.03E-06 | 1 | 0.966658386 | 1 | 0.956878334 | ENSG0000014 | 13 | CNKSR1 |
| SAT | Heart_Failure | ENSG0000024 | 0.004519909 | 0.179777727 | 0.003369441 | 0.076055212 | 0.00444264 | 0.187284677 | 0.003369066 | 0.076035069 | 7.73E-05 | 0.000416146 | 1 | 0.714462461 | 1 | 0.7171788 | ENSG0000014 | 13 | CNKSR1 |
| SAT | Heart_Failure | ENSG0000024 | 0.001378772 | 0.392758722 | 0.001613304 | 0.076055379 | 0.001404535 | 0.383924767 | 0.001613132 | 0.076035222 | -2.58E-05 | 0.025287364 | 1 | 0.863144683 | 1 | 0.854967426 | ENSG0000014 | 3 | IL22RA1 |
| SAT | Heart_Failure | ENSG0000024 | -0.00185324 | 0.021162824 | 0.000803981 | 0.076054664 | -0.00296773 | 0.000261247 | 0.000812852 | 0.07603326 | 0.00111449 | 2.31E-20 | 1 | 0.315614662 | 0.857152676 | 0.027247586 | ENSG0000014 | 3 | IL22RA1 |
| SAT | Heart_Failure | ENSG0000024 | -0.00187521 | 0.451430009 | 0.002490204 | 0.076055405 | -0.00182268 | 0.464157194 | 0.002489929 | 0.076035257 | -5.25E-05 | 0.001535051 | 1 | 0.887019168 | 1 | 0.892844363 | ENSG0000014 | 3 | IL22RA1 |
| SAT | Heart_Failure | ENSG0000024 | 0.003986225 | 0.113478809 | 0.002518542 | 0.076055102 | 0.003911645 | 0.120352367 | 0.002518282 | 0.076034964 | 7.46E-05 | 0.000108508 | 1 | 0.641850091 | 1 | 0.64481576 | ENSG0000014 | 3 | IL22RA1 |
| SAT | Heart_Failure | ENSG0000024 | 0.000345619 | 0.829490031 | 0.001604876 | 0.076055486 | 0.000321075 | 0.841415094 | 0.001604705 | 0.076035334 | 2.45E-05 | 0.033731175 | 1 | 0.976669362 | 1 | 0.972791643 | ENSG0000014 | 7 | ZNF593 |
| SAT | Heart_Failure | ENSG0000024 | -0.00156546 | 0.046675809 | 0.000786968 | 0.076054875 | -0.00320595 | 0.004626258 | 0.001132051 | 0.076034089 | 0.001640481 | 0.043836891 | 1 | 0.475002578 | 1 | 0.163212403 | ENSG0000014 | 7 | ZNF593 |
| SAT | Heart_Failure | ENSG0000024 | -0.00151312 | 0.312805325 | 0.001499094 | 0.076055334 | -0.00155408 | 0.299833002 | 0.001498931 | 0.076035172 | 4.10E-05 | 6.81E-05 | 1 | 0.822137009 | 1 | 0.810520426 | ENSG0000014 | 7 | C1orf216 |
| SAT | Heart_Failure | ENSG0000024 | 0.001090304 | 0.65970917 | 0.002476191 | 0.076055463 | 0.001218726 | 0.62255967 | 0.002475947 | 0.076035302 | -0.00012842 | 3.39E-10 | 1 | 0.962994646 | 1 | 0.94798204 | ENSG0000014 | 7 | C1orf216 |
| SAT | Heart_Failure | ENSG0000024 | -0.00344137 | 0.248862409 | 0.002984403 | 0.076055285 | -0.00329537 | 0.269510632 | 0.002984417 | 0.07603515 | -0.000146 | 0.0031006 | 1 | 0.765246076 | 1 | 0.780780192 | ENSG0000014 | 6 | EVA1B |
| SAT | Heart_Failure | ENSG0000024 | -0.00062995 | 0.700472048 | 0.001637568 | 0.07605547 | -0.00068782 | 0.674433755 | 0.00163739 | 0.076035313 | 5.79E-05 | 2.73E-07 | 1 | 0.966658386 | 1 | 0.956878334 | ENSG0000014 | 14 | NBPF3 |
| SAT | Heart_Failure | ENSG0000024 | -0.00265468 | 0.3430565 | 0.002799873 | 0.076055353 | -0.00274866 | 0.32619484 | 0.002799592 | 0.07603519 | 9.40E-05 | 2.81E-05 | 1 | 0.847011103 | 1 | 0.830490914 | ENSG0000014 | 14 | NBPF3 |
| SAT | Heart_Failure | ENSG0000024 | -0.00298023 | 0.427512908 | 0.003756008 | 0.076055395 | -0.0034424 | 0.359872874 | 0.003759683 | 0.076035209 | 0.000462172 | 0.009061574 | 1 | 0.878482333 | 1 | 0.848283315 | ENSG0000014 | 12 | HSPG2 |
| SAT | Heart_Failure | ENSG0000024 | -0.00141833 | 0.718366379 | 0.00393278 | 0.076055473 | -0.00150007 | 0.702857905 | 0.003932381 | 0.076035317 | 8.17E-05 | 0.008441446 | 1 | 0.966658386 | 1 | 0.956878334 | ENSG0000014 | 12 | HSPG2 |
| SAT | Heart_Failure | ENSG0000024 | -0.00207271 | 0.422516949 | 0.002584224 | 0.076055393 | -0.00201999 | 0.434366231 | 0.002583963 | 0.076035245 | -5.27E-05 | 0.010227458 | 1 | 0.876565321 | 1 | 0.877927942 | ENSG0000014 | 12 | HSPG2 |
| SAT | Heart_Failure | ENSG0000024 | 0.001126033 | 0.476072058 | 0.001580098 | 0.076055414 | 0.001095862 | 0.487922578 | 0.001579923 | 0.076035265 | 3.02E-05 | 0.003707439 | 1 | 0.895715287 | 1 | 0.894608312 | ENSG0000014 | 12 | HSPG2 |
| SAT | Heart_Failure | ENSG0000024 | -0.00227641 | 0.50875931 | 0.00344508 | 0.076055425 | -0.00239931 | 0.486105142 | 0.003444724 | 0.076035264 | 0.0001229 | 2.88E-06 | 1 | 0.901555828 | 1 | 0.894608312 | ENSG0000014 | 12 | HSPG2 |
| SAT | Heart_Failure | ENSG0000024 | -0.00143851 | 0.541468274 | 0.00235591 | 0.076055435 | -0.00138618 | 0.556235914 | 0.002355674 | 0.076035286 | -5.23E-05 | 0.005635171 | 1 | 0.913266518 | 1 | 0.915531921 | ENSG0000014 | 7 | RPS8 |
| SAT | Heart_Failure | ENSG0000025 | 0.000608554 | 0.738582959 | 0.001823488 | 0.076055476 | 0.000631038 | 0.729267522 | 0.001823282 | 0.076035321 | -2.25E-05 | 0.049044484 | 1 | 0.966658386 | 1 | 0.958028875 | ENSG0000014 | 7 | RPS8 |
| SAT | Heart_Failure | ENSG0000025 | 0.002767674 | 0.122189081 | 0.001790618 | 0.07605512 | 0.002795213 | 0.1184778 | 0.001790429 | 0.07603496 | -2.75E-05 | 0.035989811 | 1 | 0.646585619 | 1 | 0.64481576 | ENSG0000014 | 8 | CYP4B1 |
| SAT | Heart_Failure | ENSG0000025 | -0.0023183 | 0.214471098 | 0.00186754 | 0.076055253 | -0.00236858 | 0.20464893 | 0.001867345 | 0.076035089 | 5.03E-05 | 0.000343545 | 1 | 0.745086527 | 1 | 0.729840368 | ENSG0000014 | 8 | CYP4B1 |
| SAT | Heart_Failure | ENSG0000025 | 0.004584433 | 0.114689734 | 0.002906207 | 0.076055105 | 0.004662234 | 0.108626861 | 0.002905907 | 0.076034938 | -7.78E-05 | 0.000498898 | 1 | 0.641850091 | 1 | 0.631382451 | ENSG0000014 | 7 | SYPL2 |
| SAT | Heart_Failure | ENSG0000025 | -8.17E-06 | 0.995285604 | 0.001382698 | 0.076055493 | -4.55E-05 | 0.973724033 | 0.001382564 | 0.07603534 | 3.74E-05 | 0.001354209 | 1 | 0.999479422 | 1 | 0.998213938 | ENSG0000014 | 4 | ZNF697 |
| SAT | Heart_Failure | ENSG0000025 | 0.002618241 | 0.329711 | 0.002686209 | 0.076055345 | 0.002570673 | 0.338522016 | 0.002685924 | 0.076035197 | 4.76E-05 | 0.014528012 | 1 | 0.83656615 | 1 | 0.8376162 | ENSG0000014 | 8 | CD53 |
| SAT | Heart_Failure | ENSG0000025 | -0.00023225 | 0.80539819 | 0.000942717 | 0.076055484 | -0.00139937 | 0.142545781 | 0.000954303 | 0.076035005 | 0.001167115 | 4.88E-15 | 1 | 0.976669362 | 1 | 0.658003792 | ENSG0000014 | 8 | TIPRL |
| SAT | Heart_Failure | ENSG0000025 | -0.00285216 | 0.453565557 | 0.003805499 | 0.076055405 | -0.00275189 | 0.469549921 | 0.003805091 | 0.076035259 | -0.00010027 | 0.000209605 | 1 | 0.887019168 | 1 | 0.894229386 | ENSG0000014 | 9 | NME7 |
| SAT | Heart_Failure | ENSG0000025 | -0.00086462 | 0.772092443 | 0.002985156 | 0.07605548 | -0.00080495 | 0.787405405 | 0.002984823 | 0.076035329 | -5.97E-05 | 0.002015511 | 1 | 0.971467105 | 1 | 0.969773699 | ENSG0000014 | 3 | XCL2 |
| SAT | Heart_Failure | ENSG0000025 | -0.00402707 | 0.188507265 | 0.003062391 | 0.076055223 | -0.00350746 | 0.252328275 | 0.003064047 | 0.076035136 | -0.00051962 | 3.74E-06 | 1 | 0.722191019 | 1 | 0.765423527 | ENSG0000014 | 3 | XCL2 |
| SAT | Heart_Failure | ENSG0000025 | -0.00138904 | 0.280395952 | 0.001286824 | 0.076055311 | -0.00135425 | 0.292568189 | 0.001286689 | 0.076035167 | -3.48E-05 | 0.000233756 | 1 | 0.795640478 | 1 | 0.806652294 | ENSG0000014 | 3 | XCL2 |
| SAT | Heart_Failure | ENSG0000025 | -0.00307688 | 0.436560312 | 0.003954786 | 0.076055399 | -0.00315594 | 0.424821856 | 0.003954395 | 0.076035241 | 7.91E-05 | 0.014893807 | 1 | 0.880339889 | 1 | 0.873390936 | ENSG0000014 | 7 | RRNAD1 |
| SAT | Heart_Failure | ENSG0000025 | 0.00329502 | 0.18691832 | 0.002496695 | 0.076055221 | 0.003238617 | 0.194529252 | 0.002496432 | 0.076035078 | 5.64E-05 | 0.002115744 | 1 | 0.720656885 | 1 | 0.723506187 | ENSG0000014 | 11 | TUFT1 |
| SAT | Heart_Failure | ENSG0000025 | 0.00393107 | 0.123982085 | 0.002555506 | 0.076055124 | 0.00387731 | 0.129170772 | 0.002555262 | 0.076034981 | 5.38E-05 | 0.014592736 | 1 | 0.647388543 | 1 | 0.649215182 | ENSG0000014 | 8 | SNX27 |
| SAT | Heart_Failure | ENSG0000025 | -0.00226903 | 0.273061473 | 0.002070209 | 0.076055306 | -0.00230188 | 0.266126748 | 0.002069989 | 0.076035147 | 3.29E-05 | 0.028696365 | 1 | 0.786649253 | 1 | 0.774729353 | ENSG0000014 | 8 | SNX27 |
| SAT | Heart_Failure | ENSG0000025 | 0.009201957 | 0.006831396 | 0.003401877 | 0.076054351 | 0.008967328 | 0.008417502 | 0.003403343 | 0.076034257 | 0.000234629 | 0.039901767 | 1 | 0.181474985 | 1 | 0.209225934 | ENSG0000014 | 15 | CERS2 |
| SAT | Heart_Failure | ENSG0000025 | 0.004530616 | 0.210340601 | 0.00361688 | 0.076055248 | 0.004452267 | 0.218288077 | 0.003616511 | 0.076035104 | 7.83E-05 | 0.005422262 | 1 | 0.739879495 | 1 | 0.742028061 | ENSG0000014 | 16 | ARNT |
| SAT | Heart_Failure | ENSG0000025 | 0.004257711 | 0.278947264 | 0.003932536 | 0.07605531 | 0.004311156 | 0.272903693 | 0.003932097 | 0.076035153 | -5.34E-05 | 0.035530561 | 1 | 0.793905695 | 1 | 0.783093703 | ENSG0000014 | 16 | ARNT |
| SAT | Heart_Failure | ENSG0000025 | -0.00767345 | 0.037753571 | 0.003693551 | 0.076054819 | -0.00762027 | 0.039080182 | 0.003693161 | 0.076034676 | -5.32E-05 | 0.048356133 | 1 | 0.439253432 | 1 | 0.443745602 | ENSG0000014 | 17 | POGZ |
| SAT | Heart_Failure | ENSG0000025 | 0.003642286 | 0.152434648 | 0.002545297 | 0.076055173 | 0.003601727 | 0.157009062 | 0.002545021 | 0.076035028 | 4.06E-05 | 0.022084029 | 1 | 0.677064618 | 1 | 0.669469928 | ENSG0000014 | 17 | POGZ |
| SAT | Heart_Failure | ENSG0000025 | 0.002142539 | 0.326005424 | 0.002181383 | 0.076055342 | 0.00220462 | 0.312134671 | 0.002181164 | 0.076035181 | -6.21E-05 | 0.000381647 | 1 | 0.833010855 | 1 | 0.821155716 | ENSG0000014 | 17 | POGZ |
| SAT | Heart_Failure | ENSG0000025 | 0.00018571 | 0.890560904 | 0.00134969 | 0.07605549 | 0.000133215 | 0.921367966 | 0.001349546 | 0.076035339 | 5.25E-05 | 7.76E-08 | 1 | 0.979270459 | 1 | 0.984731983 | ENSG0000014 | 17 | POGZ |
| SAT | Heart_Failure | ENSG0000025 | -0.00108245 | 0.316591085 | 0.001080837 | 0.076055336 | -0.00111339 | 0.302903568 | 0.001080724 | 0.076035175 | 3.09E-05 | 0.000134781 | 1 | 0.824393135 | 1 | 0.810625291 | ENSG0000014 | 17 | POGZ |
| SAT | Heart_Failure | ENSG0000025 | -0.0071407 | 0.038949814 | 0.00345843 | 0.076054828 | -0.00702096 | 0.042326153 | 0.003458082 | 0.076034697 | -0.00011974 | 1.33E-05 | 1 | 0.445276442 | 1 | 0.45628996 | ENSG0000014 | 11 | HORMAD1 |
| SAT | Heart_Failure | ENSG0000025 | -0.00382955 | 0.249725213 | 0.0033271 | 0.076055286 | -0.00417007 | 0.210144957 | 0.003327621 | 0.076035095 | 0.000340516 | 2.09E-05 | 1 | 0.766457153 | 1 | 0.733958243 | ENSG0000014 | 7 | GABPB2 |
| SAT | Heart_Failure | ENSG0000025 | -0.00124421 | 0.262942751 | 0.00111143 | 0.076055297 | -0.00121544 | 0.274090946 | 0.001111316 | 0.076035153 | -2.88E-05 | 0.000771081 | 1 | 0.775544307 | 1 | 0.784122993 | ENSG0000014 | 11 | EIF2D |
| SAT | Heart_Failure | ENSG0000025 | -0.00129434 | 0.222636488 | 0.00106133 | 0.076055261 | -0.00127175 | 0.230766744 | 0.001061215 | 0.076035116 | -2.26E-05 | 0.002385439 | 1 | 0.751090303 | 1 | 0.750242469 | ENSG0000014 | 11 | EIF2D |
| SAT | Heart_Failure | ENSG0000025 | 0.00107234 | 0.762035422 | 0.003541307 | 0.076055479 | 0.001135954 | 0.748357426 | 0.003540936 | 0.076035324 | -6.36E-05 | 0.015601295 | 1 | 0.969898847 | 1 | 0.962415493 | ENSG0000014 | 11 | EIF2D |
| SAT | Heart_Failure | ENSG0000025 | 0.000494204 | 0.864407886 | 0.002894044 | 0.076055488 | 0.000588569 | 0.838828146 | 0.002893754 | 0.076035334 | -9.44E-05 | 4.87E-05 | 1 | 0.978183841 | 1 | 0.972791643 | ENSG0000014 | 14 | KCNN3 |
| SAT | Heart_Failure | ENSG0000025 | -1.07E-05 | 0.997524715 | 0.003442131 | 0.076055493 | 0.000127526 | 0.970443443 | 0.003441788 | 0.07603534 | -0.0001382 | 7.64E-07 | 1 | 0.999479422 | 1 | 0.99755193 | ENSG0000014 | 6 | RIT1 |
| SAT | Heart_Failure | ENSG0000025 | 0.002212032 | 0.308727949 | 0.002173142 | 0.076055331 | 0.00217144 | 0.317636796 | 0.002172902 | 0.076035184 | 4.06E-05 | 0.005029594 | 1 | 0.816225947 | 1 | 0.823487262 | ENSG0000014 | 10 | FLG |
| SAT | Heart_Failure | ENSG0000025 | -0.00165716 | 0.112726522 | 0.00104483 | 0.0760551 | -0.00164246 | 0.115913725 | 0.001044714 | 0.076034954 | -1.47E-05 | 0.034043526 | 1 | 0.641850091 | 1 | 0.64481576 | ENSG0000014 | 10 | FLG |
| SAT | Heart_Failure | ENSG0000025 | 0.004997109 | 0.056934786 | 0.002624786 | 0.076054927 | 0.004956829 | 0.058937317 | 0.002624514 | 0.076034784 | 4.03E-05 | 0.043265726 | 1 | 0.503931718 | 1 | 0.503576399 | ENSG0000014 | 11 | CEP170 |
| SAT | Heart_Failure | ENSG0000025 | -0.00113067 | 0.422418621 | 0.0014094 | 0.076055393 | -0.00114912 | 0.414836473 | 0.001409244 | 0.076035236 | 1.84E-05 | 0.049015254 | 1 | 0.876565321 | 1 | 0.867451005 | ENSG0000014 | 10 | SNAP47 |
| SAT | Heart_Failure | ENSG0000025 | -0.00383433 | 0.155127452 | 0.002697092 | 0.076055178 | -0.00392012 | 0.146055471 | 0.002696817 | 0.07603501 | 8.58E-05 | 5.10E-05 | 1 | 0.677064618 | 1 | 0.663722991 | ENSG0000014 | 3 | CNIH3 |
| SAT | Heart_Failure | ENSG0000025 | 0.001991757 | 0.429048942 | 0.002518594 | 0.076055395 | 0.002049782 | 0.415676603 | 0.002518328 | 0.076035237 | -5.80E-05 | 0.001714866 | 1 | 0.878482333 | 1 | 0.867451005 | ENSG0000014 | 12 | PYCR2 |
| SAT | Heart_Failure | ENSG0000025 | 0.003819625 | 0.166170777 | 0.002758618 | 0.076055194 | 0.003869391 | 0.160677532 | 0.002758337 | 0.076035033 | -4.98E-05 | 0.020873122 | 1 | 0.690134582 | 1 | 0.676259259 | ENSG0000014 | 12 | PYCR2 |
| SAT | Heart_Failure | ENSG0000025 | 0.000350448 | 0.917508206 | 0.003383582 | 0.076055491 | 0.000259307 | 0.938906367 | 0.003383244 | 0.076035339 | 9.11E-05 | 0.000873432 | 1 | 0.986998171 | 1 | 0.988983601 | ENSG0000014 | 12 | PYCR2 |
| SAT | Heart_Failure | ENSG0000025 | -0.00694004 | 0.057596585 | 0.003655036 | 0.07605493 | -0.00687409 | 0.059983124 | 0.003654639 | 0.076034788 | -6.59E-05 | 0.009016661 | 1 | 0.503931718 | 1 | 0.504627254 | ENSG0000014 | 12 | PYCR2 |
| SAT | Heart_Failure | ENSG0000025 | -0.00148456 | 0.344828038 | 0.001571512 | 0.076055354 | -0.00145923 | 0.353072688 | 0.001571346 | 0.076035206 | -2.53E-05 | 0.027462989 | 1 | 0.847011103 | 1 | 0.843366096 | ENSG0000014 | 12 | GALM |
| SAT | Heart_Failure | ENSG0000025 | 0.000364244 | 0.904791432 | 0.003045237 | 0.076055491 | 0.002784787 | 0.385853245 | 0.003211378 | 0.076035223 | -0.00242054 | 0.017724763 | 1 | 0.981517198 | 1 | 0.857132361 | ENSG0000014 | 11 | WDPCP |
| SAT | Heart_Failure | ENSG0000025 | 0.001729474 | 0.162817204 | 0.001239179 | 0.076055189 | 0.001691901 | 0.172100999 | 0.001239047 | 0.076035049 | 3.76E-05 | 2.47E-05 | 1 | 0.686431329 | 1 | 0.697976981 | ENSG0000014 | 11 | WDPCP |
| SAT | Heart_Failure | ENSG0000025 | 0.00157004 | 0.584426551 | 0.002870638 | 0.076055446 | 0.001611659 | 0.574463235 | 0.002870321 | 0.076035291 | -4.16E-05 | 0.029241196 | 1 | 0.93857245 | 1 | 0.927160988 | ENSG0000014 | 8 | REG3G |
| SAT | Heart_Failure | ENSG0000025 | -0.00409494 | 0.111816075 | 0.002575293 | 0.076055098 | -0.00396847 | 0.123331965 | 0.002575355 | 0.07603497 | -0.00012647 | 0.005457426 | 1 | 0.641378571 | 1 | 0.647747917 | ENSG0000014 | 2 | CIAO1 |
| SAT | Heart_Failure | ENSG0000025 | -0.00198188 | 0.443935308 | 0.002588783 | 0.076055402 | -0.00203324 | 0.432169548 | 0.002588513 | 0.076035244 | 5.14E-05 | 0.008060496 | 1 | 0.882758633 | 1 | 0.876305924 | ENSG0000014 | 7 | ZNF514 |
| SAT | Heart_Failure | ENSG0000025 | -0.00342499 | 0.233778363 | 0.002876488 | 0.076055272 | -0.00349462 | 0.224357954 | 0.002876179 | 0.07603511 | 6.96E-05 | 0.000587779 | 1 | 0.754244409 | 1 | 0.747837725 | ENSG0000014 | 7 | ZNF514 |
| SAT | Heart_Failure | ENSG0000025 | 0.000196091 | 0.943820305 | 0.002782656 | 0.076055492 | 0.000245243 | 0.929763879 | 0.002782368 | 0.076035339 | -4.92E-05 | 0.02021672 | 1 | 0.992414898 | 1 | 0.988475359 | ENSG0000014 | 9 | RALB |
| SAT | Heart_Failure | ENSG0000025 | 0.002088279 | 0.195874201 | 0.001614569 | 0.076055232 | 0.002061534 | 0.201611111 | 0.001614387 | 0.076035086 | 2.67E-05 | 0.008627773 | 1 | 0.726878652 | 1 | 0.728906397 | ENSG0000014 | 8 | FBLN7 |
| SAT | Heart_Failure | ENSG0000025 | -0.00035746 | 0.756260987 | 0.001151638 | 0.076055478 | -0.00038084 | 0.740847002 | 0.001151517 | 0.076035323 | 2.34E-05 | 0.006084875 | 1 | 0.967625883 | 1 | 0.960758504 | ENSG0000014 | 8 | FBLN7 |
| SAT | Heart_Failure | ENSG0000025 | 0.00222517 | 0.569948432 | 0.003916672 | 0.076055443 | 0.002143094 | 0.584222232 | 0.003916272 | 0.076035293 | 8.21E-05 | 0.007178363 | 1 | 0.92942386 | 1 | 0.933194299 | ENSG0000014 | 8 | FBLN7 |
| SAT | Heart_Failure | ENSG0000025 | 0.002716093 | 0.093590794 | 0.001619849 | 0.076055054 | 0.002666978 | 0.099639159 | 0.001619679 | 0.076034917 | 4.91E-05 | 4.20E-05 | 1 | 0.604759516 | 1 | 0.613289124 | ENSG0000014 | 8 | FBLN7 |
| SAT | Heart_Failure | ENSG0000026 | -0.00026278 | 0.799698096 | 0.001035622 | 0.076055483 | -0.0002976 | 0.773813455 | 0.001035523 | 0.076035327 | 3.48E-05 | 9.28E-05 | 1 | 0.976669362 | 1 | 0.965963214 | ENSG0000014 | 8 | FBLN7 |
| SAT | Heart_Failure | ENSG0000026 | 0.003382233 | 0.132575355 | 0.002248787 | 0.07605514 | 0.003323418 | 0.139399172 | 0.002248539 | 0.076034999 | 5.88E-05 | 8.38E-05 | 1 | 0.657069095 | 1 | 0.653383835 | ENSG0000014 | 8 | FBLN7 |
| SAT | Heart_Failure | ENSG0000026 | 0.000686745 | 0.722637804 | 0.001934842 | 0.076055473 | 0.000831547 | 0.667347811 | 0.00193477 | 0.076035311 | -0.0001448 | 6.05E-08 | 1 | 0.966658386 | 1 | 0.956878334 | ENSG0000014 | 12 | LIPT1 |
| SAT | Heart_Failure | ENSG0000026 | -0.00024453 | 0.929638882 | 0.002769378 | 0.076055492 | -0.0001974 | 0.94317001 | 0.002769082 | 0.076035339 | -4.71E-05 | 0.017577168 | 1 | 0.988739396 | 1 | 0.989303326 | ENSG0000014 | 12 | LIPT1 |
| SAT | Heart_Failure | ENSG0000026 | -0.00344002 | 0.109568761 | 0.00214983 | 0.076055093 | -0.0033885 | 0.11494756 | 0.002149594 | 0.076034952 | -5.15E-05 | 0.000351024 | 1 | 0.640599412 | 1 | 0.64481576 | ENSG0000014 | 12 | LIPT1 |
| SAT | Heart_Failure | ENSG0000026 | 0.002934696 | 0.351137684 | 0.003147513 | 0.076055357 | 0.002831126 | 0.368345231 | 0.003147171 | 0.076035214 | 0.00010357 | 1.90E-06 | 1 | 0.848363544 | 1 | 0.849290727 | ENSG0000014 | 12 | LIPT1 |
| SAT | Heart_Failure | ENSG0000026 | -0.00104902 | 0.227286924 | 0.000868844 | 0.076055266 | -0.0010254 | 0.237876498 | 0.000868755 | 0.076035123 | -2.36E-05 | 0.000474724 | 1 | 0.751217604 | 1 | 0.752162449 | ENSG0000014 | 7 | FAHD2B |
| SAT | Heart_Failure | ENSG0000026 | 0.003778109 | 0.086481857 | 0.002203932 | 0.076055034 | 0.003822315 | 0.082831211 | 0.002203707 | 0.076034871 | -4.42E-05 | 0.010362772 | 1 | 0.592308253 | 1 | 0.5844499 | ENSG0000014 | 5 | LYG1 |
| SAT | Heart_Failure | ENSG0000026 | 0.003606566 | 0.196219739 | 0.002790598 | 0.076055232 | 0.003554367 | 0.202727757 | 0.002790321 | 0.076035087 | 5.22E-05 | 0.02151707 | 1 | 0.726878652 | 1 | 0.729735526 | ENSG0000014 | 5 | LYG1 |
| SAT | Heart_Failure | ENSG0000026 | 0.001311639 | 0.572276253 | 0.002322708 | 0.076055443 | 0.001357789 | 0.558797251 | 0.002322479 | 0.076035287 | -4.61E-05 | 0.015839081 | 1 | 0.931368247 | 1 | 0.916248766 | ENSG0000014 | 5 | LYG1 |
| SAT | Heart_Failure | ENSG0000026 | 0.000128449 | 0.91177299 | 0.001159261 | 0.076055491 | 0.000293376 | 0.800557093 | 0.001161305 | 0.07603533 | -0.00016493 | 0.020927977 | 1 | 0.984491733 | 1 | 0.972791643 | ENSG0000014 | 5 | LYG1 |

| | | | | | | | | | | | | | | | | | | | | |
|---|---|---|---|---|---|---|---|---|---|---|---|---|---|---|---|---|---|---|---|---|
| SAT | Heart_Failure | ENSG0000026 | 0.00136642 | 0.588716795 | 0.002527146 | 0.076055447 | 0.001542896 | 0.541558831 | 0.002527441 | 0.076035282 | -0.00017648 | 0.001763099 | | 1 | 0.941136084 | | 1 | 0.91214298 | ENSG0000014 | 5 | LYG1 |
| SAT | Heart_Failure | ENSG0000026 | -0.00533224 | 0.044146358 | 0.00264929 | 0.076054861 | -0.00541315 | 0.041006731 | 0.002649003 | 0.076034689 | 8.09E-05 | 1.23E-05 | | 1 | 0.464244235 | | 1 | 0.453733817 | ENSG0000014 | 5 | LYG1 |
| SAT | Heart_Failure | ENSG0000026 | -0.00271878 | 0.18426754 | 0.002047689 | 0.076055218 | -0.00277823 | 0.174814646 | 0.002047489 | 0.076035053 | 5.94E-05 | 0.000488214 | | 1 | 0.720580417 | | 1 | 0.700325828 | ENSG0000014 | 14 | AFF3 |
| SAT | Heart_Failure | ENSG0000026 | 0.001247156 | 0.513409145 | 0.001908311 | 0.076055426 | 0.001294743 | 0.497423708 | 0.001908105 | 0.076035268 | -4.76E-05 | 0.000354731 | | 1 | 0.905155107 | | 1 | 0.894608312 | ENSG0000014 | 12 | GPR17 |
| SAT | Heart_Failure | ENSG0000026 | 0.00056926 | 0.786873048 | 0.002105458 | 0.076055482 | 0.000612161 | 0.771218419 | 0.002105227 | 0.076035327 | -4.29E-05 | 0.002683679 | | 1 | 0.974020771 | | 1 | 0.965963214 | ENSG0000014 | 12 | GPR17 |
| SAT | Heart_Failure | ENSG0000026 | -0.00093683 | 0.496813648 | 0.001378676 | 0.076055421 | -0.00091701 | 0.505914993 | 0.001378525 | 0.076035271 | -1.98E-05 | 0.033968826 | | 1 | 0.899087467 | | 1 | 0.900571724 | ENSG0000014 | 12 | GPR17 |
| SAT | Heart_Failure | ENSG0000026 | -0.00218926 | 0.40491411 | 0.002628546 | 0.076055385 | -0.00222674 | 0.396868519 | 0.002628267 | 0.076035228 | 3.75E-05 | 0.049443045 | | 1 | 0.870429337 | | 1 | 0.862334842 | ENSG0000014 | 12 | GPR17 |
| SAT | Heart_Failure | ENSG0000026 | 0.000113049 | 0.964780783 | 0.002560259 | 0.076055493 | 5.34E-05 | 0.983368814 | 0.002559983 | 0.07603534 | 5.97E-05 | 0.000902 | | 1 | 0.996432082 | | 1 | 0.999768899 | ENSG0000014 | 12 | GPR17 |
| SAT | Heart_Failure | ENSG0000026 | -0.00234883 | 0.033777649 | 0.001106517 | 0.07605479 | -0.00237438 | 0.03187018 | 0.001106399 | 0.076034622 | 2.55E-05 | 0.001273811 | | 1 | 0.411793489 | | 1 | 0.401948008 | ENSG0000014 | 7 | PHOSPHO2 |
| SAT | Heart_Failure | ENSG0000026 | 0.001795019 | 0.144724998 | 0.001230796 | 0.076055161 | 0.001817202 | 0.139782826 | 0.001230664 | 0.076035 | -2.22E-05 | 0.011894628 | | 1 | 0.669735853 | | 1 | 0.654247435 | ENSG0000014 | 5 | PTH2R |
| SAT | Heart_Failure | ENSG0000026 | -0.00617274 | 0.00646516 | 0.002266708 | 0.076054336 | -0.00609879 | 0.007126764 | 0.002266471 | 0.07603421 | -7.40E-05 | 1.03E-05 | | 1 | 0.179416801 | | 1 | 0.194857595 | ENSG0000014 | 5 | PTH2R |
| SAT | Heart_Failure | ENSG0000026 | 0.000483484 | 0.662490825 | 0.001107695 | 0.076055463 | 0.000527504 | 0.633884159 | 0.001107581 | 0.076035305 | -4.40E-05 | 1.71E-07 | | 1 | 0.962994646 | | 1 | 0.94798204 | ENSG0000014 | 14 | NBEAL1 |
| SAT | Heart_Failure | ENSG0000026 | -0.00212321 | 0.191580284 | 0.001625834 | 0.076055227 | -0.00214995 | 0.186001384 | 0.001625667 | 0.076035067 | 2.67E-05 | 0.033551617 | | 1 | 0.722912434 | | 1 | 0.715224819 | ENSG0000014 | 14 | NBEAL1 |
| SAT | Heart_Failure | ENSG0000026 | -0.00056137 | 0.804692864 | 0.00227019 | 0.076055483 | -0.00050832 | 0.822806662 | 0.002269933 | 0.076035332 | -5.30E-05 | 0.000186946 | | 1 | 0.976669362 | | 1 | 0.972791643 | ENSG0000014 | 5 | ACKR3 |
| SAT | Heart_Failure | ENSG0000026 | -0.00284257 | 0.453867078 | 0.003795235 | 0.076055405 | -0.00296804 | 0.434140418 | 0.003794827 | 0.076035245 | 0.000125466 | 3.29E-06 | | 1 | 0.887019168 | | 1 | 0.877927942 | ENSG0000014 | 5 | ACKR3 |
| SAT | Heart_Failure | ENSG0000026 | 0.003070984 | 0.191970063 | 0.00235365 | 0.076055227 | 0.003027624 | 0.198276503 | 0.002353419 | 0.076035082 | 4.34E-05 | 0.025553806 | | 1 | 0.722912434 | | 1 | 0.725655076 | ENSG0000014 | 11 | ANKMY1 |
| SAT | Heart_Failure | ENSG0000026 | 0.012192873 | 0.002002968 | 0.003946158 | 0.076054003 | 0.012108484 | 0.00214975 | 0.00394575 | 0.076033871 | 8.44E-05 | 0.005037603 | | 1 | 0.09524259 | | 1 | 0.111957631 | ENSG0000014 | 6 | TAMM41 |
| SAT | Heart_Failure | ENSG0000026 | -0.00421222 | 0.255777843 | 0.003706545 | 0.076055291 | -0.00411944 | 0.26635011 | 0.003706174 | 0.076035147 | -9.28E-05 | 0.001825662 | | 1 | 0.770621765 | | 1 | 0.774729353 | ENSG0000014 | 9 | RQCD1 |
| SAT | Heart_Failure | ENSG0000026 | 0.00180614 | 0.590728025 | 0.003358501 | 0.076055448 | 0.001873632 | 0.576890666 | 0.003358168 | 0.076035292 | -6.75E-05 | 0.014074178 | | 1 | 0.941376309 | | 1 | 0.927910133 | ENSG0000014 | 9 | RQCD1 |
| SAT | Heart_Failure | ENSG0000026 | 0.008077257 | 0.010353342 | 0.003150491 | 0.076054467 | 0.00799544 | 0.011146781 | 0.003150201 | 0.076034335 | 8.18E-05 | 0.003842215 | | 1 | 0.230796464 | | 1 | 0.243469768 | ENSG0000014 | 9 | RQCD1 |
| SAT | Heart_Failure | ENSG0000026 | 0.000100621 | 0.969653633 | 0.002644938 | 0.076055493 | 4.15E-05 | 0.987477463 | 0.002644657 | 0.07603534 | 5.91E-05 | 0.001941495 | | 1 | 0.996689715 | | 1 | 0.999768899 | ENSG0000014 | 10 | CTDSPL |
| SAT | Heart_Failure | ENSG0000026 | -0.0010282 | 0.64383596 | 0.002223891 | 0.07605546 | -0.00094175 | 0.671922842 | 0.002223668 | 0.076035312 | -8.65E-05 | 1.30E-06 | | 1 | 0.960732494 | | 1 | 0.956878334 | ENSG0000014 | 10 | CTDSPL |
| SAT | Heart_Failure | ENSG0000026 | 0.004085797 | 0.043086138 | 0.002019793 | 0.076054854 | 0.004154062 | 0.039698366 | 0.002019594 | 0.07603468 | -6.83E-05 | 4.39E-05 | | 1 | 0.463882417 | | 1 | 0.447595661 | ENSG0000014 | 10 | CTDSPL |
| SAT | Heart_Failure | ENSG0000026 | 0.003663808 | 0.330401116 | 0.003764287 | 0.076055345 | 0.003798799 | 0.312845061 | 0.003763902 | 0.076035181 | -0.00013499 | 4.34E-06 | | 1 | 0.837101206 | | 1 | 0.821155716 | ENSG0000014 | 5 | ZNF660 |
| SAT | Heart_Failure | ENSG0000026 | -0.00078834 | 0.631496275 | 0.001643672 | 0.076055457 | -0.00073832 | 0.653258009 | 0.001643494 | 0.076035309 | -5.00E-05 | 1.11E-05 | | 1 | 0.959428651 | | 1 | 0.954716939 | ENSG0000014 | 8 | TAGLN3 |
| SAT | Heart_Failure | ENSG0000026 | 0.001284697 | 0.62208765 | 0.002606435 | 0.076055455 | 0.001228677 | 0.637319434 | 0.002606165 | 0.076035305 | 5.60E-05 | 0.004730994 | | 1 | 0.954216727 | | 1 | 0.94798204 | ENSG0000014 | 8 | TAGLN3 |
| SAT | Heart_Failure | ENSG0000026 | 0.000867285 | 0.72464175 | 0.002462044 | 0.076055474 | 0.000818728 | 0.73945457 | 0.002461778 | 0.076035323 | 4.86E-05 | 0.004696492 | | 1 | 0.966658386 | | 1 | 0.960499534 | ENSG0000014 | 4 | PLA1A |
| SAT | Heart_Failure | ENSG0000026 | -0.00541483 | 0.071107874 | 0.003000252 | 0.076054985 | -0.00553908 | 0.064836906 | 0.002999957 | 0.076034808 | 0.000124247 | 6.20E-07 | | 1 | 0.545104986 | | 1 | 0.514983161 | ENSG0000014 | 7 | ADPRH |
| SAT | Heart_Failure | ENSG0000026 | -0.00041395 | 0.845822042 | 0.002128795 | 0.076055487 | -0.00047443 | 0.823623969 | 0.002128561 | 0.076035332 | 6.05E-05 | 2.33E-05 | | 1 | 0.976669362 | | 1 | 0.972791643 | ENSG0000014 | 7 | ADPRH |
| SAT | Heart_Failure | ENSG0000027 | -0.00317593 | 0.010640662 | 0.001243369 | 0.076054475 | -0.00319935 | 0.010070605 | 0.001243237 | 0.076034307 | 2.34E-05 | 0.009358768 | | 1 | 0.232334342 | | 1 | 0.230558242 | ENSG0000014 | 7 | ADPRH |
| SAT | Heart_Failure | ENSG0000027 | -0.00244123 | 0.527530556 | 0.003864042 | 0.076055431 | -0.00236328 | 0.540758646 | 0.003863668 | 0.076035282 | -7.80E-05 | 0.016865746 | | 1 | 0.906724424 | | 1 | 0.911336308 | ENSG0000014 | 8 | IGSF11 |
| SAT | Heart_Failure | ENSG0000027 | 0.0019837 | 0.498099779 | 0.002928044 | 0.076055421 | 0.002041687 | 0.48557433 | 0.002927713 | 0.076035264 | -5.80E-05 | 0.001557332 | | 1 | 0.899263643 | | 1 | 0.894608312 | ENSG0000014 | 8 | IGSF11 |
| SAT | Heart_Failure | ENSG0000027 | 0.001807825 | 0.142788718 | 0.0012336 | 0.076055158 | 0.00177061 | 0.151152762 | 0.001233468 | 0.076035019 | 3.72E-05 | 2.10E-05 | | 1 | 0.669181587 | | 1 | 0.665121824 | ENSG0000014 | 8 | IGSF11 |
| SAT | Heart_Failure | ENSG0000027 | -0.00023985 | 0.940369299 | 0.003206362 | 0.076055492 | -4.27E-05 | 0.989383146 | 0.003206121 | 0.07603534 | -0.00019719 | 9.30E-09 | | 1 | 0.992074492 | | 1 | 0.999768899 | ENSG0000014 | 10 | AMT |
| SAT | Heart_Failure | ENSG0000027 | 0.0061009 | 0.118857041 | 0.003911857 | 0.076055113 | 0.005997261 | 0.125214813 | 0.003911469 | 0.076034973 | 0.000103639 | 0.001135611 | | 1 | 0.646149939 | | 1 | 0.647874406 | ENSG0000014 | 10 | AMT |
| SAT | Heart_Failure | ENSG0000027 | 0.000678947 | 0.835415647 | 0.003267924 | 0.076055486 | 0.000732119 | 0.822714304 | 0.003267581 | 0.076035332 | -5.32E-05 | 0.028556977 | | 1 | 0.976669362 | | 1 | 0.972791643 | ENSG0000014 | 7 | NICN1 |
| SAT | Heart_Failure | ENSG0000027 | 0.001768638 | 0.478506653 | 0.002495584 | 0.076055415 | 0.001698646 | 0.496039753 | 0.002495313 | 0.076035268 | 7.00E-05 | 4.55E-05 | | 1 | 0.897131616 | | 1 | 0.894608312 | ENSG0000014 | 7 | NICN1 |
| SAT | Heart_Failure | ENSG0000027 | 0.003064279 | 0.116479387 | 0.00195211 | 0.076055108 | 0.003030361 | 0.120536648 | 0.001951888 | 0.076034964 | 3.39E-05 | 0.004818869 | | 1 | 0.642397685 | | 1 | 0.64481576 | ENSG0000014 | 8 | ILDR1 |
| SAT | Heart_Failure | ENSG0000027 | -0.00178957 | 0.253574571 | 0.001567447 | 0.07605529 | -0.00174911 | 0.26441753 | 0.001567285 | 0.076035146 | -4.05E-05 | 0.000659329 | | 1 | 0.768323136 | | 1 | 0.774729353 | ENSG0000014 | 8 | ILDR1 |
| SAT | Heart_Failure | ENSG0000027 | -0.00192066 | 0.536518573 | 0.003107428 | 0.076055433 | -0.00197046 | 0.525964877 | 0.003107109 | 0.076035277 | 4.98E-05 | 0.037683793 | | 1 | 0.911137391 | | 1 | 0.905399141 | ENSG0000014 | 17 | MUC4 |
| SAT | Heart_Failure | ENSG0000027 | 0.001756534 | 0.483648152 | 0.002507731 | 0.076055416 | 0.001721672 | 0.49232161 | 0.002507455 | 0.076035267 | 3.49E-05 | 0.038137262 | | 1 | 0.897349195 | | 1 | 0.894608312 | ENSG0000014 | 17 | MUC4 |
| SAT | Heart_Failure | ENSG0000027 | 0.00153247 | 0.143462064 | 0.001047469 | 0.076055159 | -0.00036548 | 0.739248592 | 0.001098031 | 0.076035323 | 0.001897948 | 8.67E-09 | | 1 | 0.669181587 | | 1 | 0.960499534 | ENSG0000014 | 17 | MUC4 |
| SAT | Heart_Failure | ENSG0000027 | -0.00398131 | 0.180638149 | 0.0029738 | 0.076055213 | -0.00391056 | 0.188462202 | 0.002973487 | 0.07603507 | -7.07E-05 | 0.001231891 | | 1 | 0.714462461 | | 1 | 0.717336988 | ENSG0000014 | 9 | SLC26A1 |
| SAT | Heart_Failure | ENSG0000027 | 0.001163121 | 0.761453848 | 0.003831446 | 0.076055479 | 0.001071649 | 0.779686635 | 0.003831046 | 0.076035328 | 9.15E-05 | 0.0014457 | | 1 | 0.969898847 | | 1 | 0.966957854 | ENSG0000014 | 9 | SLC26A1 |
| SAT | Heart_Failure | ENSG0000027 | 0.003480565 | 0.371351531 | 0.003893487 | 0.076055368 | 0.003594266 | 0.355876992 | 0.003893055 | 0.076035207 | -0.0001137 | 9.23E-06 | | 1 | 0.856926425 | | 1 | 0.844885969 | ENSG0000014 | 9 | SLC26A1 |
| SAT | Heart_Failure | ENSG0000027 | -0.00121044 | 0.434958657 | 0.001550388 | 0.076055398 | -0.00129213 | 0.40466903 | 0.001550597 | 0.076035232 | 8.17E-05 | 0.022654285 | | 1 | 0.880339889 | | 1 | 0.867451005 | ENSG0000014 | 6 | CENPC |
| SAT | Heart_Failure | ENSG0000027 | 0.00669432 | 0.01111805 | 0.002636622 | 0.076054487 | 0.006633634 | 0.01186242 | 0.002636349 | 0.076034352 | 6.07E-05 | 0.002415645 | | 1 | 0.236383009 | | 1 | 0.246278972 | ENSG0000014 | 6 | CENPC |
| SAT | Heart_Failure | ENSG0000027 | -0.00034941 | 0.904934326 | 0.002925628 | 0.076055491 | -0.00040348 | 0.890297563 | 0.002925326 | 0.076035337 | 5.41E-05 | 0.015680415 | | 1 | 0.981517198 | | 1 | 0.977805317 | ENSG0000014 | 2 | DDIT4L |
| SAT | Heart_Failure | ENSG0000027 | 0.005218725 | 0.039691827 | 0.002537122 | 0.076054833 | 0.006040518 | 0.018621154 | 0.002567115 | 0.076034476 | -0.00082179 | 0.03674464 | | 1 | 0.448303633 | | 1 | 0.304660546 | ENSG0000014 | 7 | USP53 |
| SAT | Heart_Failure | ENSG0000027 | -0.0017129 | 0.211360385 | 0.001370497 | 0.076055249 | -0.00255439 | 0.069227371 | 0.001405879 | 0.076034825 | 0.000841498 | 0.007406069 | | 1 | 0.740890409 | | 1 | 0.529994954 | ENSG0000014 | 7 | USP53 |
| SAT | Heart_Failure | ENSG0000027 | -0.00053294 | 0.386069511 | 0.000614858 | 0.076055376 | -0.00066188 | 0.282876975 | 0.000616344 | 0.07603516 | 0.00012894 | 0.003338373 | | 1 | 0.859117766 | | 1 | 0.795303647 | ENSG0000014 | 5 | FAM105A |
| SAT | Heart_Failure | ENSG0000027 | 0.003597382 | 0.235721529 | 0.003033845 | 0.076055274 | 0.003671959 | 0.226106808 | 0.003033549 | 0.076035112 | -7.46E-05 | 0.003258345 | | 1 | 0.755348658 | | 1 | 0.747837725 | ENSG0000014 | 5 | FAM105A |
| SAT | Heart_Failure | ENSG0000027 | 0.000218128 | 0.886733276 | 0.001531377 | 0.07605549 | 0.000243652 | 0.873571276 | 0.001531206 | 0.076035336 | -2.55E-05 | 0.010168368 | | 1 | 0.979270459 | | 1 | 0.974665278 | ENSG0000014 | 10 | BHMT |
| SAT | Heart_Failure | ENSG0000027 | -0.00202092 | 0.346518355 | 0.002146796 | 0.076055355 | -0.00196181 | 0.36075937 | 0.002146588 | 0.07603521 | -5.91E-05 | 0.00115111 | | 1 | 0.847011103 | | 1 | 0.848283315 | ENSG0000014 | 10 | RASA1 |
| SAT | Heart_Failure | ENSG0000027 | -0.00071097 | 0.673371411 | 0.001686654 | 0.076055465 | -0.00129347 | 0.443461927 | 0.001687814 | 0.076035248 | 0.000582507 | 1.53E-17 | | 1 | 0.966658386 | | 1 | 0.882352081 | ENSG0000014 | 10 | GTF2H2 |
| SAT | Heart_Failure | ENSG0000027 | -0.00340328 | 0.114161931 | 0.002154298 | 0.076055104 | -0.00337214 | 0.117472448 | 0.002154066 | 0.076034958 | -3.11E-05 | 0.041583023 | | 1 | 0.641850091 | | 1 | 0.64481576 | ENSG0000014 | 10 | TSLP |
| SAT | Heart_Failure | ENSG0000027 | 0.004113872 | 0.078682738 | 0.002339575 | 0.07605501 | 0.004174866 | 0.074321595 | 0.002339342 | 0.076034843 | -6.10E-05 | 0.001360556 | | 1 | 0.580653917 | | 1 | 0.55673323 | ENSG0000014 | 4 | COMMD10 |
| SAT | HTN | ENSG0000000 | -0.00042987 | 0.882755228 | 0.002914785 | 0.240335803 | -0.0004705 | 0.871760622 | 0.002914708 | 0.240312213 | 4.06E-05 | 0.037520217 | | 1 | 0.957468 | | 1 | 0.955645373 | ENSG0000014 | 4 | COMMD10 |
| SAT | HTN | ENSG0000000 | 0.017101783 | 6.11E-05 | 0.004266163 | 0.240327888 | 0.017019524 | 6.62E-05 | 0.004266076 | 0.240304376 | 8.23E-05 | 0.010881207 | | 0.200344429 | 0.001500008 | | 0.21727313 | 0.001621441 | ENSG0000014 | 4 | COMMD10 |
| SAT | HTN | ENSG0000000 | -0.00306316 | 0.585452442 | 0.005615967 | 0.240335667 | -0.00289707 | 0.605949012 | 0.005615953 | 0.240312094 | -0.00016609 | 0.002167501 | | 1 | 0.813238553 | | 1 | 0.83080598 | ENSG0000014 | 4 | COMMD10 |
| SAT | HTN | ENSG0000000 | -0.00368166 | 0.598414582 | 0.006990286 | 0.240335677 | -0.00381299 | 0.585424312 | 0.006990162 | 0.240312079 | 0.000131329 | 0.017792548 | | 1 | 0.822882751 | | 1 | 0.814247906 | ENSG0000014 | 13 | TBC1D7 |
| SAT | HTN | ENSG0000000 | -0.01034357 | 0.047148722 | 0.005210951 | 0.24033387 | -0.01365952 | 0.011266158 | 0.005389768 | 0.240309058 | 0.003315954 | 0.016098864 | | 1 | 0.205985296 | | 1 | 0.083456744 | ENSG0000014 | 6 | GFOD1 |
| SAT | HTN | ENSG0000001 | 0.014871827 | 5.87E-08 | 0.002742418 | 0.24032131 | 0.014929688 | 5.21E-08 | 0.002742348 | 0.24029761 | -5.79E-05 | 0.002200884 | | 0.000192501 | 4.70E-06 | | 0.000170892 | 4.07E-06 | ENSG0000014 | 6 | GFOD1 |
| SAT | HTN | ENSG0000001 | -0.00353103 | 0.480340558 | 0.005003193 | 0.240335568 | -0.00361401 | 0.470077196 | 0.005003096 | 0.240311968 | 8.30E-05 | 0.031742696 | | 1 | 0.746918185 | | 1 | 0.736442251 | ENSG0000014 | 6 | GFOD1 |
| SAT | HTN | ENSG0000002 | -0.00291527 | 0.012985012 | 0.001173531 | 0.24033277 | -0.0028965 | 0.013577924 | 0.001173505 | 0.240309221 | -1.88E-05 | 0.027464015 | | 1 | 0.091228745 | | 1 | 0.094584223 | ENSG0000014 | 4 | PCDHB18 |
| SAT | HTN | ENSG0000003 | 0.001813124 | 0.571457821 | 0.00320393 | 0.240335655 | 0.001138442 | 0.723714048 | 0.003220489 | 0.240312164 | 0.000674682 | 0.039509659 | | 1 | 0.805147909 | | 1 | 0.88865024 | ENSG0000014 | 6 | ZMAT2 |
| SAT | HTN | ENSG0000003 | 0.000193631 | 0.952436681 | 0.003246279 | 0.240335812 | 0.000251912 | 0.938145044 | 0.003246218 | 0.240312222 | -5.83E-05 | 0.020566696 | | 1 | 0.982687029 | | 1 | 0.978139775 | ENSG0000014 | 6 | ZMAT2 |
| SAT | HTN | ENSG0000003 | -0.00611153 | 0.092820431 | 0.003636285 | 0.24033442 | -0.00617934 | 0.089246071 | 0.00363621 | 0.240310801 | 6.78E-05 | 0.013513363 | | 1 | 0.311394513 | | 1 | 0.304120181 | ENSG0000014 | 6 | ZMAT2 |
| SAT | HTN | ENSG0000004 | 0.002766751 | 0.307283837 | 0.002710007 | 0.240335299 | 0.002716825 | 0.316083961 | 0.002709946 | 0.24031173 | 4.99E-05 | 0.01141056 | | 1 | 0.601695266 | | 1 | 0.604641264 | ENSG0000014 | 9 | DOK3 |
| SAT | HTN | ENSG0000004 | 0.004185375 | 0.372332265 | 0.004691525 | 0.240335421 | 0.004244203 | 0.365635274 | 0.004691385 | 0.240311822 | -5.88E-05 | 0.042933271 | | 1 | 0.659719927 | | 1 | 0.649864212 | ENSG0000014 | 3 | ABT1 |
| SAT | HTN | ENSG0000004 | -0.00058669 | 0.866494451 | 0.003489833 | 0.240335799 | -0.00052149 | 0.881209974 | 0.003489756 | 0.240312214 | -6.52E-05 | 0.010959546 | | 1 | 0.952737363 | | 1 | 0.957809595 | ENSG0000014 | 3 | ABT1 |
| SAT | HTN | ENSG0000005 | -0.0012382 | 0.826343669 | 0.00564375 | 0.24033579 | -0.00134653 | 0.81142182 | 0.005643661 | 0.240312197 | 0.000108332 | 0.018623609 | | 1 | 0.935230623 | | 1 | 0.926770342 | ENSG0000014 | 3 | ABT1 |
| SAT | HTN | ENSG0000005 | 0.00844675 | 0.221660243 | 0.006911516 | 0.240335077 | 0.008346648 | 0.227172488 | 0.00691133 | 0.240311506 | 0.000100102 | 0.02944585 | | 1 | 0.515379933 | | 1 | 0.517461433 | ENSG0000014 | 3 | ABT1 |
| SAT | HTN | ENSG0000006 | 0.005837984 | 0.367915923 | 0.006483876 | 0.240335414 | 0.005714833 | 0.378102039 | 0.006483816 | 0.240311842 | 0.000123151 | 0.033405439 | | 1 | 0.655337755 | | 1 | 0.66057124 | ENSG0000014 | 9 | ANO7 |
| SAT | HTN | ENSG0000006 | 0.010235483 | 7.98E-07 | 0.002073684 | 0.240323798 | 0.010163766 | 9.52E-07 | 0.002073662 | 0.240300379 | 7.17E-05 | 7.21E-05 | | 0.002618615 | 3.91E-05 | | 0.003123699 | 4.53E-05 | ENSG0000014 | 9 | ANO7 |
| SAT | HTN | ENSG0000006 | 0.007144086 | 0.045684113 | 0.003575072 | 0.240333844 | 0.00893268 | 0.014446493 | 0.003651993 | 0.240309275 | -0.00178859 | 0.016572303 | | 1 | 0.203102407 | | 1 | 0.099073573 | ENSG0000014 | 9 | ANO7 |
| SAT | HTN | ENSG0000006 | -0.00695833 | 0.000109572 | 0.001798767 | 0.240328433 | -0.00699796 | 0.000100095 | 0.001798778 | 0.240304762 | 3.96E-05 | 0.036470282 | | 0.359505355 | 0.002462365 | | 0.328412008 | 0.002296587 | ENSG0000014 | 9 | ANO7 |
| SAT | HTN | ENSG0000007 | 0.004831697 | 0.437295324 | 0.006220248 | 0.240335516 | 0.004964574 | 0.424791564 | 0.006220205 | 0.240311911 | -0.00013288 | 0.020141242 | | 1 | 0.710984122 | | 1 | 0.697917436 | ENSG0000014 | 9 | IRAK1BP1 |
| SAT | HTN | ENSG0000007 | 6.13E-05 | 0.988614397 | 0.004296836 | 0.240335813 | -8.80E-08 | 0.999983652 | 0.004296716 | 0.240312225 | 6.14E-05 | 0.028062472 | | 1 | 0.99730223 | | 1 | 0.999983652 | ENSG0000014 | 9 | IRAK1BP1 |
| SAT | HTN | ENSG0000007 | 0.016237927 | 0.008585138 | 0.006178444 | 0.240332407 | 0.016352953 | 0.008125345 | 0.006178323 | 0.240308771 | -0.00011503 | 0.015296663 | | 1 | 0.069550213 | | 1 | 0.067346705 | ENSG0000014 | 7 | MMS22L |
| SAT | HTN | ENSG0000007 | -0.01378377 | 0.016762904 | 0.005762747 | 0.240332992 | -0.01370215 | 0.017417704 | 0.005762585 | 0.240309437 | -8.16E-05 | 0.028562707 | | 1 | 0.109125177 | | 1 | 0.111398611 | ENSG0000014 | 7 | MMS22L |
| SAT | HTN | ENSG0000007 | 0.019655101 | 0.003524734 | 0.006736166 | 0.240331614 | 0.019531652 | 0.003737122 | 0.006736091 | 0.240308079 | 0.000123449 | 0.035221289 | | 1 | 0.035150923 | | 1 | 0.037382609 | ENSG0000014 | 7 | MMS22L |
| SAT | HTN | ENSG0000007 | -0.00325443 | 0.269592317 | 0.002947841 | 0.240335212 | -0.00329704 | 0.263358811 | 0.002947765 | 0.240311608 | 4.26E-05 | 0.034100105 | | 1 | 0.563396428 | | 1 | 0.553542766 | ENSG0000014 | 7 | MMS22L |
| SAT | HTN | ENSG0000007 | 0.002980051 | 0.651478731 | 0.006597278 | 0.240335713 | 0.002869697 | 0.663568982 | 0.006597138 | 0.240312132 | 0.000110354 | 0.024593433 | | 1 | 0.847542314 | | 1 | 0.859631339 | ENSG0000014 | 7 | ABRACL |
| SAT | HTN | ENSG0000008 | -0.00396921 | 0.453614738 | 0.005296495 | 0.240335536 | -0.00387109 | 0.464844907 | 0.005296384 | 0.240311962 | -9.81E-05 | 0.013321908 | | 1 | 0.726359177 | | 1 | 0.734100068 | ENSG0000014 | 7 | ABRACL |
| SAT | HTN | ENSG0000008 | 0.000354391 | 0.864206757 | 0.002072197 | 0.240335799 | 0.000320873 | 0.876939204 | 0.002072149 | 0.240312214 | 3.35E-05 | 0.023953185 | | 1 | 0.952456288 | | 1 | 0.956781838 | ENSG0000014 | 7 | ABRACL |
| SAT | HTN | ENSG0000008 | -0.01102256 | 0.091733938 | 0.00653644 | 0.240334411 | -0.01155393 | 0.077362923 | 0.00654169 | 0.240310687 | 0.000531372 | 0.049012885 | | 1 | 0.308696461 | | 1 | 0.282344552 | ENSG0000014 | 6 | SLC18B1 |
| SAT | HTN | ENSG0000008 | -0.01054116 | 0.017016241 | 0.004417244 | 0.240333005 | -0.01012442 | 0.021979213 | 0.004419727 | 0.240309638 | -0.00041674 | 0.006949305 | | 1 | 0.109471152 | | 1 | 0.129468218 | ENSG0000014 | 6 | SLC18B1 |
| SAT | HTN | ENSG0000008 | 0.008589795 | 0.103341983 | 0.005273481 | 0.240334505 | 0.008497633 | 0.107087047 | 0.005273355 | 0.240310945 | 9.22E-05 | 0.013543861 | | 1 | 0.332597644 | | 1 | 0.34237775 | ENSG0000014 | 6 | DYNLT1 |
| SAT | HTN | ENSG0000008 | 0.015799944 | 0.00369282 | 0.005442084 | 0.240331656 | 0.015609337 | 0.00412717 | 0.005442039 | 0.240308168 | 0.000190607 | 0.000101144 | | 1 | 0.036384812 | | 1 | 0.039725318 | ENSG0000014 | 6 | DYNLT1 |
| SAT | HTN | ENSG0000008 | -0.00646656 | 0.267610529 | 0.005833174 | 0.240335207 | -0.00662443 | 0.256100507 | 0.005833122 | 0.240311589 | 0.000157869 | 0.002541464 | | 1 | 0.561046104 | | 1 | 0.548573946 | ENSG0000014 | 6 | DYNLT1 |
| SAT | HTN | ENSG0000008 | -0.01116111 | 2.21E-05 | 0.002630919 | 0.240326938 | -0.01112487 | 2.35E-05 | 0.002630854 | 0.240303407 | -3.62E-05 | 0.047550759 | | 0.072607824 | 0.00066164 | | 0.077161004 | 0.000682841 | ENSG0000014 | 6 | DYNLT1 |
| SAT | HTN | ENSG0000008 | 0.001243983 | 0.735912845 | 0.003688378 | 0.240335757 | 0.001340835 | 0.716205772 | 0.003688328 | 0.24031216 | -9.69E-05 | 0.001839383 | | 1 | 0.893569851 | | 1 | 0.88553283 | ENSG0000014 | 6 | DYNLT1 |
| SAT | HTN | ENSG0000008 | -0.00126992 | 0.637037008 | 0.002691393 | 0.240335704 | -0.00122737 | 0.648358289 | 0.002691332 | 0.240312123 | -4.26E-05 | 0.029250455 | | 1 | 0.840834873 | | 1 | 0.849876055 | ENSG0000014 | 8 | PNLDC1 |
| SAT | HTN | ENSG0000009 | 0.005184095 | 9.29E-05 | 0.001326367 | 0.240328279 | 0.005156261 | 0.000101286 | 0.001326362 | 0.240304773 | 2.78E-05 | 0.027850052 | | 0.304759692 | 0.002131187 | | 0.332320906 | 0.002307784 | ENSG0000014 | 8 | PNLDC1 |
| SAT | HTN | ENSG0000009 | 0.01937856 | 0.000198023 | 0.005207116 | 0.240328983 | 0.019496423 | 0.000180944 | 0.00520701 | 0.240305312 | -0.00011786 | 0.002874605 | | 0.649715088 | 0.00389051 | | 0.593676249 | 0.003619977 | ENSG0000014 | 8 | PNLDC1 |
| SAT | HTN | ENSG0000009 | -0.00470589 | 0.162947364 | 0.003372844 | 0.240334853 | -0.00477015 | 0.157269899 | 0.003372763 | 0.240311239 | 6.43E-05 | 0.006917792 | | 1 | 0.433572431 | | 1 | 0.427154418 | ENSG0000014 | 10 | WTAP |
| SAT | HTN | ENSG0000009 | 0.007839287 | 0.014465619 | 0.003205601 | 0.240332864 | 0.00776559 | 0.015412382 | 0.003205544 | 0.240309331 | 7.37E-05 | 0.003655263 | | 1 | 0.098878534 | | 1 | 0.102989871 | ENSG0000014 | 10 | WTAP |
| SAT | HTN | ENSG0000009 | 0.015358875 | 0.008718704 | 0.00585568 | 0.24033242 | 0.01525799 | 0.009168717 | 0.005855601 | 0.240308877 | 0.000100885 | 0.040767126 | | 1 | 0.070125913 | | 1 | 0.073193579 | ENSG0000014 | 8 | VIP |
| SAT | HTN | ENSG0000009 | -0.00222392 | 0.228932246 | 0.001848465 | 0.240335099 | -0.00225706 | 0.22205892 | 0.001848424 | 0.24031149 | 3.31E-05 | 0.013943144 | | 1 | 0.520170844 | | 1 | 0.513081208 | ENSG0000014 | 9 | GNA12 |
| SAT | HTN | ENSG0000009 | -0.0017012 | 0.789428112 | 0.006370265 | 0.240335778 | -0.0018175 | 0.77540169 | 0.006370121 | 0.240312185 | 0.0001163 | 0.012301635 | | 1 | 0.918290338 | | 1 | 0.911861271 | ENSG0000014 | 9 | GNA12 |
| SAT | HTN | ENSG0000010 | -0.00542303 | 0.113419325 | 0.003425766 | 0.240334577 | -0.00536103 | 0.117595269 | 0.003425686 | 0.240311018 | -6.20E-05 | 0.011645649 | | 1 | 0.355763675 | | 1 | 0.365369392 | ENSG0000014 | 9 | GNA12 |
| SAT | HTN | ENSG0000010 | -0.00144753 | 0.692731768 | 0.003663226 | 0.240335736 | -0.00137723 | 0.706942028 | 0.003663183 | 0.240312156 | -7.03E-05 | 0.02640529 | | 1 | 0.869846415 | | 1 | 0.880236222 | ENSG0000014 | 14 | SDK1 |
| SAT | HTN | ENSG0000010 | -0.00851534 | 0.004765005 | 0.003016932 | 0.240331884 | -0.0085603 | 0.004547328 | 0.003016869 | 0.240308255 | 4.50E-05 | 0.048606944 | | 1 | 0.043792664 | | 1 | 0.042219581 | ENSG0000014 | 14 | SDK1 |
| SAT | HTN | ENSG0000010 | -0.00011622 | 0.980071619 | 0.004652511 | 0.240335813 | -0.00020474 | 0.96489853 | 0.004652392 | 0.240312224 | 8.85E-05 | 0.005439012 | | 1 | 0.99356442 | | 1 | 0.988156741 | ENSG0000014 | 14 | SDK1 |
| SAT | HTN | ENSG0000010 | 0.004880635 | 0.430320343 | 0.00618864 | 0.240335507 | 0.004965177 | 0.42236486 | 0.006188479 | 0.240311908 | -8.45E-05 | 0.044297364 | | 1 | 0.705948972 | | 1 | 0.696721522 | ENSG0000014 | 10 | WASH2P |
| SAT | HTN | ENSG0000010 | 0.004255445 | 0.393332418 | 0.004985348 | 0.240335454 | 0.004180015 | 0.401763545 | 0.00498525 | 0.240311879 | 7.54E-05 | 0.04798446 | | 1 | 0.676956485 | | 1 | 0.681743181 | ENSG0000014 | 10 | WASH2P |
| SAT | HTN | ENSG0000010 | 0.01639055 | 0.003220515 | 0.005563887 | 0.240331533 | 0.021845417 | 0.000180824 | 0.005834109 | 0.240305311 | -0.00545487 | 0.001893153 | | 1 | 0.032917472 | | 0.593283879 | 0.003619977 | ENSG0000014 | 10 | WASH2P |
| SAT | HTN | ENSG0000010 | 0.008590265 | 0.128500536 | 0.005651326 | 0.240334674 | 0.008693481 | 0.123968106 | 0.005651239 | 0.240311058 | -0.00010322 | 0.025895021 | | 1 | 0.382239581 | | 1 | 0.374529795 | ENSG0000014 | 10 | WASH2P |
| SAT | HTN | ENSG0000010 | 0.006863974 | 0.124633079 | 0.004469848 | 0.24033465 | 0.006779161 | 0.129353793 | 0.004469803 | 0.240311091 | 8.48E-05 | 0.031562667 | | 1 | 0.374127295 | | 1 | 0.384081261 | ENSG0000014 | 10 | WASH2P |
| SAT | HTN | ENSG0000010 | -0.00357769 | 0.157611782 | 0.002531708 | 0.240334828 | -0.00337198 | 0.183001775 | 0.002532335 | 0.240311351 | -0.0002057 | 0.000850277 | | 1 | 0.427022509 | | 1 | 0.463293845 | ENSG0000014 | 10 | WASH2P |
| SAT | HTN | ENSG0000010 | 0.001422427 | 0.74329366 | 0.004343354 | 0.24033576 | 0.001561518 | 0.719203923 | 0.004343305 | 0.240312162 | -0.00013909 | 0.00022607 | | 1 | 0.896532567 | | 1 | 0.886108926 | ENSG0000014 | 11 | CCZ1B |
| SAT | HTN | ENSG0000010 | -0.00025138 | 0.951541513 | 0.004136556 | 0.240335812 | -0.00019241 | 0.962899086 | 0.004136444 | 0.240312224 | -5.90E-05 | 0.032212432 | | 1 | 0.982072256 | | 1 | 0.988156741 | ENSG0000014 | 11 | CCZ1B |
| SAT | HTN | ENSG0000010 | 0.001867621 | 0.703999397 | 0.00491573 | 0.240335742 | 0.001962538 | 0.689713184 | 0.004915629 | 0.240312147 | -9.49E-05 | 0.010647664 | | 1 | 0.876281552 | | 1 | 0.870030357 | ENSG0000014 | 10 | POMZP3 |
| SAT | HTN | ENSG0000010 | -0.00167542 | 0.789094132 | 0.006263543 | 0.240335778 | -0.00157715 | 0.801192815 | 0.006263394 | 0.240312194 | -9.83E-05 | 0.027453626 | | 1 | 0.918290338 | | 1 | 0.923833778 | ENSG0000014 | 10 | POMZP3 |
| SAT | HTN | ENSG0000010 | -0.01252661 | 0.050953761 | 0.006417738 | 0.240333934 | -0.01241143 | 0.05311781 | 0.006417606 | 0.240310381 | -0.00011518 | 0.017279185 | | 1 | 0.219395395 | | 1 | 0.227405237 | ENSG0000014 | 9 | PSPH |
| SAT | HTN | ENSG0000010 | -0.00639437 | 0.069284981 | 0.003520041 | 0.240334186 | -0.00645936 | 0.06649913 | 0.003519994 | 0.240310565 | 6.50E-05 | 0.029351589 | | 1 | 0.266812233 | | 1 | 0.262480731 | ENSG0000014 | 9 | PSPH |
| SAT | HTN | ENSG0000010 | -0.0054656 | 0.332065346 | 0.005634858 | 0.240335349 | -0.00536108 | 0.341385078 | 0.005634743 | 0.240311779 | -0.00010452 | 0.014091454 | | 1 | 0.62016399 | | 1 | 0.627146943 | ENSG0000014 | 9 | PSPH |
| SAT | HTN | ENSG0000010 | 0.007501647 | 0.11633597 | 0.004777076 | 0.240334597 | 0.007571142 | 0.11298269 | 0.004776954 | 0.240310987 | -6.95E-05 | 0.034546914 | | 1 | 0.362335772 | | 1 | 0.355413428 | ENSG0000014 | 9 | PSPH |
| SAT | HTN | ENSG0000010 | -0.00410905 | 0.54887457 | 0.006854728 | 0.240335636 | -0.00394255 | 0.565180019 | 0.00685465 | 0.240312062 | -0.0001665 | 0.005110295 | | 1 | 0.791241216 | | 1 | 0.803251455 | ENSG0000014 | 9 | PSPH |
| SAT | HTN | ENSG0000010 | 0.002564077 | 0.540281101 | 0.004187011 | 0.240335628 | 0.002627646 | 0.530275394 | 0.00418691 | 0.240312031 | -6.36E-05 | 0.031217871 | | 1 | 0.786663493 | | 1 | 0.779696379 | ENSG0000014 | 9 | GIGYF1 |
| SAT | HTN | ENSG0000010 | 0.002821638 | 0.485722739 | 0.004047516 | 0.240335574 | 0.002923603 | 0.470093142 | 0.004047472 | 0.240311968 | -0.00010196 | 0.004040779 | | 1 | 0.74889864 | | 1 | 0.736442251 | ENSG0000014 | 11 | TRIM4 |
| SAT | HTN | ENSG0000010 | -0.00655508 | 0.036317939 | 0.003131375 | 0.240333652 | -0.0062683 | 0.045456804 | 0.003133513 | 0.240310252 | -0.00028678 | 0.016645311 | | 1 | 0.176751156 | | 1 | 0.203193151 | ENSG0000014 | 11 | TRIM4 |
| SAT | HTN | ENSG0000010 | -0.00669032 | 0.276525806 | 0.006148302 | 0.240335229 | -0.00656967 | 0.285275333 | 0.006148237 | 0.240311662 | -0.00012066 | 0.025279438 | | 1 | 0.571695759 | | 1 | 0.577147307 | ENSG0000014 | 11 | TRIM4 |
| SAT | HTN | ENSG0000010 | 0.001868131 | 0.467039959 | 0.002568569 | 0.240335552 | 0.001915914 | 0.455712934 | 0.002568507 | 0.240311951 | -4.78E-05 | 0.008525323 | | 1 | 0.738333035 | | 1 | 0.727942617 | ENSG0000014 | 11 | TRIM4 |
| SAT | HTN | ENSG0000010 | -0.00055297 | 0.927425188 | 0.006070947 | 0.240335809 | -0.0006586 | 0.913609187 | 0.00607079 | 0.24031222 | 0.000105632 | 0.010701658 | | 1 | 0.975926571 | | 1 | 0.972372648 | ENSG0000014 | 15 | NCAPG2 |
| SAT | HTN | ENSG0000010 | 0.004273889 | 0.348710571 | 0.004560789 | 0.24033538 | 0.005231865 | 0.252507408 | 0.004572183 | 0.24031158 | -0.00095798 | 0.003275046 | | 1 | 0.636328912 | | 1 | 0.544334301 | ENSG0000014 | 12 | CCDC25 |

| | | | | | | | | | | | | | | | | | | | |
|---|---|---|---|---|---|---|---|---|---|---|---|---|---|---|---|---|---|---|---|
| SAT | HTN | ENSG000001( | -0.01064562 | 0.133704085 | 0.007098703 | 0.240334704 | -0.01051644 | 0.138478888 | 0.007098594 | 0.240311143 | -0.00012918 | 0.02651812 | 1 | 0.386164703 | 1 | 0.393375959 | ENSG0000014 | 9 | CHMP7 |
| SAT | HTN | ENSG000001( | 0.000354225 | 0.938421127 | 0.004585174 | 0.24033581 | 0.000459369 | 0.920195839 | 0.004585108 | 0.24031222 | -0.00010514 | 0.005830213 | 1 | 0.978294984 | 1 | 0.974491801 | ENSG0000014 | 9 | CHMP7 |
| SAT | HTN | ENSG000001( | -0.00760929 | 0.146649617 | 0.00524247 | 0.240334774 | -0.00751116 | 0.15192388 | 0.005242397 | 0.240311213 | -9.81E-05 | 0.025495028 | 1 | 0.407415235 | 1 | 0.417123222 | ENSG0000014 | 9 | CHMP7 |
| SAT | HTN | ENSG000001( | -0.00734031 | 8.21E-05 | 0.001863879 | 0.240328164 | -0.00731066 | 8.77E-05 | 0.001863834 | 0.240304638 | -2.96E-05 | 0.023637343 | 0.26940511 | 0.00191236 | 0.287714717 | 0.002055105 | ENSG0000014 | 9 | RGS20 |
| SAT | HTN | ENSG000001( | -0.00975206 | 0.09778707 | 0.005890059 | 0.240334461 | -0.0096586 | 0.101038519 | 0.005889956 | 0.240310899 | -9.35E-05 | 0.045534435 | 1 | 0.321804792 | 1 | 0.329208389 | ENSG0000014 | 9 | RGS20 |
| SAT | HTN | ENSG000001( | 0.009830934 | 0.067195249 | 0.005371009 | 0.240334161 | 0.009681463 | 0.071460006 | 0.00537099 | 0.240310623 | 0.000149471 | 0.003514154 | 1 | 0.262909169 | 1 | 0.272311592 | ENSG0000014 | 11 | ADHFE1 |
| SAT | HTN | ENSG000001( | 0.001503255 | 0.608329579 | 0.002933411 | 0.240335684 | 0.001400731 | 0.632997785 | 0.002933394 | 0.240312113 | 0.000102524 | 0.000169867 | 1 | 0.829908253 | 1 | 0.840496048 | ENSG0000014 | 11 | ADHFE1 |
| SAT | HTN | ENSG000001( | -0.00025147 | 0.909681166 | 0.002216773 | 0.240335807 | -0.00021428 | 0.92299254 | 0.002216719 | 0.240312221 | -3.72E-05 | 0.017469118 | 1 | 0.968235044 | 1 | 0.974491801 | ENSG0000014 | 11 | ADHFE1 |
| SAT | HTN | ENSG000001( | -0.00776503 | 0.009776174 | 0.003005442 | 0.24033221 | -0.0053918 | 0.079807647 | 0.00307785 | 0.240310712 | -0.00237323 | 0.000353559 | 1 | 0.0752949 | 1 | 0.285677612 | ENSG0000014 | 11 | ADHFE1 |
| SAT | HTN | ENSG0000011 | 0.003125422 | 0.122475723 | 0.002023624 | 0.240334637 | 0.003159698 | 0.118420145 | 0.002023577 | 0.240311023 | -3.43E-05 | 0.018587332 | 1 | 0.372076709 | 1 | 0.366889987 | ENSG0000014 | 11 | ADHFE1 |
| SAT | HTN | ENSG0000011 | 0.002643322 | 0.496185548 | 0.003884358 | 0.240335585 | 0.002553533 | 0.510922753 | 0.003884279 | 0.240312012 | 8.98E-05 | 0.00237583 | 1 | 0.757316081 | 1 | 0.763012086 | ENSG0000014 | 11 | ADHFE1 |
| SAT | HTN | ENSG0000011 | -0.00752402 | 0.23505601 | 0.006336353 | 0.240335118 | -0.00794992 | 0.209635708 | 0.006336775 | 0.240311449 | 0.000425906 | 1.01E-05 | 1 | 0.526060442 | 1 | 0.499064033 | ENSG0000014 | 11 | ADHFE1 |
| SAT | HTN | ENSG0000011 | -0.0032002 | 0.603180075 | 0.006156209 | 0.24033568 | -0.00310661 | 0.613811916 | 0.006156092 | 0.2403121 | -9.36E-05 | 0.049760203 | 1 | 0.826982828 | 1 | 0.834690078 | ENSG0000014 | 11 | ADHFE1 |
| SAT | HTN | ENSG0000011 | 0.000409416 | 0.947782133 | 0.006251368 | 0.240335811 | 0.000302902 | 0.961353965 | 0.00625125 | 0.240312224 | 0.000106514 | 0.028654173 | 1 | 0.981389706 | 1 | 0.98809363 | ENSG0000014 | 11 | ADHFE1 |
| SAT | HTN | ENSG0000011 | -0.00330635 | 0.037269651 | 0.001587451 | 0.240333674 | -0.00305183 | 0.054907307 | 0.001589817 | 0.240310408 | -0.00025452 | 0.003859375 | 1 | 0.178482982 | 1 | 0.23245274 | ENSG0000014 | 5 | SLC39A4 |
| SAT | HTN | ENSG0000011 | 0.007157672 | 0.264317617 | 0.006412274 | 0.240335199 | 0.007043087 | 0.272031406 | 0.006412147 | 0.24031163 | 0.000114585 | 0.019268231 | 1 | 0.556219915 | 1 | 0.564538294 | ENSG0000014 | 11 | NAPRT1 |
| SAT | HTN | ENSG0000011 | 0.002088999 | 0.513752202 | 0.003199046 | 0.240335603 | 0.002174818 | 0.496627686 | 0.003199174 | 0.240311998 | -8.58E-05 | 0.044732497 | 1 | 0.765147969 | 1 | 0.755559631 | ENSG0000014 | 11 | NAPRT1 |
| SAT | HTN | ENSG0000011 | 0.001644154 | 0.602493845 | 0.00315687 | 0.24033568 | 0.001692165 | 0.591930673 | 0.003156784 | 0.240312084 | -4.80E-05 | 0.021858179 | 1 | 0.826759642 | 1 | 0.81946183 | ENSG0000014 | 12 | AK3 |
| SAT | HTN | ENSG0000011 | -0.00207179 | 0.764279649 | 0.006909044 | 0.240335769 | -0.00265527 | 0.700917515 | 0.006913291 | 0.240312153 | 0.000583487 | 0.020472279 | 1 | 0.905600735 | 1 | 0.87708252 | ENSG0000014 | 12 | AK3 |
| SAT | HTN | ENSG0000011 | -0.00064314 | 0.835403678 | 0.003095348 | 0.240335792 | -0.00069731 | 0.821760868 | 0.003095311 | 0.2403122 | 5.42E-05 | 0.042721007 | 1 | 0.940381331 | 1 | 0.934090633 | ENSG0000014 | 10 | C9orf72 |
| SAT | HTN | ENSG0000011 | 0.001796483 | 0.554290433 | 0.003037967 | 0.240335641 | 0.001874375 | 0.537237461 | 0.003037903 | 0.240312038 | -7.79E-05 | 0.000608516 | 1 | 0.795829275 | 1 | 0.782997282 | ENSG0000014 | 10 | C9orf72 |
| SAT | HTN | ENSG0000011 | 0.005079039 | 0.035208683 | 0.002419551 | 0.240333626 | 0.005557821 | 0.021623074 | 0.002419672 | 0.240309624 | -0.00047878 | 0.015143752 | 1 | 0.174237842 | 1 | 0.128291693 | ENSG0000014 | 8 | FBXO10 |
| SAT | HTN | ENSG0000011 | 0.004690239 | 0.423843411 | 0.005864471 | 0.240335498 | 0.002481564 | 0.677173876 | 0.005960672 | 0.24031214 | 0.002208674 | 0.038661337 | 1 | 0.700922495 | 1 | 0.865678672 | ENSG0000014 | 8 | FBXO10 |
| SAT | HTN | ENSG0000011 | 0.001476657 | 0.823064333 | 0.00660385 | 0.240335789 | 0.001588764 | 0.80987514 | 0.006603692 | 0.240312197 | -0.00011211 | 0.016915922 | 1 | 0.933773886 | 1 | 0.925531291 | ENSG0000014 | 12 | SIGMAR1 |
| SAT | HTN | ENSG0000011 | 0.011060152 | 0.074718493 | 0.006205925 | 0.240334247 | 0.01117678 | 0.071700489 | 0.006205809 | 0.240310626 | -0.00011663 | 0.015932305 | 1 | 0.276027247 | 1 | 0.27259479 | ENSG0000014 | 12 | SIGMAR1 |
| SAT | HTN | ENSG0000011 | 0.003747708 | 0.003345313 | 0.001277293 | 0.240331568 | 0.00372186 | 0.003569222 | 0.001277263 | 0.240308038 | 2.58E-05 | 0.004342903 | 1 | 0.033793366 | 1 | 0.036032666 | ENSG0000014 | 12 | SIGMAR1 |
| SAT | HTN | ENSG0000011 | 0.000521401 | 0.888360526 | 0.003714232 | 0.240335804 | 0.000365524 | 0.92160918 | 0.003714409 | 0.240312221 | 0.000155877 | 0.002561085 | 1 | 0.958471189 | 1 | 0.974491801 | ENSG0000014 | 12 | SIGMAR1 |
| SAT | HTN | ENSG0000011 | 0.00379255 | 0.546731258 | 0.006292955 | 0.240335634 | 0.003888314 | 0.536644505 | 0.006292834 | 0.240312037 | -9.58E-05 | 0.048885031 | 1 | 0.789827969 | 1 | 0.782991949 | ENSG0000014 | 12 | SIGMAR1 |
| SAT | HTN | ENSG0000011 | 0.002342778 | 0.597672313 | 0.004439172 | 0.240335676 | 0.002438643 | 0.582759032 | 0.004439066 | 0.240312077 | -9.59E-05 | 0.00239686 | 1 | 0.822882751 | 1 | 0.812928219 | ENSG0000014 | 11 | HIATL1 |
| SAT | HTN | ENSG0000011 | 0.00095575 | 0.63061031 | 0.001987551 | 0.240335699 | 3.38E-05 | 0.986647087 | 0.002016936 | 0.240312225 | 0.000921995 | 0.007287743 | 1 | 0.837963781 | 1 | 0.994833771 | ENSG0000014 | 11 | HIATL1 |
| SAT | HTN | ENSG0000011 | 0.00112311 | 0.69124501 | 0.002827808 | 0.240335736 | 0.001048644 | 0.710768242 | 0.002827869 | 0.240312158 | 7.45E-05 | 0.026622258 | 1 | 0.869622269 | 1 | 0.882008549 | ENSG0000014 | 11 | HIATL1 |
| SAT | HTN | ENSG0000011 | -0.01807888 | 0.000366391 | 0.005073771 | 0.240329552 | -0.0181981 | 0.000334844 | 0.005073687 | 0.240305881 | 0.000119221 | 0.00366228 | 1 | 0.006327001 | 1 | 0.005782235 | ENSG0000014 | 11 | ALAD |
| SAT | HTN | ENSG0000011 | 0.001844147 | 0.362543947 | 0.002025357 | 0.240335404 | 0.001882845 | 0.352554038 | 0.002025332 | 0.240311799 | -3.87E-05 | 0.025106386 | 1 | 0.650299517 | 1 | 0.636902944 | ENSG0000014 | 11 | WDR31 |
| SAT | HTN | ENSG0000011 | 0.004767022 | 0.467135467 | 0.006555777 | 0.240335553 | 0.004570517 | 0.485688991 | 0.006555698 | 0.240311986 | 0.000196505 | 0.000491664 | 1 | 0.738333035 | 1 | 0.747907685 | ENSG0000014 | 11 | WDR31 |
| SAT | HTN | ENSG0000011 | -0.00356461 | 0.362426417 | 0.003913923 | 0.240335404 | -0.00365401 | 0.350503087 | 0.003913831 | 0.240311796 | 8.94E-05 | 0.001403435 | 1 | 0.650299517 | 1 | 0.635605707 | ENSG0000014 | 11 | WDR31 |
| SAT | HTN | ENSG0000011 | 0.00465855 | 0.103398231 | 0.002860462 | 0.240334505 | 0.00461348 | 0.106770257 | 0.002860391 | 0.240310943 | 4.51E-05 | 0.023338029 | 1 | 0.332597644 | 1 | 0.34176899 | ENSG0000014 | 11 | WDR31 |
| SAT | HTN | ENSG0000011 | -0.00975456 | 0.10438276 | 0.006006614 | 0.240334513 | -0.00961711 | 0.109351548 | 0.006006497 | 0.240310961 | -0.00013746 | 0.002861268 | 1 | 0.334126667 | 1 | 0.347994595 | ENSG0000014 | 8 | POLE3 |
| SAT | HTN | ENSG0000011 | -0.00563276 | 0.14350401 | 0.003850493 | 0.240334758 | -0.00554094 | 0.150152774 | 0.003850567 | 0.240311204 | -9.18E-05 | 0.04141884 | 1 | 0.401395273 | 1 | 0.414759254 | ENSG0000014 | 8 | POLE3 |
| SAT | HTN | ENSG0000011 | -0.00562319 | 0.138679178 | 0.003797581 | 0.240334732 | -0.00554188 | 0.144470066 | 0.003797506 | 0.240311175 | -8.13E-05 | 0.005053763 | 1 | 0.391877031 | 1 | 0.404787607 | ENSG0000014 | 8 | POLE3 |
| SAT | HTN | ENSG0000011 | 0.001804769 | 0.771302364 | 0.006208955 | 0.240335772 | 0.001513423 | 0.807437308 | 0.006209335 | 0.240312196 | 0.000291346 | 0.001577151 | 1 | 0.909321975 | 1 | 0.925323719 | ENSG0000014 | 8 | POLE3 |
| SAT | HTN | ENSG0000011 | 0.002547944 | 0.705014148 | 0.006730606 | 0.240335743 | 0.002419993 | 0.719176819 | 0.006730442 | 0.240312162 | 0.000127951 | 0.006685459 | 1 | 0.876817876 | 1 | 0.886108926 | ENSG0000014 | 5 | SURF1 |
| SAT | HTN | ENSG0000011 | -0.00179715 | 0.515468792 | 0.002763377 | 0.240335605 | -0.00186641 | 0.499410635 | 0.002763335 | 0.240312 | 6.93E-05 | 0.002256125 | 1 | 0.765896967 | 1 | 0.756843553 | ENSG0000014 | 12 | HMCN2 |
| SAT | HTN | ENSG0000011 | -0.00312928 | 0.526325068 | 0.004938688 | 0.240335615 | -0.00321241 | 0.51538975 | 0.0049386 | 0.240312017 | 8.31E-05 | 0.033026259 | 1 | 0.776122494 | 1 | 0.76520719 | ENSG0000014 | 2 | LCN9 |
| SAT | HTN | ENSG0000011 | -0.01309396 | 0.059757797 | 0.006955327 | 0.240334065 | -0.01319263 | 0.057852322 | 0.006955129 | 0.240310451 | 9.87E-05 | 0.027565247 | 1 | 0.245081663 | 1 | 0.240880037 | ENSG0000014 | 14 | SEC16A |
| SAT | HTN | ENSG0000012 | 0.003934062 | 0.420897455 | 0.004887847 | 0.240335494 | 0.004017559 | 0.411095407 | 0.004887779 | 0.240311892 | -8.35E-05 | 0.041712215 | 1 | 0.698776883 | 1 | 0.687814395 | ENSG0000014 | 14 | SEC16A |
| SAT | HTN | ENSG0000012 | 0.042946388 | 3.76E-10 | 0.006856271 | 0.240316464 | 0.042853366 | 4.10E-10 | 0.006856083 | 0.24029296 | 9.30E-05 | 0.038628102 | 1.23E-06 | 7.71E-08 | 1.34E-06 | 8.40E-08 | ENSG0000014 | 13 | RSU1 |
| SAT | HTN | ENSG000001: | -0.00289023 | 0.428576853 | 0.003650987 | 0.240335504 | -0.00295691 | 0.417992947 | 0.00365092 | 0.240311902 | 6.67E-05 | 0.019826293 | 1 | 0.705195915 | 1 | 0.692294225 | ENSG0000014 | 10 | C10orf11 |
| SAT | HTN | ENSG000001: | -0.00906254 | 0.037193541 | 0.004349381 | 0.240333672 | -0.0093347 | 0.031876755 | 0.004349898 | 0.240309954 | 0.000272161 | 0.000643215 | 1 | 0.178436214 | 1 | 0.164445963 | ENSG0000014 | 10 | C10orf11 |
| SAT | HTN | ENSG000001: | -0.0066189 | 0.019665138 | 0.00283746 | 0.24033313 | -0.00667555 | 0.018637887 | 0.002837396 | 0.240309496 | 5.67E-05 | 0.006173289 | 1 | 0.119705596 | 1 | 0.114982479 | ENSG0000014 | 10 | C10orf11 |
| SAT | HTN | ENSG000001: | 0.012505724 | 0.062738045 | 0.006719722 | 0.240334105 | 0.012690886 | 0.058940505 | 0.006719585 | 0.240310466 | -0.00018516 | 0.000282141 | 1 | 0.251407587 | 1 | 0.243426139 | ENSG0000014 | 10 | C10orf11 |
| SAT | HTN | ENSG000001: | -0.00638155 | 0.208471698 | 0.005073661 | 0.240335033 | -0.00645794 | 0.203063251 | 0.00507351 | 0.240311426 | 7.64E-05 | 0.015740731 | 1 | 0.499334127 | 1 | 0.489890094 | ENSG0000014 | 13 | LRRC27 |
| SAT | HTN | ENSG000001: | 0.001094304 | 0.857971928 | 0.006114914 | 0.240335798 | 0.001223326 | 0.841435165 | 0.006114849 | 0.240312206 | -0.00012902 | 0.016202322 | 1 | 0.951893842 | 1 | 0.94371963 | ENSG0000014 | 14 | MTG1 |
| SAT | HTN | ENSG000001: | 0.006986376 | 0.002591739 | 0.002319179 | 0.240331338 | 0.007426189 | 0.001391382 | 0.002323261 | 0.240307187 | -0.00043981 | 0.001631017 | 1 | 0.028064341 | 1 | 0.017448468 | ENSG0000014 | 14 | MTG1 |
| SAT | HTN | ENSG000001: | -0.00510993 | 0.058226859 | 0.002697971 | 0.240334044 | -0.00441358 | 0.104049623 | 0.00271516 | 0.240310922 | -0.00069635 | 0.022945075 | 1 | 0.240304811 | 1 | 0.334692955 | ENSG0000014 | 8 | GSTO1 |
| SAT | HTN | ENSG000001: | -0.01331679 | 2.92E-07 | 0.002596565 | 0.240322842 | -0.01318405 | 3.83E-07 | 0.002596601 | 0.240299512 | -0.00013274 | 4.99E-06 | 0.000958002 | 1.71E-05 | 0.00125572 | 2.13E-05 | ENSG0000014 | 9 | PDCD11 |
| SAT | HTN | ENSG000001: | 0.02943827 | 2.95E-07 | 0.005742404 | 0.240322853 | 0.029707316 | 2.31E-07 | 0.005743393 | 0.240299032 | -0.00026905 | 0.026066505 | 0.000968997 | 1.71E-05 | 0.000758423 | 1.38E-05 | ENSG0000014 | 9 | PDCD11 |
| SAT | HTN | ENSG000001: | -0.00964716 | 3.92E-05 | 0.00234605 | 0.240327474 | -0.0107226 | 6.15E-06 | 0.002371534 | 0.240302144 | 0.001075443 | 0.001973279 | 0.128670306 | 0.001037664 | 0.020162759 | 0.000224031 | ENSG0000014 | 9 | PDCD11 |
| SAT | HTN | ENSG000001: | -0.00295282 | 0.311499833 | 0.002917572 | 0.240335308 | -0.00291079 | 0.318425726 | 0.002917498 | 0.240311735 | -4.20E-05 | 0.035800167 | 1 | 0.60449332 | 1 | 0.606006269 | ENSG0000014 | 10 | ZNF215 |
| SAT | HTN | ENSG000001: | 0.000286088 | 0.961759366 | 0.005966886 | 0.240335812 | 0.000180547 | 0.975860694 | 0.005966781 | 0.240312225 | 0.00010554 | 0.025869885 | 1 | 0.986748469 | 1 | 0.990655611 | ENSG0000014 | 10 | ZNF215 |
| SAT | HTN | ENSG000001: | 0.001075404 | 0.824813798 | 0.004858212 | 0.240335789 | 0.001293526 | 0.790050914 | 0.004858411 | 0.24031219 | -0.00021812 | 0.000819718 | 1 | 0.934466185 | 1 | 0.919969807 | ENSG0000014 | 8 | EIF3M |
| SAT | HTN | ENSG000001: | -0.00136583 | 0.773253789 | 0.00474048 | 0.240335772 | -0.00096224 | 0.839223796 | 0.004742765 | 0.240312205 | -0.00040359 | 0.009000775 | 1 | 0.909987691 | 1 | 0.942977149 | ENSG0000014 | 8 | EIF3M |
| SAT | HTN | ENSG000001: | 0.006033382 | 0.00041035 | 0.001707548 | 0.240329656 | 0.006005631 | 0.000436206 | 0.001707516 | 0.240306125 | 2.78E-05 | 0.036686616 | 1 | 0.006904399 | 1 | 0.006981431 | ENSG0000014 | 8 | PTPRJ |
| SAT | HTN | ENSG000001: | -0.00835748 | 0.076785784 | 0.004722668 | 0.240334269 | -0.00843207 | 0.074181714 | 0.004722539 | 0.240310653 | 7.46E-05 | 0.016793724 | 1 | 0.278996851 | 1 | 0.276576499 | ENSG0000014 | 9 | C11orf49 |
| SAT | HTN | ENSG000001: | -0.01449624 | 0.02227038 | 0.006342058 | 0.240333237 | -0.01432345 | 0.023914285 | 0.006341991 | 0.24030971 | -0.00017279 | 0.001903905 | 1 | 0.130247978 | 1 | 0.135984003 | ENSG0000014 | 9 | C11orf49 |
| SAT | HTN | ENSG000001: | 0.004096104 | 0.446810462 | 0.005384341 | 0.240335528 | 0.003977377 | 0.460088092 | 0.005384275 | 0.240311956 | 0.000118727 | 0.01031363 | 1 | 0.7196446 | 1 | 0.731635082 | ENSG0000014 | 9 | C11orf49 |
| SAT | HTN | ENSG000001: | 0.00800784 | 0.236885345 | 0.006770188 | 0.240335123 | 0.008094137 | 0.231856781 | 0.006769988 | 0.24031152 | -8.63E-05 | 0.041877111 | 1 | 0.52728685 | 1 | 0.519577173 | ENSG0000014 | 7 | CCDC82 |
| SAT | HTN | ENSG000001: | -0.00041664 | 0.931282814 | 0.004831645 | 0.24033581 | -0.00026962 | 0.955499125 | 0.004831616 | 0.240312224 | -0.00014702 | 0.001043833 | 1 | 0.976204842 | 1 | 0.98569618 | ENSG0000014 | 10 | CAPN5 |
| SAT | HTN | ENSG000001: | -0.01020323 | 0.034979887 | 0.004823377 | 0.240333606 | -0.00999691 | 0.038237715 | 0.004824057 | 0.240310108 | -0.00020632 | 0.028196862 | 1 | 0.172046825 | 1 | 0.181823106 | ENSG0000014 | 10 | RPS3 |
| SAT | HTN | ENSG000001: | 0.002687657 | 0.619761602 | 0.005416604 | 0.240335692 | 0.002609786 | 0.629931309 | 0.005416473 | 0.240312111 | 7.79E-05 | 0.041809053 | 1 | 0.833771344 | 1 | 0.839506103 | ENSG0000014 | 10 | RPS3 |
| SAT | HTN | ENSG000001: | 0.005609413 | 0.01173242 | 0.002225868 | 0.240332681 | 0.005641633 | 0.011256826 | 0.002225819 | 0.240309057 | -3.22E-05 | 0.048436893 | 1 | 0.084864703 | 1 | 0.083456744 | ENSG0000014 | 12 | TTC12 |
| SAT | HTN | ENSG000001: | 0.001703197 | 0.758900113 | 0.005549216 | 0.240335767 | 0.001574001 | 0.776679073 | 0.005549105 | 0.240312186 | 0.000129195 | 0.002287971 | 1 | 0.902979276 | 1 | 0.912575524 | ENSG0000014 | 12 | NPAT |
| SAT | HTN | ENSG000001: | 0.004397454 | 0.301533569 | 0.00425635 | 0.240335287 | 0.004472006 | 0.293401187 | 0.004256254 | 0.240311681 | -7.46E-05 | 0.016185797 | 1 | 0.595982915 | 1 | 0.58377762 | ENSG0000014 | 12 | NPAT |
| SAT | HTN | ENSG000001: | -0.00280933 | 0.083341821 | 0.001622368 | 0.240334335 | -0.00278225 | 0.086352224 | 0.001622335 | 0.240310775 | -2.71E-05 | 0.027154161 | 1 | 0.293903036 | 1 | 0.299568267 | ENSG0000014 | 10 | GLB1L2 |
| SAT | HTN | ENSG000001: | -0.00771945 | 0.103576876 | 0.004744989 | 0.240334508 | -0.00759262 | 0.109563906 | 0.004744917 | 0.240310963 | -0.00012682 | 0.001182169 | 1 | 0.332804614 | 1 | 0.348332534 | ENSG0000014 | 10 | GLB1L2 |
| SAT | HTN | ENSG000001: | 0.002369733 | 0.603455953 | 0.004562115 | 0.24033568 | 0.002433243 | 0.593776184 | 0.004561998 | 0.240312085 | -6.35E-05 | 0.042585673 | 1 | 0.826982828 | 1 | 0.820640533 | ENSG0000014 | 10 | GLB1L2 |
| SAT | HTN | ENSG000001: | 0.008578797 | 0.070392094 | 0.004741282 | 0.240334199 | 0.008502706 | 0.07291431 | 0.004741187 | 0.240310639 | 7.61E-05 | 0.035151525 | 1 | 0.268554024 | 1 | 0.27407569 | ENSG0000014 | 10 | SLX4IP |
| SAT | HTN | ENSG000001: | -0.00287041 | 0.137127646 | 0.001930885 | 0.240334723 | -0.0028401 | 0.141316928 | 0.001930849 | 0.240311158 | -3.03E-05 | 0.044863916 | 1 | 0.390551916 | 1 | 0.398676562 | ENSG0000014 | 7 | P4HA3 |
| SAT | HTN | ENSG000001: | 0.003581509 | 0.104042069 | 0.002203234 | 0.24033451 | 0.003629242 | 0.099502776 | 0.002203183 | 0.240310887 | -4.77E-05 | 0.002578906 | 1 | 0.333361356 | 1 | 0.325816974 | ENSG0000014 | 7 | P4HA3 |
| SAT | HTN | ENSG000001: | -0.0019353 | 0.685058088 | 0.004771795 | 0.240335732 | -0.00203665 | 0.669514752 | 0.004771739 | 0.240312136 | 0.000101345 | 0.013998702 | 1 | 0.866100648 | 1 | 0.86204999 | ENSG0000014 | 7 | GRIK4 |
| SAT | HTN | ENSG000001: | -0.00377457 | 0.518709303 | 0.005848998 | 0.240335608 | -0.00364648 | 0.532998722 | 0.005849016 | 0.240312034 | -0.00012809 | 0.032073296 | 1 | 0.768692512 | 1 | 0.780736132 | ENSG0000014 | 2 | MTA2 |
| SAT | HTN | ENSG000001: | 0.004585077 | 0.434413862 | 0.005865808 | 0.240335512 | 0.004481989 | 0.444805393 | 0.005865676 | 0.240311937 | 0.000103088 | 0.015906183 | 1 | 0.709110389 | 1 | 0.717505651 | ENSG0000014 | 2 | MTA2 |
| SAT | HTN | ENSG000001: | -0.00768477 | 0.207426029 | 0.006095753 | 0.24033503 | -0.00756648 | 0.214497917 | 0.00609564 | 0.240311466 | -0.00011829 | 0.012946494 | 1 | 0.498217277 | 1 | 0.504492951 | ENSG0000014 | 12 | FADS1 |
| SAT | HTN | ENSG000001: | -0.00941552 | 0.037133033 | 0.004517343 | 0.240333671 | -0.009528 | 0.03492994 | 0.004517405 | 0.240310032 | 0.000112483 | 0.026358706 | 1 | 0.178436214 | 1 | 0.173459202 | ENSG0000014 | 4 | ROM1 |
| SAT | HTN | ENSG000001: | 0.006291679 | 0.015455327 | 0.002598215 | 0.240332921 | 0.006424487 | 0.013424883 | 0.002598583 | 0.240309211 | -0.00013281 | 0.00889516 | 1 | 0.103207817 | 1 | 0.09371711 | ENSG0000014 | 4 | ROM1 |
| SAT | HTN | ENSG000001: | -0.00061588 | 0.851114494 | 0.003281202 | 0.240335796 | -0.00054837 | 0.867270478 | 0.00328119 | 0.240312212 | -6.75E-05 | 0.031163956 | 1 | 0.948538319 | 1 | 0.954550298 | ENSG0000014 | 4 | ROM1 |
| SAT | HTN | ENSG000001: | -0.00445807 | 0.235759569 | 0.00376001 | 0.24033512 | -0.00451493 | 0.229826251 | 0.003759909 | 0.240311514 | 5.69E-05 | 0.024219062 | 1 | 0.526207273 | 1 | 0.518610682 | ENSG0000014 | 4 | ROM1 |
| SAT | HTN | ENSG000001: | -0.00066501 | 0.914849788 | 0.006219536 | 0.240335808 | -0.00158098 | 0.799492724 | 0.006224221 | 0.240312194 | 0.000915962 | 0.000237393 | 1 | 0.969141477 | 1 | 0.923639306 | ENSG0000014 | 7 | EML3 |
| SAT | HTN | ENSG000001: | 0.006301742 | 0.196133441 | 0.004875064 | 0.240334989 | 0.006388796 | 0.190013907 | 0.004874935 | 0.240311378 | -8.71E-05 | 0.00785647 | 1 | 0.480722934 | 1 | 0.472131871 | ENSG0000014 | 7 | EML3 |
| SAT | HTN | ENSG000001: | 0.003276577 | 0.545993581 | 0.00542682 | 0.240335634 | 0.003379043 | 0.533499058 | 0.005426664 | 0.240312034 | -0.00010247 | 0.003140329 | 1 | 0.789733191 | 1 | 0.780736132 | ENSG0000014 | 7 | EML3 |
| SAT | HTN | ENSG000001: | -0.00926788 | 0.085359663 | 0.005387039 | 0.240334354 | -0.00915563 | 0.08921203 | 0.005387017 | 0.240310801 | -0.00011226 | 0.02785383 | 1 | 0.296748329 | 1 | 0.304120181 | ENSG0000014 | 10 | CCDC15 |
| SAT | HTN | ENSG000001: | -0.00565699 | 0.330389827 | 0.005811998 | 0.240335346 | -0.00526678 | 0.364951577 | 0.005813409 | 0.240311821 | -0.00039021 | 0.005443216 | 1 | 0.62016399 | 1 | 0.64900061 | ENSG0000014 | 5 | SCN2B |
| SAT | HTN | ENSG000001: | -0.00714605 | 0.28854189 | 0.00673313 | 0.240335258 | -0.0072365 | 0.28246962 | 0.006732946 | 0.240311656 | 9.04E-05 | 0.041925854 | 1 | 0.581565115 | 1 | 0.575181454 | ENSG0000014 | 11 | SIDT2 |
| SAT | HTN | ENSG000001: | 0.003303097 | 0.131708045 | 0.00219125 | 0.240334693 | 0.003266079 | 0.136083137 | 0.002191206 | 0.24031113 | 3.70E-05 | 0.026877119 | 1 | 0.384460941 | 1 | 0.389946525 | ENSG0000014 | 11 | SIDT2 |
| SAT | HTN | ENSG000001: | 0.011305697 | 0.010257947 | 0.00440421 | 0.240332563 | 0.011391714 | 0.009693056 | 0.004404125 | 0.240308926 | -8.60E-05 | 0.011533321 | 1 | 0.077370859 | 1 | 0.075362365 | ENSG0000014 | 11 | SIDT2 |
| SAT | HTN | ENSG000001: | 0.003271525 | 0.362980243 | 0.003596261 | 0.240335405 | 0.003216559 | 0.371088332 | 0.003596182 | 0.240311831 | 5.50E-05 | 0.037306465 | 1 | 0.650299517 | 1 | 0.653687263 | ENSG0000014 | 11 | SIDT2 |
| SAT | HTN | ENSG000001: | -0.00196092 | 0.768652648 | 0.006666652 | 0.24033577 | -0.0102114 | 0.157427028 | 0.007222762 | 0.24031124 | 0.008250485 | 0.002999739 | 1 | 0.907788321 | 1 | 0.427227526 | ENSG0000014 | 12 | TAGLN |
| SAT | HTN | ENSG000001: | -0.00409904 | 0.450104196 | 0.005427452 | 0.240335532 | -0.0040196 | 0.458923395 | 0.005427337 | 0.240311955 | -7.94E-05 | 0.049935675 | 1 | 0.722523685 | 1 | 0.730581106 | ENSG0000014 | 12 | TAGLN |
| SAT | HTN | ENSG000001: | -0.01554784 | 0.005807158 | 0.005636364 | 0.240332061 | -0.01543386 | 0.006175575 | 0.005636247 | 0.240308528 | -0.00011398 | 0.007046903 | 1 | 0.051081192 | 1 | 0.05390513 | ENSG0000014 | 12 | TAGLN |
| SAT | HTN | ENSG000001: | 0.002137309 | 0.756156003 | 0.006882716 | 0.240335766 | 0.002009944 | 0.770260596 | 0.006882562 | 0.240312183 | 0.000127365 | 0.011404293 | 1 | 0.901979395 | 1 | 0.91001726 | ENSG0000014 | 7 | CABLES2 |
| SAT | HTN | ENSG000001: | 0.003450045 | 0.575458058 | 0.006160446 | 0.240335659 | 0.003348471 | 0.586749848 | 0.006160348 | 0.24031208 | 0.000101574 | 0.042832872 | 1 | 0.808945111 | 1 | 0.815726353 | ENSG0000014 | 7 | CABLES2 |
| SAT | HTN | ENSG000001: | 0.002452337 | 0.342345397 | 0.002582652 | 0.240335369 | 0.002406007 | 0.351527424 | 0.002582574 | 0.240311797 | 4.63E-05 | 0.003246801 | 1 | 0.629901559 | 1 | 0.636161875 | ENSG0000014 | 9 | TRPT1 |
| SAT | HTN | ENSG000001: | -0.00605575 | 0.342500573 | 0.006379599 | 0.240335369 | -0.00595161 | 0.350858027 | 0.006379504 | 0.240311796 | -0.00010414 | 0.0485929 | 1 | 0.629901559 | 1 | 0.635651677 | ENSG0000014 | 9 | TRPT1 |
| SAT | HTN | ENSG000001: | 0.006101329 | 0.036391032 | 0.002915757 | 0.240333654 | 0.006052388 | 0.037914126 | 0.002915707 | 0.2403101 | 4.89E-05 | 0.035456566 | 1 | 0.176751156 | 1 | 0.180808499 | ENSG0000014 | 6 | TBX6 |
| SAT | HTN | ENSG000001: | 0.016017911 | 0.002574615 | 0.005313715 | 0.240331332 | 0.015911073 | 0.00274977 | 0.005313566 | 0.240307804 | 0.000106839 | 0.001912395 | 1 | 0.027977934 | 1 | 0.029292189 | ENSG0000014 | 6 | TBX6 |
| SAT | HTN | ENSG000001: | -0.00197218 | 0.750694238 | 0.006207199 | 0.240335764 | -0.00901214 | 0.178841364 | 0.0067038 | 0.240311334 | 0.007039961 | 0.005445612 | 1 | 0.899394965 | 1 | 0.458067006 | ENSG0000015 | 14 | MPP7 |
| SAT | HTN | ENSG000001: | 0.00307253 | 0.010665728 | 0.001203275 | 0.240332598 | 0.00372299 | 0.002599218 | 0.001236233 | 0.240307753 | -0.00065046 | 0.021914997 | 1 | 0.079172522 | 1 | 0.028332336 | ENSG0000015 | 18 | C10ORF68 |
| SAT | HTN | ENSG000001: | 4.68E-05 | 0.989532033 | 0.003564792 | 0.240335813 | -6.54E-06 | 0.998536639 | 0.003564706 | 0.240312225 | 5.33E-05 | 0.034199185 | 1 | 0.99730223 | 1 | 0.999787914 | ENSG0000015 | 7 | CWC15 |
| SAT | HTN | ENSG000001: | -0.00189339 | 0.702000694 | 0.004948459 | 0.240335741 | -0.00196488 | 0.691309764 | 0.00494833 | 0.240312148 | 7.15E-05 | 0.033597497 | 1 | 0.875941799 | 1 | 0.871374312 | ENSG0000015 | 7 | CWC15 |
| SAT | HTN | ENSG000001: | 0.021826371 | 6.98E-07 | 0.004398758 | 0.240323671 | 0.021719004 | 7.91E-07 | 0.004398679 | 0.240300203 | 0.000107366 | 0.001887866 | 0.002290671 | 3.52E-05 | 0.002595531 | 3.82E-05 | ENSG0000015 | 7 | CWC15 |
| SAT | HTN | ENSG000001: | 0.003022618 | 0.093892374 | 0.001804315 | 0.240334429 | 0.002998564 | 0.096527711 | 0.001804268 | 0.240310863 | 2.41E-05 | 0.049546175 | 1 | 0.313707617 | 1 | 0.319658873 | ENSG0000015 | 9 | FCGR1A |
| SAT | HTN | ENSG000001: | 0.00023744 | 0.955331764 | 0.004239042 | 0.240335812 | -0.00066284 | 0.876366422 | 0.00426051 | 0.240312213 | 0.000900278 | 0.035952713 | 1 | 0.983048498 | 1 | 0.956781838 | ENSG0000015 | 10 | CNDP1 |
| SAT | HTN | ENSG000001: | -0.00315507 | 0.229602039 | 0.002626189 | 0.240335102 | -0.00334988 | 0.20238383 | 0.002627791 | 0.240311424 | 0.000194817 | 0.041071298 | 1 | 0.52039215 | 1 | 0.488970062 | ENSG0000015 | 10 | CNDP1 |
| SAT | HTN | ENSG000001: | 0.008149541 | 0.175056802 | 0.006009387 | 0.240334906 | 0.007972455 | 0.184612169 | 0.006009287 | 0.240311357 | 0.000177086 | 0.000242675 | 1 | 0.450479504 | 1 | 0.465896151 | ENSG0000015 | 10 | CNDP1 |
| SAT | HTN | ENSG000001: | -0.00247142 | 0.435595541 | 0.003169901 | 0.240335514 | -0.00252431 | 0.42582833 | 0.003169843 | 0.240311913 | 5.29E-05 | 0.034099864 | 1 | 0.710685714 | 1 | 0.699140398 | ENSG0000015 | 9 | FREM2 |
| SAT | HTN | ENSG000001: | 0.000394575 | 0.948112896 | 0.006063223 | 0.240335811 | 0.0002561 | 0.966241123 | 0.006063107 | 0.240312225 | 0.000137965 | 0.003236353 | 1 | 0.981389706 | 1 | 0.988536677 | ENSG0000015 | 7 | PRSS53 |
| SAT | HTN | ENSG000001: | 0.00431473 | 0.432801259 | 0.005500621 | 0.24033551 | 0.00392811 | 0.475383066 | 0.005503494 | 0.240311974 | 0.00038662 | 0.03758682 | 1 | 0.708539279 | 1 | 0.74149608 | ENSG0000015 | 7 | PRSS53 |
| SAT | HTN | ENSG000001: | 0.00278857 | 0.382817522 | 0.003195272 | 0.240335438 | 0.002723554 | 0.394002257 | 0.003195225 | 0.240311867 | 6.50E-05 | 0.013973407 | 1 | 0.668098026 | 1 | 0.673994476 | ENSG0000015 | 7 | PRSS53 |
| SAT | HTN | ENSG000001: | 0.014863412 | 0.007461169 | 0.005555296 | 0.240332283 | 0.014703729 | 0.008128404 | 0.005555495 | 0.240308771 | 0.000159682 | 0.027258915 | 1 | 0.062290319 | 1 | 0.067346705 | ENSG0000015 | 4 | TMEM86A |
| SAT | HTN | ENSG0000014 | 0.006385491 | 0.215499343 | 0.005155467 | 0.240335057 | 0.006457672 | 0.210345174 | 0.005155338 | 0.240311452 | -7.22E-05 | 0.044034966 | 1 | 0.507575983 | 1 | 0.499379533 | ENSG0000015 | 4 | TMEM86A |
| SAT | HTN | ENSG0000014 | -8.32E-05 | 0.939649665 | 0.001098565 | 0.240335811 | -0.00010617 | 0.923005037 | 0.001098559 | 0.240312221 | 2.30E-05 | 0.024876473 | 1 | 0.978294984 | 1 | 0.974491801 | ENSG0000015 | 9 | UBE3B |
| SAT | HTN | ENSG0000014 | 0.004895145 | 0.117157907 | 0.003124259 | 0.240334603 | 0.004968202 | 0.111789071 | 0.003124251 | 0.240310978 | -7.31E-05 | 0.015380892 | 1 | 0.363550738 | 1 | 0.352908944 | ENSG0000015 | 9 | UBE3B |
| SAT | HTN | ENSG0000014 | 0.009598789 | 0.167345225 | 0.006951682 | 0.240334873 | 0.00972055 | 0.162017632 | 0.006951592 | 0.240311261 | -0.00012176 | 0.039678555 | 1 | 0.439214192 | 1 | 0.434297263 | ENSG0000015 | 9 | UBE3B |
| SAT | HTN | ENSG0000014 | -0.01258385 | 0.00027754 | 0.003461442 | 0.240329295 | -0.01264913 | 0.000257874 | 0.003461402 | 0.24030564 | 6.53E-05 | 0.030013277 | 0.910609609 | 0.005001434 | 0.846086136 | 0.004726738 | ENSG0000015 | 9 | UBE3B |
| SAT | HTN | ENSG0000014 | 0.009351153 | 0.164740108 | 0.006730805 | 0.240334861 | 0.009224884 | 0.170511941 | 0.006730728 | 0.240311299 | 0.000126269 | 0.030367877 | 1 | 0.435897688 | 1 | 0.443378165 | ENSG0000015 | 3 | PLBD2 |
| SAT | HTN | ENSG0000014 | -0.00049217 | 0.935281185 | 0.006061023 | 0.24033581 | -0.00039953 | 0.947442337 | 0.006060887 | 0.240312223 | -9.26E-05 | 0.036264939 | 1 | 0.97727948 | 1 | 0.981538823 | ENSG0000015 | 3 | PLBD2 |
| SAT | HTN | ENSG0000014 | 0.024014294 | 8.90E-05 | 0.006128116 | 0.24032824 | 0.024103573 | 8.38E-05 | 0.006127982 | 0.240304596 | -8.93E-05 | 0.049211472 | 0.292130826 | 0.002057259 | 0.274874119 | 0.001977512 | ENSG0000015 | 3 | PLBD2 |
| SAT | HTN | ENSG0000014 | -0.00202019 | 0.766652939 | 0.006807568 | 0.24033577 | -0.00192119 | 0.777774227 | 0.006807411 | 0.240312186 | -9.90E-05 | 0.04431138 | 1 | 0.906772997 | 1 | 0.91334189 | ENSG0000015 | 12 | DLG5 |

| | | | | | | | | | | | | | | | | | | | |
|---|---|---|---|---|---|---|---|---|---|---|---|---|---|---|---|---|---|---|---|
| SAT | HTN | ENSG0000014 | -0.00725319 | 0.074054473 | 0.004060499 | 0.24033424 | -0.00713133 | 0.079040448 | 0.004060463 | 0.240310704 | -0.00012186 | 0.000803533 | 1 | 0.276027247 | 1 | 0.285677612 | ENSG0000015 | 12 | DLG5 |
| SAT | HTN | ENSG0000014 | -0.00320216 | 0.482705575 | 0.004561753 | 0.24033557 | -0.00329908 | 0.469550703 | 0.004561706 | 0.240311967 | 9.69E-05 | 0.015936813 | 1 | 0.747733474 | 1 | 0.736442251 | ENSG0000015 | 12 | DLG5 |
| SAT | HTN | ENSG0000014 | 0.001498048 | 0.72664361 | 0.004285109 | 0.240335753 | 0.00155429 | 0.716807578 | 0.004284992 | 0.240312161 | -5.62E-05 | 0.046098375 | 1 | 0.886948544 | 1 | 0.88553283 | ENSG0000015 | 19 | MAGI1 |
| SAT | HTN | ENSG0000014 | 0.000378991 | 0.91242541 | 0.003446 | 0.240335807 | 0.000316005 | 0.926933401 | 0.003445934 | 0.240312221 | 6.30E-05 | 0.018098521 | 1 | 0.969071845 | 1 | 0.975149159 | ENSG0000015 | 19 | MAGI1 |
| SAT | HTN | ENSG0000014 | 0.003479129 | 0.610465556 | 0.006829732 | 0.240335685 | 0.003334726 | 0.625352804 | 0.006829558 | 0.240312108 | 0.000144403 | 0.002082056 | 1 | 0.830870282 | 1 | 0.837966728 | ENSG0000015 | 9 | EXT2 |
| SAT | HTN | ENSG0000014 | -0.02150946 | 0.001223545 | 0.006652369 | 0.240330657 | -0.02133111 | 0.001343634 | 0.006652433 | 0.240307155 | -0.00017835 | 0.013168597 | 1 | 0.015930364 | 1 | 0.016955623 | ENSG0000015 | 9 | EXT2 |
| SAT | HTN | ENSG0000014 | -0.00579121 | 0.318079372 | 0.005800412 | 0.240335322 | -0.00567091 | 0.328230212 | 0.005800337 | 0.240311754 | -0.0001203 | 0.014724879 | 1 | 0.610360453 | 1 | 0.615105129 | ENSG0000015 | 9 | EXT2 |
| SAT | HTN | ENSG0000014 | 0.010519818 | 0.012418749 | 0.004207884 | 0.240332731 | 0.01182034 | 0.005283333 | 0.00423783 | 0.240308389 | -0.00130052 | 0.00996956 | 1 | 0.088578075 | 1 | 0.048151705 | ENSG0000015 | 8 | EPS8 |
| SAT | HTN | ENSG0000014 | 0.011553027 | 0.01120433 | 0.004555124 | 0.240332641 | 0.011626737 | 0.010694974 | 0.004555003 | 0.240309012 | -7.37E-05 | 0.015936943 | 1 | 0.082725873 | 1 | 0.08048213 | ENSG0000015 | 9 | VTI1A |
| SAT | HTN | ENSG0000014 | 0.013438086 | 0.000692174 | 0.003960903 | 0.240330137 | 0.014008239 | 0.000411778 | 0.003965596 | 0.240306072 | -0.00057015 | 0.003780788 | 1 | 0.010414304 | 1 | 0.006721602 | ENSG0000015 | 9 | VTI1A |
| SAT | HTN | ENSG0000014 | -0.00469043 | 0.136386862 | 0.00314924 | 0.240334719 | -0.00463596 | 0.140989087 | 0.003149171 | 0.240311157 | -5.45E-05 | 0.018964357 | 1 | 0.389587189 | 1 | 0.398436861 | ENSG0000015 | 9 | VTI1A |
| SAT | HTN | ENSG0000014 | -0.01456273 | 0.024787128 | 0.006487575 | 0.240333328 | -0.01445596 | 0.025861491 | 0.00648746 | 0.240309777 | -0.00010677 | 0.037623859 | 1 | 0.139829894 | 1 | 0.142607652 | ENSG0000015 | 7 | QDPR |
| SAT | HTN | ENSG0000014 | -0.01094581 | 0.076191625 | 0.006172818 | 0.240334263 | -0.01105266 | 0.073361246 | 0.00617267 | 0.240310644 | 0.000106851 | 0.014594412 | 1 | 0.278675024 | 1 | 0.274769689 | ENSG0000015 | 7 | QDPR |
| SAT | HTN | ENSG0000014 | -0.02868272 | 8.67E-06 | 0.006448388 | 0.240326056 | -0.0285809 | 9.32E-06 | 0.006448241 | 0.240302537 | -0.00010182 | 0.029352455 | 0.028437867 | 0.00030253 | 0.030588524 | 0.000321984 | ENSG0000015 | 8 | DPYSL4 |
| SAT | HTN | ENSG0000014 | -0.01077814 | 0.001552548 | 0.003405742 | 0.240330874 | -0.00889629 | 0.010833411 | 0.003491429 | 0.240309024 | -0.00188185 | 0.014456946 | 1 | 0.019007131 | 1 | 0.081337348 | ENSG0000015 | 8 | DPYSL4 |
| SAT | HTN | ENSG0000014 | -0.00888496 | 0.156883324 | 0.006276315 | 0.240334825 | -0.0090073 | 0.151241245 | 0.006276153 | 0.24031121 | 0.00012234 | 0.004415502 | 1 | 0.425752014 | 1 | 0.415945118 | ENSG0000015 | 8 | DPYSL4 |
| SAT | HTN | ENSG0000014 | -0.00670333 | 0.163334657 | 0.004808892 | 0.240334855 | -0.00661994 | 0.168627669 | 0.004808808 | 0.240311291 | -8.34E-05 | 0.028954153 | 1 | 0.433927943 | 1 | 0.442141749 | ENSG0000015 | 8 | DPYSL4 |
| SAT | HTN | ENSG0000014 | 0.06581016 | 1.37E-22 | 0.006728725 | 0.240288645 | 0.065679101 | 1.66E-22 | 0.006728585 | 0.240265246 | 0.000131059 | 0.008974326 | 4.50E-19 | 2.25E-19 | 5.45E-19 | 2.72E-19 | ENSG0000015 | 8 | DPYSL4 |
| SAT | HTN | ENSG0000014 | -0.02124635 | 0.001485414 | 0.006686424 | 0.240330834 | -0.02134374 | 0.001412147 | 0.006686267 | 0.2403072 | 9.74E-05 | 0.042409743 | 1 | 0.018460765 | 1 | 0.017483974 | ENSG0000015 | 7 | KIN |
| SAT | HTN | ENSG0000014 | 0.007779319 | 0.074786932 | 0.004366056 | 0.240334248 | 0.007719771 | 0.077031378 | 0.004365941 | 0.240310684 | 5.95E-05 | 0.042373001 | 1 | 0.276027247 | 1 | 0.282212646 | ENSG0000015 | 7 | KIN |
| SAT | HTN | ENSG0000015 | 0.000295479 | 0.955004169 | 0.005236776 | 0.240335812 | 0.000428245 | 0.934823312 | 0.005236678 | 0.240312222 | -0.00013277 | 0.001153421 | 1 | 0.983048498 | 1 | 0.977239819 | ENSG0000015 | 7 | CENPU |
| SAT | HTN | ENSG0000015 | -0.01222355 | 0.028704634 | 0.005587842 | 0.240333453 | -0.01249598 | 0.025345126 | 0.005588271 | 0.24030976 | 0.000272432 | 0.002146352 | 1 | 0.154393286 | 1 | 0.140973783 | ENSG0000015 | 7 | CENPU |
| SAT | HTN | ENSG0000015 | 0.006719179 | 0.259130502 | 0.005954344 | 0.240335185 | 0.00662027 | 0.266197008 | 0.005954206 | 0.240311616 | 9.89E-05 | 0.020957768 | 1 | 0.550295907 | 1 | 0.55733039 | ENSG0000015 | 7 | CENPU |
| SAT | HTN | ENSG0000015 | -0.00575054 | 0.267715534 | 0.005188425 | 0.240335208 | -0.00561133 | 0.2794646 | 0.005188351 | 0.240311649 | -0.00013921 | 0.001305522 | 1 | 0.561046104 | 1 | 0.572417553 | ENSG0000015 | 7 | CENPU |
| SAT | HTN | ENSG0000015 | 0.006555744 | 0.257305456 | 0.005787284 | 0.240335181 | 0.006459539 | 0.264346364 | 0.005787193 | 0.240311611 | 9.62E-05 | 0.041486655 | 1 | 0.549440772 | 1 | 0.554552698 | ENSG0000015 | 10 | CENPJ |
| SAT | HTN | ENSG0000015 | 0.00352052 | 0.484765584 | 0.005038969 | 0.240335573 | 0.003621472 | 0.472324291 | 0.005038899 | 0.240311971 | -0.00010095 | 0.017074248 | 1 | 0.748376275 | 1 | 0.738432275 | ENSG0000015 | 10 | CENPJ |
| SAT | HTN | ENSG0000015 | -0.02335406 | 6.13E-05 | 0.005826962 | 0.240327891 | -0.0234512 | 5.71E-05 | 0.005826808 | 0.240304237 | 9.71E-05 | 0.013484311 | 0.201001009 | 0.001500008 | 0.1871961 | 0.001428978 | ENSG0000015 | 12 | PARP8 |
| SAT | HTN | ENSG0000015 | 0.003468851 | 0.346083145 | 0.003681592 | 0.240335376 | 0.004042309 | 0.273638027 | 0.003692536 | 0.240311634 | -0.00057346 | 0.045274705 | 1 | 0.633986113 | 1 | 0.566796949 | ENSG0000015 | 12 | PARP8 |
| SAT | HTN | ENSG0000015 | -0.00932267 | 0.093036593 | 0.005550544 | 0.240334422 | -0.00969723 | 0.080725376 | 0.005552393 | 0.240310721 | 0.000374562 | 0.014684542 | 1 | 0.311800881 | 1 | 0.287266767 | ENSG0000015 | 12 | PARP8 |
| SAT | HTN | ENSG0000015 | -0.00501152 | 0.471634494 | 0.006962154 | 0.240335558 | -0.0048807 | 0.483273937 | 0.006962014 | 0.240311983 | -0.00013082 | 0.013552308 | 1 | 0.740044368 | 1 | 0.747074465 | ENSG0000015 | 12 | PARP8 |
| SAT | HTN | ENSG0000015 | 0.004121935 | 0.297231411 | 0.003954312 | 0.240335277 | 0.007050446 | 0.09086856 | 0.004169796 | 0.240310816 | -0.00292851 | 0.026942648 | 1 | 0.590682168 | 1 | 0.308633276 | ENSG0000015 | 12 | PARP8 |
| SAT | HTN | ENSG0000015 | -0.00608428 | 0.019850194 | 0.002612202 | 0.240333138 | -0.00602376 | 0.021108088 | 0.002612151 | 0.240309603 | -6.05E-05 | 0.002527771 | 1 | 0.120385374 | 1 | 0.125919337 | ENSG0000015 | 17 | RABGAP1L |
| SAT | HTN | ENSG0000015 | -0.00068679 | 0.8939951 | 0.005154109 | 0.240335805 | -0.00055084 | 0.91488789 | 0.005154021 | 0.24031222 | -0.00013595 | 0.000977916 | 1 | 0.960795963 | 1 | 0.972383274 | ENSG0000015 | 17 | RABGAP1L |
| SAT | HTN | ENSG0000015 | 0.007035023 | 0.249660496 | 0.006111163 | 0.24033516 | 0.006001694 | 0.326529827 | 0.006117133 | 0.240311751 | 0.001033329 | 0.000189558 | 1 | 0.541398603 | 1 | 0.615105129 | ENSG0000015 | 5 | TMEM56 |
| SAT | HTN | ENSG0000015 | 0.021930058 | 0.001019806 | 0.006675766 | 0.240330491 | 0.021832095 | 0.001073954 | 0.006675625 | 0.240306951 | 9.80E-05 | 0.048994719 | 1 | 0.013826371 | 1 | 0.014323756 | ENSG0000015 | 4 | ARL14EP |
| SAT | HTN | ENSG0000015 | -0.0115873 | 0.017082589 | 0.004858544 | 0.240333008 | -0.01148461 | 0.018087774 | 0.004858487 | 0.24030947 | -0.00010269 | 0.01452011 | 1 | 0.109682924 | 1 | 0.113472249 | ENSG0000015 | 4 | ARL14EP |
| SAT | HTN | ENSG0000015 | 0.00524905 | 0.020606568 | 0.002267287 | 0.24033317 | 0.005532728 | 0.014782197 | 0.002269669 | 0.240309295 | -0.00028368 | 0.007670508 | 1 | 0.122927543 | 1 | 0.10024166 | ENSG0000015 | 4 | ARL14EP |
| SAT | HTN | ENSG0000015 | 0.003218186 | 0.557079153 | 0.005480694 | 0.240335643 | 0.002736563 | 0.617721545 | 0.005483188 | 0.240312103 | 0.000481623 | 0.005649956 | 1 | 0.797697485 | 1 | 0.835426376 | ENSG0000015 | 8 | EPG5 |
| SAT | HTN | ENSG0000015 | -0.00965979 | 0.126209885 | 0.006316796 | 0.24033466 | -0.00980118 | 0.120750838 | 0.006316688 | 0.240311038 | 0.000141391 | 0.00522347 | 1 | 0.376792204 | 1 | 0.369887656 | ENSG0000015 | 8 | HAUS1 |
| SAT | HTN | ENSG0000015 | -0.00626375 | 0.407145293 | 0.007556441 | 0.240335474 | -0.00657021 | 0.384643521 | 0.007557412 | 0.240311853 | 0.000306462 | 0.03142072 | 1 | 0.687516061 | 1 | 0.667766115 | ENSG0000015 | 8 | HAUS1 |
| SAT | HTN | ENSG0000015 | -0.00065707 | 0.897377671 | 0.005094556 | 0.240335805 | -0.00052427 | 0.918033996 | 0.005094472 | 0.24031222 | -0.00013279 | 0.001226221 | 1 | 0.962469823 | 1 | 0.974136703 | ENSG0000015 | 8 | HAUS1 |
| SAT | HTN | ENSG0000015 | -0.00946158 | 0.099790804 | 0.005748671 | 0.240334477 | -0.00955301 | 0.096550425 | 0.005748537 | 0.240310863 | 9.14E-05 | 0.026980044 | 1 | 0.325460864 | 1 | 0.319658873 | ENSG0000015 | 4 | SPC25 |
| SAT | HTN | ENSG0000015 | 0.026162674 | 1.46E-05 | 0.006035764 | 0.240326547 | 0.026253204 | 1.36E-05 | 0.006035606 | 0.240302895 | -9.05E-05 | 0.027472726 | 0.047918603 | 0.000460756 | 0.044732652 | 0.000430122 | ENSG0000015 | 4 | SPC25 |
| SAT | HTN | ENSG0000015 | 0.005517006 | 0.264384405 | 0.004943158 | 0.240335199 | 0.005609926 | 0.256412203 | 0.004943043 | 0.24031159 | -9.29E-05 | 0.009040161 | 1 | 0.556219915 | 1 | 0.548573946 | ENSG0000015 | 4 | SPC25 |
| SAT | HTN | ENSG0000015 | 0.009334868 | 0.029912592 | 0.004299302 | 0.240333488 | 0.009408738 | 0.028634282 | 0.004299189 | 0.240309863 | -7.39E-05 | 0.011003618 | 1 | 0.158807793 | 1 | 0.153011528 | ENSG0000015 | 4 | PDE3B |
| SAT | HTN | ENSG0000015 | 0.002464679 | 0.70102588 | 0.006419501 | 0.240335741 | 0.00260989 | 0.684328253 | 0.006419379 | 0.240312144 | -0.00014521 | 0.003518716 | 1 | 0.875548501 | 1 | 0.867573802 | ENSG0000015 | 9 | TGOLN2 |
| SAT | HTN | ENSG0000015 | -0.01611573 | 0.015014578 | 0.006626436 | 0.240332896 | -0.01595016 | 0.016081033 | 0.006626353 | 0.240309368 | -0.00016557 | 0.003456911 | 1 | 0.101155706 | 1 | 0.106374736 | ENSG0000015 | 9 | TGOLN2 |
| SAT | HTN | ENSG0000015 | -0.00236054 | 0.583278314 | 0.004302825 | 0.240335665 | -0.00242469 | 0.57307615 | 0.004302704 | 0.240312069 | 6.41E-05 | 0.021139912 | 1 | 0.812626814 | 1 | 0.806980304 | ENSG0000015 | 9 | TGOLN2 |
| SAT | HTN | ENSG0000015 | 0.00405236 | 0.25471873 | 0.003557935 | 0.240335174 | 0.004105487 | 0.248532557 | 0.003557853 | 0.240311569 | -5.31E-05 | 0.039188389 | 1 | 0.548740745 | 1 | 0.537886095 | ENSG0000015 | 11 | ATG10 |
| SAT | HTN | ENSG0000016 | 0.004957215 | 0.33737532 | 0.005167183 | 0.240335359 | 0.005111914 | 0.322509293 | 0.005167122 | 0.240311743 | -0.0001547 | 0.000538625 | 1 | 0.625030166 | 1 | 0.611295778 | ENSG0000015 | 11 | ATG10 |
| SAT | HTN | ENSG0000016 | 0.003240746 | 0.476194516 | 0.004548817 | 0.240335563 | 0.003312147 | 0.46651882 | 0.004548682 | 0.240311964 | -7.14E-05 | 0.01160616 | 1 | 0.743997242 | 1 | 0.734476126 | ENSG0000015 | 11 | ATG10 |
| SAT | HTN | ENSG0000016 | 0.001889022 | 0.174965878 | 0.001392651 | 0.240334906 | 0.001908542 | 0.170540458 | 0.001392617 | 0.240311299 | -1.95E-05 | 0.045191396 | 1 | 0.450479504 | 1 | 0.443378165 | ENSG0000015 | 11 | ATG10 |
| SAT | HTN | ENSG0000016 | -0.00502478 | 0.472687505 | 0.006997189 | 0.240335559 | -0.00515391 | 0.461381403 | 0.006997128 | 0.240311958 | 0.00012913 | 0.040127141 | 1 | 0.740564567 | 1 | 0.732029465 | ENSG0000015 | 11 | ATG10 |
| SAT | HTN | ENSG0000016 | 0.014865409 | 0.000868645 | 0.004464138 | 0.240330345 | 0.014940592 | 0.000817375 | 0.004464057 | 0.240306701 | -7.52E-05 | 0.032512136 | 1 | 0.01203601 | 1 | 0.011411947 | ENSG0000015 | 10 | POC5 |
| SAT | HTN | ENSG0000016 | -0.00413586 | 0.4341675 | 0.005288278 | 0.240335512 | -0.00404747 | 0.444043987 | 0.005288163 | 0.240311937 | -8.84E-05 | 0.02363982 | 1 | 0.709110389 | 1 | 0.716629769 | ENSG0000015 | 8 | ZNF773 |
| SAT | HTN | ENSG0000016 | 0.006282195 | 0.202839842 | 0.004932999 | 0.240335014 | 0.006204206 | 0.208493779 | 0.0049329 | 0.240311445 | 7.80E-05 | 0.037920898 | 1 | 0.490794632 | 1 | 0.497295906 | ENSG0000015 | 8 | ZNF773 |
| SAT | HTN | ENSG0000016 | 0.001719831 | 0.642310133 | 0.00370277 | 0.240335707 | 0.001610227 | 0.66365321 | 0.003702734 | 0.240312132 | 0.000109604 | 0.000832185 | 1 | 0.844302676 | 1 | 0.859631339 | ENSG0000015 | 8 | ZNF773 |
| SAT | HTN | ENSG0000016 | 0.005930031 | 0.041574184 | 0.002910069 | 0.240333765 | 0.005981646 | 0.039828101 | 0.002910022 | 0.240310142 | -5.16E-05 | 0.029009833 | 1 | 0.191311216 | 1 | 0.185366123 | ENSG0000015 | 8 | ZNF773 |
| SAT | HTN | ENSG0000016 | 0.002021525 | 0.569155815 | 0.003550923 | 0.240335654 | 0.001952975 | 0.582320183 | 0.003550869 | 0.240312076 | 6.85E-05 | 0.018780317 | 1 | 0.804220599 | 1 | 0.812928219 | ENSG0000015 | 13 | NMT2 |
| SAT | HTN | ENSG0000016 | -0.0024906 | 0.624590389 | 0.00508956 | 0.240335695 | -0.00258386 | 0.611670793 | 0.005089441 | 0.240312098 | 9.33E-05 | 0.01051008 | 1 | 0.834397828 | 1 | 0.833930915 | ENSG0000015 | 13 | NMT2 |
| SAT | HTN | ENSG0000016 | -0.00700315 | 0.285326333 | 0.006554611 | 0.24033525 | -0.00674252 | 0.303653883 | 0.006554853 | 0.240311704 | -0.00026063 | 0.002429728 | 1 | 0.57930427 | 1 | 0.595866261 | ENSG0000015 | 13 | NMT2 |
| SAT | HTN | ENSG0000016 | 0.00474276 | 0.124089336 | 0.003084047 | 0.240334647 | 0.004699333 | 0.127560451 | 0.003083966 | 0.24031108 | 4.34E-05 | 0.03584692 | 1 | 0.374127295 | 1 | 0.381499044 | ENSG0000015 | 13 | NMT2 |
| SAT | HTN | ENSG0000016 | -0.00420445 | 0.436973483 | 0.005408936 | 0.240335515 | -0.00406013 | 0.452863639 | 0.005408818 | 0.240311948 | -0.00014432 | 0.000298163 | 1 | 0.710902942 | 1 | 0.725510546 | ENSG0000015 | 13 | NMT2 |
| SAT | HTN | ENSG0000016 | -0.00414913 | 0.163068692 | 0.002974657 | 0.240334854 | -0.00410579 | 0.167498563 | 0.00297459 | 0.240311286 | -4.33E-05 | 0.044645618 | 1 | 0.433572431 | 1 | 0.440923826 | ENSG0000015 | 13 | NMT2 |
| SAT | HTN | ENSG0000016 | -0.00701991 | 0.068110028 | 0.003848039 | 0.240334172 | -0.00707727 | 0.065882473 | 0.00384796 | 0.240310557 | 5.74E-05 | 0.049014818 | 1 | 0.264460356 | 1 | 0.262366273 | ENSG0000015 | 9 | CCDC50 |
| SAT | HTN | ENSG0000016 | 0.00220197 | 0.529708766 | 0.003503805 | 0.240335619 | 0.002256641 | 0.519530488 | 0.003503721 | 0.240312021 | -5.47E-05 | 0.027563595 | 1 | 0.778612574 | 1 | 0.769056242 | ENSG0000015 | 9 | CCDC50 |
| SAT | HTN | ENSG0000016 | -0.01483846 | 0.008480565 | 0.00563702 | 0.240332396 | -0.01406196 | 0.012744898 | 0.005645494 | 0.240309166 | -0.00077651 | 0.013435599 | 1 | 0.069215759 | 1 | 0.089955233 | ENSG0000015 | 9 | CCDC50 |
| SAT | HTN | ENSG0000016 | 0.000375629 | 0.886121121 | 0.002622848 | 0.240335803 | 0.000631314 | 0.809856932 | 0.002623798 | 0.240312197 | -0.00025568 | 0.000675904 | 1 | 0.958390933 | 1 | 0.925531291 | ENSG0000015 | 9 | CCDC50 |
| SAT | HTN | ENSG0000016 | 0.009159019 | 0.030896712 | 0.004243417 | 0.240333516 | 0.009232905 | 0.029565935 | 0.004243329 | 0.240309891 | -7.39E-05 | 0.020878548 | 1 | 0.16193628 | 1 | 0.156286405 | ENSG0000015 | 9 | PLEKHH2 |
| SAT | HTN | ENSG0000016 | 0.00202574 | 0.734833008 | 0.005980859 | 0.240335757 | 0.001908133 | 0.749690301 | 0.00598071 | 0.240312175 | 0.000117606 | 0.004716249 | 1 | 0.893035719 | 1 | 0.898697069 | ENSG0000015 | 9 | PLEKHH2 |
| SAT | HTN | ENSG0000016 | -0.00615999 | 0.274860758 | 0.005641339 | 0.240335225 | -0.00780662 | 0.168852061 | 0.005673815 | 0.240311292 | 0.001646629 | 0.006836306 | 1 | 0.570049397 | 1 | 0.442141749 | ENSG0000015 | 9 | PLEKHH2 |
| SAT | HTN | ENSG0000016 | 0.007313905 | 0.022711195 | 0.003210295 | 0.240333253 | 0.007243168 | 0.024055416 | 0.003210268 | 0.240309715 | 7.07E-05 | 0.014381242 | 1 | 0.13136175 | 1 | 0.136079 | ENSG0000015 | 10 | SLC30A6 |
| SAT | HTN | ENSG0000016 | -0.0048633 | 0.13060005 | 0.00321702 | 0.240334686 | -0.0048078 | 0.135041159 | 0.003216961 | 0.240311124 | -5.55E-05 | 0.027894164 | 1 | 0.382588181 | 1 | 0.388719394 | ENSG0000015 | 13 | RASGRP3 |
| SAT | HTN | ENSG0000016 | 0.014513033 | 0.000407774 | 0.004105496 | 0.24032965 | 0.014610602 | 0.000372526 | 0.004105435 | 0.24030598 | -9.76E-05 | 0.004062189 | 1 | 0.00689643 | 1 | 0.006267988 | ENSG0000015 | 13 | RASGRP3 |
| SAT | HTN | ENSG0000016 | 0.020202921 | 0.002375352 | 0.006648322 | 0.240331259 | 0.020100828 | 0.002498579 | 0.006648134 | 0.240307717 | 0.000102093 | 0.016988116 | 1 | 0.02652693 | 1 | 0.027695394 | ENSG0000015 | 13 | RASGRP3 |
| SAT | HTN | ENSG0000016 | -0.00121071 | 0.657936615 | 0.002734446 | 0.240335717 | -0.00230091 | 0.405770124 | 0.002767643 | 0.240311885 | 0.001090196 | 0.010904111 | 1 | 0.852573121 | 1 | 0.683130653 | ENSG0000015 | 13 | RASGRP3 |
| SAT | HTN | ENSG0000016 | -0.00153916 | 0.553770565 | 0.002599409 | 0.24033564 | -0.00149831 | 0.564335894 | 0.002599362 | 0.240312062 | -4.09E-05 | 0.045293019 | 1 | 0.79549966 | 1 | 0.803251455 | ENSG0000015 | 13 | RASGRP3 |
| SAT | HTN | ENSG0000016 | -0.00604255 | 0.010372334 | 0.002357449 | 0.240332573 | -0.00607605 | 0.009953665 | 0.002357393 | 0.240308949 | 3.35E-05 | 0.045491311 | 1 | 0.077784342 | 1 | 0.076584716 | ENSG0000015 | 13 | RASGRP3 |
| SAT | HTN | ENSG0000016 | 0.005934216 | 0.385587075 | 0.006839422 | 0.240335442 | 0.005775967 | 0.398388912 | 0.006839489 | 0.240311874 | 0.000158249 | 0.032839239 | 1 | 0.669725354 | 1 | 0.678667716 | ENSG0000015 | 7 | GPR180 |
| SAT | HTN | ENSG0000016 | 0.002044439 | 0.720499893 | 0.005714059 | 0.24033575 | 0.001941262 | 0.734053472 | 0.005713986 | 0.240312168 | 0.000103176 | 0.034464527 | 1 | 0.885985829 | 1 | 0.892066631 | ENSG0000015 | 7 | GPR180 |
| SAT | HTN | ENSG0000016 | -0.00206565 | 0.752869286 | 0.006560578 | 0.240335764 | -0.00193074 | 0.768530005 | 0.006560473 | 0.240312183 | -0.00013492 | 0.011475747 | 1 | 0.900432066 | 1 | 0.908993132 | ENSG0000015 | 7 | GPR180 |
| SAT | HTN | ENSG0000016 | -0.00333886 | 0.156623457 | 0.002357081 | 0.240334824 | -0.00290319 | 0.219337594 | 0.002363596 | 0.240311481 | -0.00043567 | 0.013791021 | 1 | 0.425751089 | 1 | 0.509427202 | ENSG0000015 | 7 | GPR180 |
| SAT | HTN | ENSG0000016 | 0.001333673 | 0.749584114 | 0.004178332 | 0.240335763 | 0.001049454 | 0.801746634 | 0.004179638 | 0.240312194 | 0.000284219 | 0.011440342 | 1 | 0.899241283 | 1 | 0.923833778 | ENSG0000015 | 7 | GPR180 |
| SAT | HTN | ENSG0000016 | 0.023878767 | 0.000731681 | 0.007070066 | 0.240330188 | 0.024064028 | 0.00066502 | 0.007070106 | 0.240306512 | -0.00018526 | 0.012354556 | 1 | 0.010622319 | 1 | 0.009917863 | ENSG0000015 | 7 | GPR180 |
| SAT | HTN | ENSG0000016 | 0.007109728 | 0.002888918 | 0.00238635 | 0.240331436 | 0.007155712 | 0.002712043 | 0.002386322 | 0.240307791 | -4.60E-05 | 0.0262235 | 1 | 0.030282878 | 1 | 0.028984413 | ENSG0000015 | 7 | IFIT5 |
| SAT | HTN | ENSG0000016 | 6.42E-05 | 0.981195796 | 0.002723406 | 0.240335813 | 0.00011905 | 0.965131773 | 0.002723342 | 0.240312224 | -5.49E-05 | 0.004906177 | 1 | 0.993612162 | 1 | 0.988156741 | ENSG0000015 | 11 | PRDM8 |
| SAT | HTN | ENSG0000016 | 0.008922424 | 0.119075841 | 0.005724394 | 0.240334615 | 0.009035527 | 0.114464553 | 0.005724329 | 0.240310997 | -0.0001131 | 0.023168684 | 1 | 0.366076286 | 1 | 0.358450758 | ENSG0000015 | 11 | PRDM8 |
| SAT | HTN | ENSG0000016 | 0.00653602 | 0.217330285 | 0.005298073 | 0.240335063 | 0.007982753 | 0.132615193 | 0.005308136 | 0.24031111 | -0.00144673 | 1.23E-05 | 1 | 0.509681988 | 1 | 0.386764842 | ENSG0000015 | 10 | ZNF117 |
| SAT | HTN | ENSG0000016 | -2.18E-05 | 0.997009858 | 0.005809287 | 0.240335813 | -0.00014787 | 0.97969272 | 0.005809165 | 0.240312225 | 0.000126097 | 0.003788634 | 1 | 0.998836441 | 1 | 0.992702845 | ENSG0000015 | 14 | RAD17 |
| SAT | HTN | ENSG0000016 | 0.001382316 | 0.822306771 | 0.00615514 | 0.240335789 | 0.001603014 | 0.794534024 | 0.006155313 | 0.240312192 | -0.0002207 | 0.003912839 | 1 | 0.93323712 | 1 | 0.922042681 | ENSG0000015 | 14 | RAD17 |
| SAT | HTN | ENSG0000016 | 0.002911188 | 0.658200506 | 0.006580453 | 0.240335717 | 0.002755644 | 0.675384837 | 0.006580337 | 0.240312139 | 0.000155544 | 0.002868488 | 1 | 0.852573121 | 1 | 0.864587456 | ENSG0000015 | 14 | RAD17 |
| SAT | HTN | ENSG0000016 | 0.002757947 | 0.110078907 | 0.001726046 | 0.240334554 | 0.001800269 | 0.303977221 | 0.001751332 | 0.240311704 | 0.000957678 | 0.001262215 | 1 | 0.348282442 | 1 | 0.596144208 | ENSG0000015 | 9 | GPR125 |
| SAT | HTN | ENSG0000016 | 0.002476842 | 0.21497655 | 0.001997455 | 0.240335055 | 0.002415856 | 0.226480383 | 0.001997439 | 0.240311504 | 6.10E-05 | 0.000750024 | 1 | 0.507575983 | 1 | 0.517461433 | ENSG0000015 | 5 | CENPH |
| SAT | HTN | ENSG0000016 | 0.010281047 | 0.040480518 | 0.00501804 | 0.240333743 | 0.006843354 | 0.181847219 | 0.005125759 | 0.240311346 | 0.003437693 | 0.001019304 | 1 | 0.188077921 | 1 | 0.461796228 | ENSG0000015 | 5 | CENPH |
| SAT | HTN | ENSG0000016 | -0.00739019 | 0.289323768 | 0.006974458 | 0.24033526 | -0.0012492 | 0.862708448 | 0.00722381 | 0.240312211 | -0.00614099 | 0.001107909 | 1 | 0.581565115 | 1 | 0.952725149 | ENSG0000015 | 6 | CARHSP1 |
| SAT | HTN | ENSG0000016 | -0.00034898 | 0.896296 | 0.002677433 | 0.240335805 | -0.00019625 | 0.941574026 | 0.002677719 | 0.240312223 | -0.00015273 | 0.001228349 | 1 | 0.961971599 | 1 | 0.9791773 | ENSG0000015 | 6 | CARHSP1 |
| SAT | HTN | ENSG0000016 | 0.009925984 | 0.141865735 | 0.006757514 | 0.240334749 | 0.009834675 | 0.14555736 | 0.006757338 | 0.240311181 | 9.13E-05 | 0.046214573 | 1 | 0.39809709 | 1 | 0.406791907 | ENSG0000015 | 8 | SYCP2L |
| SAT | HTN | ENSG0000016 | 0.001280693 | 0.784369945 | 0.004680403 | 0.240335776 | 0.001361039 | 0.771205636 | 0.004680353 | 0.240312184 | -8.03E-05 | 0.049972045 | 1 | 0.916412446 | 1 | 0.91001726 | ENSG0000015 | 11 | RANBP2 |
| SAT | HTN | ENSG0000016 | -0.01508431 | 0.014305902 | 0.006158123 | 0.240332854 | -0.02253689 | 0.000402906 | 0.006369596 | 0.240306052 | 0.007452579 | 4.76E-06 | 1 | 0.098401814 | 1 | 0.00660968 | ENSG0000015 | 12 | MERTK |
| SAT | HTN | ENSG0000016 | -0.00281945 | 0.682088873 | 0.006883167 | 0.240335731 | -0.00267665 | 0.697368268 | 0.006883058 | 0.240312151 | -0.00014279 | 0.01083944 | 1 | 0.865403555 | 1 | 0.875982116 | ENSG0000015 | 7 | PLA2R1 |
| SAT | HTN | ENSG0000016 | 0.010411117 | 0.003745229 | 0.003591436 | 0.240331669 | 0.010356987 | 0.003928514 | 0.003591363 | 0.240308124 | 5.41E-05 | 0.047198652 | 1 | 0.036790708 | 1 | 0.038361473 | ENSG0000015 | 7 | PLA2R1 |
| SAT | HTN | ENSG0000016 | -0.01384207 | 0.015161731 | 0.005699839 | 0.240332905 | -0.01394062 | 0.014451455 | 0.005699702 | 0.240309275 | 9.85E-05 | 0.014418942 | 1 | 0.101937783 | 1 | 0.099073573 | ENSG0000015 | 7 | PLA2R1 |
| SAT | HTN | ENSG0000016 | -0.02053979 | 3.56E-09 | 0.003479268 | 0.240318626 | -0.02058796 | 3.27E-09 | 0.00347917 | 0.240294958 | 4.82E-05 | 0.032999576 | 1.17E-05 | 3.83E-07 | 1.07E-05 | 3.83E-07 | ENSG0000015 | 11 | SLC25A27 |
| SAT | HTN | ENSG0000016 | 0.004915038 | 0.000794335 | 0.001465084 | 0.240330263 | 0.004106623 | 0.006290732 | 0.001503024 | 0.240308544 | 0.000808415 | 0.01608431 | 1 | 0.011331359 | 1 | 0.054602888 | ENSG0000015 | 9 | NMRAL1 |
| SAT | HTN | ENSG0000016 | 0.000460617 | 0.883026948 | 0.003130612 | 0.240335803 | 0.000605183 | 0.846746146 | 0.003131253 | 0.240312207 | -0.00014457 | 0.040382344 | 1 | 0.957468 | 1 | 0.946566986 | ENSG0000015 | 9 | NMRAL1 |
| SAT | HTN | ENSG0000016 | -0.00641841 | 0.13493413 | 0.004293462 | 0.240334711 | -0.00635104 | 0.139069071 | 0.004293364 | 0.240311146 | -6.74E-05 | 0.030049926 | 1 | 0.386992028 | 1 | 0.394186122 | ENSG0000015 | 9 | NMRAL1 |
| SAT | HTN | ENSG0000017 | 0.007053393 | 0.225315805 | 0.005817158 | 0.240335088 | 0.007154732 | 0.218711845 | 0.005817034 | 0.240311479 | -0.00010134 | 0.01925266 | 1 | 0.516604581 | 1 | 0.509292807 | ENSG0000015 | 11 | C16orf89 |
| SAT | HTN | ENSG0000017 | -0.00226954 | 0.485155848 | 0.003251323 | 0.240335573 | -0.00217005 | 0.504498907 | 0.003251361 | 0.240312006 | -9.95E-05 | 0.005455818 | 1 | 0.748376275 | 1 | 0.758328059 | ENSG0000015 | 8 | ADPRHL1 |
| SAT | HTN | ENSG0000017 | 0.008012193 | 0.032332016 | 0.003743512 | 0.240333554 | 0.007948973 | 0.033717122 | 0.003743433 | 0.240310002 | 6.32E-05 | 0.024067799 | 1 | 0.1664956 | 1 | 0.170718945 | ENSG0000015 | 5 | LURAP1L |
| SAT | HTN | ENSG0000017 | -0.00795093 | 0.224828075 | 0.006550494 | 0.240335087 | -0.00760105 | 0.245930186 | 0.006550978 | 0.240311561 | -0.00034988 | 0.000652704 | 1 | 0.51644005 | 1 | 0.536079363 | ENSG0000015 | 5 | LURAP1L |
| SAT | HTN | ENSG0000017 | -0.00463247 | 0.437028721 | 0.005960297 | 0.240335515 | -0.00474171 | 0.426287374 | 0.005960201 | 0.240311913 | 0.000109238 | 0.02436939 | 1 | 0.710902942 | 1 | 0.699305573 | ENSG0000015 | 5 | LURAP1L |
| SAT | HTN | ENSG0000017 | 0.017460223 | 6.48E-05 | 0.004370858 | 0.240327943 | 0.01733567 | 7.30E-05 | 0.004370792 | 0.240304468 | 0.000124552 | 0.000526317 | 0.212571166 | 0.001574601 | 0.239584078 | 0.001774697 | ENSG0000015 | 5 | LURAP1L |
| SAT | HTN | ENSG0000017 | 0.01006824 | 0.113091921 | 0.006354405 | 0.240334575 | 0.009980769 | 0.116246326 | 0.006354229 | 0.240311009 | 8.75E-05 | 0.034520165 | 1 | 0.355076165 | 1 | 0.362072419 | ENSG0000015 | 5 | LURAP1L |
| SAT | HTN | ENSG0000017 | 0.004057021 | 0.173875342 | 0.002983414 | 0.240334901 | 0.004006202 | 0.179319863 | 0.002983348 | 0.240311336 | 5.08E-05 | 0.020387207 | 1 | 0.449200786 | 1 | 0.458229807 | ENSG0000015 | 4 | CFDP1 |
| SAT | HTN | ENSG0000017 | -0.00020637 | 0.899976452 | 0.001641902 | 0.240335806 | -0.00017604 | 0.914615842 | 0.001641863 | 0.24031222 | -3.03E-05 | 0.009725393 | 1 | 0.963714993 | 1 | 0.972383274 | ENSG0000015 | 7 | JAZF1 |
| SAT | HTN | ENSG0000017 | 0.005648262 | 0.388121638 | 0.006544692 | 0.240335446 | 0.005463863 | 0.403799437 | 0.00654468 | 0.240311882 | 0.000184399 | 0.003769443 | 1 | 0.672703167 | 1 | 0.682217278 | ENSG0000015 | 7 | JAZF1 |
| SAT | HTN | ENSG0000017 | 0.00311272 | 0.033917814 | 0.001467535 | 0.240333595 | 0.003416288 | 0.020048434 | 0.001469088 | 0.240309559 | -0.00030357 | 1.12E-05 | 1 | 0.170681517 | 1 | 0.121363303 | ENSG0000015 | 7 | JAZF1 |
| SAT | HTN | ENSG0000017 | 0.014522648 | 0.018542064 | 0.006167726 | 0.240333079 | -0.00389654 | 0.645408689 | 0.008467984 | 0.240312121 | 0.018419192 | 0.001501758 | 1 | 0.11478587 | 1 | 0.847979199 | ENSG0000015 | 9 | KCNJ16 |
| SAT | HTN | ENSG0000017 | 0.012305424 | 0.003611276 | 0.004228269 | 0.240331636 | -0.00717485 | 0.389672448 | 0.008340788 | 0.24031186 | 0.019480272 | 0.006739185 | 1 | 0.035796367 | 1 | 0.670742545 | ENSG0000015 | 9 | KCNJ16 |
| SAT | HTN | ENSG0000017 | -0.00751766 | 0.029436959 | 0.003452284 | 0.240333475 | -0.0037244 | 0.309732324 | 0.003666522 | 0.240311717 | -0.00379327 | 0.002137077 | 1 | 0.157300752 | 1 | 0.602376784 | ENSG0000015 | 5 | C6orf57 |
| SAT | HTN | ENSG0000017 | -0.01593444 | 0.002389868 | 0.005246833 | 0.240331265 | -0.01618065 | 0.002043549 | 0.005246905 | 0.240307535 | 0.000246205 | 2.98E-05 | 1 | 0.026580189 | 1 | 0.023608743 | ENSG0000015 | 10 | ROBO3 |
| SAT | HTN | ENSG0000017 | -0.00133379 | 0.81249953 | 0.005623007 | 0.240335786 | -0.00051181 | 0.927513927 | 0.005625963 | 0.240312221 | -0.00082198 | 1.62E-05 | 1 | 0.928207158 | 1 | 0.975149159 | ENSG0000015 | 10 | CC2D1B |
| SAT | HTN | ENSG0000017 | -0.00666549 | 0.046955705 | 0.003355047 | 0.240333867 | -0.00687727 | 0.040404068 | 0.003355422 | 0.240310154 | 0.000211784 | 0.000438195 | 1 | 0.205689811 | 1 | 0.18777018 | ENSG0000015 | 10 | CC2D1B |
| SAT | HTN | ENSG0000017 | 0.00903478 | 0.084109268 | 0.005230492 | 0.240334342 | 0.008918972 | 0.088152262 | 0.005230364 | 0.240310791 | 0.000115808 | 0.001573014 | 1 | 0.294571399 | 1 | 0.301592879 | ENSG0000015 | 10 | CC2D1B |
| SAT | HTN | ENSG0000017 | 0.00140228 | 0.522067682 | 0.002190508 | 0.240335611 | 0.001432367 | 0.513165962 | 0.002190444 | 0.240312015 | -3.01E-05 | 0.028600918 | 1 | 0.771578407 | 1 | 0.76520719 | ENSG0000015 | 12 | CEP112 |
| SAT | HTN | ENSG0000017 | -0.02320981 | 3.33E-09 | 0.00392405 | 0.24031856 | -0.0233707 | 2.59E-09 | 0.00392423 | 0.240294735 | 0.000160894 | 0.003002944 | 1.09E-05 | 3.76E-07 | 8.51E-06 | 3.41E-07 | ENSG0000015 | 12 | CEP112 |
| SAT | HTN | ENSG0000017 | -0.00161599 | 0.614494306 | 0.003208444 | 0.240335688 | -0.00337981 | 0.296077721 | 0.003234633 | 0.240311687 | 0.001763818 | 1.86E-05 | 1 | 0.832284161 | 1 | 0.586298316 | ENSG0000015 | 12 | CEP112 |

| | | | | | | | | | | | | | | | | |
|---|---|---|---|---|---|---|---|---|---|---|---|---|---|---|---|---|
| SAT | HTN | ENSG0000017 | -0.02702284 | 2.85E-05 | 0.006456298 | 0.240327174 | -0.02690375 | 3.09E-05 | 0.006456219 | 0.240303662 | -0.00011909 | 0.031252231 | 0.093373483 | 0.000819066 | 0.101233222 | 0.00084361 | ENSG0000015 | 12 | CEP112 |
| SAT | HTN | ENSG0000017 | -0.0055849 | 0.080354943 | 0.003193863 | 0.240334305 | -0.0053973 | 0.091102212 | 0.003194395 | 0.240310818 | -0.0001876 | 0.004636537 | 1 | 0.287194519 | 1 | 0.309106886 | ENSG0000015 | 12 | CEP112 |
| SAT | HTN | ENSG0000017 | 9.77E-05 | 0.939889305 | 0.001295785 | 0.240335811 | 0.000122976 | 0.924388954 | 0.001295752 | 0.240312221 | -2.53E-05 | 0.00471477 | 1 | 0.978294984 | 1 | 0.974588739 | ENSG0000015 | 13 | ABCA10 |
| SAT | HTN | ENSG0000017 | 0.005420809 | 0.120998393 | 0.003495945 | 0.240334628 | 0.010312241 | 0.004814434 | 0.003657851 | 0.240308306 | -0.00489143 | 5.56E-06 | 1 | 0.369642206 | 1 | 0.044371228 | ENSG0000015 | 11 | ABCA5 |
| SAT | HTN | ENSG0000017 | -0.00020262 | 0.956016854 | 0.003673868 | 0.240335812 | -0.00012242 | 0.973417275 | 0.003673807 | 0.240312225 | -8.02E-05 | 0.006740002 | 1 | 0.983048498 | 1 | 0.990013044 | ENSG0000015 | 11 | ABCA5 |
| SAT | HTN | ENSG0000017 | 0.009060645 | 0.062579707 | 0.004865637 | 0.240334103 | 0.009187759 | 0.05898317 | 0.004865565 | 0.240310467 | -0.00012711 | 0.001618117 | 1 | 0.251314587 | 1 | 0.243426139 | ENSG0000015 | 11 | ABCA5 |
| SAT | HTN | ENSG0000017 | -0.00842289 | 0.082883956 | 0.004856944 | 0.24033433 | -0.01145407 | 0.020663183 | 0.004949706 | 0.240309585 | 0.003031173 | 0.001504269 | 1 | 0.293674147 | 1 | 0.124168322 | ENSG0000015 | 7 | DISP1 |
| SAT | HTN | ENSG0000017 | 0.002460508 | 0.68455287 | 0.00605651 | 0.240335732 | 0.002820025 | 0.641518656 | 0.006057056 | 0.240312119 | -0.00035952 | 0.000375048 | 1 | 0.866100648 | 1 | 0.845498554 | ENSG0000015 | 10 | TDH |
| SAT | HTN | ENSG0000017 | -0.00072316 | 0.877572219 | 0.004694367 | 0.240335802 | -0.00065021 | 0.88983504 | 0.004694244 | 0.240312216 | -7.29E-05 | 0.021204485 | 1 | 0.957442803 | 1 | 0.960935133 | ENSG0000015 | 5 | ATP5G3 |
| SAT | HTN | ENSG0000017 | 0.023842265 | 0.000190109 | 0.006388878 | 0.240328945 | 0.023940506 | 0.000178765 | 0.006388725 | 0.2403053 | -9.82E-05 | 0.030498067 | 0.623746086 | 0.003780279 | 0.586527662 | 0.003619977 | ENSG0000015 | 5 | ATP5G3 |
| SAT | HTN | ENSG0000017 | -0.00413751 | 0.204524101 | 0.003261041 | 0.240335019 | -0.00477086 | 0.145336355 | 0.003276224 | 0.24031118 | 0.000633348 | 0.045524636 | 1 | 0.493414393 | 1 | 0.40652053 | ENSG0000015 | 5 | ATP5G3 |
| SAT | HTN | ENSG0000017 | 0.002386153 | 0.611831234 | 0.004702136 | 0.240335686 | 0.002916869 | 0.535411147 | 0.004706393 | 0.240312036 | -0.00053072 | 0.009805137 | 1 | 0.830870282 | 1 | 0.782451496 | ENSG0000015 | 11 | RABGEF1 |
| SAT | HTN | ENSG0000017 | 0.001131402 | 0.655557351 | 0.00253646 | 0.240335715 | 0.002991646 | 0.256123894 | 0.002634415 | 0.240311589 | -0.00186024 | 0.008997723 | 1 | 0.850824236 | 1 | 0.548573946 | ENSG0000015 | 11 | RABGEF1 |
| SAT | HTN | ENSG0000017 | -0.00558225 | 0.30636778 | 0.005457413 | 0.240335297 | -0.00548622 | 0.314751325 | 0.005457279 | 0.240311727 | -9.60E-05 | 0.011834762 | 1 | 0.601393141 | 1 | 0.604641264 | ENSG0000015 | 11 | RABGEF1 |
| SAT | HTN | ENSG0000017 | -0.00351987 | 0.220740801 | 0.002874392 | 0.240335074 | -0.00359289 | 0.211313119 | 0.002874391 | 0.240311455 | 7.30E-05 | 0.01007029 | 1 | 0.51391887 | 1 | 0.50095256 | ENSG0000015 | 11 | RABGEF1 |
| SAT | HTN | ENSG0000017 | -0.00320243 | 0.447812798 | 0.004218906 | 0.240335529 | -0.01425261 | 0.011306139 | 0.005626546 | 0.240309061 | 0.011050178 | 0.002996709 | 1 | 0.720446806 | 1 | 0.083456744 | ENSG0000015 | 9 | MRPL39 |
| SAT | HTN | ENSG0000017 | -0.01408143 | 0.037413877 | 0.006765935 | 0.240333677 | -0.01421733 | 0.035610713 | 0.006765803 | 0.240310048 | 0.000135896 | 0.009230929 | 1 | 0.178682575 | 1 | 0.175170538 | ENSG0000015 | 9 | MRPL39 |
| SAT | HTN | ENSG0000017 | 0.010515428 | 0.000243919 | 0.002866341 | 0.240329176 | 0.010463225 | 0.000261805 | 0.002866281 | 0.240305654 | 5.22E-05 | 0.014707737 | 0.800297117 | 0.004573126 | 0.858981653 | 0.004769464 | ENSG0000015 | 9 | MRPL39 |
| SAT | HTN | ENSG0000017 | 0.003764961 | 0.504954401 | 0.00564703 | 0.240335594 | 0.003850094 | 0.495360297 | 0.005646884 | 0.240311996 | -8.51E-05 | 0.026742144 | 1 | 0.762947945 | 1 | 0.754680724 | ENSG0000015 | 13 | TSEN2 |
| SAT | HTN | ENSG0000018 | 0.003850685 | 0.370488347 | 0.004299751 | 0.240335418 | 0.003734258 | 0.385122289 | 0.004299674 | 0.240311853 | 0.000116427 | 0.000589748 | 1 | 0.6577772 | 1 | 0.668210592 | ENSG0000015 | 13 | TSEN2 |
| SAT | HTN | ENSG0000018 | 0.019970138 | 0.000126722 | 0.005210361 | 0.240328568 | 0.01984662 | 0.000139471 | 0.005210269 | 0.24030507 | 0.000123518 | 0.002742251 | 0.415774597 | 0.002809288 | 0.457603234 | 0.002990871 | ENSG0000015 | 9 | SLFN13 |
| SAT | HTN | ENSG0000018 | -0.01028192 | 0.057548947 | 0.005414036 | 0.240334035 | -0.01019966 | 0.05956959 | 0.005413918 | 0.240310475 | -8.23E-05 | 0.039165575 | 1 | 0.238708085 | 1 | 0.245399526 | ENSG0000015 | 4 | C17orf50 |
| SAT | HTN | ENSG0000018 | 0.000475335 | 0.903667001 | 0.003927401 | 0.240335806 | -0.00278667 | 0.5001712 | 0.004133177 | 0.240312001 | 0.003262009 | 0.011352423 | 1 | 0.965816745 | 1 | 0.757052571 | ENSG0000015 | 8 | FLCN |
| SAT | HTN | ENSG0000018 | 0.000710357 | 0.913591382 | 0.006546484 | 0.240335808 | 0.00057955 | 0.929455403 | 0.006546363 | 0.240312222 | 0.000130807 | 0.010698025 | 1 | 0.969071845 | 1 | 0.975149159 | ENSG0000015 | 8 | FLCN |
| SAT | HTN | ENSG0000018 | 0.010252302 | 0.057524319 | 0.005397909 | 0.240334034 | 0.01040243 | 0.053974151 | 0.005398115 | 0.240310394 | -0.00015013 | 0.035364169 | 1 | 0.238708085 | 1 | 0.22939014 | ENSG0000015 | 8 | FLCN |
| SAT | HTN | ENSG0000018 | 0.00398885 | 0.224662463 | 0.0032851 | 0.240335086 | 0.003911511 | 0.233766741 | 0.003285015 | 0.240311526 | 7.73E-05 | 0.000522072 | 1 | 0.51644005 | 1 | 0.522471851 | ENSG0000015 | 8 | FLCN |
| SAT | HTN | ENSG0000018 | -0.00060709 | 0.865617113 | 0.003587429 | 0.240335799 | -0.00172371 | 0.634202975 | 0.003622621 | 0.240312114 | 0.00111662 | 0.027021556 | 1 | 0.952485811 | 1 | 0.841755648 | ENSG0000015 | 8 | APCDD1 |
| SAT | HTN | ENSG0000018 | 0.010837473 | 0.08307772 | 0.006253214 | 0.240334332 | 0.011549694 | 0.06511646 | 0.006261823 | 0.240310548 | -0.00071222 | 0.033023175 | 1 | 0.293726291 | 1 | 0.260227901 | ENSG0000015 | 7 | PIEZO2 |
| SAT | HTN | ENSG0000018 | 0.011510396 | 0.000824124 | 0.003441504 | 0.240330296 | 0.010930603 | 0.001548288 | 0.003453047 | 0.240307284 | 0.000579793 | 0.041280238 | 1 | 0.011654964 | 1 | 0.018884508 | ENSG0000015 | 7 | PIEZO2 |
| SAT | HTN | ENSG0000018 | 0.010766084 | 0.111585677 | 0.006766402 | 0.240334565 | 0.01063684 | 0.115943457 | 0.006766294 | 0.240311007 | 0.000129244 | 0.018666839 | 1 | 0.352031353 | 1 | 0.361606923 | ENSG0000015 | 7 | PIEZO2 |
| SAT | HTN | ENSG0000018 | -0.00064181 | 0.683130136 | 0.001572318 | 0.240335731 | -0.00068185 | 0.664530631 | 0.001572292 | 0.240312133 | 4.00E-05 | 0.001529201 | 1 | 0.865648254 | 1 | 0.86030249 | ENSG0000015 | 12 | MPPE1 |
| SAT | HTN | ENSG0000018 | -0.00413631 | 0.099735145 | 0.002512728 | 0.240334477 | -0.00409559 | 0.103108602 | 0.002512678 | 0.240310915 | -4.07E-05 | 0.03469501 | 1 | 0.325460864 | 1 | 0.33261627 | ENSG0000015 | 13 | CYP2U1 |
| SAT | HTN | ENSG0000018 | -0.01650057 | 0.003432699 | 0.005639123 | 0.240331591 | -0.01546564 | 0.006209685 | 0.005651592 | 0.240308532 | -0.00103493 | 0.006354059 | 1 | 0.034345697 | 1 | 0.054042377 | ENSG0000015 | 11 | AK9 |
| SAT | HTN | ENSG0000018 | -0.00373106 | 0.565094351 | 0.006485507 | 0.24033565 | -0.0038743 | 0.550247272 | 0.006485374 | 0.240312049 | 0.00014324 | 0.003487958 | 1 | 0.801993972 | 1 | 0.793216739 | ENSG0000015 | 11 | AK9 |
| SAT | HTN | ENSG0000018 | 0.003449726 | 0.025881345 | 0.001548355 | 0.240333365 | 0.003479254 | 0.024634352 | 0.001548334 | 0.240309735 | -2.95E-05 | 0.023695583 | 1 | 0.144416143 | 1 | 0.138583634 | ENSG0000015 | 11 | AK9 |
| SAT | HTN | ENSG0000018 | -0.00395462 | 0.45347119 | 0.005275349 | 0.240335536 | -0.00406122 | 0.441389417 | 0.005275344 | 0.240311933 | 0.000106603 | 0.039276652 | 1 | 0.726359177 | 1 | 0.713749964 | ENSG0000015 | 11 | AK9 |
| SAT | HTN | ENSG0000018 | 0.002404878 | 0.329301256 | 0.002465218 | 0.240335344 | 0.002368277 | 0.336701743 | 0.002465148 | 0.24031177 | 3.66E-05 | 0.020990764 | 1 | 0.62016399 | 1 | 0.620977189 | ENSG0000015 | 11 | AK9 |
| SAT | HTN | ENSG0000018 | -0.01334054 | 0.000964788 | 0.004041831 | 0.240330441 | -0.01343097 | 0.000890578 | 0.004041806 | 0.24030678 | 9.04E-05 | 0.015927738 | 1 | 0.013134726 | 1 | 0.012122569 | ENSG0000015 | 6 | GTF3C6 |
| SAT | HTN | ENSG0000018 | -0.00142779 | 0.457689112 | 0.001922538 | 0.240335541 | -0.00138337 | 0.471789494 | 0.001922497 | 0.24031197 | -4.44E-05 | 0.001848641 | 1 | 0.730033046 | 1 | 0.738169447 | ENSG0000015 | 7 | AGPAT5 |
| SAT | HTN | ENSG0000018 | 0.006112771 | 0.317721334 | 0.006117956 | 0.240335321 | 0.006209765 | 0.310092773 | 0.006117823 | 0.240311717 | -9.70E-05 | 0.032409401 | 1 | 0.610330034 | 1 | 0.602376784 | ENSG0000015 | 8 | HEATR3 |
| SAT | HTN | ENSG0000018 | 0.018018177 | 6.70E-07 | 0.003625383 | 0.240323632 | 0.017946603 | 7.41E-07 | 0.003625314 | 0.240300141 | 7.16E-05 | 0.010337784 | 0.002197402 | 3.43E-05 | 0.002431272 | 3.63E-05 | ENSG0000015 | 8 | HEATR3 |
| SAT | HTN | ENSG0000018 | 0.003962589 | 0.190961453 | 0.0030301 | 0.24033497 | 0.004011897 | 0.185489286 | 0.003030034 | 0.240311361 | -4.93E-05 | 0.028312402 | 1 | 0.47377744 | 1 | 0.466710388 | ENSG0000015 | 8 | HEATR3 |
| SAT | HTN | ENSG0000018 | -0.00386853 | 0.341181657 | 0.00406429 | 0.240335367 | -0.00631578 | 0.126649989 | 0.004134851 | 0.240311075 | 0.002447246 | 0.001314449 | 1 | 0.629532484 | 1 | 0.380181715 | ENSG0000015 | 7 | SETD9 |
| SAT | HTN | ENSG0000018 | -0.00998181 | 0.000364449 | 0.002800263 | 0.240329547 | -0.00993423 | 0.000388661 | 0.002800197 | 0.240306019 | -4.76E-05 | 0.017504874 | 1 | 0.006327001 | 1 | 0.006408027 | ENSG0000015 | 7 | SETD9 |
| SAT | HTN | ENSG0000018 | -0.00569662 | 0.399401979 | 0.006760041 | 0.240335463 | -0.00583391 | 0.38813128 | 0.00675994 | 0.240311858 | 0.000137287 | 0.014073747 | 1 | 0.681058492 | 1 | 0.670742545 | ENSG0000015 | 7 | SETD9 |
| SAT | HTN | ENSG0000018 | -0.01199592 | 0.016751603 | 0.00501476 | 0.240332991 | -0.01137439 | 0.023425543 | 0.005018638 | 0.240309692 | -0.00062154 | 0.002237527 | 1 | 0.109125177 | 1 | 0.134604566 | ENSG0000015 | 7 | SETD9 |
| SAT | HTN | ENSG0000018 | 0.001536101 | 0.69717462 | 0.003947451 | 0.240335739 | 0.00147314 | 0.709003123 | 0.003947368 | 0.240312157 | 6.30E-05 | 0.032750082 | 1 | 0.873398216 | 1 | 0.880526418 | ENSG0000015 | 7 | SETD9 |
| SAT | HTN | ENSG0000018 | -0.00039704 | 0.94141522 | 0.00540248 | 0.240335811 | -0.00050562 | 0.925433502 | 0.005402433 | 0.240312221 | 0.000108588 | 0.025377171 | 1 | 0.978480835 | 1 | 0.975038588 | ENSG0000015 | 7 | SETD9 |
| SAT | HTN | ENSG0000018 | 0.002585399 | 0.391384871 | 0.003016417 | 0.240335451 | 0.002539823 | 0.399776869 | 0.003016343 | 0.240311876 | 4.56E-05 | 0.030867725 | 1 | 0.675184407 | 1 | 0.679972995 | ENSG0000015 | 7 | SETD9 |
| SAT | HTN | ENSG0000018 | -0.00287318 | 0.646471471 | 0.006264147 | 0.24033571 | -0.00268276 | 0.66844955 | 0.006264067 | 0.240312135 | -0.00019042 | 0.000357368 | 1 | 0.845722845 | 1 | 0.86204999 | ENSG0000015 | 7 | SETD9 |
| SAT | HTN | ENSG0000018 | -0.00226427 | 0.536646144 | 0.003664502 | 0.240335625 | -0.00148951 | 0.685400031 | 0.003676836 | 0.240312144 | -0.00077476 | 0.010581463 | 1 | 0.783266351 | 1 | 0.86859695 | ENSG0000015 | 11 | ZKSCAN2 |
| SAT | HTN | ENSG0000018 | -0.00326126 | 0.069396477 | 0.001796012 | 0.240334187 | -0.00329425 | 0.066618177 | 0.00179597 | 0.240310566 | 3.30E-05 | 0.010111504 | 1 | 0.266928303 | 1 | 0.262480731 | ENSG0000015 | 12 | TMBIM4 |
| SAT | HTN | ENSG0000018 | -0.0177351 | 0.000348261 | 0.00495879 | 0.240329505 | -0.01428297 | 0.006555713 | 0.005253756 | 0.240308581 | -0.00345214 | 0.046779369 | 1 | 0.00614325 | 1 | 0.056160037 | ENSG0000015 | 12 | TMBIM4 |
| SAT | HTN | ENSG0000018 | 0.006970309 | 0.074461567 | 0.003907626 | 0.240334244 | 0.007051959 | 0.071123358 | 0.003907567 | 0.240310619 | -8.16E-05 | 0.011253359 | 1 | 0.276027247 | 1 | 0.271659766 | ENSG0000015 | 12 | TMBIM4 |
| SAT | HTN | ENSG0000018 | 0.001253167 | 0.831241264 | 0.005880379 | 0.240335791 | 0.001153906 | 0.844427039 | 0.005880254 | 0.240312206 | 9.93E-05 | 0.023847583 | 1 | 0.938268032 | 1 | 0.944940352 | ENSG0000015 | 13 | TTC18 |
| SAT | HTN | ENSG0000018 | 0.000616753 | 0.586640861 | 0.00113434 | 0.240335668 | 0.001310847 | 0.266349909 | 0.00117934 | 0.240311616 | -0.00069409 | 0.031574131 | 1 | 0.813855672 | 1 | 0.55733039 | ENSG0000015 | 13 | TTC18 |
| SAT | HTN | ENSG0000018 | -0.00603527 | 0.364345986 | 0.006653246 | 0.240335408 | -0.00587388 | 0.377306419 | 0.006653157 | 0.240311841 | -0.00016139 | 0.004046684 | 1 | 0.651454594 | 1 | 0.66057124 | ENSG0000015 | 13 | TTC18 |
| SAT | HTN | ENSG0000018 | 0.003335238 | 0.144765289 | 0.002287111 | 0.240334765 | 0.000433289 | 0.869848016 | 0.0026444 | 0.240312212 | 0.002901949 | 0.02882313 | 1 | 0.403890231 | 1 | 0.955333968 | ENSG0000015 | 8 | UGT2B4 |
| SAT | HTN | ENSG0000018 | 0.004125007 | 0.129609317 | 0.002721615 | 0.24033468 | 0.004173821 | 0.125125171 | 0.002721558 | 0.240311066 | -4.88E-05 | 0.017193529 | 1 | 0.382482133 | 1 | 0.376984101 | ENSG0000015 | 8 | UGT2B4 |
| SAT | HTN | ENSG0000018 | -0.00313869 | 0.059849873 | 0.001667827 | 0.240334067 | -0.00367265 | 0.029115064 | 0.00168321 | 0.240309878 | 0.000533961 | 0.019003478 | 1 | 0.24515285 | 1 | 0.154824192 | ENSG0000015 | 16 | KCNMA1 |
| SAT | HTN | ENSG0000018 | -0.00363186 | 0.002190255 | 0.001185658 | 0.240331186 | -0.00364885 | 0.002087143 | 0.00118563 | 0.240307555 | 1.70E-05 | 0.047887935 | 1 | 0.025126664 | 1 | 0.024027772 | ENSG0000015 | 5 | DCK |
| SAT | HTN | ENSG0000018 | -0.01151159 | 4.30E-07 | 0.002277063 | 0.240323209 | -0.0115503 | 3.93E-07 | 0.002277027 | 0.240299537 | 3.87E-05 | 0.037640718 | 0.00140924 | 2.43E-05 | 0.001288415 | 2.15E-05 | ENSG0000015 | 5 | DCK |
| SAT | HTN | ENSG0000018 | 0.003791837 | 0.475553322 | 0.005314617 | 0.240335562 | 0.003712509 | 0.4848261 | 0.005314501 | 0.240311985 | 7.93E-05 | 0.043373692 | 1 | 0.743349428 | 1 | 0.747867623 | ENSG0000015 | 12 | DPY19L4 |
| SAT | HTN | ENSG0000018 | -0.00018921 | 0.958941734 | 0.00367521 | 0.240335812 | -0.00013282 | 0.971170332 | 0.003675129 | 0.240312225 | -5.64E-05 | 0.036876615 | 1 | 0.985679145 | 1 | 0.990013044 | ENSG0000015 | 10 | DRAM2 |
| SAT | HTN | ENSG0000018 | -0.0058714 | 0.174979644 | 0.004328735 | 0.240334906 | -0.00599776 | 0.165895674 | 0.004328899 | 0.240311279 | 0.000126355 | 0.026944393 | 1 | 0.450479504 | 1 | 0.439073216 | ENSG0000015 | 10 | DRAM2 |
| SAT | HTN | ENSG0000018 | 0.003402047 | 0.323955561 | 0.003449083 | 0.240335334 | 0.003452728 | 0.316787057 | 0.003448991 | 0.240311731 | -5.07E-05 | 0.028779213 | 1 | 0.616849723 | 1 | 0.604641264 | ENSG0000015 | 5 | C8orf37 |
| SAT | HTN | ENSG0000018 | -0.00107913 | 0.826904484 | 0.004934887 | 0.24033579 | -0.00097416 | 0.843508302 | 0.00493476 | 0.240312206 | -0.00010497 | 0.001848449 | 1 | 0.935512501 | 1 | 0.944556566 | ENSG0000015 | 5 | C8orf37 |
| SAT | HTN | ENSG0000019 | -0.02257348 | 2.55E-09 | 0.003788609 | 0.240318306 | -0.03182573 | 8.40E-10 | 0.005185596 | 0.240293651 | 0.009252259 | 0.008976919 | 8.37E-06 | 3.32E-07 | 2.76E-06 | 1.31E-07 | ENSG0000015 | 5 | C8orf37 |
| SAT | HTN | ENSG0000019 | 0.00358361 | 0.269425296 | 0.003244883 | 0.240335212 | 0.003527274 | 0.277014966 | 0.003244811 | 0.240311643 | 5.63E-05 | 0.017988234 | 1 | 0.563396428 | 1 | 0.570293485 | ENSG0000015 | 5 | C8orf37 |
| SAT | HTN | ENSG0000019 | -0.00266115 | 0.584126355 | 0.004861722 | 0.240335666 | -0.00276252 | 0.569877309 | 0.004861596 | 0.240312066 | 0.000101367 | 0.002165698 | 1 | 0.812965812 | 1 | 0.804893436 | ENSG0000015 | 5 | C8orf37 |
| SAT | HTN | ENSG0000019 | -0.00166684 | 0.786720068 | 0.006160437 | 0.240335777 | -0.00182537 | 0.766994381 | 0.00616036 | 0.240312182 | 0.000158525 | 0.002629187 | 1 | 0.917660669 | 1 | 0.908486846 | ENSG0000015 | 12 | ART3 |
| SAT | HTN | ENSG0000019 | 0.005052539 | 0.284889245 | 0.004724641 | 0.240335249 | 0.004954951 | 0.294285234 | 0.004724543 | 0.240311683 | 9.76E-05 | 0.006000425 | 1 | 0.578774993 | 1 | 0.58518173 | ENSG0000015 | 12 | ART3 |
| SAT | HTN | ENSG0000019 | 0.006863732 | 0.001119889 | 0.002106373 | 0.240330577 | 0.008927332 | 4.77E-05 | 0.002195311 | 0.24030407 | -0.0020636 | 0.000855582 | 1 | 0.014936405 | 0.156577668 | 0.001223263 | ENSG0000015 | 12 | ART3 |
| SAT | HTN | ENSG0000019 | 0.010999228 | 1.72E-05 | 0.002559082 | 0.240326703 | 0.010937042 | 1.92E-05 | 0.002559033 | 0.240303218 | 6.22E-05 | 0.001633234 | 0.056528272 | 0.000538364 | 0.063035388 | 0.000572975 | ENSG0000015 | 12 | ART3 |
| SAT | HTN | ENSG0000019 | 0.014406169 | 0.0048163 | 0.005110232 | 0.240331894 | 0.017284393 | 0.000894136 | 0.005203166 | 0.240306783 | -0.00287822 | 0.00332746 | 1 | 0.044140448 | 1 | 0.012122569 | ENSG0000015 | 12 | ART3 |
| SAT | HTN | ENSG0000019 | -0.00267395 | 0.595755339 | 0.005040289 | 0.240335675 | -0.00253249 | 0.615353285 | 0.005040318 | 0.240312101 | -0.00014145 | 0.00727466 | 1 | 0.820946353 | 1 | 0.834690078 | ENSG0000015 | 6 | WHAMM |
| SAT | HTN | ENSG0000019 | 0.001512789 | 0.293076241 | 0.001438835 | 0.240335268 | 0.001535715 | 0.285811051 | 0.001438801 | 0.240311664 | -2.29E-05 | 0.02594745 | 1 | 0.58529001 | 1 | 0.577147307 | ENSG0000015 | 2 | RWDD2B |
| SAT | HTN | ENSG0000019 | 0.003954048 | 0.275995058 | 0.003629693 | 0.240335228 | 0.004360355 | 0.230349024 | 0.00363526 | 0.240311516 | -0.00040631 | 0.046735318 | 1 | 0.571318478 | 1 | 0.519406797 | ENSG0000015 | 2 | RWDD2B |
| SAT | HTN | ENSG0000019 | -0.00661501 | 3.81E-06 | 0.00143139 | 0.240325281 | -0.00657324 | 4.39E-06 | 0.001431432 | 0.240301826 | -4.18E-05 | 0.019720192 | 0.012509804 | 0.000157075 | 0.014402644 | 0.000167473 | ENSG0000015 | 8 | CCT8 |
| SAT | HTN | ENSG0000019 | -0.00649895 | 0.294009833 | 0.006193209 | 0.24033527 | -0.00788798 | 0.203525627 | 0.006203323 | 0.240311428 | 0.001389027 | 0.000111218 | 1 | 0.586767799 | 1 | 0.490644807 | ENSG0000015 | 5 | C14orf2 |
| SAT | HTN | ENSG0000019 | -0.00392589 | 0.158126203 | 0.002781547 | 0.240334831 | -0.005284 | 0.062408052 | 0.002835692 | 0.240310513 | 0.001358114 | 0.013915771 | 1 | 0.427709869 | 1 | 0.251548918 | ENSG0000015 | 5 | C14orf2 |
| SAT | HTN | ENSG0000019 | 0.003190965 | 0.4556291 | 0.004277066 | 0.240335539 | 0.003998582 | 0.350517592 | 0.004283032 | 0.240311796 | -0.00080762 | 0.000444739 | 1 | 0.728518069 | 1 | 0.635605707 | ENSG0000015 | 9 | FUT6 |
| SAT | HTN | ENSG0000019 | -0.00220928 | 0.287519094 | 0.002077208 | 0.240335255 | -0.00217315 | 0.295463298 | 0.002077162 | 0.240311686 | -3.61E-05 | 0.017658918 | 1 | 0.581565115 | 1 | 0.586298316 | ENSG0000015 | 14 | TDRD9 |
| SAT | HTN | ENSG0000019 | 3.50E-05 | 0.991681433 | 0.003355749 | 0.240335813 | 9.20E-05 | 0.97813641 | 0.003355689 | 0.240312225 | -5.70E-05 | 0.031659625 | 1 | 0.99730223 | 1 | 0.991752513 | ENSG0000015 | 14 | TDRD9 |
| SAT | HTN | ENSG0000019 | 0.008741565 | 0.045391362 | 0.004368566 | 0.240333839 | 0.008805087 | 0.043842046 | 0.004368463 | 0.240310222 | -6.35E-05 | 0.041577649 | 1 | 0.202074707 | 1 | 0.19786211 | ENSG0000015 | 13 | SH3RF2 |
| SAT | HTN | ENSG0000019 | 0.00392762 | 0.283834615 | 0.003664693 | 0.240335247 | 0.003863963 | 0.291701573 | 0.003664618 | 0.240311677 | 6.37E-05 | 0.021826591 | 1 | 0.578137942 | 1 | 0.582047821 | ENSG0000015 | 9 | TYSND1 |
| SAT | HTN | ENSG0000019 | 0.01337501 | 0.028825993 | 0.006118865 | 0.240333457 | 0.012934802 | 0.034574993 | 0.006120645 | 0.240310023 | 0.000440208 | 0.005770502 | 1 | 0.15453935 | 1 | 0.173191684 | ENSG0000015 | 10 | MED19 |
| SAT | HTN | ENSG0000019 | -0.00539371 | 0.303558226 | 0.005242551 | 0.240335291 | -0.00581906 | 0.26733183 | 0.005246041 | 0.240311619 | 0.000425348 | 0.031931503 | 1 | 0.598182906 | 1 | 0.559028511 | ENSG0000015 | 10 | MED19 |
| SAT | HTN | ENSG0000019 | 0.00066005 | 0.820497282 | 0.002908918 | 0.240335788 | 0.000706887 | 0.807995123 | 0.002908841 | 0.240312196 | -4.68E-05 | 0.016470004 | 1 | 0.932182301 | 1 | 0.925531291 | ENSG0000015 | 10 | MED19 |
| SAT | HTN | ENSG0000019 | 0.001634316 | 0.770090265 | 0.005592051 | 0.240335771 | 0.001725101 | 0.757703357 | 0.005591931 | 0.240312178 | -9.08E-05 | 0.028657182 | 1 | 0.908545905 | 1 | 0.904156036 | ENSG0000015 | 10 | MED19 |
| SAT | HTN | ENSG0000019 | 0.008947716 | 0.082681535 | 0.00515618 | 0.240334328 | 0.009031324 | 0.079843453 | 0.005156046 | 0.240310712 | -8.36E-05 | 0.017507172 | 1 | 0.293273641 | 1 | 0.285677612 | ENSG0000015 | 12 | MAPK13 |
| SAT | HTN | ENSG0000020 | 0.003445203 | 0.347780078 | 0.003669385 | 0.240335379 | 0.003506772 | 0.339224187 | 0.003669322 | 0.240311775 | -6.16E-05 | 0.036340888 | 1 | 0.63533766 | 1 | 0.624576071 | ENSG0000015 | 12 | MAPK13 |
| SAT | HTN | ENSG0000020 | 0.008478151 | 0.179801478 | 0.006320524 | 0.240334926 | 0.008565544 | 0.175343852 | 0.006320347 | 0.24031132 | -8.74E-05 | 0.033312784 | 1 | 0.457488848 | 1 | 0.452759918 | ENSG0000015 | 12 | MAPK13 |
| SAT | HTN | ENSG0000020 | 0.013911042 | 0.032487658 | 0.006505458 | 0.240333558 | 0.013774435 | 0.03422475 | 0.006505293 | 0.240310014 | 0.000136607 | 0.00220894 | 1 | 0.1664956 | 1 | 0.172226081 | ENSG0000015 | 2 | WDYHV1 |
| SAT | HTN | ENSG0000020 | 0.005824058 | 0.211334668 | 0.004659598 | 0.240335043 | 0.000464853 | 0.930567143 | 0.005335086 | 0.240312222 | 0.005359205 | 0.039184111 | 1 | 0.502455829 | 1 | 0.975149159 | ENSG0000015 | 2 | WDYHV1 |
| SAT | HTN | ENSG0000020 | 0.017293243 | 0.001516718 | 0.005452729 | 0.240330853 | 0.017387823 | 0.001428342 | 0.005452643 | 0.240307211 | -9.46E-05 | 0.03346605 | 1 | 0.018699145 | 1 | 0.017618005 | ENSG0000015 | 11 | FBRS |
| SAT | HTN | ENSG0000020 | 0.014085648 | 1.69E-09 | 0.002337808 | 0.24031791 | 0.020866772 | 8.98E-15 | 0.002691407 | 0.240282585 | -0.00678112 | 3.70E-07 | 5.55E-06 | 2.47E-07 | 2.95E-11 | 5.90E-12 | ENSG0000015 | 11 | ITGAD |
| SAT | HTN | ENSG0000020 | -0.00048698 | 0.881404835 | 0.003264156 | 0.240335802 | 0.001153819 | 0.728386037 | 0.003322508 | 0.240312166 | -0.00164079 | 0.008216629 | 1 | 0.957468 | 1 | 0.889755957 | ENSG0000015 | 11 | ITGAD |
| SAT | HTN | ENSG0000020 | 0.005662249 | 0.021949776 | 0.002471256 | 0.240333224 | 0.004348718 | 0.084083831 | 0.002517391 | 0.240310754 | 0.001313531 | 0.006245134 | 1 | 0.129527365 | 1 | 0.296007563 | ENSG0000015 | 11 | VPS8 |
| SAT | HTN | ENSG0000020 | -0.00359058 | 0.012541504 | 0.001438221 | 0.240332739 | -0.0041951 | 0.004120911 | 0.001462336 | 0.240308167 | 0.000604528 | 0.022470465 | 1 | 0.089025167 | 1 | 0.039725318 | ENSG0000015 | 11 | VPS8 |
| SAT | HTN | ENSG0000020 | -0.00667726 | 0.001587437 | 0.002114246 | 0.240330894 | -0.00817953 | 0.000228991 | 0.002219859 | 0.24030553 | 0.001502266 | 0.026466432 | 1 | 0.019362006 | 0.751320646 | 0.004297476 | ENSG0000015 | 8 | MPV17L |
| SAT | HTN | ENSG0000020 | -0.00342832 | 0.02096531 | 0.001485011 | 0.240333185 | -0.00466331 | 0.002803926 | 0.001560435 | 0.240307821 | 0.00123499 | 0.010008865 | 1 | 0.12461446 | 1 | 0.029676388 | ENSG0000015 | 8 | MPV17L |
| SAT | HTN | ENSG0000020 | -0.00413941 | 0.002223461 | 0.001353342 | 0.240331199 | -0.00575169 | 8.04E-05 | 0.001458584 | 0.240304557 | 0.001612281 | 0.003048923 | 1 | 0.025330473 | 0.263683982 | 0.001918893 | ENSG0000015 | 4 | NTAN1 |
| SAT | HTN | ENSG0000020 | 0.004931397 | 0.000518198 | 0.001420664 | 0.240329871 | 0.00381193 | 0.009628859 | 0.001472418 | 0.24030892 | 0.001119467 | 0.003835826 | 1 | 0.008213566 | 1 | 0.075041061 | ENSG0000015 | 4 | NTAN1 |
| SAT | HTN | ENSG0000020 | 0.002988309 | 0.006278938 | 0.001093473 | 0.24033213 | 0.002513065 | 0.023278166 | 0.00110764 | 0.240309687 | 0.000475244 | 0.00722137 | 1 | 0.054031376 | 1 | 0.134059059 | ENSG0000015 | 4 | NTAN1 |
| SAT | HTN | ENSG0000020 | -0.00255613 | 0.514223251 | 0.003918782 | 0.240335604 | -0.0057359 | 0.15612423 | 0.004044407 | 0.240311233 | 0.00317977 | 0.001489555 | 1 | 0.765198628 | 1 | 0.425451494 | ENSG0000015 | 9 | FCHO2 |
| SAT | HTN | ENSG0000020 | -0.00331397 | 0.41290837 | 0.004047415 | 0.240335483 | -0.00669227 | 0.118250142 | 0.004283975 | 0.240311022 | 0.003378298 | 0.016155327 | 1 | 0.693677605 | 1 | 0.366889987 | ENSG0000015 | 9 | FCHO2 |
| SAT | HTN | ENSG0000020 | -0.00542304 | 0.079498626 | 0.003092507 | 0.240334297 | -0.00639676 | 0.03943187 | 0.003105743 | 0.240310133 | 0.000973725 | 0.000725258 | 1 | 0.285290525 | 1 | 0.18508722 | ENSG0000015 | 4 | TIMP4 |
| SAT | HTN | ENSG0000020 | 0.006245832 | 0.062537776 | 0.003353525 | 0.240334103 | 0.007242234 | 0.032169469 | 0.003380581 | 0.240309962 | -0.0009964 | 0.019958994 | 1 | 0.251314587 | 1 | 0.165401029 | ENSG0000015 | 4 | TIMP4 |
| SAT | HTN | ENSG0000020 | -0.00216474 | 0.079301765 | 0.001233641 | 0.240334295 | -0.00353013 | 0.007382663 | 0.001317664 | 0.240308686 | 0.001365392 | 0.003199364 | 1 | 0.285290525 | 1 | 0.061950171 | ENSG0000015 | 4 | TIMP4 |
| SAT | HTN | ENSG0000020 | -0.00411521 | 0.066504252 | 0.002242603 | 0.240334153 | -0.00508394 | 0.024563186 | 0.002261329 | 0.240309733 | 0.000968737 | 0.000882072 | 1 | 0.26155018 | 1 | 0.138473904 | ENSG0000015 | 9 | STEAP2 |
| SAT | HTN | ENSG0000020 | 0.003361238 | 0.617369831 | 0.006728104 | 0.24033569 | 0.00322686 | 0.631497816 | 0.006727951 | 0.240312112 | 0.000134378 | 0.00587947 | 1 | 0.833285913 | 1 | 0.840204515 | ENSG0000015 | 9 | STEAP2 |
| SAT | HTN | ENSG0000020 | 0.006800668 | 0.045758461 | 0.003404386 | 0.240333845 | 0.006887228 | 0.043065998 | 0.003404337 | 0.240310207 | -8.66E-05 | 0.002346016 | 1 | 0.203157659 | 1 | 0.195435048 | ENSG0000015 | 9 | STEAP2 |
| SAT | HTN | ENSG0000020 | -0.00498703 | 0.031066302 | 0.002312848 | 0.24033352 | -0.00502478 | 0.029811305 | 0.002312802 | 0.240309898 | 3.78E-05 | 0.033275051 | 1 | 0.16204855 | 1 | 0.156999826 | ENSG0000015 | 9 | STEAP2 |
| SAT | HTN | ENSG0000020 | -0.00676456 | 0.008306592 | 0.002562956 | 0.240332378 | -0.00672847 | 0.008656533 | 0.002562895 | 0.240308826 | -3.61E-05 | 0.047548693 | 1 | 0.068649697 | 1 | 0.069955871 | ENSG0000015 | 9 | STEAP2 |
| SAT | HTN | ENSG0000020 | -0.00025982 | 0.955747357 | 0.004682208 | 0.240335812 | -0.00037087 | 0.936867598 | 0.0046823 | 0.240312222 | 0.000111053 | 0.043051949 | 1 | 0.983048498 | 1 | 0.978139775 | ENSG0000015 | 9 | STEAP2 |
| SAT | HTN | ENSG0000020 | -0.00575552 | 0.299700915 | 0.005549755 | 0.240335283 | -0.0058543 | 0.291478234 | 0.005549707 | 0.240311677 | 9.88E-05 | 0.047584261 | 1 | 0.59311359 | 1 | 0.582047821 | ENSG0000015 | 9 | STEAP2 |
| SAT | HTN | ENSG0000020 | -0.00077979 | 0.784578653 | 0.002852649 | 0.240335777 | -0.00196645 | 0.493421688 | 0.002871243 | 0.240311994 | 0.001186656 | 0.000290296 | 1 | 0.916412446 | 1 | 0.753334834 | ENSG0000015 | 9 | STEAP2 |
| SAT | HTN | ENSG0000020 | 0.002279651 | 0.124359195 | 0.001483439 | 0.240334649 | 0.00148092 | 0.327303594 | 0.001511821 | 0.240311752 | 0.000798731 | 0.006217556 | 1 | 0.374127295 | 1 | 0.615105129 | ENSG0000015 | 12 | CLEC18A |
| SAT | HTN | ENSG0000020 | 0.002731237 | 0.30918606 | 0.00268576 | 0.240335303 | 0.004110303 | 0.133926602 | 0.002742399 | 0.240311118 | -0.00137907 | 0.01298375 | 1 | 0.602619487 | 1 | 0.387527756 | ENSG0000015 | 12 | CLEC18A |
| SAT | HTN | ENSG0000021 | -0.00441723 | 0.156257143 | 0.003115611 | 0.240334822 | -0.00437191 | 0.160539165 | 0.003115535 | 0.240311254 | -4.53E-05 | 0.038985245 | 1 | 0.425460321 | 1 | 0.432099261 | ENSG0000015 | 10 | CACNA2D3 |
| SAT | HTN | ENSG0000021 | 0.007836285 | 0.11644039 | 0.004991601 | 0.240334598 | 0.007748828 | 0.120569219 | 0.004991543 | 0.240311037 | 8.75E-05 | 0.042463927 | 1 | 0.362335772 | 1 | 0.369887656 | ENSG0000015 | 15 | DYRK1A |
| SAT | HTN | ENSG0000021 | -0.00281757 | 0.401163232 | 0.00335606 | 0.240335466 | -0.00289336 | 0.388618552 | 0.003356078 | 0.240311859 | 7.58E-05 | 0.030233882 | 1 | 0.681924009 | 1 | 0.670742545 | ENSG0000015 | 15 | DYRK1A |
| SAT | HTN | ENSG0000021 | 0.000270573 | 0.929177366 | 0.003044252 | 0.240335809 | -0.00038537 | 0.899378249 | 0.003047694 | 0.240312218 | 0.000655944 | 9.22E-06 | 1 | 0.976204842 | 1 | 0.965911631 | ENSG0000015 | 9 | MX1 |
| SAT | HTN | ENSG0000021 | -0.00117611 | 0.585380511 | 0.002155846 | 0.240335667 | -0.00122772 | 0.569023083 | 0.002155817 | 0.240312065 | 5.16E-05 | 0.004780984 | 1 | 0.813238553 | 1 | 0.804544406 | ENSG0000015 | 9 | MX1 |
| SAT | HTN | ENSG0000021 | 0.029362665 | 2.63E-09 | 0.004932121 | 0.240318335 | 0.027571394 | 2.75E-08 | 0.004961769 | 0.240296999 | 0.001791271 | 0.000983486 | 8.63E-06 | 3.32E-07 | 9.02E-05 | 2.65E-06 | ENSG0000015 | 7 | C9orf43 |
| SAT | HTN | ENSG0000021 | 0.005146118 | 0.084025224 | 0.002978427 | 0.240334341 | 0.005188932 | 0.081471222 | 0.002978351 | 0.240310729 | -4.28E-05 | 0.036289286 | 1 | 0.294571399 | 1 | 0.289293376 | ENSG0000015 | 8 | GAREML |

| | | | | | | | | | | | | | | | | | | | |
|---|---|---|---|---|---|---|---|---|---|---|---|---|---|---|---|---|---|---|---|
| SAT | HTN | ENSG0000021 | 0.023083505 | 0.000427939 | 0.006553625 | 0.240329695 | 0.04350156 | 3.54E-08 | 0.007891823 | 0.240297242 | -0.02041806 | 3.43E-06 | 1 | 0.007020346 | 0.000116308 | 2.92E-06 | ENSG0000015 | 8 | GAREML |
| SAT | HTN | ENSG0000021 | -0.00137144 | 0.263060354 | 0.001225386 | 0.240335196 | -0.00134436 | 0.272590526 | 0.001225357 | 0.240311632 | -2.71E-05 | 0.001701067 | 1 | 0.555406063 | 1 | 0.565119463 | ENSG0000015 | 11 | SPPL3 |
| SAT | HTN | ENSG0000021 | 0.002035825 | 0.069769537 | 0.001122651 | 0.240334192 | 0.002063362 | 0.06606818 | 0.001122634 | 0.240310559 | -2.75E-05 | 0.002776776 | 1 | 0.267422724 | 1 | 0.262433049 | ENSG0000015 | 11 | SPPL3 |
| SAT | HTN | ENSG0000021 | -0.00302833 | 0.528912194 | 0.004809416 | 0.240335618 | -0.00309782 | 0.519491524 | 0.004809307 | 0.240312021 | 6.95E-05 | 0.046718587 | 1 | 0.778537869 | 1 | 0.769056242 | ENSG0000015 | 10 | FAM213B |
| SAT | HTN | ENSG0000021 | -0.00251217 | 0.566166627 | 0.004378847 | 0.240335651 | -0.0023893 | 0.585303936 | 0.004378779 | 0.240312079 | -0.00012287 | 0.000610856 | 1 | 0.802415854 | 1 | 0.814247906 | ENSG0000015 | 10 | FAM213B |
| SAT | HTN | ENSG0000021 | -0.009128 | 0.00274761 | 0.003048086 | 0.24033139 | -0.0090654 | 0.002938107 | 0.00304806 | 0.240307663 | -6.26E-05 | 0.02229834 | 1 | 0.029080345 | 1 | 0.030700413 | ENSG0000015 | 7 | C12orf43 |
| SAT | HTN | ENSG0000021 | 0.000069658 | 0.916320114 | 0.006629659 | 0.240335808 | 0.003098055 | 0.642581486 | 0.006675508 | 0.240312119 | -0.00240148 | 0.002185217 | 1 | 0.969508641 | 1 | 0.845691879 | ENSG0000015 | 7 | C12orf43 |
| SAT | HTN | ENSG0000021 | -0.00198867 | 0.253242752 | 0.001740618 | 0.24033517 | -0.00192229 | 0.269496949 | 0.00174085 | 0.240311624 | -6.64E-05 | 0.045844309 | 1 | 0.54654986 | 1 | 0.560697202 | ENSG0000015 | 5 | PEX10 |
| SAT | HTN | ENSG0000021 | 0.006744047 | 0.132810923 | 0.004486724 | 0.240334699 | 0.006856076 | 0.126489086 | 0.004486676 | 0.240311074 | -0.00011203 | 0.004347113 | 1 | 0.385638593 | 1 | 0.380046421 | ENSG0000015 | 5 | PEX10 |
| SAT | HTN | ENSG0000021 | -0.00484995 | 0.00601754 | 0.001765651 | 0.240332092 | -0.00575553 | 0.001304625 | 0.001790222 | 0.240307128 | 0.000905573 | 0.002228425 | 1 | 0.05264946 | 1 | 0.016590986 | ENSG0000015 | 5 | PEX10 |
| SAT | HTN | ENSG0000021 | 0.004702185 | 0.008422006 | 0.001784729 | 0.24033239 | 0.006301656 | 0.000737134 | 0.001866934 | 0.240306607 | -0.00159947 | 0.003526739 | 1 | 0.069215759 | 1 | 0.010515378 | ENSG0000015 | 7 | LDLRAP1 |
| SAT | HTN | ENSG0000021 | -0.00082275 | 0.844957771 | 0.004207232 | 0.240335795 | -0.00073704 | 0.860933133 | 0.004207182 | 0.24031221 | -8.57E-05 | 0.018293391 | 1 | 0.944567784 | 1 | 0.952725149 | ENSG0000015 | 7 | LDLRAP1 |
| SAT | HTN | ENSG0000021 | -0.00714554 | 0.047063557 | 0.003598438 | 0.240333869 | -0.00667052 | 0.064236131 | 0.003604629 | 0.240310537 | -0.00047502 | 0.026523176 | 1 | 0.205887374 | 1 | 0.257651278 | ENSG0000015 | 8 | KRTCAP3 |
| SAT | HTN | ENSG0000022 | -0.00285197 | 0.39958476 | 0.003385672 | 0.240335463 | -0.0029022 | 0.391323812 | 0.003385593 | 0.240311863 | 5.02E-05 | 0.03848866 | 1 | 0.681058492 | 1 | 0.671162273 | ENSG0000015 | 9 | PAFAH2 |
| SAT | HTN | ENSG0000022 | -0.00109929 | 0.820855511 | 0.004854548 | 0.240335788 | -0.00117572 | 0.808629769 | 0.004854457 | 0.240312196 | 7.64E-05 | 0.042976447 | 1 | 0.932182301 | 1 | 0.925531291 | ENSG0000015 | 9 | PAFAH2 |
| SAT | HTN | ENSG0000022 | -0.0054206 | 0.085379586 | 0.003150972 | 0.240334354 | -0.01057611 | 0.003886077 | 0.003663003 | 0.240308114 | 0.005155511 | 0.005786158 | 1 | 0.296748329 | 1 | 0.038072244 | ENSG0000015 | 6 | TRIM63 |
| SAT | HTN | ENSG0000022 | 0.002963755 | 0.03449891 | 0.001401835 | 0.240333609 | 0.002920192 | 0.037238646 | 0.00140182 | 0.240310085 | 4.36E-05 | 0.000370714 | 1 | 0.172091642 | 1 | 0.179701644 | ENSG0000015 | 5 | WDR66 |
| SAT | HTN | ENSG0000022 | 0.000587105 | 0.909804523 | 0.005182539 | 0.240335807 | 0.000476897 | 0.926680886 | 0.005182446 | 0.240312221 | 0.000110208 | 0.006992322 | 1 | 0.968235044 | 1 | 0.975149159 | ENSG0000015 | 7 | HPD |
| SAT | HTN | ENSG0000022 | -0.00552148 | 0.272344872 | 0.005030141 | 0.24033521 | -0.0053665 | 0.286023018 | 0.005030053 | 0.240311664 | -0.00015498 | 9.97E-05 | 1 | 0.567410704 | 1 | 0.577147307 | ENSG0000015 | 7 | CNNM4 |
| SAT | HTN | ENSG0000022 | -0.00349419 | 0.403562492 | 0.004183271 | 0.240335469 | -0.00339083 | 0.417606712 | 0.004183212 | 0.240311901 | -0.00010336 | 0.00309129 | 1 | 0.683930028 | 1 | 0.692294225 | ENSG0000015 | 7 | CNNM4 |
| SAT | HTN | ENSG0000022 | -0.00176297 | 0.744605682 | 0.005411874 | 0.240335761 | -0.00188313 | 0.727863943 | 0.005411777 | 0.240312166 | 0.000120153 | 0.004958318 | 1 | 0.896532567 | 1 | 0.889755957 | ENSG0000015 | 11 | EYA3 |
| SAT | HTN | ENSG0000022 | 0.003076085 | 0.006198272 | 0.001123842 | 0.240332119 | 0.003060231 | 0.006467988 | 0.001123815 | 0.240308569 | 1.59E-05 | 0.045307749 | 1 | 0.053800349 | 1 | 0.055699396 | ENSG0000015 | 11 | EYA3 |
| SAT | HTN | ENSG0000022 | -0.00084248 | 0.711555738 | 0.002278406 | 0.240335746 | -0.00212079 | 0.361289106 | 0.002323107 | 0.240311814 | 0.001278313 | 0.004877598 | 1 | 0.880322163 | 1 | 0.645706614 | ENSG0000015 | 11 | EYA3 |
| SAT | HTN | ENSG0000022 | -0.00399249 | 0.028777999 | 0.001825957 | 0.240333455 | -0.00335596 | 0.068875893 | 0.001844708 | 0.240310593 | -0.00063653 | 0.015492186 | 1 | 0.154534558 | 1 | 0.266192155 | ENSG0000015 | 11 | EYA3 |
| SAT | HTN | ENSG0000022 | -0.00810792 | 0.038024334 | 0.003908189 | 0.240333691 | -0.00899004 | 0.021816553 | 0.003919696 | 0.240309631 | 0.000882114 | 0.003562649 | 1 | 0.179766342 | 1 | 0.128741203 | ENSG0000015 | 11 | EYA3 |
| SAT | HTN | ENSG0000022 | -0.01403672 | 1.24E-11 | 0.002071658 | 0.240313173 | -0.01406991 | 1.11E-11 | 0.002071616 | 0.240289479 | 3.32E-05 | 0.035426547 | 4.07E-08 | 4.99E-09 | 3.64E-08 | 4.04E-09 | ENSG0000015 | 8 | MRAS |
| SAT | HTN | ENSG0000023 | -0.00259047 | 0.615507875 | 0.005157972 | 0.240335689 | -0.0025076 | 0.626846469 | 0.005157863 | 0.240312109 | -8.29E-05 | 0.031353605 | 1 | 0.832284161 | 1 | 0.837966728 | ENSG0000015 | 8 | MRAS |
| SAT | HTN | ENSG0000023 | -0.01211438 | 0.040057657 | 0.005900359 | 0.240333734 | -0.01220865 | 0.038528839 | 0.005900211 | 0.240310114 | 9.43E-05 | 0.021485351 | 1 | 0.186954726 | 1 | 0.182677922 | ENSG0000015 | 5 | ABHD3 |
| SAT | HTN | ENSG0000023 | 0.006437259 | 0.222976601 | 0.005282275 | 0.240335081 | 0.007696184 | 0.14641832 | 0.005299301 | 0.240311491 | -0.00125892 | 0.003245619 | 1 | 0.515564644 | 1 | 0.40780858 | ENSG0000015 | 5 | ABHD3 |
| SAT | HTN | ENSG0000023 | 0.007278392 | 0.052512685 | 0.003753854 | 0.240333959 | 0.007353141 | 0.050129881 | 0.003753788 | 0.240310333 | -7.47E-05 | 0.012092717 | 1 | 0.223758598 | 1 | 0.218427807 | ENSG0000015 | 5 | ABHD3 |
| SAT | HTN | ENSG0000023 | 0.002160921 | 0.222057877 | 0.001769686 | 0.240335078 | 0.002307092 | 0.192525264 | 0.0017704 | 0.240311388 | -0.00014617 | 0.00606597 | 1 | 0.515379933 | 1 | 0.474949011 | ENSG0000015 | 5 | ABHD3 |
| SAT | HTN | ENSG0000023 | 0.007155254 | 0.018768697 | 0.003044658 | 0.240333089 | 0.007210643 | 0.017868645 | 0.003044602 | 0.240309459 | -5.54E-05 | 0.020502392 | 1 | 0.115418861 | 1 | 0.112744278 | ENSG0000015 | 6 | MITD1 |
| SAT | HTN | ENSG0000023 | 0.005439027 | 0.018163934 | 0.00230246 | 0.240333061 | 0.006676282 | 0.005039457 | 0.002380549 | 0.240308347 | -0.00123726 | 0.040907822 | 1 | 0.113515937 | 1 | 0.046258571 | ENSG0000015 | 6 | MITD1 |
| SAT | HTN | ENSG0000023 | 0.003934965 | 0.409944591 | 0.004775491 | 0.240335479 | 0.003859902 | 0.418920169 | 0.004775352 | 0.240311903 | 7.51E-05 | 0.013210676 | 1 | 0.690214399 | 1 | 0.692780784 | ENSG0000015 | 6 | MITD1 |
| SAT | HTN | ENSG0000023 | 0.004714631 | 0.484880429 | 0.006749891 | 0.240335573 | 0.00221503 | 0.743704287 | 0.006774786 | 0.240312173 | 0.002499601 | 1.87E-05 | 1 | 0.748376275 | 1 | 0.896382412 | ENSG0000015 | 6 | MITD1 |
| SAT | HTN | ENSG0000023 | 0.004579998 | 0.13511748 | 0.003065133 | 0.240334712 | 0.005397338 | 0.079178316 | 0.00307457 | 0.240310706 | -0.00081734 | 0.000756361 | 1 | 0.387179433 | 1 | 0.285677612 | ENSG0000015 | 4 | TSPAN33 |
| SAT | HTN | ENSG0000023 | 0.000754433 | 0.611399184 | 0.001484878 | 0.240335686 | -0.00322223 | 0.124388886 | 0.00209697 | 0.240311061 | 0.003976663 | 0.007240861 | 1 | 0.830870282 | 1 | 0.375110233 | ENSG0000015 | 4 | TSPAN33 |
| SAT | HTN | ENSG0000023 | -0.0019967 | 0.299720309 | 0.001925399 | 0.240335283 | -0.00235712 | 0.222301539 | 0.001931377 | 0.240311491 | 0.000360411 | 0.018532547 | 1 | 0.59311359 | 1 | 0.513280331 | ENSG0000015 | 8 | ZFAND2B |
| SAT | HTN | ENSG0000023 | -0.00553563 | 0.361468386 | 0.006065965 | 0.240335403 | -0.00543951 | 0.369853781 | 0.00606582 | 0.240311829 | -9.61E-05 | 0.025815955 | 1 | 0.650299517 | 1 | 0.653687263 | ENSG0000015 | 8 | ZFAND2B |
| SAT | HTN | ENSG0000023 | -0.00127411 | 0.680149365 | 0.003090564 | 0.24033573 | -0.00197482 | 0.524544359 | 0.003103329 | 0.240312026 | 0.000700709 | 0.013237124 | 1 | 0.864026219 | 1 | 0.773149166 | ENSG0000015 | 8 | ZFAND2B |
| SAT | HTN | ENSG0000023 | 0.002650244 | 0.479178761 | 0.003745269 | 0.240335566 | 0.001565287 | 0.679107292 | 0.003783795 | 0.240312141 | 0.001084957 | 0.044455946 | 1 | 0.746582289 | 1 | 0.865678672 | ENSG0000015 | 10 | COPG2 |
| SAT | HTN | ENSG0000023 | -0.0024229 | 0.49882704 | 0.003582389 | 0.240335588 | -0.00253941 | 0.478407209 | 0.003582349 | 0.240311978 | 0.000116508 | 0.000191788 | 1 | 0.759114805 | 1 | 0.743207411 | ENSG0000015 | 10 | COPG2 |
| SAT | HTN | ENSG0000023 | 0.008323166 | 0.002460521 | 0.002748579 | 0.240331291 | 0.007018203 | 0.011869186 | 0.002789407 | 0.240309104 | 0.001304963 | 0.006641287 | 1 | 0.027273547 | 1 | 0.085400878 | ENSG0000015 | 6 | RNF166 |
| SAT | HTN | ENSG0000023 | -0.00144952 | 0.542625223 | 0.002380759 | 0.240335631 | -0.00239348 | 0.32133982 | 0.002413486 | 0.24031174 | 0.000943955 | 0.017368335 | 1 | 0.788531091 | 1 | 0.610136545 | ENSG0000015 | 13 | F11R |
| SAT | HTN | ENSG0000023 | -0.00195395 | 0.506747511 | 0.002943096 | 0.240335596 | -0.00218694 | 0.457760108 | 0.002945208 | 0.240311954 | 0.000232983 | 0.043271253 | 1 | 0.763819444 | 1 | 0.729437064 | ENSG0000015 | 9 | NIT1 |
| SAT | HTN | ENSG0000023 | -0.0002391 | 0.932903556 | 0.002839915 | 0.24033581 | -0.00018741 | 0.947382612 | 0.002839855 | 0.240312223 | -5.17E-05 | 0.014679116 | 1 | 0.976204842 | 1 | 0.981538823 | ENSG0000015 | 9 | NIT1 |
| SAT | HTN | ENSG0000023 | 0.00236603 | 0.590181434 | 0.004393143 | 0.24033567 | 0.002300757 | 0.600467399 | 0.004393031 | 0.24031209 | 6.53E-05 | 0.030876861 | 1 | 0.816351301 | 1 | 0.825747189 | ENSG0000015 | 9 | NIT1 |
| SAT | HTN | ENSG0000023 | -0.00121857 | 0.595324195 | 0.002294263 | 0.240335674 | -0.00012227 | 0.95867994 | 0.002359866 | 0.240312224 | -0.0010963 | 0.047437403 | 1 | 0.820696926 | 1 | 0.98710697 | ENSG0000015 | 9 | NIT1 |
| SAT | HTN | ENSG0000023 | 0.004976981 | 0.178268486 | 0.003697313 | 0.24033492 | 0.00679545 | 0.073102614 | 0.003791695 | 0.240310641 | -0.00181847 | 0.030706214 | 1 | 0.455174244 | 1 | 0.274427548 | ENSG0000015 | 7 | ZNF276 |
| SAT | HTN | ENSG0000023 | -0.00564296 | 0.222944658 | 0.004630175 | 0.240335081 | -0.00553294 | 0.232088699 | 0.004630083 | 0.240311521 | -0.00011003 | 0.001857568 | 1 | 0.515564644 | 1 | 0.519577173 | ENSG0000015 | 7 | ZNF276 |
| SAT | HTN | ENSG0000023 | 0.004002437 | 0.143039125 | 0.002732842 | 0.240334755 | 0.004055807 | 0.137774739 | 0.002732782 | 0.240311139 | -5.34E-05 | 0.008009935 | 1 | 0.400778284 | 1 | 0.392735811 | ENSG0000015 | 5 | CDA |
| SAT | HTN | ENSG0000023 | 0.009977545 | 3.80E-05 | 0.002422011 | 0.240327444 | 0.018590358 | 3.04E-07 | 0.003630397 | 0.240299294 | -0.00861281 | 0.001449444 | 0.124582329 | 0.001029606 | 0.00999886 | 1.72E-05 | ENSG0000015 | 5 | CDA |
| SAT | HTN | ENSG0000023 | -0.00167716 | 0.256337072 | 0.001477556 | 0.240335178 | -0.00265045 | 0.086158886 | 0.001544533 | 0.240310773 | 0.000973288 | 0.030606516 | 1 | 0.54929286 | 1 | 0.299456891 | ENSG0000015 | 5 | CDA |
| SAT | HTN | ENSG0000023 | 0.006275009 | 0.022571888 | 0.002751459 | 0.240333248 | 0.006224391 | 0.023682333 | 0.002751415 | 0.240309702 | 5.06E-05 | 0.023003063 | 1 | 0.130898125 | 1 | 0.135133452 | ENSG0000015 | 10 | B4GALT3 |
| SAT | HTN | ENSG0000023 | 0.013133775 | 0.04140651 | 0.006439888 | 0.240333762 | 0.01323697 | 0.03983027 | 0.006439748 | 0.240310142 | -0.00010319 | 0.030682581 | 1 | 0.191175896 | 1 | 0.185366123 | ENSG0000015 | 10 | B4GALT3 |
| SAT | HTN | ENSG0000023 | -0.01970398 | 0.002549568 | 0.006530095 | 0.240331323 | -0.02460153 | 0.000234573 | 0.006687802 | 0.240305552 | 0.004897546 | 0.000703005 | 1 | 0.027883779 | 0.769635591 | 0.004323795 | ENSG0000015 | 8 | NDUFS2 |
| SAT | HTN | ENSG0000023 | 0.008582762 | 0.125360913 | 0.005599922 | 0.240334655 | 0.00868645 | 0.120853266 | 0.005599809 | 0.240311039 | -0.00010369 | 0.014968621 | 1 | 0.3756248 | 1 | 0.369887656 | ENSG0000015 | 6 | FCER1G |
| SAT | HTN | ENSG0000023 | -0.00035958 | 0.791742931 | 0.001361795 | 0.240335779 | -0.00130962 | 0.352014838 | 0.001407154 | 0.240311798 | 0.000950044 | 0.007389526 | 1 | 0.918363088 | 1 | 0.636570061 | ENSG0000015 | 6 | FCER1G |
| SAT | HTN | ENSG0000023 | 0.027192557 | 0.000104662 | 0.007009199 | 0.240328391 | 0.027326064 | 9.67E-05 | 0.007009026 | 0.24030473 | -0.00013351 | 0.006673134 | 0.343397295 | 0.002368257 | 0.317354766 | 0.002234893 | ENSG0000015 | 6 | FCER1G |
| SAT | HTN | ENSG0000024 | -0.00218417 | 0.532665686 | 0.003500597 | 0.240335621 | -0.00452317 | 0.211629688 | 0.003621148 | 0.240311456 | 0.002338999 | 0.011650786 | 1 | 0.78125888 | 1 | 0.500979081 | ENSG0000015 | 3 | MIS18A |
| SAT | HTN | ENSG0000024 | -0.02492052 | 8.07E-05 | 0.006321471 | 0.240328149 | -0.02504725 | 7.43E-05 | 0.006321447 | 0.240304483 | 0.000126724 | 0.035519106 | 0.264933018 | 0.001905993 | 0.243622707 | 0.001791343 | ENSG0000015 | 3 | MIS18A |
| SAT | HTN | ENSG0000024 | -0.00086367 | 0.638742473 | 0.001839713 | 0.240335705 | -0.0007612 | 0.679124601 | 0.001840174 | 0.240312141 | -0.00010246 | 0.022883036 | 1 | 0.84165223 | 1 | 0.865678672 | ENSG0000015 | 3 | MIS18A |
| SAT | HTN | ENSG0000024 | -0.01233923 | 0.046050236 | 0.006185273 | 0.240333851 | -0.01224 | 0.047824284 | 0.006185156 | 0.240310294 | -9.92E-05 | 0.038230578 | 1 | 0.203626447 | 1 | 0.21061943 | ENSG0000015 | 3 | MIS18A |
| SAT | HTN | ENSG0000024 | -0.00069984 | 0.686203097 | 0.001732223 | 0.240335733 | -0.00066851 | 0.699547781 | 0.001732189 | 0.240312152 | -3.13E-05 | 0.017620172 | 1 | 0.866935834 | 1 | 0.876659689 | ENSG0000015 | 15 | IFNAR2 |
| SAT | HTN | ENSG0000024 | 0.000307121 | 0.946069224 | 0.00454027 | 0.240335811 | 0.000229251 | 0.959729117 | 0.004540201 | 0.240312224 | 7.79E-05 | 0.036711859 | 1 | 0.980743483 | 1 | 0.98710697 | ENSG0000015 | 8 | MRPL10 |
| SAT | HTN | ENSG0000024 | 0.005644768 | 0.330340305 | 0.005798846 | 0.240335346 | 0.005752159 | 0.321215948 | 0.005798762 | 0.24031174 | -0.00010739 | 0.026184929 | 1 | 0.62016399 | 1 | 0.610136545 | ENSG0000015 | 8 | MRPL10 |
| SAT | HTN | ENSG0000024 | 0.006562627 | 0.322758802 | 0.006636924 | 0.240335331 | 0.006660716 | 0.31556717 | 0.006636758 | 0.240311729 | -9.81E-05 | 0.032927904 | 1 | 0.61568118 | 1 | 0.604641264 | ENSG0000015 | 10 | IFNGR2 |
| SAT | HTN | ENSG0000024 | -0.00328258 | 0.496721794 | 0.004829761 | 0.240335586 | -0.00335226 | 0.487617621 | 0.004829622 | 0.240311988 | 6.97E-05 | 0.023110814 | 1 | 0.757316081 | 1 | 0.75005786 | ENSG0000015 | 10 | IFNGR2 |
| SAT | HTN | ENSG0000024 | -0.00510139 | 0.11899525 | 0.003272205 | 0.240334615 | -0.00502946 | 0.124280199 | 0.003272136 | 0.24031106 | -7.19E-05 | 0.003236465 | 1 | 0.366076286 | 1 | 0.375110233 | ENSG0000015 | 10 | IFNGR2 |
| SAT | HTN | ENSG0000024 | 0.000189488 | 0.892256133 | 0.001398952 | 0.240335804 | 0.005429881 | 0.006975728 | 0.002012549 | 0.240308636 | -0.00524039 | 0.00292541 | 1 | 0.959835565 | 1 | 0.05914047 | ENSG0000015 | 14 | TNNI1 |
| SAT | HTN | ENSG0000024 | -0.00255542 | 0.561549388 | 0.004401777 | 0.240335647 | -0.00243262 | 0.580501444 | 0.004401711 | 0.240312075 | -0.0001228 | 0.000730864 | 1 | 0.799324746 | 1 | 0.811169181 | ENSG0000015 | 8 | CBR1 |
| SAT | HTN | ENSG0000024 | 0.001558965 | 0.76886822 | 0.005305203 | 0.240335771 | 0.001318693 | 0.803713582 | 0.005305669 | 0.240312195 | 0.000240272 | 0.006185482 | 1 | 0.907788321 | 1 | 0.923833778 | ENSG0000015 | 5 | CBR3 |
| SAT | HTN | ENSG0000024 | -0.00259976 | 0.482218798 | 0.003699463 | 0.24033557 | -0.00254849 | 0.490887832 | 0.003699367 | 0.240311991 | -5.13E-05 | 0.040994357 | 1 | 0.747733474 | 1 | 0.75109625 | ENSG0000015 | 7 | CHAF1B |
| SAT | HTN | ENSG0000024 | 0.001707872 | 0.722783906 | 0.004814421 | 0.240335751 | 0.00162158 | 0.736250554 | 0.00481434 | 0.240312169 | 8.63E-05 | 0.025558546 | 1 | 0.885985829 | 1 | 0.893357273 | ENSG0000015 | 11 | SIM2 |
| SAT | HTN | ENSG0000024 | -0.00076862 | 0.874112346 | 0.004851236 | 0.240335801 | -0.0012404 | 0.798311822 | 0.00485419 | 0.240312193 | 0.000471788 | 0.007354069 | 1 | 0.956418386 | 1 | 0.922924978 | ENSG0000015 | 10 | HLCS |
| SAT | HTN | ENSG0000025 | 0.007916776 | 0.2182045 | 0.006429513 | 0.240335066 | 0.007807838 | 0.224596602 | 0.006429393 | 0.240311498 | 0.000108938 | 0.029639078 | 1 | 0.509681988 | 1 | 0.516432023 | ENSG0000015 | 10 | HLCS |
| SAT | HTN | ENSG0000025 | 0.00670309 | 0.133695282 | 0.00446965 | 0.240334704 | 0.006792711 | 0.128569664 | 0.004469568 | 0.240311086 | -8.96E-05 | 0.010592386 | 1 | 0.386416703 | 1 | 0.382792259 | ENSG0000015 | 8 | THEM4 |
| SAT | HTN | ENSG0000025 | 0.003783112 | 0.539078306 | 0.006159336 | 0.240335627 | 0.003899869 | 0.526620948 | 0.006159246 | 0.240312028 | -0.00011676 | 0.022370341 | 1 | 0.786082216 | 1 | 0.775861396 | ENSG0000015 | 8 | THEM4 |
| SAT | HTN | ENSG0000025 | -0.00062695 | 0.828542083 | 0.002894866 | 0.24033579 | 0.000447542 | 0.877656526 | 0.002907235 | 0.240312214 | -0.00107449 | 6.66E-05 | 1 | 0.935875598 | 1 | 0.956781838 | ENSG0000015 | 12 | MED8 |
| SAT | HTN | ENSG0000025 | -0.01223496 | 0.046578105 | 0.006147848 | 0.24033386 | -0.01212794 | 0.048524854 | 0.006147741 | 0.240310306 | -0.00010702 | 0.028527946 | 1 | 0.205131225 | 1 | 0.212847655 | ENSG0000015 | 9 | RSPRY1 |
| SAT | HTN | ENSG0000025 | 0.00048128 | 0.861217603 | 0.002752935 | 0.240335798 | -0.000298 | 0.913978082 | 0.002758661 | 0.24031222 | 0.000779277 | 1.45E-05 | 1 | 0.952246908 | 1 | 0.972372648 | ENSG0000015 | 9 | RSPRY1 |
| SAT | HTN | ENSG0000025 | 0.009897276 | 0.08597394 | 0.005764177 | 0.240334359 | 0.009422248 | 0.102207427 | 0.005765495 | 0.240310908 | 0.000475028 | 0.000468243 | 1 | 0.298497879 | 1 | 0.330732247 | ENSG0000015 | 9 | RSPRY1 |
| SAT | HTN | ENSG0000025 | -0.00160273 | 0.467211237 | 0.002204501 | 0.240335553 | -0.00075313 | 0.733834481 | 0.002214884 | 0.240312168 | -0.0008496 | 7.95E-05 | 1 | 0.738333035 | 1 | 0.892066631 | ENSG0000015 | 10 | CCDC17 |
| SAT | HTN | ENSG0000025 | -0.00930284 | 0.042957804 | 0.004595983 | 0.240333793 | -0.00936639 | 0.041549869 | 0.004595862 | 0.240310177 | 6.35E-05 | 0.040509429 | 1 | 0.194632687 | 1 | 0.192216555 | ENSG0000015 | 10 | CCDC17 |
| SAT | HTN | ENSG0000025 | -0.00555743 | 0.228890473 | 0.004618789 | 0.240335099 | -0.00548553 | 0.234960607 | 0.004618697 | 0.24031153 | -7.19E-05 | 0.041774051 | 1 | 0.520170844 | 1 | 0.524037868 | ENSG0000015 | 10 | CCDC17 |
| SAT | HTN | ENSG0000025 | -0.008053 | 0.08729884 | 0.004709838 | 0.240334372 | -0.00824851 | 0.07993503 | 0.004710562 | 0.240310713 | 0.000195508 | 0.039294169 | 1 | 0.300939215 | 1 | 0.28569372 | ENSG0000015 | 10 | CCDC17 |
| SAT | HTN | ENSG0000025 | 0.009267912 | 0.075512426 | 0.005214475 | 0.240334255 | 0.00915376 | 0.079177791 | 0.00521439 | 0.240310706 | 0.000114152 | 0.006762228 | 1 | 0.277442632 | 1 | 0.285677612 | ENSG0000015 | 7 | ATP6V0D1 |
| SAT | HTN | ENSG0000025 | 0.006829683 | 0.207078229 | 0.005413329 | 0.240335028 | 0.002346387 | 0.681082465 | 0.005709155 | 0.240312142 | 0.004483296 | 0.013490498 | 1 | 0.497746278 | 1 | 0.865857127 | ENSG0000015 | 9 | ZFYVE28 |
| SAT | HTN | ENSG0000026 | 0.007010052 | 0.274201681 | 0.006411014 | 0.240335224 | 0.007401907 | 0.248368918 | 0.006412344 | 0.240311568 | -0.00039186 | 0.006984958 | 1 | 0.569402351 | 1 | 0.537886095 | ENSG0000015 | 9 | ZFYVE28 |
| SAT | HTN | ENSG0000026 | -0.01248255 | 0.033785494 | 0.005880702 | 0.240333591 | -0.0123519 | 0.035690143 | 0.005880608 | 0.24031005 | -0.00013065 | 0.006210233 | 1 | 0.17053878 | 1 | 0.17529844 | ENSG0000015 | 10 | C16orf86 |
| SAT | HTN | ENSG0000026 | -0.00162829 | 0.78987768 | 0.006110598 | 0.240335778 | -0.00151739 | 0.803884619 | 0.006110535 | 0.240312195 | -0.0001109 | 0.039233597 | 1 | 0.918290338 | 1 | 0.923833778 | ENSG0000015 | 7 | ZNF890P |
| SAT | HTN | ENSG0000026 | 2.20E-05 | 0.992111392 | 0.002221164 | 0.240335813 | -2.75E-05 | 0.990131063 | 0.002221157 | 0.240312225 | 4.94E-05 | 0.020499234 | 1 | 0.99730223 | 1 | 0.996509208 | ENSG0000015 | 7 | ZNF890P |
| SAT | HTN | ENSG0000026 | -0.00017341 | 0.97189989 | 0.004922958 | 0.240335813 | -0.00010102 | 0.98362874 | 0.004922843 | 0.240312225 | -7.24E-05 | 0.040230453 | 1 | 0.988749111 | 1 | 0.994404109 | ENSG0000015 | 7 | ZNF890P |
| SAT | HTN | ENSG0000026 | -0.00278589 | 0.466015027 | 0.003821633 | 0.240335551 | -0.00273065 | 0.474891967 | 0.003821531 | 0.240311974 | -5.52E-05 | 0.031122756 | 1 | 0.737932097 | 1 | 0.74149608 | ENSG0000015 | 7 | ZNF890P |
| SAT | HTN | ENSG0000026 | -0.0025876 | 0.399491951 | 0.003071232 | 0.240335463 | -0.00200965 | 0.51383727 | 0.00307816 | 0.240312015 | -0.00057795 | 0.005599596 | 1 | 0.681058492 | 1 | 0.76520719 | ENSG0000015 | 10 | ZNF235 |
| SAT | HTN | ENSG0000026 | 0.003708001 | 0.29649383 | 0.003551784 | 0.240335276 | 0.003767809 | 0.28875978 | 0.003551697 | 0.24031167 | -5.98E-05 | 0.016843728 | 1 | 0.59028899 | 1 | 0.579178516 | ENSG0000016 | 7 | DFFA |
| SAT | HTN | ENSG0000026 | 0.005893124 | 0.301195139 | 0.005700035 | 0.240335286 | 0.005753698 | 0.31276576 | 0.00569991 | 0.240311723 | 0.000139426 | 0.000874918 | 1 | 0.595672845 | 1 | 0.603663737 | ENSG0000016 | 7 | DFFA |
| SAT | HTN | ENSG0000026 | -0.02132914 | 1.43E-06 | 0.004424036 | 0.24032435 | -0.02150221 | 1.17E-06 | 0.004424475 | 0.240300576 | 0.000173074 | 0.000289536 | 0.004683446 | 6.60E-05 | 0.003847323 | 5.42E-05 | ENSG0000016 | 7 | DFFA |
| SAT | HTN | ENSG0000026 | 0.009465266 | 0.006311998 | 0.003465697 | 0.240332135 | 0.009549892 | 0.00586003 | 0.003465726 | 0.240308481 | -8.46E-05 | 0.022832943 | 1 | 0.05407223 | 1 | 0.051964215 | ENSG0000016 | 7 | DFFA |
| SAT | HTN | ENSG0000026 | 0.004803437 | 0.220179253 | 0.003917804 | 0.240335072 | 0.004877399 | 0.213149 | 0.003917732 | 0.240311461 | -7.40E-05 | 0.015841141 | 1 | 0.513073955 | 1 | 0.503485868 | ENSG0000016 | 15 | ATAD3B |
| SAT | HTN | ENSG0000026 | -0.0024543 | 0.233655203 | 0.002060746 | 0.240335114 | -0.00207912 | 0.313908045 | 0.002064553 | 0.240311725 | -0.00037522 | 0.003124367 | 1 | 0.524724656 | 1 | 0.603711779 | ENSG0000016 | 15 | ATAD3B |
| SAT | HTN | ENSG0000026 | 0.003043721 | 0.247788532 | 0.002633572 | 0.240335155 | 0.002982789 | 0.257370864 | 0.00263351 | 0.240311593 | 6.09E-05 | 0.001238047 | 1 | 0.539579113 | 1 | 0.548573946 | ENSG0000016 | 15 | ATAD3B |
| SAT | HTN | ENSG0000026 | 0.002748242 | 0.590447299 | 0.005106469 | 0.24033567 | 0.00285258 | 0.576412986 | 0.005106365 | 0.240312072 | -0.00010434 | 0.00701969 | 1 | 0.816374879 | 1 | 0.808901201 | ENSG0000016 | 15 | ATAD3B |
| SAT | HTN | ENSG0000026 | 0.008718424 | 6.75E-05 | 0.002187881 | 0.240327982 | 0.008750856 | 6.34E-05 | 0.00218783 | 0.240304336 | -3.24E-05 | 0.038592064 | 0.221555214 | 0.001629082 | 0.208043723 | 0.001564239 | ENSG0000016 | 6 | FAM86C2P |
| SAT | HTN | ENSG0000026 | 0.007907418 | 0.105835289 | 0.004889567 | 0.240334524 | 0.007980777 | 0.102628203 | 0.004889443 | 0.240310912 | -7.34E-05 | 0.029813748 | 1 | 0.338116439 | 1 | 0.331746931 | ENSG0000016 | 14 | ABCG1 |
| SAT | HTN | ENSG0000026 | 0.002317095 | 0.578874291 | 0.004174718 | 0.240335661 | 0.002423794 | 0.561527448 | 0.004174819 | 0.240312059 | -0.0001067 | 0.034605919 | 1 | 0.81131968 | 1 | 0.801717772 | ENSG0000016 | 14 | ABCG1 |
| SAT | HTN | ENSG0000026 | 0.00865063 | 0.122437106 | 0.005600464 | 0.240334637 | 0.008440828 | 0.131765518 | 0.005600416 | 0.240311105 | 0.000209802 | 3.05E-05 | 1 | 0.372076709 | 1 | 0.386342332 | ENSG0000016 | 14 | ABCG1 |
| SAT | HTN | ENSG0000026 | 0.001787377 | 0.692375289 | 0.004517754 | 0.240335736 | 0.001693451 | 0.707772651 | 0.004517685 | 0.240312156 | 9.39E-05 | 0.01138122 | 1 | 0.869846415 | 1 | 0.880273494 | ENSG0000016 | 14 | ABCG1 |
| SAT | HTN | ENSG0000026 | 0.017764284 | 0.002812303 | 0.005946089 | 0.240331411 | 0.015147871 | 0.011277879 | 0.005977901 | 0.240309059 | 0.002616413 | 2.34E-05 | 1 | 0.029669341 | 1 | 0.083456744 | ENSG0000016 | 13 | NDUFV3 |
| SAT | HTN | ENSG0000027 | 0.012630248 | 0.060773888 | 0.006735625 | 0.240334079 | 0.012533503 | 0.062769724 | 0.006735461 | 0.240310518 | 9.67E-05 | 0.040879505 | 1 | 0.247326914 | 1 | 0.252696275 | ENSG0000016 | 13 | NDUFV3 |
| SAT | HTN | ENSG0000027 | 0.000287264 | 0.887856598 | 0.002037087 | 0.240335804 | 8.37E-05 | 0.967256736 | 0.002037822 | 0.240312225 | 0.000203613 | 0.000483675 | 1 | 0.958390933 | 1 | 0.989225546 | ENSG0000016 | 13 | NDUFV3 |
| SAT | HTN | ENSG0000027 | 0.009753943 | 1.62E-07 | 0.001861975 | 0.24032228 | 0.01143513 | 4.68E-09 | 0.001951999 | 0.240295302 | -0.00168119 | 0.00413752 | 0.000531387 | 1.06E-05 | 1.54E-05 | 5.12E-07 | ENSG0000016 | 13 | NDUFV3 |
| SAT | HTN | ENSG0000027 | -0.02378584 | 6.81E-06 | 0.005286248 | 0.240325828 | -0.02367666 | 7.50E-06 | 0.005286133 | 0.240302332 | -0.00010918 | 0.005029115 | 0.022345974 | 0.000251078 | 0.024609466 | 0.000261923 | ENSG0000016 | 13 | NDUFV3 |
| SAT | HTN | ENSG0000027 | 0.003730155 | 0.175911693 | 0.002756029 | 0.24033491 | 0.003518198 | 0.201861573 | 0.002756631 | 0.240311422 | 0.000211956 | 0.000879404 | 1 | 0.451616796 | 1 | 0.488427597 | ENSG0000016 | 13 | CBS |
| SAT | HTN | ENSG0000027 | -0.00151916 | 0.74584533 | 0.004686996 | 0.240335762 | -0.00160609 | 0.731841618 | 0.00468690 1 | 0.240312165 | 8.69E-05 | 0.014506975 | 1 | 0.89736653 | 1 | 0.891634737 | ENSG0000016 | 13 | CBS |
| SAT | HTN | ENSG0000027 | -0.00216485 | 0.072259974 | 0.001204378 | 0.24033422 | -0.00175266 | 0.148825126 | 0.001214008 | 0.240311198 | -0.00041219 | 0.007084583 | 1 | 0.272198594 | 1 | 0.412411518 | ENSG0000016 | 13 | CBS |
| SAT | HTN | ENSG0000027 | -0.00070363 | 0.772722753 | 0.002436255 | 0.240335772 | 0.000433514 | 0.862297899 | 0.002499364 | 0.240312211 | -0.00113714 | 0.041790449 | 1 | 0.90968904 | 1 | 0.952725149 | ENSG0000016 | 7 | U2AF1 |
| SAT | HTN | ENSG0000027 | 0.001058233 | 0.332947513 | 0.001092996 | 0.240335351 | 0.000872334 | 0.425962102 | 0.00109573 | 0.240311913 | 0.000185899 | 0.017312787 | 1 | 0.620682267 | 1 | 0.699140398 | ENSG0000016 | 7 | U2AF1 |
| SAT | HTN | ENSG0000027 | 0.005616097 | 0.138787294 | 0.003793828 | 0.240334733 | 0.005691857 | 0.133530043 | 0.003793741 | 0.240311115 | -7.58E-05 | 0.005708418 | 1 | 0.391877031 | 1 | 0.387527756 | ENSG0000016 | 7 | U2AF1 |
| SAT | HTN | ENSG0000027 | -0.0053821 | 0.159900996 | 0.003829569 | 0.240334839 | -0.00532705 | 0.164206583 | 0.003829478 | 0.240311271 | -5.51E-05 | 0.042750861 | 1 | 0.430734949 | 1 | 0.436686865 | ENSG0000016 | 14 | PDXK |
| SAT | HTN | ENSG0000027 | 0.001449574 | 0.330986626 | 0.001491133 | 0.240335347 | 0.00190655 | 0.205060079 | 0.001504457 | 0.240311433 | -0.00045698 | 0.022545551 | 1 | 0.62016399 | 1 | 0.493618576 | ENSG0000016 | 14 | PDXK |
| SAT | Obesity | ENSG000000( | 0.004697743 | 0.257426386 | 0.004148131 | 0.183453989 | 0.004359361 | 0.293523485 | 0.004150094 | 0.183420853 | 0.000338382 | 0.015116507 | 1 | 0.550238419 | 1 | 0.59812588 | ENSG0000016 | 5 | CSTB |
| SAT | Obesity | ENSG000000( | 0.008336993 | 0.129229894 | 0.005495173 | 0.183453604 | 0.007930819 | 0.149124234 | 0.005497458 | 0.183420484 | 0.000406174 | 0.020154919 | 1 | 0.37959112 | 1 | 0.414396534 | ENSG0000016 | 12 | C21orf33 |
| SAT | Obesity | ENSG000000( | 0.00130929 | 0.607878062 | 0.002551699 | 0.183454375 | 0.00127098 | 0.618395829 | 0.002551526 | 0.183421176 | 3.83E-05 | 0.02599928 | 1 | 0.823471479 | 1 | 0.828495448 | ENSG0000016 | 12 | C21orf33 |
| SAT | Obesity | ENSG000000( | -0.01456622 | 0.011529505 | 0.005765978 | 0.183452062 | -0.0144391 | 0.012269998 | 0.0057657 4 | 0.1834189 | -0.00012713 | 0.025969505 | 1 | 0.085007432 | 1 | 0.094213678 | ENSG0000016 | 12 | ICOSLG |
| SAT | Obesity | ENSG000000( | 0.000654453 | 0.741108822 | 0.001980878 | 0.183454433 | 0.000696385 | 0.725159277 | 0.001980775 | 0.183421223 | -4.19E-05 | 0.015622728 | 1 | 0.894949594 | 1 | 0.889105974 | ENSG0000016 | 12 | ICOSLG |
| SAT | Obesity | ENSG000000( | 0.002661207 | 0.483786135 | 0.003800494 | 0.183454289 | 0.002721818 | 0.473858258 | 0.003800275 | 0.183421076 | -6.06E-05 | 0.049358916 | 1 | 0.744513279 | 1 | 0.742320719 | ENSG0000016 | 11 | C21orf2 |

| | | | | | | | | | | | | | | | | | | | |
|---|---|---|---|---|---|---|---|---|---|---|---|---|---|---|---|---|---|---|---|
| SAT | Obesity | ENSG0000000( | 0.01049366 | 0.004979989 | 0.003736608 | 0.183451493 | 0.010393316 | 0.005408519 | 0.003736383 | 0.183418346 | 0.000100344 | 0.000540925 | 1 | 0.049363573 | 1 | 0.055454223 | ENSG000001( | 11 | C21orf2 |
| SAT | Obesity | ENSG0000000( | -0.025437 | 5.70E-11 | 0.003882597 | 0.183438251 | -0.02534974 | 6.61E-11 | 0.003882366 | 0.183405159 | -8.73E-05 | 0.004121427 | 1.87E-07 | 5.20E-09 | 2.17E-07 | 6.57E-09 | ENSG000001( | 11 | C21orf2 |
| SAT | Obesity | ENSG0000000( | -0.00118169 | 0.728721313 | 0.003407136 | 0.183454429 | -0.00112949 | 0.740247203 | 0.003406932 | 0.183421229 | -5.22E-05 | 0.049652889 | 1 | 0.888823282 | 1 | 0.897054301 | ENSG000001( | 7 | ZNF66 |
| SAT | Obesity | ENSG0000000( | -0.00313992 | 0.585492692 | 0.005757308 | 0.183454362 | -0.0030587 | 0.595204759 | 0.00575691 | 0.183421164 | -8.12E-05 | 0.031036368 | 1 | 0.80802013 | 1 | 0.817137678 | ENSG000001( | 7 | ZNF66 |
| SAT | Obesity | ENSG0000000( | -0.00898662 | 0.113765504 | 0.005682358 | 0.183453529 | -0.00909018 | 0.109640068 | 0.005682014 | 0.183420303 | 0.000103555 | 0.018672947 | 1 | 0.350483211 | 1 | 0.347900449 | ENSG000001( | 7 | ZNF66 |
| SAT | Obesity | ENSG0000000( | -0.00620678 | 0.206831625 | 0.004916931 | 0.183453872 | -0.00608318 | 0.215996611 | 0.004916715 | 0.183420692 | -0.00012359 | 0.009214537 | 1 | 0.493814259 | 1 | 0.508175311 | ENSG000001( | 6 | LRRC3 |
| SAT | Obesity | ENSG0000001 | 0.000621668 | 0.746414558 | 0.001922462 | 0.183454435 | 0.000688399 | 0.720291531 | 0.001922528 | 0.183421222 | -6.67E-05 | 0.02827519 | 1 | 0.89640782 | 1 | 0.885797449 | ENSG000001( | 8 | FAM207A |
| SAT | Obesity | ENSG0000001 | -0.01875501 | 3.11E-09 | 0.003164967 | 0.183441201 | -0.01678029 | 1.51E-07 | 0.003195462 | 0.183410849 | -0.00197472 | 8.02E-06 | 1.02E-05 | 2.22E-07 | 0.000495766 | 8.70E-06 | ENSG000001( | 9 | FTCD |
| SAT | Obesity | ENSG0000001 | 0.007969577 | 0.155269606 | 0.00560778 | 0.183453711 | 0.007864009 | 0.160792165 | 0.005607479 | 0.183420527 | 0.000105568 | 0.028244923 | 1 | 0.417573423 | 1 | 0.428934236 | ENSG000001( | 10 | SPATC1L |
| SAT | Obesity | ENSG0000001 | 0.001425657 | 0.552660862 | 0.002400993 | 0.183454341 | 0.001478056 | 0.538130639 | 0.002400834 | 0.183421127 | -5.24E-05 | 0.001624676 | 1 | 0.793558114 | 1 | 0.786461749 | ENSG000001( | 10 | LSS |
| SAT | Obesity | ENSG0000001 | -0.00120415 | 0.839248369 | 0.005936026 | 0.183454459 | -0.00110705 | 0.852046422 | 0.005935675 | 0.183421257 | -9.71E-05 | 0.038782505 | 1 | 0.940107169 | 1 | 0.94476658 | ENSG000001( | 10 | LSS |
| SAT | Obesity | ENSG0000001 | -0.00518927 | 0.236132923 | 0.004380207 | 0.183453944 | -0.00528448 | 0.227618621 | 0.004379941 | 0.18342072 | 9.52E-05 | 0.004973449 | 1 | 0.525379039 | 1 | 0.522317542 | ENSG000001( | 10 | LSS |
| SAT | Obesity | ENSG0000001 | -0.00120997 | 0.763277071 | 0.00401743 | 0.18345444 | -0.00127453 | 0.751037531 | 0.00401716 | 0.183421232 | 6.46E-05 | 0.018550253 | 1 | 0.904538582 | 1 | 0.901114525 | ENSG000001( | 12 | PCNT |
| SAT | Obesity | ENSG0000002 | -0.00249418 | 0.015193314 | 0.001027366 | 0.183452246 | -0.0024749 | 0.015990869 | 0.001027299 | 0.183419077 | -1.93E-05 | 0.008946622 | 1 | 0.102994344 | 1 | 0.112106928 | ENSG000001( | 12 | PCNT |
| SAT | Obesity | ENSG0000002 | 0.004473692 | 0.367921546 | 0.004968702 | 0.183454168 | 0.004554025 | 0.359351547 | 0.004968377 | 0.183420953 | -8.03E-05 | 0.022213273 | 1 | 0.664730502 | 1 | 0.660152534 | ENSG000001( | 5 | S100B |
| SAT | Obesity | ENSG0000002 | 0.009860906 | 0.074546529 | 0.005529744 | 0.183453272 | 0.009749107 | 0.07787684 | 0.005529388 | 0.183420095 | 0.000111798 | 0.005165775 | 1 | 0.274343367 | 1 | 0.287417225 | ENSG000001( | 10 | ZNF208 |
| SAT | Obesity | ENSG0000003 | 0.005724676 | 0.041239296 | 0.002804672 | 0.183452899 | 0.004221479 | 0.134269426 | 0.002819058 | 0.183420423 | 0.001503197 | 1.61E-07 | 1 | 0.192979684 | 1 | 0.392603841 | ENSG000001( | 10 | ZNF208 |
| SAT | Obesity | ENSG0000003 | 0.002698844 | 0.565721913 | 0.004698841 | 0.183454349 | 0.002788113 | 0.552914382 | 0.004698547 | 0.183421137 | -8.93E-05 | 0.010911606 | 1 | 0.797234594 | 1 | 0.793924694 | ENSG000001( | 10 | ZNF208 |
| SAT | Obesity | ENSG0000003 | -0.00814994 | 0.016752093 | 0.003407007 | 0.183452311 | -0.00822308 | 0.015791569 | 0.003406826 | 0.183419068 | 7.31E-05 | 0.013249172 | 1 | 0.10970782 | 1 | 0.111772978 | ENSG000001( | 10 | ZNF208 |
| SAT | Obesity | ENSG0000003 | -0.00537311 | 0.058658788 | 0.002841794 | 0.183453123 | -0.00531981 | 0.061192955 | 0.002841622 | 0.183419946 | -5.33E-05 | 0.015106772 | 1 | 0.24059281 | 1 | 0.250029993 | ENSG000001( | 10 | ZNF208 |
| SAT | Obesity | ENSG0000003 | -0.01376581 | 1.53E-05 | 0.003183298 | 0.183447406 | -0.01384339 | 1.37E-05 | 0.003183101 | 0.183414122 | 7.76E-05 | 0.001317823 | 0.050195442 | 0.000418295 | 0.044874705 | 0.000439948 | ENSG000001( | 10 | ZNF208 |
| SAT | Obesity | ENSG0000003 | 0.002266411 | 0.55690386 | 0.00385807 | 0.183454344 | 0.002190793 | 0.570114877 | 0.003857827 | 0.183421148 | 7.56E-05 | 0.008138809 | 1 | 0.794232732 | 1 | 0.801777501 | ENSG000001( | 14 | ADAMTS13 |
| SAT | Obesity | ENSG0000003 | 0.002050599 | 0.526700345 | 0.00323923 | 0.183454323 | 0.002005957 | 0.535710681 | 0.003239005 | 0.183421125 | 4.46E-05 | 0.033975179 | 1 | 0.778775949 | 1 | 0.78622818 | ENSG000001( | 14 | ADAMTS13 |
| SAT | Obesity | ENSG0000004 | 0.001102984 | 0.843104727 | 0.00557277 | 0.183454459 | 0.001180397 | 0.832240346 | 0.005572399 | 0.183421253 | -7.74E-05 | 0.04455791 | 1 | 0.941066406 | 1 | 0.936326075 | ENSG000001( | 11 | FCN2 |
| SAT | Obesity | ENSG0000004 | -0.00093 | 0.639872215 | 0.001987705 | 0.183454391 | -0.00089368 | 0.652974281 | 0.001987571 | 0.183421194 | -3.63E-05 | 0.007444062 | 1 | 0.842174268 | 1 | 0.849151255 | ENSG000001( | 11 | FCN2 |
| SAT | Obesity | ENSG0000004 | 0.001680481 | 0.774710539 | 0.005871312 | 0.183454443 | -0.00759091 | 0.273259663 | 0.00692862 | 0.183420817 | 0.009271393 | 0.011747354 | 1 | 0.909817544 | 1 | 0.576940124 | ENSG000001( | 11 | RDH13 |
| SAT | Obesity | ENSG0000004 | 0.003223347 | 0.002720929 | 0.001075295 | 0.183451078 | 0.003202284 | 0.002899481 | 0.001075236 | 0.183417918 | 2.11E-05 | 0.020514544 | 1 | 0.030997801 | 1 | 0.035691396 | ENSG000001( | 10 | VPS11 |
| SAT | Obesity | ENSG0000004 | 0.005029471 | 0.347117149 | 0.005349381 | 0.18345414 | 0.004948468 | 0.354908779 | 0.005349055 | 0.183420947 | 8.10E-05 | 0.048249256 | 1 | 0.644898849 | 1 | 0.656771407 | ENSG000001( | 10 | VPS11 |
| SAT | Obesity | ENSG0000004 | 0.000584358 | 0.80547717 | 0.002372881 | 0.183454451 | 0.000547895 | 0.81738205 | 0.002372728 | 0.18342125 | 3.65E-05 | 0.033531092 | 1 | 0.924045663 | 1 | 0.930870102 | ENSG000001( | 10 | FDPS |
| SAT | Obesity | ENSG0000004 | -0.00537526 | 0.190803523 | 0.004108878 | 0.183453827 | -0.00530703 | 0.196463981 | 0.004108584 | 0.18342064 | -6.82E-05 | 0.007211556 | 1 | 0.467532754 | 1 | 0.48140278 | ENSG000001( | 10 | FDPS |
| SAT | Obesity | ENSG0000004 | 0.00753509 | 0.013654681 | 0.003055306 | 0.183452175 | 0.007621112 | 0.012612244 | 0.003055112 | 0.183418918 | -8.60E-05 | 0.000133526 | 1 | 0.096553901 | 1 | 0.096011077 | ENSG000001( | 10 | GBAP1 |
| SAT | Obesity | ENSG0000004 | -0.00168872 | 0.140847933 | 0.001146729 | 0.183453654 | -0.00171759 | 0.134156933 | 0.001146657 | 0.183420422 | 2.89E-05 | 0.000748728 | 1 | 0.396214067 | 1 | 0.392603841 | ENSG000001( | 10 | GBAP1 |
| SAT | Obesity | ENSG0000005 | 0.01029324 | 0.004466849 | 0.003620334 | 0.183451419 | 0.010533917 | 0.003623772 | 0.003620904 | 0.183418072 | -0.00024068 | 0.002865163 | 1 | 0.045656481 | 1 | 0.042161685 | ENSG000001( | 10 | GBAP1 |
| SAT | Obesity | ENSG0000005 | -0.00016147 | 0.973930535 | 0.004941034 | 0.183454474 | -0.000295 | 0.952387969 | 0.004940755 | 0.183421269 | 0.000133536 | 0.001066626 | 1 | 0.993452012 | 1 | 0.987557383 | ENSG000001( | 15 | PMF1 |
| SAT | Obesity | ENSG0000005 | -0.01031533 | 0.081167781 | 0.005914913 | 0.183453324 | -0.01016184 | 0.0857756 | 0.005914514 | 0.183420155 | -0.0001535 | 0.000133796 | 1 | 0.287792178 | 1 | 0.300351913 | ENSG000001( | 15 | PMF1 |
| SAT | Obesity | ENSG0000005 | -0.00421479 | 0.486052539 | 0.006050498 | 0.183454291 | -0.00433351 | 0.473823052 | 0.006050087 | 0.183421076 | 0.000118729 | 0.003520714 | 1 | 0.746950062 | 1 | 0.742320719 | ENSG000001( | 13 | STAG3L2 |
| SAT | Obesity | ENSG0000005 | -0.00395354 | 0.136285295 | 0.002653789 | 0.183453635 | -0.00390485 | 0.141151715 | 0.002653623 | 0.183420452 | -4.87E-05 | 0.013896427 | 1 | 0.389540014 | 1 | 0.401725489 | ENSG000001( | 13 | STAG3L2 |
| SAT | Obesity | ENSG0000005 | -0.00012994 | 0.942856327 | 0.001812713 | 0.183454472 | -0.00018121 | 0.920369634 | 0.001812633 | 0.183421266 | 5.13E-05 | 0.003386069 | 1 | 0.981335116 | 1 | 0.974203713 | ENSG000001( | 13 | STAG3L2 |
| SAT | Obesity | ENSG0000005 | 0.003256158 | 0.054626144 | 0.001694289 | 0.183453078 | 0.003227529 | 0.056771481 | 0.001694178 | 0.183419899 | 2.86E-05 | 0.016494871 | 1 | 0.229192299 | 1 | 0.239958924 | ENSG000001( | 16 | FCRL3 |
| SAT | Obesity | ENSG0000006( | 0.004971341 | 0.05762742 | 0.002618527 | 0.183453112 | 0.004918031 | 0.060344302 | 0.002618378 | 0.183419937 | 5.33E-05 | 0.012858439 | 1 | 0.237234083 | 1 | 0.24841864 | ENSG000001( | 16 | FCRL3 |
| SAT | Obesity | ENSG0000006( | -0.00629075 | 0.185287513 | 0.004748984 | 0.183453811 | -0.00619217 | 0.192244565 | 0.00474871 | 0.183420628 | -9.86E-05 | 0.010399574 | 1 | 0.460203128 | 1 | 0.475323599 | ENSG000001( | 16 | FCRL3 |
| SAT | Obesity | ENSG0000006( | -0.00878133 | 0.002221421 | 0.002870721 | 0.183450937 | -0.00899387 | 0.001731465 | 0.002870856 | 0.183417561 | 0.000212539 | 8.18E-06 | 1 | 0.026030297 | 1 | 0.022999738 | ENSG000001( | 9 | LY6K |
| SAT | Obesity | ENSG0000006( | 0.017974582 | 4.15E-23 | 0.001815486 | 0.183417429 | 0.017877663 | 7.04E-23 | 0.001815392 | 0.183384625 | 9.69E-05 | 8.09E-10 | 1.36E-19 | 1.36E-20 | 2.31E-19 | 2.31E-20 | ENSG000001( | 10 | CPSF4 |
| SAT | Obesity | ENSG0000006( | -0.00724023 | 0.020716082 | 0.003130063 | 0.183452452 | -0.00381469 | 0.232847341 | 0.003197405 | 0.183420732 | -0.00342555 | 1.63E-07 | 1 | 0.127045729 | 1 | 0.525427871 | ENSG000001( | 6 | LY6E |
| SAT | Obesity | ENSG0000006( | -0.00292699 | 0.356727223 | 0.00317592 | 0.183454153 | -0.00300045 | 0.344771469 | 0.003175822 | 0.183420933 | 7.35E-05 | 0.034123421 | 1 | 0.654232542 | 1 | 0.648774721 | ENSG000001( | 6 | LY6E |
| SAT | Obesity | ENSG0000007 | -0.0051249 | 0.292688294 | 0.004870449 | 0.183454056 | -0.00501421 | 0.303213812 | 0.004870208 | 0.18342087 | -0.00011069 | 0.012049829 | 1 | 0.592053202 | 1 | 0.60366779 | ENSG000001( | 9 | VPS28 |
| SAT | Obesity | ENSG0000007 | -0.00230331 | 0.433397283 | 0.002940172 | 0.183454242 | -0.00225563 | 0.442947454 | 0.002939981 | 0.183421048 | -4.77E-05 | 0.023127011 | 1 | 0.714561047 | 1 | 0.722357979 | ENSG000001( | 9 | VPS28 |
| SAT | Obesity | ENSG0000007 | 0.008653131 | 0.022541473 | 0.003793361 | 0.183452507 | 0.008724229 | 0.021448654 | 0.003793143 | 0.183419271 | -7.11E-05 | 0.02159153 | 1 | 0.133740638 | 1 | 0.134299683 | ENSG000001( | 9 | VPS28 |
| SAT | Obesity | ENSG0000007 | 0.00426925 | 0.290031455 | 0.004035003 | 0.183454051 | 0.004207293 | 0.297055925 | 0.004034733 | 0.183420859 | 6.20E-05 | 0.02519226 | 1 | 0.588792914 | 1 | 0.601026164 | ENSG000001( | 9 | VPS28 |
| SAT | Obesity | ENSG0000007 | -0.00208464 | 0.679417463 | 0.005044395 | 0.18345441 | -0.00199826 | 0.691984758 | 0.005044041 | 0.183421211 | -8.64E-05 | 0.007089299 | 1 | 0.858646871 | 1 | 0.868937924 | ENSG000001( | 10 | MUM1 |
| SAT | Obesity | ENSG0000007 | -0.00388011 | 0.498716804 | 0.005735476 | 0.183454301 | -0.00406274 | 0.478703519 | 0.00573518 | 0.183421081 | 0.000182632 | 0.000303682 | 1 | 0.75749331 | 1 | 0.743208721 | ENSG000001( | 10 | C5orf45 |
| SAT | Obesity | ENSG0000007 | 0.027520054 | 3.05E-06 | 0.005896393 | 0.18344624 | 0.027327284 | 3.57E-06 | 0.005896086 | 0.183413152 | 0.00019277 | 0.000177994 | 0.010017246 | 0.00011008 | 0.011724467 | 0.000160609 | ENSG000001( | 7 | MAML1 |
| SAT | Obesity | ENSG0000007 | 0.006230368 | 0.310199767 | 0.006139479 | 0.183454085 | 0.00635 | 0.300971042 | 0.006139095 | 0.183420866 | -0.00011963 | 0.009183529 | 1 | 0.607417315 | 1 | 0.601026164 | ENSG000001( | 7 | MAML1 |
| SAT | Obesity | ENSG0000007 | -0.00190016 | 0.461575717 | 0.002580845 | 0.183454269 | -0.00197005 | 0.445234036 | 0.002580674 | 0.18342105 | 6.99E-05 | 9.63E-05 | 1 | 0.731904575 | 1 | 0.724809903 | ENSG000001( | 7 | MAML1 |
| SAT | Obesity | ENSG0000007 | -0.00410387 | 0.477343472 | 0.005775355 | 0.183454283 | -0.00420611 | 0.466410932 | 0.005774992 | 0.18342107 | 0.000102244 | 0.017211379 | 1 | 0.739803464 | 1 | 0.737265369 | ENSG000001( | 6 | LRWD1 |
| SAT | Obesity | ENSG0000007 | -0.00111476 | 0.803547539 | 0.004481295 | 0.183454451 | -0.00098195 | 0.826549372 | 0.004481158 | 0.183421252 | -0.00013281 | 0.006725095 | 1 | 0.922904531 | 1 | 0.935786229 | ENSG000001( | 6 | LRWD1 |
| SAT | Obesity | ENSG0000008 | -0.00066514 | 0.849533985 | 0.003506048 | 0.183454461 | -0.00076107 | 0.828141785 | 0.003505842 | 0.183421252 | 9.59E-05 | 0.000579002 | 1 | 0.94293674 | 1 | 0.936326075 | ENSG000001( | 6 | LRWD1 |
| SAT | Obesity | ENSG0000008 | -0.00472843 | 0.307789147 | 0.004636294 | 0.183454081 | -0.0046005 | 0.32103103 | 0.004636004 | 0.183420898 | -0.00012793 | 0.00022402 | 1 | 0.606520234 | 1 | 0.6232561 | ENSG000001( | 6 | LRWD1 |
| SAT | Obesity | ENSG0000008 | 0.005210585 | 0.004069354 | 0.001813798 | 0.183451355 | 0.005179793 | 0.00429089 | 0.001813679 | 0.183418188 | 3.08E-05 | 0.016284193 | 1 | 0.042520858 | 1 | 0.047531447 | ENSG000001( | 6 | CCDC116 |
| SAT | Obesity | ENSG0000008 | 0.001393233 | 0.704766369 | 0.0036771 | 0.18345442 | 0.00131444 | 0.720727568 | 0.00367689 | 0.183421222 | 7.88E-05 | 0.008963767 | 1 | 0.874485947 | 1 | 0.885797449 | ENSG000001( | 6 | CCDC116 |
| SAT | Obesity | ENSG0000008 | -0.0061674 | 0.110724772 | 0.003866836 | 0.183453513 | -0.00578545 | 0.134812952 | 0.003868852 | 0.183420425 | -0.00038195 | 0.004738515 | 1 | 0.343697234 | 1 | 0.39281238 | ENSG000001( | 13 | DVL3 |
| SAT | Obesity | ENSG0000008 | 0.009650409 | 0.036558572 | 0.004615951 | 0.183452822 | 0.009584407 | 0.03784809 | 0.004615651 | 0.183419641 | 6.60E-05 | 0.044996789 | 1 | 0.1809418 | 1 | 0.189433997 | ENSG000001( | 7 | ABCF3 |
| SAT | Obesity | ENSG0000008 | 0.017678615 | 0.000206583 | 0.004764027 | 0.183449269 | 0.017474778 | 0.000244244 | 0.004763794 | 0.183416185 | 0.000203837 | 2.45E-06 | 0.677799119 | 0.003786587 | 0.80136467 | 0.00471391 | ENSG000001( | 7 | PCYT1A |
| SAT | Obesity | ENSG0000008 | -0.00959637 | 0.026314773 | 0.004319691 | 0.183452609 | -0.0095237 | 0.027463747 | 0.004319393 | 0.183419433 | -7.27E-05 | 0.010006983 | 1 | 0.146997736 | 1 | 0.155047129 | ENSG000001( | 4 | FBXO27 |
| SAT | Obesity | ENSG0000008 | -0.00171214 | 0.737437562 | 0.005107076 | 0.183454432 | -0.00187584 | 0.71338062 | 0.005106826 | 0.183421219 | 0.000163697 | 0.000439161 | 1 | 0.892158054 | 1 | 0.882946041 | ENSG000001( | 4 | FBXO27 |
| SAT | Obesity | ENSG0000008 | 0.005848349 | 0.311721724 | 0.005781189 | 0.183454087 | 0.005996527 | 0.299596808 | 0.005780902 | 0.183420864 | -0.00014818 | 0.004591228 | 1 | 0.608768261 | 1 | 0.601026164 | ENSG000001( | 4 | FBXO27 |
| SAT | Obesity | ENSG0000008 | -0.00435282 | 0.177606724 | 0.003228704 | 0.183453787 | -0.00428214 | 0.184726144 | 0.003228527 | 0.183420605 | -7.07E-05 | 0.009642076 | 1 | 0.451027601 | 1 | 0.462308526 | ENSG000001( | 4 | FBXO27 |
| SAT | Obesity | ENSG0000008 | -0.00185059 | 0.646835149 | 0.004039141 | 0.183454395 | -0.00195581 | 0.628213674 | 0.004038912 | 0.183421182 | 0.000105228 | 0.001580753 | 1 | 0.844125734 | 1 | 0.833806256 | ENSG000001( | 4 | FBXO27 |
| SAT | Obesity | ENSG0000009 | -0.00101829 | 0.843139807 | 0.005146026 | 0.183454459 | -0.00109316 | 0.83176206 | 0.005145657 | 0.183421253 | 7.49E-05 | 0.017858272 | 1 | 0.941066406 | 1 | 0.936326075 | ENSG000001( | 16 | DMKN |
| SAT | Obesity | ENSG0000009 | 0.006949071 | 0.028559622 | 0.003173789 | 0.183452662 | 0.006993878 | 0.027539791 | 0.003173573 | 0.183419435 | -4.48E-05 | 0.034699205 | 1 | 0.154014794 | 1 | 0.155047129 | ENSG000001( | 16 | DMKN |
| SAT | Obesity | ENSG0000009 | 0.002157476 | 0.636000535 | 0.004558384 | 0.183454389 | 0.002342059 | 0.607375933 | 0.004558104 | 0.18342117 | -0.00018458 | 1.02E-07 | 1 | 0.840744539 | 1 | 0.822933669 | ENSG000001( | 16 | DMKN |
| SAT | Obesity | ENSG0000009 | -0.00077829 | 0.781517757 | 0.00280625 | 0.183454445 | -0.00087868 | 0.754177817 | 0.002806084 | 0.183421233 | 0.000100394 | 6.35E-06 | 1 | 0.913071213 | 1 | 0.901441682 | ENSG000001( | 6 | COX7A1 |
| SAT | Obesity | ENSG0000009 | -0.00115047 | 0.477051109 | 0.001617981 | 0.183454283 | -0.00119615 | 0.459706309 | 0.001617878 | 0.183421064 | 4.57E-05 | 0.0001195 | 1 | 0.739803464 | 1 | 0.732714066 | ENSG000001( | 10 | LRRC56 |
| SAT | Obesity | ENSG0000009 | 0.006271329 | 0.246476992 | 0.005411214 | 0.183453966 | 0.006137621 | 0.256679874 | 0.005411053 | 0.183420784 | 0.000133708 | 0.024853559 | 1 | 0.534848552 | 1 | 0.55551891 | ENSG000001( | 12 | PRPSAP1 |
| SAT | Obesity | ENSG0000009 | -0.00292678 | 0.32448049 | 0.002970461 | 0.183454107 | -0.00285969 | 0.335664292 | 0.002970282 | 0.18342092 | -6.71E-05 | 0.003628206 | 1 | 0.622949379 | 1 | 0.638443212 | ENSG000001( | 12 | PRPSAP1 |
| SAT | Obesity | ENSG0000009 | 0.007618727 | 0.001399586 | 0.002384761 | 0.183450616 | 0.00767829 | 0.001282303 | 0.002384614 | 0.183417352 | -5.96E-05 | 0.001046471 | 1 | 0.018078898 | 1 | 0.018134645 | ENSG000001( | 8 | ZNF577 |
| SAT | Obesity | ENSG0000009 | -0.00326945 | 0.227199967 | 0.002707386 | 0.183453923 | -0.00331673 | 0.220520541 | 0.00270721 | 0.183420703 | 4.73E-05 | 0.014771354 | 1 | 0.514945339 | 1 | 0.512776681 | ENSG000001( | 5 | CCDC155 |
| SAT | Obesity | ENSG000001( | -0.00199197 | 0.50650129 | 0.002998617 | 0.183454307 | -0.00192228 | 0.52146022 | 0.002998423 | 0.183421115 | -6.97E-05 | 0.001234051 | 1 | 0.763007683 | 1 | 0.775353375 | ENSG000001( | 5 | CCDC155 |
| SAT | Obesity | ENSG000001( | -0.00432434 | 0.177544326 | 0.003207118 | 0.183453787 | -0.00425533 | 0.184538797 | 0.003206948 | 0.183420605 | -6.90E-05 | 0.012581077 | 1 | 0.451027601 | 1 | 0.462308526 | ENSG000001( | 5 | ALDH16A1 |
| SAT | Obesity | ENSG000001( | 0.001657064 | 0.770544773 | 0.005681442 | 0.183454442 | 0.001769058 | 0.755501214 | 0.005681091 | 0.183421234 | -0.00011199 | 0.009478954 | 1 | 0.90745061 | 1 | 0.90236603 | ENSG000001( | 11 | SIGLEC11 |
| SAT | Obesity | ENSG000001( | 0.00285951 | 0.520159122 | 0.004446443 | 0.183454318 | 0.002929762 | 0.509933887 | 0.004446167 | 0.183421106 | -7.03E-05 | 0.036034922 | 1 | 0.772585821 | 1 | 0.767826106 | ENSG000001( | 2 | ASB16 |
| SAT | Obesity | ENSG000001( | -0.0033981 | 0.331843635 | 0.003501721 | 0.183454118 | -0.00351627 | 0.315282182 | 0.003501557 | 0.183420889 | 0.00011817 | 0.000313109 | 1 | 0.630089167 | 1 | 0.615584597 | ENSG000001( | 2 | ASB16 |
| SAT | Obesity | ENSG000001( | -0.00633256 | 0.125960004 | 0.004138304 | 0.183453589 | -0.00627456 | 0.129437615 | 0.004138004 | 0.183420401 | -5.80E-05 | 0.01948704 | 1 | 0.374273307 | 1 | 0.383362075 | ENSG000001( | 13 | DBF4B |
| SAT | Obesity | ENSG000001( | -0.00029429 | 0.947469108 | 0.004466765 | 0.183454472 | -0.00038367 | 0.931547029 | 0.004466488 | 0.183421267 | 8.94E-05 | 0.008163865 | 1 | 0.983182517 | 1 | 0.979468354 | ENSG000001( | 13 | DBF4B |
| SAT | Obesity | ENSG000001( | -0.00252038 | 0.133472304 | 0.001679634 | 0.183453623 | -0.0024899 | 0.138206158 | 0.001679519 | 0.18342044 | -3.05E-05 | 0.00623008 | 1 | 0.386967743 | 1 | 0.398466084 | ENSG000001( | 16 | LARP4 |
| SAT | Obesity | ENSG000001( | 0.013028985 | 0.000605855 | 0.003799643 | 0.18345003 | 0.013094019 | 0.00056832 | 0.003799418 | 0.183416781 | -6.50E-05 | 0.030395222 | 1 | 0.009245634 | 1 | 0.009661437 | ENSG000001( | 16 | LARP4 |
| SAT | Obesity | ENSG000001( | 0.008676641 | 0.033147066 | 0.00407296 | 0.183452759 | 0.008537038 | 0.036067858 | 0.004072689 | 0.18341961 | 0.000139603 | 6.73E-07 | 1 | 0.168954102 | 1 | 0.184615666 | ENSG000001( | 7 | SPC24 |
| SAT | Obesity | ENSG000001( | -0.01231388 | 0.017646533 | 0.005189249 | 0.183452346 | -0.01222614 | 0.018462611 | 0.005188891 | 0.183419172 | -8.77E-05 | 0.009792174 | 1 | 0.113720642 | 1 | 0.123121596 | ENSG000001( | 7 | SPC24 |
| SAT | Obesity | ENSG000001( | 0.005080731 | 0.105387923 | 0.003137651 | 0.183453483 | 0.004998411 | 0.111127443 | 0.003137441 | 0.183420311 | 8.23E-05 | 0.00013872 | 1 | 0.333987547 | 1 | 0.350248935 | ENSG000001( | 6 | IP6K3 |
| SAT | Obesity | ENSG000001( | -0.00495557 | 0.108365063 | 0.003086449 | 0.1834535 | -0.00499744 | 0.105390648 | 0.003086236 | 0.183420279 | 4.19E-05 | 0.039661044 | 1 | 0.337971267 | 1 | 0.340341256 | ENSG000001( | 6 | IP6K3 |
| SAT | Obesity | ENSG000001( | 0.000702231 | 0.87701023 | 0.004537536 | 0.183454465 | 0.000626828 | 0.890120567 | 0.004537246 | 0.183421263 | 7.54E-05 | 0.022877315 | 1 | 0.956287991 | 1 | 0.962898024 | ENSG000001( | 6 | IP6K3 |
| SAT | Obesity | ENSG000001( | -0.00103449 | 0.634711758 | 0.002177396 | 0.183454389 | -0.00107897 | 0.620205009 | 0.002177264 | 0.183421177 | 4.45E-05 | 0.008393502 | 1 | 0.840052149 | 1 | 0.830073312 | ENSG000001( | 6 | IP6K3 |
| SAT | Obesity | ENSG000001( | 0.002940278 | 0.587381962 | 0.005418528 | 0.183454363 | 0.003053648 | 0.573029886 | 0.005418159 | 0.18342115 | -0.00011337 | 0.001768017 | 1 | 0.808728584 | 1 | 0.802779912 | ENSG000001( | 7 | LEMD2 |
| SAT | Obesity | ENSG000001( | -0.00171693 | 0.609006252 | 0.003356713 | 0.183454375 | -0.00176247 | 0.59951715 | 0.003356473 | 0.183421166 | 4.55E-05 | 0.028083867 | 1 | 0.824225687 | 1 | 0.81924855 | ENSG000001( | 7 | LEMD2 |
| SAT | Obesity | ENSG000001( | 0.005541483 | 0.144345326 | 0.003796053 | 0.183453669 | 0.005604443 | 0.139815822 | 0.003795814 | 0.183420446 | -6.30E-05 | 0.02595296 | 1 | 0.401443831 | 1 | 0.400118009 | ENSG000001( | 7 | LEMD2 |
| SAT | Obesity | ENSG000001( | -0.00358362 | 0.444314666 | 0.004684911 | 0.183454253 | -0.00347713 | 0.457950292 | 0.004684724 | 0.183421062 | -0.00010649 | 0.023822247 | 1 | 0.720967567 | 1 | 0.731563013 | ENSG000001( | 7 | LEMD2 |
| SAT | Obesity | ENSG000001( | 0.007241431 | 0.024498316 | 0.003219516 | 0.183452562 | 0.007190854 | 0.025505945 | 0.003219315 | 0.183419385 | 5.06E-05 | 0.036674761 | 1 | 0.141512278 | 1 | 0.14864122 | ENSG000001( | 7 | LEMD2 |
| SAT | Obesity | ENSG000001( | 0.007615504 | 0.12764081 | 0.004998775 | 0.183453597 | 0.007739358 | 0.121539973 | 0.004998472 | 0.183420364 | -0.00012385 | 0.001376607 | 1 | 0.376521527 | 1 | 0.369554001 | ENSG000001( | 7 | LEMD2 |
| SAT | Obesity | ENSG000001( | -0.00040868 | 0.914252558 | 0.003795467 | 0.18345447 | -0.00032526 | 0.931703972 | 0.003795248 | 0.183421267 | -8.34E-05 | 0.006690992 | 1 | 0.969823034 | 1 | 0.979468354 | ENSG000001( | 13 | ADCY10P1 |
| SAT | Obesity | ENSG000001( | -0.00122294 | 0.654635769 | 0.002733848 | 0.183454398 | -0.0011608 | 0.671109384 | 0.002733691 | 0.183421202 | -6.21E-05 | 0.005056063 | 1 | 0.849292194 | 1 | 0.859449605 | ENSG000001( | 13 | ADCY10P1 |
| SAT | Obesity | ENSG000001( | 0.01056273 | 0.035717915 | 0.00502956 | 0.183452807 | 0.010653447 | 0.034150917 | 0.005029264 | 0.183419574 | -9.07E-05 | 0.023859331 | 1 | 0.177830774 | 1 | 0.178269465 | ENSG000001( | 13 | ADCY10P1 |
| SAT | Obesity | ENSG000001( | -0.00056768 | 0.772048755 | 0.001959576 | 0.183454442 | -0.00053858 | 0.783421235 | 0.001959441 | 0.183421242 | -2.91E-05 | 0.023932253 | 1 | 0.908895574 | 1 | 0.91311015 | ENSG000001( | 13 | ADCY10P1 |
| SAT | Obesity | ENSG000001( | 0.002478032 | 0.330242807 | 0.002545158 | 0.183454116 | 0.002517887 | 0.322491956 | 0.002544989 | 0.1834209 | -3.99E-05 | 0.02371252 | 1 | 0.628809457 | 1 | 0.62535231 | ENSG000001( | 13 | ADCY10P1 |
| SAT | Obesity | ENSG000001( | -0.00227308 | 0.108906433 | 0.001417898 | 0.183453503 | -0.00222258 | 0.116993606 | 0.001417894 | 0.183420342 | -5.05E-05 | 0.006919655 | 1 | 0.339015187 | 1 | 0.36350002 | ENSG000001( | 13 | ADCY10P1 |
| SAT | Obesity | ENSG000001( | 0.003275384 | 0.501354022 | 0.004871464 | 0.183454303 | 0.00913971 | 0.073552254 | 0.005107717 | 0.18342006 | -0.00586433 | 0.000135796 | 1 | 0.758937763 | 1 | 0.276431783 | ENSG000001( | 10 | SCIMP |
| SAT | Obesity | ENSG000001( | -0.01110409 | 0.024786241 | 0.004946748 | 0.183452569 | -0.01099695 | 0.026203286 | 0.004946469 | 0.183419402 | -0.00010714 | 0.008533045 | 1 | 0.142182373 | 1 | 0.15056564 | ENSG000001( | 8 | RPL26 |
| SAT | Obesity | ENSG000001( | -0.00073276 | 0.76366104 | 0.002437031 | 0.18345444 | -0.00067814 | 0.780793808 | 0.002436863 | 0.183421241 | -5.46E-05 | 0.000686659 | 1 | 0.904538582 | 1 | 0.91293214 | ENSG000001( | 13 | WDR90 |
| SAT | Obesity | ENSG000001( | 0.000547969 | 0.836353118 | 0.002652826 | 0.183454458 | 0.000623199 | 0.814260135 | 0.002652657 | 0.183421249 | -7.52E-05 | 0.000108469 | 1 | 0.938145156 | 1 | 0.928924722 | ENSG000001( | 13 | WDR90 |
| SAT | Obesity | ENSG000001( | 0.002516501 | 0.467266565 | 0.0034618 | 0.183454274 | 0.002588536 | 0.454587669 | 0.003461585 | 0.183421059 | -7.20E-05 | 0.005639775 | 1 | 0.73389258 | 1 | 0.730054891 | ENSG000001( | 13 | WDR90 |
| SAT | Obesity | ENSG000001( | 0.000312826 | 0.88776237 | 0.002216482 | 0.183454467 | 0.000551566 | 0.803518989 | 0.002216942 | 0.183421247 | -0.00023874 | 1.03E-05 | 1 | 0.960985924 | 1 | 0.923968271 | ENSG000001( | 13 | WDR90 |
| SAT | Obesity | ENSG000001( | 0.00494367 | 0.142207831 | 0.003368492 | 0.18345366 | 0.004883668 | 0.147084733 | 0.00336826 | 0.183420476 | 6.00E-05 | 0.006586431 | 1 | 0.398449097 | 1 | 0.412113586 | ENSG000001( | 7 | JMJD8 |
| SAT | Obesity | ENSG000001( | -0.02393534 | 5.28E-05 | 0.005920641 | 0.183448297 | -0.02404272 | 4.89E-05 | 0.005920308 | 0.183415037 | 0.000107379 | 0.028777244 | 0.173396153 | 0.00120414 | 0.16030374 | 0.001292772 | ENSG000001( | 5 | HS3ST6 |
| SAT | Obesity | ENSG000001( | -0.00710888 | 0.161504344 | 0.005077653 | 0.183453733 | -0.00702521 | 0.16646676 | 0.00507731 | 0.183420547 | -8.37E-05 | 0.014923519 | 1 | 0.425835921 | 1 | 0.436829777 | ENSG000001( | 10 | AMDHD2 |
| SAT | Obesity | ENSG000001( | -0.00481988 | 0.307686416 | 0.004724957 | 0.183454081 | -0.00490849 | 0.298850161 | 0.004724681 | 0.183420862 | 8.86E-05 | 0.019263367 | 1 | 0.606520234 | 1 | 0.601026164 | ENSG000001( | 8 | C11orf48 |
| SAT | Obesity | ENSG000001( | 0.007528426 | 0.047703087 | 0.003802218 | 0.183452992 | 0.007639676 | 0.044498003 | 0.003802018 | 0.183419744 | -0.00011125 | 0.000766859 | 1 | 0.210507537 | 1 | 0.207089289 | ENSG000001( | 8 | C11orf48 |
| SAT | Obesity | ENSG000001( | 0.003147943 | 0.384692982 | 0.003621312 | 0.183454188 | 0.003211913 | 0.375074931 | 0.003621063 | 0.183420973 | -6.40E-05 | 0.00758623 | 1 | 0.677497409 | 1 | 0.670638065 | ENSG000001( | 8 | C11orf48 |
| SAT | Obesity | ENSG000001( | 0.002696312 | 0.530927013 | 0.004303139 | 0.183454326 | 0.002784791 | 0.517507076 | 0.004302873 | 0.183421112 | -8.85E-05 | 0.006704467 | 1 | 0.781854367 | 1 | 0.773197048 | ENSG000001( | 9 | TTC9C |
| SAT | Obesity | ENSG000001( | -0.0077018 | 0.061375808 | 0.004116889 | 0.183453151 | -0.007778 | 0.058836708 | 0.004116617 | 0.183419921 | 7.62E-05 | 0.008151385 | 1 | 0.246479835 | 1 | 0.245276282 | ENSG000001( | 6 | RPL29 |
| SAT | Obesity | ENSG000001( | -0.00094313 | 0.863432106 | 0.005483063 | 0.183454463 | -0.00081638 | 0.881632074 | 0.005482708 | 0.183421262 | -0.00012675 | 0.001283487 | 1 | 0.950006955 | 1 | 0.959414539 | ENSG000001( | 6 | RPL29 |
| SAT | Obesity | ENSG000001( | 0.003459458 | 0.098130027 | 0.002091582 | 0.1834534 | 0.00341215 | 0.102791437 | 0.002091455 | 0.183420264 | 4.73E-05 | 0.003266446 | 1 | 0.319058996 | 1 | 0.334582049 | ENSG000001( | 6 | RPL29 |
| SAT | Obesity | ENSG000001( | -0.00344613 | 0.267652878 | 0.003108861 | 0.18345401 | -0.00338707 | 0.275915268 | 0.00310871 | 0.183420822 | -5.91E-05 | 0.037455434 | 1 | 0.563881016 | 1 | 0.579934654 | ENSG000001( | 6 | RPL29 |
| SAT | Obesity | ENSG000001( | -0.00806143 | 0.008896072 | 0.003081549 | 0.183451887 | -0.00815661 | 0.008119631 | 0.003081378 | 0.183418622 | 9.52E-05 | 0.000229218 | 1 | 0.072606992 | 1 | 0.072001372 | ENSG000001( | 7 | ZFPL1 |
| SAT | Obesity | ENSG000001( | 0.004264535 | 0.357533058 | 0.004634977 | 0.183454154 | 0.00434721 | 0.348263012 | 0.004634726 | 0.183420938 | -8.27E-05 | 0.036990786 | 1 | 0.655187068 | 1 | 0.650436499 | ENSG000001( | 11 | TPCN2 |
| SAT | Obesity | ENSG000001( | 0.000642912 | 0.85840754 | 0.003603729 | 0.183454462 | 0.000758809 | 0.833219958 | 0.003603529 | 0.183421253 | -0.0001159 | 0.000118953 | 1 | 0.947247673 | 1 | 0.936326075 | ENSG000001( | 11 | TPCN2 |
| SAT | Obesity | ENSG000001( | -0.02046223 | 0.000771691 | 0.006084923 | 0.1834502 | -0.02059519 | 0.000712265 | 0.006084546 | 0.18341694 | 0.000132958 | 0.003896118 | 1 | 0.011203176 | 1 | 0.01139972 | ENSG000001( | 11 | TPCN2 |
| SAT | Obesity | ENSG000001( | -0.00444157 | 0.288210637 | 0.004182049 | 0.183454048 | -0.00435499 | 0.297679412 | 0.004181768 | 0.18342086 | -8.66E-05 | 0.002449324 | 1 | 0.587064948 | 1 | 0.601026164 | ENSG000001( | 11 | TPCN2 |
| SAT | Obesity | ENSG000001( | 0.0019267 | 0.698258853 | 0.00496992 | 0.183454417 | 0.001815926 | 0.71481233 | 0.004969683 | 0.18342122 | 0.000110773 | 0.015795859 | 1 | 0.870105316 | 1 | 0.884017812 | ENSG000001( | 11 | TPCN2 |

| | | | | | | | | | | | | | | | | | | | |
|---|---|---|---|---|---|---|---|---|---|---|---|---|---|---|---|---|---|---|---|
| SAT | Obesity | ENSG000001( | 0.002344826 | 0.562421311 | 0.004048043 | 0.183454347 | 0.002283815 | 0.572609077 | 0.004047784 | 0.18342115 | 6.10E-05 | 0.03856076 | 1 | 0.797231826 | 1 | 0.802779912 | ENSG000001( | 11 | TPCN2 |
| SAT | Obesity | ENSG000001( | -0.00488869 | 0.375555041 | 0.005516986 | 0.183454177 | -0.00499134 | 0.365411517 | 0.005516657 | 0.183420961 | 0.000104446 | 0.015915428 | 1 | 0.670275034 | 1 | 0.665460489 | ENSG000001( | 7 | CYP4A22 |
| SAT | Obesity | ENSG000001( | 0.003368802 | 0.568396931 | 0.005905885 | 0.183454351 | 0.003256211 | 0.581371198 | 0.00590553 | 0.183421155 | 0.000112591 | 0.014374251 | 1 | 0.797993295 | 1 | 0.807911436 | ENSG000001( | 7 | CYP4A22 |
| SAT | Obesity | ENSG000001( | 0.004618086 | 0.360933685 | 0.005054888 | 0.183454159 | 0.004719764 | 0.350426609 | 0.005054562 | 0.183420941 | -0.00010168 | 0.005424454 | 1 | 0.65717171 | 1 | 0.65249005 | ENSG000001( | 9 | CMPK1 |
| SAT | Obesity | ENSG000001( | -0.00968961 | 0.106525636 | 0.006003434 | 0.183453489 | -0.00956535 | 0.111071909 | 0.006003118 | 0.183420311 | -0.00012426 | 0.017301951 | 1 | 0.335175838 | 1 | 0.350248935 | ENSG000001( | 9 | CMPK1 |
| SAT | Obesity | ENSG000001( | -0.00343005 | 0.34943359 | 0.003665803 | 0.183454143 | -0.00337398 | 0.357335686 | 0.003665562 | 0.18342095 | -5.61E-05 | 0.029249186 | 1 | 0.647953427 | 1 | 0.658105737 | ENSG000001( | 7 | SLC1A7 |
| SAT | Obesity | ENSG000001( | -0.00775586 | 0.028587324 | 0.003542885 | 0.183452663 | -0.00763783 | 0.031089335 | 0.0035427 | 0.183419514 | -0.00011804 | 0.000134358 | 1 | 0.154014794 | 1 | 0.166130468 | ENSG000001( | 7 | SLC1A7 |
| SAT | Obesity | ENSG000001( | -0.01098643 | 0.041211661 | 0.005381811 | 0.183452899 | -0.01085711 | 0.043646446 | 0.005381531 | 0.183419732 | -0.00012932 | 0.005986407 | 1 | 0.192979684 | 1 | 0.204577127 | ENSG000001( | 7 | SLC1A7 |
| SAT | Obesity | ENSG000001( | 0.004004074 | 0.294975205 | 0.003823342 | 0.18345406 | 0.003951498 | 0.301329061 | 0.003823086 | 0.183420866 | 5.26E-05 | 0.045333607 | 1 | 0.593750704 | 1 | 0.601253497 | ENSG000001( | 9 | C1orf123 |
| SAT | Obesity | ENSG000001( | 0.000672399 | 0.764903804 | 0.002248467 | 0.18345444 | 0.00070481 | 0.75391393 | 0.00224832 | 0.183421233 | -3.24E-05 | 0.042902946 | 1 | 0.904704175 | 1 | 0.901441682 | ENSG000001( | 9 | C1orf123 |
| SAT | Obesity | ENSG000001( | 0.005846298 | 0.27135466 | 0.00531506 | 0.183454017 | 0.005706747 | 0.282928473 | 0.005314706 | 0.183420834 | 0.000139551 | 0.000144609 | 1 | 0.567681645 | 1 | 0.587152637 | ENSG000001( | 9 | C1orf123 |
| SAT | Obesity | ENSG000001( | 0.00018105 | 0.963830114 | 0.003992467 | 0.183454473 | 0.001072812 | 0.788658515 | 0.004002235 | 0.183421243 | -0.00089176 | 0.001726095 | 1 | 0.990703823 | 1 | 0.917518993 | ENSG000001( | 9 | C1orf123 |
| SAT | Obesity | ENSG000001( | 0.005150484 | 0.19942671 | 0.004013817 | 0.183453852 | 0.005225666 | 0.192919278 | 0.004013589 | 0.18342063 | -7.52E-05 | 0.022396327 | 1 | 0.481470961 | 1 | 0.476274003 | ENSG000001( | 8 | USP24 |
| SAT | Obesity | ENSG000001( | 0.002311427 | 0.306905653 | 0.002262251 | 0.183454079 | 0.002367106 | 0.295368462 | 0.002262103 | 0.183420856 | -5.57E-05 | 0.000506421 | 1 | 0.606475433 | 1 | 0.599693023 | ENSG000001( | 8 | USP24 |
| SAT | Obesity | ENSG000001( | 0.001791442 | 0.40123763 | 0.002134158 | 0.183454208 | 0.001836136 | 0.389568319 | 0.002134046 | 0.18342099 | -4.47E-05 | 0.016010894 | 1 | 0.691650463 | 1 | 0.683150002 | ENSG000001( | 8 | USP24 |
| SAT | Obesity | ENSG000001( | -0.00780806 | 0.088952916 | 0.004590411 | 0.18345338 | -0.0077064 | 0.093172482 | 0.004590155 | 0.183420205 | -0.00010166 | 0.007703539 | 1 | 0.30212683 | 1 | 0.316069472 | ENSG000001( | 6 | ZSWIM5 |
| SAT | Obesity | ENSG000001( | -0.00580268 | 0.00037613 | 0.001631655 | 0.183449694 | -0.00575347 | 0.000421317 | 0.00163155 | 0.18341657 | -4.92E-05 | 2.42E-05 | 1 | 0.006201415 | 1 | 0.007477175 | ENSG000001( | 6 | ZSWIM5 |
| SAT | Obesity | ENSG000001( | -0.0016647 | 0.673259366 | 0.003947789 | 0.183454407 | -0.0015717 | 0.690522104 | 0.003947532 | 0.18342121 | -9.30E-05 | 0.000927342 | 1 | 0.855855862 | 1 | 0.868937924 | ENSG000001( | 7 | SLC25A34 |
| SAT | Obesity | ENSG000001( | 0.006143666 | 0.233391798 | 0.005155515 | 0.183453937 | 0.006247719 | 0.225542801 | 0.005155215 | 0.183420715 | -0.00010405 | 0.012019644 | 1 | 0.522565341 | 1 | 0.518492259 | ENSG000001( | 10 | SYNC |
| SAT | Obesity | ENSG000001( | 0.007894562 | 0.00104283 | 0.002407812 | 0.183450411 | 0.00785795 | 0.001099647 | 0.002407661 | 0.183417244 | 3.66E-05 | 0.042288609 | 1 | 0.01443682 | 1 | 0.016106882 | ENSG000001( | 7 | OMA1 |
| SAT | Obesity | ENSG000001( | 0.000673915 | 0.871228551 | 0.00415745 | 0.183454464 | 0.000535187 | 0.897565373 | 0.004157191 | 0.183421264 | 0.000138729 | 9.12E-06 | 1 | 0.955362338 | 1 | 0.963222823 | ENSG000001( | 7 | OMA1 |
| SAT | Obesity | ENSG000001( | -0.00819876 | 0.04687267 | 0.004125257 | 0.183452981 | -0.00811468 | 0.049163816 | 0.004125048 | 0.183419808 | -8.41E-05 | 0.022033066 | 1 | 0.208104508 | 1 | 0.221531933 | ENSG000001( | 7 | OMA1 |
| SAT | Obesity | ENSG000001( | 0.003184155 | 0.236508638 | 0.002689865 | 0.183453944 | 0.003220261 | 0.231202985 | 0.002689676 | 0.183420728 | -3.61E-05 | 0.03496254 | 1 | 0.525379039 | 1 | 0.524775447 | ENSG000001( | 14 | NTNG1 |
| SAT | Obesity | ENSG000001( | -0.00220933 | 0.461680484 | 0.003001462 | 0.183454269 | -0.00227266 | 0.448912238 | 0.003001276 | 0.183421053 | 6.33E-05 | 0.005195499 | 1 | 0.731904575 | 1 | 0.726631007 | ENSG000001( | 9 | EXTL2 |
| SAT | Obesity | ENSG000001( | 0.006536232 | 0.149895577 | 0.004539357 | 0.1834536 | 0.006612537 | 0.145171011 | 0.00453907 | 0.183420468 | -7.63E-05 | 0.022795275 | 1 | 0.409839489 | 1 | 0.40919767 | ENSG000001( | 11 | ARPC5 |
| SAT | Obesity | ENSG000001( | 0.005441395 | 0.034177716 | 0.00256829 | 0.183452777 | 0.005334367 | 0.037792042 | 0.002568169 | 0.18341964 | 0.000107028 | 7.27E-06 | 1 | 0.172215733 | 1 | 0.189433997 | ENSG000001( | 8 | TRIM58 |
| SAT | Obesity | ENSG000001( | 0.004098258 | 0.332561974 | 0.004229511 | 0.183454119 | 0.004026444 | 0.34107147 | 0.00422923 | 0.183420928 | 7.18E-05 | 0.014721322 | 1 | 0.630349991 | 1 | 0.64461722 | ENSG000001( | 8 | PEA15 |
| SAT | Obesity | ENSG0000011 | -0.00606103 | 0.021260828 | 0.002631421 | 0.183452469 | -0.00290834 | 0.28047356 | 0.002694759 | 0.18342083 | -0.00315269 | 6.01E-08 | 1 | 0.129189682 | 1 | 0.584646601 | ENSG000001( | 8 | PEA15 |
| SAT | Obesity | ENSG0000011 | 0.005064743 | 0.237693235 | 0.004289343 | 0.183453947 | 0.004993599 | 0.244317588 | 0.004289061 | 0.183420758 | 7.11E-05 | 0.018189256 | 1 | 0.526060746 | 1 | 0.539991654 | ENSG000001( | 6 | C1orf115 |
| SAT | Obesity | ENSG0000011 | 0.001499683 | 0.397244275 | 0.001771515 | 0.183454203 | 0.001532397 | 0.38699722 | 0.0017714 | 0.183420987 | -3.27E-05 | 0.009996582 | 1 | 0.688878681 | 1 | 0.680459742 | ENSG000001( | 6 | C1orf115 |
| SAT | Obesity | ENSG0000011 | -0.00520118 | 0.106085352 | 0.00321846 | 0.183453487 | -0.00515712 | 0.109054133 | 0.003218246 | 0.1834203 | -4.41E-05 | 0.047877805 | 1 | 0.334678885 | 1 | 0.346730908 | ENSG000001( | 6 | C1orf115 |
| SAT | Obesity | ENSG0000011 | -0.00449279 | 0.186845029 | 0.0034037 | 0.183453816 | -0.00459925 | 0.176589209 | 0.003403488 | 0.18342058 | 0.000106459 | 3.00E-05 | 1 | 0.461882921 | 1 | 0.452214424 | ENSG000001( | 7 | ACP6 |
| SAT | Obesity | ENSG0000011 | 0.004563011 | 0.410858666 | 0.005548514 | 0.183454218 | 0.004130831 | 0.456590887 | 0.005548666 | 0.183421061 | 0.00043218 | 3.89E-07 | 1 | 0.698355005 | 1 | 0.731541927 | ENSG000001( | 8 | PM20D1 |
| SAT | Obesity | ENSG0000011 | 0.001842816 | 0.617110699 | 0.003686003 | 0.18345438 | 0.002105717 | 0.567944119 | 0.003687239 | 0.183421147 | -0.0002629 | 0.014536923 | 1 | 0.829471612 | 1 | 0.800784122 | ENSG000001( | 8 | PM20D1 |
| SAT | Obesity | ENSG0000011 | -0.00085137 | 0.874482205 | 0.005389527 | 0.183454465 | -0.0007344 | 0.89160566 | 0.005389202 | 0.183421263 | -0.00011697 | 0.005248512 | 1 | 0.955754868 | 1 | 0.962922373 | ENSG000001( | 8 | PM20D1 |
| SAT | Obesity | ENSG0000011 | 0.000813373 | 0.549756009 | 0.001359868 | 0.183454339 | 0.000859171 | 0.527531111 | 0.001359922 | 0.183421119 | -4.58E-05 | 0.036408398 | 1 | 0.793070817 | 1 | 0.779574758 | ENSG000001( | 8 | B3GALNT2 |
| SAT | Obesity | ENSG0000011 | -0.00015164 | 0.93307592 | 0.001805749 | 0.18345471 | -0.0001263 | 0.944234957 | 0.001805625 | 0.183421268 | -2.53E-05 | 0.035543449 | 1 | 0.978715503 | 1 | 0.983503141 | ENSG000001( | 8 | B3GALNT2 |
| SAT | Obesity | ENSG0000011 | 0.003409344 | 0.014239553 | 0.001390904 | 0.183452203 | 0.003787027 | 0.006552622 | 0.001392916 | 0.183418477 | -0.00037768 | 9.91E-07 | 1 | 0.099045337 | 1 | 0.063606962 | ENSG000001( | 8 | B3GALNT2 |
| SAT | Obesity | ENSG0000011 | 0.008505501 | 0.129721484 | 0.005613437 | 0.183453606 | 0.008403143 | 0.134377968 | 0.005613092 | 0.183420423 | 0.000102358 | 0.016591086 | 1 | 0.380265236 | 1 | 0.392603841 | ENSG000001( | 8 | B3GALNT2 |
| SAT | Obesity | ENSG0000011 | -0.00865048 | 0.002011295 | 0.002800798 | 0.183450868 | -0.00851226 | 0.002371971 | 0.002800795 | 0.183417779 | -0.00013822 | 0.000219142 | 1 | 0.023996583 | 1 | 0.030048015 | ENSG000001( | 8 | B3GALNT2 |
| SAT | Obesity | ENSG0000011 | 0.001220878 | 0.658634437 | 0.00276342 | 0.1834544 | 0.001270745 | 0.645605272 | 0.002763232 | 0.183421219 | -4.99E-05 | 0.006948736 | 1 | 0.852457431 | 1 | 0.846276827 | ENSG000001( | 14 | MRPL55 |
| SAT | Obesity | ENSG0000011 | 0.003703626 | 0.310280835 | 0.003650209 | 0.183454085 | 0.003613666 | 0.32215113 | 0.003649991 | 0.1834209 | 9.00E-05 | 0.001660531 | 1 | 0.607417315 | 1 | 0.625060826 | ENSG000001( | 9 | C1orf145 |
| SAT | Obesity | ENSG0000011 | 0.002919108 | 0.375036698 | 0.003290696 | 0.183454177 | 0.00302815 | 0.357435057 | 0.003290522 | 0.183420961 | -0.00010904 | 0.000130185 | 1 | 0.670275034 | 1 | 0.658105737 | ENSG000001( | 11 | CLHC1 |
| SAT | Obesity | ENSG0000011 | 0.005459574 | 0.367122489 | 0.006053566 | 0.183454167 | 0.004244637 | 0.483443594 | 0.006057059 | 0.183421085 | 0.001214937 | 3.93E-08 | 1 | 0.664384383 | 1 | 0.747492193 | ENSG000001( | 5 | H3F3A |
| SAT | Obesity | ENSG0000011 | 0.001306971 | 0.645512157 | 0.002841204 | 0.183454394 | 0.001391987 | 0.624170352 | 0.002841086 | 0.183421179 | -8.50E-05 | 0.002502384 | 1 | 0.844125734 | 1 | 0.831978382 | ENSG000001( | 5 | H3F3A |
| SAT | Obesity | ENSG0000011 | -0.00079664 | 0.768790034 | 0.002710038 | 0.183454441 | -0.00086813 | 0.748700366 | 0.002709897 | 0.183421231 | 7.15E-05 | 0.00267955 | 1 | 0.907300451 | 1 | 0.900879844 | ENSG000001( | 5 | H3F3A |
| SAT | Obesity | ENSG0000011 | -0.00573636 | 0.006596863 | 0.002111636 | 0.183451685 | -0.00478832 | 0.02380765 | 0.002118514 | 0.183419339 | -0.00094804 | 4.15E-08 | 1 | 0.05946238 | 1 | 0.143326422 | ENSG000001( | 5 | H3F3A |
| SAT | Obesity | ENSG0000011 | 0.020884891 | 1.09E-08 | 0.00365393 | 0.183442126 | 0.017307355 | 3.78E-06 | 0.003743771 | 0.183413193 | 0.003577535 | 1.18E-05 | 3.59E-05 | 6.64E-07 | 0.012414489 | 0.000167763 | ENSG000001( | 11 | RPRD2 |
| SAT | Obesity | ENSG0000011 | -0.00462787 | 0.124274912 | 0.003010822 | 0.183453581 | -0.00469437 | 0.118934634 | 0.003010633 | 0.183420351 | 6.65E-05 | 0.003023648 | 1 | 0.370341494 | 1 | 0.367443065 | ENSG000001( | 6 | CTSS |
| SAT | Obesity | ENSG0000011 | 0.003950692 | 0.441928672 | 0.005137834 | 0.183454251 | 0.001989036 | 0.703268182 | 0.005221764 | 0.183421215 | 0.001961656 | 0.035902753 | 1 | 0.720521812 | 1 | 0.876674335 | ENSG000001( | 6 | CTSS |
| SAT | Obesity | ENSG0000011 | 0.008914544 | 0.122958926 | 0.005779378 | 0.183453575 | 0.009066424 | 0.116682042 | 0.005779002 | 0.18342034 | -0.00015188 | 0.000232282 | 1 | 0.367513287 | 1 | 0.363219906 | ENSG000001( | 6 | CTSS |
| SAT | Obesity | ENSG0000011 | 0.002284977 | 0.49978935 | 0.003386045 | 0.183454302 | 0.002217572 | 0.512495058 | 0.003385823 | 0.183421108 | 6.74E-05 | 0.004879313 | 1 | 0.757767494 | 1 | 0.769450316 | ENSG000001( | 14 | PACRGL |
| SAT | Obesity | ENSG0000011 | -0.01186208 | 0.000212246 | 0.003202502 | 0.183449288 | -0.01179997 | 0.000228853 | 0.003202286 | 0.183416139 | -6.21E-05 | 0.004014116 | 0.696379361 | 0.0038474 | 0.750867837 | 0.004550714 | ENSG000001( | 14 | PACRGL |
| SAT | Obesity | ENSG0000011 | -0.00431493 | 0.350232011 | 0.004619141 | 0.183454144 | -0.00423659 | 0.359015878 | 0.004618835 | 0.183420952 | -7.83E-05 | 0.014777589 | 1 | 0.648116881 | 1 | 0.660152534 | ENSG000001( | 14 | PACRGL |
| SAT | Obesity | ENSG0000011 | -0.0044882 | 0.330126843 | 0.004608674 | 0.183454116 | -0.00460946 | 0.317197799 | 0.004608384 | 0.183420892 | 0.000121261 | 0.000393097 | 1 | 0.628809457 | 1 | 0.617275195 | ENSG000001( | 14 | PACRGL |
| SAT | Obesity | ENSG0000011 | -0.00038173 | 0.842369271 | 0.001919578 | 0.183454459 | -0.00033262 | 0.862427119 | 0.001919468 | 0.183421259 | -4.91E-05 | 0.001707755 | 1 | 0.941066406 | 1 | 0.949538046 | ENSG000001( | 14 | PACRGL |
| SAT | Obesity | ENSG0000011 | -0.00888974 | 0.101800416 | 0.005433193 | 0.183453462 | -0.00872734 | 0.108187533 | 0.005432869 | 0.183420295 | -0.0001624 | 0.000134426 | 1 | 0.325294064 | 1 | 0.345600181 | ENSG000001( | 7 | LYSMD1 |
| SAT | Obesity | ENSG0000011 | -0.00066829 | 0.550082447 | 0.001118216 | 0.183454339 | -0.00069525 | 0.534079433 | 0.001118143 | 0.183421124 | 2.70E-05 | 0.000670711 | 1 | 0.793070817 | 1 | 0.784735611 | ENSG000001( | 7 | LYSMD1 |
| SAT | Obesity | ENSG0000011 | 0.000232479 | 0.927581384 | 0.002557857 | 0.183454471 | 0.000158182 | 0.950686188 | 0.002557707 | 0.183421269 | 7.43E-05 | 0.000257084 | 1 | 0.976869566 | 1 | 0.987089045 | ENSG000001( | 8 | ERCC3 |
| SAT | Obesity | ENSG0000011 | 0.007299605 | 0.1305093 | 0.004827465 | 0.18345361 | 0.007206225 | 0.135480362 | 0.0048272 | 0.183420428 | 9.34E-05 | 0.021695231 | 1 | 0.381300991 | 1 | 0.394070094 | ENSG000001( | 8 | ERCC3 |
| SAT | Obesity | ENSG0000011 | 0.003604394 | 0.267657511 | 0.003251671 | 0.18345401 | 0.00340115 | 0.295579101 | 0.003251696 | 0.183420857 | 0.000203244 | 8.10E-06 | 1 | 0.563881016 | 1 | 0.599749554 | ENSG000001( | 7 | GNPDA2 |
| SAT | Obesity | ENSG0000011 | 0.018662524 | 0.000684476 | 0.005495856 | 0.183450115 | 0.018763661 | 0.000639387 | 0.005495523 | 0.183416864 | -0.00010114 | 0.017863371 | 1 | 0.010208026 | 1 | 0.010446545 | ENSG000001( | 7 | GNPDA2 |
| SAT | Obesity | ENSG0000011 | -0.00200571 | 0.715753633 | 0.005508062 | 0.183454424 | -0.00184756 | 0.737287408 | 0.005507729 | 0.183421228 | -0.00015815 | 0.000210123 | 1 | 0.882189207 | 1 | 0.896583401 | ENSG000001( | 7 | GNPDA2 |
| SAT | Obesity | ENSG0000011 | -0.00181736 | 0.585133879 | 0.003329091 | 0.183454361 | -0.00176386 | 0.596205392 | 0.003328874 | 0.183421164 | -5.35E-05 | 0.023629612 | 1 | 0.808006842 | 1 | 0.817137678 | ENSG000001( | 7 | GNPDA2 |
| SAT | Obesity | ENSG0000011 | -0.01036429 | 0.007651139 | 0.003885977 | 0.183451785 | -0.01025109 | 0.008336543 | 0.003885729 | 0.18341864 | -0.0001132 | 6.42E-05 | 1 | 0.065715675 | 1 | 0.07313422 | ENSG000001( | 7 | GNPDA2 |
| SAT | Obesity | ENSG0000011 | -0.01358158 | 0.006416845 | 0.004982794 | 0.183451666 | -0.01438343 | 0.003917062 | 0.004985973 | 0.183418125 | 0.000801849 | 2.52E-05 | 1 | 0.058320407 | 1 | 0.044436554 | ENSG000001( | 7 | GNPDA2 |
| SAT | Obesity | ENSG0000011 | 0.00218821 | 0.208529702 | 0.001739961 | 0.183453876 | 0.00128826 | 0.465621506 | 0.001765653 | 0.183421069 | 0.00089995 | 0.00279215 | 1 | 0.496146448 | 1 | 0.736953286 | ENSG000001( | 12 | ANTXR2 |
| SAT | Obesity | ENSG0000011 | 0.005863627 | 0.017849419 | 0.002475425 | 0.183452353 | 0.00579988 | 0.01912816 | 0.002475374 | 0.183419195 | 6.37E-05 | 0.029254258 | 1 | 0.114606541 | 1 | 0.125770525 | ENSG000001( | 12 | ANTXR2 |
| SAT | Obesity | ENSG0000011 | -0.01257849 | 1.71E-07 | 0.002405883 | 0.183444142 | -0.01253659 | 1.88E-07 | 0.002405729 | 0.183411007 | -4.19E-05 | 0.016293763 | 0.000561766 | 8.14E-06 | 0.000616035 | 1.06E-05 | ENSG000001( | 12 | ANTXR2 |
| SAT | Obesity | ENSG0000011 | 0.006889473 | 0.120956479 | 0.004442602 | 0.183453565 | 0.006760695 | 0.128040605 | 0.004442341 | 0.183420395 | 0.000128777 | 0.000263307 | 1 | 0.364377321 | 1 | 0.381612058 | ENSG000001( | 12 | ANTXR2 |
| SAT | Obesity | ENSG0000011 | -0.01537978 | 0.000886847 | 0.004626635 | 0.183450297 | -0.01546001 | 0.00083258 | 0.00462632 | 0.18341705 | 8.02E-05 | 0.009954404 | 1 | 0.012434808 | 1 | 0.012764933 | ENSG000001( | 12 | ANTXR2 |
| SAT | Obesity | ENSG0000011 | -0.00032301 | 0.855445102 | 0.001773098 | 0.183454462 | -0.00027649 | 0.876078182 | 0.001773003 | 0.183421261 | -4.65E-05 | 0.002386303 | 1 | 0.945022013 | 1 | 0.958314845 | ENSG000001( | 10 | DCST2 |
| SAT | Obesity | ENSG0000011 | -0.00531015 | 0.355059894 | 0.005741814 | 0.183454151 | -0.00557979 | 0.331133809 | 0.005741503 | 0.183420913 | 0.00026964 | 3.78E-08 | 1 | 0.652999726 | 1 | 0.632292088 | ENSG000001( | 6 | POGLUT1 |
| SAT | Obesity | ENSG0000011 | 0.007431693 | 0.065547857 | 0.004035657 | 0.183453192 | 0.007492934 | 0.063339238 | 0.004035387 | 0.183419967 | -6.12E-05 | 0.026695312 | 1 | 0.257251818 | 1 | 0.256563012 | ENSG000001( | 6 | POGLUT1 |
| SAT | Obesity | ENSG0000011 | -0.00475664 | 0.305608411 | 0.004642975 | 0.183454077 | -0.00483247 | 0.297930453 | 0.004642664 | 0.183420861 | 7.58E-05 | 0.017277938 | 1 | 0.605495891 | 1 | 0.601026164 | ENSG000001( | 6 | POGLUT1 |
| SAT | Obesity | ENSG0000011 | 0.002356296 | 0.3773798 | 0.002669312 | 0.18345418 | 0.002412412 | 0.366095745 | 0.002669154 | 0.183420961 | -5.61E-05 | 0.007824232 | 1 | 0.671085989 | 1 | 0.665462682 | ENSG000001( | 6 | POGLUT1 |
| SAT | Obesity | ENSG0000011 | -0.00324444 | 0.282322315 | 0.003017756 | 0.183454037 | -0.00331894 | 0.271389438 | 0.003017571 | 0.183420813 | 7.45E-05 | 0.001303288 | 1 | 0.579661774 | 1 | 0.574631165 | ENSG000001( | 8 | SLC22A15 |
| SAT | Obesity | ENSG0000011 | 0.007367155 | 0.031510264 | 0.003425668 | 0.183452726 | 0.007289022 | 0.033345497 | 0.003425446 | 0.183419559 | 7.81E-05 | 0.001443792 | 1 | 0.163843387 | 1 | 0.175894815 | ENSG000001( | 12 | IGFN1 |
| SAT | Obesity | ENSG0000011 | -0.00124538 | 0.64613365 | 0.002712424 | 0.183454394 | -0.00119745 | 0.658853846 | 0.002712258 | 0.183421197 | -4.79E-05 | 0.021232371 | 1 | 0.844125734 | 1 | 0.852666776 | ENSG000001( | 12 | IGFN1 |
| SAT | Obesity | ENSG0000011 | -0.00030119 | 0.912535361 | 0.002742021 | 0.18345447 | -0.00023877 | 0.930604981 | 0.002741852 | 0.183421267 | -6.24E-05 | 0.002777145 | 1 | 0.968941268 | 1 | 0.979254311 | ENSG000001( | 11 | LMOD1 |
| SAT | Obesity | ENSG0000011 | -0.003468 | 0.470550951 | 0.004806092 | 0.183454277 | -0.00389419 | 0.417934709 | 0.004807587 | 0.183421022 | 0.000426198 | 0.001759534 | 1 | 0.735593387 | 1 | 0.703737593 | ENSG000001( | 7 | TMEM183A |
| SAT | Obesity | ENSG0000011 | -0.01066801 | 0.041668388 | 0.005237577 | 0.183452906 | -0.01076103 | 0.039907182 | 0.005237238 | 0.183419675 | 9.30E-05 | 0.013526459 | 1 | 0.19344648 | 1 | 0.195087821 | ENSG000001( | 7 | IGFBP7 |
| SAT | Obesity | ENSG0000011 | -0.00655979 | 0.212222694 | 0.005258438 | 0.183453886 | -0.00640691 | 0.223041945 | 0.005258113 | 0.183420709 | -0.00015288 | 0.000124291 | 1 | 0.501426545 | 1 | 0.516186585 | ENSG000001( | 7 | IGFBP7 |
| SAT | Obesity | ENSG0000011 | 0.001331696 | 0.79841986 | 0.005214307 | 0.183454449 | 0.001438385 | 0.78264647 | 0.005213957 | 0.183421241 | -0.00010669 | 0.002732245 | 1 | 0.919633097 | 1 | 0.91293214 | ENSG000001( | 7 | IGFBP7 |
| SAT | Obesity | ENSG0000011 | 3.23E-05 | 0.992360006 | 0.003371891 | 0.183454474 | 0.000111077 | 0.973720333 | 0.003371819 | 0.18342127 | -7.88E-05 | 0.046777729 | 1 | 0.996612544 | 1 | 0.992266083 | ENSG000001( | 10 | CCT3 |
| SAT | Obesity | ENSG0000011 | 0.000413135 | 0.797748493 | 0.001612156 | 0.183454449 | 0.000458408 | 0.776137423 | 0.001612091 | 0.18342124 | -4.53E-05 | 0.005122237 | 1 | 0.919633097 | 1 | 0.911862059 | ENSG000001( | 8 | SRGAP2 |
| SAT | Obesity | ENSG0000011 | -0.00270946 | 0.415049797 | 0.003324329 | 0.183454223 | -0.00261099 | 0.432179649 | 0.003324124 | 0.183421037 | -9.85E-05 | 9.41E-05 | 1 | 0.700759618 | 1 | 0.715518881 | ENSG000001( | 9 | CCDC141 |
| SAT | Obesity | ENSG0000011 | 0.013042436 | 0.016433067 | 0.005436249 | 0.183452298 | 0.012717123 | 0.019321355 | 0.005436341 | 0.183419202 | 0.000325313 | 4.33E-05 | 1 | 0.108703412 | 1 | 0.126415995 | ENSG000001( | 11 | GLB1L |
| SAT | Obesity | ENSG0000011 | -0.0132622 | 0.024406976 | 0.005892559 | 0.183452559 | -0.0134188 | 0.022763247 | 0.005892176 | 0.18341931 | 0.000156599 | 0.000210112 | 1 | 0.141482842 | 1 | 0.139600399 | ENSG000001( | 11 | GLB1L |
| SAT | Obesity | ENSG0000011 | 0.001566711 | 0.517247348 | 0.002419276 | 0.183454316 | 0.001458763 | 0.546502864 | 0.002419139 | 0.183421133 | 0.000107947 | 6.01E-08 | 1 | 0.771001245 | 1 | 0.788841592 | ENSG000001( | 11 | GLB1L |
| SAT | Obesity | ENSG0000011 | -0.00145905 | 0.788921443 | 0.005450097 | 0.183454447 | -0.00134689 | 0.804794862 | 0.005449758 | 0.183421247 | -0.00011217 | 0.006095172 | 1 | 0.914498218 | 1 | 0.923968271 | ENSG000001( | 11 | GLB1L |
| SAT | Obesity | ENSG0000011 | -0.00027844 | 0.942853609 | 0.003884259 | 0.183454472 | -0.00020218 | 0.958485261 | 0.003884005 | 0.183421269 | -7.63E-05 | 0.005644587 | 1 | 0.981335116 | 1 | 0.990485083 | ENSG000001( | 7 | NUAK2 |
| SAT | Obesity | ENSG0000011 | -0.00464785 | 0.033149546 | 0.002181809 | 0.183452759 | -0.00471952 | 0.030523638 | 0.002181694 | 0.183419502 | 7.17E-05 | 0.000168532 | 1 | 0.168954102 | 1 | 0.164446725 | ENSG000001( | 7 | NUAK2 |
| SAT | Obesity | ENSG0000011 | 0.003525746 | 0.056919944 | 0.001851826 | 0.183453104 | 0.003495443 | 0.059068339 | 0.001851705 | 0.183419923 | 3.03E-05 | 0.021567415 | 1 | 0.235503577 | 1 | 0.245320532 | ENSG000001( | 7 | NUAK2 |
| SAT | Obesity | ENSG0000012 | 0.005907306 | 0.167406861 | 0.004278839 | 0.183453754 | 0.00603249 | 0.158564175 | 0.004278607 | 0.183420519 | -0.00012518 | 0.000603885 | 1 | 0.433856169 | 1 | 0.426084405 | ENSG000001( | 7 | NUAK2 |
| SAT | Obesity | ENSG0000012 | -0.02153914 | 0.000159867 | 0.005705356 | 0.183449087 | -0.02142411 | 0.000173122 | 0.005704962 | 0.18341594 | -0.00011503 | 0.002037209 | 0.524522634 | 0.002997272 | 0.568012556 | 0.003641106 | ENSG000001( | 7 | NUAK2 |
| SAT | Obesity | ENSG0000012 | -0.00464797 | 0.006946425 | 0.001721846 | 0.18345172 | -0.00456783 | 0.007986883 | 0.001721996 | 0.183418611 | -8.01E-05 | 0.013353246 | 1 | 0.061597897 | 1 | 0.071403169 | ENSG000001( | 7 | NUAK2 |
| SAT | Obesity | ENSG0000012 | 0.002460161 | 0.252269576 | 0.002148864 | 0.183453979 | 0.00250416 | 0.243859211 | 0.002148762 | 0.183420757 | -4.40E-05 | 0.013228657 | 1 | 0.542731309 | 1 | 0.539515895 | ENSG000001( | 7 | SPTA1 |
| SAT | Obesity | ENSG0000012 | 0.001140865 | 0.78781633 | 0.004238809 | 0.183454447 | 0.00107315 | 0.800121796 | 0.004238522 | 0.183421246 | 6.77E-05 | 0.018011038 | 1 | 0.914498218 | 1 | 0.92223015 | ENSG000001( | 7 | SPTA1 |
| SAT | Obesity | ENSG0000012 | -0.00164605 | 0.705902456 | 0.004361954 | 0.18345442 | -0.00155923 | 0.720729334 | 0.004361686 | 0.183421222 | -8.68E-05 | 0.009084514 | 1 | 0.875173806 | 1 | 0.885797449 | ENSG000001( | 7 | SPTA1 |
| SAT | Obesity | ENSG0000012 | 0.008208412 | 7.43E-05 | 0.002071681 | 0.18344854 | 0.00823833 | 6.98E-05 | 0.002071544 | 0.183415293 | -2.99E-05 | 0.038796461 | 0.243690948 | 0.001562121 | 0.229108206 | 0.001660204 | ENSG000001( | 7 | SPTA1 |
| SAT | Obesity | ENSG0000012 | 0.019184731 | 0.001392009 | 0.006002128 | 0.183450612 | 0.019050009 | 0.001503146 | 0.006001716 | 0.183417463 | 0.000134721 | 0.000655871 | 1 | 0.018078898 | 1 | 0.020513962 | ENSG000001( | 8 | PRKCI |
| SAT | Obesity | ENSG0000012 | 0.007297807 | 0.022783163 | 0.003204927 | 0.183452514 | 0.007468327 | 0.019808004 | 0.003205328 | 0.183419218 | -0.00017052 | 0.010433487 | 1 | 0.134930607 | 1 | 0.127967724 | ENSG000001( | 13 | EFHB |
| SAT | Obesity | ENSG0000012 | 0.007959234 | 0.163206458 | 0.005708121 | 0.183453739 | 0.007866337 | 0.168147936 | 0.005707766 | 0.183420552 | 9.29E-05 | 0.030586068 | 1 | 0.428041877 | 1 | 0.437851886 | ENSG000001( | 13 | EFHB |
| SAT | Obesity | ENSG0000012 | 0.001778195 | 0.578038773 | 0.003196742 | 0.183454357 | 0.001718551 | 0.590834902 | 0.00319655 | 0.183421161 | 5.96E-05 | 0.016440285 | 1 | 0.804985236 | 1 | 0.815193151 | ENSG000001( | 13 | EFHB |
| SAT | Obesity | ENSG0000012 | 0.003554745 | 0.553416844 | 0.005998069 | 0.183454341 | 0.020816038 | 0.007169301 | 0.007741506 | 0.183418538 | -0.01726129 | 0.0004214 | 1 | 0.793948695 | 1 | 0.067399648 | ENSG000001( | 13 | EFHB |
| SAT | Obesity | ENSG0000012 | 0.000436351 | 0.908750033 | 0.003807088 | 0.183454469 | 8.41E-05 | 0.982381909 | 0.00380739 | 0.18342127 | 0.000352273 | 5.11E-07 | 1 | 0.967797137 | 1 | 0.995364986 | ENSG000001( | 13 | EFHB |
| SAT | Obesity | ENSG0000012 | -0.00476522 | 0.43304436 | 0.006078134 | 0.183454242 | -0.00523854 | 0.389077718 | 0.006082188 | 0.18342099 | 0.000473322 | 0.04546734 | 1 | 0.714441885 | 1 | 0.682866142 | ENSG000001( | 13 | EFHB |
| SAT | Obesity | ENSG0000012 | 0.006115728 | 0.01379911 | 0.002483585 | 0.183452182 | 0.006059563 | 0.014687708 | 0.00248343 | 0.18341902 | 5.62E-05 | 0.002424586 | 1 | 0.097156393 | 1 | 0.107328215 | ENSG000001( | 12 | ICA1L |
| SAT | Obesity | ENSG0000012 | 0.010103861 | 0.08604269 | 0.005885781 | 0.18345336 | 0.010274203 | 0.080863248 | 0.005885425 | 0.183420118 | -0.00017034 | 0.000198255 | 1 | 0.294990664 | 1 | 0.293162781 | ENSG000001( | 12 | ICA1L |
| SAT | Obesity | ENSG0000012 | 0.004506026 | 0.245434753 | 0.003879452 | 0.183453964 | 0.004442884 | 0.25207923 | 0.003879181 | 0.183420774 | 6.31E-05 | 0.011305211 | 1 | 0.534004366 | 1 | 0.549184565 | ENSG000001( | 12 | ICA1L |
| SAT | Obesity | ENSG0000012 | 0.009672778 | 0.016920488 | 0.004049832 | 0.183452318 | 0.009773183 | 0.015806968 | 0.004049632 | 0.183419069 | -0.00010041 | 0.006396529 | 1 | 0.110151033 | 1 | 0.111772978 | ENSG000001( | 12 | ICA1L |
| SAT | Obesity | ENSG0000012 | -0.00211258 | 0.556820223 | 0.003595447 | 0.183454344 | -0.00218567 | 0.543229226 | 0.003595216 | 0.183421131 | 7.31E-05 | 0.005039547 | 1 | 0.794232732 | 1 | 0.788643845 | ENSG000001( | 7 | SPICE1 |
| SAT | Obesity | ENSG0000012 | -0.00370129 | 0.40464972 | 0.004441483 | 0.183454212 | -0.00381137 | 0.390787304 | 0.004441169 | 0.183420992 | 0.000110078 | 8.43E-05 | 1 | 0.694017632 | 1 | 0.684921551 | ENSG000001( | 7 | SPICE1 |
| SAT | Obesity | ENSG0000012 | -0.00362795 | 0.497926389 | 0.005352884 | 0.1834543 | -0.00344377 | 0.519976169 | 0.005352607 | 0.183421114 | -0.00018418 | 9.54E-05 | 1 | 0.756691284 | 1 | 0.774417527 | ENSG000001( | 13 | CADPS |
| SAT | Obesity | ENSG0000012 | -0.00379205 | 0.3753422 | 0.004277493 | 0.183454177 | -0.00387155 | 0.365381458 | 0.004277207 | 0.183420961 | 7.95E-05 | 0.00673703 | 1 | 0.670275034 | 1 | 0.665460489 | ENSG000001( | 11 | WDFY3 |
| SAT | Obesity | ENSG0000012 | -0.00043195 | 0.76190854 | 0.00142568 | 0.183454439 | -0.00040876 | 0.774318389 | 0.001425589 | 0.183421239 | -2.32E-05 | 0.026643784 | 1 | 0.904421823 | 1 | 0.911862059 | ENSG000001( | 8 | PTPN13 |
| SAT | Obesity | ENSG0000012 | 0.000987218 | 0.463129136 | 0.001345525 | 0.183454271 | 0.001017561 | 0.449468251 | 0.001345442 | 0.183421054 | -3.03E-05 | 0.002903381 | 1 | 0.733008537 | 1 | 0.727172255 | ENSG000001( | 11 | RPL9 |
| SAT | Obesity | ENSG0000012 | -0.0020142 | 0.293131247 | 0.001915951 | 0.183454056 | -0.0019706 | 0.303677892 | 0.001915845 | 0.18342087 | -4.36E-05 | 0.006782926 | 1 | 0.592218978 | 1 | 0.604225085 | ENSG000001( | 11 | RPL9 |
| SAT | Obesity | ENSG0000012 | 0.002455639 | 0.226496086 | 0.002030401 | 0.183453921 | 0.003147436 | 0.121745278 | 0.002033895 | 0.183420365 | -0.0006918 | 1.54E-08 | 1 | 0.514945339 | 1 | 0.369554001 | ENSG000001( | 11 | RPL9 |
| SAT | Obesity | ENSG0000012 | 0.000499373 | 0.911637811 | 0.004499955 | 0.183454469 | 0.000586281 | 0.896333769 | 0.004499672 | 0.183421264 | -8.69E-05 | 0.009202248 | 1 | 0.968941268 | 1 | 0.963222823 | ENSG000001( | 6 | TTC14 |
| SAT | Obesity | ENSG0000012 | 0.002786548 | 0.238098204 | 0.002361972 | 0.183453948 | 0.004328916 | 0.068574193 | 0.002376933 | 0.183420017 | -0.00154237 | 8.89E-09 | 1 | 0.526060746 | 1 | 0.265839301 | ENSG000001( | 6 | TTC14 |
| SAT | Obesity | ENSG0000012 | -0.02294977 | 5.75E-24 | 0.002273026 | 0.183415949 | -0.02280671 | 1.09E-23 | 0.002272965 | 0.183383227 | -0.00014306 | 1.88E-08 | 1.89E-20 | 2.36E-21 | 3.57E-20 | 3.96E-21 | ENSG000001( | 9 | RCHY1 |
| SAT | Obesity | ENSG0000012 | 0.004009335 | 0.425356298 | 0.005029499 | 0.183454234 | 0.004562894 | 0.364350436 | 0.005030158 | 0.183420959 | -0.00055356 | 1.73E-07 | 1 | 0.708423357 | 1 | 0.664705409 | ENSG000001( | 9 | RCHY1 |
| SAT | Obesity | ENSG0000012 | 0.002366725 | 0.249153883 | 0.002053721 | 0.183453972 | 0.000665508 | 0.748531376 | 0.00207597 | 0.183421231 | 0.001701217 | 2.28E-08 | 1 | 0.538484566 | 1 | 0.900879844 | ENSG000001( | 9 | RCHY1 |
| SAT | Obesity | ENSG0000012 | 0.033750509 | 5.61E-10 | 0.005442512 | 0.183439939 | 0.027951843 | 6.71E-07 | 0.005624569 | 0.183411937 | 0.005798666 | 4.49E-05 | 1.84E-06 | 4.60E-08 | 0.002202038 | 3.34E-05 | ENSG000001( | 9 | RCHY1 |
| SAT | Obesity | ENSG0000012 | 0.00401678 | 0.416483162 | 0.004943496 | 0.183454225 | 0.003936861 | 0.425788212 | 0.004943195 | 0.18342103 | 7.99E-05 | 0.035775776 | 1 | 0.700759618 | 1 | 0.710947136 | ENSG000001( | 9 | RCHY1 |

| | | | | | | | | | | | | | | | | | | | |
|---|---|---|---|---|---|---|---|---|---|---|---|---|---|---|---|---|---|---|---|
| SAT | Obesity | ENSG0000012 | -0.00044387 | 0.919183444 | 0.00437472 | 0.18345447 | -0.0002733 | 0.950183562 | 0.004374521 | 0.183421269 | -0.00017057 | 4.00E-05 | 1 | 0.972851897 | 1 | 0.986879476 | ENSG000001 | 9 | SNRK |
| SAT | Obesity | ENSG0000012 | -0.00469229 | 0.066192003 | 0.00255415 | 0.183453198 | -0.00465088 | 0.068602303 | 0.00255398 | 0.183420017 | -4.14E-05 | 0.019422246 | 1 | 0.25885097 | 1 | 0.265839301 | ENSG000001 | 9 | SNRK |
| SAT | Obesity | ENSG0000012 | 0.000483224 | 0.887391659 | 0.003412468 | 0.183454467 | 0.000434061 | 0.89877629 | 0.003412227 | 0.183421264 | 4.92E-05 | 0.022654506 | 1 | 0.960985924 | 1 | 0.963222823 | ENSG000001 | 9 | SNRK |
| SAT | Obesity | ENSG0000012 | 0.003143771 | 0.547263953 | 0.005223378 | 0.183454337 | 0.003024483 | 0.562547539 | 0.005223072 | 0.183421143 | 0.000119288 | 0.004166924 | 1 | 0.792029636 | 1 | 0.797062812 | ENSG000001 | 4 | KIAA1143 |
| SAT | Obesity | ENSG0000012 | -0.00421551 | 0.321655583 | 0.00425353 | 0.183454103 | -0.00401153 | 0.345627769 | 0.004253535 | 0.183420934 | -0.00020398 | 0.000394337 | 1 | 0.619974849 | 1 | 0.648774721 | ENSG000001 | 4 | KIAA1143 |
| SAT | Obesity | ENSG0000012 | -0.00119946 | 0.772713536 | 0.004152861 | 0.183454443 | -0.00086996 | 0.834142417 | 0.004154695 | 0.183421254 | -0.0003295 | 0.015012341 | 1 | 0.909351905 | 1 | 0.936326075 | ENSG000001 | 10 | NMNAT3 |
| SAT | Obesity | ENSG0000012 | 0.007465631 | 0.177892275 | 0.005541282 | 0.183453788 | 0.007567645 | 0.172012529 | 0.005540946 | 0.183420565 | -0.00010201 | 0.017335025 | 1 | 0.451403367 | 1 | 0.443997285 | ENSG000001 | 10 | NMNAT3 |
| SAT | Obesity | ENSG0000012 | 0.006171326 | 0.055304854 | 0.003220139 | 0.183453086 | 0.006220942 | 0.053358051 | 0.003219922 | 0.183419859 | -4.96E-05 | 0.023434998 | 1 | 0.231153156 | 1 | 0.230862672 | ENSG000001 | 10 | TPRA1 |
| SAT | Obesity | ENSG0000012 | -0.00162797 | 0.693748435 | 0.004134287 | 0.183454415 | -0.00168976 | 0.682724889 | 0.004134002 | 0.183421207 | 6.18E-05 | 0.023082109 | 1 | 0.867449929 | 1 | 0.86636807 | ENSG000001 | 10 | TPRA1 |
| SAT | Obesity | ENSG0000012 | -0.00817559 | 0.1409271 | 0.005552748 | 0.183453655 | -0.0079772 | 0.150804865 | 0.005552463 | 0.18342049 | -0.00019838 | 5.30E-05 | 1 | 0.396214067 | 1 | 0.414511276 | ENSG000001 | 10 | TPRA1 |
| SAT | Obesity | ENSG0000012 | -0.0090931 | 0.098411782 | 0.00550228 | 0.183453442 | -0.00887409 | 0.106775352 | 0.005502078 | 0.183420287 | -0.00021901 | 0.000123193 | 1 | 0.319375923 | 1 | 0.343124319 | ENSG000001 | 8 | SENP2 |
| SAT | Obesity | ENSG0000012 | -0.0010977 | 0.812425876 | 0.00462585 | 0.183454453 | -0.00116707 | 0.800802712 | 0.004625564 | 0.183421246 | 6.94E-05 | 0.047810737 | 1 | 0.927799965 | 1 | 0.92223015 | ENSG000001 | 8 | SENP2 |
| SAT | Obesity | ENSG0000012 | -0.00209134 | 0.222104997 | 0.001712875 | 0.183453911 | -0.00205107 | 0.231107366 | 0.001712774 | 0.183420728 | -4.03E-05 | 0.003142738 | 1 | 0.509598947 | 1 | 0.524775447 | ENSG000001 | 8 | SENP2 |
| SAT | Obesity | ENSG0000012 | -0.00419046 | 0.370392758 | 0.004678218 | 0.183454171 | -0.00432467 | 0.355242884 | 0.004678004 | 0.183420947 | 0.000134207 | 0.002462916 | 1 | 0.666625694 | 1 | 0.657019111 | ENSG000001 | 7 | IFT122 |
| SAT | Obesity | ENSG0000012 | 0.000198165 | 0.973848942 | 0.006045063 | 0.183454474 | 0.000118547 | 0.984352883 | 0.006044628 | 0.18342127 | 7.96E-05 | 0.030674107 | 1 | 0.993452012 | 1 | 0.995364986 | ENSG000001 | 7 | IFT122 |
| SAT | Obesity | ENSG0000012 | -0.00344298 | 0.41563258 | 0.004229591 | 0.183454224 | -0.0032595 | 0.440898783 | 0.004229392 | 0.183421046 | -0.00018349 | 3.21E-06 | 1 | 0.700759618 | 1 | 0.721131061 | ENSG000001 | 12 | GNL3 |
| SAT | Obesity | ENSG0000012 | 0.001619688 | 0.556878886 | 0.002756991 | 0.183454344 | 0.001685505 | 0.540939738 | 0.002756827 | 0.183421129 | -6.58E-05 | 0.002481973 | 1 | 0.794232732 | 1 | 0.788109805 | ENSG000001 | 11 | UVSSA |
| SAT | Obesity | ENSG0000012 | 0.006431554 | 0.127725834 | 0.004222579 | 0.183453597 | 0.006767379 | 0.109045181 | 0.004223007 | 0.1834203 | -0.00033583 | 4.96E-05 | 1 | 0.376521527 | 1 | 0.346730908 | ENSG000001 | 11 | UVSSA |
| SAT | Obesity | ENSG0000012 | -0.00324469 | 0.305309706 | 0.003165196 | 0.183454077 | -0.00343044 | 0.278519574 | 0.003165626 | 0.183420826 | 0.000185749 | 0.005814934 | 1 | 0.605495891 | 1 | 0.581682192 | ENSG000001 | 11 | UVSSA |
| SAT | Obesity | ENSG0000012 | 0.00320619 | 0.335871911 | 0.003331609 | 0.183454124 | 0.003258227 | 0.328053276 | 0.003331372 | 0.183420909 | -5.20E-05 | 0.01223909 | 1 | 0.633626214 | 1 | 0.630177282 | ENSG000001 | 11 | UVSSA |
| SAT | Obesity | ENSG0000012 | 0.006059552 | 0.001872435 | 0.001948553 | 0.183450819 | 0.006090088 | 0.0017743 | 0.00194843 | 0.183417578 | -3.05E-05 | 0.035391409 | 1 | 0.022586247 | 1 | 0.023473705 | ENSG000001 | 7 | ARHGEF3 |
| SAT | Obesity | ENSG0000012 | -0.00404376 | 0.394441049 | 0.004748474 | 0.1834542 | -0.00419392 | 0.377098299 | 0.004748254 | 0.183420975 | 0.000150162 | 0.000761497 | 1 | 0.687020457 | 1 | 0.671441012 | ENSG000001 | 6 | LRPAP1 |
| SAT | Obesity | ENSG0000012 | -0.00084334 | 0.862189626 | 0.004858318 | 0.183454463 | -0.0009861 | 0.839147878 | 0.004858024 | 0.183421255 | 0.000142756 | 0.000148108 | 1 | 0.949276565 | 1 | 0.937115108 | ENSG000001 | 7 | EXO5 |
| SAT | Obesity | ENSG0000012 | 0.005010247 | 0.388618696 | 0.005811503 | 0.183454193 | 0.004923121 | 0.396888647 | 0.005811105 | 0.183420999 | 8.71E-05 | 0.023753011 | 1 | 0.679668413 | 1 | 0.691351433 | ENSG000001 | 7 | METAP1 |
| SAT | Obesity | ENSG0000012 | -0.00494416 | 0.374011821 | 0.005561576 | 0.183454175 | -0.00481426 | 0.386672422 | 0.00556132 | 0.183420987 | -0.0001299 | 0.013211367 | 1 | 0.669832306 | 1 | 0.680459742 | ENSG000001 | 7 | METAP1 |
| SAT | Obesity | ENSG0000012 | 0.012217475 | 0.013516465 | 0.004946601 | 0.183452168 | 0.012316742 | 0.012770815 | 0.004946271 | 0.183418927 | -9.93E-05 | 0.00362326 | 1 | 0.095990306 | 1 | 0.096541971 | ENSG000001 | 12 | DNAJB14 |
| SAT | Obesity | ENSG0000012 | -0.00124729 | 0.760654691 | 0.004094564 | 0.183454439 | -0.00115665 | 0.77755913 | 0.004094315 | 0.18342124 | -9.06E-05 | 0.003966951 | 1 | 0.904420994 | 1 | 0.912114423 | ENSG000001 | 12 | DNAJB14 |
| SAT | Obesity | ENSG0000012 | -0.00541265 | 0.280463979 | 0.00501506 | 0.183454034 | -0.0053218 | 0.288586877 | 0.005014757 | 0.183420845 | -9.08E-05 | 0.019557905 | 1 | 0.579036375 | 1 | 0.594383895 | ENSG000001 | 12 | DNAJB14 |
| SAT | Obesity | ENSG0000012 | 0.004712836 | 0.044510898 | 0.002345567 | 0.183452948 | 0.004761829 | 0.042329051 | 0.00234541 | 0.183419712 | -4.90E-05 | 0.002422248 | 1 | 0.202271825 | 1 | 0.200986421 | ENSG000001 | 10 | BDH2 |
| SAT | Obesity | ENSG0000012 | -0.00160326 | 0.259010356 | 0.0014204 | 0.183453993 | -0.00157791 | 0.266586991 | 0.001420311 | 0.183420804 | -2.53E-05 | 0.016780211 | 1 | 0.552544198 | 1 | 0.569447863 | ENSG000001 | 7 | CDC25A |
| SAT | Obesity | ENSG0000012 | -0.00048769 | 0.828281903 | 0.002248414 | 0.183454456 | -0.00045279 | 0.84038897 | 0.002248266 | 0.183421255 | -3.49E-05 | 0.027168094 | 1 | 0.936525064 | 1 | 0.937310206 | ENSG000001 | 9 | ZNF589 |
| SAT | Obesity | ENSG0000012 | 0.010670227 | 0.036618517 | 0.005105377 | 0.183452823 | 0.010792259 | 0.034511786 | 0.005105024 | 0.183419581 | -0.00012203 | 0.000255463 | 1 | 0.1809418 | 1 | 0.179450348 | ENSG000001 | 13 | ATRIP |
| SAT | Obesity | ENSG0000012 | 0.000454782 | 0.791729304 | 0.001722229 | 0.183454448 | 0.000490468 | 0.77579584 | 0.001722134 | 0.18342124 | -3.57E-05 | 0.013827267 | 1 | 0.915154651 | 1 | 0.911862059 | ENSG000001 | 8 | APEH |
| SAT | Obesity | ENSG0000012 | -0.00080412 | 0.881120242 | 0.005376969 | 0.183454466 | -0.00070082 | 0.896293348 | 0.005376665 | 0.183421264 | -0.0001033 | 0.019577574 | 1 | 0.958494761 | 1 | 0.963222823 | ENSG000001 | 8 | APEH |
| SAT | Obesity | ENSG0000012 | -0.00387452 | 0.35098977 | 0.004154215 | 0.183454145 | -0.00374058 | 0.367864744 | 0.004153981 | 0.183420964 | -0.00013394 | 9.85E-05 | 1 | 0.648421979 | 1 | 0.66613537 | ENSG000001 | 11 | RNF123 |
| SAT | Obesity | ENSG0000013 | 0.005669705 | 0.126375998 | 0.003709198 | 0.183453591 | 0.005727923 | 0.122502167 | 0.003708934 | 0.183420369 | -5.82E-05 | 0.011529163 | 1 | 0.374273307 | 1 | 0.371126141 | ENSG000001 | 7 | HSPA4L |
| SAT | Obesity | ENSG0000013 | -0.00509557 | 0.391347132 | 0.005944594 | 0.183454196 | -0.0052005 | 0.381635372 | 0.005944186 | 0.183420981 | 0.000104922 | 0.007456784 | 1 | 0.683347493 | 1 | 0.676469831 | ENSG000001 | 7 | HSPA4L |
| SAT | Obesity | ENSG0000013 | -0.01396346 | 1.62E-08 | 0.002472035 | 0.183442414 | -0.01400295 | 1.47E-08 | 0.002471887 | 0.183409143 | 3.95E-05 | 0.04085933 | 5.31E-05 | 9.66E-07 | 4.83E-05 | 1.03E-06 | ENSG000001 | 10 | C4orf29 |
| SAT | Obesity | ENSG0000013 | 0.001275406 | 0.337521219 | 0.001329829 | 0.183454126 | 0.001249712 | 0.347311855 | 0.001329739 | 0.183420936 | 2.57E-05 | 0.00437624 | 1 | 0.6350426 | 1 | 0.650045748 | ENSG000001 | 8 | MST1R |
| SAT | Obesity | ENSG0000013 | -0.00116604 | 0.45738621 | 0.00156903 | 0.183454265 | -0.00119029 | 0.448053983 | 0.001568925 | 0.183421053 | 2.42E-05 | 0.025559263 | 1 | 0.731029138 | 1 | 0.726019131 | ENSG000001 | 9 | RAD54L2 |
| SAT | Obesity | ENSG0000013 | 0.006757143 | 0.25072304 | 0.005882957 | 0.183453975 | 0.006660299 | 0.257548389 | 0.005882586 | 0.183420786 | 9.68E-05 | 0.025945876 | 1 | 0.540132826 | 1 | 0.556825651 | ENSG000001 | 9 | RAD54L2 |
| SAT | Obesity | ENSG0000013 | 0.009715726 | 0.001797798 | 0.003112252 | 0.18345079 | 0.009807601 | 0.001624761 | 0.003112099 | 0.183417517 | -9.19E-05 | 0.001187549 | 1 | 0.021927791 | 1 | 0.021937612 | ENSG000001 | 8 | TEX264 |
| SAT | Obesity | ENSG0000013 | 0.005081272 | 0.008422551 | 0.001928629 | 0.18345185 | 0.005131015 | 0.007799945 | 0.001928503 | 0.183418595 | -4.97E-05 | 0.000296791 | 1 | 0.069783813 | 1 | 0.071403169 | ENSG000001 | 8 | TEX264 |
| SAT | Obesity | ENSG0000013 | 0.008710218 | 0.141741598 | 0.005927985 | 0.183453658 | 0.008603551 | 0.146658954 | 0.005927611 | 0.183420474 | 0.000106667 | 0.014799016 | 1 | 0.39770483 | 1 | 0.411271818 | ENSG000001 | 8 | TEX264 |
| SAT | Obesity | ENSG0000013 | -0.00841742 | 0.126506887 | 0.005508688 | 0.183453592 | -0.00852174 | 0.121851792 | 0.005508379 | 0.183420366 | 0.000104324 | 0.02261741 | 1 | 0.374273307 | 1 | 0.369554001 | ENSG000001 | 10 | GRM2 |
| SAT | Obesity | ENSG0000013 | -0.00901987 | 0.045003909 | 0.004499533 | 0.183452955 | -0.0089136 | 0.047581543 | 0.004499337 | 0.183419787 | -0.00010627 | 0.01465771 | 1 | 0.203947272 | 1 | 0.216826448 | ENSG000001 | 3 | PRSS12 |
| SAT | Obesity | ENSG0000013 | 0.008158012 | 0.05099271 | 0.004180282 | 0.183453034 | 0.008078926 | 0.053270005 | 0.004180058 | 0.183419858 | 7.91E-05 | 0.027794692 | 1 | 0.219563101 | 1 | 0.230862672 | ENSG000001 | 10 | ANXA5 |
| SAT | Obesity | ENSG0000013 | -0.0045263 | 0.435407171 | 0.005803152 | 0.183454244 | -0.00462607 | 0.425327182 | 0.005802794 | 0.18342103 | 9.98E-05 | 0.023799422 | 1 | 0.716072224 | 1 | 0.710538943 | ENSG000001 | 10 | ANXA5 |
| SAT | Obesity | ENSG0000013 | -0.00773895 | 0.121502498 | 0.004997705 | 0.183453568 | -0.00762412 | 0.127107533 | 0.004997425 | 0.183420391 | -0.00011482 | 0.005603207 | 1 | 0.364910204 | 1 | 0.379817682 | ENSG000001 | 10 | ANXA5 |
| SAT | Obesity | ENSG0000013 | -0.00039273 | 0.939031308 | 0.00513463 | 0.183454472 | -0.00047177 | 0.926788268 | 0.005134303 | 0.183421267 | 7.90E-05 | 0.036082422 | 1 | 0.981045593 | 1 | 0.976804467 | ENSG000001 | 8 | CEP44 |
| SAT | Obesity | ENSG0000013 | -0.0100233 | 0.076277198 | 0.005654222 | 0.183453286 | -0.00983743 | 0.081871882 | 0.00565392 | 0.183420126 | -0.00018588 | 0.000129222 | 1 | 0.277764135 | 1 | 0.295121602 | ENSG000001 | 8 | CEP44 |
| SAT | Obesity | ENSG0000013 | 0.026691431 | 5.69E-07 | 0.005336648 | 0.183445019 | 0.026821135 | 5.01E-07 | 0.005336327 | 0.183411723 | -0.0001297 | 0.001940128 | 0.001866738 | 2.49E-05 | 0.001642551 | 2.65E-05 | ENSG000001 | 8 | CEP44 |
| SAT | Obesity | ENSG0000013 | 0.005666708 | 0.15186374 | 0.003954488 | 0.183453698 | 0.005498496 | 0.16438303 | 0.00395438 | 0.183420539 | 0.000168212 | 0.000153287 | 1 | 0.411789199 | 1 | 0.433205398 | ENSG000001 | 10 | ARFIP1 |
| SAT | Obesity | ENSG0000013 | -0.00077711 | 0.732691749 | 0.002275253 | 0.18345443 | -0.00064281 | 0.777562951 | 0.002275478 | 0.18342124 | -0.00013429 | 0.002427315 | 1 | 0.889784069 | 1 | 0.912114423 | ENSG000001 | 10 | ARFIP1 |
| SAT | Obesity | ENSG0000013 | -0.00362058 | 0.093221922 | 0.002156849 | 0.183453409 | -0.0036575 | 0.089910563 | 0.002156706 | 0.183420183 | 3.69E-05 | 0.013936293 | 1 | 0.310854834 | 1 | 0.309870332 | ENSG000001 | 10 | ARFIP1 |
| SAT | Obesity | ENSG0000013 | -0.00329323 | 0.482423621 | 0.004688473 | 0.183454288 | -0.00283866 | 0.545091157 | 0.004690971 | 0.183421132 | -0.00045457 | 0.006031573 | 1 | 0.743462612 | 1 | 0.788841592 | ENSG000001 | 5 | LSM6 |
| SAT | Obesity | ENSG0000013 | -0.0038484 | 0.457984145 | 0.005185322 | 0.183454266 | -0.00336323 | 0.516816055 | 0.005188081 | 0.183421111 | -0.00048516 | 0.008011604 | 1 | 0.731214589 | 1 | 0.772996237 | ENSG000001 | 5 | LSM6 |
| SAT | Obesity | ENSG0000013 | -0.00161439 | 0.430549878 | 0.002048063 | 0.183454239 | -0.00164282 | 0.422444087 | 0.002047918 | 0.183421027 | 2.84E-05 | 0.02796653 | 1 | 0.71201318 | 1 | 0.708004523 | ENSG000001 | 6 | ITGA2 |
| SAT | Obesity | ENSG0000013 | 0.000432426 | 0.895499168 | 0.003292184 | 0.183454468 | 0.000382823 | 0.907422552 | 0.003291962 | 0.183421265 | 4.96E-05 | 0.026475437 | 1 | 0.962747126 | 1 | 0.966723909 | ENSG000001 | 6 | ITGA2 |
| SAT | Obesity | ENSG0000013 | 0.003744454 | 0.491651445 | 0.005445012 | 0.183454295 | 0.002595136 | 0.633885256 | 0.005448924 | 0.183421184 | 0.001149318 | 1.54E-07 | 1 | 0.752031883 | 1 | 0.837394946 | ENSG000001 | 6 | ITGA2 |
| SAT | Obesity | ENSG0000013 | -0.00164235 | 0.700413339 | 0.004268483 | 0.183454418 | -0.00158146 | 0.710993973 | 0.004268196 | 0.183421218 | -6.09E-05 | 0.035635504 | 1 | 0.871446033 | 1 | 0.882288663 | ENSG000001 | 6 | ITGA2 |
| SAT | Obesity | ENSG0000013 | 0.006604472 | 0.164446202 | 0.004750481 | 0.183453743 | 0.006681755 | 0.159534289 | 0.004750147 | 0.183420522 | -7.73E-05 | 0.010829695 | 1 | 0.429576424 | 1 | 0.427990191 | ENSG000001 | 9 | ELOVL7 |
| SAT | Obesity | ENSG0000013 | 0.00012995 | 0.978615331 | 0.004847991 | 0.183454474 | 0.00025716 | 0.957693616 | 0.004847673 | 0.183421269 | -0.00012721 | 0.000205165 | 1 | 0.993452012 | 1 | 0.990485083 | ENSG000001 | 6 | LMBRD2 |
| SAT | Obesity | ENSG0000013 | 0.007651469 | 0.081943418 | 0.004398601 | 0.18345333 | 0.007731883 | 0.078764936 | 0.00439836 | 0.183420102 | -8.04E-05 | 0.030690754 | 1 | 0.288332309 | 1 | 0.289069078 | ENSG000001 | 6 | LMBRD2 |
| SAT | Obesity | ENSG0000013 | -0.00311994 | 0.50242266 | 0.004651882 | 0.183454304 | -0.00322494 | 0.48812412 | 0.004651606 | 0.183421089 | 0.000105001 | 0.004240476 | 1 | 0.760004033 | 1 | 0.750604943 | ENSG000001 | 8 | WDR41 |
| SAT | Obesity | ENSG0000013 | 0.007452941 | 0.14042533 | 0.005055544 | 0.183453653 | 0.007526927 | 0.136500955 | 0.005055188 | 0.183420432 | -7.40E-05 | 0.02121526 | 1 | 0.396084299 | 1 | 0.395635718 | ENSG000001 | 8 | WDR41 |
| SAT | Obesity | ENSG0000013 | 0.004399074 | 0.387226253 | 0.005087638 | 0.183454191 | 0.004696378 | 0.356054209 | 0.005088659 | 0.183420948 | -0.0002973 | 0.015465315 | 1 | 0.67818082 | 1 | 0.657778074 | ENSG000001 | 13 | CASP3 |
| SAT | Obesity | ENSG0000013 | -0.01529482 | 0.004917379 | 0.005438333 | 0.183451484 | -0.01520463 | 0.005173944 | 0.005437971 | 0.183418316 | -9.02E-05 | 0.016330597 | 1 | 0.049039272 | 1 | 0.053857262 | ENSG000001 | 8 | ERAP1 |
| SAT | Obesity | ENSG0000013 | 0.011334339 | 0.054543275 | 0.00589562 | 0.183453077 | 0.011226074 | 0.056875803 | 0.005895216 | 0.1834199 | 0.000108264 | 0.005435928 | 1 | 0.229137624 | 1 | 0.239958924 | ENSG000001 | 8 | ERAP1 |
| SAT | Obesity | ENSG0000013 | -0.00084797 | 0.806094279 | 0.003454505 | 0.183454451 | -0.00078083 | 0.821164115 | 0.003454275 | 0.183421251 | -6.71E-05 | 0.00505132 | 1 | 0.924430385 | 1 | 0.932801998 | ENSG000001 | 12 | ERAP2 |
| SAT | Obesity | ENSG0000013 | -0.00313495 | 0.102196636 | 0.001918223 | 0.183453464 | -0.003168 | 0.098610321 | 0.001918105 | 0.183420239 | 3.31E-05 | 0.023707816 | 1 | 0.325540933 | 1 | 0.327348022 | ENSG000001 | 8 | KIAA1430 |
| SAT | Obesity | ENSG0000013 | 0.000187695 | 0.943180287 | 0.002633461 | 0.183454472 | 0.000137271 | 0.958425563 | 0.00263328 | 0.183421269 | 5.04E-05 | 0.003780128 | 1 | 0.981335116 | 1 | 0.990485083 | ENSG000001 | 6 | UBLCP1 |
| SAT | Obesity | ENSG0000013 | -0.00785727 | 0.158843582 | 0.00557659 | 0.183453724 | -0.00798104 | 0.15235933 | 0.005576276 | 0.183420496 | 0.000123775 | 0.007442463 | 1 | 0.421996593 | 1 | 0.415882664 | ENSG000001 | 11 | FAM170A |
| SAT | Obesity | ENSG0000013 | -0.00295207 | 0.30168057 | 0.002858217 | 0.183454071 | -0.00299402 | 0.294832195 | 0.002858029 | 0.183420855 | 4.19E-05 | 0.037253254 | 1 | 0.601710607 | 1 | 0.599223798 | ENSG000001 | 11 | KLKB1 |
| SAT | Obesity | ENSG0000013 | 0.006587331 | 0.003875183 | 0.002280805 | 0.183451321 | 0.006534016 | 0.004170656 | 0.002280657 | 0.183418168 | 5.33E-05 | 0.001104738 | 1 | 0.040751521 | 1 | 0.046543954 | ENSG000001 | 5 | NSA2 |
| SAT | Obesity | ENSG0000013 | 0.004869484 | 0.274746227 | 0.004458422 | 0.183454023 | 0.004941123 | 0.267713678 | 0.004458111 | 0.183420806 | -7.16E-05 | 0.012857791 | 1 | 0.570894471 | 1 | 0.569985864 | ENSG000001 | 14 | DCBLD1 |
| SAT | Obesity | ENSG0000013 | 0.005469789 | 0.249606013 | 0.004750934 | 0.183453973 | 0.005546664 | 0.24298375 | 0.004750635 | 0.183420755 | -7.69E-05 | 0.029359423 | 1 | 0.538787717 | 1 | 0.53813209 | ENSG000001 | 14 | DCBLD1 |
| SAT | Obesity | ENSG0000013 | 0.000560704 | 0.88283181 | 0.003804468 | 0.183454466 | 0.000459201 | 0.903922033 | 0.00380422 | 0.183421265 | 0.000101503 | 0.000171217 | 1 | 0.958494761 | 1 | 0.96423859 | ENSG000001 | 10 | DAGLB |
| SAT | Obesity | ENSG0000013 | -0.00530594 | 0.282322297 | 0.004935211 | 0.183454037 | -0.0051684 | 0.294954719 | 0.004934905 | 0.183420856 | -0.00013754 | 0.000212004 | 1 | 0.579661774 | 1 | 0.599223798 | ENSG000001 | 10 | DAGLB |
| SAT | Obesity | ENSG0000013 | -0.01076316 | 0.074122679 | 0.006026878 | 0.183453269 | -0.01085603 | 0.071643237 | 0.006026501 | 0.183420044 | 9.29E-05 | 0.039236223 | 1 | 0.273561877 | 1 | 0.273327281 | ENSG000001 | 5 | STK17A |
| SAT | Obesity | ENSG0000013 | 0.003732043 | 0.488882977 | 0.005392447 | 0.183454293 | 0.003630464 | 0.500764238 | 0.005392138 | 0.183421099 | 0.000101579 | 0.020857486 | 1 | 0.749894833 | 1 | 0.761367956 | ENSG000001 | 7 | FAM183B |
| SAT | Obesity | ENSG0000013 | -0.000648 | 0.77440562 | 0.002260847 | 0.183454443 | -0.00068223 | 0.762818781 | 0.002260685 | 0.183421236 | 3.42E-05 | 0.013809689 | 1 | 0.909817544 | 1 | 0.904912194 | ENSG000001 | 7 | FAM183B |
| SAT | Obesity | ENSG0000013 | -0.00458914 | 0.41100991 | 0.005582088 | 0.183454219 | -0.0044385 | 0.426511026 | 0.005581774 | 0.183421031 | -0.00015064 | 0.001102506 | 1 | 0.698355005 | 1 | 0.711073464 | ENSG000001 | 11 | GALNT10 |
| SAT | Obesity | ENSG0000013 | -0.0021336 | 0.403201484 | 0.002552407 | 0.18345421 | -0.00221478 | 0.385518991 | 0.002552256 | 0.183420986 | 8.12E-05 | 5.94E-05 | 1 | 0.692257493 | 1 | 0.680080079 | ENSG000001 | 11 | GALNT10 |
| SAT | Obesity | ENSG0000013 | 0.007563408 | 0.103966401 | 0.004651759 | 0.183453475 | 0.007475789 | 0.108011191 | 0.004651437 | 0.183420294 | 8.76E-05 | 0.003878564 | 1 | 0.330216614 | 1 | 0.345600181 | ENSG000001 | 5 | SLU7 |
| SAT | Obesity | ENSG0000013 | -0.00467428 | 0.35951488 | 0.005101301 | 0.183454157 | -0.00459327 | 0.36786923 | 0.005100959 | 0.183420964 | -8.10E-05 | 0.020430736 | 1 | 0.655680001 | 1 | 0.66613537 | ENSG000001 | 5 | SLU7 |
| SAT | Obesity | ENSG0000013 | 0.006464235 | 0.126981096 | 0.004235737 | 0.183453594 | 0.006557352 | 0.121577812 | 0.004235503 | 0.183420364 | -9.31E-05 | 0.008863576 | 1 | 0.374999979 | 1 | 0.369554001 | ENSG000001 | 3 | CAMLG |
| SAT | Obesity | ENSG0000013 | 0.003457947 | 0.003448426 | 0.001182339 | 0.183451241 | 0.003482088 | 0.003226864 | 0.001182264 | 0.183417992 | -2.41E-05 | 0.005083897 | 1 | 0.037340878 | 1 | 0.038781468 | ENSG000001 | 3 | CAMLG |
| SAT | Obesity | ENSG0000013 | 0.011806211 | 1.67E-06 | 0.00246512 | 0.183445804 | 0.009808909 | 8.76E-05 | 0.002500657 | 0.183415455 | 0.001997302 | 2.13E-06 | 0.005492674 | 6.24E-05 | 0.287531101 | 0.001973489 | ENSG000001 | 4 | RELL2 |
| SAT | Obesity | ENSG0000013 | -0.01226853 | 0.003566133 | 0.004209909 | 0.183451264 | -0.012364 | 0.003313348 | 0.004209623 | 0.18341801 | 9.55E-05 | 0.000751389 | 1 | 0.038236873 | 1 | 0.039245833 | ENSG000001 | 10 | KIAA1324L |
| SAT | Obesity | ENSG0000013 | 0.02246155 | 3.57E-05 | 0.005433677 | 0.183448015 | 0.01361106 | 0.020379116 | 0.005868604 | 0.183419237 | 0.00885049 | 6.61E-05 | 0.117107381 | 0.000867462 | 1 | 0.13047531 | ENSG000001 | 8 | INTS4L1 |
| SAT | Obesity | ENSG0000013 | 0.009315128 | 0.008038518 | 0.003514534 | 0.183451819 | 0.006185409 | 0.084918119 | 0.00359024 | 0.183420148 | 0.003129719 | 2.05E-05 | 1 | 0.067813996 | 1 | 0.298677775 | ENSG000001 | 8 | INTS4L1 |
| SAT | Obesity | ENSG0000013 | -0.0015797 | 0.133653898 | 0.001053237 | 0.183453624 | -0.00027333 | 0.800578372 | 0.001082054 | 0.183421246 | -0.00130637 | 1.46E-07 | 1 | 0.386967743 | 1 | 0.92223015 | ENSG000001 | 16 | CTSB |
| SAT | Obesity | ENSG0000013 | -0.00347809 | 0.473861043 | 0.004856226 | 0.18345428 | -0.0033896 | 0.485156075 | 0.004855926 | 0.183421086 | -8.85E-05 | 0.016198832 | 1 | 0.737541785 | 1 | 0.748543071 | ENSG000001 | 16 | CTSB |
| SAT | Obesity | ENSG0000013 | -0.0006054 | 0.846197078 | 0.003121053 | 0.18345446 | -0.00065113 | 0.834731414 | 0.003120851 | 0.183421254 | 4.57E-05 | 0.040531258 | 1 | 0.942344807 | 1 | 0.936326075 | ENSG000001 | 16 | CTSB |
| SAT | Obesity | ENSG0000013 | 0.005556998 | 0.129807091 | 0.003668311 | 0.183453607 | 0.005614368 | 0.125868974 | 0.003668083 | 0.183420385 | -5.74E-05 | 0.038012269 | 1 | 0.380265236 | 1 | 0.378183246 | ENSG000001 | 16 | CTSB |
| SAT | Obesity | ENSG0000013 | 0.006518386 | 0.217681869 | 0.005287819 | 0.1834539 | 0.006432441 | 0.223782632 | 0.005287519 | 0.183420711 | 8.59E-05 | 0.048097895 | 1 | 0.505890452 | 1 | 0.516186585 | ENSG000001 | 3 | DEFB1 |
| SAT | Obesity | ENSG0000013 | -0.00667453 | 0.106212699 | 0.004131673 | 0.183453488 | -0.00659788 | 0.110264926 | 0.004131405 | 0.183420306 | -7.67E-05 | 0.009409686 | 1 | 0.334758757 | 1 | 0.34859618 | ENSG000001 | 3 | DEFB1 |
| SAT | Obesity | ENSG0000013 | 0.004194911 | 0.261448433 | 0.003735532 | 0.183453997 | 0.004280686 | 0.251792105 | 0.0037353 | 0.183420774 | -8.58E-05 | 0.002360754 | 1 | 0.555218322 | 1 | 0.548923519 | ENSG000001 | 3 | DEFB1 |
| SAT | Obesity | ENSG0000013 | -0.00464872 | 0.07977452 | 0.002653378 | 0.183453314 | -0.00460798 | 0.082429919 | 0.002653203 | 0.18342013 | -4.07E-05 | 0.028057924 | 1 | 0.285743367 | 1 | 0.295578034 | ENSG000001 | 13 | SUN1 |
| SAT | Obesity | ENSG0000013 | 5.45E-05 | 0.992720505 | 0.005976614 | 0.183454474 | -0.00010052 | 0.986581119 | 0.005976369 | 0.18342127 | 0.000155044 | 0.009161302 | 1 | 0.996669516 | 1 | 0.995364986 | ENSG000001 | 13 | SUN1 |
| SAT | Obesity | ENSG0000013 | 0.005400361 | 0.062527326 | 0.002899457 | 0.183453163 | 0.005451873 | 0.060051727 | 0.002899288 | 0.183419934 | -5.15E-05 | 0.02680978 | 1 | 0.250185557 | 1 | 0.24752477 | ENSG000001 | 13 | OXR1 |
| SAT | Obesity | ENSG0000013 | -0.00294223 | 0.047734717 | 0.00148618 | 0.183452993 | -0.0029641 | 0.046090384 | 0.001486083 | 0.183419767 | 2.19E-05 | 0.037452511 | 1 | 0.210507537 | 1 | 0.211796289 | ENSG000001 | 13 | OXR1 |
| SAT | Obesity | ENSG0000013 | -0.00033692 | 0.951531892 | 0.005543005 | 0.183454473 | -0.00022827 | 0.967149067 | 0.005542653 | 0.18342127 | -0.00010865 | 0.00769603 | 1 | 0.985117699 | 1 | 0.992119797 | ENSG000001 | 10 | GPER1 |
| SAT | Obesity | ENSG0000013 | 0.004075177 | 0.346870264 | 0.004332169 | 0.18345414 | 0.004016618 | 0.353811115 | 0.004331879 | 0.183420945 | 5.86E-05 | 0.048037908 | 1 | 0.644805289 | 1 | 0.656220616 | ENSG000001 | 10 | GPER1 |
| SAT | Obesity | ENSG0000013 | 0.002143637 | 0.577792234 | 0.003851218 | 0.183454357 | 0.002065394 | 0.591731969 | 0.003850989 | 0.183421161 | 7.82E-05 | 0.009793004 | 1 | 0.804985236 | 1 | 0.81608768 | ENSG000001 | 10 | GPER1 |
| SAT | Obesity | ENSG0000013 | 0.001265443 | 0.750831838 | 0.003985107 | 0.183454436 | 0.001200016 | 0.76330409 | 0.003984844 | 0.183421236 | 6.54E-05 | 0.018660739 | 1 | 0.899291443 | 1 | 0.904912194 | ENSG000001 | 11 | ALDH7A1 |
| SAT | Obesity | ENSG0000013 | -0.00372014 | 0.018565872 | 0.001580254 | 0.183452379 | -0.00375242 | 0.017562245 | 0.001580147 | 0.183419139 | 3.23E-05 | 0.002517579 | 1 | 0.117823264 | 1 | 0.118319765 | ENSG000001 | 13 | YWHAZ |
| SAT | Obesity | ENSG0000013 | -0.0076027 | 0.040525861 | 0.003711617 | 0.183452888 | -0.0087744 | 0.018662881 | 0.003730285 | 0.183419179 | 0.001171691 | 0.001837796 | 1 | 0.192172349 | 1 | 0.123670158 | ENSG000001 | 13 | YWHAZ |
| SAT | Obesity | ENSG0000013 | 0.004533994 | 0.189948381 | 0.003459127 | 0.183453825 | 0.004609457 | 0.182665875 | 0.003459012 | 0.183420599 | -7.55E-05 | 0.041819124 | 1 | 0.46712282 | 1 | 0.459606393 | ENSG000001 | 13 | YWHAZ |
| SAT | Obesity | ENSG0000013 | -0.0047855 | 0.0374498 | 0.002299803 | 0.183452837 | -0.0050197 | 0.029152041 | 0.002301103 | 0.183419472 | 0.000234193 | 0.004943402 | 1 | 0.183666357 | 1 | 0.161406237 | ENSG000001 | 13 | YWHAZ |
| SAT | Obesity | ENSG0000013 | -0.0036509 | 0.305999719 | 0.003566537 | 0.183454078 | -0.00388309 | 0.276308851 | 0.003566885 | 0.183420822 | 0.000232193 | 0.000791345 | 1 | 0.605655297 | 1 | 0.580018771 | ENSG000001 | 13 | YWHAZ |
| SAT | Obesity | ENSG0000013 | -0.00504281 | 0.219416157 | 0.00410624 | 0.183453904 | -0.00514876 | 0.209860202 | 0.004106031 | 0.183420676 | 0.000105957 | 0.003720129 | 1 | 0.506545751 | 1 | 0.501023309 | ENSG000001 | 9 | INTS8 |
| SAT | Obesity | ENSG0000013 | 0.001901064 | 0.717797684 | 0.005260219 | 0.183454425 | 0.00172823 | 0.74248382 | 0.005259915 | 0.183421229 | 0.000172834 | 4.72E-05 | 1 | 0.883049944 | 1 | 0.897931963 | ENSG000001 | 9 | INTS8 |
| SAT | Obesity | ENSG0000013 | -0.00020276 | 0.879592023 | 0.001338454 | 0.183454465 | -0.00017631 | 0.895193275 | 0.001338371 | 0.183421264 | -2.64E-05 | 0.008389228 | 1 | 0.957347365 | 1 | 0.963222823 | ENSG000001 | 9 | INTS8 |
| SAT | Obesity | ENSG0000013 | -0.00124954 | 0.647051576 | 0.002729077 | 0.183454395 | -0.00132909 | 0.626244588 | 0.002729033 | 0.183421181 | 7.96E-05 | 0.017040428 | 1 | 0.844125734 | 1 | 0.832877378 | ENSG000001 | 5 | RPP25L |
| SAT | Obesity | ENSG0000013 | 0.002908681 | 0.625552286 | 0.00596045 | 0.183454384 | 0.003154202 | 0.596697964 | 0.005960824 | 0.183421164 | -0.00024552 | 0.018636347 | 1 | 0.833307775 | 1 | 0.817137678 | ENSG000001 | 5 | RPP25L |
| SAT | Obesity | ENSG0000013 | 0.001386338 | 0.729903751 | 0.004015418 | 0.183454429 | 0.002830266 | 0.485196869 | 0.004055002 | 0.183421086 | -0.00144393 | 0.010988908 | 1 | 0.8889108 | 1 | 0.748543071 | ENSG000001 | 5 | RPP25L |
| SAT | Obesity | ENSG0000013 | -0.00567989 | 0.040648464 | 0.002774598 | 0.18345289 | -0.00573583 | 0.038698075 | 0.002774433 | 0.183419655 | 5.59E-05 | 0.010810773 | 1 | 0.192172349 | 1 | 0.191795143 | ENSG000001 | 13 | NUDT2 |
| SAT | Obesity | ENSG0000013 | -0.00883495 | 0.096019568 | 0.00530797 | 0.183453427 | -0.0090333 | 0.088767106 | 0.005307646 | 0.183420175 | 0.000198353 | 1.23E-06 | 1 | 0.315355558 | 1 | 0.307220331 | ENSG000001 | 13 | NUDT2 |
| SAT | Obesity | ENSG0000013 | -0.00720428 | 0.002893817 | 0.002418507 | 0.18345112 | -0.00724746 | 0.002727848 | 0.002418348 | 0.183417876 | 4.32E-05 | 0.011224809 | 1 | 0.032627535 | 1 | 0.034160566 | ENSG000001 | 13 | NUDT2 |
| SAT | Obesity | ENSG0000013 | -0.00386439 | 0.167114301 | 0.002797169 | 0.183453753 | -0.00397982 | 0.154770374 | 0.002797012 | 0.183420505 | 0.000115429 | 6.73E-07 | 1 | 0.433440333 | 1 | 0.418287971 | ENSG000001 | 11 | CCDC171 |
| SAT | Obesity | ENSG0000014 | -5.98E-05 | 0.977729618 | 0.002140789 | 0.183454474 | -1.73E-05 | 0.993561087 | 0.002140641 | 0.18342127 | -4.25E-05 | 0.00245062 | 1 | 0.993452012 | 1 | 0.997818772 | ENSG000001 | 11 | CCDC171 |
| SAT | Obesity | ENSG0000014 | 0.001348097 | 0.622146287 | 0.002735526 | 0.183454382 | 0.001441713 | 0.598155357 | 0.00273541 | 0.183421165 | -9.36E-05 | 0.000506259 | 1 | 0.832018977 | 1 | 0.817909964 | ENSG000001 | 11 | METTL2B |
| SAT | Obesity | ENSG0000014 | 0.00672156 | 0.072705319 | 0.003745269 | 0.183453257 | 0.006795693 | 0.069588842 | 0.003745047 | 0.183420026 | -7.41E-05 | 0.012217483 | 1 | 0.269848588 | 1 | 0.268612932 | ENSG000001 | 11 | METTL2B |
| SAT | Obesity | ENSG0000014 | -0.00095546 | 0.875255008 | 0.00608622 | 0.183454465 | -0.00079135 | 0.896542359 | 0.00608589 | 0.183421264 | -0.00016411 | 0.001539633 | 1 | 0.955789527 | 1 | 0.963222823 | ENSG000001 | 11 | METTL2B |
| SAT | Obesity | ENSG0000014 | -0.00242944 | 0.505644189 | 0.003649806 | 0.183454307 | -0.00236405 | 0.517137074 | 0.003649553 | 0.183421112 | -6.54E-05 | 0.005928178 | 1 | 0.762495381 | 1 | 0.772996237 | ENSG000001 | 11 | METTL2B |

| | | | | | | | | | | | | | | | | | | | |
|---|---|---|---|---|---|---|---|---|---|---|---|---|---|---|---|---|---|---|---|
| SAT | Obesity | ENSG000014 | 0.001903053 | 0.304850877 | 0.001854669 | 0.183454076 | 0.001929169 | 0.29822879 | 0.001854541 | 0.183420861 | -2.61E-05 | 0.032262203 | 1 | 0.605495891 | 1 | 0.601026164 | ENSG000016 | 11 | METTL2B |
| SAT | Obesity | ENSG000014 | 0.006021814 | 0.091304649 | 0.003566245 | 0.183453396 | 0.006101516 | 0.087081271 | 0.003566047 | 0.183420164 | -7.97E-05 | 0.007765053 | 1 | 0.30630936 | 1 | 0.302983724 | ENSG000016 | 10 | C9orf64 |
| SAT | Obesity | ENSG000014 | -0.00363162 | 0.361154341 | 0.00397694 | 0.183454159 | -0.0037137 | 0.350375397 | 0.003976712 | 0.183420941 | 8.21E-05 | 0.011226847 | 1 | 0.657208759 | 1 | 0.65249005 | ENSG000016 | 6 | WBSCR27 |
| SAT | Obesity | ENSG000014 | 0.008552864 | 0.146669558 | 0.005892843 | 0.183453678 | 0.008394379 | 0.154279315 | 0.005892529 | 0.183420503 | 0.000158485 | 0.001809493 | 1 | 0.405752798 | 1 | 0.417993751 | ENSG000016 | 6 | WBSCR27 |
| SAT | Obesity | ENSG000014 | 0.006599171 | 0.063235459 | 0.003552647 | 0.18345317 | 0.006661608 | 0.060760868 | 0.00355241 | 0.183419941 | -6.24E-05 | 0.011247861 | 1 | 0.250993039 | 1 | 0.249243536 | ENSG000016 | 6 | WBSCR27 |
| SAT | Obesity | ENSG000014 | -0.00251818 | 0.635077825 | 0.005305982 | 0.183454389 | -0.00242409 | 0.647751535 | 0.005305642 | 0.183421191 | -9.41E-05 | 0.014789055 | 1 | 0.840197719 | 1 | 0.846421941 | ENSG000016 | 6 | WBSCR27 |
| SAT | Obesity | ENSG000014 | 0.002705394 | 0.424291136 | 0.003385977 | 0.183454233 | 0.002641645 | 0.43526054 | 0.003385761 | 0.18342104 | 6.37E-05 | 0.00984779 | 1 | 0.707008237 | 1 | 0.717633082 | ENSG000016 | 6 | WBSCR27 |
| SAT | Obesity | ENSG000014 | -0.00263935 | 0.078528172 | 0.001500231 | 0.183453304 | -0.00261221 | 0.081629281 | 0.00150014 | 0.183420124 | -2.71E-05 | 0.019838314 | 1 | 0.282822099 | 1 | 0.294809859 | ENSG000016 | 6 | WBSCR27 |
| SAT | Obesity | ENSG000014 | -0.00107161 | 0.685724018 | 0.002646664 | 0.183454412 | -0.00101916 | 0.700165415 | 0.002646504 | 0.183421214 | -5.19E-05 | 0.011293197 | 1 | 0.862676574 | 1 | 0.876141391 | ENSG000016 | 8 | NCF1C |
| SAT | Obesity | ENSG000014 | 0.001566454 | 0.679198843 | 0.00378776 | 0.183454409 | 0.001894448 | 0.617099016 | 0.003789152 | 0.183421176 | -0.00032799 | 0.004227974 | 1 | 0.858646871 | 1 | 0.828495448 | ENSG000016 | 2 | OR1Q1 |
| SAT | Obesity | ENSG000014 | -0.00130295 | 0.207110328 | 0.001032816 | 0.183453873 | -0.0013226 | 0.200317073 | 0.001032758 | 0.18342065 | 1.97E-05 | 0.022470045 | 1 | 0.493814259 | 1 | 0.487105125 | ENSG000016 | 2 | OR1Q1 |
| SAT | Obesity | ENSG000014 | 0.003841006 | 0.519291506 | 0.005960238 | 0.183454317 | 0.003986897 | 0.50352215 | 0.005959856 | 0.183421101 | -0.00014589 | 0.000755024 | 1 | 0.772402624 | 1 | 0.763780016 | ENSG000016 | 13 | C9orf89 |
| SAT | Obesity | ENSG000014 | 0.002103381 | 0.309294596 | 0.002068823 | 0.183454083 | 0.002149054 | 0.298876463 | 0.002068692 | 0.183420862 | -4.57E-05 | 0.002727251 | 1 | 0.60688449 | 1 | 0.601026164 | ENSG000016 | 13 | C9orf89 |
| SAT | Obesity | ENSG000014 | -0.0005648 | 0.756504952 | 0.001821516 | 0.183454438 | -0.00010057 | 0.956069377 | 0.001825718 | 0.183421269 | -0.00046423 | 0.000234652 | 1 | 0.901627438 | 1 | 0.989546885 | ENSG000016 | 6 | ZNF367 |
| SAT | Obesity | ENSG000014 | 0.0056392 | 0.068099543 | 0.003091071 | 0.183453216 | 0.005698773 | 0.065222371 | 0.003090885 | 0.183419985 | -5.96E-05 | 0.013516019 | 1 | 0.261549209 | 1 | 0.260334062 | ENSG000016 | 6 | ZNF367 |
| SAT | Obesity | ENSG000014 | 0.007849306 | 0.027222515 | 0.003554426 | 0.183452631 | 0.007951255 | 0.025279334 | 0.003554247 | 0.183419379 | -0.00010195 | 0.001379404 | 1 | 0.149609837 | 1 | 0.147845799 | ENSG000016 | 6 | ZNF367 |
| SAT | Obesity | ENSG000014 | -0.00431 | 0.280538184 | 0.00399402 | 0.183454034 | -0.00439921 | 0.270677054 | 0.00399381 | 0.183420812 | 8.92E-05 | 0.010431695 | 1 | 0.579036375 | 1 | 0.574073313 | ENSG000016 | 12 | NOL6 |
| SAT | Obesity | ENSG000014 | -0.00258457 | 0.616609134 | 0.005162303 | 0.183454379 | -0.00267942 | 0.603710829 | 0.005161944 | 0.183421168 | 9.49E-05 | 0.004525275 | 1 | 0.829137118 | 1 | 0.821898436 | ENSG000016 | 12 | NOL6 |
| SAT | Obesity | ENSG000014 | 0.000244325 | 0.852926329 | 0.001317924 | 0.183454461 | 0.000276697 | 0.833700938 | 0.00131787 | 0.183421254 | -3.24E-05 | 0.01298772 | 1 | 0.943648754 | 1 | 0.936326075 | ENSG000016 | 12 | NOL6 |
| SAT | Obesity | ENSG000014 | -0.00985915 | 0.00859581 | 0.003751959 | 0.183451864 | -0.0097987 | 0.009006959 | 0.003751702 | 0.18341692 | -6.05E-05 | 0.01496342 | 1 | 0.070683838 | 1 | 0.077624322 | ENSG000016 | 11 | VCP |
| SAT | Obesity | ENSG000014 | 0.003951432 | 0.190262885 | 0.003016812 | 0.183453826 | 0.003866542 | 0.199933729 | 0.00301663 | 0.183420649 | 8.49E-05 | 0.00028158 | 1 | 0.46712282 | 1 | 0.48663395 | ENSG000016 | 11 | VCP |
| SAT | Obesity | ENSG000014 | -0.02024897 | 0.000761508 | 0.00601495 | 0.1834509 | -0.02014599 | 0.000809544 | 0.006014568 | 0.18341703 | -0.00010299 | 0.019377351 | 1 | 0.011104481 | 1 | 0.012532499 | ENSG000016 | 10 | CFL2 |
| SAT | Obesity | ENSG000014 | 0.001007295 | 0.680766107 | 0.002448348 | 0.18345441 | 0.000964594 | 0.693580334 | 0.002448201 | 0.183421212 | 4.27E-05 | 0.024808384 | 1 | 0.859735796 | 1 | 0.869649775 | ENSG000016 | 10 | CFL2 |
| SAT | Obesity | ENSG000014 | -0.00193702 | 0.733914796 | 0.005698419 | 0.18345443 | -0.00204138 | 0.720148979 | 0.00569803 | 0.183421222 | 0.000104353 | 0.005990139 | 1 | 0.88986491 | 1 | 0.885797449 | ENSG000016 | 9 | CRYL1 |
| SAT | Obesity | ENSG000014 | 0.008553206 | 0.152609595 | 0.005979688 | 0.183453701 | 0.008449717 | 0.157608122 | 0.005979289 | 0.183420515 | 0.000103489 | 0.012178732 | 1 | 0.412938483 | 1 | 0.424210212 | ENSG000016 | 8 | REEP3 |
| SAT | Obesity | ENSG000014 | -0.00123458 | 0.54740986 | 0.002052008 | 0.183454337 | -0.0011916 | 0.56141664 | 0.00205187 | 0.183421143 | -4.30E-05 | 0.002028137 | 1 | 0.792029636 | 1 | 0.796027656 | ENSG000016 | 6 | HEPACAM |
| SAT | Obesity | ENSG000014 | -0.00194699 | 0.431436529 | 0.002474767 | 0.18345424 | -0.00202475 | 0.413235893 | 0.002474606 | 0.183421017 | 7.78E-05 | 1.04E-05 | 1 | 0.71240224 | 1 | 0.701318401 | ENSG000016 | 6 | RPL36AL |
| SAT | Obesity | ENSG000014 | -0.01466988 | 9.49E-06 | 0.003312596 | 0.183447061 | -0.01475924 | 8.36E-06 | 0.003312402 | 0.183413767 | 8.94E-05 | 0.000764915 | 0.031138176 | 0.000275559 | 0.027435231 | 0.00029764 | ENSG000016 | 6 | RPL36AL |
| SAT | Obesity | ENSG000014 | -0.00384254 | 0.31535314 | 0.003827025 | 0.183454093 | -0.00377906 | 0.323381035 | 0.003826764 | 0.183420902 | -6.35E-05 | 0.013359918 | 1 | 0.613321667 | 1 | 0.626705952 | ENSG000016 | 13 | AKR1E2 |
| SAT | Obesity | ENSG000014 | 0.0075942 | 0.03926252 | 0.003683947 | 0.183452868 | 0.00951102 | 0.010359296 | 0.003710011 | 0.183418786 | -0.00191682 | 1.44E-05 | 1 | 0.189487109 | 1 | 0.083928965 | ENSG000016 | 15 | ATP5C1 |
| SAT | Obesity | ENSG000014 | -0.01008728 | 0.011415652 | 0.003987507 | 0.183452055 | -0.00997962 | 0.012318704 | 0.003987237 | 0.183418903 | -0.00010766 | 6.33E-05 | 1 | 0.084739265 | 1 | 0.094213678 | ENSG000016 | 7 | TAF3 |
| SAT | Obesity | ENSG000014 | 0.011640342 | 0.000788459 | 0.003467653 | 0.183450215 | 0.012643625 | 0.000270544 | 0.003471605 | 0.183416257 | -0.00100328 | 5.66E-09 | 1 | 0.011353235 | 0.887656266 | 0.005101473 | ENSG000016 | 7 | TAF3 |
| SAT | Obesity | ENSG000014 | 0.005128378 | 0.342582713 | 0.005403552 | 0.183454134 | 0.004953537 | 0.35925916 | 0.005403199 | 0.183420953 | 0.000174842 | 4.97E-06 | 1 | 0.641194456 | 1 | 0.660152534 | ENSG000016 | 7 | PDZD8 |
| SAT | Obesity | ENSG000014 | 0.008070042 | 0.166235933 | 0.00582926 | 0.18345375 | 0.008179773 | 0.160522934 | 0.005828884 | 0.183420526 | -0.00010973 | 0.009116725 | 1 | 0.43218708 | 1 | 0.428934236 | ENSG000016 | 7 | PDZD8 |
| SAT | Obesity | ENSG000014 | -0.00434736 | 0.504326952 | 0.006510969 | 0.183454306 | -0.00444948 | 0.494334479 | 0.006510504 | 0.183421094 | 0.000102126 | 0.011248586 | 1 | 0.761480317 | 1 | 0.755382565 | ENSG000016 | 7 | PDZD8 |
| SAT | Obesity | ENSG000014 | -0.00278094 | 0.622302521 | 0.005645545 | 0.183454382 | -0.00265316 | 0.638364542 | 0.005645184 | 0.183421187 | -0.00012778 | 0.001927595 | 1 | 0.832018977 | 1 | 0.840143029 | ENSG000016 | 7 | PDZD8 |
| SAT | Obesity | ENSG000014 | -0.00389497 | 0.467026201 | 0.005355192 | 0.183454274 | -0.00401592 | 0.453277198 | 0.005354822 | 0.183421058 | 0.000120946 | 0.000582249 | 1 | 0.733866362 | 1 | 0.728306801 | ENSG000016 | 10 | CLEC1B |
| SAT | Obesity | ENSG000014 | 9.24E-05 | 0.986706117 | 0.005546247 | 0.183454474 | 0.000229374 | 0.967010165 | 0.00554601 | 0.18342127 | -0.00013696 | 0.011617802 | 1 | 0.994152558 | 1 | 0.992119797 | ENSG000016 | 3 | SNAPC4 |
| SAT | Obesity | ENSG000014 | -0.00937699 | 0.001660852 | 0.002981535 | 0.183450735 | -0.00745492 | 0.014723654 | 0.003056406 | 0.183419022 | -0.00192207 | 0.004319295 | 1 | 0.020719605 | 1 | 0.107351796 | ENSG000016 | 3 | SNAPC4 |
| SAT | Obesity | ENSG000014 | -0.00334379 | 0.542861323 | 0.005495202 | 0.183454334 | -0.00346077 | 0.528811033 | 0.005494837 | 0.18342112 | 0.000116984 | 0.002181429 | 1 | 0.790819813 | 1 | 0.780841134 | ENSG000016 | 3 | SNAPC4 |
| SAT | Obesity | ENSG000014 | -0.00166067 | 0.693254609 | 0.00421016 | 0.183454415 | -0.00155063 | 0.712629036 | 0.004209914 | 0.183421219 | -0.00011004 | 0.001105952 | 1 | 0.867162933 | 1 | 0.882648497 | ENSG000016 | 3 | SNAPC4 |
| SAT | Obesity | ENSG000014 | -0.00472162 | 0.360804447 | 0.005166828 | 0.183454158 | -0.00460632 | 0.372620103 | 0.005166485 | 0.18342097 | -0.0001153 | 0.001308371 | 1 | 0.65717171 | 1 | 0.669643631 | ENSG000016 | 3 | SNAPC4 |
| SAT | Obesity | ENSG000014 | -0.00127602 | 0.507168523 | 0.001923882 | 0.183454308 | -0.00131762 | 0.493398033 | 0.001923772 | 0.18342109 | 4.16E-05 | 0.007837695 | 1 | 0.763662195 | 1 | 0.755129058 | ENSG000016 | 3 | SNAPC4 |
| SAT | Obesity | ENSG000014 | -0.00032549 | 0.955653728 | 0.005853278 | 0.183454473 | -0.00047698 | 0.935048672 | 0.005852903 | 0.183421268 | 0.000151488 | 0.000376596 | 1 | 0.986058193 | 1 | 0.979844999 | ENSG000016 | 3 | SNAPC4 |
| SAT | Obesity | ENSG000015 | -0.00172727 | 0.447907501 | 0.002275987 | 0.183454256 | -0.0003815 | 0.87892448 | 0.002351099 | 0.183421261 | -0.00136912 | 0.02038019 | 1 | 0.723220724 | 1 | 0.958406391 | ENSG000016 | 10 | STOX1 |
| SAT | Obesity | ENSG000015 | 0.00269348 | 0.339870969 | 0.002822108 | 0.18345413 | 0.002747344 | 0.330269856 | 0.002821923 | 0.183420912 | -5.39E-05 | 0.006975767 | 1 | 0.638034522 | 1 | 0.632292088 | ENSG000016 | 10 | STOX1 |
| SAT | Obesity | ENSG000015 | -0.02352787 | 1.50E-06 | 0.004890781 | 0.183445727 | -0.02377864 | 1.16E-06 | 0.004890954 | 0.183412337 | 0.000250763 | 0.001252572 | 0.004937235 | 5.67E-05 | 0.003818547 | 5.62E-05 | ENSG000016 | 10 | STOX1 |
| SAT | Obesity | ENSG000015 | 0.008436753 | 0.01641345 | 0.003515902 | 0.183452298 | 0.008361935 | 0.017385482 | 0.003515692 | 0.183419132 | 7.48E-05 | 0.006566792 | 1 | 0.108703412 | 1 | 0.118319765 | ENSG000016 | 7 | BMS1 |
| SAT | Obesity | ENSG000015 | -0.00166136 | 0.674541626 | 0.003956346 | 0.183454407 | -0.00157189 | 0.691125867 | 0.003956162 | 0.183421211 | -8.95E-05 | 0.016043125 | 1 | 0.856857118 | 1 | 0.868937924 | ENSG000016 | 5 | SLC39A2 |
| SAT | Obesity | ENSG000015 | 0.00112608 | 0.551752403 | 0.001892137 | 0.18345434 | 0.001093986 | 0.563118439 | 0.001892001 | 0.183421144 | 3.21E-05 | 0.005158104 | 1 | 0.793295194 | 1 | 0.797062812 | ENSG000016 | 15 | CASP7 |
| SAT | Obesity | ENSG000015 | 0.006993095 | 0.179747067 | 0.005212751 | 0.183453794 | 0.006882203 | 0.186719473 | 0.005212416 | 0.183420611 | 0.000110891 | 0.003250322 | 1 | 0.452624689 | 1 | 0.466081875 | ENSG000016 | 15 | CASP7 |
| SAT | Obesity | ENSG000015 | 0.005976272 | 0.000142848 | 0.001571374 | 0.183449007 | 0.005947517 | 0.000153636 | 0.001571277 | 0.183415855 | 2.88E-05 | 0.016516826 | 0.468683054 | 0.002709151 | 0.504078468 | 0.003273237 | ENSG000016 | 4 | TRUB1 |
| SAT | Obesity | ENSG000015 | 0.002899045 | 0.523320603 | 0.00454228 | 0.18345432 | 0.002995448 | 0.509577699 | 0.004542026 | 0.183421106 | -9.64E-05 | 0.010764878 | 1 | 0.775526151 | 1 | 0.767790037 | ENSG000016 | 8 | HSPA12A |
| SAT | Obesity | ENSG000015 | 0.002130938 | 0.535023064 | 0.003435018 | 0.183454329 | 0.0020731 | 0.54613995 | 0.003434816 | 0.183421133 | 5.78E-05 | 0.034102871 | 1 | 0.784632613 | 1 | 0.788841592 | ENSG000016 | 8 | HSPA12A |
| SAT | Obesity | ENSG000015 | -0.00593367 | 0.178525652 | 0.004410632 | 0.18345379 | -0.00579259 | 0.189049669 | 0.004410384 | 0.183420618 | -0.00014108 | 0.000109748 | 1 | 0.45217888 | 1 | 0.469903003 | ENSG000016 | 8 | HSPA12A |
| SAT | Obesity | ENSG000015 | 0.010005882 | 0.001587738 | 0.003168255 | 0.183450704 | 0.00993509 | 0.001712666 | 0.003168055 | 0.183417553 | 7.08E-05 | 0.002440989 | 1 | 0.019959267 | 1 | 0.022842502 | ENSG000016 | 11 | PACSIN3 |
| SAT | Obesity | ENSG000015 | -0.02231995 | 1.21E-05 | 0.005100491 | 0.183447236 | -0.02244065 | 1.08E-05 | 0.005100145 | 0.183413954 | 0.000120694 | 0.00048148 | 0.039660702 | 0.000336108 | 0.035524439 | 0.000355244 | ENSG000016 | 11 | PACSIN3 |
| SAT | Obesity | ENSG000015 | 0.000841108 | 0.813327121 | 0.003561961 | 0.183454453 | 0.000784707 | 0.825625575 | 0.003561744 | 0.183421252 | 5.64E-05 | 0.039219293 | 1 | 0.927860321 | 1 | 0.935559334 | ENSG000016 | 11 | PACSIN3 |
| SAT | Obesity | ENSG000015 | 0.000705455 | 0.826728483 | 0.003222737 | 0.183454456 | 0.001235901 | 0.702188417 | 0.003232227 | 0.183421215 | -0.00053045 | 0.034767449 | 1 | 0.9363121 | 1 | 0.876674335 | ENSG000016 | 4 | PSMC3 |
| SAT | Obesity | ENSG000015 | -0.00618397 | 0.203150148 | 0.004859207 | 0.183453862 | -0.00662108 | 0.173138957 | 0.00486062 | 0.183420569 | 0.000437111 | 0.001127777 | 1 | 0.487947024 | 1 | 0.445195076 | ENSG000016 | 5 | RAPSN |
| SAT | Obesity | ENSG000015 | -0.00055477 | 0.621977447 | 0.00112517 | 0.183454382 | -0.00057167 | 0.611380398 | 0.001125101 | 0.183421173 | 1.69E-05 | 0.048496138 | 1 | 0.832018977 | 1 | 0.82447147 | ENSG000016 | 5 | RAPSN |
| SAT | Obesity | ENSG000015 | -7.95E-05 | 0.968095754 | 0.001988486 | 0.183454474 | -3.81E-05 | 0.984719192 | 0.001988381 | 0.18342127 | -4.14E-05 | 0.016143758 | 1 | 0.992600678 | 1 | 0.995364986 | ENSG000016 | 5 | RAPSN |
| SAT | Obesity | ENSG000015 | 0.005168608 | 0.135404292 | 0.003461597 | 0.183453631 | 0.00755332 | 0.038520216 | 0.003650214 | 0.183419652 | -0.00238471 | 0.039657799 | 1 | 0.38936151 | 1 | 0.191782742 | ENSG000016 | 8 | TC2N |
| SAT | Obesity | ENSG000015 | -0.00344271 | 0.132314526 | 0.002287455 | 0.183453618 | -0.00338805 | 0.13854494 | 0.002287316 | 0.183420441 | -5.47E-05 | 0.001931022 | 1 | 0.384861667 | 1 | 0.398742061 | ENSG000016 | 8 | CPSF2 |
| SAT | Obesity | ENSG000015 | 0.008939065 | 0.026540407 | 0.00402984 | 0.183452614 | 0.008874345 | 0.027644804 | 0.004029586 | 0.183419437 | 6.47E-05 | 0.029942143 | 1 | 0.14734192 | 1 | 0.155261977 | ENSG000016 | 8 | CPSF2 |
| SAT | Obesity | ENSG000015 | 0.006660376 | 0.14010927 | 0.004514326 | 0.183453651 | 0.006792319 | 0.132401223 | 0.004514061 | 0.183420415 | -0.00013194 | 0.000252847 | 1 | 0.396084299 | 1 | 0.389207047 | ENSG000016 | 8 | CPSF2 |
| SAT | Obesity | ENSG000015 | 0.020296208 | 3.37E-05 | 0.004894456 | 0.183447974 | 0.020498389 | 2.82E-05 | 0.004894936 | 0.183414643 | -0.00020218 | 0.03355512 | 0.110651336 | 0.000831965 | 0.092484676 | 0.000818448 | ENSG000016 | 8 | CPSF2 |
| SAT | Obesity | ENSG000015 | 0.00513515 | 0.028582514 | 0.00234567 | 0.183452663 | 0.005186885 | 0.027007793 | 0.002345509 | 0.183419422 | -5.17E-05 | 0.00078685 | 1 | 0.154014794 | 1 | 0.153308944 | ENSG000016 | 10 | PTER |
| SAT | Obesity | ENSG000015 | 0.01592239 | 0.002916291 | 0.005349472 | 0.183451125 | 0.015441913 | 0.003927644 | 0.00535447 | 0.183418127 | 0.000480477 | 0.047259533 | 1 | 0.032768325 | 1 | 0.044436554 | ENSG000016 | 8 | C1QL3 |
| SAT | Obesity | ENSG000015 | 0.000267185 | 0.930737937 | 0.003074042 | 0.183454471 | 0.000215974 | 0.943984739 | 0.003073821 | 0.183421268 | 5.12E-05 | 0.00607147 | 1 | 0.977387556 | 1 | 0.983503141 | ENSG000016 | 6 | CEP57 |
| SAT | Obesity | ENSG000015 | -0.0112766 | 0.040156339 | 0.005495038 | 0.183452882 | -0.01140813 | 0.037875252 | 0.005494696 | 0.183419641 | 0.00013153 | 0.001484163 | 1 | 0.191780129 | 1 | 0.189433997 | ENSG000016 | 6 | CEP57 |
| SAT | Obesity | ENSG000015 | 0.009461962 | 0.105459036 | 0.005844512 | 0.183453483 | 0.009332624 | 0.110284314 | 0.005844142 | 0.183420306 | 0.000129338 | 0.002683228 | 1 | 0.333987547 | 1 | 0.34859618 | ENSG000016 | 6 | CEP57 |
| SAT | Obesity | ENSG000015 | 0.003422508 | 0.426293008 | 0.004302055 | 0.183454235 | 0.003364329 | 0.434167212 | 0.004301764 | 0.183421039 | 5.82E-05 | 0.045127801 | 1 | 0.708903881 | 1 | 0.717272217 | ENSG000016 | 6 | CEP57 |
| SAT | Obesity | ENSG000015 | 0.00048565 | 0.774600367 | 0.001695925 | 0.183454443 | 0.000518044 | 0.75999883 | 0.001695821 | 0.183421235 | -3.24E-05 | 0.011747079 | 1 | 0.909817544 | 1 | 0.904912194 | ENSG000016 | 6 | CEP57 |
| SAT | Obesity | ENSG000015 | -0.00631846 | 0.018973393 | 0.00269322 | 0.183452394 | -0.006352 | 0.018340415 | 0.00269303 | 0.183419168 | 3.35E-05 | 0.049134431 | 1 | 0.119714811 | 1 | 0.122805922 | ENSG000016 | 6 | CEP57 |
| SAT | Obesity | ENSG000015 | 0.008361886 | 0.011741537 | 0.003318435 | 0.183452074 | 0.008419267 | 0.01117183 | 0.003318216 | 0.183418837 | -5.74E-05 | 0.013331985 | 1 | 0.085991032 | 1 | 0.087690846 | ENSG000016 | 6 | CEP57 |
| SAT | Obesity | ENSG000015 | -0.0096398 | 0.014248521 | 0.003933095 | 0.183452203 | -0.00958052 | 0.014849637 | 0.003932834 | 0.183419027 | -5.93E-05 | 0.029463762 | 1 | 0.099045337 | 1 | 0.107919055 | ENSG000016 | 13 | ZFYVE19 |
| SAT | Obesity | ENSG000015 | -0.00103966 | 0.757841364 | 0.003372046 | 0.183454438 | -0.00109392 | 0.745612654 | 0.003371816 | 0.18342123 | 5.43E-05 | 0.015907675 | 1 | 0.902205195 | 1 | 0.898760093 | ENSG000016 | 2 | AVPR1A |
| SAT | Obesity | ENSG000015 | -0.00320023 | 0.504752629 | 0.004797727 | 0.183454306 | -0.00367889 | 0.44338913 | 0.004799702 | 0.183421048 | 0.000478659 | 0.00164774 | 1 | 0.761772481 | 1 | 0.722682432 | ENSG000016 | 8 | TRMT61A |
| SAT | Obesity | ENSG000015 | 0.000934622 | 0.865784309 | 0.005529792 | 0.183454463 | 0.000780787 | 0.887708285 | 0.005529467 | 0.183421263 | 0.000153835 | 0.000477649 | 1 | 0.951956541 | 1 | 0.962898024 | ENSG000016 | 8 | TRMT61A |
| SAT | Obesity | ENSG000015 | -0.01155989 | 0.00021588 | 0.00312455 | 0.1834493 | -0.01162871 | 0.000197743 | 0.00312439 | 0.183416035 | 6.88E-05 | 0.013238182 | 0.70830134 | 0.003891766 | 0.648793464 | 0.004054959 | ENSG000016 | 9 | GABRB3 |
| SAT | Obesity | ENSG000015 | -0.00525798 | 0.269931681 | 0.004766022 | 0.183454014 | -0.00517504 | 0.277527178 | 0.004765703 | 0.183420825 | -8.29E-05 | 0.011191826 | 1 | 0.566269722 | 1 | 0.580588871 | ENSG000016 | 8 | TBATA |
| SAT | Obesity | ENSG000015 | 0.005384886 | 0.19589504 | 0.004163559 | 0.183453842 | 0.005323749 | 0.200989736 | 0.004163282 | 0.183420652 | 6.11E-05 | 0.033481901 | 1 | 0.476192171 | 1 | 0.48811793 | ENSG000016 | 12 | SGPL1 |
| SAT | Obesity | ENSG000015 | 0.002776322 | 0.439055073 | 0.003587951 | 0.183454248 | 0.002707683 | 0.450427417 | 0.003587745 | 0.183421055 | 6.86E-05 | 0.018741821 | 1 | 0.717400247 | 1 | 0.727311113 | ENSG000016 | 12 | SGPL1 |
| SAT | Obesity | ENSG000015 | 0.001557874 | 0.726803615 | 0.004458958 | 0.183454428 | 0.001680881 | 0.706181576 | 0.004458695 | 0.183421216 | -0.00012301 | 0.000525851 | 1 | 0.887859509 | 1 | 0.879167935 | ENSG000016 | 4 | PCBD1 |
| SAT | Obesity | ENSG000015 | -0.00759265 | 0.13139448 | 0.005032819 | 0.183453614 | -0.00775737 | 0.123206483 | 0.005032493 | 0.183420372 | 0.000164715 | 5.56E-06 | 1 | 0.382864378 | 1 | 0.372571864 | ENSG000016 | 4 | PCBD1 |
| SAT | Obesity | ENSG000015 | -0.01118268 | 0.034321151 | 0.005284109 | 0.183452781 | -0.01108375 | 0.035931853 | 0.005283753 | 0.183419607 | -9.89E-05 | 0.005845221 | 1 | 0.172711189 | 1 | 0.184495162 | ENSG000016 | 4 | PCBD1 |
| SAT | Obesity | ENSG000015 | 0.005961845 | 0.168232727 | 0.004326741 | 0.183453756 | 0.006044212 | 0.162403999 | 0.00432646 | 0.183420533 | -8.24E-05 | 0.008002389 | 1 | 0.434800182 | 1 | 0.430422843 | ENSG000016 | 6 | COX11 |
| SAT | Obesity | ENSG000015 | 0.002979316 | 0.352737934 | 0.003206004 | 0.183454148 | 0.002927948 | 0.361067965 | 0.003205784 | 0.183420955 | 5.14E-05 | 0.015287923 | 1 | 0.650001169 | 1 | 0.6621934 | ENSG000016 | 6 | COX11 |
| SAT | Obesity | ENSG000015 | 0.008650149 | 0.021544632 | 0.003763701 | 0.183452477 | 0.008721382 | 0.020482828 | 0.003763448 | 0.183419241 | -7.12E-05 | 0.005436472 | 1 | 0.129941061 | 1 | 0.13047531 | ENSG000016 | 11 | FAM227B |
| SAT | Obesity | ENSG000015 | -0.00260914 | 0.581455962 | 0.004733052 | 0.183454359 | -0.00271028 | 0.566871565 | 0.004732752 | 0.183421146 | 0.000101142 | 0.003691237 | 1 | 0.806390405 | 1 | 0.800506675 | ENSG000016 | 11 | FAM227B |
| SAT | Obesity | ENSG000015 | -0.00049682 | 0.847564622 | 0.002584531 | 0.18345446 | -0.00046325 | 0.85773952 | 0.002584345 | 0.183421258 | -3.36E-05 | 0.033362568 | 1 | 0.942344807 | 1 | 0.947556688 | ENSG000016 | 6 | MYRFL |
| SAT | Obesity | ENSG000015 | 0.004839115 | 0.323392777 | 0.004900316 | 0.183454105 | 0.004914055 | 0.31592491 | 0.0049 | 0.18342089 | -7.49E-05 | 0.033835249 | 1 | 0.622290867 | 1 | 0.615584597 | ENSG000016 | 6 | MYRFL |
| SAT | Obesity | ENSG000015 | 0.005118838 | 0.377850538 | 0.005804563 | 0.18345418 | 0.005218739 | 0.368586876 | 0.005804246 | 0.183420965 | -9.99E-05 | 0.042329881 | 1 | 0.671085989 | 1 | 0.667026933 | ENSG000016 | 11 | C10orf32 |
| SAT | Obesity | ENSG000015 | -0.01210608 | 0.012935407 | 0.004870579 | 0.183452139 | -0.01217431 | 0.012429271 | 0.004870258 | 0.183418909 | 6.82E-05 | 0.046234913 | 1 | 0.09286886 | 1 | 0.094838226 | ENSG000016 | 11 | C10orf32 |
| SAT | Obesity | ENSG000015 | 0.001025495 | 0.741958551 | 0.003114535 | 0.183454433 | 0.001078673 | 0.729074176 | 0.003114333 | 0.183421225 | -5.32E-05 | 0.01655326 | 1 | 0.895159095 | 1 | 0.891559411 | ENSG000016 | 10 | C2 |
| SAT | Obesity | ENSG000016 | 0.01275909 | 0.00479807 | 0.004524018 | 0.183451468 | 0.012898403 | 0.00435504 | 0.004523781 | 0.183418198 | -0.00013931 | 0.000431394 | 1 | 0.047995329 | 1 | 0.047949285 | ENSG000016 | 10 | C2 |
| SAT | Obesity | ENSG000016 | 0.001925734 | 0.74531773 | 0.005928625 | 0.183454434 | 0.001841726 | 0.756051421 | 0.005928233 | 0.183421234 | 8.40E-05 | 0.04256594 | 1 | 0.895746327 | 1 | 0.902694582 | ENSG000016 | 10 | C2 |
| SAT | Obesity | ENSG000016 | 0.001452151 | 0.715428258 | 0.003983125 | 0.183454424 | 0.001523171 | 0.702140009 | 0.00398284 | 0.183421215 | -7.10E-05 | 0.003874576 | 1 | 0.882123626 | 1 | 0.876674335 | ENSG000016 | 9 | NUDT13 |
| SAT | Obesity | ENSG000016 | 0.001505744 | 0.216879581 | 0.001219354 | 0.183453898 | 0.001524472 | 0.211186256 | 0.001219273 | 0.183420679 | -1.87E-05 | 0.027140202 | 1 | 0.505890452 | 1 | 0.502449821 | ENSG000016 | 9 | NUDT13 |
| SAT | Obesity | ENSG000016 | -0.00485818 | 0.286750528 | 0.004560483 | 0.183454045 | -0.00478459 | 0.294082006 | 0.004560188 | 0.183420854 | -7.36E-05 | 0.024564332 | 1 | 0.585820974 | 1 | 0.598191606 | ENSG000016 | 9 | NUDT13 |
| SAT | Obesity | ENSG000016 | 0.000550338 | 0.845193217 | 0.002820221 | 0.18345446 | 0.000608873 | 0.829058791 | 0.002820043 | 0.183421253 | -5.85E-05 | 0.005102947 | 1 | 0.941763438 | 1 | 0.936326075 | ENSG000016 | 9 | NUDT13 |
| SAT | Obesity | ENSG000016 | -0.0004486 | 0.809391679 | 0.001859772 | 0.183454452 | -0.00048856 | 0.792767255 | 0.001859646 | 0.183421244 | 4.00E-05 | 0.001593135 | 1 | 0.925742147 | 1 | 0.919712977 | ENSG000016 | 13 | USP54 |
| SAT | Obesity | ENSG000016 | 0.004045638 | 0.316253237 | 0.004036805 | 0.183454094 | 0.003951773 | 0.32759628 | 0.004036672 | 0.183420908 | 9.39E-05 | 0.030162089 | 1 | 0.614343914 | 1 | 0.630037159 | ENSG000016 | 10 | CYB5R2 |
| SAT | Obesity | ENSG000016 | -0.00234386 | 0.69320219 | 0.005941154 | 0.183454415 | -0.00242909 | 0.682623112 | 0.005940743 | 0.183421207 | 8.52E-05 | 0.0279661 | 1 | 0.867162933 | 1 | 0.86636807 | ENSG000016 | 10 | CYB5R2 |
| SAT | Obesity | ENSG000016 | -0.01171281 | 0.055881837 | 0.006126071 | 0.183453092 | -0.01183624 | 0.053334702 | 0.006125765 | 0.183419859 | 0.000123433 | 0.025524977 | 1 | 0.232463336 | 1 | 0.230862672 | ENSG000016 | 4 | TUB |
| SAT | Obesity | ENSG000016 | 0.002162016 | 0.580100763 | 0.003907932 | 0.183454358 | 0.002225423 | 0.569019487 | 0.003907703 | 0.183421148 | -6.34E-05 | 0.042380114 | 1 | 0.806390405 | 1 | 0.801026154 | ENSG000016 | 11 | XRRA1 |
| SAT | Obesity | ENSG000016 | 0.012047544 | 0.005734184 | 0.004360914 | 0.183451589 | 0.012131913 | 0.005400709 | 0.004360672 | 0.183418345 | -8.44E-05 | 0.020680062 | 1 | 0.054062808 | 1 | 0.055454223 | ENSG000016 | 11 | XRRA1 |
| SAT | Obesity | ENSG000016 | 0.001091163 | 0.55561042 | 0.001851392 | 0.183454343 | 0.001126626 | 0.542813489 | 0.001851285 | 0.18342113 | -3.55E-05 | 0.017801829 | 1 | 0.794232732 | 1 | 0.788389136 | ENSG000016 | 11 | TRIM66 |
| SAT | Obesity | ENSG000016 | 0.010461365 | 0.023849703 | 0.004629847 | 0.183452544 | 0.010580811 | 0.022284497 | 0.004629557 | 0.183419296 | -0.00011945 | 0.000579469 | 1 | 0.140084245 | 1 | 0.137177177 | ENSG000016 | 11 | TRIM66 |
| SAT | Obesity | ENSG000016 | -0.01103483 | 0.006718127 | 0.00407112 | 0.183451697 | -0.01129126 | 0.005563891 | 0.004072671 | 0.183418365 | 0.00025643 | 0.040349074 | 1 | 0.060224517 | 1 | 0.056517422 | ENSG000016 | 11 | TRIM66 |
| SAT | Obesity | ENSG000016 | 0.004336278 | 0.180872304 | 0.003240682 | 0.183453797 | 0.004194709 | 0.195508057 | 0.003240515 | 0.183420637 | 0.00014157 | 7.24E-07 | 1 | 0.453456712 | 1 | 0.479418486 | ENSG000016 | 10 | CDYL2 |
| SAT | Obesity | ENSG000016 | -0.01017679 | 0.032532918 | 0.004760389 | 0.183452747 | -0.01026483 | 0.031050422 | 0.004760097 | 0.183419513 | 8.80E-05 | 0.015485459 | 1 | 0.16809528 | 1 | 0.166130468 | ENSG000016 | 10 | CDYL2 |
| SAT | Obesity | ENSG000016 | -0.00371802 | 0.144414275 | 0.002547376 | 0.183453669 | -0.00366559 | 0.150136606 | 0.00254723 | 0.183420488 | -5.24E-05 | 0.011476037 | 1 | 0.401443831 | 1 | 0.414396534 | ENSG000016 | 10 | CDYL2 |
| SAT | Obesity | ENSG000016 | 0.000132786 | 0.976264773 | 0.004463088 | 0.183454474 | 5.04E-05 | 0.990983953 | 0.004462799 | 0.18342127 | 8.24E-05 | 0.010275817 | 1 | 0.993452012 | 1 | 0.996450613 | ENSG000016 | 10 | CDYL2 |
| SAT | Obesity | ENSG000016 | 0.016030447 | 0.005200773 | 0.005736755 | 0.183451523 | 0.016427474 | 0.004189299 | 0.005736735 | 0.183418171 | -0.00039703 | 1.64E-07 | 1 | 0.05087712 | 1 | 0.046593526 | ENSG000016 | 10 | ZSCAN21 |
| SAT | Obesity | ENSG000016 | -0.00064973 | 0.809829963 | 0.002699932 | 0.183454452 | -0.00071031 | 0.792473006 | 0.002699748 | 0.183421244 | 6.06E-05 | 0.000799772 | 1 | 0.925742147 | 1 | 0.919712977 | ENSG000016 | 10 | ZSCAN21 |
| SAT | Obesity | ENSG000016 | -0.00749497 | 0.115782585 | 0.00473471 | 0.183453527 | -0.00736591 | 0.119750961 | 0.004734416 | 0.183420355 | -0.00012906 | 0.000285244 | 1 | 0.350423113 | 1 | 0.368420209 | ENSG000016 | 11 | RIMKLB |
| SAT | Obesity | ENSG000016 | -3.51E-05 | 0.992195832 | 0.00359313 | 0.183454474 | -0.00011436 | 0.97460829 | 0.003592911 | 0.18342127 | 7.92E-05 | 0.004099402 | 1 | 0.996612544 | 1 | 0.992763055 | ENSG000016 | 11 | RIMKLB |
| SAT | Obesity | ENSG000016 | 0.004593176 | 0.2967648 | 0.004402135 | 0.183454063 | 0.004506794 | 0.30591159 | 0.004401856 | 0.183420874 | 8.64E-05 | 0.007585142 | 1 | 0.596760583 | 1 | 0.606176583 | ENSG000016 | 11 | TK2 |
| SAT | Obesity | ENSG000016 | 0.001563518 | 0.563480831 | 0.002706549 | 0.183454348 | 0.001514649 | 0.57571359 | 0.002706388 | 0.183421152 | 4.89E-05 | 0.022218939 | 1 | 0.797231826 | 1 | 0.80345227 | ENSG000016 | 8 | TMED3 |
| SAT | Obesity | ENSG000016 | 0.000290494 | 0.945520542 | 0.004251145 | 0.183454472 | 0.000357138 | 0.933044757 | 0.004250894 | 0.183421268 | -6.66E-05 | 0.047812876 | 1 | 0.982191461 | 1 | 0.979844999 | ENSG000016 | 8 | TMED3 |
| SAT | Obesity | ENSG000016 | 0.002388564 | 0.359257272 | 0.002605378 | 0.183454156 | 0.002315267 | 0.374164451 | 0.002605227 | 0.183420972 | 7.33E-05 | 0.000472694 | 1 | 0.655574588 | 1 | 0.669643631 | ENSG000016 | 8 | TMED3 |
| SAT | Obesity | ENSG000016 | 0.003637757 | 0.23565102 | 0.003067433 | 0.183453942 | 0.003691953 | 0.228715425 | 0.003067231 | 0.183420723 | -5.42E-05 | 0.011978898 | 1 | 0.525001456 | 1 | 0.523302168 | ENSG000016 | 4 | GALR1 |
| SAT | Obesity | ENSG000016 | 0.023477296 | 6.77E-05 | 0.005892422 | 0.183448474 | 0.024608233 | 3.11E-05 | 0.005907977 | 0.183414713 | -0.00113094 | 0.009461325 | 0.222069572 | 0.000144201 | 0.102058163 | 0.000895247 | ENSG000016 | 7 | FAM96B |
| SAT | Obesity | ENSG000016 | -0.00938474 | 0.057243893 | 0.004935576 | 0.183453107 | -0.00874917 | 0.076713737 | 0.004942801 | 0.183420086 | -0.00063557 | 0.0209356 | 1 | 0.236545607 | 1 | 0.284725986 | ENSG000016 | 9 | CHRFAM7A |
| SAT | Obesity | ENSG000016 | -0.00804234 | 0.05597258 | 0.004207885 | 0.183453093 | -0.00812753 | 0.053405903 | 0.004207606 | 0.18341986 | 8.52E-05 | 0.003630987 | 1 | 0.232463336 | 1 | 0.230862672 | ENSG000016 | 8 | SPDYE6 |

| | | | | | | | | | | | | | | | | | | | |
|---|---|---|---|---|---|---|---|---|---|---|---|---|---|---|---|---|---|---|---|
| SAT | Obesity | ENSG0000016 | 0.000779337 | 0.82704928 | 0.003566963 | 0.183454456 | 0.000591607 | 0.868293646 | 0.003567626 | 0.18342126 | 0.000187731 | 0.025194097 | 1 | 0.936352204 | 1 | 0.954321831 | ENSG0000016 | 9 | ATF7IP2 |
| SAT | Obesity | ENSG0000016 | 0.000167582 | 0.941804557 | 0.002295575 | 0.183454472 | 0.000414806 | 0.856649745 | 0.002296307 | 0.183421258 | -0.00024722 | 0.000168493 | 1 | 0.981335116 | 1 | 0.947314554 | ENSG0000016 | 9 | ATF7IP2 |
| SAT | Obesity | ENSG0000016 | 0.001215968 | 0.201420157 | 0.000951821 | 0.183453857 | 0.001198306 | 0.208019943 | 0.000951769 | 0.183420671 | 1.77E-05 | 0.029395043 | 1 | 0.485212581 | 1 | 0.497821614 | ENSG0000016 | 9 | ATF7IP2 |
| SAT | Obesity | ENSG0000016 | 0.002172785 | 0.641923384 | 0.004672541 | 0.183454392 | 0.002250892 | 0.629979498 | 0.004672266 | 0.183421182 | -7.81E-05 | 0.035973905 | 1 | 0.843249571 | 1 | 0.834462145 | ENSG0000016 | 9 | ATF7IP2 |
| SAT | Obesity | ENSG0000016 | 0.000751188 | 0.844463459 | 0.003828929 | 0.18345446 | 0.000804373 | 0.833596044 | 0.00382867 | 0.183421254 | -5.32E-05 | 0.039787446 | 1 | 0.941763438 | 1 | 0.936326075 | ENSG0000016 | 5 | ZNF592 |
| SAT | Obesity | ENSG0000016 | 0.006967433 | 0.060758087 | 0.003715456 | 0.183453145 | 0.007057464 | 0.05748532 | 0.003715223 | 0.183419906 | -9.00E-05 | 0.001183097 | 1 | 0.244597894 | 1 | 0.24057313 | ENSG0000016 | 12 | CATSPER2 |
| SAT | Obesity | ENSG0000016 | -0.00050026 | 0.89105048 | 0.003652228 | 0.183454467 | -0.00057113 | 0.875727662 | 0.003652007 | 0.183421261 | 7.09E-05 | 0.012350405 | 1 | 0.962747126 | 1 | 0.958314845 | ENSG0000016 | 12 | CATSPER2 |
| SAT | Obesity | ENSG0000016 | -0.0002915 | 0.833609827 | 0.001387597 | 0.183454457 | -0.00031878 | 0.818284947 | 0.00138751 | 0.18342125 | 2.73E-05 | 0.00903385 | 1 | 0.936669124 | 1 | 0.931572835 | ENSG0000016 | 7 | STRCP1 |
| SAT | Obesity | ENSG0000016 | -0.02246971 | 1.10E-12 | 0.003157126 | 0.183435329 | -0.02251641 | 9.88E-13 | 0.003156923 | 0.183402046 | 4.67E-05 | 0.040786092 | 3.62E-09 | 1.29E-10 | 3.24E-09 | 1.20E-10 | ENSG0000016 | 6 | ZNF667-AS1 |
| SAT | Obesity | ENSG0000016 | 0.002069338 | 0.692650761 | 0.005235377 | 0.183454415 | 0.001942402 | 0.710608242 | 0.005235028 | 0.183421218 | 0.000126936 | 0.00042805 | 1 | 0.867162933 | 1 | 0.882143641 | ENSG0000016 | 3 | SAA3P |
| SAT | Obesity | ENSG0000016 | 0.007691907 | 0.119351352 | 0.004938609 | 0.183453557 | 0.006276508 | 0.206350645 | 0.004966906 | 0.183420667 | 0.001415399 | 0.007981441 | 1 | 0.361914776 | 1 | 0.495716632 | ENSG0000016 | 7 | LDHC |
| SAT | Obesity | ENSG0000016 | 0.000718703 | 0.861674006 | 0.004124701 | 0.183454463 | 0.000639338 | 0.876811046 | 0.004124411 | 0.183421261 | 7.94E-05 | 0.002478913 | 1 | 0.94902733 | 1 | 0.958314845 | ENSG0000016 | 7 | LDHC |
| SAT | Obesity | ENSG0000016 | -0.0121597 | 0.029200535 | 0.005575863 | 0.183452676 | -0.01207529 | 0.030328709 | 0.005575502 | 0.183419497 | -8.44E-05 | 0.034725846 | 1 | 0.156547313 | 1 | 0.163665286 | ENSG0000016 | 7 | LDHC |
| SAT | Obesity | ENSG0000016 | 0.007353221 | 0.222009604 | 0.006021291 | 0.18345391 | 0.007202548 | 0.231598974 | 0.006020931 | 0.183420729 | 0.000150673 | 0.001393261 | 1 | 0.509598947 | 1 | 0.524775447 | ENSG0000016 | 11 | FAM111A |
| SAT | Obesity | ENSG0000016 | -0.0076189 | 0.159402851 | 0.005414667 | 0.183453726 | -0.00752113 | 0.164801955 | 0.005414376 | 0.183420541 | -9.78E-05 | 0.03503692 | 1 | 0.423139768 | 1 | 0.433943657 | ENSG0000016 | 11 | FAM111A |
| SAT | Obesity | ENSG0000016 | 0.005114922 | 0.068773861 | 0.002810541 | 0.183453222 | 0.005034277 | 0.07324507 | 0.002810397 | 0.183420057 | 8.06E-05 | 0.001188733 | 1 | 0.262380278 | 1 | 0.275592976 | ENSG0000016 | 11 | FAM111A |
| SAT | Obesity | ENSG0000016 | -0.01093167 | 0.000103656 | 0.00281606 | 0.183448778 | -0.0108218 | 0.000121501 | 0.002815896 | 0.183415688 | -0.00010987 | 1.06E-06 | 0.340096136 | 0.002024382 | 0.398643746 | 0.002622656 | ENSG0000016 | 11 | FAM111A |
| SAT | Obesity | ENSG0000016 | 0.003592767 | 0.041485567 | 0.001762327 | 0.183452903 | 0.003621919 | 0.039849499 | 0.001762224 | 0.183419674 | -2.92E-05 | 0.040014133 | 1 | 0.193338752 | 1 | 0.195087821 | ENSG0000016 | 4 | PEX11A |
| SAT | Obesity | ENSG0000016 | 0.003781989 | 0.381450098 | 0.004321146 | 0.183454185 | 0.003865205 | 0.37103377 | 0.004320888 | 0.183420968 | -8.32E-05 | 0.014295096 | 1 | 0.674683434 | 1 | 0.668682157 | ENSG0000016 | 12 | ANKDD1A |
| SAT | Obesity | ENSG0000016 | 0.007065398 | 0.224908779 | 0.005821947 | 0.183453917 | 0.006932063 | 0.233748074 | 0.005821541 | 0.183420734 | 0.000133335 | 0.000370702 | 1 | 0.513161129 | 1 | 0.527090757 | ENSG0000016 | 12 | ANKDD1A |
| SAT | Obesity | ENSG0000016 | -0.000898 | 0.713855482 | 0.002448971 | 0.183454423 | -0.00095033 | 0.697960794 | 0.00244882 | 0.183421213 | 5.23E-05 | 0.004835085 | 1 | 0.881706793 | 1 | 0.8747171 | ENSG0000016 | 5 | DCTN5 |
| SAT | Obesity | ENSG0000016 | 0.012341178 | 0.024214558 | 0.005475927 | 0.183452554 | 0.012431014 | 0.023192772 | 0.005475611 | 0.183419322 | -8.98E-05 | 0.043069598 | 1 | 0.141366488 | 1 | 0.141441422 | ENSG0000016 | 5 | DCTN5 |
| SAT | Obesity | ENSG0000016 | 0.001029718 | 0.667065617 | 0.002393696 | 0.183454404 | -0.00036729 | 0.879498172 | 0.002422673 | 0.183421262 | 0.001397008 | 0.000194773 | 1 | 0.854600118 | 1 | 0.958406391 | ENSG0000016 | 5 | TMEM194A |
| SAT | Obesity | ENSG0000016 | -0.00192799 | 0.396744705 | 0.002275047 | 0.183454203 | -0.00188203 | 0.408068407 | 0.002274912 | 0.183421012 | -4.60E-05 | 0.010772229 | 1 | 0.688610432 | 1 | 0.698420679 | ENSG0000016 | 5 | TMEM194A |
| SAT | Obesity | ENSG0000016 | 0.003764042 | 0.290143562 | 0.003558342 | 0.183454051 | 0.003811674 | 0.284050024 | 0.0035581 | 0.183420836 | -4.76E-05 | 0.046189464 | 1 | 0.588792914 | 1 | 0.588363718 | ENSG0000016 | 5 | TMEM194A |
| SAT | Obesity | ENSG0000016 | -0.00723276 | 0.155793205 | 0.005095774 | 0.183453713 | -0.0073275 | 0.150425563 | 0.005095507 | 0.183420489 | 9.47E-05 | 0.033124952 | 1 | 0.417953806 | 1 | 0.414396534 | ENSG0000016 | 4 | MRPL16 |
| SAT | Obesity | ENSG0000016 | -0.00246542 | 0.499430311 | 0.003650388 | 0.183454302 | -0.00238979 | 0.512654188 | 0.003650139 | 0.183421108 | -7.56E-05 | 0.001888188 | 1 | 0.7576285 | 1 | 0.769450316 | ENSG0000016 | 4 | MRPL16 |
| SAT | Obesity | ENSG0000016 | -0.00864468 | 0.116802554 | 0.005511995 | 0.183453544 | -0.00854447 | 0.121081096 | 0.005511656 | 0.183420362 | -0.00010021 | 0.016966604 | 1 | 0.357489907 | 1 | 0.369554001 | ENSG0000016 | 4 | MRPL16 |
| SAT | Obesity | ENSG0000016 | 0.002891858 | 0.581290479 | 0.005243613 | 0.183454359 | 0.00277936 | 0.596054867 | 0.005243258 | 0.183421164 | 0.000112498 | 0.00148478 | 1 | 0.806390405 | 1 | 0.817137678 | ENSG0000016 | 12 | MTMR10 |
| SAT | Obesity | ENSG0000016 | 0.00097147 | 0.789724514 | 0.003642986 | 0.183454447 | 0.000905203 | 0.803752784 | 0.003642766 | 0.183421247 | 6.63E-05 | 0.019206907 | 1 | 0.914498218 | 1 | 0.923968271 | ENSG0000016 | 12 | MTMR10 |
| SAT | Obesity | ENSG0000016 | 0.0049791 | 0.173404831 | 0.003657481 | 0.183453774 | 0.00478217 | 0.191164424 | 0.003658487 | 0.183420624 | 0.00019693 | 0.046408698 | 1 | 0.442843988 | 1 | 0.473366395 | ENSG0000016 | 8 | YWHAB |
| SAT | Obesity | ENSG0000016 | 0.014173675 | 0.01588945 | 0.005877648 | 0.183452276 | 0.01427255 | 0.015164156 | 0.005877242 | 0.183419041 | -9.89E-05 | 0.010329659 | 1 | 0.106394462 | 1 | 0.108395637 | ENSG0000016 | 8 | YWHAB |
| SAT | Obesity | ENSG0000016 | -0.01486671 | 0.002099463 | 0.004833436 | 0.183450898 | -0.01473201 | 0.002302946 | 0.004833155 | 0.183417759 | -0.0001347 | 0.000511312 | 1 | 0.024689388 | 1 | 0.029515494 | ENSG0000016 | 4 | NYAP1 |
| SAT | Obesity | ENSG0000016 | 0.00621506 | 0.119909638 | 0.003996423 | 0.18345356 | 0.006138784 | 0.124496298 | 0.003996151 | 0.183420378 | 7.63E-05 | 0.004289427 | 1 | 0.362555014 | 1 | 0.375654748 | ENSG0000016 | 6 | MS4A14 |
| SAT | Obesity | ENSG0000016 | 0.001590586 | 0.665587453 | 0.003680105 | 0.183454403 | 0.001657066 | 0.652490265 | 0.003679869 | 0.183421194 | -6.65E-05 | 0.012544425 | 1 | 0.854600118 | 1 | 0.849060146 | ENSG0000016 | 6 | MS4A14 |
| SAT | Obesity | ENSG0000016 | 0.002521412 | 0.547036828 | 0.004186955 | 0.183454337 | 0.002464432 | 0.556104637 | 0.004186671 | 0.183421139 | 5.70E-05 | 0.043418797 | 1 | 0.792029636 | 1 | 0.795394048 | ENSG0000016 | 8 | DIS3L |
| SAT | Obesity | ENSG0000016 | 0.000280366 | 0.896662496 | 0.002158678 | 0.183454468 | 0.000331944 | 0.877781663 | 0.002158534 | 0.183421261 | -5.16E-05 | 0.000574102 | 1 | 0.962747126 | 1 | 0.958406391 | ENSG0000016 | 9 | CCNDBP1 |
| SAT | Obesity | ENSG0000016 | -0.0077155 | 0.158585531 | 0.005472581 | 0.183453723 | -0.00785594 | 0.151118009 | 0.005472247 | 0.183420491 | 0.000140434 | 0.00083947 | 1 | 0.421652454 | 1 | 0.414563702 | ENSG0000016 | 9 | CCNDBP1 |
| SAT | Obesity | ENSG0000016 | 0.036794767 | 2.77E-09 | 0.006189141 | 0.183441115 | 0.036937091 | 2.40E-09 | 0.006188923 | 0.183407809 | -0.00014232 | 0.029269954 | 9.07E-06 | 2.02E-07 | 7.87E-06 | 1.92E-07 | ENSG0000016 | 3 | CNPY4 |
| SAT | Obesity | ENSG0000016 | -0.00058029 | 0.781185093 | 0.002089067 | 0.183454445 | -0.0005436 | 0.794688693 | 0.002088957 | 0.183421245 | -3.67E-05 | 0.043340705 | 1 | 0.913071213 | 1 | 0.920289292 | ENSG0000016 | 2 | GNGT2 |
| SAT | Obesity | ENSG0000016 | 0.003566291 | 0.134697859 | 0.002384151 | 0.183453628 | 0.00362468 | 0.128405748 | 0.002383997 | 0.183420397 | -5.84E-05 | 0.000683772 | 1 | 0.388010251 | 1 | 0.382304228 | ENSG0000016 | 7 | PIP5KL1 |
| SAT | Obesity | ENSG0000016 | 0.007914714 | 0.114255858 | 0.005011366 | 0.183453531 | 0.008028279 | 0.109134536 | 0.005011103 | 0.1834203 | -0.00011356 | 0.009336659 | 1 | 0.351663667 | 1 | 0.346730908 | ENSG0000016 | 7 | PIP5KL1 |
| SAT | Obesity | ENSG0000016 | 0.005848491 | 9.98E-06 | 0.001323863 | 0.183447097 | 0.005885123 | 8.76E-06 | 0.001323782 | 0.183413801 | -3.66E-05 | 0.000359714 | 0.032730066 | 0.000282156 | 0.028751567 | 0.000299495 | ENSG0000016 | 7 | PIP5KL1 |
| SAT | Obesity | ENSG0000016 | -0.01277118 | 0.005898114 | 0.004638322 | 0.183451608 | -0.0111413 | 0.01650507 | 0.004646935 | 0.183419098 | -0.00162987 | 1.79E-08 | 1 | 0.055219818 | 1 | 0.114488656 | ENSG0000016 | 7 | PIP5KL1 |
| SAT | Obesity | ENSG0000016 | -0.00944634 | 0.06324206 | 0.005085542 | 0.18345317 | -0.0095234 | 0.061102346 | 0.005085228 | 0.183419945 | 7.71E-05 | 0.04539636 | 1 | 0.250993039 | 1 | 0.250029993 | ENSG0000016 | 14 | CERCAM |
| SAT | Obesity | ENSG0000016 | -0.01042045 | 0.053094943 | 0.005387604 | 0.18345306 | -0.01025127 | 0.057070022 | 0.005387527 | 0.183419902 | -0.00016918 | 0.010877869 | 1 | 0.225653508 | 1 | 0.240368091 | ENSG0000016 | 14 | CERCAM |
| SAT | Obesity | ENSG0000016 | 0.001801587 | 0.570065526 | 0.003172057 | 0.183454352 | 0.001968033 | 0.535006334 | 0.00317229 | 0.183421125 | -0.00016645 | 0.003751301 | 1 | 0.799651556 | 1 | 0.78574565 | ENSG0000016 | 6 | ENDOG |
| SAT | Obesity | ENSG0000016 | 0.011907041 | 0.038725569 | 0.005760273 | 0.183452859 | 0.011777784 | 0.040877357 | 0.005759935 | 0.18341969 | 0.000129257 | 0.004869859 | 1 | 0.188234951 | 1 | 0.196654851 | ENSG0000016 | 6 | ENDOG |
| SAT | Obesity | ENSG0000016 | 0.001460962 | 0.333624406 | 0.001511069 | 0.183454121 | 5.83E-05 | 0.969652081 | 0.001533169 | 0.18342127 | 0.001402633 | 6.99E-08 | 1 | 0.631217486 | 1 | 0.992266083 | ENSG0000016 | 6 | ENDOG |
| SAT | Obesity | ENSG0000016 | 0.002111555 | 0.227260241 | 0.001748775 | 0.183453923 | 0.002067665 | 0.237043956 | 0.001748688 | 0.183420742 | 4.39E-05 | 0.005325143 | 1 | 0.514945339 | 1 | 0.530519249 | ENSG0000016 | 12 | KATNAL2 |
| SAT | Obesity | ENSG0000016 | 0.005143829 | 0.241690996 | 0.004393536 | 0.183453956 | 7.39E-05 | 0.986870558 | 0.004487895 | 0.18342127 | 0.005069976 | 3.26E-08 | 1 | 0.530497704 | 1 | 0.995364986 | ENSG0000016 | 7 | HDHD2 |
| SAT | Obesity | ENSG0000016 | 0.009741332 | 0.083669447 | 0.00563153 | 0.183453343 | 0.010666039 | 0.058908171 | 0.005646746 | 0.183419922 | -0.00092471 | 0.028123921 | 1 | 0.291421926 | 1 | 0.245276282 | ENSG0000016 | 7 | HDHD2 |
| SAT | Obesity | ENSG0000016 | -0.00028619 | 0.934704933 | 0.003493188 | 0.183454472 | -0.0003353 | 0.923527167 | 0.003492958 | 0.183421267 | 4.91E-05 | 0.045193123 | 1 | 0.979129279 | 1 | 0.975006702 | ENSG0000016 | 2 | CCL23 |
| SAT | Obesity | ENSG0000016 | -0.00464188 | 0.447025523 | 0.006104651 | 0.183454256 | 0.004319216 | 0.494532688 | 0.006322797 | 0.183421094 | -0.00896109 | 5.46E-08 | 1 | 0.722507754 | 1 | 0.755382565 | ENSG0000016 | 2 | CCL23 |
| SAT | Obesity | ENSG0000016 | -0.00158071 | 0.500036971 | 0.002343763 | 0.183454302 | -0.00137884 | 0.55635592 | 0.002343918 | 0.183421139 | -0.00020187 | 1.14E-06 | 1 | 0.75779275 | 1 | 0.795394048 | ENSG0000016 | 6 | RNF214 |
| SAT | Obesity | ENSG0000016 | 0.009017789 | 0.136534903 | 0.006056996 | 0.183453636 | 0.001617069 | 0.796196427 | 0.006261085 | 0.183421245 | 0.007400721 | 3.14E-06 | 1 | 0.389540014 | 1 | 0.921453431 | ENSG0000016 | 6 | RNF214 |
| SAT | Obesity | ENSG0000016 | 0.02517499 | 9.75E-06 | 0.005692367 | 0.183447081 | 0.023295629 | 4.43E-05 | 0.005704671 | 0.183414968 | 0.001879361 | 8.65E-07 | 0.032005176 | 0.000278795 | 0.145510061 | 0.001202562 | ENSG0000016 | 6 | RNF214 |
| SAT | Obesity | ENSG0000016 | 0.007636315 | 0.149348126 | 0.005296229 | 0.183453688 | 0.007715089 | 0.145170379 | 0.00529589 | 0.183420468 | -7.88E-05 | 0.041705197 | 1 | 0.409366084 | 1 | 0.40919767 | ENSG0000016 | 14 | MYO5B |
| SAT | Obesity | ENSG0000016 | 0.002046201 | 0.583572058 | 0.003732752 | 0.183454361 | 0.001933613 | 0.604426349 | 0.003732511 | 0.183421169 | 0.000112588 | 2.83E-05 | 1 | 0.806765832 | 1 | 0.822274447 | ENSG0000016 | 14 | MYO5B |
| SAT | Obesity | ENSG0000016 | -0.00158983 | 0.768122292 | 0.005392327 | 0.183454441 | -0.00692451 | 0.214396951 | 0.005577237 | 0.183420688 | 0.005334682 | 0.000183451 | 1 | 0.90720275 | 1 | 0.506069351 | ENSG0000016 | 9 | ART5 |
| SAT | Obesity | ENSG0000016 | -0.01469969 | 0.014718471 | 0.006026336 | 0.183452225 | -0.0144734 | 0.016313689 | 0.006025991 | 0.18341909 | -0.00022629 | 3.93E-06 | 1 | 0.101452318 | 1 | 0.113641644 | ENSG0000016 | 9 | ART5 |
| SAT | Obesity | ENSG0000016 | 0.003575394 | 0.170518215 | 0.002608743 | 0.183453764 | 0.003621377 | 0.165059946 | 0.002608581 | 0.183420542 | -4.60E-05 | 0.019525648 | 1 | 0.437770159 | 1 | 0.433943657 | ENSG0000016 | 9 | ART5 |
| SAT | Obesity | ENSG0000016 | 0.000782438 | 0.875387848 | 0.004989445 | 0.183454465 | 0.000685417 | 0.890728642 | 0.004989118 | 0.183421263 | 9.70E-05 | 0.005873244 | 1 | 0.955789527 | 1 | 0.962898024 | ENSG0000016 | 10 | RRM1 |
| SAT | Obesity | ENSG0000016 | 0.005636284 | 0.064240636 | 0.0030458 | 0.18345318 | 0.00556658 | 0.067587357 | 0.003045588 | 0.183420008 | 6.97E-05 | 0.000380883 | 1 | 0.253944006 | 1 | 0.265533656 | ENSG0000016 | 10 | RRM1 |
| SAT | Obesity | ENSG0000016 | 0.004589355 | 0.000346132 | 0.001282623 | 0.183449635 | 0.003778689 | 0.004081271 | 0.00131578 | 0.183418153 | 0.000810666 | 0.005827625 | 1 | 0.005794173 | 1 | 0.04585839 | ENSG0000016 | 10 | RRM1 |
| SAT | Obesity | ENSG0000016 | 0.016386935 | 1.78E-13 | 0.002225174 | 0.183433976 | 0.016424187 | 1.57E-13 | 0.002225037 | 0.18340068 | -3.73E-05 | 0.029353052 | 5.85E-10 | 2.34E-11 | 5.14E-10 | 2.14E-11 | ENSG0000016 | 10 | RRM1 |
| SAT | Obesity | ENSG0000016 | -0.00030932 | 0.895999776 | 0.002366375 | 0.183454468 | -0.00035759 | 0.879877658 | 0.002366222 | 0.183421262 | 4.83E-05 | 0.004603858 | 1 | 0.962747126 | 1 | 0.958459029 | ENSG0000016 | 10 | RRM1 |
| SAT | Obesity | ENSG0000016 | 0.004163771 | 0.387564979 | 0.004818943 | 0.183454192 | 0.004285268 | 0.373837649 | 0.004818651 | 0.183420971 | -0.0001215 | 0.001136794 | 1 | 0.678187037 | 1 | 0.669643631 | ENSG0000016 | 7 | OR51E2 |
| SAT | Obesity | ENSG0000016 | -0.00897906 | 0.016878672 | 0.003757953 | 0.183452316 | -0.00892411 | 0.017555097 | 0.003757712 | 0.183419139 | -5.49E-05 | 0.043750329 | 1 | 0.110097259 | 1 | 0.118319765 | ENSG0000016 | 7 | OR51E2 |
| SAT | Obesity | ENSG0000016 | 0.00333197 | 0.153183283 | 0.002332692 | 0.183453703 | 0.003367635 | 0.14880656 | 0.002332542 | 0.183420482 | -3.57E-05 | 0.033897997 | 1 | 0.413998642 | 1 | 0.414396534 | ENSG0000016 | 8 | TRIM68 |
| SAT | Obesity | ENSG0000017 | -0.00298945 | 0.369660558 | 0.003332315 | 0.18345417 | -0.002916 | 0.381511201 | 0.003332127 | 0.183420981 | -7.35E-05 | 0.007389141 | 1 | 0.665673046 | 1 | 0.676469831 | ENSG0000016 | 10 | ZNF646 |
| SAT | Obesity | ENSG0000017 | -0.00176813 | 0.554151364 | 0.002988964 | 0.183454342 | -0.00171926 | 0.565129848 | 0.002988772 | 0.183421145 | -4.89E-05 | 0.023716296 | 1 | 0.794232732 | 1 | 0.798875928 | ENSG0000016 | 10 | ZNF646 |
| SAT | Obesity | ENSG0000017 | -0.00110201 | 0.785586048 | 0.004050794 | 0.183454446 | -0.00115766 | 0.775027269 | 0.004050506 | 0.183421239 | 5.57E-05 | 0.02779951 | 1 | 0.914334098 | 1 | 0.911862059 | ENSG0000016 | 10 | ZNF646 |
| SAT | Obesity | ENSG0000017 | -0.00489331 | 0.426990688 | 0.006160121 | 0.183454236 | -0.00498379 | 0.418460891 | 0.00615971 | 0.183421023 | 9.05E-05 | 0.033246911 | 1 | 0.70970438 | 1 | 0.703737593 | ENSG0000016 | 5 | CA4 |
| SAT | Obesity | ENSG0000017 | -0.00802653 | 0.114832405 | 0.005090253 | 0.183453534 | -0.00792125 | 0.119647473 | 0.005089933 | 0.183420355 | -0.00010528 | 0.005339533 | 1 | 0.353106955 | 1 | 0.368420209 | ENSG0000016 | 7 | RAB8A |
| SAT | Obesity | ENSG0000017 | -0.00361079 | 0.157448827 | 0.002554127 | 0.183453719 | -0.00356453 | 0.162812135 | 0.00255398 | 0.183420534 | -4.63E-05 | 0.025195918 | 1 | 0.420045334 | 1 | 0.430795656 | ENSG0000016 | 6 | JSRP1 |
| SAT | Obesity | ENSG0000017 | -0.00261829 | 0.357687043 | 0.002846645 | 0.183454154 | -0.00251965 | 0.376072982 | 0.002846563 | 0.183420974 | -9.86E-05 | 0.001792487 | 1 | 0.655187068 | 1 | 0.671325056 | ENSG0000016 | 9 | PROCA1 |
| SAT | Obesity | ENSG0000017 | -0.00053283 | 0.870861778 | 0.003277685 | 0.183454464 | -0.00062664 | 0.848373355 | 0.003277483 | 0.183421256 | 9.38E-05 | 0.000154068 | 1 | 0.955298393 | 1 | 0.942286045 | ENSG0000016 | 8 | ZNF641 |
| SAT | Obesity | ENSG0000017 | 0.005593634 | 0.329403193 | 0.005735192 | 0.183454115 | 0.005850517 | 0.307693615 | 0.005735382 | 0.183420877 | -0.00025688 | 0.004418673 | 1 | 0.628809457 | 1 | 0.607792143 | ENSG0000016 | 8 | ZNF641 |
| SAT | Obesity | ENSG0000017 | 0.009124035 | 0.080307627 | 0.005216991 | 0.183453318 | 0.009037197 | 0.083209827 | 0.005216692 | 0.183420136 | 8.68E-05 | 0.040869247 | 1 | 0.286713084 | 1 | 0.29736676 | ENSG0000016 | 8 | ZNF641 |
| SAT | Obesity | ENSG0000017 | 0.000580738 | 0.879376402 | 0.003826692 | 0.183454465 | 0.000474973 | 0.901213886 | 0.003826476 | 0.183421264 | 0.000105765 | 0.000838971 | 1 | 0.957347365 | 1 | 0.963222823 | ENSG0000016 | 8 | ZNF641 |
| SAT | Obesity | ENSG0000017 | 0.000399596 | 0.935015895 | 0.004900863 | 0.183454472 | 0.000470412 | 0.923527085 | 0.004900542 | 0.183421267 | -7.08E-05 | 0.041161114 | 1 | 0.979129279 | 1 | 0.975006702 | ENSG0000016 | 3 | CACNB3 |
| SAT | Obesity | ENSG0000017 | 0.00938636 | 0.119004083 | 0.006020872 | 0.183453555 | 0.00951018 | 0.114192607 | 0.006020517 | 0.183420327 | -0.00012382 | 0.009617074 | 1 | 0.361529998 | 1 | 0.357164865 | ENSG0000016 | 10 | C19orf55 |
| SAT | Obesity | ENSG0000017 | -0.00066717 | 0.718346585 | 0.001849805 | 0.183454425 | -0.00070615 | 0.702636894 | 0.001849695 | 0.183421215 | 3.90E-05 | 0.007427362 | 1 | 0.883393983 | 1 | 0.876674335 | ENSG0000016 | 10 | C19orf55 |
| SAT | Obesity | ENSG0000017 | -0.00750625 | 0.177212499 | 0.005562701 | 0.183453786 | -0.00759232 | 0.172267157 | 0.005562316 | 0.183420566 | 8.61E-05 | 0.017942156 | 1 | 0.450724192 | 1 | 0.443997285 | ENSG0000016 | 10 | C19orf55 |
| SAT | Obesity | ENSG0000017 | -0.00017015 | 0.948051291 | 0.002611481 | 0.183454473 | -0.00021427 | 0.934604938 | 0.002611316 | 0.183421268 | 4.41E-05 | 0.022687499 | 1 | 0.983182517 | 1 | 0.979844999 | ENSG0000016 | 10 | C19orf55 |
| SAT | Obesity | ENSG0000017 | 0.00367347 | 0.157673687 | 0.002599875 | 0.183453719 | 0.003724109 | 0.151999928 | 0.002599718 | 0.183420495 | -5.06E-05 | 0.012122153 | 1 | 0.420249688 | 1 | 0.415593137 | ENSG0000016 | 7 | C19orf33 |
| SAT | Obesity | ENSG0000017 | -0.00039372 | 0.87665025 | 0.002536589 | 0.183454465 | -0.00033812 | 0.893950892 | 0.002536435 | 0.183421263 | -5.56E-05 | 0.004600575 | 1 | 0.956287991 | 1 | 0.963222823 | ENSG0000016 | 7 | C19orf33 |
| SAT | Obesity | ENSG0000017 | -0.01136234 | 2.67E-15 | 0.001437265 | 0.183430853 | -0.01134203 | 2.98E-15 | 0.001437172 | 0.183397734 | -2.03E-05 | 0.048425759 | 8.77E-12 | 4.18E-13 | 9.78E-12 | 4.89E-13 | ENSG0000016 | 7 | C19orf33 |
| SAT | Obesity | ENSG0000017 | 0.003088926 | 0.589788586 | 0.005729329 | 0.183454364 | 0.002882232 | 0.614902596 | 0.00572908 | 0.183421175 | 0.000206694 | 0.000200894 | 1 | 0.811021102 | 1 | 0.827520679 | ENSG0000016 | 7 | C19orf33 |
| SAT | Obesity | ENSG0000017 | 0.004611824 | 0.120004494 | 0.002966266 | 0.18345356 | 0.004696401 | 0.11334076 | 0.002966104 | 0.183420323 | -8.46E-05 | 0.000761468 | 1 | 0.362555014 | 1 | 0.354838771 | ENSG0000016 | 9 | DNAAF3 |
| SAT | Obesity | ENSG0000017 | 0.001726681 | 0.178882722 | 0.001284537 | 0.183453791 | 0.002069567 | 0.10750849 | 0.001285848 | 0.183420291 | -0.00034289 | 1.52E-08 | 1 | 0.452218367 | 1 | 0.345142226 | ENSG0000016 | 9 | DNAAF3 |
| SAT | Obesity | ENSG0000017 | 0.026720038 | 7.47E-07 | 0.005399254 | 0.183445217 | 0.010674639 | 0.149844836 | 0.007412522 | 0.183420486 | 0.016045398 | 0.001582624 | 0.002450336 | 3.10E-05 | 1 | 0.414396534 | ENSG0000016 | 9 | DNAAF3 |
| SAT | Obesity | ENSG0000017 | 0.003256139 | 0.170431961 | 0.002375324 | 0.183453764 | 0.003304152 | 0.164194102 | 0.002375199 | 0.183420539 | -4.80E-05 | 0.020319108 | 1 | 0.437770159 | 1 | 0.433055345 | ENSG0000016 | 11 | CHAF1A |
| SAT | Obesity | ENSG0000017 | 0.01794911 | 1.24E-06 | 0.003700967 | 0.183445584 | -0.00071532 | 0.921949518 | 0.007300811 | 0.183421267 | 0.018664432 | 0.003019659 | 0.004055228 | 4.77E-05 | 1 | 0.974898873 | ENSG0000016 | 7 | UBXN6 |
| SAT | Obesity | ENSG0000017 | 0.009722045 | 0.067246242 | 0.005312509 | 0.183453208 | 0.009844504 | 0.063858083 | 0.005312241 | 0.183419972 | -0.00012246 | 0.009994687 | 1 | 0.26053725 | 1 | 0.257054802 | ENSG0000016 | 10 | FAM57A |
| SAT | Obesity | ENSG0000017 | 0.000142568 | 0.962371826 | 0.003021962 | 0.183454473 | 0.006659987 | 0.037980283 | 0.003209519 | 0.183419643 | -0.00651742 | 1.70E-09 | 1 | 0.989966035 | 1 | 0.18967018 | ENSG0000016 | 10 | FAM57A |
| SAT | Obesity | ENSG0000017 | -0.00055846 | 0.903237594 | 0.004593605 | 0.183454469 | -0.00079762 | 0.862147028 | 0.004593484 | 0.183421259 | 0.000239165 | 4.42E-06 | 1 | 0.964060685 | 1 | 0.949538046 | ENSG0000016 | 10 | FAM57A |
| SAT | Obesity | ENSG0000017 | -0.00479551 | 0.329973336 | 0.004922674 | 0.183454115 | -0.00391271 | 0.426934342 | 0.004925059 | 0.183421032 | -0.0008828 | 1.23E-07 | 1 | 0.628809457 | 1 | 0.711073464 | ENSG0000016 | 4 | MFSD3 |
| SAT | Obesity | ENSG0000017 | -0.00390816 | 0.306405489 | 0.003821062 | 0.183454079 | -0.00396344 | 0.299583138 | 0.00382081 | 0.183420864 | 5.53E-05 | 0.039080795 | 1 | 0.605977341 | 1 | 0.601026164 | ENSG0000016 | 9 | SERPINF2 |
| SAT | Obesity | ENSG0000017 | 0.000603298 | 0.869894474 | 0.003683305 | 0.183454464 | 0.000662583 | 0.85723161 | 0.003683077 | 0.183421258 | -5.93E-05 | 0.033349209 | 1 | 0.954875801 | 1 | 0.947314554 | ENSG0000016 | 9 | SERPINF2 |
| SAT | Obesity | ENSG0000017 | -0.00404822 | 0.168196559 | 0.0029377 | 0.183453756 | -0.00428014 | 0.145153394 | 0.002937906 | 0.183420468 | 0.000231921 | 1.07E-05 | 1 | 0.434800182 | 1 | 0.40919767 | ENSG0000016 | 9 | SERPINF2 |
| SAT | Obesity | ENSG0000017 | 0.001853491 | 0.217902897 | 0.001504304 | 0.1834539 | 0.001873524 | 0.212938326 | 0.0015042 | 0.183420684 | -2.00E-05 | 0.042103575 | 1 | 0.505890452 | 1 | 0.503707333 | ENSG0000016 | 9 | SERPINF2 |
| SAT | Obesity | ENSG0000017 | 0.002006929 | 0.661180801 | 0.004579064 | 0.183454401 | 0.001868012 | 0.683293489 | 0.004578762 | 0.183421207 | 0.000138917 | 1.39E-05 | 1 | 0.852929209 | 1 | 0.86636807 | ENSG0000016 | 9 | SERPINF2 |
| SAT | Obesity | ENSG0000017 | -0.00642587 | 0.000805341 | 0.001917613 | 0.18345023 | -0.00638942 | 0.000861686 | 0.001917477 | 0.183417074 | -3.65E-05 | 0.002321311 | 1 | 0.011499836 | 1 | 0.013149728 | ENSG0000016 | 8 | SRR |
| SAT | Obesity | ENSG0000017 | 0.000634061 | 0.853582205 | 0.003435716 | 0.183454461 | 0.00042552 | 0.901433042 | 0.003435735 | 0.183421264 | 0.00020854 | 1.21E-05 | 1 | 0.943648754 | 1 | 0.963222823 | ENSG0000016 | 8 | SRR |
| SAT | Obesity | ENSG0000017 | 0.010953294 | 0.003078692 | 0.003700707 | 0.183451163 | 0.010888814 | 0.003255678 | 0.003700506 | 0.183417998 | 6.45E-05 | 0.040448546 | 1 | 0.034241321 | 1 | 0.038984968 | ENSG0000016 | 8 | SRR |
| SAT | Obesity | ENSG0000017 | -0.00500508 | 0.187060903 | 0.003793664 | 0.183453816 | -0.00505694 | 0.18250282 | 0.0037934 | 0.183420599 | 5.19E-05 | 0.034972997 | 1 | 0.461882921 | 1 | 0.459606393 | ENSG0000016 | 8 | SRR |
| SAT | Obesity | ENSG0000017 | -0.00459228 | 0.10199488 | 0.002808287 | 0.183453463 | -0.00672315 | 0.017561695 | 0.002831108 | 0.183419139 | 0.002130864 | 3.51E-09 | 1 | 0.325530352 | 1 | 0.118319765 | ENSG0000016 | 9 | C19orf48 |
| SAT | Obesity | ENSG0000017 | 0.001658471 | 0.312891872 | 0.001643395 | 0.183454089 | 0.001685225 | 0.305114813 | 0.001643276 | 0.183420873 | -2.68E-05 | 0.006766376 | 1 | 0.609761023 | 1 | 0.605981659 | ENSG0000016 | 9 | C19orf48 |
| SAT | Obesity | ENSG0000017 | -0.0005497 | 0.922520011 | 0.005651887 | 0.183454471 | -0.00033919 | 0.952141808 | 0.005651581 | 0.183421269 | -0.00021051 | 1.23E-05 | 1 | 0.97398945 | 1 | 0.987557383 | ENSG0000016 | 9 | KLK11 |
| SAT | Obesity | ENSG0000017 | 0.000981705 | 0.873287526 | 0.006155503 | 0.183454464 | 0.000888388 | 0.885236791 | 0.006155082 | 0.183421262 | 9.33E-05 | 0.022281308 | 1 | 0.95575129 | 1 | 0.961975704 | ENSG0000016 | 14 | ZNF83 |
| SAT | Obesity | ENSG0000017 | 0.001340823 | 0.237210995 | 0.00113438 | 0.183453946 | 0.001363739 | 0.229259666 | 0.001134304 | 0.183420724 | -2.29E-05 | 0.003490241 | 1 | 0.526060746 | 1 | 0.523816827 | ENSG0000016 | 7 | SOAT2 |
| SAT | Obesity | ENSG0000017 | 0.006738117 | 0.210691845 | 0.005383315 | 0.183453882 | 0.00899886 | 0.101660132 | 0.005497632 | 0.183420258 | -0.00226074 | 0.043101616 | 1 | 0.499840884 | 1 | 0.333213679 | ENSG0000016 | 4 | ZNF558 |
| SAT | Obesity | ENSG0000017 | -0.00145969 | 0.633405833 | 0.003060542 | 0.183454388 | 0.004078363 | 0.202804791 | 0.003202225 | 0.183420657 | -0.00553805 | 4.27E-09 | 1 | 0.839339474 | 1 | 0.491434652 | ENSG0000016 | 5 | NUDT8 |
| SAT | Obesity | ENSG0000017 | -0.00621036 | 0.223542272 | 0.00510232 | 0.183453914 | -0.0061062 | 0.231377173 | 0.005102019 | 0.183420729 | -0.00010416 | 0.009890662 | 1 | 0.511109543 | 1 | 0.524775447 | ENSG0000016 | 5 | NUDT8 |
| SAT | Obesity | ENSG0000017 | 0.002448807 | 0.565359389 | 0.004259537 | 0.183454349 | 0.002606322 | 0.540595212 | 0.004259297 | 0.183421129 | -0.00015752 | 7.78E-06 | 1 | 0.797234594 | 1 | 0.787957747 | ENSG0000016 | 5 | NUDT8 |
| SAT | Obesity | ENSG0000017 | -0.0027819 | 0.512981867 | 0.004252357 | 0.183454312 | -0.00499639 | 0.24891635 | 0.004333434 | 0.183420768 | 0.002214491 | 0.008098685 | 1 | 0.767484499 | 1 | 0.545189948 | ENSG0000016 | 5 | NUDT8 |
| SAT | Obesity | ENSG0000017 | 0.004004393 | 0.450061882 | 0.005301638 | 0.183454258 | 0.004462329 | 0.39998457 | 0.005301885 | 0.183421002 | -0.00045794 | 1.85E-07 | 1 | 0.725628027 | 1 | 0.692729827 | ENSG0000016 | 5 | NUDT8 |
| SAT | Obesity | ENSG0000017 | -0.00090929 | 0.489766172 | 0.001316507 | 0.183454294 | -0.00088972 | 0.499130951 | 0.001316424 | 0.183421098 | -1.96E-05 | 0.046757284 | 1 | 0.75041911 | 1 | 0.759577297 | ENSG0000016 | 5 | NUDT8 |
| SAT | Obesity | ENSG0000017 | -0.00356514 | 0.430418324 | 0.004521556 | 0.183454239 | -0.00346865 | 0.442969904 | 0.004521253 | 0.183421048 | -9.65E-05 | 0.001896409 | 1 | 0.71201318 | 1 | 0.722357979 | ENSG0000016 | 9 | TBX10 |
| SAT | Obesity | ENSG0000017 | 0.00576352 | 0.160750534 | 0.004109303 | 0.183453731 | 0.005848224 | 0.154660586 | 0.004109026 | 0.183420505 | -8.47E-05 | 0.002392725 | 1 | 0.424997988 | 1 | 0.418287971 | ENSG0000016 | 9 | TBX10 |
| SAT | Obesity | ENSG0000017 | -0.00589285 | 0.134544437 | 0.003937963 | 0.183453628 | -0.00578352 | 0.141913529 | 0.00393784 | 0.183420455 | -0.00010933 | 0.010696252 | 1 | 0.388010251 | 1 | 0.40243586 | ENSG0000016 | 9 | ZNF232 |
| SAT | Obesity | ENSG0000017 | 0.000848717 | 0.806660784 | 0.003467885 | 0.183454451 | 0.000754878 | 0.827675584 | 0.003467771 | 0.183421252 | 9.38E-05 | 0.01180312 | 1 | 0.924756825 | 1 | 0.936326075 | ENSG0000016 | 9 | ZNF232 |
| SAT | Obesity | ENSG0000017 | 0.001157963 | 0.407561417 | 0.00139818 | 0.183454215 | 0.001132053 | 0.418106026 | 0.001398094 | 0.183421022 | 2.59E-05 | 0.015076898 | 1 | 0.695625456 | 1 | 0.703737593 | ENSG0000016 | 9 | ZNF232 |
| SAT | Obesity | ENSG0000017 | -0.00658105 | 0.076889032 | 0.003720138 | 0.183453291 | -0.00652452 | 0.07943833 | 0.003719884 | 0.183420108 | -5.65E-05 | 0.023163588 | 1 | 0.279062958 | 1 | 0.290241828 | ENSG0000016 | 9 | ZNF232 |
| SAT | Obesity | ENSG0000017 | -0.00853899 | 0.000120495 | 0.002220717 | 0.183448885 | -0.00561691 | 0.014883452 | 0.00230654 | 0.183419029 | -0.00292207 | 2.83E-06 | 0.39534337 | 0.00231195 | 1 | 0.107919055 | ENSG0000016 | 10 | TMC8 |

| | | | | | | | | | | | | | | | | | | | |
|---|---|---|---|---|---|---|---|---|---|---|---|---|---|---|---|---|---|---|---|
| SAT | Obesity | ENSG0000017 | 0.009081307 | 0.057320178 | 0.004777458 | 0.183453108 | 0.009168118 | 0.054964261 | 0.004777143 | 0.183419878 | -8.68E-05 | 0.00958842 | 1 | 0.236562896 | 1 | 0.235120913 | ENSG0000016 | 4 | TMEM99 |
| SAT | Obesity | ENSG0000017 | -0.00270995 | 0.281473417 | 0.00251616 | 0.183454036 | -0.00280382 | 0.265119699 | 0.00251605 | 0.183420801 | 9.39E-05 | 0.000109383 | 1 | 0.579661774 | 1 | 0.567932065 | ENSG0000016 | 4 | TMEM99 |
| SAT | Obesity | ENSG0000017 | -0.00167656 | 0.649865537 | 0.003693283 | 0.183454396 | -0.00104463 | 0.024939948 | 0.004925498 | 0.18341937 | 0.009368071 | 0.004049351 | 1 | 0.846029897 | 1 | 0.146382772 | ENSG0000016 | 11 | ITFG3 |
| SAT | Obesity | ENSG0000017 | -0.00024196 | 0.890095177 | 0.001750983 | 0.183454467 | -0.00021242 | 0.903437291 | 0.001750883 | 0.183421265 | -2.95E-05 | 0.03625148 | 1 | 0.962241277 | 1 | 0.96423859 | ENSG0000016 | 9 | ABCA3 |
| SAT | Obesity | ENSG0000017 | 0.003860629 | 0.514556356 | 0.005923391 | 0.183454314 | 0.003683957 | 0.533960625 | 0.005923028 | 0.183421124 | 0.000176672 | 9.57E-05 | 1 | 0.769062939 | 1 | 0.784735611 | ENSG0000016 | 9 | ABCA3 |
| SAT | Obesity | ENSG0000017 | -0.00492976 | 0.17272245 | 0.003615493 | 0.183453771 | -0.00498717 | 0.16774808 | 0.003615256 | 0.183420551 | 5.74E-05 | 0.024800812 | 1 | 0.442045521 | 1 | 0.437505128 | ENSG0000016 | 9 | ABCA3 |
| SAT | Obesity | ENSG0000017 | -0.00200088 | 0.667843555 | 0.004662865 | 0.183454405 | -0.00211868 | 0.649540304 | 0.004662575 | 0.183421192 | 0.000117799 | 0.000817229 | 1 | 0.854600118 | 1 | 0.847588987 | ENSG0000016 | 9 | ABCA3 |
| SAT | Obesity | ENSG0000017 | -0.00529886 | 0.038325714 | 0.002558154 | 0.183452852 | -0.00534508 | 0.036658194 | 0.002557998 | 0.18341962 | 4.62E-05 | 0.01894154 | 1 | 0.187401887 | 1 | 0.186062873 | ENSG0000016 | 9 | ABCA3 |
| SAT | Obesity | ENSG0000017 | -3.40E-06 | 0.999414195 | 0.004629844 | 0.183454474 | 9.37E-05 | 0.983853444 | 0.004629583 | 0.18342127 | -9.71E-05 | 0.011101271 | 1 | 0.99968511 | 1 | 0.995364986 | ENSG0000016 | 9 | ABCA3 |
| SAT | Obesity | ENSG0000017 | -0.00494143 | 0.18528491 | 0.003730339 | 0.183453811 | -0.00499929 | 0.180159048 | 0.003730067 | 0.183420591 | 5.79E-05 | 0.009380035 | 1 | 0.460203128 | 1 | 0.457509161 | ENSG0000016 | 6 | VPS37C |
| SAT | Obesity | ENSG0000017 | -0.00474738 | 0.042503892 | 0.002340273 | 0.183452919 | -0.00470802 | 0.044235158 | 0.002340131 | 0.18341974 | -3.94E-05 | 0.029696018 | 1 | 0.195589441 | 1 | 0.206451713 | ENSG0000016 | 6 | VPS37C |
| SAT | Obesity | ENSG0000018 | 0.003506382 | 0.351616965 | 0.0037644 | 0.183454146 | 0.00338162 | 0.368989297 | 0.003764177 | 0.183420965 | 0.000124762 | 2.74E-05 | 1 | 0.649215117 | 1 | 0.667026933 | ENSG0000016 | 10 | BSCL2 |
| SAT | Obesity | ENSG0000018 | -0.00885799 | 0.142757087 | 0.006043924 | 0.183453662 | -0.00911087 | 0.131684996 | 0.006043722 | 0.183420411 | 0.000252876 | 9.36E-05 | 1 | 0.39896593 | 1 | 0.388541792 | ENSG0000016 | 10 | BSCL2 |
| SAT | Obesity | ENSG0000018 | 0.003908739 | 0.512887 | 0.005973473 | 0.183454312 | 0.004002962 | 0.502752124 | 0.005973088 | 0.1834211 | -9.42E-05 | 0.02886737 | 1 | 0.767484499 | 1 | 0.763656037 | ENSG0000016 | 10 | BSCL2 |
| SAT | Obesity | ENSG0000018 | -0.00335221 | 0.498915892 | 0.004957449 | 0.183454301 | -0.00344503 | 0.487076195 | 0.004957109 | 0.183421088 | 9.28E-05 | 0.004748875 | 1 | 0.75749331 | 1 | 0.750011464 | ENSG0000016 | 5 | HRASLS5 |
| SAT | Obesity | ENSG0000018 | 0.004644349 | 0.308584714 | 0.004561345 | 0.183454082 | 0.004499602 | 0.323877727 | 0.004561078 | 0.183420902 | 0.000144748 | 6.92E-05 | 1 | 0.60688449 | 1 | 0.626706567 | ENSG0000016 | 9 | TRANK1 |
| SAT | Obesity | ENSG0000018 | 0.006392619 | 0.008578046 | 0.002432097 | 0.183451863 | 0.006348159 | 0.009046891 | 0.002431977 | 0.183418695 | 4.45E-05 | 0.043355611 | 1 | 0.070683838 | 1 | 0.07770379 | ENSG0000016 | 9 | TRANK1 |
| SAT | Obesity | ENSG0000018 | -0.00257571 | 0.586837054 | 0.004739765 | 0.183454362 | -0.00249705 | 0.5982883 | 0.004739464 | 0.183421165 | -7.87E-05 | 0.023798229 | 1 | 0.808391759 | 1 | 0.817909964 | ENSG0000016 | 9 | TRANK1 |
| SAT | Obesity | ENSG0000018 | -5.59E-05 | 0.987031634 | 0.003437833 | 0.183454474 | -0.00268518 | 0.457979405 | 0.003617968 | 0.183421062 | 0.0026293 | 0.019793606 | 1 | 0.994152558 | 1 | 0.731563013 | ENSG0000016 | 9 | TRANK1 |
| SAT | Obesity | ENSG0000018 | -0.00250173 | 0.231281259 | 0.002089887 | 0.183453932 | -0.00201001 | 0.339106575 | 0.002102671 | 0.183420925 | -0.00049172 | 0.034986251 | 1 | 0.520105422 | 1 | 0.641968257 | ENSG0000016 | 12 | TTC21A |
| SAT | Obesity | ENSG0000018 | -0.01918522 | 9.98E-09 | 0.003347546 | 0.183442059 | -0.01832876 | 4.55E-08 | 0.003351943 | 0.18340997 | -0.00085646 | 1.35E-06 | 3.28E-05 | 6.30E-07 | 0.000149321 | 2.93E-06 | ENSG0000016 | 9 | RPSA |
| SAT | Obesity | ENSG0000018 | 0.008307364 | 0.137276507 | 0.005590374 | 0.183453639 | 0.008768275 | 0.116921965 | 0.005592626 | 0.183420341 | -0.00046091 | 0.008704532 | 1 | 0.390599089 | 1 | 0.36350002 | ENSG0000016 | 9 | RPSA |
| SAT | Obesity | ENSG0000018 | 0.000107756 | 0.977675609 | 0.003850742 | 0.183454474 | 4.84E-05 | 0.989970046 | 0.003850473 | 0.18342127 | 5.94E-05 | 0.016131811 | 1 | 0.993452012 | 1 | 0.996265811 | ENSG0000016 | 9 | RPSA |
| SAT | Obesity | ENSG0000018 | -0.0058725 | 0.305491942 | 0.005730792 | 0.183454077 | -0.00601989 | 0.293484858 | 0.005730451 | 0.183420853 | 0.000147384 | 0.001105519 | 1 | 0.605495891 | 1 | 0.59812588 | ENSG0000016 | 9 | RPSA |
| SAT | Obesity | ENSG0000018 | -0.01592268 | 0.004619446 | 0.00562152 | 0.183451442 | -0.01582868 | 0.004864296 | 0.005621184 | 0.183418273 | -9.40E-05 | 0.032396593 | 1 | 0.046951916 | 1 | 0.051649693 | ENSG0000016 | 9 | RPSA |
| SAT | Obesity | ENSG0000018 | 0.003897432 | 0.027373919 | 0.001766621 | 0.183452634 | 0.00397366 | 0.02449853 | 0.001766678 | 0.183419358 | -7.62E-05 | 0.005968693 | 1 | 0.149689712 | 1 | 0.14496253 | ENSG0000016 | 9 | RPSA |
| SAT | Obesity | ENSG0000018 | -0.01064683 | 0.008386093 | 0.004038816 | 0.183451847 | -0.01078196 | 0.007591105 | 0.004038571 | 0.183418576 | 0.000135128 | 1.59E-05 | 1 | 0.069748174 | 1 | 0.070158914 | ENSG0000016 | 9 | RPSA |
| SAT | Obesity | ENSG0000018 | -0.00684383 | 0.147542502 | 0.004725524 | 0.183453681 | -0.00670679 | 0.155819491 | 0.004725504 | 0.183420509 | -0.00013704 | 0.027144216 | 1 | 0.406454198 | 1 | 0.420430716 | ENSG0000016 | 6 | ULK4 |
| SAT | Obesity | ENSG0000018 | -0.00429004 | 0.415214471 | 0.005265454 | 0.183454223 | -0.00406838 | 0.439701429 | 0.005265166 | 0.183421045 | -0.00022166 | 6.16E-07 | 1 | 0.700759618 | 1 | 0.719531366 | ENSG0000016 | 6 | ULK4 |
| SAT | Obesity | ENSG0000018 | 0.002235522 | 0.436956518 | 0.002875848 | 0.183454246 | 0.002159547 | 0.452667072 | 0.002875652 | 0.183421057 | 7.60E-05 | 7.65E-05 | 1 | 0.717185761 | 1 | 0.727870316 | ENSG0000016 | 6 | ULK4 |
| SAT | Obesity | ENSG0000018 | 0.000170593 | 0.977023795 | 0.005923281 | 0.183454474 | 1.78E-05 | 0.997601554 | 0.005922933 | 0.18342127 | 0.000152788 | 0.001225821 | 1 | 0.993452012 | 1 | 0.998819255 | ENSG0000016 | 6 | ULK4 |
| SAT | Obesity | ENSG0000018 | 0.00026854 | 0.845319514 | 0.001376464 | 0.18345446 | 0.000223427 | 0.871046986 | 0.001376385 | 0.18342126 | 4.51E-05 | 6.08E-05 | 1 | 0.941763438 | 1 | 0.955501559 | ENSG0000016 | 6 | ULK4 |
| SAT | Obesity | ENSG0000018 | -0.00212624 | 0.333789192 | 0.002199917 | 0.183454121 | -0.00209114 | 0.341802296 | 0.002199781 | 0.183420929 | -3.51E-05 | 0.035070714 | 1 | 0.631217486 | 1 | 0.645626559 | ENSG0000016 | 6 | ULK4 |
| SAT | Obesity | ENSG0000018 | -0.00619069 | 0.20988234 | 0.004937183 | 0.18345388 | -0.00447578 | 0.365688894 | 0.004947919 | 0.183420961 | -0.0017149 | 2.48E-07 | 1 | 0.498280722 | 1 | 0.665460489 | ENSG0000016 | 10 | LTBP3 |
| SAT | Obesity | ENSG0000018 | 0.000849684 | 0.435225397 | 0.001088944 | 0.183454244 | 0.000826028 | 0.448091661 | 0.001088884 | 0.183421053 | 2.37E-05 | 0.00909537 | 1 | 0.716072224 | 1 | 0.726019131 | ENSG0000016 | 10 | LTBP3 |
| SAT | Obesity | ENSG0000018 | 0.009998839 | 0.078234675 | 0.005677849 | 0.183453302 | 0.009894691 | 0.081370615 | 0.005677495 | 0.183420122 | 0.000104147 | 0.014194038 | 1 | 0.282822099 | 1 | 0.294351696 | ENSG0000016 | 10 | LTBP3 |
| SAT | Obesity | ENSG0000018 | 0.006102996 | 0.186373901 | 0.004618638 | 0.183453814 | 0.005961406 | 0.196779819 | 0.004618444 | 0.183420641 | 0.00014159 | 0.001840031 | 1 | 0.461852544 | 1 | 0.481816855 | ENSG0000016 | 10 | LTBP3 |
| SAT | Obesity | ENSG0000018 | 0.006567255 | 3.63E-05 | 0.001590154 | 0.183448027 | 0.006590148 | 3.40E-05 | 0.001590042 | 0.183414778 | -2.29E-05 | 0.021744578 | 0.119069587 | 0.000875512 | 0.111687929 | 0.000954598 | ENSG0000016 | 10 | LTBP3 |
| SAT | Obesity | ENSG0000018 | -0.02701938 | 2.23E-14 | 0.00353793 | 0.18343243 | -0.02712934 | 1.74E-14 | 0.003537764 | 0.183399048 | 0.000109958 | 0.000918754 | 7.30E-11 | 3.32E-12 | 5.72E-11 | 2.60E-12 | ENSG0000016 | 10 | LTBP3 |
| SAT | Obesity | ENSG0000018 | 0.001465773 | 0.383730748 | 0.001682778 | 0.183454187 | 0.001512041 | 0.368869053 | 0.001682673 | 0.183420965 | -4.63E-05 | 0.000269925 | 1 | 0.67678409 | 1 | 0.667026933 | ENSG0000016 | 10 | NAALADL1 |
| SAT | Obesity | ENSG0000018 | 0.004403796 | 0.396086921 | 0.005189293 | 0.183454202 | 0.004276162 | 0.409893091 | 0.005188997 | 0.183421014 | 0.000127634 | 0.002690022 | 1 | 0.6883269 | 1 | 0.700812523 | ENSG0000016 | 10 | NAALADL1 |
| SAT | Obesity | ENSG0000018 | -0.00575938 | 0.282258554 | 0.005356266 | 0.183454037 | -0.00557729 | 0.297722936 | 0.005355932 | 0.18342086 | -0.00018209 | 5.62E-06 | 1 | 0.579661774 | 1 | 0.601026164 | ENSG0000016 | 10 | NAALADL1 |
| SAT | Obesity | ENSG0000018 | 0.01068395 | 0.015755874 | 0.004424856 | 0.1834527 | 0.010760146 | 0.0150207 | 0.004424606 | 0.183419035 | -7.62E-05 | 0.036674568 | 1 | 0.105715792 | 1 | 0.108076574 | ENSG0000016 | 9 | KIAA1586 |
| SAT | Obesity | ENSG0000018 | 0.00020241 | 0.966626224 | 0.004837701 | 0.183454473 | 0.000108095 | 0.982172042 | 0.004837371 | 0.18342127 | 9.43E-05 | 0.00358309 | 1 | 0.991713771 | 1 | 0.995364986 | ENSG0000016 | 9 | KIAA1586 |
| SAT | Obesity | ENSG0000018 | 0.00722149 | 0.208968383 | 0.005747723 | 0.183453877 | 0.007313359 | 0.203204474 | 0.005747343 | 0.183420658 | -9.19E-05 | 0.022235712 | 1 | 0.49647014 | 1 | 0.491652405 | ENSG0000016 | 9 | KIAA1586 |
| SAT | Obesity | ENSG0000018 | 0.003368676 | 0.343813805 | 0.003558495 | 0.183454135 | 0.00104653 | 0.772514221 | 0.003620111 | 0.183421239 | 0.002322146 | 0.000496469 | 1 | 0.642232408 | 1 | 0.911785759 | ENSG0000016 | 9 | RNF187 |
| SAT | Obesity | ENSG0000018 | 0.004293473 | 0.079861831 | 0.002451323 | 0.183453315 | 0.004332721 | 0.077125421 | 0.002451164 | 0.183420089 | -3.92E-05 | 0.024806888 | 1 | 0.285743367 | 1 | 0.285607794 | ENSG0000016 | 9 | RNF187 |
| SAT | Obesity | ENSG0000018 | -0.00588537 | 0.32005083 | 0.005918788 | 0.1834541 | -0.00607169 | 0.304943676 | 0.005918456 | 0.183420872 | 0.000186314 | 0.000145148 | 1 | 0.618425661 | 1 | 0.605981659 | ENSG0000016 | 5 | UBTD2 |
| SAT | Obesity | ENSG0000018 | 0.006801274 | 0.269646511 | 0.006161254 | 0.183454013 | 0.006892332 | 0.263255473 | 0.006160848 | 0.183420797 | -9.11E-05 | 0.035778292 | 1 | 0.5660334 | 1 | 0.566387677 | ENSG0000016 | 14 | POLR2J3 |
| SAT | Obesity | ENSG0000018 | 0.005108586 | 0.244552352 | 0.004390008 | 0.183453962 | 0.005641413 | 0.199098386 | 0.004393191 | 0.183420647 | -0.00053283 | 0.002663827 | 1 | 0.533140377 | 1 | 0.485479182 | ENSG0000016 | 14 | POLR2J3 |
| SAT | Obesity | ENSG0000018 | -0.0100868 | 0.082880577 | 0.005816348 | 0.183453337 | -0.01108799 | 0.057206723 | 0.005830469 | 0.183419903 | 0.001001195 | 0.015360997 | 1 | 0.290088201 | 1 | 0.24057313 | ENSG0000016 | 8 | PDHB |
| SAT | Obesity | ENSG0000018 | -0.00896154 | 0.009524419 | 0.003456527 | 0.183451933 | -0.00904953 | 0.008838122 | 0.003456311 | 0.183418679 | 8.80E-05 | 0.000687242 | 1 | 0.075158099 | 1 | 0.077122017 | ENSG0000016 | 8 | PDHB |
| SAT | Obesity | ENSG0000018 | 0.004454268 | 0.346314681 | 0.004729714 | 0.183454139 | 0.004328314 | 0.3600982 | 0.004729473 | 0.183420954 | 0.000125954 | 0.002707356 | 1 | 0.644591556 | 1 | 0.660784225 | ENSG0000016 | 11 | PXK |
| SAT | Obesity | ENSG0000018 | 0.000168395 | 0.973355794 | 0.005041794 | 0.183454474 | 0.001327714 | 0.793205326 | 0.005064698 | 0.183421244 | -0.00115932 | 0.017032286 | 1 | 0.993452012 | 1 | 0.919712977 | ENSG0000016 | 9 | CX3CR1 |
| SAT | Obesity | ENSG0000018 | 0.010203665 | 0.012293542 | 0.004075575 | 0.183452105 | 0.010148191 | 0.012768507 | 0.004075299 | 0.183418927 | 5.55E-05 | 0.044085853 | 1 | 0.08923697 | 1 | 0.096541971 | ENSG0000016 | 9 | CX3CR1 |
| SAT | Obesity | ENSG0000018 | -0.00120676 | 0.669522193 | 0.002827442 | 0.183454405 | -0.00125448 | 0.657251451 | 0.002827251 | 0.183421196 | 4.77E-05 | 0.01242219 | 1 | 0.855746909 | 1 | 0.852011857 | ENSG0000016 | 9 | CX3CR1 |
| SAT | Obesity | ENSG0000018 | 0.00147324 | 0.576858894 | 0.002640314 | 0.183454356 | 0.001428057 | 0.588575484 | 0.00264014 | 0.18342116 | 4.52E-05 | 0.015141329 | 1 | 0.804708347 | 1 | 0.813760607 | ENSG0000016 | 7 | SCN11A |
| SAT | Obesity | ENSG0000018 | -8.91E-05 | 0.958789734 | 0.001724891 | 0.183454473 | -0.0012603 | 0.941751407 | 0.001724786 | 0.183421268 | 3.69E-05 | 0.005658839 | 1 | 0.987453658 | 1 | 0.982135449 | ENSG0000016 | 7 | SCN11A |
| SAT | Obesity | ENSG0000018 | -0.00211302 | 0.700020909 | 0.005484206 | 0.183454418 | -0.00195366 | 0.721650893 | 0.005483909 | 0.183421222 | -0.00015936 | 0.000647478 | 1 | 0.871308271 | 1 | 0.885797449 | ENSG0000016 | 7 | SCN11A |
| SAT | Obesity | ENSG0000018 | 0.011497658 | 0.01053333 | 0.004495099 | 0.183452001 | 0.011588893 | 0.009929558 | 0.00449481 | 0.183418758 | -9.12E-05 | 0.005280183 | 1 | 0.07998762 | 1 | 0.081872378 | ENSG0000016 | 7 | SCN11A |
| SAT | Obesity | ENSG0000018 | 0.006550838 | 0.041170946 | 0.003208349 | 0.183452898 | 0.00714347 | 0.026477216 | 0.003219017 | 0.183419409 | -0.00059263 | 0.025557467 | 1 | 0.192979684 | 1 | 0.151209505 | ENSG0000016 | 13 | SEPT2 |
| SAT | Obesity | ENSG0000018 | -0.01115556 | 0.010183381 | 0.004341447 | 0.183451978 | -0.00692536 | 0.132143603 | 0.004599413 | 0.183420413 | -0.0042302 | 0.005378197 | 1 | 0.078431158 | 1 | 0.388845886 | ENSG0000016 | 13 | SEPT2 |
| SAT | Obesity | ENSG0000018 | 0.007625298 | 0.025821797 | 0.003421122 | 0.183452596 | 0.007704946 | 0.024303782 | 0.003420924 | 0.183419353 | -7.96E-05 | 0.003979322 | 1 | 0.145319582 | 1 | 0.144457803 | ENSG0000016 | 9 | TAP1 |
| SAT | Obesity | ENSG0000018 | -0.00248497 | 0.460479932 | 0.003366911 | 0.183454268 | -0.00243382 | 0.469736589 | 0.003366704 | 0.183421073 | -5.12E-05 | 0.046266934 | 1 | 0.731904575 | 1 | 0.738833053 | ENSG0000016 | 9 | TAP1 |
| SAT | Obesity | ENSG0000018 | 0.00896584 | 0.081568778 | 0.005147874 | 0.183453327 | 0.008881725 | 0.084451228 | 0.005147553 | 0.183420145 | 8.41E-05 | 0.029251168 | 1 | 0.287792178 | 1 | 0.298677775 | ENSG0000016 | 8 | MLKL |
| SAT | Obesity | ENSG0000018 | -0.00364152 | 0.299789394 | 0.00351198 | 0.183454068 | -0.00357587 | 0.308559013 | 0.003511779 | 0.183420878 | -6.56E-05 | 0.021858725 | 1 | 0.599396101 | 1 | 0.609083345 | ENSG0000016 | 8 | MLKL |
| SAT | Obesity | ENSG0000018 | 0.000909071 | 0.641288339 | 0.00195122 | 0.183454392 | 0.000871451 | 0.655131432 | 0.001951103 | 0.183421195 | 3.76E-05 | 0.013710359 | 1 | 0.843249571 | 1 | 0.850607926 | ENSG0000016 | 10 | CMAHP |
| SAT | Obesity | ENSG0000018 | -0.00668531 | 1.66E-11 | 0.000992885 | 0.183437339 | -0.00560012 | 5.80E-08 | 0.001032274 | 0.183410148 | -0.00108519 | 0.000123658 | 5.45E-08 | 1.65E-09 | 0.000190203 | 3.66E-06 | ENSG0000016 | 10 | CMAHP |
| SAT | Obesity | ENSG0000018 | -0.00280057 | 0.42772562 | 0.003531216 | 0.183454236 | -0.00274803 | 0.436413578 | 0.003530979 | 0.183421041 | -5.25E-05 | 0.028683946 | 1 | 0.709992059 | 1 | 0.717711418 | ENSG0000016 | 10 | CMAHP |
| SAT | Obesity | ENSG0000018 | -0.00458252 | 0.294617467 | 0.004372436 | 0.183454059 | -0.00467964 | 0.284473374 | 0.004372169 | 0.183420837 | 9.71E-05 | 0.003763416 | 1 | 0.593750704 | 1 | 0.588868858 | ENSG0000016 | 10 | CMAHP |
| SAT | Obesity | ENSG0000018 | -0.00046946 | 0.929182949 | 0.005282319 | 0.183454471 | -0.00099474 | 0.85064725 | 0.005282921 | 0.183421257 | 0.000525281 | 8.71E-07 | 1 | 0.977131172 | 1 | 0.943853103 | ENSG0000016 | 10 | CMAHP |
| SAT | Obesity | ENSG0000018 | -0.00233144 | 0.689011428 | 0.005825735 | 0.183454414 | -0.00215429 | 0.711525576 | 0.005825415 | 0.183421219 | -0.00017716 | 0.000317085 | 1 | 0.864822684 | 1 | 0.882614523 | ENSG0000016 | 10 | CMAHP |
| SAT | Obesity | ENSG0000018 | -0.00889639 | 6.22E-06 | 0.001968676 | 0.183446755 | -0.00893291 | 5.69E-06 | 0.001968567 | 0.183413488 | 3.65E-05 | 0.027358486 | 0.020393468 | 0.00020165 | 0.018657521 | 0.000222113 | ENSG0000016 | 8 | FNTA |
| SAT | Obesity | ENSG0000018 | 0.005375562 | 0.007261374 | 0.002002351 | 0.18345175 | 0.002969232 | 0.19963753 | 0.002315035 | 0.183420648 | 0.002406329 | 0.038402083 | 1 | 0.063194226 | 1 | 0.486273745 | ENSG0000016 | 8 | FNTA |
| SAT | Obesity | ENSG0000018 | -0.00828604 | 0.175135377 | 0.006111156 | 0.183453779 | -0.00819104 | 0.180104942 | 0.006110737 | 0.183420591 | -9.50E-05 | 0.018571663 | 1 | 0.446826727 | 1 | 0.457509161 | ENSG0000016 | 7 | TMEM223 |
| SAT | Obesity | ENSG0000018 | 0.001720602 | 0.700662796 | 0.004475768 | 0.183454418 | 0.001960613 | 0.661365296 | 0.004475987 | 0.183421198 | -0.00024001 | 0.00132145 | 1 | 0.871446033 | 1 | 0.854643378 | ENSG0000016 | 4 | ZSWIM1 |
| SAT | Obesity | ENSG0000018 | 0.001850096 | 0.671607271 | 0.004364017 | 0.183454406 | 0.001726974 | 0.692290074 | 0.004363807 | 0.183421211 | 0.000123122 | 0.002216069 | 1 | 0.855855862 | 1 | 0.868937924 | ENSG0000016 | 6 | NDUFS5 |
| SAT | Obesity | ENSG0000018 | 9.40E-05 | 0.968531392 | 0.002382471 | 0.183454474 | 0.000136475 | 0.954316193 | 0.002382325 | 0.183421269 | -4.25E-05 | 0.018852939 | 1 | 0.992734289 | 1 | 0.988489312 | ENSG0000016 | 6 | NDUFS5 |
| SAT | Obesity | ENSG0000018 | -0.00841149 | 0.013903545 | 0.003419644 | 0.183452187 | -0.00834945 | 0.014616508 | 0.003419449 | 0.183419017 | -6.20E-05 | 0.02620649 | 1 | 0.097265525 | 1 | 0.107046342 | ENSG0000016 | 6 | NDUFS5 |
| SAT | Obesity | ENSG0000018 | 0.011410306 | 0.005756579 | 0.004132154 | 0.183451592 | 0.010660933 | 0.009917109 | 0.004134202 | 0.183418757 | 0.000749373 | 1.15E-07 | 1 | 0.054118445 | 1 | 0.081872378 | ENSG0000016 | 6 | NDUFS5 |
| SAT | Obesity | ENSG0000018 | -0.00152028 | 0.674569319 | 0.00362069 | 0.183454407 | -0.00059868 | 0.868964409 | 0.00362896 | 0.18342126 | -0.00092159 | 0.000222847 | 1 | 0.856857118 | 1 | 0.954321831 | ENSG0000016 | 10 | ZNF30 |
| SAT | Obesity | ENSG0000018 | 0.004389296 | 0.002644468 | 0.001460024 | 0.183451058 | 0.003336434 | 0.023552678 | 0.001473458 | 0.183419332 | 0.001052862 | 1.30E-07 | 1 | 0.030231706 | 1 | 0.142476911 | ENSG0000016 | 10 | ZNF30 |
| SAT | Obesity | ENSG0000018 | -7.46E-05 | 0.942709838 | 0.001038341 | 0.183454472 | -9.68E-05 | 0.925719092 | 0.001038275 | 0.183421267 | 2.22E-05 | 0.003182193 | 1 | 0.981335116 | 1 | 0.976304835 | ENSG0000016 | 10 | ZNF30 |
| SAT | Obesity | ENSG0000018 | -0.00024723 | 0.894821475 | 0.001870069 | 0.183454467 | -0.0002862 | 0.878355891 | 0.001869948 | 0.183421261 | 3.90E-05 | 0.003837363 | 1 | 0.962747126 | 1 | 0.958406391 | ENSG0000016 | 10 | ZNF30 |
| SAT | Obesity | ENSG0000018 | -0.00192645 | 0.487835624 | 0.002776846 | 0.183454292 | -0.00197241 | 0.4774843 | 0.002776649 | 0.183421079 | 4.60E-05 | 0.007931827 | 1 | 0.748926431 | 1 | 0.742751548 | ENSG0000016 | 10 | ZNF30 |
| SAT | Obesity | ENSG0000018 | -0.00455126 | 0.11362776 | 0.003018647 | 0.183453615 | -0.00459349 | 0.128056957 | 0.003018434 | 0.183420395 | 4.22E-05 | 0.027655709 | 1 | 0.383203799 | 1 | 0.381612058 | ENSG0000016 | 3 | FAM84B |
| SAT | Obesity | ENSG0000018 | 0.009022316 | 0.052457061 | 0.004652193 | 0.183453052 | 0.008906109 | 0.055555471 | 0.004651897 | 0.183419885 | 0.000116207 | 0.000661999 | 1 | 0.224103668 | 1 | 0.236724026 | ENSG0000016 | 13 | SLC16A4 |
| SAT | Obesity | ENSG0000018 | -0.00376663 | 0.241723276 | 0.003217439 | 0.183453956 | -0.00368772 | 0.251696643 | 0.003217234 | 0.183420774 | -7.89E-05 | 0.000748509 | 1 | 0.530497704 | 1 | 0.548923519 | ENSG0000016 | 13 | SLC16A4 |
| SAT | Obesity | ENSG0000018 | 0.005641784 | 0.321250424 | 0.005687898 | 0.183454102 | 0.005712516 | 0.315186859 | 0.005687497 | 0.183420889 | -7.07E-05 | 0.049534288 | 1 | 0.619468819 | 1 | 0.615584597 | ENSG0000016 | 13 | SLC16A4 |
| SAT | Obesity | ENSG0000018 | 0.000863995 | 0.853402152 | 0.004675817 | 0.183454461 | 0.000962956 | 0.836825557 | 0.004675549 | 0.183421254 | -9.90E-05 | 0.009330774 | 1 | 0.943648754 | 1 | 0.937115108 | ENSG0000016 | 13 | SLC16A4 |
| SAT | Obesity | ENSG0000018 | -0.01123999 | 0.046000682 | 0.005632971 | 0.183452969 | -0.01141162 | 0.042767435 | 0.005632645 | 0.183419719 | 0.000171636 | 0.00016425 | 1 | 0.206468178 | 1 | 0.201899213 | ENSG0000016 | 13 | SLC16A4 |
| SAT | Obesity | ENSG0000018 | -0.00499959 | 0.097701957 | 0.003018889 | 0.183453437 | -0.00494887 | 0.101127502 | 0.003018683 | 0.183420254 | -5.07E-05 | 0.011964934 | 1 | 0.318015993 | 1 | 0.331799333 | ENSG0000016 | 12 | LRP1B |
| SAT | Obesity | ENSG0000018 | -0.0067881 | 0.116067575 | 0.004319511 | 0.183453541 | -0.00668692 | 0.121580326 | 0.004319221 | 0.183420364 | -0.00010119 | 0.000583172 | 1 | 0.356237338 | 1 | 0.369554001 | ENSG0000016 | 12 | LRP1B |
| SAT | Obesity | ENSG0000018 | -0.00254048 | 0.42026734 | 0.003152118 | 0.183454229 | -0.00248578 | 0.430312192 | 0.003151909 | 0.183421035 | -5.47E-05 | 0.01278529 | 1 | 0.703340943 | 1 | 0.714674593 | ENSG0000016 | 12 | LRP1B |
| SAT | Obesity | ENSG0000018 | 0.00143817 | 0.612653325 | 0.002840606 | 0.183454377 | 0.001374283 | 0.628506935 | 0.00284042 | 0.18342118 | 6.39E-05 | 0.002435118 | 1 | 0.826188064 | 1 | 0.83385817 | ENSG0000016 | 12 | LRP1B |
| SAT | Obesity | ENSG0000018 | -0.00136156 | 0.576261929 | 0.002436345 | 0.183454356 | -0.00132726 | 0.585882449 | 0.002436176 | 0.183421182 | -3.43E-05 | 0.030825304 | 1 | 0.804708347 | 1 | 0.812070604 | ENSG0000016 | 12 | LRP1B |
| SAT | Obesity | ENSG0000019 | -0.00401527 | 0.445503836 | 0.005262938 | 0.183454254 | -0.00411352 | 0.434418074 | 0.005262586 | 0.183421039 | 9.83E-05 | 0.006699369 | 1 | 0.722137485 | 1 | 0.717325467 | ENSG0000016 | 8 | NPNT |
| SAT | Obesity | ENSG0000019 | -0.00455688 | 0.284593311 | 0.004258532 | 0.183454041 | -0.00463718 | 0.276158904 | 0.004258243 | 0.183420822 | 8.03E-05 | 0.004999735 | 1 | 0.582865575 | 1 | 0.580018771 | ENSG0000016 | 11 | GSTM4 |
| SAT | Obesity | ENSG0000019 | -0.00159981 | 0.766715171 | 0.005392476 | 0.183454441 | -0.00178271 | 0.740939928 | 0.005392187 | 0.183421229 | 0.000182896 | 8.01E-05 | 1 | 0.906519811 | 1 | 0.897054301 | ENSG0000016 | 11 | GSTM4 |
| SAT | Obesity | ENSG0000019 | 0.004978364 | 0.071802267 | 0.002765184 | 0.183453249 | 0.00503683 | 0.068512525 | 0.00276502 | 0.183420016 | -5.85E-05 | 0.007661246 | 1 | 0.268623988 | 1 | 0.265839301 | ENSG0000016 | 7 | TCTN2 |
| SAT | Obesity | ENSG0000019 | 0.00777086 | 0.060293721 | 0.00413642 | 0.18345314 | 0.007666727 | 0.0637994 | 0.004136165 | 0.183419972 | 0.000104133 | 0.000920503 | 1 | 0.243144411 | 1 | 0.257054802 | ENSG0000016 | 7 | TCTN2 |
| SAT | Obesity | ENSG0000019 | 0.003348394 | 0.36368424 | 0.003686165 | 0.183454162 | 0.003802931 | 0.302352838 | 0.003687147 | 0.183420868 | -0.00045454 | 3.98E-06 | 1 | 0.660713174 | 1 | 0.602319163 | ENSG0000016 | 7 | TCTN2 |
| SAT | Obesity | ENSG0000019 | -0.00970751 | 1.40E-07 | 0.001843861 | 0.183443997 | -0.00742193 | 0.000112433 | 0.001921736 | 0.183415633 | -0.00228558 | 2.48E-05 | 0.000460727 | 6.88E-06 | 0.368891926 | 0.002475785 | ENSG0000016 | 7 | TCTN2 |
| SAT | Obesity | ENSG0000019 | 0.025093694 | 3.96E-29 | 0.002239842 | 0.183407042 | 0.025056674 | 4.73E-29 | 0.002239707 | 0.183373979 | 3.70E-05 | 0.032057273 | 1.30E-25 | 3.25E-26 | 1.55E-25 | 3.88E-26 | ENSG0000016 | 7 | TCTN2 |
| SAT | Obesity | ENSG0000019 | -0.00344625 | 0.475551692 | 0.004830228 | 0.183454282 | -0.00330512 | 0.493784126 | 0.00482991 | 0.183421093 | -0.00014113 | 2.93E-05 | 1 | 0.738771355 | 1 | 0.755129058 | ENSG0000016 | 7 | TCTN2 |
| SAT | Obesity | ENSG0000019 | 0.001342356 | 0.774508069 | 0.004685633 | 0.183454443 | 0.001234967 | 0.792103327 | 0.004685352 | 0.183421244 | 0.000107389 | 0.003267634 | 1 | 0.909817544 | 1 | 0.919712977 | ENSG0000016 | 7 | TCTN2 |
| SAT | Obesity | ENSG0000019 | -0.01749439 | 9.20E-05 | 0.004473433 | 0.183448693 | -0.01331688 | 0.003458675 | 0.004554738 | 0.18341804 | -0.00417752 | 1.15E-06 | 0.301950166 | 0.00184116 | 1 | 0.040673517 | ENSG0000016 | 7 | TCTN2 |
| SAT | Obesity | ENSG0000019 | -0.0025879 | 0.557668002 | 0.004414024 | 0.183454344 | -0.00248758 | 0.573038099 | 0.004413868 | 0.18342115 | -0.0001004 | 0.030260791 | 1 | 0.794232732 | 1 | 0.802779912 | ENSG0000016 | 7 | TCTN2 |
| SAT | Obesity | ENSG0000019 | -0.00461965 | 0.000244758 | 0.001259546 | 0.183449389 | -0.00459975 | 0.000260086 | 0.001259465 | 0.183416229 | -1.99E-05 | 0.03042095 | 0.803050333 | 0.004279083 | 0.853342504 | 0.004934546 | ENSG0000016 | 7 | TCTN2 |
| SAT | Obesity | ENSG0000019 | -0.00499939 | 6.61E-05 | 0.001253025 | 0.183448457 | -0.00495164 | 7.76E-05 | 0.00125301 | 0.183415368 | -4.78E-05 | 0.002334033 | 0.216946595 | 0.00142728 | 0.254532644 | 0.00181809 | ENSG0000016 | 17 | PPIP5K1 |
| SAT | Obesity | ENSG0000019 | -0.0079265 | 0.011852073 | 0.003149775 | 0.18345208 | -0.00788317 | 0.012316671 | 0.003149554 | 0.183418903 | -4.33E-05 | 0.030012306 | 1 | 0.086607243 | 1 | 0.094213678 | ENSG0000016 | 17 | PPIP5K1 |
| SAT | Obesity | ENSG0000019 | 0.010813001 | 0.004145572 | 0.005422594 | 0.183452971 | 0.009107576 | 0.093565028 | 0.005431237 | 0.183420207 | 0.001705425 | 6.01E-08 | 1 | 0.20655337 | 1 | 0.316481295 | ENSG0000016 | 10 | ADAL |
| SAT | Obesity | ENSG0000019 | 0.006975662 | 0.004177575 | 0.002435257 | 0.183451373 | 0.004522051 | 0.068541578 | 0.002482686 | 0.183420016 | 0.00245361 | 3.99E-07 | 1 | 0.043375388 | 1 | 0.265839301 | ENSG0000016 | 10 | ADAL |
| SAT | Obesity | ENSG0000019 | 0.003571896 | 0.340157401 | 0.003744694 | 0.18345413 | 0.004632561 | 0.21667539 | 0.003749791 | 0.183420693 | -0.00106066 | 1.49E-07 | 1 | 0.638111167 | 1 | 0.508520712 | ENSG0000016 | 12 | ZNF507 |
| SAT | Obesity | ENSG0000019 | -0.00100119 | 0.581914925 | 0.001818401 | 0.18345436 | -0.00096095 | 0.597157189 | 0.001818286 | 0.183421165 | -4.02E-05 | 0.002825685 | 1 | 0.806390405 | 1 | 0.817137678 | ENSG0000016 | 12 | ZNF507 |
| SAT | Obesity | ENSG0000019 | -0.00459892 | 0.117480898 | 0.002937785 | 0.183453548 | -0.0045467 | 0.121682611 | 0.002937613 | 0.183420365 | -5.22E-05 | 0.025885684 | 1 | 0.358562629 | 1 | 0.369554001 | ENSG0000016 | 12 | ZNF507 |
| SAT | Obesity | ENSG0000019 | -0.00564336 | 0.014295474 | 0.002303635 | 0.183452206 | -0.00570411 | 0.013275962 | 0.002303491 | 0.183418953 | 6.08E-05 | 0.000400141 | 1 | 0.099161625 | 1 | 0.09922194 | ENSG0000016 | 10 | GFM1 |
| SAT | Obesity | ENSG0000019 | 0.003083179 | 0.42037537 | 0.003826372 | 0.183454229 | 0.003142829 | 0.411410689 | 0.003826119 | 0.183421015 | -5.97E-05 | 0.025695756 | 1 | 0.703340943 | 1 | 0.701318401 | ENSG0000016 | 10 | GFM1 |
| SAT | Obesity | ENSG0000019 | -0.01098717 | 0.004712029 | 0.003887749 | 0.183451455 | -0.01103679 | 0.004524819 | 0.003887477 | 0.183418224 | 4.96E-05 | 0.047792054 | 1 | 0.047569744 | 1 | 0.049651945 | ENSG0000016 | 10 | GFM1 |
| SAT | Obesity | ENSG0000019 | -0.01271874 | 6.23E-05 | 0.003176619 | 0.183448415 | -0.0126644 | 6.69E-05 | 0.003176418 | 0.183415263 | -5.43E-05 | 0.020687286 | 0.204492663 | 0.00134116 | 0.219571995 | 0.001602715 | ENSG0000016 | 10 | GFM1 |
| SAT | Obesity | ENSG0000019 | -0.00096085 | 0.764550408 | 0.003208051 | 0.18345444 | -0.00105067 | 0.743266117 | 0.00320785 | 0.18342123 | 8.98E-05 | 0.000187156 | 1 | 0.904612293 | 1 | 0.898125261 | ENSG0000016 | 8 | ATOH8 |
| SAT | Obesity | ENSG0000019 | 0.00103775 | 0.780553966 | 0.00372491 | 0.183454445 | 0.00097439 | 0.793627301 | 0.003724682 | 0.183421244 | 6.34E-05 | 0.025907612 | 1 | 0.913071213 | 1 | 0.919712977 | ENSG0000016 | 10 | TNIP2 |
| SAT | Obesity | ENSG0000019 | -0.00152283 | 0.796866521 | 0.005916106 | 0.183454449 | -0.00168733 | 0.77546989 | 0.005915742 | 0.183420239 | 0.0001645 | 0.000261585 | 1 | 0.919310497 | 1 | 0.911862059 | ENSG0000016 | 6 | TMEM129 |
| SAT | Obesity | ENSG0000019 | 0.007745693 | 0.149679386 | 0.005376462 | 0.18345369 | 0.007659955 | 0.154210901 | 0.005376099 | 0.183420503 | 8.57E-05 | 0.018400718 | 1 | 0.409839489 | 1 | 0.417993751 | ENSG0000016 | 6 | TMEM129 |
| SAT | Obesity | ENSG0000019 | -0.00158001 | 0.122989817 | 0.001024416 | 0.183453575 | 0.000758679 | 0.526497447 | 0.0011986 | 0.183421119 | -0.00233868 | 0.00016584 | 1 | 0.367513287 | 1 | 0.778826927 | ENSG0000016 | 11 | CEP120 |
| SAT | Obesity | ENSG0000019 | 0.000124149 | 0.978423742 | 0.004590422 | 0.183454474 | -0.00037752 | 0.934496715 | 0.004593313 | 0.183421268 | 0.000501667 | 0.00399056 | 1 | 0.993452012 | 1 | 0.979844999 | ENSG0000016 | 11 | CEP120 |
| SAT | Obesity | ENSG0000019 | 0.003617949 | 0.110704867 | 0.002268256 | 0.183453512 | 0.003337317 | 0.141311642 | 0.002268854 | 0.183420452 | 0.000280632 | 3.31E-06 | 1 | 0.343697234 | 1 | 0.401725489 | ENSG0000016 | 12 | LGALS9 |

| | | | | | | | | | | | | | | | | | | | |
|---|---|---|---|---|---|---|---|---|---|---|---|---|---|---|---|---|---|---|---|
| SAT | Obesity | ENSG0000019 | -0.00562743 | 0.038589098 | 0.002720475 | 0.183452857 | -0.00568549 | 0.036615417 | 0.002720288 | 0.183419619 | 5.81E-05 | 0.001301049 | 1 | 0.188234951 | 1 | 0.186062873 | ENSG0000016 | 12 | LGALS9 |
| SAT | Obesity | ENSG0000019 | 0.001922947 | 0.667775827 | 0.004480276 | 0.183454404 | 0.00199297 | 0.656419492 | 0.004479971 | 0.183421195 | -7.00E-05 | 0.019471032 | 1 | 0.854600118 | 1 | 0.851269705 | ENSG0000016 | 12 | LGALS9 |
| SAT | Obesity | ENSG0000019 | 8.47E-05 | 0.98619511 | 0.004895479 | 0.183454474 | 0.000166115 | 0.972929496 | 0.004895171 | 0.18342127 | -8.14E-05 | 0.025056669 | 1 | 0.994152558 | 1 | 0.992266083 | ENSG0000016 | 12 | LGALS9 |
| SAT | Obesity | ENSG0000020 | 0.006035288 | 0.181189195 | 0.0045137 | 0.183453798 | 0.006124978 | 0.174760779 | 0.004513396 | 0.183420574 | -8.97E-05 | 0.003638936 | 1 | 0.453456712 | 1 | 0.448311272 | ENSG0000016 | 3 | OR7E102P |
| SAT | Obesity | ENSG0000020 | 0.002516112 | 0.415357905 | 0.003089142 | 0.183454223 | 0.002589832 | 0.401801062 | 0.003088982 | 0.183421005 | -7.37E-05 | 0.006601977 | 1 | 0.700759618 | 1 | 0.692729827 | ENSG0000016 | 3 | OR7E102P |
| SAT | Obesity | ENSG0000020 | -0.00916516 | 0.097658673 | 0.005533463 | 0.183453437 | -0.00900329 | 0.103700414 | 0.00553308 | 0.183422027 | -0.00016186 | 6.82E-06 | 1 | 0.318015993 | 1 | 0.336539128 | ENSG0000016 | 10 | CPLX1 |
| SAT | Obesity | ENSG0000020 | 0.003032414 | 0.21775636 | 0.002460341 | 0.1834539 | 0.003072628 | 0.211683369 | 0.002460166 | 0.183420681 | -4.02E-05 | 0.008262393 | 1 | 0.505890452 | 1 | 0.502914061 | ENSG0000016 | 10 | CPLX1 |
| SAT | Obesity | ENSG0000020 | 0.002458604 | 0.665910403 | 0.005694275 | 0.183454404 | 0.002300909 | 0.686138681 | 0.005693897 | 0.183421208 | 0.000157695 | 6.78E-05 | 1 | 0.854600118 | 1 | 0.86636807 | ENSG0000016 | 10 | CPLX1 |
| SAT | Obesity | ENSG0000020 | 7.17E-05 | 0.985256887 | 0.003882101 | 0.183454474 | -0.00250037 | 0.522634459 | 0.003911172 | 0.183421116 | 0.002572111 | 7.81E-08 | 1 | 0.994152558 | 1 | 0.776262408 | ENSG0000016 | 10 | CPLX1 |
| SAT | Obesity | ENSG0000020 | -0.00059838 | 0.730147701 | 0.001734782 | 0.183454429 | -0.00165277 | 0.345087629 | 0.00175052 | 0.183420933 | 0.001054397 | 7.48E-06 | 1 | 0.8889108 | 1 | 0.648774721 | ENSG0000016 | 5 | IRS1 |
| SAT | Obesity | ENSG0000020 | -0.01001784 | 0.003165553 | 0.003394495 | 0.183451182 | -0.0058329 | 0.11941829 | 0.003745706 | 0.183420354 | -0.00418494 | 0.008249264 | 1 | 0.034736391 | 1 | 0.368420209 | ENSG0000016 | 5 | IRS1 |
| SAT | Obesity | ENSG0000020 | 0.01380219 | 0.000714009 | 0.004078463 | 0.183450145 | 0.005015805 | 0.282762654 | 0.004669622 | 0.183420834 | 0.008786384 | 0.000112145 | 1 | 0.010600293 | 1 | 0.587152637 | ENSG0000016 | 5 | IRS1 |
| SAT | Obesity | ENSG0000020 | 0.000765944 | 0.834465484 | 0.003665188 | 0.183454458 | 0.002766467 | 0.46318396 | 0.003771008 | 0.183421067 | -0.00200052 | 0.024340891 | 1 | 0.936988793 | 1 | 0.735223306 | ENSG0000016 | 5 | IRS1 |
| SAT | Obesity | ENSG0000020 | -0.0041781 | 0.041152679 | 0.002046087 | 0.183452898 | 0.001854319 | 0.431195463 | 0.002355743 | 0.183421036 | -0.00603242 | 2.40E-07 | 1 | 0.192979684 | 1 | 0.714882423 | ENSG0000016 | 5 | IRS1 |
| SAT | Obesity | ENSG0000020 | -0.00289156 | 0.311609271 | 0.002857689 | 0.183454087 | -7.44E-05 | 0.979601763 | 0.002908737 | 0.18342127 | -0.00281719 | 2.22E-07 | 1 | 0.608768261 | 1 | 0.994983894 | ENSG0000016 | 10 | UPF3A |
| SAT | Obesity | ENSG0000020 | -0.00535032 | 2.10E-07 | 0.001030888 | 0.183444293 | -0.00426634 | 6.31E-05 | 0.001066332 | 0.18341522 | -0.00108398 | 7.15E-05 | 0.000690205 | 9.86E-06 | 0.207024255 | 0.001544957 | ENSG0000016 | 7 | C8orf46 |
| SAT | Obesity | ENSG0000020 | 0.010126793 | 2.86E-06 | 0.002163634 | 0.183446194 | 0.008249118 | 0.000181971 | 0.002203971 | 0.183415976 | 0.001877675 | 8.05E-06 | 0.009394045 | 0.000104378 | 0.597045615 | 0.00377877 | ENSG0000016 | 7 | C8orf46 |
| SAT | Obesity | ENSG0000020 | -0.00230937 | 0.446993564 | 0.003036893 | 0.183454256 | -0.00065799 | 0.829496979 | 0.003055472 | 0.183421253 | -0.00165138 | 1.10E-06 | 1 | 0.722507754 | 1 | 0.936326075 | ENSG0000016 | 7 | C8orf46 |
| SAT | Obesity | ENSG0000020 | -0.00135479 | 0.749456476 | 0.004242262 | 0.183454436 | 0.001386245 | 0.752263407 | 0.004391625 | 0.183421233 | -0.00274104 | 0.015922188 | 1 | 0.898416769 | 1 | 0.901441682 | ENSG0000016 | 7 | C8orf46 |
| SAT | Obesity | ENSG0000020 | -0.00053519 | 0.756531648 | 0.001726209 | 0.183454438 | -0.00161319 | 0.354314707 | 0.001741629 | 0.183420946 | 0.001077998 | 3.51E-06 | 1 | 0.901627438 | 1 | 0.656608804 | ENSG0000016 | 7 | C8orf46 |
| SAT | Obesity | ENSG0000020 | -0.01144288 | 0.00083082 | 0.003423614 | 0.183450251 | -0.0065237 | 0.071773772 | 0.003623162 | 0.183420045 | -0.00491918 | 3.39E-05 | 1 | 0.011749661 | 1 | 0.27339388 | ENSG0000016 | 7 | C8orf46 |
| SAT | Obesity | ENSG0000020 | 0.008923711 | 0.060153375 | 0.004747483 | 0.183453139 | 0.00290362 | 0.55456837 | 0.004913654 | 0.183421138 | 0.006020092 | 2.08E-06 | 1 | 0.243144411 | 1 | 0.794558438 | ENSG0000016 | 7 | C8orf46 |
| SAT | Obesity | ENSG0000020 | 0.001391575 | 0.269047937 | 0.001259049 | 0.183454012 | 0.000101478 | 0.936815494 | 0.001280112 | 0.183421268 | 0.001290096 | 2.64E-08 | 1 | 0.5655005 | 1 | 0.980364798 | ENSG0000016 | 7 | C8orf46 |
| SAT | Obesity | ENSG0000020 | 0.004798001 | 0.009529341 | 0.001850748 | 0.183451934 | 0.002050477 | 0.291344937 | 0.001943254 | 0.183420849 | 0.002747524 | 3.59E-06 | 1 | 0.075158099 | 1 | 0.597439211 | ENSG0000016 | 7 | PARM1 |
| SAT | Obesity | ENSG0000020 | 0.00635109 | 1.04E-06 | 0.001300143 | 0.183445455 | 0.004670615 | 0.000629435 | 0.001366226 | 0.183416853 | 0.001680475 | 6.32E-05 | 0.003396026 | 4.14E-05 | 1 | 0.010377772 | ENSG0000016 | 7 | PARM1 |
| SAT | Obesity | ENSG0000020 | 0.006442758 | 5.37E-08 | 0.001184642 | 0.183443294 | 0.004687221 | 0.000241626 | 0.001276823 | 0.183416177 | 0.001755537 | 0.00022979 | 0.000176309 | 2.84E-06 | 0.792776436 | 0.004690985 | ENSG0000016 | 10 | AFAP1L2 |
| SAT | Obesity | ENSG0000020 | 0.00043641 | 0.725665429 | 0.001243696 | 0.183454428 | -0.00064199 | 0.618449814 | 0.001289002 | 0.183421176 | 0.001078396 | 0.001474517 | 1 | 0.887405245 | 1 | 0.828495448 | ENSG0000016 | 10 | AFAP1L2 |
| SAT | Obesity | ENSG0000020 | 0.014218964 | 3.40E-05 | 0.003430609 | 0.183447981 | 0.010231213 | 0.003855244 | 0.003540477 | 0.183418114 | 0.003987752 | 5.34E-06 | 0.111641196 | 0.000833143 | 1 | 0.043920336 | ENSG0000016 | 12 | RP11-231C14.4 |
| SAT | Obesity | ENSG0000020 | 0.026707702 | 4.78E-14 | 0.003543013 | 0.183432997 | 0.022714046 | 1.39E-09 | 0.003750173 | 0.183407407 | 0.003993657 | 0.001165501 | 1.57E-10 | 6.81E-12 | 4.56E-06 | 1.17E-07 | ENSG0000016 | 12 | RP11-231C14.4 |
| SAT | Obesity | ENSG0000020 | -0.00646371 | 0.001842252 | 0.002075314 | 0.183450807 | -0.0055859 | 0.007889514 | 0.002102513 | 0.183418603 | -0.00087781 | 0.009446654 | 1 | 0.022304168 | 1 | 0.071403169 | ENSG0000016 | 12 | RP11-231C14.4 |
| SAT | Obesity | ENSG0000020 | 0.000898058 | 0.740123366 | 0.00270752 | 0.183454433 | -2.93E-05 | 0.991389299 | 0.00271904 | 0.18342127 | 0.000927402 | 0.000242421 | 1 | 0.894088647 | 1 | 0.996552785 | ENSG0000016 | 13 | RGS14 |
| SAT | Obesity | ENSG0000020 | -0.00815987 | 0.011289628 | 0.003180057 | 0.183451985 | -0.00510323 | 0.116449416 | 0.003250767 | 0.183420339 | -0.00305664 | 6.06E-06 | 1 | 0.07900256 | 1 | 0.362840014 | ENSG0000016 | 13 | RGS14 |
| SAT | Obesity | ENSG0000020 | 0.000621544 | 0.832381203 | 0.00293668 | 0.183454457 | 0.002689765 | 0.363546525 | 0.002960246 | 0.183420958 | -0.00206822 | 3.45E-08 | 1 | 0.936669124 | 1 | 0.664705409 | ENSG0000016 | 13 | RGS14 |
| SAT | Obesity | ENSG0000020 | 0.00603426 | 2.31E-08 | 0.001080036 | 0.183442675 | 0.004463343 | 0.000109397 | 0.001153683 | 0.183415614 | 0.001570918 | 0.000108348 | 7.58E-05 | 1.35E-06 | 0.358933116 | 0.002425224 | ENSG0000016 | 13 | RGS14 |
| SAT | Obesity | ENSG0000020 | 0.003254238 | 0.097398344 | 0.00196322 | 0.183453436 | 0.001998297 | 0.312751281 | 0.001979557 | 0.183420885 | 0.001255941 | 8.54E-07 | 1 | 0.31765802 | 1 | 0.613351437 | ENSG0000016 | 9 | THBS3 |
| SAT | Obesity | ENSG0000020 | -0.00477589 | 0.081991479 | 0.002745947 | 0.183453331 | -0.00340521 | 0.217197386 | 0.002759454 | 0.183420695 | -0.00137068 | 6.30E-07 | 1 | 0.288332309 | 1 | 0.508767414 | ENSG0000016 | 9 | THBS3 |
| SAT | Obesity | ENSG0000020 | -0.00280546 | 0.600661764 | 0.005359568 | 0.18345437 | -0.00266759 | 0.61865707 | 0.00535925 | 0.183421177 | -0.00013787 | 0.001090785 | 1 | 0.818086861 | 1 | 0.828495448 | ENSG0000016 | 9 | THBS3 |
| SAT | Obesity | ENSG0000020 | -0.00399512 | 0.497586104 | 0.005889961 | 0.1834543 | -0.00420907 | 0.474817443 | 0.005889585 | 0.183421077 | 0.00021395 | 6.67E-07 | 1 | 0.756691284 | 1 | 0.742537751 | ENSG0000016 | 10 | SH3TC2 |
| SAT | Obesity | ENSG0000020 | 0.014353267 | 2.14E-07 | 0.002767244 | 0.183444305 | 0.012733339 | 4.93E-06 | 0.002787582 | 0.183413385 | 0.001619928 | 1.65E-06 | 0.000702043 | 9.89E-06 | 0.016168856 | 0.000199916 | ENSG0000016 | 8 | CXCL11 |
| SAT | Obesity | ENSG0000020 | -0.00381553 | 0.431075978 | 0.004846029 | 0.18345424 | -0.00372067 | 0.442593429 | 0.004845742 | 0.183421047 | -9.49E-05 | 0.013240757 | 1 | 0.712165298 | 1 | 0.722357979 | ENSG0000016 | 10 | STK32A |
| SAT | Obesity | ENSG0000020 | -0.00360989 | 0.37185999 | 0.004042447 | 0.183454173 | -0.00356105 | 0.378329436 | 0.004042155 | 0.183420977 | -4.88E-05 | 0.045861799 | 1 | 0.666706353 | 1 | 0.673155574 | ENSG0000016 | 9 | P2RY12 |
| SAT | Obesity | ENSG0000020 | -0.00079498 | 0.789653108 | 0.002980133 | 0.183454447 | -0.00070527 | 0.812913275 | 0.002979967 | 0.183421249 | -8.97E-05 | 0.00030527 | 1 | 0.914498218 | 1 | 0.928356581 | ENSG0000016 | 9 | P2RY12 |
| SAT | Obesity | ENSG0000020 | -0.00316091 | 0.410759466 | 0.003842783 | 0.183454218 | -0.00323927 | 0.399230894 | 0.003842572 | 0.183421002 | 7.84E-05 | 0.015856972 | 1 | 0.698355005 | 1 | 0.692729827 | ENSG0000016 | 4 | KIAA1024 |
| SAT | Obesity | ENSG0000020 | 0.000253408 | 0.900400101 | 0.002024738 | 0.183454468 | 0.000216045 | 0.915019832 | 0.002024616 | 0.183421266 | 3.74E-05 | 0.016843733 | 1 | 0.963816005 | 1 | 0.970950863 | ENSG0000016 | 2 | RNASE2 |
| SAT | Obesity | ENSG0000020 | -0.00662583 | 0.003139993 | 0.002243225 | 0.183451176 | -0.00659211 | 0.003294321 | 0.002243081 | 0.183418006 | -3.37E-05 | 0.036797944 | 1 | 0.034571535 | 1 | 0.039204918 | ENSG0000016 | 4 | CD52 |
| SAT | Obesity | ENSG0000020 | -0.00265658 | 0.516919351 | 0.004099009 | 0.183454315 | -0.00278761 | 0.496450727 | 0.004098915 | 0.183421095 | 0.000131037 | 0.005960253 | 1 | 0.771001245 | 1 | 0.756387058 | ENSG0000016 | 4 | CD52 |
| SAT | Obesity | ENSG0000020 | -0.00983929 | 0.042761131 | 0.004856414 | 0.183452922 | -0.00997553 | 0.03995703 | 0.004856168 | 0.183419675 | 0.000136238 | 0.001686578 | 1 | 0.195948701 | 1 | 0.195087821 | ENSG0000016 | 4 | CD52 |
| SAT | Obesity | ENSG0000020 | 0.001427796 | 0.271524835 | 0.001298517 | 0.183454017 | 0.00085163 | 0.519876486 | 0.00132336 | 0.183421114 | 0.000576166 | 0.02430067 | 1 | 0.567681645 | 1 | 0.774417527 | ENSG0000016 | 4 | CD52 |
| SAT | Obesity | ENSG0000020 | -0.00329924 | 0.160553345 | 0.002351198 | 0.18345373 | -0.00069651 | 0.771722828 | 0.002400759 | 0.183421238 | -0.00260272 | 8.70E-08 | 1 | 0.424818971 | 1 | 0.911785759 | ENSG0000016 | 4 | CD52 |
| SAT | Obesity | ENSG0000021 | -0.01023562 | 0.000174913 | 0.002727484 | 0.183449151 | -0.01018785 | 0.000187365 | 0.002727306 | 0.183415997 | -4.78E-05 | 0.013387633 | 0.573888544 | 0.00326073 | 0.614743135 | 0.003866309 | ENSG0000016 | 15 | METTL15 |
| SAT | Obesity | ENSG0000021 | 0.001877062 | 0.667491024 | 0.004369385 | 0.183454404 | 0.001763682 | 0.686457944 | 0.004369152 | 0.183421209 | 0.000113381 | 0.002659549 | 1 | 0.854600118 | 1 | 0.86636807 | ENSG0000016 | 15 | METTL15 |
| SAT | Obesity | ENSG0000021 | 0.0098064 | 0.070193843 | 0.00541592 | 0.183453235 | 0.00971026 | 0.072969777 | 0.005415579 | 0.183420055 | 9.61E-05 | 0.01644907 | 1 | 0.26532949 | 1 | 0.275592976 | ENSG0000016 | 15 | METTL15 |
| SAT | Obesity | ENSG0000021 | 0.005236968 | 0.049413063 | 0.002665103 | 0.183453015 | 0.004546446 | 0.088371491 | 0.002668016 | 0.183420173 | 0.000690522 | 1.00E-07 | 1 | 0.21559077 | 1 | 0.306497741 | ENSG0000016 | 10 | INO80E |
| SAT | Obesity | ENSG0000021 | 0.001008728 | 0.592960915 | 0.001887059 | 0.183454366 | 0.000968459 | 0.607784946 | 0.001886958 | 0.18342117 | 4.03E-05 | 0.013034632 | 1 | 0.812973298 | 1 | 0.823005533 | ENSG0000016 | 10 | INO80E |
| SAT | Obesity | ENSG0000021 | 0.012919381 | 0.002777038 | 0.004318823 | 0.183451092 | 0.010280986 | 0.017963181 | 0.004344597 | 0.183419154 | 0.002638395 | 2.97E-08 | 1 | 0.031527553 | 1 | 0.120772946 | ENSG0000016 | 9 | C15orf40 |
| SAT | Obesity | ENSG0000021 | -0.00278573 | 0.413758806 | 0.003408469 | 0.183454222 | -0.00283064 | 0.406239363 | 0.003408227 | 0.18342101 | 4.49E-05 | 0.034415496 | 1 | 0.7004864 | 1 | 0.696744041 | ENSG0000016 | 9 | C15orf40 |
| SAT | Obesity | ENSG0000021 | 0.001726794 | 0.371420056 | 0.001931931 | 0.183454172 | 0.000688745 | 0.729334184 | 0.001990525 | 0.183421225 | 0.001038049 | 0.030609161 | 1 | 0.666706353 | 1 | 0.891559411 | ENSG0000016 | 9 | C15orf40 |
| SAT | Obesity | ENSG0000021 | -0.02804322 | 1.02E-06 | 0.005736602 | 0.18344541 | -0.0149923 | 0.030007583 | 0.006908904 | 0.183419491 | -0.01305092 | 0.000701207 | 0.003335249 | 4.12E-05 | 1 | 0.162878683 | ENSG0000016 | 9 | C15orf40 |
| SAT | Obesity | ENSG0000021 | -0.00480357 | 0.013760014 | 0.001949916 | 0.18345218 | -0.00341044 | 0.089835162 | 0.002010549 | 0.183420183 | -0.00139314 | 0.004525288 | 1 | 0.097089474 | 1 | 0.309870332 | ENSG0000016 | 9 | C15orf40 |
| SAT | Obesity | ENSG0000021 | 0.01032299 | 0.066484018 | 0.005625147 | 0.183453201 | 0.010420851 | 0.063930728 | 0.005624784 | 0.183419973 | -9.79E-05 | 0.016008144 | 1 | 0.259683409 | 1 | 0.257054802 | ENSG0000016 | 9 | C15orf40 |
| SAT | Obesity | ENSG0000021 | -7.28E-05 | 0.978388152 | 0.002688813 | 0.183454474 | -0.00013054 | 0.961276774 | 0.002688639 | 0.183421269 | 5.77E-05 | 0.002865989 | 1 | 0.993452012 | 1 | 0.9914961 | ENSG0000016 | 9 | C15orf40 |
| SAT | Obesity | ENSG0000021 | -0.00268849 | 0.012228296 | 0.001073036 | 0.183452101 | -0.00266426 | 0.013025433 | 0.001072966 | 0.18341894 | -2.42E-05 | 0.001380902 | 1 | 0.089157865 | 1 | 0.097795071 | ENSG0000016 | 3 | FAM103A1 |
| SAT | Obesity | ENSG0000021 | -0.00169509 | 0.084584457 | 0.000982836 | 0.18345335 | -0.00167723 | 0.087894684 | 0.000982781 | 0.183420169 | -1.79E-05 | 0.028224658 | 1 | 0.29243583 | 1 | 0.305166621 | ENSG0000016 | 7 | RGPD8 |
| SAT | Obesity | ENSG0000021 | 0.009673476 | 0.049864908 | 0.004932613 | 0.18345302 | 0.009755405 | 0.047944978 | 0.004932301 | 0.183419792 | -8.19E-05 | 0.024561409 | 1 | 0.216697698 | 1 | 0.218179573 | ENSG0000016 | 3 | LRRC45 |
| SAT | Obesity | ENSG0000021 | -0.00758467 | 0.024494284 | 0.003372025 | 0.183452562 | -0.00763346 | 0.02357969 | 0.003371793 | 0.183419333 | 4.88E-05 | 0.028864797 | 1 | 0.141512278 | 1 | 0.142476911 | ENSG0000016 | 3 | LRRC45 |
| SAT | Obesity | ENSG0000021 | -0.00629154 | 0.110696292 | 0.003938819 | 0.183453456 | -0.00612898 | 0.109783688 | 0.003832603 | 0.183420304 | -0.00016256 | 2.63E-07 | 1 | 0.323906407 | 1 | 0.348019594 | ENSG0000016 | 3 | LRRC45 |
| SAT | Obesity | ENSG0000021 | 0.002145725 | 0.421315315 | 0.002668341 | 0.18345423 | 0.00220889 | 0.407752412 | 0.002668206 | 0.183421011 | -6.32E-05 | 0.007905186 | 1 | 0.704195389 | 1 | 0.698244083 | ENSG0000016 | 12 | HNRNPF |
| SAT | Obesity | ENSG0000021 | 0.008126751 | 0.067073856 | 0.004437979 | 0.183453207 | 0.008539886 | 0.054425264 | 0.0044399 | 0.183419872 | -0.00041313 | 0.0040273 | 1 | 0.26053725 | 1 | 0.234035768 | ENSG0000016 | 14 | CTNND2 |
| SAT | Obesity | ENSG0000021 | 0.018539281 | 0.001399453 | 0.005802987 | 0.183450616 | 0.02253179 | 0.000115136 | 0.005842867 | 0.18341565 | -0.00399251 | 5.94E-09 | 1 | 0.018078898 | 0.377760746 | 0.002501727 | ENSG0000016 | 14 | CTNND2 |
| SAT | Obesity | ENSG0000021 | -0.0020376 | 0.254614858 | 0.0017886 | 0.183453984 | -0.00200327 | 0.262675513 | 0.00178849 | 0.183420796 | -3.43E-05 | 0.011912981 | 1 | 0.546364519 | 1 | 0.565510733 | ENSG0000016 | 13 | MUC3A |
| SAT | Obesity | ENSG0000021 | 0.003271343 | 0.581484861 | 0.005934759 | 0.183454359 | 0.003153694 | 0.595120623 | 0.005934347 | 0.183421163 | 0.000117649 | 0.002236165 | 1 | 0.806390405 | 1 | 0.817137678 | ENSG0000016 | 9 | TPST1 |
| SAT | Obesity | ENSG0000021 | 0.002448342 | 0.53311077 | 0.003928253 | 0.183454327 | 0.002546239 | 0.516843534 | 0.003928052 | 0.183421111 | -9.79E-05 | 0.005035672 | 1 | 0.784366115 | 1 | 0.772996237 | ENSG0000016 | 9 | TPST1 |
| SAT | Obesity | ENSG0000021 | 0.005590088 | 0.000298355 | 0.001545611 | 0.18344953 | 0.004326528 | 0.005765072 | 0.001567092 | 0.183418389 | 0.00126356 | 1.11E-06 | 0.978902294 | 0.005121427 | 1 | 0.058022089 | ENSG0000016 | 5 | OTUD3 |
| SAT | Obesity | ENSG0000021 | 0.001029218 | 0.817309663 | 0.004455358 | 0.183454454 | 0.001124155 | 0.800786854 | 0.004455114 | 0.183421246 | -9.49E-05 | 0.011618438 | 1 | 0.930695292 | 1 | 0.92223015 | ENSG0000016 | 5 | OTUD3 |
| SAT | Obesity | ENSG0000021 | 0.002061424 | 0.165191903 | 0.001485366 | 0.183453746 | 0.002087931 | 0.159796351 | 0.00148527 | 0.183420523 | -2.65E-05 | 0.012577231 | 1 | 0.430496136 | 1 | 0.428342998 | ENSG0000016 | 5 | OTUD3 |
| SAT | Obesity | ENSG0000021 | -0.00649931 | 3.18E-05 | 0.001562245 | 0.183447932 | -0.00433663 | 0.007961925 | 0.001634188 | 0.183418609 | -0.00216269 | 6.60E-06 | 0.10432488 | 0.00079034 | 1 | 0.071403169 | ENSG0000016 | 9 | ZFPM2 |
| SAT | Obesity | ENSG0000021 | 0.002070762 | 0.511050292 | 0.003150868 | 0.183454311 | 0.003172072 | 0.314874421 | 0.003156141 | 0.183420888 | -0.00110131 | 4.07E-09 | 1 | 0.766692277 | 1 | 0.615584597 | ENSG0000016 | 12 | MAP3K2 |
| SAT | Obesity | ENSG0000022 | -0.00065006 | 0.826438758 | 0.002964624 | 0.183454456 | -0.00069433 | 0.814818441 | 0.00296443 | 0.183421249 | 4.43E-05 | 0.033931411 | 1 | 0.9363121 | 1 | 0.929234443 | ENSG0000016 | 7 | PUSL1 |
| SAT | Obesity | ENSG0000022 | -0.00015987 | 0.954252922 | 0.002786799 | 0.183454473 | -0.00011 | 0.968512447 | 0.002786613 | 0.18342127 | -4.99E-05 | 0.009527564 | 1 | 0.985800956 | 1 | 0.992119797 | ENSG0000017 | 11 | UBE2E3 |
| SAT | Obesity | ENSG0000022 | 0.008997311 | 0.034249493 | 0.004249772 | 0.18345278 | 0.008907832 | 0.036065097 | 0.004249517 | 0.18341961 | 8.95E-05 | 0.006953198 | 1 | 0.172615341 | 1 | 0.184615666 | ENSG0000017 | 10 | AC018720.10 |
| SAT | Obesity | ENSG0000022 | -7.60E-05 | 0.974736329 | 0.002399598 | 0.183454474 | -2.85E-05 | 0.990519996 | 0.002399435 | 0.18342127 | -4.75E-05 | 0.003212942 | 1 | 0.993452012 | 1 | 0.996289426 | ENSG0000017 | 14 | HNRNPA3 |
| SAT | Obesity | ENSG0000022 | -0.00708099 | 0.041289771 | 0.003470031 | 0.1834529 | -0.00712321 | 0.040079821 | 0.003469776 | 0.183419677 | 4.22E-05 | 0.037858111 | 1 | 0.192979684 | 1 | 0.195229009 | ENSG0000017 | 14 | HNRNPA3 |
| SAT | Obesity | ENSG0000022 | 0.014512026 | 0.007233012 | 0.005402975 | 0.183451747 | 0.014623306 | 0.006795891 | 0.005402641 | 0.183418501 | -0.00011128 | 0.006790253 | 1 | 0.063115725 | 1 | 0.06519684 | ENSG0000017 | 6 | CHRNB1 |
| SAT | Obesity | ENSG0000022 | 0.003556655 | 0.365403331 | 0.003929494 | 0.183454164 | 0.001485878 | 0.709554352 | 0.003989418 | 0.183421218 | 0.002070777 | 0.002723549 | 1 | 0.663101952 | 1 | 0.881168747 | ENSG0000017 | 6 | CHRNB1 |
| SAT | Obesity | ENSG0000022 | 0.015904982 | 8.10E-09 | 0.00275817 | 0.183441906 | 0.011035607 | 0.000578937 | 0.003206797 | 0.183416794 | 0.004869375 | 0.002919755 | 2.66E-05 | 5.54E-07 | 1 | 0.009691288 | ENSG0000017 | 6 | CHRNB1 |
| SAT | Obesity | ENSG0000022 | -0.00058635 | 0.921116949 | 0.005921162 | 0.18345447 | -0.00047966 | 0.935431789 | 0.005920767 | 0.183421268 | -0.0001067 | 0.00888414 | 1 | 0.973380029 | 1 | 0.979933493 | ENSG0000017 | 6 | CHRNB1 |
| SAT | Obesity | ENSG0000022 | 0.002315376 | 0.059271311 | 0.001227548 | 0.183453129 | 0.002274424 | 0.06389517 | 0.001227485 | 0.183419973 | 4.10E-05 | 0.0015416 | 1 | 0.241576612 | 1 | 0.257054802 | ENSG0000017 | 4 | ANKK1 |
| SAT | Obesity | ENSG0000022 | 0.000384383 | 0.708551225 | 0.001028304 | 0.183454421 | 1.33E-05 | 0.989776543 | 0.00103634 | 0.18342127 | 0.000371104 | 0.004175906 | 1 | 0.87726663 | 1 | 0.996265811 | ENSG0000017 | 4 | ANKK1 |
| SAT | Obesity | ENSG0000022 | 0.000154659 | 0.950203504 | 0.002476481 | 0.183454473 | 9.82E-05 | 0.96835344 | 0.002476336 | 0.18342127 | 5.64E-05 | 0.004227805 | 1 | 0.98440723 | 1 | 0.992119797 | ENSG0000017 | 4 | ANKK1 |
| SAT | Obesity | ENSG0000022 | -0.00022934 | 0.847215991 | 0.001190297 | 0.18345446 | 0.000469882 | 0.701966114 | 0.001227911 | 0.183421215 | -0.00069922 | 0.020604111 | 1 | 0.942344807 | 1 | 0.876674335 | ENSG0000017 | 4 | ANKK1 |
| SAT | Obesity | ENSG0000022 | 0.001296345 | 0.778794176 | 0.004615119 | 0.183454444 | 0.001379002 | 0.765079828 | 0.004614871 | 0.183421236 | -8.27E-05 | 0.037073343 | 1 | 0.913071213 | 1 | 0.906138181 | ENSG0000017 | 9 | PDCD6IP |
| SAT | Obesity | ENSG0000022 | 0.031905543 | 1.41E-09 | 0.00527 | 0.18344062 | 0.027332415 | 3.56E-07 | 0.005368377 | 0.183411474 | 0.004573127 | 8.16E-06 | 4.63E-06 | 1.08E-07 | 0.001166758 | 1.91E-05 | ENSG0000017 | 8 | FAM161A |
| SAT | Obesity | ENSG0000022 | -0.00084892 | 0.851604988 | 0.004537959 | 0.183454461 | -0.00093647 | 0.836496957 | 0.004537695 | 0.183421254 | 8.76E-05 | 0.016229958 | 1 | 0.943639299 | 1 | 0.937115108 | ENSG0000017 | 8 | FAM161A |
| SAT | Obesity | ENSG0000022 | -0.01073466 | 7.61E-06 | 0.00239835 | 0.183446902 | -0.01067946 | 8.47E-06 | 0.002398202 | 0.183413776 | -5.52E-05 | 0.002389275 | 0.024976216 | 0.000233423 | 0.027775013 | 0.00029764 | ENSG0000017 | 10 | CRTAP |
| SAT | Obesity | ENSG0000022 | -0.00321169 | 0.288015005 | 0.003022807 | 0.183454047 | -0.00314639 | 0.297900353 | 0.003022621 | 0.183420861 | -6.53E-05 | 0.004593069 | 1 | 0.587064948 | 1 | 0.601026164 | ENSG0000017 | 10 | CRTAP |
| SAT | Obesity | ENSG0000022 | 0.000646591 | 0.833460965 | 0.003075126 | 0.183454457 | 0.000582217 | 0.849824066 | 0.003074932 | 0.183421257 | 6.44E-05 | 0.004710042 | 1 | 0.936669124 | 1 | 0.943577922 | ENSG0000017 | 8 | CNGB3 |
| SAT | Obesity | ENSG0000022 | -0.01096889 | 0.008244574 | 0.004151899 | 0.183451836 | -0.01104636 | 0.007797741 | 0.004151649 | 0.183418595 | 7.75E-05 | 0.016681289 | 1 | 0.068830653 | 1 | 0.071403169 | ENSG0000017 | 10 | NFRKB |
| SAT | Obesity | ENSG0000022 | -0.00191758 | 0.648459025 | 0.004206111 | 0.183454396 | -0.00182237 | 0.664801971 | 0.004205847 | 0.183421199 | -9.52E-05 | 0.00237637 | 1 | 0.845625621 | 1 | 0.856049947 | ENSG0000017 | 10 | NFRKB |
| SAT | Obesity | ENSG0000022 | -0.01476772 | 0.002629017 | 0.004909321 | 0.183451054 | -0.01469596 | 0.002756575 | 0.00490901 | 0.183417883 | -7.18E-05 | 0.04678054 | 1 | 0.030160152 | 1 | 0.034389064 | ENSG0000017 | 8 | OR2A20P |
| SAT | Obesity | ENSG0000022 | 0.001388772 | 0.543574756 | 0.002286358 | 0.183454335 | 0.001423221 | 0.533594301 | 0.002286192 | 0.183421124 | -3.44E-05 | 0.011678423 | 1 | 0.790819813 | 1 | 0.784725639 | ENSG0000017 | 8 | OR2A20P |
| SAT | Obesity | ENSG0000022 | -0.00124449 | 0.565564974 | 0.002165846 | 0.183454349 | 0.001706192 | 0.448003788 | 0.002248695 | 0.183421053 | -0.00295068 | 1.10E-06 | 1 | 0.797234594 | 1 | 0.726019131 | ENSG0000017 | 8 | OR2A20P |
| SAT | Obesity | ENSG0000022 | -0.00411726 | 0.349837658 | 0.004403937 | 0.183454144 | -0.00399619 | 0.364160371 | 0.004403678 | 0.183420959 | -0.00012107 | 0.000536149 | 1 | 0.647953427 | 1 | 0.664705409 | ENSG0000017 | 9 | MGMT |
| SAT | Obesity | ENSG0000022 | -0.00222893 | 0.542786916 | 0.003662364 | 0.183454334 | -0.00214271 | 0.558484061 | 0.003662159 | 0.183421141 | -8.62E-05 | 0.004687439 | 1 | 0.790819813 | 1 | 0.795455172 | ENSG0000017 | 5 | METTL7B |
| SAT | Obesity | ENSG0000022 | -0.00121277 | 0.633777408 | 0.002545595 | 0.183454388 | -0.00126001 | 0.620594281 | 0.002545426 | 0.183421178 | 4.72E-05 | 0.00754048 | 1 | 0.839492804 | 1 | 0.830073312 | ENSG0000017 | 5 | METTL7B |
| SAT | Obesity | ENSG0000022 | 0.000773861 | 0.610292471 | 0.001518397 | 0.183454376 | 0.001583737 | 0.299346082 | 0.001525997 | 0.183420863 | -0.00080988 | 1.32E-07 | 1 | 0.824707644 | 1 | 0.601026164 | ENSG0000017 | 5 | KRT75 |
| SAT | Obesity | ENSG0000022 | -0.00280092 | 0.554356983 | 0.004737333 | 0.183454342 | -0.00289077 | 0.541697821 | 0.004737049 | 0.183421129 | 8.99E-05 | 0.015340484 | 1 | 0.794232732 | 1 | 0.788255443 | ENSG0000017 | 5 | KRT75 |
| SAT | Obesity | ENSG0000022 | 0.001813521 | 0.065300516 | 0.000983899 | 0.18345319 | 0.001793111 | 0.0683685 | 0.000983834 | 0.183420015 | 2.04E-05 | 0.003250551 | 1 | 0.256588015 | 1 | 0.265839301 | ENSG0000017 | 6 | SPATA24 |
| SAT | Obesity | ENSG0000022 | -0.00559045 | 0.029881713 | 0.002574272 | 0.183452691 | -0.00494416 | 0.056231485 | 0.002589595 | 0.183419893 | -0.00064628 | 0.022574366 | 1 | 0.157820771 | 1 | 0.238744104 | ENSG0000017 | 6 | KRT72 |
| SAT | Obesity | ENSG0000022 | -0.00135459 | 0.480528294 | 0.001920163 | 0.183454286 | -0.00131351 | 0.493906418 | 0.001920035 | 0.183421093 | -4.11E-05 | 0.001979048 | 1 | 0.741872665 | 1 | 0.755129058 | ENSG0000017 | 7 | HSD17B13 |
| SAT | Obesity | ENSG0000022 | 0.000214626 | 0.928949834 | 0.002407037 | 0.183454471 | 0.000117577 | 0.961044675 | 0.002407249 | 0.183421269 | 9.70E-05 | 0.033009519 | 1 | 0.977131172 | 1 | 0.9914961 | ENSG0000017 | 7 | HSD17B13 |
| SAT | Obesity | ENSG0000022 | 0.006181456 | 0.001940812 | 0.001994558 | 0.183450844 | 0.004232268 | 0.037428194 | 0.0020337 | 0.183419633 | 0.001949188 | 9.67E-07 | 1 | 0.023240164 | 1 | 0.188635799 | ENSG0000017 | 10 | ARL6IP1 |
| SAT | Obesity | ENSG0000022 | 0.002593278 | 0.66645316 | 0.00601659 | 0.183454404 | 0.002477764 | 0.680450797 | 0.006016206 | 0.183421206 | 0.000115514 | 0.00842259 | 1 | 0.854600118 | 1 | 0.86636807 | ENSG0000017 | 10 | ARL6IP1 |
| SAT | Obesity | ENSG0000022 | 0.000176028 | 0.936216843 | 0.002199645 | 0.183454472 | 0.000224158 | 0.918825575 | 0.002199503 | 0.183421266 | -4.81E-05 | 0.002316467 | 1 | 0.979192687 | 1 | 0.974044172 | ENSG0000017 | 12 | SMAGP |
| SAT | Obesity | ENSG0000022 | -0.00084635 | 0.59646873 | 0.001598436 | 0.183454368 | 0.00041628 | 0.796568408 | 0.001614797 | 0.183421245 | -0.00126263 | 4.18E-08 | 1 | 0.81491093 | 1 | 0.921558867 | ENSG0000017 | 12 | SMAGP |
| SAT | Obesity | ENSG0000022 | -0.00757919 | 0.038682976 | 0.003665784 | 0.183452858 | -0.00748643 | 0.041117038 | 0.003665589 | 0.183419694 | -9.28E-05 | 0.003323235 | 1 | 0.188234951 | 1 | 0.197229533 | ENSG0000017 | 7 | EMB |
| SAT | Obesity | ENSG0000022 | -0.0005778 | 0.750060057 | 0.001813776 | 0.183454436 | -0.00061731 | 0.733581655 | 0.001813663 | 0.183421226 | 3.95E-05 | 0.003853362 | 1 | 0.898811924 | 1 | 0.893959582 | ENSG0000017 | 7 | EMB |
| SAT | Obesity | ENSG0000022 | -0.01273748 | 0.016795632 | 0.005326906 | 0.183452313 | -0.01280098 | 0.016250241 | 0.005326519 | 0.183419087 | 6.35E-05 | 0.046185214 | 1 | 0.109773844 | 1 | 0.113440509 | ENSG0000017 | 7 | EMB |
| SAT | Obesity | ENSG0000023 | -0.01441269 | 0.005264515 | 0.0051651 | 0.183451531 | -0.01452697 | 0.004912768 | 0.005164758 | 0.18341828 | 0.000114283 | 0.001531206 | 1 | 0.050941439 | 1 | 0.051828914 | ENSG0000017 | 9 | ARMC10 |
| SAT | Obesity | ENSG0000023 | 0.002683012 | 0.561723409 | 0.004623619 | 0.183454347 | 0.0038129 | 0.41105582 | 0.004638351 | 0.183421015 | -0.00112989 | 0.002559772 | 1 | 0.797151602 | 1 | 0.701318401 | ENSG0000017 | 9 | ARMC10 |
| SAT | Obesity | ENSG0000023 | 0.003437945 | 0.185659514 | 0.002597555 | 0.183453812 | 0.003364065 | 0.195261101 | 0.002597386 | 0.183420636 | 7.39E-05 | 6.55E-05 | 1 | 0.460778263 | 1 | 0.479171034 | ENSG0000017 | 9 | ARMC10 |
| SAT | Obesity | ENSG0000023 | 0.005929717 | 0.071174869 | 0.003286319 | 0.183453243 | 0.00599853 | 0.067939508 | 0.003286124 | 0.183420011 | -6.88E-05 | 0.007990662 | 1 | 0.268419248 | 1 | 0.265839301 | ENSG0000017 | 9 | ARMC10 |
| SAT | Obesity | ENSG0000023 | -0.00122067 | 0.661237743 | 0.002785623 | 0.183454402 | -0.00127252 | 0.647779791 | 0.002785431 | 0.183421191 | 5.18E-05 | 0.004570002 | 1 | 0.852929209 | 1 | 0.846421941 | ENSG0000017 | 9 | ARMC10 |
| SAT | Obesity | ENSG0000023 | -0.00319263 | 0.026649206 | 0.001440311 | 0.183452617 | -0.00181986 | 0.216517721 | 0.001472566 | 0.183420693 | -0.00137277 | 7.82E-06 | 1 | 0.147637149 | 1 | 0.508514418 | ENSG0000017 | 9 | ARMC10 |
| SAT | Obesity | ENSG0000023 | 0.004727982 | 0.237470758 | 0.004002241 | 0.183453947 | 0.004802734 | 0.230112908 | 0.004002047 | 0.183420726 | -7.48E-05 | 0.041775968 | 1 | 0.526060746 | 1 | 0.523921586 | ENSG0000017 | 9 | ARMC10 |
| SAT | Obesity | ENSG0000023 | -0.00486647 | 0.015747722 | 0.002015335 | 0.18345227 | -0.00237379 | 0.254593308 | 0.002083618 | 0.183420478 | -0.00249268 | 2.53E-06 | 1 | 0.105715792 | 1 | 0.55282637 | ENSG0000017 | 12 | RNF34 |
| SAT | Obesity | ENSG0000023 | 0.00683004 | 0.001725553 | 0.002179459 | 0.183450762 | 0.008023035 | 0.000260188 | 0.002196861 | 0.18341623 | -0.001193 | 1.72E-05 | 1 | 0.02144523 | 0.853676482 | 0.004934546 | ENSG0000017 | 12 | RNF34 |
| SAT | Obesity | ENSG0000023 | 0.00103678 | 0.804280355 | 0.004183745 | 0.183454451 | 0.000973357 | 0.816018884 | 0.004183448 | 0.18342125 | 6.34E-05 | 0.015098201 | 1 | 0.92331835 | 1 | 0.929961083 | ENSG0000017 | 2 | TMEM133 |

| | | | | | | | | | | | | | | | | | | |
|---|---|---|---|---|---|---|---|---|---|---|---|---|---|---|---|---|---|---|
| SAT | Obesity | ENSG0000023 | 0.012327609 | 0.036949923 | 0.005908774 | 0.183452829 | 0.009476136 | 0.110066302 | 0.005930375 | 0.183420305 | 0.002851473 | 2.53E-08 | 1 | 0.181758169 | 1 | 0.348578702 | ENSG0000017 | 10 | RASA4B |
| SAT | Obesity | ENSG0000023 | 0.01076893 | 9.32E-10 | 0.001759432 | 0.183440314 | 0.008473329 | 5.63E-05 | 0.002103628 | 0.183415139 | 0.002295602 | 0.046537445 | 3.06E-06 | 7.46E-08 | 0.184626332 | 0.001429286 | ENSG0000017 | 10 | RASA4B |
| SAT | Obesity | ENSG0000023 | 0.000539471 | 0.899531647 | 0.004272934 | 0.183454468 | 0.000465714 | 0.913203268 | 0.004272637 | 0.183421266 | 7.38E-05 | 0.007772631 | 1 | 0.963553161 | 1 | 0.969964365 | ENSG0000017 | 10 | RASA4B |
| SAT | Obesity | ENSG0000023 | -0.00297347 | 0.25485478 | 0.002611427 | 0.183453984 | -0.00286769 | 0.272127313 | 0.002611316 | 0.183420814 | -0.00010578 | 3.47E-05 | 1 | 0.546521917 | 1 | 0.575660681 | ENSG0000017 | 7 | KCNS3 |
| SAT | Obesity | ENSG0000023 | 0.000444175 | 0.889547612 | 0.003198337 | 0.183454467 | 0.000505736 | 0.874350904 | 0.003198138 | 0.183421261 | -6.16E-05 | 0.010237554 | 1 | 0.962025309 | 1 | 0.957844846 | ENSG0000017 | 7 | KCNS3 |
| SAT | Obesity | ENSG0000023 | 0.003480651 | 0.401686352 | 0.004150481 | 0.183454208 | 0.003545773 | 0.392904975 | 0.0041502 | 0.183420994 | -6.51E-05 | 0.020587802 | 1 | 0.691650463 | 1 | 0.68753132 | ENSG0000017 | 7 | KCNS3 |
| SAT | Obesity | ENSG0000023 | 0.00376453 | 0.478395573 | 0.005310487 | 0.183454284 | 0.003890871 | 0.463727047 | 0.005310144 | 0.183421067 | -0.00012634 | 0.000953228 | 1 | 0.74073425 | 1 | 0.735373824 | ENSG0000017 | 7 | KCNS3 |
| SAT | Obesity | ENSG0000023 | 0.003307483 | 0.22160137 | 0.002705989 | 0.183453909 | 0.00241407 | 0.374273206 | 0.002717022 | 0.183420972 | 0.000893414 | 0.000302745 | 1 | 0.50915553 | 1 | 0.669643631 | ENSG0000017 | 7 | KCNS3 |
| SAT | Obesity | ENSG0000023 | -0.00087628 | 0.789257243 | 0.003278582 | 0.183454447 | -0.00225401 | 0.496160156 | 0.003312067 | 0.183421095 | 0.001377724 | 0.003502404 | 1 | 0.914498218 | 1 | 0.756387058 | ENSG0000017 | 15 | AKAP13 |
| SAT | Obesity | ENSG0000023 | -0.00875403 | 0.005245661 | 0.003135892 | 0.183451528 | -0.00886482 | 0.004698227 | 0.003135726 | 0.18341825 | 0.000110792 | 4.59E-05 | 1 | 0.050941439 | 1 | 0.050822526 | ENSG0000017 | 15 | AKAP13 |
| SAT | Obesity | ENSG0000023 | -0.00146309 | 0.521985282 | 0.002285047 | 0.183454319 | -0.00149982 | 0.511566057 | 0.002284914 | 0.183421107 | 3.67E-05 | 0.045589511 | 1 | 0.774246705 | 1 | 0.768872301 | ENSG0000017 | 15 | AKAP13 |
| SAT | Obesity | ENSG0000023 | -0.00828476 | 0.019372291 | 0.003543076 | 0.183452407 | -0.00821235 | 0.020448403 | 0.003542824 | 0.183419239 | -7.24E-05 | 0.001008226 | 1 | 0.121067591 | 1 | 0.13047531 | ENSG0000017 | 15 | AKAP13 |
| SAT | Obesity | ENSG0000023 | -0.00117076 | 0.688785409 | 0.002923212 | 0.183454413 | -0.00111371 | 0.703193067 | 0.002923026 | 0.183421215 | -5.70E-05 | 0.007615711 | 1 | 0.864822684 | 1 | 0.876674335 | ENSG0000017 | 2 | GPR27 |
| SAT | Obesity | ENSG0000023 | 0.000515479 | 0.82197508 | 0.002290963 | 0.183454455 | 0.000470087 | 0.837411166 | 0.002290805 | 0.183421254 | 4.54E-05 | 0.002611854 | 1 | 0.933051052 | 1 | 0.937115108 | ENSG0000017 | 2 | GPR27 |
| SAT | Obesity | ENSG0000023 | 0.008033854 | 0.000116029 | 0.002084334 | 0.183448859 | 0.00630703 | 0.002836556 | 0.002112956 | 0.183417903 | 0.001726824 | 6.80E-07 | 0.38069216 | 0.002239366 | 1 | 0.035252799 | ENSG0000017 | 2 | GPR27 |
| SAT | Obesity | ENSG0000023 | 0.00908808 | 0.017676784 | 0.00383087 | 0.183452347 | 0.009027217 | 0.018443979 | 0.003830627 | 0.183419171 | 6.09E-05 | 0.030316254 | 1 | 0.113720642 | 1 | 0.123121596 | ENSG0000017 | 2 | GPR27 |
| SAT | Obesity | ENSG0000023 | -0.00836877 | 3.08E-05 | 0.002008254 | 0.18344791 | -0.0061124 | 0.003086069 | 0.002065664 | 0.183417961 | -0.00225637 | 3.18E-06 | 0.10119758 | 0.000772501 | 1 | 0.037501449 | ENSG0000017 | 2 | GPR27 |
| SAT | Obesity | ENSG0000023 | -0.00047153 | 0.884192547 | 0.003237294 | 0.183454466 | 0.003698993 | 0.265211987 | 0.00331999 | 0.183420801 | -0.00417053 | 1.57E-08 | 1 | 0.959020082 | 1 | 0.567932065 | ENSG0000017 | 2 | GPR27 |
| SAT | Obesity | ENSG0000023 | 0.006277797 | 0.105852915 | 0.003882083 | 0.183453486 | 0.007264532 | 0.061742252 | 0.003888631 | 0.183419951 | -0.00098674 | 2.04E-05 | 1 | 0.334504825 | 1 | 0.251335397 | ENSG0000017 | 7 | LILRA3 |
| SAT | Obesity | ENSG0000023 | -0.00017544 | 0.930389515 | 0.002008409 | 0.183454471 | -0.00022424 | 0.911096159 | 0.00200828 | 0.183421265 | 4.88E-05 | 0.00084612 | 1 | 0.977387556 | 1 | 0.968667044 | ENSG0000017 | 7 | LILRA3 |
| SAT | Obesity | ENSG0000023 | -0.00724115 | 0.126208994 | 0.004735173 | 0.18345359 | -0.00734123 | 0.121035782 | 0.004734923 | 0.183420362 | 0.000100079 | 0.015128926 | 1 | 0.374273307 | 1 | 0.369554001 | ENSG0000017 | 7 | LILRA3 |
| SAT | Obesity | ENSG0000023 | 0.003249719 | 0.031711818 | 0.001512883 | 0.18345273 | 0.003215342 | 0.033551467 | 0.001512798 | 0.183419563 | 3.44E-05 | 0.005900939 | 1 | 0.164370418 | 1 | 0.176545003 | ENSG0000017 | 7 | LILRA3 |
| SAT | Obesity | ENSG0000023 | 0.000598491 | 0.88264581 | 0.004054374 | 0.183454466 | 0.000668494 | 0.869028796 | 0.004054124 | 0.18342126 | -7.00E-05 | 0.022946507 | 1 | 0.958494761 | 1 | 0.954321831 | ENSG0000017 | 8 | PAQR8 |
| SAT | Obesity | ENSG0000023 | 0.000222704 | 0.925824765 | 0.002392115 | 0.183454471 | 0.000307695 | 0.897645021 | 0.002391964 | 0.183421264 | -8.50E-05 | 1.80E-06 | 1 | 0.976102523 | 1 | 0.963222823 | ENSG0000017 | 8 | PAQR8 |
| SAT | Obesity | ENSG0000023 | 0.005025548 | 0.000102329 | 0.001293566 | 0.183448769 | 0.003227087 | 0.017007339 | 0.001352193 | 0.183419118 | 0.001798461 | 5.06E-06 | 0.335743054 | 0.002010437 | 1 | 0.11722916 | ENSG0000017 | 8 | PAQR8 |
| SAT | Obesity | ENSG0000023 | 0.000862337 | 0.819787747 | 0.003785202 | 0.183454454 | 0.000917998 | 0.808361294 | 0.003784923 | 0.183421248 | -5.57E-05 | 0.011113377 | 1 | 0.932313205 | 1 | 0.924767575 | ENSG0000017 | 8 | PAQR8 |
| SAT | Obesity | ENSG0000023 | 0.004525005 | 0.060322935 | 0.002408929 | 0.18345314 | 0.004443585 | 0.065076256 | 0.002408789 | 0.183419984 | 8.14E-05 | 2.64E-05 | 1 | 0.243144411 | 1 | 0.260067232 | ENSG0000017 | 9 | PRKCDBP |
| SAT | Obesity | ENSG0000023 | 0.008934761 | 0.113003202 | 0.00563764 | 0.183453525 | 0.009100503 | 0.106454023 | 0.005637285 | 0.183420285 | -0.00016574 | 7.56E-05 | 1 | 0.349776893 | 1 | 0.342763151 | ENSG0000017 | 9 | PRKCDBP |
| SAT | Obesity | ENSG0000023 | 0.006955223 | 0.22376162 | 0.005716991 | 0.183453915 | 9.19E-05 | 0.987482117 | 0.005855007 | 0.18342127 | 0.006863361 | 5.93E-08 | 1 | 0.511254788 | 1 | 0.99567573 | ENSG0000017 | 9 | PRKCDBP |
| SAT | Obesity | ENSG0000023 | 0.001231728 | 0.681608533 | 0.003002242 | 0.18345441 | 0.001291019 | 0.667163132 | 0.003002055 | 0.1834212 | -5.93E-05 | 0.008856384 | 1 | 0.860468487 | 1 | 0.857743823 | ENSG0000017 | 9 | PRKCDBP |
| SAT | Obesity | ENSG0000023 | 0.004648721 | 9.63E-05 | 0.001192045 | 0.183448726 | 0.003205245 | 0.009262706 | 0.001231738 | 0.183418711 | 0.001443476 | 3.35E-06 | 0.315930883 | 0.001914733 | 1 | 0.078523369 | ENSG0000017 | 10 | TSNARE1 |
| SAT | Obesity | ENSG0000023 | 0.035129419 | 1.03E-08 | 0.006135616 | 0.183442084 | 0.03522512 | 9.39E-09 | 0.006135211 | 0.183408812 | -9.57E-05 | 0.026660096 | 3.39E-05 | 6.39E-07 | 3.08E-05 | 6.85E-07 | ENSG0000017 | 10 | TSNARE1 |
| SAT | Obesity | ENSG0000023 | 0.001749813 | 0.550146047 | 0.002928353 | 0.183454339 | 0.001808309 | 0.536869097 | 0.002928175 | 0.183421126 | -5.85E-05 | 0.009737847 | 1 | 0.793070817 | 1 | 0.78622818 | ENSG0000017 | 11 | FAM86JP |
| SAT | Obesity | ENSG0000024 | -0.00235312 | 0.069226008 | 0.001295098 | 0.183453226 | -0.00340294 | 0.009839979 | 0.001318247 | 0.183418752 | 0.001049816 | 2.07E-05 | 1 | 0.262882561 | 1 | 0.081872378 | ENSG0000017 | 11 | FAM86JP |
| SAT | Obesity | ENSG0000024 | 0.011318795 | 0.000221051 | 0.003064363 | 0.183449317 | 0.008075804 | 0.01084637 | 0.003169941 | 0.183418817 | 0.003242992 | 6.54E-05 | 0.72526703 | 0.003941669 | 1 | 0.086166924 | ENSG0000017 | 11 | FAM86JP |
| SAT | Obesity | ENSG0000024 | 0.006854391 | 0.243931719 | 0.005882502 | 0.183453961 | 0.006649733 | 0.258269869 | 0.005882162 | 0.183420787 | 0.000204659 | 1.70E-05 | 1 | 0.532140939 | 1 | 0.557489106 | ENSG0000017 | 6 | TRMT61B |
| SAT | Obesity | ENSG0000024 | 0.001623785 | 0.220372546 | 0.001324955 | 0.183453906 | 0.001649426 | 0.213142027 | 0.001324868 | 0.183420684 | -2.56E-05 | 0.005923836 | 1 | 0.507398121 | 1 | 0.503707333 | ENSG0000017 | 14 | MFN1 |
| SAT | Obesity | ENSG0000024 | 0.001437145 | 0.282039282 | 0.001335946 | 0.183454037 | 0.002054613 | 0.1263783 | 0.001344164 | 0.183420387 | -0.00061747 | 3.63E-05 | 1 | 0.579661774 | 1 | 0.37867367 | ENSG0000017 | 14 | MFN1 |
| SAT | Obesity | ENSG0000024 | 0.004638032 | 0.303971848 | 0.004511905 | 0.183454075 | 0.00473754 | 0.293683207 | 0.004511611 | 0.183420853 | -9.95E-05 | 0.002048191 | 1 | 0.604809967 | 1 | 0.59812588 | ENSG0000017 | 14 | MFN1 |
| SAT | Obesity | ENSG0000024 | 0.004093814 | 0.449636474 | 0.005414942 | 0.183454258 | 0.004214783 | 0.436328425 | 0.005414613 | 0.183421041 | -0.00012097 | 0.003824301 | 1 | 0.72529856 | 1 | 0.717711418 | ENSG0000017 | 13 | ATP6V0E2 |
| SAT | Obesity | ENSG0000024 | -0.00203041 | 0.451488868 | 0.002696651 | 0.18345426 | -0.00211688 | 0.432421945 | 0.002696475 | 0.183421037 | 8.65E-05 | 6.23E-06 | 1 | 0.726500723 | 1 | 0.715518881 | ENSG0000017 | 8 | GAA |
| SAT | Obesity | ENSG0000024 | -0.00373442 | 0.042287245 | 0.00183899 | 0.183452915 | -0.00369719 | 0.044371018 | 0.001838875 | 0.183419742 | -3.72E-05 | 0.006540349 | 1 | 0.194865803 | 1 | 0.206791634 | ENSG0000017 | 8 | GAA |
| SAT | Obesity | ENSG0000024 | 0.001504016 | 0.549261611 | 0.002511435 | 0.183454339 | 0.001463236 | 0.560117729 | 0.002511273 | 0.183421142 | 4.08E-05 | 0.024186752 | 1 | 0.792840892 | 1 | 0.795863471 | ENSG0000017 | 8 | GAA |
| SAT | Obesity | ENSG0000024 | 0.001617965 | 0.19593387 | 0.001251109 | 0.183453842 | 0.002225676 | 0.078505417 | 0.001265 | 0.1834201 | -0.00060771 | 0.001206477 | 1 | 0.476192171 | 1 | 0.288439275 | ENSG0000017 | 8 | GAA |
| SAT | Obesity | ENSG0000024 | 0.01054527 | 0.069495018 | 0.005809451 | 0.183453229 | 0.010673383 | 0.066156329 | 0.005809064 | 0.183419994 | -0.00012811 | 0.00140404 | 1 | 0.262990952 | 1 | 0.26214845 | ENSG0000017 | 8 | GAA |
| SAT | Obesity | ENSG0000024 | -0.00326545 | 0.43994049 | 0.004228243 | 0.183454249 | -0.00333738 | 0.429901205 | 0.004227945 | 0.183421035 | 7.19E-05 | 0.007264561 | 1 | 0.718489173 | 1 | 0.714541973 | ENSG0000017 | 6 | CHST11 |
| SAT | Obesity | ENSG0000024 | -0.00046331 | 0.871502432 | 0.00286435 | 0.183454464 | -0.00039985 | 0.888973403 | 0.002864173 | 0.183421263 | -6.35E-05 | 0.003267445 | 1 | 0.955362338 | 1 | 0.962898024 | ENSG0000017 | 6 | CHST11 |
| SAT | Obesity | ENSG0000024 | -0.00663566 | 5.97E-08 | 0.001224332 | 0.183443371 | -0.00366357 | 0.037545095 | 0.001761508 | 0.183419635 | -0.00297209 | 0.018944066 | 0.000195832 | 3.11E-06 | 1 | 0.18893475 | ENSG0000017 | 6 | CHST11 |
| SAT | Obesity | ENSG0000024 | -0.0002612 | 0.926338614 | 0.002825258 | 0.183454471 | -0.00019149 | 0.945960314 | 0.002825111 | 0.183421268 | -6.97E-05 | 0.004923554 | 1 | 0.976330547 | 1 | 0.984050663 | ENSG0000017 | 8 | PGAM1 |
| SAT | Obesity | ENSG0000024 | -0.00596287 | 0.121778522 | 0.00385359 | 0.183453569 | -0.00584641 | 0.129212269 | 0.003853373 | 0.1834204 | -0.00011646 | 0.00025378 | 1 | 0.36522425 | 1 | 0.383362075 | ENSG0000017 | 8 | PGAM1 |
| SAT | Obesity | ENSG0000024 | -0.00470577 | 0.310972203 | 0.004644534 | 0.183454086 | -0.00495984 | 0.285594838 | 0.004644754 | 0.183420839 | 0.000254065 | 0.000998651 | 1 | 0.608407751 | 1 | 0.589702117 | ENSG0000017 | 5 | LURAP1 |
| SAT | Obesity | ENSG0000024 | 0.010265439 | 0.021262551 | 0.004456845 | 0.183452469 | 0.010201512 | 0.022073874 | 0.004456557 | 0.18341929 | 6.39E-05 | 0.045480534 | 1 | 0.129189682 | 1 | 0.136607693 | ENSG0000017 | 5 | LURAP1 |
| SAT | Obesity | ENSG0000024 | 0.001362019 | 0.59303452 | 0.002548477 | 0.183454366 | 0.001401078 | 0.582451751 | 0.002548307 | 0.183421156 | -3.91E-05 | 0.026100588 | 1 | 0.812973298 | 1 | 0.809070362 | ENSG0000017 | 5 | LURAP1 |
| SAT | Obesity | ENSG0000024 | -0.00699881 | 4.55E-06 | 0.001526614 | 0.18344653 | -0.00636387 | 3.32E-05 | 0.001533448 | 0.183414761 | -0.00063494 | 1.37E-05 | 0.014932386 | 0.000153942 | 0.109084671 | 0.000940385 | ENSG0000017 | 5 | LURAP1 |
| SAT | Obesity | ENSG0000024 | 0.002645087 | 0.544004973 | 0.004359303 | 0.183454335 | 0.00275335 | 0.527622622 | 0.004359051 | 0.183421119 | -0.00010826 | 0.002174066 | 1 | 0.790819813 | 1 | 0.779574758 | ENSG0000017 | 5 | LURAP1 |
| SAT | Obesity | ENSG0000024 | -0.02591645 | 9.27E-06 | 0.005845588 | 0.183447044 | -0.02658876 | 5.52E-06 | 0.005851536 | 0.183413467 | 0.000672312 | 0.014624395 | 0.030425754 | 0.000271659 | 0.01812343 | 0.000218355 | ENSG0000017 | 5 | LURAP1 |
| SAT | Obesity | ENSG0000024 | 0.003197604 | 0.018310471 | 0.001355326 | 0.18345237 | 0.003227211 | 0.017252331 | 0.001355234 | 0.183419127 | -2.96E-05 | 0.001236225 | 1 | 0.116653699 | 1 | 0.118173061 | ENSG0000017 | 5 | LURAP1 |
| SAT | Obesity | ENSG0000024 | -0.00639041 | 0.180226917 | 0.00476875 | 0.183453795 | -0.00628518 | 0.187480428 | 0.004768464 | 0.183420614 | -0.00010523 | 0.004648292 | 1 | 0.452624689 | 1 | 0.46706856 | ENSG0000017 | 8 | MRPL36 |
| SAT | Obesity | ENSG0000024 | -9.99E-05 | 0.986971229 | 0.006119282 | 0.183454474 | 1.95E-05 | 0.99745519 | 0.006118914 | 0.18342127 | -0.00011944 | 0.012319232 | 1 | 0.994152558 | 1 | 0.998819255 | ENSG0000017 | 8 | MRPL36 |
| SAT | Obesity | ENSG0000024 | -0.00461192 | 0.455174938 | 0.006175445 | 0.183454263 | -0.00841407 | 0.180981178 | 0.006289754 | 0.183420594 | 0.003802146 | 0.001485893 | 1 | 0.730067807 | 1 | 0.458532235 | ENSG0000017 | 8 | MRPL36 |
| SAT | Obesity | ENSG0000024 | 0.006605857 | 0.117109094 | 0.004215535 | 0.183453546 | 0.006462966 | 0.125222103 | 0.004215287 | 0.183420382 | 0.00014289 | 1.99E-05 | 1 | 0.35809407 | 1 | 0.376584526 | ENSG0000017 | 8 | MRPL36 |
| SAT | Obesity | ENSG0000025 | -0.00025317 | 0.863000817 | 0.001467152 | 0.183454463 | 0.00074081 | 0.617936947 | 0.001485254 | 0.183421176 | -0.00099398 | 1.84E-05 | 1 | 0.94985095 | 1 | 0.828495448 | ENSG0000017 | 8 | MRPL36 |
| SAT | Obesity | ENSG0000025 | -0.0027106 | 0.400103733 | 0.003221391 | 0.183454206 | -0.00263637 | 0.413101237 | 0.003221183 | 0.183421017 | -7.42E-05 | 0.00140843 | 1 | 0.691279804 | 1 | 0.701318401 | ENSG0000017 | 8 | MRPL36 |
| SAT | Obesity | ENSG0000025 | 0.004768924 | 0.396879709 | 0.00562899 | 0.183454203 | 0.004676982 | 0.406016971 | 0.005628652 | 0.183421009 | 9.19E-05 | 0.036309025 | 1 | 0.688610432 | 1 | 0.696726821 | ENSG0000017 | 6 | KRT27 |
| SAT | Obesity | ENSG0000025 | 0.007946618 | 0.019286537 | 0.003396058 | 0.183452404 | 0.007862159 | 0.020601335 | 0.003395859 | 0.183419244 | 8.45E-05 | 0.001804213 | 1 | 0.120761697 | 1 | 0.130740771 | ENSG0000017 | 6 | KRT27 |
| SAT | Obesity | ENSG0000025 | 0.007897194 | 0.04357116 | 0.003912992 | 0.183452934 | 0.008020935 | 0.040370441 | 0.003912758 | 0.183419682 | -0.00012374 | 5.67E-05 | 1 | 0.199104425 | 1 | 0.195361972 | ENSG0000017 | 4 | CDK5R2 |
| SAT | Obesity | ENSG0000025 | -0.00152879 | 0.687122484 | 0.003795761 | 0.183454413 | -0.00146602 | 0.699311454 | 0.003795513 | 0.183421214 | -6.28E-05 | 0.019588988 | 1 | 0.863266708 | 1 | 0.87540667 | ENSG0000017 | 6 | ZNF561 |
| SAT | Obesity | ENSG0000025 | -0.01537993 | 0.004342341 | 0.005392356 | 0.183451399 | -0.01522267 | 0.004755415 | 0.00539206 | 0.183418258 | -0.00015725 | 0.000546724 | 1 | 0.04466213 | 1 | 0.050822526 | ENSG0000017 | 6 | ZNF561 |
| SAT | Obesity | ENSG0000025 | -0.0003387 | 0.871902563 | 0.00210059 | 0.183454464 | -0.000295 | 0.888309257 | 0.002100456 | 0.183421263 | -4.37E-05 | 0.004543097 | 1 | 0.955463635 | 1 | 0.962898024 | ENSG0000017 | 10 | COL24A1 |
| SAT | Obesity | ENSG0000025 | -0.00366278 | 0.14834496 | 0.002534087 | 0.183453684 | -0.00234709 | 0.356374728 | 0.002544836 | 0.183420949 | -0.00131569 | 2.51E-08 | 1 | 0.407296916 | 1 | 0.657825701 | ENSG0000017 | 10 | COL24A1 |
| SAT | Obesity | ENSG0000025 | 0.007193198 | 0.002840258 | 0.002410159 | 0.183451107 | 0.006309343 | 0.008989069 | 0.002415078 | 0.183418691 | 0.000883855 | 1.95E-08 | 1 | 0.03213409 | 1 | 0.077624322 | ENSG0000017 | 10 | COL24A1 |
| SAT | Obesity | ENSG0000025 | -0.00054824 | 0.913477328 | 0.005045785 | 0.18345447 | -0.00114751 | 0.820131128 | 0.005046736 | 0.183421251 | 0.000599267 | 4.99E-07 | 1 | 0.96957309 | 1 | 0.932801998 | ENSG0000017 | 7 | ETFDH |
| SAT | Obesity | ENSG0000025 | -0.00321556 | 0.095654373 | 0.00192976 | 0.183453425 | -0.00216318 | 0.264536518 | 0.001938792 | 0.1834208 | -0.00105238 | 2.44E-08 | 1 | 0.315355558 | 1 | 0.567283867 | ENSG0000017 | 7 | ETFDH |
| SAT | Obesity | ENSG0000025 | 0.008183389 | 0.137501356 | 0.005510096 | 0.18345364 | 0.008014121 | 0.145798847 | 0.00550975 | 0.183420471 | 0.000169268 | 3.62E-05 | 1 | 0.390599089 | 1 | 0.410262452 | ENSG0000017 | 9 | TBCA |
| SAT | Obesity | ENSG0000025 | -0.00190393 | 0.557409672 | 0.003245182 | 0.183454344 | -0.0019654 | 0.54473455 | 0.003245 | 0.183421132 | 6.15E-05 | 0.022790795 | 1 | 0.794232732 | 1 | 0.788841592 | ENSG0000017 | 9 | TBCA |
| SAT | Obesity | ENSG0000025 | -0.01189436 | 0.026406404 | 0.005357364 | 0.183452611 | -0.01197999 | 0.02533243 | 0.005357054 | 0.18341938 | 8.56E-05 | 0.048332164 | 1 | 0.147095776 | 1 | 0.147892708 | ENSG0000017 | 9 | TBCA |
| SAT | Obesity | ENSG0000025 | 0.02056763 | 2.56E-11 | 0.003083638 | 0.183437659 | 0.020655418 | 2.11E-11 | 0.003083541 | 0.183404313 | -8.78E-05 | 0.008955652 | 8.41E-08 | 2.47E-09 | 6.91E-08 | 2.16E-09 | ENSG0000017 | 8 | BCL2L1 |
| SAT | Obesity | ENSG0000025 | -0.00972302 | 0.00927465 | 0.003737077 | 0.183451915 | -0.00963368 | 0.00993703 | 0.003736847 | 0.183418758 | -8.93E-05 | 0.001594597 | 1 | 0.074401287 | 1 | 0.081872378 | ENSG0000017 | 13 | ZNF584 |
| SAT | Obesity | ENSG0000025 | -0.00160438 | 0.464425117 | 0.002193029 | 0.183454272 | -0.00155965 | 0.476940107 | 0.002192879 | 0.183421079 | -4.47E-05 | 0.00226843 | 1 | 0.733866362 | 1 | 0.742751548 | ENSG0000017 | 13 | ZNF584 |
| SAT | Obesity | ENSG0000025 | 0.011154741 | 0.030506376 | 0.005155975 | 0.183452705 | 0.011071927 | 0.031751552 | 0.005155655 | 0.183419527 | 8.28E-05 | 0.032863994 | 1 | 0.159635439 | 1 | 0.168844153 | ENSG0000017 | 13 | ZNF584 |
| SAT | Obesity | ENSG0000025 | -0.00535333 | 0.194212538 | 0.004123593 | 0.183453837 | -0.00568387 | 0.168135026 | 0.004124057 | 0.183420552 | 0.000330542 | 7.02E-05 | 1 | 0.474115579 | 1 | 0.437851886 | ENSG0000017 | 13 | ZNF584 |
| SAT | Obesity | ENSG0000025 | -0.00191526 | 0.721048554 | 0.005363992 | 0.183454426 | -0.00179354 | 0.738089536 | 0.005363696 | 0.183421228 | -0.00012172 | 0.00680437 | 1 | 0.884077949 | 1 | 0.896583401 | ENSG0000017 | 9 | RGS19 |
| SAT | Obesity | ENSG0000025 | -8.10E-05 | 0.985838179 | 0.004565659 | 0.183454474 | -0.00020785 | 0.963687234 | 0.004565394 | 0.183421269 | 0.000126808 | 0.000565874 | 1 | 0.994152558 | 1 | 0.991742114 | ENSG0000017 | 9 | RGS19 |
| SAT | Obesity | ENSG0000025 | -0.00884999 | 0.0179408 | 0.00373915 | 0.183452357 | -0.00878939 | 0.018733334 | 0.00373889 | 0.183419182 | -6.06E-05 | 0.011984527 | 1 | 0.114968288 | 1 | 0.123670158 | ENSG0000017 | 11 | GPHN |
| SAT | Obesity | ENSG0000025 | -0.00465979 | 0.412733204 | 0.005688955 | 0.183454221 | -0.00454926 | 0.423874605 | 0.005688578 | 0.183421028 | -0.00011053 | 0.005142837 | 1 | 0.699592222 | 1 | 0.709195604 | ENSG0000017 | 11 | GPHN |
| SAT | Obesity | ENSG0000025 | 0.014021695 | 0.003089194 | 0.004739084 | 0.183451165 | 0.010104495 | 0.043205136 | 0.004997953 | 0.183419725 | 0.0039172 | 0.01368519 | 1 | 0.034242044 | 1 | 0.2030889 | ENSG0000017 | 14 | CAMTA1 |
| SAT | Obesity | ENSG0000025 | 0.000707612 | 0.876926449 | 0.004569168 | 0.183454465 | 0.000563373 | 0.901865441 | 0.004568929 | 0.183421264 | 0.000144239 | 0.000306052 | 1 | 0.956287991 | 1 | 0.963222823 | ENSG0000017 | 4 | LGALS4 |
| SAT | Obesity | ENSG0000026 | 0.003172937 | 0.048977258 | 0.001611614 | 0.183453009 | 0.00313974 | 0.051379614 | 0.001611528 | 0.183419836 | 3.32E-05 | 0.016770327 | 1 | 0.21425918 | 1 | 0.226581335 | ENSG0000017 | 11 | LRRC34 |
| SAT | Obesity | ENSG0000026 | 0.014668504 | 0.008949547 | 0.005611547 | 0.183451891 | 0.015013648 | 0.007472084 | 0.005612476 | 0.183418566 | -0.00034514 | 0.006610506 | 1 | 0.07286219 | 1 | 0.069450161 | ENSG0000017 | 13 | SPATA5L1 |
| SAT | Obesity | ENSG0000026 | 0.010507702 | 0.041257584 | 0.005148469 | 0.1834529 | 0.010620581 | 0.039114978 | 0.005148172 | 0.183419662 | -0.00011288 | 0.007011441 | 1 | 0.192979684 | 1 | 0.192986833 | ENSG0000017 | 13 | SPATA5L1 |
| SAT | Obesity | ENSG0000026 | 0.000387389 | 0.942268725 | 0.00534929 | 0.183454472 | 0.000483843 | 0.927926052 | 0.005349014 | 0.183421267 | -9.65E-05 | 0.041046839 | 1 | 0.981335116 | 1 | 0.977375723 | ENSG0000017 | 13 | SPATA5L1 |
| SAT | Obesity | ENSG0000026 | -0.00924768 | 0.031906376 | 0.004310093 | 0.183452734 | -0.0091778 | 0.033212315 | 0.004309811 | 0.183419556 | -6.99E-05 | 0.021565602 | 1 | 0.165118011 | 1 | 0.175474405 | ENSG0000017 | 11 | SYCE1 |
| SAT | Obesity | ENSG0000026 | 0.003119815 | 0.245903035 | 0.002688662 | 0.183453965 | 0.003679196 | 0.172136031 | 0.002694642 | 0.183420566 | -0.00055938 | 0.002244045 | 1 | 0.534004366 | 1 | 0.443997285 | ENSG0000017 | 7 | METTL18 |
| SAT | Obesity | ENSG0000026 | -0.00796094 | 0.010441302 | 0.003108685 | 0.183451995 | -0.00788891 | 0.01115328 | 0.003108479 | 0.183418836 | -7.20E-05 | 0.000854979 | 1 | 0.079855276 | 1 | 0.087690846 | ENSG0000017 | 7 | METTL18 |
| SAT | Obesity | ENSG0000026 | 0.007297029 | 0.143644428 | 0.004989914 | 0.183453666 | 0.007182152 | 0.150030579 | 0.004989595 | 0.183420487 | 0.000114877 | 0.001585131 | 1 | 0.400717148 | 1 | 0.414396534 | ENSG0000017 | 7 | METTL18 |
| SAT | Obesity | ENSG0000026 | 0.003361227 | 0.385469544 | 0.003872991 | 0.183454189 | 0.003173384 | 0.412565847 | 0.003872874 | 0.183421016 | 0.000187843 | 9.98E-06 | 1 | 0.677819359 | 1 | 0.701318401 | ENSG0000017 | 4 | C3AR1 |
| SAT | Obesity | ENSG0000026 | -0.00574736 | 0.157219944 | 0.004063212 | 0.183453718 | -0.00579717 | 0.153624228 | 0.004062922 | 0.183420501 | 4.98E-05 | 0.048052336 | 1 | 0.420045334 | 1 | 0.417598255 | ENSG0000017 | 4 | C3AR1 |
| SAT | Obesity | ENSG0000026 | 0.000662116 | 0.84694154 | 0.003430254 | 0.18345446 | 0.0007326 | 0.830872266 | 0.003430048 | 0.183421253 | -7.05E-05 | 0.00835294 | 1 | 0.942344807 | 1 | 0.936326075 | ENSG0000017 | 5 | PTEN |
| SAT | Obesity | ENSG0000026 | 0.002828509 | 0.466113989 | 0.003880959 | 0.183454273 | 0.002884207 | 0.457350346 | 0.003880704 | 0.183421061 | -5.57E-05 | 0.041045778 | 1 | 0.733866362 | 1 | 0.731563013 | ENSG0000017 | 5 | PTEN |
| SAT | Obesity | ENSG0000026 | 0.000266048 | 0.882788533 | 0.001804507 | 0.183454466 | 0.000854814 | 0.636316437 | 0.001807769 | 0.183421186 | -0.00058877 | 1.21E-07 | 1 | 0.958494761 | 1 | 0.838966415 | ENSG0000017 | 9 | TVP23B |
| SAT | Obesity | ENSG0000026 | -0.01134862 | 0.022279313 | 0.004965314 | 0.1834525 | -0.01143567 | 0.021264587 | 0.004964991 | 0.183419265 | 8.71E-05 | 0.014448492 | 1 | 0.132906229 | 1 | 0.133679732 | ENSG0000017 | 9 | TVP23B |
| SAT | Obesity | ENSG0000026 | 0.00175893 | 0.445497142 | 0.002305451 | 0.183454254 | 0.001700572 | 0.460709881 | 0.002305301 | 0.183421065 | 5.84E-05 | 0.000358301 | 1 | 0.722137485 | 1 | 0.732714066 | ENSG0000017 | 11 | ATPAF2 |
| SAT | Obesity | ENSG0000026 | -0.00159259 | 0.721739324 | 0.004471859 | 0.183454426 | -0.00147674 | 0.74121128 | 0.004471581 | 0.183421229 | -0.00011585 | 0.000581686 | 1 | 0.884077949 | 1 | 0.897054301 | ENSG0000017 | 11 | ATPAF2 |
| SAT | Obesity | ENSG0000026 | 0.00225433 | 0.493162957 | 0.003289614 | 0.183454297 | 0.002188446 | 0.505862858 | 0.003289443 | 0.183421103 | 6.59E-05 | 0.022545292 | 1 | 0.752324516 | 1 | 0.765560903 | ENSG0000017 | 13 | LRRC48 |
| SAT | Obesity | ENSG0000026 | 0.002617535 | 0.218807038 | 0.002128583 | 0.183453903 | 0.002583796 | 0.224772331 | 0.002128443 | 0.183420713 | 3.37E-05 | 0.024798877 | 1 | 0.506281943 | 1 | 0.517165511 | ENSG0000017 | 10 | C20orf196 |
| SAT | Obesity | ENSG0000026 | 0.001255333 | 0.685161306 | 0.003096296 | 0.183454412 | 0.00119936 | 0.69847974 | 0.003096132 | 0.183421213 | 5.60E-05 | 0.037029299 | 1 | 0.86231772 | 1 | 0.875033229 | ENSG0000017 | 3 | ZNF556 |
| SAT | Obesity | ENSG0000026 | 0.00877536 | 0.073519172 | 0.004903537 | 0.183453264 | 0.008612459 | 0.079009584 | 0.004903289 | 0.183420104 | 0.000162902 | 0.000199345 | 1 | 0.271914102 | 1 | 0.289319694 | ENSG0000017 | 3 | ZNF556 |
| SAT | Obesity | ENSG0000026 | 0.001848876 | 0.593204555 | 0.003461029 | 0.183454366 | 0.001783076 | 0.606401311 | 0.003460823 | 0.18342117 | 6.58E-05 | 0.015677195 | 1 | 0.812973298 | 1 | 0.822933669 | ENSG0000017 | 3 | ZNF556 |
| SAT | Obesity | ENSG0000026 | -0.0011198 | 0.779100098 | 0.003992255 | 0.183454444 | -0.00119528 | 0.764620284 | 0.003991984 | 0.183421236 | 7.55E-05 | 0.004996313 | 1 | 0.913071213 | 1 | 0.906001861 | ENSG0000017 | 7 | EIF2AK3 |
| SAT | Obesity | ENSG0000026 | 0.011309931 | 0.004236423 | 0.003954504 | 0.183451382 | 0.011209713 | 0.004585172 | 0.003954278 | 0.183418233 | 0.000100218 | 0.001874525 | 1 | 0.043847645 | 1 | 0.050146494 | ENSG0000017 | 7 | EIF2AK3 |
| SAT | Obesity | ENSG0000026 | -0.0268707 | 8.66E-09 | 0.004668921 | 0.183441955 | -0.02675474 | 1.00E-08 | 0.00466866 | 0.183408859 | -0.00011596 | 0.002704859 | 2.84E-05 | 5.57E-07 | 3.28E-05 | 7.14E-07 | ENSG0000017 | 4 | NME6 |
| SAT | Obesity | ENSG0000026 | 0.026243341 | 4.63E-07 | 0.005206023 | 0.183444869 | 0.023598735 | 6.52E-06 | 0.005233744 | 0.183413587 | 0.002644606 | 1.08E-06 | 0.001520332 | 2.05E-05 | 0.021379575 | 0.000245156 | ENSG0000017 | 4 | NME6 |
| SAT | Obesity | ENSG0000027 | 0.004438871 | 0.386107437 | 0.005121583 | 0.18345419 | 0.004522792 | 0.377161166 | 0.005121264 | 0.183420975 | -8.39E-05 | 0.02877519 | 1 | 0.677819359 | 1 | 0.671441012 | ENSG0000017 | 4 | NME6 |
| SAT | Obesity | ENSG0000027 | -0.00344619 | 0.558924411 | 0.005896561 | 0.183454345 | -0.00356899 | 0.544975545 | 0.005896173 | 0.183421132 | 0.000122805 | 0.003050799 | 1 | 0.794898566 | 1 | 0.788841592 | ENSG0000017 | 17 | CD8B |
| SAT | Obesity | ENSG0000027 | 0.000352958 | 0.843113143 | 0.001783401 | 0.183454459 | 7.13E-05 | 0.968098725 | 0.00178397 | 0.18342127 | 0.000281612 | 3.45E-08 | 1 | 0.941066406 | 1 | 0.992119797 | ENSG0000017 | 8 | SLFN12 |
| SAT | Obesity | ENSG0000027 | -0.01101851 | 0.067757329 | 0.006032182 | 0.183453213 | -0.01091229 | 0.070431015 | 0.006031779 | 0.183420033 | -0.00010622 | 0.01050169 | 1 | 0.261235952 | 1 | 0.270590351 | ENSG0000017 | 8 | SLFN12 |
| SAT | Obesity | ENSG0000027 | -0.00101446 | 0.760914484 | 0.003333977 | 0.183454439 | -0.00106576 | 0.749206504 | 0.003333766 | 0.183421232 | 5.13E-05 | 0.036911885 | 1 | 0.904420994 | 1 | 0.900889371 | ENSG0000017 | 3 | MTBP |
| SAT | Obesity | ENSG0000027 | -0.00332148 | 0.041543377 | 0.001629716 | 0.183452904 | -0.00092007 | 0.590213026 | 0.001708483 | 0.183421161 | -0.00240141 | 2.88E-06 | 1 | 0.193338752 | 1 | 0.815020596 | ENSG0000017 | 6 | TEFM |
| SAT | Obesity | ENSG0000027 | 0.006489497 | 0.01506005 | 0.002669542 | 0.18345224 | 0.006432889 | 0.015957665 | 0.00266937 | 0.183419075 | 5.66E-05 | 0.003234065 | 1 | 0.102604462 | 1 | 0.112106928 | ENSG0000017 | 6 | TEFM |
| SAT | Obesity | ENSG0000027 | -0.05120648 | 1.83E-28 | 0.004626909 | 0.183408188 | -0.05112997 | 2.18E-28 | 0.004626618 | 0.183375124 | -7.65E-05 | 0.024636876 | 5.99E-25 | 1.20E-25 | 7.15E-25 | 1.43E-25 | ENSG0000017 | 10 | ISG20 |
| SAT | Obesity | ENSG0000027 | -0.03114348 | 1.90E-09 | 0.005185445 | 0.18344084 | -0.03130869 | 1.56E-09 | 0.005185331 | 0.183407493 | 0.000165216 | 0.006620156 | 6.25E-06 | 1.42E-07 | 5.13E-06 | 1.28E-07 | ENSG0000017 | 3 | C1QTNF4 |
| SAT | Obesity | ENSG0000027 | -0.00265973 | 0.438956191 | 0.003436534 | 0.183454248 | -0.00270818 | 0.430634462 | 0.003436303 | 0.183421035 | 4.84E-05 | 0.039097197 | 1 | 0.717400247 | 1 | 0.714674593 | ENSG0000017 | 3 | C1QTNF4 |
| SAT | Obesity | ENSG0000027 | -0.00113598 | 0.63777103 | 0.00241279 | 0.18345439 | -0.00138146 | 0.567015395 | 0.002413221 | 0.183421146 | 0.000245471 | 1.17E-05 | 1 | 0.841561068 | 1 | 0.800506675 | ENSG0000017 | 3 | C1QTNF4 |
| SAT | Obesity | ENSG0000027 | 0.003649226 | 0.355335961 | 0.003948141 | 0.183454151 | 0.002436128 | 0.542000695 | 0.003995023 | 0.18342113 | 0.001213098 | 0.047651216 | 1 | 0.653141338 | 1 | 0.788255443 | ENSG0000017 | 3 | C1QTNF4 |
| SAT | Obesity | ENSG0000027 | -0.00234644 | 0.026061464 | 0.001054439 | 0.183452602 | -0.00172106 | 0.105381325 | 0.001062833 | 0.183420279 | -0.00062539 | 3.08E-06 | 1 | 0.146166945 | 1 | 0.340341256 | ENSG0000017 | 3 | C1QTNF4 |
| SAT | Obesity | ENSG0000027 | -0.00503672 | 0.018186356 | 0.002132568 | 0.183452366 | -0.00251839 | 0.249684347 | 0.002187774 | 0.183420769 | -0.00251833 | 2.63E-07 | 1 | 0.116221806 | 1 | 0.546117316 | ENSG0000017 | 3 | C1QTNF4 |
| SAT | Obesity | ENSG0000027 | 0.00630067 | 4.51E-11 | 0.000956608 | 0.183438077 | 0.006008114 | 3.73E-10 | 0.000958971 | 0.183406436 | 0.000292556 | 1.92E-05 | 1.48E-07 | 4.23E-09 | 1.22E-06 | 3.40E-08 | ENSG0000017 | 9 | CLEC12A |

| | | | | | | | | | | | | | | | | | | |
|---|---|---|---|---|---|---|---|---|---|---|---|---|---|---|---|---|---|---|---|
| SAT | Obesity | ENSG0000027 | 0.000969001 | 0.770460924 | 0.003321086 | 0.183454442 | 0.001029666 | 0.75651613 | 0.003320871 | 0.183421234 | -6.07E-05 | 0.01095314 | 1 | 0.90745061 | 1 | 0.902920853 | ENSG0000017 | 9 | CLEC12A |
| SAT | Obesity | ENSG0000027 | 0.001797596 | 0.479204547 | 0.002540473 | 0.183454285 | 0.001751938 | 0.490414288 | 0.002540327 | 0.18342109 | 4.57E-05 | 0.027062214 | 1 | 0.741636848 | 1 | 0.752696629 | ENSG0000017 | 9 | CLEC12A |
| SAT | Obesity | ENSG0000027 | -0.00297288 | 0.212277193 | 0.002383393 | 0.183453886 | -0.00301678 | 0.205572077 | 0.002383228 | 0.183420665 | 4.39E-05 | 0.004680667 | 1 | 0.501426545 | 1 | 0.494851053 | ENSG0000017 | 9 | CLEC12A |
| SAT | Obesity | ENSG0000027 | -0.00231416 | 0.592810825 | 0.004327416 | 0.183454366 | -0.00225498 | 0.60227903 | 0.004327121 | 0.183421168 | -5.92E-05 | 0.040543218 | 1 | 0.812973298 | 1 | 0.820971125 | ENSG0000017 | 9 | CLEC12A |
| SAT | Obesity | ENSG0000027 | -0.01322107 | 8.03E-05 | 0.003352544 | 0.183448596 | -0.01316384 | 8.61E-05 | 0.003352327 | 0.183415443 | -5.72E-05 | 0.016751441 | 0.263401104 | 0.001656611 | 0.282504754 | 0.001973489 | ENSG0000017 | 10 | SUCLG2 |
| SAT | Obesity | ENSG0000027 | -0.00100483 | 0.873313735 | 0.006301844 | 0.183454484 | -0.00111208 | 0.859917242 | 0.006301451 | 0.183421258 | 0.000107243 | 0.023237506 | 1 | 0.95575129 | 1 | 0.948758084 | ENSG0000017 | 10 | SUCLG2 |
| SAT | Obesity | ENSG0000027 | 0.001546645 | 0.308887073 | 0.001519953 | 0.183454083 | 0.001526637 | 0.31515294 | 0.001519844 | 0.183420889 | 2.00E-05 | 0.032762744 | 1 | 0.60688449 | 1 | 0.615584597 | ENSG0000017 | 12 | IL16 |
| SAT | Obesity | ENSG0000027 | 0.001498465 | 0.241663895 | 0.001279821 | 0.183453956 | 0.001522912 | 0.234039031 | 0.001279736 | 0.183420735 | -2.44E-05 | 0.006157816 | 1 | 0.530497704 | 1 | 0.527090757 | ENSG0000017 | 12 | IL16 |
| SAT | Obesity | ENSG0000027 | -0.00024877 | 0.848911209 | 0.001305848 | 0.18345446 | 0.000735457 | 0.576684025 | 0.001317468 | 0.183421152 | -0.00098423 | 2.04E-08 | 1 | 0.942883439 | 1 | 0.804464408 | ENSG0000017 | 14 | GTPBP2 |
| SAT | Stroke | ENSG0000002 | 0.001136542 | 0.307639929 | 0.001114052 | 0.057490515 | 0.001180344 | 0.289452676 | 0.001114242 | 0.057488926 | -4.38E-05 | 0.040660719 | 1 | 0.871327204 | 1 | 0.865823277 | ENSG0000017 | 14 | GTPBP2 |
| SAT | Stroke | ENSG0000004 | -0.00056374 | 0.863511055 | 0.003279322 | 0.057490634 | -0.00469791 | 0.224818157 | 0.003870357 | 0.057488885 | 0.004134169 | 0.044320871 | 1 | 0.985453834 | 1 | 0.846254871 | ENSG0000017 | 16 | AFF1 |
| SAT | Stroke | ENSG0000009 | -0.0072541 | 0.001328798 | 0.002260051 | 0.057489422 | -0.00721283 | 0.001416155 | 0.002260115 | 0.057487857 | -4.13E-05 | 0.046433953 | 1 | 0.36331564 | 1 | 0.38845011 | ENSG0000017 | 16 | AFF1 |
| SAT | Stroke | ENSG0000012 | 0.001572011 | 0.621755984 | 0.003186314 | 0.057490609 | 0.001520956 | 0.633125282 | 0.003186363 | 0.057489032 | 5.11E-05 | 0.035366279 | 1 | 0.94794674 | 1 | 0.950783463 | ENSG0000017 | 16 | AFF1 |
| SAT | Stroke | ENSG0000016 | 0.001403052 | 0.481390475 | 0.001992777 | 0.057490579 | 0.001303719 | 0.513078128 | 0.001993293 | 0.057489008 | 9.93E-05 | 0.032695256 | 1 | 0.923200221 | 1 | 0.930997953 | ENSG0000017 | 10 | NIPAL4 |
| SAT | Stroke | ENSG0000016 | -0.00347589 | 0.119491588 | 0.002232551 | 0.057490352 | -0.00355089 | 0.111767395 | 0.00223284 | 0.05748876 | 7.50E-05 | 0.047135356 | 1 | 0.771457664 | 1 | 0.769465514 | ENSG0000017 | 6 | PDE3A |
| SAT | Stroke | ENSG0000016 | -6.07E-05 | 0.984773951 | 0.003183168 | 0.057490638 | -0.0033062 | 0.342315294 | 0.003481672 | 0.057488952 | 0.003245453 | 0.021404895 | 1 | 0.998676328 | 1 | 0.892510712 | ENSG0000017 | 14 | MRPL52 |
| SAT | Stroke | ENSG0000017 | 0.00232425 | 0.312186851 | 0.002299769 | 0.057490517 | 0.00253826 | 0.270173122 | 0.002301932 | 0.057488915 | -0.00021401 | 0.033232244 | 1 | 0.875457313 | 1 | 0.858951562 | ENSG0000017 | 14 | MRPL52 |
| SAT | Stroke | ENSG0000018 | 0.001314012 | 0.453914615 | 0.001754577 | 0.057490572 | 0.000810071 | 0.647535471 | 0.001771853 | 0.057489034 | 0.000503941 | 0.041318426 | 1 | 0.909770222 | 1 | 0.955863681 | ENSG0000017 | 8 | SMPDL3A |
| SAT | Stroke | ENSG0000018 | -0.00079592 | 0.688859858 | 0.001987807 | 0.057490619 | -0.00170733 | 0.398549516 | 0.002022392 | 0.057488975 | 0.00091141 | 0.014431837 | 1 | 0.955101857 | 1 | 0.907572399 | ENSG0000017 | 8 | SMPDL3A |
| SAT | Stroke | ENSG0000018 | -0.00152885 | 0.58720764 | 0.002816153 | 0.057490603 | -0.00148262 | 0.598567968 | 0.002816197 | 0.057489026 | -4.62E-05 | 0.032261515 | 1 | 0.943546012 | 1 | 0.946914901 | ENSG0000017 | 3 | RAD9A |
| SAT | Stroke | ENSG0000018 | 0.001885006 | 0.564011525 | 0.003267509 | 0.057490598 | 0.00200874 | 0.538757705 | 0.003267875 | 0.057489014 | -0.00012373 | 0.016944184 | 1 | 0.937689169 | 1 | 0.93449293 | ENSG0000017 | 3 | RAD9A |
| SAT | Stroke | ENSG0000019 | 0.000442505 | 0.882452002 | 0.002992694 | 0.057490635 | 0.00026682 | 0.92898013 | 0.002993669 | 0.057489058 | 0.000175685 | 0.024291091 | 1 | 0.986976745 | 1 | 0.995096555 | ENSG0000017 | 3 | RAD9A |
| SAT | Stroke | ENSG0000020 | -0.00224018 | 0.326816277 | 0.002284617 | 0.057490524 | -0.00294047 | 0.201317925 | 0.002301184 | 0.057488866 | 0.000700294 | 0.011143515 | 1 | 0.882615609 | 1 | 0.833541007 | ENSG0000017 | 3 | RAD9A |
| SAT | Stroke | ENSG0000020 | -0.00202326 | 0.286022787 | 0.001896416 | 0.057490503 | -0.00381891 | 0.068005591 | 0.002092584 | 0.057488666 | 0.001795646 | 0.04238155 | 1 | 0.864235017 | 1 | 0.732839136 | ENSG0000017 | 3 | RAD9A |
| SAT | Stroke | ENSG0000020 | 0.000519108 | 0.720973427 | 0.001453439 | 0.057490623 | 0.000836023 | 0.567301024 | 0.001461498 | 0.05748902 | -0.00031691 | 0.03890885 | 1 | 0.960236691 | 1 | 0.938376016 | ENSG0000017 | 3 | RAD9A |
| SAT | Stroke | ENSG0000020 | -0.00027768 | 0.881060401 | 0.00185584 | 0.057490635 | -0.00089489 | 0.633343004 | 0.001875983 | 0.057489032 | 0.000617214 | 0.024465266 | 1 | 0.986976745 | 1 | 0.950783463 | ENSG0000017 | 3 | RAD9A |
| SAT | Stroke | ENSG0000020 | -0.0002711 | 0.722015516 | 0.000762021 | 0.057490623 | -0.000648 | 0.544203488 | 0.000766401 | 0.057489015 | 0.000193697 | 0.01805239 | 1 | 0.960236691 | 1 | 0.937286953 | ENSG0000017 | 8 | EFEMP2 |
| SAT | Stroke | ENSG0000020 | -8.55E-05 | 0.906239289 | 0.000725538 | 0.057490636 | -0.00037924 | 0.608045792 | 0.000739444 | 0.057489028 | 0.000293779 | 0.039636211 | 1 | 0.992541054 | 1 | 0.94961806 | ENSG0000017 | 8 | EFEMP2 |
| SAT | Stroke | ENSG0000022 | -0.00102564 | 0.591564046 | 0.001911466 | 0.057490604 | -0.00135228 | 0.480590022 | 0.001917164 | 0.057489 | 0.000326642 | 0.027349051 | 1 | 0.943546012 | 1 | 0.92815772 | ENSG0000017 | 8 | EFEMP2 |
| SAT | Stroke | ENSG0000022 | 0.000377981 | 0.597154205 | 0.000715198 | 0.057490605 | -0.00078822 | 0.368593977 | 0.000876667 | 0.057488963 | 0.001166201 | 0.021437365 | 1 | 0.943916331 | 1 | 0.905937727 | ENSG0000017 | 8 | AGAP5 |
| SAT | Stroke | ENSG0000022 | -0.0016553 | 0.269517534 | 0.001499129 | 0.057490494 | -0.00140462 | 0.350279963 | 0.0015038 | 0.057488956 | -0.00025068 | 0.034674415 | 1 | 0.858043425 | 1 | 0.898540262 | ENSG0000017 | 8 | AGAP5 |
| SAT | Stroke | ENSG0000023 | -0.00039611 | 0.674021298 | 0.000941695 | 0.057490617 | -0.00054492 | 0.564041566 | 0.000944653 | 0.057489019 | 0.000148812 | 0.046825824 | 1 | 0.955009236 | 1 | 0.938376016 | ENSG0000017 | 13 | FAM21C |
| SAT | Stroke | ENSG0000023 | -0.00123842 | 0.639995212 | 0.002647856 | 0.057490612 | -0.00166087 | 0.531754498 | 0.002655989 | 0.057489013 | 0.000422453 | 0.042418219 | 1 | 0.951392057 | 1 | 0.933410978 | ENSG0000017 | 13 | FAM21C |
| SAT | Stroke | ENSG0000023 | -0.00077617 | 0.671764511 | 0.001831776 | 0.057490617 | -0.00136661 | 0.460248715 | 0.001850683 | 0.057488994 | 0.00059044 | 0.025340838 | 1 | 0.955009236 | 1 | 0.92077807 | ENSG0000017 | 15 | ZFAND4 |
| SAT | Stroke | ENSG0000023 | -0.00064658 | 0.630535788 | 0.001344316 | 0.05749061 | -0.00114629 | 0.400803257 | 0.001364329 | 0.057488975 | 0.000499713 | 0.013928826 | 1 | 0.949661187 | 1 | 0.907572399 | ENSG0000017 | 9 | ZNF738 |
| SAT | Stroke | ENSG0000024 | -0.00432294 | 0.105510862 | 0.00267061 | 0.057490329 | -0.00438178 | 0.100862248 | 0.002670691 | 0.05748741 | 5.88E-05 | 0.01901498 | 1 | 0.749315451 | 1 | 0.758998867 | ENSG0000017 | 9 | ZNF738 |
| SAT | Stroke | ENSG0000027 | -5.21E-05 | 0.971646174 | 0.001466424 | 0.057490638 | -0.00017978 | 0.902505633 | 0.001467607 | 0.057489057 | 0.000127655 | 0.031676526 | 1 | 0.997892448 | 1 | 0.991426711 | ENSG0000017 | 11 | FUT10 |
| SAT | T2DM | ENSG0000000 | -0.00390275 | 0.216073418 | 0.003154916 | 0.081436703 | -0.00395008 | 0.210515677 | 0.003154639 | 0.081418869 | 4.73E-05 | 0.022963663 | 1 | 0.580276647 | 1 | 0.579977552 | ENSG0000017 | 11 | FUT10 |
| SAT | T2DM | ENSG0000000 | 0.001191819 | 0.482411864 | 0.001696711 | 0.081436876 | 0.001150779 | 0.497581862 | 0.001696564 | 0.081419054 | 4.10E-05 | 0.000307082 | 1 | 0.800198851 | 1 | 0.81497549 | ENSG0000017 | 11 | FUT10 |
| SAT | T2DM | ENSG0000000 | 0.000173968 | 0.894931117 | 0.001317264 | 0.081436956 | 0.000198723 | 0.880077342 | 0.001317168 | 0.081419127 | -2.48E-05 | 0.028115552 | 1 | 0.970180416 | 1 | 0.966700287 | ENSG0000017 | 6 | RP11-344H11.4 |
| SAT | T2DM | ENSG0000000 | 0.003510866 | 0.157439326 | 0.002483392 | 0.081436624 | 0.003437026 | 0.166323429 | 0.002483191 | 0.081418811 | 7.38E-05 | 8.60E-05 | 1 | 0.507474184 | 1 | 0.526744374 | ENSG0000017 | 12 | FAM192A |
| SAT | T2DM | ENSG0000000 | -0.00151324 | 0.504082897 | 0.002265063 | 0.081436884 | -0.00146359 | 0.518142194 | 0.002264881 | 0.081419061 | -4.96E-05 | 0.00419252 | 1 | 0.810314363 | 1 | 0.826056628 | ENSG0000017 | 12 | RAB43 |
| SAT | T2DM | ENSG0000000 | 0.002165345 | 0.486449019 | 0.003111269 | 0.081436877 | 0.002098326 | 0.500003962 | 0.003111009 | 0.081419055 | 6.70E-05 | 0.002604218 | 1 | 0.801645999 | 1 | 0.815984012 | ENSG0000017 | 12 | RAB43 |
| SAT | T2DM | ENSG0000000 | -0.00205444 | 0.550436902 | 0.003440666 | 0.081436899 | -0.00198461 | 0.564038326 | 0.003440399 | 0.081419075 | -6.98E-05 | 0.010945292 | 1 | 0.83552458 | 1 | 0.84223981 | ENSG0000017 | 12 | RAB43 |
| SAT | T2DM | ENSG0000000 | -0.00254677 | 0.435951408 | 0.003269083 | 0.081436857 | -0.00244761 | 0.454001667 | 0.003268877 | 0.081419037 | -9.92E-05 | 0.001659278 | 1 | 0.772274669 | 1 | 0.794600418 | ENSG0000017 | 12 | RAB43 |
| SAT | T2DM | ENSG0000001 | -0.00043021 | 0.736436775 | 0.001278187 | 0.081436939 | -0.00037597 | 0.768654505 | 0.001278206 | 0.081419116 | -5.42E-05 | 0.007171937 | 1 | 0.923995816 | 1 | 0.937978714 | ENSG0000017 | 3 | HOXC5 |
| SAT | T2DM | ENSG0000001 | -0.00278558 | 0.185686937 | 0.002104787 | 0.081436666 | -0.00100728 | 0.635491566 | 0.002125013 | 0.081419093 | -0.0017783 | 1.48E-09 | 1 | 0.542563333 | 1 | 0.884482576 | ENSG0000017 | 3 | HOXC5 |
| SAT | T2DM | ENSG0000001 | 0.009656686 | 1.46E-09 | 0.001596364 | 0.081430844 | 0.009709795 | 1.18E-09 | 0.001596227 | 0.081412949 | -5.31E-05 | 1.38E-06 | 4.78E-06 | 3.68E-07 | 3.87E-06 | 6.46E-07 | ENSG0000017 | 13 | SNX32 |
| SAT | T2DM | ENSG0000001 | 0.001836644 | 0.419497065 | 0.00227506 | 0.08143685 | 0.00188506 | 0.407308424 | 0.002274884 | 0.081419016 | -4.84E-05 | 0.007985757 | 1 | 0.76380126 | 1 | 0.761035842 | ENSG0000017 | 13 | SNX32 |
| SAT | T2DM | ENSG0000001 | -0.00056276 | 0.846780158 | 0.002912386 | 0.081436952 | -0.0006703 | 0.817958209 | 0.002912154 | 0.081419122 | 0.000107541 | 1.74E-06 | 1 | 0.963201596 | 1 | 0.957271954 | ENSG0000017 | 13 | SNX32 |
| SAT | T2DM | ENSG0000001 | -0.00126472 | 0.578153932 | 0.002274343 | 0.081436907 | -0.00123148 | 0.588155284 | 0.002274144 | 0.081419082 | -3.32E-05 | 0.02722525 | 1 | 0.842939695 | 1 | 0.853229568 | ENSG0000017 | 13 | SNX32 |
| SAT | T2DM | ENSG0000001 | -0.00151163 | 0.571437099 | 0.002671031 | 0.081436905 | -0.00155529 | 0.560345277 | 0.002670803 | 0.081419074 | 4.37E-05 | 0.018661049 | 1 | 0.840736467 | 1 | 0.84223981 | ENSG0000017 | 13 | SNX32 |
| SAT | T2DM | ENSG0000002 | -0.00059048 | 0.865845096 | 0.003495263 | 0.081436954 | -0.00066902 | 0.848195094 | 0.003494993 | 0.081419125 | 7.85E-05 | 0.005057962 | 1 | 0.965942795 | 1 | 0.965561588 | ENSG0000017 | 13 | SNX32 |
| SAT | T2DM | ENSG0000002 | -0.00359347 | 0.276784049 | 0.003304121 | 0.081436761 | -0.00351371 | 0.28754581 | 0.003303841 | 0.081418942 | -7.98E-05 | 0.000612986 | 1 | 0.63926506 | 1 | 0.653237213 | ENSG0000017 | 13 | SNX32 |
| SAT | T2DM | ENSG0000002 | 0.000277684 | 0.888506642 | 0.00198071 | 0.081436955 | 0.000310143 | 0.875563914 | 0.00198054 | 0.081419127 | -3.25E-05 | 0.01768525 | 1 | 0.969612316 | 1 | 0.966700287 | ENSG0000017 | 7 | NADSYN1 |
| SAT | T2DM | ENSG0000002 | -0.00136248 | 0.590610265 | 0.002532719 | 0.08143691 | -0.0014296 | 0.572421681 | 0.002532563 | 0.081419078 | 6.71E-05 | 0.00677407 | 1 | 0.846620707 | 1 | 0.847525062 | ENSG0000017 | 7 | NADSYN1 |
| SAT | T2DM | ENSG0000002 | 0.006322987 | 0.085385282 | 0.003675593 | 0.081436464 | 0.006237623 | 0.089663176 | 0.003675286 | 0.08141865 | 8.54E-05 | 0.001192388 | 1 | 0.402357222 | 1 | 0.421804746 | ENSG0000017 | 7 | NADSYN1 |
| SAT | T2DM | ENSG0000002 | -0.00128996 | 0.330616076 | 0.001325921 | 0.0814368 | -0.00116608 | 0.379195503 | 0.001326021 | 0.081419002 | -0.00012388 | 1.15E-06 | 1 | 0.693131851 | 1 | 0.739239717 | ENSG0000017 | 7 | NADSYN1 |
| SAT | T2DM | ENSG0000003 | 0.00532108 | 0.004329525 | 0.001865009 | 0.081435598 | 0.00443273 | 0.018044911 | 0.00187454 | 0.081418197 | 0.000888349 | 3.20E-06 | 1 | 0.069633188 | 1 | 0.185291928 | ENSG0000017 | 4 | RNASEH2C |
| SAT | T2DM | ENSG0000003 | -0.00139953 | 0.65419715 | 0.003124366 | 0.081436925 | -0.00133676 | 0.668734263 | 0.003124112 | 0.0814191 | -6.28E-05 | 0.007106508 | 1 | 0.882995924 | 1 | 0.896403813 | ENSG0000017 | 4 | RNASEH2C |
| SAT | T2DM | ENSG0000003 | -0.00650845 | 0.004070223 | 0.002265638 | 0.081435579 | -0.006551 | 0.003832212 | 0.002265474 | 0.081417734 | 4.25E-05 | 0.029618303 | 1 | 0.068134697 | 1 | 0.072792717 | ENSG0000017 | 4 | RNASEH2C |
| SAT | T2DM | ENSG0000003 | 0.00036665 | 0.846154288 | 0.001889676 | 0.081436952 | 0.000402664 | 0.831246552 | 0.001889526 | 0.081419123 | -3.60E-05 | 0.013947082 | 1 | 0.963201596 | 1 | 0.961544037 | ENSG0000017 | 4 | RNASEH2C |
| SAT | T2DM | ENSG0000003 | -0.00503134 | 0.01745549 | 0.002116695 | 0.081436014 | -0.00506914 | 0.016619393 | 0.002116523 | 0.081418172 | 3.78E-05 | 0.018021267 | 1 | 0.172324494 | 1 | 0.178781076 | ENSG0000017 | 4 | RNASEH2C |
| SAT | T2DM | ENSG0000003 | -0.00289863 | 0.258506441 | 0.002565318 | 0.081436745 | -0.00296445 | 0.247811479 | 0.002565107 | 0.081418908 | 6.58E-05 | 0.000543359 | 1 | 0.622274126 | 1 | 0.620663712 | ENSG0000017 | 4 | RNASEH2C |
| SAT | T2DM | ENSG0000004 | 0.002762004 | 0.456102639 | 0.003705993 | 0.081436866 | 0.00283617 | 0.444058624 | 0.003705677 | 0.081419033 | -7.42E-05 | 0.003942381 | 1 | 0.786411516 | 1 | 0.790106478 | ENSG0000017 | 3 | MYEOV |
| SAT | T2DM | ENSG0000004 | 0.011311338 | 0.003753828 | 0.003902946 | 0.081435555 | 0.001403515 | 0.760582798 | 0.004605982 | 0.081419115 | 0.009907823 | 5.12E-05 | 1 | 0.064822685 | 1 | 0.932999239 | ENSG0000017 | 3 | MYEOV |
| SAT | T2DM | ENSG0000004 | -0.0001153 | 0.871875969 | 0.000714949 | 0.081436954 | -0.00012793 | 0.857976087 | 0.000714897 | 0.081419126 | 1.26E-05 | 0.039795933 | 1 | 0.966468088 | 1 | 0.965566132 | ENSG0000017 | 3 | MYEOV |
| SAT | T2DM | ENSG0000004 | 0.009583775 | 0.007045391 | 0.003556524 | 0.081435745 | 0.009493453 | 0.007596231 | 0.00355624 | 0.08141794 | 9.03E-05 | 0.000939287 | 1 | 0.09396718 | 1 | 0.108835078 | ENSG0000017 | 10 | ANKRD13D |
| SAT | T2DM | ENSG0000004 | -0.00419496 | 0.19966623 | 0.003270913 | 0.081436684 | -0.00411647 | 0.20818605 | 0.003270747 | 0.081418866 | -7.85E-05 | 0.026670145 | 1 | 0.56039769 | 1 | 0.579977552 | ENSG0000017 | 10 | ANKRD13D |
| SAT | T2DM | ENSG0000004 | 0.000816965 | 0.76482662 | 0.002730963 | 0.081436946 | 0.000871466 | 0.749624543 | 0.002730717 | 0.081419114 | -5.45E-05 | 0.001272464 | 1 | 0.93459819 | 1 | 0.928470414 | ENSG0000017 | 10 | ANKRD13D |
| SAT | T2DM | ENSG0000004 | 0.005145588 | 0.01131007 | 0.002031437 | 0.081435886 | 0.005175572 | 0.010836103 | 0.002031269 | 0.081418046 | -3.00E-05 | 0.044409669 | 1 | 0.131589853 | 1 | 0.138879898 | ENSG0000017 | 8 | MRGPRF |
| SAT | T2DM | ENSG0000004 | -0.00096736 | 0.204596149 | 0.000762564 | 0.081436689 | -0.00098066 | 0.198407109 | 0.000762502 | 0.081418855 | 1.33E-05 | 0.019366256 | 1 | 0.565931149 | 1 | 0.566556767 | ENSG0000017 | 8 | MRGPRF |
| SAT | T2DM | ENSG0000004 | -0.00193577 | 0.42054271 | 0.002403243 | 0.08143685 | -0.0019966 | 0.40605333 | 0.002403056 | 0.081419016 | 6.08E-05 | 0.001404621 | 1 | 0.764009208 | 1 | 0.759884747 | ENSG0000017 | 8 | MRGPRF |
| SAT | T2DM | ENSG0000004 | -0.00775128 | 0.029867639 | 0.00356898 | 0.08143617 | -0.00782116 | 0.028408598 | 0.003568691 | 0.081418328 | 6.99E-05 | 0.009565673 | 1 | 0.240185597 | 1 | 0.247237694 | ENSG0000017 | 13 | ADH6 |
| SAT | T2DM | ENSG0000005 | 6.74E-05 | 0.977656149 | 0.002407602 | 0.081436958 | -7.78E-05 | 0.974209217 | 0.002407937 | 0.081419131 | 0.000145279 | 0.006788347 | 1 | 0.995203291 | 1 | 0.996688631 | ENSG0000017 | 13 | ADH6 |
| SAT | T2DM | ENSG0000005 | -0.00132705 | 0.686255826 | 0.003285256 | 0.081436931 | -0.00140399 | 0.669092636 | 0.003285006 | 0.0814191 | 7.69E-05 | 0.004091552 | 1 | 0.895709047 | 1 | 0.896403813 | ENSG0000017 | 7 | DCAKD |
| SAT | T2DM | ENSG0000005 | 0.007144346 | 0.023867609 | 0.003162249 | 0.081436105 | 0.007205193 | 0.022685602 | 0.003161981 | 0.081418263 | -6.08E-05 | 0.006246606 | 1 | 0.212275376 | 1 | 0.215743361 | ENSG0000017 | 2 | TRMT112 |
| SAT | T2DM | ENSG0000005 | 0.006687534 | 0.096466831 | 0.004023225 | 0.081436497 | 0.006622092 | 0.099741775 | 0.004022873 | 0.081418678 | 6.54E-05 | 0.014459357 | 1 | 0.420328914 | 1 | 0.440431278 | ENSG0000017 | 2 | TRMT112 |
| SAT | T2DM | ENSG0000005 | 0.005097369 | 7.22E-06 | 0.001135978 | 0.081433594 | 0.005059899 | 8.41E-06 | 0.001135894 | 0.081415816 | 3.75E-05 | 8.53E-05 | 0.02368204 | 0.000526268 | 0.027589044 | 0.000689726 | ENSG0000017 | 2 | TRMT112 |
| SAT | T2DM | ENSG0000005 | 0.001767891 | 0.316431837 | 0.001764682 | 0.08143679 | 0.00181757 | 0.302985334 | 0.00176454 | 0.081418954 | -4.97E-05 | 0.000210649 | 1 | 0.677242569 | 1 | 0.671232195 | ENSG0000017 | 2 | TRMT112 |
| SAT | T2DM | ENSG0000005 | -2.47E-05 | 0.983651481 | 0.001205416 | 0.081436958 | -5.24E-05 | 0.965326623 | 0.001205341 | 0.081419131 | 2.77E-05 | 0.017901725 | 1 | 0.995937959 | 1 | 0.99427016 | ENSG0000017 | 2 | TRMT112 |
| SAT | T2DM | ENSG0000006 | 0.000858738 | 0.738670166 | 0.00257404 | 0.08143694 | 0.000917281 | 0.721550973 | 0.002573835 | 0.08141911 | -5.85E-05 | 0.003284289 | 1 | 0.924676388 | 1 | 0.920812547 | ENSG0000017 | 2 | RHOD |
| SAT | T2DM | ENSG0000006 | -0.00118684 | 0.495363794 | 0.001740736 | 0.081436881 | -0.00122401 | 0.481923591 | 0.001740602 | 0.081419048 | 3.72E-05 | 0.008415053 | 1 | 0.806594843 | 1 | 0.80426821 | ENSG0000017 | 2 | RHOD |
| SAT | T2DM | ENSG0000006 | 0.003311768 | 0.223465512 | 0.002720437 | 0.081436711 | 0.003265421 | 0.229974852 | 0.00272022 | 0.08141889 | 4.63E-05 | 0.027309797 | 1 | 0.587734143 | 1 | 0.605344964 | ENSG0000017 | 2 | RHOD |
| SAT | T2DM | ENSG0000006 | -0.00297391 | 0.119262075 | 0.001908943 | 0.081436553 | -0.00316503 | 0.097327132 | 0.001908994 | 0.081418672 | 0.000191112 | 1.36E-09 | 1 | 0.454470229 | 1 | 0.435054932 | ENSG0000017 | 9 | PARP14 |
| SAT | T2DM | ENSG0000006 | 0.00035905 | 0.766130631 | 0.001207133 | 0.081436944 | 0.00029587 | 0.806364621 | 0.001207046 | 0.081419121 | 6.32E-05 | 1.89E-09 | 1 | 0.934630346 | 1 | 0.952410029 | ENSG0000017 | 9 | PARP14 |
| SAT | T2DM | ENSG0000006 | 0.002547712 | 0.334681633 | 0.002640861 | 0.081436803 | 0.00249414 | 0.344903863 | 0.00264064 | 0.081418982 | 5.36E-05 | 0.00476236 | 1 | 0.695221474 | 1 | 0.713247438 | ENSG0000017 | 9 | PARP14 |
| SAT | T2DM | ENSG0000006 | -0.00163126 | 0.433125126 | 0.002081075 | 0.081436856 | -1.82E-05 | 0.993185805 | 0.002125729 | 0.081419131 | -0.00161311 | 0.000205187 | 1 | 0.772274669 | 1 | 0.998405122 | ENSG0000017 | 8 | PARP15 |
| SAT | T2DM | ENSG0000006 | 0.000463622 | 0.697165454 | 0.001191372 | 0.081436933 | 0.000430409 | 0.717873496 | 0.001191272 | 0.081419109 | 3.32E-05 | 9.53E-05 | 1 | 0.900770568 | 1 | 0.918559531 | ENSG0000017 | 8 | PARP15 |
| SAT | T2DM | ENSG0000006 | 0.001469721 | 0.636411206 | 0.003109055 | 0.081436921 | 0.001415475 | 0.648866133 | 0.003108813 | 0.081419096 | 5.42E-05 | 0.028232128 | 1 | 0.873655621 | 1 | 0.891013292 | ENSG0000017 | 8 | PARP15 |
| SAT | T2DM | ENSG0000006 | 0.002289087 | 0.100778477 | 0.001394854 | 0.081436508 | 0.002272803 | 0.103193137 | 0.001394726 | 0.081418687 | 1.63E-05 | 0.049227379 | 1 | 0.425727224 | 1 | 0.440431278 | ENSG0000017 | 8 | PARP15 |
| SAT | T2DM | ENSG0000006 | 0.001844892 | 0.449417498 | 0.002439085 | 0.081436863 | 0.001796276 | 0.46141712 | 0.002438883 | 0.08141904 | 4.86E-05 | 0.006315974 | 1 | 0.78183394 | 1 | 0.797039301 | ENSG0000017 | 8 | PARP15 |
| SAT | T2DM | ENSG0000007 | -0.00580492 | 0.108891186 | 0.003620832 | 0.081436529 | -0.00571904 | 0.114202003 | 0.003620589 | 0.081418714 | -8.59E-05 | 0.009874925 | 1 | 0.436210685 | 1 | 0.455762334 | ENSG0000017 | 18 | AHSA2 |
| SAT | T2DM | ENSG0000007 | 0.002435843 | 0.398675936 | 0.002886107 | 0.081436839 | 0.002395005 | 0.406587925 | 0.002885848 | 0.081419016 | 4.08E-05 | 0.024181955 | 1 | 0.750003279 | 1 | 0.760122496 | ENSG0000017 | 18 | AHSA2 |
| SAT | T2DM | ENSG0000007 | -0.0004636 | 0.81135765 | 0.0019424 | 0.081436949 | -0.00049795 | 0.797656064 | 0.001942232 | 0.08141912 | 3.44E-05 | 0.009070043 | 1 | 0.951539016 | 1 | 0.952160336 | ENSG0000017 | 5 | KIAA1919 |
| SAT | T2DM | ENSG0000007 | 0.001632947 | 0.542620369 | 0.002681993 | 0.081436896 | 0.001593752 | 0.552316442 | 0.002681762 | 0.081419072 | 3.92E-05 | 0.031988678 | 1 | 0.832700048 | 1 | 0.842050121 | ENSG0000017 | 12 | IQCB1 |
| SAT | T2DM | ENSG0000007 | -0.00087199 | 0.74888351 | 0.002724013 | 0.081436941 | -0.00079353 | 0.770795649 | 0.002723787 | 0.081419117 | -7.85E-05 | 7.49E-05 | 1 | 0.928253417 | 1 | 0.938518235 | ENSG0000017 | 12 | IQCB1 |
| SAT | T2DM | ENSG0000007 | -0.00244185 | 0.46666095 | 0.003354543 | 0.0814367 | -0.00237329 | 0.479227157 | 0.003354247 | 0.081419047 | -6.86E-05 | 0.001580089 | 1 | 0.79334291 | 1 | 0.80363893 | ENSG0000017 | 12 | IQCB1 |
| SAT | T2DM | ENSG0000007 | 0.002453045 | 0.520058684 | 0.003813485 | 0.081436889 | 0.00231449 | 0.543873916 | 0.003813213 | 0.081419069 | 0.000138555 | 3.20E-05 | 1 | 0.819948363 | 1 | 0.840136685 | ENSG0000017 | 8 | ZNF483 |
| SAT | T2DM | ENSG0000007 | 0.002164617 | 0.57298794 | 0.00384031 | 0.081436905 | 0.002043727 | 0.594573127 | 0.003839996 | 0.081419084 | 0.00012089 | 2.33E-05 | 1 | 0.840775238 | 1 | 0.854809759 | ENSG0000017 | 8 | ZNF483 |
| SAT | T2DM | ENSG0000007 | 0.000700951 | 0.772267791 | 0.002421999 | 0.081436944 | 0.001302053 | 0.591497196 | 0.002426179 | 0.081419083 | -0.0006011 | 4.23E-05 | 1 | 0.936021656 | 1 | 0.854809759 | ENSG0000017 | 9 | MZT2A |
| SAT | T2DM | ENSG0000008 | -0.00115589 | 0.707005154 | 0.003083119 | 0.081436935 | -0.00107724 | 0.7267689 | 0.003082868 | 0.08141911 | -8.17E-05 | 0.000402543 | 1 | 0.906221055 | 1 | 0.921455409 | ENSG0000017 | 7 | EHBP1L1 |
| SAT | T2DM | ENSG0000008 | 0.001928844 | 0.109807138 | 0.001206234 | 0.081436531 | 0.001908109 | 0.11364895 | 0.001206133 | 0.081418713 | 2.07E-05 | 0.016403969 | 1 | 0.436210685 | 1 | 0.455762334 | ENSG0000017 | 7 | EHBP1L1 |
| SAT | T2DM | ENSG0000008 | 0.001372751 | 0.394610843 | 0.001612561 | 0.081436837 | 0.001345214 | 0.404122141 | 0.00161242 | 0.081419015 | 2.75E-05 | 0.010908966 | 1 | 0.74451879 | 1 | 0.758538184 | ENSG0000017 | 7 | EHBP1L1 |
| SAT | T2DM | ENSG0000008 | -0.00196735 | 0.444202956 | 0.002571315 | 0.081436861 | -0.00170239 | 0.508140045 | 0.002572604 | 0.081419058 | -0.00026497 | 0.003193497 | 1 | 0.779791277 | 1 | 0.817738235 | ENSG0000017 | 5 | VEGFB |
| SAT | T2DM | ENSG0000008 | 0.008066249 | 0.008596882 | 0.003069708 | 0.081435805 | 0.00799963 | 0.009155298 | 0.003069449 | 0.081417996 | 6.66E-05 | 0.0021672 | 1 | 0.109327018 | 1 | 0.124126174 | ENSG0000017 | 5 | VEGFB |
| SAT | T2DM | ENSG0000008 | 0.005328774 | 0.092545396 | 0.003167884 | 0.081436486 | 0.005191806 | 0.101213829 | 0.003167666 | 0.081418682 | 0.000136968 | 1.59E-06 | 1 | 0.414244809 | 1 | 0.440431278 | ENSG0000017 | 5 | VEGFB |
| SAT | T2DM | ENSG0000008 | 0.001496241 | 0.350235243 | 0.001601742 | 0.081436813 | 0.001577953 | 0.324539087 | 0.0016017 | 0.081418969 | -8.17E-05 | 8.02E-05 | 1 | 0.708896874 | 1 | 0.693504571 | ENSG0000017 | 5 | TNFRSF10D |
| SAT | T2DM | ENSG0000008 | 0.004095361 | 0.333326273 | 0.004233208 | 0.081436802 | 0.003962451 | 0.349216344 | 0.004232885 | 0.081418984 | 0.000132909 | 0.000113022 | 1 | 0.695221474 | 1 | 0.717008026 | ENSG0000017 | 11 | MST1 |
| SAT | T2DM | ENSG0000008 | -0.00426988 | 0.124463802 | 0.002779316 | 0.081436564 | -0.00432013 | 0.120066987 | 0.002779119 | 0.081418727 | 5.02E-05 | 0.039976227 | 1 | 0.463429598 | 1 | 0.467258033 | ENSG0000017 | 11 | MST1 |
| SAT | T2DM | ENSG0000008 | 0.000249047 | 0.941531054 | 0.003395525 | 0.081436958 | 0.000161752 | 0.962003197 | 0.0033929 | 0.081419131 | 8.73E-05 | 0.004138365 | 1 | 0.982870948 | 1 | 0.993672317 | ENSG0000017 | 9 | TNFRSF10C |
| SAT | T2DM | ENSG0000008 | 0.004906895 | 0.135805023 | 0.003289689 | 0.081436587 | 0.004812729 | 0.143442459 | 0.00328942 | 0.081418773 | 9.42E-05 | 0.000125815 | 1 | 0.478599658 | 1 | 0.501004535 | ENSG0000017 | 8 | GMPPB |
| SAT | T2DM | ENSG0000008 | -0.00418997 | 0.021985308 | 0.001829181 | 0.081436082 | -0.00412618 | 0.0240762 | 0.001829048 | 0.081418281 | -6.38E-05 | 5.08E-05 | 1 | 0.20149105 | 1 | 0.223974689 | ENSG0000017 | 7 | AP000769.1 |
| SAT | T2DM | ENSG0000008 | -0.00079899 | 0.709789807 | 0.002147024 | 0.081436935 | -0.00070073 | 0.744123363 | 0.002146866 | 0.081419113 | -9.83E-05 | 5.66E-08 | 1 | 0.907710061 | 1 | 0.927432137 | ENSG0000017 | 7 | AP000769.1 |
| SAT | T2DM | ENSG0000008 | -0.00227744 | 0.548894448 | 0.003799422 | 0.081436898 | -0.00221806 | 0.5593271 | 0.003799073 | 0.081419074 | -5.94E-05 | 0.008198562 | 1 | 0.83552458 | 1 | 0.842199272 | ENSG0000017 | 7 | AP000769.1 |
| SAT | T2DM | ENSG0000008 | 0.004565758 | 0.203465232 | 0.00359016 | 0.081436688 | 0.004639165 | 0.196259987 | 0.003589901 | 0.081418852 | -7.34E-05 | 0.018123938 | 1 | 0.565257769 | 1 | 0.564850016 | ENSG0000017 | 11 | HAP1 |
| SAT | T2DM | ENSG0000009 | -0.00110648 | 0.680818057 | 0.002689892 | 0.08143693 | -0.00103697 | 0.699843277 | 0.002689711 | 0.081419106 | -6.95E-05 | 0.004820659 | 1 | 0.892786811 | 1 | 0.909380512 | ENSG0000017 | 15 | RNF213 |
| SAT | T2DM | ENSG0000009 | 0.001859299 | 0.016034849 | 0.000772093 | 0.081435989 | 0.00184462 | 0.016882279 | 0.000772043 | 0.081418177 | 1.47E-05 | 0.046281474 | 1 | 0.162066172 | 1 | 0.179453096 | ENSG0000017 | 15 | RNF213 |
| SAT | T2DM | ENSG0000009 | -0.00185698 | 0.216955299 | 0.001504036 | 0.081436704 | -0.00189784 | 0.20697585 | 0.001503926 | 0.081418865 | 4.09E-05 | 0.001508697 | 1 | 0.581494104 | 1 | 0.579921232 | ENSG0000017 | 4 | GOLIM4 |
| SAT | T2DM | ENSG0000009 | -0.01019111 | 4.98E-09 | 0.001742702 | 0.081431244 | -0.01021837 | 4.52E-09 | 0.001742547 | 0.081413386 | 2.73E-05 | 0.014226059 | 1.63E-05 | 1.17E-06 | 1.48E-05 | 1.85E-06 | ENSG0000017 | 9 | HOXB2 |
| SAT | T2DM | ENSG0000009 | 0.000186728 | 0.900358484 | 0.001491337 | 0.081436956 | 0.000229713 | 0.877575629 | 0.001491225 | 0.081419127 | -4.30E-05 | 0.000506731 | 1 | 0.972055342 | 1 | 0.966700287 | ENSG0000017 | 9 | HOXB2 |
| SAT | T2DM | ENSG0000009 | 0.004221912 | 0.146062534 | 0.002904486 | 0.081436605 | 0.004156064 | 0.152424152 | 0.00290426 | 0.081418789 | 6.58E-05 | 0.004350513 | 1 | 0.492023791 | 1 | 0.512250482 | ENSG0000017 | 9 | HOXB2 |
| SAT | T2DM | ENSG0000009 | 0.002208713 | 0.466200182 | 0.003031131 | 0.0814367 | 0.00230518 | 0.446918645 | 0.003030887 | 0.081419034 | -9.65E-05 | 2.77E-05 | 1 | 0.79334291 | 1 | 0.791227469 | ENSG0000017 | 5 | NRROS |
| SAT | T2DM | ENSG0000009 | 0.002503272 | 0.17975424 | 0.001866005 | 0.081436658 | 0.002447611 | 0.189592497 | 0.001865859 | 0.081418843 | 5.57E-05 | 0.000162396 | 1 | 0.533249243 | 1 | 0.559400165 | ENSG0000017 | 7 | CEP19 |
| SAT | T2DM | ENSG0000009 | 0.004087484 | 0.230468858 | 0.003408641 | 0.081436718 | 0.004026824 | 0.237426619 | 0.003408389 | 0.081418896 | 6.07E-05 | 0.034566716 | 1 | 0.5943147 | 1 | 0.613383258 | ENSG0000017 | 7 | CEP19 |
| SAT | T2DM | ENSG0000009 | -0.00412879 | 0.000124452 | 0.001075987 | 0.081434498 | -0.00414476 | 0.000116993 | 0.001075898 | 0.081416651 | 1.60E-05 | 0.041875036 | 0.408327732 | 0.00600482 | 0.383854135 | 0.006618175 | ENSG0000017 | 7 | CEP19 |
| SAT | T2DM | ENSG0000009 | -0.00141315 | 0.474366507 | 0.001975342 | 0.081436873 | -0.00137395 | 0.486675374 | 0.001975185 | 0.08141905 | -3.92E-05 | 0.010193968 | 1 | 0.796607042 | 1 | 0.807270931 | ENSG0000017 | 7 | CEP19 |

| | | | | | | | | | | | | | | | | | | | |
|---|---|---|---|---|---|---|---|---|---|---|---|---|---|---|---|---|---|---|---|
| SAT | T2DM | ENSG000001( | -0.00052302 | 0.79310828 | 0.001994161 | 0.081436947 | -0.00047064 | 0.813408564 | 0.001993994 | 0.081419122 | -5.24E-05 | 0.000250998 | 1 | 0.94681813 | 1 | 0.955415085 | ENSG0000017 | 6 | CTSF |
| SAT | T2DM | ENSG000001( | -0.00661899 | 0.079641563 | 0.003776294 | 0.081436445 | -0.00655258 | 0.082683551 | 0.003775994 | 0.081418628 | -6.64E-05 | 0.02331988 | 1 | 0.394965061 | 1 | 0.412914353 | ENSG0000017 | 6 | CTSF |
| SAT | T2DM | ENSG000001( | 0.002767069 | 0.109341172 | 0.001728161 | 0.08143653 | 0.002720292 | 0.115434761 | 0.001728014 | 0.081418717 | 4.68E-05 | 9.97E-05 | 1 | 0.436210685 | 1 | 0.456640392 | ENSG0000017 | 6 | CTSF |
| SAT | T2DM | ENSG000001( | -0.0063078 | 0.032856623 | 0.002956083 | 0.081436197 | -0.00624875 | 0.034512961 | 0.002955844 | 0.081418384 | -5.90E-05 | 0.007979325 | 1 | 0.252465059 | 1 | 0.272172417 | ENSG0000017 | 6 | CTSF |
| SAT | T2DM | ENSG000001( | 0.001562925 | 0.5020967 | 0.002328571 | 0.081436883 | 0.001480339 | 0.524926264 | 0.002328418 | 0.081419063 | 8.26E-05 | 0.000145728 | 1 | 0.810314363 | 1 | 0.827311811 | ENSG0000017 | 6 | CTSF |
| SAT | T2DM | ENSG000001( | 0.000765623 | 0.761861352 | 0.002526494 | 0.081436943 | 0.000845671 | 0.737815619 | 0.002526294 | 0.081419112 | -8.00E-05 | 5.08E-05 | 1 | 0.93459819 | 1 | 0.925041991 | ENSG0000017 | 6 | CTSF |
| SAT | T2DM | ENSG000001( | -0.00285267 | 0.29427786 | 0.002719982 | 0.081436775 | -0.00278154 | 0.306442454 | 0.002719752 | 0.081418956 | -7.11E-05 | 0.000222667 | 1 | 0.651501794 | 1 | 0.674696375 | ENSG0000017 | 3 | TLR1 |
| SAT | T2DM | ENSG000001( | 0.00092278 | 0.733307547 | 0.002708251 | 0.081436939 | 0.000876361 | 0.746228125 | 0.002708018 | 0.081419113 | 4.64E-05 | 0.012270425 | 1 | 0.922893004 | 1 | 0.927766001 | ENSG0000017 | 8 | CTU2 |
| SAT | T2DM | ENSG000001( | -0.00349658 | 0.310871884 | 0.003450363 | 0.081436787 | -0.00342387 | 0.320998023 | 0.00345006 | 0.081418966 | -7.27E-05 | 0.001322424 | 1 | 0.671033323 | 1 | 0.691873008 | ENSG0000017 | 11 | AGAP8 |
| SAT | T2DM | ENSG000001( | 0.00339697 | 0.125861596 | 0.002219328 | 0.081436567 | 0.003357329 | 0.130306254 | 0.002219138 | 0.081418748 | 3.96E-05 | 0.009241096 | 1 | 0.464512818 | 1 | 0.480064491 | ENSG0000017 | 11 | AGAP8 |
| SAT | T2DM | ENSG000001( | -0.00158877 | 0.598544604 | 0.003017631 | 0.081436912 | -0.00166194 | 0.581778854 | 0.003017379 | 0.08141908 | 7.32E-05 | 0.000768178 | 1 | 0.850140626 | 1 | 0.851769934 | ENSG0000017 | 4 | SNX31 |
| SAT | T2DM | ENSG000001( | -0.00752834 | 0.006070312 | 0.002743592 | 0.0814357 | -0.00756838 | 0.005801349 | 0.002743343 | 0.081417859 | 4.00E-05 | 0.017423724 | 1 | 0.085847824 | 1 | 0.091952785 | ENSG0000017 | 4 | SNX31 |
| SAT | T2DM | ENSG000001( | 0.00321104 | 0.014791751 | 0.001317379 | 0.081435966 | 0.003187387 | 0.015533534 | 0.001317263 | 0.081418153 | 2.37E-05 | 0.006868315 | 1 | 0.15455967 | 1 | 0.171731497 | ENSG0000017 | 4 | SNX31 |
| SAT | T2DM | ENSG000001( | -0.00174237 | 0.600190474 | 0.003324335 | 0.081436913 | -0.00166004 | 0.617497663 | 0.003324071 | 0.081419089 | -8.23E-05 | 0.001372817 | 1 | 0.851004731 | 1 | 0.869905467 | ENSG0000017 | 4 | SNX31 |
| SAT | T2DM | ENSG000001( | -0.00361263 | 0.069506876 | 0.001990305 | 0.081436408 | -0.00355765 | 0.073838535 | 0.001990166 | 0.081418597 | -5.50E-05 | 0.001817539 | 1 | 0.366054671 | 1 | 0.395413696 | ENSG0000017 | 4 | SNX31 |
| SAT | T2DM | ENSG000001( | 0.005852071 | 0.084875162 | 0.00339629 | 0.081436462 | 0.005790824 | 0.08816031 | 0.003396007 | 0.081418645 | 6.12E-05 | 0.012113422 | 1 | 0.401115612 | 1 | 0.421383296 | ENSG0000017 | 4 | SNX31 |
| SAT | T2DM | ENSG000001( | -0.00044745 | 0.912792246 | 0.004085639 | 0.081436956 | -0.0005497 | 0.892963548 | 0.0040853 | 0.081419128 | 0.000102251 | 0.000570904 | 1 | 0.976164068 | 1 | 0.972100523 | ENSG0000017 | 13 | STAG3L3 |
| SAT | T2DM | ENSG000001( | 0.00568754 | 0.026653688 | 0.002565932 | 0.081436137 | 0.005646308 | 0.027759904 | 0.002565723 | 0.081418322 | 4.12E-05 | 0.031788339 | 1 | 0.225970935 | 1 | 0.242234694 | ENSG0000017 | 13 | STAG3L3 |
| SAT | T2DM | ENSG000001( | -0.00542258 | 0.031648798 | 0.002523509 | 0.081436187 | -0.00537536 | 0.033149247 | 0.002523316 | 0.081418373 | -4.72E-05 | 0.021364391 | 1 | 0.248186538 | 1 | 0.267755401 | ENSG0000017 | 13 | STAG3L3 |
| SAT | T2DM | ENSG000001( | 0.003709085 | 0.363441036 | 0.00408117 | 0.08143682 | 0.002866122 | 0.483547871 | 0.004090906 | 0.081419049 | 0.000842962 | 0.003471375 | 1 | 0.721005279 | 1 | 0.805749399 | ENSG0000017 | 13 | STAG3L3 |
| SAT | T2DM | ENSG000001( | 0.001881927 | 0.103880525 | 0.001157163 | 0.081436516 | 0.001849736 | 0.10991478 | 0.001157113 | 0.081418704 | 3.22E-05 | 0.01534278 | 1 | 0.429259448 | 1 | 0.452205306 | ENSG0000017 | 13 | STAG3L3 |
| SAT | T2DM | ENSG000001( | 0.002314448 | 0.343086375 | 0.00244118 | 0.081436808 | 0.002369632 | 0.331663372 | 0.002440983 | 0.081418973 | -5.52E-05 | 0.002742976 | 1 | 0.703910998 | 1 | 0.700003408 | ENSG0000017 | 9 | LIG4 |
| SAT | T2DM | ENSG000001( | 0.00084371 | 0.362022864 | 0.00092561 | 0.08143682 | 0.000814999 | 0.378568664 | 0.00092557 | 0.081419001 | 2.87E-05 | 0.007104311 | 1 | 0.720313533 | 1 | 0.738661557 | ENSG0000017 | 8 | SNAPC5 |
| SAT | T2DM | ENSG000001( | -0.00427582 | 0.193680168 | 0.003289669 | 0.081436676 | -0.00420703 | 0.200912787 | 0.00328942 | 0.081418858 | -6.88E-05 | 0.010748714 | 1 | 0.553257876 | 1 | 0.570731475 | ENSG0000017 | 6 | TMEM81 |
| SAT | T2DM | ENSG000001( | 0.004204164 | 0.23983732 | 0.00357681 | 0.081436728 | 0.004154326 | 0.245414671 | 0.003576505 | 0.081418905 | 4.98E-05 | 0.044703556 | 1 | 0.60584911 | 1 | 0.620663712 | ENSG0000017 | 13 | ZNF266 |
| SAT | T2DM | ENSG000001( | 0.002979638 | 0.119488651 | 0.001913795 | 0.081436553 | 0.003015025 | 0.115133491 | 0.001913651 | 0.081418716 | -3.54E-05 | 0.024893237 | 1 | 0.45480541 | 1 | 0.456639917 | ENSG0000017 | 13 | ZNF266 |
| SAT | T2DM | ENSG000001( | -0.00142557 | 0.583766914 | 0.002601928 | 0.081436908 | -0.00152419 | 0.55798635 | 0.002601744 | 0.081419074 | 9.86E-05 | 1.75E-05 | 1 | 0.845251212 | 1 | 0.842050121 | ENSG0000017 | 13 | ZNF266 |
| SAT | T2DM | ENSG000001( | 0.002817609 | 0.024997846 | 0.001257051 | 0.081436119 | 0.0028405 | 0.023831578 | 0.001256947 | 0.081418278 | -2.29E-05 | 0.012343964 | 1 | 0.216409969 | 1 | 0.223866198 | ENSG0000017 | 13 | ZNF266 |
| SAT | T2DM | ENSG000001( | 0.001362185 | 0.082026278 | 0.000783292 | 0.081436453 | 0.00134752 | 0.085348308 | 0.000783229 | 0.081418636 | 1.47E-05 | 0.01486962 | 1 | 0.395495266 | 1 | 0.415329589 | ENSG0000017 | 13 | ZNF266 |
| SAT | T2DM | ENSG000001( | 0.005471884 | 0.055000378 | 0.002851606 | 0.081436343 | 0.005421621 | 0.0572498 | 0.002851379 | 0.081418527 | 5.03E-05 | 0.021891621 | 1 | 0.325754403 | 1 | 0.345923743 | ENSG0000017 | 13 | ZNF266 |
| SAT | T2DM | ENSG000001( | -0.00233174 | 0.113601067 | 0.00147372 | 0.08143654 | -0.00215099 | 0.144486289 | 0.001473996 | 0.081418775 | -0.00018076 | 4.75E-07 | 1 | 0.443192746 | 1 | 0.501004535 | ENSG0000017 | 7 | SH3PXD2B |
| SAT | T2DM | ENSG000001( | 0.003249445 | 0.146864588 | 0.002239918 | 0.081436607 | 0.003195329 | 0.153677204 | 0.002239719 | 0.081418791 | 5.41E-05 | 0.000166222 | 1 | 0.492391981 | 1 | 0.513457135 | ENSG0000017 | 7 | KIAA1551 |
| SAT | T2DM | ENSG000001( | 0.002378202 | 0.297778571 | 0.002284075 | 0.081436777 | 0.002427146 | 0.287909598 | 0.0022839 | 0.081418942 | -4.89E-05 | 0.008203362 | 1 | 0.654394836 | 1 | 0.653237213 | ENSG0000017 | 12 | HRAS |
| SAT | T2DM | ENSG000001( | -0.00442286 | 0.128268729 | 0.002907924 | 0.081436572 | -0.00437735 | 0.132212223 | 0.002907695 | 0.081418752 | -4.55E-05 | 0.045549739 | 1 | 0.468889004 | 1 | 0.485221815 | ENSG0000017 | 11 | SRP72 |
| SAT | T2DM | ENSG000001( | -0.00528458 | 0.169311862 | 0.003844954 | 0.081436643 | -0.00537862 | 0.161819277 | 0.00384467 | 0.081418804 | 9.40E-05 | 0.003790121 | 1 | 0.523170556 | 1 | 0.520626624 | ENSG0000017 | 7 | BTC |
| SAT | T2DM | ENSG000001( | -0.00168391 | 0.505393656 | 0.002528288 | 0.081436884 | -0.00159308 | 0.528598819 | 0.002528107 | 0.081419065 | -9.08E-05 | 3.51E-05 | 1 | 0.810314363 | 1 | 0.82995678 | ENSG0000017 | 7 | BTC |
| SAT | T2DM | ENSG000001( | -0.00462803 | 0.090897337 | 0.002737367 | 0.081436481 | -0.0046682 | 0.088100572 | 0.002737134 | 0.081418645 | 4.02E-05 | 0.035003359 | 1 | 0.413867045 | 1 | 0.421383296 | ENSG0000017 | 7 | BTC |
| SAT | T2DM | ENSG000001( | 0.005508281 | 0.130850279 | 0.003646033 | 0.081436577 | 0.005570517 | 0.126523543 | 0.003645726 | 0.081418741 | -6.22E-05 | 0.016433704 | 1 | 0.47229897 | 1 | 0.47645145 | ENSG0000017 | 10 | YIF1A |
| SAT | T2DM | ENSG000001( | 0.000765019 | 0.743204578 | 0.00233513 | 0.08143694 | 0.00081546 | 0.726908407 | 0.002334947 | 0.081419111 | -5.04E-05 | 0.00592017 | 1 | 0.927272852 | 1 | 0.921455409 | ENSG0000017 | 8 | RSRC1 |
| SAT | T2DM | ENSG000001( | -0.00418772 | 0.030714194 | 0.001938077 | 0.081436178 | -0.00416064 | 0.031796168 | 0.001937914 | 0.081418361 | -2.71E-05 | 0.047948651 | 1 | 0.244489871 | 1 | 0.263779676 | ENSG0000017 | 8 | RSRC1 |
| SAT | T2DM | ENSG000001( | -0.00392481 | 0.035726848 | 0.00186893 | 0.081436221 | -0.0039579 | 0.034181784 | 0.001868761 | 0.081418381 | 3.31E-05 | 0.004170815 | 1 | 0.263415251 | 1 | 0.271550686 | ENSG0000017 | 7 | PTDSS2 |
| SAT | T2DM | ENSG000001( | 0.002672621 | 0.264415786 | 0.002394787 | 0.08143675 | 0.002755733 | 0.249811487 | 0.002394606 | 0.08141891 | -8.31E-05 | 2.40E-05 | 1 | 0.627688499 | 1 | 0.621386978 | ENSG0000017 | 7 | PTDSS2 |
| SAT | T2DM | ENSG000001( | 0.008719664 | 0.031160282 | 0.004046199 | 0.081436182 | 0.008638351 | 0.032753128 | 0.004045872 | 0.081418369 | 8.13E-05 | 0.007799825 | 1 | 0.245761744 | 1 | 0.267755401 | ENSG0000017 | 8 | FBXW8 |
| SAT | T2DM | ENSG000001( | 0.003170459 | 0.426865773 | 0.003990172 | 0.081436853 | 0.003260053 | 0.41388345 | 0.003989885 | 0.081419019 | -8.96E-05 | 0.009638767 | 1 | 0.771499322 | 1 | 0.766308941 | ENSG0000017 | 7 | PACS1 |
| SAT | T2DM | ENSG000001( | -0.00101483 | 0.666664892 | 0.002356081 | 0.081436927 | -0.00095958 | 0.683781448 | 0.002355914 | 0.081419103 | -5.53E-05 | 0.00744263 | 1 | 0.888630991 | 1 | 0.901372745 | ENSG0000017 | 7 | PACS1 |
| SAT | T2DM | ENSG000001( | -0.00200807 | 0.574746527 | 0.003578962 | 0.081436906 | -0.00189761 | 0.59593963 | 0.003578708 | 0.081419084 | -0.00011046 | 0.000433923 | 1 | 0.841779887 | 1 | 0.854809759 | ENSG0000017 | 7 | PACS1 |
| SAT | T2DM | ENSG000001( | 0.000591495 | 0.692398841 | 0.001495178 | 0.081436932 | 0.000617115 | 0.679774276 | 0.001495052 | 0.081419102 | -2.56E-05 | 0.01539197 | 1 | 0.900268043 | 1 | 0.90041962 | ENSG0000017 | 6 | MARCKSL1 |
| SAT | T2DM | ENSG000001( | -0.00773121 | 0.019057002 | 0.003297706 | 0.08143604 | -0.00781009 | 0.017859152 | 0.003297439 | 0.081418194 | 7.89E-05 | 0.001565284 | 1 | 0.181761693 | 1 | 0.184263769 | ENSG0000017 | 2 | ZNF408 |
| SAT | T2DM | ENSG000001( | 0.005347363 | 0.130241569 | 0.003533922 | 0.081436576 | 0.005297411 | 0.133836193 | 0.003533617 | 0.081418755 | 5.00E-05 | 0.038123582 | 1 | 0.472179657 | 1 | 0.486825442 | ENSG0000017 | 7 | ARHGAP1 |
| SAT | T2DM | ENSG000001( | -0.00134706 | 0.422130484 | 0.001678093 | 0.081436851 | -0.00137714 | 0.411799749 | 0.001677946 | 0.081419018 | 3.01E-05 | 0.006325256 | 1 | 0.766045419 | 1 | 0.765504236 | ENSG0000017 | 7 | ARHGAP1 |
| SAT | T2DM | ENSG000001( | 0.003696298 | 0.163838832 | 0.002654856 | 0.081436634 | 0.004361966 | 0.101209925 | 0.002661327 | 0.081418682 | -0.00066567 | 0.000447192 | 1 | 0.515393296 | 1 | 0.440431278 | ENSG0000017 | 7 | ARHGAP1 |
| SAT | T2DM | ENSG000001( | 0.004437406 | 0.282885469 | 0.004132197 | 0.081436766 | 0.004571569 | 0.268548387 | 0.004131883 | 0.081418926 | -0.00013416 | 7.54E-05 | 1 | 0.641542051 | 1 | 0.639410201 | ENSG0000017 | 7 | DOLK |
| SAT | T2DM | ENSG000001( | -0.00115292 | 0.39918291 | 0.001367508 | 0.08143684 | -0.00117715 | 0.389308102 | 0.001367393 | 0.081419007 | 2.42E-05 | 0.011878918 | 1 | 0.750125503 | 1 | 0.749514821 | ENSG0000017 | 12 | CCNE2 |
| SAT | T2DM | ENSG000001( | 0.001115034 | 0.642871421 | 0.002404706 | 0.081436922 | 0.000205306 | 0.932847643 | 0.002436502 | 0.08141913 | 0.000909727 | 0.020929364 | 1 | 0.877329554 | 1 | 0.983506786 | ENSG0000017 | 3 | BANF1 |
| SAT | T2DM | ENSG000001( | 0.002572068 | 0.335214987 | 0.002669051 | 0.081436803 | 0.002626601 | 0.32503298 | 0.002668851 | 0.081418969 | -5.45E-05 | 0.014019711 | 1 | 0.695221474 | 1 | 0.693840734 | ENSG0000017 | 3 | BANF1 |
| SAT | T2DM | ENSG000001( | 0.000614063 | 0.477427052 | 0.000864332 | 0.081436874 | 0.000639448 | 0.459379235 | 0.00086427 | 0.081419039 | -2.54E-05 | 0.000745648 | 1 | 0.796607042 | 1 | 0.795972372 | ENSG0000017 | 3 | BANF1 |
| SAT | T2DM | ENSG000001( | -0.00964626 | 0.001572363 | 0.003051645 | 0.081435289 | -0.00958312 | 0.001686433 | 0.003051418 | 0.081417483 | -6.31E-05 | 0.013540319 | 1 | 0.035825853 | 1 | 0.043914172 | ENSG0000017 | 12 | CHRNA7 |
| SAT | T2DM | ENSG000001( | 0.00307485 | 0.241422055 | 0.002624844 | 0.081436729 | 0.003121092 | 0.234378419 | 0.002624626 | 0.081418895 | -4.62E-05 | 0.015538495 | 1 | 0.607040297 | 1 | 0.610313962 | ENSG0000017 | 8 | SCUBE2 |
| SAT | T2DM | ENSG000001( | 0.001938109 | 0.388611205 | 0.002248023 | 0.081436834 | 0.001894994 | 0.399210895 | 0.002247831 | 0.081419012 | 4.31E-05 | 0.005531182 | 1 | 0.739604086 | 1 | 0.756241886 | ENSG0000017 | 11 | SMAD2 |
| SAT | T2DM | ENSG000001( | 0.005341395 | 0.098385777 | 0.003231855 | 0.081436502 | 0.00538593 | 0.095583039 | 0.003231575 | 0.081418667 | -4.45E-05 | 0.040642342 | 1 | 0.421965667 | 1 | 0.432044519 | ENSG0000017 | 11 | SMAD2 |
| SAT | T2DM | ENSG000001( | 0.00113986 | 0.476533195 | 0.001601175 | 0.081436874 | 0.001092808 | 0.494885609 | 0.001601045 | 0.081419053 | 4.71E-05 | 8.90E-05 | 1 | 0.796607042 | 1 | 0.813894578 | ENSG0000017 | 11 | SMAD2 |
| SAT | T2DM | ENSG000001( | -0.00089804 | 0.745278243 | 0.002764282 | 0.081436941 | -0.00095766 | 0.728988931 | 0.002764055 | 0.081419111 | 5.96E-05 | 0.003479054 | 1 | 0.927287795 | 1 | 0.922641931 | ENSG0000017 | 8 | EIF3F |
| SAT | T2DM | ENSG000001( | -0.0066965 | 0.014632067 | 0.002742928 | 0.081435962 | -0.00659582 | 0.016180166 | 0.002742735 | 0.081418165 | -0.00010068 | 3.12E-05 | 1 | 0.153379588 | 1 | 0.176369183 | ENSG0000017 | 8 | EIF3F |
| SAT | T2DM | ENSG000001( | 0.001571814 | 0.48829834 | 0.00226807 | 0.081436878 | 0.001341945 | 0.554357686 | 0.0022697 | 0.081419072 | 0.000229869 | 0.012762135 | 1 | 0.80306108 | 1 | 0.842050121 | ENSG0000017 | 8 | EIF3F |
| SAT | T2DM | ENSG000001( | 0.011228556 | 0.003015359 | 0.003785524 | 0.081435488 | 0.01117446 | 0.003155794 | 0.003785187 | 0.081417675 | 5.41E-05 | 0.025276807 | 1 | 0.054659637 | 1 | 0.064643525 | ENSG0000017 | 8 | EIF3F |
| SAT | T2DM | ENSG000001( | -0.00428473 | 0.180610167 | 0.003200231 | 0.081436659 | -0.00417852 | 0.191643138 | 0.003200127 | 0.081418846 | -0.00010621 | 0.007405457 | 1 | 0.534339005 | 1 | 0.560852203 | ENSG0000017 | 8 | EIF3F |
| SAT | T2DM | ENSG000001( | 0.002207998 | 0.195984618 | 0.001707552 | 0.081436679 | 0.002144773 | 0.209067627 | 0.001707439 | 0.081418867 | 6.32E-05 | 6.79E-05 | 1 | 0.55576969 | 1 | 0.579977552 | ENSG0000017 | 8 | EIF3F |
| SAT | T2DM | ENSG000001( | 0.001822364 | 0.576964502 | 0.003266913 | 0.081436906 | 0.001590591 | 0.62635686 | 0.00326703 | 0.081419091 | 0.000231773 | 3.16E-05 | 1 | 0.842502133 | 1 | 0.876644981 | ENSG0000017 | 11 | ZNF25 |
| SAT | T2DM | ENSG000001( | 0.00064898 | 0.615012552 | 0.001290395 | 0.081436916 | 0.00066697 | 0.605214606 | 0.001290286 | 0.081419086 | -1.80E-05 | 0.04827489 | 1 | 0.859969008 | 1 | 0.858870727 | ENSG0000017 | 11 | ZNF25 |
| SAT | T2DM | ENSG0000011 | -0.00207506 | 0.235587094 | 0.001749493 | 0.081436723 | -0.00057075 | 0.750044753 | 0.00179153 | 0.081419114 | -0.00150431 | 0.000100075 | 1 | 0.600737047 | 1 | 0.928640315 | ENSG0000017 | 8 | CLTB |
| SAT | T2DM | ENSG0000011 | 0.001530319 | 0.591559521 | 0.002851999 | 0.08143691 | 0.001477512 | 0.604384804 | 0.002851759 | 0.081419086 | 5.28E-05 | 0.009099436 | 1 | 0.846620707 | 1 | 0.858807511 | ENSG0000017 | 8 | CLTB |
| SAT | T2DM | ENSG0000011 | 0.002304389 | 0.308135397 | 0.002261104 | 0.081436785 | 0.002250251 | 0.319600384 | 0.002260922 | 0.081418965 | 5.41E-05 | 0.001609863 | 1 | 0.66864566 | 1 | 0.690783176 | ENSG0000017 | 8 | CLTB |
| SAT | T2DM | ENSG0000011 | -0.00036388 | 0.921413397 | 0.00368843 | 0.081436957 | -0.00058669 | 0.873620364 | 0.003688453 | 0.081419127 | 0.000222815 | 7.22E-05 | 1 | 0.978648294 | 1 | 0.966700287 | ENSG0000017 | 10 | LPL |
| SAT | T2DM | ENSG0000011 | -0.00190701 | 0.564387266 | 0.003308849 | 0.081436903 | -0.00184596 | 0.576890148 | 0.003308573 | 0.081419079 | -6.11E-05 | 0.010274263 | 1 | 0.837959912 | 1 | 0.849979671 | ENSG0000017 | 10 | LPL |
| SAT | T2DM | ENSG0000011 | 0.00734804 | 0.040316832 | 0.003583547 | 0.081436256 | 0.007406397 | 0.038740109 | 0.003583262 | 0.081418417 | -5.84E-05 | 0.035574423 | 1 | 0.276735412 | 1 | 0.283087523 | ENSG0000017 | 10 | LPL |
| SAT | T2DM | ENSG0000011 | -0.00243897 | 0.226180923 | 0.002015246 | 0.081436714 | -0.00248197 | 0.218063403 | 0.002015082 | 0.081418877 | 4.30E-05 | 0.004294011 | 1 | 0.591314429 | 1 | 0.591783312 | ENSG0000017 | 10 | LPL |
| SAT | T2DM | ENSG0000011 | -0.00033317 | 0.712506874 | 0.000904144 | 0.081436936 | -0.00029083 | 0.747713421 | 0.000904162 | 0.081419114 | -4.23E-05 | 0.003597898 | 1 | 0.908107919 | 1 | 0.927854665 | ENSG0000017 | 7 | TBC1D10C |
| SAT | T2DM | ENSG0000011 | 4.90E-05 | 0.978711103 | 0.001836357 | 0.081436958 | 7.80E-05 | 0.966129844 | 0.001836201 | 0.081419131 | -2.90E-05 | 0.023752474 | 1 | 0.995203291 | 1 | 0.994313682 | ENSG0000017 | 7 | TBC1D10C |
| SAT | T2DM | ENSG0000011 | -0.00044598 | 0.629360142 | 0.000924068 | 0.081436919 | -0.00011903 | 0.897650951 | 0.000925388 | 0.081419128 | -0.00032695 | 1.81E-10 | 1 | 0.868347614 | 1 | 0.972878089 | ENSG0000017 | 7 | TBC1D10C |
| SAT | T2DM | ENSG0000011 | 0.004988234 | 0.181422233 | 0.003732617 | 0.08143666 | 0.004919268 | 0.187497649 | 0.003732317 | 0.081418841 | 6.90E-05 | 0.015522537 | 1 | 0.535187534 | 1 | 0.557313872 | ENSG0000017 | 9 | LRRC25 |
| SAT | T2DM | ENSG0000011 | 0.003953718 | 0.041597075 | 0.001940442 | 0.081436265 | 0.003909584 | 0.043911346 | 0.001940297 | 0.081418453 | 4.41E-05 | 0.006093114 | 1 | 0.28140207 | 1 | 0.303311846 | ENSG0000017 | 8 | PAAF1 |
| SAT | T2DM | ENSG0000011 | 0.00166483 | 0.371310548 | 0.001862181 | 0.081436825 | 0.001845278 | 0.321713667 | 0.001862143 | 0.081418967 | -0.00018045 | 4.15E-13 | 1 | 0.725888698 | 1 | 0.691873008 | ENSG0000017 | 8 | PAAF1 |
| SAT | T2DM | ENSG0000011 | 0.000294276 | 0.872771889 | 0.001837631 | 0.081436954 | 0.000333508 | 0.855972237 | 0.00183747 | 0.081419125 | -3.92E-05 | 0.001287083 | 1 | 0.966468088 | 1 | 0.965566132 | ENSG0000017 | 8 | PAAF1 |
| SAT | T2DM | ENSG0000011 | -0.00143101 | 0.555477546 | 0.002427201 | 0.0814369 | -0.00151448 | 0.532620637 | 0.002427009 | 0.081419066 | 8.35E-05 | 1.01E-05 | 1 | 0.835948344 | 1 | 0.83186484 | ENSG0000017 | 8 | PAAF1 |
| SAT | T2DM | ENSG0000011 | 0.005236458 | 0.161489437 | 0.003740108 | 0.081436631 | 0.005390557 | 0.14948782 | 0.003739949 | 0.081418784 | -0.0001541 | 0.000369587 | 1 | 0.512811064 | 1 | 0.509843595 | ENSG0000017 | 8 | PAAF1 |
| SAT | T2DM | ENSG0000011 | 0.004798402 | 0.232829823 | 0.004021786 | 0.081436721 | 0.004276004 | 0.28795438 | 0.004024016 | 0.081418942 | 0.000522399 | 0.000364194 | 1 | 0.598143699 | 1 | 0.653237213 | ENSG0000017 | 9 | MRPL48 |
| SAT | T2DM | ENSG0000011 | -0.00077948 | 0.6799084 | 0.001889249 | 0.08143693 | -0.00070536 | 0.708866868 | 0.001889133 | 0.081419108 | -7.41E-05 | 5.78E-05 | 1 | 0.892786811 | 1 | 0.912941362 | ENSG0000017 | 9 | MRPL48 |
| SAT | T2DM | ENSG0000011 | 0.000121882 | 0.946069253 | 0.001801818 | 0.081436958 | 7.78E-05 | 0.96557705 | 0.001801688 | 0.081419131 | 4.41E-05 | 0.00457127 | 1 | 0.984788457 | 1 | 0.99427016 | ENSG0000017 | 9 | MRPL48 |
| SAT | T2DM | ENSG0000011 | -0.00541746 | 0.103612652 | 0.00332852 | 0.081436516 | -0.00534938 | 0.107995516 | 0.003328235 | 0.081418699 | -6.81E-05 | 0.002897655 | 1 | 0.429233725 | 1 | 0.450218948 | ENSG0000017 | 15 | RAB6A |
| SAT | T2DM | ENSG0000011 | 0.001860454 | 0.292778978 | 0.001768415 | 0.081436773 | 0.00191549 | 0.278695358 | 0.001768271 | 0.081418935 | -5.50E-05 | 3.17E-05 | 1 | 0.649937638 | 1 | 0.644397089 | ENSG0000017 | 15 | RAB6A |
| SAT | T2DM | ENSG0000011 | -0.00189425 | 0.177247647 | 0.001403896 | 0.081436654 | -0.00125896 | 0.371386951 | 0.001408425 | 0.081418997 | -0.00063529 | 3.08E-08 | 1 | 0.530526907 | 1 | 0.734641671 | ENSG0000017 | 7 | RMI2 |
| SAT | T2DM | ENSG0000011 | -0.00075265 | 0.756767055 | 0.00243001 | 0.081436942 | -0.00338619 | 0.173785069 | 0.002489585 | 0.081418822 | 0.002633548 | 1.21E-06 | 1 | 0.93284139 | 1 | 0.532948952 | ENSG0000017 | 7 | RMI2 |
| SAT | T2DM | ENSG0000011 | -0.00027238 | 0.943512019 | 0.003844136 | 0.081436958 | -0.00018947 | 0.960685639 | 0.003843812 | 0.08141913 | -8.29E-05 | 0.00240749 | 1 | 0.98331034 | 1 | 0.993672317 | ENSG0000017 | 7 | RMI2 |
| SAT | T2DM | ENSG0000011 | 0.00239346 | 0.359918194 | 0.002614313 | 0.081436818 | 0.002353182 | 0.368019589 | 0.002614094 | 0.081418995 | 4.03E-05 | 0.032287418 | 1 | 0.71830389 | 1 | 0.732878736 | ENSG0000017 | 7 | RMI2 |
| SAT | T2DM | ENSG0000011 | -0.00168582 | 0.428518586 | 0.002129289 | 0.081436854 | -0.00164509 | 0.439718378 | 0.002129104 | 0.081419031 | -4.07E-05 | 0.004577104 | 1 | 0.771792681 | 1 | 0.786806474 | ENSG0000017 | 13 | TOM1L2 |
| SAT | T2DM | ENSG0000011 | -0.00088131 | 0.489829728 | 0.001276192 | 0.081436879 | -0.00083087 | 0.514981752 | 0.001276094 | 0.08141906 | -5.04E-05 | 9.88E-07 | 1 | 0.80382562 | 1 | 0.824105482 | ENSG0000017 | 7 | BAK1P1 |
| SAT | T2DM | ENSG0000011 | 0.002256976 | 0.532124743 | 0.003612512 | 0.081436893 | 0.0023932 | 0.507633338 | 0.003612226 | 0.081419058 | -0.00013622 | 1.32E-06 | 1 | 0.827049399 | 1 | 0.817738235 | ENSG0000017 | 10 | TTLL11 |
| SAT | T2DM | ENSG0000011 | 0.000637359 | 0.5707192 | 0.001124103 | 0.081436905 | 0.000619383 | 0.581601281 | 0.001124011 | 0.08141908 | 1.80E-05 | 0.02974585 | 1 | 0.840736467 | 1 | 0.851769934 | ENSG0000017 | 7 | CTDNEP1 |
| SAT | T2DM | ENSG0000011 | 0.002052223 | 0.005777786 | 0.00074352 | 0.081435685 | 0.002036605 | 0.006155928 | 0.000743457 | 0.081417877 | 1.56E-05 | 0.003069502 | 1 | 0.083144374 | 1 | 0.095234728 | ENSG0000017 | 7 | CTDNEP1 |
| SAT | T2DM | ENSG0000011 | -0.00207971 | 0.547938226 | 0.003461268 | 0.081436898 | -0.00200103 | 0.563155009 | 0.003461024 | 0.081419075 | -7.87E-05 | 0.010120177 | 1 | 0.835027823 | 1 | 0.84223981 | ENSG0000017 | 8 | ETV4 |
| SAT | T2DM | ENSG0000011 | -0.0045331 | 0.126526263 | 0.002966793 | 0.081436568 | -0.00459501 | 0.121397835 | 0.002966558 | 0.08141873 | 6.19E-05 | 0.007328224 | 1 | 0.464874211 | 1 | 0.469700823 | ENSG0000017 | 8 | ETV4 |
| SAT | T2DM | ENSG0000011 | -0.00203997 | 0.607884206 | 0.003975805 | 0.081436914 | -0.00211027 | 0.595541988 | 0.003975481 | 0.081419084 | 7.03E-05 | 0.018225932 | 1 | 0.855260755 | 1 | 0.854809759 | ENSG0000017 | 7 | SWI5 |
| SAT | T2DM | ENSG0000011 | 0.00161285 | 0.455683358 | 0.002162073 | 0.081436865 | 0.001526809 | 0.480072083 | 0.002162045 | 0.081419048 | 8.60E-05 | 0.004234481 | 1 | 0.786411516 | 1 | 0.80363893 | ENSG0000017 | 7 | HOXD8 |
| SAT | T2DM | ENSG0000011 | 0.003052195 | 0.403595472 | 0.00365437 | 0.081436842 | 0.00311837 | 0.393441055 | 0.003654078 | 0.081419009 | -6.62E-05 | 0.018700037 | 1 | 0.753240469 | 1 | 0.753672849 | ENSG0000017 | 4 | AC127496.1 |
| SAT | T2DM | ENSG0000011 | 0.003624923 | 0.101508492 | 0.002213576 | 0.08143651 | 0.003686567 | 0.095798647 | 0.002213389 | 0.081418667 | -6.16E-05 | 8.77E-05 | 1 | 0.425727224 | 1 | 0.432044519 | ENSG0000017 | 6 | ORAI3 |
| SAT | T2DM | ENSG0000011 | -0.00576183 | 0.025763638 | 0.002584053 | 0.081436128 | -0.00568715 | 0.027732923 | 0.002583836 | 0.081418321 | -7.47E-05 | 4.85E-05 | 1 | 0.220131498 | 1 | 0.242234694 | ENSG0000017 | 12 | ZNF613 |
| SAT | T2DM | ENSG0000011 | -0.00085888 | 0.601830072 | 0.001646082 | 0.081436913 | -0.00092936 | 0.572338621 | 0.001646018 | 0.081419078 | 7.05E-05 | 0.00031209 | 1 | 0.851489636 | 1 | 0.847525062 | ENSG0000017 | 12 | ZNF613 |
| SAT | T2DM | ENSG0000011 | -0.00712228 | 8.48E-06 | 0.001599492 | 0.081433645 | -0.00709451 | 9.17E-06 | 0.00159936 | 0.081415843 | -2.78E-05 | 0.017239456 | 0.027810273 | 0.000596732 | 0.030097647 | 0.000734089 | ENSG0000017 | 12 | ZNF613 |
| SAT | T2DM | ENSG0000011 | 0.002906816 | 0.418489148 | 0.003592894 | 0.081436849 | 0.00296399 | 0.409357615 | 0.003592605 | 0.081419017 | -5.72E-05 | 0.03739799 | 1 | 0.763661232 | 1 | 0.761827756 | ENSG0000017 | 12 | ZNF613 |
| SAT | T2DM | ENSG0000011 | -0.00413588 | 0.000451391 | 0.001178957 | 0.081434902 | -0.00407721 | 0.000543105 | 0.001178872 | 0.081417132 | -5.87E-05 | 5.40E-09 | 1 | 0.014727922 | 1 | 0.019368778 | ENSG0000017 | 8 | TMEM39A |
| SAT | T2DM | ENSG0000011 | 0.001126722 | 0.55318148 | 0.001900037 | 0.0814369 | 0.001093243 | 0.56500147 | 0.001899875 | 0.081419076 | 3.35E-05 | 0.011001557 | 1 | 0.835948344 | 1 | 0.84223981 | ENSG0000017 | 11 | TCP11L1 |
| SAT | T2DM | ENSG0000011 | 0.00842396 | 0.0272825 | 0.003816135 | 0.081436144 | 0.008266059 | 0.030293482 | 0.003815859 | 0.081418347 | 0.000157901 | 1.50E-06 | 1 | 0.227073047 | 1 | 0.256527252 | ENSG0000017 | 11 | TCP11L1 |
| SAT | T2DM | ENSG0000011 | -0.0020267 | 0.373702515 | 0.002278315 | 0.081436826 | -0.00209055 | 0.358795367 | 0.002278124 | 0.08141899 | 6.39E-05 | 8.88E-05 | 1 | 0.72766644 | 1 | 0.725902637 | ENSG0000017 | 11 | TCP11L1 |
| SAT | T2DM | ENSG0000011 | 0.006004917 | 0.060226932 | 0.003195578 | 0.081436368 | 0.005649331 | 0.077171053 | 0.003196507 | 0.081418609 | 0.000355586 | 8.27E-05 | 1 | 0.342468915 | 1 | 0.404470007 | ENSG0000017 | 11 | TCP11L1 |
| SAT | T2DM | ENSG0000011 | 0.00172283 | 0.619161971 | 0.003466188 | 0.081436917 | 0.001654229 | 0.633157232 | 0.003465892 | 0.081419093 | 6.86E-05 | 0.004198373 | 1 | 0.862986588 | 1 | 0.883243571 | ENSG0000017 | 14 | CCDC57 |
| SAT | T2DM | ENSG0000011 | 0.003577017 | 0.031694654 | 0.001665087 | 0.081436187 | 0.003538532 | 0.033560849 | 0.001664945 | 0.081418376 | 3.85E-05 | 0.000878125 | 1 | 0.248186538 | 1 | 0.267915193 | ENSG0000017 | 14 | CCDC57 |
| SAT | T2DM | ENSG0000011 | 0.003861002 | 0.118603045 | 0.002473944 | 0.081436551 | 0.003775092 | 0.12699646 | 0.002473757 | 0.081418742 | 8.59E-05 | 2.29E-05 | 1 | 0.453748792 | 1 | 0.476745292 | ENSG0000017 | 7 | BNIP3 |
| SAT | T2DM | ENSG0000011 | -0.00028354 | 0.92462137 | 0.002996795 | 0.081436957 | -0.00022248 | 0.940815166 | 0.002996551 | 0.08141913 | -6.11E-05 | 0.006554908 | 1 | 0.978648294 | 1 | 0.986202735 | ENSG0000017 | 7 | BNIP3 |
| SAT | T2DM | ENSG0000011 | -0.0032281 | 0.424856546 | 0.004045117 | 0.081436852 | -0.00317252 | 0.432833623 | 0.004044762 | 0.081419028 | -5.56E-05 | 0.036308435 | 1 | 0.770140513 | 1 | 0.781577939 | ENSG0000017 | 13 | CIDEA |
| SAT | T2DM | ENSG0000011 | -0.00289211 | 0.097722537 | 0.001746441 | 0.0814365 | -0.00284249 | 0.103584479 | 0.001746297 | 0.081418688 | -4.96E-05 | 0.000103642 | 1 | 0.421965667 | 1 | 0.441377502 | ENSG0000017 | 5 | CYCSP34 |
| SAT | T2DM | ENSG0000011 | -0.00198005 | 0.281571683 | 0.001838829 | 0.081436765 | -0.00195039 | 0.288801379 | 0.00183868 | 0.081418943 | -2.97E-05 | 0.031321321 | 1 | 0.641415693 | 1 | 0.653487811 | ENSG0000017 | 15 | ZNF135 |
| SAT | T2DM | ENSG0000011 | -0.00581706 | 0.096175056 | 0.003496481 | 0.081436496 | -0.00575852 | 0.099542364 | 0.003496202 | 0.081418678 | -5.85E-05 | 0.029112458 | 1 | 0.420328914 | 1 | 0.440431278 | ENSG0000017 | 15 | ZNF135 |
| SAT | T2DM | ENSG0000011 | 0.00887519 | 0.001737748 | 0.002833935 | 0.081435319 | 0.008916566 | 0.001651815 | 0.0028337 | 0.081417477 | -4.14E-05 | 0.045914723 | 1 | 0.038414593 | 1 | 0.04335685 | ENSG0000017 | 4 | CDC26 |
| SAT | T2DM | ENSG0000011 | -0.00299821 | 0.181009326 | 0.002241393 | 0.081436659 | -0.00291671 | 0.193139722 | 0.002241301 | 0.081418848 | -8.15E-05 | 0.001872883 | 1 | 0.535037478 | 1 | 0.561285588 | ENSG0000017 | 4 | CDC26 |
| SAT | T2DM | ENSG0000011 | -0.00080684 | 0.451635855 | 0.001071942 | 0.081436864 | -0.00077213 | 0.471305457 | 0.001071877 | 0.081419044 | -3.47E-05 | 0.001063928 | 1 | 0.783398902 | 1 | 0.80363893 | ENSG0000017 | 10 | CRLF3 |
| SAT | T2DM | ENSG0000011 | -0.00150792 | 0.495155499 | 0.002210596 | 0.081436881 | -0.00147084 | 0.505786195 | 0.002210418 | 0.081419057 | -3.71E-05 | 0.028072159 | 1 | 0.806594843 | 1 | 0.817738235 | ENSG0000017 | 10 | CRLF3 |
| SAT | T2DM | ENSG0000011 | 0.000981992 | 0.802091683 | 0.003917922 | 0.081436948 | 0.000863495 | 0.825548173 | 0.003917589 | 0.081419123 | 0.000118498 | 1.59E-05 | 1 | 0.948256967 | 1 | 0.959754419 | ENSG0000017 | 10 | CRLF3 |

| | | | | | | | | | | | | | | | | | | |
|---|---|---|---|---|---|---|---|---|---|---|---|---|---|---|---|---|---|---|
| SAT | T2DM | ENSG0000011 | -0.00240329 | 0.135162333 | 0.001608573 | 0.081436585 | -0.00246955 | 0.124695809 | 0.001608451 | 0.081418737 | 6.63E-05 | 5.16E-07 | 1 | 0.477360189 | 1 | 0.474800718 | ENSG0000017 | 10 CRLF3 |
| SAT | T2DM | ENSG0000011 | -0.00293287 | 0.017685111 | 0.001236375 | 0.081436018 | -0.00291585 | 0.018344446 | 0.001236264 | 0.081418201 | -1.70E-05 | 0.028226976 | 1 | 0.172693001 | 1 | 0.187482528 | ENSG0000017 | 6 RNPEP |
| SAT | T2DM | ENSG0000011 | 0.000745712 | 0.836958196 | 0.003623727 | 0.081436951 | 0.000804915 | 0.824203722 | 0.003623434 | 0.081419123 | -5.92E-05 | 0.030419774 | 1 | 0.959823323 | 1 | 0.959280742 | ENSG0000017 | 6 RNPEP |
| SAT | T2DM | ENSG0000011 | 0.005890171 | 0.036475986 | 0.002816125 | 0.081436227 | 0.005841101 | 0.038049497 | 0.002815902 | 0.081418412 | 4.91E-05 | 0.02501078 | 1 | 0.266450175 | 1 | 0.281806774 | ENSG0000017 | 10 SPRYD4 |
| SAT | T2DM | ENSG0000011 | -0.00567711 | 0.16085853 | 0.004048739 | 0.08143663 | -0.00573838 | 0.156351634 | 0.004048379 | 0.081418795 | 6.13E-05 | 0.018729246 | 1 | 0.512133793 | 1 | 0.51775889 | ENSG0000017 | 10 SPRYD4 |
| SAT | T2DM | ENSG0000011 | 0.001487186 | 0.227070508 | 0.001231174 | 0.081436715 | 0.001466617 | 0.233522853 | 0.00123107 | 0.081418894 | 2.06E-05 | 0.017992328 | 1 | 0.591955891 | 1 | 0.610313962 | ENSG0000017 | 10 SPRYD4 |
| SAT | T2DM | ENSG0000012 | 0.002737168 | 0.303786377 | 0.002661713 | 0.081436782 | 0.002813182 | 0.290516027 | 0.002661499 | 0.081418944 | -7.60E-05 | 0.000192882 | 1 | 0.66271483 | 1 | 0.655417523 | ENSG0000017 | 10 SPRYD4 |
| SAT | T2DM | ENSG0000012 | -0.00311336 | 0.411802341 | 0.003793423 | 0.081436846 | -0.00305098 | 0.421194568 | 0.00379309 | 0.081419023 | -6.24E-05 | 0.012129469 | 1 | 0.757945834 | 1 | 0.770311805 | ENSG0000017 | 10 SPRYD4 |
| SAT | T2DM | ENSG0000012 | -0.00158025 | 0.43560328 | 0.002026903 | 0.081436857 | -0.00160648 | 0.427980941 | 0.00202672 | 0.081419026 | 2.62E-05 | 0.036149631 | 1 | 0.772274669 | 1 | 0.777953168 | ENSG0000017 | 5 VPS37D |
| SAT | T2DM | ENSG0000012 | -0.00280262 | 0.014364199 | 0.001144845 | 0.081435957 | -0.00276007 | 0.015921946 | 0.001144922 | 0.08141816 | -4.25E-05 | 0.047651654 | 1 | 0.151551916 | 1 | 0.174133018 | ENSG0000017 | 5 VPS37D |
| SAT | T2DM | ENSG0000012 | 0.001332922 | 0.636211218 | 0.002818 | 0.081436921 | 0.001289671 | 0.647172155 | 0.002817754 | 0.081419096 | 4.33E-05 | 0.021836832 | 1 | 0.873655621 | 1 | 0.890302658 | ENSG0000017 | 5 VPS37D |
| SAT | T2DM | ENSG0000012 | -0.0010437 | 0.747263525 | 0.003238801 | 0.081436941 | -0.00098551 | 0.760892151 | 0.00323852 | 0.081419115 | -5.82E-05 | 0.007696742 | 1 | 0.927647229 | 1 | 0.932999239 | ENSG0000017 | 5 VPS37D |
| SAT | T2DM | ENSG0000012 | -0.00358124 | 0.092856478 | 0.002131028 | 0.081436486 | -0.00349416 | 0.101112888 | 0.002131252 | 0.081418682 | -8.71E-05 | 0.048476268 | 1 | 0.414991889 | 1 | 0.440431278 | ENSG0000017 | 6 ZNF575 |
| SAT | T2DM | ENSG0000012 | -0.00127094 | 0.737520511 | 0.003792284 | 0.08143694 | -0.00141943 | 0.708162671 | 0.003791979 | 0.081419107 | 0.000148491 | 2.71E-07 | 1 | 0.924295185 | 1 | 0.912941362 | ENSG0000017 | 6 ZNF575 |
| SAT | T2DM | ENSG0000012 | 0.007920118 | 0.049325853 | 0.004029018 | 0.081436313 | 0.008019387 | 0.046531298 | 0.004028736 | 0.081418469 | -9.93E-05 | 0.005652669 | 1 | 0.30826309 | 1 | 0.310935212 | ENSG0000017 | 6 PLA2G16 |
| SAT | T2DM | ENSG0000012 | 0.000706114 | 0.780324044 | 0.002531809 | 0.081436945 | 0.000594454 | 0.814379259 | 0.002531957 | 0.081419122 | 0.00011166 | 0.016153765 | 1 | 0.940919952 | 1 | 0.955647609 | ENSG0000017 | 6 PLA2G16 |
| SAT | T2DM | ENSG0000012 | 0.004633477 | 0.077676925 | 0.002626195 | 0.081436438 | 0.004681773 | 0.074608118 | 0.002625976 | 0.0814186 | -4.83E-05 | 0.010911646 | 1 | 0.389096167 | 1 | 0.395620568 | ENSG0000017 | 6 PHLDB3 |
| SAT | T2DM | ENSG0000012 | 0.000432086 | 0.811005302 | 0.001806914 | 0.081436949 | 0.000480594 | 0.790244161 | 0.001806786 | 0.081419119 | -4.85E-05 | 0.002306069 | 1 | 0.951539016 | 1 | 0.947957612 | ENSG0000017 | 11 ACSF3 |
| SAT | T2DM | ENSG0000012 | 0.003466706 | 0.035828958 | 0.001651702 | 0.081436222 | 0.003435122 | 0.037533502 | 0.001651565 | 0.081418408 | 3.16E-05 | 0.008719313 | 1 | 0.263575813 | 1 | 0.280518039 | ENSG0000017 | 10 RUFY1 |
| SAT | T2DM | ENSG0000012 | -0.00148625 | 0.286870204 | 0.001395524 | 0.081436769 | -0.00151679 | 0.277043985 | 0.001395415 | 0.081418933 | 3.05E-05 | 0.005482056 | 1 | 0.644230759 | 1 | 0.64327119 | ENSG0000017 | 10 RUFY1 |
| SAT | T2DM | ENSG0000012 | 0.00526685 | 0.178150935 | 0.003911592 | 0.081436655 | 0.005405066 | 0.166997335 | 0.003911276 | 0.081418812 | -0.00013822 | 3.21E-06 | 1 | 0.530526907 | 1 | 0.527859591 | ENSG0000017 | 7 MAMSTR |
| SAT | T2DM | ENSG0000012 | 0.007993065 | 0.012798624 | 0.003210926 | 0.081435923 | 0.007928236 | 0.013536519 | 0.003210667 | 0.081418112 | 6.48E-05 | 0.007867527 | 1 | 0.143532419 | 1 | 0.157494043 | ENSG0000017 | 10 ANKLE2 |
| SAT | T2DM | ENSG0000012 | -0.00087637 | 0.797572088 | 0.003416788 | 0.081436947 | -0.00082634 | 0.808883856 | 0.003416504 | 0.081419121 | -5.00E-05 | 0.043154338 | 1 | 0.94681813 | 1 | 0.953285895 | ENSG0000017 | 4 TOB2P1 |
| SAT | T2DM | ENSG0000012 | -0.00077477 | 0.44468803 | 0.001013703 | 0.081436861 | -0.00075912 | 0.453904536 | 0.001013622 | 0.081419037 | -1.57E-05 | 0.046114548 | 1 | 0.780081459 | 1 | 0.794600418 | ENSG0000017 | 4 TOB2P1 |
| SAT | T2DM | ENSG0000012 | 0.004791799 | 0.045051524 | 0.002390904 | 0.081436287 | 0.00471993 | 0.048350237 | 0.002390707 | 0.08141848 | 7.19E-05 | 4.08E-05 | 1 | 0.292287338 | 1 | 0.319189392 | ENSG0000017 | 4 TOB2P1 |
| SAT | T2DM | ENSG0000012 | -0.00217723 | 0.257446997 | 0.001922589 | 0.081436744 | -0.00222216 | 0.247725573 | 0.001922462 | 0.081418908 | 4.49E-05 | 0.012016424 | 1 | 0.622005594 | 1 | 0.620663712 | ENSG0000017 | 4 TOB2P1 |
| SAT | T2DM | ENSG0000012 | -0.00494541 | 0.094037011 | 0.002953401 | 0.08143649 | -0.00498815 | 0.091199999 | 0.002953135 | 0.081418654 | 4.27E-05 | 0.020259311 | 1 | 0.417503971 | 1 | 0.424593011 | ENSG0000017 | 4 TOB2P1 |
| SAT | T2DM | ENSG0000012 | 0.001041597 | 0.49200482 | 0.001515878 | 0.08143688 | 0.00101952 | 0.501192044 | 0.001515754 | 0.081419055 | 2.21E-05 | 0.049869732 | 1 | 0.803918235 | 1 | 0.816084912 | ENSG0000017 | 7 THAP4 |
| SAT | T2DM | ENSG0000012 | 0.004049777 | 0.25523309 | 0.003559519 | 0.081436742 | 0.004130389 | 0.245861842 | 0.003559266 | 0.081418906 | -8.06E-05 | 0.009857967 | 1 | 0.620408004 | 1 | 0.620663712 | ENSG0000017 | 13 SHMT1 |
| SAT | T2DM | ENSG0000012 | -0.0052016 | 0.099769933 | 0.003160199 | 0.081436506 | -0.00515809 | 0.102607197 | 0.003159923 | 0.081418686 | -4.35E-05 | 0.038196975 | 1 | 0.425727224 | 1 | 0.440431278 | ENSG0000017 | 13 SHMT1 |
| SAT | T2DM | ENSG0000012 | -0.00110476 | 0.38580675 | 0.001273864 | 0.081436833 | -0.00106679 | 0.402306748 | 0.001273769 | 0.081419014 | -3.80E-05 | 0.000372207 | 1 | 0.736376933 | 1 | 0.757591271 | ENSG0000017 | 13 SHMT1 |
| SAT | T2DM | ENSG0000012 | -0.00282912 | 0.233964978 | 0.002376995 | 0.081436722 | -0.0028727 | 0.22679979 | 0.00237679 | 0.081418887 | 4.36E-05 | 0.007242907 | 1 | 0.598969104 | 1 | 0.601281476 | ENSG0000017 | 3 HCG4 |
| SAT | T2DM | ENSG0000012 | -0.00295839 | 0.446271953 | 0.003884203 | 0.081436861 | -0.00290055 | 0.455171199 | 0.00388386 | 0.081419038 | -5.78E-05 | 0.022197446 | 1 | 0.780081459 | 1 | 0.794600418 | ENSG0000017 | 3 HCG4 |
| SAT | T2DM | ENSG0000012 | 0.004561033 | 0.039346314 | 0.0022135 | 0.081436249 | 0.004524676 | 0.040925603 | 0.002213326 | 0.081418433 | 3.64E-05 | 0.035569603 | 1 | 0.274087595 | 1 | 0.289545565 | ENSG0000017 | 3 HCG4 |
| SAT | T2DM | ENSG0000012 | 0.001087728 | 0.420408181 | 0.001350018 | 0.08143685 | 0.001661 | 0.21934769 | 0.001352313 | 0.081418879 | -0.00057327 | 1.73E-12 | 1 | 0.764009208 | 1 | 0.592816944 | ENSG0000017 | 3 HCG4 |
| SAT | T2DM | ENSG0000012 | 0.001659229 | 0.579207211 | 0.002992063 | 0.081436907 | 0.001759919 | 0.556368836 | 0.002991819 | 0.081419073 | -0.00010069 | 6.66E-06 | 1 | 0.842939695 | 1 | 0.842050121 | ENSG0000017 | 3 HCG4 |
| SAT | T2DM | ENSG0000012 | 0.000257654 | 0.8696856 | 0.001570509 | 0.081436954 | 0.001227113 | 0.437484074 | 0.001580417 | 0.08141903 | -0.00096946 | 5.33E-08 | 1 | 0.966468088 | 1 | 0.786081735 | ENSG0000017 | 3 HCG4 |
| SAT | T2DM | ENSG0000012 | 0.000173242 | 0.908750619 | 0.001511515 | 0.081436956 | 0.000286279 | 0.84977336 | 0.001511444 | 0.081419125 | -0.00011304 | 2.42E-11 | 1 | 0.974472349 | 1 | 0.965561588 | ENSG0000017 | 10 TMEM80 |
| SAT | T2DM | ENSG0000012 | -0.00080717 | 0.809193293 | 0.003342774 | 0.081436949 | -0.00047067 | 0.888037989 | 0.003343149 | 0.081419128 | -0.0003365 | 1.74E-06 | 1 | 0.951539016 | 1 | 0.971217547 | ENSG0000017 | 10 TMEM80 |
| SAT | T2DM | ENSG0000012 | -0.00093281 | 0.410227117 | 0.001132741 | 0.081436845 | -0.00094996 | 0.401631542 | 0.001132644 | 0.081419013 | 1.72E-05 | 0.026831831 | 1 | 0.757820042 | 1 | 0.757591271 | ENSG0000017 | 6 TALDO1 |
| SAT | T2DM | ENSG0000012 | 0.0016677 | 0.222026326 | 0.00136567 | 0.081436709 | 0.000622486 | 0.652035211 | 0.001380428 | 0.081419097 | 0.001045215 | 2.39E-07 | 1 | 0.587001107 | 1 | 0.891013292 | ENSG0000017 | 1 SPACA4 |
| SAT | T2DM | ENSG0000012 | 0.007316809 | 0.043213081 | 0.003619227 | 0.081436275 | 0.002532316 | 0.49836327 | 0.003740122 | 0.081419054 | 0.004784493 | 4.09E-07 | 1 | 0.286428522 | 1 | 0.81497549 | ENSG0000017 | 6 FBXO39 |
| SAT | T2DM | ENSG0000012 | -0.00409536 | 0.124430671 | 0.002665485 | 0.081436564 | -0.00405228 | 0.12841004 | 0.002665267 | 0.081418745 | -4.31E-05 | 0.029656214 | 1 | 0.463429598 | 1 | 0.478221727 | ENSG0000017 | 6 ZBTB38 |
| SAT | T2DM | ENSG0000012 | -0.00109748 | 0.518143333 | 0.001698335 | 0.081436889 | -0.00106109 | 0.532077055 | 0.001698189 | 0.081419066 | -3.64E-05 | 0.001796671 | 1 | 0.818675014 | 1 | 0.831703104 | ENSG0000017 | 6 ZBTB38 |
| SAT | T2DM | ENSG0000012 | -0.00066667 | 0.796970828 | 0.002591316 | 0.081436947 | -0.00063391 | 0.806727499 | 0.002591081 | 0.081419121 | -3.28E-05 | 0.039748042 | 1 | 0.94681813 | 1 | 0.952410029 | ENSG0000017 | 4 TIMM22 |
| SAT | T2DM | ENSG0000012 | -0.00164621 | 0.560489963 | 0.00282799 | 0.081436902 | -0.00149278 | 0.597589586 | 0.002827935 | 0.081419084 | -0.00015343 | 5.26E-05 | 1 | 0.837416925 | 1 | 0.855769266 | ENSG0000017 | 7 SAMD9L |
| SAT | T2DM | ENSG0000012 | -0.00210668 | 0.445202003 | 0.002759457 | 0.081436861 | -0.00174572 | 0.527147884 | 0.002760621 | 0.081419064 | -0.00036095 | 5.99E-05 | 1 | 0.780081459 | 1 | 0.828775497 | ENSG0000017 | 7 SAMD9L |
| SAT | T2DM | ENSG0000012 | 0.005212753 | 0.157073039 | 0.003683959 | 0.081436624 | 0.005290724 | 0.150927174 | 0.003683664 | 0.081418786 | -7.80E-05 | 0.00586716 | 1 | 0.507474184 | 1 | 0.5109822 | ENSG0000017 | 7 SAMD9L |
| SAT | T2DM | ENSG0000012 | 0.003459211 | 0.000501109 | 0.000993973 | 0.081434934 | 0.003432358 | 0.000553525 | 0.000993894 | 0.081417138 | 2.69E-05 | 0.000514398 | 1 | 0.015422559 | 1 | 0.019528141 | ENSG0000017 | 15 ARIH2 |
| SAT | T2DM | ENSG0000012 | 0.000995294 | 0.717319368 | 0.002749098 | 0.081436937 | 0.000939716 | 0.732458626 | 0.002748857 | 0.081419111 | 5.56E-05 | 0.002206355 | 1 | 0.911865496 | 1 | 0.923670747 | ENSG0000017 | 9 SAMD12 |
| SAT | T2DM | ENSG0000012 | -0.01415563 | 8.55E-06 | 0.003180309 | 0.081433648 | -0.01420936 | 7.89E-06 | 0.003180062 | 0.081415795 | 5.37E-05 | 0.034136514 | 0.02804641 | 0.000596732 | 0.025878571 | 0.000663553 | ENSG0000017 | 6 RP5-1051J4.6 |
| SAT | T2DM | ENSG0000012 | 0.00038549 | 0.916837006 | 0.003691765 | 0.081436957 | 0.000453702 | 0.902182891 | 0.003691503 | 0.081419128 | -6.82E-05 | 0.035226446 | 1 | 0.977304164 | 1 | 0.972878089 | ENSG0000017 | 6 RP5-1051J4.6 |
| SAT | T2DM | ENSG0000012 | 0.002344123 | 0.384815122 | 0.002697311 | 0.081436832 | 0.002404458 | 0.372657633 | 0.002697071 | 0.081418998 | -6.03E-05 | 0.000519058 | 1 | 0.734911767 | 1 | 0.735400932 | ENSG0000017 | 13 PIDD |
| SAT | T2DM | ENSG0000012 | -0.00173125 | 0.636055244 | 0.00365843 | 0.081436921 | -0.00157721 | 0.666364905 | 0.00365822 | 0.0814191 | -0.00015403 | 3.72E-05 | 1 | 0.873655621 | 1 | 0.896403813 | ENSG0000017 | 11 GBA |
| SAT | T2DM | ENSG0000012 | 0.00472836 | 0.230218154 | 0.003940962 | 0.081436718 | 0.004783715 | 0.224765946 | 0.003940607 | 0.081418885 | -5.54E-05 | 0.024940281 | 1 | 0.594292496 | 1 | 0.599071543 | ENSG0000017 | 11 GBA |
| SAT | T2DM | ENSG0000012 | 0.000780117 | 0.808435218 | 0.003217702 | 0.081436949 | 0.000721111 | 0.822659957 | 0.003217446 | 0.081419123 | 5.90E-05 | 0.017922323 | 1 | 0.951539016 | 1 | 0.958398582 | ENSG0000017 | 11 GBA |
| SAT | T2DM | ENSG0000012 | 0.003769325 | 0.180182803 | 0.002812521 | 0.081436658 | 0.003860193 | 0.169879139 | 0.002812334 | 0.081418816 | -9.09E-05 | 0.000499183 | 1 | 0.533555754 | 1 | 0.531756789 | ENSG0000017 | 11 GBA |
| SAT | T2DM | ENSG0000012 | 0.00025728 | 0.70489417 | 0.000679336 | 0.081436934 | 0.000269245 | 0.691831763 | 0.000679275 | 0.081419105 | -1.20E-05 | 0.005480855 | 1 | 0.906221055 | 1 | 0.904075912 | ENSG0000017 | 13 ACAD9 |
| SAT | T2DM | ENSG0000012 | -0.00523766 | 0.092020593 | 0.003108701 | 0.081436484 | -0.00530903 | 0.087650217 | 0.003108459 | 0.081418644 | 7.14E-05 | 0.003956707 | 1 | 0.414244809 | 1 | 0.420439126 | ENSG0000017 | 9 IL17RA |
| SAT | T2DM | ENSG0000012 | 0.001005989 | 0.632696464 | 0.002104865 | 0.08143692 | 0.00112834 | 0.591958705 | 0.002105112 | 0.081419083 | -0.00012235 | 0.00631771 | 1 | 0.871484928 | 1 | 0.854809759 | ENSG0000017 | 9 IL17RA |
| SAT | T2DM | ENSG0000012 | 0.003014821 | 0.375731476 | 0.003403542 | 0.081436827 | 0.002936861 | 0.388164375 | 0.003403274 | 0.081419006 | 7.80E-05 | 0.003488351 | 1 | 0.728082648 | 1 | 0.748716823 | ENSG0000017 | 9 IL17RA |
| SAT | T2DM | ENSG0000012 | -0.00080789 | 0.79777712 | 0.003153034 | 0.081436947 | -0.0008705 | 0.782467401 | 0.003152768 | 0.081419118 | 6.26E-05 | 0.004931082 | 1 | 0.94681813 | 1 | 0.944893464 | ENSG0000017 | 11 AGTRAP |
| SAT | T2DM | ENSG0000012 | 0.000291347 | 0.895363044 | 0.002215206 | 0.081436956 | 0.000319281 | 0.885386411 | 0.002215006 | 0.081419127 | -2.79E-05 | 0.041405673 | 1 | 0.970180416 | 1 | 0.968806494 | ENSG0000017 | 11 AGTRAP |
| SAT | T2DM | ENSG0000012 | 0.001959545 | 0.130445268 | 0.001295695 | 0.081436576 | 0.001979163 | 0.126608117 | 0.001295588 | 0.081418741 | -1.96E-05 | 0.038983996 | 1 | 0.47229897 | 1 | 0.47645145 | ENSG0000017 | 11 AGTRAP |
| SAT | T2DM | ENSG0000012 | -0.00270514 | 0.402341577 | 0.003230226 | 0.081436841 | -0.00280193 | 0.385679985 | 0.003229966 | 0.081419005 | 9.68E-05 | 8.66E-05 | 1 | 0.752518444 | 1 | 0.746964998 | ENSG0000017 | 12 EFCAB4A |
| SAT | T2DM | ENSG0000012 | -0.00073403 | 0.593227981 | 0.001374162 | 0.081436911 | -0.00070912 | 0.605799663 | 0.001374052 | 0.081419086 | -2.49E-05 | 0.017104239 | 1 | 0.846620707 | 1 | 0.85932931 | ENSG0000017 | 10 NAALADL2 |
| SAT | T2DM | ENSG0000012 | -0.00017101 | 0.963092066 | 0.003695552 | 0.081436958 | -9.23E-05 | 0.980066457 | 0.003695312 | 0.081419131 | -7.87E-05 | 0.023966843 | 1 | 0.9918095 | 1 | 0.997085517 | ENSG0000017 | 13 CD151 |
| SAT | T2DM | ENSG0000012 | 0.007084352 | 0.004013598 | 0.00246232 | 0.081435575 | 0.006993672 | 0.004506719 | 0.002462265 | 0.081417783 | 9.07E-05 | 0.005219207 | 1 | 0.068072202 | 1 | 0.079766896 | ENSG0000017 | 4 PCDHB9 |
| SAT | T2DM | ENSG0000012 | 0.008865954 | 0.007025839 | 0.003289012 | 0.081435744 | 0.008933702 | 0.006598677 | 0.003288731 | 0.081417898 | -6.77E-05 | 0.003055815 | 1 | 0.09396718 | 1 | 0.099770777 | ENSG0000017 | 9 ODF3B |
| SAT | T2DM | ENSG0000012 | 0.002497582 | 0.216122543 | 0.002019217 | 0.081436703 | 0.002457257 | 0.223597195 | 0.002019077 | 0.081418883 | 4.03E-05 | 0.026139345 | 1 | 0.580276647 | 1 | 0.597775996 | ENSG0000017 | 7 DPY19L2 |
| SAT | T2DM | ENSG0000012 | -0.00136179 | 0.582571862 | 0.002477646 | 0.081436908 | -0.00131982 | 0.594217058 | 0.00247744 | 0.081419083 | -4.20E-05 | 0.020019925 | 1 | 0.845212395 | 1 | 0.854809759 | ENSG0000017 | 7 DPY19L2 |
| SAT | T2DM | ENSG0000012 | 0.000152131 | 0.916592924 | 0.001452648 | 0.081436957 | 0.000180502 | 0.901103048 | 0.001452522 | 0.081419128 | -2.84E-05 | 0.003567418 | 1 | 0.977304164 | 1 | 0.972878089 | ENSG0000017 | 7 DPY19L2 |
| SAT | T2DM | ENSG0000012 | 0.006337841 | 0.061910584 | 0.003394775 | 0.081436376 | 0.006420696 | 0.058556392 | 0.003394475 | 0.081418533 | -8.29E-05 | 0.000170474 | 1 | 0.345456849 | 1 | 0.349315498 | ENSG0000017 | 7 DPY19L2 |
| SAT | T2DM | ENSG0000012 | 0.000544588 | 0.839070593 | 0.002681611 | 0.081436952 | 0.000604643 | 0.821598074 | 0.002681463 | 0.081419122 | -6.01E-05 | 0.031604788 | 1 | 0.959889764 | 1 | 0.958012298 | ENSG0000017 | 7 DPY19L2 |
| SAT | T2DM | ENSG0000012 | -0.00040825 | 0.909078587 | 0.003574844 | 0.081436956 | -0.00031448 | 0.929895644 | 0.003574574 | 0.08141913 | -9.38E-05 | 0.001479331 | 1 | 0.974472349 | 1 | 0.982921265 | ENSG0000017 | 4 GPR150 |
| SAT | T2DM | ENSG0000012 | 0.000336368 | 0.903073237 | 0.00276209 | 0.081436956 | 0.000433313 | 0.875331339 | 0.002761882 | 0.081419127 | -9.69E-05 | 2.05E-05 | 1 | 0.97231278 | 1 | 0.966700287 | ENSG0000017 | 7 STAP2 |
| SAT | T2DM | ENSG0000012 | -0.0011915 | 0.653670059 | 0.002655631 | 0.081436925 | -0.00115287 | 0.66417278 | 0.002655403 | 0.081419099 | -3.86E-05 | 0.03408026 | 1 | 0.882995924 | 1 | 0.895996588 | ENSG0000017 | 12 DALRD3 |
| SAT | T2DM | ENSG0000013 | -0.00444412 | 0.071508065 | 0.002465885 | 0.081436416 | -0.00440726 | 0.073864717 | 0.002465662 | 0.081418597 | -3.69E-05 | 0.015077233 | 1 | 0.373553186 | 1 | 0.395413696 | ENSG0000017 | 12 DALRD3 |
| SAT | T2DM | ENSG0000013 | -0.00388006 | 0.238443594 | 0.003291291 | 0.081436726 | -0.00392701 | 0.232770148 | 0.003291007 | 0.081418893 | 4.69E-05 | 0.035906551 | 1 | 0.60584911 | 1 | 0.610313962 | ENSG0000017 | 10 SH2B1 |
| SAT | T2DM | ENSG0000013 | -0.00096706 | 0.274098294 | 0.000884226 | 0.081436759 | -0.00098082 | 0.26728617 | 0.000884149 | 0.081418925 | 1.38E-05 | 0.020719095 | 1 | 0.6356488 | 1 | 0.638593145 | ENSG0000017 | 10 PLEC |
| SAT | T2DM | ENSG0000013 | 4.68E-05 | 0.974234288 | 0.001448905 | 0.081436958 | 2.40E-05 | 0.986790099 | 0.001448778 | 0.081419131 | 2.28E-05 | 0.016801211 | 1 | 0.995031844 | 1 | 0.997162029 | ENSG0000017 | 10 AP000345.1 |
| SAT | T2DM | ENSG0000013 | 5.38E-06 | 0.996650523 | 0.001282518 | 0.081436958 | 3.34E-05 | 0.979232997 | 0.001282411 | 0.081419131 | -2.80E-05 | 0.002388432 | 1 | 0.998171663 | 1 | 0.997085517 | ENSG0000017 | 13 WDR6 |
| SAT | T2DM | ENSG0000013 | 0.000844881 | 0.82571304 | 0.003836824 | 0.08143695 | 0.000753267 | 0.844342541 | 0.003836506 | 0.081419124 | 9.16E-05 | 0.001027634 | 1 | 0.955051074 | 1 | 0.96477102 | ENSG0000017 | 10 GEN1 |
| SAT | T2DM | ENSG0000013 | -0.00553654 | 0.130679649 | 0.003663111 | 0.081436577 | -0.00560618 | 0.125879327 | 0.003662835 | 0.081418739 | 6.96E-05 | 0.021547517 | 1 | 0.47229897 | 1 | 0.47645145 | ENSG0000017 | 10 GEN1 |
| SAT | T2DM | ENSG0000013 | -0.0023189 | 0.192759909 | 0.001780401 | 0.081436675 | -0.00236199 | 0.184588018 | 0.001780269 | 0.081418837 | 4.31E-05 | 0.004042511 | 1 | 0.552241504 | 1 | 0.550575716 | ENSG0000017 | 10 GEN1 |
| SAT | T2DM | ENSG0000013 | -0.00677584 | 0.065498954 | 0.003678838 | 0.081436391 | -0.00670999 | 0.068137785 | 0.003678526 | 0.081418575 | -6.58E-05 | 0.010540232 | 1 | 0.357574154 | 1 | 0.380219613 | ENSG0000017 | 10 GEN1 |
| SAT | T2DM | ENSG0000013 | 0.002220091 | 0.390382134 | 0.002584731 | 0.081436835 | 0.002163458 | 0.402551801 | 0.00258455 | 0.081419014 | 5.66E-05 | 0.013753029 | 1 | 0.741307668 | 1 | 0.757591271 | ENSG0000017 | 10 GEN1 |
| SAT | T2DM | ENSG0000013 | 0.002523547 | 0.53397372 | 0.004057463 | 0.081436894 | 0.002432878 | 0.548738836 | 0.004057154 | 0.081419071 | 9.07E-05 | 0.00613802 | 1 | 0.828750699 | 1 | 0.841179809 | ENSG0000017 | 7 ZFAND2A |
| SAT | T2DM | ENSG0000013 | 0.00018911 | 0.945720578 | 0.002777687 | 0.081436958 | 6.99E-05 | 0.979928621 | 0.002777487 | 0.081419131 | 0.000119233 | 6.81E-07 | 1 | 0.984737929 | 1 | 0.997085517 | ENSG0000017 | 7 ZFAND2A |
| SAT | T2DM | ENSG0000013 | -0.00302773 | 0.436189907 | 0.003888472 | 0.081436857 | -0.00297088 | 0.444814205 | 0.003888134 | 0.081419033 | -5.69E-05 | 0.029447956 | 1 | 0.772274669 | 1 | 0.790845546 | ENSG0000017 | 7 ZFAND2A |
| SAT | T2DM | ENSG0000013 | 0.00184828 | 0.200973134 | 0.00144534 | 0.081436685 | 0.001822241 | 0.207351776 | 0.001445211 | 0.081418865 | 2.60E-05 | 0.004464298 | 1 | 0.561663418 | 1 | 0.579977552 | ENSG0000017 | 8 ZNF223 |
| SAT | T2DM | ENSG0000013 | -0.0033876 | 0.308016242 | 0.003323146 | 0.081436785 | -0.00332638 | 0.316804585 | 0.003322897 | 0.081418963 | -6.12E-05 | 0.026353298 | 1 | 0.66864566 | 1 | 0.68742915 | ENSG0000017 | 8 ZNF223 |
| SAT | T2DM | ENSG0000013 | 0.006085935 | 0.074688731 | 0.003414509 | 0.081436428 | 0.005983027 | 0.079708822 | 0.003414226 | 0.081418618 | 0.000102908 | 3.55E-05 | 1 | 0.382299104 | 1 | 0.409271745 | ENSG0000017 | 8 FAM220A |
| SAT | T2DM | ENSG0000013 | 0.000674141 | 0.627194772 | 0.001388038 | 0.081436919 | 0.000630784 | 0.649486334 | 0.001387937 | 0.081419096 | 4.34E-05 | 0.000256454 | 1 | 0.866453072 | 1 | 0.891013292 | ENSG0000017 | 8 FAM220A |
| SAT | T2DM | ENSG0000013 | 0.002553511 | 0.471754096 | 0.003548374 | 0.081436872 | 0.002690874 | 0.448211982 | 0.003548093 | 0.081419035 | -0.00013736 | 7.13E-07 | 1 | 0.79607871 | 1 | 0.791227469 | ENSG0000017 | 13 P4HTM |
| SAT | T2DM | ENSG0000013 | 0.004248608 | 0.106159392 | 0.002629574 | 0.081436522 | 0.004164504 | 0.113241698 | 0.002629451 | 0.081418712 | 8.41E-05 | 0.00433314 | 1 | 0.433220108 | 1 | 0.455325994 | ENSG0000017 | 13 P4HTM |
| SAT | T2DM | ENSG0000013 | 0.000967501 | 0.748580513 | 0.003018613 | 0.081436941 | 0.001011302 | 0.73758533 | 0.003018336 | 0.081419112 | -4.38E-05 | 0.014718704 | 1 | 0.92822852 | 1 | 0.925041991 | ENSG0000017 | 5 CA8 |
| SAT | T2DM | ENSG0000013 | 0.001245196 | 0.514444171 | 0.001910004 | 0.081436887 | 0.001289415 | 0.499594578 | 0.001909882 | 0.081419055 | -4.42E-05 | 0.015307618 | 1 | 0.816039871 | 1 | 0.815958247 | ENSG0000017 | 7 GPR35 |
| SAT | T2DM | ENSG0000013 | -0.00284676 | 0.132058072 | 0.001890234 | 0.081436579 | -0.00281436 | 0.136479367 | 0.001890059 | 0.08141876 | -3.24E-05 | 0.003422569 | 1 | 0.472458584 | 1 | 0.492616944 | ENSG0000017 | 7 GPR35 |
| SAT | T2DM | ENSG0000013 | -0.0016354 | 0.121457193 | 0.001055991 | 0.081436558 | -0.00165307 | 0.117455314 | 0.001055905 | 0.081418721 | 1.77E-05 | 0.025142601 | 1 | 0.459102591 | 1 | 0.462451956 | ENSG0000017 | 7 GPR35 |
| SAT | T2DM | ENSG0000013 | -0.00140889 | 0.434687856 | 0.0018035 | 0.081436856 | -0.00137229 | 0.44667462 | 0.001803341 | 0.081419034 | -3.66E-05 | 0.001692439 | 1 | 0.772274669 | 1 | 0.791227469 | ENSG0000017 | 10 ZNF713 |
| SAT | T2DM | ENSG0000013 | 0.002101841 | 0.185705158 | 0.001588221 | 0.081436666 | 0.002064941 | 0.193512861 | 0.0015881 | 0.081418848 | 3.69E-05 | 0.00444526 | 1 | 0.542563333 | 1 | 0.561375505 | ENSG0000017 | 10 ZNF713 |
| SAT | T2DM | ENSG0000013 | -0.01079696 | 0.00053458 | 0.003117962 | 0.081434955 | -0.01046809 | 0.000791863 | 0.003119545 | 0.081417249 | -0.00032887 | 0.002669781 | 1 | 0.015863016 | 1 | 0.024351274 | ENSG0000017 | 10 ZNF713 |
| SAT | T2DM | ENSG0000013 | -0.01087329 | 0.001614993 | 0.003448331 | 0.081435297 | -0.0105124 | 0.002311502 | 0.003450081 | 0.08141758 | -0.00036089 | 0.002878277 | 1 | 0.036293105 | 1 | 0.051945464 | ENSG0000017 | 9 PARP10 |
| SAT | T2DM | ENSG0000013 | -0.00205619 | 0.131068337 | 0.001361802 | 0.081436577 | -0.00208432 | 0.125844883 | 0.001361681 | 0.081418739 | 2.81E-05 | 0.0011499 | 1 | 0.472458584 | 1 | 0.47645145 | ENSG0000017 | 4 KCTD12 |
| SAT | T2DM | ENSG0000013 | -0.00086726 | 0.40910458 | 0.001050627 | 0.081436845 | -0.00088992 | 0.396933788 | 0.001050536 | 0.081419011 | 2.27E-05 | 0.001371603 | 1 | 0.756926483 | 1 | 0.756180206 | ENSG0000017 | 4 KCTD12 |
| SAT | T2DM | ENSG0000013 | 0.002297928 | 0.293767012 | 0.002188722 | 0.081436774 | 0.002237976 | 0.306501032 | 0.002188532 | 0.081418956 | 6.00E-05 | 4.46E-05 | 1 | 0.651249707 | 1 | 0.674696375 | ENSG0000017 | 5 RPP25 |
| SAT | T2DM | ENSG0000013 | -0.00011604 | 0.974431431 | 0.003620425 | 0.081436958 | -0.00061593 | 0.86500336 | 0.003622933 | 0.081419126 | 0.000499892 | 0.000568556 | 1 | 0.995031844 | 1 | 0.966241215 | ENSG0000017 | 5 RPP25 |
| SAT | T2DM | ENSG0000013 | -3.70E-05 | 0.989591989 | 0.002837805 | 0.081436958 | 2.99E-05 | 0.991606033 | 0.002837559 | 0.081419131 | -6.69E-05 | 0.000451497 | 1 | 0.996461007 | 1 | 0.998267105 | ENSG0000017 | 2 STX19 |
| SAT | T2DM | ENSG0000013 | 0.002752511 | 0.38357471 | 0.003158979 | 0.081436832 | 0.002811374 | 0.373443642 | 0.003158698 | 0.081418999 | -5.89E-05 | 0.003556003 | 1 | 0.734252407 | 1 | 0.735400932 | ENSG0000017 | 13 MPI |
| SAT | T2DM | ENSG0000013 | 0.000977835 | 0.475122512 | 0.001369192 | 0.081436873 | 0.001022064 | 0.455346363 | 0.001369083 | 0.081419038 | -4.42E-05 | 3.01E-05 | 1 | 0.796607042 | 1 | 0.794600418 | ENSG0000017 | 7 TRIM73 |
| SAT | T2DM | ENSG0000013 | 0.00132051 | 0.673679841 | 0.003135833 | 0.081436929 | 0.00147789 | 0.637411296 | 0.00313563 | 0.081419094 | -0.00015738 | 1.18E-07 | 1 | 0.891627092 | 1 | 0.884719251 | ENSG0000017 | 7 TRIM73 |
| SAT | T2DM | ENSG0000013 | 0.001487511 | 0.249974547 | 0.001293024 | 0.081436737 | 0.001516307 | 0.240883963 | 0.001292915 | 0.081418901 | -2.88E-05 | 0.001737024 | 1 | 0.615333003 | 1 | 0.618903903 | ENSG0000017 | 11 EFCAB13 |
| SAT | T2DM | ENSG0000013 | -0.00032109 | 0.920658703 | 0.003223641 | 0.081436957 | -0.00022929 | 0.943291443 | 0.003223368 | 0.08141913 | -9.18E-05 | 5.44E-05 | 1 | 0.978648294 | 1 | 0.98735976 | ENSG0000017 | 11 EFCAB13 |
| SAT | T2DM | ENSG0000013 | 0.007795714 | 0.038332478 | 0.003763705 | 0.081436241 | 0.007897047 | 0.03587364 | 0.003763433 | 0.081418395 | -0.00010133 | 0.00182526 | 1 | 0.271246148 | 1 | 0.27605566 | ENSG0000017 | 12 HYI |
| SAT | T2DM | ENSG0000013 | -0.00278779 | 0.367415308 | 0.003092991 | 0.081436823 | -0.00284067 | 0.358360313 | 0.003092748 | 0.08141899 | 5.29E-05 | 0.029497334 | 1 | 0.723275188 | 1 | 0.725902637 | ENSG0000017 | 12 HYI |
| SAT | T2DM | ENSG0000013 | -0.00620692 | 0.066560168 | 0.003383186 | 0.081436396 | -0.00591849 | 0.080280691 | 0.003383804 | 0.08141862 | -0.00028843 | 0.000415968 | 1 | 0.358786109 | 1 | 0.409385619 | ENSG0000017 | 4 TUFM |
| SAT | T2DM | ENSG0000013 | 0.006129203 | 0.090105321 | 0.003616378 | 0.081436478 | 0.006184749 | 0.08720134 | 0.003616067 | 0.081418642 | -5.55E-05 | 0.024871188 | 1 | 0.41272218 | 1 | 0.419512605 | ENSG0000017 | 4 TUFM |
| SAT | T2DM | ENSG0000013 | -0.00327335 | 0.355933895 | 0.003545886 | 0.081436816 | -0.00321213 | 0.36496138 | 0.003545577 | 0.081418994 | -6.12E-05 | 0.009779098 | 1 | 0.715575434 | 1 | 0.730228601 | ENSG0000017 | 4 TUFM |
| SAT | T2DM | ENSG0000013 | -0.00143128 | 0.564597586 | 0.002484746 | 0.081436903 | -0.00138417 | 0.577452132 | 0.002484544 | 0.081419079 | -4.71E-05 | 0.011467007 | 1 | 0.837959912 | 1 | 0.849979671 | ENSG0000017 | 11 RMI1 |
| SAT | T2DM | ENSG0000013 | -0.00579299 | 0.101437398 | 0.003536781 | 0.08143651 | -0.00775344 | 0.033223927 | 0.003641185 | 0.081418373 | 0.001960449 | 0.02379487 | 1 | 0.425727224 | 1 | 0.267755401 | ENSG0000017 | 11 RMI1 |
| SAT | T2DM | ENSG0000013 | -0.00062339 | 0.793051095 | 0.002376172 | 0.081436947 | -0.0005432 | 0.819163912 | 0.002375986 | 0.081419122 | -8.02E-05 | 1.99E-05 | 1 | 0.94681813 | 1 | 0.957271954 | ENSG0000017 | 9 CTC1 |
| SAT | T2DM | ENSG0000013 | -0.00054224 | 0.795616401 | 0.002093404 | 0.081436947 | -0.00057644 | 0.783021571 | 0.002093231 | 0.081419118 | 3.42E-05 | 0.026021047 | 1 | 0.94681813 | 1 | 0.945214781 | ENSG0000017 | 9 CTC1 |
| SAT | T2DM | ENSG0000013 | 0.000962369 | 0.7558501 | 0.00309508 | 0.081436942 | 0.00087222 | 0.778072492 | 0.003094824 | 0.081419118 | 9.01E-05 | 6.85E-05 | 1 | 0.932309841 | 1 | 0.941773638 | ENSG0000017 | 9 CTC1 |
| SAT | T2DM | ENSG0000013 | 0.001027259 | 0.791232849 | 0.003880693 | 0.081436947 | -0.0026906 | 0.522183363 | 0.004204161 | 0.081419062 | 0.003717854 | 0.021586999 | 1 | 0.94681813 | 1 | 0.826644476 | ENSG0000017 | 5 C3orf38 |
| SAT | T2DM | ENSG0000013 | -0.00862158 | 0.001092398 | 0.00264012 | 0.081435176 | -0.00856078 | 0.001183505 | 0.002639896 | 0.081417374 | -6.08E-05 | 0.001082603 | 1 | 0.027029242 | 1 | 0.033188714 | ENSG0000017 | 5 C3orf38 |
| SAT | T2DM | ENSG0000013 | -0.00362434 | 0.221518038 | 0.002964691 | 0.081436709 | -0.00358552 | 0.226465925 | 0.002964427 | 0.081418886 | -3.88E-05 | 0.039410407 | 1 | 0.586129584 | 1 | 0.601160762 | ENSG0000017 | 7 KLHDC7A |

| | | | | | | | | | | | | | | | | | | | |
|---|---|---|---|---|---|---|---|---|---|---|---|---|---|---|---|---|---|---|---|
| SAT | T2DM | ENSG0000013 | -0.00050922 | 0.860016577 | 0.002887519 | 0.081436953 | -0.00056095 | 0.845954494 | 0.002887281 | 0.081419125 | 5.17E-05 | 0.015058379 | 1 | 0.965007451 | 1 | 0.96477102 | ENSG0000017 | 11 | FUCA1 |
| SAT | T2DM | ENSG0000013 | 0.001361311 | 0.607101876 | 0.002647358 | 0.081436914 | 0.00142593 | 0.590115565 | 0.002647137 | 0.081419082 | -6.46E-05 | 0.000694919 | 1 | 0.854526494 | 1 | 0.854809759 | ENSG0000017 | 11 | FUCA1 |
| SAT | T2DM | ENSG0000013 | -0.00160695 | 0.529093492 | 0.002553177 | 0.081436892 | -0.00166754 | 0.513643909 | 0.002552973 | 0.08141906 | 6.06E-05 | 0.002155631 | 1 | 0.823749329 | 1 | 0.82328562 | ENSG0000017 | 11 | FUCA1 |
| SAT | T2DM | ENSG0000013 | -0.00488257 | 0.122240781 | 0.003159342 | 0.081436559 | -0.00483395 | 0.125974268 | 0.003159085 | 0.08141874 | -4.86E-05 | 0.039251513 | 1 | 0.459418101 | 1 | 0.47645145 | ENSG0000017 | 11 | FUCA1 |
| SAT | T2DM | ENSG0000013 | 0.000219607 | 0.930806173 | 0.002529138 | 0.081436957 | 0.000159235 | 0.949793948 | 0.002528924 | 0.08141913 | 6.04E-05 | 0.000749565 | 1 | 0.980452059 | 1 | 0.989619036 | ENSG0000017 | 11 | FUCA1 |
| SAT | T2DM | ENSG0000013 | -0.00189397 | 0.330950389 | 0.001948118 | 0.0814368 | -0.00185891 | 0.339939511 | 0.001947957 | 0.081418979 | -3.51E-05 | 0.013220015 | 1 | 0.693389672 | 1 | 0.707255253 | ENSG0000017 | 11 | FUCA1 |
| SAT | T2DM | ENSG0000013 | -0.00134477 | 0.681903002 | 0.003280982 | 0.08143693 | -0.00122444 | 0.708983417 | 0.003280716 | 0.081419108 | -0.00012033 | 1.02E-06 | 1 | 0.892786811 | 1 | 0.912941362 | ENSG0000017 | 11 | FUCA1 |
| SAT | T2DM | ENSG0000013 | 0.001006544 | 0.801636484 | 0.004006465 | 0.081436948 | 0.00093213 | 0.816013311 | 0.004006134 | 0.081419122 | 7.44E-05 | 0.01109924 | 1 | 0.948256967 | 1 | 0.95619274 | ENSG0000017 | 11 | CDH4 |
| SAT | T2DM | ENSG0000013 | -0.00098644 | 0.783256838 | 0.003586029 | 0.081436946 | -0.00105904 | 0.767729485 | 0.003585755 | 0.081419116 | 7.26E-05 | 0.012877893 | 1 | 0.942378322 | 1 | 0.937978714 | ENSG0000017 | 9 | CTGLF12P |
| SAT | T2DM | ENSG0000013 | -1.91E-05 | 0.989872977 | 0.001503377 | 0.081436958 | -4.10E-05 | 0.978245395 | 0.00150324 | 0.081419131 | 2.19E-05 | 0.01678826 | 1 | 0.996461007 | 1 | 0.99701704 | ENSG0000017 | 12 | NSUN7 |
| SAT | T2DM | ENSG0000013 | 0.002128049 | 0.150415917 | 0.001479799 | 0.081436613 | 0.002094016 | 0.157012947 | 0.00147967 | 0.081418796 | 3.40E-05 | 0.000522074 | 1 | 0.496972349 | 1 | 0.51775889 | ENSG0000017 | 12 | NSUN7 |
| SAT | T2DM | ENSG0000013 | -0.00502053 | 0.176397552 | 0.003713594 | 0.081436653 | -0.00484244 | 0.192209261 | 0.003713316 | 0.081418847 | -0.00017809 | 6.86E-09 | 1 | 0.530526907 | 1 | 0.560852203 | ENSG0000017 | 12 | HLA-DQB1 |
| SAT | T2DM | ENSG0000013 | -0.00685343 | 0.070996351 | 0.003795854 | 0.081436414 | -0.00677388 | 0.074310343 | 0.003795524 | 0.081418599 | -7.96E-05 | 0.001767733 | 1 | 0.373189908 | 1 | 0.395620568 | ENSG0000017 | 12 | HLA-DQB1 |
| SAT | T2DM | ENSG0000013 | -3.86E-05 | 0.9818333 | 0.001697285 | 0.081436958 | -8.75E-05 | 0.958865745 | 0.001697153 | 0.08141913 | 4.89E-05 | 0.000307274 | 1 | 0.995935491 | 1 | 0.993672317 | ENSG0000017 | 13 | ARID3B |
| SAT | T2DM | ENSG0000013 | -0.00018059 | 0.953442013 | 0.00309317 | 0.081436958 | -0.00024944 | 0.935720184 | 0.003092897 | 0.08141913 | 6.88E-05 | 0.000590639 | 1 | 0.988996775 | 1 | 0.984952815 | ENSG0000017 | 13 | ARID3B |
| SAT | T2DM | ENSG0000013 | 0.003938397 | 0.008570613 | 0.001498211 | 0.081435804 | 0.003964673 | 0.008133198 | 0.001498081 | 0.081417961 | -2.63E-05 | 0.009710353 | 1 | 0.109327018 | 1 | 0.115021654 | ENSG0000017 | 6 | LINC00174 |
| SAT | T2DM | ENSG0000013 | 0.001772997 | 0.349328074 | 0.001894441 | 0.081436812 | 0.001743405 | 0.35739188 | 0.00189429 | 0.081418989 | 2.96E-05 | 0.043258213 | 1 | 0.708371701 | 1 | 0.725119681 | ENSG0000017 | 6 | LINC00174 |
| SAT | T2DM | ENSG0000013 | -0.00286751 | 0.157065145 | 0.002026491 | 0.081436624 | -0.00281844 | 0.164251564 | 0.002026322 | 0.081418808 | -4.91E-05 | 0.000832784 | 1 | 0.507474184 | 1 | 0.523492072 | ENSG0000017 | 6 | LINC00174 |
| SAT | T2DM | ENSG0000013 | 0.001743186 | 0.287587672 | 0.001639212 | 0.081436769 | 0.000644013 | 0.698528281 | 0.001662793 | 0.081419106 | 0.001099173 | 8.69E-05 | 1 | 0.644566475 | 1 | 0.908954249 | ENSG0000017 | 6 | GEMIN4 |
| SAT | T2DM | ENSG0000013 | 0.001553902 | 0.50124565 | 0.002310526 | 0.081436883 | 0.001516549 | 0.511556029 | 0.002310342 | 0.081419059 | 3.74E-05 | 0.036720583 | 1 | 0.810314363 | 1 | 0.821142531 | ENSG0000017 | 6 | GEMIN4 |
| SAT | T2DM | ENSG0000013 | 0.001919888 | 0.468203953 | 0.002646643 | 0.08143687 | 0.001847279 | 0.485164569 | 0.002646453 | 0.081419049 | 7.26E-05 | 0.001609611 | 1 | 0.79334291 | 1 | 0.806395618 | ENSG0000017 | 11 | LBX2 |
| SAT | T2DM | ENSG0000013 | 0.006017389 | 0.095838551 | 0.003613233 | 0.081436495 | -0.00331391 | 0.395732901 | 0.003902086 | 0.08141901 | 0.009331304 | 2.46E-10 | 1 | 0.420328914 | 1 | 0.755715818 | ENSG0000017 | 11 | LBX2 |
| SAT | T2DM | ENSG0000013 | 0.006087638 | 0.00918323 | 0.002336756 | 0.081435824 | 0.00499607 | 0.036343647 | 0.002386961 | 0.081418399 | 0.001091568 | 0.025353363 | 1 | 0.114563415 | 1 | 0.276092282 | ENSG0000017 | 6 | FCER1A |
| SAT | T2DM | ENSG0000013 | -0.00142096 | 0.674922021 | 0.003388061 | 0.081436929 | -0.00046658 | 0.890718955 | 0.00339594 | 0.081419128 | -0.00095438 | 5.47E-05 | 1 | 0.892191439 | 1 | 0.971822875 | ENSG0000017 | 6 | FCER1A |
| SAT | T2DM | ENSG0000013 | 0.001254617 | 0.073262163 | 0.000700435 | 0.081436422 | 0.001860896 | 0.009707149 | 0.000719576 | 0.081418013 | -0.00060628 | 0.000242606 | 1 | 0.37912505 | 1 | 0.129468108 | ENSG0000017 | 6 | FCER1A |
| SAT | T2DM | ENSG0000013 | 0.007147416 | 0.016053492 | 0.002968559 | 0.08143599 | 0.007094266 | 0.016848213 | 0.002968298 | 0.081418176 | 5.31E-05 | 0.006170277 | 1 | 0.162066172 | 1 | 0.179453096 | ENSG0000017 | 9 | KIAA1875 |
| SAT | T2DM | ENSG0000013 | -0.00411067 | 0.057903821 | 0.002167582 | 0.081436357 | -0.00415061 | 0.055490504 | 0.002167397 | 0.081418518 | 3.99E-05 | 0.008164628 | 1 | 0.333873157 | 1 | 0.341997131 | ENSG0000017 | 9 | KIAA1875 |
| SAT | T2DM | ENSG0000013 | 0.000445042 | 0.871279855 | 0.002746611 | 0.081436954 | 0.000499131 | 0.855785771 | 0.002746379 | 0.081419125 | -5.41E-05 | 0.005265412 | 1 | 0.966468088 | 1 | 0.965566132 | ENSG0000017 | 9 | KIAA1875 |
| SAT | T2DM | ENSG0000013 | -0.00687605 | 0.005646764 | 0.002484453 | 0.081435678 | -0.00679484 | 0.006235207 | 0.002484254 | 0.081417881 | -8.12E-05 | 1.90E-05 | 1 | 0.082656666 | 1 | 0.095436475 | ENSG0000017 | 7 | ATOH7 |
| SAT | T2DM | ENSG0000013 | 0.001244482 | 0.48589762 | 0.00178587 | 0.081436877 | 0.001270077 | 0.476934427 | 0.001785716 | 0.081419046 | -2.56E-05 | 0.035425146 | 1 | 0.801645999 | 1 | 0.80363893 | ENSG0000017 | 6 | B3GNT3 |
| SAT | T2DM | ENSG0000013 | -0.00041188 | 0.917451636 | 0.003973993 | 0.081436957 | -0.00051936 | 0.89601489 | 0.003973752 | 0.081419128 | 0.000107476 | 0.006160403 | 1 | 0.977324291 | 1 | 0.9725607 | ENSG0000017 | 10 | SSC5D |
| SAT | T2DM | ENSG0000013 | -0.00176571 | 0.073972253 | 0.000988203 | 0.081436425 | -0.00177958 | 0.071707144 | 0.000988119 | 0.081418589 | 1.39E-05 | 0.044296531 | 1 | 0.379223381 | 1 | 0.390815844 | ENSG0000017 | 10 | RP11-1319K7.1 |
| SAT | T2DM | ENSG0000013 | 0.001431988 | 0.697578568 | 0.003685073 | 0.081436933 | 0.001488366 | 0.686268759 | 0.003684766 | 0.081419104 | -5.64E-05 | 0.034514614 | 1 | 0.900770568 | 1 | 0.902825425 | ENSG0000017 | 10 | RP11-1319K7.1 |
| SAT | T2DM | ENSG0000013 | 0.001378984 | 0.632133142 | 0.002880524 | 0.08143692 | 0.00133196 | 0.643764698 | 0.002880275 | 0.081419095 | 4.70E-05 | 0.016338723 | 1 | 0.871484928 | 1 | 0.888950368 | ENSG0000017 | 7 | CRIPAK |
| SAT | T2DM | ENSG0000013 | 0.001688984 | 0.509508456 | 0.002560604 | 0.081436886 | 0.001638196 | 0.522290155 | 0.002560403 | 0.081419062 | 5.08E-05 | 0.011563607 | 1 | 0.812130745 | 1 | 0.826644476 | ENSG0000017 | 7 | CRIPAK |
| SAT | T2DM | ENSG0000013 | 0.007750453 | 0.00067346 | 0.002279422 | 0.081435026 | 0.007705973 | 0.00072242 | 0.002279235 | 0.081417221 | 4.45E-05 | 0.008182761 | 1 | 0.018725617 | 1 | 0.023237829 | ENSG0000017 | 7 | CRIPAK |
| SAT | T2DM | ENSG0000013 | 0.002055401 | 0.060898897 | 0.001096662 | 0.081436371 | 0.002033288 | 0.063707847 | 0.001096571 | 0.081418556 | 2.21E-05 | 0.005323779 | 1 | 0.34323074 | 1 | 0.366069085 | ENSG0000017 | 7 | CRIPAK |
| SAT | T2DM | ENSG0000013 | 0.000551151 | 0.59975465 | 0.001050304 | 0.081436912 | 0.000519876 | 0.620586391 | 0.001050213 | 0.08141909 | 3.13E-05 | 1.15E-05 | 1 | 0.850754434 | 1 | 0.87257824 | ENSG0000018 | 4 | TMEM86B |
| SAT | T2DM | ENSG0000013 | 0.003170958 | 0.148828592 | 0.002196435 | 0.08143661 | 0.003220947 | 0.142496702 | 0.002196262 | 0.081418772 | -5.00E-05 | 0.003477451 | 1 | 0.493635361 | 1 | 0.500269293 | ENSG0000018 | 4 | TMEM86B |
| SAT | T2DM | ENSG0000013 | 0.005214399 | 0.050529547 | 0.002666586 | 0.081436319 | 0.005157969 | 0.053059068 | 0.002666382 | 0.081418506 | 5.64E-05 | 0.009040917 | 1 | 0.311148486 | 1 | 0.332861953 | ENSG0000018 | 4 | TMEM86B |
| SAT | T2DM | ENSG0000013 | -0.00287405 | 0.24412855 | 0.002467565 | 0.081436732 | -0.00381035 | 0.124419573 | 0.002479909 | 0.081418736 | 0.0009363 | 0.000178503 | 1 | 0.61016229 | 1 | 0.474800718 | ENSG0000018 | 11 | TDRD6 |
| SAT | T2DM | ENSG0000013 | -0.00480574 | 0.03667407 | 0.002300077 | 0.081436229 | -0.00485583 | 0.034749639 | 0.002299959 | 0.08141838 | 5.01E-05 | 0.043588674 | 1 | 0.266450175 | 1 | 0.272172417 | ENSG0000018 | 14 | LYNX1 |
| SAT | T2DM | ENSG0000013 | -0.00246071 | 0.107473648 | 0.001528717 | 0.081436525 | -0.00264264 | 0.08403993 | 0.001529556 | 0.081418632 | 0.00018193 | 0.001047647 | 1 | 0.435334617 | 1 | 0.414575198 | ENSG0000018 | 7 | FAHD1 |
| SAT | T2DM | ENSG0000013 | -0.00077277 | 0.825161575 | 0.003498102 | 0.08143695 | -0.0008829 | 0.800722112 | 0.003497832 | 0.08141912 | 0.000110127 | 8.82E-05 | 1 | 0.955051074 | 1 | 0.952160336 | ENSG0000018 | 7 | FAHD1 |
| SAT | T2DM | ENSG0000013 | 0.002443029 | 0.178189775 | 0.001814555 | 0.081436655 | 0.002359415 | 0.193492646 | 0.001814491 | 0.081418848 | 8.36E-05 | 0.000155509 | 1 | 0.530526907 | 1 | 0.561375505 | ENSG0000018 | 7 | FAHD1 |
| SAT | T2DM | ENSG0000013 | -0.00106635 | 0.219303927 | 0.000868095 | 0.081436706 | -0.00103077 | 0.235076024 | 0.0008681 | 0.081418895 | -3.56E-05 | 0.006842311 | 1 | 0.584513552 | 1 | 0.611160407 | ENSG0000018 | 7 | FAHD1 |
| SAT | T2DM | ENSG0000013 | 0.002330904 | 0.556427527 | 0.003963067 | 0.081436901 | 0.002544108 | 0.520921628 | 0.003963243 | 0.081419062 | -0.0002132 | 0.002171355 | 1 | 0.835948344 | 1 | 0.826644476 | ENSG0000018 | 5 | RP1-278E11.3 |
| SAT | T2DM | ENSG0000013 | -0.00192898 | 0.295841774 | 0.001845215 | 0.081436776 | -0.00198779 | 0.28132279 | 0.00184507 | 0.081418937 | 5.88E-05 | 5.15E-05 | 1 | 0.653201117 | 1 | 0.647765649 | ENSG0000018 | 5 | RP1-278E11.3 |
| SAT | T2DM | ENSG0000013 | 0.00309971 | 0.379810511 | 0.003529432 | 0.08143683 | 0.003006197 | 0.394315043 | 0.003529151 | 0.08141901 | 9.35E-05 | 0.000606725 | 1 | 0.732172907 | 1 | 0.754765468 | ENSG0000018 | 6 | HCLS1 |
| SAT | T2DM | ENSG0000013 | -0.00092498 | 0.565130977 | 0.001607995 | 0.081436903 | -0.00095895 | 0.55089925 | 0.001607858 | 0.081419071 | 3.40E-05 | 0.002453302 | 1 | 0.837959912 | 1 | 0.842050121 | ENSG0000018 | 7 | MCFD2 |
| SAT | T2DM | ENSG0000013 | -0.00574744 | 0.075517186 | 0.00323378 | 0.081436431 | -0.00585413 | 0.070241947 | 0.0032337 | 0.081418583 | 0.000106689 | 0.011269687 | 1 | 0.382364025 | 1 | 0.386976108 | ENSG0000018 | 7 | MCFD2 |
| SAT | T2DM | ENSG0000013 | -0.00153085 | 0.432022564 | 0.001948304 | 0.081436855 | -0.00157052 | 0.420151044 | 0.001948148 | 0.081419022 | 3.97E-05 | 0.008068386 | 1 | 0.772274669 | 1 | 0.769260924 | ENSG0000018 | 6 | GLIPR1L2 |
| SAT | T2DM | ENSG0000014 | 0.007254543 | 0.015633641 | 0.003001011 | 0.081435982 | 0.007303191 | 0.014942245 | 0.003000755 | 0.081418141 | -4.86E-05 | 0.019717312 | 1 | 0.159794321 | 1 | 0.166753416 | ENSG0000018 | 3 | BHLHA15 |
| SAT | T2DM | ENSG0000014 | -0.0006678 | 0.769898875 | 0.002283031 | 0.081436944 | -0.00070178 | 0.758525855 | 0.002282827 | 0.081419115 | 3.40E-05 | 0.019224328 | 1 | 0.935400227 | 1 | 0.932894921 | ENSG0000018 | 8 | RNF182 |
| SAT | T2DM | ENSG0000014 | 0.001189239 | 0.513168433 | 0.001818651 | 0.081436887 | 0.001234175 | 0.497351376 | 0.001818537 | 0.081419054 | -4.49E-05 | 0.010446215 | 1 | 0.81574885 | 1 | 0.81497549 | ENSG0000018 | 8 | RNF182 |
| SAT | T2DM | ENSG0000014 | -0.00780003 | 0.053911549 | 0.004046603 | 0.081436338 | -0.00769578 | 0.057181299 | 0.004046307 | 0.081418526 | -0.00010425 | 0.002478655 | 1 | 0.322192699 | 1 | 0.345923743 | ENSG0000018 | 3 | TSPYL5 |
| SAT | T2DM | ENSG0000014 | 0.007213607 | 0.026060408 | 0.003241608 | 0.081436131 | 0.007161191 | 0.027152614 | 0.00324135 | 0.081418315 | 5.24E-05 | 0.036110204 | 1 | 0.221604234 | 1 | 0.240777641 | ENSG0000018 | 3 | TSPYL5 |
| SAT | T2DM | ENSG0000014 | -0.00495139 | 0.166066208 | 0.00357512 | 0.081436638 | -0.00500766 | 0.161272195 | 0.003574839 | 0.081418803 | 5.63E-05 | 0.046044114 | 1 | 0.518423625 | 1 | 0.520626624 | ENSG0000018 | 9 | ZBTB12P1 |
| SAT | T2DM | ENSG0000014 | -0.0032624 | 0.222345476 | 0.002673406 | 0.08143671 | -0.00330995 | 0.215640428 | 0.002673181 | 0.081418875 | 4.75E-05 | 0.012389255 | 1 | 0.587161187 | 1 | 0.587638077 | ENSG0000018 | 9 | ZBTB12P1 |
| SAT | T2DM | ENSG0000014 | 0.000181965 | 0.924244678 | 0.001913645 | 0.081436957 | 0.000226136 | 0.905924139 | 0.001913473 | 0.081419129 | -4.42E-05 | 0.000225824 | 1 | 0.978648294 | 1 | 0.975470314 | ENSG0000018 | 10 | SLC36A4 |
| SAT | T2DM | ENSG0000014 | -0.01032613 | 0.000128161 | 0.002696119 | 0.081434507 | -0.01026109 | 0.000141143 | 0.002695897 | 0.08141671 | -6.50E-05 | 0.00104193 | 0.420494863 | 0.006094128 | 0.463090023 | 0.007718167 | ENSG0000018 | 8 | OR51E1 |
| SAT | T2DM | ENSG0000014 | -0.01649626 | 6.72E-06 | 0.003663866 | 0.081433571 | -0.01643338 | 7.27E-06 | 0.003663559 | 0.081415769 | -6.29E-05 | 0.016413767 | 0.022048263 | 0.000501097 | 0.023857506 | 0.000627829 | ENSG0000018 | 7 | PSMG4 |
| SAT | T2DM | ENSG0000014 | 0.003923258 | 0.289670348 | 0.00370522 | 0.081436771 | 0.003975843 | 0.283213393 | 0.003704904 | 0.081418938 | -5.26E-05 | 0.041688795 | 1 | 0.646536335 | 1 | 0.649244222 | ENSG0000018 | 7 | PSMG4 |
| SAT | T2DM | ENSG0000014 | -0.00061695 | 0.75946359 | 0.00201495 | 0.081436943 | -0.00070883 | 0.724981231 | 0.002014806 | 0.08141911 | 9.19E-05 | 1.55E-07 | 1 | 0.934086217 | 1 | 0.921192527 | ENSG0000018 | 1 | BHLHE22 |
| SAT | T2DM | ENSG0000014 | 0.006836584 | 0.078487563 | 0.003885453 | 0.081436441 | 0.006759602 | 0.081882853 | 0.003885124 | 0.081418625 | 7.70E-05 | 0.004859156 | 1 | 0.391645528 | 1 | 0.411433676 | ENSG0000018 | 1 | BHLHE22 |
| SAT | T2DM | ENSG0000014 | 0.004662432 | 0.077842448 | 0.002644079 | 0.081436439 | 0.004579259 | 0.083269874 | 0.002643877 | 0.08141863 | 8.32E-05 | 0.000104948 | 1 | 0.389330903 | 1 | 0.414575198 | ENSG0000018 | 1 | BHLHE22 |
| SAT | T2DM | ENSG0000014 | 0.005172921 | 0.146882367 | 0.003565972 | 0.081436607 | 0.005230425 | 0.142406975 | 0.003565659 | 0.081418771 | -5.75E-05 | 0.014673835 | 1 | 0.492391981 | 1 | 0.500269293 | ENSG0000018 | 3 | MAP6D1 |
| SAT | T2DM | ENSG0000014 | -0.00319684 | 0.255273028 | 0.002810073 | 0.081436742 | -0.00314119 | 0.263598491 | 0.002809829 | 0.081418922 | -5.57E-05 | 0.003156272 | 1 | 0.620408004 | 1 | 0.63733725 | ENSG0000018 | 3 | MAP6D1 |
| SAT | T2DM | ENSG0000014 | -0.00167328 | 0.222623748 | 0.001372007 | 0.08143671 | -0.0016535 | 0.228101495 | 0.001371893 | 0.081418888 | -1.98E-05 | 0.049840799 | 1 | 0.587161187 | 1 | 0.602834606 | ENSG0000018 | 11 | ZNF443 |
| SAT | T2DM | ENSG0000014 | 0.00185597 | 0.234220469 | 0.001560217 | 0.081436722 | 0.001905931 | 0.221834482 | 0.001560107 | 0.081418882 | -5.00E-05 | 0.000310033 | 1 | 0.598969104 | 1 | 0.596100684 | ENSG0000018 | 11 | ZNF443 |
| SAT | T2DM | ENSG0000014 | 0.000445449 | 0.850427955 | 0.002362218 | 0.081436952 | 0.000498904 | 0.832715925 | 0.002362016 | 0.081419123 | -5.35E-05 | 0.001071594 | 1 | 0.963534946 | 1 | 0.961544037 | ENSG0000018 | 11 | ZNF443 |
| SAT | T2DM | ENSG0000014 | 0.005743679 | 0.027806956 | 0.002610756 | 0.08143615 | 0.005795099 | 0.026427623 | 0.002610551 | 0.081418308 | -5.14E-05 | 0.011976399 | 1 | 0.230185673 | 1 | 0.236264392 | ENSG0000018 | 9 | PDIA3P1 |
| SAT | T2DM | ENSG0000014 | 0.001680209 | 0.390620019 | 0.001957159 | 0.081436835 | 0.001715754 | 0.380635693 | 0.001957001 | 0.081419002 | -3.55E-05 | 0.016735151 | 1 | 0.741307668 | 1 | 0.740726993 | ENSG0000018 | 6 | KCTD2 |
| SAT | T2DM | ENSG0000014 | -0.00155693 | 0.138996865 | 0.00105231 | 0.081436593 | -0.0015404 | 0.143206747 | 0.001052216 | 0.081418773 | -1.65E-05 | 0.01273825 | 1 | 0.483935715 | 1 | 0.501004535 | ENSG0000018 | 7 | ST20 |
| SAT | T2DM | ENSG0000014 | -0.00303742 | 0.443379793 | 0.003962719 | 0.08143686 | -0.00290759 | 0.463073302 | 0.003962389 | 0.081419041 | -0.00012983 | 5.85E-06 | 1 | 0.779180021 | 1 | 0.797402301 | ENSG0000018 | 7 | ST20 |
| SAT | T2DM | ENSG0000014 | -0.00466928 | 0.152171868 | 0.003260887 | 0.081436616 | -0.0046162 | 0.156848553 | 0.003260594 | 0.081418796 | -5.31E-05 | 0.009707257 | 1 | 0.499775674 | 1 | 0.51775889 | ENSG0000018 | 7 | ST20 |
| SAT | T2DM | ENSG0000014 | -0.00252572 | 0.066361443 | 0.001375682 | 0.081436395 | -0.00250137 | 0.068999615 | 0.001375569 | 0.081418578 | -2.44E-05 | 0.016122604 | 1 | 0.358786109 | 1 | 0.38370803 | ENSG0000018 | 5 | LRRC57 |
| SAT | T2DM | ENSG0000014 | 0.004396616 | 0.089510328 | 0.002589306 | 0.081436477 | 0.004345553 | 0.093267161 | 0.002589085 | 0.08141866 | 5.11E-05 | 0.004238583 | 1 | 0.411321268 | 1 | 0.428129221 | ENSG0000018 | 5 | LRRC57 |
| SAT | T2DM | ENSG0000014 | -0.00030467 | 0.897439062 | 0.002363646 | 0.081436956 | -0.00021822 | 0.926435047 | 0.002363482 | 0.081419129 | -8.64E-05 | 4.42E-05 | 1 | 0.970180416 | 1 | 0.981878481 | ENSG0000018 | 5 | LRRC57 |
| SAT | T2DM | ENSG0000014 | 0.004667496 | 0.252218047 | 0.004076487 | 0.081436739 | 0.004737239 | 0.245161473 | 0.004076154 | 0.081418905 | -6.97E-05 | 0.021502891 | 1 | 0.617791928 | 1 | 0.620663712 | ENSG0000018 | 5 | MRPL14 |
| SAT | T2DM | ENSG0000014 | -0.00144437 | 0.099312329 | 0.000876334 | 0.081436504 | -0.00142539 | 0.103815782 | 0.000876281 | 0.081418689 | -1.90E-05 | 0.028259417 | 1 | 0.424828882 | 1 | 0.441789338 | ENSG0000018 | 6 | FKRP |
| SAT | T2DM | ENSG0000014 | 0.004076864 | 0.10217109 | 0.002494378 | 0.081436512 | 0.0041172 | 0.098793 | 0.002494159 | 0.081418676 | -4.03E-05 | 0.013993427 | 1 | 0.425727224 | 1 | 0.439213865 | ENSG0000018 | 6 | FKRP |
| SAT | T2DM | ENSG0000014 | 0.001147181 | 0.585266849 | 0.002102188 | 0.081436909 | 0.001087473 | 0.604917498 | 0.002102038 | 0.081419086 | 5.97E-05 | 0.00117083 | 1 | 0.845534946 | 1 | 0.858820558 | ENSG0000018 | 6 | FKRP |
| SAT | T2DM | ENSG0000014 | -0.00215973 | 0.281627047 | 0.002005932 | 0.081436765 | -0.00220188 | 0.272305781 | 0.002005773 | 0.08141893 | 4.21E-05 | 0.006588118 | 1 | 0.641415693 | 1 | 0.64192524 | ENSG0000018 | 6 | FKRP |
| SAT | T2DM | ENSG0000014 | 0.001634986 | 0.315359087 | 0.001628405 | 0.08143679 | 0.001599092 | 0.326062427 | 0.001628276 | 0.08141897 | 3.59E-05 | 0.004520353 | 1 | 0.675828325 | 1 | 0.695122755 | ENSG0000018 | 9 | HIGD1A |
| SAT | T2DM | ENSG0000014 | 0.006445889 | 0.00052482 | 0.001858794 | 0.081434949 | 0.006406391 | 0.000567287 | 0.001858641 | 0.081417146 | 3.95E-05 | 0.004053583 | 1 | 0.015797562 | 1 | 0.019800723 | ENSG0000018 | 9 | HIGD1A |
| SAT | T2DM | ENSG0000014 | -0.00274143 | 0.49047126 | 0.003975622 | 0.081436879 | -0.00281661 | 0.478615629 | 0.003975281 | 0.081419047 | 7.52E-05 | 0.005898374 | 1 | 0.80382562 | 1 | 0.80363893 | ENSG0000018 | 5 | ADIPOQ |
| SAT | T2DM | ENSG0000014 | -0.00988966 | 0.010653092 | 0.003872395 | 0.081435868 | -0.0098063 | 0.011325915 | 0.003872198 | 0.081418059 | -8.34E-05 | 0.046507488 | 1 | 0.127894796 | 1 | 0.141833307 | ENSG0000018 | 5 | ADIPOQ |
| SAT | T2DM | ENSG0000014 | 6.83E-05 | 0.969881927 | 0.001808694 | 0.081436958 | 1.48E-05 | 0.993449141 | 0.001808569 | 0.081419131 | 5.34E-05 | 0.000944816 | 1 | 0.9941737 | 1 | 0.998405122 | ENSG0000018 | 5 | ADIPOQ |
| SAT | T2DM | ENSG0000014 | 0.000897334 | 0.348790459 | 0.000957729 | 0.081436812 | 0.000920139 | 0.336639764 | 0.000957653 | 0.081418977 | -2.28E-05 | 0.002062651 | 1 | 0.708371701 | 1 | 0.704365099 | ENSG0000018 | 5 | ADIPOQ |
| SAT | T2DM | ENSG0000014 | 0.001538162 | 0.336196299 | 0.001599405 | 0.081436804 | 0.001575159 | 0.324664133 | 0.001599277 | 0.081418969 | -3.70E-05 | 0.00268406 | 1 | 0.696147596 | 1 | 0.693504571 | ENSG0000018 | 5 | ADIPOQ |
| SAT | T2DM | ENSG0000014 | -0.00314076 | 0.217107674 | 0.002544656 | 0.081436704 | -0.00303471 | 0.233004889 | 0.002544495 | 0.081418893 | -0.00010605 | 1.45E-05 | 1 | 0.581494104 | 1 | 0.610313962 | ENSG0000018 | 5 | ADIPOQ |
| SAT | T2DM | ENSG0000014 | -0.00137808 | 0.402283613 | 0.001645366 | 0.081436841 | -0.00140522 | 0.393037887 | 0.001645227 | 0.081419009 | 2.71E-05 | 0.019155489 | 1 | 0.752518444 | 1 | 0.753672849 | ENSG0000018 | 5 | ADIPOQ |
| SAT | T2DM | ENSG0000014 | -0.00191639 | 0.467694332 | 0.0026388 | 0.08143687 | -0.00186469 | 0.479759972 | 0.002638626 | 0.081419047 | -5.17E-05 | 0.036255771 | 1 | 0.79334291 | 1 | 0.80363893 | ENSG0000018 | 5 | ADIPOQ |
| SAT | T2DM | ENSG0000014 | 0.000118529 | 0.964345458 | 0.002651579 | 0.081436958 | 0.00019878 | 0.94023598 | 0.002651349 | 0.08141913 | -8.03E-05 | 6.53E-06 | 1 | 0.992060403 | 1 | 0.986202735 | ENSG0000018 | 6 | F2R |
| SAT | T2DM | ENSG0000014 | 0.001470929 | 0.523502347 | 0.002305689 | 0.08143689 | 0.001973878 | 0.392480374 | 0.002308282 | 0.081419009 | -0.00050295 | 1.14E-05 | 1 | 0.821144329 | 1 | 0.753498015 | ENSG0000018 | 6 | F2R |
| SAT | T2DM | ENSG0000014 | -0.00216038 | 0.23860656 | 0.001833191 | 0.081436726 | -0.0021075 | 0.250255905 | 0.00183304 | 0.08141891 | -5.29E-05 | 9.12E-05 | 1 | 0.60584911 | 1 | 0.621566711 | ENSG0000018 | 6 | F2R |
| SAT | T2DM | ENSG0000014 | -0.00538412 | 0.153954568 | 0.003776463 | 0.081436619 | -0.00531023 | 0.15965079 | 0.003776168 | 0.0814188 | -7.39E-05 | 0.013458689 | 1 | 0.504116703 | 1 | 0.519656987 | ENSG0000018 | 6 | F2R |
| SAT | T2DM | ENSG0000014 | -0.00380004 | 0.290259255 | 0.003593237 | 0.081436772 | -0.0038796 | 0.280238227 | 0.003592934 | 0.081418936 | 7.96E-05 | 0.001784204 | 1 | 0.646746051 | 1 | 0.646143093 | ENSG0000018 | 9 | HLA-V |
| SAT | T2DM | ENSG0000014 | 0.001314965 | 0.734326849 | 0.003874659 | 0.081436939 | 0.001426838 | 0.712665032 | 0.003874334 | 0.081419108 | -0.00011187 | 5.70E-05 | 1 | 0.923192618 | 1 | 0.915526221 | ENSG0000018 | 9 | HLA-V |
| SAT | T2DM | ENSG0000014 | -0.00040688 | 0.925028395 | 0.004323833 | 0.081436957 | -0.0004824 | 0.911157371 | 0.00432344 | 0.081419129 | 7.55E-05 | 0.004196027 | 1 | 0.978648294 | 1 | 0.978562139 | ENSG0000018 | 9 | HLA-V |
| SAT | T2DM | ENSG0000014 | -0.00138569 | 0.466726853 | 0.001903898 | 0.08143687 | -7.79E-05 | 0.967871723 | 0.001933552 | 0.081419131 | -0.00130781 | 0.0001118 | 1 | 0.79334291 | 1 | 0.995169891 | ENSG0000018 | 9 | HLA-V |
| SAT | T2DM | ENSG0000014 | -0.00013842 | 0.97058475 | 0.003753717 | 0.081436958 | -6.99E-05 | 0.985142734 | 0.003753404 | 0.081419131 | -6.85E-05 | 0.011770028 | 1 | 0.994419011 | 1 | 0.997162029 | ENSG0000018 | 9 | HLA-V |
| SAT | T2DM | ENSG0000014 | 0.007359933 | 0.038668907 | 0.003559481 | 0.081436244 | 0.007303247 | 0.040174712 | 0.003559168 | 0.081418427 | 5.67E-05 | 0.015083981 | 1 | 0.272844483 | 1 | 0.2890641 | ENSG0000018 | 9 | HLA-V |
| SAT | T2DM | ENSG0000014 | 0.002906456 | 0.359876055 | 0.003174365 | 0.081436818 | 0.003062242 | 0.334800294 | 0.003174982 | 0.081418975 | -0.00015579 | 0.046522885 | 1 | 0.71830389 | 1 | 0.702779133 | ENSG0000018 | 9 | HLA-V |
| SAT | T2DM | ENSG0000014 | -0.00069428 | 0.81648996 | 0.002991796 | 0.081436949 | -0.00065412 | 0.826916236 | 0.002991532 | 0.081419123 | -4.02E-05 | 0.038697129 | 1 | 0.95232974 | 1 | 0.960053847 | ENSG0000018 | 9 | HLA-V |
| SAT | T2DM | ENSG0000014 | -0.00545684 | 0.005914443 | 0.001982505 | 0.081435692 | -0.00351054 | 0.084095897 | 0.002032264 | 0.081418632 | -0.00194631 | 1.39E-05 | 1 | 0.084370819 | 1 | 0.414575198 | ENSG0000018 | 9 | HLA-V |
| SAT | T2DM | ENSG0000014 | 0.002398935 | 0.529248273 | 0.003812951 | 0.081436892 | 0.002499882 | 0.512032163 | 0.003812673 | 0.081419059 | -0.00010095 | 0.001911564 | 1 | 0.823749329 | 1 | 0.821219305 | ENSG0000018 | 4 | HIST3H2A |
| SAT | T2DM | ENSG0000014 | -0.00161284 | 0.638724182 | 0.003435358 | 0.081436922 | -0.00155259 | 0.651282609 | 0.003435066 | 0.081419097 | -6.03E-05 | 0.012117551 | 1 | 0.874326778 | 1 | 0.891013292 | ENSG0000018 | 4 | HIST3H2A |
| SAT | T2DM | ENSG0000014 | 0.006888877 | 0.009463648 | 0.002654832 | 0.081435833 | 0.006932081 | 0.009019349 | 0.002654617 | 0.081417992 | -4.32E-05 | 0.031133029 | 1 | 0.116626425 | 1 | 0.123302012 | ENSG0000018 | 10 | POLR2A |
| SAT | T2DM | ENSG0000014 | 0.00104136 | 0.790360808 | 0.003917211 | 0.081436947 | 0.000959983 | 0.806388288 | 0.003916891 | 0.081419121 | 8.14E-05 | 0.005317451 | 1 | 0.94681813 | 1 | 0.952410029 | ENSG0000018 | 5 | TMEM132E |
| SAT | T2DM | ENSG0000014 | 0.001005512 | 0.496946514 | 0.001480212 | 0.081436881 | 0.001034021 | 0.484792179 | 0.001480098 | 0.081419049 | -2.85E-05 | 0.016946764 | 1 | 0.807969034 | 1 | 0.806185068 | ENSG0000018 | 5 | TMEM132E |
| SAT | T2DM | ENSG0000014 | -0.01067829 | 0.000562712 | 0.003096053 | 0.08143497 | -0.01074647 | 0.000518027 | 0.003095824 | 0.081417118 | 6.82E-05 | 0.009006468 | 1 | 0.016484453 | 1 | 0.018677439 | ENSG0000018 | 6 | ZNF322 |
| SAT | T2DM | ENSG0000014 | 0.004403461 | 0.257910725 | 0.003892238 | 0.081436745 | 0.004294704 | 0.269813006 | 0.003891912 | 0.081418927 | 0.000108758 | 9.86E-05 | 1 | 0.622209623 | 1 | 0.640098679 | ENSG0000018 | 6 | ZNF322 |
| SAT | T2DM | ENSG0000014 | 0.001113835 | 0.65448267 | 0.002488773 | 0.081436925 | 0.001178285 | 0.635872714 | 0.002488575 | 0.081419093 | -6.44E-05 | 0.000848033 | 1 | 0.882995924 | 1 | 0.884482576 | ENSG0000018 | 6 | ZNF322 |
| SAT | T2DM | ENSG0000014 | 0.007409183 | 0.020303209 | 0.003192644 | 0.081436058 | 0.007504038 | 0.018743342 | 0.003192386 | 0.081418208 | -9.49E-05 | 9.44E-05 | 1 | 0.189785835 | 1 | 0.18980526 | ENSG0000018 | 6 | ZNF322 |
| SAT | T2DM | ENSG0000014 | -0.00070292 | 0.820654614 | 0.003100623 | 0.08143695 | -0.00063827 | 0.83689684 | 0.003100434 | 0.081419124 | -6.47E-05 | 0.034569943 | 1 | 0.953693013 | 1 | 0.963362294 | ENSG0000018 | 6 | ZNF322 |
| SAT | T2DM | ENSG0000015 | -0.00069667 | 0.710453958 | 0.001876564 | 0.081436935 | -0.00066002 | 0.725028952 | 0.001876405 | 0.08141911 | -3.67E-05 | 0.005491126 | 1 | 0.907710061 | 1 | 0.921192527 | ENSG0000018 | 8 | OGFOD3 |
| SAT | T2DM | ENSG0000015 | 0.001870133 | 0.539552887 | 0.003048354 | 0.081436896 | 0.00195053 | 0.52222871 | 0.003048113 | 0.081419062 | -8.04E-05 | 0.000721469 | 1 | 0.831736309 | 1 | 0.826644476 | ENSG0000018 | 11 | GPR135 |
| SAT | T2DM | ENSG0000015 | -0.00840472 | 0.009768844 | 0.003252706 | 0.081435843 | -0.00853558 | 0.008687971 | 0.003252759 | 0.08141798 | 0.000130863 | 0.011313822 | 1 | 0.11916337 | 1 | 0.121125172 | ENSG0000018 | 11 | GPR135 |
| SAT | T2DM | ENSG0000015 | -0.00215085 | 0.333901714 | 0.002225891 | 0.081436802 | -0.00218878 | 0.325409196 | 0.002225723 | 0.081418969 | 3.79E-05 | 0.038827213 | 1 | 0.695221474 | 1 | 0.694192179 | ENSG0000018 | 11 | GPR135 |
| SAT | T2DM | ENSG0000015 | -0.00323438 | 0.218892877 | 0.002630688 | 0.081436706 | -0.00317758 | 0.227061185 | 0.002630519 | 0.081418887 | -5.68E-05 | 0.023047398 | 1 | 0.583892301 | 1 | 0.601281476 | ENSG0000018 | 13 | SLX1B |
| SAT | T2DM | ENSG0000015 | -0.0007181 | 0.568146695 | 0.001258103 | 0.081436904 | -0.00074016 | 0.556284491 | 0.001257989 | 0.081419073 | 2.21E-05 | 0.003994074 | 1 | 0.840032807 | 1 | 0.842050121 | ENSG0000018 | 10 | ANKRD62 |
| SAT | T2DM | ENSG0000015 | -0.00249552 | 0.089249924 | 0.001468497 | 0.081436476 | -0.00252921 | 0.084986474 | 0.001468368 | 0.081418635 | 3.37E-05 | 0.000481435 | 1 | 0.410699859 | 1 | 0.414941399 | ENSG0000018 | 4 | C3orf58 |
| SAT | T2DM | ENSG0000015 | -0.01742123 | 2.80E-07 | 0.003391869 | 0.08143255 | -0.01752012 | 2.40E-07 | 0.003391574 | 0.081414672 | 9.89E-05 | 1.55E-05 | 0.00092031 | 3.54E-05 | 0.000785911 | 4.37E-05 | ENSG0000018 | 4 | C3orf58 |
| SAT | T2DM | ENSG0000015 | -0.00120324 | 0.245341095 | 0.001035722 | 0.081436733 | -0.00118814 | 0.251278293 | 0.001035637 | 0.081418911 | -1.51E-05 | 0.049446608 | 1 | 0.61016229 | 1 | 0.622908325 | ENSG0000018 | 6 | AMIGO1 |
| SAT | T2DM | ENSG0000015 | -0.00480968 | 0.235313745 | 0.004052703 | 0.081436723 | -0.00473469 | 0.242657004 | 0.004052377 | 0.081418903 | -7.50E-05 | 0.015033319 | 1 | 0.600737047 | 1 | 0.620663712 | ENSG0000018 | 6 | AMIGO1 |

| | | | | | | | | | | | | | | | | | | | |
|---|---|---|---|---|---|---|---|---|---|---|---|---|---|---|---|---|---|---|---|
| SAT | T2DM | ENSG0000015 | 0.000305893 | 0.850839832 | 0.001626685 | 0.081436953 | 0.000262093 | 0.871988581 | 0.001626561 | 0.081419127 | 4.38E-05 | 0.000949664 | 1 | 0.963534946 | 1 | 0.966383771 | ENSG0000018 | 6 | GPR3 |
| SAT | T2DM | ENSG0000015 | 0.001659821 | 0.209360644 | 0.001322222 | 0.081436695 | 0.001686164 | 0.20218906 | 0.001322128 | 0.081418859 | -2.63E-05 | 0.022502293 | 1 | 0.571950268 | 1 | 0.573364137 | ENSG0000018 | 5 | ODF3L2 |
| SAT | T2DM | ENSG0000015 | -0.0024161 | 0.55504431 | 0.00409356 | 0.0814369 | -0.0022763 | 0.578144141 | 0.004093339 | 0.081419079 | -0.0001398 | 0.001189443 | 1 | 0.835948344 | 1 | 0.849979671 | ENSG0000018 | 5 | ODF3L2 |
| SAT | T2DM | ENSG0000015 | -0.00168677 | 0.267303931 | 0.001520583 | 0.081436753 | -0.00165878 | 0.275285806 | 0.001520461 | 0.081418932 | -2.80E-05 | 0.016398213 | 1 | 0.630625715 | 1 | 0.64327119 | ENSG0000018 | 5 | ODF3L2 |
| SAT | T2DM | ENSG0000015 | 0.001994787 | 0.092965266 | 0.0011874 | 0.081436487 | 0.002018148 | 0.089176329 | 0.001187312 | 0.081418648 | -2.34E-05 | 0.019994617 | 1 | 0.414991889 | 1 | 0.421804746 | ENSG0000018 | 5 | RELL1 |
| SAT | T2DM | ENSG0000015 | 0.003051969 | 0.309037474 | 0.003000231 | 0.081436785 | 0.003163848 | 0.291601221 | 0.002999998 | 0.081418945 | -0.00011188 | 3.15E-06 | 1 | 0.669717274 | 1 | 0.657104124 | ENSG0000018 | 5 | RELL1 |
| SAT | T2DM | ENSG0000015 | 0.001221659 | 0.731283657 | 0.003557345 | 0.081436939 | 0.000856263 | 0.809956209 | 0.003560604 | 0.081419121 | 0.000365395 | 0.02335785 | 1 | 0.92246416 | 1 | 0.953864437 | ENSG0000018 | 5 | RELL1 |
| SAT | T2DM | ENSG0000015 | 0.005872931 | 0.033693748 | 0.002765393 | 0.081436205 | 0.00581314 | 0.035535155 | 0.002765248 | 0.081418392 | 5.98E-05 | 0.042637663 | 1 | 0.256395884 | 1 | 0.274331399 | ENSG0000018 | 14 | CLDN7 |
| SAT | T2DM | ENSG0000015 | 0.000714081 | 0.565619941 | 0.001242929 | 0.081436903 | 0.000734599 | 0.554470961 | 0.001242821 | 0.081419081 | -2.05E-05 | 0.013617143 | 1 | 0.837959912 | 1 | 0.842050121 | ENSG0000018 | 3 | ADO |
| SAT | T2DM | ENSG0000015 | -0.00281361 | 0.141009547 | 0.001911362 | 0.081436596 | -0.00212984 | 0.267636889 | 0.001921331 | 0.081418926 | -0.00068377 | 0.000535394 | 1 | 0.485469384 | 1 | 0.638630278 | ENSG0000018 | 3 | ADO |
| SAT | T2DM | ENSG0000015 | 0.000523052 | 0.875554117 | 0.00333989 | 0.081436954 | 0.000572239 | 0.863949438 | 0.003339612 | 0.081419126 | -4.92E-05 | 0.042519759 | 1 | 0.967171747 | 1 | 0.966241215 | ENSG0000018 | 13 | PNMAL1 |
| SAT | T2DM | ENSG0000015 | -0.00030065 | 0.781748834 | 0.001085224 | 0.081436946 | -0.00032187 | 0.766760421 | 0.001085144 | 0.081419116 | 2.12E-05 | 0.020598694 | 1 | 0.941848527 | 1 | 0.937659687 | ENSG0000018 | 13 | PNMAL1 |
| SAT | T2DM | ENSG0000015 | 0.000497294 | 0.898173359 | 0.003886041 | 0.081436956 | 0.000419987 | 0.913928605 | 0.003885724 | 0.081419129 | 7.73E-05 | 0.007608618 | 1 | 0.970287017 | 1 | 0.979654273 | ENSG0000018 | 7 | TRPC2 |
| SAT | T2DM | ENSG0000015 | -0.0039238 | 0.028450422 | 0.001790853 | 0.081436156 | -0.00394912 | 0.027429245 | 0.001790693 | 0.081418318 | 2.53E-05 | 0.026322033 | 1 | 0.233364583 | 1 | 0.241274407 | ENSG0000018 | 7 | TRPC2 |
| SAT | T2DM | ENSG0000015 | -0.00234641 | 0.550565115 | 0.003930897 | 0.081436899 | -0.00242245 | 0.537687596 | 0.00393055 | 0.081419067 | 7.60E-05 | 0.003078061 | 1 | 0.83552458 | 1 | 0.836091471 | ENSG0000018 | 6 | PTCHD3 |
| SAT | T2DM | ENSG0000015 | -0.00253798 | 0.369513701 | 0.002828198 | 0.081436824 | -0.00246316 | 0.383758556 | 0.002827994 | 0.081419004 | -7.48E-05 | 0.002217915 | 1 | 0.723805643 | 1 | 0.745584853 | ENSG0000018 | 4 | NOP10 |
| SAT | T2DM | ENSG0000015 | -0.00307383 | 0.08547485 | 0.001787352 | 0.081436464 | -0.0030467 | 0.088242513 | 0.001787187 | 0.081418645 | -2.71E-05 | 0.009396773 | 1 | 0.402357222 | 1 | 0.421383296 | ENSG0000018 | 4 | NOP10 |
| SAT | T2DM | ENSG0000015 | 0.004135665 | 0.255921413 | 0.003640276 | 0.081436743 | 0.004066794 | 0.263884432 | 0.003639976 | 0.081418922 | 6.89E-05 | 0.010030618 | 1 | 0.62099686 | 1 | 0.637558779 | ENSG0000018 | 11 | TDRKH |
| SAT | T2DM | ENSG0000015 | -0.00263337 | 0.24020905 | 0.002242184 | 0.081436728 | -0.00266611 | 0.234372374 | 0.002241988 | 0.081418895 | 3.27E-05 | 0.027911955 | 1 | 0.60584911 | 1 | 0.610313962 | ENSG0000018 | 11 | TDRKH |
| SAT | T2DM | ENSG0000015 | 0.001612682 | 0.48021816 | 0.002284406 | 0.081436875 | 0.00158388 | 0.488054285 | 0.002284199 | 0.081419051 | 2.88E-05 | 0.040351307 | 1 | 0.798174155 | 1 | 0.807924374 | ENSG0000018 | 11 | TDRKH |
| SAT | T2DM | ENSG0000015 | -0.00016734 | 0.963701085 | 0.003677049 | 0.081436958 | -0.0002571 | 0.944253648 | 0.003676765 | 0.08141913 | 8.98E-05 | 0.002319576 | 1 | 0.992060403 | 1 | 0.987598412 | ENSG0000018 | 11 | TDRKH |
| SAT | T2DM | ENSG0000015 | 0.004341759 | 0.036637498 | 0.002077609 | 0.081436229 | 0.00425384 | 0.04059844 | 0.002077463 | 0.08141843 | 8.79E-05 | 1.68E-06 | 1 | 0.266450175 | 1 | 0.289075371 | ENSG0000018 | 5 | ZNF708 |
| SAT | T2DM | ENSG0000015 | 0.004170214 | 0.018036729 | 0.0017634 | 0.081436024 | 0.004198059 | 0.017272827 | 0.001763256 | 0.081418184 | -2.78E-05 | 0.034498967 | 1 | 0.175084346 | 1 | 0.179453096 | ENSG0000018 | 5 | ZNF708 |
| SAT | T2DM | ENSG0000015 | -0.00399951 | 0.169725934 | 0.002912788 | 0.081436643 | -0.0040502 | 0.164342834 | 0.002912525 | 0.081418808 | 5.07E-05 | 0.005319719 | 1 | 0.523374802 | 1 | 0.523492072 | ENSG0000018 | 5 | ZNF708 |
| SAT | T2DM | ENSG0000015 | 0.008297923 | 0.002240692 | 0.002714987 | 0.081435397 | 0.008349819 | 0.002100165 | 0.002714765 | 0.08141755 | -5.19E-05 | 0.010379881 | 1 | 0.044750749 | 1 | 0.049971446 | ENSG0000018 | 5 | ZNF708 |
| SAT | T2DM | ENSG0000015 | -0.00792525 | 0.043131303 | 0.003918656 | 0.081436275 | -0.00786642 | 0.044687235 | 0.003918334 | 0.081418457 | -5.88E-05 | 0.042036055 | 1 | 0.286428522 | 1 | 0.306542588 | ENSG0000018 | 5 | ZNF708 |
| SAT | T2DM | ENSG0000015 | 0.00490351 | 0.098232781 | 0.00296556 | 0.081436502 | 0.005001287 | 0.091683571 | 0.002965331 | 0.081418656 | -9.78E-05 | 4.33E-05 | 1 | 0.421965667 | 1 | 0.424878243 | ENSG0000018 | 11 | ERCC6L2 |
| SAT | T2DM | ENSG0000015 | -0.00123438 | 0.603613902 | 0.002377416 | 0.081436913 | -0.00116057 | 0.625408796 | 0.002377251 | 0.081419091 | -7.38E-05 | 0.000548614 | 1 | 0.852910082 | 1 | 0.876644981 | ENSG0000018 | 11 | ERCC6L2 |
| SAT | T2DM | ENSG0000015 | -0.00438669 | 0.045038569 | 0.002188641 | 0.081436287 | -0.00443736 | 0.042601724 | 0.002188479 | 0.081418444 | 5.07E-05 | 0.006041824 | 1 | 0.292287338 | 1 | 0.296764878 | ENSG0000018 | 11 | ERCC6L2 |
| SAT | T2DM | ENSG0000015 | -0.00047864 | 0.886263066 | 0.003346341 | 0.081436955 | -0.0005983 | 0.858090107 | 0.003346061 | 0.081419126 | 0.000119654 | 6.60E-07 | 1 | 0.969599573 | 1 | 0.965566132 | ENSG0000018 | 7 | MRPL41 |
| SAT | T2DM | ENSG0000015 | -0.0016579 | 0.151985124 | 0.0011573 | 0.081436615 | -0.00167599 | 0.147530462 | 0.001157202 | 0.08141878 | 1.81E-05 | 0.025503576 | 1 | 0.499700102 | 1 | 0.506773649 | ENSG0000018 | 7 | FAM153B |
| SAT | T2DM | ENSG0000015 | 0.003719904 | 0.137178047 | 0.002502653 | 0.081436589 | 0.003754187 | 0.133559858 | 0.002502436 | 0.081418755 | -3.43E-05 | 0.042639473 | 1 | 0.480267343 | 1 | 0.486825442 | ENSG0000018 | 11 | BACE2 |
| SAT | T2DM | ENSG0000015 | -0.00155972 | 0.620048765 | 0.003145989 | 0.081436917 | -0.00163636 | 0.602934252 | 0.003145727 | 0.081419086 | 7.66E-05 | 0.000774243 | 1 | 0.863241392 | 1 | 0.858233093 | ENSG0000018 | 11 | BACE2 |
| SAT | T2DM | ENSG0000015 | -0.00498071 | 0.182573693 | 0.003736819 | 0.081436662 | -0.00486801 | 0.192637005 | 0.003736523 | 0.081418847 | -0.0001127 | 9.98E-05 | 1 | 0.537241514 | 1 | 0.560852203 | ENSG0000018 | 11 | BACE2 |
| SAT | T2DM | ENSG0000015 | -0.00430266 | 0.264654294 | 0.003857302 | 0.08143675 | -0.00411853 | 0.28561134 | 0.003857021 | 0.0814189 | -0.00018414 | 2.32E-08 | 1 | 0.627688499 | 1 | 0.651854486 | ENSG0000018 | 11 | BACE2 |
| SAT | T2DM | ENSG0000015 | -0.00254248 | 0.219596047 | 0.002071092 | 0.081436707 | -0.00251263 | 0.225019639 | 0.00207092 | 0.081418885 | -2.99E-05 | 0.046477809 | 1 | 0.58481707 | 1 | 0.599260906 | ENSG0000018 | 11 | BACE2 |
| SAT | T2DM | ENSG0000016 | 0.004731339 | 0.115718894 | 0.003007839 | 0.081436545 | 0.004846522 | 0.10709036 | 0.003007622 | 0.081418697 | -0.00011518 | 9.56E-06 | 1 | 0.447728409 | 1 | 0.448503105 | ENSG0000018 | 11 | BACE2 |
| SAT | T2DM | ENSG0000016 | 0.001090884 | 0.680352575 | 0.002647891 | 0.08143693 | 0.001155859 | 0.662430873 | 0.002647653 | 0.081419099 | -6.50E-05 | 7.95E-05 | 1 | 0.892786811 | 1 | 0.895728881 | ENSG0000018 | 9 | YBEY |
| SAT | T2DM | ENSG0000016 | 0.000193255 | 0.81157954 | 0.000810671 | 0.081436949 | 0.000213868 | 0.791904685 | 0.000810602 | 0.081419119 | -2.06E-05 | 0.000279552 | 1 | 0.951539016 | 1 | 0.948608715 | ENSG0000018 | 9 | YBEY |
| SAT | T2DM | ENSG0000016 | -0.00376609 | 0.214301917 | 0.003032708 | 0.081436701 | -0.00368373 | 0.224454614 | 0.003032453 | 0.081418884 | -8.24E-05 | 0.000134861 | 1 | 0.576804422 | 1 | 0.598728122 | ENSG0000018 | 14 | CACNB4 |
| SAT | T2DM | ENSG0000016 | 0.00063652 | 0.606728611 | 0.001236563 | 0.081436914 | 0.000616063 | 0.618309323 | 0.001236457 | 0.081419089 | 2.05E-05 | 0.015858551 | 1 | 0.854526494 | 1 | 0.87067506 | ENSG0000018 | 9 | NPLOC4 |
| SAT | T2DM | ENSG0000016 | 0.000516054 | 0.736768077 | 0.00153525 | 0.08143694 | 0.000553909 | 0.718233773 | 0.001535137 | 0.081419109 | -3.79E-05 | 0.003736177 | 1 | 0.924058127 | 1 | 0.918559531 | ENSG0000018 | 9 | NPLOC4 |
| SAT | T2DM | ENSG0000016 | -0.00055874 | 0.890890724 | 0.004073103 | 0.081436955 | -0.00068861 | 0.86573754 | 0.004072821 | 0.081419126 | 0.000129876 | 0.000390422 | 1 | 0.970180416 | 1 | 0.966241215 | ENSG0000018 | 4 | CAPN12 |
| SAT | T2DM | ENSG0000016 | -0.00076508 | 0.824969223 | 0.003459443 | 0.08143695 | -0.00087272 | 0.800814939 | 0.003459151 | 0.08141912 | 0.000107634 | 1.04E-05 | 1 | 0.955051074 | 1 | 0.952160336 | ENSG0000018 | 7 | FES |
| SAT | T2DM | ENSG0000016 | 0.003598193 | 0.147447466 | 0.002483896 | 0.081436608 | 0.003910715 | 0.115888524 | 0.002487305 | 0.081418718 | -0.00031252 | 0.02088501 | 1 | 0.492642703 | 1 | 0.457557457 | ENSG0000018 | 7 | FES |
| SAT | T2DM | ENSG0000016 | 0.001856793 | 0.521976466 | 0.00289987 | 0.08143689 | 0.001922878 | 0.50724066 | 0.002899653 | 0.081419057 | -6.61E-05 | 0.006155199 | 1 | 0.821144329 | 1 | 0.817738235 | ENSG0000018 | 7 | FES |
| SAT | T2DM | ENSG0000016 | -0.00316991 | 0.266342437 | 0.00285185 | 0.081436752 | -0.00323238 | 0.256994885 | 0.002851623 | 0.081418916 | 6.25E-05 | 0.004834734 | 1 | 0.630350538 | 1 | 0.629723837 | ENSG0000018 | 4 | GLRX5 |
| SAT | T2DM | ENSG0000016 | 0.003612786 | 0.003342325 | 0.001231193 | 0.081435519 | 0.003632437 | 0.003172084 | 0.001231099 | 0.081417676 | -1.97E-05 | 0.049294353 | 1 | 0.059276589 | 1 | 0.064643525 | ENSG0000018 | 6 | CAV3 |
| SAT | T2DM | ENSG0000016 | 0.004470581 | 0.25656993 | 0.003940446 | 0.081436743 | 0.004412831 | 0.262722394 | 0.003940094 | 0.081418921 | 5.78E-05 | 0.020913932 | 1 | 0.62099686 | 1 | 0.636156586 | ENSG0000018 | 9 | MXRA7 |
| SAT | T2DM | ENSG0000016 | -0.00280012 | 0.389352187 | 0.003252957 | 0.081436835 | -0.00274999 | 0.397856731 | 0.003252666 | 0.081419011 | -5.01E-05 | 0.014784164 | 1 | 0.740559146 | 1 | 0.756180206 | ENSG0000018 | 9 | MXRA7 |
| SAT | T2DM | ENSG0000016 | 0.001096569 | 0.582076703 | 0.001992479 | 0.081436908 | 0.001048894 | 0.598566016 | 0.001992337 | 0.081419085 | 4.77E-05 | 0.006153649 | 1 | 0.845212395 | 1 | 0.855952281 | ENSG0000018 | 9 | MXRA7 |
| SAT | T2DM | ENSG0000016 | -4.89E-05 | 0.987324677 | 0.003078336 | 0.081436958 | 5.15E-05 | 0.986645136 | 0.003078083 | 0.081419131 | -0.00010043 | 1.00E-05 | 1 | 0.996461007 | 1 | 0.997162029 | ENSG0000018 | 8 | MFSD5 |
| SAT | T2DM | ENSG0000016 | 0.001688814 | 0.556449614 | 0.002871529 | 0.081436901 | 0.00163296 | 0.569547521 | 0.002871298 | 0.081419077 | 5.59E-05 | 0.010651034 | 1 | 0.835948344 | 1 | 0.845822679 | ENSG0000018 | 10 | TRAK1 |
| SAT | T2DM | ENSG0000016 | -0.00196821 | 0.436343201 | 0.002474285 | 0.081436853 | -0.00200615 | 0.417439534 | 0.002474065 | 0.081419021 | 3.79E-05 | 0.016945467 | 1 | 0.77145714 | 1 | 0.768154297 | ENSG0000018 | 9 | TTC3 |
| SAT | T2DM | ENSG0000016 | 0.000200645 | 0.949561777 | 0.003171894 | 0.081436958 | 0.000146275 | 0.963214666 | 0.003171617 | 0.081419131 | 5.44E-05 | 0.009790298 | 1 | 0.987171163 | 1 | 0.993672317 | ENSG0000018 | 3 | BRICD5 |
| SAT | T2DM | ENSG0000016 | 0.000465029 | 0.86361175 | 0.002707145 | 0.081436953 | -0.00010572 | 0.96885971 | 0.002708145 | 0.081419131 | 0.000570751 | 9.54E-12 | 1 | 0.965214867 | 1 | 0.995561763 | ENSG0000018 | 13 | GRID1 |
| SAT | T2DM | ENSG0000016 | -0.00126993 | 0.555738488 | 0.002155402 | 0.0814369 | -0.00133035 | 0.537062157 | 0.00215525 | 0.081419067 | 6.04E-05 | 0.001549101 | 1 | 0.835948344 | 1 | 0.835911261 | ENSG0000018 | 8 | TMEM198B |
| SAT | T2DM | ENSG0000016 | -0.00113631 | 0.502350747 | 0.001693973 | 0.081436883 | -0.00110132 | 0.515570848 | 0.001693844 | 0.08141906 | -3.50E-05 | 0.010989074 | 1 | 0.810314363 | 1 | 0.824105482 | ENSG0000018 | 8 | TMEM198B |
| SAT | T2DM | ENSG0000016 | 0.000210333 | 0.943402079 | 0.002962661 | 0.081436958 | 0.000141456 | 0.961915298 | 0.002962412 | 0.081419131 | 6.89E-05 | 0.001168224 | 1 | 0.98331034 | 1 | 0.993672317 | ENSG0000018 | 8 | ACBD3 |
| SAT | T2DM | ENSG0000016 | 0.002228667 | 0.559145025 | 0.003815477 | 0.081436901 | 0.002457855 | 0.519449008 | 0.003815387 | 0.081419062 | -0.00022919 | 4.64E-06 | 1 | 0.837033211 | 1 | 0.826644476 | ENSG0000018 | 8 | RRP7B |
| SAT | T2DM | ENSG0000016 | 0.001535651 | 0.392330133 | 0.001795243 | 0.081436836 | 0.001507728 | 0.400954296 | 0.001795087 | 0.081419013 | 2.79E-05 | 0.020429552 | 1 | 0.742778516 | 1 | 0.757591271 | ENSG0000018 | 2 | GPIHBP1 |
| SAT | T2DM | ENSG0000016 | -0.00899702 | 0.004269912 | 0.003148546 | 0.081435594 | -0.00891303 | 0.004639404 | 0.003148287 | 0.081417792 | -8.40E-05 | 0.000299019 | 1 | 0.06901272 | 1 | 0.0804112 | ENSG0000018 | 9 | ALG12 |
| SAT | T2DM | ENSG0000016 | -0.00098771 | 0.679309982 | 0.00238921 | 0.08143693 | -0.00106667 | 0.655244267 | 0.002389018 | 0.081419098 | 7.90E-05 | 1.54E-05 | 1 | 0.892786811 | 1 | 0.891871358 | ENSG0000018 | 9 | ALG12 |
| SAT | T2DM | ENSG0000016 | -0.00158735 | 0.478543909 | 0.00223997 | 0.081436874 | -0.00164516 | 0.462635094 | 0.002239789 | 0.081419041 | 5.78E-05 | 0.000653631 | 1 | 0.796855602 | 1 | 0.797402301 | ENSG0000018 | 9 | ALG12 |
| SAT | T2DM | ENSG0000016 | 0.001469042 | 0.03255195 | 0.000687248 | 0.081436195 | 0.001481192 | 0.031129127 | 0.000687193 | 0.081418355 | -1.22E-05 | 0.021374064 | 1 | 0.252459754 | 1 | 0.262336831 | ENSG0000018 | 10 | COL18A1 |
| SAT | T2DM | ENSG0000016 | 0.002844008 | 0.100675937 | 0.001732473 | 0.081436508 | 0.002780936 | 0.108427365 | 0.00173234 | 0.0814187 | 6.31E-05 | 7.84E-06 | 1 | 0.425727224 | 1 | 0.450887431 | ENSG0000018 | 10 | COL18A1 |
| SAT | T2DM | ENSG0000016 | 0.001740398 | 0.393487229 | 0.002039582 | 0.081436837 | 0.001808268 | 0.375261242 | 0.00203941 | 0.081419 | -6.79E-05 | 2.61E-06 | 1 | 0.743169868 | 1 | 0.736823541 | ENSG0000018 | 9 | SLC25A18 |
| SAT | T2DM | ENSG0000016 | 0.002907597 | 0.056788469 | 0.001526346 | 0.081436352 | 0.002880653 | 0.059100947 | 0.001526218 | 0.081418536 | 2.69E-05 | 0.013626404 | 1 | 0.330946654 | 1 | 0.351641584 | ENSG0000018 | 9 | SLC25A18 |
| SAT | T2DM | ENSG0000016 | 0.000319103 | 0.777559343 | 0.001129563 | 0.081436945 | 0.000356892 | 0.752018989 | 0.001129481 | 0.081419114 | -3.78E-05 | 8.39E-05 | 1 | 0.939432466 | 1 | 0.929871654 | ENSG0000018 | 10 | C11orf54 |
| SAT | T2DM | ENSG0000016 | 0.002252423 | 0.565411593 | 0.003918466 | 0.081436903 | 0.003267524 | 0.405580107 | 0.003928741 | 0.081419015 | -0.0010151 | 0.000461137 | 1 | 0.837959912 | 1 | 0.759868776 | ENSG0000018 | 10 | C11orf54 |
| SAT | T2DM | ENSG0000016 | -0.00041948 | 0.625778302 | 0.000860166 | 0.081436919 | -0.00037372 | 0.663958674 | 0.000860207 | 0.081419099 | -4.58E-05 | 0.002674368 | 1 | 0.866359035 | 1 | 0.895996588 | ENSG0000018 | 10 | C11orf54 |
| SAT | T2DM | ENSG0000016 | -0.00169433 | 0.605610251 | 0.003281362 | 0.081436914 | -0.00116746 | 0.722384921 | 0.003286094 | 0.08141911 | -0.00052687 | 0.003956989 | 1 | 0.853892236 | 1 | 0.920812547 | ENSG0000018 | 10 | C11orf54 |
| SAT | T2DM | ENSG0000016 | -0.00022821 | 0.934981515 | 0.002797407 | 0.081436957 | -0.00032751 | 0.906791971 | 0.002797167 | 0.081419129 | 9.93E-05 | 2.54E-07 | 1 | 0.981839134 | 1 | 0.975470314 | ENSG0000018 | 10 | C11orf54 |
| SAT | T2DM | ENSG0000016 | 0.001111966 | 0.639187577 | 0.002371762 | 0.081436922 | 0.000911881 | 0.700673535 | 0.002372148 | 0.081419106 | 0.000200084 | 0.00030033 | 1 | 0.874326778 | 1 | 0.909587128 | ENSG0000018 | 10 | C11orf54 |
| SAT | T2DM | ENSG0000016 | -0.00481824 | 0.001600321 | 0.001526758 | 0.081435294 | -0.00469757 | 0.002098839 | 0.001527219 | 0.08141755 | -0.00012068 | 0.005845953 | 1 | 0.036211407 | 1 | 0.049971446 | ENSG0000018 | 10 | C11orf54 |
| SAT | T2DM | ENSG0000016 | -0.00026951 | 0.670197108 | 0.000632839 | 0.081436928 | -0.00029024 | 0.646475171 | 0.000632793 | 0.081419096 | 2.07E-05 | 0.000129756 | 1 | 0.889125844 | 1 | 0.890183286 | ENSG0000018 | 12 | CEP63 |
| SAT | T2DM | ENSG0000016 | -0.00808681 | 0.000625091 | 0.002364212 | 0.08143503 | -0.00803222 | 0.000679613 | 0.00236402 | 0.081417202 | -5.46E-05 | 0.002039068 | 1 | 0.017680364 | 1 | 0.022298088 | ENSG0000018 | 12 | CEP63 |
| SAT | T2DM | ENSG0000016 | 0.002070852 | 0.505177963 | 0.003107683 | 0.081436884 | 0.002151111 | 0.488783828 | 0.003107442 | 0.081419051 | -8.03E-05 | 0.001270742 | 1 | 0.810314363 | 1 | 0.808724024 | ENSG0000018 | 12 | CEP63 |
| SAT | T2DM | ENSG0000016 | -0.00407991 | 0.124673184 | 0.002657135 | 0.081436564 | -0.00419662 | 0.114262258 | 0.002657224 | 0.081418714 | 0.000116713 | 0.009437878 | 1 | 0.463429598 | 1 | 0.455762334 | ENSG0000018 | 12 | CEP63 |
| SAT | T2DM | ENSG0000016 | 4.26E-05 | 0.973147314 | 0.001264565 | 0.081436958 | 6.85E-05 | 0.956818689 | 0.001264459 | 0.08141913 | -2.59E-05 | 0.004606789 | 1 | 0.995031844 | 1 | 0.993142081 | ENSG0000018 | 12 | CEP63 |
| SAT | T2DM | ENSG0000016 | 0.002112884 | 0.117570734 | 0.001350038 | 0.081436549 | 0.002153399 | 0.110673837 | 0.001349946 | 0.081418706 | -4.05E-05 | 0.000909048 | 1 | 0.451697397 | 1 | 0.452205306 | ENSG0000018 | 2 | ODF3L1 |
| SAT | T2DM | ENSG0000016 | 0.004530854 | 0.066615132 | 0.002470118 | 0.081436396 | 0.004598991 | 0.062603472 | 0.002469918 | 0.081418552 | -6.81E-05 | 0.000252133 | 1 | 0.358786109 | 1 | 0.362901045 | ENSG0000018 | 4 | HMGN4 |
| SAT | T2DM | ENSG0000016 | -0.00783393 | 0.00024875 | 0.002138331 | 0.081434715 | -0.00787714 | 0.000229539 | 0.002138146 | 0.081416863 | 4.32E-05 | 0.002742951 | 0.816147259 | 0.009170194 | 0.753118457 | 0.009909453 | ENSG0000018 | 4 | HMGN4 |
| SAT | T2DM | ENSG0000016 | -0.0014394 | 0.652270206 | 0.003194321 | 0.081436924 | -0.00149327 | 0.640133055 | 0.003194073 | 0.081419094 | 5.39E-05 | 0.035155628 | 1 | 0.882995924 | 1 | 0.885818875 | ENSG0000018 | 4 | HMGN4 |
| SAT | T2DM | ENSG0000016 | -0.00048826 | 0.888466997 | 0.00348149 | 0.081436955 | -0.00057221 | 0.869438106 | 0.003481193 | 0.081419126 | 8.40E-05 | 0.000529197 | 1 | 0.969612316 | 1 | 0.966336865 | ENSG0000018 | 4 | HMGN4 |
| SAT | T2DM | ENSG0000016 | -0.00712977 | 0.029181936 | 0.003268999 | 0.081436163 | -0.00708126 | 0.030282614 | 0.003268706 | 0.081418347 | -4.85E-05 | 0.018245245 | 1 | 0.236994883 | 1 | 0.256527252 | ENSG0000018 | 4 | HMGN4 |
| SAT | T2DM | ENSG0000016 | 0.002781477 | 0.237725217 | 0.0023558 | 0.081436725 | 0.002818107 | 0.231562065 | 0.002355595 | 0.081418892 | -3.66E-05 | 0.020463651 | 1 | 0.604632897 | 1 | 0.608290741 | ENSG0000018 | 4 | HMGN4 |
| SAT | T2DM | ENSG0000016 | -0.00347061 | 0.205760531 | 0.002742891 | 0.081436691 | -0.00352948 | 0.198133168 | 0.002742645 | 0.081418854 | 5.89E-05 | 0.000684305 | 1 | 0.568086176 | 1 | 0.566556767 | ENSG0000018 | 4 | HMGN4 |
| SAT | T2DM | ENSG0000016 | -0.00085445 | 0.830998839 | 0.004003603 | 0.081436951 | -0.00091948 | 0.818339805 | 0.004003291 | 0.081419122 | 6.50E-05 | 0.040856735 | 1 | 0.95668325 | 1 | 0.957271954 | ENSG0000018 | 4 | HMGN4 |
| SAT | T2DM | ENSG0000016 | 0.006972839 | 0.004228602 | 0.002437546 | 0.081435591 | 0.00692952 | 0.00446827 | 0.002437334 | 0.08141778 | 4.33E-05 | 0.008356084 | 1 | 0.068881061 | 1 | 0.079676052 | ENSG0000018 | 8 | ZNF320 |
| SAT | T2DM | ENSG0000016 | -0.00439151 | 0.222530053 | 0.003600104 | 0.08143671 | -0.0042851 | 0.23399622 | 0.003599842 | 0.081418894 | -0.00010641 | 0.000537109 | 1 | 0.587161187 | 1 | 0.610313962 | ENSG0000018 | 8 | ZNF320 |
| SAT | T2DM | ENSG0000016 | 0.004848067 | 0.187881413 | 0.003681496 | 0.081436669 | 0.004990351 | 0.175218653 | 0.003681213 | 0.081418824 | -0.00014228 | 1.89E-06 | 1 | 0.54503883 | 1 | 0.535281564 | ENSG0000018 | 8 | ZNF320 |
| SAT | T2DM | ENSG0000016 | 0.000845283 | 0.65103309 | 0.001868739 | 0.081436924 | 0.000810297 | 0.664552806 | 0.001868611 | 0.081419099 | 3.50E-05 | 0.037552773 | 1 | 0.882256298 | 1 | 0.895996588 | ENSG0000018 | 8 | ZNF320 |
| SAT | T2DM | ENSG0000016 | 0.001165529 | 0.663376838 | 0.002677801 | 0.081436927 | 0.001210189 | 0.65129043 | 0.002677583 | 0.081419097 | -4.47E-05 | 0.026012984 | 1 | 0.887596281 | 1 | 0.891013292 | ENSG0000018 | 11 | LYSMD4 |
| SAT | T2DM | ENSG0000016 | -0.00081067 | 0.66508543 | 0.001872648 | 0.081436927 | -0.00075986 | 0.68488944 | 0.001872501 | 0.081419103 | -5.08E-05 | 0.0005434 | 1 | 0.888806281 | 1 | 0.901372745 | ENSG0000018 | 11 | LYSMD4 |
| SAT | T2DM | ENSG0000016 | -0.00595616 | 0.006715353 | 0.002197315 | 0.081435731 | -0.00600739 | 0.006253838 | 0.002197145 | 0.081417882 | 5.12E-05 | 0.003592929 | 1 | 0.090914049 | 1 | 0.095436475 | ENSG0000018 | 11 | LYSMD4 |
| SAT | T2DM | ENSG0000016 | 0.005733924 | 0.138443568 | 0.003870051 | 0.081436592 | 0.005630564 | 0.145660017 | 0.003869708 | 0.081418777 | 0.00010336 | 3.30E-05 | 1 | 0.483226964 | 1 | 0.503061344 | ENSG0000018 | 8 | SMDT1 |
| SAT | T2DM | ENSG0000016 | -0.0010901 | 0.605930655 | 0.002113049 | 0.081436914 | -0.0010455 | 0.620724661 | 0.002112878 | 0.08141909 | -4.46E-05 | 0.00525494 | 1 | 0.85397701 | 1 | 0.87257824 | ENSG0000018 | 8 | SMDT1 |
| SAT | T2DM | ENSG0000016 | -0.01003345 | 0.000986441 | 0.003045621 | 0.081435145 | -0.0099846 | 0.001043225 | 0.003045366 | 0.081417335 | -4.88E-05 | 0.024218892 | 1 | 0.024896248 | 1 | 0.029763665 | ENSG0000018 | 8 | GDPGP1 |
| SAT | T2DM | ENSG0000016 | 0.002688211 | 0.122431179 | 0.001740335 | 0.08143656 | 0.002775003 | 0.110863593 | 0.001740549 | 0.081418706 | -8.68E-05 | 0.020783506 | 1 | 0.459607206 | 1 | 0.452417225 | ENSG0000018 | 8 | GDPGP1 |
| SAT | T2DM | ENSG0000016 | -0.0063016 | 0.05887613 | 0.003336558 | 0.081436362 | -0.00637242 | 0.056130478 | 0.003336298 | 0.081418521 | 6.93E-05 | 0.009205317 | 1 | 0.337124925 | 1 | 0.342949902 | ENSG0000018 | 8 | GDPGP1 |
| SAT | T2DM | ENSG0000016 | -0.00396334 | 0.328367687 | 0.004054951 | 0.081436799 | -0.00390724 | 0.335217721 | 0.004054586 | 0.081418976 | -5.61E-05 | 0.02566798 | 1 | 0.692334629 | 1 | 0.702779133 | ENSG0000018 | 7 | C21orf67 |
| SAT | T2DM | ENSG0000016 | 0.003498932 | 0.027935528 | 0.001591728 | 0.081436151 | 0.002177445 | 0.176489796 | 0.00161096 | 0.081418826 | 0.001321487 | 1.15E-07 | 1 | 0.230185673 | 1 | 0.537662971 | ENSG0000018 | 7 | PTTG1IP |
| SAT | T2DM | ENSG0000016 | 0.000455385 | 0.763448687 | 0.00151313 | 0.081436943 | 0.000501205 | 0.740445942 | 0.001513011 | 0.081419113 | -4.58E-05 | 0.000114621 | 1 | 0.93459819 | 1 | 0.925595787 | ENSG0000018 | 10 | PLGLB1 |
| SAT | T2DM | ENSG0000016 | 0.002233736 | 0.509727268 | 0.003388232 | 0.081436886 | 0.002151641 | 0.525377061 | 0.003387989 | 0.081419064 | 8.21E-05 | 0.005344007 | 1 | 0.812130745 | 1 | 0.827311811 | ENSG0000018 | 7 | ZNF623 |
| SAT | T2DM | ENSG0000016 | -0.00030745 | 0.851966748 | 0.001647538 | 0.081436953 | -0.00036074 | 0.826672942 | 0.001647451 | 0.081419123 | 5.33E-05 | 0.002268386 | 1 | 0.963534946 | 1 | 0.960053847 | ENSG0000018 | 7 | ZNF623 |
| SAT | T2DM | ENSG0000016 | -0.01895392 | 1.17E-06 | 0.003900012 | 0.08143011 | -0.01905231 | 1.03E-06 | 0.003899687 | 0.081415143 | 9.84E-05 | 0.000512455 | 0.003853313 | 9.88E-05 | 0.003384238 | 0.000120866 | ENSG0000018 | 6 | C10orf107 |
| SAT | T2DM | ENSG0000016 | -0.00087026 | 0.802882613 | 0.003486367 | 0.081436948 | -0.000928 | 0.790083338 | 0.003486067 | 0.081419119 | 5.77E-05 | 0.015118091 | 1 | 0.948256967 | 1 | 0.947957612 | ENSG0000018 | 5 | FHL3 |
| SAT | T2DM | ENSG0000016 | 0.003989177 | 0.187799294 | 0.003028714 | 0.081436669 | 0.004056652 | 0.180404724 | 0.003028455 | 0.081418831 | -6.75E-05 | 0.001347256 | 1 | 0.54503883 | 1 | 0.543907656 | ENSG0000018 | 11 | NPIPA1 |
| SAT | T2DM | ENSG0000016 | 0.026394479 | 3.24E-12 | 0.00378835 | 0.081428847 | 0.01966209 | 2.08E-06 | 0.004143324 | 0.081415368 | 0.006732389 | 6.06E-05 | 1.06E-08 | 1.18E-09 | 0.006826591 | 0.000206866 | ENSG0000018 | 11 | NPIPA1 |
| SAT | T2DM | ENSG0000016 | 0.001114276 | 0.765927585 | 0.003742871 | 0.081436944 | 0.001172609 | 0.754038019 | 0.003742535 | 0.081419114 | -5.83E-05 | 0.013099727 | 1 | 0.934630346 | 1 | 0.930424498 | ENSG0000018 | 11 | NPIPA1 |
| SAT | T2DM | ENSG0000016 | 0.001863236 | 0.488820182 | 0.002691811 | 0.081436878 | 0.001913657 | 0.477096895 | 0.002691576 | 0.081419046 | -5.04E-05 | 0.004947296 | 1 | 0.803516541 | 1 | 0.80363893 | ENSG0000018 | 11 | NPIPA1 |
| SAT | T2DM | ENSG0000016 | 0.002329028 | 0.336298061 | 0.00242227 | 0.081436804 | 0.002267714 | 0.349155665 | 0.002422078 | 0.081418984 | 6.13E-05 | 0.001143966 | 1 | 0.696147596 | 1 | 0.717008026 | ENSG0000018 | 11 | NPIPA1 |
| SAT | T2DM | ENSG0000016 | 0.000904182 | 0.509902266 | 0.001372071 | 0.081436886 | 0.001153331 | 0.401856573 | 0.00137578 | 0.081419013 | -0.00024915 | 0.015539948 | 1 | 0.812130745 | 1 | 0.757591271 | ENSG0000018 | 7 | SF3A3 |
| SAT | T2DM | ENSG0000016 | -0.00275474 | 0.49518675 | 0.004038713 | 0.081436881 | -0.0028213 | 0.484785407 | 0.004038353 | 0.081419049 | 6.66E-05 | 0.009634717 | 1 | 0.806594843 | 1 | 0.806185068 | ENSG0000018 | 7 | SF3A3 |
| SAT | T2DM | ENSG0000016 | 0.005457378 | 0.162582496 | 0.003908063 | 0.081436633 | 0.005522289 | 0.157605097 | 0.003907719 | 0.081418797 | -6.49E-05 | 0.011165285 | 1 | 0.513507272 | 1 | 0.51775889 | ENSG0000018 | 7 | SF3A3 |
| SAT | T2DM | ENSG0000016 | 0.000509558 | 0.874014925 | 0.003213643 | 0.081436954 | 0.000569329 | 0.859371984 | 0.003213393 | 0.081419126 | -5.98E-05 | 0.019649158 | 1 | 0.966670474 | 1 | 0.965566132 | ENSG0000018 | 10 | OR7E38P |
| SAT | T2DM | ENSG0000016 | 0.001432303 | 0.36695901 | 0.001587594 | 0.081436822 | 0.001404838 | 0.376177667 | 0.001587456 | 0.081419 | 2.75E-05 | 0.010311603 | 1 | 0.723118627 | 1 | 0.737979921 | ENSG0000018 | 10 | OR7E38P |
| SAT | T2DM | ENSG0000016 | 0.001504715 | 0.548042944 | 0.002504954 | 0.081436898 | 0.001468529 | 0.557674211 | 0.002504741 | 0.081419073 | 3.62E-05 | 0.038330744 | 1 | 0.835027823 | 1 | 0.842050121 | ENSG0000018 | 9 | RP11-958N24.1 |
| SAT | T2DM | ENSG0000016 | 0.003099357 | 0.265511526 | 0.002783538 | 0.081436751 | 0.003047704 | 0.273517673 | 0.002783294 | 0.081418931 | 5.17E-05 | 0.005133206 | 1 | 0.628984344 | 1 | 0.64192524 | ENSG0000018 | 9 | RP11-958N24.1 |
| SAT | T2DM | ENSG0000016 | -0.00103605 | 0.704760144 | 0.002734331 | 0.081436934 | -0.000961 | 0.725223807 | 0.002734111 | 0.08141911 | -7.50E-05 | 0.000316868 | 1 | 0.906221055 | 1 | 0.921192527 | ENSG0000018 | 9 | RP11-958N24.1 |
| SAT | T2DM | ENSG0000016 | -0.00049533 | 0.895502171 | 0.003771218 | 0.081436956 | -0.00037029 | 0.921777268 | 0.003770913 | 0.081419129 | -0.00012505 | 1.11E-05 | 1 | 0.970180416 | 1 | 0.979654273 | ENSG0000018 | 8 | MX2 |
| SAT | T2DM | ENSG0000016 | 0.00607931 | 0.139635569 | 0.004115562 | 0.081436594 | 0.00617057 | 0.133768363 | 0.004115337 | 0.081418755 | -9.13E-05 | 0.034235626 | 1 | 0.484402728 | 1 | 0.486825442 | ENSG0000018 | 8 | MX2 |
| SAT | T2DM | ENSG0000016 | 0.004255612 | 0.002187311 | 0.001389106 | 0.08143539 | 0.004292774 | 0.001998061 | 0.001389006 | 0.081417535 | -3.72E-05 | 0.002026596 | 1 | 0.044524014 | 1 | 0.049345351 | ENSG0000018 | 8 | MX2 |
| SAT | T2DM | ENSG0000016 | -0.00265996 | 0.093639359 | 0.001585304 | 0.081436488 | -0.0026363 | 0.096293356 | 0.001585171 | 0.081418669 | -2.37E-05 | 0.037060007 | 1 | 0.416229576 | 1 | 0.432792465 | ENSG0000018 | 12 | PI4KAP2 |
| SAT | T2DM | ENSG0000016 | 0.001664867 | 0.330192436 | 0.001709787 | 0.0814368 | 0.001639581 | 0.337548231 | 0.001709639 | 0.081418977 | 2.53E-05 | 0.027972853 | 1 | 0.692789976 | 1 | 0.704513834 | ENSG0000018 | 12 | PI4KAP2 |
| SAT | T2DM | ENSG0000016 | -0.00205396 | 0.391379518 | 0.00239635 | 0.081436836 | -0.00201509 | 0.400364834 | 0.002396153 | 0.081419013 | -3.89E-05 | 0.027927352 | 1 | 0.741834893 | 1 | 0.757591271 | ENSG0000018 | 12 | PI4KAP2 |

| | | | | | | | | | | | | | | | | | | |
|---|---|---|---|---|---|---|---|---|---|---|---|---|---|---|---|---|---|---|
| SAT | T2DM | ENSG0000016 | 0.00261298 | 0.43294711 | 0.003332206 | 0.081436856 | 0.002711557 | 0.415759516 | 0.003331967 | 0.08141902 | -9.86E-05 | 0.000673861 | 1 | 0.772274669 | 1 | 0.766754833 | ENSG0000018 | 12 | PI4KAP2 |
| SAT | T2DM | ENSG0000016 | 0.000629243 | 0.474772451 | 0.000880385 | 0.081436873 | 0.000648161 | 0.461558876 | 0.000880315 | 0.08141904 | -1.89E-05 | 0.005649685 | 1 | 0.796607042 | 1 | 0.797039301 | ENSG0000018 | 12 | PI4KAP2 |
| SAT | T2DM | ENSG0000016 | 0.001425511 | 0.445000256 | 0.001866396 | 0.081436861 | 0.001389158 | 0.456661903 | 0.001866257 | 0.081419038 | 3.64E-05 | 0.019729425 | 1 | 0.780081459 | 1 | 0.794600418 | ENSG0000018 | 7 | PSMG1 |
| SAT | T2DM | ENSG0000016 | 3.55E-05 | 0.990820269 | 0.003084041 | 0.081436958 | 0.000790952 | 0.797954584 | 0.003089714 | 0.08141912 | -0.00075547 | 8.79E-05 | 1 | 0.996461007 | 1 | 0.952160336 | ENSG0000018 | 7 | PSMG1 |
| SAT | T2DM | ENSG0000016 | -0.0002009 | 0.86030076 | 0.001141557 | 0.081436953 | -0.00017595 | 0.877493798 | 0.001141457 | 0.081419127 | -2.50E-05 | 0.001138882 | 1 | 0.965007451 | 1 | 0.966700287 | ENSG0000018 | 7 | PSMG1 |
| SAT | T2DM | ENSG0000016 | 0.001454301 | 0.667151557 | 0.003381616 | 0.081436928 | 0.001378635 | 0.683480621 | 0.003381341 | 0.081419103 | 7.57E-05 | 0.002838185 | 1 | 0.888630991 | 1 | 0.901372745 | ENSG0000018 | 7 | PSMG1 |
| SAT | T2DM | ENSG0000016 | -0.00206867 | 0.133321502 | 0.00137807 | 0.081436582 | -0.0019692 | 0.153156832 | 0.001378534 | 0.08141879 | -9.95E-05 | 0.015606922 | 1 | 0.474948803 | 1 | 0.512250482 | ENSG0000018 | 2 | AC131971.1 |
| SAT | T2DM | ENSG0000016 | -0.00350715 | 0.359886688 | 0.003830514 | 0.081436818 | -0.00361705 | 0.344993503 | 0.003830215 | 0.081418982 | 0.000109898 | 0.000295688 | 1 | 0.71830389 | 1 | 0.713247438 | ENSG0000018 | 2 | AC131971.1 |
| SAT | T2DM | ENSG0000016 | 0.001420842 | 0.15732216 | 0.001004741 | 0.081436624 | 0.000724662 | 0.477163958 | 0.001019398 | 0.081419046 | 0.00069618 | 5.65E-05 | 1 | 0.507474184 | 1 | 0.80363893 | ENSG0000018 | 12 | PCBP3 |
| SAT | T2DM | ENSG0000016 | -0.00149282 | 0.199180125 | 0.001162729 | 0.081436683 | -0.00152677 | 0.189121485 | 0.001162649 | 0.081418843 | 3.40E-05 | 0.001264203 | 1 | 0.559511977 | 1 | 0.55901585 | ENSG0000018 | 10 | RP11-347C12.2 |
| SAT | T2DM | ENSG0000016 | 0.012743118 | 1.29E-05 | 0.002920983 | 0.081433778 | 0.011064621 | 0.000208443 | 0.002983514 | 0.081416833 | 0.001678497 | 0.005855984 | 0.04217545 | 0.00082697 | 0.68390041 | 0.009368499 | ENSG0000018 | 10 | RP11-347C12.2 |
| SAT | T2DM | ENSG0000016 | 0.002175362 | 0.459971985 | 0.002944081 | 0.081436867 | 0.002235317 | 0.447665096 | 0.002943863 | 0.081419035 | -6.00E-05 | 0.01601185 | 1 | 0.789726889 | 1 | 0.791227469 | ENSG0000018 | 10 | RP11-347C12.2 |
| SAT | T2DM | ENSG0000016 | -0.00918696 | 0.012914705 | 0.003695299 | 0.081435926 | -0.00912898 | 0.013487487 | 0.003694985 | 0.081418111 | -5.80E-05 | 0.024556454 | 1 | 0.143739766 | 1 | 0.157494043 | ENSG0000018 | 4 | MRPL54 |
| SAT | T2DM | ENSG0000016 | -0.00274741 | 0.498582351 | 0.004059872 | 0.081436882 | 0.001711667 | 0.683950816 | 0.004204763 | 0.081419103 | -0.00445907 | 4.73E-05 | 1 | 0.809826086 | 1 | 0.901372745 | ENSG0000018 | 9 | HMCES |
| SAT | T2DM | ENSG0000016 | 0.000168626 | 0.913841408 | 0.001558548 | 0.081436956 | 0.000251425 | 0.871847222 | 0.00155862 | 0.081419127 | -8.28E-05 | 0.002608551 | 1 | 0.976649401 | 1 | 0.966383771 | ENSG0000018 | 8 | DGCR6 |
| SAT | T2DM | ENSG0000016 | 0.001324104 | 0.303505306 | 0.001286854 | 0.081436781 | 0.001348681 | 0.294581587 | 0.001286756 | 0.081418947 | -2.46E-05 | 0.019094661 | 1 | 0.662542189 | 1 | 0.661548383 | ENSG0000018 | 8 | DGCR6 |
| SAT | T2DM | ENSG0000016 | 0.001639097 | 0.664984959 | 0.003785082 | 0.081436927 | 0.000371277 | 0.922027551 | 0.003793184 | 0.081419129 | 0.00126782 | 6.01E-07 | 1 | 0.888056281 | 1 | 0.979654273 | ENSG0000018 | 8 | DGCR6 |
| SAT | T2DM | ENSG0000016 | -0.00564925 | 0.108676958 | 0.003521602 | 0.081436528 | -0.00559235 | 0.112254204 | 0.003521311 | 0.081418709 | -5.69E-05 | 0.027131898 | 1 | 0.436210685 | 1 | 0.453578872 | ENSG0000018 | 8 | DGCR6 |
| SAT | T2DM | ENSG0000016 | 0.002093878 | 0.398889278 | 0.002482052 | 0.081436839 | 0.002029165 | 0.413583956 | 0.002481844 | 0.081419019 | 6.47E-05 | 0.000270956 | 1 | 0.750003279 | 1 | 0.766308941 | ENSG0000018 | 8 | DGCR6 |
| SAT | T2DM | ENSG0000016 | 0.004913811 | 0.071297999 | 0.002724478 | 0.081436415 | 0.004987915 | 0.067114852 | 0.002724284 | 0.081418571 | -7.41E-05 | 0.001896244 | 1 | 0.373189908 | 1 | 0.377708111 | ENSG0000018 | 5 | NDUFB1 |
| SAT | T2DM | ENSG0000016 | -0.00256251 | 0.474703833 | 0.003584696 | 0.081436873 | -0.00771813 | 0.037369012 | 0.003707583 | 0.081418407 | 0.005155622 | 5.41E-08 | 1 | 0.796607042 | 1 | 0.280518039 | ENSG0000018 | 6 | ALYREF |
| SAT | T2DM | ENSG0000016 | 0.003557657 | 0.084689577 | 0.002063481 | 0.081436462 | 0.00360978 | 0.080204281 | 0.002063314 | 0.081418619 | -5.21E-05 | 0.000881775 | 1 | 0.401115612 | 1 | 0.409385619 | ENSG0000018 | 6 | ALYREF |
| SAT | T2DM | ENSG0000016 | -0.01085724 | 0.006733186 | 0.004006698 | 0.081435731 | -0.01075235 | 0.007279334 | 0.004006393 | 0.081417927 | -0.00010488 | 0.001303515 | 1 | 0.090914049 | 1 | 0.10521364 | ENSG0000018 | 6 | ALYREF |
| SAT | T2DM | ENSG0000016 | -0.00022824 | 0.895314865 | 0.001734553 | 0.081436956 | -0.00017613 | 0.919112196 | 0.001734413 | 0.081419129 | -5.21E-05 | 8.13E-05 | 1 | 0.970180416 | 1 | 0.979654273 | ENSG0000018 | 4 | FAM101B |
| SAT | T2DM | ENSG0000016 | -0.00222831 | 0.197647282 | 0.001729673 | 0.081436681 | -0.00225233 | 0.192819654 | 0.00172952 | 0.081418848 | 2.40E-05 | 0.032275566 | 1 | 0.558130589 | 1 | 0.560852203 | ENSG0000018 | 8 | KREMEN1 |
| SAT | T2DM | ENSG0000016 | 0.002322095 | 0.006473546 | 0.000852837 | 0.081435719 | 0.001470023 | 0.092903917 | 0.000874869 | 0.081418659 | 0.000852072 | 1.31E-05 | 1 | 0.088498767 | 1 | 0.428114821 | ENSG0000018 | 5 | ZNF703 |
| SAT | T2DM | ENSG0000016 | 0.000559944 | 0.889642011 | 0.004035418 | 0.081436955 | 0.003226321 | 0.431954286 | 0.004105505 | 0.081419028 | -0.00266638 | 0.000433365 | 1 | 0.969612316 | 1 | 0.781358298 | ENSG0000018 | 5 | ZNF703 |
| SAT | T2DM | ENSG0000016 | 0.002363501 | 0.110253568 | 0.00147991 | 0.081436532 | 0.002397776 | 0.105158128 | 0.00147979 | 0.081418692 | -3.43E-05 | 0.002276897 | 1 | 0.436277088 | 1 | 0.444046099 | ENSG0000018 | 9 | NPIPA5 |
| SAT | T2DM | ENSG0000016 | -0.00195298 | 0.62340504 | 0.003977305 | 0.081436918 | -0.00188728 | 0.635108258 | 0.003976996 | 0.081419093 | -6.57E-05 | 0.038083856 | 1 | 0.864876358 | 1 | 0.884482576 | ENSG0000018 | 9 | NPIPA5 |
| SAT | T2DM | ENSG0000016 | -0.00499536 | 0.131843727 | 0.00331505 | 0.081436579 | -0.00493928 | 0.136205948 | 0.003314793 | 0.08141876 | -5.61E-05 | 0.033561644 | 1 | 0.472458584 | 1 | 0.492171492 | ENSG0000018 | 6 | FAM162B |
| SAT | T2DM | ENSG0000016 | 0.002317418 | 0.534460197 | 0.003730476 | 0.081436894 | 0.00239747 | 0.520408205 | 0.003730212 | 0.081419062 | -8.01E-05 | 0.014936239 | 1 | 0.828750699 | 1 | 0.826644476 | ENSG0000018 | 6 | FAM162B |
| SAT | T2DM | ENSG0000016 | -0.00316019 | 0.229989075 | 0.002632642 | 0.081436718 | -0.00311041 | 0.237372406 | 0.002632413 | 0.081418898 | -4.98E-05 | 0.004757015 | 1 | 0.594168627 | 1 | 0.613383258 | ENSG0000018 | 10 | NUDT14 |
| SAT | T2DM | ENSG0000016 | 0.000252487 | 0.889805331 | 0.001822346 | 0.081436955 | 0.000333205 | 0.85494108 | 0.001822609 | 0.081419125 | -8.07E-05 | 0.049266777 | 1 | 0.969612316 | 1 | 0.965566132 | ENSG0000018 | 10 | NUDT14 |
| SAT | T2DM | ENSG0000016 | -0.00476248 | 0.01764823 | 0.002007008 | 0.081436017 | -0.00479194 | 0.016949456 | 0.002006836 | 0.081418178 | 2.95E-05 | 0.03408761 | 1 | 0.172693001 | 1 | 0.179453096 | ENSG0000018 | 10 | NUDT14 |
| SAT | T2DM | ENSG0000017 | 0.001249167 | 0.572894263 | 0.002215642 | 0.081436905 | 0.001300551 | 0.557183896 | 0.002215476 | 0.081419073 | -5.14E-05 | 0.005241065 | 1 | 0.840775238 | 1 | 0.842050121 | ENSG0000018 | 12 | MAATS1 |
| SAT | T2DM | ENSG0000017 | -0.00343204 | 0.084179945 | 0.001987358 | 0.08143646 | -0.00337928 | 0.089032463 | 0.001987193 | 0.081418648 | -5.28E-05 | 0.000246465 | 1 | 0.401115612 | 1 | 0.421804746 | ENSG0000018 | 12 | MAATS1 |
| SAT | T2DM | ENSG0000017 | 0.000199242 | 0.961203247 | 0.004095946 | 0.081436958 | 0.00013496 | 0.973712432 | 0.004095595 | 0.081419131 | 6.43E-05 | 0.022796977 | 1 | 0.99110869 | 1 | 0.996491107 | ENSG0000018 | 12 | MAATS1 |
| SAT | T2DM | ENSG0000017 | 0.005439587 | 0.045602754 | 0.002721077 | 0.081436291 | 0.005393357 | 0.047454706 | 0.002720861 | 0.081418474 | 4.62E-05 | 0.028795229 | 1 | 0.293843465 | 1 | 0.314543215 | ENSG0000018 | 12 | FAM3B |
| SAT | T2DM | ENSG0000017 | -0.00420796 | 0.078543967 | 0.002391972 | 0.081436441 | -0.00416631 | 0.081520742 | 0.002391774 | 0.081418624 | -4.16E-05 | 0.01683456 | 1 | 0.391645528 | 1 | 0.411433676 | ENSG0000018 | 12 | FAM3B |
| SAT | T2DM | ENSG0000017 | -0.01111007 | 0.000242929 | 0.003027573 | 0.081434708 | -0.01105536 | 0.000260389 | 0.003027333 | 0.081416903 | -5.47E-05 | 0.020238047 | 0.797048645 | 0.009057371 | 0.854336845 | 0.011095284 | ENSG0000018 | 12 | FAM3B |
| SAT | T2DM | ENSG0000017 | 0.000994479 | 0.68278898 | 0.00243351 | 0.081436931 | 0.000941016 | 0.698961161 | 0.002433306 | 0.081419106 | 5.35E-05 | 0.002130407 | 1 | 0.893590205 | 1 | 0.908954249 | ENSG0000018 | 12 | FAM3B |
| SAT | T2DM | ENSG0000017 | -0.0037947 | 0.164305223 | 0.002728547 | 0.081436635 | -0.00374492 | 0.169875827 | 0.00272833 | 0.081418816 | -4.98E-05 | 0.018085574 | 1 | 0.516146667 | 1 | 0.531756789 | ENSG0000018 | 8 | NPIPA7 |
| SAT | T2DM | ENSG0000017 | -0.00194881 | 0.571917394 | 0.00344781 | 0.081436905 | -0.00202623 | 0.556712825 | 0.003447538 | 0.081419073 | 7.74E-05 | 0.004065669 | 1 | 0.840736467 | 1 | 0.842050121 | ENSG0000018 | 8 | NPIPA7 |
| SAT | T2DM | ENSG0000017 | 0.0048086 | 0.027337353 | 0.002179117 | 0.081436145 | 0.004761981 | 0.028855798 | 0.00217894 | 0.081418333 | 4.66E-05 | 0.004260912 | 1 | 0.227073047 | 1 | 0.249804413 | ENSG0000018 | 5 | TTC32 |
| SAT | T2DM | ENSG0000017 | 0.005860018 | 0.124337411 | 0.003813074 | 0.081436564 | 0.006091248 | 0.110167618 | 0.003813124 | 0.081418705 | -0.00023123 | 0.000108713 | 1 | 0.463429598 | 1 | 0.452205306 | ENSG0000018 | 5 | TTC32 |
| SAT | T2DM | ENSG0000017 | -0.00082733 | 0.811526693 | 0.003469522 | 0.081436949 | -0.00095119 | 0.783947867 | 0.003469257 | 0.081419118 | 0.000123862 | 1.16E-05 | 1 | 0.951539016 | 1 | 0.945615795 | ENSG0000018 | 9 | PP2D1 |
| SAT | T2DM | ENSG0000017 | -0.00118498 | 0.655634067 | 0.002657207 | 0.081436925 | -0.00112784 | 0.671214778 | 0.002656986 | 0.081419101 | -5.71E-05 | 0.003090087 | 1 | 0.883423151 | 1 | 0.898343777 | ENSG0000018 | 9 | PP2D1 |
| SAT | T2DM | ENSG0000017 | -0.00154784 | 0.542957533 | 0.00254434 | 0.081436897 | -0.00161531 | 0.525485466 | 0.002544148 | 0.081419064 | 6.75E-05 | 0.001253169 | 1 | 0.832700048 | 1 | 0.827311811 | ENSG0000018 | 4 | TREML3P |
| SAT | T2DM | ENSG0000017 | 0.001508782 | 0.297192846 | 0.001447311 | 0.081436777 | 0.001545045 | 0.285695444 | 0.001447195 | 0.08141894 | -3.63E-05 | 0.00115466 | 1 | 0.654394836 | 1 | 0.651854486 | ENSG0000018 | 10 | EIF3C |
| SAT | T2DM | ENSG0000017 | -0.00278566 | 0.145385388 | 0.00191319 | 0.081436604 | -0.0028131 | 0.141425737 | 0.00191302 | 0.08141877 | 2.74E-05 | 0.024583206 | 1 | 0.491255879 | 1 | 0.499917672 | ENSG0000018 | 10 | EIF3C |
| SAT | T2DM | ENSG0000017 | 0.006758461 | 0.038312717 | 0.003262594 | 0.081436241 | 0.006831282 | 0.036261336 | 0.003262327 | 0.081418398 | -7.28E-05 | 0.002685691 | 1 | 0.271246148 | 1 | 0.276092282 | ENSG0000018 | 10 | EIF3C |
| SAT | T2DM | ENSG0000017 | -0.00047842 | 0.668672778 | 0.001117878 | 0.081436928 | -0.00045449 | 0.684301884 | 0.001117788 | 0.081419103 | -2.39E-05 | 0.004608752 | 1 | 0.888630991 | 1 | 0.901372745 | ENSG0000018 | 10 | EIF3C |
| SAT | T2DM | ENSG0000017 | -0.00503563 | 0.122208977 | 0.003258102 | 0.081436559 | -0.00493622 | 0.129725151 | 0.003257825 | 0.081418747 | -9.94E-05 | 1.22E-05 | 1 | 0.459418101 | 1 | 0.479851431 | ENSG0000018 | 10 | EIF3C |
| SAT | T2DM | ENSG0000017 | -0.00270321 | 0.120155474 | 0.001739379 | 0.081436555 | -0.00274087 | 0.115049616 | 0.001739242 | 0.081418716 | 3.77E-05 | 0.005697345 | 1 | 0.456565492 | 1 | 0.456639917 | ENSG0000018 | 12 | RP13-1039J1.2 |
| SAT | T2DM | ENSG0000017 | -0.00162446 | 0.660540106 | 0.003698942 | 0.081436926 | -0.00168043 | 0.649583695 | 0.003698615 | 0.081419096 | 5.60E-05 | 0.020120085 | 1 | 0.885563559 | 1 | 0.891013292 | ENSG0000018 | 12 | RP13-1039J1.2 |
| SAT | T2DM | ENSG0000017 | 0.003530079 | 0.042084761 | 0.001736657 | 0.081436268 | 0.003471316 | 0.045607358 | 0.001736514 | 0.081418463 | 5.88E-05 | 4.08E-06 | 1 | 0.28326473 | 1 | 0.308531429 | ENSG0000018 | 7 | NIPSNAP1 |
| SAT | T2DM | ENSG0000017 | -0.00848124 | 0.001509733 | 0.002673087 | 0.081435276 | -0.00856443 | 0.001354533 | 0.002672887 | 0.081417415 | 8.32E-05 | 0.000188244 | 1 | 0.034843648 | 1 | 0.037035189 | ENSG0000018 | 7 | NIPSNAP1 |
| SAT | T2DM | ENSG0000017 | -0.0026759 | 0.121658647 | 0.001728781 | 0.081436558 | -0.00262151 | 0.129390747 | 0.001728644 | 0.081418747 | -5.44E-05 | 5.46E-05 | 1 | 0.459236339 | 1 | 0.479366489 | ENSG0000018 | 10 | CRELD2 |
| SAT | T2DM | ENSG0000017 | 8.02E-05 | 0.931743225 | 0.000935797 | 0.081436957 | 0.000147208 | 0.875011044 | 0.000935864 | 0.081419127 | -6.71E-05 | 0.000171372 | 1 | 0.980452059 | 1 | 0.966700287 | ENSG0000018 | 10 | CRELD2 |
| SAT | T2DM | ENSG0000017 | -0.00358813 | 0.000173865 | 0.000955746 | 0.081434603 | -0.00357062 | 0.000186796 | 0.000955666 | 0.081416798 | -1.75E-05 | 0.010338429 | 0.570449786 | 0.007408439 | 0.612876884 | 0.008632069 | ENSG0000018 | 10 | CRELD2 |
| SAT | T2DM | ENSG0000017 | 0.00379343 | 0.133489698 | 0.002528137 | 0.081436582 | 0.003836658 | 0.129089643 | 0.002527936 | 0.081418746 | -4.32E-05 | 0.026661186 | 1 | 0.475032212 | 1 | 0.479121176 | ENSG0000018 | 10 | CRELD2 |
| SAT | T2DM | ENSG0000017 | 0.0035121 | 0.07494876 | 0.001972224 | 0.081436428 | 0.003545845 | 0.072173874 | 0.001972079 | 0.081418591 | -3.37E-05 | 0.043765174 | 1 | 0.382364025 | 1 | 0.39204956 | ENSG0000018 | 7 | GOLGA6L4 |
| SAT | T2DM | ENSG0000017 | 0.000413672 | 0.628212651 | 0.000854264 | 0.081436919 | 0.000581776 | 0.496285332 | 0.000855117 | 0.081419054 | -0.0001681 | 2.93E-05 | 1 | 0.867129031 | 1 | 0.814437198 | ENSG0000018 | 4 | OAF |
| SAT | T2DM | ENSG0000017 | 0.002498658 | 0.486460586 | 0.003590284 | 0.081436877 | -0.01337601 | 0.00665231 | 0.004928924 | 0.0814179 | 0.015874664 | 2.60E-06 | 1 | 0.801645999 | 1 | 0.100085232 | ENSG0000018 | 4 | OAF |
| SAT | T2DM | ENSG0000017 | 0.000656245 | 0.67779975 | 0.001579533 | 0.08143693 | 0.000694765 | 0.660019766 | 0.00157942 | 0.081419099 | -3.85E-05 | 0.005124663 | 1 | 0.892786811 | 1 | 0.894350146 | ENSG0000018 | 8 | TM2D3 |
| SAT | T2DM | ENSG0000017 | -0.00176572 | 0.206271153 | 0.001397054 | 0.081436691 | -0.00174599 | 0.211345883 | 0.001396932 | 0.08141887 | -1.97E-05 | 0.0338845 | 1 | 0.568086176 | 1 | 0.58173309 | ENSG0000018 | 8 | TM2D3 |
| SAT | T2DM | ENSG0000017 | -0.00032644 | 0.934418838 | 0.003967132 | 0.081436957 | -0.00026406 | 0.9469266 | 0.003966809 | 0.08141913 | -6.24E-05 | 0.035423832 | 1 | 0.981839134 | 1 | 0.988503397 | ENSG0000018 | 5 | TACSTD2 |
| SAT | T2DM | ENSG0000017 | 0.008143767 | 0.000937244 | 0.002461291 | 0.081435129 | -0.00010848 | 0.982172551 | 0.004854927 | 0.081419131 | 0.008252252 | 0.048620796 | 1 | 0.023837978 | 1 | 0.997162029 | ENSG0000018 | 5 | TACSTD2 |
| SAT | T2DM | ENSG0000017 | -0.00718147 | 0.000352105 | 0.002009576 | 0.081434824 | -0.0053438 | 0.012282422 | 0.002134161 | 0.081418083 | -0.00183768 | 0.010600941 | 1 | 0.012289954 | 1 | 0.150368007 | ENSG0000018 | 15 | MROH7 |
| SAT | T2DM | ENSG0000017 | -0.00741655 | 0.015170295 | 0.00305422 | 0.081435973 | -0.00750992 | 0.013933481 | 0.003054078 | 0.081418121 | 9.34E-05 | 0.006519173 | 1 | 0.156521191 | 1 | 0.160406141 | ENSG0000018 | 9 | CSF1 |
| SAT | T2DM | ENSG0000017 | 0.000242162 | 0.941023309 | 0.003273182 | 0.081436958 | 0.000688287 | 0.833524958 | 0.003274705 | 0.081419124 | -0.00044612 | 5.82E-05 | 1 | 0.982870948 | 1 | 0.961601754 | ENSG0000018 | 9 | CSF1 |
| SAT | T2DM | ENSG0000017 | 0.002031157 | 0.406845221 | 0.002448773 | 0.081436843 | 0.002073445 | 0.397108734 | 0.002448574 | 0.081419011 | -4.23E-05 | 0.021501236 | 1 | 0.756247299 | 1 | 0.756180206 | ENSG0000018 | 9 | CSF1 |
| SAT | T2DM | ENSG0000017 | -0.00047037 | 0.8096747 | 0.001952998 | 0.081436949 | -0.00056142 | 0.77376826 | 0.001953099 | 0.081419117 | 9.10E-05 | 0.009410815 | 1 | 0.951539016 | 1 | 0.939923606 | ENSG0000018 | 9 | CSF1 |
| SAT | T2DM | ENSG0000017 | -0.00140596 | 0.644244633 | 0.003044706 | 0.081436923 | -0.00146155 | 0.631176741 | 0.003044447 | 0.081419092 | 5.56E-05 | 0.009141526 | 1 | 0.877766855 | 1 | 0.881139272 | ENSG0000018 | 9 | CSF1 |
| SAT | T2DM | ENSG0000017 | -0.01013955 | 9.04E-06 | 0.002284246 | 0.081433666 | -0.01026764 | 6.96E-06 | 0.002284213 | 0.081415755 | 0.00012809 | 4.94E-05 | 0.029672588 | 0.000618179 | 0.022826296 | 0.000616927 | ENSG0000018 | 14 | PLA2G6 |
| SAT | T2DM | ENSG0000017 | 0.001815589 | 0.359676919 | 0.001982124 | 0.081436818 | 0.00753832 | 0.008850615 | 0.002879659 | 0.081417986 | -0.00572273 | 0.006157627 | 1 | 0.71830389 | 1 | 0.12252687 | ENSG0000018 | 14 | PLA2G6 |
| SAT | T2DM | ENSG0000017 | 0.000862784 | 0.732392645 | 0.002523168 | 0.081436939 | 0.000819074 | 0.745446084 | 0.002522944 | 0.081419113 | 4.37E-05 | 0.006963399 | 1 | 0.922803482 | 1 | 0.927496626 | ENSG0000018 | 2 | C21orf128 |
| SAT | T2DM | ENSG0000017 | -0.00054552 | 0.770220902 | 0.001867653 | 0.081436944 | -0.00155108 | 0.410040894 | 0.001882784 | 0.081419017 | 0.001005564 | 2.75E-05 | 1 | 0.935400227 | 1 | 0.762666765 | ENSG0000018 | 2 | C21orf128 |
| SAT | T2DM | ENSG0000017 | -0.00086181 | 0.43031405 | 0.001092754 | 0.081436854 | -0.00083687 | 0.443733005 | 0.001092655 | 0.081419033 | -2.49E-05 | 0.000159287 | 1 | 0.772274669 | 1 | 0.789955502 | ENSG0000018 | 2 | C21orf128 |
| SAT | T2DM | ENSG0000017 | 0.004697195 | 0.211367036 | 0.003758307 | 0.081436697 | 0.004869364 | 0.195072047 | 0.003758033 | 0.08141885 | -0.00017217 | 8.81E-08 | 1 | 0.575258443 | 1 | 0.563293473 | ENSG0000018 | 2 | C21orf128 |
| SAT | T2DM | ENSG0000017 | -0.00176078 | 0.343598243 | 0.001859169 | 0.081436809 | -0.00167339 | 0.368130489 | 0.001859366 | 0.081418996 | -8.74E-05 | 0.023463802 | 1 | 0.704151053 | 1 | 0.732878736 | ENSG0000018 | 2 | C21orf128 |
| SAT | T2DM | ENSG0000017 | -0.00168605 | 0.025397744 | 0.00075428 | 0.081436123 | -0.00167432 | 0.026423022 | 0.000754215 | 0.081418307 | -1.17E-05 | 0.024172387 | 1 | 0.217571794 | 1 | 0.236264392 | ENSG0000018 | 8 | A3GALT2 |
| SAT | T2DM | ENSG0000017 | -0.00224569 | 0.393609184 | 0.00263241 | 0.081436837 | -0.00220033 | 0.403194454 | 0.002632197 | 0.081419014 | -4.54E-05 | 0.023091499 | 1 | 0.743169868 | 1 | 0.757663806 | ENSG0000018 | 8 | A3GALT2 |
| SAT | T2DM | ENSG0000017 | 0.004434607 | 0.02931988 | 0.002035003 | 0.081436165 | 0.007415485 | 0.000496071 | 0.002129117 | 0.081417104 | -0.00298088 | 1.98E-06 | 1 | 0.237037613 | 1 | 0.018287726 | ENSG0000018 | 8 | A3GALT2 |
| SAT | T2DM | ENSG0000017 | 0.000674759 | 0.752368862 | 0.002138578 | 0.081436942 | 0.000726461 | 0.734067849 | 0.002138413 | 0.081419112 | -5.17E-05 | 0.002693688 | 1 | 0.929413492 | 1 | 0.924370544 | ENSG0000018 | 8 | A3GALT2 |
| SAT | T2DM | ENSG0000017 | -0.00298388 | 0.379129247 | 0.003392686 | 0.081436829 | -0.00290718 | 0.3914661 | 0.003392422 | 0.081419008 | -7.67E-05 | 0.004594088 | 1 | 0.731289276 | 1 | 0.752431326 | ENSG0000018 | 8 | A3GALT2 |
| SAT | T2DM | ENSG0000017 | -0.00231375 | 0.091352547 | 0.001370455 | 0.081436482 | -0.00234509 | 0.08702247 | 0.001370338 | 0.081418642 | 3.13E-05 | 0.00101075 | 1 | 0.413867045 | 1 | 0.419266847 | ENSG0000018 | 10 | SS18L1 |
| SAT | T2DM | ENSG0000017 | 0.002748417 | 0.331859328 | 0.00283232 | 0.081436801 | 0.002796241 | 0.323478572 | 0.002832108 | 0.081418968 | -4.78E-05 | 0.041578661 | 1 | 0.693964599 | 1 | 0.691873008 | ENSG0000018 | 10 | SS18L1 |
| SAT | T2DM | ENSG0000017 | -0.00632848 | 0.025196162 | 0.002827247 | 0.081436121 | -0.00804861 | 0.005212355 | 0.002881068 | 0.081417827 | 0.001720136 | 0.00197264 | 1 | 0.216409969 | 1 | 0.085938379 | ENSG0000018 | 10 | SS18L1 |
| SAT | T2DM | ENSG0000017 | 0.000252976 | 0.942796552 | 0.003525527 | 0.081436958 | 0.000444027 | 0.899777448 | 0.003525632 | 0.081419128 | -0.00019105 | 0.001164342 | 1 | 0.983253492 | 1 | 0.972878089 | ENSG0000018 | 8 | TOP1MT |
| SAT | T2DM | ENSG0000017 | -0.00090448 | 0.76347823 | 0.00300575 | 0.081436943 | -0.00081694 | 0.785763868 | 0.003005491 | 0.081419119 | -8.75E-05 | 2.14E-05 | 1 | 0.93459819 | 1 | 0.946500713 | ENSG0000018 | 11 | WDR27 |
| SAT | T2DM | ENSG0000017 | -0.00100166 | 0.652488836 | 0.002224384 | 0.081436925 | -0.00095414 | 0.667938935 | 0.002224206 | 0.0814191 | -4.75E-05 | 0.005237015 | 1 | 0.882995924 | 1 | 0.896403813 | ENSG0000018 | 15 | TXNRD2 |
| SAT | T2DM | ENSG0000017 | 0.007593656 | 0.044122567 | 0.00377244 | 0.081436281 | 0.007526779 | 0.046002113 | 0.003772105 | 0.081418466 | 6.69E-05 | 0.005850532 | 1 | 0.290112507 | 1 | 0.309334779 | ENSG0000018 | 15 | TXNRD2 |
| SAT | T2DM | ENSG0000017 | 0.004146753 | 0.129139449 | 0.002732609 | 0.081436574 | 0.004199147 | 0.12433963 | 0.002732372 | 0.081418736 | -5.24E-05 | 0.004460899 | 1 | 0.470262523 | 1 | 0.474800718 | ENSG0000018 | 6 | HDDC3 |
| SAT | T2DM | ENSG0000017 | 0.00274338 | 0.080244848 | 0.001568299 | 0.081436447 | 0.002714565 | 0.083444703 | 0.001568167 | 0.08141863 | 2.88E-05 | 0.009827981 | 1 | 0.395495266 | 1 | 0.414575198 | ENSG0000018 | 6 | HDDC3 |
| SAT | T2DM | ENSG0000017 | -0.00505253 | 0.013224886 | 0.002039233 | 0.081435933 | -0.00508978 | 0.012555533 | 0.002039058 | 0.08141809 | 3.72E-05 | 0.008582451 | 1 | 0.145107 | 1 | 0.150896353 | ENSG0000018 | 6 | HDDC3 |
| SAT | T2DM | ENSG0000017 | 0.006077134 | 0.102247197 | 0.003719043 | 0.081436512 | 0.006154388 | 0.097931081 | 0.003718729 | 0.081418673 | -7.73E-05 | 0.003467349 | 1 | 0.425727224 | 1 | 0.436565051 | ENSG0000018 | 5 | LPAR5 |
| SAT | T2DM | ENSG0000017 | -0.00055885 | 0.808553151 | 0.002306491 | 0.081436949 | -0.00062675 | 0.785817721 | 0.00230637 | 0.081419119 | 6.79E-05 | 0.005904999 | 1 | 0.951539016 | 1 | 0.946500713 | ENSG0000018 | 5 | LPAR5 |
| SAT | T2DM | ENSG0000017 | 0.002395613 | 0.313738351 | 0.002377993 | 0.081436789 | 0.000337664 | 0.890447327 | 0.002451489 | 0.081419128 | 0.00205795 | 0.000564144 | 1 | 0.674437649 | 1 | 0.971822875 | ENSG0000018 | 4 | C8orf12 |
| SAT | T2DM | ENSG0000017 | -0.00016691 | 0.935089271 | 0.00204945 | 0.081436957 | -0.00012247 | 0.952343693 | 0.002049279 | 0.08141913 | -4.44E-05 | 0.002575011 | 1 | 0.981839134 | 1 | 0.99107822 | ENSG0000018 | 4 | C8orf12 |
| SAT | T2DM | ENSG0000017 | -0.00194632 | 0.47703614 | 0.002737137 | 0.081436874 | -0.00122621 | 0.654433946 | 0.002739451 | 0.081419097 | -0.00072011 | 1.74E-09 | 1 | 0.796607042 | 1 | 0.891871358 | ENSG0000018 | 4 | C8orf12 |
| SAT | T2DM | ENSG0000017 | 0.004359471 | 0.177928991 | 0.003236043 | 0.081436655 | 0.004414738 | 0.172461931 | 0.003235811 | 0.08141882 | -5.53E-05 | 0.049244619 | 1 | 0.530526907 | 1 | 0.532948952 | ENSG0000018 | 4 | C8orf12 |
| SAT | T2DM | ENSG0000017 | -0.00170089 | 0.24932633 | 0.001476485 | 0.081436737 | 0.000417683 | 0.785324372 | 0.001533416 | 0.081419118 | -0.00211858 | 3.21E-07 | 1 | 0.615333003 | 1 | 0.946500713 | ENSG0000018 | 8 | GSTT1 |
| SAT | T2DM | ENSG0000017 | -0.00057267 | 0.788782817 | 0.002137688 | 0.081436946 | -0.00054003 | 0.800542819 | 0.0021375 | 0.08141912 | -3.26E-05 | 0.020201763 | 1 | 0.946250977 | 1 | 0.952160336 | ENSG0000018 | 8 | GSTT1 |
| SAT | T2DM | ENSG0000017 | -0.00751874 | 0.024832541 | 0.003350589 | 0.081436117 | -0.00747266 | 0.025718441 | 0.003350303 | 0.0814183 | -4.61E-05 | 0.047676976 | 1 | 0.21611556 | 1 | 0.233772911 | ENSG0000018 | 8 | GSTT1 |
| SAT | T2DM | ENSG0000017 | 0.001630088 | 0.521162378 | 0.002540839 | 0.08143689 | 0.001684601 | 0.507292538 | 0.002540647 | 0.081419057 | -5.45E-05 | 0.009503794 | 1 | 0.82089955 | 1 | 0.817738235 | ENSG0000018 | 8 | GSTT1 |
| SAT | T2DM | ENSG0000017 | -0.00086321 | 0.679668323 | 0.002090512 | 0.08143693 | -0.00089672 | 0.66793846 | 0.002090349 | 0.0814191 | 3.35E-05 | 0.043608176 | 1 | 0.892786811 | 1 | 0.896403813 | ENSG0000018 | 8 | ATL3 |
| SAT | T2DM | ENSG0000017 | 0.001746498 | 0.296574905 | 0.001673199 | 0.081436776 | 0.001686349 | 0.313493268 | 0.001673098 | 0.081418961 | 6.01E-05 | 0.000270413 | 1 | 0.654379465 | 1 | 0.682529139 | ENSG0000018 | 8 | ATL3 |
| SAT | T2DM | ENSG0000017 | 0.003738576 | 0.165584441 | 0.002696346 | 0.081436637 | 0.003780858 | 0.160816142 | 0.002696118 | 0.081418802 | -4.23E-05 | 0.026583206 | 1 | 0.518060238 | 1 | 0.520626624 | ENSG0000018 | 8 | ATL3 |
| SAT | T2DM | ENSG0000017 | 0.004660296 | 0.135432718 | 0.00312139 | 0.081436586 | 0.004721225 | 0.130368016 | 0.003121152 | 0.081418749 | -6.09E-05 | 0.016812267 | 1 | 0.477800803 | 1 | 0.480064491 | ENSG0000018 | 8 | ATL3 |
| SAT | T2DM | ENSG0000017 | -0.0002676 | 0.715329623 | 0.000733749 | 0.081436936 | -0.00028188 | 0.700833971 | 0.000733693 | 0.081419106 | 1.43E-05 | 0.018917995 | 1 | 0.910394295 | 1 | 0.909587128 | ENSG0000018 | 9 | BTBD6 |
| SAT | T2DM | ENSG0000017 | -0.00086938 | 0.825296506 | 0.003938504 | 0.08143695 | -0.00093561 | 0.812211334 | 0.00393819 | 0.081419121 | 6.62E-05 | 0.02926178 | 1 | 0.955051074 | 1 | 0.95480666 | ENSG0000018 | 9 | BTBD6 |
| SAT | T2DM | ENSG0000017 | -0.00209123 | 0.051818993 | 0.001075384 | 0.081436326 | -0.00206365 | 0.05496573 | 0.001075294 | 0.081418515 | -2.76E-05 | 0.00032921 | 1 | 0.315548759 | 1 | 0.341997131 | ENSG0000018 | 8 | NUTM2A |
| SAT | T2DM | ENSG0000017 | -0.00073621 | 0.759428854 | 0.0024041 | 0.081436943 | -0.0007742 | 0.747408214 | 0.002403897 | 0.081419114 | 3.80E-05 | 0.025094018 | 1 | 0.934086217 | 1 | 0.927854665 | ENSG0000018 | 8 | NUTM2A |
| SAT | T2DM | ENSG0000017 | 0.002365867 | 0.551312677 | 0.003970944 | 0.081436899 | 0.002437052 | 0.539366304 | 0.003970615 | 0.081419068 | -7.12E-05 | 0.014063621 | 1 | 0.83588581 | 1 | 0.836431746 | ENSG0000018 | 9 | AQP12A |
| SAT | T2DM | ENSG0000017 | 0.001512163 | 0.625806435 | 0.003100995 | 0.081436919 | 0.001461275 | 0.637450872 | 0.003100742 | 0.081419094 | 5.09E-05 | 0.02837002 | 1 | 0.866359035 | 1 | 0.884719251 | ENSG0000018 | 9 | AQP12A |
| SAT | T2DM | ENSG0000017 | -0.00244209 | 0.342448859 | 0.002572411 | 0.081436808 | -0.00237326 | 0.356185843 | 0.002572196 | 0.081418989 | -6.88E-05 | 0.000193171 | 1 | 0.703113082 | 1 | 0.724067999 | ENSG0000018 | 9 | AQP12A |
| SAT | T2DM | ENSG0000017 | -0.00142337 | 0.546319024 | 0.002359356 | 0.081436898 | -0.00146707 | 0.534033183 | 0.002359154 | 0.081419066 | 4.37E-05 | 0.007688419 | 1 | 0.835027823 | 1 | 0.832950936 | ENSG0000018 | 11 | FAM227A |
| SAT | T2DM | ENSG0000017 | -0.01041069 | 0.003614541 | 0.003577568 | 0.081435543 | -0.01047233 | 0.003417695 | 0.003577286 | 0.081417699 | 6.16E-05 | 0.028182767 | 1 | 0.062747674 | 1 | 0.066764938 | ENSG0000018 | 11 | FAM227A |
| SAT | T2DM | ENSG0000017 | 0.000629071 | 0.744590871 | 0.001930968 | 0.081436941 | 0.000668501 | 0.729171145 | 0.001930808 | 0.081419111 | -3.94E-05 | 0.005259662 | 1 | 0.927287795 | 1 | 0.922641931 | ENSG0000018 | 2 | NOC4L |
| SAT | T2DM | ENSG0000017 | 0.004892086 | 0.064139219 | 0.002642635 | 0.081436386 | 0.004849187 | 0.066486401 | 0.002642415 | 0.081418568 | 4.29E-05 | 0.024802603 | 1 | 0.351320163 | 1 | 0.375381853 | ENSG0000018 | 2 | NOC4L |
| SAT | T2DM | ENSG0000017 | 0.002481779 | 0.136909017 | 0.001668532 | 0.081436589 | 0.002452718 | 0.141533679 | 0.001668395 | 0.08141877 | 2.91E-05 | 0.019642706 | 1 | 0.479912911 | 1 | 0.499917672 | ENSG0000018 | 2 | NOC4L |
| SAT | T2DM | ENSG0000017 | 0.00345427 | 0.293334473 | 0.003287164 | 0.081436774 | 0.003539577 | 0.281534232 | 0.00328688 | 0.081418937 | -8.53E-05 | 0.000136963 | 1 | 0.650730497 | 1 | 0.647765649 | ENSG0000018 | 2 | NOC4L |
| SAT | T2DM | ENSG0000017 | -0.00120903 | 0.664141686 | 0.002784486 | 0.081436927 | -0.00116082 | 0.676736397 | 0.002784258 | 0.081419102 | -4.82E-05 | 0.019882308 | 1 | 0.887596281 | 1 | 0.898778511 | ENSG0000018 | 5 | NDUFA6 |
| SAT | T2DM | ENSG0000017 | -0.00111386 | 0.56245426 | 0.001923106 | 0.081436902 | -0.00117085 | 0.54261365 | 0.001923001 | 0.081419069 | 5.70E-05 | 0.004629377 | 1 | 0.837959912 | 1 | 0.839771408 | ENSG0000018 | 5 | NDUFA6 |
| SAT | T2DM | ENSG0000017 | -0.01107677 | 0.004240772 | 0.003873422 | 0.081435592 | -0.01101346 | 0.00446131 | 0.003873112 | 0.08141778 | -6.33E-05 | 0.03264365 | 1 | 0.068881061 | 1 | 0.079676052 | ENSG0000018 | 5 | NDUFA6 |
| SAT | T2DM | ENSG0000017 | -0.00169726 | 0.493765258 | 0.002480173 | 0.08143688 | -0.00172673 | 0.486255111 | 0.002479946 | 0.08141905 | 2.95E-05 | 0.047760351 | 1 | 0.805591154 | 1 | 0.807270931 | ENSG0000018 | 5 | NDUFA6 |

| | | | | | | | | | | | | | | | | | | | |
|---|---|---|---|---|---|---|---|---|---|---|---|---|---|---|---|---|---|---|---|
| SAT | T2DM | ENSG0000017 | 0.001456626 | 0.202875434 | 0.001143884 | 0.081436687 | 0.001484617 | 0.194296944 | 0.001143796 | 0.081418849 | -2.80E-05 | 0.002497228 | 1 | 0.565054583 | 1 | 0.562655141 | ENSG0000018 | 4 | TMEM106A |
| SAT | T2DM | ENSG0000018 | 9.99E-05 | 0.968173492 | 0.00250291 | 0.081436958 | 3.92E-05 | 0.987516514 | 0.002502714 | 0.081419131 | 6.07E-05 | 0.002082894 | 1 | 0.993613755 | 1 | 0.997162029 | ENSG0000018 | 4 | TMEM106A |
| SAT | T2DM | ENSG0000018 | 0.00713194 | 0.072447198 | 0.003970347 | 0.081436419 | 0.007194753 | 0.069944529 | 0.003970015 | 0.081418582 | -6.28E-05 | 0.027877401 | 1 | 0.376702465 | 1 | 0.38680588 | ENSG0000018 | 8 | SIVA1 |
| SAT | T2DM | ENSG0000018 | -0.0070157 | 0.006021496 | 0.002554301 | 0.081435698 | -0.00705829 | 0.005718218 | 0.002554087 | 0.081417855 | 4.26E-05 | 0.020374412 | 1 | 0.085526101 | 1 | 0.091112401 | ENSG0000018 | 8 | SIVA1 |
| SAT | T2DM | ENSG0000018 | -0.00148967 | 0.651272403 | 0.003295763 | 0.081436924 | -0.00154294 | 0.639640922 | 0.003295473 | 0.081419094 | 5.33E-05 | 0.013851652 | 1 | 0.882256298 | 1 | 0.885701439 | ENSG0000018 | 8 | SIVA1 |
| SAT | T2DM | ENSG0000018 | -0.00243983 | 0.421153644 | 0.003033022 | 0.0814368 | -0.00253665 | 0.402924952 | 0.003032785 | 0.081419014 | 9.68E-05 | 0.008956127 | 1 | 0.764695687 | 1 | 0.757591271 | ENSG0000018 | 7 | MAFF |
| SAT | T2DM | ENSG0000018 | -0.0001802 | 0.844147918 | 0.000916636 | 0.081436952 | -0.0001501 | 0.863989132 | 0.000916563 | 0.081419126 | -2.32E-05 | 0.000988265 | 1 | 0.962017825 | 1 | 0.966241215 | ENSG0000018 | 6 | CIB1 |
| SAT | T2DM | ENSG0000018 | -0.00259392 | 0.256535415 | 0.002286158 | 0.081436743 | -0.00456074 | 0.057995663 | 0.002405791 | 0.08141853 | 0.001966824 | 0.008729333 | 1 | 0.62099686 | 1 | 0.347397154 | ENSG0000018 | 6 | CIB1 |
| SAT | T2DM | ENSG0000018 | 0.000181324 | 0.94051202 | 0.002429753 | 0.081436957 | 0.000224078 | 0.926515019 | 0.002429541 | 0.081419129 | -4.28E-05 | 0.008366668 | 1 | 0.982870948 | 1 | 0.981878481 | ENSG0000018 | 6 | CIB1 |
| SAT | T2DM | ENSG0000018 | -0.00030809 | 0.824445885 | 0.001388861 | 0.08143695 | -0.00093989 | 0.501183585 | 0.001397339 | 0.081419055 | 0.000631799 | 4.62E-05 | 1 | 0.955051074 | 1 | 0.816084912 | ENSG0000018 | 6 | FLRT2 |
| SAT | T2DM | ENSG0000018 | 0.000949592 | 0.81563181 | 0.004072583 | 0.081436949 | 0.001038755 | 0.798662341 | 0.004072292 | 0.08141912 | -8.92E-05 | 0.011990564 | 1 | 0.952005681 | 1 | 0.952160336 | ENSG0000018 | 8 | ANO9 |
| SAT | T2DM | ENSG0000018 | -0.00169189 | 0.447221342 | 0.002226001 | 0.081436862 | -0.00119211 | 0.592751894 | 0.00222886 | 0.081419083 | -0.00049977 | 2.13E-05 | 1 | 0.780081459 | 1 | 0.854809759 | ENSG0000018 | 8 | ANO9 |
| SAT | T2DM | ENSG0000018 | -0.00029359 | 0.937047373 | 0.003717259 | 0.081436957 | -0.00065767 | 0.859621661 | 0.003718696 | 0.081419126 | 0.000364079 | 0.001875084 | 1 | 0.982567092 | 1 | 0.965566132 | ENSG0000018 | 8 | ANO9 |
| SAT | T2DM | ENSG0000018 | 0.000423866 | 0.868527038 | 0.002560664 | 0.081436954 | 0.000390283 | 0.878849294 | 0.002560436 | 0.081419127 | 3.36E-05 | 0.040294552 | 1 | 0.966468088 | 1 | 0.966700287 | ENSG0000018 | 5 | DDX51 |
| SAT | T2DM | ENSG0000018 | -0.00221986 | 0.186847363 | 0.001681756 | 0.081436667 | -0.00226564 | 0.177883524 | 0.001681612 | 0.081418828 | 4.58E-05 | 8.19E-05 | 1 | 0.544446003 | 1 | 0.539691192 | ENSG0000018 | 6 | GP1BA |
| SAT | T2DM | ENSG0000018 | 0.002527509 | 0.507164899 | 0.003810741 | 0.081436885 | 0.002465891 | 0.517541228 | 0.003810441 | 0.081419061 | 6.16E-05 | 0.038864524 | 1 | 0.810627063 | 1 | 0.825499645 | ENSG0000018 | 6 | KLHL33 |
| SAT | T2DM | ENSG0000018 | -0.00240743 | 0.519204523 | 0.00373492 | 0.081436889 | -0.00247861 | 0.506890443 | 0.003734605 | 0.081419057 | 7.12E-05 | 0.006931088 | 1 | 0.819389148 | 1 | 0.817738235 | ENSG0000018 | 3 | NUPR1L |
| SAT | T2DM | ENSG0000018 | -0.00614205 | 0.022089299 | 0.002683483 | 0.081436083 | -0.00619677 | 0.020921064 | 0.002683267 | 0.08141824 | 5.47E-05 | 0.007404145 | 1 | 0.201880195 | 1 | 0.204291699 | ENSG0000018 | 3 | NUPR1L |
| SAT | T2DM | ENSG0000018 | -0.00355193 | 0.310413577 | 0.003501661 | 0.081436786 | -0.003479 | 0.320417592 | 0.003501404 | 0.081418966 | -7.29E-05 | 0.014182865 | 1 | 0.670485151 | 1 | 0.691183511 | ENSG0000018 | 3 | NUPR1L |
| SAT | T2DM | ENSG0000018 | 0.00041429 | 0.828483316 | 0.001912277 | 0.081436951 | 0.000371532 | 0.845937448 | 0.001912112 | 0.081419125 | 4.28E-05 | 0.000984087 | 1 | 0.956121618 | 1 | 0.96477102 | ENSG0000018 | 3 | NUPR1L |
| SAT | T2DM | ENSG0000018 | 0.000382188 | 0.85478465 | 0.002088261 | 0.081436953 | -0.00116261 | 0.581386976 | 0.002108619 | 0.08141908 | 0.001544797 | 1.48E-07 | 1 | 0.963534946 | 1 | 0.851769934 | ENSG0000018 | 3 | NUPR1L |
| SAT | T2DM | ENSG0000018 | 0.000160055 | 0.839354994 | 0.000789544 | 0.081436952 | 0.000135253 | 0.863973952 | 0.000789486 | 0.081419126 | 2.48E-05 | 0.000197333 | 1 | 0.959889764 | 1 | 0.966241215 | ENSG0000018 | 8 | CCDC137 |
| SAT | T2DM | ENSG0000018 | -0.00010326 | 0.959250484 | 0.002020905 | 0.081436958 | -7.21E-05 | 0.971543574 | 0.002020728 | 0.081419131 | -3.12E-05 | 0.019726981 | 1 | 0.990651822 | 1 | 0.996397533 | ENSG0000018 | 8 | CCDC137 |
| SAT | T2DM | ENSG0000018 | 0.001095162 | 0.763517145 | 0.003640039 | 0.081436943 | 0.001485665 | 0.683561255 | 0.003644832 | 0.081419103 | -0.0003905 | 0.044635917 | 1 | 0.93459819 | 1 | 0.901372745 | ENSG0000018 | 8 | CCDC137 |
| SAT | T2DM | ENSG0000018 | -0.00051673 | 0.572363778 | 0.000915255 | 0.081436905 | -0.00054116 | 0.554313332 | 0.000915184 | 0.081419072 | 2.44E-05 | 0.00089414 | 1 | 0.840736467 | 1 | 0.842050121 | ENSG0000018 | 8 | CCDC137 |
| SAT | T2DM | ENSG0000018 | -0.00506215 | 0.1230442 | 0.003282584 | 0.081436561 | -0.00370228 | 0.260405557 | 0.003289644 | 0.081418919 | -0.00135987 | 7.26E-10 | 1 | 0.460845874 | 1 | 0.633351098 | ENSG0000018 | 8 | CCDC137 |
| SAT | T2DM | ENSG0000018 | 0.000265558 | 0.8511417 | 0.001415088 | 0.081436953 | 0.0002481 | 0.860812087 | 0.00141496 | 0.081419126 | 1.75E-05 | 0.046507572 | 1 | 0.963534946 | 1 | 0.965732208 | ENSG0000018 | 17 | CDK10 |
| SAT | T2DM | ENSG0000018 | -7.67E-05 | 0.923218609 | 0.000796135 | 0.081436957 | -8.90E-05 | 0.910937362 | 0.000796069 | 0.081419129 | 1.23E-05 | 0.035608906 | 1 | 0.978648294 | 1 | 0.978562139 | ENSG0000018 | 17 | CDK10 |
| SAT | T2DM | ENSG0000018 | -0.0014382 | 0.713138433 | 0.003911934 | 0.081436936 | -0.00158795 | 0.684777158 | 0.003911659 | 0.081419103 | 0.000149749 | 1.56E-05 | 1 | 0.908310248 | 1 | 0.901372745 | ENSG0000018 | 17 | CDK10 |
| SAT | T2DM | ENSG0000018 | 0.001278638 | 0.156002503 | 0.000901308 | 0.081436622 | 0.001295059 | 0.150726501 | 0.000901242 | 0.081418786 | -1.64E-05 | 0.03068842 | 1 | 0.506776449 | 1 | 0.51088187 | ENSG0000018 | 17 | CDK10 |
| SAT | T2DM | ENSG0000018 | 0.002540702 | 0.40802635 | 0.003070808 | 0.081436844 | 0.002479227 | 0.419434857 | 0.00307062 | 0.081419022 | 6.15E-05 | 0.041133496 | 1 | 0.756247299 | 1 | 0.768824902 | ENSG0000018 | 11 | GAS2L1 |
| SAT | T2DM | ENSG0000018 | -0.00265109 | 0.440432255 | 0.003436439 | 0.081436859 | -0.00150803 | 0.662223143 | 0.003452092 | 0.081419099 | -0.00114306 | 0.000581214 | 1 | 0.776495555 | 1 | 0.895728881 | ENSG0000018 | 11 | GAS2L1 |
| SAT | T2DM | ENSG0000018 | 4.98E-05 | 0.972341156 | 0.001435018 | 0.081436958 | 2.30E-05 | 0.987212445 | 0.001434891 | 0.081419131 | 2.68E-05 | 0.003744225 | 1 | 0.995031844 | 1 | 0.997162029 | ENSG0000018 | 11 | GAS2L1 |
| SAT | T2DM | ENSG0000018 | 0.00118919 | 0.26077697 | 0.001057473 | 0.081436747 | 0.001203209 | 0.255155673 | 0.001057378 | 0.081418915 | -1.40E-05 | 0.034905589 | 1 | 0.625445349 | 1 | 0.62709046 | ENSG0000018 | 9 | ATP6V0A2 |
| SAT | T2DM | ENSG0000018 | -0.00282917 | 0.229181031 | 0.002352794 | 0.081436717 | -0.00288529 | 0.220047078 | 0.002352638 | 0.0814188 | 5.61E-05 | 0.010170348 | 1 | 0.593149171 | 1 | 0.593241137 | ENSG0000018 | 9 | ATP6V0A2 |
| SAT | T2DM | ENSG0000018 | -0.00196481 | 0.079143594 | 0.001119117 | 0.081436443 | -0.00193378 | 0.083972196 | 0.001119025 | 0.081418632 | -3.10E-05 | 0.000186019 | 1 | 0.393439592 | 1 | 0.414575198 | ENSG0000018 | 9 | ATP6V0A2 |
| SAT | T2DM | ENSG0000018 | -0.01387295 | 7.45E-05 | 0.003502045 | 0.081434336 | -0.01395176 | 6.77E-05 | 0.003501798 | 0.081416479 | 7.88E-05 | 0.010910868 | 0.244521641 | 0.003943897 | 0.22220762 | 0.004114956 | ENSG0000018 | 9 | ATP6V0A2 |
| SAT | T2DM | ENSG0000018 | 0.00195811 | 0.570439158 | 0.003450991 | 0.081436905 | 0.001895131 | 0.582870301 | 0.00345073 | 0.081419072 | 6.30E-05 | 0.026248128 | 1 | 0.840736467 | 1 | 0.85184742 | ENSG0000018 | 9 | ATP6V0A2 |
| SAT | T2DM | ENSG0000018 | 8.06E-06 | 0.998194947 | 0.003561298 | 0.081436958 | 8.18E-05 | 0.981669797 | 0.003561006 | 0.081419131 | -7.38E-05 | 0.005190955 | 1 | 0.998511991 | 1 | 0.997162029 | ENSG0000018 | 9 | ATP6V0A2 |
| SAT | T2DM | ENSG0000018 | -0.00068132 | 0.53425762 | 0.001096217 | 0.081436894 | -0.00073198 | 0.504334837 | 0.001096298 | 0.081419056 | 5.07E-05 | 0.015834902 | 1 | 0.828750699 | 1 | 0.817738235 | ENSG0000018 | 12 | SP140L |
| SAT | T2DM | ENSG0000018 | 0.007733516 | 0.043038125 | 0.003822144 | 0.081436274 | 0.007831105 | 0.040457497 | 0.003821817 | 0.081418429 | -9.76E-05 | 0.000237095 | 1 | 0.286428522 | 1 | 0.289075371 | ENSG0000018 | 12 | SP140L |
| SAT | T2DM | ENSG0000018 | 0.00740093 | 0.000453374 | 0.002110382 | 0.081434903 | 0.007364308 | 0.000483352 | 0.002110213 | 0.081417096 | 3.66E-05 | 0.024188753 | 1 | 0.014727922 | 1 | 0.018228475 | ENSG0000018 | 12 | SP140L |
| SAT | T2DM | ENSG0000018 | 0.000345533 | 0.913537701 | 0.003182367 | 0.081436956 | 0.000395138 | 0.901176633 | 0.003182099 | 0.081419128 | -4.96E-05 | 0.028778981 | 1 | 0.976642944 | 1 | 0.972878089 | ENSG0000018 | 12 | SP140L |
| SAT | T2DM | ENSG0000018 | -0.00134784 | 0.448839787 | 0.001779681 | 0.081436863 | -0.00131692 | 0.459280856 | 0.001779543 | 0.081419039 | -3.09E-05 | 0.029418485 | 1 | 0.78183394 | 1 | 0.795972372 | ENSG0000018 | 11 | TARSL2 |
| SAT | T2DM | ENSG0000018 | 0.004573745 | 0.053206528 | 0.002365838 | 0.081436334 | 0.001791297 | 0.456712247 | 0.002406778 | 0.081419038 | 0.002782448 | 3.48E-10 | 1 | 0.320901873 | 1 | 0.794600418 | ENSG0000018 | 9 | SH3BGR |
| SAT | T2DM | ENSG0000018 | 0.006959711 | 0.068559877 | 0.00382126 | 0.081436404 | 0.006876286 | 0.071919203 | 0.003820937 | 0.08141859 | 8.34E-05 | 0.001998821 | 1 | 0.363988605 | 1 | 0.391321567 | ENSG0000018 | 9 | SH3BGR |
| SAT | T2DM | ENSG0000018 | -0.0008339 | 0.60895105 | 0.001630069 | 0.081436915 | -0.00079209 | 0.626990418 | 0.001629932 | 0.081419091 | -4.18E-05 | 0.000335321 | 1 | 0.855383565 | 1 | 0.876877903 | ENSG0000018 | 12 | MUC1 |
| SAT | T2DM | ENSG0000018 | 0.002707245 | 0.491464854 | 0.003935056 | 0.081436879 | 0.002589622 | 0.510448623 | 0.00393476 | 0.081419059 | 0.000117623 | 0.000301356 | 1 | 0.80382562 | 1 | 0.819765996 | ENSG0000018 | 12 | MUC1 |
| SAT | T2DM | ENSG0000018 | -0.00108958 | 0.790253953 | 0.004096475 | 0.081436947 | -0.00101904 | 0.803529644 | 0.004096127 | 0.081419121 | -7.05E-05 | 0.013487173 | 1 | 0.94681813 | 1 | 0.952314593 | ENSG0000018 | 12 | MUC1 |
| SAT | T2DM | ENSG0000018 | -0.00310994 | 0.286711871 | 0.002919132 | 0.081436769 | -0.00257556 | 0.377952126 | 0.002921212 | 0.081419001 | -0.00053437 | 6.33E-06 | 1 | 0.644230759 | 1 | 0.738661557 | ENSG0000018 | 5 | SP1 |
| SAT | T2DM | ENSG0000018 | -0.00052576 | 0.819020551 | 0.002297832 | 0.08143695 | -0.00059816 | 0.794606032 | 0.002297645 | 0.08141912 | 7.24E-05 | 2.46E-05 | 1 | 0.953693013 | 1 | 0.950456578 | ENSG0000018 | 13 | PCGF3 |
| SAT | T2DM | ENSG0000018 | -0.00175616 | 0.576550836 | 0.003144811 | 0.081436906 | -0.00184582 | 0.557215703 | 0.003144594 | 0.081419073 | 8.97E-05 | 0.001516843 | 1 | 0.842502133 | 1 | 0.842050121 | ENSG0000018 | 13 | PCGF3 |
| SAT | T2DM | ENSG0000018 | -0.00102489 | 0.54722092 | 0.001702674 | 0.081436898 | -0.00106579 | 0.531312396 | 0.001702528 | 0.081419065 | 4.09E-05 | 0.000483132 | 1 | 0.835027823 | 1 | 0.831380066 | ENSG0000018 | 13 | PCGF3 |
| SAT | T2DM | ENSG0000018 | 0.003211859 | 0.087541823 | 0.001879915 | 0.08143647 | 0.00318025 | 0.090676102 | 0.001879752 | 0.081418653 | 3.16E-05 | 0.012840513 | 1 | 0.407026915 | 1 | 0.423536093 | ENSG0000018 | 13 | PCGF3 |
| SAT | T2DM | ENSG0000018 | 0.002215943 | 0.206942423 | 0.001755871 | 0.081436692 | 0.002191827 | 0.21188847 | 0.001755722 | 0.0814187 | 2.41E-05 | 0.049714557 | 1 | 0.568658367 | 1 | 0.582737696 | ENSG0000018 | 10 | P4HB |
| SAT | T2DM | ENSG0000018 | 0.000436445 | 0.703589495 | 0.001147091 | 0.081436934 | 0.000414435 | 0.717859839 | 0.001147 | 0.081419109 | 2.20E-05 | 0.013425262 | 1 | 0.905285151 | 1 | 0.918559531 | ENSG0000018 | 13 | PSMD13 |
| SAT | T2DM | ENSG0000018 | -0.00061555 | 0.85428653 | 0.003351726 | 0.081436953 | -0.00050271 | 0.880765719 | 0.003351458 | 0.081419127 | -0.00011284 | 1.13E-05 | 1 | 0.963534946 | 1 | 0.967132638 | ENSG0000018 | 13 | PSMD13 |
| SAT | T2DM | ENSG0000018 | -0.0032287 | 0.375913443 | 0.003646391 | 0.081436827 | -0.00309416 | 0.396095337 | 0.003646124 | 0.081419011 | -0.00013454 | 1.47E-05 | 1 | 0.728082648 | 1 | 0.755715818 | ENSG0000018 | 10 | PMEL |
| SAT | T2DM | ENSG0000018 | 0.005560851 | 0.061227984 | 0.002970779 | 0.081436373 | 0.005641715 | 0.057537008 | 0.002970553 | 0.081418528 | -8.09E-05 | 0.00092598 | 1 | 0.343400026 | 1 | 0.346763912 | ENSG0000018 | 10 | PMEL |
| SAT | T2DM | ENSG0000018 | 0.001129336 | 0.770949315 | 0.003879099 | 0.081436944 | 0.001200858 | 0.756866511 | 0.003878762 | 0.081419115 | -7.15E-05 | 0.005894258 | 1 | 0.935460319 | 1 | 0.932511837 | ENSG0000018 | 10 | PMEL |
| SAT | T2DM | ENSG0000018 | 3.64E-05 | 0.972257872 | 0.001045472 | 0.081436958 | 1.50E-05 | 0.988541083 | 0.001045384 | 0.081419131 | 2.13E-05 | 0.004251667 | 1 | 0.995031844 | 1 | 0.997162029 | ENSG0000018 | 8 | EP400NL |
| SAT | T2DM | ENSG0000018 | -0.00638381 | 0.026997319 | 0.002886561 | 0.08143614 | -0.0019532 | 0.523016321 | 0.003058076 | 0.081419063 | -0.00443061 | 1.16E-05 | 1 | 0.226852733 | 1 | 0.82668068 | ENSG0000018 | 8 | EP400NL |
| SAT | T2DM | ENSG0000018 | -0.00017501 | 0.938670723 | 0.002274657 | 0.081436957 | -0.00011685 | 0.959026365 | 0.002274486 | 0.08141913 | -5.82E-05 | 0.001922585 | 1 | 0.982870948 | 1 | 0.993627317 | ENSG0000018 | 8 | EP400NL |
| SAT | T2DM | ENSG0000018 | -0.00322871 | 0.329224353 | 0.003309193 | 0.081436799 | -0.00334522 | 0.312031415 | 0.003308924 | 0.08141896 | 0.000116514 | 2.66E-06 | 1 | 0.692334629 | 1 | 0.68070151 | ENSG0000018 | 8 | EP400NL |
| SAT | T2DM | ENSG0000018 | 0.002269078 | 0.450282909 | 0.003005624 | 0.081436863 | 0.002212866 | 0.461544279 | 0.003005358 | 0.08141904 | 5.62E-05 | 0.00363898 | 1 | 0.782095408 | 1 | 0.797039301 | ENSG0000018 | 8 | EP400NL |
| SAT | T2DM | ENSG0000018 | -0.00392925 | 0.092473129 | 0.002335369 | 0.081436485 | -0.0038748 | 0.097054339 | 0.002335191 | 0.081418671 | -5.45E-05 | 0.004063689 | 1 | 0.414244809 | 1 | 0.434658078 | ENSG0000018 | 8 | EP400NL |
| SAT | T2DM | ENSG0000018 | -0.00158158 | 0.491438272 | 0.002298734 | 0.081436879 | -0.00162812 | 0.478743456 | 0.002298548 | 0.081419047 | 4.65E-05 | 0.007367498 | 1 | 0.80382562 | 1 | 0.80363893 | ENSG0000018 | 8 | EP400NL |
| SAT | T2DM | ENSG0000018 | -0.00194185 | 0.134353952 | 0.001297026 | 0.081436584 | -0.0019665 | 0.129447946 | 0.001296922 | 0.081418747 | 2.47E-05 | 0.012945689 | 1 | 0.475922014 | 1 | 0.479366489 | ENSG0000018 | 12 | MYBL1 |
| SAT | T2DM | ENSG0000018 | -0.00226893 | 0.220292551 | 0.00185105 | 0.081436707 | -0.00229365 | 0.215267143 | 0.001850888 | 0.081418874 | 2.47E-05 | 0.043336139 | 1 | 0.585721118 | 1 | 0.587596919 | ENSG0000018 | 12 | MYBL1 |
| SAT | T2DM | ENSG0000018 | -0.00013901 | 0.833258237 | 0.000660305 | 0.081436951 | 0.000816314 | 0.234374662 | 0.000686459 | 0.081418895 | -0.00095532 | 3.71E-07 | 1 | 0.957925815 | 1 | 0.610313962 | ENSG0000018 | 12 | MYBL1 |
| SAT | T2DM | ENSG0000018 | 0.000340317 | 0.868636464 | 0.002057652 | 0.081436954 | 0.000311807 | 0.879542976 | 0.002057471 | 0.081419127 | 2.85E-05 | 0.034030352 | 1 | 0.966468088 | 1 | 0.966700287 | ENSG0000018 | 12 | MYBL1 |
| SAT | T2DM | ENSG0000018 | 0.001228485 | 0.600826153 | 0.002347962 | 0.081436913 | 0.001261706 | 0.590985703 | 0.002347759 | 0.081419083 | -3.32E-05 | 0.036495765 | 1 | 0.851317034 | 1 | 0.854809759 | ENSG0000018 | 5 | RP11-645C24.2 |
| SAT | T2DM | ENSG0000018 | -0.00039272 | 0.895138625 | 0.002979516 | 0.081436956 | -0.00044501 | 0.881262011 | 0.002979285 | 0.081419127 | 5.23E-05 | 0.027453582 | 1 | 0.970180416 | 1 | 0.96735385 | ENSG0000018 | 9 | SRL |
| SAT | T2DM | ENSG0000018 | 0.002281178 | 0.432698813 | 0.002907506 | 0.081436856 | 0.00220428 | 0.448334397 | 0.002907271 | 0.081419035 | 7.69E-05 | 0.000495722 | 1 | 0.772274669 | 1 | 0.791227469 | ENSG0000018 | 9 | SRL |
| SAT | T2DM | ENSG0000018 | -0.00315164 | 0.369282768 | 0.003510336 | 0.081436824 | -0.00348157 | 0.321339851 | 0.003510672 | 0.081418967 | 0.000329922 | 3.53E-06 | 1 | 0.723805643 | 1 | 0.691873008 | ENSG0000018 | 9 | SRL |
| SAT | T2DM | ENSG0000018 | 0.004446271 | 0.250948267 | 0.003872888 | 0.081436738 | 0.004530738 | 0.242022909 | 0.003872602 | 0.081418902 | -8.45E-05 | 0.009746385 | 1 | 0.615827421 | 1 | 0.620663712 | ENSG0000018 | 6 | C11orf87 |
| SAT | T2DM | ENSG0000018 | -0.00251323 | 0.233234371 | 0.002108291 | 0.081436721 | -0.00244555 | 0.246025446 | 0.002108126 | 0.081418906 | -6.77E-05 | 4.48E-05 | 1 | 0.598312722 | 1 | 0.620663712 | ENSG0000018 | 6 | C11orf87 |
| SAT | T2DM | ENSG0000018 | 0.003411874 | 0.389832893 | 0.003967659 | 0.081436835 | 0.00332537 | 0.401924707 | 0.003967325 | 0.081419014 | 8.65E-05 | 0.002126723 | 1 | 0.741043872 | 1 | 0.757591271 | ENSG0000018 | 10 | KCNIP4 |
| SAT | T2DM | ENSG0000018 | 0.006174338 | 0.038009589 | 0.002975933 | 0.081436239 | 0.006272252 | 0.035066155 | 0.002976017 | 0.081418389 | -9.79E-05 | 0.047416766 | 1 | 0.27051944 | 1 | 0.272172417 | ENSG0000018 | 10 | KCNIP4 |
| SAT | T2DM | ENSG0000018 | 0.000573025 | 0.843456774 | 0.002901777 | 0.081436952 | 0.000505729 | 0.861634377 | 0.002901581 | 0.081419126 | 6.73E-05 | 0.011339102 | 1 | 0.961656125 | 1 | 0.965732208 | ENSG0000018 | 7 | DMWD |
| SAT | T2DM | ENSG0000018 | 0.002714211 | 0.183030678 | 0.002038487 | 0.081436662 | 0.002745629 | 0.177978016 | 0.002038315 | 0.081418828 | -3.14E-05 | 0.030174672 | 1 | 0.537621892 | 1 | 0.539691192 | ENSG0000018 | 11 | ARL17A |
| SAT | T2DM | ENSG0000018 | 0.005281354 | 0.000857185 | 0.001584252 | 0.08143501 | 0.005316135 | 0.000791175 | 0.001584123 | 0.081417249 | -3.48E-05 | 0.003538979 | 1 | 0.022145067 | 1 | 0.024351274 | ENSG0000018 | 12 | NPIPB4 |
| SAT | T2DM | ENSG0000018 | -0.00115148 | 0.612569457 | 0.002273812 | 0.081436916 | -0.00119477 | 0.599244861 | 0.002273638 | 0.081419085 | 4.33E-05 | 0.018858968 | 1 | 0.857440439 | 1 | 0.855952281 | ENSG0000018 | 12 | NPIPB4 |
| SAT | T2DM | ENSG0000018 | -0.00143688 | 0.600929678 | 0.002747042 | 0.081436913 | -0.00186115 | 0.498288974 | 0.002748357 | 0.081419054 | 0.000424266 | 6.73E-06 | 1 | 0.851317044 | 1 | 0.81497549 | ENSG0000018 | 9 | ZNF829 |
| SAT | T2DM | ENSG0000018 | -0.00307547 | 0.255580193 | 0.002705137 | 0.081436742 | -0.00311639 | 0.249268876 | 0.002704897 | 0.081418909 | 4.09E-05 | 0.018215602 | 1 | 0.620694754 | 1 | 0.621386978 | ENSG0000018 | 9 | ZNF829 |
| SAT | T2DM | ENSG0000018 | 0.000907988 | 0.349661534 | 0.000970854 | 0.081436812 | 0.000302262 | 0.757695324 | 0.000979751 | 0.081419115 | 0.000605726 | 4.85E-06 | 1 | 0.708609939 | 1 | 0.932832405 | ENSG0000018 | 9 | ZNF829 |
| SAT | T2DM | ENSG0000018 | 0.000683556 | 0.602355772 | 0.001311968 | 0.081436913 | 0.000700948 | 0.593119775 | 0.001311849 | 0.081419083 | -1.74E-05 | 0.030548228 | 1 | 0.851866072 | 1 | 0.854809759 | ENSG0000018 | 8 | TRIM69 |
| SAT | T2DM | ENSG0000018 | 0.000402784 | 0.892149273 | 0.002970711 | 0.081436955 | 0.000457929 | 0.877483362 | 0.002970473 | 0.081419127 | -5.51E-05 | 0.01590327 | 1 | 0.970180416 | 1 | 0.966700287 | ENSG0000018 | 7 | LINC00839 |
| SAT | T2DM | ENSG0000018 | -5.55E-05 | 0.960931671 | 0.001132488 | 0.081436958 | -3.43E-05 | 0.97584007 | 0.00113239 | 0.081419131 | -2.12E-05 | 0.005889096 | 1 | 0.99110869 | 1 | 0.99701704 | ENSG0000018 | 9 | SETD4 |
| SAT | T2DM | ENSG0000018 | 0.001407153 | 0.673510345 | 0.003339745 | 0.081436929 | 0.001455923 | 0.66285407 | 0.003339454 | 0.081419099 | -4.88E-05 | 0.029924657 | 1 | 0.891627092 | 1 | 0.895728881 | ENSG0000018 | 10 | SDHAP3 |
| SAT | T2DM | ENSG0000018 | -0.00032112 | 0.808581758 | 0.001325525 | 0.081436949 | -0.00035422 | 0.789275921 | 0.001325424 | 0.081419119 | 3.31E-05 | 0.002257226 | 1 | 0.951539016 | 1 | 0.947957612 | ENSG0000018 | 7 | PLK5 |
| SAT | T2DM | ENSG0000018 | 0.005548166 | 0.0443861 | 0.002759689 | 0.081436283 | 0.005503108 | 0.046123722 | 0.002759467 | 0.081418466 | 4.51E-05 | 0.031993564 | 1 | 0.29068023 | 1 | 0.309334779 | ENSG0000018 | 4 | KRT73 |
| SAT | T2DM | ENSG0000018 | -9.87E-05 | 0.978509884 | 0.00366589 | 0.081436958 | -0.00015235 | 0.966848534 | 0.003665578 | 0.081419131 | 5.36E-05 | 0.0359364 | 1 | 0.995203291 | 1 | 0.994741311 | ENSG0000018 | 7 | KRT5 |
| SAT | T2DM | ENSG0000018 | 0.006035353 | 0.051071845 | 0.003093656 | 0.081436322 | 0.005985071 | 0.053017128 | 0.003093402 | 0.081418505 | 5.03E-05 | 0.0278216 | 1 | 0.312624484 | 1 | 0.332861953 | ENSG0000018 | 10 | C2orf76 |
| SAT | T2DM | ENSG0000018 | -0.0029574 | 0.166856601 | 0.002139355 | 0.081436639 | -0.00291053 | 0.173646173 | 0.002139179 | 0.081418822 | -4.69E-05 | 0.002879509 | 1 | 0.519037734 | 1 | 0.532948952 | ENSG0000018 | 10 | C2orf76 |
| SAT | T2DM | ENSG0000018 | 0.009591563 | 0.005016725 | 0.003418269 | 0.081435643 | 0.009508518 | 0.005404608 | 0.003418011 | 0.081417838 | 8.30E-05 | 0.003182614 | 1 | 0.074817614 | 1 | 0.088387036 | ENSG0000018 | 12 | WWOX |
| SAT | T2DM | ENSG0000018 | 0.003501074 | 0.367977333 | 0.003888918 | 0.081436823 | 0.003701161 | 0.341247293 | 0.003888981 | 0.08141898 | -0.00020009 | 0.001175698 | 1 | 0.723275188 | 1 | 0.708179865 | ENSG0000018 | 4 | CYP4Z1 |
| SAT | T2DM | ENSG0000018 | 0.000405387 | 0.896281308 | 0.003109738 | 0.081436956 | 0.00050781 | 0.870275187 | 0.003109501 | 0.081419126 | -0.00010242 | 5.07E-05 | 1 | 0.970180416 | 1 | 0.966383771 | ENSG0000018 | 6 | ZNF749 |
| SAT | T2DM | ENSG0000018 | 0.004718668 | 0.207763107 | 0.003745742 | 0.081436693 | 0.004591059 | 0.220286804 | 0.003745457 | 0.0814188 | 0.000127609 | 2.94E-05 | 1 | 0.569276333 | 1 | 0.593399838 | ENSG0000018 | 6 | ZNF749 |
| SAT | T2DM | ENSG0000018 | 0.000151379 | 0.939897938 | 0.002007731 | 0.081436957 | 0.000204195 | 0.918984184 | 0.002007557 | 0.081419129 | -5.28E-05 | 9.05E-05 | 1 | 0.982870948 | 1 | 0.979654273 | ENSG0000018 | 6 | ZNF749 |
| SAT | T2DM | ENSG0000018 | -0.00405494 | 0.158073336 | 0.002872618 | 0.081436625 | -0.00399214 | 0.164577271 | 0.002872371 | 0.081418808 | -6.28E-05 | 0.001373969 | 1 | 0.507474184 | 1 | 0.523492072 | ENSG0000018 | 6 | ZNF749 |
| SAT | T2DM | ENSG0000018 | 0.000720643 | 0.593842865 | 0.001351351 | 0.081436911 | 0.000676876 | 0.616426193 | 0.001351259 | 0.081419089 | 4.38E-05 | 0.000374651 | 1 | 0.8467616 | 1 | 0.869142389 | ENSG0000018 | 6 | ZNF749 |
| SAT | T2DM | ENSG0000019 | -0.0022844 | 0.014092932 | 0.000930547 | 0.081435951 | -0.00230012 | 0.013436131 | 0.000930468 | 0.08141811 | 1.57E-05 | 0.016138231 | 1 | 0.151493517 | 1 | 0.157494043 | ENSG0000018 | 6 | ZNF749 |
| SAT | T2DM | ENSG0000019 | -0.00201848 | 0.285241538 | 0.001888863 | 0.081436768 | -0.00206853 | 0.273425374 | 0.001888707 | 0.08141893 | 5.01E-05 | 0.000304716 | 1 | 0.643214767 | 1 | 0.64192524 | ENSG0000018 | 7 | NUDT17 |
| SAT | T2DM | ENSG0000019 | -0.00391482 | 0.311873278 | 0.003871074 | 0.081436788 | -0.00397987 | 0.303863868 | 0.003870776 | 0.081418954 | 6.51E-05 | 0.037380571 | 1 | 0.67252715 | 1 | 0.671707395 | ENSG0000018 | 10 | CCDC30 |
| SAT | T2DM | ENSG0000019 | -0.00303666 | 0.060850272 | 0.00161991 | 0.081436371 | -0.00300722 | 0.063371851 | 0.001619767 | 0.081418555 | -2.94E-05 | 0.005152701 | 1 | 0.34323074 | 1 | 0.365339457 | ENSG0000018 | 9 | GLDN |
| SAT | T2DM | ENSG0000019 | 0.00706177 | 1.39E-07 | 0.001340879 | 0.081432323 | 0.002446034 | 0.113882837 | 0.001547162 | 0.081418713 | 0.004615737 | 2.25E-09 | 0.00045635 | 1.90E-05 | 1 | 0.455762334 | ENSG0000018 | 9 | GLDN |
| SAT | T2DM | ENSG0000019 | 0.004861411 | 0.057073266 | 0.002554934 | 0.081436353 | 0.004899223 | 0.055146992 | 0.002554711 | 0.081418516 | -3.78E-05 | 0.026034814 | 1 | 0.331288318 | 1 | 0.341997131 | ENSG0000018 | 10 | ZNF197 |
| SAT | T2DM | ENSG0000019 | -0.00298855 | 0.290962205 | 0.002830032 | 0.081436772 | -0.00308148 | 0.276179156 | 0.002829787 | 0.081418933 | 9.29E-05 | 1.37E-06 | 1 | 0.647523626 | 1 | 0.64327119 | ENSG0000018 | 10 | ZNF197 |
| SAT | T2DM | ENSG0000019 | 0.000971289 | 0.645722217 | 0.002112815 | 0.081436923 | 0.000935558 | 0.657881794 | 0.002112632 | 0.081419098 | 3.57E-05 | 0.012566334 | 1 | 0.878338086 | 1 | 0.892315075 | ENSG0000018 | 8 | FAM228A |
| SAT | T2DM | ENSG0000019 | 0.002653023 | 0.409285912 | 0.003215194 | 0.081436845 | 0.002596866 | 0.419231751 | 0.003214916 | 0.081419022 | 5.62E-05 | 0.009783611 | 1 | 0.756926483 | 1 | 0.768824902 | ENSG0000018 | 8 | FAM228A |
| SAT | T2DM | ENSG0000019 | -0.0004315 | 0.90422326 | 0.003586023 | 0.081436956 | -0.00054145 | 0.879975924 | 0.003585762 | 0.081419127 | 0.00010995 | 0.000337938 | 1 | 0.973026079 | 1 | 0.966700287 | ENSG0000018 | 8 | FAM228A |
| SAT | T2DM | ENSG0000019 | -0.01341148 | 1.52E-08 | 0.002369952 | 0.081431607 | 0.000612058 | 0.891276763 | 0.004477742 | 0.081419128 | -0.01402354 | 0.000223263 | 5.00E-05 | 2.76E-06 | 1 | 0.971822875 | ENSG0000018 | 8 | FAM228A |
| SAT | T2DM | ENSG0000019 | -0.00054645 | 0.766277242 | 0.001838375 | 0.081436944 | -0.00051539 | 0.779190095 | 0.001838229 | 0.081419118 | -3.11E-05 | 0.029980513 | 1 | 0.934630346 | 1 | 0.941975941 | ENSG0000018 | 8 | RPS23 |
| SAT | T2DM | ENSG0000019 | 0.002149266 | 0.434518877 | 0.00275024 | 0.081436856 | 0.002082965 | 0.448788272 | 0.002750017 | 0.081419035 | 6.63E-05 | 0.001340404 | 1 | 0.772274669 | 1 | 0.791227469 | ENSG0000018 | 8 | RPS23 |
| SAT | T2DM | ENSG0000019 | 0.004887867 | 0.012279908 | 0.001952019 | 0.081435911 | 0.005103516 | 0.008938257 | 0.001952066 | 0.081417989 | -0.00021565 | 1.59E-11 | 1 | 0.138932334 | 1 | 0.1231999 | ENSG0000018 | 8 | RPS23 |
| SAT | T2DM | ENSG0000019 | 0.001797213 | 0.472704561 | 0.002502783 | 0.081436872 | 0.001757397 | 0.482534797 | 0.002502587 | 0.081419048 | 3.98E-05 | 0.044588361 | 1 | 0.796581235 | 1 | 0.804878835 | ENSG0000018 | 8 | RPS23 |
| SAT | T2DM | ENSG0000019 | -0.00195799 | 0.480877941 | 0.002777716 | 0.081436875 | -0.00202048 | 0.466956053 | 0.002777517 | 0.081419042 | 6.25E-05 | 0.009732265 | 1 | 0.798461803 | 1 | 0.800114089 | ENSG0000018 | 9 | BTN3A2 |
| SAT | T2DM | ENSG0000019 | 0.001187177 | 0.554253656 | 0.002007405 | 0.0814369 | 0.00121552 | 0.544799872 | 0.002007228 | 0.08141907 | -2.83E-05 | 0.030899743 | 1 | 0.835948344 | 1 | 0.840498821 | ENSG0000018 | 9 | BTN3A2 |
| SAT | T2DM | ENSG0000019 | 0.00395216 | 0.060988577 | 0.002109413 | 0.081436372 | 0.0039155 | 0.063403623 | 0.002109241 | 0.081418555 | 3.67E-05 | 0.019707556 | 1 | 0.34323074 | 1 | 0.365339457 | ENSG0000018 | 9 | BTN3A2 |
| SAT | T2DM | ENSG0000019 | -0.00335057 | 0.006283357 | 0.001226135 | 0.081435711 | -0.00146661 | 0.251084042 | 0.001277845 | 0.081418911 | -0.00188396 | 1.70E-07 | 1 | 0.086986048 | 1 | 0.622908325 | ENSG0000018 | 9 | BTN3A2 |
| SAT | T2DM | ENSG0000019 | 0.002446279 | 0.10055924 | 0.001489679 | 0.081436508 | 0.002421589 | 0.104012536 | 0.00148956 | 0.081418689 | 2.47E-05 | 0.031678259 | 1 | 0.425727224 | 1 | 0.442053277 | ENSG0000018 | 9 | BTN3A2 |
| SAT | T2DM | ENSG0000019 | -0.00249176 | 0.437791144 | 0.003211318 | 0.081436858 | -0.00240532 | 0.453811698 | 0.003211043 | 0.081419037 | -8.64E-05 | 9.92E-05 | 1 | 0.77433571 | 1 | 0.794600418 | ENSG0000018 | 9 | BTN3A2 |
| SAT | T2DM | ENSG0000019 | 0.004307661 | 0.287408469 | 0.004049219 | 0.081436769 | 0.004405531 | 0.276562874 | 0.004048925 | 0.081418933 | -9.79E-05 | 0.00479201 | 1 | 0.644566475 | 1 | 0.64327119 | ENSG0000018 | 10 | TMEM222 |
| SAT | T2DM | ENSG0000019 | 0.0012468 | 0.96854754 | 0.003162058 | 0.081436958 | 4.64E-05 | 0.988280126 | 0.003161874 | 0.081419131 | 7.82E-05 | 0.014525177 | 1 | 0.993613755 | 1 | 0.997162029 | ENSG0000018 | 10 | SEPT10 |
| SAT | T2DM | ENSG0000019 | -0.00738694 | 0.017752542 | 0.003115868 | 0.081436019 | -0.00748352 | 0.016308975 | 0.003115619 | 0.081418167 | 9.66E-05 | 5.92E-05 | 1 | 0.172837064 | 1 | 0.177184592 | ENSG0000018 | 10 | SEPT10 |
| SAT | T2DM | ENSG0000019 | -0.00420454 | 0.004492361 | 0.001479765 | 0.081435609 | -0.00417347 | 0.004793786 | 0.001479647 | 0.081417802 | -3.11E-05 | 0.006349906 | 1 | 0.071190619 | 1 | 0.081103506 | ENSG0000018 | 7 | SMYD4 |

| | | | | | | | | | | | | | | | | | | | |
|---|---|---|---|---|---|---|---|---|---|---|---|---|---|---|---|---|---|---|---|
| SAT | T2DM | ENSG0000019 | -0.00466989 | 0.116447756 | 0.00297471 | 0.081436547 | -0.00139949 | 0.644021648 | 0.003028641 | 0.081419095 | -0.0032704 | 1.00E-08 | 1 | 0.4484332 | 1 | 0.888950368 | ENSG0000018 | 7 | SMYD4 |
| SAT | T2DM | ENSG0000019 | 0.001820316 | 0.588967498 | 0.003368877 | 0.08143691 | 0.001770403 | 0.599192638 | 0.003368589 | 0.081419085 | 4.99E-05 | 0.033010396 | 1 | 0.846620707 | 1 | 0.855952281 | ENSG0000018 | 7 | SMYD4 |
| SAT | T2DM | ENSG0000019 | 0.00258206 | 0.487957873 | 0.003722903 | 0.081436878 | 0.002244658 | 0.546890907 | 0.003726031 | 0.08141907 | 0.000337402 | 0.037599247 | 1 | 0.802903602 | 1 | 0.841095699 | ENSG0000018 | 7 | SMYD4 |
| SAT | T2DM | ENSG0000019 | 0.000499041 | 0.681633973 | 0.001216478 | 0.08143693 | 0.000466139 | 0.70156019 | 0.001216389 | 0.081419106 | 3.29E-05 | 0.001599812 | 1 | 0.892786811 | 1 | 0.909657964 | ENSG0000018 | 8 | CEACAM19 |
| SAT | T2DM | ENSG0000019 | -0.00189391 | 0.023028815 | 0.000833234 | 0.081436095 | -0.00187212 | 0.024648155 | 0.000833208 | 0.081418287 | -2.18E-05 | 0.036606843 | 1 | 0.20700696 | 1 | 0.227804495 | ENSG0000018 | 8 | CEACAM19 |
| SAT | T2DM | ENSG0000019 | -1.30E-05 | 0.995044486 | 0.002094728 | 0.081436958 | 1.51E-05 | 0.994259478 | 0.0020945 | 0.081419131 | -2.81E-05 | 0.033770903 | 1 | 0.99758396 | 1 | 0.998405122 | ENSG0000018 | 8 | CEACAM19 |
| SAT | T2DM | ENSG0000019 | -0.00690825 | 0.055333876 | 0.00360509 | 0.081436345 | -0.00774823 | 0.031883475 | 0.003610758 | 0.081418362 | 0.000839979 | 5.94E-05 | 1 | 0.326529581 | 1 | 0.263779676 | ENSG0000018 | 8 | CEACAM19 |
| SAT | T2DM | ENSG0000019 | 0.001440851 | 0.373531663 | 0.001619155 | 0.081436826 | -0.00032165 | 0.845495427 | 0.001650578 | 0.081419125 | 0.001762498 | 4.17E-08 | 1 | 0.72766644 | 1 | 0.96477102 | ENSG0000018 | 13 | NF2 |
| SAT | T2DM | ENSG0000019 | -0.00300384 | 0.22762303 | 0.0024897 | 0.081436715 | -0.00216065 | 0.386119647 | 0.002493025 | 0.081419005 | -0.00084319 | 2.99E-10 | 1 | 0.591955891 | 1 | 0.746964998 | ENSG0000018 | 13 | NF2 |
| SAT | T2DM | ENSG0000019 | 0.001328858 | 0.271769382 | 0.001209154 | 0.081436757 | 0.001361317 | 0.260193363 | 0.001209054 | 0.081418919 | -3.25E-05 | 0.000249697 | 1 | 0.634354473 | 1 | 0.633304468 | ENSG0000018 | 7 | C6orf1 |
| SAT | T2DM | ENSG0000019 | 0.002358014 | 0.227380475 | 0.001953396 | 0.081436715 | 0.002401053 | 0.218973104 | 0.001953243 | 0.081418878 | -4.30E-05 | 0.00529635 | 1 | 0.591955891 | 1 | 0.592333986 | ENSG0000018 | 7 | C6orf1 |
| SAT | T2DM | ENSG0000019 | 0.000218902 | 0.931402173 | 0.002542976 | 0.081436957 | 0.000271181 | 0.915068189 | 0.002542762 | 0.081419129 | -5.23E-05 | 0.003961607 | 1 | 0.980452059 | 1 | 0.979654273 | ENSG0000018 | 8 | C17orf58 |
| SAT | T2DM | ENSG0000019 | -0.00221779 | 0.390875424 | 0.002584743 | 0.081436835 | -0.002276 | 0.378519387 | 0.002584514 | 0.081419001 | 5.82E-05 | 0.000483851 | 1 | 0.741307668 | 1 | 0.738661557 | ENSG0000018 | 8 | C17orf58 |
| SAT | T2DM | ENSG0000019 | -0.00751736 | 0.029065012 | 0.003444208 | 0.081436162 | -0.00758277 | 0.027680739 | 0.003443914 | 0.081418321 | 6.54E-05 | 0.006376426 | 1 | 0.236875227 | 1 | 0.242234694 | ENSG0000018 | 8 | C17orf58 |
| SAT | T2DM | ENSG0000019 | 0.00226256 | 0.288861749 | 0.002133237 | 0.08143677 | 0.002197486 | 0.302916131 | 0.002133065 | 0.081418954 | 6.51E-05 | 5.63E-05 | 1 | 0.645569425 | 1 | 0.671232195 | ENSG0000018 | 8 | CYP27C1 |
| SAT | T2DM | ENSG0000019 | 0.005084311 | 0.18902139 | 0.003870866 | 0.08143667 | 0.005010243 | 0.195512964 | 0.003870578 | 0.081418851 | 7.41E-05 | 0.021870324 | 1 | 0.547377916 | 1 | 0.563318267 | ENSG0000018 | 6 | MST1L |
| SAT | T2DM | ENSG0000019 | 0.004093778 | 0.09827155 | 0.002476134 | 0.081436502 | 0.004040203 | 0.102724518 | 0.002475934 | 0.081418686 | 5.36E-05 | 0.00441697 | 1 | 0.421965667 | 1 | 0.440431278 | ENSG0000018 | 6 | MST1L |
| SAT | T2DM | ENSG0000019 | 0.00417232 | 0.288907089 | 0.003934208 | 0.08143677 | 0.004102424 | 0.297022523 | 0.003933893 | 0.081418949 | 6.99E-05 | 0.020122367 | 1 | 0.645569425 | 1 | 0.66437884 | ENSG0000018 | 6 | MST1L |
| SAT | T2DM | ENSG0000019 | -0.00264283 | 0.113912066 | 0.001671776 | 0.08143654 | -0.0026715 | 0.110014683 | 0.001671642 | 0.081418704 | 2.87E-05 | 0.025723022 | 1 | 0.443351708 | 1 | 0.452205306 | ENSG0000018 | 6 | MST1L |
| SAT | T2DM | ENSG0000019 | 0.004563166 | 0.200149419 | 0.003561835 | 0.081436684 | 0.003847549 | 0.280157731 | 0.003562656 | 0.081418936 | 0.000715617 | 1.26E-14 | 1 | 0.561024613 | 1 | 0.646143093 | ENSG0000018 | 6 | MST1L |
| SAT | T2DM | ENSG0000019 | 0.002134817 | 0.550155579 | 0.003572751 | 0.081436899 | 0.002074305 | 0.561482896 | 0.003572442 | 0.081419075 | 6.05E-05 | 0.012408345 | 1 | 0.83552458 | 1 | 0.84223981 | ENSG0000018 | 7 | FOXI2 |
| SAT | T2DM | ENSG0000019 | 0.01172958 | 0.000536664 | 0.00338831 | 0.081434956 | 0.011664832 | 0.000575495 | 0.003388053 | 0.08141715 | 6.47E-05 | 0.019573323 | 1 | 0.015863016 | 1 | 0.019875797 | ENSG0000018 | 7 | FOXI2 |
| SAT | T2DM | ENSG0000019 | -0.00480153 | 1.79E-12 | 0.000681068 | 0.081428653 | -0.00285877 | 0.00033098 | 0.000796362 | 0.081416978 | -0.00194276 | 2.52E-06 | 5.88E-09 | 7.35E-10 | 1 | 0.013406744 | ENSG0000018 | 7 | FOXI2 |
| SAT | T2DM | ENSG0000019 | -0.00192609 | 0.584165728 | 0.003519185 | 0.081436908 | -0.00179162 | 0.61066862 | 0.003519051 | 0.081419088 | -0.00013447 | 0.001367534 | 1 | 0.845302694 | 1 | 0.863994715 | ENSG0000018 | 7 | FOXI2 |
| SAT | T2DM | ENSG0000019 | -0.00235764 | 0.117985272 | 0.001508128 | 0.08143655 | -0.0026075 | 0.083891624 | 0.001508496 | 0.081418632 | 0.000249859 | 4.76E-10 | 1 | 0.452230931 | 1 | 0.414575198 | ENSG0000018 | 11 | HYAL3 |
| SAT | T2DM | ENSG0000019 | 0.00102311 | 0.571488137 | 0.001808057 | 0.081436905 | 0.000983383 | 0.586485 | 0.001807898 | 0.081419081 | 3.97E-05 | 0.00092437 | 1 | 0.840736467 | 1 | 0.853229568 | ENSG0000018 | 11 | HYAL3 |
| SAT | T2DM | ENSG0000019 | -0.00855912 | 0.034593647 | 0.004050523 | 0.081436212 | -0.00848341 | 0.036211183 | 0.004050221 | 0.081418398 | -7.57E-05 | 0.025292101 | 1 | 0.257958536 | 1 | 0.276092282 | ENSG0000018 | 6 | TRABD2A |
| SAT | T2DM | ENSG0000019 | 0.002085105 | 0.21817771 | 0.001693294 | 0.081436705 | 0.002120059 | 0.210518845 | 0.001693147 | 0.081418869 | -3.50E-05 | 0.002105074 | 1 | 0.582458151 | 1 | 0.579977552 | ENSG0000018 | 6 | TRABD2A |
| SAT | T2DM | ENSG0000019 | -0.00243505 | 0.417197858 | 0.003001447 | 0.081436848 | -0.00238929 | 0.425966209 | 0.003001188 | 0.081419025 | -4.58E-05 | 0.025467427 | 1 | 0.763620456 | 1 | 0.776441739 | ENSG0000018 | 6 | TRABD2A |
| SAT | T2DM | ENSG0000020 | -0.00036472 | 0.906879267 | 0.003117913 | 0.081436956 | -0.00029313 | 0.925091158 | 0.003117662 | 0.081419129 | -7.16E-05 | 0.002562097 | 1 | 0.974472349 | 1 | 0.981878481 | ENSG0000018 | 6 | TRABD2A |
| SAT | T2DM | ENSG0000020 | 0.000552485 | 0.855140497 | 0.00302626 | 0.081436953 | 0.000605127 | 0.841499926 | 0.003026008 | 0.081419124 | -5.26E-05 | 0.015766408 | 1 | 0.963534946 | 1 | 0.96477102 | ENSG0000018 | 6 | TRABD2A |
| SAT | T2DM | ENSG0000020 | -0.00051264 | 0.744297707 | 0.00157171 | 0.081436941 | -0.00054974 | 0.726489673 | 0.001571588 | 0.08141911 | 3.71E-05 | 0.002981239 | 1 | 0.927287795 | 1 | 0.921455409 | ENSG0000018 | 6 | QRFPR |
| SAT | T2DM | ENSG0000020 | -0.0062653 | 0.002288322 | 0.002054177 | 0.081435404 | -0.00620847 | 0.002506473 | 0.002054033 | 0.081417604 | -5.68E-05 | 0.001730483 | 1 | 0.045228813 | 1 | 0.054078961 | ENSG0000018 | 6 | QRFPR |
| SAT | T2DM | ENSG0000020 | -0.00356081 | 0.239733445 | 0.003028788 | 0.081436727 | -0.00348678 | 0.249605201 | 0.003028535 | 0.081418909 | -7.40E-05 | 0.000676157 | 1 | 0.60584911 | 1 | 0.621386978 | ENSG0000018 | 5 | TMEM17 |
| SAT | T2DM | ENSG0000020 | -0.00626172 | 0.088770589 | 0.003679206 | 0.081436474 | -0.00617543 | 0.093227474 | 0.003678881 | 0.0841866 | -8.63E-05 | 0.000305785 | 1 | 0.410282965 | 1 | 0.428129221 | ENSG0000018 | 14 | RHD |
| SAT | T2DM | ENSG0000020 | 0.003499474 | 0.343266564 | 0.003692473 | 0.081436808 | 0.003409898 | 0.355721717 | 0.003692169 | 0.081418988 | 8.96E-05 | 0.000941446 | 1 | 0.703910998 | 1 | 0.724067999 | ENSG0000018 | 14 | RHD |
| SAT | T2DM | ENSG0000020 | -0.00043297 | 0.791252974 | 0.001635812 | 0.081436947 | -0.0039907 | 0.807247224 | 0.001635664 | 0.081419121 | -3.39E-05 | 0.000847164 | 1 | 0.94681813 | 1 | 0.952410029 | ENSG0000018 | 2 | C17orf82 |
| SAT | T2DM | ENSG0000020 | 0.001314617 | 0.59810692 | 0.002493936 | 0.081436912 | 0.001360483 | 0.58536762 | 0.00249373 | 0.081419081 | -4.59E-05 | 0.012191499 | 1 | 0.84988688 | 1 | 0.853216864 | ENSG0000018 | 8 | CYP4A11 |
| SAT | T2DM | ENSG0000020 | -0.00158641 | 0.626993022 | 0.003264464 | 0.081436919 | -0.00304812 | 0.352351894 | 0.003277412 | 0.081418986 | 0.001461706 | 7.26E-07 | 1 | 0.866453072 | 1 | 0.720881299 | ENSG0000018 | 8 | CYP4A11 |
| SAT | T2DM | ENSG0000020 | -0.00216211 | 0.426523393 | 0.002719103 | 0.081436853 | -0.00435807 | 0.111525408 | 0.002738555 | 0.081418708 | 0.002195964 | 2.26E-11 | 1 | 0.77145714 | 1 | 0.45286493 | ENSG0000018 | 5 | TMEM216 |
| SAT | T2DM | ENSG0000020 | 0.011874764 | 0.001713981 | 0.003786841 | 0.081435315 | 0.011762554 | 0.001893709 | 0.003786516 | 0.081417519 | 0.00011221 | 1.57E-05 | 1 | 0.038255586 | 1 | 0.047650044 | ENSG0000018 | 5 | TMEM216 |
| SAT | T2DM | ENSG0000020 | -0.00151993 | 0.556041193 | 0.002581705 | 0.0814369 | -0.0026847 | 0.30198569 | 0.002600983 | 0.081418953 | 0.001164764 | 0.00025406 | 1 | 0.835948344 | 1 | 0.670829416 | ENSG0000018 | 5 | TMEM216 |
| SAT | T2DM | ENSG0000020 | 0.008647889 | 0.001431037 | 0.002712353 | 0.08143526 | -0.00057627 | 0.852781729 | 0.003105394 | 0.081419125 | 0.009224159 | 1.07E-09 | 1 | 0.03377865 | 1 | 0.965561588 | ENSG0000018 | 5 | TMEM216 |
| SAT | T2DM | ENSG0000020 | 0.00867025 | 0.00037541 | 0.002437643 | 0.081434844 | 0.006909595 | 0.005866451 | 0.002507869 | 0.081417863 | 0.001760655 | 0.002861264 | 1 | 0.012965483 | 1 | 0.092094861 | ENSG0000018 | 5 | TMEM216 |
| SAT | T2DM | ENSG0000020 | 0.001222942 | 0.70002394 | 0.003174095 | 0.081436934 | 0.001363262 | 0.667538459 | 0.003173853 | 0.0814191 | -0.00014032 | 5.96E-08 | 1 | 0.902046837 | 1 | 0.896403813 | ENSG0000018 | 5 | TMEM216 |
| SAT | T2DM | ENSG0000020 | -0.00030116 | 0.824861155 | 0.001360901 | 0.08143695 | 0.003075668 | 0.049626088 | 0.001566678 | 0.081418487 | -0.00337683 | 1.37E-05 | 1 | 0.955051074 | 1 | 0.324677987 | ENSG0000018 | 2 | C3orf70 |
| SAT | T2DM | ENSG0000020 | 0.003060123 | 0.07689251 | 0.001729847 | 0.081436435 | 0.004050044 | 0.019881184 | 0.001739268 | 0.081418225 | -0.00098992 | 5.94E-08 | 1 | 0.387125475 | 1 | 0.195966491 | ENSG0000018 | 9 | PLCD1 |
| SAT | T2DM | ENSG0000020 | -0.00568614 | 0.002766223 | 0.001900065 | 0.081435462 | -0.00496624 | 0.010230045 | 0.00193392 | 0.081418029 | -0.0007199 | 0.046339867 | 1 | 0.050988644 | 1 | 0.133193565 | ENSG0000018 | 9 | PLCD1 |
| SAT | T2DM | ENSG0000020 | 0.005372573 | 0.000187889 | 0.001438518 | 0.081434627 | 0.003748757 | 0.010516772 | 0.001465292 | 0.081418037 | 0.001623815 | 6.33E-09 | 0.616462728 | 0.007553524 | 1 | 0.136385495 | ENSG0000018 | 4 | HEATR4 |
| SAT | T2DM | ENSG0000020 | 0.005454754 | 0.053192903 | 0.002821391 | 0.081436334 | 0.003295674 | 0.259138543 | 0.002920581 | 0.081418918 | 0.00215908 | 0.000281412 | 1 | 0.320901873 | 1 | 0.631674265 | ENSG0000018 | 10 | MRPS21 |
| SAT | T2DM | ENSG0000020 | 0.000900405 | 0.683532146 | 0.002208781 | 0.081436931 | -0.00163063 | 0.465150872 | 0.002232541 | 0.081419042 | 0.002531035 | 9.03E-15 | 1 | 0.89384973 | 1 | 0.799014258 | ENSG0000018 | 10 | MRPS21 |
| SAT | T2DM | ENSG0000020 | -5.14E-05 | 0.964247975 | 0.001147715 | 0.081436958 | -0.00047079 | 0.684317479 | 0.001157929 | 0.081419103 | 0.000419345 | 0.00660857 | 1 | 0.992060403 | 1 | 0.901372745 | ENSG0000018 | 10 | MRPS21 |
| SAT | T2DM | ENSG0000020 | -0.00257622 | 0.257862817 | 0.002276906 | 0.081436744 | -0.00086863 | 0.71846866 | 0.002409481 | 0.081419109 | -0.00170759 | 0.030432367 | 1 | 0.622209623 | 1 | 0.918559531 | ENSG0000018 | 1 | H1FNT |
| SAT | T2DM | ENSG0000020 | 0.004708289 | 0.001375216 | 0.001471423 | 0.081435247 | 0.002008444 | 0.188697938 | 0.00152798 | 0.081418842 | 0.002699846 | 5.89E-11 | 1 | 0.032934918 | 1 | 0.558266847 | ENSG0000018 | 1 | H1FNT |
| SAT | T2DM | ENSG0000020 | 0.009607239 | 0.002337094 | 0.003156446 | 0.08143541 | 0.004337524 | 0.184264656 | 0.003266848 | 0.081418836 | 0.005269715 | 4.13E-10 | 1 | 0.045916193 | 1 | 0.550111317 | ENSG0000018 | 1 | H1FNT |
| SAT | T2DM | ENSG0000020 | 0.000623056 | 0.456747651 | 0.000837201 | 0.081436866 | 8.37E-05 | 0.9216273 | 0.000851189 | 0.081419129 | 0.000539313 | 0.000468734 | 1 | 0.786668754 | 1 | 0.979654273 | ENSG0000018 | 1 | H1FNT |
| SAT | T2DM | ENSG0000020 | -0.00094988 | 0.294942822 | 0.00090694 | 0.081436775 | -0.00164738 | 0.070886672 | 0.000912064 | 0.081418586 | 0.0006975 | 8.38E-13 | 1 | 0.652072776 | 1 | 0.387631954 | ENSG0000018 | 11 | WDR86 |
| SAT | T2DM | ENSG0000020 | -0.00062786 | 0.609941474 | 0.001230726 | 0.081436915 | -0.00154039 | 0.233208513 | 0.001292129 | 0.081418893 | 0.00091253 | 0.020559185 | 1 | 0.855586993 | 1 | 0.610313962 | ENSG0000018 | 10 | ZNF385C |
| SAT | T2DM | ENSG0000020 | 0.000592815 | 0.492851432 | 0.000864437 | 0.08143688 | -0.00095532 | 0.292899835 | 0.00090829 | 0.081418946 | 0.001548138 | 2.91E-08 | 1 | 0.80490072 | 1 | 0.658673309 | ENSG0000018 | 10 | DHRS4L2 |
| SAT | T2DM | ENSG0000020 | 0.000852857 | 0.278991834 | 0.000787794 | 0.081436763 | -0.00042797 | 0.614205556 | 0.000849011 | 0.081419088 | 0.001280826 | 5.26E-05 | 1 | 0.63926506 | 1 | 0.867485217 | ENSG0000018 | 10 | DHRS4L2 |
| SAT | T2DM | ENSG0000020 | 0.003098635 | 0.000179015 | 0.000826974 | 0.081434612 | 0.001675333 | 0.050612035 | 0.000857053 | 0.081418264 | 0.001423302 | 2.69E-10 | 0.58734888 | 0.007434796 | 1 | 0.326243786 | ENSG0000018 | 10 | TRPV2 |
| SAT | T2DM | ENSG0000020 | 0.001516835 | 0.017171609 | 0.000636518 | 0.081436009 | 0.001198233 | 0.063096336 | 0.000644726 | 0.081418554 | 0.000318602 | 0.001974024 | 1 | 0.171768447 | 1 | 0.365113014 | ENSG0000018 | 10 | TRPV2 |
| SAT | T2DM | ENSG0000020 | 0.001244035 | 0.585509112 | 0.002281144 | 0.081436909 | -0.00197368 | 0.40181267 | 0.002354131 | 0.081419013 | 0.003217713 | 3.34E-08 | 1 | 0.845534946 | 1 | 0.757591271 | ENSG0000018 | 9 | KBTBD12 |
| SAT | T2DM | ENSG0000020 | 0.003273725 | 0.164676557 | 0.002356019 | 0.081436636 | -0.00024501 | 0.921727794 | 0.00249358 | 0.081419129 | 0.003518738 | 1.68E-05 | 1 | 0.516146667 | 1 | 0.979654273 | ENSG0000018 | 9 | KBTBD12 |
| SAT | T2DM | ENSG0000020 | 0.004830308 | 0.000465236 | 0.001380074 | 0.081434911 | 0.004299143 | 0.002105722 | 0.001398133 | 0.081417551 | 0.000531165 | 0.018184564 | 1 | 0.014911936 | 1 | 0.049971446 | ENSG0000018 | 10 | FANCA |
| SAT | T2DM | ENSG0000020 | 0.00284377 | 0.17872044 | 0.002114787 | 0.081436656 | 0.004309662 | 0.046197513 | 0.00216176 | 0.081418467 | -0.00146589 | 0.001104181 | 1 | 0.530902204 | 1 | 0.309334779 | ENSG0000018 | 10 | FANCA |
| SAT | T2DM | ENSG0000020 | -0.00055055 | 0.777922388 | 0.001952111 | 0.081436945 | 0.000926027 | 0.637924135 | 0.001967744 | 0.081419094 | -0.00147658 | 3.13E-09 | 1 | 0.939432466 | 1 | 0.884889906 | ENSG0000018 | 10 | FANCA |
| SAT | T2DM | ENSG0000020 | 0.001174122 | 0.102045629 | 0.000718109 | 0.081436512 | -3.77E-06 | 0.996079761 | 0.000766978 | 0.081419131 | 0.001177891 | 1.25E-05 | 1 | 0.425727224 | 1 | 0.998405122 | ENSG0000018 | 10 | FANCA |
| SAT | T2DM | ENSG0000020 | 0.010244766 | 0.004115918 | 0.00357066 | 0.081435583 | 0.010322513 | 0.003838202 | 0.003570351 | 0.081417734 | -7.77E-05 | 0.0013598 | 1 | 0.068549877 | 1 | 0.072792717 | ENSG0000018 | 10 | FANCA |
| SAT | T2DM | ENSG0000020 | 0.003423686 | 0.008724968 | 0.001305426 | 0.081435809 | 0.002354833 | 0.073608893 | 0.001316254 | 0.081418596 | 0.001068853 | 2.90E-10 | 1 | 0.11010239 | 1 | 0.395413696 | ENSG0000018 | 10 | FANCA |
| SAT | T2DM | ENSG0000020 | -0.00178135 | 0.329280621 | 0.001825968 | 0.081436799 | -0.00128394 | 0.484100055 | 0.001834921 | 0.081419049 | -0.00049741 | 0.006576872 | 1 | 0.692334629 | 1 | 0.806185068 | ENSG0000018 | 10 | FANCA |
| SAT | T2DM | ENSG0000020 | 2.64E-05 | 0.991535653 | 0.002488484 | 0.081436958 | -1.74E-05 | 0.994418751 | 0.00248827 | 0.081419131 | 4.38E-05 | 0.010052448 | 1 | 0.996461007 | 1 | 0.998405122 | ENSG0000018 | 5 | HSPE1P11 |
| SAT | T2DM | ENSG0000020 | 0.003376442 | 0.38862464 | 0.003916463 | 0.081436834 | 0.0033146 | 0.397495033 | 0.003916137 | 0.081419011 | 6.30E-05 | 0.026562218 | 1 | 0.739604086 | 1 | 0.756180206 | ENSG0000018 | 5 | HSPE1P11 |
| SAT | T2DM | ENSG0000020 | 0.002474154 | 0.17880126 | 0.001840263 | 0.081436656 | 0.001176047 | 0.525801959 | 0.001853713 | 0.081419064 | 0.001298108 | 7.41E-09 | 1 | 0.530902204 | 1 | 0.827413059 | ENSG0000018 | 12 | DNAH17 |
| SAT | T2DM | ENSG0000020 | -0.00401516 | 0.212753954 | 0.003222364 | 0.081436699 | -0.0039607 | 0.218988087 | 0.003222112 | 0.081418878 | -5.45E-05 | 0.032207396 | 1 | 0.575921597 | 1 | 0.592332986 | ENSG0000018 | 9 | MCRS1 |
| SAT | T2DM | ENSG0000020 | -0.00045206 | 0.851336847 | 0.002412129 | 0.081436953 | -0.00051204 | 0.831876859 | 0.002411942 | 0.081419123 | 6.00E-05 | 0.001807536 | 1 | 0.963534946 | 1 | 0.961544037 | ENSG0000018 | 6 | FANCM |
| SAT | T2DM | ENSG0000020 | 0.001677212 | 0.565705945 | 0.002920002 | 0.081436903 | 0.001634978 | 0.575499084 | 0.002919752 | 0.081419079 | 4.22E-05 | 0.035949601 | 1 | 0.837959912 | 1 | 0.84981421 | ENSG0000018 | 6 | FANCM |
| SAT | T2DM | ENSG0000020 | -0.00227911 | 0.366055394 | 0.002521455 | 0.081436822 | -0.00231489 | 0.358537658 | 0.00252124 | 0.08141899 | 3.58E-05 | 0.040131813 | 1 | 0.721771482 | 1 | 0.725902637 | ENSG0000018 | 6 | FANCM |
| SAT | T2DM | ENSG0000020 | -0.00496231 | 0.075495073 | 0.002791819 | 0.08143643 | -0.00490275 | 0.079046212 | 0.0027916 | 0.081418616 | -5.96E-05 | 0.006670282 | 1 | 0.382364025 | 1 | 0.407783994 | ENSG0000018 | 5 | ZFP69 |
| SAT | T2DM | ENSG0000020 | -0.00198898 | 0.464538195 | 0.002719437 | 0.081436869 | -0.00193418 | 0.476895597 | 0.002719197 | 0.081419046 | -5.48E-05 | 0.002025457 | 1 | 0.792954767 | 1 | 0.80363893 | ENSG0000018 | 4 | TMEM220 |
| SAT | T2DM | ENSG0000020 | 0.003449187 | 0.033425819 | 0.001621669 | 0.081436202 | 0.003417118 | 0.035089586 | 0.00162154 | 0.081418389 | 3.21E-05 | 0.010644288 | 1 | 0.25564129 | 1 | 0.272172417 | ENSG0000018 | 2 | HIST1H1C |
| SAT | T2DM | ENSG0000020 | 0.004402675 | 0.071037792 | 0.002523457 | 0.08143645 | 0.004446486 | 0.078035545 | 0.002523253 | 0.081418612 | -4.38E-05 | 0.0217247 | 1 | 0.395495266 | 1 | 0.406404163 | ENSG0000018 | 8 | P2RX2 |
| SAT | T2DM | ENSG0000020 | 0.000758551 | 0.700329599 | 0.001981719 | 0.081436944 | 0.000655379 | 0.740843185 | 0.001981571 | 0.081419113 | -7.68E-05 | 3.49E-06 | 1 | 0.935400227 | 1 | 0.925595787 | ENSG0000018 | 7 | TTC24 |
| SAT | T2DM | ENSG0000020 | 0.000590573 | 0.817255551 | 0.00255532 | 0.081436949 | 0.00049539 | 0.846269733 | 0.002555135 | 0.081419125 | 9.52E-05 | 1.49E-05 | 1 | 0.952848981 | 1 | 0.96477102 | ENSG0000018 | 7 | TTC24 |
| SAT | T2DM | ENSG0000020 | -0.00202818 | 0.131949943 | 0.001346324 | 0.081436579 | -0.00205883 | 0.126179715 | 0.001346216 | 0.08141874 | 3.06E-05 | 0.002990723 | 1 | 0.472458584 | 1 | 0.47645145 | ENSG0000018 | 7 | LDLRAD2 |
| SAT | T2DM | ENSG0000020 | 0.002341195 | 0.095648826 | 0.001405003 | 0.081436494 | 0.002302577 | 0.101217854 | 0.001404883 | 0.081418682 | 3.86E-05 | 7.98E-05 | 1 | 0.420328914 | 1 | 0.440431278 | ENSG0000018 | 7 | LDLRAD2 |
| SAT | T2DM | ENSG0000020 | -0.00425639 | 0.187655445 | 0.003230539 | 0.081436668 | -0.00439035 | 0.174112438 | 0.003230316 | 0.081418822 | 0.000133961 | 3.92E-06 | 1 | 0.54503883 | 1 | 0.532948952 | ENSG0000018 | 7 | LDLRAD2 |
| SAT | T2DM | ENSG0000020 | 0.002237008 | 0.177930204 | 0.001660539 | 0.081436655 | 0.001116341 | 0.504158239 | 0.001671265 | 0.081419056 | 0.001120667 | 4.11E-09 | 1 | 0.530526907 | 1 | 0.817738235 | ENSG0000018 | 7 | LDLRAD2 |
| SAT | T2DM | ENSG0000020 | 0.00550573 | 1.82E-10 | 0.000863484 | 0.081430165 | 0.004371075 | 6.79E-07 | 0.000879964 | 0.081415008 | 0.001134655 | 2.43E-11 | 5.96E-07 | 5.42E-08 | 0.002227894 | 0.000101268 | ENSG0000018 | 6 | PMS2CL |
| SAT | T2DM | ENSG0000020 | -0.00077433 | 0.620397197 | 0.001563396 | 0.081436917 | 0.000946645 | 0.553157765 | 0.001596272 | 0.081419072 | -0.00172098 | 1.00E-07 | 1 | 0.863241392 | 1 | 0.842050121 | ENSG0000018 | 5 | ZSCAN23 |
| SAT | T2DM | ENSG0000021 | -0.0033294 | 0.066389997 | 0.001813611 | 0.081436395 | -0.0032855 | 0.070028251 | 0.001813458 | 0.081418582 | -4.39E-05 | 0.000591347 | 1 | 0.358786109 | 1 | 0.38680588 | ENSG0000018 | 5 | ZSCAN23 |
| SAT | T2DM | ENSG0000021 | 0.00057414 | 0.872211951 | 0.003569424 | 0.081436954 | 0.00066352 | 0.852520731 | 0.003569167 | 0.081419125 | -8.94E-05 | 0.00390302 | 1 | 0.966468088 | 1 | 0.965561588 | ENSG0000018 | 5 | ZSCAN23 |
| SAT | T2DM | ENSG0000021 | 0.004390328 | 0.1401703 | 0.002976171 | 0.081436595 | 0.004436417 | 0.13602182 | 0.002975918 | 0.08141876 | -4.61E-05 | 0.026207892 | 1 | 0.484614073 | 1 | 0.492171492 | ENSG0000018 | 5 | ZSCAN23 |
| SAT | T2DM | ENSG0000021 | 0.001263607 | 0.668153955 | 0.002947648 | 0.081436928 | 0.001393071 | 0.636471613 | 0.002947437 | 0.081419094 | -0.00012946 | 4.26E-07 | 1 | 0.888630991 | 1 | 0.884482576 | ENSG0000018 | 5 | ZSCAN23 |
| SAT | T2DM | ENSG0000021 | 0.001054228 | 0.551902499 | 0.001772074 | 0.081436899 | 0.00070615 | 0.690584919 | 0.001773971 | 0.081419104 | 0.000348079 | 5.28E-05 | 1 | 0.835918602 | 1 | 0.904075912 | ENSG0000018 | 5 | ZSCAN23 |
| SAT | T2DM | ENSG0000021 | -0.00145986 | 0.244708422 | 0.001254928 | 0.081436732 | -0.00149914 | 0.232208405 | 0.001254836 | 0.081418892 | 3.93E-05 | 0.000224888 | 1 | 0.61016229 | 1 | 0.609500622 | ENSG0000018 | 5 | ZSCAN23 |
| SAT | T2DM | ENSG0000021 | 0.008889284 | 0.001960715 | 0.00287109 | 0.081435356 | 0.007720366 | 0.007515601 | 0.002888168 | 0.081417937 | 0.001168918 | 0.000220585 | 1 | 0.041237855 | 1 | 0.108152135 | ENSG0000018 | 5 | ZSCAN23 |
| SAT | T2DM | ENSG0000021 | 0.002398678 | 0.166508568 | 0.00173376 | 0.081436639 | 0.002429986 | 0.161008297 | 0.001733611 | 0.081418803 | -3.13E-05 | 0.00853016 | 1 | 0.519037734 | 1 | 0.520626624 | ENSG0000018 | 5 | ZSCAN23 |
| SAT | T2DM | ENSG0000021 | -0.00217452 | 0.337392398 | 0.002266706 | 0.081436805 | -0.00220948 | 0.329639921 | 0.002266501 | 0.081418972 | 3.50E-05 | 0.011803494 | 1 | 0.696216641 | 1 | 0.698223745 | ENSG0000018 | 5 | ZSCAN23 |
| SAT | T2DM | ENSG0000021 | 0.004997626 | 0.000104214 | 0.001284702 | 0.0814343 | 0.002928322 | 0.026946831 | 0.001323659 | 0.081418313 | 0.002069303 | 9.10E-11 | 0.328801711 | 0.005087718 | 1 | 0.2402515 | ENSG0000018 | 6 | TMEM221 |
| SAT | T2DM | ENSG0000021 | -0.00181391 | 0.264773204 | 0.00162656 | 0.081436751 | -0.00184645 | 0.256261461 | 0.001626441 | 0.081418916 | 3.25E-05 | 0.018718935 | 1 | 0.627688499 | 1 | 0.628866009 | ENSG0000018 | 7 | TREML4 |
| SAT | T2DM | ENSG0000021 | -0.01328907 | 0.000495011 | 0.003814901 | 0.081434931 | -0.0036076 | 0.432260642 | 0.004593735 | 0.081419028 | -0.00968148 | 0.000155352 | 1 | 0.015422559 | 1 | 0.781403397 | ENSG0000018 | 7 | TREML4 |
| SAT | T2DM | ENSG0000021 | 0.00226023 | 0.081298129 | 0.001296593 | 0.081436451 | 0.001553301 | 0.245266586 | 0.001336835 | 0.081418905 | 0.000706928 | 0.030167642 | 1 | 0.395495266 | 1 | 0.620663712 | ENSG0000018 | 7 | TREML4 |
| SAT | T2DM | ENSG0000021 | 0.002888057 | 0.439986035 | 0.003739953 | 0.081436859 | 0.002949653 | 0.430256875 | 0.003739642 | 0.081419027 | -6.16E-05 | 0.023224664 | 1 | 0.776495555 | 1 | 0.780817802 | ENSG0000018 | 7 | TREML4 |
| SAT | T2DM | ENSG0000021 | -0.00088967 | 0.212303957 | 0.000713304 | 0.081436698 | -0.00086987 | 0.222616135 | 0.000713243 | 0.081418882 | -1.98E-05 | 8.08E-05 | 1 | 0.575921597 | 1 | 0.596734918 | ENSG0000018 | 8 | PRSS45 |
| SAT | T2DM | ENSG0000021 | -0.00051882 | 0.427252402 | 0.000653503 | 0.081436853 | -0.00050069 | 0.443543005 | 0.000653453 | 0.081419033 | -1.81E-05 | 0.000717382 | 1 | 0.771499796 | 1 | 0.789555502 | ENSG0000018 | 6 | LAMTOR4 |
| SAT | T2DM | ENSG0000021 | 0.000758981 | 0.76588483 | 0.00254895 | 0.081436944 | 0.000899395 | 0.724181465 | 0.002548757 | 0.08141911 | -0.00014041 | 1.73E-11 | 1 | 0.934630346 | 1 | 0.921192527 | ENSG0000018 | 8 | ZNF793 |
| SAT | T2DM | ENSG0000021 | -0.00451489 | 0.003932405 | 0.001565741 | 0.081435569 | -0.00454564 | 0.003691053 | 0.001565605 | 0.081417722 | 3.08E-05 | 0.003572988 | 1 | 0.067550897 | 1 | 0.071237315 | ENSG0000018 | 9 | AGAP4 |
| SAT | T2DM | ENSG0000021 | -0.00049987 | 0.467468513 | 0.000687952 | 0.08143687 | -0.0005136 | 0.455285334 | 0.000687894 | 0.081419038 | 1.37E-05 | 0.004241054 | 1 | 0.79334291 | 1 | 0.794600418 | ENSG0000018 | 9 | AGAP4 |
| SAT | T2DM | ENSG0000021 | -0.0029504 | 0.096345911 | 0.001774318 | 0.081436496 | -0.00290228 | 0.101875849 | 0.001774196 | 0.081418684 | -4.81E-05 | 0.002542572 | 1 | 0.420328914 | 1 | 0.440431278 | ENSG0000018 | 9 | CENPP |
| SAT | T2DM | ENSG0000021 | -0.01028091 | 0.007720923 | 0.003859131 | 0.081435772 | -0.00907052 | 0.01957536 | 0.003885593 | 0.08141822 | -0.00121039 | 0.007977549 | 1 | 0.100525186 | 1 | 0.195966491 | ENSG0000018 | 9 | CENPP |
| SAT | T2DM | ENSG0000021 | 0.004435368 | 0.260994775 | 0.003945901 | 0.081436747 | 0.004375656 | 0.267426693 | 0.003945554 | 0.081418925 | 5.97E-05 | 0.020259644 | 1 | 0.625510487 | 1 | 0.638593145 | ENSG0000018 | 9 | CENPP |
| SAT | T2DM | ENSG0000021 | 0.001649287 | 0.108565572 | 0.0010278 | 0.081436528 | 0.000564156 | 0.588236904 | 0.001042046 | 0.081419082 | 0.001085131 | 3.07E-10 | 1 | 0.436210685 | 1 | 0.853229568 | ENSG0000018 | 9 | ZNF559 |
| SAT | T2DM | ENSG0000021 | -0.00254718 | 0.014214627 | 0.001038901 | 0.081435954 | -0.00105198 | 0.333019269 | 0.001086698 | 0.081418974 | -0.0014952 | 2.79E-06 | 1 | 0.151551916 | 1 | 0.701740984 | ENSG0000018 | 9 | ZNF559 |
| SAT | T2DM | ENSG0000021 | -0.00039737 | 0.871103763 | 0.002449052 | 0.081436954 | -0.00035354 | 0.885209233 | 0.002448875 | 0.081419127 | -4.38E-05 | 0.038174586 | 1 | 0.966468088 | 1 | 0.968806494 | ENSG0000018 | 9 | ZNF559 |
| SAT | T2DM | ENSG0000021 | 0.000988111 | 0.430042674 | 0.001252169 | 0.081436854 | 0.000959859 | 0.443306396 | 0.001252062 | 0.081419033 | 2.83E-05 | 0.001118646 | 1 | 0.772274669 | 1 | 0.789955502 | ENSG0000018 | 9 | ZNF559 |
| SAT | T2DM | ENSG0000021 | 0.003335252 | 0.111328172 | 0.002094673 | 0.081436535 | 0.004067576 | 0.05254423 | 0.002098146 | 0.081418303 | -0.00073232 | 4.21E-09 | 1 | 0.437446384 | 1 | 0.332371916 | ENSG0000018 | 9 | ZNF559 |
| SAT | T2DM | ENSG0000021 | 0.000454798 | 0.778488232 | 0.001616829 | 0.081436945 | 0.000495874 | 0.759058948 | 0.001616715 | 0.081419115 | -4.11E-05 | 0.003932652 | 1 | 0.939432466 | 1 | 0.932998601 | ENSG0000018 | 10 | ZP3 |
| SAT | T2DM | ENSG0000022 | 0.001424743 | 0.70009969 | 0.003698843 | 0.081436934 | 0.001503845 | 0.68430185 | 0.003698585 | 0.081419103 | -7.91E-05 | 0.01639872 | 1 | 0.902046837 | 1 | 0.901372745 | ENSG0000018 | 10 | ZP3 |
| SAT | T2DM | ENSG0000022 | -0.00328208 | 0.095844827 | 0.001970814 | 0.081436495 | -0.0033209 | 0.091954553 | 0.001970648 | 0.081418656 | 3.88E-05 | 0.00600424 | 1 | 0.420328914 | 1 | 0.425532987 | ENSG0000018 | 7 | AC018892.9 |
| SAT | T2DM | ENSG0000022 | -0.00090162 | 0.626601516 | 0.001853215 | 0.081436919 | -0.00086725 | 0.639778242 | 0.001853057 | 0.081419094 | -3.44E-05 | 0.007475566 | 1 | 0.866453072 | 1 | 0.885701439 | ENSG0000018 | 4 | SELL |
| SAT | T2DM | ENSG0000022 | 0.005887497 | 0.08201443 | 0.003385333 | 0.081436453 | 0.005827608 | 0.085146407 | 0.003385036 | 0.081418636 | 5.99E-05 | 0.007544119 | 1 | 0.395495266 | 1 | 0.415104547 | ENSG0000018 | 4 | SELL |
| SAT | T2DM | ENSG0000022 | 0.002904687 | 0.438817279 | 0.003751892 | 0.081436858 | 0.002977323 | 0.427417615 | 0.003751577 | 0.081419026 | -7.26E-05 | 0.006600372 | 1 | 0.775314751 | 1 | 0.777953168 | ENSG0000018 | 4 | SELL |
| SAT | T2DM | ENSG0000022 | 0.003986465 | 0.307270262 | 0.003904594 | 0.081436784 | 0.004063265 | 0.298010478 | 0.003904318 | 0.08141895 | -7.68E-05 | 0.025453179 | 1 | 0.667651477 | 1 | 0.664773075 | ENSG0000018 | 9 | SERPINA5 |
| SAT | T2DM | ENSG0000022 | -0.00898481 | 0.000481053 | 0.002573624 | 0.081434922 | -0.0090243 | 0.000453644 | 0.002573403 | 0.081417076 | 3.95E-05 | 0.025640364 | 1 | 0.015176308 | 1 | 0.017307053 | ENSG0000018 | 9 | SERPINA5 |
| SAT | T2DM | ENSG0000022 | 0.001618368 | 0.535686189 | 0.002613011 | 0.081436894 | 0.003080055 | 0.245604952 | 0.002652722 | 0.081418906 | -0.00146169 | 0.001447602 | 1 | 0.828930489 | 1 | 0.620663712 | ENSG0000018 | 10 | C19orf54 |
| SAT | T2DM | ENSG0000022 | 0.004647019 | 0.011291585 | 0.001834191 | 0.081435886 | 0.001527378 | 0.473770219 | 0.002132142 | 0.081419045 | 0.00311964 | 0.004119519 | 1 | 0.131589853 | 1 | 0.80363893 | ENSG0000018 | 10 | SRSF10 |

| | | | | | | | | | | | | | | | | | | | |
|---|---|---|---|---|---|---|---|---|---|---|---|---|---|---|---|---|---|---|---|
| SAT | T2DM | ENSG0000022 | 0.001194064 | 0.143390595 | 0.000816017 | 0.081436601 | 0.001162292 | 0.154317138 | 0.000815959 | 0.081418792 | 3.18E-05 | 8.17E-06 | 1 | 0.488540542 | 1 | 0.514547286 | ENSG0000018 | 10 | SRSF10 |
| SAT | T2DM | ENSG0000022 | 0.00180599 | 0.00825561 | 0.000683713 | 0.081435792 | 0.001146993 | 0.095995262 | 0.000689054 | 0.081418668 | 0.000658997 | 2.09E-14 | 1 | 0.105807246 | 1 | 0.432044519 | ENSG0000018 | 13 | NPIPP1 |
| SAT | T2DM | ENSG0000022 | -0.00245562 | 0.001916615 | 0.000791401 | 0.081435349 | -0.000915 | 0.262402834 | 0.000816427 | 0.081418921 | -0.00154062 | 1.74E-14 | 1 | 0.04093133 | 1 | 0.636156586 | ENSG0000018 | 10 | ASAH2 |
| SAT | T2DM | ENSG0000022 | 0.007278909 | 0.037726636 | 0.00350315 | 0.081436237 | 0.004057193 | 0.255536174 | 0.003568312 | 0.081418915 | 0.003221716 | 2.22E-06 | 1 | 0.269351866 | 1 | 0.62755553 | ENSG0000018 | 7 | ZNF177 |
| SAT | T2DM | ENSG0000022 | -0.00562161 | 0.062399123 | 0.003016773 | 0.081436378 | -0.00566857 | 0.060222628 | 0.003016536 | 0.081418541 | 4.70E-05 | 0.048360112 | 1 | 0.346391409 | 1 | 0.35334636 | ENSG0000018 | 7 | ZNF177 |
| SAT | T2DM | ENSG0000022 | -0.00058023 | 0.858588204 | 0.003256594 | 0.081436953 | -0.00063794 | 0.844685018 | 0.003256363 | 0.081419124 | 5.77E-05 | 0.043493726 | 1 | 0.964974089 | 1 | 0.96477102 | ENSG0000018 | 7 | ZNF177 |
| SAT | T2DM | ENSG0000022 | 0.007537057 | 0.00633059 | 0.002760665 | 0.081435713 | 0.007435898 | 0.007065963 | 0.002760446 | 0.081417919 | 0.000101159 | 2.15E-06 | 1 | 0.087271699 | 1 | 0.103497438 | ENSG0000018 | 7 | ZNF177 |
| SAT | T2DM | ENSG0000022 | -0.00125873 | 0.407705085 | 0.001520314 | 0.081436844 | -0.00122828 | 0.419097565 | 0.001520175 | 0.081419022 | -3.04E-05 | 0.000852847 | 1 | 0.756247299 | 1 | 0.768824902 | ENSG0000018 | 5 | FAM154B |
| SAT | T2DM | ENSG0000022 | 0.001946445 | 0.506208932 | 0.002928074 | 0.081436885 | 0.001994306 | 0.495776053 | 0.002927845 | 0.081419053 | -4.79E-05 | 0.038965496 | 1 | 0.810627063 | 1 | 0.814419437 | ENSG0000018 | 5 | HILS1 |
| SAT | T2DM | ENSG0000022 | -0.00545699 | 0.025028065 | 0.002435093 | 0.081436119 | -0.00541548 | 0.026142267 | 0.002434912 | 0.081418304 | -4.15E-05 | 0.041254361 | 1 | 0.216409969 | 1 | 0.235639496 | ENSG0000018 | 5 | HILS1 |
| SAT | T2DM | ENSG0000022 | 0.002362471 | 0.221447279 | 0.001932191 | 0.081436709 | 0.00232839 | 0.228146649 | 0.001932036 | 0.081418888 | 3.41E-05 | 0.021588454 | 1 | 0.586129584 | 1 | 0.602834606 | ENSG0000018 | 12 | RHCE |
| SAT | T2DM | ENSG0000022 | 0.005369819 | 0.142984936 | 0.003665986 | 0.0814366 | 0.005276595 | 0.15001959 | 0.003665665 | 0.081418785 | 9.32E-05 | 0.00011916 | 1 | 0.488540542 | 1 | 0.51082416 | ENSG0000018 | 10 | PARVB |
| SAT | T2DM | ENSG0000022 | -0.00710483 | 0.024728686 | 0.003163858 | 0.081436116 | -0.00705106 | 0.025827639 | 0.003163611 | 0.081418301 | -5.38E-05 | 0.031805552 | 1 | 0.21611556 | 1 | 0.234089731 | ENSG0000018 | 10 | PARVB |
| SAT | T2DM | ENSG0000022 | 0.00270996 | 0.109346375 | 0.001692518 | 0.08143653 | 0.002681437 | 0.113098016 | 0.001692374 | 0.081418711 | 2.85E-05 | 0.015271243 | 1 | 0.436210685 | 1 | 0.455306244 | ENSG0000018 | 4 | ZBED6CL |
| SAT | T2DM | ENSG0000022 | -0.0004759 | 0.637323024 | 0.001009442 | 0.081436921 | -0.00022554 | 0.82406391 | 0.001014473 | 0.081419123 | -0.00025036 | 0.014114501 | 1 | 0.874187643 | 1 | 0.959280742 | ENSG0000018 | 9 | FSIP2 |
| SAT | T2DM | ENSG0000022 | 0.00389933 | 0.134194769 | 0.002603431 | 0.081436584 | 0.003860173 | 0.138114874 | 0.002603212 | 0.081418764 | 3.92E-05 | 0.034250811 | 1 | 0.475922014 | 1 | 0.49579311 | ENSG0000018 | 6 | TMEM198 |
| SAT | T2DM | ENSG0000022 | -0.00342274 | 0.277263741 | 0.003150277 | 0.081436761 | -0.00349835 | 0.266751797 | 0.00315003 | 0.081418925 | 7.56E-05 | 0.002385289 | 1 | 0.63926506 | 1 | 0.638375379 | ENSG0000018 | 9 | MTF1 |
| SAT | T2DM | ENSG0000022 | -9.60E-05 | 0.883274556 | 0.000654199 | 0.081436955 | -0.00010589 | 0.871405546 | 0.000654144 | 0.081419127 | 9.84E-06 | 0.032821941 | 1 | 0.969020186 | 1 | 0.966383771 | ENSG0000018 | 9 | MTF1 |
| SAT | T2DM | ENSG0000022 | 0.001334259 | 0.435625784 | 0.001711463 | 0.081436857 | 0.000581225 | 0.735661272 | 0.001721613 | 0.081419112 | 0.000753035 | 6.40E-05 | 1 | 0.772274669 | 1 | 0.92442683 | ENSG0000018 | 8 | ZDHHC11 |
| SAT | T2DM | ENSG0000022 | 0.003127206 | 0.014324407 | 0.001276915 | 0.081435956 | 0.003161393 | 0.013286035 | 0.001276806 | 0.081418107 | -3.42E-05 | 0.000112973 | 1 | 0.151551916 | 1 | 0.157369966 | ENSG0000018 | 8 | FAM26F |
| SAT | T2DM | ENSG0000022 | -0.00172278 | 0.656749097 | 0.00387659 | 0.081436925 | -0.00202307 | 0.601925271 | 0.003878334 | 0.081419085 | 0.000300291 | 0.020595442 | 1 | 0.884093538 | 1 | 0.858221553 | ENSG0000018 | 8 | FAM26F |
| SAT | T2DM | ENSG0000022 | -0.00050479 | 0.588100195 | 0.000932052 | 0.081436909 | -0.00053102 | 0.568826683 | 0.00093198 | 0.081419077 | 2.62E-05 | 0.000558332 | 1 | 0.846296816 | 1 | 0.845271624 | ENSG0000018 | 9 | ZNF563 |
| SAT | T2DM | ENSG0000022 | 0.000781882 | 0.555503335 | 0.001326272 | 0.0814369 | -0.00045773 | 0.734982957 | 0.001352212 | 0.081419112 | 0.001239613 | 2.73E-06 | 1 | 0.835948344 | 1 | 0.92442683 | ENSG0000018 | 12 | FBF1 |
| SAT | T2DM | ENSG0000022 | 0.002527176 | 0.227551921 | 0.002094303 | 0.081436715 | 0.002559974 | 0.221536131 | 0.002094125 | 0.081418881 | -3.28E-05 | 0.025313334 | 1 | 0.591955891 | 1 | 0.595786923 | ENSG0000018 | 12 | FBF1 |
| SAT | T2DM | ENSG0000022 | 0.000486537 | 0.903175431 | 0.003999449 | 0.081436956 | 0.00039606 | 0.921109019 | 0.003999118 | 0.081419129 | 9.05E-05 | 0.001895663 | 1 | 0.97231278 | 1 | 0.979654273 | ENSG0000018 | 12 | FBF1 |
| SAT | T2DM | ENSG0000022 | -0.00530321 | 0.091441358 | 0.003141999 | 0.081436482 | -0.00523842 | 0.0954423 | 0.003141735 | 0.081418666 | -6.48E-05 | 0.003734136 | 1 | 0.413867045 | 1 | 0.432044519 | ENSG0000018 | 12 | FBF1 |
| SAT | T2DM | ENSG0000022 | 0.000311064 | 0.831554522 | 0.001462396 | 0.081436951 | 0.000347564 | 0.812123483 | 0.001462273 | 0.081419121 | -3.65E-05 | 0.000497552 | 1 | 0.956754735 | 1 | 0.95480666 | ENSG0000018 | 7 | MSL1 |
| SAT | T2DM | ENSG0000022 | 5.85E-05 | 0.973121966 | 0.001736772 | 0.081436958 | 3.20E-05 | 0.985278878 | 0.001736633 | 0.081419131 | 2.65E-05 | 0.046573849 | 1 | 0.995031844 | 1 | 0.997162029 | ENSG0000018 | 9 | LRRK2 |
| SAT | T2DM | ENSG0000022 | -0.00531696 | 0.110625407 | 0.003332698 | 0.081436533 | -0.00525232 | 0.114999267 | 0.003332439 | 0.081418716 | -6.46E-05 | 0.014746415 | 1 | 0.436277088 | 1 | 0.456639917 | ENSG0000018 | 9 | LRRK2 |
| SAT | T2DM | ENSG0000022 | -0.00174633 | 0.10038607 | 0.001062901 | 0.081436507 | -0.00079551 | 0.458773502 | 0.001073753 | 0.081419039 | -0.00095082 | 5.23E-10 | 1 | 0.425727224 | 1 | 0.795972372 | ENSG0000018 | 12 | PTPLAD2 |
| SAT | T2DM | ENSG0000022 | -0.00399542 | 0.013285089 | 0.001613629 | 0.081435934 | -0.00402266 | 0.012662692 | 0.001613499 | 0.081418092 | 2.72E-05 | 0.025721566 | 1 | 0.145107 | 1 | 0.151628804 | ENSG0000018 | 12 | PTPLAD2 |
| SAT | T2DM | ENSG0000022 | -0.00033087 | 0.927091353 | 0.003615864 | 0.081436957 | -0.00026571 | 0.941416625 | 0.003615554 | 0.08141913 | -6.52E-05 | 0.008769647 | 1 | 0.978695859 | 1 | 0.986380169 | ENSG0000018 | 12 | PTPLAD2 |
| SAT | T2DM | ENSG0000022 | 0.000859345 | 0.68768034 | 0.002137647 | 0.081436931 | 0.000895557 | 0.675228851 | 0.002137456 | 0.081419102 | -3.62E-05 | 0.007553126 | 1 | 0.896416049 | 1 | 0.898778511 | ENSG0000018 | 5 | UTS2B |
| SAT | T2DM | ENSG0000022 | 4.18E-05 | 0.985337155 | 0.002274988 | 0.081436958 | -0.00131712 | 0.563741193 | 0.0022815 | 0.081419075 | 0.001358935 | 1.22E-14 | 1 | 0.996461007 | 1 | 0.84223981 | ENSG0000018 | 11 | ADAT2 |
| SAT | T2DM | ENSG0000022 | -0.00096131 | 0.693262707 | 0.002437209 | 0.081436932 | -0.00091068 | 0.708640701 | 0.002437034 | 0.081419108 | -5.06E-05 | 0.016505543 | 1 | 0.900268043 | 1 | 0.912941362 | ENSG0000018 | 11 | ADAT2 |
| SAT | T2DM | ENSG0000023 | -0.00999579 | 0.003610869 | 0.003434615 | 0.081435543 | -0.01008074 | 0.003332494 | 0.00343432 | 0.081417691 | 8.49E-05 | 0.000372067 | 1 | 0.062747674 | 1 | 0.065866952 | ENSG0000018 | 11 | ADAT2 |
| SAT | T2DM | ENSG0000023 | -0.00114536 | 0.507228698 | 0.001727121 | 0.081436885 | -0.00118808 | 0.491483089 | 0.001726976 | 0.081419052 | 4.27E-05 | 0.000502991 | 1 | 0.810627063 | 1 | 0.811177221 | ENSG0000018 | 9 | AC138783.12 |
| SAT | T2DM | ENSG0000023 | 0.002715431 | 0.213986949 | 0.002185143 | 0.0814367 | 0.002754633 | 0.207411364 | 0.002184973 | 0.081418865 | -3.92E-05 | 0.023759113 | 1 | 0.576804422 | 1 | 0.579977552 | ENSG0000018 | 9 | AC138783.12 |
| SAT | T2DM | ENSG0000023 | -0.00055441 | 0.764664305 | 0.001851984 | 0.081436943 | -0.00059625 | 0.747466452 | 0.001851821 | 0.081419114 | 4.18E-05 | 0.000574614 | 1 | 0.93459819 | 1 | 0.927854665 | ENSG0000018 | 9 | AC138783.12 |
| SAT | T2DM | ENSG0000023 | -0.00043812 | 0.670619838 | 0.001030145 | 0.081436928 | -0.00031822 | 0.757470961 | 0.001030499 | 0.081419115 | -0.0001199 | 0.000110138 | 1 | 0.889125844 | 1 | 0.932832405 | ENSG0000018 | 6 | FAM35DP |
| SAT | T2DM | ENSG0000023 | 0.000403881 | 0.915125665 | 0.003789618 | 0.081436957 | 0.000472561 | 0.900753338 | 0.003789285 | 0.081419128 | -6.87E-05 | 0.005827824 | 1 | 0.977304164 | 1 | 0.972878089 | ENSG0000018 | 9 | ANKDD1B |
| SAT | T2DM | ENSG0000023 | 0.00075796 | 0.668895219 | 0.001772321 | 0.081436928 | 0.000789464 | 0.655976039 | 0.001772182 | 0.081419098 | -3.15E-05 | 0.023564864 | 1 | 0.888630991 | 1 | 0.891942554 | ENSG0000018 | 9 | ANKDD1B |
| SAT | T2DM | ENSG0000023 | 0.001468482 | 0.27323122 | 0.00134028 | 0.081436758 | 0.002197974 | 0.112686458 | 0.001385654 | 0.08141871 | -0.00072949 | 0.038366465 | 1 | 0.634986223 | 1 | 0.4547654 | ENSG0000018 | 9 | ANKDD1B |
| SAT | T2DM | ENSG0000023 | -0.00068393 | 0.75977819 | 0.002236746 | 0.081436943 | -0.00071392 | 0.749571094 | 0.002236553 | 0.081419114 | 3.00E-05 | 0.049849077 | 1 | 0.934086217 | 1 | 0.928470414 | ENSG0000018 | 6 | ALKBH2 |
| SAT | T2DM | ENSG0000023 | -0.00419142 | 0.131608034 | 0.002779834 | 0.081436578 | -0.00424221 | 0.126960348 | 0.002779585 | 0.081418742 | 5.08E-05 | 0.003967916 | 1 | 0.472458584 | 1 | 0.476745292 | ENSG0000018 | 6 | ALKBH2 |
| SAT | T2DM | ENSG0000023 | 0.001785213 | 0.649576422 | 0.003929148 | 0.081436924 | 0.000401761 | 0.918850397 | 0.003943401 | 0.081419129 | 0.001383451 | 4.71E-05 | 1 | 0.881625728 | 1 | 0.979654273 | ENSG0000018 | 6 | ALKBH2 |
| SAT | T2DM | ENSG0000023 | 0.009025723 | 1.22E-14 | 0.001170015 | 0.081427015 | 0.006404531 | 4.68E-06 | 0.001398775 | 0.081415628 | 0.002621192 | 0.000628984 | 4.00E-11 | 5.72E-12 | 0.015356883 | 0.000430718 | ENSG0000018 | 6 | ALKBH2 |
| SAT | T2DM | ENSG0000023 | 0.003348955 | 0.060521735 | 0.001784227 | 0.08143637 | 0.004356753 | 0.014913948 | 0.00178961 | 0.081418141 | -0.0010078 | 9.67E-13 | 1 | 0.342956501 | 1 | 0.166753416 | ENSG0000018 | 5 | VSTM1 |
| SAT | T2DM | ENSG0000023 | 6.80E-05 | 0.968780982 | 0.001736283 | 0.081436958 | 0.00012058 | 0.944630363 | 0.001736174 | 0.08141913 | -5.26E-05 | 0.001719646 | 1 | 0.993613755 | 1 | 0.987677572 | ENSG0000018 | 8 | CLDN4 |
| SAT | T2DM | ENSG0000023 | 0.004746812 | 3.98E-08 | 0.000864329 | 0.081431918 | -0.00056142 | 0.645547911 | 0.001220586 | 0.081419096 | 0.005308228 | 7.33E-10 | 0.000130525 | 6.53E-06 | 1 | 0.889933905 | ENSG0000018 | 9 | ZNF573 |
| SAT | T2DM | ENSG0000023 | 0.001594217 | 0.455289866 | 0.00213523 | 0.081436865 | 0.001529931 | 0.473635671 | 0.002135055 | 0.081419045 | 6.43E-05 | 5.53E-05 | 1 | 0.786213711 | 1 | 0.80363893 | ENSG0000018 | 9 | ZNF573 |
| SAT | T2DM | ENSG0000023 | 0.000289591 | 0.796112129 | 0.001120786 | 0.081436947 | -0.00021615 | 0.847532449 | 0.001124199 | 0.081419125 | 0.000505738 | 1.37E-08 | 1 | 0.94681813 | 1 | 0.965539571 | ENSG0000018 | 9 | FAM47E |
| SAT | T2DM | ENSG0000023 | 0.002904765 | 0.170804089 | 0.002120844 | 0.081436645 | 0.002848382 | 0.179222573 | 0.002120666 | 0.081418829 | 5.64E-05 | 0.000184635 | 1 | 0.524235936 | 1 | 0.541962453 | ENSG0000018 | 9 | FAM47E |
| SAT | T2DM | ENSG0000023 | -0.00894951 | 0.024064466 | 0.003966798 | 0.081436108 | -0.00887191 | 0.025305211 | 0.003966486 | 0.081418295 | -7.76E-05 | 0.012601856 | 1 | 0.212818095 | 1 | 0.232403611 | ENSG0000018 | 9 | FAM47E |
| SAT | T2DM | ENSG0000023 | 0.006887001 | 0.02861959 | 0.003151407 | 0.08143616 | 0.005655124 | 0.073591457 | 0.00316079 | 0.081418596 | 0.001231876 | 6.61E-07 | 1 | 0.236149844 | 1 | 0.395413696 | ENSG0000018 | 11 | ZNF33A |
| SAT | T2DM | ENSG0000023 | 0.002055456 | 0.253232447 | 0.001799032 | 0.08143674 | 0.000735335 | 0.683947652 | 0.001806355 | 0.081419103 | 0.00132012 | 1.07E-15 | 1 | 0.618206663 | 1 | 0.901372745 | ENSG0000018 | 11 | ZNF33A |
| SAT | T2DM | ENSG0000023 | 0.00186506 | 0.392287505 | 0.00218014 | 0.081436836 | -0.00074834 | 0.734022583 | 0.002202434 | 0.081419112 | 0.002613402 | 9.03E-17 | 1 | 0.742778516 | 1 | 0.924370544 | ENSG0000018 | 11 | ZNF33A |
| SAT | T2DM | ENSG0000023 | 0.000973403 | 0.640652714 | 0.002085328 | 0.081436922 | 0.00091666 | 0.660221847 | 0.002085179 | 0.081419099 | 5.67E-05 | 0.00180437 | 1 | 0.875315948 | 1 | 0.894350146 | ENSG0000018 | 5 | KRT77 |
| SAT | T2DM | ENSG0000023 | -0.0034082 | 0.148005535 | 0.002355987 | 0.081436609 | -0.00337865 | 0.151515558 | 0.002355774 | 0.081418787 | -2.96E-05 | 0.041980625 | 1 | 0.493001178 | 1 | 0.511442948 | ENSG0000018 | 5 | KRT77 |
| SAT | T2DM | ENSG0000023 | 0.000870118 | 0.490799434 | 0.0012628 | 0.081436879 | 0.000843226 | 0.504263183 | 0.001262698 | 0.081419056 | 2.69E-05 | 0.004697495 | 1 | 0.80382562 | 1 | 0.817738235 | ENSG0000018 | 7 | ZNF600 |
| SAT | T2DM | ENSG0000023 | 0.00623999 | 9.62E-05 | 0.001599952 | 0.081434416 | 0.004396489 | 0.006772984 | 0.001623629 | 0.081417906 | 0.001843501 | 2.92E-11 | 0.315506995 | 0.005008048 | 1 | 0.100552768 | ENSG0000018 | 7 | ZNF600 |
| SAT | T2DM | ENSG0000023 | 0.013959332 | 0.000159101 | 0.00369642 | 0.081434575 | 0.013909923 | 0.000167623 | 0.003696083 | 0.081416764 | 4.94E-05 | 0.027055031 | 0.522008791 | 0.006960117 | 0.549971522 | 0.00820853 | ENSG0000018 | 7 | ZNF600 |
| SAT | T2DM | ENSG0000023 | 0.002269822 | 0.101452258 | 0.001385849 | 0.08143651 | 0.00102071 | 0.467485129 | 0.001404819 | 0.081419043 | 0.001249113 | 6.37E-08 | 1 | 0.425727224 | 1 | 0.800114089 | ENSG0000018 | 7 | ZNF600 |
| SAT | T2DM | ENSG0000023 | 0.001569904 | 0.359477273 | 0.001713191 | 0.081436818 | 0.001755709 | 0.305767191 | 0.001714317 | 0.081418956 | -0.00018581 | 0.005620646 | 1 | 0.71830389 | 1 | 0.674208436 | ENSG0000018 | 7 | ZNF600 |
| SAT | T2DM | ENSG0000023 | -0.00398988 | 0.269392172 | 0.003612497 | 0.081436755 | -0.0039135 | 0.278638569 | 0.003612294 | 0.081418935 | -7.64E-05 | 0.040516972 | 1 | 0.632695575 | 1 | 0.644397089 | ENSG0000018 | 7 | ZNF600 |
| SAT | T2DM | ENSG0000023 | -0.00365047 | 0.027228761 | 0.001653121 | 0.081436144 | -0.0036035 | 0.029258549 | 0.001652986 | 0.081418337 | -4.70E-05 | 0.000138903 | 1 | 0.227073047 | 1 | 0.251961415 | ENSG0000018 | 8 | C22orf43 |
| SAT | T2DM | ENSG0000023 | -0.00074614 | 0.569399276 | 0.001311471 | 0.081436904 | -0.00076857 | 0.557818147 | 0.001311362 | 0.081419074 | 2.24E-05 | 0.017109251 | 1 | 0.840736467 | 1 | 0.842050121 | ENSG0000018 | 9 | ZKSCAN3 |
| SAT | T2DM | ENSG0000023 | -0.00086822 | 0.515621483 | 0.001335502 | 0.081436888 | 0.000400005 | 0.770892731 | 0.001373607 | 0.081419117 | -0.00126823 | 8.15E-05 | 1 | 0.816877879 | 1 | 0.938518235 | ENSG0000018 | 9 | ZKSCAN3 |
| SAT | T2DM | ENSG0000023 | -0.0011142 | 0.278930864 | 0.001029069 | 0.081436763 | -0.00109218 | 0.288516869 | 0.001029014 | 0.081418943 | -2.20E-05 | 0.042673446 | 1 | 0.63926506 | 1 | 0.65329458 | ENSG0000018 | 9 | ZKSCAN3 |
| SAT | T2DM | ENSG0000023 | -0.00168254 | 0.434352861 | 0.002152228 | 0.081436856 | -6.84E-05 | 0.975259098 | 0.002207039 | 0.081419131 | -0.00161409 | 0.000983787 | 1 | 0.772274669 | 1 | 0.99701704 | ENSG0000018 | 9 | ZKSCAN3 |
| SAT | T2DM | ENSG0000023 | 0.000772309 | 0.764806833 | 0.002581463 | 0.081436943 | 0.001091493 | 0.672940808 | 0.002585767 | 0.081419101 | -0.00031918 | 0.03813576 | 1 | 0.93459819 | 1 | 0.898778511 | ENSG0000018 | 6 | RRP7A |
| SAT | T2DM | ENSG0000023 | -0.00212814 | 0.271838583 | 0.001936717 | 0.081436757 | -0.00209175 | 0.280080991 | 0.001936559 | 0.081418936 | -3.64E-05 | 0.011361075 | 1 | 0.634354473 | 1 | 0.646143093 | ENSG0000018 | 6 | RRP7A |
| SAT | T2DM | ENSG0000023 | -0.00198816 | 0.375230157 | 0.002242153 | 0.081436827 | -0.00194215 | 0.386345505 | 0.002241983 | 0.081419006 | -4.60E-05 | 0.012016202 | 1 | 0.728082648 | 1 | 0.746964998 | ENSG0000018 | 6 | RRP7A |
| SAT | T2DM | ENSG0000023 | 0.002845351 | 0.425817474 | 0.003572902 | 0.081436852 | 0.002780775 | 0.436357613 | 0.003572611 | 0.08141903 | 6.46E-05 | 0.015786499 | 1 | 0.771328212 | 1 | 0.784487303 | ENSG0000018 | 6 | RRP7A |
| SAT | T2DM | ENSG0000023 | -0.0014269 | 0.369734914 | 0.001590804 | 0.081436824 | -0.00139185 | 0.381570907 | 0.001590673 | 0.081419003 | -3.51E-05 | 0.002769309 | 1 | 0.723806834 | 1 | 0.742106784 | ENSG0000018 | 7 | SLC35E2B |
| SAT | T2DM | ENSG0000023 | 0.000445067 | 0.604833355 | 0.000860094 | 0.081436914 | -0.00066706 | 0.458102908 | 0.000899029 | 0.081419039 | 0.001112124 | 2.19E-05 | 1 | 0.853560529 | 1 | 0.795256953 | ENSG0000018 | 7 | SLC35E2B |
| SAT | T2DM | ENSG0000023 | 0.004187635 | 0.263956971 | 0.003748704 | 0.08143675 | 0.004253354 | 0.256496119 | 0.003748396 | 0.081418916 | -6.57E-05 | 0.018050807 | 1 | 0.627688499 | 1 | 0.628971425 | ENSG0000018 | 7 | TMEM194B |
| SAT | T2DM | ENSG0000023 | 0.004934119 | 0.194277099 | 0.003801231 | 0.081436677 | 0.000520736 | 0.893585851 | 0.003892824 | 0.081419128 | 0.004413382 | 1.56E-07 | 1 | 0.553799446 | 1 | 0.972100523 | ENSG0000018 | 7 | TMEM194B |
| SAT | T2DM | ENSG0000023 | 0.004392815 | 0.177789492 | 0.003259746 | 0.081436655 | 0.004461645 | 0.171056251 | 0.003259483 | 0.081418818 | -6.88E-05 | 0.005522802 | 1 | 0.530526907 | 1 | 0.531976833 | ENSG0000018 | 10 | KIAA0408 |
| SAT | T2DM | ENSG0000023 | -0.00115705 | 0.562177221 | 0.00199625 | 0.081436902 | -0.00111769 | 0.575521846 | 0.001996089 | 0.081419079 | -3.94E-05 | 0.008968519 | 1 | 0.837959912 | 1 | 0.84981421 | ENSG0000018 | 5 | OR7E12P |
| SAT | T2DM | ENSG0000023 | 0.00146518 | 0.064555474 | 0.000792705 | 0.081436387 | 0.000399178 | 0.626003752 | 0.000819063 | 0.081419091 | 0.001066002 | 2.42E-07 | 1 | 0.353010849 | 1 | 0.876644981 | ENSG0000019 | 7 | BLOC1S2 |
| SAT | T2DM | ENSG0000023 | 0.006043307 | 0.020446433 | 0.002607053 | 0.08143606 | 0.006075187 | 0.019780066 | 0.002606815 | 0.081418223 | -3.19E-05 | 0.043994014 | 1 | 0.190581663 | 1 | 0.195966491 | ENSG0000019 | 7 | BLOC1S2 |
| SAT | T2DM | ENSG0000023 | 0.013320553 | 0.001095669 | 0.00408011 | 0.081435177 | 0.013394316 | 0.001026754 | 0.004079763 | 0.08141733 | -7.38E-05 | 0.010026356 | 1 | 0.027029242 | 1 | 0.029550704 | ENSG0000019 | 7 | BLOC1S2 |
| SAT | T2DM | ENSG0000023 | -0.00096241 | 0.621198317 | 0.001947602 | 0.081436918 | -0.00092085 | 0.636320842 | 0.001947445 | 0.081419094 | -4.16E-05 | 0.005033029 | 1 | 0.863989689 | 1 | 0.884482576 | ENSG0000019 | 6 | HLA-DRB1 |
| SAT | T2DM | ENSG0000024 | 0.000431847 | 0.832164297 | 0.002037717 | 0.081436951 | -0.0037654 | 0.074027384 | 0.002107759 | 0.081418598 | 0.004197248 | 7.44E-15 | 1 | 0.957003525 | 1 | 0.395576296 | ENSG0000019 | 6 | HLA-DRB1 |
| SAT | T2DM | ENSG0000024 | -0.00767506 | 0.037004009 | 0.003679804 | 0.081436231 | -0.00775949 | 0.034962023 | 0.003679568 | 0.081418388 | 8.44E-05 | 0.016114346 | 1 | 0.266835505 | 1 | 0.272172417 | ENSG0000019 | 6 | HLA-DRB1 |
| SAT | T2DM | ENSG0000024 | -0.00023885 | 0.951300393 | 0.003910882 | 0.081436958 | -0.00030469 | 0.937895404 | 0.003910581 | 0.08141913 | 6.58E-05 | 0.037057914 | 1 | 0.988605339 | 1 | 0.985647128 | ENSG0000019 | 6 | HLA-DRB1 |
| SAT | T2DM | ENSG0000024 | -0.00063911 | 0.468188339 | 0.000881003 | 0.08143687 | -0.00062618 | 0.477193904 | 0.000880927 | 0.081419046 | -1.29E-05 | 0.034087623 | 1 | 0.79334291 | 1 | 0.80363893 | ENSG0000019 | 6 | HLA-DRB1 |
| SAT | T2DM | ENSG0000024 | -0.00227038 | 0.328368501 | 0.002322862 | 0.081436799 | -0.0023141 | 0.319098592 | 0.002322663 | 0.081418965 | 4.37E-05 | 0.006409963 | 1 | 0.692334629 | 1 | 0.690153251 | ENSG0000019 | 6 | WDSUB1 |
| SAT | T2DM | ENSG0000024 | -0.00203859 | 0.056960991 | 0.001070903 | 0.081436353 | -0.0019379 | 0.070412904 | 0.001071107 | 0.081418584 | -0.00010069 | 0.000122383 | 1 | 0.331288318 | 1 | 0.386976108 | ENSG0000019 | 6 | WDSUB1 |
| SAT | T2DM | ENSG0000024 | 0.000517469 | 0.88572031 | 0.003600495 | 0.081436955 | 0.000596432 | 0.86841992 | 0.003600209 | 0.081419126 | -7.90E-05 | 0.004611415 | 1 | 0.969439538 | 1 | 0.966241215 | ENSG0000019 | 6 | WDSUB1 |
| SAT | T2DM | ENSG0000024 | -0.00192691 | 0.282488777 | 0.001792896 | 0.081436765 | -0.0019648 | 0.273089832 | 0.001792744 | 0.08141893 | 3.79E-05 | 0.00248837 | 1 | 0.641415693 | 1 | 0.64192524 | ENSG0000019 | 6 | WDSUB1 |
| SAT | T2DM | ENSG0000024 | 0.000569306 | 0.572346372 | 0.001008336 | 0.081436905 | 0.000585829 | 0.561218956 | 0.001008255 | 0.081419074 | -1.65E-05 | 0.031549695 | 1 | 0.840736467 | 1 | 0.84223981 | ENSG0000019 | 6 | WDSUB1 |
| SAT | T2DM | ENSG0000024 | -0.0004257 | 0.893456972 | 0.003178512 | 0.081436955 | -0.00050918 | 0.872720242 | 0.003178304 | 0.081419127 | 8.35E-05 | 0.005178962 | 1 | 0.970180416 | 1 | 0.966383771 | ENSG0000019 | 6 | WDSUB1 |
| SAT | T2DM | ENSG0000024 | 0.004311019 | 0.102856204 | 0.002642908 | 0.081436514 | 0.004224966 | 0.10988218 | 0.002642708 | 0.081418704 | 8.61E-05 | 7.31E-05 | 1 | 0.42676853 | 1 | 0.452205306 | ENSG0000019 | 6 | WDSUB1 |
| SAT | T2DM | ENSG0000024 | -0.00124297 | 0.712702802 | 0.003375542 | 0.081436936 | -0.00114774 | 0.73382465 | 0.003375289 | 0.081419112 | -9.52E-05 | 0.000705727 | 1 | 0.908107919 | 1 | 0.924370544 | ENSG0000019 | 6 | ZNF79 |
| SAT | T2DM | ENSG0000024 | 0.000703925 | 0.855422774 | 0.003863392 | 0.081436953 | 0.000759858 | 0.844063588 | 0.003863062 | 0.081419124 | -5.59E-05 | 0.036655809 | 1 | 0.963534946 | 1 | 0.96477102 | ENSG0000019 | 6 | ZNF79 |
| SAT | T2DM | ENSG0000024 | 0.000417464 | 0.87129915 | 0.002576799 | 0.081436954 | 0.000460257 | 0.858227092 | 0.002576574 | 0.081419126 | -4.28E-05 | 0.012633087 | 1 | 0.966468088 | 1 | 0.965566132 | ENSG0000019 | 6 | ZNF79 |
| SAT | T2DM | ENSG0000024 | 0.000259531 | 0.920673767 | 0.002606128 | 0.081436957 | 0.000212707 | 0.934945197 | 0.002605919 | 0.08141913 | 4.68E-05 | 0.01885137 | 1 | 0.978648294 | 1 | 0.984768922 | ENSG0000019 | 6 | ZNF79 |
| SAT | T2DM | ENSG0000024 | -0.00429946 | 0.023995071 | 0.001904763 | 0.081436107 | -0.00425853 | 0.025358273 | 0.001904607 | 0.081418296 | -4.09E-05 | 0.003997663 | 1 | 0.212777913 | 1 | 0.232403611 | ENSG0000019 | 6 | ZNF79 |
| SAT | T2DM | ENSG0000024 | -0.00415237 | 3.41E-07 | 0.000814315 | 0.081432613 | -0.00159731 | 0.172716075 | 0.001171451 | 0.08141882 | -0.00255506 | 0.002415015 | 0.001119471 | 4.15E-05 | 1 | 0.532948952 | ENSG0000019 | 6 | ZNF79 |
| SAT | T2DM | ENSG0000024 | -0.00362339 | 0.053710496 | 0.001878214 | 0.081436336 | -0.00359142 | 0.055840729 | 0.0018780 | 0.08141852 | -3.20E-05 | 0.049802168 | 1 | 0.321968593 | 1 | 0.342399143 | ENSG0000019 | 2 | HRCT1 |
| SAT | T2DM | ENSG0000024 | -0.00944216 | 0.000228651 | 0.002562263 | 0.081434689 | -0.00930267 | 0.000282444 | 0.002562071 | 0.081416928 | -0.00013949 | 4.35E-11 | 0.750203982 | 0.008723302 | 0.926699671 | 0.011730376 | ENSG0000019 | 2 | HRCT1 |
| SAT | T2DM | ENSG0000024 | -0.00365486 | 0.216426715 | 0.0029568 | 0.081436703 | -0.00370561 | 0.210076727 | 0.002956553 | 0.081418868 | 5.08E-05 | 0.017145591 | 1 | 0.580618195 | 1 | 0.579977552 | ENSG0000019 | 2 | HRCT1 |
| SAT | T2DM | ENSG0000024 | -0.00363381 | 0.000342139 | 0.00101471 | 0.081434815 | -0.00300401 | 0.003205274 | 0.001019226 | 0.081417679 | -0.0006298 | 8.30E-11 | 1 | 0.012070532 | 1 | 0.064916684 | ENSG0000019 | 7 | RNF216P1 |
| SAT | T2DM | ENSG0000024 | -0.00193547 | 0.618496 | 0.003886621 | 0.081436917 | -0.00257641 | 0.507825414 | 0.003890513 | 0.081419058 | 0.000640934 | 0.000469764 | 1 | 0.862424724 | 1 | 0.817738235 | ENSG0000019 | 7 | RNF216P1 |
| SAT | T2DM | ENSG0000024 | 0.006253936 | 0.127693256 | 0.004105614 | 0.081436571 | 0.004195252 | 0.315741639 | 0.004181669 | 0.081418963 | 0.002058684 | 0.009719749 | 1 | 0.467990294 | 1 | 0.685604445 | ENSG0000019 | 7 | RNF216P1 |
| SAT | T2DM | ENSG0000025 | 0.003963733 | 0.03567044 | 0.00188689 | 0.081436221 | 0.003989643 | 0.034465235 | 0.00188672 | 0.081418384 | -2.59E-05 | 0.02839214 | 1 | 0.263415251 | 1 | 0.272172417 | ENSG0000019 | 8 | SIRPB2 |
| SAT | T2DM | ENSG0000025 | -0.0012936 | 0.365932451 | 0.001430779 | 0.081436822 | -0.00132267 | 0.355222373 | 0.001430673 | 0.081418988 | 2.91E-05 | 0.015992378 | 1 | 0.721771482 | 1 | 0.724067999 | ENSG0000019 | 8 | SIRPB2 |
| SAT | T2DM | ENSG0000025 | 0.006598195 | 0.017607048 | 0.002779606 | 0.081436017 | 0.00655233 | 0.018399687 | 0.002779375 | 0.081418202 | 4.59E-05 | 0.02277168 | 1 | 0.172693001 | 1 | 0.187482528 | ENSG0000019 | 11 | SRGAP3 |
| SAT | T2DM | ENSG0000025 | -0.00140125 | 0.619748769 | 0.00282393 | 0.081436917 | -0.00160616 | 0.569725121 | 0.002825478 | 0.081419077 | 0.000204909 | 0.045497907 | 1 | 0.863241392 | 1 | 0.845822679 | ENSG0000019 | 11 | SRGAP3 |
| SAT | T2DM | ENSG0000025 | 0.008966145 | 0.022790501 | 0.003937812 | 0.081436092 | 0.009042289 | 0.021649681 | 0.00393748 | 0.08141825 | -7.61E-05 | 0.006520465 | 1 | 0.206365356 | 1 | 0.208306757 | ENSG0000019 | 8 | XPNPEP3 |
| SAT | T2DM | ENSG0000025 | -0.00322185 | 0.309289887 | 0.003168885 | 0.081436786 | -0.00283573 | 0.371128755 | 0.003170671 | 0.081418997 | -0.00038612 | 0.000896035 | 1 | 0.669821861 | 1 | 0.734641671 | ENSG0000019 | 8 | XPNPEP3 |
| SAT | T2DM | ENSG0000025 | -0.00059312 | 0.778750268 | 0.002111142 | 0.081436945 | -0.0005652 | 0.788893879 | 0.002110954 | 0.081419119 | -2.79E-05 | 0.039432093 | 1 | 0.939432466 | 1 | 0.947957612 | ENSG0000019 | 7 | ZNF107 |
| SAT | T2DM | ENSG0000025 | 0.005928468 | 0.147425524 | 0.004092303 | 0.081436608 | 0.005850432 | 0.152796139 | 0.004091993 | 0.081418789 | 7.80E-05 | 0.020195381 | 1 | 0.492642703 | 1 | 0.512250482 | ENSG0000019 | 7 | ZNF107 |
| SAT | T2DM | ENSG0000025 | -0.00373346 | 0.148420834 | 0.002583514 | 0.081436609 | -0.00379354 | 0.141972968 | 0.002583301 | 0.081418771 | 6.01E-05 | 0.001556816 | 1 | 0.493635361 | 1 | 0.499917672 | ENSG0000019 | 7 | ZNF107 |
| SAT | T2DM | ENSG0000025 | 0.00389033 | 0.172950831 | 0.002854691 | 0.081436648 | 0.003939692 | 0.167527235 | 0.002854446 | 0.081418813 | -4.94E-05 | 0.01212853 | 1 | 0.526970477 | 1 | 0.529024888 | ENSG0000019 | 3 | SFTA2 |
| SAT | T2DM | ENSG0000025 | -0.00313093 | 0.40284439 | 0.003742657 | 0.081436841 | -0.00320193 | 0.392224027 | 0.00374236 | 0.081419009 | 7.10E-05 | 0.014878461 | 1 | 0.752695013 | 1 | 0.75344674 | ENSG0000019 | 3 | SFTA2 |
| SAT | T2DM | ENSG0000025 | 0.009751325 | 0.000178306 | 0.002601775 | 0.081434611 | 0.009791969 | 0.000167324 | 0.00260157 | 0.081416764 | -4.06E-05 | 0.046437954 | 0.585021353 | 0.007434796 | 0.548990588 | 0.00820853 | ENSG0000019 | 3 | SFTA2 |
| SAT | T2DM | ENSG0000025 | -0.00028302 | 0.910715539 | 0.002523863 | 0.081436956 | -0.0002186 | 0.930972531 | 0.00252365 | 0.08141913 | -6.44E-05 | 0.000287591 | 1 | 0.975126969 | 1 | 0.983109389 | ENSG0000019 | 3 | SFTA2 |
| SAT | T2DM | ENSG0000025 | -0.00097369 | 0.383978581 | 0.001118422 | 0.081436832 | -0.00099867 | 0.37185808 | 0.001118331 | 0.081418998 | 2.50E-05 | 0.002899835 | 1 | 0.734596925 | 1 | 0.734979735 | ENSG0000019 | 3 | ZNF471 |
| SAT | T2DM | ENSG0000025 | 0.001142691 | 0.497703569 | 0.001685116 | 0.081436882 | 0.001769135 | 0.295812373 | 0.001692213 | 0.081418948 | -0.00062644 | 6.48E-05 | 1 | 0.808799113 | 1 | 0.662977391 | ENSG0000019 | 3 | HLA-DRB9 |
| SAT | T2DM | ENSG0000025 | -0.00396398 | 0.249575038 | 0.003442796 | 0.081436737 | -0.00416615 | 0.226322477 | 0.003443415 | 0.081418886 | 0.000202167 | 0.014631232 | 1 | 0.615333003 | 1 | 0.601160762 | ENSG0000019 | 3 | HLA-DRB9 |
| SAT | T2DM | ENSG0000025 | 0.00084767 | 0.596827909 | 0.001602497 | 0.081436912 | 0.000401027 | 0.802783052 | 0.001605734 | 0.08141912 | 0.000446642 | 1.96E-05 | 1 | 0.848804669 | 1 | 0.952314593 | ENSG0000019 | 3 | HLA-DRB9 |
| SAT | T2DM | ENSG0000025 | -0.00094337 | 0.778592751 | 0.003355367 | 0.081436945 | -0.00125517 | 0.708394 | 0.003355931 | 0.081419108 | 0.000311792 | 7.99E-05 | 1 | 0.939432466 | 1 | 0.912941362 | ENSG0000019 | 7 | ZNF565 |
| SAT | T2DM | ENSG0000025 | 0.000656615 | 0.608874958 | 0.001283252 | 0.081436915 | 0.001159612 | 0.368402777 | 0.001289217 | 0.081418996 | -0.000503 | 5.98E-05 | 1 | 0.855383565 | 1 | 0.732878736 | ENSG0000019 | 7 | ZNF565 |
| SAT | T2DM | ENSG0000025 | 0.000541725 | 0.882474954 | 0.003664452 | 0.081436955 | 0.000488212 | 0.894003654 | 0.003664153 | 0.081419128 | 5.35E-05 | 0.049322988 | 1 | 0.969020186 | 1 | 0.972150687 | ENSG0000019 | 7 | ZNF565 |

| | | | | | | | | | | | | | | | | | |
|---|---|---|---|---|---|---|---|---|---|---|---|---|---|---|---|---|---|
| SAT | T2DM | ENSG0000025 | 0.008952537 | 1.27E-05 | 0.002050555 | 0.081433773 | 0.009006153 | 1.12E-05 | 0.002050451 | 0.081415908 | -5.36E-05 | 0.015920674 | 0.041544541 | 0.00082697 | 0.036810281 | 0.000856053 | ENSG0000019 | 6 | ZNF781 |
| SAT | T2DM | ENSG0000025 | -0.00232166 | 0.35010984 | 0.002484711 | 0.081436813 | -0.00225658 | 0.363742165 | 0.002484511 | 0.081418993 | -6.51E-05 | 0.000593755 | 1 | 0.708896874 | 1 | 0.729039733 | ENSG0000019 | 8 | EVL |
| SAT | T2DM | ENSG0000025 | 0.004082613 | 0.233653528 | 0.003427882 | 0.081436721 | 0.004023224 | 0.240487277 | 0.003427605 | 0.081418901 | 5.94E-05 | 0.021850775 | 1 | 0.598919708 | 1 | 0.618368932 | ENSG0000019 | 4 | ZNF775 |
| SAT | T2DM | ENSG0000025 | -0.00097645 | 0.707907517 | 0.002606177 | 0.081436935 | -0.00092622 | 0.722273522 | 0.002605962 | 0.08141911 | -5.02E-05 | 0.008707082 | 1 | 0.906221055 | 1 | 0.920812547 | ENSG0000019 | 6 | ZNF605 |
| SAT | T2DM | ENSG0000025 | 0.001028255 | 0.734793611 | 0.003035383 | 0.081436939 | 0.000934017 | 0.758285433 | 0.00303515 | 0.081419115 | 9.42E-05 | 0.000122429 | 1 | 0.923192618 | 1 | 0.932894921 | ENSG0000019 | 6 | ZNF605 |
| SAT | T2DM | ENSG0000025 | 0.00035943 | 0.76042191 | 0.001178742 | 0.081436943 | 0.000342704 | 0.771233344 | 0.001178641 | 0.081419117 | 1.67E-05 | 0.037837353 | 1 | 0.934086217 | 1 | 0.938518235 | ENSG0000019 | 6 | ZNF605 |
| SAT | T2DM | ENSG0000025 | 0.000499416 | 0.812471914 | 0.002105123 | 0.081436949 | 0.000462541 | 0.826073294 | 0.002104941 | 0.081419123 | 3.69E-05 | 0.009560102 | 1 | 0.951539016 | 1 | 0.959754419 | ENSG0000019 | 8 | RFX8 |
| SAT | T2DM | ENSG0000025 | -0.00339026 | 0.172809515 | 0.002486928 | 0.081436648 | -0.00335494 | 0.177289478 | 0.002486707 | 0.081418827 | -3.53E-05 | 0.026255508 | 1 | 0.526970477 | 1 | 0.539255458 | ENSG0000019 | 9 | TPK1 |
| SAT | T2DM | ENSG0000025 | 5.71E-06 | 0.998699494 | 0.003501242 | 0.081436958 | -6.29E-05 | 0.985675577 | 0.003500974 | 0.081419131 | 6.86E-05 | 0.015533724 | 1 | 0.998699494 | 1 | 0.997162029 | ENSG0000019 | 9 | TPK1 |
| SAT | T2DM | ENSG0000025 | -0.00682093 | 0.071316694 | 0.003782137 | 0.081436415 | -0.00674212 | 0.074623725 | 0.003781814 | 0.0814186 | -7.88E-05 | 0.00256164 | 1 | 0.373189908 | 1 | 0.395620568 | ENSG0000019 | 10 | MYO18A |
| SAT | T2DM | ENSG0000025 | -0.00111349 | 0.493509615 | 0.001626161 | 0.08143688 | -0.00108683 | 0.503879171 | 0.001626026 | 0.081419056 | -2.67E-05 | 0.024951295 | 1 | 0.805574651 | 1 | 0.817738235 | ENSG0000019 | 10 | MYO18A |
| SAT | T2DM | ENSG0000025 | 0.000616545 | 0.655483268 | 0.001381898 | 0.081436925 | 0.000640265 | 0.643106762 | 0.001381788 | 0.081419095 | -2.37E-05 | 0.026550662 | 1 | 0.883423151 | 1 | 0.888435068 | ENSG0000019 | 10 | MYO18A |
| SAT | T2DM | ENSG0000025 | -0.00036512 | 0.77584294 | 0.001282273 | 0.081436945 | -0.00038863 | 0.761811936 | 0.001282165 | 0.081419116 | 2.35E-05 | 0.010759166 | 1 | 0.939125227 | 1 | 0.932999239 | ENSG0000019 | 10 | MYO18A |
| SAT | T2DM | ENSG0000025 | 0.00411271 | 0.124715571 | 0.0026788 | 0.081436565 | 0.004068525 | 0.128785869 | 0.002678582 | 0.081418745 | 4.42E-05 | 0.027598091 | 1 | 0.463429598 | 1 | 0.478535034 | ENSG0000019 | 10 | MYO18A |
| SAT | T2DM | ENSG0000025 | -0.00175576 | 0.563285201 | 0.003037813 | 0.081436903 | -0.00183913 | 0.544876865 | 0.003037593 | 0.08141907 | 8.34E-05 | 0.001441917 | 1 | 0.837959912 | 1 | 0.840498821 | ENSG0000019 | 10 | MYO18A |
| SAT | T2DM | ENSG0000025 | 0.00385587 | 0.341405375 | 0.004052867 | 0.081436807 | 0.003753271 | 0.354367885 | 0.004052547 | 0.081418988 | 0.000102599 | 0.001163511 | 1 | 0.702729632 | 1 | 0.723060342 | ENSG0000019 | 10 | SULF2 |
| SAT | T2DM | ENSG0000025 | 0.005978443 | 0.003484514 | 0.002046414 | 0.081435532 | 0.005925845 | 0.003780377 | 0.002046257 | 0.08141773 | 5.26E-05 | 0.001478665 | 1 | 0.06146607 | 1 | 0.072534597 | ENSG0000019 | 7 | ZNF502 |
| SAT | T2DM | ENSG0000026 | 0.001929843 | 0.071727677 | 0.001071631 | 0.081436416 | 0.001883336 | 0.078820371 | 0.001071553 | 0.081418615 | 4.65E-05 | 4.55E-07 | 1 | 0.373553186 | 1 | 0.407783994 | ENSG0000019 | 5 | TRAPPC4 |
| SAT | T2DM | ENSG0000026 | 0.00231793 | 0.319198609 | 0.002326989 | 0.081436793 | 0.002272771 | 0.328675633 | 0.002326786 | 0.081418971 | 4.52E-05 | 0.003519136 | 1 | 0.680500738 | 1 | 0.696630976 | ENSG0000019 | 3 | FAM180B |
| SAT | T2DM | ENSG0000026 | -0.00399058 | 0.046242196 | 0.002002116 | 0.081436295 | -0.0038977 | 0.051557452 | 0.002002089 | 0.081418498 | -9.29E-05 | 0.000784941 | 1 | 0.295751746 | 1 | 0.328466017 | ENSG0000019 | 10 | ZNF33B |
| SAT | T2DM | ENSG0000026 | 0.005898798 | 0.039548641 | 0.002865667 | 0.08143625 | 0.005975805 | 0.037025827 | 0.002865427 | 0.081418404 | -7.70E-05 | 0.000177825 | 1 | 0.274913329 | 1 | 0.279268363 | ENSG0000019 | 10 | ZNF33B |
| SAT | T2DM | ENSG0000026 | 0.001818544 | 0.413658552 | 0.002224592 | 0.081436847 | 0.001865005 | 0.401789067 | 0.002224399 | 0.081419013 | -4.65E-05 | 0.001842338 | 1 | 0.760343815 | 1 | 0.757591271 | ENSG0000019 | 10 | ZNF33B |
| SAT | T2DM | ENSG0000026 | -0.00144529 | 0.657220937 | 0.003256967 | 0.081436926 | -0.00153175 | 0.638113233 | 0.003256688 | 0.081419094 | 8.65E-05 | 0.000121453 | 1 | 0.884109018 | 1 | 0.884889906 | ENSG0000019 | 10 | ZNF33B |
| SAT | T2DM | ENSG0000026 | 0.008615086 | 0.009419035 | 0.003318001 | 0.081435832 | 0.008549713 | 0.009967116 | 0.003317728 | 0.081418021 | 6.54E-05 | 0.007345328 | 1 | 0.116618318 | 1 | 0.131863332 | ENSG0000019 | 12 | NF1 |
| SAT | T2DM | ENSG0000026 | -0.00727628 | 0.004722129 | 0.002575295 | 0.081435624 | -0.00733332 | 0.004403692 | 0.002575163 | 0.081417776 | 5.70E-05 | 0.040160364 | 1 | 0.073427987 | 1 | 0.079676052 | ENSG0000019 | 12 | NF1 |
| SAT | T2DM | ENSG0000026 | 5.90E-05 | 0.979355131 | 0.002280578 | 0.081436958 | 9.82E-05 | 0.965642443 | 0.002280398 | 0.081419131 | -3.92E-05 | 0.027996293 | 1 | 0.995203291 | 1 | 0.99427016 | ENSG0000019 | 8 | DAPK1 |
| SAT | T2DM | ENSG0000026 | -0.00236122 | 0.049023471 | 0.001199568 | 0.081436311 | -0.00186971 | 0.119739381 | 0.001201713 | 0.081418726 | -0.00049151 | 2.92E-11 | 1 | 0.306958031 | 1 | 0.466585403 | ENSG0000019 | 8 | DAPK1 |
| SAT | T2DM | ENSG0000026 | -0.00711624 | 0.031125102 | 0.003301471 | 0.081436182 | -0.00719702 | 0.029248448 | 0.003301193 | 0.081418337 | 8.08E-05 | 0.000617769 | 1 | 0.245761744 | 1 | 0.251961415 | ENSG0000019 | 8 | DAPK1 |
| SAT | T2DM | ENSG0000026 | 9.10E-05 | 0.952684927 | 0.001533018 | 0.081436958 | 5.20E-05 | 0.972913989 | 0.00153289 | 0.081419131 | 3.89E-05 | 0.000394179 | 1 | 0.988645274 | 1 | 0.996397533 | ENSG0000019 | 7 | HLA-DQA1 |
| SAT | T2DM | ENSG0000026 | 0.005548827 | 0.004953996 | 0.001974656 | 0.081435639 | 0.005586931 | 0.004661389 | 0.001974487 | 0.081417793 | -3.81E-05 | 0.005714515 | 1 | 0.074555423 | 1 | 0.0804112 | ENSG0000019 | 7 | HLA-DQA1 |
| SAT | T2DM | ENSG0000026 | 0.005413489 | 0.012963479 | 0.002178659 | 0.081435927 | 0.005446393 | 0.012416176 | 0.00217847 | 0.081418087 | -3.29E-05 | 0.026088756 | 1 | 0.143739766 | 1 | 0.150879531 | ENSG0000019 | 7 | HLA-DQA1 |
| SAT | T2DM | ENSG0000026 | -1.06E-05 | 0.994083803 | 0.001426618 | 0.081436958 | 1.45E-05 | 0.991877709 | 0.001426495 | 0.081419131 | -2.51E-05 | 0.009442139 | 1 | 0.99758396 | 1 | 0.998267105 | ENSG0000019 | 7 | HLA-DQA1 |
| SAT | T2DM | ENSG0000026 | -0.00473197 | 0.081907159 | 0.002719945 | 0.081436453 | -0.004781 | 0.078764576 | 0.002719718 | 0.081418615 | 4.90E-05 | 0.01300018 | 1 | 0.395495266 | 1 | 0.407783994 | ENSG0000019 | 7 | HLA-DQA1 |
| SAT | T2DM | ENSG0000026 | 0.002591628 | 0.162411356 | 0.001855126 | 0.081436632 | 0.002616141 | 0.15843643 | 0.001854956 | 0.081418799 | -2.45E-05 | 0.027587218 | 1 | 0.513507272 | 1 | 0.51775889 | ENSG0000019 | 7 | HLA-DQA1 |
| SAT | T2DM | ENSG0000026 | 0.002220074 | 0.08130692 | 0.001273594 | 0.081436451 | 0.00224817 | 0.077503037 | 0.001273487 | 0.08141861 | -2.81E-05 | 0.002079455 | 1 | 0.395495266 | 1 | 0.405562145 | ENSG0000019 | 7 | HLA-DQA1 |
| SAT | T2DM | ENSG0000026 | -0.00091429 | 0.69668465 | 0.002345548 | 0.081436933 | -0.00087038 | 0.710556328 | 0.002345356 | 0.081419108 | -4.39E-05 | 0.011387352 | 1 | 0.900770568 | 1 | 0.913890753 | ENSG0000019 | 7 | HLA-DQA1 |
| SAT | T2DM | ENSG0000026 | 0.000218637 | 0.848723892 | 0.001146215 | 0.081436952 | 0.000251966 | 0.825994398 | 0.001146122 | 0.081419123 | -3.33E-05 | 0.000130503 | 1 | 0.963201596 | 1 | 0.959754419 | ENSG0000019 | 4 | GM2A |
| SAT | T2DM | ENSG0000026 | -0.00038784 | 0.799823274 | 0.001529496 | 0.081436948 | -0.00029053 | 0.849348818 | 0.001529533 | 0.081419125 | -9.73E-05 | 0.000101782 | 1 | 0.948056417 | 1 | 0.965561588 | ENSG0000019 | 4 | GM2A |
| SAT | T2DM | ENSG0000026 | -0.00069007 | 0.768944012 | 0.002349133 | 0.081436944 | -0.0006432 | 0.784218403 | 0.002348922 | 0.081419118 | -4.69E-05 | 0.001420492 | 1 | 0.935400227 | 1 | 0.945615795 | ENSG0000019 | 4 | GM2A |
| SAT | T2DM | ENSG0000026 | -0.00109432 | 0.764563675 | 0.003653887 | 0.081436943 | 0.000696531 | 0.850205807 | 0.003688159 | 0.081419125 | -0.00179085 | 0.000385826 | 1 | 0.93459819 | 1 | 0.965561588 | ENSG0000019 | 4 | GM2A |
| SAT | T2DM | ENSG0000026 | 0.000179027 | 0.880451926 | 0.001190366 | 0.081436955 | 0.00030092 | 0.800468105 | 0.001190625 | 0.08141912 | -0.00012189 | 6.29E-05 | 1 | 0.968084038 | 1 | 0.952160336 | ENSG0000019 | 4 | GM2A |
| SAT | T2DM | ENSG0000026 | 0.004704605 | 0.198348146 | 0.003657545 | 0.081436682 | 0.005091175 | 0.164093049 | 0.003658929 | 0.081418807 | -0.00038657 | 0.000716803 | 1 | 0.558130589 | 1 | 0.523492072 | ENSG0000019 | 4 | GM2A |
| SAT | T2DM | ENSG0000026 | -2.48E-05 | 0.983424905 | 0.001193379 | 0.081436958 | 5.52E-05 | 0.96308001 | 0.001193437 | 0.081419131 | -8.00E-05 | 0.000166145 | 1 | 0.995937959 | 1 | 0.993672317 | ENSG0000019 | 2 | HIST1H2AG |
| SAT | T2DM | ENSG0000026 | 0.005177476 | 0.112252218 | 0.003260059 | 0.081436537 | 0.005062717 | 0.120409553 | 0.003259834 | 0.081418728 | 0.000114759 | 8.91E-05 | 1 | 0.44002333 | 1 | 0.467258033 | ENSG0000019 | 9 | ZNF239 |
| SAT | T2DM | ENSG0000026 | 0.000447482 | 0.877663687 | 0.00290702 | 0.081436954 | 0.000500643 | 0.863254259 | 0.002906775 | 0.081419126 | -5.32E-05 | 0.009833282 | 1 | 0.96728739 | 1 | 0.966241215 | ENSG0000019 | 9 | ZNF239 |
| SAT | T2DM | ENSG0000026 | -0.0012556 | 0.642011524 | 0.002700857 | 0.081436922 | -0.00121174 | 0.653659446 | 0.00270064 | 0.081419097 | -4.39E-05 | 0.032764931 | 1 | 0.87658752 | 1 | 0.891871358 | ENSG0000019 | 9 | ZNF239 |
| SAT | T2DM | ENSG0000026 | 0.002925368 | 0.161611289 | 0.002090034 | 0.081436631 | 0.002959562 | 0.15673459 | 0.002089876 | 0.081418796 | -3.42E-05 | 0.047945964 | 1 | 0.512811064 | 1 | 0.51775889 | ENSG0000019 | 9 | ZNF239 |
| SAT | T2DM | ENSG0000026 | -0.00541202 | 0.164707577 | 0.003895182 | 0.081436636 | -0.0053518 | 0.169422648 | 0.003894874 | 0.081418815 | -6.02E-05 | 0.047648494 | 1 | 0.516146667 | 1 | 0.531756789 | ENSG0000019 | 5 | C6orf106 |
| SAT | T2DM | ENSG0000026 | -0.00170707 | 0.523075708 | 0.002673094 | 0.08143689 | -0.0016617 | 0.534144703 | 0.002672868 | 0.081419066 | -4.54E-05 | 0.015925651 | 1 | 0.821144329 | 1 | 0.832950936 | ENSG0000019 | 5 | C6orf106 |
| SAT | T2DM | ENSG0000026 | 0.000320194 | 0.903092585 | 0.002629809 | 0.081436956 | 0.000260941 | 0.920954289 | 0.00262961 | 0.081419129 | 5.93E-05 | 0.006090278 | 1 | 0.97231278 | 1 | 0.979654273 | ENSG0000019 | 5 | C6orf106 |
| SAT | T2DM | ENSG0000026 | 0.002678099 | 0.22407679 | 0.002202818 | 0.081436711 | 0.002621336 | 0.234009488 | 0.002202627 | 0.081418894 | 5.68E-05 | 0.000124437 | 1 | 0.588156759 | 1 | 0.610313962 | ENSG0000019 | 5 | C6orf106 |
| SAT | T2DM | ENSG0000026 | 0.009734587 | 0.004916828 | 0.003461253 | 0.081435637 | 0.008164478 | 0.018955608 | 0.003479561 | 0.081418211 | 0.001570109 | 1.30E-05 | 1 | 0.07434153 | 1 | 0.191364154 | ENSG0000019 | 8 | ADA |
| SAT | T2DM | ENSG0000027 | -0.00012111 | 0.968115373 | 0.003029865 | 0.081436958 | -7.34E-05 | 0.980675088 | 0.003029593 | 0.081419131 | -4.77E-05 | 0.011856462 | 1 | 0.993613755 | 1 | 0.997085517 | ENSG0000019 | 3 | KRT39 |
| SAT | T2DM | ENSG0000027 | -0.00052352 | 0.681716526 | 0.0012765 | 0.08143693 | -0.00054724 | 0.668113583 | 0.001276392 | 0.0814191 | 2.37E-05 | 0.008788106 | 1 | 0.892786811 | 1 | 0.896403813 | ENSG0000019 | 6 | KIAA1211L |
| SAT | T2DM | ENSG0000027 | 0.009364953 | 0.004763613 | 0.003317832 | 0.081435627 | 0.009548328 | 0.004001812 | 0.003317658 | 0.081417747 | -0.00018338 | 2.46E-07 | 1 | 0.073507661 | 1 | 0.075459453 | ENSG0000019 | 6 | KIAA1211L |
| SAT | T2DM | ENSG0000027 | 0.002242403 | 0.579601327 | 0.004047896 | 0.081436907 | 0.002109437 | 0.602256213 | 0.004047588 | 0.081419086 | 0.000132966 | 5.46E-05 | 1 | 0.842939695 | 1 | 0.858221553 | ENSG0000019 | 8 | ANKRD36B |
| SAT | T2DM | ENSG0000027 | -0.00098234 | 0.40743396 | 0.001185798 | 0.081436844 | -0.00110616 | 0.351046255 | 0.001186154 | 0.081418986 | 0.000123826 | 0.000266385 | 1 | 0.756247299 | 1 | 0.71896552 | ENSG0000019 | 8 | ANKRD36B |
| SAT | T2DM | ENSG0000027 | 0.004569461 | 0.254667495 | 0.004011512 | 0.081436742 | 0.004637083 | 0.247663902 | 0.004011166 | 0.081418908 | -6.76E-05 | 0.013477546 | 1 | 0.620408004 | 1 | 0.620663712 | ENSG0000019 | 8 | ANKRD36B |
| SAT | T2DM | ENSG0000027 | -0.00161947 | 0.465104382 | 0.00221703 | 0.081436869 | -0.00165821 | 0.454459684 | 0.002216848 | 0.081419037 | 3.87E-05 | 0.018348154 | 1 | 0.792954767 | 1 | 0.794600418 | ENSG0000019 | 8 | ANKRD36B |
| SAT | T2DM | ENSG0000027 | 0.002266882 | 0.382347403 | 0.002594933 | 0.081436831 | 0.002226393 | 0.39086435 | 0.002594708 | 0.081419008 | 4.05E-05 | 0.019961126 | 1 | 0.732758077 | 1 | 0.751715083 | ENSG0000019 | 8 | ANKRD36B |
| SAT | T2DM | ENSG0000027 | 0.001481391 | 0.507142845 | 0.002233386 | 0.081436885 | 0.001516506 | 0.497089894 | 0.00223319 | 0.081419054 | -3.51E-05 | 0.016250733 | 1 | 0.810627063 | 1 | 0.81497549 | ENSG0000019 | 8 | ANKRD36B |
| SAT | T2DM | ENSG0000027 | 0.001433733 | 0.091578017 | 0.000849804 | 0.081436483 | 0.001420959 | 0.094477196 | 0.000849732 | 0.081418664 | 1.28E-05 | 0.036300289 | 1 | 0.413867045 | 1 | 0.429887299 | ENSG0000019 | 8 | ANKRD36B |
| SAT | T2DM | ENSG0000027 | 0.004734072 | 0.007658772 | 0.001775212 | 0.08143577 | 0.004707035 | 0.008007716 | 0.001775064 | 0.081417956 | 2.70E-05 | 0.035559055 | 1 | 0.100113265 | 1 | 0.114231808 | ENSG0000019 | 8 | AP2A1 |
| SAT | T2DM | ENSG0000027 | -0.00410764 | 0.181923857 | 0.003077211 | 0.081436661 | -0.0040552 | 0.187529082 | 0.003076958 | 0.081418841 | -5.24E-05 | 0.020642634 | 1 | 0.535809853 | 1 | 0.557313872 | ENSG0000019 | 8 | AP2A1 |
| SAT | T2DM | ENSG0000027 | -0.01280102 | 0.000205406 | 0.003448271 | 0.081434655 | -0.01290776 | 0.00018156 | 0.003448129 | 0.08141679 | 0.000106741 | 0.008105044 | 0.673938506 | 0.008023077 | 0.595698457 | 0.008583862 | ENSG0000019 | 2 | PCDHB16 |
| SAT | T2DM | ENSG0000027 | -0.00245842 | 0.282124118 | 0.002285711 | 0.081436765 | -0.00249503 | 0.27497851 | 0.002285514 | 0.081418932 | 3.66E-05 | 0.019444958 | 1 | 0.641415693 | 1 | 0.64327119 | ENSG0000019 | 2 | PCDHB16 |
| SAT | T2DM | ENSG0000027 | 0.008612197 | 0.029838037 | 0.003964662 | 0.08143617 | 0.008502415 | 0.031975399 | 0.00396434 | 0.081418362 | 0.000109782 | 0.000220357 | 1 | 0.240185597 | 1 | 0.263779676 | ENSG0000019 | 2 | PCDHB16 |
| SAT | T2DM | ENSG0000027 | 0.002592251 | 0.106134644 | 0.001604298 | 0.081436522 | 0.002408323 | 0.133373623 | 0.001604551 | 0.081418754 | 0.000183927 | 7.08E-07 | 1 | 0.433220108 | 1 | 0.486825442 | ENSG0000019 | 3 | HIST1H3E |
| SAT | T2DM | ENSG0000027 | 0.001631005 | 0.549970486 | 0.002728325 | 0.081436899 | 0.001580461 | 0.562369716 | 0.002728105 | 0.081419075 | 5.05E-05 | 0.01461579 | 1 | 0.83552458 | 1 | 0.84223981 | ENSG0000019 | 3 | HIST1H3E |
| SAT | T2DM | ENSG0000027 | -0.00245244 | 0.417887209 | 0.003027361 | 0.081436849 | -0.00251417 | 0.406229151 | 0.003027118 | 0.081419016 | 6.17E-05 | 0.007690153 | 1 | 0.763620456 | 1 | 0.759884747 | ENSG0000019 | 3 | HIST1H3E |
| SAT | T2DM | ENSG0000027 | -0.00349002 | 6.42E-07 | 0.000701059 | 0.081432817 | -0.00323397 | 4.73E-06 | 0.000706624 | 0.081415631 | -0.00025605 | 0.004054741 | 0.002106314 | 6.58E-05 | 0.01550583 | 0.000430718 | ENSG0000019 | 3 | HIST1H3E |
| SAT | T2DM | ENSG0000027 | -0.0008708 | 0.539191676 | 0.001418157 | 0.081436895 | 0.000498255 | 0.731983125 | 0.001454805 | 0.081419111 | -0.00136905 | 2.55E-05 | 1 | 0.831736309 | 1 | 0.923670747 | ENSG0000019 | 3 | HIST1H3E |
| SAT | T2DM | ENSG0000027 | 0.009633511 | 0.005668318 | 0.003482343 | 0.081435679 | 0.009735531 | 0.005175601 | 0.003482063 | 0.081417825 | -0.00010202 | 0.000129231 | 1 | 0.082656666 | 1 | 0.08576336 | ENSG0000019 | 6 | ZNF585A |
| SAT | T2DM | ENSG0000027 | 7.00E-05 | 0.912442136 | 0.000636239 | 0.081436956 | -0.00024906 | 0.696160393 | 0.000637792 | 0.081419105 | 0.000319024 | 2.27E-12 | 1 | 0.976107808 | 1 | 0.907470103 | ENSG0000019 | 6 | FUT11 |
| SAT | T2DM | ENSG0000027 | 0.003551298 | 0.107818005 | 0.002208407 | 0.081436526 | 0.003598086 | 0.103228178 | 0.002208222 | 0.081418687 | -4.68E-05 | 0.003359428 | 1 | 0.435653784 | 1 | 0.440431278 | ENSG0000019 | 6 | FUT11 |
| SAT | T2DM | ENSG0000027 | -0.00325524 | 0.142820798 | 0.002221445 | 0.0814366 | -0.00321021 | 0.148400363 | 0.002221282 | 0.081418782 | -4.50E-05 | 0.016712812 | 1 | 0.488540542 | 1 | 0.507874308 | ENSG0000019 | 6 | FUT11 |
| SAT | T2DM | ENSG0000027 | -0.00113742 | 0.514595091 | 0.001745306 | 0.081436887 | -0.00169805 | 0.330936091 | 0.001746546 | 0.081418973 | 0.00056063 | 2.22E-15 | 1 | 0.816039871 | 1 | 0.700003408 | ENSG0000019 | 2 | WDR5B |
| SAT | T2DM | ENSG0000027 | -0.00415791 | 0.062154623 | 0.002229209 | 0.081436377 | -0.00410351 | 0.065630124 | 0.002229021 | 0.081418565 | -5.44E-05 | 0.00058205 | 1 | 0.346229741 | 1 | 0.37355836 | ENSG0000019 | 5 | ZNF138 |
| SAT | T2DM | ENSG0000027 | 0.003744004 | 0.347572759 | 0.003985914 | 0.081436811 | 0.003794991 | 0.341003711 | 0.003985561 | 0.081418979 | -5.10E-05 | 0.047619586 | 1 | 0.708371701 | 1 | 0.708179865 | ENSG0000019 | 6 | ZNF100 |
| SAT | T2DM | ENSG0000027 | -0.00458871 | 0.273464046 | 0.004190137 | 0.081436758 | -0.0046567 | 0.266379832 | 0.004189796 | 0.081418924 | 6.80E-05 | 0.03044191 | 1 | 0.634986223 | 1 | 0.638375379 | ENSG0000019 | 7 | ZNF763 |
| SAT | T2DM | ENSG0000027 | -0.00057402 | 0.570034138 | 0.001010592 | 0.081436905 | -0.00058803 | 0.560621319 | 0.001010501 | 0.081419074 | 1.40E-05 | 0.023662764 | 1 | 0.840736467 | 1 | 0.84223981 | ENSG0000019 | 7 | ZNF763 |
| SAT | T2DM | ENSG0000027 | -0.00058051 | 0.503630151 | 0.000867997 | 0.081436884 | 2.44E-05 | 0.977795164 | 0.000875701 | 0.081419131 | -0.00060488 | 2.13E-07 | 1 | 0.810314363 | 1 | 0.99701704 | ENSG0000019 | 7 | ZNF763 |
| SAT | T2DM | ENSG0000027 | 0.005992552 | 0.013312163 | 0.002420924 | 0.081435934 | 0.006043067 | 0.012547379 | 0.002420739 | 0.08141809 | -5.05E-05 | 0.010363148 | 1 | 0.145107 | 1 | 0.150896353 | ENSG0000019 | 7 | RP5-874C20.3 |
| VAT | CAD | ENSG0000005 | 0.000842682 | 0.543536932 | 0.00138719 | 0.138947791 | 0.000865581 | 0.532646462 | 0.001387212 | 0.138943761 | -2.29E-05 | 0.033550653 | 1 | 0.84421043 | 1 | 0.841020729 | ENSG0000019 | 7 | RP5-874C20.3 |
| VAT | CAD | ENSG0000007 | -0.00794774 | 0.116224314 | 0.005059604 | 0.138947193 | -0.00857148 | 0.090862625 | 0.005069276 | 0.138943057 | 0.000623732 | 0.047133318 | 1 | 0.535009893 | 1 | 0.519153608 | ENSG0000019 | 7 | RP5-874C20.3 |
| VAT | CAD | ENSG0000009 | 0.019464309 | 6.84E-05 | 0.004888315 | 0.138943376 | 0.020058784 | 4.21E-05 | 0.004897574 | 0.138939089 | -0.00059447 | 0.049184682 | 0.188795807 | 0.017163255 | 0.116207024 | 0.0124879 | ENSG0000019 | 7 | RP5-874C20.3 |
| VAT | CAD | ENSG0000010 | 0.005695742 | 0.090105006 | 0.003360622 | 0.138947077 | 0.006322662 | 0.061038047 | 0.003375284 | 0.138942872 | -0.00062692 | 0.046429039 | 1 | 0.513821934 | 1 | 0.434398716 | ENSG0000019 | 7 | RP5-874C20.3 |
| VAT | CAD | ENSG0000012 | -0.00769739 | 0.104965746 | 0.004747827 | 0.138947147 | -0.00761061 | 0.108952653 | 0.004747959 | 0.138943139 | -8.68E-05 | 0.046586359 | 1 | 0.526589031 | 1 | 0.535978172 | ENSG0000019 | 7 | RP5-874C20.3 |
| VAT | CAD | ENSG0000012 | -0.00465482 | 0.001615201 | 0.001476238 | 0.138945062 | -0.00467946 | 0.001525524 | 0.001476263 | 0.138941007 | 2.46E-05 | 0.035876406 | 1 | 0.080942347 | 1 | 0.081258362 | ENSG0000019 | 7 | RP5-874C20.3 |
| VAT | CAD | ENSG0000012 | 0.003432641 | 0.068168941 | 0.001882041 | 0.138946948 | 0.00358708 | 0.056843375 | 0.001883458 | 0.138942838 | -0.00015444 | 0.036281088 | 1 | 0.461578079 | 1 | 0.422852497 | ENSG0000019 | 7 | RP5-874C20.3 |
| VAT | CAD | ENSG0000013 | 0.001556356 | 0.761091544 | 0.005118786 | 0.13894787 | 0.001638886 | 0.748843068 | 0.005118873 | 0.138943843 | -8.25E-05 | 0.041720942 | 1 | 0.918627635 | 1 | 0.916133253 | ENSG0000019 | 8 | IGF2R |
| VAT | CAD | ENSG0000014 | 0.005631779 | 0.164963179 | 0.004055802 | 0.138947347 | 0.006491574 | 0.111463306 | 0.004078511 | 0.13894315 | -0.0008598 | 0.045735161 | 1 | 0.616100643 | 1 | 0.535978172 | ENSG0000019 | 8 | SLC25A29 |
| VAT | CAD | ENSG0000018 | -0.00925455 | 0.003120185 | 0.003131132 | 0.138945406 | -0.01173758 | 0.000338054 | 0.003274746 | 0.138940209 | 0.002483025 | 0.009646373 | 1 | 0.10764638 | 0.933027817 | 0.038876159 | ENSG0000019 | 7 | ZNF772 |
| VAT | CAD | ENSG0000018 | 0.00288358 | 0.462314356 | 0.003923014 | 0.138947742 | 0.002962195 | 0.450209344 | 0.003923091 | 0.138943709 | -7.86E-05 | 0.015409428 | 1 | 0.811043869 | 1 | 0.799828966 | ENSG0000019 | 7 | ZNF257 |
| VAT | CAD | ENSG0000018 | -0.00431956 | 0.270632518 | 0.003921137 | 0.13894755 | -0.00439784 | 0.26206154 | 0.003921274 | 0.138943513 | 7.83E-05 | 0.044884596 | 1 | 0.697428338 | 1 | 0.696035802 | ENSG0000019 | 7 | ZNF257 |
| VAT | CAD | ENSG0000020 | -0.00228513 | 0.352785665 | 0.002459239 | 0.13894765 | -0.00223982 | 0.36242283 | 0.002459293 | 0.138943636 | -4.53E-05 | 0.03124847 | 1 | 0.749857359 | 1 | 0.756460842 | ENSG0000019 | 7 | ZNF257 |
| VAT | CAD | ENSG0000023 | -0.00308331 | 0.332250414 | 0.00318001 | 0.138947628 | -0.00302447 | 0.34156971 | 0.003180073 | 0.138943614 | -5.88E-05 | 0.025249008 | 1 | 0.743723555 | 1 | 0.747632686 | ENSG0000019 | 2 | AL133458.1 |
| VAT | CAD | ENSG0000023 | -0.00306018 | 0.295511794 | 0.002925298 | 0.138947584 | -0.00300732 | 0.30394165 | 0.002925353 | 0.138943571 | -5.29E-05 | 0.026425113 | 1 | 0.712469718 | 1 | 0.724807082 | ENSG0000019 | 2 | AL133458.1 |
| VAT | CAD | ENSG0000025 | -0.0031856 | 0.220334954 | 0.002599134 | 0.138947468 | -0.00313763 | 0.227374271 | 0.002599198 | 0.138943457 | -4.80E-05 | 0.036178862 | 1 | 0.675304527 | 1 | 0.678432654 | ENSG0000019 | 6 | SND1 |
| VAT | CAD | ENSG0000026 | -0.00067018 | 0.840320627 | 0.003326209 | 0.138947885 | -0.00061439 | 0.853457225 | 0.003326277 | 0.138943862 | -5.58E-05 | 0.044580493 | 1 | 0.949931696 | 1 | 0.959970096 | ENSG0000019 | 6 | SND1 |
| VAT | Heart_Failure | ENSG0000000 | -0.00428398 | 0.106751299 | 0.002655952 | 0.076055087 | -0.00439307 | 0.098123341 | 0.002655991 | 0.07604845 | 0.000109097 | 0.000145843 | 1 | 0.637447692 | 1 | 0.624888429 | ENSG0000019 | 6 | SND1 |
| VAT | Heart_Failure | ENSG0000000 | -0.00328287 | 0.114022649 | 0.002077281 | 0.076055103 | -0.00332631 | 0.109311877 | 0.002077261 | 0.076048477 | 4.34E-05 | 0.010763657 | 1 | 0.655403408 | 1 | 0.628112053 | ENSG0000019 | 6 | SULT1A2 |
| VAT | Heart_Failure | ENSG0000000 | 0.003055538 | 0.199853015 | 0.002383466 | 0.076055237 | 0.003094109 | 0.194226771 | 0.002383425 | 0.076048614 | -3.86E-05 | 0.024758821 | 1 | 0.756191037 | 1 | 0.738152304 | ENSG0000019 | 6 | SULT1A2 |
| VAT | Heart_Failure | ENSG0000000 | 0.004378696 | 0.13547362 | 0.002933086 | 0.076055145 | 0.003583264 | 0.22502103 | 0.002953354 | 0.076048647 | 0.000795432 | 0.021688884 | 1 | 0.688213472 | 1 | 0.76157815 | ENSG0000019 | 6 | SULT1A2 |
| VAT | Heart_Failure | ENSG0000000 | 0.002386657 | 0.50427979 | 0.003574063 | 0.076055423 | 0.000331232 | 0.928895076 | 0.003711905 | 0.076048876 | 0.002055425 | 0.04037544 | 1 | 0.904941123 | 1 | 0.990517646 | ENSG0000019 | 6 | SULT1A2 |
| VAT | Heart_Failure | ENSG0000001 | -0.00025851 | 0.91794659 | 0.002509296 | 0.076055491 | -0.00020134 | 0.9360489 | 0.002509266 | 0.076048876 | -5.72E-05 | 0.00423219 | 1 | 0.984985763 | 1 | 0.990517646 | ENSG0000019 | 6 | SULT1A2 |
| VAT | Heart_Failure | ENSG0000001 | 0.000129369 | 0.950804221 | 0.00209685 | 0.076055492 | 9.26E-05 | 0.96476985 | 0.002096817 | 0.076048877 | 3.68E-05 | 0.018088972 | 1 | 0.992630426 | 1 | 0.993760632 | ENSG0000019 | 6 | SULT1A2 |
| VAT | Heart_Failure | ENSG0000001 | -0.00213374 | 0.423235704 | 0.002664442 | 0.076055393 | -0.00590873 | 0.062587744 | 0.003173132 | 0.076048336 | 0.00377499 | 0.02849475 | 1 | 0.87173921 | 1 | 0.543610282 | ENSG0000019 | 6 | SULT1A2 |
| VAT | Heart_Failure | ENSG0000002 | 0.000351197 | 0.58777763 | 0.000647894 | 0.076055447 | 0.000362775 | 0.575523756 | 0.000647888 | 0.076048828 | -1.16E-05 | 0.03282663 | 1 | 0.929665478 | 1 | 0.92458997 | ENSG0000019 | 7 | NEK5 |
| VAT | Heart_Failure | ENSG0000003 | -0.00173071 | 0.542871458 | 0.002844336 | 0.076055435 | -0.00091708 | 0.747992104 | 0.00285438 | 0.076048861 | -0.00081363 | 0.00072423 | 1 | 0.921324909 | 1 | 0.960126407 | ENSG0000019 | 7 | NEK5 |
| VAT | Heart_Failure | ENSG0000004 | 0.000267104 | 0.828124342 | 0.00123028 | 0.076055486 | 0.000242013 | 0.844049865 | 0.001230265 | 0.076048871 | 2.51E-05 | 0.010277241 | 1 | 0.972185844 | 1 | 0.969484287 | ENSG0000019 | 7 | NEK5 |
| VAT | Heart_Failure | ENSG0000005 | 0.002620645 | 0.368279248 | 0.002912792 | 0.076055367 | 0.00273684 | 0.34743619 | 0.00291285 | 0.076048739 | -0.00011619 | 0.000393092 | 1 | 0.834524405 | 1 | 0.821053473 | ENSG0000019 | 7 | NEK5 |
| VAT | Heart_Failure | ENSG0000005 | -0.0029879 | 0.420904974 | 0.003712355 | 0.076055392 | -0.00592475 | 0.135356517 | 0.003967526 | 0.076048529 | 0.002936849 | 0.035970945 | 1 | 0.870783317 | 1 | 0.670707336 | ENSG0000019 | 7 | PIWIL2 |
| VAT | Heart_Failure | ENSG0000005 | -0.00279518 | 0.251616732 | 0.002438156 | 0.076055288 | -0.00284204 | 0.243751557 | 0.00243813 | 0.076048665 | 4.69E-05 | 0.01783083 | 1 | 0.790959203 | 1 | 0.775956515 | ENSG0000019 | 7 | PIWIL2 |
| VAT | Heart_Failure | ENSG0000005 | 0.00056881 | 0.836977381 | 0.002764414 | 0.076055486 | 0.000520335 | 0.850696286 | 0.002764357 | 0.076048871 | 4.85E-05 | 0.00921759 | 1 | 0.972185844 | 1 | 0.972627071 | ENSG0000019 | 7 | PIWIL2 |
| VAT | Heart_Failure | ENSG0000005 | 0.002463484 | 0.016379215 | 0.001026297 | 0.076054594 | 0.002492713 | 0.01514613 | 0.001026283 | 0.076047956 | -2.92E-05 | 0.000245513 | 1 | 0.282541459 | 1 | 0.274406299 | ENSG0000019 | 7 | PIWIL2 |
| VAT | Heart_Failure | ENSG0000005 | 0.003394117 | 0.354235376 | 0.003663746 | 0.076055359 | 0.003467761 | 0.343883518 | 0.003663693 | 0.076048737 | -7.36E-05 | 0.008435496 | 1 | 0.826718867 | 1 | 0.81750087 | ENSG0000019 | 7 | PIWIL2 |
| VAT | Heart_Failure | ENSG0000006 | -0.00014478 | 0.961415367 | 0.00299267 | 0.076055493 | -8.63E-05 | 0.976995826 | 0.002992618 | 0.076048877 | -5.85E-05 | 0.006810982 | 1 | 0.993060803 | 1 | 0.995646729 | ENSG0000019 | 8 | ENTPD4 |
| VAT | Heart_Failure | ENSG0000006 | -0.00284525 | 0.094127135 | 0.001699647 | 0.076055056 | -0.00291745 | 0.086087687 | 0.001699739 | 0.076048417 | 7.22E-05 | 0.002331743 | 1 | 0.635185556 | 1 | 0.607677788 | ENSG0000019 | 8 | ENTPD4 |
| VAT | Heart_Failure | ENSG0000006 | -0.00526142 | 0.111467328 | 0.003305668 | 0.076055098 | -0.00533971 | 0.106237578 | 0.003305624 | 0.07604847 | 7.83E-05 | 0.002358473 | 1 | 0.64371215 | 1 | 0.628112053 | ENSG0000019 | 8 | ENTPD4 |
| VAT | Heart_Failure | ENSG0000006 | 0.000333038 | 0.758602183 | 0.001083696 | 0.076055478 | 0.0003141 | 0.771933977 | 0.001083678 | 0.076048864 | 1.89E-05 | 0.016779635 | 1 | 0.964199078 | 1 | 0.965780945 | ENSG0000019 | 7 | C1D |
| VAT | Heart_Failure | ENSG0000006 | -0.00266542 | 0.08465042 | 0.001545779 | 0.076055029 | -0.0027028 | 0.080380223 | 0.001545792 | 0.0760484 | 3.74E-05 | 0.018200657 | 1 | 0.623489698 | 1 | 0.600796054 | ENSG0000019 | 7 | C1D |
| VAT | Heart_Failure | ENSG0000007 | 0.002865728 | 0.2906763 | 0.002712113 | 0.076055319 | 0.007128524 | 0.01460563 | 0.002919108 | 0.076047946 | -0.0042628 | 7.91E-05 | 1 | 0.795899392 | 1 | 0.274406299 | ENSG0000019 | 8 | TBC1D9B |
| VAT | Heart_Failure | ENSG0000007 | -0.00374511 | 0.126704654 | 0.002452226 | 0.076055129 | -0.00378908 | 0.122302191 | 0.002452187 | 0.076048504 | 4.40E-05 | 0.016160134 | 1 | 0.673805094 | 1 | 0.652670563 | ENSG0000019 | 8 | TBC1D9B |
| VAT | Heart_Failure | ENSG0000007 | -0.00450758 | 0.233630408 | 0.003784512 | 0.076055272 | -0.00441874 | 0.242969093 | 0.003784466 | 0.076048664 | -8.88E-05 | 0.0030417 | 1 | 0.779051326 | 1 | 0.774790476 | ENSG0000019 | 8 | TBC1D9B |
| VAT | Heart_Failure | ENSG0000008 | -5.44E-05 | 0.977808252 | 0.001955512 | 0.076055493 | -8.40E-05 | 0.965716444 | 0.001955479 | 0.076048877 | 2.97E-05 | 0.037062363 | 1 | 0.994659842 | 1 | 0.993760632 | ENSG0000019 | 7 | C6orf141 |
| VAT | Heart_Failure | ENSG0000008 | 0.004526803 | 0.059890851 | 0.002405826 | 0.07605494 | 0.004491504 | 0.061908115 | 0.002405788 | 0.076048333 | 3.53E-05 | 0.048703225 | 1 | 0.546800541 | 1 | 0.54177348 | ENSG0000019 | 7 | C6orf141 |

| | | | | | | | | | | | | | | | | | | | |
|---|---|---|---|---|---|---|---|---|---|---|---|---|---|---|---|---|---|---|---|
| VAT | Heart_Failure | ENSG0000008 | 0.000722575 | 0.42911941 | 0.000913843 | 0.076055395 | -0.00064475 | 0.542115414 | 0.001057623 | 0.076048819 | 0.001367321 | 0.010232842 | 1 | 0.872618112 | 1 | 0.913136234 | ENSG0000019 | 7 | C6orf141 |
| VAT | Heart_Failure | ENSG0000008 | 0.011513686 | 8.62E-05 | 0.002932242 | 0.076053087 | 0.011575077 | 7.90E-05 | 0.002932205 | 0.076046445 | -6.14E-05 | 0.007869365 | 0.237839757 | 0.011325703 | 0.217930737 | 0.011470039 | ENSG0000019 | 3 | ZNF165 |
| VAT | Heart_Failure | ENSG0000008 | 0.000481076 | 0.868431371 | 0.002904147 | 0.076055489 | 0.000434798 | 0.880987342 | 0.002904116 | 0.076048873 | 4.63E-05 | 0.049017606 | 1 | 0.982255997 | 1 | 0.982211765 | ENSG0000019 | 3 | ZNF165 |
| VAT | Heart_Failure | ENSG0000008 | 0.00319082 | 0.121169147 | 0.002058743 | 0.076055118 | 0.003224474 | 0.117287917 | 0.002058707 | 0.076048494 | -3.37E-05 | 0.023501174 | 1 | 0.669281111 | 1 | 0.64556656 | ENSG0000019 | 3 | ZNF165 |
| VAT | Heart_Failure | ENSG0000008 | 0.003546614 | 0.245366437 | 0.003053006 | 0.076055282 | 0.003488009 | 0.253248059 | 0.00305297 | 0.076048673 | 5.86E-05 | 0.015725493 | 1 | 0.782288072 | 1 | 0.784151852 | ENSG0000019 | 3 | ZNF165 |
| VAT | Heart_Failure | ENSG0000009 | 0.001853942 | 0.510136343 | 0.002814863 | 0.076055425 | 0.00190648 | 0.498219454 | 0.002814846 | 0.076048805 | -5.25E-05 | 0.031129929 | 1 | 0.906664036 | 1 | 0.899741745 | ENSG0000019 | 3 | ZNF165 |
| VAT | Heart_Failure | ENSG0000009 | 0.000508077 | 0.746857202 | 0.001574035 | 0.076055477 | 0.00046257 | 0.768852868 | 0.001574032 | 0.076048863 | 4.55E-05 | 0.001421894 | 1 | 0.963913289 | 1 | 0.965438542 | ENSG0000019 | 6 | ZNF720 |
| VAT | Heart_Failure | ENSG0000009 | 0.000627294 | 0.42149114 | 0.000780374 | 0.076055392 | 0.000600395 | 0.441693872 | 0.000780406 | 0.076048784 | 2.69E-05 | 0.008421544 | 1 | 0.870783317 | 1 | 0.875281919 | ENSG0000019 | 6 | ZNF720 |
| VAT | Heart_Failure | ENSG0000009 | 0.003768421 | 0.29742702 | 0.003616638 | 0.076055324 | 0.004700609 | 0.194529268 | 0.003623384 | 0.076048614 | -0.00093219 | 3.06E-05 | 1 | 0.802442401 | 1 | 0.738152304 | ENSG0000019 | 8 | MRPL21 |
| VAT | Heart_Failure | ENSG0000009 | -0.00282667 | 0.339558353 | 0.002959747 | 0.076055351 | -0.0027778 | 0.34796908 | 0.002959709 | 0.076048739 | -4.89E-05 | 0.035504472 | 1 | 0.815649307 | 1 | 0.821053473 | ENSG0000019 | 8 | MRPL21 |
| VAT | Heart_Failure | ENSG0000009 | 0.003500782 | 0.198696321 | 0.002723758 | 0.076055235 | 0.003438497 | 0.206796068 | 0.002723721 | 0.076048628 | 6.23E-05 | 0.003090576 | 1 | 0.756191037 | 1 | 0.748456354 | ENSG0000019 | 5 | ZNF517 |
| VAT | Heart_Failure | ENSG0000010 | -0.00171521 | 0.471646042 | 0.00238288 | 0.076055412 | -0.0016742 | 0.482304133 | 0.00238285 | 0.0760488 | -4.10E-05 | 0.027720015 | 1 | 0.884782233 | 1 | 0.895800408 | ENSG0000019 | 5 | ZNF517 |
| VAT | Heart_Failure | ENSG0000010 | -0.00111752 | 0.673561653 | 0.00265276 | 0.076055465 | -0.00105658 | 0.690415902 | 0.002652774 | 0.076048852 | -6.09E-05 | 0.02000967 | 1 | 0.963913289 | 1 | 0.960065682 | ENSG0000019 | 8 | DCHS2 |
| VAT | Heart_Failure | ENSG0000010 | -0.00029191 | 0.910274223 | 0.00259034 | 0.076055491 | -0.00034592 | 0.893761594 | 0.002590297 | 0.076048874 | 5.40E-05 | 0.004265211 | 1 | 0.984985763 | 1 | 0.98412197 | ENSG0000019 | 7 | VEPH1 |
| VAT | Heart_Failure | ENSG0000010 | -0.00251734 | 0.186895512 | 0.001907336 | 0.076055221 | -0.00254922 | 0.181371342 | 0.001907317 | 0.076048598 | 3.19E-05 | 0.042032156 | 1 | 0.756191037 | 1 | 0.732920798 | ENSG0000019 | 7 | VEPH1 |
| VAT | Heart_Failure | ENSG0000010 | 0.000488432 | 0.894177371 | 0.003671858 | 0.07605549 | 0.000549797 | 0.880974115 | 0.003671814 | 0.076048873 | -6.14E-05 | 0.035588827 | 1 | 0.984985763 | 1 | 0.982211765 | ENSG0000019 | 8 | MAP3K5 |
| VAT | Heart_Failure | ENSG0000010 | 0.001839993 | 0.091461075 | 0.001090209 | 0.076055048 | 0.001855169 | 0.088812664 | 0.001090187 | 0.076048425 | -1.52E-05 | 0.042023012 | 1 | 0.623489698 | 1 | 0.608245538 | ENSG0000019 | 5 | STMN3 |
| VAT | Heart_Failure | ENSG0000010 | -0.0039859 | 0.288145197 | 0.003752488 | 0.076055317 | -0.00390205 | 0.298399944 | 0.003752434 | 0.076048708 | -8.38E-05 | 0.003281005 | 1 | 0.795233108 | 1 | 0.800488833 | ENSG0000019 | 4 | PCDHB11 |
| VAT | Heart_Failure | ENSG0000010 | -0.0044377 | 0.2203561 | 0.003620885 | 0.076055259 | -0.00449411 | 0.214536845 | 0.003620811 | 0.076048636 | 5.64E-05 | 0.021773026 | 1 | 0.772901704 | 1 | 0.756570334 | ENSG0000019 | 4 | PCDHB11 |
| VAT | Heart_Failure | ENSG0000010 | 0.00169236 | 0.318545309 | 0.001696682 | 0.076055338 | 0.001725689 | 0.309099003 | 0.00169665 | 0.076048715 | -3.33E-05 | 0.005041728 | 1 | 0.811008798 | 1 | 0.807108087 | ENSG0000019 | 8 | RPF2 |
| VAT | Heart_Failure | ENSG0000010 | 0.00056559 | 0.848389329 | 0.002958509 | 0.076055487 | 0.000653523 | 0.8251769 | 0.002958567 | 0.076048869 | -8.79E-05 | 0.008123251 | 1 | 0.977595833 | 1 | 0.96872144 | ENSG0000019 | 10 | MYO5A |
| VAT | Heart_Failure | ENSG0000010 | 0.001002551 | 0.793361528 | 0.003827287 | 0.076055482 | 0.000919861 | 0.810063169 | 0.003827257 | 0.076048868 | 8.27E-05 | 0.010470969 | 1 | 0.972185844 | 1 | 0.96824489 | ENSG0000019 | 5 | PIGN |
| VAT | Heart_Failure | ENSG0000010 | 0.001249969 | 0.703990996 | 0.003289923 | 0.07605547 | 0.001201224 | 0.715015422 | 0.003289865 | 0.076048856 | 4.87E-05 | 0.039116043 | 1 | 0.963913289 | 1 | 0.960065682 | ENSG0000019 | 5 | PIGN |
| VAT | Heart_Failure | ENSG0000010 | 0.005530453 | 0.117435213 | 0.003532407 | 0.07605511 | 0.005603451 | 0.112675194 | 0.003532435 | 0.076048484 | -7.30E-05 | 0.041800898 | 1 | 0.661746357 | 1 | 0.637261341 | ENSG0000019 | 5 | PIGN |
| VAT | Heart_Failure | ENSG0000010 | -0.00233214 | 0.343363831 | 0.002461252 | 0.076055353 | -0.00227792 | 0.35469636 | 0.002461235 | 0.076048743 | -5.42E-05 | 0.010032391 | 1 | 0.818147112 | 1 | 0.822657104 | ENSG0000019 | 9 | HHLA3 |
| VAT | Heart_Failure | ENSG0000010 | 0.000440091 | 0.765091116 | 0.001472851 | 0.076055479 | 0.000199491 | 0.892619631 | 0.001477817 | 0.076048874 | 0.0002406 | 0.04828452 | 1 | 0.967686555 | 1 | 0.98412197 | ENSG0000019 | 9 | HHLA3 |
| VAT | Heart_Failure | ENSG0000010 | 0.003034656 | 0.240210964 | 0.002583871 | 0.076055278 | 0.002897393 | 0.262223089 | 0.002584296 | 0.076048681 | 0.000137264 | 0.009176403 | 1 | 0.779051326 | 1 | 0.791877836 | ENSG0000019 | 9 | HHLA3 |
| VAT | Heart_Failure | ENSG0000010 | 0.000934149 | 0.607879284 | 0.001820586 | 0.076055452 | 0.001227783 | 0.501005477 | 0.001824589 | 0.076048806 | -0.00029363 | 0.016084579 | 1 | 0.938337149 | 1 | 0.901880257 | ENSG0000019 | 7 | BCO2 |
| VAT | Heart_Failure | ENSG0000010 | -0.00054718 | 0.858671506 | 0.003072899 | 0.076055488 | -0.00106869 | 0.728509749 | 0.003078827 | 0.076048858 | 0.00052151 | 0.006920138 | 1 | 0.979476883 | 1 | 0.960065682 | ENSG0000019 | 7 | BCO2 |
| VAT | Heart_Failure | ENSG0000010 | -0.00264742 | 0.099243188 | 0.001605924 | 0.076055069 | -0.00261315 | 0.103695254 | 0.001605917 | 0.076048464 | -3.43E-05 | 0.016029741 | 1 | 0.637447692 | 1 | 0.624888429 | ENSG0000019 | 8 | ENTPD6 |
| VAT | Heart_Failure | ENSG0000010 | -0.00265557 | 0.381558933 | 0.003034842 | 0.076055374 | -0.00253809 | 0.403016181 | 0.00303509 | 0.076048768 | -0.00011749 | 0.014473262 | 1 | 0.837245697 | 1 | 0.852871973 | ENSG0000019 | 5 | MPEG1 |
| VAT | Heart_Failure | ENSG0000010 | -0.00154994 | 0.533033623 | 0.002486338 | 0.076055432 | -0.00107105 | 0.667985626 | 0.002497114 | 0.076048848 | -0.00047889 | 0.039761517 | 1 | 0.9158182 | 1 | 0.957156395 | ENSG0000019 | 5 | MPEG1 |
| VAT | Heart_Failure | ENSG0000010 | -0.00019196 | 0.889864466 | 0.001386215 | 0.07605549 | 0.000386659 | 0.784696261 | 0.001415275 | 0.076048865 | -0.00057862 | 0.042784455 | 1 | 0.984985763 | 1 | 0.965780945 | ENSG0000019 | 10 | DNAH10 |
| VAT | Heart_Failure | ENSG0000010 | 0.00252431 | 0.346847464 | 0.002683373 | 0.076055355 | 0.002162909 | 0.420677422 | 0.002686018 | 0.076048776 | 0.000361401 | 0.002996362 | 1 | 0.820677452 | 1 | 0.871425441 | ENSG0000019 | 5 | RPS26 |
| VAT | Heart_Failure | ENSG0000010 | 0.000466686 | 0.903069459 | 0.003832054 | 0.076055491 | 0.000336029 | 0.930125814 | 0.003832161 | 0.076048876 | 0.000130657 | 0.004353568 | 1 | 0.984985763 | 1 | 0.990517646 | ENSG0000019 | 5 | RPS26 |
| VAT | Heart_Failure | ENSG0000010 | -0.00188169 | 0.064209841 | 0.00101673 | 0.076054958 | -0.0018637 | 0.066795835 | 0.001016719 | 0.076048352 | -1.80E-05 | 0.027329389 | 1 | 0.564941346 | 1 | 0.560354119 | ENSG0000019 | 5 | RPS26 |
| VAT | Heart_Failure | ENSG0000010 | -0.00207265 | 0.284424316 | 0.001936272 | 0.076055314 | -0.00204248 | 0.291486613 | 0.001936241 | 0.076048703 | -3.02E-05 | 0.035237518 | 1 | 0.795233108 | 1 | 0.79496349 | ENSG0000019 | 5 | RPS26 |
| VAT | Heart_Failure | ENSG0000010 | 0.009931808 | 0.006380826 | 0.003641289 | 0.076054332 | 0.009994086 | 0.006056989 | 0.003641241 | 0.076047701 | -6.23E-05 | 0.028669851 | 1 | 0.195678662 | 1 | 0.192152742 | ENSG0000019 | 4 | C14orf178 |
| VAT | Heart_Failure | ENSG0000010 | -0.00179596 | 0.54184263 | 0.002944057 | 0.076055435 | -0.00185543 | 0.528540522 | 0.00294402 | 0.076048815 | 5.95E-05 | 0.009940097 | 1 | 0.920870181 | 1 | 0.908819144 | ENSG0000019 | 10 | ZNF780A |
| VAT | Heart_Failure | ENSG0000011 | -0.00044307 | 0.705062607 | 0.001170611 | 0.076055471 | -0.00042459 | 0.716821654 | 0.001170591 | 0.076048856 | -1.85E-05 | 0.028974617 | 1 | 0.963913289 | 1 | 0.960065682 | ENSG0000019 | 10 | ZNF780A |
| VAT | Heart_Failure | ENSG0000011 | -0.00149873 | 0.663254001 | 0.003441999 | 0.076055463 | -0.00143316 | 0.67713474 | 0.00344198 | 0.07604885 | -6.56E-05 | 0.02892263 | 1 | 0.963584692 | 1 | 0.958291901 | ENSG0000019 | 9 | CCDC180 |
| VAT | Heart_Failure | ENSG0000011 | -0.00400936 | 0.201535813 | 0.003139205 | 0.076055238 | -0.00395132 | 0.208130504 | 0.00313915 | 0.07604863 | -5.80E-05 | 0.010291215 | 1 | 0.756841892 | 1 | 0.749921919 | ENSG0000019 | 9 | CCDC180 |
| VAT | Heart_Failure | ENSG0000011 | 0.001485798 | 0.346599451 | 0.00157861 | 0.076055355 | 0.001453424 | 0.357206943 | 0.001578606 | 0.076048745 | 3.24E-05 | 0.023009848 | 1 | 0.820677452 | 1 | 0.826396615 | ENSG0000019 | 9 | CCDC180 |
| VAT | Heart_Failure | ENSG0000011 | -0.00282505 | 0.39874248 | 0.003347726 | 0.076055382 | -0.00291527 | 0.383860974 | 0.00334779 | 0.076048758 | 9.02E-05 | 0.0159753 | 1 | 0.850486278 | 1 | 0.838615487 | ENSG0000019 | 9 | CCDC180 |
| VAT | Heart_Failure | ENSG0000011 | -0.00831038 | 0.009032593 | 0.003183043 | 0.076054429 | -0.00825811 | 0.009475041 | 0.003183015 | 0.076047826 | -5.23E-05 | 0.047919758 | 1 | 0.232990241 | 1 | 0.239368781 | ENSG0000019 | 6 | OCLN |
| VAT | Heart_Failure | ENSG0000011 | -0.00057276 | 0.526753373 | 0.000904883 | 0.07605543 | -0.00059209 | 0.512893972 | 0.000904871 | 0.07604881 | 1.93E-05 | 0.00681293 | 1 | 0.913789635 | 1 | 0.904747387 | ENSG0000019 | 6 | OCLN |
| VAT | Heart_Failure | ENSG0000011 | -0.00539011 | 0.148973332 | 0.003734913 | 0.076055168 | -0.00532188 | 0.154180939 | 0.003734859 | 0.07604856 | -6.82E-05 | 0.016494543 | 1 | 0.721315241 | 1 | 0.704981382 | ENSG0000019 | 6 | OCLN |
| VAT | Heart_Failure | ENSG0000011 | 0.004043443 | 0.03380545 | 0.001905136 | 0.07605479 | 0.004007995 | 0.035396087 | 0.001905118 | 0.076048186 | 3.54E-05 | 0.023908318 | 1 | 0.438042449 | 1 | 0.430366522 | ENSG0000019 | 8 | SLC22A20 |
| VAT | Heart_Failure | ENSG0000011 | -0.00387118 | 0.056148887 | 0.002026919 | 0.076054924 | -0.00382521 | 0.059132116 | 0.002026909 | 0.076048321 | -4.60E-05 | 0.009714032 | 1 | 0.533809522 | 1 | 0.532804147 | ENSG0000019 | 8 | SLC22A20 |
| VAT | Heart_Failure | ENSG0000011 | 0.007659161 | 0.01365409 | 0.003105595 | 0.076054544 | 0.007717379 | 0.012954894 | 0.003105565 | 0.076047913 | -5.82E-05 | 0.023049122 | 1 | 0.266586081 | 1 | 0.264855602 | ENSG0000019 | 8 | SLC22A20 |
| VAT | Heart_Failure | ENSG0000011 | 0.000525542 | 0.857501719 | 0.002926905 | 0.076055488 | 0.000474652 | 0.871171019 | 0.002926854 | 0.076048873 | 5.09E-05 | 0.016107457 | 1 | 0.978786081 | 1 | 0.980677945 | ENSG0000019 | 6 | HIST1H2BK |
| VAT | Heart_Failure | ENSG0000011 | 0.000719262 | 0.834048674 | 0.003433036 | 0.076055486 | 0.000648263 | 0.850224702 | 0.00343302 | 0.076048871 | 7.10E-05 | 0.018775124 | 1 | 0.972185844 | 1 | 0.97249075 | ENSG0000019 | 6 | HIST1H2BK |
| VAT | Heart_Failure | ENSG0000011 | -0.0027164 | 0.073386648 | 0.001517184 | 0.076054993 | -0.00273828 | 0.071094582 | 0.001517157 | 0.076048368 | 2.19E-05 | 0.043770991 | 1 | 0.603545069 | 1 | 0.580042752 | ENSG0000019 | 6 | HIST1H2BK |
| VAT | Heart_Failure | ENSG0000011 | -0.00063113 | 0.529851034 | 0.001004609 | 0.076055431 | -0.00061267 | 0.541952612 | 0.001004595 | 0.076048819 | -1.85E-05 | 0.017426579 | 1 | 0.915695819 | 1 | 0.913136234 | ENSG0000019 | 6 | HIST1H2BK |
| VAT | Heart_Failure | ENSG0000011 | -0.00482917 | 0.177741745 | 0.003583156 | 0.07605521 | -0.00495406 | 0.166822946 | 0.003583444 | 0.076048579 | 0.000124891 | 0.026917407 | 1 | 0.74593461 | 1 | 0.71714634 | ENSG0000019 | 6 | HIST1H2BK |
| VAT | Heart_Failure | ENSG0000011 | 0.002880365 | 0.321392993 | 0.002904764 | 0.07605534 | 0.00283267 | 0.329464635 | 0.002904725 | 0.076048728 | 4.77E-05 | 0.034593985 | 1 | 0.811008798 | 1 | 0.812752882 | ENSG0000019 | 6 | HIST1H2BK |
| VAT | Heart_Failure | ENSG0000012 | -0.00547844 | 0.142787352 | 0.003738295 | 0.076055158 | -0.00556959 | 0.136260782 | 0.003738319 | 0.07604853 | 9.11E-05 | 0.01478487 | 1 | 0.707715645 | 1 | 0.672705156 | ENSG0000019 | 6 | HIST1H2BK |
| VAT | Heart_Failure | ENSG0000012 | 0.004431789 | 0.124904008 | 0.002888074 | 0.076055126 | 0.004496057 | 0.119523374 | 0.002888049 | 0.076048499 | -6.43E-05 | 0.007612436 | 1 | 0.67041525 | 1 | 0.64556656 | ENSG0000019 | 6 | HIST1H2BK |
| VAT | Heart_Failure | ENSG0000012 | 0.00090999 | 0.30874787 | 0.000894028 | 0.076055331 | -0.00033533 | 0.744908808 | 0.001030651 | 0.07604886 | 0.001245322 | 0.015173847 | 1 | 0.811008798 | 1 | 0.960065682 | ENSG0000019 | 6 | HIST1H2BK |
| VAT | Heart_Failure | ENSG0000012 | -0.0010063 | 0.557436159 | 0.001715319 | 0.076055439 | -0.00098041 | 0.567614569 | 0.001715295 | 0.076048826 | -2.59E-05 | 0.049692879 | 1 | 0.92326913 | 1 | 0.921911445 | ENSG0000019 | 2 | HIST1H4K |
| VAT | Heart_Failure | ENSG0000012 | 0.002093042 | 0.341355945 | 0.002199751 | 0.076055352 | 0.002047643 | 0.351926733 | 0.002199734 | 0.076048742 | 4.54E-05 | 0.014967998 | 1 | 0.818052084 | 1 | 0.822453669 | ENSG0000019 | 5 | HRNR |
| VAT | Heart_Failure | ENSG0000012 | -0.00129876 | 0.234388608 | 0.001092193 | 0.076055272 | -0.00131669 | 0.22798776 | 0.00109218 | 0.07604865 | 1.79E-05 | 0.039046225 | 1 | 0.779051326 | 1 | 0.763693205 | ENSG0000019 | 5 | HRNR |
| VAT | Heart_Failure | ENSG0000012 | 0.000642057 | 0.756039393 | 0.002066578 | 0.076055478 | -0.0008286 | 0.700746762 | 0.002156061 | 0.076048854 | 0.001470659 | 0.016788868 | 1 | 0.964199078 | 1 | 0.960065682 | ENSG0000019 | 5 | HRNR |
| VAT | Heart_Failure | ENSG0000012 | -0.00037785 | 0.677159542 | 0.000907542 | 0.076055466 | -0.00039305 | 0.664941945 | 0.000907531 | 0.076048848 | 1.52E-05 | 0.034841866 | 1 | 0.963913289 | 1 | 0.957156395 | ENSG0000019 | 6 | ZNF311 |
| VAT | Heart_Failure | ENSG0000012 | -0.00518956 | 0.088421915 | 0.003045902 | 0.07605504 | -0.00513379 | 0.091897717 | 0.003045902 | 0.076048433 | -5.58E-05 | 0.048866352 | 1 | 0.623489698 | 1 | 0.616607764 | ENSG0000019 | 6 | ZNF311 |
| VAT | Heart_Failure | ENSG0000012 | 0.002692817 | 0.165155147 | 0.00194015 | 0.076055192 | 0.002757257 | 0.15528422 | 0.001940208 | 0.076048562 | -6.44E-05 | 0.006147637 | 1 | 0.729325131 | 1 | 0.704981382 | ENSG0000019 | 6 | ZNF311 |
| VAT | Heart_Failure | ENSG0000012 | -0.002415 | 0.456469801 | 0.003243039 | 0.076055406 | -0.00236416 | 0.465999109 | 0.003242991 | 0.076048794 | -5.08E-05 | 0.03869371 | 1 | 0.881202785 | 1 | 0.884473968 | ENSG0000019 | 6 | ZNF311 |
| VAT | Heart_Failure | ENSG0000012 | -0.00036362 | 0.720741017 | 0.00101721 | 0.076055473 | -0.00038463 | 0.705336551 | 0.001017194 | 0.076048855 | 2.10E-05 | 0.005439213 | 1 | 0.963913289 | 1 | 0.960065682 | ENSG0000019 | 6 | ZNF311 |
| VAT | Heart_Failure | ENSG0000012 | 0.001973233 | 0.021197605 | 0.000856269 | 0.076054664 | 0.001954263 | 0.022471415 | 0.000856265 | 0.076048064 | 1.90E-05 | 0.011453374 | 1 | 0.33431651 | 1 | 0.351455777 | ENSG0000019 | 6 | ZNF311 |
| VAT | Heart_Failure | ENSG0000012 | 0.005444101 | 0.046304648 | 0.002732145 | 0.076054873 | 0.005490687 | 0.044463972 | 0.002732099 | 0.076048246 | -4.66E-05 | 0.019764189 | 1 | 0.497279488 | 1 | 0.477344364 | ENSG0000019 | 6 | ZNF311 |
| VAT | Heart_Failure | ENSG0000012 | -0.00890391 | 0.011704046 | 0.00353196 | 0.076054501 | -0.00883957 | 0.012323117 | 0.003531925 | 0.076047899 | -6.43E-05 | 0.026157866 | 1 | 0.256374348 | 1 | 0.258531674 | ENSG0000019 | 6 | ZNF311 |
| VAT | Heart_Failure | ENSG0000012 | -0.00156775 | 0.260199316 | 0.001392417 | 0.076055295 | -0.00141163 | 0.311029061 | 0.001393426 | 0.076048717 | -0.00015612 | 0.004224574 | 1 | 0.79317431 | 1 | 0.809810347 | ENSG0000019 | 6 | ZNF311 |
| VAT | Heart_Failure | ENSG0000012 | -0.00336904 | 0.087773276 | 0.001973356 | 0.076055038 | -0.00331216 | 0.093282816 | 0.001973482 | 0.076048437 | -5.69E-05 | 0.049345969 | 1 | 0.623489698 | 1 | 0.617411444 | ENSG0000019 | 6 | ZNF311 |
| VAT | Heart_Failure | ENSG0000012 | -0.00054621 | 0.811865225 | 0.002294793 | 0.076055484 | -0.0005928 | 0.796154776 | 0.00229476 | 0.076048866 | 4.66E-05 | 0.007848029 | 1 | 0.972185844 | 1 | 0.965780945 | ENSG0000019 | 6 | ZNF311 |
| VAT | Heart_Failure | ENSG0000012 | -0.00038379 | 0.891362157 | 0.002810011 | 0.07605549 | 0.004542758 | 0.13026796 | 0.003002387 | 0.07604852 | -0.00492655 | 3.20E-06 | 1 | 0.984985763 | 1 | 0.665314041 | ENSG0000019 | 4 | ZNF71 |
| VAT | Heart_Failure | ENSG0000012 | 0.000673555 | 0.550141423 | 0.001127198 | 0.076055437 | 0.000692236 | 0.539128165 | 0.001127178 | 0.076048818 | -1.87E-05 | 0.020892138 | 1 | 0.92326913 | 1 | 0.911760867 | ENSG0000019 | 4 | ZNF71 |
| VAT | Heart_Failure | ENSG0000012 | 0.002031161 | 0.055964522 | 0.001062703 | 0.076054923 | 0.002053798 | 0.053281189 | 0.001062691 | 0.076048294 | -2.26E-05 | 0.007951464 | 1 | 0.533809522 | 1 | 0.508844578 | ENSG0000019 | 8 | RPL12 |
| VAT | Heart_Failure | ENSG0000012 | -0.00325575 | 0.252545405 | 0.002845462 | 0.076055289 | -0.00321449 | 0.258599237 | 0.00284541 | 0.076048678 | -4.13E-05 | 0.042027249 | 1 | 0.791175162 | 1 | 0.786050543 | ENSG0000019 | 8 | RPL12 |
| VAT | Heart_Failure | ENSG0000012 | -0.00085005 | 0.794385669 | 0.003261638 | 0.076055482 | -0.00091934 | 0.778043675 | 0.003261593 | 0.076048864 | 6.93E-05 | 0.005768306 | 1 | 0.972185844 | 1 | 0.965780945 | ENSG0000019 | 8 | RPL12 |
| VAT | Heart_Failure | ENSG0000012 | -0.00365105 | 0.164631652 | 0.002627291 | 0.076055192 | -0.00361436 | 0.168906934 | 0.002627239 | 0.076048581 | -3.67E-05 | 0.043843908 | 1 | 0.729325131 | 1 | 0.71714634 | ENSG0000019 | 8 | RPL12 |
| VAT | Heart_Failure | ENSG0000013 | 0.003326197 | 0.380485651 | 0.003792687 | 0.076055373 | 0.003242996 | 0.392514107 | 0.003792678 | 0.076048763 | 8.32E-05 | 0.015268594 | 1 | 0.837245697 | 1 | 0.842989349 | ENSG0000019 | 8 | RPL12 |
| VAT | Heart_Failure | ENSG0000013 | -0.00123468 | 0.713680528 | 0.003365 | 0.076055472 | -0.00117167 | 0.727688922 | 0.00336493 | 0.076048858 | -6.30E-05 | 0.005655131 | 1 | 0.963913289 | 1 | 0.960065682 | ENSG0000019 | 8 | RPL12 |
| VAT | Heart_Failure | ENSG0000013 | 0.00402012 | 0.068153136 | 0.002204017 | 0.076054974 | 0.003984191 | 0.070650844 | 0.002203988 | 0.076048367 | 3.59E-05 | 0.036084939 | 1 | 0.586322251 | 1 | 0.580042752 | ENSG0000019 | 7 | VPS13A |
| VAT | Heart_Failure | ENSG0000013 | 0.00186371 | 0.525541646 | 0.002935778 | 0.07605543 | 0.001802991 | 0.539121545 | 0.002935786 | 0.076048818 | 6.07E-05 | 0.03197183 | 1 | 0.91226097 | 1 | 0.911760867 | ENSG0000019 | 7 | VPS13A |
| VAT | Heart_Failure | ENSG0000013 | 0.000564953 | 0.703350347 | 0.001483588 | 0.07605547 | 0.000531925 | 0.719944488 | 0.001483616 | 0.076048857 | 3.30E-05 | 0.046296509 | 1 | 0.963913289 | 1 | 0.960065682 | ENSG0000019 | 7 | VPS13A |
| VAT | Heart_Failure | ENSG0000013 | -0.00980512 | 0.012945566 | 0.003945291 | 0.076054529 | -0.0097331 | 0.013623142 | 0.003945223 | 0.076047927 | -7.20E-05 | 0.011547335 | 1 | 0.266586081 | 1 | 0.267212404 | ENSG0000019 | 7 | VPS13A |
| VAT | Heart_Failure | ENSG0000013 | -0.00266196 | 0.126439888 | 0.001741786 | 0.076055128 | -0.0026099 | 0.134047541 | 0.001741868 | 0.076048526 | -5.21E-05 | 0.026316321 | 1 | 0.673695158 | 1 | 0.6690257 | ENSG0000019 | 8 | MRPL42 |
| VAT | Heart_Failure | ENSG0000013 | -0.00040848 | 0.739698359 | 0.001229436 | 0.076055476 | -0.00038516 | 0.754064688 | 0.001229414 | 0.076048862 | -2.33E-05 | 0.007541337 | 1 | 0.963913289 | 1 | 0.960414647 | ENSG0000019 | 8 | MRPL42 |
| VAT | Heart_Failure | ENSG0000013 | -0.00347729 | 0.358517856 | 0.003787093 | 0.076055361 | -0.00341027 | 0.367849369 | 0.003787047 | 0.07604875 | -6.70E-05 | 0.025425483 | 1 | 0.82873678 | 1 | 0.830566839 | ENSG0000019 | 6 | ZNF335 |
| VAT | Heart_Failure | ENSG0000013 | 0.002668523 | 0.321153964 | 0.002689798 | 0.076055339 | 0.002596021 | 0.334476045 | 0.002689792 | 0.076048731 | 7.25E-05 | 0.002932067 | 1 | 0.811008798 | 1 | 0.812752882 | ENSG0000019 | 6 | ZNF335 |
| VAT | Heart_Failure | ENSG0000013 | 0.004543061 | 0.054520364 | 0.002362875 | 0.076054916 | 0.0044935 | 0.057207276 | 0.002362851 | 0.076048312 | 4.96E-05 | 0.010085067 | 1 | 0.529845795 | 1 | 0.526306936 | ENSG0000019 | 6 | ZNF335 |
| VAT | Heart_Failure | ENSG0000013 | -0.00186435 | 0.505154964 | 0.002797645 | 0.076055424 | -0.00190615 | 0.495647987 | 0.002797599 | 0.076048804 | 4.18E-05 | 0.04166447 | 1 | 0.904963929 | 1 | 0.898557334 | ENSG0000019 | 6 | ZNF335 |
| VAT | Heart_Failure | ENSG0000013 | 0.00439196 | 0.000893267 | 0.001322015 | 0.07605377 | 0.004411477 | 0.000846964 | 0.001321992 | 0.076047139 | -1.95E-05 | 0.039910828 | 1 | 0.0632158 | 1 | 0.058440502 | ENSG0000019 | 3 | ZNF560 |
| VAT | Heart_Failure | ENSG0000013 | -5.37E-05 | 0.987837901 | 0.003521621 | 0.076055493 | -0.00010504 | 0.976203808 | 0.003521545 | 0.076048877 | 5.14E-05 | 0.0283649 | 1 | 0.996216992 | 1 | 0.995646729 | ENSG0000019 | 7 | ZNF273 |
| VAT | Heart_Failure | ENSG0000013 | -0.00463411 | 0.069339311 | 0.002551531 | 0.076054978 | -0.00451383 | 0.076944282 | 0.002552059 | 0.076048389 | -0.00012028 | 0.035061161 | 1 | 0.592493737 | 1 | 0.597746085 | ENSG0000019 | 7 | ZNF273 |
| VAT | Heart_Failure | ENSG0000013 | 0.003113204 | 0.082775733 | 0.001794554 | 0.076055023 | 0.004194915 | 0.024473449 | 0.001864721 | 0.076048087 | -0.00108171 | 0.032880719 | 1 | 0.619367186 | 1 | 0.371135825 | ENSG0000019 | 7 | ZNF667 |
| VAT | Heart_Failure | ENSG0000013 | -0.00133296 | 0.387958692 | 0.001543986 | 0.076055377 | -0.00129048 | 0.403260118 | 0.001543979 | 0.076048768 | -4.25E-05 | 0.001885728 | 1 | 0.840475658 | 1 | 0.852871973 | ENSG0000019 | 7 | ZNF667 |
| VAT | Heart_Failure | ENSG0000013 | 0.001260433 | 0.730161839 | 0.003654378 | 0.076055474 | 0.001334407 | 0.714992762 | 0.003654317 | 0.076048856 | -7.40E-05 | 0.005689938 | 1 | 0.963913289 | 1 | 0.960065682 | ENSG0000019 | 7 | SIRPA |
| VAT | Heart_Failure | ENSG0000013 | -0.00208229 | 0.552790746 | 0.003507993 | 0.076055438 | -0.00114048 | 0.745628526 | 0.003515555 | 0.07604886 | -0.00094181 | 5.20E-05 | 1 | 0.92326913 | 1 | 0.960126407 | ENSG0000019 | 6 | ZNF649 |
| VAT | Heart_Failure | ENSG0000014 | 0.001430842 | 0.690829499 | 0.003597522 | 0.076055468 | 0.001276884 | 0.722642082 | 0.003597565 | 0.076048857 | 0.000153958 | 4.88E-05 | 1 | 0.963913289 | 1 | 0.960065682 | ENSG0000019 | 6 | ZNF649 |
| VAT | Heart_Failure | ENSG0000014 | 0.005042632 | 0.173251203 | 0.003702827 | 0.076055204 | 0.004960896 | 0.180319658 | 0.003702787 | 0.076048597 | 8.17E-05 | 0.006262165 | 1 | 0.73906232 | 1 | 0.732359213 | ENSG0000019 | 6 | ZNF649 |
| VAT | Heart_Failure | ENSG0000014 | -0.00480164 | 0.133006664 | 0.003196083 | 0.076055141 | -0.00486099 | 0.128274691 | 0.003196038 | 0.076048516 | 5.93E-05 | 0.015393348 | 1 | 0.68290901 | 1 | 0.662992784 | ENSG0000019 | 4 | ADH4 |
| VAT | Heart_Failure | ENSG0000014 | 0.002378367 | 0.545783072 | 0.003937098 | 0.076055436 | 0.00245534 | 0.532857842 | 0.003937048 | 0.076048816 | -7.70E-05 | 0.013082381 | 1 | 0.921324909 | 1 | 0.908819144 | ENSG0000019 | 4 | ADH4 |
| VAT | Heart_Failure | ENSG0000014 | -0.00644593 | 0.076003002 | 0.003632807 | 0.076055002 | -0.00654173 | 0.071740743 | 0.003632752 | 0.076048371 | 9.58E-05 | 0.000466029 | 1 | 0.612031969 | 1 | 0.580042752 | ENSG0000019 | 4 | ADH4 |
| VAT | Heart_Failure | ENSG0000014 | 0.001261504 | 0.567588824 | 0.002206944 | 0.076055442 | 0.001225589 | 0.57865964 | 0.0022069 | 0.076048829 | 3.59E-05 | 0.017929929 | 1 | 0.92639479 | 1 | 0.92586219 | ENSG0000019 | 7 | ZNF248 |
| VAT | Heart_Failure | ENSG0000014 | -0.00012532 | 0.920598727 | 0.00125723 | 0.076055491 | -0.00010755 | 0.931826477 | 0.001257208 | 0.076048876 | -1.78E-05 | 0.048520403 | 1 | 0.984985763 | 1 | 0.990517646 | ENSG0000019 | 7 | ZNF248 |
| VAT | Heart_Failure | ENSG0000014 | -0.00289646 | 0.108028675 | 0.001802265 | 0.07605509 | -0.00326202 | 0.070960601 | 0.00180648 | 0.076048368 | 0.000365566 | 0.003320703 | 1 | 0.639208475 | 1 | 0.580042752 | ENSG0000019 | 9 | ZNF544 |
| VAT | Heart_Failure | ENSG0000014 | 0.006005852 | 0.045337157 | 0.003000649 | 0.076054868 | 0.006632701 | 0.027936802 | 0.003017361 | 0.076048123 | -0.00062685 | 0.048976941 | 1 | 0.49654981 | 1 | 0.387205367 | ENSG0000019 | 9 | ZNF544 |
| VAT | Heart_Failure | ENSG0000014 | 0.005283993 | 0.080776818 | 0.003025995 | 0.076055017 | 0.005328429 | 0.078253114 | 0.00302594 | 0.076048393 | -4.44E-05 | 0.039952539 | 1 | 0.619367186 | 1 | 0.597746085 | ENSG0000019 | 9 | ZNF544 |
| VAT | Heart_Failure | ENSG0000014 | 0.00763122 | 0.010300266 | 0.002974447 | 0.076054466 | 0.007581163 | 0.010810133 | 0.00297442 | 0.076047863 | 5.01E-05 | 0.041868456 | 1 | 0.240921465 | 1 | 0.25004753 | ENSG0000019 | 4 | ZNF251 |
| VAT | Heart_Failure | ENSG0000014 | 0.0035739 | 0.353174736 | 0.003849319 | 0.076055358 | 0.003515527 | 0.361085113 | 0.003849256 | 0.076048747 | 5.84E-05 | 0.039623089 | 1 | 0.826718867 | 1 | 0.829804255 | ENSG0000019 | 4 | ZNF251 |
| VAT | Heart_Failure | ENSG0000014 | 0.003914233 | 0.212093188 | 0.003136827 | 0.07605525 | 0.004435444 | 0.158172954 | 0.003142929 | 0.076048566 | -0.00052121 | 0.008463065 | 1 | 0.766257518 | 1 | 0.704981382 | ENSG0000019 | 8 | DDRGK1 |
| VAT | Heart_Failure | ENSG0000014 | 0.000201189 | 0.774025462 | 0.000700728 | 0.07605548 | 0.000440301 | 0.533155806 | 0.00070652 | 0.076048816 | -0.00023911 | 0.008253724 | 1 | 0.969984771 | 1 | 0.908819144 | ENSG0000019 | 7 | ZNF334 |
| VAT | Heart_Failure | ENSG0000014 | -0.00359919 | 0.088610285 | 0.002113716 | 0.07605504 | -0.00323655 | 0.126594512 | 0.002118615 | 0.076048513 | -0.00036264 | 0.012577881 | 1 | 0.623489698 | 1 | 0.658005371 | ENSG0000019 | 5 | SULT1C2 |
| VAT | Heart_Failure | ENSG0000014 | 0.000763752 | 0.694794844 | 0.001946576 | 0.076055469 | 0.000136044 | 0.944588474 | 0.001957357 | 0.076048876 | 0.000627708 | 0.002305549 | 1 | 0.963913289 | 1 | 0.991197882 | ENSG0000019 | 5 | SULT1C2 |
| VAT | Heart_Failure | ENSG0000014 | -0.00012823 | 0.954150816 | 0.002230333 | 0.076055493 | -6.78E-05 | 0.975737525 | 0.002230303 | 0.076048877 | -6.04E-05 | 0.000472087 | 1 | 0.993060803 | 1 | 0.995646729 | ENSG0000019 | 5 | SULT1C2 |
| VAT | Heart_Failure | ENSG0000014 | 0.001740086 | 0.493191857 | 0.002539376 | 0.07605542 | 0.001780384 | 0.48322681 | 0.002539334 | 0.0760488 | -4.03E-05 | 0.030237419 | 1 | 0.896935503 | 1 | 0.896227848 | ENSG0000019 | 7 | RPS6KL1 |
| VAT | Heart_Failure | ENSG0000014 | -0.00346258 | 0.21245516 | 0.002777074 | 0.07605525 | -0.00341987 | 0.218142181 | 0.00277703 | 0.07604864 | -4.27E-05 | 0.038188897 | 1 | 0.766257518 | 1 | 0.761153501 | ENSG0000019 | 7 | RPS6KL1 |
| VAT | Heart_Failure | ENSG0000014 | 0.002732058 | 0.035146205 | 0.001296857 | 0.0760548 | 0.00269561 | 0.037658258 | 0.001296864 | 0.076048203 | 3.64E-05 | 0.004236213 | 1 | 0.442938475 | 1 | 0.442284224 | ENSG0000019 | 7 | RPS6KL1 |
| VAT | Heart_Failure | ENSG0000015 | 0.002331606 | 0.349469667 | 0.002492046 | 0.076055356 | 0.002376188 | 0.340331246 | 0.002492038 | 0.076048735 | -4.46E-05 | 0.0461725 | 1 | 0.825801611 | 1 | 0.812752882 | ENSG0000019 | 6 | QRICH1 |
| VAT | Heart_Failure | ENSG0000015 | -0.00370117 | 0.260112549 | 0.003286636 | 0.076055295 | -0.00377591 | 0.250607456 | 0.003286611 | 0.076048671 | 7.47E-05 | 0.007127241 | 1 | 0.79317431 | 1 | 0.781555457 | ENSG0000019 | 6 | QRICH1 |
| VAT | Heart_Failure | ENSG0000015 | -0.00135783 | 0.257078876 | 0.001198096 | 0.076055293 | -0.00138125 | 0.248957725 | 0.001198078 | 0.076048669 | 2.34E-05 | 0.010472047 | 1 | 0.79317431 | 1 | 0.780278252 | ENSG0000019 | 7 | RP11-98J23.2 |
| VAT | Heart_Failure | ENSG0000015 | -0.00313857 | 0.276428046 | 0.002883711 | 0.076055308 | -0.00318592 | 0.269240747 | 0.00288367 | 0.076048686 | 4.73E-05 | 0.032173747 | 1 | 0.795233108 | 1 | 0.791877836 | ENSG0000019 | 8 | TMEM116 |
| VAT | Heart_Failure | ENSG0000015 | -0.00505814 | 0.055751587 | 0.00264412 | 0.076054922 | -0.00509672 | 0.053905926 | 0.002644078 | 0.076048297 | 3.86E-05 | 0.049750838 | 1 | 0.533809522 | 1 | 0.512041201 | ENSG0000019 | 6 | ZKSCAN8 |
| VAT | Heart_Failure | ENSG0000015 | 0.002104822 | 0.138077333 | 0.001419308 | 0.07605515 | 0.002071654 | 0.144391284 | 0.001419297 | 0.076048544 | 3.32E-05 | 0.005412736 | 1 | 0.694159268 | 1 | 0.696882186 | ENSG0000019 | 6 | ZKSCAN8 |
| VAT | Heart_Failure | ENSG0000015 | 0.001717925 | 0.442985997 | 0.002239327 | 0.076055401 | 0.001748982 | 0.43477597 | 0.002239283 | 0.076048782 | -3.11E-05 | 0.043128626 | 1 | 0.873500102 | 1 | 0.875281919 | ENSG0000019 | 6 | ZKSCAN8 |
| VAT | Heart_Failure | ENSG0000015 | 0.001914622 | 0.32150221 | 0.001931276 | 0.07605534 | 0.001944372 | 0.314033033 | 0.001931246 | 0.076048719 | -2.97E-05 | 0.038365881 | 1 | 0.811008798 | 1 | 0.812343568 | ENSG0000019 | 6 | ZKSCAN8 |

| | | | | | | | | | | | | | | | | | | | |
|---|---|---|---|---|---|---|---|---|---|---|---|---|---|---|---|---|---|---|---|
| VAT | Heart_Failure | ENSG0000015 | -0.00086601 | 0.681516662 | 0.002110178 | 0.076055467 | -0.00093595 | 0.657373408 | 0.002110176 | 0.076048846 | 6.99E-05 | 0.00034965 | 1 | 0.963913289 | 1 | 0.956411867 | ENSG0000019 | 6 | ZKSCAN8 |
| VAT | Heart_Failure | ENSG0000015 | -0.00236143 | 0.258316929 | 0.002089056 | 0.076055294 | -0.00231901 | 0.266964635 | 0.002089049 | 0.076048685 | -4.24E-05 | 0.023348512 | 1 | 0.79317431 | 1 | 0.791877836 | ENSG0000019 | 5 | MYL4 |
| VAT | Heart_Failure | ENSG0000016 | 0.000247167 | 0.849614103 | 0.001303548 | 0.076055487 | 0.000292154 | 0.822667017 | 0.001303584 | 0.076048869 | -4.50E-05 | 0.003708286 | 1 | 0.977595833 | 1 | 0.968476262 | ENSG0000019 | 5 | MYL4 |
| VAT | Heart_Failure | ENSG0000016 | -0.00313892 | 0.268264934 | 0.002835348 | 0.076055302 | -0.00309501 | 0.275011836 | 0.002835306 | 0.076048691 | -4.39E-05 | 0.040213529 | 1 | 0.795233108 | 1 | 0.791877836 | ENSG0000019 | 8 | ZNF568 |
| VAT | Heart_Failure | ENSG0000016 | -0.0077662 | 0.003248761 | 0.002638711 | 0.076054141 | -0.00781718 | 0.003051361 | 0.002638684 | 0.076047507 | 5.10E-05 | 0.018379231 | 1 | 0.122857446 | 1 | 0.126139004 | ENSG0000019 | 8 | ZNF568 |
| VAT | Heart_Failure | ENSG0000016 | -0.00386393 | 0.309900026 | 0.003805207 | 0.076055332 | -0.00293498 | 0.442483805 | 0.003821551 | 0.076048785 | -0.00092896 | 0.008841797 | 1 | 0.811008798 | 1 | 0.875576038 | ENSG0000019 | 8 | ZNF587 |
| VAT | Heart_Failure | ENSG0000016 | -0.00063415 | 0.729680932 | 0.001835201 | 0.076055474 | -0.00068638 | 0.708399125 | 0.001835199 | 0.076048855 | 5.22E-05 | 0.002023527 | 1 | 0.963913289 | 1 | 0.960065682 | ENSG0000019 | 8 | ZNF587 |
| VAT | Heart_Failure | ENSG0000016 | 0.001030365 | 0.748130743 | 0.003208806 | 0.076055477 | 0.001428624 | 0.656787915 | 0.00321507 | 0.076048846 | -0.00039826 | 0.049565777 | 1 | 0.963913289 | 1 | 0.956411867 | ENSG0000019 | 8 | ZNF587 |
| VAT | Heart_Failure | ENSG0000016 | -0.00042324 | 0.881054145 | 0.002828528 | 0.07605549 | -0.00047463 | 0.866738895 | 0.002828508 | 0.076048872 | 5.14E-05 | 0.033335558 | 1 | 0.984137328 | 1 | 0.980029484 | ENSG0000019 | 8 | ZNF587 |
| VAT | Heart_Failure | ENSG0000016 | 0.001082667 | 0.743708718 | 0.003311452 | 0.076055476 | 0.001148806 | 0.728647856 | 0.003311397 | 0.076048858 | -6.61E-05 | 0.006506445 | 1 | 0.963913289 | 1 | 0.960065682 | ENSG0000019 | 3 | FLVCR1-AS1 |
| VAT | Heart_Failure | ENSG0000016 | 0.003105304 | 0.327153103 | 0.003169113 | 0.076055343 | 0.00338262 | 0.285997343 | 0.00317038 | 0.076048699 | -0.00027732 | 0.003309335 | 1 | 0.811008798 | 1 | 0.794840503 | ENSG0000019 | 3 | FLVCR1-AS1 |
| VAT | Heart_Failure | ENSG0000016 | -0.0009209 | 0.784393459 | 0.003365881 | 0.076055481 | -0.00086172 | 0.797934861 | 0.003365839 | 0.076048867 | -5.92E-05 | 0.025634954 | 1 | 0.972185844 | 1 | 0.965780945 | ENSG0000019 | 6 | SH3BGRL2 |
| VAT | Heart_Failure | ENSG0000016 | 0.000399348 | 0.862415591 | 0.00230437 | 0.076055488 | 0.000369267 | 0.87268427 | 0.00230432 | 0.076048873 | 3.01E-05 | 0.048508059 | 1 | 0.980340622 | 1 | 0.980677945 | ENSG0000019 | 6 | SH3BGRL2 |
| VAT | Heart_Failure | ENSG0000016 | 0.000326055 | 0.876236547 | 0.002093558 | 0.076055489 | 0.000282986 | 0.892477553 | 0.002093559 | 0.076048874 | 4.31E-05 | 0.028584552 | 1 | 0.983439565 | 1 | 0.98412197 | ENSG0000019 | 6 | SH3BGRL2 |
| VAT | Heart_Failure | ENSG0000016 | 0.002599518 | 0.28654724 | 0.002439195 | 0.076055316 | 0.002664707 | 0.274628294 | 0.002439164 | 0.076048691 | -6.52E-05 | 0.000696378 | 1 | 0.795233108 | 1 | 0.791877836 | ENSG0000019 | 6 | SH3BGRL2 |
| VAT | Heart_Failure | ENSG0000016 | 0.000708833 | 0.194525197 | 0.000546387 | 0.07605523 | 0.000719552 | 0.187859255 | 0.000546382 | 0.076048606 | -1.07E-05 | 0.017942213 | 1 | 0.756191037 | 1 | 0.738152304 | ENSG0000019 | 5 | HLA-DRB5 |
| VAT | Heart_Failure | ENSG0000016 | -0.00206321 | 0.334364968 | 0.002137246 | 0.076055348 | -0.00190713 | 0.372355301 | 0.002137871 | 0.076048753 | -0.00015608 | 0.004818291 | 1 | 0.813847772 | 1 | 0.834172589 | ENSG0000019 | 5 | HLA-DRB5 |
| VAT | Heart_Failure | ENSG0000016 | 0.000879646 | 0.57612314 | 0.001573445 | 0.076055444 | 0.000849439 | 0.589289543 | 0.001573424 | 0.076048831 | 3.02E-05 | 0.013328313 | 1 | 0.92639479 | 1 | 0.927008193 | ENSG0000019 | 5 | HLA-DRB5 |
| VAT | Heart_Failure | ENSG0000016 | 0.004652333 | 0.208914971 | 0.003702447 | 0.076055247 | 0.00472431 | 0.201956709 | 0.003702442 | 0.076048623 | -7.20E-05 | 0.034024401 | 1 | 0.765744116 | 1 | 0.74232065 | ENSG0000019 | 5 | HLA-DRB5 |
| VAT | Heart_Failure | ENSG0000016 | 0.00191214 | 0.498633322 | 0.002825925 | 0.076055422 | 0.0018633 | 0.509661463 | 0.002825899 | 0.076048809 | 4.88E-05 | 0.036369604 | 1 | 0.900611917 | 1 | 0.904026759 | ENSG0000019 | 5 | HLA-DRB5 |
| VAT | Heart_Failure | ENSG0000016 | -0.00038466 | 0.738921632 | 0.001154164 | 0.076055476 | -0.00040094 | 0.728299283 | 0.001154142 | 0.076048858 | 1.63E-05 | 0.044213061 | 1 | 0.963913289 | 1 | 0.960065682 | ENSG0000019 | 5 | HLA-DRB5 |
| VAT | Heart_Failure | ENSG0000016 | 0.001783223 | 0.445019072 | 0.002334839 | 0.076055402 | 0.001752742 | 0.452828982 | 0.002334787 | 0.076048789 | 3.05E-05 | 0.044842936 | 1 | 0.873500102 | 1 | 0.879776385 | ENSG0000019 | 9 | CNGA1 |
| VAT | Heart_Failure | ENSG0000016 | -0.00677248 | 0.050148983 | 0.003457647 | 0.076054894 | -0.00660339 | 0.056216098 | 0.00345843 | 0.076048308 | -0.00016909 | 0.035287955 | 1 | 0.51300547 | 1 | 0.520659166 | ENSG0000019 | 9 | CNGA1 |
| VAT | Heart_Failure | ENSG0000016 | 0.001390843 | 0.088417382 | 0.000816314 | 0.07605504 | 0.001476896 | 0.070714252 | 0.000817178 | 0.076048367 | -8.61E-05 | 0.024813048 | 1 | 0.623489698 | 1 | 0.580042752 | ENSG0000019 | 5 | SLC34A3 |
| VAT | Heart_Failure | ENSG0000016 | 0.001957556 | 0.552071027 | 0.003291891 | 0.076055438 | 0.002115926 | 0.520467835 | 0.003292631 | 0.076048812 | -0.00015837 | 0.037775967 | 1 | 0.92326913 | 1 | 0.90770989 | ENSG0000019 | 5 | SLC34A3 |
| VAT | Heart_Failure | ENSG0000016 | -0.00315403 | 0.416418905 | 0.003881169 | 0.07605539 | -0.00323044 | 0.405213823 | 0.003881126 | 0.076048769 | 7.64E-05 | 0.014523502 | 1 | 0.869982193 | 1 | 0.854385142 | ENSG0000019 | 8 | CCDC69 |
| VAT | Heart_Failure | ENSG0000016 | -0.00478446 | 0.027161939 | 0.00216571 | 0.076054731 | -0.00482159 | 0.025990079 | 0.002165679 | 0.076048103 | 3.71E-05 | 0.024717519 | 1 | 0.390557318 | 1 | 0.383743978 | ENSG0000019 | 8 | CCDC69 |
| VAT | Heart_Failure | ENSG0000016 | -0.00285496 | 0.191139729 | 0.002184001 | 0.076055226 | -0.00290466 | 0.18352475 | 0.002183986 | 0.076048601 | 4.97E-05 | 0.007736116 | 1 | 0.756191037 | 1 | 0.733556367 | ENSG0000019 | 8 | CCDC69 |
| VAT | Heart_Failure | ENSG0000016 | -0.00027918 | 0.856177938 | 0.001540377 | 0.076055488 | -0.00030663 | 0.842215285 | 0.00154036 | 0.076048871 | 2.74E-05 | 0.02786895 | 1 | 0.978786081 | 1 | 0.969450344 | ENSG0000019 | 8 | CCDC69 |
| VAT | Heart_Failure | ENSG0000016 | -0.00054039 | 0.727838691 | 0.001552824 | 0.076055474 | -4.40E-05 | 0.977592311 | 0.001565566 | 0.076048877 | -0.00049641 | 0.012997909 | 1 | 0.963913289 | 1 | 0.995646729 | ENSG0000019 | 6 | ZNF534 |
| VAT | Heart_Failure | ENSG0000016 | -0.00216943 | 0.151888885 | 0.001514021 | 0.076055173 | -0.00219615 | 0.146899924 | 0.00151399 | 0.076048548 | 2.67E-05 | 0.01024431 | 1 | 0.721315241 | 1 | 0.701115705 | ENSG0000019 | 9 | NCOA6 |
| VAT | Heart_Failure | ENSG0000016 | 0.001270389 | 0.715676948 | 0.003487753 | 0.076055472 | 0.001212467 | 0.728111211 | 0.003487714 | 0.076048858 | 5.79E-05 | 0.038978503 | 1 | 0.963913289 | 1 | 0.960065682 | ENSG0000019 | 9 | NCOA6 |
| VAT | Heart_Failure | ENSG0000016 | -0.0011713 | 0.327487019 | 0.001196191 | 0.076055343 | -0.00119564 | 0.317530702 | 0.001196183 | 0.076048721 | 2.43E-05 | 0.01771503 | 1 | 0.811008798 | 1 | 0.812752882 | ENSG0000019 | 9 | NCOA6 |
| VAT | Heart_Failure | ENSG0000016 | 0.00223421 | 0.352714121 | 0.002404087 | 0.076055358 | 0.003373528 | 0.166303856 | 0.002437203 | 0.076048578 | -0.00113932 | 0.004499322 | 1 | 0.826718867 | 1 | 0.71714634 | ENSG0000019 | 6 | TTC37 |
| VAT | Heart_Failure | ENSG0000016 | 0.004855461 | 0.14408407 | 0.003323946 | 0.07605516 | 0.005433324 | 0.102768578 | 0.003330099 | 0.076048461 | -0.00057786 | 0.004765187 | 1 | 0.709748879 | 1 | 0.624888429 | ENSG0000019 | 6 | TTC37 |
| VAT | Heart_Failure | ENSG0000016 | -0.00184916 | 0.098142893 | 0.001118044 | 0.076055066 | -0.00182468 | 0.102673177 | 0.001118042 | 0.076048461 | -2.45E-05 | 0.016656203 | 1 | 0.637447692 | 1 | 0.624888429 | ENSG0000019 | 6 | RPL10A |
| VAT | Heart_Failure | ENSG0000016 | -4.43E-05 | 0.98721423 | 0.002765977 | 0.076055493 | -4.72E-06 | 0.998637732 | 0.002765925 | 0.076048877 | -3.96E-05 | 0.041990423 | 1 | 0.996216992 | 1 | 0.999361908 | ENSG0000019 | 6 | RPL10A |
| VAT | Heart_Failure | ENSG0000016 | 0.001549936 | 0.532500201 | 0.002483097 | 0.076055432 | 0.00158265 | 0.52387478 | 0.002483038 | 0.076048813 | -3.27E-05 | 0.035261875 | 1 | 0.9158182 | 1 | 0.908734352 | ENSG0000019 | 6 | COLGALT2 |
| VAT | Heart_Failure | ENSG0000016 | 0.002410424 | 0.200703049 | 0.001883802 | 0.076055237 | 0.002437098 | 0.19575676 | 0.001883766 | 0.076048616 | -2.67E-05 | 0.04303569 | 1 | 0.756749202 | 1 | 0.738152304 | ENSG0000019 | 6 | COLGALT2 |
| VAT | Heart_Failure | ENSG0000016 | 0.000415632 | 0.65223467 | 0.000922274 | 0.076055461 | 0.00043176 | 0.639675632 | 0.000922264 | 0.076048843 | -1.61E-05 | 0.030354581 | 1 | 0.96297588 | 1 | 0.953349011 | ENSG0000019 | 6 | COLGALT2 |
| VAT | Heart_Failure | ENSG0000016 | 0.005336788 | 0.095861273 | 0.003204775 | 0.07605506 | 0.005391476 | 0.092501657 | 0.003204729 | 0.076048435 | -5.47E-05 | 0.025616072 | 1 | 0.637447692 | 1 | 0.616677715 | ENSG0000019 | 6 | COLGALT2 |
| VAT | Heart_Failure | ENSG0000016 | 0.000852053 | 0.790070935 | 0.003200577 | 0.076055482 | 0.000901727 | 0.778140931 | 0.003200534 | 0.076048864 | -4.97E-05 | 0.046227939 | 1 | 0.972185844 | 1 | 0.965780945 | ENSG0000019 | 4 | GRIN3A |
| VAT | Heart_Failure | ENSG0000016 | 0.002120555 | 0.103508755 | 0.001302489 | 0.076055079 | 0.00190051 | 0.145391888 | 0.001305291 | 0.076048546 | 0.000220045 | 0.01080457 | 1 | 0.637447692 | 1 | 0.697881062 | ENSG0000019 | 7 | SCAMP5 |
| VAT | Heart_Failure | ENSG0000016 | -0.00085279 | 0.589054911 | 0.001578645 | 0.076055447 | -0.00116168 | 0.463864187 | 0.001585917 | 0.076048793 | 0.000308889 | 0.04269435 | 1 | 0.930570288 | 1 | 0.881725315 | ENSG0000019 | 7 | SCAMP5 |
| VAT | Heart_Failure | ENSG0000016 | -0.00245537 | 0.43935366 | 0.003175239 | 0.0760554 | -0.0029488 | 0.353920272 | 0.00318097 | 0.076048743 | 0.000493433 | 0.010633646 | 1 | 0.873500102 | 1 | 0.822657104 | ENSG0000019 | 7 | SCAMP5 |
| VAT | Heart_Failure | ENSG0000016 | 0.001384358 | 0.431636676 | 0.001760386 | 0.076055397 | 0.001348989 | 0.443498298 | 0.001760393 | 0.076048785 | 3.54E-05 | 0.039893657 | 1 | 0.872618112 | 1 | 0.875576038 | ENSG0000019 | 5 | DENND4B |
| VAT | Heart_Failure | ENSG0000017 | 0.001686312 | 0.438537551 | 0.002176826 | 0.076055399 | 0.001735885 | 0.425196772 | 0.002176825 | 0.076048778 | -4.96E-05 | 0.014631501 | 1 | 0.872855704 | 1 | 0.875281919 | ENSG0000019 | 8 | TYW1 |
| VAT | Heart_Failure | ENSG0000017 | 0.001097521 | 0.648917122 | 0.002410718 | 0.076055461 | 0.001142623 | 0.635514669 | 0.002410702 | 0.076048842 | -4.51E-05 | 0.029153448 | 1 | 0.961874179 | 1 | 0.950579326 | ENSG0000019 | 8 | TYW1 |
| VAT | Heart_Failure | ENSG0000017 | 0.002253506 | 0.321093376 | 0.002271188 | 0.076055339 | 0.002178294 | 0.337534193 | 0.002271304 | 0.076048733 | 7.52E-05 | 0.016116527 | 1 | 0.811008798 | 1 | 0.812752882 | ENSG0000019 | 8 | TYW1 |
| VAT | Heart_Failure | ENSG0000017 | -0.00090854 | 0.656913051 | 0.002045442 | 0.076055462 | -0.00086511 | 0.672331463 | 0.002045422 | 0.076048849 | -4.34E-05 | 0.01003947 | 1 | 0.963584692 | 1 | 0.957156395 | ENSG0000019 | 5 | ITPRIPL1 |
| VAT | Heart_Failure | ENSG0000017 | -0.00216985 | 0.125246126 | 0.001415311 | 0.076055126 | -0.00219744 | 0.120512224 | 0.001415302 | 0.076048501 | 2.76E-05 | 0.024036768 | 1 | 0.67041525 | 1 | 0.648517413 | ENSG0000019 | 5 | ITPRIPL1 |
| VAT | Heart_Failure | ENSG0000017 | -0.00110786 | 0.462244951 | 0.001506969 | 0.076055409 | -0.00114059 | 0.449117698 | 0.001506953 | 0.076048787 | 3.27E-05 | 0.007430459 | 1 | 0.88303338 | 1 | 0.877588101 | ENSG0000019 | 5 | ITPRIPL1 |
| VAT | Heart_Failure | ENSG0000017 | 0.0015911 | 0.07095052 | 0.000881106 | 0.076054984 | 0.001553202 | 0.077968452 | 0.000881199 | 0.076048392 | 3.79E-05 | 0.01250337 | 1 | 0.597905872 | 1 | 0.597746085 | ENSG0000019 | 5 | ITPRIPL1 |
| VAT | Heart_Failure | ENSG0000017 | 0.002522865 | 0.481246207 | 0.003582087 | 0.076055416 | 0.002460923 | 0.492073954 | 0.003582054 | 0.076048803 | 6.19E-05 | 0.036481728 | 1 | 0.892788619 | 1 | 0.897302672 | ENSG0000019 | 5 | ITPRIPL1 |
| VAT | Heart_Failure | ENSG0000017 | 0.001266704 | 0.57057415 | 0.002233233 | 0.076055443 | 0.001214786 | 0.586468929 | 0.002233225 | 0.076048831 | 5.19E-05 | 0.009087508 | 1 | 0.92639479 | 1 | 0.927008193 | ENSG0000019 | 6 | C1orf174 |
| VAT | Heart_Failure | ENSG0000017 | 0.001300163 | 0.581010646 | 0.002355755 | 0.076055445 | 0.00133657 | 0.570461736 | 0.002355721 | 0.076048827 | -3.64E-05 | 0.043153639 | 1 | 0.92639479 | 1 | 0.922350742 | ENSG0000019 | 6 | C1orf174 |
| VAT | Heart_Failure | ENSG0000017 | -0.00246055 | 0.315039372 | 0.002449027 | 0.076055335 | -0.00242633 | 0.321806297 | 0.002448971 | 0.076048724 | -3.42E-05 | 0.029490186 | 1 | 0.811008798 | 1 | 0.812752882 | ENSG0000019 | 7 | C9orf114 |
| VAT | Heart_Failure | ENSG0000017 | -0.00147247 | 0.314434778 | 0.001463745 | 0.076055335 | -0.00139281 | 0.341438865 | 0.001464076 | 0.076048736 | -7.97E-05 | 0.019027579 | 1 | 0.811008798 | 1 | 0.814313543 | ENSG0000019 | 5 | KIAA0753 |
| VAT | Heart_Failure | ENSG0000017 | -0.00223366 | 0.569085941 | 0.003922844 | 0.076055442 | -0.00228564 | 0.560121749 | 0.003922759 | 0.076048824 | 5.20E-05 | 0.045514117 | 1 | 0.92639479 | 1 | 0.921646963 | ENSG0000019 | 5 | KIAA0753 |
| VAT | Heart_Failure | ENSG0000017 | 0.007512204 | 0.026188892 | 0.003378692 | 0.076054721 | 0.00744421 | 0.027574682 | 0.003378677 | 0.076048119 | 6.80E-05 | 0.022564296 | 1 | 0.383927688 | 1 | 0.387205367 | ENSG0000019 | 5 | KIAA0753 |
| VAT | Heart_Failure | ENSG0000017 | 0.002480854 | 0.360706037 | 0.002714219 | 0.076055363 | 0.002528763 | 0.351502951 | 0.002714201 | 0.076048741 | -4.79E-05 | 0.039520082 | 1 | 0.829623884 | 1 | 0.822453669 | ENSG0000019 | 6 | ATG9A |
| VAT | Heart_Failure | ENSG0000017 | 0.002385224 | 0.114220666 | 0.001510108 | 0.076055104 | 0.002361108 | 0.117923261 | 0.001510091 | 0.076048495 | 2.41E-05 | 0.048700548 | 1 | 0.655403408 | 1 | 0.64556656 | ENSG0000019 | 3 | L3MBTL3 |
| VAT | Heart_Failure | ENSG0000017 | -0.00243951 | 0.194895154 | 0.001881999 | 0.076055231 | -0.0024094 | 0.200457253 | 0.001881974 | 0.076048621 | -3.01E-05 | 0.040647804 | 1 | 0.756191037 | 1 | 0.74232065 | ENSG0000019 | 3 | L3MBTL3 |
| VAT | Heart_Failure | ENSG0000017 | 0.001032605 | 0.657603917 | 0.002329758 | 0.076055462 | 0.000601514 | 0.796908204 | 0.002337333 | 0.076048866 | 0.000431091 | 0.022747866 | 1 | 0.963584692 | 1 | 0.965780945 | ENSG0000019 | 3 | L3MBTL3 |
| VAT | Heart_Failure | ENSG0000017 | -0.00095117 | 0.789785826 | 0.003567928 | 0.076055482 | -0.00089113 | 0.802768661 | 0.003567872 | 0.076048867 | -6.00E-05 | 0.023521093 | 1 | 0.972185844 | 1 | 0.966005801 | ENSG0000019 | 3 | L3MBTL3 |
| VAT | Heart_Failure | ENSG0000017 | 0.005097956 | 0.081011266 | 0.002921713 | 0.076055018 | 0.00515886 | 0.077447442 | 0.002921714 | 0.07604839 | -6.09E-05 | 0.026533015 | 1 | 0.619367186 | 1 | 0.597746085 | ENSG0000019 | 3 | L3MBTL3 |
| VAT | Heart_Failure | ENSG0000017 | -0.00206846 | 0.068962831 | 0.001137352 | 0.076054977 | -0.00203553 | 0.073502371 | 0.001137359 | 0.076048377 | -3.29E-05 | 0.003635574 | 1 | 0.591109981 | 1 | 0.584037962 | ENSG0000019 | 7 | TGM2 |
| VAT | Heart_Failure | ENSG0000017 | -0.00390277 | 0.090552279 | 0.002305926 | 0.076055046 | -0.00393497 | 0.087917474 | 0.002305883 | 0.076048422 | 3.22E-05 | 0.048167577 | 1 | 0.623489698 | 1 | 0.608245538 | ENSG0000019 | 7 | TGM2 |
| VAT | Heart_Failure | ENSG0000017 | -0.00030111 | 0.885944964 | 0.002099217 | 0.07605549 | -0.00035666 | 0.865087025 | 0.002099195 | 0.076048872 | 5.56E-05 | 0.001104066 | 1 | 0.98475518 | 1 | 0.980029484 | ENSG0000019 | 5 | PJA2 |
| VAT | Heart_Failure | ENSG0000017 | -0.01516232 | 4.21E-25 | 0.001464968 | 0.076038778 | -0.01773778 | 1.19E-22 | 0.001810986 | 0.076033908 | 0.002575457 | 0.015569498 | 1.16E-21 | 1.16E-21 | 3.30E-19 | 3.30E-19 | ENSG0000020 | 2 | AL590822.1 |
| VAT | Heart_Failure | ENSG0000017 | -2.21E-06 | 0.999310141 | 0.002555273 | 0.076055493 | -5.71E-05 | 0.982158767 | 0.002555247 | 0.076048877 | 5.49E-05 | 0.008317687 | 1 | 0.999535911 | 1 | 0.995646729 | ENSG0000020 | 2 | AL590822.1 |
| VAT | Heart_Failure | ENSG0000018 | 0.005174543 | 0.108804191 | 0.003226841 | 0.076055092 | 0.005231276 | 0.104978198 | 0.003226819 | 0.076048467 | -5.67E-05 | 0.040272245 | 1 | 0.64079958 | 1 | 0.626491505 | ENSG0000020 | 2 | AC020907.2 |
| VAT | Heart_Failure | ENSG0000018 | 0.004017421 | 0.197624928 | 0.003118269 | 0.076055234 | 0.004081812 | 0.190530321 | 0.003118235 | 0.076048609 | -6.44E-05 | 0.010457508 | 1 | 0.756191037 | 1 | 0.738152304 | ENSG0000020 | 3 | AC099048.1 |
| VAT | Heart_Failure | ENSG0000018 | -0.00043048 | 0.678087354 | 0.001037111 | 0.076055466 | -0.00041504 | 0.689013465 | 0.001037095 | 0.076048852 | -1.54E-05 | 0.049901513 | 1 | 0.963913289 | 1 | 0.960065682 | ENSG0000020 | 8 | EFCAB2 |
| VAT | Heart_Failure | ENSG0000018 | -0.00434225 | 0.052096433 | 0.002235577 | 0.076054904 | -0.00438734 | 0.049700392 | 0.002235542 | 0.076048276 | 4.51E-05 | 0.006947846 | 1 | 0.518983358 | 1 | 0.500631689 | ENSG0000020 | 1 | CHML |
| VAT | Heart_Failure | ENSG0000018 | -0.00251995 | 0.276686111 | 0.002316563 | 0.076055308 | -0.00510097 | 0.035252822 | 0.002422747 | 0.076048185 | 0.002581023 | 0.000276334 | 1 | 0.795233108 | 1 | 0.430366522 | ENSG0000020 | 1 | CHML |
| VAT | Heart_Failure | ENSG0000018 | 6.53E-05 | 0.950049784 | 0.001042537 | 0.076055492 | -0.00047779 | 0.648263807 | 0.001047386 | 0.076048844 | 0.000543101 | 7.86E-08 | 1 | 0.992630426 | 1 | 0.955604921 | ENSG0000020 | 1 | CHML |
| VAT | Heart_Failure | ENSG0000018 | -0.00051959 | 0.727153126 | 0.001489149 | 0.076055474 | -0.00054635 | 0.713702792 | 0.001489142 | 0.076048856 | 2.68E-05 | 0.041244034 | 1 | 0.963913289 | 1 | 0.960065682 | ENSG0000020 | 6 | TATDN3 |
| VAT | Heart_Failure | ENSG0000018 | 0.00635959 | 0.081698976 | 0.003653014 | 0.07605502 | 0.006143149 | 0.092760269 | 0.003654424 | 0.076048436 | 0.000216442 | 0.04314758 | 1 | 0.619367186 | 1 | 0.616911666 | ENSG0000020 | 6 | TATDN3 |
| VAT | Heart_Failure | ENSG0000018 | 0.001043072 | 0.436024546 | 0.001339119 | 0.076055398 | 0.001018494 | 0.44690745 | 0.001339099 | 0.076048787 | 2.46E-05 | 0.016282075 | 1 | 0.872618112 | 1 | 0.877588101 | ENSG0000020 | 6 | TATDN3 |
| VAT | Heart_Failure | ENSG0000018 | -0.00241749 | 0.327583578 | 0.00246936 | 0.076055343 | -0.00236865 | 0.337442727 | 0.002469324 | 0.076048733 | -4.88E-05 | 0.009114527 | 1 | 0.811008798 | 1 | 0.812752882 | ENSG0000020 | 3 | RAET1G |
| VAT | Heart_Failure | ENSG0000018 | -0.00056346 | 0.792056896 | 0.002137221 | 0.076055482 | -0.00060426 | 0.777381437 | 0.002137199 | 0.076048864 | 4.08E-05 | 0.019597587 | 1 | 0.972185844 | 1 | 0.965780945 | ENSG0000020 | 7 | ECT2L |
| VAT | Heart_Failure | ENSG0000018 | 0.00390685 | 0.052650486 | 0.002016146 | 0.076054907 | 0.003944862 | 0.050389175 | 0.00201613 | 0.076048279 | -3.80E-05 | 0.025649096 | 1 | 0.518983358 | 1 | 0.502072644 | ENSG0000020 | 3 | TMEM244 |
| VAT | Heart_Failure | ENSG0000018 | 0.000677097 | 0.850491302 | 0.003592187 | 0.076055487 | 0.000610576 | 0.865030014 | 0.003592158 | 0.076048872 | 6.65E-05 | 0.028076577 | 1 | 0.977595833 | 1 | 0.980029484 | ENSG0000020 | 3 | FAM229B |
| VAT | Heart_Failure | ENSG0000018 | -0.0050008 | 0.012934037 | 0.002011918 | 0.076054529 | -0.0050433 | 0.012185806 | 0.002011907 | 0.076047896 | 4.25E-05 | 0.015831174 | 1 | 0.266586081 | 1 | 0.258531674 | ENSG0000020 | 3 | FAM229B |
| VAT | Heart_Failure | ENSG0000018 | -0.00028961 | 0.844102343 | 0.001472738 | 0.076055487 | -0.00031859 | 0.828732505 | 0.001472713 | 0.07604887 | 2.90E-05 | 0.007128909 | 1 | 0.975350458 | 1 | 0.96872144 | ENSG0000020 | 9 | METTL10 |
| VAT | Heart_Failure | ENSG0000018 | 0.005776716 | 0.119419336 | 0.003709638 | 0.076055115 | 0.005849407 | 0.114833864 | 0.003709585 | 0.076048489 | -7.27E-05 | 0.010367535 | 1 | 0.663876855 | 1 | 0.641581914 | ENSG0000020 | 9 | METTL10 |
| VAT | Heart_Failure | ENSG0000018 | 0.00249531 | 0.063867153 | 0.001346553 | 0.076054957 | 0.002518174 | 0.061467119 | 0.001346527 | 0.076048331 | -2.29E-05 | 0.015872043 | 1 | 0.564941346 | 1 | 0.540284228 | ENSG0000020 | 9 | METTL10 |
| VAT | Heart_Failure | ENSG0000018 | 0.000768773 | 0.493696353 | 0.001123212 | 0.07605542 | 0.000747045 | 0.505982144 | 0.001123194 | 0.076048808 | 2.17E-05 | 0.009891709 | 1 | 0.897038797 | 1 | 0.902059208 | ENSG0000020 | 4 | C6orf164 |
| VAT | Heart_Failure | ENSG0000018 | -0.00333761 | 0.347003836 | 0.00354907 | 0.076055355 | -0.00341919 | 0.335338458 | 0.003549023 | 0.076048732 | 8.16E-05 | 0.003171177 | 1 | 0.820677452 | 1 | 0.812752882 | ENSG0000020 | 4 | C6orf164 |
| VAT | Heart_Failure | ENSG0000018 | 0.000404582 | 0.697471975 | 0.001040765 | 0.076055469 | 0.003934038 | 0.043085275 | 0.001944763 | 0.076048238 | -0.00352946 | 0.031685967 | 1 | 0.963913289 | 1 | 0.477344364 | ENSG0000020 | 6 | C6orf163 |
| VAT | Heart_Failure | ENSG0000018 | -0.00568667 | 0.038180915 | 0.002743323 | 0.076054822 | -0.00577465 | 0.03529232 | 0.002743307 | 0.076048185 | 8.80E-05 | 0.000231712 | 1 | 0.462190028 | 1 | 0.430366522 | ENSG0000020 | 6 | C6orf163 |
| VAT | Heart_Failure | ENSG0000018 | -0.00118021 | 0.383434261 | 0.001354088 | 0.076055374 | -0.001157 | 0.392849697 | 0.001354069 | 0.076048763 | -2.32E-05 | 0.024858885 | 1 | 0.837245697 | 1 | 0.842989349 | ENSG0000020 | 5 | OOEP |
| VAT | Heart_Failure | ENSG0000018 | 0.003910861 | 0.177834805 | 0.002902409 | 0.07605521 | 0.003997706 | 0.168392933 | 0.002902382 | 0.076048581 | -8.68E-05 | 0.000297648 | 1 | 0.74593461 | 1 | 0.71714634 | ENSG0000020 | 5 | OOEP |
| VAT | Heart_Failure | ENSG0000018 | 0.005350857 | 0.04375745 | 0.002653653 | 0.076054858 | 0.005292922 | 0.046091399 | 0.002653678 | 0.076048256 | 5.79E-05 | 0.03410465 | 1 | 0.491334625 | 1 | 0.483462092 | ENSG0000020 | 4 | FOXO6 |
| VAT | Heart_Failure | ENSG0000018 | -0.00104584 | 0.718468363 | 0.002901027 | 0.076055473 | -0.00113532 | 0.695540685 | 0.002901044 | 0.076048853 | 8.95E-05 | 0.001943095 | 1 | 0.963913289 | 1 | 0.960065682 | ENSG0000020 | 4 | FOXO6 |
| VAT | Heart_Failure | ENSG0000018 | -0.0004444 | 0.787468957 | 0.001648378 | 0.076055482 | -0.00048497 | 0.768592515 | 0.001648351 | 0.076048863 | 4.06E-05 | 0.000799235 | 1 | 0.972185844 | 1 | 0.965438542 | ENSG0000020 | 4 | FOXO6 |
| VAT | Heart_Failure | ENSG0000018 | -0.00269991 | 0.436020441 | 0.003466167 | 0.076055398 | -0.00275065 | 0.42743784 | 0.003466104 | 0.076048779 | 5.07E-05 | 0.039724603 | 1 | 0.872618112 | 1 | 0.875281919 | ENSG0000020 | 8 | INPP5B |
| VAT | Heart_Failure | ENSG0000018 | -0.00511139 | 0.124402211 | 0.003326519 | 0.076055125 | -0.00577439 | 0.08328539 | 0.003334063 | 0.076048409 | 0.000662998 | 0.003396207 | 1 | 0.67041525 | 1 | 0.604015714 | ENSG0000020 | 8 | CLPSL1 |
| VAT | Heart_Failure | ENSG0000018 | 0.001017882 | 0.444262602 | 0.001330537 | 0.076055402 | 0.001041639 | 0.433698346 | 0.001330524 | 0.076048781 | -2.38E-05 | 0.028455437 | 1 | 0.873500102 | 1 | 0.875281919 | ENSG0000020 | 8 | CLPSL1 |
| VAT | Heart_Failure | ENSG0000018 | 0.001750148 | 0.504235804 | 0.002620609 | 0.076055423 | 0.001610804 | 0.538874265 | 0.002621254 | 0.076048818 | 0.000139344 | 0.027140757 | 1 | 0.904941123 | 1 | 0.911760867 | ENSG0000020 | 8 | CLPSL1 |
| VAT | Heart_Failure | ENSG0000018 | -0.00367637 | 0.290400592 | 0.00347731 | 0.076055319 | -0.003539 | 0.308824868 | 0.003477473 | 0.076048715 | -0.00013737 | 0.003268244 | 1 | 0.795899392 | 1 | 0.807108087 | ENSG0000020 | 8 | CLPSL1 |
| VAT | Heart_Failure | ENSG0000018 | -0.00584498 | 0.000308823 | 0.001620091 | 0.076053462 | -0.00581165 | 0.000334166 | 0.001620067 | 0.076046869 | -3.33E-05 | 0.006225832 | 0.852352241 | 0.032782778 | 0.922297947 | 0.036010503 | ENSG0000020 | 5 | ASAH2B |
| VAT | Heart_Failure | ENSG0000018 | -0.00340501 | 0.239553495 | 0.002895158 | 0.076055277 | -0.0034481 | 0.233651055 | 0.002895108 | 0.076048655 | 4.31E-05 | 0.039997109 | 1 | 0.779051326 | 1 | 0.76771061 | ENSG0000020 | 5 | ASAH2B |
| VAT | Heart_Failure | ENSG0000019 | 0.00059036 | 0.644080728 | 0.001277833 | 0.07605546 | 0.000614185 | 0.630767223 | 0.001277829 | 0.076048841 | -2.38E-05 | 0.037514777 | 1 | 0.960091406 | 1 | 0.950282497 | ENSG0000020 | 5 | ASAH2B |
| VAT | Heart_Failure | ENSG0000019 | -0.00992109 | 1.39E-07 | 0.00188383 | 0.076051164 | -0.00671724 | 0.002291595 | 0.002202664 | 0.076047425 | -0.00320385 | 0.005006287 | 0.000384045 | 4.27E-05 | 1 | 0.110580144 | ENSG0000020 | 5 | AGAP6 |
| VAT | Heart_Failure | ENSG0000019 | -0.00147707 | 0.609920777 | 0.002895151 | 0.076055452 | -0.00154456 | 0.593688801 | 0.002895151 | 0.076048832 | 6.75E-05 | 0.01241343 | 1 | 0.93938691 | 1 | 0.929058336 | ENSG0000020 | 5 | AGAP6 |
| VAT | Heart_Failure | ENSG0000019 | 0.002767743 | 0.37820113 | 0.003140821 | 0.076055372 | 0.00281674 | 0.369811494 | 0.003140783 | 0.076048751 | -4.90E-05 | 0.049532981 | 1 | 0.837245697 | 1 | 0.830566839 | ENSG0000020 | 9 | C10orf128 |
| VAT | Heart_Failure | ENSG0000019 | -0.00023789 | 0.850924921 | 0.001265768 | 0.076055488 | -0.00021476 | 0.86527049 | 0.001265754 | 0.076048872 | -2.31E-05 | 0.023251937 | 1 | 0.977595833 | 1 | 0.980029484 | ENSG0000020 | 9 | C10orf128 |
| VAT | Heart_Failure | ENSG0000019 | -0.00028549 | 0.899274956 | 0.002255464 | 0.076055491 | -0.00032063 | 0.886955374 | 0.002255431 | 0.076048874 | 3.51E-05 | 0.040285597 | 1 | 0.984985763 | 1 | 0.98412197 | ENSG0000020 | 6 | AGAP7 |
| VAT | Heart_Failure | ENSG0000019 | 0.000994932 | 0.67824014 | 0.002398201 | 0.076055466 | 0.001044864 | 0.66305832 | 0.002398154 | 0.076048847 | -4.99E-05 | 0.002690723 | 1 | 0.963913289 | 1 | 0.957156395 | ENSG0000020 | 6 | AGAP7 |
| VAT | Heart_Failure | ENSG0000019 | 0.002080813 | 0.260863021 | 0.001850672 | 0.076055296 | 0.001977661 | 0.285277866 | 0.001850808 | 0.076048699 | 0.000103152 | 0.000262941 | 1 | 0.79317431 | 1 | 0.794840503 | ENSG0000020 | 6 | AGAP7 |
| VAT | Heart_Failure | ENSG0000019 | 0.003966641 | 0.273870701 | 0.003625174 | 0.076055306 | 0.003871903 | 0.285490468 | 0.003625147 | 0.076048699 | 9.47E-05 | 0.002108197 | 1 | 0.795233108 | 1 | 0.794840503 | ENSG0000020 | 6 | AGAP7 |
| VAT | Heart_Failure | ENSG0000019 | 0.001118879 | 0.644871864 | 0.002427605 | 0.07605546 | 0.000689582 | 0.776688648 | 0.002431211 | 0.076048864 | 0.000429297 | 0.001388128 | 1 | 0.960521503 | 1 | 0.965780945 | ENSG0000020 | 6 | AGAP7 |
| VAT | Heart_Failure | ENSG0000019 | 0.002949307 | 0.417024073 | 0.003633961 | 0.07605539 | 0.003008548 | 0.407722903 | 0.003633916 | 0.07604877 | -5.92E-05 | 0.038814507 | 1 | 0.869982193 | 1 | 0.855404372 | ENSG0000020 | 6 | AGAP7 |
| VAT | Heart_Failure | ENSG0000019 | -0.00183962 | 0.00474411 | 0.00065144 | 0.076054248 | 2.07E-05 | 0.982219161 | 0.000930664 | 0.076048877 | -0.00186036 | 0.005127668 | 1 | 0.1596798 | 1 | 0.995646729 | ENSG0000020 | 6 | LRRC37A5P |
| VAT | Heart_Failure | ENSG0000020 | 0.002174044 | 0.367355939 | 0.002411751 | 0.076055366 | 0.002137575 | 0.375440307 | 0.002411715 | 0.076048754 | 3.65E-05 | 0.046568517 | 1 | 0.833801308 | 1 | 0.834984084 | ENSG0000020 | 6 | BMS1P1 |
| VAT | Heart_Failure | ENSG0000020 | 0.00149338 | 0.454680353 | 0.00199747 | 0.076055406 | 0.001545876 | 0.438981934 | 0.001997477 | 0.076048783 | -5.25E-05 | 0.006696255 | 1 | 0.879409792 | 1 | 0.875281919 | ENSG0000020 | 6 | BMS1P1 |
| VAT | Heart_Failure | ENSG0000020 | 0.000503333 | 0.890587169 | 0.003658969 | 0.07605549 | 0.000560996 | 0.878143127 | 0.003658904 | 0.076048873 | -5.77E-05 | 0.028131012 | 1 | 0.984985763 | 1 | 0.982211765 | ENSG0000020 | 6 | DAXX |
| VAT | Heart_Failure | ENSG0000020 | -0.00010189 | 0.96300525 | 0.002196806 | 0.076055493 | -0.00014564 | 0.94713998 | 0.002196792 | 0.076048876 | 4.38E-05 | 0.020366641 | 1 | 0.993060803 | 1 | 0.991197882 | ENSG0000020 | 6 | DAXX |
| VAT | Heart_Failure | ENSG0000020 | 0.00617409 | 0.001048165 | 0.001883902 | 0.076053817 | 0.002929597 | 0.162006056 | 0.002095025 | 0.076048572 | 0.003244494 | 0.000401 | 1 | 0.070559418 | 1 | 0.70956483 | ENSG0000020 | 4 | RING1 |
| VAT | Heart_Failure | ENSG0000020 | -0.00046058 | 0.683776689 | 0.00113078 | 0.076055467 | -0.00158942 | 0.191003119 | 0.001215504 | 0.07604861 | 0.001128832 | 0.011370312 | 1 | 0.963913289 | 1 | 0.738152304 | ENSG0000020 | 4 | RING1 |
| VAT | Heart_Failure | ENSG0000020 | -0.00878317 | 0.001683277 | 0.002796213 | 0.076053953 | -0.00590952 | 0.04015136 | 0.002879609 | 0.07604822 | -0.00287365 | 2.99E-05 | 1 | 0.089726808 | 1 | 0.461626679 | ENSG0000020 | 4 | RING1 |
| VAT | Heart_Failure | ENSG0000020 | -0.00162863 | 0.110495393 | 0.001020461 | 0.076055095 | -0.00229934 | 0.026864179 | 0.001038782 | 0.076048112 | 0.000670709 | 0.000563409 | 1 | 0.643497633 | 1 | 0.385375739 | ENSG0000020 | 3 | HSD17B8 |
| VAT | Heart_Failure | ENSG0000020 | 0.00190086 | 0.118436055 | 0.001217428 | 0.076055113 | 0.003147682 | 0.011645211 | 0.00124773 | 0.076047884 | -0.00124682 | 5.18E-06 | 1 | 0.663876855 | 1 | 0.257959522 | ENSG0000020 | 3 | HSD17B8 |
| VAT | Heart_Failure | ENSG0000020 | 0.005381983 | 0.014244082 | 0.00219578 | 0.076054555 | 0.004405685 | 0.0462442 | 0.002210399 | 0.076048257 | 0.000976297 | 0.000125643 | 1 | 0.271128728 | 1 | 0.483462092 | ENSG0000020 | 3 | HSD17B8 |
| VAT | Heart_Failure | ENSG0000020 | 0.002451008 | 0.002310812 | 0.000804377 | 0.076054044 | 0.002793396 | 0.000577761 | 0.000811593 | 0.076047028 | -0.00034239 | 0.001564142 | 1 | 0.11189196 | 1 | 0.04577203 | ENSG0000020 | 3 | HSD17B8 |

| | | | | | | | | | | | | | | | | | | |
|---|---|---|---|---|---|---|---|---|---|---|---|---|---|---|---|---|---|---|
| VAT | Heart_Failure | ENSG000002( | -0.00225057 | 0.030322233 | 0.001039109 | 0.076054761 | -0.00138177 | 0.217358975 | 0.00112013 | 0.076048639 | -0.00086879 | 0.037836783 | 1 | 0.412262874 | 1 | 0.759489439 | ENSG00000002( | 3 | HSD17B8 |
| VAT | Heart_Failure | ENSG00000002( | -0.00017332 | 0.830964101 | 0.000811947 | 0.076055486 | -0.00100091 | 0.22504943 | 0.000825011 | 0.076048647 | 0.000827591 | 1.59E-08 | 1 | 0.97185844 | 1 | 0.76157815 | ENSG00000002( | 3 | HSD17B8 |
| VAT | Heart_Failure | ENSG00000002( | -0.00076573 | 0.206817501 | 0.000606583 | 0.076055244 | -0.0013053 | 0.034123178 | 0.000616109 | 0.076048176 | 0.000539569 | 5.96E-07 | 1 | 0.762104543 | 1 | 0.425960325 | ENSG00000002( | 3 | HSD17B8 |
| VAT | Heart_Failure | ENSG00000002( | 0.000461683 | 0.869093651 | 0.002801304 | 0.076055489 | 0.00092835 | 0.740858866 | 0.002807088 | 0.07604886 | -0.00046667 | 0.010346567 | 1 | 0.982255997 | 1 | 0.960065682 | ENSG00000002( | 3 | HSD17B8 |
| VAT | Heart_Failure | ENSG00000002( | -0.0023776 | 0.235473808 | 0.002004084 | 0.076055273 | -0.00334696 | 0.103264376 | 0.002054317 | 0.076048463 | 0.000969361 | 0.031951599 | 1 | 0.779051326 | 1 | 0.624888429 | ENSG00000002( | 3 | HSD17B8 |
| VAT | Heart_Failure | ENSG00000002( | -0.00447521 | 0.177405961 | 0.003317954 | 0.076055209 | -0.00534796 | 0.10932579 | 0.0033399 | 0.076048417 | 0.000872747 | 0.022869298 | 1 | 0.74593461 | 1 | 0.628112053 | ENSG00000002( | 4 | OXLD1 |
| VAT | Heart_Failure | ENSG00000002( | -0.00139531 | 0.195195901 | 0.001077156 | 0.076055231 | -0.00224826 | 0.039506148 | 0.001091981 | 0.076048215 | 0.000852951 | 2.05E-06 | 1 | 0.756191037 | 1 | 0.460071599 | ENSG00000002( | 4 | OXLD1 |
| VAT | Heart_Failure | ENSG00000002( | 0.001344123 | 0.68605477 | 0.003325266 | 0.07605468 | -4.13E-05 | 0.990168043 | 0.003350981 | 0.076048877 | 0.001385416 | 0.000854894 | 1 | 0.963913289 | 1 | 0.998073832 | ENSG00000002( | 4 | OXLD1 |
| VAT | Heart_Failure | ENSG00000002( | -0.00517536 | 0.009653969 | 0.001999756 | 0.076054448 | -0.00627649 | 0.001924175 | 0.002023559 | 0.076047375 | 0.001101138 | 0.000383293 | 1 | 0.237475638 | 1 | 0.098346714 | ENSG00000002( | 4 | OXLD1 |
| VAT | Heart_Failure | ENSG00000002( | -0.00248658 | 0.389183651 | 0.002887678 | 0.076055377 | -0.00243732 | 0.398640158 | 0.002887642 | 0.076048766 | -4.93E-05 | 0.029571503 | 1 | 0.840717801 | 1 | 0.848955891 | ENSG00000002( | 4 | OXLD1 |
| VAT | Heart_Failure | ENSG00000002( | -3.23E-05 | 0.987915184 | 0.002131049 | 0.076055493 | -0.00057001 | 0.789873796 | 0.002139064 | 0.076048866 | 0.000537728 | 0.003852156 | 1 | 0.996216992 | 1 | 0.965780945 | ENSG00000002( | 4 | OXLD1 |
| VAT | Heart_Failure | ENSG00000002( | -0.0021432 | 0.238821558 | 0.001819448 | 0.076055276 | -0.00211105 | 0.245937358 | 0.001819436 | 0.076048667 | -3.22E-05 | 0.038800136 | 1 | 0.779051326 | 1 | 0.778245128 | ENSG00000002( | 4 | OXLD1 |
| VAT | Heart_Failure | ENSG00000002( | -0.00740571 | 0.00947721 | 0.002854549 | 0.076054443 | -0.00733646 | 0.010166593 | 0.00285452 | 0.076047846 | -6.92E-05 | 0.002919402 | 1 | 0.237475638 | 1 | 0.241894809 | ENSG00000002( | 6 | COL11A2 |
| VAT | Heart_Failure | ENSG00000002( | 5.67E-06 | 0.997040468 | 0.001529569 | 0.076055493 | -3.24E-05 | 0.983118548 | 0.001529545 | 0.076048877 | 3.80E-05 | 0.000882072 | 1 | 0.998849979 | 1 | 0.995646729 | ENSG00000002( | 8 | HLA-DOA |
| VAT | Heart_Failure | ENSG00000002( | -0.0018857 | 0.243501125 | 0.001616845 | 0.076055281 | -0.00209443 | 0.196038275 | 0.001619916 | 0.076048616 | 0.00020873 | 0.038453361 | 1 | 0.779655573 | 1 | 0.738152304 | ENSG00000002( | 8 | HLA-DOA |
| VAT | Heart_Failure | ENSG00000002( | 0.004543518 | 0.183115513 | 0.003413035 | 0.076055216 | 0.00447625 | 0.189679735 | 0.003413002 | 0.076048608 | 6.73E-05 | 0.01636313 | 1 | 0.754326591 | 1 | 0.738152304 | ENSG00000002( | 8 | HLA-DOA |
| VAT | Heart_Failure | ENSG00000002( | -0.00374396 | 0.037343946 | 0.001798263 | 0.076054816 | -0.00475161 | 0.008869418 | 0.00181563 | 0.076047808 | 0.001007642 | 6.00E-05 | 1 | 0.458085743 | 1 | 0.236280118 | ENSG00000002( | 8 | BRD2 |
| VAT | Heart_Failure | ENSG00000002( | 0.000272435 | 0.821212652 | 0.001205539 | 0.076055485 | -0.00112566 | 0.366855994 | 0.001247436 | 0.07604875 | 0.001398095 | 1.31E-05 | 1 | 0.972185844 | 1 | 0.830566839 | ENSG00000002( | 10 | HLA-DMA |
| VAT | Heart_Failure | ENSG00000021 | 0.00214345 | 0.262074782 | 0.001911232 | 0.076055297 | 0.00211326 | 0.268846747 | 0.001911201 | 0.076048686 | 3.02E-05 | 0.032086639 | 1 | 0.79317431 | 1 | 0.791877836 | ENSG00000002( | 10 | HLA-DMA |
| VAT | Heart_Failure | ENSG00000021 | -0.00610693 | 0.056282091 | 0.003199272 | 0.076054924 | -0.00623541 | 0.051304446 | 0.003199403 | 0.076048284 | 0.000128481 | 0.002007785 | 1 | 0.533809522 | 1 | 0.50391556 | ENSG00000002( | 9 | TAP2 |
| VAT | Heart_Failure | ENSG00000021 | -0.0003854 | 0.659714866 | 0.000875291 | 0.076055463 | -0.00040078 | 0.64703653 | 0.000875282 | 0.076048844 | 1.54E-05 | 0.032081517 | 1 | 0.963584692 | 1 | 0.955604921 | ENSG00000002( | 9 | TAP2 |
| VAT | Heart_Failure | ENSG00000021 | 0.002010939 | 0.106756667 | 0.001246748 | 0.076055087 | 0.001974865 | 0.113194152 | 0.001246757 | 0.076048485 | 3.61E-05 | 0.0040912 | 1 | 0.637447692 | 1 | 0.638887238 | ENSG00000002( | 9 | TAP2 |
| VAT | Heart_Failure | ENSG00000021 | 0.008801328 | 8.49E-06 | 0.001976725 | 0.076052399 | 0.007146307 | 0.000376644 | 0.002009673 | 0.076046903 | 0.001655022 | 5.07E-06 | 0.02343312 | 0.00146457 | 1 | 0.036010503 | ENSG00000002( | 9 | TAP2 |
| VAT | Heart_Failure | ENSG00000021 | 0.000273187 | 0.832755016 | 0.001293683 | 0.076055486 | -0.00083363 | 0.525100161 | 0.00131176 | 0.076048814 | 0.001106815 | 3.54E-07 | 1 | 0.972185844 | 1 | 0.908819144 | ENSG00000002( | 9 | TAP2 |
| VAT | Heart_Failure | ENSG00000021 | -0.00014682 | 0.894203848 | 0.00110404 | 0.07605549 | -2.36E-05 | 0.982939174 | 0.001104852 | 0.076048877 | -0.0001232 | 0.004718322 | 1 | 0.984985763 | 1 | 0.995646729 | ENSG00000002( | 9 | HLA-DRA |
| VAT | Heart_Failure | ENSG00000021 | 8.47E-05 | 0.965715651 | 0.001970372 | 0.076055493 | 0.000486717 | 0.805486193 | 0.001976488 | 0.076048867 | -0.00040203 | 0.010176729 | 1 | 0.993060803 | 1 | 0.967412756 | ENSG00000002( | 9 | HLA-DRA |
| VAT | Heart_Failure | ENSG00000021 | 0.000872839 | 0.754017678 | 0.002785541 | 0.076055478 | 0.000766624 | 0.783165626 | 0.002785735 | 0.076048865 | 0.000106215 | 0.011332097 | 1 | 0.963913289 | 1 | 0.965780945 | ENSG00000002( | 9 | HLA-DRA |
| VAT | Heart_Failure | ENSG00000021 | -0.00158124 | 0.592084796 | 0.002951086 | 0.076055448 | -0.00154158 | 0.601400507 | 0.002951026 | 0.076048834 | -3.97E-05 | 0.047055919 | 1 | 0.930611638 | 1 | 0.931462064 | ENSG00000002( | 5 | NOTCH4 |
| VAT | Heart_Failure | ENSG00000021 | -0.00999539 | 0.00274095 | 0.003336908 | 0.076054093 | -0.01004956 | 0.002598041 | 0.003336842 | 0.07604746 | 5.42E-05 | 0.018917656 | 1 | 0.122016507 | 1 | 0.117595502 | ENSG00000002( | 5 | NOTCH4 |
| VAT | Heart_Failure | ENSG00000021 | 0.000717099 | 0.579223534 | 0.001293189 | 0.076055445 | 0.000692517 | 0.592293875 | 0.001293179 | 0.076048832 | 2.46E-05 | 0.025714316 | 1 | 0.92639479 | 1 | 0.928824486 | ENSG00000002( | 5 | NOTCH4 |
| VAT | Heart_Failure | ENSG00000022 | -0.00109498 | 0.774230598 | 0.003817302 | 0.07605548 | -0.0013806 | 0.717682762 | 0.00381848 | 0.076048856 | 0.000285624 | 0.004806155 | 1 | 0.969984771 | 1 | 0.960065682 | ENSG00000002( | 9 | AGER |
| VAT | Heart_Failure | ENSG00000022 | 0.000246765 | 0.866580329 | 0.001468805 | 0.076055489 | 0.000371078 | 0.800678963 | 0.001469799 | 0.076048867 | -0.00012431 | 0.025797257 | 1 | 0.982255997 | 1 | 0.965780945 | ENSG00000002( | 9 | AGER |
| VAT | Heart_Failure | ENSG00000022 | -0.00172311 | 0.07105548 | 0.000954566 | 0.076054984 | -0.00210376 | 0.029541774 | 0.000966133 | 0.076048138 | 0.000380645 | 0.012888366 | 1 | 0.597905872 | 1 | 0.398269521 | ENSG00000002( | 9 | AGER |
| VAT | Heart_Failure | ENSG00000022 | 0.000977323 | 0.313315797 | 0.000969288 | 0.076055334 | 0.001400761 | 0.154241565 | 0.00098319 | 0.0760486 | -0.00042344 | 0.010282099 | 1 | 0.811008798 | 1 | 0.704981382 | ENSG00000002( | 9 | AGER |
| VAT | Heart_Failure | ENSG00000022 | -0.00193605 | 0.463121409 | 0.002638681 | 0.076055409 | -0.00199153 | 0.450396794 | 0.002638649 | 0.076048788 | 5.55E-05 | 0.007669361 | 1 | 0.88303338 | 1 | 0.877892056 | ENSG00000002( | 9 | AGER |
| VAT | Heart_Failure | ENSG00000022 | -0.00127298 | 0.230414728 | 0.001061446 | 0.076055269 | -0.0023164 | 0.033606226 | 0.00109019 | 0.076048172 | 0.00104342 | 2.76E-05 | 1 | 0.777367927 | 1 | 0.423530522 | ENSG00000002( | 9 | AGER |
| VAT | Heart_Failure | ENSG00000022 | 0.002897904 | 0.192316423 | 0.00222273 | 0.076055228 | 0.002388427 | 0.283870578 | 0.002228705 | 0.076048698 | 0.000509477 | 0.001939133 | 1 | 0.756191037 | 1 | 0.794840503 | ENSG00000002( | 9 | AGER |
| VAT | Heart_Failure | ENSG00000022 | -0.00212994 | 0.573275921 | 0.003781627 | 0.07605544 | -0.0037802 | 0.326128479 | 0.003849713 | 0.076048726 | 0.001650258 | 0.022217298 | 1 | 0.92639479 | 1 | 0.812752882 | ENSG00000002( | 9 | AGER |
| VAT | Heart_Failure | ENSG00000022 | -0.00144827 | 0.289613276 | 0.00136762 | 0.076055318 | -0.00218958 | 0.114199169 | 0.001386163 | 0.076048487 | 0.000741311 | 0.00105582 | 1 | 0.795899392 | 1 | 0.640561135 | ENSG00000002( | 9 | AGER |
| VAT | Heart_Failure | ENSG00000022 | 0.001698068 | 0.353788591 | 0.001831262 | 0.076055359 | 0.001669096 | 0.362055069 | 0.001831239 | 0.07604874 | 2.90E-05 | 0.04466479 | 1 | 0.826718867 | 1 | 0.830566839 | ENSG00000002( | 9 | AGER |
| VAT | Heart_Failure | ENSG00000022 | -0.00536276 | 0.069768284 | 0.002957272 | 0.07605498 | -0.00542118 | 0.066772984 | 0.002957215 | 0.076048352 | 5.84E-05 | 0.004708581 | 1 | 0.592493737 | 1 | 0.560354119 | ENSG00000002( | 9 | RNF5 |
| VAT | Heart_Failure | ENSG00000023 | 0.001811253 | 0.408528396 | 0.002191508 | 0.076055386 | 0.00173637 | 0.428196038 | 0.002191604 | 0.076048779 | 7.49E-05 | 0.009551307 | 1 | 0.861373852 | 1 | 0.875281919 | ENSG00000002( | 6 | PRRT1 |
| VAT | Heart_Failure | ENSG00000023 | 0.00067001 | 0.675680804 | 0.001601499 | 0.076055466 | 0.000992515 | 0.537218903 | 0.00160855 | 0.076048817 | -0.0003225 | 0.032914649 | 1 | 0.963913289 | 1 | 0.911760867 | ENSG00000002( | 6 | PRRT1 |
| VAT | Heart_Failure | ENSG00000023 | 0.001882338 | 0.154085019 | 0.001320706 | 0.076055176 | 0.001912061 | 0.147679278 | 0.001320685 | 0.07604855 | -2.97E-05 | 0.002681129 | 1 | 0.721315241 | 1 | 0.701115705 | ENSG00000002( | 6 | PRRT1 |
| VAT | Heart_Failure | ENSG00000023 | -0.00082165 | 0.461018081 | 0.001114601 | 0.076055408 | -0.00133146 | 0.238533064 | 0.00112963 | 0.076048666 | 0.000509809 | 0.005593581 | 1 | 0.88303338 | 1 | 0.771971926 | ENSG00000002( | 6 | PRRT1 |
| VAT | Heart_Failure | ENSG00000023 | -0.00411982 | 0.079929042 | 0.002352701 | 0.076055014 | -0.00407838 | 0.083006706 | 0.00235268 | 0.076048408 | -4.14E-05 | 0.033177004 | 1 | 0.619367186 | 1 | 0.604015714 | ENSG00000002( | 6 | PRRT1 |
| VAT | Heart_Failure | ENSG00000023 | 0.001403965 | 0.525293053 | 0.002210245 | 0.07605543 | 0.001449511 | 0.511938886 | 0.002210219 | 0.07604881 | -4.55E-05 | 0.009844868 | 1 | 0.91226097 | 1 | 0.904578313 | ENSG00000002( | 8 | MRPL38 |
| VAT | Heart_Failure | ENSG00000023 | -0.00356832 | 0.148097759 | 0.002467229 | 0.076055167 | -0.00294581 | 0.2331994 | 0.00247099 | 0.076048655 | -0.0006225 | 6.65E-06 | 1 | 0.719389945 | 1 | 0.76771061 | ENSG00000002( | 8 | MRPL38 |
| VAT | Heart_Failure | ENSG00000023 | -0.00306321 | 0.239293108 | 0.002603093 | 0.076055277 | -0.00315198 | 0.225975388 | 0.002603238 | 0.07604868 | 8.88E-05 | 0.015399667 | 1 | 0.779051326 | 1 | 0.76157815 | ENSG00000002( | 8 | MRPL38 |
| VAT | Heart_Failure | ENSG00000023 | -0.0010109 | 0.279603792 | 0.000934971 | 0.076055311 | -0.00135822 | 0.147230152 | 0.000937102 | 0.076048549 | 0.000347322 | 5.13E-08 | 1 | 0.795233108 | 1 | 0.701115705 | ENSG00000002( | 8 | MRPL38 |
| VAT | Heart_Failure | ENSG00000023 | -0.00056757 | 0.801088118 | 0.002252805 | 0.076055483 | -0.00060559 | 0.788069548 | 0.002252782 | 0.076048866 | 3.80E-05 | 0.038282934 | 1 | 0.972185844 | 1 | 0.965780945 | ENSG00000002( | 4 | CYP21A1P |
| VAT | Heart_Failure | ENSG00000023 | -0.00667944 | 0.089309807 | 0.003931271 | 0.076055042 | -0.00676078 | 0.085475187 | 0.003931213 | 0.076048415 | 8.13E-05 | 0.006523962 | 1 | 0.623489698 | 1 | 0.607677788 | ENSG00000002( | 4 | CYP21A1P |
| VAT | Heart_Failure | ENSG00000023 | 0.002839716 | 0.231442338 | 0.002373048 | 0.07605527 | 0.002317326 | 0.331116556 | 0.002384403 | 0.076048729 | 0.00052239 | 0.025245851 | 1 | 0.777647581 | 1 | 0.812752882 | ENSG00000002( | 4 | CYP21A1P |
| VAT | Heart_Failure | ENSG00000023 | -0.00053027 | 0.566506382 | 0.000925106 | 0.076055442 | -0.00042382 | 0.647350195 | 0.0009265 | 0.076048844 | -0.00010645 | 0.038896577 | 1 | 0.92639479 | 1 | 0.955604921 | ENSG00000002( | 4 | CYP21A1P |
| VAT | Heart_Failure | ENSG00000023 | 0.004450919 | 0.034600015 | 0.002106431 | 0.076054796 | 0.004513059 | 0.032150278 | 0.002106403 | 0.0760416 | -6.21E-05 | 0.000154604 | 1 | 0.440073928 | 1 | 0.414648442 | ENSG00000002( | 4 | CYP21A1P |
| VAT | Heart_Failure | ENSG00000023 | -0.0007881 | 0.627648812 | 0.001624825 | 0.076055456 | -0.00107785 | 0.507497086 | 0.001626349 | 0.076048808 | 0.000289744 | 5.73E-05 | 1 | 0.952138605 | 1 | 0.903672231 | ENSG00000002( | 4 | CYP21A1P |
| VAT | Heart_Failure | ENSG00000023 | -0.00195317 | 0.096088593 | 0.001173694 | 0.07605061 | -0.00192689 | 0.10064377 | 0.001173687 | 0.076048456 | -2.63E-05 | 0.009881258 | 1 | 0.637447692 | 1 | 0.624888429 | ENSG00000002( | 4 | CYP21A1P |
| VAT | Heart_Failure | ENSG00000023 | 0.012914392 | 0.000409686 | 0.003654545 | 0.076053544 | 0.011893973 | 0.001178173 | 0.003666304 | 0.076047234 | 0.001020419 | 0.000548825 | 1 | 0.035335395 | 1 | 0.073737866 | ENSG00000002( | 4 | CYP21A1P |
| VAT | Heart_Failure | ENSG00000023 | -0.0035127 | 0.104578567 | 0.002164257 | 0.076055082 | -0.00347591 | 0.108259148 | 0.002164234 | 0.076048474 | -3.68E-05 | 0.036750567 | 1 | 0.637447692 | 1 | 0.628112053 | ENSG00000002( | 4 | CYP21A1P |
| VAT | Heart_Failure | ENSG00000023 | -0.00147033 | 0.695053553 | 0.00375079 | 0.076055469 | -0.00154915 | 0.679588391 | 0.003750745 | 0.07604885 | 7.88E-05 | 0.008132989 | 1 | 0.963913289 | 1 | 0.960065682 | ENSG00000002( | 10 | STK19 |
| VAT | Heart_Failure | ENSG00000023 | 0.002577026 | 0.324437361 | 0.002615258 | 0.076055341 | 0.002110264 | 0.420873229 | 0.002621746 | 0.076048776 | 0.000466762 | 0.012058354 | 1 | 0.811008798 | 1 | 0.871425441 | ENSG00000002( | 10 | STK19 |
| VAT | Heart_Failure | ENSG00000023 | 0.005091028 | 0.060575302 | 0.002712919 | 0.076054943 | 0.0051344 | 0.058410486 | 0.002712869 | 0.076048318 | -4.34E-05 | 0.024561517 | 1 | 0.546800541 | 1 | 0.532055913 | ENSG00000002( | 10 | STK19 |
| VAT | Heart_Failure | ENSG00000023 | 0.001792013 | 0.281379403 | 0.001663543 | 0.076055312 | 0.001817268 | 0.274645615 | 0.001663513 | 0.076048691 | -2.53E-05 | 0.033275244 | 1 | 0.795233108 | 1 | 0.791877836 | ENSG00000002( | 10 | STK19 |
| VAT | Heart_Failure | ENSG00000024 | 0.001659515 | 0.233526614 | 0.001392999 | 0.076055272 | 0.002974086 | 0.036931232 | 0.001425375 | 0.076048197 | -0.00131457 | 1.38E-05 | 1 | 0.779051326 | 1 | 0.441256279 | ENSG00000002( | 10 | STK19 |
| VAT | Heart_Failure | ENSG00000024 | -0.00338135 | 0.383111058 | 0.003876903 | 0.076055374 | -0.00331516 | 0.392486949 | 0.003876847 | 0.076048763 | -6.62E-05 | 0.025177905 | 1 | 0.837245697 | 1 | 0.842989349 | ENSG00000002( | 5 | DXO |
| VAT | Heart_Failure | ENSG00000024 | -0.00216523 | 0.492905683 | 0.003157721 | 0.07605542 | -0.00222819 | 0.480415823 | 0.00315771 | 0.076048799 | 6.30E-05 | 0.02634287 | 1 | 0.896935503 | 1 | 0.892894055 | ENSG00000002( | 5 | DXO |
| VAT | Heart_Failure | ENSG00000024 | 0.001239584 | 0.295738764 | 0.001185505 | 0.076055322 | -0.00039894 | 0.770278936 | 0.001366178 | 0.076048864 | 0.001638522 | 0.015826767 | 1 | 0.799450526 | 1 | 0.965780945 | ENSG00000002( | 5 | DXO |
| VAT | Heart_Failure | ENSG00000024 | -0.00049733 | 0.578193495 | 0.000894428 | 0.076055445 | -0.0005124 | 0.566717743 | 0.000894415 | 0.076048826 | 1.51E-05 | 0.024719704 | 1 | 0.92639479 | 1 | 0.92172808 | ENSG00000002( | 5 | DXO |
| VAT | Heart_Failure | ENSG00000024 | -0.000988 | 0.1546881 | 0.000694226 | 0.076055177 | -0.00153666 | 0.028338944 | 0.000700848 | 0.076048127 | 0.000548657 | 1.24E-08 | 1 | 0.721315241 | 1 | 0.387205367 | ENSG00000002( | 7 | SKIV2L |
| VAT | Heart_Failure | ENSG00000024 | 0.001598375 | 0.600340844 | 0.003050853 | 0.07605545 | 0.001903071 | 0.532959813 | 0.003052266 | 0.076048816 | -0.0003047 | 0.001707657 | 1 | 0.93401394 | 1 | 0.908819144 | ENSG00000002( | 7 | SKIV2L |
| VAT | Heart_Failure | ENSG00000024 | -0.00381833 | 0.326022568 | 0.003887692 | 0.076055342 | -0.00376509 | 0.332803068 | 0.003887615 | 0.07604873 | -5.32E-05 | 0.047663801 | 1 | 0.811008798 | 1 | 0.812752882 | ENSG00000002( | 7 | SKIV2L |
| VAT | Heart_Failure | ENSG00000024 | 0.007709962 | 0.003343134 | 0.002627528 | 0.076054149 | 0.00775741 | 0.003153274 | 0.002627491 | 0.076047516 | -4.74E-05 | 0.019384552 | 1 | 0.124689861 | 1 | 0.126139004 | ENSG00000002( | 7 | SKIV2L |
| VAT | Heart_Failure | ENSG00000024 | -0.00057677 | 0.566536211 | 0.001006296 | 0.076055442 | -0.00085948 | 0.395626027 | 0.001011797 | 0.076048764 | 0.000282711 | 0.007525991 | 1 | 0.92639479 | 1 | 0.845800026 | ENSG00000002( | 7 | SKIV2L |
| VAT | Heart_Failure | ENSG00000024 | -0.00265183 | 0.007596889 | 0.000993383 | 0.076054381 | -0.00449359 | 0.000752122 | 0.001333466 | 0.076047105 | 0.001841765 | 0.038424729 | 1 | 0.21841057 | 1 | 0.05462779 | ENSG00000002( | 7 | SKIV2L |
| VAT | Heart_Failure | ENSG00000024 | -0.00167323 | 0.381159559 | 0.001910513 | 0.076055373 | -0.00171219 | 0.370143917 | 0.00191049 | 0.076048751 | 3.90E-05 | 0.010194068 | 1 | 0.837245697 | 1 | 0.830566839 | ENSG00000002( | 7 | SKIV2L |
| VAT | Heart_Failure | ENSG00000024 | -0.00402998 | 0.00153618 | 0.001272179 | 0.076053927 | -0.0045749 | 0.000346877 | 0.001278784 | 0.076046879 | 0.000544918 | 2.92E-05 | 1 | 0.085321306 | 0.957379669 | 0.036010503 | ENSG00000002( | 7 | SKIV2L |
| VAT | Heart_Failure | ENSG00000024 | -0.00168395 | 0.085286813 | 0.000978581 | 0.076055031 | -0.00229781 | 0.024817065 | 0.001023868 | 0.076048091 | 0.000613865 | 0.041601692 | 1 | 0.623489698 | 1 | 0.372255978 | ENSG00000002( | 7 | SKIV2L |
| VAT | Heart_Failure | ENSG00000025 | -0.0025423 | 0.186141149 | 0.00192295 | 0.07605522 | -0.00248857 | 0.195614091 | 0.001922941 | 0.076048615 | -5.37E-05 | 0.001519364 | 1 | 0.756191037 | 1 | 0.738152304 | ENSG00000002( | 7 | NELFE |
| VAT | Heart_Failure | ENSG00000025 | -0.00215019 | 0.401664087 | 0.002563859 | 0.076055383 | -0.00219373 | 0.392196523 | 0.002563837 | 0.076048763 | 4.35E-05 | 0.042031438 | 1 | 0.85407772 | 1 | 0.842989349 | ENSG00000002( | 7 | NELFE |
| VAT | Heart_Failure | ENSG00000025 | -0.00048608 | 0.762726021 | 0.001610054 | 0.076055479 | -0.00045911 | 0.775524899 | 0.001610038 | 0.076048864 | -2.70E-05 | 0.041638675 | 1 | 0.966718282 | 1 | 0.965780945 | ENSG00000002( | 7 | NELFE |
| VAT | Heart_Failure | ENSG00000025 | 0.001068123 | 0.616355724 | 0.002131888 | 0.076055454 | 0.001159776 | 0.586498803 | 0.002132267 | 0.076048831 | -9.17E-05 | 0.041061496 | 1 | 0.945604112 | 1 | 0.927008193 | ENSG00000002( | 7 | EHMT2 |
| VAT | Heart_Failure | ENSG00000025 | -0.0024452 | 0.395163121 | 0.002875723 | 0.07605538 | -0.0023862 | 0.406659347 | 0.00287568 | 0.076048769 | -5.90E-05 | 0.006690927 | 1 | 0.846538141 | 1 | 0.855404372 | ENSG00000002( | 7 | EHMT2 |
| VAT | Heart_Failure | ENSG00000025 | 0.002011623 | 0.59602464 | 0.003794612 | 0.076055449 | 0.001911578 | 0.614427564 | 0.003794592 | 0.076048837 | 0.000100045 | 0.002570181 | 1 | 0.931133617 | 1 | 0.942122265 | ENSG00000002( | 7 | EHMT2 |
| VAT | Heart_Failure | ENSG00000025 | -0.00573907 | 0.110333986 | 0.00359434 | 0.076055095 | -0.00567415 | 0.114419072 | 0.003594325 | 0.076048488 | -6.49E-05 | 0.041342636 | 1 | 0.643497633 | 1 | 0.640561135 | ENSG00000002( | 7 | EHMT2 |
| VAT | Heart_Failure | ENSG00000025 | 9.61E-05 | 0.939890866 | 0.001275062 | 0.07605492 | 6.85E-05 | 0.957154058 | 0.001275054 | 0.076048876 | 2.76E-05 | 0.01189036 | 1 | 0.990975896 | 1 | 0.992070179 | ENSG00000002( | 8 | C6orf48 |
| VAT | Heart_Failure | ENSG00000025 | -0.00196786 | 0.422359639 | 0.002452668 | 0.076055393 | -0.00476463 | 0.08086156 | 0.002729334 | 0.076048401 | 0.002796771 | 0.019526646 | 1 | 0.871415448 | 1 | 0.600796054 | ENSG00000002( | 8 | C6orf48 |
| VAT | Heart_Failure | ENSG00000025 | 0.002016204 | 0.090631016 | 0.001191552 | 0.076055046 | 0.001994685 | 0.094124084 | 0.001191542 | 0.076048439 | 2.15E-05 | 0.030284149 | 1 | 0.623489698 | 1 | 0.620005898 | ENSG00000002( | 8 | C6orf48 |
| VAT | Heart_Failure | ENSG00000025 | -0.00086613 | 0.624875049 | 0.001771397 | 0.076055456 | -0.00089681 | 0.612660558 | 0.001771368 | 0.076048837 | 3.07E-05 | 0.018785643 | 1 | 0.952138605 | 1 | 0.940457808 | ENSG00000002( | 8 | C6orf48 |
| VAT | Heart_Failure | ENSG00000025 | 0.001071026 | 0.527792768 | 0.001696327 | 0.07605543 | 0.00102866 | 0.544242429 | 0.00169631 | 0.076048819 | 4.24E-05 | 0.00235111 | 1 | 0.915017613 | 1 | 0.913136234 | ENSG00000002( | 10 | MSH5 |
| VAT | Heart_Failure | ENSG00000026 | 0.000120071 | 0.972232984 | 0.003449528 | 0.076055493 | 3.84E-05 | 0.991115311 | 0.003449506 | 0.076048877 | 8.17E-05 | 0.006117612 | 1 | 0.994659842 | 1 | 0.998073832 | ENSG00000002( | 10 | MSH5 |
| VAT | Heart_Failure | ENSG00000026 | 0.002899172 | 0.423906643 | 0.003625493 | 0.076055393 | 0.003020762 | 0.404736113 | 0.003625523 | 0.076048768 | -0.00012159 | 0.000993985 | 1 | 0.871819921 | 1 | 0.85403033 | ENSG00000002( | 10 | MSH5 |
| VAT | Heart_Failure | ENSG00000026 | -0.0011297 | 0.71858697 | 0.003135011 | 0.076055473 | -0.00117906 | 0.706844009 | 0.003134975 | 0.076048855 | 4.94E-05 | 0.049189596 | 1 | 0.963913289 | 1 | 0.960065682 | ENSG00000002( | 10 | MSH5 |
| VAT | Heart_Failure | ENSG00000026 | -0.0072111 | 0.03374685 | 0.003396515 | 0.07605479 | -0.00730234 | 0.031558052 | 0.003396486 | 0.076048155 | 9.12E-05 | 0.001392214 | 1 | 0.438042449 | 1 | 0.409322718 | ENSG00000002( | 7 | ABHD16A |
| VAT | Heart_Failure | ENSG00000026 | 0.001419462 | 0.6127765 | 0.002804627 | 0.076055453 | 0.001368937 | 0.62547514 | 0.00280459 | 0.07604884 | 5.05E-05 | 0.020515992 | 1 | 0.942733077 | 1 | 0.947481552 | ENSG00000002( | 7 | ABHD16A |
| VAT | Heart_Failure | ENSG00000026 | -0.00034047 | 0.875051609 | 0.002165244 | 0.076055489 | -0.00037801 | 0.861409089 | 0.002165226 | 0.076048872 | 3.75E-05 | 0.039319966 | 1 | 0.983439565 | 1 | 0.979196493 | ENSG00000002( | 7 | ABHD16A |
| VAT | Heart_Failure | ENSG00000026 | -0.00501843 | 0.169085998 | 0.003649371 | 0.076055198 | -0.00529647 | 0.1469277 | 0.003651547 | 0.076048548 | 0.000278038 | 0.033236825 | 1 | 0.733769426 | 1 | 0.701115705 | ENSG00000002( | 7 | ABHD16A |
| VAT | Heart_Failure | ENSG00000026 | -0.00126249 | 0.381077186 | 0.001441341 | 0.076055373 | -0.00129898 | 0.367462852 | 0.001441326 | 0.07604875 | 3.65E-05 | 0.001852337 | 1 | 0.837245697 | 1 | 0.830566839 | ENSG00000002( | 8 | LY6G5C |
| VAT | Heart_Failure | ENSG00000026 | -0.00187858 | 0.445237966 | 0.00246087 | 0.076055402 | -0.00183195 | 0.456611842 | 0.002460849 | 0.07604879 | -4.66E-05 | 0.023248351 | 1 | 0.873500102 | 1 | 0.880469391 | ENSG00000002( | 8 | LY6G5C |
| VAT | Heart_Failure | ENSG00000026 | 0.0035269 | 0.249324562 | 0.003061564 | 0.076055286 | 0.003464816 | 0.25774906 | 0.003061525 | 0.076048677 | 6.21E-05 | 0.00990027 | 1 | 0.789536793 | 1 | 0.786050543 | ENSG00000002( | 8 | LY6G5C |
| VAT | Heart_Failure | ENSG00000026 | 0.003333746 | 0.309266746 | 0.003278784 | 0.076055332 | 0.00339146 | 0.300955793 | 0.003278715 | 0.076048719 | -5.77E-05 | 0.00850064 | 1 | 0.811008798 | 1 | 0.801314322 | ENSG00000002( | 8 | LY6G5C |
| VAT | Heart_Failure | ENSG00000027 | -0.00249989 | 0.134314203 | 0.001669593 | 0.076055143 | -0.00252476 | 0.130475142 | 0.00166956 | 0.07604852 | 2.49E-05 | 0.031044979 | 1 | 0.686494815 | 1 | 0.665314041 | ENSG00000002( | 8 | LY6G5C |
| VAT | Heart_Failure | ENSG00000027 | 0.001709834 | 0.122095181 | 0.001105943 | 0.07605512 | 0.001690534 | 0.126363331 | 0.001105933 | 0.076048512 | 1.93E-05 | 0.035531773 | 1 | 0.669945723 | 1 | 0.658005371 | ENSG00000002( | 8 | LY6G5C |
| VAT | Heart_Failure | ENSG00000027 | -0.00179716 | 0.549783387 | 0.003004855 | 0.076055437 | -0.00186183 | 0.535513649 | 0.003004826 | 0.076048817 | 6.47E-05 | 0.00862114 | 1 | 0.92326913 | 1 | 0.910019404 | ENSG00000002( | 6 | GPANK1 |
| VAT | Heart_Failure | ENSG00000027 | 0.003788891 | 0.257962993 | 0.003349385 | 0.076055293 | 0.006234355 | 0.065761245 | 0.003388141 | 0.076048348 | -0.00244546 | 1.79E-06 | 1 | 0.79317431 | 1 | 0.560188382 | ENSG00000002( | 6 | GPANK1 |
| VAT | Heart_Failure | ENSG00000027 | 0.000124339 | 0.970483362 | 0.00336033 | 0.076055493 | 0.000197261 | 0.953188649 | 0.003360315 | 0.076048876 | -7.29E-05 | 0.014014867 | 1 | 0.994628325 | 1 | 0.992070179 | ENSG00000002( | 6 | GPANK1 |
| VAT | Heart_Failure | ENSG00000027 | 0.003105033 | 0.436239216 | 0.003988174 | 0.076055398 | -0.00311169 | 0.478960569 | 0.004395199 | 0.076048799 | 0.006216728 | 0.000766102 | 1 | 0.872618112 | 1 | 0.891945927 | ENSG00000002( | 6 | GPANK1 |
| VAT | Heart_Failure | ENSG00000027 | -0.00430799 | 0.220872207 | 0.003518981 | 0.076055259 | -0.00448785 | 0.202255481 | 0.003519457 | 0.076048623 | 0.000179863 | 0.006878619 | 1 | 0.772901704 | 1 | 0.74232065 | ENSG00000002( | 6 | GPANK1 |
| VAT | Heart_Failure | ENSG00000027 | 0.002824148 | 0.000350861 | 0.00079007 | 0.076053499 | 0.003118532 | 8.58E-05 | 0.000794011 | 0.076046469 | -0.00029438 | 0.000208211 | 0.968377101 | 0.034584896 | 0.236867022 | 0.011843351 | ENSG00000002( | 3 | C6orf47 |
| VAT | Heart_Failure | ENSG00000027 | 0.003204933 | 0.320406901 | 0.003225502 | 0.076055339 | 0.003263762 | 0.311600938 | 0.003225477 | 0.076048717 | -5.88E-05 | 0.036048398 | 1 | 0.811008798 | 1 | 0.809810347 | ENSG00000002( | 8 | BAG6 |
| VAT | Heart_Failure | ENSG00000027 | -0.00076868 | 0.545223811 | 0.001270692 | 0.076055436 | -0.00099932 | 0.433513731 | 0.001275956 | 0.076048781 | 0.000230637 | 0.04753125 | 1 | 0.921324909 | 1 | 0.875281919 | ENSG00000002( | 8 | BAG6 |
| VAT | Heart_Failure | ENSG00000027 | 0.00440934 | 0.195708218 | 0.003407849 | 0.076055232 | 0.00446548 | 0.19002317 | 0.003407812 | 0.076048609 | -5.66E-05 | 0.039788271 | 1 | 0.756191037 | 1 | 0.738152304 | ENSG00000002( | 8 | BAG6 |
| VAT | HTN | ENSG00000000( | 0.000427465 | 0.872139307 | 0.002656024 | 0.240335801 | 0.000498242 | 0.851194099 | 0.002655944 | 0.240302367 | -7.08E-05 | 0.002705902 | 1 | 0.953686405 | 1 | 0.945771221 | ENSG00000002( | 8 | BAG6 |
| VAT | HTN | ENSG00000000( | -0.01437498 | 0.000647766 | 0.004214543 | 0.240330076 | -0.0149199 | 0.000411253 | 0.004223274 | 0.24029623 | 0.000544915 | 0.048344661 | 1 | 0.01103602 | 1 | 0.00822505 | ENSG00000002( | 8 | BAG6 |
| VAT | HTN | ENSG00000000( | -0.00033816 | 0.931163534 | 0.003914792 | 0.24033581 | -0.00050493 | 0.897377879 | 0.003914946 | 0.240302377 | 0.000166764 | 0.00392035 | 1 | 0.973508158 | 1 | 0.964471552 | ENSG00000002( | 8 | BAG6 |
| VAT | HTN | ENSG00000000( | 0.00939445 | 0.050140359 | 0.004796098 | 0.240333921 | 0.00930933 | 0.05224934 | 0.004795958 | 0.240300527 | 8.51E-05 | 0.047884744 | 1 | 0.226864576 | 1 | 0.229630856 | ENSG00000002( | 6 | NFKBIL1 |
| VAT | HTN | ENSG00000000( | 0.007681419 | 0.140687673 | 0.005213972 | 0.240334743 | 0.005391271 | 0.304451734 | 0.005249868 | 0.240301865 | 0.002290147 | 0.000200486 | 1 | 0.408304917 | 1 | 0.604088272 | ENSG00000002( | 6 | NFKBIL1 |
| VAT | HTN | ENSG00000000( | 0.009399045 | 0.07642743 | 0.005304783 | 0.240334265 | 0.00948333 | 0.073812892 | 0.005304542 | 0.240300809 | -8.43E-05 | 0.022471877 | 1 | 0.292971817 | 1 | 0.285327146 | ENSG00000002( | 5 | MICB |
| VAT | HTN | ENSG00000000( | -0.01251419 | 0.048874336 | 0.006353377 | 0.2403339 | -0.01984619 | 0.00263143 | 0.006598199 | 0.240297923 | 0.007332 | 3.90E-05 | 1 | 0.222963912 | 1 | 0.030644506 | ENSG00000002( | 5 | MICB |
| VAT | HTN | ENSG00000001 | 0.025914169 | 6.26E-09 | 0.004460472 | 0.240319168 | 0.026018476 | 5.44E-09 | 0.004460302 | 0.240285605 | -0.00010431 | 0.003327953 | 1.73E-05 | 8.23E-07 | 1.50E-05 | 6.52E-07 | ENSG00000002( | 5 | MICB |
| VAT | HTN | ENSG00000001 | -0.00031399 | 0.945481985 | 0.004591704 | 0.240335811 | -0.00020611 | 0.964195288 | 0.004591516 | 0.240302384 | -0.00010788 | 0.001925112 | 1 | 0.976968793 | 1 | 0.985245375 | ENSG00000002( | 5 | MICB |
| VAT | HTN | ENSG00000001 | 0.005079416 | 0.239547286 | 0.004318789 | 0.240335131 | 0.005316382 | 0.218318715 | 0.004318706 | 0.240301637 | -0.00023697 | 4.95E-08 | 1 | 0.549661061 | 1 | 0.517020606 | ENSG00000002( | 5 | MICB |
| VAT | HTN | ENSG00000003 | -0.00355023 | 0.021691389 | 0.001546444 | 0.240333214 | -0.0036557 | 0.018110111 | 0.001546817 | 0.24029963 | 0.000105469 | 0.006248271 | 1 | 0.133634452 | 1 | 0.116241644 | ENSG00000002( | 5 | MICB |
| VAT | HTN | ENSG00000003 | -0.01788202 | 0.004921881 | 0.006358922 | 0.240331913 | -0.01803668 | 0.004561216 | 0.006358771 | 0.240298417 | 0.000154661 | 0.011071013 | 1 | 0.049397792 | 1 | 0.046861116 | ENSG00000002( | 5 | MICB |
| VAT | HTN | ENSG00000003 | 0.005403638 | 0.437224999 | 0.006955488 | 0.240335516 | 0.005283831 | 0.447441304 | 0.006955262 | 0.2403021 | 0.000119807 | 0.045497108 | 1 | 0.716423851 | 1 | 0.721401667 | ENSG00000002( | 5 | MICB |
| VAT | HTN | ENSG00000003 | 0.0053913 | 0.850902759 | 0.002868218 | 0.240335796 | 0.000463049 | 0.871741004 | 0.002868106 | 0.240302372 | 7.61E-05 | 0.000696798 | 1 | 0.942297189 | 1 | 0.956043422 | ENSG00000002( | 5 | MICB |
| VAT | HTN | ENSG00000004 | -0.00131555 | 0.326405241 | 0.001340511 | 0.240335338 | -0.00134065 | 0.317241544 | 0.001340457 | 0.240301891 | 2.51E-05 | 0.015464539 | 1 | 0.627494074 | 1 | 0.616610324 | ENSG00000002( | 5 | MICB |
| VAT | HTN | ENSG00000005 | -0.0054212 | 0.41136818 | 0.006599233 | 0.240335481 | -0.01894994 | 0.007211288 | 0.007052615 | 0.240298825 | 0.013528738 | 5.48E-08 | 1 | 0.700167128 | 1 | 0.065470903 | ENSG00000002( | 5 | MICB |

| | | | | | | | | | | | | | | | | | | | |
|---|---|---|---|---|---|---|---|---|---|---|---|---|---|---|---|---|---|---|---|
| VAT | HTN | ENSG0000005 | 0.001358538 | 0.782198603 | 0.004914135 | 0.240335776 | 0.00128752 | 0.793309474 | 0.004913904 | 0.240302351 | 7.10E-05 | 0.031867244 | 1 | 0.904048637 | 1 | 0.918168712 | ENSG000002 | 8 | MICA |
| VAT | HTN | ENSG0000005 | 0.004369065 | 0.287663266 | 0.004109111 | 0.240335256 | 0.004292235 | 0.296207605 | 0.004108968 | 0.240301847 | 7.68E-05 | 0.024631393 | 1 | 0.59321241 | 1 | 0.596304149 | ENSG000002 | 8 | MICA |
| VAT | HTN | ENSG0000005 | 0.003719797 | 0.563713137 | 0.006443043 | 0.240335649 | 0.003625162 | 0.57365836 | 0.006442759 | 0.240302229 | 9.46E-05 | 0.03971606 | 1 | 0.792605307 | 1 | 0.801328599 | ENSG000002 | 8 | MICA |
| VAT | HTN | ENSG0000005 | -2.52E-06 | 0.998897878 | 0.001824398 | 0.240335813 | 2.73E-05 | 0.988053784 | 0.001824326 | 0.240302385 | -2.98E-05 | 0.035231162 | 1 | 0.999622242 | 1 | 0.997085354 | ENSG000002 | 8 | MICA |
| VAT | HTN | ENSG0000005 | -0.00147366 | 0.820991038 | 0.006512828 | 0.240335788 | -0.00134354 | 0.836557397 | 0.006512565 | 0.240302364 | -0.00013013 | 0.008835937 | 1 | 0.926718801 | 1 | 0.939635856 | ENSG000002 | 8 | MICA |
| VAT | HTN | ENSG0000006 | 0.007498404 | 0.009074973 | 0.002873803 | 0.240332456 | 0.007287529 | 0.011223149 | 0.002873991 | 0.240299214 | 0.000210876 | 7.76E-06 | 1 | 0.075215989 | 1 | 0.086044142 | ENSG000002 | 8 | MICA |
| VAT | HTN | ENSG0000006 | 0.012686668 | 0.004667781 | 0.004484313 | 0.240331866 | 0.012754888 | 0.004448854 | 0.004484115 | 0.240298395 | -6.82E-05 | 0.032886065 | 1 | 0.047539021 | 1 | 0.046161039 | ENSG000002 | 10 | HLA-C |
| VAT | HTN | ENSG0000006 | 0.015153473 | 5.29E-07 | 0.003021293 | 0.240323407 | 0.015011965 | 6.75E-07 | 0.003021378 | 0.240290211 | 0.000141507 | 0.000789119 | 0.001459982 | 3.29E-05 | 0.001862089 | 3.88E-05 | ENSG000002 | 10 | HLA-C |
| VAT | HTN | ENSG0000006 | -0.00351307 | 0.546485956 | 0.005825646 | 0.240335634 | -0.00366723 | 0.529006102 | 0.005825394 | 0.240302189 | 0.000154161 | 0.000259053 | 1 | 0.783126292 | 1 | 0.771700234 | ENSG000002 | 10 | HLA-C |
| VAT | HTN | ENSG0000006 | -0.00381386 | 0.459550252 | 0.005156737 | 0.240335544 | -0.0039384 | 0.445004 | 0.005156522 | 0.240302097 | 0.000124541 | 0.001237824 | 1 | 0.733862963 | 1 | 0.720405231 | ENSG000002 | 10 | HLA-C |
| VAT | HTN | ENSG0000006 | 0.008157435 | 0.042384649 | 0.004018977 | 0.240333782 | 0.008238171 | 0.040375324 | 0.004018828 | 0.240300313 | -8.07E-05 | 0.012707189 | 1 | 0.204075679 | 1 | 0.197548996 | ENSG000002 | 10 | HLA-C |
| VAT | HTN | ENSG0000007 | -0.00166556 | 0.721233376 | 0.004667901 | 0.240335751 | -0.00156899 | 0.736769345 | 0.004667728 | 0.240302329 | -9.66E-05 | 0.010078054 | 1 | 0.869815229 | 1 | 0.880677087 | ENSG000002 | 10 | HLA-C |
| VAT | HTN | ENSG0000007 | -0.00173466 | 0.641480128 | 0.003725402 | 0.240335706 | -0.00125128 | 0.737142494 | 0.003728051 | 0.240302329 | -0.00048338 | 0.001024608 | 1 | 0.830433937 | 1 | 0.880741681 | ENSG000002 | 10 | HLA-C |
| VAT | HTN | ENSG0000007 | 0.002163427 | 0.67023116 | 0.005080481 | 0.240335724 | 0.002078583 | 0.682432261 | 0.005080288 | 0.240302302 | 8.48E-05 | 0.035269405 | 1 | 0.843623709 | 1 | 0.856249354 | ENSG000002 | 10 | HLA-C |
| VAT | HTN | ENSG0000007 | 0.001395311 | 0.829377658 | 0.006474764 | 0.24033579 | 0.00159263 | 0.805694478 | 0.00647452 | 0.240302355 | -0.00019732 | 0.000137526 | 1 | 0.931521652 | 1 | 0.924385659 | ENSG000002 | 10 | HLA-C |
| VAT | HTN | ENSG0000007 | -0.00415298 | 0.484542026 | 0.005941173 | 0.240335572 | -0.00428827 | 0.470407029 | 0.005940928 | 0.240302128 | 0.000135288 | 0.00247593 | 1 | 0.748369329 | 1 | 0.73755282 | ENSG000002 | 10 | HLA-C |
| VAT | HTN | ENSG0000007 | -0.00168901 | 0.770768416 | 0.005796802 | 0.240335772 | -0.00179905 | 0.756282728 | 0.005796536 | 0.240302337 | 0.000110033 | 0.005669728 | 1 | 0.899121229 | 1 | 0.892265229 | ENSG000002 | 10 | HLA-C |
| VAT | HTN | ENSG0000007 | 0.005561026 | 0.374079145 | 0.006256362 | 0.240335424 | 0.005689183 | 0.363145936 | 0.006256058 | 0.240301977 | -0.00012816 | 0.001593395 | 1 | 0.671507939 | 1 | 0.662009765 | ENSG000002 | 10 | HLA-C |
| VAT | HTN | ENSG0000007 | 0.001583707 | 0.679049465 | 0.003827591 | 0.240335729 | 0.001511161 | 0.692972212 | 0.003827421 | 0.240302308 | 7.25E-05 | 0.007433587 | 1 | 0.849581379 | 1 | 0.861145117 | ENSG000002 | 10 | HLA-C |
| VAT | HTN | ENSG0000007 | 0.012253474 | 0.076230395 | 0.006911178 | 0.240334263 | 0.012635207 | 0.067554381 | 0.006912145 | 0.240300737 | -0.00038173 | 0.00699368 | 1 | 0.292622935 | 1 | 0.269826471 | ENSG000002 | 11 | POU5F1 |
| VAT | HTN | ENSG0000007 | -0.00455039 | 0.493700142 | 0.006648373 | 0.240335582 | -0.00441774 | 0.506363928 | 0.006648115 | 0.240302167 | -0.00013264 | 0.010798206 | 1 | 0.752792925 | 1 | 0.76099237 | ENSG000002 | 11 | POU5F1 |
| VAT | HTN | ENSG0000007 | -0.00735259 | 0.274431916 | 0.006727504 | 0.240335224 | -0.00724116 | 0.281752302 | 0.006727247 | 0.240301813 | -0.00011143 | 0.036534633 | 1 | 0.583987732 | 1 | 0.58432047 | ENSG000002 | 11 | POU5F1 |
| VAT | HTN | ENSG0000008 | 0.003198166 | 0.548908087 | 0.005335635 | 0.240335636 | 0.003119667 | 0.558743273 | 0.005335413 | 0.240302216 | 7.85E-05 | 0.049075898 | 1 | 0.783206863 | 1 | 0.792055179 | ENSG000002 | 11 | POU5F1 |
| VAT | HTN | ENSG0000008 | -0.00112648 | 0.759297724 | 0.003676451 | 0.240335767 | -0.00145371 | 0.692555815 | 0.003676668 | 0.240302308 | 0.000327238 | 2.85E-08 | 1 | 0.89291083 | 1 | 0.861126259 | ENSG000002 | 11 | POU5F1 |
| VAT | HTN | ENSG0000008 | -0.00966699 | 0.06738625 | 0.005285129 | 0.240334163 | -0.00957816 | 0.069932114 | 0.005284926 | 0.240300765 | -8.88E-05 | 0.033073979 | 1 | 0.269986697 | 1 | 0.275732335 | ENSG000002 | 11 | POU5F1 |
| VAT | HTN | ENSG0000008 | -0.00592714 | 0.246443817 | 0.005113874 | 0.240335151 | -0.0060194 | 0.239150454 | 0.005113692 | 0.240301701 | 9.23E-05 | 0.028525972 | 1 | 0.555714288 | 1 | 0.545951408 | ENSG000002 | 11 | POU5F1 |
| VAT | HTN | ENSG0000008 | -0.00479327 | 0.264934177 | 0.004299648 | 0.2403352 | -0.00662301 | 0.124835602 | 0.004315249 | 0.240301223 | 0.001829737 | 7.69E-07 | 1 | 0.572277862 | 1 | 0.382404287 | ENSG000002 | 11 | POU5F1 |
| VAT | HTN | ENSG0000008 | -0.00494491 | 0.372170835 | 0.005541046 | 0.240335421 | -0.00484522 | 0.381867726 | 0.005540805 | 0.240302008 | -9.97E-05 | 0.012424636 | 1 | 0.670600511 | 1 | 0.673023578 | ENSG000002 | 11 | POU5F1 |
| VAT | HTN | ENSG0000008 | 0.010581729 | 0.033014035 | 0.004963477 | 0.240333572 | 0.010688519 | 0.031277585 | 0.004963256 | 0.240300098 | -0.00010679 | 0.002457411 | 1 | 0.17572191 | 1 | 0.171622533 | ENSG000002 | 11 | POU5F1 |
| VAT | HTN | ENSG0000008 | 0.003260487 | 0.54799156 | 0.005427154 | 0.240335635 | 0.003139202 | 0.562962556 | 0.005426948 | 0.24030222 | 0.000121284 | 0.004924983 | 1 | 0.783206863 | 1 | 0.794767593 | ENSG000002 | 9 | CCHCR1 |
| VAT | HTN | ENSG0000008 | -0.0015696 | 0.635397372 | 0.003310397 | 0.240335703 | -0.0014912 | 0.652368048 | 0.003310283 | 0.240302285 | -7.84E-05 | 0.004818715 | 1 | 0.827629557 | 1 | 0.843737494 | ENSG000002 | 9 | CCHCR1 |
| VAT | HTN | ENSG0000009 | -0.00945507 | 0.058814937 | 0.005003799 | 0.240334052 | -0.00956276 | 0.05598442 | 0.005003638 | 0.240300584 | 0.000107689 | 0.01294064 | 1 | 0.251992588 | 1 | 0.242158595 | ENSG000002 | 9 | CCHCR1 |
| VAT | HTN | ENSG0000009 | 0.011175921 | 6.49E-05 | 0.002798024 | 0.240327945 | 0.011233713 | 5.95E-05 | 0.002797944 | 0.240294436 | -5.78E-05 | 0.022657891 | 0.179171607 | 0.001794776 | 0.164102622 | 0.001676064 | ENSG000002 | 9 | CCHCR1 |
| VAT | HTN | ENSG0000009 | 0.002179864 | 0.74323268 | 0.006654537 | 0.24033576 | 0.002285829 | 0.731212015 | 0.006654254 | 0.240302327 | -0.00010596 | 0.030295058 | 1 | 0.880557529 | 1 | 0.878114224 | ENSG000002 | 9 | CCHCR1 |
| VAT | HTN | ENSG0000009 | -0.00071808 | 0.751770371 | 0.002270201 | 0.240335764 | -0.00067769 | 0.765303975 | 0.002270136 | 0.240302341 | -4.04E-05 | 0.049158227 | 1 | 0.887082609 | 1 | 0.898827402 | ENSG000002 | 9 | CCHCR1 |
| VAT | HTN | ENSG0000009 | 0.002486031 | 0.665718663 | 0.00575428 | 0.240335721 | 0.002578737 | 0.654037133 | 0.00575404 | 0.240302286 | -9.27E-05 | 0.031053106 | 1 | 0.842665958 | 1 | 0.843796725 | ENSG000002 | 9 | CCHCR1 |
| VAT | HTN | ENSG0000009 | 0.00279801 | 0.508498527 | 0.004231857 | 0.240335598 | 0.002694224 | 0.524334532 | 0.004231687 | 0.240302185 | 0.000103786 | 0.001373846 | 1 | 0.765243148 | 1 | 0.769652481 | ENSG000002 | 3 | PSORS1C2 |
| VAT | HTN | ENSG0000009 | 0.009862716 | 0.016793209 | 0.004124567 | 0.240332993 | 0.004435554 | 0.321145576 | 0.00447084 | 0.240301899 | 0.005427161 | 0.001662983 | 1 | 0.110883392 | 1 | 0.619471487 | ENSG000002 | 3 | PSORS1C2 |
| VAT | HTN | ENSG0000009 | -0.00155055 | 0.263680437 | 0.001387223 | 0.240335197 | -0.00159687 | 0.249689088 | 0.001387245 | 0.240301731 | 4.63E-05 | 0.010703792 | 1 | 0.571238623 | 1 | 0.556098166 | ENSG000002 | 3 | PSORS1C2 |
| VAT | HTN | ENSG0000009 | 0.00154668 | 0.733263118 | 0.004538535 | 0.240335756 | 0.001623825 | 0.720494658 | 0.004538385 | 0.240302322 | -7.71E-05 | 0.046747179 | 1 | 0.875348705 | 1 | 0.872943483 | ENSG000002 | 3 | PSORS1C2 |
| VAT | HTN | ENSG0000009 | 0.020932359 | 0.001130343 | 0.00642902 | 0.240330585 | 0.021988685 | 0.000640344 | 0.006440841 | 0.240296637 | -0.00105633 | 0.007867766 | 1 | 0.0168635 | 1 | 0.01096636 | ENSG000002 | 3 | PSORS1C2 |
| VAT | HTN | ENSG0000009 | -0.0212417 | 0.001200846 | 0.006558716 | 0.24033064 | -0.02115237 | 0.00125882 | 0.006558416 | 0.240297255 | -8.93E-05 | 0.048166383 | 1 | 0.01768665 | 1 | 0.018581684 | ENSG000002 | 3 | PSORS1C2 |
| VAT | HTN | ENSG0000009 | 0.005478246 | 0.257873393 | 0.004841865 | 0.240335182 | 0.005399397 | 0.264768533 | 0.004841673 | 0.240301771 | 7.88E-05 | 0.035117179 | 1 | 0.567568234 | 1 | 0.569128623 | ENSG000002 | 3 | PSORS1C2 |
| VAT | HTN | ENSG0000009 | 0.001340323 | 0.530145962 | 0.002135008 | 0.240335619 | 0.000791546 | 0.71161417 | 0.002141113 | 0.240302317 | 0.000548777 | 0.000789965 | 1 | 0.775412218 | 1 | 0.869767456 | ENSG000002 | 6 | PSORS1C1 |
| VAT | HTN | ENSG0000009 | -0.00276412 | 0.506195237 | 0.004157984 | 0.240335595 | -0.00373373 | 0.370144228 | 0.004166164 | 0.240301989 | 0.000969613 | 0.000260767 | 1 | 0.763441997 | 1 | 0.665877875 | ENSG000002 | 6 | PSORS1C1 |
| VAT | HTN | ENSG0000009 | -0.00494597 | 0.449776485 | 0.006544124 | 0.240335532 | -0.00453108 | 0.488763721 | 0.006545182 | 0.240302148 | -0.00041489 | 0.003188993 | 1 | 0.72510695 | 1 | 0.748605922 | ENSG000002 | 6 | PSORS1C1 |
| VAT | HTN | ENSG0000009 | -0.00301762 | 0.261001848 | 0.002684653 | 0.24033519 | -0.0064426 | 0.021970359 | 0.002812273 | 0.240299797 | 0.003424975 | 4.39E-05 | 1 | 0.570001694 | 1 | 0.135655909 | ENSG000002 | 6 | PSORS1C1 |
| VAT | HTN | ENSG0000009 | -0.00300472 | 0.511598736 | 0.004577918 | 0.240335601 | -0.00292247 | 0.523205614 | 0.004577719 | 0.240302184 | -8.22E-05 | 0.012672818 | 1 | 0.767398104 | 1 | 0.769652481 | ENSG000002 | 6 | PSORS1C1 |
| VAT | HTN | ENSG0000009 | -0.00229533 | 0.404957901 | 0.002756164 | 0.240335471 | -0.00245157 | 0.373771472 | 0.002756337 | 0.240301995 | 0.00015624 | 0.000493889 | 1 | 0.695156362 | 1 | 0.668408979 | ENSG000002 | 6 | PSORS1C1 |
| VAT | HTN | ENSG0000001 | -0.0060149 | 0.122337621 | 0.003893043 | 0.240334636 | -0.00613764 | 0.114896227 | 0.003893042 | 0.240301159 | 0.000122735 | 0.007496077 | 1 | 0.378109558 | 1 | 0.366605303 | ENSG000002 | 6 | PSORS1C1 |
| VAT | HTN | ENSG0000001 | -0.00096015 | 0.79423704 | 0.00368137 | 0.24033578 | -0.0010181 | 0.782113932 | 0.003681224 | 0.240302347 | 5.80E-05 | 0.041911372 | 1 | 0.911852841 | 1 | 0.912852444 | ENSG000002 | 5 | DHX16 |
| VAT | HTN | ENSG0000001 | -0.0009837 | 0.869490161 | 0.00598698 | 0.2403358 | -0.00111309 | 0.852503465 | 0.005986785 | 0.240302368 | 0.000129396 | 0.011886871 | 1 | 0.95263744 | 1 | 0.94611263 | ENSG000002 | 5 | DHX16 |
| VAT | HTN | ENSG0000001 | -0.01299247 | 0.05205007 | 0.00668776 | 0.240333952 | -0.01285663 | 0.054542834 | 0.006687434 | 0.240300562 | -0.00013585 | 0.001587154 | 1 | 0.229853111 | 1 | 0.238570877 | ENSG000002 | 5 | DHX16 |
| VAT | HTN | ENSG0000001 | 0.00768675 | 0.219970833 | 0.006266676 | 0.240335071 | 0.007588526 | 0.225900549 | 0.006266392 | 0.240301662 | 9.82E-05 | 0.024328223 | 1 | 0.523830457 | 1 | 0.526616698 | ENSG000002 | 5 | DHX16 |
| VAT | HTN | ENSG0000001 | -7.24E-05 | 0.976882858 | 0.002498505 | 0.240335813 | -2.30E-05 | 0.992661261 | 0.002498409 | 0.240302385 | -4.94E-05 | 0.012294079 | 1 | 0.98866831 | 1 | 0.997393679 | ENSG000002 | 5 | DHX16 |
| VAT | HTN | ENSG0000001 | -0.00529079 | 0.261878214 | 0.004715648 | 0.240335193 | -0.00514151 | 0.275567863 | 0.00471555 | 0.240301799 | -0.00014928 | 0.001348023 | 1 | 0.570001694 | 1 | 0.580170048 | ENSG000002 | 5 | DHX16 |
| VAT | HTN | ENSG0000001 | 0.003102247 | 0.299601911 | 0.002990727 | 0.240335283 | 0.003026091 | 0.311602331 | 0.002990603 | 0.24030188 | 7.62E-05 | 0.000661311 | 1 | 0.601382744 | 1 | 0.610862846 | ENSG000002 | 5 | DHX16 |
| VAT | HTN | ENSG0000001 | -0.00013131 | 0.967847266 | 0.00325769 | 0.240335813 | -0.00028939 | 0.929216094 | 0.003257794 | 0.240302381 | 0.000158081 | 0.000662684 | 1 | 0.985704227 | 1 | 0.973666067 | ENSG000002 | 8 | ABCF1 |
| VAT | HTN | ENSG0000001 | -0.00304802 | 0.126182067 | 0.001993034 | 0.24033466 | -0.00307526 | 0.122812703 | 0.001992941 | 0.240301211 | 2.72E-05 | 0.042660468 | 1 | 0.388252513 | 1 | 0.379153311 | ENSG000002 | 8 | ABCF1 |
| VAT | HTN | ENSG0000001 | -0.00985223 | 0.108331967 | 0.006135639 | 0.240334542 | -0.00998909 | 0.103504322 | 0.006135426 | 0.240301078 | 0.000136855 | 0.007722973 | 1 | 0.353658534 | 1 | 0.344320158 | ENSG000002 | 8 | ABCF1 |
| VAT | HTN | ENSG0000001 | 0.004729315 | 0.406637955 | 0.005699169 | 0.240335474 | 0.004598555 | 0.419716791 | 0.005698945 | 0.240302064 | 0.00013076 | 0.003150389 | 1 | 0.696665428 | 1 | 0.703433516 | ENSG000002 | 8 | ABCF1 |
| VAT | HTN | ENSG0000001 | -0.00762928 | 0.20628294 | 0.006036516 | 0.240335026 | -0.00752502 | 0.212531219 | 0.006036255 | 0.240301618 | -0.00010426 | 0.017358183 | 1 | 0.50538674 | 1 | 0.51251931 | ENSG000002 | 8 | ABCF1 |
| VAT | HTN | ENSG0000001 | -0.00547881 | 0.138572224 | 0.003699068 | 0.240334731 | -0.00554434 | 0.133895764 | 0.003698896 | 0.240301277 | 6.55E-05 | 0.009189635 | 1 | 0.406439256 | 1 | 0.400381698 | ENSG000002 | 8 | ABCF1 |
| VAT | HTN | ENSG0000001 | 0.005436448 | 0.303266233 | 0.005280897 | 0.240335291 | 0.005555148 | 0.292811749 | 0.005280687 | 0.240301839 | -0.0001187 | 0.003517113 | 1 | 0.605216777 | 1 | 0.592989502 | ENSG000002 | 8 | ABCF1 |
| VAT | HTN | ENSG0000001 | 0.001895595 | 0.242634718 | 0.001622347 | 0.24033514 | 0.001868487 | 0.249420983 | 0.001622291 | 0.240301731 | 2.71E-05 | 0.046664695 | 1 | 0.552590633 | 1 | 0.556098166 | ENSG000002 | 5 | HLA-E |
| VAT | HTN | ENSG0000001 | -0.00032846 | 0.937596548 | 0.004195418 | 0.24033581 | -0.00039238 | 0.925482885 | 0.004195241 | 0.24030238 | 6.39E-05 | 0.039638041 | 1 | 0.976192895 | 1 | 0.973079147 | ENSG000002 | 5 | HLA-E |
| VAT | HTN | ENSG0000001 | -0.01050929 | 0.013081437 | 0.004234963 | 0.240332776 | -0.01059897 | 0.012321052 | 0.004234805 | 0.240299296 | 8.97E-05 | 0.008397951 | 1 | 0.094022827 | 1 | 0.090201866 | ENSG000002 | 5 | HLA-E |
| VAT | HTN | ENSG0000001 | 9.25E-05 | 0.980174018 | 0.003722267 | 0.240335813 | 0.000165471 | 0.964540943 | 0.003722125 | 0.240302384 | -7.30E-05 | 0.013334509 | 1 | 0.990945161 | 1 | 0.985245375 | ENSG000002 | 5 | HLA-E |
| VAT | HTN | ENSG0000001 | 0.002689386 | 0.173870393 | 0.001977673 | 0.240334901 | 0.002637424 | 0.182318979 | 0.001977597 | 0.240301508 | 5.20E-05 | 0.000906274 | 1 | 0.458916465 | 1 | 0.472932691 | ENSG000002 | 5 | HLA-E |
| VAT | HTN | ENSG0000001 | -0.00574328 | 0.193706036 | 0.004418943 | 0.24033498 | -0.00567409 | 0.199109155 | 0.004418747 | 0.240301572 | -6.92E-05 | 0.027558906 | 1 | 0.489138755 | 1 | 0.493820082 | ENSG000002 | 5 | HLA-E |
| VAT | HTN | ENSG0000001 | -0.00138845 | 0.783301896 | 0.005048563 | 0.240335776 | -0.00147398 | 0.770306548 | 0.005048327 | 0.240302343 | 8.55E-05 | 0.012120644 | 1 | 0.904415117 | 1 | 0.903546992 | ENSG000002 | 5 | HLA-E |
| VAT | HTN | ENSG0000001 | 0.014299976 | 0.038332943 | 0.006903924 | 0.240333697 | 0.014443919 | 0.036419101 | 0.006903623 | 0.240300226 | -0.00014394 | 0.003908486 | 1 | 0.19180587 | 1 | 0.186988481 | ENSG000002 | 6 | TRIM31 |
| VAT | HTN | ENSG0000001 | 0.000826608 | 0.896123139 | 0.006331234 | 0.240335805 | 0.000728385 | 0.908404598 | 0.006330964 | 0.240302378 | 9.82E-05 | 0.034114541 | 1 | 0.959290439 | 1 | 0.96802961 | ENSG000002 | 6 | TRIM31 |
| VAT | HTN | ENSG0000001 | -0.00271213 | 0.557214123 | 0.004620441 | 0.240335643 | -0.00257639 | 0.577100119 | 0.004620288 | 0.240302231 | -0.00013574 | 0.00056661 | 1 | 0.788268056 | 1 | 0.80282073 | ENSG000002 | 6 | TRIM31 |
| VAT | HTN | ENSG0000001 | -0.0035144 | 0.460173531 | 0.004758439 | 0.240335544 | -0.00344348 | 0.469257723 | 0.004758239 | 0.240302127 | -7.09E-05 | 0.044279733 | 1 | 0.733862963 | 1 | 0.73755282 | ENSG000002 | 4 | RNF39 |
| VAT | HTN | ENSG0000001 | 0.004270556 | 0.536113996 | 0.006902461 | 0.240335625 | 0.004732043 | 0.49315907 | 0.006905134 | 0.240302153 | -0.00046149 | 0.026981985 | 1 | 0.777236628 | 1 | 0.749901432 | ENSG000002 | 4 | RNF39 |
| VAT | HTN | ENSG0000001 | -0.00209474 | 0.590255472 | 0.003890199 | 0.24033567 | -0.00218646 | 0.574078181 | 0.003890114 | 0.240302229 | 9.17E-05 | 0.015867256 | 1 | 0.807604154 | 1 | 0.801328599 | ENSG000002 | 4 | RNF39 |
| VAT | HTN | ENSG0000001 | 0.003909603 | 0.540488292 | 0.006387455 | 0.240335629 | 0.003997912 | 0.531361078 | 0.006387154 | 0.240302192 | -8.83E-05 | 0.039440253 | 1 | 0.778573949 | 1 | 0.773365062 | ENSG000002 | 4 | RNF39 |
| VAT | HTN | ENSG0000001 | 0.002025062 | 0.666944971 | 0.004705672 | 0.240335722 | 0.001927909 | 0.682013211 | 0.004705458 | 0.240302302 | 9.72E-05 | 0.00287391 | 1 | 0.842887455 | 1 | 0.856249354 | ENSG000002 | 4 | RNF39 |
| VAT | HTN | ENSG0000001 | 0.001715488 | 0.475814554 | 0.002405842 | 0.240335563 | 0.001675176 | 0.486223116 | 0.002405725 | 0.240302146 | 4.03E-05 | 0.009829773 | 1 | 0.741910088 | 1 | 0.746371412 | ENSG000002 | 4 | RNF39 |
| VAT | HTN | ENSG0000001 | -0.00384299 | 0.023223415 | 0.001693135 | 0.240333273 | -0.00388233 | 0.021845433 | 0.001693084 | 0.240299792 | 3.93E-05 | 0.008712286 | 1 | 0.139902718 | 1 | 0.135186986 | ENSG000002 | 6 | HLA-J |
| VAT | HTN | ENSG0000001 | 0.005456065 | 0.194559937 | 0.004206004 | 0.240334983 | 0.005560455 | 0.186141611 | 0.004205834 | 0.240301523 | -0.00010439 | 0.001154556 | 1 | 0.489950206 | 1 | 0.477318114 | ENSG000002 | 6 | HLA-J |
| VAT | HTN | ENSG0000001 | 0.004869576 | 0.276155766 | 0.004471616 | 0.240335228 | 0.004730079 | 0.290139991 | 0.004471553 | 0.240301833 | 0.000139497 | 0.003058341 | 1 | 0.584877028 | 1 | 0.59182601 | ENSG000002 | 6 | HLA-J |
| VAT | HTN | ENSG0000001 | -0.00700173 | 0.00748941 | 0.00261818 | 0.240332286 | -0.00775657 | 0.00315018 | 0.002626936 | 0.240298085 | 0.00075484 | 0.00049121 | 1 | 0.066465499 | 1 | 0.035927668 | ENSG000002 | 6 | HLA-J |
| VAT | HTN | ENSG0000001 | 0.001629219 | 0.661766686 | 0.003724137 | 0.240335719 | 0.001567394 | 0.673834997 | 0.003723992 | 0.240302297 | 6.18E-05 | 0.034067941 | 1 | 0.840330084 | 1 | 0.850381614 | ENSG000002 | 6 | HLA-J |
| VAT | HTN | ENSG0000001 | 0.003078085 | 0.56829902 | 0.00539486 | 0.240335653 | 0.003360297 | 0.533393018 | 0.00539516 | 0.240302193 | -0.00028221 | 0.000951944 | 1 | 0.794096482 | 1 | 0.774008796 | ENSG000002 | 6 | HLA-J |
| VAT | HTN | ENSG0000001 | 0.006355327 | 0.150457492 | 0.004419807 | 0.240334794 | 0.007843448 | 0.077228372 | 0.004438843 | 0.240300845 | -0.00148812 | 0.000324997 | 1 | 0.423737427 | 1 | 0.293595464 | ENSG000002 | 6 | HLA-J |
| VAT | HTN | ENSG0000001 | 0.004388593 | 0.496552546 | 0.00645453 | 0.240335585 | 0.004189371 | 0.516288239 | 0.00645434 | 0.240302177 | 0.000199223 | 0.000568913 | 1 | 0.754348741 | 1 | 0.768034541 | ENSG000002 | 6 | HLA-J |
| VAT | HTN | ENSG0000001 | 0.006082695 | 0.319719806 | 0.006113047 | 0.240335325 | 0.005076983 | 0.406510797 | 0.006116477 | 0.240302045 | 0.001005712 | 3.60E-06 | 1 | 0.622518701 | 1 | 0.690868103 | ENSG000002 | 6 | HLA-J |
| VAT | HTN | ENSG0000001 | 0.002717014 | 0.568952277 | 0.004770076 | 0.240335653 | 0.00210114 | 0.659892887 | 0.004774653 | 0.240302289 | 0.000615874 | 0.004434994 | 1 | 0.794096482 | 1 | 0.846845836 | ENSG000002 | 6 | HLA-J |
| VAT | HTN | ENSG0000001 | -0.0040514 | 0.552015464 | 0.006812012 | 0.240335639 | -0.00427501 | 0.530287587 | 0.006812025 | 0.240302191 | 0.000223608 | 0.006047697 | 1 | 0.785341587 | 1 | 0.772344982 | ENSG000002 | 6 | HCG9 |
| VAT | HTN | ENSG0000001 | 0.001752896 | 0.319323796 | 0.001760201 | 0.240335324 | 0.001725354 | 0.326966061 | 0.001760127 | 0.240301911 | 2.75E-05 | 0.03312299 | 1 | 0.622410789 | 1 | 0.624860924 | ENSG000002 | 6 | HCG9 |
| VAT | HTN | ENSG0000001 | 0.00020064 | 0.973878611 | 0.006127492 | 0.240335813 | 0.000313247 | 0.959227101 | 0.006127252 | 0.240302383 | -0.00011261 | 0.018195734 | 1 | 0.988563798 | 1 | 0.983460383 | ENSG000002 | 6 | HCG9 |
| VAT | HTN | ENSG0000001 | -0.00035098 | 0.925762948 | 0.003766844 | 0.240335809 | -0.00027333 | 0.942151965 | 0.003766675 | 0.240302382 | -7.77E-05 | 0.003457333 | 1 | 0.971153834 | 1 | 0.978587755 | ENSG000002 | 6 | HCG9 |
| VAT | HTN | ENSG0000001 | -0.00690311 | 0.199721401 | 0.00538318 | 0.240335002 | -0.00699913 | 0.193518155 | 0.005382937 | 0.240301551 | 9.60E-05 | 0.010563929 | 1 | 0.49587241 | 1 | 0.485554643 | ENSG000002 | 6 | HCG9 |
| VAT | HTN | ENSG0000011 | 0.004027444 | 0.340038748 | 0.004221243 | 0.240335364 | 0.004998984 | 0.236927985 | 0.00422675 | 0.240301695 | -0.00097154 | 1.14E-05 | 1 | 0.642812975 | 1 | 0.542174105 | ENSG000002 | 6 | HCG9 |
| VAT | HTN | ENSG0000011 | -0.01158404 | 0.022776773 | 0.005087045 | 0.240333256 | -0.00796675 | 0.123012267 | 0.005165653 | 0.240301212 | -0.00361729 | 5.80E-05 | 1 | 0.138162403 | 1 | 0.379345092 | ENSG000002 | 6 | HCG9 |
| VAT | HTN | ENSG0000011 | 0.001367014 | 0.705030854 | 0.003611296 | 0.240335743 | 0.001445158 | 0.689019913 | 0.003611218 | 0.240302306 | -7.81E-05 | 0.027087465 | 1 | 0.861817357 | 1 | 0.860491307 | ENSG000002 | 6 | HCG9 |
| VAT | HTN | ENSG0000011 | -0.00650331 | 0.287841691 | 0.006118631 | 0.240335256 | -0.0063851 | 0.296678392 | 0.006118438 | 0.240301848 | -0.0001182 | 0.026743843 | 1 | 0.59321241 | 1 | 0.59681659 | ENSG000002 | 6 | HCG9 |
| VAT | HTN | ENSG0000011 | -1.66E-05 | 0.994198379 | 0.00228339 | 0.240335813 | 1.57E-05 | 0.994507306 | 0.00228329 | 0.240302385 | -3.23E-05 | 0.047235928 | 1 | 0.997230388 | 1 | 0.997760874 | ENSG000002 | 7 | HLA-G |
| VAT | HTN | ENSG0000011 | 0.005775761 | 0.300663894 | 0.005580381 | 0.240335285 | 0.005859381 | 0.293699176 | 0.005580138 | 0.240301841 | -8.36E-05 | 0.037545981 | 1 | 0.603075834 | 1 | 0.592989502 | ENSG000002 | 7 | HLA-G |
| VAT | HTN | ENSG0000011 | 0.002597335 | 0.535210216 | 0.004188759 | 0.240335624 | 0.0026675 | 0.524222518 | 0.004188582 | 0.240302185 | -7.02E-05 | 0.023547409 | 1 | 0.777236628 | 1 | 0.769652481 | ENSG000002 | 7 | HLA-G |
| VAT | HTN | ENSG0000011 | -0.00285072 | 0.510387901 | 0.004330863 | 0.2403356 | -0.00277103 | 0.522265574 | 0.004330696 | 0.240302183 | -7.97E-05 | 0.019373197 | 1 | 0.767061751 | 1 | 0.769595828 | ENSG000002 | 7 | HLA-G |
| VAT | HTN | ENSG0000011 | 0.005453946 | 0.281298252 | 0.005062095 | 0.240335241 | 0.005358734 | 0.289763101 | 0.005061892 | 0.240301832 | 9.52E-05 | 0.013926794 | 1 | 0.586749061 | 1 | 0.59182601 | ENSG000002 | 7 | HLA-G |
| VAT | HTN | ENSG0000011 | -0.0010823 | 0.87284843 | 0.006762635 | 0.240335801 | -0.00093494 | 0.890038048 | 0.006762396 | 0.240302375 | -0.00014736 | 0.008471544 | 1 | 0.954083829 | 1 | 0.961073949 | ENSG000002 | 7 | HLA-G |
| VAT | HTN | ENSG0000011 | -0.00118001 | 0.620014004 | 0.002379863 | 0.240335692 | -0.00112695 | 0.635816519 | 0.002379761 | 0.240302274 | -5.31E-05 | 0.002163839 | 1 | 0.821428835 | 1 | 0.832512881 | ENSG000002 | 7 | HLA-G |
| VAT | HTN | ENSG0000011 | 0.000567756 | 0.768852663 | 0.001931956 | 0.240335771 | 0.000527538 | 0.784802116 | 0.001931902 | 0.240302348 | 4.02E-05 | 0.022604337 | 1 | 0.898405313 | 1 | 0.913561299 | ENSG000002 | 7 | HLA-G |
| VAT | HTN | ENSG0000011 | -0.00388692 | 0.366405524 | 0.004303365 | 0.240335411 | -0.00379715 | 0.377558408 | 0.004303189 | 0.240302001 | -8.98E-05 | 0.005767942 | 1 | 0.667014755 | 1 | 0.669772956 | ENSG000002 | 9 | HLA-F |
| VAT | HTN | ENSG0000011 | -0.00309573 | 0.450498207 | 0.004102563 | 0.240335533 | -0.00332347 | 0.41790453 | 0.004102732 | 0.240302061 | 0.000227736 | 0.000191538 | 1 | 0.725846499 | 1 | 0.702429993 | ENSG000002 | 9 | HLA-F |
| VAT | HTN | ENSG0000011 | 0.027687316 | 3.01E-08 | 0.00499663 | 0.240320671 | 0.027607456 | 3.29E-08 | 0.004996429 | 0.24028733 | 7.99E-05 | 0.035688624 | 8.30E-05 | 3.19E-06 | 9.08E-05 | 3.24E-06 | ENSG000002 | 9 | HLA-F |
| VAT | HTN | ENSG0000011 | -0.0021929 | 0.718916171 | 0.006092954 | 0.240335749 | -0.00208392 | 0.732326179 | 0.006092737 | 0.240302327 | -0.00010899 | 0.029578225 | 1 | 0.86836264 | 1 | 0.878114224 | ENSG000002 | 9 | HLA-F |
| VAT | HTN | ENSG0000011 | 0.004321756 | 0.536108878 | 0.006985128 | 0.240335625 | 0.004528502 | 0.516772548 | 0.006984883 | 0.240302177 | -0.00020675 | 0.00037212 | 1 | 0.777236628 | 1 | 0.768034541 | ENSG000002 | 9 | HLA-F |
| VAT | HTN | ENSG0000011 | 0.005477176 | 0.29012408 | 0.005177646 | 0.240335261 | 0.00540039 | 0.296918374 | 0.005177421 | 0.240301848 | 7.68E-05 | 0.039684028 | 1 | 0.59321241 | 1 | 0.596864321 | ENSG000002 | 9 | HLA-F |
| VAT | HTN | ENSG0000011 | 0.003209815 | 0.635920999 | 0.006780203 | 0.240335703 | 0.003346741 | 0.62157425 | 0.006779985 | 0.240302265 | -0.00013693 | 0.01939499 | 1 | 0.827897149 | 1 | 0.826686941 | ENSG000002 | 9 | HLA-F |
| VAT | HTN | ENSG0000011 | 0.004845368 | 0.380118401 | 0.005520661 | 0.240335433 | 0.004963973 | 0.368549346 | 0.005520465 | 0.240301986 | -0.00011861 | 0.009198381 | 1 | 0.673481092 | 1 | 0.664239406 | ENSG000002 | 5 | ZFP57 |
| VAT | HTN | ENSG0000011 | 0.0134173 | 0.039075949 | 0.006502546 | 0.240333714 | 0.013542726 | 0.037274195 | 0.006502317 | 0.240300246 | -0.00012543 | 0.020125598 | 1 | 0.192933127 | 1 | 0.189809556 | ENSG000002 | 5 | ZFP57 |
| VAT | HTN | ENSG0000011 | -0.00645524 | 0.326478801 | 0.006578708 | 0.240335339 | -0.00656363 | 0.318401278 | 0.00657843 | 0.240301894 | 0.000108387 | 0.025858613 | 1 | 0.627494074 | 1 | 0.616693002 | ENSG000002 | 5 | ZFP57 |
| VAT | HTN | ENSG0000011 | 0.010754131 | 0.030034439 | 0.004956638 | 0.240333492 | 0.010835223 | 0.028808133 | 0.004956397 | 0.240300028 | -8.11E-05 | 0.011703822 | 1 | 0.167127118 | 1 | 0.164277785 | ENSG000002 | 5 | ZFP57 |
| VAT | HTN | ENSG0000011 | 0.002954879 | 0.539874907 | 0.004820336 | 0.240335628 | 0.003119169 | 0.517565822 | 0.004820207 | 0.240302178 | -0.00016429 | 0.000226788 | 1 | 0.778497687 | 1 | 0.768034541 | ENSG000002 | 5 | ZFP57 |
| VAT | HTN | ENSG0000011 | -0.0147576 | 0.003376498 | 0.005034628 | 0.240331576 | -0.01485142 | 0.003178075 | 0.00503441 | 0.240298093 | 9.38E-05 | 0.010154132 | 1 | 0.037128021 | 1 | 0.03594872 | ENSG000002 | 5 | ZFP57 |
| VAT | HTN | ENSG0000011 | 0.00489329 | 0.422031643 | 0.006094503 | 0.240335495 | 0.004780334 | 0.432808079 | 0.006094286 | 0.240302081 | 0.000112956 | 0.024196686 | 1 | 0.706371944 | 1 | 0.711889332 | ENSG000002 | 9 | CRHR1-IT1 |
| VAT | HTN | ENSG0000011 | -0.00012453 | 0.977205489 | 0.004358375 | 0.240335813 | -4.95E-05 | 0.990936923 | 0.004358213 | 0.240302385 | -7.50E-05 | 0.032676404 | 1 | 0.98866831 | 1 | 0.997393679 | ENSG000002 | 9 | CRHR1-IT1 |
| VAT | HTN | ENSG0000011 | 0.006022248 | 0.362796936 | 0.006617496 | 0.240335405 | 0.0091636 | 0.17482335 | 0.006753488 | 0.240301477 | -0.00314135 | 0.020037731 | 1 | 0.665980483 | 1 | 0.461734397 | ENSG000002 | 2 | OR2H2 |
| VAT | HTN | ENSG0000011 | -0.00790787 | 0.051628001 | 0.004063178 | 0.240333945 | -0.00796923 | 0.049831793 | 0.004063012 | 0.240300488 | 6.14E-05 | 0.046738959 | 1 | 0.229793907 | 1 | 0.224365004 | ENSG000002 | 6 | FAM153C |
| VAT | HTN | ENSG0000011 | -0.00483766 | 0.427948172 | 0.006102701 | 0.240335503 | -0.00505604 | 0.407377815 | 0.006102512 | 0.240302046 | 0.000218377 | 4.78E-05 | 1 | 0.710672055 | 1 | 0.69191555 | ENSG000002 | 6 | FAM153C |
| VAT | HTN | ENSG0000011 | -0.0001378 | 0.959251316 | 0.002697017 | 0.240335812 | -0.0001823 | 0.946105974 | 0.002696898 | 0.240302383 | 4.45E-05 | 0.021050607 | 1 | 0.98228229 | 1 | 0.9791881 | ENSG000002 | 6 | FAM153C |
| VAT | HTN | ENSG0000011 | -0.00441669 | 0.434077262 | 0.005646245 | 0.240335512 | -0.004319 | 0.444292202 | 0.005646003 | 0.240302096 | -9.77E-05 | 0.017789713 | 1 | 0.713724511 | 1 | 0.720050779 | ENSG000002 | 6 | FAM153C |

| | | | | | | | | | | | | | | | | | | | |
|---|---|---|---|---|---|---|---|---|---|---|---|---|---|---|---|---|---|---|---|
| VAT | HTN | ENSG0000011 | -0.00927838 | 0.129085254 | 0.006113362 | 0.240334677 | -0.00942393 | 0.123177287 | 0.006113177 | 0.240301213 | 0.000145546 | 0.007292169 | | 1 | 0.391942027 | | 1 | 0.379430035 | ENSG000002 | 6 | GABBR1 |
| VAT | HTN | ENSG0000011 | 0.003669897 | 0.485138274 | 0.005257259 | 0.240335573 | 0.00376297 | 0.474118807 | 0.005257056 | 0.240302132 | -9.31E-05 | 0.024595489 | | 1 | 0.748439173 | | 1 | 0.740140219 | ENSG000002 | 4 | C6orf100 |
| VAT | HTN | ENSG0000011 | 0.011374051 | 0.037713602 | 0.005473663 | 0.240333684 | 0.011291417 | 0.039117338 | 0.005473416 | 0.240300286 | 8.26E-05 | 0.030616059 | | 1 | 0.189598436 | | 1 | 0.193604012 | ENSG000002 | 4 | C6orf100 |
| VAT | HTN | ENSG0000011 | -0.00029925 | 0.913907159 | 0.00276794 | 0.240335808 | 2.68E-05 | 0.992279471 | 0.002769441 | 0.240302385 | -0.00032604 | 0.000760369 | | 1 | 0.966800981 | | 1 | 0.997393679 | ENSG000002 | 4 | C6orf100 |
| VAT | HTN | ENSG0000011 | -0.00602248 | 0.186058351 | 0.004554442 | 0.240334951 | -0.00612683 | 0.178534878 | 0.004554309 | 0.240301492 | 0.000104346 | 0.010769649 | | 1 | 0.477250044 | | 1 | 0.46743134 | ENSG000002 | 4 | C6orf100 |
| VAT | HTN | ENSG0000011 | -0.00874472 | 0.007808064 | 0.003287157 | 0.240332323 | -0.00870228 | 0.008109568 | 0.003286998 | 0.240298928 | -4.24E-05 | 0.046028598 | | 1 | 0.067768104 | | 1 | 0.070885362 | ENSG000002 | 4 | C6orf100 |
| VAT | HTN | ENSG0000011 | -0.00356833 | 0.390155493 | 0.004152422 | 0.240335449 | -0.00366617 | 0.377275116 | 0.004152283 | 0.240302 | 9.78E-05 | 0.005565291 | | 1 | 0.682680345 | | 1 | 0.669772956 | ENSG000002 | 4 | C6orf100 |
| VAT | HTN | ENSG0000011 | -0.00765934 | 0.159089796 | 0.005439328 | 0.240334835 | -0.00754845 | 0.165192211 | 0.005439065 | 0.240301435 | -0.0001109 | 0.001720057 | | 1 | 0.43652375 | | 1 | 0.449635604 | ENSG000002 | 4 | C6orf100 |
| VAT | HTN | ENSG0000011 | -0.00473035 | 0.060158099 | 0.002516631 | 0.240334071 | -0.00467711 | 0.063092103 | 0.002516542 | 0.240300681 | -5.32E-05 | 0.010159904 | | 1 | 0.252880519 | | 1 | 0.260935385 | ENSG000002 | 6 | TRIM27 |
| VAT | HTN | ENSG0000011 | 0.008307396 | 0.154398073 | 0.005833152 | 0.240334813 | 0.008395044 | 0.150079187 | 0.005832913 | 0.240301363 | -8.76E-05 | 0.047181202 | | 1 | 0.430829979 | | 1 | 0.423969862 | ENSG000002 | 6 | TRIM27 |
| VAT | HTN | ENSG0000011 | 0.003704994 | 0.523764923 | 0.005811271 | 0.240335613 | 0.00380251 | 0.512878783 | 0.005811017 | 0.240302174 | -9.75E-05 | 0.019733152 | | 1 | 0.772678586 | | 1 | 0.76521159 | ENSG000002 | 6 | TRIM27 |
| VAT | HTN | ENSG0000011 | 0.000420526 | 0.949516605 | 0.006641933 | 0.240335811 | 0.000573488 | 0.931190799 | 0.00664169 | 0.240302381 | -0.00015296 | 0.004615603 | | 1 | 0.979347123 | | 1 | 0.974255726 | ENSG000002 | 6 | TRIM27 |
| VAT | HTN | ENSG0000012 | 0.000490433 | 0.889828467 | 0.003540486 | 0.240335804 | 0.000418683 | 0.905862614 | 0.003540396 | 0.240302378 | 7.17E-05 | 0.030682481 | | 1 | 0.958223397 | | 1 | 0.967584999 | ENSG000002 | 6 | TRIM27 |
| VAT | HTN | ENSG0000012 | -0.00924399 | 0.036175059 | 0.00441249 | 0.240333649 | -0.0091559 | 0.037981726 | 0.004412359 | 0.240300261 | -8.81E-05 | 0.025396762 | | 1 | 0.185702856 | | 1 | 0.191644541 | ENSG000002 | 6 | TRIM27 |
| VAT | HTN | ENSG0000012 | 0.002771032 | 0.613329739 | 0.005483646 | 0.240335687 | 0.002676321 | 0.625495837 | 0.005483403 | 0.240302267 | 9.47E-05 | 0.015070341 | | 1 | 0.819907808 | | 1 | 0.827116213 | ENSG000002 | 3 | CNN2P12 |
| VAT | HTN | ENSG0000012 | 0.013321105 | 5.21E-05 | 0.003292517 | 0.24032774 | 0.013377832 | 4.84E-05 | 0.003292379 | 0.240294243 | -5.67E-05 | 0.020541572 | 0.143896838 | 0.001530817 | 0.133575816 | 0.001451911 | ENSG000002 | 5 | MALRD1 |
| VAT | HTN | ENSG0000012 | -0.01059005 | 0.094686396 | 0.00633686 | 0.240334436 | -0.01069391 | 0.091481236 | 0.006336608 | 0.24030098 | 0.00010386 | 0.034016844 | | 1 | 0.329967743 | | 1 | 0.324534976 | ENSG000002 | 5 | MALRD1 |
| VAT | HTN | ENSG0000012 | -0.01256389 | 0.058673988 | 0.006645335 | 0.24033405 | -0.01270576 | 0.055874316 | 0.006645205 | 0.240300582 | 0.000141872 | 0.032799649 | | 1 | 0.251851022 | | 1 | 0.242158595 | ENSG000002 | 4 | RP11-15J10.1 |
| VAT | HTN | ENSG0000012 | 0.001722663 | 0.604789677 | 0.003328654 | 0.240335681 | 0.001667465 | 0.616399556 | 0.00332854 | 0.240302261 | 5.52E-05 | 0.048498374 | | 1 | 0.81487424 | | 1 | 0.824255221 | ENSG000002 | 4 | RP11-15J10.1 |
| VAT | HTN | ENSG0000012 | -0.00067643 | 0.851192621 | 0.003605777 | 0.240335796 | -0.00062226 | 0.862980315 | 0.003605623 | 0.24030237 | -5.42E-05 | 0.03971432 | | 1 | 0.942297189 | | 1 | 0.949828955 | ENSG000002 | 4 | RP11-15J10.1 |
| VAT | HTN | ENSG0000012 | -0.00643071 | 0.330662179 | 0.006610638 | 0.240335347 | -0.00632645 | 0.338542648 | 0.006610364 | 0.240301933 | -0.00010426 | 0.035281734 | | 1 | 0.630703258 | | 1 | 0.637283706 | ENSG000002 | 4 | RP11-15J10.1 |
| VAT | HTN | ENSG0000012 | -0.00530173 | 0.28405763 | 0.004949106 | 0.240335247 | -0.0053935 | 0.275784843 | 0.004948896 | 0.240301799 | 9.18E-05 | 0.011871724 | | 1 | 0.589701944 | | 1 | 0.580170048 | ENSG000002 | 4 | RP11-15J10.1 |
| VAT | HTN | ENSG0000012 | 0.002696957 | 0.32772721 | 0.002755644 | 0.240335341 | 0.002752825 | 0.317789324 | 0.002755547 | 0.240301893 | -5.59E-05 | 0.014655093 | | 1 | 0.627864285 | | 1 | 0.616693002 | ENSG000002 | 3 | ZNF204P |
| VAT | HTN | ENSG0000012 | -0.0060509 | 0.036774325 | 0.00289757 | 0.240333663 | -0.00609776 | 0.035333162 | 0.002897451 | 0.240300201 | 4.69E-05 | 0.03292148 | | 1 | 0.18691922 | | 1 | 0.183949871 | ENSG000002 | 3 | ZNF204P |
| VAT | HTN | ENSG0000012 | 0.014454048 | 0.012155542 | 0.005764083 | 0.240332712 | 0.014550091 | 0.011590379 | 0.005763814 | 0.240299242 | -9.60E-05 | 0.013932582 | | 1 | 0.089779421 | | 1 | 0.08740286 | ENSG000002 | 3 | ZNF204P |
| VAT | HTN | ENSG0000012 | -0.0071221 | 0.016552971 | 0.002971877 | 0.240332981 | -0.00718892 | 0.015560983 | 0.002971781 | 0.240299499 | 6.68E-05 | 0.009085121 | | 1 | 0.110087228 | | 1 | 0.105524112 | ENSG000002 | 3 | ZNF204P |
| VAT | HTN | ENSG0000012 | -0.01922515 | 0.003412163 | 0.006566069 | 0.240331585 | -0.01907151 | 0.00367685 | 0.006565864 | 0.240298224 | -0.00015364 | 0.007395161 | | 1 | 0.037371304 | | 1 | 0.039953174 | ENSG000002 | 3 | ZNF204P |
| VAT | HTN | ENSG0000012 | -0.00039488 | 0.871613095 | 0.002443387 | 0.2403358 | -0.00048875 | 0.84145359 | 0.002443315 | 0.240302365 | 9.39E-05 | 1.79E-05 | | 1 | 0.953488761 | | 1 | 0.940295692 | ENSG000002 | 3 | ZNF204P |
| VAT | HTN | ENSG0000012 | -0.00073706 | 0.835881581 | 0.003557869 | 0.240335792 | -0.00084218 | 0.812875504 | 0.003557752 | 0.240302357 | 0.000105121 | 0.000566303 | | 1 | 0.934399823 | | 1 | 0.928203638 | ENSG000002 | 2 | AC104135.3 |
| VAT | HTN | ENSG0000012 | -0.01750487 | 1.62E-05 | 0.004059497 | 0.240326643 | -0.01736898 | 1.88E-05 | 0.004059756 | 0.240293359 | -0.00013588 | 0.040102766 | 0.04464555 | 0.00055118 | 0.051980452 | 0.000641734 | ENSG000002 | 2 | AC104135.3 |
| VAT | HTN | ENSG0000012 | 0.005294769 | 0.358388734 | 0.005764956 | 0.240335397 | 0.005381761 | 0.350526972 | 0.00576472 | 0.240301955 | -8.70E-05 | 0.046612275 | | 1 | 0.662083605 | | 1 | 0.650170996 | ENSG000002 | 2 | AC104135.3 |
| VAT | HTN | ENSG0000012 | 0.006681546 | 0.163400512 | 0.004794011 | 0.240334855 | 0.006792434 | 0.156512614 | 0.004793868 | 0.240301395 | -0.00011089 | 0.009370476 | | 1 | 0.441724348 | | 1 | 0.433565668 | ENSG000002 | 2 | AC104135.3 |
| VAT | HTN | ENSG0000012 | -0.00661435 | 0.199786275 | 0.005158749 | 0.24033500 | -0.00670868 | 0.193434968 | 0.005158597 | 0.240301551 | 9.43E-05 | 0.040900412 | | 1 | 0.49587241 | | 1 | 0.485554643 | ENSG000002 | 2 | AC104135.3 |
| VAT | HTN | ENSG0000012 | 0.004935495 | 0.474048648 | 0.006894038 | 0.240335561 | 0.00480517 | 0.48578452 | 0.006893781 | 0.240302145 | 0.000130325 | 0.018689189 | | 1 | 0.741580968 | | 1 | 0.746371412 | ENSG000002 | 2 | AC104135.3 |
| VAT | HTN | ENSG0000012 | 0.003812377 | 0.432478309 | 0.004856788 | 0.240335509 | 0.003902956 | 0.421603632 | 0.00485658 | 0.240302066 | -9.06E-05 | 0.010789813 | | 1 | 0.713571923 | | 1 | 0.704374107 | ENSG000002 | 2 | AC104135.3 |
| VAT | HTN | ENSG0000012 | 0.001217019 | 0.649975831 | 0.002681863 | 0.240335712 | 0.001111282 | 0.67861266 | 0.00268194 | 0.2403023 | 0.000105737 | 0.005000299 | | 1 | 0.835367368 | | 1 | 0.853678642 | ENSG000002 | 4 | MROH6 |
| VAT | HTN | ENSG0000012 | 0.004275023 | 0.161248238 | 0.003051652 | 0.240334845 | 0.004333829 | 0.155552393 | 0.003051579 | 0.24030139 | -5.88E-05 | 0.043630622 | | 1 | 0.439333798 | | 1 | 0.432828844 | ENSG000002 | 4 | MROH6 |
| VAT | HTN | ENSG0000012 | -0.00937413 | 0.060356479 | 0.00499106 | 0.240334074 | -0.00945271 | 0.058224141 | 0.004990841 | 0.240300616 | 7.86E-05 | 0.028255045 | | 1 | 0.252880519 | | 1 | 0.248759487 | ENSG000002 | 6 | ZNF155 |
| VAT | HTN | ENSG0000012 | 0.000984699 | 0.630473605 | 0.002046934 | 0.240335699 | 0.001018224 | 0.618864793 | 0.002046845 | 0.240302263 | -3.35E-05 | 0.023940323 | | 1 | 0.825823207 | | 1 | 0.825554802 | ENSG000002 | 6 | ZNF155 |
| VAT | HTN | ENSG0000012 | 0.004830661 | 0.050984366 | 0.002475211 | 0.240333935 | 0.005068328 | 0.04073654 | 0.002476938 | 0.24030032 | -0.00023767 | 0.014282636 | | 1 | 0.229180536 | | 1 | 0.198159466 | ENSG000002 | 2 | CD177P1 |
| VAT | HTN | ENSG0000012 | 0.007079439 | 0.06037091 | 0.00376951 | 0.240334074 | 0.007135162 | 0.058364979 | 0.003769333 | 0.240300618 | -5.57E-05 | 0.027818783 | | 1 | 0.252880519 | | 1 | 0.2489758 | ENSG000002 | 2 | CD177P1 |
| VAT | HTN | ENSG0000012 | -0.00400677 | 0.253368491 | 0.003507923 | 0.24033517 | -0.00382612 | 0.275420693 | 0.003508056 | 0.240301798 | -0.00018066 | 0.000445014 | | 1 | 0.563738197 | | 1 | 0.580170048 | ENSG000002 | 5 | ZNF783 |
| VAT | HTN | ENSG0000012 | -0.00194237 | 0.637604116 | 0.004123487 | 0.240335704 | -0.00107115 | 0.79545375 | 0.004130917 | 0.240302352 | -0.00087123 | 0.000556148 | | 1 | 0.828194673 | | 1 | 0.918168712 | ENSG000002 | 5 | ZNF783 |
| VAT | HTN | ENSG0000012 | 0.006109848 | 0.217211425 | 0.004951339 | 0.240335062 | 0.005679163 | 0.25147728 | 0.004952312 | 0.240301736 | 0.000430685 | 0.000161682 | | 1 | 0.519951026 | | 1 | 0.558388812 | ENSG000002 | 4 | ZNF425 |
| VAT | HTN | ENSG0000012 | -0.0109964 | 0.030326225 | 0.005077268 | 0.2403335 | -0.01108522 | 0.02900733 | 0.00507706 | 0.240300034 | 8.88E-05 | 0.020694249 | | 1 | 0.168253907 | | 1 | 0.164732983 | ENSG000002 | 4 | ZNF425 |
| VAT | HTN | ENSG0000012 | -0.00764449 | 0.082135603 | 0.004397351 | 0.240334323 | -0.00754892 | 0.08602806 | 0.00439725 | 0.240300931 | -9.56E-05 | 0.024330063 | | 1 | 0.303472909 | | 1 | 0.314034154 | ENSG000002 | 4 | ZNF425 |
| VAT | HTN | ENSG0000012 | -0.0072254 | 0.239343732 | 0.006140758 | 0.240335131 | -0.00841614 | 0.171617776 | 0.006156536 | 0.240301463 | 0.001190744 | 0.007648572 | | 1 | 0.549661061 | | 1 | 0.456168919 | ENSG000002 | 4 | ZNF425 |
| VAT | HTN | ENSG0000012 | -0.00080905 | 0.857145933 | 0.004494495 | 0.240335797 | -0.00190283 | 0.673557443 | 0.004516874 | 0.240302297 | 0.00109378 | 0.015570232 | | 1 | 0.945910746 | | 1 | 0.850381614 | ENSG000002 | 5 | C12orf73 |
| VAT | HTN | ENSG0000012 | 0.001360259 | 0.742627474 | 0.004142381 | 0.24033576 | 0.0012929 | 0.754943968 | 0.004142221 | 0.240302337 | 6.74E-05 | 0.038549873 | | 1 | 0.880434634 | | 1 | 0.892265229 | ENSG000002 | 3 | ARHGEF34P |
| VAT | HTN | ENSG0000012 | -0.00120895 | 0.756109562 | 0.003892394 | 0.240335766 | -0.00445672 | 0.276750075 | 0.004097568 | 0.240301600 | 0.003247767 | 0.011246662 | | 1 | 0.890491808 | | 1 | 0.581087867 | ENSG000002 | 3 | ARHGEF34P |
| VAT | HTN | ENSG0000012 | -0.0033251 | 0.03401232 | 0.001568492 | 0.240333597 | -0.00329571 | 0.035617605 | 0.001568434 | 0.240300207 | -2.94E-05 | 0.020071473 | | 1 | 0.178807628 | | 1 | 0.184436381 | ENSG000002 | 3 | ARHGEF34P |
| VAT | HTN | ENSG0000012 | -0.00317732 | 0.092584961 | 0.001889105 | 0.240334418 | -0.00314372 | 0.096073888 | 0.001889034 | 0.240301019 | -3.36E-05 | 0.026678076 | | 1 | 0.32589131 | | 1 | 0.333838771 | ENSG000002 | 5 | TRIM13 |
| VAT | HTN | ENSG0000012 | -0.00386467 | 0.388975412 | 0.004486095 | 0.240335447 | -0.00378439 | 0.39888398 | 0.004485914 | 0.240302034 | -8.03E-05 | 0.019205252 | | 1 | 0.682680345 | | 1 | 0.68668204 | ENSG000002 | 6 | SPIRE2 |
| VAT | HTN | ENSG0000012 | -0.00120166 | 0.717037952 | 0.003315642 | 0.240335749 | -0.00110771 | 0.738303037 | 0.003315502 | 0.24030233 | -9.39E-05 | 0.000124054 | | 1 | 0.867520033 | | 1 | 0.881179801 | ENSG000002 | 6 | SPIRE2 |
| VAT | HTN | ENSG0000013 | 0.009643055 | 0.000229056 | 0.002617099 | 0.240329118 | 0.009688663 | 0.000213751 | 0.002616992 | 0.240295626 | -4.56E-05 | 0.021787159 | 0.632193253 | 0.005057546 | 0.589951873 | 0.004835671 | ENSG000002 | 6 | SPIRE2 |
| VAT | HTN | ENSG0000013 | 0.004378075 | 0.369356605 | 0.0048771 | 0.240335416 | 0.004538191 | 0.352089672 | 0.00487692 | 0.240301958 | -0.00016012 | 4.85E-05 | | 1 | 0.668640123 | | 1 | 0.651755529 | ENSG000002 | 6 | SPIRE2 |
| VAT | HTN | ENSG0000013 | 0.007031341 | 0.033405599 | 0.003305475 | 0.240333582 | 0.006925391 | 0.036153028 | 0.003305348 | 0.24030022 | 0.00010595 | 4.74E-05 | | 1 | 0.176610754 | | 1 | 0.186465689 | ENSG000002 | 6 | SPIRE2 |
| VAT | HTN | ENSG0000013 | 0.006896631 | 0.315019086 | 0.006864046 | 0.240335315 | 0.007036241 | 0.305304342 | 0.006863783 | 0.240301867 | -0.00013961 | 0.010196752 | | 1 | 0.618387395 | | 1 | 0.604212978 | ENSG000002 | 6 | SPIRE2 |
| VAT | HTN | ENSG0000013 | -0.02296525 | 0.000165273 | 0.006096507 | 0.240328815 | -0.02311843 | 0.000149318 | 0.006096261 | 0.240295293 | 0.000153188 | 0.001051882 | 0.456152124 | 0.003801268 | 0.412116568 | 0.003552729 | ENSG000002 | 7 | PKHD1L1 |
| VAT | HTN | ENSG0000013 | 0.005883347 | 0.262272038 | 0.005248123 | 0.240335194 | 0.005964783 | 0.255705811 | 0.005247915 | 0.240301748 | -8.14E-05 | 0.045026568 | | 1 | 0.570001694 | | 1 | 0.562797479 | ENSG000002 | 7 | PKHD1L1 |
| VAT | HTN | ENSG0000013 | -0.00372606 | 0.31983026 | 0.003745507 | 0.240335325 | -0.00343298 | 0.359613507 | 0.003747376 | 0.240301971 | -0.00029308 | 0.020329204 | | 1 | 0.622518701 | | 1 | 0.657742398 | ENSG000002 | 4 | SLFN12L |
| VAT | HTN | ENSG0000013 | 0.02497382 | 1.05E-05 | 0.005668137 | 0.240326239 | 0.024846442 | 1.17E-05 | 0.005667903 | 0.240292908 | 0.000127377 | 0.002699129 | 0.029067997 | 0.000398192 | 0.032209103 | 0.000435258 | ENSG000002 | 4 | SLFN12L |
| VAT | HTN | ENSG0000013 | 0.000691692 | 0.874576243 | 0.004382002 | 0.240335801 | 0.000772065 | 0.860138427 | 0.004381803 | 0.240302369 | -8.04E-05 | 0.008385419 | | 1 | 0.954460432 | | 1 | 0.949732956 | ENSG000002 | 3 | FLJ00104 |
| VAT | HTN | ENSG0000013 | -0.0092884 | 0.075106793 | 0.005218743 | 0.240334251 | -0.00941686 | 0.071157968 | 0.005218622 | 0.240300779 | 0.000128458 | 0.010692665 | | 1 | 0.290259707 | | 1 | 0.278181293 | ENSG000002 | 3 | FLJ00104 |
| VAT | HTN | ENSG0000013 | 0.010619409 | 0.04129088 | 0.005204055 | 0.24033376 | 0.010730292 | 0.039210036 | 0.005203874 | 0.240300288 | -0.00011088 | 0.010700553 | | 1 | 0.200638783 | | 1 | 0.193604012 | ENSG000002 | 3 | CLLU1OS |
| VAT | HTN | ENSG0000013 | 0.004650136 | 0.252019166 | 0.004059623 | 0.240335166 | 0.004713891 | 0.245556368 | 0.004059458 | 0.24030172 | -6.38E-05 | 0.039077566 | | 1 | 0.562761245 | | 1 | 0.551175422 | ENSG000002 | 5 | SYCE1L |
| VAT | HTN | ENSG0000013 | -0.00511798 | 0.205707311 | 0.004044365 | 0.240335024 | -0.00518855 | 0.199507889 | 0.004044216 | 0.240301573 | 7.06E-05 | 0.031402284 | | 1 | 0.505148194 | | 1 | 0.494292435 | ENSG000002 | 5 | SYCE1L |
| VAT | HTN | ENSG0000013 | -0.00534829 | 0.10076015 | 0.003258805 | 0.240334485 | -0.00529715 | 0.104044569 | 0.003258665 | 0.240301082 | -5.11E-05 | 0.030660116 | | 1 | 0.33914392 | | 1 | 0.344320158 | ENSG000002 | 6 | TMEM231 |
| VAT | HTN | ENSG0000013 | -0.00394057 | 0.574206945 | 0.007013351 | 0.240335658 | -0.00378535 | 0.589364451 | 0.007013047 | 0.240302241 | -0.00015522 | 0.00219823 | | 1 | 0.797991524 | | 1 | 0.809098734 | ENSG000002 | 5 | FAM205A |
| VAT | HTN | ENSG0000013 | -0.00120865 | 0.566223037 | 0.00210704 | 0.240335651 | -0.00288216 | 0.180706145 | 0.002153141 | 0.240301501 | 0.001673517 | 0.000163125 | | 1 | 0.79306004 | | 1 | 0.469962518 | ENSG000002 | 4 | TMEM170B |
| VAT | HTN | ENSG0000013 | 0.015147241 | 0.007077869 | 0.005624318 | 0.240332236 | 0.014889446 | 0.00811586 | 0.005624555 | 0.240298929 | 0.000257795 | 0.002137501 | | 1 | 0.064259602 | | 1 | 0.070885362 | ENSG000002 | 4 | TMEM170B |
| VAT | HTN | ENSG0000013 | 0.002230812 | 0.307380113 | 0.002185493 | 0.2403353 | 0.002263062 | 0.300418035 | 0.002185397 | 0.240301856 | -3.23E-05 | 0.037702368 | | 1 | 0.609460569 | | 1 | 0.599966554 | ENSG000002 | 4 | TMEM170B |
| VAT | HTN | ENSG0000013 | -0.00266616 | 0.135614788 | 0.001786579 | 0.240334715 | -0.00262211 | 0.142179601 | 0.001786516 | 0.240301322 | -4.40E-05 | 0.00285392 | | 1 | 0.402902923 | | 1 | 0.412928922 | ENSG000002 | 4 | TMEM170B |
| VAT | HTN | ENSG0000013 | -0.00370523 | 0.542594527 | 0.006085176 | 0.240335631 | -0.0038681 | 0.524983046 | 0.006084949 | 0.240302186 | 0.00016287 | 0.000871609 | | 1 | 0.780738853 | | 1 | 0.769900748 | ENSG000002 | 4 | TMEM170B |
| VAT | HTN | ENSG0000013 | -0.00024601 | 0.970850051 | 0.006732096 | 0.240335813 | -4.83E-05 | 0.994278036 | 0.006731839 | 0.240302385 | -0.00019773 | 0.000207508 | | 1 | 0.987235058 | | 1 | 0.997760874 | ENSG000002 | 4 | TMEM170B |
| VAT | HTN | ENSG0000013 | -0.00194768 | 0.765766763 | 0.006537657 | 0.24033577 | -0.00159524 | 0.807226266 | 0.00653772 | 0.240302355 | -0.00035244 | 1.83E-05 | | 1 | 0.896317331 | | 1 | 0.924888243 | ENSG000002 | 4 | TMEM170B |
| VAT | HTN | ENSG0000013 | -0.00211513 | 0.544537063 | 0.003490507 | 0.240335632 | -0.00205329 | 0.556348087 | 0.00349036 | 0.240302214 | -6.18E-05 | 0.0166438 | | 1 | 0.780738853 | | 1 | 0.791790437 | ENSG000002 | 9 | NT5M |
| VAT | HTN | ENSG0000013 | 0.000380644 | 0.908284367 | 0.003304115 | 0.240335807 | 0.000293829 | 0.929136338 | 0.003303986 | 0.240302381 | 8.68E-05 | 0.000792584 | | 1 | 0.964197726 | | 1 | 0.973666067 | ENSG000002 | 9 | NT5M |
| VAT | HTN | ENSG0000013 | 0.009407759 | 0.146045193 | 0.006471838 | 0.240334771 | 0.009593395 | 0.138241412 | 0.006471642 | 0.240301301 | -0.00018564 | 0.001227144 | | 1 | 0.417271979 | | 1 | 0.407127621 | ENSG000002 | 9 | NT5M |
| VAT | HTN | ENSG0000013 | 0.000459626 | 0.820790656 | 0.002028999 | 0.240335788 | 0.000502259 | 0.804481967 | 0.002028912 | 0.240302355 | -4.26E-05 | 0.004138857 | | 1 | 0.926718801 | | 1 | 0.924383942 | ENSG000002 | 5 | TECPR1 |
| VAT | HTN | ENSG0000013 | 0.001741713 | 0.772446437 | 0.006023012 | 0.240335772 | 0.001572813 | 0.793980717 | 0.006022754 | 0.240302351 | 0.000168901 | 0.000128406 | | 1 | 0.900698 | | 1 | 0.918168712 | ENSG000002 | 7 | ATP6AP1L |
| VAT | HTN | ENSG0000013 | 0.003031462 | 0.36673902 | 0.003358591 | 0.240335412 | 0.00307924 | 0.359213894 | 0.003358445 | 0.24030197 | -4.78E-05 | 0.048688433 | | 1 | 0.667014755 | | 1 | 0.65744718 | ENSG000002 | 7 | ATP6AP1L |
| VAT | HTN | ENSG0000013 | -0.01225057 | 0.000914821 | 0.004535686 | 0.240332216 | -0.01180958 | 0.009235234 | 0.004536508 | 0.240299043 | -0.00044099 | 1.38E-05 | | 1 | 0.062986484 | | 1 | 0.076483154 | ENSG000002 | 7 | ATP6AP1L |
| VAT | HTN | ENSG0000013 | 0.006507568 | 0.147226884 | 0.00448984 | 0.240334777 | 0.006401791 | 0.153902901 | 0.004489705 | 0.240301382 | 0.000105777 | 0.007983692 | | 1 | 0.418982277 | | 1 | 0.429931182 | ENSG000002 | 4 | CCDC85C |
| VAT | HTN | ENSG0000013 | -0.00428711 | 0.308725263 | 0.004211712 | 0.240335302 | -0.00419137 | 0.319634972 | 0.004211547 | 0.240301896 | -9.57E-05 | 0.003542618 | | 1 | 0.611674165 | | 1 | 0.617799211 | ENSG000002 | 9 | AC004980.7 |
| VAT | HTN | ENSG0000013 | -0.0052031 | 0.082027762 | 0.002991933 | 0.240334322 | -0.00513444 | 0.086131831 | 0.002991804 | 0.240300932 | -6.87E-05 | 0.001478231 | | 1 | 0.303472909 | | 1 | 0.314034154 | ENSG000002 | 9 | AC004980.7 |
| VAT | HTN | ENSG0000013 | -0.00590385 | 0.337769739 | 0.006158935 | 0.24033536 | -0.0056589 | 0.358226914 | 0.006159353 | 0.240301969 | -0.00024494 | 0.016417308 | | 1 | 0.640277802 | | 1 | 0.656234077 | ENSG000002 | 9 | AC004980.7 |
| VAT | HTN | ENSG0000013 | 0.006494722 | 0.11437372 | 0.004113607 | 0.240334584 | 0.006408136 | 0.11926796 | 0.004113432 | 0.240301188 | 8.66E-05 | 0.00414006 | | 1 | 0.36551286 | | 1 | 0.374067693 | ENSG000002 | 9 | AC004980.7 |
| VAT | HTN | ENSG0000013 | 0.008488347 | 0.15453684 | 0.005962215 | 0.240334814 | 0.008586278 | 0.149819412 | 0.005961981 | 0.240301362 | -9.79E-05 | 0.034998525 | | 1 | 0.430829979 | | 1 | 0.42366965 | ENSG000002 | 8 | RGL3 |
| VAT | HTN | ENSG0000013 | -0.00371599 | 0.237597276 | 0.003146437 | 0.240335125 | -0.00299991 | 0.3413978 | 0.00315313 | 0.240301938 | -0.00071608 | 0.00059899 | | 1 | 0.549204878 | | 1 | 0.639686306 | ENSG000002 | 8 | RGL3 |
| VAT | HTN | ENSG0000013 | 0.009662708 | 0.002245386 | 0.003162179 | 0.240331208 | 0.009614262 | 0.002361738 | 0.003162034 | 0.240297826 | 4.84E-05 | 0.025927094 | | 1 | 0.027915609 | | 1 | 0.02884247 | ENSG000002 | 8 | RGL3 |
| VAT | HTN | ENSG0000013 | -0.00497153 | 0.473831103 | 0.006940959 | 0.24033556 | -0.00508631 | 0.463665018 | 0.006940681 | 0.24030212 | 0.000114778 | 0.031223773 | | 1 | 0.741580968 | | 1 | 0.733361289 | ENSG000002 | 8 | NAP1L4 |
| VAT | HTN | ENSG0000013 | -0.00078407 | 0.700105316 | 0.002035609 | 0.24033574 | -0.0008357 | 0.681398891 | 0.002035534 | 0.240302302 | 5.16E-05 | 0.001658964 | | 1 | 0.860778235 | | 1 | 0.856249354 | ENSG000002 | 8 | CPT1B |
| VAT | HTN | ENSG0000013 | 0.003401973 | 0.34059866 | 0.003569811 | 0.240335365 | 0.003457406 | 0.332769165 | 0.003569664 | 0.240301922 | -5.54E-05 | 0.039177246 | | 1 | 0.642908166 | | 1 | 0.631637966 | ENSG000002 | 8 | CPT1B |
| VAT | HTN | ENSG0000013 | -0.00423801 | 0.361021543 | 0.004639706 | 0.240335402 | -0.00412747 | 0.3736625 | 0.004639514 | 0.240301994 | -0.00011053 | 0.00149603 | | 1 | 0.665223012 | | 1 | 0.668408979 | ENSG000002 | 8 | CPT1B |
| VAT | HTN | ENSG0000013 | -0.00232036 | 0.693883896 | 0.005895365 | 0.240335737 | -0.0021757 | 0.712079761 | 0.005895187 | 0.240302318 | -0.00014465 | 0.005655797 | | 1 | 0.858028474 | | 1 | 0.869767456 | ENSG000002 | 8 | CPT1B |
| VAT | HTN | ENSG0000013 | -0.01042314 | 0.078768456 | 0.005929376 | 0.240334289 | -0.01029113 | 0.08262202 | 0.005929179 | 0.240300899 | -0.00013201 | 0.008845076 | | 1 | 0.297402103 | | 1 | 0.306698747 | ENSG000002 | 6 | RP11-497H16.6 |
| VAT | HTN | ENSG0000013 | -0.01012407 | 0.108532166 | 0.006308504 | 0.240334543 | -0.01001949 | 0.11221449 | 0.00630823 | 0.240301141 | -0.00010458 | 0.021365655 | | 1 | 0.353658534 | | 1 | 0.362660414 | ENSG000002 | 6 | RP11-497H16.6 |
| VAT | HTN | ENSG0000013 | 0.00037064 | 0.892579674 | 0.002744654 | 0.240335804 | 0.000442707 | 0.871854895 | 0.002744571 | 0.240302372 | -7.21E-05 | 0.003020984 | | 1 | 0.959290439 | | 1 | 0.956043422 | ENSG000002 | 8 | SMN2 |
| VAT | HTN | ENSG0000013 | -0.0091143 | 0.0874382 | 0.005332895 | 0.240334373 | -0.00903527 | 0.090203831 | 0.005332661 | 0.240300969 | -7.90E-05 | 0.038688236 | | 1 | 0.315463309 | | 1 | 0.32217795 | ENSG000002 | 8 | SMN2 |
| VAT | HTN | ENSG0000013 | 0.00016439 | 0.906135688 | 0.001394148 | 0.240335807 | 0.00019406 | 0.889291472 | 0.001394097 | 0.240302375 | -2.97E-05 | 0.008936233 | | 1 | 0.963884879 | | 1 | 0.960643625 | ENSG000002 | 8 | SMN2 |
| VAT | HTN | ENSG0000013 | 0.010971407 | 0.015239733 | 0.004521234 | 0.240332909 | 0.007206738 | 0.128305718 | 0.0047372 | 0.240301244 | 0.003764669 | 0.008024307 | | 1 | 0.105417704 | | 1 | 0.390434159 | ENSG000002 | 8 | SMN2 |
| VAT | HTN | ENSG0000014 | 0.003138911 | 0.114706338 | 0.001989939 | 0.240334586 | 0.003089364 | 0.120531771 | 0.001989867 | 0.240301196 | 4.95E-05 | 0.002213235 | | 1 | 0.365576781 | | 1 | 0.376532767 | ENSG000002 | 5 | SERF1B |
| VAT | HTN | ENSG0000014 | 0.000988229 | 0.598214639 | 0.001875303 | 0.240335676 | 0.001021718 | 0.585858665 | 0.001875237 | 0.240302238 | -3.35E-05 | 0.031032934 | | 1 | 0.81054119 | | 1 | 0.807829017 | ENSG000002 | 5 | SERF1B |
| VAT | HTN | ENSG0000014 | 0.008287685 | 0.051833652 | 0.004262078 | 0.240333949 | 0.008394324 | 0.048885549 | 0.004261958 | 0.240300472 | -0.00010664 | 0.005978118 | | 1 | 0.229793907 | | 1 | 0.221550272 | ENSG000002 | 4 | POM121B |
| VAT | HTN | ENSG0000014 | 1.74E-05 | 0.994918358 | 0.002734858 | 0.240335813 | -4.82E-05 | 0.9859397 | 0.002734766 | 0.240302385 | 6.56E-05 | 0.004620682 | | 1 | 0.997448117 | | 1 | 0.996044499 | ENSG000002 | 4 | POM121B |
| VAT | HTN | ENSG0000014 | -0.00373788 | 0.415323613 | 0.004588829 | 0.240335486 | -0.00367006 | 0.423818148 | 0.004588634 | 0.240302069 | -6.78E-05 | 0.044436139 | | 1 | 0.701684662 | | 1 | 0.704889231 | ENSG000002 | 4 | POM121B |
| VAT | HTN | ENSG0000014 | -0.01086496 | 0.000605586 | 0.003168438 | 0.240330014 | -0.01092071 | 0.000567152 | 0.00316829 | 0.240296526 | 5.57E-05 | 0.009214846 | | 1 | 0.010578596 | | 1 | 0.010098967 | ENSG000002 | 4 | POM121B |
| VAT | HTN | ENSG0000014 | -0.00280835 | 0.324814807 | 0.002852243 | 0.240335335 | -0.00274332 | 0.336127103 | 0.002852136 | 0.240301929 | -6.50E-05 | 0.004267197 | | 1 | 0.626300732 | | 1 | 0.635876512 | ENSG000002 | 4 | POM121B |
| VAT | HTN | ENSG0000014 | 0.012178569 | 0.034783237 | 0.005769431 | 0.240333616 | 0.012294314 | 0.033086614 | 0.00576917 | 0.240300145 | -0.00011575 | 0.004188466 | | 1 | 0.18113535 | | 1 | 0.176632601 | ENSG000002 | 9 | STAG3L1 |
| VAT | HTN | ENSG0000014 | -0.02671043 | 2.14E-07 | 0.005149977 | 0.240322547 | -0.02660741 | 2.38E-07 | 0.00514983 | 0.240289221 | -0.00010302 | 0.026886975 | 0.000591608 | 1.64E-05 | 0.000658012 | 1.88E-05 | ENSG000002 | 9 | STAG3L1 |
| VAT | HTN | ENSG0000014 | 0.004241125 | 0.544525813 | 0.006998744 | 0.240335632 | 0.004456652 | 0.524253229 | 0.006998474 | 0.240302185 | -0.00021553 | 9.27E-05 | | 1 | 0.780738853 | | 1 | 0.769652481 | ENSG000002 | 6 | CDPF1 |
| VAT | HTN | ENSG0000014 | -0.00654957 | 0.351206661 | 0.007025538 | 0.240335385 | -0.00666436 | 0.342811718 | 0.007025284 | 0.240301941 | 0.000114788 | 0.045504677 | | 1 | 0.655395798 | | 1 | 0.641030042 | ENSG000002 | 6 | CDPF1 |
| VAT | HTN | ENSG0000014 | 0.013156465 | 0.04162127 | 0.006457818 | 0.240333766 | 0.013003722 | 0.044039184 | 0.006457553 | 0.240300385 | 0.000152743 | 0.001694539 | | 1 | 0.20153457 | | 1 | 0.209205076 | ENSG000002 | 6 | LIN52 |
| VAT | HTN | ENSG0000014 | -0.00980289 | 0.031018806 | 0.004545028 | 0.240333519 | -0.00991416 | 0.029153321 | 0.004544834 | 0.240300038 | 0.000111276 | 0.000881549 | | 1 | 0.169864891 | | 1 | 0.165136315 | ENSG000002 | 5 | LINC00634 |
| VAT | HTN | ENSG0000014 | -0.00411081 | 0.195936406 | 0.003178742 | 0.240334989 | -0.00416132 | 0.190480478 | 0.003178615 | 0.24030154 | 5.05E-05 | 0.038814327 | | 1 | 0.491622256 | | 1 | 0.482167409 | ENSG000002 | 3 | C17orf107 |
| VAT | HTN | ENSG0000014 | 0.001935969 | 0.386357292 | 0.002234902 | 0.240335443 | 0.001980536 | 0.375497429 | 0.002234804 | 0.240301997 | -4.46E-05 | 0.005378083 | | 1 | 0.680067683 | | 1 | 0.668627681 | ENSG000002 | 3 | C17orf107 |
| VAT | HTN | ENSG0000014 | -0.00087303 | 0.737731322 | 0.002607153 | 0.240335758 | -0.00080844 | 0.756485738 | 0.00260705 | 0.240302337 | -6.46E-05 | 0.001357132 | | 1 | 0.878176701 | | 1 | 0.892265229 | ENSG000002 | 3 | C17orf107 |
| VAT | HTN | ENSG0000014 | 0.002588395 | 0.419138356 | 0.003203785 | 0.240335491 | 0.002061217 | 0.520948699 | 0.003211198 | 0.240302182 | 0.000527178 | 0.017211757 | | 1 | 0.704520013 | | 1 | 0.768793407 | ENSG000002 | 3 | C17orf107 |
| VAT | HTN | ENSG0000014 | 4.52E-05 | 0.99417445 | 0.006195834 | 0.240335813 | 0.000201988 | 0.973992102 | 0.006195595 | 0.240302384 | -0.00015675 | 0.001287689 | | 1 | 0.997230388 | | 1 | 0.989761439 | ENSG000002 | 2 | CATSPER2P1 |
| VAT | HTN | ENSG0000014 | 0.024144966 | 3.91E-05 | 0.005870643 | 0.240327471 | 0.023751049 | 5.24E-05 | 0.005872228 | 0.240294318 | 0.000393917 | 0.009972471 | 0.107888249 | 0.001198758 | 0.144656132 | 0.001530313 | ENSG000002 | 2 | CATSPER2P1 |
| VAT | HTN | ENSG0000014 | 0.002249956 | 0.693834757 | 0.005715529 | 0.240335737 | 0.002085045 | 0.715247554 | 0.005715305 | 0.240302319 | 0.000164911 | 0.00021101 | | 1 | 0.858028474 | | 1 | 0.870791023 | ENSG000002 | 2 | CATSPER2P1 |
| VAT | HTN | ENSG0000014 | -0.00382954 | 0.573285871 | 0.006799371 | 0.240335657 | -0.00506016 | 0.457679007 | 0.006813422 | 0.240302113 | 0.00123062 | 0.005648931 | | 1 | 0.797624595 | | 1 | 0.728905978 | ENSG000002 | 2 | CATSPER2P1 |
| VAT | HTN | ENSG0000014 | -0.00246564 | 0.534939457 | 0.003973735 | 0.240335623 | -0.00251994 | 0.525965627 | 0.003973553 | 0.240302186 | 5.43E-05 | 0.047350043 | | 1 | 0.777236628 | | 1 | 0.770114128 | ENSG000002 | 2 | CATSPER2P1 |
| VAT | HTN | ENSG0000014 | 0.002541749 | 0.705079799 | 0.006715809 | 0.240335743 | 0.002663959 | 0.691601969 | 0.006715599 | 0.240302307 | -0.00012221 | 0.037763131 | | 1 | 0.861817357 | | 1 | 0.861126259 | ENSG000002 | 2 | CATSPER2P1 |
| VAT | HTN | ENSG0000014 | 0.004579621 | 0.464621533 | 0.006262656 | 0.24033555 | 0.0047298 | 0.450088238 | 0.006262409 | 0.240302103 | -0.00015018 | 0.001986285 | | 1 | 0.737028926 | | 1 | 0.723286064 | ENSG000002 | 6 | TPTE2P6 |
| VAT | HTN | ENSG0000014 | 0.005440835 | 0.096542095 | 0.003273949 | 0.240334451 | 0.005386497 | 0.099903794 | 0.003273823 | 0.24030105 | 5.43E-05 | 0.035111025 | | 1 | 0.333487087 | | 1 | 0.33915679 | ENSG000002 | 3 | LINC00487 |

| | | | | | | | | | | | | | | | | | | | |
|---|---|---|---|---|---|---|---|---|---|---|---|---|---|---|---|---|---|---|---|
| VAT | HTN | ENSG0000014 | 0.004738387 | 0.370176398 | 0.00528753 | 0.240335417 | 0.004835646 | 0.360417794 | 0.005287343 | 0.240301972 | -9.73E-05 | 0.026142706 | 1 | 0.66884126 | 1 | 0.658776895 | ENSG0000020 | 5 | PDPK2 |
| VAT | HTN | ENSG0000014 | 0.008389141 | 0.170407501 | 0.006119451 | 0.240334886 | 0.008480814 | 0.165766951 | 0.006119194 | 0.240301438 | -9.17E-05 | 0.04385992 | 1 | 0.454626907 | 1 | 0.450561028 | ENSG0000020 | 7 | RNPS1 |
| VAT | HTN | ENSG0000014 | 0.000966462 | 0.823264941 | 0.00432716 | 0.240335789 | 0.000876646 | 0.839447462 | 0.004326969 | 0.240302365 | 8.98E-05 | 0.00354412 | 1 | 0.928190865 | 1 | 0.94007326 | ENSG0000020 | 7 | RNPS1 |
| VAT | HTN | ENSG0000014 | -0.01267788 | 0.027273612 | 0.005742871 | 0.24033341 | -0.01275798 | 0.026308351 | 0.005742606 | 0.240299951 | 8.01E-05 | 0.041393603 | 1 | 0.15617255 | 1 | 0.15378118 | ENSG0000020 | 5 | NYNRIN |
| VAT | HTN | ENSG0000014 | -0.00394422 | 0.444063233 | 0.00515348 | 0.240335524 | -0.00383858 | 0.456343778 | 0.005153279 | 0.240302111 | -0.00010564 | 0.008768552 | 1 | 0.721374057 | 1 | 0.728497638 | ENSG0000020 | 5 | NYNRIN |
| VAT | HTN | ENSG0000014 | 0.004007247 | 0.417416355 | 0.004941652 | 0.240335489 | 0.003877929 | 0.432590594 | 0.004941504 | 0.240302081 | 0.000129318 | 0.003233546 | 1 | 0.702909786 | 1 | 0.711889332 | ENSG0000020 | 5 | NYNRIN |
| VAT | HTN | ENSG0000014 | -0.01632534 | 0.000485476 | 0.004679534 | 0.240329811 | -0.01624424 | 0.000517647 | 0.004679343 | 0.240296442 | -8.11E-05 | 0.021847921 | 1 | 0.009115068 | 1 | 0.009771271 | ENSG0000020 | 2 | IFITM5 |
| VAT | HTN | ENSG0000014 | -0.01919498 | 5.32E-06 | 0.004217048 | 0.240325596 | -0.01909674 | 5.94E-06 | 0.004216946 | 0.240292272 | -9.82E-05 | 0.014180058 | 0.014686681 | 0.000225949 | 0.016393759 | 0.000252212 | ENSG0000020 | 2 | IFITM5 |
| VAT | HTN | ENSG0000014 | -0.00323109 | 0.239452676 | 0.002746693 | 0.240335131 | -0.00328619 | 0.231521597 | 0.002746619 | 0.240301679 | 5.51E-05 | 0.030008282 | 1 | 0.549661061 | 1 | 0.534280609 | ENSG0000020 | 2 | IFITM5 |
| VAT | HTN | ENSG0000014 | 0.004369627 | 0.271480488 | 0.003973613 | 0.240335217 | 0.004433165 | 0.264552793 | 0.003973452 | 0.240301771 | -6.35E-05 | 0.035651443 | 1 | 0.579737555 | 1 | 0.569128623 | ENSG0000020 | 1 | CTA-373H7.7 |
| VAT | HTN | ENSG0000014 | -0.013079 | 0.00049983 | 0.003757392 | 0.240329838 | -0.01213279 | 0.001274589 | 0.003766006 | 0.240297267 | -0.00094621 | 0.000249255 | 1 | 0.009321154 | 1 | 0.018581684 | ENSG0000020 | 6 | ZDHHC11B |
| VAT | HTN | ENSG0000014 | 0.006704518 | 0.184606514 | 0.005053506 | 0.240334945 | 0.006594582 | 0.191894341 | 0.00505333 | 0.240301545 | 0.000109936 | 0.008868708 | 1 | 0.474407803 | 1 | 0.484121007 | ENSG0000020 | 3 | RP11-503N18.1 |
| VAT | HTN | ENSG0000014 | 0.005941854 | 0.133956995 | 0.003964722 | 0.240334706 | 0.006049 | 0.127068659 | 0.003964565 | 0.240301237 | -0.00010715 | 0.000485215 | 1 | 0.400743043 | 1 | 0.387952984 | ENSG0000020 | 4 | P2RX6P |
| VAT | HTN | ENSG0000014 | -0.00241254 | 0.59338885 | 0.004518443 | 0.240335673 | -0.00255847 | 0.571260696 | 0.004518698 | 0.240302227 | 0.000145928 | 0.041950834 | 1 | 0.808777562 | 1 | 0.801076492 | ENSG0000020 | 4 | P2RX6P |
| VAT | HTN | ENSG0000014 | 0.000736061 | 0.893189836 | 0.005481983 | 0.240335804 | 0.000818158 | 0.881355353 | 0.005481747 | 0.240302374 | -8.21E-05 | 0.040224543 | 1 | 0.959290439 | 1 | 0.959100799 | ENSG0000020 | 4 | HBA1 |
| VAT | HTN | ENSG0000014 | 0.010829674 | 0.001366551 | 0.003382537 | 0.240330758 | 0.010890964 | 0.001282493 | 0.003382405 | 0.240297272 | -6.13E-05 | 0.020932534 | 1 | 0.019243274 | 1 | 0.018581684 | ENSG0000020 | 4 | HBA1 |
| VAT | HTN | ENSG0000014 | 0.001494034 | 0.688051805 | 0.003721126 | 0.240335734 | 0.001410767 | 0.70458886 | 0.003721029 | 0.240302314 | 8.33E-05 | 0.016346621 | 1 | 0.855718693 | 1 | 0.868203529 | ENSG0000020 | 6 | HLA-H |
| VAT | HTN | ENSG0000014 | 0.012778606 | 0.036964434 | 0.006125413 | 0.240333667 | 0.012624727 | 0.03929477 | 0.006125261 | 0.24030029 | 0.000153878 | 0.007836077 | 1 | 0.187472424 | 1 | 0.193604012 | ENSG0000020 | 6 | HLA-H |
| VAT | HTN | ENSG0000014 | -0.0033317 | 0.431117522 | 0.004231904 | 0.240335508 | -0.00325935 | 0.44117098 | 0.004231721 | 0.240302092 | -7.24E-05 | 0.018142294 | 1 | 0.713571923 | 1 | 0.717128893 | ENSG0000020 | 6 | HLA-H |
| VAT | HTN | ENSG0000014 | -0.01399074 | 0.001968088 | 0.004520399 | 0.240331089 | -0.01410957 | 0.001800158 | 0.004520299 | 0.24029758 | 0.000118833 | 0.006999627 | 1 | 0.0258663 | 1 | 0.024236277 | ENSG0000020 | 6 | HLA-H |
| VAT | HTN | ENSG0000014 | 0.007490868 | 0.001156702 | 0.002305332 | 0.240330606 | 0.007556028 | 0.001046632 | 0.002305282 | 0.240297087 | -6.52E-05 | 0.00402222 | 1 | 0.01716397 | 1 | 0.016048355 | ENSG0000020 | 6 | HLA-H |
| VAT | HTN | ENSG0000015 | 0.001864831 | 0.213729101 | 0.00149981 | 0.240335051 | 0.001826495 | 0.223276027 | 0.001499752 | 0.240301653 | 3.83E-05 | 0.001106708 | 1 | 0.513843482 | 1 | 0.523569953 | ENSG0000020 | 6 | HLA-H |
| VAT | HTN | ENSG0000015 | -0.00621461 | 0.077555382 | 0.003520916 | 0.240334277 | -0.0050599 | 0.152412565 | 0.003535758 | 0.240301375 | -0.00115471 | 0.000401024 | 1 | 0.294028647 | 1 | 0.427064649 | ENSG0000020 | 6 | HLA-H |
| VAT | HTN | ENSG0000015 | -0.00953449 | 0.111821846 | 0.00599631 | 0.240334566 | -0.00945352 | 0.114881502 | 0.005996034 | 0.240301159 | -8.10E-05 | 0.048779958 | 1 | 0.361931749 | 1 | 0.366605303 | ENSG0000020 | 6 | HLA-H |
| VAT | HTN | ENSG0000015 | -0.00262055 | 0.400705817 | 0.003118355 | 0.240335465 | -0.00266679 | 0.392425812 | 0.003118227 | 0.240302024 | 4.62E-05 | 0.049092311 | 1 | 0.692212916 | 1 | 0.681708076 | ENSG0000020 | 6 | HLA-H |
| VAT | HTN | ENSG0000015 | 0.009254341 | 0.005510473 | 0.003334196 | 0.240332014 | 0.008523796 | 0.010852081 | 0.003346029 | 0.240299185 | 0.000730545 | 0.010061598 | 1 | 0.053552481 | 1 | 0.084874033 | ENSG0000020 | 6 | HLA-H |
| VAT | HTN | ENSG0000015 | 0.002629079 | 0.348098766 | 0.002802007 | 0.240335379 | 0.002688104 | 0.337367187 | 0.002801914 | 0.240301931 | -5.90E-05 | 0.013666969 | 1 | 0.650475689 | 1 | 0.636028046 | ENSG0000020 | 6 | HLA-H |
| VAT | HTN | ENSG0000015 | -0.00875799 | 0.010306691 | 0.00341392 | 0.240332568 | -0.00914269 | 0.007446369 | 0.00341629 | 0.240298853 | 0.0003847 | 0.003953178 | 1 | 0.081742724 | 1 | 0.066988633 | ENSG0000020 | 6 | HLA-H |
| VAT | HTN | ENSG0000015 | 0.00421815 | 0.133695673 | 0.002812683 | 0.240334704 | 0.005180326 | 0.067918199 | 0.002837675 | 0.240300741 | -0.00096218 | 0.010759592 | 1 | 0.400651528 | 1 | 0.270496723 | ENSG0000020 | 6 | HCG27 |
| VAT | HTN | ENSG0000015 | -0.00445862 | 0.407568876 | 0.005383636 | 0.240335475 | -0.00463384 | 0.389373261 | 0.005383463 | 0.240302019 | 0.000175224 | 0.000171174 | 1 | 0.696936974 | 1 | 0.679740797 | ENSG0000020 | 6 | HCG27 |
| VAT | HTN | ENSG0000015 | -0.00122492 | 0.803947625 | 0.004934389 | 0.240335783 | -0.00131454 | 0.789922525 | 0.004934231 | 0.24030235 | 8.96E-05 | 0.03603581 | 1 | 0.918797285 | 1 | 0.915481504 | ENSG0000020 | 6 | HCG27 |
| VAT | HTN | ENSG0000015 | 0.010410386 | 0.117730602 | 0.006654669 | 0.240334606 | 0.010244518 | 0.123687883 | 0.006654533 | 0.240301216 | 0.000165869 | 0.011743179 | 1 | 0.369484015 | 1 | 0.379887847 | ENSG0000020 | 6 | HCG27 |
| VAT | HTN | ENSG0000015 | -0.00023655 | 0.958628746 | 0.004560069 | 0.240335812 | -0.00039629 | 0.930745142 | 0.004559909 | 0.240302381 | 0.000159739 | 2.46E-05 | 1 | 0.98228229 | 1 | 0.974255726 | ENSG0000020 | 6 | HCG27 |
| VAT | HTN | ENSG0000015 | -0.01529805 | 0.002611524 | 0.005082195 | 0.240331345 | -0.01538528 | 0.002466677 | 0.005081987 | 0.240297865 | 8.72E-05 | 0.023582377 | 1 | 0.030626438 | 1 | 0.029600124 | ENSG0000020 | 6 | HCG27 |
| VAT | HTN | ENSG0000015 | 0.0037668 | 0.524096558 | 0.005912936 | 0.240335613 | 0.003663602 | 0.535510556 | 0.005912692 | 0.240302195 | 0.000103198 | 0.020225575 | 1 | 0.772678586 | 1 | 0.776265302 | ENSG0000020 | 6 | HCG27 |
| VAT | HTN | ENSG0000015 | 0.012397515 | 0.002214721 | 0.004051696 | 0.240331196 | 0.012466744 | 0.00209083 | 0.004051546 | 0.240297716 | -6.92E-05 | 0.034152496 | 1 | 0.02785549 | 1 | 0.026978653 | ENSG0000020 | 6 | HCG27 |
| VAT | HTN | ENSG0000015 | -0.00565304 | 0.353319398 | 0.006090516 | 0.240335388 | -0.00585002 | 0.336776925 | 0.006090257 | 0.24030193 | 0.000196981 | 1.10E-05 | 1 | 0.657128126 | 1 | 0.636028046 | ENSG0000020 | 6 | HCG27 |
| VAT | HTN | ENSG0000015 | -0.00353573 | 0.316930431 | 0.003532954 | 0.240335319 | -0.00358376 | 0.310376594 | 0.003532775 | 0.240301877 | 4.80E-05 | 0.027362596 | 1 | 0.620374461 | 1 | 0.60927411 | ENSG0000020 | 6 | HCG27 |
| VAT | HTN | ENSG0000015 | -0.00536231 | 0.181661176 | 0.004014726 | 0.240334934 | -0.00530171 | 0.186628578 | 0.004014559 | 0.240301525 | -6.06E-05 | 0.043948465 | 1 | 0.470059822 | 1 | 0.477318114 | ENSG0000020 | 6 | HCG27 |
| VAT | HTN | ENSG0000015 | -0.0085896 | 0.148096386 | 0.005939059 | 0.240334782 | -0.00869895 | 0.142989213 | 0.005938852 | 0.240301327 | 0.000109342 | 0.027344849 | 1 | 0.418982277 | 1 | 0.412957063 | ENSG0000020 | 6 | HCG27 |
| VAT | HTN | ENSG0000015 | -0.00742436 | 0.038890442 | 0.003594722 | 0.24033371 | -0.00732467 | 0.041585913 | 0.003594672 | 0.240300337 | -9.97E-05 | 0.008577025 | 1 | 0.192789179 | 1 | 0.201363369 | ENSG0000020 | 6 | HCG27 |
| VAT | HTN | ENSG0000015 | 0.000555598 | 0.886287849 | 0.003885218 | 0.240335803 | 0.000471615 | 0.903381223 | 0.003885081 | 0.240302378 | 8.40E-05 | 0.009080453 | 1 | 0.957399007 | 1 | 0.967157554 | ENSG0000020 | 7 | HLA-A |
| VAT | HTN | ENSG0000015 | 0.000492383 | 0.870134722 | 0.00301174 | 0.2403358 | 0.000443499 | 0.882924441 | 0.003011611 | 0.240302374 | 4.89E-05 | 0.026240405 | 1 | 0.95263744 | 1 | 0.959100799 | ENSG0000020 | 7 | HLA-A |
| VAT | HTN | ENSG0000015 | 0.005002016 | 0.230572598 | 0.004172217 | 0.240335104 | 0.004895842 | 0.240623352 | 0.004172243 | 0.240301706 | 0.000106174 | 0.038465633 | 1 | 0.53748342 | 1 | 0.54683908 | ENSG0000020 | 7 | HLA-A |
| VAT | HTN | ENSG0000015 | 0.009816742 | 0.137588849 | 0.006611347 | 0.240334726 | 0.009680271 | 0.143128578 | 0.006611116 | 0.240301327 | 0.000136472 | 0.012999254 | 1 | 0.404531314 | 1 | 0.412957063 | ENSG0000020 | 7 | HLA-A |
| VAT | HTN | ENSG0000016 | -0.00917917 | 0.081780108 | 0.00527401 | 0.240334319 | -0.00932359 | 0.07707632 | 0.005273787 | 0.240300843 | 0.000144423 | 0.000222186 | 1 | 0.3029706 | 1 | 0.293421577 | ENSG0000020 | 7 | HLA-A |
| VAT | HTN | ENSG0000016 | -0.02705811 | 5.52E-05 | 0.006709757 | 0.240327793 | -0.02692813 | 5.98E-05 | 0.006709484 | 0.240294442 | -0.00012999 | 0.010648576 | 0.152239551 | 0.001602522 | 0.165182137 | 0.001676064 | ENSG0000020 | 7 | HLA-A |
| VAT | HTN | ENSG0000016 | -0.00253147 | 0.591365216 | 0.004715333 | 0.240335671 | -0.00263965 | 0.575605635 | 0.004715219 | 0.24030223 | 0.000108183 | 0.0159615 | 1 | 0.807604154 | 1 | 0.802143866 | ENSG0000020 | 7 | HLA-A |
| VAT | HTN | ENSG0000016 | -0.00217521 | 0.614334062 | 0.004316769 | 0.240335688 | -0.002262 | 0.600262827 | 0.004316609 | 0.240302249 | 8.68E-05 | 0.012486258 | 1 | 0.819907808 | 1 | 0.814916578 | ENSG0000020 | 7 | HLA-A |
| VAT | HTN | ENSG0000016 | 0.00288508 | 0.585027304 | 0.005283482 | 0.240335666 | 0.002793168 | 0.597024294 | 0.005283237 | 0.240302247 | 9.19E-05 | 0.010582881 | 1 | 0.804121195 | 1 | 0.814290189 | ENSG0000020 | 7 | HLA-A |
| VAT | HTN | ENSG0000016 | -0.00276433 | 0.647921408 | 0.006053476 | 0.240335711 | -0.00259252 | 0.668445039 | 0.006053287 | 0.240302294 | -0.00017181 | 0.00117605 | 1 | 0.835218069 | 1 | 0.849405298 | ENSG0000020 | 7 | HLA-A |
| VAT | HTN | ENSG0000016 | -0.00516906 | 0.081138226 | 0.002963702 | 0.240334313 | -0.00510891 | 0.084732709 | 0.002963635 | 0.240300919 | -6.02E-05 | 0.035977967 | 1 | 0.301756109 | 1 | 0.311050019 | ENSG0000020 | 3 | LNP1 |
| VAT | HTN | ENSG0000016 | -0.0037361 | 0.286513305 | 0.003505435 | 0.240335253 | -0.00243522 | 0.493413406 | 0.003555631 | 0.240302153 | -0.00130088 | 0.029265981 | 1 | 0.592342114 | 1 | 0.749901432 | ENSG0000020 | 7 | METTL6 |
| VAT | HTN | ENSG0000016 | -0.01373413 | 2.85E-05 | 0.003281437 | 0.240327174 | -0.01359099 | 3.45E-05 | 0.003281518 | 0.240293926 | -0.00014314 | 0.00143558 | 0.078580679 | 0.000913729 | 0.095173587 | 0.001119689 | ENSG0000020 | 7 | METTL6 |
| VAT | HTN | ENSG0000016 | 0.000186476 | 0.953248746 | 0.003180678 | 0.240335812 | 0.0001377 | 0.965466743 | 0.003180538 | 0.240302384 | 4.88E-05 | 0.031523803 | 1 | 0.981080063 | 1 | 0.985461617 | ENSG0000021 | 2 | TAS2R15 |
| VAT | HTN | ENSG0000016 | 0.000536465 | 0.6834937 | 0.001315831 | 0.240335731 | 0.000558055 | 0.671473953 | 0.001315776 | 0.240302296 | -2.16E-05 | 0.027647681 | 1 | 0.852822158 | 1 | 0.850122986 | ENSG0000021 | 5 | TAS2R14 |
| VAT | HTN | ENSG0000016 | 0.000933932 | 0.336277241 | 0.00097128 | 0.240335357 | 0.000961821 | 0.322029978 | 0.000971246 | 0.240301901 | -2.79E-05 | 0.000531952 | 1 | 0.638628342 | 1 | 0.620239176 | ENSG0000021 | 5 | TAS2R14 |
| VAT | HTN | ENSG0000016 | -0.00275821 | 0.289617929 | 0.002604633 | 0.24033526 | -0.00270773 | 0.298522456 | 0.002604569 | 0.240301852 | -5.05E-05 | 0.041565546 | 1 | 0.59321241 | 1 | 0.597477868 | ENSG0000021 | 5 | TAS2R14 |
| VAT | HTN | ENSG0000016 | -0.01566213 | 0.011542947 | 0.006200794 | 0.240332667 | -0.01554734 | 0.012161671 | 0.006200514 | 0.240299284 | -0.00011479 | 0.007848089 | 1 | 0.087283652 | 1 | 0.089985214 | ENSG0000021 | 5 | TAS2R14 |
| VAT | HTN | ENSG0000016 | 0.003921936 | 0.301936462 | 0.003799255 | 0.240335288 | 0.004245928 | 0.263878767 | 0.003800265 | 0.240301769 | -0.00032399 | 0.000995275 | 1 | 0.60414959 | 1 | 0.569128623 | ENSG0000021 | 4 | AC084018.1 |
| VAT | HTN | ENSG0000016 | 0.002450425 | 0.613303423 | 0.004848831 | 0.240335687 | 0.002358428 | 0.626677363 | 0.004848652 | 0.240302268 | 9.20E-05 | 0.018606524 | 1 | 0.819907808 | 1 | 0.827573934 | ENSG0000021 | 4 | AC084018.1 |
| VAT | HTN | ENSG0000016 | -0.00644771 | 0.047327809 | 0.003250901 | 0.240333873 | -0.00656446 | 0.043471303 | 0.003251086 | 0.240300374 | 0.000116752 | 0.024049038 | 1 | 0.21807219 | 1 | 0.207579233 | ENSG0000021 | 4 | AC084018.1 |
| VAT | HTN | ENSG0000016 | -0.01283996 | 0.016518027 | 0.005356069 | 0.240332979 | -0.01266191 | 0.018073572 | 0.005355876 | 0.240299629 | -0.00017805 | 4.68E-05 | 1 | 0.110087228 | 1 | 0.116241644 | ENSG0000021 | 4 | AC084018.1 |
| VAT | HTN | ENSG0000016 | 0.004198837 | 0.547497431 | 0.006980438 | 0.240335635 | 0.004310011 | 0.536926064 | 0.006980132 | 0.240302197 | -0.00011117 | 0.026358195 | 1 | 0.783206863 | 1 | 0.776342689 | ENSG0000021 | 4 | AC084018.1 |
| VAT | HTN | ENSG0000016 | 0.003820767 | 0.566348311 | 0.00666291 | 0.240335651 | 0.003985736 | 0.549695264 | 0.006662689 | 0.240302208 | -0.00016497 | 0.003770942 | 1 | 0.79306004 | 1 | 0.786610816 | ENSG0000021 | 4 | AC084018.1 |
| VAT | HTN | ENSG0000016 | -0.00647011 | 0.089010989 | 0.003804513 | 0.240334387 | -0.00569935 | 0.136014769 | 0.003823019 | 0.240301289 | -0.00077076 | 0.041635273 | 1 | 0.317403528 | 1 | 0.403617533 | ENSG0000021 | 4 | AC084018.1 |
| VAT | HTN | ENSG0000016 | -0.00728918 | 0.107807858 | 0.004532712 | 0.240334538 | -0.00719453 | 0.112445035 | 0.004532553 | 0.240301142 | -9.47E-05 | 0.011846842 | 1 | 0.35296523 | 1 | 0.362980465 | ENSG0000021 | 1 | OR2A42 |
| VAT | HTN | ENSG0000016 | -0.02213756 | 0.000316158 | 0.006146396 | 0.240329416 | -0.02152984 | 0.000461586 | 0.006147631 | 0.240296337 | -0.00060772 | 2.08E-05 | 0.872594828 | 0.006610567 | 1 | 0.008847074 | ENSG0000021 | 3 | MAP10 |
| VAT | HTN | ENSG0000016 | -0.00386408 | 0.000748506 | 0.001451107 | 0.240332316 | -0.00358066 | 0.013702203 | 0.001452607 | 0.240299388 | -0.00028342 | 3.21E-05 | 1 | 0.067627177 | 1 | 0.096721436 | ENSG0000021 | 6 | NUP62 |
| VAT | HTN | ENSG0000016 | 0.002662181 | 0.690740388 | 0.006691406 | 0.240335735 | 0.002569022 | 0.701018794 | 0.006691107 | 0.240302312 | 9.32E-05 | 0.047955411 | 1 | 0.857708065 | 1 | 0.866075144 | ENSG0000021 | 5 | FAM106A |
| VAT | HTN | ENSG0000016 | -0.0003443 | 0.783322884 | 0.001252037 | 0.240335776 | -0.00032032 | 0.798067605 | 0.001251977 | 0.240302353 | -2.40E-05 | 0.003569189 | 1 | 0.904415117 | 1 | 0.92007794 | ENSG0000021 | 5 | DARC |
| VAT | HTN | ENSG0000016 | -0.00344096 | 0.556520343 | 0.0058518 | 0.240335643 | -0.00304631 | 0.602733475 | 0.005852962 | 0.240302251 | -0.00039465 | 0.003590541 | 1 | 0.788086792 | 1 | 0.816595742 | ENSG0000021 | 5 | DARC |
| VAT | HTN | ENSG0000016 | -0.00107995 | 0.781263102 | 0.00388929 | 0.240335775 | -0.00093575 | 0.809871593 | 0.003889381 | 0.240302356 | -0.0001442 | 0.006491937 | 1 | 0.903724293 | 1 | 0.92710311 | ENSG0000021 | 2 | AC007256.5 |
| VAT | HTN | ENSG0000016 | -0.00156271 | 0.761476193 | 0.00514824 | 0.240335768 | 0.009359907 | 0.185891458 | 0.007075651 | 0.240301522 | -0.01092262 | 0.024443169 | 1 | 0.893569002 | 1 | 0.47726551 | ENSG0000021 | 2 | AC007256.5 |
| VAT | HTN | ENSG0000016 | -0.0007307 | 0.81964567 | 0.003204794 | 0.240335788 | -0.00066424 | 0.835795787 | 0.003204661 | 0.240302364 | -6.65E-05 | 0.005514049 | 1 | 0.926718801 | 1 | 0.939506392 | ENSG0000021 | 6 | TCTEX1D2 |
| VAT | HTN | ENSG0000016 | 0.013874298 | 0.022256386 | 0.006069324 | 0.240333236 | 0.013755022 | 0.023426857 | 0.006069085 | 0.240299852 | 0.000119276 | 0.01127014 | 1 | 0.135901828 | 1 | 0.142418781 | ENSG0000021 | 2 | AC022498.1 |
| VAT | HTN | ENSG0000016 | -0.00783008 | 0.029776779 | 0.003603261 | 0.240333484 | -0.00777057 | 0.03103513 | 0.003603112 | 0.240300091 | -5.95E-05 | 0.028381517 | 1 | 0.166662296 | 1 | 0.171622533 | ENSG0000021 | 2 | AC022498.1 |
| VAT | HTN | ENSG0000016 | 0.003020404 | 0.596796142 | 0.005709501 | 0.240335675 | 0.003118399 | 0.584929308 | 0.005709271 | 0.240302238 | -9.80E-05 | 0.025154251 | 1 | 0.810316927 | 1 | 0.807829017 | ENSG0000021 | 2 | LINGO4 |
| VAT | HTN | ENSG0000016 | -0.01729796 | 0.00041986 | 0.004904023 | 0.240329677 | -0.01720898 | 0.000449318 | 0.004903812 | 0.240296312 | -8.90E-05 | 0.012194668 | 1 | 0.00816066 | 1 | 0.008733218 | ENSG0000021 | 2 | LINGO4 |
| VAT | HTN | ENSG0000016 | -0.01559524 | 9.68E-09 | 0.002718634 | 0.240319585 | -0.01803648 | 9.29E-11 | 0.002784199 | 0.240281691 | 0.002441248 | 4.96E-05 | 2.67E-05 | 1.11E-06 | 2.57E-07 | 3.66E-08 | ENSG0000021 | 3 | ASIC3 |
| VAT | HTN | ENSG0000016 | -0.00431632 | 0.117626516 | 0.002758356 | 0.240334606 | -0.00436837 | 0.113251538 | 0.002758244 | 0.240301148 | 5.20E-05 | 0.013658266 | 1 | 0.369484015 | 1 | 0.364132457 | ENSG0000021 | 4 | DNLZ |
| VAT | HTN | ENSG0000016 | 0.001469499 | 0.69803842 | 0.00378766 | 0.240335739 | 0.001521509 | 0.687888709 | 0.003787466 | 0.240302305 | -5.20E-05 | 0.023014288 | 1 | 0.860778235 | 1 | 0.859471632 | ENSG0000021 | 4 | DNLZ |
| VAT | HTN | ENSG0000016 | 0.003067231 | 0.418697877 | 0.003792872 | 0.240335491 | 0.003132279 | 0.408884276 | 0.003792751 | 0.240302048 | -6.50E-05 | 0.0481588 | 1 | 0.704520013 | 1 | 0.692521557 | ENSG0000021 | 4 | DNLZ |
| VAT | HTN | ENSG0000016 | -0.00405812 | 0.143150083 | 0.002771634 | 0.240334756 | -0.0040104 | 0.147896855 | 0.002771522 | 0.240301352 | -4.77E-05 | 0.024127969 | 1 | 0.411985639 | 1 | 0.420521528 | ENSG0000021 | 4 | DNLZ |
| VAT | HTN | ENSG0000016 | -0.00891367 | 0.03237582 | 0.00416576 | 0.240333555 | -0.0077206 | 0.065040316 | 0.004184642 | 0.240300706 | -0.00119307 | 0.00286117 | 1 | 0.174179193 | 1 | 0.263213011 | ENSG0000021 | 4 | DNLZ |
| VAT | HTN | ENSG0000016 | 0.001838647 | 0.686225963 | 0.004551307 | 0.240335733 | 0.001748788 | 0.700789644 | 0.004551113 | 0.240302312 | 8.99E-05 | 0.007232644 | 1 | 0.854300251 | 1 | 0.866075144 | ENSG0000021 | 4 | DNLZ |
| VAT | HTN | ENSG0000016 | -0.00144721 | 0.574181223 | 0.002575543 | 0.240335658 | -0.0013664 | 0.595732591 | 0.002575459 | 0.240302246 | -8.08E-05 | 0.000245644 | 1 | 0.797991524 | 1 | 0.814290189 | ENSG0000021 | 4 | DNLZ |
| VAT | HTN | ENSG0000016 | -0.00130517 | 0.458254328 | 0.001759638 | 0.240335542 | -0.00127027 | 0.470342289 | 0.001759564 | 0.240302128 | -3.49E-05 | 0.007317487 | 1 | 0.733206925 | 1 | 0.73755282 | ENSG0000021 | 7 | NOTCH2NL |
| VAT | HTN | ENSG0000016 | 0.009744755 | 0.106581352 | 0.006038561 | 0.240334529 | 0.009646456 | 0.110146161 | 0.006038321 | 0.240301126 | 9.83E-05 | 0.03476774 | 1 | 0.350613267 | 1 | 0.358494581 | ENSG0000021 | 2 | SUPT4H1 |
| VAT | HTN | ENSG0000016 | -0.00222096 | 0.657418725 | 0.005008021 | 0.240335716 | -0.00229818 | 0.646293859 | 0.005007823 | 0.240302281 | 7.72E-05 | 0.046379806 | 1 | 0.837990221 | 1 | 0.839816879 | ENSG0000021 | 3 | MARCKSL1P1 |
| VAT | HTN | ENSG0000016 | 0.02005027 | 0.002059877 | 0.006506707 | 0.24033113 | 0.009136039 | 0.236004636 | 0.007709517 | 0.240301903 | 0.010914232 | 0.008315492 | 1 | 0.026691367 | 1 | 0.541007306 | ENSG0000021 | 3 | MARCKSL1P1 |
| VAT | HTN | ENSG0000016 | -0.0046567 | 0.21662735 | 0.003768934 | 0.24033506 | -0.00397527 | 0.291891783 | 0.003771666 | 0.240301837 | -0.00068143 | 5.72E-06 | 1 | 0.519003026 | 1 | 0.592804504 | ENSG0000021 | 4 | CTD-3035D6.1 |
| VAT | HTN | ENSG0000016 | -0.00066419 | 0.739947897 | 0.002001054 | 0.240335759 | -0.00060542 | 0.762224514 | 0.00200971 | 0.24030234 | -5.88E-05 | 8.87E-05 | 1 | 0.879146016 | 1 | 0.896852841 | ENSG0000021 | 6 | MXD3 |
| VAT | HTN | ENSG0000016 | 0.003283147 | 0.3425071 | 0.003458769 | 0.240335369 | 0.003155676 | 0.361560877 | 0.00345867 | 0.240301974 | 0.000127471 | 4.56E-05 | 1 | 0.643512318 | 1 | 0.660428868 | ENSG0000021 | 8 | GSTM2 |
| VAT | HTN | ENSG0000016 | -0.00208584 | 0.736540081 | 0.006199766 | 0.240335758 | -0.00621418 | 0.321182499 | 0.006264094 | 0.240301899 | 0.004128339 | 4.33E-06 | 1 | 0.877742066 | 1 | 0.619471487 | ENSG0000021 | 8 | GSTM2 |
| VAT | HTN | ENSG0000016 | 0.002712599 | 0.606167626 | 0.005261545 | 0.240335682 | 0.003238817 | 0.538316433 | 0.005263277 | 0.240302198 | -0.00052622 | 0.000399581 | 1 | 0.815710701 | 1 | 0.777474283 | ENSG0000021 | 3 | LCAT |
| VAT | HTN | ENSG0000016 | -0.00591802 | 0.181641212 | 0.004430583 | 0.240334933 | -0.00582037 | 0.188934544 | 0.004430386 | 0.240301534 | -9.76E-05 | 0.001839397 | 1 | 0.470059822 | 1 | 0.48016514 | ENSG0000021 | 2 | PTPRCAP |
| VAT | HTN | ENSG0000016 | 0.005556469 | 0.19078067 | 0.004247172 | 0.240334969 | 0.005644744 | 0.183809402 | 0.004246982 | 0.240301514 | -8.83E-05 | 0.003204568 | 1 | 0.483521258 | 1 | 0.475904267 | ENSG0000021 | 2 | PTPRCAP |
| VAT | HTN | ENSG0000016 | -0.00366932 | 0.025213724 | 0.001639464 | 0.240333343 | -0.0036152 | 0.027440908 | 0.001639403 | 0.240299987 | -5.41E-05 | 4.37E-05 | 1 | 0.147535207 | 1 | 0.158114629 | ENSG0000021 | 2 | PTPRCAP |
| VAT | HTN | ENSG0000016 | -0.00262618 | 0.65725505 | 0.005918729 | 0.240335716 | -0.00279467 | 0.636790466 | 0.005918513 | 0.240302275 | 0.000168492 | 0.000465494 | 1 | 0.837990221 | 1 | 0.833353099 | ENSG0000021 | 2 | PTPRCAP |
| VAT | HTN | ENSG0000016 | 0.016646259 | 0.006548415 | 0.006122224 | 0.240332167 | 0.01676672 | 0.006167932 | 0.006122079 | 0.240298686 | -0.00012046 | 0.040133879 | 1 | 0.06063534 | 1 | 0.058100659 | ENSG0000021 | 2 | PTPRCAP |
| VAT | HTN | ENSG0000016 | 0.011264499 | 0.048009169 | 0.005696932 | 0.240333885 | 0.01139965 | 0.045382533 | 0.005696702 | 0.24030041 | -0.00013515 | 0.001915366 | 1 | 0.220474719 | 1 | 0.213136987 | ENSG0000021 | 2 | PTPRCAP |
| VAT | HTN | ENSG0000016 | 0.00481416 | 0.39746812 | 0.005689471 | 0.24033546 | 0.004942494 | 0.384988176 | 0.005689247 | 0.240302013 | -0.00012833 | 0.003767096 | 1 | 0.69081361 | 1 | 0.675933439 | ENSG0000021 | 5 | RP11-54C4.1 |
| VAT | HTN | ENSG0000016 | -0.00827423 | 0.052702551 | 0.004270892 | 0.240333962 | -0.0084123 | 0.048867693 | 0.004270746 | 0.240300471 | 0.000138064 | 0.000122488 | 1 | 0.232362685 | 1 | 0.221550272 | ENSG0000021 | 2 | RP11-75L1.2 |
| VAT | HTN | ENSG0000016 | 0.007767576 | 0.000794181 | 0.002315337 | 0.240330263 | 0.007202816 | 0.001907316 | 0.00232026 | 0.240297633 | 0.00056476 | 0.000232943 | 1 | 0.01289377 | 1 | 0.025341822 | ENSG0000021 | 2 | RP11-75L1.2 |
| VAT | HTN | ENSG0000016 | -0.00666649 | 0.051203283 | 0.003419105 | 0.240333938 | -0.00674423 | 0.048602321 | 0.003418964 | 0.240300467 | 7.59E-05 | 0.00313021 | 1 | 0.229790343 | 1 | 0.221550272 | ENSG0000021 | 2 | RP11-75L1.2 |
| VAT | HTN | ENSG0000016 | -0.00388575 | 0.305789362 | 0.003794311 | 0.240335296 | -0.00382338 | 0.313597448 | 0.003794157 | 0.240301884 | -6.24E-05 | 0.030571288 | 1 | 0.607969384 | 1 | 0.612903088 | ENSG0000021 | 2 | RP11-75L1.2 |
| VAT | HTN | ENSG0000016 | -0.0174188 | 5.34E-10 | 0.002805443 | 0.240316802 | -0.01750783 | 4.35E-10 | 0.002805345 | 0.240283179 | 8.90E-05 | 0.000148554 | 1.47E-06 | 1.23E-07 | 1.20E-06 | 1.00E-07 | ENSG0000021 | 4 | SIPA1 |
| VAT | HTN | ENSG0000016 | -0.00239525 | 0.457230195 | 0.003221956 | 0.240335541 | -0.00245818 | 0.445474787 | 0.003221803 | 0.240302098 | 6.29E-05 | 0.003406147 | 1 | 0.73258513 | 1 | 0.720517323 | ENSG0000021 | 4 | SIPA1 |
| VAT | HTN | ENSG0000016 | -0.00601818 | 0.171907481 | 0.004405364 | 0.240334893 | -0.00590819 | 0.179867473 | 0.004405264 | 0.240301498 | -0.00010999 | 0.009806566 | 1 | 0.45577776 | 1 | 0.468776417 | ENSG0000021 | 4 | SIPA1 |
| VAT | HTN | ENSG0000016 | 0.004132237 | 0.279150468 | 0.003818258 | 0.240335236 | 0.005867585 | 0.131989777 | 0.003895352 | 0.240301266 | -0.00173535 | 0.02467544 | 1 | 0.58589756 | 1 | 0.395969331 | ENSG0000021 | 4 | ERV3-1 |
| VAT | HTN | ENSG0000016 | -0.00792752 | 0.241841368 | 0.006773354 | 0.240335138 | -0.00853154 | 0.208081349 | 0.006777191 | 0.240301603 | 0.000604017 | 0.01243452 | 1 | 0.552094438 | 1 | 0.5074681 | ENSG0000021 | 4 | ERV3-1 |
| VAT | HTN | ENSG0000017 | -0.01344031 | 0.042817426 | 0.006635579 | 0.24033379 | -0.01332085 | 0.044691132 | 0.006635358 | 0.240300397 | -0.00011946 | 0.034556256 | 1 | 0.204346659 | 1 | 0.211211515 | ENSG0000021 | 4 | ERV3-1 |
| VAT | HTN | ENSG0000017 | 0.011592837 | 0.068298924 | 0.006359091 | 0.240334174 | 0.011461539 | 0.071475301 | 0.006358868 | 0.240300783 | 0.000131297 | 0.012706193 | 1 | 0.271760451 | 1 | 0.278635048 | ENSG0000021 | 4 | ERV3-1 |
| VAT | HTN | ENSG0000017 | 0.005786894 | 0.29380511 | 0.005512315 | 0.24033527 | 0.00587843 | 0.286219139 | 0.005512129 | 0.240301824 | -9.15E-05 | 0.049775321 | 1 | 0.594939182 | 1 | 0.590747324 | ENSG0000021 | 3 | ARL2 |
| VAT | HTN | ENSG0000017 | -0.00760943 | 0.074415732 | 0.004265249 | 0.240334244 | -0.00754433 | 0.076916821 | 0.004265066 | 0.240300842 | -6.51E-05 | 0.036552122 | 1 | 0.288060897 | 1 | 0.293421577 | ENSG0000021 | 5 | CCL14 |
| VAT | HTN | ENSG0000017 | 0.005053555 | 0.41379378 | 0.006183713 | 0.240335484 | 0.000918024 | 0.883693236 | 0.006275412 | 0.240302374 | 0.00413553 | 0.000113309 | 1 | 0.70100582 | 1 | 0.959100799 | ENSG0000021 | 5 | CCL14 |
| VAT | HTN | ENSG0000017 | -0.01483567 | 0.013840784 | 0.006027383 | 0.240332825 | -0.01513554 | 0.012037471 | 0.006027567 | 0.240299275 | 0.000299872 | 0.000441696 | 1 | 0.098709469 | 1 | 0.089310271 | ENSG0000021 | 5 | CCL14 |
| VAT | HTN | ENSG0000017 | -0.0046338 | 0.476058973 | 0.006502158 | 0.240335563 | -0.00447679 | 0.491131149 | 0.006502131 | 0.240302151 | -0.00015701 | 0.034552226 | 1 | 0.741910088 | 1 | 0.749321156 | ENSG0000021 | 5 | CCL14 |
| VAT | HTN | ENSG0000017 | -0.02205549 | 0.000712534 | 0.006516169 | 0.240330163 | -0.02177911 | 0.000831422 | 0.006516522 | 0.240296877 | -0.00027638 | 0.006950945 | 1 | 0.011705911 | 1 | 0.013419446 | ENSG0000021 | 5 | SRA1 |
| VAT | HTN | ENSG0000017 | 4.02E-05 | 0.989933694 | 0.003184621 | 0.240335813 | 9.15E-05 | 0.977087166 | 0.003184472 | 0.240302384 | -5.13E-05 | 0.017000679 | 1 | 0.996068901 | 1 | 0.991091723 | ENSG0000021 | 5 | SRA1 |
| VAT | HTN | ENSG0000017 | 0.000472997 | 0.813852852 | 0.002008834 | 0.240335786 | 0.000522978 | 0.794594905 | 0.00200876 | 0.240302351 | -5.00E-05 | 0.002111272 | 1 | 0.923869473 | 1 | 0.918168712 | ENSG0000021 | 5 | SRA1 |
| VAT | HTN | ENSG0000017 | -1.82E-05 | 0.993335826 | 0.00218413 | 0.240335813 | 4.41E-05 | 0.983880349 | 0.00218405 | 0.240302385 | -6.24E-05 | 0.000454757 | 1 | 0.997230388 | 1 | 0.995421467 | ENSG0000021 | 5 | SRA1 |
| VAT | HTN | ENSG0000017 | 0.004003969 | 0.135003116 | 0.002678844 | 0.240334712 | 0.003956517 | 0.1396754 | 0.002678746 | 0.240301309 | 4.75E-05 | 0.029048613 | 1 | 0.40220992 | 1 | 0.410547502 | ENSG0000021 | 4 | ZBTB9 |
| VAT | HTN | ENSG0000017 | -0.01916446 | 7.92E-09 | 0.00332121 | 0.240319393 | -0.01911062 | 8.70E-09 | 0.00332106 | 0.240286057 | -5.38E-05 | 0.021818334 | 2.18E-05 | 9.93E-07 | 2.40E-05 | 1.00E-06 | ENSG0000021 | 4 | ZBTB9 |
| VAT | HTN | ENSG0000017 | 0.000766827 | 0.624431221 | 0.001566295 | 0.240335695 | 0.000660508 | 0.673267769 | 0.001566418 | 0.240302297 | 0.000106319 | 8.09E-05 | 1 | 0.825002475 | 1 | 0.850381614 | ENSG0000021 | 4 | ZBTB9 |
| VAT | HTN | ENSG0000017 | 0.013379949 | 0.017855387 | 0.005648858 | 0.240333046 | 0.013458818 | 0.017187301 | 0.005648595 | 0.240299585 | -7.89E-05 | 0.039790877 | 1 | 0.114606667 | 1 | 0.1118796 | ENSG0000021 | 6 | ATF6B |

| | | | | | | | | | | | | | | | | | | | |
|---|---|---|---|---|---|---|---|---|---|---|---|---|---|---|---|---|---|---|---|
| VAT | HTN | ENSG0000017 | -0.00099466 | 0.718138623 | 0.00275568 | 0.240335749 | -0.00095261 | 0.729564775 | 0.002755562 | 0.240302326 | -4.20E-05 | 0.036689386 | 1 | 0.86780324 | 1 | 0.878043729 | ENSG0000021 | 6 | ATF6B |
| VAT | HTN | ENSG0000017 | -0.00060575 | 0.711591755 | 0.001638394 | 0.240335746 | 0.002778239 | 0.21749675 | 0.002252846 | 0.240301635 | -0.00338399 | 0.028648091 | 1 | 0.864869003 | 1 | 0.517020606 | ENSG0000021 | 6 | ATF6B |
| VAT | HTN | ENSG0000017 | -0.00640021 | 0.09572193 | 0.003841755 | 0.240334445 | -0.00629583 | 0.101243916 | 0.003841603 | 0.24030106 | -0.00010438 | 0.000444013 | 1 | 0.332536044 | 1 | 0.342746569 | ENSG0000021 | 6 | ATF6B |
| VAT | HTN | ENSG0000017 | 0.004538348 | 0.070370302 | 0.002508035 | 0.240334198 | 0.005042081 | 0.044581296 | 0.00251026 | 0.240300395 | -0.00050373 | 4.48E-06 | 1 | 0.27741042 | 1 | 0.211053818 | ENSG0000021 | 6 | ATF6B |
| VAT | HTN | ENSG0000017 | -0.01870619 | 0.002761551 | 0.006249728 | 0.240331395 | -0.01883074 | 0.002585544 | 0.006249496 | 0.240297908 | 0.000124558 | 0.01354134 | 1 | 0.031626067 | 1 | 0.030627042 | ENSG0000021 | 6 | ATF6B |
| VAT | HTN | ENSG0000017 | -0.0019482 | 0.65335348 | 0.004337928 | 0.240335714 | -0.00188583 | 0.663743928 | 0.004337722 | 0.240302292 | -6.24E-05 | 0.030087326 | 1 | 0.837554855 | 1 | 0.847332674 | ENSG0000021 | 6 | ATF6B |
| VAT | HTN | ENSG0000017 | 0.00263213 | 0.626328092 | 0.005405874 | 0.240335696 | 0.002744414 | 0.611669736 | 0.00540567 | 0.240302258 | -0.00011228 | 0.00926801 | 1 | 0.825696865 | 1 | 0.821992369 | ENSG0000021 | 6 | ATF6B |
| VAT | HTN | ENSG0000017 | 0.005016913 | 0.062860578 | 0.002697003 | 0.240334107 | -0.00850489 | 0.032410428 | 0.003975517 | 0.240300128 | 0.013521807 | 3.67E-06 | 1 | 0.25933512 | 1 | 0.175054366 | ENSG0000021 | 6 | ATF6B |
| VAT | HTN | ENSG0000017 | 0.002573561 | 0.563726166 | 0.004457802 | 0.240335649 | 0.002677758 | 0.548031106 | 0.004457628 | 0.240302207 | -0.0001042 | 0.002825937 | 1 | 0.792605307 | 1 | 0.785748494 | ENSG0000021 | 5 | TREX1 |
| VAT | HTN | ENSG0000017 | -0.00413982 | 0.071974018 | 0.002300812 | 0.240334217 | -0.00152251 | 0.523983486 | 0.002389317 | 0.240302185 | -0.00261731 | 4.93E-05 | 1 | 0.282170866 | 1 | 0.769652481 | ENSG0000021 | 5 | TREX1 |
| VAT | HTN | ENSG0000017 | -0.00847791 | 0.139312576 | 0.005734677 | 0.240334735 | -0.00806345 | 0.159725602 | 0.005735037 | 0.24030141 | -0.00041446 | 8.74E-06 | 1 | 0.40653371 | 1 | 0.439962736 | ENSG0000021 | 5 | TREX1 |
| VAT | HTN | ENSG0000017 | -0.0003953 | 0.918085948 | 0.003843613 | 0.240335808 | -0.00030403 | 0.936950593 | 0.003843468 | 0.240302382 | -9.13E-05 | 0.002836757 | 1 | 0.968477978 | 1 | 0.975842882 | ENSG0000021 | 4 | CLIC1 |
| VAT | HTN | ENSG0000017 | -0.00174001 | 0.51683373 | 0.002684237 | 0.240335606 | -0.00165731 | 0.53697036 | 0.002684327 | 0.240302197 | -8.27E-05 | 0.031922478 | 1 | 0.769926105 | 1 | 0.776342689 | ENSG0000021 | 4 | CLIC1 |
| VAT | HTN | ENSG0000017 | 0.006090919 | 0.289069325 | 0.005745247 | 0.240335259 | 0.006190407 | 0.281249901 | 0.005745066 | 0.240301812 | -9.95E-05 | 0.047601182 | 1 | 0.59321241 | 1 | 0.584169217 | ENSG0000021 | 4 | CLIC1 |
| VAT | HTN | ENSG0000017 | -0.002406 | 0.539222312 | 0.003918629 | 0.240335627 | -0.00292968 | 0.454940612 | 0.003920861 | 0.240302109 | 0.000523686 | 0.000186359 | 1 | 0.778154916 | 1 | 0.727062009 | ENSG0000021 | 4 | CLIC1 |
| VAT | HTN | ENSG0000017 | 0.008546549 | 0.014581547 | 0.003498931 | 0.240332871 | 0.006689157 | 0.057899247 | 0.003527172 | 0.240300611 | 0.001857392 | 3.29E-05 | 1 | 0.102351368 | 1 | 0.247754918 | ENSG0000021 | 6 | ATP6V1G2 |
| VAT | HTN | ENSG0000017 | -0.00075831 | 0.909162752 | 0.00664632 | 0.240335807 | -0.00065409 | 0.92159992 | 0.006646033 | 0.24030238 | -0.00010422 | 0.030944679 | 1 | 0.964369407 | 1 | 0.972406143 | ENSG0000021 | 6 | ATP6V1G2 |
| VAT | HTN | ENSG0000017 | 0.000781837 | 0.811739974 | 0.003282519 | 0.240335785 | -5.87E-05 | 0.985761253 | 0.003287648 | 0.240302385 | 0.000840511 | 7.45E-06 | 1 | 0.923115916 | 1 | 0.996044499 | ENSG0000021 | 6 | ATP6V1G2 |
| VAT | HTN | ENSG0000017 | 0.00080549 | 0.756892811 | 0.002602015 | 0.240335766 | 0.001028699 | 0.692645035 | 0.002602534 | 0.240302308 | -0.00022321 | 0.000218498 | 1 | 0.890841859 | 1 | 0.861126259 | ENSG0000021 | 6 | ATP6V1G2 |
| VAT | HTN | ENSG0000017 | 0.012299711 | 0.062657864 | 0.006607008 | 0.240334104 | 0.012405685 | 0.060417451 | 0.006606714 | 0.240300646 | -0.00010597 | 0.023439592 | 1 | 0.259273918 | 1 | 0.253180669 | ENSG0000021 | 6 | ATP6V1G2 |
| VAT | HTN | ENSG0000017 | -0.0004336 | 0.924345382 | 0.004566013 | 0.240335809 | -0.00088539 | 0.846266771 | 0.004566607 | 0.240302366 | 0.000451796 | 7.34E-07 | 1 | 0.970404433 | 1 | 0.942573159 | ENSG0000021 | 6 | ATP6V1G2 |
| VAT | HTN | ENSG0000017 | -0.00023116 | 0.879204412 | 0.00152102 | 0.240335802 | -0.0001817 | 0.90490676 | 0.001520963 | 0.240302378 | -4.95E-05 | 5.14E-05 | 1 | 0.955786877 | 1 | 0.967289953 | ENSG0000021 | 6 | ATP6V1G2 |
| VAT | HTN | ENSG0000017 | -0.00057945 | 0.899826106 | 0.004603181 | 0.240335806 | -0.0007113 | 0.87719696 | 0.004603199 | 0.240302373 | 0.000131849 | 0.018023928 | 1 | 0.959290439 | 1 | 0.9580782 | ENSG0000021 | 6 | ATP6V1G2 |
| VAT | HTN | ENSG0000017 | 0.023924886 | 6.79E-05 | 0.006006033 | 0.240327988 | 0.023690121 | 8.00E-05 | 0.006005851 | 0.240294712 | 0.000234765 | 9.42E-06 | 0.187471523 | 0.001837956 | 0.220732361 | 0.002143033 | ENSG0000021 | 6 | ATP6V1G2 |
| VAT | HTN | ENSG0000017 | -0.00641871 | 0.147748138 | 0.004434236 | 0.24033478 | -0.00846384 | 0.059998534 | 0.004500108 | 0.240300664 | 0.002045136 | 0.007823842 | 1 | 0.418982277 | 1 | 0.252944336 | ENSG0000021 | 6 | ATP6V1G2 |
| VAT | HTN | ENSG0000017 | -0.00136183 | 0.695866371 | 0.003483762 | 0.240335738 | -0.00129848 | 0.709343294 | 0.003483622 | 0.240302316 | -6.33E-05 | 0.017726044 | 1 | 0.859324915 | 1 | 0.869462276 | ENSG0000021 | 4 | MT1P1 |
| VAT | HTN | ENSG0000017 | -0.00176426 | 0.65979111 | 0.004007852 | 0.240335718 | -0.00182818 | 0.648266056 | 0.004007658 | 0.240302282 | 6.39E-05 | 0.014604492 | 1 | 0.838794779 | 1 | 0.840256636 | ENSG0000021 | 4 | RP11-3J10.4 |
| VAT | HTN | ENSG0000017 | -0.00592945 | 0.152222611 | 0.004141464 | 0.240334802 | -0.00715768 | 0.084935941 | 0.004154817 | 0.240300921 | 0.001228229 | 0.000261621 | 1 | 0.426965859 | 1 | 0.311318987 | ENSG0000021 | 4 | RP11-3J10.4 |
| VAT | HTN | ENSG0000017 | 0.003171009 | 0.617101642 | 0.006342493 | 0.24033569 | 0.00328156 | 0.60486712 | 0.006342227 | 0.240302253 | -0.00011055 | 0.018972032 | 1 | 0.821428835 | 1 | 0.817356612 | ENSG0000021 | 6 | SLC23A3 |
| VAT | HTN | ENSG0000017 | 0.002428223 | 0.655996761 | 0.005451202 | 0.240335716 | 0.001896104 | 0.728060191 | 0.005453162 | 0.240302325 | 0.000532119 | 0.000860824 | 1 | 0.837990221 | 1 | 0.877487391 | ENSG0000021 | 6 | SLC23A3 |
| VAT | HTN | ENSG0000017 | -0.00643555 | 0.001457171 | 0.002021788 | 0.240330816 | -0.00637175 | 0.001623851 | 0.002021748 | 0.240297487 | -6.38E-05 | 0.001526866 | 1 | 0.020312084 | 1 | 0.022409137 | ENSG0000021 | 6 | SLC23A3 |
| VAT | HTN | ENSG0000017 | 0.006097784 | 0.102243874 | 0.003731644 | 0.240334496 | 0.005990542 | 0.108407625 | 0.003731507 | 0.240301114 | 0.000107242 | 0.000395373 | 1 | 0.342052233 | 1 | 0.354088812 | ENSG0000021 | 6 | SLC23A3 |
| VAT | HTN | ENSG0000017 | -0.00585013 | 0.210212989 | 0.004668967 | 0.240335039 | -0.0057769 | 0.215959092 | 0.004668781 | 0.24030163 | -7.32E-05 | 0.041914948 | 1 | 0.508766538 | 1 | 0.515497803 | ENSG0000021 | 6 | SLC23A3 |
| VAT | HTN | ENSG0000018 | -8.57E-05 | 0.963481191 | 0.001870894 | 0.240335812 | -0.00013102 | 0.944168619 | 0.001870848 | 0.240302382 | 4.54E-05 | 0.010820104 | 1 | 0.983071382 | 1 | 0.978965976 | ENSG0000021 | 4 | LTB4R |
| VAT | HTN | ENSG0000018 | 0.006821626 | 0.277451683 | 0.00628106 | 0.240335232 | 0.006664304 | 0.288663224 | 0.0062808 | 0.24030183 | 0.000157322 | 0.00082313 | 1 | 0.585762318 | 1 | 0.59182601 | ENSG0000021 | 6 | TSPAN4 |
| VAT | HTN | ENSG0000018 | -0.00157215 | 0.805245247 | 0.006376222 | 0.240335783 | -0.00139575 | 0.826721755 | 0.006375982 | 0.240302361 | -0.0001764 | 0.000532716 | 1 | 0.918978716 | 1 | 0.936295463 | ENSG0000021 | 3 | CPSF1P1 |
| VAT | HTN | ENSG0000018 | 0.009147231 | 0.09890578 | 0.005543156 | 0.24033447 | 0.009251129 | 0.095119962 | 0.00554295 | 0.240301011 | -0.0001039 | 0.020124048 | 1 | 0.336272805 | 1 | 0.332830936 | ENSG0000021 | 7 | CPNE1 |
| VAT | HTN | ENSG0000018 | -0.00023437 | 0.952973161 | 0.003974068 | 0.240335812 | -0.00031732 | 0.936355063 | 0.003973902 | 0.240302382 | 8.30E-05 | 0.005203164 | 1 | 0.981080063 | 1 | 0.975590779 | ENSG0000021 | 7 | CPNE1 |
| VAT | HTN | ENSG0000018 | 0.000209659 | 0.93868973 | 0.002725788 | 0.24033581 | 0.000163248 | 0.952241153 | 0.00272568 | 0.240302383 | 4.64E-05 | 0.026885729 | 1 | 0.976192895 | 1 | 0.98139865 | ENSG0000021 | 7 | CPNE1 |
| VAT | HTN | ENSG0000018 | -0.00496063 | 0.228351489 | 0.004118015 | 0.240335098 | -0.00911412 | 0.0343212 | 0.004306661 | 0.240300176 | 0.004153482 | 0.000994363 | 1 | 0.534110263 | 1 | 0.180088428 | ENSG0000021 | 7 | CPNE1 |
| VAT | HTN | ENSG0000018 | 0.005823806 | 0.109711614 | 0.003641031 | 0.240334552 | 0.005889836 | 0.105733485 | 0.003640929 | 0.240301094 | -6.60E-05 | 0.04662793 | 1 | 0.356659663 | 1 | 0.348224683 | ENSG0000021 | 7 | CPNE1 |
| VAT | HTN | ENSG0000018 | 0.005431294 | 0.402937984 | 0.006493762 | 0.240335468 | 0.004907611 | 0.449967074 | 0.006496098 | 0.240302103 | 0.000523684 | 0.005913346 | 1 | 0.693767209 | 1 | 0.723286064 | ENSG0000021 | 4 | ARL16 |
| VAT | HTN | ENSG0000018 | 0.026665426 | 1.17E-09 | 0.004382104 | 0.240317553 | 0.026755312 | 1.02E-09 | 0.004381897 | 0.240284001 | -8.99E-05 | 0.0023042 | 3.22E-06 | 1.89E-07 | 2.82E-06 | 1.76E-07 | ENSG0000021 | 4 | ARL16 |
| VAT | HTN | ENSG0000018 | 0.003377409 | 0.573364927 | 0.005997852 | 0.240335657 | 0.003686567 | 0.538803069 | 0.005998081 | 0.240302198 | -0.00030916 | 0.000449573 | 1 | 0.797624595 | 1 | 0.777770121 | ENSG0000021 | 4 | ARL16 |
| VAT | HTN | ENSG0000018 | -0.00742771 | 0.021982968 | 0.003242593 | 0.240333226 | -0.00734971 | 0.023409675 | 0.003242489 | 0.240299851 | -7.80E-05 | 0.005407018 | 1 | 0.13482887 | 1 | 0.142418781 | ENSG0000021 | 4 | ARL16 |
| VAT | HTN | ENSG0000018 | -0.05754077 | 2.09E-19 | 0.00638713 | 0.240295792 | -0.05772693 | 1.60E-19 | 0.006386854 | 0.240262105 | 0.000186153 | 7.08E-05 | 5.77E-16 | 5.77E-16 | 4.40E-16 | 4.40E-16 | ENSG0000021 | 4 | ARL16 |
| VAT | HTN | ENSG0000018 | -0.00422109 | 0.065131199 | 0.002288644 | 0.240334136 | -0.00428129 | 0.061384494 | 0.002288579 | 0.240300659 | 6.02E-05 | 0.003683121 | 1 | 0.263580805 | 1 | 0.255210403 | ENSG0000021 | 3 | WEE2 |
| VAT | HTN | ENSG0000018 | -0.00466071 | 0.390172684 | 0.005423815 | 0.240335449 | -0.00424372 | 0.434018602 | 0.005424439 | 0.240302083 | -0.00041699 | 6.29E-05 | 1 | 0.682680345 | 1 | 0.712632987 | ENSG0000021 | 4 | AC024560.3 |
| VAT | HTN | ENSG0000018 | -0.00035434 | 0.827702074 | 0.00162801 | 0.24033579 | -0.0003843 | 0.813382154 | 0.001627942 | 0.240302357 | 3.00E-05 | 0.012851533 | 1 | 0.930911869 | 1 | 0.928203638 | ENSG0000021 | 4 | AC024560.3 |
| VAT | HTN | ENSG0000018 | -0.00591997 | 0.098579265 | 0.003583987 | 0.240334468 | -0.00584573 | 0.102864502 | 0.003583866 | 0.240301073 | -7.42E-05 | 0.014218356 | 1 | 0.336272805 | 1 | 0.344320158 | ENSG0000021 | 4 | AC024560.3 |
| VAT | HTN | ENSG0000018 | 0.004894115 | 0.000164036 | 0.001298578 | 0.240328808 | 0.00485425 | 0.000185348 | 0.001298545 | 0.240295494 | 3.99E-05 | 0.001109221 | 0.452738877 | 0.003801268 | 0.511559856 | 0.004298822 | ENSG0000021 | 4 | C17orf67 |
| VAT | HTN | ENSG0000018 | -0.00059851 | 0.895528726 | 0.004557955 | 0.240335805 | -0.00049802 | 0.912990065 | 0.004557778 | 0.240302379 | -0.00010049 | 0.005023952 | 1 | 0.959290439 | 1 | 0.968801453 | ENSG0000021 | 4 | FIS1 |
| VAT | HTN | ENSG0000018 | -0.00125784 | 0.661912175 | 0.002876553 | 0.240335719 | -0.00121155 | 0.673609318 | 0.002876435 | 0.240302297 | -4.63E-05 | 0.033275451 | 1 | 0.840330084 | 1 | 0.850381614 | ENSG0000021 | 8 | RP11-108K14.4 |
| VAT | HTN | ENSG0000018 | -0.00144663 | 0.635415591 | 0.003051199 | 0.240335703 | -0.00149388 | 0.624399173 | 0.003051058 | 0.240302267 | 4.72E-05 | 0.023593944 | 1 | 0.827629557 | 1 | 0.826939404 | ENSG0000021 | 3 | MBLAC1 |
| VAT | HTN | ENSG0000018 | 0.002812206 | 0.579409088 | 0.005073908 | 0.240335662 | 0.002736756 | 0.58961033 | 0.005073693 | 0.240302241 | 7.54E-05 | 0.043845893 | 1 | 0.800563876 | 1 | 0.809098734 | ENSG0000021 | 6 | SOGA3 |
| VAT | HTN | ENSG0000018 | -0.00134314 | 0.566214866 | 0.002341445 | 0.240335651 | -0.00129909 | 0.578997877 | 0.002341343 | 0.240302233 | -4.40E-05 | 0.009211404 | 1 | 0.79306004 | 1 | 0.805054983 | ENSG0000021 | 6 | SOGA3 |
| VAT | HTN | ENSG0000018 | 0.005809986 | 0.362898989 | 0.006385612 | 0.240335405 | 0.005644017 | 0.376753444 | 0.006385395 | 0.240302 | 0.000165969 | 0.002052332 | 1 | 0.665980483 | 1 | 0.669772956 | ENSG0000021 | 5 | VSTM5 |
| VAT | HTN | ENSG0000018 | -0.00236698 | 0.160477455 | 0.001686517 | 0.240334842 | -0.00233677 | 0.165868376 | 0.001686459 | 0.240301438 | -3.02E-05 | 0.033382288 | 1 | 0.438197474 | 1 | 0.450561028 | ENSG0000021 | 5 | VSTM5 |
| VAT | HTN | ENSG0000018 | 0.003115626 | 0.547378574 | 0.005178096 | 0.240335635 | 0.003017489 | 0.560058845 | 0.005177976 | 0.240302217 | 9.81E-05 | 0.048961777 | 1 | 0.783206863 | 1 | 0.793105393 | ENSG0000021 | 5 | VSTM5 |
| VAT | HTN | ENSG0000018 | -0.00552062 | 0.03540402 | 0.002624226 | 0.240333631 | -0.00557909 | 0.033500013 | 0.002624163 | 0.240300156 | 5.85E-05 | 0.01984352 | 1 | 0.183674992 | 1 | 0.177661645 | ENSG0000021 | 5 | VSTM5 |
| VAT | HTN | ENSG0000018 | 0.006323494 | 0.365974146 | 0.006994699 | 0.24033541 | 0.006510823 | 0.351924986 | 0.006994396 | 0.240301957 | -0.00018733 | 0.00022602 | 1 | 0.667014755 | 1 | 0.651755529 | ENSG0000021 | 5 | VSTM5 |
| VAT | HTN | ENSG0000018 | -0.00567205 | 0.407549316 | 0.006848522 | 0.240335475 | -0.01284978 | 0.070070255 | 0.007093601 | 0.240300767 | 0.00717773 | 0.000104811 | 1 | 0.696936974 | 1 | 0.275849438 | ENSG0000021 | 5 | VSTM5 |
| VAT | HTN | ENSG0000018 | 0.006020036 | 0.157023781 | 0.004253977 | 0.240334826 | 0.005950792 | 0.161830701 | 0.004253777 | 0.24030142 | 6.92E-05 | 0.015161346 | 1 | 0.435044054 | 1 | 0.4435479 | ENSG0000021 | 5 | VSTM5 |
| VAT | HTN | ENSG0000018 | 0.005549037 | 0.102498141 | 0.003398345 | 0.240334498 | 0.005605844 | 0.09901519 | 0.003398202 | 0.240301043 | -5.68E-05 | 0.024296637 | 1 | 0.342135622 | 1 | 0.337802132 | ENSG0000021 | 2 | LRRC37A4P |
| VAT | HTN | ENSG0000018 | -0.0014555 | 0.825312675 | 0.006594419 | 0.240335789 | -0.00135606 | 0.837067945 | 0.006594153 | 0.240302364 | -9.94E-05 | 0.048527852 | 1 | 0.930119634 | 1 | 0.939635856 | ENSG0000021 | 2 | LRRC37A4P |
| VAT | HTN | ENSG0000018 | -0.00408079 | 0.088229706 | 0.002393688 | 0.24033438 | -0.00404597 | 0.090963095 | 0.002393581 | 0.240300976 | -3.48E-05 | 0.038783635 | 1 | 0.316251933 | 1 | 0.323528535 | ENSG0000021 | 3 | AS3MT |
| VAT | HTN | ENSG0000018 | -0.00947165 | 0.045868679 | 0.004743879 | 0.240333847 | -0.00939483 | 0.04764671 | 0.00474364 | 0.24030045 | -7.68E-05 | 0.008885855 | 1 | 0.214201949 | 1 | 0.218810182 | ENSG0000021 | 3 | AS3MT |
| VAT | HTN | ENSG0000018 | -0.00161722 | 0.630078355 | 0.003357894 | 0.240335699 | -0.00169563 | 0.613569386 | 0.003357785 | 0.240302259 | 7.84E-05 | 0.006724205 | 1 | 0.825823207 | 1 | 0.823173728 | ENSG0000021 | 4 | SEC14L6 |
| VAT | HTN | ENSG0000018 | -0.00303053 | 0.441785567 | 0.003939941 | 0.240335522 | -0.00295808 | 0.452759422 | 0.003939783 | 0.240302107 | -7.25E-05 | 0.01664264 | 1 | 0.720643123 | 1 | 0.724217571 | ENSG0000021 | 4 | PPME1 |
| VAT | HTN | ENSG0000018 | 0.012426135 | 0.011649359 | 0.004925921 | 0.240332675 | 0.012511484 | 0.011084863 | 0.004925739 | 0.240299203 | -8.53E-05 | 0.031990576 | 1 | 0.087847622 | 1 | 0.085698103 | ENSG0000021 | 4 | PPME1 |
| VAT | HTN | ENSG0000018 | 0.008450482 | 0.017374739 | 0.00355258 | 0.240333023 | 0.00756262 | 0.033653066 | 0.003560203 | 0.24030016 | 0.000887862 | 0.000174169 | 1 | 0.112568733 | 1 | 0.17793575 | ENSG0000021 | 4 | PPME1 |
| VAT | HTN | ENSG0000018 | 0.001429409 | 0.571867313 | 0.002528567 | 0.240335656 | 0.00133013 | 0.598864565 | 0.002528599 | 0.240302248 | 9.93E-05 | 0.002182857 | 1 | 0.796745978 | 1 | 0.814290189 | ENSG0000021 | 4 | PPME1 |
| VAT | HTN | ENSG0000018 | -0.00144136 | 0.424429104 | 0.00180449 | 0.240335499 | -0.00138895 | 0.441449996 | 0.001804418 | 0.240302093 | -5.24E-05 | 0.000145157 | 1 | 0.708665655 | 1 | 0.717128893 | ENSG0000021 | 3 | STARD10 |
| VAT | HTN | ENSG0000018 | 0.00135825 | 0.462859334 | 0.001850106 | 0.240335548 | 0.011377274 | 0.000998015 | 0.003456976 | 0.240297043 | -0.01001902 | 0.000601844 | 1 | 0.735882351 | 1 | 0.015650693 | ENSG0000021 | 9 | NUTM2D |
| VAT | HTN | ENSG0000018 | 0.00252072 | 0.625150655 | 0.005159448 | 0.240335696 | 0.002658608 | 0.606338458 | 0.005159266 | 0.240302254 | -0.00013789 | 0.001233547 | 1 | 0.825288349 | 1 | 0.817934576 | ENSG0000021 | 4 | TMEM249 |
| VAT | HTN | ENSG0000018 | 0.000371754 | 0.942532256 | 0.005156984 | 0.240335811 | 0.000179469 | 0.972237682 | 0.005156881 | 0.240302384 | 0.000192285 | 0.000179491 | 1 | 0.976968793 | 1 | 0.989080723 | ENSG0000021 | 4 | KRT8P26 |
| VAT | HTN | ENSG0000018 | -0.00329962 | 0.159109744 | 0.002343354 | 0.240334836 | -0.00321919 | 0.169526521 | 0.002343397 | 0.240301454 | -8.04E-05 | 0.009707582 | 1 | 0.43652375 | 1 | 0.454706703 | ENSG0000021 | 4 | KRT8P26 |
| VAT | HTN | ENSG0000018 | 0.002847698 | 0.688295471 | 0.00709849 | 0.240335734 | -0.00249882 | 0.728593159 | 0.007201257 | 0.240302325 | 0.005346518 | 1.08E-05 | 1 | 0.855718693 | 1 | 0.87774645 | ENSG0000021 | 4 | KRT8P26 |
| VAT | HTN | ENSG0000018 | -0.00990635 | 0.096727978 | 0.005964334 | 0.240334453 | -0.01000637 | 0.0933927 | 0.005964088 | 0.240300997 | 0.00010002 | 0.025966556 | 1 | 0.333711525 | 1 | 0.329200323 | ENSG0000021 | 4 | KRT8P26 |
| VAT | HTN | ENSG0000018 | 0.003895354 | 0.51006494 | 0.005913367 | 0.240335599 | 0.002917714 | 0.622503306 | 0.005926628 | 0.240302265 | 0.00097764 | 0.015097635 | 1 | 0.767061751 | 1 | 0.826686941 | ENSG0000021 | 6 | ARHGEF33 |
| VAT | HTN | ENSG0000018 | 0.000197694 | 0.974486427 | 0.006181414 | 0.240335813 | 0.000609247 | 0.921488403 | 0.006181542 | 0.24030238 | -0.00041155 | 7.15E-07 | 1 | 0.98866831 | 1 | 0.972406143 | ENSG0000021 | 6 | IFRD2 |
| VAT | HTN | ENSG0000018 | 0.009245964 | 0.031412859 | 0.004296832 | 0.24033353 | 0.009168276 | 0.032858985 | 0.004296673 | 0.240300139 | 7.77E-05 | 0.02475004 | 1 | 0.171134257 | 1 | 0.17644124 | ENSG0000021 | 4 | SEPT7P2 |
| VAT | HTN | ENSG0000018 | -0.0018567 | 0.766651496 | 0.006256614 | 0.24033577 | -0.0017394 | 0.780998026 | 0.006256413 | 0.240302347 | -0.0001173 | 0.030300362 | 1 | 0.896662855 | 1 | 0.912597186 | ENSG0000021 | 4 | SEPT7P2 |
| VAT | HTN | ENSG0000018 | -0.00870358 | 0.114402302 | 0.005513082 | 0.240334584 | -0.00860492 | 0.118551035 | 0.005512836 | 0.240301183 | -9.87E-05 | 0.011236197 | 1 | 0.36551286 | 1 | 0.372666123 | ENSG0000021 | 4 | DDX12P |
| VAT | HTN | ENSG0000018 | -0.00047557 | 0.908622747 | 0.004143461 | 0.240335807 | -0.00054483 | 0.895381684 | 0.004143291 | 0.240302376 | 6.93E-05 | 0.026583819 | 1 | 0.964197726 | 1 | 0.962700992 | ENSG0000021 | 4 | SMTNL1 |
| VAT | HTN | ENSG0000018 | 0.000165189 | 0.954359026 | 0.002886223 | 0.240335812 | -4.94E-06 | 0.998634087 | 0.002886523 | 0.240302385 | 0.000170131 | 0.001545377 | 1 | 0.981382605 | 1 | 0.998848822 | ENSG0000021 | 4 | SMTNL1 |
| VAT | HTN | ENSG0000018 | -0.00245766 | 0.711636777 | 0.006648455 | 0.240335746 | -0.00227325 | 0.732400188 | 0.006648192 | 0.240302327 | -0.00018441 | 0.000343129 | 1 | 0.864869003 | 1 | 0.878114224 | ENSG0000021 | 3 | RPL23AP3 |
| VAT | HTN | ENSG0000018 | -0.00225379 | 0.321103442 | 0.002271526 | 0.240335328 | -0.00219913 | 0.33296584 | 0.002271459 | 0.240301923 | -5.47E-05 | 0.007257627 | 1 | 0.624075039 | 1 | 0.631637966 | ENSG0000021 | 6 | PARGP1 |
| VAT | HTN | ENSG0000019 | 0.002099858 | 0.26149278 | 0.00187008 | 0.240335192 | 0.002045239 | 0.27408512 | 0.00187001 | 0.240301795 | 5.46E-05 | 0.000285217 | 1 | 0.570001694 | 1 | 0.579436835 | ENSG0000021 | 6 | PARGP1 |
| VAT | HTN | ENSG0000019 | -0.00312733 | 0.640054763 | 0.006687728 | 0.240335706 | -0.00322493 | 0.629636597 | 0.006687408 | 0.24030227 | 9.76E-05 | 0.026535127 | 1 | 0.829366735 | 1 | 0.829101626 | ENSG0000021 | 3 | UBXN2B |
| VAT | HTN | ENSG0000019 | 0.009046637 | 0.140238963 | 0.006133712 | 0.240334741 | 0.01011916 | 0.09930162 | 0.006139328 | 0.240301045 | -0.00107252 | 8.24E-05 | 1 | 0.408123093 | 1 | 0.338361074 | ENSG0000021 | 4 | RP11-313J2.1 |
| VAT | HTN | ENSG0000019 | 0.001045309 | 0.653346821 | 0.002327471 | 0.240335714 | 0.00174032 | 0.45878199 | 0.002349072 | 0.240302114 | -0.00069501 | 0.029360385 | 1 | 0.837554855 | 1 | 0.728992166 | ENSG0000021 | 5 | RP11-54D18.2 |
| VAT | HTN | ENSG0000019 | -0.00533901 | 0.176857777 | 0.003953372 | 0.240334914 | -0.00544224 | 0.168620419 | 0.003953243 | 0.240301445 | 0.000103231 | 0.002413729 | 1 | 0.462241918 | 1 | 0.45395764 | ENSG0000021 | 5 | VPS16 |
| VAT | HTN | ENSG0000019 | -0.00018811 | 0.958692025 | 0.00365118 | 0.240335812 | -0.00027575 | 0.939796815 | 0.003651062 | 0.240302382 | 8.66E-05 | 0.000602663 | 1 | 0.98228229 | 1 | 0.977875185 | ENSG0000021 | 2 | HMGB3P7 |
| VAT | HTN | ENSG0000019 | -0.01088356 | 0.01068144 | 0.004263111 | 0.240332599 | -0.01082013 | 0.011142815 | 0.004262918 | 0.240299208 | -6.34E-05 | 0.031977592 | 1 | 0.083044439 | 1 | 0.085905504 | ENSG0000021 | 2 | HMGB3P7 |
| VAT | HTN | ENSG0000019 | 0.017659445 | 7.96E-08 | 0.003289737 | 0.240321602 | 0.017547845 | 9.62E-08 | 0.003289893 | 0.240288356 | 0.0001116 | 0.026340207 | 0.000219786 | 6.66E-06 | 0.000265476 | 8.30E-06 | ENSG0000021 | 3 | ZNF663P |
| VAT | HTN | ENSG0000019 | 0.004644433 | 0.028490525 | 0.002120289 | 0.240333447 | 0.006047241 | 0.006704325 | 0.00223047 | 0.24029876 | -0.00140281 | 0.042893902 | 1 | 0.16246663 | 1 | 0.061679788 | ENSG0000021 | 3 | ZNF663P |
| VAT | HTN | ENSG0000019 | -0.00734675 | 0.037069266 | 0.003523612 | 0.240333669 | -0.00728457 | 0.038691987 | 0.003523449 | 0.240300277 | -6.22E-05 | 0.009830012 | 1 | 0.187472424 | 1 | 0.193570362 | ENSG0000021 | 3 | ZNF663P |
| VAT | HTN | ENSG0000019 | 0.027863242 | 1.53E-05 | 0.006444141 | 0.240326593 | 0.027713692 | 1.70E-05 | 0.006443927 | 0.240293264 | 0.00014955 | 0.006326786 | 0.042334961 | 0.000549805 | 0.046990952 | 0.000594822 | ENSG0000021 | 4 | AP000354.2 |
| VAT | HTN | ENSG0000019 | 0.002544655 | 0.499046409 | 0.003764331 | 0.240335588 | 0.002641906 | 0.482770441 | 0.003764181 | 0.240302142 | -9.73E-05 | 0.000817392 | 1 | 0.755550241 | 1 | 0.74386019 | ENSG0000021 | 3 | AP005482.1 |
| VAT | HTN | ENSG0000019 | -0.00342134 | 0.593849942 | 0.006415831 | 0.240335673 | -0.00329559 | 0.607417408 | 0.006415569 | 0.240302255 | -0.00012575 | 0.009595994 | 1 | 0.808777562 | 1 | 0.818845598 | ENSG0000021 | 2 | SLC35E2 |
| VAT | HTN | ENSG0000019 | -0.00815336 | 0.004352562 | 0.002859399 | 0.240331803 | -0.00810164 | 0.004605358 | 0.002859308 | 0.240298426 | -5.17E-05 | 0.037144118 | 1 | 0.045161918 | 1 | 0.047053283 | ENSG0000021 | 5 | CROCCP2 |
| VAT | HTN | ENSG0000019 | 0.001881919 | 0.70518472 | 0.004974264 | 0.240335743 | 0.001802549 | 0.717061351 | 0.00497407 | 0.24030232 | 7.94E-05 | 0.041203891 | 1 | 0.861817357 | 1 | 0.872614342 | ENSG0000021 | 5 | CROCCP2 |
| VAT | HTN | ENSG0000019 | 0.00304547 | 0.528903433 | 0.004836527 | 0.240335618 | 0.002923091 | 0.545578216 | 0.004836354 | 0.240302205 | 0.000122379 | 0.002096408 | 1 | 0.774633714 | 1 | 0.78353797 | ENSG0000021 | 5 | CROCCP2 |
| VAT | HTN | ENSG0000019 | -0.00264927 | 0.279871088 | 0.002451645 | 0.240335237 | -0.00257432 | 0.293683608 | 0.002451556 | 0.240301841 | -7.50E-05 | 0.000168678 | 1 | 0.586749061 | 1 | 0.592989502 | ENSG0000021 | 5 | ATAD3C |
| VAT | HTN | ENSG0000019 | -0.00773731 | 0.253565126 | 0.006776804 | 0.24033517 | -0.00601358 | 0.378136058 | 0.006823251 | 0.240302002 | -0.00172373 | 0.030926826 | 1 | 0.563738197 | 1 | 0.669868754 | ENSG0000021 | 2 | RP1-152L7.5 |
| VAT | HTN | ENSG0000019 | -0.00127793 | 0.813000816 | 0.005402194 | 0.240335786 | -0.00135789 | 0.801527662 | 0.005401944 | 0.240302354 | 8.00E-05 | 0.030023692 | 1 | 0.923295638 | 1 | 0.923295638 | ENSG0000021 | 1 | AL022393.7 |
| VAT | HTN | ENSG0000019 | 0.001859277 | 0.631036646 | 0.003871327 | 0.2403357 | 0.001942929 | 0.61573738 | 0.003871141 | 0.240302261 | -8.37E-05 | 0.000955662 | 1 | 0.825823207 | 1 | 0.823768865 | ENSG0000021 | 1 | AL022393.7 |
| VAT | HTN | ENSG0000019 | 0.004886912 | 0.281597719 | 0.004538626 | 0.240335242 | 0.004804547 | 0.289765362 | 0.004538424 | 0.240301832 | 8.24E-05 | 0.01052396 | 1 | 0.586749061 | 1 | 0.59182601 | ENSG0000021 | 1 | AL022393.7 |
| VAT | HTN | ENSG0000019 | -0.01029089 | 0.003587202 | 0.003533516 | 0.24033163 | -0.01039659 | 0.003259267 | 0.003533637 | 0.240298116 | 0.000105707 | 0.037997993 | 1 | 0.03843057 | 1 | 0.036419338 | ENSG0000021 | 1 | AL022393.7 |
| VAT | HTN | ENSG0000019 | -0.00438911 | 0.395388272 | 0.005164343 | 0.240335457 | -0.00429569 | 0.405505733 | 0.005164153 | 0.240302044 | -9.34E-05 | 0.024981871 | 1 | 0.688065342 | 1 | 0.690868103 | ENSG0000021 | 1 | AL022393.7 |
| VAT | HTN | ENSG0000019 | -0.00602214 | 0.035506578 | 0.002864222 | 0.240333633 | -0.0059436 | 0.037969209 | 0.002864118 | 0.240300261 | -7.85E-05 | 0.000742213 | 1 | 0.183861456 | 1 | 0.191644541 | ENSG0000021 | 1 | AL022393.7 |
| VAT | HTN | ENSG0000020 | -0.00615576 | 0.284128533 | 0.005747181 | 0.240335248 | -0.00633377 | 0.270416033 | 0.005746968 | 0.240301786 | 0.000178009 | 0.000125485 | 1 | 0.589701944 | 1 | 0.575885996 | ENSG0000021 | 2 | RP11-350J20.9 |
| VAT | HTN | ENSG0000020 | -0.00627924 | 0.117241341 | 0.004008553 | 0.240334603 | -0.0061857 | 0.122784924 | 0.004008399 | 0.24030121 | -9.35E-05 | 0.003091358 | 1 | 0.369484015 | 1 | 0.379153311 | ENSG0000021 | 3 | PAM16 |
| VAT | HTN | ENSG0000020 | -0.00482383 | 0.139340902 | 0.003263195 | 0.240334736 | -0.00475138 | 0.145363712 | 0.00326307 | 0.240301339 | -7.24E-05 | 0.004939647 | 1 | 0.40653371 | 1 | 0.41661874 | ENSG0000021 | 3 | PAM16 |
| VAT | HTN | ENSG0000020 | -0.00025551 | 0.923156692 | 0.002648876 | 0.240335809 | -0.00018895 | 0.943131677 | 0.002648799 | 0.240302382 | -6.66E-05 | 0.005236667 | 1 | 0.970079269 | 1 | 0.978587755 | ENSG0000021 | 2 | AC016773.1 |
| VAT | HTN | ENSG0000020 | 0.005001029 | 0.020841019 | 0.002164142 | 0.24033318 | 0.006046108 | 0.006411267 | 0.002217956 | 0.24029872 | -0.00104508 | 0.031630859 | 1 | 0.129552281 | 1 | 0.05937952 | ENSG0000021 | 4 | AP000350.4 |
| VAT | HTN | ENSG0000020 | -0.01002882 | 5.66E-08 | 0.001847115 | 0.240321275 | -0.01185149 | 2.65E-09 | 0.001991068 | 0.240284914 | 0.001822668 | 0.014248766 | 0.000156089 | 5.34E-06 | 7.30E-06 | 4.06E-07 | ENSG0000021 | 1 | AP000350.4 |
| VAT | HTN | ENSG0000020 | -0.00360157 | 0.000931258 | 0.001087913 | 0.240330408 | -0.00413257 | 0.000224101 | 0.001119876 | 0.24029567 | 0.000530998 | 0.045881742 | 1 | 0.014521318 | 0.618518387 | 0.005028605 | ENSG0000021 | 1 | AP000350.4 |
| VAT | HTN | ENSG0000020 | -0.00254277 | 0.072780324 | 0.001417207 | 0.240334226 | -0.00411424 | 0.00847294 | 0.001562789 | 0.240299167 | 0.001571478 | 0.017076411 | 1 | 0.283719908 | 1 | 0.072176896 | ENSG0000021 | 3 | AC011298.2 |
| VAT | HTN | ENSG0000020 | 0.004332237 | 0.002686284 | 0.001443337 | 0.24033137 | 0.003735061 | 0.010869582 | 0.001466528 | 0.240299186 | 0.000597176 | 0.021786046 | 1 | 0.031021521 | 1 | 0.084874033 | ENSG0000021 | 3 | AC011298.2 |
| VAT | HTN | ENSG0000020 | 0.000250821 | 0.943871352 | 0.003562546 | 0.240335811 | -0.0016959 | 0.642355561 | 0.003651751 | 0.240302278 | 0.001946724 | 0.015378186 | 1 | 0.976968793 | 1 | 0.837994779 | ENSG0000021 | 3 | AC011298.2 |
| VAT | HTN | ENSG0000020 | -0.00913294 | 0.121515264 | 0.005898126 | 0.240334631 | -0.01102181 | 0.063387544 | 0.005936982 | 0.240300685 | 0.001888877 | 0.005594544 | 1 | 0.376410917 | 1 | 0.261118836 | ENSG0000021 | 3 | RP11-181C21.4 |
| VAT | HTN | ENSG0000020 | 0.003473645 | 0.069661425 | 0.001914793 | 0.24033419 | 0.002677528 | 0.167776795 | 0.001941103 | 0.240301446 | 0.000796117 | 0.012653765 | 1 | 0.276243583 | 1 | 0.452652937 | ENSG0000021 | 1 | RP1-265C24.5 |
| VAT | HTN | ENSG0000020 | 0.002204295 | 0.535207092 | 0.00355487 | 0.240335624 | 0.000580154 | 0.871870034 | 0.003597102 | 0.240302372 | 0.001624141 | 0.003211848 | 1 | 0.777236628 | 1 | 0.956043422 | ENSG0000021 | 1 | RP1-265C24.5 |
| VAT | HTN | ENSG0000020 | -0.00186904 | 0.076221151 | 0.001054137 | 0.240334263 | -0.00276715 | 0.014347312 | 0.001130161 | 0.240299428 | 0.000898113 | 0.02760349 | 1 | 0.292622935 | 1 | 0.100249572 | ENSG0000021 | 1 | RP1-265C24.5 |
| VAT | HTN | ENSG0000020 | 0.002615879 | 0.610335882 | 0.005133257 | 0.240335685 | 0.002727138 | 0.595221985 | 0.005133057 | 0.240302246 | -0.00011123 | 0.005706794 | 1 | 0.819230676 | 1 | 0.814079622 | ENSG0000021 | 1 | RP1-265C24.5 |

| | | | | | | | | | | | | | | | | | | | |
|---|---|---|---|---|---|---|---|---|---|---|---|---|---|---|---|---|---|---|---|
| VAT | HTN | ENSG000002( | -0.00450302 | 0.131139267 | 0.002982871 | 0.240334689 | -0.00651429 | 0.038714072 | 0.003151232 | 0.240300277 | 0.002011262 | 0.047939319 | 1 | 0.397304475 | 1 | 0.193570362 | ENSG0000021 | 1 | RP1-265C24.5 |
| VAT | HTN | ENSG000002( | 0.001468889 | 0.649717037 | 0.003234329 | 0.240335712 | 0.001532506 | 0.635613697 | 0.003234222 | 0.240301867 | -6.36E-05 | 0.021450448 | 1 | 0.835367368 | 1 | 0.832512881 | ENSG0000021 | 1 | RP1-265C24.5 |
| VAT | HTN | ENSG000002( | -0.00836853 | 0.099113023 | 0.005074383 | 0.240334472 | -0.00823092 | 0.104780221 | 0.0050742 | 0.240301087 | -0.00013762 | 0.000876758 | 1 | 0.336272805 | 1 | 0.345925131 | ENSG0000021 | 6 | NBPF1 |
| VAT | HTN | ENSG000002( | -0.00773945 | 0.11269989 | 0.004879309 | 0.240334573 | -0.00766441 | 0.116213047 | 0.004879079 | 0.240301168 | -7.50E-05 | 0.021349466 | 1 | 0.363803153 | 1 | 0.368868078 | ENSG0000021 | 6 | NBPF1 |
| VAT | HTN | ENSG000002( | 0.004083121 | 0.520287618 | 0.006351072 | 0.24033561 | 0.008029803 | 0.213333156 | 0.006452471 | 0.240301621 | -0.00394668 | 0.00054785 | 1 | 0.77245499 | 1 | 0.513786658 | ENSG0000021 | 6 | NBPF1 |
| VAT | HTN | ENSG000002( | 0.003526801 | 0.529333038 | 0.005606775 | 0.240335618 | 0.003353297 | 0.54977256 | 0.00560657 | 0.240302208 | 0.000173504 | 0.000443699 | 1 | 0.774633714 | 1 | 0.786610816 | ENSG0000021 | 6 | NBPF1 |
| VAT | HTN | ENSG000002( | 0.017365668 | 0.004838097 | 0.006163211 | 0.240331898 | 0.017507615 | 0.004500833 | 0.006163009 | 0.240298405 | -0.00014195 | 0.007305415 | 1 | 0.048912633 | 1 | 0.046525467 | ENSG0000021 | 6 | NBPF1 |
| VAT | HTN | ENSG000002( | 0.005691825 | 0.350282548 | 0.006093755 | 0.240335383 | 0.005782446 | 0.342644232 | 0.006093491 | 0.240301941 | -9.06E-05 | 0.039909109 | 1 | 0.654113554 | 1 | 0.641030042 | ENSG0000021 | 4 | FAM228B |
| VAT | HTN | ENSG000002( | -0.00471739 | 0.005422116 | 0.001696391 | 0.240332 | -0.00468492 | 0.005748297 | 0.001696317 | 0.240298623 | -3.25E-05 | 0.008084191 | 1 | 0.053256368 | 1 | 0.055667722 | ENSG0000021 | 1 | RP1-199J3.5 |
| VAT | HTN | ENSG000002( | 0.004686333 | 0.321308199 | 0.004725201 | 0.240335328 | 0.004776426 | 0.312077554 | 0.004725049 | 0.240301881 | -9.01E-05 | 0.0274413 | 1 | 0.624075039 | 1 | 0.610875211 | ENSG0000021 | 1 | RP1-199J3.5 |
| VAT | HTN | ENSG000002( | -0.00145155 | 0.61353613 | 0.002874173 | 0.240335688 | -0.00208117 | 0.469839095 | 0.002879556 | 0.240302127 | 0.000629624 | 0.000443699 | 1 | 0.819907808 | 1 | 0.73755282 | ENSG0000021 | 1 | RP1-199J3.5 |
| VAT | HTN | ENSG000002( | 0.003588121 | 0.310646819 | 0.003539046 | 0.240335306 | 0.003533592 | 0.318036889 | 0.003538896 | 0.240301893 | 5.45E-05 | 0.036681037 | 1 | 0.613294148 | 1 | 0.616693002 | ENSG0000021 | 1 | RP1-199J3.5 |
| VAT | HTN | ENSG000002( | -0.00347257 | 0.310026532 | 0.003420687 | 0.240335305 | -0.00354698 | 0.299757307 | 0.003420574 | 0.240301855 | 7.44E-05 | 0.010997481 | 1 | 0.612581159 | 1 | 0.599080498 | ENSG0000021 | 2 | ZSCAN12P1 |
| VAT | HTN | ENSG000002( | 0.001871417 | 0.588003688 | 0.003454509 | 0.240335669 | 0.001938712 | 0.574641971 | 0.003454406 | 0.240302229 | -6.73E-05 | 0.029127825 | 1 | 0.806605456 | 1 | 0.801420839 | ENSG0000021 | 2 | ZSCAN12P1 |
| VAT | HTN | ENSG000002( | 0.011588071 | 0.085625026 | 0.006741386 | 0.240334356 | 0.01169174 | 0.082847466 | 0.00674108 | 0.240300901 | -0.00010367 | 0.027145441 | 1 | 0.31229706 | 1 | 0.306924839 | ENSG0000021 | 2 | ZSCAN12P1 |
| VAT | HTN | ENSG0000021 | -0.00464157 | 0.173922689 | 0.003413646 | 0.240334902 | -0.00458789 | 0.178935871 | 0.003413507 | 0.240301494 | -5.37E-05 | 0.037499779 | 1 | 0.458916465 | 1 | 0.46743134 | ENSG0000021 | 2 | ZSCAN12P1 |
| VAT | HTN | ENSG0000021 | -0.00170791 | 0.721056445 | 0.004783418 | 0.240335751 | -0.00203083 | 0.671295962 | 0.004785513 | 0.240302296 | 0.000322923 | 0.034139207 | 1 | 0.869815229 | 1 | 0.850122986 | ENSG0000021 | 2 | ZSCAN12P1 |
| VAT | HTN | ENSG0000021 | 0.000284484 | 0.854926947 | 0.001555953 | 0.240335797 | 0.000259357 | 0.867611877 | 0.001555897 | 0.240302371 | 2.51E-05 | 0.048847385 | 1 | 0.944984744 | 1 | 0.953267827 | ENSG0000022 | 1 | ACTBP8 |
| VAT | HTN | ENSG0000021 | -0.00038518 | 0.883261301 | 0.002623216 | 0.240335803 | -0.00122066 | 0.642465241 | 0.00262929 | 0.240302278 | 0.000835478 | 4.05E-06 | 1 | 0.956145095 | 1 | 0.837994779 | ENSG0000022 | 1 | RPL35P2 |
| VAT | HTN | ENSG0000021 | -0.00199763 | 0.36740385 | 0.002216274 | 0.240335413 | -0.0020506 | 0.354828166 | 0.002216232 | 0.240301963 | 5.30E-05 | 0.017710741 | 1 | 0.667567233 | 1 | 0.6537555 | ENSG0000022 | 1 | RPL35P2 |
| VAT | HTN | ENSG0000021 | -0.00495811 | 0.415417659 | 0.006088066 | 0.240335486 | -0.0052923 | 0.384799144 | 0.006089486 | 0.240302012 | 0.000334197 | 0.025748006 | 1 | 0.701684662 | 1 | 0.675933439 | ENSG0000022 | 1 | RPL35P2 |
| VAT | HTN | ENSG0000021 | -0.00287536 | 0.242659365 | 0.002461008 | 0.24033514 | -0.00283004 | 0.250143769 | 0.002460899 | 0.240301733 | -4.53E-05 | 0.009523232 | 1 | 0.552590633 | 1 | 0.556098166 | ENSG0000022 | 1 | RPL35P2 |
| VAT | HTN | ENSG0000021 | -0.00223361 | 0.745028741 | 0.006868378 | 0.240335761 | -0.00196941 | 0.774306587 | 0.006868136 | 0.240302344 | -0.00026419 | 3.34E-06 | 1 | 0.881766434 | 1 | 0.907082419 | ENSG0000022 | 6 | PPP3R1 |
| VAT | HTN | ENSG0000021 | -0.02156212 | 0.000904797 | 0.006497374 | 0.240330382 | -0.02175491 | 0.000813093 | 0.006497261 | 0.240296856 | 0.000192786 | 0.003753613 | 1 | 0.01418886 | 1 | 0.013200804 | ENSG0000022 | 6 | PPP3R1 |
| VAT | HTN | ENSG0000021 | 0.01707219 | 0.000311983 | 0.004735478 | 0.240329403 | 0.017156351 | 0.000291116 | 0.00473526 | 0.240295912 | -8.42E-05 | 0.009791397 | 0.86107294 | 0.006573076 | 0.803478988 | 0.006228519 | ENSG0000022 | 5 | FANCG |
| VAT | HTN | ENSG0000021 | 0.0025189 | 0.2738068 | 0.002301755 | 0.240335223 | 0.003121343 | 0.176113631 | 0.002307285 | 0.240301482 | -0.00060244 | 0.000199644 | 1 | 0.583904713 | 1 | 0.463882578 | ENSG0000022 | 5 | FANCG |
| VAT | HTN | ENSG0000021 | 0.001108461 | 0.732893569 | 0.003247958 | 0.240335756 | 0.001164168 | 0.720010025 | 0.003247825 | 0.240302321 | -5.57E-05 | 0.022728013 | 1 | 0.875286132 | 1 | 0.872943483 | ENSG0000022 | 5 | FANCG |
| VAT | HTN | ENSG0000021 | 0.000532998 | 0.816649465 | 0.002298825 | 0.240335787 | 0.000483997 | 0.833240059 | 0.002298749 | 0.240302363 | 4.90E-05 | 0.012373129 | 1 | 0.925513978 | 1 | 0.939506392 | ENSG0000022 | 5 | AP4M1 |
| VAT | HTN | ENSG0000022 | -0.01273254 | 0.060606419 | 0.006785767 | 0.240334077 | -0.01323256 | 0.051236827 | 0.006787682 | 0.240300511 | 0.000500013 | 0.005487428 | 1 | 0.252880519 | 1 | 0.227450279 | ENSG0000022 | 4 | ZNF880 |
| VAT | HTN | ENSG0000022 | 0.000751682 | 0.878460173 | 0.00491547 | 0.240335802 | 0.00087736 | 0.85833472 | 0.004915333 | 0.240302369 | -0.00012568 | 0.005120393 | 1 | 0.955786877 | 1 | 0.949120123 | ENSG0000022 | 3 | TIGD1 |
| VAT | HTN | ENSG0000022 | 0.004597802 | 0.117778725 | 0.002939455 | 0.240334607 | 0.004539018 | 0.12253127 | 0.002939325 | 0.240301209 | 5.88E-05 | 0.004926472 | 1 | 0.369484015 | 1 | 0.379153311 | ENSG0000022 | 3 | TIGD1 |
| VAT | HTN | ENSG0000022 | -0.00734349 | 0.267270402 | 0.006619503 | 0.240335206 | -0.00714214 | 0.280605369 | 0.006619458 | 0.240301811 | -0.00020135 | 0.006579369 | 1 | 0.573612994 | 1 | 0.584169217 | ENSG0000022 | 3 | TIGD1 |
| VAT | HTN | ENSG0000022 | -0.00254308 | 0.461560368 | 0.003453952 | 0.240335546 | -0.00247901 | 0.472903132 | 0.003453795 | 0.240302131 | -6.41E-05 | 0.007628621 | 1 | 0.734663562 | 1 | 0.739078508 | ENSG0000022 | 3 | TIGD1 |
| VAT | HTN | ENSG0000022 | 0.003126207 | 0.171811682 | 0.002287902 | 0.240334893 | 0.002645503 | 0.250205792 | 0.002300736 | 0.240301733 | 0.000480703 | 0.048970577 | 1 | 0.45577776 | 1 | 0.556098166 | ENSG0000022 | 3 | XKR9 |
| VAT | HTN | ENSG0000022 | 4.75E-05 | 0.988476015 | 0.003286995 | 0.240335813 | 0.000149583 | 0.963701956 | 0.003286922 | 0.240302384 | -0.00010211 | 0.001374072 | 1 | 0.995476191 | 1 | 0.985245375 | ENSG0000022 | 3 | XKR9 |
| VAT | HTN | ENSG0000022 | -0.00111456 | 0.865820699 | 0.006596191 | 0.240335799 | -0.00122871 | 0.852223003 | 0.006595932 | 0.240302368 | 0.000114154 | 0.026235555 | 1 | 0.950651672 | 1 | 0.94611263 | ENSG0000022 | 3 | XKR9 |
| VAT | HTN | ENSG0000022 | 0.003162116 | 0.291232904 | 0.002996068 | 0.240335264 | 0.003978951 | 0.185288948 | 0.003003782 | 0.240301519 | -0.00081683 | 0.000179621 | 1 | 0.59321241 | 1 | 0.477049902 | ENSG0000022 | 6 | SLC12A8 |
| VAT | HTN | ENSG0000022 | 0.001664884 | 0.256911553 | 0.001468511 | 0.240335179 | 0.001506968 | 0.305345484 | 0.001470157 | 0.240301867 | 0.000157916 | 0.02755063 | 1 | 0.566122845 | 1 | 0.604212978 | ENSG0000022 | 6 | SLC12A8 |
| VAT | HTN | ENSG0000022 | 0.00439329 | 0.318975857 | 0.00440843 | 0.240335324 | 0.004267775 | 0.332983058 | 0.004408295 | 0.240301923 | 0.000125516 | 0.001219847 | 1 | 0.622410789 | 1 | 0.631637966 | ENSG0000022 | 3 | UBA52 |
| VAT | HTN | ENSG0000022 | 0.005245407 | 0.05392571 | 0.002721441 | 0.240333981 | 0.005302861 | 0.051341132 | 0.002721339 | 0.240300512 | -5.75E-05 | 0.008445705 | 1 | 0.236976525 | 1 | 0.227450279 | ENSG0000022 | 3 | UBA52 |
| VAT | HTN | ENSG0000022 | 0.010382196 | 0.084373828 | 0.006015677 | 0.240334344 | 0.010222129 | 0.0892613 | 0.006015456 | 0.240300961 | 0.000160067 | 0.001011233 | 1 | 0.309258654 | 1 | 0.321201027 | ENSG0000022 | 8 | PPT2 |
| VAT | HTN | ENSG0000022 | -0.0104576 | 0.055029183 | 0.005450497 | 0.240333998 | -0.01065125 | 0.05067879 | 0.005450453 | 0.240300502 | 0.000193647 | 0.001370539 | 1 | 0.239559218 | 1 | 0.226332458 | ENSG0000022 | 8 | PPT2 |
| VAT | HTN | ENSG0000022 | 0.002615101 | 0.200498839 | 0.002042835 | 0.240335005 | 0.002508335 | 0.219546431 | 0.002043057 | 0.240301641 | 0.000106766 | 0.005594471 | 1 | 0.497193887 | 1 | 0.517020606 | ENSG0000022 | 8 | PPT2 |
| VAT | HTN | ENSG0000022 | 0.004782018 | 0.327808853 | 0.004886901 | 0.240335341 | 0.009675759 | 0.07202152 | 0.005378452 | 0.240300789 | -0.00489374 | 0.029419384 | 1 | 0.627864285 | 1 | 0.27997098 | ENSG0000022 | 4 | TIAF1 |
| VAT | HTN | ENSG0000022 | -0.00068268 | 0.72625818 | 0.001949905 | 0.240335753 | -0.00071799 | 0.712698728 | 0.001949832 | 0.240302318 | 3.53E-05 | 0.023815894 | 1 | 0.873302353 | 1 | 0.869990486 | ENSG0000022 | 4 | BTBD19 |
| VAT | HTN | ENSG0000022 | -0.00752902 | 0.288648083 | 0.007095533 | 0.240335258 | -0.00737926 | 0.298329346 | 0.007095279 | 0.240301851 | -0.00014976 | 0.010179021 | 1 | 0.59321241 | 1 | 0.597477868 | ENSG0000022 | 4 | BTBD19 |
| VAT | HTN | ENSG0000022 | -0.00073887 | 0.684660984 | 0.001819382 | 0.240335732 | -0.00079155 | 0.663502869 | 0.001819312 | 0.240302291 | 5.27E-05 | 0.000234959 | 1 | 0.853121587 | 1 | 0.847332674 | ENSG0000022 | 3 | URAHP |
| VAT | HTN | ENSG0000022 | 0.004393562 | 0.469880674 | 0.006079592 | 0.240335556 | 0.00428714 | 0.480685626 | 0.006079319 | 0.24030214 | 0.000106421 | 0.012636454 | 1 | 0.740158428 | 1 | 0.743661619 | ENSG0000022 | 3 | URAHP |
| VAT | HTN | ENSG0000022 | -0.00637612 | 0.054092468 | 0.00331038 | 0.240333984 | -0.00642902 | 0.052116881 | 0.003310221 | 0.240300525 | 5.29E-05 | 0.015126787 | 1 | 0.236976525 | 1 | 0.229414021 | ENSG0000022 | 3 | URAHP |
| VAT | HTN | ENSG0000022 | -0.00458572 | 0.239148421 | 0.003895711 | 0.24033513 | -0.00471889 | 0.225787575 | 0.003895779 | 0.240301661 | 0.000133173 | 0.009517643 | 1 | 0.549661061 | 1 | 0.526616698 | ENSG0000022 | 3 | URAHP |
| VAT | HTN | ENSG0000022 | 0.006730569 | 0.079011877 | 0.003831911 | 0.240334292 | 0.006651911 | 0.082564498 | 0.003831746 | 0.240300899 | 7.87E-05 | 0.004713849 | 1 | 0.297599038 | 1 | 0.306698747 | ENSG0000022 | 3 | URAHP |
| VAT | HTN | ENSG0000022 | -0.00218171 | 0.656119633 | 0.004899661 | 0.240335716 | -0.00104919 | 0.830858768 | 0.004911929 | 0.240302362 | -0.00113252 | 0.001281953 | 1 | 0.837990221 | 1 | 0.939438836 | ENSG0000022 | 2 | RPS10P7 |
| VAT | HTN | ENSG0000022 | 0.003410427 | 0.414815607 | 0.004182268 | 0.240335485 | 0.003479807 | 0.405371172 | 0.00418212 | 0.240302043 | -6.94E-05 | 0.044843908 | 1 | 0.701684662 | 1 | 0.690868103 | ENSG0000022 | 1 | GS1-124K5.3 |
| VAT | HTN | ENSG0000022 | 0.003062774 | 0.435670316 | 0.003929021 | 0.240335514 | 0.003154366 | 0.422051474 | 0.003928872 | 0.240302067 | -9.16E-05 | 0.003498443 | 1 | 0.715318306 | 1 | 0.704580121 | ENSG0000022 | 1 | EXOSC6 |
| VAT | HTN | ENSG0000022 | -0.00529377 | 0.153339724 | 0.003707544 | 0.240334808 | -0.0052283 | 0.158471817 | 0.003707402 | 0.240301404 | -6.55E-05 | 0.024853407 | 1 | 0.429662578 | 1 | 0.437382215 | ENSG0000022 | 1 | EXOSC6 |
| VAT | HTN | ENSG0000022 | -0.00081557 | 0.880263996 | 0.005414216 | 0.240335802 | -0.00105347 | 0.845721874 | 0.005414065 | 0.240302366 | 0.0002379 | 1.54E-06 | 1 | 0.955786877 | 1 | 0.942573159 | ENSG0000022 | 1 | EXOSC6 |
| VAT | HTN | ENSG0000022 | 0.005763311 | 0.400336113 | 0.006852749 | 0.240335465 | 0.005636578 | 0.41075917 | 0.00685249 | 0.240302051 | 0.000126734 | 0.02025869 | 1 | 0.692212916 | 1 | 0.69398935 | ENSG0000022 | 4 | RP11-632K20.7 |
| VAT | HTN | ENSG0000022 | 0.002251613 | 0.664420259 | 0.005190217 | 0.240335721 | 0.00039005 | 0.940303675 | 0.005208428 | 0.240302382 | 0.001861564 | 2.27E-05 | 1 | 0.842351822 | 1 | 0.977875185 | ENSG0000022 | 5 | CKMT1A |
| VAT | HTN | ENSG0000022 | 0.007344656 | 0.094007816 | 0.004385843 | 0.24033443 | 0.008063247 | 0.066399267 | 0.00439241 | 0.240300723 | -0.00071859 | 0.003442236 | 1 | 0.328017159 | 1 | 0.266756879 | ENSG0000022 | 2 | AC093107.7 |
| VAT | HTN | ENSG0000022 | 0.027443794 | 4.55E-06 | 0.005986222 | 0.240325448 | 0.027344422 | 4.92E-06 | 0.005985969 | 0.240292095 | 9.94E-05 | 0.02419142 | 0.012563585 | 0.000199422 | 0.01358799 | 0.000219161 | ENSG0000022 | 2 | TSSC2 |
| VAT | HTN | ENSG0000022 | 0.01055131 | 0.066215086 | 0.005743869 | 0.240334149 | 0.009497868 | 0.099441383 | 0.00576477 | 0.240301046 | 0.001053441 | 0.03334604 | 1 | 0.26718368 | 1 | 0.338419502 | ENSG0000022 | 2 | TSSC2 |
| VAT | HTN | ENSG0000022 | 0.002267275 | 0.642503177 | 0.004884241 | 0.240335707 | 0.002148061 | 0.660072454 | 0.004884028 | 0.240302289 | 0.000119214 | 0.000705358 | 1 | 0.830978804 | 1 | 0.846845836 | ENSG0000022 | 2 | TSSC2 |
| VAT | HTN | ENSG0000022 | 0.005175886 | 0.138799763 | 0.003496563 | 0.240334733 | 0.005121339 | 0.142993487 | 0.003496424 | 0.240301327 | 5.45E-05 | 0.041466148 | 1 | 0.40653371 | 1 | 0.412957063 | ENSG0000022 | 2 | TSSC2 |
| VAT | HTN | ENSG0000022 | 0.005312546 | 0.228958019 | 0.004415903 | 0.2403351 | 0.005435135 | 0.218375147 | 0.004415715 | 0.240301638 | -0.00012259 | 0.000168455 | 1 | 0.534622786 | 1 | 0.517020606 | ENSG0000022 | 1 | RP11-465B22.5 |
| VAT | HTN | ENSG0000022 | -0.00236072 | 0.155499061 | 0.001662041 | 0.240334818 | -0.00261848 | 0.115969944 | 0.001665786 | 0.240301166 | 0.000257758 | 0.022966865 | 1 | 0.431768017 | 1 | 0.368752358 | ENSG0000022 | 1 | RP11-74C1.2 |
| VAT | HTN | ENSG0000022 | 0.004110256 | 0.522366394 | 0.006425263 | 0.240335612 | 0.00434875 | 0.498507771 | 0.006425077 | 0.240302159 | -0.00023849 | 3.89E-05 | 1 | 0.772678586 | 1 | 0.754734749 | ENSG0000022 | 1 | RP11-74C1.2 |
| VAT | HTN | ENSG0000022 | -0.00794641 | 0.047224078 | 0.004004661 | 0.240333871 | -0.00787698 | 0.049180974 | 0.004004516 | 0.240300477 | -6.94E-05 | 0.033301338 | 1 | 0.21807219 | 1 | 0.222523752 | ENSG0000022 | 6 | HLA-DPB1 |
| VAT | HTN | ENSG0000022 | -0.01139414 | 0.104281574 | 0.007014177 | 0.240334512 | -0.01129511 | 0.107311227 | 0.007013856 | 0.240301106 | -9.90E-05 | 0.040377097 | 1 | 0.346052657 | 1 | 0.351756515 | ENSG0000022 | 6 | HLA-DPB1 |
| VAT | HTN | ENSG0000022 | -0.00535162 | 0.376358427 | 0.006049581 | 0.240335427 | -0.00578942 | 0.338756124 | 0.006051909 | 0.240301933 | 0.000437798 | 0.016396846 | 1 | 0.672329618 | 1 | 0.637283706 | ENSG0000022 | 6 | HLA-DPB1 |
| VAT | HTN | ENSG0000022 | 0.003159374 | 0.086384816 | 0.001842429 | 0.240334363 | 0.002911905 | 0.114800874 | 0.001846508 | 0.240301158 | 0.000247469 | 0.046959232 | 1 | 0.314113273 | 1 | 0.366605303 | ENSG0000022 | 6 | HLA-DPB1 |
| VAT | HTN | ENSG0000022 | -0.00283005 | 0.540153286 | 0.004619876 | 0.240335628 | -0.00784124 | 0.102379388 | 0.004800475 | 0.240301069 | 0.005011185 | 0.000123805 | 1 | 0.778497687 | 1 | 0.344320158 | ENSG0000022 | 6 | HLA-DPB1 |
| VAT | HTN | ENSG0000022 | 0.00270712 | 0.372837117 | 0.003037706 | 0.240335422 | 0.002460968 | 0.418105976 | 0.003039312 | 0.240302061 | 0.000246152 | 0.019141443 | 1 | 0.670600511 | 1 | 0.702429993 | ENSG0000022 | 2 | AC112715.2 |
| VAT | HTN | ENSG0000022 | 0.003394712 | 0.377578613 | 0.003847278 | 0.240335429 | 0.003457367 | 0.368820555 | 0.003847137 | 0.240301987 | -6.27E-05 | 0.045451481 | 1 | 0.673387034 | 1 | 0.664239406 | ENSG0000022 | 1 | SDHDP6 |
| VAT | HTN | ENSG0000022 | 0.009007688 | 5.90E-05 | 0.002242488 | 0.240327856 | 0.008955716 | 6.50E-05 | 0.002242438 | 0.240294519 | 5.20E-05 | 0.016895311 | 0.162827818 | 0.001690495 | 0.179533751 | 0.001777562 | ENSG0000022 | 2 | MSL3P1 |
| VAT | HTN | ENSG0000022 | 0.005201828 | 0.390315067 | 0.006055348 | 0.240335449 | 0.005083667 | 0.401149707 | 0.006055081 | 0.240302037 | 0.000118161 | 0.006268026 | 1 | 0.682680345 | 1 | 0.687928367 | ENSG0000022 | 2 | MSL3P1 |
| VAT | HTN | ENSG0000022 | -0.00379238 | 0.364427887 | 0.004181414 | 0.240335408 | -0.00386934 | 0.354756377 | 0.00418124 | 0.240301962 | 7.70E-05 | 0.013781169 | 1 | 0.667014755 | 1 | 0.6537555 | ENSG0000022 | 1 | RP11-479O9.2 |
| VAT | HTN | ENSG0000022 | 0.001186644 | 0.543785539 | 0.001954614 | 0.240335632 | 0.000570236 | 0.773208007 | 0.00197874 | 0.240302344 | 0.000616408 | 0.045996696 | 1 | 0.780738853 | 1 | 0.906180085 | ENSG0000022 | 1 | RP11-479O9.2 |
| VAT | HTN | ENSG0000022 | 0.005838158 | 0.011991111 | 0.002323723 | 0.2403327 | 0.005798236 | 0.01258385 | 0.002323622 | 0.240299314 | 3.99E-05 | 0.017244781 | 1 | 0.089447203 | 1 | 0.090807843 | ENSG0000022 | 1 | RP11-479O9.2 |
| VAT | HTN | ENSG0000022 | 0.00724784 | 0.016881635 | 0.003033478 | 0.240332998 | 0.007158156 | 0.018391535 | 0.003033471 | 0.240299644 | 9.60E-05 | 0.006355731 | 1 | 0.11097825 | 1 | 0.11723011 | ENSG0000022 | 4 | C4B |
| VAT | HTN | ENSG0000022 | 0.001133382 | 0.807392891 | 0.004648993 | 0.240335784 | 0.000414136 | 0.929191071 | 0.004660404 | 0.240302381 | 0.000719246 | 0.029540705 | 1 | 0.920447905 | 1 | 0.973666067 | ENSG0000022 | 4 | C4B |
| VAT | HTN | ENSG0000022 | -0.0109071 | 0.107271091 | 0.006772138 | 0.240334534 | -0.01109185 | 0.101441386 | 0.006771958 | 0.240301062 | 0.000184753 | 0.00329264 | 1 | 0.351624954 | 1 | 0.342746569 | ENSG0000022 | 4 | C4B |
| VAT | HTN | ENSG0000022 | -0.00033307 | 0.780921476 | 0.001197592 | 0.240335775 | -0.00093837 | 0.447739513 | 0.001236018 | 0.240302025 | 0.000605299 | 0.048010277 | 1 | 0.903724293 | 1 | 0.721401667 | ENSG0000022 | 4 | C4B |
| VAT | HTN | ENSG0000022 | -0.00141066 | 0.383459861 | 0.00161858 | 0.240335439 | -0.00138664 | 0.391591095 | 0.001618511 | 0.240302023 | -2.40E-05 | 0.042329491 | 1 | 0.67626148 | 1 | 0.68145739 | ENSG0000022 | 4 | C4B |
| VAT | HTN | ENSG0000022 | 0.011265955 | 0.011045019 | 0.00443318 | 0.240332628 | 0.011147013 | 0.011919533 | 0.004433045 | 0.240299267 | 0.000118943 | 0.002417147 | 1 | 0.084345828 | 1 | 0.088913277 | ENSG0000022 | 4 | C4B |
| VAT | HTN | ENSG0000024 | -0.0120793 | 0.000356163 | 0.003382965 | 0.240329526 | -0.01214352 | 0.000331028 | 0.003382832 | 0.24029603 | 6.42E-05 | 0.014859072 | 0.983010846 | 0.007252285 | 0.91363702 | 0.006869451 | ENSG0000022 | 4 | C4B |
| VAT | HTN | ENSG0000024 | -0.00619436 | 0.195572146 | 0.004785982 | 0.240334987 | -0.00631563 | 0.186949594 | 0.004785804 | 0.240301526 | 0.000121266 | 0.001642443 | 1 | 0.491154798 | 1 | 0.477318114 | ENSG0000022 | 3 | LINC00422 |
| VAT | HTN | ENSG0000024 | 0.00263214 | 0.626404357 | 0.00540709 | 0.240335697 | 0.002776372 | 0.607607168 | 0.005406842 | 0.240302255 | -0.00014423 | 0.000104148 | 1 | 0.825696865 | 1 | 0.818845598 | ENSG0000022 | 3 | LINC00422 |
| VAT | HTN | ENSG0000024 | 0.003287095 | 0.319134215 | 0.003299502 | 0.240335324 | 0.004164795 | 0.207982238 | 0.003307664 | 0.240301603 | -0.0008777 | 0.000193649 | 1 | 0.622410789 | 1 | 0.5074681 | ENSG0000022 | 1 | RPL36P4 |
| VAT | HTN | ENSG0000024 | -0.01355751 | 0.015724176 | 0.005613261 | 0.240332936 | -0.01368083 | 0.014797557 | 0.005613096 | 0.240299455 | 0.000123319 | 0.014381652 | 1 | 0.107157347 | 1 | 0.102874703 | ENSG0000022 | 1 | RPL36P4 |
| VAT | HTN | ENSG0000024 | -0.00429827 | 0.492230272 | 0.006258708 | 0.240335581 | -0.00420747 | 0.501399331 | 0.006258417 | 0.240302162 | -9.08E-05 | 0.03227046 | 1 | 0.75257754 | 1 | 0.756622282 | ENSG0000022 | 1 | RPL36P4 |
| VAT | HTN | ENSG0000024 | -0.01002271 | 0.097345629 | 0.006045555 | 0.240334458 | -0.01012457 | 0.093976708 | 0.006045291 | 0.240301002 | 0.000101868 | 0.019469877 | 1 | 0.334303787 | 1 | 0.330414924 | ENSG0000022 | 1 | RPL36P4 |
| VAT | HTN | ENSG0000024 | -0.00088023 | 0.642158324 | 0.001894248 | 0.240335707 | -0.00082973 | 0.661353468 | 0.001894169 | 0.24030229 | -5.05E-05 | 0.000363914 | 1 | 0.830922164 | 1 | 0.847332674 | ENSG0000022 | 1 | RPL36P4 |
| VAT | HTN | ENSG0000024 | 0.001027784 | 0.404941161 | 0.001234086 | 0.240335471 | 0.000657662 | 0.597575229 | 0.001245829 | 0.240302247 | 0.000370122 | 0.030688922 | 1 | 0.695156362 | 1 | 0.814290189 | ENSG0000022 | 1 | RPL36P4 |
| VAT | HTN | ENSG0000024 | 0.001424705 | 0.626642989 | 0.002928734 | 0.240335697 | 0.00134824 | 0.645252279 | 0.002928613 | 0.24030228 | 7.65E-05 | 0.000506179 | 1 | 0.825696865 | 1 | 0.839493846 | ENSG0000022 | 3 | HLA-DPB2 |
| VAT | HTN | ENSG0000024 | 0.005576873 | 0.23248718 | 0.004670834 | 0.24033511 | 0.005648223 | 0.226547556 | 0.004670648 | 0.240301664 | -7.14E-05 | 0.047946958 | 1 | 0.541032561 | 1 | 0.526616698 | ENSG0000022 | 3 | HLA-DPB2 |
| VAT | HTN | ENSG0000024 | 0.004335963 | 0.42064181 | 0.005384223 | 0.240335494 | 0.004419945 | 0.411680024 | 0.005383995 | 0.240302052 | -8.40E-05 | 0.03460002 | 1 | 0.705324046 | 1 | 0.694521313 | ENSG0000022 | 3 | HLA-DPB2 |
| VAT | HTN | ENSG0000024 | -0.01034276 | 0.060551014 | 0.005510953 | 0.240334076 | -0.01053076 | 0.056014985 | 0.005510822 | 0.240300584 | 0.000188001 | 0.000372032 | 1 | 0.252880519 | 1 | 0.242158595 | ENSG0000022 | 3 | HLA-DPB2 |
| VAT | HTN | ENSG0000024 | 0.000659006 | 0.709010277 | 0.001765891 | 0.240335745 | 0.004227084 | 0.074539171 | 0.002370381 | 0.240300817 | -0.00356808 | 0.024052089 | 1 | 0.863959543 | 1 | 0.287731625 | ENSG0000022 | 3 | HLA-DPB2 |
| VAT | HTN | ENSG0000024 | 0.006885142 | 0.042631154 | 0.003396194 | 0.240333786 | 0.00679507 | 0.045407445 | 0.003396065 | 0.240300411 | 9.01E-05 | 0.000834468 | 1 | 0.204284466 | 1 | 0.213136987 | ENSG0000022 | 3 | HLA-DPB2 |
| VAT | HTN | ENSG0000024 | -0.01429867 | 0.042868375 | 0.007061089 | 0.240333791 | -0.0144447 | 0.040780644 | 0.007060805 | 0.240300321 | 0.000146032 | 0.007070562 | 1 | 0.204346659 | 1 | 0.198159466 | ENSG0000022 | 3 | HLA-DPB2 |
| VAT | HTN | ENSG0000024 | 0.003042501 | 0.02473298 | 0.001354899 | 0.240333326 | 0.003067757 | 0.023557212 | 0.001354848 | 0.240299856 | -2.53E-05 | 0.01887388 | 1 | 0.146802202 | 1 | 0.142896495 | ENSG0000022 | 1 | RP4-564F22.5 |
| VAT | HTN | ENSG0000024 | 0.000815537 | 0.839576458 | 0.00402863 | 0.240335793 | 0.000957639 | 0.812101541 | 0.004028502 | 0.240302357 | -0.0001421 | 5.08E-05 | 1 | 0.937501866 | 1 | 0.928116047 | ENSG0000022 | 1 | RP4-564F22.5 |
| VAT | HTN | ENSG0000024 | 0.001564729 | 0.48900096 | 0.002261499 | 0.240335577 | 0.000968364 | 0.670112594 | 0.002273188 | 0.240302295 | 0.000596366 | 0.010076301 | 1 | 0.75021826 | 1 | 0.850122986 | ENSG0000022 | 1 | RP4-564F22.5 |
| VAT | HTN | ENSG0000024 | -0.01151489 | 0.044695028 | 0.005735882 | 0.240333826 | -0.01138281 | 0.047194975 | 0.005735708 | 0.240300443 | -0.00013208 | 0.00923444 | 1 | 0.210868848 | 1 | 0.217096886 | ENSG0000022 | 1 | RP4-564F22.5 |
| VAT | HTN | ENSG0000024 | -0.00231351 | 0.602456379 | 0.004441608 | 0.24033568 | -0.00242514 | 0.585050091 | 0.004441461 | 0.240302238 | 0.000111633 | 0.003211431 | 1 | 0.814289719 | 1 | 0.807829017 | ENSG0000022 | 1 | RP4-564F22.5 |
| VAT | HTN | ENSG0000024 | -0.00412421 | 0.284910705 | 0.00385673 | 0.240335249 | -0.00404774 | 0.293916536 | 0.003856574 | 0.240301842 | -7.65E-05 | 0.009288009 | 1 | 0.589912638 | 1 | 0.592989502 | ENSG0000022 | 1 | RP4-564F22.5 |
| VAT | HTN | ENSG0000025 | 0.008736098 | 0.176635232 | 0.006465485 | 0.240334913 | 0.00861217 | 0.182835145 | 0.006465214 | 0.240301511 | 0.000123927 | 0.009998627 | 1 | 0.462067857 | 1 | 0.473826292 | ENSG0000022 | 1 | RASAL2-AS1 |
| VAT | HTN | ENSG0000025 | -0.00207976 | 0.515221157 | 0.003196038 | 0.240335605 | -0.00215099 | 0.500920425 | 0.003195912 | 0.240302161 | 7.12E-05 | 0.004150408 | 1 | 0.769486144 | 1 | 0.756313114 | ENSG0000022 | 1 | AC069213.1 |
| VAT | HTN | ENSG0000025 | -0.01474598 | 1.60E-05 | 0.003418254 | 0.240326635 | -0.01466861 | 1.78E-05 | 0.00341815 | 0.240293303 | -7.74E-05 | 0.010198147 | 0.044280439 | 0.00055118 | 0.04901568 | 0.000612696 | ENSG0000022 | 1 | AC069213.1 |
| VAT | HTN | ENSG0000025 | 0.001378567 | 0.780527244 | 0.004947624 | 0.240335775 | 0.001090908 | 0.825539895 | 0.004949102 | 0.240302361 | 0.000287659 | 0.032157171 | 1 | 0.903724293 | 1 | 0.935340768 | ENSG0000022 | 1 | RPL32P1 |
| VAT | HTN | ENSG0000025 | -0.00200205 | 0.437381953 | 0.002577897 | 0.240335516 | -0.00195847 | 0.44740199 | 0.00257777 | 0.2403021 | -4.36E-05 | 0.008042042 | 1 | 0.716423851 | 1 | 0.721401667 | ENSG0000022 | 1 | RPL32P1 |
| VAT | HTN | ENSG0000025 | 0.004360874 | 0.327122922 | 0.004450206 | 0.24033534 | 0.00444432 | 0.317936716 | 0.00445007 | 0.240301893 | -8.34E-05 | 0.033987954 | 1 | 0.627864285 | 1 | 0.616693002 | ENSG0000022 | 1 | RPL32P1 |
| VAT | HTN | ENSG0000025 | -0.00633937 | 0.18304446 | 0.004761284 | 0.240334939 | -0.00641276 | 0.178009145 | 0.004761086 | 0.24030149 | 7.34E-05 | 0.039251093 | 1 | 0.472152066 | 1 | 0.466576677 | ENSG0000022 | 1 | RPL32P1 |
| VAT | HTN | ENSG0000025 | -0.00257874 | 0.575440302 | 0.004604415 | 0.240335659 | -0.00249662 | 0.587649233 | 0.004604222 | 0.24030224 | -8.21E-05 | 0.016323709 | 1 | 0.798499363 | 1 | 0.808584778 | ENSG0000022 | 1 | RPL32P1 |
| VAT | HTN | ENSG0000025 | -0.00084232 | 0.69149008 | 0.002122596 | 0.240335736 | 0.000438643 | 0.837706928 | 0.002141525 | 0.240302364 | -0.00128096 | 7.07E-06 | 1 | 0.857708065 | 1 | 0.939635856 | ENSG0000022 | 1 | RPL32P1 |
| VAT | HTN | ENSG0000025 | -0.01087727 | 0.004803311 | 0.003857256 | 0.240331891 | -0.01093901 | 0.004567261 | 0.003857084 | 0.240298418 | 6.17E-05 | 0.024231017 | 1 | 0.048739475 | 1 | 0.046861116 | ENSG0000022 | 1 | RPL32P1 |
| VAT | HTN | ENSG0000025 | 0.007583604 | 0.113865259 | 0.004796535 | 0.24033458 | 0.0074165 | 0.122038507 | 0.004796362 | 0.240301206 | 0.000167104 | 1.92E-05 | 1 | 0.36506767 | 1 | 0.379153311 | ENSG0000022 | 1 | RPL32P1 |
| VAT | HTN | ENSG0000025 | 0.002896912 | 0.46010396 | 0.003921762 | 0.240335544 | 0.002788974 | 0.476973842 | 0.003921622 | 0.240302135 | 0.000107938 | 0.000830801 | 1 | 0.733862963 | 1 | 0.740826001 | ENSG0000022 | 1 | RPL32P1 |
| VAT | HTN | ENSG0000025 | 0.000581121 | 0.814033628 | 0.002470484 | 0.240335786 | -0.00027406 | 0.911929086 | 0.002477848 | 0.240302379 | 0.000855186 | 9.47E-06 | 1 | 0.923869473 | 1 | 0.968393726 | ENSG0000022 | 1 | RPL32P1 |
| VAT | HTN | ENSG0000025 | -0.00038203 | 0.940478905 | 0.005116423 | 0.240335811 | 0.000524633 | 0.918391751 | 0.005120374 | 0.24030238 | -0.00090667 | 1.57E-05 | 1 | 0.976202248 | 1 | 0.971917651 | ENSG0000022 | 1 | RP11-134G8.8 |
| VAT | HTN | ENSG0000025 | -0.00244671 | 0.495494561 | 0.003589669 | 0.240335584 | -0.00042781 | 0.905883927 | 0.003618431 | 0.240302378 | -0.00201889 | 1.01E-05 | 1 | 0.753479333 | 1 | 0.967584999 | ENSG0000022 | 1 | RP11-134G8.8 |
| VAT | HTN | ENSG0000025 | 0.008829996 | 0.060317738 | 0.004700637 | 0.240334073 | 0.008927607 | 0.057525664 | 0.004700473 | 0.240300606 | -9.76E-05 | 0.01276557 | 1 | 0.252880519 | 1 | 0.246538561 | ENSG0000022 | 2 | LINC00240 |
| VAT | HTN | ENSG0000025 | 0.00040574 | 0.804883525 | 0.00164246 | 0.240335783 | 0.000345914 | 0.83319096 | 0.001642433 | 0.240302363 | 5.98E-05 | 0.000401868 | 1 | 0.918978716 | 1 | 0.939506392 | ENSG0000022 | 2 | LINC00240 |
| VAT | HTN | ENSG0000025 | -0.00682086 | 0.289108363 | 0.006434283 | 0.240335259 | -0.00667625 | 0.299434373 | 0.006434021 | 0.240301854 | -0.00014461 | 0.003043665 | 1 | 0.59321241 | 1 | 0.598868745 | ENSG0000022 | 2 | LINC00240 |
| VAT | HTN | ENSG0000025 | -0.00086305 | 0.737350677 | 0.002573465 | 0.240335758 | 0.0004518945 | 0.201384255 | 0.003537004 | 0.24030158 | -0.00538199 | 0.026563554 | 1 | 0.878176701 | 1 | 0.497601204 | ENSG0000022 | 2 | LINC00240 |
| VAT | HTN | ENSG0000025 | 0.007757967 | 0.250101677 | 0.006745449 | 0.240335161 | 0.007524254 | 0.264640571 | 0.006745238 | 0.240301771 | 0.000233713 | 7.43E-05 | 1 | 0.56074787 | 1 | 0.569128623 | ENSG0000022 | 3 | RP4-758J18.2 |
| VAT | HTN | ENSG0000025 | -0.02343764 | 0.00024427 | 0.006389373 | 0.240329177 | -0.02331879 | 0.000262543 | 0.00638918 | 0.240295816 | -0.00011885 | 0.035642981 | 0.674184667 | 0.005267068 | 0.724618892 | 0.005750944 | ENSG0000022 | 2 | LINC00162 |

| | | | | | | | | | | | | | | | | | | | |
|---|---|---|---|---|---|---|---|---|---|---|---|---|---|---|---|---|---|---|---|
| VAT | HTN | ENSG0000025 | -0.00158706 | 0.700162006 | 0.004121139 | 0.24033574 | -0.00167821 | 0.683834547 | 0.004120967 | 0.240302303 | 9.11E-05 | 0.002940624 | 1 | 0.860778235 | 1 | 0.856733249 | ENSG0000022 | 2 | LINC00162 |
| VAT | HTN | ENSG0000025 | -0.02148579 | 0.00124762 | 0.006656517 | 0.240330675 | -0.02128996 | 0.001381589 | 0.006656257 | 0.24029734 | -0.00019583 | 0.000156425 | 1 | 0.018219212 | 1 | 0.019554794 | ENSG0000022 | 2 | LINC00162 |
| VAT | HTN | ENSG0000025 | -0.00334874 | 0.113885241 | 0.002118153 | 0.240334581 | -0.00340708 | 0.107710628 | 0.002118079 | 0.240301109 | 5.83E-05 | 0.000952668 | 1 | 0.36506767 | 1 | 0.352646896 | ENSG0000022 | 1 | AL049840.1 |
| VAT | HTN | ENSG0000025 | -0.01006836 | 0.060901451 | 0.005372043 | 0.240334081 | -0.00993298 | 0.064447329 | 0.005371858 | 0.240300699 | -0.00013538 | 0.002587326 | 1 | 0.253144583 | 1 | 0.26197345 | ENSG0000022 | 1 | AL049840.1 |
| VAT | HTN | ENSG0000025 | 0.011430503 | 0.000150248 | 0.003015416 | 0.240328727 | 0.011369371 | 0.000162907 | 0.003015308 | 0.240295374 | 6.11E-05 | 0.013538506 | 0.41468463 | 0.003519708 | 0.449623483 | 0.003842936 | ENSG0000022 | 1 | AL049840.1 |
| VAT | HTN | ENSG0000026 | -0.01975808 | 0.001272407 | 0.006131955 | 0.240330693 | -0.01988988 | 0.001179756 | 0.006131756 | 0.240297196 | 0.000131797 | 0.012804985 | 1 | 0.018405959 | 1 | 0.017696346 | ENSG0000022 | 2 | LINC00092 |
| VAT | HTN | ENSG0000026 | 0.006151996 | 0.339798637 | 0.006444822 | 0.240335364 | 0.006317255 | 0.326976237 | 0.006444708 | 0.240301911 | -0.00016526 | 0.011828163 | 1 | 0.64279934 | 1 | 0.624860924 | ENSG0000022 | 2 | LINC00092 |
| VAT | HTN | ENSG0000026 | 0.002345985 | 0.614547213 | 0.004658489 | 0.240335688 | 0.002271452 | 0.625823285 | 0.004658297 | 0.240302268 | 7.45E-05 | 0.033559458 | 1 | 0.819907808 | 1 | 0.827116213 | ENSG0000022 | 3 | RP11-640M9.2 |
| VAT | HTN | ENSG0000026 | 0.008910713 | 0.000101634 | 0.002292623 | 0.240328363 | 0.008962444 | 9.25E-05 | 0.002292541 | 0.240294848 | -5.17E-05 | 0.006400555 | 0.280510026 | 0.002597315 | 0.255413501 | 0.00243251 | ENSG0000022 | 1 | AP000350.6 |
| VAT | HTN | ENSG0000026 | -0.00387472 | 0.004470225 | 0.001362931 | 0.240331827 | -0.00380091 | 0.005295542 | 0.001363067 | 0.24029855 | -7.38E-05 | 0.003244148 | 1 | 0.046036649 | 1 | 0.052198915 | ENSG0000022 | 1 | AP000350.6 |
| VAT | HTN | ENSG0000026 | 0.000291304 | 0.958693826 | 0.005624418 | 0.240335812 | 0.000180897 | 0.974341243 | 0.005624198 | 0.240302384 | 0.000110407 | 0.011703554 | 1 | 0.98228229 | 1 | 0.989761439 | ENSG0000022 | 1 | OSTCP2 |
| VAT | HTN | ENSG0000026 | -0.01017361 | 9.36E-05 | 0.002604213 | 0.240328287 | -0.01023207 | 8.53E-05 | 0.002604155 | 0.240294772 | 5.85E-05 | 0.020871239 | 0.258385761 | 0.002437602 | 0.235331206 | 0.0022628 | ENSG0000022 | 2 | RP11-395L14.5 |
| VAT | HTN | ENSG0000026 | 0.008163651 | 0.157696218 | 0.005778085 | 0.240334829 | 0.008311586 | 0.1502868 | 0.005777868 | 0.240301364 | -0.00014794 | 0.001385681 | 1 | 0.435044054 | 1 | 0.424122257 | ENSG0000022 | 2 | GBP1P1 |
| VAT | HTN | ENSG0000026 | 0.000131907 | 0.976866731 | 0.004548941 | 0.240335813 | 0.000203235 | 0.964362987 | 0.004548764 | 0.240302384 | -7.13E-05 | 0.045140435 | 1 | 0.98866831 | 1 | 0.985245375 | ENSG0000022 | 2 | GBP1P1 |
| VAT | HTN | ENSG0000026 | 0.006022034 | 0.137225481 | 0.004051953 | 0.240334724 | 0.00608275 | 0.133289891 | 0.004051773 | 0.240301273 | -6.07E-05 | 0.034440728 | 1 | 0.404207393 | 1 | 0.399435503 | ENSG0000022 | 2 | GBP1P1 |
| VAT | HTN | ENSG0000026 | -0.00164086 | 0.558056842 | 0.002801395 | 0.240335644 | -0.00157549 | 0.573829977 | 0.002801271 | 0.240302229 | -6.54E-05 | 0.000984172 | 1 | 0.788352234 | 1 | 0.801328599 | ENSG0000022 | 1 | AC002064.5 |
| VAT | HTN | ENSG0000026 | 0.010134529 | 0.031150182 | 0.004702459 | 0.240333523 | 0.010045806 | 0.032649665 | 0.004702275 | 0.240300134 | 8.87E-05 | 0.015501713 | 1 | 0.170246542 | 1 | 0.176002101 | ENSG0000022 | 1 | AC002064.5 |
| VAT | HTN | ENSG0000026 | 0.003466214 | 0.471368639 | 0.004812485 | 0.240335558 | 0.003351608 | 0.48614159 | 0.004812353 | 0.240302146 | 0.000114606 | 0.009501462 | 1 | 0.741297688 | 1 | 0.746371412 | ENSG0000022 | 1 | AC002064.5 |
| VAT | HTN | ENSG0000026 | 0.005509053 | 0.147925775 | 0.003807489 | 0.240334781 | 0.005586675 | 0.142280944 | 0.003807317 | 0.240301323 | -7.76E-05 | 0.003534071 | 1 | 0.418982277 | 1 | 0.412928922 | ENSG0000022 | 1 | AC002064.5 |
| VAT | HTN | ENSG0000026 | -0.01721144 | 0.000817018 | 0.00514237 | 0.240330289 | -0.01732967 | 0.000751499 | 0.005142201 | 0.240296784 | 0.000118237 | 0.007329852 | 1 | 0.013186962 | 1 | 0.012346048 | ENSG0000022 | 1 | AC002064.5 |
| VAT | HTN | ENSG0000026 | -0.00110883 | 0.80006679 | 0.004378218 | 0.240335782 | -0.00102645 | 0.814635021 | 0.004378099 | 0.240302358 | -8.24E-05 | 0.040662054 | 1 | 0.916259063 | 1 | 0.928320668 | ENSG0000022 | 3 | FAM225B |
| VAT | HTN | ENSG0000026 | 0.000397199 | 0.945358304 | 0.005795411 | 0.240335811 | 0.000210519 | 0.971022369 | 0.005795261 | 0.240302384 | 0.00018668 | 0.000557188 | 1 | 0.976968793 | 1 | 0.988573124 | ENSG0000022 | 3 | FAM225B |
| VAT | HTN | ENSG0000026 | -0.00172599 | 0.559348114 | 0.002956417 | 0.240335645 | -0.00167157 | 0.571782858 | 0.002956295 | 0.240302227 | -5.44E-05 | 0.014352648 | 1 | 0.788860907 | 1 | 0.801076492 | ENSG0000022 | 3 | FAM225B |
| VAT | HTN | ENSG0000026 | -0.00255995 | 0.486515388 | 0.003678819 | 0.240335575 | -0.00263126 | 0.474438183 | 0.003678656 | 0.240302132 | 7.13E-05 | 0.006444524 | 1 | 0.749738957 | 1 | 0.740220116 | ENSG0000022 | 1 | AC007000.10 |
| VAT | HTN | ENSG0000026 | 0.003646384 | 0.390065857 | 0.004242454 | 0.240335449 | 0.003728905 | 0.379408425 | 0.004242273 | 0.240302004 | -8.25E-05 | 0.007971207 | 1 | 0.682680345 | 1 | 0.671045748 | ENSG0000022 | 1 | AC007000.10 |
| VAT | HTN | ENSG0000026 | -0.00882605 | 0.023161095 | 0.003886802 | 0.24033327 | -0.0087699 | 0.024043853 | 0.003886618 | 0.240299874 | -5.61E-05 | 0.029767978 | 1 | 0.139878824 | 1 | 0.144666325 | ENSG0000022 | 5 | RP11-592B15.4 |
| VAT | HTN | ENSG0000026 | 0.011376911 | 0.099663462 | 0.006909794 | 0.240334476 | 0.011238119 | 0.103849981 | 0.006909508 | 0.24030108 | 0.000138792 | 0.007245228 | 1 | 0.336272805 | 1 | 0.344320158 | ENSG0000022 | 5 | RP11-592B15.4 |
| VAT | HTN | ENSG0000026 | 0.014323374 | 0.000131702 | 0.003746353 | 0.240328604 | 0.014209688 | 0.000148836 | 0.003746259 | 0.24029529 | 0.000113686 | 0.001261807 | 0.363496717 | 0.003188568 | 0.410786292 | 0.003552729 | ENSG0000022 | 5 | RP11-592B15.4 |
| VAT | HTN | ENSG0000026 | 0.00330664 | 0.602857839 | 0.00635531 | 0.24033568 | 0.003578631 | 0.573385456 | 0.006355536 | 0.240302228 | -0.00027199 | 0.003165026 | 1 | 0.814433497 | 1 | 0.801328599 | ENSG0000022 | 1 | MKRN7P |
| VAT | HTN | ENSG0000026 | 0.002985966 | 0.202686765 | 0.002343891 | 0.240335013 | 0.00261204 | 0.266508841 | 0.002350776 | 0.240301776 | 0.000373926 | 0.039808654 | 1 | 0.501009921 | 1 | 0.572019261 | ENSG0000022 | 1 | HLA-S |
| VAT | HTN | ENSG0000027 | 0.005210723 | 0.070458226 | 0.002880512 | 0.240334199 | 0.005265322 | 0.067552371 | 0.002880397 | 0.240300737 | -5.46E-05 | 0.013835907 | 1 | 0.27741042 | 1 | 0.269826471 | ENSG0000022 | 1 | HLA-S |
| VAT | HTN | ENSG0000027 | -0.00304947 | 0.120872186 | 0.00196597 | 0.240334627 | -0.00310349 | 0.114414477 | 0.001965901 | 0.240301156 | 5.40E-05 | 0.000931766 | 1 | 0.37568382 | 1 | 0.366605303 | ENSG0000022 | 1 | HLA-S |
| VAT | HTN | ENSG0000027 | 0.000782695 | 0.792260677 | 0.002971776 | 0.240335779 | 0.000889933 | 0.764579303 | 0.002971665 | 0.240302341 | -0.00010724 | 6.92E-06 | 1 | 0.909962326 | 1 | 0.898738874 | ENSG0000022 | 1 | HLA-S |
| VAT | HTN | ENSG0000027 | 0.005368727 | 0.210327036 | 0.004285833 | 0.240335039 | 0.005301224 | 0.216098173 | 0.004285647 | 0.24030163 | 6.75E-05 | 0.029020826 | 1 | 0.508766538 | 1 | 0.515497803 | ENSG0000022 | 2 | AC002398.5 |
| VAT | HTN | ENSG0000027 | -0.00372587 | 0.161803549 | 0.002663172 | 0.240334848 | -0.00377233 | 0.156617743 | 0.00266306 | 0.240301395 | 4.65E-05 | 0.018433582 | 1 | 0.440412026 | 1 | 0.433565668 | ENSG0000022 | 4 | LINC00115 |
| VAT | HTN | ENSG0000027 | -0.0028445 | 0.042835951 | 0.001404474 | 0.24033379 | -0.00229953 | 0.103272066 | 0.001411448 | 0.240301076 | -0.00054497 | 0.000112273 | 1 | 0.204346659 | 1 | 0.344320158 | ENSG0000022 | 4 | LINC00115 |
| VAT | HTN | ENSG0000027 | -0.00330544 | 0.63067222 | 0.006875154 | 0.240335699 | -0.00348977 | 0.611729103 | 0.006874938 | 0.240302258 | 0.000184323 | 0.00213303 | 1 | 0.825823207 | 1 | 0.821992369 | ENSG0000022 | 4 | LINC00115 |
| VAT | HTN | ENSG0000027 | -0.00010307 | 0.969639383 | 0.002708057 | 0.240335813 | -0.00031431 | 0.907619543 | 0.002708558 | 0.240302378 | 0.000211235 | 0.000550252 | 1 | 0.987165141 | 1 | 0.96802961 | ENSG0000022 | 3 | RP11-384K6.2 |
| VAT | HTN | ENSG0000027 | -0.00026957 | 0.937565448 | 0.003441443 | 0.24033581 | -0.00112085 | 0.745222593 | 0.003449352 | 0.240302333 | 0.000851282 | 0.000327376 | 1 | 0.976192895 | 1 | 0.884511252 | ENSG0000022 | 3 | RP11-384K6.2 |
| VAT | HTN | ENSG0000027 | -0.0064296 | 0.179319382 | 0.004788008 | 0.240334924 | -0.00635005 | 0.184742945 | 0.004787812 | 0.240301517 | -7.96E-05 | 0.02807208 | 1 | 0.467789692 | 1 | 0.476781711 | ENSG0000022 | 1 | RP11-460N20.5 |
| VAT | HTN | ENSG0000027 | 0.012272877 | 0.045849153 | 0.00614632 | 0.240333847 | 0.012426078 | 0.043199098 | 0.006146092 | 0.240300369 | -0.0001532 | 0.002008523 | 1 | 0.214201949 | 1 | 0.20699568 | ENSG0000022 | 1 | RP11-460N20.5 |
| VAT | HTN | ENSG0000027 | 0.000641707 | 0.776343417 | 0.002258835 | 0.240335774 | 0.000186905 | 0.934325035 | 0.002268134 | 0.240302381 | 0.000454802 | 0.027936873 | 1 | 0.902839726 | 1 | 0.974609668 | ENSG0000022 | 1 | RP11-460N20.5 |
| VAT | HTN | ENSG0000027 | 0.006788488 | 0.262460967 | 0.006057933 | 0.240335194 | 0.006914072 | 0.253717662 | 0.00605771 | 0.240301742 | -0.00012558 | 0.010334262 | 1 | 0.570001694 | 1 | 0.559962186 | ENSG0000022 | 2 | CCT8P1 |
| VAT | HTN | ENSG0000027 | -0.00101007 | 0.657431618 | 0.002277683 | 0.240335716 | -0.00097065 | 0.669979342 | 0.002277587 | 0.240302295 | -3.94E-05 | 0.019624319 | 1 | 0.837990221 | 1 | 0.850122986 | ENSG0000022 | 2 | CCT8P1 |
| VAT | HTN | ENSG0000027 | 0.002246199 | 0.332948505 | 0.002319992 | 0.240335351 | 0.002864551 | 0.218091587 | 0.00232584 | 0.240301637 | -0.00061835 | 0.000215367 | 1 | 0.634625603 | 1 | 0.517020606 | ENSG0000022 | 2 | CCT8P1 |
| VAT | Obesity | ENSG0000000 | -0.00159468 | 0.49287072 | 0.002325448 | 0.183454296 | -0.00154504 | 0.506404442 | 0.002325306 | 0.183417379 | -4.96E-05 | 0.016112667 | 1 | 0.766182814 | 1 | 0.783011911 | ENSG0000022 | 2 | AC010536.1 |
| VAT | Obesity | ENSG0000000 | 0.000708704 | 0.729413134 | 0.002048832 | 0.183454429 | 0.000768348 | 0.70762726 | 0.002048683 | 0.183417493 | -5.96E-05 | 0.000103856 | 1 | 0.883749012 | 1 | 0.876422561 | ENSG0000022 | 2 | AC010536.1 |
| VAT | Obesity | ENSG0000000 | -0.00017637 | 0.958954928 | 0.003426964 | 0.183454473 | -0.00033955 | 0.921074824 | 0.003426997 | 0.183417542 | 0.000163178 | 0.001341853 | 1 | 0.98546132 | 1 | 0.973721371 | ENSG0000022 | 1 | RP1-137D17.1 |
| VAT | Obesity | ENSG0000000 | 0.002765582 | 0.421207102 | 0.003438369 | 0.18345423 | 0.002839188 | 0.408920157 | 0.003438122 | 0.183417288 | -7.36E-05 | 0.004835485 | 1 | 0.71233554 | 1 | 0.72254778 | ENSG0000022 | 4 | RP11-419C5.2 |
| VAT | Obesity | ENSG0000000 | 0.005035704 | 0.356841113 | 0.005465263 | 0.183454153 | 0.004840221 | 0.375783039 | 0.005464892 | 0.18341725 | 0.000195483 | 9.57E-06 | 1 | 0.668015266 | 1 | 0.699838858 | ENSG0000022 | 4 | RP11-419C5.2 |
| VAT | Obesity | ENSG0000000 | -0.00145077 | 0.455920643 | 0.001945818 | 0.183454264 | -0.00142153 | 0.465018214 | 0.001945677 | 0.183417344 | -2.92E-05 | 0.044463116 | 1 | 0.739580277 | 1 | 0.76239704 | ENSG0000022 | 4 | RP11-419C5.2 |
| VAT | Obesity | ENSG0000000 | -0.00447322 | 0.217639789 | 0.003628419 | 0.1834539 | -0.00440946 | 0.224240572 | 0.003628197 | 0.183416988 | -6.38E-05 | 0.047897885 | 1 | 0.521961016 | 1 | 0.533537912 | ENSG0000022 | 6 | GTF2H2B |
| VAT | Obesity | ENSG0000000 | -0.00470542 | 0.30259372 | 0.004564435 | 0.183454072 | -0.00727291 | 0.113528799 | 0.004595741 | 0.1834166 | 0.002567484 | 1.94E-06 | 1 | 0.611389947 | 1 | 0.36994036 | ENSG0000022 | 6 | GTF2H2B |
| VAT | Obesity | ENSG0000000 | 0.011407537 | 0.040235531 | 0.005561049 | 0.183452884 | 0.003620645 | 0.530717659 | 0.00577537 | 0.183417398 | 0.007786892 | 6.05E-07 | 1 | 0.193804653 | 1 | 0.786996417 | ENSG0000022 | 2 | AC009961.3 |
| VAT | Obesity | ENSG0000001 | -0.00114973 | 0.80714216 | 0.004709805 | 0.183454452 | -0.00122135 | 0.795372801 | 0.004709437 | 0.183417521 | 7.16E-05 | 0.022904179 | 1 | 0.921269427 | 1 | 0.918573915 | ENSG0000022 | 1 | PSPHP1 |
| VAT | Obesity | ENSG0000001 | 0.001427245 | 0.616769817 | 0.002852015 | 0.183454379 | 0.001379669 | 0.628536763 | 0.002851806 | 0.183417458 | 4.76E-05 | 0.024497353 | 1 | 0.831570896 | 1 | 0.842487235 | ENSG0000022 | 1 | PSPHP1 |
| VAT | Obesity | ENSG0000001 | 0.002018167 | 0.590057941 | 0.003746001 | 0.183454364 | 0.002078055 | 0.579046958 | 0.003745743 | 0.18341743 | -5.99E-05 | 0.04513677 | 1 | 0.817872626 | 1 | 0.816782844 | ENSG0000022 | 1 | PSPHP1 |
| VAT | Obesity | ENSG0000001 | 0.001207149 | 0.769506206 | 0.004119666 | 0.183454442 | 0.00112213 | 0.785314863 | 0.004119425 | 0.183417518 | 8.50E-05 | 0.02438535 | 1 | 0.902991976 | 1 | 0.913850934 | ENSG0000022 | 3 | ZNF192P1 |
| VAT | Obesity | ENSG0000001 | 0.003153053 | 0.419433109 | 0.003905166 | 0.183454228 | 0.003239871 | 0.406712353 | 0.0039049 | 0.183417286 | -8.68E-05 | 0.005872346 | 1 | 0.712286006 | 1 | 0.721556426 | ENSG0000022 | 3 | ZNF192P1 |
| VAT | Obesity | ENSG0000001 | -0.00350155 | 0.383834192 | 0.004020822 | 0.183454187 | -0.00340144 | 0.397543892 | 0.004020535 | 0.183417276 | -0.00010011 | 0.00109913 | 1 | 0.691502852 | 1 | 0.715434904 | ENSG0000022 | 3 | ZNF192P1 |
| VAT | Obesity | ENSG0000001 | -0.00171614 | 0.618640033 | 0.003447592 | 0.18345438 | -0.00164825 | 0.63256495 | 0.003447351 | 0.18341746 | -6.79E-05 | 0.012158 | 1 | 0.831754083 | 1 | 0.84382758 | ENSG0000022 | 3 | ZNF192P1 |
| VAT | Obesity | ENSG0000001 | 0.006989675 | 0.034527444 | 0.003306588 | 0.183452785 | 0.006833962 | 0.038774682 | 0.003306901 | 0.183415932 | 0.000155713 | 0.017166657 | 1 | 0.174534333 | 1 | 0.194932828 | ENSG0000022 | 3 | ZNF192P1 |
| VAT | Obesity | ENSG0000001 | -0.00056748 | 0.907906421 | 0.004905595 | 0.183454469 | 0.00323329 | 0.51752932 | 0.004996128 | 0.183417388 | -0.00380077 | 6.24E-05 | 1 | 0.967125327 | 1 | 0.784425829 | ENSG0000022 | 3 | ZNF192P1 |
| VAT | Obesity | ENSG0000001 | 0.003076039 | 0.415769575 | 0.003779924 | 0.183454224 | 0.003263047 | 0.387972841 | 0.003779736 | 0.183417264 | -0.00018701 | 9.45E-07 | 1 | 0.710460915 | 1 | 0.708846095 | ENSG0000022 | 3 | ZNF192P1 |
| VAT | Obesity | ENSG0000002 | -0.00246475 | 0.014505407 | 0.001008281 | 0.183452215 | -0.00243779 | 0.015609554 | 0.001008215 | 0.183415337 | -2.70E-05 | 0.001422334 | 1 | 0.098365905 | 1 | 0.108519823 | ENSG0000022 | 2 | CTA-217C2.1 |
| VAT | Obesity | ENSG0000002 | -0.0032781 | 0.418588206 | 0.004052678 | 0.183454227 | -0.00337984 | 0.404259762 | 0.004052385 | 0.183417283 | 0.00010174 | 0.000883043 | 1 | 0.712286006 | 1 | 0.720669275 | ENSG0000022 | 2 | CTA-217C2.1 |
| VAT | Obesity | ENSG0000002 | -0.00352096 | 0.059575746 | 0.001868948 | 0.183453133 | -0.00440635 | 0.018875314 | 0.001876646 | 0.183415462 | 0.000885392 | 2.60E-07 | 1 | 0.248007631 | 1 | 0.120957701 | ENSG0000022 | 2 | CTA-217C2.1 |
| VAT | Obesity | ENSG0000003 | 0.003701988 | 0.402943185 | 0.004426218 | 0.18345421 | 0.002465896 | 0.578774775 | 0.004441651 | 0.18341743 | 0.001236091 | 0.000986316 | 1 | 0.706963646 | 1 | 0.816782844 | ENSG0000022 | 2 | CTA-217C2.1 |
| VAT | Obesity | ENSG0000003 | -0.00380075 | 0.004997916 | 0.001353939 | 0.183451495 | -0.00386726 | 0.004294372 | 0.001354224 | 0.183414464 | 6.65E-05 | 0.049128678 | 1 | 0.044071084 | 1 | 0.043575249 | ENSG0000022 | 1 | AC093375.1 |
| VAT | Obesity | ENSG0000003 | -0.00678374 | 0.011829289 | 0.002694946 | 0.183452079 | -0.00685051 | 0.011017352 | 0.002694764 | 0.183415104 | 6.68E-05 | 0.002297039 | 1 | 0.084112755 | 1 | 0.084466367 | ENSG0000022 | 1 | AC093375.1 |
| VAT | Obesity | ENSG0000003 | 0.034124219 | 9.38E-08 | 0.006392272 | 0.183443702 | 0.033988019 | 1.05E-07 | 0.00639182 | 0.183406861 | 0.0001362 | 0.005605716 | 0.000258986 | 4.89E-06 | 0.000290608 | 6.46E-06 | ENSG0000022 | 3 | TMEM185B |
| VAT | Obesity | ENSG0000003 | -0.02607395 | 2.82E-06 | 0.005567445 | 0.183446184 | -0.02622576 | 2.47E-06 | 0.005567143 | 0.183409159 | 0.000151808 | 0.004787049 | 0.007793827 | 0.000108248 | 0.006811713 | 0.000111667 | ENSG0000022 | 3 | TMEM185B |
| VAT | Obesity | ENSG0000003 | 0.01496502 | 0.013987275 | 0.006089285 | 0.183452191 | 0.014799805 | 0.015073352 | 0.0060889 | 0.183415313 | 0.000165215 | 0.001685018 | 1 | 0.095320688 | 1 | 0.107059606 | ENSG0000022 | 3 | TMEM185B |
| VAT | Obesity | ENSG0000003 | -0.00136467 | 0.54389782 | 0.002248478 | 0.183454335 | -0.00132516 | 0.555594673 | 0.002248334 | 0.183417415 | -3.95E-05 | 0.040273163 | 1 | 0.79046238 | 1 | 0.805802048 | ENSG0000022 | 3 | TMEM185B |
| VAT | Obesity | ENSG0000004 | -0.00078909 | 0.680185367 | 0.001914278 | 0.18345441 | -0.00074517 | 0.697057637 | 0.001914147 | 0.183417489 | -4.39E-05 | 0.004235479 | 1 | 0.862732905 | 1 | 0.870280156 | ENSG0000022 | 3 | AC012309.5 |
| VAT | Obesity | ENSG0000004 | -0.00164174 | 0.172512427 | 0.001203465 | 0.183453771 | -0.0016674 | 0.165869197 | 0.001203376 | 0.18341682 | 2.57E-05 | 0.003722598 | 1 | 0.459620793 | 1 | 0.456484191 | ENSG0000022 | 1 | BSN-AS2 |
| VAT | Obesity | ENSG0000004 | -0.01628048 | 0.012621234 | 0.006527099 | 0.183452122 | -0.01613937 | 0.013404387 | 0.006526628 | 0.183415235 | -0.00014111 | 0.004122831 | 1 | 0.0877446 | 1 | 0.098393908 | ENSG0000022 | 3 | WDR46 |
| VAT | Obesity | ENSG0000005 | 0.003067899 | 0.595354659 | 0.005776576 | 0.183454367 | -0.00207707 | 0.736524364 | 0.006173305 | 0.183417503 | 0.005144967 | 0.018212016 | 1 | 0.818796889 | 1 | 0.891658005 | ENSG0000022 | 3 | WDR46 |
| VAT | Obesity | ENSG0000005 | -0.00227807 | 0.663169211 | 0.005230416 | 0.183454402 | -0.00211319 | 0.686181207 | 0.005230108 | 0.183417484 | -0.00016488 | 0.000556155 | 1 | 0.854105003 | 1 | 0.869882588 | ENSG0000022 | 2 | RP11-460N20.4 |
| VAT | Obesity | ENSG0000005 | -0.00477473 | 0.366465432 | 0.005286953 | 0.183454166 | -0.00466499 | 0.377547542 | 0.005286556 | 0.183417252 | -0.00010974 | 0.003694234 | 1 | 0.674746226 | 1 | 0.700657728 | ENSG0000022 | 2 | RP11-460N20.4 |
| VAT | Obesity | ENSG0000005 | 0.003272499 | 0.040446169 | 0.001596988 | 0.183452887 | 0.003305304 | 0.038466315 | 0.001596876 | 0.183415927 | -3.28E-05 | 0.008231604 | 1 | 0.194479835 | 1 | 0.193735457 | ENSG0000022 | 2 | RP11-460N20.4 |
| VAT | Obesity | ENSG0000006 | -0.00356446 | 0.545188986 | 0.005891819 | 0.183454336 | -0.00345335 | 0.557765358 | 0.005891431 | 0.183417416 | -0.00011112 | 0.023752192 | 1 | 0.791125973 | 1 | 0.808055518 | ENSG0000022 | 4 | ZNF717 |
| VAT | Obesity | ENSG0000006 | -0.00279566 | 0.298663802 | 0.00268993 | 0.183454066 | -0.00274124 | 0.308131487 | 0.002689728 | 0.183417153 | -5.44E-05 | 0.004744087 | 1 | 0.608797706 | 1 | 0.629957707 | ENSG0000022 | 4 | ZNF717 |
| VAT | Obesity | ENSG0000006 | -0.00175325 | 0.737380877 | 0.005228519 | 0.183454432 | -0.00049727 | 0.924775502 | 0.005266533 | 0.183417543 | -0.00125598 | 0.048276975 | 1 | 0.887171413 | 1 | 0.974118595 | ENSG0000022 | 4 | ZNF717 |
| VAT | Obesity | ENSG0000006 | 0.02639805 | 1.87E-23 | 0.00264497 | 0.183416829 | 0.026202429 | 3.92E-23 | 0.002644965 | 0.183380463 | 0.000195621 | 1.16E-07 | 5.15E-20 | 8.58E-21 | 1.08E-19 | 1.80E-20 | ENSG0000022 | 3 | HCG4B |
| VAT | Obesity | ENSG0000006 | -0.00864577 | 0.090016951 | 0.005099814 | 0.183453388 | -0.0087782 | 0.085177571 | 0.005099432 | 0.183416426 | 0.000132426 | 0.000307738 | 1 | 0.314092016 | 1 | 0.311377611 | ENSG0000022 | 3 | HCG4B |
| VAT | Obesity | ENSG0000006 | -0.00190295 | 0.412670653 | 0.002322928 | 0.18345422 | -0.00186042 | 0.423161796 | 0.002322775 | 0.183417304 | -4.25E-05 | 0.027777932 | 1 | 0.708533979 | 1 | 0.728768041 | ENSG0000022 | 3 | HCG4B |
| VAT | Obesity | ENSG0000006 | -0.00766678 | 0.005714364 | 0.002774051 | 0.183451587 | -0.00488643 | 0.083993926 | 0.002827842 | 0.183416418 | -0.00278035 | 4.36E-07 | 1 | 0.048828617 | 1 | 0.308446194 | ENSG0000022 | 3 | HCG4B |
| VAT | Obesity | ENSG0000006 | 0.003569633 | 0.177177842 | 0.002645157 | 0.183453786 | 0.003464488 | 0.190272657 | 0.002645102 | 0.183416898 | 0.000105146 | 0.001628445 | 1 | 0.4663773 | 1 | 0.500365103 | ENSG0000022 | 3 | HCG4B |
| VAT | Obesity | ENSG0000006 | -0.00177504 | 0.694474035 | 0.00451905 | 0.183454416 | -0.00188257 | 0.676959735 | 0.004518718 | 0.18341748 | 0.000107526 | 0.001212288 | 1 | 0.866149932 | 1 | 0.865004105 | ENSG0000022 | 3 | HCG4B |
| VAT | Obesity | ENSG0000006 | 0.004316396 | 0.473090917 | 0.006016221 | 0.18345428 | 0.004219981 | 0.482999459 | 0.006015764 | 0.18341736 | 9.64E-05 | 0.022603185 | 1 | 0.755994642 | 1 | 0.775358255 | ENSG0000022 | 3 | HCG4B |
| VAT | Obesity | ENSG0000006 | -0.00146299 | 0.686776852 | 0.00362816 | 0.183454413 | -0.00176335 | 0.627178194 | 0.003630514 | 0.183417457 | 0.000300356 | 0.032534397 | 1 | 0.863333733 | 1 | 0.842487235 | ENSG0000022 | 1 | RP11-202G18.1 |
| VAT | Obesity | ENSG0000007 | -0.01220357 | 0.002824998 | 0.004086683 | 0.183451103 | -0.01213357 | 0.00298535 | 0.0040864 | 0.183414214 | -7.00E-05 | 0.030746761 | 1 | 0.029202227 | 1 | 0.033090625 | ENSG0000022 | 1 | RP11-202G18.1 |
| VAT | Obesity | ENSG0000007 | 0.002436998 | 0.454963812 | 0.003261656 | 0.183454263 | 0.003179336 | 0.330010104 | 0.003263889 | 0.183417187 | -0.00074234 | 9.44E-09 | 1 | 0.739081884 | 1 | 0.650677127 | ENSG0000022 | 1 | RPS3AP53 |
| VAT | Obesity | ENSG0000007 | -0.00034397 | 0.938350964 | 0.004447406 | 0.183454472 | -0.00044948 | 0.919492962 | 0.004447099 | 0.183417542 | 0.000105505 | 0.002869694 | 1 | 0.979148832 | 1 | 0.973721371 | ENSG0000022 | 1 | AC005884.1 |
| VAT | Obesity | ENSG0000007 | -0.00101618 | 0.726295483 | 0.0029029 | 0.183454428 | -0.00107161 | 0.711995508 | 0.002902699 | 0.183417495 | 5.54E-05 | 0.016490523 | 1 | 0.882200818 | 1 | 0.877671998 | ENSG0000022 | 1 | AP000344.4 |
| VAT | Obesity | ENSG0000007 | 0.009663071 | 0.077838135 | 0.005479877 | 0.183453299 | 0.009766315 | 0.074691947 | 0.005479443 | 0.183416345 | -0.00010324 | 0.004106909 | 1 | 0.286642319 | 1 | 0.291599575 | ENSG0000022 | 1 | HCG4P5 |
| VAT | Obesity | ENSG0000007 | -0.00189825 | 0.676798656 | 0.00455395 | 0.183454408 | -0.00199604 | 0.661138395 | 0.004553616 | 0.183417473 | 9.78E-05 | 0.003662308 | 1 | 0.860416532 | 1 | 0.855080586 | ENSG0000022 | 1 | HCG4P5 |
| VAT | Obesity | ENSG0000007 | 0.009780495 | 0.000141643 | 0.002570222 | 0.183449001 | 0.009746773 | 0.000149162 | 0.002570019 | 0.18341211 | 3.37E-05 | 0.046399732 | 0.390934604 | 0.002874519 | 0.41168648 | 0.003293492 | ENSG0000022 | 1 | HCG4P5 |
| VAT | Obesity | ENSG0000007 | 0.000107122 | 0.983124662 | 0.005064459 | 0.183454474 | 1.34E-05 | 0.997882508 | 0.005064097 | 0.183417546 | 9.37E-05 | 0.015650315 | 1 | 0.99502166 | 1 | 0.999330814 | ENSG0000022 | 1 | HCG4P5 |
| VAT | Obesity | ENSG0000007 | -0.00122134 | 0.802083179 | 0.004872658 | 0.18345445 | -0.00131277 | 0.787595119 | 0.004872307 | 0.183417519 | 9.14E-05 | 0.013056043 | 1 | 0.918961541 | 1 | 0.915268433 | ENSG0000022 | 1 | HCG4P5 |
| VAT | Obesity | ENSG0000008 | 0.012321992 | 0.028111249 | 0.00561178 | 0.183452652 | 0.012404202 | 0.027067548 | 0.005611367 | 0.183415699 | -8.22E-05 | 0.046974784 | 1 | 0.152430349 | 1 | 0.155231297 | ENSG0000022 | 1 | HCG4P5 |
| VAT | Obesity | ENSG0000008 | -0.00347903 | 0.425549634 | 0.004366084 | 0.183454234 | -0.00356436 | 0.414259395 | 0.004365831 | 0.183417294 | 8.53E-05 | 0.034572117 | 1 | 0.714174021 | 1 | 0.72386593 | ENSG0000022 | 1 | HCG4P5 |
| VAT | Obesity | ENSG0000008 | 0.006341945 | 0.004068008 | 0.002207542 | 0.183451354 | 0.006290028 | 0.004378496 | 0.002207387 | 0.183414477 | 5.19E-05 | 0.002451653 | 1 | 0.037931426 | 1 | 0.0439491 | ENSG0000022 | 3 | SLC9A7P1 |
| VAT | Obesity | ENSG0000008 | 0.000858329 | 0.84304168 | 0.004334897 | 0.183454459 | 0.00093005 | 0.830105568 | 0.004334557 | 0.183417529 | -7.17E-05 | 0.012930433 | 1 | 0.938891035 | 1 | 0.936897537 | ENSG0000022 | 3 | SLC9A7P1 |
| VAT | Obesity | ENSG0000008 | 0.006747231 | 0.013782104 | 0.002739545 | 0.183452181 | 0.006696175 | 0.014507531 | 0.002739334 | 0.183415288 | 5.11E-05 | 0.006577721 | 1 | 0.094623402 | 1 | 0.104818812 | ENSG0000022 | 3 | RP11-958N24.2 |
| VAT | Obesity | ENSG0000008 | 0.013859775 | 1.66E-05 | 0.003218386 | 0.183447464 | 0.013535305 | 2.61E-05 | 0.003218472 | 0.183410862 | 0.00032447 | 2.34E-10 | 0.045798762 | 0.000487221 | 0.071910214 | 0.00080798 | ENSG0000022 | 3 | RP11-958N24.2 |
| VAT | Obesity | ENSG0000008 | 0.008955991 | 0.016749783 | 0.003743888 | 0.183452311 | 0.008859399 | 0.017955869 | 0.003743616 | 0.183415429 | 9.66E-05 | 0.000552363 | 1 | 0.108012621 | 1 | 0.119020171 | ENSG0000022 | 3 | RP11-958N24.2 |
| VAT | Obesity | ENSG0000008 | 0.014924507 | 0.001073733 | 0.004563402 | 0.183450431 | 0.014842102 | 0.001143441 | 0.004563087 | 0.183413548 | 8.24E-05 | 0.022976729 | 1 | 0.014179435 | 1 | 0.016436964 | ENSG0000022 | 3 | RP11-958N24.2 |
| VAT | Obesity | ENSG0000008 | -0.00484586 | 0.130401265 | 0.003203827 | 0.183453609 | -0.0047948 | 0.134473352 | 0.003203589 | 0.183416699 | -5.11E-05 | 0.028726353 | 1 | 0.387831349 | 1 | 0.405181715 | ENSG0000022 | 2 | LINC00948 |
| VAT | Obesity | ENSG0000008 | -0.00412152 | 0.273444553 | 0.003763374 | 0.183454021 | -0.00601033 | 0.111536658 | 0.003776931 | 0.183416589 | 0.001888813 | 5.68E-09 | 1 | 0.578319515 | 1 | 0.366914394 | ENSG0000022 | 1 | AL353791.1 |
| VAT | Obesity | ENSG0000008 | -0.00606974 | 0.038204594 | 0.002928481 | 0.18345285 | -0.00615997 | 0.03541163 | 0.002928262 | 0.183415874 | 9.02E-05 | 1.95E-05 | 1 | 0.186351781 | 1 | 0.184211409 | ENSG0000022 | 1 | AL353791.1 |
| VAT | Obesity | ENSG0000008 | 0.012266138 | 0.004760622 | 0.004345359 | 0.183451462 | 0.012381224 | 0.004378799 | 0.004345031 | 0.183414477 | -0.00011509 | 0.000191069 | 1 | 0.042384891 | 1 | 0.0439491 | ENSG0000022 | 1 | RP11-399H11.3 |
| VAT | Obesity | ENSG0000008 | -0.00347825 | 0.521341186 | 0.005423924 | 0.183454319 | -0.00356596 | 0.510862362 | 0.00542355 | 0.183417383 | 8.77E-05 | 0.041822516 | 1 | 0.778563922 | 1 | 0.784425829 | ENSG0000022 | 1 | TRIM60P18 |
| VAT | Obesity | ENSG0000009 | -0.000333 | 0.93941657 | 0.004381445 | 0.183454472 | -0.00051142 | 0.907073694 | 0.00438117 | 0.183417541 | 0.000178412 | 2.79E-06 | 1 | 0.979890299 | 1 | 0.970734158 | ENSG0000022 | 1 | HCG20 |
| VAT | Obesity | ENSG0000009 | -0.00346992 | 0.551396781 | 0.005825251 | 0.18345434 | -0.00336891 | 0.563013961 | 0.00582482 | 0.18341742 | -0.00010101 | 0.017222544 | 1 | 0.793904838 | 1 | 0.809555852 | ENSG0000022 | 1 | HCG20 |
| VAT | Obesity | ENSG0000009 | -0.0014471 | 0.670389314 | 0.00340002 | 0.183454406 | -0.00152708 | 0.653311431 | 0.003399797 | 0.18341747 | 8.00E-05 | 0.005022909 | 1 | 0.857647266 | 1 | 0.852951538 | ENSG0000022 | 1 | HCG20 |
| VAT | Obesity | ENSG0000009 | -0.02953061 | 6.03E-11 | 0.004513151 | 0.183438292 | -0.02943088 | 6.96E-11 | 0.004512839 | 0.183401473 | -9.97E-05 | 0.005154988 | 1.66E-07 | 4.89E-09 | 1.92E-07 | 5.82E-09 | ENSG0000022 | 1 | HCG20 |
| VAT | Obesity | ENSG0000009 | 0.003411091 | 0.086093404 | 0.001987377 | 0.183453361 | 0.003454021 | 0.082196949 | 0.00198726 | 0.183416404 | -4.29E-05 | 0.018160715 | 1 | 0.303083921 | 1 | 0.305457316 | ENSG0000022 | 5 | POLR2J2 |
| VAT | Obesity | ENSG0000009 | 0.000359167 | 0.922769297 | 0.003704819 | 0.183454471 | 0.000223074 | 0.951983434 | 0.003704554 | 0.183417545 | 0.000136093 | 1.56E-06 | 1 | 0.974681691 | 1 | 0.984072763 | ENSG0000022 | 5 | POLR2J2 |
| VAT | Obesity | ENSG0000009 | 0.000978568 | 0.272404275 | 0.000891598 | 0.183454019 | 0.000959805 | 0.281668867 | 0.000891532 | 0.183417108 | 1.88E-05 | 0.003989629 | 1 | 0.576561196 | 1 | 0.600777491 | ENSG0000022 | 5 | POLR2J2 |
| VAT | Obesity | ENSG0000009 | -0.00472705 | 0.234374034 | 0.003975091 | 0.18345394 | -0.00465718 | 0.241331996 | 0.003974834 | 0.183417027 | -6.99E-05 | 0.038746562 | 1 | 0.529788971 | 1 | 0.547310032 | ENSG0000022 | 5 | POLR2J2 |
| VAT | Obesity | ENSG0000009 | -0.01055598 | 0.052646512 | 0.005447366 | 0.183453055 | -0.01064656 | 0.050632257 | 0.005446957 | 0.183416102 | 9.06E-05 | 0.020196689 | 1 | 0.231376391 | 1 | 0.236113017 | ENSG0000022 | 1 | TOP3BP1 |
| VAT | Obesity | ENSG0000009 | 0.024937978 | 9.41E-06 | 0.005628991 | 0.183447055 | 0.026003821 | 4.00E-06 | 0.005639176 | 0.18340951 | -0.00106584 | 0.002206137 | 0.025980893 | 0.000309296 | 0.011048227 | 0.000172629 | ENSG0000022 | 1 | TOP3BP1 |
| VAT | Obesity | ENSG0000009 | 0.010907708 | 0.046085147 | 0.005468571 | 0.18345297 | 0.010788666 | 0.048504135 | 0.005468348 | 0.183416075 | 0.000119043 | 0.046461811 | 1 | 0.212642286 | 1 | 0.229164841 | ENSG0000022 | 1 | TOP3BP1 |
| VAT | Obesity | ENSG0000009 | -0.01246746 | 0.006789148 | 0.004605596 | 0.183451704 | -0.01239126 | 0.007131169 | 0.004605272 | 0.18341481 | -7.62E-05 | 0.033573221 | 1 | 0.055111911 | 1 | 0.062273965 | ENSG0000022 | 6 | RP11-222A11.1 |
| VAT | Obesity | ENSG0000009 | 8.07E-05 | 0.988718483 | 0.005704963 | 0.183454474 | -0.00017044 | 0.976167361 | 0.005705126 | 0.183417546 | 0.000251103 | 0.006167864 | 1 | 0.997756129 | 1 | 0.993659205 | ENSG0000022 | 6 | RP11-222A11.1 |

| | | | | | | | | | | | | | | | | | | | |
|---|---|---|---|---|---|---|---|---|---|---|---|---|---|---|---|---|---|---|---|
| VAT | Obesity | ENSG0000009 | -0.00745839 | 0.048875457 | 0.003786596 | 0.183453008 | -0.00751822 | 0.047075176 | 0.003786317 | 0.183416056 | 5.98E-05 | 0.031160501 | 1 | 0.219637062 | 1 | 0.223627344 | ENSG0000022 | 6 | RP11-222A11.1 |
| VAT | Obesity | ENSG0000009 | -0.00257737 | 0.238532841 | 0.002186689 | 0.183453949 | -0.00253122 | 0.247010008 | 0.002186532 | 0.18341704 | -4.61E-05 | 0.005278907 | 1 | 0.533077442 | 1 | 0.554266359 | ENSG0000022 | 6 | RP11-222A11.1 |
| VAT | Obesity | ENSG0000009 | -0.00733863 | 0.201181159 | 0.005741403 | 0.183453857 | -0.00722918 | 0.207948919 | 0.005740964 | 0.183416947 | -0.00010946 | 0.006189079 | 1 | 0.501589882 | 1 | 0.520051841 | ENSG0000022 | 6 | RP11-222A11.1 |
| VAT | Obesity | ENSG0000009 | -0.00813013 | 0.055117023 | 0.004238953 | 0.183453084 | -0.00826893 | 0.051076795 | 0.004238653 | 0.183416108 | 0.000138799 | 2.47E-05 | 1 | 0.236951687 | 1 | 0.237726734 | ENSG0000022 | 6 | RP11-222A11.1 |
| VAT | Obesity | ENSG0000009 | 0.01122281 | 0.049708992 | 0.005718732 | 0.183453018 | 0.011140203 | 0.051395219 | 0.005718291 | 0.183416112 | 8.26E-05 | 0.035120938 | 1 | 0.222001323 | 1 | 0.238404712 | ENSG0000022 | 6 | RP11-222A11.1 |
| VAT | Obesity | ENSG0000010 | -0.00183093 | 0.56995105 | 0.003222767 | 0.183454352 | -0.00190677 | 0.554053812 | 0.003222542 | 0.183417414 | 7.58E-05 | 0.002813871 | 1 | 0.806905296 | 1 | 0.805615291 | ENSG0000022 | 4 | BOD1L2 |
| VAT | Obesity | ENSG0000010 | 5.67E-05 | 0.991365587 | 0.005240826 | 0.183454474 | -7.37E-05 | 0.988780569 | 0.005240489 | 0.183417546 | 0.000130407 | 0.003574943 | 1 | 0.997766076 | 1 | 0.996361581 | ENSG0000022 | 4 | BOD1L2 |
| VAT | Obesity | ENSG0000010 | -0.00105764 | 0.799813848 | 0.004170708 | 0.18345445 | -0.00096874 | 0.816313429 | 0.004170384 | 0.183417526 | -8.89E-05 | 0.001647911 | 1 | 0.918637629 | 1 | 0.929465786 | ENSG0000022 | 1 | HLA-U |
| VAT | Obesity | ENSG0000010 | -0.00323316 | 0.139347989 | 0.002187187 | 0.183453648 | -0.00318074 | 0.145846381 | 0.002187034 | 0.183416747 | -5.24E-05 | 0.002081187 | 1 | 0.401503968 | 1 | 0.421062772 | ENSG0000022 | 1 | HLA-U |
| VAT | Obesity | ENSG0000010 | -0.00207869 | 0.573629114 | 0.003694044 | 0.183454354 | -0.00199678 | 0.588796547 | 0.003693773 | 0.183417436 | -8.19E-05 | 0.002699349 | 1 | 0.806905296 | 1 | 0.823637411 | ENSG0000022 | 1 | HLA-U |
| VAT | Obesity | ENSG0000010 | 0.011299341 | 0.023518711 | 0.004988872 | 0.183452535 | 0.011191464 | 0.024868794 | 0.004988525 | 0.183415644 | 0.000107876 | 0.005949108 | 1 | 0.134392636 | 1 | 0.146976171 | ENSG0000022 | 1 | HLA-U |
| VAT | Obesity | ENSG0000010 | -0.00113742 | 0.776853752 | 0.004013178 | 0.183454444 | -0.00124084 | 0.757158529 | 0.004012894 | 0.18341751 | 0.000103424 | 0.000892638 | 1 | 0.905454542 | 1 | 0.900448937 | ENSG0000022 | 1 | HLA-U |
| VAT | Obesity | ENSG0000010 | 0.001186333 | 0.795818043 | 0.004584629 | 0.183454449 | 0.00108585 | 0.812764544 | 0.004584331 | 0.183417525 | 0.000100483 | 0.009329516 | 1 | 0.915954045 | 1 | 0.928683673 | ENSG0000022 | 1 | HLA-U |
| VAT | Obesity | ENSG0000010 | 0.001000042 | 0.7362316 | 0.002968821 | 0.183454431 | 0.001088427 | 0.713885085 | 0.002968624 | 0.183417495 | -8.84E-05 | 0.00031413 | 1 | 0.886561612 | 1 | 0.878823744 | ENSG0000022 | 1 | RP11-452F19.3 |
| VAT | Obesity | ENSG0000010 | -3.49E-05 | 0.992910346 | 0.003932787 | 0.183454474 | 3.41E-05 | 0.993087731 | 0.003932511 | 0.183417546 | -6.90E-05 | 0.024478588 | 1 | 0.997766076 | 1 | 0.997787455 | ENSG0000022 | 1 | RP11-452F19.3 |
| VAT | Obesity | ENSG0000010 | -0.00107755 | 0.720972142 | 0.003017005 | 0.183454426 | -0.0010034 | 0.739430312 | 0.003016774 | 0.183417504 | -7.42E-05 | 0.000390223 | 1 | 0.880089833 | 1 | 0.892750508 | ENSG0000022 | 1 | RP11-452F19.3 |
| VAT | Obesity | ENSG0000010 | -0.00098332 | 0.814092016 | 0.004181659 | 0.183454453 | -0.0011113 | 0.790415075 | 0.004181412 | 0.183417519 | 0.000127982 | 0.000789635 | 1 | 0.92437246 | 1 | 0.917009082 | ENSG0000022 | 1 | RP11-452F19.3 |
| VAT | Obesity | ENSG0000010 | 0.008637711 | 0.005740178 | 0.003127025 | 0.18345159 | 0.00821851 | 0.008681617 | 0.003131631 | 0.183414943 | 0.000419202 | 0.01688208 | 1 | 0.048897816 | 1 | 0.071955746 | ENSG0000022 | 1 | RP11-452F19.3 |
| VAT | Obesity | ENSG0000010 | -0.00138286 | 0.642630838 | 0.002980152 | 0.183454393 | -0.00143653 | 0.629761182 | 0.002979958 | 0.183417458 | 5.37E-05 | 0.032618479 | 1 | 0.84339568 | 1 | 0.842487235 | ENSG0000022 | 2 | AC007952.5 |
| VAT | Obesity | ENSG0000010 | -0.0031354 | 0.569694924 | 0.005515212 | 0.183454352 | -0.0030439 | 0.580981395 | 0.005514795 | 0.183417431 | -9.15E-05 | 0.019402989 | 1 | 0.806905296 | 1 | 0.816866353 | ENSG0000022 | 2 | AC007952.5 |
| VAT | Obesity | ENSG0000010 | -0.00497622 | 0.011053124 | 0.001958352 | 0.183452033 | -0.00491394 | 0.012094349 | 0.001958222 | 0.183415166 | -6.23E-05 | 0.000130459 | 1 | 0.079444329 | 1 | 0.090106525 | ENSG0000022 | 2 | AC007952.5 |
| VAT | Obesity | ENSG0000010 | 0.002908693 | 0.072897844 | 0.001621821 | 0.183453258 | 0.002951695 | 0.068743757 | 0.001621718 | 0.183416294 | -4.30E-05 | 0.002048278 | 1 | 0.279186061 | 1 | 0.277387091 | ENSG0000022 | 4 | AC073218.2 |
| VAT | Obesity | ENSG0000010 | 0.005919491 | 0.294477472 | 0.005646478 | 0.183454059 | 0.00584182 | 0.300820905 | 0.00564604 | 0.183417141 | 7.77E-05 | 0.04250389 | 1 | 0.603383685 | 1 | 0.623322597 | ENSG0000022 | 4 | AC073218.2 |
| VAT | Obesity | ENSG0000010 | 0.008426757 | 0.131150593 | 0.005582176 | 0.183453613 | 0.008269939 | 0.138449801 | 0.005581796 | 0.183416716 | 0.000156818 | 0.000483206 | 1 | 0.387974943 | 1 | 0.412910374 | ENSG0000022 | 9 | C19orf24 |
| VAT | Obesity | ENSG0000010 | -0.00468394 | 0.311079333 | 0.00462401 | 0.183454086 | -0.00454173 | 0.325965555 | 0.004623687 | 0.183417181 | -0.00014222 | 8.84E-05 | 1 | 0.619458427 | 1 | 0.646775652 | ENSG0000022 | 1 | CCT6P1 |
| VAT | Obesity | ENSG0000010 | -0.00232517 | 0.526684324 | 0.003672811 | 0.183454323 | -0.00239376 | 0.514530868 | 0.003672543 | 0.183417385 | 6.86E-05 | 0.011969298 | 1 | 0.779823873 | 1 | 0.784425829 | ENSG0000022 | 2 | CTSLP3 |
| VAT | Obesity | ENSG0000010 | -0.00131635 | 0.753001529 | 0.00418308 | 0.183454437 | -0.0014018 | 0.737524132 | 0.004182792 | 0.183417504 | 8.54E-05 | 0.010283407 | 1 | 0.893787648 | 1 | 0.892400966 | ENSG0000022 | 2 | AC073052.1 |
| VAT | Obesity | ENSG0000010 | 0.005782985 | 0.001339036 | 0.001802958 | 0.183450585 | 0.005819632 | 0.001246349 | 0.001802818 | 0.183413608 | -3.66E-05 | 0.00291629 | 1 | 0.016875521 | 1 | 0.01764063 | ENSG0000022 | 2 | AC073052.1 |
| VAT | Obesity | ENSG0000010 | -0.00072339 | 0.82431215 | 0.003258491 | 0.183454455 | -0.00065723 | 0.840142232 | 0.003258264 | 0.183417531 | -6.62E-05 | 0.009571067 | 1 | 0.927365584 | 1 | 0.939543177 | ENSG0000022 | 2 | HNRNPCP7 |
| VAT | Obesity | ENSG0000010 | 0.001418548 | 0.412667665 | 0.001731607 | 0.18345422 | 0.001389155 | 0.42238555 | 0.001731487 | 0.183417303 | 2.94E-05 | 0.03229319 | 1 | 0.708533979 | 1 | 0.728159973 | ENSG0000022 | 2 | HNRNPCP7 |
| VAT | Obesity | ENSG0000010 | -0.01899107 | 0.001625312 | 0.00602634 | 0.18345072 | -0.01914295 | 0.001489429 | 0.006025948 | 0.183413732 | 0.000151883 | 0.002844318 | 1 | 0.019588911 | 1 | 0.020451858 | ENSG0000022 | 2 | PROB1 |
| VAT | Obesity | ENSG0000010 | -0.0002506 | 0.948350671 | 0.003868664 | 0.183454473 | -0.00019544 | 0.959706385 | 0.003868373 | 0.183417545 | -5.52E-05 | 0.045279631 | 1 | 0.982080274 | 1 | 0.98754495 | ENSG0000022 | 2 | PROB1 |
| VAT | Obesity | ENSG0000010 | 0.005406987 | 0.101858249 | 0.003305178 | 0.183453463 | 0.005488419 | 0.09677903 | 0.003304929 | 0.183416504 | -8.14E-05 | 0.000541467 | 1 | 0.342005798 | 1 | 0.339834761 | ENSG0000022 | 2 | PROB1 |
| VAT | Obesity | ENSG0000010 | -0.01048761 | 0.022859797 | 0.004608363 | 0.183452516 | -0.01038651 | 0.024199704 | 0.004608137 | 0.183415626 | -0.00010109 | 0.030633182 | 1 | 0.132827449 | 1 | 0.143946518 | ENSG0000022 | 4 | ARL17B |
| VAT | Obesity | ENSG0000010 | -0.00355236 | 0.393791628 | 0.004165715 | 0.183454199 | -0.0034436 | 0.408399828 | 0.00416541 | 0.183417288 | -0.00010876 | 0.00042946 | 1 | 0.701655839 | 1 | 0.72254778 | ENSG0000022 | 4 | ARL17B |
| VAT | Obesity | ENSG0000010 | -0.00261882 | 0.639556306 | 0.005591941 | 0.183454391 | -0.00251293 | 0.653130143 | 0.005591507 | 0.18341747 | -0.00010589 | 0.005320154 | 1 | 0.841849822 | 1 | 0.852951538 | ENSG0000022 | 4 | ARL17B |
| VAT | Obesity | ENSG0000010 | 0.01044811 | 0.023211563 | 0.00460281 | 0.183452526 | 0.010550471 | 0.021890686 | 0.004602637 | 0.18341556 | -0.00010236 | 0.047688758 | 1 | 0.133602008 | 1 | 0.13459386 | ENSG0000022 | 2 | DHFR |
| VAT | Obesity | ENSG0000010 | -0.00442349 | 0.282920189 | 0.004119532 | 0.183454038 | -0.00450127 | 0.274504312 | 0.004119217 | 0.183417095 | 7.78E-05 | 0.006830894 | 1 | 0.588439881 | 1 | 0.593290447 | ENSG0000022 | 3 | MRPL45P2 |
| VAT | Obesity | ENSG0000010 | 0.002532648 | 0.588758366 | 0.004684566 | 0.183454364 | 0.002631383 | 0.574284754 | 0.004684238 | 0.183417427 | -9.87E-05 | 0.007007251 | 1 | 0.816569392 | 1 | 0.814504585 | ENSG0000022 | 2 | HCG22 |
| VAT | Obesity | ENSG0000010 | -0.00424404 | 0.275942651 | 0.003895473 | 0.183454025 | -0.00430872 | 0.268659919 | 0.003895219 | 0.183417084 | 6.47E-05 | 0.048987925 | 1 | 0.580931898 | 1 | 0.584363319 | ENSG0000022 | 2 | HCG22 |
| VAT | Obesity | ENSG0000010 | -0.00052536 | 0.893235876 | 0.003914426 | 0.183454467 | -0.00074571 | 0.848908168 | 0.003914253 | 0.183417532 | 0.000220353 | 1.17E-07 | 1 | 0.961627151 | 1 | 0.944752639 | ENSG0000022 | 2 | HCG22 |
| VAT | Obesity | ENSG0000010 | -0.00386302 | 0.313224995 | 0.003830536 | 0.18345409 | -0.00393019 | 0.30485441 | 0.003830292 | 0.183417148 | 6.72E-05 | 0.04121847 | 1 | 0.619765744 | 1 | 0.626973302 | ENSG0000022 | 2 | HCG22 |
| VAT | Obesity | ENSG0000010 | -0.00406936 | 0.075815584 | 0.002291946 | 0.183453283 | -0.00509481 | 0.026721063 | 0.002299543 | 0.183415691 | 0.001025454 | 6.38E-08 | 1 | 0.283034564 | 1 | 0.154937256 | ENSG0000022 | 2 | HCG22 |
| VAT | Obesity | ENSG0000010 | -0.00951641 | 0.087171641 | 0.005563484 | 0.183453368 | -0.00969094 | 0.081510106 | 0.005563131 | 0.183416399 | 0.000174528 | 0.000277662 | 1 | 0.306098891 | 1 | 0.304422046 | ENSG0000022 | 2 | HCG22 |
| VAT | Obesity | ENSG0000010 | -0.00480274 | 0.408638306 | 0.005812391 | 0.183454216 | -0.0049493 | 0.394459271 | 0.005812048 | 0.183417272 | 0.000146564 | 0.005682342 | 1 | 0.708044263 | 1 | 0.714375058 | ENSG0000022 | 1 | OR1H1P |
| VAT | Obesity | ENSG0000010 | -0.00198291 | 0.258306593 | 0.001754163 | 0.183453991 | -0.00152304 | 0.385967813 | 0.001756774 | 0.183417262 | -0.00045987 | 3.32E-06 | 1 | 0.557845225 | 1 | 0.707997923 | ENSG0000022 | 1 | OR1H1P |
| VAT | Obesity | ENSG0000010 | -0.00557586 | 0.237704314 | 0.00472232 | 0.183453947 | -0.00522538 | 0.268509872 | 0.004722432 | 0.183417083 | -0.00035048 | 2.47E-06 | 1 | 0.532768171 | 1 | 0.584363319 | ENSG0000022 | 4 | OR2A9P |
| VAT | Obesity | ENSG0000010 | 0.009993365 | 0.009802877 | 0.003869324 | 0.183451953 | 0.012038339 | 0.001948608 | 0.003885877 | 0.183413919 | -0.00204497 | 1.70E-08 | 1 | 0.072731024 | 1 | 0.023938382 | ENSG0000022 | 4 | OR2A9P |
| VAT | Obesity | ENSG0000010 | 0.002366986 | 0.131499311 | 0.001569392 | 0.183453614 | 0.002314372 | 0.140272509 | 0.0015693 | 0.183416724 | 5.26E-05 | 0.000246898 | 1 | 0.387974943 | 1 | 0.416292606 | ENSG0000022 | 3 | RP11-697E2.7 |
| VAT | Obesity | ENSG0000010 | 0.000263813 | 0.902663956 | 0.00215715 | 0.183454468 | 0.001568143 | 0.476426064 | 0.002202253 | 0.183417354 | -0.00130433 | 0.003340946 | 1 | 0.965567705 | 1 | 0.772080125 | ENSG0000022 | 3 | RP11-697E2.7 |
| VAT | Obesity | ENSG0000010 | -0.00798379 | 0.135825918 | 0.005352791 | 0.183453633 | -0.00841237 | 0.11624057 | 0.005355624 | 0.183416614 | 0.000428576 | 0.024136819 | 1 | 0.395299075 | 1 | 0.375232716 | ENSG0000022 | 2 | ANKRD20A8P |
| VAT | Obesity | ENSG0000010 | -0.00075717 | 0.794720965 | 0.002910108 | 0.183454449 | 0.003850171 | 0.229511396 | 0.003204151 | 0.183417 | -0.00460734 | 0.000593639 | 1 | 0.915072951 | 1 | 0.539107619 | ENSG0000022 | 2 | ANKRD20A8P |
| VAT | Obesity | ENSG0000010 | -0.01153207 | 4.85E-05 | 0.002838393 | 0.183448235 | -0.01145581 | 5.43E-05 | 0.002838205 | 0.183411389 | -7.63E-05 | 0.001135891 | 0.133797875 | 0.001205386 | 0.149901575 | 0.001441361 | ENSG0000022 | 2 | HSPA8P8 |
| VAT | Obesity | ENSG0000010 | 0.002392757 | 0.427058737 | 0.003012655 | 0.183454236 | 0.002442487 | 0.417480146 | 0.003012436 | 0.183417298 | -4.97E-05 | 0.027404761 | 1 | 0.715653985 | 1 | 0.725782338 | ENSG0000022 | 3 | ADAM1A |
| VAT | Obesity | ENSG0000010 | 0.010396396 | 0.052604148 | 0.005364051 | 0.183453054 | 0.010516266 | 0.049920941 | 0.005363675 | 0.183416093 | -0.00011987 | 0.004262775 | 1 | 0.231376391 | 1 | 0.234322784 | ENSG0000022 | 3 | RP11-561C5.4 |
| VAT | Obesity | ENSG0000010 | -0.00891714 | 0.051613983 | 0.004581481 | 0.183453042 | -0.00880041 | 0.054732223 | 0.004581161 | 0.183416151 | -0.00011673 | 0.001140458 | 1 | 0.22939548 | 1 | 0.245627538 | ENSG0000022 | 3 | RP11-561C5.4 |
| VAT | Obesity | ENSG0000010 | -0.00442845 | 0.441894619 | 0.005758727 | 0.183454251 | -0.00456268 | 0.428149802 | 0.005758323 | 0.183417309 | 0.000134226 | 0.002850105 | 1 | 0.727515266 | 1 | 0.732605984 | ENSG0000022 | 3 | RP11-561C5.4 |
| VAT | Obesity | ENSG0000011 | -0.00163899 | 0.657352355 | 0.003694979 | 0.1834544 | -0.0005381 | 0.884358706 | 0.003699683 | 0.183417538 | -0.00110088 | 1.33E-08 | 1 | 0.852180602 | 1 | 0.960056703 | ENSG0000022 | 3 | RP11-561C5.4 |
| VAT | Obesity | ENSG0000011 | -0.01061548 | 0.017142106 | 0.004453449 | 0.183452326 | -0.0063405 | 0.160881772 | 0.004522098 | 0.183416803 | -0.00427498 | 5.66E-08 | 1 | 0.108763707 | 1 | 0.449882159 | ENSG0000022 | 3 | RP11-561C5.4 |
| VAT | Obesity | ENSG0000011 | 0.000938425 | 0.576746237 | 0.001681331 | 0.183454356 | 0.000975992 | 0.561557485 | 0.001681208 | 0.183417419 | -3.76E-05 | 0.002672632 | 1 | 0.806947626 | 1 | 0.809555852 | ENSG0000022 | 1 | RP4-580N22.2 |
| VAT | Obesity | ENSG0000011 | -0.01572363 | 0.001134082 | 0.004830637 | 0.183450469 | -0.01580394 | 0.001068617 | 0.004830307 | 0.1834135 | 8.03E-05 | 0.038546761 | 1 | 0.014764466 | 1 | 0.015746258 | ENSG0000022 | 1 | RP4-580N22.2 |
| VAT | Obesity | ENSG0000011 | 0.001906222 | 0.571833244 | 0.003371732 | 0.183454353 | 0.001988534 | 0.555322386 | 0.003371518 | 0.183417415 | -8.23E-05 | 0.004539902 | 1 | 0.806905296 | 1 | 0.805802048 | ENSG0000022 | 1 | MICD |
| VAT | Obesity | ENSG0000011 | -0.00059002 | 0.876320886 | 0.003791031 | 0.183454465 | -0.0004543 | 0.904607251 | 0.003790766 | 0.183417541 | -0.00013572 | 4.77E-06 | 1 | 0.951543054 | 1 | 0.970729398 | ENSG0000022 | 1 | MICD |
| VAT | Obesity | ENSG0000011 | -0.00350131 | 0.31040811 | 0.003451714 | 0.183454085 | -0.00355584 | 0.30289773 | 0.00345147 | 0.183417145 | 5.45E-05 | 0.041424256 | 1 | 0.618575006 | 1 | 0.624344835 | ENSG0000022 | 1 | MICD |
| VAT | Obesity | ENSG0000011 | 0.019542063 | 1.51E-10 | 0.003051066 | 0.183438968 | 0.016884885 | 6.22E-08 | 0.003119694 | 0.183406476 | 0.002657178 | 4.60E-05 | 4.15E-07 | 1.12E-08 | 0.000171758 | 3.90E-06 | ENSG0000022 | 1 | MICD |
| VAT | Obesity | ENSG0000011 | 0.004518367 | 0.075121775 | 0.002538802 | 0.183453277 | 0.004475424 | 0.077910617 | 0.002538608 | 0.183416371 | 4.29E-05 | 0.015286847 | 1 | 0.282737093 | 1 | 0.297007324 | ENSG0000022 | 1 | MICD |
| VAT | Obesity | ENSG0000011 | -0.01701182 | 0.00032436 | 0.004732011 | 0.183449589 | -0.01685235 | 0.000368778 | 0.004731812 | 0.183412752 | -0.00015948 | 0.001805306 | 0.895232629 | 0.005526127 | 1 | 0.006696239 | ENSG0000022 | 1 | MICD |
| VAT | Obesity | ENSG0000011 | -0.00903046 | 0.005477992 | 0.003251285 | 0.183451558 | -0.00893253 | 0.006005828 | 0.003251178 | 0.183414693 | -9.79E-05 | 0.009603069 | 1 | 0.047544835 | 1 | 0.054526597 | ENSG0000022 | 1 | MICD |
| VAT | Obesity | ENSG0000011 | 0.000425708 | 0.929924979 | 0.004840927 | 0.183454471 | 0.00052281 | 0.913991692 | 0.004840604 | 0.183417542 | -9.71E-05 | 0.01491921 | 1 | 0.976170888 | 1 | 0.972419923 | ENSG0000022 | 1 | MICD |
| VAT | Obesity | ENSG0000011 | 0.000330534 | 0.873924815 | 0.002083087 | 0.183454465 | 0.000364709 | 0.861005977 | 0.002082933 | 0.183417534 | -3.42E-05 | 0.024896422 | 1 | 0.951116912 | 1 | 0.950288579 | ENSG0000022 | 1 | MICD |
| VAT | Obesity | ENSG0000011 | -0.00987246 | 0.000463978 | 0.002820092 | 0.183449842 | -0.01119687 | 7.77E-05 | 0.002833565 | 0.183411645 | 0.00132441 | 2.06E-06 | 1 | 0.00731759 | 0.214367329 | 0.001981293 | ENSG0000022 | 1 | MICD |
| VAT | Obesity | ENSG0000011 | -0.00357207 | 0.034676652 | 0.001691225 | 0.183452788 | -0.00363406 | 0.031642968 | 0.001691128 | 0.183415801 | 6.20E-05 | 7.78E-05 | 1 | 0.17455266 | 1 | 0.172598009 | ENSG0000022 | 1 | MICD |
| VAT | Obesity | ENSG0000011 | 0.005273545 | 0.168299968 | 0.003827819 | 0.183453757 | 0.005327005 | 0.163994266 | 0.003827518 | 0.183416814 | -5.35E-05 | 0.034956084 | 1 | 0.450948512 | 1 | 0.455295507 | ENSG0000022 | 3 | HLA-DRB6 |
| VAT | Obesity | ENSG0000011 | -0.01810916 | 0.000256284 | 0.004953373 | 0.183449422 | -0.01796903 | 0.000285796 | 0.004953049 | 0.183412572 | -0.00014013 | 0.000703086 | 0.707344985 | 0.004684404 | 0.788795859 | 0.005533516 | ENSG0000022 | 3 | HLA-DRB6 |
| VAT | Obesity | ENSG0000011 | -0.01094535 | 0.000283795 | 0.00301551 | 0.183449494 | -0.01083171 | 0.000327891 | 0.003015316 | 0.183412669 | -0.00011364 | 9.45E-06 | 0.783274966 | 0.005022832 | 0.904978408 | 0.006073681 | ENSG0000022 | 3 | HLA-DRB6 |
| VAT | Obesity | ENSG0000011 | -0.00549511 | 0.368899894 | 0.006115618 | 0.183454169 | -0.00532989 | 0.383438883 | 0.006115213 | 0.183417259 | -0.00016522 | 0.001104598 | 1 | 0.676520736 | 1 | 0.707013865 | ENSG0000022 | 3 | HLA-DRB6 |
| VAT | Obesity | ENSG0000011 | 0.010988886 | 0.060493906 | 0.00585393 | 0.183453142 | 0.01113751 | 0.057085482 | 0.005853652 | 0.183416178 | -0.00014862 | 0.013825785 | 1 | 0.25037186 | 1 | 0.251902356 | ENSG0000022 | 1 | AJ006998.2 |
| VAT | Obesity | ENSG0000011 | -0.00453332 | 0.31729381 | 0.004533157 | 0.183454096 | -0.00463756 | 0.306258422 | 0.004532822 | 0.18341715 | 0.000104238 | 0.001696926 | 1 | 0.623838113 | 1 | 0.628923546 | ENSG0000022 | 1 | AJ006998.2 |
| VAT | Obesity | ENSG0000011 | 0.002073587 | 0.484299282 | 0.002964787 | 0.183454289 | 0.0021393 | 0.470529956 | 0.002964589 | 0.183417349 | -6.57E-05 | 0.007091169 | 1 | 0.764245865 | 1 | 0.76685518 | ENSG0000022 | 2 | RP11-305L7.6 |
| VAT | Obesity | ENSG0000011 | 0.002399533 | 0.292448299 | 0.002279263 | 0.183454055 | 0.002537527 | 0.265594164 | 0.00227935 | 0.183417078 | -0.00013799 | 0.000282492 | 1 | 0.601010652 | 1 | 0.579936623 | ENSG0000022 | 2 | RP11-305L7.6 |
| VAT | Obesity | ENSG0000011 | 3.68E-05 | 0.994863009 | 0.005712069 | 0.183454474 | 0.000669243 | 0.906742828 | 0.005712797 | 0.183417541 | -0.00063247 | 2.18E-07 | 1 | 0.997766076 | 1 | 0.970734158 | ENSG0000022 | 1 | MRPS18AP1 |
| VAT | Obesity | ENSG0000011 | 0.009837497 | 0.015584117 | 0.004067568 | 0.183452263 | 0.009634957 | 0.017858473 | 0.004067876 | 0.183415426 | 0.000202541 | 0.007968358 | 1 | 0.10325657 | 1 | 0.119020171 | ENSG0000022 | 1 | MRPS18AP1 |
| VAT | Obesity | ENSG0000011 | 0.001507045 | 0.632794424 | 0.003154148 | 0.183454388 | 0.001582329 | 0.615880439 | 0.00315395 | 0.183417451 | -7.53E-05 | 0.005979287 | 1 | 0.840477676 | 1 | 0.839253732 | ENSG0000022 | 1 | MRPS18AP1 |
| VAT | Obesity | ENSG0000011 | 0.004964015 | 0.252620196 | 0.004339138 | 0.183453979 | 0.005055899 | 0.24390711 | 0.004338792 | 0.183417033 | -9.19E-05 | 0.001140291 | 1 | 0.549867303 | 1 | 0.550886762 | ENSG0000022 | 1 | AC093899.3 |
| VAT | Obesity | ENSG0000011 | -0.00010875 | 0.945337394 | 0.001586078 | 0.183454472 | -6.07E-05 | 0.969454674 | 0.001585991 | 0.183417545 | -4.80E-05 | 0.001526511 | 1 | 0.981611028 | 1 | 0.990920808 | ENSG0000022 | 1 | XXbac-BPG248L24.10 |
| VAT | Obesity | ENSG0000011 | 0.011112002 | 0.037220345 | 0.005333732 | 0.183452833 | 0.01100033 | 0.039156212 | 0.005333372 | 0.183415938 | 0.000111672 | 0.00990284 | 1 | 0.182790308 | 1 | 0.196136378 | ENSG0000023 | 3 | RP11-206L10.8 |
| VAT | Obesity | ENSG0000011 | -0.00338888 | 0.516335864 | 0.005221662 | 0.183454315 | -0.00328147 | 0.529690509 | 0.005221284 | 0.183417397 | -0.00010741 | 0.00614392 | 1 | 0.776929212 | 1 | 0.786996417 | ENSG0000023 | 2 | GS1-124K5.2 |
| VAT | Obesity | ENSG0000011 | 0.005887536 | 0.284425526 | 0.005500151 | 0.183454041 | 0.00578469 | 0.292886817 | 0.005499743 | 0.183417128 | 0.000102847 | 0.010054801 | 1 | 0.59068055 | 1 | 0.615229003 | ENSG0000023 | 2 | GS1-124K5.2 |
| VAT | Obesity | ENSG0000011 | -0.0043255 | 0.223930379 | 0.003556735 | 0.183453915 | -0.00441006 | 0.214973021 | 0.00355648 | 0.183416965 | 8.46E-05 | 0.001753774 | 1 | 0.52199987 | 1 | 0.526433789 | ENSG0000023 | 1 | PHBP9 |
| VAT | Obesity | ENSG0000011 | 0.008741162 | 0.125255755 | 0.005701684 | 0.183453586 | 0.00865386 | 0.129042804 | 0.005701244 | 0.183416675 | 8.73E-05 | 0.025178229 | 1 | 0.379063468 | 1 | 0.395731266 | ENSG0000023 | 1 | PHBP9 |
| VAT | Obesity | ENSG0000011 | -0.01075317 | 0.04412347 | 0.005342073 | 0.183452943 | -0.01094649 | 0.040439396 | 0.005341739 | 0.183415959 | 0.000193322 | 3.59E-05 | 1 | 0.206758534 | 1 | 0.201467029 | ENSG0000023 | 1 | PHBP9 |
| VAT | Obesity | ENSG0000011 | -0.00592623 | 0.012065138 | 0.002360818 | 0.183452092 | -0.00597075 | 0.011429585 | 0.002360641 | 0.183415128 | 4.45E-05 | 0.008478976 | 1 | 0.085384052 | 1 | 0.087142688 | ENSG0000023 | 1 | PHBP9 |
| VAT | Obesity | ENSG0000011 | -0.00999564 | 0.063290562 | 0.00538225 | 0.18345317 | -0.01011987 | 0.060061253 | 0.005381912 | 0.18341621 | 0.000124227 | 0.008012798 | 1 | 0.258023563 | 1 | 0.25801405 | ENSG0000023 | 1 | PHBP9 |
| VAT | Obesity | ENSG0000011 | -0.00373999 | 0.548689246 | 0.006236173 | 0.183454338 | -0.00361788 | 0.561789947 | 0.00623574 | 0.183417419 | -0.00012211 | 0.013058701 | 1 | 0.793647904 | 1 | 0.809555852 | ENSG0000023 | 3 | HERC2P4 |
| VAT | Obesity | ENSG0000011 | -0.00329427 | 0.505026167 | 0.004941874 | 0.183454306 | -0.00321172 | 0.515726799 | 0.004941514 | 0.183417386 | -8.25E-05 | 0.024894074 | 1 | 0.770249732 | 1 | 0.784425829 | ENSG0000023 | 3 | HERC2P4 |
| VAT | Obesity | ENSG0000011 | 0.016749774 | 0.000474739 | 0.004792986 | 0.183449858 | 0.01663217 | 0.000519791 | 0.004792624 | 0.183412995 | 0.000117604 | 0.00052198 | 1 | 0.007444768 | 1 | 0.0087477 | ENSG0000023 | 3 | HERC2P4 |
| VAT | Obesity | ENSG0000011 | 0.006049173 | 0.10662206 | 0.003748945 | 0.18345349 | 0.006133798 | 0.101789555 | 0.00374871 | 0.183416534 | -8.46E-05 | 0.0098324 | 1 | 0.351165734 | 1 | 0.351173964 | ENSG0000023 | 1 | RP11-458F8.2 |
| VAT | Obesity | ENSG0000011 | 0.005119082 | 0.114783792 | 0.003245978 | 0.183453534 | 0.004941884 | 0.127902234 | 0.003246052 | 0.18341667 | 0.000177198 | 0.000507231 | 1 | 0.365551911 | 1 | 0.395731266 | ENSG0000023 | 1 | RP11-458F8.2 |
| VAT | Obesity | ENSG0000011 | -0.00012711 | 0.964769197 | 0.002877777 | 0.183454473 | -7.37E-05 | 0.97956526 | 0.00287755 | 0.183417546 | -5.34E-05 | 0.004738359 | 1 | 0.985729121 | 1 | 0.994336196 | ENSG0000023 | 1 | CTD-2192J16.11 |
| VAT | Obesity | ENSG0000011 | -0.00519509 | 0.350053012 | 0.00555929 | 0.183454144 | -0.00509781 | 0.359113088 | 0.005558873 | 0.183417228 | -9.73E-05 | 0.014716978 | 1 | 0.661582235 | 1 | 0.683932408 | ENSG0000023 | 1 | RP4-737E23.2 |
| VAT | Obesity | ENSG0000011 | -0.00015061 | 0.935018976 | 0.001847231 | 0.183454472 | -0.00010571 | 0.954359914 | 0.00184711 | 0.183417545 | -4.49E-05 | 0.00378911 | 1 | 0.977307539 | 1 | 0.984913318 | ENSG0000023 | 1 | RP4-737E23.2 |
| VAT | Obesity | ENSG0000011 | 0.001839852 | 0.612779993 | 0.003635285 | 0.183454377 | 0.001713999 | 0.637269752 | 0.003635051 | 0.183417462 | 0.000125854 | 4.79E-05 | 1 | 0.83120188 | 1 | 0.845380113 | ENSG0000023 | 1 | RP4-737E23.2 |
| VAT | Obesity | ENSG0000011 | -0.01085387 | 0.022643061 | 0.004761698 | 0.18345251 | -0.01076847 | 0.02371921 | 0.004761325 | 0.183415613 | -8.54E-05 | 0.006908361 | 1 | 0.132124418 | 1 | 0.141699174 | ENSG0000023 | 1 | RP4-737E23.2 |
| VAT | Obesity | ENSG0000011 | -0.00436225 | 0.047720506 | 0.002203317 | 0.183452992 | -0.0042924 | 0.051381444 | 0.002203168 | 0.183416111 | -6.98E-05 | 0.000100302 | 1 | 0.216270272 | 1 | 0.238404712 | ENSG0000023 | 1 | RP4-737E23.2 |
| VAT | Obesity | ENSG0000011 | -0.00270031 | 0.597015939 | 0.005107488 | 0.183454368 | -0.00257868 | 0.613615595 | 0.00510712 | 0.18341745 | -0.00012163 | 0.001613477 | 1 | 0.819937013 | 1 | 0.839253732 | ENSG0000023 | 5 | RP11-420G6.4 |
| VAT | Obesity | ENSG0000011 | 0.011322848 | 7.00E-07 | 0.002282117 | 0.183445169 | 0.011394059 | 5.94E-07 | 0.002281946 | 0.183408124 | -7.12E-05 | 1.70E-05 | 0.001930744 | 2.93E-05 | 0.001640224 | 2.98E-05 | ENSG0000023 | 5 | RP11-420G6.4 |
| VAT | Obesity | ENSG0000011 | 0.004203309 | 0.408682101 | 0.005087422 | 0.183454216 | 0.004317057 | 0.396083263 | 0.005087043 | 0.183417274 | -0.00011375 | 0.00197143 | 1 | 0.708044263 | 1 | 0.715434904 | ENSG0000023 | 5 | RP11-420G6.4 |
| VAT | Obesity | ENSG0000011 | -0.00407622 | 0.028359531 | 0.00185935 | 0.183452657 | -0.00411453 | 0.026895115 | 0.001859216 | 0.183415695 | 3.83E-05 | 0.006646339 | 1 | 0.153475108 | 1 | 0.155231297 | ENSG0000023 | 5 | RP11-420G6.4 |
| VAT | Obesity | ENSG0000011 | -0.00074029 | 0.882480255 | 0.005007855 | 0.183454466 | -0.00066666 | 0.89408785 | 0.005007488 | 0.183417539 | -7.36E-05 | 0.046750583 | 1 | 0.95522585 | 1 | 0.968028006 | ENSG0000023 | 5 | RP11-420G6.4 |
| VAT | Obesity | ENSG0000011 | -0.00582578 | 0.128016002 | 0.003827777 | 0.183453599 | -0.00576716 | 0.131870553 | 0.003827503 | 0.183416688 | -5.86E-05 | 0.04463034 | 1 | 0.383214931 | 1 | 0.399591076 | ENSG0000023 | 5 | RP11-420G6.4 |
| VAT | Obesity | ENSG0000012 | 0.004068801 | 0.396631636 | 0.004800078 | 0.183454203 | 0.003977707 | 0.40725308 | 0.004799716 | 0.183417286 | 9.11E-05 | 0.007511774 | 1 | 0.701928581 | 1 | 0.721556426 | ENSG0000023 | 1 | U73166.2 |
| VAT | Obesity | ENSG0000012 | -0.00890387 | 0.103329066 | 0.005466095 | 0.183453471 | -0.00877341 | 0.108461189 | 0.005465783 | 0.183416572 | -0.00013046 | 0.010514865 | 1 | 0.344846701 | 1 | 0.365064489 | ENSG0000023 | 1 | U73166.2 |
| VAT | Obesity | ENSG0000012 | -0.00092363 | 0.616367556 | 0.001843552 | 0.183454379 | -0.00089463 | 0.627455637 | 0.001843412 | 0.183417457 | -2.90E-05 | 0.026096865 | 1 | 0.831570896 | 1 | 0.842487235 | ENSG0000023 | 1 | U73166.2 |
| VAT | Obesity | ENSG0000012 | 0.004181378 | 0.146985415 | 0.002883183 | 0.183453679 | 0.004234835 | 0.141857304 | 0.002882974 | 0.183416731 | -5.35E-05 | 0.013536574 | 1 | 0.413958925 | 1 | 0.417278146 | ENSG0000023 | 1 | HCG4P7 |
| VAT | Obesity | ENSG0000012 | 0.005857508 | 0.001473888 | 0.001842103 | 0.183450652 | 0.005940957 | 0.001259209 | 0.001842079 | 0.183413615 | -8.34E-05 | 0.000650606 | 1 | 0.018160406 | 1 | 0.017731721 | ENSG0000023 | 1 | HCG4P7 |
| VAT | Obesity | ENSG0000012 | 0.004286197 | 0.141283475 | 0.002913737 | 0.183453656 | 0.004204949 | 0.148953419 | 0.002913547 | 0.183416759 | 8.12E-05 | 0.000908523 | 1 | 0.405767317 | 1 | 0.426597721 | ENSG0000023 | 1 | HCG4P7 |
| VAT | Obesity | ENSG0000012 | 0.007166296 | 0.023193541 | 0.003156626 | 0.183452526 | 0.007218639 | 0.022196808 | 0.003156392 | 0.183415569 | -5.23E-05 | 0.022933629 | 1 | 0.133602008 | 1 | 0.135971568 | ENSG0000023 | 1 | HCG4P7 |
| VAT | Obesity | ENSG0000012 | -0.00193443 | 0.695549177 | 0.004943153 | 0.183454416 | -0.00184837 | 0.70844157 | 0.004942822 | 0.183417493 | -8.61E-05 | 0.033556986 | 1 | 0.866149932 | 1 | 0.876422561 | ENSG0000023 | 1 | HCG4P7 |
| VAT | Obesity | ENSG0000012 | -0.00776983 | 0.072961343 | 0.004333241 | 0.183453259 | -0.00788227 | 0.068887828 | 0.004332922 | 0.183416295 | 0.000112442 | 0.00043245 | 1 | 0.279186061 | 1 | 0.277410173 | ENSG0000023 | 1 | HCG4P7 |
| VAT | Obesity | ENSG0000012 | 0.001848246 | 0.642185939 | 0.00397776 | 0.183454392 | 0.001720504 | 0.665336126 | 0.00397751 | 0.183417475 | 0.000127742 | 0.000222091 | 1 | 0.843374123 | 1 | 0.856496133 | ENSG0000023 | 1 | HCG4P7 |
| VAT | Obesity | ENSG0000012 | -0.00113752 | 0.568630572 | 0.001995399 | 0.183454351 | -0.00120137 | 0.547103657 | 0.001995278 | 0.183417409 | 6.39E-05 | 0.0003372 | 1 | 0.806905296 | 1 | 0.799791363 | ENSG0000023 | 3 | NOS2P3 |
| VAT | Obesity | ENSG0000012 | 0.007588087 | 0.002774936 | 0.002536427 | 0.183451091 | 0.007546505 | 0.002925596 | 0.002536244 | 0.1834142 | 4.16E-05 | 0.029855662 | 1 | 0.029101822 | 1 | 0.032690866 | ENSG0000023 | 3 | NOS2P3 |
| VAT | Obesity | ENSG0000012 | 0.007946237 | 0.115283421 | 0.005045595 | 0.183453537 | 0.008044282 | 0.110837624 | 0.005045201 | 0.183416585 | -9.80E-05 | 0.003771209 | 1 | 0.366186259 | 1 | 0.366800769 | ENSG0000023 | 2 | RP11-517P14.2 |
| VAT | Obesity | ENSG0000012 | 0.005902935 | 0.023283538 | 0.002601836 | 0.183452528 | 0.005830119 | 0.025032172 | 0.002601672 | 0.183415648 | 7.28E-05 | 0.001168548 | 1 | 0.133602008 | 1 | 0.147625628 | ENSG0000023 | 2 | AC112229.4 |
| VAT | Obesity | ENSG0000012 | -0.00071676 | 0.759365663 | 0.002339951 | 0.183454439 | -0.00075356 | 0.74740237 | 0.002339767 | 0.183417507 | 3.68E-05 | 0.017406526 | 1 | 0.897964537 | 1 | 0.89626095 | ENSG0000023 | 2 | AC112229.4 |
| VAT | Obesity | ENSG0000012 | 0.00354889 | 0.389399529 | 0.004123223 | 0.183454194 | 0.003479787 | 0.398664623 | 0.004122925 | 0.183417277 | 6.91E-05 | 0.025861016 | 1 | 0.696527997 | 1 | 0.716012529 | ENSG0000023 | 2 | AC112229.4 |
| VAT | Obesity | ENSG0000012 | 0.01401068 | 0.004364492 | 0.004915072 | 0.183451403 | 0.013920304 | 0.004620509 | 0.004914705 | 0.183414514 | 9.04E-05 | 0.010695075 | 1 | 0.040287616 | 1 | 0.044966202 | ENSG0000023 | 6 | AC159540.1 |

| | | | | | | | | | | | | | | | | | | | |
|---|---|---|---|---|---|---|---|---|---|---|---|---|---|---|---|---|---|---|---|
| VAT | Obesity | ENSG0000012 | 0.005131385 | 0.016427912 | 0.002138722 | 0.183452298 | 0.005078731 | 0.017558846 | 0.002138593 | 0.183415415 | 5.27E-05 | 0.00591379 | 1 | 0.106434359 | 1 | 0.117913417 | ENSG0000023 | 6 | AC159540.1 |
| VAT | Obesity | ENSG0000012 | 0.003476615 | 0.162038787 | 0.002486405 | 0.183453735 | 0.003436244 | 0.166935432 | 0.002486212 | 0.183416824 | 4.04E-05 | 0.016087417 | 1 | 0.441391512 | 1 | 0.457993828 | ENSG0000023 | 2 | AP000354.4 |
| VAT | Obesity | ENSG0000012 | 0.001047666 | 0.533896721 | 0.001684163 | 0.183454328 | 0.001075973 | 0.522870969 | 0.001684032 | 0.183417392 | -2.83E-05 | 0.012630371 | 1 | 0.783986954 | 1 | 0.784841324 | ENSG0000023 | 2 | HLA-K |
| VAT | Obesity | ENSG0000012 | 0.003723603 | 0.231901659 | 0.003114745 | 0.183453934 | 0.003662893 | 0.239571419 | 0.003114545 | 0.183417023 | 6.07E-05 | 0.022484094 | 1 | 0.52715189 | 1 | 0.546616798 | ENSG0000023 | 2 | HLA-K |
| VAT | Obesity | ENSG0000012 | -0.03634146 | 1.53E-24 | 0.003553866 | 0.183414956 | -0.03602194 | 3.86E-24 | 0.003553989 | 0.183378729 | -0.00031952 | 3.71E-08 | 4.21E-21 | 8.43E-22 | 1.07E-20 | 2.13E-21 | ENSG0000023 | 2 | HLA-K |
| VAT | Obesity | ENSG0000012 | 0.001681958 | 0.246202583 | 0.001450434 | 0.183453966 | 0.00083714 | 0.566887709 | 0.001461891 | 0.183417422 | 0.000844818 | 4.30E-06 | 1 | 0.543180758 | 1 | 0.811940881 | ENSG0000023 | 2 | HLA-K |
| VAT | Obesity | ENSG0000012 | 0.008710428 | 0.00391321 | 0.00301912 | 0.183451328 | 0.008563181 | 0.004562759 | 0.003019035 | 0.183414506 | 0.000147247 | 4.91E-05 | 1 | 0.036733053 | 1 | 0.044966202 | ENSG0000023 | 2 | HLA-K |
| VAT | Obesity | ENSG0000012 | -0.0013383 | 0.74976842 | 0.004196025 | 0.183454436 | -0.00117192 | 0.780006683 | 0.004195763 | 0.183417517 | -0.00016638 | 5.77E-06 | 1 | 0.892799597 | 1 | 0.911107144 | ENSG0000023 | 2 | HLA-K |
| VAT | Obesity | ENSG0000012 | 0.016017362 | 0.008777804 | 0.006112098 | 0.183451878 | 0.015835343 | 0.009570391 | 0.006111703 | 0.183415009 | 0.000182018 | 0.000430687 | 1 | 0.06692469 | 1 | 0.076785697 | ENSG0000023 | 2 | HLA-K |
| VAT | Obesity | ENSG0000012 | 0.005020226 | 0.094263768 | 0.003000148 | 0.183453416 | 0.005138433 | 0.086743851 | 0.002999961 | 0.183416437 | -0.00011821 | 6.87E-06 | 1 | 0.323190063 | 1 | 0.316168943 | ENSG0000023 | 2 | HLA-K |
| VAT | Obesity | ENSG0000012 | -0.00389702 | 0.388166031 | 0.00451593 | 0.183454193 | -0.0040481 | 0.370008307 | 0.004515659 | 0.183417242 | 0.000151086 | 0.000215602 | 1 | 0.695473688 | 1 | 0.692818993 | ENSG0000023 | 2 | HLA-K |
| VAT | Obesity | ENSG0000012 | -0.00786874 | 0.01774991 | 0.003319022 | 0.18345235 | -0.00779589 | 0.018823674 | 0.003318796 | 0.183415461 | -7.29E-05 | 0.006398864 | 1 | 0.110802893 | 1 | 0.120957701 | ENSG0000023 | 2 | HLA-K |
| VAT | Obesity | ENSG0000012 | -0.0054523 | 0.222267442 | 0.004467187 | 0.183453911 | -0.00532348 | 0.233356504 | 0.004466911 | 0.183417009 | -0.00012882 | 0.001059089 | 1 | 0.52199987 | 1 | 0.540524906 | ENSG0000023 | 1 | AC104135.4 |
| VAT | Obesity | ENSG0000012 | -0.01051141 | 0.080559233 | 0.006015269 | 0.18345332 | -0.0106646 | 0.076221978 | 0.00601485 | 0.183416358 | 0.000153189 | 0.001227498 | 1 | 0.291025502 | 1 | 0.293816565 | ENSG0000023 | 1 | AC104135.4 |
| VAT | Obesity | ENSG0000012 | 0.001489668 | 0.504398662 | 0.002231427 | 0.183454306 | 0.001522804 | 0.494930923 | 0.002231255 | 0.18341737 | -3.31E-05 | 0.02979099 | 1 | 0.770249732 | 1 | 0.775358255 | ENSG0000023 | 1 | AC104135.4 |
| VAT | Obesity | ENSG0000012 | 0.000216697 | 0.970943897 | 0.0059492 | 0.183454474 | 0.002636031 | 0.659609135 | 0.005984816 | 0.183417473 | -0.00241933 | 0.000233116 | 1 | 0.988370418 | 1 | 0.854766662 | ENSG0000023 | 1 | AC104135.4 |
| VAT | Obesity | ENSG0000012 | 0.005189585 | 0.218263134 | 0.004215195 | 0.183453901 | 0.005257479 | 0.212268553 | 0.004214907 | 0.183416958 | -6.79E-05 | 0.045506162 | 1 | 0.521961016 | 1 | 0.524026123 | ENSG0000023 | 1 | PPIHP1 |
| VAT | Obesity | ENSG0000012 | -0.00206806 | 0.671163929 | 0.004871163 | 0.183454406 | -0.00226388 | 0.64208813 | 0.004870846 | 0.183417464 | 0.00019582 | 1.90E-06 | 1 | 0.857647266 | 1 | 0.848972395 | ENSG0000023 | 1 | RP11-314C16.1 |
| VAT | Obesity | ENSG0000012 | 0.005381446 | 0.005816089 | 0.001951223 | 0.183451599 | 0.005431133 | 0.005375979 | 0.001951112 | 0.183414618 | -4.97E-05 | 0.006512404 | 1 | 0.049240505 | 1 | 0.05044591 | ENSG0000023 | 1 | RP11-314C16.1 |
| VAT | Obesity | ENSG0000012 | 0.000875894 | 0.820011835 | 0.003849578 | 0.183454455 | 0.001086107 | 0.777827159 | 0.00384937 | 0.183417516 | -0.00021021 | 1.62E-08 | 1 | 0.927365584 | 1 | 0.910459344 | ENSG0000023 | 1 | RP11-314C16.1 |
| VAT | Obesity | ENSG0000012 | -0.00441439 | 0.223530238 | 0.003626686 | 0.183453914 | -0.00447219 | 0.217494093 | 0.003626431 | 0.183416971 | 5.78E-05 | 0.040648538 | 1 | 0.52199987 | 1 | 0.529248865 | ENSG0000023 | 1 | RP11-314C16.1 |
| VAT | Obesity | ENSG0000012 | 0.004280894 | 0.228246247 | 0.003552933 | 0.183453925 | 0.004339465 | 0.221912451 | 0.003552686 | 0.183416982 | -5.86E-05 | 0.037174176 | 1 | 0.524101338 | 1 | 0.532716487 | ENSG0000023 | 2 | RP4-631H13.6 |
| VAT | Obesity | ENSG0000012 | -0.00197187 | 0.233164899 | 0.001653913 | 0.183453937 | -0.00193645 | 0.241634794 | 0.0016538 | 0.183417028 | -3.54E-05 | 0.007723255 | 1 | 0.527920526 | 1 | 0.547546824 | ENSG0000023 | 4 | CDC20P1 |
| VAT | Obesity | ENSG0000012 | -0.02034942 | 1.68E-05 | 0.004728849 | 0.183447475 | -0.0202641 | 1.82E-05 | 0.004728568 | 0.183410605 | -8.53E-05 | 0.046947536 | 0.046462655 | 0.000489081 | 0.050340929 | 0.000606517 | ENSG0000023 | 2 | AC007238.1 |
| VAT | Obesity | ENSG0000012 | 0.002788405 | 0.608453413 | 0.005443098 | 0.183454375 | 0.002689762 | 0.621171921 | 0.005442765 | 0.183417454 | 9.86E-05 | 0.041280945 | 1 | 0.828767095 | 1 | 0.840409069 | ENSG0000023 | 2 | AP000688.8 |
| VAT | Obesity | ENSG0000012 | -0.00090599 | 0.750469226 | 0.002848837 | 0.183454436 | -0.00085377 | 0.764394664 | 0.002848622 | 0.183417512 | -5.22E-05 | 0.009633996 | 1 | 0.892799597 | 1 | 0.904298874 | ENSG0000023 | 1 | HLA-T |
| VAT | Obesity | ENSG0000013 | 0.005004423 | 0.241113184 | 0.00426922 | 0.183453955 | 0.005088836 | 0.233236483 | 0.004268926 | 0.183417009 | -8.44E-05 | 0.01342419 | 1 | 0.535635762 | 1 | 0.540524906 | ENSG0000023 | 1 | HLA-T |
| VAT | Obesity | ENSG0000013 | 0.000501048 | 0.747379539 | 0.001555583 | 0.183454435 | 0.000528733 | 0.733916856 | 0.001555462 | 0.183417502 | -2.77E-05 | 0.008714259 | 1 | 0.892799597 | 1 | 0.890236789 | ENSG0000023 | 1 | HLA-T |
| VAT | Obesity | ENSG0000013 | 0.006055934 | 0.313502022 | 0.006008456 | 0.18345409 | 0.00620419 | 0.301768807 | 0.006008035 | 0.183417143 | -0.00014826 | 0.001652689 | 1 | 0.619817751 | 1 | 0.623730655 | ENSG0000023 | 1 | HLA-T |
| VAT | Obesity | ENSG0000013 | 0.008752042 | 0.001207561 | 0.002703661 | 0.183450513 | 0.008814609 | 0.001112362 | 0.00270348 | 0.183413528 | -6.26E-05 | 0.004787193 | 1 | 0.015501716 | 1 | 0.016244016 | ENSG0000023 | 1 | HLA-T |
| VAT | Obesity | ENSG0000013 | 0.005469992 | 0.231872985 | 0.004575295 | 0.183453934 | 0.005368762 | 0.240589175 | 0.004574934 | 0.183417026 | 0.00010123 | 0.000749821 | 1 | 0.52715189 | 1 | 0.546973742 | ENSG0000023 | 1 | HLA-T |
| VAT | Obesity | ENSG0000013 | -0.00989404 | 0.002549521 | 0.003278977 | 0.183451033 | -0.0096078 | 0.003403407 | 0.00328051 | 0.183414304 | -0.00028624 | 0.009583504 | 1 | 0.027812961 | 1 | 0.036161216 | ENSG0000023 | 7 | ZNF826P |
| VAT | Obesity | ENSG0000013 | 0.002804536 | 0.551423744 | 0.004708533 | 0.18345434 | 0.002693495 | 0.567264742 | 0.004708209 | 0.183417422 | 0.000111041 | 0.003114253 | 1 | 0.793904838 | 1 | 0.81202149 | ENSG0000023 | 7 | ZNF826P |
| VAT | Obesity | ENSG0000013 | 0.013248535 | 0.000112175 | 0.003429903 | 0.183448834 | 0.013137927 | 0.000127808 | 0.003429662 | 0.183412 | 0.000110608 | 3.52E-05 | 0.309603817 | 0.00240003 | 0.352748922 | 0.00291528 | ENSG0000023 | 7 | ZNF826P |
| VAT | Obesity | ENSG0000013 | 0.019366818 | 9.50E-05 | 0.004961938 | 0.183448716 | 0.019289287 | 0.000101199 | 0.004961579 | 0.183411834 | 7.75E-05 | 0.037341893 | 0.262175373 | 0.002112498 | 0.279309184 | 0.002428776 | ENSG0000023 | 7 | ZNF826P |
| VAT | Obesity | ENSG0000013 | -0.0033654 | 0.463671466 | 0.004592421 | 0.183454271 | -0.00347555 | 0.44913706 | 0.004592098 | 0.18341733 | 0.000110157 | 0.00210723 | 1 | 0.746635499 | 1 | 0.747200025 | ENSG0000023 | 2 | AP001604.3 |
| VAT | Obesity | ENSG0000013 | 0.001277897 | 0.800503487 | 0.005057059 | 0.18345445 | 0.00118459 | 0.814782415 | 0.005056709 | 0.183417525 | 9.33E-05 | 0.019819908 | 1 | 0.918665124 | 1 | 0.928683673 | ENSG0000023 | 1 | WASF5P |
| VAT | Obesity | ENSG0000013 | 0.001870959 | 0.506650155 | 0.002817441 | 0.183454307 | 0.001938535 | 0.491395771 | 0.002817266 | 0.183417367 | -6.76E-05 | 0.006285105 | 1 | 0.771793347 | 1 | 0.775358255 | ENSG0000023 | 1 | WASF5P |
| VAT | Obesity | ENSG0000013 | -0.00153673 | 0.661624952 | 0.003511149 | 0.183454402 | -0.00165487 | 0.637404353 | 0.003511059 | 0.183417462 | 0.000118144 | 0.005995617 | 1 | 0.854099711 | 1 | 0.845380113 | ENSG0000023 | 1 | WASF5P |
| VAT | Obesity | ENSG0000013 | -0.00056278 | 0.901969192 | 0.004568948 | 0.183454468 | -0.00066325 | 0.884573984 | 0.0045687 | 0.183417538 | 0.000100477 | 0.022360129 | 1 | 0.965567705 | 1 | 0.960056703 | ENSG0000023 | 2 | CTC-490G23.2 |
| VAT | Obesity | ENSG0000013 | 0.001571905 | 0.730067519 | 0.004555773 | 0.183454429 | 0.001690376 | 0.710589284 | 0.004555473 | 0.183417494 | -0.00011847 | 0.001835667 | 1 | 0.884153731 | 1 | 0.876721691 | ENSG0000023 | 2 | CTC-490G23.2 |
| VAT | Obesity | ENSG0000013 | 0.002161746 | 0.523576317 | 0.003389151 | 0.18345432 | 0.002222665 | 0.511910147 | 0.003388897 | 0.183417383 | -6.09E-05 | 0.011916828 | 1 | 0.779013819 | 1 | 0.784425829 | ENSG0000023 | 2 | AC024937.6 |
| VAT | Obesity | ENSG0000013 | -0.0145125 | 0.018086609 | 0.006139355 | 0.183452362 | -0.01441321 | 0.018882579 | 0.0061389 | 0.183415463 | -9.93E-05 | 0.025856832 | 1 | 0.111926101 | 1 | 0.120957701 | ENSG0000023 | 1 | AC024937.4 |
| VAT | Obesity | ENSG0000013 | -0.00364986 | 0.119150025 | 0.002342126 | 0.183453556 | -0.00369627 | 0.114497875 | 0.002341937 | 0.183416605 | 4.64E-05 | 0.001907523 | 1 | 0.372684435 | 1 | 0.371781334 | ENSG0000023 | 2 | RPS18 |
| VAT | Obesity | ENSG0000013 | 0.003788917 | 0.047678747 | 0.001913377 | 0.183452992 | 0.003816042 | 0.046092236 | 0.001913232 | 0.183416043 | -2.71E-05 | 0.045478187 | 1 | 0.216270272 | 1 | 0.220094413 | ENSG0000023 | 2 | RPS18 |
| VAT | Obesity | ENSG0000013 | 0.016095568 | 0.001780487 | 0.005151217 | 0.183450784 | 0.016020355 | 0.001869425 | 0.005150832 | 0.18341389 | 7.52E-05 | 0.042291767 | 1 | 0.020647668 | 1 | 0.023559873 | ENSG0000023 | 2 | RPS18 |
| VAT | Obesity | ENSG0000013 | 0.011312132 | 0.05496147 | 0.005894236 | 0.183453082 | 0.011414305 | 0.052788085 | 0.005893824 | 0.183416129 | -0.00010217 | 0.027176735 | 1 | 0.236651571 | 1 | 0.241837823 | ENSG0000023 | 2 | RPS18 |
| VAT | Obesity | ENSG0000013 | -0.00568792 | 0.200676297 | 0.004444977 | 0.183453855 | -0.00562547 | 0.205630338 | 0.004444643 | 0.183416941 | -6.25E-05 | 0.049173058 | 1 | 0.501589882 | 1 | 0.517356182 | ENSG0000023 | 2 | RPS18 |
| VAT | Obesity | ENSG0000013 | -0.00397784 | 0.487014649 | 0.00572297 | 0.183454291 | -0.0037035 | 0.517539465 | 0.005722847 | 0.183417388 | -0.00027434 | 0.00013889 | 1 | 0.765031548 | 1 | 0.784425829 | ENSG0000023 | 2 | RPS18 |
| VAT | Obesity | ENSG0000013 | 7.32E-05 | 0.980894616 | 0.003055914 | 0.183454474 | 0.000146995 | 0.961632389 | 0.003055691 | 0.183417545 | -7.38E-05 | 0.001123279 | 1 | 0.99502166 | 1 | 0.98754495 | ENSG0000023 | 2 | RPS18 |
| VAT | Obesity | ENSG0000013 | -0.00383176 | 0.107503798 | 0.002380687 | 0.183453495 | -0.00378974 | 0.111389487 | 0.002380519 | 0.183416588 | -4.20E-05 | 0.022393784 | 1 | 0.353035267 | 1 | 0.366914394 | ENSG0000023 | 1 | Z95114.3 |
| VAT | Obesity | ENSG0000013 | 0.003959891 | 0.330941246 | 0.004073037 | 0.183454117 | 0.003897228 | 0.338615844 | 0.004072744 | 0.1834172 | 6.27E-05 | 0.042313294 | 1 | 0.640950731 | 1 | 0.660896608 | ENSG0000023 | 2 | AP002856.5 |
| VAT | Obesity | ENSG0000013 | -0.03654725 | 1.25E-17 | 0.004275348 | 0.183426856 | -0.03665955 | 9.92E-18 | 0.004275084 | 0.183389759 | 0.000112302 | 0.003075767 | 3.46E-14 | 2.66E-15 | 2.74E-14 | 2.11E-15 | ENSG0000023 | 2 | AP002856.5 |
| VAT | Obesity | ENSG0000013 | 0.00042483 | 0.933618513 | 0.005100426 | 0.183454471 | 0.000507074 | 0.920801026 | 0.005100074 | 0.183417542 | -8.22E-05 | 0.042579869 | 1 | 0.977307539 | 1 | 0.973721371 | ENSG0000023 | 1 | RP5-855F14.1 |
| VAT | Obesity | ENSG0000013 | 0.005180458 | 0.360501599 | 0.005665352 | 0.183454158 | 0.005319266 | 0.347746423 | 0.005664999 | 0.183417213 | -0.00013881 | 0.005161692 | 1 | 0.670499496 | 1 | 0.670656147 | ENSG0000023 | 1 | RP5-855F14.1 |
| VAT | Obesity | ENSG0000013 | -0.00382927 | 0.324951742 | 0.003890213 | 0.183454108 | -0.00393176 | 0.312138873 | 0.003889964 | 0.18341716 | 0.000102494 | 0.002129471 | 1 | 0.633061352 | 1 | 0.634391229 | ENSG0000023 | 4 | DBIL5P |
| VAT | Obesity | ENSG0000013 | 0.000957202 | 0.855936316 | 0.0052724 | 0.183454462 | 0.000851036 | 0.871758511 | 0.00527201 | 0.183417536 | 0.000106166 | 0.005817521 | 1 | 0.944116239 | 1 | 0.955921132 | ENSG0000023 | 4 | DBIL5P |
| VAT | Obesity | ENSG0000013 | 0.002151237 | 0.181146555 | 0.00160872 | 0.183453798 | 0.002186664 | 0.174035495 | 0.001608607 | 0.183416848 | -3.54E-05 | 0.004529744 | 1 | 0.473899992 | 1 | 0.470919574 | ENSG0000023 | 4 | DBIL5P |
| VAT | Obesity | ENSG0000013 | -0.0110046 | 0.00558056 | 0.00397067 | 0.183451571 | -0.01068138 | 0.007152481 | 0.003971256 | 0.183414812 | -0.00032323 | 0.000255719 | 1 | 0.048283214 | 1 | 0.062273965 | ENSG0000023 | 4 | DBIL5P |
| VAT | Obesity | ENSG0000013 | -0.00385601 | 0.32657802 | 0.003930574 | 0.18345411 | -0.00397632 | 0.311681461 | 0.003930332 | 0.183417159 | 0.000120307 | 0.000547326 | 1 | 0.634310581 | 1 | 0.633928395 | ENSG0000023 | 4 | DBIL5P |
| VAT | Obesity | ENSG0000013 | 0.000817266 | 0.861373728 | 0.004680098 | 0.183454463 | 0.000734683 | 0.875252198 | 0.004679785 | 0.183417537 | 8.26E-05 | 0.031848307 | 1 | 0.944533766 | 1 | 0.956332568 | ENSG0000023 | 4 | DBIL5P |
| VAT | Obesity | ENSG0000013 | -0.003725 | 0.312422511 | 0.003687565 | 0.183454088 | -0.00366866 | 0.319764549 | 0.003687305 | 0.183417172 | -5.63E-05 | 0.049112525 | 1 | 0.619458427 | 1 | 0.641269508 | ENSG0000023 | 4 | DOC2GP |
| VAT | Obesity | ENSG0000013 | 0.016127751 | 0.003126383 | 0.0054577 | 0.183451173 | 0.008345427 | 0.149229867 | 0.005786357 | 0.18341676 | 0.007782324 | 5.23E-05 | 1 | 0.031840649 | 1 | 0.426812885 | ENSG0000023 | 4 | DOC2GP |
| VAT | Obesity | ENSG0000013 | 0.006194203 | 0.026562002 | 0.002792821 | 0.183452615 | 0.002436847 | 0.401075493 | 0.002902035 | 0.18341728 | 0.003757356 | 1.95E-06 | 1 | 0.147671738 | 1 | 0.718344166 | ENSG0000023 | 4 | DOC2GP |
| VAT | Obesity | ENSG0000013 | -0.00284393 | 0.048940867 | 0.001444272 | 0.183453008 | -0.00110327 | 0.457327937 | 0.001484381 | 0.183417337 | -0.00174066 | 3.98E-07 | 1 | 0.219637062 | 1 | 0.755285959 | ENSG0000023 | 4 | DOC2GP |
| VAT | Obesity | ENSG0000013 | -0.00201362 | 0.442070259 | 0.002619502 | 0.183454251 | -0.00196603 | 0.452898453 | 0.002619306 | 0.183417333 | -4.76E-05 | 0.011606719 | 1 | 0.727515266 | 1 | 0.750751641 | ENSG0000023 | 4 | DOC2GP |
| VAT | Obesity | ENSG0000013 | 0.024469577 | 6.96E-08 | 0.004537951 | 0.183443484 | 0.024583661 | 6.05E-08 | 0.004537804 | 0.183406454 | -0.00011408 | 0.031684423 | 0.000192178 | 3.70E-06 | 0.000166866 | 3.88E-06 | ENSG0000023 | 4 | DOC2GP |
| VAT | Obesity | ENSG0000013 | -0.02128729 | 0.000100368 | 0.005472687 | 0.183448755 | -0.02107747 | 0.000117355 | 0.005472366 | 0.18341194 | -0.00020982 | 2.70E-05 | 0.277016792 | 0.002198546 | 0.32389977 | 0.002744913 | ENSG0000023 | 2 | RP13-93L13.2 |
| VAT | Obesity | ENSG0000013 | 0.001460895 | 0.49545392 | 0.002143142 | 0.183454298 | 0.001493785 | 0.485766857 | 0.002142985 | 0.183417362 | -3.29E-05 | 0.038281884 | 1 | 0.766551816 | 1 | 0.775358255 | ENSG0000023 | 1 | RP4-782L23.1 |
| VAT | Obesity | ENSG0000013 | 0.001442475 | 0.28907056 | 0.001360615 | 0.183454049 | 0.001462632 | 0.282349404 | 0.001360515 | 0.183417109 | -2.02E-05 | 0.044446889 | 1 | 0.59807702 | 1 | 0.601299657 | ENSG0000023 | 2 | CYP21A2 |
| VAT | Obesity | ENSG0000013 | -0.00194799 | 0.748511413 | 0.006076019 | 0.183454435 | -0.00204368 | 0.736587048 | 0.006075586 | 0.183417503 | 9.57E-05 | 0.03987224 | 1 | 0.892799597 | 1 | 0.891658005 | ENSG0000023 | 2 | CYP21A2 |
| VAT | Obesity | ENSG0000013 | -0.00422555 | 0.277235467 | 0.00388895 | 0.183454028 | -0.00753053 | 0.059540541 | 0.003996706 | 0.183416204 | 0.003304988 | 0.000345202 | 1 | 0.582299881 | 1 | 0.257361325 | ENSG0000023 | 7 | TAPBP |
| VAT | Obesity | ENSG0000013 | -3.61E-05 | 0.990779578 | 0.003125289 | 0.183454474 | 3.80E-05 | 0.990292539 | 0.003125064 | 0.183417546 | -7.41E-05 | 0.001732599 | 1 | 0.997766076 | 1 | 0.99714778 | ENSG0000023 | 7 | TAPBP |
| VAT | Obesity | ENSG0000013 | 0.00537429 | 0.18588029 | 0.004062609 | 0.183453813 | 0.005464522 | 0.178569679 | 0.004062316 | 0.183416862 | -9.02E-05 | 0.003324074 | 1 | 0.481717936 | 1 | 0.480362878 | ENSG0000023 | 7 | TAPBP |
| VAT | Obesity | ENSG0000013 | -0.00699959 | 0.177500868 | 0.005190681 | 0.183453787 | -0.00681933 | 0.188897837 | 0.005190349 | 0.183416894 | -0.00018026 | 4.97E-05 | 1 | 0.466573709 | 1 | 0.497954184 | ENSG0000023 | 7 | TAPBP |
| VAT | Obesity | ENSG0000013 | -0.00099827 | 0.808113508 | 0.004110466 | 0.183454452 | -0.00031366 | 0.939351998 | 0.004122577 | 0.183417544 | -0.00068461 | 0.033001942 | 1 | 0.921269427 | 1 | 0.980488158 | ENSG0000023 | 7 | TAPBP |
| VAT | Obesity | ENSG0000013 | -0.00522546 | 0.015674044 | 0.002162474 | 0.183452267 | -0.00525425 | 0.015101812 | 0.0021623 | 0.183415315 | 2.88E-05 | 0.035311518 | 1 | 0.103365207 | 1 | 0.107059606 | ENSG0000023 | 7 | TAPBP |
| VAT | Obesity | ENSG0000013 | 0.003181976 | 0.514169022 | 0.004877636 | 0.183454313 | 0.003061757 | 0.530160811 | 0.004877265 | 0.183417397 | 0.00012022 | 0.000441463 | 1 | 0.776929212 | 1 | 0.786996417 | ENSG0000023 | 7 | TAPBP |
| VAT | Obesity | ENSG0000013 | -0.00022747 | 0.967157413 | 0.005524533 | 0.183454473 | -0.00014519 | 0.979031025 | 0.005524121 | 0.183417546 | -8.23E-05 | 0.038970576 | 1 | 0.985729121 | 1 | 0.994336196 | ENSG0000023 | 7 | TAPBP |
| VAT | Obesity | ENSG0000013 | -0.00543617 | 0.08520846 | 0.003158293 | 0.183453354 | -0.00536096 | 0.089599222 | 0.003158117 | 0.183416457 | -7.52E-05 | 0.011986007 | 1 | 0.301506858 | 1 | 0.322837926 | ENSG0000023 | 7 | TAPBP |
| VAT | Obesity | ENSG0000013 | -0.00168163 | 0.168268153 | 0.001220525 | 0.183453757 | -0.00165227 | 0.175792555 | 0.001220443 | 0.183416853 | -2.94E-05 | 0.00316227 | 1 | 0.450948512 | 1 | 0.474743104 | ENSG0000023 | 3 | RP11-395L14.4 |
| VAT | Obesity | ENSG0000014 | -0.01138792 | 0.022808393 | 0.005002081 | 0.183452515 | -0.01152394 | 0.021223989 | 0.005001747 | 0.18341554 | 0.000136023 | 0.000960731 | 1 | 0.132808366 | 1 | 0.132256412 | ENSG0000023 | 2 | SCAND3 |
| VAT | Obesity | ENSG0000014 | -0.00133863 | 0.414800111 | 0.001641529 | 0.183454223 | -0.00126674 | 0.44027109 | 0.001641421 | 0.183417321 | -7.19E-05 | 1.56E-07 | 1 | 0.709763365 | 1 | 0.742750586 | ENSG0000023 | 2 | SCAND3 |
| VAT | Obesity | ENSG0000014 | 0.003303608 | 0.536777271 | 0.00534829 | 0.18345433 | 0.00339236 | 0.525860818 | 0.005347878 | 0.183417394 | -8.88E-05 | 0.015607071 | 1 | 0.78620379 | 1 | 0.785965627 | ENSG0000023 | 2 | SCAND3 |
| VAT | Obesity | ENSG0000014 | 0.000944135 | 0.800237595 | 0.003731171 | 0.18345445 | 0.001048185 | 0.778754109 | 0.003730952 | 0.183417516 | -0.00010405 | 0.002267505 | 1 | 0.918665124 | 1 | 0.910529706 | ENSG0000023 | 2 | SCAND3 |
| VAT | Obesity | ENSG0000014 | -0.00912286 | 0.071238146 | 0.005057126 | 0.183453244 | -0.00899648 | 0.075224238 | 0.005056762 | 0.18341635 | -0.00012638 | 0.000922361 | 1 | 0.274916222 | 1 | 0.291599575 | ENSG0000023 | 2 | AC009784.3 |
| VAT | Obesity | ENSG0000014 | 0.001527903 | 0.581731303 | 0.002773678 | 0.183454359 | 0.001483049 | 0.59283787 | 0.002773463 | 0.183417438 | 4.49E-05 | 0.016967413 | 1 | 0.810488842 | 1 | 0.825129864 | ENSG0000023 | 1 | RP11-307E17.8 |
| VAT | Obesity | ENSG0000014 | -0.0007541 | 0.762642007 | 0.002496918 | 0.18345444 | -0.00070841 | 0.776615967 | 0.002496747 | 0.183417516 | -4.57E-05 | 0.021870136 | 1 | 0.899221841 | 1 | 0.909787806 | ENSG0000023 | 1 | MRPL35P2 |
| VAT | Obesity | ENSG0000014 | -0.00464993 | 0.357194312 | 0.005050295 | 0.183454154 | -0.00457593 | 0.36486108 | 0.005049908 | 0.183417236 | -7.40E-05 | 0.034581094 | 1 | 0.668015266 | 1 | 0.688411445 | ENSG0000023 | 1 | MRPL35P2 |
| VAT | Obesity | ENSG0000014 | -0.03721244 | 1.54E-16 | 0.004508587 | 0.183428727 | -0.03709417 | 1.91E-16 | 0.004508325 | 0.183391964 | -0.00011827 | 0.004264177 | 4.25E-13 | 3.04E-14 | 5.27E-13 | 3.76E-14 | ENSG0000023 | 1 | MRPL35P2 |
| VAT | Obesity | ENSG0000014 | -0.00451902 | 0.37293082 | 0.005071862 | 0.183454174 | -0.00439728 | 0.385914418 | 0.005071533 | 0.183417262 | -0.00012174 | 0.004541638 | 1 | 0.68300535 | 1 | 0.707997923 | ENSG0000023 | 3 | TMA7 |
| VAT | Obesity | ENSG0000014 | -0.00256689 | 0.67532553 | 0.006128423 | 0.183454408 | -0.00245087 | 0.689196378 | 0.006127997 | 0.183417486 | -0.00011603 | 0.016188677 | 1 | 0.860416532 | 1 | 0.869882588 | ENSG0000023 | 1 | PHBP15 |
| VAT | Obesity | ENSG0000014 | 0.001417706 | 0.246818305 | 0.001224152 | 0.183453967 | 0.001390544 | 0.255959865 | 0.001224077 | 0.183417058 | 2.72E-05 | 0.013088206 | 1 | 0.543398386 | 1 | 0.566137648 | ENSG0000023 | 2 | LINC00493 |
| VAT | Obesity | ENSG0000014 | 0.005386238 | 0.350361261 | 0.005767531 | 0.183454144 | 0.007436393 | 0.198620171 | 0.005784851 | 0.183416922 | -0.00205016 | 6.55E-06 | 1 | 0.661582235 | 1 | 0.51015583 | ENSG0000023 | 2 | LINC00493 |
| VAT | Obesity | ENSG0000014 | -0.00045097 | 0.941562512 | 0.006151858 | 0.183454472 | -0.0005741 | 0.9256435 | 0.006151443 | 0.183417543 | 0.000123129 | 0.014003747 | 1 | 0.981016434 | 1 | 0.974118595 | ENSG0000023 | 2 | LINC00493 |
| VAT | Obesity | ENSG0000014 | -0.00257041 | 0.164673984 | 0.001849846 | 0.183453744 | -0.00254261 | 0.169257116 | 0.001849711 | 0.183416832 | -2.78E-05 | 0.042803476 | 1 | 0.445152005 | 1 | 0.463442103 | ENSG0000023 | 2 | LINC00493 |
| VAT | Obesity | ENSG0000014 | -0.00076052 | 0.737319089 | 0.002267473 | 0.183454432 | -0.00082245 | 0.716798243 | 0.002267316 | 0.183417496 | 6.19E-05 | 0.000565463 | 1 | 0.887171413 | 1 | 0.880054782 | ENSG0000023 | 1 | RP4-730K3.3 |
| VAT | Obesity | ENSG0000014 | 0.001481703 | 0.515392917 | 0.002277917 | 0.183454314 | 0.001426902 | 0.53101807 | 0.002277748 | 0.183417398 | 5.48E-05 | 0.000867379 | 1 | 0.776929212 | 1 | 0.786996417 | ENSG0000023 | 1 | RP4-756H11.1 |
| VAT | Obesity | ENSG0000014 | 0.000479156 | 0.863294031 | 0.002782835 | 0.183454463 | 0.000420885 | 0.879775237 | 0.002782634 | 0.183417537 | 5.83E-05 | 0.005345829 | 1 | 0.94551251 | 1 | 0.958352832 | ENSG0000023 | 1 | RP11-458F8.1 |
| VAT | Obesity | ENSG0000014 | 0.00832192 | 0.001119516 | 0.002553794 | 0.18345046 | 0.008914991 | 0.000524442 | 0.002570662 | 0.183413001 | -0.00059307 | 0.045299527 | 1 | 0.014643911 | 1 | 0.008772485 | ENSG0000023 | 1 | RP11-458F8.1 |
| VAT | Obesity | ENSG0000014 | -0.00219022 | 0.120944287 | 0.001412294 | 0.183453565 | -0.00221664 | 0.116499969 | 0.001412198 | 0.183416615 | 2.64E-05 | 0.020642362 | 1 | 0.373803171 | 1 | 0.37563074 | ENSG0000023 | 2 | GS1-124K5.12 |
| VAT | Obesity | ENSG0000014 | 0.002793007 | 0.621609441 | 0.005658774 | 0.183454382 | 0.002905514 | 0.607611066 | 0.0056584 | 0.183417446 | -0.00011251 | 0.016739602 | 1 | 0.833933632 | 1 | 0.834474842 | ENSG0000023 | 2 | GTF2IP1 |
| VAT | Obesity | ENSG0000014 | 0.005366383 | 0.13816419 | 0.003619418 | 0.183453643 | 0.005306171 | 0.142614533 | 0.003619177 | 0.183416734 | 6.02E-05 | 0.044243695 | 1 | 0.399279871 | 1 | 0.418027797 | ENSG0000023 | 2 | GTF2IP1 |
| VAT | Obesity | ENSG0000014 | 0.002709921 | 0.526530976 | 0.004278974 | 0.183454322 | 0.002784302 | 0.515213139 | 0.004278655 | 0.183417386 | -7.44E-05 | 0.015819364 | 1 | 0.779823873 | 1 | 0.784425829 | ENSG0000023 | 2 | GTF2IP1 |
| VAT | Obesity | ENSG0000014 | -0.00989854 | 0.056820429 | 0.005196917 | 0.183453103 | -0.01000739 | 0.05413257 | 0.005196543 | 0.183416144 | 0.000108855 | 0.005682233 | 1 | 0.24109463 | 1 | 0.24496821 | ENSG0000023 | 4 | CTD-2012J19.1 |
| VAT | Obesity | ENSG0000014 | -0.00532208 | 0.326494144 | 0.005424046 | 0.18345411 | -0.00520361 | 0.337344104 | 0.005423666 | 0.183417198 | -0.00011847 | 0.005163116 | 1 | 0.634310581 | 1 | 0.659397824 | ENSG0000023 | 4 | CTD-2012J19.1 |
| VAT | Obesity | ENSG0000014 | 0.007904673 | 0.123983135 | 0.005138675 | 0.18345358 | 0.007513225 | 0.143811154 | 0.005139894 | 0.183416739 | 0.000391448 | 0.003367741 | 1 | 0.37657343 | 1 | 0.419132824 | ENSG0000023 | 4 | CTD-2012J19.1 |
| VAT | Obesity | ENSG0000014 | -0.00272878 | 0.576238575 | 0.004882515 | 0.183454356 | -0.00160751 | 0.742662741 | 0.004896039 | 0.183417505 | -0.00112126 | 0.002454073 | 1 | 0.806947626 | 1 | 0.893915902 | ENSG0000023 | 4 | CTD-2012J19.1 |
| VAT | Obesity | ENSG0000014 | -0.0037451 | 0.398192343 | 0.004432837 | 0.183454204 | -0.0040256 | 0.363853694 | 0.004433245 | 0.183417234 | 0.000280492 | 0.001269576 | 1 | 0.703592104 | 1 | 0.687833011 | ENSG0000023 | 4 | CTD-2012J19.1 |
| VAT | Obesity | ENSG0000014 | -0.00843471 | 0.132300678 | 0.005604111 | 0.183453618 | -0.00851877 | 0.128460822 | 0.005603707 | 0.183416673 | 8.41E-05 | 0.046996467 | 1 | 0.388871002 | 1 | 0.395731266 | ENSG0000023 | 4 | HLA-DQB2 |
| VAT | Obesity | ENSG0000014 | -0.00311973 | 0.183470604 | 0.002345406 | 0.183453805 | -0.00303367 | 0.195840911 | 0.002345331 | 0.183416914 | -8.61E-05 | 0.001770315 | 1 | 0.477265662 | 1 | 0.508008379 | ENSG0000023 | 4 | HLA-DQB2 |
| VAT | Obesity | ENSG0000014 | -0.00317741 | 0.525274896 | 0.005001941 | 0.183454322 | -0.00332708 | 0.505919812 | 0.005001587 | 0.183417379 | 0.000149667 | 0.000108619 | 1 | 0.779823873 | 1 | 0.782701055 | ENSG0000023 | 4 | HLA-DQB2 |
| VAT | Obesity | ENSG0000014 | 0.005609101 | 0.346006214 | 0.005952153 | 0.183454138 | 0.004099303 | 0.491888798 | 0.005964286 | 0.183417368 | 0.001509798 | 0.000105952 | 1 | 0.657697762 | 1 | 0.775358255 | ENSG0000023 | 4 | HLA-DQB2 |
| VAT | Obesity | ENSG0000014 | -0.00177919 | 0.77196279 | 0.006139198 | 0.183454442 | -0.00191801 | 0.754705337 | 0.006138799 | 0.183417509 | 0.000138822 | 0.007366051 | 1 | 0.904559068 | 1 | 0.898976285 | ENSG0000023 | 4 | HLA-DQB2 |
| VAT | Obesity | ENSG0000014 | -0.0014806 | 0.513435554 | 0.002265659 | 0.183454313 | -0.00151857 | 0.50266798 | 0.002265507 | 0.183417376 | 3.80E-05 | 0.040742958 | 1 | 0.776692113 | 1 | 0.778980138 | ENSG0000023 | 3 | WI2-3658N16.1 |
| VAT | Obesity | ENSG0000014 | -0.01526274 | 0.001632529 | 0.00484524 | 0.183450723 | -0.01517069 | 0.001740586 | 0.004844889 | 0.18341384 | -9.21E-05 | 0.011445396 | 1 | 0.019590343 | 1 | 0.022554068 | ENSG0000023 | 4 | GOLGA8N |
| VAT | Obesity | ENSG0000014 | 0.002901396 | 0.404276868 | 0.003478859 | 0.183454211 | 0.002814375 | 0.418483462 | 0.003478593 | 0.183417299 | 8.70E-05 | 0.000310791 | 1 | 0.707119945 | 1 | 0.725782338 | ENSG0000023 | 4 | GOLGA8N |
| VAT | Obesity | ENSG0000014 | 0.001110876 | 0.597128042 | 0.002101802 | 0.183454369 | 0.001043718 | 0.619460783 | 0.002101664 | 0.183417453 | 6.72E-05 | 0.000135703 | 1 | 0.819937013 | 1 | 0.8401532 | ENSG0000023 | 4 | GOLGA8N |
| VAT | Obesity | ENSG0000014 | 0.001235689 | 0.793162115 | 0.004712652 | 0.183454448 | 0.00131591 | 0.780052964 | 0.004712297 | 0.183417517 | -8.02E-05 | 0.016653457 | 1 | 0.914040683 | 1 | 0.911107144 | ENSG0000023 | 2 | LINC00665 |
| VAT | Obesity | ENSG0000014 | 0.005446036 | 0.320776365 | 0.005485174 | 0.183454101 | 0.005581823 | 0.308825426 | 0.005484793 | 0.183417155 | -0.00013579 | 0.001724604 | 1 | 0.628794579 | 1 | 0.630590092 | ENSG0000023 | 2 | LINC00665 |
| VAT | Obesity | ENSG0000014 | -0.00296436 | 0.580111615 | 0.00535836 | 0.183454358 | -0.00287771 | 0.591206254 | 0.005357969 | 0.183417437 | -8.67E-05 | 0.029428811 | 1 | 0.809049044 | 1 | 0.824941855 | ENSG0000023 | 2 | LINC00665 |
| VAT | Obesity | ENSG0000014 | -0.00491528 | 0.194452122 | 0.003788207 | 0.183453838 | -0.00503253 | 0.183989614 | 0.003787924 | 0.183416879 | 0.000117248 | 1.71E-05 | 1 | 0.492826316 | 1 | 0.489908029 | ENSG0000023 | 5 | LYRM9 |
| VAT | Obesity | ENSG0000014 | -0.00617684 | 0.131574111 | 0.00409625 | 0.183453615 | -0.00607375 | 0.138110647 | 0.004095956 | 0.183416715 | -0.00010309 | 0.000919601 | 1 | 0.387974943 | 1 | 0.412910374 | ENSG0000023 | 1 | AC000078.5 |
| VAT | Obesity | ENSG0000014 | -0.00172958 | 0.639430653 | 0.003691778 | 0.183454391 | -0.00163776 | 0.657296328 | 0.003691573 | 0.183417472 | -9.18E-05 | 0.008814613 | 1 | 0.841849822 | 1 | 0.854766662 | ENSG0000023 | 1 | AC000078.5 |
| VAT | Obesity | ENSG0000014 | -0.01227195 | 0.000191001 | 0.003289486 | 0.183449213 | -0.01085074 | 0.00099774 | 0.003296911 | 0.183413452 | -0.00142122 | 3.12E-10 | 0.527162594 | 0.003738742 | 1 | 0.015047886 | ENSG0000023 | 1 | AC000078.5 |
| VAT | Obesity | ENSG0000014 | 0.001219264 | 0.574588471 | 0.002172187 | 0.183454355 | 0.001186404 | 0.584915019 | 0.002172026 | 0.183417433 | 3.29E-05 | 0.037555099 | 1 | 0.806905296 | 1 | 0.820551558 | ENSG0000023 | 1 | AC000078.5 |
| VAT | Obesity | ENSG0000014 | 0.005169868 | 0.364913739 | 0.005705988 | 0.183454164 | 0.005090123 | 0.372321168 | 0.005705554 | 0.183417245 | 7.97E-05 | 0.046094969 | 1 | 0.674137831 | 1 | 0.695586169 | ENSG0000023 | 1 | AC067959.1 |

| | | | | | | | | | | | | | | | | | | | |
|---|---|---|---|---|---|---|---|---|---|---|---|---|---|---|---|---|---|---|---|
| VAT | Obesity | ENSG0000014 | -0.007366 | 0.095928295 | 0.004424221 | 0.183453426 | -0.00747969 | 0.090888033 | 0.004423925 | 0.183416466 | 0.000113688 | 0.001798702 | 1 | 0.326866783 | 1 | 0.32624827 | ENSG0000023 | 1 | AC067959.1 |
| VAT | Obesity | ENSG0000014 | 0.006961019 | 0.04496242 | 0.003471811 | 0.183452955 | 0.007065824 | 0.041817167 | 0.003471569 | 0.18341598 | -0.00010481 | 0.000128084 | 1 | 0.209268597 | 1 | 0.205736949 | ENSG0000023 | 1 | MICF |
| VAT | Obesity | ENSG0000014 | -0.00292131 | 0.378645712 | 0.003318186 | 0.183454181 | -0.00286273 | 0.388246864 | 0.003317944 | 0.183417265 | -5.86E-05 | 0.017548774 | 1 | 0.687088866 | 1 | 0.708846095 | ENSG0000023 | 1 | MICF |
| VAT | Obesity | ENSG0000014 | 0.007275592 | 0.000440618 | 0.002070163 | 0.183449805 | 0.007243889 | 0.000466278 | 0.002070014 | 0.183412918 | 3.17E-05 | 0.043250346 | 1 | 0.007102966 | 1 | 0.00819516 | ENSG0000023 | 1 | MICF |
| VAT | Obesity | ENSG0000014 | -0.00173899 | 0.146346795 | 0.001197189 | 0.183453677 | -0.0017632 | 0.140779954 | 0.001197099 | 0.183416726 | 2.42E-05 | 0.005073206 | 1 | 0.413425951 | 1 | 0.417278146 | ENSG0000023 | 1 | MICF |
| VAT | Obesity | ENSG0000014 | -0.0035393 | 0.460942682 | 0.004800382 | 0.183454269 | -0.00345566 | 0.47157152 | 0.004800028 | 0.18341735 | -8.36E-05 | 0.01747579 | 1 | 0.744619218 | 1 | 0.766963697 | ENSG0000023 | 1 | MICF |
| VAT | Obesity | ENSG0000014 | 0.001529172 | 0.638711333 | 0.003257011 | 0.183454391 | 0.001454417 | 0.655181392 | 0.003256819 | 0.183417471 | 7.48E-05 | 0.012151615 | 1 | 0.841849822 | 1 | 0.854180748 | ENSG0000023 | 1 | MICF |
| VAT | Obesity | ENSG0000014 | 0.000493479 | 0.926687547 | 0.005363125 | 0.183454471 | 0.000353092 | 0.947504776 | 0.005362826 | 0.183417544 | 0.000140387 | 0.005748913 | 1 | 0.976170888 | 1 | 0.981754039 | ENSG0000023 | 1 | MICF |
| VAT | Obesity | ENSG0000014 | -0.01007165 | 0.01093075 | 0.003957561 | 0.183452026 | -0.01019455 | 0.009992262 | 0.003957347 | 0.183415038 | 0.000122895 | 0.001320375 | 1 | 0.079183388 | 1 | 0.079124756 | ENSG0000023 | 1 | MICF |
| VAT | Obesity | ENSG0000015 | -0.0034656 | 0.444097181 | 0.004528461 | 0.183454253 | -0.00407333 | 0.368756504 | 0.00453194 | 0.183417241 | 0.000607733 | 0.001288949 | 1 | 0.729116989 | 1 | 0.69094905 | ENSG0000023 | 4 | RASA4DP |
| VAT | Obesity | ENSG0000015 | 0.005315696 | 0.028050238 | 0.002419977 | 0.18345265 | 0.005271185 | 0.029381521 | 0.002419822 | 0.183415753 | 4.45E-05 | 0.031531697 | 1 | 0.152430349 | 1 | 0.163493946 | ENSG0000023 | 4 | RASA4DP |
| VAT | Obesity | ENSG0000015 | 0.001468017 | 0.574461927 | 0.00261449 | 0.183454355 | 0.001420708 | 0.586828471 | 0.002614297 | 0.183417434 | 4.73E-05 | 0.013349137 | 1 | 0.806905296 | 1 | 0.822155624 | ENSG0000023 | 6 | USP32P2 |
| VAT | Obesity | ENSG0000015 | 0.00188829 | 0.681662846 | 0.004603393 | 0.183454411 | 0.001348609 | 0.769747785 | 0.0046074 | 0.183417514 | 0.000539682 | 0.007816451 | 1 | 0.863333733 | 1 | 0.907738428 | ENSG0000023 | 3 | RP11-458D21.1 |
| VAT | Obesity | ENSG0000015 | 0.004078411 | 0.00189479 | 0.001312964 | 0.183450827 | 0.004053569 | 0.002018203 | 0.001312873 | 0.183413943 | 2.48E-05 | 0.016289057 | 1 | 0.021521074 | 1 | 0.024430883 | ENSG0000023 | 2 | PMS2P9 |
| VAT | Obesity | ENSG0000015 | 0.002557211 | 0.671270974 | 0.006025408 | 0.183454406 | 0.002414686 | 0.688585874 | 0.006025039 | 0.183417485 | 0.000142525 | 0.007608845 | 1 | 0.857647266 | 1 | 0.869882588 | ENSG0000023 | 2 | PMS2P9 |
| VAT | Obesity | ENSG0000015 | -0.00387022 | 0.209207486 | 0.003081998 | 0.183453878 | -0.00285009 | 0.357105742 | 0.003094909 | 0.183417226 | -0.00102013 | 0.000356735 | 1 | 0.509182242 | 1 | 0.683029694 | ENSG0000023 | 2 | PMS2P9 |
| VAT | Obesity | ENSG0000015 | -0.00024745 | 0.894405482 | 0.001864299 | 0.183454467 | -0.00020775 | 0.911264313 | 0.001864165 | 0.183417541 | -3.97E-05 | 0.004896964 | 1 | 0.962401221 | 1 | 0.971531422 | ENSG0000023 | 2 | PMS2P9 |
| VAT | Obesity | ENSG0000015 | -0.00068372 | 0.522124261 | 0.001068187 | 0.183454319 | -0.00072976 | 0.494475415 | 0.001068142 | 0.18341737 | 4.60E-05 | 7.29E-05 | 1 | 0.778563922 | 1 | 0.775358255 | ENSG0000023 | 2 | PMS2P9 |
| VAT | Obesity | ENSG0000015 | 0.000404495 | 0.875309555 | 0.002577751 | 0.183454465 | 0.000353774 | 0.890831405 | 0.002577545 | 0.183417539 | 5.07E-05 | 0.002243048 | 1 | 0.951543054 | 1 | 0.965708829 | ENSG0000023 | 1 | RP11-500B12.1 |
| VAT | Obesity | ENSG0000015 | -0.01436483 | 0.006213033 | 0.005249663 | 0.183451644 | -0.01422216 | 0.006741471 | 0.00524926 | 0.183414772 | -0.00014267 | 7.93E-05 | 1 | 0.051797881 | 1 | 0.059827845 | ENSG0000023 | 1 | RP11-500B12.1 |
| VAT | Obesity | ENSG0000015 | -0.00253083 | 0.674220245 | 0.006020582 | 0.183454407 | -0.00242862 | 0.686642106 | 0.006020143 | 0.183417485 | -0.00010221 | 0.022584317 | 1 | 0.859911219 | 1 | 0.869882588 | ENSG0000023 | 1 | RP11-153K11.3 |
| VAT | Obesity | ENSG0000015 | 0.004793221 | 0.023901107 | 0.002122094 | 0.183452546 | -0.00082623 | 0.742655609 | 0.002516395 | 0.183417505 | 0.005619451 | 3.26E-05 | 1 | 0.136014549 | 1 | 0.893915902 | ENSG0000023 | 3 | RP11-62H7.2 |
| VAT | Obesity | ENSG0000015 | -0.00147451 | 0.708196648 | 0.00393961 | 0.183454421 | -0.00158747 | 0.686962588 | 0.003939318 | 0.183417485 | 0.000112954 | 8.15E-05 | 1 | 0.872628294 | 1 | 0.869882588 | ENSG0000023 | 3 | RP11-62H7.2 |
| VAT | Obesity | ENSG0000015 | 0.009450462 | 0.001202623 | 0.002918362 | 0.18345051 | 0.008905953 | 0.002358135 | 0.002928635 | 0.183414051 | 0.00054451 | 0.028448951 | 1 | 0.015501716 | 1 | 0.027933277 | ENSG0000023 | 3 | RP11-62H7.2 |
| VAT | Obesity | ENSG0000015 | 0.007955296 | 0.062435412 | 0.004269706 | 0.183453162 | 0.008022346 | 0.060238688 | 0.00426937 | 0.183416212 | -6.71E-05 | 0.017780972 | 1 | 0.255670231 | 1 | 0.25801405 | ENSG0000023 | 1 | EBAG9P1 |
| VAT | Obesity | ENSG0000015 | -0.00296742 | 0.108892891 | 0.001850946 | 0.183453503 | 1.95E-05 | 0.992089784 | 0.001969938 | 0.183417546 | -0.00298695 | 9.57E-06 | 1 | 0.355343638 | 1 | 0.99714778 | ENSG0000023 | 1 | AP001628.7 |
| VAT | Obesity | ENSG0000015 | 0.010834643 | 0.062904 | 0.005825473 | 0.183453167 | 0.010701631 | 0.06619059 | 0.005825174 | 0.18341627 | 0.000133012 | 0.021535162 | 1 | 0.257207467 | 1 | 0.27337482 | ENSG0000023 | 1 | AP001628.7 |
| VAT | Obesity | ENSG0000015 | -0.00315557 | 0.208656183 | 0.002509855 | 0.183453877 | -0.0032304 | 0.198034715 | 0.002509687 | 0.18341692 | 7.48E-05 | 0.000287325 | 1 | 0.508805001 | 1 | 0.509865497 | ENSG0000023 | 2 | RP11-535M15.2 |
| VAT | Obesity | ENSG0000015 | 0.003289751 | 0.231078224 | 0.002746982 | 0.183453932 | 0.003243993 | 0.237595634 | 0.002746774 | 0.183417019 | 4.58E-05 | 0.018001946 | 1 | 0.52708752 | 1 | 0.544132965 | ENSG0000023 | 3 | RP11-255H23.2 |
| VAT | Obesity | ENSG0000015 | 0.000152358 | 0.943372205 | 0.002144913 | 0.183454472 | -0.00017876 | 0.933639291 | 0.002146873 | 0.183417543 | 0.000331121 | 0.000611648 | 1 | 0.981243858 | 1 | 0.977187882 | ENSG0000023 | 3 | RP11-255H23.2 |
| VAT | Obesity | ENSG0000015 | 0.006899516 | 0.083938167 | 0.003992122 | 0.183453345 | 0.006745851 | 0.091047535 | 0.003991862 | 0.183416467 | 0.000153664 | 4.70E-06 | 1 | 0.298928183 | 1 | 0.326336482 | ENSG0000023 | 3 | RP11-255H23.2 |
| VAT | Obesity | ENSG0000015 | 0.008166974 | 0.066386415 | 0.004448704 | 0.1834532 | 0.008085203 | 0.069131994 | 0.004448383 | 0.183416298 | 8.18E-05 | 0.014685892 | 1 | 0.264946219 | 1 | 0.277735522 | ENSG0000023 | 1 | PAIP1P1 |
| VAT | Obesity | ENSG0000015 | -0.00235082 | 0.060506533 | 0.001252374 | 0.183453142 | -0.0023308 | 0.062711208 | 0.001252285 | 0.183416237 | -2.00E-05 | 0.037576711 | 1 | 0.25037186 | 1 | 0.263845936 | ENSG0000023 | 1 | PAIP1P1 |
| VAT | Obesity | ENSG0000015 | -0.00697618 | 0.049216215 | 0.003547117 | 0.183453012 | -0.00690583 | 0.05153339 | 0.003546879 | 0.183416113 | -7.04E-05 | 0.015386357 | 1 | 0.22051421 | 1 | 0.238644559 | ENSG0000023 | 1 | PAIP1P1 |
| VAT | Obesity | ENSG0000015 | 0.003362976 | 0.491800349 | 0.004891966 | 0.183454295 | 0.00346174 | 0.479140041 | 0.004891627 | 0.183417357 | -9.88E-05 | 0.010658139 | 1 | 0.766182814 | 1 | 0.774198351 | ENSG0000023 | 1 | PAIP1P1 |
| VAT | Obesity | ENSG0000015 | 0.010759815 | 0.050381404 | 0.005498914 | 0.183453027 | 0.010339075 | 0.060191252 | 0.00550128 | 0.183416211 | 0.00042074 | 0.018845449 | 1 | 0.224640832 | 1 | 0.25801405 | ENSG0000023 | 1 | ZDHHC20P1 |
| VAT | Obesity | ENSG0000015 | -0.00447376 | 0.173127332 | 0.003284154 | 0.183453773 | -0.004555 | 0.165429517 | 0.003283965 | 0.183416819 | 8.12E-05 | 0.007691291 | 1 | 0.460092631 | 1 | 0.456129337 | ENSG0000023 | 1 | ZDHHC20P1 |
| VAT | Obesity | ENSG0000015 | -0.0143296 | 0.000448894 | 0.004083023 | 0.183449818 | -0.0144806 | 0.000390016 | 0.004082752 | 0.183412792 | 0.000150996 | 8.83E-06 | 1 | 0.007148695 | 1 | 0.007035575 | ENSG0000023 | 1 | ZDHHC20P1 |
| VAT | Obesity | ENSG0000015 | 0.002485334 | 0.44397803 | 0.003246708 | 0.183454253 | 0.002549036 | 0.432363281 | 0.00324654 | 0.183417313 | -6.37E-05 | 0.047741035 | 1 | 0.729116989 | 1 | 0.737081891 | ENSG0000023 | 1 | ZDHHC20P1 |
| VAT | Obesity | ENSG0000015 | 0.005464432 | 0.305387359 | 0.005331414 | 0.183454077 | 0.005267237 | 0.323136203 | 0.005331025 | 0.183417177 | 0.000197196 | 5.96E-07 | 1 | 0.613888646 | 1 | 0.643939293 | ENSG0000023 | 1 | ZDHHC20P1 |
| VAT | Obesity | ENSG0000015 | 0.002751706 | 0.368792855 | 0.003061746 | 0.183454169 | 0.002853313 | 0.351340381 | 0.003061516 | 0.183417218 | -0.00010161 | 3.50E-06 | 1 | 0.676520736 | 1 | 0.674808247 | ENSG0000023 | 1 | AL133493.2 |
| VAT | Obesity | ENSG0000015 | -0.00118581 | 0.735848657 | 0.003515 | 0.183454431 | -0.00113208 | 0.747381433 | 0.003514746 | 0.183417507 | -5.37E-05 | 0.042002543 | 1 | 0.886561612 | 1 | 0.89626095 | ENSG0000023 | 4 | TRIM26 |
| VAT | Obesity | ENSG0000015 | -0.00034633 | 0.946922587 | 0.00520231 | 0.183454472 | -0.00048287 | 0.926043453 | 0.005201968 | 0.183417543 | 0.00013654 | 0.001669454 | 1 | 0.981782998 | 1 | 0.974118595 | ENSG0000023 | 4 | TRIM26 |
| VAT | Obesity | ENSG0000015 | 0.007752502 | 0.157212667 | 0.005480695 | 0.183453718 | 0.007883865 | 0.150272453 | 0.005480341 | 0.183416764 | -0.00013136 | 0.004748116 | 1 | 0.432609133 | 1 | 0.42846278 | ENSG0000023 | 4 | TRIM26 |
| VAT | Obesity | ENSG0000015 | -0.00070092 | 0.86733301 | 0.004195956 | 0.183454464 | -0.0007723 | 0.853955581 | 0.004195625 | 0.183417533 | 7.14E-05 | 0.009885258 | 1 | 0.948025973 | 1 | 0.947822399 | ENSG0000023 | 4 | TRIM26 |
| VAT | Obesity | ENSG0000015 | -0.00182745 | 0.591039824 | 0.003400976 | 0.183454365 | -0.00197501 | 0.561404393 | 0.00340075 | 0.183417419 | 0.000147563 | 1.58E-07 | 1 | 0.818499706 | 1 | 0.809555852 | ENSG0000023 | 4 | TRIM26 |
| VAT | Obesity | ENSG0000015 | 0.00054481 | 0.849842529 | 0.002877729 | 0.183454461 | 0.000497486 | 0.862740367 | 0.002877522 | 0.183417535 | 4.73E-05 | 0.030547482 | 1 | 0.941875299 | 1 | 0.950564236 | ENSG0000023 | 4 | TRIM26 |
| VAT | Obesity | ENSG0000015 | -0.00057131 | 0.875701118 | 0.003652353 | 0.183454465 | -0.00069083 | 0.849974372 | 0.003652259 | 0.183417533 | 0.000119524 | 0.007492431 | 1 | 0.951543054 | 1 | 0.945557947 | ENSG0000023 | 4 | TRIM26 |
| VAT | Obesity | ENSG0000016 | -0.00455741 | 0.323827036 | 0.004619195 | 0.183454106 | -0.0046505 | 0.314006885 | 0.004618856 | 0.183417163 | 9.31E-05 | 0.006419931 | 1 | 0.633061352 | 1 | 0.636904895 | ENSG0000023 | 4 | TRIM26 |
| VAT | Obesity | ENSG0000016 | 0.000507825 | 0.867662328 | 0.003047648 | 0.183454464 | 0.000451326 | 0.882262548 | 0.003047407 | 0.183417538 | 5.65E-05 | 0.004764056 | 1 | 0.948025973 | 1 | 0.959812626 | ENSG0000023 | 4 | TRIM26 |
| VAT | Obesity | ENSG0000016 | -0.0047497 | 0.030430221 | 0.002194411 | 0.183452703 | -0.00478874 | 0.029081619 | 0.002194268 | 0.183415746 | 3.90E-05 | 0.034717749 | 1 | 0.160281317 | 1 | 0.162152058 | ENSG0000023 | 4 | TRIM26 |
| VAT | Obesity | ENSG0000016 | -0.00185731 | 0.518445238 | 0.002876237 | 0.183454316 | -0.00195056 | 0.497677763 | 0.002876301 | 0.183417372 | 9.32E-05 | 0.038492184 | 1 | 0.778044231 | 1 | 0.776039901 | ENSG0000023 | 4 | TRIM26 |
| VAT | Obesity | ENSG0000016 | 0.000583089 | 0.608293144 | 0.001137708 | 0.183454375 | 0.000553623 | 0.626509026 | 0.001137628 | 0.183417457 | 2.95E-05 | 0.00082346 | 1 | 0.828767095 | 1 | 0.842487235 | ENSG0000023 | 4 | TRIM26 |
| VAT | Obesity | ENSG0000016 | -0.00302522 | 0.068090022 | 0.001658186 | 0.183453216 | -0.00300112 | 0.070293261 | 0.001658057 | 0.183416308 | -2.41E-05 | 0.030767519 | 1 | 0.26846923 | 1 | 0.281173044 | ENSG0000023 | 1 | RP3-460G2.2 |
| VAT | Obesity | ENSG0000016 | 0.007416922 | 0.206563529 | 0.005872125 | 0.183453871 | 0.007532895 | 0.199522475 | 0.005871702 | 0.183416924 | -0.00011597 | 0.009192209 | 1 | 0.507220054 | 1 | 0.510412237 | ENSG0000023 | 1 | RP3-460G2.2 |
| VAT | Obesity | ENSG0000016 | -0.00174282 | 0.607142692 | 0.003389661 | 0.183454374 | -0.0018041 | 0.594535092 | 0.003389398 | 0.183417439 | 6.13E-05 | 0.007940157 | 1 | 0.827969162 | 1 | 0.825553026 | ENSG0000023 | 1 | RP3-460G2.2 |
| VAT | Obesity | ENSG0000016 | -0.00641262 | 0.1268456 | 0.004200421 | 0.183453593 | -0.00651328 | 0.12097426 | 0.004200217 | 0.183416637 | 0.000100651 | 0.018988011 | 1 | 0.381781739 | 1 | 0.382826475 | ENSG0000023 | 1 | RP3-460G2.2 |
| VAT | Obesity | ENSG0000016 | -0.00809562 | 0.066541042 | 0.004412346 | 0.183453202 | -0.00801251 | 0.069361851 | 0.004412028 | 0.1834163 | -8.31E-05 | 0.012594739 | 1 | 0.264946219 | 1 | 0.278253938 | ENSG0000023 | 2 | RP11-353N4.5 |
| VAT | Obesity | ENSG0000016 | -0.0124099 | 0.00264616 | 0.004128209 | 0.183451058 | -0.01249006 | 0.002480624 | 0.004127977 | 0.183414086 | 8.02E-05 | 0.039604275 | 1 | 0.028230105 | 1 | 0.029010686 | ENSG0000023 | 2 | RP11-353N4.5 |
| VAT | Obesity | ENSG0000016 | 0.001441229 | 0.733127207 | 0.004226864 | 0.18345443 | 0.001362772 | 0.747128651 | 0.004226595 | 0.183417507 | 7.85E-05 | 0.030904245 | 1 | 0.884914569 | 1 | 0.89626095 | ENSG0000023 | 1 | RP11-797H7.1 |
| VAT | Obesity | ENSG0000016 | -0.0053758 | 0.154875485 | 0.003779061 | 0.183453709 | -0.00548029 | 0.146984259 | 0.003778808 | 0.183416751 | 0.000104489 | 0.000735359 | 1 | 0.428742567 | 1 | 0.423904445 | ENSG0000023 | 2 | AP000347.4 |
| VAT | Obesity | ENSG0000016 | -0.00102351 | 0.444504124 | 0.0013386 | 0.183454253 | -0.00104683 | 0.434159219 | 0.001338499 | 0.183417315 | 2.33E-05 | 0.014564022 | 1 | 0.729116989 | 1 | 0.737402735 | ENSG0000023 | 4 | AC006547.14 |
| VAT | Obesity | ENSG0000016 | 0.007377056 | 0.110611157 | 0.004623794 | 0.183453512 | 0.007243394 | 0.11719295 | 0.004623439 | 0.183416618 | 0.000133662 | 2.64E-05 | 1 | 0.357478681 | 1 | 0.376596898 | ENSG0000023 | 4 | AC006547.14 |
| VAT | Obesity | ENSG0000016 | 0.014745951 | 0.011004346 | 0.005799623 | 0.18345203 | 0.014839909 | 0.010499081 | 0.00579921 | 0.183415071 | -9.40E-05 | 0.035240638 | 1 | 0.079444329 | 1 | 0.081397369 | ENSG0000023 | 5 | ZNF736 |
| VAT | Obesity | ENSG0000016 | -0.00549298 | 0.299948102 | 0.005299314 | 0.183454068 | -0.00526777 | 0.320170066 | 0.005298982 | 0.183417173 | -0.00022521 | 1.09E-06 | 1 | 0.608848729 | 1 | 0.641269508 | ENSG0000023 | 5 | ZNF736 |
| VAT | Obesity | ENSG0000016 | -0.004068 | 0.429831303 | 0.005152748 | 0.183454239 | -0.00415614 | 0.419871032 | 0.005152368 | 0.1834173 | 8.81E-05 | 0.019571121 | 1 | 0.718120094 | 1 | 0.726092763 | ENSG0000023 | 5 | ZNF736 |
| VAT | Obesity | ENSG0000016 | -0.01664324 | 0.006053411 | 0.006063363 | 0.183451626 | -0.01653341 | 0.006392063 | 0.006062916 | 0.183414736 | -0.00010983 | 0.013578423 | 1 | 0.050937236 | 1 | 0.057279527 | ENSG0000023 | 5 | ZNF736 |
| VAT | Obesity | ENSG0000016 | -0.00875057 | 0.106324283 | 0.005418508 | 0.183453488 | -0.00884309 | 0.102651277 | 0.005418105 | 0.183416539 | 9.25E-05 | 0.018531122 | 1 | 0.35060337 | 1 | 0.353263747 | ENSG0000023 | 6 | PINLYP |
| VAT | Obesity | ENSG0000016 | 0.010363641 | 0.017784667 | 0.004372698 | 0.183452351 | 0.010460442 | 0.016741948 | 0.004372483 | 0.183415383 | -9.68E-05 | 0.029196115 | 1 | 0.110802893 | 1 | 0.113642834 | ENSG0000023 | 2 | CCT6P3 |
| VAT | Obesity | ENSG0000016 | 0.007059964 | 0.119435871 | 0.004533905 | 0.183453558 | 0.007128645 | 0.115856107 | 0.004533578 | 0.183416612 | -6.87E-05 | 0.045381016 | 1 | 0.372684435 | 1 | 0.37486853 | ENSG0000023 | 2 | CCT6P3 |
| VAT | Obesity | ENSG0000016 | 0.007351956 | 0.225286689 | 0.006063012 | 0.183453918 | 0.007441192 | 0.219671048 | 0.006062541 | 0.183416977 | -8.92E-05 | 0.030040518 | 1 | 0.52272452 | 1 | 0.531115411 | ENSG0000023 | 2 | CCT6P3 |
| VAT | Obesity | ENSG0000016 | 0.000511639 | 0.843951838 | 0.002599247 | 0.183454459 | 0.000470962 | 0.856207315 | 0.002599068 | 0.183417534 | 4.07E-05 | 0.048931983 | 1 | 0.938891035 | 1 | 0.948422961 | ENSG0000023 | 5 | HLA-B |
| VAT | Obesity | ENSG0000016 | -0.00484311 | 0.114472757 | 0.003068356 | 0.183453532 | -0.00327104 | 0.293244355 | 0.003112221 | 0.183417129 | -0.00157207 | 0.002622003 | 1 | 0.365254115 | 1 | 0.615229003 | ENSG0000023 | 5 | HLA-B |
| VAT | Obesity | ENSG0000016 | 0.005705534 | 0.04702461 | 0.002872756 | 0.183452983 | 0.005859642 | 0.041375915 | 0.002872728 | 0.183415974 | -0.00015411 | 6.92E-05 | 1 | 0.21488067 | 1 | 0.204683628 | ENSG0000023 | 5 | HLA-B |
| VAT | Obesity | ENSG0000016 | 0.021204101 | 1.71E-05 | 0.004931925 | 0.183447487 | 0.021509946 | 1.30E-05 | 0.004933608 | 0.183410362 | -0.00030585 | 0.037106966 | 0.04728667 | 0.000492569 | 0.035924051 | 0.000466546 | ENSG0000023 | 5 | HLA-B |
| VAT | Obesity | ENSG0000016 | -0.00131453 | 0.122134295 | 0.000850346 | 0.183453571 | -0.00129448 | 0.127911134 | 0.000850289 | 0.18341667 | -2.01E-05 | 0.004161552 | 1 | 0.374700593 | 1 | 0.395731266 | ENSG0000023 | 5 | HLA-B |
| VAT | Obesity | ENSG0000016 | -0.01434498 | 0.003400937 | 0.004897606 | 0.183451231 | -0.01445093 | 0.003169532 | 0.004897269 | 0.183414255 | 0.000105946 | 0.006823023 | 1 | 0.033404223 | 1 | 0.034440579 | ENSG0000023 | 5 | HLA-B |
| VAT | Obesity | ENSG0000016 | -0.00040585 | 0.930280836 | 0.004638708 | 0.183454471 | -0.00028584 | 0.95086337 | 0.004638519 | 0.183417545 | -0.00012001 | 0.018011067 | 1 | 0.976170888 | 1 | 0.983283215 | ENSG0000023 | 5 | HLA-B |
| VAT | Obesity | ENSG0000016 | -0.01312716 | 0.015600721 | 0.005428633 | 0.183452264 | -0.0130423 | 0.016275658 | 0.005428224 | 0.183415364 | -8.49E-05 | 0.027715651 | 1 | 0.10325657 | 1 | 0.111743324 | ENSG0000023 | 5 | HLA-B |
| VAT | Obesity | ENSG0000016 | -0.00395829 | 0.23398755 | 0.00332587 | 0.183453939 | -0.00361125 | 0.27767733 | 0.003326649 | 0.183417101 | -0.00034703 | 5.49E-05 | 1 | 0.529348883 | 1 | 0.597807668 | ENSG0000023 | 5 | HLA-B |
| VAT | Obesity | ENSG0000016 | -0.0017244 | 0.710881854 | 0.004652084 | 0.183454422 | -0.00182971 | 0.694073593 | 0.004651817 | 0.183417488 | 0.000105316 | 0.014921987 | 1 | 0.873568084 | 1 | 0.870280156 | ENSG0000023 | 1 | EIF3EP1 |
| VAT | Obesity | ENSG0000016 | -0.00159003 | 0.782682784 | 0.005764625 | 0.183454445 | -0.00169369 | 0.768891937 | 0.005764281 | 0.183417513 | 0.000103662 | 0.047235045 | 1 | 0.909245983 | 1 | 0.907673971 | ENSG0000023 | 1 | EIF3EP1 |
| VAT | Obesity | ENSG0000016 | 0.007034236 | 0.226137001 | 0.005811635 | 0.18345392 | 0.00694366 | 0.232135412 | 0.005811189 | 0.183417006 | 9.06E-05 | 0.024232608 | 1 | 0.52272452 | 1 | 0.540524906 | ENSG0000023 | 1 | EIF3EP1 |
| VAT | Obesity | ENSG0000016 | -0.00884946 | 0.059141187 | 0.004689334 | 0.183453128 | -0.00875945 | 0.061751889 | 0.004689021 | 0.183416227 | -9.00E-05 | 0.019536891 | 1 | 0.246943534 | 1 | 0.2614037 | ENSG0000023 | 1 | EIF3EP1 |
| VAT | Obesity | ENSG0000016 | -0.01073646 | 0.078956306 | 0.006111449 | 0.183453308 | -0.01057151 | 0.083645909 | 0.006110993 | 0.183416415 | -0.00016495 | 0.000182557 | 1 | 0.289017777 | 1 | 0.308446194 | ENSG0000023 | 1 | RP11-534L20.4 |
| VAT | Obesity | ENSG0000016 | 0.004408539 | 0.271222177 | 0.004006839 | 0.183454017 | 0.004504691 | 0.260872722 | 0.004006548 | 0.183417068 | -9.62E-05 | 0.001373782 | 1 | 0.575086695 | 1 | 0.574169626 | ENSG0000023 | 1 | RP11-439E19.7 |
| VAT | Obesity | ENSG0000016 | -0.0108008 | 0.065420819 | 0.005862428 | 0.183453191 | -0.01091646 | 0.062568975 | 0.005861983 | 0.183416235 | 0.000115661 | 0.005026397 | 1 | 0.264360266 | 1 | 0.263845936 | ENSG0000023 | 1 | AC010492.2 |
| VAT | Obesity | ENSG0000016 | -0.01852984 | 5.76E-06 | 0.004085926 | 0.1834467 | -0.01861344 | 5.22E-06 | 0.004085645 | 0.183409702 | 8.36E-05 | 0.010697325 | 0.015899387 | 0.000201258 | 0.014406061 | 0.000218274 | ENSG0000023 | 3 | ZSCAN31 |
| VAT | Obesity | ENSG0000016 | -0.00155105 | 0.453832972 | 0.00207072 | 0.183454262 | -0.00151133 | 0.465448918 | 0.002070588 | 0.183417345 | -3.97E-05 | 0.025493763 | 1 | 0.738859748 | 1 | 0.76239704 | ENSG0000023 | 3 | ZSCAN31 |
| VAT | Obesity | ENSG0000016 | 0.000535298 | 0.924006795 | 0.005611801 | 0.183454471 | 0.00090707 | 0.8716425 | 0.005614008 | 0.183417536 | -0.00037177 | 0.035069902 | 1 | 0.975615438 | 1 | 0.955921132 | ENSG0000023 | 3 | ZSCAN31 |
| VAT | Obesity | ENSG0000016 | -0.00098292 | 0.751771061 | 0.003107503 | 0.183454436 | -0.00231017 | 0.459100503 | 0.003120457 | 0.183417339 | 0.001327247 | 3.89E-06 | 1 | 0.892809005 | 1 | 0.756487993 | ENSG0000023 | 3 | ZSCAN31 |
| VAT | Obesity | ENSG0000016 | 0.001161492 | 0.749204877 | 0.003633203 | 0.183454435 | 0.001225079 | 0.735953845 | 0.003632915 | 0.183417503 | -6.36E-05 | 0.007452253 | 1 | 0.892799597 | 1 | 0.891658005 | ENSG0000023 | 3 | ZSCAN31 |
| VAT | Obesity | ENSG0000016 | -0.01577978 | 6.98E-05 | 0.003967809 | 0.183448496 | -0.01566342 | 7.89E-05 | 0.003967549 | 0.183411656 | -0.00011636 | 0.000452062 | 0.192689484 | 0.001632962 | 0.217643783 | 0.001981293 | ENSG0000023 | 3 | ZSCAN31 |
| VAT | Obesity | ENSG0000016 | -0.01949216 | 0.000291702 | 0.00538073 | 0.183449514 | -0.019023 | 0.000408121 | 0.005381633 | 0.183412824 | -0.00046916 | 0.00016504 | 0.805097424 | 0.005063506 | 1 | 0.007220609 | ENSG0000023 | 3 | ZSCAN31 |
| VAT | Obesity | ENSG0000016 | -0.00052439 | 0.679790783 | 0.001270471 | 0.18345441 | -0.0002649 | 0.834995898 | 0.001271741 | 0.18341753 | -0.00025948 | 1.34E-05 | 1 | 0.862732905 | 1 | 0.938350439 | ENSG0000023 | 3 | ZSCAN31 |
| VAT | Obesity | ENSG0000016 | 0.002839975 | 0.125595641 | 0.001854125 | 0.183453587 | 0.002812026 | 0.129331823 | 0.001853988 | 0.183416677 | 2.79E-05 | 0.038316805 | 1 | 0.37967576 | 1 | 0.396177394 | ENSG0000023 | 3 | ZSCAN31 |
| VAT | Obesity | ENSG0000016 | -5.58E-05 | 0.959396582 | 0.001096281 | 0.183454473 | -2.87E-05 | 0.979082738 | 0.001096195 | 0.183417546 | -2.71E-05 | 0.000206081 | 1 | 0.98546132 | 1 | 0.994336196 | ENSG0000023 | 3 | AP000351.10 |
| VAT | Obesity | ENSG0000016 | 0.007895591 | 0.123184287 | 0.005121861 | 0.183453576 | 0.008271184 | 0.106396092 | 0.005122717 | 0.183416561 | -0.00037559 | 0.001540757 | 1 | 0.37609362 | 1 | 0.361633427 | ENSG0000023 | 3 | AP000351.10 |
| VAT | Obesity | ENSG0000016 | 0.001422238 | 0.676274038 | 0.003406124 | 0.183454408 | 0.001557526 | 0.64747228 | 0.003406091 | 0.183417467 | -0.00013529 | 0.003204259 | 1 | 0.860416532 | 1 | 0.851774782 | ENSG0000023 | 1 | RP11-108M9.5 |
| VAT | Obesity | ENSG0000016 | -0.00085829 | 0.661858114 | 0.001962473 | 0.183454402 | -0.00090708 | 0.643909286 | 0.001962352 | 0.183417465 | 4.88E-05 | 0.004754403 | 1 | 0.854099711 | 1 | 0.849655254 | ENSG0000023 | 1 | RP11-108M9.5 |
| VAT | Obesity | ENSG0000016 | -0.0021153 | 0.450882156 | 0.002805646 | 0.183454259 | -0.00205958 | 0.462866755 | 0.002805444 | 0.183417342 | -5.57E-05 | 0.00867852 | 1 | 0.735047106 | 1 | 0.76223881 | ENSG0000023 | 1 | RP11-108M9.5 |
| VAT | Obesity | ENSG0000016 | -0.00762055 | 0.015692037 | 0.003154191 | 0.183452268 | -0.00755229 | 0.016641578 | 0.003153962 | 0.183415379 | -6.83E-05 | 0.003812318 | 1 | 0.103365207 | 1 | 0.113409271 | ENSG0000023 | 1 | HLA-W |
| VAT | Obesity | ENSG0000016 | -0.04070794 | 3.80E-16 | 0.004997839 | 0.1834294 | -0.04059158 | 4.58E-16 | 0.004997485 | 0.183392616 | -0.00011636 | 0.002453135 | 1.05E-12 | 6.55E-14 | 1.26E-12 | 7.90E-14 | ENSG0000023 | 1 | HLA-W |
| VAT | Obesity | ENSG0000016 | 0.000349264 | 0.928854855 | 0.003911758 | 0.183454471 | 0.003710105 | 0.358129064 | 0.004037389 | 0.183417227 | -0.00336084 | 0.00078543 | 1 | 0.976170888 | 1 | 0.683932408 | ENSG0000023 | 1 | HLA-W |
| VAT | Obesity | ENSG0000016 | -0.0006008 | 0.724748841 | 0.001706228 | 0.183454427 | -0.00069448 | 0.684008255 | 0.001706344 | 0.183417483 | 9.37E-05 | 0.002794144 | 1 | 0.881580785 | 1 | 0.869882588 | ENSG0000023 | 1 | HLA-W |
| VAT | Obesity | ENSG0000016 | -0.0027487 | 0.521961746 | 0.004292668 | 0.183454319 | -0.00265588 | 0.536083583 | 0.004292349 | 0.183417401 | -9.28E-05 | 0.002913821 | 1 | 0.778563922 | 1 | 0.791224967 | ENSG0000023 | 1 | HLA-W |
| VAT | Obesity | ENSG0000016 | -0.0021624 | 0.35469415 | 0.002336409 | 0.18345415 | -0.00244395 | 0.295779234 | 0.00233753 | 0.183417133 | 0.000281553 | 0.000406946 | 1 | 0.66550364 | 1 | 0.617860951 | ENSG0000023 | 1 | HLA-W |
| VAT | Obesity | ENSG0000016 | 0.002396313 | 0.10405532 | 0.001474195 | 0.183453475 | 0.002352781 | 0.110472824 | 0.001474105 | 0.183416583 | 4.35E-05 | 0.000908266 | 1 | 0.346851066 | 1 | 0.366472348 | ENSG0000023 | 1 | HLA-W |
| VAT | Obesity | ENSG0000016 | -0.0036926 | 0.123395524 | 0.002396732 | 0.183453577 | -0.00363863 | 0.128948169 | 0.002396568 | 0.183416675 | -5.40E-05 | 0.005006085 | 1 | 0.376322261 | 1 | 0.395731266 | ENSG0000023 | 1 | HLA-W |
| VAT | Obesity | ENSG0000016 | -0.00036657 | 0.879324064 | 0.002414431 | 0.183454465 | -0.00041513 | 0.863476458 | 0.00241426 | 0.183417535 | 4.86E-05 | 0.009360772 | 1 | 0.953234256 | 1 | 0.95099562 | ENSG0000023 | 1 | HLA-W |
| VAT | Obesity | ENSG0000016 | 7.29E-05 | 0.981223477 | 0.003098267 | 0.183454474 | 0.000147056 | 0.962140714 | 0.003098044 | 0.183417545 | -7.41E-05 | 0.001579714 | 1 | 0.99502166 | 1 | 0.98754495 | ENSG0000023 | 1 | AC006946.12 |
| VAT | Obesity | ENSG0000016 | -0.00135499 | 0.78798245 | 0.005038409 | 0.183454447 | -0.00152009 | 0.762866594 | 0.005038108 | 0.183417512 | 0.000165105 | 0.000297172 | 1 | 0.911115024 | 1 | 0.903653133 | ENSG0000023 | 5 | DNM1P51 |
| VAT | Obesity | ENSG0000016 | -0.00081108 | 0.818314996 | 0.003530853 | 0.183454454 | -0.00086484 | 0.806489273 | 0.003530572 | 0.183417523 | 5.38E-05 | 0.019667479 | 1 | 0.926394336 | 1 | 0.925919465 | ENSG0000023 | 5 | DNM1P51 |
| VAT | Obesity | ENSG0000016 | 0.011991797 | 0.033022884 | 0.005625169 | 0.183452756 | 0.01215746 | 0.030663431 | 0.005624757 | 0.18341578 | -0.00016566 | 7.39E-05 | 1 | 0.16909677 | 1 | 0.168587788 | ENSG0000023 | 1 | SSR4P1 |
| VAT | Obesity | ENSG0000016 | 0.008829659 | 0.026749336 | 0.003986009 | 0.183452619 | 0.008723553 | 0.028618763 | 0.003985714 | 0.183415736 | 0.000106106 | 0.000271352 | 1 | 0.147671738 | 1 | 0.160694428 | ENSG0000023 | 1 | SSR4P1 |
| VAT | Obesity | ENSG0000016 | 0.007407744 | 0.031284782 | 0.003439962 | 0.183452721 | 0.007348787 | 0.032642496 | 0.003439704 | 0.183415821 | 5.90E-05 | 0.016431887 | 1 | 0.163240261 | 1 | 0.17527877 | ENSG0000023 | 1 | SSR4P1 |
| VAT | Obesity | ENSG0000016 | 0.002961998 | 0.188951339 | 0.002254717 | 0.183453822 | 0.003017047 | 0.180835503 | 0.002254575 | 0.183416869 | -5.50E-05 | 0.00479081 | 1 | 0.485121578 | 1 | 0.483914845 | ENSG0000023 | 1 | RP11-667F9.1 |
| VAT | Obesity | ENSG0000016 | 0.001404958 | 0.361765958 | 0.001540513 | 0.18345416 | 0.001454573 | 0.345025926 | 0.0015404 | 0.183417209 | -4.96E-05 | 1.28E-05 | 1 | 0.671468759 | 1 | 0.667792116 | ENSG0000023 | 1 | RP11-667F9.1 |
| VAT | Obesity | ENSG0000016 | 0.001647674 | 0.723822961 | 0.004662952 | 0.183454427 | 0.001746602 | 0.707960627 | 0.004662622 | 0.183417493 | -9.89E-05 | 0.005975644 | 1 | 0.881231307 | 1 | 0.876422561 | ENSG0000023 | 1 | RP11-667F9.1 |
| VAT | Obesity | ENSG0000016 | -0.00215776 | 0.682162111 | 0.005269058 | 0.183454411 | -0.00204446 | 0.697987193 | 0.005268691 | 0.183417489 | -0.0001133 | 0.006253453 | 1 | 0.863333733 | 1 | 0.870280156 | ENSG0000023 | 2 | RP11-122G18.5 |
| VAT | Obesity | ENSG0000016 | -0.00036051 | 0.934471144 | 0.004384688 | 0.183454472 | -0.00046981 | 0.914666506 | 0.004384376 | 0.183417542 | 0.000109293 | 0.001170339 | 1 | 0.977307539 | 1 | 0.972419923 | ENSG0000023 | 2 | RP11-122G18.5 |
| VAT | Obesity | ENSG0000016 | 0.006163013 | 2.26E-05 | 0.001454423 | 0.183447687 | 0.006216213 | 1.92E-05 | 0.001454327 | 0.183410642 | -5.32E-05 | 1.39E-05 | 0.062413418 | 0.000630439 | 0.052932954 | 0.000626206 | ENSG0000023 | 2 | RP11-122G18.5 |
| VAT | Obesity | ENSG0000016 | 0.003637454 | 0.04614954 | 0.001824174 | 0.183452971 | 0.003675399 | 0.043908994 | 0.001824052 | 0.183416012 | -3.79E-05 | 0.011590442 | 1 | 0.212642286 | 1 | 0.212985632 | ENSG0000023 | 2 | RP11-122G18.5 |
| VAT | Obesity | ENSG0000016 | 0.008821622 | 0.047139402 | 0.004444029 | 0.183452985 | 0.00888875 | 0.045467973 | 0.004443699 | 0.183416034 | -6.71E-05 | 0.038284348 | 1 | 0.215024497 | 1 | 0.218780414 | ENSG0000023 | 2 | RP11-122G18.5 |
| VAT | Obesity | ENSG0000016 | -0.00902475 | 0.00623076 | 0.003299248 | 0.183451646 | -0.00828895 | 0.012051762 | 0.003301535 | 0.183415164 | -0.0007358 | 2.13E-08 | 1 | 0.051797881 | 1 | 0.090106525 | ENSG0000023 | 4 | POU5F1P3 |
| VAT | Obesity | ENSG0000016 | 0.002191569 | 0.469167393 | 0.003027719 | 0.183454276 | 0.002043417 | 0.499712269 | 0.00302754 | 0.183417374 | 0.000148152 | 7.04E-08 | 1 | 0.751667156 | 1 | 0.776858792 | ENSG0000023 | 3 | LINC00484 |
| VAT | Obesity | ENSG0000016 | 0.000680317 | 0.900267719 | 0.005428502 | 0.183454468 | -0.00397789 | 0.468293433 | 0.005484784 | 0.183417347 | 0.004658207 | 3.32E-09 | 1 | 0.965087305 | 1 | 0.76491497 | ENSG0000023 | 3 | LINC00484 |

| | | | | | | | | | | | | | | | | | | | |
|---|---|---|---|---|---|---|---|---|---|---|---|---|---|---|---|---|---|---|---|
| VAT | Obesity | ENSG0000016 | -0.00033027 | 0.942856332 | 0.004607577 | 0.183454472 | 0.000341085 | 0.941006599 | 0.004608957 | 0.183417544 | -0.00067136 | 2.60E-07 | 1 | 0.981243858 | 1 | 0.980488158 | ENSG0000023 | 3 | LINC00484 |
| VAT | Obesity | ENSG0000016 | -0.01275551 | 0.001004704 | 0.003877966 | 0.183450385 | -0.01269284 | 0.00106302 | 0.003877675 | 0.183413497 | -6.27E-05 | 0.023437973 | 1 | 0.013660016 | 1 | 0.015746258 | ENSG0000023 | 1 | NPM1P26 |
| VAT | Obesity | ENSG0000016 | -0.00057156 | 0.873725058 | 0.003596355 | 0.183454465 | -0.00066142 | 0.854070401 | 0.003596101 | 0.183417533 | 8.99E-05 | 0.001315534 | 1 | 0.951116912 | 1 | 0.947822399 | ENSG0000023 | 1 | NPM1P26 |
| VAT | Obesity | ENSG0000016 | 0.010834544 | 0.000666363 | 0.003183747 | 0.183450097 | 0.010253168 | 0.001290601 | 0.003186113 | 0.183413632 | 0.000581376 | 8.68E-06 | 1 | 0.009679794 | 1 | 0.017990194 | ENSG0000023 | 1 | IFITM4P |
| VAT | Obesity | ENSG0000016 | 0.001885573 | 0.520093971 | 0.002931546 | 0.183454318 | 0.001834148 | 0.531507725 | 0.002931324 | 0.183417398 | 5.14E-05 | 0.012951141 | 1 | 0.778563922 | 1 | 0.786996417 | ENSG0000023 | 1 | IFITM4P |
| VAT | Obesity | ENSG0000016 | 0.005548237 | 0.135744115 | 0.003719079 | 0.183453633 | 0.005604952 | 0.131761334 | 0.003718797 | 0.183416688 | -5.67E-05 | 0.030599515 | 1 | 0.395299075 | 1 | 0.399591076 | ENSG0000023 | 1 | IFITM4P |
| VAT | Obesity | ENSG0000016 | 0.016333557 | 0.003917953 | 0.00566212 | 0.183451329 | 0.018508377 | 0.001310122 | 0.005759082 | 0.183413643 | -0.00217482 | 0.039347935 | 1 | 0.036733053 | 1 | 0.018170536 | ENSG0000023 | 1 | IFITM4P |
| VAT | Obesity | ENSG0000016 | -0.00255786 | 0.074838477 | 0.001435823 | 0.183453274 | -0.00251882 | 0.07936529 | 0.001435728 | 0.183416383 | -3.90E-05 | 0.000936016 | 1 | 0.282737093 | 1 | 0.298885799 | ENSG0000023 | 1 | IFITM4P |
| VAT | Obesity | ENSG0000016 | -0.00812616 | 0.116664775 | 0.005179428 | 0.183453544 | -0.00821075 | 0.112883342 | 0.005179082 | 0.183416596 | 8.46E-05 | 0.047062715 | 1 | 0.367994033 | 1 | 0.368271896 | ENSG0000023 | 1 | IFITM4P |
| VAT | Obesity | ENSG0000016 | 0.000777563 | 0.856386516 | 0.0042965 | 0.183454462 | 0.000874233 | 0.838756975 | 0.004296326 | 0.18341753 | -9.67E-05 | 0.04021504 | 1 | 0.944116239 | 1 | 0.939543177 | ENSG0000023 | 1 | IFITM4P |
| VAT | Obesity | ENSG0000016 | 0.006203682 | 0.213556145 | 0.004987492 | 0.183453889 | 0.006278946 | 0.208020736 | 0.004987136 | 0.183416947 | -7.53E-05 | 0.048914184 | 1 | 0.516511799 | 1 | 0.520051841 | ENSG0000023 | 1 | IFITM4P |
| VAT | Obesity | ENSG0000016 | -0.00265852 | 0.424428083 | 0.003328297 | 0.183454233 | -0.00259656 | 0.43527175 | 0.00332806 | 0.183417316 | -6.20E-05 | 0.015087773 | 1 | 0.714174021 | 1 | 0.738383547 | ENSG0000023 | 1 | IFITM4P |
| VAT | Obesity | ENSG0000016 | 0.012991767 | 0.00909089 | 0.004980307 | 0.183451902 | 0.013112841 | 0.008460695 | 0.004979962 | 0.183414926 | -0.00012107 | 0.002171881 | 1 | 0.067950356 | 1 | 0.070977262 | ENSG0000023 | 1 | IFITM4P |
| VAT | Obesity | ENSG0000016 | 0.010514241 | 0.004916799 | 0.003738466 | 0.183451484 | 0.010381771 | 0.005483251 | 0.003738223 | 0.183414631 | 0.00013247 | 2.48E-05 | 1 | 0.043494762 | 1 | 0.05044591 | ENSG0000023 | 2 | B3GALT4 |
| VAT | Obesity | ENSG0000016 | -0.00659058 | 0.074902545 | 0.003700358 | 0.183453275 | -0.00651847 | 0.078124176 | 0.003700144 | 0.183416373 | -7.21E-05 | 0.034796787 | 1 | 0.282737093 | 1 | 0.297410657 | ENSG0000023 | 2 | B3GALT4 |
| VAT | Obesity | ENSG0000016 | 0.007252847 | 0.000346074 | 0.002026985 | 0.183449635 | 0.006408825 | 0.001604268 | 0.002031228 | 0.183413784 | 0.000844023 | 3.24E-10 | 0.955165045 | 0.005824177 | 1 | 0.021287397 | ENSG0000023 | 2 | B3GALT4 |
| VAT | Obesity | ENSG0000016 | 0.000552322 | 0.853600045 | 0.002993177 | 0.183454461 | 0.000480669 | 0.872408599 | 0.00299296 | 0.183417536 | 7.17E-05 | 0.001464557 | 1 | 0.943884665 | 1 | 0.956254064 | ENSG0000023 | 4 | AP001468.1 |
| VAT | Obesity | ENSG0000016 | 0.002347803 | 0.479702913 | 0.003321826 | 0.183454285 | 0.002417449 | 0.466736869 | 0.003321588 | 0.183417346 | -6.96E-05 | 0.005864357 | 1 | 0.762547611 | 1 | 0.763147961 | ENSG0000023 | 4 | AP001468.1 |
| VAT | Obesity | ENSG0000016 | -0.01951368 | 1.93E-15 | 0.002455737 | 0.18343061 | -0.01955501 | 1.68E-15 | 0.002455575 | 0.183393581 | 4.13E-05 | 0.044136993 | 5.32E-12 | 3.01E-13 | 4.63E-12 | 2.72E-13 | ENSG0000023 | 3 | ZNF815P |
| VAT | Obesity | ENSG0000016 | 0.001397652 | 0.620245426 | 0.002820673 | 0.183454381 | 0.001337236 | 0.635413955 | 0.002820453 | 0.183417461 | 6.04E-05 | 0.001412668 | 1 | 0.833030354 | 1 | 0.844363271 | ENSG0000023 | 1 | MCCD1P1 |
| VAT | Obesity | ENSG0000016 | -0.00275965 | 0.408970411 | 0.003342164 | 0.183454216 | 0.000413015 | 0.90358392 | 0.003409539 | 0.18341754 | -0.00317267 | 2.70E-06 | 1 | 0.708044263 | 1 | 0.970385844 | ENSG0000023 | 1 | MCCD1P1 |
| VAT | Obesity | ENSG0000017 | 0.041569515 | 4.76E-19 | 0.004661227 | 0.183424415 | 0.041701703 | 3.66E-19 | 0.004660919 | 0.183387297 | -0.00013219 | 0.000716823 | 1.31E-15 | 1.19E-16 | 1.01E-15 | 9.19E-17 | ENSG0000023 | 1 | MCCD1P1 |
| VAT | Obesity | ENSG0000017 | -0.0028686 | 0.130823724 | 0.001898652 | 0.183453611 | -0.00281995 | 0.137459691 | 0.001898547 | 0.183416712 | -4.87E-05 | 0.00731291 | 1 | 0.387974943 | 1 | 0.412354078 | ENSG0000023 | 1 | MCCD1P1 |
| VAT | Obesity | ENSG0000017 | 0.019927458 | 0.000641584 | 0.005837975 | 0.18345007 | 0.019824786 | 0.000683629 | 0.005837543 | 0.183413187 | 0.000102673 | 0.015779413 | 1 | 0.00936916 | 1 | 0.010986495 | ENSG0000023 | 4 | RP3-470B24.5 |
| VAT | Obesity | ENSG0000017 | -0.00623057 | 0.065855172 | 0.003387263 | 0.183453195 | -0.00616023 | 0.068950499 | 0.003387069 | 0.183416296 | -7.03E-05 | 0.025988671 | 1 | 0.264689498 | 1 | 0.277410173 | ENSG0000023 | 4 | RP3-470B24.5 |
| VAT | Obesity | ENSG0000017 | -0.00085539 | 0.86014088 | 0.004854813 | 0.183454462 | -0.0007659 | 0.874635284 | 0.004854426 | 0.183417537 | -8.95E-05 | 0.004441027 | 1 | 0.944533766 | 1 | 0.956332568 | ENSG0000023 | 2 | ZBTB22 |
| VAT | Obesity | ENSG0000017 | 0.002936518 | 0.57788226 | 0.00527694 | 0.183454357 | 0.00278631 | 0.597488432 | 0.005276941 | 0.183417441 | 0.000150208 | 0.044911111 | 1 | 0.80716348 | 1 | 0.827015081 | ENSG0000023 | 2 | ZBTB22 |
| VAT | Obesity | ENSG0000017 | 0.002347496 | 0.531514516 | 0.003751814 | 0.183454326 | 0.002417895 | 0.519250934 | 0.003751577 | 0.183417389 | -7.04E-05 | 0.029905003 | 1 | 0.783174974 | 1 | 0.784425829 | ENSG0000023 | 2 | ZBTB22 |
| VAT | Obesity | ENSG0000017 | 0.000523041 | 0.903252893 | 0.004302975 | 0.183454469 | 0.00041962 | 0.922309096 | 0.004302666 | 0.183417542 | 0.000103421 | 0.00156358 | 1 | 0.965567705 | 1 | 0.974118595 | ENSG0000023 | 2 | ZBTB22 |
| VAT | Obesity | ENSG0000017 | -0.00969804 | 0.089129513 | 0.005704693 | 0.183453382 | -0.00938138 | 0.100080555 | 0.005704817 | 0.183416524 | -0.00031666 | 0.000384305 | 1 | 0.311783847 | 1 | 0.347887069 | ENSG0000023 | 2 | ZBTB22 |
| VAT | Obesity | ENSG0000017 | 0.004723106 | 0.162884267 | 0.003384675 | 0.183453738 | 0.004792132 | 0.156795362 | 0.003384431 | 0.183416788 | -6.90E-05 | 0.00702394 | 1 | 0.441873282 | 1 | 0.441136798 | ENSG0000023 | 2 | RP11-231P20.2 |
| VAT | Obesity | ENSG0000017 | -0.00098343 | 0.576021919 | 0.001758618 | 0.183454356 | -0.00094218 | 0.592105005 | 0.0017585 | 0.183417438 | -4.12E-05 | 0.003925141 | 1 | 0.806947626 | 1 | 0.824941855 | ENSG0000023 | 1 | AC104135.2 |
| VAT | Obesity | ENSG0000017 | -0.00205087 | 0.324733346 | 0.002082568 | 0.183454108 | -0.00209075 | 0.315377216 | 0.002082416 | 0.183417165 | 3.99E-05 | 0.010392891 | 1 | 0.633061352 | 1 | 0.637753261 | ENSG0000023 | 1 | AC104135.2 |
| VAT | Obesity | ENSG0000017 | -0.0047103 | 0.01376668 | 0.001912187 | 0.18345218 | -0.00466423 | 0.014712826 | 0.001912057 | 0.183415297 | -4.61E-05 | 0.002903867 | 1 | 0.094623402 | 1 | 0.105748437 | ENSG0000023 | 1 | AC104135.2 |
| VAT | Obesity | ENSG0000017 | 0.002592094 | 0.239202574 | 0.002202319 | 0.18345395 | 0.002646312 | 0.22948896 | 0.002202181 | 0.183417 | -5.42E-05 | 0.004723148 | 1 | 0.533709866 | 1 | 0.539107619 | ENSG0000023 | 1 | AC104135.2 |
| VAT | Obesity | ENSG0000017 | 0.00351516 | 0.07397229 | 0.001967306 | 0.183453267 | 0.003545307 | 0.071505508 | 0.001967148 | 0.183416318 | -3.01E-05 | 0.016029261 | 1 | 0.281216969 | 1 | 0.283556326 | ENSG0000023 | 2 | DDX11L2 |
| VAT | Obesity | ENSG0000017 | 0.008753089 | 0.002608842 | 0.002907579 | 0.183451048 | 0.008799469 | 0.002473166 | 0.002907357 | 0.183414084 | -4.64E-05 | 0.022152655 | 1 | 0.028230105 | 1 | 0.029010686 | ENSG0000023 | 2 | DDX11L2 |
| VAT | Obesity | ENSG0000017 | 0.017589656 | 0.001873237 | 0.005656485 | 0.183450819 | 0.017679161 | 0.001774017 | 0.005656091 | 0.183413854 | -8.95E-05 | 0.04452364 | 1 | 0.021380487 | 1 | 0.02256353 | ENSG0000023 | 2 | DDX11L2 |
| VAT | Obesity | ENSG0000017 | -0.00349871 | 0.22824993 | 0.002903778 | 0.183453925 | -0.0034567 | 0.233848918 | 0.002903559 | 0.18341701 | -4.20E-05 | 0.042575421 | 1 | 0.524101338 | 1 | 0.540555288 | ENSG0000023 | 1 | NDUFAF4P3 |
| VAT | Obesity | ENSG0000017 | -0.0064489 | 0.247344574 | 0.005574661 | 0.183453968 | -0.0063238 | 0.256603094 | 0.005574296 | 0.18341706 | -0.0001251 | 0.007450393 | 1 | 0.543960975 | 1 | 0.566579631 | ENSG0000023 | 2 | CTSLP1 |
| VAT | Obesity | ENSG0000017 | -0.00980171 | 0.031471058 | 0.004556663 | 0.183452725 | -0.00971221 | 0.033041644 | 0.004556338 | 0.183415829 | -8.95E-05 | 0.01020918 | 1 | 0.163578381 | 1 | 0.176392531 | ENSG0000023 | 2 | CTSLP1 |
| VAT | Obesity | ENSG0000017 | 0.0028874 | 0.637428151 | 0.006126485 | 0.18345439 | 0.003006359 | 0.623602044 | 0.006126024 | 0.183417455 | -0.00011896 | 0.006493659 | 1 | 0.841638558 | 1 | 0.841508821 | ENSG0000023 | 1 | RP1-156L9.1 |
| VAT | Obesity | ENSG0000017 | 0.000869241 | 0.870179958 | 0.005318739 | 0.183454464 | 0.000964046 | 0.856158598 | 0.005318397 | 0.183417534 | -9.48E-05 | 0.036644715 | 1 | 0.948684727 | 1 | 0.948422961 | ENSG0000023 | 1 | AL035610.2 |
| VAT | Obesity | ENSG0000017 | -0.01916252 | 1.96E-15 | 0.002412239 | 0.183430623 | -0.0191183 | 2.27E-15 | 0.002412061 | 0.183393806 | -4.42E-05 | 0.012162849 | 5.42E-12 | 3.01E-13 | 6.25E-12 | 3.47E-13 | ENSG0000023 | 1 | RANP1 |
| VAT | Obesity | ENSG0000017 | -0.00188596 | 0.574901771 | 0.003362691 | 0.183454355 | -0.00180507 | 0.591383697 | 0.003362454 | 0.183417437 | -8.09E-05 | 0.001962932 | 1 | 0.806905296 | 1 | 0.824941855 | ENSG0000023 | 1 | AP000343.2 |
| VAT | Obesity | ENSG0000017 | 0.00271995 | 0.44733443 | 0.003579508 | 0.183454256 | 0.002582031 | 0.470671259 | 0.003579255 | 0.183417349 | 0.000137918 | 6.98E-07 | 1 | 0.731423594 | 1 | 0.76685518 | ENSG0000023 | 1 | AP000343.2 |
| VAT | Obesity | ENSG0000017 | 0.002910228 | 0.184941909 | 0.00219525 | 0.18345381 | 0.003462898 | 0.115003644 | 0.002197132 | 0.183416607 | -0.00055267 | 9.02E-09 | 1 | 0.480635362 | 1 | 0.37298479 | ENSG0000023 | 4 | CCDC163P |
| VAT | Obesity | ENSG0000017 | -0.01350837 | 0.001062118 | 0.004126516 | 0.183450423 | -0.01308316 | 0.001526428 | 0.004127661 | 0.183413749 | -0.00042521 | 0.000176251 | 1 | 0.014093491 | 1 | 0.020753408 | ENSG0000023 | 4 | CCDC163P |
| VAT | Obesity | ENSG0000017 | 0.002401574 | 0.167055662 | 0.001738094 | 0.183453752 | 0.002445929 | 0.1593251 | 0.001737972 | 0.183416797 | -4.44E-05 | 0.001066368 | 1 | 0.449389499 | 1 | 0.446887475 | ENSG0000023 | 4 | CCDC163P |
| VAT | Obesity | ENSG0000017 | 0.009233168 | 9.17E-05 | 0.002360494 | 0.183448691 | -0.00206327 | 0.553260256 | 0.003480067 | 0.183417413 | 0.011296438 | 9.99E-06 | 0.253151054 | 0.002075009 | 1 | 0.805509891 | ENSG0000023 | 4 | CCDC163P |
| VAT | Obesity | ENSG0000017 | 0.007224752 | 0.064122306 | 0.00390246 | 0.183453179 | 0.007313744 | 0.060894234 | 0.003902189 | 0.183416218 | -8.90E-05 | 0.003921276 | 1 | 0.259497897 | 1 | 0.258964693 | ENSG0000023 | 4 | CCDC163P |
| VAT | Obesity | ENSG0000017 | 0.001137912 | 0.572078676 | 0.002014028 | 0.183454353 | 0.004614818 | 0.027349022 | 0.002091459 | 0.183415706 | -0.00347691 | 7.35E-10 | 1 | 0.806905296 | 1 | 0.155231297 | ENSG0000023 | 4 | CCDC163P |
| VAT | Obesity | ENSG0000017 | 0.020374379 | 4.93E-05 | 0.005019835 | 0.183448247 | 0.020836133 | 3.32E-05 | 0.00501999 | 0.183411035 | -0.00046175 | 1.57E-08 | 0.13617451 | 0.001215844 | 0.091525667 | 0.000984147 | ENSG0000023 | 1 | AC068282.3 |
| VAT | Obesity | ENSG0000017 | -0.00413987 | 0.218638864 | 0.003365319 | 0.183453902 | -0.00402811 | 0.231296098 | 0.00336509 | 0.183417005 | -0.00011176 | 3.80E-05 | 1 | 0.521961016 | 1 | 0.540524906 | ENSG0000023 | 1 | AC068282.3 |
| VAT | Obesity | ENSG0000017 | 0.001436482 | 0.541038006 | 0.002350094 | 0.183454333 | 0.001514691 | 0.519236535 | 0.002350096 | 0.183417389 | -7.82E-05 | 0.019463744 | 1 | 0.788540277 | 1 | 0.784425829 | ENSG0000023 | 1 | AC068282.3 |
| VAT | Obesity | ENSG0000017 | 0.001695326 | 0.644250179 | 0.003671405 | 0.183454394 | 0.001631656 | 0.65671605 | 0.003671165 | 0.183417471 | 6.37E-05 | 0.038446083 | 1 | 0.844163628 | 1 | 0.854766662 | ENSG0000023 | 1 | AC068282.3 |
| VAT | Obesity | ENSG0000017 | -0.01109916 | 0.001646544 | 0.003526285 | 0.183450729 | -0.01074919 | 0.002321423 | 0.003529287 | 0.18341404 | -0.00034997 | 0.022914586 | 1 | 0.019672995 | 1 | 0.027736481 | ENSG0000023 | 4 | ARHGEF38 |
| VAT | Obesity | ENSG0000017 | 0.000635563 | 0.900398482 | 0.005078086 | 0.183454468 | 0.000746966 | 0.883048119 | 0.005077728 | 0.183417538 | -0.0001114 | 0.00480771 | 1 | 0.965087305 | 1 | 0.960056703 | ENSG0000023 | 1 | AC108448.2 |
| VAT | Obesity | ENSG0000017 | -0.00095131 | 0.781543294 | 0.003430524 | 0.183454445 | -0.00157739 | 0.645830855 | 0.003432384 | 0.183417466 | 0.00062608 | 3.61E-07 | 1 | 0.90900105 | 1 | 0.850570266 | ENSG0000023 | 1 | AC108448.2 |
| VAT | Obesity | ENSG0000017 | -0.01103668 | 0.001730967 | 0.003522829 | 0.183450764 | -0.01511439 | 0.000208654 | 0.004075795 | 0.183412349 | 0.004077709 | 0.04673641 | 1 | 0.0201581 | 0.575885908 | 0.004290423 | ENSG0000023 | 1 | AC108448.2 |
| VAT | Obesity | ENSG0000017 | 0.000271785 | 0.929317262 | 0.003063958 | 0.183454471 | -0.00204804 | 0.507277459 | 0.003088667 | 0.18341738 | 0.002319829 | 3.36E-09 | 1 | 0.976170888 | 1 | 0.783244156 | ENSG0000023 | 1 | LINC00421 |
| VAT | Obesity | ENSG0000017 | -0.00697202 | 0.230850149 | 0.005818876 | 0.183453931 | -0.00687706 | 0.237229935 | 0.005818452 | 0.183417018 | -9.50E-05 | 0.028327219 | 1 | 0.52708752 | 1 | 0.544132965 | ENSG0000023 | 1 | DDX10P1 |
| VAT | Obesity | ENSG0000017 | 0.013336825 | 0.000552538 | 0.003861353 | 0.183449965 | 0.013230121 | 0.00061136 | 0.003861066 | 0.183413109 | 0.000106704 | 0.000123983 | 1 | 0.008472254 | 1 | 0.009984341 | ENSG0000023 | 3 | CDKN2AIPNL |
| VAT | Obesity | ENSG0000017 | 0.001872141 | 0.514723837 | 0.002873584 | 0.183454314 | 0.000949958 | 0.741340734 | 0.002877974 | 0.183417505 | 0.000922183 | 1.89E-08 | 1 | 0.776929212 | 1 | 0.893915902 | ENSG0000023 | 4 | ANO7P1 |
| VAT | Obesity | ENSG0000017 | -0.00119543 | 0.667261421 | 0.002780662 | 0.183454404 | -0.00110832 | 0.690180946 | 0.002780461 | 0.183417486 | -8.71E-05 | 3.36E-05 | 1 | 0.857608118 | 1 | 0.869882588 | ENSG0000023 | 1 | GS1-124K5.4 |
| VAT | Obesity | ENSG0000017 | 6.61E-05 | 0.976854278 | 0.002277958 | 0.183454474 | 0.000371732 | 0.870393226 | 0.002278339 | 0.183417536 | -0.00030564 | 6.98E-09 | 1 | 0.992659365 | 1 | 0.955921132 | ENSG0000023 | 1 | GS1-124K5.4 |
| VAT | Obesity | ENSG0000017 | 0.001353332 | 0.23164131 | 0.001131413 | 0.183453933 | 0.001370469 | 0.22574989 | 0.001131328 | 0.183416991 | -1.71E-05 | 0.034325484 | 1 | 0.52715189 | 1 | 0.535743505 | ENSG0000023 | 1 | GS1-124K5.4 |
| VAT | Obesity | ENSG0000017 | 0.007126894 | 0.122184976 | 0.004610877 | 0.183453571 | 0.007058934 | 0.125759684 | 0.004610543 | 0.18341666 | 6.80E-05 | 0.049018624 | 1 | 0.374700593 | 1 | 0.391460914 | ENSG0000023 | 1 | GS1-124K5.4 |
| VAT | Obesity | ENSG0000017 | 0.006844987 | 0.190061221 | 0.005223586 | 0.183453825 | 0.006663656 | 0.202038379 | 0.005223253 | 0.183416931 | 0.000181331 | 5.24E-05 | 1 | 0.487064968 | 1 | 0.512836579 | ENSG0000023 | 4 | RP11-206L10.9 |
| VAT | Obesity | ENSG0000017 | -0.01482307 | 0.000208853 | 0.003997496 | 0.183449277 | -0.01536972 | 0.000120833 | 0.003997888 | 0.18341196 | 0.000546649 | 7.64E-12 | 0.576433352 | 0.004003009 | 0.333499937 | 0.00280252 | ENSG0000023 | 4 | RP11-206L10.9 |
| VAT | Obesity | ENSG0000017 | -0.00658149 | 0.063048748 | 0.003540623 | 0.183453168 | -0.00649854 | 0.066427452 | 0.003540414 | 0.183416273 | -8.29E-05 | 0.010150775 | 1 | 0.257417965 | 1 | 0.27337482 | ENSG0000023 | 3 | AC008268.2 |
| VAT | Obesity | ENSG0000017 | -0.00047706 | 0.720209656 | 0.001331914 | 0.183454426 | -0.00044409 | 0.738798573 | 0.001331823 | 0.183417504 | -3.30E-05 | 0.002118256 | 1 | 0.879937428 | 1 | 0.892750508 | ENSG0000023 | 3 | AC008268.2 |
| VAT | Obesity | ENSG0000017 | 0.001256748 | 0.755131214 | 0.004029578 | 0.183454437 | 0.001144895 | 0.776310305 | 0.004029465 | 0.183417516 | 0.000111853 | 0.021286123 | 1 | 0.894874259 | 1 | 0.909787806 | ENSG0000023 | 3 | AC008268.2 |
| VAT | Obesity | ENSG0000017 | 0.007328578 | 0.161529182 | 0.005234889 | 0.183453733 | 0.007579606 | 0.147673785 | 0.00523526 | 0.183416754 | -0.00025103 | 0.009659893 | 1 | 0.441056458 | 1 | 0.424562132 | ENSG0000023 | 3 | AC008268.2 |
| VAT | Obesity | ENSG0000017 | -0.01001756 | 0.001020217 | 0.003049568 | 0.183450395 | -0.00993194 | 0.001125781 | 0.003049351 | 0.183413537 | -8.56E-05 | 0.000247907 | 1 | 0.013782824 | 1 | 0.016353444 | ENSG0000023 | 3 | HLA-DQA2 |
| VAT | Obesity | ENSG0000017 | -0.00283201 | 0.173201538 | 0.002079315 | 0.183453773 | -0.00289181 | 0.164273748 | 0.002079181 | 0.183416815 | 5.98E-05 | 0.000742223 | 1 | 0.460092631 | 1 | 0.455295507 | ENSG0000023 | 3 | HLA-DQA2 |
| VAT | Obesity | ENSG0000017 | 0.000956637 | 0.78554272 | 0.003515714 | 0.183454446 | 0.001007131 | 0.774504963 | 0.003515446 | 0.183417515 | -5.05E-05 | 0.040366754 | 1 | 0.909319831 | 1 | 0.909523293 | ENSG0000023 | 3 | HLA-DQA2 |
| VAT | Obesity | ENSG0000017 | -0.01091788 | 0.001859885 | 0.003508589 | 0.183450814 | -0.01097532 | 0.001757854 | 0.003508312 | 0.183413847 | 5.74E-05 | 0.012673928 | 1 | 0.021380487 | 1 | 0.02256353 | ENSG0000023 | 1 | HCG4P3 |
| VAT | Obesity | ENSG0000017 | 0.006541163 | 0.071190923 | 0.003625396 | 0.183453244 | 0.005039155 | 0.165889001 | 0.003636969 | 0.183416821 | 0.001502009 | 3.35E-07 | 1 | 0.274916222 | 1 | 0.456484191 | ENSG0000023 | 1 | HCG4P3 |
| VAT | Obesity | ENSG0000017 | -0.00991319 | 5.48E-05 | 0.002457409 | 0.183448323 | -0.00703892 | 0.005446824 | 0.002532569 | 0.183414627 | -0.00287427 | 2.78E-06 | 0.151367782 | 0.001339538 | 1 | 0.05044591 | ENSG0000023 | 1 | HCG4P3 |
| VAT | Obesity | ENSG0000017 | 0.003349393 | 0.206088352 | 0.002649005 | 0.18345387 | 0.003396773 | 0.199715973 | 0.002648839 | 0.183416925 | -4.74E-05 | 0.039713964 | 1 | 0.507220054 | 1 | 0.510412237 | ENSG0000023 | 1 | HCG4P3 |
| VAT | Obesity | ENSG0000017 | -0.00231595 | 0.407080221 | 0.00279352 | 0.183454214 | -0.00236824 | 0.396535068 | 0.002793312 | 0.183417274 | 5.23E-05 | 0.009822572 | 1 | 0.708044263 | 1 | 0.715434904 | ENSG0000023 | 1 | HCG4P3 |
| VAT | Obesity | ENSG0000017 | -0.00029154 | 0.869161241 | 0.001769877 | 0.183454464 | -0.00021874 | 0.901633174 | 0.001769786 | 0.18341754 | -7.28E-05 | 3.34E-05 | 1 | 0.948684727 | 1 | 0.969573834 | ENSG0000023 | 1 | HCG4P3 |
| VAT | Obesity | ENSG0000017 | -0.04022362 | 1.98E-13 | 0.005472469 | 0.183434055 | -0.04038699 | 1.58E-13 | 0.005472089 | 0.183396961 | 0.000163369 | 0.000177277 | 5.47E-10 | 2.38E-11 | 4.36E-10 | 1.98E-11 | ENSG0000023 | 1 | HCG4P3 |
| VAT | Obesity | ENSG0000017 | -0.00486688 | 0.02914508 | 0.002230951 | 0.183452675 | -0.00128908 | 0.5890798 | 0.002386432 | 0.183417436 | -0.0035778 | 2.45E-05 | 1 | 0.155892287 | 1 | 0.823637411 | ENSG0000023 | 1 | HCG4P3 |
| VAT | Obesity | ENSG0000017 | 0.004957814 | 0.225192113 | 0.004087777 | 0.183453918 | 0.005082944 | 0.213670734 | 0.00408749 | 0.183416962 | -0.00012513 | 8.77E-05 | 1 | 0.52272452 | 1 | 0.525139114 | ENSG0000023 | 1 | HCG4P3 |
| VAT | Obesity | ENSG0000017 | -0.00444714 | 0.47091429 | 0.006168084 | 0.183454278 | -0.00422081 | 0.493771779 | 0.006167863 | 0.183417369 | -0.00022634 | 0.001256315 | 1 | 0.753462863 | 1 | 0.775358255 | ENSG0000023 | 1 | HCG4P3 |
| VAT | Obesity | ENSG0000017 | -0.01271944 | 0.02098737 | 0.0055105 | 0.18345246 | -0.01258445 | 0.022378533 | 0.005510101 | 0.183415575 | -0.00013499 | 0.001082373 | 1 | 0.125108299 | 1 | 0.136056182 | ENSG0000023 | 1 | HCG4P3 |
| VAT | Obesity | ENSG0000017 | 0.003399296 | 0.333368281 | 0.003514019 | 0.18345412 | 0.003458076 | 0.325040286 | 0.003513755 | 0.18341718 | -5.88E-05 | 0.018833151 | 1 | 0.642540807 | 1 | 0.64633371 | ENSG0000023 | 4 | IRGM |
| VAT | Obesity | ENSG0000017 | -0.00073589 | 0.82652082 | 0.003357698 | 0.183454456 | -0.00068132 | 0.839192802 | 0.003357464 | 0.18341753 | -5.46E-05 | 0.038438677 | 1 | 0.928070571 | 1 | 0.939543177 | ENSG0000023 | 1 | AC079922.3 |
| VAT | Obesity | ENSG0000018 | -0.00142732 | 0.206294748 | 0.001129367 | 0.18345387 | -0.00140621 | 0.21305171 | 0.001129287 | 0.18341696 | -2.11E-05 | 0.01641728 | 1 | 0.507220054 | 1 | 0.525074761 | ENSG0000023 | 1 | AC079922.3 |
| VAT | Obesity | ENSG0000018 | -0.00308198 | 0.606401831 | 0.005981909 | 0.183454374 | -0.00184202 | 0.759298032 | 0.006011759 | 0.183417511 | -0.00123996 | 0.040185533 | 1 | 0.827729503 | 1 | 0.90093259 | ENSG0000023 | 1 | AC079922.3 |
| VAT | Obesity | ENSG0000018 | 0.008918028 | 0.104834198 | 0.005498646 | 0.18345348 | 0.008801494 | 0.109425925 | 0.005498247 | 0.183416578 | 0.000116534 | 0.00472263 | 1 | 0.347941332 | 1 | 0.365676336 | ENSG0000023 | 3 | LINC00674 |
| VAT | Obesity | ENSG0000018 | 0.001980662 | 0.722796437 | 0.00558367 | 0.183454427 | 0.002136108 | 0.702023967 | 0.005583283 | 0.183417491 | -0.00015545 | 0.000442321 | 1 | 0.88098326 | 1 | 0.872786554 | ENSG0000023 | 1 | RPL23AP32 |
| VAT | Obesity | ENSG0000018 | 0.002899808 | 0.42119178 | 0.00360513 | 0.18345423 | -0.00080683 | 0.830542633 | 0.003770145 | 0.183417529 | 0.003706641 | 0.000789798 | 1 | 0.71233554 | 1 | 0.936897537 | ENSG0000023 | 1 | RP3-393E18.2 |
| VAT | Obesity | ENSG0000018 | 9.49E-06 | 0.998354567 | 0.004604146 | 0.183454474 | 0.000123741 | 0.978557302 | 0.004603869 | 0.183417546 | -0.00011425 | 0.005778378 | 1 | 0.999228023 | 1 | 0.994336196 | ENSG0000023 | 1 | RP3-393E18.2 |
| VAT | Obesity | ENSG0000018 | -0.00143816 | 0.534926891 | 0.002317726 | 0.183454329 | -0.00149448 | 0.519028804 | 0.002317585 | 0.183417389 | 5.63E-05 | 0.006414356 | 1 | 0.784524453 | 1 | 0.784425829 | ENSG0000023 | 1 | RP3-393E18.2 |
| VAT | Obesity | ENSG0000018 | -0.01155953 | 0.00115808 | 0.003557844 | 0.183450484 | -0.01294381 | 0.000294877 | 0.00357585 | 0.183412594 | 0.001384279 | 0.000131487 | 1 | 0.015006104 | 0.813859235 | 0.005574378 | ENSG0000023 | 2 | PTENP1 |
| VAT | Obesity | ENSG0000018 | 0.011531291 | 0.042586673 | 0.005686752 | 0.18345292 | 0.0111183 | 0.050644531 | 0.00568861 | 0.183416102 | 0.000412991 | 0.012970795 | 1 | 0.201957418 | 1 | 0.236113017 | ENSG0000023 | 2 | PTENP1 |
| VAT | Obesity | ENSG0000018 | 0.002667916 | 0.486797561 | 0.003836455 | 0.183454291 | 0.002719473 | 0.478383459 | 0.003836157 | 0.183417356 | -5.16E-05 | 0.047298357 | 1 | 0.765031548 | 1 | 0.77393807 | ENSG0000023 | 2 | OR2I1P |
| VAT | Obesity | ENSG0000018 | 0.003577587 | 0.076091176 | 0.002016867 | 0.183453285 | 0.003635077 | 0.071473047 | 0.002016726 | 0.183416318 | -5.75E-05 | 0.000307844 | 1 | 0.283034564 | 1 | 0.283556326 | ENSG0000023 | 2 | OR2I1P |
| VAT | Obesity | ENSG0000018 | -0.01430314 | 0.006447745 | 0.005250573 | 0.183451669 | -0.01395446 | 0.007868248 | 0.005250613 | 0.183414877 | -0.00034867 | 6.20E-06 | 1 | 0.052963619 | 1 | 0.066819583 | ENSG0000023 | 2 | OR2I1P |
| VAT | Obesity | ENSG0000018 | -0.00369773 | 0.192637506 | 0.002838257 | 0.183453833 | -0.00360313 | 0.20423992 | 0.002838079 | 0.183416937 | -9.46E-05 | 0.000130192 | 1 | 0.490311549 | 1 | 0.515267074 | ENSG0000023 | 2 | OR2I1P |
| VAT | Obesity | ENSG0000018 | -0.00587553 | 0.131364118 | 0.003894311 | 0.183453614 | -0.00600596 | 0.122989842 | 0.003894036 | 0.183416647 | 0.000130427 | 1.65E-05 | 1 | 0.387974943 | 1 | 0.385025715 | ENSG0000023 | 1 | DDX39BP2 |
| VAT | Obesity | ENSG0000018 | 0.00939725 | 0.092997879 | 0.005594285 | 0.183453408 | 0.009269741 | 0.097495324 | 0.005593877 | 0.183416508 | 0.000127509 | 0.002241433 | 1 | 0.319981445 | 1 | 0.341914986 | ENSG0000023 | 1 | DDX39BP2 |
| VAT | Obesity | ENSG0000018 | 0.020499536 | 0.000249891 | 0.005597298 | 0.183449404 | 0.020748871 | 0.000209858 | 0.005597406 | 0.183412353 | -0.00024933 | 0.004077566 | 0.689699497 | 0.004597997 | 0.57920706 | 0.004290423 | ENSG0000023 | 1 | DDX39BP2 |
| VAT | Obesity | ENSG0000018 | 0.000573067 | 0.687611313 | 0.00142519 | 0.183454413 | 0.000523688 | 0.713262628 | 0.001425086 | 0.183417495 | 4.94E-05 | 3.23E-06 | 1 | 0.863613457 | 1 | 0.878449288 | ENSG0000023 | 1 | RP5-1185K9.1 |
| VAT | Obesity | ENSG0000018 | -0.00347031 | 0.268742376 | 0.003137817 | 0.183454012 | -0.003393 | 0.279521364 | 0.003137614 | 0.183417104 | -7.73E-05 | 0.003589176 | 1 | 0.57160411 | 1 | 0.59920191 | ENSG0000023 | 1 | RP5-1185K9.1 |
| VAT | Obesity | ENSG0000018 | -0.0034168 | 0.219190624 | 0.00278086 | 0.183453903 | -0.0014123 | 0.615361698 | 0.002810916 | 0.183417451 | -0.0020045 | 1.13E-06 | 1 | 0.521961016 | 1 | 0.839253732 | ENSG0000023 | 1 | RP5-1185K9.1 |
| VAT | Obesity | ENSG0000018 | 0.00062332 | 0.58348755 | 0.001136827 | 0.18345436 | 0.000593218 | 0.601776353 | 0.001136764 | 0.183417443 | 3.01E-05 | 0.00528383 | 1 | 0.810955212 | 1 | 0.829207556 | ENSG0000023 | 1 | RP5-1185K9.1 |
| VAT | Obesity | ENSG0000018 | 0.012641372 | 0.002703049 | 0.004214284 | 0.183451073 | 0.01255758 | 0.002882733 | 0.004213969 | 0.18341419 | 8.38E-05 | 0.005558763 | 1 | 0.0286939 | 1 | 0.032474874 | ENSG0000023 | 3 | LRRC37A2 |
| VAT | Obesity | ENSG0000018 | -0.01226423 | 0.004349358 | 0.004300736 | 0.1834514 | -0.01218577 | 0.00460266 | 0.004300432 | 0.183414512 | -7.85E-05 | 0.018367712 | 1 | 0.040282648 | 1 | 0.044966202 | ENSG0000023 | 3 | LRRC37A2 |
| VAT | Obesity | ENSG0000018 | -0.00475758 | 0.126219861 | 0.003111189 | 0.18345359 | -0.00470356 | 0.1305531 | 0.003110967 | 0.183416682 | -5.40E-05 | 0.023358835 | 1 | 0.381145314 | 1 | 0.398993348 | ENSG0000023 | 3 | LRRC37A2 |
| VAT | Obesity | ENSG0000018 | 0.012445078 | 0.000967381 | 0.00377139 | 0.183450358 | 0.01237416 | 0.001033511 | 0.003771159 | 0.183413477 | 7.09E-05 | 0.034276884 | 1 | 0.013283447 | 1 | 0.015502666 | ENSG0000023 | 3 | GOLGA2P5 |
| VAT | Obesity | ENSG0000018 | 0.016113282 | 0.000286 | 0.00444175 | 0.1834495 | 0.016017532 | 0.000310513 | 0.004441426 | 0.183412631 | 9.58E-05 | 0.003752329 | 0.789359804 | 0.005022832 | 0.85701644 | 0.005830044 | ENSG0000023 | 1 | RP11-108M9.4 |
| VAT | Obesity | ENSG0000018 | 0.001087396 | 0.853152522 | 0.005874715 | 0.183454461 | 0.000470516 | 0.936189918 | 0.005877075 | 0.183417544 | 0.00061688 | 0.000924093 | 1 | 0.943767921 | 1 | 0.979114882 | ENSG0000023 | 1 | RP11-108M9.4 |
| VAT | Obesity | ENSG0000018 | 0.000417338 | 0.940495364 | 0.005590801 | 0.183454472 | 0.000323866 | 0.953802622 | 0.005590435 | 0.183417545 | 9.35E-05 | 0.046396255 | 1 | 0.980274624 | 1 | 0.984913318 | ENSG0000023 | 2 | OR2W3 |
| VAT | Obesity | ENSG0000018 | -0.00326169 | 0.090513357 | 0.001926914 | 0.183453391 | -0.00321051 | 0.095663269 | 0.001926779 | 0.183416497 | -5.12E-05 | 0.00068267 | 1 | 0.314235052 | 1 | 0.337231331 | ENSG0000023 | 2 | OR2W3 |
| VAT | Obesity | ENSG0000018 | 0.005747992 | 0.134336245 | 0.003839106 | 0.183453627 | 0.005686353 | 0.138534423 | 0.003838828 | 0.183416717 | 6.16E-05 | 0.032529207 | 1 | 0.392762749 | 1 | 0.412910374 | ENSG0000023 | 1 | SLC35E1P1 |
| VAT | Obesity | ENSG0000018 | -0.00347799 | 0.449672986 | 0.004600759 | 0.183454258 | -0.00355554 | 0.439602012 | 0.004600457 | 0.18341732 | 7.75E-05 | 0.044018086 | 1 | 0.733942898 | 1 | 0.74253461 | ENSG0000023 | 1 | SLC35E1P1 |
| VAT | Obesity | ENSG0000018 | -0.00524269 | 0.326984798 | 0.00534856 | 0.183454111 | -0.00531874 | 0.319979281 | 0.005348145 | 0.183417172 | 7.60E-05 | 0.036775214 | 1 | 0.634654038 | 1 | 0.641269508 | ENSG0000023 | 1 | RPL23AP1 |
| VAT | Obesity | ENSG0000018 | 0.004402548 | 0.472183029 | 0.006123732 | 0.183454279 | 0.004594176 | 0.453086086 | 0.006123277 | 0.183417333 | -0.00019163 | 1.64E-05 | 1 | 0.755055133 | 1 | 0.750751641 | ENSG0000023 | 1 | RPL23AP1 |
| VAT | Obesity | ENSG0000018 | -0.00657268 | 0.221433984 | 0.005375437 | 0.183453909 | -0.00667235 | 0.214474824 | 0.005375051 | 0.183416964 | 9.97E-05 | 0.014805122 | 1 | 0.52199987 | 1 | 0.526178235 | ENSG0000023 | 1 | RPL23AP1 |
| VAT | Obesity | ENSG0000018 | -0.00163607 | 0.660388533 | 0.003723615 | 0.183454401 | -0.00168681 | 0.650521706 | 0.003723324 | 0.183417468 | 5.07E-05 | 0.04178097 | 1 | 0.853311026 | 1 | 0.85253557 | ENSG0000023 | 1 | RPL23AP1 |
| VAT | Obesity | ENSG0000018 | -0.00551299 | 0.271005322 | 0.005008385 | 0.183454016 | -0.00560557 | 0.263007486 | 0.005008044 | 0.183417073 | 9.26E-05 | 0.022318278 | 1 | 0.575086695 | 1 | 0.577946387 | ENSG0000023 | 1 | RPL23AP1 |
| VAT | Obesity | ENSG0000018 | -0.01456904 | 0.000270665 | 0.004000399 | 0.183449461 | -0.01462871 | 0.000255125 | 0.0040001 | 0.183412492 | 5.97E-05 | 0.039285853 | 0.747034477 | 0.004882578 | 0.704144699 | 0.004995804 | ENSG0000023 | 1 | RPL23AP1 |
| VAT | Obesity | ENSG0000018 | 0.002035438 | 0.374527036 | 0.002292092 | 0.183454176 | 0.001999205 | 0.383054647 | 0.002291923 | 0.183417258 | 3.62E-05 | 0.031401165 | 1 | 0.684277319 | 1 | 0.707013865 | ENSG0000023 | 1 | RPL23AP1 |

| | | | | | | | | | | | | | | | | | | | |
|---|---|---|---|---|---|---|---|---|---|---|---|---|---|---|---|---|---|---|---|
| VAT | Obesity | ENSG0000018 | 0.000181132 | 0.894911945 | 0.001371265 | 0.183454468 | 0.000156443 | 0.90916269 | 0.001371165 | 0.183417541 | 2.47E-05 | 0.016452257 | 1 | 0.962570916 | 1 | 0.971175886 | ENSG0000023 | 1 | RPL23AP1 |
| VAT | Obesity | ENSG0000018 | 0.00614909 | 0.04163268 | 0.003018434 | 0.183452905 | 0.006207132 | 0.039732068 | 0.003018256 | 0.183415948 | -5.80E-05 | 0.035254226 | 1 | 0.199144188 | 1 | 0.198660338 | ENSG0000023 | 1 | RPL23AP1 |
| VAT | Obesity | ENSG0000018 | -0.00373383 | 0.499056019 | 0.005523604 | 0.183454301 | -0.00384267 | 0.486597778 | 0.005523218 | 0.183417363 | 0.000108847 | 0.011814941 | 1 | 0.768206699 | 1 | 0.775358255 | ENSG0000023 | 1 | RPL21P10 |
| VAT | Obesity | ENSG0000018 | -0.00334836 | 0.466264653 | 0.004595791 | 0.183454273 | -0.00324316 | 0.480355833 | 0.004595463 | 0.183417358 | -0.0001052 | 0.002778882 | 1 | 0.749063121 | 1 | 0.774858036 | ENSG0000023 | 4 | ASB14 |
| VAT | Obesity | ENSG0000018 | -0.00020947 | 0.961256459 | 0.004312228 | 0.183454473 | -0.00012935 | 0.97606763 | 0.004311933 | 0.183417546 | -8.01E-05 | 0.020724201 | 1 | 0.985729121 | 1 | 0.993659205 | ENSG0000023 | 4 | ASB14 |
| VAT | Obesity | ENSG0000018 | -0.00201486 | 0.282533641 | 0.001874904 | 0.183454038 | -0.00195317 | 0.297494374 | 0.001874762 | 0.183417136 | -6.17E-05 | 3.42E-06 | 1 | 0.588079072 | 1 | 0.619387123 | ENSG0000023 | 2 | AC004951.5 |
| VAT | Obesity | ENSG0000018 | -0.0028017 | 0.192435069 | 0.002149514 | 0.183453832 | -0.00287313 | 0.18130832 | 0.002149358 | 0.183416871 | 7.14E-05 | 9.55E-06 | 1 | 0.490311549 | 1 | 0.48442494 | ENSG0000023 | 2 | AC004951.5 |
| VAT | Obesity | ENSG0000018 | 0.002894936 | 0.406003192 | 0.003483894 | 0.183454213 | 0.002799486 | 0.421654051 | 0.003483873 | 0.183417302 | 9.54E-05 | 0.046577886 | 1 | 0.708044263 | 1 | 0.727353238 | ENSG0000023 | 1 | RP11-168J18.6 |
| VAT | Obesity | ENSG0000018 | 0.00195577 | 0.341662887 | 0.002056788 | 0.183454132 | 0.001872486 | 0.362584107 | 0.002056653 | 0.183417233 | 8.33E-05 | 1.31E-06 | 1 | 0.651685949 | 1 | 0.686859082 | ENSG0000023 | 1 | RP11-168J18.6 |
| VAT | Obesity | ENSG0000018 | 0.002365581 | 0.512683145 | 0.003613416 | 0.183454312 | 0.002502982 | 0.488510897 | 0.003613476 | 0.183417365 | -0.0001374 | 0.013005181 | 1 | 0.776196094 | 1 | 0.775358255 | ENSG0000023 | 1 | RP11-168J18.6 |
| VAT | Obesity | ENSG0000018 | -0.00584322 | 7.70E-11 | 0.00089805 | 0.183438473 | -0.00482813 | 2.76E-07 | 0.000939412 | 0.183407564 | -0.00101509 | 0.000234339 | 2.12E-07 | 5.90E-09 | 0.000760503 | 1.52E-05 | ENSG0000023 | 1 | RP11-302B13.1 |
| VAT | Obesity | ENSG0000018 | 0.005322825 | 0.212440906 | 0.004268904 | 0.183453886 | 0.005238348 | 0.219758623 | 0.004268635 | 0.183416977 | 8.45E-05 | 0.022489196 | 1 | 0.515160001 | 1 | 0.531115411 | ENSG0000023 | 3 | NME1 |
| VAT | Obesity | ENSG0000018 | -0.00331296 | 0.11577571 | 0.002106465 | 0.183453539 | -0.00334502 | 0.112266959 | 0.002106315 | 0.183416593 | 3.21E-05 | 0.049110542 | 1 | 0.367288459 | 1 | 0.367944097 | ENSG0000023 | 6 | TNFSF12 |
| VAT | Obesity | ENSG0000018 | 0.015111198 | 0.00081952 | 0.004516012 | 0.183450242 | 0.015239998 | 0.000738525 | 0.00451571 | 0.183413242 | -0.0001288 | 0.000526472 | 1 | 0.011599357 | 1 | 0.011600152 | ENSG0000023 | 6 | TNFSF12 |
| VAT | Obesity | ENSG0000018 | -0.00359959 | 0.300062521 | 0.003473498 | 0.183454068 | -0.00352628 | 0.309981167 | 0.003473273 | 0.183417156 | -7.33E-05 | 0.012577931 | 1 | 0.608848729 | 1 | 0.631867076 | ENSG0000023 | 6 | TNFSF12 |
| VAT | Obesity | ENSG0000018 | 0.006671717 | 0.016870452 | 0.002792066 | 0.183452316 | 0.006751908 | 0.015588571 | 0.002791871 | 0.183415336 | -8.02E-05 | 0.000267316 | 1 | 0.108150968 | 1 | 0.108519823 | ENSG0000023 | 3 | APOBEC3G |
| VAT | Obesity | ENSG0000018 | 0.005937849 | 0.188451914 | 0.004514876 | 0.18345382 | 0.005751485 | 0.202677284 | 0.004514642 | 0.183416933 | 0.000186364 | 2.95E-05 | 1 | 0.484397268 | 1 | 0.512836579 | ENSG0000024 | 8 | LY6G5B |
| VAT | Obesity | ENSG0000018 | 0.002744828 | 0.180932199 | 0.002051606 | 0.183453798 | 0.002837234 | 0.166680822 | 0.002051581 | 0.183416823 | -9.24E-05 | 0.000713836 | 1 | 0.4737883 | 1 | 0.457993828 | ENSG0000024 | 8 | LY6G5B |
| VAT | Obesity | ENSG0000018 | -0.00147923 | 0.486603626 | 0.002126185 | 0.183454291 | -0.00155036 | 0.465863817 | 0.00212603 | 0.183417345 | 7.11E-05 | 6.53E-06 | 1 | 0.765031548 | 1 | 0.762624042 | ENSG0000024 | 8 | LY6G5B |
| VAT | Obesity | ENSG0000018 | -0.00067635 | 0.904131055 | 0.005615453 | 0.183454469 | -0.00058049 | 0.917659981 | 0.005615043 | 0.183417542 | -9.59E-05 | 0.021493831 | 1 | 0.965567705 | 1 | 0.973721371 | ENSG0000024 | 8 | LY6G5B |
| VAT | Obesity | ENSG0000018 | 0.005003325 | 0.412819224 | 0.00610949 | 0.183454221 | 0.005141316 | 0.400019511 | 0.006109071 | 0.183417278 | -0.00013799 | 0.004863672 | 1 | 0.708533979 | 1 | 0.716918084 | ENSG0000024 | 8 | LY6G5B |
| VAT | Obesity | ENSG0000018 | -0.00749889 | 0.150971983 | 0.005221674 | 0.183453695 | -0.00770167 | 0.140199862 | 0.005221296 | 0.183416724 | 0.000202778 | 2.56E-07 | 1 | 0.420095411 | 1 | 0.416292606 | ENSG0000024 | 8 | LY6G5B |
| VAT | Obesity | ENSG0000018 | 0.001043067 | 0.741065001 | 0.003156567 | 0.183454433 | 0.001090043 | 0.729830676 | 0.003156334 | 0.183417501 | -4.70E-05 | 0.04212248 | 1 | 0.889278002 | 1 | 0.887801393 | ENSG0000024 | 8 | LY6G5B |
| VAT | Obesity | ENSG0000018 | -0.00215221 | 0.434479136 | 0.002753775 | 0.183454243 | -0.00210376 | 0.444861451 | 0.002753581 | 0.183417325 | -4.84E-05 | 0.024001606 | 1 | 0.722822432 | 1 | 0.745369224 | ENSG0000024 | 8 | LY6G5B |
| VAT | Obesity | ENSG0000018 | 0.004450056 | 0.000986268 | 0.00135078 | 0.183450372 | 0.003533143 | 0.009633654 | 0.001364823 | 0.183415013 | 0.000916913 | 2.96E-06 | 1 | 0.013475737 | 1 | 0.077028846 | ENSG0000024 | 8 | LY6G5B |
| VAT | Obesity | ENSG0000018 | 0.005795366 | 0.252836631 | 0.005068148 | 0.18345398 | 0.005586585 | 0.270375632 | 0.005068581 | 0.183417087 | 0.000208781 | 0.032641023 | 1 | 0.54990473 | 1 | 0.586203255 | ENSG0000024 | 3 | PSMB9 |
| VAT | Obesity | ENSG0000018 | 0.001408292 | 0.754114204 | 0.004496185 | 0.183454437 | 0.001496046 | 0.739316327 | 0.004495884 | 0.183417504 | -8.78E-05 | 0.0175122 | 1 | 0.89443713 | 1 | 0.892750508 | ENSG0000024 | 3 | PSMB9 |
| VAT | Obesity | ENSG0000018 | -0.00391977 | 0.244954202 | 0.003371283 | 0.183453963 | -0.00332657 | 0.324872268 | 0.003378956 | 0.18341718 | -0.0005932 | 0.010742166 | 1 | 0.541424131 | 1 | 0.64633371 | ENSG0000024 | 3 | PSMB9 |
| VAT | Obesity | ENSG0000018 | 0.002163831 | 0.653929815 | 0.004826642 | 0.183454398 | 0.002321568 | 0.630497901 | 0.004826278 | 0.183417459 | -0.00015774 | 4.40E-06 | 1 | 0.849804294 | 1 | 0.842487235 | ENSG0000024 | 3 | PSMB9 |
| VAT | Obesity | ENSG0000018 | 0.00370651 | 0.228122349 | 0.003075403 | 0.183453925 | 0.003657061 | 0.234354599 | 0.003075182 | 0.183417012 | 4.94E-05 | 0.03385797 | 1 | 0.524101338 | 1 | 0.541270873 | ENSG0000024 | 3 | PSMB9 |
| VAT | Obesity | ENSG0000018 | -0.00525417 | 0.017123791 | 0.002203886 | 0.183452326 | -0.00521305 | 0.018004067 | 0.002203744 | 0.183415431 | -4.11E-05 | 0.02800946 | 1 | 0.108763707 | 1 | 0.119020171 | ENSG0000024 | 3 | PSMB9 |
| VAT | Obesity | ENSG0000018 | -0.0190162 | 1.16E-08 | 0.00333331 | 0.183442173 | -0.01526222 | 1.03E-05 | 0.003459473 | 0.18341019 | -0.00375398 | 5.11E-05 | 3.21E-05 | 6.99E-07 | 0.02831071 | 0.000403836 | ENSG0000024 | 3 | PSMB9 |
| VAT | Obesity | ENSG0000019 | 0.000869113 | 0.595495668 | 0.001637089 | 0.183454368 | 0.000812975 | 0.619449352 | 0.001636979 | 0.183417453 | 5.61E-05 | 2.61E-05 | 1 | 0.818796889 | 1 | 0.8401532 | ENSG0000024 | 3 | PSMB9 |
| VAT | Obesity | ENSG0000019 | -0.00669949 | 0.111350044 | 0.004207806 | 0.183453516 | -0.00658609 | 0.117513602 | 0.004207561 | 0.18341662 | -0.0001134 | 0.003362521 | 1 | 0.359025843 | 1 | 0.377136675 | ENSG0000024 | 3 | PSMB9 |
| VAT | Obesity | ENSG0000019 | 0.001201558 | 0.805950959 | 0.00489128 | 0.183454451 | 0.001374226 | 0.778730604 | 0.004890939 | 0.183417516 | -0.00017267 | 6.81E-06 | 1 | 0.921063356 | 1 | 0.910529706 | ENSG0000024 | 2 | TCAM1P |
| VAT | Obesity | ENSG0000019 | -0.00496303 | 0.396742241 | 0.005856401 | 0.183454203 | -0.00504914 | 0.388564113 | 0.005855946 | 0.183417265 | 8.61E-05 | 0.030167861 | 1 | 0.701928581 | 1 | 0.708846095 | ENSG0000024 | 4 | PPIL3 |
| VAT | Obesity | ENSG0000019 | -0.0016725 | 0.411771013 | 0.002037695 | 0.183454219 | -0.00033222 | 0.87166831 | 0.002056587 | 0.183417536 | -0.00134028 | 1.64E-06 | 1 | 0.708533979 | 1 | 0.955921132 | ENSG0000024 | 4 | PPIL3 |
| VAT | Obesity | ENSG0000019 | 0.003913178 | 0.258242042 | 0.003461283 | 0.183453991 | 0.003843771 | 0.266751612 | 0.003461062 | 0.18341708 | 6.94E-05 | 0.019432123 | 1 | 0.557845225 | 1 | 0.582003517 | ENSG0000024 | 2 | RPL23AP7 |
| VAT | Obesity | ENSG0000019 | 0.012959803 | 0.000221918 | 0.003509583 | 0.18344932 | 0.013017935 | 0.000207675 | 0.00350933 | 0.183412346 | -5.81E-05 | 0.02932811 | 0.612494646 | 0.004195169 | 0.57318292 | 0.004290423 | ENSG0000024 | 1 | RP1-228P16.1 |
| VAT | Obesity | ENSG0000019 | 0.003925196 | 0.219535805 | 0.003197025 | 0.183453904 | 0.003838895 | 0.22981093 | 0.003196821 | 0.183417001 | 8.63E-05 | 0.00166081 | 1 | 0.521961016 | 1 | 0.539352182 | ENSG0000024 | 1 | RP1-228P16.1 |
| VAT | Obesity | ENSG0000019 | -0.00499442 | 0.226013974 | 0.00412526 | 0.18345392 | 0.002139408 | 0.674686674 | 0.005097168 | 0.183417479 | -0.00713383 | 0.017202557 | 1 | 0.52272452 | 1 | 0.862964608 | ENSG0000024 | 1 | RP1-228P16.1 |
| VAT | Obesity | ENSG0000019 | 0.000160996 | 0.95770016 | 0.003035381 | 0.183454473 | 0.00022515 | 0.940867353 | 0.003035192 | 0.183417544 | -6.42E-05 | 0.015538038 | 1 | 0.985127657 | 1 | 0.980488158 | ENSG0000024 | 1 | RP1-228P16.1 |
| VAT | Obesity | ENSG0000019 | 0.004115572 | 0.400347822 | 0.004893659 | 0.183454207 | 0.003700724 | 0.449673639 | 0.004895397 | 0.18341733 | 0.000414848 | 0.00498776 | 1 | 0.705593862 | 1 | 0.747200025 | ENSG0000024 | 1 | RPL7AP10 |
| VAT | Obesity | ENSG0000019 | 0.010408986 | 0.039294531 | 0.005050222 | 0.183452868 | -0.00183554 | 0.752099836 | 0.005811018 | 0.183417508 | 0.012244523 | 2.06E-05 | 1 | 0.190602646 | 1 | 0.89822395 | ENSG0000024 | 1 | RPL7AP10 |
| VAT | Obesity | ENSG0000019 | -0.00542324 | 0.045225655 | 0.002708161 | 0.183452958 | -0.00547625 | 0.043148153 | 0.002707962 | 0.183416 | 5.30E-05 | 0.00836982 | 1 | 0.209588906 | 1 | 0.210230467 | ENSG0000024 | 1 | RPL7AP10 |
| VAT | Obesity | ENSG0000019 | -0.0103243 | 2.66E-08 | 0.00185593 | 0.183442777 | -0.00799086 | 4.26E-05 | 0.001952418 | 0.183411215 | -0.00233345 | 0.000119915 | 7.33E-05 | 1.53E-06 | 0.117648929 | 0.001170502 | ENSG0000024 | 1 | RPL7AP10 |
| VAT | Obesity | ENSG0000019 | 0.009428414 | 0.077085792 | 0.005333251 | 0.183453293 | 0.009308697 | 0.080894514 | 0.005332899 | 0.183416395 | 0.000119717 | 0.006893134 | 1 | 0.284920982 | 1 | 0.303354427 | ENSG0000024 | 1 | RPL7AP10 |
| VAT | Obesity | ENSG0000019 | -0.00535803 | 0.300041422 | 0.00517012 | 0.183454068 | -0.00547498 | 0.289579096 | 0.005169727 | 0.183417122 | 0.00011695 | 0.001274401 | 1 | 0.608848729 | 1 | 0.611488195 | ENSG0000024 | 1 | L1TD1 |
| VAT | Obesity | ENSG0000019 | 0.06296627 | 6.18E-29 | 0.005640208 | 0.183407376 | 0.062849037 | 7.74E-29 | 0.005639844 | 0.183370625 | 0.000117233 | 0.013833941 | 1.71E-25 | 8.53E-26 | 2.14E-25 | 1.07E-25 | ENSG0000024 | 2 | ARHGEF25 |
| VAT | Obesity | ENSG0000019 | -0.00479685 | 0.145589168 | 0.003296145 | 0.183453674 | -0.00473402 | 0.150909054 | 0.003295914 | 0.183416766 | -6.28E-05 | 0.014801183 | 1 | 0.412455621 | 1 | 0.42939071 | ENSG0000024 | 1 | LINC00886 |
| VAT | Obesity | ENSG0000019 | -2.78E-05 | 0.996049969 | 0.005616404 | 0.183454474 | 9.52E-05 | 0.986475474 | 0.005615997 | 0.183417546 | -0.000123 | 0.003527849 | 1 | 0.998582606 | 1 | 0.996361581 | ENSG0000024 | 2 | HIST2H2BB |
| VAT | Obesity | ENSG0000019 | -0.00240833 | 0.394817478 | 0.002830287 | 0.1834542 | 0.011248937 | 0.035503456 | 0.005350076 | 0.183415875 | -0.01365727 | 0.002629737 | 1 | 0.701928581 | 1 | 0.184211409 | ENSG0000024 | 1 | MIF |
| VAT | Obesity | ENSG0000019 | -0.01212772 | 0.000280042 | 0.003338101 | 0.183449485 | -0.00870082 | 0.010999238 | 0.003421839 | 0.183415103 | -0.0034269 | 5.46E-06 | 0.772915908 | 0.005018934 | 1 | 0.084466367 | ENSG0000024 | 1 | MIF |
| VAT | Obesity | ENSG0000019 | -0.00519673 | 0.016794935 | 0.002173295 | 0.183452313 | -0.0052868 | 0.014992832 | 0.00217335 | 0.18341531 | 9.01E-05 | 0.008984204 | 1 | 0.108051331 | 1 | 0.107059606 | ENSG0000024 | 1 | MIF |
| VAT | Obesity | ENSG0000019 | -0.00192484 | 0.492533824 | 0.002804718 | 0.183454296 | -0.00187216 | 0.504423715 | 0.002804541 | 0.183417378 | -5.27E-05 | 0.030599067 | 1 | 0.766182814 | 1 | 0.78126232 | ENSG0000024 | 1 | MIF |
| VAT | Obesity | ENSG0000019 | 0.038906975 | 7.15E-13 | 0.005421214 | 0.183435007 | 0.038775569 | 8.50E-13 | 0.005420844 | 0.183398211 | 0.000131406 | 0.002444105 | 1.97E-09 | 8.22E-11 | 2.35E-09 | 9.77E-11 | ENSG0000024 | 1 | RP11-313M3.1 |
| VAT | Obesity | ENSG0000019 | -0.00441939 | 4.52E-05 | 0.001083433 | 0.183448185 | -0.00443337 | 4.27E-05 | 0.001083347 | 0.183411217 | 1.40E-05 | 0.046986894 | 0.12482401 | 0.001145174 | 0.117906757 | 0.001170502 | ENSG0000024 | 1 | RP11-313M3.1 |
| VAT | Obesity | ENSG0000019 | 0.014004748 | 0.000394452 | 0.003951909 | 0.183449727 | 0.009871607 | 0.015238396 | 0.004067962 | 0.183415321 | 0.00413314 | 1.89E-05 | 1 | 0.006519084 | 1 | 0.107453986 | ENSG0000024 | 1 | AC004985.12 |
| VAT | Obesity | ENSG0000019 | 0.001897307 | 0.10014979 | 0.001153988 | 0.183453452 | 0.001869574 | 0.10519499 | 0.001153932 | 0.183416554 | 2.77E-05 | 0.01927757 | 1 | 0.337913718 | 1 | 0.359264719 | ENSG0000024 | 1 | AC004985.12 |
| VAT | Obesity | ENSG0000019 | 0.002604284 | 0.490574267 | 0.00377763 | 0.183454294 | 0.002127962 | 0.573778417 | 0.00378307 | 0.183417426 | 0.000476322 | 0.023158132 | 1 | 0.766182814 | 1 | 0.814504585 | ENSG0000024 | 2 | HLA-DOB |
| VAT | Obesity | ENSG0000019 | 0.003853234 | 0.362792738 | 0.004234057 | 0.183454161 | 0.003773762 | 0.372743458 | 0.004233774 | 0.183417246 | 7.95E-05 | 0.022633463 | 1 | 0.672794162 | 1 | 0.695586169 | ENSG0000024 | 2 | HLA-DOB |
| VAT | Obesity | ENSG0000019 | -0.00060424 | 0.778300556 | 0.002146235 | 0.183454444 | -0.00053076 | 0.804665932 | 0.002146091 | 0.183417523 | -7.35E-05 | 3.05E-05 | 1 | 0.90675793 | 1 | 0.924210558 | ENSG0000024 | 2 | HLA-DOB |
| VAT | Obesity | ENSG0000019 | 0.009787347 | 0.023400297 | 0.004317613 | 0.183452532 | 0.009868379 | 0.022267808 | 0.0043173 | 0.183415571 | -8.10E-05 | 0.012544791 | 1 | 0.133993401 | 1 | 0.135971568 | ENSG0000024 | 2 | HLA-DOB |
| VAT | Obesity | ENSG0000019 | -0.00930964 | 0.116596899 | 0.005932647 | 0.183453543 | -0.00600216 | 0.314979262 | 0.005973312 | 0.183417164 | -0.00330748 | 2.36E-06 | 1 | 0.367994033 | 1 | 0.637753261 | ENSG0000024 | 2 | HLA-DOB |
| VAT | Obesity | ENSG0000019 | -0.00634794 | 0.304427033 | 0.006181124 | 0.183454075 | -0.00613141 | 0.321185081 | 0.006180697 | 0.183417174 | -0.00021652 | 9.82E-06 | 1 | 0.613298257 | 1 | 0.641712662 | ENSG0000024 | 2 | HLA-DOB |
| VAT | Obesity | ENSG0000019 | -0.0023274 | 0.42549021 | 0.00292045 | 0.183454234 | -0.00248738 | 0.394457359 | 0.00292096 | 0.183417272 | 0.000159976 | 0.019565543 | 1 | 0.714174021 | 1 | 0.714375058 | ENSG0000024 | 3 | TDGF1 |
| VAT | Obesity | ENSG0000019 | 0.001610482 | 0.713518485 | 0.00438662 | 0.183454423 | 0.001514638 | 0.729862812 | 0.004386336 | 0.183417501 | 9.58E-05 | 0.010091317 | 1 | 0.875776046 | 1 | 0.887801393 | ENSG0000024 | 3 | TDGF1 |
| VAT | Obesity | ENSG0000019 | 0.0015757 | 0.759162083 | 0.005139581 | 0.183454439 | 0.001401302 | 0.785109645 | 0.005139251 | 0.183417518 | 0.000174398 | 6.91E-05 | 1 | 0.897964537 | 1 | 0.913850934 | ENSG0000024 | 3 | TDGF1 |
| VAT | Obesity | ENSG0000019 | -0.00038479 | 0.935170004 | 0.004730553 | 0.183454472 | -0.00046819 | 0.921154529 | 0.004730185 | 0.183417542 | 8.34E-05 | 0.009369588 | 1 | 0.977307539 | 1 | 0.973721371 | ENSG0000024 | 3 | TDGF1 |
| VAT | Obesity | ENSG0000019 | 0.001904865 | 0.246760947 | 0.001644601 | 0.183453967 | 0.001931221 | 0.24024648 | 0.001644471 | 0.183417025 | -2.64E-05 | 0.01551751 | 1 | 0.543398386 | 1 | 0.546644917 | ENSG0000024 | 2 | CRCP |
| VAT | Obesity | ENSG0000019 | 0.00868444 | 0.01039125 | 0.003388996 | 0.183451992 | 0.008749844 | 0.009822039 | 0.00338873 | 0.183415026 | -6.54E-05 | 0.003632478 | 1 | 0.075863144 | 1 | 0.078123421 | ENSG0000024 | 2 | CRCP |
| VAT | Obesity | ENSG0000019 | -0.00107142 | 0.290640509 | 0.001013909 | 0.183454052 | 0.001059174 | 0.464612119 | 0.001448395 | 0.183417344 | -0.00213059 | 0.039429287 | 1 | 0.599975921 | 1 | 0.76239704 | ENSG0000024 | 5 | ARHGAP8 |
| VAT | Obesity | ENSG0000019 | 0.003635418 | 0.360439292 | 0.00397518 | 0.183454158 | 0.003579261 | 0.367870961 | 0.003974883 | 0.18341724 | 5.62E-05 | 0.049767141 | 1 | 0.670499496 | 1 | 0.690690966 | ENSG0000024 | 5 | ARHGAP8 |
| VAT | Obesity | ENSG0000019 | -0.00399251 | 0.183356506 | 0.003000776 | 0.183453805 | -0.00412277 | 0.169467793 | 0.003000739 | 0.183416833 | 0.000130257 | 0.001095397 | 1 | 0.477265662 | 1 | 0.463559076 | ENSG0000024 | 4 | GUSBP2 |
| VAT | Obesity | ENSG0000019 | -0.00751007 | 0.096731479 | 0.004521652 | 0.183453431 | -0.00743755 | 0.099973481 | 0.004521347 | 0.183416523 | -7.25E-05 | 0.048521881 | 1 | 0.328791727 | 1 | 0.347887069 | ENSG0000024 | 4 | GUSBP2 |
| VAT | Obesity | ENSG0000019 | -0.0062235 | 0.263023132 | 0.005560292 | 0.183454001 | -0.00603012 | 0.278112427 | 0.005559911 | 0.183417101 | -0.00019338 | 1.44E-05 | 1 | 0.564497547 | 1 | 0.59781176 | ENSG0000024 | 4 | GUSBP2 |
| VAT | Obesity | ENSG0000019 | -0.00378401 | 0.131231039 | 0.002507185 | 0.183453613 | -0.00368681 | 0.14140095 | 0.002507015 | 0.183416729 | -9.72E-05 | 1.64E-06 | 1 | 0.387974943 | 1 | 0.417278146 | ENSG0000024 | 4 | GUSBP2 |
| VAT | Obesity | ENSG0000019 | 0.002136507 | 0.684678417 | 0.005261198 | 0.183454412 | 0.002278414 | 0.664951589 | 0.005260866 | 0.183417475 | -0.00014191 | 0.001879556 | 1 | 0.863333733 | 1 | 0.856400553 | ENSG0000024 | 4 | GUSBP2 |
| VAT | Obesity | ENSG0000019 | 0.005406454 | 0.039719469 | 0.002628754 | 0.183452875 | 0.005348045 | 0.041892813 | 0.002628563 | 0.183415982 | 5.84E-05 | 0.002848644 | 1 | 0.191653384 | 1 | 0.205736949 | ENSG0000024 | 4 | GUSBP2 |
| VAT | Obesity | ENSG0000019 | 0.00049458 | 0.927182193 | 0.005411708 | 0.183454471 | 0.000594976 | 0.912448503 | 0.005411299 | 0.183417541 | -0.0001004 | 0.0089835 | 1 | 0.976170888 | 1 | 0.971531422 | ENSG0000024 | 4 | GUSBP2 |
| VAT | Obesity | ENSG0000019 | 0.004634456 | 0.068754188 | 0.002546354 | 0.183453222 | 0.004685874 | 0.065715303 | 0.002546166 | 0.183416266 | -5.14E-05 | 0.005724496 | 1 | 0.269094893 | 1 | 0.272854254 | ENSG0000024 | 4 | GUSBP2 |
| VAT | Obesity | ENSG0000019 | 0.00101243 | 0.789410359 | 0.003790785 | 0.183454447 | -0.00112951 | 0.770853572 | 0.003878021 | 0.183417514 | 0.002141937 | 0.008971998 | 1 | 0.911620331 | 1 | 0.908047742 | ENSG0000024 | 1 | RP11-381E24.1 |
| VAT | Obesity | ENSG0000020 | 0.005086854 | 0.371235968 | 0.005688966 | 0.183454172 | 0.004928001 | 0.386331092 | 0.005688622 | 0.183417262 | 0.000158853 | 0.001829361 | 1 | 0.68035277 | 1 | 0.707997923 | ENSG0000024 | 2 | INMT |
| VAT | Obesity | ENSG0000020 | -0.0120197 | 0.016888792 | 0.005030997 | 0.183452317 | -0.0121826 | 0.015449646 | 0.005030658 | 0.18341533 | 0.000162907 | 7.40E-05 | 1 | 0.108150968 | 1 | 0.108225945 | ENSG0000024 | 2 | INMT |
| VAT | Obesity | ENSG0000020 | -0.00908989 | 0.009590089 | 0.003509231 | 0.183451938 | -0.00901439 | 0.010201175 | 0.003508986 | 0.183415052 | -7.55E-05 | 0.006033237 | 1 | 0.071344059 | 1 | 0.080066313 | ENSG0000024 | 2 | INMT |
| VAT | Obesity | ENSG0000020 | -0.00750385 | 0.008622901 | 0.002856798 | 0.183451866 | -0.00745613 | 0.009050733 | 0.002856598 | 0.183414971 | -4.77E-05 | 0.033221443 | 1 | 0.066293054 | 1 | 0.073687385 | ENSG0000024 | 2 | RP11-80H8.4 |
| VAT | Obesity | ENSG0000020 | 0.002879959 | 0.394829178 | 0.003384631 | 0.1834542 | 0.001876034 | 0.580251072 | 0.003392354 | 0.18341743 | 0.001003925 | 1.75E-05 | 1 | 0.701928581 | 1 | 0.816782844 | ENSG0000024 | 2 | RP11-80H8.4 |
| VAT | Obesity | ENSG0000020 | -0.00044105 | 0.915997379 | 0.004181445 | 0.18345447 | -0.00519295 | 0.223831255 | 0.004269098 | 0.183416987 | 0.004751906 | 3.63E-08 | 1 | 0.972156914 | 1 | 0.533483819 | ENSG0000024 | 3 | TMEFF1 |
| VAT | Obesity | ENSG0000020 | -0.00208258 | 0.542372825 | 0.003418378 | 0.183454334 | -0.00219698 | 0.520394308 | 0.003418162 | 0.18341739 | 0.000114408 | 0.00011539 | 1 | 0.789547633 | 1 | 0.784425829 | ENSG0000024 | 2 | ATP5O |
| VAT | Obesity | ENSG0000020 | 0.000423464 | 0.857734858 | 0.002362312 | 0.183454462 | -0.00317587 | 0.194398887 | 0.002447352 | 0.18341691 | 0.003599338 | 1.91E-08 | 1 | 0.944116239 | 1 | 0.505693617 | ENSG0000024 | 2 | ATP5O |
| VAT | Obesity | ENSG0000020 | 0.005638796 | 0.204235654 | 0.004441473 | 0.183453865 | -0.00170906 | 0.71572942 | 0.004693008 | 0.183417496 | 0.00734786 | 1.28E-06 | 1 | 0.505551932 | 1 | 0.879525022 | ENSG0000024 | 2 | ATP5O |
| VAT | Obesity | ENSG0000020 | -0.0046807 | 0.007803568 | 0.001759354 | 0.183451799 | -0.00159977 | 0.397607963 | 0.001891196 | 0.183417276 | -0.00308093 | 9.07E-06 | 1 | 0.06153671 | 1 | 0.715434904 | ENSG0000024 | 2 | ATP5O |
| VAT | Obesity | ENSG0000020 | -0.00791718 | 6.21E-07 | 0.001588327 | 0.183445083 | -0.00694358 | 1.75E-05 | 0.001616797 | 0.183410576 | -0.0009736 | 0.001305824 | 0.001714273 | 2.68E-05 | 0.048301174 | 0.000589039 | ENSG0000024 | 5 | PLEKHO2 |
| VAT | Obesity | ENSG0000020 | 0.00279926 | 0.368424197 | 0.003112261 | 0.183454168 | -0.00248914 | 0.446973028 | 0.003273148 | 0.183417327 | 0.005288397 | 1.86E-07 | 1 | 0.676520736 | 1 | 0.74540517 | ENSG0000024 | 5 | PLEKHO2 |
| VAT | Obesity | ENSG0000020 | -0.0011332 | 0.549784019 | 0.001894725 | 0.183454339 | -0.00238978 | 0.218423663 | 0.001941754 | 0.183416974 | 0.001256574 | 0.003154294 | 1 | 0.793904838 | 1 | 0.529803765 | ENSG0000024 | 5 | PLEKHO2 |
| VAT | Obesity | ENSG0000020 | -0.02385117 | 1.69E-12 | 0.003379184 | 0.183435644 | -0.01973324 | 1.37E-07 | 0.00374496 | 0.183407053 | -0.00411793 | 0.010769324 | 4.66E-09 | 1.86E-10 | 0.00037817 | 7.88E-06 | ENSG0000024 | 5 | PLEKHO2 |
| VAT | Obesity | ENSG0000020 | -0.00017416 | 0.9593533 | 0.00341716 | 0.183454473 | 0.001520556 | 0.658448389 | 0.00343973 | 0.183417472 | -0.00169471 | 1.91E-05 | 1 | 0.98546132 | 1 | 0.854766662 | ENSG0000024 | 5 | PLEKHO2 |
| VAT | Obesity | ENSG0000020 | 0.000224928 | 0.857389484 | 0.0012517 | 0.183454462 | -0.00059905 | 0.635246406 | 0.001262862 | 0.183417461 | 0.000823974 | 1.00E-06 | 1 | 0.944116239 | 1 | 0.844363271 | ENSG0000024 | 6 | PI4KA |
| VAT | Obesity | ENSG0000020 | -0.00298427 | 0.157705406 | 0.002112256 | 0.18345372 | -0.00321577 | 0.128398807 | 0.002115011 | 0.183416672 | 0.000231501 | 0.038761966 | 1 | 0.433532788 | 1 | 0.395731266 | ENSG0000024 | 4 | LINC00969 |
| VAT | Obesity | ENSG0000020 | 0.004396101 | 0.006554414 | 0.001616995 | 0.18345168 | 0.00173033 | 0.320872782 | 0.001743112 | 0.183417174 | 0.00266577 | 4.26E-05 | 1 | 0.053526038 | 1 | 0.641712662 | ENSG0000024 | 4 | LINC00969 |
| VAT | Obesity | ENSG0000020 | 0.003372572 | 0.000399013 | 0.000952499 | 0.183449735 | 0.002241651 | 0.022242512 | 0.00098051 | 0.183415571 | 0.001130921 | 1.22E-06 | 1 | 0.006555209 | 1 | 0.135971568 | ENSG0000024 | 4 | LINC00969 |
| VAT | Obesity | ENSG0000020 | 0.005729579 | 3.87E-06 | 0.001240664 | 0.183446413 | 0.003457349 | 0.011508964 | 0.001368238 | 0.183415133 | 0.00227223 | 8.25E-05 | 0.010688541 | 0.000140639 | 1 | 0.087265774 | ENSG0000024 | 3 | AMACR |
| VAT | Obesity | ENSG0000020 | -0.00127559 | 0.176517637 | 0.000943792 | 0.183453784 | -0.00079783 | 0.40524168 | 0.000958584 | 0.183417284 | -0.00047776 | 0.004523263 | 1 | 0.465763555 | 1 | 0.720669275 | ENSG0000024 | 3 | AMACR |
| VAT | Obesity | ENSG0000020 | 0.001044658 | 0.810735086 | 0.00436223 | 0.183454452 | 0.000176006 | 0.967881123 | 0.004371079 | 0.183417545 | 0.000868652 | 0.002287783 | 1 | 0.923114207 | 1 | 0.990920808 | ENSG0000024 | 3 | AMACR |
| VAT | Obesity | ENSG0000020 | 0.009781839 | 0.001708198 | 0.003118425 | 0.183450755 | 0.00686076 | 0.031840969 | 0.003196392 | 0.183415805 | 0.002921079 | 3.25E-05 | 1 | 0.020002655 | 1 | 0.172994243 | ENSG0000024 | 3 | AMACR |
| VAT | Obesity | ENSG0000020 | 0.011197748 | 0.030095309 | 0.005163012 | 0.183452696 | 0.008583677 | 0.098593633 | 0.005196832 | 0.183416515 | 0.002614071 | 1.17E-05 | 1 | 0.159430046 | 1 | 0.344453707 | ENSG0000024 | 3 | AMACR |
| VAT | Obesity | ENSG0000020 | -0.02649797 | 4.38E-06 | 0.005769663 | 0.183446502 | -0.01890885 | 0.001756312 | 0.006043805 | 0.183413847 | -0.00758911 | 2.52E-05 | 0.012082985 | 0.000156922 | 1 | 0.02256353 | ENSG0000024 | 1 | RP11-299L17.1 |
| VAT | Obesity | ENSG0000020 | -0.0088971 | 0.085669031 | 0.005176637 | 0.183453358 | -0.00634171 | 0.224096108 | 0.005216464 | 0.183416987 | -0.0025554 | 7.93E-05 | 1 | 0.301975129 | 1 | 0.533537912 | ENSG0000024 | 5 | RP11-568K15.1 |
| VAT | Obesity | ENSG0000020 | 0.004931849 | 0.113098862 | 0.003112716 | 0.183453525 | 0.0030823 | 0.327767604 | 0.003149635 | 0.183417184 | 0.001849549 | 0.000127787 | 1 | 0.362126285 | 1 | 0.648715527 | ENSG0000024 | 1 | BGLAP |
| VAT | Obesity | ENSG0000020 | 0.005374648 | 5.75E-09 | 0.000922861 | 0.183441654 | 0.004078199 | 3.77E-05 | 0.000989502 | 0.183411126 | 0.001296449 | 0.00028386 | 1.59E-05 | 3.69E-07 | 0.103922864 | 0.001105562 | ENSG0000024 | 1 | BGLAP |
| VAT | Obesity | ENSG0000020 | -0.00748345 | 0.095907975 | 0.004494491 | 0.183453426 | -0.00738224 | 0.100461842 | 0.004494179 | 0.183416526 | -0.00010121 | 0.004316032 | 1 | 0.326866783 | 1 | 0.348773187 | ENSG0000024 | 2 | STAG3L5P |
| VAT | Obesity | ENSG0000020 | 0.014907423 | 1.14E-08 | 0.002611382 | 0.183442156 | 0.011580548 | 2.70E-05 | 0.002758823 | 0.183410887 | 0.003326875 | 0.000186927 | 3.15E-05 | 6.99E-07 | 0.074453131 | 0.000827257 | ENSG0000024 | 2 | EIF6 |
| VAT | Obesity | ENSG0000020 | -0.01141776 | 0.000575485 | 0.003316285 | 0.183449993 | -0.01034131 | 0.001891382 | 0.003328611 | 0.183413898 | -0.00107644 | 0.000205117 | 1 | 0.008727132 | 1 | 0.023620883 | ENSG0000024 | 2 | EIF6 |
| VAT | Obesity | ENSG0000020 | -0.00256912 | 0.36421688 | 0.002831422 | 0.183454163 | -0.00251705 | 0.373989703 | 0.002831241 | 0.183417247 | -5.21E-05 | 0.031786662 | 1 | 0.673301132 | 1 | 0.697440257 | ENSG0000024 | 2 | EIF6 |
| VAT | Obesity | ENSG0000020 | 0.005732036 | 0.22631644 | 0.004737597 | 0.183453921 | 0.005644583 | 0.23344409 | 0.004737235 | 0.183417009 | 8.75E-05 | 0.007898097 | 1 | 0.52272452 | 1 | 0.540524906 | ENSG0000024 | 1 | UPK3BP1 |
| VAT | Obesity | ENSG0000020 | 0.003922609 | 0.480436037 | 0.005559237 | 0.183454286 | 0.006103429 | 0.279863972 | 0.005648049 | 0.183417105 | -0.00218082 | 0.029325911 | 1 | 0.762547611 | 1 | 0.59924326 | ENSG0000024 | 1 | UPK3BP1 |
| VAT | Obesity | ENSG0000020 | -0.01615507 | 0.004685479 | 0.005712722 | 0.183451451 | -0.01367359 | 0.017902638 | 0.005775213 | 0.183415427 | -0.00248148 | 0.003548413 | 1 | 0.042123525 | 1 | 0.119020171 | ENSG0000024 | 1 | UPK3BP1 |
| VAT | Obesity | ENSG0000020 | -0.00375573 | 0.486357886 | 0.005395288 | 0.183454291 | -0.00354555 | 0.51105318 | 0.005394938 | 0.183417383 | -0.00021019 | 4.17E-06 | 1 | 0.765031548 | 1 | 0.784425829 | ENSG0000024 | 3 | GTF2A1L |
| VAT | Obesity | ENSG0000020 | -0.00207822 | 0.54423783 | 0.003427049 | 0.183454335 | -0.00220468 | 0.520017121 | 0.003427039 | 0.18341739 | 0.000126458 | 0.008252021 | 1 | 0.79046238 | 1 | 0.784425829 | ENSG0000024 | 1 | LINC00960 |
| VAT | Obesity | ENSG0000020 | 0.000425228 | 0.824139495 | 0.001913511 | 0.183454455 | 0.000386882 | 0.83976398 | 0.001913406 | 0.183417531 | 3.83E-05 | 0.036123091 | 1 | 0.927365584 | 1 | 0.939543177 | ENSG0000024 | 1 | LINC00960 |
| VAT | Obesity | ENSG0000020 | -0.00709187 | 0.006074731 | 0.002584755 | 0.183451629 | -0.00717925 | 0.005474262 | 0.002584575 | 0.18341463 | 8.74E-05 | 2.07E-05 | 1 | 0.05096127 | 1 | 0.05044591 | ENSG0000024 | 1 | LINC00960 |
| VAT | Obesity | ENSG0000020 | -0.00661769 | 0.109640183 | 0.004136534 | 0.183453507 | -0.00653557 | 0.114092795 | 0.004136269 | 0.183416602 | -8.21E-05 | 0.02009526 | 1 | 0.355833781 | 1 | 0.370902372 | ENSG0000024 | 2 | MBL1P |
| VAT | Obesity | ENSG0000020 | -0.00254377 | 0.312034919 | 0.002516185 | 0.183454088 | -0.0034345 | 0.173056338 | 0.002520828 | 0.183416844 | 0.000890736 | 1.42E-08 | 1 | 0.619458427 | 1 | 0.470490411 | ENSG0000024 | 3 | GNG10 |
| VAT | Obesity | ENSG0000020 | -0.01879857 | 3.87E-20 | 0.002045077 | 0.183422541 | -0.01884251 | 3.15E-20 | 0.002044932 | 0.183385464 | 4.39E-05 | 0.005765058 | 1.07E-16 | 1.19E-17 | 8.68E-17 | 9.65E-18 | ENSG0000024 | 2 | ZNF702P |
| VAT | Obesity | ENSG0000020 | -0.00270091 | 0.366933405 | 0.002993579 | 0.183454166 | -0.0028066 | 0.348449607 | 0.002993386 | 0.183417214 | 0.000105697 | 3.37E-05 | 1 | 0.675157466 | 1 | 0.670656147 | ENSG0000024 | 2 | ZNF702P |
| VAT | Obesity | ENSG0000020 | -0.00049009 | 0.871255769 | 0.003024084 | 0.183454464 | -0.00039056 | 0.897233834 | 0.003023895 | 0.18341754 | -9.95E-05 | 0.000168095 | 1 | 0.949335145 | 1 | 0.968028006 | ENSG0000024 | 2 | ZNF702P |
| VAT | Obesity | ENSG0000020 | 0.009369088 | 0.052132317 | 0.004824343 | 0.183453048 | 0.009287123 | 0.054205519 | 0.00482399 | 0.183416145 | 8.20E-05 | 0.021689438 | 1 | 0.229848555 | 1 | 0.24496821 | ENSG0000024 | 1 | EIF4EBP3 |

| | | | | | | | | | | | | | | | | | | | |
|---|---|---|---|---|---|---|---|---|---|---|---|---|---|---|---|---|---|---|---|
| VAT | Obesity | ENSG0000020 | -0.01002136 | 0.047450334 | 0.005055514 | 0.183452989 | -0.00989432 | 0.05031703 | 0.005055181 | 0.183416098 | -0.00012704 | 0.002591761 | 1 | 0.215754403 | 1 | 0.23538136 | ENSG0000024 | 1 | EIF4EBP3 |
| VAT | Obesity | ENSG0000020 | -0.0007653 | 0.784484859 | 0.002798376 | 0.183454446 | 0.000831184 | 0.768606325 | 0.002825249 | 0.183417513 | -0.00159648 | 4.40E-05 | 1 | 0.909319831 | 1 | 0.907673971 | ENSG0000024 | 1 | EIF4EBP3 |
| VAT | Obesity | ENSG0000020 | -0.00226856 | 0.226513958 | 0.001875793 | 0.183453921 | -0.00113196 | 0.559764142 | 0.001940965 | 0.183417417 | -0.00113661 | 0.022861248 | 1 | 0.52272452 | 1 | 0.809555852 | ENSG0000024 | 1 | STON1 |
| VAT | Obesity | ENSG0000020 | -0.00907852 | 0.039477611 | 0.004408807 | 0.183452871 | -0.00705131 | 0.111138452 | 0.004426155 | 0.183416587 | -0.00202722 | 3.15E-07 | 1 | 0.191154746 | 1 | 0.366914394 | ENSG0000024 | 3 | PALM2 |
| VAT | Obesity | ENSG0000020 | -0.0002387 | 0.965102883 | 0.005455903 | 0.183454473 | -8.12E-05 | 0.988119418 | 0.005455514 | 0.183417546 | -0.00015747 | 0.000164573 | 1 | 0.985729121 | 1 | 0.996361581 | ENSG0000024 | 3 | PALM2 |
| VAT | Obesity | ENSG0000021 | -0.01183542 | 7.48E-05 | 0.002988435 | 0.183448545 | -0.01177004 | 8.19E-05 | 0.002988221 | 0.183411683 | -6.54E-05 | 0.0040622 | 0.206537765 | 0.001721148 | 0.226036704 | 0.002018185 | ENSG0000024 | 3 | AOX2P |
| VAT | Obesity | ENSG0000021 | -0.00743342 | 0.055522083 | 0.003882141 | 0.183453088 | -0.00736779 | 0.057697205 | 0.003881877 | 0.183416185 | -6.56E-05 | 0.036147161 | 1 | 0.237582866 | 1 | 0.252768707 | ENSG0000024 | 3 | AOX2P |
| VAT | Obesity | ENSG0000021 | -0.00525299 | 0.081431782 | 0.003014728 | 0.183453326 | -0.00519544 | 0.084805824 | 0.003014537 | 0.183416423 | -5.76E-05 | 0.027234439 | 1 | 0.293409555 | 1 | 0.310429805 | ENSG0000024 | 4 | IL10RB |
| VAT | Obesity | ENSG0000021 | 0.000330963 | 0.948410326 | 0.005115089 | 0.183454473 | 0.000251231 | 0.96082436 | 0.005114719 | 0.183417545 | 7.97E-05 | 0.038432291 | 1 | 0.982080274 | 1 | 0.98754495 | ENSG0000024 | 4 | IL10RB |
| VAT | Obesity | ENSG0000021 | 0.005236009 | 0.293546583 | 0.004984899 | 0.183454057 | 0.005367348 | 0.281592103 | 0.004984763 | 0.183417108 | -0.00013134 | 0.029775961 | 1 | 0.602370683 | 1 | 0.600777491 | ENSG0000024 | 4 | IL10RB |
| VAT | Obesity | ENSG0000021 | 0.00331496 | 0.42858028 | 0.004187543 | 0.183454237 | 0.002927342 | 0.484691796 | 0.004189237 | 0.183417361 | 0.000387617 | 0.003593713 | 1 | 0.71733267 | 1 | 0.775358255 | ENSG0000024 | 2 | TTC4 |
| VAT | Obesity | ENSG0000021 | 0.000402739 | 0.767504915 | 0.00136226 | 0.183454441 | 0.000358576 | 0.792366515 | 0.001362169 | 0.18341752 | 4.42E-05 | 8.38E-05 | 1 | 0.901410028 | 1 | 0.918062321 | ENSG0000024 | 1 | HLA-L |
| VAT | Obesity | ENSG0000021 | 0.008193612 | 0.00035941 | 0.002296256 | 0.183449661 | 0.007284576 | 0.00155021 | 0.002301506 | 0.18341376 | 0.000909036 | 1.01E-08 | 0.991971782 | 0.005975734 | 1 | 0.020868421 | ENSG0000024 | 1 | HLA-L |
| VAT | Obesity | ENSG0000021 | 0.002973158 | 0.439500479 | 0.003846072 | 0.183454248 | 0.003032046 | 0.43045901 | 0.003845787 | 0.183417311 | -5.89E-05 | 0.035797359 | 1 | 0.726033105 | 1 | 0.735057673 | ENSG0000024 | 1 | HLA-L |
| VAT | Obesity | ENSG0000021 | 0.008572563 | 0.012385967 | 0.003427706 | 0.18345211 | -8.44E-05 | 0.983378242 | 0.004050701 | 0.183417546 | 0.008656954 | 6.08E-05 | 1 | 0.086985415 | 1 | 0.996238962 | ENSG0000024 | 1 | HLA-L |
| VAT | Obesity | ENSG0000021 | -0.00051202 | 0.791902051 | 0.001940623 | 0.183454448 | -0.00056414 | 0.771269497 | 0.001940527 | 0.183417514 | 5.21E-05 | 0.007876598 | 1 | 0.913008119 | 1 | 0.908150091 | ENSG0000024 | 1 | HLA-L |
| VAT | Obesity | ENSG0000021 | 0.005252967 | 0.002783653 | 0.001756442 | 0.183451093 | 0.005209813 | 0.00301453 | 0.001756353 | 0.183414221 | 4.32E-05 | 0.013927948 | 1 | 0.029101822 | 1 | 0.033147822 | ENSG0000024 | 1 | HLA-L |
| VAT | Obesity | ENSG0000021 | -0.00468035 | 0.020070182 | 0.002013019 | 0.183452431 | -0.00389515 | 0.056339424 | 0.002041052 | 0.18341617 | -0.0007852 | 0.020288104 | 1 | 0.120683447 | 1 | 0.24999487 | ENSG0000024 | 1 | HLA-L |
| VAT | Obesity | ENSG0000021 | 0.009722559 | 0.004841748 | 0.003450909 | 0.183451474 | 0.009568048 | 0.005564308 | 0.003451152 | 0.183414641 | 0.000154511 | 0.015479844 | 1 | 0.04296857 | 1 | 0.051021564 | ENSG0000024 | 1 | HLA-L |
| VAT | Obesity | ENSG0000021 | 0.014886186 | 0.003764004 | 0.005137942 | 0.183451301 | 0.014773845 | 0.004032406 | 0.005137604 | 0.183414421 | 0.000112342 | 0.008675888 | 1 | 0.036051294 | 1 | 0.041068046 | ENSG0000024 | 1 | ST13P15 |
| VAT | Obesity | ENSG0000021 | -0.0089568 | 0.028768807 | 0.004096135 | 0.183452667 | -0.00902544 | 0.027554953 | 0.004095822 | 0.183415711 | 6.86E-05 | 0.016324546 | 1 | 0.154178459 | 1 | 0.15580997 | ENSG0000024 | 6 | JMJD7 |
| VAT | Obesity | ENSG0000021 | -0.00140426 | 0.721660041 | 0.003941883 | 0.183454426 | -0.00147217 | 0.708779198 | 0.003941585 | 0.183417493 | 6.79E-05 | 0.015298619 | 1 | 0.880540103 | 1 | 0.876447308 | ENSG0000024 | 6 | JMJD7 |
| VAT | Obesity | ENSG0000021 | -0.00336476 | 0.123868793 | 0.0021867 | 0.183453579 | -0.00341386 | 0.118454217 | 0.002186556 | 0.183416625 | 4.91E-05 | 0.007308581 | 1 | 0.37657343 | 1 | 0.37883388 | ENSG0000024 | 3 | RN7SL832P |
| VAT | Obesity | ENSG0000021 | -0.00906925 | 0.104228913 | 0.005582137 | 0.183453476 | -0.00897397 | 0.10789248 | 0.005581725 | 0.183416569 | -9.53E-05 | 0.019898525 | 1 | 0.347010616 | 1 | 0.363593705 | ENSG0000024 | 3 | RN7SL832P |
| VAT | Obesity | ENSG0000021 | -0.00462327 | 0.221783643 | 0.003783981 | 0.18345391 | -0.00447248 | 0.237248695 | 0.003784167 | 0.183417018 | -0.00015079 | 0.02124728 | 1 | 0.52199987 | 1 | 0.544132965 | ENSG0000024 | 3 | RN7SL832P |
| VAT | Obesity | ENSG0000021 | 0.001049872 | 0.827503176 | 0.004818027 | 0.183454456 | 0.001198095 | 0.803604385 | 0.004817716 | 0.183417523 | -0.00014822 | 0.000303767 | 1 | 0.928795757 | 1 | 0.92407402 | ENSG0000024 | 2 | RP5-839B4.8 |
| VAT | Obesity | ENSG0000021 | -0.01188475 | 0.048076582 | 0.00601243 | 0.183452997 | -0.0116319 | 0.053019071 | 0.00601203 | 0.183416131 | -0.00025285 | 3.15E-07 | 1 | 0.216635313 | 1 | 0.242272577 | ENSG0000024 | 2 | RP5-839B4.8 |
| VAT | Obesity | ENSG0000021 | 0.001156462 | 0.622523918 | 0.002349211 | 0.183454383 | 0.001113859 | 0.63537574 | 0.002349048 | 0.183417461 | 4.26E-05 | 0.022107016 | 1 | 0.833933632 | 1 | 0.844363271 | ENSG0000024 | 2 | FAM86DP |
| VAT | Obesity | ENSG0000021 | 0.00609358 | 3.22E-06 | 0.001308694 | 0.183446279 | 0.004908492 | 0.00023558 | 0.001334745 | 0.183412435 | 0.001185088 | 6.62E-06 | 0.008890984 | 0.000120148 | 0.650200703 | 0.004745991 | ENSG0000024 | 2 | FAM86DP |
| VAT | Obesity | ENSG0000021 | -0.00177837 | 0.397425836 | 0.002101529 | 0.183454203 | 0.000453291 | 0.833012594 | 0.002149935 | 0.183417529 | -0.00223166 | 9.16E-07 | 1 | 0.702687577 | 1 | 0.93758435 | ENSG0000024 | 6 | TMEM199 |
| VAT | Obesity | ENSG0000021 | 0.00492702 | 0.083075198 | 0.002842863 | 0.183453339 | 0.0049904 | 0.079167577 | 0.002842658 | 0.183416381 | -6.34E-05 | 0.003136426 | 1 | 0.297357955 | 1 | 0.298885799 | ENSG0000024 | 6 | TMEM199 |
| VAT | Obesity | ENSG0000021 | -0.00165893 | 0.351137717 | 0.001779231 | 0.183454145 | -0.00159944 | 0.368647554 | 0.001779114 | 0.183417241 | -5.95E-05 | 6.02E-05 | 1 | 0.661582235 | 1 | 0.69094905 | ENSG0000024 | 1 | PDCL3P4 |
| VAT | Obesity | ENSG0000021 | -0.01204895 | 0.001346548 | 0.003758382 | 0.183450589 | -0.00879315 | 0.021147769 | 0.003814247 | 0.183415537 | -0.00325581 | 6.04E-07 | 1 | 0.01689306 | 1 | 0.132256412 | ENSG0000024 | 1 | P2RY11 |
| VAT | Obesity | ENSG0000022 | -0.00784704 | 0.186624892 | 0.005941884 | 0.183453815 | -0.00864753 | 0.145672693 | 0.005943358 | 0.183416746 | 0.000800487 | 3.43E-07 | 1 | 0.482741052 | 1 | 0.421062772 | ENSG0000024 | 1 | P2RY11 |
| VAT | Obesity | ENSG0000022 | 0.003875688 | 0.068824242 | 0.002129995 | 0.183453223 | 0.003834302 | 0.071820465 | 0.002129862 | 0.183416321 | 4.14E-05 | 0.02595129 | 1 | 0.269094893 | 1 | 0.283582953 | ENSG0000024 | 1 | P2RY11 |
| VAT | Obesity | ENSG0000022 | -0.00662018 | 0.221893957 | 0.005419675 | 0.18345391 | -0.00649354 | 0.230827018 | 0.005419268 | 0.183417003 | -0.00012664 | 0.00106084 | 1 | 0.52199987 | 1 | 0.540207072 | ENSG0000024 | 1 | P2RY11 |
| VAT | Obesity | ENSG0000022 | 0.000218478 | 0.949622701 | 0.003457988 | 0.183454473 | 0.000304828 | 0.92975104 | 0.003457739 | 0.183417543 | -8.63E-05 | 0.000965017 | 1 | 0.982080274 | 1 | 0.976079449 | ENSG0000024 | 1 | P2RY11 |
| VAT | Obesity | ENSG0000022 | -0.02224963 | 0.000125171 | 0.005800516 | 0.183448913 | -0.02207068 | 0.000141769 | 0.005800302 | 0.183412074 | -0.00017896 | 0.006188773 | 0.345471582 | 0.002637188 | 0.39128379 | 0.003155514 | ENSG0000024 | 1 | P2RY11 |
| VAT | Obesity | ENSG0000022 | -0.00150055 | 0.726241582 | 0.004285702 | 0.183454428 | -0.00141421 | 0.741400152 | 0.004285472 | 0.183417505 | -8.63E-05 | 0.037344505 | 1 | 0.882200818 | 1 | 0.893915902 | ENSG0000024 | 2 | GS1-259H13.2 |
| VAT | Obesity | ENSG0000022 | -0.00040369 | 0.869649636 | 0.002459961 | 0.183454464 | -0.00035765 | 0.88439654 | 0.002459804 | 0.183417538 | -4.60E-05 | 0.029566676 | 1 | 0.948684727 | 1 | 0.960056703 | ENSG0000024 | 2 | GS1-259H13.2 |
| VAT | Obesity | ENSG0000022 | -0.00278188 | 0.357484981 | 0.003023224 | 0.183454154 | -0.00273899 | 0.364907951 | 0.003022994 | 0.183417236 | -4.29E-05 | 0.043748823 | 1 | 0.668015266 | 1 | 0.688411445 | ENSG0000024 | 2 | GS1-259H13.2 |
| VAT | Obesity | ENSG0000022 | 0.002299169 | 0.250977122 | 0.002002795 | 0.183453976 | 0.001291349 | 0.521399569 | 0.002013989 | 0.183417391 | 0.00100782 | 2.46E-06 | 1 | 0.549323438 | 1 | 0.784425829 | ENSG0000024 | 2 | GS1-259H13.2 |
| VAT | Obesity | ENSG0000022 | 0.014273928 | 0.000128122 | 0.003726804 | 0.183448929 | 0.002444052 | 0.614001843 | 0.004845756 | 0.18341745 | 0.011829876 | 0.000133919 | 0.353616871 | 0.002678916 | 1 | 0.839253732 | ENSG0000024 | 1 | C7orf13 |
| VAT | Obesity | ENSG0000022 | 0.001931546 | 0.054092077 | 0.001002826 | 0.183453072 | 0.001907719 | 0.057113505 | 0.001002772 | 0.183416178 | 2.38E-05 | 0.014090989 | 1 | 0.236224894 | 1 | 0.251902356 | ENSG0000024 | 1 | AOX3P |
| VAT | Obesity | ENSG0000022 | 0.000739088 | 0.61883744 | 0.001485606 | 0.183454381 | -0.00032391 | 0.829530804 | 0.001504449 | 0.183417529 | 0.001063001 | 8.16E-06 | 1 | 0.831754083 | 1 | 0.936897537 | ENSG0000024 | 1 | AOX3P |
| VAT | Obesity | ENSG0000022 | 0.00032938 | 0.893337686 | 0.002456557 | 0.183454467 | 0.000236042 | 0.923447531 | 0.002456421 | 0.183417543 | 9.33E-05 | 6.72E-05 | 1 | 0.961627151 | 1 | 0.974118595 | ENSG0000024 | 2 | CFHR1 |
| VAT | Obesity | ENSG0000022 | -0.00486642 | 0.334732609 | 0.005044874 | 0.183454122 | -0.0046247 | 0.359312316 | 0.005045067 | 0.183417228 | -0.00024172 | 0.004026963 | 1 | 0.64291023 | 1 | 0.683932408 | ENSG0000024 | 2 | CFHR1 |
| VAT | Obesity | ENSG0000022 | -0.00042915 | 0.776016082 | 0.001508372 | 0.183454444 | 0.000709543 | 0.642809086 | 0.00152993 | 0.183417465 | -0.0011387 | 9.26E-06 | 1 | 0.904860323 | 1 | 0.849283426 | ENSG0000024 | 2 | CFHR1 |
| VAT | Obesity | ENSG0000022 | 0.00734849 | 0.000401411 | 0.002076326 | 0.183449739 | 0.006041627 | 0.003900377 | 0.002093337 | 0.183414398 | 0.001306863 | 1.07E-06 | 1 | 0.006555585 | 1 | 0.040018734 | ENSG0000024 | 5 | LILRA6 |
| VAT | Obesity | ENSG0000022 | -0.02894543 | 3.98E-08 | 0.005270455 | 0.183443073 | -0.02878792 | 4.70E-08 | 0.005270076 | 0.183406269 | -0.00015751 | 7.42E-05 | 0.000109728 | 2.24E-06 | 0.000129621 | 3.09E-06 | ENSG0000024 | 5 | LILRA6 |
| VAT | Obesity | ENSG0000022 | -0.00153721 | 0.485628365 | 0.002204584 | 0.18345429 | -0.00150402 | 0.495067821 | 0.002204425 | 0.18341737 | -3.32E-05 | 0.046473865 | 1 | 0.765003071 | 1 | 0.775358255 | ENSG0000024 | 5 | LILRA6 |
| VAT | Obesity | ENSG0000022 | -0.01914943 | 0.001863007 | 0.006154877 | 0.183450815 | -0.01943018 | 0.001593722 | 0.006154496 | 0.183413779 | 0.000280749 | 2.39E-07 | 1 | 0.021380487 | 1 | 0.021249624 | ENSG0000024 | 2 | RP4-800G7.2 |
| VAT | Obesity | ENSG0000022 | -0.00094101 | 0.833588914 | 0.00447883 | 0.183454457 | -0.00086148 | 0.847460661 | 0.004478505 | 0.183417532 | -7.95E-05 | 0.018217698 | 1 | 0.933349047 | 1 | 0.944145183 | ENSG0000024 | 1 | CCDC13 |
| VAT | Obesity | ENSG0000022 | 0.000481394 | 0.77070857 | 0.00165173 | 0.183454442 | 0.002038434 | 0.229510966 | 0.001696403 | 0.183417 | -0.00155704 | 5.86E-05 | 1 | 0.904018552 | 1 | 0.539107619 | ENSG0000024 | 1 | CCDC13 |
| VAT | Obesity | ENSG0000022 | 0.000269125 | 0.963640039 | 0.005903632 | 0.183454473 | 0.000186142 | 0.97484494 | 0.005903186 | 0.183417546 | 8.30E-05 | 0.046894855 | 1 | 0.985729121 | 1 | 0.993586204 | ENSG0000024 | 1 | CCDC13 |
| VAT | Obesity | ENSG0000022 | 0.001157831 | 0.367758307 | 0.001285506 | 0.183454167 | 0.000853667 | 0.507109764 | 0.001286912 | 0.18341738 | 0.000304164 | 1.29E-06 | 1 | 0.676224469 | 1 | 0.783244156 | ENSG0000024 | 1 | CCDC13 |
| VAT | Obesity | ENSG0000022 | 0.005226518 | 0.130882315 | 0.003459817 | 0.183453612 | 0.006316933 | 0.068606738 | 0.003468927 | 0.183416293 | -0.00109042 | 2.06E-05 | 1 | 0.387974943 | 1 | 0.277239528 | ENSG0000024 | 1 | OR2AE1 |
| VAT | Obesity | ENSG0000022 | -0.00237868 | 0.651484674 | 0.005266045 | 0.183454397 | -0.00260596 | 0.620674874 | 0.005265686 | 0.183417453 | 0.000227281 | 8.24E-08 | 1 | 0.848236169 | 1 | 0.840409069 | ENSG0000024 | 1 | CTA-211A9.5 |
| VAT | Obesity | ENSG0000022 | -0.00742798 | 0.119550477 | 0.004771725 | 0.183453558 | -0.00756955 | 0.112649551 | 0.004771533 | 0.183416595 | 0.000141571 | 0.006962282 | 1 | 0.372684435 | 1 | 0.367944097 | ENSG0000024 | 1 | CTA-211A9.5 |
| VAT | Obesity | ENSG0000022 | 0.001520923 | 0.395052746 | 0.001788288 | 0.183454201 | 0.001355336 | 0.448549725 | 0.00178843 | 0.183417329 | 0.000165586 | 1.06E-06 | 1 | 0.701928581 | 1 | 0.747131709 | ENSG0000024 | 1 | CTAGE8 |
| VAT | Obesity | ENSG0000022 | -0.01715758 | 0.00355177 | 0.005885027 | 0.183451261 | -0.01191549 | 0.046704336 | 0.005990764 | 0.183416051 | -0.00524209 | 3.08E-06 | 1 | 0.034455189 | 1 | 0.222632069 | ENSG0000024 | 3 | ASLP1 |
| VAT | Obesity | ENSG0000022 | 0.002570862 | 0.439828751 | 0.00332805 | 0.183454249 | 0.002516804 | 0.449473568 | 0.003327814 | 0.18341733 | 5.41E-05 | 0.034901133 | 1 | 0.726033105 | 1 | 0.747200025 | ENSG0000024 | 3 | C4A |
| VAT | Obesity | ENSG0000022 | -0.01469889 | 0.017686187 | 0.006196492 | 0.183452347 | -0.01455863 | 0.018790783 | 0.006196049 | 0.18341546 | -0.00014026 | 0.002978122 | 1 | 0.110689061 | 1 | 0.120957701 | ENSG0000024 | 3 | C4A |
| VAT | Obesity | ENSG0000022 | -0.00585254 | 0.005960705 | 0.002128239 | 0.183451616 | -0.00433856 | 0.044289832 | 0.002157047 | 0.183416017 | -0.00151398 | 1.76E-05 | 1 | 0.050310539 | 1 | 0.214456031 | ENSG0000024 | 3 | C4A |
| VAT | Obesity | ENSG0000022 | -0.00151592 | 0.319045968 | 0.001521368 | 0.183454099 | -0.00148376 | 0.329389458 | 0.001521263 | 0.183417187 | -3.22E-05 | 0.008373212 | 1 | 0.626292228 | 1 | 0.650296785 | ENSG0000024 | 3 | C4A |
| VAT | Obesity | ENSG0000022 | 0.006576188 | 0.04354673 | 0.003258066 | 0.183452934 | 0.006660618 | 0.040905055 | 0.003257828 | 0.183415966 | -8.44E-05 | 0.000506537 | 1 | 0.205215702 | 1 | 0.203053868 | ENSG0000024 | 3 | C4A |
| VAT | Obesity | ENSG0000022 | 0.003339077 | 0.145853872 | 0.002295946 | 0.183453675 | 9.13E-05 | 0.96973808 | 0.002405317 | 0.183417545 | 0.003247826 | 6.05E-06 | 1 | 0.412455621 | 1 | 0.990920808 | ENSG0000024 | 3 | C4A |
| VAT | Obesity | ENSG0000022 | -0.02124974 | 6.79E-07 | 0.004277871 | 0.183445148 | -0.01391001 | 0.003132945 | 0.004708239 | 0.183414247 | -0.00733972 | 0.000190878 | 0.001873842 | 2.88E-05 | 1 | 0.03417758 | ENSG0000024 | 3 | C4A |
| VAT | Obesity | ENSG0000022 | -0.00103265 | 0.867993346 | 0.006213009 | 0.183454464 | -0.0008268 | 0.894126624 | 0.006212598 | 0.183417539 | -0.00020585 | 6.58E-05 | 1 | 0.948025973 | 1 | 0.968028006 | ENSG0000024 | 3 | C4A |
| VAT | Obesity | ENSG0000022 | 0.000536887 | 0.73604818 | 0.001592703 | 0.183454431 | 0.000505346 | 0.751008197 | 0.001592592 | 0.183417508 | 3.15E-05 | 0.011230133 | 1 | 0.886561612 | 1 | 0.897653804 | ENSG0000024 | 3 | C4A |
| VAT | Obesity | ENSG0000022 | -0.0082344 | 0.121893724 | 0.005323246 | 0.18345357 | -0.00831483 | 0.118264223 | 0.005322841 | 0.183416624 | 8.04E-05 | 0.031703144 | 1 | 0.374639953 | 1 | 0.37866503 | ENSG0000024 | 3 | C4A |
| VAT | Obesity | ENSG0000022 | 0.007474284 | 6.80E-05 | 0.001876402 | 0.183448477 | 0.005851993 | 0.002134658 | 0.001905662 | 0.183413982 | 0.001622292 | 1.16E-06 | 0.187604572 | 0.001603458 | 1 | 0.025615891 | ENSG0000024 | 2 | CRYBB2 |
| VAT | Obesity | ENSG0000022 | -0.0015036 | 0.597719346 | 0.00284943 | 0.183454369 | -0.00142601 | 0.616730065 | 0.002849233 | 0.183417451 | -7.76E-05 | 0.000610253 | 1 | 0.820340823 | 1 | 0.839253732 | ENSG0000024 | 2 | RP11-33B1.1 |
| VAT | Obesity | ENSG0000022 | -0.00537163 | 0.357283822 | 0.005835215 | 0.183454154 | -0.00573691 | 0.325557738 | 0.005835515 | 0.183417181 | 0.000365276 | 0.000332089 | 1 | 0.668015266 | 1 | 0.646431191 | ENSG0000024 | 2 | RP11-33B1.1 |
| VAT | Obesity | ENSG0000022 | -0.00240024 | 0.135150375 | 0.001606479 | 0.18345363 | -0.0003451 | 0.835723755 | 0.001664182 | 0.18341753 | -0.00205514 | 2.31E-06 | 1 | 0.394724904 | 1 | 0.938551038 | ENSG0000024 | 2 | RP11-33B1.1 |
| VAT | Obesity | ENSG0000022 | 0.005190599 | 0.376841483 | 0.005873501 | 0.183454179 | 0.005054623 | 0.389437675 | 0.005873109 | 0.183417266 | 0.000135976 | 0.004889258 | 1 | 0.685170491 | 1 | 0.709470616 | ENSG0000024 | 2 | RP11-33B1.1 |
| VAT | Obesity | ENSG0000022 | -0.00531319 | 0.067464481 | 0.002905653 | 0.18345321 | -0.00315906 | 0.282034199 | 0.002936577 | 0.183417109 | -0.00215414 | 4.54E-07 | 1 | 0.266383359 | 1 | 0.601092193 | ENSG0000024 | 3 | RAB30-AS1 |
| VAT | Obesity | ENSG0000023 | -0.00637316 | 0.024811283 | 0.002839666 | 0.18345257 | -0.00237792 | 0.426733716 | 0.002991874 | 0.183417307 | -0.00399524 | 2.27E-05 | 1 | 0.139753347 | 1 | 0.73108942 | ENSG0000024 | 3 | RAB30-AS1 |
| VAT | Obesity | ENSG0000023 | 0.002026367 | 0.554902369 | 0.003432016 | 0.183454342 | 0.001952198 | 0.569455213 | 0.003431805 | 0.183417424 | 7.42E-05 | 0.014640442 | 1 | 0.795474689 | 1 | 0.812246196 | ENSG0000024 | 2 | RP11-10L7.1 |
| VAT | Obesity | ENSG0000023 | 0.008774748 | 0.066620535 | 0.004783887 | 0.183453202 | 0.008954583 | 0.061214812 | 0.004783572 | 0.183416222 | -0.00017984 | 6.58E-06 | 1 | 0.264946219 | 1 | 0.259777168 | ENSG0000024 | 2 | RP11-10L7.1 |
| VAT | Obesity | ENSG0000023 | 0.001071664 | 0.749380662 | 0.00335465 | 0.183454436 | 0.001005778 | 0.764300813 | 0.003354404 | 0.183417512 | 6.59E-05 | 0.007889972 | 1 | 0.892799597 | 1 | 0.904298874 | ENSG0000024 | 3 | RP11-736K20.6 |
| VAT | Obesity | ENSG0000023 | 0.007566621 | 0.00239821 | 0.002492371 | 0.18345099 | 0.006426216 | 0.010253329 | 0.002503223 | 0.183415055 | 0.001140406 | 1.27E-06 | 1 | 0.026266109 | 1 | 0.080167672 | ENSG0000024 | 3 | RP11-736K20.6 |
| VAT | Obesity | ENSG0000023 | -0.00404815 | 0.27386216 | 0.003699604 | 0.183454022 | -0.00412102 | 0.26528756 | 0.003699359 | 0.183417077 | 7.29E-05 | 0.017324169 | 1 | 0.578759235 | 1 | 0.579936623 | ENSG0000024 | 3 | RP11-736K20.6 |
| VAT | Obesity | ENSG0000023 | -0.00386254 | 0.025966391 | 0.001734632 | 0.1834526 | -0.00256502 | 0.144541989 | 0.001757966 | 0.183416741 | -0.00129752 | 5.93E-06 | 1 | 0.145075383 | 1 | 0.420375017 | ENSG0000024 | 3 | RP11-736K20.6 |
| VAT | Obesity | ENSG0000023 | -0.00310708 | 0.396101183 | 0.00366139 | 0.183454202 | -0.00304715 | 0.405244037 | 0.003661146 | 0.183417284 | -5.99E-05 | 0.047093499 | 1 | 0.701928581 | 1 | 0.720669275 | ENSG0000024 | 2 | AC093162.5 |
| VAT | Obesity | ENSG0000023 | 0.007469391 | 0.029872823 | 0.003439296 | 0.183452691 | 0.007592465 | 0.02726455 | 0.003439059 | 0.183415704 | -0.00012307 | 7.31E-06 | 1 | 0.158861254 | 1 | 0.155231297 | ENSG0000024 | 2 | AC093162.5 |
| VAT | Obesity | ENSG0000023 | -0.00579948 | 0.008420245 | 0.00220115 | 0.18345185 | -0.00286404 | 0.210150085 | 0.002285467 | 0.183416953 | -0.00293543 | 1.87E-06 | 1 | 0.065097695 | 1 | 0.521595534 | ENSG0000024 | 2 | AC093162.5 |
| VAT | Obesity | ENSG0000023 | 0.006644786 | 0.102533083 | 0.004069819 | 0.183453466 | 0.006584236 | 0.105675607 | 0.004069511 | 0.183416557 | 6.06E-05 | 0.035876612 | 1 | 0.343436055 | 1 | 0.360079845 | ENSG0000024 | 2 | CTD-2260A17.2 |
| VAT | Obesity | ENSG0000023 | -0.00279401 | 0.555519462 | 0.004739553 | 0.183454343 | -0.00294953 | 0.533706391 | 0.004739272 | 0.1834174 | 0.000155513 | 0.000313356 | 1 | 0.795474689 | 1 | 0.788982131 | ENSG0000024 | 2 | CTD-2260A17.2 |
| VAT | Obesity | ENSG0000023 | 0.004022962 | 0.348080129 | 0.004287406 | 0.183454141 | 0.003896832 | 0.363367891 | 0.004287099 | 0.183417234 | 0.00012613 | 0.00010696 | 1 | 0.660100296 | 1 | 0.687385454 | ENSG0000024 | 2 | RP11-434C1.1 |
| VAT | Obesity | ENSG0000023 | -0.00245645 | 0.682427299 | 0.00600373 | 0.183454411 | -0.00264131 | 0.659955572 | 0.006003313 | 0.183417473 | 0.00018486 | 9.90E-05 | 1 | 0.863333733 | 1 | 0.854766662 | ENSG0000024 | 2 | RP11-434C1.1 |
| VAT | Obesity | ENSG0000023 | 0.005114993 | 0.260303964 | 0.004543937 | 0.183453995 | 0.002853358 | 0.531465106 | 0.004559744 | 0.183417398 | 0.002261635 | 4.14E-09 | 1 | 0.561280421 | 1 | 0.786996417 | ENSG0000024 | 2 | RP11-434C1.1 |
| VAT | Obesity | ENSG0000023 | 0.00098432 | 0.772152596 | 0.003399359 | 0.183454442 | 0.000881267 | 0.795431759 | 0.003399112 | 0.183417521 | 0.000103053 | 4.86E-05 | 1 | 0.904559068 | 1 | 0.918573915 | ENSG0000024 | 4 | TWF2 |
| VAT | Obesity | ENSG0000023 | 0.013159031 | 0.000608689 | 0.003838987 | 0.183450033 | 0.013618858 | 0.000396691 | 0.003844637 | 0.183412804 | -0.00045983 | 0.032791561 | 1 | 0.009032155 | 1 | 0.007109524 | ENSG0000024 | 4 | TWF2 |
| VAT | Obesity | ENSG0000023 | 0.018410362 | 0.000442649 | 0.005240218 | 0.183449809 | 0.018270716 | 0.000488716 | 0.005239832 | 0.183412951 | 0.000139646 | 0.000262429 | 1 | 0.007102966 | 1 | 0.008430344 | ENSG0000024 | 6 | BCKDHA |
| VAT | Obesity | ENSG0000023 | -0.00782382 | 0.145345522 | 0.005372862 | 0.183453673 | -0.00764927 | 0.154512043 | 0.005372526 | 0.18341678 | -0.00017454 | 0.000189663 | 1 | 0.412285344 | 1 | 0.436939795 | ENSG0000024 | 6 | BCKDHA |
| VAT | Obesity | ENSG0000023 | 0.003080289 | 0.617671951 | 0.006171034 | 0.18345438 | 0.002970997 | 0.630178092 | 0.006170595 | 0.183417458 | 0.000109292 | 0.021299329 | 1 | 0.831570896 | 1 | 0.842487235 | ENSG0000024 | 1 | RP11-499E18.1 |
| VAT | Obesity | ENSG0000023 | 0.00602959 | 0.230569491 | 0.005029291 | 0.183453931 | 0.003950284 | 0.433850462 | 0.00504751 | 0.183417315 | 0.002079306 | 1.69E-06 | 1 | 0.52708752 | 1 | 0.737402735 | ENSG0000024 | 1 | RP11-455B3.1 |
| VAT | Obesity | ENSG0000023 | -0.00507194 | 0.012579291 | 0.002032458 | 0.18345212 | -0.0033167 | 0.109598089 | 0.002072929 | 0.183416579 | -0.00175525 | 1.74E-05 | 1 | 0.0877446 | 1 | 0.365676336 | ENSG0000024 | 1 | RP11-455B3.1 |
| VAT | Obesity | ENSG0000023 | -0.01741927 | 4.53E-08 | 0.003185141 | 0.183443169 | -0.01750575 | 3.88E-08 | 0.003184915 | 0.183406129 | 8.65E-05 | 0.000465278 | 0.000125033 | 2.45E-06 | 0.000107003 | 2.63E-06 | ENSG0000024 | 1 | RP11-455B3.1 |
| VAT | Obesity | ENSG0000023 | 0.002039848 | 0.633392371 | 0.00427679 | 0.183454388 | 0.00194474 | 0.649286779 | 0.004276471 | 0.183417468 | 9.51E-05 | 0.002113914 | 1 | 0.840565826 | 1 | 0.852367106 | ENSG0000024 | 1 | RP11-455B3.1 |
| VAT | Obesity | ENSG0000023 | 0.001558216 | 0.686913361 | 0.003866085 | 0.183454413 | 0.001680213 | 0.663827405 | 0.003865801 | 0.183417475 | -0.000122 | 2.00E-05 | 1 | 0.863333733 | 1 | 0.855751349 | ENSG0000024 | 1 | RP11-455B3.1 |
| VAT | Obesity | ENSG0000023 | -0.00780598 | 0.165267946 | 0.005625638 | 0.183453746 | -0.00765565 | 0.173535956 | 0.005625305 | 0.183416846 | -0.00015032 | 0.003327906 | 1 | 0.446320479 | 1 | 0.470490411 | ENSG0000024 | 1 | RP11-834C11.3 |
| VAT | Obesity | ENSG0000023 | -0.0005459 | 0.903003014 | 0.004479426 | 0.183454468 | -0.00061729 | 0.890385214 | 0.004479094 | 0.183417539 | 7.14E-05 | 0.028621005 | 1 | 0.965567705 | 1 | 0.965604397 | ENSG0000024 | 1 | RP11-834C11.3 |
| VAT | Obesity | ENSG0000023 | -0.00288197 | 0.411025858 | 0.003505667 | 0.183454219 | -0.00281776 | 0.421496477 | 0.00350543 | 0.183417302 | -6.42E-05 | 0.024624122 | 1 | 0.708533979 | 1 | 0.727353238 | ENSG0000024 | 1 | RP11-834C11.3 |
| VAT | Obesity | ENSG0000023 | -0.02520588 | 4.04E-05 | 0.006140368 | 0.183448105 | -0.02512247 | 4.28E-05 | 0.006139898 | 0.183411219 | -8.34E-05 | 0.049858233 | 0.111634994 | 0.001043318 | 0.118220734 | 0.001170502 | ENSG0000024 | 1 | TNXA |
| VAT | Obesity | ENSG0000023 | 0.009330705 | 0.078087442 | 0.005295831 | 0.183453301 | 0.00893113 | 0.091826269 | 0.005297717 | 0.183416472 | 0.000399575 | 0.012524771 | 1 | 0.286642319 | 1 | 0.327866111 | ENSG0000024 | 1 | TNXA |
| VAT | Obesity | ENSG0000023 | -0.00350528 | 0.297839489 | 0.00336697 | 0.183454064 | 0.00368412 | 0.319111031 | 0.003697848 | 0.183417171 | -0.0071894 | 2.60E-06 | 1 | 0.608015525 | 1 | 0.641269508 | ENSG0000024 | 1 | TNXA |
| VAT | Obesity | ENSG0000023 | 0.000766425 | 0.63475925 | 0.001613392 | 0.183454389 | 0.000255415 | 0.874485204 | 0.001616916 | 0.183417537 | 0.00051101 | 2.83E-06 | 1 | 0.840660043 | 1 | 0.956332568 | ENSG0000024 | 1 | TNXA |
| VAT | Obesity | ENSG0000023 | 0.000490688 | 0.733216304 | 0.001439599 | 0.18345443 | 0.000854084 | 0.55356847 | 0.001441682 | 0.183417413 | -0.0003634 | 5.75E-06 | 1 | 0.884914569 | 1 | 0.805509891 | ENSG0000024 | 1 | RP11-177B4.1 |
| VAT | Obesity | ENSG0000023 | 0.008014927 | 2.53E-07 | 0.001554647 | 0.183444428 | 0.006747713 | 1.93E-05 | 0.001579257 | 0.183410647 | 0.001267214 | 5.37E-06 | 0.000698563 | 1.20E-05 | 0.053308014 | 0.000626206 | ENSG0000024 | 1 | RP11-177B4.1 |
| VAT | Obesity | ENSG0000023 | 0.002068475 | 0.436636072 | 0.002659095 | 0.183454245 | 0.001618063 | 0.543056747 | 0.002660424 | 0.183417406 | 0.000450412 | 1.02E-06 | 1 | 0.725099615 | 1 | 0.797218136 | ENSG0000024 | 1 | RP11-452C8.1 |
| VAT | Obesity | ENSG0000023 | -0.00409312 | 0.001390058 | 0.00128041 | 0.183450611 | -0.00275924 | 0.034895089 | 0.001307956 | 0.183415864 | -0.00133388 | 6.21E-07 | 1 | 0.017360006 | 1 | 0.183099707 | ENSG0000024 | 1 | RP11-452C8.1 |
| VAT | Obesity | ENSG0000023 | -0.00308001 | 0.552357461 | 0.005183173 | 0.183454341 | -0.00315857 | 0.542235921 | 0.005182779 | 0.183417406 | 7.86E-05 | 0.030436386 | 1 | 0.793904838 | 1 | 0.796907844 | ENSG0000024 | 1 | RP11-452C8.1 |
| VAT | Obesity | ENSG0000023 | 0.007721647 | 0.174548396 | 0.005687153 | 0.183453777 | 0.009424962 | 0.098531763 | 0.005705125 | 0.183416515 | -0.00170331 | 0.000211035 | 1 | 0.46322459 | 1 | 0.344453707 | ENSG0000024 | 2 | SRP14-AS1 |
| VAT | Obesity | ENSG0000023 | -0.00982309 | 0.063861239 | 0.005300746 | 0.183453176 | -0.00991971 | 0.061273528 | 0.005300348 | 0.183416222 | 9.66E-05 | 0.010943256 | 1 | 0.2592015 | 1 | 0.259777168 | ENSG0000024 | 2 | RP11-247C2.2 |
| VAT | Obesity | ENSG0000023 | 0.001703677 | 0.562016435 | 0.002938136 | 0.183454347 | 0.001750773 | 0.551228287 | 0.002937929 | 0.183417412 | -4.71E-05 | 0.038918898 | 1 | 0.802465266 | 1 | 0.803268253 | ENSG0000024 | 1 | RP11-311D14.1 |
| VAT | Obesity | ENSG0000023 | 0.004732189 | 0.005678275 | 0.001710956 | 0.183451583 | 0.003527459 | 0.04169598 | 0.001732078 | 0.183415979 | 0.00120473 | 8.59E-06 | 1 | 0.048670928 | 1 | 0.205736949 | ENSG0000024 | 1 | RP11-666A20.4 |
| VAT | Obesity | ENSG0000023 | 0.004628896 | 0.00075157 | 0.001373534 | 0.183450181 | 0.00260112 | 0.071125433 | 0.00144132 | 0.183416315 | 0.002027776 | 3.52E-06 | 1 | 0.010692438 | 1 | 0.283270125 | ENSG0000024 | 1 | CTD-2248H3.1 |
| VAT | Obesity | ENSG0000023 | 0.016457482 | 3.47E-05 | 0.003975163 | 0.183447995 | 0.016517246 | 3.25E-05 | 0.003974862 | 0.18341102 | -5.98E-05 | 0.033529241 | 0.095840376 | 0.000921542 | 0.089627377 | 0.000974211 | ENSG0000024 | 1 | CTD-2248H3.1 |
| VAT | Obesity | ENSG0000023 | 0.006603415 | 0.104912619 | 0.004072429 | 0.18345348 | 0.005396353 | 0.186205966 | 0.004082306 | 0.183416886 | 0.001207062 | 3.07E-05 | 1 | 0.347941332 | 1 | 0.493687287 | ENSG0000024 | 1 | CTD-2248H3.1 |
| VAT | Obesity | ENSG0000023 | 0.004061363 | 0.000107112 | 0.001048386 | 0.183448802 | 0.00284328 | 0.008595219 | 0.001082017 | 0.183414936 | 0.001218083 | 5.51E-06 | 0.295630061 | 0.00230961 | 1 | 0.071670101 | ENSG0000024 | 4 | FMN1 |
| VAT | Obesity | ENSG0000023 | -0.00168248 | 0.702842662 | 0.004410327 | 0.183454419 | -0.00178868 | 0.685043716 | 0.004410059 | 0.183417484 | 0.000106199 | 0.00697231 | 1 | 0.871233847 | 1 | 0.869882588 | ENSG0000024 | 4 | FMN1 |
| VAT | Obesity | ENSG0000024 | -0.00135581 | 0.531764457 | 0.002168197 | 0.183454326 | -0.00285365 | 0.198326784 | 0.002218435 | 0.183416921 | 0.001497843 | 0.001453611 | 1 | 0.783174974 | 1 | 0.510141588 | ENSG0000024 | 4 | FMN1 |
| VAT | Obesity | ENSG0000024 | 0.00769467 | 1.20E-06 | 0.001584496 | 0.18344556 | 0.005532217 | 0.00190308 | 0.001781728 | 0.183413902 | 0.002162453 | 0.007976922 | 0.003303224 | 4.79E-05 | 1 | 0.023659912 | ENSG0000024 | 4 | FMN1 |
| VAT | Obesity | ENSG0000024 | -0.00072925 | 0.611900819 | 0.001437341 | 0.183454377 | -0.00075744 | 0.598186572 | 0.001437242 | 0.183417441 | 2.82E-05 | 0.013939462 | 1 | 0.83120188 | 1 | 0.827151773 | ENSG0000024 | 1 | CTD-2334D19.1 |
| VAT | Obesity | ENSG0000024 | 0.000948228 | 0.748819467 | 0.002961392 | 0.183454435 | 0.000876836 | 0.76714643 | 0.002961186 | 0.183417513 | 7.14E-05 | 0.002243639 | 1 | 0.892799597 | 1 | 0.906388761 | ENSG0000024 | 1 | CTD-2334D19.1 |

| | | | | | | | | | | | | | | | | | | | |
|---|---|---|---|---|---|---|---|---|---|---|---|---|---|---|---|---|---|---|---|
| VAT | Obesity | ENSG0000024 | 0.011488922 | 0.056954239 | 0.006035155 | 0.183453104 | 0.011378891 | 0.059353864 | 0.00603473 | 0.183416202 | 0.000110031 | 0.019005106 | 1 | 0.24109463 | 1 | 0.257169016 | ENSG0000024 | 1 | CTD-2334D19.1 |
| VAT | Obesity | ENSG0000024 | -0.00186733 | 0.375954474 | 0.002109085 | 0.183454178 | -0.00190033 | 0.367545592 | 0.002108939 | 0.183417239 | 3.30E-05 | 0.048661508 | 1 | 0.685170491 | 1 | 0.690555366 | ENSG0000024 | 1 | RP11-259O2.1 |
| VAT | Obesity | ENSG0000024 | 0.003116542 | 0.579581492 | 0.005625563 | 0.183454358 | 0.003320252 | 0.555025996 | 0.005625201 | 0.183417414 | -0.00020371 | 2.13E-05 | 1 | 0.808718361 | 1 | 0.805802048 | ENSG0000024 | 1 | RP11-259O2.1 |
| VAT | Obesity | ENSG0000024 | -0.00172428 | 0.712065584 | 0.004671784 | 0.183454423 | -0.00186353 | 0.689960071 | 0.004671551 | 0.183417486 | 0.000139247 | 0.003086943 | 1 | 0.874633294 | 1 | 0.869882588 | ENSG0000024 | 3 | C1orf213 |
| VAT | Obesity | ENSG0000024 | 0.000877603 | 0.530659365 | 0.001399685 | 0.183454326 | 0.001813214 | 0.199726527 | 0.001413996 | 0.183416925 | -0.00093561 | 3.49E-06 | 1 | 0.782382397 | 1 | 0.510412237 | ENSG0000024 | 1 | RP11-1026M7.2 |
| VAT | Obesity | ENSG0000024 | 0.003714074 | 0.042140206 | 0.001827668 | 0.183452913 | 0.003136788 | 0.086826544 | 0.001831825 | 0.183416438 | 0.000577287 | 4.61E-06 | 1 | 0.200529258 | 1 | 0.316168943 | ENSG0000024 | 3 | TMEM150C |
| VAT | Obesity | ENSG0000024 | -0.00870428 | 0.11225813 | 0.005480844 | 0.183453521 | -0.00861749 | 0.115855178 | 0.005480417 | 0.183416612 | -8.68E-05 | 0.018817887 | 1 | 0.361110068 | 1 | 0.37486853 | ENSG0000024 | 3 | TMEM150C |
| VAT | Obesity | ENSG0000024 | 0.007658149 | 0.147898593 | 0.005292446 | 0.183453683 | 0.007499091 | 0.15646953 | 0.005292052 | 0.183416787 | 0.000159058 | 3.24E-05 | 1 | 0.415682399 | 1 | 0.440669287 | ENSG0000024 | 1 | CTD-2587M2.1 |
| VAT | Obesity | ENSG0000024 | -0.0013487 | 0.415980014 | 0.001658068 | 0.183454224 | -0.0013093 | 0.429697556 | 0.001657947 | 0.18341731 | -3.94E-05 | 0.001417932 | 1 | 0.710460915 | 1 | 0.734343811 | ENSG0000024 | 2 | SERBP1P5 |
| VAT | Obesity | ENSG0000024 | 0.003023415 | 0.547837206 | 0.005030602 | 0.183454338 | 0.002908738 | 0.563096401 | 0.005030252 | 0.18341742 | 0.000114677 | 0.003886356 | 1 | 0.793647904 | 1 | 0.809555852 | ENSG0000024 | 1 | RP11-496H1.1 |
| VAT | Obesity | ENSG0000024 | -0.01685131 | 9.54E-05 | 0.004318705 | 0.183448719 | -0.01677681 | 0.00010237 | 0.004318426 | 0.183411842 | -7.45E-05 | 0.041368108 | 0.263413849 | 0.002112498 | 0.282541352 | 0.002435701 | ENSG0000024 | 1 | RP11-259O2.3 |
| VAT | Obesity | ENSG0000024 | 0.003647492 | 0.544445284 | 0.006017921 | 0.183454335 | 0.003798856 | 0.527843729 | 0.0060175 | 0.183417395 | -0.00015136 | 0.00132643 | 1 | 0.79046238 | 1 | 0.786996417 | ENSG0000024 | 1 | RP11-259O2.3 |
| VAT | Obesity | ENSG0000024 | 0.002476306 | 0.643298191 | 0.005347324 | 0.183454393 | 0.002389797 | 0.654912607 | 0.005346921 | 0.183417471 | 8.65E-05 | 0.022859916 | 1 | 0.843870251 | 1 | 0.854180748 | ENSG0000024 | 1 | CTD-2127H9.1 |
| VAT | Obesity | ENSG0000024 | 0.000748415 | 0.773348082 | 0.002598679 | 0.183454443 | 0.000798806 | 0.758530354 | 0.002598492 | 0.18341751 | -5.04E-05 | 0.010673541 | 1 | 0.904681739 | 1 | 0.900448937 | ENSG0000024 | 1 | RP11-352E6.2 |
| VAT | Obesity | ENSG0000024 | 0.00467412 | 0.146953702 | 0.00322269 | 0.183453679 | 0.004600789 | 0.153371889 | 0.003222464 | 0.183416776 | 7.33E-05 | 0.003472205 | 1 | 0.413958925 | 1 | 0.434702786 | ENSG0000024 | 1 | AC011330.5 |
| VAT | Obesity | ENSG0000024 | -0.00057571 | 0.822350216 | 0.002564137 | 0.183454455 | -0.00064852 | 0.800317102 | 0.002563955 | 0.183417522 | 7.28E-05 | 0.00022869 | 1 | 0.927365584 | 1 | 0.921516563 | ENSG0000024 | 1 | AC011330.5 |
| VAT | Obesity | ENSG0000024 | 0.004519757 | 0.338064454 | 0.004717921 | 0.183454127 | 0.004442671 | 0.3463346 | 0.004717595 | 0.183417211 | 7.71E-05 | 0.039602438 | 1 | 0.646762125 | 1 | 0.669386202 | ENSG0000024 | 1 | AC011330.5 |
| VAT | Obesity | ENSG0000024 | -0.00805573 | 1.87E-07 | 0.001545514 | 0.183444205 | -0.00463343 | 0.025525384 | 0.002074644 | 0.183415661 | -0.0034223 | 0.013418272 | 0.000514893 | 9.19E-06 | 1 | 0.149575496 | ENSG0000024 | 2 | RP11-177C12.1 |
| VAT | Obesity | ENSG0000024 | -0.00918707 | 0.002000069 | 0.002972931 | 0.183450864 | -0.00923884 | 0.001884532 | 0.002972725 | 0.183413896 | 5.18E-05 | 0.028614783 | 1 | 0.022348948 | 1 | 0.023620883 | ENSG0000024 | 2 | RP11-177C12.1 |
| VAT | Obesity | ENSG0000024 | -0.01046835 | 0.090417649 | 0.006182573 | 0.18345339 | -0.01056533 | 0.087449792 | 0.006182135 | 0.183416442 | 9.70E-05 | 0.042381986 | 1 | 0.314235052 | 1 | 0.317028233 | ENSG0000024 | 1 | RP11-122C5.1 |
| VAT | Obesity | ENSG0000024 | -0.00082143 | 0.881691052 | 0.005519408 | 0.183454466 | -0.00071696 | 0.89663939 | 0.005519023 | 0.18341754 | -0.00010447 | 0.015994078 | 1 | 0.95522585 | 1 | 0.968028006 | ENSG0000025 | 1 | RP11-584P21.2 |
| VAT | Obesity | ENSG0000024 | -0.00846676 | 0.016349574 | 0.00352631 | 0.183452295 | -0.00838236 | 0.017443057 | 0.003526088 | 0.18341541 | -8.44E-05 | 0.005898904 | 1 | 0.106176055 | 1 | 0.117421555 | ENSG0000025 | 1 | RP11-584P21.2 |
| VAT | Obesity | ENSG0000024 | 0.002881922 | 0.504032014 | 0.004313224 | 0.183454305 | 0.002973665 | 0.490522751 | 0.004312921 | 0.183417366 | -9.17E-05 | 0.00650003 | 1 | 0.770249732 | 1 | 0.775358255 | ENSG0000025 | 3 | RP11-848G14.5 |
| VAT | Obesity | ENSG0000024 | -0.00764477 | 0.066007257 | 0.004158432 | 0.183453197 | -0.00773187 | 0.062966661 | 0.00415819 | 0.183416239 | 8.71E-05 | 0.023062076 | 1 | 0.264689498 | 1 | 0.264517478 | ENSG0000025 | 1 | RP11-173M11.2 |
| VAT | Obesity | ENSG0000024 | 0.003296222 | 0.396710558 | 0.003889295 | 0.183454203 | 0.003193849 | 0.411509003 | 0.003889048 | 0.183417291 | 0.000102374 | 0.00223037 | 1 | 0.701928581 | 1 | 0.722625422 | ENSG0000025 | 1 | CTD-2005D20.1 |
| VAT | Obesity | ENSG0000024 | 0.007395383 | 0.028506343 | 0.003376497 | 0.183452661 | 0.007447733 | 0.02739045 | 0.003376256 | 0.183415707 | -5.23E-05 | 0.043588147 | 1 | 0.153667004 | 1 | 0.155231297 | ENSG0000025 | 1 | CTD-2005D20.1 |
| VAT | Obesity | ENSG0000024 | -0.00321612 | 0.363130639 | 0.003536464 | 0.183454161 | -0.00327552 | 0.354298042 | 0.003536188 | 0.183417222 | 5.94E-05 | 0.012223512 | 1 | 0.672794162 | 1 | 0.679543152 | ENSG0000025 | 1 | CTD-2005D20.1 |
| VAT | Obesity | ENSG0000024 | -0.00427209 | 0.473318385 | 0.005957531 | 0.18345428 | -0.00415376 | 0.485628662 | 0.005957099 | 0.183417362 | -0.00011833 | 0.008047146 | 1 | 0.755994642 | 1 | 0.775358255 | ENSG0000025 | 1 | RP11-119J18.1 |
| VAT | Obesity | ENSG0000025 | 0.000434 | 0.876729705 | 0.002797899 | 0.183454465 | 0.000362484 | 0.896910853 | 0.002797704 | 0.18341754 | 7.15E-05 | 0.001156551 | 1 | 0.951543054 | 1 | 0.968028006 | ENSG0000025 | 1 | KLHL2P1 |
| VAT | Obesity | ENSG0000025 | -0.00329713 | 0.030384614 | 0.00152289 | 0.183452702 | -0.00102778 | 0.518901843 | 0.001593355 | 0.183417389 | -0.00226935 | 1.31E-06 | 1 | 0.160281317 | 1 | 0.784425829 | ENSG0000025 | 1 | ENPP7P9 |
| VAT | Obesity | ENSG0000025 | -0.00498393 | 0.044563194 | 0.002481098 | 0.183452949 | -0.0050161 | 0.04318865 | 0.002480902 | 0.183416001 | 3.22E-05 | 0.049822255 | 1 | 0.208112376 | 1 | 0.210230467 | ENSG0000025 | 2 | CDK3 |
| VAT | Obesity | ENSG0000025 | 0.00192376 | 0.692685567 | 0.004867648 | 0.183454415 | 0.001832599 | 0.706536269 | 0.004867302 | 0.183417492 | 9.12E-05 | 0.014849397 | 1 | 0.866149932 | 1 | 0.876422518 | ENSG0000025 | 2 | CDK3 |
| VAT | Obesity | ENSG0000025 | 0.001301825 | 0.744207844 | 0.003989823 | 0.183454434 | 0.00121667 | 0.760394159 | 0.003989562 | 0.183417511 | 8.52E-05 | 0.010980484 | 1 | 0.892204507 | 1 | 0.901498229 | ENSG0000025 | 2 | CDK3 |
| VAT | Obesity | ENSG0000025 | -0.00123716 | 0.768016584 | 0.004194172 | 0.183454441 | -0.00129623 | 0.757261531 | 0.004193855 | 0.18341751 | 5.91E-05 | 0.046875769 | 1 | 0.901627295 | 1 | 0.900448937 | ENSG0000025 | 2 | CDK3 |
| VAT | Obesity | ENSG0000025 | -0.0230568 | 4.83E-09 | 0.003939343 | 0.183441526 | -0.02298448 | 5.38E-09 | 0.00393907 | 0.183404679 | -7.23E-05 | 0.020108523 | 1.33E-05 | 3.18E-07 | 1.49E-05 | 3.91E-07 | ENSG0000025 | 2 | CDK3 |
| VAT | Obesity | ENSG0000025 | 0.004539381 | 0.173117329 | 0.003332249 | 0.183453773 | 0.004470277 | 0.179723085 | 0.003332024 | 0.183416866 | 6.91E-05 | 0.010629946 | 1 | 0.460092631 | 1 | 0.482525015 | ENSG0000025 | 2 | CDK3 |
| VAT | Obesity | ENSG0000025 | 0.000854237 | 0.705056055 | 0.002256875 | 0.18345442 | 0.000900754 | 0.689784646 | 0.002256695 | 0.183417486 | -4.65E-05 | 0.00137197 | 1 | 0.871233847 | 1 | 0.869882588 | ENSG0000025 | 2 | RP11-680H20.2 |
| VAT | Obesity | ENSG0000025 | 0.007991683 | 0.003200161 | 0.002711028 | 0.183451189 | 0.008117002 | 0.002752625 | 0.002710992 | 0.183414158 | -0.00012532 | 0.000471787 | 1 | 0.032235198 | 1 | 0.031393572 | ENSG0000025 | 2 | RP11-680H20.2 |
| VAT | Obesity | ENSG0000025 | -0.00259766 | 0.505127542 | 0.003897784 | 0.183454306 | -0.00246527 | 0.527048674 | 0.003897543 | 0.183417395 | -0.00013239 | 0.000115252 | 1 | 0.770249732 | 1 | 0.7867249 | ENSG0000025 | 1 | STK19P |
| VAT | Obesity | ENSG0000025 | -0.00034733 | 0.931337419 | 0.004031064 | 0.183454471 | -0.00027689 | 0.945234081 | 0.004030772 | 0.183417544 | -7.04E-05 | 0.019904455 | 1 | 0.976259505 | 1 | 0.981754039 | ENSG0000025 | 1 | STK19P |
| VAT | Obesity | ENSG0000025 | -0.00281154 | 0.130239027 | 0.001858052 | 0.183453609 | -0.00139866 | 0.455599476 | 0.001874598 | 0.183417336 | -0.00141287 | 1.57E-08 | 1 | 0.387831349 | 1 | 0.752966798 | ENSG0000025 | 1 | STK19P |
| VAT | Obesity | ENSG0000025 | -0.0017939 | 0.669267138 | 0.004199654 | 0.183454405 | -0.00193425 | 0.645081863 | 0.004199369 | 0.183417466 | 0.000140352 | 3.64E-05 | 1 | 0.857647266 | 1 | 0.850251166 | ENSG0000025 | 1 | STK19P |
| VAT | Obesity | ENSG0000025 | -0.00352292 | 0.524155836 | 0.005530902 | 0.183454321 | -0.0036228 | 0.512429369 | 0.005530481 | 0.183417384 | 9.99E-05 | 0.009860957 | 1 | 0.779212983 | 1 | 0.784425829 | ENSG0000025 | 1 | STK19P |
| VAT | Obesity | ENSG0000025 | 0.001648237 | 0.188022468 | 0.001252028 | 0.183453819 | 0.001631003 | 0.192647893 | 0.001251933 | 0.183416905 | 1.72E-05 | 0.048294936 | 1 | 0.484270267 | 1 | 0.50208516 | ENSG0000025 | 1 | STK19P |
| VAT | Obesity | ENSG0000025 | 0.000660776 | 0.847470948 | 0.003435343 | 0.18345446 | 0.000529293 | 0.877544342 | 0.003435113 | 0.183417537 | 0.000131483 | 3.16E-06 | 1 | 0.940876837 | 1 | 0.957699638 | ENSG0000025 | 1 | STK19P |
| VAT | Obesity | ENSG0000025 | 0.007094833 | 0.00103713 | 0.002162877 | 0.183450407 | 0.006169819 | 0.004452802 | 0.00216928 | 0.183414489 | 0.000925014 | 4.64E-08 | 1 | 0.013828397 | 1 | 0.044318348 | ENSG0000025 | 1 | STK19P |
| VAT | Obesity | ENSG0000025 | 0.008905588 | 0.046771787 | 0.004478847 | 0.18345298 | 0.009912568 | 0.026997748 | 0.004482167 | 0.183415698 | -0.00100698 | 4.37E-08 | 1 | 0.214079822 | 1 | 0.155231297 | ENSG0000025 | 1 | STK19P |
| VAT | Obesity | ENSG0000025 | -0.00446298 | 0.155522286 | 0.003142287 | 0.183453712 | -0.00220013 | 0.487299535 | 0.003167424 | 0.183417364 | -0.00226285 | 1.63E-08 | 1 | 0.429671181 | 1 | 0.775358255 | ENSG0000025 | 1 | AC064853.3 |
| VAT | Obesity | ENSG0000025 | -0.00725816 | 0.218139756 | 0.005893799 | 0.183453901 | -0.00736902 | 0.21115926 | 0.005893399 | 0.183416955 | 0.00011086 | 0.020267004 | 1 | 0.521961016 | 1 | 0.522690187 | ENSG0000025 | 1 | AC064853.3 |
| VAT | Obesity | ENSG0000025 | 0.00051542 | 0.876558794 | 0.003318162 | 0.183454465 | 0.000378281 | 0.90924583 | 0.00331855 | 0.183417541 | 0.000137139 | 0.047440485 | 1 | 0.951543054 | 1 | 0.971175886 | ENSG0000025 | 2 | ST3GAL1P1 |
| VAT | Obesity | ENSG0000025 | 0.006511484 | 0.247565561 | 0.005631388 | 0.183453969 | 0.006672329 | 0.236045186 | 0.005630984 | 0.183417015 | -0.00016084 | 0.000173898 | 1 | 0.544013495 | 1 | 0.543810279 | ENSG0000025 | 2 | ST3GAL1P1 |
| VAT | Obesity | ENSG0000025 | -0.00217279 | 0.496497994 | 0.00319523 | 0.183454299 | -0.00209661 | 0.51168439 | 0.003194996 | 0.183417383 | -7.62E-05 | 0.001216431 | 1 | 0.766551816 | 1 | 0.784425829 | ENSG0000025 | 2 | ST3GAL1P1 |
| VAT | Obesity | ENSG0000025 | 0.001508672 | 0.689390346 | 0.003774673 | 0.183454414 | 0.001583954 | 0.67473827 | 0.003774427 | 0.183417479 | -7.53E-05 | 0.018174415 | 1 | 0.86447858 | 1 | 0.862964608 | ENSG0000025 | 2 | ST3GAL1P1 |
| VAT | Obesity | ENSG0000025 | 0.011020701 | 0.062033635 | 0.005905867 | 0.183453158 | 0.010838109 | 0.066468434 | 0.005905504 | 0.183416273 | 0.000182592 | 0.000458603 | 1 | 0.254402428 | 1 | 0.27337482 | ENSG0000025 | 2 | ST3GAL1P1 |
| VAT | Obesity | ENSG0000025 | -0.00814855 | 0.023899593 | 0.003607555 | 0.183452546 | -0.00822193 | 0.022652388 | 0.003607291 | 0.183415583 | 7.34E-05 | 0.006173274 | 1 | 0.136014549 | 1 | 0.136806542 | ENSG0000025 | 2 | RP5-1063M23.1 |
| VAT | Obesity | ENSG0000025 | 0.001234921 | 0.749910331 | 0.003874167 | 0.183454436 | 0.001359257 | 0.725688344 | 0.003873995 | 0.1834175 | -0.00012434 | 0.002511643 | 1 | 0.892799597 | 1 | 0.885846895 | ENSG0000025 | 1 | RP11-122F24.1 |
| VAT | Obesity | ENSG0000025 | 0.012985928 | 0.004520732 | 0.004573555 | 0.183451427 | 0.012873274 | 0.004879288 | 0.004573246 | 0.183414551 | 0.000112653 | 0.002357482 | 1 | 0.041315295 | 1 | 0.046598047 | ENSG0000025 | 1 | AC141928.1 |
| VAT | Obesity | ENSG0000025 | 0.000581717 | 0.70904724 | 0.001558993 | 0.183454421 | 0.000559476 | 0.719672296 | 0.001558875 | 0.183417497 | 2.22E-05 | 0.0416578 | 1 | 0.872868145 | 1 | 0.881622519 | ENSG0000025 | 1 | AC141928.1 |
| VAT | Obesity | ENSG0000025 | -0.0013394 | 0.818199976 | 0.005827017 | 0.183454454 | -0.00116378 | 0.841688024 | 0.005826607 | 0.183417531 | -0.00017563 | 0.000107708 | 1 | 0.926394336 | 1 | 0.940035744 | ENSG0000025 | 1 | AC141928.1 |
| VAT | Obesity | ENSG0000025 | -0.00615368 | 0.12302276 | 0.003990163 | 0.183453575 | -0.00625063 | 0.117208962 | 0.003989935 | 0.183416618 | 9.70E-05 | 0.009284871 | 1 | 0.37609362 | 1 | 0.376596898 | ENSG0000025 | 1 | ORAOV1P1 |
| VAT | Obesity | ENSG0000025 | 0.003832891 | 0.529468808 | 0.006095394 | 0.183454325 | 0.003935841 | 0.518437445 | 0.006094932 | 0.183417388 | -0.00010295 | 0.016829314 | 1 | 0.781416001 | 1 | 0.784425829 | ENSG0000025 | 1 | ORAOV1P1 |
| VAT | Obesity | ENSG0000025 | 0.003172045 | 0.206222312 | 0.002509483 | 0.18345387 | 0.003378217 | 0.178465032 | 0.002510756 | 0.183416862 | -0.00020617 | 0.01847197 | 1 | 0.507220054 | 1 | 0.480362878 | ENSG0000025 | 4 | RP11-37B2.1 |
| VAT | Obesity | ENSG0000026 | -0.00530063 | 0.347483713 | 0.005642079 | 0.18345414 | -0.00513457 | 0.36277408 | 0.005641805 | 0.183417233 | -0.00016607 | 0.00391675 | 1 | 0.660045114 | 1 | 0.686859082 | ENSG0000025 | 2 | AC005355.2 |
| VAT | Obesity | ENSG0000026 | -0.00150547 | 0.775815068 | 0.005286486 | 0.183454443 | -0.00137361 | 0.794977929 | 0.005286127 | 0.183417521 | -0.00013186 | 0.002039665 | 1 | 0.904860323 | 1 | 0.918573915 | ENSG0000025 | 2 | AC005355.2 |
| VAT | Obesity | ENSG0000026 | -0.00530722 | 8.74E-06 | 0.001193642 | 0.183447002 | -0.00523471 | 1.16E-05 | 0.001193725 | 0.183410279 | -7.25E-05 | 0.000956916 | 0.024123505 | 0.000294189 | 0.031990896 | 0.000420933 | ENSG0000025 | 1 | RP11-469N6.1 |
| VAT | Obesity | ENSG0000026 | -0.00569771 | 0.005769493 | 0.002063927 | 0.183451593 | -0.00573753 | 0.005434588 | 0.002063791 | 0.183414625 | 3.98E-05 | 0.021541507 | 1 | 0.048996306 | 1 | 0.05044591 | ENSG0000025 | 1 | RP11-328N19.1 |
| VAT | Obesity | ENSG0000026 | -0.00063413 | 0.897508614 | 0.004923035 | 0.183454468 | -0.00071586 | 0.884378523 | 0.004922692 | 0.183417538 | 8.17E-05 | 0.034646667 | 1 | 0.963797845 | 1 | 0.960056703 | ENSG0000025 | 1 | RP11-332J15.3 |
| VAT | Obesity | ENSG0000026 | 0.000840935 | 0.666381571 | 0.001950583 | 0.183454404 | 0.001273369 | 0.51434102 | 0.00195274 | 0.183417385 | -0.00043243 | 6.43E-06 | 1 | 0.857042468 | 1 | 0.784425829 | ENSG0000025 | 1 | RP11-542G1.3 |
| VAT | Obesity | ENSG0000026 | 0.005164812 | 0.307153365 | 0.005057519 | 0.18345408 | 0.005304505 | 0.294221285 | 0.005057173 | 0.18341713 | -0.00013969 | 0.000573405 | 1 | 0.615029354 | 1 | 0.616533352 | ENSG0000025 | 1 | RP11-542G1.3 |
| VAT | Obesity | ENSG0000026 | 0.004901052 | 0.143970658 | 0.003354207 | 0.183453667 | 0.004957498 | 0.139383782 | 0.003353985 | 0.18341672 | -5.64E-05 | 0.042404161 | 1 | 0.410919356 | 1 | 0.414546593 | ENSG0000025 | 1 | RP11-1348G14.4 |
| VAT | Obesity | ENSG0000026 | -0.00262714 | 0.440179006 | 0.003403515 | 0.183454249 | -0.00268205 | 0.430647919 | 0.003403248 | 0.183417311 | 5.49E-05 | 0.015580944 | 1 | 0.726069693 | 1 | 0.735057673 | ENSG0000025 | 1 | RP11-1348G14.4 |
| VAT | Obesity | ENSG0000026 | 0.000903265 | 0.80275952 | 0.003616277 | 0.183454451 | 0.000799269 | 0.825064992 | 0.003616025 | 0.183417528 | 0.000103995 | 0.000249459 | 1 | 0.918961541 | 1 | 0.934468454 | ENSG0000025 | 1 | RP11-1348G14.4 |
| VAT | Obesity | ENSG0000026 | 0.001565559 | 0.727034781 | 0.004484906 | 0.183454428 | 0.001628767 | 0.716459879 | 0.00448456 | 0.183417496 | -6.32E-05 | 0.039388589 | 1 | 0.882348289 | 1 | 0.880030826 | ENSG0000025 | 1 | RP11-1348G14.4 |
| VAT | Obesity | ENSG0000026 | -0.00758608 | 0.162980798 | 0.00543759 | 0.183453738 | -0.00751667 | 0.166829752 | 0.00543715 | 0.183416824 | -6.94E-05 | 0.04246985 | 1 | 0.441873282 | 1 | 0.457993828 | ENSG0000025 | 1 | RP11-1348G14.4 |
| VAT | Obesity | ENSG0000026 | -0.01216523 | 0.003883482 | 0.004213083 | 0.183451323 | -0.01227428 | 0.003573702 | 0.004212839 | 0.183414338 | 0.000109053 | 0.005118287 | 1 | 0.036721461 | 1 | 0.037380051 | ENSG0000025 | 1 | RP11-1348G14.4 |
| VAT | Obesity | ENSG0000026 | 0.003099854 | 0.3523702 | 0.003333167 | 0.183454147 | 0.003178385 | 0.340268726 | 0.003332915 | 0.183417202 | -7.85E-05 | 0.000856992 | 1 | 0.663398193 | 1 | 0.661367382 | ENSG0000025 | 1 | RP11-1348G14.4 |
| VAT | Obesity | ENSG0000026 | -0.00632373 | 0.160165421 | 0.004502422 | 0.183453728 | -0.00642481 | 0.153563484 | 0.004502132 | 0.183416776 | 0.000101079 | 0.008560405 | 1 | 0.438548177 | 1 | 0.434702786 | ENSG0000025 | 2 | RP11-369K16.1 |
| VAT | Obesity | ENSG0000026 | 0.000799948 | 0.892324312 | 0.005909616 | 0.183454467 | 0.000901856 | 0.878699037 | 0.00590921 | 0.183417537 | -0.00010191 | 0.030852141 | 1 | 0.961627151 | 1 | 0.958352832 | ENSG0000025 | 1 | CTD-2245E15.3 |
| VAT | Obesity | ENSG0000026 | -0.00639077 | 0.207978459 | 0.005075483 | 0.183453875 | -0.00653523 | 0.197857529 | 0.005075195 | 0.183416919 | 0.000144464 | 0.002420433 | 1 | 0.508805001 | 1 | 0.509865497 | ENSG0000025 | 1 | CTD-2245E15.3 |
| VAT | Obesity | ENSG0000026 | 0.00251852 | 0.434238423 | 0.003220778 | 0.183454243 | 0.002466495 | 0.443757062 | 0.003220535 | 0.183417324 | 5.20E-05 | 0.022840918 | 1 | 0.722822432 | 1 | 0.744875937 | ENSG0000025 | 1 | RP11-539L10.3 |
| VAT | Obesity | ENSG0000026 | 0.008672078 | 0.01958134 | 0.003715093 | 0.183452414 | 0.008785919 | 0.018025519 | 0.003714817 | 0.183415432 | -0.00011384 | 2.41E-05 | 1 | 0.118518637 | 1 | 0.119020171 | ENSG0000025 | 1 | RP11-539L10.3 |
| VAT | Obesity | ENSG0000026 | 0.000680365 | 0.84154312 | 0.003403182 | 0.183454459 | 0.000731524 | 0.82979104 | 0.003402916 | 0.183417529 | -5.12E-05 | 0.024726788 | 1 | 0.938448085 | 1 | 0.936897537 | ENSG0000025 | 1 | RP11-539L10.3 |
| VAT | Obesity | ENSG0000026 | 0.007150479 | 0.121427946 | 0.004616755 | 0.183453567 | 0.007073037 | 0.125487717 | 0.004616433 | 0.183416659 | 7.74E-05 | 0.0325302 | 1 | 0.374041441 | 1 | 0.391351525 | ENSG0000025 | 1 | RP11-539L10.3 |
| VAT | Obesity | ENSG0000026 | -0.00085702 | 0.848976927 | 0.004500596 | 0.18345446 | -0.00093773 | 0.834940803 | 0.004500314 | 0.18341753 | 8.07E-05 | 0.03957968 | 1 | 0.941412743 | 1 | 0.938350439 | ENSG0000025 | 1 | RP11-539L10.3 |
| VAT | Obesity | ENSG0000026 | 0.003688551 | 0.488558663 | 0.00532563 | 0.183454293 | 0.003495346 | 0.511593154 | 0.005325354 | 0.183417383 | 0.000193204 | 0.00023685 | 1 | 0.76550139 | 1 | 0.784425829 | ENSG0000025 | 1 | MSNP1 |
| VAT | Obesity | ENSG0000026 | -0.00185185 | 0.408307761 | 0.002239573 | 0.183454216 | -0.00180952 | 0.419070436 | 0.002239408 | 0.183417299 | -4.23E-05 | 0.010022167 | 1 | 0.708044263 | 1 | 0.725782338 | ENSG0000025 | 1 | MSNP1 |
| VAT | Obesity | ENSG0000026 | -0.00323624 | 0.459077715 | 0.004371124 | 0.183454267 | -0.00334987 | 0.44342745 | 0.004370803 | 0.183417324 | 0.00011363 | 0.000415296 | 1 | 0.742704861 | 1 | 0.744875937 | ENSG0000025 | 1 | RP11-673E1.1 |
| VAT | Obesity | ENSG0000026 | 0.001742326 | 0.595248997 | 0.003279701 | 0.183454367 | 0.001657162 | 0.613343879 | 0.003279519 | 0.18341745 | 8.52E-05 | 0.006206908 | 1 | 0.818796889 | 1 | 0.839253732 | ENSG0000025 | 1 | RP11-673E1.1 |
| VAT | Obesity | ENSG0000026 | -0.00267574 | 0.528219451 | 0.004242314 | 0.183454324 | -0.00276822 | 0.514033192 | 0.00424202 | 0.183417385 | 9.25E-05 | 0.006103254 | 1 | 0.780452723 | 1 | 0.784425829 | ENSG0000025 | 3 | RP11-774O3.3 |
| VAT | Obesity | ENSG0000026 | -0.0021688 | 0.333008901 | 0.002240325 | 0.18345412 | -0.00212344 | 0.343184216 | 0.002240172 | 0.183417206 | -4.54E-05 | 0.011534262 | 1 | 0.642540807 | 1 | 0.665851594 | ENSG0000025 | 3 | RP11-774O3.3 |
| VAT | Obesity | ENSG0000026 | 0.002231364 | 0.455905627 | 0.002992684 | 0.183454264 | 0.002162362 | 0.469931735 | 0.002992513 | 0.183417349 | 6.90E-05 | 0.013312895 | 1 | 0.739580277 | 1 | 0.766555312 | ENSG0000025 | 2 | RP11-20I20.1 |
| VAT | Obesity | ENSG0000026 | 0.002075553 | 0.311745021 | 0.002051817 | 0.183454087 | 0.001315953 | 0.522500412 | 0.002057796 | 0.183417391 | 0.0007596 | 1.89E-06 | 1 | 0.619458427 | 1 | 0.784841324 | ENSG0000025 | 2 | RP11-20I20.1 |
| VAT | Obesity | ENSG0000026 | -0.02756286 | 9.16E-06 | 0.006213181 | 0.183447035 | -0.02737299 | 1.05E-05 | 0.006212746 | 0.18341021 | -0.00018988 | 8.81E-05 | 0.02527726 | 0.000304545 | 0.029076197 | 0.000403836 | ENSG0000025 | 2 | GUSBP3 |
| VAT | Obesity | ENSG0000027 | -0.00756927 | 0.127121643 | 0.004961655 | 0.183453594 | -0.00747505 | 0.131894011 | 0.00496128 | 0.183416688 | -9.42E-05 | 0.007263759 | 1 | 0.381801917 | 1 | 0.399591076 | ENSG0000025 | 1 | TMEM200B |
| VAT | Obesity | ENSG0000027 | -0.0038224 | 0.422516241 | 0.004765704 | 0.183454231 | -0.0038994 | 0.413198528 | 0.004765373 | 0.183417293 | 7.70E-05 | 0.040862377 | 1 | 0.712802461 | 1 | 0.72386593 | ENSG0000025 | 1 | TMEM200B |
| VAT | Obesity | ENSG0000027 | 0.002231735 | 0.624150692 | 0.004554779 | 0.183454383 | 0.002409314 | 0.596820541 | 0.004554653 | 0.18341744 | -0.00017758 | 0.001248435 | 1 | 0.835148305 | 1 | 0.827015081 | ENSG0000025 | 1 | PCDHGB6 |
| VAT | Obesity | ENSG0000027 | -0.00364934 | 0.311928222 | 0.003608971 | 0.183454088 | -0.0035909 | 0.319705268 | 0.003608706 | 0.183417172 | -5.84E-05 | 0.028148664 | 1 | 0.619458427 | 1 | 0.641269508 | ENSG0000025 | 2 | RP11-261C10.7 |
| VAT | Obesity | ENSG0000027 | 0.001691692 | 0.62702604 | 0.00348144 | 0.183454385 | 0.00174593 | 0.61599453 | 0.003481168 | 0.183417451 | -5.42E-05 | 0.020328875 | 1 | 0.836438797 | 1 | 0.839253732 | ENSG0000025 | 2 | RP11-261C10.7 |
| VAT | Obesity | ENSG0000027 | -0.00447584 | 0.253641809 | 0.00392085 | 0.183453982 | -0.00455561 | 0.245246669 | 0.003920583 | 0.183417036 | 7.98E-05 | 0.011745557 | 1 | 0.549998458 | 1 | 0.552555761 | ENSG0000025 | 2 | RP11-261C10.7 |
| VAT | Obesity | ENSG0000027 | 0.000378149 | 0.946045411 | 0.005587823 | 0.183454472 | 0.000478344 | 0.931775961 | 0.005587432 | 0.183417543 | -0.0001002 | 0.022093966 | 1 | 0.981611028 | 1 | 0.976719199 | ENSG0000025 | 2 | RP11-261C10.7 |
| VAT | Obesity | ENSG0000027 | 0.000703047 | 0.886013888 | 0.00490441 | 0.183454466 | -0.00027919 | 0.954649519 | 0.004909307 | 0.183417545 | 0.000982235 | 1.95E-05 | 1 | 0.957634941 | 1 | 0.984913318 | ENSG0000025 | 1 | RP11-497H16.8 |
| VAT | Obesity | ENSG0000027 | 0.007038933 | 0.253676825 | 0.006166575 | 0.183453982 | 0.007138915 | 0.246962465 | 0.006166143 | 0.183417039 | -1.00E-04 | 0.038159579 | 1 | 0.549998458 | 1 | 0.554266359 | ENSG0000025 | 1 | KB-1460A1.1 |
| VAT | Obesity | ENSG0000027 | -0.00097405 | 0.708120979 | 0.002601767 | 0.183454421 | -0.00090833 | 0.726980211 | 0.002601591 | 0.1834175 | -6.57E-05 | 0.001866501 | 1 | 0.872628294 | 1 | 0.886980127 | ENSG0000025 | 1 | AC133633.2 |
| VAT | Obesity | ENSG0000027 | -0.01167558 | 0.059926101 | 0.006205985 | 0.183453136 | -0.00109163 | 0.873194997 | 0.006839704 | 0.183417536 | -0.01058395 | 0.000233842 | 1 | 0.24909042 | 1 | 0.956332568 | ENSG0000025 | 1 | PCDHGA5 |
| VAT | Obesity | ENSG0000027 | -0.00289797 | 0.605559267 | 0.005611625 | 0.183454373 | -0.00300877 | 0.591819993 | 0.005611271 | 0.183417437 | 0.000110796 | 0.022789579 | 1 | 0.827343939 | 1 | 0.824941855 | ENSG0000025 | 1 | FAM86HP |
| VAT | Obesity | ENSG0000027 | 0.000939457 | 0.708707825 | 0.002514659 | 0.183454421 | 6.16E-05 | 0.980497064 | 0.002521458 | 0.183417546 | 0.000877818 | 3.19E-06 | 1 | 0.872839624 | 1 | 0.99462398 | ENSG0000025 | 1 | FAM86HP |
| VAT | Obesity | ENSG0000027 | -0.00246357 | 0.500613705 | 0.003657721 | 0.183454303 | -0.0025178 | 0.491199875 | 0.003657451 | 0.183417367 | 5.42E-05 | 0.043155256 | 1 | 0.768636987 | 1 | 0.775358255 | ENSG0000025 | 3 | KB-1732A1.1 |
| VAT | Obesity | ENSG0000027 | -0.00074845 | 0.882624098 | 0.005069324 | 0.183454466 | -0.0006596 | 0.896468306 | 0.005068979 | 0.18341754 | -8.89E-05 | 0.029748195 | 1 | 0.95522585 | 1 | 0.968028006 | ENSG0000025 | 1 | RP11-267M23.4 |
| VAT | Obesity | ENSG0000027 | 0.00089686 | 0.811081351 | 0.003752058 | 0.183454453 | 0.000839753 | 0.822891194 | 0.003751778 | 0.183417527 | 5.71E-05 | 0.035132232 | 1 | 0.923127641 | 1 | 0.933105873 | ENSG0000025 | 1 | PCDHGA6 |
| VAT | Obesity | ENSG0000027 | -0.00203256 | 0.098331107 | 0.001229616 | 0.183453441 | -0.00163535 | 0.185693542 | 0.00123569 | 0.183416884 | -0.00039721 | 0.001310479 | 1 | 0.333396805 | 1 | 0.492802092 | ENSG0000025 | 1 | PCDHGA6 |
| VAT | Obesity | ENSG0000027 | -0.01522522 | 0.000624548 | 0.004450848 | 0.183450051 | -0.01047159 | 0.021645912 | 0.00455974 | 0.183415553 | -0.00475363 | 1.67E-06 | 1 | 0.00916889 | 1 | 0.13365261 | ENSG0000025 | 1 | PCDHGA6 |
| VAT | Obesity | ENSG0000027 | 0.00082394 | 0.891657765 | 0.006049177 | 0.183454467 | 0.000950678 | 0.875110415 | 0.006048702 | 0.183417537 | -0.00012674 | 0.001624163 | 1 | 0.961627151 | 1 | 0.956332568 | ENSG0000025 | 1 | PCDHGA6 |
| VAT | Obesity | ENSG0000027 | -0.02077428 | 0.000195936 | 0.005578145 | 0.183449231 | -0.02068604 | 0.000208345 | 0.005577697 | 0.183412348 | -8.82E-05 | 0.013088252 | 0.540783532 | 0.003808335 | 0.575030975 | 0.004290423 | ENSG0000025 | 1 | RP11-350F16.1 |
| VAT | Obesity | ENSG0000027 | 0.007044907 | 1.08E-11 | 0.001036676 | 0.183437019 | 0.00669793 | 1.25E-10 | 0.001041098 | 0.183401905 | 0.000346977 | 0.000343545 | 2.98E-08 | 1.03E-09 | 3.44E-07 | 1.01E-08 | ENSG0000025 | 1 | TPTE2P1 |
| VAT | Obesity | ENSG0000027 | 0.000287906 | 0.924799305 | 0.003050175 | 0.183454471 | 0.000374634 | 0.902244839 | 0.003050115 | 0.18341754 | -8.67E-05 | 0.02540391 | 1 | 0.975705689 | 1 | 0.969573834 | ENSG0000025 | 1 | TPTE2P1 |
| VAT | Obesity | ENSG0000027 | 0.00783017 | 0.193295363 | 0.006019069 | 0.183453834 | 0.00771467 | 0.199918921 | 0.006018696 | 0.183416925 | 0.0001155 | 0.029054225 | 1 | 0.490645638 | 1 | 0.510431287 | ENSG0000025 | 1 | COX6B1P4 |
| VAT | Obesity | ENSG0000027 | -0.01661551 | 7.37E-05 | 0.004191653 | 0.183448535 | -0.01652107 | 8.09E-05 | 0.004191353 | 0.183411674 | -9.44E-05 | 0.00311467 | 0.203494252 | 0.001710036 | 0.223329467 | 0.002011977 | ENSG0000025 | 1 | COX6B1P4 |
| VAT | Obesity | ENSG0000027 | -0.00604186 | 0.261418717 | 0.005379892 | 0.183453997 | -0.0059412 | 0.269415494 | 0.005379522 | 0.183417085 | -0.00010066 | 0.019174472 | 1 | 0.563244075 | 1 | 0.585237719 | ENSG0000025 | 1 | COX6B1P4 |
| VAT | Obesity | ENSG0000027 | 0.001431053 | 0.738800673 | 0.004291757 | 0.183454432 | 0.001351753 | 0.752770888 | 0.004291443 | 0.183417509 | 7.93E-05 | 0.01240157 | 1 | 0.888105338 | 1 | 0.898390751 | ENSG0000025 | 1 | COX6B1P4 |
| VAT | Obesity | ENSG0000027 | -0.00686952 | 0.171087653 | 0.005018943 | 0.183453766 | -0.006963 | 0.165310056 | 0.005018616 | 0.183416819 | 9.35E-05 | 0.027279158 | 1 | 0.457560003 | 1 | 0.456129337 | ENSG0000025 | 2 | RP11-1415C14.3 |
| VAT | Obesity | ENSG0000027 | 0.002207319 | 0.264343012 | 0.001977554 | 0.183454003 | 0.001337658 | 0.500531499 | 0.001985674 | 0.183417375 | 0.000869661 | 1.67E-06 | 1 | 0.565471432 | 1 | 0.776886587 | ENSG0000025 | 2 | RP11-1415C14.3 |
| VAT | Obesity | ENSG0000027 | -0.00186561 | 0.223177159 | 0.001531542 | 0.183453913 | -0.00027096 | 0.86214587 | 0.001560444 | 0.183417535 | -0.00159465 | 1.03E-07 | 1 | 0.52199987 | 1 | 0.950288579 | ENSG0000025 | 2 | RP11-1415C14.3 |
| VAT | Stroke | ENSG0000019 | 0.002822017 | 0.074174397 | 0.001580484 | 0.057490262 | 0.008307609 | 0.005442403 | 0.002988753 | 0.057488797 | -0.00548559 | 0.030579464 | 1 | 0.73651102 | 1 | 0.551358311 | ENSG0000025 | 2 | CD8BP |
| VAT | T2DM | ENSG0000000 | 0.003011447 | 0.273191547 | 0.002748314 | 0.081436758 | 0.002925737 | 0.287068745 | 0.002748267 | 0.081424463 | 8.57E-05 | 0.003930626 | 1 | 0.652447775 | 1 | 0.663927047 | ENSG0000025 | 2 | CD8BP |
| VAT | T2DM | ENSG0000000 | 0.00323482 | 0.187323195 | 0.002453332 | 0.081436668 | 0.003568463 | 0.14663084 | 0.002458402 | 0.0814243 | -0.00033364 | 0.037831661 | 1 | 0.570024276 | 1 | 0.514898084 | ENSG0000025 | 2 | CD8BP |
| VAT | T2DM | ENSG0000000 | -0.00536298 | 0.012596792 | 0.002149508 | 0.081435918 | -0.00541876 | 0.011701337 | 0.002149418 | 0.08142359 | 5.58E-05 | 0.00155036 | 1 | 0.14522599 | 1 | 0.148828067 | ENSG0000025 | 1 | CTD-3107M8.2 |
| VAT | T2DM | ENSG0000000 | 0.001407328 | 0.301450946 | 0.001361938 | 0.08143678 | 0.001430793 | 0.293440056 | 0.001361873 | 0.081424468 | -2.35E-05 | 0.022319663 | 1 | 0.675668349 | 1 | 0.66767894 | ENSG0000025 | 1 | CTD-3107M8.2 |

| | | | | | | | | | | | | | | | | | | | |
|---|---|---|---|---|---|---|---|---|---|---|---|---|---|---|---|---|---|---|---|
| VAT | T2DM | ENSG000000( | 0.001194897 | 0.742302189 | 0.003634043 | 0.08143694 | 0.001113779 | 0.759225721 | 0.003633888 | 0.081424637 | 8.11E-05 | 0.005963098 | 1 | 0.916157236 | 1 | 0.915051408 | ENSG0000025 | 1 | RP11-578O24.2 |
| VAT | T2DM | ENSG000000( | 0.001398367 | 0.60360006 | 0.002693152 | 0.081436913 | 0.001329389 | 0.621559169 | 0.002693021 | 0.081424612 | 6.90E-05 | 0.000505236 | 1 | 0.857656828 | 1 | 0.868609269 | ENSG0000025 | 1 | RP11-578O24.2 |
| VAT | T2DM | ENSG000000( | -0.00123455 | 0.608869982 | 0.002412701 | 0.081436915 | -0.0011786 | 0.625183471 | 0.002412613 | 0.081424613 | -5.59E-05 | 0.008672454 | 1 | 0.859136518 | 1 | 0.869014343 | ENSG0000025 | 1 | RP11-578O24.2 |
| VAT | T2DM | ENSG0000001 | -0.0078997 | 0.001514383 | 0.002490502 | 0.081435277 | -0.0078557 | 0.001608371 | 0.002490391 | 0.08142299 | -4.40E-05 | 0.025166033 | 1 | 0.039062596 | 1 | 0.045352953 | ENSG0000025 | 1 | RP11-578O24.2 |
| VAT | T2DM | ENSG0000001 | 0.006542228 | 0.087007357 | 0.003822727 | 0.081436469 | 0.006606345 | 0.08394236 | 0.003822543 | 0.081424153 | -6.41E-05 | 0.023098172 | 1 | 0.406073085 | 1 | 0.410615651 | ENSG0000025 | 2 | SIGLEC12 |
| VAT | T2DM | ENSG0000001 | 0.017073235 | 4.85E-11 | 0.002596439 | 0.081429733 | 0.017150923 | 3.96E-11 | 0.002596324 | 0.081417361 | -7.77E-05 | 0.000172445 | 1.34E-07 | 1.49E-08 | 1.09E-07 | 2.18E-08 | ENSG0000025 | 2 | SIGLEC12 |
| VAT | T2DM | ENSG0000001 | 0.002801618 | 0.294558161 | 0.002672852 | 0.081436775 | 0.002844422 | 0.287220614 | 0.002672726 | 0.081424463 | -4.28E-05 | 0.034503447 | 1 | 0.671884731 | 1 | 0.663927047 | ENSG0000025 | 1 | AP003068.23 |
| VAT | T2DM | ENSG0000001 | 0.000777957 | 0.734343739 | 0.00229247 | 0.081436939 | 0.000843856 | 0.712787025 | 0.002292366 | 0.08142463 | -6.59E-05 | 0.000209645 | 1 | 0.912967891 | 1 | 0.903325484 | ENSG0000025 | 1 | AP003068.23 |
| VAT | T2DM | ENSG0000001 | -0.00251364 | 0.361926756 | 0.002757094 | 0.08143682 | -0.00869792 | 0.008071068 | 0.003283357 | 0.08142348 | 0.006184274 | 0.000524525 | 1 | 0.719681446 | 1 | 0.117539869 | ENSG0000025 | 1 | AP003068.23 |
| VAT | T2DM | ENSG0000001 | 0.00250068 | 0.319880029 | 0.002513991 | 0.081436793 | 0.002583005 | 0.304196071 | 0.002513928 | 0.081424476 | -8.23E-05 | 0.001135974 | 1 | 0.690842541 | 1 | 0.676208846 | ENSG0000025 | 1 | AP003068.23 |
| VAT | T2DM | ENSG0000002 | 0.002975294 | 0.12593176 | 0.001944197 | 0.081436567 | 0.002930831 | 0.131673566 | 0.001944118 | 0.081424273 | 4.45E-05 | 0.006404199 | 1 | 0.480735348 | 1 | 0.491745938 | ENSG0000025 | 1 | AP003068.23 |
| VAT | T2DM | ENSG0000002 | -0.0011607 | 0.350322209 | 0.00124277 | 0.081436813 | -0.00171239 | 0.170007699 | 0.001247931 | 0.081424338 | 0.000551683 | 1.42E-06 | 1 | 0.710948012 | 1 | 0.54753664 | ENSG0000025 | 1 | AP003068.23 |
| VAT | T2DM | ENSG0000003 | 0.001652204 | 0.610264621 | 0.003241548 | 0.081436915 | 0.001708211 | 0.598196721 | 0.003241413 | 0.081424606 | -5.60E-05 | 0.035738214 | 1 | 0.85979089 | 1 | 0.856779751 | ENSG0000025 | 3 | RP11-231C14.3 |
| VAT | T2DM | ENSG0000003 | -0.00442855 | 0.013469469 | 0.001792123 | 0.081435938 | -0.00446286 | 0.012761323 | 0.001792045 | 0.081423616 | 3.43E-05 | 0.016863553 | 1 | 0.149821016 | 1 | 0.156538892 | ENSG0000025 | 1 | AP000783.1 |
| VAT | T2DM | ENSG0000003 | -0.00155859 | 0.700275465 | 0.004048828 | 0.081436934 | -0.00165425 | 0.682839685 | 0.004048672 | 0.081424625 | 9.57E-05 | 0.00608033 | 1 | 0.901290186 | 1 | 0.89223498 | ENSG0000025 | 1 | AP000783.1 |
| VAT | T2DM | ENSG0000003 | 0.000210523 | 0.888039638 | 0.001495348 | 0.081436955 | 0.000237339 | 0.87388571 | 0.001495288 | 0.081424648 | -2.68E-05 | 0.033681046 | 1 | 0.969128142 | 1 | 0.96520345 | ENSG0000025 | 1 | AP000783.1 |
| VAT | T2DM | ENSG0000005 | -0.00203739 | 0.499068539 | 0.003014082 | 0.081436882 | -0.00191922 | 0.524281611 | 0.003014046 | 0.081424585 | -0.00011817 | 0.000494403 | 1 | 0.797584926 | 1 | 0.820071559 | ENSG0000025 | 2 | PKD1P5 |
| VAT | T2DM | ENSG0000005 | -0.00400791 | 0.296794349 | 0.003841447 | 0.081436777 | -0.00966445 | 0.018567375 | 0.004105347 | 0.081423726 | 0.005656538 | 9.47E-05 | 1 | 0.674754862 | 1 | 0.195735137 | ENSG0000025 | 2 | PKD1P5 |
| VAT | T2DM | ENSG0000005 | 0.004922159 | 0.191892327 | 0.00377176 | 0.081436674 | 0.00482004 | 0.201256875 | 0.003771606 | 0.08142438 | 0.00010212 | 0.001165042 | 1 | 0.572736771 | 1 | 0.581634659 | ENSG0000025 | 2 | MEX3A |
| VAT | T2DM | ENSG0000005 | 0.004839923 | 0.049517465 | 0.002464176 | 0.081436314 | 0.00480069 | 0.051382175 | 0.002464067 | 0.081424018 | 3.92E-05 | 0.043843032 | 1 | 0.314179779 | 1 | 0.328275005 | ENSG0000025 | 1 | CTD-2005H7.1 |
| VAT | T2DM | ENSG0000005 | 0.002839783 | 0.414098407 | 0.003477129 | 0.081436847 | 0.002917464 | 0.401428437 | 0.003477006 | 0.081424535 | -7.77E-05 | 0.012636288 | 1 | 0.757898941 | 1 | 0.754215443 | ENSG0000025 | 1 | CTD-2005H7.1 |
| VAT | T2DM | ENSG0000005 | -0.00090988 | 0.460571697 | 0.001233051 | 0.081436867 | -0.00093107 | 0.450168816 | 0.001232992 | 0.081424557 | 2.12E-05 | 0.02134458 | 1 | 0.78904441 | 1 | 0.786370844 | ENSG0000025 | 1 | CTD-2005H7.1 |
| VAT | T2DM | ENSG0000005 | -0.00014219 | 0.946653464 | 0.002125145 | 0.081436958 | -0.00019204 | 0.92799457 | 0.002125114 | 0.081424651 | 4.98E-05 | 0.033564374 | 1 | 0.987266065 | 1 | 0.984723188 | ENSG0000025 | 1 | RP11-680E19.1 |
| VAT | T2DM | ENSG0000006 | 0.00481181 | 0.219431541 | 0.003918276 | 0.081436706 | 0.004891175 | 0.211904145 | 0.003918117 | 0.081424392 | -7.94E-05 | 0.015712631 | 1 | 0.601421106 | 1 | 0.593897083 | ENSG0000025 | 1 | RP11-109L13.1 |
| VAT | T2DM | ENSG0000006 | -0.00042566 | 0.808762179 | 0.001758756 | 0.081436949 | -0.00056244 | 0.749129626 | 0.001758794 | 0.081424635 | 0.00013678 | 2.50E-08 | 1 | 0.941055486 | 1 | 0.911563685 | ENSG0000025 | 1 | RP11-109L13.1 |
| VAT | T2DM | ENSG0000006 | -0.00234332 | 0.142597388 | 0.001598235 | 0.081436599 | -0.00231546 | 0.147385471 | 0.001598157 | 0.081424302 | -2.79E-05 | 0.01681068 | 1 | 0.507766954 | 1 | 0.515568947 | ENSG0000025 | 1 | RP11-109L13.1 |
| VAT | T2DM | ENSG0000006 | -0.01592862 | 2.64E-06 | 0.003391064 | 0.081433271 | -0.01598076 | 2.44E-06 | 0.003390896 | 0.081420941 | 5.21E-05 | 0.033802743 | 0.007280315 | 0.000260011 | 0.006744149 | 0.000293224 | ENSG0000025 | 1 | RP11-109L13.1 |
| VAT | T2DM | ENSG0000006 | -0.0036689 | 0.046640138 | 0.00184408 | 0.081436297 | -0.00197905 | 0.292437007 | 0.00187981 | 0.081424467 | -0.00168986 | 3.76E-06 | 1 | 0.303391075 | 1 | 0.666746052 | ENSG0000025 | 2 | RP13-870H17.3 |
| VAT | T2DM | ENSG0000006 | -0.00097584 | 0.384185324 | 0.00112138 | 0.081436832 | -0.00099796 | 0.373473153 | 0.001121325 | 0.08142452 | 2.21E-05 | 0.006925309 | 1 | 0.739952195 | 1 | 0.733070343 | ENSG0000025 | 2 | RP13-870H17.3 |
| VAT | T2DM | ENSG0000006 | 0.000420144 | 0.730174536 | 0.001218185 | 0.081436939 | 0.000399529 | 0.742921484 | 0.001218123 | 0.081424635 | 2.06E-05 | 0.015762112 | 1 | 0.910550482 | 1 | 0.911123584 | ENSG0000025 | 2 | RP11-632K20.2 |
| VAT | T2DM | ENSG0000006 | 0.00310273 | 0.368737646 | 0.003451922 | 0.081436823 | 0.003055877 | 0.375986314 | 0.003451734 | 0.081424522 | 4.69E-05 | 0.036860443 | 1 | 0.727345349 | 1 | 0.735451613 | ENSG0000025 | 2 | RP11-632K20.2 |
| VAT | T2DM | ENSG0000006 | 0.002121807 | 0.36442938 | 0.002339476 | 0.081436821 | 0.002161191 | 0.355573952 | 0.002339375 | 0.08142451 | -3.94E-05 | 0.036778113 | 1 | 0.722575494 | 1 | 0.715294539 | ENSG0000025 | 2 | RP11-632K20.2 |
| VAT | T2DM | ENSG0000007 | -0.00540457 | 0.054131182 | 0.002806416 | 0.081436339 | -0.00202189 | 0.503253409 | 0.003020537 | 0.081424578 | -0.00338268 | 0.002469897 | 1 | 0.330306509 | 1 | 0.809900529 | ENSG0000025 | 3 | DPP3 |
| VAT | T2DM | ENSG0000007 | -0.00323699 | 0.393731356 | 0.003795403 | 0.081436837 | -0.0031758 | 0.402711972 | 0.003795226 | 0.081424535 | -6.12E-05 | 0.033758117 | 1 | 0.747385518 | 1 | 0.755809114 | ENSG0000025 | 3 | DPP3 |
| VAT | T2DM | ENSG0000007 | 0.005552158 | 0.060463808 | 0.002957365 | 0.081436369 | 0.005500071 | 0.062904577 | 0.002957235 | 0.081424074 | 5.21E-05 | 0.026395629 | 1 | 0.351326547 | 1 | 0.367054194 | ENSG0000025 | 3 | DPP3 |
| VAT | T2DM | ENSG0000007 | -0.00501126 | 0.168816516 | 0.003641857 | 0.081436642 | -0.00495488 | 0.173637144 | 0.003641659 | 0.081424343 | -5.64E-05 | 0.017031113 | 1 | 0.546218791 | 1 | 0.548422551 | ENSG0000025 | 3 | DPP3 |
| VAT | T2DM | ENSG0000007 | -0.00088872 | 0.791874325 | 0.003367918 | 0.081436947 | -0.00096819 | 0.773739522 | 0.00336776 | 0.081424639 | 7.95E-05 | 0.001882256 | 1 | 0.932507176 | 1 | 0.923905803 | ENSG0000025 | 3 | DPP3 |
| VAT | T2DM | ENSG0000007 | 0.003382497 | 0.382108616 | 0.003870052 | 0.081436831 | 0.003316295 | 0.391472486 | 0.003869878 | 0.08142453 | 6.62E-05 | 0.028842148 | 1 | 0.739046798 | 1 | 0.748625 | ENSG0000025 | 3 | DPP3 |
| VAT | T2DM | ENSG0000007 | -0.00018739 | 0.939201245 | 0.002456865 | 0.081436957 | -0.00014135 | 0.954118486 | 0.002456733 | 0.081424652 | -4.60E-05 | 0.0046453 | 1 | 0.986373964 | 1 | 0.988501134 | ENSG0000025 | 2 | BRK1 |
| VAT | T2DM | ENSG0000007 | 0.01201365 | 0.000253171 | 0.003283263 | 0.081434721 | 0.011965055 | 0.000268002 | 0.003283098 | 0.081422433 | 4.86E-05 | 0.036867782 | 0.698751413 | 0.009841569 | 0.739685085 | 0.012455159 | ENSG0000025 | 2 | BRK1 |
| VAT | T2DM | ENSG0000007 | 0.000985382 | 0.761014711 | 0.003239802 | 0.081436943 | 0.000932399 | 0.773493405 | 0.003239649 | 0.081424639 | 5.30E-05 | 0.029362302 | 1 | 0.920907272 | 1 | 0.923905803 | ENSG0000025 | 1 | RP11-680E19.2 |
| VAT | T2DM | ENSG0000007 | -0.00385012 | 0.325535155 | 0.003916116 | 0.081436797 | -0.00377811 | 0.334644811 | 0.003915943 | 0.081424497 | -7.20E-05 | 0.020261296 | 1 | 0.693804654 | 1 | 0.702906909 | ENSG0000025 | 2 | RP11-110I1.12 |
| VAT | T2DM | ENSG0000008 | 0.003275556 | 0.12587417 | 0.002140076 | 0.081436567 | 0.003185229 | 0.136674441 | 0.00214019 | 0.081424282 | 9.03E-05 | 0.00849112 | 1 | 0.480735348 | 1 | 0.502180434 | ENSG0000025 | 1 | RP11-655M14.4 |
| VAT | T2DM | ENSG0000008 | 0.003565478 | 0.116907864 | 0.002274064 | 0.081436548 | 0.003620305 | 0.111368859 | 0.002273956 | 0.081424229 | -5.48E-05 | 0.001361443 | 1 | 0.462271782 | 1 | 0.455952462 | ENSG0000025 | 1 | RP11-655M14.4 |
| VAT | T2DM | ENSG0000008 | 0.00506311 | 0.041972436 | 0.002489484 | 0.081436267 | 0.005023336 | 0.043600304 | 0.002489365 | 0.081423972 | 3.98E-05 | 0.031835919 | 1 | 0.284233402 | 1 | 0.296396158 | ENSG0000025 | 1 | RP11-655M14.4 |
| VAT | T2DM | ENSG0000008 | 0.003638262 | 0.237916929 | 0.003082722 | 0.081436726 | 0.003690678 | 0.231201364 | 0.003082574 | 0.081424413 | -5.24E-05 | 0.02238195 | 1 | 0.624786607 | 1 | 0.618894184 | ENSG0000025 | 1 | RP11-655M14.4 |
| VAT | T2DM | ENSG0000008 | 0.010558659 | 0.007768534 | 0.00396647 | 0.081435774 | 0.010631431 | 0.007352666 | 0.003966292 | 0.081423452 | -7.28E-05 | 0.019291362 | 1 | 0.106355675 | 1 | 0.112740874 | ENSG0000025 | 1 | RP11-655M14.4 |
| VAT | T2DM | ENSG0000008 | 0.010170732 | 4.66E-06 | 0.002220986 | 0.081433454 | 0.010205169 | 4.33E-06 | 0.002220882 | 0.081421124 | -3.44E-05 | 0.041032465 | 0.012874837 | 0.00044396 | 0.011941327 | 0.000497555 | ENSG0000025 | 1 | RP11-655M14.4 |
| VAT | T2DM | ENSG0000008 | 0.002701213 | 0.280474219 | 0.002502848 | 0.081436764 | 0.001912137 | 0.4465226 | 0.002511919 | 0.081424556 | 0.000789076 | 0.000250158 | 1 | 0.657696555 | 1 | 0.785418349 | ENSG0000025 | 1 | AP003068.18 |
| VAT | T2DM | ENSG0000008 | -0.00669526 | 0.020133594 | 0.002881099 | 0.081436056 | -0.00662458 | 0.021481677 | 0.002880982 | 0.081423769 | -7.07E-05 | 0.003257251 | 1 | 0.193619235 | 1 | 0.210994407 | ENSG0000025 | 1 | AP003068.18 |
| VAT | T2DM | ENSG0000008 | -0.0044233 | 0.086994524 | 0.002584498 | 0.081436469 | -0.00432958 | 0.093880891 | 0.002584399 | 0.081424184 | -9.37E-05 | 2.59E-05 | 1 | 0.406073085 | 1 | 0.429776321 | ENSG0000025 | 1 | AP003068.18 |
| VAT | T2DM | ENSG0000008 | 0.003619295 | 0.210328016 | 0.002889275 | 0.081436696 | 0.003689681 | 0.201570747 | 0.002889129 | 0.08142438 | -7.04E-05 | 0.000605488 | 1 | 0.592033071 | 1 | 0.581634659 | ENSG0000025 | 1 | AP003068.18 |
| VAT | T2DM | ENSG0000008 | -0.00026387 | 0.935550444 | 0.003263094 | 0.081436957 | -0.00031069 | 0.924141295 | 0.003262932 | 0.081424651 | 4.68E-05 | 0.046344485 | 1 | 0.985917994 | 1 | 0.982969566 | ENSG0000025 | 1 | AP003068.18 |
| VAT | T2DM | ENSG0000008 | -0.00264335 | 0.3586009 | 0.002879354 | 0.081436818 | -0.00259767 | 0.366942699 | 0.002879209 | 0.081424516 | -4.57E-05 | 0.025864266 | 1 | 0.716682464 | 1 | 0.72593143 | ENSG0000025 | 1 | AP003068.18 |
| VAT | T2DM | ENSG0000008 | 0.004263845 | 0.086806709 | 0.002489848 | 0.081436468 | 0.004304727 | 0.083812859 | 0.002489746 | 0.081424153 | -4.09E-05 | 0.048318988 | 1 | 0.406073085 | 1 | 0.410615651 | ENSG0000025 | 2 | OR8G5 |
| VAT | T2DM | ENSG0000009 | -0.00124632 | 0.668735615 | 0.002912748 | 0.081436928 | -0.00130612 | 0.653842093 | 0.002912637 | 0.081424619 | 5.98E-05 | 0.017743568 | 1 | 0.884384426 | 1 | 0.875674196 | ENSG0000025 | 1 | RP11-326C3.12 |
| VAT | T2DM | ENSG0000009 | -0.00254124 | 0.260859104 | 0.002260159 | 0.081436747 | -0.002595 | 0.250889452 | 0.002260069 | 0.081424432 | 5.38E-05 | 0.004816432 | 1 | 0.643030277 | 1 | 0.632237235 | ENSG0000025 | 1 | RP11-326C3.12 |
| VAT | T2DM | ENSG0000009 | -0.00023324 | 0.694008754 | 0.000592854 | 0.081436933 | -0.00024341 | 0.681375777 | 0.000592825 | 0.081424624 | 1.02E-05 | 0.01768714 | 1 | 0.898615554 | 1 | 0.891626351 | ENSG0000025 | 1 | RP11-326C3.12 |
| VAT | T2DM | ENSG0000009 | 0.005683481 | 0.128845349 | 0.003742398 | 0.081436573 | 0.007158179 | 0.056233452 | 0.003749256 | 0.081424043 | -0.0014747 | 1.84E-10 | 1 | 0.485394797 | 1 | 0.344116711 | ENSG0000025 | 1 | TAS2R43 |
| VAT | T2DM | ENSG0000009 | -0.0005461 | 0.790704162 | 0.002057676 | 0.081436947 | -0.00049931 | 0.808261282 | 0.002057576 | 0.081424643 | -4.68E-05 | 0.001957353 | 1 | 0.932507176 | 1 | 0.938393612 | ENSG0000025 | 2 | C8orf49 |
| VAT | T2DM | ENSG0000009 | -0.00277585 | 0.464293827 | 0.003793207 | 0.081436869 | -0.00292547 | 0.440588167 | 0.0037934 | 0.081424553 | 0.000149622 | 0.013081096 | 1 | 0.78904441 | 1 | 0.781760596 | ENSG0000025 | 2 | C8orf49 |
| VAT | T2DM | ENSG0000009 | 0.000676311 | 0.650370335 | 0.001492144 | 0.081436924 | 0.00070188 | 0.638066657 | 0.001492078 | 0.081424616 | -2.56E-05 | 0.031085065 | 1 | 0.874341025 | 1 | 0.87456323 | ENSG0000025 | 2 | C8orf49 |
| VAT | T2DM | ENSG0000009 | 0.001042775 | 0.711399028 | 0.002818478 | 0.081436936 | 0.000973352 | 0.729821945 | 0.002818349 | 0.081424633 | 6.94E-05 | 0.001438091 | 1 | 0.903987715 | 1 | 0.908589792 | ENSG0000025 | 1 | EBLN2 |
| VAT | T2DM | ENSG000001( | 0.001797505 | 0.51707366 | 0.002774513 | 0.081436888 | 0.001837071 | 0.50786973 | 0.002774367 | 0.081424579 | -3.96E-05 | 0.034899971 | 1 | 0.812597159 | 1 | 0.811718027 | ENSG0000025 | 1 | RP11-326C3.4 |
| VAT | T2DM | ENSG000001( | -0.00010302 | 0.973292842 | 0.003077257 | 0.081436958 | -0.00016015 | 0.958491445 | 0.003077121 | 0.081424652 | 5.71E-05 | 0.019380934 | 1 | 0.992835034 | 1 | 0.989894944 | ENSG0000025 | 1 | RP4-809F18.1 |
| VAT | T2DM | ENSG000001( | 0.000502895 | 0.729510668 | 0.001454393 | 0.081436938 | 0.000528895 | 0.716104795 | 0.001454328 | 0.08142463 | -2.60E-05 | 0.023649859 | 1 | 0.910527923 | 1 | 0.906212395 | ENSG0000025 | 1 | RP4-809F18.1 |
| VAT | T2DM | ENSG000001( | -0.00045939 | 0.903363106 | 0.003783683 | 0.081436956 | -0.00052837 | 0.888936682 | 0.003783529 | 0.081424649 | 6.90E-05 | 0.029315878 | 1 | 0.974557156 | 1 | 0.971063504 | ENSG0000025 | 1 | RP4-809F18.1 |
| VAT | T2DM | ENSG000001( | -0.00304506 | 0.089800189 | 0.001794952 | 0.081436477 | -0.00300738 | 0.09382814 | 0.001794869 | 0.081424183 | -3.77E-05 | 0.006095819 | 1 | 0.410345234 | 1 | 0.429776321 | ENSG0000025 | 1 | TAS2R20 |
| VAT | T2DM | ENSG000001( | 0.003987281 | 0.099636909 | 0.002421495 | 0.081436505 | 0.003935264 | 0.104120006 | 0.002421399 | 0.081424211 | 5.20E-05 | 0.011125663 | 1 | 0.430095137 | 1 | 0.442883689 | ENSG0000025 | 1 | TAS2R20 |
| VAT | T2DM | ENSG000001( | 0.000413476 | 0.622367206 | 0.000839548 | 0.081436918 | 0.000394528 | 0.638391102 | 0.000839511 | 0.081424616 | 1.89E-05 | 0.004052298 | 1 | 0.868321617 | 1 | 0.87456323 | ENSG0000025 | 1 | TAS2R20 |
| VAT | T2DM | ENSG000001( | -0.00905171 | 0.028474204 | 0.004131884 | 0.081436156 | -0.00898399 | 0.02967362 | 0.004131662 | 0.081423862 | -6.77E-05 | 0.012938545 | 1 | 0.23459344 | 1 | 0.251532736 | ENSG0000025 | 3 | DYX1C1 |
| VAT | T2DM | ENSG000001( | -0.00150739 | 0.426671314 | 0.001896317 | 0.081436853 | -0.00156133 | 0.410322057 | 0.001896367 | 0.081424539 | 5.39E-05 | 0.045942914 | 1 | 0.769178855 | 1 | 0.762700254 | ENSG0000025 | 3 | DYX1C1 |
| VAT | T2DM | ENSG000001( | -0.00139737 | 0.228410699 | 0.001160157 | 0.081436716 | -0.00141372 | 0.222987956 | 0.001160096 | 0.081424404 | 1.64E-05 | 0.036624674 | 1 | 0.612920192 | 1 | 0.604564596 | ENSG0000025 | 3 | DYX1C1 |
| VAT | T2DM | ENSG000001( | 0.003811948 | 0.120636109 | 0.002455975 | 0.081436556 | 0.003860315 | 0.115978946 | 0.002455858 | 0.08142424 | -4.84E-05 | 0.008485788 | 1 | 0.472283245 | 1 | 0.466853188 | ENSG0000025 | 2 | C21orf119 |
| VAT | T2DM | ENSG000001( | 0.000574128 | 0.872291849 | 0.003571597 | 0.081436954 | 0.00049134 | 0.890577015 | 0.003571452 | 0.081424649 | 8.28E-05 | 0.005633044 | 1 | 0.965652689 | 1 | 0.971465036 | ENSG0000025 | 2 | C21orf119 |
| VAT | T2DM | ENSG000001( | 0.001271156 | 0.63378716 | 0.002668235 | 0.08143692 | 0.001196251 | 0.653900187 | 0.002668113 | 0.081424619 | 7.49E-05 | 0.000277302 | 1 | 0.871141713 | 1 | 0.875674196 | ENSG0000025 | 2 | ZNF432 |
| VAT | T2DM | ENSG000001( | -0.00520118 | 0.171031695 | 0.003799535 | 0.081436645 | -0.00513949 | 0.176144843 | 0.003799369 | 0.081424347 | -6.17E-05 | 0.041190215 | 1 | 0.546218791 | 1 | 0.549954486 | ENSG0000025 | 2 | ZNF432 |
| VAT | T2DM | ENSG000001( | 0.000751376 | 0.707919968 | 0.002005533 | 0.081436935 | 0.000806603 | 0.687529983 | 0.002005429 | 0.081424625 | -5.52E-05 | 7.09E-05 | 1 | 0.902964787 | 1 | 0.893821363 | ENSG0000025 | 2 | ZNF432 |
| VAT | T2DM | ENSG000001( | -0.00748099 | 0.043728707 | 0.003709544 | 0.081436279 | -0.00761636 | 0.040047096 | 0.003709384 | 0.081423948 | 0.00013537 | 6.05E-06 | 1 | 0.291524711 | 1 | 0.286347109 | ENSG0000025 | 2 | ZNF432 |
| VAT | T2DM | ENSG000001( | -0.00205413 | 0.343118404 | 0.002166749 | 0.081436808 | -0.00199206 | 0.357878092 | 0.002166654 | 0.081424511 | -6.21E-05 | 0.000298859 | 1 | 0.705138034 | 1 | 0.718056961 | ENSG0000025 | 2 | hsa-mir-8072 |
| VAT | T2DM | ENSG000001( | 0.006244035 | 0.109094397 | 0.003896969 | 0.081436529 | 0.006301427 | 0.105860282 | 0.003896778 | 0.081424216 | -5.74E-05 | 0.04344395 | 1 | 0.446736699 | 1 | 0.446749813 | ENSG0000025 | 2 | hsa-mir-8072 |
| VAT | T2DM | ENSG000001( | 0.002289592 | 0.046717829 | 0.001151211 | 0.081436297 | 0.002267194 | 0.048897916 | 0.00115116 | 0.081424004 | 2.24E-05 | 0.014008964 | 1 | 0.303391075 | 1 | 0.318297756 | ENSG0000025 | 2 | hsa-mir-8072 |
| VAT | T2DM | ENSG000001( | -0.0108005 | 0.007025968 | 0.00400668 | 0.081435744 | -0.0108722 | 0.00665536 | 0.00400652 | 0.081423422 | 7.17E-05 | 0.033917593 | 1 | 0.099957076 | 1 | 0.104368145 | ENSG0000025 | 2 | hsa-mir-8072 |
| VAT | T2DM | ENSG000001( | 0.000859209 | 0.7792161 | 0.003064872 | 0.081436945 | 0.00099437 | 0.745598308 | 0.003064796 | 0.081424635 | -0.00013516 | 1.20E-05 | 1 | 0.928876022 | 1 | 0.911123584 | ENSG0000025 | 1 | XXbac-BPG248L24.13 |
| VAT | T2DM | ENSG000001( | -0.00663449 | 0.016611213 | 0.002769898 | 0.081436 | -0.00659049 | 0.0173393 | 0.002769765 | 0.081423706 | -4.40E-05 | 0.032065262 | 1 | 0.173007353 | 1 | 0.189906624 | ENSG0000025 | 1 | XXbac-BPG248L24.13 |
| VAT | T2DM | ENSG000001( | 0.011027362 | 0.003018834 | 0.003718139 | 0.081435488 | 0.011113765 | 0.002796996 | 0.003717941 | 0.081423159 | -8.64E-05 | 0.000536644 | 1 | 0.060817379 | 1 | 0.064871495 | ENSG0000025 | 1 | ZNF10 |
| VAT | T2DM | ENSG000001( | 0.005032578 | 0.203823897 | 0.003960371 | 0.081436689 | 0.00492779 | 0.213380766 | 0.003960213 | 0.081424394 | 0.000104788 | 0.00171814 | 1 | 0.582793504 | 1 | 0.59427943 | ENSG0000025 | 2 | HMBS |
| VAT | T2DM | ENSG000001( | 0.003662275 | 0.282029779 | 0.003404323 | 0.081436765 | 0.003612445 | 0.28860522 | 0.003404154 | 0.081424464 | 4.98E-05 | 0.04154252 | 1 | 0.658546692 | 1 | 0.664345628 | ENSG0000025 | 2 | HMBS |
| VAT | T2DM | ENSG000001( | 0.004631994 | 0.075155178 | 0.002602946 | 0.081436429 | 0.00457096 | 0.07907076 | 0.002602894 | 0.081424137 | 6.10E-05 | 0.025999856 | 1 | 0.379905296 | 1 | 0.401383586 | ENSG0000025 | 1 | TAS2R64P |
| VAT | T2DM | ENSG000001( | 0.003252992 | 0.400429752 | 0.003868674 | 0.08143684 | 0.003195455 | 0.4087905 | 0.003868474 | 0.081424538 | 5.75E-05 | 0.031554282 | 1 | 0.750041208 | 1 | 0.762700254 | ENSG0000025 | 1 | TAS2R64P |
| VAT | T2DM | ENSG000001( | -0.00351515 | 0.200728376 | 0.002747324 | 0.081436685 | -0.00346355 | 0.207394974 | 0.002747187 | 0.081424387 | -5.16E-05 | 0.009052216 | 1 | 0.581584201 | 1 | 0.588899308 | ENSG0000025 | 1 | RP1-102E24.8 |
| VAT | T2DM | ENSG000001( | 0.004140814 | 0.283942446 | 0.00386448 | 0.081436767 | 0.004011388 | 0.2992468 | 0.003864345 | 0.081424472 | 0.000129426 | 0.000206198 | 1 | 0.659508298 | 1 | 0.673122386 | ENSG0000025 | 1 | RP1-102E24.8 |
| VAT | T2DM | ENSG000001( | 0.002200625 | 0.034950954 | 0.001043478 | 0.081436215 | 0.002179867 | 0.036698497 | 0.001043444 | 0.081423923 | 2.08E-05 | 0.031314759 | 1 | 0.260715228 | 1 | 0.279029897 | ENSG0000025 | 1 | TAS2R31 |
| VAT | T2DM | ENSG000001( | 0.002101759 | 0.575894055 | 0.003757216 | 0.081436906 | 0.002033493 | 0.58834076 | 0.003757083 | 0.081424604 | 6.83E-05 | 0.0425042 | 1 | 0.844236594 | 1 | 0.852399212 | ENSG0000025 | 1 | RP13-977J11.2 |
| VAT | T2DM | ENSG000001( | 0.000940017 | 0.791652507 | 0.00355844 | 0.081436947 | 0.000559868 | 0.875049537 | 0.003560414 | 0.081424648 | 0.000380149 | 0.002624976 | 1 | 0.932507176 | 1 | 0.966054689 | ENSG0000025 | 1 | RP13-977J11.2 |
| VAT | T2DM | ENSG000001( | -0.0014157 | 0.464371656 | 0.001934897 | 0.081436869 | 0.000588004 | 0.782556608 | 0.002130533 | 0.08142464 | -0.00200371 | 0.024706588 | 1 | 0.78904441 | 1 | 0.92833376 | ENSG0000025 | 3 | RP11-705C15.2 |
| VAT | T2DM | ENSG000001( | 0.001436645 | 0.433350469 | 0.001833691 | 0.081436856 | 0.001463787 | 0.424688964 | 0.0018336 | 0.081424546 | -2.71E-05 | 0.038856726 | 1 | 0.772141571 | 1 | 0.771653418 | ENSG0000025 | 3 | CYP2B7P |
| VAT | T2DM | ENSG000001( | 0.005392142 | 0.123191834 | 0.003497947 | 0.081436561 | 0.005456778 | 0.118744053 | 0.003497778 | 0.081424246 | -6.46E-05 | 0.012640106 | 1 | 0.475900889 | 1 | 0.472623853 | ENSG0000025 | 3 | CYP2B7P |
| VAT | T2DM | ENSG000001( | 0.001746464 | 0.088288669 | 0.001024621 | 0.081436473 | 0.001728225 | 0.091646381 | 0.001024571 | 0.081424177 | 1.82E-05 | 0.015356525 | 1 | 0.407340768 | 1 | 0.426549767 | ENSG0000025 | 1 | CLEC12B |
| VAT | T2DM | ENSG000001( | -0.00181055 | 0.028435214 | 0.000826267 | 0.081436156 | -0.00182366 | 0.027298961 | 0.000826223 | 0.081423838 | 1.31E-05 | 0.016845733 | 1 | 0.23459344 | 1 | 0.237681802 | ENSG0000025 | 1 | RP11-500M8.6 |
| VAT | T2DM | ENSG000001( | 0.005085925 | 0.153900246 | 0.003566838 | 0.081436619 | 0.005173406 | 0.146924294 | 0.003566676 | 0.081424301 | -8.75E-05 | 0.001622599 | 1 | 0.525049048 | 1 | 0.515261817 | ENSG0000025 | 1 | AP001877.1 |
| VAT | T2DM | ENSG000001( | 0.005233827 | 0.070468736 | 0.002893392 | 0.081436412 | 0.0051907 | 0.072802761 | 0.002893255 | 0.081424115 | 4.31E-05 | 0.047424913 | 1 | 0.369646805 | 1 | 0.382830909 | ENSG0000025 | 1 | AP001877.1 |
| VAT | T2DM | ENSG0000011 | 0.000542608 | 0.825230627 | 0.00245721 | 0.08143695 | 0.001091826 | 0.657216737 | 0.002460401 | 0.08142462 | -0.00054922 | 2.03E-05 | 1 | 0.94546971 | 1 | 0.876362712 | ENSG0000025 | 1 | AP001877.1 |
| VAT | T2DM | ENSG0000011 | -0.00504284 | 0.08857459 | 0.002961203 | 0.081436474 | -0.00319532 | 0.287941926 | 0.003006939 | 0.081424464 | -0.00184752 | 0.000418956 | 1 | 0.407340768 | 1 | 0.664345628 | ENSG0000025 | 1 | AP001877.1 |
| VAT | T2DM | ENSG0000011 | -0.00211096 | 0.553391492 | 0.00356169 | 0.0814369 | -0.00199954 | 0.574509372 | 0.003561557 | 0.0814246 | -0.00011142 | 0.000334008 | 1 | 0.830087238 | 1 | 0.848256597 | ENSG0000025 | 1 | RP11-476M19.2 |
| VAT | T2DM | ENSG0000011 | -0.00062542 | 0.793917409 | 0.00239417 | 0.081436947 | -0.00065982 | 0.782847404 | 0.002394048 | 0.08142464 | 3.44E-05 | 0.039914423 | 1 | 0.933577731 | 1 | 0.92833376 | ENSG0000025 | 1 | RP11-476M19.2 |
| VAT | T2DM | ENSG0000011 | 0.00287182 | 0.23887767 | 0.002438296 | 0.081436727 | 0.00291784 | 0.231412608 | 0.002438178 | 0.081424413 | -4.60E-05 | 0.010729271 | 1 | 0.626118109 | 1 | 0.618894184 | ENSG0000025 | 2 | KIAA1147 |
| VAT | T2DM | ENSG0000011 | -0.00059633 | 0.835844144 | 0.002877889 | 0.081436951 | -0.00024203 | 0.933101741 | 0.002883257 | 0.081424651 | -0.0003543 | 0.048245684 | 1 | 0.94740445 | 1 | 0.987645456 | ENSG0000025 | 2 | KIAA1147 |
| VAT | T2DM | ENSG0000011 | 0.000604886 | 0.76560202 | 0.002028917 | 0.081436944 | -0.00152465 | 0.462380114 | 0.002074542 | 0.081424562 | 0.002129538 | 8.95E-07 | 1 | 0.922331547 | 1 | 0.791032387 | ENSG0000025 | 1 | GATC |
| VAT | T2DM | ENSG0000011 | 0.003067444 | 0.069230028 | 0.001688266 | 0.081436407 | 0.003044286 | 0.071342519 | 0.00168818 | 0.081424109 | 2.32E-05 | 0.048656225 | 1 | 0.369583901 | 1 | 0.380126161 | ENSG0000025 | 1 | GATC |
| VAT | T2DM | ENSG0000011 | 0.000874335 | 0.781026378 | 0.003145296 | 0.081436946 | 0.001010046 | 0.748108092 | 0.003145237 | 0.081424635 | -0.00013571 | 5.12E-05 | 1 | 0.92915207 | 1 | 0.911563685 | ENSG0000025 | 1 | GATC |
| VAT | T2DM | ENSG0000011 | 0.003410039 | 0.114683569 | 0.002161687 | 0.081436543 | 0.003542146 | 0.101294339 | 0.002161672 | 0.081424204 | -0.00013211 | 1.81E-07 | 1 | 0.457521084 | 1 | 0.440271456 | ENSG0000025 | 1 | GATC |
| VAT | T2DM | ENSG0000011 | -0.00352938 | 0.272851934 | 0.003218711 | 0.081436757 | -0.00345297 | 0.283350041 | 0.003218576 | 0.08142446 | -7.64E-05 | 0.003749378 | 1 | 0.652447775 | 1 | 0.659909226 | ENSG0000025 | 1 | RP11-263K4.3 |
| VAT | T2DM | ENSG0000011 | 0.000153314 | 0.911878683 | 0.001385331 | 0.081436956 | 0.000185183 | 0.89365495 | 0.001385263 | 0.081424649 | -3.19E-05 | 0.001553566 | 1 | 0.978106206 | 1 | 0.973390089 | ENSG0000025 | 1 | RP11-263K4.3 |
| VAT | T2DM | ENSG0000011 | 0.001167274 | 0.629526536 | 0.002419754 | 0.08143692 | 0.001121075 | 0.643133357 | 0.002419643 | 0.081424617 | 4.62E-05 | 0.01371185 | 1 | 0.869869063 | 1 | 0.87456323 | ENSG0000025 | 1 | RP11-486A14.1 |
| VAT | T2DM | ENSG0000011 | -0.00378841 | 0.043374283 | 0.001875366 | 0.081436276 | -0.00436626 | 0.020497725 | 0.001884348 | 0.081423755 | 0.000577842 | 0.001810977 | 1 | 0.290565585 | 1 | 0.205722619 | ENSG0000025 | 1 | RP11-486A14.1 |
| VAT | T2DM | ENSG0000011 | -0.00153049 | 0.666089534 | 0.003546741 | 0.081436927 | -0.00145344 | 0.681943256 | 0.003546593 | 0.081424624 | -7.71E-05 | 0.008227738 | 1 | 0.88342485 | 1 | 0.891626351 | ENSG0000025 | 1 | RP11-834C11.10 |
| VAT | T2DM | ENSG0000011 | -0.00870426 | 0.032297934 | 0.004066062 | 0.081436193 | -0.00861422 | 0.034120326 | 0.004065896 | 0.081423902 | -9.00E-05 | 0.007750246 | 1 | 0.252282367 | 1 | 0.269834096 | ENSG0000025 | 1 | RP11-834C11.10 |
| VAT | T2DM | ENSG0000011 | -0.00320917 | 0.346700741 | 0.003410345 | 0.08143681 | -0.00329968 | 0.333253026 | 0.003410232 | 0.081424496 | 9.05E-05 | 0.003981996 | 1 | 0.706225056 | 1 | 0.702906909 | ENSG0000025 | 2 | RP3-473L9.4 |
| VAT | T2DM | ENSG0000011 | 0.004921712 | 0.03988646 | 0.002395077 | 0.081436253 | 0.004971831 | 0.037899936 | 0.002394977 | 0.081423932 | -5.01E-05 | 0.010927396 | 1 | 0.282273412 | 1 | 0.281394741 | ENSG0000025 | 1 | RP11-350F4.2 |
| VAT | T2DM | ENSG0000011 | 0.000907244 | 0.818193895 | 0.003946793 | 0.08143695 | 0.000810509 | 0.837285729 | 0.003946642 | 0.081424645 | 9.67E-05 | 0.004548599 | 1 | 0.943325602 | 1 | 0.949823878 | ENSG0000025 | 1 | RP11-350F4.2 |
| VAT | T2DM | ENSG0000011 | -0.00214424 | 0.306625422 | 0.002097409 | 0.081436784 | -0.00209307 | 0.318295005 | 0.002097331 | 0.081424486 | -5.12E-05 | 0.005401807 | 1 | 0.677179801 | 1 | 0.688782388 | ENSG0000025 | 1 | RP11-350F4.2 |
| VAT | T2DM | ENSG0000011 | 0.003002241 | 0.298093575 | 0.002885296 | 0.081436777 | 0.003061733 | 0.288595706 | 0.002885138 | 0.081424464 | -5.95E-05 | 0.001486897 | 1 | 0.674852962 | 1 | 0.664345628 | ENSG0000025 | 1 | RP11-350F4.2 |
| VAT | T2DM | ENSG0000011 | -0.00354945 | 0.000763721 | 0.001054613 | 0.081435065 | -0.00352895 | 0.000819006 | 0.00105458 | 0.081422781 | -2.05E-05 | 0.039827099 | 1 | 0.023683941 | 1 | 0.027234428 | ENSG0000025 | 1 | RP11-1136G11.6 |
| VAT | T2DM | ENSG0000011 | 0.009116489 | 0.010177565 | 0.003547621 | 0.081435855 | 0.009052684 | 0.010715003 | 0.003547473 | 0.081423564 | 6.38E-05 | 0.028722932 | 1 | 0.128265203 | 1 | 0.141499556 | ENSG0000025 | 1 | RP3-462E2.3 |
| VAT | T2DM | ENSG0000011 | -0.00031511 | 0.929983791 | 0.003586242 | 0.081436957 | -0.00023512 | 0.947724203 | 0.003586054 | 0.081424652 | -8.00E-05 | 0.001044988 | 1 | 0.983954728 | 1 | 0.988242509 | ENSG0000025 | 1 | RP3-462E2.3 |
| VAT | T2DM | ENSG0000011 | -0.00186149 | 0.600273755 | 0.003552416 | 0.081436913 | -0.00199461 | 0.574456283 | 0.003552285 | 0.0814246 | 0.00013312 | 2.06E-05 | 1 | 0.854879031 | 1 | 0.848256597 | ENSG0000025 | 2 | TEN1 |

| | | | | | | | | | | | | | | | | | | | |
|---|---|---|---|---|---|---|---|---|---|---|---|---|---|---|---|---|---|---|---|
| VAT | T2DM | ENSG0000011 | -0.0031749 | 0.443852029 | 0.004146372 | 0.08143686 | -0.00310773 | 0.453532994 | 0.004146187 | 0.081424559 | -6.72E-05 | 0.038851906 | 1 | 0.779859126 | 1 | 0.789415267 | ENSG0000025 | 2 | TEN1 |
| VAT | T2DM | ENSG0000011 | 0.003478306 | 0.107061602 | 0.002158367 | 0.081436524 | 0.003387609 | 0.116544147 | 0.002158471 | 0.081424241 | 9.07E-05 | 0.007581345 | 1 | 0.442145471 | 1 | 0.466853188 | ENSG0000025 | 2 | TEN1 |
| VAT | T2DM | ENSG0000011 | -0.00124806 | 0.30956467 | 0.001228244 | 0.081436786 | -0.0012153 | 0.32241845 | 0.001228194 | 0.081424489 | -3.28E-05 | 0.00139309 | 1 | 0.680925546 | 1 | 0.693593042 | ENSG0000025 | 1 | RP11-121E16.1 |
| VAT | T2DM | ENSG0000011 | -0.0036859 | 0.12728446 | 0.002417143 | 0.08143657 | -0.00375191 | 0.120598463 | 0.002417047 | 0.08142425 | 6.60E-05 | 0.001314252 | 1 | 0.482731692 | 1 | 0.473820696 | ENSG0000025 | 1 | RP11-121E16.1 |
| VAT | T2DM | ENSG0000011 | 0.000547389 | 0.862744696 | 0.003166268 | 0.081436953 | 0.000600851 | 0.849484194 | 0.003166096 | 0.081424646 | -5.35E-05 | 0.009426539 | 1 | 0.959377663 | 1 | 0.95585823 | ENSG0000025 | 1 | RP11-121E16.1 |
| VAT | T2DM | ENSG0000011 | -0.00284519 | 0.052115227 | 0.001464944 | 0.081436328 | -0.00280877 | 0.055187639 | 0.001464883 | 0.081424038 | -3.64E-05 | 0.00252178 | 1 | 0.323231522 | 1 | 0.342287379 | ENSG0000025 | 1 | RP11-493L12.4 |
| VAT | T2DM | ENSG0000011 | 0.002116288 | 0.163170338 | 0.001517607 | 0.081436633 | 0.002164564 | 0.153761802 | 0.001517532 | 0.081424313 | -4.83E-05 | 1.18E-05 | 1 | 0.541286218 | 1 | 0.52392045 | ENSG0000025 | 2 | RP11-511H9.3 |
| VAT | T2DM | ENSG0000011 | 0.000297538 | 0.816640744 | 0.001283221 | 0.081436949 | 0.000322769 | 0.801395759 | 0.001283166 | 0.081424642 | -2.52E-05 | 0.014966219 | 1 | 0.943325602 | 1 | 0.935824336 | ENSG0000025 | 2 | RP11-511H9.3 |
| VAT | T2DM | ENSG0000011 | 0.00072507 | 0.830280441 | 0.003382773 | 0.081436951 | 0.000783994 | 0.816714927 | 0.003382605 | 0.081424644 | -5.89E-05 | 0.015517103 | 1 | 0.946239005 | 1 | 0.941969577 | ENSG0000025 | 3 | CHURC1 |
| VAT | T2DM | ENSG0000011 | -0.00200144 | 0.431588383 | 0.002544813 | 0.081436855 | -0.00194338 | 0.445047268 | 0.002544695 | 0.081424555 | -5.81E-05 | 0.002743166 | 1 | 0.771992181 | 1 | 0.783873938 | ENSG0000025 | 3 | CHURC1 |
| VAT | T2DM | ENSG0000012 | 0.000502482 | 0.874916828 | 0.00319206 | 0.081436954 | 0.000451634 | 0.887479861 | 0.003191899 | 0.081424649 | 5.08E-05 | 0.024972447 | 1 | 0.966352796 | 1 | 0.971063504 | ENSG0000025 | 3 | CHURC1 |
| VAT | T2DM | ENSG0000012 | -0.00641921 | 0.077388727 | 0.003634791 | 0.081436437 | -0.00631053 | 0.082528308 | 0.00363467 | 0.081424149 | -0.00010868 | 0.001132571 | 1 | 0.386243918 | 1 | 0.407209212 | ENSG0000025 | 2 | RP11-488C13.5 |
| VAT | T2DM | ENSG0000012 | -0.00676306 | 0.080406651 | 0.003868287 | 0.081436448 | -0.00683946 | 0.077039828 | 0.003868187 | 0.08142413 | 7.64E-05 | 0.048312528 | 1 | 0.393299377 | 1 | 0.39522291 | ENSG0000025 | 5 | ZNF578 |
| VAT | T2DM | ENSG0000012 | 0.003114038 | 0.321930847 | 0.003143909 | 0.081436794 | 0.00025265 | 0.941290973 | 0.003430531 | 0.081424652 | 0.002861388 | 0.037192836 | 1 | 0.69146112 | 1 | 0.987645456 | ENSG0000025 | 1 | RP11-796G6.2 |
| VAT | T2DM | ENSG0000012 | 0.000411779 | 0.742245468 | 0.001252059 | 0.08143694 | 0.000438413 | 0.726215937 | 0.001252022 | 0.081424632 | -2.66E-05 | 0.026181673 | 1 | 0.916157236 | 1 | 0.908355795 | ENSG0000025 | 1 | RP11-72M17.1 |
| VAT | T2DM | ENSG0000012 | 0.003277193 | 0.052020246 | 0.001686692 | 0.081436328 | 0.003246296 | 0.054262085 | 0.001686613 | 0.081424033 | 3.09E-05 | 0.015694153 | 1 | 0.323231522 | 1 | 0.33883112 | ENSG0000025 | 1 | RP11-1070N10.3 |
| VAT | T2DM | ENSG0000012 | -0.00542078 | 0.156119475 | 0.003822169 | 0.081436622 | -0.00535456 | 0.161222292 | 0.003822027 | 0.081424325 | -6.62E-05 | 0.047351018 | 1 | 0.527404835 | 1 | 0.537053745 | ENSG0000025 | 1 | RP5-1021I20.2 |
| VAT | T2DM | ENSG0000012 | 0.000532894 | 0.707907435 | 0.001422308 | 0.081436935 | 0.000501158 | 0.724562385 | 0.001422258 | 0.081424632 | 3.17E-05 | 0.01273773 | 1 | 0.902964787 | 1 | 0.907346725 | ENSG0000025 | 1 | RP5-1021I20.2 |
| VAT | T2DM | ENSG0000012 | 0.001173489 | 0.606178071 | 0.002276246 | 0.081436914 | 0.00111504 | 0.624219963 | 0.002276156 | 0.081424612 | 5.84E-05 | 0.002466533 | 1 | 0.857656828 | 1 | 0.86880842 | ENSG0000025 | 1 | LINC00930 |
| VAT | T2DM | ENSG0000012 | 0.003312824 | 0.109690787 | 0.002071049 | 0.081436531 | 0.003253136 | 0.116223249 | 0.002070969 | 0.08142424 | 5.97E-05 | 0.000772136 | 1 | 0.448335641 | 1 | 0.466853188 | ENSG0000025 | 2 | C20orf141 |
| VAT | T2DM | ENSG0000012 | 0.002217068 | 0.348140098 | 0.002363098 | 0.081436811 | 0.002420094 | 0.305807035 | 0.002363233 | 0.081424477 | -0.00020303 | 1.37E-07 | 1 | 0.708603739 | 1 | 0.676208846 | ENSG0000025 | 2 | C20orf141 |
| VAT | T2DM | ENSG0000012 | 0.000893699 | 0.354110315 | 0.000964444 | 0.081436815 | 0.000277318 | 0.775426427 | 0.000972077 | 0.081424639 | 0.000616381 | 4.50E-07 | 1 | 0.714271445 | 1 | 0.925281859 | ENSG0000025 | 1 | RP11-7F17.3 |
| VAT | T2DM | ENSG0000012 | -0.00069777 | 0.728170578 | 0.00200762 | 0.081436938 | -0.0007739 | 0.69988229 | 0.002007616 | 0.081424628 | 7.61E-05 | 0.001760162 | 1 | 0.910304385 | 1 | 0.897664737 | ENSG0000025 | 1 | RP11-7F17.3 |
| VAT | T2DM | ENSG0000012 | 0.013517766 | 0.000882614 | 0.004064862 | 0.08143511 | 0.013392092 | 0.000985229 | 0.004064699 | 0.081422839 | 0.000125674 | 0.000233202 | 1 | 0.026057806 | 1 | 0.030900358 | ENSG0000025 | 1 | RP11-7F17.3 |
| VAT | T2DM | ENSG0000012 | 0.003819584 | 0.176685583 | 0.002827158 | 0.081436653 | 0.003872348 | 0.17076146 | 0.00282702 | 0.081424339 | -5.28E-05 | 0.010732118 | 1 | 0.556533092 | 1 | 0.54753664 | ENSG0000025 | 1 | RP11-7F17.3 |
| VAT | T2DM | ENSG0000012 | 0.000446274 | 0.880582029 | 0.002970577 | 0.081436955 | 0.000528798 | 0.858708455 | 0.002970464 | 0.081424647 | -8.25E-05 | 0.001337185 | 1 | 0.968646917 | 1 | 0.959900123 | ENSG0000025 | 1 | RP11-7F17.3 |
| VAT | T2DM | ENSG0000012 | 0.002845046 | 0.476843721 | 0.003999285 | 0.081436874 | 0.002777889 | 0.4872893 | 0.003999107 | 0.081424572 | 6.72E-05 | 0.033263901 | 1 | 0.791889203 | 1 | 0.799594809 | ENSG0000025 | 1 | RP11-7F17.3 |
| VAT | T2DM | ENSG0000012 | 0.00071577 | 0.852354237 | 0.003845838 | 0.081436953 | 0.000772309 | 0.840833656 | 0.003845634 | 0.081424646 | -5.65E-05 | 0.028831699 | 1 | 0.95477787 | 1 | 0.95149688 | ENSG0000025 | 2 | DUXA |
| VAT | T2DM | ENSG0000012 | 0.000479873 | 0.882231465 | 0.003239299 | 0.081436955 | 0.000409385 | 0.899426435 | 0.003239168 | 0.08142465 | 7.05E-05 | 0.009358827 | 1 | 0.968732296 | 1 | 0.975350965 | ENSG0000025 | 2 | DUXA |
| VAT | T2DM | ENSG0000012 | 0.001309012 | 0.594321243 | 0.002457847 | 0.081436911 | 0.00127057 | 0.605177671 | 0.002457726 | 0.081424608 | 3.84E-05 | 0.031171013 | 1 | 0.851009022 | 1 | 0.858231221 | ENSG0000025 | 1 | RP3-449M8.6 |
| VAT | T2DM | ENSG0000012 | -0.0001654 | 0.939914321 | 0.00219426 | 0.081436957 | -0.00013502 | 0.950933709 | 0.002194143 | 0.081424652 | -3.04E-05 | 0.039346969 | 1 | 0.986373964 | 1 | 0.988354266 | ENSG0000025 | 1 | RP3-449M8.6 |
| VAT | T2DM | ENSG0000012 | 0.002786993 | 0.276248901 | 0.002559726 | 0.08143676 | 0.002849413 | 0.265621514 | 0.002559652 | 0.081424445 | -6.24E-05 | 0.011516184 | 1 | 0.653996814 | 1 | 0.646388519 | ENSG0000025 | 1 | DUXAP1 |
| VAT | T2DM | ENSG0000012 | 0.001215507 | 0.675927688 | 0.002907725 | 0.081436929 | 0.004535851 | 0.144283668 | 0.003106693 | 0.081424296 | -0.00332034 | 0.002416609 | 1 | 0.888785335 | 1 | 0.514898084 | ENSG0000025 | 1 | DUXAP1 |
| VAT | T2DM | ENSG0000012 | 0.001919957 | 0.635935863 | 0.004055771 | 0.081436921 | 0.001994111 | 0.622937843 | 0.004055616 | 0.081424612 | -7.42E-05 | 0.034511727 | 1 | 0.871491053 | 1 | 0.868736298 | ENSG0000025 | 1 | DUXAP1 |
| VAT | T2DM | ENSG0000012 | -0.00226838 | 0.530837178 | 0.003619396 | 0.081436893 | -0.00234369 | 0.51726994 | 0.003619262 | 0.081424582 | 7.53E-05 | 0.017519425 | 1 | 0.819413094 | 1 | 0.817922279 | ENSG0000025 | 1 | LINC00924 |
| VAT | T2DM | ENSG0000012 | 0.00398808 | 0.175421958 | 0.002943258 | 0.081436652 | 0.003943453 | 0.180282758 | 0.002943124 | 0.081424352 | 4.46E-05 | 0.049595361 | 1 | 0.553330975 | 1 | 0.556577643 | ENSG0000025 | 2 | PRKXP1 |
| VAT | T2DM | ENSG0000012 | 0.002062784 | 0.479880626 | 0.002919743 | 0.081436875 | 0.002007016 | 0.491814301 | 0.002919608 | 0.081424574 | 5.58E-05 | 0.012549749 | 1 | 0.791889203 | 1 | 0.800277918 | ENSG0000025 | 2 | PRKXP1 |
| VAT | T2DM | ENSG0000013 | -0.00194594 | 0.490943871 | 0.002825073 | 0.081436879 | -0.00189655 | 0.501992498 | 0.002824943 | 0.081424577 | -4.94E-05 | 0.022545803 | 1 | 0.793793254 | 1 | 0.809302075 | ENSG0000025 | 2 | PRKXP1 |
| VAT | T2DM | ENSG0000013 | 4.65E-06 | 0.997080154 | 0.001269548 | 0.081436958 | 3.82E-05 | 0.976003753 | 0.001269501 | 0.081424652 | -3.35E-05 | 0.002484159 | 1 | 0.998501236 | 1 | 0.991815301 | ENSG0000025 | 2 | PRKXP1 |
| VAT | T2DM | ENSG0000013 | 0.003878961 | 0.334147315 | 0.004016341 | 0.081436803 | 0.003943327 | 0.32616515 | 0.004016146 | 0.081424491 | -6.44E-05 | 0.029139667 | 1 | 0.701928909 | 1 | 0.697301172 | ENSG0000025 | 2 | PRKXP1 |
| VAT | T2DM | ENSG0000013 | -0.00012309 | 0.936453322 | 0.001543821 | 0.081436957 | -9.87E-05 | 0.949047028 | 0.001543746 | 0.081424652 | -2.44E-05 | 0.031591138 | 1 | 0.986373964 | 1 | 0.988242509 | ENSG0000025 | 2 | PRKXP1 |
| VAT | T2DM | ENSG0000013 | -0.00121593 | 0.756695444 | 0.003924577 | 0.081436942 | -0.0012863 | 0.743088158 | 0.003924441 | 0.081424635 | 7.04E-05 | 0.047355935 | 1 | 0.919012884 | 1 | 0.911123584 | ENSG0000025 | 2 | PRKXP1 |
| VAT | T2DM | ENSG0000013 | 0.00984628 | 0.000288913 | 0.002716163 | 0.081434762 | 0.009891253 | 0.000270764 | 0.002716033 | 0.081422436 | -4.50E-05 | 0.025973238 | 0.797400319 | 0.010923292 | 0.747309516 | 0.012455159 | ENSG0000025 | 2 | PRKXP1 |
| VAT | T2DM | ENSG0000013 | -0.0060412 | 0.078431317 | 0.003432764 | 0.081436441 | -0.00610056 | 0.07553044 | 0.003432614 | 0.081424125 | 5.94E-05 | 0.030304962 | 1 | 0.390036819 | 1 | 0.389652366 | ENSG0000025 | 2 | PRKXP1 |
| VAT | T2DM | ENSG0000013 | 0.000214818 | 0.924957732 | 0.002280667 | 0.081436957 | 0.000171896 | 0.939916947 | 0.002280564 | 0.081424652 | 4.29E-05 | 0.015528943 | 1 | 0.982255998 | 1 | 0.987645456 | ENSG0000025 | 2 | PRKXP1 |
| VAT | T2DM | ENSG0000013 | 0.005143768 | 0.075413323 | 0.002893096 | 0.08143643 | 0.005037096 | 0.081676122 | 0.002893168 | 0.081424146 | 0.000106672 | 0.006564032 | 1 | 0.380513292 | 1 | 0.407209212 | ENSG0000025 | 3 | ITGB3 |
| VAT | T2DM | ENSG0000013 | -0.0007506 | 0.804845598 | 0.003037867 | 0.081436948 | -0.00084659 | 0.780484164 | 0.003037779 | 0.08142464 | 9.60E-05 | 0.001049554 | 1 | 0.940065109 | 1 | 0.927707275 | ENSG0000025 | 3 | ITGB3 |
| VAT | T2DM | ENSG0000013 | -0.00150258 | 0.428307162 | 0.001896972 | 0.081436854 | -0.00153686 | 0.417824018 | 0.001896878 | 0.081424543 | 3.43E-05 | 0.012828389 | 1 | 0.771120526 | 1 | 0.764717698 | ENSG0000025 | 2 | CSPG4P12 |
| VAT | T2DM | ENSG0000013 | -0.00664441 | 0.103624196 | 0.004082499 | 0.081436516 | -0.00654324 | 0.108972068 | 0.004082297 | 0.081424223 | -0.00010117 | 0.000607177 | 1 | 0.435867474 | 1 | 0.450918902 | ENSG0000025 | 2 | RP11-307C19.1 |
| VAT | T2DM | ENSG0000013 | -0.00027768 | 0.789462796 | 0.00103998 | 0.081436947 | -0.00025347 | 0.807433088 | 0.001039936 | 0.081424643 | -2.42E-05 | 0.004846335 | 1 | 0.932507176 | 1 | 0.938342488 | ENSG0000025 | 1 | CTD-2054N24.2 |
| VAT | T2DM | ENSG0000013 | 0.004430485 | 0.180792699 | 0.003310482 | 0.081436659 | 0.007476905 | 0.031494864 | 0.003476385 | 0.08142388 | -0.00304642 | 0.004120717 | 1 | 0.559898669 | 1 | 0.259967574 | ENSG0000025 | 1 | CTD-2054N24.2 |
| VAT | T2DM | ENSG0000013 | -0.00702294 | 0.013516461 | 0.002843444 | 0.081435939 | -0.00707599 | 0.012824275 | 0.002843338 | 0.081423618 | 5.30E-05 | 0.033239142 | 1 | 0.149821016 | 1 | 0.156615037 | ENSG0000025 | 1 | CTD-2054N24.2 |
| VAT | T2DM | ENSG0000013 | 0.00145101 | 0.526997583 | 0.002293738 | 0.081436892 | 0.001715199 | 0.455040428 | 0.002295997 | 0.081424559 | -0.00026419 | 0.012417404 | 1 | 0.81918483 | 1 | 0.789881498 | ENSG0000025 | 1 | CTD-2054N24.2 |
| VAT | T2DM | ENSG0000013 | -0.0003122 | 0.903937073 | 0.002586823 | 0.081436956 | -0.0003871 | 0.881041887 | 0.002586721 | 0.081424649 | 7.49E-05 | 0.000654558 | 1 | 0.974557156 | 1 | 0.96786083 | ENSG0000025 | 2 | CTD-2651B20.4 |
| VAT | T2DM | ENSG0000013 | 0.000395891 | 0.777187951 | 0.00139898 | 0.081436945 | 0.000425427 | 0.761041588 | 0.00139891 | 0.081424637 | -2.95E-05 | 0.003186137 | 1 | 0.928588201 | 1 | 0.915917135 | ENSG0000025 | 2 | CTD-2651B20.4 |
| VAT | T2DM | ENSG0000013 | 0.002475384 | 0.020666583 | 0.001069729 | 0.081436064 | 0.002504671 | 0.019206196 | 0.001069681 | 0.081423736 | -2.93E-05 | 0.000423818 | 1 | 0.19668886 | 1 | 0.196330006 | ENSG0000025 | 2 | CTD-2651B20.4 |
| VAT | T2DM | ENSG0000013 | 0.000720347 | 0.7828412 | 0.002613565 | 0.081436946 | 0.000670186 | 0.79761487 | 0.00261347 | 0.081424641 | 5.02E-05 | 0.030688398 | 1 | 0.929809975 | 1 | 0.935209552 | ENSG0000025 | 1 | CTD-2651B20.1 |
| VAT | T2DM | ENSG0000013 | 0.006919188 | 0.000887476 | 0.002081594 | 0.081435112 | 0.006881096 | 0.000947059 | 0.00208151 | 0.081422826 | 3.81E-05 | 0.028423028 | 1 | 0.026057806 | 1 | 0.030693597 | ENSG0000025 | 1 | CTD-2651B20.1 |
| VAT | T2DM | ENSG0000013 | 0.021960061 | 1.43E-09 | 0.003628296 | 0.081430837 | 0.015733453 | 4.31E-05 | 0.003846608 | 0.081421857 | 0.006226607 | 1.10E-06 | 3.94E-06 | 3.58E-07 | 0.118967945 | 0.003109431 | ENSG0000025 | 1 | RP11-643G16.4 |
| VAT | T2DM | ENSG0000013 | 0.005699843 | 0.00214449 | 0.001856946 | 0.081435384 | 0.003508408 | 0.069019937 | 0.001929506 | 0.0814241 | 0.002191435 | 2.95E-05 | 1 | 0.048514701 | 1 | 0.375985973 | ENSG0000025 | 3 | RP11-89K11.1 |
| VAT | T2DM | ENSG0000013 | 0.00096635 | 0.314326552 | 0.000960408 | 0.081436789 | 0.002050532 | 0.037768626 | 0.000987084 | 0.081423931 | -0.00108418 | 2.03E-06 | 1 | 0.685828291 | 1 | 0.281394741 | ENSG0000025 | 1 | IQCH-AS1 |
| VAT | T2DM | ENSG0000013 | -0.01165115 | 0.001734564 | 0.003719685 | 0.081435319 | -0.01298886 | 0.000541869 | 0.003754897 | 0.081422653 | 0.001337719 | 0.009406965 | 1 | 0.042758635 | 1 | 0.020487111 | ENSG0000025 | 1 | CTD-3110H11.1 |
| VAT | T2DM | ENSG0000013 | 0.002970284 | 0.306730574 | 0.002906039 | 0.081436784 | 0.002916294 | 0.315584784 | 0.00290591 | 0.081424484 | 5.40E-05 | 0.01909354 | 1 | 0.677179801 | 1 | 0.685837798 | ENSG0000025 | 1 | CTD-3110H11.1 |
| VAT | T2DM | ENSG0000013 | 0.003678751 | 0.304840951 | 0.003585148 | 0.081436782 | 0.00386801 | 0.28066334 | 0.003585369 | 0.081424458 | -0.00018926 | 0.001447271 | 1 | 0.675668349 | 1 | 0.657179108 | ENSG0000025 | 1 | CSPG4P11 |
| VAT | T2DM | ENSG0000013 | -0.00105591 | 0.753164886 | 0.003357768 | 0.081436942 | -0.00112094 | 0.738491097 | 0.0033576 | 0.081424634 | 6.50E-05 | 0.006824431 | 1 | 0.918605915 | 1 | 0.910293329 | ENSG0000025 | 1 | CSPG4P11 |
| VAT | T2DM | ENSG0000013 | 0.001529028 | 0.65953147 | 0.003470643 | 0.081436926 | 0.001600576 | 0.64465783 | 0.003470486 | 0.081424617 | -7.15E-05 | 0.008139614 | 1 | 0.879375293 | 1 | 0.87456323 | ENSG0000025 | 1 | CSPG4P11 |
| VAT | T2DM | ENSG0000013 | -0.00437795 | 0.137723431 | 0.002949458 | 0.08143659 | -0.00401023 | 0.174379328 | 0.002952462 | 0.081424344 | -0.00036772 | 0.007696744 | 1 | 0.500466051 | 1 | 0.548422551 | ENSG0000025 | 1 | CASC7 |
| VAT | T2DM | ENSG0000013 | 0.005234071 | 0.004266936 | 0.001831545 | 0.081435594 | 0.004989943 | 0.006554584 | 0.001835431 | 0.081423417 | 0.000244128 | 0.044420246 | 1 | 0.074865219 | 1 | 0.103375158 | ENSG0000025 | 1 | CASC7 |
| VAT | T2DM | ENSG0000013 | -0.00287314 | 0.266584226 | 0.002586171 | 0.081436752 | -0.00449829 | 0.090565491 | 0.002657898 | 0.081424174 | 0.00162515 | 0.00814005 | 1 | 0.646548738 | 1 | 0.422945439 | ENSG0000025 | 2 | SLC22A31 |
| VAT | T2DM | ENSG0000013 | 2.75E-05 | 0.991834142 | 0.002688826 | 0.081436958 | -4.93E-05 | 0.985381925 | 0.002688872 | 0.081424652 | 7.68E-05 | 0.035780118 | 1 | 0.998465252 | 1 | 0.995445662 | ENSG0000025 | 1 | AL022344.7 |
| VAT | T2DM | ENSG0000013 | 0.002196934 | 0.524444416 | 0.003451531 | 0.081436891 | 0.002297009 | 0.505711371 | 0.003451395 | 0.081424578 | -0.00010008 | 0.000650875 | 1 | 0.81721896 | 1 | 0.810353632 | ENSG0000025 | 1 | RP11-410E4.1 |
| VAT | T2DM | ENSG0000013 | 0.00320914 | 0.322431327 | 0.003243276 | 0.081436795 | 0.00313609 | 0.333544339 | 0.003243112 | 0.081424496 | 7.30E-05 | 0.001421575 | 1 | 0.69146112 | 1 | 0.702906909 | ENSG0000025 | 1 | CDC37P1 |
| VAT | T2DM | ENSG0000013 | 0.00141086 | 0.626423214 | 0.002898426 | 0.081436919 | 0.001483716 | 0.608704998 | 0.002898318 | 0.081424609 | -7.29E-05 | 0.003994958 | 1 | 0.869127163 | 1 | 0.859788021 | ENSG0000025 | 1 | CDC37P1 |
| VAT | T2DM | ENSG0000013 | 0.000952254 | 0.240639335 | 0.00081154 | 0.081436728 | 0.000967791 | 0.233031813 | 0.000811505 | 0.081424415 | -1.55E-05 | 0.018675514 | 1 | 0.628348691 | 1 | 0.619918077 | ENSG0000025 | 1 | CDC37P1 |
| VAT | T2DM | ENSG0000013 | 0.004494922 | 0.21561317 | 0.003629975 | 0.081436702 | 0.0051359 | 0.157991458 | 0.003637681 | 0.081424319 | -0.00064098 | 0.007782155 | 1 | 0.597482277 | 1 | 0.531380313 | ENSG0000025 | 1 | CDC37P1 |
| VAT | T2DM | ENSG0000014 | 0.007898043 | 0.033868149 | 0.003722605 | 0.081436206 | 0.007797217 | 0.036206984 | 0.003722531 | 0.081423919 | 0.000100825 | 0.010157306 | 1 | 0.256802447 | 1 | 0.276817942 | ENSG0000025 | 1 | CDC37P1 |
| VAT | T2DM | ENSG0000014 | -0.00010629 | 0.922433611 | 0.001091624 | 0.081436957 | -8.54E-05 | 0.937661328 | 0.001091579 | 0.081424651 | -2.09E-05 | 0.020659319 | 1 | 0.981085459 | 1 | 0.987645456 | ENSG0000026 | 1 | CTA-445C9.15 |
| VAT | T2DM | ENSG0000014 | 0.001510485 | 0.54264113 | 0.002480987 | 0.081436896 | 0.001581636 | 0.523783593 | 0.002480902 | 0.081424585 | -7.12E-05 | 0.001627726 | 1 | 0.823132089 | 1 | 0.820071559 | ENSG0000026 | 1 | CTA-445C9.15 |
| VAT | T2DM | ENSG0000014 | -3.02E-05 | 0.992173465 | 0.003079621 | 0.081436958 | -0.0001413 | 0.963402196 | 0.003079507 | 0.081424652 | 0.000111093 | 3.90E-05 | 1 | 0.998465252 | 1 | 0.989894944 | ENSG0000026 | 1 | CTA-445C9.15 |
| VAT | T2DM | ENSG0000014 | -0.00144565 | 0.667192466 | 0.003361944 | 0.081436928 | -0.00134208 | 0.689734975 | 0.003361786 | 0.081424626 | -0.00010358 | 4.61E-05 | 1 | 0.883734796 | 1 | 0.894473859 | ENSG0000026 | 1 | CTA-445C9.15 |
| VAT | T2DM | ENSG0000014 | 0.016580856 | 1.25E-06 | 0.003420891 | 0.081433033 | 0.016661083 | 1.11E-06 | 0.003420748 | 0.081420689 | -8.02E-05 | 0.004451763 | 0.003461556 | 0.000138462 | 0.003071798 | 0.00015359 | ENSG0000026 | 1 | NSFP1 |
| VAT | T2DM | ENSG0000014 | -0.00134869 | 0.407275907 | 0.001627485 | 0.081436844 | -0.00137627 | 0.397735418 | 0.001627423 | 0.081424533 | 2.76E-05 | 0.049996109 | 1 | 0.753911136 | 1 | 0.752171241 | ENSG0000026 | 1 | NSFP1 |
| VAT | T2DM | ENSG0000014 | -0.00249993 | 0.00213136 | 0.000813963 | 0.081435382 | -0.00251443 | 0.002006842 | 0.000813934 | 0.081423058 | 1.45E-05 | 0.045519437 | 1 | 0.048514701 | 1 | 0.05225363 | ENSG0000026 | 1 | RP11-254F7.2 |
| VAT | T2DM | ENSG0000014 | 0.002560597 | 0.504448944 | 0.003836062 | 0.081436884 | 0.003315993 | 0.38878533 | 0.00384765 | 0.081424528 | -0.0007554 | 0.012404482 | 1 | 0.802929115 | 1 | 0.74719619 | ENSG0000026 | 1 | RP11-254F7.2 |
| VAT | T2DM | ENSG0000014 | 0.000689195 | 0.349444235 | 0.00073658 | 0.081436812 | 0.000701636 | 0.34078975 | 0.000736543 | 0.081424501 | -1.24E-05 | 0.016933911 | 1 | 0.710210669 | 1 | 0.704514894 | ENSG0000026 | 1 | RP11-2C24.5 |
| VAT | T2DM | ENSG0000014 | 0.004939559 | 0.001051545 | 0.001507627 | 0.081435165 | 0.004915812 | 0.001111163 | 0.00150756 | 0.081422876 | 2.37E-05 | 0.044191146 | 1 | 0.02961495 | 1 | 0.034458528 | ENSG0000026 | 1 | RP11-2C24.5 |
| VAT | T2DM | ENSG0000014 | -0.00343159 | 0.043301895 | 0.001698139 | 0.081436276 | -0.00385264 | 0.024210494 | 0.001709412 | 0.081423804 | 0.000421052 | 0.032674076 | 1 | 0.290565585 | 1 | 0.221996552 | ENSG0000026 | 1 | RP11-92G12.3 |
| VAT | T2DM | ENSG0000014 | 0.004153843 | 0.292678123 | 0.003947523 | 0.081436773 | 0.004092772 | 0.299807163 | 0.00394732 | 0.081424473 | 6.11E-05 | 0.026344079 | 1 | 0.668507183 | 1 | 0.67328541 | ENSG0000026 | 1 | RP11-33B1.4 |
| VAT | T2DM | ENSG0000014 | 0.000984297 | 0.674688836 | 0.002345118 | 0.081436929 | 0.001032212 | 0.65981099 | 0.002345006 | 0.08142462 | -4.79E-05 | 0.006206335 | 1 | 0.888785335 | 1 | 0.877628112 | ENSG0000026 | 1 | CTD-3126B10.4 |
| VAT | T2DM | ENSG0000014 | -0.00181512 | 0.629499168 | 0.003762441 | 0.08143692 | -0.00175399 | 0.641069252 | 0.003762284 | 0.081424616 | -6.11E-05 | 0.048018448 | 1 | 0.869869063 | 1 | 0.87456323 | ENSG0000026 | 1 | CTD-3126B10.4 |
| VAT | T2DM | ENSG0000014 | 0.003955328 | 0.100053009 | 0.002405039 | 0.081436506 | 0.003895544 | 0.105273811 | 0.002404939 | 0.081424214 | 5.98E-05 | 0.002504899 | 1 | 0.430095137 | 1 | 0.445637604 | ENSG0000026 | 1 | RP5-1085F17.3 |
| VAT | T2DM | ENSG0000014 | 0.000173274 | 0.961681827 | 0.003606628 | 0.081436958 | 0.000271004 | 0.940100252 | 0.003606467 | 0.081424652 | -9.77E-05 | 0.000565826 | 1 | 0.991412214 | 1 | 0.987645456 | ENSG0000026 | 1 | RP5-1085F17.3 |
| VAT | T2DM | ENSG0000014 | 0.002744364 | 0.38698116 | 0.003172287 | 0.081436833 | 0.002868018 | 0.365995337 | 0.003172583 | 0.081424516 | -0.00012365 | 0.033937114 | 1 | 0.741456409 | 1 | 0.7256222 | ENSG0000026 | 1 | RP11-44F21.5 |
| VAT | T2DM | ENSG0000014 | 0.001090525 | 0.725427068 | 0.003105005 | 0.081436938 | 0.001902347 | 0.542328023 | 0.003122201 | 0.08142459 | -0.00081182 | 0.013734411 | 1 | 0.908074099 | 1 | 0.82887997 | ENSG0000026 | 1 | RP11-44F21.5 |
| VAT | T2DM | ENSG0000014 | 0.000812458 | 0.589690176 | 0.001506545 | 0.08143691 | 0.000785174 | 0.602229245 | 0.00150648 | 0.081424607 | 2.73E-05 | 0.024974992 | 1 | 0.849919041 | 1 | 0.856779751 | ENSG0000026 | 1 | RP11-44F21.5 |
| VAT | T2DM | ENSG0000014 | -0.00117451 | 0.611623103 | 0.002313132 | 0.081436915 | -0.00120787 | 0.601528015 | 0.002313012 | 0.081424607 | 3.34E-05 | 0.036364968 | 1 | 0.860549903 | 1 | 0.856779751 | ENSG0000026 | 1 | RP11-817O13.8 |
| VAT | T2DM | ENSG0000014 | 0.000950371 | 0.496621757 | 0.001397984 | 0.081436881 | 0.000910326 | 0.51491938 | 0.001397927 | 0.081424582 | 4.00E-05 | 0.000531353 | 1 | 0.79758205 | 1 | 0.817290281 | ENSG0000026 | 1 | RP11-973H7.1 |
| VAT | T2DM | ENSG0000014 | 0.002180107 | 0.365116637 | 0.002407204 | 0.081436821 | 0.002232547 | 0.353678869 | 0.002407115 | 0.081424509 | -5.24E-05 | 0.013440789 | 1 | 0.722899512 | 1 | 0.714084622 | ENSG0000026 | 1 | RP11-973H7.1 |
| VAT | T2DM | ENSG0000014 | 0.003365455 | 0.355916751 | 0.003645524 | 0.081436816 | 0.003421032 | 0.348007177 | 0.003645358 | 0.081424505 | -5.56E-05 | 0.049305073 | 1 | 0.714526689 | 1 | 0.709379474 | ENSG0000026 | 1 | RP11-973H7.1 |
| VAT | T2DM | ENSG0000014 | -0.00084169 | 0.658742008 | 0.001905788 | 0.081436926 | -0.00088728 | 0.641506767 | 0.001905703 | 0.081424616 | 4.56E-05 | 0.002389325 | 1 | 0.878747192 | 1 | 0.87456323 | ENSG0000026 | 1 | RP11-973H7.1 |
| VAT | T2DM | ENSG0000014 | 0.002058839 | 0.530078679 | 0.003279 | 0.081436893 | 0.002106962 | 0.520488049 | 0.00327884 | 0.081424583 | -4.81E-05 | 0.044700966 | 1 | 0.819413094 | 1 | 0.817953302 | ENSG0000026 | 1 | RP11-973H7.1 |
| VAT | T2DM | ENSG0000014 | 0.005003039 | 0.015132383 | 0.002059539 | 0.081435972 | 0.00504713 | 0.014256967 | 0.002059438 | 0.081423649 | -4.41E-05 | 0.00346196 | 1 | 0.163254046 | 1 | 0.168816097 | ENSG0000026 | 1 | RP11-973H7.1 |
| VAT | T2DM | ENSG0000014 | 0.002451144 | 0.202263435 | 0.001922268 | 0.081436687 | 0.003232918 | 0.097350654 | 0.00195008 | 0.081424193 | -0.00078177 | 0.017502405 | 1 | 0.582793504 | 1 | 0.435146086 | ENSG0000026 | 1 | RP11-973H7.1 |
| VAT | T2DM | ENSG0000014 | 0.012388368 | 0.001869701 | 0.003983139 | 0.081435342 | 0.01232627 | 0.001969823 | 0.003982947 | 0.081423052 | 6.21E-05 | 0.034389995 | 1 | 0.045266446 | 1 | 0.05224565 | ENSG0000026 | 1 | RP11-973H7.1 |
| VAT | T2DM | ENSG0000014 | -0.00046743 | 0.848984209 | 0.002454818 | 0.081436952 | -0.00041079 | 0.86709734 | 0.002454743 | 0.081424648 | -5.66E-05 | 0.015125004 | 1 | 0.954545261 | 1 | 0.962279315 | ENSG0000026 | 1 | RP11-973H7.1 |
| VAT | T2DM | ENSG0000014 | -0.00139315 | 0.561095187 | 0.002396964 | 0.081436902 | -0.00143761 | 0.548650271 | 0.002396883 | 0.081424592 | 4.45E-05 | 0.042755251 | 1 | 0.835470159 | 1 | 0.830883001 | ENSG0000026 | 1 | RP11-973H7.1 |
| VAT | T2DM | ENSG0000014 | -0.00541128 | 0.013359331 | 0.002187211 | 0.081435936 | -0.00472719 | 0.03105628 | 0.002192212 | 0.081423876 | -0.00068408 | 5.38E-06 | 1 | 0.149821016 | 1 | 0.258654894 | ENSG0000026 | 1 | RP11-645C24.5 |
| VAT | T2DM | ENSG0000014 | -0.00458994 | 0.042150814 | 0.002258789 | 0.081436268 | -0.00462965 | 0.040394009 | 0.002258692 | 0.08142395 | 3.97E-05 | 0.029155814 | 1 | 0.284233402 | 1 | 0.287161671 | ENSG0000026 | 1 | RP11-540B6.2 |
| VAT | T2DM | ENSG0000014 | -0.0006687 | 0.643380798 | 0.001444342 | 0.081436923 | -0.00069169 | 0.631994583 | 0.00144427 | 0.081424614 | 2.30E-05 | 0.028326264 | 1 | 0.874341025 | 1 | 0.87433837 | ENSG0000026 | 1 | LINC00562 |
| VAT | T2DM | ENSG0000014 | -0.00016719 | 0.964851068 | 0.003794036 | 0.081436958 | -0.0002286 | 0.951952206 | 0.003793844 | 0.081424652 | 6.14E-05 | 0.02184679 | 1 | 0.991412214 | 1 | 0.988354266 | ENSG0000026 | 1 | LINC00562 |
| VAT | T2DM | ENSG0000014 | -0.00085536 | 0.771224145 | 0.002941678 | 0.081436944 | -0.00090361 | 0.758700405 | 0.002941557 | 0.081424637 | 4.82E-05 | 0.048484614 | 1 | 0.924263413 | 1 | 0.915051408 | ENSG0000026 | 1 | LINC00562 |
| VAT | T2DM | ENSG0000014 | 0.00226419 | 0.326560874 | 0.002307889 | 0.081436798 | 0.002308221 | 0.317219177 | 0.002307783 | 0.081424485 | -4.40E-05 | 0.013779158 | 1 | 0.694146198 | 1 | 0.687858086 | ENSG0000026 | 2 | RP11-384K6.6 |
| VAT | T2DM | ENSG0000014 | 0.001565631 | 0.477970535 | 0.002206447 | 0.081436874 | 0.001600196 | 0.468285839 | 0.002206341 | 0.081424565 | -3.46E-05 | 0.035512565 | 1 | 0.791889203 | 1 | 0.792925716 | ENSG0000026 | 1 | CTD-2196E14.9 |
| VAT | T2DM | ENSG0000014 | -0.00576418 | 0.028413238 | 0.002630197 | 0.081436156 | -0.00562956 | 0.03233537 | 0.002630331 | 0.081423887 | -0.00013463 | 0.001265765 | 1 | 0.23459344 | 1 | 0.261717366 | ENSG0000026 | 1 | CTD-2196E14.9 |
| VAT | T2DM | ENSG0000014 | 0.006238411 | 0.080189557 | 0.00356564 | 0.081436447 | 0.006151342 | 0.084487543 | 0.00356553 | 0.081424155 | 8.71E-05 | 0.00974549 | 1 | 0.393299377 | 1 | 0.410615651 | ENSG0000026 | 1 | BMS1P8 |
| VAT | T2DM | ENSG0000014 | -0.00091754 | 0.656969378 | 0.002066068 | 0.081436925 | -0.00095364 | 0.644373702 | 0.002065981 | 0.081424617 | 3.61E-05 | 0.032722772 | 1 | 0.877230519 | 1 | 0.87456323 | ENSG0000026 | 1 | BMS1P8 |
| VAT | T2DM | ENSG0000015 | -3.08E-05 | 0.984767593 | 0.001613633 | 0.081436958 | -3.00E-06 | 0.998518465 | 0.001613568 | 0.081424652 | -2.78E-05 | 0.039808062 | 1 | 0.997544462 | 1 | 0.999423969 | ENSG0000026 | 1 | RP11-252A24.7 |
| VAT | T2DM | ENSG0000015 | -0.00021552 | 0.893452871 | 0.001609089 | 0.081436955 | -0.00024585 | 0.878560247 | 0.001609025 | 0.081424649 | 3.03E-05 | 0.02570244 | 1 | 0.970456484 | 1 | 0.96786083 | ENSG0000026 | 1 | RP11-252A24.7 |
| VAT | T2DM | ENSG0000015 | -0.00377805 | 0.137527084 | 0.002544031 | 0.08143659 | -0.00371026 | 0.144711824 | 0.002543941 | 0.081424297 | -6.78E-05 | 0.002841469 | 1 | 0.500466051 | 1 | 0.514898084 | ENSG0000026 | 1 | RP11-252A24.7 |
| VAT | T2DM | ENSG0000015 | 0.000724234 | 0.406795551 | 0.000873048 | 0.081436843 | 0.00070818 | 0.417254215 | 0.000873009 | 0.081424543 | 1.61E-05 | 0.018946069 | 1 | 0.753911136 | 1 | 0.764717698 | ENSG0000026 | 1 | RP11-252A24.7 |
| VAT | T2DM | ENSG0000015 | -0.00411253 | 0.238713455 | 0.003490488 | 0.081436726 | -0.00406439 | 0.244230376 | 0.003490306 | 0.081424426 | -4.81E-05 | 0.044167806 | 1 | 0.626118109 | 1 | 0.630010905 | ENSG0000026 | 1 | RP3-522J7.6 |
| VAT | T2DM | ENSG0000015 | 0.004356187 | 0.122492997 | 0.00282064 | 0.08143656 | 0.003669306 | 0.195682548 | 0.002835735 | 0.081424373 | 0.000686881 | 0.01956945 | 1 | 0.475900889 | 1 | 0.578867987 | ENSG0000026 | 1 | RP3-522J7.6 |
| VAT | T2DM | ENSG0000015 | -0.00023165 | 0.938122931 | 0.002983992 | 0.081436957 | -0.0002893 | 0.922762708 | 0.002983854 | 0.081424651 | 5.77E-05 | 0.01170366 | 1 | 0.986373964 | 1 | 0.982591477 | ENSG0000026 | 1 | AC026150.8 |
| VAT | T2DM | ENSG0000015 | -0.0020952 | 0.423824826 | 0.002619646 | 0.081436852 | -0.00214736 | 0.412357223 | 0.002619517 | 0.08142454 | 5.22E-05 | 0.00614633 | 1 | 0.767766503 | 1 | 0.762700254 | ENSG0000026 | 1 | AC026150.8 |
| VAT | T2DM | ENSG0000015 | -0.00051383 | 0.647322623 | 0.001123173 | 0.081436923 | -0.00267711 | 0.044906897 | 0.001334864 | 0.08142398 | 0.002163276 | 0.002714437 | 1 | 0.874341025 | 1 | 0.302300087 | ENSG0000026 | 1 | RP11-184E9.2 |
| VAT | T2DM | ENSG0000015 | 0.005680904 | 0.003422462 | 0.001940851 | 0.081435527 | 0.005315005 | 0.006356302 | 0.001947728 | 0.081423408 | 0.000365899 | 0.026794868 | 1 | 0.06550978 | 1 | 0.10224671 | ENSG0000026 | 2 | MIR940 |
| VAT | T2DM | ENSG0000015 | 0.002347293 | 0.152080321 | 0.001638915 | 0.081436616 | 0.002379273 | 0.146555706 | 0.001638835 | 0.0814243 | -3.20E-05 | 0.007886697 | 1 | 0.522716918 | 1 | 0.514898084 | ENSG0000026 | 2 | MIR940 |

| | | | | | | | | | | | | | | | | | | | |
|---|---|---|---|---|---|---|---|---|---|---|---|---|---|---|---|---|---|---|---|
| VAT | T2DM | ENSG0000015 | -0.0001176 | 0.970064916 | 0.003133845 | 0.081436958 | -6.37E-05 | 0.983789762 | 0.003133726 | 0.081424652 | -5.39E-05 | 0.046904301 | 1 | 0.991412214 | 1 | 0.995445662 | ENSG0000026 | 1 | RP11-797A18.4 |
| VAT | T2DM | ENSG0000015 | -0.00289186 | 0.083118535 | 0.00166882 | 0.081436457 | -0.00294416 | 0.077683621 | 0.001668752 | 0.081424132 | 5.23E-05 | 0.000180811 | 1 | 0.398276316 | 1 | 0.397049619 | ENSG0000026 | 1 | RP11-797A18.4 |
| VAT | T2DM | ENSG0000015 | 0.001486424 | 0.297134664 | 0.001425691 | 0.081436777 | 0.00133113 | 0.350926341 | 0.001427034 | 0.081424507 | 0.000155293 | 0.015776379 | 1 | 0.674852962 | 1 | 0.712174044 | ENSG0000026 | 1 | RP11-797A18.4 |
| VAT | T2DM | ENSG0000015 | 0.003376144 | 0.39167812 | 0.003941428 | 0.081436836 | 0.001160672 | 0.77373692 | 0.004037255 | 0.081424639 | 0.002215472 | 0.011410718 | 1 | 0.746568793 | 1 | 0.923905803 | ENSG0000026 | 2 | RP11-566E18.3 |
| VAT | T2DM | ENSG0000015 | 0.000927458 | 0.688972821 | 0.002317199 | 0.081436932 | 0.0009646 | 0.677190455 | 0.002317078 | 0.081424624 | -3.71E-05 | 0.019410776 | 1 | 0.895575687 | 1 | 0.89117459 | ENSG0000026 | 2 | RP11-566E18.3 |
| VAT | T2DM | ENSG0000015 | -2.89E-05 | 0.98845638 | 0.001998437 | 0.081436958 | 1.25E-05 | 0.994995927 | 0.001998341 | 0.081424652 | -4.14E-05 | 0.00529775 | 1 | 0.997544462 | 1 | 0.99870972 | ENSG0000026 | 2 | RP11-566E18.3 |
| VAT | T2DM | ENSG0000015 | -0.0014064 | 0.634508181 | 0.002958399 | 0.081436921 | -0.00145204 | 0.623538626 | 0.002958261 | 0.081424612 | 4.56E-05 | 0.041884394 | 1 | 0.871259726 | 1 | 0.868736298 | ENSG0000026 | 2 | RP11-578F21.9 |
| VAT | T2DM | ENSG0000015 | -0.00689886 | 0.060139628 | 0.003670052 | 0.081436368 | -0.00681806 | 0.063193997 | 0.00366991 | 0.081424076 | -8.08E-05 | 0.010269617 | 1 | 0.350180111 | 1 | 0.367965046 | ENSG0000026 | 1 | RP11-211G23.2 |
| VAT | T2DM | ENSG0000015 | 0.005749829 | 0.115891449 | 0.003657052 | 0.081436545 | 0.005406701 | 0.13947296 | 0.003658716 | 0.081424288 | 0.000343128 | 0.003965891 | 1 | 0.460230789 | 1 | 0.507843495 | ENSG0000026 | 1 | RP11-211G23.2 |
| VAT | T2DM | ENSG0000015 | 0.00456567 | 0.036533834 | 0.002183547 | 0.081436228 | 0.004495132 | 0.039523132 | 0.002183477 | 0.081423944 | 7.05E-05 | 0.000492984 | 1 | 0.26817389 | 1 | 0.285285693 | ENSG0000026 | 1 | RP11-211G23.2 |
| VAT | T2DM | ENSG0000015 | 0.005825499 | 0.031933821 | 0.002715541 | 0.081436189 | 0.005731145 | 0.034808475 | 0.002715429 | 0.081423908 | 9.44E-05 | 2.53E-05 | 1 | 0.250390184 | 1 | 0.271107773 | ENSG0000026 | 1 | AC008088.4 |
| VAT | T2DM | ENSG0000015 | -0.01026761 | 4.01E-08 | 0.001870037 | 0.081431921 | -0.01030653 | 3.56E-08 | 0.001869956 | 0.081419577 | 3.89E-05 | 0.009701792 | 0.000110618 | 6.51E-06 | 9.82E-05 | 8.92E-06 | ENSG0000026 | 1 | RP11-196G11.2 |
| VAT | T2DM | ENSG0000015 | -0.00239561 | 0.245444527 | 0.00206254 | 0.081436733 | -0.00243423 | 0.237900238 | 0.002062463 | 0.08142442 | 3.86E-05 | 0.031871953 | 1 | 0.631339139 | 1 | 0.62355618 | ENSG0000026 | 1 | RP11-196G11.2 |
| VAT | T2DM | ENSG0000015 | -0.00585029 | 0.098913438 | 0.003545315 | 0.081436503 | -0.00595606 | 0.092945968 | 0.003545142 | 0.081424181 | 0.000105766 | 4.99E-05 | 1 | 0.429246993 | 1 | 0.428877415 | ENSG0000026 | 1 | RP11-384P7.7 |
| VAT | T2DM | ENSG0000015 | -0.00240276 | 0.237899982 | 0.002035801 | 0.081436726 | -0.00237084 | 0.244168928 | 0.0020357 | 0.081424426 | -3.19E-05 | 0.029192535 | 1 | 0.624786607 | 1 | 0.630010905 | ENSG0000026 | 1 | RP11-384P7.7 |
| VAT | T2DM | ENSG0000015 | 0.004743428 | 0.170045429 | 0.003457158 | 0.081436644 | 0.004663301 | 0.177357659 | 0.003457018 | 0.081424348 | 8.01E-05 | 0.005471758 | 1 | 0.546218791 | 1 | 0.552491128 | ENSG0000026 | 1 | RP11-384P7.7 |
| VAT | T2DM | ENSG0000015 | -0.00165615 | 0.64949237 | 0.003644143 | 0.081436924 | -0.00154581 | 0.671415402 | 0.003643997 | 0.081424622 | -0.00011034 | 0.000320318 | 1 | 0.874341025 | 1 | 0.888954186 | ENSG0000026 | 1 | RP11-384P7.7 |
| VAT | T2DM | ENSG0000015 | -0.00166825 | 0.460734795 | 0.002261605 | 0.081436867 | -0.00173575 | 0.442772996 | 0.002261511 | 0.081424554 | 6.75E-05 | 0.000314763 | 1 | 0.78904441 | 1 | 0.783615427 | ENSG0000026 | 1 | RP11-105C19.2 |
| VAT | T2DM | ENSG0000015 | -0.00397344 | 0.147828203 | 0.002745512 | 0.081436608 | -0.00392011 | 0.153322285 | 0.002745373 | 0.081424312 | -5.33E-05 | 0.006185292 | 1 | 0.514322353 | 1 | 0.52392045 | ENSG0000026 | 1 | CTC-471C19.1 |
| VAT | T2DM | ENSG0000015 | -0.0014724 | 0.615510877 | 0.002931771 | 0.081436916 | -0.00152901 | 0.601980288 | 0.002931634 | 0.081424607 | 5.66E-05 | 0.011277565 | 1 | 0.863595144 | 1 | 0.856779751 | ENSG0000026 | 1 | RP11-1000B6.3 |
| VAT | T2DM | ENSG0000016 | -0.00284725 | 0.03478645 | 0.001348872 | 0.081436214 | -0.00281118 | 0.037151262 | 0.001348865 | 0.081423927 | -3.61E-05 | 0.024498894 | 1 | 0.260715228 | 1 | 0.280157059 | ENSG0000026 | 1 | RP11-1000B6.3 |
| VAT | T2DM | ENSG0000016 | -0.00558579 | 0.019713059 | 0.002395512 | 0.08143605 | -0.00563068 | 0.018743537 | 0.002395423 | 0.081423729 | 4.49E-05 | 0.03256998 | 1 | 0.190905412 | 1 | 0.195735137 | ENSG0000026 | 1 | ADAMTS7P3 |
| VAT | T2DM | ENSG0000016 | 0.000591178 | 0.841293091 | 0.002952351 | 0.081436952 | 0.000653058 | 0.824929849 | 0.002952223 | 0.081424644 | -6.19E-05 | 0.008831858 | 1 | 0.951626611 | 1 | 0.946021651 | ENSG0000026 | 1 | ADAMTS7P3 |
| VAT | T2DM | ENSG0000016 | 9.21E-05 | 0.917568067 | 0.000889983 | 0.081436957 | 7.77E-05 | 0.930421117 | 0.000889938 | 0.081424651 | 1.44E-05 | 0.022944512 | 1 | 0.979877489 | 1 | 0.986539486 | ENSG0000026 | 1 | ADAMTS7P3 |
| VAT | T2DM | ENSG0000016 | -0.00199149 | 0.517293201 | 0.003075542 | 0.081436888 | -0.00206214 | 0.502518745 | 0.003075381 | 0.081424577 | 7.07E-05 | 0.000736278 | 1 | 0.812597159 | 1 | 0.809662427 | ENSG0000026 | 1 | ADAMTS7P3 |
| VAT | T2DM | ENSG0000016 | -0.00108985 | 0.75710313 | 0.003523761 | 0.081436942 | -0.00095127 | 0.78718352 | 0.003523629 | 0.08142464 | -0.00013858 | 6.93E-06 | 1 | 0.919012884 | 1 | 0.931258686 | ENSG0000026 | 1 | CTD-2568A17.1 |
| VAT | T2DM | ENSG0000016 | -0.00140428 | 0.486873236 | 0.002019705 | 0.081436878 | -0.00144139 | 0.475415911 | 0.002019616 | 0.081424567 | 3.71E-05 | 0.020416351 | 1 | 0.792496248 | 1 | 0.795723417 | ENSG0000026 | 1 | RP3-465N24.5 |
| VAT | T2DM | ENSG0000016 | -0.00150275 | 0.70938157 | 0.004032196 | 0.081436935 | -0.00142132 | 0.724456663 | 0.004031998 | 0.081424632 | -8.14E-05 | 0.005491258 | 1 | 0.903452435 | 1 | 0.907346725 | ENSG0000026 | 1 | CTA-14H9.5 |
| VAT | T2DM | ENSG0000016 | 0.001259682 | 0.751280949 | 0.003974376 | 0.081436942 | 0.001335227 | 0.736891896 | 0.003974217 | 0.081424634 | -7.55E-05 | 0.024264775 | 1 | 0.918605915 | 1 | 0.910293329 | ENSG0000026 | 1 | CTA-14H9.5 |
| VAT | T2DM | ENSG0000016 | 0.004100464 | 0.297751371 | 0.00393795 | 0.081436777 | 0.004162858 | 0.290438025 | 0.003937766 | 0.081424466 | -6.24E-05 | 0.037497759 | 1 | 0.674852962 | 1 | 0.665135964 | ENSG0000026 | 1 | LINC01003 |
| VAT | T2DM | ENSG0000016 | -0.0014121 | 0.413062045 | 0.001725191 | 0.081436846 | -0.00146036 | 0.397265968 | 0.001725141 | 0.081424533 | 4.83E-05 | 0.003847907 | 1 | 0.757898941 | 1 | 0.752171241 | ENSG0000026 | 4 | RP11-296I10.6 |
| VAT | T2DM | ENSG0000016 | 0.001969231 | 0.254621183 | 0.001728612 | 0.081436742 | 0.001923676 | 0.265754761 | 0.001728536 | 0.081424446 | 4.56E-05 | 0.000892921 | 1 | 0.635542337 | 1 | 0.646388519 | ENSG0000026 | 4 | RP11-296I10.6 |
| VAT | T2DM | ENSG0000016 | -0.00094783 | 0.72190228 | 0.002663049 | 0.081436937 | -0.0011067 | 0.677744734 | 0.002663274 | 0.081424624 | 0.000158878 | 0.000865341 | 1 | 0.907308877 | 1 | 0.89117459 | ENSG0000026 | 4 | RP11-296I10.6 |
| VAT | T2DM | ENSG0000016 | -0.00158597 | 0.295544551 | 0.00151617 | 0.081436776 | -0.00154959 | 0.30674619 | 0.001516125 | 0.081424478 | -3.64E-05 | 0.011629727 | 1 | 0.673522043 | 1 | 0.676208846 | ENSG0000026 | 1 | RP11-457M11.5 |
| VAT | T2DM | ENSG0000016 | 0.00253156 | 0.483083009 | 0.003609537 | 0.081436876 | 0.002614225 | 0.468886783 | 0.003609351 | 0.081424565 | -8.27E-05 | 0.001005936 | 1 | 0.791923137 | 1 | 0.793004278 | ENSG0000026 | 1 | RP11-457M11.5 |
| VAT | T2DM | ENSG0000016 | 0.003809567 | 0.330858573 | 0.003917748 | 0.0814368 | 0.003712342 | 0.343330214 | 0.003917598 | 0.081424502 | 9.72E-05 | 0.004074001 | 1 | 0.699746867 | 1 | 0.705801938 | ENSG0000026 | 1 | RP11-457M11.5 |
| VAT | T2DM | ENSG0000016 | 0.003659734 | 0.169574952 | 0.00266439 | 0.081436643 | 0.003720535 | 0.162577066 | 0.002664265 | 0.081424327 | -6.08E-05 | 0.002547826 | 1 | 0.546218791 | 1 | 0.53867071 | ENSG0000026 | 1 | RP11-457M11.5 |
| VAT | T2DM | ENSG0000016 | -0.00155784 | 0.689413274 | 0.003897989 | 0.081436932 | -0.00163667 | 0.674560821 | 0.00389779 | 0.081424623 | 7.88E-05 | 0.003991965 | 1 | 0.895575687 | 1 | 0.890807592 | ENSG0000026 | 1 | RP11-457M11.5 |
| VAT | T2DM | ENSG0000016 | 0.001558537 | 0.668656612 | 0.003641504 | 0.081436928 | 0.001616328 | 0.657125216 | 0.003641314 | 0.08142462 | -5.78E-05 | 0.01991875 | 1 | 0.884384426 | 1 | 0.876362712 | ENSG0000026 | 1 | RP11-457M11.5 |
| VAT | T2DM | ENSG0000016 | -0.00113163 | 0.343371565 | 0.001194298 | 0.081436808 | -0.00115373 | 0.334005092 | 0.001194237 | 0.081424497 | 2.21E-05 | 0.008276895 | 1 | 0.705138034 | 1 | 0.702906909 | ENSG0000026 | 1 | RP11-457M11.5 |
| VAT | T2DM | ENSG0000016 | -0.00234867 | 0.546521793 | 0.003895102 | 0.081436898 | -0.00240723 | 0.536544613 | 0.003894904 | 0.081424589 | 5.86E-05 | 0.032690894 | 1 | 0.826107649 | 1 | 0.825591361 | ENSG0000026 | 1 | RP11-457M11.5 |
| VAT | T2DM | ENSG0000016 | -0.00018605 | 0.798513902 | 0.000728817 | 0.081436948 | -0.00017547 | 0.809726115 | 0.000728778 | 0.081424643 | -1.06E-05 | 0.027348893 | 1 | 0.935837949 | 1 | 0.938393612 | ENSG0000026 | 1 | RP11-457M11.5 |
| VAT | T2DM | ENSG0000016 | 0.000710252 | 0.703407827 | 0.001865527 | 0.081436934 | 0.000750497 | 0.687451241 | 0.001865439 | 0.081424625 | -4.02E-05 | 0.00388331 | 1 | 0.90214015 | 1 | 0.893821363 | ENSG0000026 | 1 | RP11-457M11.5 |
| VAT | T2DM | ENSG0000016 | 0.001588272 | 0.541678582 | 0.002602542 | 0.081436896 | 0.001538764 | 0.554336711 | 0.002602452 | 0.081424594 | 4.95E-05 | 0.035685922 | 1 | 0.823132089 | 1 | 0.83604881 | ENSG0000026 | 1 | RP11-457M11.5 |
| VAT | T2DM | ENSG0000016 | -0.01418581 | 1.97E-05 | 0.003323469 | 0.081433914 | -0.01413532 | 2.11E-05 | 0.003323316 | 0.08142163 | -5.05E-05 | 0.047457523 | 0.054355121 | 0.001469057 | 0.058128376 | 0.001816512 | ENSG0000026 | 2 | RP11-923I11.6 |
| VAT | T2DM | ENSG0000016 | -0.00850975 | 0.002873282 | 0.002854668 | 0.081435473 | -0.00846073 | 0.003037125 | 0.002854528 | 0.081423185 | -4.90E-05 | 0.017699187 | 1 | 0.059181022 | 1 | 0.067059723 | ENSG0000026 | 2 | RP11-923I11.6 |
| VAT | T2DM | ENSG0000016 | -0.00076646 | 0.379471809 | 0.000872099 | 0.081436829 | -0.00075426 | 0.387080759 | 0.000872053 | 0.081424527 | -1.22E-05 | 0.037804389 | 1 | 0.737965059 | 1 | 0.745528886 | ENSG0000026 | 1 | CTC-462L7.1 |
| VAT | T2DM | ENSG0000016 | 0.00127656 | 0.411099975 | 0.001553068 | 0.081436846 | 0.00114383 | 0.461656174 | 0.001553856 | 0.081424562 | 0.00013273 | 0.01235733 | 1 | 0.755416731 | 1 | 0.791032387 | ENSG0000026 | 1 | CTC-462L7.1 |
| VAT | T2DM | ENSG0000016 | 0.001609267 | 0.100701901 | 0.000980385 | 0.081436508 | 0.001576294 | 0.107859906 | 0.00098035 | 0.08142422 | 3.30E-05 | 0.000137387 | 1 | 0.430384653 | 1 | 0.449687827 | ENSG0000026 | 1 | CTC-462L7.1 |
| VAT | T2DM | ENSG0000016 | 0.002306021 | 0.14796929 | 0.001593939 | 0.081436609 | 0.002333018 | 0.143264506 | 0.00159387 | 0.081424294 | -2.70E-05 | 0.036559113 | 1 | 0.514322353 | 1 | 0.514828934 | ENSG0000026 | 2 | C15orf65 |
| VAT | T2DM | ENSG0000016 | -0.00103935 | 0.517434118 | 0.001605657 | 0.081436888 | -0.00107542 | 0.502984411 | 0.001605582 | 0.081424578 | 3.61E-05 | 0.003322278 | 1 | 0.812597159 | 1 | 0.809900529 | ENSG0000026 | 2 | C15orf65 |
| VAT | T2DM | ENSG0000016 | 0.00127151 | 0.509579942 | 0.001928013 | 0.081436886 | 0.001320783 | 0.493309429 | 0.001927994 | 0.081424574 | -4.93E-05 | 0.026331136 | 1 | 0.808707681 | 1 | 0.800431525 | ENSG0000026 | 1 | RP11-475B2.1 |
| VAT | T2DM | ENSG0000016 | 0.000641396 | 0.604333649 | 0.001237788 | 0.081436914 | 0.000605587 | 0.624651899 | 0.001237741 | 0.081424612 | 3.58E-05 | 0.000747848 | 1 | 0.857656828 | 1 | 0.868971392 | ENSG0000026 | 1 | RP11-61A14.2 |
| VAT | T2DM | ENSG0000016 | -0.00310929 | 0.199763949 | 0.002424918 | 0.081436684 | -0.00375047 | 0.123640727 | 0.00243589 | 0.081424256 | 0.000641181 | 0.005898573 | 1 | 0.58063629 | 1 | 0.477270497 | ENSG0000026 | 1 | AP001505.9 |
| VAT | T2DM | ENSG0000016 | 0.002730498 | 0.465387613 | 0.003740384 | 0.081436869 | 0.002805403 | 0.453214627 | 0.003740203 | 0.081424558 | -7.49E-05 | 0.006475088 | 1 | 0.78904441 | 1 | 0.789415267 | ENSG0000026 | 2 | CTC-523E23.1 |
| VAT | T2DM | ENSG0000016 | -0.00023508 | 0.838982892 | 0.001156926 | 0.081436952 | -0.00020943 | 0.856348239 | 0.001156887 | 0.081424647 | -2.57E-05 | 0.015316788 | 1 | 0.949791953 | 1 | 0.958443285 | ENSG0000026 | 2 | RP11-1223D19.1 |
| VAT | T2DM | ENSG0000016 | -0.00080401 | 0.432488227 | 0.001024294 | 0.081436855 | -0.00078323 | 0.444456238 | 0.001024245 | 0.081424555 | -2.08E-05 | 0.006076585 | 1 | 0.772100587 | 1 | 0.783833365 | ENSG0000026 | 1 | RP4-555D20.2 |
| VAT | T2DM | ENSG0000016 | -0.00589674 | 0.093859251 | 0.003519632 | 0.081436489 | -0.00582729 | 0.097776139 | 0.003519455 | 0.081424194 | -6.95E-05 | 0.005579196 | 1 | 0.41970912 | 1 | 0.435146086 | ENSG0000026 | 1 | GOLGA8H |
| VAT | T2DM | ENSG0000016 | 0.003942099 | 0.262083724 | 0.003515083 | 0.081436748 | 0.003873574 | 0.270446145 | 0.003514922 | 0.08142445 | 6.85E-05 | 0.011467204 | 1 | 0.643030277 | 1 | 0.64741007 | ENSG0000026 | 1 | GOLGA8H |
| VAT | T2DM | ENSG0000016 | -0.00081799 | 0.779020869 | 0.002915196 | 0.081436945 | -0.00090689 | 0.755719945 | 0.002915063 | 0.081424636 | 8.89E-05 | 8.16E-05 | 1 | 0.928876022 | 1 | 0.91441782 | ENSG0000026 | 2 | LINC00662 |
| VAT | T2DM | ENSG0000016 | -0.00148976 | 0.60600716 | 0.002888363 | 0.081436914 | -0.00141869 | 0.623289073 | 0.002888252 | 0.081424612 | -7.11E-05 | 0.00427833 | 1 | 0.857656828 | 1 | 0.868736298 | ENSG0000026 | 2 | LINC00662 |
| VAT | T2DM | ENSG0000016 | 0.002100063 | 0.50986352 | 0.003186497 | 0.081436886 | 0.002145214 | 0.500784079 | 0.003186323 | 0.081424577 | -4.52E-05 | 0.028441162 | 1 | 0.808707681 | 1 | 0.808756032 | ENSG0000026 | 1 | RP11-388M20.9 |
| VAT | T2DM | ENSG0000016 | 0.000599676 | 0.535252364 | 0.000967205 | 0.081436894 | 0.000624477 | 0.5184895 | 0.000967167 | 0.081424583 | -2.48E-05 | 0.00254639 | 1 | 0.81975639 | 1 | 0.817922279 | ENSG0000026 | 1 | RP11-433P17.3 |
| VAT | T2DM | ENSG0000016 | 0.001143921 | 0.345645111 | 0.001212975 | 0.08143681 | 0.001171148 | 0.33426609 | 0.001212924 | 0.081424497 | -2.72E-05 | 0.006212294 | 1 | 0.706225056 | 1 | 0.702906909 | ENSG0000026 | 1 | RP11-81A22.5 |
| VAT | T2DM | ENSG0000016 | -0.0036077 | 0.100091201 | 0.002193914 | 0.081436507 | -0.0033157 | 0.130985437 | 0.002195491 | 0.081424271 | -0.00029201 | 0.00083868 | 1 | 0.430095137 | 1 | 0.491745938 | ENSG0000026 | 1 | RP11-81A22.5 |
| VAT | T2DM | ENSG0000016 | -0.00219746 | 0.380443652 | 0.002505427 | 0.08143683 | -0.00214116 | 0.39274938 | 0.002505322 | 0.08142453 | -5.63E-05 | 0.006221489 | 1 | 0.737965059 | 1 | 0.749646119 | ENSG0000026 | 1 | RP11-473M20.9 |
| VAT | T2DM | ENSG0000016 | -0.00251625 | 0.211382618 | 0.002013366 | 0.081436697 | -0.00256558 | 0.202552197 | 0.002013296 | 0.081424381 | 4.93E-05 | 0.006729624 | 1 | 0.592343831 | 1 | 0.583144839 | ENSG0000026 | 1 | RP11-473M20.9 |
| VAT | T2DM | ENSG0000016 | 0.007896848 | 0.02865843 | 0.003608899 | 0.081436158 | 0.006074343 | 0.095738584 | 0.003646334 | 0.081424189 | 0.001822505 | 0.000492461 | 1 | 0.235408535 | 1 | 0.433697593 | ENSG0000026 | 1 | RP11-473M20.9 |
| VAT | T2DM | ENSG0000016 | -3.77E-06 | 0.99901808 | 0.003062774 | 0.081436958 | 0.000301223 | 0.921679797 | 0.003063764 | 0.081424651 | -0.00030499 | 0.000423314 | 1 | 0.99901808 | 1 | 0.982577635 | ENSG0000026 | 1 | RP11-473M20.9 |
| VAT | T2DM | ENSG0000016 | 0.002557739 | 0.137133852 | 0.001720585 | 0.081436589 | 0.002530254 | 0.141388139 | 0.001720508 | 0.081424291 | 2.75E-05 | 0.041501669 | 1 | 0.500466051 | 1 | 0.510969582 | ENSG0000026 | 1 | RP11-95P2.1 |
| VAT | T2DM | ENSG0000016 | 0.000411289 | 0.845950137 | 0.0021169 | 0.081436952 | -2.38E-05 | 0.991052104 | 0.002118529 | 0.081424652 | 0.000435048 | 5.80E-07 | 1 | 0.954545261 | 1 | 0.997194242 | ENSG0000026 | 1 | FAM187B2P |
| VAT | T2DM | ENSG0000016 | 0.001244554 | 0.548895085 | 0.002076275 | 0.081436898 | 0.001279964 | 0.537562931 | 0.002076168 | 0.081424589 | -3.54E-05 | 0.01460368 | 1 | 0.828182675 | 1 | 0.825843697 | ENSG0000026 | 1 | RP11-259G18.2 |
| VAT | T2DM | ENSG0000016 | 0.001683358 | 0.387827015 | 0.001949311 | 0.081436834 | 0.001653468 | 0.396285033 | 0.001949211 | 0.081424532 | 2.99E-05 | 0.028442418 | 1 | 0.741792098 | 1 | 0.752171241 | ENSG0000026 | 1 | RP11-259G18.2 |
| VAT | T2DM | ENSG0000016 | -4.31E-05 | 0.963961582 | 0.000954345 | 0.081436958 | -1.86E-05 | 0.984487626 | 0.000954304 | 0.081424652 | -2.46E-05 | 0.001434993 | 1 | 0.991412214 | 1 | 0.995445662 | ENSG0000026 | 1 | RP11-259G18.3 |
| VAT | T2DM | ENSG0000016 | -0.00046429 | 0.785472348 | 0.001705721 | 0.081436946 | -0.00043825 | 0.797225034 | 0.001705633 | 0.081424641 | -2.60E-05 | 0.025933654 | 1 | 0.931230103 | 1 | 0.935209552 | ENSG0000026 | 1 | RP11-259G18.3 |
| VAT | T2DM | ENSG0000016 | 0.000282614 | 0.932085379 | 0.003316227 | 0.081436957 | 0.000343502 | 0.917497147 | 0.003316074 | 0.081424651 | -6.09E-05 | 0.016322825 | 1 | 0.984451223 | 1 | 0.981889153 | ENSG0000026 | 1 | RP11-497H17.1 |
| VAT | T2DM | ENSG0000016 | -0.00061983 | 0.77944814 | 0.00221337 | 0.081436945 | -0.00058652 | 0.791009251 | 0.002213267 | 0.081424641 | -3.33E-05 | 0.048734495 | 1 | 0.928876022 | 1 | 0.933384153 | ENSG0000026 | 1 | RP11-497H17.1 |
| VAT | T2DM | ENSG0000016 | 0.00302009 | 0.361822525 | 0.003311871 | 0.081436819 | 0.003122042 | 0.345822024 | 0.003311721 | 0.081424504 | -0.00010195 | 7.69E-05 | 1 | 0.719681446 | 1 | 0.707749931 | ENSG0000026 | 1 | RP11-46I8.4 |
| VAT | T2DM | ENSG0000016 | -0.01271065 | 3.18E-07 | 0.002486052 | 0.08143259 | -0.0128006 | 2.62E-07 | 0.002485951 | 0.081420222 | 9.00E-05 | 1.71E-05 | 0.000876472 | 4.17E-05 | 0.000722378 | 5.16E-05 | ENSG0000026 | 1 | RP11-46I8.4 |
| VAT | T2DM | ENSG0000016 | 0.003669403 | 0.006478 | 0.001347775 | 0.08143572 | 0.003271726 | 0.01542019 | 0.001350633 | 0.081423672 | 0.000397677 | 8.51E-06 | 1 | 0.094101469 | 1 | 0.176596367 | ENSG0000026 | 1 | RP11-46I8.4 |
| VAT | T2DM | ENSG0000016 | 0.000669817 | 0.410303239 | 0.000813516 | 0.081436845 | 0.000629589 | 0.439074856 | 0.000813678 | 0.081424552 | 4.02E-05 | 0.034929538 | 1 | 0.754454991 | 1 | 0.781680226 | ENSG0000026 | 1 | RP11-46I8.4 |
| VAT | T2DM | ENSG0000016 | 0.001374189 | 0.653057973 | 0.003057026 | 0.081436925 | 0.001426674 | 0.640707611 | 0.003056875 | 0.081424616 | -5.25E-05 | 0.017489161 | 1 | 0.875814904 | 1 | 0.87456323 | ENSG0000026 | 1 | RP11-46I8.4 |
| VAT | T2DM | ENSG0000017 | 0.002964985 | 0.423140309 | 0.003701671 | 0.081436851 | 0.002898199 | 0.433641691 | 0.003701518 | 0.08142455 | 6.68E-05 | 0.029442439 | 1 | 0.767766503 | 1 | 0.778186649 | ENSG0000026 | 3 | RP11-1055B8.4 |
| VAT | T2DM | ENSG0000017 | 0.001677081 | 0.184012621 | 0.001262384 | 0.081436663 | 0.001725587 | 0.171636213 | 0.001262347 | 0.08142434 | -4.85E-05 | 7.06E-05 | 1 | 0.56430537 | 1 | 0.54753664 | ENSG0000026 | 3 | ALOX12P2 |
| VAT | T2DM | ENSG0000017 | 0.002250944 | 0.48299017 | 0.003208749 | 0.081436876 | 0.002314482 | 0.470705557 | 0.003208622 | 0.081424566 | -6.35E-05 | 0.019311248 | 1 | 0.791923137 | 1 | 0.794121331 | ENSG0000026 | 1 | RP11-166P13.3 |
| VAT | T2DM | ENSG0000017 | -0.00047018 | 0.766849793 | 0.001585781 | 0.081436944 | -0.00049588 | 0.754493259 | 0.001585705 | 0.081424636 | 2.57E-05 | 0.029917997 | 1 | 0.923431688 | 1 | 0.913932577 | ENSG0000026 | 2 | ROCK1P1 |
| VAT | T2DM | ENSG0000017 | 0.003360085 | 0.240170428 | 0.002860713 | 0.081436728 | 0.003296785 | 0.249120811 | 0.002860578 | 0.081424431 | 6.33E-05 | 0.003412218 | 1 | 0.628313159 | 1 | 0.630351056 | ENSG0000026 | 2 | ROCK1P1 |
| VAT | T2DM | ENSG0000017 | -0.00112451 | 0.595218114 | 0.002116569 | 0.081436911 | -0.00108595 | 0.607887297 | 0.002116482 | 0.081424608 | -3.86E-05 | 0.027163342 | 1 | 0.851009022 | 1 | 0.859788021 | ENSG0000026 | 2 | ROCK1P1 |
| VAT | T2DM | ENSG0000017 | 0.004779485 | 0.033714063 | 0.002250778 | 0.081436205 | 0.00483901 | 0.0315533 | 0.002250671 | 0.08142388 | -5.95E-05 | 0.000402677 | 1 | 0.256548047 | 1 | 0.259967574 | ENSG0000026 | 1 | RP11-692N5.1 |
| VAT | T2DM | ENSG0000017 | -0.00032109 | 0.783635157 | 0.001169353 | 0.081436946 | -0.0002938 | 0.801609463 | 0.001169303 | 0.081424642 | -2.73E-05 | 0.003943006 | 1 | 0.930250767 | 1 | 0.935824336 | ENSG0000026 | 1 | RP11-692N5.1 |
| VAT | T2DM | ENSG0000017 | 0.00290314 | 0.047445197 | 0.001464524 | 0.081436302 | 0.002937073 | 0.044904145 | 0.001464468 | 0.08142398 | -3.39E-05 | 0.007323264 | 1 | 0.306671528 | 1 | 0.302300087 | ENSG0000026 | 1 | LINC00672 |
| VAT | T2DM | ENSG0000017 | -0.00289694 | 0.026823582 | 0.001308416 | 0.081436139 | -0.00287518 | 0.027979846 | 0.001308343 | 0.081423846 | -2.18E-05 | 0.008878256 | 1 | 0.225024577 | 1 | 0.242843944 | ENSG0000026 | 1 | MAGOH2 |
| VAT | T2DM | ENSG0000017 | 0.000917958 | 0.441866303 | 0.001193632 | 0.08143686 | 0.000936778 | 0.432540478 | 0.001193573 | 0.08142455 | -1.88E-05 | 0.026803448 | 1 | 0.777976538 | 1 | 0.776799597 | ENSG0000026 | 1 | RP11-173M1.8 |
| VAT | T2DM | ENSG0000017 | -0.00694942 | 0.088639796 | 0.004081603 | 0.081436474 | -0.00687926 | 0.091892087 | 0.004081428 | 0.081424178 | -7.02E-05 | 0.033440642 | 1 | 0.407340768 | 1 | 0.426973333 | ENSG0000026 | 1 | RP11-173M1.8 |
| VAT | T2DM | ENSG0000017 | -0.00096171 | 0.291516453 | 0.000911748 | 0.081436772 | -0.00100656 | 0.269634098 | 0.000911814 | 0.081424449 | 4.48E-05 | 0.004288086 | 1 | 0.667050516 | 1 | 0.647121836 | ENSG0000026 | 1 | RP11-173M1.8 |
| VAT | T2DM | ENSG0000017 | -0.00347599 | 0.393628035 | 0.00407475 | 0.081436837 | -0.00336287 | 0.409181938 | 0.004074548 | 0.081424539 | -0.00011313 | 0.000112611 | 1 | 0.747385518 | 1 | 0.762700254 | ENSG0000026 | 1 | RP11-173M1.8 |
| VAT | T2DM | ENSG0000017 | -0.00320846 | 0.177709679 | 0.002380443 | 0.081436655 | -0.00325991 | 0.170838652 | 0.002380336 | 0.081424339 | 5.14E-05 | 0.005844479 | 1 | 0.556533092 | 1 | 0.54753664 | ENSG0000026 | 1 | RP11-173M1.8 |
| VAT | T2DM | ENSG0000017 | 0.000437248 | 0.764561353 | 0.001459945 | 0.081436943 | 0.000679247 | 0.642042642 | 0.001461233 | 0.081424616 | -0.000242 | 0.000152812 | 1 | 0.922285548 | 1 | 0.87456323 | ENSG0000026 | 2 | SUZ12P |
| VAT | T2DM | ENSG0000017 | -0.00424676 | 0.121695886 | 0.002743919 | 0.081436558 | -0.00409246 | 0.135959736 | 0.002744757 | 0.081424281 | -0.0001543 | 0.041575459 | 1 | 0.475078708 | 1 | 0.501669614 | ENSG0000026 | 2 | SUZ12P |
| VAT | T2DM | ENSG0000017 | -0.00092766 | 0.300727738 | 0.0008964 | 0.081436779 | -0.0009163 | 0.306657027 | 0.000896349 | 0.081424478 | -1.14E-05 | 0.038707192 | 1 | 0.675668349 | 1 | 0.676208846 | ENSG0000026 | 2 | SUZ12P |
| VAT | T2DM | ENSG0000017 | 0.004749447 | 0.002484329 | 0.001569931 | 0.081435429 | 0.000821434 | 0.722620311 | 0.002314166 | 0.081424631 | 0.003928013 | 0.020877472 | 1 | 0.053310623 | 1 | 0.907346725 | ENSG0000026 | 2 | RP11-271K11.5 |
| VAT | T2DM | ENSG0000017 | -0.00146658 | 0.571954344 | 0.00259491 | 0.081436905 | -0.00140515 | 0.58814545 | 0.002594794 | 0.081424603 | -6.14E-05 | 0.00249068 | 1 | 0.842366056 | 1 | 0.852399212 | ENSG0000026 | 2 | RP11-271K11.5 |
| VAT | T2DM | ENSG0000017 | -0.00538447 | 5.81E-05 | 0.001339297 | 0.081434257 | -0.0043243 | 0.001876123 | 0.001390813 | 0.081423037 | -0.00106017 | 0.004735931 | 0.16039721 | 0.003341609 | 1 | 0.050765682 | ENSG0000026 | 1 | STRADA |
| VAT | T2DM | ENSG0000017 | -0.00727644 | 0.029275378 | 0.003338177 | 0.081436164 | -0.00707737 | 0.03400582 | 0.003338367 | 0.081423901 | -0.00019907 | 0.000243782 | 1 | 0.239053383 | 1 | 0.269701333 | ENSG0000026 | 1 | STRADA |
| VAT | T2DM | ENSG0000017 | -0.001225 | 0.584027037 | 0.002237387 | 0.081436908 | -0.00117074 | 0.600774704 | 0.002237289 | 0.081424607 | -5.43E-05 | 0.002302337 | 1 | 0.848256181 | 1 | 0.856779751 | ENSG0000026 | 2 | SNORA76 |
| VAT | T2DM | ENSG0000017 | -0.00107241 | 0.6604396 | 0.002441151 | 0.081436926 | -0.00112517 | 0.644845551 | 0.002441051 | 0.081424617 | 5.28E-05 | 0.009633378 | 1 | 0.880097732 | 1 | 0.87456323 | ENSG0000026 | 2 | SNORA76 |
| VAT | T2DM | ENSG0000017 | -0.00330756 | 0.0653984 | 0.00179512 | 0.081436391 | -0.00334763 | 0.062189343 | 0.001795027 | 0.081424071 | 4.01E-05 | 0.001248105 | 1 | 0.364645623 | 1 | 0.365363373 | ENSG0000026 | 2 | SNORA76 |
| VAT | T2DM | ENSG0000017 | 0.002241179 | 0.229362655 | 0.001864536 | 0.081436717 | 0.002209538 | 0.235983313 | 0.001864451 | 0.081424418 | 3.16E-05 | 0.028222783 | 1 | 0.612920192 | 1 | 0.622076357 | ENSG0000026 | 2 | MYO15B |
| VAT | T2DM | ENSG0000017 | 0.006560894 | 0.004024123 | 0.002281036 | 0.081435576 | 0.006261813 | 0.006076929 | 0.002282324 | 0.081423395 | 0.00029908 | 0.000246118 | 1 | 0.072120646 | 1 | 0.100155459 | ENSG0000026 | 2 | MYO15B |
| VAT | T2DM | ENSG0000017 | 0.00660909 | 0.001174829 | 0.002036737 | 0.081435199 | 0.005744515 | 0.005144059 | 0.002053164 | 0.081423344 | 0.000864575 | 0.000898258 | 1 | 0.032752796 | 1 | 0.090430593 | ENSG0000026 | 1 | LINC00663 |
| VAT | T2DM | ENSG0000017 | -0.00320875 | 0.185908655 | 0.002425762 | 0.081436666 | -0.00317226 | 0.190940703 | 0.002425639 | 0.081424367 | -3.65E-05 | 0.033273468 | 1 | 0.568225789 | 1 | 0.573647304 | ENSG0000026 | 1 | LINC00663 |
| VAT | T2DM | ENSG0000017 | -3.10E-05 | 0.987072486 | 0.001910772 | 0.081436958 | -0.00043239 | 0.821247186 | 0.001913745 | 0.081424644 | 0.000401434 | 0.00023679 | 1 | 0.997544462 | 1 | 0.94480438 | ENSG0000026 | 1 | LINC00663 |
| VAT | T2DM | ENSG0000017 | 0.001028527 | 0.577977488 | 0.001848732 | 0.081436907 | 0.0010603 | 0.566268951 | 0.001848645 | 0.081424598 | -3.18E-05 | 0.023162141 | 1 | 0.844236594 | 1 | 0.843444309 | ENSG0000026 | 2 | CTD-2085J24.4 |
| VAT | T2DM | ENSG0000017 | 0.004058434 | 0.271548597 | 0.003691148 | 0.081436756 | 0.004134885 | 0.262607517 | 0.003691036 | 0.081424443 | -7.65E-05 | 0.029170325 | 1 | 0.651395653 | 1 | 0.643691606 | ENSG0000026 | 2 | CTD-2085J24.4 |
| VAT | T2DM | ENSG0000017 | -0.0017164 | 0.022517789 | 0.000752304 | 0.081436089 | -0.00170597 | 0.023343737 | 0.000752266 | 0.081423793 | -1.04E-05 | 0.049138952 | 1 | 0.207163659 | 1 | 0.220256845 | ENSG0000026 | 1 | DHX40P1 |
| VAT | T2DM | ENSG0000017 | -0.00218871 | 0.429743986 | 0.002771814 | 0.081436854 | -0.00225476 | 0.41593442 | 0.002771687 | 0.081424542 | 6.60E-05 | 0.002040576 | 1 | 0.771654721 | 1 | 0.764134655 | ENSG0000026 | 1 | DHX40P1 |
| VAT | T2DM | ENSG0000017 | -0.00396177 | 0.136075901 | 0.002657894 | 0.081436587 | -0.004144 | 0.119012166 | 0.002658223 | 0.081424246 | 0.000182231 | 0.000600568 | 1 | 0.500073799 | 1 | 0.472623853 | ENSG0000026 | 1 | DHX40P1 |
| VAT | T2DM | ENSG0000017 | -0.00395104 | 0.093252178 | 0.002353923 | 0.081436488 | -0.0038931 | 0.098141375 | 0.002353842 | 0.081424195 | -5.79E-05 | 0.006563354 | 1 | 0.41970912 | 1 | 0.435146086 | ENSG0000026 | 1 | RP11-703I16.3 |
| VAT | T2DM | ENSG0000017 | 0.001096445 | 0.215578693 | 0.000885392 | 0.081436702 | 0.001118444 | 0.206491537 | 0.000885354 | 0.081424386 | -2.20E-05 | 0.0019736 | 1 | 0.597482277 | 1 | 0.588001967 | ENSG0000026 | 1 | RP11-703I16.3 |
| VAT | T2DM | ENSG0000017 | -0.0026326 | 0.056890062 | 0.001382554 | 0.081436352 | -0.00267016 | 0.053433851 | 0.0013825 | 0.081424029 | 3.76E-05 | 0.001499277 | 1 | 0.336223918 | 1 | 0.335175972 | ENSG0000026 | 1 | RP11-703I16.3 |
| VAT | T2DM | ENSG0000017 | -0.00653649 | 0.005082287 | 0.002332973 | 0.081435647 | -0.00657131 | 0.004849728 | 0.002332846 | 0.081423327 | 3.48E-05 | 0.022294609 | 1 | 0.081154081 | 1 | 0.087485285 | ENSG0000026 | 2 | AC005336.4 |
| VAT | T2DM | ENSG0000017 | -0.00145564 | 0.373020615 | 0.001634031 | 0.081436826 | 0.000545116 | 0.746157267 | 0.00168396 | 0.081424635 | -0.00200076 | 9.12E-07 | 1 | 0.731205182 | 1 | 0.911123584 | ENSG0000026 | 1 | RP11-15A1.3 |
| VAT | T2DM | ENSG0000017 | 0.004558974 | 0.265272641 | 0.004092375 | 0.081436751 | 0.004618806 | 0.259027563 | 0.004092169 | 0.08142444 | -5.98E-05 | 0.039851278 | 1 | 0.645637115 | 1 | 0.639459815 | ENSG0000026 | 1 | RP11-15A1.3 |
| VAT | T2DM | ENSG0000017 | 0.000745082 | 0.46005057 | 0.001008552 | 0.081436867 | 0.000728381 | 0.47014802 | 0.001008505 | 0.081424565 | 1.67E-05 | 0.02943191 | 1 | 0.78904441 | 1 | 0.794121331 | ENSG0000026 | 1 | RP11-686D22.7 |
| VAT | T2DM | ENSG0000017 | 0.002295443 | 0.534116595 | 0.003691998 | 0.081436894 | 0.002359265 | 0.522789484 | 0.003691821 | 0.081424584 | -6.38E-05 | 0.019978109 | 1 | 0.81975639 | 1 | 0.820071559 | ENSG0000026 | 1 | RP11-686D22.7 |
| VAT | T2DM | ENSG0000017 | -0.01121443 | 0.005720051 | 0.004058164 | 0.081435682 | -0.0112711 | 0.005477625 | 0.004057959 | 0.081423363 | 5.67E-05 | 0.048840188 | 1 | 0.087222876 | 1 | 0.093902135 | ENSG0000026 | 1 | RP11-686D22.7 |
| VAT | T2DM | ENSG0000017 | -0.00200344 | 0.088699928 | 0.001176902 | 0.081436474 | -0.00194986 | 0.09755678 | 0.001176872 | 0.081424194 | -5.36E-05 | 4.82E-06 | 1 | 0.407340768 | 1 | 0.435146086 | ENSG0000026 | 1 | RP11-861E21.1 |

| | | | | | | | | | | | | | | | | | | | |
|---|---|---|---|---|---|---|---|---|---|---|---|---|---|---|---|---|---|---|---|
| VAT | T2DM | ENSG0000017 | -0.00199038 | 0.281647002 | 0.001848713 | 0.081436765 | -0.00203074 | 0.271980043 | 0.001848625 | 0.081424451 | 4.04E-05 | 0.003467286 | 1 | 0.658209759 | 1 | 0.648137145 | ENSG0000026 | 1 | RP11-861E21.1 |
| VAT | T2DM | ENSG0000017 | -0.00012934 | 0.945583595 | 0.001894974 | 0.081436958 | -0.0001665 | 0.929982779 | 0.001894886 | 0.081424651 | 3.72E-05 | 0.009585429 | 1 | 0.987266065 | 1 | 0.986453678 | ENSG0000026 | 1 | RP11-861E21.1 |
| VAT | T2DM | ENSG0000017 | 0.009704183 | 0.000603449 | 0.00282914 | 0.081434992 | 0.009638938 | 0.000656413 | 0.002829001 | 0.081422713 | 6.52E-05 | 0.001493045 | 1 | 0.019594341 | 1 | 0.023226937 | ENSG0000026 | 1 | CTD-2528L19.6 |
| VAT | T2DM | ENSG0000017 | -0.00167486 | 0.683048695 | 0.00410197 | 0.081436931 | -0.00156062 | 0.703603213 | 0.004101927 | 0.081424628 | -0.00011424 | 0.014499371 | 1 | 0.891775969 | 1 | 0.90021629 | ENSG0000026 | 1 | RP11-178C3.2 |
| VAT | T2DM | ENSG0000017 | 0.00499844 | 0.172478345 | 0.003663779 | 0.081436647 | 0.005056614 | 0.167516674 | 0.003663604 | 0.081424334 | -5.82E-05 | 0.03372965 | 1 | 0.547323832 | 1 | 0.54753664 | ENSG0000026 | 1 | RP11-178C3.2 |
| VAT | T2DM | ENSG0000017 | 0.00187995 | 0.399803503 | 0.002232791 | 0.08143684 | 0.001917499 | 0.39043559 | 0.002232692 | 0.081424529 | -3.75E-05 | 0.032428773 | 1 | 0.750041208 | 1 | 0.748625 | ENSG0000026 | 1 | RP11-773H22.2 |
| VAT | T2DM | ENSG0000018 | -0.00216514 | 0.467228848 | 0.0029782 | 0.08143687 | -0.00210507 | 0.479655445 | 0.002978072 | 0.081424569 | -6.01E-05 | 0.012409597 | 1 | 0.789199279 | 1 | 0.798561663 | ENSG0000026 | 1 | EIF5AP3 |
| VAT | T2DM | ENSG0000018 | -0.00328495 | 0.32521635 | 0.003339056 | 0.081436797 | -0.003223 | 0.334405001 | 0.003338927 | 0.081424497 | -6.20E-05 | 0.030438402 | 1 | 0.693804654 | 1 | 0.702906909 | ENSG0000026 | 1 | EIF5AP3 |
| VAT | T2DM | ENSG0000018 | 0.000623792 | 0.866532917 | 0.003711633 | 0.081436954 | 0.000683663 | 0.85385447 | 0.003711471 | 0.081424647 | -5.99E-05 | 0.043422307 | 1 | 0.962039763 | 1 | 0.958443285 | ENSG0000026 | 1 | RP11-1094M14.8 |
| VAT | T2DM | ENSG0000018 | -0.00389573 | 0.325116247 | 0.003959074 | 0.081436797 | -0.00400972 | 0.311141203 | 0.003958918 | 0.081424481 | 0.00011399 | 0.000716584 | 1 | 0.693804654 | 1 | 0.681547397 | ENSG0000026 | 1 | CTC-296K1.4 |
| VAT | T2DM | ENSG0000018 | -0.00386533 | 0.253611722 | 0.003385824 | 0.081436741 | -0.0039208 | 0.246840845 | 0.003385671 | 0.081424428 | 5.55E-05 | 0.035887114 | 1 | 0.635542337 | 1 | 0.630010905 | ENSG0000026 | 1 | CTD-2554C21.1 |
| VAT | T2DM | ENSG0000018 | 0.001526359 | 0.618139523 | 0.003061971 | 0.081436917 | 0.001585533 | 0.604575337 | 0.003061864 | 0.081424608 | -5.92E-05 | 0.032191808 | 1 | 0.864706073 | 1 | 0.858231221 | ENSG0000026 | 1 | CTC-296K1.3 |
| VAT | T2DM | ENSG0000018 | -0.00062577 | 0.791395621 | 0.002365854 | 0.081436947 | -0.0017561 | 0.46020231 | 0.002377878 | 0.081424561 | 0.001130331 | 2.63E-06 | 1 | 0.932507176 | 1 | 0.791032387 | ENSG0000026 | 1 | ZNF285 |
| VAT | T2DM | ENSG0000018 | -0.00388772 | 0.133206433 | 0.002589089 | 0.081436582 | -0.00395992 | 0.126132575 | 0.002588971 | 0.081424262 | 7.22E-05 | 0.000299657 | 1 | 0.495484846 | 1 | 0.482945681 | ENSG0000026 | 1 | ZNF285 |
| VAT | T2DM | ENSG0000018 | 0.004514159 | 0.275011384 | 0.004135377 | 0.081436759 | 0.004594121 | 0.266576886 | 0.004135191 | 0.081424446 | -8.00E-05 | 0.013673609 | 1 | 0.653812443 | 1 | 0.647099564 | ENSG0000026 | 1 | S1PR2 |
| VAT | T2DM | ENSG0000018 | -0.00447101 | 0.229194608 | 0.003718286 | 0.081436717 | -0.0045888 | 0.217137675 | 0.003718105 | 0.081424398 | 0.000117791 | 1.57E-05 | 1 | 0.612920192 | 1 | 0.598701283 | ENSG0000026 | 1 | RP11-686D22.4 |
| VAT | T2DM | ENSG0000018 | -0.00262856 | 0.356970054 | 0.00285355 | 0.081436817 | -0.00259276 | 0.363530117 | 0.002853391 | 0.081424515 | -3.58E-05 | 0.045969101 | 1 | 0.714526689 | 1 | 0.72339086 | ENSG0000026 | 1 | CTC-507E2.2 |
| VAT | T2DM | ENSG0000018 | -0.00038743 | 0.682668156 | 0.000947673 | 0.08143693 | -0.00040764 | 0.667069499 | 0.000947628 | 0.081424622 | 2.02E-05 | 0.003940563 | 1 | 0.891700952 | 1 | 0.885149913 | ENSG0000026 | 5 | CYP4F24P |
| VAT | T2DM | ENSG0000018 | -0.00139636 | 0.663984629 | 0.003214327 | 0.081436927 | -0.00133714 | 0.677399667 | 0.003214172 | 0.081424624 | -5.92E-05 | 0.01252282 | 1 | 0.882329117 | 1 | 0.89117459 | ENSG0000026 | 5 | CYP4F24P |
| VAT | T2DM | ENSG0000018 | -0.00129299 | 0.484390093 | 0.001849077 | 0.081436877 | 0.000176993 | 0.924557192 | 0.001869083 | 0.081424651 | -0.00146998 | 7.82E-08 | 1 | 0.792012238 | 1 | 0.982969566 | ENSG0000026 | 1 | RP11-686D22.3 |
| VAT | T2DM | ENSG0000018 | 0.002390643 | 0.393574311 | 0.002802123 | 0.081436837 | 0.002341326 | 0.403382674 | 0.002801983 | 0.081424536 | 4.93E-05 | 0.013882666 | 1 | 0.747385518 | 1 | 0.756008271 | ENSG0000026 | 1 | RP11-686D22.3 |
| VAT | T2DM | ENSG0000018 | 0.004253959 | 0.170735285 | 0.003105426 | 0.081436645 | 0.004202366 | 0.175963573 | 0.003105296 | 0.081424346 | 5.16E-05 | 0.042021078 | 1 | 0.546218791 | 1 | 0.549954486 | ENSG0000026 | 1 | RP11-686D22.3 |
| VAT | T2DM | ENSG0000018 | -0.00138546 | 0.35086044 | 0.001485071 | 0.081436813 | -0.00142276 | 0.3380373 | 0.001485055 | 0.081424499 | 3.73E-05 | 0.027008054 | 1 | 0.71151713 | 1 | 0.703807164 | ENSG0000026 | 1 | RP11-95O2.1 |
| VAT | T2DM | ENSG0000018 | -7.97E-05 | 0.986357846 | 0.004661487 | 0.081436958 | 1.35E-05 | 0.997690137 | 0.004661282 | 0.081424652 | -9.32E-05 | 0.011864667 | 1 | 0.997544462 | 1 | 0.999423969 | ENSG0000026 | 1 | RP1-193H18.3 |
| VAT | T2DM | ENSG0000018 | -0.00632724 | 0.117449146 | 0.004041485 | 0.081436549 | -0.00640946 | 0.112742098 | 0.0040413 | 0.081424232 | 8.22E-05 | 0.008381607 | 1 | 0.4637477 | 1 | 0.458589157 | ENSG0000026 | 1 | AC004156.3 |
| VAT | T2DM | ENSG0000018 | -0.00225508 | 0.078410497 | 0.001281305 | 0.081436441 | -0.00222435 | 0.082549352 | 0.001281247 | 0.081424149 | -3.07E-05 | 0.001986397 | 1 | 0.390036819 | 1 | 0.407209212 | ENSG0000026 | 1 | RP11-49K24.5 |
| VAT | T2DM | ENSG0000018 | -0.00462595 | 0.070001638 | 0.00255309 | 0.08143641 | -0.0046847 | 0.066506175 | 0.002552969 | 0.08142409 | 5.87E-05 | 0.002264717 | 1 | 0.369646805 | 1 | 0.374084884 | ENSG0000026 | 1 | CTD-2325M2.1 |
| VAT | T2DM | ENSG0000018 | 8.58E-06 | 0.997349668 | 0.002584473 | 0.081436958 | 4.72E-05 | 0.985429442 | 0.002584339 | 0.081424652 | -3.86E-05 | 0.029990485 | 1 | 0.998501236 | 1 | 0.995445662 | ENSG0000026 | 1 | PRED62 |
| VAT | T2DM | ENSG0000018 | 0.002201326 | 0.235357383 | 0.001855041 | 0.081436723 | 0.002143171 | 0.247943154 | 0.00185498 | 0.081424429 | 5.82E-05 | 0.000681527 | 1 | 0.620857424 | 1 | 0.630010905 | ENSG0000026 | 1 | AC004466.1 |
| VAT | T2DM | ENSG0000018 | -0.00333955 | 0.169347973 | 0.002429994 | 0.081436643 | -0.00441302 | 0.074415158 | 0.002473591 | 0.081424121 | 0.001073474 | 0.020514624 | 1 | 0.546218791 | 1 | 0.386790651 | ENSG0000026 | 1 | AC004466.1 |
| VAT | T2DM | ENSG0000018 | -0.00066815 | 0.552120377 | 0.001123722 | 0.081436899 | -0.0006473 | 0.564577429 | 0.001123669 | 0.081424597 | -2.09E-05 | 0.013783736 | 1 | 0.829533065 | 1 | 0.842744027 | ENSG0000026 | 1 | CTC-429P9.2 |
| VAT | T2DM | ENSG0000018 | -0.00264463 | 0.203977727 | 0.00208189 | 0.081436689 | -0.00270801 | 0.193328933 | 0.002081812 | 0.08142437 | 6.34E-05 | 0.000505954 | 1 | 0.582793504 | 1 | 0.574986912 | ENSG0000026 | 1 | CTC-429P9.2 |
| VAT | T2DM | ENSG0000018 | -0.00392025 | 0.074819434 | 0.002200444 | 0.081436428 | -0.00404505 | 0.066091244 | 0.002201019 | 0.081424088 | 0.000124798 | 0.02888537 | 1 | 0.379598598 | 1 | 0.374084884 | ENSG0000026 | 1 | VN1R84P |
| VAT | T2DM | ENSG0000018 | 0.001595951 | 0.631708313 | 0.003329586 | 0.08143692 | 0.001536764 | 0.644391304 | 0.003329441 | 0.081424617 | 5.92E-05 | 0.026618279 | 1 | 0.870017438 | 1 | 0.87456323 | ENSG0000026 | 1 | VN1R84P |
| VAT | T2DM | ENSG0000018 | -0.00058028 | 0.75376862 | 0.001849952 | 0.081436942 | -0.00062911 | 0.733793418 | 0.001849864 | 0.081424633 | 4.88E-05 | 0.000421937 | 1 | 0.918605915 | 1 | 0.908589792 | ENSG0000026 | 1 | CTD-2331H12.5 |
| VAT | T2DM | ENSG0000018 | 0.003742074 | 0.05853645 | 0.001978193 | 0.08143636 | 0.003704727 | 0.061086421 | 0.001978098 | 0.081424066 | 3.73E-05 | 0.010961929 | 1 | 0.343016143 | 1 | 0.362577469 | ENSG0000026 | 1 | CTD-2331H12.5 |
| VAT | T2DM | ENSG0000018 | 0.000406942 | 0.655421745 | 0.000911929 | 0.081436925 | 0.000386261 | 0.671868997 | 0.000911885 | 0.081424623 | 2.07E-05 | 0.002470927 | 1 | 0.876436054 | 1 | 0.888954186 | ENSG0000026 | 1 | CTD-2331H12.5 |
| VAT | T2DM | ENSG0000018 | -0.00414699 | 0.212872641 | 0.003329023 | 0.081436699 | -0.00410216 | 0.21783506 | 0.003328846 | 0.081424399 | -4.48E-05 | 0.044216591 | 1 | 0.594063185 | 1 | 0.598987824 | ENSG0000026 | 1 | CTD-2331H12.5 |
| VAT | T2DM | ENSG0000018 | 0.000744383 | 0.710746156 | 0.00200721 | 0.081436935 | 0.000793818 | 0.692476476 | 0.002007141 | 0.081424626 | -4.94E-05 | 0.006847204 | 1 | 0.903574108 | 1 | 0.894473859 | ENSG0000026 | 1 | RP3-477O4.16 |
| VAT | T2DM | ENSG0000018 | -0.0046872 | 0.102124556 | 0.002867416 | 0.081436512 | -0.00458385 | 0.109894606 | 0.002867293 | 0.081424225 | -0.00010335 | 8.16E-06 | 1 | 0.432306403 | 1 | 0.452700168 | ENSG0000026 | 1 | RP3-477O4.16 |
| VAT | T2DM | ENSG0000018 | -0.00202216 | 0.054213351 | 0.0010504 | 0.081436339 | -0.00200431 | 0.05636207 | 0.001050351 | 0.081424044 | -1.78E-05 | 0.026250087 | 1 | 0.330306509 | 1 | 0.344116711 | ENSG0000026 | 1 | RP3-477O4.16 |
| VAT | T2DM | ENSG0000018 | -0.0012392 | 0.364822822 | 0.001367443 | 0.081436821 | -0.00126679 | 0.354223928 | 0.001367387 | 0.081424509 | 2.76E-05 | 0.015106765 | 1 | 0.722836316 | 1 | 0.714662311 | ENSG0000026 | 1 | RP3-477O4.16 |
| VAT | T2DM | ENSG0000018 | -0.00405596 | 0.091375513 | 0.00240255 | 0.081436482 | -0.00414212 | 0.084711796 | 0.002402649 | 0.081424156 | 8.62E-05 | 0.018953536 | 1 | 0.414115625 | 1 | 0.410615651 | ENSG0000026 | 1 | CTD-2017D11.1 |
| VAT | T2DM | ENSG0000018 | -9.37E-05 | 0.875358105 | 0.000597244 | 0.081436954 | 0.00092382 | 0.139215517 | 0.000624742 | 0.081424287 | -0.0010175 | 2.92E-08 | 1 | 0.966383933 | 1 | 0.507843495 | ENSG0000026 | 1 | CTD-2017D11.1 |
| VAT | T2DM | ENSG0000018 | -0.00286265 | 0.041048663 | 0.00140117 | 0.081436261 | -0.00284017 | 0.042653241 | 0.001401106 | 0.081423966 | -2.25E-05 | 0.035701906 | 1 | 0.284233402 | 1 | 0.294307366 | ENSG0000026 | 1 | CTD-2545M3.8 |
| VAT | T2DM | ENSG0000018 | 0.000918154 | 0.620934681 | 0.001856634 | 0.081436918 | 0.000970936 | 0.600989992 | 0.001856553 | 0.081424607 | -5.28E-05 | 0.000403935 | 1 | 0.867297429 | 1 | 0.856779751 | ENSG0000026 | 1 | LINC00664 |
| VAT | T2DM | ENSG0000018 | -0.0032691 | 0.317061473 | 0.003267418 | 0.081436791 | -0.00334899 | 0.305359717 | 0.003267277 | 0.081424477 | 7.99E-05 | 0.002369291 | 1 | 0.688402356 | 1 | 0.676208846 | ENSG0000026 | 1 | LINC00664 |
| VAT | T2DM | ENSG0000018 | 0.001157563 | 0.756528169 | 0.003733553 | 0.081436942 | 0.001228384 | 0.742134858 | 0.003733375 | 0.081424634 | -7.08E-05 | 0.010934031 | 1 | 0.919012884 | 1 | 0.911123584 | ENSG0000026 | 1 | CTB-61M7.2 |
| VAT | T2DM | ENSG0000018 | -0.00915993 | 0.008331577 | 0.003471855 | 0.081435795 | -0.00921434 | 0.007951463 | 0.003471691 | 0.081423475 | 5.44E-05 | 0.037463263 | 1 | 0.111226029 | 1 | 0.117236996 | ENSG0000026 | 1 | AC011298.1 |
| VAT | T2DM | ENSG0000018 | 0.00100247 | 0.264383714 | 0.000898197 | 0.08143675 | 0.000457822 | 0.613934576 | 0.000907539 | 0.08142461 | 0.000544648 | 2.93E-05 | 1 | 0.645180415 | 1 | 0.864265654 | ENSG0000026 | 1 | AC011298.1 |
| VAT | T2DM | ENSG0000018 | -0.0001688 | 0.902424451 | 0.001376806 | 0.081436956 | -0.00013994 | 0.91903927 | 0.001376748 | 0.081424651 | -2.89E-05 | 0.010114281 | 1 | 0.974557156 | 1 | 0.981949202 | ENSG0000026 | 1 | AC011298.1 |
| VAT | T2DM | ENSG0000018 | 0.002568062 | 0.227280244 | 0.002126942 | 0.081436715 | 0.002533382 | 0.233597443 | 0.002126847 | 0.081424415 | 3.47E-05 | 0.037107477 | 1 | 0.612591282 | 1 | 0.619918077 | ENSG0000026 | 1 | CTD-2192J16.26 |
| VAT | T2DM | ENSG0000018 | -0.00203482 | 0.190410753 | 0.001554048 | 0.081436672 | -0.0020744 | 0.181922305 | 0.001554022 | 0.081424355 | 3.96E-05 | 0.019131007 | 1 | 0.572476773 | 1 | 0.557702654 | ENSG0000026 | 2 | CTC-523E23.5 |
| VAT | T2DM | ENSG0000018 | 0.000435232 | 0.90372513 | 0.003598234 | 0.081436956 | 0.000547297 | 0.879108544 | 0.003598286 | 0.081424649 | -0.00011207 | 0.020398391 | 1 | 0.974557156 | 1 | 0.96786083 | ENSG0000026 | 1 | RP4-614O4.12 |
| VAT | T2DM | ENSG0000018 | 0.002853342 | 0.339735988 | 0.002988769 | 0.081436806 | 0.002912705 | 0.329763388 | 0.002988642 | 0.081424494 | -5.94E-05 | 0.014488453 | 1 | 0.703429352 | 1 | 0.701227106 | ENSG0000026 | 1 | RP4-614O4.12 |
| VAT | T2DM | ENSG0000018 | -0.00516951 | 0.1072146 | 0.003209194 | 0.081436525 | -0.00511227 | 0.111141239 | 0.003209031 | 0.081424228 | -5.72E-05 | 0.01151296 | 1 | 0.442145471 | 1 | 0.45579468 | ENSG0000026 | 1 | LRRC3DN |
| VAT | T2DM | ENSG0000018 | 0.005373912 | 0.008592795 | 0.002044981 | 0.081435804 | 0.005324738 | 0.009216485 | 0.002044886 | 0.08142352 | 4.92E-05 | 0.001665263 | 1 | 0.112933871 | 1 | 0.128472216 | ENSG0000026 | 1 | LRRC3DN |
| VAT | T2DM | ENSG0000018 | -0.00208916 | 0.386393292 | 0.002411929 | 0.081436833 | -0.00212824 | 0.377547845 | 0.002411815 | 0.081424522 | 3.91E-05 | 0.031571917 | 1 | 0.741393167 | 1 | 0.736939216 | ENSG0000026 | 1 | LRRC3DN |
| VAT | T2DM | ENSG0000018 | -0.00624689 | 0.004829273 | 0.00221661 | 0.081435631 | -0.00322215 | 0.161314893 | 0.002300442 | 0.081424325 | -0.00302475 | 9.06E-07 | 1 | 0.079821197 | 1 | 0.537053745 | ENSG0000026 | 1 | LRRC3DN |
| VAT | T2DM | ENSG0000019 | -0.00118973 | 0.274432778 | 0.001088583 | 0.081436759 | -0.00121733 | 0.263433189 | 0.001088536 | 0.081424444 | 2.76E-05 | 0.00163439 | 1 | 0.653524131 | 1 | 0.643814606 | ENSG0000026 | 1 | AC006967.1 |
| VAT | T2DM | ENSG0000019 | -0.00150884 | 0.642466044 | 0.003250042 | 0.081436922 | -0.0013929 | 0.668216617 | 0.003249899 | 0.081424622 | -0.00011595 | 7.07E-06 | 1 | 0.874341025 | 1 | 0.885820299 | ENSG0000026 | 1 | CTC-360G5.9 |
| VAT | T2DM | ENSG0000019 | -0.00140056 | 0.54279 | 0.002301282 | 0.081436897 | -0.0014655 | 0.524228335 | 0.002301193 | 0.081424585 | 6.49E-05 | 0.00104336 | 1 | 0.823132089 | 1 | 0.820071559 | ENSG0000026 | 1 | CTC-360G5.9 |
| VAT | T2DM | ENSG0000019 | -0.00200209 | 0.604694697 | 0.00386757 | 0.081436914 | -0.00206753 | 0.592923726 | 0.003867398 | 0.081424605 | 6.54E-05 | 0.031808019 | 1 | 0.857656828 | 1 | 0.855446673 | ENSG0000026 | 1 | CTC-360G5.9 |
| VAT | T2DM | ENSG0000019 | 0.009418255 | 0.002533324 | 0.003119301 | 0.081435435 | 0.009479185 | 0.00237378 | 0.00311918 | 0.081423109 | -6.09E-05 | 0.022895975 | 1 | 0.053784425 | 1 | 0.057470467 | ENSG0000026 | 1 | AL031663.2 |
| VAT | T2DM | ENSG0000019 | -0.00401246 | 0.059041092 | 0.002125363 | 0.081436363 | -0.00406524 | 0.055774369 | 0.002125282 | 0.081424041 | 5.28E-05 | 0.004045911 | 1 | 0.345240286 | 1 | 0.344116711 | ENSG0000026 | 1 | AC104057.1 |
| VAT | T2DM | ENSG0000019 | -0.02069391 | 4.52E-14 | 0.0027426 | 0.081427445 | -0.01658716 | 9.85E-07 | 0.003388762 | 0.081420649 | -0.00410674 | 0.039105977 | 1.25E-10 | 2.49E-11 | 0.002717469 | 0.000143025 | ENSG0000026 | 1 | AC104057.1 |
| VAT | T2DM | ENSG0000019 | 1.54E-05 | 0.995050177 | 0.002481596 | 0.081436958 | 5.65E-05 | 0.981836342 | 0.002481468 | 0.081424652 | -4.11E-05 | 0.01697325 | 1 | 0.998465252 | 1 | 0.994447084 | ENSG0000026 | 1 | AC104057.1 |
| VAT | T2DM | ENSG0000019 | 0.000653283 | 0.746190843 | 0.002018383 | 0.081436941 | 0.000693913 | 0.730989771 | 0.002018307 | 0.081424633 | -4.06E-05 | 0.020935035 | 1 | 0.9165495 | 1 | 0.908589792 | ENSG0000026 | 2 | SNHG8 |
| VAT | T2DM | ENSG0000019 | 0.001372001 | 0.473720853 | 0.001915029 | 0.081436873 | 0.001271147 | 0.506852671 | 0.001915108 | 0.081424579 | 0.000100854 | 0.000564267 | 1 | 0.791889203 | 1 | 0.811434671 | ENSG0000026 | 2 | SNHG8 |
| VAT | T2DM | ENSG0000019 | 0.005584532 | 0.086108555 | 0.003253828 | 0.081436466 | 0.005386844 | 0.0979434 | 0.003255068 | 0.081424195 | 0.000197688 | 0.044290426 | 1 | 0.406073085 | 1 | 0.435146086 | ENSG0000026 | 1 | RP11-646I6.5 |
| VAT | T2DM | ENSG0000019 | -0.00399213 | 0.001218534 | 0.001234224 | 0.08143521 | -0.00202099 | 0.119574862 | 0.001298367 | 0.081424248 | -0.00197114 | 1.02E-06 | 1 | 0.033298544 | 1 | 0.472896409 | ENSG0000026 | 1 | RP11-646I6.5 |
| VAT | T2DM | ENSG0000019 | -0.00484889 | 0.163837137 | 0.003482686 | 0.081436634 | -0.00478199 | 0.169709862 | 0.003482523 | 0.081424337 | -6.69E-05 | 0.011288162 | 1 | 0.542275761 | 1 | 0.54753664 | ENSG0000026 | 1 | RP11-646I6.5 |
| VAT | T2DM | ENSG0000019 | 0.000343948 | 0.920295167 | 0.003437348 | 0.081436957 | 0.000287603 | 0.933315506 | 0.003437174 | 0.081424651 | 5.63E-05 | 0.02052491 | 1 | 0.980360785 | 1 | 0.987645456 | ENSG0000026 | 1 | RP11-849F2.8 |
| VAT | T2DM | ENSG0000019 | 0.001039151 | 0.588996446 | 0.001923317 | 0.08143691 | 0.001011594 | 0.598895243 | 0.001923218 | 0.081424606 | 2.76E-05 | 0.04081533 | 1 | 0.849780549 | 1 | 0.856779751 | ENSG0000026 | 1 | RP11-73M18.9 |
| VAT | T2DM | ENSG0000019 | 0.002855569 | 0.444505916 | 0.003734688 | 0.081436861 | 0.002941045 | 0.430971382 | 0.003734513 | 0.081424549 | -8.55E-05 | 0.002491034 | 1 | 0.779859126 | 1 | 0.776708652 | ENSG0000026 | 1 | RP11-73M18.9 |
| VAT | T2DM | ENSG0000019 | -0.00052134 | 0.760840071 | 0.001712806 | 0.081436943 | -0.0005639 | 0.741975339 | 0.001712736 | 0.081424634 | 4.26E-05 | 0.002845089 | 1 | 0.920907272 | 1 | 0.911123584 | ENSG0000026 | 1 | RP11-73M18.9 |
| VAT | T2DM | ENSG0000019 | 0.000407483 | 0.828501814 | 0.001881065 | 0.081436951 | 0.007104649 | 0.045780827 | 0.003556924 | 0.081423986 | -0.00669717 | 0.026527642 | 1 | 0.945851136 | 1 | 0.305944511 | ENSG0000026 | 1 | RP11-497H16.9 |
| VAT | T2DM | ENSG0000019 | -0.00537273 | 0.015498578 | 0.002219655 | 0.081435979 | -0.002682 | 0.238506809 | 0.002275331 | 0.08142442 | -0.00269073 | 7.82E-08 | 1 | 0.165798744 | 1 | 0.623960942 | ENSG0000026 | 1 | RP11-497H16.9 |
| VAT | T2DM | ENSG0000019 | -0.00268444 | 0.106793233 | 0.00166448 | 0.081436524 | -0.00272099 | 0.102091083 | 0.001664417 | 0.081424206 | 3.66E-05 | 0.011383282 | 1 | 0.442145471 | 1 | 0.441647944 | ENSG0000026 | 1 | CTD-3074O7.12 |
| VAT | T2DM | ENSG0000019 | 0.002782361 | 0.289713914 | 0.002627967 | 0.081436771 | -0.00069185 | 0.798142397 | 0.002705153 | 0.081424642 | 0.00347421 | 6.37E-08 | 1 | 0.665288122 | 1 | 0.935209552 | ENSG0000026 | 1 | CTD-3074O7.12 |
| VAT | T2DM | ENSG0000019 | 0.00189035 | 0.451738033 | 0.002512022 | 0.081436864 | 0.001302178 | 0.604719304 | 0.002515673 | 0.081424608 | 0.000588172 | 2.30E-05 | 1 | 0.78446312 | 1 | 0.858231221 | ENSG0000026 | 1 | CTD-3074O7.12 |
| VAT | T2DM | ENSG0000019 | 0.00171559 | 0.229310339 | 0.001427115 | 0.081436717 | 0.001749274 | 0.220277029 | 0.001427054 | 0.081424401 | -3.37E-05 | 0.003681719 | 1 | 0.612920192 | 1 | 0.601178278 | ENSG0000026 | 1 | CTD-3074O7.12 |
| VAT | T2DM | ENSG0000019 | -0.00038218 | 0.925905592 | 0.004109588 | 0.081436957 | -0.00024743 | 0.951987252 | 0.004109404 | 0.081424652 | -0.00013475 | 3.06E-05 | 1 | 0.98250651 | 1 | 0.988354266 | ENSG0000027 | 1 | RP3-462E2.5 |
| VAT | T2DM | ENSG0000019 | -0.00276266 | 0.359949419 | 0.003017773 | 0.081436818 | -0.0026959 | 0.371659672 | 0.003017681 | 0.081424519 | -6.68E-05 | 0.01973634 | 1 | 0.717818205 | 1 | 0.731049227 | ENSG0000027 | 1 | RP11-463O9.9 |
| VAT | T2DM | ENSG0000019 | 0.001948454 | 0.535514772 | 0.003144645 | 0.081436894 | 0.001888496 | 0.548123374 | 0.00314448 | 0.081424592 | 6.00E-05 | 0.00519223 | 1 | 0.81975639 | 1 | 0.830883001 | ENSG0000027 | 1 | RP11-463O9.9 |
| VAT | T2DM | ENSG0000019 | -0.00180813 | 0.422346656 | 0.002253519 | 0.081436851 | -0.0017684 | 0.432587312 | 0.002253397 | 0.08142455 | -3.97E-05 | 0.007045031 | 1 | 0.767538154 | 1 | 0.776799597 | ENSG0000027 | 1 | RP11-463O9.9 |
| VAT | T2DM | ENSG0000019 | -0.00105981 | 0.532177826 | 0.001696551 | 0.081436893 | -0.00109173 | 0.519878472 | 0.001696461 | 0.081424583 | 3.19E-05 | 0.004920231 | 1 | 0.81975639 | 1 | 0.817922279 | ENSG0000027 | 1 | RP11-326C3.13 |
| VAT | T2DM | ENSG0000019 | -0.00078156 | 0.639241351 | 0.001667286 | 0.081436922 | -0.00074778 | 0.653777582 | 0.001667215 | 0.081424619 | -3.38E-05 | 0.012694987 | 1 | 0.87299478 | 1 | 0.875674196 | ENSG0000027 | 1 | RP11-326C3.13 |
| VAT | T2DM | ENSG0000019 | -0.00291347 | 0.085344766 | 0.001693397 | 0.081436464 | -0.00288487 | 0.088440881 | 0.001693315 | 0.081424167 | -2.86E-05 | 0.021579832 | 1 | 0.404227064 | 1 | 0.417476427 | ENSG0000027 | 1 | AP001172.2 |
| VAT | T2DM | ENSG0000020 | -0.0044571 | 0.182767822 | 0.003345462 | 0.081436662 | -0.00452866 | 0.175823169 | 0.003345318 | 0.081424346 | 7.16E-05 | 0.008087502 | 1 | 0.562893955 | 1 | 0.549954486 | ENSG0000027 | 1 | RP5-935K16.1 |
| VAT | T2DM | ENSG0000020 | -0.00187076 | 0.2250304 | 0.001541922 | 0.081436712 | -0.0018232 | 0.237022084 | 0.001541868 | 0.081424419 | -4.76E-05 | 0.000610411 | 1 | 0.610120125 | 1 | 0.623029477 | ENSG0000027 | 1 | AP000473.8 |
| VAT | T2DM | ENSG0000020 | 0.004524843 | 0.044370541 | 0.002250518 | 0.081436283 | 0.003841721 | 0.088547851 | 0.002255707 | 0.081424168 | 0.000683122 | 1.10E-05 | 1 | 0.294381476 | 1 | 0.417476427 | ENSG0000027 | 1 | AP000473.8 |
| VAT | T2DM | ENSG0000020 | 0.004301517 | 0.121878181 | 0.00278066 | 0.081436559 | 0.001705896 | 0.547927961 | 0.002839051 | 0.081424592 | 0.002595621 | 6.09E-06 | 1 | 0.475118335 | 1 | 0.830883001 | ENSG0000027 | 1 | RP11-526I2.5 |
| VAT | T2DM | ENSG0000020 | 0.003852932 | 0.090086692 | 0.002273191 | 0.081436478 | 0.003791548 | 0.095315364 | 0.002273104 | 0.081424188 | 6.14E-05 | 0.001660287 | 1 | 0.410675884 | 1 | 0.433697593 | ENSG0000027 | 1 | RP11-526I2.5 |
| VAT | T2DM | ENSG0000020 | -0.00015054 | 0.923644597 | 0.001570671 | 0.081436957 | -0.00262856 | 0.106235706 | 0.00162724 | 0.081424216 | 0.002478022 | 5.89E-09 | 1 | 0.9816169 | 1 | 0.447649694 | ENSG0000027 | 1 | RP11-526I2.5 |
| VAT | T2DM | ENSG0000020 | 0.008198813 | 0.005505604 | 0.0029536 | 0.081435671 | 0.002221032 | 0.476681939 | 0.003120961 | 0.081424568 | 0.005977781 | 3.12E-09 | 1 | 0.086337881 | 1 | 0.795911768 | ENSG0000027 | 1 | RP11-526I2.5 |
| VAT | T2DM | ENSG0000020 | -0.00101042 | 0.387843682 | 0.001170101 | 0.081436834 | 0.000902502 | 0.473027346 | 0.001257733 | 0.081424566 | -0.00191293 | 3.39E-05 | 1 | 0.741792098 | 1 | 0.795041781 | ENSG0000027 | 1 | RP11-526I2.5 |
| VAT | T2DM | ENSG0000020 | -0.00046854 | 0.820866094 | 0.002069233 | 0.08143695 | -0.00371021 | 0.088229315 | 0.002176311 | 0.081424167 | 0.003241672 | 1.55E-06 | 1 | 0.943325602 | 1 | 0.417476427 | ENSG0000027 | 1 | RP11-526I2.5 |
| VAT | T2DM | ENSG0000020 | -0.01000401 | 8.48E-06 | 0.002246737 | 0.081433645 | -0.00603231 | 0.015397448 | 0.002489706 | 0.081423672 | -0.0039717 | 0.000214469 | 0.02341086 | 0.000719735 | 1 | 0.176596367 | ENSG0000027 | 1 | RP11-526I2.5 |
| VAT | T2DM | ENSG0000020 | 5.27E-05 | 0.949552195 | 0.000832356 | 0.081436958 | -0.00034931 | 0.677451231 | 0.000839799 | 0.081424624 | 0.000401972 | 0.000333023 | 1 | 0.987266065 | 1 | 0.89117459 | ENSG0000027 | 1 | RP11-526I2.5 |
| VAT | T2DM | ENSG0000020 | -0.00100313 | 0.475082455 | 0.001404484 | 0.081436873 | -0.00122364 | 0.384256624 | 0.001406355 | 0.081424526 | 0.000220511 | 0.003096543 | 1 | 0.791889203 | 1 | 0.742680871 | ENSG0000027 | 1 | RP11-526I2.5 |
| VAT | T2DM | ENSG0000020 | -0.00228998 | 0.033193888 | 0.001075243 | 0.0814362 | -0.00392544 | 0.000707124 | 0.001159033 | 0.081422736 | 0.001635456 | 0.000158144 | 1 | 0.255105869 | 1 | 0.024704568 | ENSG0000027 | 1 | RP1-170O19.21 |
| VAT | T2DM | ENSG0000020 | 0.000132143 | 0.834712245 | 0.000633287 | 0.081436951 | -0.00069193 | 0.288497487 | 0.00065189 | 0.081424464 | 0.000824075 | 1.02E-07 | 1 | 0.947236318 | 1 | 0.664345628 | ENSG0000027 | 1 | RP1-170O19.21 |
| VAT | T2DM | ENSG0000020 | 0.002185612 | 0.008064836 | 0.00082496 | 0.081435785 | 0.000443295 | 0.62605056 | 0.000909708 | 0.081424613 | 0.001742317 | 5.55E-06 | 1 | 0.109112491 | 1 | 0.869014343 | ENSG0000027 | 1 | RP11-453E17.3 |
| VAT | T2DM | ENSG0000020 | 0.00263825 | 0.001688851 | 0.000840173 | 0.081435311 | 0.002239813 | 0.008696988 | 0.000853668 | 0.081423502 | 0.000398437 | 0.008552134 | 1 | 0.042374798 | 1 | 0.122996468 | ENSG0000027 | 1 | RP11-453E17.3 |
| VAT | T2DM | ENSG0000020 | 0.010752177 | 0.000207822 | 0.002898675 | 0.081434659 | 0.010150218 | 0.000474877 | 0.002904573 | 0.081422612 | 0.00060196 | 0.001391707 | 0.573588307 | 0.008962317 | 1 | 0.018940823 | ENSG0000027 | 1 | RP11-381K20.5 |
| VAT | T2DM | ENSG0000020 | 0.004228911 | 0.041426594 | 0.002073768 | 0.081436263 | 0.001285603 | 0.545316619 | 0.00212569 | 0.081424591 | 0.002943308 | 3.09E-10 | 1 | 0.284233402 | 1 | 0.830754257 | ENSG0000027 | 1 | RP1-90J20.12 |
| VAT | T2DM | ENSG0000020 | 0.000502393 | 0.883662767 | 0.003433339 | 0.081436955 | -0.00207202 | 0.548804243 | 0.003455939 | 0.081424592 | 0.002574412 | 8.75E-11 | 1 | 0.968732296 | 1 | 0.830883001 | ENSG0000027 | 1 | RP1-90J20.12 |
| VAT | T2DM | ENSG0000020 | -0.00037935 | 0.921229403 | 0.003836255 | 0.081436957 | 0.005054395 | 0.20846543 | 0.004018447 | 0.081424388 | -0.00543374 | 5.66E-06 | 1 | 0.980545052 | 1 | 0.590967513 | ENSG0000027 | 1 | RP11-536O18.2 |
| VAT | T2DM | ENSG0000020 | 0.004662651 | 2.87E-05 | 0.001114595 | 0.081434034 | 0.003817344 | 0.000729002 | 0.001129908 | 0.081422745 | 0.000845307 | 5.41E-06 | 0.079321181 | 0.00198303 | 1 | 0.025150583 | ENSG0000027 | 1 | RP11-536O18.2 |
| VAT | T2DM | ENSG0000020 | -0.00177096 | 0.606777085 | 0.003440898 | 0.081436914 | -0.0008112 | 0.815022881 | 0.003467396 | 0.081424643 | -0.00095976 | 0.025590741 | 1 | 0.85794301 | 1 | 0.940454387 | ENSG0000027 | 1 | RP11-536O18.2 |
| VAT | T2DM | ENSG0000020 | 0.003703441 | 0.073501969 | 0.002069303 | 0.081436423 | 0.001676183 | 0.423412292 | 0.002093878 | 0.081424545 | 0.002027258 | 2.64E-10 | 1 | 0.374965392 | 1 | 0.770348006 | ENSG0000027 | 1 | HCG17 |
| VAT | T2DM | ENSG0000020 | 0.001071933 | 0.080654872 | 0.000613619 | 0.081436448 | -0.00013631 | 0.835851443 | 0.000657873 | 0.081424645 | 0.001208246 | 3.56E-07 | 1 | 0.393299377 | 1 | 0.948971609 | ENSG0000027 | 1 | HCG17 |
| VAT | T2DM | ENSG0000020 | -0.00753366 | 0.011694555 | 0.002988077 | 0.081435896 | -0.00747361 | 0.012375742 | 0.002987943 | 0.081423607 | -6.00E-05 | 0.010361636 | 1 | 0.137935783 | 1 | 0.152486826 | ENSG0000027 | 1 | CTC-325H20.7 |
| VAT | T2DM | ENSG0000020 | 0.006657489 | 0.000125966 | 0.001736323 | 0.081434502 | 0.003834783 | 0.036568494 | 0.001834337 | 0.081423922 | 0.002822706 | 1.85E-06 | 0.347666071 | 0.005892645 | 1 | 0.278809514 | ENSG0000027 | 1 | MTRNR2L6 |
| VAT | T2DM | ENSG0000020 | -0.00046129 | 0.834300785 | 0.002205154 | 0.081436951 | 9.48E-05 | 0.96583012 | 0.002213377 | 0.081424652 | -0.00055611 | 0.003871099 | 1 | 0.947211093 | 1 | 0.989894944 | ENSG0000027 | 1 | MTRNR2L6 |
| VAT | T2DM | ENSG0000020 | 1.94E-05 | 0.992916267 | 0.002183564 | 0.081436958 | -6.88E-05 | 0.974857315 | 0.002183501 | 0.081424652 | 8.82E-05 | 2.84E-05 | 1 | 0.998465252 | 1 | 0.991745739 | ENSG0000027 | 1 | RP13-685P2.7 |
| VAT | T2DM | ENSG0000020 | -0.0008833 | 0.755808081 | 0.002840279 | 0.081436942 | -0.00083 | 0.770101696 | 0.002840128 | 0.081424638 | -5.33E-05 | 0.004978779 | 1 | 0.919012884 | 1 | 0.923597644 | ENSG0000027 | 1 | AP000436.4 |
| VAT | T2DM | ENSG0000020 | 0.000480529 | 0.893450717 | 0.003587667 | 0.081436955 | 0.000605605 | 0.865947404 | 0.003587528 | 0.081424648 | -0.00012508 | 4.89E-05 | 1 | 0.970456484 | 1 | 0.961389717 | ENSG0000027 | 1 | AP000436.4 |
| VAT | T2DM | ENSG0000020 | 0.002715497 | 0.233413722 | 0.002278842 | 0.081436721 | 0.002777785 | 0.222875091 | 0.00227889 | 0.081424404 | -6.23E-05 | 0.049438828 | 1 | 0.618342254 | 1 | 0.604564596 | ENSG0000027 | 1 | AP000436.4 |
| VAT | T2DM | ENSG0000020 | -0.0002798 | 0.825902791 | 0.001272055 | 0.08143695 | -0.00030651 | 0.809584212 | 0.001272017 | 0.081424643 | 2.67E-05 | 0.028669208 | 1 | 0.94584718 | 1 | 0.938393612 | ENSG0000027 | 1 | AP000436.4 |
| VAT | T2DM | ENSG0000020 | -0.00462944 | 0.000664372 | 0.001360042 | 0.081435022 | -0.00465748 | 0.000615596 | 0.00135998 | 0.081422693 | 2.80E-05 | 0.007784065 | 1 | 0.021076643 | 1 | 0.022653918 | ENSG0000027 | 1 | RP11-103J8.2 |
| VAT | T2DM | ENSG0000020 | 0.013928655 | 5.94E-05 | 0.003468976 | 0.081434264 | 0.013875109 | 6.34E-05 | 0.003468801 | 0.081421979 | 5.35E-05 | 0.02901971 | 0.163950978 | 0.003345938 | 0.174870981 | 0.004163595 | ENSG0000027 | 1 | RP11-274B21.8 |
| VAT | T2DM | ENSG0000020 | 0.003444884 | 0.094485984 | 0.002060093 | 0.081436491 | 0.003404671 | 0.098380854 | 0.002059994 | 0.081424196 | 4.02E-05 | 0.008123184 | 1 | 0.419937705 | 1 | 0.435146086 | ENSG0000027 | 1 | RP11-274B21.8 |
| VAT | T2DM | ENSG0000020 | -4.48E-05 | 0.982064053 | 0.001991202 | 0.081436958 | -0.00012045 | 0.951763361 | 0.001991124 | 0.081424652 | 7.57E-05 | 8.87E-06 | 1 | 0.996872669 | 1 | 0.988354266 | ENSG0000027 | 1 | CTC-523E23.8 |
| VAT | T2DM | ENSG0000020 | 5.25E-05 | 0.977449258 | 0.001857271 | 0.081436958 | 7.80E-05 | 0.966513296 | 0.001857171 | 0.081424652 | -2.55E-05 | 0.035384534 | 1 | 0.993284224 | 1 | 0.989894944 | ENSG0000027 | 1 | CTC-523E23.8 |
| VAT | T2DM | ENSG0000020 | -0.0008461 | 0.79195537 | 0.003207676 | 0.081436947 | -0.00090368 | 0.778145685 | 0.00320752 | 0.081424639 | 5.76E-05 | 0.014509441 | 1 | 0.932507176 | 1 | 0.926363331 | ENSG0000027 | 1 | RP11-27I1.6 |

| | | | | | | | | | | | | | | | | | | | |
|---|---|---|---|---|---|---|---|---|---|---|---|---|---|---|---|---|---|---|---|
| VAT | T2DM | ENSG0000002( | 0.002569223 | 0.444697789 | 0.003361603 | 0.081436861 | 0.002657631 | 0.429168307 | 0.003361467 | 0.081424548 | -8.84E-05 | 0.001741651 | 1 | 0.779859126 | 1 | 0.774674827 | ENSG0000027 | 1 | RP11-460N20.7 |
| VAT | T2DM | ENSG0000002( | -0.00152076 | 0.413780228 | 0.001860802 | 0.081436847 | -0.00097749 | 0.602857726 | 0.001878719 | 0.081424607 | -0.00054327 | 0.036548813 | 1 | 0.757898941 | 1 | 0.857232006 | ENSG0000027 | 1 | RP11-502I4.3 |
| VAT | T2DM | ENSG0000002( | -0.00053725 | 0.854583333 | 0.002931414 | 0.081436953 | 0.001056365 | 0.719638739 | 0.002942998 | 0.081424631 | -0.00159362 | 1.43E-09 | 1 | 0.955830337 | 1 | 0.906651719 | ENSG0000027 | 1 | RP11-326C3.15 |
| VAT | T2DM | ENSG0000021 | -0.00387726 | 0.051031502 | 0.001987098 | 0.081436322 | -0.00384765 | 0.052818636 | 0.001987004 | 0.081424026 | -2.96E-05 | 0.048606546 | 1 | 0.31865825 | 1 | 0.332694881 | ENSG0000027 | 1 | RP11-326C3.15 |
| VAT | T2DM | ENSG0000021 | -0.00214808 | 0.517021117 | 0.003315225 | 0.081436888 | -0.00206699 | 0.532965115 | 0.003315217 | 0.081424588 | -8.11E-05 | 0.043058137 | 1 | 0.812597159 | 1 | 0.824080513 | ENSG0000027 | 1 | RP11-326C3.15 |
| VAT | T2DM | ENSG0000021 | 0.003332952 | 0.000233342 | 0.000905716 | 0.081434695 | 0.003317361 | 0.000249452 | 0.000905678 | 0.081422411 | 1.56E-05 | 0.035740654 | 0.644022967 | 0.009333666 | 0.688488117 | 0.011907029 | ENSG0000027 | 1 | RP5-933K21.3 |
| VAT | T2DM | ENSG0000021 | 0.00190465 | 0.375200394 | 0.002147839 | 0.081436827 | 0.001858906 | 0.386752891 | 0.002147729 | 0.081424527 | 4.57E-05 | 0.002252748 | 1 | 0.733387742 | 1 | 0.745528886 | ENSG0000027 | 1 | RP5-933K21.3 |
| VAT | T2DM | ENSG0000021 | 0.003805713 | 0.128798131 | 0.002505636 | 0.081436573 | 0.003845256 | 0.124852474 | 0.002505504 | 0.081424259 | -3.95E-05 | 0.019595713 | 1 | 0.485394797 | 1 | 0.480487243 | ENSG0000027 | 1 | AC003973.6 |
| VAT | T2DM | ENSG0000021 | 0.000749456 | 0.62356314 | 0.001526987 | 0.081436918 | 0.000368224 | 0.809874487 | 0.001530514 | 0.081424643 | 0.000381233 | 0.00030306 | 1 | 0.868407764 | 1 | 0.938393612 | ENSG0000027 | 1 | RP11-230C9.4 |
| VAT | T2DM | ENSG0000021 | 0.000671524 | 0.60270237 | 0.001290105 | 0.081436913 | 0.000633626 | 0.623317264 | 0.001290074 | 0.081424612 | 3.79E-05 | 0.003557245 | 1 | 0.857011097 | 1 | 0.868736298 | ENSG0000027 | 1 | SRXN1 |
| VAT | T2DM | ENSG0000021 | 0.000539832 | 0.814041454 | 0.002295052 | 0.081436949 | 0.000623751 | 0.785809888 | 0.002295264 | 0.08142464 | -8.39E-05 | 0.046109343 | 1 | 0.943325602 | 1 | 0.930032286 | ENSG0000027 | 1 | RP11-10C24.2 |
| VAT | T2DM | ENSG0000021 | -0.00102719 | 0.12055286 | 0.000661656 | 0.081436556 | -0.00099929 | 0.130956261 | 0.000661632 | 0.081424271 | -2.79E-05 | 1.63E-06 | 1 | 0.472283245 | 1 | 0.491745938 | ENSG0000027 | 1 | RP11-10C24.2 |
| VAT | T2DM | ENSG0000021 | -0.00425993 | 0.117779081 | 0.002723449 | 0.08143655 | -0.00430834 | 0.113644925 | 0.002723307 | 0.081424234 | 4.84E-05 | 0.009721501 | 1 | 0.464386092 | 1 | 0.459912014 | ENSG0000027 | 1 | RP11-697E2.9 |
| VAT | T2DM | ENSG0000021 | -0.0044738 | 0.087955834 | 0.002621953 | 0.081436472 | -0.00452268 | 0.084525037 | 0.002621819 | 0.081424155 | 4.89E-05 | 0.007987139 | 1 | 0.407340768 | 1 | 0.410615651 | ENSG0000027 | 1 | RP11-326I11.4 |
| VAT | T2DM | ENSG0000021 | -0.00329559 | 0.39719688 | 0.003892556 | 0.081436839 | -0.00319005 | 0.412468338 | 0.003892397 | 0.08142454 | -0.00010554 | 0.001152212 | 1 | 0.749325624 | 1 | 0.762700254 | ENSG0000027 | 1 | RP11-326I11.4 |
| VAT | T2DM | ENSG0000021 | 0.002293354 | 0.008405585 | 0.000870231 | 0.081435798 | 0.001370902 | 0.122448007 | 0.000887556 | 0.081424254 | 0.000922451 | 1.32E-07 | 1 | 0.111535642 | 1 | 0.475526929 | ENSG0000027 | 1 | RP11-561N12.7 |
| VAT | T2DM | ENSG0000021 | -0.0024709 | 0.077033225 | 0.001397435 | 0.081436436 | -0.00080273 | 0.574461463 | 0.00142964 | 0.0814246 | -0.00166817 | 3.39E-08 | 1 | 0.385865156 | 1 | 0.848256597 | ENSG0000027 | 1 | RP11-561N12.7 |
| VAT | T2DM | ENSG0000021 | 0.003181664 | 0.202937487 | 0.002498895 | 0.081436688 | 0.00544102 | 0.031917297 | 0.002536073 | 0.081423883 | -0.00225936 | 1.90E-07 | 1 | 0.582793504 | 1 | 0.260626449 | ENSG0000027 | 1 | RP11-230C9.2 |
| VAT | T2DM | ENSG0000022 | -0.0079089 | 0.045259562 | 0.003950029 | 0.081436288 | -0.00863957 | 0.028771171 | 0.003951118 | 0.081423854 | 0.000730668 | 3.14E-12 | 1 | 0.2966907 | 1 | 0.246610035 | ENSG0000027 | 1 | RP11-274B21.10 |
| VAT | T2DM | ENSG0000022 | -0.00267069 | 0.458546782 | 0.003602996 | 0.081436867 | -0.00261632 | 0.46772441 | 0.003602814 | 0.081424564 | -5.44E-05 | 0.033118991 | 1 | 0.78904441 | 1 | 0.792857737 | ENSG0000027 | 1 | RP11-274B21.10 |
| VAT | T2DM | ENSG0000022 | -0.00633331 | 0.060693873 | 0.003376465 | 0.08143637 | -0.00644163 | 0.056409685 | 0.003376362 | 0.081424044 | 0.000108318 | 0.000747228 | 1 | 0.351922455 | 1 | 0.344116711 | ENSG0000027 | 1 | RP11-274B21.10 |
| VAT | T2DM | ENSG0000022 | 0.00996774 | 5.73E-05 | 0.002477285 | 0.081434253 | 0.002828991 | 0.379679035 | 0.003220293 | 0.081424524 | 0.007138749 | 0.000521806 | 0.15817682 | 0.003341609 | 1 | 0.73818881 | ENSG0000027 | 1 | RP11-274B21.10 |
| VAT | T2DM | ENSG0000022 | 0.000960621 | 0.149601502 | 0.000666662 | 0.081436611 | 0.000937611 | 0.159586076 | 0.000666642 | 0.081424322 | 2.30E-05 | 0.000345503 | 1 | 0.516979293 | 1 | 0.534944655 | ENSG0000027 | 1 | RP11-274B21.10 |
| VAT | T2DM | ENSG0000022 | 0.002705594 | 0.006160378 | 0.000987756 | 0.081435705 | 0.001697759 | 0.089651362 | 0.001000304 | 0.081424171 | 0.001007835 | 1.97E-10 | 1 | 0.091412061 | 1 | 0.421529404 | ENSG0000027 | 1 | XXbac-BPG248L24.12 |
| VAT | T2DM | ENSG0000022 | -0.00111266 | 0.560892661 | 0.001913377 | 0.081436902 | -0.00115204 | 0.547099353 | 0.001913322 | 0.081424592 | 3.94E-05 | 0.034008628 | 1 | 0.835470159 | 1 | 0.830883001 | ENSG0000027 | 1 | XXbac-BPG248L24.12 |
| VAT | T2DM | ENSG0000022 | -0.00331573 | 0.000946916 | 0.001002984 | 0.081435132 | -0.00225293 | 0.026793233 | 0.001017344 | 0.081423833 | -0.00106279 | 4.82E-10 | 1 | 0.027223824 | 1 | 0.235964845 | ENSG0000027 | 1 | XXbac-BPG248L24.12 |
| VAT | T2DM | ENSG0000022 | -0.00133553 | 0.333111198 | 0.00137987 | 0.081436802 | -0.00224072 | 0.107255671 | 0.001391187 | 0.081424219 | 0.000905191 | 3.62E-07 | 1 | 0.701286733 | 1 | 0.448599281 | ENSG0000027 | 1 | XXbac-BPG248L24.12 |
| VAT | T2DM | ENSG0000022 | 0.000846333 | 0.718143406 | 0.002344793 | 0.081436937 | 0.000889346 | 0.704461703 | 0.002344675 | 0.081424628 | -4.30E-05 | 0.010052231 | 1 | 0.905268686 | 1 | 0.90021629 | ENSG0000027 | 1 | XXbac-BPG248L24.12 |
| VAT | T2DM | ENSG0000022 | 0.010238103 | 0.012361952 | 0.004092541 | 0.081435913 | 0.010125945 | 0.013348786 | 0.00409239 | 0.081423629 | 0.000112158 | 0.001821976 | 1 | 0.143357087 | 1 | 0.158804523 | ENSG0000027 | 1 | XXbac-BPG248L24.12 |
| VAT | T2DM | ENSG0000022 | 0.001906435 | 0.627307611 | 0.003926582 | 0.081436919 | 0.00183672 | 0.63993548 | 0.003926384 | 0.081424616 | 6.97E-05 | 0.011917076 | 1 | 0.869127163 | 1 | 0.87456323 | ENSG0000027 | 1 | RP11-89C3.4 |
| VAT | T2DM | ENSG0000022 | -0.01165866 | 1.70E-06 | 0.002435828 | 0.08143313 | -0.01169676 | 1.57E-06 | 0.0024357 | 0.081420799 | 3.81E-05 | 0.021773828 | 0.004689663 | 0.000173691 | 0.00433226 | 0.000200258 | ENSG0000027 | 1 | RP11-89C3.4 |
| VAT | T2DM | ENSG0000022 | 0.004002258 | 0.081842607 | 0.00230002 | 0.081436452 | 0.004625303 | 0.044893046 | 0.002306125 | 0.08142398 | -0.00062305 | 0.000249232 | 1 | 0.395539673 | 1 | 0.302300087 | ENSG0000027 | 1 | RP11-435I10.5 |
| VAT | T2DM | ENSG0000022 | 0.002243152 | 0.521797687 | 0.003501766 | 0.08143689 | 0.002184368 | 0.53274809 | 0.003501617 | 0.081424587 | 5.88E-05 | 0.038080244 | 1 | 0.814571049 | 1 | 0.824080513 | ENSG0000027 | 1 | RP11-435I10.5 |
| VAT | T2DM | ENSG0000022 | 0.002444204 | 0.532223259 | 0.003913129 | 0.081436893 | 0.005207965 | 0.191077484 | 0.003983452 | 0.081424367 | -0.00276376 | 0.000214789 | 1 | 0.81975639 | 1 | 0.573647304 | ENSG0000027 | 1 | RP11-435I10.5 |
| VAT | T2DM | ENSG0000022 | 0.001562077 | 0.480092737 | 0.002212091 | 0.081436875 | 0.001518384 | 0.492439865 | 0.002211989 | 0.081424574 | 4.37E-05 | 0.009719616 | 1 | 0.791889203 | 1 | 0.800277918 | ENSG0000027 | 1 | RP11-435I10.5 |
| VAT | T2DM | ENSG0000022 | 0.001197683 | 0.764973101 | 0.004006209 | 0.081436943 | 0.001103153 | 0.783029345 | 0.004006021 | 0.08142464 | 9.45E-05 | 0.001891292 | 1 | 0.922331547 | 1 | 0.92833376 | ENSG0000027 | 1 | RP11-85G21.3 |
| VAT | T2DM | ENSG0000022 | -0.00103294 | 0.465450473 | 0.001415179 | 0.081436869 | -0.00046141 | 0.747685457 | 0.001434322 | 0.081424635 | -0.00057153 | 0.014674552 | 1 | 0.78904441 | 1 | 0.911489338 | ENSG0000027 | 1 | RP11-110I1.14 |
| VAT | T2DM | ENSG0000022 | 0.002480347 | 0.014222207 | 0.001011722 | 0.081435954 | 0.002503244 | 0.013348278 | 0.001011678 | 0.081423629 | -2.29E-05 | 0.004838565 | 1 | 0.157013171 | 1 | 0.158804523 | ENSG0000027 | 1 | RP11-110I1.14 |
| VAT | T2DM | ENSG0000022 | 0.002922732 | 0.177312798 | 0.002166468 | 0.081436654 | 0.002978038 | 0.169234119 | 0.002166364 | 0.081424337 | -5.53E-05 | 0.00063215 | 1 | 0.556533092 | 1 | 0.54753664 | ENSG0000027 | 1 | RP1-153G14.4 |
| VAT | T2DM | ENSG0000022 | -0.00320771 | 0.25948403 | 0.00284469 | 0.081436746 | 0.003866736 | 0.216809347 | 0.003130812 | 0.081424398 | -0.00707444 | 6.36E-08 | 1 | 0.643030277 | 1 | 0.598393798 | ENSG0000027 | 1 | RP1-153G14.4 |
| VAT | T2DM | ENSG0000022 | 0.000409807 | 0.718063968 | 0.00113505 | 0.081436937 | 0.000387297 | 0.732930358 | 0.001135001 | 0.081424633 | 2.25E-05 | 0.013337845 | 1 | 0.905268686 | 1 | 0.908589792 | ENSG0000027 | 1 | RP1-153G14.4 |
| VAT | T2DM | ENSG0000022 | -0.00146602 | 0.722636087 | 0.004130351 | 0.081436937 | -0.00135497 | 0.742862209 | 0.004130179 | 0.081424634 | -0.00011105 | 0.001062702 | 1 | 0.907404731 | 1 | 0.911123584 | ENSG0000027 | 1 | RP1-79C4.4 |
| VAT | T2DM | ENSG0000022 | -0.00010709 | 0.931610349 | 0.001247805 | 0.081436957 | -0.00129164 | 0.308087597 | 0.001267256 | 0.081424479 | 0.001184558 | 9.16E-08 | 1 | 0.984451223 | 1 | 0.677547225 | ENSG0000027 | 1 | RP1-79C4.4 |
| VAT | T2DM | ENSG0000022 | 0.001105035 | 0.559792998 | 0.001894943 | 0.081436902 | 0.001144878 | 0.545709227 | 0.001894858 | 0.081424591 | -3.98E-05 | 0.007619293 | 1 | 0.835470159 | 1 | 0.830754257 | ENSG0000027 | 1 | RP1-79C4.4 |
| VAT | T2DM | ENSG0000022 | -0.00266433 | 0.012628347 | 0.001068256 | 0.081435919 | -0.0009658 | 0.382814925 | 0.001106654 | 0.081424525 | -0.00169853 | 4.31E-09 | 1 | 0.14522599 | 1 | 0.741436359 | ENSG0000027 | 1 | XXbac-BPG299F13.14 |
| VAT | T2DM | ENSG0000022 | 0.001149761 | 0.76844638 | 0.003905334 | 0.081436944 | 0.0010493 | 0.788164481 | 0.003905172 | 0.08142464 | 0.000100461 | 0.001797436 | 1 | 0.924118506 | 1 | 0.932019695 | ENSG0000027 | 1 | XXbac-BPG299F13.14 |
| VAT | T2DM | ENSG0000022 | -0.00307771 | 0.110955295 | 0.001930912 | 0.081436534 | -0.00200252 | 0.304818933 | 0.001951483 | 0.081424477 | -0.00107519 | 0.000149759 | 1 | 0.451011213 | 1 | 0.676208846 | ENSG0000027 | 1 | XXbac-BPG299F13.14 |
| VAT | T2DM | ENSG0000022 | -0.00041739 | 0.787134516 | 0.0015457 | 0.081436946 | -0.00044925 | 0.77131226 | 0.00154563 | 0.081424638 | 3.19E-05 | 0.007563562 | 1 | 0.932399685 | 1 | 0.923905803 | ENSG0000027 | 1 | XXbac-BPG299F13.14 |
| VAT | T2DM | ENSG0000022 | -0.00035396 | 0.920307367 | 0.003537958 | 0.081436957 | -0.0004031 | 0.909284463 | 0.003537776 | 0.08142465 | 4.91E-05 | 0.04554298 | 1 | 0.980360785 | 1 | 0.9774361 | ENSG0000027 | 1 | RP1-224A6.9 |
| VAT | T2DM | ENSG0000023 | 0.004239089 | 0.18263903 | 0.003180887 | 0.081436662 | 0.004337694 | 0.172653083 | 0.003180757 | 0.081424342 | -9.86E-05 | 0.000203853 | 1 | 0.562893955 | 1 | 0.548422551 | ENSG0000027 | 1 | RP11-493E12.1 |
| VAT | T2DM | ENSG0000023 | 0.000794786 | 0.721606089 | 0.002230583 | 0.081436937 | 0.000762604 | 0.732424358 | 0.002230473 | 0.081424633 | 3.22E-05 | 0.04700481 | 1 | 0.907308877 | 1 | 0.908589792 | ENSG0000027 | 1 | RP11-661A12.14 |
| VAT | T2DM | ENSG0000023 | 0.000557903 | 0.736376369 | 0.00165719 | 0.081436939 | -6.17E-05 | 0.970410356 | 0.001664433 | 0.081424652 | 0.000619643 | 7.47E-05 | 1 | 0.913437653 | 1 | 0.990507612 | ENSG0000027 | 1 | RP11-23P13.7 |
| VAT | T2DM | ENSG0000023 | -0.00131808 | 0.592133411 | 0.002460263 | 0.08143691 | -0.0013723 | 0.576974652 | 0.002460161 | 0.0814246 | 5.42E-05 | 0.00749146 | 1 | 0.849919041 | 1 | 0.848358775 | ENSG0000027 | 1 | RP11-23P13.7 |
| VAT | T2DM | ENSG0000023 | -5.49E-05 | 0.962068626 | 0.001153361 | 0.081436958 | 0.00052905 | 0.650829131 | 0.001168884 | 0.081424618 | -0.0005839 | 0.002163452 | 1 | 0.991412214 | 1 | 0.875384211 | ENSG0000027 | 1 | RP11-23P13.7 |
| VAT | T2DM | ENSG0000023 | 0.001367579 | 0.549871954 | 0.002287104 | 0.081436899 | 0.001411717 | 0.537051822 | 0.002287004 | 0.081424589 | -4.41E-05 | 0.015629712 | 1 | 0.828182675 | 1 | 0.825773275 | ENSG0000027 | 1 | RP11-91J19.4 |
| VAT | T2DM | ENSG0000023 | -0.00150883 | 0.679250847 | 0.003649056 | 0.08143693 | -0.00141964 | 0.697231103 | 0.003648878 | 0.081424627 | -8.92E-05 | 0.00085475 | 1 | 0.890278989 | 1 | 0.896718473 | ENSG0000027 | 1 | CTD-3025N20.3 |
| VAT | T2DM | ENSG0000023 | -0.00079589 | 0.586621775 | 0.001463737 | 0.081436909 | 0.000958349 | 0.528301873 | 0.00151974 | 0.081424586 | -0.00175424 | 1.81E-05 | 1 | 0.848256181 | 1 | 0.822106477 | ENSG0000027 | 1 | CTD-3025N20.3 |
| VAT | T2DM | ENSG0000023 | -0.0019834 | 0.486649407 | 0.00285116 | 0.081436878 | -0.00203039 | 0.476365303 | 0.002851029 | 0.081424568 | 4.70E-05 | 0.032281124 | 1 | 0.792496248 | 1 | 0.795911768 | ENSG0000027 | 1 | CTD-3025N20.3 |
| VAT | T2DM | ENSG0000023 | 0.000778851 | 0.796568415 | 0.003021253 | 0.081436947 | -0.00016109 | 0.957625287 | 0.003031835 | 0.081424652 | 0.000939945 | 0.000238301 | 1 | 0.934673346 | 1 | 0.989894944 | ENSG0000027 | 1 | CTD-3025N20.3 |
| VAT | T2DM | ENSG0000023 | 0.0046417 | 0.069046285 | 0.002553021 | 0.081436406 | 0.005474668 | 0.032258668 | 0.002556826 | 0.081423886 | -0.00083297 | 5.71E-09 | 1 | 0.369583901 | 1 | 0.261717366 | ENSG0000027 | 1 | CTD-3025N20.3 |
| VAT | T2DM | ENSG0000023 | 0.001075237 | 0.59102788 | 0.002001008 | 0.08143691 | 0.001106686 | 0.580201385 | 0.002000912 | 0.081424601 | -3.14E-05 | 0.033909238 | 1 | 0.849919041 | 1 | 0.849509852 | ENSG0000027 | 1 | CTD-3025N20.3 |
| VAT | T2DM | ENSG0000023 | 0.010067435 | 0.00386394 | 0.003484657 | 0.081435564 | 0.00998189 | 0.004174693 | 0.003484489 | 0.081423281 | 8.55E-05 | 0.000857727 | 1 | 0.070440121 | 1 | 0.082893182 | ENSG0000027 | 1 | GTF2H5 |
| VAT | T2DM | ENSG0000023 | -0.00163047 | 0.648070057 | 0.003572102 | 0.081436924 | -0.00147846 | 0.678942823 | 0.003571965 | 0.081424624 | -0.00015201 | 8.06E-07 | 1 | 0.874341025 | 1 | 0.891475829 | ENSG0000027 | 1 | GTF2H5 |
| VAT | T2DM | ENSG0000023 | -0.00596888 | 0.004834091 | 0.002118202 | 0.081435631 | -0.00600842 | 0.004558538 | 0.002118106 | 0.081423308 | 3.95E-05 | 0.016635964 | 1 | 0.079821197 | 1 | 0.086954128 | ENSG0000027 | 1 | GTF2H5 |
| VAT | T2DM | ENSG0000023 | 0.004956943 | 1.35E-08 | 0.000872691 | 0.081431567 | 0.002739032 | 0.012179632 | 0.001092595 | 0.081423602 | 0.00221791 | 0.000742261 | 3.72E-05 | 2.67E-06 | 1 | 0.151924604 | ENSG0000027 | 1 | RP11-421M1.8 |
| VAT | T2DM | ENSG0000023 | 0.003334249 | 0.354096432 | 0.003598084 | 0.081436815 | 0.003269493 | 0.363496278 | 0.003597895 | 0.081424514 | 6.48E-05 | 0.007945074 | 1 | 0.714271445 | 1 | 0.72339086 | ENSG0000027 | 1 | RP11-250B2.6 |
| VAT | T2DM | ENSG0000023 | -0.00023135 | 0.811010246 | 0.000967485 | 0.081436949 | -0.00044285 | 0.647881032 | 0.000969659 | 0.081424618 | 0.000211503 | 0.001348407 | 1 | 0.942876276 | 1 | 0.874829573 | ENSG0000027 | 1 | RP11-477N3.1 |
| VAT | T2DM | ENSG0000023 | -0.00331291 | 0.406800506 | 0.003993687 | 0.081436843 | -0.00337608 | 0.397887685 | 0.003993473 | 0.081424533 | 6.32E-05 | 0.017078021 | 1 | 0.753911136 | 1 | 0.752171241 | ENSG0000027 | 1 | RP11-477N3.1 |
| VAT | T2DM | ENSG0000023 | 0.002215532 | 0.340978648 | 0.002326666 | 0.081436807 | 0.002251272 | 0.333221773 | 0.002326546 | 0.081424496 | -3.57E-05 | 0.025799191 | 1 | 0.703529273 | 1 | 0.702906909 | ENSG0000027 | 1 | CASC15 |
| VAT | T2DM | ENSG0000023 | 0.002601858 | 0.289338882 | 0.002455569 | 0.081436771 | 0.001350855 | 0.584024187 | 0.002467241 | 0.081424602 | 0.001251003 | 2.23E-07 | 1 | 0.665288122 | 1 | 0.850991929 | ENSG0000027 | 1 | CASC15 |
| VAT | T2DM | ENSG0000023 | 0.003419245 | 0.122261897 | 0.0022126 | 0.081436559 | 0.003367375 | 0.128019238 | 0.002212522 | 0.081424265 | 5.19E-05 | 0.00915303 | 1 | 0.475900889 | 1 | 0.486377366 | ENSG0000027 | 1 | XXbac-BPG181B23.7 |
| VAT | T2DM | ENSG0000023 | -0.00038999 | 0.846494213 | 0.0020145 | 0.081436952 | -0.00042664 | 0.8322655 | 0.002014405 | 0.081424645 | 3.67E-05 | 0.015641444 | 1 | 0.954545261 | 1 | 0.9486742 | ENSG0000027 | 1 | XXbac-BPG181B23.7 |
| VAT | T2DM | ENSG0000023 | 0.002564194 | 0.01311676 | 0.001033702 | 0.08143593 | 0.001665023 | 0.112819579 | 0.001050058 | 0.081424232 | 0.000899171 | 1.18E-06 | 1 | 0.148234846 | 1 | 0.458589157 | ENSG0000027 | 1 | XXbac-BPG181B23.7 |
| VAT | T2DM | ENSG0000023 | 0.001427789 | 0.395768026 | 0.001681326 | 0.081436838 | 0.001183052 | 0.482052898 | 0.001682849 | 0.08142457 | 0.000244737 | 0.001020844 | 1 | 0.748694591 | 1 | 0.798935611 | ENSG0000027 | 1 | XXbac-BPG181B23.7 |
| VAT | T2DM | ENSG0000023 | -0.00046774 | 0.88295778 | 0.003177133 | 0.081436955 | -0.00041372 | 0.896389941 | 0.003176993 | 0.08142465 | -5.40E-05 | 0.032188188 | 1 | 0.968732296 | 1 | 0.973968942 | ENSG0000027 | 1 | XXbac-BPG181B23.7 |
| VAT | T2DM | ENSG0000023 | -4.55E-05 | 0.985357467 | 0.002478835 | 0.081436958 | 3.54E-05 | 0.988595044 | 0.00247878 | 0.081424652 | -8.09E-05 | 0.001605462 | 1 | 0.997544462 | 1 | 0.996594905 | ENSG0000027 | 1 | XXbac-BPG181B23.7 |
| VAT | T2DM | ENSG0000023 | -0.00378005 | 0.031869902 | 0.001761403 | 0.081436189 | -0.00371057 | 0.035162912 | 0.0017615 | 0.081423911 | -6.95E-05 | 0.014525897 | 1 | 0.250390184 | 1 | 0.271847722 | ENSG0000027 | 1 | XXbac-BPG181B23.7 |
| VAT | T2DM | ENSG0000023 | 0.000219542 | 0.948601775 | 0.003405718 | 0.081436958 | 0.000392827 | 0.908181479 | 0.003406038 | 0.08142465 | -0.00017329 | 0.00571287 | 1 | 0.987266065 | 1 | 0.9774361 | ENSG0000027 | 1 | XXbac-BPG181B23.7 |
| VAT | T2DM | ENSG0000023 | -0.00028962 | 0.8213778 | 0.001282792 | 0.08143695 | -0.00031618 | 0.80530561 | 0.001282733 | 0.081424642 | 2.66E-05 | 0.006587661 | 1 | 0.943325602 | 1 | 0.937428716 | ENSG0000027 | 1 | XXbac-BPG181B23.7 |
| VAT | T2DM | ENSG0000023 | -0.00066256 | 0.436513978 | 0.000851517 | 0.081436857 | 0.000210507 | 0.808774593 | 0.000869838 | 0.081424643 | -0.00087307 | 9.21E-07 | 1 | 0.77527579 | 1 | 0.938393612 | ENSG0000027 | 1 | XXbac-BPG181B23.7 |
| VAT | T2DM | ENSG0000023 | 0.000425629 | 0.744382145 | 0.001305385 | 0.081436941 | 0.000398947 | 0.759890517 | 0.001305348 | 0.081424637 | 2.67E-05 | 0.035121961 | 1 | 0.916313203 | 1 | 0.915051408 | ENSG0000027 | 1 | XXbac-BPG181B23.7 |
| VAT | T2DM | ENSG0000023 | 0.000460435 | 0.614153098 | 0.000913281 | 0.081436916 | -0.00110679 | 0.248130556 | 0.000958339 | 0.08142443 | 0.001567224 | 6.94E-08 | 1 | 0.862627253 | 1 | 0.630010905 | ENSG0000027 | 1 | XXbac-BPG181B23.7 |
| VAT | T2DM | ENSG0000023 | -0.00217134 | 0.422347441 | 0.002706202 | 0.081436851 | -0.00333387 | 0.219099161 | 0.002712825 | 0.0814244 | 0.001162532 | 1.52E-09 | 1 | 0.767538154 | 1 | 0.601043397 | ENSG0000027 | 1 | XXbac-BPG170G13.32 |
| VAT | T2DM | ENSG0000023 | 0.001466922 | 0.035356595 | 0.000697121 | 0.081436218 | 0.000452901 | 0.529036908 | 0.000719488 | 0.081424586 | 0.001014021 | 1.27E-08 | 1 | 0.262323122 | 1 | 0.822106477 | ENSG0000027 | 1 | XXbac-BPG170G13.32 |
| VAT | T2DM | ENSG0000023 | 0.002293496 | 0.489053323 | 0.003315183 | 0.081436878 | 0.002355524 | 0.477363104 | 0.003315069 | 0.081424568 | -6.20E-05 | 0.039205507 | 1 | 0.793269357 | 1 | 0.796085901 | ENSG0000027 | 1 | XXbac-BPG170G13.32 |
| VAT | T2DM | ENSG0000024 | 0.004383191 | 3.19E-05 | 0.00105369 | 0.081434067 | 0.001495504 | 0.206865909 | 0.001184809 | 0.081424386 | 0.002887686 | 9.88E-08 | 0.087909583 | 0.002093085 | 1 | 0.588001967 | ENSG0000027 | 1 | XXbac-BPG170G13.32 |
| VAT | T2DM | ENSG0000024 | 0.003382977 | 0.040780491 | 0.001653653 | 0.081436259 | 0.003405826 | 0.039428616 | 0.001653562 | 0.081423944 | -2.28E-05 | 0.031136337 | 1 | 0.284233402 | 1 | 0.285285693 | ENSG0000027 | 1 | XXbac-BPG170G13.32 |
| VAT | T2DM | ENSG0000024 | 0.001773149 | 0.635575373 | 0.003741655 | 0.081436921 | 0.001838393 | 0.623178552 | 0.003741507 | 0.081424612 | -6.52E-05 | 0.040431895 | 1 | 0.871429722 | 1 | 0.868736298 | ENSG0000027 | 1 | XXbac-BPG170G13.32 |
| VAT | T2DM | ENSG0000024 | 0.00107668 | 0.247281554 | 0.000930596 | 0.081436735 | 0.001355244 | 0.149435509 | 0.000940142 | 0.081424305 | -0.00027856 | 0.037811675 | 1 | 0.632482181 | 1 | 0.519448368 | ENSG0000027 | 1 | RP11-222K16.2 |
| VAT | T2DM | ENSG0000024 | -0.00170467 | 0.160573187 | 0.00121489 | 0.081436629 | -0.00208815 | 0.086371706 | 0.001217679 | 0.081424161 | 0.000383476 | 4.64E-06 | 1 | 0.536539946 | 1 | 0.413170734 | ENSG0000027 | 1 | RP11-722E23.2 |
| VAT | T2DM | ENSG0000024 | 0.00183277 | 0.614920762 | 0.003643228 | 0.081436916 | 0.00188445 | 0.604964745 | 0.003643036 | 0.081424608 | -5.17E-05 | 0.036307225 | 1 | 0.863266176 | 1 | 0.858231221 | ENSG0000027 | 1 | RP11-722E23.2 |
| VAT | T2DM | ENSG0000024 | -0.00494337 | 0.203593352 | 0.003888186 | 0.081436688 | -0.0048485 | 0.212382761 | 0.003887997 | 0.081424393 | -9.49E-05 | 0.000871992 | 1 | 0.582793504 | 1 | 0.593897083 | ENSG0000027 | 1 | RP11-446N19.1 |
| VAT | T2DM | ENSG0000024 | 0.001374894 | 0.696027873 | 0.003519159 | 0.081436933 | 0.001305157 | 0.710719415 | 0.003518984 | 0.081424629 | 6.97E-05 | 0.006001235 | 1 | 0.899282267 | 1 | 0.902661061 | ENSG0000027 | 1 | RP11-1017G21.6 |
| VAT | T2DM | ENSG0000024 | 0.001314183 | 0.423940634 | 0.001643542 | 0.081436852 | 0.001293538 | 0.431231966 | 0.001643452 | 0.081424549 | 2.06E-05 | 0.049147888 | 1 | 0.767766503 | 1 | 0.776708652 | ENSG0000027 | 2 | U91328.19 |
| VAT | T2DM | ENSG0000024 | 0.000748008 | 0.497536203 | 0.001102651 | 0.081436882 | 0.000769963 | 0.48497948 | 0.001102599 | 0.081424571 | -2.20E-05 | 0.007745901 | 1 | 0.79758205 | 1 | 0.798935611 | ENSG0000027 | 2 | U91328.19 |
| VAT | T2DM | ENSG0000024 | 0.008152138 | 0.02186867 | 0.003555773 | 0.08143608 | 0.008095999 | 0.022788176 | 0.003555595 | 0.081423786 | 5.61E-05 | 0.026428083 | 1 | 0.203910572 | 1 | 0.218753557 | ENSG0000027 | 2 | U91328.19 |
| VAT | T2DM | ENSG0000024 | 0.005791509 | 0.039410613 | 0.002811577 | 0.081436249 | 0.005853936 | 0.037328552 | 0.002811476 | 0.081423928 | -6.24E-05 | 0.012570768 | 1 | 0.280343536 | 1 | 0.280464735 | ENSG0000027 | 2 | U91328.19 |
| VAT | T2DM | ENSG0000024 | 0.009108743 | 1.35E-07 | 0.001727873 | 0.081432314 | 0.00914322 | 1.21E-07 | 0.001727793 | 0.081419973 | -3.45E-05 | 0.009554029 | 0.000373314 | 2.07E-05 | 0.00033434 | 2.79E-05 | ENSG0000027 | 2 | U91328.19 |
| VAT | T2DM | ENSG0000024 | -0.00044932 | 0.833914779 | 0.002142826 | 0.081436951 | -0.00049058 | 0.818905823 | 0.002142731 | 0.081424644 | 4.13E-05 | 0.015144048 | 1 | 0.947162465 | 1 | 0.943313886 | ENSG0000027 | 2 | U91328.19 |
| VAT | T2DM | ENSG0000024 | -0.00215612 | 0.205974845 | 0.001704829 | 0.081436691 | -0.00220325 | 0.196212426 | 0.001704748 | 0.081424373 | 4.71E-05 | 0.000231237 | 1 | 0.586072755 | 1 | 0.579193901 | ENSG0000027 | 2 | U91328.19 |
| VAT | T2DM | ENSG0000024 | 0.007096342 | 0.024585345 | 0.003156928 | 0.081436114 | 0.007295069 | 0.020898663 | 0.003158288 | 0.081423761 | -0.00019873 | 0.048016411 | 1 | 0.215414452 | 1 | 0.20766569 | ENSG0000027 | 1 | XXbac-BPGBPG55C20.1 |
| VAT | T2DM | ENSG0000024 | 0.002989366 | 0.271564087 | 0.002718918 | 0.081436756 | 0.003042658 | 0.263089219 | 0.002718793 | 0.081424443 | -5.33E-05 | 0.01115488 | 1 | 0.651395653 | 1 | 0.643814606 | ENSG0000027 | 1 | XXbac-BPGBPG55C20.1 |
| VAT | T2DM | ENSG0000024 | -0.00095303 | 0.761071297 | 0.003134186 | 0.081436943 | -0.0008861 | 0.77738099 | 0.003134035 | 0.081424639 | -6.69E-05 | 0.003817568 | 1 | 0.920907272 | 1 | 0.926363331 | ENSG0000027 | 1 | XXbac-BPGBPG55C20.1 |
| VAT | T2DM | ENSG0000024 | -0.00464523 | 6.21E-06 | 0.001027913 | 0.081433546 | -0.00097083 | 0.481685121 | 0.001379807 | 0.08142457 | -0.0036744 | 6.56E-05 | 0.017145536 | 0.000571518 | 1 | 0.798935611 | ENSG0000027 | 1 | XXbac-BPGBPG55C20.1 |
| VAT | T2DM | ENSG0000024 | -0.00457422 | 0.070711243 | 0.002530927 | 0.081436413 | -0.00452954 | 0.073493387 | 0.002530814 | 0.081424117 | -4.47E-05 | 0.02466982 | 1 | 0.369646805 | 1 | 0.38344376 | ENSG0000027 | 1 | RP11-638I8.1 |
| VAT | T2DM | ENSG0000024 | 0.006350871 | 0.024190648 | 0.002817482 | 0.081436109 | 0.006294803 | 0.02546364 | 0.002817346 | 0.081423818 | 5.61E-05 | 0.007104918 | 1 | 0.214682275 | 1 | 0.230425072 | ENSG0000027 | 1 | RP11-638I8.1 |
| VAT | T2DM | ENSG0000024 | -0.00074608 | 0.750374086 | 0.002345087 | 0.081436942 | -0.00069547 | 0.766791545 | 0.002344998 | 0.081424638 | -5.06E-05 | 0.013161912 | 1 | 0.918605915 | 1 | 0.920903057 | ENSG0000027 | 1 | DOC2B |
| VAT | T2DM | ENSG0000024 | -0.00600207 | 0.036393192 | 0.002868355 | 0.081436227 | -0.005947 | 0.038134707 | 0.002868224 | 0.081423934 | -5.51E-05 | 0.012817662 | 1 | 0.267853895 | 1 | 0.281907017 | ENSG0000027 | 1 | DOC2B |
| VAT | T2DM | ENSG0000024 | -5.77E-05 | 0.980419253 | 0.002351377 | 0.081436958 | -0.00010111 | 0.96570104 | 0.002351252 | 0.081424652 | 4.34E-05 | 0.005818714 | 1 | 0.995569219 | 1 | 0.989894944 | ENSG0000027 | 1 | CTA-228A9.3 |
| VAT | T2DM | ENSG0000025 | -0.00502448 | 0.178805695 | 0.003737223 | 0.081436656 | -0.00508197 | 0.173864724 | 0.00373704 | 0.081424343 | 5.75E-05 | 0.034944115 | 1 | 0.556533092 | 1 | 0.548422551 | ENSG0000027 | 1 | CTA-228A9.3 |
| VAT | T2DM | ENSG0000025 | -0.00180664 | 0.331504182 | 0.001860429 | 0.081436801 | -0.00184361 | 0.321684441 | 0.001860345 | 0.081424488 | 3.70E-05 | 0.010601283 | 1 | 0.700039437 | 1 | 0.693593042 | ENSG0000027 | 1 | RP5-1159O4.2 |
| VAT | T2DM | ENSG0000025 | -0.00197873 | 0.050690945 | 0.001012609 | 0.08143632 | -0.00056962 | 0.59081006 | 0.001059443 | 0.081424604 | -0.00140911 | 6.19E-06 | 1 | 0.31865825 | 1 | 0.854057999 | ENSG0000027 | 1 | RP5-1159O4.2 |
| VAT | T2DM | ENSG0000025 | -0.00324705 | 0.31579566 | 0.003236894 | 0.08143679 | -0.00331286 | 0.306065004 | 0.003236744 | 0.081424477 | 6.58E-05 | 0.007865755 | 1 | 0.687360522 | 1 | 0.676208846 | ENSG0000027 | 1 | KB-208E9.1 |
| VAT | T2DM | ENSG0000025 | -0.00195464 | 0.57399358 | 0.003476898 | 0.081436906 | -0.00187146 | 0.590382202 | 0.003476731 | 0.081424604 | -8.32E-05 | 0.001244088 | 1 | 0.844018264 | 1 | 0.854057999 | ENSG0000027 | 1 | KB-208E9.1 |
| VAT | T2DM | ENSG0000025 | 0.003138027 | 0.275898927 | 0.002880035 | 0.08143676 | 0.002956429 | 0.304786021 | 0.002880878 | 0.081424477 | 0.000181598 | 0.020151553 | 1 | 0.653996814 | 1 | 0.676208846 | ENSG0000027 | 1 | KB-208E9.1 |
| VAT | T2DM | ENSG0000025 | 0.00330169 | 0.136107581 | 0.002215236 | 0.081436587 | 0.003261688 | 0.140899347 | 0.002215139 | 0.08142429 | 4.00E-05 | 0.024127619 | 1 | 0.500073799 | 1 | 0.510969582 | ENSG0000027 | 1 | KB-208E9.1 |
| VAT | T2DM | ENSG0000025 | 0.004287639 | 0.168370849 | 0.003112715 | 0.081436641 | 0.00436108 | 0.161181532 | 0.003112587 | 0.081424324 | -7.34E-05 | 0.004507983 | 1 | 0.546218791 | 1 | 0.537053745 | ENSG0000027 | 1 | KB-226F1.2 |
| VAT | T2DM | ENSG0000025 | 0.000972739 | 0.431120137 | 0.001235574 | 0.081436855 | 0.001614176 | 0.195362171 | 0.001246583 | 0.081424372 | -0.00064144 | 0.000111681 | 1 | 0.771654721 | 1 | 0.578540335 | ENSG0000027 | 1 | KB-226F1.2 |
| VAT | T2DM | ENSG0000025 | 0.003065582 | 0.272226235 | 0.002792092 | 0.081436757 | 0.002991685 | 0.283931611 | 0.002791974 | 0.081424461 | 7.39E-05 | 0.00116735 | 1 | 0.651643026 | 1 | 0.660194815 | ENSG0000027 | 1 | KB-226F1.2 |
| VAT | T2DM | ENSG0000025 | 0.004107672 | 0.071964894 | 0.002282872 | 0.081436417 | 0.004067669 | 0.074766926 | 0.002282777 | 0.081424122 | 4.00E-05 | 0.033341222 | 1 | 0.369875435 | 1 | 0.38716082 | ENSG0000027 | 1 | RP11-474N24.6 |
| VAT | T2DM | ENSG0000025 | 0.000197024 | 0.891027315 | 0.001438082 | 0.081436955 | -0.00022933 | 0.873670794 | 0.00144236 | 0.081424648 | 0.000426355 | 0.000148736 | 1 | 0.969128142 | 1 | 0.96520345 | ENSG0000027 | 1 | RP11-474N24.6 |
| VAT | T2DM | ENSG0000025 | -0.00256445 | 0.437594028 | 0.003303587 | 0.081436858 | -0.00250606 | 0.448081205 | 0.003303447 | 0.081424556 | -5.84E-05 | 0.02980634 | 1 | 0.775765788 | 1 | 0.786370844 | ENSG0000027 | 1 | RP11-474N24.6 |
| VAT | T2DM | ENSG0000025 | 0.00087283 | 0.769473771 | 0.002978297 | 0.081436944 | 0.001319255 | 0.658042961 | 0.002980578 | 0.08142462 | -0.00044642 | 0.000259597 | 1 | 0.924118506 | 1 | 0.876362712 | ENSG0000027 | 1 | RP11-474N24.6 |
| VAT | T2DM | ENSG0000025 | 0.001313279 | 0.529680676 | 0.002089566 | 0.081436892 | 0.002330359 | 0.268562814 | 0.002106292 | 0.081424448 | -0.00101708 | 0.000132877 | 1 | 0.819413094 | 1 | 0.647121836 | ENSG0000027 | 1 | U91328.22 |
| VAT | T2DM | ENSG0000025 | -0.00796995 | 0.041969474 | 0.003918696 | 0.081436267 | -0.00803785 | 0.040243737 | 0.003918527 | 0.081423949 | 6.79E-05 | 0.032058714 | 1 | 0.284233402 | 1 | 0.287009601 | ENSG0000027 | 1 | U91328.22 |
| VAT | T2DM | ENSG0000025 | 0.00232377 | 0.066319603 | 0.00126549 | 0.081436395 | 0.002303736 | 0.068680919 | 0.00126543 | 0.081424099 | 2.00E-05 | 0.034145869 | 1 | 0.366008125 | 1 | 0.375985973 | ENSG0000027 | 1 | U91328.22 |
| VAT | T2DM | ENSG0000025 | -0.00525093 | 0.013466167 | 0.002124846 | 0.081435938 | -0.00520419 | 0.014312669 | 0.002124743 | 0.08142365 | -4.67E-05 | 0.002614345 | 1 | 0.149821016 | 1 | 0.168816097 | ENSG0000027 | 1 | U91328.22 |

| | | | | | | | | | | | | | | | | | | | |
|---|---|---|---|---|---|---|---|---|---|---|---|---|---|---|---|---|---|---|---|
| VAT | T2DM | ENSG0000025 | 0.003046963 | 0.224766705 | 0.002509954 | 0.081436712 | 0.003090192 | 0.21823975 | 0.002509854 | 0.081424399 | -4.32E-05 | 0.041604753 | 1 | 0.610120125 | 1 | 0.599344984 | ENSG0000027 | 1 | RP11-347I19.8 |
| VAT | T2DM | ENSG0000025 | -0.00347229 | 0.376531222 | 0.003926564 | 0.081436828 | -0.00357706 | 0.362283619 | 0.003926417 | 0.081424514 | 0.000104771 | 0.002278001 | 1 | 0.734435458 | 1 | 0.721986899 | ENSG0000027 | 1 | RP11-347I19.8 |
| VAT | T2DM | ENSG0000025 | -0.00737047 | 0.057152938 | 0.003874821 | 0.081436354 | -0.00730998 | 0.059212126 | 0.003874645 | 0.081424058 | -6.05E-05 | 0.044854139 | 1 | 0.33656546 | 1 | 0.354502101 | ENSG0000027 | 1 | RP11-347I19.8 |
| VAT | T2DM | ENSG0000025 | 0.000175971 | 0.937277428 | 0.002236191 | 0.081436957 | 0.00020916 | 0.925475738 | 0.002236086 | 0.081424651 | -3.32E-05 | 0.04938787 | 1 | 0.986373964 | 1 | 0.982969566 | ENSG0000027 | 1 | RP11-347I19.8 |
| VAT | T2DM | ENSG0000025 | -0.00038148 | 0.757021033 | 0.001232991 | 0.081436942 | -0.00041528 | 0.736249741 | 0.00123294 | 0.081424634 | 3.38E-05 | 0.001006744 | 1 | 0.919012884 | 1 | 0.910008637 | ENSG0000027 | 1 | RP11-347I19.8 |
| VAT | T2DM | ENSG0000026 | 0.001700231 | 0.055830688 | 0.000889075 | 0.081436347 | 0.001682727 | 0.058391508 | 0.000889038 | 0.081424054 | 1.75E-05 | 0.016219119 | 1 | 0.334382936 | 1 | 0.351112337 | ENSG0000027 | 1 | RP11-347I19.8 |
| VAT | T2DM | ENSG0000026 | -0.00998512 | 0.00330097 | 0.003398332 | 0.081435516 | -0.01004882 | 0.003105534 | 0.003398191 | 0.081423191 | 6.37E-05 | 0.02338843 | 1 | 0.064614724 | 1 | 0.068025986 | ENSG0000027 | 1 | RP11-347I19.8 |
| VAT | T2DM | ENSG0000026 | 0.001479692 | 0.673749406 | 0.00351464 | 0.081436929 | 0.001389759 | 0.692521446 | 0.003514499 | 0.081424626 | 8.99E-05 | 0.002329325 | 1 | 0.888036466 | 1 | 0.894473859 | ENSG0000027 | 1 | RP11-347I19.8 |
| VAT | T2DM | ENSG0000026 | -0.00165383 | 0.638011069 | 0.003515183 | 0.081436921 | -0.0015914 | 0.65073576 | 0.003515034 | 0.081424618 | -6.24E-05 | 0.029580443 | 1 | 0.87299478 | 1 | 0.875384211 | ENSG0000027 | 1 | RP11-408A13.4 |
| VAT | T2DM | ENSG0000026 | 0.005589215 | 0.039290897 | 0.00271172 | 0.081436248 | 0.005536838 | 0.041160572 | 0.002711592 | 0.081423956 | 5.24E-05 | 0.010295175 | 1 | 0.280343536 | 1 | 0.290139674 | ENSG0000027 | 1 | RP11-299H21.1 |
| VAT | T2DM | ENSG0000026 | -0.00227915 | 0.132724076 | 0.001515947 | 0.081436581 | -0.00232561 | 0.124996319 | 0.001515903 | 0.081424259 | 4.65E-05 | 0.001609401 | 1 | 0.49502493 | 1 | 0.480487243 | ENSG0000027 | 1 | RP11-299H21.1 |
| VAT | T2DM | ENSG0000026 | -0.00239246 | 0.065011947 | 0.001296602 | 0.081436389 | -0.00201111 | 0.121307266 | 0.001298069 | 0.081424251 | -0.00038135 | 2.13E-09 | 1 | 0.364130175 | 1 | 0.474232369 | ENSG0000027 | 1 | CTA-246H3.12 |
| VAT | T2DM | ENSG0000026 | 0.000773833 | 0.818037017 | 0.00336346 | 0.08143695 | 0.000862966 | 0.797501482 | 0.003363314 | 0.081424641 | -8.91E-05 | 0.000933893 | 1 | 0.943325602 | 1 | 0.935209552 | ENSG0000027 | 1 | RP11-508N22.12 |
| VAT | T2DM | ENSG0000026 | 0.005132821 | 0.01834703 | 0.002176264 | 0.081436029 | 0.005171408 | 0.017483696 | 0.002176168 | 0.081423709 | -3.86E-05 | 0.025011363 | 1 | 0.184809504 | 1 | 0.189980318 | ENSG0000027 | 1 | RP11-352G9.1 |
| VAT | T2DM | ENSG0000026 | 0.01131335 | 0.000216965 | 0.003058964 | 0.081434673 | 0.011217579 | 0.000245298 | 0.003058939 | 0.081422405 | 9.58E-05 | 0.006951596 | 0.598823748 | 0.009212673 | 0.677023394 | 0.011907029 | ENSG0000027 | 1 | RP11-479G22.8 |
| VAT | T2DM | ENSG0000026 | 0.001092246 | 0.608876614 | 0.002134636 | 0.081436915 | 0.001047869 | 0.623487928 | 0.00213453 | 0.081424612 | 4.44E-05 | 0.003790212 | 1 | 0.859136518 | 1 | 0.868736298 | ENSG0000027 | 1 | RP11-479G22.8 |
| VAT | T2DM | ENSG0000026 | -0.00181541 | 0.606263939 | 0.003522237 | 0.081436914 | -0.00176691 | 0.615898045 | 0.003522044 | 0.08142461 | -4.85E-05 | 0.032955204 | 1 | 0.857656828 | 1 | 0.865252117 | ENSG0000027 | 1 | RP11-479G22.8 |
| VAT | T2DM | ENSG0000026 | 0.002243214 | 0.478896724 | 0.003168029 | 0.081436875 | 0.002181836 | 0.49099019 | 0.003167888 | 0.081424573 | 6.14E-05 | 0.013725781 | 1 | 0.791889203 | 1 | 0.800277918 | ENSG0000027 | 1 | RP11-109E12.1 |
| VAT | T2DM | ENSG0000026 | 0.002052466 | 0.168081643 | 0.001489025 | 0.081436641 | 0.002090568 | 0.160303759 | 0.001488952 | 0.081424323 | -3.81E-05 | 0.000475729 | 1 | 0.546218791 | 1 | 0.536288938 | ENSG0000027 | 1 | RP11-109E12.1 |
| VAT | T2DM | ENSG0000026 | -0.00130381 | 0.653687936 | 0.002906101 | 0.081436925 | -0.00136651 | 0.638181362 | 0.00290596 | 0.081424616 | 6.27E-05 | 0.003307367 | 1 | 0.875814904 | 1 | 0.87456323 | ENSG0000027 | 1 | RP11-458F8.4 |
| VAT | T2DM | ENSG0000026 | 0.00122808 | 0.537118512 | 0.001989835 | 0.081436895 | 0.001170066 | 0.556505483 | 0.001989771 | 0.081424595 | 5.80E-05 | 0.001804954 | 1 | 0.819766464 | 1 | 0.837489168 | ENSG0000027 | 1 | RP11-458F8.4 |
| VAT | T2DM | ENSG0000027 | 0.009692532 | 0.003879311 | 0.003356342 | 0.081435565 | 0.009749351 | 0.003673889 | 0.003356176 | 0.081423242 | -5.68E-05 | 0.019727384 | 1 | 0.070440121 | 1 | 0.076597393 | ENSG0000027 | 1 | RP11-458F8.4 |
| VAT | T2DM | ENSG0000027 | 0.000831425 | 0.63034284 | 0.001727655 | 0.08143692 | 0.000807584 | 0.640164377 | 0.001727565 | 0.081424616 | 2.38E-05 | 0.04575407 | 1 | 0.869869063 | 1 | 0.87456323 | ENSG0000027 | 1 | RP11-458F8.4 |
| VAT | T2DM | ENSG0000027 | 0.001163035 | 0.758997419 | 0.003790885 | 0.081436943 | 0.001227967 | 0.745983267 | 0.003790718 | 0.081424635 | -6.49E-05 | 0.031020392 | 1 | 0.919996872 | 1 | 0.911123584 | ENSG0000027 | 1 | RP11-458F8.4 |
| VAT | T2DM | ENSG0000027 | -0.00034415 | 0.763627867 | 0.001144404 | 0.081436943 | -0.00037266 | 0.744689469 | 0.001144357 | 0.081424635 | 2.85E-05 | 0.00268612 | 1 | 0.921562271 | 1 | 0.911123584 | ENSG0000027 | 1 | AC108004.2 |
| VAT | T2DM | ENSG0000027 | -0.00608699 | 0.065042093 | 0.003299229 | 0.08143639 | -0.00600251 | 0.068842005 | 0.00329906 | 0.081424099 | -8.45E-05 | 0.000232692 | 1 | 0.364130175 | 1 | 0.375985973 | ENSG0000027 | 1 | AC108004.2 |
| VAT | T2DM | ENSG0000027 | -0.00132235 | 0.581570106 | 0.002399508 | 0.081436908 | -0.00126697 | 0.597474558 | 0.002399391 | 0.081424606 | -5.54E-05 | 0.001629193 | 1 | 0.846589395 | 1 | 0.856424723 | ENSG0000027 | 1 | OR2A13P |
| VAT | T2DM | ENSG0000027 | 0.000736125 | 0.750481057 | 0.002314819 | 0.081436942 | 0.000771804 | 0.738805008 | 0.002314697 | 0.081424634 | -3.57E-05 | 0.022164697 | 1 | 0.918605915 | 1 | 0.910293329 | ENSG0000027 | 1 | RP11-212P7.2 |
| VAT | T2DM | ENSG0000027 | -0.00508831 | 0.050954538 | 0.002606892 | 0.081436322 | -0.00513424 | 0.048887943 | 0.00260678 | 0.081424004 | 4.59E-05 | 0.028887917 | 1 | 0.31865825 | 1 | 0.318297756 | ENSG0000027 | 1 | AP000350.5 |
| VAT | T2DM | ENSG0000027 | 0.004510049 | 0.009130118 | 0.001729875 | 0.081435822 | 0.004539494 | 0.008683278 | 0.001729799 | 0.081423502 | -2.94E-05 | 0.033936642 | 1 | 0.11830576 | 1 | 0.122996468 | ENSG0000027 | 1 | AP000350.5 |
| VAT | T2DM | ENSG0000027 | 0.014415064 | 0.000477617 | 0.004126809 | 0.081434919 | 0.020804375 | 4.77E-06 | 0.004547796 | 0.081421156 | -0.00638931 | 0.000831369 | 1 | 0.016477795 | 0.013171662 | 0.000526866 | ENSG0000027 | 1 | AP000350.5 |
| VAT | T2DM | ENSG0000027 | -0.00073143 | 0.661769233 | 0.001671944 | 0.081436926 | -0.00129027 | 0.44152012 | 0.001676483 | 0.081424554 | 0.000558842 | 7.79E-06 | 1 | 0.880657224 | 1 | 0.782153743 | ENSG0000027 | 1 | MICE |
| VAT | T2DM | ENSG0000027 | -0.00339357 | 3.31E-05 | 0.00081754 | 0.081434079 | -0.0029815 | 0.00028465 | 0.000821598 | 0.081422452 | -0.00041206 | 5.24E-07 | 0.091399934 | 0.00212558 | 0.785633404 | 0.012671507 | ENSG0000027 | 1 | MICE |
| VAT | T2DM | ENSG0000027 | 0.001784746 | 0.546548031 | 0.002960066 | 0.081436898 | 0.004695658 | 0.121512226 | 0.00303247 | 0.081424252 | -0.00291091 | 1.02E-05 | 1 | 0.826107649 | 1 | 0.474361731 | ENSG0000027 | 1 | MICE |
| VAT | T2DM | ENSG0000027 | -0.00350608 | 0.344563733 | 0.003709402 | 0.081436809 | -0.00344851 | 0.352516524 | 0.003709195 | 0.081424508 | -5.76E-05 | 0.013482984 | 1 | 0.706010321 | 1 | 0.71278393 | ENSG0000027 | 1 | MICE |
| VAT | T2DM | ENSG0000027 | 0.000224544 | 0.74467422 | 0.000689484 | 0.081436941 | -5.41E-05 | 0.937743576 | 0.000692433 | 0.081424651 | 0.000278628 | 1.51E-05 | 1 | 0.916313203 | 1 | 0.987645456 | ENSG0000027 | 1 | MICE |
| VAT | T2DM | ENSG0000027 | 0.002396875 | 0.250036184 | 0.002083763 | 0.081436737 | 0.002352636 | 0.258862814 | 0.002083667 | 0.081424439 | 4.42E-05 | 0.005782985 | 1 | 0.633538232 | 1 | 0.639459815 | ENSG0000027 | 1 | MICE |
| VAT | T2DM | ENSG0000027 | -0.00569257 | 0.041106593 | 0.002787118 | 0.081436261 | -0.005651 | 0.042597857 | 0.002786987 | 0.081423965 | -4.16E-05 | 0.048641937 | 1 | 0.284233402 | 1 | 0.294307366 | ENSG0000027 | 1 | MICE |
| VAT | T2DM | ENSG0000027 | 0.001814388 | 0.586710525 | 0.003337668 | 0.081436909 | 0.001743932 | 0.601307846 | 0.003337535 | 0.081424607 | 7.05E-05 | 0.01235161 | 1 | 0.848256181 | 1 | 0.856779751 | ENSG0000027 | 1 | RP11-166O4.6 |
| VAT | T2DM | ENSG0000027 | -0.00072662 | 0.475443729 | 0.001018175 | 0.081436873 | 0.000228897 | 0.825369614 | 0.001037402 | 0.081424644 | -0.00095552 | 1.61E-06 | 1 | 0.791889203 | 1 | 0.946021651 | ENSG0000027 | 1 | RP11-166O4.6 |

Table S9 Cell-type common and unique associated genes identified from TWAS analysis, based on estimated CTS expression profiles of all cell types in the target tissue

| | | *Summary of trait-associated genes (p(bonferroni)<0.05)* | | | | *Summary of trait-associated genes (FDR<=0.05)* | | | | | |
|---|---|---|---|---|---|---|---|---|---|---|---|
| Tissue | Disease/Disorder | Cell type specific TWAS gene number | Cell type common TWAS gene number | Total TWAS gene number | Proportion of cell type common TWAS gene | Cell type specific TWAS gene number | Cell type common TWAS gene number | Total TWAS gene number | Proportion of cell type common TWAS gene | Case size | Sample size |
| Frontal cortex | Anxiety | 0 | 0 | 0 | | 0 | 0 | 0 | 0.000 | 7163 | 479906 |
| Frontal cortex | Bipolar | 1 | 0 | 1 | 0.000 | 186 | 0 | 186 | 0.000 | 1603 | 479906 |
| Frontal cortex | Depression | 0 | 2 | 2 | 1.000 | 0 | 0 | 0 | 0.000 | 27875 | 479906 |
| Frontal cortex | Psychosis | 0 | 0 | 0 | | 0 | 2 | 2 | 0.000 | 1070 | 479906 |
| Pancreas | Prediabetes | 5 | 0 | 5 | 0.000 | 1367 | 772 | 2139 | 0.361 | 1039 | 479906 |
| Pancreas | T1DM | 3153 | 29 | 3182 | 0.009 | 3576 | 70 | 3646 | 0.019 | 3965 | 479906 |
| Pancreas | T2DM | 16 | 27 | 43 | 0.628 | 2970 | 235 | 3205 | 0.073 | 43371 | 479962 |
| Pancreas | T2DM related hospitalization | 46 | 43 | 89 | 0.483 | 3123 | 235 | 3358 | 0.070 | 38553 | 479906 |
| Pancreas | T2DM requires insulin | 2 | 10 | 12 | 0.833 | 3128 | 20 | 3148 | 0.006 | 5627 | 479906 |
| Subcutaneous adipose | Obesity | 1857 | 2457 | 4314 | 0.570 | 106 | 6145 | 6251 | 0.983 | 122160 | 479906 |
| Subcutaneous adipose | CAD | 6 | 11 | 17 | 0.647 | 605 | 46 | 651 | 0.071 | 80164 | 479962 |
| Subcutaneous adipose | T2DM | 2323 | 988 | 3311 | 0.298 | 104 | 6148 | 6252 | 0.983 | 43371 | 479962 |
| Subcutaneous adipose | Hypertension | 69 | 138 | 207 | 0.667 | 2160 | 3567 | 5727 | 0.623 | 194236 | 479962 |
| Subcutaneous adipose | Heart failure | 4732 | 46 | 4778 | 0.010 | 1524 | 4552 | 6076 | 0.749 | 39744 | 479962 |
| Subcutaneous adipose | Stroke | 1 | 2 | 3 | 0.667 | 2 | 2 | 4 | 0.500 | 29348 | 479962 |
| Visceral adipose | Obesity | 1856 | 2207 | 4063 | 0.543 | 125 | 4694 | 4819 | 0.974 | 122160 | 479962 |
| Visceral adipose | CAD | 20 | 11 | 31 | 0.355 | 781 | 870 | 1651 | 0.527 | 80164 | 479962 |
| Visceral adipose | T2DM | 2559 | 851 | 3410 | 0.250 | 90 | 4738 | 4828 | 0.981 | 43371 | 479962 |
| Visceral adipose | Hypertension | 1560 | 280 | 1840 | 0.152 | 391 | 4371 | 4762 | 0.918 | 194236 | 479962 |
| Visceral adipose | Heart failure | 1796 | 56 | 1852 | 0.030 | 1982 | 2258 | 4240 | 0.533 | 39744 | 479962 |
| Visceral adipose | Stroke | 3 | 0 | 3 | 0.000 | 806 | 0 | 806 | 0.000 | 29348 | 479962 |

Table S10 Identified trait-associated genes from CTS TWAS analysis

a. Identified trait-associated genes from CTS TWAS analysis in frontal cortex

| disorder | celltype | gene | Estimates | Pvalues | Pvalues_BH | Pvalue_Bonferroni |
|---|---|---|---|---|---|---|
| Bipolar | Ex | *SCYL3* | 0.027141259 | 0.001944181 | 0.036916749 | 1 |
| Bipolar | Ex | *HS3ST1* | 0.100104921 | 0.000349965 | 0.017785524 | 0.999850749 |
| Bipolar | Ex | *LRRC23* | 0.023913402 | 9.74E-05 | 0.017785524 | 0.278253741 |
| Bipolar | Ex | *TTC27* | 0.096481997 | 0.000254131 | 0.017785524 | 0.726052582 |
| Bipolar | Ex | *MIPEP* | 0.096386215 | 0.000484428 | 0.019268993 | 1 |
| Bipolar | Ex | *DAPK2* | 0.109473131 | 0.001524137 | 0.032255251 | 1 |
| Bipolar | Ex | *CNTLN* | 0.067101237 | 0.000881344 | 0.022684674 | 1 |
| Bipolar | Ex | *HSPA5* | 0.08921208 | 0.000252358 | 0.017785524 | 0.720986834 |
| Bipolar | Ex | *SEC61A1* | 0.028233956 | 0.001544632 | 0.032448629 | 1 |
| Bipolar | Ex | *POLR3E* | 0.068081267 | 0.002891056 | 0.046403074 | 1 |
| Bipolar | Ex | *TM7SF3* | 0.074207345 | 0.003004127 | 0.047267494 | 1 |
| Bipolar | Ex | *CNN2* | 0.51795813 | 0.000830444 | 0.022173639 | 1 |
| Bipolar | Ex | *THUMPD1* | 0.068140106 | 0.001015879 | 0.024863087 | 1 |
| Bipolar | Ex | *ZXDC* | 0.083688634 | 0.000533128 | 0.019627556 | 1 |
| Bipolar | Ex | *SIRT6* | 0.101995073 | 0.001198255 | 0.027608192 | 1 |
| Bipolar | Ex | *SAR1A* | 0.065256008 | 0.000757159 | 0.021374262 | 1 |
| Bipolar | Ex | *BZW1* | 0.077678083 | 0.002798199 | 0.045423038 | 1 |
| Bipolar | Ex | *PIBF1* | 0.07365563 | 0.000268893 | 0.017785524 | 0.768226125 |
| Bipolar | Ex | *TDRD3* | 0.069803176 | 8.57E-05 | 0.017785524 | 0.244783206 |
| Bipolar | Ex | *AGBL5* | 0.055402727 | 0.001649448 | 0.034148349 | 1 |
| Bipolar | Ex | *CD59* | 0.089714863 | 0.000385965 | 0.017785524 | 1 |
| Bipolar | Ex | *COQ9* | 0.061369046 | 0.002110635 | 0.03816509 | 1 |
| Bipolar | Ex | *ANAPC5* | 0.069349178 | 0.000294314 | 0.017785524 | 0.840855155 |
| Bipolar | Ex | *NOS1* | 0.075982365 | 0.000591722 | 0.019814124 | 1 |
| Bipolar | Ex | *TOX4* | 0.090542364 | 0.000850148 | 0.022340816 | 1 |
| Bipolar | Ex | *NUP50* | 0.095825454 | 0.001074835 | 0.025590029 | 1 |
| Bipolar | Ex | *LRRFIP2* | 0.029712671 | 0.002818444 | 0.045493189 | 1 |
| Bipolar | Ex | *IFT74* | 0.003835244 | 0.002143996 | 0.038524515 | 1 |
| Bipolar | Ex | *SH3GLB1* | 0.092761915 | 0.000314031 | 0.017785524 | 0.897187562 |
| Bipolar | Ex | *MAPK1* | 0.068642355 | 0.000519466 | 0.019627556 | 1 |
| Bipolar | Ex | *TXN2* | 0.104890464 | 0.000359287 | 0.017785524 | 1 |
| Bipolar | Ex | *COCH* | 0.03222192 | 0.001964069 | 0.036916749 | 1 |
| Bipolar | Ex | *GSTZ1* | 0.168281563 | 0.00062136 | 0.019946344 | 1 |
| Bipolar | Ex | *PLTP* | 0.160106249 | 0.000744799 | 0.021374262 | 1 |
| Bipolar | Ex | *MIB1* | 0.072834656 | 0.000283396 | 0.017785524 | 0.809662824 |
| Bipolar | Ex | *GGA2* | 0.026268798 | 0.002783907 | 0.045423038 | 1 |
| Bipolar | Ex | *C16orf62* | 0.078618965 | 0.000282072 | 0.017785524 | 0.805879132 |
| Bipolar | Ex | *AKAP8* | 0.089840368 | 0.002071635 | 0.037977054 | 1 |
| Bipolar | Ex | *PLA2G4C* | 0.00798525 | 0.002492841 | 0.042220698 | 1 |
| Bipolar | Ex | *RPA3* | 0.101612217 | 0.001266052 | 0.028936885 | 1 |
| Bipolar | Ex | *CDK5RAP3* | 0.04789584 | 0.001354497 | 0.029712702 | 1 |
| Bipolar | Ex | *EZH1* | 0.088657948 | 0.000314276 | 0.017785524 | 0.897885754 |
| Bipolar | Ex | *SLAIN2* | 0.084587842 | 0.000771069 | 0.021374262 | 1 |
| Bipolar | Ex | *LAMTOR3* | 0.08507558 | 0.000100579 | 0.017785524 | 0.287354044 |
| Bipolar | Ex | *MTCH2* | 0.088978841 | 0.000369774 | 0.017785524 | 1 |

| | | | | | | |
|---|---|---|---|---|---|---|
| Bipolar | Ex | *TSPAN11* | 0.025285227 | 0.003131499 | 0.048889038 | 1 |
| Bipolar | Ex | *ALDH2* | 0.111958062 | 0.000273746 | 0.017785524 | 0.782092437 |
| Bipolar | Ex | *DDX55* | 0.006418404 | 0.001033249 | 0.025016877 | 1 |
| Bipolar | Ex | *SPSB2* | 0.015717161 | 0.001065481 | 0.025580492 | 1 |
| Bipolar | Ex | *NUDT12* | 0.041237775 | 0.002500812 | 0.042220698 | 1 |
| Bipolar | Ex | *ARSB* | 0.046080327 | 0.001834948 | 0.035907173 | 1 |
| Bipolar | Ex | *SLC27A6* | 0.092439543 | 0.00060615 | 0.019814124 | 1 |
| Bipolar | Ex | *ACTR8* | 0.175078832 | 0.00030816 | 0.017785524 | 0.880414456 |
| Bipolar | Ex | *TFG* | 0.034998372 | 0.000304446 | 0.017785524 | 0.86980309 |
| Bipolar | Ex | *ABCC5* | 0.017175059 | 0.000151509 | 0.017785524 | 0.432859798 |
| Bipolar | Ex | *NKTR* | 0.039812138 | 0.001196746 | 0.027608192 | 1 |
| Bipolar | Ex | *EPB41L5* | 0.083281089 | 0.000288846 | 0.017785524 | 0.825232363 |
| Bipolar | Ex | *TTC31* | 0.175935511 | 0.001091151 | 0.025763787 | 1 |
| Bipolar | Ex | *COX7A2L* | 0.073856939 | 0.000871573 | 0.02263713 | 1 |
| Bipolar | Ex | *THADA* | 0.067236538 | 0.000473523 | 0.019268993 | 1 |
| Bipolar | Ex | *ARMC2* | 0.048790082 | 0.00199552 | 0.037262758 | 1 |
| Bipolar | Ex | *DNAL1* | 0.006796564 | 0.000542976 | 0.019627556 | 1 |
| Bipolar | Ex | *CRIPT* | 0.110097355 | 0.001792646 | 0.035588271 | 1 |
| Bipolar | Ex | *CRHR1* | 0.026704615 | 0.001916577 | 0.036916749 | 1 |
| Bipolar | Ex | *ZSCAN18* | 0.089169923 | 0.000603034 | 0.019814124 | 1 |
| Bipolar | Ex | *FLT3* | 0.10508716 | 0.00023961 | 0.017785524 | 0.684566052 |
| Bipolar | Ex | *NQO2* | 0.07944372 | 0.000562472 | 0.019627556 | 1 |
| Bipolar | Ex | *GLO1* | 0.084905566 | 0.001310929 | 0.029260342 | 1 |
| Bipolar | Ex | *BBS2* | 0.072794748 | 0.00039399 | 0.017867153 | 1 |
| Bipolar | Ex | *MIF4GD* | 0.09284618 | 0.002336129 | 0.040450422 | 1 |
| Bipolar | Ex | *PCNXL4* | 0.055244581 | 0.000435895 | 0.018048587 | 1 |
| Bipolar | Ex | *POT1* | 0.045588472 | 0.000316037 | 0.017785524 | 0.902917046 |
| Bipolar | Ex | *ORMDL1* | 0.055217599 | 0.000673753 | 0.020593961 | 1 |
| Bipolar | Ex | *NDUFA10* | 0.07823106 | 0.000374433 | 0.017785524 | 1 |
| Bipolar | Ex | *PNPLA7* | 0.091927315 | 0.000434478 | 0.018048587 | 1 |
| Bipolar | Ex | *ATPIF1* | 0.049497135 | 0.00059642 | 0.019814124 | 1 |
| Bipolar | Ex | *PPFIA1* | 0.042877336 | 0.00301109 | 0.047267494 | 1 |
| Bipolar | Ex | *SNRPA1* | 0.095458655 | 0.000125363 | 0.017785524 | 0.358162572 |
| Bipolar | Ex | *C1QTNF6* | 0.201866358 | 0.000417104 | 0.018048587 | 1 |
| Bipolar | Ex | *LARS* | 0.084731397 | 0.000280605 | 0.017785524 | 0.801689144 |
| Bipolar | Ex | *WDR36* | 0.055361015 | 0.00171756 | 0.034815981 | 1 |
| Bipolar | Ex | *CCDC64* | 0.011232023 | 0.000750787 | 0.021374262 | 1 |
| Bipolar | Ex | *PWP1* | 0.066378203 | 0.001950972 | 0.036916749 | 1 |
| Bipolar | Ex | *NUPL2* | 0.060666103 | 0.002949858 | 0.047082369 | 1 |
| Bipolar | Ex | *XPA* | 0.091935539 | 0.000261206 | 0.017785524 | 0.746266786 |
| Bipolar | Ex | *CPEB2* | 0.064782168 | 0.000492347 | 0.019268993 | 1 |
| Bipolar | Ex | *NARS2* | 0.084885456 | 0.000762955 | 0.021374262 | 1 |
| Bipolar | Ex | *PPP2R1B* | 0.106139082 | 0.000289573 | 0.017785524 | 0.82731083 |
| Bipolar | Ex | *MAPKBP1* | 0.061315766 | 0.002395418 | 0.040980292 | 1 |
| Bipolar | Ex | *RPS24* | 0.039562745 | 0.000408819 | 0.017969172 | 1 |
| Bipolar | Ex | *DISP2* | 0.08258394 | 0.001018194 | 0.024863087 | 1 |
| Bipolar | Ex | *SCAPER* | 0.054086865 | 0.000577386 | 0.019814124 | 1 |
| Bipolar | Ex | *DEF8* | 0.084094877 | 0.000383374 | 0.017785524 | 1 |

| | | | | | | |
|---|---|---|---|---|---|---|
| Bipolar | Ex | *FBXO15* | 0.150368226 | 0.001953981 | 0.036916749 | 1 |
| Bipolar | Ex | *ZNF385B* | 0.055008804 | 0.001685745 | 0.034648726 | 1 |
| Bipolar | Ex | *SCD5* | 0.030230211 | 0.002793724 | 0.045423038 | 1 |
| Bipolar | Ex | *GTF2H2* | 0.095484945 | 0.00029895 | 0.017785524 | 0.854101009 |
| Bipolar | Ex | *C7orf50* | 0.013379487 | 0.001457322 | 0.031459942 | 1 |
| Bipolar | Ex | *GBAS* | 0.081642271 | 9.15E-05 | 0.017785524 | 0.26142673 |
| Bipolar | Ex | *NTRK2* | 0.045855516 | 0.000186156 | 0.017785524 | 0.531846302 |
| Bipolar | Ex | *UGCG* | 0.086479669 | 0.000773263 | 0.021374262 | 1 |
| Bipolar | Ex | *CACNA1B* | 0.061681866 | 0.000900376 | 0.022967637 | 1 |
| Bipolar | Ex | *ADD3* | 0.054544478 | 0.000174752 | 0.017785524 | 0.499265881 |
| Bipolar | Ex | *PPP1R1C* | 0.126958766 | 0.001464534 | 0.031459942 | 1 |
| Bipolar | Ex | *NBAS* | 0.032031615 | 0.002533954 | 0.042336302 | 1 |
| Bipolar | Ex | *GRID2* | 0.051183446 | 0.000302675 | 0.017785524 | 0.864743807 |
| Bipolar | Ex | *NMRAL1* | 0.072871299 | 0.000684783 | 0.020593961 | 1 |
| Bipolar | Ex | *RPIA* | 0.106106099 | 0.00096705 | 0.024024889 | 1 |
| Bipolar | Ex | *ANKH* | 0.056180455 | 6.88E-05 | 0.017785524 | 0.196485395 |
| Bipolar | Ex | *AGPAT5* | 0.082089537 | 0.000275737 | 0.017785524 | 0.787779563 |
| Bipolar | Ex | *FAM126B* | 0.072355301 | 0.000333375 | 0.017785524 | 0.952453598 |
| Bipolar | Ex | *NMNAT2* | 0.070866523 | 0.000372573 | 0.017785524 | 1 |
| Bipolar | Ex | *DYRK1A* | 0.069469653 | 0.000334181 | 0.017785524 | 0.954756487 |
| Bipolar | Ex | *RADIL* | 0.134966366 | 0.001353803 | 0.029712702 | 1 |
| Bipolar | Ex | *CDA* | 0.325855322 | 0.000674062 | 0.020593961 | 1 |
| Bipolar | Ex | *JAK1* | 0.081417443 | 0.000259488 | 0.017785524 | 0.74135734 |
| Bipolar | Ex | *ZNF496* | 0.113499486 | 0.000227774 | 0.017785524 | 0.650748975 |
| Bipolar | Ex | *PPP1R21* | 0.008303401 | 0.00274856 | 0.045390957 | 1 |
| Bipolar | Ex | *GABRB1* | 0.059323896 | 0.000337325 | 0.017785524 | 0.963738874 |
| Bipolar | Ex | *ADORA1* | 0.03059968 | 0.002333218 | 0.040450422 | 1 |
| Bipolar | Ex | *HPS3* | 0.120526185 | 0.000273919 | 0.017785524 | 0.782585362 |
| Bipolar | Ex | *LRPAP1* | 0.046350286 | 0.001514412 | 0.032255251 | 1 |
| Bipolar | Ex | *WDR41* | 0.082374125 | 0.000610306 | 0.019814124 | 1 |
| Bipolar | Ex | *INTS1* | 0.078919481 | 0.001279412 | 0.029010159 | 1 |
| Bipolar | Ex | *YWHAZ* | 0.027268685 | 2.33E-05 | 0.017785524 | 0.066694858 |
| Bipolar | Ex | *CCDC171* | 0.016986638 | 0.002236563 | 0.039408386 | 1 |
| Bipolar | Ex | *GABRB3* | 0.054555523 | 0.002083059 | 0.037977054 | 1 |
| Bipolar | Ex | *DCHS1* | 0.081688452 | 0.001784529 | 0.035588271 | 1 |
| Bipolar | Ex | *CRK* | 0.097178487 | 0.000402305 | 0.01795913 | 1 |
| Bipolar | Ex | *ING5* | 0.095483126 | 0.000542557 | 0.019627556 | 1 |
| Bipolar | Ex | *PTPN9* | 0.086552303 | 0.000232363 | 0.017785524 | 0.66386013 |
| Bipolar | Ex | *CNTROB* | 0.131029358 | 0.000759637 | 0.021374262 | 1 |
| Bipolar | Ex | *MGMT* | 0.102037103 | 0.000758561 | 0.021374262 | 1 |
| Bipolar | Ex | *EIF2AK3* | 0.097631548 | 0.000322153 | 0.017785524 | 0.920389816 |
| Bipolar | Ex | *FUT9* | 0.062582622 | 0.000295001 | 0.017785524 | 0.842818135 |
| Bipolar | Ex | *GXYLT2* | 0.082007876 | 0.000192866 | 0.017785524 | 0.55101816 |
| Bipolar | Ex | *PACS1* | 0.026509846 | 0.001834214 | 0.035907173 | 1 |
| Bipolar | Ex | *LRRC37A3* | 0.071894219 | 0.000258223 | 0.017785524 | 0.737743364 |
| Bipolar | Ex | *PLEC* | 0.065986922 | 0.002641062 | 0.043869266 | 1 |
| Bipolar | Ex | *EFCAB13* | 0.117430785 | 0.000193719 | 0.017785524 | 0.553454873 |
| Bipolar | Ex | *LRRC3B* | 0.065131992 | 0.000956915 | 0.024024889 | 1 |
| Bipolar | Ex | *CCDC66* | 0.066485001 | 0.000485877 | 0.019268993 | 1 |
| Bipolar | Ex | *NME9* | 0.087499911 | 6.65E-05 | 0.017785524 | 0.189982597 |

| | | | | | | |
|---|---|---|---|---|---|---|
| Bipolar | Ex | *ERCC6L2* | 0.041414669 | 0.003185609 | 0.049463506 | 1 |
| Bipolar | Ex | *C12orf60* | 0.073863876 | 0.000959891 | 0.024024889 | 1 |
| Bipolar | Ex | *SMDT1* | 0.095007563 | 0.000353988 | 0.017785524 | 1 |
| Bipolar | Ex | *ZNF623* | 0.010667589 | 0.000132344 | 0.017785524 | 0.378106927 |
| Bipolar | Ex | *ACSM5* | 0.076415806 | 0.002080731 | 0.037977054 | 1 |
| Bipolar | Ex | *KCTD16* | 0.058667499 | 0.000340094 | 0.017785524 | 0.971647275 |
| Bipolar | Ex | *TM2D3* | 0.061862318 | 0.003009502 | 0.047267494 | 1 |
| Bipolar | Ex | *TXNRD2* | 0.106776798 | 0.001362396 | 0.029712702 | 1 |
| Bipolar | Ex | *CHRM5* | 0.08461714 | 0.000679139 | 0.020593961 | 1 |
| Bipolar | Ex | *GSAP* | 0.083861341 | 0.000299412 | 0.017785524 | 0.855419845 |
| Bipolar | Ex | *NBPF12* | 0.092534241 | 0.000385157 | 0.017785524 | 1 |
| Bipolar | Ex | *ISG15* | 0.113102001 | 0.000348952 | 0.017785524 | 0.996956921 |
| Bipolar | Ex | *TMEM220* | 0.15028626 | 0.000324361 | 0.017785524 | 0.926699434 |
| Bipolar | Ex | *UROS* | 0.056937689 | 0.000793 | 0.021573372 | 1 |
| Bipolar | Ex | *APOD* | 0.051244742 | 0.002086943 | 0.037977054 | 1 |
| Bipolar | Ex | *FAM47E* | 0.361722939 | 0.001646514 | 0.034148349 | 1 |
| Bipolar | Ex | *OR2L13* | 0.076867029 | 0.001793739 | 0.035588271 | 1 |
| Bipolar | Ex | *GTF2IRD2* | 0.100104685 | 0.00055076 | 0.019627556 | 1 |
| Bipolar | Ex | *ZNF124* | 0.079406911 | 0.000203051 | 0.017785524 | 0.580118082 |
| Bipolar | Ex | *MYO18A* | 0.090569031 | 0.002512257 | 0.042220698 | 1 |
| Bipolar | Ex | *MYO6* | 0.048803713 | 0.000563339 | 0.019627556 | 1 |
| Bipolar | Ex | *SLC39A10* | 0.055583284 | 0.001718255 | 0.034815981 | 1 |
| Bipolar | Ex | *PARVA* | 0.06918228 | 0.000778062 | 0.021374262 | 1 |
| Bipolar | Ex | *PRMT6* | 0.141695773 | 0.00187245 | 0.036391759 | 1 |
| Bipolar | Ex | *ZNF814* | 0.093145595 | 0.000431188 | 0.018048587 | 1 |
| Bipolar | Ex | *DPF3* | 0.071020405 | 0.001141723 | 0.026736912 | 1 |
| Bipolar | Ex | *CSNK1E* | 0.051574822 | 0.000642662 | 0.020400957 | 1 |
| Bipolar | Ex | *PGM5P2* | 0.107340433 | 0.000318251 | 0.017785524 | 0.909241805 |
| Bipolar | Ex | *LINC01103* | 0.109029541 | 0.000800412 | 0.021573372 | 1 |
| Bipolar | Ex | *RP4-669L17.1(* | 0.131358897 | 0.002230447 | 0.039408386 | 1 |
| Bipolar | Ex | *FAM200B* | 0.092173628 | 0.000512969 | 0.019627556 | 1 |
| Bipolar | Ex | *LRRC37A2* | 0.023790525 | 0.000852345 | 0.022340816 | 1 |
| Bipolar | Ex | *AC007405.6* | 0.123102328 | 0.001295014 | 0.029132721 | 1 |
| Bipolar | Ex | *MRPL20* | 0.093141622 | 0.000655639 | 0.02058419 | 1 |
| Bipolar | Ex | *NEAT1* | 0.027171008 | 0.002236689 | 0.039408386 | 1 |
| Bipolar | Ex | *LINC00290* | 0.128420856 | 0.000286072 | 0.017785524 | 0.817308685 |
| Bipolar | Ex | *AP006621.5* | 0.11497268 | 0.000539088 | 0.019627556 | 1 |
| Bipolar | Ex | *HMBS* | 0.032824608 | 0.003230179 | 0.04988444 | 1 |
| Bipolar | Ex | *PWAR6* | 0.061004539 | 0.000239215 | 0.017785524 | 0.683438221 |
| Bipolar | Ex | *LINC01003* | 0.150607602 | 0.000302919 | 0.017785524 | 0.865438351 |
| Bipolar | Ex | *CTC-498M16.4* | 0.054336635 | 0.002394209 | 0.040980292 | 1 |
| Bipolar | Ex | *RP11-301G7.1* | 0.165188597 | 0.002248361 | 0.039408386 | 1 |
| Bipolar | Inhib | *TSSC1* | -0.058200782 | 1.51E-05 | 0.043049224 | 0.043049224 |
| Depression | Astros | *ZSCAN9* | -1.544015402 | 2.16E-07 | 0.000617702 | 0.000617702 |
| Depression | Astros | *BTN3A2* | 0.904951231 | 1.04E-05 | 0.014797886 | 0.029595772 |
| Depression | Ex | *ZSCAN9* | -0.050278755 | 1.10E-07 | 0.000314214 | 0.000314214 |
| Depression | Ex | *BTN3A2* | 0.028083903 | 1.26E-05 | 0.017941799 | 0.035883599 |
| Depression | Inhib | *ZSCAN9* | -0.067792998 | 5.92E-08 | 0.000169066 | 0.000169066 |
| Depression | Inhib | *BTN3A2* | 0.035683921 | 2.52E-05 | 0.036023585 | 0.07204717 |
| Depression | Micro_Macro | *ZSCAN9* | -2.028384466 | 2.88E-07 | 0.000823229 | 0.000823229 |

| | | | | | | |
|---|---|---|---|---|---|---|
| Depression | Micro_Macro | *BTN3A2* | 1.191006204 | 2.07E-05 | 0.029509561 | 0.059019123 |
| Depression | Oligos | *ZSCAN9* | -0.112933231 | 1.33E-07 | 0.000378634 | 0.000378634 |
| Depression | OPCs | *ZSCAN9* | -0.425614173 | 3.19E-08 | 9.12E-05 | 9.12E-05 |

b. Identified trait-associated genes from CTS TWAS analysis in pancreas

| disorder | celltype | gene | Estimates | Pvalues | Pvalues_BH | Pvalue_Bonferroni |
|---|---|---|---|---|---|---|
| Prediabetes | duct_epithelial_cell | *STPG1* | -0.164458875 | 7.44E-06 | 0.008963481 | 0.029218456 |
| Prediabetes | duct_epithelial_cell | *M6PR* | -0.162374367 | 0.002265684 | 0.013326143 | 1 |
| Prediabetes | duct_epithelial_cell | *NDUFAF7* | -0.234139996 | 0.002443751 | 0.013915212 | 1 |
| Prediabetes | duct_epithelial_cell | *PLXND1* | -0.11676646 | 0.004978932 | 0.021832837 | 1 |
| Prediabetes | duct_epithelial_cell | *KDM1A* | -0.130241815 | 0.009347995 | 0.033088535 | 1 |
| Prediabetes | duct_epithelial_cell | *RECQL* | -0.092816137 | 0.01369914 | 0.044117967 | 1 |
| Prediabetes | duct_epithelial_cell | *SLC25A13* | -0.26018646 | 0.001380988 | 0.010271223 | 1 |
| Prediabetes | duct_epithelial_cell | *KMT2E* | -0.140269826 | 0.00058333 | 0.008963481 | 1 |
| Prediabetes | duct_epithelial_cell | *IBTK* | -0.107859807 | 0.000842593 | 0.008963481 | 1 |
| Prediabetes | duct_epithelial_cell | *GGNBP2* | -0.082247068 | 0.006086741 | 0.024776105 | 1 |
| Prediabetes | duct_epithelial_cell | *SYNRG* | -0.136021584 | 0.002066413 | 0.012568013 | 1 |
| Prediabetes | duct_epithelial_cell | *VPS41* | -0.071307492 | 0.012878112 | 0.042129976 | 1 |
| Prediabetes | duct_epithelial_cell | *CCDC124* | -0.134728403 | 0.004266027 | 0.019612585 | 1 |
| Prediabetes | duct_epithelial_cell | *BAIAP3* | -0.064871685 | 0.007128577 | 0.027648747 | 1 |
| Prediabetes | duct_epithelial_cell | *CYB561* | -0.091110785 | 0.004242887 | 0.019551648 | 1 |
| Prediabetes | duct_epithelial_cell | *MED24* | -0.200540851 | 0.005435313 | 0.023196885 | 1 |
| Prediabetes | duct_epithelial_cell | *RPS20* | -0.082678623 | 0.000891397 | 0.00905104 | 1 |
| Prediabetes | duct_epithelial_cell | *REV3L* | -0.13111087 | 0.004712464 | 0.020968597 | 1 |
| Prediabetes | duct_epithelial_cell | *ETV7* | -0.361470073 | 0.005971416 | 0.024567217 | 1 |
| Prediabetes | duct_epithelial_cell | *FUZ* | -0.24765069 | 0.006176398 | 0.024966119 | 1 |
| Prediabetes | duct_epithelial_cell | *LRRC23* | -0.068835012 | 0.007142713 | 0.027676253 | 1 |
| Prediabetes | duct_epithelial_cell | *RABGAP1* | -0.193667678 | 0.000202707 | 0.008963481 | 0.796435181 |
| Prediabetes | duct_epithelial_cell | *PPP5C* | -0.264869766 | 0.000715204 | 0.008963481 | 1 |
| Prediabetes | duct_epithelial_cell | *ERCC1* | -0.165407848 | 0.000627605 | 0.008963481 | 1 |
| Prediabetes | duct_epithelial_cell | *EXTL3* | -0.104751838 | 0.005046085 | 0.02202293 | 1 |
| Prediabetes | duct_epithelial_cell | *MAN2B2* | -0.220189247 | 0.009889357 | 0.034568759 | 1 |
| Prediabetes | duct_epithelial_cell | *UFL1* | -0.145404715 | 0.001047051 | 0.009336398 | 1 |
| Prediabetes | duct_epithelial_cell | *MDH1* | -0.126607466 | 0.002110454 | 0.012737289 | 1 |
| Prediabetes | duct_epithelial_cell | *NUDCD3* | -0.111108683 | 0.009473597 | 0.033412712 | 1 |
| Prediabetes | duct_epithelial_cell | *RNH1* | -0.097655502 | 0.003746436 | 0.018194991 | 1 |
| Prediabetes | duct_epithelial_cell | *NDUFS1* | -0.101281622 | 0.004095261 | 0.019223751 | 1 |
| Prediabetes | duct_epithelial_cell | *RNASET2* | -0.085633389 | 0.0046819 | 0.02090362 | 1 |
| Prediabetes | duct_epithelial_cell | *AGPAT4* | -0.08036806 | 0.007345992 | 0.028324243 | 1 |
| Prediabetes | duct_epithelial_cell | *VEZT* | -0.104723807 | 0.006088516 | 0.024776105 | 1 |
| Prediabetes | duct_epithelial_cell | *RABEP1* | -0.091026879 | 0.005188664 | 0.022377892 | 1 |
| Prediabetes | duct_epithelial_cell | *ARID4A* | -0.108047355 | 0.00541767 | 0.02316216 | 1 |
| Prediabetes | duct_epithelial_cell | *ABCF2* | -0.269688271 | 0.000365826 | 0.008963481 | 1 |
| Prediabetes | duct_epithelial_cell | *UBA6* | -0.185232982 | 0.000469162 | 0.008963481 | 1 |
| Prediabetes | duct_epithelial_cell | *FAM136A* | -0.194861329 | 0.000364529 | 0.008963481 | 1 |
| Prediabetes | duct_epithelial_cell | *DAPK2* | -0.069331892 | 0.015344246 | 0.04807619 | 1 |
| Prediabetes | duct_epithelial_cell | *RPL26L1* | -0.106098795 | 0.003546532 | 0.017534673 | 1 |
| Prediabetes | duct_epithelial_cell | *NSUN2* | -0.147770331 | 0.008430261 | 0.030897851 | 1 |
| Prediabetes | duct_epithelial_cell | *AGA* | -0.146276957 | 0.004528701 | 0.020499155 | 1 |
| Prediabetes | duct_epithelial_cell | *DNAH5* | -0.168907936 | 0.00118627 | 0.009719245 | 1 |
| Prediabetes | duct_epithelial_cell | *PHLPP2* | -0.206399117 | 0.004242991 | 0.019551648 | 1 |
| Prediabetes | duct_epithelial_cell | *INPP4A* | -0.282310192 | 0.000857473 | 0.008963481 | 1 |

| | | | | | | |
|---|---|---|---|---|---|---|
| Prediabetes | duct_epithelial_cell | *PSMA4* | -0.170337862 | 0.000344864 | 0.008963481 | 1 |
| Prediabetes | duct_epithelial_cell | *CNTLN* | -0.310803401 | 0.000950248 | 0.009132479 | 1 |
| Prediabetes | duct_epithelial_cell | *WDR37* | -0.107116305 | 0.002331512 | 0.013496739 | 1 |
| Prediabetes | duct_epithelial_cell | *YTHDC2* | -0.1222248 | 0.001399497 | 0.010278575 | 1 |
| Prediabetes | duct_epithelial_cell | *DTNBP1* | -0.22472068 | 0.0032736 | 0.016769199 | 1 |
| Prediabetes | duct_epithelial_cell | *GOPC* | -0.186129871 | 0.000793832 | 0.008963481 | 1 |
| Prediabetes | duct_epithelial_cell | *ZNF800* | -0.156231503 | 0.00065039 | 0.008963481 | 1 |
| Prediabetes | duct_epithelial_cell | *H6PD* | -0.244342683 | 0.00616527 | 0.024966119 | 1 |
| Prediabetes | duct_epithelial_cell | *VAMP3* | -0.122819369 | 0.002777039 | 0.01511602 | 1 |
| Prediabetes | duct_epithelial_cell | *RCN1* | -0.0806333 | 0.005355881 | 0.022972989 | 1 |
| Prediabetes | duct_epithelial_cell | *JKAMP* | -0.087350648 | 0.012438877 | 0.041069201 | 1 |
| Prediabetes | duct_epithelial_cell | *BCAR1* | -0.128404414 | 0.005238708 | 0.022568952 | 1 |
| Prediabetes | duct_epithelial_cell | *HEBP2* | -0.154416261 | 0.000581163 | 0.008963481 | 1 |
| Prediabetes | duct_epithelial_cell | *RRP12* | -0.174505075 | 0.000986317 | 0.009283441 | 1 |
| Prediabetes | duct_epithelial_cell | *FNIP2* | -0.098793072 | 0.001572717 | 0.010878879 | 1 |
| Prediabetes | duct_epithelial_cell | *MCF2L2* | -0.089162453 | 0.00899366 | 0.032207801 | 1 |
| Prediabetes | duct_epithelial_cell | *LY75* | -0.223537603 | 0.00128572 | 0.010042933 | 1 |
| Prediabetes | duct_epithelial_cell | *KIF1B* | -0.165598061 | 0.000617782 | 0.008963481 | 1 |
| Prediabetes | duct_epithelial_cell | *TBC1D22A* | -0.139957163 | 0.001635012 | 0.011037642 | 1 |
| Prediabetes | duct_epithelial_cell | *SYNE2* | -0.193843821 | 0.002912767 | 0.015486146 | 1 |
| Prediabetes | duct_epithelial_cell | *ATP9A* | -0.19453204 | 0.001366217 | 0.010271223 | 1 |
| Prediabetes | duct_epithelial_cell | *USP36* | -0.167385445 | 0.000751437 | 0.008963481 | 1 |
| Prediabetes | duct_epithelial_cell | *MCOLN3* | -0.075975211 | 0.007552374 | 0.028893162 | 1 |
| Prediabetes | duct_epithelial_cell | *GDI2* | -0.115252629 | 0.000361547 | 0.008963481 | 1 |
| Prediabetes | duct_epithelial_cell | *RIMBP2* | -0.053537323 | 0.016146299 | 0.049755927 | 1 |
| Prediabetes | duct_epithelial_cell | *MPC1* | -0.16272113 | 0.000447158 | 0.008963481 | 1 |
| Prediabetes | duct_epithelial_cell | *EIF4B* | -0.088954365 | 0.000963846 | 0.009147222 | 1 |
| Prediabetes | duct_epithelial_cell | *HAGH* | -0.085028847 | 0.004163819 | 0.019406459 | 1 |
| Prediabetes | duct_epithelial_cell | *RNF4* | -0.24012995 | 0.000126159 | 0.008963481 | 0.495676775 |
| Prediabetes | duct_epithelial_cell | *ST3GAL6* | -0.086481589 | 0.000365966 | 0.008963481 | 1 |
| Prediabetes | duct_epithelial_cell | *HIPK2* | -0.140475053 | 0.001105045 | 0.009564575 | 1 |
| Prediabetes | duct_epithelial_cell | *PMS1* | -0.158265121 | 0.006378496 | 0.02552048 | 1 |
| Prediabetes | duct_epithelial_cell | *IPO5* | -0.132641939 | 0.003283817 | 0.016799632 | 1 |
| Prediabetes | duct_epithelial_cell | *OAT* | -0.087474003 | 0.010969958 | 0.037414245 | 1 |
| Prediabetes | duct_epithelial_cell | *GSTO2* | -0.135042175 | 0.001007982 | 0.009336398 | 1 |
| Prediabetes | duct_epithelial_cell | *ATXN3* | -0.164324015 | 0.000206372 | 0.008963481 | 0.810836149 |
| Prediabetes | duct_epithelial_cell | *EML1* | -0.305050405 | 0.000639555 | 0.008963481 | 1 |
| Prediabetes | duct_epithelial_cell | *STOML1* | -0.264961023 | 0.00022401 | 0.008963481 | 0.880136968 |
| Prediabetes | duct_epithelial_cell | *REEP1* | -0.062988974 | 0.013869891 | 0.044485554 | 1 |
| Prediabetes | duct_epithelial_cell | *POLR1A* | -0.05592608 | 0.002300459 | 0.013430167 | 1 |
| Prediabetes | duct_epithelial_cell | *LAPTM4A* | -0.126826638 | 0.000450758 | 0.008963481 | 1 |
| Prediabetes | duct_epithelial_cell | *KIF2A* | -0.128941797 | 0.011449081 | 0.038546222 | 1 |
| Prediabetes | duct_epithelial_cell | *MAST4* | -0.159706485 | 0.000614616 | 0.008963481 | 1 |
| Prediabetes | duct_epithelial_cell | *RAB27A* | -0.068266437 | 0.005966214 | 0.024567217 | 1 |
| Prediabetes | duct_epithelial_cell | *PTPN3* | -0.201621016 | 0.0054376 | 0.023196885 | 1 |
| Prediabetes | duct_epithelial_cell | *FGF10* | -0.260384319 | 0.001584846 | 0.010911694 | 1 |
| Prediabetes | duct_epithelial_cell | *DGCR2* | -0.175237706 | 0.000868203 | 0.009000449 | 1 |
| Prediabetes | duct_epithelial_cell | *ZXDC* | -0.212925266 | 0.000584427 | 0.008963481 | 1 |

| | | | | | | |
|---|---|---|---|---|---|---|
| Prediabetes | duct_epithelial_cell | *WIPI1* | -0.217414107 | 0.004092969 | 0.019223751 | 1 |
| Prediabetes | duct_epithelial_cell | *ASNS* | -0.116844339 | 0.006335171 | 0.025438743 | 1 |
| Prediabetes | duct_epithelial_cell | *RPL31* | -0.081781741 | 0.001111068 | 0.009564575 | 1 |
| Prediabetes | duct_epithelial_cell | *SEL1L* | -0.128277264 | 0.000153946 | 0.008963481 | 0.604852939 |
| Prediabetes | duct_epithelial_cell | *CYBRD1* | -0.141352674 | 0.004616346 | 0.020788435 | 1 |
| Prediabetes | duct_epithelial_cell | *TRNT1* | -0.225478528 | 0.000621891 | 0.008963481 | 1 |
| Prediabetes | duct_epithelial_cell | *CRMP1* | -0.054384437 | 0.004768806 | 0.021123605 | 1 |
| Prediabetes | duct_epithelial_cell | *DERL2* | -0.170819322 | 0.000340585 | 0.008963481 | 1 |
| Prediabetes | duct_epithelial_cell | *NDE1* | -0.195809056 | 0.002605065 | 0.014586734 | 1 |
| Prediabetes | duct_epithelial_cell | *SMARCE1* | -0.110385989 | 0.001246296 | 0.009849499 | 1 |
| Prediabetes | duct_epithelial_cell | *MRPS34* | -0.077548227 | 0.013290539 | 0.043155809 | 1 |
| Prediabetes | duct_epithelial_cell | *ZZEF1* | -0.151108079 | 0.000903609 | 0.009057947 | 1 |
| Prediabetes | duct_epithelial_cell | *ANO8* | -0.133126966 | 0.011581419 | 0.038858577 | 1 |
| Prediabetes | duct_epithelial_cell | *GRAMD4* | -0.055853377 | 0.011001638 | 0.037424619 | 1 |
| Prediabetes | duct_epithelial_cell | *CACNG5* | -1.570507843 | 0.001915189 | 0.012108933 | 1 |
| Prediabetes | duct_epithelial_cell | *DLG1* | -0.172336987 | 0.001085616 | 0.009519336 | 1 |
| Prediabetes | duct_epithelial_cell | *MKRN2* | -0.152360163 | 0.007018059 | 0.027368761 | 1 |
| Prediabetes | duct_epithelial_cell | *TPD52* | -0.090423779 | 0.00086624 | 0.009000449 | 1 |
| Prediabetes | duct_epithelial_cell | *ACACB* | -0.062167899 | 0.006159876 | 0.024966119 | 1 |
| Prediabetes | duct_epithelial_cell | *GPATCH1* | -0.11665869 | 0.003861838 | 0.018571801 | 1 |
| Prediabetes | duct_epithelial_cell | *XAB2* | -0.193627554 | 0.002105953 | 0.012729677 | 1 |
| Prediabetes | duct_epithelial_cell | *APBB1IP* | 0.053622004 | 0.008648281 | 0.031435931 | 1 |
| Prediabetes | duct_epithelial_cell | *N4BP2* | -0.118378118 | 0.001845517 | 0.011790302 | 1 |
| Prediabetes | duct_epithelial_cell | *MLLT10* | -0.148618985 | 0.001684224 | 0.011177897 | 1 |
| Prediabetes | duct_epithelial_cell | *PCM1* | -0.093461398 | 0.002769476 | 0.01511602 | 1 |
| Prediabetes | duct_epithelial_cell | *TP73* | -1.490527174 | 0.006966218 | 0.027335463 | 1 |
| Prediabetes | duct_epithelial_cell | *TOLLIP* | -0.091782514 | 0.002192467 | 0.012987238 | 1 |
| Prediabetes | duct_epithelial_cell | *FKBP7* | -0.083420566 | 0.013830565 | 0.044431963 | 1 |
| Prediabetes | duct_epithelial_cell | *LXN* | -0.048530363 | 0.006172969 | 0.024966119 | 1 |
| Prediabetes | duct_epithelial_cell | *SAR1A* | -0.107489185 | 0.0012267 | 0.009805258 | 1 |
| Prediabetes | duct_epithelial_cell | *FDFT1* | -0.155768917 | 4.25E-05 | 0.008963481 | 0.166857771 |
| Prediabetes | duct_epithelial_cell | *DDX1* | -0.081890501 | 0.01040756 | 0.035995866 | 1 |
| Prediabetes | duct_epithelial_cell | *RABL2B* | -0.275458599 | 0.009493529 | 0.033452982 | 1 |
| Prediabetes | duct_epithelial_cell | *RIF1* | -0.146864888 | 0.004244331 | 0.019551648 | 1 |
| Prediabetes | duct_epithelial_cell | *COL5A3* | -0.181774095 | 0.003232258 | 0.016709924 | 1 |
| Prediabetes | duct_epithelial_cell | *IGSF9B* | -0.165361571 | 0.008047815 | 0.030163992 | 1 |
| Prediabetes | duct_epithelial_cell | *ZNF506* | -0.173530921 | 0.000662868 | 0.008963481 | 1 |
| Prediabetes | duct_epithelial_cell | *ME2* | -0.184551431 | 0.00068626 | 0.008963481 | 1 |
| Prediabetes | duct_epithelial_cell | *BCKDHB* | -0.170625034 | 0.01161572 | 0.038940413 | 1 |
| Prediabetes | duct_epithelial_cell | *PIBF1* | -0.149861283 | 0.006860949 | 0.027010691 | 1 |
| Prediabetes | duct_epithelial_cell | *CHMP2B* | -0.157538202 | 0.00053169 | 0.008963481 | 1 |
| Prediabetes | duct_epithelial_cell | *NCOA1* | -0.188866175 | 0.000445589 | 0.008963481 | 1 |
| Prediabetes | duct_epithelial_cell | *SLC25A24* | -0.14042076 | 0.004476117 | 0.020307924 | 1 |
| Prediabetes | duct_epithelial_cell | *CPNE3* | -0.097504302 | 0.001730522 | 0.011369938 | 1 |
| Prediabetes | duct_epithelial_cell | *RRN3* | -0.135098676 | 0.000281907 | 0.008963481 | 1 |
| Prediabetes | duct_epithelial_cell | *EPDR1* | -0.233292159 | 0.001419028 | 0.010342775 | 1 |
| Prediabetes | duct_epithelial_cell | *TMED2* | -0.092568383 | 0.00144746 | 0.010415881 | 1 |
| Prediabetes | duct_epithelial_cell | *PGS1* | -0.19131308 | 0.001668738 | 0.01113209 | 1 |
| Prediabetes | duct_epithelial_cell | *PSMC5* | -0.06758318 | 0.006215766 | 0.025099428 | 1 |
| Prediabetes | duct_epithelial_cell | *UIMC1* | -0.135915766 | 0.005555777 | 0.023430359 | 1 |

| | | | | | | |
|---|---|---|---|---|---|---|
| Prediabetes | duct_epithelial_cell | *SF3B2* | -0.08355709 | 0.003984934 | 0.018886376 | 1 |
| Prediabetes | duct_epithelial_cell | *ZNF343* | -0.150041072 | 0.013865715 | 0.044485554 | 1 |
| Prediabetes | duct_epithelial_cell | *MAVS* | -0.15824973 | 0.009298589 | 0.032973065 | 1 |
| Prediabetes | duct_epithelial_cell | *SLC23A2* | -0.113955874 | 0.009020709 | 0.03224965 | 1 |
| Prediabetes | duct_epithelial_cell | *TMEM230* | -0.096568265 | 0.002771139 | 0.01511602 | 1 |
| Prediabetes | duct_epithelial_cell | *GRAMD1A* | -0.141360827 | 0.000504704 | 0.008963481 | 1 |
| Prediabetes | duct_epithelial_cell | *DDX24* | -0.087216868 | 0.001350942 | 0.010256616 | 1 |
| Prediabetes | duct_epithelial_cell | *GPATCH2L* | -0.147930189 | 0.000610286 | 0.008963481 | 1 |
| Prediabetes | duct_epithelial_cell | *STRN4* | -0.158215128 | 0.001502966 | 0.010620779 | 1 |
| Prediabetes | duct_epithelial_cell | *MUL1* | -0.213072598 | 0.000189546 | 0.008963481 | 0.744727995 |
| Prediabetes | duct_epithelial_cell | *SPG21* | -0.177621691 | 0.000537844 | 0.008963481 | 1 |
| Prediabetes | duct_epithelial_cell | *GLG1* | -0.1586374 | 0.000421181 | 0.008963481 | 1 |
| Prediabetes | duct_epithelial_cell | *NRCAM* | -0.054828482 | 0.009214671 | 0.032764203 | 1 |
| Prediabetes | duct_epithelial_cell | *TMEM101* | -0.107513116 | 0.014697362 | 0.046474444 | 1 |
| Prediabetes | duct_epithelial_cell | *SCFD1* | -0.10968843 | 0.002141718 | 0.012886383 | 1 |
| Prediabetes | duct_epithelial_cell | *NUP50* | -0.146004979 | 0.000265869 | 0.008963481 | 1 |
| Prediabetes | duct_epithelial_cell | *COMT* | -0.158048846 | 0.006971274 | 0.027335463 | 1 |
| Prediabetes | duct_epithelial_cell | *SUCO* | -0.072853694 | 0.003956007 | 0.018817373 | 1 |
| Prediabetes | duct_epithelial_cell | *MSH2* | -0.411825349 | 0.001347345 | 0.010256616 | 1 |
| Prediabetes | duct_epithelial_cell | *TMEM38B* | -0.067969129 | 0.014384978 | 0.045690039 | 1 |
| Prediabetes | duct_epithelial_cell | *PSMD5* | -0.243318537 | 0.000558668 | 0.008963481 | 1 |
| Prediabetes | duct_epithelial_cell | *CRAT* | -0.157428029 | 0.005080291 | 0.022048127 | 1 |
| Prediabetes | duct_epithelial_cell | *IKZF5* | -0.206582954 | 2.46E-05 | 0.008963481 | 0.096573773 |
| Prediabetes | duct_epithelial_cell | *SRPK1* | -0.140181168 | 0.003778997 | 0.018285321 | 1 |
| Prediabetes | duct_epithelial_cell | *EFHC1* | -0.071337772 | 0.004045816 | 0.019105782 | 1 |
| Prediabetes | duct_epithelial_cell | *ABL1* | -0.21585135 | 0.000378841 | 0.008963481 | 1 |
| Prediabetes | duct_epithelial_cell | *SH3GLB1* | -0.11650065 | 0.000306923 | 0.008963481 | 1 |
| Prediabetes | duct_epithelial_cell | *ABLIM1* | -0.09967574 | 0.003517105 | 0.017534673 | 1 |
| Prediabetes | duct_epithelial_cell | *HNRNPM* | -0.089585044 | 0.001726967 | 0.011365586 | 1 |
| Prediabetes | duct_epithelial_cell | *TIMM13* | -0.118308666 | 0.00092166 | 0.009057947 | 1 |
| Prediabetes | duct_epithelial_cell | *POLR2E* | -0.191801406 | 0.000444909 | 0.008963481 | 1 |
| Prediabetes | duct_epithelial_cell | *ARVCF* | -0.169136747 | 0.000444135 | 0.008963481 | 1 |
| Prediabetes | duct_epithelial_cell | *TRMT2A* | -0.106678145 | 0.001376619 | 0.010271223 | 1 |
| Prediabetes | duct_epithelial_cell | *MED15* | -0.244447595 | 0.002404615 | 0.013772204 | 1 |
| Prediabetes | duct_epithelial_cell | *CABIN1* | -0.101106483 | 0.011066599 | 0.037577086 | 1 |
| Prediabetes | duct_epithelial_cell | *SPECC1L* | -0.126971762 | 0.001424865 | 0.010342775 | 1 |
| Prediabetes | duct_epithelial_cell | *SLC25A1* | -0.191077659 | 0.000835983 | 0.008963481 | 1 |
| Prediabetes | duct_epithelial_cell | *HPS4* | -0.099834592 | 0.003249361 | 0.016734297 | 1 |
| Prediabetes | duct_epithelial_cell | *CBY1* | -0.160606658 | 0.002796245 | 0.015172844 | 1 |
| Prediabetes | duct_epithelial_cell | *XBP1* | -0.076627491 | 0.00089887 | 0.009057947 | 1 |
| Prediabetes | duct_epithelial_cell | *TIMP3* | -0.068954928 | 0.000449074 | 0.008963481 | 1 |
| Prediabetes | duct_epithelial_cell | *MIOX* | -0.130748461 | 0.013065457 | 0.042604802 | 1 |
| Prediabetes | duct_epithelial_cell | *TOM1* | -0.166106802 | 0.008738289 | 0.031543827 | 1 |
| Prediabetes | duct_epithelial_cell | *TTLL12* | -0.330541617 | 0.005337434 | 0.022918885 | 1 |
| Prediabetes | duct_epithelial_cell | *RBFOX2* | -0.120260998 | 0.002333918 | 0.013496739 | 1 |
| Prediabetes | duct_epithelial_cell | *SAMM50* | -0.223997728 | 0.003392811 | 0.017190742 | 1 |
| Prediabetes | duct_epithelial_cell | *SGSM3* | -0.184422713 | 0.002883339 | 0.015391907 | 1 |
| Prediabetes | duct_epithelial_cell | *IFT27* | -0.095034928 | 0.002968044 | 0.01569508 | 1 |
| Prediabetes | duct_epithelial_cell | *ST13* | -0.097621489 | 0.000915212 | 0.009057947 | 1 |
| Prediabetes | duct_epithelial_cell | *POLR3H* | -0.15512586 | 0.002643769 | 0.014712986 | 1 |

| | | | | | | |
|---|---|---|---|---|---|---|
| Prediabetes | duct_epithelial_cell | *PMM1* | -0.195340569 | 0.001416299 | 0.010342775 | 1 |
| Prediabetes | duct_epithelial_cell | *PRMT5* | -0.25336405 | 0.000480426 | 0.008963481 | 1 |
| Prediabetes | duct_epithelial_cell | *VCPKMT* | -0.258287756 | 0.000199508 | 0.008963481 | 0.783867646 |
| Prediabetes | duct_epithelial_cell | *SOS2* | -0.109031558 | 0.015872917 | 0.049339155 | 1 |
| Prediabetes | duct_epithelial_cell | *CDKL1* | -0.122507496 | 0.005532339 | 0.023421841 | 1 |
| Prediabetes | duct_epithelial_cell | *NIN* | -0.271533375 | 0.00104677 | 0.009336398 | 1 |
| Prediabetes | duct_epithelial_cell | *TRIM9* | -0.057688366 | 0.014149619 | 0.045198253 | 1 |
| Prediabetes | duct_epithelial_cell | *GSTZ1* | -0.112790312 | 0.012348 | 0.040906653 | 1 |
| Prediabetes | duct_epithelial_cell | *TMED8* | -0.083616202 | 0.011575621 | 0.038858577 | 1 |
| Prediabetes | duct_epithelial_cell | *DAAM1* | -0.097229095 | 0.001970717 | 0.01235437 | 1 |
| Prediabetes | duct_epithelial_cell | *DHRS7* | -0.166855972 | 0.001044915 | 0.009336398 | 1 |
| Prediabetes | duct_epithelial_cell | *GSKIP* | -0.245284434 | 0.00066521 | 0.008963481 | 1 |
| Prediabetes | duct_epithelial_cell | *TRIP11* | -0.193966711 | 0.000411487 | 0.008963481 | 1 |
| Prediabetes | duct_epithelial_cell | *NELFCD* | -0.159672569 | 0.000878478 | 0.009011853 | 1 |
| Prediabetes | duct_epithelial_cell | *MTG2* | -0.242883035 | 5.48E-05 | 0.008963481 | 0.215365873 |
| Prediabetes | duct_epithelial_cell | *POFUT1* | -0.28011742 | 0.000832973 | 0.008963481 | 1 |
| Prediabetes | duct_epithelial_cell | *IDH3B* | -0.193293238 | 0.000655778 | 0.008963481 | 1 |
| Prediabetes | duct_epithelial_cell | *MAPRE1* | -0.09530991 | 0.00749572 | 0.028732375 | 1 |
| Prediabetes | duct_epithelial_cell | *TTI1* | -0.331958544 | 0.000565488 | 0.008963481 | 1 |
| Prediabetes | duct_epithelial_cell | *CHMP4B* | -0.1372382 | 0.000433459 | 0.008963481 | 1 |
| Prediabetes | duct_epithelial_cell | *DNTTIP1* | -0.160701424 | 0.002541624 | 0.014327176 | 1 |
| Prediabetes | duct_epithelial_cell | *VAPA* | -0.114607585 | 0.000621925 | 0.008963481 | 1 |
| Prediabetes | duct_epithelial_cell | *LIPG* | -0.332084603 | 0.000654721 | 0.008963481 | 1 |
| Prediabetes | duct_epithelial_cell | *RIOK3* | -0.111227811 | 0.001593448 | 0.010911694 | 1 |
| Prediabetes | duct_epithelial_cell | *NALCN* | -0.076613837 | 0.010109162 | 0.035211789 | 1 |
| Prediabetes | duct_epithelial_cell | *FGF14* | -0.050433785 | 0.004073315 | 0.019166532 | 1 |
| Prediabetes | duct_epithelial_cell | *FNDC3A* | -0.087969239 | 0.001637807 | 0.011037642 | 1 |
| Prediabetes | duct_epithelial_cell | *DNAJC3* | -0.117830284 | 0.000834139 | 0.008963481 | 1 |
| Prediabetes | duct_epithelial_cell | *UGGT2* | -0.141312993 | 0.002315183 | 0.013483149 | 1 |
| Prediabetes | duct_epithelial_cell | *ARHGEF7* | -0.116417247 | 0.001872424 | 0.011928428 | 1 |
| Prediabetes | duct_epithelial_cell | *CLN5* | -0.174107709 | 0.000364759 | 0.008963481 | 1 |
| Prediabetes | duct_epithelial_cell | *MGRN1* | -0.125443297 | 0.000286915 | 0.008963481 | 1 |
| Prediabetes | duct_epithelial_cell | *LONP2* | -0.111642562 | 0.002128091 | 0.012824032 | 1 |
| Prediabetes | duct_epithelial_cell | *USB1* | -0.162646285 | 0.006849417 | 0.026992335 | 1 |
| Prediabetes | duct_epithelial_cell | *CYB5B* | -0.195690772 | 0.000801056 | 0.008963481 | 1 |
| Prediabetes | duct_epithelial_cell | *NME3* | -0.112567537 | 0.000315507 | 0.008963481 | 1 |
| Prediabetes | duct_epithelial_cell | *NPRL3* | -0.107165788 | 0.002669544 | 0.014746113 | 1 |
| Prediabetes | duct_epithelial_cell | *NAGPA* | -0.185205291 | 0.006791376 | 0.026844381 | 1 |
| Prediabetes | duct_epithelial_cell | *NME4* | -0.124759247 | 0.001187206 | 0.009719245 | 1 |
| Prediabetes | duct_epithelial_cell | *NOMO3* | -0.111845941 | 0.000961836 | 0.009147222 | 1 |
| Prediabetes | duct_epithelial_cell | *LMF1* | -0.229140116 | 0.010324925 | 0.035836244 | 1 |
| Prediabetes | duct_epithelial_cell | *HAGHL* | -0.076437516 | 0.01113412 | 0.037693769 | 1 |
| Prediabetes | duct_epithelial_cell | *METRN* | -0.082378936 | 0.002874033 | 0.015391043 | 1 |
| Prediabetes | duct_epithelial_cell | *HMOX2* | -0.202333873 | 0.000452245 | 0.008963481 | 1 |
| Prediabetes | duct_epithelial_cell | *RBL2* | -0.080386147 | 0.015975802 | 0.049463299 | 1 |
| Prediabetes | duct_epithelial_cell | *KAT8* | -0.150123335 | 0.006444824 | 0.02571346 | 1 |
| Prediabetes | duct_epithelial_cell | *RNF40* | -0.118806742 | 0.0072169 | 0.027881219 | 1 |
| Prediabetes | duct_epithelial_cell | *AP3B2* | -0.086043789 | 0.00965663 | 0.033906075 | 1 |
| Prediabetes | duct_epithelial_cell | *RPAP1* | -0.224221797 | 0.001048637 | 0.009336398 | 1 |
| Prediabetes | duct_epithelial_cell | *DMXL2* | -0.088927341 | 0.00431652 | 0.019789508 | 1 |

| | | | | | | |
|---|---|---|---|---|---|---|
| Prediabetes | duct_epithelial_cell | *DNAJC17* | -0.189360626 | 0.012604367 | 0.04147618 | 1 |
| Prediabetes | duct_epithelial_cell | *ZDHHC2* | -0.21165749 | 0.000391329 | 0.008963481 | 1 |
| Prediabetes | duct_epithelial_cell | *NBN* | -0.126828828 | 0.006446363 | 0.02571346 | 1 |
| Prediabetes | duct_epithelial_cell | *RAB2A* | -0.056812572 | 0.011218154 | 0.037931263 | 1 |
| Prediabetes | duct_epithelial_cell | *EIF3E* | -0.085497064 | 0.001601177 | 0.01094091 | 1 |
| Prediabetes | duct_epithelial_cell | *ESRP1* | -0.222827131 | 0.000899696 | 0.009057947 | 1 |
| Prediabetes | duct_epithelial_cell | *ARMC1* | -0.118371804 | 0.001290794 | 0.010044933 | 1 |
| Prediabetes | duct_epithelial_cell | *CHRAC1* | -0.164503012 | 0.001386851 | 0.010271223 | 1 |
| Prediabetes | duct_epithelial_cell | *INTS10* | -0.116519051 | 0.001242493 | 0.009849499 | 1 |
| Prediabetes | duct_epithelial_cell | *LEPROTL1* | -0.172074824 | 0.001668704 | 0.01113209 | 1 |
| Prediabetes | duct_epithelial_cell | *KLHDC4* | -0.105122124 | 0.004630558 | 0.020792527 | 1 |
| Prediabetes | duct_epithelial_cell | *ASAH1* | -0.099424083 | 0.001311151 | 0.010116188 | 1 |
| Prediabetes | duct_epithelial_cell | *MAN2B1* | -0.096305189 | 0.008704182 | 0.031543827 | 1 |
| Prediabetes | duct_epithelial_cell | *HNRNPL* | -0.108216198 | 0.001920047 | 0.012108933 | 1 |
| Prediabetes | duct_epithelial_cell | *LIN7B* | -0.080640554 | 0.00129109 | 0.010044933 | 1 |
| Prediabetes | duct_epithelial_cell | *ERCC2* | -0.144387207 | 0.010796063 | 0.036981459 | 1 |
| Prediabetes | duct_epithelial_cell | *NUMBL* | -0.104816808 | 0.004268491 | 0.019612585 | 1 |
| Prediabetes | duct_epithelial_cell | *PLIN3* | -0.045181725 | 0.012199694 | 0.040552114 | 1 |
| Prediabetes | duct_epithelial_cell | *ZNF175* | -0.141808458 | 0.014039268 | 0.044918798 | 1 |
| Prediabetes | duct_epithelial_cell | *PLA2G4C* | -0.073031667 | 0.016047468 | 0.049607005 | 1 |
| Prediabetes | duct_epithelial_cell | *GCDH* | -0.187481739 | 0.001122237 | 0.009564575 | 1 |
| Prediabetes | duct_epithelial_cell | *PIK3R2* | -0.238318037 | 0.001017636 | 0.009336398 | 1 |
| Prediabetes | duct_epithelial_cell | *UPK1A* | -0.722949664 | 0.002875293 | 0.015391043 | 1 |
| Prediabetes | duct_epithelial_cell | *COPE* | -0.106853279 | 0.000764238 | 0.008963481 | 1 |
| Prediabetes | duct_epithelial_cell | *SIPA1L3* | -0.211269271 | 0.000544795 | 0.008963481 | 1 |
| Prediabetes | duct_epithelial_cell | *PMPCB* | -0.202506536 | 0.000690321 | 0.008963481 | 1 |
| Prediabetes | duct_epithelial_cell | *TSPAN12* | -0.124529689 | 0.003493719 | 0.017450459 | 1 |
| Prediabetes | duct_epithelial_cell | *CHCHD2* | -0.071513831 | 0.002887207 | 0.015391907 | 1 |
| Prediabetes | duct_epithelial_cell | *EIF3B* | -0.115584871 | 0.001109091 | 0.009564575 | 1 |
| Prediabetes | duct_epithelial_cell | *C1GALT1* | -0.151708782 | 0.000615999 | 0.008963481 | 1 |
| Prediabetes | duct_epithelial_cell | *RPA3* | -0.121464597 | 0.000444789 | 0.008963481 | 1 |
| Prediabetes | duct_epithelial_cell | *TMEM106B* | -0.110913799 | 0.005078551 | 0.022048127 | 1 |
| Prediabetes | duct_epithelial_cell | *ACTR3C* | -0.241691309 | 0.011826672 | 0.03954638 | 1 |
| Prediabetes | duct_epithelial_cell | *TSPAN13* | -0.07349659 | 0.002818512 | 0.015228566 | 1 |
| Prediabetes | duct_epithelial_cell | *COA1* | -0.192862989 | 0.000493702 | 0.008963481 | 1 |
| Prediabetes | duct_epithelial_cell | *BLVRA* | -0.171432337 | 0.000349637 | 0.008963481 | 1 |
| Prediabetes | duct_epithelial_cell | *TMEM248* | -0.104887499 | 0.002339343 | 0.013496739 | 1 |
| Prediabetes | duct_epithelial_cell | *RHEB* | -0.102118637 | 0.000681382 | 0.008963481 | 1 |
| Prediabetes | duct_epithelial_cell | *SPIN1* | -0.198275344 | 0.000281449 | 0.008963481 | 1 |
| Prediabetes | duct_epithelial_cell | *NMRK1* | -0.135740724 | 0.000793494 | 0.008963481 | 1 |
| Prediabetes | duct_epithelial_cell | *SUSD1* | -0.116012425 | 0.002200136 | 0.012987238 | 1 |
| Prediabetes | duct_epithelial_cell | *DNM1* | -0.081468799 | 0.005467628 | 0.023274443 | 1 |
| Prediabetes | duct_epithelial_cell | *NPDC1* | -0.050959021 | 0.005864987 | 0.02428191 | 1 |
| Prediabetes | duct_epithelial_cell | *HPS1* | -0.189705927 | 0.001776158 | 0.01151572 | 1 |
| Prediabetes | duct_epithelial_cell | *ATE1* | -0.158562958 | 0.003745175 | 0.018194991 | 1 |
| Prediabetes | duct_epithelial_cell | *PLEKHA1* | -0.118522765 | 0.000118976 | 0.008963481 | 0.467457682 |
| Prediabetes | duct_epithelial_cell | *SPOCK2* | -0.65536316 | 0.000698866 | 0.008963481 | 1 |
| Prediabetes | duct_epithelial_cell | *BMPR1A* | -0.293794992 | 0.001184207 | 0.009719245 | 1 |
| Prediabetes | duct_epithelial_cell | *LIPA* | -0.206818286 | 0.000838229 | 0.008963481 | 1 |
| Prediabetes | duct_epithelial_cell | *TNKS2* | -0.127714501 | 0.000674747 | 0.008963481 | 1 |

| | | | | | | |
|---|---|---|---|---|---|---|
| Prediabetes | duct_epithelial_cell | *ACBD5* | -0.195915416 | 0.000742108 | 0.008963481 | 1 |
| Prediabetes | duct_epithelial_cell | *GTPBP4* | -0.162780104 | 0.000844938 | 0.008963481 | 1 |
| Prediabetes | duct_epithelial_cell | *XPNPEP1* | -0.20477485 | 0.003887107 | 0.01867047 | 1 |
| Prediabetes | duct_epithelial_cell | *SMC3* | -0.126954288 | 0.00113691 | 0.00963464 | 1 |
| Prediabetes | duct_epithelial_cell | *CWC25* | -0.160392519 | 0.004039963 | 0.019101102 | 1 |
| Prediabetes | duct_epithelial_cell | *TRIM37* | -0.107022542 | 0.002562474 | 0.014403376 | 1 |
| Prediabetes | duct_epithelial_cell | *MED13* | -0.08618318 | 0.00187321 | 0.011928428 | 1 |
| Prediabetes | duct_epithelial_cell | *CCDC47* | -0.144444913 | 0.000482087 | 0.008963481 | 1 |
| Prediabetes | duct_epithelial_cell | *DRG2* | -0.184193579 | 0.000518696 | 0.008963481 | 1 |
| Prediabetes | duct_epithelial_cell | *SMARCD2* | -0.198927944 | 0.004680611 | 0.02090362 | 1 |
| Prediabetes | duct_epithelial_cell | *C17orf75* | -0.072273951 | 0.014930271 | 0.046928828 | 1 |
| Prediabetes | duct_epithelial_cell | *NAGLU* | -0.107981825 | 1.51E-05 | 0.008963481 | 0.059205576 |
| Prediabetes | duct_epithelial_cell | *RND2* | -0.419276645 | 0.000368551 | 0.008963481 | 1 |
| Prediabetes | duct_epithelial_cell | *MPP2* | -0.070562521 | 0.007275707 | 0.028080799 | 1 |
| Prediabetes | duct_epithelial_cell | *HLF* | -0.06249945 | 0.009874087 | 0.034546116 | 1 |
| Prediabetes | duct_epithelial_cell | *EFNB3* | -0.100512923 | 0.003418556 | 0.017241986 | 1 |
| Prediabetes | duct_epithelial_cell | *RANGRF* | -0.175348894 | 0.001352234 | 0.010256616 | 1 |
| Prediabetes | duct_epithelial_cell | *DPH1* | -0.247810079 | 0.001120451 | 0.009564575 | 1 |
| Prediabetes | duct_epithelial_cell | *OCIAD1* | -0.100749352 | 0.000874323 | 0.009011853 | 1 |
| Prediabetes | duct_epithelial_cell | *UBE2D3* | -0.078236769 | 0.00529699 | 0.022770103 | 1 |
| Prediabetes | duct_epithelial_cell | *TRIM2* | -0.104343162 | 0.002249011 | 0.01324792 | 1 |
| Prediabetes | duct_epithelial_cell | *ST3GAL4* | -0.100671419 | 0.009320625 | 0.033021401 | 1 |
| Prediabetes | duct_epithelial_cell | *PRPF19* | -0.119585103 | 0.004116851 | 0.019282694 | 1 |
| Prediabetes | duct_epithelial_cell | *MDK* | -0.160028007 | 0.002898229 | 0.01542973 | 1 |
| Prediabetes | duct_epithelial_cell | *MADD* | -0.091906058 | 0.006039165 | 0.02471654 | 1 |
| Prediabetes | duct_epithelial_cell | *NAA40* | -0.183999389 | 0.008675447 | 0.031502618 | 1 |
| Prediabetes | duct_epithelial_cell | *NUP98* | -0.167143219 | 0.000705297 | 0.008963481 | 1 |
| Prediabetes | duct_epithelial_cell | *GTF2H1* | -0.123133921 | 0.001591647 | 0.010911694 | 1 |
| Prediabetes | duct_epithelial_cell | *METAP2* | -0.10371939 | 0.000639856 | 0.008963481 | 1 |
| Prediabetes | duct_epithelial_cell | *ITFG2* | -0.2595012 | 0.00119087 | 0.009719245 | 1 |
| Prediabetes | duct_epithelial_cell | *CREBL2* | -0.149128842 | 0.004972854 | 0.021830553 | 1 |
| Prediabetes | duct_epithelial_cell | *LTBR* | -0.103694089 | 0.015007454 | 0.047096075 | 1 |
| Prediabetes | duct_epithelial_cell | *CAND1* | -0.103332583 | 0.001916969 | 0.012108933 | 1 |
| Prediabetes | duct_epithelial_cell | *KRR1* | -0.124381751 | 9.13E-05 | 0.008963481 | 0.358668134 |
| Prediabetes | duct_epithelial_cell | *ATN1* | -0.121778877 | 0.011959987 | 0.039890314 | 1 |
| Prediabetes | duct_epithelial_cell | *GOLT1B* | -0.11488142 | 0.001743014 | 0.011413834 | 1 |
| Prediabetes | duct_epithelial_cell | *C2CD5* | -0.269758313 | 0.000253938 | 0.008963481 | 0.997723735 |
| Prediabetes | duct_epithelial_cell | *SRSF9* | -0.087856859 | 0.008120756 | 0.030300524 | 1 |
| Prediabetes | duct_epithelial_cell | *RSPH4A* | -0.052072841 | 0.010418972 | 0.036003641 | 1 |
| Prediabetes | duct_epithelial_cell | *SMIM8* | -0.202385421 | 0.003477246 | 0.017403948 | 1 |
| Prediabetes | duct_epithelial_cell | *NEDD9* | -0.127419771 | 0.001030973 | 0.009336398 | 1 |
| Prediabetes | duct_epithelial_cell | *CEP85L* | -0.08671947 | 0.012359217 | 0.04090932 | 1 |
| Prediabetes | duct_epithelial_cell | *RNF8* | -0.151405584 | 0.002864374 | 0.015391043 | 1 |
| Prediabetes | duct_epithelial_cell | *GLP1R* | -0.048447344 | 0.011822573 | 0.03954638 | 1 |
| Prediabetes | duct_epithelial_cell | *FBXL4* | -0.217031287 | 0.000658973 | 0.008963481 | 1 |
| Prediabetes | duct_epithelial_cell | *ASCC3* | -0.093229828 | 0.015418314 | 0.048231336 | 1 |
| Prediabetes | duct_epithelial_cell | *C6orf62* | -0.170259197 | 0.000484421 | 0.008963481 | 1 |
| Prediabetes | duct_epithelial_cell | *B3GAT2* | -0.354106693 | 0.000581964 | 0.008963481 | 1 |
| Prediabetes | duct_epithelial_cell | *HBS1L* | -0.143145962 | 0.00101661 | 0.009336398 | 1 |
| Prediabetes | duct_epithelial_cell | *PEX7* | -0.21485091 | 0.003532712 | 0.017534673 | 1 |

| | | | | | | |
|---|---|---|---|---|---|---|
| Prediabetes | duct_epithelial_cell | *PHACTR2* | -0.117421093 | 0.000925919 | 0.009057947 | 1 |
| Prediabetes | duct_epithelial_cell | *TFEB* | -0.173281403 | 0.001192933 | 0.009719245 | 1 |
| Prediabetes | duct_epithelial_cell | *DUSP22* | -0.250273745 | 0.004342217 | 0.019814833 | 1 |
| Prediabetes | duct_epithelial_cell | *FBRSL1* | -0.248106112 | 0.000195644 | 0.008963481 | 0.768687046 |
| Prediabetes | duct_epithelial_cell | *SEMA5A* | -0.099719233 | 0.002992631 | 0.015740359 | 1 |
| Prediabetes | duct_epithelial_cell | *GHR* | -1.014259622 | 0.0005803 | 0.008963481 | 1 |
| Prediabetes | duct_epithelial_cell | *BRD8* | -0.207830839 | 0.000646606 | 0.008963481 | 1 |
| Prediabetes | duct_epithelial_cell | *NNT* | -0.092916216 | 0.000784445 | 0.008963481 | 1 |
| Prediabetes | duct_epithelial_cell | *IK* | -0.138143581 | 0.000601177 | 0.008963481 | 1 |
| Prediabetes | duct_epithelial_cell | *PDE8B* | -0.082596462 | 0.010721337 | 0.036757533 | 1 |
| Prediabetes | duct_epithelial_cell | *GOLPH3* | -0.163401718 | 0.000392002 | 0.008963481 | 1 |
| Prediabetes | duct_epithelial_cell | *RAD50* | -0.182629289 | 0.000447926 | 0.008963481 | 1 |
| Prediabetes | duct_epithelial_cell | *SMAD5* | -0.110391943 | 0.001994551 | 0.012380078 | 1 |
| Prediabetes | duct_epithelial_cell | *HMGXB3* | -0.120639871 | 0.002039034 | 0.012486647 | 1 |
| Prediabetes | duct_epithelial_cell | *STC2* | -0.129480977 | 0.00322394 | 0.016688879 | 1 |
| Prediabetes | duct_epithelial_cell | *ACTR8* | -0.307787274 | 0.000913432 | 0.009057947 | 1 |
| Prediabetes | duct_epithelial_cell | *TIMMDC1* | -0.117125874 | 0.004136965 | 0.019327151 | 1 |
| Prediabetes | duct_epithelial_cell | *RBP1* | -0.154546837 | 0.001248422 | 0.009849499 | 1 |
| Prediabetes | duct_epithelial_cell | *RNF7* | -0.080372355 | 0.003542376 | 0.017534673 | 1 |
| Prediabetes | duct_epithelial_cell | *WNT5A* | -0.087320811 | 0.016121831 | 0.049752675 | 1 |
| Prediabetes | duct_epithelial_cell | *CBLB* | -0.141219456 | 0.003310076 | 0.016870817 | 1 |
| Prediabetes | duct_epithelial_cell | *NCBP2* | -0.131144214 | 0.004823325 | 0.021317033 | 1 |
| Prediabetes | duct_epithelial_cell | *ATP6V1A* | -0.168462236 | 0.000535618 | 0.008963481 | 1 |
| Prediabetes | duct_epithelial_cell | *PODXL2* | -0.054768955 | 0.008629967 | 0.031424597 | 1 |
| Prediabetes | duct_epithelial_cell | *PLSCR4* | -0.268129764 | 0.008375313 | 0.030753836 | 1 |
| Prediabetes | duct_epithelial_cell | *COMMD2* | -0.196629601 | 0.000263706 | 0.008963481 | 1 |
| Prediabetes | duct_epithelial_cell | *ARHGEF26* | -0.294436456 | 0.005084144 | 0.022048127 | 1 |
| Prediabetes | duct_epithelial_cell | *PLCH1* | -0.280575543 | 0.00105043 | 0.009336398 | 1 |
| Prediabetes | duct_epithelial_cell | *NKTR* | -0.100074748 | 0.000832859 | 0.008963481 | 1 |
| Prediabetes | duct_epithelial_cell | *NEK4* | -0.186633014 | 0.00549884 | 0.023344611 | 1 |
| Prediabetes | duct_epithelial_cell | *ACTR3* | -0.130137059 | 0.000448402 | 0.008963481 | 1 |
| Prediabetes | duct_epithelial_cell | *POMC* | -0.108930966 | 0.002928534 | 0.015548931 | 1 |
| Prediabetes | duct_epithelial_cell | *RTN4* | -0.12992592 | 0.000286964 | 0.008963481 | 1 |
| Prediabetes | duct_epithelial_cell | *HTRA2* | -0.142161501 | 0.014541765 | 0.046039156 | 1 |
| Prediabetes | duct_epithelial_cell | *WDR75* | -0.212354078 | 0.000672205 | 0.008963481 | 1 |
| Prediabetes | duct_epithelial_cell | *OTX1* | -0.155963914 | 0.000200663 | 0.008963481 | 0.788404783 |
| Prediabetes | duct_epithelial_cell | *SF3B1* | -0.119880018 | 0.000706886 | 0.008963481 | 1 |
| Prediabetes | duct_epithelial_cell | *CNPPD1* | -0.078083175 | 0.007582956 | 0.028953774 | 1 |
| Prediabetes | duct_epithelial_cell | *HDLBP* | -0.086972235 | 0.002384312 | 0.013695852 | 1 |
| Prediabetes | duct_epithelial_cell | *CEBPZ* | -0.096346095 | 0.00144422 | 0.010411636 | 1 |
| Prediabetes | duct_epithelial_cell | *WIPF1* | -0.18287194 | 0.002011457 | 0.012406615 | 1 |
| Prediabetes | duct_epithelial_cell | *COX7A2L* | -0.094899996 | 0.001048316 | 0.009336398 | 1 |
| Prediabetes | duct_epithelial_cell | *PNO1* | -0.112240749 | 0.011084714 | 0.037577086 | 1 |
| Prediabetes | duct_epithelial_cell | *ARID3A* | -0.144939642 | 0.001594124 | 0.010911694 | 1 |
| Prediabetes | duct_epithelial_cell | *ALMS1* | -0.112861521 | 0.001616946 | 0.011022922 | 1 |
| Prediabetes | duct_epithelial_cell | *MARK1* | -0.13807993 | 0.00343633 | 0.017287247 | 1 |
| Prediabetes | duct_epithelial_cell | *SCP2* | -0.124981087 | 0.000740503 | 0.008963481 | 1 |
| Prediabetes | duct_epithelial_cell | *FAM20B* | -0.12803453 | 0.007817989 | 0.029507089 | 1 |
| Prediabetes | duct_epithelial_cell | *WRAP73* | -0.304373543 | 0.001053248 | 0.009336398 | 1 |
| Prediabetes | duct_epithelial_cell | *PARK7* | -0.087910162 | 0.001985305 | 0.012369325 | 1 |

| | | | | | | |
|---|---|---|---|---|---|---|
| Prediabetes | duct_epithelial_cell | *EDEM3* | -0.096950478 | 0.003547993 | 0.017534673 | 1 |
| Prediabetes | duct_epithelial_cell | *FBXO6* | -0.250885965 | 0.013550783 | 0.043806482 | 1 |
| Prediabetes | duct_epithelial_cell | *SMG7* | -0.16966424 | 0.001142719 | 0.00963464 | 1 |
| Prediabetes | duct_epithelial_cell | *BCAS2* | -0.107558989 | 0.005022583 | 0.021950756 | 1 |
| Prediabetes | duct_epithelial_cell | *PLEKHM2* | -0.119554036 | 0.00340418 | 0.017213674 | 1 |
| Prediabetes | duct_epithelial_cell | *UAP1* | -0.119576022 | 0.000100104 | 0.008963481 | 0.393307251 |
| Prediabetes | duct_epithelial_cell | *CTBS* | -0.071601187 | 0.004117633 | 0.019282694 | 1 |
| Prediabetes | duct_epithelial_cell | *CD46* | -0.118236156 | 0.000688703 | 0.008963481 | 1 |
| Prediabetes | duct_epithelial_cell | *CCDC181* | -0.061393609 | 0.013620451 | 0.043936578 | 1 |
| Prediabetes | duct_epithelial_cell | *NSUN4* | -0.130128999 | 0.002037273 | 0.012486647 | 1 |
| Prediabetes | duct_epithelial_cell | *TMED5* | -0.176022849 | 0.000507513 | 0.008963481 | 1 |
| Prediabetes | duct_epithelial_cell | *DR1* | -0.120358487 | 0.00308209 | 0.01610824 | 1 |
| Prediabetes | duct_epithelial_cell | *PTBP2* | -0.093341381 | 0.009149229 | 0.032620074 | 1 |
| Prediabetes | duct_epithelial_cell | *NSL1* | -0.129039203 | 0.001986534 | 0.012369325 | 1 |
| Prediabetes | duct_epithelial_cell | *ESYT2* | -0.125722257 | 0.007034123 | 0.027390555 | 1 |
| Prediabetes | duct_epithelial_cell | *TTR* | -0.02676366 | 0.006306624 | 0.025388037 | 1 |
| Prediabetes | duct_epithelial_cell | *SPCS2* | -0.115608782 | 0.000852545 | 0.008963481 | 1 |
| Prediabetes | duct_epithelial_cell | *ELOVL4* | -0.078079708 | 0.010453606 | 0.036028262 | 1 |
| Prediabetes | duct_epithelial_cell | *FILIP1* | -0.26639268 | 3.39E-05 | 0.008963481 | 0.133315594 |
| Prediabetes | duct_epithelial_cell | *RNF146* | -0.120244883 | 0.002396768 | 0.013747304 | 1 |
| Prediabetes | duct_epithelial_cell | *MED28* | -0.095654541 | 0.011308542 | 0.038171186 | 1 |
| Prediabetes | duct_epithelial_cell | *ARMC2* | -0.072757969 | 0.002705713 | 0.014880018 | 1 |
| Prediabetes | duct_epithelial_cell | *PKD2* | -0.250635749 | 0.000679972 | 0.008963481 | 1 |
| Prediabetes | duct_epithelial_cell | *CCDC92* | -0.199547415 | 0.001323328 | 0.010174863 | 1 |
| Prediabetes | duct_epithelial_cell | *RBM18* | -0.242813993 | 0.000388079 | 0.008963481 | 1 |
| Prediabetes | duct_epithelial_cell | *DCAF4* | -0.076574054 | 0.008275733 | 0.030617094 | 1 |
| Prediabetes | duct_epithelial_cell | *IFI27L2* | -0.082540793 | 0.004625128 | 0.020791908 | 1 |
| Prediabetes | duct_epithelial_cell | *ZC2HC1C* | -0.161587115 | 0.000173404 | 0.008963481 | 0.681303087 |
| Prediabetes | duct_epithelial_cell | *AFTPH* | -0.097004478 | 0.008002729 | 0.030059963 | 1 |
| Prediabetes | duct_epithelial_cell | *EPCAM* | -0.128007344 | 0.002691328 | 0.014830613 | 1 |
| Prediabetes | duct_epithelial_cell | *SLC17A5* | -0.185098432 | 3.87E-06 | 0.008963481 | 0.015204514 |
| Prediabetes | duct_epithelial_cell | *HELLS* | -0.191029292 | 0.014239606 | 0.045368317 | 1 |
| Prediabetes | duct_epithelial_cell | *RCL1* | -0.158181529 | 0.001215219 | 0.009784005 | 1 |
| Prediabetes | duct_epithelial_cell | *CD274* | -0.28743252 | 0.002546017 | 0.014331379 | 1 |
| Prediabetes | duct_epithelial_cell | *SLC10A7* | -0.08948795 | 0.003720799 | 0.01813774 | 1 |
| Prediabetes | duct_epithelial_cell | *IQSEC3* | -0.060341458 | 0.012511135 | 0.041203899 | 1 |
| Prediabetes | duct_epithelial_cell | *DNAJC15* | -0.125672476 | 0.001194807 | 0.009719245 | 1 |
| Prediabetes | duct_epithelial_cell | *PROSER1* | -0.23513474 | 0.002653217 | 0.014744682 | 1 |
| Prediabetes | duct_epithelial_cell | *WBP4* | -0.178617954 | 0.000464167 | 0.008963481 | 1 |
| Prediabetes | duct_epithelial_cell | *KBTBD7* | -0.146397585 | 0.002750025 | 0.015090572 | 1 |
| Prediabetes | duct_epithelial_cell | *FAM53C* | -0.094417924 | 0.010344159 | 0.035853926 | 1 |
| Prediabetes | duct_epithelial_cell | *SIL1* | -0.060613187 | 0.014021417 | 0.044898247 | 1 |
| Prediabetes | duct_epithelial_cell | *UTP20* | -0.10724264 | 0.00609786 | 0.024776105 | 1 |
| Prediabetes | duct_epithelial_cell | *PPP3CC* | -0.16026733 | 0.000855341 | 0.008963481 | 1 |
| Prediabetes | duct_epithelial_cell | *EPHX2* | -0.219092994 | 0.000619483 | 0.008963481 | 1 |
| Prediabetes | duct_epithelial_cell | *IAPP* | -0.026628457 | 0.008337048 | 0.030749039 | 1 |
| Prediabetes | duct_epithelial_cell | *ZNF211* | -0.045002347 | 0.010448593 | 0.036028262 | 1 |
| Prediabetes | duct_epithelial_cell | *POLK* | -0.092868652 | 0.011079887 | 0.037577086 | 1 |
| Prediabetes | duct_epithelial_cell | *UBL3* | -0.098355002 | 0.001372276 | 0.010271223 | 1 |
| Prediabetes | duct_epithelial_cell | *NAA60* | -0.220047845 | 0.000192148 | 0.008963481 | 0.754949697 |

| | | | | | | |
|---|---|---|---|---|---|---|
| Prediabetes | duct_epithelial_cell | *KLHL7* | -0.2467194 | 0.002764132 | 0.01511602 | 1 |
| Prediabetes | duct_epithelial_cell | *CBX3* | -0.188284312 | 0.000451995 | 0.008963481 | 1 |
| Prediabetes | duct_epithelial_cell | *INHBA* | -0.062222592 | 0.006734108 | 0.026644824 | 1 |
| Prediabetes | duct_epithelial_cell | *BHLHE41* | -0.184640755 | 0.000492884 | 0.008963481 | 1 |
| Prediabetes | duct_epithelial_cell | *USP45* | -0.17532423 | 0.001541362 | 0.010718604 | 1 |
| Prediabetes | duct_epithelial_cell | *ARFGEF2* | -0.149032086 | 0.001168334 | 0.009719245 | 1 |
| Prediabetes | duct_epithelial_cell | *ZNFX1* | -0.274161595 | 0.001466662 | 0.01048303 | 1 |
| Prediabetes | duct_epithelial_cell | *BCAS4* | -0.140446551 | 0.010583647 | 0.036348905 | 1 |
| Prediabetes | duct_epithelial_cell | *PEPD* | -0.129463033 | 0.002050211 | 0.012527651 | 1 |
| Prediabetes | duct_epithelial_cell | *SNRNP27* | -0.094273429 | 0.006093513 | 0.024776105 | 1 |
| Prediabetes | duct_epithelial_cell | *USP22* | -0.129182657 | 0.000743592 | 0.008963481 | 1 |
| Prediabetes | duct_epithelial_cell | *MRS2* | -0.249102474 | 0.000477541 | 0.008963481 | 1 |
| Prediabetes | duct_epithelial_cell | *SNRPC* | -0.162046574 | 0.000199209 | 0.008963481 | 0.78269333 |
| Prediabetes | duct_epithelial_cell | *SERPINB6* | -0.115965993 | 0.001028465 | 0.009336398 | 1 |
| Prediabetes | duct_epithelial_cell | *GLO1* | -0.165354504 | 0.000783274 | 0.008963481 | 1 |
| Prediabetes | duct_epithelial_cell | *ATXN1* | -0.203531134 | 0.000296402 | 0.008963481 | 1 |
| Prediabetes | duct_epithelial_cell | *LRRFIP1* | -0.12550133 | 0.002079562 | 0.012608949 | 1 |
| Prediabetes | duct_epithelial_cell | *SH3TC1* | -0.166447795 | 0.00198967 | 0.012369325 | 1 |
| Prediabetes | duct_epithelial_cell | *MT1G* | -0.022136699 | 0.011323344 | 0.038188343 | 1 |
| Prediabetes | duct_epithelial_cell | *TM9SF2* | -0.100439392 | 0.012796104 | 0.041896577 | 1 |
| Prediabetes | duct_epithelial_cell | *MRPS7* | -0.119489427 | 0.003599122 | 0.017765011 | 1 |
| Prediabetes | duct_epithelial_cell | *PPDPF* | -0.144802707 | 0.000526025 | 0.008963481 | 1 |
| Prediabetes | duct_epithelial_cell | *CHCHD5* | -0.110970536 | 0.003045754 | 0.015976995 | 1 |
| Prediabetes | duct_epithelial_cell | *NAPB* | -0.070813719 | 0.00597835 | 0.024570017 | 1 |
| Prediabetes | duct_epithelial_cell | *RBCK1* | -0.083124111 | 0.015466624 | 0.048305536 | 1 |
| Prediabetes | duct_epithelial_cell | *TUBGCP3* | -0.193240371 | 0.000917366 | 0.009057947 | 1 |
| Prediabetes | duct_epithelial_cell | *MCF2L* | -0.095117888 | 0.000954055 | 0.009132479 | 1 |
| Prediabetes | duct_epithelial_cell | *NR1D1* | -0.087574846 | 0.006005665 | 0.024630748 | 1 |
| Prediabetes | duct_epithelial_cell | *PRMT1* | -0.08865452 | 0.0024385 | 0.013905466 | 1 |
| Prediabetes | duct_epithelial_cell | *BECN1* | -0.128977006 | 0.005884928 | 0.024287691 | 1 |
| Prediabetes | duct_epithelial_cell | *ZNF384* | -0.294094964 | 0.000389421 | 0.008963481 | 1 |
| Prediabetes | duct_epithelial_cell | *HSPA2* | -0.096833387 | 0.00935706 | 0.03309081 | 1 |
| Prediabetes | duct_epithelial_cell | *ABHD8* | -0.093228832 | 0.009933762 | 0.034693111 | 1 |
| Prediabetes | duct_epithelial_cell | *YEATS4* | -0.105904353 | 0.007648475 | 0.029024613 | 1 |
| Prediabetes | duct_epithelial_cell | *IDUA* | -0.290135448 | 0.000588465 | 0.008963481 | 1 |
| Prediabetes | duct_epithelial_cell | *UBR4* | -0.138896284 | 0.001402985 | 0.010284193 | 1 |
| Prediabetes | duct_epithelial_cell | *WDR24* | -0.472765615 | 0.000531211 | 0.008963481 | 1 |
| Prediabetes | duct_epithelial_cell | *KDM4B* | -0.18488678 | 0.007201356 | 0.027848551 | 1 |
| Prediabetes | duct_epithelial_cell | *METTL25* | -0.30310594 | 0.001025825 | 0.009336398 | 1 |
| Prediabetes | duct_epithelial_cell | *POR* | -0.206938688 | 0.001521971 | 0.010704482 | 1 |
| Prediabetes | duct_epithelial_cell | *RBM48* | -0.257488573 | 0.000320137 | 0.008963481 | 1 |
| Prediabetes | duct_epithelial_cell | *A4GALT* | 0.042714311 | 0.003671633 | 0.017966065 | 1 |
| Prediabetes | duct_epithelial_cell | *NAA38* | -0.102924548 | 0.00067299 | 0.008963481 | 1 |
| Prediabetes | duct_epithelial_cell | *PRKRIP1* | -0.159710061 | 0.001753254 | 0.011442746 | 1 |
| Prediabetes | duct_epithelial_cell | *VGF* | -0.037232182 | 0.005486065 | 0.02332765 | 1 |
| Prediabetes | duct_epithelial_cell | *MTX2* | -0.183102013 | 0.000517805 | 0.008963481 | 1 |
| Prediabetes | duct_epithelial_cell | *PSMG2* | -0.127688187 | 0.002272092 | 0.013326143 | 1 |
| Prediabetes | duct_epithelial_cell | *TWSG1* | -0.114004901 | 0.009625142 | 0.033856026 | 1 |
| Prediabetes | duct_epithelial_cell | *TMOD2* | -0.12288885 | 0.002489217 | 0.014112749 | 1 |
| Prediabetes | duct_epithelial_cell | *ELL3* | -0.082245558 | 0.00828732 | 0.03063112 | 1 |

| | | | | | | |
|---|---|---|---|---|---|---|
| Prediabetes | duct_epithelial_cell | *SUMF2* | -0.120628635 | 0.005451489 | 0.023230913 | 1 |
| Prediabetes | duct_epithelial_cell | *DCTD* | -0.134059596 | 0.011023952 | 0.037468087 | 1 |
| Prediabetes | duct_epithelial_cell | *MTUS1* | -0.096895839 | 0.008069381 | 0.030194855 | 1 |
| Prediabetes | duct_epithelial_cell | *HEATR5A* | -0.219317982 | 0.00339527 | 0.017190742 | 1 |
| Prediabetes | duct_epithelial_cell | *MIS18BP1* | -0.067613258 | 0.003064682 | 0.016054848 | 1 |
| Prediabetes | duct_epithelial_cell | *RNASE1* | -0.081754656 | 0.005502944 | 0.023344611 | 1 |
| Prediabetes | duct_epithelial_cell | *TEP1* | -0.122495008 | 0.003729525 | 0.018157748 | 1 |
| Prediabetes | duct_epithelial_cell | *REEP5* | -0.102923082 | 0.000959773 | 0.009147222 | 1 |
| Prediabetes | duct_epithelial_cell | *MAU2* | -0.112773158 | 0.004236888 | 0.019551648 | 1 |
| Prediabetes | duct_epithelial_cell | *SHC2* | -0.226686607 | 0.000624333 | 0.008963481 | 1 |
| Prediabetes | duct_epithelial_cell | *CBFA2T3* | -0.089366442 | 0.010143777 | 0.035301061 | 1 |
| Prediabetes | duct_epithelial_cell | *PHF10* | -0.1920159 | 0.001819057 | 0.011681295 | 1 |
| Prediabetes | duct_epithelial_cell | *ECSIT* | -0.135040705 | 0.005603603 | 0.023493587 | 1 |
| Prediabetes | duct_epithelial_cell | *CDC16* | -0.137968737 | 0.002980367 | 0.015696866 | 1 |
| Prediabetes | duct_epithelial_cell | *DPP6* | -0.046239447 | 0.01069353 | 0.036694217 | 1 |
| Prediabetes | duct_epithelial_cell | *TULP4* | -0.149023921 | 9.82E-05 | 0.008963481 | 0.385941856 |
| Prediabetes | duct_epithelial_cell | *NDUFA10* | -0.097902729 | 0.001339897 | 0.010242129 | 1 |
| Prediabetes | duct_epithelial_cell | *UBAC1* | -0.164900678 | 0.000407138 | 0.008963481 | 1 |
| Prediabetes | duct_epithelial_cell | *ATXN10* | -0.130084444 | 0.001368768 | 0.010271223 | 1 |
| Prediabetes | duct_epithelial_cell | *PAK4* | -0.264187352 | 0.001011945 | 0.009336398 | 1 |
| Prediabetes | duct_epithelial_cell | *ADRM1* | -0.162040327 | 0.000419224 | 0.008963481 | 1 |
| Prediabetes | duct_epithelial_cell | *UCK1* | -0.225727602 | 0.000365321 | 0.008963481 | 1 |
| Prediabetes | duct_epithelial_cell | *MAP3K10* | -0.196908053 | 0.000198603 | 0.008963481 | 0.780311971 |
| Prediabetes | duct_epithelial_cell | *DNMT1* | -0.209815417 | 0.001528433 | 0.010707166 | 1 |
| Prediabetes | duct_epithelial_cell | *HSD17B3* | -0.946365795 | 0.00267224 | 0.014746113 | 1 |
| Prediabetes | duct_epithelial_cell | *SYNE1* | -0.19621394 | 0.000556383 | 0.008963481 | 1 |
| Prediabetes | duct_epithelial_cell | *PDLIM4* | -0.11287976 | 0.001470673 | 0.01048303 | 1 |
| Prediabetes | duct_epithelial_cell | *MGAT1* | -0.144171763 | 0.000721303 | 0.008963481 | 1 |
| Prediabetes | duct_epithelial_cell | *DIAPH1* | -0.137347286 | 0.013795947 | 0.044357017 | 1 |
| Prediabetes | duct_epithelial_cell | *PPFIA1* | -0.141004895 | 0.008374299 | 0.030753836 | 1 |
| Prediabetes | duct_epithelial_cell | *TRAF7* | -0.113576685 | 0.012682412 | 0.041663208 | 1 |
| Prediabetes | duct_epithelial_cell | *CHD1L* | -0.103753551 | 0.004383035 | 0.019977896 | 1 |
| Prediabetes | duct_epithelial_cell | *PIAS3* | -0.114208649 | 0.003317779 | 0.016885431 | 1 |
| Prediabetes | duct_epithelial_cell | *ZNF304* | -0.351583002 | 0.00097082 | 0.00916911 | 1 |
| Prediabetes | duct_epithelial_cell | *GCH1* | -0.091783632 | 0.005608421 | 0.023493587 | 1 |
| Prediabetes | duct_epithelial_cell | *ENOSF1* | -0.227894872 | 0.000737412 | 0.008963481 | 1 |
| Prediabetes | duct_epithelial_cell | *IMMT* | -0.111869616 | 0.000787538 | 0.008963481 | 1 |
| Prediabetes | duct_epithelial_cell | *TMEM128* | -0.154818237 | 0.00122696 | 0.009805258 | 1 |
| Prediabetes | duct_epithelial_cell | *SEC61G* | -0.075844807 | 0.001642054 | 0.011047314 | 1 |
| Prediabetes | duct_epithelial_cell | *FIGNL1* | -0.145058724 | 0.00480373 | 0.021254343 | 1 |
| Prediabetes | duct_epithelial_cell | *EIF5A* | -0.075638628 | 0.007442076 | 0.028573059 | 1 |
| Prediabetes | duct_epithelial_cell | *ANKEF1* | -0.318085255 | 0.000373891 | 0.008963481 | 1 |
| Prediabetes | duct_epithelial_cell | *BTBD3* | -0.094600875 | 0.002606232 | 0.014586734 | 1 |
| Prediabetes | duct_epithelial_cell | *SERINC3* | -0.0679166 | 0.012423876 | 0.041069201 | 1 |
| Prediabetes | duct_epithelial_cell | *TRPC4* | -0.178246749 | 0.008389602 | 0.030777541 | 1 |
| Prediabetes | duct_epithelial_cell | *RFC3* | -0.136773454 | 0.005154454 | 0.022303801 | 1 |
| Prediabetes | duct_epithelial_cell | *CNDP2* | -0.191974445 | 0.00624151 | 0.02517751 | 1 |
| Prediabetes | duct_epithelial_cell | *SLC2A11* | -0.115113094 | 0.004244733 | 0.019551648 | 1 |
| Prediabetes | duct_epithelial_cell | *FAM83F* | -0.262707264 | 0.000464301 | 0.008963481 | 1 |
| Prediabetes | duct_epithelial_cell | *AMPD3* | -0.06346267 | 0.005880761 | 0.024287691 | 1 |

| | | | | | | |
|---|---|---|---|---|---|---|
| Prediabetes | duct_epithelial_cell | *HSD17B4* | -0.190597673 | 0.001373348 | 0.010271223 | 1 |
| Prediabetes | duct_epithelial_cell | *COX16* | -0.116552112 | 0.001117119 | 0.009564575 | 1 |
| Prediabetes | duct_epithelial_cell | *ELP3* | -0.166332806 | 0.001987419 | 0.012369325 | 1 |
| Prediabetes | duct_epithelial_cell | *MRPS36* | -0.08611608 | 0.00578419 | 0.024023343 | 1 |
| Prediabetes | duct_epithelial_cell | *THUMPD3* | -0.102703325 | 0.00698305 | 0.027354339 | 1 |
| Prediabetes | duct_epithelial_cell | *SORT1* | -0.229934628 | 0.000838807 | 0.008963481 | 1 |
| Prediabetes | duct_epithelial_cell | *CEPT1* | -0.153273453 | 0.0014728 | 0.01048303 | 1 |
| Prediabetes | duct_epithelial_cell | *NAPG* | -0.213030694 | 0.001001065 | 0.009336398 | 1 |
| Prediabetes | duct_epithelial_cell | *SPIRE1* | -0.127863303 | 0.001160705 | 0.009685284 | 1 |
| Prediabetes | duct_epithelial_cell | *FKBP11* | -0.045759637 | 0.015791623 | 0.049125325 | 1 |
| Prediabetes | duct_epithelial_cell | *KIDINS220* | -0.160671919 | 0.000495376 | 0.008963481 | 1 |
| Prediabetes | duct_epithelial_cell | *LPIN1* | -0.067115746 | 0.003529514 | 0.017534673 | 1 |
| Prediabetes | duct_epithelial_cell | *IAH1* | -0.149518099 | 0.001747589 | 0.011424754 | 1 |
| Prediabetes | duct_epithelial_cell | *KLRC1* | -2.519034006 | 0.012458399 | 0.041099117 | 1 |
| Prediabetes | duct_epithelial_cell | *PRPF38A* | -0.198391661 | 0.003720401 | 0.01813774 | 1 |
| Prediabetes | duct_epithelial_cell | *TIMM10* | -0.142239198 | 0.001734479 | 0.011376911 | 1 |
| Prediabetes | duct_epithelial_cell | *TMEM165* | -0.172927456 | 0.000543098 | 0.008963481 | 1 |
| Prediabetes | duct_epithelial_cell | *TMED7* | -0.130622728 | 0.000639974 | 0.008963481 | 1 |
| Prediabetes | duct_epithelial_cell | *NREP* | -0.297461669 | 0.000918949 | 0.009057947 | 1 |
| Prediabetes | duct_epithelial_cell | *WDR36* | -0.308290061 | 0.000754225 | 0.008963481 | 1 |
| Prediabetes | duct_epithelial_cell | *OSTF1* | -0.192212289 | 0.002197431 | 0.012987238 | 1 |
| Prediabetes | duct_epithelial_cell | *MSI1* | -0.086147691 | 0.00980452 | 0.034333298 | 1 |
| Prediabetes | duct_epithelial_cell | *PNPLA8* | -0.090774193 | 0.000835417 | 0.008963481 | 1 |
| Prediabetes | duct_epithelial_cell | *MTO1* | -0.242091058 | 0.000421409 | 0.008963481 | 1 |
| Prediabetes | duct_epithelial_cell | *LCA5* | -0.287859243 | 0.000298641 | 0.008963481 | 1 |
| Prediabetes | duct_epithelial_cell | *LMO2* | -0.100333563 | 0.005064372 | 0.022038603 | 1 |
| Prediabetes | duct_epithelial_cell | *TFCP2* | -0.244216691 | 0.000457235 | 0.008963481 | 1 |
| Prediabetes | duct_epithelial_cell | *PKIB* | -0.053316925 | 0.008250103 | 0.030579862 | 1 |
| Prediabetes | duct_epithelial_cell | *GNS* | -0.168843916 | 0.000515597 | 0.008963481 | 1 |
| Prediabetes | duct_epithelial_cell | *MPHOSPH6* | -0.169520192 | 0.007653219 | 0.029024613 | 1 |
| Prediabetes | duct_epithelial_cell | *NTPCR* | -0.111611056 | 0.008906456 | 0.031928346 | 1 |
| Prediabetes | duct_epithelial_cell | *GLUL* | -0.175995498 | 0.001424925 | 0.010342775 | 1 |
| Prediabetes | duct_epithelial_cell | *USP37* | -0.115725068 | 0.005736309 | 0.023925645 | 1 |
| Prediabetes | duct_epithelial_cell | *ARMC9* | -0.264403627 | 0.000681585 | 0.008963481 | 1 |
| Prediabetes | duct_epithelial_cell | *COX5B* | -0.108604305 | 0.000779692 | 0.008963481 | 1 |
| Prediabetes | duct_epithelial_cell | *REV1* | -0.098851544 | 6.54E-05 | 0.008963481 | 0.257104008 |
| Prediabetes | duct_epithelial_cell | *TSGA10* | -0.055336696 | 5.49E-05 | 0.008963481 | 0.215687485 |
| Prediabetes | duct_epithelial_cell | *TGFBRAP1* | -0.105225866 | 0.003253705 | 0.016734297 | 1 |
| Prediabetes | duct_epithelial_cell | *MRPS9* | -0.168158819 | 0.000337709 | 0.008963481 | 1 |
| Prediabetes | duct_epithelial_cell | *ANKRD36* | -0.360400764 | 0.001496041 | 0.010590892 | 1 |
| Prediabetes | duct_epithelial_cell | *DRAM1* | -0.288015862 | 0.000371721 | 0.008963481 | 1 |
| Prediabetes | duct_epithelial_cell | *VPS36* | -0.272479498 | 0.00010542 | 0.008963481 | 0.414195525 |
| Prediabetes | duct_epithelial_cell | *RNASEH2B* | -0.122899881 | 0.013006794 | 0.042515553 | 1 |
| Prediabetes | duct_epithelial_cell | *SUCLA2* | -0.143814672 | 0.0010983 | 0.00956008 | 1 |
| Prediabetes | duct_epithelial_cell | *KDELR2* | -0.089811616 | 0.00147133 | 0.01048303 | 1 |
| Prediabetes | duct_epithelial_cell | *ALPK3* | -0.073490838 | 0.004672466 | 0.02090362 | 1 |
| Prediabetes | duct_epithelial_cell | *CIB2* | -0.049087877 | 0.003984021 | 0.018886376 | 1 |
| Prediabetes | duct_epithelial_cell | *TEX2* | -0.193549235 | 0.007385182 | 0.028419569 | 1 |
| Prediabetes | duct_epithelial_cell | *MRPL47* | -0.129907848 | 0.00136096 | 0.010271223 | 1 |
| Prediabetes | duct_epithelial_cell | *SMPD4* | -0.20104017 | 0.000602906 | 0.008963481 | 1 |

| | | | | | | |
|---|---|---|---|---|---|---|
| Prediabetes | duct_epithelial_cell | *CCDC115* | -0.120081894 | 0.005184733 | 0.022377892 | 1 |
| Prediabetes | duct_epithelial_cell | *NIPSNAP3A* | -0.129616948 | 0.001999885 | 0.012393611 | 1 |
| Prediabetes | duct_epithelial_cell | *CDK9* | -0.245990833 | 0.000294426 | 0.008963481 | 1 |
| Prediabetes | duct_epithelial_cell | *TXN* | -0.141158932 | 0.000756596 | 0.008963481 | 1 |
| Prediabetes | duct_epithelial_cell | *TOR1A* | -0.158761342 | 0.003762956 | 0.018230151 | 1 |
| Prediabetes | duct_epithelial_cell | *STX17* | -0.09893476 | 0.008736081 | 0.031543827 | 1 |
| Prediabetes | duct_epithelial_cell | *ATP6V1G1* | -0.058864788 | 0.011397686 | 0.038406096 | 1 |
| Prediabetes | duct_epithelial_cell | *ARPC5L* | -0.133397864 | 0.00071638 | 0.008963481 | 1 |
| Prediabetes | duct_epithelial_cell | *APTX* | -0.160011829 | 0.001348162 | 0.010256616 | 1 |
| Prediabetes | duct_epithelial_cell | *TMEM8B* | -0.199748107 | 0.001212594 | 0.009782919 | 1 |
| Prediabetes | duct_epithelial_cell | *PPIL1* | -0.148516613 | 0.002057412 | 0.012532667 | 1 |
| Prediabetes | duct_epithelial_cell | *KIAA0319* | -0.118563161 | 0.01470291 | 0.046474444 | 1 |
| Prediabetes | duct_epithelial_cell | *RIPK1* | -0.193615976 | 0.001787304 | 0.011538034 | 1 |
| Prediabetes | duct_epithelial_cell | *MTCH1* | -0.12612543 | 0.000803092 | 0.008963481 | 1 |
| Prediabetes | duct_epithelial_cell | *VARS2* | -0.026393901 | 0.01305541 | 0.042604802 | 1 |
| Prediabetes | duct_epithelial_cell | *ANKRD42* | -0.129181347 | 0.002167093 | 0.012920349 | 1 |
| Prediabetes | duct_epithelial_cell | *CREBZF* | -0.18178001 | 0.000712659 | 0.008963481 | 1 |
| Prediabetes | duct_epithelial_cell | *NARS2* | -0.265102806 | 0.008903916 | 0.031928346 | 1 |
| Prediabetes | duct_epithelial_cell | *GGH* | -0.058017474 | 0.005010184 | 0.02192095 | 1 |
| Prediabetes | duct_epithelial_cell | *FDX1* | -0.139764556 | 0.002017659 | 0.012425367 | 1 |
| Prediabetes | duct_epithelial_cell | *C11orf1* | -0.079753966 | 0.010946098 | 0.037397582 | 1 |
| Prediabetes | duct_epithelial_cell | *ALKBH8* | -0.462941761 | 0.00036501 | 0.008963481 | 1 |
| Prediabetes | duct_epithelial_cell | *TUBGCP4* | -0.153861066 | 0.000585026 | 0.008963481 | 1 |
| Prediabetes | duct_epithelial_cell | *ADAM10* | -0.186685308 | 0.00094626 | 0.009132479 | 1 |
| Prediabetes | duct_epithelial_cell | *ZNF280D* | -0.119627058 | 0.001534267 | 0.010707166 | 1 |
| Prediabetes | duct_epithelial_cell | *RSL24D1* | -0.09296825 | 0.002272465 | 0.013326143 | 1 |
| Prediabetes | duct_epithelial_cell | *SPTBN5* | 0.069400509 | 0.008138526 | 0.030336239 | 1 |
| Prediabetes | duct_epithelial_cell | *RTCA* | -0.159426119 | 0.00044396 | 0.008963481 | 1 |
| Prediabetes | duct_epithelial_cell | *ACTR2* | -0.139826548 | 0.000105159 | 0.008963481 | 0.413169196 |
| Prediabetes | duct_epithelial_cell | *ATRAID* | -0.092753994 | 0.002944703 | 0.015613682 | 1 |
| Prediabetes | duct_epithelial_cell | *LRPPRC* | -0.145170784 | 0.000256043 | 0.008963481 | 1 |
| Prediabetes | duct_epithelial_cell | *NCOA4* | -0.137741042 | 0.001650168 | 0.011064011 | 1 |
| Prediabetes | duct_epithelial_cell | *ASCC1* | -0.168158637 | 0.008362578 | 0.030753836 | 1 |
| Prediabetes | duct_epithelial_cell | *RPS24* | -0.067231703 | 0.001175238 | 0.009719245 | 1 |
| Prediabetes | duct_epithelial_cell | *USP8* | -0.119886812 | 0.001189828 | 0.009719245 | 1 |
| Prediabetes | duct_epithelial_cell | *SPPL2A* | -0.1648329 | 0.000302668 | 0.008963481 | 1 |
| Prediabetes | duct_epithelial_cell | *RAP1GDS1* | -0.214295626 | 0.001001728 | 0.009336398 | 1 |
| Prediabetes | duct_epithelial_cell | *G3BP2* | -0.109028753 | 0.001227871 | 0.009805258 | 1 |
| Prediabetes | duct_epithelial_cell | *SCARB2* | -0.066039481 | 0.00762018 | 0.028976426 | 1 |
| Prediabetes | duct_epithelial_cell | *CNOT6L* | -0.129539463 | 0.002072397 | 0.012584925 | 1 |
| Prediabetes | duct_epithelial_cell | *USO1* | -0.125622942 | 0.00201066 | 0.012406615 | 1 |
| Prediabetes | duct_epithelial_cell | *ZCRB1* | -0.116338748 | 0.000837853 | 0.008963481 | 1 |
| Prediabetes | duct_epithelial_cell | *PEX5* | -0.126681235 | 0.001230337 | 0.009805258 | 1 |
| Prediabetes | duct_epithelial_cell | *SNRPF* | -0.074260064 | 0.010357395 | 0.035853926 | 1 |
| Prediabetes | duct_epithelial_cell | *GLTP* | -0.211007215 | 0.000436422 | 0.008963481 | 1 |
| Prediabetes | duct_epithelial_cell | *MTMR6* | -0.110851495 | 0.002661964 | 0.014746113 | 1 |
| Prediabetes | duct_epithelial_cell | *KANSL2* | -0.309491233 | 0.000109234 | 0.008963481 | 0.429179406 |
| Prediabetes | duct_epithelial_cell | *WDFY2* | -0.137974265 | 0.003623943 | 0.017842699 | 1 |
| Prediabetes | duct_epithelial_cell | *TMX1* | -0.150377878 | 0.00072821 | 0.008963481 | 1 |
| Prediabetes | duct_epithelial_cell | *NIPA2* | -0.206677394 | 0.000416407 | 0.008963481 | 1 |

| | | | | | | |
|---|---|---|---|---|---|---|
| Prediabetes | duct_epithelial_cell | *SERF2* | -0.077048268 | 0.000579687 | 0.008963481 | 1 |
| Prediabetes | duct_epithelial_cell | *BNIP2* | -0.196080806 | 0.000482247 | 0.008963481 | 1 |
| Prediabetes | duct_epithelial_cell | *GTF2A2* | -0.129986746 | 0.000426728 | 0.008963481 | 1 |
| Prediabetes | duct_epithelial_cell | *SRP14* | -0.097196157 | 0.000627007 | 0.008963481 | 1 |
| Prediabetes | duct_epithelial_cell | *TLE3* | -0.184647696 | 0.000637982 | 0.008963481 | 1 |
| Prediabetes | duct_epithelial_cell | *SCAPER* | -0.081061745 | 0.002458614 | 0.013979585 | 1 |
| Prediabetes | duct_epithelial_cell | *TSPAN3* | -0.175546807 | 0.000101853 | 0.008963481 | 0.400180227 |
| Prediabetes | duct_epithelial_cell | *DNAJA4* | -0.049405829 | 0.013557822 | 0.043806482 | 1 |
| Prediabetes | duct_epithelial_cell | *IGF1R* | -0.079427489 | 0.01048557 | 0.036075136 | 1 |
| Prediabetes | duct_epithelial_cell | *CELF6* | -0.090086065 | 0.008613512 | 0.031393773 | 1 |
| Prediabetes | duct_epithelial_cell | *POLG* | -0.291401793 | 0.000701664 | 0.008963481 | 1 |
| Prediabetes | duct_epithelial_cell | *PMM2* | -0.189301452 | 0.000234422 | 0.008963481 | 0.921043976 |
| Prediabetes | duct_epithelial_cell | *UQCRC2* | -0.210070113 | 3.10E-05 | 0.008963481 | 0.121653861 |
| Prediabetes | duct_epithelial_cell | *ARHGAP17* | -0.240732424 | 0.00046718 | 0.008963481 | 1 |
| Prediabetes | duct_epithelial_cell | *KIFC3* | -0.123346633 | 0.014053429 | 0.044927521 | 1 |
| Prediabetes | duct_epithelial_cell | *NUDT7* | -0.112265132 | 0.006892725 | 0.027108625 | 1 |
| Prediabetes | duct_epithelial_cell | *GCSH* | -0.164423071 | 0.000430816 | 0.008963481 | 1 |
| Prediabetes | duct_epithelial_cell | *MAP1LC3B* | -0.114345137 | 0.000901125 | 0.009057947 | 1 |
| Prediabetes | duct_epithelial_cell | *RPS2* | -0.05612199 | 0.00322193 | 0.016688879 | 1 |
| Prediabetes | duct_epithelial_cell | *PDPK1* | -0.106198924 | 0.01613258 | 0.049752675 | 1 |
| Prediabetes | duct_epithelial_cell | *DEF8* | -0.119442539 | 0.001082456 | 0.009514476 | 1 |
| Prediabetes | duct_epithelial_cell | *NPEPPS* | -0.148193654 | 0.002412653 | 0.013798128 | 1 |
| Prediabetes | duct_epithelial_cell | *PTRH2* | -0.227946226 | 0.000805243 | 0.008963481 | 1 |
| Prediabetes | duct_epithelial_cell | *GNAL* | -0.079684266 | 0.006342491 | 0.025438743 | 1 |
| Prediabetes | duct_epithelial_cell | *RPRD1A* | -0.158051661 | 0.000798318 | 0.008963481 | 1 |
| Prediabetes | duct_epithelial_cell | *SAT2* | -0.167974119 | 0.000414794 | 0.008963481 | 1 |
| Prediabetes | duct_epithelial_cell | *ARHGDIA* | -0.119849738 | 0.000108516 | 0.008963481 | 0.426360458 |
| Prediabetes | duct_epithelial_cell | *TTYH2* | -0.74231099 | 0.000747452 | 0.008963481 | 1 |
| Prediabetes | duct_epithelial_cell | *NARF* | -0.127255107 | 0.000954448 | 0.009132479 | 1 |
| Prediabetes | duct_epithelial_cell | *WDR45B* | -0.082203435 | 0.005601787 | 0.023493587 | 1 |
| Prediabetes | duct_epithelial_cell | *PMAIP1* | -0.234069167 | 0.000545651 | 0.008963481 | 1 |
| Prediabetes | duct_epithelial_cell | *TXNL4A* | -0.077223005 | 0.009549759 | 0.033620969 | 1 |
| Prediabetes | duct_epithelial_cell | *SH3GL1* | -0.096164714 | 0.015667478 | 0.048855176 | 1 |
| Prediabetes | duct_epithelial_cell | *SIRT3* | -0.274788895 | 0.006021091 | 0.024668265 | 1 |
| Prediabetes | duct_epithelial_cell | *COL6A1* | -0.062592997 | 0.004389817 | 0.019985619 | 1 |
| Prediabetes | duct_epithelial_cell | *AKT1* | -0.108826516 | 0.006071159 | 0.024776105 | 1 |
| Prediabetes | duct_epithelial_cell | *RERE* | -0.194743587 | 0.000425484 | 0.008963481 | 1 |
| Prediabetes | duct_epithelial_cell | *ZNF593* | -0.187042777 | 0.000272561 | 0.008963481 | 1 |
| Prediabetes | duct_epithelial_cell | *CELA3A* | -0.021126512 | 0.007667907 | 0.029052273 | 1 |
| Prediabetes | duct_epithelial_cell | *PRKACB* | -0.142054552 | 0.000861872 | 0.00898221 | 1 |
| Prediabetes | duct_epithelial_cell | *PIGK* | -0.162655812 | 0.000921004 | 0.009057947 | 1 |
| Prediabetes | duct_epithelial_cell | *RPS8* | -0.063507356 | 0.001030226 | 0.009336398 | 1 |
| Prediabetes | duct_epithelial_cell | *ALDH9A1* | -0.179408 | 0.00098765 | 0.009283441 | 1 |
| Prediabetes | duct_epithelial_cell | *MGST3* | -0.139450995 | 0.000536555 | 0.008963481 | 1 |
| Prediabetes | duct_epithelial_cell | *LYPLAL1* | -0.172263257 | 0.003135698 | 0.016318092 | 1 |
| Prediabetes | duct_epithelial_cell | *SNX27* | -0.165524071 | 0.000336547 | 0.008963481 | 1 |
| Prediabetes | duct_epithelial_cell | *CTSK* | -0.129729817 | 0.00916723 | 0.03265462 | 1 |
| Prediabetes | duct_epithelial_cell | *TAF1A* | -0.253987928 | 0.009241052 | 0.032821135 | 1 |
| Prediabetes | duct_epithelial_cell | *KCNN3* | -0.325682697 | 0.00216364 | 0.012919364 | 1 |
| Prediabetes | duct_epithelial_cell | *SRP9* | -0.129534998 | 0.000184375 | 0.008963481 | 0.724410987 |

| | | | | | | |
|---|---|---|---|---|---|---|
| Prediabetes | duct_epithelial_cell | *ARL8A* | -0.145182598 | 0.001770309 | 0.011496768 | 1 |
| Prediabetes | duct_epithelial_cell | *GALM* | -0.388820836 | 0.007045832 | 0.027408984 | 1 |
| Prediabetes | duct_epithelial_cell | *EML4* | -0.170631341 | 0.001095252 | 0.00956008 | 1 |
| Prediabetes | duct_epithelial_cell | *WDPCP* | -0.222308166 | 0.003083074 | 0.01610824 | 1 |
| Prediabetes | duct_epithelial_cell | *CIAO1* | -0.116586745 | 0.008158468 | 0.030354755 | 1 |
| Prediabetes | duct_epithelial_cell | *TEX261* | -0.248262295 | 0.001121215 | 0.009564575 | 1 |
| Prediabetes | duct_epithelial_cell | *LIPT1* | -0.239909816 | 0.000186944 | 0.008963481 | 0.734501109 |
| Prediabetes | duct_epithelial_cell | *FAM171B* | -0.074692951 | 0.003528474 | 0.017534673 | 1 |
| Prediabetes | duct_epithelial_cell | *METTL21A* | -0.154860802 | 0.007197914 | 0.027848551 | 1 |
| Prediabetes | duct_epithelial_cell | *HES6* | -0.143148077 | 0.001392393 | 0.010271223 | 1 |
| Prediabetes | duct_epithelial_cell | *RAB5A* | -0.182355308 | 3.81E-05 | 0.008963481 | 0.149882245 |
| Prediabetes | duct_epithelial_cell | *UBA3* | -0.149155146 | 0.001532543 | 0.010707166 | 1 |
| Prediabetes | duct_epithelial_cell | *ARL6IP5* | -0.16750958 | 0.000409312 | 0.008963481 | 1 |
| Prediabetes | duct_epithelial_cell | *TAGLN3* | -0.032748112 | 0.013571003 | 0.043813042 | 1 |
| Prediabetes | duct_epithelial_cell | *EIF2A* | -0.090771106 | 0.003446252 | 0.017314992 | 1 |
| Prediabetes | duct_epithelial_cell | *TRPC1* | -0.113198538 | 0.004559141 | 0.020613192 | 1 |
| Prediabetes | duct_epithelial_cell | *PYURF* | -0.090732767 | 0.002286299 | 0.013387283 | 1 |
| Prediabetes | duct_epithelial_cell | *TIFA* | -0.155589208 | 0.003683872 | 0.018002406 | 1 |
| Prediabetes | duct_epithelial_cell | *SSBP2* | -0.126840682 | 0.001713762 | 0.011297601 | 1 |
| Prediabetes | duct_epithelial_cell | *GTF2H2* | -0.283294475 | 0.000375491 | 0.008963481 | 1 |
| Prediabetes | duct_epithelial_cell | *BTF3* | -0.080968435 | 0.002329872 | 0.013496739 | 1 |
| Prediabetes | duct_epithelial_cell | *COMMD10* | -0.306404129 | 0.000831504 | 0.008963481 | 1 |
| Prediabetes | duct_epithelial_cell | *ATG12* | -0.146026038 | 0.000322557 | 0.008963481 | 1 |
| Prediabetes | duct_epithelial_cell | *PCYOX1L* | -0.273354857 | 0.001082456 | 0.009514476 | 1 |
| Prediabetes | duct_epithelial_cell | *TBC1D7* | -0.321830108 | 0.00050891 | 0.008963481 | 1 |
| Prediabetes | duct_epithelial_cell | *ZMAT2* | -0.178456727 | 0.000553373 | 0.008963481 | 1 |
| Prediabetes | duct_epithelial_cell | *SLC17A4* | 0.056017371 | 0.010315937 | 0.035836244 | 1 |
| Prediabetes | duct_epithelial_cell | *TRIM7* | -0.180386043 | 3.39E-05 | 0.008963481 | 0.133280029 |
| Prediabetes | duct_epithelial_cell | *RPL7L1* | -0.105350859 | 0.001038685 | 0.009336398 | 1 |
| Prediabetes | duct_epithelial_cell | *IRAK1BP1* | -0.193707439 | 0.000776559 | 0.008963481 | 1 |
| Prediabetes | duct_epithelial_cell | *FAXC* | -0.149368289 | 0.007625763 | 0.028976426 | 1 |
| Prediabetes | duct_epithelial_cell | *DYNLT1* | -0.113630004 | 0.00506512 | 0.022038603 | 1 |
| Prediabetes | duct_epithelial_cell | *GIGYF1* | -0.147092628 | 0.006315443 | 0.02539752 | 1 |
| Prediabetes | duct_epithelial_cell | *TMEM209* | -0.098666868 | 0.003272818 | 0.016769199 | 1 |
| Prediabetes | duct_epithelial_cell | *CCDC25* | -0.130302257 | 0.003100228 | 0.016176354 | 1 |
| Prediabetes | duct_epithelial_cell | *LACTB2* | -0.181925343 | 0.001159285 | 0.009685284 | 1 |
| Prediabetes | duct_epithelial_cell | *TERF1* | -0.17172447 | 0.00047312 | 0.008963481 | 1 |
| Prediabetes | duct_epithelial_cell | *LRP12* | -0.10847017 | 0.005763637 | 0.023988697 | 1 |
| Prediabetes | duct_epithelial_cell | *ZNF7* | -0.270953351 | 0.000391569 | 0.008963481 | 1 |
| Prediabetes | duct_epithelial_cell | *AK3* | -0.171320279 | 0.0003998 | 0.008963481 | 1 |
| Prediabetes | duct_epithelial_cell | *C9orf72* | -0.142808306 | 0.000313285 | 0.008963481 | 1 |
| Prediabetes | duct_epithelial_cell | *FBXO10* | -0.087693237 | 0.012748265 | 0.041783516 | 1 |
| Prediabetes | duct_epithelial_cell | *ZNF462* | -0.074457408 | 0.005644776 | 0.023593961 | 1 |
| Prediabetes | duct_epithelial_cell | *SURF6* | -0.214690984 | 0.000452619 | 0.008963481 | 1 |
| Prediabetes | duct_epithelial_cell | *GPR107* | -0.162547162 | 0.002617285 | 0.014627755 | 1 |
| Prediabetes | duct_epithelial_cell | *GLUD1* | -0.163881823 | 0.000485686 | 0.008963481 | 1 |
| Prediabetes | duct_epithelial_cell | *DNAJB12* | -0.182836181 | 0.000633833 | 0.008963481 | 1 |
| Prediabetes | duct_epithelial_cell | *PTPRJ* | -0.100979401 | 0.004605872 | 0.02077666 | 1 |
| Prediabetes | duct_epithelial_cell | *CAPN5* | 0.029597336 | 0.005579115 | 0.023475689 | 1 |
| Prediabetes | duct_epithelial_cell | *RPS3* | -0.060998165 | 0.001522984 | 0.010704482 | 1 |

| | | | | | | |
|---|---|---|---|---|---|---|
| Prediabetes | duct_epithelial_cell | *AASDHPPT* | -0.144472043 | 0.000852528 | 0.008963481 | 1 |
| Prediabetes | duct_epithelial_cell | *DUSP15* | -0.29415639 | 3.13E-05 | 0.008963481 | 0.123054651 |
| Prediabetes | duct_epithelial_cell | *YTHDF1* | -0.203834667 | 0.000445791 | 0.008963481 | 1 |
| Prediabetes | duct_epithelial_cell | *TMEM218* | -0.249010295 | 0.000710159 | 0.008963481 | 1 |
| Prediabetes | duct_epithelial_cell | *MIA2* | -0.087825044 | 0.007021561 | 0.027368761 | 1 |
| Prediabetes | duct_epithelial_cell | *DLG5* | -0.131205561 | 0.00138142 | 0.010271223 | 1 |
| Prediabetes | duct_epithelial_cell | *SLC2A13* | -0.081351214 | 0.000541399 | 0.008963481 | 1 |
| Prediabetes | duct_epithelial_cell | *MAGI1* | -0.089013954 | 0.014871882 | 0.046782724 | 1 |
| Prediabetes | duct_epithelial_cell | *CSNK1G3* | -0.106838798 | 0.001369922 | 0.010271223 | 1 |
| Prediabetes | duct_epithelial_cell | *AKAP6* | -0.139383094 | 0.00846561 | 0.030969628 | 1 |
| Prediabetes | duct_epithelial_cell | *TMEM18* | -0.172831927 | 0.000629404 | 0.008963481 | 1 |
| Prediabetes | duct_epithelial_cell | *NDUFC2* | -0.089735063 | 0.00121008 | 0.009782723 | 1 |
| Prediabetes | duct_epithelial_cell | *QDPR* | -0.058277782 | 0.002799774 | 0.015172844 | 1 |
| Prediabetes | duct_epithelial_cell | *ZNF827* | -0.120285376 | 0.001031711 | 0.009336398 | 1 |
| Prediabetes | duct_epithelial_cell | *KIN* | -0.215469291 | 0.000815151 | 0.008963481 | 1 |
| Prediabetes | duct_epithelial_cell | *BICD1* | -0.104327935 | 0.008759347 | 0.031544891 | 1 |
| Prediabetes | duct_epithelial_cell | *GUF1* | -0.114590481 | 0.004326807 | 0.019802187 | 1 |
| Prediabetes | duct_epithelial_cell | *CACUL1* | -0.136445603 | 0.000843516 | 0.008963481 | 1 |
| Prediabetes | duct_epithelial_cell | *RABGAP1L* | -0.132485617 | 0.001227156 | 0.009805258 | 1 |
| Prediabetes | duct_epithelial_cell | *ARL14EP* | -0.166919752 | 0.000524243 | 0.008963481 | 1 |
| Prediabetes | duct_epithelial_cell | *EPG5* | -0.254858506 | 9.59E-05 | 0.008963481 | 0.376712789 |
| Prediabetes | duct_epithelial_cell | *PDK1* | -0.145616719 | 0.001630428 | 0.011037642 | 1 |
| Prediabetes | duct_epithelial_cell | *PDE3B* | -0.044442537 | 0.000756898 | 0.008963481 | 1 |
| Prediabetes | duct_epithelial_cell | *TGOLN2* | -0.147700476 | 0.000114902 | 0.008963481 | 0.451449662 |
| Prediabetes | duct_epithelial_cell | *USP12* | -0.078550035 | 0.006942265 | 0.027276159 | 1 |
| Prediabetes | duct_epithelial_cell | *IGSF10* | -0.308015012 | 0.002532526 | 0.014316973 | 1 |
| Prediabetes | duct_epithelial_cell | *GPR180* | -0.092873286 | 0.006480487 | 0.025823362 | 1 |
| Prediabetes | duct_epithelial_cell | *IFIT5* | -0.16497803 | 0.000100703 | 0.008963481 | 0.395661704 |
| Prediabetes | duct_epithelial_cell | *MED21* | -0.147497615 | 0.002813473 | 0.015226082 | 1 |
| Prediabetes | duct_epithelial_cell | *SRP19* | -0.110040747 | 0.000815016 | 0.008963481 | 1 |
| Prediabetes | duct_epithelial_cell | *CAST* | -0.135807213 | 0.00346965 | 0.017388079 | 1 |
| Prediabetes | duct_epithelial_cell | *PLA2R1* | -0.911524241 | 0.002707868 | 0.014880018 | 1 |
| Prediabetes | duct_epithelial_cell | *RPIA* | -0.176744819 | 0.003412686 | 0.017234503 | 1 |
| Prediabetes | duct_epithelial_cell | *GTF2E1* | -0.291184747 | 0.000518724 | 0.008963481 | 1 |
| Prediabetes | duct_epithelial_cell | *JAZF1* | -0.089684011 | 0.000924598 | 0.009057947 | 1 |
| Prediabetes | duct_epithelial_cell | *CMIP* | -0.142020976 | 0.00087653 | 0.009011853 | 1 |
| Prediabetes | duct_epithelial_cell | *NUS1* | -0.135445457 | 0.000669901 | 0.008963481 | 1 |
| Prediabetes | duct_epithelial_cell | *PITPNC1* | -0.176329857 | 0.001382705 | 0.010271223 | 1 |
| Prediabetes | duct_epithelial_cell | *CEP112* | -0.195443182 | 0.010986253 | 0.037414245 | 1 |
| Prediabetes | duct_epithelial_cell | *TSEN2* | -0.139458002 | 0.000944767 | 0.009132479 | 1 |
| Prediabetes | duct_epithelial_cell | *FLCN* | -0.175685394 | 0.001647928 | 0.011064011 | 1 |
| Prediabetes | duct_epithelial_cell | *RAB6B* | -0.213711569 | 0.001782585 | 0.011538034 | 1 |
| Prediabetes | duct_epithelial_cell | *ANKRD40* | -0.129122182 | 0.004225348 | 0.019551648 | 1 |
| Prediabetes | duct_epithelial_cell | *ATP6V1C1* | -0.156088773 | 0.00084114 | 0.008963481 | 1 |
| Prediabetes | duct_epithelial_cell | *MARVELD1* | -0.154381251 | 0.009067511 | 0.0323875 | 1 |
| Prediabetes | duct_epithelial_cell | *DBI* | -0.087865585 | 0.008697543 | 0.031543827 | 1 |
| Prediabetes | duct_epithelial_cell | *LRGUK* | -0.127134631 | 0.001890349 | 0.011998678 | 1 |
| Prediabetes | duct_epithelial_cell | *MIER3* | -0.150570086 | 0.003615199 | 0.017821977 | 1 |
| Prediabetes | duct_epithelial_cell | *RMND1* | -0.081758818 | 0.015937062 | 0.049382268 | 1 |
| Prediabetes | duct_epithelial_cell | *PSD3* | -0.147561569 | 0.003341513 | 0.016984223 | 1 |

| | | | | | | |
|---|---|---|---|---|---|---|
| Prediabetes | duct_epithelial_cell | *CCT8* | -0.053367215 | 0.007995756 | 0.030059963 | 1 |
| Prediabetes | duct_epithelial_cell | *ZDHHC5* | -0.156433876 | 0.014186996 | 0.045268501 | 1 |
| Prediabetes | duct_epithelial_cell | *MED19* | -0.17717621 | 0.002367143 | 0.013617139 | 1 |
| Prediabetes | duct_epithelial_cell | *MAPK13* | -0.065530849 | 0.006720995 | 0.026639575 | 1 |
| Prediabetes | duct_epithelial_cell | *EXOG* | -0.152309923 | 5.97E-05 | 0.008963481 | 0.234492697 |
| Prediabetes | duct_epithelial_cell | *ZFYVE9* | -0.191988088 | 0.002055671 | 0.012532667 | 1 |
| Prediabetes | duct_epithelial_cell | *GATAD1* | -0.162160476 | 0.000767435 | 0.008963481 | 1 |
| Prediabetes | duct_epithelial_cell | *DHRS4* | -0.127722182 | 0.004704391 | 0.020956406 | 1 |
| Prediabetes | duct_epithelial_cell | *SLC38A10* | -0.133825572 | 0.000381616 | 0.008963481 | 1 |
| Prediabetes | duct_epithelial_cell | *SLC37A3* | -0.186839085 | 0.002321381 | 0.013492166 | 1 |
| Prediabetes | duct_epithelial_cell | *SPPL3* | -0.183026451 | 0.00033123 | 0.008963481 | 1 |
| Prediabetes | duct_epithelial_cell | *TNFRSF14* | -0.3649251 | 0.001257353 | 0.009900082 | 1 |
| Prediabetes | duct_epithelial_cell | *PEX10* | -0.167704139 | 0.010103431 | 0.035211789 | 1 |
| Prediabetes | duct_epithelial_cell | *LDLRAP1* | -0.114515345 | 0.001245236 | 0.009849499 | 1 |
| Prediabetes | duct_epithelial_cell | *XKR8* | -0.148169753 | 0.014481187 | 0.045921375 | 1 |
| Prediabetes | duct_epithelial_cell | *FAIM* | -0.171216144 | 0.01048424 | 0.036075136 | 1 |
| Prediabetes | duct_epithelial_cell | *PINK1* | -0.137537024 | 0.000855375 | 0.008963481 | 1 |
| Prediabetes | duct_epithelial_cell | *CCAR2* | -0.269580265 | 0.000436691 | 0.008963481 | 1 |
| Prediabetes | duct_epithelial_cell | *EPB41* | -0.122738064 | 0.006537581 | 0.025971847 | 1 |
| Prediabetes | duct_epithelial_cell | *ADPGK* | -0.144967692 | 0.004181316 | 0.019464918 | 1 |
| Prediabetes | duct_epithelial_cell | *THEM4* | -0.128069246 | 0.007885631 | 0.029705317 | 1 |
| Prediabetes | duct_epithelial_cell | *UBR1* | -0.261397202 | 0.000473534 | 0.008963481 | 1 |
| Prediabetes | duct_epithelial_cell | *TEPP* | -2.666297899 | 0.000746848 | 0.008963481 | 1 |
| Prediabetes | duct_epithelial_cell | *ZDHHC1* | -0.156271372 | 0.008347905 | 0.030749039 | 1 |
| Prediabetes | duct_epithelial_cell | *PDXK* | -0.176541048 | 3.85E-05 | 0.008963481 | 0.151459877 |
| Prediabetes | duct_epithelial_cell | *FTCD* | -0.045147495 | 0.013066629 | 0.042604802 | 1 |
| Prediabetes | duct_epithelial_cell | *LSS* | -0.060246412 | 0.008527303 | 0.031108424 | 1 |
| Prediabetes | duct_epithelial_cell | *SLC25A44* | -0.132883869 | 0.005797474 | 0.024053088 | 1 |
| Prediabetes | duct_epithelial_cell | *VPS28* | -0.08612628 | 0.005862665 | 0.02428191 | 1 |
| Prediabetes | duct_epithelial_cell | *TONSL* | -0.590473693 | 0.000363393 | 0.008963481 | 1 |
| Prediabetes | duct_epithelial_cell | *ZNF333* | -0.225965268 | 0.000255804 | 0.008963481 | 1 |
| Prediabetes | duct_epithelial_cell | *RPL8* | -0.062706068 | 0.001620845 | 0.011022922 | 1 |
| Prediabetes | duct_epithelial_cell | *MAML1* | -0.111587023 | 0.013204202 | 0.042946447 | 1 |
| Prediabetes | duct_epithelial_cell | *RPL26* | -0.07282838 | 0.000423271 | 0.008963481 | 1 |
| Prediabetes | duct_epithelial_cell | *ZNF75A* | -0.165516343 | 0.001276278 | 0.010008972 | 1 |
| Prediabetes | duct_epithelial_cell | *ZFPL1* | -0.109537513 | 0.003121707 | 0.016266825 | 1 |
| Prediabetes | duct_epithelial_cell | *CTRC* | -0.024651373 | 0.011238161 | 0.037966239 | 1 |
| Prediabetes | duct_epithelial_cell | *TM2D1* | -0.210755893 | 0.001010207 | 0.009336398 | 1 |
| Prediabetes | duct_epithelial_cell | *ZNF496* | -0.268674385 | 0.000564219 | 0.008963481 | 1 |
| Prediabetes | duct_epithelial_cell | *FLVCR1* | -0.106647452 | 0.001985901 | 0.012369325 | 1 |
| Prediabetes | duct_epithelial_cell | *ACP6* | -0.068125732 | 0.013161123 | 0.042877323 | 1 |
| Prediabetes | duct_epithelial_cell | *SMC6* | -0.268419434 | 0.000470125 | 0.008963481 | 1 |
| Prediabetes | duct_epithelial_cell | *MSX1* | -0.088784909 | 0.010505241 | 0.036111192 | 1 |
| Prediabetes | duct_epithelial_cell | *PACRGL* | -0.11509856 | 0.008776383 | 0.031577296 | 1 |
| Prediabetes | duct_epithelial_cell | *DCAF16* | -0.121021175 | 0.002635355 | 0.014686963 | 1 |
| Prediabetes | duct_epithelial_cell | *NPPC* | -0.807586342 | 0.000529857 | 0.008963481 | 1 |
| Prediabetes | duct_epithelial_cell | *EOGT* | -0.210808379 | 0.000592195 | 0.008963481 | 1 |
| Prediabetes | duct_epithelial_cell | *ARPC2* | -0.162060637 | 0.000754906 | 0.008963481 | 1 |
| Prediabetes | duct_epithelial_cell | *CWC22* | -0.207313159 | 0.000705984 | 0.008963481 | 1 |
| Prediabetes | duct_epithelial_cell | *HDAC11* | -0.119523229 | 0.013752872 | 0.044254737 | 1 |

| | | | | | | |
|---|---|---|---|---|---|---|
| Prediabetes | duct_epithelial_cell | *SUCLG1* | -0.208861733 | 0.000389337 | 0.008963481 | 1 |
| Prediabetes | duct_epithelial_cell | *ATXN7* | -0.155373713 | 0.000163277 | 0.008963481 | 0.641513494 |
| Prediabetes | duct_epithelial_cell | *PSMD6* | -0.106606381 | 0.001313122 | 0.010116188 | 1 |
| Prediabetes | duct_epithelial_cell | *IL17RC* | -0.11279427 | 0.000780268 | 0.008963481 | 1 |
| Prediabetes | duct_epithelial_cell | *TTC14* | -0.238563814 | 0.000955319 | 0.009132479 | 1 |
| Prediabetes | duct_epithelial_cell | *NMNAT3* | -0.092616384 | 0.014309074 | 0.045485722 | 1 |
| Prediabetes | duct_epithelial_cell | *SMIM12* | -0.168127 | 0.001260913 | 0.009908255 | 1 |
| Prediabetes | duct_epithelial_cell | *TPRA1* | -0.230970703 | 0.008487885 | 0.031022233 | 1 |
| Prediabetes | duct_epithelial_cell | *MEAF6* | -0.136981516 | 0.0004609 | 0.008963481 | 1 |
| Prediabetes | duct_epithelial_cell | *DNALI1* | -0.0845404 | 0.003967294 | 0.018848246 | 1 |
| Prediabetes | duct_epithelial_cell | *RPN1* | -0.137739996 | 0.001632351 | 0.011037642 | 1 |
| Prediabetes | duct_epithelial_cell | *RFC4* | -0.148145338 | 0.001494047 | 0.010590892 | 1 |
| Prediabetes | duct_epithelial_cell | *TKT* | -0.128975155 | 0.000578756 | 0.008963481 | 1 |
| Prediabetes | duct_epithelial_cell | *LRPAP1* | -0.055069928 | 0.008649082 | 0.031435931 | 1 |
| Prediabetes | duct_epithelial_cell | *PIGX* | -0.147252343 | 0.001560935 | 0.010816429 | 1 |
| Prediabetes | duct_epithelial_cell | *SAP30* | -0.126239913 | 0.000735611 | 0.008963481 | 1 |
| Prediabetes | duct_epithelial_cell | *ARFIP1* | -0.095633201 | 0.00558063 | 0.023475689 | 1 |
| Prediabetes | duct_epithelial_cell | *STARD4* | -0.12179822 | 0.001940316 | 0.012217151 | 1 |
| Prediabetes | duct_epithelial_cell | *CMBL* | -0.125906315 | 0.006403342 | 0.025593825 | 1 |
| Prediabetes | duct_epithelial_cell | *PRRC1* | -0.166869126 | 0.000745085 | 0.008963481 | 1 |
| Prediabetes | duct_epithelial_cell | *AGGF1* | -0.182726473 | 0.000297872 | 0.008963481 | 1 |
| Prediabetes | duct_epithelial_cell | *RICTOR* | -0.153434528 | 0.000694731 | 0.008963481 | 1 |
| Prediabetes | duct_epithelial_cell | *UTP15* | -0.310663861 | 0.000684237 | 0.008963481 | 1 |
| Prediabetes | duct_epithelial_cell | *NSA2* | -0.072750475 | 0.004864296 | 0.021463151 | 1 |
| Prediabetes | duct_epithelial_cell | *PDSS2* | -0.179010791 | 0.005748243 | 0.023949997 | 1 |
| Prediabetes | duct_epithelial_cell | *SAP30L* | -0.124888315 | 0.008741784 | 0.031543827 | 1 |
| Prediabetes | duct_epithelial_cell | *COG5* | -0.204202388 | 0.000816802 | 0.008963481 | 1 |
| Prediabetes | duct_epithelial_cell | *SLU7* | -0.138554056 | 0.000588539 | 0.008963481 | 1 |
| Prediabetes | duct_epithelial_cell | *CAMLG* | -0.08093184 | 0.003800493 | 0.018364621 | 1 |
| Prediabetes | duct_epithelial_cell | *BRI3* | -0.109918667 | 0.000683199 | 0.008963481 | 1 |
| Prediabetes | duct_epithelial_cell | *LMTK2* | -0.228287191 | 2.09E-05 | 0.008963481 | 0.082073335 |
| Prediabetes | duct_epithelial_cell | *PEX2* | -0.162366651 | 0.000582088 | 0.008963481 | 1 |
| Prediabetes | duct_epithelial_cell | *SUN1* | -0.190062919 | 0.000131961 | 0.008963481 | 0.518474291 |
| Prediabetes | duct_epithelial_cell | *INTS1* | -0.267241769 | 0.001133512 | 0.00963464 | 1 |
| Prediabetes | duct_epithelial_cell | *YWHAZ* | -0.141373177 | 0.000818388 | 0.008963481 | 1 |
| Prediabetes | duct_epithelial_cell | *DCAF13* | -0.125798439 | 0.002632425 | 0.014686963 | 1 |
| Prediabetes | duct_epithelial_cell | *GEM* | -0.082561519 | 0.002201451 | 0.012987238 | 1 |
| Prediabetes | duct_epithelial_cell | *RPP25L* | -0.246161 | 0.003671863 | 0.017966065 | 1 |
| Prediabetes | duct_epithelial_cell | *FAM219A* | -0.179258977 | 0.00170255 | 0.011242555 | 1 |
| Prediabetes | duct_epithelial_cell | *NUDT2* | -0.068607724 | 0.004573735 | 0.020655409 | 1 |
| Prediabetes | duct_epithelial_cell | *NOL6* | -0.220414953 | 0.001885738 | 0.01198878 | 1 |
| Prediabetes | duct_epithelial_cell | *WRN* | -0.20106498 | 0.000608382 | 0.008963481 | 1 |
| Prediabetes | duct_epithelial_cell | *CFL2* | -0.116659965 | 0.007604553 | 0.028976426 | 1 |
| Prediabetes | duct_epithelial_cell | *PGM2L1* | -0.082745331 | 0.01185783 | 0.03961685 | 1 |
| Prediabetes | duct_epithelial_cell | *PHYHIPL* | -0.058623592 | 0.011952227 | 0.039890314 | 1 |
| Prediabetes | duct_epithelial_cell | *INPPL1* | -0.156320714 | 0.003755917 | 0.018218516 | 1 |
| Prediabetes | duct_epithelial_cell | *OXGR1* | -0.168361994 | 0.009409001 | 0.033244573 | 1 |
| Prediabetes | duct_epithelial_cell | *VDAC2* | -0.128847762 | 0.000602711 | 0.008963481 | 1 |
| Prediabetes | duct_epithelial_cell | *TSC1* | -0.185714005 | 0.000917899 | 0.009057947 | 1 |
| Prediabetes | duct_epithelial_cell | *ZMYND19* | -0.222218255 | 0.014297858 | 0.045485722 | 1 |

| | | | | | | |
|---|---|---|---|---|---|---|
| Prediabetes | duct_epithelial_cell | *METTL3* | -0.175318055 | 0.013446583 | 0.043518636 | 1 |
| Prediabetes | duct_epithelial_cell | *TRUB1* | -0.179089329 | 0.000437193 | 0.008963481 | 1 |
| Prediabetes | duct_epithelial_cell | *IFI27L1* | -0.114281239 | 0.000620101 | 0.008963481 | 1 |
| Prediabetes | duct_epithelial_cell | *HTRA1* | -0.104921447 | 0.00541091 | 0.023158459 | 1 |
| Prediabetes | duct_epithelial_cell | *CEP57* | -0.108565508 | 0.002201116 | 0.012987238 | 1 |
| Prediabetes | duct_epithelial_cell | *BAG5* | -0.123620824 | 0.000937571 | 0.009132479 | 1 |
| Prediabetes | duct_epithelial_cell | *LARP6* | -0.120713266 | 0.00975896 | 0.034204242 | 1 |
| Prediabetes | duct_epithelial_cell | *ZNF319* | -0.348617431 | 0.001434034 | 0.010357204 | 1 |
| Prediabetes | duct_epithelial_cell | *FRS2* | -0.071157748 | 0.005608802 | 0.023493587 | 1 |
| Prediabetes | duct_epithelial_cell | *PCBD1* | -0.081176109 | 0.015775277 | 0.049113362 | 1 |
| Prediabetes | duct_epithelial_cell | *COX11* | -0.149958224 | 0.000244982 | 0.008963481 | 0.962534711 |
| Prediabetes | duct_epithelial_cell | *RIC3* | -0.147876157 | 0.000552689 | 0.008963481 | 1 |
| Prediabetes | duct_epithelial_cell | *ZNF143* | -0.27512056 | 0.000442446 | 0.008963481 | 1 |
| Prediabetes | duct_epithelial_cell | *COG1* | -0.172713956 | 0.000473164 | 0.008963481 | 1 |
| Prediabetes | duct_epithelial_cell | *B2M* | -0.083576763 | 0.000450933 | 0.008963481 | 1 |
| Prediabetes | duct_epithelial_cell | *SAAL1* | -0.257790314 | 0.007880787 | 0.029705317 | 1 |
| Prediabetes | duct_epithelial_cell | *GREM1* | -0.087485057 | 0.004941443 | 0.021716922 | 1 |
| Prediabetes | duct_epithelial_cell | *TSC22D4* | -0.078299879 | 0.003258592 | 0.016735957 | 1 |
| Prediabetes | duct_epithelial_cell | *NAT16* | -0.046510287 | 0.012432312 | 0.041069201 | 1 |
| Prediabetes | duct_epithelial_cell | *ENDOG* | -0.150185973 | 0.000184413 | 0.008963481 | 0.72455921 |
| Prediabetes | duct_epithelial_cell | *ZNF23* | -0.26352581 | 0.000432757 | 0.008963481 | 1 |
| Prediabetes | duct_epithelial_cell | *ZNF180* | -0.354151904 | 0.00875497 | 0.031544891 | 1 |
| Prediabetes | duct_epithelial_cell | *GPX4* | -0.083222928 | 0.00393959 | 0.018783824 | 1 |
| Prediabetes | duct_epithelial_cell | *ZNF641* | -0.195396018 | 0.005166129 | 0.022329724 | 1 |
| Prediabetes | duct_epithelial_cell | *RAB4B* | -0.087101122 | 0.008222247 | 0.03050539 | 1 |
| Prediabetes | duct_epithelial_cell | *UBXN6* | -0.168155859 | 0.000121302 | 0.008963481 | 0.476594996 |
| Prediabetes | duct_epithelial_cell | *NDUFV1* | -0.152101374 | 0.00700786 | 0.027368761 | 1 |
| Prediabetes | duct_epithelial_cell | *HID1* | -0.160893277 | 0.006074633 | 0.024776105 | 1 |
| Prediabetes | duct_epithelial_cell | *SRP68* | -0.166636226 | 0.000751921 | 0.008963481 | 1 |
| Prediabetes | duct_epithelial_cell | *TMEM68* | -0.185692168 | 0.006248145 | 0.025178422 | 1 |
| Prediabetes | duct_epithelial_cell | *MLST8* | -0.087547677 | 0.00528809 | 0.022756741 | 1 |
| Prediabetes | duct_epithelial_cell | *C2CD3* | -0.228783758 | 0.000641601 | 0.008963481 | 1 |
| Prediabetes | duct_epithelial_cell | *RPSA* | -0.050909815 | 0.009191503 | 0.032711427 | 1 |
| Prediabetes | duct_epithelial_cell | *ENTPD3* | -0.049232213 | 0.002316397 | 0.013483149 | 1 |
| Prediabetes | duct_epithelial_cell | *LTBP3* | -0.060240753 | 0.002835799 | 0.015283753 | 1 |
| Prediabetes | duct_epithelial_cell | *SAC3D1* | -0.257561408 | 0.00706191 | 0.027444355 | 1 |
| Prediabetes | duct_epithelial_cell | *LMBRD1* | -0.141208374 | 0.002845871 | 0.015317022 | 1 |
| Prediabetes | duct_epithelial_cell | *TTC39C* | -0.107592707 | 0.005557927 | 0.023430359 | 1 |
| Prediabetes | duct_epithelial_cell | *POLR2J3* | -0.110225931 | 0.004192052 | 0.019468837 | 1 |
| Prediabetes | duct_epithelial_cell | *NT5DC2* | -0.084928779 | 0.015511651 | 0.048407686 | 1 |
| Prediabetes | duct_epithelial_cell | *COA6* | -0.23628923 | 0.002336969 | 0.013496739 | 1 |
| Prediabetes | duct_epithelial_cell | *PXK* | -0.126308174 | 0.001041199 | 0.009336398 | 1 |
| Prediabetes | duct_epithelial_cell | *ING5* | -0.15759742 | 0.001161051 | 0.009685284 | 1 |
| Prediabetes | duct_epithelial_cell | *HEXIM2* | -0.348886295 | 0.00083581 | 0.008963481 | 1 |
| Prediabetes | duct_epithelial_cell | *TRAPPC11* | -0.177506894 | 0.00924739 | 0.032821135 | 1 |
| Prediabetes | duct_epithelial_cell | *COL3A1* | -0.061215889 | 0.004619064 | 0.020788435 | 1 |
| Prediabetes | duct_epithelial_cell | *ING2* | -0.073910496 | 0.008040879 | 0.030163992 | 1 |
| Prediabetes | duct_epithelial_cell | *DNAJC21* | -0.167875611 | 0.000472711 | 0.008963481 | 1 |
| Prediabetes | duct_epithelial_cell | *CNNM3* | -0.193373796 | 0.000857793 | 0.008963481 | 1 |
| Prediabetes | duct_epithelial_cell | *PPIP5K1* | -0.231313243 | 7.31E-05 | 0.008963481 | 0.287196018 |

| | | | | | | |
|---|---|---|---|---|---|---|
| Prediabetes | duct_epithelial_cell | *ZNF507* | -0.195843431 | 0.001099811 | 0.00956008 | 1 |
| Prediabetes | duct_epithelial_cell | *FSTL5* | -0.036560605 | 0.008053456 | 0.030163992 | 1 |
| Prediabetes | duct_epithelial_cell | *CTRB1* | -0.023284854 | 0.012065744 | 0.040194196 | 1 |
| Prediabetes | duct_epithelial_cell | *MFF* | -0.172909339 | 0.000828517 | 0.008963481 | 1 |
| Prediabetes | duct_epithelial_cell | *FAM110B* | -0.228949776 | 0.005557863 | 0.023430359 | 1 |
| Prediabetes | duct_epithelial_cell | *PRELID1* | -0.113493205 | 0.000521613 | 0.008963481 | 1 |
| Prediabetes | duct_epithelial_cell | *NMD3* | -0.129903275 | 0.001422801 | 0.010342775 | 1 |
| Prediabetes | duct_epithelial_cell | *SDC2* | -0.087570558 | 0.000918384 | 0.009057947 | 1 |
| Prediabetes | duct_epithelial_cell | *HINT1* | -0.073624894 | 0.002297606 | 0.013430167 | 1 |
| Prediabetes | duct_epithelial_cell | *MT1E* | -0.033594698 | 0.007574673 | 0.028950283 | 1 |
| Prediabetes | duct_epithelial_cell | *LIMS1* | -0.114485216 | 0.001507201 | 0.010631588 | 1 |
| Prediabetes | duct_epithelial_cell | *AVEN* | -0.093164295 | 0.006821378 | 0.026908829 | 1 |
| Prediabetes | duct_epithelial_cell | *TM4SF4* | -0.09556379 | 0.001668822 | 0.01113209 | 1 |
| Prediabetes | duct_epithelial_cell | *OTUD3* | -0.123718459 | 0.000608381 | 0.008963481 | 1 |
| Prediabetes | duct_epithelial_cell | *MAP3K2* | -0.08467784 | 0.002767489 | 0.01511602 | 1 |
| Prediabetes | duct_epithelial_cell | *UBE2E1* | -0.219647965 | 0.000417126 | 0.008963481 | 1 |
| Prediabetes | duct_epithelial_cell | *USP47* | -0.139944764 | 0.000701718 | 0.008963481 | 1 |
| Prediabetes | duct_epithelial_cell | *CRTAP* | -0.180894839 | 0.000603333 | 0.008963481 | 1 |
| Prediabetes | duct_epithelial_cell | *NFRKB* | -0.220607952 | 0.014743286 | 0.046564606 | 1 |
| Prediabetes | duct_epithelial_cell | *MGMT* | -0.219606191 | 0.000314296 | 0.008963481 | 1 |
| Prediabetes | duct_epithelial_cell | *NUDT9* | -0.04584725 | 0.014835108 | 0.046765612 | 1 |
| Prediabetes | duct_epithelial_cell | *IRX1* | -0.104834469 | 0.014816513 | 0.046758297 | 1 |
| Prediabetes | duct_epithelial_cell | *HSPA4* | -0.106219591 | 0.005880378 | 0.024287691 | 1 |
| Prediabetes | duct_epithelial_cell | *ACYP2* | -0.110791022 | 0.00364911 | 0.01792169 | 1 |
| Prediabetes | duct_epithelial_cell | *FOXN2* | -0.114596171 | 0.001808249 | 0.011646905 | 1 |
| Prediabetes | duct_epithelial_cell | *MTSS1* | -0.100597245 | 0.001832078 | 0.011742633 | 1 |
| Prediabetes | duct_epithelial_cell | *RPS9* | -0.068128483 | 0.000501439 | 0.008963481 | 1 |
| Prediabetes | duct_epithelial_cell | *NDUFA3* | -0.121627704 | 0.001203577 | 0.009750216 | 1 |
| Prediabetes | duct_epithelial_cell | *ZNF160* | -0.122997604 | 0.013440285 | 0.043518636 | 1 |
| Prediabetes | duct_epithelial_cell | *TSNARE1* | -0.108454232 | 0.007723929 | 0.029236335 | 1 |
| Prediabetes | duct_epithelial_cell | *TRMT61B* | -0.052964683 | 0.008119489 | 0.030300524 | 1 |
| Prediabetes | duct_epithelial_cell | *MFN1* | -0.087706012 | 0.010156574 | 0.035314317 | 1 |
| Prediabetes | duct_epithelial_cell | *JAGN1* | -0.193770374 | 0.000369284 | 0.008963481 | 1 |
| Prediabetes | duct_epithelial_cell | *TRIM8* | -0.141131263 | 0.010807308 | 0.036987729 | 1 |
| Prediabetes | duct_epithelial_cell | *ZNF440* | -0.138038706 | 0.013179916 | 0.042902975 | 1 |
| Prediabetes | duct_epithelial_cell | *EXOSC1* | -0.132014291 | 0.003495421 | 0.017450459 | 1 |
| Prediabetes | duct_epithelial_cell | *MRPL36* | -0.138213612 | 0.000110908 | 0.008963481 | 0.435759048 |
| Prediabetes | duct_epithelial_cell | *PTGER4* | -0.11194702 | 0.008270571 | 0.030617094 | 1 |
| Prediabetes | duct_epithelial_cell | *VAT1L* | -0.086136201 | 0.008091894 | 0.030250286 | 1 |
| Prediabetes | duct_epithelial_cell | *PWWP2B* | -0.225853417 | 0.008215471 | 0.03050539 | 1 |
| Prediabetes | duct_epithelial_cell | *PTEN* | -0.065101982 | 0.01333212 | 0.043255079 | 1 |
| Prediabetes | duct_epithelial_cell | *TVP23B* | -0.088735651 | 0.004335579 | 0.019807548 | 1 |
| Prediabetes | duct_epithelial_cell | *SRGAP2C* | -0.213257492 | 0.000280726 | 0.008963481 | 1 |
| Prediabetes | duct_epithelial_cell | *ZNF554* | -0.116119081 | 0.016017696 | 0.049553959 | 1 |
| Prediabetes | duct_epithelial_cell | *REG1B* | -0.022233531 | 0.0153736 | 0.048129782 | 1 |
| Prediabetes | duct_epithelial_cell | *NME6* | -0.14183716 | 0.010853144 | 0.037112275 | 1 |
| Prediabetes | duct_epithelial_cell | *TEFM* | -0.174245321 | 0.003944173 | 0.018783824 | 1 |
| Prediabetes | duct_epithelial_cell | *ZNF131* | -0.222152084 | 0.000284682 | 0.008963481 | 1 |
| Prediabetes | duct_epithelial_cell | *BSG* | -0.093402152 | 0.001158534 | 0.009685284 | 1 |
| Prediabetes | duct_epithelial_cell | *HINFP* | -0.364686052 | 0.000134235 | 0.008963481 | 0.527408959 |

| | | | | | | |
|---|---|---|---|---|---|---|
| Prediabetes | duct_epithelial_cell | *CERS6* | -0.076423316 | 0.007737566 | 0.029259765 | 1 |
| Prediabetes | duct_epithelial_cell | *SUCLG2* | -0.225368258 | 0.002538645 | 0.014327176 | 1 |
| Prediabetes | duct_epithelial_cell | *ZNF24* | -0.101927445 | 0.00064402 | 0.008963481 | 1 |
| Prediabetes | duct_epithelial_cell | *AFF1* | -0.126792907 | 0.003372574 | 0.01711995 | 1 |
| Prediabetes | duct_epithelial_cell | *DCP2* | -0.185226625 | 0.002028995 | 0.012475623 | 1 |
| Prediabetes | duct_epithelial_cell | *DMXL1* | -0.133559714 | 0.00178841 | 0.011538034 | 1 |
| Prediabetes | duct_epithelial_cell | *LCLAT1* | -0.20314786 | 0.000109949 | 0.008963481 | 0.43198872 |
| Prediabetes | duct_epithelial_cell | *TADA2B* | -0.207267083 | 0.00038188 | 0.008963481 | 1 |
| Prediabetes | duct_epithelial_cell | *IQCB1* | -0.123704163 | 0.003424296 | 0.017248792 | 1 |
| Prediabetes | duct_epithelial_cell | *ZNF483* | -0.057332834 | 0.015092026 | 0.04732368 | 1 |
| Prediabetes | duct_epithelial_cell | *NDNF* | -0.981340384 | 0.01364685 | 0.043985621 | 1 |
| Prediabetes | duct_epithelial_cell | *ARV1* | -0.207251323 | 0.004004415 | 0.018955838 | 1 |
| Prediabetes | duct_epithelial_cell | *PPP1R14B* | -0.106738254 | 0.005050308 | 0.02202293 | 1 |
| Prediabetes | duct_epithelial_cell | *NUDT18* | -0.307950246 | 0.000520668 | 0.008963481 | 1 |
| Prediabetes | duct_epithelial_cell | *UQCRH* | -0.121635038 | 0.000159599 | 0.008963481 | 0.627064688 |
| Prediabetes | duct_epithelial_cell | *DPY19L1* | -0.281800646 | 0.000681131 | 0.008963481 | 1 |
| Prediabetes | duct_epithelial_cell | *RALGAPA1* | -0.079822667 | 0.003809449 | 0.018364621 | 1 |
| Prediabetes | duct_epithelial_cell | *GOLT1A* | -0.208139746 | 0.001055068 | 0.009336398 | 1 |
| Prediabetes | duct_epithelial_cell | *ANGEL2* | -0.171700631 | 0.000663102 | 0.008963481 | 1 |
| Prediabetes | duct_epithelial_cell | *BRSK2* | -0.062102753 | 0.012304973 | 0.04084842 | 1 |
| Prediabetes | duct_epithelial_cell | *BRMS1* | -0.168680399 | 0.000621164 | 0.008963481 | 1 |
| Prediabetes | duct_epithelial_cell | *HRAS* | -0.159515369 | 0.000580988 | 0.008963481 | 1 |
| Prediabetes | duct_epithelial_cell | *SRP72* | -0.089909186 | 0.008350528 | 0.030749039 | 1 |
| Prediabetes | duct_epithelial_cell | *DENND6A* | -0.10023822 | 0.006367417 | 0.025502123 | 1 |
| Prediabetes | duct_epithelial_cell | *NDUFA11* | -0.111835039 | 8.36E-05 | 0.008963481 | 0.328420668 |
| Prediabetes | duct_epithelial_cell | *DHX36* | -0.115401647 | 0.000815484 | 0.008963481 | 1 |
| Prediabetes | duct_epithelial_cell | *ZNF519* | -0.146658252 | 0.003461492 | 0.017369351 | 1 |
| Prediabetes | duct_epithelial_cell | *TMEM9B* | -0.113130191 | 0.001768875 | 0.011496768 | 1 |
| Prediabetes | duct_epithelial_cell | *EIF3F* | -0.101095721 | 0.001193479 | 0.009719245 | 1 |
| Prediabetes | duct_epithelial_cell | *ZNF25* | -0.092353868 | 0.009126644 | 0.032569106 | 1 |
| Prediabetes | duct_epithelial_cell | *PPP2R2D* | -0.145598735 | 0.001312901 | 0.010116188 | 1 |
| Prediabetes | duct_epithelial_cell | *UCP2* | -0.079365879 | 0.014249046 | 0.045368317 | 1 |
| Prediabetes | duct_epithelial_cell | *SWI5* | -0.054609484 | 0.012193819 | 0.040552114 | 1 |
| Prediabetes | duct_epithelial_cell | *ZDHHC21* | -0.12114135 | 0.006151944 | 0.024966119 | 1 |
| Prediabetes | duct_epithelial_cell | *BNIP3* | -0.058046686 | 0.011560438 | 0.038854544 | 1 |
| Prediabetes | duct_epithelial_cell | *SMIM19* | -0.120468558 | 0.003639838 | 0.017898526 | 1 |
| Prediabetes | duct_epithelial_cell | *SLC35G1* | -0.117483007 | 0.006510299 | 0.02591587 | 1 |
| Prediabetes | duct_epithelial_cell | *COX8A* | -0.078282582 | 0.001835732 | 0.011746889 | 1 |
| Prediabetes | duct_epithelial_cell | *WDR25* | -0.123885848 | 0.002962269 | 0.015685656 | 1 |
| Prediabetes | duct_epithelial_cell | *KBTBD11* | -0.167558431 | 0.003045015 | 0.015976995 | 1 |
| Prediabetes | duct_epithelial_cell | *ANKLE2* | -0.10251994 | 0.003667146 | 0.017966065 | 1 |
| Prediabetes | duct_epithelial_cell | *DPP7* | -0.089810906 | 0.001491365 | 0.010590892 | 1 |
| Prediabetes | duct_epithelial_cell | *DEAF1* | -0.190621236 | 0.000554927 | 0.008963481 | 1 |
| Prediabetes | duct_epithelial_cell | *MAN1B1* | -0.173941181 | 0.000891513 | 0.00905104 | 1 |
| Prediabetes | duct_epithelial_cell | *PPFIA3* | -0.09689483 | 0.00750322 | 0.028733091 | 1 |
| Prediabetes | duct_epithelial_cell | *PAWR* | -0.134882025 | 0.001399602 | 0.010278575 | 1 |
| Prediabetes | duct_epithelial_cell | *AGTRAP* | -0.241511837 | 0.0003732 | 0.008963481 | 1 |
| Prediabetes | duct_epithelial_cell | *THAP5* | -0.101355693 | 0.002040323 | 0.012486647 | 1 |
| Prediabetes | duct_epithelial_cell | *CHID1* | -0.067101953 | 0.001902429 | 0.012055874 | 1 |
| Prediabetes | duct_epithelial_cell | *ZBTB41* | -0.110240362 | 0.002777747 | 0.01511602 | 1 |

| | | | | | | |
|---|---|---|---|---|---|---|
| Prediabetes | duct_epithelial_cell | *BET1L* | -0.164499957 | 0.001533212 | 0.010707166 | 1 |
| Prediabetes | duct_epithelial_cell | *IMP3* | -0.119714047 | 0.012750938 | 0.041783516 | 1 |
| Prediabetes | duct_epithelial_cell | *NT5DC1* | -0.120199479 | 0.007625156 | 0.028976426 | 1 |
| Prediabetes | duct_epithelial_cell | *ZNF713* | -0.289358536 | 0.0158944 | 0.049366875 | 1 |
| Prediabetes | duct_epithelial_cell | *RPP25* | -0.155112296 | 0.000873917 | 0.009011853 | 1 |
| Prediabetes | duct_epithelial_cell | *SPTY2D1* | -0.16587181 | 0.000498814 | 0.008963481 | 1 |
| Prediabetes | duct_epithelial_cell | *CALR* | -0.084051316 | 0.003922717 | 0.018769071 | 1 |
| Prediabetes | duct_epithelial_cell | *ELMOD2* | -0.192133204 | 0.000758186 | 0.008963481 | 1 |
| Prediabetes | duct_epithelial_cell | *RPRML* | -0.054122682 | 0.004272938 | 0.019612585 | 1 |
| Prediabetes | duct_epithelial_cell | *CCDC66* | -0.135488381 | 0.002093546 | 0.012674181 | 1 |
| Prediabetes | duct_epithelial_cell | *MCFD2* | -0.131896568 | 0.000428794 | 0.008963481 | 1 |
| Prediabetes | duct_epithelial_cell | *PPA1* | -0.092658636 | 0.0006387 | 0.008963481 | 1 |
| Prediabetes | duct_epithelial_cell | *MAP6D1* | -0.099034887 | 0.00973565 | 0.034153006 | 1 |
| Prediabetes | duct_epithelial_cell | *D2HGDH* | -0.167248949 | 0.000255908 | 0.008963481 | 1 |
| Prediabetes | duct_epithelial_cell | *LRRC57* | -0.110736551 | 0.010351001 | 0.035853926 | 1 |
| Prediabetes | duct_epithelial_cell | *ZFP82* | -0.165851748 | 0.001140393 | 0.00963464 | 1 |
| Prediabetes | duct_epithelial_cell | *RPH3AL* | -0.05356274 | 0.008854455 | 0.031799959 | 1 |
| Prediabetes | duct_epithelial_cell | *HIGD1A* | -0.141886668 | 0.000675387 | 0.008963481 | 1 |
| Prediabetes | duct_epithelial_cell | *ZNF707* | -0.391152384 | 0.012344831 | 0.040906653 | 1 |
| Prediabetes | duct_epithelial_cell | *OGFOD3* | -0.062220368 | 0.001020121 | 0.009336398 | 1 |
| Prediabetes | duct_epithelial_cell | *ZNF467* | -0.168478756 | 0.01166782 | 0.039081727 | 1 |
| Prediabetes | duct_epithelial_cell | *TMEM45A* | -0.066276288 | 0.014194653 | 0.045268501 | 1 |
| Prediabetes | duct_epithelial_cell | *GPR135* | -0.162590161 | 0.004868675 | 0.021463151 | 1 |
| Prediabetes | duct_epithelial_cell | *RFX7* | -0.174807969 | 0.001590742 | 0.010911694 | 1 |
| Prediabetes | duct_epithelial_cell | *CCDC149* | -0.164495084 | 0.001311631 | 0.010116188 | 1 |
| Prediabetes | duct_epithelial_cell | *ZNF708* | -0.177840771 | 0.001624401 | 0.011022922 | 1 |
| Prediabetes | duct_epithelial_cell | *IST1* | -0.175188755 | 0.00046551 | 0.008963481 | 1 |
| Prediabetes | duct_epithelial_cell | *ERCC6L2* | -0.176727979 | 0.000496899 | 0.008963481 | 1 |
| Prediabetes | duct_epithelial_cell | *MRPL41* | -0.138758995 | 0.000483238 | 0.008963481 | 1 |
| Prediabetes | duct_epithelial_cell | *TP53TG1* | -0.081972063 | 0.001188086 | 0.009719245 | 1 |
| Prediabetes | duct_epithelial_cell | *ARL6IP4* | -0.075640282 | 0.002604268 | 0.014586734 | 1 |
| Prediabetes | duct_epithelial_cell | *FIGN* | -0.097822575 | 0.008834133 | 0.031756002 | 1 |
| Prediabetes | duct_epithelial_cell | *NPLOC4* | -0.138911845 | 0.001188496 | 0.009719245 | 1 |
| Prediabetes | duct_epithelial_cell | *SATB1* | -0.192115572 | 0.00214785 | 0.012899732 | 1 |
| Prediabetes | duct_epithelial_cell | *C11orf54* | -0.051761733 | 0.015915127 | 0.049373666 | 1 |
| Prediabetes | duct_epithelial_cell | *CEP63* | -0.096187912 | 0.000756684 | 0.008963481 | 1 |
| Prediabetes | duct_epithelial_cell | *GPC6* | -0.071548359 | 0.013235277 | 0.043011913 | 1 |
| Prediabetes | duct_epithelial_cell | *FANCF* | -0.300807458 | 0.000112048 | 0.008963481 | 0.440236997 |
| Prediabetes | duct_epithelial_cell | *CALN1* | -0.18888279 | 0.003207596 | 0.016648141 | 1 |
| Prediabetes | duct_epithelial_cell | *BOLA2* | -0.12211825 | 0.004648867 | 0.02085091 | 1 |
| Prediabetes | duct_epithelial_cell | *MRPL54* | -0.083433327 | 0.002786604 | 0.015143249 | 1 |
| Prediabetes | duct_epithelial_cell | *TRMT12* | -0.265775403 | 0.000286227 | 0.008963481 | 1 |
| Prediabetes | duct_epithelial_cell | *CBX6* | -0.095864343 | 0.003154143 | 0.016392366 | 1 |
| Prediabetes | duct_epithelial_cell | *UQCR10* | -0.10146397 | 0.000219713 | 0.008963481 | 0.863252581 |
| Prediabetes | duct_epithelial_cell | *CRELD2* | -0.058024167 | 0.002979804 | 0.015696866 | 1 |
| Prediabetes | duct_epithelial_cell | *UBE2F* | -0.135925873 | 0.000202302 | 0.008963481 | 0.794844999 |
| Prediabetes | duct_epithelial_cell | *TM2D3* | -0.124221803 | 0.001054425 | 0.009336398 | 1 |
| Prediabetes | duct_epithelial_cell | *COPB2* | -0.125759586 | 0.003254009 | 0.016734297 | 1 |
| Prediabetes | duct_epithelial_cell | *CCR10* | -0.058904138 | 0.005636673 | 0.023585185 | 1 |
| Prediabetes | duct_epithelial_cell | *ZFP1* | -0.264804864 | 0.000813536 | 0.008963481 | 1 |

| | | | | | | |
|---|---|---|---|---|---|---|
| Prediabetes | duct_epithelial_cell | *XPOT* | -0.08509942 | 0.00406066 | 0.019152863 | 1 |
| Prediabetes | duct_epithelial_cell | *ZBTB40* | -0.107314748 | 0.015455539 | 0.048305536 | 1 |
| Prediabetes | duct_epithelial_cell | *UBE2G2* | -0.060157651 | 0.012071558 | 0.040194196 | 1 |
| Prediabetes | duct_epithelial_cell | *TMED9* | -0.103331766 | 0.004933777 | 0.021707514 | 1 |
| Prediabetes | duct_epithelial_cell | *BTBD6* | -0.231473828 | 0.000993433 | 0.009315508 | 1 |
| Prediabetes | duct_epithelial_cell | *LCN12* | -0.079522196 | 0.010099724 | 0.035211789 | 1 |
| Prediabetes | duct_epithelial_cell | *SIVA1* | -0.093388398 | 0.001329578 | 0.010202955 | 1 |
| Prediabetes | duct_epithelial_cell | *BRI3BP* | -0.208459967 | 0.001463452 | 0.01048303 | 1 |
| Prediabetes | duct_epithelial_cell | *CIB1* | -0.155511083 | 0.000340913 | 0.008963481 | 1 |
| Prediabetes | duct_epithelial_cell | *ZNF445* | -0.257009492 | 0.007765693 | 0.029337893 | 1 |
| Prediabetes | duct_epithelial_cell | *RBM11* | -0.085418793 | 0.00742177 | 0.02853242 | 1 |
| Prediabetes | duct_epithelial_cell | *ATP6V0A2* | -0.142867627 | 0.001142302 | 0.00963464 | 1 |
| Prediabetes | duct_epithelial_cell | *RAD51D* | -0.162509714 | 0.002506838 | 0.014192172 | 1 |
| Prediabetes | duct_epithelial_cell | *GPRIN3* | -0.073499791 | 0.008221605 | 0.03050539 | 1 |
| Prediabetes | duct_epithelial_cell | *P4HB* | -0.074890008 | 0.001690262 | 0.011199058 | 1 |
| Prediabetes | duct_epithelial_cell | *DRG1* | -0.223091209 | 1.21E-05 | 0.008963481 | 0.047710491 |
| Prediabetes | duct_epithelial_cell | *MORF4L1* | -0.089749355 | 0.000890195 | 0.00905104 | 1 |
| Prediabetes | duct_epithelial_cell | *DMWD* | -0.115974165 | 0.000371853 | 0.008963481 | 1 |
| Prediabetes | duct_epithelial_cell | *ZNF529* | -0.16396428 | 0.000474032 | 0.008963481 | 1 |
| Prediabetes | duct_epithelial_cell | *AIDA* | -0.084791539 | 0.014509093 | 0.045972762 | 1 |
| Prediabetes | duct_epithelial_cell | *WWOX* | -0.223476664 | 0.000232543 | 0.008963481 | 0.913662547 |
| Prediabetes | duct_epithelial_cell | *POLR1D* | -0.108784167 | 0.000723912 | 0.008963481 | 1 |
| Prediabetes | duct_epithelial_cell | *TOR3A* | -0.173020805 | 0.000267127 | 0.008963481 | 1 |
| Prediabetes | duct_epithelial_cell | *ZNF555* | -0.086519477 | 0.004329366 | 0.019802187 | 1 |
| Prediabetes | duct_epithelial_cell | *ZNF197* | -0.175925496 | 0.001087855 | 0.009519336 | 1 |
| Prediabetes | duct_epithelial_cell | *RPS23* | -0.065564808 | 0.001393373 | 0.010271223 | 1 |
| Prediabetes | duct_epithelial_cell | *ZNF396* | -0.114832741 | 0.004519013 | 0.020478893 | 1 |
| Prediabetes | duct_epithelial_cell | *FAM86B1* | -0.845942662 | 0.000386516 | 0.008963481 | 1 |
| Prediabetes | duct_epithelial_cell | *RTN4RL2* | -0.135798555 | 0.005396469 | 0.023121839 | 1 |
| Prediabetes | duct_epithelial_cell | *TMEM232* | -0.128785419 | 0.004768783 | 0.021123605 | 1 |
| Prediabetes | duct_epithelial_cell | *FAM183A* | -0.0491613 | 0.015717205 | 0.048971372 | 1 |
| Prediabetes | duct_epithelial_cell | *BCAM* | -0.065841093 | 0.004688817 | 0.020910738 | 1 |
| Prediabetes | duct_epithelial_cell | *CHP1* | -0.165807034 | 0.003895868 | 0.018689701 | 1 |
| Prediabetes | duct_epithelial_cell | *DHRS4L2* | -0.167106057 | 0.000704352 | 0.008963481 | 1 |
| Prediabetes | duct_epithelial_cell | *KBTBD12* | -0.095333655 | 0.001116307 | 0.009564575 | 1 |
| Prediabetes | duct_epithelial_cell | *KLHL17* | -0.129079284 | 0.010437066 | 0.036028262 | 1 |
| Prediabetes | duct_epithelial_cell | *RINL* | -0.177033969 | 0.001338608 | 0.010242129 | 1 |
| Prediabetes | duct_epithelial_cell | *ZNF490* | -0.122346001 | 0.005538022 | 0.023421841 | 1 |
| Prediabetes | duct_epithelial_cell | *FAM166A* | -0.238735576 | 0.000232479 | 0.008963481 | 0.913410247 |
| Prediabetes | duct_epithelial_cell | *LAMTOR4* | -0.069420079 | 0.00814577 | 0.030336239 | 1 |
| Prediabetes | duct_epithelial_cell | *ZNF559* | -0.029144201 | 0.006522925 | 0.025939852 | 1 |
| Prediabetes | duct_epithelial_cell | *FOCAD* | -0.095304909 | 0.006802162 | 0.026859993 | 1 |
| Prediabetes | duct_epithelial_cell | *FBLL1* | -0.081557978 | 0.009651274 | 0.033906075 | 1 |
| Prediabetes | duct_epithelial_cell | *UROS* | -0.211604115 | 0.000440334 | 0.008963481 | 1 |
| Prediabetes | duct_epithelial_cell | *FAM221A* | -0.079112087 | 0.008460389 | 0.030969628 | 1 |
| Prediabetes | duct_epithelial_cell | *MTF1* | -0.100743472 | 0.002883203 | 0.015391907 | 1 |
| Prediabetes | duct_epithelial_cell | *RPL14* | -0.070414022 | 0.000542522 | 0.008963481 | 1 |
| Prediabetes | duct_epithelial_cell | *ZNF567* | -0.164606362 | 0.000926774 | 0.009057947 | 1 |
| Prediabetes | duct_epithelial_cell | *ARID2* | -0.127183932 | 0.00063895 | 0.008963481 | 1 |
| Prediabetes | duct_epithelial_cell | *PNRC2* | -0.119324592 | 0.001944038 | 0.012221002 | 1 |

| | | | | | | |
|---|---|---|---|---|---|---|
| Prediabetes | duct_epithelial_cell | *SPATA41* | -0.181947269 | 0.007379223 | 0.028419569 | 1 |
| Prediabetes | duct_epithelial_cell | *ZNF681* | -0.221684816 | 0.002464574 | 0.013993224 | 1 |
| Prediabetes | duct_epithelial_cell | *MPHOSPH8* | -0.095873125 | 0.001201183 | 0.009750216 | 1 |
| Prediabetes | duct_epithelial_cell | *LCOR* | -0.142174782 | 0.000264682 | 0.008963481 | 1 |
| Prediabetes | duct_epithelial_cell | *XPNPEP3* | -0.14187082 | 0.001111016 | 0.009564575 | 1 |
| Prediabetes | duct_epithelial_cell | *ZNF471* | -0.324026754 | 0.001388528 | 0.010271223 | 1 |
| Prediabetes | duct_epithelial_cell | *ZNF493* | -0.212402748 | 0.000492332 | 0.008963481 | 1 |
| Prediabetes | duct_epithelial_cell | *GTF2IRD2* | -0.168105658 | 0.004656867 | 0.020862975 | 1 |
| Prediabetes | duct_epithelial_cell | *LONP1* | -0.121219682 | 0.00499096 | 0.021861184 | 1 |
| Prediabetes | duct_epithelial_cell | *ZNF34* | -0.223533372 | 0.004223145 | 0.019551648 | 1 |
| Prediabetes | duct_epithelial_cell | *ZNF124* | -0.352449188 | 0.002175049 | 0.012948135 | 1 |
| Prediabetes | duct_epithelial_cell | *XRCC6* | -0.093031318 | 0.002422972 | 0.013837002 | 1 |
| Prediabetes | duct_epithelial_cell | *ZNF502* | -0.29594826 | 0.000447311 | 0.008963481 | 1 |
| Prediabetes | duct_epithelial_cell | *ZNF33B* | -0.15118933 | 0.014432504 | 0.045803964 | 1 |
| Prediabetes | duct_epithelial_cell | *AMZ2* | -0.11771932 | 0.002153786 | 0.012899732 | 1 |
| Prediabetes | duct_epithelial_cell | *ZNF431* | -0.175048916 | 0.002669524 | 0.014746113 | 1 |
| Prediabetes | duct_epithelial_cell | *FAM3C* | -0.140059529 | 0.000706042 | 0.008963481 | 1 |
| Prediabetes | duct_epithelial_cell | *AP2A1* | -0.145618433 | 0.000885005 | 0.00905104 | 1 |
| Prediabetes | duct_epithelial_cell | *ZNF429* | -0.246461849 | 0.000442139 | 0.008963481 | 1 |
| Prediabetes | duct_epithelial_cell | *ZNF682* | -0.208893105 | 0.001151958 | 0.009685284 | 1 |
| Prediabetes | duct_epithelial_cell | *SND1* | -0.074418783 | 0.011138322 | 0.037693769 | 1 |
| Prediabetes | duct_epithelial_cell | *PIWIL2* | -0.063667119 | 0.001759013 | 0.011461296 | 1 |
| Prediabetes | duct_epithelial_cell | *GTF2E2* | -0.136639352 | 0.004068263 | 0.019165715 | 1 |
| Prediabetes | duct_epithelial_cell | *OGDHL* | -0.037402247 | 0.003536487 | 0.017534673 | 1 |
| Prediabetes | duct_epithelial_cell | *RPS26* | -0.044933798 | 0.013396309 | 0.043427474 | 1 |
| Prediabetes | duct_epithelial_cell | *LHFPL5* | -0.119905092 | 0.008524026 | 0.031108424 | 1 |
| Prediabetes | duct_epithelial_cell | *ATAD3A* | -0.12787455 | 0.005142936 | 0.022278498 | 1 |
| Prediabetes | duct_epithelial_cell | *SLC9A8* | -0.262842784 | 0.003248872 | 0.016734297 | 1 |
| Prediabetes | duct_epithelial_cell | *RPL12* | -0.054493815 | 0.002003084 | 0.012393885 | 1 |
| Prediabetes | duct_epithelial_cell | *ENTPD7* | -0.091558492 | 0.008742995 | 0.031543827 | 1 |
| Prediabetes | duct_epithelial_cell | *ZNF273* | -0.194000289 | 0.012309628 | 0.04084842 | 1 |
| Prediabetes | duct_epithelial_cell | *ZNF84* | -0.159275135 | 0.001385709 | 0.010271223 | 1 |
| Prediabetes | duct_epithelial_cell | *ZNF667* | -0.160782503 | 0.002153655 | 0.012899732 | 1 |
| Prediabetes | duct_epithelial_cell | *CD2AP* | -0.216567958 | 0.00071169 | 0.008963481 | 1 |
| Prediabetes | duct_epithelial_cell | *SFI1* | -0.078957448 | 0.007008366 | 0.027368761 | 1 |
| Prediabetes | duct_epithelial_cell | *SVIP* | -0.09084218 | 0.001295536 | 0.010059608 | 1 |
| Prediabetes | duct_epithelial_cell | *DDRGK1* | -0.115019782 | 0.010989066 | 0.037414245 | 1 |
| Prediabetes | duct_epithelial_cell | *HSD17B11* | -0.106129459 | 0.00799701 | 0.030059963 | 1 |
| Prediabetes | duct_epithelial_cell | *HELZ* | -0.15448286 | 0.000840524 | 0.008963481 | 1 |
| Prediabetes | duct_epithelial_cell | *ZNF583* | -0.152153939 | 0.002198765 | 0.012987238 | 1 |
| Prediabetes | duct_epithelial_cell | *ZNF587* | -0.215482759 | 0.000623038 | 0.008963481 | 1 |
| Prediabetes | duct_epithelial_cell | *ZNF627* | -0.169840243 | 0.001698885 | 0.01123724 | 1 |
| Prediabetes | duct_epithelial_cell | *NCOA6* | -0.212762132 | 0.003310613 | 0.016870817 | 1 |
| Prediabetes | duct_epithelial_cell | *PAPSS2* | -0.062678376 | 0.002869966 | 0.015391043 | 1 |
| Prediabetes | duct_epithelial_cell | *CEP290* | -0.159797365 | 0.001068597 | 0.009434874 | 1 |
| Prediabetes | duct_epithelial_cell | *ECI2* | -0.102126597 | 0.005995852 | 0.024616198 | 1 |
| Prediabetes | duct_epithelial_cell | *DENND4B* | -0.10180204 | 0.004722025 | 0.020987373 | 1 |
| Prediabetes | duct_epithelial_cell | *TSEN15* | -0.184651935 | 0.000465799 | 0.008963481 | 1 |
| Prediabetes | duct_epithelial_cell | *CCDC152* | -0.109685723 | 0.001623866 | 0.011022922 | 1 |
| Prediabetes | duct_epithelial_cell | *MAP3K3* | -0.201602828 | 0.001541343 | 0.010718604 | 1 |

| | | | | | | |
|---|---|---|---|---|---|---|
| Prediabetes | duct_epithelial_cell | *C1orf174* | -0.127139443 | 0.002821684 | 0.015228566 | 1 |
| Prediabetes | duct_epithelial_cell | *TBKBP1* | -0.049382619 | 0.010987813 | 0.037414245 | 1 |
| Prediabetes | duct_epithelial_cell | *CCDC167* | -0.138051792 | 0.016090905 | 0.049702174 | 1 |
| Prediabetes | duct_epithelial_cell | *SOWAHA* | -0.115329767 | 0.004126077 | 0.019299233 | 1 |
| Prediabetes | duct_epithelial_cell | *PJA2* | -0.061691875 | 0.00759873 | 0.028976426 | 1 |
| Prediabetes | duct_epithelial_cell | *EFCAB2* | -0.081707432 | 0.005507879 | 0.023344611 | 1 |
| Prediabetes | duct_epithelial_cell | *SAMD5* | -0.07731194 | 0.004412237 | 0.020064443 | 1 |
| Prediabetes | duct_epithelial_cell | *PCMTD2* | -0.153903808 | 0.00095146 | 0.009132479 | 1 |
| Prediabetes | duct_epithelial_cell | *NOTCH4* | -0.031744308 | 0.007005839 | 0.027368761 | 1 |
| Prediabetes | duct_epithelial_cell | *MRPL38* | -0.117660115 | 0.006345118 | 0.025438743 | 1 |
| Prediabetes | duct_epithelial_cell | *ABCF1* | -0.105180199 | 0.003926744 | 0.018769071 | 1 |
| Prediabetes | duct_epithelial_cell | *ASPDH* | -0.185896287 | 0.004741392 | 0.021049638 | 1 |
| Prediabetes | duct_epithelial_cell | *SARNP* | -0.228017037 | 2.29E-05 | 0.008963481 | 0.089950414 |
| Prediabetes | duct_epithelial_cell | *LCMT1* | -0.19422821 | 0.000387537 | 0.008963481 | 1 |
| Prediabetes | duct_epithelial_cell | *LIN52* | -0.180276067 | 0.00216242 | 0.012919364 | 1 |
| Prediabetes | duct_epithelial_cell | *ZNF254* | -0.174111134 | 0.000313569 | 0.008963481 | 1 |
| Prediabetes | duct_epithelial_cell | *SUPT4H1* | -0.198859505 | 0.000789137 | 0.008963481 | 1 |
| Prediabetes | duct_epithelial_cell | *C8orf82* | -0.104390078 | 0.004872774 | 0.021463151 | 1 |
| Prediabetes | duct_epithelial_cell | *LEPROT* | -0.158370173 | 0.000674577 | 0.008963481 | 1 |
| Prediabetes | duct_epithelial_cell | *PPP1CB* | -0.109359547 | 0.000835863 | 0.008963481 | 1 |
| Prediabetes | duct_epithelial_cell | *NUDT19* | -0.103879393 | 0.002663968 | 0.014746113 | 1 |
| Prediabetes | duct_epithelial_cell | *ZNF90* | -0.50979216 | 0.000262568 | 0.008963481 | 1 |
| Prediabetes | duct_epithelial_cell | *REPIN1* | -0.169830974 | 0.000692775 | 0.008963481 | 1 |
| Prediabetes | duct_epithelial_cell | *MEG3* | -0.060536668 | 0.003900812 | 0.018690598 | 1 |
| Prediabetes | duct_epithelial_cell | *EML6* | -0.127754589 | 0.005778837 | 0.024023343 | 1 |
| Prediabetes | duct_epithelial_cell | *IFRD2* | -0.23584343 | 0.000334034 | 0.008963481 | 1 |
| Prediabetes | duct_epithelial_cell | *C5orf49* | -0.102297948 | 0.004192068 | 0.019468837 | 1 |
| Prediabetes | duct_epithelial_cell | *VPS16* | -0.10601946 | 0.009000806 | 0.032207801 | 1 |
| Prediabetes | duct_epithelial_cell | *ATAD3C* | -0.079969868 | 0.009455938 | 0.033380395 | 1 |
| Prediabetes | duct_epithelial_cell | *PAM16* | -0.142555843 | 0.000514842 | 0.008963481 | 1 |
| Prediabetes | duct_epithelial_cell | *CELA3B* | -0.024184468 | 0.012744221 | 0.041783516 | 1 |
| Prediabetes | duct_epithelial_cell | *VAMP2* | -0.074823997 | 0.001431675 | 0.010357204 | 1 |
| Prediabetes | duct_epithelial_cell | *PPP3R1* | -0.139969392 | 0.000943564 | 0.009132479 | 1 |
| Prediabetes | duct_epithelial_cell | *CEBPD* | -0.121383927 | 0.000553481 | 0.008963481 | 1 |
| Prediabetes | duct_epithelial_cell | *UBA52* | -0.063406228 | 0.006073164 | 0.024776105 | 1 |
| Prediabetes | duct_epithelial_cell | *ARL17B* | -0.076538514 | 0.015921719 | 0.049373666 | 1 |
| Prediabetes | duct_epithelial_cell | *HSBP1* | -0.108109729 | 0.001578445 | 0.010899314 | 1 |
| Prediabetes | duct_epithelial_cell | *RPS18* | -0.059257462 | 0.000966218 | 0.009147642 | 1 |
| Prediabetes | duct_epithelial_cell | *SNHG15* | -0.112690205 | 0.003293789 | 0.016828736 | 1 |
| Prediabetes | duct_epithelial_cell | *RPS28* | -0.061265523 | 0.001819535 | 0.011681295 | 1 |
| Prediabetes | duct_epithelial_cell | *ZNF879* | -0.398628588 | 7.42E-05 | 0.008963481 | 0.291378099 |
| Prediabetes | duct_epithelial_cell | *PINLYP* | 0.104427179 | 0.008308274 | 0.030679707 | 1 |
| Prediabetes | duct_epithelial_cell | *ANKRD65* | -0.081665187 | 0.005712848 | 0.023853116 | 1 |
| Prediabetes | duct_epithelial_cell | *ZNF737* | -0.298947187 | 2.12E-05 | 0.008963481 | 0.083265099 |
| Prediabetes | duct_epithelial_cell | *TXNDC5* | -0.089590881 | 0.005946999 | 0.024518112 | 1 |
| Prediabetes | duct_epithelial_cell | *NME1* | -0.067449697 | 0.008742093 | 0.031543827 | 1 |
| Prediabetes | duct_epithelial_cell | *CDRT4* | -0.087503633 | 0.007083016 | 0.027499182 | 1 |
| Prediabetes | duct_epithelial_cell | *SMKR1* | -0.07463146 | 0.006726001 | 0.026639575 | 1 |
| Prediabetes | duct_epithelial_cell | *LINC00886* | -0.058633573 | 0.014854539 | 0.046765612 | 1 |
| Prediabetes | duct_epithelial_cell | *MIF* | -0.063283251 | 0.001460945 | 0.01048303 | 1 |

| | | | | | | |
|---|---|---|---|---|---|---|
| Prediabetes | duct_epithelial_cell | *NSUN6* | -0.192380817 | 0.000662442 | 0.008963481 | 1 |
| Prediabetes | duct_epithelial_cell | *CRCP* | -0.091151813 | 0.007446885 | 0.028573059 | 1 |
| Prediabetes | duct_epithelial_cell | *PI4KA* | -0.13563189 | 0.0010099 | 0.009336398 | 1 |
| Prediabetes | duct_epithelial_cell | *DECR2* | -0.181681809 | 0.014956726 | 0.046974403 | 1 |
| Prediabetes | duct_epithelial_cell | *GNG10* | -0.048210212 | 0.012477539 | 0.041127729 | 1 |
| Prediabetes | duct_epithelial_cell | *PSG4* | 0.20815524 | 0.002345215 | 0.013510775 | 1 |
| Prediabetes | duct_epithelial_cell | *MICAL3* | -0.199460315 | 0.006570216 | 0.026075129 | 1 |
| Prediabetes | duct_epithelial_cell | *VSIG8* | -0.467056533 | 0.000528266 | 0.008963481 | 1 |
| Prediabetes | duct_epithelial_cell | *C4orf48* | -0.082691657 | 0.001426771 | 0.010342775 | 1 |
| Prediabetes | duct_epithelial_cell | *IL10RB* | -0.123917794 | 0.001283013 | 0.010041748 | 1 |
| Prediabetes | duct_epithelial_cell | *TMEM141* | -0.166034899 | 5.69E-05 | 0.008963481 | 0.223444659 |
| Prediabetes | duct_epithelial_cell | *NEAT1* | -0.054627875 | 0.003812854 | 0.018364621 | 1 |
| Prediabetes | duct_epithelial_cell | *LINC01091* | -0.048004845 | 0.011504318 | 0.038699029 | 1 |
| Prediabetes | duct_epithelial_cell | *PDCD6* | -0.126003121 | 0.004161554 | 0.019406459 | 1 |
| Prediabetes | duct_epithelial_cell | *ATP6V1E2* | -0.097437558 | 0.012636188 | 0.041546093 | 1 |
| Prediabetes | duct_epithelial_cell | *ZNF550* | -0.161837938 | 0.001241003 | 0.009849499 | 1 |
| Prediabetes | duct_epithelial_cell | *ALG11* | -0.370308369 | 0.000187071 | 0.008963481 | 0.735003271 |
| Prediabetes | duct_epithelial_cell | *SNHG9* | -0.094512024 | 0.000391153 | 0.008963481 | 1 |
| Prediabetes | duct_epithelial_cell | *ZNF432* | -0.159142287 | 0.000676356 | 0.008963481 | 1 |
| Prediabetes | duct_epithelial_cell | *ZNF10* | -0.166713698 | 0.001681537 | 0.011177897 | 1 |
| Prediabetes | duct_epithelial_cell | *GATC* | -0.150631505 | 0.004437547 | 0.020156211 | 1 |
| Prediabetes | duct_epithelial_cell | *TEN1* | -0.095114243 | 0.00394324 | 0.018783824 | 1 |
| Prediabetes | duct_epithelial_cell | *CHURC1* | -0.062963335 | 0.01484694 | 0.046765612 | 1 |
| Prediabetes | duct_epithelial_cell | *TMEM179* | -0.04699401 | 0.013882688 | 0.044490278 | 1 |
| Prediabetes | duct_epithelial_cell | *TMEM178B* | -0.076751892 | 0.001971542 | 0.01235437 | 1 |
| Prediabetes | duct_epithelial_cell | *LINC01003* | -0.186899669 | 0.000410644 | 0.008963481 | 1 |
| Prediabetes | duct_epithelial_cell | *MRPL12* | -0.186844707 | 0.000579441 | 0.008963481 | 1 |
| Prediabetes | duct_epithelial_cell | *LINC00667* | -0.119642634 | 0.001680254 | 0.011177897 | 1 |
| Prediabetes | duct_epithelial_cell | *FBXO17* | -0.103000842 | 0.003814083 | 0.018364621 | 1 |
| Prediabetes | duct_epithelial_cell | *SNHG8* | -0.062797446 | 0.00297207 | 0.015695248 | 1 |
| Prediabetes | duct_epithelial_cell | *GTF2H5* | -0.138211901 | 0.001544681 | 0.010722704 | 1 |
| Prediabetes | PP_cell | *GCLC* | -0.310359378 | 0.002550512 | 0.01537007 | 1 |
| Prediabetes | PP_cell | *LASP1* | -0.307808579 | 0.001461864 | 0.011646409 | 1 |
| Prediabetes | PP_cell | *SLC25A13* | -0.411728505 | 0.00307958 | 0.016741022 | 1 |
| Prediabetes | PP_cell | *PON1* | -0.084080162 | 0.01463689 | 0.039524633 | 1 |
| Prediabetes | PP_cell | *IBTK* | -0.309129654 | 0.001280359 | 0.011111559 | 1 |
| Prediabetes | PP_cell | *SYNRG* | -0.361715777 | 0.001077152 | 0.010267243 | 1 |
| Prediabetes | PP_cell | *IL32* | -0.176244993 | 0.000839134 | 0.009444968 | 1 |
| Prediabetes | PP_cell | *MED24* | -0.443513327 | 0.001989727 | 0.013597804 | 1 |
| Prediabetes | PP_cell | *ETV7* | -0.156593721 | 0.006028735 | 0.024356489 | 1 |
| Prediabetes | PP_cell | *GIPR* | -0.147509042 | 0.011824748 | 0.035704358 | 1 |
| Prediabetes | PP_cell | *TTC19* | -0.223754562 | 0.016563731 | 0.04234151 | 1 |
| Prediabetes | PP_cell | *LARS2* | -0.248445647 | 0.015084481 | 0.040072295 | 1 |
| Prediabetes | PP_cell | *PPP5C* | -0.420817798 | 0.001783552 | 0.012957043 | 1 |
| Prediabetes | PP_cell | *ERCC1* | -0.299060671 | 0.000220517 | 0.008519859 | 0.866412642 |
| Prediabetes | PP_cell | *EXTL3* | -0.292673616 | 0.003553067 | 0.018224541 | 1 |
| Prediabetes | PP_cell | *ALOX5* | -0.132694136 | 0.009036808 | 0.030982214 | 1 |
| Prediabetes | PP_cell | *MAN2B2* | -0.448612726 | 0.000327958 | 0.008519859 | 1 |
| Prediabetes | PP_cell | *DDX11* | -0.243064383 | 0.00691085 | 0.026361874 | 1 |
| Prediabetes | PP_cell | *MDH1* | -0.271411813 | 0.000737584 | 0.009084543 | 1 |

| | | | | | | |
|---|---|---|---|---|---|---|
| Prediabetes | PP_cell | *DPEP1* | -0.084119265 | 0.010387016 | 0.033588961 | 1 |
| Prediabetes | PP_cell | *TTC27* | -0.268240799 | 0.01460159 | 0.039474478 | 1 |
| Prediabetes | PP_cell | *RNASET2* | -0.255099557 | 0.000406965 | 0.008519859 | 1 |
| Prediabetes | PP_cell | *CD44* | -0.109458713 | 0.001115776 | 0.010487757 | 1 |
| Prediabetes | PP_cell | *AGPAT4* | -0.37446771 | 0.002095823 | 0.01395676 | 1 |
| Prediabetes | PP_cell | *SH2D2A* | -0.249213869 | 0.015781821 | 0.04125534 | 1 |
| Prediabetes | PP_cell | *VEZT* | -0.284942902 | 0.005936981 | 0.024153447 | 1 |
| Prediabetes | PP_cell | *BAK1* | -0.137668222 | 0.014265308 | 0.039085352 | 1 |
| Prediabetes | PP_cell | *ABCF2* | -0.448939198 | 0.00404678 | 0.019357575 | 1 |
| Prediabetes | PP_cell | *APBA2* | -0.174459108 | 0.012949847 | 0.037800852 | 1 |
| Prediabetes | PP_cell | *ASTE1* | -0.187613116 | 0.009973314 | 0.032763505 | 1 |
| Prediabetes | PP_cell | *SH3YL1* | -0.128795719 | 0.003154772 | 0.017002881 | 1 |
| Prediabetes | PP_cell | *FAM136A* | -0.282815814 | 0.000306634 | 0.008519859 | 1 |
| Prediabetes | PP_cell | *MYOM2* | -0.383366355 | 0.00434788 | 0.020119092 | 1 |
| Prediabetes | PP_cell | *RPL26L1* | -0.268461711 | 0.004437193 | 0.020248234 | 1 |
| Prediabetes | PP_cell | *NSUN2* | -0.412329336 | 0.000357579 | 0.008519859 | 1 |
| Prediabetes | PP_cell | *AGA* | -0.385339396 | 0.000273321 | 0.008519859 | 1 |
| Prediabetes | PP_cell | *SPATA7* | -0.266154052 | 0.0068627 | 0.026305899 | 1 |
| Prediabetes | PP_cell | *CAPG* | -0.077468341 | 0.003986957 | 0.019154283 | 1 |
| Prediabetes | PP_cell | *ADRB1* | -0.07591933 | 0.014391632 | 0.039235459 | 1 |
| Prediabetes | PP_cell | *PREX2* | -0.250074336 | 0.017354859 | 0.043822135 | 1 |
| Prediabetes | PP_cell | *WDR37* | -0.379193475 | 0.001291416 | 0.011111559 | 1 |
| Prediabetes | PP_cell | *DTNBP1* | -0.4024696 | 0.002044997 | 0.01385309 | 1 |
| Prediabetes | PP_cell | *GOPC* | -0.364496815 | 0.002787585 | 0.0160564 | 1 |
| Prediabetes | PP_cell | *MRPS10* | -0.18061212 | 0.007662043 | 0.027998993 | 1 |
| Prediabetes | PP_cell | *ERCC8* | -0.436224737 | 0.000552092 | 0.008519859 | 1 |
| Prediabetes | PP_cell | *CYBA* | -0.299113375 | 0.001684573 | 0.012559178 | 1 |
| Prediabetes | PP_cell | *THOC3* | -0.185376532 | 0.00155739 | 0.011998012 | 1 |
| Prediabetes | PP_cell | *HEBP2* | -0.264225639 | 0.001590245 | 0.012147101 | 1 |
| Prediabetes | PP_cell | *FNIP2* | -0.298704297 | 0.000438465 | 0.008519859 | 1 |
| Prediabetes | PP_cell | *MRTO4* | -0.221284652 | 0.013426633 | 0.03803406 | 1 |
| Prediabetes | PP_cell | *MCF2L2* | -0.226007096 | 0.001996486 | 0.013597804 | 1 |
| Prediabetes | PP_cell | *LAMA3* | -0.104958868 | 0.011785733 | 0.035704358 | 1 |
| Prediabetes | PP_cell | *ANAPC4* | -0.336764711 | 0.001468982 | 0.011646409 | 1 |
| Prediabetes | PP_cell | *LY75* | -0.244884609 | 0.002674002 | 0.015685035 | 1 |
| Prediabetes | PP_cell | *OPN3* | -0.192135395 | 0.017028308 | 0.04320283 | 1 |
| Prediabetes | PP_cell | *TBC1D22A* | -0.326875084 | 0.000415136 | 0.008519859 | 1 |
| Prediabetes | PP_cell | *SYNE2* | -0.290185791 | 0.008400742 | 0.029688131 | 1 |
| Prediabetes | PP_cell | *ATP9A* | -0.157708789 | 0.001616795 | 0.012263296 | 1 |
| Prediabetes | PP_cell | *CYFIP2* | -0.238549108 | 0.016512417 | 0.042268461 | 1 |
| Prediabetes | PP_cell | *MCOLN3* | -0.044675175 | 0.012125513 | 0.036255474 | 1 |
| Prediabetes | PP_cell | *ITIH4* | -0.080301654 | 0.014854281 | 0.039783551 | 1 |
| Prediabetes | PP_cell | *GDI2* | -0.324186778 | 0.000365435 | 0.008519859 | 1 |
| Prediabetes | PP_cell | *LAMC2* | -0.068818347 | 0.018809749 | 0.046626816 | 1 |
| Prediabetes | PP_cell | *RASGRF1* | -0.127372816 | 0.016153869 | 0.041700755 | 1 |
| Prediabetes | PP_cell | *CROCC* | -0.236071812 | 0.01056552 | 0.033914974 | 1 |
| Prediabetes | PP_cell | *GNA15* | -0.353067775 | 0.01951053 | 0.047834311 | 1 |
| Prediabetes | PP_cell | *RIMBP2* | -0.146077979 | 0.003735878 | 0.018556591 | 1 |
| Prediabetes | PP_cell | *MPC1* | -0.293368281 | 0.001752767 | 0.012855972 | 1 |
| Prediabetes | PP_cell | *BCAT1* | -0.06999088 | 0.009895836 | 0.032720527 | 1 |

| | | | | | | |
|---|---|---|---|---|---|---|
| Prediabetes | PP_cell | *EIF4B* | -0.191751446 | 0.009598874 | 0.032069706 | 1 |
| Prediabetes | PP_cell | *SPHK2* | -0.344827305 | 0.002073459 | 0.013902083 | 1 |
| Prediabetes | PP_cell | *RNF4* | -0.342645052 | 0.003770429 | 0.018659548 | 1 |
| Prediabetes | PP_cell | *TM7SF3* | -0.229298926 | 0.002379587 | 0.014840315 | 1 |
| Prediabetes | PP_cell | *ST3GAL6* | -0.29339817 | 7.45E-05 | 0.008519859 | 0.29254693 |
| Prediabetes | PP_cell | *TMEM161A* | -0.476991096 | 0.001744811 | 0.012855972 | 1 |
| Prediabetes | PP_cell | *SNX24* | -0.322963858 | 0.003632551 | 0.01839612 | 1 |
| Prediabetes | PP_cell | *PMS1* | -0.185645027 | 0.004085492 | 0.019452089 | 1 |
| Prediabetes | PP_cell | *IPO5* | -0.266884985 | 0.00941047 | 0.031819051 | 1 |
| Prediabetes | PP_cell | *OAT* | -0.242410285 | 0.014299957 | 0.039125719 | 1 |
| Prediabetes | PP_cell | *GSTO2* | -0.281038498 | 0.000197392 | 0.008519859 | 0.775554376 |
| Prediabetes | PP_cell | *CTNNA2* | -0.166281946 | 0.009487346 | 0.031941544 | 1 |
| Prediabetes | PP_cell | *STOML1* | -0.188219376 | 0.001820141 | 0.013073736 | 1 |
| Prediabetes | PP_cell | *HEATR6* | -0.326245823 | 0.019092341 | 0.047089646 | 1 |
| Prediabetes | PP_cell | *REEP1* | -0.234526692 | 0.003383886 | 0.017633008 | 1 |
| Prediabetes | PP_cell | *POLR1A* | -0.194939492 | 0.000677728 | 0.008780065 | 1 |
| Prediabetes | PP_cell | *MAST4* | -0.355698335 | 0.000335739 | 0.008519859 | 1 |
| Prediabetes | PP_cell | *CLEC2D* | -0.527568422 | 0.001624262 | 0.012296195 | 1 |
| Prediabetes | PP_cell | *PLA2G10* | -0.297950135 | 0.004989624 | 0.021710114 | 1 |
| Prediabetes | PP_cell | *RAB27A* | -0.251793687 | 0.000674282 | 0.008780065 | 1 |
| Prediabetes | PP_cell | *PTPN3* | -0.291322523 | 0.000451904 | 0.008519859 | 1 |
| Prediabetes | PP_cell | *FGF10* | -0.235557421 | 0.003197257 | 0.017184708 | 1 |
| Prediabetes | PP_cell | *DGCR2* | -0.397603139 | 0.000294413 | 0.008519859 | 1 |
| Prediabetes | PP_cell | *ASNS* | -0.168962699 | 0.004119429 | 0.019523809 | 1 |
| Prediabetes | PP_cell | *RPL31* | -0.224284809 | 0.000991738 | 0.009839747 | 1 |
| Prediabetes | PP_cell | *SEL1L* | -0.277307253 | 0.002238474 | 0.014348148 | 1 |
| Prediabetes | PP_cell | *CPSF1* | -0.18108201 | 0.011857614 | 0.035727429 | 1 |
| Prediabetes | PP_cell | *ALDH3A2* | -0.165234225 | 0.00535022 | 0.022774663 | 1 |
| Prediabetes | PP_cell | *P4HA2* | -0.191693543 | 0.012266119 | 0.036372141 | 1 |
| Prediabetes | PP_cell | *TRNT1* | -0.456140227 | 0.002338926 | 0.014690752 | 1 |
| Prediabetes | PP_cell | *CRMP1* | -0.306656127 | 0.000208254 | 0.008519859 | 0.818228967 |
| Prediabetes | PP_cell | *DERL2* | -0.233224915 | 0.017093826 | 0.043330092 | 1 |
| Prediabetes | PP_cell | *NDE1* | -0.288730809 | 0.000179717 | 0.008519859 | 0.70610884 |
| Prediabetes | PP_cell | *SCARB1* | -0.21814856 | 0.005594518 | 0.023253761 | 1 |
| Prediabetes | PP_cell | *NLE1* | -0.068772371 | 0.01860953 | 0.046247213 | 1 |
| Prediabetes | PP_cell | *MRPS34* | -0.232265107 | 0.002476991 | 0.015206402 | 1 |
| Prediabetes | PP_cell | *IPCEF1* | -0.137634446 | 0.007610695 | 0.027900502 | 1 |
| Prediabetes | PP_cell | *ANO8* | -0.177808691 | 0.003596562 | 0.018323322 | 1 |
| Prediabetes | PP_cell | *GRAMD4* | -0.157466037 | 0.001129151 | 0.010537852 | 1 |
| Prediabetes | PP_cell | *TIMM21* | -0.359339612 | 0.007359007 | 0.027276923 | 1 |
| Prediabetes | PP_cell | *CACNG5* | -2.297660509 | 0.003086167 | 0.016741022 | 1 |
| Prediabetes | PP_cell | *EXOSC7* | -0.317867499 | 0.009918592 | 0.032720527 | 1 |
| Prediabetes | PP_cell | *MKRN2* | -0.258456177 | 0.001458715 | 0.011646409 | 1 |
| Prediabetes | PP_cell | *TPD52* | -0.285799682 | 0.00018572 | 0.008519859 | 0.729693382 |
| Prediabetes | PP_cell | *ACACB* | -0.266007197 | 0.000444686 | 0.008519859 | 1 |
| Prediabetes | PP_cell | *GPATCH1* | -0.284550211 | 0.020586376 | 0.049760752 | 1 |
| Prediabetes | PP_cell | *ICAM3* | -0.10276832 | 0.017564789 | 0.044153587 | 1 |
| Prediabetes | PP_cell | *ACTL6B* | -0.168967643 | 0.002651394 | 0.01561651 | 1 |
| Prediabetes | PP_cell | *APBB1IP* | 0.105745311 | 0.00583552 | 0.023907984 | 1 |
| Prediabetes | PP_cell | *NAALAD2* | -0.191712784 | 0.010044151 | 0.032886224 | 1 |

| | | | | | | |
|---|---|---|---|---|---|---|
| Prediabetes | PP_cell | *MCCC1* | -0.177702535 | 0.002084353 | 0.013904889 | 1 |
| Prediabetes | PP_cell | *N4BP2* | -0.317923778 | 0.008904829 | 0.030690414 | 1 |
| Prediabetes | PP_cell | *VDAC3* | -0.212593639 | 0.015151172 | 0.040167986 | 1 |
| Prediabetes | PP_cell | *TP73* | -0.790487395 | 0.013474365 | 0.038114313 | 1 |
| Prediabetes | PP_cell | *TOLLIP* | -0.28145024 | 0.001952558 | 0.013500582 | 1 |
| Prediabetes | PP_cell | *CLUL1* | -0.398687505 | 0.009589116 | 0.032064372 | 1 |
| Prediabetes | PP_cell | *FKBP7* | -0.36173626 | 0.001996929 | 0.013597804 | 1 |
| Prediabetes | PP_cell | *LXN* | -0.188149204 | 0.000509639 | 0.008519859 | 1 |
| Prediabetes | PP_cell | *FDFT1* | -0.322151141 | 1.37E-05 | 0.008519859 | 0.05389867 |
| Prediabetes | PP_cell | *SCGN* | -0.141184438 | 0.006298117 | 0.024961783 | 1 |
| Prediabetes | PP_cell | *SCTR* | -0.063648034 | 0.015914497 | 0.041381906 | 1 |
| Prediabetes | PP_cell | *ZNF506* | -0.28986405 | 0.014653138 | 0.039533432 | 1 |
| Prediabetes | PP_cell | *CADPS2* | -0.276327265 | 0.001790714 | 0.012963792 | 1 |
| Prediabetes | PP_cell | *ME2* | -0.396750669 | 0.000397697 | 0.008519859 | 1 |
| Prediabetes | PP_cell | *CCNT2* | -0.19736883 | 0.011344067 | 0.035126529 | 1 |
| Prediabetes | PP_cell | *PIBF1* | -0.386475421 | 0.001341174 | 0.011283666 | 1 |
| Prediabetes | PP_cell | *CHMP2B* | -0.236848796 | 0.00650413 | 0.02547829 | 1 |
| Prediabetes | PP_cell | *PPIE* | -0.238507941 | 0.001017928 | 0.009973668 | 1 |
| Prediabetes | PP_cell | *CD82* | -0.124755258 | 0.013187659 | 0.037909761 | 1 |
| Prediabetes | PP_cell | *SLC25A24* | -0.29932633 | 0.003916228 | 0.019030442 | 1 |
| Prediabetes | PP_cell | *CPNE3* | -0.285913008 | 0.000980242 | 0.009817609 | 1 |
| Prediabetes | PP_cell | *DDHD2* | -0.193868528 | 0.001271548 | 0.011102024 | 1 |
| Prediabetes | PP_cell | *TTC39A* | -0.194885217 | 0.002956015 | 0.016546556 | 1 |
| Prediabetes | PP_cell | *EPDR1* | -0.304301477 | 0.001231681 | 0.010954577 | 1 |
| Prediabetes | PP_cell | *ITPKC* | -0.385471051 | 0.015220091 | 0.040296319 | 1 |
| Prediabetes | PP_cell | *TMED2* | -0.213329086 | 0.007975892 | 0.028855691 | 1 |
| Prediabetes | PP_cell | *ACOX3* | -0.350538184 | 0.000274513 | 0.008519859 | 1 |
| Prediabetes | PP_cell | *TRIP6* | -0.087666525 | 0.008205125 | 0.029383885 | 1 |
| Prediabetes | PP_cell | *PGS1* | -0.312563009 | 0.018295563 | 0.045524551 | 1 |
| Prediabetes | PP_cell | *UIMC1* | -0.25979263 | 0.017032625 | 0.04320283 | 1 |
| Prediabetes | PP_cell | *METTL2A* | -0.226464234 | 0.001256623 | 0.011020698 | 1 |
| Prediabetes | PP_cell | *SLC15A1* | -0.130517283 | 0.015844621 | 0.041316373 | 1 |
| Prediabetes | PP_cell | *TGDS* | -0.167121861 | 0.00590921 | 0.024134393 | 1 |
| Prediabetes | PP_cell | *DOCK3* | -0.209066765 | 0.003687713 | 0.018410449 | 1 |
| Prediabetes | PP_cell | *ARHGAP28* | -0.167322302 | 0.013411814 | 0.03803406 | 1 |
| Prediabetes | PP_cell | *PPP1R13B* | -0.361066171 | 0.010912777 | 0.034411155 | 1 |
| Prediabetes | PP_cell | *ZNF343* | -0.281759904 | 0.002165435 | 0.014132881 | 1 |
| Prediabetes | PP_cell | *DZANK1* | -0.128672742 | 0.011735607 | 0.035704358 | 1 |
| Prediabetes | PP_cell | *GRAMD1A* | -0.357679412 | 0.003326051 | 0.017447335 | 1 |
| Prediabetes | PP_cell | *CDIP1* | -0.242474182 | 0.01090254 | 0.03440649 | 1 |
| Prediabetes | PP_cell | *DDX24* | -0.194064007 | 0.011776749 | 0.035704358 | 1 |
| Prediabetes | PP_cell | *SPTLC1* | -0.177523666 | 0.011351509 | 0.035126529 | 1 |
| Prediabetes | PP_cell | *LYZ* | -0.068657907 | 0.014264203 | 0.039085352 | 1 |
| Prediabetes | PP_cell | *NRCAM* | -0.158484166 | 0.001514742 | 0.011846392 | 1 |
| Prediabetes | PP_cell | *PITPNM3* | -0.18093891 | 0.013606499 | 0.038249975 | 1 |
| Prediabetes | PP_cell | *SCFD1* | -0.33244441 | 0.000544593 | 0.008519859 | 1 |
| Prediabetes | PP_cell | *EZR* | -0.295557335 | 0.000674484 | 0.008780065 | 1 |
| Prediabetes | PP_cell | *NUP50* | -0.336172076 | 0.002969031 | 0.016546556 | 1 |
| Prediabetes | PP_cell | *COMT* | -0.321052577 | 0.000412019 | 0.008519859 | 1 |
| Prediabetes | PP_cell | *SUCO* | -0.280149653 | 0.000536111 | 0.008519859 | 1 |

| | | | | | | |
|---|---|---|---|---|---|---|
| Prediabetes | PP_cell | *TMEM38B* | -0.233764614 | 0.006308743 | 0.024961783 | 1 |
| Prediabetes | PP_cell | *CRAT* | -0.366032805 | 0.00507661 | 0.02194376 | 1 |
| Prediabetes | PP_cell | *CWF19L1* | -0.123808478 | 0.003248605 | 0.017225057 | 1 |
| Prediabetes | PP_cell | *SEMA4G* | -0.197843779 | 0.008627452 | 0.030190258 | 1 |
| Prediabetes | PP_cell | *TLL2* | -0.264595681 | 0.013342421 | 0.037957281 | 1 |
| Prediabetes | PP_cell | *SRPK1* | -0.253864562 | 0.001001166 | 0.009851066 | 1 |
| Prediabetes | PP_cell | *ITPR3* | -0.103932585 | 0.006065396 | 0.024391958 | 1 |
| Prediabetes | PP_cell | *IFT74* | -0.248011756 | 0.005009553 | 0.021772712 | 1 |
| Prediabetes | PP_cell | *SYDE2* | -0.064145737 | 0.010932982 | 0.034447222 | 1 |
| Prediabetes | PP_cell | *PCSK5* | -0.094761427 | 0.018171393 | 0.045272926 | 1 |
| Prediabetes | PP_cell | *ABLIM1* | -0.267012467 | 0.000282458 | 0.008519859 | 1 |
| Prediabetes | PP_cell | *MZF1* | -0.165905183 | 0.007109768 | 0.026721477 | 1 |
| Prediabetes | PP_cell | *HSD3B7* | -0.121775134 | 0.004679686 | 0.020822749 | 1 |
| Prediabetes | PP_cell | *TIMM13* | -0.233751647 | 0.011259602 | 0.034971522 | 1 |
| Prediabetes | PP_cell | *CEP170B* | -0.114728497 | 0.00447544 | 0.020304855 | 1 |
| Prediabetes | PP_cell | *RASSF7* | -0.359168877 | 0.002739909 | 0.015948298 | 1 |
| Prediabetes | PP_cell | *ARVCF* | -0.44323773 | 0.000107581 | 0.008519859 | 0.422685569 |
| Prediabetes | PP_cell | *MED15* | -0.339688636 | 0.011791438 | 0.035704358 | 1 |
| Prediabetes | PP_cell | *SPECC1L* | -0.334261875 | 0.002441793 | 0.015060919 | 1 |
| Prediabetes | PP_cell | *SLC25A1* | -0.306548181 | 0.007202874 | 0.02687568 | 1 |
| Prediabetes | PP_cell | *GCAT* | -0.092866132 | 0.009799171 | 0.032572709 | 1 |
| Prediabetes | PP_cell | *HSCB* | -0.200083773 | 0.003014218 | 0.016609903 | 1 |
| Prediabetes | PP_cell | *CBY1* | -0.14443253 | 0.003176641 | 0.017097291 | 1 |
| Prediabetes | PP_cell | *XBP1* | -0.152652828 | 0.000400662 | 0.008519859 | 1 |
| Prediabetes | PP_cell | *MIOX* | -0.159518422 | 0.010579896 | 0.033933396 | 1 |
| Prediabetes | PP_cell | *LMF2* | -0.138991987 | 0.009097421 | 0.031136291 | 1 |
| Prediabetes | PP_cell | *TOM1* | -0.36855483 | 0.003377895 | 0.017625163 | 1 |
| Prediabetes | PP_cell | *CHKB* | -0.228556845 | 0.002948763 | 0.016546556 | 1 |
| Prediabetes | PP_cell | *BIK* | -0.086221471 | 0.014558649 | 0.03942173 | 1 |
| Prediabetes | PP_cell | *ARSA* | -0.095245384 | 0.014524667 | 0.039384002 | 1 |
| Prediabetes | PP_cell | *RBFOX2* | -0.311298539 | 0.001415966 | 0.011542178 | 1 |
| Prediabetes | PP_cell | *SYNGR1* | -0.106497989 | 0.002966058 | 0.016546556 | 1 |
| Prediabetes | PP_cell | *SGSM3* | -0.146331145 | 0.002252957 | 0.014369919 | 1 |
| Prediabetes | PP_cell | *ST13* | -0.242494127 | 0.002573629 | 0.015428508 | 1 |
| Prediabetes | PP_cell | *PMM1* | -0.204820024 | 0.001295264 | 0.011111559 | 1 |
| Prediabetes | PP_cell | *PRMT5* | -0.514178486 | 0.000524167 | 0.008519859 | 1 |
| Prediabetes | PP_cell | *VCPKMT* | -0.402524411 | 0.008980998 | 0.030871689 | 1 |
| Prediabetes | PP_cell | *TRIM9* | -0.309900005 | 0.000764815 | 0.009244451 | 1 |
| Prediabetes | PP_cell | *ATP6V1D* | -0.270710946 | 0.002502245 | 0.015266026 | 1 |
| Prediabetes | PP_cell | *GSTZ1* | -0.146848184 | 0.002944614 | 0.016546556 | 1 |
| Prediabetes | PP_cell | *TMED8* | -0.281398712 | 0.005683363 | 0.02351138 | 1 |
| Prediabetes | PP_cell | *DAAM1* | -0.340563602 | 0.000488959 | 0.008519859 | 1 |
| Prediabetes | PP_cell | *DHRS7* | -0.301288969 | 0.0008769 | 0.009444968 | 1 |
| Prediabetes | PP_cell | *MTHFD1* | -0.400081564 | 0.007342337 | 0.027240832 | 1 |
| Prediabetes | PP_cell | *ABHD12* | -0.238170784 | 0.009537989 | 0.032025881 | 1 |
| Prediabetes | PP_cell | *CD40* | -0.468945074 | 0.001661943 | 0.012440588 | 1 |
| Prediabetes | PP_cell | *SALL4* | -0.173240307 | 0.001915713 | 0.01336916 | 1 |
| Prediabetes | PP_cell | *NELFCD* | -0.282015766 | 0.000448823 | 0.008519859 | 1 |
| Prediabetes | PP_cell | *MTG2* | -0.362027201 | 0.004570974 | 0.020408359 | 1 |
| Prediabetes | PP_cell | *ARFRP1* | -0.233185053 | 0.002053898 | 0.013865574 | 1 |

| | | | | | | |
|---|---|---|---|---|---|---|
| Prediabetes | PP_cell | *POFUT1* | -0.383791922 | 0.007455143 | 0.027529377 | 1 |
| Prediabetes | PP_cell | *MROH8* | -0.102567708 | 0.011153749 | 0.034835517 | 1 |
| Prediabetes | PP_cell | *NOP56* | -0.230439719 | 0.012862018 | 0.03760809 | 1 |
| Prediabetes | PP_cell | *MANBAL* | -0.245674698 | 0.007189393 | 0.026854533 | 1 |
| Prediabetes | PP_cell | *IDH3B* | -0.277367105 | 0.01179104 | 0.035704358 | 1 |
| Prediabetes | PP_cell | *MAPRE1* | -0.23190292 | 0.010848986 | 0.034320181 | 1 |
| Prediabetes | PP_cell | *TTI1* | -0.434075326 | 0.000407387 | 0.008519859 | 1 |
| Prediabetes | PP_cell | *DNTTIP1* | -0.361922951 | 0.002137477 | 0.014043725 | 1 |
| Prediabetes | PP_cell | *APMAP* | -0.117152447 | 0.016430437 | 0.04220961 | 1 |
| Prediabetes | PP_cell | *MYOM1* | -0.116575722 | 0.010776116 | 0.034287057 | 1 |
| Prediabetes | PP_cell | *RIOK3* | -0.257412695 | 0.003233517 | 0.017219313 | 1 |
| Prediabetes | PP_cell | *FNDC3A* | -0.323588052 | 0.000277993 | 0.008519859 | 1 |
| Prediabetes | PP_cell | *KLF5* | -0.073695551 | 0.007545863 | 0.027760015 | 1 |
| Prediabetes | PP_cell | *DNAJC3* | -0.222329671 | 0.008757114 | 0.030367193 | 1 |
| Prediabetes | PP_cell | *UGGT2* | -0.199911203 | 0.000982536 | 0.009817609 | 1 |
| Prediabetes | PP_cell | *PARP4* | -0.25867269 | 0.008442889 | 0.029750772 | 1 |
| Prediabetes | PP_cell | *CLN5* | -0.266369906 | 0.006841064 | 0.026292348 | 1 |
| Prediabetes | PP_cell | *OLFM4* | -0.056047132 | 0.015463535 | 0.040748644 | 1 |
| Prediabetes | PP_cell | *DHODH* | -0.194259005 | 0.004359301 | 0.020119092 | 1 |
| Prediabetes | PP_cell | *USB1* | -0.41656957 | 0.001133438 | 0.010552788 | 1 |
| Prediabetes | PP_cell | *CYB5B* | -0.308015023 | 0.010703721 | 0.03417498 | 1 |
| Prediabetes | PP_cell | *NME3* | -0.342628326 | 0.000203613 | 0.008519859 | 0.799993617 |
| Prediabetes | PP_cell | *VAC14* | -0.296090572 | 0.001521564 | 0.011846392 | 1 |
| Prediabetes | PP_cell | *SMPD3* | -0.226082527 | 0.006636268 | 0.025799843 | 1 |
| Prediabetes | PP_cell | *HAGHL* | -0.10435504 | 0.006811317 | 0.026269771 | 1 |
| Prediabetes | PP_cell | *METRN* | -0.251347379 | 0.001651477 | 0.012440588 | 1 |
| Prediabetes | PP_cell | *RHBDL1* | -0.154255959 | 0.005508753 | 0.023090348 | 1 |
| Prediabetes | PP_cell | *PIEZO1* | -0.140417157 | 0.005730147 | 0.023599316 | 1 |
| Prediabetes | PP_cell | *AQP8* | -0.063669948 | 0.017388681 | 0.043823045 | 1 |
| Prediabetes | PP_cell | *HMOX2* | -0.36077933 | 0.000123217 | 0.008519859 | 0.48412085 |
| Prediabetes | PP_cell | *KNOP1* | -0.356964594 | 0.008512244 | 0.029941457 | 1 |
| Prediabetes | PP_cell | *TRIP4* | -0.355025479 | 0.002759598 | 0.015968275 | 1 |
| Prediabetes | PP_cell | *AP3B2* | -0.183828207 | 0.003212524 | 0.017212803 | 1 |
| Prediabetes | PP_cell | *RPAP1* | -0.185950461 | 0.003151182 | 0.017002881 | 1 |
| Prediabetes | PP_cell | *HOMER2* | -0.170719819 | 0.001752987 | 0.012855972 | 1 |
| Prediabetes | PP_cell | *DMXL2* | -0.308817223 | 0.000431449 | 0.008519859 | 1 |
| Prediabetes | PP_cell | *ZDHHC2* | -0.310680991 | 0.000344216 | 0.008519859 | 1 |
| Prediabetes | PP_cell | *TRIM35* | -0.173827189 | 0.003857941 | 0.018923659 | 1 |
| Prediabetes | PP_cell | *RIPK2* | -0.245968617 | 0.004089444 | 0.019452089 | 1 |
| Prediabetes | PP_cell | *LAPTM4B* | -0.300481618 | 0.001582029 | 0.012117789 | 1 |
| Prediabetes | PP_cell | *EIF3E* | -0.258385461 | 0.000692565 | 0.008780065 | 1 |
| Prediabetes | PP_cell | *ESRP1* | -0.373616014 | 0.000454995 | 0.008519859 | 1 |
| Prediabetes | PP_cell | *ARMC1* | -0.322638589 | 0.000483752 | 0.008519859 | 1 |
| Prediabetes | PP_cell | *SNX16* | -0.447724455 | 0.000379186 | 0.008519859 | 1 |
| Prediabetes | PP_cell | *SH2D4A* | -0.16039282 | 0.011407667 | 0.035181102 | 1 |
| Prediabetes | PP_cell | *INTS10* | -0.322627476 | 0.001765808 | 0.012915503 | 1 |
| Prediabetes | PP_cell | *LEPROTL1* | -0.321185981 | 0.002959937 | 0.016546556 | 1 |
| Prediabetes | PP_cell | *KLHDC4* | -0.280849655 | 0.000462674 | 0.008519859 | 1 |
| Prediabetes | PP_cell | *MCM4* | -0.51086888 | 0.001791636 | 0.012963792 | 1 |
| Prediabetes | PP_cell | *FGL1* | -0.065254341 | 0.013609943 | 0.038249975 | 1 |

| | | | | | | |
|---|---|---|---|---|---|---|
| Prediabetes | PP_cell | *ECH1* | -0.188681179 | 0.005334837 | 0.022733811 | 1 |
| Prediabetes | PP_cell | *HNRNPL* | -0.222626991 | 0.011070906 | 0.034770255 | 1 |
| Prediabetes | PP_cell | *SARS2* | -0.085921164 | 0.006848418 | 0.026292348 | 1 |
| Prediabetes | PP_cell | *LIN7B* | -0.391077269 | 5.08E-05 | 0.008519859 | 0.199679214 |
| Prediabetes | PP_cell | *PPP1R13L* | -0.082622123 | 0.009563991 | 0.032025881 | 1 |
| Prediabetes | PP_cell | *ERCC2* | -0.414515046 | 0.00793464 | 0.028742703 | 1 |
| Prediabetes | PP_cell | *KLC3* | -0.066491919 | 0.008857513 | 0.030634833 | 1 |
| Prediabetes | PP_cell | *AMH* | -0.575951526 | 0.00068866 | 0.008780065 | 1 |
| Prediabetes | PP_cell | *NUMBL* | -0.467065587 | 0.00024079 | 0.008519859 | 0.946063805 |
| Prediabetes | PP_cell | *MYH14* | -0.158284479 | 0.004818082 | 0.021151112 | 1 |
| Prediabetes | PP_cell | *CAPS* | -0.155128695 | 0.007033827 | 0.026624187 | 1 |
| Prediabetes | PP_cell | *PPP2R1A* | -0.233404856 | 0.009916788 | 0.032720527 | 1 |
| Prediabetes | PP_cell | *GCDH* | -0.17379527 | 0.002110496 | 0.013980215 | 1 |
| Prediabetes | PP_cell | *PIK3R2* | -0.207362199 | 0.001538719 | 0.011877462 | 1 |
| Prediabetes | PP_cell | *UPK1A* | -1.185796777 | 0.003064594 | 0.016741022 | 1 |
| Prediabetes | PP_cell | *COPE* | -0.220884486 | 0.005804137 | 0.023804231 | 1 |
| Prediabetes | PP_cell | *SIPA1L3* | -0.451097791 | 0.00039351 | 0.008519859 | 1 |
| Prediabetes | PP_cell | *ETHE1* | -0.253605618 | 0.002283056 | 0.014499386 | 1 |
| Prediabetes | PP_cell | *SMG9* | -0.217055774 | 0.019162768 | 0.047174509 | 1 |
| Prediabetes | PP_cell | *ITGB8* | -0.33584716 | 0.00013013 | 0.008519859 | 0.51127946 |
| Prediabetes | PP_cell | *DNAH11* | -0.37201856 | 0.001313156 | 0.011188791 | 1 |
| Prediabetes | PP_cell | *ANKRD7* | -0.127165284 | 0.007667833 | 0.027998993 | 1 |
| Prediabetes | PP_cell | *TSPAN12* | -0.478800411 | 0.000526694 | 0.008519859 | 1 |
| Prediabetes | PP_cell | *ABHD11* | -0.182627179 | 0.002211798 | 0.014319626 | 1 |
| Prediabetes | PP_cell | *CHCHD2* | -0.172733171 | 0.006794409 | 0.026255163 | 1 |
| Prediabetes | PP_cell | *AGFG2* | -0.312322628 | 0.002228365 | 0.014336949 | 1 |
| Prediabetes | PP_cell | *C1GALT1* | -0.266127371 | 0.013870059 | 0.038431214 | 1 |
| Prediabetes | PP_cell | *RPA3* | -0.364143587 | 0.000226449 | 0.008519859 | 0.889719406 |
| Prediabetes | PP_cell | *CLDN15* | -0.095388926 | 0.002611521 | 0.015570055 | 1 |
| Prediabetes | PP_cell | *TMEM106B* | -0.254841269 | 0.002229543 | 0.014336949 | 1 |
| Prediabetes | PP_cell | *TSPAN13* | -0.297059492 | 0.000295504 | 0.008519859 | 1 |
| Prediabetes | PP_cell | *AGR2* | -0.047701311 | 0.015756623 | 0.04125534 | 1 |
| Prediabetes | PP_cell | *COA1* | -0.369280137 | 6.84E-05 | 0.008519859 | 0.268893334 |
| Prediabetes | PP_cell | *BLVRA* | -0.401359023 | 0.000122988 | 0.008519859 | 0.483220597 |
| Prediabetes | PP_cell | *TMEM248* | -0.237427349 | 0.005438664 | 0.022930133 | 1 |
| Prediabetes | PP_cell | *RHEB* | -0.252455844 | 0.001463371 | 0.011646409 | 1 |
| Prediabetes | PP_cell | *LIMK1* | -0.251119037 | 0.002858251 | 0.016251909 | 1 |
| Prediabetes | PP_cell | *SLC1A1* | -0.074524886 | 0.010221436 | 0.033269422 | 1 |
| Prediabetes | PP_cell | *NMRK1* | -0.336524663 | 0.003325573 | 0.017447335 | 1 |
| Prediabetes | PP_cell | *SUSD1* | -0.344670386 | 0.004467023 | 0.020304368 | 1 |
| Prediabetes | PP_cell | *AK1* | -0.171097017 | 0.007068186 | 0.026651539 | 1 |
| Prediabetes | PP_cell | *PLGRKT* | -0.165309724 | 0.008135956 | 0.029272821 | 1 |
| Prediabetes | PP_cell | *NPDC1* | -0.228747749 | 0.00528597 | 0.022550027 | 1 |
| Prediabetes | PP_cell | *HPS1* | -0.326108717 | 0.000448262 | 0.008519859 | 1 |
| Prediabetes | PP_cell | *CXCL12* | -0.096651629 | 0.014151843 | 0.03888293 | 1 |
| Prediabetes | PP_cell | *PLEKHA1* | -0.3177807 | 0.00027081 | 0.008519859 | 1 |
| Prediabetes | PP_cell | *PALD1* | -0.33209836 | 0.003664374 | 0.01839612 | 1 |
| Prediabetes | PP_cell | *SPOCK2* | -0.114480672 | 0.011685278 | 0.035704358 | 1 |
| Prediabetes | PP_cell | *BMPR1A* | -0.353669726 | 0.000453175 | 0.008519859 | 1 |
| Prediabetes | PP_cell | *LIPA* | -0.447307775 | 0.000766557 | 0.009244451 | 1 |

| | | | | | | |
|---|---|---|---|---|---|---|
| Prediabetes | PP_cell | *NPM3* | -0.141022387 | 0.005078946 | 0.02194376 | 1 |
| Prediabetes | PP_cell | *NEURL1* | -0.137321209 | 0.002643768 | 0.01561651 | 1 |
| Prediabetes | PP_cell | *SORCS1* | -0.244162706 | 0.011903217 | 0.035755154 | 1 |
| Prediabetes | PP_cell | *XPNPEP1* | -0.328404977 | 0.011307839 | 0.0350936 | 1 |
| Prediabetes | PP_cell | *SMC3* | -0.208823223 | 0.017858991 | 0.044721463 | 1 |
| Prediabetes | PP_cell | *PBLD* | -0.155837529 | 0.003320447 | 0.017447335 | 1 |
| Prediabetes | PP_cell | *DHRS11* | -0.377673774 | 0.000319617 | 0.008519859 | 1 |
| Prediabetes | PP_cell | *TRIM37* | -0.405759511 | 0.000422199 | 0.008519859 | 1 |
| Prediabetes | PP_cell | *TRIM16L* | -0.099267943 | 0.017483984 | 0.044034984 | 1 |
| Prediabetes | PP_cell | *CDK5RAP3* | -0.1521252 | 0.014745575 | 0.039627472 | 1 |
| Prediabetes | PP_cell | *MED13* | -0.221379956 | 0.004557361 | 0.020393929 | 1 |
| Prediabetes | PP_cell | *RAI1* | -0.442751762 | 0.00391846 | 0.019030442 | 1 |
| Prediabetes | PP_cell | *DRG2* | -0.440335564 | 0.000427172 | 0.008519859 | 1 |
| Prediabetes | PP_cell | *SMARCD2* | -0.435987055 | 0.000507614 | 0.008519859 | 1 |
| Prediabetes | PP_cell | *SYNGR2* | -0.249735942 | 0.003224389 | 0.017212803 | 1 |
| Prediabetes | PP_cell | *C17orf75* | -0.126916483 | 0.002238589 | 0.014348148 | 1 |
| Prediabetes | PP_cell | *NAGLU* | -0.27979974 | 5.40E-06 | 0.008519859 | 0.021207864 |
| Prediabetes | PP_cell | *CNTNAP1* | 0.089855303 | 0.006255484 | 0.024851158 | 1 |
| Prediabetes | PP_cell | *RND2* | -0.749490895 | 0.00092272 | 0.009590919 | 1 |
| Prediabetes | PP_cell | *ABCC3* | -0.066716891 | 0.003023629 | 0.016638427 | 1 |
| Prediabetes | PP_cell | *MPP2* | -0.296082608 | 0.000487017 | 0.008519859 | 1 |
| Prediabetes | PP_cell | *EFNB3* | -0.392524314 | 0.000260677 | 0.008519859 | 1 |
| Prediabetes | PP_cell | *RANGRF* | -0.397813118 | 0.000303274 | 0.008519859 | 1 |
| Prediabetes | PP_cell | *AADAT* | -0.350372903 | 0.005411891 | 0.022930133 | 1 |
| Prediabetes | PP_cell | *TRIM2* | -0.333897613 | 0.003716545 | 0.01850736 | 1 |
| Prediabetes | PP_cell | *SH3D19* | -0.503987442 | 0.000551443 | 0.008519859 | 1 |
| Prediabetes | PP_cell | *GLRB* | -0.43525092 | 0.001656584 | 0.012440588 | 1 |
| Prediabetes | PP_cell | *FAM149A* | -0.241984065 | 0.008764672 | 0.030367193 | 1 |
| Prediabetes | PP_cell | *HTATIP2* | -0.160506945 | 0.007061896 | 0.026651539 | 1 |
| Prediabetes | PP_cell | *B3GAT1* | -0.156913475 | 0.016062956 | 0.041521635 | 1 |
| Prediabetes | PP_cell | *P2RX3* | -1.2737616 | 0.013163535 | 0.037909761 | 1 |
| Prediabetes | PP_cell | *VWA5A* | -0.227470519 | 0.018794243 | 0.046626816 | 1 |
| Prediabetes | PP_cell | *PRPF19* | -0.316919067 | 0.001208127 | 0.010887001 | 1 |
| Prediabetes | PP_cell | *CEP164* | -0.247336284 | 0.004692494 | 0.020832552 | 1 |
| Prediabetes | PP_cell | *MDK* | -0.311496476 | 0.001213937 | 0.010904418 | 1 |
| Prediabetes | PP_cell | *MADD* | -0.260473001 | 0.000872098 | 0.009444968 | 1 |
| Prediabetes | PP_cell | *GTF2H1* | -0.248045241 | 0.00054315 | 0.008519859 | 1 |
| Prediabetes | PP_cell | *CORO1C* | -0.436583708 | 0.000562584 | 0.008521817 | 1 |
| Prediabetes | PP_cell | *KRT18* | -0.058084582 | 0.004384554 | 0.02017203 | 1 |
| Prediabetes | PP_cell | *ACSS3* | -0.154540449 | 0.01331179 | 0.0379275 | 1 |
| Prediabetes | PP_cell | *WNT5B* | -0.144517721 | 0.013299896 | 0.0379275 | 1 |
| Prediabetes | PP_cell | *ITFG2* | -0.219596888 | 0.001732089 | 0.012837572 | 1 |
| Prediabetes | PP_cell | *GSG1* | -0.453136907 | 0.020606534 | 0.049760752 | 1 |
| Prediabetes | PP_cell | *RASAL1* | -0.450887432 | 0.009953037 | 0.032751739 | 1 |
| Prediabetes | PP_cell | *MDM1* | -0.186168201 | 0.002575999 | 0.015428508 | 1 |
| Prediabetes | PP_cell | *NUP107* | -0.380535396 | 0.002806354 | 0.01609659 | 1 |
| Prediabetes | PP_cell | *GNB3* | -0.21983803 | 0.00572842 | 0.023599316 | 1 |
| Prediabetes | PP_cell | *CDCA3* | -0.169194397 | 0.002065873 | 0.013874899 | 1 |
| Prediabetes | PP_cell | *SPSB2* | -0.292903565 | 0.011890804 | 0.035745194 | 1 |
| Prediabetes | PP_cell | *NT5DC3* | -0.118235082 | 0.015248858 | 0.040345295 | 1 |

| | | | | | | |
|---|---|---|---|---|---|---|
| Prediabetes | PP_cell | *GOLT1B* | -0.325642111 | 0.003413954 | 0.017742628 | 1 |
| Prediabetes | PP_cell | *C2CD5* | -0.385196147 | 0.000330946 | 0.008519859 | 1 |
| Prediabetes | PP_cell | *FRK* | -0.464978896 | 0.00365639 | 0.01839612 | 1 |
| Prediabetes | PP_cell | *RSPH4A* | -0.216014335 | 0.003563403 | 0.018246003 | 1 |
| Prediabetes | PP_cell | *NEDD9* | -0.307647661 | 0.000151421 | 0.008519859 | 0.594932644 |
| Prediabetes | PP_cell | *CEP85L* | -0.286918981 | 0.001346929 | 0.01130353 | 1 |
| Prediabetes | PP_cell | *TPD52L1* | -0.155614269 | 0.000632307 | 0.008645299 | 1 |
| Prediabetes | PP_cell | *RNF8* | -0.376731091 | 0.001470252 | 0.011646409 | 1 |
| Prediabetes | PP_cell | *GLP1R* | -0.061191289 | 0.008413229 | 0.029699528 | 1 |
| Prediabetes | PP_cell | *BACH2* | -0.270976555 | 0.000633488 | 0.008645299 | 1 |
| Prediabetes | PP_cell | *WASF1* | -0.356474975 | 0.013796302 | 0.038362117 | 1 |
| Prediabetes | PP_cell | *ALDH5A1* | -0.3559123 | 0.002932687 | 0.016546556 | 1 |
| Prediabetes | PP_cell | *C6orf62* | -0.306848467 | 0.002084495 | 0.013904889 | 1 |
| Prediabetes | PP_cell | *HBS1L* | -0.30591142 | 0.006979969 | 0.026522533 | 1 |
| Prediabetes | PP_cell | *PEX7* | -0.38988709 | 0.000213475 | 0.008519859 | 0.838741516 |
| Prediabetes | PP_cell | *PHACTR2* | -0.241436825 | 0.01054762 | 0.033892939 | 1 |
| Prediabetes | PP_cell | *EPM2A* | -0.227163789 | 0.00601026 | 0.024326947 | 1 |
| Prediabetes | PP_cell | *DNPH1* | -0.15973435 | 0.006796004 | 0.026255163 | 1 |
| Prediabetes | PP_cell | *FBRSL1* | -0.295627519 | 0.000342918 | 0.008519859 | 1 |
| Prediabetes | PP_cell | *NUDT12* | -0.128428856 | 0.004283333 | 0.019996719 | 1 |
| Prediabetes | PP_cell | *SEMA5A* | -0.194259471 | 0.000974321 | 0.009804971 | 1 |
| Prediabetes | PP_cell | *DAP* | -0.187761349 | 0.003341229 | 0.017480279 | 1 |
| Prediabetes | PP_cell | *BRD8* | -0.349897233 | 0.015683741 | 0.041136895 | 1 |
| Prediabetes | PP_cell | *NNT* | -0.22665756 | 0.00619662 | 0.024667192 | 1 |
| Prediabetes | PP_cell | *TMCO6* | -0.122091573 | 0.012284508 | 0.036372141 | 1 |
| Prediabetes | PP_cell | *PDE8B* | -0.103220049 | 0.005445979 | 0.022930133 | 1 |
| Prediabetes | PP_cell | *CCNG1* | -0.227717063 | 0.002129579 | 0.014043725 | 1 |
| Prediabetes | PP_cell | *GOLPH3* | -0.336913283 | 0.001383221 | 0.011450342 | 1 |
| Prediabetes | PP_cell | *RAD50* | -0.353195508 | 0.000392589 | 0.008519859 | 1 |
| Prediabetes | PP_cell | *PPWD1* | -0.339609205 | 0.010716105 | 0.03417498 | 1 |
| Prediabetes | PP_cell | *WWC1* | -0.086937355 | 0.010307769 | 0.033442795 | 1 |
| Prediabetes | PP_cell | *STC2* | -0.142849469 | 0.003707229 | 0.018484393 | 1 |
| Prediabetes | PP_cell | *EHHADH* | -0.203933258 | 0.011254861 | 0.034971522 | 1 |
| Prediabetes | PP_cell | *ACTR8* | -0.480678292 | 0.000327662 | 0.008519859 | 1 |
| Prediabetes | PP_cell | *CLDN16* | -0.887894319 | 0.01577841 | 0.04125534 | 1 |
| Prediabetes | PP_cell | *FAM162A* | -0.194763134 | 0.01083973 | 0.034318532 | 1 |
| Prediabetes | PP_cell | *RBP1* | -0.169731152 | 0.000606946 | 0.008645299 | 1 |
| Prediabetes | PP_cell | *RNF7* | -0.163863191 | 0.016582227 | 0.042361229 | 1 |
| Prediabetes | PP_cell | *SERPINI2* | -0.087936952 | 0.00618356 | 0.024659802 | 1 |
| Prediabetes | PP_cell | *WNT5A* | -0.374705933 | 0.001387041 | 0.011450342 | 1 |
| Prediabetes | PP_cell | *TUSC2* | -0.233457578 | 0.002182733 | 0.014198607 | 1 |
| Prediabetes | PP_cell | *C3orf14* | -0.151535275 | 0.01113663 | 0.034835517 | 1 |
| Prediabetes | PP_cell | *NCBP2* | -0.271502852 | 0.011185062 | 0.034877863 | 1 |
| Prediabetes | PP_cell | *C3orf52* | -0.057103388 | 0.012239315 | 0.036372141 | 1 |
| Prediabetes | PP_cell | *SLC41A3* | -0.2226307 | 0.002956249 | 0.016546556 | 1 |
| Prediabetes | PP_cell | *ATP6V1A* | -0.329781466 | 0.000868156 | 0.009444968 | 1 |
| Prediabetes | PP_cell | *GORASP1* | -0.206807264 | 0.016250549 | 0.041846259 | 1 |
| Prediabetes | PP_cell | *PLCH1* | -0.248492516 | 0.002791174 | 0.0160564 | 1 |
| Prediabetes | PP_cell | *NEK4* | -0.428852987 | 0.002588015 | 0.015476883 | 1 |
| Prediabetes | PP_cell | *KANSL3* | -0.202481095 | 0.005156078 | 0.022091856 | 1 |

| | | | | | | |
|---|---|---|---|---|---|---|
| Prediabetes | PP_cell | *ACTR1B* | -0.229496628 | 0.000778307 | 0.009253454 | 1 |
| Prediabetes | PP_cell | *TFCP2L1* | -0.069212572 | 0.012973952 | 0.037843101 | 1 |
| Prediabetes | PP_cell | *TANC1* | -0.155024948 | 0.020643936 | 0.049760752 | 1 |
| Prediabetes | PP_cell | *REEP6* | -0.102686463 | 0.010894327 | 0.03440649 | 1 |
| Prediabetes | PP_cell | *INO80B* | -0.0766001 | 0.009871076 | 0.032714383 | 1 |
| Prediabetes | PP_cell | *MOGS* | -0.220841454 | 0.014736229 | 0.039627472 | 1 |
| Prediabetes | PP_cell | *GALNT3* | -0.062576002 | 0.009953061 | 0.032751739 | 1 |
| Prediabetes | PP_cell | *MRPL19* | -0.202335696 | 0.017646696 | 0.044302791 | 1 |
| Prediabetes | PP_cell | *OTX1* | -0.559118173 | 5.86E-05 | 0.008519859 | 0.230154106 |
| Prediabetes | PP_cell | *IL1R2* | -0.093361489 | 0.010474255 | 0.033815404 | 1 |
| Prediabetes | PP_cell | *IL1RL2* | -0.195282627 | 0.004769279 | 0.021054489 | 1 |
| Prediabetes | PP_cell | *CNPPD1* | -0.273560158 | 0.000842227 | 0.009444968 | 1 |
| Prediabetes | PP_cell | *HDLBP* | -0.257194415 | 0.000609843 | 0.008645299 | 1 |
| Prediabetes | PP_cell | *TAF1B* | -0.445116684 | 0.000542934 | 0.008519859 | 1 |
| Prediabetes | PP_cell | *COX7A2L* | -0.258760452 | 0.000655318 | 0.008752256 | 1 |
| Prediabetes | PP_cell | *PNO1* | -0.276825389 | 0.016436931 | 0.04220961 | 1 |
| Prediabetes | PP_cell | *ARID3A* | -0.305872463 | 0.000482599 | 0.008519859 | 1 |
| Prediabetes | PP_cell | *ALMS1* | -0.31840867 | 0.000153071 | 0.008519859 | 0.601414632 |
| Prediabetes | PP_cell | *MARK1* | -0.413206314 | 0.000707302 | 0.008816433 | 1 |
| Prediabetes | PP_cell | *MORN1* | -0.351578841 | 0.010144394 | 0.033119439 | 1 |
| Prediabetes | PP_cell | *RALGPS2* | -0.269460845 | 0.000660846 | 0.008757086 | 1 |
| Prediabetes | PP_cell | *PARK7* | -0.183161558 | 0.012110092 | 0.036238044 | 1 |
| Prediabetes | PP_cell | *MECR* | -0.188994706 | 0.004470165 | 0.020304368 | 1 |
| Prediabetes | PP_cell | *FBXO6* | -0.12921114 | 0.005146543 | 0.022075073 | 1 |
| Prediabetes | PP_cell | *SMG7* | -0.312212383 | 0.009408117 | 0.031819051 | 1 |
| Prediabetes | PP_cell | *TMEM9* | -0.131622849 | 0.001592201 | 0.012147101 | 1 |
| Prediabetes | PP_cell | *WARS2* | -0.288996003 | 0.00933574 | 0.031675407 | 1 |
| Prediabetes | PP_cell | *HPCAL4* | -0.187535738 | 0.008783918 | 0.03040706 | 1 |
| Prediabetes | PP_cell | *BMP8B* | -0.143773441 | 0.006116917 | 0.024498846 | 1 |
| Prediabetes | PP_cell | *UAP1* | -0.32923226 | 2.24E-05 | 0.008519859 | 0.087859594 |
| Prediabetes | PP_cell | *CTBS* | -0.273684998 | 0.000898395 | 0.009499809 | 1 |
| Prediabetes | PP_cell | *CD46* | -0.276929818 | 0.000865497 | 0.009444968 | 1 |
| Prediabetes | PP_cell | *TMED5* | -0.328761004 | 0.001390128 | 0.011450342 | 1 |
| Prediabetes | PP_cell | *DR1* | -0.234394815 | 0.020720655 | 0.049888972 | 1 |
| Prediabetes | PP_cell | *PTBP2* | -0.233003318 | 0.00129272 | 0.011111559 | 1 |
| Prediabetes | PP_cell | *NSL1* | -0.339369538 | 0.000436914 | 0.008519859 | 1 |
| Prediabetes | PP_cell | *RPA2* | -0.170693176 | 0.016476398 | 0.042228159 | 1 |
| Prediabetes | PP_cell | *DDX59* | -0.385354543 | 0.00109452 | 0.010322435 | 1 |
| Prediabetes | PP_cell | *TTR* | -0.152619639 | 0.000432983 | 0.008519859 | 1 |
| Prediabetes | PP_cell | *SPCS2* | -0.187706286 | 0.018041629 | 0.045006706 | 1 |
| Prediabetes | PP_cell | *ELOVL4* | -0.562676716 | 0.003930044 | 0.019039633 | 1 |
| Prediabetes | PP_cell | *FILIP1* | -0.597271781 | 1.59E-05 | 0.008519859 | 0.0624835 |
| Prediabetes | PP_cell | *RNF146* | -0.468450371 | 0.000471809 | 0.008519859 | 1 |
| Prediabetes | PP_cell | *VAMP8* | -0.145685492 | 0.002132742 | 0.014043725 | 1 |
| Prediabetes | PP_cell | *SPP1* | -0.058060458 | 0.006390517 | 0.025183893 | 1 |
| Prediabetes | PP_cell | *DNAH7* | -0.103234328 | 0.00603178 | 0.024356489 | 1 |
| Prediabetes | PP_cell | *PIGZ* | -0.055753688 | 0.0112369 | 0.03495628 | 1 |
| Prediabetes | PP_cell | *NEK6* | -0.193363932 | 0.010548621 | 0.033892939 | 1 |
| Prediabetes | PP_cell | *RBM18* | -0.385584814 | 0.000321473 | 0.008519859 | 1 |
| Prediabetes | PP_cell | *DCAF4* | -0.15179742 | 0.001120235 | 0.01050454 | 1 |

| | | | | | | |
|---|---|---|---|---|---|---|
| Prediabetes | PP_cell | *IFI27L2* | -0.257136221 | 0.000845896 | 0.009444968 | 1 |
| Prediabetes | PP_cell | *KLHL29* | -0.083800621 | 0.016779871 | 0.042754938 | 1 |
| Prediabetes | PP_cell | *BCL11A* | -0.087818692 | 0.013759062 | 0.038295695 | 1 |
| Prediabetes | PP_cell | *CRIPT* | -0.269816249 | 0.017508437 | 0.044068322 | 1 |
| Prediabetes | PP_cell | *EPCAM* | -0.168583341 | 0.000434379 | 0.008519859 | 1 |
| Prediabetes | PP_cell | *PRLHR* | -0.089567443 | 0.013065597 | 0.037909761 | 1 |
| Prediabetes | PP_cell | *HOXB3* | -0.262003406 | 0.013232145 | 0.037909761 | 1 |
| Prediabetes | PP_cell | *RCL1* | -0.4641047 | 0.000795176 | 0.009253454 | 1 |
| Prediabetes | PP_cell | *CD274* | -0.088400578 | 0.012361645 | 0.036545451 | 1 |
| Prediabetes | PP_cell | *PLEKHG1* | -0.138342582 | 0.002295479 | 0.014546671 | 1 |
| Prediabetes | PP_cell | *PCDHB10* | -0.119717573 | 0.010292223 | 0.033419952 | 1 |
| Prediabetes | PP_cell | *SLC10A7* | -0.14765659 | 0.001733549 | 0.012837572 | 1 |
| Prediabetes | PP_cell | *MRC1* | -0.074072004 | 0.008711748 | 0.030237155 | 1 |
| Prediabetes | PP_cell | *IQSEC3* | -0.107917533 | 0.005444708 | 0.022930133 | 1 |
| Prediabetes | PP_cell | *CCDC77* | -0.111698114 | 0.017106978 | 0.043335473 | 1 |
| Prediabetes | PP_cell | *MTRF1* | -0.154576282 | 0.003820991 | 0.018812875 | 1 |
| Prediabetes | PP_cell | *KBTBD7* | -0.454834477 | 0.003404494 | 0.017716895 | 1 |
| Prediabetes | PP_cell | *SIL1* | -0.182079965 | 0.00148573 | 0.01172175 | 1 |
| Prediabetes | PP_cell | *PLS1* | -0.281972785 | 0.001671687 | 0.012486802 | 1 |
| Prediabetes | PP_cell | *UTP20* | -0.384069236 | 0.000543573 | 0.008519859 | 1 |
| Prediabetes | PP_cell | *APAF1* | -0.112497449 | 0.011597521 | 0.035654664 | 1 |
| Prediabetes | PP_cell | *PPP3CC* | -0.34199254 | 0.00645507 | 0.025367051 | 1 |
| Prediabetes | PP_cell | *EPHX2* | -0.205305355 | 0.001779892 | 0.012957043 | 1 |
| Prediabetes | PP_cell | *CRISPLD1* | -0.343408441 | 0.020602334 | 0.049760752 | 1 |
| Prediabetes | PP_cell | *TRIM6* | -0.172744032 | 0.003282689 | 0.017358932 | 1 |
| Prediabetes | PP_cell | *IAPP* | -0.05831874 | 0.003045342 | 0.016712144 | 1 |
| Prediabetes | PP_cell | *ZNF211* | -0.17696113 | 0.001023076 | 0.00999917 | 1 |
| Prediabetes | PP_cell | *CAT* | -0.37881541 | 0.001218386 | 0.010904418 | 1 |
| Prediabetes | PP_cell | *CSMD2* | -0.349004462 | 0.009153502 | 0.031273138 | 1 |
| Prediabetes | PP_cell | *UBL3* | -0.211810842 | 0.014359859 | 0.039207704 | 1 |
| Prediabetes | PP_cell | *NBPF14* | -0.247829932 | 0.010615485 | 0.033992045 | 1 |
| Prediabetes | PP_cell | *PMS2* | -0.265150945 | 0.002429438 | 0.015031909 | 1 |
| Prediabetes | PP_cell | *EEPD1* | -0.335480292 | 0.001250266 | 0.010991254 | 1 |
| Prediabetes | PP_cell | *NPY* | -0.066599985 | 0.009026382 | 0.030973499 | 1 |
| Prediabetes | PP_cell | *INHBA* | -0.068214581 | 0.006358509 | 0.025082914 | 1 |
| Prediabetes | PP_cell | *BHLHE41* | -0.357552003 | 0.000391784 | 0.008519859 | 1 |
| Prediabetes | PP_cell | *EBPL* | -0.242097199 | 0.008143343 | 0.029272821 | 1 |
| Prediabetes | PP_cell | *IKZF4* | -0.292121916 | 0.003247201 | 0.017225057 | 1 |
| Prediabetes | PP_cell | *SARDH* | -0.118259188 | 0.019137249 | 0.047170798 | 1 |
| Prediabetes | PP_cell | *USP45* | -0.481806499 | 0.000364619 | 0.008519859 | 1 |
| Prediabetes | PP_cell | *C4BPA* | -0.136442783 | 0.00594395 | 0.024153447 | 1 |
| Prediabetes | PP_cell | *ARHGAP40* | -3.331089861 | 0.014686139 | 0.039551271 | 1 |
| Prediabetes | PP_cell | *MATN4* | -0.149820255 | 0.018805821 | 0.046626816 | 1 |
| Prediabetes | PP_cell | *PARD6B* | -0.0448849 | 0.013630612 | 0.038253338 | 1 |
| Prediabetes | PP_cell | *ARFGEF2* | -0.33445742 | 0.004202649 | 0.019870287 | 1 |
| Prediabetes | PP_cell | *BCAS4* | -0.389115343 | 0.000953619 | 0.009706653 | 1 |
| Prediabetes | PP_cell | *PEPD* | -0.263146712 | 0.012157923 | 0.036270675 | 1 |
| Prediabetes | PP_cell | *SNRNP27* | -0.239088945 | 0.013543274 | 0.038233958 | 1 |
| Prediabetes | PP_cell | *USP22* | -0.209586685 | 0.014432815 | 0.039243274 | 1 |
| Prediabetes | PP_cell | *BTN2A2* | -0.142513863 | 0.015430817 | 0.040717045 | 1 |

| | | | | | | |
|---|---|---|---|---|---|---|
| Prediabetes | PP_cell | *MRS2* | -0.334430786 | 0.000404512 | 0.008519859 | 1 |
| Prediabetes | PP_cell | *ABCC10* | -0.380086894 | 0.000882177 | 0.009444968 | 1 |
| Prediabetes | PP_cell | *MOCS1* | -0.106549901 | 0.014972805 | 0.0399258 | 1 |
| Prediabetes | PP_cell | *KCNK17* | -0.053352014 | 0.015206813 | 0.040288313 | 1 |
| Prediabetes | PP_cell | *ATXN1* | -0.369203794 | 0.000383366 | 0.008519859 | 1 |
| Prediabetes | PP_cell | *LRRFIP1* | -0.262683756 | 0.000219944 | 0.008519859 | 0.864158377 |
| Prediabetes | PP_cell | *MT1G* | -0.085210155 | 0.007184007 | 0.026854533 | 1 |
| Prediabetes | PP_cell | *CLYBL* | -0.271210977 | 0.002344571 | 0.014691894 | 1 |
| Prediabetes | PP_cell | *TM9SF2* | -0.23046581 | 0.012202292 | 0.036320307 | 1 |
| Prediabetes | PP_cell | *GRK4* | -0.198510596 | 0.00251123 | 0.015273409 | 1 |
| Prediabetes | PP_cell | *HS3ST3B1* | -0.20885188 | 0.009819355 | 0.032612212 | 1 |
| Prediabetes | PP_cell | *PPDPF* | -0.210856808 | 0.004775886 | 0.021059995 | 1 |
| Prediabetes | PP_cell | *CHCHD5* | -0.267202481 | 0.002690497 | 0.0157471 | 1 |
| Prediabetes | PP_cell | *INSIG2* | -0.488152457 | 7.08E-06 | 0.008519859 | 0.027830768 |
| Prediabetes | PP_cell | *PSD4* | -0.226361874 | 0.009767791 | 0.032495895 | 1 |
| Prediabetes | PP_cell | *CLPP* | -0.225811564 | 0.016252836 | 0.041846259 | 1 |
| Prediabetes | PP_cell | *SH2D3A* | -0.110237056 | 0.000148586 | 0.008519859 | 0.583792537 |
| Prediabetes | PP_cell | *GPR108* | -0.249560356 | 0.020638304 | 0.049760752 | 1 |
| Prediabetes | PP_cell | *SYMPK* | -0.235175896 | 0.002554506 | 0.01537007 | 1 |
| Prediabetes | PP_cell | *NAPB* | -0.305051973 | 0.001951316 | 0.013500582 | 1 |
| Prediabetes | PP_cell | *PSMF1* | -0.187053995 | 0.018222588 | 0.045371704 | 1 |
| Prediabetes | PP_cell | *RBCK1* | -0.207869112 | 0.001185696 | 0.010758887 | 1 |
| Prediabetes | PP_cell | *MGME1* | -0.158019594 | 0.005417527 | 0.022930133 | 1 |
| Prediabetes | PP_cell | *ITPA* | -0.292531724 | 0.010808186 | 0.034287057 | 1 |
| Prediabetes | PP_cell | *BANF2* | -0.899269185 | 0.013232421 | 0.037909761 | 1 |
| Prediabetes | PP_cell | *MCF2L* | -0.321930099 | 0.00017152 | 0.008519859 | 0.673900154 |
| Prediabetes | PP_cell | *PROZ* | -0.989144219 | 0.008901993 | 0.030690414 | 1 |
| Prediabetes | PP_cell | *ASL* | -0.386659509 | 0.000480029 | 0.008519859 | 1 |
| Prediabetes | PP_cell | *TRAP1* | -0.136715399 | 0.005121164 | 0.021990223 | 1 |
| Prediabetes | PP_cell | *ZNF384* | -0.52518175 | 0.000836953 | 0.009444968 | 1 |
| Prediabetes | PP_cell | *RHOJ* | -0.113635562 | 0.011146018 | 0.034835517 | 1 |
| Prediabetes | PP_cell | *HSPA2* | -0.441736366 | 0.002306066 | 0.014586494 | 1 |
| Prediabetes | PP_cell | *PLEKHG3* | -0.423242231 | 0.000529916 | 0.008519859 | 1 |
| Prediabetes | PP_cell | *FGD3* | -0.388706495 | 0.01400367 | 0.038637936 | 1 |
| Prediabetes | PP_cell | *HELB* | -0.358462077 | 0.003896857 | 0.019001519 | 1 |
| Prediabetes | PP_cell | *YEATS4* | -0.278941842 | 0.002552976 | 0.01537007 | 1 |
| Prediabetes | PP_cell | *IDUA* | -0.619283644 | 0.000225196 | 0.008519859 | 0.884794244 |
| Prediabetes | PP_cell | *EMC1* | -0.300684986 | 0.020392828 | 0.049458901 | 1 |
| Prediabetes | PP_cell | *SYNGR3* | -0.134279625 | 0.009966074 | 0.032763505 | 1 |
| Prediabetes | PP_cell | *KDM4B* | -0.386097008 | 0.001145207 | 0.010597251 | 1 |
| Prediabetes | PP_cell | *METTL25* | -0.436352252 | 0.000868525 | 0.009444968 | 1 |
| Prediabetes | PP_cell | *POR* | -0.368745887 | 0.000513368 | 0.008519859 | 1 |
| Prediabetes | PP_cell | *RBM48* | -0.430697239 | 0.000273154 | 0.008519859 | 1 |
| Prediabetes | PP_cell | *SRD5A3* | -0.174627433 | 0.01003712 | 0.032886224 | 1 |
| Prediabetes | PP_cell | *GALR3* | -0.620859684 | 0.005548935 | 0.023167203 | 1 |
| Prediabetes | PP_cell | *NAA38* | -0.247874094 | 0.004981721 | 0.021699757 | 1 |
| Prediabetes | PP_cell | *PRKRIP1* | -0.346291777 | 0.005882747 | 0.024051313 | 1 |
| Prediabetes | PP_cell | *VGF* | -0.216642868 | 0.001956775 | 0.013500582 | 1 |
| Prediabetes | PP_cell | *MTX2* | -0.374100835 | 0.0003961 | 0.008519859 | 1 |
| Prediabetes | PP_cell | *GAD1* | -0.179348613 | 0.005944613 | 0.024153447 | 1 |

| | | | | | | |
|---|---|---|---|---|---|---|
| Prediabetes | PP_cell | *TWSG1* | -0.271396161 | 0.020523778 | 0.049653894 | 1 |
| Prediabetes | PP_cell | *EIF2AK4* | -0.256169797 | 0.008476512 | 0.029842488 | 1 |
| Prediabetes | PP_cell | *MYO5C* | -0.236751273 | 0.001216425 | 0.010904418 | 1 |
| Prediabetes | PP_cell | *CGNL1* | -0.075908317 | 0.011757254 | 0.035704358 | 1 |
| Prediabetes | PP_cell | *ANAPC13* | -0.078665962 | 0.011761286 | 0.035704358 | 1 |
| Prediabetes | PP_cell | *SUMF2* | -0.297621482 | 0.001871098 | 0.013293931 | 1 |
| Prediabetes | PP_cell | *DCTD* | -0.262085737 | 0.020319825 | 0.049330421 | 1 |
| Prediabetes | PP_cell | *ATP1B2* | -0.200169995 | 0.008872583 | 0.030659964 | 1 |
| Prediabetes | PP_cell | *AP1M2* | -0.089137914 | 0.006832429 | 0.026292348 | 1 |
| Prediabetes | PP_cell | *DTD2* | -0.328988204 | 0.013966602 | 0.03856274 | 1 |
| Prediabetes | PP_cell | *MIS18BP1* | -0.160830995 | 0.017945004 | 0.044810578 | 1 |
| Prediabetes | PP_cell | *RNASE1* | -0.080819374 | 0.006187355 | 0.024659802 | 1 |
| Prediabetes | PP_cell | *EPB41L4A* | -0.762465209 | 0.002340649 | 0.014690752 | 1 |
| Prediabetes | PP_cell | *REEP5* | -0.214299465 | 0.00694077 | 0.026424694 | 1 |
| Prediabetes | PP_cell | *SHC2* | -0.387841768 | 0.00045849 | 0.008519859 | 1 |
| Prediabetes | PP_cell | *SYT5* | -0.144149583 | 0.008157601 | 0.029297271 | 1 |
| Prediabetes | PP_cell | *CBFA2T3* | -0.204383492 | 0.002786767 | 0.0160564 | 1 |
| Prediabetes | PP_cell | *ERMARD* | -0.172997914 | 0.016452976 | 0.042210376 | 1 |
| Prediabetes | PP_cell | *ECSIT* | -0.335625185 | 0.002004042 | 0.013622633 | 1 |
| Prediabetes | PP_cell | *CDC16* | -0.345756722 | 0.00052801 | 0.008519859 | 1 |
| Prediabetes | PP_cell | *DPP6* | -0.288698471 | 0.000338156 | 0.008519859 | 1 |
| Prediabetes | PP_cell | *TULP4* | -0.233262722 | 0.009875097 | 0.032714383 | 1 |
| Prediabetes | PP_cell | *RSPH3* | -0.367293097 | 0.001232355 | 0.010954577 | 1 |
| Prediabetes | PP_cell | *NDUFA10* | -0.192031423 | 0.011803725 | 0.035704358 | 1 |
| Prediabetes | PP_cell | *SCO2* | -0.310397023 | 0.00109556 | 0.010322435 | 1 |
| Prediabetes | PP_cell | *ATXN10* | -0.264104968 | 0.00733195 | 0.027228007 | 1 |
| Prediabetes | PP_cell | *PNPLA7* | -0.357464676 | 0.001958598 | 0.013500582 | 1 |
| Prediabetes | PP_cell | *PAK4* | -0.343391394 | 0.000493822 | 0.008519859 | 1 |
| Prediabetes | PP_cell | *LAMA5* | -0.19966642 | 0.020327297 | 0.049330421 | 1 |
| Prediabetes | PP_cell | *UCK1* | -0.418520684 | 0.000997532 | 0.009851066 | 1 |
| Prediabetes | PP_cell | *MAP3K10* | -0.260907284 | 0.00021518 | 0.008519859 | 0.845444122 |
| Prediabetes | PP_cell | *SMPDL3B* | -0.062299962 | 0.017951604 | 0.044810578 | 1 |
| Prediabetes | PP_cell | *CLIP1* | -0.208732887 | 0.013606112 | 0.038249975 | 1 |
| Prediabetes | PP_cell | *ZNF317* | -0.210737847 | 0.009371859 | 0.031770521 | 1 |
| Prediabetes | PP_cell | *LRP3* | -0.31362986 | 0.003093403 | 0.016741022 | 1 |
| Prediabetes | PP_cell | *HSD17B3* | -0.956499823 | 0.003794583 | 0.018729796 | 1 |
| Prediabetes | PP_cell | *C1QL1* | -0.089005845 | 0.009935449 | 0.03274864 | 1 |
| Prediabetes | PP_cell | *F12* | -0.09843622 | 0.014179314 | 0.038931185 | 1 |
| Prediabetes | PP_cell | *NFATC1* | -0.297962519 | 0.007688077 | 0.028046848 | 1 |
| Prediabetes | PP_cell | *PDLIM4* | -0.32972699 | 0.000341733 | 0.008519859 | 1 |
| Prediabetes | PP_cell | *MGAT1* | -0.278291153 | 0.007445349 | 0.027521593 | 1 |
| Prediabetes | PP_cell | *DIAPH1* | -0.298209865 | 0.000907618 | 0.009499809 | 1 |
| Prediabetes | PP_cell | *EXOC4* | -0.27896846 | 0.015442428 | 0.040720336 | 1 |
| Prediabetes | PP_cell | *ANO1* | -0.193536576 | 0.005494079 | 0.023062216 | 1 |
| Prediabetes | PP_cell | *CHD1L* | -0.422657426 | 0.0005118 | 0.008519859 | 1 |
| Prediabetes | PP_cell | *PIAS3* | -0.372153898 | 0.002216704 | 0.014319626 | 1 |
| Prediabetes | PP_cell | *ZNF304* | -0.364558411 | 0.00117995 | 0.010731532 | 1 |
| Prediabetes | PP_cell | *RHPN2* | -0.101642126 | 0.012555383 | 0.037006826 | 1 |
| Prediabetes | PP_cell | *GCH1* | -0.28699103 | 0.00062764 | 0.008645299 | 1 |
| Prediabetes | PP_cell | *CCT6B* | -0.217739613 | 0.00267472 | 0.015685035 | 1 |

| | | | | | | |
|---|---|---|---|---|---|---|
| Prediabetes | PP_cell | *PPARG* | -0.483943604 | 0.002031877 | 0.01378799 | 1 |
| Prediabetes | PP_cell | *ENOSF1* | -0.360507494 | 0.00039087 | 0.008519859 | 1 |
| Prediabetes | PP_cell | *IMMT* | -0.244181402 | 0.005712199 | 0.02357482 | 1 |
| Prediabetes | PP_cell | *TMEM128* | -0.282535799 | 0.000555124 | 0.008519859 | 1 |
| Prediabetes | PP_cell | *SEC61G* | -0.185143511 | 0.007369158 | 0.027288805 | 1 |
| Prediabetes | PP_cell | *FIGNL1* | -0.23912218 | 0.001836165 | 0.013116896 | 1 |
| Prediabetes | PP_cell | *EIF5A* | -0.192333583 | 0.011085221 | 0.034787408 | 1 |
| Prediabetes | PP_cell | *RBM38* | -0.312418988 | 0.007937346 | 0.028742703 | 1 |
| Prediabetes | PP_cell | *POSTN* | -0.322576992 | 0.01360328 | 0.038249975 | 1 |
| Prediabetes | PP_cell | *EPHB2* | -0.127890515 | 0.004525217 | 0.020366067 | 1 |
| Prediabetes | PP_cell | *BTBD2* | -0.111070654 | 0.005058121 | 0.02194376 | 1 |
| Prediabetes | PP_cell | *CNDP2* | -0.352745396 | 0.000325842 | 0.008519859 | 1 |
| Prediabetes | PP_cell | *SLC2A11* | -0.416527486 | 0.003333856 | 0.017464961 | 1 |
| Prediabetes | PP_cell | *FAM83F* | -0.374379498 | 0.000325751 | 0.008519859 | 1 |
| Prediabetes | PP_cell | *GIMAP6* | -0.11661617 | 0.019157307 | 0.047174509 | 1 |
| Prediabetes | PP_cell | *AGAP3* | -0.349827352 | 0.004359349 | 0.020119092 | 1 |
| Prediabetes | PP_cell | *SFTPD* | -0.1111293 | 0.006306808 | 0.024961783 | 1 |
| Prediabetes | PP_cell | *IMPA1* | -0.17680396 | 0.004506623 | 0.020366067 | 1 |
| Prediabetes | PP_cell | *RRAS2* | -0.229966979 | 0.006006046 | 0.024326947 | 1 |
| Prediabetes | PP_cell | *HSD17B4* | -0.385705249 | 0.000580681 | 0.008616393 | 1 |
| Prediabetes | PP_cell | *COX16* | -0.261328751 | 0.003004335 | 0.016581525 | 1 |
| Prediabetes | PP_cell | *ELP3* | -0.276097657 | 0.001089453 | 0.010314364 | 1 |
| Prediabetes | PP_cell | *PEBP4* | -0.098726502 | 0.011017839 | 0.034659 | 1 |
| Prediabetes | PP_cell | *MRO* | -0.526458532 | 0.013752427 | 0.038295695 | 1 |
| Prediabetes | PP_cell | *MRPS36* | -0.249864687 | 0.001250468 | 0.010991254 | 1 |
| Prediabetes | PP_cell | *THUMPD3* | -0.235607971 | 0.014978526 | 0.0399258 | 1 |
| Prediabetes | PP_cell | *REG4* | -0.079606943 | 0.016007494 | 0.041435884 | 1 |
| Prediabetes | PP_cell | *SORT1* | -0.261347776 | 0.00048077 | 0.008519859 | 1 |
| Prediabetes | PP_cell | *NOTCH2* | -0.085821515 | 0.003234373 | 0.017219313 | 1 |
| Prediabetes | PP_cell | *CEPT1* | -0.43136282 | 0.003256774 | 0.017245099 | 1 |
| Prediabetes | PP_cell | *NAPG* | -0.25990262 | 0.018109604 | 0.045147611 | 1 |
| Prediabetes | PP_cell | *FKBP11* | -0.107314963 | 0.002644318 | 0.01561651 | 1 |
| Prediabetes | PP_cell | *IAH1* | -0.322089517 | 0.000718094 | 0.008872296 | 1 |
| Prediabetes | PP_cell | *ECHDC3* | -0.256264416 | 0.003871046 | 0.018940647 | 1 |
| Prediabetes | PP_cell | *KLRC1* | -2.036010899 | 0.013148518 | 0.037909761 | 1 |
| Prediabetes | PP_cell | *DSC2* | -0.186966466 | 0.001399383 | 0.011478447 | 1 |
| Prediabetes | PP_cell | *TPGS2* | -0.254052268 | 0.005118055 | 0.021990223 | 1 |
| Prediabetes | PP_cell | *DAGLA* | -0.161910591 | 0.004491878 | 0.020343144 | 1 |
| Prediabetes | PP_cell | *TIMM10* | -0.279586755 | 0.00427037 | 0.019996719 | 1 |
| Prediabetes | PP_cell | *CLOCK* | -0.22381984 | 0.00683702 | 0.026292348 | 1 |
| Prediabetes | PP_cell | *GGACT* | -0.180692985 | 0.014019703 | 0.038655027 | 1 |
| Prediabetes | PP_cell | *CLDN10* | -0.033197074 | 0.009535225 | 0.032025881 | 1 |
| Prediabetes | PP_cell | *CARS2* | -0.230272285 | 0.001174871 | 0.010731532 | 1 |
| Prediabetes | PP_cell | *TMED7* | -0.29205269 | 0.001239937 | 0.010970459 | 1 |
| Prediabetes | PP_cell | *OSTF1* | -0.404893675 | 0.001281635 | 0.011111559 | 1 |
| Prediabetes | PP_cell | *GOLM1* | -0.174341109 | 0.00079156 | 0.009253454 | 1 |
| Prediabetes | PP_cell | *MSI1* | -0.162172815 | 0.003866296 | 0.018940647 | 1 |
| Prediabetes | PP_cell | *PNPLA8* | -0.211319594 | 0.008991996 | 0.030882477 | 1 |
| Prediabetes | PP_cell | *TES* | -0.18217552 | 0.002986413 | 0.016581525 | 1 |
| Prediabetes | PP_cell | *LCA5* | -0.450702175 | 0.000288409 | 0.008519859 | 1 |

| | | | | | | |
|---|---|---|---|---|---|---|
| Prediabetes | PP_cell | *TFCP2* | -0.359720199 | 0.000437654 | 0.008519859 | 1 |
| Prediabetes | PP_cell | *KRT7* | -0.034494 | 0.008704881 | 0.030237155 | 1 |
| Prediabetes | PP_cell | *PKIB* | -0.314337192 | 0.001824128 | 0.013078464 | 1 |
| Prediabetes | PP_cell | *GNS* | -0.35620279 | 0.000624192 | 0.008645299 | 1 |
| Prediabetes | PP_cell | *MPHOSPH6* | -0.354546438 | 0.010168019 | 0.03312616 | 1 |
| Prediabetes | PP_cell | *NTPCR* | -0.238140602 | 0.000974889 | 0.009804971 | 1 |
| Prediabetes | PP_cell | *GLUL* | -0.304679958 | 0.006050416 | 0.024381623 | 1 |
| Prediabetes | PP_cell | *USP37* | -0.317943032 | 0.000386056 | 0.008519859 | 1 |
| Prediabetes | PP_cell | *ARMC9* | -0.610322997 | 0.00048935 | 0.008519859 | 1 |
| Prediabetes | PP_cell | *COX5B* | -0.211540397 | 0.004494235 | 0.020343144 | 1 |
| Prediabetes | PP_cell | *REV1* | -0.259789458 | 0.000331745 | 0.008519859 | 1 |
| Prediabetes | PP_cell | *TSGA10* | -0.102626104 | 8.35E-05 | 0.008519859 | 0.32811416 |
| Prediabetes | PP_cell | *MRPS9* | -0.411680737 | 0.000136406 | 0.008519859 | 0.53594097 |
| Prediabetes | PP_cell | *ANKRD36* | -0.434328033 | 0.020761 | 0.049920422 | 1 |
| Prediabetes | PP_cell | *USP44* | -0.099462587 | 0.013443808 | 0.038055275 | 1 |
| Prediabetes | PP_cell | *DRAM1* | -0.480655922 | 0.001659595 | 0.012440588 | 1 |
| Prediabetes | PP_cell | *VPS36* | -0.34145479 | 0.000195196 | 0.008519859 | 0.766926611 |
| Prediabetes | PP_cell | *RNASEH2B* | -0.301291745 | 0.000430402 | 0.008519859 | 1 |
| Prediabetes | PP_cell | *SUCLA2* | -0.310354246 | 0.003596191 | 0.018323322 | 1 |
| Prediabetes | PP_cell | *LMO7* | -0.210705725 | 0.000847891 | 0.009444968 | 1 |
| Prediabetes | PP_cell | *C7orf25* | -0.18715331 | 0.008305017 | 0.029556532 | 1 |
| Prediabetes | PP_cell | *RAPGEF5* | -0.296967453 | 0.000688468 | 0.008780065 | 1 |
| Prediabetes | PP_cell | *KDELR2* | -0.271391625 | 0.00108821 | 0.010314364 | 1 |
| Prediabetes | PP_cell | *TBRG4* | -0.213390992 | 0.009253677 | 0.031505802 | 1 |
| Prediabetes | PP_cell | *ALPK3* | -0.324532525 | 0.003819784 | 0.018812875 | 1 |
| Prediabetes | PP_cell | *CIB2* | -0.171157778 | 0.00188078 | 0.013314564 | 1 |
| Prediabetes | PP_cell | *TEX2* | -0.389082684 | 0.000930365 | 0.009619487 | 1 |
| Prediabetes | PP_cell | *BLK* | -0.995188266 | 0.010370858 | 0.033564334 | 1 |
| Prediabetes | PP_cell | *SMPD4* | -0.373054933 | 0.006549752 | 0.025605945 | 1 |
| Prediabetes | PP_cell | *CCDC115* | -0.292953871 | 0.000250602 | 0.008519859 | 0.984616854 |
| Prediabetes | PP_cell | *HS6ST1* | -0.42920877 | 0.002109916 | 0.013980215 | 1 |
| Prediabetes | PP_cell | *NIPSNAP3A* | -0.229480753 | 0.00045224 | 0.008519859 | 1 |
| Prediabetes | PP_cell | *TXN* | -0.163606356 | 0.01926451 | 0.047346586 | 1 |
| Prediabetes | PP_cell | *SLC2A8* | -0.43328336 | 0.000329395 | 0.008519859 | 1 |
| Prediabetes | PP_cell | *STX17* | -0.252353166 | 0.010228929 | 0.033269422 | 1 |
| Prediabetes | PP_cell | *FPGS* | -0.506400174 | 0.001533683 | 0.011877462 | 1 |
| Prediabetes | PP_cell | *POLR1E* | -0.183405355 | 0.004533917 | 0.020368512 | 1 |
| Prediabetes | PP_cell | *TMEM8B* | -0.179009062 | 0.001836025 | 0.013116896 | 1 |
| Prediabetes | PP_cell | *ARHGEF39* | -0.272211858 | 0.005527183 | 0.023127052 | 1 |
| Prediabetes | PP_cell | *PPIL1* | -0.142262164 | 0.00363576 | 0.01839612 | 1 |
| Prediabetes | PP_cell | *GMPR* | -0.214939456 | 0.002065566 | 0.013874899 | 1 |
| Prediabetes | PP_cell | *KIAA0319* | -0.588450243 | 0.002061703 | 0.013874899 | 1 |
| Prediabetes | PP_cell | *LRRC1* | -0.400654304 | 0.000909119 | 0.009499809 | 1 |
| Prediabetes | PP_cell | *UQCC2* | -0.182504479 | 0.008181022 | 0.029327769 | 1 |
| Prediabetes | PP_cell | *HMGA1* | -0.169049237 | 0.000843082 | 0.009444968 | 1 |
| Prediabetes | PP_cell | *VARS2* | -0.095951919 | 0.001232245 | 0.010954577 | 1 |
| Prediabetes | PP_cell | *ANKRD42* | -0.296048997 | 0.000301256 | 0.008519859 | 1 |
| Prediabetes | PP_cell | *GGH* | -0.200720004 | 0.00049582 | 0.008519859 | 1 |
| Prediabetes | PP_cell | *SDCBP* | -0.200982614 | 0.015884512 | 0.041331289 | 1 |
| Prediabetes | PP_cell | *NEK1* | -0.153367746 | 0.012247018 | 0.036372141 | 1 |

| | | | | | | |
|---|---|---|---|---|---|---|
| Prediabetes | PP_cell | *MMP7* | -0.038886499 | 0.015951835 | 0.041414773 | 1 |
| Prediabetes | PP_cell | *FDX1* | -0.296213433 | 0.000788268 | 0.009253454 | 1 |
| Prediabetes | PP_cell | *C11orf1* | -0.25247705 | 0.00128533 | 0.011111559 | 1 |
| Prediabetes | PP_cell | *ALKBH8* | -0.511253277 | 0.000694986 | 0.008780065 | 1 |
| Prediabetes | PP_cell | *TUBGCP4* | -0.388055417 | 0.000143528 | 0.008519859 | 0.563921771 |
| Prediabetes | PP_cell | *ADAM10* | -0.267284245 | 0.011205889 | 0.034915098 | 1 |
| Prediabetes | PP_cell | *RSL24D1* | -0.237608838 | 0.002886258 | 0.01636379 | 1 |
| Prediabetes | PP_cell | *SPTBN5* | 0.218967228 | 0.014961464 | 0.0399258 | 1 |
| Prediabetes | PP_cell | *TTLL7* | -0.239414745 | 0.007008471 | 0.026553791 | 1 |
| Prediabetes | PP_cell | *SLC44A5* | -0.152446985 | 0.018759897 | 0.046591426 | 1 |
| Prediabetes | PP_cell | *RTCA* | -0.396522183 | 0.000338414 | 0.008519859 | 1 |
| Prediabetes | PP_cell | *DYNC2LI1* | -0.222904203 | 0.008236265 | 0.029391721 | 1 |
| Prediabetes | PP_cell | *DUSP5* | -0.31275067 | 0.000187241 | 0.008519859 | 0.735670836 |
| Prediabetes | PP_cell | *DNAJC13* | -0.438756548 | 0.014687021 | 0.039551271 | 1 |
| Prediabetes | PP_cell | *ATIC* | -0.136629144 | 0.00836638 | 0.029640674 | 1 |
| Prediabetes | PP_cell | *SMARCAL1* | -0.574994939 | 0.000622996 | 0.008645299 | 1 |
| Prediabetes | PP_cell | *BARD1* | -0.187251421 | 0.003986479 | 0.019154283 | 1 |
| Prediabetes | PP_cell | *FASTKD1* | -0.150526295 | 0.015626215 | 0.041056337 | 1 |
| Prediabetes | PP_cell | *WDR12* | -0.181001756 | 0.01049818 | 0.03386482 | 1 |
| Prediabetes | PP_cell | *SLC35A5* | -0.185586887 | 0.004033538 | 0.019326551 | 1 |
| Prediabetes | PP_cell | *HCN4* | -0.112117044 | 0.00274871 | 0.015953783 | 1 |
| Prediabetes | PP_cell | *SEMA7A* | -0.155311667 | 0.013081038 | 0.037909761 | 1 |
| Prediabetes | PP_cell | *AP1AR* | -0.226164809 | 0.019274108 | 0.047346586 | 1 |
| Prediabetes | PP_cell | *RAP1GDS1* | -0.437340783 | 0.000479041 | 0.008519859 | 1 |
| Prediabetes | PP_cell | *G3BP2* | -0.257035523 | 0.003928767 | 0.019039633 | 1 |
| Prediabetes | PP_cell | *SCARB2* | -0.183161389 | 0.00705153 | 0.026639866 | 1 |
| Prediabetes | PP_cell | *USO1* | -0.318493621 | 0.001520488 | 0.011846392 | 1 |
| Prediabetes | PP_cell | *SHROOM3* | -0.12105196 | 0.00432586 | 0.020113968 | 1 |
| Prediabetes | PP_cell | *EGF* | -0.186938821 | 0.011855443 | 0.035727429 | 1 |
| Prediabetes | PP_cell | *RGS3* | -0.340189816 | 0.000103923 | 0.008519859 | 0.408313501 |
| Prediabetes | PP_cell | *ETV6* | -0.407977323 | 0.000193221 | 0.008519859 | 0.759164103 |
| Prediabetes | PP_cell | *ALG10* | -0.933591803 | 0.003937237 | 0.019050989 | 1 |
| Prediabetes | PP_cell | *ZCRB1* | -0.264224854 | 0.00151762 | 0.011846392 | 1 |
| Prediabetes | PP_cell | *KLRG1* | -0.446491932 | 0.009231118 | 0.031483562 | 1 |
| Prediabetes | PP_cell | *PEX5* | -0.39726327 | 0.000188792 | 0.008519859 | 0.741762592 |
| Prediabetes | PP_cell | *SLC38A4* | -0.072476051 | 0.012263606 | 0.036372141 | 1 |
| Prediabetes | PP_cell | *LRIG3* | -0.339823661 | 0.004435743 | 0.020248234 | 1 |
| Prediabetes | PP_cell | *GLIPR1* | -0.491838776 | 0.001449614 | 0.011646409 | 1 |
| Prediabetes | PP_cell | *ASCL1* | -0.101464113 | 0.014328544 | 0.039167161 | 1 |
| Prediabetes | PP_cell | *PDX1* | -0.164006468 | 0.002456293 | 0.015103998 | 1 |
| Prediabetes | PP_cell | *KANSL2* | -0.499952812 | 0.000131917 | 0.008519859 | 0.518300599 |
| Prediabetes | PP_cell | *GALNT6* | -0.124013646 | 0.016756366 | 0.042722754 | 1 |
| Prediabetes | PP_cell | *RAB20* | -0.226804016 | 0.006603977 | 0.0257511 | 1 |
| Prediabetes | PP_cell | *GRTP1* | -0.217646128 | 0.008656233 | 0.030204563 | 1 |
| Prediabetes | PP_cell | *SERF2* | -0.17976053 | 0.004285375 | 0.019996719 | 1 |
| Prediabetes | PP_cell | *GTF2A2* | -0.20334701 | 0.016073914 | 0.041521635 | 1 |
| Prediabetes | PP_cell | *SCAPER* | -0.259132937 | 0.000773885 | 0.009253454 | 1 |
| Prediabetes | PP_cell | *DNAJA4* | -0.239889075 | 0.000969322 | 0.009804971 | 1 |
| Prediabetes | PP_cell | *TPM1* | -0.140876543 | 0.000946976 | 0.009678612 | 1 |
| Prediabetes | PP_cell | *IGF1R* | -0.264099427 | 0.004839423 | 0.021221086 | 1 |

| | | | | | | |
|---|---|---|---|---|---|---|
| Prediabetes | PP_cell | *ULK3* | -0.235084022 | 0.019031552 | 0.047015043 | 1 |
| Prediabetes | PP_cell | *CELF6* | -0.519821334 | 0.000363775 | 0.008519859 | 1 |
| Prediabetes | PP_cell | *WDR93* | -0.279365799 | 0.010357984 | 0.033559662 | 1 |
| Prediabetes | PP_cell | *PMM2* | -0.406586306 | 0.001041662 | 0.010105406 | 1 |
| Prediabetes | PP_cell | *SLC5A2* | -0.262115674 | 0.010805915 | 0.034287057 | 1 |
| Prediabetes | PP_cell | *TXNL4B* | -0.205943778 | 0.002048746 | 0.013854598 | 1 |
| Prediabetes | PP_cell | *MARVELD3* | -0.133904233 | 0.006076494 | 0.024411598 | 1 |
| Prediabetes | PP_cell | *KIFC3* | -0.338456759 | 0.001945111 | 0.013500582 | 1 |
| Prediabetes | PP_cell | *NUDT7* | -0.179389721 | 0.002754074 | 0.015959817 | 1 |
| Prediabetes | PP_cell | *GCSH* | -0.325088915 | 0.000682152 | 0.008780065 | 1 |
| Prediabetes | PP_cell | *OSGIN1* | -0.305420493 | 0.002280827 | 0.014499386 | 1 |
| Prediabetes | PP_cell | *PDPK1* | -0.2111952 | 0.001299485 | 0.011123481 | 1 |
| Prediabetes | PP_cell | *DEF8* | -0.236504613 | 0.011979388 | 0.035956468 | 1 |
| Prediabetes | PP_cell | *GALNS* | -0.260756624 | 0.009097598 | 0.031136291 | 1 |
| Prediabetes | PP_cell | *GAS8* | -0.279080447 | 0.000165131 | 0.008519859 | 0.648801445 |
| Prediabetes | PP_cell | *TOM1L1* | -0.118849663 | 0.006084096 | 0.024417172 | 1 |
| Prediabetes | PP_cell | *NPEPPS* | -0.290089685 | 0.000238225 | 0.008519859 | 0.935985278 |
| Prediabetes | PP_cell | *SPACA3* | -0.075995408 | 0.003606784 | 0.018332544 | 1 |
| Prediabetes | PP_cell | *PTRH2* | -0.409788845 | 0.001156989 | 0.010645921 | 1 |
| Prediabetes | PP_cell | *RPRD1A* | -0.353888359 | 0.000614561 | 0.008645299 | 1 |
| Prediabetes | PP_cell | *NPC1* | -0.305719803 | 0.008348965 | 0.029619945 | 1 |
| Prediabetes | PP_cell | *SLC14A1* | -0.10449589 | 0.015958759 | 0.041414773 | 1 |
| Prediabetes | PP_cell | *ZMYND15* | -0.241334916 | 0.001242518 | 0.010970459 | 1 |
| Prediabetes | PP_cell | *TMC6* | -0.120058438 | 0.012081085 | 0.036178797 | 1 |
| Prediabetes | PP_cell | *TTYH2* | -0.725635234 | 0.000679185 | 0.008780065 | 1 |
| Prediabetes | PP_cell | *TBCD* | -0.290839761 | 0.006732848 | 0.026062425 | 1 |
| Prediabetes | PP_cell | *NARF* | -0.284280331 | 0.003223195 | 0.017212803 | 1 |
| Prediabetes | PP_cell | *WDR45B* | -0.193692343 | 0.014505071 | 0.039384002 | 1 |
| Prediabetes | PP_cell | *TXNL4A* | -0.201133919 | 0.015130226 | 0.040139539 | 1 |
| Prediabetes | PP_cell | *AKT1* | -0.224979661 | 0.016524397 | 0.042268461 | 1 |
| Prediabetes | PP_cell | *CBLC* | -0.069573313 | 0.009302328 | 0.031644023 | 1 |
| Prediabetes | PP_cell | *CTU1* | -0.276649924 | 0.006703993 | 0.025976319 | 1 |
| Prediabetes | PP_cell | *CNKSR1* | -0.140490129 | 0.010687112 | 0.034165713 | 1 |
| Prediabetes | PP_cell | *ZNF593* | -0.266485801 | 0.000175343 | 0.008519859 | 0.688920821 |
| Prediabetes | PP_cell | *CELA3A* | -0.061872008 | 0.001311786 | 0.011188791 | 1 |
| Prediabetes | PP_cell | *PIGK* | -0.289270914 | 0.000483821 | 0.008519859 | 1 |
| Prediabetes | PP_cell | *SLC44A3* | -0.104359352 | 0.012862586 | 0.03760809 | 1 |
| Prediabetes | PP_cell | *CELSR2* | -0.360002925 | 0.001878072 | 0.013314564 | 1 |
| Prediabetes | PP_cell | *ALDH9A1* | -0.25712671 | 0.016249609 | 0.041846259 | 1 |
| Prediabetes | PP_cell | *ATP1B1* | -0.105043585 | 0.001177494 | 0.010731532 | 1 |
| Prediabetes | PP_cell | *NME7* | -0.067992954 | 0.007574306 | 0.027812568 | 1 |
| Prediabetes | PP_cell | *LYPLAL1* | -0.289295559 | 0.000884639 | 0.009444968 | 1 |
| Prediabetes | PP_cell | *CTSK* | -0.211253633 | 0.003426203 | 0.017782763 | 1 |
| Prediabetes | PP_cell | *TAF1A* | -0.106027334 | 0.015064858 | 0.040047244 | 1 |
| Prediabetes | PP_cell | *SLC27A3* | -0.311486931 | 0.001388543 | 0.011450342 | 1 |
| Prediabetes | PP_cell | *PSEN2* | -0.329490744 | 0.002629683 | 0.015607286 | 1 |
| Prediabetes | PP_cell | *LBR* | -0.192939516 | 0.001001009 | 0.009851066 | 1 |
| Prediabetes | PP_cell | *ARL8A* | -0.310199605 | 0.00086872 | 0.009444968 | 1 |
| Prediabetes | PP_cell | *GALM* | -0.240097097 | 0.003045532 | 0.016712144 | 1 |
| Prediabetes | PP_cell | *EML4* | -0.35863383 | 0.000789191 | 0.009253454 | 1 |

| | | | | | | |
|---|---|---|---|---|---|---|
| Prediabetes | PP_cell | *WDPCP* | -0.560782527 | 0.001032266 | 0.010051656 | 1 |
| Prediabetes | PP_cell | *REG3G* | -0.04819581 | 0.013880751 | 0.038433735 | 1 |
| Prediabetes | PP_cell | *CIAO1* | -0.273035719 | 0.006904831 | 0.026361874 | 1 |
| Prediabetes | PP_cell | *THNSL2* | -0.062091569 | 0.015853746 | 0.041316373 | 1 |
| Prediabetes | PP_cell | *RALB* | -0.247232557 | 0.018833202 | 0.046655517 | 1 |
| Prediabetes | PP_cell | *LIPT1* | -0.186022711 | 0.001995667 | 0.013597804 | 1 |
| Prediabetes | PP_cell | *FAM171B* | -0.281435087 | 0.000277372 | 0.008519859 | 1 |
| Prediabetes | PP_cell | *SUMF1* | -0.340810275 | 0.001768527 | 0.012915503 | 1 |
| Prediabetes | PP_cell | *HES6* | -0.425624453 | 0.000455695 | 0.008519859 | 1 |
| Prediabetes | PP_cell | *ESPNL* | -0.99887796 | 0.007167599 | 0.02684604 | 1 |
| Prediabetes | PP_cell | *RAB5A* | -0.224240319 | 0.016596614 | 0.042370433 | 1 |
| Prediabetes | PP_cell | *CNTN4* | -0.084894457 | 0.011749662 | 0.035704358 | 1 |
| Prediabetes | PP_cell | *POMGNT2* | -0.291489712 | 0.01407532 | 0.038726844 | 1 |
| Prediabetes | PP_cell | *IL17RD* | -0.27877186 | 0.011056746 | 0.034753564 | 1 |
| Prediabetes | PP_cell | *UBA3* | -0.392517841 | 0.000505737 | 0.008519859 | 1 |
| Prediabetes | PP_cell | *ARL6IP5* | -0.294555028 | 0.000581152 | 0.008616393 | 1 |
| Prediabetes | PP_cell | *TAGLN3* | -0.160482654 | 0.002102517 | 0.013977644 | 1 |
| Prediabetes | PP_cell | *EIF2A* | -0.278105009 | 0.001146305 | 0.010597251 | 1 |
| Prediabetes | PP_cell | *ALDH1L1* | -0.090343043 | 0.014031775 | 0.038661181 | 1 |
| Prediabetes | PP_cell | *NCEH1* | -0.167322195 | 0.001428433 | 0.011619697 | 1 |
| Prediabetes | PP_cell | *DGKQ* | -0.105477348 | 0.008580926 | 0.030075341 | 1 |
| Prediabetes | PP_cell | *SCD5* | -0.06575034 | 0.007987197 | 0.028870005 | 1 |
| Prediabetes | PP_cell | *TBCK* | -0.126456628 | 0.015632548 | 0.041056337 | 1 |
| Prediabetes | PP_cell | *TIFA* | -0.35975142 | 0.000704358 | 0.008813447 | 1 |
| Prediabetes | PP_cell | *RNF175* | -3.862678103 | 0.016958059 | 0.043069304 | 1 |
| Prediabetes | PP_cell | *IQGAP2* | -0.206179484 | 0.000273488 | 0.008519859 | 1 |
| Prediabetes | PP_cell | *GTF2H2* | -0.152424224 | 0.002962743 | 0.016546556 | 1 |
| Prediabetes | PP_cell | *SPATA9* | -0.47485228 | 0.019532231 | 0.047834311 | 1 |
| Prediabetes | PP_cell | *COMMD10* | -0.347487822 | 0.00048687 | 0.008519859 | 1 |
| Prediabetes | PP_cell | *PCYOX1L* | -0.737934691 | 0.001438344 | 0.011628097 | 1 |
| Prediabetes | PP_cell | *TBC1D7* | -0.536792689 | 0.000101822 | 0.008519859 | 0.400056853 |
| Prediabetes | PP_cell | *ZMAT2* | -0.350819238 | 0.001045602 | 0.01010918 | 1 |
| Prediabetes | PP_cell | *DCDC2* | -0.408223141 | 0.000896013 | 0.009499809 | 1 |
| Prediabetes | PP_cell | *TRIM7* | -0.270197613 | 0.000126089 | 0.008519859 | 0.495403559 |
| Prediabetes | PP_cell | *ANO7* | -0.213763091 | 0.009615895 | 0.032072031 | 1 |
| Prediabetes | PP_cell | *IRAK1BP1* | -0.404714968 | 0.000302141 | 0.008519859 | 1 |
| Prediabetes | PP_cell | *FAXC* | -0.062877977 | 0.013545856 | 0.038233958 | 1 |
| Prediabetes | PP_cell | *ABRACL* | -0.189033192 | 0.001710053 | 0.012724999 | 1 |
| Prediabetes | PP_cell | *DYNLT1* | -0.222168027 | 0.005088461 | 0.02194376 | 1 |
| Prediabetes | PP_cell | *TMEM209* | -0.353541486 | 0.007105466 | 0.026721477 | 1 |
| Prediabetes | PP_cell | *CSGALNACT1* | -0.299333163 | 0.014893342 | 0.039860996 | 1 |
| Prediabetes | PP_cell | *BIN3* | -0.459132641 | 0.00250145 | 0.015266026 | 1 |
| Prediabetes | PP_cell | *CHMP7* | -0.246193173 | 0.014660272 | 0.039533432 | 1 |
| Prediabetes | PP_cell | *LACTB2* | -0.372593974 | 0.000416591 | 0.008519859 | 1 |
| Prediabetes | PP_cell | *AK3* | -0.250518757 | 0.008049518 | 0.029041834 | 1 |
| Prediabetes | PP_cell | *C9orf72* | -0.292228477 | 0.00754487 | 0.027760015 | 1 |
| Prediabetes | PP_cell | *FBXO10* | -0.199046959 | 0.002951589 | 0.016546556 | 1 |
| Prediabetes | PP_cell | *ZNF462* | -0.415670125 | 0.000293799 | 0.008519859 | 1 |
| Prediabetes | PP_cell | *SURF4* | -0.19998983 | 0.002501929 | 0.015266026 | 1 |
| Prediabetes | PP_cell | *SURF6* | -0.424684247 | 0.000390544 | 0.008519859 | 1 |

| | | | | | | |
|---|---|---|---|---|---|---|
| Prediabetes | PP_cell | *SH3GLB2* | -0.307153756 | 0.001434733 | 0.011622813 | 1 |
| Prediabetes | PP_cell | *LCN2* | -0.030665584 | 0.01122134 | 0.034935536 | 1 |
| Prediabetes | PP_cell | *GPR107* | -0.367557482 | 0.002204352 | 0.014315539 | 1 |
| Prediabetes | PP_cell | *SEC16A* | -0.211527797 | 0.013852733 | 0.038410295 | 1 |
| Prediabetes | PP_cell | *MSRB2* | -0.222825492 | 0.001842975 | 0.01314165 | 1 |
| Prediabetes | PP_cell | *FRA10AC1* | -0.313357357 | 0.002434937 | 0.015042244 | 1 |
| Prediabetes | PP_cell | *DNAJB12* | -0.374483801 | 0.000522352 | 0.008519859 | 1 |
| Prediabetes | PP_cell | *PAOX* | -0.285546073 | 0.019235389 | 0.047323634 | 1 |
| Prediabetes | PP_cell | *APIP* | -0.119454102 | 0.010783468 | 0.034287057 | 1 |
| Prediabetes | PP_cell | *PTPRJ* | -0.155109098 | 0.000431342 | 0.008519859 | 1 |
| Prediabetes | PP_cell | *CCDC82* | -0.127970832 | 0.017373572 | 0.043823045 | 1 |
| Prediabetes | PP_cell | *AASDHPPT* | -0.356523541 | 0.001340456 | 0.011283666 | 1 |
| Prediabetes | PP_cell | *EML3* | -0.493359688 | 0.004704302 | 0.020861403 | 1 |
| Prediabetes | PP_cell | *CHEK1* | -0.184943468 | 0.01181261 | 0.035704358 | 1 |
| Prediabetes | PP_cell | *SIDT2* | -0.247758505 | 0.004856462 | 0.021272062 | 1 |
| Prediabetes | PP_cell | *DUSP15* | -0.123103171 | 0.003668888 | 0.01839612 | 1 |
| Prediabetes | PP_cell | *YTHDF1* | -0.25997527 | 0.020621253 | 0.049760752 | 1 |
| Prediabetes | PP_cell | *TRPT1* | -0.338505805 | 0.001054731 | 0.01015696 | 1 |
| Prediabetes | PP_cell | *TBX6* | -0.190696479 | 0.017236256 | 0.043597099 | 1 |
| Prediabetes | PP_cell | *MPP7* | -0.318660246 | 0.000987008 | 0.009817609 | 1 |
| Prediabetes | PP_cell | *MIA2* | -0.257545009 | 0.00117673 | 0.010731532 | 1 |
| Prediabetes | PP_cell | *LYPD1* | -0.104409732 | 0.013817565 | 0.038387898 | 1 |
| Prediabetes | PP_cell | *MTMR12* | -0.328132852 | 0.003971587 | 0.019146462 | 1 |
| Prediabetes | PP_cell | *IL18* | -0.104126402 | 0.013926864 | 0.038507143 | 1 |
| Prediabetes | PP_cell | *DLG5* | -0.367969635 | 0.000374609 | 0.008519859 | 1 |
| Prediabetes | PP_cell | *AKAP6* | -0.140833989 | 0.005556339 | 0.023167203 | 1 |
| Prediabetes | PP_cell | *NDUFC2* | -0.171102689 | 0.017193608 | 0.043526859 | 1 |
| Prediabetes | PP_cell | *C4orf33* | -0.291553649 | 0.006342776 | 0.025045996 | 1 |
| Prediabetes | PP_cell | *QDPR* | -0.219380519 | 0.002748972 | 0.015953783 | 1 |
| Prediabetes | PP_cell | *TEX9* | -0.359937031 | 0.001450779 | 0.011646409 | 1 |
| Prediabetes | PP_cell | *KIN* | -0.343922135 | 0.006186513 | 0.024659802 | 1 |
| Prediabetes | PP_cell | *CENPU* | -0.188423329 | 0.01961114 | 0.047918016 | 1 |
| Prediabetes | PP_cell | *BICD1* | -0.375919609 | 0.00117086 | 0.010731532 | 1 |
| Prediabetes | PP_cell | *GUF1* | -0.365794573 | 0.000507523 | 0.008519859 | 1 |
| Prediabetes | PP_cell | *CCL28* | -0.415091535 | 0.002352817 | 0.014710984 | 1 |
| Prediabetes | PP_cell | *RABGAP1L* | -0.380893852 | 0.001062539 | 0.010207131 | 1 |
| Prediabetes | PP_cell | *ARL14EP* | -0.360570845 | 0.001374207 | 0.011450342 | 1 |
| Prediabetes | PP_cell | *EPG5* | -0.408282468 | 0.003216578 | 0.017212803 | 1 |
| Prediabetes | PP_cell | *PDE3B* | -0.169156288 | 9.33E-05 | 0.008519859 | 0.366636485 |
| Prediabetes | PP_cell | *POC5* | -0.061504661 | 0.0131064 | 0.037909761 | 1 |
| Prediabetes | PP_cell | *GUCY1A2* | -0.502741247 | 0.000479283 | 0.008519859 | 1 |
| Prediabetes | PP_cell | *DCLRE1C* | -0.210934378 | 0.004106289 | 0.019508595 | 1 |
| Prediabetes | PP_cell | *ZSCAN1* | -0.210972561 | 0.004447152 | 0.020270141 | 1 |
| Prediabetes | PP_cell | *GRIA4* | -0.059831002 | 0.014771914 | 0.039644024 | 1 |
| Prediabetes | PP_cell | *IGSF10* | -0.583258352 | 0.000358665 | 0.008519859 | 1 |
| Prediabetes | PP_cell | *GPD1L* | -0.565802736 | 0.001465288 | 0.011646409 | 1 |
| Prediabetes | PP_cell | *CCNO* | -0.080736213 | 0.011868275 | 0.035732147 | 1 |
| Prediabetes | PP_cell | *CATSPER3* | -0.527749895 | 0.009452663 | 0.031889469 | 1 |
| Prediabetes | PP_cell | *GPR180* | -0.270531572 | 0.013963479 | 0.03856274 | 1 |
| Prediabetes | PP_cell | *MED21* | -0.263816175 | 0.017893685 | 0.044751296 | 1 |

| | | | | | | |
|---|---|---|---|---|---|---|
| Prediabetes | PP_cell | *ACMSD* | -0.090615338 | 0.013762668 | 0.038295695 | 1 |
| Prediabetes | PP_cell | *TMEM87B* | -0.238862106 | 0.000975756 | 0.009804971 | 1 |
| Prediabetes | PP_cell | *PLA2R1* | -0.539307759 | 0.004517298 | 0.020366067 | 1 |
| Prediabetes | PP_cell | *CMTM7* | -0.106692683 | 0.006930605 | 0.026411589 | 1 |
| Prediabetes | PP_cell | *PTPRD* | -0.075495885 | 0.008218136 | 0.029383885 | 1 |
| Prediabetes | PP_cell | *ZDHHC7* | -0.276175894 | 0.003770853 | 0.018659548 | 1 |
| Prediabetes | PP_cell | *JAZF1* | -0.219994578 | 0.000149033 | 0.008519859 | 0.585551619 |
| Prediabetes | PP_cell | *CMIP* | -0.351533685 | 0.000354964 | 0.008519859 | 1 |
| Prediabetes | PP_cell | *PID1* | -0.140902876 | 0.004431305 | 0.020248234 | 1 |
| Prediabetes | PP_cell | *KCTD15* | -0.666437728 | 0.002720523 | 0.015858955 | 1 |
| Prediabetes | PP_cell | *DDAH1* | -0.309675597 | 6.94E-05 | 0.008519859 | 0.272530487 |
| Prediabetes | PP_cell | *NUS1* | -0.346735218 | 0.000937887 | 0.009671806 | 1 |
| Prediabetes | PP_cell | *CC2D1B* | -0.117188846 | 0.009465088 | 0.031893938 | 1 |
| Prediabetes | PP_cell | *MRPL39* | -0.211639291 | 0.01254574 | 0.037006165 | 1 |
| Prediabetes | PP_cell | *TSEN2* | -0.534920306 | 0.001047197 | 0.01010918 | 1 |
| Prediabetes | PP_cell | *FLCN* | -0.309139233 | 0.000629628 | 0.008645299 | 1 |
| Prediabetes | PP_cell | *MPPE1* | -0.164889163 | 0.011534459 | 0.035488558 | 1 |
| Prediabetes | PP_cell | *RAB6B* | -0.511676383 | 0.000455536 | 0.008519859 | 1 |
| Prediabetes | PP_cell | *TRMT44* | -0.234556895 | 0.002784569 | 0.0160564 | 1 |
| Prediabetes | PP_cell | *PPM1J* | -0.172692997 | 0.011825637 | 0.035704358 | 1 |
| Prediabetes | PP_cell | *HEATR3* | -0.072633152 | 0.011753122 | 0.035704358 | 1 |
| Prediabetes | PP_cell | *LRGUK* | -0.520137979 | 0.000791182 | 0.009253454 | 1 |
| Prediabetes | PP_cell | *MIER3* | -0.375007597 | 0.000766562 | 0.009244451 | 1 |
| Prediabetes | PP_cell | *TTN* | -0.498062171 | 0.006532535 | 0.025564074 | 1 |
| Prediabetes | PP_cell | *RMND1* | -0.288329219 | 0.003316746 | 0.017447335 | 1 |
| Prediabetes | PP_cell | *PSD3* | -0.153167535 | 0.002954978 | 0.016546556 | 1 |
| Prediabetes | PP_cell | *MMP16* | -0.105538858 | 0.012716241 | 0.03734089 | 1 |
| Prediabetes | PP_cell | *ADK* | -0.266034979 | 0.000796047 | 0.009253454 | 1 |
| Prediabetes | PP_cell | *BATF* | -0.972956858 | 0.013489719 | 0.038130291 | 1 |
| Prediabetes | PP_cell | *DRAM2* | -0.243644746 | 0.002319217 | 0.014602893 | 1 |
| Prediabetes | PP_cell | *CCT8* | -0.154500585 | 0.011125559 | 0.034830535 | 1 |
| Prediabetes | PP_cell | *MAPK13* | -0.196616504 | 0.000221736 | 0.008519859 | 0.87120178 |
| Prediabetes | PP_cell | *TBC1D31* | -0.125779826 | 0.013409756 | 0.03803406 | 1 |
| Prediabetes | PP_cell | *MPV17L* | -0.194616532 | 0.000757876 | 0.009244451 | 1 |
| Prediabetes | PP_cell | *EXOG* | -0.291099061 | 0.005791194 | 0.023775968 | 1 |
| Prediabetes | PP_cell | *ZFYVE9* | -0.130975297 | 0.004237115 | 0.019985143 | 1 |
| Prediabetes | PP_cell | *SLC6A1* | -1.613361554 | 0.001858509 | 0.013228408 | 1 |
| Prediabetes | PP_cell | *STEAP2* | -0.337583067 | 0.009563151 | 0.032025881 | 1 |
| Prediabetes | PP_cell | *GATAD1* | -0.295788786 | 0.004301182 | 0.020046669 | 1 |
| Prediabetes | PP_cell | *SLC38A10* | -0.247237672 | 0.015857748 | 0.041316373 | 1 |
| Prediabetes | PP_cell | *AP3S2* | -0.308857626 | 0.003004847 | 0.016581525 | 1 |
| Prediabetes | PP_cell | *TNFRSF14* | -0.279252126 | 0.001395449 | 0.011470127 | 1 |
| Prediabetes | PP_cell | *PEX10* | -0.30088293 | 0.000857935 | 0.009444968 | 1 |
| Prediabetes | PP_cell | *LDLRAP1* | -0.381630874 | 0.000253836 | 0.008519859 | 0.997322368 |
| Prediabetes | PP_cell | *KRTCAP3* | -0.046766981 | 0.012907031 | 0.037703885 | 1 |
| Prediabetes | PP_cell | *PAFAH2* | -0.211415269 | 0.003434021 | 0.017799828 | 1 |
| Prediabetes | PP_cell | *EXTL1* | -0.340215285 | 0.019521622 | 0.047834311 | 1 |
| Prediabetes | PP_cell | *TRIM63* | -1.280646068 | 0.00107618 | 0.010267243 | 1 |
| Prediabetes | PP_cell | *XKR8* | -0.129205977 | 0.007163195 | 0.02684604 | 1 |
| Prediabetes | PP_cell | *CNNM4* | -0.229262963 | 0.000134786 | 0.008519859 | 0.529573471 |

| | | | | | | |
|---|---|---|---|---|---|---|
| Prediabetes | PP_cell | *ESYT3* | -0.138213229 | 0.013753474 | 0.038295695 | 1 |
| Prediabetes | PP_cell | *FAIM* | -0.317807693 | 0.014535931 | 0.039387361 | 1 |
| Prediabetes | PP_cell | *COPG2* | -0.081465041 | 0.015834263 | 0.041316373 | 1 |
| Prediabetes | PP_cell | *NPM2* | -0.097453425 | 0.00744603 | 0.027521593 | 1 |
| Prediabetes | PP_cell | *CDA* | -0.792218731 | 0.019967792 | 0.048728853 | 1 |
| Prediabetes | PP_cell | *EPB41* | -0.281890343 | 0.000907231 | 0.009499809 | 1 |
| Prediabetes | PP_cell | *MIS18A* | -0.154795982 | 0.011123797 | 0.034830535 | 1 |
| Prediabetes | PP_cell | *ALG8* | -0.584012133 | 0.0018963 | 0.013352263 | 1 |
| Prediabetes | PP_cell | *GJD2* | -0.177844821 | 0.011765844 | 0.035704358 | 1 |
| Prediabetes | PP_cell | *CLDN14* | -0.326176842 | 0.001483955 | 0.01172175 | 1 |
| Prediabetes | PP_cell | *ADPGK* | -0.375532532 | 0.004391203 | 0.020178992 | 1 |
| Prediabetes | PP_cell | *UBR1* | -0.40511296 | 0.000366305 | 0.008519859 | 1 |
| Prediabetes | PP_cell | *TEPP* | -0.577057245 | 0.010550029 | 0.033892939 | 1 |
| Prediabetes | PP_cell | *NPR2* | -0.256872709 | 0.016845717 | 0.042867114 | 1 |
| Prediabetes | PP_cell | *TMPRSS3* | -0.20872093 | 0.014498569 | 0.039384002 | 1 |
| Prediabetes | PP_cell | *SLC37A1* | -0.181535588 | 0.010157527 | 0.033119439 | 1 |
| Prediabetes | PP_cell | *NDUFV3* | -0.110299167 | 0.013834885 | 0.038387898 | 1 |
| Prediabetes | PP_cell | *PDXK* | -0.30789584 | 3.10E-05 | 0.008519859 | 0.121938146 |
| Prediabetes | PP_cell | *ZNF66* | -0.329129594 | 0.006101038 | 0.024460181 | 1 |
| Prediabetes | PP_cell | *LSS* | -0.220312242 | 0.001945669 | 0.013500582 | 1 |
| Prediabetes | PP_cell | *VAV2* | -0.456537987 | 0.000964809 | 0.009795175 | 1 |
| Prediabetes | PP_cell | *RDH13* | -0.094185674 | 0.003724614 | 0.018524062 | 1 |
| Prediabetes | PP_cell | *MPZL3* | -0.104895773 | 0.008440712 | 0.029750772 | 1 |
| Prediabetes | PP_cell | *S100A1* | -0.151637094 | 0.014070044 | 0.038726844 | 1 |
| Prediabetes | PP_cell | *SLC25A44* | -0.356895751 | 0.000877867 | 0.009444968 | 1 |
| Prediabetes | PP_cell | *CCR5* | -9.022961597 | 0.016390775 | 0.042146174 | 1 |
| Prediabetes | PP_cell | *TONSL* | -0.17788178 | 0.006400585 | 0.025198295 | 1 |
| Prediabetes | PP_cell | *PGAP3* | -0.618969884 | 0.002156857 | 0.0141015 | 1 |
| Prediabetes | PP_cell | *PRPSAP1* | -0.30359402 | 0.002410339 | 0.014960855 | 1 |
| Prediabetes | PP_cell | *SIGLEC11* | -1.329819892 | 0.006188487 | 0.024659802 | 1 |
| Prediabetes | PP_cell | *SCIMP* | -0.147090444 | 0.014422992 | 0.039243274 | 1 |
| Prediabetes | PP_cell | *AMDHD2* | -0.210147196 | 0.013912168 | 0.038493597 | 1 |
| Prediabetes | PP_cell | *ZFPL1* | -0.3010667 | 0.002568318 | 0.015428508 | 1 |
| Prediabetes | PP_cell | *TPCN2* | -0.167148687 | 0.00518166 | 0.022153146 | 1 |
| Prediabetes | PP_cell | *PDZK1IP1* | -0.046409204 | 0.015684161 | 0.041136895 | 1 |
| Prediabetes | PP_cell | *CTRC* | -0.073549283 | 0.001904738 | 0.01336916 | 1 |
| Prediabetes | PP_cell | *OMA1* | -0.13040884 | 0.01320191 | 0.037909761 | 1 |
| Prediabetes | PP_cell | *TM2D1* | -0.28473267 | 0.013629498 | 0.038253338 | 1 |
| Prediabetes | PP_cell | *FUBP1* | -0.241503782 | 0.005942614 | 0.024153447 | 1 |
| Prediabetes | PP_cell | *VCAM1* | -0.118164699 | 0.002143678 | 0.014060951 | 1 |
| Prediabetes | PP_cell | *ZNF496* | -0.490267354 | 0.000781362 | 0.009253454 | 1 |
| Prediabetes | PP_cell | *ACP6* | -0.194651299 | 0.001349289 | 0.01130353 | 1 |
| Prediabetes | PP_cell | *MSX1* | -0.285315577 | 0.000847244 | 0.009444968 | 1 |
| Prediabetes | PP_cell | *PACRGL* | -0.340697929 | 0.004405183 | 0.020219583 | 1 |
| Prediabetes | PP_cell | *TDRD10* | -1.26222529 | 0.00856096 | 0.030058993 | 1 |
| Prediabetes | PP_cell | *NPPC* | -0.154195465 | 0.013264094 | 0.037909824 | 1 |
| Prediabetes | PP_cell | *NIPAL1* | -0.150469557 | 0.003794061 | 0.018729796 | 1 |
| Prediabetes | PP_cell | *DAPL1* | -0.102930129 | 0.008284771 | 0.029511212 | 1 |
| Prediabetes | PP_cell | *ATP1A1* | -0.236638661 | 0.000506557 | 0.008519859 | 1 |
| Prediabetes | PP_cell | *SRGAP2* | -0.309264559 | 0.004467048 | 0.020304368 | 1 |

| | | | | | | |
|---|---|---|---|---|---|---|
| Prediabetes | PP_cell | *EOMES* | -0.455604047 | 0.006459421 | 0.025367051 | 1 |
| Prediabetes | PP_cell | *HDAC11* | -0.15812647 | 0.002485587 | 0.015235369 | 1 |
| Prediabetes | PP_cell | *NUAK2* | -0.186235272 | 0.003082635 | 0.016741022 | 1 |
| Prediabetes | PP_cell | *CADPS* | -0.153608526 | 0.003464121 | 0.017885059 | 1 |
| Prediabetes | PP_cell | *ATXN7* | -0.283611523 | 0.007265138 | 0.027056614 | 1 |
| Prediabetes | PP_cell | *PSMD6* | -0.251308327 | 0.005630342 | 0.023335038 | 1 |
| Prediabetes | PP_cell | *IL17RC* | -0.23030658 | 0.000183305 | 0.008519859 | 0.720205294 |
| Prediabetes | PP_cell | *CRELD1* | -0.137865727 | 0.010534855 | 0.033892939 | 1 |
| Prediabetes | PP_cell | *CXCL1* | -0.221771946 | 0.004337518 | 0.020119092 | 1 |
| Prediabetes | PP_cell | *CCDC158* | -0.203022522 | 0.004901525 | 0.02139788 | 1 |
| Prediabetes | PP_cell | *TM4SF18* | -0.09794602 | 0.012700337 | 0.03734089 | 1 |
| Prediabetes | PP_cell | *NMNAT3* | -0.348317376 | 0.002407321 | 0.014960855 | 1 |
| Prediabetes | PP_cell | *TPRA1* | -0.372691547 | 0.013699561 | 0.038295695 | 1 |
| Prediabetes | PP_cell | *MEAF6* | -0.26328916 | 0.003854313 | 0.018923659 | 1 |
| Prediabetes | PP_cell | *RPN1* | -0.271597938 | 0.003657198 | 0.01839612 | 1 |
| Prediabetes | PP_cell | *RFC4* | -0.214867017 | 0.000917452 | 0.009561452 | 1 |
| Prediabetes | PP_cell | *TKT* | -0.279544447 | 0.000486055 | 0.008519859 | 1 |
| Prediabetes | PP_cell | *UVSSA* | -0.18290714 | 0.020494107 | 0.04961266 | 1 |
| Prediabetes | PP_cell | *ARHGEF3* | -0.293718433 | 0.015767817 | 0.04125534 | 1 |
| Prediabetes | PP_cell | *LRPAP1* | -0.171049701 | 0.00835299 | 0.029619945 | 1 |
| Prediabetes | PP_cell | *PIGX* | -0.360191215 | 0.001265876 | 0.011077124 | 1 |
| Prediabetes | PP_cell | *EXO5* | -0.258864672 | 0.016860222 | 0.042870225 | 1 |
| Prediabetes | PP_cell | *ERMAP* | -0.205523871 | 0.006996912 | 0.026553791 | 1 |
| Prediabetes | PP_cell | *CEP44* | -0.286665771 | 0.012713146 | 0.03734089 | 1 |
| Prediabetes | PP_cell | *ARFIP1* | -0.337688285 | 0.001748197 | 0.012855972 | 1 |
| Prediabetes | PP_cell | *ITGA2* | -0.149877493 | 0.003077722 | 0.016741022 | 1 |
| Prediabetes | PP_cell | *STARD4* | -0.330214984 | 0.010360873 | 0.033559662 | 1 |
| Prediabetes | PP_cell | *PRRC1* | -0.269180389 | 0.012407859 | 0.036654496 | 1 |
| Prediabetes | PP_cell | *WDR41* | -0.202203401 | 0.002903362 | 0.016437045 | 1 |
| Prediabetes | PP_cell | *SERINC5* | -0.449747069 | 0.000331056 | 0.008519859 | 1 |
| Prediabetes | PP_cell | *UTP15* | -0.483716884 | 0.00032872 | 0.008519859 | 1 |
| Prediabetes | PP_cell | *NSA2* | -0.167302686 | 0.016235772 | 0.041846259 | 1 |
| Prediabetes | PP_cell | *DACT2* | -0.200616588 | 0.010057319 | 0.03290192 | 1 |
| Prediabetes | PP_cell | *PDSS2* | -0.403758484 | 0.001138454 | 0.01057443 | 1 |
| Prediabetes | PP_cell | *STK17A* | -0.360150283 | 6.23E-05 | 0.008519859 | 0.244854228 |
| Prediabetes | PP_cell | *GALNT10* | -0.267793385 | 0.013247672 | 0.037909761 | 1 |
| Prediabetes | PP_cell | *COG5* | -0.358450935 | 0.016009079 | 0.041435884 | 1 |
| Prediabetes | PP_cell | *SLU7* | -0.233126552 | 0.015560117 | 0.040920818 | 1 |
| Prediabetes | PP_cell | *STEAP1* | -0.29221506 | 0.020442655 | 0.049518615 | 1 |
| Prediabetes | PP_cell | *CHMP4C* | -0.091079561 | 0.004896852 | 0.02139788 | 1 |
| Prediabetes | PP_cell | *LMTK2* | -0.348728802 | 4.43E-05 | 0.008519859 | 0.174119503 |
| Prediabetes | PP_cell | *PEX2* | -0.341241313 | 0.001662334 | 0.012440588 | 1 |
| Prediabetes | PP_cell | *CSMD3* | -4.605035106 | 0.013038594 | 0.037891004 | 1 |
| Prediabetes | PP_cell | *OSGIN2* | -0.359510925 | 0.000364308 | 0.008519859 | 1 |
| Prediabetes | PP_cell | *DEFB1* | -0.057381196 | 0.004796994 | 0.0211057 | 1 |
| Prediabetes | PP_cell | *INTS1* | -0.417212517 | 0.002880069 | 0.016352298 | 1 |
| Prediabetes | PP_cell | *YWHAZ* | -0.184220581 | 0.015877222 | 0.041331289 | 1 |
| Prediabetes | PP_cell | *GEM* | -0.415870445 | 0.000168679 | 0.008519859 | 0.662738319 |
| Prediabetes | PP_cell | *RPP25L* | -0.539517802 | 0.000274721 | 0.008519859 | 1 |
| Prediabetes | PP_cell | *FAM219A* | -0.341915373 | 0.016791907 | 0.042757876 | 1 |

| | | | | | | |
|---|---|---|---|---|---|---|
| Prediabetes | PP_cell | *NUDT2* | -0.195598348 | 0.008328022 | 0.029611582 | 1 |
| Prediabetes | PP_cell | *METTL2B* | -0.153641149 | 0.017699065 | 0.044405892 | 1 |
| Prediabetes | PP_cell | *MAMDC2* | -0.365499288 | 0.005100566 | 0.021949753 | 1 |
| Prediabetes | PP_cell | *PRSS37* | -37.8743355 | 0.01316267 | 0.037909761 | 1 |
| Prediabetes | PP_cell | *C9orf64* | -0.436639778 | 0.000488876 | 0.008519859 | 1 |
| Prediabetes | PP_cell | *CLDN3* | -0.089804988 | 0.003752864 | 0.018617428 | 1 |
| Prediabetes | PP_cell | *WNK2* | -0.084836682 | 0.00975793 | 0.032490598 | 1 |
| Prediabetes | PP_cell | *NOL6* | -0.50169353 | 0.000478078 | 0.008519859 | 1 |
| Prediabetes | PP_cell | *ARHGAP12* | -0.37020719 | 0.001126194 | 0.010535274 | 1 |
| Prediabetes | PP_cell | *WRN* | -0.400546003 | 0.003668699 | 0.01839612 | 1 |
| Prediabetes | PP_cell | *CFL2* | -0.273901896 | 0.006333101 | 0.025032951 | 1 |
| Prediabetes | PP_cell | *INPPL1* | -0.475530472 | 0.000601927 | 0.008645299 | 1 |
| Prediabetes | PP_cell | *NGB* | 0.787414796 | 0.01928087 | 0.047346586 | 1 |
| Prediabetes | PP_cell | *VDAC2* | -0.26008519 | 0.001292528 | 0.011111559 | 1 |
| Prediabetes | PP_cell | *SNAPC4* | -0.19971427 | 0.00349204 | 0.018005545 | 1 |
| Prediabetes | PP_cell | *TSC1* | -0.268670285 | 0.000605958 | 0.008645299 | 1 |
| Prediabetes | PP_cell | *ZMYND19* | -0.444326973 | 0.005562669 | 0.023167203 | 1 |
| Prediabetes | PP_cell | *METTL3* | -0.456932798 | 0.004367927 | 0.020119092 | 1 |
| Prediabetes | PP_cell | *SLC39A13* | -0.433827166 | 0.000461916 | 0.008519859 | 1 |
| Prediabetes | PP_cell | *IFI27L1* | -0.285779778 | 0.00013096 | 0.008519859 | 0.514542325 |
| Prediabetes | PP_cell | *LIPC* | -0.094870198 | 0.013647396 | 0.038273105 | 1 |
| Prediabetes | PP_cell | *CEP57* | -0.306417807 | 0.000643907 | 0.008645299 | 1 |
| Prediabetes | PP_cell | *BTRC* | -0.23508889 | 0.002814977 | 0.016099046 | 1 |
| Prediabetes | PP_cell | *ZNF319* | -0.164338941 | 0.005093588 | 0.02194376 | 1 |
| Prediabetes | PP_cell | *SGPL1* | -0.159809436 | 0.007296213 | 0.027120926 | 1 |
| Prediabetes | PP_cell | *FRS2* | -0.260415618 | 0.000482711 | 0.008519859 | 1 |
| Prediabetes | PP_cell | *PCBD1* | -0.158147072 | 0.001894882 | 0.013352263 | 1 |
| Prediabetes | PP_cell | *COX11* | -0.35653798 | 0.000277124 | 0.008519859 | 1 |
| Prediabetes | PP_cell | *ZNF202* | -0.637329125 | 0.000709084 | 0.008816433 | 1 |
| Prediabetes | PP_cell | *WBP1L* | -0.173361979 | 0.006613098 | 0.0257511 | 1 |
| Prediabetes | PP_cell | *TUB* | -0.16215936 | 0.01889641 | 0.046769771 | 1 |
| Prediabetes | PP_cell | *RIC3* | -0.315314883 | 0.000384659 | 0.008519859 | 1 |
| Prediabetes | PP_cell | *XRRA1* | -0.179568741 | 0.007006991 | 0.026553791 | 1 |
| Prediabetes | PP_cell | *TRIM66* | -0.126975138 | 0.010792866 | 0.034287057 | 1 |
| Prediabetes | PP_cell | *ZNF143* | -0.417111425 | 0.000317448 | 0.008519859 | 1 |
| Prediabetes | PP_cell | *SPDYE6* | -0.219822995 | 0.014425703 | 0.039243274 | 1 |
| Prediabetes | PP_cell | *ATF7IP2* | -0.264305748 | 0.00070006 | 0.008813447 | 1 |
| Prediabetes | PP_cell | *COG1* | -0.404596174 | 0.000287696 | 0.008519859 | 1 |
| Prediabetes | PP_cell | *AP1G1* | -0.262501941 | 0.004319847 | 0.020109809 | 1 |
| Prediabetes | PP_cell | *SLFN5* | -0.343706423 | 0.004363902 | 0.020119092 | 1 |
| Prediabetes | PP_cell | *SCNN1G* | -0.586121492 | 0.014601478 | 0.039474478 | 1 |
| Prediabetes | PP_cell | *STAT6* | -0.372197198 | 0.004344272 | 0.020119092 | 1 |
| Prediabetes | PP_cell | *TSC22D4* | -0.262348898 | 0.00054131 | 0.008519859 | 1 |
| Prediabetes | PP_cell | *URM1* | -0.263560283 | 0.00284568 | 0.016227394 | 1 |
| Prediabetes | PP_cell | *CERCAM* | -0.119223131 | 0.015360873 | 0.040614315 | 1 |
| Prediabetes | PP_cell | *DOLPP1* | -0.273008735 | 0.012643981 | 0.037212134 | 1 |
| Prediabetes | PP_cell | *ENDOG* | -0.292355447 | 0.005089073 | 0.02194376 | 1 |
| Prediabetes | PP_cell | *PRR15L* | -0.105693211 | 0.007045085 | 0.026639866 | 1 |
| Prediabetes | PP_cell | *ACAA2* | -0.273830278 | 0.000524944 | 0.008519859 | 1 |
| Prediabetes | PP_cell | *ZNF229* | -0.102310396 | 0.008178491 | 0.029327769 | 1 |

| | | | | | | |
|---|---|---|---|---|---|---|
| Prediabetes | PP_cell | *PROCA1* | -0.393074975 | 0.011786213 | 0.035704358 | 1 |
| Prediabetes | PP_cell | *TP53I13* | -0.153200371 | 0.003963576 | 0.019131314 | 1 |
| Prediabetes | PP_cell | *ZNF528* | -0.390569661 | 0.00033076 | 0.008519859 | 1 |
| Prediabetes | PP_cell | *TMC4* | -0.127254241 | 0.002136966 | 0.014043725 | 1 |
| Prediabetes | PP_cell | *C19orf33* | -0.130776385 | 0.005566261 | 0.023167203 | 1 |
| Prediabetes | PP_cell | *UBXN6* | -0.248215259 | 0.010749299 | 0.034253037 | 1 |
| Prediabetes | PP_cell | *KIFC2* | -0.160300493 | 0.020030538 | 0.048760833 | 1 |
| Prediabetes | PP_cell | *TRPV3* | -0.309097212 | 0.018939122 | 0.046829333 | 1 |
| Prediabetes | PP_cell | *NDUFV1* | -0.289941953 | 0.005600866 | 0.023253761 | 1 |
| Prediabetes | PP_cell | *HID1* | -0.364442654 | 0.002284327 | 0.014499386 | 1 |
| Prediabetes | PP_cell | *TMEM68* | -0.35673063 | 0.000536699 | 0.008519859 | 1 |
| Prediabetes | PP_cell | *MLST8* | -0.286633832 | 0.000483953 | 0.008519859 | 1 |
| Prediabetes | PP_cell | *ECI1* | -0.338697643 | 0.000877231 | 0.009444968 | 1 |
| Prediabetes | PP_cell | *ABCA3* | -0.193294244 | 0.009504488 | 0.031971859 | 1 |
| Prediabetes | PP_cell | *RPSA* | -0.125459286 | 0.019339298 | 0.047460401 | 1 |
| Prediabetes | PP_cell | *ENTPD3* | -0.118431387 | 0.000607852 | 0.008645299 | 1 |
| Prediabetes | PP_cell | *LTBP3* | -0.200222739 | 0.00107925 | 0.010267243 | 1 |
| Prediabetes | PP_cell | *SAC3D1* | -0.402900251 | 0.0165193 | 0.042268461 | 1 |
| Prediabetes | PP_cell | *CCDC88B* | -0.072426581 | 0.01851558 | 0.046042858 | 1 |
| Prediabetes | PP_cell | *PBK* | -0.145464863 | 0.01597193 | 0.041421592 | 1 |
| Prediabetes | PP_cell | *FAM83B* | -0.079298279 | 0.009863997 | 0.032714383 | 1 |
| Prediabetes | PP_cell | *LMBRD1* | -0.228701962 | 0.017380805 | 0.043823045 | 1 |
| Prediabetes | PP_cell | *TTC39C* | -0.119787564 | 0.001991362 | 0.013597804 | 1 |
| Prediabetes | PP_cell | *NT5DC2* | -0.128230722 | 0.00401655 | 0.019268653 | 1 |
| Prediabetes | PP_cell | *COA6* | -0.423803049 | 0.000471311 | 0.008519859 | 1 |
| Prediabetes | PP_cell | *ING5* | -0.432807523 | 0.001433935 | 0.011622813 | 1 |
| Prediabetes | PP_cell | *HEXIM2* | -0.218026761 | 0.004243163 | 0.019989673 | 1 |
| Prediabetes | PP_cell | *SERINC2* | -0.067814196 | 0.005755578 | 0.023679231 | 1 |
| Prediabetes | PP_cell | *TRAPPC11* | -0.252763817 | 0.001408848 | 0.011508037 | 1 |
| Prediabetes | PP_cell | *COL3A1* | -0.216284184 | 0.020013435 | 0.048749401 | 1 |
| Prediabetes | PP_cell | *ING2* | -0.205323839 | 0.000908126 | 0.009499809 | 1 |
| Prediabetes | PP_cell | *ZSWIM1* | -0.171875728 | 0.014287272 | 0.039118253 | 1 |
| Prediabetes | PP_cell | *ZNF30* | -0.150556245 | 0.005604813 | 0.023253761 | 1 |
| Prediabetes | PP_cell | *SLC16A4* | -0.219516354 | 0.003202462 | 0.01718917 | 1 |
| Prediabetes | PP_cell | *PPIP5K1* | -0.076699727 | 0.006497716 | 0.02547829 | 1 |
| Prediabetes | PP_cell | *NSG1* | -0.050641351 | 0.01313256 | 0.037909761 | 1 |
| Prediabetes | PP_cell | *FSTL5* | -0.170634554 | 0.001497323 | 0.011789544 | 1 |
| Prediabetes | PP_cell | *CTRB1* | -0.071501293 | 0.001910056 | 0.01336916 | 1 |
| Prediabetes | PP_cell | *CTRB2* | -0.063450389 | 0.013570538 | 0.038249975 | 1 |
| Prediabetes | PP_cell | *MFF* | -0.273768845 | 0.004073197 | 0.019421835 | 1 |
| Prediabetes | PP_cell | *XPO6* | -0.319600997 | 0.003442558 | 0.017820568 | 1 |
| Prediabetes | PP_cell | *PRELID1* | -0.175858792 | 0.020724239 | 0.049888972 | 1 |
| Prediabetes | PP_cell | *CXCL10* | -11.68687263 | 0.013196877 | 0.037909761 | 1 |
| Prediabetes | PP_cell | *NMD3* | -0.329751793 | 0.000366803 | 0.008519859 | 1 |
| Prediabetes | PP_cell | *SNUPN* | -0.492024992 | 0.000386875 | 0.008519859 | 1 |
| Prediabetes | PP_cell | *CRADD* | -0.293434534 | 0.017282497 | 0.043667479 | 1 |
| Prediabetes | PP_cell | *SDC2* | -0.171363683 | 0.000456459 | 0.008519859 | 1 |
| Prediabetes | PP_cell | *CLIC4* | -0.320547605 | 0.007109625 | 0.026721477 | 1 |
| Prediabetes | PP_cell | *AGPAT2* | -0.205501339 | 0.002693373 | 0.0157471 | 1 |
| Prediabetes | PP_cell | *MT1E* | -0.077974438 | 0.001806423 | 0.013030484 | 1 |

| | | | | | | |
|---|---|---|---|---|---|---|
| Prediabetes | PP_cell | *LDB2* | -0.179985895 | 0.002797347 | 0.016068387 | 1 |
| Prediabetes | PP_cell | *RAC3* | -0.295080514 | 0.002655085 | 0.01561651 | 1 |
| Prediabetes | PP_cell | *AVEN* | -0.461623465 | 0.000386561 | 0.008519859 | 1 |
| Prediabetes | PP_cell | *CTNND2* | -0.063311151 | 0.009193363 | 0.031382034 | 1 |
| Prediabetes | PP_cell | *WNT10B* | -4.015588291 | 0.013833006 | 0.038387898 | 1 |
| Prediabetes | PP_cell | *OTUD3* | -0.315658055 | 0.000119774 | 0.008519859 | 0.470590606 |
| Prediabetes | PP_cell | *PUSL1* | -0.263004343 | 0.005512534 | 0.023090348 | 1 |
| Prediabetes | PP_cell | *MYRIP* | -0.186058609 | 0.009328488 | 0.031675407 | 1 |
| Prediabetes | PP_cell | *CNTROB* | -0.184636575 | 0.003987835 | 0.019154283 | 1 |
| Prediabetes | PP_cell | *KCNAB3* | 0.096110556 | 0.019084687 | 0.047089646 | 1 |
| Prediabetes | PP_cell | *FAM153A* | -0.187697283 | 0.019552533 | 0.047834311 | 1 |
| Prediabetes | PP_cell | *ZNF778* | -0.288916333 | 0.002518858 | 0.015295049 | 1 |
| Prediabetes | PP_cell | *CST2* | -0.240898465 | 0.006889779 | 0.026358267 | 1 |
| Prediabetes | PP_cell | *KRT8* | -0.062509399 | 0.005443143 | 0.022930133 | 1 |
| Prediabetes | PP_cell | *MGMT* | -0.328520434 | 0.007218334 | 0.026907811 | 1 |
| Prediabetes | PP_cell | *NPAS2* | -0.391847029 | 0.000159895 | 0.008519859 | 0.628228698 |
| Prediabetes | PP_cell | *NUDT9* | -0.147212533 | 0.019624586 | 0.047921067 | 1 |
| Prediabetes | PP_cell | *SMAGP* | -0.179589961 | 0.010898286 | 0.03440649 | 1 |
| Prediabetes | PP_cell | *IRX1* | -0.098017876 | 0.013026637 | 0.037884276 | 1 |
| Prediabetes | PP_cell | *CDH2* | -0.071633646 | 0.010255096 | 0.033326942 | 1 |
| Prediabetes | PP_cell | *ACYP2* | -0.298513281 | 0.000811023 | 0.009399738 | 1 |
| Prediabetes | PP_cell | *FOXN2* | -0.450304209 | 0.000282936 | 0.008519859 | 1 |
| Prediabetes | PP_cell | *MTSS1* | -0.290899007 | 0.000904106 | 0.009499809 | 1 |
| Prediabetes | PP_cell | *NDUFA3* | -0.293381719 | 0.0023158 | 0.014602893 | 1 |
| Prediabetes | PP_cell | *TSNARE1* | -0.124466854 | 0.004524633 | 0.020366067 | 1 |
| Prediabetes | PP_cell | *SOX7* | -0.150071677 | 0.007859387 | 0.02853931 | 1 |
| Prediabetes | PP_cell | *TRMT61B* | -0.188475982 | 0.002605176 | 0.01555583 | 1 |
| Prediabetes | PP_cell | *MFN1* | -0.262204811 | 0.000430193 | 0.008519859 | 1 |
| Prediabetes | PP_cell | *KCNMB3* | -0.371294607 | 0.020427738 | 0.049513007 | 1 |
| Prediabetes | PP_cell | *ATP6V0E2* | -0.110230108 | 0.00806951 | 0.029087251 | 1 |
| Prediabetes | PP_cell | *JAGN1* | -0.306334231 | 0.000317842 | 0.008519859 | 1 |
| Prediabetes | PP_cell | *MORN4* | -0.126817217 | 0.00496816 | 0.021664708 | 1 |
| Prediabetes | PP_cell | *TRIM8* | -0.250832799 | 0.012316123 | 0.036438288 | 1 |
| Prediabetes | PP_cell | *EXOSC1* | -0.289141454 | 0.009726676 | 0.032414002 | 1 |
| Prediabetes | PP_cell | *ZNF561* | -0.360720586 | 0.000742939 | 0.009121901 | 1 |
| Prediabetes | PP_cell | *PTGER4* | -0.183978045 | 0.004055674 | 0.019361778 | 1 |
| Prediabetes | PP_cell | *NMUR1* | -0.738748245 | 0.013148248 | 0.037909761 | 1 |
| Prediabetes | PP_cell | *PIK3CD* | -0.439054917 | 0.002213942 | 0.014319626 | 1 |
| Prediabetes | PP_cell | *ENC1* | -0.130333789 | 0.006612558 | 0.0257511 | 1 |
| Prediabetes | PP_cell | *RGS19* | -0.201869107 | 0.003677966 | 0.018402303 | 1 |
| Prediabetes | PP_cell | *ANO5* | -0.141121718 | 0.010521928 | 0.033892939 | 1 |
| Prediabetes | PP_cell | *GPHN* | -0.391237954 | 0.003520403 | 0.018080605 | 1 |
| Prediabetes | PP_cell | *VAT1L* | -0.113435176 | 0.005684859 | 0.02351138 | 1 |
| Prediabetes | PP_cell | *LGALS4* | -0.040610148 | 0.00822659 | 0.029383885 | 1 |
| Prediabetes | PP_cell | *SPATA5L1* | -0.26055312 | 0.000415994 | 0.008519859 | 1 |
| Prediabetes | PP_cell | *CTPS1* | -0.182683602 | 0.008022575 | 0.028971229 | 1 |
| Prediabetes | PP_cell | *PWWP2B* | -0.191875441 | 0.001981505 | 0.013597804 | 1 |
| Prediabetes | PP_cell | *TVP23B* | -0.218860082 | 0.000878229 | 0.009444968 | 1 |
| Prediabetes | PP_cell | *SRGAP2C* | -0.356095137 | 0.01007632 | 0.032936658 | 1 |
| Prediabetes | PP_cell | *ZNF556* | -0.108404035 | 0.01550937 | 0.040814678 | 1 |

| | | | | | | |
|---|---|---|---|---|---|---|
| Prediabetes | PP_cell | *GAP43* | -0.050858358 | 0.020098499 | 0.0488355 | 1 |
| Prediabetes | PP_cell | *REG1B* | -0.071398039 | 0.002395532 | 0.014916081 | 1 |
| Prediabetes | PP_cell | *NME6* | -0.367550701 | 0.000233004 | 0.008519859 | 0.915474431 |
| Prediabetes | PP_cell | *SNTB1* | -0.170842698 | 0.007278849 | 0.027082005 | 1 |
| Prediabetes | PP_cell | *BSG* | -0.213695118 | 0.008402444 | 0.029688131 | 1 |
| Prediabetes | PP_cell | *CERS6* | -0.267488654 | 0.000817689 | 0.009434491 | 1 |
| Prediabetes | PP_cell | *SPTLC3* | -0.206741395 | 0.014399779 | 0.039235459 | 1 |
| Prediabetes | PP_cell | *SUCLG2* | -0.319253647 | 0.000456015 | 0.008519859 | 1 |
| Prediabetes | PP_cell | *CSDC2* | -0.230242075 | 0.004767041 | 0.021054489 | 1 |
| Prediabetes | PP_cell | *AFF1* | -0.297820549 | 0.006672937 | 0.025881509 | 1 |
| Prediabetes | PP_cell | *SMPDL3A* | -0.235342473 | 0.006303815 | 0.024961783 | 1 |
| Prediabetes | PP_cell | *EFEMP2* | -0.522640654 | 0.002625461 | 0.015605802 | 1 |
| Prediabetes | PP_cell | *AGAP5* | -1.352947627 | 0.000369745 | 0.008519859 | 1 |
| Prediabetes | PP_cell | *FUT10* | -0.176240984 | 0.001381014 | 0.011450342 | 1 |
| Prediabetes | PP_cell | *NAA16* | -0.241689542 | 0.009110295 | 0.031152611 | 1 |
| Prediabetes | PP_cell | *DMXL1* | -0.345988444 | 0.000566349 | 0.008521817 | 1 |
| Prediabetes | PP_cell | *NADSYN1* | -0.240339329 | 0.014956299 | 0.0399258 | 1 |
| Prediabetes | PP_cell | *TADA2B* | -0.388126826 | 0.000458619 | 0.008519859 | 1 |
| Prediabetes | PP_cell | *NOC3L* | -0.164121882 | 0.020051332 | 0.048781228 | 1 |
| Prediabetes | PP_cell | *RHOD* | -0.153819341 | 0.012587613 | 0.037074012 | 1 |
| Prediabetes | PP_cell | *VANGL1* | -0.374279332 | 0.000929091 | 0.009619487 | 1 |
| Prediabetes | PP_cell | *IQCB1* | -0.303649518 | 0.000553102 | 0.008519859 | 1 |
| Prediabetes | PP_cell | *ARV1* | -0.357433982 | 0.000260851 | 0.008519859 | 1 |
| Prediabetes | PP_cell | *PPP1R14B* | -0.207718308 | 0.007646146 | 0.027971794 | 1 |
| Prediabetes | PP_cell | *TNFRSF10D* | -0.174301203 | 0.013135207 | 0.037909761 | 1 |
| Prediabetes | PP_cell | *GMPPB* | -0.243468503 | 0.013418014 | 0.03803406 | 1 |
| Prediabetes | PP_cell | *NUDT18* | -0.556404892 | 0.000293757 | 0.008519859 | 1 |
| Prediabetes | PP_cell | *PC* | -0.098910189 | 0.014978431 | 0.0399258 | 1 |
| Prediabetes | PP_cell | *UQCRH* | -0.18471351 | 0.011620578 | 0.035697617 | 1 |
| Prediabetes | PP_cell | *HEG1* | -0.360624051 | 0.000771684 | 0.009253454 | 1 |
| Prediabetes | PP_cell | *DPY19L1* | -0.444172832 | 0.004806888 | 0.021125572 | 1 |
| Prediabetes | PP_cell | *GPR160* | -0.187271611 | 0.007113939 | 0.026721477 | 1 |
| Prediabetes | PP_cell | *C9orf131* | -0.148639134 | 0.01495222 | 0.0399258 | 1 |
| Prediabetes | PP_cell | *CTU2* | -0.315047317 | 0.002974218 | 0.016551989 | 1 |
| Prediabetes | PP_cell | *ADCY6* | -0.103400906 | 0.006203734 | 0.024670519 | 1 |
| Prediabetes | PP_cell | *RALGAPA1* | -0.265836952 | 0.001807486 | 0.013030484 | 1 |
| Prediabetes | PP_cell | *GOLT1A* | -0.120735275 | 0.003451399 | 0.017842824 | 1 |
| Prediabetes | PP_cell | *BRSK2* | -0.343341303 | 0.000513594 | 0.008519859 | 1 |
| Prediabetes | PP_cell | *BRMS1* | -0.304120603 | 0.00455465 | 0.020393929 | 1 |
| Prediabetes | PP_cell | *HRAS* | -0.290533708 | 0.002624223 | 0.015605802 | 1 |
| Prediabetes | PP_cell | *YIF1A* | -0.151542013 | 0.008635328 | 0.030190258 | 1 |
| Prediabetes | PP_cell | *NDUFA11* | -0.24359127 | 0.001561556 | 0.012006559 | 1 |
| Prediabetes | PP_cell | *PPM1E* | -0.051328568 | 0.015800094 | 0.041275644 | 1 |
| Prediabetes | PP_cell | *ZNF408* | -0.089997387 | 0.01890313 | 0.046769771 | 1 |
| Prediabetes | PP_cell | *MED16* | -0.214022498 | 0.008636765 | 0.030190258 | 1 |
| Prediabetes | PP_cell | *CHST1* | -0.135087139 | 0.013210361 | 0.037909761 | 1 |
| Prediabetes | PP_cell | *DOLK* | -0.324293339 | 6.74E-05 | 0.008519859 | 0.264886874 |
| Prediabetes | PP_cell | *ZNF519* | -0.351954981 | 0.000420381 | 0.008519859 | 1 |
| Prediabetes | PP_cell | *CLCF1* | -0.256706861 | 0.000704045 | 0.008813447 | 1 |
| Prediabetes | PP_cell | *UCP2* | -0.134880975 | 0.005559512 | 0.023167203 | 1 |

| | | | | | | |
|---|---|---|---|---|---|---|
| Prediabetes | PP_cell | *PAAF1* | -0.116057516 | 0.007190371 | 0.026854533 | 1 |
| Prediabetes | PP_cell | *TMEM70* | -0.193432202 | 0.006638748 | 0.025799843 | 1 |
| Prediabetes | PP_cell | *RPS6KB2* | -0.264852417 | 0.003077266 | 0.016741022 | 1 |
| Prediabetes | PP_cell | *RMI2* | -0.24928134 | 0.007788232 | 0.028333299 | 1 |
| Prediabetes | PP_cell | *SWI5* | -0.197454512 | 0.002506648 | 0.015269176 | 1 |
| Prediabetes | PP_cell | *ZDHHC21* | -0.349355081 | 0.00083736 | 0.009444968 | 1 |
| Prediabetes | PP_cell | *CHMP6* | -0.318742194 | 0.00098574 | 0.009817609 | 1 |
| Prediabetes | PP_cell | *DLEU1* | -0.299139103 | 0.005194847 | 0.022185385 | 1 |
| Prediabetes | PP_cell | *CCDC57* | -0.138064722 | 0.00221956 | 0.014319626 | 1 |
| Prediabetes | PP_cell | *BNIP3* | -0.221024262 | 0.001522634 | 0.011846392 | 1 |
| Prediabetes | PP_cell | *SMIM19* | -0.26674229 | 0.001468116 | 0.011646409 | 1 |
| Prediabetes | PP_cell | *SLC35G1* | -0.431421079 | 0.000564821 | 0.008521817 | 1 |
| Prediabetes | PP_cell | *COX8A* | -0.173861628 | 0.01204574 | 0.036128026 | 1 |
| Prediabetes | PP_cell | *CRLF3* | -0.205244173 | 0.00443697 | 0.020248234 | 1 |
| Prediabetes | PP_cell | *RNPEP* | -0.36158207 | 0.00364278 | 0.01839612 | 1 |
| Prediabetes | PP_cell | *DMRTA1* | -0.0594169 | 0.017837935 | 0.044697223 | 1 |
| Prediabetes | PP_cell | *WDR25* | -0.091224602 | 0.006153538 | 0.024620417 | 1 |
| Prediabetes | PP_cell | *PHLDB3* | -0.07517216 | 0.020083603 | 0.048829502 | 1 |
| Prediabetes | PP_cell | *PRR15* | -0.146474463 | 0.020007668 | 0.048749401 | 1 |
| Prediabetes | PP_cell | *RMDN1* | -0.211150297 | 0.008693136 | 0.030237155 | 1 |
| Prediabetes | PP_cell | *LRRC37A* | -0.245304333 | 0.013270031 | 0.037909824 | 1 |
| Prediabetes | PP_cell | *ACSF3* | -0.456122792 | 0.002998143 | 0.016581525 | 1 |
| Prediabetes | PP_cell | *PFN4* | 0.134334978 | 0.013583527 | 0.038249975 | 1 |
| Prediabetes | PP_cell | *VSIG10* | -0.269855445 | 0.001193745 | 0.01078212 | 1 |
| Prediabetes | PP_cell | *WSB2* | -0.192174364 | 0.006903254 | 0.026361874 | 1 |
| Prediabetes | PP_cell | *MUC20* | -0.122044027 | 0.004900667 | 0.02139788 | 1 |
| Prediabetes | PP_cell | *SHMT1* | -0.062133028 | 0.019544975 | 0.047834311 | 1 |
| Prediabetes | PP_cell | *DPP7* | -0.275929502 | 0.0004134 | 0.008519859 | 1 |
| Prediabetes | PP_cell | *DEAF1* | -0.351310413 | 0.000339618 | 0.008519859 | 1 |
| Prediabetes | PP_cell | *DSCAML1* | -0.244888081 | 0.003243018 | 0.017225057 | 1 |
| Prediabetes | PP_cell | *KCNA2* | -0.439138774 | 0.013178963 | 0.037909761 | 1 |
| Prediabetes | PP_cell | *PPFIA3* | -0.311104396 | 0.000767038 | 0.009244451 | 1 |
| Prediabetes | PP_cell | *PAWR* | -0.327859823 | 0.000402328 | 0.008519859 | 1 |
| Prediabetes | PP_cell | *ACOT4* | -0.322118078 | 0.00307745 | 0.016741022 | 1 |
| Prediabetes | PP_cell | *RBM44* | -0.164445325 | 0.013302963 | 0.0379275 | 1 |
| Prediabetes | PP_cell | *IL17RA* | -0.216634485 | 0.004715687 | 0.020888312 | 1 |
| Prediabetes | PP_cell | *AGTRAP* | -0.161338583 | 0.00158219 | 0.012117789 | 1 |
| Prediabetes | PP_cell | *THAP5* | -0.357862331 | 0.00064102 | 0.008645299 | 1 |
| Prediabetes | PP_cell | *CD151* | -0.292466464 | 0.000526236 | 0.008519859 | 1 |
| Prediabetes | PP_cell | *CHID1* | -0.240878205 | 0.000420914 | 0.008519859 | 1 |
| Prediabetes | PP_cell | *ZBTB41* | -0.370163284 | 0.000557841 | 0.008521817 | 1 |
| Prediabetes | PP_cell | *BET1L* | -0.225269502 | 0.000906624 | 0.009499809 | 1 |
| Prediabetes | PP_cell | *GPR150* | -0.081567086 | 0.009569454 | 0.032025881 | 1 |
| Prediabetes | PP_cell | *ALS2CL* | -0.119380449 | 0.01135421 | 0.035126529 | 1 |
| Prediabetes | PP_cell | *STAP2* | -0.138332858 | 0.011522819 | 0.035480529 | 1 |
| Prediabetes | PP_cell | *DALRD3* | -0.254700523 | 0.005785771 | 0.023775968 | 1 |
| Prediabetes | PP_cell | *ZNF114* | -0.179168624 | 0.014729203 | 0.039627472 | 1 |
| Prediabetes | PP_cell | *WDR6* | -0.181002824 | 0.00646282 | 0.025367051 | 1 |
| Prediabetes | PP_cell | *NT5DC1* | -0.265656263 | 0.000866474 | 0.009444968 | 1 |
| Prediabetes | PP_cell | *P4HTM* | -0.164799882 | 0.003619864 | 0.018375251 | 1 |

| | | | | | | |
|---|---|---|---|---|---|---|
| Prediabetes | PP_cell | *ERBB4* | -0.231897798 | 0.016868763 | 0.042870225 | 1 |
| Prediabetes | PP_cell | *ERN1* | -0.239032914 | 0.013201016 | 0.037909761 | 1 |
| Prediabetes | PP_cell | *RPP25* | -0.267667257 | 0.000524685 | 0.008519859 | 1 |
| Prediabetes | PP_cell | *GAK* | -0.255043595 | 0.005460648 | 0.022946401 | 1 |
| Prediabetes | PP_cell | *IGSF22* | -0.126734175 | 0.014200578 | 0.03896234 | 1 |
| Prediabetes | PP_cell | *FARSA* | -0.155300604 | 0.003353594 | 0.017521636 | 1 |
| Prediabetes | PP_cell | *SPTY2D1* | -0.349864234 | 0.001537386 | 0.011877462 | 1 |
| Prediabetes | PP_cell | *FUCA1* | -0.230628762 | 0.000640853 | 0.008645299 | 1 |
| Prediabetes | PP_cell | *HLA-DQB1* | -0.071420427 | 0.014784146 | 0.039649766 | 1 |
| Prediabetes | PP_cell | *ELMOD2* | -0.345621146 | 0.000284041 | 0.008519859 | 1 |
| Prediabetes | PP_cell | *DNHD1* | -0.138093488 | 0.005426013 | 0.022930133 | 1 |
| Prediabetes | PP_cell | *ZBTB42* | -0.488388859 | 0.005400502 | 0.022930133 | 1 |
| Prediabetes | PP_cell | *RPRML* | -0.195358505 | 0.001405397 | 0.011503757 | 1 |
| Prediabetes | PP_cell | *SYCN* | -0.047537571 | 0.015120751 | 0.040139539 | 1 |
| Prediabetes | PP_cell | *CITED4* | -0.210016555 | 0.001651687 | 0.012440588 | 1 |
| Prediabetes | PP_cell | *SSC5D* | -0.088384791 | 0.01227556 | 0.036372141 | 1 |
| Prediabetes | PP_cell | *ANKRD18A* | -0.0659255 | 0.004541312 | 0.020368512 | 1 |
| Prediabetes | PP_cell | *TH* | -0.345294388 | 0.016673652 | 0.042539466 | 1 |
| Prediabetes | PP_cell | *ZNF816* | -0.386000238 | 0.002649621 | 0.01561651 | 1 |
| Prediabetes | PP_cell | *MCFD2* | -0.261127072 | 0.002355105 | 0.014710984 | 1 |
| Prediabetes | PP_cell | *TSPYL5* | -0.164471733 | 0.002178426 | 0.014194086 | 1 |
| Prediabetes | PP_cell | *CHST13* | -5.572960075 | 0.013018676 | 0.037884276 | 1 |
| Prediabetes | PP_cell | *PPA1* | -0.253761425 | 0.000257345 | 0.008519859 | 1 |
| Prediabetes | PP_cell | *MAP6D1* | -0.357448059 | 0.005410479 | 0.022930133 | 1 |
| Prediabetes | PP_cell | *D2HGDH* | -0.123087411 | 0.002554351 | 0.01537007 | 1 |
| Prediabetes | PP_cell | *ZFP82* | -0.191469917 | 0.00188739 | 0.013337331 | 1 |
| Prediabetes | PP_cell | *RPH3AL* | -0.277364043 | 0.000384165 | 0.008519859 | 1 |
| Prediabetes | PP_cell | *HIGD1A* | -0.275909789 | 0.000299729 | 0.008519859 | 1 |
| Prediabetes | PP_cell | *OGFOD3* | -0.213439009 | 0.000661963 | 0.008757086 | 1 |
| Prediabetes | PP_cell | *ZNF467* | -0.408175768 | 0.001454921 | 0.011646409 | 1 |
| Prediabetes | PP_cell | *ZNF678* | -0.124871061 | 0.011378254 | 0.035145567 | 1 |
| Prediabetes | PP_cell | *TMEM45A* | -0.533494159 | 0.002077887 | 0.013904889 | 1 |
| Prediabetes | PP_cell | *ACBD4* | -0.856876819 | 0.008579359 | 0.030075341 | 1 |
| Prediabetes | PP_cell | *SGSH* | -0.248819769 | 0.013709599 | 0.038295695 | 1 |
| Prediabetes | PP_cell | *GPR135* | -0.192250421 | 0.003897996 | 0.019001519 | 1 |
| Prediabetes | PP_cell | *GPR88* | -1.714165873 | 0.001378662 | 0.011450342 | 1 |
| Prediabetes | PP_cell | *ZNF329* | -0.2859448 | 0.008272779 | 0.029495235 | 1 |
| Prediabetes | PP_cell | *IDH2* | -0.29872328 | 0.000630091 | 0.008645299 | 1 |
| Prediabetes | PP_cell | *ERCC6L2* | -0.432303062 | 0.0009484 | 0.009678612 | 1 |
| Prediabetes | PP_cell | *TP53TG1* | -0.313419883 | 0.000205044 | 0.008519859 | 0.805618528 |
| Prediabetes | PP_cell | *ARL6IP4* | -0.168764453 | 0.012758333 | 0.037436514 | 1 |
| Prediabetes | PP_cell | *FAM153B* | -0.169921046 | 0.01774465 | 0.04449185 | 1 |
| Prediabetes | PP_cell | *BACE2* | -0.36226316 | 0.001002908 | 0.009851066 | 1 |
| Prediabetes | PP_cell | *ZNF804B* | -4.933063687 | 0.011812815 | 0.035704358 | 1 |
| Prediabetes | PP_cell | *NPLOC4* | -0.316691278 | 0.002113578 | 0.013980215 | 1 |
| Prediabetes | PP_cell | *SATB1* | -0.374587456 | 0.001241372 | 0.010970459 | 1 |
| Prediabetes | PP_cell | *ACTL10* | -0.121210327 | 0.00990717 | 0.032720527 | 1 |
| Prediabetes | PP_cell | *SKA2* | -0.270106281 | 0.008100505 | 0.029172212 | 1 |
| Prediabetes | PP_cell | *ANXA2* | -0.256035159 | 0.000220395 | 0.008519859 | 0.86593054 |
| Prediabetes | PP_cell | *GPIHBP1* | -0.602707173 | 0.013425666 | 0.03803406 | 1 |

| | | | | | | |
|---|---|---|---|---|---|---|
| Prediabetes | PP_cell | *ALG12* | -0.164218941 | 0.009546314 | 0.032025881 | 1 |
| Prediabetes | PP_cell | *SPATA13* | -0.412994517 | 0.001598629 | 0.012172507 | 1 |
| Prediabetes | PP_cell | *SLC25A21* | -0.281562994 | 0.008541674 | 0.0300181 | 1 |
| Prediabetes | PP_cell | *LYSMD4* | -0.30448249 | 0.004536495 | 0.020368512 | 1 |
| Prediabetes | PP_cell | *BEGAIN* | -0.274996781 | 0.000752448 | 0.009209872 | 1 |
| Prediabetes | PP_cell | *GPC6* | -0.342337475 | 0.000947968 | 0.009678612 | 1 |
| Prediabetes | PP_cell | *FANCF* | -0.12152023 | 0.003509454 | 0.018071617 | 1 |
| Prediabetes | PP_cell | *CALN1* | -0.47891947 | 0.000844758 | 0.009444968 | 1 |
| Prediabetes | PP_cell | *MX2* | -0.163292999 | 0.017627527 | 0.044282962 | 1 |
| Prediabetes | PP_cell | *SFXN4* | -0.204578156 | 0.002857963 | 0.016251909 | 1 |
| Prediabetes | PP_cell | *MRPL54* | -0.255500176 | 0.000684671 | 0.008780065 | 1 |
| Prediabetes | PP_cell | *MACC1* | -0.054556201 | 0.010812335 | 0.034287057 | 1 |
| Prediabetes | PP_cell | *KREMEN1* | -0.16584698 | 0.004049867 | 0.019357575 | 1 |
| Prediabetes | PP_cell | *NUDT14* | -0.214168861 | 0.002157038 | 0.0141015 | 1 |
| Prediabetes | PP_cell | *TMEM236* | -0.092816174 | 0.013351225 | 0.037957281 | 1 |
| Prediabetes | PP_cell | *UQCR10* | -0.191410944 | 0.005061405 | 0.02194376 | 1 |
| Prediabetes | PP_cell | *CRELD2* | -0.19159081 | 0.000366723 | 0.008519859 | 1 |
| Prediabetes | PP_cell | *UBE2F* | -0.355172659 | 0.000178739 | 0.008519859 | 0.702263708 |
| Prediabetes | PP_cell | *OAF* | -0.2652699 | 0.015036635 | 0.039999281 | 1 |
| Prediabetes | PP_cell | *TM2D3* | -0.341436114 | 0.000620978 | 0.008645299 | 1 |
| Prediabetes | PP_cell | *COPB2* | -0.279256213 | 0.004566755 | 0.020408359 | 1 |
| Prediabetes | PP_cell | *LRRC19* | 0.113067768 | 0.013707441 | 0.038295695 | 1 |
| Prediabetes | PP_cell | *CCR10* | -0.279505051 | 0.000657143 | 0.008752256 | 1 |
| Prediabetes | PP_cell | *NCMAP* | -0.158087084 | 0.004784829 | 0.021075778 | 1 |
| Prediabetes | PP_cell | *WDR27* | -0.209693003 | 0.004606387 | 0.020543126 | 1 |
| Prediabetes | PP_cell | *ZFP1* | -0.419712727 | 0.008647702 | 0.030201619 | 1 |
| Prediabetes | PP_cell | *XPOT* | -0.306241166 | 0.000714306 | 0.008853338 | 1 |
| Prediabetes | PP_cell | *C14orf180* | -6.923210078 | 0.013247502 | 0.037909761 | 1 |
| Prediabetes | PP_cell | *ZBTB40* | -0.258973739 | 0.001939602 | 0.013500582 | 1 |
| Prediabetes | PP_cell | *LCN12* | -0.16416116 | 0.00423522 | 0.019985143 | 1 |
| Prediabetes | PP_cell | *MUC6* | -0.043352977 | 0.011363761 | 0.035128417 | 1 |
| Prediabetes | PP_cell | *BRI3BP* | -0.376111411 | 0.000425792 | 0.008519859 | 1 |
| Prediabetes | PP_cell | *UBOX5* | -0.351931069 | 0.016069157 | 0.041521635 | 1 |
| Prediabetes | PP_cell | *CIB1* | -0.274908828 | 0.000478996 | 0.008519859 | 1 |
| Prediabetes | PP_cell | *INPP5J* | -0.135873718 | 0.010156611 | 0.033119439 | 1 |
| Prediabetes | PP_cell | *MFSD6L* | -0.109969973 | 0.01071127 | 0.03417498 | 1 |
| Prediabetes | PP_cell | *ZNF445* | -0.298529505 | 0.00181706 | 0.013073736 | 1 |
| Prediabetes | PP_cell | *ZNF74* | -0.102351057 | 0.012536746 | 0.037006165 | 1 |
| Prediabetes | PP_cell | *RBM11* | -0.174098218 | 0.001734984 | 0.012837572 | 1 |
| Prediabetes | PP_cell | *ATP6V0A2* | -0.365894472 | 0.003517251 | 0.018080605 | 1 |
| Prediabetes | PP_cell | *RAD51D* | -0.153624858 | 0.00411127 | 0.019508673 | 1 |
| Prediabetes | PP_cell | *GPRIN3* | -0.310648509 | 0.000552992 | 0.008519859 | 1 |
| Prediabetes | PP_cell | *MUC1* | -0.044783848 | 0.0121436 | 0.036255474 | 1 |
| Prediabetes | PP_cell | *IRF7* | -0.368515236 | 0.000670063 | 0.008780065 | 1 |
| Prediabetes | PP_cell | *FAM131C* | -0.240365054 | 0.017536222 | 0.044109998 | 1 |
| Prediabetes | PP_cell | *LSAMP* | -0.107736753 | 0.00433322 | 0.020119092 | 1 |
| Prediabetes | PP_cell | *PDIA2* | -0.044771549 | 0.012068011 | 0.036167213 | 1 |
| Prediabetes | PP_cell | *P4HB* | -0.226266767 | 0.000531491 | 0.008519859 | 1 |
| Prediabetes | PP_cell | *DRG1* | -0.452411573 | 6.26E-05 | 0.008519859 | 0.245894704 |
| Prediabetes | PP_cell | *DMWD* | -0.257258252 | 0.004425708 | 0.020248234 | 1 |

| | | | | | | |
|---|---|---|---|---|---|---|
| Prediabetes | PP_cell | *TRIM69* | -0.118929909 | 0.017946385 | 0.044810578 | 1 |
| Prediabetes | PP_cell | *ZNF529* | -0.467498586 | 0.000846766 | 0.009444968 | 1 |
| Prediabetes | PP_cell | *WWOX* | -0.22722799 | 0.000644712 | 0.008645299 | 1 |
| Prediabetes | PP_cell | *POLR1D* | -0.25437502 | 0.001192171 | 0.01078212 | 1 |
| Prediabetes | PP_cell | *ZNF749* | -0.081339055 | 0.019590277 | 0.047896825 | 1 |
| Prediabetes | PP_cell | *TOR3A* | -0.303901719 | 0.000223052 | 0.008519859 | 0.876370609 |
| Prediabetes | PP_cell | *ZNF555* | -0.37590253 | 0.001616129 | 0.012263296 | 1 |
| Prediabetes | PP_cell | *FAM86B1* | -0.142729488 | 0.01327664 | 0.037909824 | 1 |
| Prediabetes | PP_cell | *BCR* | -0.363475072 | 0.001913211 | 0.01336916 | 1 |
| Prediabetes | PP_cell | *RTN4RL2* | -0.098920378 | 0.009319059 | 0.031673516 | 1 |
| Prediabetes | PP_cell | *TMEM232* | -0.108117799 | 0.008953715 | 0.030831853 | 1 |
| Prediabetes | PP_cell | *ESPN* | -1.444212284 | 0.005079544 | 0.02194376 | 1 |
| Prediabetes | PP_cell | *TMEM216* | -0.121883265 | 0.016000807 | 0.041435884 | 1 |
| Prediabetes | PP_cell | *BCAM* | -0.240198137 | 0.001519585 | 0.011846392 | 1 |
| Prediabetes | PP_cell | *CHP1* | -0.251142402 | 0.014399983 | 0.039235459 | 1 |
| Prediabetes | PP_cell | *RGPD1* | -7.445059191 | 0.012850262 | 0.03760809 | 1 |
| Prediabetes | PP_cell | *KBTBD12* | -0.296052579 | 0.000289883 | 0.008519859 | 1 |
| Prediabetes | PP_cell | *PALM3* | -0.082681691 | 0.013660847 | 0.0382835 | 1 |
| Prediabetes | PP_cell | *KLHL17* | -0.152463551 | 0.007745748 | 0.028204861 | 1 |
| Prediabetes | PP_cell | *ZNF490* | -0.289095379 | 0.001152687 | 0.010631239 | 1 |
| Prediabetes | PP_cell | *NRN1L* | -0.177541082 | 0.013002879 | 0.037884276 | 1 |
| Prediabetes | PP_cell | *FAM166A* | -0.709812695 | 7.55E-05 | 0.008519859 | 0.296662989 |
| Prediabetes | PP_cell | *ZNF559* | -0.113562292 | 0.00129381 | 0.011111559 | 1 |
| Prediabetes | PP_cell | *FOCAD* | -0.399747432 | 0.007151599 | 0.026837281 | 1 |
| Prediabetes | PP_cell | *SERPINA5* | -0.060892448 | 0.004687528 | 0.020832552 | 1 |
| Prediabetes | PP_cell | *FBLL1* | -0.391839069 | 0.000508309 | 0.008519859 | 1 |
| Prediabetes | PP_cell | *UROS* | -0.350112935 | 0.001325157 | 0.011220999 | 1 |
| Prediabetes | PP_cell | *FAM221A* | -0.325237538 | 6.83E-05 | 0.008519859 | 0.268484702 |
| Prediabetes | PP_cell | *TMEM198* | -0.088191063 | 0.008889756 | 0.030690414 | 1 |
| Prediabetes | PP_cell | *RPL14* | -0.165302293 | 0.004271221 | 0.019996719 | 1 |
| Prediabetes | PP_cell | *ZNF567* | -0.421996115 | 0.00057238 | 0.008550883 | 1 |
| Prediabetes | PP_cell | *ARID2* | -0.350311722 | 0.000615805 | 0.008645299 | 1 |
| Prediabetes | PP_cell | *CLDN4* | -0.062250096 | 0.003670796 | 0.01839612 | 1 |
| Prediabetes | PP_cell | *FAM47E* | -0.127123674 | 0.011709312 | 0.035704358 | 1 |
| Prediabetes | PP_cell | *S100A14* | -0.060073418 | 0.009455646 | 0.031889469 | 1 |
| Prediabetes | PP_cell | *CXCL17* | -0.057192794 | 0.011681833 | 0.035704358 | 1 |
| Prediabetes | PP_cell | *SPATA41* | -0.348867005 | 0.003566539 | 0.018246003 | 1 |
| Prediabetes | PP_cell | *PTPRT* | -0.087824998 | 0.011166279 | 0.034846952 | 1 |
| Prediabetes | PP_cell | *ZNF676* | -0.189903571 | 0.014519698 | 0.039384002 | 1 |
| Prediabetes | PP_cell | *HLA-DRB1* | -0.060957105 | 0.003303562 | 0.017445616 | 1 |
| Prediabetes | PP_cell | *ZNF681* | -0.068323681 | 0.012828353 | 0.03760809 | 1 |
| Prediabetes | PP_cell | *HRCT1* | -0.429167325 | 0.015419017 | 0.04071325 | 1 |
| Prediabetes | PP_cell | *XPNPEP3* | -0.294321215 | 0.000245896 | 0.008519859 | 0.966124909 |
| Prediabetes | PP_cell | *ZNF471* | -0.647708062 | 0.000504666 | 0.008519859 | 1 |
| Prediabetes | PP_cell | *ZNF493* | -0.439581745 | 0.000685426 | 0.008780065 | 1 |
| Prediabetes | PP_cell | *SUPT3H* | -0.604789145 | 8.70E-05 | 0.008519859 | 0.34177395 |
| Prediabetes | PP_cell | *LONP1* | -0.321572535 | 0.002810832 | 0.016098772 | 1 |
| Prediabetes | PP_cell | *EVL* | -0.170599806 | 0.004280279 | 0.019996719 | 1 |
| Prediabetes | PP_cell | *ZNF124* | -0.387521297 | 0.000619129 | 0.008645299 | 1 |
| Prediabetes | PP_cell | *PIK3R4* | -0.259950141 | 0.002416086 | 0.014972873 | 1 |

| | | | | | | |
|---|---|---|---|---|---|---|
| Prediabetes | PP_cell | *C20orf96* | -0.289378016 | 0.003909832 | 0.019030442 | 1 |
| Prediabetes | PP_cell | *SULT1A1* | -0.182957949 | 0.006661717 | 0.025863524 | 1 |
| Prediabetes | PP_cell | *MYO18A* | -0.226521112 | 0.003600307 | 0.018323322 | 1 |
| Prediabetes | PP_cell | *XRCC2* | -0.187205949 | 0.013346672 | 0.037957281 | 1 |
| Prediabetes | PP_cell | *MYO6* | -0.308048209 | 0.003307962 | 0.017445616 | 1 |
| Prediabetes | PP_cell | *MMP1* | -0.102672766 | 0.00867729 | 0.030231024 | 1 |
| Prediabetes | PP_cell | *TRAPPC4* | -0.181458406 | 0.003661631 | 0.01839612 | 1 |
| Prediabetes | PP_cell | *ZNF33B* | -0.129156208 | 0.004256294 | 0.019996719 | 1 |
| Prediabetes | PP_cell | *AMZ2* | -0.31094535 | 0.011889119 | 0.035745194 | 1 |
| Prediabetes | PP_cell | *MVB12B* | -0.253341533 | 0.004646164 | 0.020697029 | 1 |
| Prediabetes | PP_cell | *ANKRD36B* | -0.127798036 | 0.01313877 | 0.037909761 | 1 |
| Prediabetes | PP_cell | *FAM3C* | -0.221571319 | 0.013741899 | 0.038295695 | 1 |
| Prediabetes | PP_cell | *AP2A1* | -0.302175683 | 0.00822521 | 0.029383885 | 1 |
| Prediabetes | PP_cell | *ZNF100* | -0.241346073 | 0.00307228 | 0.016741022 | 1 |
| Prediabetes | PP_cell | *ARRDC1* | -0.132935723 | 0.008699043 | 0.030237155 | 1 |
| Prediabetes | PP_cell | *ACSL5* | -0.182773217 | 0.002456466 | 0.015103998 | 1 |
| Prediabetes | PP_cell | *SND1* | -0.250834075 | 0.00197736 | 0.013597804 | 1 |
| Prediabetes | PP_cell | *PIWIL2* | -0.17114521 | 0.000182282 | 0.008519859 | 0.716185733 |
| Prediabetes | PP_cell | *C6orf141* | -0.118933249 | 0.020807903 | 0.049972037 | 1 |
| Prediabetes | PP_cell | *ZNF165* | -0.11532065 | 0.010592047 | 0.033944658 | 1 |
| Prediabetes | PP_cell | *SVIL* | -0.432113804 | 0.001069798 | 0.010251798 | 1 |
| Prediabetes | PP_cell | *ZNF98* | -0.109986339 | 0.010628311 | 0.034005404 | 1 |
| Prediabetes | PP_cell | *OGDHL* | -0.087141662 | 0.000865244 | 0.009444968 | 1 |
| Prediabetes | PP_cell | *ATG7* | -0.405740728 | 0.015394833 | 0.040676731 | 1 |
| Prediabetes | PP_cell | *HHLA3* | -0.152321703 | 0.007554297 | 0.027765046 | 1 |
| Prediabetes | PP_cell | *ENTPD6* | -0.229231553 | 0.00133483 | 0.011278593 | 1 |
| Prediabetes | PP_cell | *DPP4* | -0.053929453 | 0.01301277 | 0.037884276 | 1 |
| Prediabetes | PP_cell | *NMB* | -0.193613007 | 0.000434908 | 0.008519859 | 1 |
| Prediabetes | PP_cell | *LHFPL5* | -0.132409929 | 0.005881061 | 0.024051313 | 1 |
| Prediabetes | PP_cell | *FAM118B* | -0.20317648 | 0.011831783 | 0.035704358 | 1 |
| Prediabetes | PP_cell | *SLC9A8* | -0.364478549 | 0.000449813 | 0.008519859 | 1 |
| Prediabetes | PP_cell | *ZNF181* | -0.33189706 | 0.016458716 | 0.042210376 | 1 |
| Prediabetes | PP_cell | *TEAD4* | -0.225139083 | 0.001908512 | 0.01336916 | 1 |
| Prediabetes | PP_cell | *RPL12* | -0.138260711 | 0.008976711 | 0.030871689 | 1 |
| Prediabetes | PP_cell | *LEKR1* | -0.113130343 | 0.013747469 | 0.038295695 | 1 |
| Prediabetes | PP_cell | *ENTPD7* | -0.342917828 | 0.000641537 | 0.008645299 | 1 |
| Prediabetes | PP_cell | *ZNF84* | -0.393450578 | 0.00426035 | 0.019996719 | 1 |
| Prediabetes | PP_cell | *ZNF667* | -0.404897418 | 0.01083096 | 0.034318421 | 1 |
| Prediabetes | PP_cell | *CD2AP* | -0.324510859 | 0.002988872 | 0.016581525 | 1 |
| Prediabetes | PP_cell | *SFI1* | -0.294498796 | 9.55E-05 | 0.008519859 | 0.375243518 |
| Prediabetes | PP_cell | *SOWAHC* | -0.466748411 | 0.000170713 | 0.008519859 | 0.670730702 |
| Prediabetes | PP_cell | *DDRGK1* | -0.249714675 | 0.015727444 | 0.041222901 | 1 |
| Prediabetes | PP_cell | *ZNF607* | -0.108249041 | 0.014608269 | 0.039474478 | 1 |
| Prediabetes | PP_cell | *ZNF587* | -0.362761388 | 0.000406213 | 0.008519859 | 1 |
| Prediabetes | PP_cell | *HLA-DRB5* | -0.062072966 | 0.019998896 | 0.048749401 | 1 |
| Prediabetes | PP_cell | *CNGA1* | -0.136845787 | 0.008373945 | 0.029640748 | 1 |
| Prediabetes | PP_cell | *ZNF789* | -0.151240036 | 0.010466629 | 0.033815404 | 1 |
| Prediabetes | PP_cell | *NCOA6* | -0.417563974 | 0.000945544 | 0.009678612 | 1 |
| Prediabetes | PP_cell | *STK39* | -0.104887294 | 0.006952629 | 0.026444221 | 1 |
| Prediabetes | PP_cell | *PAPSS2* | -0.181512185 | 0.000500478 | 0.008519859 | 1 |

| | | | | | | |
|---|---|---|---|---|---|---|
| Prediabetes | PP_cell | *ECI2* | -0.249377433 | 0.014819076 | 0.039716338 | 1 |
| Prediabetes | PP_cell | *MSRB1* | -0.310308578 | 0.014996084 | 0.039945502 | 1 |
| Prediabetes | PP_cell | *COLGALT2* | -0.07281752 | 0.007910789 | 0.028699438 | 1 |
| Prediabetes | PP_cell | *DENND4B* | -0.287488249 | 0.000606775 | 0.008645299 | 1 |
| Prediabetes | PP_cell | *TSEN15* | -0.235459414 | 0.000586859 | 0.008635842 | 1 |
| Prediabetes | PP_cell | *CCDC152* | -0.47339769 | 0.000692814 | 0.008780065 | 1 |
| Prediabetes | PP_cell | *TYW1* | -0.170945324 | 0.006012081 | 0.024326947 | 1 |
| Prediabetes | PP_cell | *ITPRIPL1* | -0.117904558 | 0.007540472 | 0.027760015 | 1 |
| Prediabetes | PP_cell | *C1orf174* | -0.294234582 | 0.000371572 | 0.008519859 | 1 |
| Prediabetes | PP_cell | *TBKBP1* | -0.152773023 | 0.000619401 | 0.008645299 | 1 |
| Prediabetes | PP_cell | *CCDC167* | -0.367446457 | 0.000293892 | 0.008519859 | 1 |
| Prediabetes | PP_cell | *INF2* | -0.162496537 | 0.003681397 | 0.018402303 | 1 |
| Prediabetes | PP_cell | *CHML* | -0.263835129 | 0.000641208 | 0.008645299 | 1 |
| Prediabetes | PP_cell | *RAET1G* | -0.395899469 | 0.011017824 | 0.034659 | 1 |
| Prediabetes | PP_cell | *PCMTD2* | -0.290314721 | 0.019038161 | 0.047015043 | 1 |
| Prediabetes | PP_cell | *HSD17B8* | -0.101811398 | 0.012260785 | 0.036372141 | 1 |
| Prediabetes | PP_cell | *NOTCH4* | -0.136880051 | 0.000124953 | 0.008519859 | 0.490939425 |
| Prediabetes | PP_cell | *RNF5* | -0.119910628 | 0.009419766 | 0.031823094 | 1 |
| Prediabetes | PP_cell | *MRPL38* | -0.301881699 | 0.001316875 | 0.011188791 | 1 |
| Prediabetes | PP_cell | *LAYN* | -0.557090453 | 0.006040087 | 0.024364991 | 1 |
| Prediabetes | PP_cell | *PSORS1C1* | -0.059719248 | 0.015948725 | 0.041414773 | 1 |
| Prediabetes | PP_cell | *DHX16* | -0.315007647 | 0.006888534 | 0.026358267 | 1 |
| Prediabetes | PP_cell | *LILRB3* | -0.176627494 | 0.005984293 | 0.024289554 | 1 |
| Prediabetes | PP_cell | *ZNF783* | -0.684750635 | 0.000594922 | 0.008645299 | 1 |
| Prediabetes | PP_cell | *ZNF425* | -0.167063765 | 0.012190208 | 0.036311846 | 1 |
| Prediabetes | PP_cell | *PABPN1L* | -6.033700647 | 0.012864666 | 0.03760809 | 1 |
| Prediabetes | PP_cell | *SLC35B4* | -0.52662041 | 0.009381198 | 0.031774765 | 1 |
| Prediabetes | PP_cell | *SARNP* | -0.268195297 | 0.006813158 | 0.026269771 | 1 |
| Prediabetes | PP_cell | *MT1A* | -0.038513302 | 0.011407129 | 0.035181102 | 1 |
| Prediabetes | PP_cell | *ATP6AP1L* | -0.219091041 | 0.011678681 | 0.035704358 | 1 |
| Prediabetes | PP_cell | *RGL3* | -0.149744404 | 0.007636647 | 0.027963081 | 1 |
| Prediabetes | PP_cell | *CPT1B* | -0.091415287 | 0.005449417 | 0.022930133 | 1 |
| Prediabetes | PP_cell | *LCMT1* | -0.337613982 | 0.000266334 | 0.008519859 | 1 |
| Prediabetes | PP_cell | *LIN52* | -0.637132408 | 0.000448384 | 0.008519859 | 1 |
| Prediabetes | PP_cell | *GOLGA8O* | -0.480015042 | 0.00088266 | 0.009444968 | 1 |
| Prediabetes | PP_cell | *MAP10* | -0.14299727 | 0.012189319 | 0.036311846 | 1 |
| Prediabetes | PP_cell | *CCL14* | -0.138939073 | 0.00300287 | 0.016581525 | 1 |
| Prediabetes | PP_cell | *ZBTB9* | -0.170110144 | 0.005694609 | 0.02352694 | 1 |
| Prediabetes | PP_cell | *PPP1CB* | -0.205816893 | 0.012138719 | 0.036255474 | 1 |
| Prediabetes | PP_cell | *NUDT19* | -0.427393565 | 0.00016569 | 0.008519859 | 0.650997891 |
| Prediabetes | PP_cell | *AP1G2* | -0.192061496 | 0.002522574 | 0.015295049 | 1 |
| Prediabetes | PP_cell | *ZNF90* | -0.766230902 | 0.000268245 | 0.008519859 | 1 |
| Prediabetes | PP_cell | *MBLAC1* | -0.193829479 | 0.020797291 | 0.049972037 | 1 |
| Prediabetes | PP_cell | *SEC14L6* | -0.041212018 | 0.017878324 | 0.04474136 | 1 |
| Prediabetes | PP_cell | *MEG3* | -0.113466272 | 0.00153561 | 0.011877462 | 1 |
| Prediabetes | PP_cell | *IFRD2* | -0.446496766 | 0.00027966 | 0.008519859 | 1 |
| Prediabetes | PP_cell | *UBXN2B* | -0.110827294 | 0.002640466 | 0.01561651 | 1 |
| Prediabetes | PP_cell | *C5orf49* | -0.214909364 | 0.001318506 | 0.011188791 | 1 |
| Prediabetes | PP_cell | *ATAD3C* | -0.352885794 | 0.000681617 | 0.008780065 | 1 |
| Prediabetes | PP_cell | *PAM16* | -0.250664248 | 0.000355091 | 0.008519859 | 1 |

| | | | | | | |
|---|---|---|---|---|---|---|
| Prediabetes | PP_cell | *CELA3B* | -0.06486107 | 0.002697327 | 0.0157471 | 1 |
| Prediabetes | PP_cell | *PPP3R1* | -0.326220175 | 0.004273627 | 0.019996719 | 1 |
| Prediabetes | PP_cell | *TRIM16* | -0.570983367 | 0.003826009 | 0.018814004 | 1 |
| Prediabetes | PP_cell | *SLC12A8* | -0.458526644 | 0.002822577 | 0.016119051 | 1 |
| Prediabetes | PP_cell | *LINC00365* | -0.187417408 | 0.013190305 | 0.037909761 | 1 |
| Prediabetes | PP_cell | *LINC00937* | -0.098222817 | 0.007612456 | 0.027900502 | 1 |
| Prediabetes | PP_cell | *CYP4F26P* | -3.288813334 | 0.015030876 | 0.039999281 | 1 |
| Prediabetes | PP_cell | *EPPK1* | -0.159428801 | 0.007541211 | 0.027760015 | 1 |
| Prediabetes | PP_cell | *ARL17B* | -0.251713217 | 0.00054477 | 0.008519859 | 1 |
| Prediabetes | PP_cell | *PSMG3-AS1* | -0.2052873 | 0.014118264 | 0.038817816 | 1 |
| Prediabetes | PP_cell | *HSBP1* | -0.210643389 | 0.011770858 | 0.035704358 | 1 |
| Prediabetes | PP_cell | *RPS18* | -0.126684111 | 0.013704449 | 0.038295695 | 1 |
| Prediabetes | PP_cell | *LYRM9* | -0.183034221 | 0.016358397 | 0.042090466 | 1 |
| Prediabetes | PP_cell | *ANKRD65* | -0.266799519 | 0.008679205 | 0.030231024 | 1 |
| Prediabetes | PP_cell | *C12orf75* | -0.20705527 | 0.000398645 | 0.008519859 | 1 |
| Prediabetes | PP_cell | *ZNF737* | -0.387625001 | 0.003092271 | 0.016741022 | 1 |
| Prediabetes | PP_cell | *SHISA9* | -0.497211049 | 0.014335041 | 0.039167161 | 1 |
| Prediabetes | PP_cell | *LRRC37A2* | -0.215781001 | 0.000568266 | 0.008521817 | 1 |
| Prediabetes | PP_cell | *TXNDC5* | -0.258555654 | 0.010029828 | 0.032886224 | 1 |
| Prediabetes | PP_cell | *RBM14* | -0.175074005 | 0.005175754 | 0.022152003 | 1 |
| Prediabetes | PP_cell | *NME1* | -0.194104127 | 0.007736093 | 0.028195833 | 1 |
| Prediabetes | PP_cell | *CDRT4* | -0.108049849 | 0.008336241 | 0.029614005 | 1 |
| Prediabetes | PP_cell | *AMY2B* | -0.046764078 | 0.01146595 | 0.035333113 | 1 |
| Prediabetes | PP_cell | *SMKR1* | -0.289298138 | 0.000777907 | 0.009253454 | 1 |
| Prediabetes | PP_cell | *LINC00886* | -0.116715917 | 0.003949085 | 0.019084817 | 1 |
| Prediabetes | PP_cell | *MIF* | -0.16074376 | 0.005932165 | 0.024153447 | 1 |
| Prediabetes | PP_cell | *PI4KA* | -0.3792914 | 0.000450719 | 0.008519859 | 1 |
| Prediabetes | PP_cell | *EIF4EBP3* | -0.208789862 | 0.006062176 | 0.024391958 | 1 |
| Prediabetes | PP_cell | *MICAL3* | -0.298766761 | 0.000586279 | 0.008635842 | 1 |
| Prediabetes | PP_cell | *VSIG8* | -0.17975039 | 0.009607354 | 0.032070769 | 1 |
| Prediabetes | PP_cell | *C4orf48* | -0.25357959 | 0.000621435 | 0.008645299 | 1 |
| Prediabetes | PP_cell | *IL10RB* | -0.34012149 | 0.000339996 | 0.008519859 | 1 |
| Prediabetes | PP_cell | *RN7SL832P* | -0.30342327 | 0.007848541 | 0.02852629 | 1 |
| Prediabetes | PP_cell | *GSTA1* | -0.043817022 | 0.015477337 | 0.040757678 | 1 |
| Prediabetes | PP_cell | *GSTA2* | -0.063601218 | 0.006573107 | 0.025671708 | 1 |
| Prediabetes | PP_cell | *TMEM141* | -0.209941149 | 0.000137547 | 0.008519859 | 0.540421437 |
| Prediabetes | PP_cell | *UBAP1L* | -0.315375029 | 0.002243737 | 0.014351084 | 1 |
| Prediabetes | PP_cell | *BCKDHA* | -0.350912841 | 0.000879476 | 0.009444968 | 1 |
| Prediabetes | PP_cell | *PCDHAC1* | -0.123907945 | 0.020735223 | 0.049888972 | 1 |
| Prediabetes | PP_cell | *FMN1* | -0.099383935 | 0.004522907 | 0.020366067 | 1 |
| Prediabetes | PP_cell | *PDCD6* | -0.264957419 | 0.002246352 | 0.014351084 | 1 |
| Prediabetes | PP_cell | *PCDHA10* | -0.200184246 | 0.004189246 | 0.01983078 | 1 |
| Prediabetes | PP_cell | *ATP6V1E2* | -0.151462661 | 0.002551777 | 0.01537007 | 1 |
| Prediabetes | PP_cell | *SNHG18* | -0.179021821 | 0.002309188 | 0.014586494 | 1 |
| Prediabetes | PP_cell | *ZNF550* | -0.466705094 | 0.000208444 | 0.008519859 | 0.818975429 |
| Prediabetes | PP_cell | *LINC01099* | -0.233971357 | 0.003575527 | 0.018268201 | 1 |
| Prediabetes | PP_cell | *LYN* | -0.35923479 | 0.001359517 | 0.011364982 | 1 |
| Prediabetes | PP_cell | *AP5B1* | -0.328967384 | 0.009250355 | 0.031505802 | 1 |
| Prediabetes | PP_cell | *PABPC4L* | -1.046294151 | 0.00545094 | 0.022930133 | 1 |
| Prediabetes | PP_cell | *NPIPA2* | -0.749586497 | 0.010007339 | 0.032847816 | 1 |

| | | | | | | |
|---|---|---|---|---|---|---|
| Prediabetes | PP_cell | *EID3* | -0.300707455 | 0.001033563 | 0.010051656 | 1 |
| Prediabetes | PP_cell | *SNHG9* | -0.287264474 | 0.000863944 | 0.009444968 | 1 |
| Prediabetes | PP_cell | *ZNF10* | -0.378669196 | 0.017243546 | 0.043597099 | 1 |
| Prediabetes | PP_cell | *LIMS3* | -1.323072494 | 0.006852472 | 0.026292348 | 1 |
| Prediabetes | PP_cell | *GATC* | -0.358596656 | 0.001753831 | 0.012855972 | 1 |
| Prediabetes | PP_cell | *TEN1* | -0.238555467 | 0.011743224 | 0.035704358 | 1 |
| Prediabetes | PP_cell | *LINC00638* | -0.575327495 | 0.004731078 | 0.020932887 | 1 |
| Prediabetes | PP_cell | *TMEM179* | -0.17042538 | 0.001784108 | 0.012957043 | 1 |
| Prediabetes | PP_cell | *SH3RF3-AS1* | -0.345715596 | 0.011321368 | 0.035107856 | 1 |
| Prediabetes | PP_cell | *TMEM178B* | -0.339257613 | 0.003112631 | 0.016821908 | 1 |
| Prediabetes | PP_cell | *MRPL12* | -0.367908002 | 0.003885811 | 0.018989244 | 1 |
| Prediabetes | PP_cell | *LINC00667* | -0.252446741 | 0.014760542 | 0.03964058 | 1 |
| Prediabetes | PP_cell | *FSBP* | -0.117408563 | 0.011114787 | 0.034830535 | 1 |
| Prediabetes | PP_cell | *ZNF285* | -0.394216429 | 0.009891861 | 0.032720527 | 1 |
| Prediabetes | PP_cell | *FBXO17* | -0.293593331 | 0.000562949 | 0.008521817 | 1 |
| Prediabetes | PP_cell | *SNHG8* | -0.235732351 | 0.000790385 | 0.009253454 | 1 |
| Prediabetes | PP_cell | *GTF2H5* | -0.305227523 | 0.000818824 | 0.009434491 | 1 |
| T1DM | acinar_cell | *CEP68* | 0.2918121 | 0.000375486 | 0.046102602 | 1 |
| T1DM | acinar_cell | *UQCC2* | 0.420967841 | 3.74E-05 | 0.005069852 | 0.147025694 |
| T1DM | acinar_cell | *HLA-DQB1* | -0.313913978 | 1.25E-28 | 8.21E-26 | 4.92E-25 |
| T1DM | acinar_cell | *BTN3A2* | -0.073687797 | 2.38E-05 | 0.003335122 | 0.093383413 |
| T1DM | acinar_cell | *ZKSCAN3* | -0.467317287 | 2.51E-09 | 5.58E-07 | 9.88E-06 |
| T1DM | acinar_cell | *HLA-DRB1* | -0.113206073 | 1.05E-08 | 2.07E-06 | 4.13E-05 |
| T1DM | acinar_cell | *SFTA2* | -0.206287569 | 2.56E-09 | 5.58E-07 | 1.00E-05 |
| T1DM | acinar_cell | *RPS26* | 0.230137041 | 0.000410187 | 0.048388528 | 1 |
| T1DM | acinar_cell | *HLA-DRB5* | -0.204406187 | 1.05E-23 | 5.17E-21 | 4.13E-20 |
| T1DM | acinar_cell | *HSD17B8* | 0.64987386 | 5.90E-28 | 3.31E-25 | 2.32E-24 |
| T1DM | acinar_cell | *HLA-DMA* | 0.212329234 | 1.96E-29 | 1.92E-26 | 7.69E-26 |
| T1DM | acinar_cell | *NOTCH4* | -0.423133901 | 5.86E-29 | 4.61E-26 | 2.30E-25 |
| T1DM | acinar_cell | *AGER* | 0.37811136 | 4.65E-06 | 0.000676734 | 0.018271809 |
| T1DM | acinar_cell | *RNF5* | -0.347499166 | 1.81E-14 | 5.93E-12 | 7.11E-11 |
| T1DM | acinar_cell | *EHMT2* | 0.457659235 | 0.000151076 | 0.019147691 | 0.593578434 |
| T1DM | acinar_cell | *MICA* | 0.102564971 | 1.91E-07 | 3.41E-05 | 0.000749455 |
| T1DM | acinar_cell | *POU5F1* | 0.147460858 | 0.000418735 | 0.048388528 | 1 |
| T1DM | acinar_cell | *CCHCR1* | 0.369408365 | 9.26E-09 | 1.91E-06 | 3.64E-05 |
| T1DM | acinar_cell | *PSORS1C1* | -0.153212321 | 4.44E-11 | 1.09E-08 | 1.74E-07 |
| T1DM | acinar_cell | *RNF39* | 0.303782609 | 1.50E-13 | 4.54E-11 | 5.90E-10 |
| T1DM | acinar_cell | *ZFP57* | 0.239327096 | 3.28E-07 | 5.39E-05 | 0.001288796 |
| T1DM | acinar_cell | *HCG27* | 0.229720492 | 2.42E-08 | 4.52E-06 | 9.49E-05 |
| T1DM | acinar_cell | *ATF6B* | 0.342432947 | 4.43E-11 | 1.09E-08 | 1.74E-07 |
| T1DM | acinar_cell | *PPT2* | 0.537468961 | 2.28E-12 | 6.40E-10 | 8.96E-09 |
| T1DM | acinar_cell | *C4B* | 1.82037135 | 6.46E-07 | 0.000101488 | 0.002537198 |
| T1DM | acinar_cell | *HLA-DQB2* | 0.950641717 | 1.53E-52 | 6.02E-49 | 6.02E-49 |
| T1DM | acinar_cell | *HLA-B* | 0.136705996 | 4.35E-05 | 0.005695233 | 0.170856997 |
| T1DM | acinar_cell | *ZSCAN31* | 0.144010099 | 3.29E-07 | 5.39E-05 | 0.001292888 |
| T1DM | acinar_cell | *B3GALT4* | -0.248056779 | 1.59E-06 | 0.000239598 | 0.00622954 |
| T1DM | acinar_cell | *HLA-DQA2* | 0.371724601 | 8.15E-33 | 1.07E-29 | 3.20E-29 |
| T1DM | acinar_cell | *PSMB9* | 0.528297116 | 7.87E-21 | 3.09E-18 | 3.09E-17 |
| T1DM | acinar_cell | *HLA-DOB* | 0.605244643 | 4.49E-20 | 1.61E-17 | 1.77E-16 |
| T1DM | acinar_cell | *HLA-DMB* | 1.01527798 | 6.03E-35 | 1.18E-31 | 2.37E-31 |

| | | | | | | |
|---|---|---|---|---|---|---|
| T1DM | acinar_cell | *C4A* | -2.28398029 | 1.72E-23 | 7.50E-21 | 6.75E-20 |
| T1DM | alpha_cell | *UQCC2* | 0.072238164 | 9.31E-05 | 0.011795073 | 0.365647278 |
| T1DM | alpha_cell | *HLA-DQB1* | -0.051911614 | 1.40E-29 | 9.14E-27 | 5.49E-26 |
| T1DM | alpha_cell | *CAMK1D* | 0.113129848 | 0.000202874 | 0.024154352 | 0.797093606 |
| T1DM | alpha_cell | *BTN3A2* | -0.01215429 | 1.54E-05 | 0.002165613 | 0.060637152 |
| T1DM | alpha_cell | *ZKSCAN3* | -0.078897783 | 7.59E-10 | 1.75E-07 | 2.98E-06 |
| T1DM | alpha_cell | *HLA-DRB1* | -0.01957382 | 1.93E-09 | 4.20E-07 | 7.56E-06 |
| T1DM | alpha_cell | *SFTA2* | -0.032123119 | 7.97E-09 | 1.57E-06 | 3.13E-05 |
| T1DM | alpha_cell | *HLA-DRB5* | -0.036470321 | 9.33E-27 | 4.58E-24 | 3.66E-23 |
| T1DM | alpha_cell | *HSD17B8* | 0.114337254 | 2.12E-30 | 2.08E-27 | 8.32E-27 |
| T1DM | alpha_cell | *HLA-DMA* | 0.034550439 | 7.84E-30 | 6.16E-27 | 3.08E-26 |
| T1DM | alpha_cell | *NOTCH4* | -0.068553376 | 3.69E-29 | 2.07E-26 | 1.45E-25 |
| T1DM | alpha_cell | *AGER* | 0.065009243 | 3.62E-06 | 0.00052681 | 0.014223861 |
| T1DM | alpha_cell | *RNF5* | -0.058890951 | 1.39E-15 | 4.57E-13 | 5.48E-12 |
| T1DM | alpha_cell | *EHMT2* | 0.076807654 | 0.000391922 | 0.043996045 | 1 |
| T1DM | alpha_cell | *SLC44A4* | -0.021575313 | 0.000186426 | 0.022889677 | 0.732469659 |
| T1DM | alpha_cell | *MICA* | 0.016782553 | 1.55E-07 | 2.65E-05 | 0.000608818 |
| T1DM | alpha_cell | *POU5F1* | 0.024848145 | 0.000227018 | 0.02623394 | 0.891953945 |
| T1DM | alpha_cell | *CCHCR1* | 0.060980108 | 9.99E-09 | 1.87E-06 | 3.93E-05 |
| T1DM | alpha_cell | *PSORS1C1* | -0.02455395 | 5.67E-11 | 1.48E-08 | 2.23E-07 |
| T1DM | alpha_cell | *TRIM31* | -0.124949522 | 5.81E-05 | 0.00761292 | 0.228387612 |
| T1DM | alpha_cell | *RNF39* | 0.051477346 | 2.71E-14 | 8.18E-12 | 1.06E-10 |
| T1DM | alpha_cell | *ZFP57* | 0.039163627 | 2.44E-07 | 3.83E-05 | 0.000957722 |
| T1DM | alpha_cell | *HCG27* | 0.039072963 | 6.87E-09 | 1.42E-06 | 2.70E-05 |
| T1DM | alpha_cell | *ATF6B* | 0.055703081 | 8.02E-11 | 1.97E-08 | 3.15E-07 |
| T1DM | alpha_cell | *PPT2* | 0.097887353 | 1.03E-13 | 2.90E-11 | 4.06E-10 |
| T1DM | alpha_cell | *C4B* | 0.302846823 | 2.83E-07 | 4.28E-05 | 0.001111755 |
| T1DM | alpha_cell | *HLA-DQB2* | 0.162495579 | 4.13E-56 | 1.62E-52 | 1.62E-52 |
| T1DM | alpha_cell | *HLA-B* | 0.023038146 | 4.03E-05 | 0.005458968 | 0.158310069 |
| T1DM | alpha_cell | *ZSCAN31* | 0.023627342 | 2.23E-07 | 3.66E-05 | 0.000877697 |
| T1DM | alpha_cell | *B3GALT4* | -0.046647473 | 1.09E-07 | 1.95E-05 | 0.000429029 |
| T1DM | alpha_cell | *HLA-DQA2* | 0.062491959 | 1.45E-34 | 1.90E-31 | 5.70E-31 |
| T1DM | alpha_cell | *PSMB9* | 0.089432151 | 9.27E-22 | 3.64E-19 | 3.64E-18 |
| T1DM | alpha_cell | *HLA-DOB* | 0.097766656 | 3.48E-20 | 1.24E-17 | 1.37E-16 |
| T1DM | alpha_cell | *HLA-DMB* | 0.177258168 | 3.35E-38 | 6.58E-35 | 1.32E-34 |
| T1DM | alpha_cell | *C4A* | -0.375616261 | 3.57E-24 | 1.56E-21 | 1.40E-20 |
| T1DM | beta_cell | *BTN2A2* | -0.019729908 | 0.000272089 | 0.031442283 | 1 |
| T1DM | beta_cell | *RHPN2* | -0.080181934 | 0.000140952 | 0.017306283 | 0.55380106 |
| T1DM | beta_cell | *UQCC2* | 0.051425045 | 2.46E-05 | 0.003335101 | 0.096717937 |
| T1DM | beta_cell | *HLA-DQB1* | -0.031759006 | 3.84E-30 | 3.77E-27 | 1.51E-26 |
| T1DM | beta_cell | *BTN3A2* | -0.007339744 | 1.56E-05 | 0.002194721 | 0.061452201 |
| T1DM | beta_cell | *ZKSCAN3* | -0.04808706 | 5.44E-10 | 1.26E-07 | 2.14E-06 |
| T1DM | beta_cell | *HLA-DRB1* | -0.011980257 | 1.24E-09 | 2.70E-07 | 4.86E-06 |
| T1DM | beta_cell | *SFTA2* | -0.020345715 | 1.70E-09 | 3.52E-07 | 6.68E-06 |
| T1DM | beta_cell | *HLA-DRB5* | -0.021873579 | 3.90E-26 | 1.92E-23 | 1.53E-22 |
| T1DM | beta_cell | *HSD17B8* | 0.069411637 | 7.71E-30 | 6.06E-27 | 3.03E-26 |
| T1DM | beta_cell | *HLA-DMA* | 0.020712566 | 3.16E-29 | 1.78E-26 | 1.24E-25 |
| T1DM | beta_cell | *NOTCH4* | -0.041715914 | 1.67E-29 | 1.09E-26 | 6.54E-26 |
| T1DM | beta_cell | *AGER* | 0.039850728 | 2.78E-06 | 0.000404766 | 0.010928678 |
| T1DM | beta_cell | *RNF5* | -0.036373338 | 5.73E-16 | 1.88E-13 | 2.25E-12 |

| | | | | | | |
|---|---|---|---|---|---|---|
| T1DM | beta_cell | *EHMT2* | 0.052458389 | 0.000129578 | 0.016423026 | 0.50911381 |
| T1DM | beta_cell | *SLC44A4* | -0.013015399 | 0.000195645 | 0.023293644 | 0.768690247 |
| T1DM | beta_cell | *MICA* | 0.009983383 | 2.42E-07 | 4.14E-05 | 0.000952655 |
| T1DM | beta_cell | *POU5F1* | 0.014645442 | 0.000327521 | 0.035745305 | 1 |
| T1DM | beta_cell | *CCHCR1* | 0.036442046 | 1.77E-08 | 3.31E-06 | 6.95E-05 |
| T1DM | beta_cell | *PSORS1C1* | -0.015064586 | 2.78E-11 | 7.29E-09 | 1.09E-07 |
| T1DM | beta_cell | *TRIM31* | -0.068930038 | 0.000293186 | 0.032912229 | 1 |
| T1DM | beta_cell | *RNF39* | 0.030459535 | 1.13E-13 | 3.43E-11 | 4.46E-10 |
| T1DM | beta_cell | *ZFP57* | 0.023571544 | 2.74E-07 | 4.48E-05 | 0.001075696 |
| T1DM | beta_cell | *HCG27* | 0.023467723 | 8.84E-09 | 1.74E-06 | 3.48E-05 |
| T1DM | beta_cell | *ATF6B* | 0.033593953 | 1.27E-10 | 3.11E-08 | 4.97E-07 |
| T1DM | beta_cell | *PPT2* | 0.059239994 | 1.41E-13 | 3.96E-11 | 5.54E-10 |
| T1DM | beta_cell | *C4B* | 0.180788363 | 4.07E-07 | 6.14E-05 | 0.00159749 |
| T1DM | beta_cell | *HLA-DQB2* | 0.098617133 | 3.02E-56 | 1.19E-52 | 1.19E-52 |
| T1DM | beta_cell | *HLA-B* | 0.013841291 | 6.91E-05 | 0.009044847 | 0.271345401 |
| T1DM | beta_cell | *ZSCAN31* | 0.01409498 | 3.25E-07 | 5.11E-05 | 0.001278307 |
| T1DM | beta_cell | *B3GALT4* | -0.028884486 | 6.38E-08 | 1.14E-05 | 0.000250539 |
| T1DM | beta_cell | *HLA-DQA2* | 0.038117607 | 1.29E-34 | 1.69E-31 | 5.08E-31 |
| T1DM | beta_cell | *PSMB9* | 0.053596119 | 5.13E-21 | 2.01E-18 | 2.01E-17 |
| T1DM | beta_cell | *HLA-DOB* | 0.058757729 | 4.12E-20 | 1.47E-17 | 1.62E-16 |
| T1DM | beta_cell | *HLA-DMB* | 0.105824216 | 6.07E-37 | 1.19E-33 | 2.39E-33 |
| T1DM | beta_cell | *C4A* | -0.225552654 | 4.36E-23 | 1.90E-20 | 1.71E-19 |
| T1DM | delta_cell | *HLA-DQB1* | -0.267478713 | 1.67E-28 | 1.31E-25 | 6.57E-25 |
| T1DM | delta_cell | *BTN3A2* | -0.063536436 | 1.22E-05 | 0.001841323 | 0.047874394 |
| T1DM | delta_cell | *ZKSCAN3* | -0.207486451 | 8.84E-05 | 0.01253115 | 0.347463168 |
| T1DM | delta_cell | *HLA-DRB1* | -0.100954905 | 4.52E-09 | 1.04E-06 | 1.77E-05 |
| T1DM | delta_cell | *SFTA2* | -0.167027428 | 1.23E-08 | 2.55E-06 | 4.84E-05 |
| T1DM | delta_cell | *HLA-DRB5* | -0.178335213 | 2.53E-25 | 1.24E-22 | 9.94E-22 |
| T1DM | delta_cell | *HSD17B8* | 0.538616806 | 2.43E-27 | 1.59E-24 | 9.56E-24 |
| T1DM | delta_cell | *HLA-DMA* | 0.176725873 | 4.12E-29 | 4.04E-26 | 1.62E-25 |
| T1DM | delta_cell | *NOTCH4* | -0.335777135 | 2.72E-26 | 1.52E-23 | 1.07E-22 |
| T1DM | delta_cell | *RNF5* | -0.272389816 | 9.34E-14 | 3.06E-11 | 3.67E-10 |
| T1DM | delta_cell | *MICA* | 0.087189029 | 2.02E-07 | 3.97E-05 | 0.000793131 |
| T1DM | delta_cell | *POU5F1* | 0.129020996 | 0.000264049 | 0.035774095 | 1 |
| T1DM | delta_cell | *CCHCR1* | 0.309306516 | 5.54E-09 | 1.21E-06 | 2.18E-05 |
| T1DM | delta_cell | *PSORS1C1* | -0.125618447 | 1.80E-10 | 4.71E-08 | 7.06E-07 |
| T1DM | delta_cell | *RNF39* | 0.233257689 | 4.41E-13 | 1.33E-10 | 1.73E-09 |
| T1DM | delta_cell | *ZFP57* | 0.202255811 | 4.01E-07 | 6.86E-05 | 0.001577147 |
| T1DM | delta_cell | *HCG27* | 0.132785248 | 4.96E-07 | 8.13E-05 | 0.001950029 |
| T1DM | delta_cell | *ATF6B* | 0.254831973 | 2.49E-09 | 6.11E-07 | 9.77E-06 |
| T1DM | delta_cell | *PPT2* | 0.464504911 | 3.90E-12 | 1.10E-09 | 1.53E-08 |
| T1DM | delta_cell | *C4B* | 1.561576925 | 2.24E-07 | 4.19E-05 | 0.000879877 |
| T1DM | delta_cell | *HLA-DQB2* | 0.786938286 | 3.38E-52 | 1.33E-48 | 1.33E-48 |
| T1DM | delta_cell | *HLA-B* | 0.095617957 | 8.93E-05 | 0.01253115 | 0.350872208 |
| T1DM | delta_cell | *ZSCAN31* | 0.121859034 | 3.68E-07 | 6.57E-05 | 0.001444568 |
| T1DM | delta_cell | *B3GALT4* | -0.213409569 | 6.06E-07 | 9.53E-05 | 0.002381902 |
| T1DM | delta_cell | *HLA-DQA2* | 0.297316379 | 8.51E-32 | 1.67E-28 | 3.35E-28 |
| T1DM | delta_cell | *PSMB9* | 0.428161294 | 1.52E-20 | 5.45E-18 | 5.99E-17 |
| T1DM | delta_cell | *HLA-DOB* | 0.533061398 | 1.08E-20 | 4.25E-18 | 4.25E-17 |
| T1DM | delta_cell | *HLA-DMB* | 0.684494405 | 1.20E-29 | 1.57E-26 | 4.72E-26 |

| | | | | | | |
|---|---|---|---|---|---|---|
| T1DM | delta_cell | *C4A* | -1.955120837 | 2.45E-21 | 1.07E-18 | 9.64E-18 |
| T1DM | duct_epithelial_cell | *BTN2A2* | -0.113589964 | 0.000372411 | 0.045725078 | 1 |
| T1DM | duct_epithelial_cell | *UQCC2* | 0.210846591 | 0.000208686 | 0.026449273 | 0.819927461 |
| T1DM | duct_epithelial_cell | *HLA-DQB1* | -0.200854382 | 2.25E-30 | 2.21E-27 | 8.86E-27 |
| T1DM | duct_epithelial_cell | *BTN3A2* | -0.046017969 | 1.67E-05 | 0.002342122 | 0.065579414 |
| T1DM | duct_epithelial_cell | *ZKSCAN3* | -0.291259675 | 8.85E-10 | 2.17E-07 | 3.48E-06 |
| T1DM | duct_epithelial_cell | *HLA-DRB1* | -0.080194801 | 2.30E-10 | 6.02E-08 | 9.03E-07 |
| T1DM | duct_epithelial_cell | *SFTA2* | -0.100032476 | 7.05E-08 | 1.54E-05 | 0.000277113 |
| T1DM | duct_epithelial_cell | *HLA-DRB5* | -0.127785976 | 4.91E-25 | 2.41E-22 | 1.93E-21 |
| T1DM | duct_epithelial_cell | *HSD17B8* | 0.419646829 | 1.54E-28 | 8.66E-26 | 6.06E-25 |
| T1DM | duct_epithelial_cell | *HLA-DMA* | 0.130533846 | 3.57E-29 | 2.81E-26 | 1.40E-25 |
| T1DM | duct_epithelial_cell | *NOTCH4* | -0.255098961 | 6.37E-29 | 4.17E-26 | 2.50E-25 |
| T1DM | duct_epithelial_cell | *AGER* | 0.243132997 | 1.75E-06 | 0.000264795 | 0.006884663 |
| T1DM | duct_epithelial_cell | *RNF5* | -0.207990136 | 1.14E-14 | 3.72E-12 | 4.47E-11 |
| T1DM | duct_epithelial_cell | *EHMT2* | 0.285117566 | 0.000204997 | 0.026449273 | 0.805431376 |
| T1DM | duct_epithelial_cell | *MICA* | 0.059005349 | 9.03E-07 | 0.000141931 | 0.003548279 |
| T1DM | duct_epithelial_cell | *CCHCR1* | 0.206334126 | 1.23E-07 | 2.53E-05 | 0.000481572 |
| T1DM | duct_epithelial_cell | *PSORS1C1* | -0.072213488 | 2.63E-09 | 6.09E-07 | 1.03E-05 |
| T1DM | duct_epithelial_cell | *RNF39* | 0.075513781 | 1.01E-05 | 0.00147337 | 0.039780979 |
| T1DM | duct_epithelial_cell | *ZFP57* | 0.150083998 | 2.23E-07 | 4.18E-05 | 0.000877126 |
| T1DM | duct_epithelial_cell | *HCG27* | 0.128880372 | 1.41E-07 | 2.77E-05 | 0.000553319 |
| T1DM | duct_epithelial_cell | *ATF6B* | 0.210857879 | 1.01E-10 | 2.83E-08 | 3.96E-07 |
| T1DM | duct_epithelial_cell | *PPT2* | 0.373847219 | 5.92E-14 | 1.79E-11 | 2.33E-10 |
| T1DM | duct_epithelial_cell | *C4B* | 1.116200311 | 6.93E-07 | 0.00011351 | 0.002724241 |
| T1DM | duct_epithelial_cell | *HLA-DQB2* | 0.62342939 | 1.75E-56 | 6.90E-53 | 6.90E-53 |
| T1DM | duct_epithelial_cell | *HLA-B* | 0.086893685 | 3.08E-05 | 0.00417461 | 0.121063692 |
| T1DM | duct_epithelial_cell | *ZSCAN31* | 0.088912833 | 3.52E-07 | 6.28E-05 | 0.001382052 |
| T1DM | duct_epithelial_cell | *B3GALT4* | -0.162350135 | 6.89E-07 | 0.00011351 | 0.002707219 |
| T1DM | duct_epithelial_cell | *HLA-DQA2* | 0.248218403 | 9.42E-36 | 1.23E-32 | 3.70E-32 |
| T1DM | duct_epithelial_cell | *PSMB9* | 0.259386952 | 2.94E-16 | 1.05E-13 | 1.15E-12 |
| T1DM | duct_epithelial_cell | *HLA-DOB* | 0.334105561 | 7.19E-18 | 2.82E-15 | 2.82E-14 |
| T1DM | duct_epithelial_cell | *HLA-DMB* | 0.660749828 | 2.48E-36 | 4.87E-33 | 9.74E-33 |
| T1DM | duct_epithelial_cell | *C4A* | -1.446395005 | 3.63E-22 | 1.58E-19 | 1.43E-18 |
| T1DM | endothelial_cell | *USH1C* | -0.172630568 | 0.00075327 | 0.047033508 | 1 |
| T1DM | endothelial_cell | *BAIAP3* | -0.181622087 | 0.000780852 | 0.047033508 | 1 |
| T1DM | endothelial_cell | *CEP68* | 0.250335056 | 0.000245964 | 0.025431367 | 0.96639196 |
| T1DM | endothelial_cell | *RNASET2* | -0.206332283 | 0.000774936 | 0.047033508 | 1 |
| T1DM | endothelial_cell | *GPATCH1* | 0.428399869 | 8.59E-05 | 0.009926625 | 0.337505243 |
| T1DM | endothelial_cell | *CTSH* | 0.089434422 | 0.000338777 | 0.032464756 | 1 |
| T1DM | endothelial_cell | *VIPR1* | 0.351193879 | 4.78E-05 | 0.007217939 | 0.187666416 |
| T1DM | endothelial_cell | *PCDHB10* | 0.274161266 | 0.000459568 | 0.03611288 | 1 |
| T1DM | endothelial_cell | *EPS8L1* | 0.182685937 | 0.000625917 | 0.043721785 | 1 |
| T1DM | endothelial_cell | *PIAS3* | 0.338420983 | 5.51E-05 | 0.00746945 | 0.216614055 |
| T1DM | endothelial_cell | *RHPN2* | -0.495529888 | 8.32E-05 | 0.009910504 | 0.327046637 |
| T1DM | endothelial_cell | *NHSL1* | 0.397635268 | 0.000433064 | 0.03585838 | 1 |
| T1DM | endothelial_cell | *SPTBN5* | 0.113622459 | 0.000551055 | 0.040850829 | 1 |
| T1DM | endothelial_cell | *IFITM3* | 0.149336654 | 0.000449672 | 0.036056347 | 1 |
| T1DM | endothelial_cell | *ARHGEF19* | 0.129014431 | 0.000294134 | 0.028891357 | 1 |
| T1DM | endothelial_cell | *SPC25* | -0.120056268 | 0.000767467 | 0.047033508 | 1 |
| T1DM | endothelial_cell | *SLC25A27* | -0.13383973 | 0.000597635 | 0.042692883 | 1 |

| | | | | | | |
|---|---|---|---|---|---|---|
| T1DM | endothelial_cell | *CDK20* | -0.244311821 | 0.000485509 | 0.037403226 | 1 |
| T1DM | endothelial_cell | *DIP2A* | 0.241843072 | 0.000140466 | 0.015330349 | 0.551892563 |
| T1DM | endothelial_cell | *SRGAP2* | 0.240968143 | 6.33E-05 | 0.008295095 | 0.248852851 |
| T1DM | endothelial_cell | *EXO5* | 0.212426875 | 0.000539531 | 0.04076569 | 1 |
| T1DM | endothelial_cell | *ZNF589* | -0.113172074 | 0.000276542 | 0.027859872 | 1 |
| T1DM | endothelial_cell | *LMBRD2* | -0.276999619 | 0.000721504 | 0.047033508 | 1 |
| T1DM | endothelial_cell | *WBP1L* | 0.465327683 | 0.000634294 | 0.043721785 | 1 |
| T1DM | endothelial_cell | *ENGASE* | 0.134862876 | 0.000802048 | 0.047033508 | 1 |
| T1DM | endothelial_cell | *CACNB3* | 0.181607691 | 0.000425801 | 0.03585838 | 1 |
| T1DM | endothelial_cell | *SMAGP* | 0.137568624 | 0.000666213 | 0.045130171 | 1 |
| T1DM | endothelial_cell | *UBE2U* | -0.478011335 | 0.000879764 | 0.049379895 | 1 |
| T1DM | endothelial_cell | *HLA-DQB1* | -0.241813964 | 1.98E-24 | 1.56E-21 | 7.78E-21 |
| T1DM | endothelial_cell | *LBX2* | 0.595874436 | 0.000429591 | 0.03585838 | 1 |
| T1DM | endothelial_cell | *TH* | 1.030000948 | 0.000118262 | 0.013275776 | 0.464652152 |
| T1DM | endothelial_cell | *ZBTB40* | 0.263963309 | 0.000570063 | 0.041477338 | 1 |
| T1DM | endothelial_cell | *RBM11* | -0.21008307 | 0.000687841 | 0.045805555 | 1 |
| T1DM | endothelial_cell | *C19orf54* | 0.309388256 | 0.00085407 | 0.048632506 | 1 |
| T1DM | endothelial_cell | *ZKSCAN3* | -0.287880507 | 2.82E-06 | 0.000502975 | 0.011065445 |
| T1DM | endothelial_cell | *HLA-DRB1* | -0.067970205 | 5.14E-05 | 0.00746945 | 0.201923325 |
| T1DM | endothelial_cell | *MVB12B* | 0.455301854 | 8.19E-05 | 0.009910504 | 0.321735518 |
| T1DM | endothelial_cell | *HLA-DRB5* | -0.084895944 | 7.04E-10 | 1.84E-07 | 2.77E-06 |
| T1DM | endothelial_cell | *HSD17B8* | 0.399795302 | 2.32E-18 | 1.14E-15 | 9.11E-15 |
| T1DM | endothelial_cell | *HLA-DMA* | 0.185961368 | 3.12E-32 | 3.07E-29 | 1.23E-28 |
| T1DM | endothelial_cell | *AGER* | 0.256243939 | 1.06E-07 | 2.31E-05 | 0.000415277 |
| T1DM | endothelial_cell | *RNF5* | -0.294741057 | 7.07E-15 | 3.09E-12 | 2.78E-11 |
| T1DM | endothelial_cell | *EHMT2* | 0.360194048 | 0.000763076 | 0.047033508 | 1 |
| T1DM | endothelial_cell | *SLC44A4* | -0.119507823 | 4.20E-05 | 0.006606389 | 0.165159727 |
| T1DM | endothelial_cell | *MICB* | 0.070487359 | 0.000438076 | 0.03585838 | 1 |
| T1DM | endothelial_cell | *MICA* | 0.090838821 | 3.55E-09 | 8.72E-07 | 1.40E-05 |
| T1DM | endothelial_cell | *HLA-C* | 0.101833135 | 3.45E-05 | 0.005652281 | 0.135654736 |
| T1DM | endothelial_cell | *POU5F1* | 0.137368208 | 6.95E-05 | 0.008805333 | 0.272965308 |
| T1DM | endothelial_cell | *CCHCR1* | 0.352108132 | 1.04E-10 | 3.14E-08 | 4.08E-07 |
| T1DM | endothelial_cell | *PSORS1C1* | -0.093096795 | 2.61E-07 | 5.13E-05 | 0.0010252 |
| T1DM | endothelial_cell | *TRIM31* | -0.675751789 | 0.000399223 | 0.035648823 | 1 |
| T1DM | endothelial_cell | *RNF39* | 0.224850273 | 3.54E-11 | 1.39E-08 | 1.39E-07 |
| T1DM | endothelial_cell | *HCG9* | 0.206906724 | 0.000828831 | 0.047889343 | 1 |
| T1DM | endothelial_cell | *ZFP57* | 0.180152183 | 3.17E-06 | 0.000541389 | 0.012451952 |
| T1DM | endothelial_cell | *HCG27* | 0.131800372 | 5.33E-05 | 0.00746945 | 0.209476479 |
| T1DM | endothelial_cell | *ATF6B* | 0.289033505 | 5.65E-11 | 1.85E-08 | 2.22E-07 |
| T1DM | endothelial_cell | *LTB4R* | 0.376217735 | 0.000794406 | 0.047033508 | 1 |
| T1DM | endothelial_cell | *PPT2* | 0.207613287 | 5.39E-10 | 1.51E-07 | 2.12E-06 |
| T1DM | endothelial_cell | *C4B* | 1.615558904 | 1.47E-07 | 3.05E-05 | 0.000579043 |
| T1DM | endothelial_cell | *HLA-DQB2* | 0.85179145 | 1.66E-55 | 6.53E-52 | 6.53E-52 |
| T1DM | endothelial_cell | *HLA-B* | 0.132562651 | 3.19E-07 | 5.96E-05 | 0.001252 |
| T1DM | endothelial_cell | *ZSCAN31* | 0.083430313 | 0.000154124 | 0.016366333 | 0.605554324 |
| T1DM | endothelial_cell | *B3GALT4* | -0.224353038 | 3.35E-08 | 7.75E-06 | 0.000131812 |
| T1DM | endothelial_cell | *HLA-DQA2* | 0.341567382 | 1.09E-35 | 2.14E-32 | 4.27E-32 |
| T1DM | endothelial_cell | *PSMB9* | 0.133193866 | 5.27E-11 | 1.85E-08 | 2.07E-07 |
| T1DM | endothelial_cell | *PCDHGC3* | 0.228195793 | 0.000353669 | 0.033084936 | 1 |
| T1DM | endothelial_cell | *HLA-DOB* | 0.510281778 | 8.91E-20 | 5.00E-17 | 3.50E-16 |

| | | | | | | |
|---|---|---|---|---|---|---|
| T1DM | endothelial_cell | *HLA-DMB* | 0.725085828 | 3.61E-35 | 4.72E-32 | 1.42E-31 |
| T1DM | endothelial_cell | *C4A* | -2.003959178 | 4.82E-23 | 3.16E-20 | 1.90E-19 |
| T1DM | endothelial_cell | *GOLGA8K* | -1.782772651 | 0.000393075 | 0.035648823 | 1 |
| T1DM | epsilon_cell | *HLA-DQB1* | -2.321486487 | 1.21E-22 | 1.19E-19 | 4.74E-19 |
| T1DM | epsilon_cell | *BTN3A2* | -0.792895094 | 1.80E-05 | 0.004413869 | 0.070621911 |
| T1DM | epsilon_cell | *HLA-DRB1* | -1.133798914 | 1.73E-09 | 6.18E-07 | 6.80E-06 |
| T1DM | epsilon_cell | *SFTA2* | -2.018488114 | 3.40E-09 | 1.03E-06 | 1.34E-05 |
| T1DM | epsilon_cell | *HLA-DRB5* | -2.224245973 | 3.13E-17 | 1.54E-14 | 1.23E-13 |
| T1DM | epsilon_cell | *HLA-DMA* | 1.458547128 | 3.99E-18 | 2.24E-15 | 1.57E-14 |
| T1DM | epsilon_cell | *NOTCH4* | -3.262658665 | 3.35E-23 | 4.39E-20 | 1.32E-19 |
| T1DM | epsilon_cell | *RNF5* | -0.926517314 | 4.78E-06 | 0.001251774 | 0.018776606 |
| T1DM | epsilon_cell | *PSORS1C1* | -1.654965124 | 8.46E-12 | 3.32E-09 | 3.32E-08 |
| T1DM | epsilon_cell | *RNF39* | 2.387380195 | 2.62E-09 | 8.59E-07 | 1.03E-05 |
| T1DM | epsilon_cell | *C4B* | 17.7296358 | 3.58E-06 | 0.001004827 | 0.014067581 |
| T1DM | epsilon_cell | *HLA-DQB2* | 8.558347809 | 2.24E-45 | 8.79E-42 | 8.79E-42 |
| T1DM | epsilon_cell | *HLA-DQA2* | 3.660692057 | 1.19E-29 | 2.33E-26 | 4.66E-26 |
| T1DM | epsilon_cell | *HLA-DOB* | 6.547150285 | 4.28E-20 | 2.80E-17 | 1.68E-16 |
| T1DM | epsilon_cell | *HLA-DMB* | 5.239849096 | 6.46E-16 | 2.82E-13 | 2.54E-12 |
| T1DM | epsilon_cell | *C4A* | -24.84005838 | 3.52E-22 | 2.77E-19 | 1.38E-18 |
| T1DM | leukocyte | *GCLC* | -0.927374905 | 9.06E-09 | 1.72E-08 | 3.56E-05 |
| T1DM | leukocyte | *NFYA* | -0.773456522 | 2.29E-09 | 8.21E-09 | 8.98E-06 |
| T1DM | leukocyte | *STPG1* | -0.665720209 | 8.04E-10 | 8.14E-09 | 3.16E-06 |
| T1DM | leukocyte | *LASP1* | -0.626746433 | 4.20E-09 | 9.96E-09 | 1.65E-05 |
| T1DM | leukocyte | *SNX11* | -0.680640514 | 2.26E-08 | 3.79E-08 | 8.89E-05 |
| T1DM | leukocyte | *TMEM176A* | -0.395563141 | 2.02E-06 | 2.62E-06 | 0.007919863 |
| T1DM | leukocyte | *M6PR* | -1.079925561 | 1.66E-06 | 2.17E-06 | 0.006533214 |
| T1DM | leukocyte | *DBNDD1* | -0.434971516 | 2.78E-07 | 3.90E-07 | 0.001093499 |
| T1DM | leukocyte | *CASP10* | -1.537979902 | 1.27E-07 | 1.85E-07 | 0.000500249 |
| T1DM | leukocyte | *NDUFAF7* | -1.358263824 | 4.48E-09 | 1.03E-08 | 1.76E-05 |
| T1DM | leukocyte | *POLDIP2* | -0.514870385 | 2.64E-08 | 4.36E-08 | 0.000103735 |
| T1DM | leukocyte | *PLXND1* | -1.354957685 | 5.23E-09 | 1.13E-08 | 2.05E-05 |
| T1DM | leukocyte | *CD38* | -3.071299585 | 0.000581045 | 0.000674026 | 1 |
| T1DM | leukocyte | *KDM1A* | -0.701262181 | 1.25E-09 | 8.14E-09 | 4.93E-06 |
| T1DM | leukocyte | *RBM6* | -0.736118875 | 1.46E-09 | 8.14E-09 | 5.72E-06 |
| T1DM | leukocyte | *RECQL* | -0.690037 | 2.43E-09 | 8.21E-09 | 9.55E-06 |
| T1DM | leukocyte | *SLC25A13* | -0.686111315 | 1.51E-09 | 8.14E-09 | 5.93E-06 |
| T1DM | leukocyte | *MEOX1* | 1.182917444 | 0.000161103 | 0.000191 | 0.632974828 |
| T1DM | leukocyte | *PON1* | -0.797918548 | 2.92E-09 | 8.49E-09 | 1.15E-05 |
| T1DM | leukocyte | *KMT2E* | -0.684586557 | 3.56E-09 | 9.10E-09 | 1.40E-05 |
| T1DM | leukocyte | *SOX8* | -0.79200015 | 0.001089193 | 0.00125203 | 1 |
| T1DM | leukocyte | *IBTK* | -0.669977221 | 3.66E-09 | 9.22E-09 | 1.44E-05 |
| T1DM | leukocyte | *ZNF195* | -0.943699462 | 8.46E-09 | 1.62E-08 | 3.33E-05 |
| T1DM | leukocyte | *GGNBP2* | -0.703491442 | 2.83E-09 | 8.43E-09 | 1.11E-05 |
| T1DM | leukocyte | *ASB4* | 0.88290214 | 0.006648967 | 0.007434204 | 1 |
| T1DM | leukocyte | *TMEM98* | -0.520262093 | 4.67E-09 | 1.06E-08 | 1.83E-05 |
| T1DM | leukocyte | *SYNRG* | -0.925506413 | 8.39E-09 | 1.61E-08 | 3.30E-05 |
| T1DM | leukocyte | *SPATA20* | -0.710685758 | 6.96E-09 | 1.39E-08 | 2.73E-05 |
| T1DM | leukocyte | *FARP2* | -1.383043417 | 1.71E-05 | 2.11E-05 | 0.067180039 |
| T1DM | leukocyte | *USH1C* | -0.733328085 | 1.19E-10 | 8.14E-09 | 4.67E-07 |
| T1DM | leukocyte | *VPS41* | -0.852656818 | 1.37E-08 | 2.45E-08 | 5.40E-05 |

| | | | | | | |
|---|---|---|---|---|---|---|
| T1DM | leukocyte | *ELAC2* | -0.733118436 | 2.05E-09 | 8.14E-09 | 8.06E-06 |
| T1DM | leukocyte | *CCDC124* | -0.590330845 | 3.07E-09 | 8.60E-09 | 1.20E-05 |
| T1DM | leukocyte | *NOS2* | -11.19298429 | 1.61E-09 | 8.14E-09 | 6.32E-06 |
| T1DM | leukocyte | *CD79B* | -2.252071228 | 0.000113337 | 0.000135145 | 0.445302712 |
| T1DM | leukocyte | *ST7L* | -0.561630236 | 0.000788689 | 0.000911132 | 1 |
| T1DM | leukocyte | *BAIAP3* | -0.702259964 | 1.41E-10 | 8.14E-09 | 5.54E-07 |
| T1DM | leukocyte | *FMO3* | -1.625708588 | 0.000589916 | 0.000684114 | 1 |
| T1DM | leukocyte | *E2F2* | -0.883931499 | 0.03878369 | 0.042082606 | 1 |
| T1DM | leukocyte | *CYB561* | -0.606468371 | 1.88E-09 | 8.14E-09 | 7.37E-06 |
| T1DM | leukocyte | *MGST1* | -0.455874056 | 1.63E-08 | 2.83E-08 | 6.40E-05 |
| T1DM | leukocyte | *ST3GAL1* | -1.240292852 | 1.59E-07 | 2.29E-07 | 0.000625842 |
| T1DM | leukocyte | *IL32* | -0.318669351 | 1.40E-09 | 8.14E-09 | 5.49E-06 |
| T1DM | leukocyte | *MED24* | -0.934243164 | 6.86E-10 | 8.14E-09 | 2.70E-06 |
| T1DM | leukocyte | *RPS20* | -0.447213483 | 3.28E-09 | 8.79E-09 | 1.29E-05 |
| T1DM | leukocyte | *REV3L* | -0.580394713 | 1.86E-09 | 8.14E-09 | 7.32E-06 |
| T1DM | leukocyte | *MLXIPL* | -0.51305447 | 3.14E-06 | 4.03E-06 | 0.012349103 |
| T1DM | leukocyte | *ETV7* | -1.256673436 | 1.93E-09 | 8.14E-09 | 7.57E-06 |
| T1DM | leukocyte | *NCAPD2* | -1.072929221 | 1.06E-07 | 1.57E-07 | 0.000416722 |
| T1DM | leukocyte | *GIPR* | -0.55459422 | 3.42E-09 | 8.93E-09 | 1.35E-05 |
| T1DM | leukocyte | *FUZ* | -1.043838607 | 1.15E-09 | 8.14E-09 | 4.53E-06 |
| T1DM | leukocyte | *LRRC23* | -0.513799381 | 1.83E-09 | 8.14E-09 | 7.19E-06 |
| T1DM | leukocyte | *TSPAN9* | -0.589524909 | 1.41E-09 | 8.14E-09 | 5.53E-06 |
| T1DM | leukocyte | *BTBD7* | -0.728493421 | 2.23E-09 | 8.21E-09 | 8.75E-06 |
| T1DM | leukocyte | *APBA3* | -1.273925654 | 1.66E-09 | 8.14E-09 | 6.50E-06 |
| T1DM | leukocyte | *TTC19* | -0.553413586 | 1.95E-09 | 8.14E-09 | 7.64E-06 |
| T1DM | leukocyte | *DPF1* | -0.682879364 | 4.19E-07 | 5.76E-07 | 0.001646206 |
| T1DM | leukocyte | *LARS2* | 0.908336856 | 0.042465715 | 0.046001597 | 1 |
| T1DM | leukocyte | *RABGAP1* | -0.66326967 | 2.53E-09 | 8.21E-09 | 9.93E-06 |
| T1DM | leukocyte | *PPP5C* | -0.669151542 | 1.51E-09 | 8.14E-09 | 5.92E-06 |
| T1DM | leukocyte | *CEP68* | -0.615539774 | 4.43E-05 | 5.39E-05 | 0.173995424 |
| T1DM | leukocyte | *ERCC1* | -0.806015113 | 9.44E-10 | 8.14E-09 | 3.71E-06 |
| T1DM | leukocyte | *CD22* | 0.302020225 | 1.62E-06 | 2.12E-06 | 0.006373764 |
| T1DM | leukocyte | *LTF* | -1.74310081 | 2.57E-09 | 8.21E-09 | 1.01E-05 |
| T1DM | leukocyte | *EXTL3* | -0.606097227 | 1.87E-09 | 8.14E-09 | 7.34E-06 |
| T1DM | leukocyte | *ALOX5* | 0.34536668 | 2.03E-08 | 3.43E-08 | 7.97E-05 |
| T1DM | leukocyte | *CALCOCO1* | -0.927024019 | 0.009819041 | 0.010894948 | 1 |
| T1DM | leukocyte | *UBR7* | -0.690614476 | 3.15E-09 | 8.69E-09 | 1.24E-05 |
| T1DM | leukocyte | *MAN2B2* | -0.950769348 | 2.26E-09 | 8.21E-09 | 8.89E-06 |
| T1DM | leukocyte | *ANGEL1* | -0.881050057 | 1.96E-08 | 3.34E-08 | 7.71E-05 |
| T1DM | leukocyte | *DDX11* | -0.727110128 | 2.74E-09 | 8.30E-09 | 1.08E-05 |
| T1DM | leukocyte | *GPRC5A* | -0.778651836 | 7.89E-08 | 1.19E-07 | 0.000309996 |
| T1DM | leukocyte | *TACC3* | 0.342134298 | 0.007225339 | 0.008069459 | 1 |
| T1DM | leukocyte | *UFL1* | -0.640424517 | 2.49E-09 | 8.21E-09 | 9.77E-06 |
| T1DM | leukocyte | *POLA2* | -1.204070075 | 1.23E-09 | 8.14E-09 | 4.85E-06 |
| T1DM | leukocyte | *ZC3H3* | -0.613796709 | 1.92E-08 | 3.28E-08 | 7.55E-05 |
| T1DM | leukocyte | *MDH1* | -0.637692425 | 2.62E-09 | 8.21E-09 | 1.03E-05 |
| T1DM | leukocyte | *MTMR11* | -2.252858438 | 2.60E-08 | 4.30E-08 | 0.000102058 |
| T1DM | leukocyte | *DPEP1* | -0.420480754 | 9.15E-10 | 8.14E-09 | 3.60E-06 |
| T1DM | leukocyte | *NUDCD3* | -0.594744968 | 2.87E-09 | 8.44E-09 | 1.13E-05 |
| T1DM | leukocyte | *TTC27* | -1.359723432 | 2.51E-08 | 4.16E-08 | 9.86E-05 |

| | | | | | | |
|---|---|---|---|---|---|---|
| T1DM | leukocyte | *SNAI2* | -1.547682763 | 5.15E-07 | 7.02E-07 | 0.002023666 |
| T1DM | leukocyte | *PLEKHB1* | -0.623559488 | 4.42E-08 | 7.00E-08 | 0.000173745 |
| T1DM | leukocyte | *OSBPL5* | -1.190554777 | 2.76E-06 | 3.56E-06 | 0.010862841 |
| T1DM | leukocyte | *NLRP2* | -1.264823029 | 6.62E-08 | 1.01E-07 | 0.00026001 |
| T1DM | leukocyte | *ZDHHC6* | -0.892234582 | 1.78E-10 | 8.14E-09 | 6.98E-07 |
| T1DM | leukocyte | *RNH1* | -0.615865074 | 4.54E-09 | 1.04E-08 | 1.78E-05 |
| T1DM | leukocyte | *NDUFS1* | -0.583612667 | 3.74E-09 | 9.32E-09 | 1.47E-05 |
| T1DM | leukocyte | *TYMP* | 0.323316881 | 9.90E-08 | 1.47E-07 | 0.000389164 |
| T1DM | leukocyte | *RNASET2* | 0.527378254 | 0.001049919 | 0.001207591 | 1 |
| T1DM | leukocyte | *CD44* | -0.35213849 | 3.05E-09 | 8.59E-09 | 1.20E-05 |
| T1DM | leukocyte | *KCNG1* | -2.037269288 | 4.38E-07 | 6.02E-07 | 0.001722741 |
| T1DM | leukocyte | *AGPAT4* | -1.247318837 | 0.001485535 | 0.001697199 | 1 |
| T1DM | leukocyte | *SH2D2A* | -1.581271871 | 0.00414375 | 0.004671677 | 1 |
| T1DM | leukocyte | *VEZT* | -0.67871326 | 3.51E-09 | 9.03E-09 | 1.38E-05 |
| T1DM | leukocyte | *RABEP1* | -1.0371908 | 2.89E-08 | 4.73E-08 | 0.000113385 |
| T1DM | leukocyte | *BAK1* | -1.237623002 | 2.16E-09 | 8.21E-09 | 8.48E-06 |
| T1DM | leukocyte | *ARID4A* | -0.857597467 | 4.37E-09 | 1.02E-08 | 1.72E-05 |
| T1DM | leukocyte | *PNPLA6* | -0.858228002 | 6.18E-10 | 8.14E-09 | 2.43E-06 |
| T1DM | leukocyte | *ABCF2* | -0.914201335 | 1.86E-09 | 8.14E-09 | 7.31E-06 |
| T1DM | leukocyte | *LRRC7* | -0.727579054 | 1.18E-05 | 1.47E-05 | 0.046399933 |
| T1DM | leukocyte | *UBA6* | -0.982919712 | 7.57E-09 | 1.49E-08 | 2.97E-05 |
| T1DM | leukocyte | *APBA2* | -1.788097264 | 1.39E-09 | 8.14E-09 | 5.48E-06 |
| T1DM | leukocyte | *ASTE1* | -0.672487845 | 5.65E-06 | 7.14E-06 | 0.022198804 |
| T1DM | leukocyte | *RNF19A* | -0.595270137 | 4.02E-10 | 8.14E-09 | 1.58E-06 |
| T1DM | leukocyte | *SH3YL1* | -0.448910206 | 1.42E-09 | 8.14E-09 | 5.59E-06 |
| T1DM | leukocyte | *FAM136A* | -0.635630978 | 2.21E-09 | 8.21E-09 | 8.67E-06 |
| T1DM | leukocyte | *DAPK2* | -0.716596692 | 1.03E-09 | 8.14E-09 | 4.06E-06 |
| T1DM | leukocyte | *TIMP2* | -0.998971121 | 7.47E-07 | 1.00E-06 | 0.002933049 |
| T1DM | leukocyte | *MYOM2* | 0.88667154 | 9.80E-09 | 1.83E-08 | 3.85E-05 |
| T1DM | leukocyte | *RPL26L1* | -0.586537164 | 2.65E-09 | 8.21E-09 | 1.04E-05 |
| T1DM | leukocyte | *NSUN2* | -0.953113529 | 8.41E-10 | 8.14E-09 | 3.30E-06 |
| T1DM | leukocyte | *MRI1* | -0.784185231 | 2.70E-09 | 8.26E-09 | 1.06E-05 |
| T1DM | leukocyte | *AGA* | -0.765492121 | 1.12E-09 | 8.14E-09 | 4.41E-06 |
| T1DM | leukocyte | *BOD1L1* | -0.546728348 | 6.30E-08 | 9.69E-08 | 0.000247627 |
| T1DM | leukocyte | *DNAH5* | -0.981592531 | 3.58E-09 | 9.11E-09 | 1.41E-05 |
| T1DM | leukocyte | *PNKP* | -0.818471808 | 0.000596629 | 0.000691695 | 1 |
| T1DM | leukocyte | *PHLPP2* | -0.614632444 | 1.68E-09 | 8.14E-09 | 6.61E-06 |
| T1DM | leukocyte | *SPDL1* | 0.940076503 | 5.47E-07 | 7.44E-07 | 0.002149098 |
| T1DM | leukocyte | *CTNS* | -0.859676041 | 1.03E-09 | 8.14E-09 | 4.06E-06 |
| T1DM | leukocyte | *INPP4A* | -0.647111331 | 1.05E-09 | 8.14E-09 | 4.14E-06 |
| T1DM | leukocyte | *PSMA4* | -0.607646465 | 1.59E-09 | 8.14E-09 | 6.25E-06 |
| T1DM | leukocyte | *LSG1* | -0.399028695 | 5.13E-06 | 6.49E-06 | 0.020139911 |
| T1DM | leukocyte | *SPATA7* | -0.871674909 | 3.98E-09 | 9.64E-09 | 1.56E-05 |
| T1DM | leukocyte | *CAPG* | 0.603492157 | 1.68E-06 | 2.19E-06 | 0.006585852 |
| T1DM | leukocyte | *TRIT1* | -1.413306361 | 5.51E-09 | 1.17E-08 | 2.17E-05 |
| T1DM | leukocyte | *CNTLN* | -0.929348908 | 2.43E-09 | 8.21E-09 | 9.53E-06 |
| T1DM | leukocyte | *PREX2* | -1.320230183 | 1.11E-09 | 8.14E-09 | 4.36E-06 |
| T1DM | leukocyte | *WDR37* | -0.903083807 | 4.58E-09 | 1.04E-08 | 1.80E-05 |
| T1DM | leukocyte | *YTHDC2* | -0.956262894 | 9.08E-09 | 1.72E-08 | 3.57E-05 |
| T1DM | leukocyte | *DTNBP1* | -0.744840923 | 2.06E-09 | 8.14E-09 | 8.09E-06 |

| | | | | | | |
|---|---|---|---|---|---|---|
| T1DM | leukocyte | *GOPC* | -0.747896372 | 1.99E-09 | 8.14E-09 | 7.81E-06 |
| T1DM | leukocyte | *ZNF800* | -0.730736503 | 2.82E-09 | 8.41E-09 | 1.11E-05 |
| T1DM | leukocyte | *LMO3* | -0.635317888 | 0.000130188 | 0.000154815 | 0.511507278 |
| T1DM | leukocyte | *MRPS10* | -0.740774219 | 1.15E-07 | 1.68E-07 | 0.000450107 |
| T1DM | leukocyte | *COL9A2* | -0.716383658 | 2.01E-09 | 8.14E-09 | 7.88E-06 |
| T1DM | leukocyte | *ERCC8* | -1.926550126 | 2.13E-06 | 2.76E-06 | 0.008366492 |
| T1DM | leukocyte | *H6PD* | -1.486829908 | 6.21E-08 | 9.56E-08 | 0.000244057 |
| T1DM | leukocyte | *VAMP3* | -0.73594355 | 6.60E-09 | 1.34E-08 | 2.59E-05 |
| T1DM | leukocyte | *RCN1* | -0.532930935 | 2.16E-09 | 8.21E-09 | 8.47E-06 |
| T1DM | leukocyte | *RFC2* | -0.711308552 | 8.88E-09 | 1.69E-08 | 3.49E-05 |
| T1DM | leukocyte | *JKAMP* | -0.632230686 | 9.78E-10 | 8.14E-09 | 3.84E-06 |
| T1DM | leukocyte | *PTGER3* | -0.577535758 | 3.18E-08 | 5.17E-08 | 0.000124969 |
| T1DM | leukocyte | *BCAR1* | -0.535164167 | 2.02E-09 | 8.14E-09 | 7.94E-06 |
| T1DM | leukocyte | *CYBA* | 0.696573967 | 1.27E-05 | 1.58E-05 | 0.05009404 |
| T1DM | leukocyte | *THOC3* | -0.537157294 | 1.87E-09 | 8.14E-09 | 7.35E-06 |
| T1DM | leukocyte | *HEBP2* | -0.569110159 | 4.53E-09 | 1.04E-08 | 1.78E-05 |
| T1DM | leukocyte | *RRP12* | -0.486708328 | 1.38E-09 | 8.14E-09 | 5.42E-06 |
| T1DM | leukocyte | *FNIP2* | -0.916188855 | 0.000103136 | 0.000123242 | 0.405221163 |
| T1DM | leukocyte | *MRTO4* | -0.77785074 | 1.44E-09 | 8.14E-09 | 5.68E-06 |
| T1DM | leukocyte | *MCF2L2* | -1.071172841 | 1.89E-09 | 8.14E-09 | 7.44E-06 |
| T1DM | leukocyte | *LAMA3* | -1.062836493 | 2.97E-09 | 8.50E-09 | 1.17E-05 |
| T1DM | leukocyte | *ANAPC4* | -1.379328203 | 0.00470011 | 0.005286783 | 1 |
| T1DM | leukocyte | *LY75* | -2.370949294 | 1.33E-08 | 2.38E-08 | 5.21E-05 |
| T1DM | leukocyte | *ARID4B* | -0.55412006 | 5.84E-09 | 1.22E-08 | 2.30E-05 |
| T1DM | leukocyte | *OPN3* | -0.83614891 | 0.002057833 | 0.002340152 | 1 |
| T1DM | leukocyte | *SDCCAG8* | -0.645622053 | 2.41E-09 | 8.21E-09 | 9.46E-06 |
| T1DM | leukocyte | *KIF1B* | -0.634113478 | 1.41E-09 | 8.14E-09 | 5.56E-06 |
| T1DM | leukocyte | *TBC1D22A* | -0.685293706 | 1.71E-09 | 8.14E-09 | 6.73E-06 |
| T1DM | leukocyte | *SYNE2* | -0.619475749 | 1.81E-09 | 8.14E-09 | 7.11E-06 |
| T1DM | leukocyte | *ATP9A* | -0.673405421 | 2.55E-09 | 8.21E-09 | 1.00E-05 |
| T1DM | leukocyte | *CHRDL2* | -2.499543449 | 9.66E-10 | 8.14E-09 | 3.79E-06 |
| T1DM | leukocyte | *GALC* | -0.871416712 | 2.00E-08 | 3.39E-08 | 7.86E-05 |
| T1DM | leukocyte | *CYFIP2* | -0.53348876 | 4.47E-09 | 1.03E-08 | 1.76E-05 |
| T1DM | leukocyte | *USP36* | -0.955645916 | 4.07E-09 | 9.78E-09 | 1.60E-05 |
| T1DM | leukocyte | *MCOLN3* | -0.652388665 | 8.86E-10 | 8.14E-09 | 3.48E-06 |
| T1DM | leukocyte | *ITIH4* | -1.334087608 | 3.61E-09 | 9.13E-09 | 1.42E-05 |
| T1DM | leukocyte | *PCGF2* | -0.641579923 | 2.53E-09 | 8.21E-09 | 9.92E-06 |
| T1DM | leukocyte | *GDI2* | -0.667777814 | 2.32E-09 | 8.21E-09 | 9.12E-06 |
| T1DM | leukocyte | *ATP11B* | -1.034514159 | 5.79E-09 | 1.21E-08 | 2.28E-05 |
| T1DM | leukocyte | *LAMC2* | -0.466295278 | 7.35E-09 | 1.46E-08 | 2.89E-05 |
| T1DM | leukocyte | *RASGRF1* | -0.512765309 | 1.92E-09 | 8.14E-09 | 7.54E-06 |
| T1DM | leukocyte | *CROCC* | -1.022594374 | 2.22E-09 | 8.21E-09 | 8.73E-06 |
| T1DM | leukocyte | *FLYWCH1* | -0.435396508 | 8.13E-08 | 1.22E-07 | 0.000319528 |
| T1DM | leukocyte | *OGFR* | -1.158052046 | 0.000108994 | 0.000130084 | 0.428237175 |
| T1DM | leukocyte | *GNA15* | 1.155596148 | 5.61E-09 | 1.18E-08 | 2.20E-05 |
| T1DM | leukocyte | *PIGV* | 0.794424204 | 7.70E-08 | 1.16E-07 | 0.000302621 |
| T1DM | leukocyte | *RIMBP2* | -0.555139539 | 1.31E-09 | 8.14E-09 | 5.15E-06 |
| T1DM | leukocyte | *MPC1* | -0.5985703 | 2.25E-09 | 8.21E-09 | 8.84E-06 |
| T1DM | leukocyte | *BCAT1* | 0.269838821 | 2.38E-08 | 3.97E-08 | 9.34E-05 |
| T1DM | leukocyte | *MRPS35* | -0.685808187 | 1.74E-09 | 8.14E-09 | 6.84E-06 |

| | | | | | | |
|---|---|---|---|---|---|---|
| T1DM | leukocyte | *ZNF112* | -0.936409927 | 1.08E-09 | 8.14E-09 | 4.23E-06 |
| T1DM | leukocyte | *EIF4B* | -0.573110968 | 5.98E-09 | 1.24E-08 | 2.35E-05 |
| T1DM | leukocyte | *SLC6A16* | -1.818980434 | 1.26E-09 | 8.14E-09 | 4.96E-06 |
| T1DM | leukocyte | *SPHK2* | -0.807084115 | 2.48E-09 | 8.21E-09 | 9.75E-06 |
| T1DM | leukocyte | *HAGH* | -0.736099141 | 4.59E-09 | 1.04E-08 | 1.80E-05 |
| T1DM | leukocyte | *RNF4* | -0.757152822 | 2.77E-09 | 8.34E-09 | 1.09E-05 |
| T1DM | leukocyte | *TM7SF3* | -1.083279335 | 2.61E-08 | 4.31E-08 | 0.000102402 |
| T1DM | leukocyte | *ATP2C2* | -0.790700904 | 0.00036977 | 0.000431618 | 1 |
| T1DM | leukocyte | *HIPK2* | -0.545520434 | 1.93E-09 | 8.14E-09 | 7.57E-06 |
| T1DM | leukocyte | *TMEM161A* | -0.741686913 | 3.98E-09 | 9.64E-09 | 1.56E-05 |
| T1DM | leukocyte | *SNX24* | -1.303483613 | 5.17E-08 | 8.09E-08 | 0.000203207 |
| T1DM | leukocyte | *ABCA7* | -0.948720739 | 3.73E-09 | 9.32E-09 | 1.47E-05 |
| T1DM | leukocyte | *BCAS1* | 3.931557661 | 7.09E-08 | 1.08E-07 | 0.000278616 |
| T1DM | leukocyte | *PMS1* | -0.656819404 | 2.33E-09 | 8.21E-09 | 9.17E-06 |
| T1DM | leukocyte | *IPO5* | -0.576756928 | 1.52E-09 | 8.14E-09 | 5.99E-06 |
| T1DM | leukocyte | *OAT* | -0.596864927 | 2.86E-09 | 8.43E-09 | 1.12E-05 |
| T1DM | leukocyte | *WDR18* | -0.710535685 | 8.90E-09 | 1.69E-08 | 3.50E-05 |
| T1DM | leukocyte | *MCM10* | 1.389628116 | 1.04E-08 | 1.93E-08 | 4.10E-05 |
| T1DM | leukocyte | *ERBB3* | -0.582269977 | 1.75E-09 | 8.14E-09 | 6.86E-06 |
| T1DM | leukocyte | *ANKRD44* | -1.316323861 | 0.004654678 | 0.00523868 | 1 |
| T1DM | leukocyte | *ADAT1* | -0.644803223 | 6.98E-09 | 1.40E-08 | 2.74E-05 |
| T1DM | leukocyte | *MYLK* | -0.610009064 | 1.51E-09 | 8.14E-09 | 5.92E-06 |
| T1DM | leukocyte | *GSTO2* | -0.455748464 | 1.98E-09 | 8.14E-09 | 7.80E-06 |
| T1DM | leukocyte | *SEC61A2* | -1.876336464 | 1.56E-09 | 8.14E-09 | 6.13E-06 |
| T1DM | leukocyte | *PRKCQ* | -1.415597888 | 2.42E-07 | 3.43E-07 | 0.000952174 |
| T1DM | leukocyte | *TBC1D1* | -0.69419579 | 1.27E-09 | 8.14E-09 | 5.01E-06 |
| T1DM | leukocyte | *CTNNA2* | -0.851261432 | 2.21E-09 | 8.21E-09 | 8.67E-06 |
| T1DM | leukocyte | *CD84* | 0.108545375 | 6.19E-10 | 8.14E-09 | 2.43E-06 |
| T1DM | leukocyte | *ATXN3* | -0.901760398 | 1.32E-07 | 1.91E-07 | 0.000518363 |
| T1DM | leukocyte | *EML1* | -1.006440537 | 2.34E-09 | 8.21E-09 | 9.20E-06 |
| T1DM | leukocyte | *STOML1* | -0.613266387 | 1.32E-09 | 8.14E-09 | 5.18E-06 |
| T1DM | leukocyte | *DNTTIP2* | -0.591096461 | 2.38E-09 | 8.21E-09 | 9.36E-06 |
| T1DM | leukocyte | *RRP15* | -0.703433129 | 7.44E-10 | 8.14E-09 | 2.92E-06 |
| T1DM | leukocyte | *HEATR6* | 1.427658153 | 0.000101765 | 0.000121678 | 0.399832964 |
| T1DM | leukocyte | *REEP1* | -0.610712681 | 4.00E-09 | 9.65E-09 | 1.57E-05 |
| T1DM | leukocyte | *POLR1A* | -0.53538599 | 1.02E-07 | 1.51E-07 | 0.000400884 |
| T1DM | leukocyte | *LAPTM4A* | -0.555888006 | 3.00E-09 | 8.53E-09 | 1.18E-05 |
| T1DM | leukocyte | *KIF2A* | -0.62519762 | 1.70E-09 | 8.14E-09 | 6.67E-06 |
| T1DM | leukocyte | *MAST4* | -0.700310057 | 2.39E-09 | 8.21E-09 | 9.38E-06 |
| T1DM | leukocyte | *ADAM7* | -5.083552021 | 0.00920897 | 0.010235373 | 1 |
| T1DM | leukocyte | *CLEC2D* | 1.689000718 | 1.19E-07 | 1.74E-07 | 0.000466203 |
| T1DM | leukocyte | *DRD4* | -2.204135485 | 2.39E-09 | 8.21E-09 | 9.41E-06 |
| T1DM | leukocyte | *PLA2G10* | -2.468363517 | 2.60E-09 | 8.21E-09 | 1.02E-05 |
| T1DM | leukocyte | *PIGB* | -0.89651222 | 0.030311755 | 0.033026868 | 1 |
| T1DM | leukocyte | *GNB5* | -0.776624579 | 1.08E-08 | 1.99E-08 | 4.25E-05 |
| T1DM | leukocyte | *RAB27A* | -0.568538632 | 1.86E-08 | 3.19E-08 | 7.31E-05 |
| T1DM | leukocyte | *PTPN3* | -1.018879496 | 2.33E-09 | 8.21E-09 | 9.17E-06 |
| T1DM | leukocyte | *SPTB* | -0.506243485 | 2.19E-07 | 3.11E-07 | 0.000859957 |
| T1DM | leukocyte | *FGF10* | -2.146455158 | 2.80E-10 | 8.14E-09 | 1.10E-06 |
| T1DM | leukocyte | *DGCR2* | -0.848006573 | 2.49E-09 | 8.21E-09 | 9.78E-06 |

| | | | | | | |
|---|---|---|---|---|---|---|
| T1DM | leukocyte | *ZXDC* | -1.390547799 | 4.94E-08 | 7.74E-08 | 0.000194012 |
| T1DM | leukocyte | *WIPI1* | -1.316016045 | 1.31E-07 | 1.91E-07 | 0.00051642 |
| T1DM | leukocyte | *GBA2* | -0.657063032 | 4.23E-09 | 9.97E-09 | 1.66E-05 |
| T1DM | leukocyte | *ASNS* | -0.671312611 | 1.39E-09 | 8.14E-09 | 5.47E-06 |
| T1DM | leukocyte | *AP3M2* | -0.895722731 | 4.16E-07 | 5.72E-07 | 0.00163287 |
| T1DM | leukocyte | *MAP4K4* | -0.576036938 | 1.68E-09 | 8.14E-09 | 6.61E-06 |
| T1DM | leukocyte | *RPL31* | -0.492262133 | 3.27E-09 | 8.79E-09 | 1.28E-05 |
| T1DM | leukocyte | *SEL1L* | -0.557082142 | 4.29E-09 | 1.00E-08 | 1.68E-05 |
| T1DM | leukocyte | *TCF3* | -0.638045291 | 5.46E-10 | 8.14E-09 | 2.15E-06 |
| T1DM | leukocyte | *CPSF1* | -0.594840829 | 2.83E-09 | 8.42E-09 | 1.11E-05 |
| T1DM | leukocyte | *CYBRD1* | 0.513187876 | 2.23E-08 | 3.74E-08 | 8.77E-05 |
| T1DM | leukocyte | *LNX1* | -0.52444978 | 1.37E-09 | 8.14E-09 | 5.37E-06 |
| T1DM | leukocyte | *ALDH3A2* | -0.46484141 | 8.04E-10 | 8.14E-09 | 3.16E-06 |
| T1DM | leukocyte | *P4HA2* | -0.500451663 | 4.96E-09 | 1.09E-08 | 1.95E-05 |
| T1DM | leukocyte | *TRNT1* | -1.819032179 | 9.69E-08 | 1.44E-07 | 0.000380555 |
| T1DM | leukocyte | *CRMP1* | -0.553432649 | 2.12E-09 | 8.16E-09 | 8.32E-06 |
| T1DM | leukocyte | *DERL2* | -0.662776429 | 1.89E-09 | 8.14E-09 | 7.42E-06 |
| T1DM | leukocyte | *SIDT1* | -0.440628894 | 0.017561574 | 0.019322157 | 1 |
| T1DM | leukocyte | *NDE1* | 1.164178375 | 0.00040435 | 0.000471003 | 1 |
| T1DM | leukocyte | *XRCC1* | -0.875678145 | 1.59E-08 | 2.77E-08 | 6.23E-05 |
| T1DM | leukocyte | *SCARB1* | -1.613197577 | 1.31E-09 | 8.14E-09 | 5.13E-06 |
| T1DM | leukocyte | *NLE1* | -0.674307647 | 7.16E-08 | 1.09E-07 | 0.000281384 |
| T1DM | leukocyte | *SMARCE1* | -0.571352381 | 2.66E-09 | 8.24E-09 | 1.05E-05 |
| T1DM | leukocyte | *MRPS34* | -0.524321566 | 5.91E-09 | 1.23E-08 | 2.32E-05 |
| T1DM | leukocyte | *TEAD2* | -0.879439731 | 1.63E-08 | 2.83E-08 | 6.39E-05 |
| T1DM | leukocyte | *IPCEF1* | -1.070613667 | 6.29E-07 | 8.48E-07 | 0.002469791 |
| T1DM | leukocyte | *ZZEF1* | -0.904294449 | 4.23E-09 | 9.97E-09 | 1.66E-05 |
| T1DM | leukocyte | *ANO8* | 1.548397214 | 3.86E-05 | 4.71E-05 | 0.151657607 |
| T1DM | leukocyte | *WSCD2* | -0.729130984 | 1.57E-09 | 8.14E-09 | 6.17E-06 |
| T1DM | leukocyte | *TIPIN* | -0.780404735 | 2.13E-07 | 3.03E-07 | 0.000837209 |
| T1DM | leukocyte | *SEMA3A* | -1.927702441 | 1.64E-09 | 8.14E-09 | 6.45E-06 |
| T1DM | leukocyte | *TTC38* | -1.179653789 | 7.49E-09 | 1.48E-08 | 2.94E-05 |
| T1DM | leukocyte | *GRAMD4* | -0.695358897 | 1.43E-05 | 1.77E-05 | 0.056250949 |
| T1DM | leukocyte | *CELSR1* | -3.304622979 | 8.45E-09 | 1.62E-08 | 3.32E-05 |
| T1DM | leukocyte | *TIMM21* | -0.988297443 | 8.80E-10 | 8.14E-09 | 3.46E-06 |
| T1DM | leukocyte | *VPS9D1* | -0.646829014 | 9.59E-09 | 1.80E-08 | 3.77E-05 |
| T1DM | leukocyte | *ZNF37A* | -0.583852226 | 1.18E-07 | 1.72E-07 | 0.000462308 |
| T1DM | leukocyte | *MARK3* | -0.712061021 | 3.56E-10 | 8.14E-09 | 1.40E-06 |
| T1DM | leukocyte | *CACNG5* | -20.6763649 | 5.15E-10 | 8.14E-09 | 2.02E-06 |
| T1DM | leukocyte | *MOCOS* | -1.572165404 | 4.02E-06 | 5.12E-06 | 0.015811256 |
| T1DM | leukocyte | *ATP12A* | 1.89464192 | 7.13E-08 | 1.08E-07 | 0.000280307 |
| T1DM | leukocyte | *DLG1* | -0.640622539 | 1.53E-09 | 8.14E-09 | 6.01E-06 |
| T1DM | leukocyte | *SEC31B* | 0.892084117 | 1.92E-05 | 2.36E-05 | 0.075366733 |
| T1DM | leukocyte | *ARHGAP15* | -2.944814635 | 0.004756255 | 0.005346874 | 1 |
| T1DM | leukocyte | *EXOSC7* | -0.565755362 | 2.09E-09 | 8.15E-09 | 8.20E-06 |
| T1DM | leukocyte | *MKRN2* | -0.816166597 | 1.38E-09 | 8.14E-09 | 5.43E-06 |
| T1DM | leukocyte | *MLH1* | -0.865680593 | 7.86E-09 | 1.54E-08 | 3.09E-05 |
| T1DM | leukocyte | *TPD52* | -0.513365199 | 3.53E-09 | 9.07E-09 | 1.39E-05 |
| T1DM | leukocyte | *ACACB* | -0.803429081 | 1.41E-09 | 8.14E-09 | 5.52E-06 |
| T1DM | leukocyte | *GPATCH1* | -0.593146505 | 9.27E-08 | 1.38E-07 | 0.000364206 |

| | | | | | | |
|---|---|---|---|---|---|---|
| T1DM | leukocyte | *ICAM3* | -0.7119138 | 4.25E-10 | 8.14E-09 | 1.67E-06 |
| T1DM | leukocyte | *XAB2* | -0.688441891 | 6.36E-10 | 8.14E-09 | 2.50E-06 |
| T1DM | leukocyte | *ACTL6B* | -0.526110001 | 1.33E-07 | 1.93E-07 | 0.000524518 |
| T1DM | leukocyte | *IL4R* | -0.574452738 | 3.39E-08 | 5.49E-08 | 0.000133129 |
| T1DM | leukocyte | *APBB1IP* | 0.288128782 | 9.55E-09 | 1.80E-08 | 3.75E-05 |
| T1DM | leukocyte | *GPR137B* | -0.809422006 | 3.54E-06 | 4.52E-06 | 0.013906469 |
| T1DM | leukocyte | *NAALAD2* | -1.712327053 | 4.84E-08 | 7.59E-08 | 0.000190085 |
| T1DM | leukocyte | *MCCC1* | -0.694394411 | 2.32E-09 | 8.21E-09 | 9.12E-06 |
| T1DM | leukocyte | *N4BP2* | -0.914154088 | 4.21E-09 | 9.96E-09 | 1.65E-05 |
| T1DM | leukocyte | *MLLT10* | -0.853472969 | 2.11E-09 | 8.16E-09 | 8.28E-06 |
| T1DM | leukocyte | *VDAC3* | -0.659481716 | 2.34E-08 | 3.91E-08 | 9.18E-05 |
| T1DM | leukocyte | *PCM1* | -0.605586664 | 1.58E-09 | 8.14E-09 | 6.22E-06 |
| T1DM | leukocyte | *MYH7B* | -0.977502542 | 0.001687633 | 0.001923618 | 1 |
| T1DM | leukocyte | *TP73* | 2.067906618 | 4.84E-09 | 1.08E-08 | 1.90E-05 |
| T1DM | leukocyte | *TOLLIP* | -0.673705683 | 3.42E-09 | 8.93E-09 | 1.34E-05 |
| T1DM | leukocyte | *CLUL1* | -1.744684504 | 3.18E-09 | 8.70E-09 | 1.25E-05 |
| T1DM | leukocyte | *FKBP7* | -0.889709405 | 1.75E-09 | 8.14E-09 | 6.87E-06 |
| T1DM | leukocyte | *LXN* | -0.47661009 | 5.75E-08 | 8.91E-08 | 0.000225975 |
| T1DM | leukocyte | *SAR1A* | -0.53026602 | 1.59E-09 | 8.14E-09 | 6.25E-06 |
| T1DM | leukocyte | *CDC14A* | -0.956621315 | 1.39E-08 | 2.48E-08 | 5.48E-05 |
| T1DM | leukocyte | *FDFT1* | -0.731788048 | 6.53E-10 | 8.14E-09 | 2.57E-06 |
| T1DM | leukocyte | *SCGN* | -0.308483665 | 9.90E-09 | 1.84E-08 | 3.89E-05 |
| T1DM | leukocyte | *DDX1* | -0.583474625 | 4.89E-09 | 1.08E-08 | 1.92E-05 |
| T1DM | leukocyte | *RABL2B* | -1.005044638 | 1.38E-09 | 8.14E-09 | 5.44E-06 |
| T1DM | leukocyte | *DCT* | -4.02948659 | 3.55E-09 | 9.10E-09 | 1.40E-05 |
| T1DM | leukocyte | *SCTR* | -0.697291801 | 8.42E-10 | 8.14E-09 | 3.31E-06 |
| T1DM | leukocyte | *RIF1* | -1.072423175 | 1.69E-08 | 2.92E-08 | 6.62E-05 |
| T1DM | leukocyte | *IGSF9B* | -1.223029995 | 1.26E-08 | 2.28E-08 | 4.95E-05 |
| T1DM | leukocyte | *COL4A4* | -1.52248225 | 1.65E-09 | 8.14E-09 | 6.48E-06 |
| T1DM | leukocyte | *ZNF506* | -0.665073647 | 1.08E-09 | 8.14E-09 | 4.26E-06 |
| T1DM | leukocyte | *CADPS2* | -0.716883951 | 1.26E-09 | 8.14E-09 | 4.94E-06 |
| T1DM | leukocyte | *C1QTNF3* | 2.14450914 | 2.20E-07 | 3.11E-07 | 0.000863126 |
| T1DM | leukocyte | *ME2* | -1.246474767 | 1.61E-07 | 2.31E-07 | 0.000633005 |
| T1DM | leukocyte | *CCNT2* | -0.748460823 | 6.47E-08 | 9.92E-08 | 0.000254033 |
| T1DM | leukocyte | *COL19A1* | -10.50840369 | 0.015954406 | 0.017573553 | 1 |
| T1DM | leukocyte | *SEMA5B* | 1.133292316 | 2.67E-09 | 8.25E-09 | 1.05E-05 |
| T1DM | leukocyte | *BCKDHB* | -1.67438068 | 1.25E-08 | 2.27E-08 | 4.93E-05 |
| T1DM | leukocyte | *DIS3* | -0.641193418 | 3.60E-09 | 9.12E-09 | 1.41E-05 |
| T1DM | leukocyte | *PIBF1* | -0.976768688 | 1.12E-08 | 2.05E-08 | 4.39E-05 |
| T1DM | leukocyte | *CHMP2B* | -0.571318825 | 2.41E-09 | 8.21E-09 | 9.45E-06 |
| T1DM | leukocyte | *PPIE* | -0.715669786 | 1.02E-08 | 1.90E-08 | 4.02E-05 |
| T1DM | leukocyte | *NCOA1* | -0.745805304 | 2.90E-09 | 8.49E-09 | 1.14E-05 |
| T1DM | leukocyte | *AGBL5* | -1.121491887 | 4.48E-08 | 7.08E-08 | 0.00017606 |
| T1DM | leukocyte | *CAD* | -0.697345087 | 1.86E-07 | 2.66E-07 | 0.000730976 |
| T1DM | leukocyte | *CD59* | -0.415745001 | 3.17E-09 | 8.70E-09 | 1.25E-05 |
| T1DM | leukocyte | *CD82* | -0.94260626 | 9.78E-07 | 1.30E-06 | 0.003842378 |
| T1DM | leukocyte | *MECOM* | -0.473439676 | 8.08E-09 | 1.57E-08 | 3.18E-05 |
| T1DM | leukocyte | *SLC25A24* | -0.646848963 | 4.16E-09 | 9.92E-09 | 1.64E-05 |
| T1DM | leukocyte | *PILRA* | 0.244282295 | 5.72E-09 | 1.20E-08 | 2.25E-05 |
| T1DM | leukocyte | *ZNF213* | -0.806103535 | 8.74E-11 | 8.14E-09 | 3.43E-07 |

| | | | | | | |
|---|---|---|---|---|---|---|
| T1DM | leukocyte | *CPNE3* | -0.508718346 | 2.02E-09 | 8.14E-09 | 7.94E-06 |
| T1DM | leukocyte | *RRN3* | -0.586753478 | 1.83E-09 | 8.14E-09 | 7.19E-06 |
| T1DM | leukocyte | *WNT11* | -1.28999027 | 4.08E-05 | 4.97E-05 | 0.160289651 |
| T1DM | leukocyte | *DDHD2* | -0.68580844 | 1.55E-07 | 2.22E-07 | 0.00060706 |
| T1DM | leukocyte | *TTC39A* | -0.668389863 | 1.40E-08 | 2.48E-08 | 5.49E-05 |
| T1DM | leukocyte | *EPS15* | -0.738837944 | 7.94E-07 | 1.06E-06 | 0.003120277 |
| T1DM | leukocyte | *ORC1* | -1.944754465 | 9.77E-07 | 1.30E-06 | 0.003839021 |
| T1DM | leukocyte | *MGST2* | -0.47676153 | 3.79E-09 | 9.41E-09 | 1.49E-05 |
| T1DM | leukocyte | *USP40* | -1.005666304 | 3.31E-09 | 8.82E-09 | 1.30E-05 |
| T1DM | leukocyte | *POMGNT1* | -0.850807468 | 1.51E-09 | 8.14E-09 | 5.91E-06 |
| T1DM | leukocyte | *NFX1* | -1.011687087 | 5.16E-09 | 1.12E-08 | 2.03E-05 |
| T1DM | leukocyte | *EIF2AK1* | -0.626379942 | 3.06E-08 | 4.99E-08 | 0.000120148 |
| T1DM | leukocyte | *EPDR1* | 1.347674301 | 0.001878883 | 0.002138508 | 1 |
| T1DM | leukocyte | *ITPKC* | -0.83783512 | 1.61E-09 | 8.14E-09 | 6.32E-06 |
| T1DM | leukocyte | *TMED2* | -0.492591038 | 2.33E-09 | 8.21E-09 | 9.17E-06 |
| T1DM | leukocyte | *ACOX3* | -1.243475797 | 2.34E-07 | 3.32E-07 | 0.000920741 |
| T1DM | leukocyte | *TRIP6* | -0.492791848 | 8.42E-08 | 1.26E-07 | 0.000330628 |
| T1DM | leukocyte | *ACHE* | 0.596116956 | 0.003069403 | 0.003473412 | 1 |
| T1DM | leukocyte | *PGS1* | -1.030300057 | 1.41E-09 | 8.14E-09 | 5.53E-06 |
| T1DM | leukocyte | *PSMC5* | -0.550932229 | 3.83E-10 | 8.14E-09 | 1.50E-06 |
| T1DM | leukocyte | *UIMC1* | -0.718225765 | 3.22E-09 | 8.75E-09 | 1.27E-05 |
| T1DM | leukocyte | *SH3BP2* | 0.869407851 | 1.73E-07 | 2.48E-07 | 0.000680678 |
| T1DM | leukocyte | *SF3B2* | -0.545925027 | 2.62E-09 | 8.21E-09 | 1.03E-05 |
| T1DM | leukocyte | *KLHL42* | -0.56829096 | 1.81E-09 | 8.14E-09 | 7.13E-06 |
| T1DM | leukocyte | *TFAP2C* | -1.517947867 | 1.35E-06 | 1.77E-06 | 0.005285234 |
| T1DM | leukocyte | *METTL2A* | -1.001350976 | 2.40E-09 | 8.21E-09 | 9.44E-06 |
| T1DM | leukocyte | *GP6* | 1.091319928 | 1.66E-08 | 2.88E-08 | 6.52E-05 |
| T1DM | leukocyte | *EDEM2* | -0.71792082 | 4.13E-08 | 6.55E-08 | 0.000162163 |
| T1DM | leukocyte | *SLC15A1* | -1.24987474 | 6.68E-08 | 1.02E-07 | 0.000262419 |
| T1DM | leukocyte | *TGDS* | -0.690034774 | 2.69E-09 | 8.26E-09 | 1.06E-05 |
| T1DM | leukocyte | *DOCK3* | -0.782571042 | 1.53E-09 | 8.14E-09 | 6.02E-06 |
| T1DM | leukocyte | *ARHGAP28* | -1.76620239 | 1.32E-09 | 8.14E-09 | 5.19E-06 |
| T1DM | leukocyte | *PPP1R13B* | -0.495043444 | 5.60E-09 | 1.18E-08 | 2.20E-05 |
| T1DM | leukocyte | *ZNF343* | -0.956905714 | 5.55E-10 | 8.14E-09 | 2.18E-06 |
| T1DM | leukocyte | *MAVS* | -1.134622602 | 9.24E-08 | 1.38E-07 | 0.00036295 |
| T1DM | leukocyte | *DYNLL1* | -0.509084571 | 1.95E-08 | 3.32E-08 | 7.68E-05 |
| T1DM | leukocyte | *RBBP9* | -0.628547831 | 6.91E-10 | 8.14E-09 | 2.72E-06 |
| T1DM | leukocyte | *SLC23A2* | -1.011765722 | 1.38E-08 | 2.46E-08 | 5.43E-05 |
| T1DM | leukocyte | *SLC8B1* | 0.223448568 | 4.67E-09 | 1.06E-08 | 1.84E-05 |
| T1DM | leukocyte | *TMEM230* | -0.624482831 | 5.62E-10 | 8.14E-09 | 2.21E-06 |
| T1DM | leukocyte | *KIF16B* | -0.785365716 | 1.40E-06 | 1.84E-06 | 0.005500812 |
| T1DM | leukocyte | *ZNF302* | -0.641518179 | 6.41E-10 | 8.14E-09 | 2.52E-06 |
| T1DM | leukocyte | *GRAMD1A* | -0.987316991 | 5.61E-09 | 1.18E-08 | 2.20E-05 |
| T1DM | leukocyte | *CDIP1* | -0.704694112 | 1.71E-09 | 8.14E-09 | 6.73E-06 |
| T1DM | leukocyte | *MLF2* | -0.661460517 | 2.83E-09 | 8.42E-09 | 1.11E-05 |
| T1DM | leukocyte | *DDX24* | -0.53446751 | 2.44E-09 | 8.21E-09 | 9.60E-06 |
| T1DM | leukocyte | *ANKRD24* | -4.893161637 | 8.48E-08 | 1.27E-07 | 0.000333256 |
| T1DM | leukocyte | *GPATCH2L* | -0.751758237 | 3.90E-09 | 9.52E-09 | 1.53E-05 |
| T1DM | leukocyte | *SLC9A1* | -0.587822794 | 6.25E-09 | 1.28E-08 | 2.46E-05 |
| T1DM | leukocyte | *SPTLC1* | -0.625976504 | 5.50E-09 | 1.17E-08 | 2.16E-05 |

| | | | | | | |
|---|---|---|---|---|---|---|
| T1DM | leukocyte | *YPEL3* | -0.664915067 | 1.05E-07 | 1.55E-07 | 0.00041294 |
| T1DM | leukocyte | *NDUFB2* | -0.481081963 | 1.32E-08 | 2.38E-08 | 5.20E-05 |
| T1DM | leukocyte | *MAEA* | -0.528920716 | 1.24E-07 | 1.81E-07 | 0.000486929 |
| T1DM | leukocyte | *STRN4* | -0.641139247 | 1.82E-09 | 8.14E-09 | 7.17E-06 |
| T1DM | leukocyte | *LYZ* | 0.169911896 | 3.34E-09 | 8.82E-09 | 1.31E-05 |
| T1DM | leukocyte | *MUL1* | -0.704758251 | 9.57E-10 | 8.14E-09 | 3.76E-06 |
| T1DM | leukocyte | *SPG21* | -0.844142401 | 2.48E-09 | 8.21E-09 | 9.75E-06 |
| T1DM | leukocyte | *USP48* | -0.640119386 | 7.27E-09 | 1.44E-08 | 2.86E-05 |
| T1DM | leukocyte | *GLG1* | -0.644947553 | 1.46E-09 | 8.14E-09 | 5.73E-06 |
| T1DM | leukocyte | *NRCAM* | -0.444685531 | 1.46E-09 | 8.14E-09 | 5.72E-06 |
| T1DM | leukocyte | *TF* | -0.905756859 | 8.03E-07 | 1.07E-06 | 0.003155084 |
| T1DM | leukocyte | *MYO15A* | -1.418241946 | 2.92E-07 | 4.08E-07 | 0.00114923 |
| T1DM | leukocyte | *PITPNM3* | -1.916509921 | 1.29E-09 | 8.14E-09 | 5.08E-06 |
| T1DM | leukocyte | *SPAG7* | -0.650063004 | 6.76E-08 | 1.03E-07 | 0.000265533 |
| T1DM | leukocyte | *TMEM101* | -0.803442045 | 3.83E-09 | 9.46E-09 | 1.51E-05 |
| T1DM | leukocyte | *PPP2R3C* | -0.771780849 | 5.51E-06 | 6.96E-06 | 0.021649383 |
| T1DM | leukocyte | *OSGEP* | -0.918432991 | 5.78E-07 | 7.83E-07 | 0.002272517 |
| T1DM | leukocyte | *SCFD1* | -0.653324243 | 3.90E-09 | 9.52E-09 | 1.53E-05 |
| T1DM | leukocyte | *RPGRIP1* | 0.953375489 | 2.97E-09 | 8.50E-09 | 1.17E-05 |
| T1DM | leukocyte | *EZR* | -0.45822211 | 1.51E-09 | 8.14E-09 | 5.95E-06 |
| T1DM | leukocyte | *NUP50* | -1.049142434 | 1.17E-08 | 2.13E-08 | 4.60E-05 |
| T1DM | leukocyte | *COMT* | -0.580815103 | 1.84E-09 | 8.14E-09 | 7.22E-06 |
| T1DM | leukocyte | *ECHDC1* | -0.547156036 | 2.16E-09 | 8.21E-09 | 8.50E-06 |
| T1DM | leukocyte | *CBX5* | -0.547851627 | 1.60E-09 | 8.14E-09 | 6.27E-06 |
| T1DM | leukocyte | *SUCO* | -0.458109599 | 2.39E-09 | 8.21E-09 | 9.40E-06 |
| T1DM | leukocyte | *MSH2* | -1.363584967 | 2.21E-09 | 8.21E-09 | 8.67E-06 |
| T1DM | leukocyte | *TMEM38B* | -0.831221567 | 4.57E-08 | 7.21E-08 | 0.000179498 |
| T1DM | leukocyte | *PSMD5* | -0.628956999 | 1.59E-09 | 8.14E-09 | 6.26E-06 |
| T1DM | leukocyte | *CRAT* | -1.050276335 | 2.55E-09 | 8.21E-09 | 1.00E-05 |
| T1DM | leukocyte | *SH2D3C* | -0.968204611 | 2.13E-09 | 8.18E-09 | 8.37E-06 |
| T1DM | leukocyte | *CWF19L1* | -0.632198764 | 5.53E-09 | 1.17E-08 | 2.17E-05 |
| T1DM | leukocyte | *SEMA4G* | -2.307657455 | 0.034205871 | 0.037177004 | 1 |
| T1DM | leukocyte | *IKZF5* | -0.957517447 | 5.01E-09 | 1.10E-08 | 1.97E-05 |
| T1DM | leukocyte | *TLL2* | -2.810672794 | 1.06E-09 | 8.14E-09 | 4.16E-06 |
| T1DM | leukocyte | *TPSD1* | -22.0801647 | 4.12E-07 | 5.68E-07 | 0.001619679 |
| T1DM | leukocyte | *SRPK1* | -0.755353107 | 3.20E-09 | 8.73E-09 | 1.26E-05 |
| T1DM | leukocyte | *PGC* | -0.345549303 | 0.000794818 | 0.000917942 | 1 |
| T1DM | leukocyte | *EFHC1* | -0.575006458 | 7.74E-09 | 1.52E-08 | 3.04E-05 |
| T1DM | leukocyte | *ITPR3* | -0.70337988 | 3.60E-09 | 9.12E-09 | 1.41E-05 |
| T1DM | leukocyte | *IFT74* | -0.863860505 | 6.39E-09 | 1.30E-08 | 2.51E-05 |
| T1DM | leukocyte | *ABL1* | -0.679101515 | 2.58E-09 | 8.21E-09 | 1.01E-05 |
| T1DM | leukocyte | *SH3GLB1* | -0.662018426 | 3.39E-09 | 8.89E-09 | 1.33E-05 |
| T1DM | leukocyte | *CDC7* | -0.89524395 | 1.22E-08 | 2.20E-08 | 4.78E-05 |
| T1DM | leukocyte | *SYDE2* | -0.734775719 | 1.93E-08 | 3.28E-08 | 7.57E-05 |
| T1DM | leukocyte | *ABLIM1* | -0.536328405 | 9.73E-09 | 1.82E-08 | 3.82E-05 |
| T1DM | leukocyte | *ERMP1* | -0.661897182 | 1.93E-09 | 8.14E-09 | 7.57E-06 |
| T1DM | leukocyte | *PALMD* | -0.460804726 | 1.86E-06 | 2.42E-06 | 0.007297552 |
| T1DM | leukocyte | *MZF1* | -0.637876342 | 1.39E-07 | 2.02E-07 | 0.00054783 |
| T1DM | leukocyte | *OCEL1* | -0.851655685 | 5.45E-09 | 1.16E-08 | 2.14E-05 |
| T1DM | leukocyte | *CATSPERG* | -0.803631596 | 0.00019733 | 0.000233246 | 0.775308517 |

| | | | | | | |
|---|---|---|---|---|---|---|
| T1DM | leukocyte | *STX1B* | 0.663187839 | 2.08E-06 | 2.70E-06 | 0.008175093 |
| T1DM | leukocyte | *HSD3B7* | -1.682672944 | 1.07E-06 | 1.42E-06 | 0.004212056 |
| T1DM | leukocyte | *HNRNPM* | -0.518121843 | 4.22E-09 | 9.97E-09 | 1.66E-05 |
| T1DM | leukocyte | *TIMM13* | -0.6377243 | 2.78E-09 | 8.34E-09 | 1.09E-05 |
| T1DM | leukocyte | *CEP170B* | -0.961531477 | 2.76E-09 | 8.33E-09 | 1.08E-05 |
| T1DM | leukocyte | *POLR2E* | -0.849208626 | 3.81E-09 | 9.44E-09 | 1.50E-05 |
| T1DM | leukocyte | *RASSF7* | -0.592841065 | 1.62E-09 | 8.14E-09 | 6.36E-06 |
| T1DM | leukocyte | *ARVCF* | -1.271759784 | 7.70E-10 | 8.14E-09 | 3.03E-06 |
| T1DM | leukocyte | *TRMT2A* | -0.657680352 | 4.86E-10 | 8.14E-09 | 1.91E-06 |
| T1DM | leukocyte | *MED15* | -1.409247926 | 3.34E-07 | 4.64E-07 | 0.001312828 |
| T1DM | leukocyte | *LZTR1* | -0.935307256 | 7.21E-07 | 9.69E-07 | 0.002832351 |
| T1DM | leukocyte | *SMARCB1* | -0.687667018 | 2.91E-09 | 8.49E-09 | 1.14E-05 |
| T1DM | leukocyte | *DDT* | -0.632730005 | 2.84E-08 | 4.66E-08 | 0.000111459 |
| T1DM | leukocyte | *CABIN1* | -0.857906927 | 3.15E-09 | 8.69E-09 | 1.24E-05 |
| T1DM | leukocyte | *SPECC1L* | -0.97016319 | 6.11E-09 | 1.26E-08 | 2.40E-05 |
| T1DM | leukocyte | *PES1* | -0.694740314 | 5.09E-10 | 8.14E-09 | 2.00E-06 |
| T1DM | leukocyte | *TOP3B* | -1.028489185 | 8.91E-09 | 1.69E-08 | 3.50E-05 |
| T1DM | leukocyte | *CARD10* | -0.466794497 | 2.77E-08 | 4.55E-08 | 0.000108714 |
| T1DM | leukocyte | *SLC25A1* | -0.725579103 | 2.97E-09 | 8.50E-09 | 1.17E-05 |
| T1DM | leukocyte | *HPS4* | -0.745772327 | 2.71E-09 | 8.26E-09 | 1.07E-05 |
| T1DM | leukocyte | *GCAT* | -0.953404026 | 1.32E-09 | 8.14E-09 | 5.19E-06 |
| T1DM | leukocyte | *CCDC134* | -1.637123669 | 3.14E-06 | 4.02E-06 | 0.012331363 |
| T1DM | leukocyte | *CYP2D6* | 1.906830427 | 8.28E-09 | 1.60E-08 | 3.25E-05 |
| T1DM | leukocyte | *DDX17* | -0.622117339 | 1.32E-08 | 2.37E-08 | 5.18E-05 |
| T1DM | leukocyte | *HSCB* | -0.980893114 | 2.66E-07 | 3.73E-07 | 0.00104535 |
| T1DM | leukocyte | *CBY1* | -0.725612589 | 2.41E-09 | 8.21E-09 | 9.48E-06 |
| T1DM | leukocyte | *XBP1* | -0.462362858 | 2.71E-09 | 8.26E-09 | 1.06E-05 |
| T1DM | leukocyte | *TIMP3* | -0.736016955 | 4.78E-09 | 1.07E-08 | 1.88E-05 |
| T1DM | leukocyte | *CYB5R3* | -0.606682764 | 6.90E-09 | 1.39E-08 | 2.71E-05 |
| T1DM | leukocyte | *MIOX* | -1.631469137 | 1.27E-09 | 8.14E-09 | 4.99E-06 |
| T1DM | leukocyte | *LMF2* | -1.111054484 | 8.48E-08 | 1.27E-07 | 0.000333232 |
| T1DM | leukocyte | *RHBDD3* | -0.741556592 | 6.53E-09 | 1.33E-08 | 2.56E-05 |
| T1DM | leukocyte | *AP1B1* | 0.704526993 | 0.009530711 | 0.010583992 | 1 |
| T1DM | leukocyte | *TOM1* | -0.983204501 | 1.74E-08 | 3.01E-08 | 6.85E-05 |
| T1DM | leukocyte | *CHKB* | -1.512887726 | 6.43E-08 | 9.87E-08 | 0.00025247 |
| T1DM | leukocyte | *BIK* | -1.03398048 | 2.43E-06 | 3.13E-06 | 0.009530081 |
| T1DM | leukocyte | *ARSA* | -0.450234341 | 0.00022882 | 0.000269494 | 0.89903246 |
| T1DM | leukocyte | *TTLL12* | -1.02127702 | 1.40E-09 | 8.14E-09 | 5.51E-06 |
| T1DM | leukocyte | *RBFOX2* | -0.620788922 | 5.89E-09 | 1.22E-08 | 2.31E-05 |
| T1DM | leukocyte | *SYNGR1* | -1.316019501 | 4.13E-06 | 5.25E-06 | 0.016226648 |
| T1DM | leukocyte | *MTMR3* | -0.764247034 | 7.14E-08 | 1.08E-07 | 0.000280393 |
| T1DM | leukocyte | *APOL4* | -1.205763427 | 0.002378622 | 0.002700262 | 1 |
| T1DM | leukocyte | *SAMM50* | -0.641295376 | 8.03E-10 | 8.14E-09 | 3.15E-06 |
| T1DM | leukocyte | *FOXRED2* | -0.773662834 | 3.13E-09 | 8.69E-09 | 1.23E-05 |
| T1DM | leukocyte | *SGSM3* | -0.94042904 | 4.88E-09 | 1.08E-08 | 1.92E-05 |
| T1DM | leukocyte | *IFT27* | -0.828425249 | 1.03E-09 | 8.14E-09 | 4.05E-06 |
| T1DM | leukocyte | *FAM118A* | -0.608920777 | 9.20E-05 | 0.00011017 | 0.361577428 |
| T1DM | leukocyte | *ST13* | -0.558036615 | 3.03E-09 | 8.56E-09 | 1.19E-05 |
| T1DM | leukocyte | *POLR3H* | -1.295607135 | 5.39E-08 | 8.40E-08 | 0.000211704 |
| T1DM | leukocyte | *PMM1* | -0.857077329 | 3.43E-09 | 8.93E-09 | 1.35E-05 |

| | | | | | | |
|---|---|---|---|---|---|---|
| T1DM | leukocyte | *DESI1* | -0.871382994 | 2.95E-09 | 8.50E-09 | 1.16E-05 |
| T1DM | leukocyte | *HDAC10* | 1.064769873 | 2.83E-05 | 3.47E-05 | 0.111145414 |
| T1DM | leukocyte | *KHNYN* | -0.996924136 | 5.94E-11 | 8.14E-09 | 2.33E-07 |
| T1DM | leukocyte | *RBM23* | -0.599101883 | 3.28E-08 | 5.33E-08 | 0.000128976 |
| T1DM | leukocyte | *PRMT5* | -0.648975846 | 1.46E-09 | 8.14E-09 | 5.75E-06 |
| T1DM | leukocyte | *VCPKMT* | -0.948726615 | 1.71E-09 | 8.14E-09 | 6.70E-06 |
| T1DM | leukocyte | *SOS2* | -0.822187331 | 4.59E-09 | 1.04E-08 | 1.80E-05 |
| T1DM | leukocyte | *CDKL1* | -1.147942641 | 3.63E-08 | 5.84E-08 | 0.000142731 |
| T1DM | leukocyte | *NIN* | -1.361746387 | 1.73E-08 | 2.98E-08 | 6.78E-05 |
| T1DM | leukocyte | *PYGL* | 1.394869731 | 1.27E-05 | 1.57E-05 | 0.049834837 |
| T1DM | leukocyte | *TRIM9* | -0.537890642 | 1.19E-09 | 8.14E-09 | 4.67E-06 |
| T1DM | leukocyte | *ATP6V1D* | -0.603233626 | 1.66E-09 | 8.14E-09 | 6.51E-06 |
| T1DM | leukocyte | *PLEK2* | -0.965235226 | 5.00E-08 | 7.83E-08 | 0.000196554 |
| T1DM | leukocyte | *GSTZ1* | -0.931646096 | 1.54E-08 | 2.70E-08 | 6.03E-05 |
| T1DM | leukocyte | *TMED8* | -0.743805592 | 3.33E-09 | 8.82E-09 | 1.31E-05 |
| T1DM | leukocyte | *DAAM1* | -0.598454395 | 1.54E-09 | 8.14E-09 | 6.07E-06 |
| T1DM | leukocyte | *DHRS7* | -0.676668248 | 5.38E-09 | 1.15E-08 | 2.11E-05 |
| T1DM | leukocyte | *MTHFD1* | -1.858983857 | 8.93E-07 | 1.19E-06 | 0.003509091 |
| T1DM | leukocyte | *TELO2* | -0.652359393 | 1.15E-05 | 1.43E-05 | 0.045110413 |
| T1DM | leukocyte | *GSKIP* | -0.671662322 | 1.28E-09 | 8.14E-09 | 5.03E-06 |
| T1DM | leukocyte | *TRIP11* | -0.574886656 | 1.59E-09 | 8.14E-09 | 6.23E-06 |
| T1DM | leukocyte | *DCAF11* | -0.814046113 | 1.35E-08 | 2.41E-08 | 5.29E-05 |
| T1DM | leukocyte | *EMC9* | -0.673526909 | 2.02E-08 | 3.41E-08 | 7.92E-05 |
| T1DM | leukocyte | *NFATC4* | -0.958380738 | 1.51E-09 | 8.14E-09 | 5.94E-06 |
| T1DM | leukocyte | *PYGB* | -0.603901223 | 5.79E-07 | 7.83E-07 | 0.002274174 |
| T1DM | leukocyte | *ABHD12* | -0.976698521 | 8.38E-07 | 1.12E-06 | 0.003291963 |
| T1DM | leukocyte | *PROCR* | -0.622388395 | 4.98E-10 | 8.14E-09 | 1.96E-06 |
| T1DM | leukocyte | *CD40* | -0.747134759 | 1.48E-09 | 8.14E-09 | 5.83E-06 |
| T1DM | leukocyte | *IFT52* | -0.658772985 | 1.02E-09 | 8.14E-09 | 4.00E-06 |
| T1DM | leukocyte | *MYBL2* | -1.494628864 | 4.49E-07 | 6.16E-07 | 0.001765925 |
| T1DM | leukocyte | *PABPC1L* | -0.320616062 | 0.019056689 | 0.020926141 | 1 |
| T1DM | leukocyte | *SALL4* | 0.93719024 | 1.71E-06 | 2.24E-06 | 0.006737105 |
| T1DM | leukocyte | *NELFCD* | -0.622943545 | 1.66E-09 | 8.14E-09 | 6.52E-06 |
| T1DM | leukocyte | *MTG2* | -0.741937959 | 3.27E-09 | 8.79E-09 | 1.29E-05 |
| T1DM | leukocyte | *DIDO1* | -0.730434328 | 6.83E-08 | 1.04E-07 | 0.000268518 |
| T1DM | leukocyte | *SLC17A9* | 0.805488738 | 5.51E-08 | 8.56E-08 | 0.000216335 |
| T1DM | leukocyte | *CDC25B* | -1.097201622 | 2.20E-07 | 3.11E-07 | 0.00086258 |
| T1DM | leukocyte | *ISM1* | 1.094897809 | 5.86E-09 | 1.22E-08 | 2.30E-05 |
| T1DM | leukocyte | *ARFRP1* | -1.006970173 | 4.23E-09 | 9.97E-09 | 1.66E-05 |
| T1DM | leukocyte | *SLC52A3* | -1.336041956 | 2.98E-06 | 3.82E-06 | 0.01169006 |
| T1DM | leukocyte | *RSPO4* | -5.784273787 | 0.0034132 | 0.003858015 | 1 |
| T1DM | leukocyte | *SIRPB1* | 0.506153589 | 3.74E-09 | 9.32E-09 | 1.47E-05 |
| T1DM | leukocyte | *PLCB4* | -0.541107573 | 9.39E-08 | 1.40E-07 | 0.000368831 |
| T1DM | leukocyte | *POFUT1* | -0.658076556 | 1.42E-09 | 8.14E-09 | 5.59E-06 |
| T1DM | leukocyte | *NOP56* | -0.872732658 | 6.05E-09 | 1.25E-08 | 2.38E-05 |
| T1DM | leukocyte | *MANBAL* | -0.613413908 | 1.91E-09 | 8.14E-09 | 7.51E-06 |
| T1DM | leukocyte | *IDH3B* | -0.797774362 | 1.96E-09 | 8.14E-09 | 7.72E-06 |
| T1DM | leukocyte | *MAPRE1* | -0.53320056 | 5.47E-09 | 1.17E-08 | 2.15E-05 |
| T1DM | leukocyte | *OXT* | 0.660475684 | 7.39E-09 | 1.46E-08 | 2.90E-05 |
| T1DM | leukocyte | *TTI1* | -1.012991175 | 2.04E-09 | 8.14E-09 | 8.03E-06 |

| | | | | | | |
|---|---|---|---|---|---|---|
| T1DM | leukocyte | *CHMP4B* | -0.546487965 | 2.07E-09 | 8.14E-09 | 8.14E-06 |
| T1DM | leukocyte | *DHX35* | -1.070558405 | 1.63E-08 | 2.83E-08 | 6.39E-05 |
| T1DM | leukocyte | *DNTTIP1* | -1.28803477 | 2.33E-08 | 3.90E-08 | 9.15E-05 |
| T1DM | leukocyte | *MAP1LC3A* | -0.420588827 | 5.93E-08 | 9.15E-08 | 0.00023296 |
| T1DM | leukocyte | *APMAP* | -0.592920563 | 2.09E-06 | 2.71E-06 | 0.008202977 |
| T1DM | leukocyte | *VAPA* | -0.526991405 | 2.61E-09 | 8.21E-09 | 1.02E-05 |
| T1DM | leukocyte | *MYOM1* | -1.59815577 | 7.16E-09 | 1.43E-08 | 2.82E-05 |
| T1DM | leukocyte | *CEP192* | -0.52684932 | 1.30E-07 | 1.89E-07 | 0.00051236 |
| T1DM | leukocyte | *LIPG* | -0.753937058 | 1.35E-09 | 8.14E-09 | 5.32E-06 |
| T1DM | leukocyte | *LAMA1* | -2.723414237 | 2.87E-05 | 3.52E-05 | 0.112955646 |
| T1DM | leukocyte | *POLI* | -0.713920354 | 0.000266553 | 0.000313091 | 1 |
| T1DM | leukocyte | *RIOK3* | -0.581863725 | 3.16E-09 | 8.69E-09 | 1.24E-05 |
| T1DM | leukocyte | *NALCN* | -0.652017529 | 1.38E-09 | 8.14E-09 | 5.40E-06 |
| T1DM | leukocyte | *FGF14* | -0.573586638 | 2.46E-07 | 3.47E-07 | 0.000967184 |
| T1DM | leukocyte | *FNDC3A* | -0.512434637 | 1.75E-09 | 8.14E-09 | 6.88E-06 |
| T1DM | leukocyte | *KLF5* | -0.497520655 | 1.20E-09 | 8.14E-09 | 4.71E-06 |
| T1DM | leukocyte | *ACP5* | 0.046549814 | 4.28E-09 | 1.00E-08 | 1.68E-05 |
| T1DM | leukocyte | *DNAJC3* | -0.564942904 | 2.81E-09 | 8.40E-09 | 1.10E-05 |
| T1DM | leukocyte | *UGGT2* | -0.614862736 | 3.16E-09 | 8.69E-09 | 1.24E-05 |
| T1DM | leukocyte | *ARHGEF7* | -0.847141508 | 3.28E-09 | 8.79E-09 | 1.29E-05 |
| T1DM | leukocyte | *PARP4* | -1.396030326 | 8.01E-07 | 1.07E-06 | 0.00314753 |
| T1DM | leukocyte | *MEDAG* | -1.023376395 | 1.75E-08 | 3.01E-08 | 6.86E-05 |
| T1DM | leukocyte | *CLN5* | -0.775091892 | 4.76E-09 | 1.07E-08 | 1.87E-05 |
| T1DM | leukocyte | *OLFM4* | -0.330828356 | 2.26E-09 | 8.21E-09 | 8.86E-06 |
| T1DM | leukocyte | *MGRN1* | -0.745156904 | 7.46E-08 | 1.13E-07 | 0.000293043 |
| T1DM | leukocyte | *LONP2* | -0.639135394 | 2.02E-09 | 8.14E-09 | 7.93E-06 |
| T1DM | leukocyte | *CBLN1* | -1.84355106 | 6.00E-07 | 8.11E-07 | 0.002359136 |
| T1DM | leukocyte | *DHODH* | -1.787013066 | 6.65E-09 | 1.35E-08 | 2.61E-05 |
| T1DM | leukocyte | *PARD6A* | -0.48961139 | 4.14E-09 | 9.90E-09 | 1.63E-05 |
| T1DM | leukocyte | *MMP15* | -1.150215044 | 1.18E-06 | 1.55E-06 | 0.004622287 |
| T1DM | leukocyte | *USB1* | -0.942119322 | 3.01E-09 | 8.54E-09 | 1.18E-05 |
| T1DM | leukocyte | *CYB5B* | -1.110285405 | 1.92E-08 | 3.28E-08 | 7.55E-05 |
| T1DM | leukocyte | *CCDC113* | -0.774750697 | 3.33E-07 | 4.62E-07 | 0.001307933 |
| T1DM | leukocyte | *NME3* | -0.681421843 | 8.49E-09 | 1.63E-08 | 3.34E-05 |
| T1DM | leukocyte | *VAC14* | -0.733572248 | 8.96E-10 | 8.14E-09 | 3.52E-06 |
| T1DM | leukocyte | *SMPD3* | 1.479924953 | 3.32E-06 | 4.24E-06 | 0.013044792 |
| T1DM | leukocyte | *NPRL3* | -0.930093219 | 3.47E-08 | 5.61E-08 | 0.000136284 |
| T1DM | leukocyte | *TAF1C* | 0.323187141 | 0.018146902 | 0.019949407 | 1 |
| T1DM | leukocyte | *NAGPA* | -1.006969774 | 0.045098712 | 0.048826905 | 1 |
| T1DM | leukocyte | *ZNF500* | -1.296024152 | 9.58E-09 | 1.80E-08 | 3.76E-05 |
| T1DM | leukocyte | *NME4* | -0.694982849 | 3.24E-09 | 8.77E-09 | 1.27E-05 |
| T1DM | leukocyte | *NOMO3* | -1.016937697 | 1.08E-09 | 8.14E-09 | 4.24E-06 |
| T1DM | leukocyte | *LMF1* | -0.692826023 | 1.55E-09 | 8.14E-09 | 6.08E-06 |
| T1DM | leukocyte | *MTHFSD* | -0.333791104 | 0.015503556 | 0.017096119 | 1 |
| T1DM | leukocyte | *HAGHL* | -1.08981238 | 1.97E-09 | 8.14E-09 | 7.75E-06 |
| T1DM | leukocyte | *METRN* | -0.538915191 | 1.19E-09 | 8.14E-09 | 4.67E-06 |
| T1DM | leukocyte | *PIEZO1* | -0.801642049 | 4.11E-09 | 9.85E-09 | 1.62E-05 |
| T1DM | leukocyte | *CLUAP1* | -0.663694807 | 5.43E-07 | 7.39E-07 | 0.002133504 |
| T1DM | leukocyte | *UBFD1* | -0.787397272 | 3.39E-07 | 4.70E-07 | 0.001331638 |
| T1DM | leukocyte | *AQP8* | -1.121701432 | 1.42E-09 | 8.14E-09 | 5.59E-06 |

| | | | | | | |
|---|---|---|---|---|---|---|
| T1DM | leukocyte | *USP31* | -0.499402352 | 8.35E-09 | 1.61E-08 | 3.28E-05 |
| T1DM | leukocyte | *HMOX2* | -0.602632662 | 3.80E-09 | 9.42E-09 | 1.49E-05 |
| T1DM | leukocyte | *RBL2* | -0.833825284 | 3.82E-09 | 9.44E-09 | 1.50E-05 |
| T1DM | leukocyte | *KAT8* | -0.786804307 | 2.02E-09 | 8.14E-09 | 7.93E-06 |
| T1DM | leukocyte | *NOMO1* | -0.909693401 | 5.23E-07 | 7.13E-07 | 0.002054376 |
| T1DM | leukocyte | *RNF40* | -1.106087019 | 1.73E-08 | 2.98E-08 | 6.78E-05 |
| T1DM | leukocyte | *KNOP1* | -0.832863319 | 1.91E-09 | 8.14E-09 | 7.51E-06 |
| T1DM | leukocyte | *TRIP4* | -0.731836379 | 1.99E-09 | 8.14E-09 | 7.81E-06 |
| T1DM | leukocyte | *AP3B2* | -0.752209404 | 1.41E-09 | 8.14E-09 | 5.55E-06 |
| T1DM | leukocyte | *IGDCC4* | 1.824997136 | 2.75E-08 | 4.53E-08 | 0.000108084 |
| T1DM | leukocyte | *RPAP1* | -1.040681207 | 1.24E-09 | 8.14E-09 | 4.88E-06 |
| T1DM | leukocyte | *HOMER2* | -0.52526979 | 6.00E-09 | 1.24E-08 | 2.36E-05 |
| T1DM | leukocyte | *CEP152* | 0.750621704 | 0.005347699 | 0.005998032 | 1 |
| T1DM | leukocyte | *ATP8B4* | 0.402129818 | 3.40E-09 | 8.90E-09 | 1.33E-05 |
| T1DM | leukocyte | *DNAJC17* | -0.679636441 | 2.23E-09 | 8.21E-09 | 8.75E-06 |
| T1DM | leukocyte | *OIP5* | -2.500260067 | 1.84E-06 | 2.40E-06 | 0.007235422 |
| T1DM | leukocyte | *SLC30A4* | -1.074729918 | 0.000842491 | 0.000972429 | 1 |
| T1DM | leukocyte | *ZDHHC2* | -0.467952473 | 1.40E-09 | 8.14E-09 | 5.49E-06 |
| T1DM | leukocyte | *TRIM35* | -1.549263575 | 3.13E-09 | 8.69E-09 | 1.23E-05 |
| T1DM | leukocyte | *RIPK2* | -0.82637981 | 8.41E-10 | 8.14E-09 | 3.30E-06 |
| T1DM | leukocyte | *NBN* | -0.947791158 | 1.85E-09 | 8.14E-09 | 7.28E-06 |
| T1DM | leukocyte | *LAPTM4B* | -0.435144531 | 1.34E-09 | 8.14E-09 | 5.28E-06 |
| T1DM | leukocyte | *RAB2A* | -0.481547505 | 1.29E-09 | 8.14E-09 | 5.06E-06 |
| T1DM | leukocyte | *EIF3E* | -0.528421688 | 2.55E-09 | 8.21E-09 | 1.00E-05 |
| T1DM | leukocyte | *ESRP1* | -0.559223147 | 1.97E-09 | 8.14E-09 | 7.75E-06 |
| T1DM | leukocyte | *ARMC1* | -0.726712453 | 5.58E-09 | 1.18E-08 | 2.19E-05 |
| T1DM | leukocyte | *SPAG1* | -0.621616281 | 3.35E-09 | 8.83E-09 | 1.31E-05 |
| T1DM | leukocyte | *CHRAC1* | -0.562017686 | 9.98E-10 | 8.14E-09 | 3.92E-06 |
| T1DM | leukocyte | *SNX16* | -1.798535432 | 1.40E-08 | 2.49E-08 | 5.52E-05 |
| T1DM | leukocyte | *GSDMD* | -0.541998437 | 4.27E-09 | 1.00E-08 | 1.68E-05 |
| T1DM | leukocyte | *SH2D4A* | -0.776786483 | 8.81E-10 | 8.14E-09 | 3.46E-06 |
| T1DM | leukocyte | *INTS10* | -0.821335537 | 5.78E-09 | 1.21E-08 | 2.27E-05 |
| T1DM | leukocyte | *ERI1* | -1.464292355 | 2.93E-08 | 4.79E-08 | 0.000114953 |
| T1DM | leukocyte | *LEPROTL1* | -0.770645142 | 9.98E-10 | 8.14E-09 | 3.92E-06 |
| T1DM | leukocyte | *ERICH1* | -0.942419758 | 7.14E-10 | 8.14E-09 | 2.81E-06 |
| T1DM | leukocyte | *ARHGEF10* | -0.713021657 | 5.51E-09 | 1.17E-08 | 2.17E-05 |
| T1DM | leukocyte | *KLHDC4* | -0.763176153 | 8.28E-08 | 1.25E-07 | 0.000325399 |
| T1DM | leukocyte | *MCM4* | -0.845170504 | 1.55E-09 | 8.14E-09 | 6.09E-06 |
| T1DM | leukocyte | *FGL1* | -1.143059221 | 2.18E-08 | 3.66E-08 | 8.56E-05 |
| T1DM | leukocyte | *ASAH1* | 0.504089888 | 2.03E-06 | 2.64E-06 | 0.007980047 |
| T1DM | leukocyte | *MAN2B1* | 0.726118819 | 7.74E-06 | 9.70E-06 | 0.030399322 |
| T1DM | leukocyte | *ECH1* | -0.645171145 | 1.77E-10 | 8.14E-09 | 6.94E-07 |
| T1DM | leukocyte | *HNRNPL* | -0.660369377 | 2.48E-09 | 8.21E-09 | 9.76E-06 |
| T1DM | leukocyte | *SARS2* | -0.79866916 | 1.75E-09 | 8.14E-09 | 6.88E-06 |
| T1DM | leukocyte | *LIN7B* | -0.847152116 | 4.41E-09 | 1.02E-08 | 1.73E-05 |
| T1DM | leukocyte | *PPP1R13L* | -0.710656797 | 3.05E-08 | 4.97E-08 | 0.000119802 |
| T1DM | leukocyte | *ERCC2* | -0.751517836 | 1.20E-09 | 8.14E-09 | 4.71E-06 |
| T1DM | leukocyte | *KLC3* | -0.702009498 | 1.15E-10 | 8.14E-09 | 4.51E-07 |
| T1DM | leukocyte | *AMH* | -0.76450466 | 2.11E-09 | 8.16E-09 | 8.31E-06 |
| T1DM | leukocyte | *TLE6* | -0.965295216 | 2.89E-08 | 4.73E-08 | 0.000113438 |

| | | | | | | |
|---|---|---|---|---|---|---|
| T1DM | leukocyte | *IL27RA* | -0.860420225 | 6.59E-08 | 1.01E-07 | 0.000258955 |
| T1DM | leukocyte | *C19orf44* | -0.584277628 | 1.27E-05 | 1.57E-05 | 0.049733767 |
| T1DM | leukocyte | *CASP14* | 1.601993487 | 0.000204312 | 0.000241136 | 0.802742263 |
| T1DM | leukocyte | *PDCD5* | -0.515902839 | 3.62E-09 | 9.14E-09 | 1.42E-05 |
| T1DM | leukocyte | *NUMBL* | -0.583743707 | 1.42E-09 | 8.14E-09 | 5.57E-06 |
| T1DM | leukocyte | *SLC1A5* | -0.832478809 | 9.89E-09 | 1.84E-08 | 3.88E-05 |
| T1DM | leukocyte | *PRKD2* | -0.807555845 | 2.40E-10 | 8.14E-09 | 9.43E-07 |
| T1DM | leukocyte | *APLP1* | -0.307418489 | 4.79E-08 | 7.52E-08 | 0.000188068 |
| T1DM | leukocyte | *PLIN3* | -0.690089049 | 2.39E-09 | 8.21E-09 | 9.38E-06 |
| T1DM | leukocyte | *MYH14* | -0.567776205 | 9.29E-10 | 8.14E-09 | 3.65E-06 |
| T1DM | leukocyte | *CARD8* | -0.926011284 | 0.006591519 | 0.007372069 | 1 |
| T1DM | leukocyte | *ZNF175* | -0.652488009 | 8.80E-10 | 8.14E-09 | 3.46E-06 |
| T1DM | leukocyte | *PLA2G4C* | -0.865846861 | 4.99E-09 | 1.10E-08 | 1.96E-05 |
| T1DM | leukocyte | *CAPS* | -0.893852658 | 2.17E-09 | 8.21E-09 | 8.51E-06 |
| T1DM | leukocyte | *RASIP1* | -0.721816361 | 4.41E-07 | 6.04E-07 | 0.001731537 |
| T1DM | leukocyte | *PPP2R1A* | -0.564491656 | 3.37E-09 | 8.87E-09 | 1.33E-05 |
| T1DM | leukocyte | *GCDH* | -0.827247337 | 3.53E-10 | 8.14E-09 | 1.39E-06 |
| T1DM | leukocyte | *PIK3R2* | -1.504096988 | 1.00E-06 | 1.33E-06 | 0.003936409 |
| T1DM | leukocyte | *UPK1A* | -10.04180692 | 3.44E-09 | 8.94E-09 | 1.35E-05 |
| T1DM | leukocyte | *COPE* | -0.556682567 | 3.05E-09 | 8.59E-09 | 1.20E-05 |
| T1DM | leukocyte | *ETV2* | -0.757282392 | 5.70E-07 | 7.72E-07 | 0.002238957 |
| T1DM | leukocyte | *PBX4* | -2.587525782 | 1.95E-08 | 3.32E-08 | 7.67E-05 |
| T1DM | leukocyte | *SIPA1L3* | -1.424773018 | 1.20E-07 | 1.75E-07 | 0.000470527 |
| T1DM | leukocyte | *ETHE1* | -0.896895225 | 1.04E-07 | 1.54E-07 | 0.000409743 |
| T1DM | leukocyte | *SMG9* | -0.633971269 | 2.28E-09 | 8.21E-09 | 8.94E-06 |
| T1DM | leukocyte | *RUNDC3B* | -3.014558977 | 4.55E-08 | 7.18E-08 | 0.000178792 |
| T1DM | leukocyte | *PMPCB* | -0.755720537 | 3.18E-09 | 8.70E-09 | 1.25E-05 |
| T1DM | leukocyte | *PON2* | -0.575262351 | 2.61E-09 | 8.21E-09 | 1.03E-05 |
| T1DM | leukocyte | *ITGB8* | -0.607092518 | 1.87E-09 | 8.14E-09 | 7.37E-06 |
| T1DM | leukocyte | *DNAH11* | -2.124811876 | 1.13E-05 | 1.40E-05 | 0.044236549 |
| T1DM | leukocyte | *ADAP1* | 1.901232543 | 4.22E-08 | 6.70E-08 | 0.000165809 |
| T1DM | leukocyte | *CAV2* | -0.414823529 | 4.42E-09 | 1.02E-08 | 1.74E-05 |
| T1DM | leukocyte | *CAV1* | -0.505585846 | 1.37E-09 | 8.14E-09 | 5.40E-06 |
| T1DM | leukocyte | *ANKRD7* | -1.200085952 | 3.82E-09 | 9.44E-09 | 1.50E-05 |
| T1DM | leukocyte | *VIPR2* | -0.550032819 | 0.000668316 | 0.00077298 | 1 |
| T1DM | leukocyte | *TSPAN12* | -0.99970462 | 1.85E-09 | 8.14E-09 | 7.28E-06 |
| T1DM | leukocyte | *CPVL* | 0.098668279 | 3.93E-09 | 9.56E-09 | 1.55E-05 |
| T1DM | leukocyte | *ABHD11* | -0.892822075 | 4.69E-09 | 1.06E-08 | 1.84E-05 |
| T1DM | leukocyte | *CHCHD2* | -0.446912014 | 1.87E-09 | 8.14E-09 | 7.34E-06 |
| T1DM | leukocyte | *EIF3B* | -0.645266648 | 4.30E-09 | 1.01E-08 | 1.69E-05 |
| T1DM | leukocyte | *WASL* | -0.538715357 | 5.11E-09 | 1.12E-08 | 2.01E-05 |
| T1DM | leukocyte | *TFR2* | 1.052623175 | 0.0002011 | 0.00023756 | 0.790122977 |
| T1DM | leukocyte | *AGFG2* | -0.668321072 | 1.04E-09 | 8.14E-09 | 4.10E-06 |
| T1DM | leukocyte | *C1GALT1* | -0.576157369 | 1.58E-09 | 8.14E-09 | 6.21E-06 |
| T1DM | leukocyte | *RPA3* | -0.705082142 | 2.06E-09 | 8.14E-09 | 8.10E-06 |
| T1DM | leukocyte | *ZNHIT1* | -0.571849553 | 1.81E-09 | 8.14E-09 | 7.12E-06 |
| T1DM | leukocyte | *CLDN15* | -0.464073616 | 1.51E-06 | 1.98E-06 | 0.005913984 |
| T1DM | leukocyte | *TMEM106B* | -0.570887999 | 4.88E-09 | 1.08E-08 | 1.92E-05 |
| T1DM | leukocyte | *ACTR3C* | -1.071140935 | 4.28E-09 | 1.00E-08 | 1.68E-05 |
| T1DM | leukocyte | *TSPAN13* | -0.446747263 | 1.86E-09 | 8.14E-09 | 7.31E-06 |

| | | | | | | |
|---|---|---|---|---|---|---|
| T1DM | leukocyte | *RARRES2* | -0.327490985 | 7.06E-09 | 1.41E-08 | 2.77E-05 |
| T1DM | leukocyte | *AGR2* | -0.475765692 | 1.99E-08 | 3.38E-08 | 7.84E-05 |
| T1DM | leukocyte | *TMEM176B* | -0.356469628 | 0.000256859 | 0.000301975 | 1 |
| T1DM | leukocyte | *COA1* | -0.804213028 | 5.86E-09 | 1.22E-08 | 2.30E-05 |
| T1DM | leukocyte | *BLVRA* | -1.009782247 | 4.34E-08 | 6.88E-08 | 0.000170619 |
| T1DM | leukocyte | *TMEM248* | -0.744739266 | 8.10E-09 | 1.57E-08 | 3.18E-05 |
| T1DM | leukocyte | *RHEB* | -0.554217614 | 2.36E-09 | 8.21E-09 | 9.26E-06 |
| T1DM | leukocyte | *TBL2* | -0.610328656 | 3.57E-08 | 5.75E-08 | 0.000140101 |
| T1DM | leukocyte | *LIMK1* | -1.616101388 | 3.04E-05 | 3.73E-05 | 0.119566792 |
| T1DM | leukocyte | *SLC1A1* | -0.724167693 | 0.000490233 | 0.000569691 | 1 |
| T1DM | leukocyte | *SPIN1* | -0.916276091 | 8.07E-09 | 1.57E-08 | 3.17E-05 |
| T1DM | leukocyte | *NMRK1* | -0.702352504 | 3.09E-09 | 8.63E-09 | 1.21E-05 |
| T1DM | leukocyte | *C5* | -0.634023015 | 4.84E-05 | 5.88E-05 | 0.190141959 |
| T1DM | leukocyte | *OGN* | -1.497367014 | 2.71E-09 | 8.26E-09 | 1.06E-05 |
| T1DM | leukocyte | *SUSD1* | -1.067679506 | 3.56E-09 | 9.10E-09 | 1.40E-05 |
| T1DM | leukocyte | *DNM1* | -0.494760625 | 1.30E-09 | 8.14E-09 | 5.10E-06 |
| T1DM | leukocyte | *AK1* | -0.494128441 | 8.50E-09 | 1.63E-08 | 3.34E-05 |
| T1DM | leukocyte | *PLGRKT* | -0.57454277 | 3.46E-09 | 8.98E-09 | 1.36E-05 |
| T1DM | leukocyte | *DOCK8* | 0.340402651 | 9.38E-09 | 1.77E-08 | 3.68E-05 |
| T1DM | leukocyte | *MPDZ* | -0.560063027 | 4.30E-09 | 1.01E-08 | 1.69E-05 |
| T1DM | leukocyte | *RAPGEF1* | -0.518694811 | 0.010249171 | 0.011368999 | 1 |
| T1DM | leukocyte | *NPDC1* | -0.46270205 | 2.54E-09 | 8.21E-09 | 9.98E-06 |
| T1DM | leukocyte | *HPS1* | -0.769256814 | 2.40E-09 | 8.21E-09 | 9.42E-06 |
| T1DM | leukocyte | *CXCL12* | -0.657954184 | 2.52E-09 | 8.21E-09 | 9.90E-06 |
| T1DM | leukocyte | *TRDMT1* | -1.332864491 | 3.46E-08 | 5.59E-08 | 0.000135907 |
| T1DM | leukocyte | *ATE1* | -1.012982674 | 4.80E-09 | 1.07E-08 | 1.89E-05 |
| T1DM | leukocyte | *NSMCE4A* | -0.537966957 | 2.21E-07 | 3.13E-07 | 0.000868838 |
| T1DM | leukocyte | *PLEKHA1* | -0.50438864 | 3.09E-09 | 8.64E-09 | 1.21E-05 |
| T1DM | leukocyte | *PALD1* | -3.616821685 | 4.64E-08 | 7.29E-08 | 0.000182124 |
| T1DM | leukocyte | *SPOCK2* | -1.052046433 | 1.41E-09 | 8.14E-09 | 5.54E-06 |
| T1DM | leukocyte | *BMPR1A* | -1.316675005 | 9.67E-09 | 1.81E-08 | 3.80E-05 |
| T1DM | leukocyte | *ACTA2* | -0.446157626 | 0.033139444 | 0.036057844 | 1 |
| T1DM | leukocyte | *LIPA* | 0.241962025 | 9.56E-09 | 1.80E-08 | 3.76E-05 |
| T1DM | leukocyte | *SFXN3* | -1.028391216 | 6.06E-10 | 8.14E-09 | 2.38E-06 |
| T1DM | leukocyte | *NPM3* | -0.751158683 | 2.06E-09 | 8.14E-09 | 8.08E-06 |
| T1DM | leukocyte | *TNKS2* | -0.701356741 | 3.55E-09 | 9.10E-09 | 1.40E-05 |
| T1DM | leukocyte | *ACBD5* | -0.586547641 | 2.21E-09 | 8.21E-09 | 8.70E-06 |
| T1DM | leukocyte | *LHPP* | -1.042708475 | 9.13E-08 | 1.37E-07 | 0.000358709 |
| T1DM | leukocyte | *GTPBP4* | -0.556186131 | 1.79E-09 | 8.14E-09 | 7.03E-06 |
| T1DM | leukocyte | *NEURL1* | -0.772837906 | 2.55E-09 | 8.21E-09 | 1.00E-05 |
| T1DM | leukocyte | *SORCS1* | -2.151901355 | 0.009396455 | 0.010440801 | 1 |
| T1DM | leukocyte | *XPNPEP1* | -0.643410458 | 1.36E-09 | 8.14E-09 | 5.33E-06 |
| T1DM | leukocyte | *SMC3* | -0.543903127 | 1.54E-09 | 8.14E-09 | 6.03E-06 |
| T1DM | leukocyte | *RPL28* | -0.377146787 | 7.78E-08 | 1.17E-07 | 0.000305539 |
| T1DM | leukocyte | *PBLD* | -0.543717517 | 2.78E-08 | 4.57E-08 | 0.000109063 |
| T1DM | leukocyte | *KRT23* | -0.259216299 | 0.000335413 | 0.000392214 | 1 |
| T1DM | leukocyte | *DHRS11* | -0.597680805 | 2.57E-09 | 8.21E-09 | 1.01E-05 |
| T1DM | leukocyte | *CWC25* | -0.770375243 | 2.84E-09 | 8.43E-09 | 1.11E-05 |
| T1DM | leukocyte | *WNT3* | -0.8470709 | 2.52E-05 | 3.10E-05 | 0.098976032 |
| T1DM | leukocyte | *RAD51C* | -0.70724229 | 2.57E-09 | 8.21E-09 | 1.01E-05 |

| | | | | | | |
|---|---|---|---|---|---|---|
| T1DM | leukocyte | *TRIM37* | -0.640323086 | 1.59E-09 | 8.14E-09 | 6.23E-06 |
| T1DM | leukocyte | *RPS6KB1* | -0.719045272 | 9.43E-09 | 1.78E-08 | 3.70E-05 |
| T1DM | leukocyte | *TRIM16L* | -0.98343874 | 3.03E-08 | 4.95E-08 | 0.000119244 |
| T1DM | leukocyte | *CDK5RAP3* | -0.889495034 | 2.43E-07 | 3.43E-07 | 0.000955056 |
| T1DM | leukocyte | *CAMTA2* | -0.968209091 | 1.86E-08 | 3.18E-08 | 7.29E-05 |
| T1DM | leukocyte | *MED13* | -0.631606293 | 2.05E-09 | 8.14E-09 | 8.04E-06 |
| T1DM | leukocyte | *RAI1* | -0.55496067 | 1.32E-09 | 8.14E-09 | 5.18E-06 |
| T1DM | leukocyte | *CCDC47* | -0.605444907 | 1.18E-09 | 8.14E-09 | 4.65E-06 |
| T1DM | leukocyte | *DRG2* | -0.713596904 | 9.90E-10 | 8.14E-09 | 3.89E-06 |
| T1DM | leukocyte | *FTSJ3* | -0.637022935 | 1.96E-09 | 8.14E-09 | 7.69E-06 |
| T1DM | leukocyte | *SMARCD2* | -1.314581104 | 8.37E-09 | 1.61E-08 | 3.29E-05 |
| T1DM | leukocyte | *SYNGR2* | -0.577531726 | 3.08E-09 | 8.62E-09 | 1.21E-05 |
| T1DM | leukocyte | *C17orf75* | -0.87537938 | 1.18E-08 | 2.15E-08 | 4.64E-05 |
| T1DM | leukocyte | *DHX58* | -1.292281247 | 4.45E-09 | 1.03E-08 | 1.75E-05 |
| T1DM | leukocyte | *NAGLU* | -0.697819221 | 2.85E-09 | 8.43E-09 | 1.12E-05 |
| T1DM | leukocyte | *HSD17B1* | -0.780805514 | 1.49E-09 | 8.14E-09 | 5.84E-06 |
| T1DM | leukocyte | *CNTNAP1* | -0.411461213 | 0.030911367 | 0.03366152 | 1 |
| T1DM | leukocyte | *RND2* | -1.600406216 | 2.80E-09 | 8.39E-09 | 1.10E-05 |
| T1DM | leukocyte | *ABCC3* | -0.494772321 | 7.48E-08 | 1.13E-07 | 0.000293864 |
| T1DM | leukocyte | *MPP2* | -0.491433441 | 2.09E-09 | 8.15E-09 | 8.21E-06 |
| T1DM | leukocyte | *HLF* | -0.442584126 | 1.79E-09 | 8.14E-09 | 7.02E-06 |
| T1DM | leukocyte | *EFNB3* | -0.638283312 | 8.65E-10 | 8.14E-09 | 3.40E-06 |
| T1DM | leukocyte | *RANGRF* | -0.813115218 | 2.96E-09 | 8.50E-09 | 1.16E-05 |
| T1DM | leukocyte | *DPH1* | -1.56220974 | 4.89E-10 | 8.14E-09 | 1.92E-06 |
| T1DM | leukocyte | *MYH3* | -1.341431393 | 0.00060557 | 0.000701647 | 1 |
| T1DM | leukocyte | *VTN* | -0.39431369 | 3.21E-06 | 4.11E-06 | 0.012607169 |
| T1DM | leukocyte | *OCIAD1* | -0.555012517 | 1.74E-09 | 8.14E-09 | 6.84E-06 |
| T1DM | leukocyte | *UBE2D3* | -0.509360781 | 1.53E-08 | 2.69E-08 | 6.01E-05 |
| T1DM | leukocyte | *MAPK10* | -0.51959695 | 1.57E-08 | 2.74E-08 | 6.15E-05 |
| T1DM | leukocyte | *ANXA10* | -0.55502603 | 3.53E-05 | 4.31E-05 | 0.138821653 |
| T1DM | leukocyte | *AADAT* | -1.91931467 | 2.00E-09 | 8.14E-09 | 7.86E-06 |
| T1DM | leukocyte | *TRIM2* | -0.619166452 | 2.66E-09 | 8.23E-09 | 1.04E-05 |
| T1DM | leukocyte | *CLNK* | -1.133241468 | 0.008458065 | 0.00942477 | 1 |
| T1DM | leukocyte | *SH3D19* | -1.263167164 | 4.10E-09 | 9.83E-09 | 1.61E-05 |
| T1DM | leukocyte | *GLRB* | -0.639181819 | 4.60E-09 | 1.04E-08 | 1.81E-05 |
| T1DM | leukocyte | *LRP2BP* | -0.770080947 | 6.82E-10 | 8.14E-09 | 2.68E-06 |
| T1DM | leukocyte | *UFSP2* | -0.768639783 | 3.94E-08 | 6.29E-08 | 0.000154982 |
| T1DM | leukocyte | *KLHL5* | -0.800973703 | 2.27E-09 | 8.21E-09 | 8.91E-06 |
| T1DM | leukocyte | *FAM149A* | -0.636351379 | 4.64E-09 | 1.05E-08 | 1.82E-05 |
| T1DM | leukocyte | *DDX25* | -0.589373772 | 3.72E-07 | 5.14E-07 | 0.001462713 |
| T1DM | leukocyte | *HTATIP2* | -0.764337363 | 9.91E-08 | 1.47E-07 | 0.000389351 |
| T1DM | leukocyte | *B3GAT1* | -1.679347149 | 2.05E-09 | 8.14E-09 | 8.06E-06 |
| T1DM | leukocyte | *P2RX3* | -13.35316837 | 1.18E-09 | 8.14E-09 | 4.62E-06 |
| T1DM | leukocyte | *VWA5A* | -0.708407094 | 3.44E-08 | 5.56E-08 | 0.000135027 |
| T1DM | leukocyte | *DTX4* | -0.611376873 | 1.66E-06 | 2.17E-06 | 0.006538647 |
| T1DM | leukocyte | *FOXRED1* | -1.008059788 | 1.31E-07 | 1.91E-07 | 0.000516061 |
| T1DM | leukocyte | *ST3GAL4* | -1.105854473 | 2.45E-09 | 8.21E-09 | 9.63E-06 |
| T1DM | leukocyte | *PRPF19* | -0.589000502 | 2.57E-09 | 8.21E-09 | 1.01E-05 |
| T1DM | leukocyte | *APOA5* | -3.570242902 | 0.023134044 | 0.025325622 | 1 |
| T1DM | leukocyte | *CEP164* | -1.074750825 | 2.02E-07 | 2.88E-07 | 0.000795459 |

| | | | | | | |
|---|---|---|---|---|---|---|
| T1DM | leukocyte | *PDHX* | -0.729669367 | 1.14E-07 | 1.67E-07 | 0.000447474 |
| T1DM | leukocyte | *SLC1A2* | -0.717970782 | 2.28E-09 | 8.21E-09 | 8.97E-06 |
| T1DM | leukocyte | *CD5* | -2.45329741 | 1.95E-06 | 2.53E-06 | 0.007661835 |
| T1DM | leukocyte | *ACCS* | 0.37712244 | 6.15E-09 | 1.26E-08 | 2.42E-05 |
| T1DM | leukocyte | *MDK* | -0.636792558 | 1.96E-09 | 8.14E-09 | 7.71E-06 |
| T1DM | leukocyte | *MADD* | -0.843806746 | 6.81E-09 | 1.37E-08 | 2.68E-05 |
| T1DM | leukocyte | *NAA40* | -0.806538312 | 3.70E-09 | 9.29E-09 | 1.46E-05 |
| T1DM | leukocyte | *CD81* | -0.393006464 | 1.32E-07 | 1.92E-07 | 0.000520134 |
| T1DM | leukocyte | *SLC35F2* | -0.687886223 | 8.34E-09 | 1.61E-08 | 3.28E-05 |
| T1DM | leukocyte | *NUP98* | -0.689252247 | 1.79E-09 | 8.14E-09 | 7.04E-06 |
| T1DM | leukocyte | *GTF2H1* | -1.073910245 | 5.51E-09 | 1.17E-08 | 2.16E-05 |
| T1DM | leukocyte | *PTPN5* | 1.479697915 | 3.06E-09 | 8.60E-09 | 1.20E-05 |
| T1DM | leukocyte | *PRDM4* | -0.531799646 | 4.89E-09 | 1.08E-08 | 1.92E-05 |
| T1DM | leukocyte | *COQ5* | -0.276771972 | 0.020999363 | 0.023014364 | 1 |
| T1DM | leukocyte | *CORO1C* | -0.772563721 | 1.84E-09 | 8.14E-09 | 7.24E-06 |
| T1DM | leukocyte | *LIN7A* | -1.553239362 | 1.27E-09 | 8.14E-09 | 5.00E-06 |
| T1DM | leukocyte | *KRT18* | -0.243224699 | 2.18E-09 | 8.21E-09 | 8.56E-06 |
| T1DM | leukocyte | *ACSS3* | 0.862504996 | 0.02133864 | 0.023379675 | 1 |
| T1DM | leukocyte | *METAP2* | -0.597121252 | 5.56E-09 | 1.18E-08 | 2.18E-05 |
| T1DM | leukocyte | *SLC6A12* | -3.526540134 | 3.38E-07 | 4.69E-07 | 0.001327357 |
| T1DM | leukocyte | *WNT5B* | -1.599497404 | 1.33E-09 | 8.14E-09 | 5.22E-06 |
| T1DM | leukocyte | *ITFG2* | -1.482914059 | 0.001871846 | 0.002131116 | 1 |
| T1DM | leukocyte | *PRR4* | -0.509972772 | 0.00093659 | 0.001079772 | 1 |
| T1DM | leukocyte | *VPS29* | -0.593175854 | 6.33E-10 | 8.14E-09 | 2.49E-06 |
| T1DM | leukocyte | *AKAP3* | -0.484465235 | 0.003320015 | 0.003754847 | 1 |
| T1DM | leukocyte | *CREBL2* | -0.815768471 | 2.44E-09 | 8.21E-09 | 9.58E-06 |
| T1DM | leukocyte | *GPRC5D* | -11.43902562 | 0.005248469 | 0.005890099 | 1 |
| T1DM | leukocyte | *GSG1* | -3.976299261 | 1.16E-06 | 1.53E-06 | 0.004548296 |
| T1DM | leukocyte | *LTBR* | -0.733798229 | 2.03E-09 | 8.14E-09 | 7.97E-06 |
| T1DM | leukocyte | *CDK2AP1* | -0.623172947 | 4.19E-09 | 9.96E-09 | 1.65E-05 |
| T1DM | leukocyte | *OAS3* | 3.142036771 | 1.87E-05 | 2.31E-05 | 0.073660165 |
| T1DM | leukocyte | *RASAL1* | 2.186982905 | 0.001018669 | 0.001172678 | 1 |
| T1DM | leukocyte | *DDX55* | -0.948220032 | 1.14E-08 | 2.09E-08 | 4.49E-05 |
| T1DM | leukocyte | *STX2* | -0.853927584 | 1.03E-08 | 1.90E-08 | 4.04E-05 |
| T1DM | leukocyte | *CAND1* | -0.721338537 | 3.59E-09 | 9.11E-09 | 1.41E-05 |
| T1DM | leukocyte | *NUP107* | -0.862448479 | 1.96E-09 | 8.14E-09 | 7.69E-06 |
| T1DM | leukocyte | *KRR1* | -0.632795939 | 4.33E-09 | 1.01E-08 | 1.70E-05 |
| T1DM | leukocyte | *GNB3* | -0.824981494 | 3.03E-09 | 8.56E-09 | 1.19E-05 |
| T1DM | leukocyte | *CDCA3* | -1.305192209 | 5.35E-09 | 1.15E-08 | 2.10E-05 |
| T1DM | leukocyte | *CHPT1* | -0.893406223 | 1.38E-07 | 2.00E-07 | 0.000543388 |
| T1DM | leukocyte | *SPSB2* | -0.707131151 | 9.62E-10 | 8.14E-09 | 3.78E-06 |
| T1DM | leukocyte | *ATN1* | -0.679386203 | 4.57E-09 | 1.04E-08 | 1.80E-05 |
| T1DM | leukocyte | *NT5DC3* | -0.796433961 | 1.13E-08 | 2.06E-08 | 4.43E-05 |
| T1DM | leukocyte | *GOLT1B* | -0.627815444 | 1.88E-09 | 8.14E-09 | 7.40E-06 |
| T1DM | leukocyte | *PRKAB1* | -0.597838642 | 9.00E-09 | 1.70E-08 | 3.53E-05 |
| T1DM | leukocyte | *C2CD5* | -0.652112905 | 1.04E-09 | 8.14E-09 | 4.08E-06 |
| T1DM | leukocyte | *SRSF9* | -0.561289117 | 1.78E-09 | 8.14E-09 | 6.98E-06 |
| T1DM | leukocyte | *BTN3A3* | -0.514180224 | 0.000249537 | 0.000293454 | 0.980430364 |
| T1DM | leukocyte | *FRK* | -0.702817349 | 1.76E-09 | 8.14E-09 | 6.92E-06 |
| T1DM | leukocyte | *RSPH4A* | -0.642683802 | 2.11E-07 | 3.00E-07 | 0.000829207 |

| | | | | | | |
|---|---|---|---|---|---|---|
| T1DM | leukocyte | *SMIM8* | -0.788594961 | 1.57E-09 | 8.14E-09 | 6.16E-06 |
| T1DM | leukocyte | *NEDD9* | -0.38600162 | 1.33E-09 | 8.14E-09 | 5.23E-06 |
| T1DM | leukocyte | *CEP85L* | -0.683764163 | 6.75E-09 | 1.36E-08 | 2.65E-05 |
| T1DM | leukocyte | *GABRR2* | -0.921128151 | 0.002045286 | 0.002326558 | 1 |
| T1DM | leukocyte | *HDDC2* | -0.535045868 | 1.79E-09 | 8.14E-09 | 7.05E-06 |
| T1DM | leukocyte | *TPD52L1* | -0.568494305 | 4.57E-09 | 1.04E-08 | 1.80E-05 |
| T1DM | leukocyte | *MTRF1L* | -0.755284948 | 3.90E-09 | 9.52E-09 | 1.53E-05 |
| T1DM | leukocyte | *MRPL18* | -0.874849123 | 4.38E-09 | 1.02E-08 | 1.72E-05 |
| T1DM | leukocyte | *RNF8* | -0.673880125 | 1.50E-09 | 8.14E-09 | 5.91E-06 |
| T1DM | leukocyte | *MDGA1* | 1.216754477 | 1.68E-09 | 8.14E-09 | 6.60E-06 |
| T1DM | leukocyte | *CD83* | 0.148730574 | 2.32E-09 | 8.21E-09 | 9.12E-06 |
| T1DM | leukocyte | *GLP1R* | -0.641893212 | 1.16E-08 | 2.11E-08 | 4.55E-05 |
| T1DM | leukocyte | *BACH2* | -0.636003639 | 2.43E-10 | 8.14E-09 | 9.56E-07 |
| T1DM | leukocyte | *FBXL4* | -0.798844116 | 1.61E-09 | 8.14E-09 | 6.34E-06 |
| T1DM | leukocyte | *ASCC3* | -0.560036129 | 1.40E-09 | 8.14E-09 | 5.51E-06 |
| T1DM | leukocyte | *WASF1* | -0.611788502 | 1.86E-09 | 8.14E-09 | 7.33E-06 |
| T1DM | leukocyte | *ALDH5A1* | -1.547208068 | 1.35E-06 | 1.78E-06 | 0.005317884 |
| T1DM | leukocyte | *C6orf62* | -0.728741224 | 3.44E-09 | 8.93E-09 | 1.35E-05 |
| T1DM | leukocyte | *B3GAT2* | -0.992691058 | 1.61E-09 | 8.14E-09 | 6.34E-06 |
| T1DM | leukocyte | *HBS1L* | -0.670750003 | 1.75E-09 | 8.14E-09 | 6.86E-06 |
| T1DM | leukocyte | *TRIM38* | 0.6938188 | 0.001669903 | 0.001904513 | 1 |
| T1DM | leukocyte | *PEX7* | -0.641834223 | 9.92E-10 | 8.14E-09 | 3.90E-06 |
| T1DM | leukocyte | *PHACTR2* | -0.516804717 | 1.69E-09 | 8.14E-09 | 6.64E-06 |
| T1DM | leukocyte | *EPM2A* | -1.10607995 | 8.43E-10 | 8.14E-09 | 3.31E-06 |
| T1DM | leukocyte | *CCR6* | 2.31035677 | 4.43E-07 | 6.07E-07 | 0.001738994 |
| T1DM | leukocyte | *TFEB* | -0.827274615 | 8.44E-10 | 8.14E-09 | 3.31E-06 |
| T1DM | leukocyte | *PRPH2* | -0.715184913 | 2.88E-06 | 3.70E-06 | 0.011327224 |
| T1DM | leukocyte | *DNPH1* | -0.590806997 | 3.33E-07 | 4.62E-07 | 0.001307445 |
| T1DM | leukocyte | *DUSP22* | -0.853706894 | 9.49E-10 | 8.14E-09 | 3.73E-06 |
| T1DM | leukocyte | *LAMA4* | -0.548091851 | 1.47E-09 | 8.14E-09 | 5.79E-06 |
| T1DM | leukocyte | *FBRSL1* | -0.794132808 | 1.91E-09 | 8.14E-09 | 7.51E-06 |
| T1DM | leukocyte | *ENPP5* | -0.569215549 | 8.89E-09 | 1.69E-08 | 3.49E-05 |
| T1DM | leukocyte | *NUDT12* | -0.84041499 | 4.27E-09 | 1.00E-08 | 1.68E-05 |
| T1DM | leukocyte | *CEP72* | -2.034120754 | 1.03E-08 | 1.91E-08 | 4.06E-05 |
| T1DM | leukocyte | *SEMA5A* | -0.783204708 | 6.45E-09 | 1.31E-08 | 2.54E-05 |
| T1DM | leukocyte | *GHR* | -1.833599424 | 1.28E-09 | 8.14E-09 | 5.05E-06 |
| T1DM | leukocyte | *DAP* | -0.52718907 | 3.89E-09 | 9.52E-09 | 1.53E-05 |
| T1DM | leukocyte | *NME5* | -0.84421973 | 1.28E-06 | 1.68E-06 | 0.005010624 |
| T1DM | leukocyte | *BRD8* | -0.780347998 | 1.62E-09 | 8.14E-09 | 6.37E-06 |
| T1DM | leukocyte | *NNT* | -0.849634978 | 1.38E-07 | 2.00E-07 | 0.000542562 |
| T1DM | leukocyte | *TMCO6* | 1.206315221 | 3.26E-07 | 4.53E-07 | 0.001280997 |
| T1DM | leukocyte | *IK* | -0.634246678 | 2.94E-09 | 8.50E-09 | 1.16E-05 |
| T1DM | leukocyte | *PDE8B* | -0.454353162 | 2.42E-09 | 8.21E-09 | 9.50E-06 |
| T1DM | leukocyte | *ITK* | 1.004559926 | 7.01E-05 | 8.44E-05 | 0.275420013 |
| T1DM | leukocyte | *BTNL8* | 5.89928522 | 2.51E-07 | 3.54E-07 | 0.00098807 |
| T1DM | leukocyte | *MSH3* | -0.730651347 | 1.25E-09 | 8.14E-09 | 4.92E-06 |
| T1DM | leukocyte | *CCNG1* | -0.98853739 | 2.85E-07 | 3.98E-07 | 0.00111893 |
| T1DM | leukocyte | *DROSHA* | -0.590306282 | 1.42E-09 | 8.14E-09 | 5.57E-06 |
| T1DM | leukocyte | *GOLPH3* | -0.6458861 | 2.64E-09 | 8.21E-09 | 1.04E-05 |
| T1DM | leukocyte | *NPR3* | -1.992184209 | 4.67E-09 | 1.06E-08 | 1.83E-05 |

| | | | | | | |
|---|---|---|---|---|---|---|
| T1DM | leukocyte | *RAD50* | -0.798745457 | 3.44E-09 | 8.93E-09 | 1.35E-05 |
| T1DM | leukocyte | *PPWD1* | 1.174385548 | 5.11E-06 | 6.47E-06 | 0.020060186 |
| T1DM | leukocyte | *WWC1* | -0.395653657 | 2.70E-09 | 8.26E-09 | 1.06E-05 |
| T1DM | leukocyte | *SMAD5* | -0.58020335 | 5.95E-09 | 1.23E-08 | 2.34E-05 |
| T1DM | leukocyte | *HMGXB3* | -0.829782326 | 1.47E-09 | 8.14E-09 | 5.78E-06 |
| T1DM | leukocyte | *BNIP1* | -0.499714839 | 2.33E-06 | 3.00E-06 | 0.009135434 |
| T1DM | leukocyte | *STC2* | -0.527886887 | 1.31E-09 | 8.14E-09 | 5.16E-06 |
| T1DM | leukocyte | *EHHADH* | -0.910323527 | 2.34E-09 | 8.21E-09 | 9.21E-06 |
| T1DM | leukocyte | *ACTR8* | -0.662496608 | 1.27E-09 | 8.14E-09 | 5.00E-06 |
| T1DM | leukocyte | *TIMMDC1* | -0.721269161 | 3.89E-09 | 9.52E-09 | 1.53E-05 |
| T1DM | leukocyte | *CLDN16* | 3.29830364 | 4.80E-08 | 7.53E-08 | 0.000188583 |
| T1DM | leukocyte | *NPHP3* | -0.705472937 | 3.25E-07 | 4.52E-07 | 0.001278001 |
| T1DM | leukocyte | *FAM162A* | -0.756754563 | 1.04E-09 | 8.14E-09 | 4.09E-06 |
| T1DM | leukocyte | *RBP1* | -0.555716931 | 2.65E-09 | 8.23E-09 | 1.04E-05 |
| T1DM | leukocyte | *RNF7* | -0.579022188 | 9.46E-09 | 1.78E-08 | 3.72E-05 |
| T1DM | leukocyte | *SERPINI2* | -0.452117383 | 2.60E-09 | 8.21E-09 | 1.02E-05 |
| T1DM | leukocyte | *WNT5A* | 1.704700511 | 9.01E-06 | 1.13E-05 | 0.035417694 |
| T1DM | leukocyte | *HYAL1* | -0.453962128 | 0.009004991 | 0.010014324 | 1 |
| T1DM | leukocyte | *TUSC2* | -0.820548727 | 2.42E-09 | 8.21E-09 | 9.50E-06 |
| T1DM | leukocyte | *CYB561D2* | -0.571201421 | 1.47E-08 | 2.60E-08 | 5.78E-05 |
| T1DM | leukocyte | *C3orf14* | -0.518012842 | 1.89E-08 | 3.23E-08 | 7.42E-05 |
| T1DM | leukocyte | *CBLB* | -1.033075737 | 2.69E-09 | 8.26E-09 | 1.06E-05 |
| T1DM | leukocyte | *IFT57* | -0.570087019 | 7.27E-09 | 1.44E-08 | 2.86E-05 |
| T1DM | leukocyte | *NCBP2* | -0.60541879 | 1.19E-09 | 8.14E-09 | 4.67E-06 |
| T1DM | leukocyte | *C3orf52* | -0.576565594 | 4.02E-09 | 9.69E-09 | 1.58E-05 |
| T1DM | leukocyte | *SLC41A3* | -0.563331735 | 1.38E-07 | 2.00E-07 | 0.000541439 |
| T1DM | leukocyte | *ATP6V1A* | -0.724241227 | 3.67E-09 | 9.23E-09 | 1.44E-05 |
| T1DM | leukocyte | *PODXL2* | -0.517361049 | 1.45E-09 | 8.14E-09 | 5.71E-06 |
| T1DM | leukocyte | *UPK1B* | -0.866056658 | 8.54E-07 | 1.14E-06 | 0.003356346 |
| T1DM | leukocyte | *NEK11* | -0.559062124 | 2.20E-09 | 8.21E-09 | 8.64E-06 |
| T1DM | leukocyte | *PLSCR4* | -1.062091189 | 2.02E-09 | 8.14E-09 | 7.95E-06 |
| T1DM | leukocyte | *COMMD2* | -0.625253808 | 7.60E-10 | 8.14E-09 | 2.98E-06 |
| T1DM | leukocyte | *GORASP1* | -1.170277154 | 1.78E-06 | 2.32E-06 | 0.006985055 |
| T1DM | leukocyte | *AADAC* | -0.439331209 | 5.42E-06 | 6.85E-06 | 0.021296182 |
| T1DM | leukocyte | *ARHGEF26* | -1.538407283 | 1.48E-08 | 2.61E-08 | 5.82E-05 |
| T1DM | leukocyte | *PLCH1* | -2.503537383 | 5.82E-08 | 9.01E-08 | 0.000228673 |
| T1DM | leukocyte | *VIPR1* | -0.881272999 | 1.63E-06 | 2.14E-06 | 0.006418135 |
| T1DM | leukocyte | *NKTR* | -0.608122022 | 4.33E-09 | 1.01E-08 | 1.70E-05 |
| T1DM | leukocyte | *NEK4* | -1.518778104 | 1.23E-07 | 1.80E-07 | 0.000484212 |
| T1DM | leukocyte | *KANSL3* | -1.254388699 | 7.66E-09 | 1.50E-08 | 3.01E-05 |
| T1DM | leukocyte | *ACTR1B* | -1.006840624 | 1.62E-08 | 2.82E-08 | 6.35E-05 |
| T1DM | leukocyte | *SLC35F5* | -0.611447774 | 1.09E-08 | 2.00E-08 | 4.28E-05 |
| T1DM | leukocyte | *ACTR3* | -0.690101965 | 2.53E-09 | 8.21E-09 | 9.94E-06 |
| T1DM | leukocyte | *POMC* | -1.01060992 | 4.91E-09 | 1.09E-08 | 1.93E-05 |
| T1DM | leukocyte | *TANC1* | -0.528431662 | 6.68E-08 | 1.02E-07 | 0.000262392 |
| T1DM | leukocyte | *REEP6* | -0.845192923 | 1.25E-09 | 8.14E-09 | 4.89E-06 |
| T1DM | leukocyte | *INO80B* | -1.936983668 | 2.99E-08 | 4.88E-08 | 0.000117467 |
| T1DM | leukocyte | *MOGS* | -1.0751636 | 9.54E-10 | 8.14E-09 | 3.75E-06 |
| T1DM | leukocyte | *RTN4* | -0.516838167 | 2.58E-09 | 8.21E-09 | 1.01E-05 |
| T1DM | leukocyte | *HTRA2* | -1.007463293 | 5.29E-09 | 1.14E-08 | 2.08E-05 |

| | | | | | | |
|---|---|---|---|---|---|---|
| T1DM | leukocyte | *GALNT3* | -0.735300174 | 1.49E-09 | 8.14E-09 | 5.86E-06 |
| T1DM | leukocyte | *MRPL19* | -0.662929421 | 7.45E-09 | 1.47E-08 | 2.93E-05 |
| T1DM | leukocyte | *WDR75* | -1.110845587 | 3.21E-09 | 8.73E-09 | 1.26E-05 |
| T1DM | leukocyte | *FANCL* | -1.440552434 | 1.97E-10 | 8.14E-09 | 7.74E-07 |
| T1DM | leukocyte | *DNAH6* | -0.376799356 | 0.000101709 | 0.000121649 | 0.399616577 |
| T1DM | leukocyte | *GGCX* | -0.794523163 | 6.28E-09 | 1.29E-08 | 2.47E-05 |
| T1DM | leukocyte | *OTX1* | -1.098775655 | 0.042427963 | 0.045973377 | 1 |
| T1DM | leukocyte | *SF3B1* | -0.679370163 | 5.26E-09 | 1.14E-08 | 2.07E-05 |
| T1DM | leukocyte | *CHMP3* | -0.56806522 | 1.56E-09 | 8.14E-09 | 6.15E-06 |
| T1DM | leukocyte | *IL1R2* | -0.702171571 | 1.25E-09 | 8.14E-09 | 4.90E-06 |
| T1DM | leukocyte | *IL1R1* | -0.611242503 | 2.29E-09 | 8.21E-09 | 9.00E-06 |
| T1DM | leukocyte | *IL1RL2* | 0.708433985 | 5.07E-09 | 1.11E-08 | 1.99E-05 |
| T1DM | leukocyte | *IL18R1* | 0.621524238 | 3.32E-08 | 5.39E-08 | 0.000130445 |
| T1DM | leukocyte | *MLPH* | -0.958596158 | 4.41E-08 | 6.98E-08 | 0.000173214 |
| T1DM | leukocyte | *CNPPD1* | -0.559659797 | 2.91E-09 | 8.49E-09 | 1.14E-05 |
| T1DM | leukocyte | *HDLBP* | -0.492686737 | 1.27E-09 | 8.14E-09 | 5.01E-06 |
| T1DM | leukocyte | *PROC* | 0.901904809 | 1.60E-08 | 2.79E-08 | 6.27E-05 |
| T1DM | leukocyte | *TAF1B* | -0.628685733 | 2.85E-09 | 8.43E-09 | 1.12E-05 |
| T1DM | leukocyte | *STRN* | -1.031685376 | 5.75E-08 | 8.91E-08 | 0.000225774 |
| T1DM | leukocyte | *CEBPZ* | -0.59261582 | 8.36E-09 | 1.61E-08 | 3.28E-05 |
| T1DM | leukocyte | *WIPF1* | 0.462125261 | 1.62E-08 | 2.82E-08 | 6.35E-05 |
| T1DM | leukocyte | *COX7A2L* | -0.520095522 | 2.50E-09 | 8.21E-09 | 9.84E-06 |
| T1DM | leukocyte | *PNO1* | -0.679576287 | 1.92E-09 | 8.14E-09 | 7.54E-06 |
| T1DM | leukocyte | *ORC4* | -0.679154909 | 2.42E-09 | 8.21E-09 | 9.51E-06 |
| T1DM | leukocyte | *C2orf42* | 1.138214995 | 2.44E-07 | 3.44E-07 | 0.000956956 |
| T1DM | leukocyte | *ARID3A* | 0.837209323 | 0.020254008 | 0.022216079 | 1 |
| T1DM | leukocyte | *SUMO1* | -0.55833012 | 1.42E-08 | 2.52E-08 | 5.59E-05 |
| T1DM | leukocyte | *GRIN3B* | 1.10065407 | 2.93E-06 | 3.76E-06 | 0.011498683 |
| T1DM | leukocyte | *ALMS1* | -0.81708416 | 1.93E-09 | 8.14E-09 | 7.58E-06 |
| T1DM | leukocyte | *MARK1* | -1.048681391 | 2.22E-09 | 8.21E-09 | 8.73E-06 |
| T1DM | leukocyte | *MORN1* | -1.192449614 | 1.12E-08 | 2.05E-08 | 4.41E-05 |
| T1DM | leukocyte | *GPX7* | -0.610405357 | 3.56E-09 | 9.10E-09 | 1.40E-05 |
| T1DM | leukocyte | *SCP2* | -0.604057624 | 5.05E-09 | 1.11E-08 | 1.99E-05 |
| T1DM | leukocyte | *RALGPS2* | -0.638920947 | 1.44E-09 | 8.14E-09 | 5.64E-06 |
| T1DM | leukocyte | *ANGPTL1* | -1.204362974 | 8.79E-10 | 8.14E-09 | 3.45E-06 |
| T1DM | leukocyte | *CEP104* | -0.496019919 | 0.005662924 | 0.006344348 | 1 |
| T1DM | leukocyte | *FAM20B* | -0.713961659 | 9.25E-09 | 1.75E-08 | 3.63E-05 |
| T1DM | leukocyte | *WRAP73* | -1.737036368 | 2.69E-06 | 3.47E-06 | 0.010576488 |
| T1DM | leukocyte | *ICMT* | -0.814761825 | 6.71E-09 | 1.36E-08 | 2.64E-05 |
| T1DM | leukocyte | *PARK7* | -0.5041593 | 4.53E-09 | 1.04E-08 | 1.78E-05 |
| T1DM | leukocyte | *MECR* | -1.631615071 | 2.05E-08 | 3.46E-08 | 8.04E-05 |
| T1DM | leukocyte | *EDEM3* | -0.73303053 | 1.86E-09 | 8.14E-09 | 7.29E-06 |
| T1DM | leukocyte | *FBXO6* | -1.712537276 | 0.00034521 | 0.00040343 | 1 |
| T1DM | leukocyte | *SMG7* | -0.655327324 | 1.70E-09 | 8.14E-09 | 6.70E-06 |
| T1DM | leukocyte | *PDC* | -6.449235299 | 2.96E-08 | 4.83E-08 | 0.000116179 |
| T1DM | leukocyte | *RGS2* | 0.423766453 | 1.03E-08 | 1.90E-08 | 4.03E-05 |
| T1DM | leukocyte | *BCAS2* | -0.580869436 | 2.03E-09 | 8.14E-09 | 7.97E-06 |
| T1DM | leukocyte | *AGMAT* | -0.649062783 | 0.000148908 | 0.000176808 | 0.585058755 |
| T1DM | leukocyte | *TNNI3K* | -1.671158808 | 6.44E-08 | 9.89E-08 | 0.000252964 |
| T1DM | leukocyte | *PLEKHM2* | -1.131056638 | 9.02E-06 | 1.13E-05 | 0.035435275 |

| | | | | | | |
|---|---|---|---|---|---|---|
| T1DM | leukocyte | *CRYZ* | -0.474765393 | 1.81E-09 | 8.14E-09 | 7.10E-06 |
| T1DM | leukocyte | *TMEM9* | -0.570126473 | 4.07E-08 | 6.47E-08 | 0.000159874 |
| T1DM | leukocyte | *WARS2* | -0.688317099 | 1.69E-09 | 8.14E-09 | 6.65E-06 |
| T1DM | leukocyte | *C1orf109* | -0.45291492 | 2.72E-07 | 3.81E-07 | 0.001067663 |
| T1DM | leukocyte | *LGALS8* | -0.806019732 | 1.68E-09 | 8.14E-09 | 6.61E-06 |
| T1DM | leukocyte | *HPCAL4* | -1.78242467 | 3.55E-09 | 9.10E-09 | 1.40E-05 |
| T1DM | leukocyte | *BMP8B* | -0.625720872 | 8.31E-10 | 8.14E-09 | 3.27E-06 |
| T1DM | leukocyte | *MYCL* | 0.370225504 | 3.99E-09 | 9.64E-09 | 1.57E-05 |
| T1DM | leukocyte | *MFAP2* | -0.392854849 | 5.21E-05 | 6.32E-05 | 0.204808367 |
| T1DM | leukocyte | *UAP1* | -0.490866594 | 1.24E-09 | 8.14E-09 | 4.85E-06 |
| T1DM | leukocyte | *CTBS* | -0.752486173 | 2.88E-09 | 8.46E-09 | 1.13E-05 |
| T1DM | leukocyte | *ZNHIT6* | -0.461768325 | 2.91E-08 | 4.77E-08 | 0.00011441 |
| T1DM | leukocyte | *GBP3* | -0.213580652 | 0.003519083 | 0.003974268 | 1 |
| T1DM | leukocyte | *CD46* | -0.513733058 | 2.00E-09 | 8.14E-09 | 7.86E-06 |
| T1DM | leukocyte | *SLC2A1* | -0.934975668 | 1.06E-09 | 8.14E-09 | 4.16E-06 |
| T1DM | leukocyte | *PRDX1* | -0.532114846 | 4.53E-09 | 1.04E-08 | 1.78E-05 |
| T1DM | leukocyte | *CCDC181* | -0.540143079 | 8.59E-10 | 8.14E-09 | 3.37E-06 |
| T1DM | leukocyte | *NSUN4* | -1.351791362 | 6.89E-05 | 8.31E-05 | 0.270847301 |
| T1DM | leukocyte | *TMED5* | -0.836309479 | 6.07E-09 | 1.25E-08 | 2.39E-05 |
| T1DM | leukocyte | *DR1* | -0.814370542 | 8.84E-09 | 1.68E-08 | 3.47E-05 |
| T1DM | leukocyte | *PTBP2* | -0.660167877 | 1.16E-08 | 2.11E-08 | 4.55E-05 |
| T1DM | leukocyte | *NSL1* | -0.961826265 | 6.28E-09 | 1.29E-08 | 2.47E-05 |
| T1DM | leukocyte | *RPA2* | -0.825335269 | 4.48E-09 | 1.03E-08 | 1.76E-05 |
| T1DM | leukocyte | *ESYT2* | -0.707297034 | 4.87E-09 | 1.08E-08 | 1.91E-05 |
| T1DM | leukocyte | *DDX59* | -0.599699301 | 5.59E-10 | 8.14E-09 | 2.20E-06 |
| T1DM | leukocyte | *TTR* | -0.25671243 | 1.71E-09 | 8.14E-09 | 6.74E-06 |
| T1DM | leukocyte | *SPCS2* | -0.572979966 | 2.97E-09 | 8.50E-09 | 1.17E-05 |
| T1DM | leukocyte | *ELOVL4* | -0.955144576 | 1.57E-09 | 8.14E-09 | 6.18E-06 |
| T1DM | leukocyte | *FILIP1* | -0.734647049 | 2.13E-09 | 8.18E-09 | 8.37E-06 |
| T1DM | leukocyte | *AKAP7* | -1.107431399 | 4.22E-07 | 5.79E-07 | 0.001656558 |
| T1DM | leukocyte | *ALDH8A1* | -0.51581511 | 0.005586636 | 0.006260665 | 1 |
| T1DM | leukocyte | *RNF146* | -0.737268098 | 1.46E-09 | 8.14E-09 | 5.74E-06 |
| T1DM | leukocyte | *MED28* | -0.864877725 | 5.19E-08 | 8.11E-08 | 0.000203891 |
| T1DM | leukocyte | *VAMP8* | -0.631521013 | 1.36E-08 | 2.43E-08 | 5.34E-05 |
| T1DM | leukocyte | *ARMC2* | -0.581251445 | 9.64E-09 | 1.81E-08 | 3.79E-05 |
| T1DM | leukocyte | *PKD2* | -0.550758043 | 1.40E-09 | 8.14E-09 | 5.50E-06 |
| T1DM | leukocyte | *SPP1* | -0.281276084 | 2.98E-08 | 4.86E-08 | 0.000116957 |
| T1DM | leukocyte | *WDR35* | -0.613151278 | 2.00E-09 | 8.14E-09 | 7.86E-06 |
| T1DM | leukocyte | *DNAH7* | -0.62522741 | 3.84E-08 | 6.14E-08 | 0.000150875 |
| T1DM | leukocyte | *NDUFB3* | -0.606870537 | 1.54E-09 | 8.14E-09 | 6.04E-06 |
| T1DM | leukocyte | *GTF3C3* | -0.884378086 | 5.18E-08 | 8.10E-08 | 0.000203557 |
| T1DM | leukocyte | *CPSF3* | -0.945941277 | 3.61E-09 | 9.13E-09 | 1.42E-05 |
| T1DM | leukocyte | *PIGZ* | 0.409517754 | 0.039395367 | 0.042734511 | 1 |
| T1DM | leukocyte | *CCDC92* | -0.691539267 | 1.73E-09 | 8.14E-09 | 6.82E-06 |
| T1DM | leukocyte | *CTNNAL1* | -0.489063694 | 5.41E-08 | 8.43E-08 | 0.000212535 |
| T1DM | leukocyte | *NEK6* | -0.724728739 | 3.94E-08 | 6.28E-08 | 0.00015478 |
| T1DM | leukocyte | *BSPRY* | -0.48445832 | 3.90E-07 | 5.38E-07 | 0.001532588 |
| T1DM | leukocyte | *RBM18* | -0.834372292 | 1.94E-09 | 8.14E-09 | 7.61E-06 |
| T1DM | leukocyte | *INVS* | -0.520997191 | 9.18E-07 | 1.22E-06 | 0.003608494 |
| T1DM | leukocyte | *DCAF4* | -0.830119211 | 3.24E-09 | 8.77E-09 | 1.27E-05 |

| | | | | | | |
|---|---|---|---|---|---|---|
| T1DM | leukocyte | *IFI27L2* | -0.550325733 | 5.84E-09 | 1.22E-08 | 2.30E-05 |
| T1DM | leukocyte | *ACOT2* | -0.9229397 | 1.94E-07 | 2.77E-07 | 0.00076177 |
| T1DM | leukocyte | *ZC2HC1C* | -1.613262069 | 3.82E-06 | 4.86E-06 | 0.015000174 |
| T1DM | leukocyte | *ESRRB* | -1.258340636 | 0.026707819 | 0.029156716 | 1 |
| T1DM | leukocyte | *KLHL29* | -1.800765483 | 3.94E-06 | 5.02E-06 | 0.015493189 |
| T1DM | leukocyte | *AFTPH* | -0.666001609 | 8.24E-10 | 8.14E-09 | 3.24E-06 |
| T1DM | leukocyte | *BCL11A* | -1.135131523 | 1.03E-09 | 8.14E-09 | 4.04E-06 |
| T1DM | leukocyte | *CRIPT* | -0.760586907 | 2.00E-09 | 8.14E-09 | 7.86E-06 |
| T1DM | leukocyte | *EPCAM* | -0.411802715 | 1.79E-09 | 8.14E-09 | 7.04E-06 |
| T1DM | leukocyte | *SLC17A5* | -1.051496825 | 0.001477508 | 0.00168852 | 1 |
| T1DM | leukocyte | *OGFRL1* | 0.791645121 | 0.000193634 | 0.000229222 | 0.760786893 |
| T1DM | leukocyte | *CUTC* | -0.714377603 | 6.30E-08 | 9.69E-08 | 0.000247651 |
| T1DM | leukocyte | *PYROXD2* | -1.136283961 | 1.48E-10 | 8.14E-09 | 5.81E-07 |
| T1DM | leukocyte | *HELLS* | -0.826200347 | 3.75E-10 | 8.14E-09 | 1.47E-06 |
| T1DM | leukocyte | *PRLHR* | -0.943197366 | 1.41E-09 | 8.14E-09 | 5.54E-06 |
| T1DM | leukocyte | *WDR11* | -0.985135042 | 9.96E-09 | 1.85E-08 | 3.91E-05 |
| T1DM | leukocyte | *KANSL1* | -0.690661849 | 1.44E-09 | 8.14E-09 | 5.65E-06 |
| T1DM | leukocyte | *HOXB3* | -0.881963993 | 1.35E-08 | 2.41E-08 | 5.30E-05 |
| T1DM | leukocyte | *RCL1* | -1.098769448 | 3.63E-09 | 9.16E-09 | 1.43E-05 |
| T1DM | leukocyte | *CD274* | -1.82906526 | 5.08E-09 | 1.11E-08 | 1.99E-05 |
| T1DM | leukocyte | *PLEKHG1* | -0.650865924 | 6.88E-09 | 1.38E-08 | 2.70E-05 |
| T1DM | leukocyte | *PCDHB10* | -1.092562999 | 4.71E-08 | 7.41E-08 | 0.000185165 |
| T1DM | leukocyte | *MRPS14* | -0.975433928 | 5.61E-08 | 8.71E-08 | 0.000220314 |
| T1DM | leukocyte | *GORAB* | -0.686051943 | 1.40E-09 | 8.14E-09 | 5.48E-06 |
| T1DM | leukocyte | *SNX19* | -0.623381116 | 6.29E-10 | 8.14E-09 | 2.47E-06 |
| T1DM | leukocyte | *KCNJ5* | 0.241209169 | 1.41E-09 | 8.14E-09 | 5.53E-06 |
| T1DM | leukocyte | *SLC10A7* | -1.180855238 | 9.58E-09 | 1.80E-08 | 3.77E-05 |
| T1DM | leukocyte | *MASTL* | -0.502945982 | 5.49E-06 | 6.94E-06 | 0.021560807 |
| T1DM | leukocyte | *MRC1* | 0.218737631 | 7.11E-09 | 1.42E-08 | 2.79E-05 |
| T1DM | leukocyte | *IQSEC3* | -0.726006765 | 1.43E-09 | 8.14E-09 | 5.61E-06 |
| T1DM | leukocyte | *MTRF1* | -1.172339474 | 6.83E-06 | 8.59E-06 | 0.026849332 |
| T1DM | leukocyte | *DNAJC15* | -0.636859472 | 3.05E-09 | 8.59E-09 | 1.20E-05 |
| T1DM | leukocyte | *PROSER1* | -0.859945719 | 1.87E-09 | 8.14E-09 | 7.35E-06 |
| T1DM | leukocyte | *WBP4* | -0.648510791 | 1.92E-09 | 8.14E-09 | 7.56E-06 |
| T1DM | leukocyte | *KBTBD7* | -1.292545291 | 2.62E-09 | 8.21E-09 | 1.03E-05 |
| T1DM | leukocyte | *FAM53C* | -1.160109504 | 4.55E-08 | 7.18E-08 | 0.00017867 |
| T1DM | leukocyte | *SIL1* | -0.628103203 | 5.45E-09 | 1.16E-08 | 2.14E-05 |
| T1DM | leukocyte | *PLS1* | -0.564630345 | 1.13E-09 | 8.14E-09 | 4.43E-06 |
| T1DM | leukocyte | *ZFP30* | -0.865036274 | 5.97E-09 | 1.24E-08 | 2.35E-05 |
| T1DM | leukocyte | *UTP20* | -1.211081444 | 8.75E-08 | 1.31E-07 | 0.000343923 |
| T1DM | leukocyte | *TMPO* | -0.844396453 | 9.80E-08 | 1.45E-07 | 0.000384857 |
| T1DM | leukocyte | *APAF1* | -0.746082671 | 0.045459882 | 0.049177279 | 1 |
| T1DM | leukocyte | *PPP3CC* | -0.793892345 | 2.81E-09 | 8.40E-09 | 1.10E-05 |
| T1DM | leukocyte | *EPHX2* | -0.771355943 | 2.15E-09 | 8.19E-09 | 8.44E-06 |
| T1DM | leukocyte | *CRISPLD1* | -3.599097146 | 1.22E-08 | 2.20E-08 | 4.78E-05 |
| T1DM | leukocyte | *TEX14* | -1.234900324 | 7.00E-05 | 8.43E-05 | 0.275141804 |
| T1DM | leukocyte | *TRIM6* | 0.870049843 | 1.41E-07 | 2.04E-07 | 0.000552916 |
| T1DM | leukocyte | *PYROXD1* | -0.553064941 | 1.12E-09 | 8.14E-09 | 4.39E-06 |
| T1DM | leukocyte | *IAPP* | -0.233266936 | 2.91E-09 | 8.49E-09 | 1.14E-05 |
| T1DM | leukocyte | *ZSCAN18* | -0.537009282 | 1.33E-08 | 2.39E-08 | 5.23E-05 |

| | | | | | | |
|---|---|---|---|---|---|---|
| T1DM | leukocyte | *ZNF211* | -0.428465581 | 0.015521656 | 0.017111276 | 1 |
| T1DM | leukocyte | *LHX4* | -1.049519594 | 0.001597407 | 0.001822361 | 1 |
| T1DM | leukocyte | *TRMT1L* | -0.688685685 | 3.48E-09 | 9.00E-09 | 1.37E-05 |
| T1DM | leukocyte | *CAT* | -1.223846817 | 6.21E-08 | 9.56E-08 | 0.000243869 |
| T1DM | leukocyte | *PILRB* | -0.358942167 | 0.000602483 | 0.000698276 | 1 |
| T1DM | leukocyte | *CPXM2* | -0.606065473 | 0.032091602 | 0.034927397 | 1 |
| T1DM | leukocyte | *CSMD2* | 0.944822679 | 4.81E-09 | 1.07E-08 | 1.89E-05 |
| T1DM | leukocyte | *ACVR2A* | -0.62011291 | 1.70E-09 | 8.14E-09 | 6.70E-06 |
| T1DM | leukocyte | *POLK* | -0.877763258 | 2.42E-09 | 8.21E-09 | 9.49E-06 |
| T1DM | leukocyte | *FLT3* | -0.631904514 | 0.033253122 | 0.036161504 | 1 |
| T1DM | leukocyte | *UBL3* | -0.563160827 | 2.30E-09 | 8.21E-09 | 9.04E-06 |
| T1DM | leukocyte | *FMOD* | -0.774870418 | 2.25E-06 | 2.92E-06 | 0.008859288 |
| T1DM | leukocyte | *LDB3* | -0.936194747 | 2.01E-07 | 2.86E-07 | 0.000788444 |
| T1DM | leukocyte | *NAA60* | -0.837210901 | 1.68E-09 | 8.14E-09 | 6.59E-06 |
| T1DM | leukocyte | *TRMT13* | -1.428911071 | 3.85E-09 | 9.49E-09 | 1.51E-05 |
| T1DM | leukocyte | *LRRC39* | -0.572150568 | 0.000138203 | 0.000164247 | 0.542999299 |
| T1DM | leukocyte | *RWDD3* | -0.66649946 | 1.15E-09 | 8.14E-09 | 4.53E-06 |
| T1DM | leukocyte | *CCDC18* | -0.681701103 | 4.25E-09 | 9.99E-09 | 1.67E-05 |
| T1DM | leukocyte | *NBPF14* | -1.871906215 | 1.72E-09 | 8.14E-09 | 6.78E-06 |
| T1DM | leukocyte | *PMS2* | -1.159530754 | 1.68E-07 | 2.41E-07 | 0.00066009 |
| T1DM | leukocyte | *EEPD1* | 0.411612184 | 1.35E-08 | 2.41E-08 | 5.29E-05 |
| T1DM | leukocyte | *KLHL7* | -0.920524022 | 1.99E-09 | 8.14E-09 | 7.82E-06 |
| T1DM | leukocyte | *CBX3* | -0.678392654 | 2.91E-09 | 8.49E-09 | 1.14E-05 |
| T1DM | leukocyte | *NPY* | -0.316919136 | 1.89E-09 | 8.14E-09 | 7.43E-06 |
| T1DM | leukocyte | *INHBA* | -0.594577152 | 1.63E-09 | 8.14E-09 | 6.39E-06 |
| T1DM | leukocyte | *CCZ1* | -0.898349552 | 1.19E-08 | 2.17E-08 | 4.69E-05 |
| T1DM | leukocyte | *POLM* | -0.541593436 | 7.41E-07 | 9.95E-07 | 0.002911234 |
| T1DM | leukocyte | *SMU1* | -0.733805027 | 1.47E-09 | 8.14E-09 | 5.76E-06 |
| T1DM | leukocyte | *SLC25A51* | -0.775629041 | 1.56E-08 | 2.74E-08 | 6.13E-05 |
| T1DM | leukocyte | *ACO1* | -0.649105572 | 1.10E-09 | 8.14E-09 | 4.32E-06 |
| T1DM | leukocyte | *PLAU* | -0.988290197 | 1.29E-09 | 8.14E-09 | 5.08E-06 |
| T1DM | leukocyte | *CHST3* | 0.73815687 | 1.13E-05 | 1.41E-05 | 0.044564189 |
| T1DM | leukocyte | *RBM19* | -0.605377558 | 1.56E-09 | 8.14E-09 | 6.13E-06 |
| T1DM | leukocyte | *BHLHE41* | -0.975437198 | 1.94E-07 | 2.76E-07 | 0.000761133 |
| T1DM | leukocyte | *ITPR2* | -1.566899116 | 5.43E-08 | 8.45E-08 | 0.000213342 |
| T1DM | leukocyte | *EBPL* | -0.576674473 | 2.53E-09 | 8.21E-09 | 9.95E-06 |
| T1DM | leukocyte | *ATP7B* | -0.535614334 | 0.006235313 | 0.006977655 | 1 |
| T1DM | leukocyte | *TSFM* | -0.829938819 | 8.00E-09 | 1.56E-08 | 3.14E-05 |
| T1DM | leukocyte | *CDK2* | -1.073078812 | 1.51E-06 | 1.98E-06 | 0.005937154 |
| T1DM | leukocyte | *IKZF4* | -0.918223597 | 2.20E-08 | 3.69E-08 | 8.63E-05 |
| T1DM | leukocyte | *SARDH* | 0.995668371 | 0.001494802 | 0.00170729 | 1 |
| T1DM | leukocyte | *USP45* | -0.56935339 | 2.19E-09 | 8.21E-09 | 8.60E-06 |
| T1DM | leukocyte | *SLC36A1* | 0.392456995 | 0.033236058 | 0.036152955 | 1 |
| T1DM | leukocyte | *C4BPA* | -0.977680196 | 2.14E-06 | 2.77E-06 | 0.008400061 |
| T1DM | leukocyte | *C4BPB* | -1.593092677 | 2.58E-07 | 3.63E-07 | 0.001014913 |
| T1DM | leukocyte | *RAB38* | -1.892221234 | 2.75E-08 | 4.54E-08 | 0.000108242 |
| T1DM | leukocyte | *CKS2* | -0.613370138 | 4.97E-09 | 1.10E-08 | 1.95E-05 |
| T1DM | leukocyte | *SNX21* | -0.754438163 | 4.07E-10 | 8.14E-09 | 1.60E-06 |
| T1DM | leukocyte | *WFDC3* | -0.441408184 | 0.040216903 | 0.043601604 | 1 |
| T1DM | leukocyte | *ARHGAP40* | -36.03515973 | 7.47E-10 | 8.14E-09 | 2.93E-06 |

| | | | | | | |
|---|---|---|---|---|---|---|
| T1DM | leukocyte | *NCOA3* | -0.683429649 | 4.98E-09 | 1.10E-08 | 1.96E-05 |
| T1DM | leukocyte | *MATN4* | -3.944183973 | 5.52E-07 | 7.49E-07 | 0.00216865 |
| T1DM | leukocyte | *VAPB* | -0.563986534 | 6.36E-09 | 1.30E-08 | 2.50E-05 |
| T1DM | leukocyte | *PARD6B* | -0.847223667 | 1.49E-09 | 8.14E-09 | 5.87E-06 |
| T1DM | leukocyte | *ARFGEF2* | -0.799855582 | 3.56E-09 | 9.10E-09 | 1.40E-05 |
| T1DM | leukocyte | *ZNFX1* | -1.520572174 | 1.18E-05 | 1.46E-05 | 0.04627059 |
| T1DM | leukocyte | *CSE1L* | -0.773301893 | 2.36E-09 | 8.21E-09 | 9.28E-06 |
| T1DM | leukocyte | *BCAS4* | -0.510033085 | 1.04E-09 | 8.14E-09 | 4.09E-06 |
| T1DM | leukocyte | *KCNK15* | -0.640028276 | 8.94E-09 | 1.70E-08 | 3.51E-05 |
| T1DM | leukocyte | *PEPD* | -0.963375637 | 4.73E-09 | 1.06E-08 | 1.86E-05 |
| T1DM | leukocyte | *PAIP2B* | -0.454848653 | 2.09E-07 | 2.98E-07 | 0.000822436 |
| T1DM | leukocyte | *SNRNP27* | -0.621392376 | 7.03E-09 | 1.40E-08 | 2.76E-05 |
| T1DM | leukocyte | *USP22* | -0.743128366 | 6.94E-09 | 1.39E-08 | 2.73E-05 |
| T1DM | leukocyte | *CRISP2* | -20.01033639 | 0.003956941 | 0.004464911 | 1 |
| T1DM | leukocyte | *BTN2A2* | -1.239075816 | 5.95E-10 | 8.14E-09 | 2.34E-06 |
| T1DM | leukocyte | *SIRT5* | 0.959444492 | 9.48E-08 | 1.41E-07 | 0.00037261 |
| T1DM | leukocyte | *MRS2* | -1.043799258 | 3.66E-09 | 9.22E-09 | 1.44E-05 |
| T1DM | leukocyte | *SNRPC* | -0.678800175 | 2.68E-09 | 8.26E-09 | 1.05E-05 |
| T1DM | leukocyte | *SERPINB6* | -0.595073248 | 2.77E-09 | 8.34E-09 | 1.09E-05 |
| T1DM | leukocyte | *ABCC10* | -0.876237264 | 8.58E-10 | 8.14E-09 | 3.37E-06 |
| T1DM | leukocyte | *PEX6* | -0.607630705 | 8.63E-08 | 1.30E-07 | 0.000339194 |
| T1DM | leukocyte | *NQO2* | -1.216838396 | 6.59E-07 | 8.87E-07 | 0.002590568 |
| T1DM | leukocyte | *UNC5CL* | -0.626507158 | 2.93E-09 | 8.50E-09 | 1.15E-05 |
| T1DM | leukocyte | *ZNF391* | -0.365307379 | 0.000652851 | 0.00075576 | 1 |
| T1DM | leukocyte | *MOCS1* | 0.757293033 | 7.35E-06 | 9.23E-06 | 0.028888596 |
| T1DM | leukocyte | *MED20* | -0.962504305 | 2.70E-08 | 4.46E-08 | 0.000106226 |
| T1DM | leukocyte | *TREM1* | 0.728997569 | 7.92E-08 | 1.19E-07 | 0.000311064 |
| T1DM | leukocyte | *GLO1* | -0.675611719 | 4.00E-09 | 9.65E-09 | 1.57E-05 |
| T1DM | leukocyte | *KCNK17* | -0.52794038 | 3.49E-09 | 9.01E-09 | 1.37E-05 |
| T1DM | leukocyte | *ATXN1* | -0.836973672 | 5.25E-09 | 1.14E-08 | 2.06E-05 |
| T1DM | leukocyte | *LRRFIP1* | -0.534386455 | 1.86E-09 | 8.14E-09 | 7.30E-06 |
| T1DM | leukocyte | *CXCL6* | -1.022669494 | 8.91E-08 | 1.33E-07 | 0.000350053 |
| T1DM | leukocyte | *SH3TC1* | -0.720502428 | 3.33E-09 | 8.82E-09 | 1.31E-05 |
| T1DM | leukocyte | *MT1G* | -0.278996329 | 0.002210192 | 0.002512686 | 1 |
| T1DM | leukocyte | *DOK4* | -0.506826053 | 1.08E-09 | 8.14E-09 | 4.25E-06 |
| T1DM | leukocyte | *CLYBL* | -1.490151154 | 2.36E-08 | 3.95E-08 | 9.29E-05 |
| T1DM | leukocyte | *ABCC4* | 0.865691151 | 1.86E-05 | 2.30E-05 | 0.073165438 |
| T1DM | leukocyte | *TM9SF2* | -0.583897192 | 2.07E-09 | 8.14E-09 | 8.13E-06 |
| T1DM | leukocyte | *KIF25* | -9.472310768 | 1.02E-09 | 8.14E-09 | 4.02E-06 |
| T1DM | leukocyte | *BMP4* | -1.201930839 | 7.46E-07 | 1.00E-06 | 0.002932216 |
| T1DM | leukocyte | *GRK4* | -0.785974194 | 1.43E-07 | 2.07E-07 | 0.000562377 |
| T1DM | leukocyte | *HS3ST3B1* | -1.600486044 | 1.36E-09 | 8.14E-09 | 5.35E-06 |
| T1DM | leukocyte | *MRPS7* | -0.552440517 | 2.34E-09 | 8.21E-09 | 9.18E-06 |
| T1DM | leukocyte | *GGA3* | -1.29090899 | 2.11E-07 | 3.01E-07 | 0.000830937 |
| T1DM | leukocyte | *DDX31* | -0.820940122 | 3.28E-05 | 4.02E-05 | 0.129040062 |
| T1DM | leukocyte | *OPRL1* | 0.945424035 | 6.29E-05 | 7.60E-05 | 0.24705324 |
| T1DM | leukocyte | *PPDPF* | -0.48723985 | 2.46E-09 | 8.21E-09 | 9.67E-06 |
| T1DM | leukocyte | *CHCHD5* | -0.588821843 | 2.53E-09 | 8.21E-09 | 9.95E-06 |
| T1DM | leukocyte | *PAX8* | 0.298013581 | 0.037124884 | 0.040316106 | 1 |
| T1DM | leukocyte | *INSIG2* | -0.58107561 | 1.12E-09 | 8.14E-09 | 4.40E-06 |

| | | | | | | |
|---|---|---|---|---|---|---|
| T1DM | leukocyte | *POLR1B* | -0.556895539 | 1.65E-05 | 2.04E-05 | 0.064913147 |
| T1DM | leukocyte | *PSD4* | -0.865587317 | 2.41E-08 | 4.02E-08 | 9.47E-05 |
| T1DM | leukocyte | *CLPP* | -0.63452978 | 5.56E-09 | 1.18E-08 | 2.19E-05 |
| T1DM | leukocyte | *SH2D3A* | -0.268998303 | 0.005664612 | 0.00634443 | 1 |
| T1DM | leukocyte | *GPR108* | -0.888032686 | 9.75E-09 | 1.82E-08 | 3.83E-05 |
| T1DM | leukocyte | *TNFSF14* | -0.957262966 | 0.012164034 | 0.013462674 | 1 |
| T1DM | leukocyte | *FOSB* | -0.452104963 | 0.008536846 | 0.009504468 | 1 |
| T1DM | leukocyte | *SYMPK* | -0.955517809 | 6.89E-06 | 8.66E-06 | 0.027069433 |
| T1DM | leukocyte | *NAPB* | -0.586862085 | 1.89E-08 | 3.24E-08 | 7.44E-05 |
| T1DM | leukocyte | *PSMF1* | -0.68894771 | 1.58E-08 | 2.76E-08 | 6.20E-05 |
| T1DM | leukocyte | *RBCK1* | -0.586531581 | 1.93E-08 | 3.28E-08 | 7.57E-05 |
| T1DM | leukocyte | *FLRT3* | -0.589869775 | 1.31E-09 | 8.14E-09 | 5.15E-06 |
| T1DM | leukocyte | *MGME1* | -2.284597473 | 1.28E-07 | 1.86E-07 | 0.000502853 |
| T1DM | leukocyte | *ITPA* | -0.873275505 | 1.93E-09 | 8.14E-09 | 7.58E-06 |
| T1DM | leukocyte | *BANF2* | -9.46542881 | 1.44E-09 | 8.14E-09 | 5.64E-06 |
| T1DM | leukocyte | *MRPS26* | -0.57415212 | 9.42E-10 | 8.14E-09 | 3.70E-06 |
| T1DM | leukocyte | *ZNF436* | -1.613626435 | 4.42E-10 | 8.14E-09 | 1.74E-06 |
| T1DM | leukocyte | *TUBGCP3* | -0.830250803 | 8.44E-10 | 8.14E-09 | 3.32E-06 |
| T1DM | leukocyte | *MCF2L* | -0.50779849 | 2.82E-09 | 8.42E-09 | 1.11E-05 |
| T1DM | leukocyte | *PROZ* | -4.812132715 | 0.001470191 | 0.001681125 | 1 |
| T1DM | leukocyte | *IGFLR1* | 0.258494843 | 5.67E-09 | 1.19E-08 | 2.23E-05 |
| T1DM | leukocyte | *NR1D1* | -0.576062672 | 1.70E-09 | 8.14E-09 | 6.66E-06 |
| T1DM | leukocyte | *PRMT1* | -0.55705311 | 7.32E-10 | 8.14E-09 | 2.88E-06 |
| T1DM | leukocyte | *PRR12* | -0.696473648 | 3.71E-09 | 9.29E-09 | 1.46E-05 |
| T1DM | leukocyte | *FLRT1* | -1.104946955 | 0.000411026 | 0.000478637 | 1 |
| T1DM | leukocyte | *ASL* | -0.710549021 | 2.11E-09 | 8.16E-09 | 8.28E-06 |
| T1DM | leukocyte | *WNK4* | -0.763297164 | 5.44E-07 | 7.40E-07 | 0.002139052 |
| T1DM | leukocyte | *BECN1* | -0.690025215 | 4.67E-09 | 1.06E-08 | 1.83E-05 |
| T1DM | leukocyte | *PRKCG* | 1.986046823 | 1.28E-07 | 1.86E-07 | 0.000501412 |
| T1DM | leukocyte | *TRAP1* | -0.503489151 | 1.25E-07 | 1.82E-07 | 0.000490411 |
| T1DM | leukocyte | *ZNF384* | -0.846942468 | 1.61E-09 | 8.14E-09 | 6.33E-06 |
| T1DM | leukocyte | *RHOJ* | -1.161755122 | 1.65E-09 | 8.14E-09 | 6.46E-06 |
| T1DM | leukocyte | *L3HYPDH* | -0.7116166 | 6.59E-07 | 8.87E-07 | 0.002589708 |
| T1DM | leukocyte | *HSPA2* | -1.091703721 | 4.68E-09 | 1.06E-08 | 1.84E-05 |
| T1DM | leukocyte | *ZBTB1* | -0.881659255 | 2.35E-09 | 8.21E-09 | 9.21E-06 |
| T1DM | leukocyte | *PLEKHG3* | -0.939214843 | 3.49E-09 | 9.01E-09 | 1.37E-05 |
| T1DM | leukocyte | *FGD3* | 2.181371269 | 9.93E-09 | 1.85E-08 | 3.90E-05 |
| T1DM | leukocyte | *BCL11B* | 1.328893616 | 3.71E-09 | 9.29E-09 | 1.46E-05 |
| T1DM | leukocyte | *ABHD8* | -1.168441893 | 4.25E-06 | 5.40E-06 | 0.016708109 |
| T1DM | leukocyte | *BEST3* | -1.489169563 | 5.11E-07 | 6.98E-07 | 0.002009576 |
| T1DM | leukocyte | *YEATS4* | -0.582904519 | 2.58E-08 | 4.27E-08 | 0.000101363 |
| T1DM | leukocyte | *TAS2R5* | -2.414345568 | 0.002622687 | 0.002973893 | 1 |
| T1DM | leukocyte | *LRRC61* | -0.982167943 | 4.47E-09 | 1.03E-08 | 1.76E-05 |
| T1DM | leukocyte | *IDUA* | -0.698805534 | 1.07E-09 | 8.14E-09 | 4.20E-06 |
| T1DM | leukocyte | *FGFRL1* | -0.587530208 | 1.08E-07 | 1.59E-07 | 0.000423097 |
| T1DM | leukocyte | *EMC1* | -0.766418705 | 2.95E-09 | 8.50E-09 | 1.16E-05 |
| T1DM | leukocyte | *UBR4* | -0.710379476 | 2.95E-09 | 8.50E-09 | 1.16E-05 |
| T1DM | leukocyte | *SYNGR3* | -0.832959109 | 2.21E-09 | 8.21E-09 | 8.69E-06 |
| T1DM | leukocyte | *WFIKKN1* | -0.718637824 | 4.54E-09 | 1.04E-08 | 1.78E-05 |
| T1DM | leukocyte | *WDR24* | 1.588911896 | 1.17E-05 | 1.45E-05 | 0.045968708 |

| | | | | | | |
|---|---|---|---|---|---|---|
| T1DM | leukocyte | *KDM4B* | -1.344575974 | 4.64E-08 | 7.29E-08 | 0.00018219 |
| T1DM | leukocyte | *METTL25* | 1.557754185 | 0.004662759 | 0.005246272 | 1 |
| T1DM | leukocyte | *PNKD* | -0.835059953 | 1.14E-09 | 8.14E-09 | 4.47E-06 |
| T1DM | leukocyte | *PTPN12* | -0.647194345 | 7.56E-09 | 1.49E-08 | 2.97E-05 |
| T1DM | leukocyte | *POR* | -0.702774868 | 3.34E-09 | 8.82E-09 | 1.31E-05 |
| T1DM | leukocyte | *STYXL1* | -0.827980516 | 0.013308711 | 0.014712978 | 1 |
| T1DM | leukocyte | *RBM48* | -1.18598941 | 4.79E-09 | 1.07E-08 | 1.88E-05 |
| T1DM | leukocyte | *SRD5A3* | -0.788069235 | 4.65E-08 | 7.32E-08 | 0.00018281 |
| T1DM | leukocyte | *PPAT* | -0.760198749 | 5.72E-09 | 1.20E-08 | 2.25E-05 |
| T1DM | leukocyte | *A4GALT* | -0.585666718 | 1.50E-08 | 2.65E-08 | 5.91E-05 |
| T1DM | leukocyte | *GALR3* | -1.900307548 | 0.033837636 | 0.03678696 | 1 |
| T1DM | leukocyte | *NAA38* | -0.545271979 | 2.00E-09 | 8.14E-09 | 7.85E-06 |
| T1DM | leukocyte | *PRKRIP1* | -0.65325984 | 1.70E-09 | 8.14E-09 | 6.69E-06 |
| T1DM | leukocyte | *VGF* | -0.347271125 | 1.87E-09 | 8.14E-09 | 7.35E-06 |
| T1DM | leukocyte | *IRF5* | 0.316567353 | 5.23E-09 | 1.13E-08 | 2.06E-05 |
| T1DM | leukocyte | *MTX2* | -0.898231771 | 4.65E-09 | 1.05E-08 | 1.83E-05 |
| T1DM | leukocyte | *GAD1* | -2.398425376 | 3.47E-09 | 8.99E-09 | 1.36E-05 |
| T1DM | leukocyte | *ORMDL1* | -0.593671414 | 2.45E-08 | 4.07E-08 | 9.61E-05 |
| T1DM | leukocyte | *PSMG2* | -0.722383735 | 4.59E-09 | 1.04E-08 | 1.80E-05 |
| T1DM | leukocyte | *TWSG1* | -0.966959666 | 2.98E-09 | 8.50E-09 | 1.17E-05 |
| T1DM | leukocyte | *ARHGAP22* | 0.646698674 | 0.000988235 | 0.001138644 | 1 |
| T1DM | leukocyte | *EIF2AK4* | -0.881546187 | 1.36E-08 | 2.43E-08 | 5.36E-05 |
| T1DM | leukocyte | *MYO5C* | -0.485255904 | 2.00E-09 | 8.14E-09 | 7.84E-06 |
| T1DM | leukocyte | *CGNL1* | -0.751293582 | 2.28E-09 | 8.21E-09 | 8.97E-06 |
| T1DM | leukocyte | *TMOD2* | -0.754508717 | 1.68E-09 | 8.14E-09 | 6.62E-06 |
| T1DM | leukocyte | *ELL3* | -1.246784691 | 0.00020467 | 0.000241486 | 0.804148848 |
| T1DM | leukocyte | *IVD* | -0.706004592 | 7.57E-06 | 9.49E-06 | 0.029729596 |
| T1DM | leukocyte | *KNSTRN* | -0.356348705 | 0.00051965 | 0.000603163 | 1 |
| T1DM | leukocyte | *CHAC1* | -0.762053101 | 8.60E-09 | 1.64E-08 | 3.38E-05 |
| T1DM | leukocyte | *ACKR4* | -1.236081227 | 1.10E-07 | 1.62E-07 | 0.000432416 |
| T1DM | leukocyte | *ANAPC13* | -0.448931838 | 7.56E-09 | 1.49E-08 | 2.97E-05 |
| T1DM | leukocyte | *SUMF2* | -0.721987837 | 8.54E-09 | 1.63E-08 | 3.35E-05 |
| T1DM | leukocyte | *TPH1* | -1.558964233 | 0.000107885 | 0.000128799 | 0.423878899 |
| T1DM | leukocyte | *DCTD* | -0.780731023 | 3.40E-09 | 8.90E-09 | 1.34E-05 |
| T1DM | leukocyte | *PLD2* | 1.259177385 | 0.001003228 | 0.00115558 | 1 |
| T1DM | leukocyte | *ATP1B2* | -0.852134324 | 0.035722058 | 0.038803419 | 1 |
| T1DM | leukocyte | *KRI1* | -0.576279489 | 2.49E-09 | 8.21E-09 | 9.79E-06 |
| T1DM | leukocyte | *AP1M2* | -0.598086078 | 2.99E-09 | 8.52E-09 | 1.18E-05 |
| T1DM | leukocyte | *CDKN2D* | -0.580398741 | 1.75E-08 | 3.02E-08 | 6.89E-05 |
| T1DM | leukocyte | *MTUS1* | -0.557948027 | 2.69E-09 | 8.26E-09 | 1.06E-05 |
| T1DM | leukocyte | *ADCY4* | -0.735275681 | 5.22E-09 | 1.13E-08 | 2.05E-05 |
| T1DM | leukocyte | *DTD2* | -0.594260782 | 3.17E-09 | 8.69E-09 | 1.24E-05 |
| T1DM | leukocyte | *HEATR5A* | -1.225300799 | 1.98E-08 | 3.36E-08 | 7.78E-05 |
| T1DM | leukocyte | *MIS18BP1* | -0.904902665 | 4.11E-05 | 5.01E-05 | 0.161526263 |
| T1DM | leukocyte | *RNASE1* | 0.12947153 | 5.60E-09 | 1.18E-08 | 2.20E-05 |
| T1DM | leukocyte | *TEP1* | 1.046267808 | 4.42E-05 | 5.39E-05 | 0.173815789 |
| T1DM | leukocyte | *EPB41L4A* | -0.798150283 | 1.37E-09 | 8.14E-09 | 5.37E-06 |
| T1DM | leukocyte | *REEP5* | -0.540459313 | 3.19E-09 | 8.72E-09 | 1.26E-05 |
| T1DM | leukocyte | *MAU2* | -1.373793109 | 3.33E-09 | 8.82E-09 | 1.31E-05 |
| T1DM | leukocyte | *SHC2* | -0.741959573 | 1.94E-09 | 8.14E-09 | 7.61E-06 |

| | | | | | | |
|---|---|---|---|---|---|---|
| T1DM | leukocyte | *SYT5* | -0.391796189 | 5.59E-07 | 7.59E-07 | 0.002197627 |
| T1DM | leukocyte | *CBFA2T3* | -0.678338071 | 1.40E-09 | 8.14E-09 | 5.52E-06 |
| T1DM | leukocyte | *ERMARD* | -1.202562165 | 2.10E-08 | 3.54E-08 | 8.25E-05 |
| T1DM | leukocyte | *PHF10* | -0.688211532 | 1.19E-09 | 8.14E-09 | 4.69E-06 |
| T1DM | leukocyte | *SH3BP4* | -0.420374959 | 2.85E-09 | 8.43E-09 | 1.12E-05 |
| T1DM | leukocyte | *DOCK6* | -0.528306097 | 3.34E-09 | 8.82E-09 | 1.31E-05 |
| T1DM | leukocyte | *ECSIT* | -0.749429189 | 8.78E-09 | 1.67E-08 | 3.45E-05 |
| T1DM | leukocyte | *CDC16* | -0.88038442 | 3.10E-09 | 8.64E-09 | 1.22E-05 |
| T1DM | leukocyte | *DPP6* | -0.374856309 | 1.45E-09 | 8.14E-09 | 5.68E-06 |
| T1DM | leukocyte | *GTPBP3* | -0.70297643 | 1.98E-09 | 8.14E-09 | 7.77E-06 |
| T1DM | leukocyte | *PLVAP* | -0.348130185 | 2.95E-09 | 8.50E-09 | 1.16E-05 |
| T1DM | leukocyte | *MRPL34* | -0.467895598 | 2.99E-07 | 4.16E-07 | 0.001174044 |
| T1DM | leukocyte | *LSM7* | -0.459881047 | 1.62E-08 | 2.83E-08 | 6.38E-05 |
| T1DM | leukocyte | *TULP4* | -0.668378228 | 4.68E-09 | 1.06E-08 | 1.84E-05 |
| T1DM | leukocyte | *QRSL1* | -0.788137119 | 4.24E-10 | 8.14E-09 | 1.67E-06 |
| T1DM | leukocyte | *RSPH3* | -0.741801306 | 1.04E-08 | 1.92E-08 | 4.08E-05 |
| T1DM | leukocyte | *STK33* | -0.576590636 | 1.15E-09 | 8.14E-09 | 4.51E-06 |
| T1DM | leukocyte | *NDUFA10* | -0.653116369 | 1.32E-09 | 8.14E-09 | 5.19E-06 |
| T1DM | leukocyte | *EPO* | -0.818115128 | 0.013822302 | 0.015263582 | 1 |
| T1DM | leukocyte | *SCO2* | 0.764377927 | 3.98E-08 | 6.34E-08 | 0.000156452 |
| T1DM | leukocyte | *TRPM4* | -1.886318447 | 7.99E-05 | 9.59E-05 | 0.313985478 |
| T1DM | leukocyte | *UBAC1* | -0.650252905 | 2.06E-09 | 8.14E-09 | 8.10E-06 |
| T1DM | leukocyte | *ATXN10* | -0.56341057 | 1.44E-09 | 8.14E-09 | 5.67E-06 |
| T1DM | leukocyte | *PNPLA7* | -1.355531389 | 1.35E-09 | 8.14E-09 | 5.32E-06 |
| T1DM | leukocyte | *PAK4* | -0.84356921 | 1.46E-09 | 8.14E-09 | 5.72E-06 |
| T1DM | leukocyte | *CEP85* | 0.890197927 | 4.13E-06 | 5.26E-06 | 0.016244863 |
| T1DM | leukocyte | *LAMA5* | -0.464097611 | 5.29E-09 | 1.14E-08 | 2.08E-05 |
| T1DM | leukocyte | *ADRM1* | -0.61982949 | 2.49E-09 | 8.21E-09 | 9.80E-06 |
| T1DM | leukocyte | *UCK1* | -1.067398991 | 1.21E-08 | 2.20E-08 | 4.77E-05 |
| T1DM | leukocyte | *MAP3K10* | -1.324589608 | 9.50E-08 | 1.41E-07 | 0.0003734 |
| T1DM | leukocyte | *SMPDL3B* | -0.583940003 | 3.77E-09 | 9.36E-09 | 1.48E-05 |
| T1DM | leukocyte | *CLIP1* | -0.723237593 | 1.52E-08 | 2.67E-08 | 5.97E-05 |
| T1DM | leukocyte | *ZNF317* | -0.82601278 | 4.08E-08 | 6.49E-08 | 0.000160437 |
| T1DM | leukocyte | *DNMT1* | -1.051454029 | 2.33E-09 | 8.21E-09 | 9.14E-06 |
| T1DM | leukocyte | *ZNF426* | -0.925874773 | 2.19E-09 | 8.21E-09 | 8.60E-06 |
| T1DM | leukocyte | *LRP3* | -1.087407927 | 9.23E-10 | 8.14E-09 | 3.63E-06 |
| T1DM | leukocyte | *PKDREJ* | 0.751750568 | 0.000295487 | 0.000346145 | 1 |
| T1DM | leukocyte | *HSD17B3* | -8.666678651 | 8.91E-10 | 8.14E-09 | 3.50E-06 |
| T1DM | leukocyte | *FBP2* | -1.563339218 | 0.000102808 | 0.000122888 | 0.403931948 |
| T1DM | leukocyte | *SYNE1* | -0.596208988 | 1.52E-09 | 8.14E-09 | 5.96E-06 |
| T1DM | leukocyte | *EPS8L1* | -1.374327981 | 0.000124253 | 0.000147892 | 0.488190807 |
| T1DM | leukocyte | *C1QL1* | -0.3042386 | 7.83E-09 | 1.53E-08 | 3.08E-05 |
| T1DM | leukocyte | *F12* | -0.773438658 | 2.03E-08 | 3.43E-08 | 7.96E-05 |
| T1DM | leukocyte | *TRAF3* | -0.554690211 | 8.04E-09 | 1.56E-08 | 3.16E-05 |
| T1DM | leukocyte | *GALNT15* | -0.84980347 | 0.000129987 | 0.000154623 | 0.510718435 |
| T1DM | leukocyte | *NAPSA* | -1.592612199 | 6.69E-05 | 8.07E-05 | 0.262689916 |
| T1DM | leukocyte | *PDLIM4* | -0.737333307 | 2.53E-09 | 8.21E-09 | 9.94E-06 |
| T1DM | leukocyte | *DIAPH1* | -0.516301896 | 3.24E-09 | 8.77E-09 | 1.27E-05 |
| T1DM | leukocyte | *EXOC4* | -1.185268711 | 9.15E-08 | 1.37E-07 | 0.000359435 |
| T1DM | leukocyte | *C1orf159* | -0.954029597 | 0.025348556 | 0.027711318 | 1 |

| | | | | | | |
|---|---|---|---|---|---|---|
| T1DM | leukocyte | *ANO1* | -1.697120807 | 8.43E-10 | 8.14E-09 | 3.31E-06 |
| T1DM | leukocyte | *PPFIA1* | -0.603381932 | 1.78E-09 | 8.14E-09 | 6.98E-06 |
| T1DM | leukocyte | *TRAF7* | -0.883187499 | 1.52E-09 | 8.14E-09 | 5.96E-06 |
| T1DM | leukocyte | *MAP1B* | -0.426821125 | 3.75E-09 | 9.33E-09 | 1.47E-05 |
| T1DM | leukocyte | *CHD1L* | -1.37720164 | 8.05E-08 | 1.21E-07 | 0.000316378 |
| T1DM | leukocyte | *PIAS3* | -0.512067195 | 2.21E-08 | 3.72E-08 | 8.70E-05 |
| T1DM | leukocyte | *RBM8A* | -0.459532185 | 3.27E-09 | 8.79E-09 | 1.28E-05 |
| T1DM | leukocyte | *ZNF304* | -2.316041831 | 3.65E-07 | 5.04E-07 | 0.001433609 |
| T1DM | leukocyte | *ZNF132* | 0.600559194 | 0.000397339 | 0.000462972 | 1 |
| T1DM | leukocyte | *RHPN2* | -0.709318416 | 1.22E-10 | 8.14E-09 | 4.79E-07 |
| T1DM | leukocyte | *GCH1* | -0.497618745 | 1.15E-09 | 8.14E-09 | 4.51E-06 |
| T1DM | leukocyte | *CC2D1A* | -0.894923933 | 5.73E-09 | 1.20E-08 | 2.25E-05 |
| T1DM | leukocyte | *CCT6B* | -1.065896884 | 1.64E-09 | 8.14E-09 | 6.46E-06 |
| T1DM | leukocyte | *PPARG* | -1.086628061 | 4.16E-09 | 9.92E-09 | 1.63E-05 |
| T1DM | leukocyte | *ENOSF1* | -1.338259605 | 3.78E-07 | 5.22E-07 | 0.001485939 |
| T1DM | leukocyte | *TRIM22* | 0.850383894 | 3.08E-06 | 3.95E-06 | 0.012095119 |
| T1DM | leukocyte | *IMMT* | -0.719143521 | 1.51E-09 | 8.14E-09 | 5.93E-06 |
| T1DM | leukocyte | *MRPL35* | -0.721634995 | 4.13E-09 | 9.87E-09 | 1.62E-05 |
| T1DM | leukocyte | *SCLY* | -1.038469436 | 1.60E-07 | 2.30E-07 | 0.000628831 |
| T1DM | leukocyte | *CARD6* | -1.259340986 | 1.43E-08 | 2.53E-08 | 5.61E-05 |
| T1DM | leukocyte | *INPP5K* | -0.741845561 | 4.42E-09 | 1.02E-08 | 1.73E-05 |
| T1DM | leukocyte | *RPA1* | -0.811029053 | 5.45E-09 | 1.16E-08 | 2.14E-05 |
| T1DM | leukocyte | *TMEM128* | -0.969506499 | 3.94E-09 | 9.57E-09 | 1.55E-05 |
| T1DM | leukocyte | *SEC61G* | -0.507926723 | 9.36E-10 | 8.14E-09 | 3.68E-06 |
| T1DM | leukocyte | *FIGNL1* | -1.938001846 | 8.28E-08 | 1.25E-07 | 0.000325453 |
| T1DM | leukocyte | *ITGB4* | -1.338945162 | 1.10E-07 | 1.62E-07 | 0.000432443 |
| T1DM | leukocyte | *TRIM47* | -0.67761824 | 1.44E-09 | 8.14E-09 | 5.67E-06 |
| T1DM | leukocyte | *EIF5A* | -0.582905374 | 3.51E-10 | 8.14E-09 | 1.38E-06 |
| T1DM | leukocyte | *PRMT7* | -0.934696497 | 0.000368915 | 0.000430748 | 1 |
| T1DM | leukocyte | *ANKEF1* | -0.60144983 | 1.38E-09 | 8.14E-09 | 5.43E-06 |
| T1DM | leukocyte | *BTBD3* | -0.56113884 | 1.71E-09 | 8.14E-09 | 6.72E-06 |
| T1DM | leukocyte | *IGHMBP2* | -0.395069438 | 0.004652627 | 0.005237872 | 1 |
| T1DM | leukocyte | *LPIN3* | -0.697575246 | 7.93E-05 | 9.53E-05 | 0.311745004 |
| T1DM | leukocyte | *RBM38* | -1.065062713 | 3.48E-10 | 8.14E-09 | 1.37E-06 |
| T1DM | leukocyte | *VSTM2L* | -0.57955753 | 2.77E-09 | 8.34E-09 | 1.09E-05 |
| T1DM | leukocyte | *SERINC3* | -0.621242634 | 1.08E-08 | 1.99E-08 | 4.26E-05 |
| T1DM | leukocyte | *BHMT2* | -0.787207509 | 4.88E-09 | 1.08E-08 | 1.92E-05 |
| T1DM | leukocyte | *WASF3* | -0.708286586 | 4.02E-09 | 9.69E-09 | 1.58E-05 |
| T1DM | leukocyte | *SCO1* | -0.988857515 | 2.82E-07 | 3.95E-07 | 0.001108414 |
| T1DM | leukocyte | *CHI3L1* | -0.608038592 | 3.74E-08 | 5.99E-08 | 0.000146891 |
| T1DM | leukocyte | *SLC41A1* | -0.7769999 | 1.68E-09 | 8.14E-09 | 6.61E-06 |
| T1DM | leukocyte | *TRPC4* | -2.095153645 | 2.52E-10 | 8.14E-09 | 9.90E-07 |
| T1DM | leukocyte | *POSTN* | 0.719376509 | 4.84E-09 | 1.08E-08 | 1.90E-05 |
| T1DM | leukocyte | *RFC3* | -0.584760397 | 1.27E-09 | 8.14E-09 | 5.00E-06 |
| T1DM | leukocyte | *EPHB2* | -1.187576366 | 0.00708222 | 0.007911869 | 1 |
| T1DM | leukocyte | *BTBD2* | -1.390142408 | 1.49E-08 | 2.63E-08 | 5.85E-05 |
| T1DM | leukocyte | *CNDP2* | -0.684341274 | 1.82E-09 | 8.14E-09 | 7.15E-06 |
| T1DM | leukocyte | *WDR74* | -0.89342377 | 1.02E-07 | 1.50E-07 | 0.000398973 |
| T1DM | leukocyte | *GSTT2B* | -0.595880347 | 3.60E-08 | 5.80E-08 | 0.000141414 |
| T1DM | leukocyte | *SLC2A11* | -1.045601703 | 2.64E-09 | 8.21E-09 | 1.04E-05 |

| | | | | | | |
|---|---|---|---|---|---|---|
| T1DM | leukocyte | *C1QTNF6* | -1.166761132 | 6.17E-10 | 8.14E-09 | 2.43E-06 |
| T1DM | leukocyte | *FAM83F* | -0.737084642 | 1.67E-09 | 8.14E-09 | 6.56E-06 |
| T1DM | leukocyte | *GIMAP6* | -1.341337865 | 3.24E-10 | 8.14E-09 | 1.27E-06 |
| T1DM | leukocyte | *AGAP3* | -1.379805846 | 4.86E-05 | 5.90E-05 | 0.190992013 |
| T1DM | leukocyte | *SFTPD* | -0.590381162 | 1.40E-06 | 1.84E-06 | 0.005487494 |
| T1DM | leukocyte | *IMPA1* | -0.602292807 | 1.52E-08 | 2.67E-08 | 5.96E-05 |
| T1DM | leukocyte | *LRRCC1* | -1.061561412 | 2.11E-09 | 8.16E-09 | 8.27E-06 |
| T1DM | leukocyte | *RRAS2* | -0.562342739 | 9.46E-09 | 1.78E-08 | 3.72E-05 |
| T1DM | leukocyte | *HSD17B4* | -1.101084264 | 5.56E-09 | 1.18E-08 | 2.19E-05 |
| T1DM | leukocyte | *RNF122* | -0.979154978 | 5.86E-05 | 7.09E-05 | 0.230244976 |
| T1DM | leukocyte | *COX16* | -0.625837899 | 3.30E-09 | 8.81E-09 | 1.30E-05 |
| T1DM | leukocyte | *ELP3* | -1.315959763 | 4.41E-09 | 1.02E-08 | 1.73E-05 |
| T1DM | leukocyte | *PEBP4* | -0.954607686 | 3.59E-09 | 9.11E-09 | 1.41E-05 |
| T1DM | leukocyte | *MRO* | -1.663946805 | 9.25E-08 | 1.38E-07 | 0.000363337 |
| T1DM | leukocyte | *MRPS36* | -0.709075995 | 5.34E-09 | 1.15E-08 | 2.10E-05 |
| T1DM | leukocyte | *THUMPD3* | -0.730917595 | 1.70E-09 | 8.14E-09 | 6.67E-06 |
| T1DM | leukocyte | *CHL1* | -0.743198746 | 5.81E-08 | 9.00E-08 | 0.000228369 |
| T1DM | leukocyte | *GSTM1* | 1.308198236 | 4.59E-07 | 6.29E-07 | 0.0018046 |
| T1DM | leukocyte | *REG4* | -0.785915645 | 1.90E-09 | 8.14E-09 | 7.48E-06 |
| T1DM | leukocyte | *GSTM5* | -0.978704005 | 6.95E-05 | 8.37E-05 | 0.273003003 |
| T1DM | leukocyte | *GSTM3* | -0.460414953 | 4.55E-08 | 7.18E-08 | 0.000178856 |
| T1DM | leukocyte | *PTPN22* | -1.79196484 | 0.007006343 | 0.00782933 | 1 |
| T1DM | leukocyte | *SORT1* | -0.819764923 | 3.28E-09 | 8.79E-09 | 1.29E-05 |
| T1DM | leukocyte | *NOTCH2* | -0.654511773 | 1.84E-05 | 2.27E-05 | 0.072306569 |
| T1DM | leukocyte | *CEPT1* | -1.087916481 | 1.54E-08 | 2.71E-08 | 6.07E-05 |
| T1DM | leukocyte | *NAPG* | -0.679881247 | 2.07E-09 | 8.14E-09 | 8.11E-06 |
| T1DM | leukocyte | *SPIRE1* | -0.702099558 | 1.27E-09 | 8.14E-09 | 4.98E-06 |
| T1DM | leukocyte | *FKBP11* | -0.413857136 | 2.95E-09 | 8.50E-09 | 1.16E-05 |
| T1DM | leukocyte | *KIDINS220* | -0.701699657 | 2.89E-09 | 8.48E-09 | 1.13E-05 |
| T1DM | leukocyte | *LPIN1* | -0.497353773 | 4.28E-09 | 1.00E-08 | 1.68E-05 |
| T1DM | leukocyte | *IAH1* | -0.798327191 | 1.85E-08 | 3.18E-08 | 7.29E-05 |
| T1DM | leukocyte | *ANO3* | 1.240171272 | 1.52E-09 | 8.14E-09 | 5.97E-06 |
| T1DM | leukocyte | *IL2RA* | 0.240264345 | 4.36E-09 | 1.02E-08 | 1.71E-05 |
| T1DM | leukocyte | *ECHDC3* | -0.676967617 | 1.54E-09 | 8.14E-09 | 6.05E-06 |
| T1DM | leukocyte | *TMEM241* | -1.226291403 | 9.38E-10 | 8.14E-09 | 3.69E-06 |
| T1DM | leukocyte | *KLRC1* | 37.32803231 | 7.20E-07 | 9.68E-07 | 0.002830109 |
| T1DM | leukocyte | *LRP4* | -0.527966138 | 0.00579062 | 0.006481865 | 1 |
| T1DM | leukocyte | *AGO4* | -1.139571516 | 8.66E-09 | 1.65E-08 | 3.40E-05 |
| T1DM | leukocyte | *PRPF38A* | -0.676160749 | 1.75E-09 | 8.14E-09 | 6.86E-06 |
| T1DM | leukocyte | *TPGS2* | -0.791118163 | 5.35E-09 | 1.15E-08 | 2.10E-05 |
| T1DM | leukocyte | *DAGLA* | -0.795549027 | 5.88E-09 | 1.22E-08 | 2.31E-05 |
| T1DM | leukocyte | *TIMM10* | -0.676008412 | 3.14E-09 | 8.69E-09 | 1.23E-05 |
| T1DM | leukocyte | *FADS2* | -0.762218488 | 5.33E-09 | 1.15E-08 | 2.09E-05 |
| T1DM | leukocyte | *TMEM165* | -0.561061562 | 1.48E-09 | 8.14E-09 | 5.80E-06 |
| T1DM | leukocyte | *CLOCK* | -0.776030933 | 3.30E-09 | 8.81E-09 | 1.30E-05 |
| T1DM | leukocyte | *GGACT* | -1.124663036 | 1.70E-09 | 8.14E-09 | 6.67E-06 |
| T1DM | leukocyte | *CLDN10* | -0.301213768 | 3.64E-08 | 5.85E-08 | 0.000142948 |
| T1DM | leukocyte | *CARS2* | -0.670875935 | 9.00E-10 | 8.14E-09 | 3.54E-06 |
| T1DM | leukocyte | *TMED7* | -0.59423691 | 2.42E-09 | 8.21E-09 | 9.49E-06 |
| T1DM | leukocyte | *NREP* | -2.319521774 | 7.36E-05 | 8.85E-05 | 0.28921711 |

| | | | | | | |
|---|---|---|---|---|---|---|
| T1DM | leukocyte | *WDR36* | -0.729362535 | 1.36E-09 | 8.14E-09 | 5.33E-06 |
| T1DM | leukocyte | *OSTF1* | -0.927172409 | 6.12E-09 | 1.26E-08 | 2.40E-05 |
| T1DM | leukocyte | *GOLM1* | -0.533467301 | 8.33E-10 | 8.14E-09 | 3.27E-06 |
| T1DM | leukocyte | *ADAM19* | -1.974730924 | 0.001100042 | 0.001264131 | 1 |
| T1DM | leukocyte | *SDS* | 0.051401582 | 3.05E-09 | 8.59E-09 | 1.20E-05 |
| T1DM | leukocyte | *MSI1* | -1.167529479 | 2.05E-09 | 8.14E-09 | 8.07E-06 |
| T1DM | leukocyte | *DTX1* | -1.737276781 | 1.30E-09 | 8.14E-09 | 5.12E-06 |
| T1DM | leukocyte | *PNPLA8* | -0.537012313 | 2.40E-09 | 8.21E-09 | 9.43E-06 |
| T1DM | leukocyte | *TES* | -0.454744793 | 3.16E-09 | 8.69E-09 | 1.24E-05 |
| T1DM | leukocyte | *MDFIC* | -0.992419129 | 0.000464355 | 0.000540098 | 1 |
| T1DM | leukocyte | *MTO1* | -0.664060128 | 1.11E-09 | 8.14E-09 | 4.37E-06 |
| T1DM | leukocyte | *LCA5* | -1.564853454 | 3.62E-08 | 5.83E-08 | 0.000142152 |
| T1DM | leukocyte | *LMO2* | -0.604658456 | 2.63E-09 | 8.21E-09 | 1.03E-05 |
| T1DM | leukocyte | *RDH5* | -0.613254535 | 1.52E-09 | 8.14E-09 | 5.95E-06 |
| T1DM | leukocyte | *TFCP2* | -1.725426222 | 4.53E-08 | 7.15E-08 | 0.000177814 |
| T1DM | leukocyte | *KRT7* | -0.330671506 | 3.59E-09 | 9.12E-09 | 1.41E-05 |
| T1DM | leukocyte | *NHSL1* | -0.856288863 | 3.88E-08 | 6.19E-08 | 0.000152379 |
| T1DM | leukocyte | *AHI1* | -0.497953911 | 6.42E-10 | 8.14E-09 | 2.52E-06 |
| T1DM | leukocyte | *HEY2* | -0.995244458 | 2.32E-09 | 8.21E-09 | 9.12E-06 |
| T1DM | leukocyte | *PKIB* | -0.483604902 | 1.66E-09 | 8.14E-09 | 6.50E-06 |
| T1DM | leukocyte | *SMPD2* | -0.898589237 | 0.001184971 | 0.001359344 | 1 |
| T1DM | leukocyte | *SEMA4F* | -0.985019473 | 7.64E-10 | 8.14E-09 | 3.00E-06 |
| T1DM | leukocyte | *GNS* | -1.070127489 | 6.28E-08 | 9.66E-08 | 0.000246739 |
| T1DM | leukocyte | *MPHOSPH6* | -0.618808816 | 1.73E-09 | 8.14E-09 | 6.79E-06 |
| T1DM | leukocyte | *AGT* | -0.464037998 | 3.02E-10 | 8.14E-09 | 1.19E-06 |
| T1DM | leukocyte | *NTPCR* | -0.853850522 | 1.19E-08 | 2.17E-08 | 4.69E-05 |
| T1DM | leukocyte | *USP37* | -0.881611045 | 1.55E-09 | 8.14E-09 | 6.10E-06 |
| T1DM | leukocyte | *ARMC9* | -1.24412781 | 0.01925393 | 0.021136823 | 1 |
| T1DM | leukocyte | *COX5B* | -0.509389415 | 5.19E-09 | 1.13E-08 | 2.04E-05 |
| T1DM | leukocyte | *REV1* | -0.630928171 | 2.51E-09 | 8.21E-09 | 9.88E-06 |
| T1DM | leukocyte | *TSGA10* | -0.581545787 | 3.69E-06 | 4.70E-06 | 0.014480895 |
| T1DM | leukocyte | *TGFBRAP1* | -0.779082225 | 1.89E-09 | 8.14E-09 | 7.45E-06 |
| T1DM | leukocyte | *GCC2* | -0.69427473 | 7.05E-09 | 1.41E-08 | 2.77E-05 |
| T1DM | leukocyte | *MRPS9* | -0.596570062 | 2.38E-09 | 8.21E-09 | 9.34E-06 |
| T1DM | leukocyte | *ANKRD36* | -0.907109209 | 1.45E-09 | 8.14E-09 | 5.71E-06 |
| T1DM | leukocyte | *USP44* | -0.856235299 | 3.67E-08 | 5.90E-08 | 0.000144312 |
| T1DM | leukocyte | *PLXNC1* | 1.281717249 | 3.14E-07 | 4.37E-07 | 0.001232249 |
| T1DM | leukocyte | *DRAM1* | -1.229875603 | 4.21E-09 | 9.96E-09 | 1.65E-05 |
| T1DM | leukocyte | *VILL* | 1.012333141 | 1.76E-08 | 3.03E-08 | 6.91E-05 |
| T1DM | leukocyte | *FLNB* | -0.550006176 | 1.37E-08 | 2.45E-08 | 5.39E-05 |
| T1DM | leukocyte | *NEK3* | -0.457167293 | 1.12E-07 | 1.65E-07 | 0.00044129 |
| T1DM | leukocyte | *VPS36* | -0.971138702 | 4.37E-09 | 1.02E-08 | 1.72E-05 |
| T1DM | leukocyte | *RNASEH2B* | -1.12069331 | 5.73E-08 | 8.89E-08 | 0.000225279 |
| T1DM | leukocyte | *CKAP2* | -0.502792408 | 1.50E-05 | 1.86E-05 | 0.058922129 |
| T1DM | leukocyte | *TBC1D4* | -0.720038652 | 1.50E-09 | 8.14E-09 | 5.89E-06 |
| T1DM | leukocyte | *SUCLA2* | -0.764691158 | 1.77E-09 | 8.14E-09 | 6.96E-06 |
| T1DM | leukocyte | *LMO7* | -0.400166239 | 1.21E-09 | 8.14E-09 | 4.76E-06 |
| T1DM | leukocyte | *SETDB2* | 0.963440934 | 7.84E-06 | 9.82E-06 | 0.030811664 |
| T1DM | leukocyte | *C7orf25* | -0.573715046 | 1.02E-08 | 1.90E-08 | 4.02E-05 |
| T1DM | leukocyte | *RAPGEF5* | -0.495418221 | 1.43E-09 | 8.14E-09 | 5.63E-06 |

| | | | | | | |
|---|---|---|---|---|---|---|
| T1DM | leukocyte | *KDELR2* | -0.585463501 | 3.01E-09 | 8.53E-09 | 1.18E-05 |
| T1DM | leukocyte | *TBRG4* | -1.403911539 | 5.74E-10 | 8.14E-09 | 2.25E-06 |
| T1DM | leukocyte | *DBNL* | -1.449670569 | 2.21E-06 | 2.85E-06 | 0.008668853 |
| T1DM | leukocyte | *ALPK3* | -0.50318591 | 1.52E-09 | 8.14E-09 | 5.96E-06 |
| T1DM | leukocyte | *CIB2* | -0.481052013 | 0.00042863 | 0.000498989 | 1 |
| T1DM | leukocyte | *NMT1* | -0.583557346 | 1.14E-09 | 8.14E-09 | 4.46E-06 |
| T1DM | leukocyte | *CHAD* | -2.323760126 | 0.001143115 | 0.001313246 | 1 |
| T1DM | leukocyte | *TEX2* | -0.496783673 | 2.12E-09 | 8.16E-09 | 8.33E-06 |
| T1DM | leukocyte | *LIMD2* | 0.621995849 | 6.71E-10 | 8.14E-09 | 2.64E-06 |
| T1DM | leukocyte | *MRPL47* | -0.607790082 | 5.29E-09 | 1.14E-08 | 2.08E-05 |
| T1DM | leukocyte | *SMPD4* | -1.343481288 | 6.51E-08 | 9.97E-08 | 0.000255654 |
| T1DM | leukocyte | *CCDC115* | -0.719397729 | 4.49E-09 | 1.03E-08 | 1.76E-05 |
| T1DM | leukocyte | *BIN1* | -1.148304324 | 5.52E-06 | 6.98E-06 | 0.021699287 |
| T1DM | leukocyte | *HS6ST1* | -0.947777999 | 2.73E-09 | 8.29E-09 | 1.07E-05 |
| T1DM | leukocyte | *GYPC* | -0.61387902 | 1.20E-07 | 1.76E-07 | 0.000472057 |
| T1DM | leukocyte | *NIPSNAP3A* | -0.9120156 | 6.88E-09 | 1.38E-08 | 2.70E-05 |
| T1DM | leukocyte | *CDK9* | -0.795613623 | 1.49E-09 | 8.14E-09 | 5.84E-06 |
| T1DM | leukocyte | *TXN* | -0.468830454 | 3.14E-09 | 8.69E-09 | 1.23E-05 |
| T1DM | leukocyte | *TOR1B* | -1.050126363 | 5.32E-09 | 1.15E-08 | 2.09E-05 |
| T1DM | leukocyte | *TOR1A* | -0.638357944 | 1.89E-09 | 8.14E-09 | 7.44E-06 |
| T1DM | leukocyte | *SLC2A8* | -0.731484353 | 1.44E-09 | 8.14E-09 | 5.66E-06 |
| T1DM | leukocyte | *ZFP37* | -0.643855079 | 1.82E-09 | 8.14E-09 | 7.14E-06 |
| T1DM | leukocyte | *STX17* | -0.624122925 | 5.52E-09 | 1.17E-08 | 2.17E-05 |
| T1DM | leukocyte | *PRPF4* | -0.835826949 | 2.87E-08 | 4.71E-08 | 0.000112724 |
| T1DM | leukocyte | *FPGS* | -0.856349297 | 1.47E-09 | 8.14E-09 | 5.78E-06 |
| T1DM | leukocyte | *ATP6V1G1* | -0.484641414 | 6.94E-09 | 1.39E-08 | 2.73E-05 |
| T1DM | leukocyte | *ARPC5L* | -0.509683016 | 2.30E-09 | 8.21E-09 | 9.04E-06 |
| T1DM | leukocyte | *DSCC1* | -0.167432513 | 0.029968735 | 0.032680311 | 1 |
| T1DM | leukocyte | *POLR1E* | -0.459301192 | 2.03E-08 | 3.43E-08 | 7.96E-05 |
| T1DM | leukocyte | *APTX* | -0.577426665 | 1.21E-09 | 8.14E-09 | 4.76E-06 |
| T1DM | leukocyte | *CD72* | -1.166367156 | 5.97E-08 | 9.21E-08 | 0.000234726 |
| T1DM | leukocyte | *TMEM8B* | -0.893065859 | 8.96E-10 | 8.14E-09 | 3.52E-06 |
| T1DM | leukocyte | *GRHPR* | -0.636870768 | 3.18E-09 | 8.70E-09 | 1.25E-05 |
| T1DM | leukocyte | *ALDH1B1* | -1.10951239 | 4.33E-07 | 5.94E-07 | 0.001700759 |
| T1DM | leukocyte | *ARHGEF39* | -2.496882617 | 2.48E-09 | 8.21E-09 | 9.75E-06 |
| T1DM | leukocyte | *PPIL1* | -1.420271484 | 1.10E-08 | 2.02E-08 | 4.32E-05 |
| T1DM | leukocyte | *GMPR* | -1.147365269 | 1.18E-09 | 8.14E-09 | 4.64E-06 |
| T1DM | leukocyte | *KIAA0319* | -0.723745213 | 2.24E-09 | 8.21E-09 | 8.79E-06 |
| T1DM | leukocyte | *LRRC1* | -0.629838126 | 1.79E-09 | 8.14E-09 | 7.03E-06 |
| T1DM | leukocyte | *RIPK1* | -0.697382316 | 1.26E-09 | 8.14E-09 | 4.93E-06 |
| T1DM | leukocyte | *TUBB2B* | -0.56029622 | 3.62E-09 | 9.14E-09 | 1.42E-05 |
| T1DM | leukocyte | *UQCC2* | -0.552409732 | 2.40E-07 | 3.39E-07 | 0.000941292 |
| T1DM | leukocyte | *HMGA1* | -0.486977004 | 1.18E-09 | 8.14E-09 | 4.63E-06 |
| T1DM | leukocyte | *TCF19* | 0.580734345 | 3.70E-08 | 5.94E-08 | 0.000145333 |
| T1DM | leukocyte | *CLPS* | -0.24406181 | 1.01E-07 | 1.49E-07 | 0.000396515 |
| T1DM | leukocyte | *MTCH1* | -0.576582108 | 2.39E-09 | 8.21E-09 | 9.37E-06 |
| T1DM | leukocyte | *FCHSD2* | 0.854942589 | 4.97E-05 | 6.03E-05 | 0.195302162 |
| T1DM | leukocyte | *ANKRD42* | -0.95763258 | 1.35E-09 | 8.14E-09 | 5.29E-06 |
| T1DM | leukocyte | *CREBZF* | -0.841673558 | 3.17E-09 | 8.70E-09 | 1.25E-05 |
| T1DM | leukocyte | *PRCP* | -0.749419895 | 8.61E-07 | 1.15E-06 | 0.003381146 |

| | | | | | | |
|---|---|---|---|---|---|---|
| T1DM | leukocyte | *NARS2* | -0.809835399 | 2.12E-09 | 8.17E-09 | 8.34E-06 |
| T1DM | leukocyte | *TTPA* | -4.666326987 | 3.69E-07 | 5.10E-07 | 0.001451555 |
| T1DM | leukocyte | *GGH* | -0.703079957 | 6.84E-07 | 9.20E-07 | 0.002687285 |
| T1DM | leukocyte | *SDCBP* | -0.791337907 | 3.54E-06 | 4.52E-06 | 0.013919224 |
| T1DM | leukocyte | *NEK1* | -0.893257388 | 4.63E-05 | 5.63E-05 | 0.181744486 |
| T1DM | leukocyte | *TRPC6* | -0.946142828 | 0.002660333 | 0.00301484 | 1 |
| T1DM | leukocyte | *MMP7* | -0.289457293 | 4.95E-09 | 1.09E-08 | 1.95E-05 |
| T1DM | leukocyte | *TRIM29* | -1.862124833 | 6.68E-06 | 8.41E-06 | 0.026256455 |
| T1DM | leukocyte | *POU2F3* | -1.022959854 | 0.002397366 | 0.002719968 | 1 |
| T1DM | leukocyte | *FDX1* | -0.650777065 | 2.94E-09 | 8.50E-09 | 1.16E-05 |
| T1DM | leukocyte | *C11orf1* | -0.659799793 | 1.37E-09 | 8.14E-09 | 5.40E-06 |
| T1DM | leukocyte | *FXYD2* | -0.365377674 | 2.26E-09 | 8.21E-09 | 8.88E-06 |
| T1DM | leukocyte | *CASP1* | 0.287798907 | 3.25E-09 | 8.77E-09 | 1.28E-05 |
| T1DM | leukocyte | *ALKBH8* | -0.952762933 | 1.37E-09 | 8.14E-09 | 5.37E-06 |
| T1DM | leukocyte | *NDUFAF1* | -0.629926609 | 3.04E-06 | 3.90E-06 | 0.011953436 |
| T1DM | leukocyte | *TUBGCP4* | -1.000056904 | 5.65E-09 | 1.19E-08 | 2.22E-05 |
| T1DM | leukocyte | *UACA* | -0.567279977 | 1.38E-08 | 2.46E-08 | 5.43E-05 |
| T1DM | leukocyte | *PLCB2* | 0.440362336 | 1.22E-08 | 2.21E-08 | 4.80E-05 |
| T1DM | leukocyte | *PAK6* | -0.382050088 | 8.13E-09 | 1.57E-08 | 3.19E-05 |
| T1DM | leukocyte | *ADAM10* | -0.784236708 | 5.85E-09 | 1.22E-08 | 2.30E-05 |
| T1DM | leukocyte | *DUOX1* | -0.77908962 | 5.88E-08 | 9.08E-08 | 0.000230866 |
| T1DM | leukocyte | *CYP19A1* | -5.397194598 | 5.86E-08 | 9.06E-08 | 0.000230252 |
| T1DM | leukocyte | *ZNF280D* | -0.7871168 | 2.99E-09 | 8.51E-09 | 1.17E-05 |
| T1DM | leukocyte | *RSL24D1* | -0.588881819 | 4.83E-09 | 1.08E-08 | 1.90E-05 |
| T1DM | leukocyte | *TTLL7* | -0.649688295 | 1.35E-09 | 8.14E-09 | 5.32E-06 |
| T1DM | leukocyte | *GIPC2* | -0.493127901 | 8.99E-05 | 0.000107744 | 0.353293538 |
| T1DM | leukocyte | *SLC44A5* | -0.720745658 | 5.20E-05 | 6.31E-05 | 0.20429396 |
| T1DM | leukocyte | *RTCA* | -1.180108035 | 4.50E-06 | 5.70E-06 | 0.01766502 |
| T1DM | leukocyte | *KHK* | -1.385902411 | 0.000606428 | 0.000702433 | 1 |
| T1DM | leukocyte | *DYNC2LI1* | -0.769022413 | 4.76E-10 | 8.14E-09 | 1.87E-06 |
| T1DM | leukocyte | *ACTR2* | -0.581383329 | 1.91E-09 | 8.14E-09 | 7.51E-06 |
| T1DM | leukocyte | *ATRAID* | -0.591341374 | 9.77E-09 | 1.83E-08 | 3.84E-05 |
| T1DM | leukocyte | *LRPPRC* | -0.697204764 | 7.34E-09 | 1.46E-08 | 2.88E-05 |
| T1DM | leukocyte | *MYOF* | -0.36302747 | 1.69E-06 | 2.21E-06 | 0.006647999 |
| T1DM | leukocyte | *BTBD16* | -4.355941166 | 0.000274456 | 0.000322181 | 1 |
| T1DM | leukocyte | *DUSP5* | -1.064242071 | 3.07E-08 | 5.01E-08 | 0.000120712 |
| T1DM | leukocyte | *CALHM2* | 0.583464455 | 6.26E-05 | 7.56E-05 | 0.245802524 |
| T1DM | leukocyte | *ARL3* | -0.523975147 | 8.34E-08 | 1.25E-07 | 0.000327828 |
| T1DM | leukocyte | *PLCE1* | -0.772692122 | 1.09E-08 | 2.00E-08 | 4.29E-05 |
| T1DM | leukocyte | *DNAJC13* | -0.614188829 | 1.80E-09 | 8.14E-09 | 7.07E-06 |
| T1DM | leukocyte | *FAM149B1* | -0.716440693 | 1.96E-09 | 8.14E-09 | 7.70E-06 |
| T1DM | leukocyte | *NCOA4* | -1.069992648 | 3.87E-07 | 5.33E-07 | 0.001519829 |
| T1DM | leukocyte | *ASCC1* | -0.790974751 | 1.97E-09 | 8.14E-09 | 7.75E-06 |
| T1DM | leukocyte | *RPS24* | -0.432504436 | 3.96E-09 | 9.61E-09 | 1.55E-05 |
| T1DM | leukocyte | *DNA2* | -0.749896745 | 5.33E-05 | 6.45E-05 | 0.209223332 |
| T1DM | leukocyte | *MYPN* | 2.950083814 | 3.62E-08 | 5.83E-08 | 0.000142351 |
| T1DM | leukocyte | *ATIC* | -0.982374999 | 3.22E-08 | 5.23E-08 | 0.000126478 |
| T1DM | leukocyte | *SMARCAL1* | -0.925686346 | 1.37E-09 | 8.14E-09 | 5.40E-06 |
| T1DM | leukocyte | *BARD1* | -0.735656872 | 3.20E-09 | 8.73E-09 | 1.26E-05 |
| T1DM | leukocyte | *STAT4* | 0.939409742 | 9.43E-08 | 1.41E-07 | 0.00037043 |

| | | | | | | |
|---|---|---|---|---|---|---|
| T1DM | leukocyte | *FASTKD1* | -0.781770053 | 8.06E-09 | 1.56E-08 | 3.17E-05 |
| T1DM | leukocyte | *FAM117B* | -0.835416809 | 1.07E-09 | 8.14E-09 | 4.21E-06 |
| T1DM | leukocyte | *WDR12* | -0.962181167 | 5.71E-10 | 8.14E-09 | 2.24E-06 |
| T1DM | leukocyte | *SLC35A5* | -0.925768277 | 1.65E-09 | 8.14E-09 | 6.49E-06 |
| T1DM | leukocyte | *USP8* | -0.762780551 | 5.58E-09 | 1.18E-08 | 2.19E-05 |
| T1DM | leukocyte | *SPPL2A* | -0.68547792 | 2.69E-09 | 8.26E-09 | 1.06E-05 |
| T1DM | leukocyte | *APH1B* | -0.939589932 | 2.97E-08 | 4.85E-08 | 0.000116566 |
| T1DM | leukocyte | *HCN4* | -0.927354092 | 1.52E-09 | 8.14E-09 | 5.95E-06 |
| T1DM | leukocyte | *SEMA7A* | -1.606622367 | 1.48E-09 | 8.14E-09 | 5.82E-06 |
| T1DM | leukocyte | *AP1AR* | -0.566887097 | 2.44E-09 | 8.21E-09 | 9.59E-06 |
| T1DM | leukocyte | *BBS7* | -0.788517893 | 4.20E-09 | 9.96E-09 | 1.65E-05 |
| T1DM | leukocyte | *RAP1GDS1* | -0.78882026 | 2.51E-09 | 8.21E-09 | 9.86E-06 |
| T1DM | leukocyte | *PRDM5* | -0.682155332 | 1.39E-09 | 8.14E-09 | 5.46E-06 |
| T1DM | leukocyte | *NUP54* | -0.541132668 | 9.52E-08 | 1.42E-07 | 0.000373948 |
| T1DM | leukocyte | *G3BP2* | -0.543708172 | 9.82E-10 | 8.14E-09 | 3.86E-06 |
| T1DM | leukocyte | *SCARB2* | -0.609728003 | 1.52E-09 | 8.14E-09 | 5.98E-06 |
| T1DM | leukocyte | *CNOT6L* | -0.785567223 | 3.30E-09 | 8.82E-09 | 1.30E-05 |
| T1DM | leukocyte | *USO1* | -0.546444731 | 1.19E-09 | 8.14E-09 | 4.69E-06 |
| T1DM | leukocyte | *SHROOM3* | -1.142444571 | 4.51E-09 | 1.04E-08 | 1.77E-05 |
| T1DM | leukocyte | *ANXA3* | -0.490314085 | 3.05E-10 | 8.14E-09 | 1.20E-06 |
| T1DM | leukocyte | *PPA2* | -0.647086612 | 1.86E-09 | 8.14E-09 | 7.31E-06 |
| T1DM | leukocyte | *EGF* | -1.385168812 | 5.32E-06 | 6.73E-06 | 0.020894676 |
| T1DM | leukocyte | *MAPK8IP3* | -0.700669425 | 3.31E-09 | 8.82E-09 | 1.30E-05 |
| T1DM | leukocyte | *RGS3* | -0.617610913 | 6.98E-10 | 8.14E-09 | 2.74E-06 |
| T1DM | leukocyte | *ETV6* | -1.180902557 | 1.47E-08 | 2.60E-08 | 5.78E-05 |
| T1DM | leukocyte | *ALG10* | -1.22162568 | 1.35E-09 | 8.14E-09 | 5.31E-06 |
| T1DM | leukocyte | *PIK3C2G* | 1.361772072 | 0.001008109 | 0.001160861 | 1 |
| T1DM | leukocyte | *SLCO1C1* | -10.71024755 | 1.86E-09 | 8.14E-09 | 7.31E-06 |
| T1DM | leukocyte | *ZCRB1* | -0.574283826 | 2.94E-09 | 8.50E-09 | 1.16E-05 |
| T1DM | leukocyte | *C1RL* | -0.739715045 | 2.50E-07 | 3.53E-07 | 0.000983231 |
| T1DM | leukocyte | *CLSTN3* | -0.867209092 | 4.72E-07 | 6.46E-07 | 0.001855819 |
| T1DM | leukocyte | *KLRG1* | -4.478588817 | 8.73E-10 | 8.14E-09 | 3.43E-06 |
| T1DM | leukocyte | *VAMP1* | 0.480687233 | 1.63E-06 | 2.13E-06 | 0.006396006 |
| T1DM | leukocyte | *TAPBPL* | -0.443629401 | 3.01E-05 | 3.68E-05 | 0.118120781 |
| T1DM | leukocyte | *PEX5* | -0.663666851 | 2.13E-09 | 8.18E-09 | 8.37E-06 |
| T1DM | leukocyte | *SLC38A4* | -0.945158837 | 4.12E-09 | 9.86E-09 | 1.62E-05 |
| T1DM | leukocyte | *LRIG3* | -0.692012879 | 8.91E-10 | 8.14E-09 | 3.50E-06 |
| T1DM | leukocyte | *GLIPR1* | -0.782811415 | 1.48E-09 | 8.14E-09 | 5.83E-06 |
| T1DM | leukocyte | *SNRPF* | -0.51158247 | 5.91E-09 | 1.23E-08 | 2.32E-05 |
| T1DM | leukocyte | *AMDHD1* | -0.720552652 | 0.000617096 | 0.000714581 | 1 |
| T1DM | leukocyte | *ASCL1* | -1.085065552 | 1.05E-09 | 8.14E-09 | 4.13E-06 |
| T1DM | leukocyte | *RITA1* | -0.615119626 | 0.000121898 | 0.000145264 | 0.478935404 |
| T1DM | leukocyte | *GLTP* | -0.539652205 | 1.51E-09 | 8.14E-09 | 5.94E-06 |
| T1DM | leukocyte | *MTMR6* | -0.791016707 | 1.05E-08 | 1.94E-08 | 4.13E-05 |
| T1DM | leukocyte | *PDX1* | -0.894394124 | 1.96E-09 | 8.14E-09 | 7.68E-06 |
| T1DM | leukocyte | *SUOX* | -0.82519582 | 6.87E-10 | 8.14E-09 | 2.70E-06 |
| T1DM | leukocyte | *CCDC65* | -0.885606273 | 1.21E-06 | 1.59E-06 | 0.004738131 |
| T1DM | leukocyte | *SLC39A5* | -0.200495872 | 0.007677996 | 0.008567693 | 1 |
| T1DM | leukocyte | *KANSL2* | -1.378641917 | 2.09E-08 | 3.53E-08 | 8.23E-05 |
| T1DM | leukocyte | *CERS5* | -0.722327252 | 2.46E-07 | 3.47E-07 | 0.000967512 |

| | | | | | | |
|---|---|---|---|---|---|---|
| T1DM | leukocyte | *GALNT6* | 2.258404025 | 0.002284606 | 0.002596534 | 1 |
| T1DM | leukocyte | *ZNF740* | -1.314172886 | 1.85E-08 | 3.17E-08 | 7.26E-05 |
| T1DM | leukocyte | *WDFY2* | -0.755611035 | 1.53E-09 | 8.14E-09 | 5.99E-06 |
| T1DM | leukocyte | *GRTP1* | -1.447074471 | 9.52E-09 | 1.79E-08 | 3.74E-05 |
| T1DM | leukocyte | *TSSK4* | -5.153966865 | 2.26E-06 | 2.92E-06 | 0.008886397 |
| T1DM | leukocyte | *TMX1* | -0.877700392 | 6.02E-09 | 1.24E-08 | 2.37E-05 |
| T1DM | leukocyte | *RAB15* | -0.88880528 | 2.08E-08 | 3.51E-08 | 8.18E-05 |
| T1DM | leukocyte | *ESR2* | -0.893370576 | 3.33E-06 | 4.26E-06 | 0.01309602 |
| T1DM | leukocyte | *PTGR2* | -0.44702253 | 9.09E-07 | 1.21E-06 | 0.003573035 |
| T1DM | leukocyte | *NIPA2* | -0.559669356 | 1.47E-09 | 8.14E-09 | 5.76E-06 |
| T1DM | leukocyte | *SERF2* | -0.474038101 | 3.56E-09 | 9.10E-09 | 1.40E-05 |
| T1DM | leukocyte | *ZSCAN29* | -0.739603113 | 0.008503333 | 0.009469839 | 1 |
| T1DM | leukocyte | *BNIP2* | -0.97755697 | 9.28E-09 | 1.75E-08 | 3.65E-05 |
| T1DM | leukocyte | *GTF2A2* | -0.581574052 | 2.27E-09 | 8.21E-09 | 8.93E-06 |
| T1DM | leukocyte | *SRP14* | -0.465596116 | 4.24E-09 | 9.99E-09 | 1.67E-05 |
| T1DM | leukocyte | *BAHD1* | -0.746989678 | 5.13E-09 | 1.12E-08 | 2.02E-05 |
| T1DM | leukocyte | *DISP2* | -0.48418537 | 2.28E-07 | 3.23E-07 | 0.000897 |
| T1DM | leukocyte | *TLE3* | -1.211267414 | 1.09E-07 | 1.60E-07 | 0.000426643 |
| T1DM | leukocyte | *SCAPER* | -0.557551166 | 2.33E-08 | 3.89E-08 | 9.14E-05 |
| T1DM | leukocyte | *TSPAN3* | -0.507274455 | 2.64E-09 | 8.21E-09 | 1.04E-05 |
| T1DM | leukocyte | *MAN2C1* | -0.584963964 | 6.99E-08 | 1.06E-07 | 0.000274464 |
| T1DM | leukocyte | *DNAJA4* | -0.576013451 | 4.07E-09 | 9.78E-09 | 1.60E-05 |
| T1DM | leukocyte | *TPM1* | -0.379773142 | 1.10E-09 | 8.14E-09 | 4.33E-06 |
| T1DM | leukocyte | *IGF1R* | -0.561414642 | 1.09E-09 | 8.14E-09 | 4.27E-06 |
| T1DM | leukocyte | *ADAMTS17* | -1.188571287 | 1.44E-05 | 1.79E-05 | 0.056678004 |
| T1DM | leukocyte | *ULK3* | -0.741631808 | 4.74E-11 | 8.14E-09 | 1.86E-07 |
| T1DM | leukocyte | *CELF6* | -0.809140264 | 1.32E-09 | 8.14E-09 | 5.18E-06 |
| T1DM | leukocyte | *POLG* | -1.035934166 | 1.41E-09 | 8.14E-09 | 5.54E-06 |
| T1DM | leukocyte | *WDR93* | -1.842823229 | 3.32E-05 | 4.06E-05 | 0.130312011 |
| T1DM | leukocyte | *UNC45A* | -0.748807753 | 5.24E-10 | 8.14E-09 | 2.06E-06 |
| T1DM | leukocyte | *IQGAP1* | -0.613015109 | 2.11E-08 | 3.56E-08 | 8.30E-05 |
| T1DM | leukocyte | *PMM2* | -0.663026326 | 1.54E-09 | 8.14E-09 | 6.07E-06 |
| T1DM | leukocyte | *SLC5A2* | -2.665152477 | 1.73E-09 | 8.14E-09 | 6.82E-06 |
| T1DM | leukocyte | *UQCRC2* | -0.684020244 | 1.91E-09 | 8.14E-09 | 7.50E-06 |
| T1DM | leukocyte | *ARHGAP17* | -1.409142371 | 4.40E-08 | 6.97E-08 | 0.000172991 |
| T1DM | leukocyte | *TXNL4B* | -1.270708595 | 2.62E-07 | 3.68E-07 | 0.001028656 |
| T1DM | leukocyte | *MARVELD3* | -0.552886766 | 1.17E-08 | 2.14E-08 | 4.61E-05 |
| T1DM | leukocyte | *KIFC3* | -0.720751776 | 4.19E-09 | 9.96E-09 | 1.65E-05 |
| T1DM | leukocyte | *NUDT7* | -0.618310575 | 1.45E-09 | 8.14E-09 | 5.68E-06 |
| T1DM | leukocyte | *GCSH* | -0.615430927 | 1.65E-09 | 8.14E-09 | 6.49E-06 |
| T1DM | leukocyte | *MAP1LC3B* | -0.526418832 | 2.53E-09 | 8.21E-09 | 9.95E-06 |
| T1DM | leukocyte | *OSGIN1* | -0.535434029 | 1.12E-09 | 8.14E-09 | 4.40E-06 |
| T1DM | leukocyte | *RPL3L* | -1.219570169 | 3.05E-06 | 3.91E-06 | 0.011965528 |
| T1DM | leukocyte | *RPS2* | -0.4031318 | 2.43E-09 | 8.21E-09 | 9.57E-06 |
| T1DM | leukocyte | *PDPK1* | -0.773181239 | 1.86E-08 | 3.19E-08 | 7.32E-05 |
| T1DM | leukocyte | *DEF8* | -0.706542051 | 4.59E-09 | 1.04E-08 | 1.80E-05 |
| T1DM | leukocyte | *GALNS* | 0.777246823 | 0.013573751 | 0.014997544 | 1 |
| T1DM | leukocyte | *GAS8* | -1.089809496 | 1.08E-08 | 1.99E-08 | 4.24E-05 |
| T1DM | leukocyte | *RANBP10* | -0.607672421 | 5.30E-08 | 8.28E-08 | 0.000208421 |
| T1DM | leukocyte | *PRPSAP2* | -0.796332379 | 5.42E-09 | 1.16E-08 | 2.13E-05 |

| | | | | | | |
|---|---|---|---|---|---|---|
| T1DM | leukocyte | *TOM1L1* | -0.638198232 | 2.28E-09 | 8.21E-09 | 8.96E-06 |
| T1DM | leukocyte | *C17orf80* | -1.000983961 | 6.43E-08 | 9.87E-08 | 0.000252592 |
| T1DM | leukocyte | *VPS53* | -0.618202305 | 5.87E-08 | 9.08E-08 | 0.000230787 |
| T1DM | leukocyte | *NPEPPS* | -0.799491303 | 3.80E-08 | 6.09E-08 | 0.000149376 |
| T1DM | leukocyte | *LRRC46* | 0.577738847 | 6.45E-05 | 7.78E-05 | 0.253283212 |
| T1DM | leukocyte | *SCRN2* | -0.981594675 | 1.12E-06 | 1.49E-06 | 0.004414723 |
| T1DM | leukocyte | *RHBDL3* | -0.746776699 | 0.00147061 | 0.001681125 | 1 |
| T1DM | leukocyte | *SPACA3* | -0.415592024 | 7.98E-05 | 9.58E-05 | 0.313549333 |
| T1DM | leukocyte | *ARSG* | -1.191098259 | 0.00325506 | 0.003682444 | 1 |
| T1DM | leukocyte | *PTRH2* | -0.739526319 | 1.85E-09 | 8.14E-09 | 7.27E-06 |
| T1DM | leukocyte | *GNAL* | -0.985627922 | 2.42E-09 | 8.21E-09 | 9.51E-06 |
| T1DM | leukocyte | *RPRD1A* | -0.649724298 | 4.06E-09 | 9.77E-09 | 1.60E-05 |
| T1DM | leukocyte | *NPC1* | -1.196236497 | 2.36E-08 | 3.94E-08 | 9.26E-05 |
| T1DM | leukocyte | *SLC14A1* | -0.789672065 | 1.03E-05 | 1.28E-05 | 0.040446031 |
| T1DM | leukocyte | *ZMYND15* | 0.170168935 | 2.95E-09 | 8.50E-09 | 1.16E-05 |
| T1DM | leukocyte | *SAT2* | -0.728611579 | 6.62E-09 | 1.34E-08 | 2.60E-05 |
| T1DM | leukocyte | *CCDC40* | -0.700106284 | 6.91E-09 | 1.39E-08 | 2.71E-05 |
| T1DM | leukocyte | *ARHGDIA* | -0.570525028 | 1.82E-08 | 3.13E-08 | 7.16E-05 |
| T1DM | leukocyte | *TMC6* | -1.099279993 | 1.28E-09 | 8.14E-09 | 5.05E-06 |
| T1DM | leukocyte | *TTYH2* | -1.218247218 | 1.47E-09 | 8.14E-09 | 5.79E-06 |
| T1DM | leukocyte | *EIF4A3* | -0.718286495 | 2.41E-09 | 8.21E-09 | 9.45E-06 |
| T1DM | leukocyte | *TBCD* | -1.051088746 | 1.94E-08 | 3.30E-08 | 7.61E-05 |
| T1DM | leukocyte | *FN3KRP* | -0.531665376 | 0.0002781 | 0.000326361 | 1 |
| T1DM | leukocyte | *NARF* | -0.866793478 | 5.97E-09 | 1.24E-08 | 2.35E-05 |
| T1DM | leukocyte | *TRIM65* | -0.909335216 | 9.35E-08 | 1.40E-07 | 0.000367416 |
| T1DM | leukocyte | *RNF157* | -0.800920192 | 5.95E-10 | 8.14E-09 | 2.34E-06 |
| T1DM | leukocyte | *WDR45B* | -0.528610169 | 1.01E-08 | 1.88E-08 | 3.99E-05 |
| T1DM | leukocyte | *TNFRSF11A* | -0.632255411 | 9.23E-10 | 8.14E-09 | 3.63E-06 |
| T1DM | leukocyte | *ZCCHC2* | -0.852550911 | 1.18E-08 | 2.15E-08 | 4.64E-05 |
| T1DM | leukocyte | *PMAIP1* | -0.632711655 | 1.39E-09 | 8.14E-09 | 5.48E-06 |
| T1DM | leukocyte | *NT5C3B* | -0.614898114 | 9.56E-08 | 1.42E-07 | 0.000375701 |
| T1DM | leukocyte | *PIP4K2B* | -0.65591991 | 3.43E-09 | 8.93E-09 | 1.35E-05 |
| T1DM | leukocyte | *TXNL4A* | -0.549625164 | 9.66E-09 | 1.81E-08 | 3.79E-05 |
| T1DM | leukocyte | *PRDM15* | -0.854106099 | 1.26E-07 | 1.84E-07 | 0.000496493 |
| T1DM | leukocyte | *MVB12A* | -0.74679532 | 4.03E-08 | 6.42E-08 | 0.000158411 |
| T1DM | leukocyte | *SH3GL1* | -0.616197479 | 1.97E-09 | 8.14E-09 | 7.75E-06 |
| T1DM | leukocyte | *DUS3L* | -0.485098131 | 4.60E-08 | 7.24E-08 | 0.000180574 |
| T1DM | leukocyte | *SIRT3* | -0.936200136 | 1.41E-09 | 8.14E-09 | 5.53E-06 |
| T1DM | leukocyte | *IFITM3* | -0.413195606 | 7.95E-09 | 1.55E-08 | 3.12E-05 |
| T1DM | leukocyte | *COL6A1* | -0.894062224 | 7.85E-10 | 8.14E-09 | 3.08E-06 |
| T1DM | leukocyte | *IFNAR1* | -0.622906832 | 9.34E-10 | 8.14E-09 | 3.67E-06 |
| T1DM | leukocyte | *SIK1* | -0.861216324 | 0.001404816 | 0.001608253 | 1 |
| T1DM | leukocyte | *TMEM50B* | -0.56751356 | 2.94E-07 | 4.10E-07 | 0.00115609 |
| T1DM | leukocyte | *URB1* | -0.432621194 | 0.006296624 | 0.007044258 | 1 |
| T1DM | leukocyte | *AKT1* | -0.893325572 | 1.62E-08 | 2.82E-08 | 6.36E-05 |
| T1DM | leukocyte | *CBLC* | -0.647233538 | 1.41E-08 | 2.51E-08 | 5.55E-05 |
| T1DM | leukocyte | *EVI5L* | -0.744166685 | 2.48E-07 | 3.49E-07 | 0.000972683 |
| T1DM | leukocyte | *CTU1* | -1.04843821 | 2.84E-09 | 8.43E-09 | 1.12E-05 |
| T1DM | leukocyte | *RERE* | -0.559371294 | 1.57E-09 | 8.14E-09 | 6.18E-06 |
| T1DM | leukocyte | *FHAD1* | -0.687031908 | 5.82E-09 | 1.21E-08 | 2.29E-05 |

| | | | | | | |
|---|---|---|---|---|---|---|
| T1DM | leukocyte | *PGD* | -0.843644383 | 6.78E-10 | 8.14E-09 | 2.66E-06 |
| T1DM | leukocyte | *CNKSR1* | -0.973202385 | 3.90E-06 | 4.97E-06 | 0.015333898 |
| T1DM | leukocyte | *ZNF593* | -0.613226057 | 3.09E-09 | 8.63E-09 | 1.21E-05 |
| T1DM | leukocyte | *FCN3* | -0.322762979 | 6.26E-07 | 8.46E-07 | 0.002461126 |
| T1DM | leukocyte | *CELA3A* | -0.297923361 | 1.46E-09 | 8.14E-09 | 5.74E-06 |
| T1DM | leukocyte | *NBPF3* | -0.606086381 | 7.89E-06 | 9.88E-06 | 0.031009085 |
| T1DM | leukocyte | *HSPG2* | -0.356265678 | 1.51E-08 | 2.65E-08 | 5.92E-05 |
| T1DM | leukocyte | *ITGB3BP* | -0.565323403 | 6.48E-07 | 8.73E-07 | 0.002545484 |
| T1DM | leukocyte | *PRKACB* | -0.729288967 | 4.97E-09 | 1.10E-08 | 1.95E-05 |
| T1DM | leukocyte | *PIGK* | -1.038648961 | 5.85E-09 | 1.22E-08 | 2.30E-05 |
| T1DM | leukocyte | *RPS8* | -0.389782115 | 2.41E-09 | 8.21E-09 | 9.47E-06 |
| T1DM | leukocyte | *CYP4B1* | 3.758109142 | 5.28E-06 | 6.69E-06 | 0.020750349 |
| T1DM | leukocyte | *SYPL2* | -2.011757738 | 2.52E-09 | 8.21E-09 | 9.90E-06 |
| T1DM | leukocyte | *SLC44A3* | -0.660134193 | 2.64E-09 | 8.21E-09 | 1.04E-05 |
| T1DM | leukocyte | *ZNF697* | -0.830164415 | 2.79E-06 | 3.60E-06 | 0.010981546 |
| T1DM | leukocyte | *CELSR2* | -0.772377673 | 1.25E-08 | 2.26E-08 | 4.90E-05 |
| T1DM | leukocyte | *ALDH9A1* | -0.586377611 | 1.60E-09 | 8.14E-09 | 6.27E-06 |
| T1DM | leukocyte | *ATP1B1* | -0.402180467 | 1.41E-09 | 8.14E-09 | 5.53E-06 |
| T1DM | leukocyte | *TIPRL* | -0.566310217 | 3.84E-09 | 9.48E-09 | 1.51E-05 |
| T1DM | leukocyte | *NME7* | -0.925910185 | 8.06E-09 | 1.56E-08 | 3.17E-05 |
| T1DM | leukocyte | *CREG1* | 0.514509891 | 6.66E-08 | 1.02E-07 | 0.000261715 |
| T1DM | leukocyte | *XCL1* | -11.3570334 | 0.009130189 | 0.010150682 | 1 |
| T1DM | leukocyte | *MGST3* | -0.588455364 | 1.32E-08 | 2.37E-08 | 5.18E-05 |
| T1DM | leukocyte | *LYPLAL1* | -0.793953041 | 1.09E-09 | 8.14E-09 | 4.29E-06 |
| T1DM | leukocyte | *SNX27* | -0.979481144 | 8.66E-09 | 1.65E-08 | 3.40E-05 |
| T1DM | leukocyte | *CTSK* | -1.871375766 | 6.67E-08 | 1.02E-07 | 0.000262165 |
| T1DM | leukocyte | *SELENBP1* | -0.804144714 | 1.35E-09 | 8.14E-09 | 5.31E-06 |
| T1DM | leukocyte | *POGZ* | -0.696166397 | 9.79E-09 | 1.83E-08 | 3.85E-05 |
| T1DM | leukocyte | *GABPB2* | -1.121707437 | 4.20E-09 | 9.96E-09 | 1.65E-05 |
| T1DM | leukocyte | *TAF1A* | 1.53448456 | 1.08E-07 | 1.59E-07 | 0.000424527 |
| T1DM | leukocyte | *SLC27A3* | -0.917548617 | 2.58E-09 | 8.21E-09 | 1.01E-05 |
| T1DM | leukocyte | *KCNN3* | -1.225344227 | 1.05E-09 | 8.14E-09 | 4.12E-06 |
| T1DM | leukocyte | *RIT1* | -0.671294931 | 0.001286449 | 0.001474032 | 1 |
| T1DM | leukocyte | *SRP9* | -0.607771246 | 4.31E-09 | 1.01E-08 | 1.69E-05 |
| T1DM | leukocyte | *CNIH4* | -0.645374166 | 2.37E-09 | 8.21E-09 | 9.31E-06 |
| T1DM | leukocyte | *PSEN2* | -1.22978948 | 1.37E-08 | 2.45E-08 | 5.38E-05 |
| T1DM | leukocyte | *PYCR2* | -0.727672405 | 2.69E-09 | 8.26E-09 | 1.06E-05 |
| T1DM | leukocyte | *LBR* | -0.729962662 | 4.33E-08 | 6.86E-08 | 0.000169938 |
| T1DM | leukocyte | *ARL8A* | -0.940378164 | 1.53E-08 | 2.68E-08 | 5.99E-05 |
| T1DM | leukocyte | *GALM* | 1.634784665 | 0.00117488 | 0.001348555 | 1 |
| T1DM | leukocyte | *EML4* | -0.977502272 | 1.72E-08 | 2.97E-08 | 6.74E-05 |
| T1DM | leukocyte | *WDPCP* | -1.569003483 | 1.46E-08 | 2.58E-08 | 5.74E-05 |
| T1DM | leukocyte | *REG3G* | -0.641340351 | 3.72E-08 | 5.96E-08 | 0.000146077 |
| T1DM | leukocyte | *CIAO1* | -0.842449792 | 7.57E-09 | 1.49E-08 | 2.97E-05 |
| T1DM | leukocyte | *ZNF514* | -0.749498629 | 2.94E-10 | 8.14E-09 | 1.15E-06 |
| T1DM | leukocyte | *TPRKB* | -0.573952464 | 2.15E-09 | 8.19E-09 | 8.44E-06 |
| T1DM | leukocyte | *TEX261* | -0.751166184 | 1.86E-09 | 8.14E-09 | 7.31E-06 |
| T1DM | leukocyte | *THNSL2* | -0.325500288 | 0.030096127 | 0.032801021 | 1 |
| T1DM | leukocyte | *RALB* | -0.724738588 | 3.07E-10 | 8.14E-09 | 1.21E-06 |
| T1DM | leukocyte | *RABL2A* | -1.009076812 | 2.54E-09 | 8.21E-09 | 9.98E-06 |

| | | | | | | |
|---|---|---|---|---|---|---|
| T1DM | leukocyte | *SLC20A1* | -0.98662473 | 3.88E-09 | 9.52E-09 | 1.53E-05 |
| T1DM | leukocyte | *LIPT1* | 1.235732788 | 0.003525153 | 0.003979979 | 1 |
| T1DM | leukocyte | *LYG1* | -2.653342919 | 3.40E-05 | 4.15E-05 | 0.133518648 |
| T1DM | leukocyte | *AFF3* | -0.796351623 | 0.019814172 | 0.021739705 | 1 |
| T1DM | leukocyte | *GULP1* | -0.647822389 | 2.77E-09 | 8.34E-09 | 1.09E-05 |
| T1DM | leukocyte | *FAM171B* | -0.599387037 | 1.48E-09 | 8.14E-09 | 5.83E-06 |
| T1DM | leukocyte | *METTL21A* | -0.688082562 | 2.09E-09 | 8.15E-09 | 8.20E-06 |
| T1DM | leukocyte | *SUMF1* | -0.919411015 | 1.17E-09 | 8.14E-09 | 4.60E-06 |
| T1DM | leukocyte | *HES6* | -1.341376355 | 1.72E-07 | 2.47E-07 | 0.000677566 |
| T1DM | leukocyte | *ESPNL* | -5.046093011 | 0.00070443 | 0.00081451 | 1 |
| T1DM | leukocyte | *TAMM41* | -0.781903276 | 2.27E-08 | 3.80E-08 | 8.91E-05 |
| T1DM | leukocyte | *RAB5A* | -0.614142007 | 2.57E-09 | 8.21E-09 | 1.01E-05 |
| T1DM | leukocyte | *CNTN4* | -0.628976017 | 2.51E-08 | 4.16E-08 | 9.86E-05 |
| T1DM | leukocyte | *POMGNT2* | -1.113722036 | 3.72E-09 | 9.31E-09 | 1.46E-05 |
| T1DM | leukocyte | *CTDSPL* | -0.626101541 | 1.33E-09 | 8.14E-09 | 5.21E-06 |
| T1DM | leukocyte | *IL17RD* | -2.763403848 | 4.54E-09 | 1.04E-08 | 1.78E-05 |
| T1DM | leukocyte | *UBA3* | -0.952327253 | 3.02E-09 | 8.56E-09 | 1.19E-05 |
| T1DM | leukocyte | *ARL6IP5* | -0.593489411 | 2.48E-09 | 8.21E-09 | 9.76E-06 |
| T1DM | leukocyte | *LIMD1* | -1.051414641 | 6.79E-09 | 1.37E-08 | 2.67E-05 |
| T1DM | leukocyte | *ZNF660* | -0.610106013 | 6.07E-05 | 7.35E-05 | 0.2385573 |
| T1DM | leukocyte | *NXPE3* | -0.602279254 | 2.13E-08 | 3.58E-08 | 8.36E-05 |
| T1DM | leukocyte | *PHLDB2* | -0.503789202 | 1.38E-09 | 8.14E-09 | 5.42E-06 |
| T1DM | leukocyte | *ABHD10* | -0.721756634 | 9.02E-09 | 1.71E-08 | 3.54E-05 |
| T1DM | leukocyte | *TAGLN3* | -0.426555323 | 5.48E-09 | 1.17E-08 | 2.15E-05 |
| T1DM | leukocyte | *EIF2A* | -0.556844577 | 1.33E-09 | 8.14E-09 | 5.21E-06 |
| T1DM | leukocyte | *ALDH1L1* | -0.958080028 | 1.43E-09 | 8.14E-09 | 5.61E-06 |
| T1DM | leukocyte | *TRPC1* | -0.809035288 | 1.43E-09 | 8.14E-09 | 5.61E-06 |
| T1DM | leukocyte | *NCEH1* | -0.420263921 | 2.25E-09 | 8.21E-09 | 8.85E-06 |
| T1DM | leukocyte | *AMT* | -0.76781445 | 4.78E-05 | 5.81E-05 | 0.187887389 |
| T1DM | leukocyte | *DGKQ* | -0.711394001 | 1.34E-08 | 2.40E-08 | 5.26E-05 |
| T1DM | leukocyte | *SCD5* | -0.569193533 | 1.40E-09 | 8.14E-09 | 5.52E-06 |
| T1DM | leukocyte | *PYURF* | -0.527277137 | 2.58E-09 | 8.21E-09 | 1.01E-05 |
| T1DM | leukocyte | *TBCK* | -0.760867176 | 4.64E-06 | 5.89E-06 | 0.018238745 |
| T1DM | leukocyte | *DDIT4L* | -1.124318584 | 9.83E-05 | 0.000117615 | 0.386131126 |
| T1DM | leukocyte | *TIFA* | -0.820768259 | 1.83E-09 | 8.14E-09 | 7.21E-06 |
| T1DM | leukocyte | *RNF175* | -12.51515212 | 0.027666121 | 0.030194497 | 1 |
| T1DM | leukocyte | *CYP4V2* | -0.709038711 | 0.002529874 | 0.002869479 | 1 |
| T1DM | leukocyte | *SSBP2* | -0.554743006 | 1.42E-09 | 8.14E-09 | 5.58E-06 |
| T1DM | leukocyte | *IQGAP2* | 0.39995143 | 2.65E-07 | 3.72E-07 | 0.001040754 |
| T1DM | leukocyte | *GTF2H2* | -1.505626254 | 4.75E-08 | 7.47E-08 | 0.000186794 |
| T1DM | leukocyte | *BTF3* | -0.476399934 | 4.40E-09 | 1.02E-08 | 1.73E-05 |
| T1DM | leukocyte | *SPATA9* | -4.563052175 | 1.14E-07 | 1.68E-07 | 0.000448466 |
| T1DM | leukocyte | *COMMD10* | -0.778840172 | 1.52E-09 | 8.14E-09 | 5.98E-06 |
| T1DM | leukocyte | *ATG12* | -0.597846265 | 4.93E-09 | 1.09E-08 | 1.94E-05 |
| T1DM | leukocyte | *PCYOX1L* | -2.444439697 | 7.00E-06 | 8.80E-06 | 0.027505667 |
| T1DM | leukocyte | *MYLK4* | 1.584193815 | 5.83E-05 | 7.06E-05 | 0.229144098 |
| T1DM | leukocyte | *ZMAT2* | -0.783047019 | 2.37E-09 | 8.21E-09 | 9.31E-06 |
| T1DM | leukocyte | *KLHL3* | -1.454709438 | 0.001167897 | 0.001340931 | 1 |
| T1DM | leukocyte | *DCDC2* | -0.515719302 | 1.48E-09 | 8.14E-09 | 5.80E-06 |
| T1DM | leukocyte | *TRIM7* | -1.757058732 | 2.17E-07 | 3.08E-07 | 0.000850946 |

| | | | | | | |
|---|---|---|---|---|---|---|
| T1DM | leukocyte | *ANO7* | -1.827546284 | 8.24E-09 | 1.59E-08 | 3.24E-05 |
| T1DM | leukocyte | *RPL7L1* | -0.597107366 | 1.59E-09 | 8.14E-09 | 6.24E-06 |
| T1DM | leukocyte | *IRAK1BP1* | -0.588296162 | 1.07E-09 | 8.14E-09 | 4.21E-06 |
| T1DM | leukocyte | *MMS22L* | -0.941067828 | 4.75E-09 | 1.07E-08 | 1.87E-05 |
| T1DM | leukocyte | *FAXC* | -1.023799346 | 1.48E-09 | 8.14E-09 | 5.81E-06 |
| T1DM | leukocyte | *SCML4* | -1.702353085 | 4.70E-05 | 5.71E-05 | 0.184529378 |
| T1DM | leukocyte | *TBC1D32* | -0.908594986 | 0.010944048 | 0.012129524 | 1 |
| T1DM | leukocyte | *ABRACL* | -0.620484218 | 4.80E-08 | 7.54E-08 | 0.00018872 |
| T1DM | leukocyte | *DYNLT1* | -0.527779679 | 2.90E-09 | 8.49E-09 | 1.14E-05 |
| T1DM | leukocyte | *WTAP* | -0.547921589 | 1.06E-09 | 8.14E-09 | 4.18E-06 |
| T1DM | leukocyte | *SLC22A3* | -4.263483528 | 5.40E-08 | 8.42E-08 | 0.00021226 |
| T1DM | leukocyte | *VWDE* | -0.614584537 | 2.12E-08 | 3.57E-08 | 8.33E-05 |
| T1DM | leukocyte | *GNA12* | -0.952734721 | 4.18E-08 | 6.64E-08 | 0.000164249 |
| T1DM | leukocyte | *SDK1* | -0.806820453 | 3.75E-09 | 9.33E-09 | 1.47E-05 |
| T1DM | leukocyte | *CCZ1B* | -0.893015149 | 3.16E-07 | 4.40E-07 | 0.001243227 |
| T1DM | leukocyte | *RBAK* | -1.314319011 | 3.43E-08 | 5.55E-08 | 0.000134795 |
| T1DM | leukocyte | *POMZP3* | -0.721343859 | 7.52E-09 | 1.48E-08 | 2.95E-05 |
| T1DM | leukocyte | *PSPH* | -0.753050091 | 4.21E-06 | 5.34E-06 | 0.016526507 |
| T1DM | leukocyte | *GIGYF1* | -1.322954824 | 7.99E-10 | 8.14E-09 | 3.14E-06 |
| T1DM | leukocyte | *TRIM4* | -0.529694492 | 3.52E-08 | 5.68E-08 | 0.000138272 |
| T1DM | leukocyte | *TMEM209* | -0.980673428 | 1.88E-09 | 8.14E-09 | 7.38E-06 |
| T1DM | leukocyte | *NCAPG2* | 0.68785511 | 3.28E-08 | 5.33E-08 | 0.000128967 |
| T1DM | leukocyte | *RAB19* | -3.641333855 | 2.78E-05 | 3.41E-05 | 0.109195222 |
| T1DM | leukocyte | *CSGALNACT1* | -0.610475074 | 3.04E-09 | 8.59E-09 | 1.20E-05 |
| T1DM | leukocyte | *CCDC25* | -0.542358735 | 1.36E-09 | 8.14E-09 | 5.35E-06 |
| T1DM | leukocyte | *BIN3* | -1.54116065 | 2.13E-07 | 3.02E-07 | 0.000835224 |
| T1DM | leukocyte | *CHMP7* | -0.970945923 | 2.50E-09 | 8.21E-09 | 9.83E-06 |
| T1DM | leukocyte | *STAR* | 0.524800681 | 7.43E-09 | 1.47E-08 | 2.92E-05 |
| T1DM | leukocyte | *ADHFE1* | 1.300599674 | 0.004154724 | 0.004682706 | 1 |
| T1DM | leukocyte | *LACTB2* | -0.604565823 | 1.18E-09 | 8.14E-09 | 4.65E-06 |
| T1DM | leukocyte | *TERF1* | -1.125603841 | 2.88E-08 | 4.73E-08 | 0.000113251 |
| T1DM | leukocyte | *LRP12* | -0.673255672 | 1.37E-09 | 8.14E-09 | 5.39E-06 |
| T1DM | leukocyte | *ZNF7* | -0.591124446 | 1.38E-09 | 8.14E-09 | 5.41E-06 |
| T1DM | leukocyte | *AK3* | -0.580907422 | 2.08E-09 | 8.15E-09 | 8.19E-06 |
| T1DM | leukocyte | *CER1* | 2.773120846 | 1.83E-09 | 8.14E-09 | 7.20E-06 |
| T1DM | leukocyte | *C9orf72* | -0.925918801 | 2.55E-09 | 8.21E-09 | 1.00E-05 |
| T1DM | leukocyte | *FBXO10* | -0.960358051 | 8.96E-09 | 1.70E-08 | 3.52E-05 |
| T1DM | leukocyte | *SIGMAR1* | -0.761513957 | 1.75E-09 | 8.14E-09 | 6.88E-06 |
| T1DM | leukocyte | *ZNF462* | -0.593076358 | 1.95E-09 | 8.14E-09 | 7.67E-06 |
| T1DM | leukocyte | *UGCG* | -0.638647299 | 5.91E-09 | 1.23E-08 | 2.32E-05 |
| T1DM | leukocyte | *ALAD* | -0.692975784 | 5.41E-08 | 8.43E-08 | 0.000212568 |
| T1DM | leukocyte | *SURF4* | -0.561393086 | 6.46E-10 | 8.14E-09 | 2.54E-06 |
| T1DM | leukocyte | *SURF1* | -0.586783476 | 7.30E-07 | 9.81E-07 | 0.002868269 |
| T1DM | leukocyte | *SURF6* | -0.925294652 | 2.63E-09 | 8.21E-09 | 1.03E-05 |
| T1DM | leukocyte | *SH3GLB2* | -1.00583602 | 2.25E-09 | 8.21E-09 | 8.85E-06 |
| T1DM | leukocyte | *LCN2* | -0.289450069 | 1.33E-09 | 8.14E-09 | 5.24E-06 |
| T1DM | leukocyte | *GPR107* | -0.868809261 | 4.66E-09 | 1.06E-08 | 1.83E-05 |
| T1DM | leukocyte | *INPP5E* | -1.125715176 | 1.49E-07 | 2.15E-07 | 0.000585428 |
| T1DM | leukocyte | *SEC16A* | -0.790262855 | 1.43E-07 | 2.06E-07 | 0.000561464 |
| T1DM | leukocyte | *MSRB2* | -0.985840416 | 8.09E-10 | 8.14E-09 | 3.18E-06 |

| | | | | | | |
|---|---|---|---|---|---|---|
| T1DM | leukocyte | *RSU1* | -0.856776778 | 7.30E-10 | 8.14E-09 | 2.87E-06 |
| T1DM | leukocyte | *GLUD1* | -0.749435595 | 2.78E-09 | 8.34E-09 | 1.09E-05 |
| T1DM | leukocyte | *FRA10AC1* | -0.511993911 | 2.79E-09 | 8.35E-09 | 1.10E-05 |
| T1DM | leukocyte | *DNAJB12* | -0.809140683 | 2.72E-09 | 8.27E-09 | 1.07E-05 |
| T1DM | leukocyte | *FUOM* | 0.74325801 | 2.92E-07 | 4.07E-07 | 0.001145381 |
| T1DM | leukocyte | *LRRC27* | -0.723996116 | 0.039475101 | 0.042809184 | 1 |
| T1DM | leukocyte | *MTG1* | -0.563273814 | 8.37E-09 | 1.61E-08 | 3.29E-05 |
| T1DM | leukocyte | *PAOX* | -1.162609314 | 0.025843155 | 0.028226272 | 1 |
| T1DM | leukocyte | *GSTO1* | -0.683677111 | 5.85E-07 | 7.91E-07 | 0.002297808 |
| T1DM | leukocyte | *ZNF215* | -0.929561629 | 0.000126676 | 0.00015073 | 0.497710493 |
| T1DM | leukocyte | *HSD17B12* | -0.344278028 | 1.08E-06 | 1.42E-06 | 0.004225724 |
| T1DM | leukocyte | *APIP* | -0.582091556 | 3.65E-08 | 5.87E-08 | 0.000143557 |
| T1DM | leukocyte | *PTPRJ* | -0.890252596 | 1.52E-09 | 8.14E-09 | 5.96E-06 |
| T1DM | leukocyte | *CCDC82* | -0.664438417 | 9.42E-10 | 8.14E-09 | 3.70E-06 |
| T1DM | leukocyte | *KLHL35* | -0.476663335 | 0.000550798 | 0.000639127 | 1 |
| T1DM | leukocyte | *CAPN5* | -0.419213584 | 0.001396948 | 0.001599711 | 1 |
| T1DM | leukocyte | *RPS3* | -0.41057529 | 3.12E-09 | 8.69E-09 | 1.23E-05 |
| T1DM | leukocyte | *TTC12* | -0.532879361 | 6.08E-05 | 7.36E-05 | 0.238983342 |
| T1DM | leukocyte | *AASDHPPT* | -0.682969127 | 2.07E-09 | 8.14E-09 | 8.14E-06 |
| T1DM | leukocyte | *GLB1L2* | -0.648801228 | 3.95E-08 | 6.30E-08 | 0.000155326 |
| T1DM | leukocyte | *FADS1* | -0.411345041 | 1.53E-05 | 1.89E-05 | 0.059964595 |
| T1DM | leukocyte | *TMC2* | -46.43375462 | 4.42E-07 | 6.06E-07 | 0.001735715 |
| T1DM | leukocyte | *EML3* | -1.751450819 | 0.000159987 | 0.000189734 | 0.628587344 |
| T1DM | leukocyte | *CHEK1* | -0.84148752 | 2.48E-09 | 8.21E-09 | 9.74E-06 |
| T1DM | leukocyte | *SIDT2* | -0.855391711 | 4.96E-10 | 8.14E-09 | 1.95E-06 |
| T1DM | leukocyte | *TAGLN* | -0.550703059 | 3.37E-09 | 8.87E-09 | 1.32E-05 |
| T1DM | leukocyte | *DUSP15* | -0.864298174 | 1.58E-09 | 8.14E-09 | 6.20E-06 |
| T1DM | leukocyte | *DSN1* | -0.791957188 | 2.99E-09 | 8.52E-09 | 1.18E-05 |
| T1DM | leukocyte | *YTHDF1* | -0.717539985 | 2.47E-09 | 8.21E-09 | 9.72E-06 |
| T1DM | leukocyte | *CABLES2* | -1.02074819 | 1.12E-11 | 3.16E-09 | 4.42E-08 |
| T1DM | leukocyte | *TRPT1* | -0.672597065 | 1.67E-09 | 8.14E-09 | 6.56E-06 |
| T1DM | leukocyte | *TBX6* | -1.048338404 | 0.000112064 | 0.000133667 | 0.440298584 |
| T1DM | leukocyte | *DOC2A* | 1.082460219 | 2.88E-06 | 3.70E-06 | 0.011296507 |
| T1DM | leukocyte | *MPP7* | -0.558236363 | 1.75E-09 | 8.14E-09 | 6.86E-06 |
| T1DM | leukocyte | *DCUN1D2* | -0.772082445 | 5.36E-08 | 8.37E-08 | 0.000210592 |
| T1DM | leukocyte | *TMCO3* | -0.729939836 | 8.30E-09 | 1.60E-08 | 3.26E-05 |
| T1DM | leukocyte | *TMEM218* | -1.020038184 | 8.80E-10 | 8.14E-09 | 3.46E-06 |
| T1DM | leukocyte | *MIA2* | -0.619865958 | 3.14E-09 | 8.69E-09 | 1.23E-05 |
| T1DM | leukocyte | *LYPD1* | -1.109811009 | 1.19E-09 | 8.14E-09 | 4.66E-06 |
| T1DM | leukocyte | *LYPD6B* | -1.811628182 | 1.22E-09 | 8.14E-09 | 4.81E-06 |
| T1DM | leukocyte | *WDR17* | -0.552105355 | 1.05E-08 | 1.94E-08 | 4.14E-05 |
| T1DM | leukocyte | *CNDP1* | -2.055968084 | 0.000292501 | 0.000342851 | 1 |
| T1DM | leukocyte | *PRSS23* | -0.529082651 | 7.94E-10 | 8.14E-09 | 3.12E-06 |
| T1DM | leukocyte | *MTMR12* | -0.552900882 | 2.56E-09 | 8.21E-09 | 1.01E-05 |
| T1DM | leukocyte | *DOCK1* | -0.690548616 | 1.88E-09 | 8.14E-09 | 7.40E-06 |
| T1DM | leukocyte | *IL18* | 0.427424727 | 6.23E-09 | 1.28E-08 | 2.45E-05 |
| T1DM | leukocyte | *PRSS53* | 0.45707225 | 4.29E-06 | 5.45E-06 | 0.016854687 |
| T1DM | leukocyte | *TMEM86A* | -0.855322592 | 1.39E-09 | 8.14E-09 | 5.48E-06 |
| T1DM | leukocyte | *DLG5* | -0.66103288 | 6.19E-10 | 8.14E-09 | 2.43E-06 |
| T1DM | leukocyte | *SLC2A13* | -0.510879391 | 3.15E-09 | 8.69E-09 | 1.24E-05 |

| | | | | | | |
|---|---|---|---|---|---|---|
| T1DM | leukocyte | *MAGI1* | -0.680299708 | 1.39E-09 | 8.14E-09 | 5.48E-06 |
| T1DM | leukocyte | *CSNK1G3* | -0.745066515 | 3.91E-09 | 9.54E-09 | 1.54E-05 |
| T1DM | leukocyte | *AKAP6* | -0.829035396 | 1.25E-09 | 8.14E-09 | 4.93E-06 |
| T1DM | leukocyte | *MBIP* | -0.711508309 | 8.54E-09 | 1.63E-08 | 3.36E-05 |
| T1DM | leukocyte | *MIPOL1* | -0.639861172 | 1.91E-09 | 8.14E-09 | 7.49E-06 |
| T1DM | leukocyte | *TMEM18* | -0.701611854 | 1.91E-09 | 8.14E-09 | 7.50E-06 |
| T1DM | leukocyte | *NDUFC2* | -0.512088293 | 2.85E-09 | 8.43E-09 | 1.12E-05 |
| T1DM | leukocyte | *VIPAS39* | -1.045855644 | 9.59E-09 | 1.80E-08 | 3.77E-05 |
| T1DM | leukocyte | *C4orf33* | -1.188035768 | 4.17E-07 | 5.74E-07 | 0.001639496 |
| T1DM | leukocyte | *QDPR* | -0.503905358 | 1.62E-09 | 8.14E-09 | 6.36E-06 |
| T1DM | leukocyte | *TEX9* | -1.14082712 | 3.58E-09 | 9.11E-09 | 1.41E-05 |
| T1DM | leukocyte | *ZNF827* | -0.633002822 | 8.36E-10 | 8.14E-09 | 3.28E-06 |
| T1DM | leukocyte | *AKR1C2* | -1.230719935 | 2.34E-06 | 3.02E-06 | 0.009195026 |
| T1DM | leukocyte | *DPYSL4* | -0.487101518 | 0.001864117 | 0.002122932 | 1 |
| T1DM | leukocyte | *KIN* | -1.054246707 | 1.78E-08 | 3.07E-08 | 7.01E-05 |
| T1DM | leukocyte | *TMEM45B* | -1.065272997 | 6.48E-09 | 1.32E-08 | 2.55E-05 |
| T1DM | leukocyte | *CENPU* | -1.127114686 | 0.000243536 | 0.000286483 | 0.956854293 |
| T1DM | leukocyte | *BICD1* | -0.59585445 | 1.37E-09 | 8.14E-09 | 5.40E-06 |
| T1DM | leukocyte | *GUF1* | -1.452225165 | 1.98E-08 | 3.36E-08 | 7.77E-05 |
| T1DM | leukocyte | *CCL28* | -0.869155054 | 2.55E-10 | 8.14E-09 | 1.00E-06 |
| T1DM | leukocyte | *CACUL1* | -0.872629659 | 7.68E-09 | 1.51E-08 | 3.02E-05 |
| T1DM | leukocyte | *KCNE4* | -0.772770658 | 8.75E-08 | 1.31E-07 | 0.000343932 |
| T1DM | leukocyte | *RABGAP1L* | -0.739138133 | 2.25E-09 | 8.21E-09 | 8.84E-06 |
| T1DM | leukocyte | *PTPN14* | -0.568091956 | 3.86E-08 | 6.17E-08 | 0.00015158 |
| T1DM | leukocyte | *ARL14EP* | -0.852253053 | 3.13E-09 | 8.69E-09 | 1.23E-05 |
| T1DM | leukocyte | *EPG5* | -1.044351721 | 3.47E-09 | 8.98E-09 | 1.36E-05 |
| T1DM | leukocyte | *HAUS1* | -0.90000936 | 1.17E-09 | 8.14E-09 | 4.60E-06 |
| T1DM | leukocyte | *SPC25* | -0.819264842 | 2.76E-09 | 8.33E-09 | 1.08E-05 |
| T1DM | leukocyte | *PDK1* | -0.825315709 | 2.14E-09 | 8.18E-09 | 8.39E-06 |
| T1DM | leukocyte | *PDE3B* | -0.43252596 | 6.18E-10 | 8.14E-09 | 2.43E-06 |
| T1DM | leukocyte | *TGOLN2* | -0.661446708 | 1.07E-09 | 8.14E-09 | 4.19E-06 |
| T1DM | leukocyte | *ATG10* | -0.519205873 | 2.12E-05 | 2.61E-05 | 0.083193278 |
| T1DM | leukocyte | *POC5* | -0.500430305 | 0.000512472 | 0.000595007 | 1 |
| T1DM | leukocyte | *GUCY1A2* | -0.699954962 | 1.35E-09 | 8.14E-09 | 5.32E-06 |
| T1DM | leukocyte | *ZNF773* | -0.52142521 | 1.65E-06 | 2.16E-06 | 0.006490617 |
| T1DM | leukocyte | *DCLRE1C* | -1.227188835 | 0.000155218 | 0.000184134 | 0.609852055 |
| T1DM | leukocyte | *ZSCAN1* | -0.720656834 | 4.41E-10 | 8.14E-09 | 1.73E-06 |
| T1DM | leukocyte | *USP12* | -0.604577631 | 5.04E-09 | 1.11E-08 | 1.98E-05 |
| T1DM | leukocyte | *CCDC50* | -0.514867413 | 5.60E-09 | 1.18E-08 | 2.20E-05 |
| T1DM | leukocyte | *PLEKHH2* | -0.571471721 | 2.13E-06 | 2.76E-06 | 0.008362166 |
| T1DM | leukocyte | *GRIA4* | -0.782479905 | 1.04E-07 | 1.54E-07 | 0.000408029 |
| T1DM | leukocyte | *IGSF10* | -1.674252986 | 2.30E-09 | 8.21E-09 | 9.05E-06 |
| T1DM | leukocyte | *SPEF2* | -0.688567635 | 5.50E-07 | 7.47E-07 | 0.002161564 |
| T1DM | leukocyte | *GPD1L* | -0.791975897 | 1.65E-09 | 8.14E-09 | 6.50E-06 |
| T1DM | leukocyte | *CCNO* | -0.741593816 | 4.45E-08 | 7.03E-08 | 0.000174664 |
| T1DM | leukocyte | *CLEC4F* | -81.76471542 | 4.32E-05 | 5.26E-05 | 0.169725237 |
| T1DM | leukocyte | *RASGRP3* | -0.713110521 | 1.55E-07 | 2.23E-07 | 0.000608136 |
| T1DM | leukocyte | *CATSPER3* | -3.856485521 | 1.29E-09 | 8.14E-09 | 5.07E-06 |
| T1DM | leukocyte | *GPR180* | -0.603127042 | 4.77E-09 | 1.07E-08 | 1.88E-05 |
| T1DM | leukocyte | *IFIT5* | -0.738765896 | 2.23E-09 | 8.21E-09 | 8.76E-06 |

| | | | | | | |
|---|---|---|---|---|---|---|
| T1DM | leukocyte | *PRDM8* | 0.759777106 | 4.75E-07 | 6.49E-07 | 0.001865387 |
| T1DM | leukocyte | *ZNF117* | -0.485231178 | 1.19E-05 | 1.48E-05 | 0.046798156 |
| T1DM | leukocyte | *MED21* | -0.983512115 | 2.30E-09 | 8.21E-09 | 9.04E-06 |
| T1DM | leukocyte | *LGI2* | -1.076678675 | 2.72E-09 | 8.27E-09 | 1.07E-05 |
| T1DM | leukocyte | *SRP19* | -0.548475199 | 2.53E-09 | 8.21E-09 | 9.95E-06 |
| T1DM | leukocyte | *DAB2* | -0.861937572 | 2.69E-09 | 8.26E-09 | 1.06E-05 |
| T1DM | leukocyte | *ACMSD* | -0.952265113 | 2.04E-09 | 8.14E-09 | 8.03E-06 |
| T1DM | leukocyte | *CAST* | -0.562592345 | 4.51E-09 | 1.04E-08 | 1.77E-05 |
| T1DM | leukocyte | *SYCP2L* | -1.588910002 | 3.91E-06 | 4.98E-06 | 0.015360016 |
| T1DM | leukocyte | *MERTK* | 0.380107993 | 6.36E-06 | 8.01E-06 | 0.024992883 |
| T1DM | leukocyte | *TMEM87B* | -0.731866867 | 4.88E-09 | 1.08E-08 | 1.92E-05 |
| T1DM | leukocyte | *PLA2R1* | -2.413418646 | 1.15E-09 | 8.14E-09 | 4.53E-06 |
| T1DM | leukocyte | *SLC25A27* | -0.956074487 | 1.19E-09 | 8.14E-09 | 4.68E-06 |
| T1DM | leukocyte | *ASAP1* | -0.608447379 | 4.88E-09 | 1.08E-08 | 1.92E-05 |
| T1DM | leukocyte | *NMRAL1* | -0.74260346 | 1.14E-07 | 1.68E-07 | 0.000448387 |
| T1DM | leukocyte | *TEX29* | -0.860354812 | 0.000902661 | 0.001040961 | 1 |
| T1DM | leukocyte | *CMTM7* | -0.561031926 | 1.72E-08 | 2.98E-08 | 6.77E-05 |
| T1DM | leukocyte | *RPIA* | -0.698418332 | 1.77E-09 | 8.14E-09 | 6.95E-06 |
| T1DM | leukocyte | *PTPRD* | -0.552974687 | 5.38E-06 | 6.81E-06 | 0.021155457 |
| T1DM | leukocyte | *GTF2E1* | -1.518177404 | 7.02E-09 | 1.40E-08 | 2.76E-05 |
| T1DM | leukocyte | *CFDP1* | -0.604960008 | 6.89E-10 | 8.14E-09 | 2.71E-06 |
| T1DM | leukocyte | *ZDHHC7* | -1.059033307 | 4.25E-08 | 6.75E-08 | 0.000167133 |
| T1DM | leukocyte | *JAZF1* | -0.796086048 | 2.92E-09 | 8.49E-09 | 1.15E-05 |
| T1DM | leukocyte | *CMIP* | -0.687480505 | 2.11E-09 | 8.16E-09 | 8.29E-06 |
| T1DM | leukocyte | *KCNJ16* | -0.330377763 | 0.030826058 | 0.033577927 | 1 |
| T1DM | leukocyte | *KCTD15* | -0.984831483 | 1.71E-09 | 8.14E-09 | 6.72E-06 |
| T1DM | leukocyte | *MCOLN2* | -2.833796373 | 0.001567334 | 0.001788573 | 1 |
| T1DM | leukocyte | *LGI4* | 0.8538153 | 1.16E-07 | 1.70E-07 | 0.000456365 |
| T1DM | leukocyte | *DDAH1* | -1.369716575 | 6.59E-09 | 1.34E-08 | 2.59E-05 |
| T1DM | leukocyte | *NUS1* | -0.743691058 | 1.26E-09 | 8.14E-09 | 4.95E-06 |
| T1DM | leukocyte | *PPP2R5E* | -0.74909321 | 2.05E-09 | 8.14E-09 | 8.04E-06 |
| T1DM | leukocyte | *THY1* | 0.599094724 | 3.17E-05 | 3.88E-05 | 0.124416555 |
| T1DM | leukocyte | *ROBO3* | -2.082277038 | 1.36E-09 | 8.14E-09 | 5.34E-06 |
| T1DM | leukocyte | *PITPNC1* | -0.752468226 | 1.86E-09 | 8.14E-09 | 7.33E-06 |
| T1DM | leukocyte | *LRRK1* | 1.187462216 | 1.16E-07 | 1.71E-07 | 0.000457497 |
| T1DM | leukocyte | *CEP112* | -0.581396439 | 1.79E-09 | 8.14E-09 | 7.04E-06 |
| T1DM | leukocyte | *ABCA5* | -0.884447747 | 4.05E-08 | 6.44E-08 | 0.000159153 |
| T1DM | leukocyte | *GBP5* | -14.24338052 | 5.95E-07 | 8.04E-07 | 0.002337231 |
| T1DM | leukocyte | *SRSF12* | -1.188681441 | 2.66E-08 | 4.40E-08 | 0.00010465 |
| T1DM | leukocyte | *PDLIM3* | -0.452909735 | 1.34E-08 | 2.40E-08 | 5.26E-05 |
| T1DM | leukocyte | *PDE1C* | -0.74056367 | 1.58E-06 | 2.07E-06 | 0.006202244 |
| T1DM | leukocyte | *RABGEF1* | -0.724142461 | 1.06E-09 | 8.14E-09 | 4.18E-06 |
| T1DM | leukocyte | *MRPL39* | -0.667018364 | 4.86E-07 | 6.63E-07 | 0.001908643 |
| T1DM | leukocyte | *TSEN2* | -1.593871377 | 5.75E-10 | 8.14E-09 | 2.26E-06 |
| T1DM | leukocyte | *C17orf50* | -0.949671583 | 0.008487781 | 0.009455201 | 1 |
| T1DM | leukocyte | *FLCN* | -1.019836835 | 3.96E-09 | 9.62E-09 | 1.56E-05 |
| T1DM | leukocyte | *MPPE1* | -0.735995006 | 8.92E-08 | 1.34E-07 | 0.00035058 |
| T1DM | leukocyte | *RAB6B* | -0.852822197 | 1.31E-09 | 8.14E-09 | 5.17E-06 |
| T1DM | leukocyte | *ACSS1* | -0.939320448 | 3.36E-07 | 4.66E-07 | 0.001319184 |
| T1DM | leukocyte | *ANKRD40* | -0.846649936 | 2.68E-08 | 4.42E-08 | 0.000105275 |

| | | | | | | |
|---|---|---|---|---|---|---|
| T1DM | leukocyte | *ATP6V1C1* | -0.635287602 | 1.63E-09 | 8.14E-09 | 6.40E-06 |
| T1DM | leukocyte | *MARVELD1* | -1.670505435 | 8.58E-06 | 1.07E-05 | 0.033706739 |
| T1DM | leukocyte | *TRMT44* | -0.631465102 | 8.50E-09 | 1.63E-08 | 3.34E-05 |
| T1DM | leukocyte | *PPM1J* | -1.780148419 | 1.56E-09 | 8.14E-09 | 6.15E-06 |
| T1DM | leukocyte | *DBI* | -0.598727931 | 1.01E-08 | 1.87E-08 | 3.96E-05 |
| T1DM | leukocyte | *HEATR3* | -0.531319223 | 4.66E-07 | 6.38E-07 | 0.001831214 |
| T1DM | leukocyte | *LRGUK* | -1.759495878 | 3.32E-06 | 4.24E-06 | 0.013034446 |
| T1DM | leukocyte | *SETD9* | -0.761680346 | 8.01E-10 | 8.14E-09 | 3.15E-06 |
| T1DM | leukocyte | *MIER3* | -1.294891159 | 6.40E-09 | 1.31E-08 | 2.52E-05 |
| T1DM | leukocyte | *TTN* | -1.576993496 | 4.92E-09 | 1.09E-08 | 1.93E-05 |
| T1DM | leukocyte | *DEPTOR* | -0.758803796 | 1.35E-08 | 2.41E-08 | 5.30E-05 |
| T1DM | leukocyte | *RMND1* | -1.059969643 | 2.62E-08 | 4.32E-08 | 0.00010277 |
| T1DM | leukocyte | *TMBIM4* | -0.557572434 | 1.67E-08 | 2.90E-08 | 6.56E-05 |
| T1DM | leukocyte | *PSD3* | -0.611964476 | 1.19E-09 | 8.14E-09 | 4.69E-06 |
| T1DM | leukocyte | *FAM161B* | -1.010579862 | 1.44E-07 | 2.07E-07 | 0.000564766 |
| T1DM | leukocyte | *MMP16* | -0.58106943 | 5.87E-09 | 1.22E-08 | 2.31E-05 |
| T1DM | leukocyte | *ADK* | -0.720284114 | 8.52E-09 | 1.63E-08 | 3.35E-05 |
| T1DM | leukocyte | *BATF* | -10.27075069 | 1.21E-09 | 8.14E-09 | 4.73E-06 |
| T1DM | leukocyte | *DRAM2* | -0.903798167 | 1.02E-06 | 1.35E-06 | 0.003989805 |
| T1DM | leukocyte | *CCT8* | -0.574899931 | 2.15E-10 | 8.14E-09 | 8.44E-07 |
| T1DM | leukocyte | *CDK20* | -0.725131754 | 5.87E-11 | 8.14E-09 | 2.31E-07 |
| T1DM | leukocyte | *SFXN2* | -0.330641445 | 0.003615941 | 0.004081308 | 1 |
| T1DM | leukocyte | *FUT6* | -0.922528662 | 0.00030969 | 0.000362566 | 1 |
| T1DM | leukocyte | *TDRD9* | -0.456925652 | 2.11E-09 | 8.16E-09 | 8.28E-06 |
| T1DM | leukocyte | *HKDC1* | -1.499349781 | 2.73E-07 | 3.83E-07 | 0.001073225 |
| T1DM | leukocyte | *ZDHHC5* | -0.790595621 | 2.12E-09 | 8.16E-09 | 8.31E-06 |
| T1DM | leukocyte | *MED19* | -0.648216687 | 2.07E-09 | 8.14E-09 | 8.14E-06 |
| T1DM | leukocyte | *MAPK13* | -0.782749944 | 3.49E-09 | 9.01E-09 | 1.37E-05 |
| T1DM | leukocyte | *TBC1D31* | 1.824311664 | 1.41E-07 | 2.04E-07 | 0.000553824 |
| T1DM | leukocyte | *COX6A2* | -0.73006683 | 1.20E-06 | 1.58E-06 | 0.004704103 |
| T1DM | leukocyte | *MPV17L* | -0.669969345 | 2.92E-09 | 8.50E-09 | 1.15E-05 |
| T1DM | leukocyte | *TATDN2* | -1.088754645 | 1.53E-08 | 2.68E-08 | 6.00E-05 |
| T1DM | leukocyte | *EXOG* | -0.821526714 | 4.68E-09 | 1.06E-08 | 1.84E-05 |
| T1DM | leukocyte | *ZFYVE9* | -0.74942969 | 1.24E-09 | 8.14E-09 | 4.89E-06 |
| T1DM | leukocyte | *SLC6A1* | -4.08395471 | 1.32E-09 | 8.14E-09 | 5.17E-06 |
| T1DM | leukocyte | *NECAP2* | -0.858115032 | 2.01E-09 | 8.14E-09 | 7.91E-06 |
| T1DM | leukocyte | *STEAP2* | -1.020131705 | 3.50E-09 | 9.02E-09 | 1.38E-05 |
| T1DM | leukocyte | *GATAD1* | -0.831064479 | 4.76E-09 | 1.07E-08 | 1.87E-05 |
| T1DM | leukocyte | *TMED6* | -0.295688815 | 8.10E-05 | 9.71E-05 | 0.318126164 |
| T1DM | leukocyte | *CLEC18A* | -2.232110515 | 2.21E-10 | 8.14E-09 | 8.69E-07 |
| T1DM | leukocyte | *DHRS4* | -0.793079699 | 1.73E-09 | 8.14E-09 | 6.79E-06 |
| T1DM | leukocyte | *DHRS1* | -0.799311279 | 3.57E-08 | 5.75E-08 | 0.00014029 |
| T1DM | leukocyte | *AASDH* | -0.535215698 | 1.92E-08 | 3.28E-08 | 7.56E-05 |
| T1DM | leukocyte | *SLC38A10* | -0.765823435 | 2.55E-09 | 8.21E-09 | 1.00E-05 |
| T1DM | leukocyte | *SLC37A3* | -0.562742194 | 6.90E-10 | 8.14E-09 | 2.71E-06 |
| T1DM | leukocyte | *AP3S2* | -0.820367673 | 4.40E-09 | 1.02E-08 | 1.73E-05 |
| T1DM | leukocyte | *SPPL3* | -0.747870526 | 2.43E-09 | 8.21E-09 | 9.57E-06 |
| T1DM | leukocyte | *TNFRSF14* | 0.55789313 | 2.51E-08 | 4.16E-08 | 9.86E-05 |
| T1DM | leukocyte | *CIB4* | -10.11900979 | 2.54E-07 | 3.58E-07 | 0.000998619 |
| T1DM | leukocyte | *C12orf43* | -0.804237185 | 1.16E-08 | 2.11E-08 | 4.55E-05 |

| | | | | | | |
|---|---|---|---|---|---|---|
| T1DM | leukocyte | *PEX10* | -1.03359249 | 8.44E-09 | 1.62E-08 | 3.31E-05 |
| T1DM | leukocyte | *LDLRAP1* | -0.558677279 | 2.69E-09 | 8.26E-09 | 1.06E-05 |
| T1DM | leukocyte | *KRTCAP3* | -0.687047654 | 1.41E-09 | 8.14E-09 | 5.54E-06 |
| T1DM | leukocyte | *PAFAH2* | -1.791147431 | 3.70E-09 | 9.28E-09 | 1.45E-05 |
| T1DM | leukocyte | *EXTL1* | -2.672320296 | 3.32E-06 | 4.24E-06 | 0.013054008 |
| T1DM | leukocyte | *TRIM63* | -8.648809509 | 7.29E-08 | 1.11E-07 | 0.000286255 |
| T1DM | leukocyte | *XKR8* | -1.215858774 | 1.26E-09 | 8.14E-09 | 4.97E-06 |
| T1DM | leukocyte | *CNNM4* | -0.68330354 | 0.000654844 | 0.000757844 | 1 |
| T1DM | leukocyte | *ESYT3* | -1.229037717 | 2.14E-07 | 3.03E-07 | 0.000839185 |
| T1DM | leukocyte | *FAIM* | -0.719311741 | 6.78E-10 | 8.14E-09 | 2.66E-06 |
| T1DM | leukocyte | *AHCYL2* | -0.740140099 | 1.94E-07 | 2.77E-07 | 0.000762157 |
| T1DM | leukocyte | *COPG2* | -0.574799968 | 5.54E-07 | 7.52E-07 | 0.002176624 |
| T1DM | leukocyte | *RNF166* | -0.873122276 | 1.38E-08 | 2.46E-08 | 5.42E-05 |
| T1DM | leukocyte | *NIT1* | -0.940991301 | 4.54E-08 | 7.17E-08 | 0.00017832 |
| T1DM | leukocyte | *NPM2* | -0.813999783 | 1.83E-09 | 8.14E-09 | 7.17E-06 |
| T1DM | leukocyte | *FGF17* | -1.495772938 | 5.48E-07 | 7.44E-07 | 0.002151799 |
| T1DM | leukocyte | *VWA5B1* | 2.119991762 | 4.72E-09 | 1.06E-08 | 1.86E-05 |
| T1DM | leukocyte | *CDA* | -8.562375664 | 4.17E-09 | 9.92E-09 | 1.64E-05 |
| T1DM | leukocyte | *PINK1* | -0.773072725 | 1.12E-08 | 2.04E-08 | 4.38E-05 |
| T1DM | leukocyte | *DMTN* | -0.689020697 | 1.98E-07 | 2.82E-07 | 0.00077636 |
| T1DM | leukocyte | *SLC5A11* | 1.196644645 | 3.71E-05 | 4.53E-05 | 0.145915766 |
| T1DM | leukocyte | *CCAR2* | -0.953578732 | 2.74E-09 | 8.30E-09 | 1.08E-05 |
| T1DM | leukocyte | *EPB41* | -0.698544675 | 7.71E-10 | 8.14E-09 | 3.03E-06 |
| T1DM | leukocyte | *MIS18A* | -0.723834314 | 1.46E-07 | 2.10E-07 | 0.00057316 |
| T1DM | leukocyte | *ALG8* | -0.675549303 | 1.32E-09 | 8.14E-09 | 5.20E-06 |
| T1DM | leukocyte | *IFNAR2* | -1.072887036 | 1.19E-06 | 1.57E-06 | 0.004655888 |
| T1DM | leukocyte | *IFNGR2* | -0.645535793 | 1.28E-08 | 2.31E-08 | 5.03E-05 |
| T1DM | leukocyte | *C1QC* | 0.045239889 | 3.33E-09 | 8.82E-09 | 1.31E-05 |
| T1DM | leukocyte | *GJD2* | -0.470053784 | 3.18E-10 | 8.14E-09 | 1.25E-06 |
| T1DM | leukocyte | *CLDN14* | -1.763723683 | 1.00E-09 | 8.14E-09 | 3.94E-06 |
| T1DM | leukocyte | *HLCS* | -0.645306452 | 2.98E-09 | 8.50E-09 | 1.17E-05 |
| T1DM | leukocyte | *ADPGK* | 0.815061513 | 3.32E-06 | 4.24E-06 | 0.013057619 |
| T1DM | leukocyte | *THEM4* | -0.585383977 | 1.01E-09 | 8.14E-09 | 3.95E-06 |
| T1DM | leukocyte | *UBR1* | -0.874529564 | 1.99E-09 | 8.14E-09 | 7.80E-06 |
| T1DM | leukocyte | *TEPP* | 2.915476562 | 1.11E-08 | 2.03E-08 | 4.36E-05 |
| T1DM | leukocyte | *ZDHHC1* | -0.705123741 | 3.26E-09 | 8.79E-09 | 1.28E-05 |
| T1DM | leukocyte | *ZFYVE28* | -0.947459523 | 3.28E-09 | 8.79E-09 | 1.29E-05 |
| T1DM | leukocyte | *NPR2* | -0.88268598 | 2.56E-09 | 8.21E-09 | 1.01E-05 |
| T1DM | leukocyte | *ATAD3B* | -1.419476669 | 2.73E-07 | 3.82E-07 | 0.001072052 |
| T1DM | leukocyte | *ANKLE1* | 0.414735895 | 1.42E-06 | 1.87E-06 | 0.005593481 |
| T1DM | leukocyte | *KALRN* | -0.995817468 | 2.77E-09 | 8.34E-09 | 1.09E-05 |
| T1DM | leukocyte | *CILP2* | -0.741750896 | 0.009761882 | 0.010837647 | 1 |
| T1DM | leukocyte | *TMPRSS3* | -2.071764332 | 1.07E-08 | 1.97E-08 | 4.20E-05 |
| T1DM | leukocyte | *RSPH1* | -0.469535847 | 8.65E-09 | 1.65E-08 | 3.40E-05 |
| T1DM | leukocyte | *SLC37A1* | -0.753261409 | 2.58E-09 | 8.21E-09 | 1.01E-05 |
| T1DM | leukocyte | *NDUFV3* | -0.504957484 | 1.06E-08 | 1.95E-08 | 4.16E-05 |
| T1DM | leukocyte | *HSF2BP* | -1.629055801 | 6.04E-06 | 7.62E-06 | 0.023736372 |
| T1DM | leukocyte | *PDXK* | -0.773565003 | 3.58E-09 | 9.11E-09 | 1.41E-05 |
| T1DM | leukocyte | *CSTB* | -0.653431393 | 1.57E-07 | 2.25E-07 | 0.000614985 |
| T1DM | leukocyte | *ICOSLG* | 0.369878109 | 2.22E-08 | 3.73E-08 | 8.72E-05 |

| | | | | | | |
|---|---|---|---|---|---|---|
| T1DM | leukocyte | *ZNF66* | -1.630134204 | 4.19E-07 | 5.76E-07 | 0.001646004 |
| T1DM | leukocyte | *FTCD* | -0.989646899 | 8.03E-10 | 8.14E-09 | 3.15E-06 |
| T1DM | leukocyte | *SPATC1L* | -0.356111404 | 0.000122695 | 0.00014617 | 0.482067789 |
| T1DM | leukocyte | *LSS* | -0.814166642 | 1.04E-09 | 8.14E-09 | 4.07E-06 |
| T1DM | leukocyte | *VAV2* | -1.035940451 | 6.11E-09 | 1.26E-08 | 2.40E-05 |
| T1DM | leukocyte | *MCM3AP* | -0.762359215 | 4.62E-05 | 5.62E-05 | 0.181558392 |
| T1DM | leukocyte | *DIP2A* | -0.513367754 | 5.25E-07 | 7.16E-07 | 0.002064421 |
| T1DM | leukocyte | *ZNF208* | -0.707787666 | 4.94E-11 | 8.14E-09 | 1.94E-07 |
| T1DM | leukocyte | *ADAMTS13* | -0.503099195 | 0.011942686 | 0.013221418 | 1 |
| T1DM | leukocyte | *SLC2A6* | -1.191517424 | 3.31E-09 | 8.82E-09 | 1.30E-05 |
| T1DM | leukocyte | *RDH13* | -0.831356515 | 2.74E-09 | 8.31E-09 | 1.08E-05 |
| T1DM | leukocyte | *MPZL3* | -0.747797127 | 6.03E-09 | 1.25E-08 | 2.37E-05 |
| T1DM | leukocyte | *S100A1* | -1.466238511 | 2.41E-08 | 4.03E-08 | 9.48E-05 |
| T1DM | leukocyte | *VPS11* | -1.395283933 | 9.19E-11 | 8.14E-09 | 3.61E-07 |
| T1DM | leukocyte | *PAQR6* | -0.824767703 | 3.01E-09 | 8.54E-09 | 1.18E-05 |
| T1DM | leukocyte | *SLC25A44* | -1.077770711 | 3.40E-09 | 8.90E-09 | 1.33E-05 |
| T1DM | leukocyte | *CCR5* | -89.25643941 | 2.40E-08 | 4.01E-08 | 9.43E-05 |
| T1DM | leukocyte | *NBEAL2* | -0.505312791 | 2.46E-08 | 4.09E-08 | 9.65E-05 |
| T1DM | leukocyte | *LY6K* | -1.12227913 | 3.88E-07 | 5.36E-07 | 0.001526357 |
| T1DM | leukocyte | *LY6E* | -0.717262565 | 8.45E-09 | 1.62E-08 | 3.32E-05 |
| T1DM | leukocyte | *VPS28* | -0.581805394 | 5.73E-09 | 1.20E-08 | 2.25E-05 |
| T1DM | leukocyte | *TONSL* | -2.90794089 | 1.45E-07 | 2.10E-07 | 0.000570999 |
| T1DM | leukocyte | *ZNF333* | -1.575251348 | 6.71E-05 | 8.09E-05 | 0.263517336 |
| T1DM | leukocyte | *RPL8* | -0.402167434 | 2.64E-09 | 8.21E-09 | 1.04E-05 |
| T1DM | leukocyte | *MAML1* | -1.132947695 | 6.62E-08 | 1.01E-07 | 0.000260042 |
| T1DM | leukocyte | *LRWD1* | -1.013345096 | 2.75E-09 | 8.32E-09 | 1.08E-05 |
| T1DM | leukocyte | *CCDC116* | -5.122986348 | 1.33E-05 | 1.64E-05 | 0.052128281 |
| T1DM | leukocyte | *U2AF1L4* | -0.531151498 | 3.54E-08 | 5.71E-08 | 0.000139195 |
| T1DM | leukocyte | *ZNF382* | -0.697691483 | 8.34E-09 | 1.61E-08 | 3.28E-05 |
| T1DM | leukocyte | *DUSP14* | -0.873841473 | 1.79E-09 | 8.14E-09 | 7.02E-06 |
| T1DM | leukocyte | *PGAP3* | -0.766499302 | 1.81E-09 | 8.14E-09 | 7.10E-06 |
| T1DM | leukocyte | *PRPSAP1* | -0.687123072 | 7.97E-10 | 8.14E-09 | 3.13E-06 |
| T1DM | leukocyte | *SIGLEC11* | -11.92426086 | 6.92E-09 | 1.39E-08 | 2.72E-05 |
| T1DM | leukocyte | *DBF4B* | -0.761765543 | 1.03E-05 | 1.28E-05 | 0.040402545 |
| T1DM | leukocyte | *SPC24* | -1.799175563 | 1.24E-07 | 1.81E-07 | 0.000486997 |
| T1DM | leukocyte | *IP6K3* | 1.25835801 | 7.20E-05 | 8.66E-05 | 0.282698564 |
| T1DM | leukocyte | *LEMD2* | -1.212984895 | 1.14E-07 | 1.68E-07 | 0.000448995 |
| T1DM | leukocyte | *CXCL16* | 0.290504583 | 8.18E-09 | 1.58E-08 | 3.21E-05 |
| T1DM | leukocyte | *RPL26* | -0.427390069 | 2.60E-09 | 8.21E-09 | 1.02E-05 |
| T1DM | leukocyte | *SNRNP25* | -0.616440611 | 9.65E-10 | 8.14E-09 | 3.79E-06 |
| T1DM | leukocyte | *WDR90* | -0.995887823 | 3.10E-08 | 5.04E-08 | 0.00012163 |
| T1DM | leukocyte | *CCDC78* | -2.174435368 | 1.77E-09 | 8.14E-09 | 6.96E-06 |
| T1DM | leukocyte | *AMDHD2* | 0.764447916 | 1.86E-08 | 3.19E-08 | 7.32E-05 |
| T1DM | leukocyte | *ZNF75A* | -1.162471298 | 1.92E-08 | 3.28E-08 | 7.55E-05 |
| T1DM | leukocyte | *ASRGL1* | -0.573572452 | 1.74E-09 | 8.14E-09 | 6.85E-06 |
| T1DM | leukocyte | *TAF6L* | -0.87434192 | 4.62E-08 | 7.27E-08 | 0.000181406 |
| T1DM | leukocyte | *ZFPL1* | -0.748410614 | 6.37E-09 | 1.30E-08 | 2.50E-05 |
| T1DM | leukocyte | *LRP5* | -1.25838486 | 1.59E-09 | 8.14E-09 | 6.24E-06 |
| T1DM | leukocyte | *TPCN2* | -0.826789137 | 2.76E-07 | 3.86E-07 | 0.001082744 |
| T1DM | leukocyte | *PDZK1IP1* | -0.498382729 | 1.66E-09 | 8.14E-09 | 6.52E-06 |

| | | | | | | |
|---|---|---|---|---|---|---|
| T1DM | leukocyte | *CTRC* | -0.352534165 | 1.61E-09 | 8.14E-09 | 6.34E-06 |
| T1DM | leukocyte | *SDC3* | -1.349565559 | 1.28E-09 | 8.14E-09 | 5.03E-06 |
| T1DM | leukocyte | *OMA1* | -0.962281013 | 1.79E-08 | 3.07E-08 | 7.03E-05 |
| T1DM | leukocyte | *TM2D1* | -0.911150899 | 6.70E-09 | 1.36E-08 | 2.63E-05 |
| T1DM | leukocyte | *FUBP1* | -0.573336037 | 1.29E-09 | 8.14E-09 | 5.05E-06 |
| T1DM | leukocyte | *SNX7* | -0.569200106 | 1.93E-09 | 8.14E-09 | 7.59E-06 |
| T1DM | leukocyte | *NTNG1* | 0.963389686 | 7.62E-07 | 1.02E-06 | 0.002994184 |
| T1DM | leukocyte | *HFM1* | -1.312321443 | 7.79E-09 | 1.53E-08 | 3.06E-05 |
| T1DM | leukocyte | *VCAM1* | -0.39411309 | 2.28E-09 | 8.21E-09 | 8.97E-06 |
| T1DM | leukocyte | *ZNF496* | -0.694005209 | 1.84E-09 | 8.14E-09 | 7.24E-06 |
| T1DM | leukocyte | *TRIM58* | -2.465359864 | 6.53E-07 | 8.80E-07 | 0.002565506 |
| T1DM | leukocyte | *PEA15* | -0.621640792 | 3.93E-09 | 9.56E-09 | 1.54E-05 |
| T1DM | leukocyte | *FLVCR1* | -0.566026402 | 6.72E-10 | 8.14E-09 | 2.64E-06 |
| T1DM | leukocyte | *TDRD5* | -2.349813364 | 6.22E-09 | 1.28E-08 | 2.44E-05 |
| T1DM | leukocyte | *C1orf115* | -0.435470873 | 1.59E-06 | 2.09E-06 | 0.006256849 |
| T1DM | leukocyte | *ACP6* | -0.892743622 | 5.52E-09 | 1.17E-08 | 2.17E-05 |
| T1DM | leukocyte | *OXER1* | -1.180044209 | 0.011204851 | 0.012411576 | 1 |
| T1DM | leukocyte | *HAAO* | -0.80793726 | 2.64E-09 | 8.21E-09 | 1.04E-05 |
| T1DM | leukocyte | *RFTN2* | -1.779208917 | 1.77E-11 | 4.65E-09 | 6.97E-08 |
| T1DM | leukocyte | *KCNJ3* | -0.645448985 | 2.27E-09 | 8.21E-09 | 8.93E-06 |
| T1DM | leukocyte | *CLHC1* | -0.520582678 | 4.73E-07 | 6.47E-07 | 0.001860335 |
| T1DM | leukocyte | *DUSP19* | -1.024821638 | 4.08E-09 | 9.80E-09 | 1.60E-05 |
| T1DM | leukocyte | *SMC6* | -1.096008518 | 5.41E-09 | 1.16E-08 | 2.12E-05 |
| T1DM | leukocyte | *NOSTRIN* | -0.628055197 | 9.87E-07 | 1.31E-06 | 0.003876199 |
| T1DM | leukocyte | *CTSS* | 0.17857312 | 1.21E-08 | 2.20E-08 | 4.76E-05 |
| T1DM | leukocyte | *MSX1* | -0.783103574 | 1.47E-09 | 8.14E-09 | 5.78E-06 |
| T1DM | leukocyte | *PACRGL* | -0.790546711 | 2.05E-09 | 8.14E-09 | 8.05E-06 |
| T1DM | leukocyte | *ERCC3* | -0.868723032 | 1.40E-08 | 2.49E-08 | 5.51E-05 |
| T1DM | leukocyte | *TDRD10* | -11.08304818 | 6.88E-08 | 1.05E-07 | 0.000270272 |
| T1DM | leukocyte | *CCNYL1* | -1.494672888 | 1.47E-08 | 2.60E-08 | 5.77E-05 |
| T1DM | leukocyte | *DCAF16* | -0.84321827 | 3.74E-09 | 9.32E-09 | 1.47E-05 |
| T1DM | leukocyte | *NPPC* | -5.333696453 | 1.05E-06 | 1.39E-06 | 0.004119592 |
| T1DM | leukocyte | *GNPDA2* | -0.449677743 | 1.08E-05 | 1.35E-05 | 0.042495826 |
| T1DM | leukocyte | *NIPAL1* | -0.607572916 | 1.65E-08 | 2.87E-08 | 6.49E-05 |
| T1DM | leukocyte | *CGGBP1* | -0.612800819 | 1.68E-08 | 2.91E-08 | 6.59E-05 |
| T1DM | leukocyte | *DAPL1* | -1.103579632 | 0.001042562 | 0.00119948 | 1 |
| T1DM | leukocyte | *EOGT* | -0.70936071 | 1.56E-09 | 8.14E-09 | 6.15E-06 |
| T1DM | leukocyte | *POGLUT1* | -0.575309602 | 2.85E-09 | 8.43E-09 | 1.12E-05 |
| T1DM | leukocyte | *IGFN1* | -0.784712747 | 1.21E-07 | 1.77E-07 | 0.00047654 |
| T1DM | leukocyte | *ATP1A1* | -0.459225589 | 1.48E-09 | 8.14E-09 | 5.83E-06 |
| T1DM | leukocyte | *ARPC2* | -0.648425735 | 5.16E-09 | 1.12E-08 | 2.03E-05 |
| T1DM | leukocyte | *CCT3* | -0.509141673 | 1.64E-09 | 8.14E-09 | 6.45E-06 |
| T1DM | leukocyte | *SRGAP2* | -0.691213924 | 0.031677071 | 0.034485788 | 1 |
| T1DM | leukocyte | *CCDC141* | -0.799478513 | 0.001983565 | 0.002257002 | 1 |
| T1DM | leukocyte | *EOMES* | -3.822267386 | 2.93E-08 | 4.79E-08 | 0.000114927 |
| T1DM | leukocyte | *CWC22* | -0.741634637 | 2.52E-09 | 8.21E-09 | 9.89E-06 |
| T1DM | leukocyte | *HDAC11* | -1.439135286 | 7.53E-06 | 9.45E-06 | 0.029602888 |
| T1DM | leukocyte | *SUCLG1* | -0.647928866 | 2.45E-09 | 8.21E-09 | 9.61E-06 |
| T1DM | leukocyte | *NUAK2* | 0.796900948 | 4.64E-05 | 5.64E-05 | 0.182325697 |
| T1DM | leukocyte | *EFHB* | -0.352931055 | 4.05E-05 | 4.95E-05 | 0.159284404 |

| | | | | | | |
|---|---|---|---|---|---|---|
| T1DM | leukocyte | *ICA1L* | -0.554429653 | 3.45E-07 | 4.78E-07 | 0.001355533 |
| T1DM | leukocyte | *RYBP* | -0.529229851 | 7.21E-10 | 8.14E-09 | 2.83E-06 |
| T1DM | leukocyte | *GTPBP8* | -1.096294442 | 2.48E-09 | 8.21E-09 | 9.75E-06 |
| T1DM | leukocyte | *CADPS* | -0.354962722 | 7.62E-09 | 1.50E-08 | 2.99E-05 |
| T1DM | leukocyte | *ATXN7* | -0.735831549 | 5.69E-09 | 1.20E-08 | 2.24E-05 |
| T1DM | leukocyte | *PSMD6* | -0.582725196 | 2.62E-09 | 8.21E-09 | 1.03E-05 |
| T1DM | leukocyte | *PRICKLE2* | -0.743854229 | 1.79E-09 | 8.14E-09 | 7.04E-06 |
| T1DM | leukocyte | *RPL9* | -0.42441974 | 1.36E-09 | 8.14E-09 | 5.34E-06 |
| T1DM | leukocyte | *IL17RC* | -0.76141085 | 9.30E-10 | 8.14E-09 | 3.65E-06 |
| T1DM | leukocyte | *CRELD1* | -0.508643347 | 2.18E-08 | 3.66E-08 | 8.56E-05 |
| T1DM | leukocyte | *TTC14* | -1.017565753 | 7.46E-09 | 1.47E-08 | 2.93E-05 |
| T1DM | leukocyte | *CXCL3* | 0.659388382 | 1.59E-07 | 2.28E-07 | 0.000623909 |
| T1DM | leukocyte | *CXCL1* | -0.48312949 | 8.17E-10 | 8.14E-09 | 3.21E-06 |
| T1DM | leukocyte | *CCDC158* | -1.720073939 | 2.54E-08 | 4.21E-08 | 9.98E-05 |
| T1DM | leukocyte | *TM4SF18* | -0.725268565 | 2.47E-09 | 8.21E-09 | 9.70E-06 |
| T1DM | leukocyte | *SNRK* | -0.487456897 | 3.47E-09 | 8.99E-09 | 1.36E-05 |
| T1DM | leukocyte | *KIAA1143* | -0.561247864 | 3.40E-10 | 8.14E-09 | 1.34E-06 |
| T1DM | leukocyte | *CCR1* | 0.629531192 | 1.14E-07 | 1.68E-07 | 0.000448812 |
| T1DM | leukocyte | *NMNAT3* | -0.645349 | 8.40E-10 | 8.14E-09 | 3.30E-06 |
| T1DM | leukocyte | *SMIM12* | -0.647377929 | 2.10E-09 | 8.16E-09 | 8.24E-06 |
| T1DM | leukocyte | *TPRA1* | -1.149066718 | 4.75E-09 | 1.07E-08 | 1.87E-05 |
| T1DM | leukocyte | *MEAF6* | -0.581410379 | 2.33E-09 | 8.21E-09 | 9.14E-06 |
| T1DM | leukocyte | *DNALI1* | -0.718075568 | 3.29E-09 | 8.81E-09 | 1.29E-05 |
| T1DM | leukocyte | *CAMK2N2* | -0.4567714 | 1.10E-09 | 8.14E-09 | 4.33E-06 |
| T1DM | leukocyte | *RPN1* | -0.70044653 | 2.08E-09 | 8.15E-09 | 8.18E-06 |
| T1DM | leukocyte | *IFT122* | -0.762874712 | 5.42E-07 | 7.37E-07 | 0.002128143 |
| T1DM | leukocyte | *RFC4* | -0.861613242 | 1.69E-09 | 8.14E-09 | 6.64E-06 |
| T1DM | leukocyte | *TKT* | -0.664289852 | 4.78E-09 | 1.07E-08 | 1.88E-05 |
| T1DM | leukocyte | *RFT1* | -1.168249153 | 1.91E-09 | 8.14E-09 | 7.50E-06 |
| T1DM | leukocyte | *UVSSA* | -0.918525299 | 1.42E-06 | 1.87E-06 | 0.005581262 |
| T1DM | leukocyte | *ARHGEF3* | -0.672614199 | 4.35E-09 | 1.01E-08 | 1.71E-05 |
| T1DM | leukocyte | *LRPAP1* | -0.54961834 | 7.96E-09 | 1.55E-08 | 3.13E-05 |
| T1DM | leukocyte | *SLC51A* | 1.22567284 | 2.55E-05 | 3.13E-05 | 0.100243754 |
| T1DM | leukocyte | *PIGX* | -0.878678282 | 3.89E-09 | 9.52E-09 | 1.53E-05 |
| T1DM | leukocyte | *EXO5* | -0.93746713 | 1.48E-06 | 1.94E-06 | 0.005811201 |
| T1DM | leukocyte | *ERMAP* | -1.612611416 | 2.07E-06 | 2.69E-06 | 0.008151201 |
| T1DM | leukocyte | *BDH2* | -0.655618981 | 2.84E-09 | 8.43E-09 | 1.12E-05 |
| T1DM | leukocyte | *CDC25A* | -1.006622252 | 1.90E-07 | 2.72E-07 | 0.000748046 |
| T1DM | leukocyte | *TEX264* | -0.759026318 | 7.55E-09 | 1.49E-08 | 2.97E-05 |
| T1DM | leukocyte | *PPM1M* | -0.433873734 | 0.005305081 | 0.005951931 | 1 |
| T1DM | leukocyte | *SAP30* | -0.66635873 | 2.64E-09 | 8.21E-09 | 1.04E-05 |
| T1DM | leukocyte | *ANXA5* | -0.431119914 | 2.97E-09 | 8.50E-09 | 1.17E-05 |
| T1DM | leukocyte | *CEP44* | -0.759600131 | 1.96E-09 | 8.14E-09 | 7.69E-06 |
| T1DM | leukocyte | *HPGD* | -1.028313552 | 8.55E-07 | 1.14E-06 | 0.003361096 |
| T1DM | leukocyte | *NPY1R* | 1.565019911 | 6.41E-06 | 8.08E-06 | 0.025203078 |
| T1DM | leukocyte | *ARFIP1* | -0.93458899 | 5.37E-09 | 1.15E-08 | 2.11E-05 |
| T1DM | leukocyte | *ITGA2* | -0.47589852 | 7.62E-09 | 1.50E-08 | 2.99E-05 |
| T1DM | leukocyte | *ELOVL7* | -0.663408608 | 5.32E-09 | 1.15E-08 | 2.09E-05 |
| T1DM | leukocyte | *LMBRD2* | -0.823179118 | 9.24E-11 | 8.14E-09 | 3.63E-07 |
| T1DM | leukocyte | *STARD4* | -0.920098998 | 4.34E-10 | 8.14E-09 | 1.71E-06 |

| | | | | | | |
|---|---|---|---|---|---|---|
| T1DM | leukocyte | *CMBL* | -0.7207857 | 1.76E-09 | 8.14E-09 | 6.90E-06 |
| T1DM | leukocyte | *PRRC1* | -0.545620757 | 1.36E-09 | 8.14E-09 | 5.34E-06 |
| T1DM | leukocyte | *AGGF1* | -0.842890282 | 5.76E-09 | 1.20E-08 | 2.26E-05 |
| T1DM | leukocyte | *WDR41* | -1.025422719 | 3.11E-09 | 8.66E-09 | 1.22E-05 |
| T1DM | leukocyte | *SERINC5* | -1.373122146 | 0.0009853 | 0.001135596 | 1 |
| T1DM | leukocyte | *RICTOR* | -0.888361773 | 6.41E-09 | 1.31E-08 | 2.52E-05 |
| T1DM | leukocyte | *UTP15* | -0.632342686 | 1.38E-09 | 8.14E-09 | 5.42E-06 |
| T1DM | leukocyte | *KLKB1* | -0.848114071 | 0.000139139 | 0.00016526 | 0.546678474 |
| T1DM | leukocyte | *NSA2* | -0.461610935 | 2.69E-09 | 8.26E-09 | 1.06E-05 |
| T1DM | leukocyte | *LEAP2* | -1.107522584 | 2.01E-09 | 8.14E-09 | 7.88E-06 |
| T1DM | leukocyte | *DACT2* | -3.022567926 | 1.70E-09 | 8.14E-09 | 6.67E-06 |
| T1DM | leukocyte | *PDSS2* | -0.705429903 | 1.01E-09 | 8.14E-09 | 3.98E-06 |
| T1DM | leukocyte | *STK17A* | -0.59352169 | 3.24E-09 | 8.77E-09 | 1.27E-05 |
| T1DM | leukocyte | *GALNT10* | -1.237170078 | 3.56E-07 | 4.92E-07 | 0.001398542 |
| T1DM | leukocyte | *SAP30L* | -0.88817672 | 3.23E-09 | 8.77E-09 | 1.27E-05 |
| T1DM | leukocyte | *HCN1* | -1.645323891 | 3.21E-06 | 4.11E-06 | 0.01260495 |
| T1DM | leukocyte | *COG5* | -1.134841567 | 1.34E-09 | 8.14E-09 | 5.26E-06 |
| T1DM | leukocyte | *SLU7* | -0.678723514 | 3.44E-09 | 8.93E-09 | 1.35E-05 |
| T1DM | leukocyte | *CAMLG* | -0.535953308 | 4.17E-09 | 9.93E-09 | 1.64E-05 |
| T1DM | leukocyte | *RELL2* | -0.919263882 | 3.03E-06 | 3.89E-06 | 0.011914213 |
| T1DM | leukocyte | *KIF6* | -1.184552513 | 1.69E-08 | 2.94E-08 | 6.66E-05 |
| T1DM | leukocyte | *STEAP1* | -0.944292908 | 4.50E-09 | 1.04E-08 | 1.77E-05 |
| T1DM | leukocyte | *FNDC1* | -3.009288302 | 0.000200623 | 0.000237067 | 0.788247653 |
| T1DM | leukocyte | *CHMP4C* | -0.661658812 | 1.59E-09 | 8.14E-09 | 6.25E-06 |
| T1DM | leukocyte | *BRI3* | -0.690602197 | 5.36E-08 | 8.37E-08 | 0.000210707 |
| T1DM | leukocyte | *LMTK2* | -0.680725818 | 3.67E-09 | 9.23E-09 | 1.44E-05 |
| T1DM | leukocyte | *CTSB* | 0.381353011 | 3.09E-08 | 5.03E-08 | 0.000121287 |
| T1DM | leukocyte | *DLC1* | -0.666017276 | 2.60E-07 | 3.66E-07 | 0.001021785 |
| T1DM | leukocyte | *PEX2* | -0.589318997 | 1.69E-09 | 8.14E-09 | 6.64E-06 |
| T1DM | leukocyte | *MED30* | -0.687593215 | 1.52E-09 | 8.14E-09 | 5.96E-06 |
| T1DM | leukocyte | *TNFRSF11B* | -1.949109847 | 1.64E-07 | 2.35E-07 | 0.000644984 |
| T1DM | leukocyte | *CSMD3* | -48.52474452 | 1.38E-09 | 8.14E-09 | 5.44E-06 |
| T1DM | leukocyte | *OSGIN2* | -0.883606786 | 4.63E-09 | 1.05E-08 | 1.82E-05 |
| T1DM | leukocyte | *DEFB1* | -0.322569237 | 1.63E-09 | 8.14E-09 | 6.41E-06 |
| T1DM | leukocyte | *SUN1* | -0.539186428 | 2.23E-09 | 8.21E-09 | 8.75E-06 |
| T1DM | leukocyte | *TMEM184A* | -0.723734865 | 2.64E-09 | 8.21E-09 | 1.04E-05 |
| T1DM | leukocyte | *INTS1* | -0.716549585 | 1.57E-09 | 8.14E-09 | 6.17E-06 |
| T1DM | leukocyte | *YWHAZ* | -0.565040566 | 3.39E-09 | 8.89E-09 | 1.33E-05 |
| T1DM | leukocyte | *DCAF13* | -0.695331153 | 2.29E-09 | 8.21E-09 | 9.00E-06 |
| T1DM | leukocyte | *INTS8* | -0.797664466 | 1.45E-06 | 1.91E-06 | 0.005703822 |
| T1DM | leukocyte | *GEM* | -0.706424102 | 9.95E-09 | 1.85E-08 | 3.91E-05 |
| T1DM | leukocyte | *RPP25L* | -1.04322884 | 3.06E-09 | 8.60E-09 | 1.20E-05 |
| T1DM | leukocyte | *FAM219A* | -1.03670416 | 2.30E-09 | 8.21E-09 | 9.04E-06 |
| T1DM | leukocyte | *NUDT2* | -0.737289681 | 4.21E-09 | 9.96E-09 | 1.65E-05 |
| T1DM | leukocyte | *NIPSNAP3B* | 1.104981851 | 4.34E-05 | 5.29E-05 | 0.170551216 |
| T1DM | leukocyte | *METTL2B* | -0.614299874 | 1.62E-06 | 2.12E-06 | 0.006375688 |
| T1DM | leukocyte | *FXN* | -1.289655315 | 9.14E-09 | 1.73E-08 | 3.59E-05 |
| T1DM | leukocyte | *MAMDC2* | -1.513500838 | 1.52E-08 | 2.67E-08 | 5.97E-05 |
| T1DM | leukocyte | *PRSS37* | -396.6281638 | 1.71E-09 | 8.14E-09 | 6.73E-06 |
| T1DM | leukocyte | *C9orf64* | -0.892561661 | 2.94E-09 | 8.50E-09 | 1.15E-05 |

| | | | | | | |
|---|---|---|---|---|---|---|
| T1DM | leukocyte | *ANKS6* | -0.627417629 | 5.47E-08 | 8.50E-08 | 0.000214784 |
| T1DM | leukocyte | *CLDN3* | -0.691309701 | 1.22E-09 | 8.14E-09 | 4.78E-06 |
| T1DM | leukocyte | *WNK2* | -1.049188584 | 3.33E-09 | 8.82E-09 | 1.31E-05 |
| T1DM | leukocyte | *NOL6* | -1.313882796 | 1.91E-09 | 8.14E-09 | 7.50E-06 |
| T1DM | leukocyte | *PIGO* | -1.111332039 | 1.86E-07 | 2.66E-07 | 0.000732407 |
| T1DM | leukocyte | *ARHGAP12* | -0.720483342 | 3.91E-09 | 9.54E-09 | 1.54E-05 |
| T1DM | leukocyte | *WRN* | -0.772178342 | 1.79E-09 | 8.14E-09 | 7.02E-06 |
| T1DM | leukocyte | *CFL2* | -0.673418684 | 3.82E-09 | 9.44E-09 | 1.50E-05 |
| T1DM | leukocyte | *PGM2L1* | -1.050318347 | 3.37E-08 | 5.47E-08 | 0.000132491 |
| T1DM | leukocyte | *PHYHIPL* | -0.707168084 | 1.63E-09 | 8.14E-09 | 6.41E-06 |
| T1DM | leukocyte | *PKNOX2* | 2.867712586 | 9.23E-08 | 1.38E-07 | 0.000362584 |
| T1DM | leukocyte | *RPL36AL* | -0.464064741 | 1.82E-08 | 3.12E-08 | 7.14E-05 |
| T1DM | leukocyte | *TMEM63C* | -0.614442898 | 4.11E-09 | 9.85E-09 | 1.62E-05 |
| T1DM | leukocyte | *NGB* | -3.07104838 | 0.012668885 | 0.014009583 | 1 |
| T1DM | leukocyte | *NOXRED1* | -2.515159685 | 4.93E-05 | 5.98E-05 | 0.193598901 |
| T1DM | leukocyte | *AKR1E2* | -1.954012379 | 0.000316292 | 0.000369965 | 1 |
| T1DM | leukocyte | *OXGR1* | -1.590820129 | 2.64E-09 | 8.21E-09 | 1.04E-05 |
| T1DM | leukocyte | *VDAC2* | -0.573940755 | 2.96E-09 | 8.50E-09 | 1.16E-05 |
| T1DM | leukocyte | *SNAPC4* | -0.721681337 | 2.24E-09 | 8.21E-09 | 8.81E-06 |
| T1DM | leukocyte | *TSC1* | -0.954052037 | 1.98E-09 | 8.14E-09 | 7.79E-06 |
| T1DM | leukocyte | *ZMYND19* | -0.695750782 | 1.41E-09 | 8.14E-09 | 5.53E-06 |
| T1DM | leukocyte | *METTL17* | -0.909159954 | 8.02E-09 | 1.56E-08 | 3.15E-05 |
| T1DM | leukocyte | *CASP7* | -0.416373449 | 5.26E-07 | 7.17E-07 | 0.002067777 |
| T1DM | leukocyte | *VWA2* | -1.864339142 | 9.00E-10 | 8.14E-09 | 3.54E-06 |
| T1DM | leukocyte | *METTL3* | -0.757929515 | 1.95E-09 | 8.14E-09 | 7.68E-06 |
| T1DM | leukocyte | *TRUB1* | -0.929191417 | 3.44E-09 | 8.93E-09 | 1.35E-05 |
| T1DM | leukocyte | *ARHGAP42* | -1.093724027 | 2.56E-09 | 8.21E-09 | 1.01E-05 |
| T1DM | leukocyte | *SLC39A13* | -0.678799849 | 1.55E-09 | 8.14E-09 | 6.08E-06 |
| T1DM | leukocyte | *PSMC3* | -0.521821634 | 1.08E-08 | 1.99E-08 | 4.24E-05 |
| T1DM | leukocyte | *CPSF2* | -0.550907257 | 5.03E-08 | 7.87E-08 | 0.000197551 |
| T1DM | leukocyte | *IFI27L1* | -0.710495904 | 2.98E-09 | 8.50E-09 | 1.17E-05 |
| T1DM | leukocyte | *C1QL3* | -1.078440123 | 0.0165531 | 0.018227895 | 1 |
| T1DM | leukocyte | *SMCO4* | -0.722737286 | 3.34E-09 | 8.82E-09 | 1.31E-05 |
| T1DM | leukocyte | *ABTB2* | -1.038430084 | 3.25E-09 | 8.77E-09 | 1.28E-05 |
| T1DM | leukocyte | *HTRA1* | 0.827705755 | 0.002331227 | 0.002647988 | 1 |
| T1DM | leukocyte | *CEP57* | -0.591801406 | 2.26E-09 | 8.21E-09 | 8.89E-06 |
| T1DM | leukocyte | *JAM3* | -0.559941983 | 6.15E-09 | 1.26E-08 | 2.42E-05 |
| T1DM | leukocyte | *IKBIP* | -0.785058713 | 8.93E-09 | 1.70E-08 | 3.51E-05 |
| T1DM | leukocyte | *ZFYVE19* | -0.641672935 | 4.91E-08 | 7.69E-08 | 0.000192769 |
| T1DM | leukocyte | *CKB* | -0.518800986 | 1.95E-10 | 8.14E-09 | 7.68E-07 |
| T1DM | leukocyte | *BTRC* | -1.142965808 | 6.71E-08 | 1.02E-07 | 0.000263763 |
| T1DM | leukocyte | *BAG5* | -0.692613358 | 3.85E-09 | 9.49E-09 | 1.51E-05 |
| T1DM | leukocyte | *LARP6* | -0.593057135 | 2.04E-09 | 8.14E-09 | 8.03E-06 |
| T1DM | leukocyte | *ZNF319* | -1.838386054 | 4.74E-09 | 1.06E-08 | 1.86E-05 |
| T1DM | leukocyte | *SGPL1* | -1.084262402 | 1.53E-09 | 8.14E-09 | 6.02E-06 |
| T1DM | leukocyte | *FRS2* | -0.676261411 | 4.63E-08 | 7.29E-08 | 0.000181927 |
| T1DM | leukocyte | *PCBD1* | -0.643575652 | 1.37E-09 | 8.14E-09 | 5.40E-06 |
| T1DM | leukocyte | *COX11* | -0.746525829 | 3.97E-09 | 9.63E-09 | 1.56E-05 |
| T1DM | leukocyte | *ZNF202* | 0.958675946 | 8.16E-08 | 1.23E-07 | 0.000320541 |
| T1DM | leukocyte | *MYRFL* | -0.425427561 | 0.001239019 | 0.001420929 | 1 |

| | | | | | | |
|---|---|---|---|---|---|---|
| T1DM | leukocyte | *WBP1L* | -1.269873911 | 9.50E-08 | 1.41E-07 | 0.000373104 |
| T1DM | leukocyte | *NUDT13* | -0.496843218 | 0.008884142 | 0.009882728 | 1 |
| T1DM | leukocyte | *CYB5R2* | -0.622498989 | 6.30E-07 | 8.50E-07 | 0.002474613 |
| T1DM | leukocyte | *TUB* | -0.900489879 | 8.05E-09 | 1.56E-08 | 3.16E-05 |
| T1DM | leukocyte | *RIC3* | -0.537385881 | 1.68E-09 | 8.14E-09 | 6.61E-06 |
| T1DM | leukocyte | *XRRA1* | -0.721151441 | 3.88E-09 | 9.51E-09 | 1.52E-05 |
| T1DM | leukocyte | *TRIM66* | -0.762488626 | 3.10E-06 | 3.98E-06 | 0.012197734 |
| T1DM | leukocyte | *CDYL2* | -0.732967295 | 2.85E-09 | 8.43E-09 | 1.12E-05 |
| T1DM | leukocyte | *ZNF143* | -0.695760131 | 1.46E-09 | 8.14E-09 | 5.74E-06 |
| T1DM | leukocyte | *CCDC68* | -0.685383115 | 2.04E-08 | 3.45E-08 | 8.02E-05 |
| T1DM | leukocyte | *ZSCAN21* | -1.039760528 | 6.40E-09 | 1.31E-08 | 2.51E-05 |
| T1DM | leukocyte | *RIMKLB* | -0.674487148 | 1.63E-09 | 8.14E-09 | 6.39E-06 |
| T1DM | leukocyte | *GALR1* | 0.919502382 | 1.30E-09 | 8.14E-09 | 5.10E-06 |
| T1DM | leukocyte | *SPDYE6* | 1.224827713 | 2.51E-06 | 3.24E-06 | 0.009867738 |
| T1DM | leukocyte | *ATF7IP2* | -0.894147729 | 2.11E-09 | 8.16E-09 | 8.28E-06 |
| T1DM | leukocyte | *COG1* | -0.669025937 | 2.17E-09 | 8.21E-09 | 8.54E-06 |
| T1DM | leukocyte | *B2M* | -0.427054752 | 2.18E-09 | 8.21E-09 | 8.55E-06 |
| T1DM | leukocyte | *ACSM1* | -1.409971703 | 0.00033983 | 0.000397261 | 1 |
| T1DM | leukocyte | *AP1G1* | -0.513592066 | 3.74E-09 | 9.32E-09 | 1.47E-05 |
| T1DM | leukocyte | *SLFN5* | -0.571267353 | 1.69E-09 | 8.14E-09 | 6.65E-06 |
| T1DM | leukocyte | *SAAL1* | -0.683155967 | 1.61E-09 | 8.14E-09 | 6.34E-06 |
| T1DM | leukocyte | *SCNN1G* | -6.301304667 | 9.82E-10 | 8.14E-09 | 3.86E-06 |
| T1DM | leukocyte | *GLYATL1* | -4.691372152 | 0.013736934 | 0.015173577 | 1 |
| T1DM | leukocyte | *DCTN5* | -0.767151976 | 4.61E-10 | 8.14E-09 | 1.81E-06 |
| T1DM | leukocyte | *STAT6* | -1.104643981 | 1.06E-08 | 1.95E-08 | 4.16E-05 |
| T1DM | leukocyte | *PATL1* | -0.607789489 | 1.44E-09 | 8.14E-09 | 5.65E-06 |
| T1DM | leukocyte | *MRPL16* | -0.871886556 | 6.31E-06 | 7.96E-06 | 0.024805887 |
| T1DM | leukocyte | *GREM1* | -1.012289981 | 1.95E-08 | 3.32E-08 | 7.66E-05 |
| T1DM | leukocyte | *TSC22D4* | -0.699361285 | 3.72E-06 | 4.74E-06 | 0.014619097 |
| T1DM | leukocyte | *MS4A14* | 0.168992939 | 1.36E-08 | 2.42E-08 | 5.33E-05 |
| T1DM | leukocyte | *DIS3L* | -1.049271799 | 4.17E-09 | 9.93E-09 | 1.64E-05 |
| T1DM | leukocyte | *MS4A8* | -1.329159585 | 0.000176926 | 0.00020957 | 0.695143636 |
| T1DM | leukocyte | *RCCD1* | -1.476200507 | 6.74E-08 | 1.03E-07 | 0.000264976 |
| T1DM | leukocyte | *MAPRE2* | -0.633531663 | 6.86E-09 | 1.38E-08 | 2.70E-05 |
| T1DM | leukocyte | *NAT16* | -0.468790641 | 2.07E-09 | 8.14E-09 | 8.13E-06 |
| T1DM | leukocyte | *GNGT2* | 0.334639325 | 7.38E-09 | 1.46E-08 | 2.90E-05 |
| T1DM | leukocyte | *SLC27A4* | -0.762271639 | 6.30E-09 | 1.29E-08 | 2.47E-05 |
| T1DM | leukocyte | *URM1* | -0.833714102 | 3.51E-08 | 5.67E-08 | 0.000137904 |
| T1DM | leukocyte | *CERCAM* | -0.606898574 | 2.11E-08 | 3.55E-08 | 8.28E-05 |
| T1DM | leukocyte | *DOLPP1* | -0.796480783 | 1.08E-08 | 1.99E-08 | 4.26E-05 |
| T1DM | leukocyte | *ENDOG* | -0.773987055 | 6.54E-09 | 1.33E-08 | 2.57E-05 |
| T1DM | leukocyte | *PRR15L* | -0.96526895 | 7.80E-09 | 1.53E-08 | 3.07E-05 |
| T1DM | leukocyte | *RNF214* | -1.027882083 | 9.73E-08 | 1.45E-07 | 0.000382428 |
| T1DM | leukocyte | *POP5* | -0.762523275 | 4.54E-10 | 8.14E-09 | 1.79E-06 |
| T1DM | leukocyte | *ENGASE* | -0.36898788 | 0.000766759 | 0.000886058 | 1 |
| T1DM | leukocyte | *ACAA2* | -0.71418931 | 1.98E-09 | 8.14E-09 | 7.77E-06 |
| T1DM | leukocyte | *FN3K* | -0.906718286 | 3.88E-09 | 9.51E-09 | 1.52E-05 |
| T1DM | leukocyte | *ZNF23* | -0.672798122 | 1.55E-09 | 8.14E-09 | 6.10E-06 |
| T1DM | leukocyte | *ZNF229* | -0.95066565 | 1.23E-09 | 8.14E-09 | 4.83E-06 |
| T1DM | leukocyte | *ZNF180* | -0.746235706 | 1.38E-09 | 8.14E-09 | 5.44E-06 |

| | | | | | | |
|---|---|---|---|---|---|---|
| T1DM | leukocyte | *CA4* | -0.44520753 | 1.11E-06 | 1.47E-06 | 0.004370751 |
| T1DM | leukocyte | *GPX4* | -0.624062389 | 1.31E-08 | 2.35E-08 | 5.13E-05 |
| T1DM | leukocyte | *ZNF641* | -1.069085779 | 3.26E-09 | 8.78E-09 | 1.28E-05 |
| T1DM | leukocyte | *CACNB3* | -0.709436575 | 1.09E-05 | 1.35E-05 | 0.042686513 |
| T1DM | leukocyte | *TP53I13* | -0.640656689 | 1.84E-08 | 3.17E-08 | 7.24E-05 |
| T1DM | leukocyte | *TUBA1C* | -0.450440317 | 3.07E-09 | 8.60E-09 | 1.21E-05 |
| T1DM | leukocyte | *ZNF528* | -1.933351434 | 9.91E-07 | 1.31E-06 | 0.003892904 |
| T1DM | leukocyte | *RAB4B* | -0.801427843 | 4.13E-09 | 9.87E-09 | 1.62E-05 |
| T1DM | leukocyte | *TMC4* | -0.701865811 | 1.46E-09 | 8.14E-09 | 5.73E-06 |
| T1DM | leukocyte | *PPP1R14A* | -0.414069883 | 0.000218351 | 0.000257473 | 0.857900361 |
| T1DM | leukocyte | *C19orf33* | -0.616282379 | 1.32E-09 | 8.14E-09 | 5.19E-06 |
| T1DM | leukocyte | *DNAAF3* | -2.507018219 | 1.19E-09 | 8.14E-09 | 4.66E-06 |
| T1DM | leukocyte | *CHAF1A* | -0.817293204 | 1.79E-06 | 2.32E-06 | 0.007015048 |
| T1DM | leukocyte | *UBXN6* | -0.845997845 | 6.43E-09 | 1.31E-08 | 2.53E-05 |
| T1DM | leukocyte | *NXN* | -0.502512618 | 4.25E-09 | 9.99E-09 | 1.67E-05 |
| T1DM | leukocyte | *KIFC2* | -2.128173952 | 6.53E-08 | 1.00E-07 | 0.000256442 |
| T1DM | leukocyte | *SLC43A2* | 0.506035206 | 2.13E-05 | 2.62E-05 | 0.083594317 |
| T1DM | leukocyte | *SERPINF2* | 0.32581462 | 4.81E-07 | 6.57E-07 | 0.001889709 |
| T1DM | leukocyte | *SRR* | -0.674335012 | 2.94E-07 | 4.10E-07 | 0.001154103 |
| T1DM | leukocyte | *TRPV3* | -3.559118859 | 2.92E-10 | 8.14E-09 | 1.15E-06 |
| T1DM | leukocyte | *C19orf48* | -0.965640788 | 1.60E-06 | 2.10E-06 | 0.006288348 |
| T1DM | leukocyte | *KLK11* | -0.384817738 | 0.007375825 | 0.008235185 | 1 |
| T1DM | leukocyte | *ZNF83* | -0.867148932 | 2.23E-09 | 8.21E-09 | 8.74E-06 |
| T1DM | leukocyte | *ZNF558* | -0.781691653 | 2.51E-07 | 3.54E-07 | 0.000987885 |
| T1DM | leukocyte | *NDUFV1* | -0.629427693 | 1.34E-09 | 8.14E-09 | 5.26E-06 |
| T1DM | leukocyte | *ZNF232* | -0.530348375 | 8.36E-07 | 1.12E-06 | 0.003285342 |
| T1DM | leukocyte | *MIS12* | -0.881090032 | 3.37E-10 | 8.14E-09 | 1.32E-06 |
| T1DM | leukocyte | *HID1* | -0.748209795 | 1.96E-09 | 8.14E-09 | 7.69E-06 |
| T1DM | leukocyte | *EVPL* | -0.771078605 | 0.03519023 | 0.038236287 | 1 |
| T1DM | leukocyte | *SRP68* | -0.58213484 | 1.49E-09 | 8.14E-09 | 5.85E-06 |
| T1DM | leukocyte | *TMC8* | 0.695268572 | 7.39E-07 | 9.92E-07 | 0.002901859 |
| T1DM | leukocyte | *TMEM68* | -0.894326359 | 4.38E-09 | 1.02E-08 | 1.72E-05 |
| T1DM | leukocyte | *GSDMA* | -2.161612623 | 0.008561189 | 0.00952887 | 1 |
| T1DM | leukocyte | *MLST8* | -0.567185197 | 2.31E-09 | 8.21E-09 | 9.09E-06 |
| T1DM | leukocyte | *ECI1* | -0.548213519 | 1.91E-09 | 8.14E-09 | 7.52E-06 |
| T1DM | leukocyte | *ABCA3* | -0.587219187 | 1.12E-07 | 1.65E-07 | 0.000440455 |
| T1DM | leukocyte | *BSCL2* | -0.505740238 | 1.54E-07 | 2.21E-07 | 0.000604571 |
| T1DM | leukocyte | *C2CD3* | -1.266670291 | 1.01E-09 | 8.14E-09 | 3.96E-06 |
| T1DM | leukocyte | *TRANK1* | -1.66235315 | 1.18E-08 | 2.14E-08 | 4.62E-05 |
| T1DM | leukocyte | *RPSA* | -0.42757119 | 5.87E-09 | 1.22E-08 | 2.31E-05 |
| T1DM | leukocyte | *ENTPD3* | -0.434627609 | 2.19E-08 | 3.68E-08 | 8.61E-05 |
| T1DM | leukocyte | *ULK4* | -0.716421144 | 1.59E-06 | 2.08E-06 | 0.006241768 |
| T1DM | leukocyte | *LTBP3* | -0.579745733 | 1.77E-09 | 8.14E-09 | 6.96E-06 |
| T1DM | leukocyte | *SAC3D1* | -0.815079802 | 1.19E-09 | 8.14E-09 | 4.67E-06 |
| T1DM | leukocyte | *PBK* | -1.563549547 | 1.96E-09 | 8.14E-09 | 7.70E-06 |
| T1DM | leukocyte | *KIAA1586* | -0.881148439 | 7.29E-10 | 8.14E-09 | 2.86E-06 |
| T1DM | leukocyte | *FAM83B* | -0.670346218 | 2.65E-07 | 3.72E-07 | 0.001040654 |
| T1DM | leukocyte | *LMBRD1* | -0.779751529 | 2.27E-09 | 8.21E-09 | 8.91E-06 |
| T1DM | leukocyte | *TTC39C* | -0.635174559 | 3.34E-09 | 8.82E-09 | 1.31E-05 |
| T1DM | leukocyte | *POLR2J3* | -0.728663486 | 3.79E-09 | 9.41E-09 | 1.49E-05 |

| | | | | | | |
|---|---|---|---|---|---|---|
| T1DM | leukocyte | *FOXI1* | 2.164820032 | 6.35E-06 | 8.00E-06 | 0.024933702 |
| T1DM | leukocyte | *COA6* | -0.745017419 | 1.83E-09 | 8.14E-09 | 7.20E-06 |
| T1DM | leukocyte | *PXK* | -0.884235251 | 3.57E-09 | 9.10E-09 | 1.40E-05 |
| T1DM | leukocyte | *ING5* | -1.120564878 | 7.23E-09 | 1.44E-08 | 2.84E-05 |
| T1DM | leukocyte | *RFWD3* | -0.581012094 | 0.00019707 | 0.000233009 | 0.774289412 |
| T1DM | leukocyte | *BMP1* | -0.823457254 | 9.78E-10 | 8.14E-09 | 3.84E-06 |
| T1DM | leukocyte | *HEXIM2* | -0.877874243 | 1.40E-09 | 8.14E-09 | 5.50E-06 |
| T1DM | leukocyte | *SERINC2* | -0.47267808 | 2.29E-09 | 8.21E-09 | 8.99E-06 |
| T1DM | leukocyte | *TRAPPC11* | -0.586180322 | 1.23E-09 | 8.14E-09 | 4.83E-06 |
| T1DM | leukocyte | *COL3A1* | 0.791397591 | 0.0023181 | 0.002633838 | 1 |
| T1DM | leukocyte | *ING2* | -0.498374237 | 8.65E-09 | 1.65E-08 | 3.40E-05 |
| T1DM | leukocyte | *ADAM9* | -1.143011285 | 7.52E-08 | 1.14E-07 | 0.000295585 |
| T1DM | leukocyte | *NDUFS5* | -0.510940929 | 1.87E-09 | 8.14E-09 | 7.35E-06 |
| T1DM | leukocyte | *ZNF30* | -0.608586418 | 0.027826941 | 0.030361581 | 1 |
| T1DM | leukocyte | *DNAJC21* | -0.725140111 | 1.90E-09 | 8.14E-09 | 7.46E-06 |
| T1DM | leukocyte | *CNNM3* | -0.675487628 | 2.08E-09 | 8.15E-09 | 8.17E-06 |
| T1DM | leukocyte | *GSTM4* | -0.797522665 | 2.88E-09 | 8.46E-09 | 1.13E-05 |
| T1DM | leukocyte | *TCTN2* | -0.461847428 | 0.003373858 | 0.003814645 | 1 |
| T1DM | leukocyte | *SHOX2* | -5.704497485 | 1.06E-07 | 1.57E-07 | 0.000416846 |
| T1DM | leukocyte | *PPIP5K1* | -1.750911227 | 1.49E-09 | 8.14E-09 | 5.85E-06 |
| T1DM | leukocyte | *ADAL* | -1.514981811 | 2.75E-06 | 3.54E-06 | 0.010810415 |
| T1DM | leukocyte | *LCMT2* | -1.091310049 | 5.46E-10 | 8.14E-09 | 2.15E-06 |
| T1DM | leukocyte | *ZNF507* | -0.837270453 | 2.47E-09 | 8.21E-09 | 9.70E-06 |
| T1DM | leukocyte | *NSG1* | -0.532583863 | 1.42E-09 | 8.14E-09 | 5.59E-06 |
| T1DM | leukocyte | *ZBTB49* | -1.838842574 | 2.93E-09 | 8.50E-09 | 1.15E-05 |
| T1DM | leukocyte | *GFM1* | -0.587539081 | 9.50E-10 | 8.14E-09 | 3.73E-06 |
| T1DM | leukocyte | *FSTL5* | -0.472525209 | 4.26E-09 | 1.00E-08 | 1.67E-05 |
| T1DM | leukocyte | *CTRB1* | -0.303120947 | 1.64E-09 | 8.14E-09 | 6.45E-06 |
| T1DM | leukocyte | *CTRB2* | -0.280536625 | 3.51E-09 | 9.03E-09 | 1.38E-05 |
| T1DM | leukocyte | *TMEM129* | -0.803392362 | 6.67E-07 | 8.98E-07 | 0.002621954 |
| T1DM | leukocyte | *PPIC* | -0.345901305 | 1.27E-07 | 1.85E-07 | 0.000499948 |
| T1DM | leukocyte | *STXBP6* | -0.747997377 | 6.56E-09 | 1.33E-08 | 2.58E-05 |
| T1DM | leukocyte | *MFF* | -0.646742639 | 3.07E-09 | 8.60E-09 | 1.21E-05 |
| T1DM | leukocyte | *CPLX1* | -0.709022912 | 5.53E-09 | 1.17E-08 | 2.17E-05 |
| T1DM | leukocyte | *ROR2* | -1.767695888 | 4.71E-07 | 6.45E-07 | 0.001852357 |
| T1DM | leukocyte | *HSPBAP1* | 1.272507284 | 0.00020351 | 0.000240262 | 0.799590298 |
| T1DM | leukocyte | *FAM110B* | -0.736690329 | 1.19E-09 | 8.14E-09 | 4.67E-06 |
| T1DM | leukocyte | *ATF5* | -0.7958717 | 4.63E-09 | 1.05E-08 | 1.82E-05 |
| T1DM | leukocyte | *XPO6* | -1.08408043 | 1.33E-08 | 2.39E-08 | 5.24E-05 |
| T1DM | leukocyte | *RGS14* | -0.905406796 | 6.57E-08 | 1.01E-07 | 0.000258253 |
| T1DM | leukocyte | *TBC1D10B* | -0.877690271 | 8.18E-10 | 8.14E-09 | 3.21E-06 |
| T1DM | leukocyte | *PRELID1* | -0.505897002 | 4.09E-09 | 9.81E-09 | 1.61E-05 |
| T1DM | leukocyte | *CXCL10* | -122.9291475 | 1.48E-09 | 8.14E-09 | 5.80E-06 |
| T1DM | leukocyte | *NMD3* | -0.657343849 | 2.02E-09 | 8.14E-09 | 7.92E-06 |
| T1DM | leukocyte | *STK32A* | -0.768463822 | 1.42E-08 | 2.52E-08 | 5.57E-05 |
| T1DM | leukocyte | *P2RY12* | -25.03357988 | 0.003043583 | 0.003445186 | 1 |
| T1DM | leukocyte | *SNUPN* | -1.689914474 | 2.35E-09 | 8.21E-09 | 9.25E-06 |
| T1DM | leukocyte | *CRADD* | -0.750630548 | 1.70E-09 | 8.14E-09 | 6.67E-06 |
| T1DM | leukocyte | *COL22A1* | -7.422228919 | 5.78E-10 | 8.14E-09 | 2.27E-06 |
| T1DM | leukocyte | *SDC2* | -0.913520992 | 0.00039496 | 0.000460337 | 1 |

| | | | | | | |
|---|---|---|---|---|---|---|
| T1DM | leukocyte | *CLIC4* | -0.415261625 | 1.72E-09 | 8.14E-09 | 6.75E-06 |
| T1DM | leukocyte | *SLC38A11* | -6.145753406 | 3.83E-11 | 8.14E-09 | 1.51E-07 |
| T1DM | leukocyte | *CCDC8* | -0.876555736 | 1.05E-05 | 1.31E-05 | 0.041366154 |
| T1DM | leukocyte | *HINT1* | -0.462850219 | 1.00E-09 | 8.14E-09 | 3.95E-06 |
| T1DM | leukocyte | *DTWD2* | -1.373437412 | 0.000289116 | 0.000339086 | 1 |
| T1DM | leukocyte | *INO80E* | -0.826576177 | 1.09E-08 | 2.00E-08 | 4.28E-05 |
| T1DM | leukocyte | *DFFB* | 0.969243852 | 3.08E-07 | 4.28E-07 | 0.00120909 |
| T1DM | leukocyte | *C15orf40* | -0.630825006 | 3.28E-08 | 5.33E-08 | 0.000128876 |
| T1DM | leukocyte | *HIC2* | -1.012016256 | 0.030049866 | 0.03275969 | 1 |
| T1DM | leukocyte | *AGPAT2* | -0.689757084 | 1.84E-09 | 8.14E-09 | 7.23E-06 |
| T1DM | leukocyte | *FASN* | -1.093853008 | 4.20E-09 | 9.96E-09 | 1.65E-05 |
| T1DM | leukocyte | *MT1E* | -0.391478586 | 9.46E-09 | 1.78E-08 | 3.72E-05 |
| T1DM | leukocyte | *LDB2* | -0.738203982 | 2.68E-08 | 4.42E-08 | 0.000105187 |
| T1DM | leukocyte | *RAC3* | -0.971972804 | 7.36E-10 | 8.14E-09 | 2.89E-06 |
| T1DM | leukocyte | *LIMS1* | -0.759933762 | 2.04E-09 | 8.14E-09 | 8.02E-06 |
| T1DM | leukocyte | *CSGALNACT2* | -1.144613342 | 9.02E-05 | 0.000108037 | 0.354360083 |
| T1DM | leukocyte | *AVEN* | -1.630302496 | 7.30E-08 | 1.11E-07 | 0.000286677 |
| T1DM | leukocyte | *CTNND2* | -0.735170865 | 3.91E-09 | 9.52E-09 | 1.53E-05 |
| T1DM | leukocyte | *WNT10B* | -42.6415921 | 1.33E-09 | 8.14E-09 | 5.23E-06 |
| T1DM | leukocyte | *CALML6* | 0.844566147 | 2.64E-09 | 8.21E-09 | 1.04E-05 |
| T1DM | leukocyte | *TM4SF4* | -0.638216158 | 9.77E-10 | 8.14E-09 | 3.84E-06 |
| T1DM | leukocyte | *OTUD3* | -0.527165678 | 1.08E-09 | 8.14E-09 | 4.24E-06 |
| T1DM | leukocyte | *MAP3K2* | -0.776354618 | 1.46E-08 | 2.59E-08 | 5.75E-05 |
| T1DM | leukocyte | *PUSL1* | -1.277245482 | 5.88E-08 | 9.09E-08 | 0.000231202 |
| T1DM | leukocyte | *IFFO2* | -0.667449827 | 6.27E-10 | 8.14E-09 | 2.46E-06 |
| T1DM | leukocyte | *MYRIP* | -0.936456158 | 1.83E-09 | 8.14E-09 | 7.19E-06 |
| T1DM | leukocyte | *KCNAB3* | 0.677647003 | 8.13E-07 | 1.09E-06 | 0.003193507 |
| T1DM | leukocyte | *FAM153A* | -1.460881044 | 1.47E-08 | 2.60E-08 | 5.78E-05 |
| T1DM | leukocyte | *ZNF778* | -0.908444075 | 1.45E-09 | 8.14E-09 | 5.70E-06 |
| T1DM | leukocyte | *NIPA1* | -0.85117852 | 6.56E-10 | 8.14E-09 | 2.58E-06 |
| T1DM | leukocyte | *FOXD4* | -6.755974902 | 8.21E-05 | 9.85E-05 | 0.322623513 |
| T1DM | leukocyte | *UBE2E1* | -1.059536232 | 9.47E-09 | 1.78E-08 | 3.72E-05 |
| T1DM | leukocyte | *ANKK1* | -19.92259845 | 2.13E-10 | 8.14E-09 | 8.39E-07 |
| T1DM | leukocyte | *USP47* | -0.752124221 | 5.05E-09 | 1.11E-08 | 1.99E-05 |
| T1DM | leukocyte | *PDCD6IP* | -0.651962696 | 2.40E-09 | 8.21E-09 | 9.44E-06 |
| T1DM | leukocyte | *ZNF212* | 0.517088953 | 0.020359557 | 0.022325621 | 1 |
| T1DM | leukocyte | *CRTAP* | -1.095990346 | 1.72E-08 | 2.97E-08 | 6.75E-05 |
| T1DM | leukocyte | *STX8* | -0.790830897 | 1.44E-07 | 2.07E-07 | 0.000564097 |
| T1DM | leukocyte | *NFRKB* | -1.143751657 | 4.51E-09 | 1.04E-08 | 1.77E-05 |
| T1DM | leukocyte | *SETMAR* | -0.690132802 | 7.86E-10 | 8.14E-09 | 3.09E-06 |
| T1DM | leukocyte | *CST5* | -2.325251065 | 0.002938181 | 0.003326833 | 1 |
| T1DM | leukocyte | *CST2* | -1.079776095 | 9.19E-06 | 1.15E-05 | 0.036103032 |
| T1DM | leukocyte | *KRT8* | -0.297138583 | 1.88E-09 | 8.14E-09 | 7.37E-06 |
| T1DM | leukocyte | *ADORA2B* | 1.318827078 | 0.000195346 | 0.000231109 | 0.767513896 |
| T1DM | leukocyte | *MGMT* | -0.862295323 | 2.54E-09 | 8.21E-09 | 9.99E-06 |
| T1DM | leukocyte | *NFXL1* | -1.148489884 | 2.71E-10 | 8.14E-09 | 1.06E-06 |
| T1DM | leukocyte | *DNAJC18* | -0.581877299 | 0.000110963 | 0.000132394 | 0.435973995 |
| T1DM | leukocyte | *KRT4* | -25.04554711 | 0.000225478 | 0.000265638 | 0.885902504 |
| T1DM | leukocyte | *SLC23A1* | -5.620035958 | 3.63E-08 | 5.84E-08 | 0.000142714 |
| T1DM | leukocyte | *NPAS2* | -0.559053705 | 5.23E-09 | 1.13E-08 | 2.06E-05 |

| | | | | | | |
|---|---|---|---|---|---|---|
| T1DM | leukocyte | *NUDT9* | -0.564079981 | 1.14E-08 | 2.09E-08 | 4.50E-05 |
| T1DM | leukocyte | *SMAGP* | -0.483008924 | 9.70E-08 | 1.44E-07 | 0.000380948 |
| T1DM | leukocyte | *IRX1* | -1.033267945 | 1.39E-09 | 8.14E-09 | 5.47E-06 |
| T1DM | leukocyte | *CDH2* | -0.573405836 | 1.45E-09 | 8.14E-09 | 5.71E-06 |
| T1DM | leukocyte | *HSPA4* | -0.682225002 | 4.38E-09 | 1.02E-08 | 1.72E-05 |
| T1DM | leukocyte | *ARMC10* | -0.885008992 | 1.29E-08 | 2.32E-08 | 5.07E-05 |
| T1DM | leukocyte | *ACYP2* | -0.618496112 | 2.34E-09 | 8.21E-09 | 9.18E-06 |
| T1DM | leukocyte | *ATF7* | -0.72739853 | 1.60E-09 | 8.14E-09 | 6.29E-06 |
| T1DM | leukocyte | *RASA4B* | -0.576394115 | 0.01234354 | 0.013657497 | 1 |
| T1DM | leukocyte | *KCNS3* | -0.933989221 | 3.74E-09 | 9.32E-09 | 1.47E-05 |
| T1DM | leukocyte | *FOXN2* | -1.400046775 | 9.46E-06 | 1.18E-05 | 0.037163016 |
| T1DM | leukocyte | *GPR27* | -0.715908341 | 6.90E-10 | 8.14E-09 | 2.71E-06 |
| T1DM | leukocyte | *MTSS1* | -0.753418915 | 1.49E-06 | 1.96E-06 | 0.005863211 |
| T1DM | leukocyte | *TMEM43* | -0.692273725 | 2.03E-09 | 8.14E-09 | 7.97E-06 |
| T1DM | leukocyte | *RPS9* | -0.417033205 | 2.64E-09 | 8.21E-09 | 1.04E-05 |
| T1DM | leukocyte | *NDUFA3* | -0.690243464 | 5.07E-09 | 1.11E-08 | 1.99E-05 |
| T1DM | leukocyte | *ZNF160* | -0.693424901 | 2.30E-09 | 8.21E-09 | 9.02E-06 |
| T1DM | leukocyte | *ZNF415* | -0.67282962 | 9.51E-09 | 1.79E-08 | 3.74E-05 |
| T1DM | leukocyte | *TSNARE1* | 0.739375099 | 7.51E-09 | 1.48E-08 | 2.95E-05 |
| T1DM | leukocyte | *SOX7* | -0.933509105 | 1.48E-09 | 8.14E-09 | 5.81E-06 |
| T1DM | leukocyte | *TRMT61B* | -0.585563048 | 8.67E-09 | 1.65E-08 | 3.41E-05 |
| T1DM | leukocyte | *INSR* | -0.492587079 | 2.71E-09 | 8.26E-09 | 1.06E-05 |
| T1DM | leukocyte | *MFN1* | -0.714173758 | 5.73E-09 | 1.20E-08 | 2.25E-05 |
| T1DM | leukocyte | *KCNMB3* | -1.247697692 | 2.35E-10 | 8.14E-09 | 9.24E-07 |
| T1DM | leukocyte | *ATP6V0E2* | -0.537928814 | 2.90E-08 | 4.74E-08 | 0.00011375 |
| T1DM | leukocyte | *JAGN1* | -0.736008556 | 2.22E-09 | 8.21E-09 | 8.73E-06 |
| T1DM | leukocyte | *MORN4* | -0.557626756 | 4.84E-11 | 8.14E-09 | 1.90E-07 |
| T1DM | leukocyte | *TRIM8* | -0.757618912 | 1.90E-09 | 8.14E-09 | 7.47E-06 |
| T1DM | leukocyte | *CDC42BPG* | -0.504663656 | 3.70E-09 | 9.29E-09 | 1.46E-05 |
| T1DM | leukocyte | *ZNF440* | -2.001839024 | 1.73E-08 | 2.98E-08 | 6.78E-05 |
| T1DM | leukocyte | *GAA* | -0.505976087 | 0.000354181 | 0.000413667 | 1 |
| T1DM | leukocyte | *EXOSC1* | -0.864383037 | 3.15E-09 | 8.69E-09 | 1.24E-05 |
| T1DM | leukocyte | *PGAM1* | -0.553547296 | 3.02E-08 | 4.94E-08 | 0.00011884 |
| T1DM | leukocyte | *ESCO2* | -2.359286127 | 2.62E-07 | 3.69E-07 | 0.001031068 |
| T1DM | leukocyte | *MRPL36* | -0.67562254 | 2.14E-09 | 8.19E-09 | 8.43E-06 |
| T1DM | leukocyte | *MCC* | -0.843156656 | 2.07E-08 | 3.49E-08 | 8.12E-05 |
| T1DM | leukocyte | *ZNF561* | -0.834568495 | 4.82E-10 | 8.14E-09 | 1.89E-06 |
| T1DM | leukocyte | *PPID* | -0.785508994 | 4.19E-06 | 5.33E-06 | 0.016468726 |
| T1DM | leukocyte | *RXFP1* | -11.34765297 | 6.79E-05 | 8.19E-05 | 0.266875675 |
| T1DM | leukocyte | *PTGER4* | 0.483787195 | 1.26E-08 | 2.28E-08 | 4.95E-05 |
| T1DM | leukocyte | *MAP6* | -0.482619959 | 9.78E-09 | 1.83E-08 | 3.84E-05 |
| T1DM | leukocyte | *ZNF584* | -0.709354483 | 0.000154925 | 0.000183842 | 0.608701012 |
| T1DM | leukocyte | *NMUR1* | -7.76197343 | 1.25E-09 | 8.14E-09 | 4.90E-06 |
| T1DM | leukocyte | *CLSTN1* | -0.61408651 | 9.41E-11 | 8.14E-09 | 3.70E-07 |
| T1DM | leukocyte | *PIK3CD* | 0.690353044 | 1.71E-08 | 2.95E-08 | 6.70E-05 |
| T1DM | leukocyte | *ENC1* | -0.578892953 | 1.62E-07 | 2.33E-07 | 0.000638018 |
| T1DM | leukocyte | *RGS19* | -1.039160853 | 1.33E-08 | 2.39E-08 | 5.22E-05 |
| T1DM | leukocyte | *ANO5* | -0.876461076 | 1.91E-09 | 8.14E-09 | 7.51E-06 |
| T1DM | leukocyte | *GPHN* | -1.062459968 | 3.72E-08 | 5.96E-08 | 0.000146063 |
| T1DM | leukocyte | *VAT1L* | -0.868673176 | 5.23E-09 | 1.13E-08 | 2.06E-05 |

| | | | | | | |
|---|---|---|---|---|---|---|
| T1DM | leukocyte | *LGALS4* | -0.398435253 | 1.60E-09 | 8.14E-09 | 6.27E-06 |
| T1DM | leukocyte | *LRRC34* | -0.857803664 | 3.42E-09 | 8.93E-09 | 1.34E-05 |
| T1DM | leukocyte | *SPATA5L1* | -1.291268643 | 2.51E-09 | 8.21E-09 | 9.88E-06 |
| T1DM | leukocyte | *CTPS1* | -0.689888869 | 3.61E-10 | 8.14E-09 | 1.42E-06 |
| T1DM | leukocyte | *METTL18* | -0.897050696 | 2.77E-09 | 8.34E-09 | 1.09E-05 |
| T1DM | leukocyte | *PWWP2B* | -0.855137219 | 1.26E-09 | 8.14E-09 | 4.96E-06 |
| T1DM | leukocyte | *PCDHB1* | 0.990678601 | 0.015787227 | 0.017399162 | 1 |
| T1DM | leukocyte | *FAM90A1* | -1.327110692 | 0.001856462 | 0.002114827 | 1 |
| T1DM | leukocyte | *PTEN* | -0.79915139 | 4.35E-10 | 8.14E-09 | 1.71E-06 |
| T1DM | leukocyte | *CYP4F11* | -1.672965796 | 7.08E-08 | 1.08E-07 | 0.00027829 |
| T1DM | leukocyte | *TLN2* | -1.398666915 | 1.36E-09 | 8.14E-09 | 5.34E-06 |
| T1DM | leukocyte | *TVP23B* | -1.008981936 | 1.16E-07 | 1.71E-07 | 0.000457128 |
| T1DM | leukocyte | *SRGAP2C* | -1.290148068 | 2.52E-09 | 8.21E-09 | 9.88E-06 |
| T1DM | leukocyte | *ATPAF2* | -0.843503905 | 8.95E-09 | 1.70E-08 | 3.52E-05 |
| T1DM | leukocyte | *ZNF556* | 0.600651235 | 2.59E-07 | 3.65E-07 | 0.001018517 |
| T1DM | leukocyte | *ZNF554* | -0.790954525 | 1.40E-09 | 8.14E-09 | 5.49E-06 |
| T1DM | leukocyte | *GAP43* | -0.560410475 | 1.49E-09 | 8.14E-09 | 5.86E-06 |
| T1DM | leukocyte | *REG1B* | -0.32925581 | 6.24E-10 | 8.14E-09 | 2.45E-06 |
| T1DM | leukocyte | *NME6* | -1.074218374 | 6.76E-08 | 1.03E-07 | 0.000265521 |
| T1DM | leukocyte | *SLFN12* | 0.758943634 | 1.33E-08 | 2.39E-08 | 5.23E-05 |
| T1DM | leukocyte | *SLC9C1* | -2.230276823 | 0.00266198 | 0.003015836 | 1 |
| T1DM | leukocyte | *TEFM* | -0.897970333 | 1.83E-09 | 8.14E-09 | 7.20E-06 |
| T1DM | leukocyte | *MBOAT1* | -1.160841082 | 1.48E-06 | 1.94E-06 | 0.005800816 |
| T1DM | leukocyte | *ZNF131* | -0.711825047 | 1.52E-09 | 8.14E-09 | 5.99E-06 |
| T1DM | leukocyte | *BSG* | -0.568012018 | 3.07E-09 | 8.60E-09 | 1.21E-05 |
| T1DM | leukocyte | *HINFP* | -1.756323128 | 5.53E-08 | 8.59E-08 | 0.000217163 |
| T1DM | leukocyte | *CERS6* | -0.660160656 | 1.45E-09 | 8.14E-09 | 5.71E-06 |
| T1DM | leukocyte | *SPTLC3* | 0.898714027 | 1.38E-08 | 2.45E-08 | 5.41E-05 |
| T1DM | leukocyte | *COPRS* | -0.470179237 | 3.81E-08 | 6.10E-08 | 0.000149782 |
| T1DM | leukocyte | *B3GALT1* | -1.131862076 | 0.005782391 | 0.006474498 | 1 |
| T1DM | leukocyte | *SUCLG2* | -1.123946559 | 6.82E-09 | 1.37E-08 | 2.68E-05 |
| T1DM | leukocyte | *CSDC2* | -1.60854965 | 2.34E-06 | 3.02E-06 | 0.009178155 |
| T1DM | leukocyte | *ARNT2* | -0.570303351 | 1.55E-08 | 2.72E-08 | 6.11E-05 |
| T1DM | leukocyte | *FUT9* | -2.027081283 | 0.000203084 | 0.000239831 | 0.797916492 |
| T1DM | leukocyte | *ZNF24* | -0.73247111 | 5.28E-10 | 8.14E-09 | 2.07E-06 |
| T1DM | leukocyte | *MANEA* | -0.774504381 | 4.54E-09 | 1.04E-08 | 1.78E-05 |
| T1DM | leukocyte | *AFF1* | -0.761706805 | 9.95E-10 | 8.14E-09 | 3.91E-06 |
| T1DM | leukocyte | *PDE3A* | -0.576581993 | 0.000897292 | 0.001035074 | 1 |
| T1DM | leukocyte | *SMPDL3A* | -0.847146548 | 0.005538362 | 0.006208338 | 1 |
| T1DM | leukocyte | *RAD9A* | -1.150689014 | 3.10E-08 | 5.05E-08 | 0.000121967 |
| T1DM | leukocyte | *EFEMP2* | -0.764641964 | 6.41E-10 | 8.14E-09 | 2.52E-06 |
| T1DM | leukocyte | *AGAP5* | 3.246831587 | 2.17E-06 | 2.81E-06 | 0.008525882 |
| T1DM | leukocyte | *ZNF738* | -0.803698492 | 1.12E-07 | 1.65E-07 | 0.000439045 |
| T1DM | leukocyte | *NAA16* | -0.690067569 | 2.32E-09 | 8.21E-09 | 9.11E-06 |
| T1DM | leukocyte | *RAB43* | 0.617674825 | 0.02368405 | 0.025913293 | 1 |
| T1DM | leukocyte | *SNX32* | -0.985425099 | 0.004235826 | 0.004772745 | 1 |
| T1DM | leukocyte | *OVOL1* | -0.695334887 | 0.023865377 | 0.026104417 | 1 |
| T1DM | leukocyte | *DMXL1* | -0.995697493 | 1.85E-08 | 3.17E-08 | 7.25E-05 |
| T1DM | leukocyte | *NADSYN1* | -1.083623089 | 3.21E-07 | 4.47E-07 | 0.001262474 |
| T1DM | leukocyte | *RNASEH2C* | -0.67718411 | 1.11E-08 | 2.03E-08 | 4.36E-05 |

| | | | | | | |
|---|---|---|---|---|---|---|
| T1DM | leukocyte | *LCLAT1* | -0.719452245 | 8.02E-10 | 8.14E-09 | 3.15E-06 |
| T1DM | leukocyte | *SH3RF3* | -1.172219171 | 1.82E-10 | 8.14E-09 | 7.13E-07 |
| T1DM | leukocyte | *DCAKD* | -0.528189285 | 1.13E-08 | 2.06E-08 | 4.43E-05 |
| T1DM | leukocyte | *TADA2B* | -0.789399874 | 2.40E-09 | 8.21E-09 | 9.42E-06 |
| T1DM | leukocyte | *EVC2* | -1.349114615 | 0.000475066 | 0.000552393 | 1 |
| T1DM | leukocyte | *TRMT112* | -0.587820695 | 3.35E-09 | 8.83E-09 | 1.32E-05 |
| T1DM | leukocyte | *NOC3L* | -0.701131673 | 1.33E-09 | 8.14E-09 | 5.22E-06 |
| T1DM | leukocyte | *RHOD* | -0.494518417 | 5.27E-10 | 8.14E-09 | 2.07E-06 |
| T1DM | leukocyte | *PARP14* | -0.679190587 | 1.34E-08 | 2.40E-08 | 5.26E-05 |
| T1DM | leukocyte | *VANGL1* | -0.723996941 | 9.46E-10 | 8.14E-09 | 3.72E-06 |
| T1DM | leukocyte | *IQCB1* | -1.441001698 | 4.09E-08 | 6.50E-08 | 0.00016078 |
| T1DM | leukocyte | *ZNF483* | -0.570238967 | 4.49E-09 | 1.03E-08 | 1.76E-05 |
| T1DM | leukocyte | *SLC2A14* | -2.810553878 | 1.95E-09 | 8.14E-09 | 7.65E-06 |
| T1DM | leukocyte | *MZT2A* | -0.495112628 | 6.34E-08 | 9.75E-08 | 0.000249255 |
| T1DM | leukocyte | *PPP1R3B* | -1.378559534 | 1.85E-07 | 2.65E-07 | 0.000727837 |
| T1DM | leukocyte | *MAP3K11* | -0.654164333 | 1.16E-09 | 8.14E-09 | 4.57E-06 |
| T1DM | leukocyte | *NDNF* | 0.80067785 | 3.89E-09 | 9.52E-09 | 1.53E-05 |
| T1DM | leukocyte | *ARV1* | -0.636110881 | 2.00E-09 | 8.14E-09 | 7.85E-06 |
| T1DM | leukocyte | *EHBP1L1* | -0.677766187 | 5.40E-08 | 8.41E-08 | 0.000212018 |
| T1DM | leukocyte | *PPP1R14B* | -0.540073468 | 6.78E-09 | 1.37E-08 | 2.66E-05 |
| T1DM | leukocyte | *ZNF417* | 0.53012987 | 8.28E-09 | 1.60E-08 | 3.25E-05 |
| T1DM | leukocyte | *PEAK1* | -0.536617321 | 1.16E-07 | 1.70E-07 | 0.000454404 |
| T1DM | leukocyte | *TNFRSF10D* | -0.859537904 | 2.10E-09 | 8.16E-09 | 8.23E-06 |
| T1DM | leukocyte | *GMPPB* | -1.0119745 | 7.84E-09 | 1.53E-08 | 3.08E-05 |
| T1DM | leukocyte | *NUDT18* | -0.722448247 | 1.45E-09 | 8.14E-09 | 5.69E-06 |
| T1DM | leukocyte | *NUDT4* | -0.62131381 | 8.33E-09 | 1.61E-08 | 3.27E-05 |
| T1DM | leukocyte | *PC* | -1.084589343 | 2.64E-09 | 8.21E-09 | 1.04E-05 |
| T1DM | leukocyte | *LRFN4* | 1.402505848 | 1.97E-05 | 2.43E-05 | 0.077542188 |
| T1DM | leukocyte | *UQCRH* | -0.573668738 | 1.32E-09 | 8.14E-09 | 5.20E-06 |
| T1DM | leukocyte | *HEG1* | -0.708180492 | 1.26E-09 | 8.14E-09 | 4.97E-06 |
| T1DM | leukocyte | *HAP1* | -0.865615007 | 2.36E-07 | 3.34E-07 | 0.000927065 |
| T1DM | leukocyte | *ENDOV* | -1.233259343 | 7.39E-09 | 1.46E-08 | 2.90E-05 |
| T1DM | leukocyte | *DPY19L1* | -0.849306362 | 1.03E-09 | 8.14E-09 | 4.04E-06 |
| T1DM | leukocyte | *TUBB8* | -12.79308011 | 0.002758072 | 0.003123801 | 1 |
| T1DM | leukocyte | *GPR160* | -0.993342065 | 8.28E-09 | 1.60E-08 | 3.25E-05 |
| T1DM | leukocyte | *XXYLT1* | -1.052349992 | 1.47E-08 | 2.60E-08 | 5.77E-05 |
| T1DM | leukocyte | *GNG5* | -0.573031916 | 2.87E-09 | 8.44E-09 | 1.13E-05 |
| T1DM | leukocyte | *C9orf131* | -1.386089586 | 6.17E-08 | 9.51E-08 | 0.000242416 |
| T1DM | leukocyte | *FAM53A* | -2.592278997 | 2.12E-06 | 2.75E-06 | 0.008330439 |
| T1DM | leukocyte | *SELP* | -4.286617657 | 1.30E-09 | 8.14E-09 | 5.12E-06 |
| T1DM | leukocyte | *CTU2* | -0.658595993 | 7.19E-09 | 1.43E-08 | 2.83E-05 |
| T1DM | leukocyte | *SNX31* | -3.603707568 | 0.000506145 | 0.000587834 | 1 |
| T1DM | leukocyte | *ADCY6* | -0.790447093 | 8.81E-10 | 8.14E-09 | 3.46E-06 |
| T1DM | leukocyte | *PODN* | -0.684368165 | 9.70E-08 | 1.44E-07 | 0.000381017 |
| T1DM | leukocyte | *SLC6A19* | 0.629354223 | 0.025848339 | 0.028226272 | 1 |
| T1DM | leukocyte | *RALGAPA1* | -0.630245875 | 3.14E-09 | 8.69E-09 | 1.23E-05 |
| T1DM | leukocyte | *CNTNAP2* | 1.508055202 | 6.84E-09 | 1.38E-08 | 2.69E-05 |
| T1DM | leukocyte | *GCSAM* | -2.377815436 | 0.045163453 | 0.048883528 | 1 |
| T1DM | leukocyte | *GOLT1A* | -0.667972163 | 2.47E-09 | 8.21E-09 | 9.70E-06 |
| T1DM | leukocyte | *NPAS4* | -1.310787295 | 4.57E-08 | 7.21E-08 | 0.000179596 |

| | | | | | | |
|---|---|---|---|---|---|---|
| T1DM | leukocyte | *ANGEL2* | -0.903447529 | 2.64E-09 | 8.21E-09 | 1.04E-05 |
| T1DM | leukocyte | *ZNF266* | -0.461389353 | 7.75E-07 | 1.04E-06 | 0.003046597 |
| T1DM | leukocyte | *BRSK2* | -0.658692389 | 1.38E-09 | 8.14E-09 | 5.43E-06 |
| T1DM | leukocyte | *BRMS1* | -0.626924627 | 1.94E-09 | 8.14E-09 | 7.63E-06 |
| T1DM | leukocyte | *HRAS* | -0.622118226 | 1.69E-09 | 8.14E-09 | 6.63E-06 |
| T1DM | leukocyte | *SRP72* | -0.520086171 | 2.15E-09 | 8.19E-09 | 8.43E-06 |
| T1DM | leukocyte | *PCP2* | -1.96939054 | 1.60E-05 | 1.98E-05 | 0.062925806 |
| T1DM | leukocyte | *BTC* | -1.044408645 | 8.13E-09 | 1.57E-08 | 3.20E-05 |
| T1DM | leukocyte | *DENND6A* | -0.668998159 | 9.40E-10 | 8.14E-09 | 3.69E-06 |
| T1DM | leukocyte | *YIF1A* | -0.53723309 | 6.85E-08 | 1.04E-07 | 0.000269116 |
| T1DM | leukocyte | *NDUFA11* | -0.593632121 | 3.88E-09 | 9.51E-09 | 1.52E-05 |
| T1DM | leukocyte | *RSRC1* | -1.087379267 | 3.71E-07 | 5.13E-07 | 0.001459606 |
| T1DM | leukocyte | *FUT1* | -1.223450591 | 1.44E-09 | 8.14E-09 | 5.66E-06 |
| T1DM | leukocyte | *DHX36* | -0.670110715 | 2.35E-09 | 8.21E-09 | 9.24E-06 |
| T1DM | leukocyte | *PSMD2* | -0.554991858 | 3.35E-09 | 8.83E-09 | 1.32E-05 |
| T1DM | leukocyte | *FAM182B* | -0.608807902 | 0.040936851 | 0.0443699 | 1 |
| T1DM | leukocyte | *PPM1E* | -0.567127017 | 5.14E-10 | 8.14E-09 | 2.02E-06 |
| T1DM | leukocyte | *ZNF408* | -0.899067093 | 0.008692758 | 0.009672571 | 1 |
| T1DM | leukocyte | *ARHGAP1* | -0.625261441 | 1.20E-08 | 2.18E-08 | 4.71E-05 |
| T1DM | leukocyte | *MED16* | -0.632732209 | 2.41E-09 | 8.21E-09 | 9.49E-06 |
| T1DM | leukocyte | *GAL3ST3* | 24.33800015 | 2.30E-07 | 3.26E-07 | 0.000904726 |
| T1DM | leukocyte | *CHST1* | -1.419265816 | 1.45E-09 | 8.14E-09 | 5.71E-06 |
| T1DM | leukocyte | *DOLK* | -1.171882198 | 7.05E-08 | 1.07E-07 | 0.000276882 |
| T1DM | leukocyte | *PHYHD1* | -0.377691429 | 0.000346944 | 0.000405336 | 1 |
| T1DM | leukocyte | *CCNE2* | -0.745227248 | 3.39E-07 | 4.70E-07 | 0.001331371 |
| T1DM | leukocyte | *ZNF519* | -1.315055264 | 3.40E-10 | 8.14E-09 | 1.34E-06 |
| T1DM | leukocyte | *CHRNA7* | -1.140754055 | 5.43E-08 | 8.45E-08 | 0.000213333 |
| T1DM | leukocyte | *TMEM9B* | -0.637873421 | 5.19E-09 | 1.13E-08 | 2.04E-05 |
| T1DM | leukocyte | *SCUBE2* | -0.372341225 | 2.57E-05 | 3.16E-05 | 0.101042448 |
| T1DM | leukocyte | *EIF3F* | -0.631204476 | 8.00E-09 | 1.56E-08 | 3.14E-05 |
| T1DM | leukocyte | *ZNF25* | -0.649268823 | 2.79E-09 | 8.35E-09 | 1.10E-05 |
| T1DM | leukocyte | *PPP2R2D* | -0.640625544 | 1.39E-09 | 8.14E-09 | 5.48E-06 |
| T1DM | leukocyte | *CLCF1* | -0.983503738 | 1.61E-09 | 8.14E-09 | 6.31E-06 |
| T1DM | leukocyte | *UCP2* | 0.607246114 | 1.56E-08 | 2.74E-08 | 6.14E-05 |
| T1DM | leukocyte | *PAAF1* | -0.440755935 | 0.006667553 | 0.007452864 | 1 |
| T1DM | leukocyte | *MRPL48* | -0.840197536 | 3.77E-09 | 9.36E-09 | 1.48E-05 |
| T1DM | leukocyte | *TMEM70* | -0.634647162 | 3.07E-09 | 8.60E-09 | 1.21E-05 |
| T1DM | leukocyte | *RPS6KB2* | -0.949389196 | 3.33E-09 | 8.82E-09 | 1.31E-05 |
| T1DM | leukocyte | *RMI2* | -2.166222728 | 3.51E-08 | 5.67E-08 | 0.000137868 |
| T1DM | leukocyte | *SFN* | -1.035184309 | 6.32E-05 | 7.64E-05 | 0.248484115 |
| T1DM | leukocyte | *SWI5* | -0.817809421 | 6.28E-07 | 8.48E-07 | 0.002467485 |
| T1DM | leukocyte | *ZDHHC21* | -1.080639887 | 8.67E-08 | 1.30E-07 | 0.000340586 |
| T1DM | leukocyte | *LRRN1* | -0.477755678 | 0.007648054 | 0.008536706 | 1 |
| T1DM | leukocyte | *TUBB6* | -0.367486946 | 3.43E-09 | 8.93E-09 | 1.35E-05 |
| T1DM | leukocyte | *CHMP6* | -0.694987979 | 4.15E-09 | 9.91E-09 | 1.63E-05 |
| T1DM | leukocyte | *DLEU1* | -0.790750147 | 1.21E-08 | 2.20E-08 | 4.76E-05 |
| T1DM | leukocyte | *TCP11L1* | 0.965026962 | 2.34E-05 | 2.87E-05 | 0.091860748 |
| T1DM | leukocyte | *CCDC57* | -0.76263322 | 7.05E-08 | 1.07E-07 | 0.000277128 |
| T1DM | leukocyte | *BNIP3* | -0.505426054 | 1.31E-09 | 8.14E-09 | 5.14E-06 |
| T1DM | leukocyte | *SMIM19* | -0.561630508 | 4.60E-09 | 1.04E-08 | 1.81E-05 |

| | | | | | | |
|---|---|---|---|---|---|---|
| T1DM | leukocyte | *SLC35G1* | -0.970975636 | 1.44E-09 | 8.14E-09 | 5.65E-06 |
| T1DM | leukocyte | *COX8A* | -0.469191012 | 3.99E-09 | 9.64E-09 | 1.57E-05 |
| T1DM | leukocyte | *CDC26* | -0.707626759 | 1.89E-09 | 8.14E-09 | 7.41E-06 |
| T1DM | leukocyte | *RNPEP* | -1.106364105 | 2.04E-08 | 3.44E-08 | 8.00E-05 |
| T1DM | leukocyte | *DMRTA1* | -0.674280912 | 3.83E-10 | 8.14E-09 | 1.51E-06 |
| T1DM | leukocyte | *CLEC14A* | -0.44412676 | 1.50E-09 | 8.14E-09 | 5.88E-06 |
| T1DM | leukocyte | *WDR25* | -1.449681932 | 4.35E-08 | 6.89E-08 | 0.000170917 |
| T1DM | leukocyte | *DIRAS1* | -0.832568618 | 2.33E-08 | 3.90E-08 | 9.15E-05 |
| T1DM | leukocyte | *PHLDB3* | -0.606836231 | 6.77E-09 | 1.37E-08 | 2.66E-05 |
| T1DM | leukocyte | *PRR15* | 1.080321791 | 9.41E-09 | 1.78E-08 | 3.70E-05 |
| T1DM | leukocyte | *KBTBD11* | -0.614094237 | 2.05E-09 | 8.14E-09 | 8.07E-06 |
| T1DM | leukocyte | *RMDN1* | -0.754627152 | 2.51E-08 | 4.17E-08 | 9.88E-05 |
| T1DM | leukocyte | *LRRC37A* | -3.755328202 | 1.48E-09 | 8.14E-09 | 5.83E-06 |
| T1DM | leukocyte | *ACSF3* | -0.918029423 | 1.43E-09 | 8.14E-09 | 5.63E-06 |
| T1DM | leukocyte | *PFN4* | 1.294524206 | 0.000796152 | 0.000919213 | 1 |
| T1DM | leukocyte | *RUFY1* | -1.396832306 | 6.46E-08 | 9.91E-08 | 0.000253791 |
| T1DM | leukocyte | *VSIG10* | -0.890108912 | 1.12E-07 | 1.65E-07 | 0.000441939 |
| T1DM | leukocyte | *WSB2* | -0.573325102 | 1.72E-08 | 2.98E-08 | 6.78E-05 |
| T1DM | leukocyte | *ANKLE2* | -0.525980131 | 3.50E-08 | 5.66E-08 | 0.000137634 |
| T1DM | leukocyte | *FUT2* | -0.21573348 | 0.004884006 | 0.005487349 | 1 |
| T1DM | leukocyte | *MUC20* | -0.715349096 | 2.31E-09 | 8.21E-09 | 9.07E-06 |
| T1DM | leukocyte | *NFATC2IP* | -0.957603721 | 3.74E-09 | 9.32E-09 | 1.47E-05 |
| T1DM | leukocyte | *DPP7* | -0.607403776 | 3.22E-09 | 8.74E-09 | 1.26E-05 |
| T1DM | leukocyte | *MTHFR* | -0.753136937 | 7.19E-10 | 8.14E-09 | 2.82E-06 |
| T1DM | leukocyte | *DEAF1* | -0.866810797 | 3.14E-09 | 8.69E-09 | 1.24E-05 |
| T1DM | leukocyte | *TMEM80* | -0.592743657 | 7.08E-10 | 8.14E-09 | 2.78E-06 |
| T1DM | leukocyte | *DSCAML1* | -0.855253313 | 9.86E-09 | 1.84E-08 | 3.87E-05 |
| T1DM | leukocyte | *PUS1* | -1.364684572 | 7.92E-09 | 1.55E-08 | 3.11E-05 |
| T1DM | leukocyte | *MAN1B1* | -1.073225896 | 8.05E-09 | 1.56E-08 | 3.16E-05 |
| T1DM | leukocyte | *KCNA2* | -2.66718292 | 0.000235968 | 0.000277747 | 0.927118888 |
| T1DM | leukocyte | *PPFIA3* | -0.573913873 | 1.07E-09 | 8.14E-09 | 4.20E-06 |
| T1DM | leukocyte | *UBE2U* | -3.373557583 | 9.96E-09 | 1.85E-08 | 3.91E-05 |
| T1DM | leukocyte | *PAWR* | -0.651535133 | 1.31E-09 | 8.14E-09 | 5.14E-06 |
| T1DM | leukocyte | *ACOT4* | -0.906380632 | 6.77E-09 | 1.37E-08 | 2.66E-05 |
| T1DM | leukocyte | *ARIH2* | -0.583247074 | 3.93E-08 | 6.27E-08 | 0.000154216 |
| T1DM | leukocyte | *RBM44* | 3.156918282 | 1.68E-06 | 2.19E-06 | 0.006594077 |
| T1DM | leukocyte | *IRX3* | -0.830163696 | 0.001264533 | 0.001449344 | 1 |
| T1DM | leukocyte | *SAMD12* | -0.832517145 | 1.82E-07 | 2.60E-07 | 0.000713753 |
| T1DM | leukocyte | *CD163* | 0.074771034 | 2.27E-09 | 8.21E-09 | 8.92E-06 |
| T1DM | leukocyte | *ACAD9* | -0.642508048 | 7.54E-06 | 9.46E-06 | 0.029622156 |
| T1DM | leukocyte | *IL17RA* | -0.730011552 | 0.001182944 | 0.001357415 | 1 |
| T1DM | leukocyte | *AGTRAP* | -0.733614478 | 2.59E-09 | 8.21E-09 | 1.02E-05 |
| T1DM | leukocyte | *THAP5* | -0.692580041 | 2.18E-09 | 8.21E-09 | 8.56E-06 |
| T1DM | leukocyte | *CD151* | -0.549370006 | 1.58E-09 | 8.14E-09 | 6.21E-06 |
| T1DM | leukocyte | *CHID1* | -0.551988032 | 5.29E-09 | 1.14E-08 | 2.08E-05 |
| T1DM | leukocyte | *ZBTB41* | -0.851291396 | 2.98E-09 | 8.50E-09 | 1.17E-05 |
| T1DM | leukocyte | *BET1L* | -0.932469077 | 9.25E-09 | 1.75E-08 | 3.63E-05 |
| T1DM | leukocyte | *RIC8A* | -0.855173953 | 3.81E-09 | 9.43E-09 | 1.50E-05 |
| T1DM | leukocyte | *IMP3* | -0.721292338 | 5.70E-09 | 1.20E-08 | 2.24E-05 |
| T1DM | leukocyte | *DPY19L2* | -0.59345691 | 1.37E-06 | 1.80E-06 | 0.005364474 |

| | | | | | | |
|---|---|---|---|---|---|---|
| T1DM | leukocyte | *GPR150* | -0.634472188 | 5.89E-07 | 7.96E-07 | 0.002313535 |
| T1DM | leukocyte | *ADAMTSL1* | -2.344221238 | 7.69E-05 | 9.24E-05 | 0.302301797 |
| T1DM | leukocyte | *ALS2CL* | -0.977716919 | 2.07E-09 | 8.14E-09 | 8.12E-06 |
| T1DM | leukocyte | *STAP2* | -0.553205679 | 3.87E-10 | 8.14E-09 | 1.52E-06 |
| T1DM | leukocyte | *DALRD3* | -0.696963633 | 7.30E-08 | 1.11E-07 | 0.000286912 |
| T1DM | leukocyte | *ZNF114* | -1.890274542 | 2.30E-09 | 8.21E-09 | 9.03E-06 |
| T1DM | leukocyte | *PLEC* | -1.0977152 | 1.62E-08 | 2.83E-08 | 6.37E-05 |
| T1DM | leukocyte | *WDR6* | -0.724924584 | 1.15E-09 | 8.14E-09 | 4.51E-06 |
| T1DM | leukocyte | *AQP11* | -0.502517447 | 0.000749763 | 0.000866673 | 1 |
| T1DM | leukocyte | *ZFAND2A* | -0.820985369 | 8.13E-09 | 1.57E-08 | 3.19E-05 |
| T1DM | leukocyte | *DNAJC22* | -0.288460124 | 0.000292202 | 0.000342603 | 1 |
| T1DM | leukocyte | *NT5DC1* | -0.650034421 | 3.71E-09 | 9.29E-09 | 1.46E-05 |
| T1DM | leukocyte | *COX14* | -0.648515859 | 1.69E-08 | 2.94E-08 | 6.66E-05 |
| T1DM | leukocyte | *P4HTM* | -0.479149414 | 7.25E-09 | 1.44E-08 | 2.85E-05 |
| T1DM | leukocyte | *ERBB4* | -5.240389496 | 3.10E-08 | 5.05E-08 | 0.00012184 |
| T1DM | leukocyte | *ERN1* | -0.815511239 | 2.50E-08 | 4.15E-08 | 9.82E-05 |
| T1DM | leukocyte | *GPR35* | 0.615219076 | 0.012453283 | 0.013775042 | 1 |
| T1DM | leukocyte | *PARP10* | -0.827767999 | 1.77E-08 | 3.06E-08 | 6.97E-05 |
| T1DM | leukocyte | *RPP25* | -1.055324878 | 3.34E-09 | 8.82E-09 | 1.31E-05 |
| T1DM | leukocyte | *ZHX2* | -0.784123203 | 8.90E-10 | 8.14E-09 | 3.50E-06 |
| T1DM | leukocyte | *TRIM73* | 0.961854668 | 9.98E-06 | 1.25E-05 | 0.039203325 |
| T1DM | leukocyte | *EFCAB13* | -0.360096781 | 0.010599358 | 0.011750812 | 1 |
| T1DM | leukocyte | *MSC* | 0.767782305 | 2.20E-05 | 2.70E-05 | 0.086247876 |
| T1DM | leukocyte | *HYI* | -0.7751287 | 3.87E-09 | 9.51E-09 | 1.52E-05 |
| T1DM | leukocyte | *GAK* | -0.784482872 | 4.70E-09 | 1.06E-08 | 1.84E-05 |
| T1DM | leukocyte | *TUFM* | -0.50519858 | 4.02E-09 | 9.69E-09 | 1.58E-05 |
| T1DM | leukocyte | *RMI1* | 0.74072301 | 0.000163302 | 0.000193548 | 0.6416118 |
| T1DM | leukocyte | *C3orf38* | -0.935367327 | 7.33E-08 | 1.11E-07 | 0.000288079 |
| T1DM | leukocyte | *KLHDC7A* | 1.072429554 | 5.94E-07 | 8.03E-07 | 0.002333029 |
| T1DM | leukocyte | *IGSF22* | -0.540452053 | 1.51E-09 | 8.14E-09 | 5.93E-06 |
| T1DM | leukocyte | *HTR1F* | -1.848151406 | 6.72E-09 | 1.36E-08 | 2.64E-05 |
| T1DM | leukocyte | *TMTC2* | -1.351268197 | 1.42E-10 | 8.14E-09 | 5.59E-07 |
| T1DM | leukocyte | *FARSA* | -0.723486535 | 4.73E-10 | 8.14E-09 | 1.86E-06 |
| T1DM | leukocyte | *SPTY2D1* | -0.696342556 | 3.86E-09 | 9.50E-09 | 1.52E-05 |
| T1DM | leukocyte | *SAMD4B* | -0.571609464 | 1.60E-08 | 2.80E-08 | 6.29E-05 |
| T1DM | leukocyte | *TCAIM* | -0.938526134 | 2.23E-09 | 8.21E-09 | 8.75E-06 |
| T1DM | leukocyte | *FUCA1* | 0.411585484 | 5.10E-08 | 7.98E-08 | 0.000200316 |
| T1DM | leukocyte | *CALR* | -0.491718943 | 1.25E-08 | 2.26E-08 | 4.90E-05 |
| T1DM | leukocyte | *CDH4* | -0.910645019 | 0.003468351 | 0.0039181 | 1 |
| T1DM | leukocyte | *SMCO3* | -15.50853889 | 3.62E-07 | 5.00E-07 | 0.0014216 |
| T1DM | leukocyte | *NSUN7* | -0.851195974 | 0.00013493 | 0.000160405 | 0.530139973 |
| T1DM | leukocyte | *HLA-DQB1* | 0.082700401 | 1.56E-05 | 1.93E-05 | 0.061228186 |
| T1DM | leukocyte | *ELMOD2* | -1.091453562 | 1.05E-08 | 1.93E-08 | 4.11E-05 |
| T1DM | leukocyte | *LINC00174* | -0.6893889 | 7.22E-10 | 8.14E-09 | 2.84E-06 |
| T1DM | leukocyte | *GEMIN4* | -0.720289916 | 1.57E-09 | 8.14E-09 | 6.19E-06 |
| T1DM | leukocyte | *LBX2* | 0.838261477 | 1.06E-09 | 8.14E-09 | 4.17E-06 |
| T1DM | leukocyte | *CDC42EP4* | -0.576076041 | 3.57E-09 | 9.10E-09 | 1.40E-05 |
| T1DM | leukocyte | *ZBTB42* | -1.544128279 | 5.56E-09 | 1.18E-08 | 2.18E-05 |
| T1DM | leukocyte | *SYCN* | -0.354662117 | 2.80E-08 | 4.60E-08 | 0.000110101 |
| T1DM | leukocyte | *ATOH7* | -0.426221027 | 0.008345461 | 0.009304573 | 1 |

| | | | | | | |
|---|---|---|---|---|---|---|
| T1DM | leukocyte | *CITED4* | -0.497293392 | 4.89E-10 | 8.14E-09 | 1.92E-06 |
| T1DM | leukocyte | *ABCA13* | -13.8842351 | 7.96E-08 | 1.20E-07 | 0.00031278 |
| T1DM | leukocyte | *ZNF154* | -0.61807159 | 7.69E-07 | 1.03E-06 | 0.003019591 |
| T1DM | leukocyte | *B3GNT3* | -1.508152706 | 5.71E-08 | 8.86E-08 | 0.000224438 |
| T1DM | leukocyte | *LINC00652* | -10.59420479 | 0.001035693 | 0.001191927 | 1 |
| T1DM | leukocyte | *PUF60* | -0.630338533 | 1.54E-09 | 8.14E-09 | 6.05E-06 |
| T1DM | leukocyte | *SSC5D* | 1.107877515 | 0.000274352 | 0.000322155 | 1 |
| T1DM | leukocyte | *TSHZ1* | -0.570770072 | 6.79E-09 | 1.37E-08 | 2.67E-05 |
| T1DM | leukocyte | *ANKRD18A* | -0.408417489 | 1.26E-05 | 1.56E-05 | 0.049340963 |
| T1DM | leukocyte | *EXOC3* | -0.858387811 | 9.14E-06 | 1.14E-05 | 0.035897352 |
| T1DM | leukocyte | *TH* | -3.170028182 | 2.83E-07 | 3.96E-07 | 0.001113246 |
| T1DM | leukocyte | *FAHD1* | -0.57809615 | 4.30E-07 | 5.91E-07 | 0.001690496 |
| T1DM | leukocyte | *ZNF816* | -1.027698138 | 9.34E-10 | 8.14E-09 | 3.67E-06 |
| T1DM | leukocyte | *CCDC66* | -1.349086329 | 2.65E-07 | 3.73E-07 | 0.001043099 |
| T1DM | leukocyte | *MCFD2* | -0.518313508 | 1.84E-09 | 8.14E-09 | 7.21E-06 |
| T1DM | leukocyte | *SERTM1* | 0.605145721 | 1.55E-08 | 2.72E-08 | 6.10E-05 |
| T1DM | leukocyte | *GLIPR1L2* | -1.049875251 | 8.33E-07 | 1.11E-06 | 0.003273277 |
| T1DM | leukocyte | *TSPYL5* | -0.522215965 | 5.30E-09 | 1.14E-08 | 2.08E-05 |
| T1DM | leukocyte | *SHISA2* | -0.973104384 | 1.39E-05 | 1.72E-05 | 0.054694721 |
| T1DM | leukocyte | *CHST13* | -58.75171988 | 1.38E-09 | 8.14E-09 | 5.41E-06 |
| T1DM | leukocyte | *PPA1* | -0.488018883 | 2.02E-09 | 8.14E-09 | 7.95E-06 |
| T1DM | leukocyte | *PSMG4* | -0.584972298 | 4.62E-10 | 8.14E-09 | 1.82E-06 |
| T1DM | leukocyte | *MAP6D1* | -1.329905443 | 3.39E-08 | 5.48E-08 | 0.000133005 |
| T1DM | leukocyte | *ZNF792* | -1.523925143 | 5.67E-09 | 1.19E-08 | 2.23E-05 |
| T1DM | leukocyte | *D2HGDH* | -1.870419055 | 7.44E-07 | 9.98E-07 | 0.002922171 |
| T1DM | leukocyte | *FAM83H* | -1.035245339 | 3.27E-09 | 8.79E-09 | 1.29E-05 |
| T1DM | leukocyte | *LRRC57* | -1.037650779 | 1.65E-08 | 2.86E-08 | 6.47E-05 |
| T1DM | leukocyte | *MRPL14* | -0.658541198 | 4.79E-09 | 1.07E-08 | 1.88E-05 |
| T1DM | leukocyte | *ZFP82* | -0.661833675 | 1.65E-09 | 8.14E-09 | 6.48E-06 |
| T1DM | leukocyte | *LSMEM1* | -0.680200155 | 0.00048699 | 0.00056609 | 1 |
| T1DM | leukocyte | *FKRP* | -0.684918346 | 2.02E-10 | 8.14E-09 | 7.93E-07 |
| T1DM | leukocyte | *RPH3AL* | -0.524884144 | 1.29E-09 | 8.14E-09 | 5.08E-06 |
| T1DM | leukocyte | *HIGD1A* | -0.532720802 | 1.54E-09 | 8.14E-09 | 6.05E-06 |
| T1DM | leukocyte | *F2R* | -0.628529172 | 3.86E-11 | 8.14E-09 | 1.52E-07 |
| T1DM | leukocyte | *ZNF707* | -0.888049193 | 1.38E-09 | 8.14E-09 | 5.44E-06 |
| T1DM | leukocyte | *SLC25A41* | -1.719019506 | 0.010417218 | 0.011552144 | 1 |
| T1DM | leukocyte | *OGFOD3* | -0.568522986 | 4.22E-09 | 9.96E-09 | 1.66E-05 |
| T1DM | leukocyte | *ZNF467* | -0.881931557 | 5.12E-09 | 1.12E-08 | 2.01E-05 |
| T1DM | leukocyte | *ZNF678* | -0.577690949 | 0.020369071 | 0.022329821 | 1 |
| T1DM | leukocyte | *TMEM45A* | -0.571222032 | 1.34E-09 | 8.14E-09 | 5.26E-06 |
| T1DM | leukocyte | *ACBD4* | 1.85384378 | 8.84E-07 | 1.18E-06 | 0.003472215 |
| T1DM | leukocyte | *SGSH* | -0.816236468 | 0.017203842 | 0.018939169 | 1 |
| T1DM | leukocyte | *GPR135* | -0.817519663 | 1.45E-09 | 8.14E-09 | 5.71E-06 |
| T1DM | leukocyte | *ANKRD62* | -0.824902595 | 1.24E-06 | 1.64E-06 | 0.00487992 |
| T1DM | leukocyte | *GPR88* | -5.938755547 | 0.004328806 | 0.004876112 | 1 |
| T1DM | leukocyte | *RFX7* | -1.503175493 | 3.18E-09 | 8.70E-09 | 1.25E-05 |
| T1DM | leukocyte | *ZNF329* | -0.855633759 | 2.84E-08 | 4.67E-08 | 0.000111689 |
| T1DM | leukocyte | *CCDC149* | -1.042342377 | 2.50E-09 | 8.21E-09 | 9.84E-06 |
| T1DM | leukocyte | *IDH2* | -0.536311326 | 2.12E-09 | 8.16E-09 | 8.32E-06 |
| T1DM | leukocyte | *TNRC18* | -0.822636389 | 2.95E-09 | 8.50E-09 | 1.16E-05 |

| | | | | | | |
|---|---|---|---|---|---|---|
| T1DM | leukocyte | *TDRKH* | -0.753431006 | 3.59E-09 | 9.11E-09 | 1.41E-05 |
| T1DM | leukocyte | *ZNF708* | -0.544717717 | 1.53E-09 | 8.14E-09 | 6.03E-06 |
| T1DM | leukocyte | *IST1* | -0.913090794 | 9.69E-09 | 1.81E-08 | 3.81E-05 |
| T1DM | leukocyte | *ERCC6L2* | -1.415661296 | 4.44E-06 | 5.64E-06 | 0.017456073 |
| T1DM | leukocyte | *MRPL41* | -0.60941996 | 3.16E-09 | 8.69E-09 | 1.24E-05 |
| T1DM | leukocyte | *TP53TG1* | -0.690204607 | 2.25E-09 | 8.21E-09 | 8.84E-06 |
| T1DM | leukocyte | *ARL6IP4* | -0.479853388 | 1.27E-08 | 2.29E-08 | 4.98E-05 |
| T1DM | leukocyte | *FAM153B* | -0.980025022 | 7.92E-09 | 1.55E-08 | 3.11E-05 |
| T1DM | leukocyte | *BACE2* | -0.421411951 | 7.96E-10 | 8.14E-09 | 3.13E-06 |
| T1DM | leukocyte | *SYNM* | -0.557260053 | 1.62E-06 | 2.12E-06 | 0.006354192 |
| T1DM | leukocyte | *FIGN* | -0.679610462 | 2.35E-09 | 8.21E-09 | 9.24E-06 |
| T1DM | leukocyte | *IZUMO1* | -2.058129123 | 0.038382992 | 0.041659331 | 1 |
| T1DM | leukocyte | *B4GALNT4* | -0.998553261 | 1.08E-05 | 1.35E-05 | 0.042524662 |
| T1DM | leukocyte | *ZNF804B* | -50.9666607 | 1.46E-09 | 8.14E-09 | 5.74E-06 |
| T1DM | leukocyte | *YBEY* | -0.515999995 | 0.000103369 | 0.000123484 | 0.406137371 |
| T1DM | leukocyte | *NPLOC4* | -0.681954049 | 2.61E-09 | 8.21E-09 | 1.02E-05 |
| T1DM | leukocyte | *CAPN12* | -1.234070078 | 5.12E-09 | 1.12E-08 | 2.01E-05 |
| T1DM | leukocyte | *GLRX5* | -0.523628126 | 1.98E-07 | 2.83E-07 | 0.000779881 |
| T1DM | leukocyte | *MXRA7* | -0.553698805 | 5.16E-09 | 1.12E-08 | 2.03E-05 |
| T1DM | leukocyte | *ADI1* | -1.217268568 | 2.80E-08 | 4.60E-08 | 0.000109997 |
| T1DM | leukocyte | *SATB1* | -0.973479559 | 3.24E-09 | 8.77E-09 | 1.27E-05 |
| T1DM | leukocyte | *EPHB3* | -1.441082421 | 1.71E-05 | 2.11E-05 | 0.066995366 |
| T1DM | leukocyte | *ACTL10* | 2.176655969 | 2.09E-05 | 2.57E-05 | 0.082119722 |
| T1DM | leukocyte | *SKA2* | -1.011616305 | 1.63E-08 | 2.84E-08 | 6.41E-05 |
| T1DM | leukocyte | *TTC3* | -0.392327144 | 2.46E-08 | 4.09E-08 | 9.66E-05 |
| T1DM | leukocyte | *BRICD5* | -1.13651574 | 2.73E-05 | 3.35E-05 | 0.107129737 |
| T1DM | leukocyte | *TSKU* | -0.775042881 | 1.10E-09 | 8.14E-09 | 4.34E-06 |
| T1DM | leukocyte | *ANXA2* | -0.402627092 | 1.08E-09 | 8.14E-09 | 4.26E-06 |
| T1DM | leukocyte | *SLC35D3* | -1.107809762 | 7.43E-08 | 1.13E-07 | 0.000292096 |
| T1DM | leukocyte | *GPIHBP1* | -6.34063553 | 1.51E-09 | 8.14E-09 | 5.93E-06 |
| T1DM | leukocyte | *VMO1* | 0.085625462 | 2.88E-09 | 8.46E-09 | 1.13E-05 |
| T1DM | leukocyte | *ALG12* | -1.078237486 | 1.48E-07 | 2.13E-07 | 0.000580843 |
| T1DM | leukocyte | *COL18A1* | -0.298960731 | 2.83E-07 | 3.96E-07 | 0.001112662 |
| T1DM | leukocyte | *C11orf54* | -0.860494952 | 5.16E-08 | 8.08E-08 | 0.000202852 |
| T1DM | leukocyte | *CEP63* | -0.806966932 | 4.08E-10 | 8.14E-09 | 1.60E-06 |
| T1DM | leukocyte | *HMGN4* | -0.794088329 | 7.98E-08 | 1.20E-07 | 0.000313669 |
| T1DM | leukocyte | *SPATA13* | -0.638372081 | 2.26E-09 | 8.21E-09 | 8.89E-06 |
| T1DM | leukocyte | *ZNF320* | 0.244607973 | 0.015239962 | 0.016814886 | 1 |
| T1DM | leukocyte | *SLC25A21* | -1.756076367 | 1.36E-09 | 8.14E-09 | 5.34E-06 |
| T1DM | leukocyte | *CAMK1D* | -1.321623506 | 6.14E-06 | 7.74E-06 | 0.024124546 |
| T1DM | leukocyte | *LYSMD4* | -0.763048284 | 1.59E-08 | 2.77E-08 | 6.23E-05 |
| T1DM | leukocyte | *BEGAIN* | -0.984384401 | 1.39E-09 | 8.14E-09 | 5.48E-06 |
| T1DM | leukocyte | *GPC6* | -0.774357202 | 4.77E-09 | 1.07E-08 | 1.88E-05 |
| T1DM | leukocyte | *FAM43B* | -3.113533839 | 0.000216961 | 0.000255911 | 0.85243981 |
| T1DM | leukocyte | *FANCF* | -1.713590854 | 1.91E-06 | 2.48E-06 | 0.007496668 |
| T1DM | leukocyte | *CALN1* | -2.707136037 | 4.61E-06 | 5.85E-06 | 0.018130366 |
| T1DM | leukocyte | *SMDT1* | -0.42744348 | 1.21E-07 | 1.76E-07 | 0.000474448 |
| T1DM | leukocyte | *POTEC* | -10.62557719 | 0.02280018 | 0.024967087 | 1 |
| T1DM | leukocyte | *PTTG1IP* | -0.503168411 | 3.04E-07 | 4.23E-07 | 0.001194564 |
| T1DM | leukocyte | *ZNF623* | -0.621345841 | 2.35E-09 | 8.21E-09 | 9.24E-06 |

| | | | | | | |
|---|---|---|---|---|---|---|
| T1DM | leukocyte | *CCDC125* | -1.204638178 | 5.78E-06 | 7.30E-06 | 0.022706929 |
| T1DM | leukocyte | *BOLA2* | -0.635312119 | 4.84E-09 | 1.08E-08 | 1.90E-05 |
| T1DM | leukocyte | *FHL3* | -0.663138977 | 3.67E-08 | 5.90E-08 | 0.000144226 |
| T1DM | leukocyte | *SF3A3* | -0.555643825 | 2.05E-08 | 3.47E-08 | 8.06E-05 |
| T1DM | leukocyte | *MX2* | 0.92707197 | 9.44E-08 | 1.41E-07 | 0.00037104 |
| T1DM | leukocyte | *PCBP3* | -0.867530282 | 1.12E-06 | 1.48E-06 | 0.004397173 |
| T1DM | leukocyte | *SFXN4* | -0.780496069 | 1.83E-10 | 8.14E-09 | 7.17E-07 |
| T1DM | leukocyte | *MRPL54* | -0.627029261 | 1.20E-09 | 8.14E-09 | 4.73E-06 |
| T1DM | leukocyte | *DGCR6* | -0.572105213 | 2.33E-06 | 3.01E-06 | 0.009164482 |
| T1DM | leukocyte | *NDUFB1* | -0.551201066 | 1.51E-07 | 2.17E-07 | 0.000592489 |
| T1DM | leukocyte | *TRMT12* | -0.773155395 | 1.06E-09 | 8.14E-09 | 4.15E-06 |
| T1DM | leukocyte | *FIGLA* | -12.27429656 | 3.21E-06 | 4.11E-06 | 0.012610721 |
| T1DM | leukocyte | *CBX6* | -0.539612781 | 2.42E-08 | 4.03E-08 | 9.49E-05 |
| T1DM | leukocyte | *MACC1* | -0.510713874 | 1.06E-09 | 8.14E-09 | 4.17E-06 |
| T1DM | leukocyte | *NPIPA5* | 0.437513684 | 0.000232943 | 0.000274268 | 0.915231297 |
| T1DM | leukocyte | *NUDT14* | -0.641265694 | 1.29E-09 | 8.14E-09 | 5.09E-06 |
| T1DM | leukocyte | *FAM3B* | -0.606676213 | 1.00E-09 | 8.14E-09 | 3.93E-06 |
| T1DM | leukocyte | *TOB2* | -0.740111397 | 2.95E-09 | 8.50E-09 | 1.16E-05 |
| T1DM | leukocyte | *TTC32* | -0.669722211 | 8.20E-10 | 8.14E-09 | 3.22E-06 |
| T1DM | leukocyte | *LRRC55* | -5.105664391 | 0.00025827 | 0.000303452 | 1 |
| T1DM | leukocyte | *TMEM236* | -0.527017375 | 4.63E-09 | 1.05E-08 | 1.82E-05 |
| T1DM | leukocyte | *UQCR10* | -0.553553402 | 8.90E-10 | 8.14E-09 | 3.50E-06 |
| T1DM | leukocyte | *EIF3C* | -0.632886614 | 5.36E-09 | 1.15E-08 | 2.11E-05 |
| T1DM | leukocyte | *NIPSNAP1* | -0.642291159 | 3.17E-09 | 8.69E-09 | 1.24E-05 |
| T1DM | leukocyte | *CRELD2* | -0.538032612 | 1.43E-09 | 8.14E-09 | 5.63E-06 |
| T1DM | leukocyte | *UBE2F* | -0.907062102 | 7.45E-08 | 1.13E-07 | 0.00029253 |
| T1DM | leukocyte | *GOLGA6L4* | -0.739917262 | 1.15E-07 | 1.69E-07 | 0.000453247 |
| T1DM | leukocyte | *PGP* | -0.907612421 | 8.91E-10 | 8.14E-09 | 3.50E-06 |
| T1DM | leukocyte | *ACOT1* | -3.700992955 | 1.72E-07 | 2.46E-07 | 0.000675416 |
| T1DM | leukocyte | *OAF* | -0.797125262 | 5.18E-09 | 1.13E-08 | 2.04E-05 |
| T1DM | leukocyte | *TM2D3* | -0.635689335 | 2.06E-09 | 8.14E-09 | 8.08E-06 |
| T1DM | leukocyte | *MROH7* | -0.660758396 | 0.001523489 | 0.001739044 | 1 |
| T1DM | leukocyte | *PLA2G6* | 0.659895741 | 6.55E-07 | 8.82E-07 | 0.002572059 |
| T1DM | leukocyte | *TOP1MT* | -1.473341942 | 9.87E-09 | 1.84E-08 | 3.88E-05 |
| T1DM | leukocyte | *COPB2* | -0.713114298 | 2.16E-09 | 8.21E-09 | 8.50E-06 |
| T1DM | leukocyte | *CCR10* | -0.897557657 | 0.000377873 | 0.000440945 | 1 |
| T1DM | leukocyte | *NCMAP* | -0.677963777 | 1.66E-09 | 8.14E-09 | 6.52E-06 |
| T1DM | leukocyte | *WDR27* | -1.199717158 | 2.60E-06 | 3.35E-06 | 0.010203169 |
| T1DM | leukocyte | *TXNRD2* | -0.698110894 | 1.36E-09 | 8.14E-09 | 5.32E-06 |
| T1DM | leukocyte | *ZFP1* | -0.888336858 | 7.81E-10 | 8.14E-09 | 3.07E-06 |
| T1DM | leukocyte | *XPOT* | -0.646628443 | 5.44E-10 | 8.14E-09 | 2.14E-06 |
| T1DM | leukocyte | *C14orf180* | -72.83102587 | 1.45E-09 | 8.14E-09 | 5.71E-06 |
| T1DM | leukocyte | *ZBTB40* | -1.006481625 | 3.46E-08 | 5.59E-08 | 0.000135907 |
| T1DM | leukocyte | *UBE2G2* | -0.632619957 | 2.78E-08 | 4.58E-08 | 0.00010932 |
| T1DM | leukocyte | *OSBP2* | -0.803728277 | 1.37E-09 | 8.14E-09 | 5.37E-06 |
| T1DM | leukocyte | *TMED9* | -0.547628764 | 3.70E-09 | 9.28E-09 | 1.45E-05 |
| T1DM | leukocyte | *BTBD6* | -0.83227309 | 2.48E-09 | 8.21E-09 | 9.73E-06 |
| T1DM | leukocyte | *CLCNKB* | 2.377364749 | 0.014628436 | 0.016144698 | 1 |
| T1DM | leukocyte | *JAG2* | -0.645578096 | 9.68E-10 | 8.14E-09 | 3.80E-06 |
| T1DM | leukocyte | *LCN12* | -1.974809814 | 8.10E-08 | 1.22E-07 | 0.000318361 |

| | | | | | | |
|---|---|---|---|---|---|---|
| T1DM | leukocyte | *FAM227A* | -0.712374698 | 2.41E-08 | 4.03E-08 | 9.49E-05 |
| T1DM | leukocyte | *MUC6* | -0.443581733 | 1.73E-09 | 8.14E-09 | 6.78E-06 |
| T1DM | leukocyte | *NOC4L* | -1.024167308 | 2.07E-09 | 8.14E-09 | 8.14E-06 |
| T1DM | leukocyte | *NDUFA6* | -0.425447512 | 5.76E-07 | 7.80E-07 | 0.002261901 |
| T1DM | leukocyte | *SIVA1* | -0.624141215 | 4.03E-09 | 9.70E-09 | 1.58E-05 |
| T1DM | leukocyte | *BRI3BP* | -0.916851669 | 1.40E-09 | 8.14E-09 | 5.50E-06 |
| T1DM | leukocyte | *UBOX5* | -0.690724211 | 1.31E-09 | 8.14E-09 | 5.14E-06 |
| T1DM | leukocyte | *MAFF* | -0.454098402 | 9.02E-10 | 8.14E-09 | 3.55E-06 |
| T1DM | leukocyte | *SEMA4B* | -0.681970215 | 1.66E-09 | 8.14E-09 | 6.53E-06 |
| T1DM | leukocyte | *CIB1* | -0.599196489 | 2.98E-09 | 8.50E-09 | 1.17E-05 |
| T1DM | leukocyte | *ANO9* | -0.563041214 | 0.001673224 | 0.001907747 | 1 |
| T1DM | leukocyte | *MFSD6L* | -1.964648896 | 1.26E-08 | 2.28E-08 | 4.96E-05 |
| T1DM | leukocyte | *AQP12B* | -1.553240922 | 0.043601042 | 0.047218438 | 1 |
| T1DM | leukocyte | *ZNF445* | -1.894316708 | 3.27E-05 | 4.00E-05 | 0.12851162 |
| T1DM | leukocyte | *ZNF74* | -1.332987256 | 2.08E-07 | 2.96E-07 | 0.000817694 |
| T1DM | leukocyte | *RBM11* | -0.626429523 | 1.70E-10 | 8.14E-09 | 6.69E-07 |
| T1DM | leukocyte | *RGPD2* | -2.04451463 | 6.86E-09 | 1.38E-08 | 2.70E-05 |
| T1DM | leukocyte | *GAS2L1* | -1.047187135 | 0.000258157 | 0.000303409 | 1 |
| T1DM | leukocyte | *ATP6V0A2* | -1.003754368 | 6.39E-09 | 1.30E-08 | 2.51E-05 |
| T1DM | leukocyte | *RAD51D* | -1.564674038 | 7.82E-09 | 1.53E-08 | 3.07E-05 |
| T1DM | leukocyte | *SH3BGR* | -0.335347502 | 7.57E-06 | 9.49E-06 | 0.029742252 |
| T1DM | leukocyte | *GPRIN3* | 0.931537779 | 0.000308574 | 0.000361367 | 1 |
| T1DM | leukocyte | *MUC1* | -0.468351725 | 1.11E-09 | 8.14E-09 | 4.38E-06 |
| T1DM | leukocyte | *IRF7* | -1.08804812 | 1.24E-08 | 2.25E-08 | 4.89E-05 |
| T1DM | leukocyte | *SV2B* | -0.525538516 | 3.18E-05 | 3.89E-05 | 0.125008666 |
| T1DM | leukocyte | *FAM131C* | -0.499682436 | 2.40E-09 | 8.21E-09 | 9.42E-06 |
| T1DM | leukocyte | *LSAMP* | -0.394272916 | 2.55E-09 | 8.21E-09 | 1.00E-05 |
| T1DM | leukocyte | *OLFML2A* | -1.984198594 | 1.04E-09 | 8.14E-09 | 4.10E-06 |
| T1DM | leukocyte | *PDIA2* | -0.365379017 | 1.41E-09 | 8.14E-09 | 5.54E-06 |
| T1DM | leukocyte | *PCGF3* | -0.708773103 | 1.27E-09 | 8.14E-09 | 4.99E-06 |
| T1DM | leukocyte | *P4HB* | -0.447280852 | 2.07E-09 | 8.14E-09 | 8.14E-06 |
| T1DM | leukocyte | *PSMD13* | -0.603124096 | 1.42E-07 | 2.05E-07 | 0.000557093 |
| T1DM | leukocyte | *BRWD1* | -0.712299421 | 9.65E-10 | 8.14E-09 | 3.79E-06 |
| T1DM | leukocyte | *DRG1* | -0.626989497 | 1.96E-09 | 8.14E-09 | 7.71E-06 |
| T1DM | leukocyte | *ADAMTSL5* | -3.977743348 | 2.04E-05 | 2.51E-05 | 0.080207453 |
| T1DM | leukocyte | *MORF4L1* | -0.490173444 | 2.51E-09 | 8.21E-09 | 9.86E-06 |
| T1DM | leukocyte | *DMWD* | -1.064976658 | 4.42E-09 | 1.02E-08 | 1.74E-05 |
| T1DM | leukocyte | *PIGP* | -0.511943405 | 6.14E-09 | 1.26E-08 | 2.41E-05 |
| T1DM | leukocyte | *NPAP1* | -13.83152141 | 0.000459629 | 0.000534759 | 1 |
| T1DM | leukocyte | *ARL17A* | -0.5427096 | 3.73E-08 | 5.98E-08 | 0.000146675 |
| T1DM | leukocyte | *TRIM69* | -0.747549685 | 5.01E-10 | 8.14E-09 | 1.97E-06 |
| T1DM | leukocyte | *IFITM1* | -1.273218022 | 9.53E-07 | 1.27E-06 | 0.003743714 |
| T1DM | leukocyte | *SETD4* | -1.42208464 | 0.000454655 | 0.000529129 | 1 |
| T1DM | leukocyte | *ZNF529* | -1.09787128 | 1.62E-08 | 2.83E-08 | 6.38E-05 |
| T1DM | leukocyte | *AIDA* | -0.606345482 | 2.20E-09 | 8.21E-09 | 8.62E-06 |
| T1DM | leukocyte | *GSAP* | 0.856388949 | 1.22E-07 | 1.77E-07 | 0.000478077 |
| T1DM | leukocyte | *WWOX* | -0.671161924 | 2.03E-09 | 8.14E-09 | 7.99E-06 |
| T1DM | leukocyte | *POLR1D* | -0.562597964 | 3.31E-09 | 8.82E-09 | 1.30E-05 |
| T1DM | leukocyte | *FFAR4* | -0.416139563 | 0.000659167 | 0.000762623 | 1 |
| T1DM | leukocyte | *EDARADD* | -0.724965936 | 9.68E-10 | 8.14E-09 | 3.80E-06 |

| | | | | | | |
|---|---|---|---|---|---|---|
| T1DM | leukocyte | *ZNF749* | -0.478209179 | 1.71E-08 | 2.96E-08 | 6.72E-05 |
| T1DM | leukocyte | *TOR3A* | -0.702890916 | 1.59E-09 | 8.14E-09 | 6.26E-06 |
| T1DM | leukocyte | *GABRA5* | 12.39546516 | 8.86E-07 | 1.18E-06 | 0.003481185 |
| T1DM | leukocyte | *ZNF555* | -0.572060488 | 1.39E-09 | 8.14E-09 | 5.46E-06 |
| T1DM | leukocyte | *NUDT17* | -0.993290942 | 7.33E-10 | 8.14E-09 | 2.88E-06 |
| T1DM | leukocyte | *CYP4X1* | 1.071628124 | 2.40E-09 | 8.21E-09 | 9.41E-06 |
| T1DM | leukocyte | *ZNF197* | -0.626993145 | 1.93E-09 | 8.14E-09 | 7.57E-06 |
| T1DM | leukocyte | *FAM228A* | -0.632363739 | 1.14E-05 | 1.42E-05 | 0.044962409 |
| T1DM | leukocyte | *RPS23* | -0.42841383 | 3.87E-09 | 9.51E-09 | 1.52E-05 |
| T1DM | leukocyte | *BTN3A2* | -0.413113184 | 6.75E-09 | 1.36E-08 | 2.65E-05 |
| T1DM | leukocyte | *ZNF396* | -1.429196622 | 1.74E-06 | 2.26E-06 | 0.006822687 |
| T1DM | leukocyte | *FAM86B1* | 2.376441206 | 3.38E-07 | 4.69E-07 | 0.001329008 |
| T1DM | leukocyte | *SMYD4* | -0.412465434 | 0.002364884 | 0.002685442 | 1 |
| T1DM | leukocyte | *CEACAM19* | -0.438637282 | 4.90E-06 | 6.21E-06 | 0.019269621 |
| T1DM | leukocyte | *ZFP91* | -0.679661964 | 9.03E-08 | 1.35E-07 | 0.000354735 |
| T1DM | leukocyte | *BCR* | -0.630127133 | 2.61E-09 | 8.21E-09 | 1.03E-05 |
| T1DM | leukocyte | *FSCN2* | -1.165849039 | 7.86E-08 | 1.19E-07 | 0.000309006 |
| T1DM | leukocyte | *RTN4RL2* | -0.720836942 | 1.42E-09 | 8.14E-09 | 5.60E-06 |
| T1DM | leukocyte | *TMEM232* | -1.088240631 | 5.71E-10 | 8.14E-09 | 2.24E-06 |
| T1DM | leukocyte | *FAM183A* | -0.578721721 | 2.02E-09 | 8.14E-09 | 7.95E-06 |
| T1DM | leukocyte | *EMID1* | -0.789457194 | 5.97E-08 | 9.21E-08 | 0.000234722 |
| T1DM | leukocyte | *RHD* | -0.607963956 | 0.01165322 | 0.012904595 | 1 |
| T1DM | leukocyte | *ESPN* | -11.98657955 | 5.90E-08 | 9.11E-08 | 0.000231764 |
| T1DM | leukocyte | *LYPD6* | -2.13923146 | 5.24E-05 | 6.35E-05 | 0.205784489 |
| T1DM | leukocyte | *AKR1C1* | -0.529949005 | 1.09E-09 | 8.14E-09 | 4.29E-06 |
| T1DM | leukocyte | *BCAM* | -0.470106931 | 2.46E-08 | 4.09E-08 | 9.67E-05 |
| T1DM | leukocyte | *CHP1* | -0.564994701 | 1.25E-09 | 8.14E-09 | 4.91E-06 |
| T1DM | leukocyte | *KCNJ11* | -1.996301841 | 7.62E-09 | 1.50E-08 | 2.99E-05 |
| T1DM | leukocyte | *ZNF385C* | -2.288150544 | 9.95E-10 | 8.14E-09 | 3.91E-06 |
| T1DM | leukocyte | *EXD3* | -0.74349333 | 1.29E-09 | 8.14E-09 | 5.08E-06 |
| T1DM | leukocyte | *C17orf97* | -0.477625245 | 2.94E-06 | 3.77E-06 | 0.011545865 |
| T1DM | leukocyte | *RGPD1* | -78.53317337 | 1.36E-09 | 8.14E-09 | 5.36E-06 |
| T1DM | leukocyte | *DHRS4L2* | -0.686534613 | 1.56E-09 | 8.14E-09 | 6.14E-06 |
| T1DM | leukocyte | *KBTBD12* | -1.417296573 | 1.50E-05 | 1.86E-05 | 0.058901902 |
| T1DM | leukocyte | *FANCA* | -0.677694132 | 0.000313482 | 0.000366787 | 1 |
| T1DM | leukocyte | *MCRS1* | -0.749310907 | 5.41E-05 | 6.55E-05 | 0.21263357 |
| T1DM | leukocyte | *ZFP69* | 1.576637873 | 0.000310662 | 0.000363595 | 1 |
| T1DM | leukocyte | *TMEM220* | -0.511189056 | 0.00039087 | 0.000455976 | 1 |
| T1DM | leukocyte | *PALM3* | -0.876139188 | 1.28E-09 | 8.14E-09 | 5.02E-06 |
| T1DM | leukocyte | *KLHL17* | -1.231708206 | 3.23E-10 | 8.14E-09 | 1.27E-06 |
| T1DM | leukocyte | *ZSCAN23* | -1.187679123 | 0.000391483 | 0.000456555 | 1 |
| T1DM | leukocyte | *RINL* | -0.852696488 | 2.00E-09 | 8.14E-09 | 7.86E-06 |
| T1DM | leukocyte | *OR7D2* | -8.351267673 | 0.000149189 | 0.000177089 | 0.586163118 |
| T1DM | leukocyte | *ZNF490* | -1.211856408 | 1.73E-09 | 8.14E-09 | 6.79E-06 |
| T1DM | leukocyte | *NRN1L* | -1.863541544 | 1.48E-09 | 8.14E-09 | 5.82E-06 |
| T1DM | leukocyte | *NWD1* | -0.985787677 | 0.008457286 | 0.00942477 | 1 |
| T1DM | leukocyte | *RAB42* | 0.391547138 | 1.94E-08 | 3.30E-08 | 7.61E-05 |
| T1DM | leukocyte | *MAPK12* | -0.840162055 | 2.63E-09 | 8.21E-09 | 1.03E-05 |
| T1DM | leukocyte | *FAM166A* | -2.830568238 | 8.86E-05 | 0.000106256 | 0.348305591 |
| T1DM | leukocyte | *LINC00265* | -1.47314832 | 1.31E-09 | 8.14E-09 | 5.14E-06 |

| | | | | | | |
|---|---|---|---|---|---|---|
| T1DM | leukocyte | *LAMTOR4* | -0.567480844 | 2.08E-08 | 3.51E-08 | 8.17E-05 |
| T1DM | leukocyte | *PRKAR1B* | -0.814942673 | 5.12E-09 | 1.12E-08 | 2.01E-05 |
| T1DM | leukocyte | *NUTM2B* | -6.985929966 | 0.001507488 | 0.001721279 | 1 |
| T1DM | leukocyte | *AGAP4* | -1.660556848 | 1.36E-09 | 8.14E-09 | 5.33E-06 |
| T1DM | leukocyte | *CENPP* | -0.602653566 | 8.02E-10 | 8.14E-09 | 3.15E-06 |
| T1DM | leukocyte | *ZNF559* | -0.453302368 | 7.97E-06 | 9.97E-06 | 0.031303278 |
| T1DM | leukocyte | *FOCAD* | -0.722901216 | 2.18E-09 | 8.21E-09 | 8.55E-06 |
| T1DM | leukocyte | *ZP3* | -0.911934845 | 1.37E-08 | 2.45E-08 | 5.39E-05 |
| T1DM | leukocyte | *INSC* | -2.033824642 | 0.005039797 | 0.005659149 | 1 |
| T1DM | leukocyte | *SERPINA5* | -0.353876933 | 7.66E-05 | 9.20E-05 | 0.300842763 |
| T1DM | leukocyte | *SRSF10* | -0.611048416 | 5.36E-09 | 1.15E-08 | 2.11E-05 |
| T1DM | leukocyte | *RALGAPA2* | -0.72405841 | 8.63E-10 | 8.14E-09 | 3.39E-06 |
| T1DM | leukocyte | *NDOR1* | -2.016893127 | 1.37E-09 | 8.14E-09 | 5.38E-06 |
| T1DM | leukocyte | *FBLL1* | -1.404326351 | 2.29E-09 | 8.21E-09 | 9.01E-06 |
| T1DM | leukocyte | *ASAH2* | -1.34006143 | 3.47E-07 | 4.80E-07 | 0.001362326 |
| T1DM | leukocyte | *ZNF177* | -0.441170011 | 0.004727881 | 0.005316498 | 1 |
| T1DM | leukocyte | *S100A16* | -0.376906997 | 1.35E-09 | 8.14E-09 | 5.29E-06 |
| T1DM | leukocyte | *PARVB* | -0.456452927 | 1.45E-08 | 2.56E-08 | 5.68E-05 |
| T1DM | leukocyte | *UROS* | -0.782771363 | 3.24E-09 | 8.77E-09 | 1.27E-05 |
| T1DM | leukocyte | *QRFP* | -5.359095261 | 0.011195809 | 0.012405058 | 1 |
| T1DM | leukocyte | *FAM221A* | -0.88679146 | 6.69E-09 | 1.36E-08 | 2.63E-05 |
| T1DM | leukocyte | *NOXA1* | -1.280522265 | 1.30E-09 | 8.14E-09 | 5.11E-06 |
| T1DM | leukocyte | *TMEM198* | -0.574005784 | 4.85E-09 | 1.08E-08 | 1.91E-05 |
| T1DM | leukocyte | *ZNF548* | -0.763810189 | 1.24E-07 | 1.80E-07 | 0.000486079 |
| T1DM | leukocyte | *MTF1* | -0.651957436 | 7.22E-09 | 1.44E-08 | 2.84E-05 |
| T1DM | leukocyte | *ZDHHC11* | -1.712496679 | 4.62E-08 | 7.27E-08 | 0.000181326 |
| T1DM | leukocyte | *ENTPD8* | -2.925042622 | 8.67E-05 | 0.000103909 | 0.340509138 |
| T1DM | leukocyte | *RPL14* | -0.420295564 | 2.56E-09 | 8.21E-09 | 1.01E-05 |
| T1DM | leukocyte | *FBF1* | 0.686963598 | 4.73E-09 | 1.06E-08 | 1.86E-05 |
| T1DM | leukocyte | *MSL1* | -0.593641456 | 5.53E-07 | 7.51E-07 | 0.002173692 |
| T1DM | leukocyte | *NOC2L* | -0.72227313 | 7.46E-08 | 1.13E-07 | 0.000293147 |
| T1DM | leukocyte | *ZNF567* | -0.664126885 | 5.70E-09 | 1.20E-08 | 2.24E-05 |
| T1DM | leukocyte | *ARID2* | -0.860123479 | 2.35E-09 | 8.21E-09 | 9.25E-06 |
| T1DM | leukocyte | *PLAC9* | -0.682448963 | 4.46E-09 | 1.03E-08 | 1.75E-05 |
| T1DM | leukocyte | *CLDN4* | -0.391143834 | 1.74E-09 | 8.14E-09 | 6.84E-06 |
| T1DM | leukocyte | *FAM47E* | -0.552985072 | 0.000224525 | 0.000264674 | 0.882158797 |
| T1DM | leukocyte | *ZNF527* | -0.904523433 | 1.05E-08 | 1.93E-08 | 4.11E-05 |
| T1DM | leukocyte | *ZNF600* | -0.646961809 | 1.69E-07 | 2.43E-07 | 0.00066591 |
| T1DM | leukocyte | *PNRC2* | -0.562242193 | 4.70E-09 | 1.06E-08 | 1.85E-05 |
| T1DM | leukocyte | *ZKSCAN3* | -2.032888682 | 1.44E-17 | 8.08E-15 | 5.66E-14 |
| T1DM | leukocyte | *RRP7A* | -0.721669286 | 4.34E-09 | 1.01E-08 | 1.70E-05 |
| T1DM | leukocyte | *S100A14* | -0.587987547 | 3.28E-09 | 8.79E-09 | 1.29E-05 |
| T1DM | leukocyte | *SLC35E2B* | -0.750128818 | 0.002632645 | 0.002984323 | 1 |
| T1DM | leukocyte | *CXCL17* | -0.572019519 | 4.21E-09 | 9.96E-09 | 1.65E-05 |
| T1DM | leukocyte | *SPATA41* | -2.701499303 | 9.75E-08 | 1.45E-07 | 0.000383189 |
| T1DM | leukocyte | *BLOC1S2* | -0.586971139 | 2.49E-07 | 3.51E-07 | 0.000977993 |
| T1DM | leukocyte | *PTPRT* | 1.473675143 | 0.018105907 | 0.01990991 | 1 |
| T1DM | leukocyte | *ZNF676* | -0.945315761 | 1.49E-10 | 8.14E-09 | 5.85E-07 |
| T1DM | leukocyte | *HLA-DRB1* | 0.090465349 | 8.32E-07 | 1.11E-06 | 0.00326993 |
| T1DM | leukocyte | *WDSUB1* | -0.880815255 | 2.20E-09 | 8.21E-09 | 8.64E-06 |

| | | | | | | |
|---|---|---|---|---|---|---|
| T1DM | leukocyte | *ZNF681* | -2.118738592 | 7.80E-05 | 9.37E-05 | 0.306489302 |
| T1DM | leukocyte | *TMEM63A* | 1.020533223 | 1.31E-06 | 1.73E-06 | 0.005143407 |
| T1DM | leukocyte | *HRCT1* | 0.867981143 | 7.46E-09 | 1.47E-08 | 2.93E-05 |
| T1DM | leukocyte | *MPHOSPH8* | -0.644145983 | 1.99E-09 | 8.14E-09 | 7.82E-06 |
| T1DM | leukocyte | *SIRPB2* | 1.343616783 | 3.68E-09 | 9.24E-09 | 1.44E-05 |
| T1DM | leukocyte | *LCOR* | -0.597636871 | 2.33E-09 | 8.21E-09 | 9.15E-06 |
| T1DM | leukocyte | *SUPT5H* | -0.572477287 | 2.87E-09 | 8.44E-09 | 1.13E-05 |
| T1DM | leukocyte | *XPNPEP3* | -0.937898242 | 2.91E-09 | 8.49E-09 | 1.14E-05 |
| T1DM | leukocyte | *ZNF107* | -0.739166785 | 3.58E-09 | 9.11E-09 | 1.41E-05 |
| T1DM | leukocyte | *SFTA2* | -1.059829121 | 3.69E-15 | 1.61E-12 | 1.45E-11 |
| T1DM | leukocyte | *PPIA* | -0.511733151 | 5.54E-09 | 1.18E-08 | 2.18E-05 |
| T1DM | leukocyte | *ZNF471* | -0.99735032 | 1.40E-09 | 8.14E-09 | 5.48E-06 |
| T1DM | leukocyte | *ZNF493* | -0.871218591 | 2.39E-09 | 8.21E-09 | 9.38E-06 |
| T1DM | leukocyte | *GTF2IRD2* | -0.887192467 | 5.29E-09 | 1.14E-08 | 2.08E-05 |
| T1DM | leukocyte | *SUPT3H* | -0.603478329 | 1.96E-09 | 8.14E-09 | 7.71E-06 |
| T1DM | leukocyte | *POM121* | -0.696603481 | 3.53E-09 | 9.06E-09 | 1.39E-05 |
| T1DM | leukocyte | *LONP1* | -0.930742609 | 2.48E-09 | 8.21E-09 | 9.75E-06 |
| T1DM | leukocyte | *ZNF34* | -0.9633072 | 1.95E-09 | 8.14E-09 | 7.64E-06 |
| T1DM | leukocyte | *ZNF781* | -0.68927756 | 6.77E-05 | 8.16E-05 | 0.265948515 |
| T1DM | leukocyte | *EVL* | -1.028015559 | 2.20E-05 | 2.70E-05 | 0.086330477 |
| T1DM | leukocyte | *ZNF124* | -1.106737065 | 1.80E-09 | 8.14E-09 | 7.09E-06 |
| T1DM | leukocyte | *XRCC6* | -0.588866821 | 5.17E-09 | 1.13E-08 | 2.03E-05 |
| T1DM | leukocyte | *PIK3R4* | -1.096823497 | 1.34E-09 | 8.14E-09 | 5.26E-06 |
| T1DM | leukocyte | *ZNF605* | -0.994502678 | 0.000394867 | 0.000460337 | 1 |
| T1DM | leukocyte | *C20orf96* | -1.151063713 | 1.17E-07 | 1.72E-07 | 0.000461238 |
| T1DM | leukocyte | *SULT1A1* | -1.303398129 | 2.05E-09 | 8.14E-09 | 8.04E-06 |
| T1DM | leukocyte | *ARL9* | 0.885813211 | 2.52E-08 | 4.17E-08 | 9.89E-05 |
| T1DM | leukocyte | *TPK1* | -1.458929996 | 5.62E-07 | 7.62E-07 | 0.002208594 |
| T1DM | leukocyte | *AFAP1* | -0.779075276 | 1.63E-09 | 8.14E-09 | 6.42E-06 |
| T1DM | leukocyte | *MYO18A* | -0.565432848 | 1.96E-08 | 3.34E-08 | 7.71E-05 |
| T1DM | leukocyte | *XRCC2* | -2.572000865 | 1.40E-09 | 8.14E-09 | 5.51E-06 |
| T1DM | leukocyte | *MYO6* | -0.500099002 | 1.61E-09 | 8.14E-09 | 6.34E-06 |
| T1DM | leukocyte | *MMP1* | 0.684384505 | 4.05E-07 | 5.57E-07 | 0.001589503 |
| T1DM | leukocyte | *ZNF502* | -1.760512374 | 3.36E-05 | 4.10E-05 | 0.131847338 |
| T1DM | leukocyte | *TRAPPC4* | -0.614631592 | 3.12E-09 | 8.69E-09 | 1.23E-05 |
| T1DM | leukocyte | *ZNF33B* | -1.170005865 | 8.88E-10 | 8.14E-09 | 3.49E-06 |
| T1DM | leukocyte | *AMZ2* | -0.799664974 | 2.21E-09 | 8.21E-09 | 8.69E-06 |
| T1DM | leukocyte | *ZNF431* | -0.679243883 | 2.04E-09 | 8.14E-09 | 8.01E-06 |
| T1DM | leukocyte | *DAPK1* | -1.024961354 | 3.38E-08 | 5.48E-08 | 0.000132938 |
| T1DM | leukocyte | *HLA-DQA1* | 0.066555411 | 3.41E-09 | 8.93E-09 | 1.34E-05 |
| T1DM | leukocyte | *GM2A* | 0.588839158 | 4.06E-07 | 5.59E-07 | 0.001593459 |
| T1DM | leukocyte | *CLPSL2* | -0.817894446 | 2.48E-08 | 4.12E-08 | 9.75E-05 |
| T1DM | leukocyte | *ADA* | 0.354788722 | 5.68E-08 | 8.81E-08 | 0.000223011 |
| T1DM | leukocyte | *PPTC7* | -0.974597009 | 1.63E-09 | 8.14E-09 | 6.41E-06 |
| T1DM | leukocyte | *ZFP28* | -0.965190186 | 1.68E-09 | 8.14E-09 | 6.59E-06 |
| T1DM | leukocyte | *ANKRD36B* | -0.752462115 | 2.75E-08 | 4.53E-08 | 0.000108104 |
| T1DM | leukocyte | *SRGAP1* | -0.782501467 | 1.94E-09 | 8.14E-09 | 7.64E-06 |
| T1DM | leukocyte | *FAM3C* | -0.544909016 | 1.41E-09 | 8.14E-09 | 5.54E-06 |
| T1DM | leukocyte | *AP2A1* | -0.810042374 | 3.22E-09 | 8.75E-09 | 1.27E-05 |
| T1DM | leukocyte | *WDR5B* | -1.7785553 | 1.01E-07 | 1.50E-07 | 0.000397066 |

| | | | | | | |
|---|---|---|---|---|---|---|
| T1DM | leukocyte | *ZNF138* | -0.524272306 | 3.97E-08 | 6.32E-08 | 0.000155995 |
| T1DM | leukocyte | *ZNF429* | -0.656236307 | 1.50E-09 | 8.14E-09 | 5.91E-06 |
| T1DM | leukocyte | *ZNF100* | -0.792125026 | 9.68E-08 | 1.44E-07 | 0.000380476 |
| T1DM | leukocyte | *ZNF763* | -1.336592122 | 3.82E-08 | 6.11E-08 | 0.000150014 |
| T1DM | leukocyte | *ARRDC1* | -0.641118996 | 1.98E-09 | 8.14E-09 | 7.79E-06 |
| T1DM | leukocyte | *ZNF682* | -1.215309605 | 2.92E-09 | 8.49E-09 | 1.15E-05 |
| T1DM | leukocyte | *ZNF772* | 0.519054911 | 0.019291008 | 0.021171612 | 1 |
| T1DM | leukocyte | *ZNF257* | -1.1587458 | 4.13E-11 | 8.14E-09 | 1.62E-07 |
| T1DM | leukocyte | *ADAM32* | 0.865413492 | 5.67E-08 | 8.80E-08 | 0.000222836 |
| T1DM | leukocyte | *ACSL5* | -1.151734907 | 1.11E-09 | 8.14E-09 | 4.38E-06 |
| T1DM | leukocyte | *SND1* | -0.644477214 | 2.71E-09 | 8.26E-09 | 1.06E-05 |
| T1DM | leukocyte | *SULT1A2* | -1.737077099 | 3.44E-05 | 4.21E-05 | 0.135291031 |
| T1DM | leukocyte | *PIWIL2* | 0.837416271 | 9.67E-07 | 1.28E-06 | 0.003801023 |
| T1DM | leukocyte | *TBC1D9B* | -0.921232162 | 2.04E-06 | 2.64E-06 | 0.00800583 |
| T1DM | leukocyte | *C6orf141* | -1.218636895 | 2.80E-08 | 4.60E-08 | 0.000109828 |
| T1DM | leukocyte | *GTF2E2* | -0.653468292 | 1.19E-09 | 8.14E-09 | 4.69E-06 |
| T1DM | leukocyte | *GUCA2A* | -0.691253013 | 0.015884495 | 0.017501453 | 1 |
| T1DM | leukocyte | *ZNF165* | -0.574226905 | 2.14E-06 | 2.77E-06 | 0.008406379 |
| T1DM | leukocyte | *SVIL* | -0.632121165 | 1.12E-09 | 8.14E-09 | 4.40E-06 |
| T1DM | leukocyte | *MRPL21* | -0.629659655 | 6.03E-09 | 1.25E-08 | 2.37E-05 |
| T1DM | leukocyte | *ZNF98* | -1.000851381 | 8.12E-10 | 8.14E-09 | 3.19E-06 |
| T1DM | leukocyte | *ZNF786* | 0.354571741 | 0.025409533 | 0.027770252 | 1 |
| T1DM | leukocyte | *ZNF517* | -1.455490872 | 0.001145532 | 0.001315637 | 1 |
| T1DM | leukocyte | *SLC22A5* | -1.013724306 | 1.27E-09 | 8.14E-09 | 4.98E-06 |
| T1DM | leukocyte | *ADARB1* | 0.601438514 | 0.045288686 | 0.049005577 | 1 |
| T1DM | leukocyte | *C5AR1* | 0.097478513 | 3.64E-09 | 9.18E-09 | 1.43E-05 |
| T1DM | leukocyte | *VEPH1* | -1.120411731 | 1.85E-06 | 2.41E-06 | 0.007268021 |
| T1DM | leukocyte | *SHPK* | -0.863260437 | 7.81E-08 | 1.18E-07 | 0.000307039 |
| T1DM | leukocyte | *OGDHL* | -0.550736228 | 5.33E-05 | 6.46E-05 | 0.209551304 |
| T1DM | leukocyte | *PCDHB11* | -1.012138485 | 2.52E-06 | 3.25E-06 | 0.009909304 |
| T1DM | leukocyte | *MIB2* | -0.630535171 | 1.97E-08 | 3.34E-08 | 7.72E-05 |
| T1DM | leukocyte | *ATG7* | -1.219554546 | 6.33E-09 | 1.30E-08 | 2.49E-05 |
| T1DM | leukocyte | *PIGN* | -0.609458884 | 7.05E-09 | 1.41E-08 | 2.77E-05 |
| T1DM | leukocyte | *HHLA3* | -0.557446489 | 2.25E-07 | 3.19E-07 | 0.000884883 |
| T1DM | leukocyte | *BCO2* | -0.736739402 | 5.56E-09 | 1.18E-08 | 2.19E-05 |
| T1DM | leukocyte | *ENTPD6* | -0.776730802 | 1.20E-09 | 8.14E-09 | 4.70E-06 |
| T1DM | leukocyte | *ENPP1* | -0.572093196 | 3.82E-09 | 9.44E-09 | 1.50E-05 |
| T1DM | leukocyte | *CCDC154* | 2.197481734 | 4.82E-05 | 5.86E-05 | 0.189421123 |
| T1DM | leukocyte | *DPP4* | -0.484015014 | 4.96E-10 | 8.14E-09 | 1.95E-06 |
| T1DM | leukocyte | *NMB* | -0.651353035 | 2.86E-09 | 8.44E-09 | 1.13E-05 |
| T1DM | leukocyte | *HSPB9* | -0.325810114 | 0.013376501 | 0.014783762 | 1 |
| T1DM | leukocyte | *RPS26* | -0.389328738 | 7.53E-07 | 1.01E-06 | 0.002956857 |
| T1DM | leukocyte | *LHFPL5* | -1.527784443 | 1.00E-07 | 1.49E-07 | 0.000394583 |
| T1DM | leukocyte | *ZNF780A* | -0.54883114 | 6.47E-08 | 9.92E-08 | 0.000254092 |
| T1DM | leukocyte | *ATAD3A* | -1.123658081 | 1.60E-09 | 8.14E-09 | 6.30E-06 |
| T1DM | leukocyte | *FAM118B* | -1.317811485 | 1.51E-07 | 2.17E-07 | 0.000591345 |
| T1DM | leukocyte | *CCDC180* | -1.146173353 | 3.39E-05 | 4.15E-05 | 0.133380022 |
| T1DM | leukocyte | *SLC9A8* | -1.808409902 | 2.84E-07 | 3.97E-07 | 0.00111488 |
| T1DM | leukocyte | *OCLN* | -0.482623609 | 9.42E-10 | 8.14E-09 | 3.70E-06 |
| T1DM | leukocyte | *ZNF181* | -0.93881104 | 2.68E-09 | 8.26E-09 | 1.05E-05 |

| | | | | | | |
|---|---|---|---|---|---|---|
| T1DM | leukocyte | *TEAD4* | -0.593513655 | 1.76E-09 | 8.14E-09 | 6.92E-06 |
| T1DM | leukocyte | *C2orf27A* | -1.105729492 | 2.04E-10 | 8.14E-09 | 8.01E-07 |
| T1DM | leukocyte | *PLCG2* | -0.733875405 | 3.99E-09 | 9.64E-09 | 1.57E-05 |
| T1DM | leukocyte | *RPL12* | -0.414863622 | 1.06E-09 | 8.14E-09 | 4.15E-06 |
| T1DM | leukocyte | *VPS13A* | -0.626678419 | 5.21E-09 | 1.13E-08 | 2.05E-05 |
| T1DM | leukocyte | *LEKR1* | -1.188586611 | 1.82E-09 | 8.14E-09 | 7.14E-06 |
| T1DM | leukocyte | *ENTPD7* | -1.08455962 | 6.65E-06 | 8.38E-06 | 0.026143031 |
| T1DM | leukocyte | *AGAP9* | -1.279393087 | 4.94E-08 | 7.74E-08 | 0.000194136 |
| T1DM | leukocyte | *ZNF273* | -1.275323418 | 1.82E-09 | 8.14E-09 | 7.16E-06 |
| T1DM | leukocyte | *ZNF84* | -1.240840118 | 5.86E-08 | 9.06E-08 | 0.000230055 |
| T1DM | leukocyte | *ZNF667* | -0.519486508 | 1.16E-09 | 8.14E-09 | 4.57E-06 |
| T1DM | leukocyte | *SIRPA* | 0.600991712 | 9.14E-05 | 0.000109403 | 0.358951 |
| T1DM | leukocyte | *CD2AP* | -0.651301966 | 2.00E-09 | 8.14E-09 | 7.85E-06 |
| T1DM | leukocyte | *SFI1* | -1.209173814 | 2.47E-07 | 3.49E-07 | 0.000971886 |
| T1DM | leukocyte | *HIBCH* | -0.629349438 | 1.08E-09 | 8.14E-09 | 4.24E-06 |
| T1DM | leukocyte | *TMEM229B* | -3.022229271 | 5.49E-09 | 1.17E-08 | 2.16E-05 |
| T1DM | leukocyte | *SOWAHC* | -1.100027751 | 4.41E-09 | 1.02E-08 | 1.73E-05 |
| T1DM | leukocyte | *SVIP* | -0.571551129 | 1.69E-09 | 8.14E-09 | 6.64E-06 |
| T1DM | leukocyte | *DDRGK1* | -0.680595772 | 7.35E-09 | 1.46E-08 | 2.89E-05 |
| T1DM | leukocyte | *ZNF607* | -0.874741251 | 7.22E-10 | 8.14E-09 | 2.83E-06 |
| T1DM | leukocyte | *HSD17B11* | -0.916680034 | 1.74E-08 | 3.01E-08 | 6.85E-05 |
| T1DM | leukocyte | *RPS6KL1* | -0.835735618 | 2.44E-08 | 4.06E-08 | 9.58E-05 |
| T1DM | leukocyte | *QRICH1* | -1.048950941 | 3.49E-09 | 9.01E-09 | 1.37E-05 |
| T1DM | leukocyte | *DDX42* | -0.558001586 | 1.01E-08 | 1.88E-08 | 3.98E-05 |
| T1DM | leukocyte | *HELZ* | -0.700441318 | 2.24E-09 | 8.21E-09 | 8.79E-06 |
| T1DM | leukocyte | *CARD11* | -1.260518257 | 0.001793794 | 0.00204403 | 1 |
| T1DM | leukocyte | *ZNF583* | -0.618922022 | 1.42E-09 | 8.14E-09 | 5.60E-06 |
| T1DM | leukocyte | *ZNF568* | -0.6674488 | 3.48E-07 | 4.82E-07 | 0.001368876 |
| T1DM | leukocyte | *ZNF587* | -0.778994969 | 1.56E-09 | 8.14E-09 | 6.14E-06 |
| T1DM | leukocyte | *TPM2* | -0.756835058 | 1.13E-09 | 8.14E-09 | 4.43E-06 |
| T1DM | leukocyte | *HLA-DRB5* | 0.063134155 | 1.89E-05 | 2.33E-05 | 0.074197122 |
| T1DM | leukocyte | *CNGA1* | -0.922110221 | 2.07E-06 | 2.68E-06 | 0.008128414 |
| T1DM | leukocyte | *ZNF789* | -1.228139976 | 1.76E-05 | 2.17E-05 | 0.06910549 |
| T1DM | leukocyte | *CCDC69* | -0.565579042 | 9.86E-05 | 0.000118003 | 0.387521615 |
| T1DM | leukocyte | *NCOA6* | -0.625755977 | 1.08E-09 | 8.14E-09 | 4.24E-06 |
| T1DM | leukocyte | *STK39* | -0.611942892 | 3.40E-09 | 8.90E-09 | 1.33E-05 |
| T1DM | leukocyte | *PAPSS2* | -0.562652278 | 1.72E-09 | 8.14E-09 | 6.74E-06 |
| T1DM | leukocyte | *CEP290* | -0.752997725 | 1.49E-09 | 8.14E-09 | 5.86E-06 |
| T1DM | leukocyte | *ECI2* | -0.699298008 | 7.54E-09 | 1.48E-08 | 2.96E-05 |
| T1DM | leukocyte | *MSRB1* | -0.889008608 | 1.03E-09 | 8.14E-09 | 4.05E-06 |
| T1DM | leukocyte | *OXCT2* | -0.697940999 | 3.29E-07 | 4.57E-07 | 0.001292963 |
| T1DM | leukocyte | *COLGALT2* | -0.675969533 | 0.000238797 | 0.000280992 | 0.938233401 |
| T1DM | leukocyte | *PNP* | -0.397380367 | 2.87E-07 | 4.01E-07 | 0.001128985 |
| T1DM | leukocyte | *DENND4B* | -0.70564589 | 5.27E-09 | 1.14E-08 | 2.07E-05 |
| T1DM | leukocyte | *CES1* | -1.196417657 | 2.36E-06 | 3.05E-06 | 0.009281499 |
| T1DM | leukocyte | *TSEN15* | -0.80315977 | 2.50E-09 | 8.21E-09 | 9.81E-06 |
| T1DM | leukocyte | *LTN1* | -0.700878543 | 4.94E-10 | 8.14E-09 | 1.94E-06 |
| T1DM | leukocyte | *CCDC152* | 0.817456262 | 8.47E-08 | 1.27E-07 | 0.000332897 |
| T1DM | leukocyte | *TYW1* | -1.005061034 | 6.07E-09 | 1.25E-08 | 2.38E-05 |
| T1DM | leukocyte | *ITPRIPL1* | -0.664971783 | 0.000194789 | 0.00023052 | 0.765327105 |

| | | | | | | |
|---|---|---|---|---|---|---|
| T1DM | leukocyte | *MAP3K3* | -0.743372568 | 8.43E-10 | 8.14E-09 | 3.31E-06 |
| T1DM | leukocyte | *C1orf174* | -0.795905279 | 1.52E-08 | 2.67E-08 | 5.96E-05 |
| T1DM | leukocyte | *KIAA0753* | -0.520975933 | 0.023437323 | 0.025650485 | 1 |
| T1DM | leukocyte | *ATG9A* | -0.796896791 | 2.57E-08 | 4.25E-08 | 0.000100861 |
| T1DM | leukocyte | *TBKBP1* | -0.686406995 | 3.86E-08 | 6.17E-08 | 0.000151697 |
| T1DM | leukocyte | *CCDC167* | -0.546747783 | 2.63E-09 | 8.21E-09 | 1.03E-05 |
| T1DM | leukocyte | *SOWAHA* | -0.623058162 | 1.42E-09 | 8.14E-09 | 5.56E-06 |
| T1DM | leukocyte | *L3MBTL3* | -0.705622217 | 0.000124033 | 0.000147675 | 0.487326252 |
| T1DM | leukocyte | *MFAP3L* | -1.126848735 | 1.42E-09 | 8.14E-09 | 5.59E-06 |
| T1DM | leukocyte | *PJA2* | -0.554612815 | 7.41E-09 | 1.47E-08 | 2.91E-05 |
| T1DM | leukocyte | *INF2* | -0.973203546 | 3.32E-09 | 8.82E-09 | 1.31E-05 |
| T1DM | leukocyte | *EFCAB2* | -0.6419391 | 5.86E-08 | 9.06E-08 | 0.000230255 |
| T1DM | leukocyte | *RAET1G* | -1.734853908 | 0.00548356 | 0.00614866 | 1 |
| T1DM | leukocyte | *SAMD5* | -0.799033804 | 3.03E-09 | 8.57E-09 | 1.19E-05 |
| T1DM | leukocyte | *SPRN* | -3.291866967 | 1.01E-05 | 1.26E-05 | 0.039567091 |
| T1DM | leukocyte | *DDO* | -1.12525612 | 0.000105153 | 0.000125576 | 0.413144243 |
| T1DM | leukocyte | *C6orf163* | -0.809883349 | 0.000295087 | 0.000345779 | 1 |
| T1DM | leukocyte | *PCMTD2* | -0.653308548 | 1.58E-09 | 8.14E-09 | 6.22E-06 |
| T1DM | leukocyte | *LIME1* | -0.596969615 | 0.001452966 | 0.001661923 | 1 |
| T1DM | leukocyte | *EFCAB7* | 1.133616264 | 0.001412727 | 0.001616838 | 1 |
| T1DM | leukocyte | *FOXO6* | -1.33765777 | 2.79E-09 | 8.35E-09 | 1.10E-05 |
| T1DM | leukocyte | *SYS1* | -0.672522851 | 2.72E-09 | 8.27E-09 | 1.07E-05 |
| T1DM | leukocyte | *INPP5B* | -0.822666445 | 6.27E-06 | 7.90E-06 | 0.02462985 |
| T1DM | leukocyte | *TRAF3IP1* | -1.464068978 | 5.36E-07 | 7.29E-07 | 0.002104783 |
| T1DM | leukocyte | *C2orf72* | -1.913781764 | 3.22E-06 | 4.11E-06 | 0.012637873 |
| T1DM | leukocyte | *CLPSL1* | -0.264189692 | 2.04E-05 | 2.51E-05 | 0.079966564 |
| T1DM | leukocyte | *HLA-DOA* | 0.128276863 | 1.69E-09 | 8.14E-09 | 6.62E-06 |
| T1DM | leukocyte | *HLA-DMA* | 0.234652493 | 3.98E-20 | 2.61E-17 | 1.56E-16 |
| T1DM | leukocyte | *NOTCH4* | -1.333638859 | 1.58E-32 | 3.10E-29 | 6.21E-29 |
| T1DM | leukocyte | *RNF5* | -0.989259939 | 2.63E-21 | 2.06E-18 | 1.03E-17 |
| T1DM | leukocyte | *MRPL38* | -0.676787839 | 2.39E-09 | 8.21E-09 | 9.41E-06 |
| T1DM | leukocyte | *SMIM5* | -0.598914548 | 3.38E-10 | 8.14E-09 | 1.33E-06 |
| T1DM | leukocyte | *LAYN* | -0.660974048 | 1.47E-09 | 8.14E-09 | 5.79E-06 |
| T1DM | leukocyte | *SLC44A4* | -0.597451968 | 4.78E-12 | 1.45E-09 | 1.88E-08 |
| T1DM | leukocyte | *VWA7* | -0.607326628 | 7.44E-05 | 8.95E-05 | 0.292467776 |
| T1DM | leukocyte | *MICB* | 0.288071595 | 0.018749904 | 0.020600776 | 1 |
| T1DM | leukocyte | *HLA-C* | -0.215660404 | 0.001060976 | 0.001219951 | 1 |
| T1DM | leukocyte | *POU5F1* | 0.554593827 | 0.001447103 | 0.001655698 | 1 |
| T1DM | leukocyte | *CCHCR1* | 1.356489238 | 2.76E-14 | 1.08E-11 | 1.08E-10 |
| T1DM | leukocyte | *PSORS1C1* | -0.761517967 | 6.49E-16 | 3.19E-13 | 2.55E-12 |
| T1DM | leukocyte | *DHX16* | -0.749017444 | 1.98E-08 | 3.36E-08 | 7.76E-05 |
| T1DM | leukocyte | *ABCF1* | -0.643072746 | 1.83E-08 | 3.14E-08 | 7.18E-05 |
| T1DM | leukocyte | *LILRB3* | 0.635032853 | 1.51E-06 | 1.98E-06 | 0.005934214 |
| T1DM | leukocyte | *LINC01123* | -1.856080657 | 3.00E-05 | 3.68E-05 | 0.118065667 |
| T1DM | leukocyte | *ZNF468* | -1.332778737 | 3.69E-07 | 5.10E-07 | 0.001451664 |
| T1DM | leukocyte | *TRIM31* | -2.273443894 | 0.004584763 | 0.005162951 | 1 |
| T1DM | leukocyte | *RNF39* | 0.449727935 | 0.002389428 | 0.002711746 | 1 |
| T1DM | leukocyte | *HLA-G* | -0.486325436 | 2.75E-05 | 3.37E-05 | 0.107891271 |
| T1DM | leukocyte | *HLA-F* | -0.37152315 | 5.12E-09 | 1.12E-08 | 2.01E-05 |
| T1DM | leukocyte | *ZFP57* | 0.627399061 | 0.000882199 | 0.001017962 | 1 |

| | | | | | | |
|---|---|---|---|---|---|---|
| T1DM | leukocyte | *ASPDH* | -1.23526857 | 1.40E-09 | 8.14E-09 | 5.48E-06 |
| T1DM | leukocyte | *KRT40* | -0.706507492 | 0.037626172 | 0.040849193 | 1 |
| T1DM | leukocyte | *ZNF425* | -1.72698858 | 7.42E-09 | 1.47E-08 | 2.92E-05 |
| T1DM | leukocyte | *PCDHA5* | -0.967710197 | 6.84E-07 | 9.20E-07 | 0.002688533 |
| T1DM | leukocyte | *PCDHA4* | -0.329033865 | 0.003444216 | 0.003891954 | 1 |
| T1DM | leukocyte | *SPIRE2* | -0.558970597 | 6.56E-08 | 1.01E-07 | 0.00025779 |
| T1DM | leukocyte | *PABPN1L* | -95.11083259 | 1.40E-09 | 8.14E-09 | 5.49E-06 |
| T1DM | leukocyte | *PKHD1L1* | -2.89010175 | 9.96E-10 | 8.14E-09 | 3.91E-06 |
| T1DM | leukocyte | *SYCE1L* | -0.997117381 | 1.42E-08 | 2.52E-08 | 5.60E-05 |
| T1DM | leukocyte | *TMEM231* | -0.70861642 | 5.15E-09 | 1.12E-08 | 2.02E-05 |
| T1DM | leukocyte | *LINC00654* | -2.000630407 | 0.000167291 | 0.000198216 | 0.657285575 |
| T1DM | leukocyte | *SAP25* | -0.347609148 | 0.001323448 | 0.001515985 | 1 |
| T1DM | leukocyte | *SARNP* | -0.632866573 | 2.43E-09 | 8.21E-09 | 9.54E-06 |
| T1DM | leukocyte | *MT1A* | -0.298857455 | 6.26E-05 | 7.57E-05 | 0.245984459 |
| T1DM | leukocyte | *ATP6AP1L* | 1.338999748 | 1.01E-07 | 1.50E-07 | 0.000396925 |
| T1DM | leukocyte | *CCDC85C* | -0.719593501 | 2.27E-09 | 8.21E-09 | 8.90E-06 |
| T1DM | leukocyte | *RGL3* | -0.926287314 | 6.91E-06 | 8.68E-06 | 0.027131039 |
| T1DM | leukocyte | *CPT1B* | -0.763841108 | 2.48E-08 | 4.11E-08 | 9.73E-05 |
| T1DM | leukocyte | *SMN2* | -0.863927166 | 3.01E-09 | 8.54E-09 | 1.18E-05 |
| T1DM | leukocyte | *SERF1B* | -5.55327236 | 1.43E-06 | 1.88E-06 | 0.005618315 |
| T1DM | leukocyte | *HMGN1* | -0.508393542 | 2.24E-09 | 8.21E-09 | 8.81E-06 |
| T1DM | leukocyte | *LCMT1* | -0.885782855 | 2.81E-09 | 8.40E-09 | 1.10E-05 |
| T1DM | leukocyte | *LIN52* | -1.409451648 | 3.38E-09 | 8.89E-09 | 1.33E-05 |
| T1DM | leukocyte | *DENND1C* | 0.297837365 | 3.00E-09 | 8.53E-09 | 1.18E-05 |
| T1DM | leukocyte | *GMNC* | -1.017993599 | 0.009813548 | 0.010891929 | 1 |
| T1DM | leukocyte | *LINC00487* | -10.18061188 | 0.018348931 | 0.02016586 | 1 |
| T1DM | leukocyte | *DEFB134* | -77.72504757 | 1.76E-06 | 2.30E-06 | 0.006922123 |
| T1DM | leukocyte | *RNPS1* | -0.629089016 | 1.35E-09 | 8.14E-09 | 5.29E-06 |
| T1DM | leukocyte | *ZDHHC11B* | -1.178164263 | 2.29E-09 | 8.21E-09 | 8.99E-06 |
| T1DM | leukocyte | *GOLGA8O* | -1.968240629 | 3.93E-08 | 6.27E-08 | 0.000154407 |
| T1DM | leukocyte | *HLA-A* | -0.328136203 | 3.47E-06 | 4.43E-06 | 0.013625702 |
| T1DM | leukocyte | *STK38L* | -1.063194235 | 1.21E-07 | 1.76E-07 | 0.000473907 |
| T1DM | leukocyte | *TAS2R14* | -0.402496643 | 0.017745705 | 0.019519282 | 1 |
| T1DM | leukocyte | *MAP10* | -0.834277038 | 7.28E-10 | 8.14E-09 | 2.86E-06 |
| T1DM | leukocyte | *NUP62* | -0.739097872 | 1.58E-09 | 8.14E-09 | 6.20E-06 |
| T1DM | leukocyte | *FAM106A* | -0.826104565 | 5.05E-07 | 6.88E-07 | 0.001982285 |
| T1DM | leukocyte | *ZNF254* | -0.801225668 | 1.87E-09 | 8.14E-09 | 7.37E-06 |
| T1DM | leukocyte | *LINGO4* | -0.791579811 | 4.76E-06 | 6.03E-06 | 0.018689744 |
| T1DM | leukocyte | *ASIC3* | -0.882974382 | 1.18E-07 | 1.73E-07 | 0.00046402 |
| T1DM | leukocyte | *ARHGEF35* | -0.48680877 | 2.01E-09 | 8.14E-09 | 7.90E-06 |
| T1DM | leukocyte | *SUPT4H1* | -0.763242679 | 3.21E-09 | 8.74E-09 | 1.26E-05 |
| T1DM | leukocyte | *MXD3* | -0.670567517 | 5.99E-08 | 9.24E-08 | 0.000235429 |
| T1DM | leukocyte | *COG8* | -0.990863675 | 1.49E-09 | 8.14E-09 | 5.85E-06 |
| T1DM | leukocyte | *ERV3-1* | -0.697061691 | 0.000191499 | 0.000226763 | 0.752398222 |
| T1DM | leukocyte | *CCL14* | -0.538940142 | 0.004050904 | 0.004568313 | 1 |
| T1DM | leukocyte | *RBMXL1* | -0.608497066 | 4.72E-07 | 6.46E-07 | 0.001856105 |
| T1DM | leukocyte | *SRA1* | -0.526504404 | 2.29E-09 | 8.21E-09 | 9.01E-06 |
| T1DM | leukocyte | *C8orf82* | -1.079889668 | 1.33E-07 | 1.93E-07 | 0.000523521 |
| T1DM | leukocyte | *ZBTB9* | 1.06867395 | 7.25E-06 | 9.11E-06 | 0.028493584 |
| T1DM | leukocyte | *LEPROT* | -0.640355938 | 3.36E-09 | 8.85E-09 | 1.32E-05 |

| | | | | | | |
|---|---|---|---|---|---|---|
| T1DM | leukocyte | *PPP1CB* | -0.524365627 | 2.40E-09 | 8.21E-09 | 9.42E-06 |
| T1DM | leukocyte | *NCKIPSD* | -0.800585386 | 1.02E-06 | 1.35E-06 | 0.004010674 |
| T1DM | leukocyte | *TREX1* | 4.69327075 | 7.79E-06 | 9.76E-06 | 0.030593115 |
| T1DM | leukocyte | *ATP6V1G2* | -1.108211799 | 8.47E-07 | 1.13E-06 | 0.003329313 |
| T1DM | leukocyte | *ZNF888* | -1.020077127 | 1.34E-09 | 8.14E-09 | 5.28E-06 |
| T1DM | leukocyte | *EMP2* | -0.735804474 | 9.33E-10 | 8.14E-09 | 3.67E-06 |
| T1DM | leukocyte | *LTB4R* | -0.919608367 | 5.82E-09 | 1.21E-08 | 2.29E-05 |
| T1DM | leukocyte | *DNASE1* | -0.661463501 | 1.17E-09 | 8.14E-09 | 4.58E-06 |
| T1DM | leukocyte | *NUDT19* | -0.885155324 | 4.22E-09 | 9.96E-09 | 1.66E-05 |
| T1DM | leukocyte | *AP1G2* | -1.90022981 | 2.88E-06 | 3.70E-06 | 0.01132152 |
| T1DM | leukocyte | *ZNF90* | -0.873466273 | 1.33E-09 | 8.14E-09 | 5.24E-06 |
| T1DM | leukocyte | *REPIN1* | -0.724084025 | 2.49E-09 | 8.21E-09 | 9.79E-06 |
| T1DM | leukocyte | *TSPAN4* | -0.601881198 | 6.07E-06 | 7.65E-06 | 0.023835033 |
| T1DM | leukocyte | *CPNE1* | -0.372613384 | 0.017482004 | 0.019239999 | 1 |
| T1DM | leukocyte | *ARL16* | -0.408844598 | 6.15E-05 | 7.44E-05 | 0.241729253 |
| T1DM | leukocyte | *WEE2* | -2.016395981 | 0.004832691 | 0.005431248 | 1 |
| T1DM | leukocyte | *FIS1* | -0.530846869 | 2.54E-09 | 8.21E-09 | 9.99E-06 |
| T1DM | leukocyte | *MBLAC1* | -0.715062642 | 4.47E-05 | 5.45E-05 | 0.175804705 |
| T1DM | leukocyte | *VSTM5* | -1.02291089 | 0.015390532 | 0.016976249 | 1 |
| T1DM | leukocyte | *SEC14L6* | -0.28134513 | 7.23E-05 | 8.70E-05 | 0.284047221 |
| T1DM | leukocyte | *MEG3* | -0.430016815 | 1.62E-09 | 8.14E-09 | 6.35E-06 |
| T1DM | leukocyte | *EML6* | -0.870737087 | 2.73E-09 | 8.29E-09 | 1.07E-05 |
| T1DM | leukocyte | *C12orf71* | -8.679765491 | 8.01E-06 | 1.00E-05 | 0.031455634 |
| T1DM | leukocyte | *IFRD2* | -0.724088873 | 1.59E-09 | 8.14E-09 | 6.26E-06 |
| T1DM | leukocyte | *ARHGEF28* | -0.88417464 | 7.14E-09 | 1.42E-08 | 2.81E-05 |
| T1DM | leukocyte | *COL28A1* | -3.112359637 | 2.91E-07 | 4.07E-07 | 0.001144887 |
| T1DM | leukocyte | *UBXN2B* | -1.033143683 | 1.08E-08 | 1.99E-08 | 4.25E-05 |
| T1DM | leukocyte | *C5orf49* | -0.592158463 | 1.38E-09 | 8.14E-09 | 5.41E-06 |
| T1DM | leukocyte | *GOLGA8B* | -0.946090804 | 1.92E-09 | 8.14E-09 | 7.54E-06 |
| T1DM | leukocyte | *VPS16* | -1.025922777 | 9.08E-08 | 1.36E-07 | 0.000356877 |
| T1DM | leukocyte | *LINC01139* | -1.635635619 | 1.96E-06 | 2.54E-06 | 0.007691201 |
| T1DM | leukocyte | *ATAD3C* | -0.647508087 | 1.28E-09 | 8.14E-09 | 5.04E-06 |
| T1DM | leukocyte | *SYCE3* | -1.30788106 | 1.16E-08 | 2.11E-08 | 4.55E-05 |
| T1DM | leukocyte | *PAM16* | -0.618698371 | 1.62E-09 | 8.14E-09 | 6.35E-06 |
| T1DM | leukocyte | *LINC00339* | -0.747168288 | 2.44E-08 | 4.06E-08 | 9.58E-05 |
| T1DM | leukocyte | *CELA3B* | -0.33972163 | 1.72E-09 | 8.14E-09 | 6.77E-06 |
| T1DM | leukocyte | *FAM228B* | -0.502706966 | 6.30E-05 | 7.60E-05 | 0.247374843 |
| T1DM | leukocyte | *VAMP2* | -0.477619859 | 3.16E-09 | 8.69E-09 | 1.24E-05 |
| T1DM | leukocyte | *PPP3R1* | -0.947243741 | 8.21E-09 | 1.59E-08 | 3.22E-05 |
| T1DM | leukocyte | *FANCG* | -1.019518634 | 7.76E-07 | 1.04E-06 | 0.003047815 |
| T1DM | leukocyte | *AP4M1* | -0.590674286 | 1.45E-09 | 8.14E-09 | 5.70E-06 |
| T1DM | leukocyte | *ZNF880* | -0.530525715 | 5.42E-08 | 8.43E-08 | 0.000212884 |
| T1DM | leukocyte | *TRIM16* | -0.977420052 | 1.69E-09 | 8.14E-09 | 6.65E-06 |
| T1DM | leukocyte | *TIGD1* | -1.040616616 | 1.41E-08 | 2.51E-08 | 5.54E-05 |
| T1DM | leukocyte | *XKR9* | -0.427325722 | 0.005234812 | 0.00587645 | 1 |
| T1DM | leukocyte | *SLC12A8* | -1.073975356 | 2.73E-10 | 8.14E-09 | 1.07E-06 |
| T1DM | leukocyte | *UBA52* | -0.478806925 | 4.80E-09 | 1.07E-08 | 1.89E-05 |
| T1DM | leukocyte | *PPT2* | -0.350511678 | 0.024053994 | 0.026303408 | 1 |
| T1DM | leukocyte | *EXOSC6* | -0.718543287 | 3.53E-09 | 9.06E-09 | 1.39E-05 |
| T1DM | leukocyte | *CKMT1A* | -0.68767838 | 8.32E-09 | 1.60E-08 | 3.27E-05 |

| | | | | | | |
|---|---|---|---|---|---|---|
| T1DM | leukocyte | *HLA-DPB1* | 0.083092708 | 9.83E-09 | 1.83E-08 | 3.86E-05 |
| T1DM | leukocyte | *C4B* | 5.464021612 | 0.000283458 | 0.000332549 | 1 |
| T1DM | leukocyte | *LINC00365* | 3.237859776 | 4.30E-07 | 5.90E-07 | 0.001688046 |
| T1DM | leukocyte | *LINC00115* | -1.864911899 | 1.85E-09 | 8.14E-09 | 7.25E-06 |
| T1DM | leukocyte | *LINC00937* | -0.83245945 | 1.24E-09 | 8.14E-09 | 4.86E-06 |
| T1DM | leukocyte | *TMEM185B* | -0.752009813 | 1.54E-09 | 8.14E-09 | 6.03E-06 |
| T1DM | leukocyte | *CYP4F26P* | 3.824068683 | 3.99E-09 | 9.64E-09 | 1.57E-05 |
| T1DM | leukocyte | *WDR46* | -0.640881331 | 6.68E-09 | 1.35E-08 | 2.62E-05 |
| T1DM | leukocyte | *ZNF717* | -0.423927112 | 5.47E-07 | 7.44E-07 | 0.002151104 |
| T1DM | leukocyte | *EPPK1* | -1.124833322 | 7.01E-06 | 8.80E-06 | 0.027536177 |
| T1DM | leukocyte | *ZNF883* | 0.784576259 | 0.000493561 | 0.000573389 | 1 |
| T1DM | leukocyte | *PROB1* | -1.324825358 | 1.98E-09 | 8.14E-09 | 7.80E-06 |
| T1DM | leukocyte | *ARL17B* | -1.5140467 | 4.18E-07 | 5.75E-07 | 0.001643118 |
| T1DM | leukocyte | *DHFR* | -0.864718329 | 4.84E-07 | 6.60E-07 | 0.001899678 |
| T1DM | leukocyte | *PSMG3-AS1* | -0.854199739 | 3.21E-09 | 8.74E-09 | 1.26E-05 |
| T1DM | leukocyte | *HSBP1* | -0.573520977 | 2.61E-09 | 8.21E-09 | 1.02E-05 |
| T1DM | leukocyte | *RPS18* | -0.386509452 | 3.15E-09 | 8.69E-09 | 1.24E-05 |
| T1DM | leukocyte | *DIRC3* | -1.97684783 | 2.71E-05 | 3.32E-05 | 0.106324467 |
| T1DM | leukocyte | *LINC00865* | -1.730354084 | 2.58E-09 | 8.21E-09 | 1.01E-05 |
| T1DM | leukocyte | *HLA-DQB2* | 1.716026932 | 2.08E-35 | 8.18E-32 | 8.18E-32 |
| T1DM | leukocyte | *GOLGA8N* | -0.756112353 | 1.21E-07 | 1.76E-07 | 0.000474311 |
| T1DM | leukocyte | *LINC00665* | -0.527902673 | 3.18E-09 | 8.70E-09 | 1.25E-05 |
| T1DM | leukocyte | *LYRM9* | -0.811470903 | 1.39E-09 | 8.14E-09 | 5.46E-06 |
| T1DM | leukocyte | *SNHG15* | -0.634235964 | 4.80E-09 | 1.07E-08 | 1.89E-05 |
| T1DM | leukocyte | *RPS28* | -0.414003762 | 2.92E-09 | 8.49E-09 | 1.15E-05 |
| T1DM | leukocyte | *TRIM26* | -0.897179124 | 4.94E-11 | 8.14E-09 | 1.94E-07 |
| T1DM | leukocyte | *ZNF879* | -0.912690472 | 2.14E-09 | 8.18E-09 | 8.39E-06 |
| T1DM | leukocyte | *PINLYP* | -1.409252819 | 5.36E-09 | 1.15E-08 | 2.11E-05 |
| T1DM | leukocyte | *MCIDAS* | 25.90276846 | 2.20E-07 | 3.12E-07 | 0.000864304 |
| T1DM | leukocyte | *HLA-B* | -0.250661303 | 0.000138256 | 0.00016426 | 0.543207933 |
| T1DM | leukocyte | *ANKRD65* | -0.630906757 | 1.51E-09 | 8.14E-09 | 5.92E-06 |
| T1DM | leukocyte | *C12orf75* | -0.389604348 | 1.37E-09 | 8.14E-09 | 5.38E-06 |
| T1DM | leukocyte | *LINC00957* | -1.049249006 | 3.43E-07 | 4.75E-07 | 0.001346664 |
| T1DM | leukocyte | *B3GALT4* | -0.677907326 | 7.99E-13 | 2.85E-10 | 3.14E-09 |
| T1DM | leukocyte | *ARHGEF38* | -0.734394458 | 0.000224919 | 0.000265059 | 0.8837071 |
| T1DM | leukocyte | *ZNF737* | -0.9345784 | 2.73E-09 | 8.28E-09 | 1.07E-05 |
| T1DM | leukocyte | *SHISA9* | -5.323278378 | 9.79E-10 | 8.14E-09 | 3.85E-06 |
| T1DM | leukocyte | *HLA-DQA2* | 0.093856908 | 4.45E-12 | 1.45E-09 | 1.75E-08 |
| T1DM | leukocyte | *LRRC37A2* | -0.815458758 | 8.49E-07 | 1.13E-06 | 0.003335952 |
| T1DM | leukocyte | *TXNDC5* | -0.686357625 | 1.21E-09 | 8.14E-09 | 4.76E-06 |
| T1DM | leukocyte | *RBM14* | -0.868122161 | 3.93E-09 | 9.56E-09 | 1.54E-05 |
| T1DM | leukocyte | *PCDHA13* | -1.478878509 | 0.000196799 | 0.000232758 | 0.773223412 |
| T1DM | leukocyte | *NME1* | -0.502153882 | 2.50E-09 | 8.21E-09 | 9.82E-06 |
| T1DM | leukocyte | *CDRT4* | -1.518388513 | 1.13E-09 | 8.14E-09 | 4.42E-06 |
| T1DM | leukocyte | *APOBEC3G* | 1.227270374 | 6.21E-07 | 8.39E-07 | 0.002441087 |
| T1DM | leukocyte | *AMY2B* | -0.570969792 | 1.47E-09 | 8.14E-09 | 5.78E-06 |
| T1DM | leukocyte | *LY6G5B* | -0.675800741 | 1.21E-06 | 1.60E-06 | 0.0047729 |
| T1DM | leukocyte | *PSMB9* | -0.197028437 | 0.029269253 | 0.0319264 | 1 |
| T1DM | leukocyte | *PCDHGC3* | -0.746483963 | 1.20E-07 | 1.75E-07 | 0.000470891 |
| T1DM | leukocyte | *SMKR1* | -0.846184709 | 7.24E-09 | 1.44E-08 | 2.85E-05 |

| | | | | | | |
|---|---|---|---|---|---|---|
| T1DM | leukocyte | *C1QTNF9* | -4.678587569 | 0.018908449 | 0.020769162 | 1 |
| T1DM | leukocyte | *LINC00886* | -0.890499914 | 1.70E-07 | 2.44E-07 | 0.000669636 |
| T1DM | leukocyte | *MIF* | -0.401870237 | 2.91E-09 | 8.49E-09 | 1.14E-05 |
| T1DM | leukocyte | *NSUN6* | -0.668871976 | 1.48E-09 | 8.14E-09 | 5.83E-06 |
| T1DM | leukocyte | *HLA-DOB* | 2.408624987 | 4.88E-27 | 6.40E-24 | 1.92E-23 |
| T1DM | leukocyte | *CRCP* | -0.779509521 | 7.53E-10 | 8.14E-09 | 2.96E-06 |
| T1DM | leukocyte | *ARHGAP8* | -0.573359258 | 0.000123091 | 0.000146598 | 0.483625865 |
| T1DM | leukocyte | *PLEKHO2* | 0.497156347 | 0.001243431 | 0.001425573 | 1 |
| T1DM | leukocyte | *C8orf58* | -1.007322632 | 3.95E-08 | 6.30E-08 | 0.000155337 |
| T1DM | leukocyte | *PWP2* | 2.118421309 | 0.008132501 | 0.009069712 | 1 |
| T1DM | leukocyte | *PI4KA* | -0.948399907 | 1.88E-07 | 2.69E-07 | 0.000738799 |
| T1DM | leukocyte | *AMACR* | -0.57576344 | 4.97E-09 | 1.10E-08 | 1.95E-05 |
| T1DM | leukocyte | *ARHGDIG* | -0.682917462 | 8.19E-06 | 1.03E-05 | 0.032195689 |
| T1DM | leukocyte | *BGLAP* | -0.87990552 | 6.69E-06 | 8.42E-06 | 0.026296636 |
| T1DM | leukocyte | *MRPL20* | -0.526207694 | 2.47E-08 | 4.10E-08 | 9.70E-05 |
| T1DM | leukocyte | *LINC00960* | -0.394394112 | 6.85E-05 | 8.26E-05 | 0.269248223 |
| T1DM | leukocyte | *HLA-DMB* | 0.087393404 | 1.62E-10 | 8.14E-09 | 6.36E-07 |
| T1DM | leukocyte | *DECR2* | -0.713280751 | 1.64E-09 | 8.14E-09 | 6.46E-06 |
| T1DM | leukocyte | *GNG10* | -0.554651578 | 1.31E-08 | 2.35E-08 | 5.13E-05 |
| T1DM | leukocyte | *EIF4EBP3* | -0.927076379 | 2.77E-10 | 8.14E-09 | 1.09E-06 |
| T1DM | leukocyte | *PSG4* | 2.246323025 | 2.98E-07 | 4.15E-07 | 0.001169801 |
| T1DM | leukocyte | *MICAL3* | -0.662489524 | 1.48E-09 | 8.14E-09 | 5.82E-06 |
| T1DM | leukocyte | *STON1* | -0.823933974 | 7.55E-07 | 1.01E-06 | 0.002968301 |
| T1DM | leukocyte | *VSIG8* | -1.809625604 | 1.45E-09 | 8.14E-09 | 5.69E-06 |
| T1DM | leukocyte | *C4orf48* | -0.486452423 | 2.21E-09 | 8.21E-09 | 8.68E-06 |
| T1DM | leukocyte | *UPK3B* | 1.63660568 | 0.004891843 | 0.005494583 | 1 |
| T1DM | leukocyte | *IL10RB* | -1.200653286 | 8.81E-07 | 1.17E-06 | 0.003461453 |
| T1DM | leukocyte | *PLA2G4B* | -0.901513519 | 6.34E-07 | 8.55E-07 | 0.002491107 |
| T1DM | leukocyte | *LEFTY1* | -5.532421282 | 1.97E-08 | 3.34E-08 | 7.73E-05 |
| T1DM | leukocyte | *TTC4* | -0.718982967 | 2.08E-09 | 8.15E-09 | 8.17E-06 |
| T1DM | leukocyte | *JMJD7* | 0.834121246 | 0.009493112 | 0.010545218 | 1 |
| T1DM | leukocyte | *APOBEC3D* | -2.640945823 | 2.23E-08 | 3.74E-08 | 8.76E-05 |
| T1DM | leukocyte | *GSTA1* | -0.400657475 | 3.46E-09 | 8.97E-09 | 1.36E-05 |
| T1DM | leukocyte | *GSTA2* | -0.532044246 | 1.44E-09 | 8.14E-09 | 5.66E-06 |
| T1DM | leukocyte | *TMEM141* | -0.773814782 | 1.26E-08 | 2.27E-08 | 4.94E-05 |
| T1DM | leukocyte | *CFHR1* | 5.160101915 | 2.38E-06 | 3.07E-06 | 0.009345148 |
| T1DM | leukocyte | *APOBEC3C* | -1.303985633 | 1.32E-07 | 1.91E-07 | 0.000517796 |
| T1DM | leukocyte | *C4A* | -9.995810018 | 3.04E-24 | 2.99E-21 | 1.19E-20 |
| T1DM | leukocyte | *NEAT1* | -0.440835924 | 5.83E-09 | 1.22E-08 | 2.29E-05 |
| T1DM | leukocyte | *LINC00920* | -5.142378259 | 5.63E-07 | 7.63E-07 | 0.002212141 |
| T1DM | leukocyte | *UBAP1L* | -0.735392418 | 2.70E-09 | 8.26E-09 | 1.06E-05 |
| T1DM | leukocyte | *TWF2* | -0.718693236 | 1.55E-06 | 2.03E-06 | 0.006088762 |
| T1DM | leukocyte | *LINC00926* | 3.995444818 | 1.05E-06 | 1.39E-06 | 0.004137932 |
| T1DM | leukocyte | *BCKDHA* | -0.684956018 | 5.42E-09 | 1.16E-08 | 2.13E-05 |
| T1DM | leukocyte | *PCDHAC1* | -1.079607217 | 1.46E-09 | 8.14E-09 | 5.72E-06 |
| T1DM | leukocyte | *TMEM150C* | -0.80810391 | 3.50E-09 | 9.01E-09 | 1.37E-05 |
| T1DM | leukocyte | *LINC01091* | -0.512391689 | 7.77E-10 | 8.14E-09 | 3.05E-06 |
| T1DM | leukocyte | *PDCD6* | -0.636605294 | 2.50E-09 | 8.21E-09 | 9.82E-06 |
| T1DM | leukocyte | *GOLGA8K* | -12.15079308 | 2.45E-07 | 3.46E-07 | 0.000962329 |
| T1DM | leukocyte | *YJEFN3* | -1.231494442 | 9.66E-10 | 8.14E-09 | 3.79E-06 |

| | | | | | | |
|---|---|---|---|---|---|---|
| T1DM | leukocyte | *PCDHA10* | -0.657722436 | 1.29E-09 | 8.14E-09 | 5.07E-06 |
| T1DM | leukocyte | *ZNF718* | -0.852271998 | 3.82E-08 | 6.11E-08 | 0.000150064 |
| T1DM | leukocyte | *CDK3* | 0.333717758 | 3.34E-06 | 4.27E-06 | 0.013128511 |
| T1DM | leukocyte | *SNHG18* | -0.702454902 | 1.82E-09 | 8.14E-09 | 7.15E-06 |
| T1DM | leukocyte | *ZNF550* | -0.779353491 | 5.33E-09 | 1.15E-08 | 2.09E-05 |
| T1DM | leukocyte | *LINC01099* | -0.850706855 | 1.52E-09 | 8.14E-09 | 5.97E-06 |
| T1DM | leukocyte | *PCDHA12* | -0.85827359 | 1.02E-05 | 1.27E-05 | 0.039904843 |
| T1DM | leukocyte | *ALG11* | -1.195140809 | 2.51E-09 | 8.21E-09 | 9.88E-06 |
| T1DM | leukocyte | *ZNF260* | -0.994361513 | 2.84E-06 | 3.66E-06 | 0.011171098 |
| T1DM | leukocyte | *LYN* | 0.68091902 | 0.021641119 | 0.023704477 | 1 |
| T1DM | leukocyte | *AP5B1* | -1.074731554 | 9.85E-08 | 1.46E-07 | 0.000386968 |
| T1DM | leukocyte | *SIGLEC12* | 1.517241578 | 3.64E-09 | 9.18E-09 | 1.43E-05 |
| T1DM | leukocyte | *PABPC4L* | -1.987353578 | 1.63E-09 | 8.14E-09 | 6.42E-06 |
| T1DM | leukocyte | *FPGT* | 0.693762549 | 0.014001887 | 0.015457549 | 1 |
| T1DM | leukocyte | *NPIPA2* | -7.612300136 | 1.01E-09 | 8.14E-09 | 3.98E-06 |
| T1DM | leukocyte | *DPP3* | -0.500085456 | 1.12E-05 | 1.40E-05 | 0.044042279 |
| T1DM | leukocyte | *BRK1* | -0.502381703 | 4.98E-09 | 1.10E-08 | 1.96E-05 |
| T1DM | leukocyte | *EID3* | -1.35176857 | 3.86E-09 | 9.50E-09 | 1.52E-05 |
| T1DM | leukocyte | *SNHG9* | -0.595375295 | 9.60E-09 | 1.80E-08 | 3.77E-05 |
| T1DM | leukocyte | *PCDHA3* | -0.894943833 | 4.34E-06 | 5.50E-06 | 0.017036123 |
| T1DM | leukocyte | *ZNF432* | -1.291980043 | 1.96E-09 | 8.14E-09 | 7.72E-06 |
| T1DM | leukocyte | *LINC01152* | -3.38676528 | 1.50E-09 | 8.14E-09 | 5.91E-06 |
| T1DM | leukocyte | *SMLR1* | 2.800222641 | 5.75E-09 | 1.20E-08 | 2.26E-05 |
| T1DM | leukocyte | *ZNF10* | -0.871952583 | 8.66E-10 | 8.14E-09 | 3.40E-06 |
| T1DM | leukocyte | *HMBS* | -0.310210226 | 0.025695217 | 0.028074668 | 1 |
| T1DM | leukocyte | *TAS2R31* | -2.549361225 | 6.55E-09 | 1.33E-08 | 2.58E-05 |
| T1DM | leukocyte | *LIMS3* | -4.870709803 | 0.008080956 | 0.009014786 | 1 |
| T1DM | leukocyte | *GATC* | -0.893469997 | 9.57E-10 | 8.14E-09 | 3.76E-06 |
| T1DM | leukocyte | *TEN1* | -1.143410848 | 0.000115213 | 0.00013734 | 0.45267212 |
| T1DM | leukocyte | *CHURC1* | -0.533078785 | 4.03E-09 | 9.70E-09 | 1.58E-05 |
| T1DM | leukocyte | *LINC00638* | -4.302582634 | 2.74E-07 | 3.84E-07 | 0.001076133 |
| T1DM | leukocyte | *SLC25A21-AS1* | -0.702478052 | 1.61E-09 | 8.14E-09 | 6.32E-06 |
| T1DM | leukocyte | *TMEM179* | -0.509779288 | 1.78E-09 | 8.14E-09 | 6.98E-06 |
| T1DM | leukocyte | *IQCH-AS1* | -1.033819116 | 4.47E-07 | 6.12E-07 | 0.001755341 |
| T1DM | leukocyte | *SLC22A31* | -0.526064224 | 0.003963945 | 0.004471531 | 1 |
| T1DM | leukocyte | *SH3RF3-AS1* | 1.546382496 | 1.73E-06 | 2.25E-06 | 0.006780538 |
| T1DM | leukocyte | *PWRN1* | -0.874586076 | 7.01E-08 | 1.07E-07 | 0.000275253 |
| T1DM | leukocyte | *TMEM178B* | -0.636623485 | 3.56E-09 | 9.10E-09 | 1.40E-05 |
| T1DM | leukocyte | *LINC01003* | -0.779717304 | 4.21E-09 | 9.96E-09 | 1.65E-05 |
| T1DM | leukocyte | *GOLGA8H* | -10.52581079 | 1.74E-07 | 2.49E-07 | 0.000682581 |
| T1DM | leukocyte | *MRPL12* | -0.83122724 | 2.42E-09 | 8.21E-09 | 9.52E-06 |
| T1DM | leukocyte | *LINC00667* | -0.718762068 | 2.23E-09 | 8.21E-09 | 8.76E-06 |
| T1DM | leukocyte | *FSBP* | -1.011489174 | 1.35E-09 | 8.14E-09 | 5.31E-06 |
| T1DM | leukocyte | *MYO15B* | -1.315655666 | 2.19E-07 | 3.11E-07 | 0.000859956 |
| T1DM | leukocyte | *LINC00663* | -0.924021006 | 8.01E-07 | 1.07E-06 | 0.00314802 |
| T1DM | leukocyte | *ZNF285* | -1.637516309 | 4.52E-09 | 1.04E-08 | 1.78E-05 |
| T1DM | leukocyte | *FBXO17* | -0.581498697 | 1.13E-09 | 8.14E-09 | 4.45E-06 |
| T1DM | leukocyte | *LINC00528* | 0.420810732 | 3.38E-08 | 5.48E-08 | 0.000132725 |
| T1DM | leukocyte | *SNHG8* | -0.489792752 | 1.80E-09 | 8.14E-09 | 7.07E-06 |
| T1DM | leukocyte | *MILR1* | 0.657393354 | 1.79E-08 | 3.07E-08 | 7.02E-05 |

| | | | | | | |
|---|---|---|---|---|---|---|
| T1DM | leukocyte | *GTF2H5* | -0.618552173 | 4.60E-09 | 1.04E-08 | 1.81E-05 |
| T1DM | mesenchymal_cell | *CEP68* | 0.348351975 | 0.000159927 | 0.019041039 | 0.628354292 |
| T1DM | mesenchymal_cell | *GPATCH1* | 0.558100378 | 0.000208894 | 0.024139529 | 0.820743997 |
| T1DM | mesenchymal_cell | *BTN2A2* | -0.212060841 | 0.000391173 | 0.042692206 | 1 |
| T1DM | mesenchymal_cell | *UQCC2* | 0.452736717 | 1.15E-05 | 0.001677589 | 0.045294908 |
| T1DM | mesenchymal_cell | *ENGASE* | 0.194384966 | 0.00043889 | 0.046605381 | 1 |
| T1DM | mesenchymal_cell | *HLA-DQB1* | -0.288176327 | 2.56E-23 | 2.01E-20 | 1.01E-19 |
| T1DM | mesenchymal_cell | *BTN3A2* | -0.085453142 | 1.27E-05 | 0.001784055 | 0.049953552 |
| T1DM | mesenchymal_cell | *ZKSCAN3* | -0.525284169 | 9.45E-09 | 2.18E-06 | 3.71E-05 |
| T1DM | mesenchymal_cell | *HLA-DRB1* | -0.098962803 | 2.84E-07 | 5.07E-05 | 0.001114982 |
| T1DM | mesenchymal_cell | *SFTA2* | -0.234955038 | 1.02E-09 | 2.50E-07 | 4.00E-06 |
| T1DM | mesenchymal_cell | *RPS26* | 0.242443253 | 0.000159785 | 0.019041039 | 0.627796376 |
| T1DM | mesenchymal_cell | *HLA-DRB5* | -0.140271059 | 7.97E-16 | 3.48E-13 | 3.13E-12 |
| T1DM | mesenchymal_cell | *HSD17B8* | 0.603100824 | 4.32E-24 | 4.24E-21 | 1.70E-20 |
| T1DM | mesenchymal_cell | *HLA-DMA* | 0.242350085 | 1.60E-28 | 2.09E-25 | 6.27E-25 |
| T1DM | mesenchymal_cell | *NOTCH4* | -0.260066779 | 2.85E-17 | 1.60E-14 | 1.12E-13 |
| T1DM | mesenchymal_cell | *AGER* | 0.476404239 | 2.14E-06 | 0.00033678 | 0.008419508 |
| T1DM | mesenchymal_cell | *RNF5* | -0.386473182 | 1.95E-13 | 6.37E-11 | 7.64E-10 |
| T1DM | mesenchymal_cell | *EHMT2* | 0.608159436 | 2.59E-05 | 0.003280272 | 0.101688444 |
| T1DM | mesenchymal_cell | *MICA* | 0.093123479 | 1.14E-05 | 0.001677589 | 0.044758602 |
| T1DM | mesenchymal_cell | *HLA-C* | 0.125137352 | 0.000283464 | 0.031820855 | 1 |
| T1DM | mesenchymal_cell | *CCHCR1* | 0.387088224 | 1.12E-07 | 2.33E-05 | 0.000441929 |
| T1DM | mesenchymal_cell | *PSORS1C1* | -0.177825285 | 3.54E-11 | 9.94E-09 | 1.39E-07 |
| T1DM | mesenchymal_cell | *RNF39* | 0.351438706 | 5.49E-14 | 1.96E-11 | 2.16E-10 |
| T1DM | mesenchymal_cell | *ZFP57* | 0.268512124 | 8.13E-07 | 0.000133152 | 0.003195642 |
| T1DM | mesenchymal_cell | *HCG27* | 0.256802562 | 1.36E-08 | 2.97E-06 | 5.35E-05 |
| T1DM | mesenchymal_cell | *ATF6B* | 0.407481973 | 1.29E-11 | 3.89E-09 | 5.06E-08 |
| T1DM | mesenchymal_cell | *PPT2* | 0.384973498 | 2.41E-07 | 4.74E-05 | 0.000947363 |
| T1DM | mesenchymal_cell | *C4B* | 2.164702984 | 2.59E-07 | 4.84E-05 | 0.001015811 |
| T1DM | mesenchymal_cell | *HLA-DQB2* | 1.081329738 | 1.25E-51 | 4.93E-48 | 4.93E-48 |
| T1DM | mesenchymal_cell | *HLA-B* | 0.160946451 | 2.41E-05 | 0.003158508 | 0.094755236 |
| T1DM | mesenchymal_cell | *ZSCAN31* | 0.15202471 | 3.22E-07 | 5.50E-05 | 0.001264232 |
| T1DM | mesenchymal_cell | *B3GALT4* | -0.254859134 | 2.27E-05 | 0.003076309 | 0.089212952 |
| T1DM | mesenchymal_cell | *HLA-DQA2* | 0.448564337 | 6.98E-33 | 1.37E-29 | 2.74E-29 |
| T1DM | mesenchymal_cell | *PSMB9* | 0.293126454 | 1.02E-09 | 2.50E-07 | 3.99E-06 |
| T1DM | mesenchymal_cell | *HLA-DOB* | 0.596012192 | 1.60E-16 | 7.86E-14 | 6.29E-13 |
| T1DM | mesenchymal_cell | *HLA-DMB* | 0.490643988 | 5.23E-14 | 1.96E-11 | 2.06E-10 |
| T1DM | mesenchymal_cell | *C4A* | -2.679420158 | 1.84E-21 | 1.20E-18 | 7.23E-18 |
| T1DM | PP_cell | *BTN2A2* | -0.417583381 | 0.000257227 | 0.037431237 | 1 |
| T1DM | PP_cell | *ENGASE* | 0.324638666 | 0.000222071 | 0.03355832 | 0.872516314 |
| T1DM | PP_cell | *HLA-DQB1* | -0.631844149 | 1.17E-28 | 1.84E-25 | 4.58E-25 |
| T1DM | PP_cell | *BTN3A2* | -0.142114363 | 8.75E-05 | 0.014318254 | 0.343638107 |
| T1DM | PP_cell | *ZKSCAN3* | -0.731676744 | 1.34E-06 | 0.000262646 | 0.00525292 |
| T1DM | PP_cell | *HLA-DRB1* | -0.248757107 | 6.63E-10 | 1.74E-07 | 2.61E-06 |
| T1DM | PP_cell | *SFTA2* | -0.370112356 | 4.77E-09 | 1.17E-06 | 1.87E-05 |
| T1DM | PP_cell | *HLA-DRB5* | -0.429361523 | 1.16E-16 | 5.69E-14 | 4.55E-13 |
| T1DM | PP_cell | *HSD17B8* | 0.438575183 | 2.76E-08 | 6.37E-06 | 0.000108281 |
| T1DM | PP_cell | *HLA-DMA* | 0.419682567 | 2.28E-27 | 2.24E-24 | 8.95E-24 |
| T1DM | PP_cell | *NOTCH4* | -0.726933956 | 9.35E-26 | 7.35E-23 | 3.68E-22 |
| T1DM | PP_cell | *RNF5* | -0.638819902 | 1.05E-12 | 4.13E-10 | 4.13E-09 |

| | | | | | | |
|---|---|---|---|---|---|---|
| T1DM | PP_cell | *SLC44A4* | -0.205907889 | 0.000319121 | 0.043525559 | 1 |
| T1DM | PP_cell | *MICA* | 0.209373819 | 4.06E-07 | 8.40E-05 | 0.001595457 |
| T1DM | PP_cell | *PSORS1C1* | -0.322838073 | 1.95E-11 | 6.38E-09 | 7.65E-08 |
| T1DM | PP_cell | *TRIM31* | -1.716230849 | 0.000321263 | 0.043525559 | 1 |
| T1DM | PP_cell | *RNF39* | 0.378700201 | 2.04E-07 | 4.45E-05 | 0.00080189 |
| T1DM | PP_cell | *ZFP57* | 0.461264161 | 2.23E-06 | 0.000416437 | 0.008745173 |
| T1DM | PP_cell | *ATF6B* | 0.586626748 | 3.85E-10 | 1.08E-07 | 1.51E-06 |
| T1DM | PP_cell | *PPT2* | 1.14728567 | 9.13E-13 | 3.99E-10 | 3.59E-09 |
| T1DM | PP_cell | *C4B* | 3.131498624 | 1.99E-05 | 0.003555647 | 0.078224229 |
| T1DM | PP_cell | *HLA-DQB2* | 1.793953017 | 9.15E-46 | 3.60E-42 | 3.60E-42 |
| T1DM | PP_cell | *HLA-B* | 0.243435054 | 0.000167764 | 0.026365863 | 0.659146581 |
| T1DM | PP_cell | *B3GALT4* | -0.397828616 | 8.10E-05 | 0.013842616 | 0.318380163 |
| T1DM | PP_cell | *HLA-DQA2* | 0.701517073 | 1.41E-28 | 1.84E-25 | 5.53E-25 |
| T1DM | PP_cell | *PSMB9* | 0.669560985 | 6.66E-12 | 2.38E-09 | 2.62E-08 |
| T1DM | PP_cell | *HLA-DOB* | 1.240414239 | 5.06E-19 | 2.84E-16 | 1.99E-15 |
| T1DM | PP_cell | *HLA-DMB* | 0.693262541 | 1.29E-10 | 3.91E-08 | 5.09E-07 |
| T1DM | PP_cell | *C4A* | -4.851704286 | 2.48E-22 | 1.63E-19 | 9.76E-19 |
| T2DM | acinar_cell | *RBM6* | -0.754793139 | 1.80E-07 | 6.44E-05 | 0.000708573 |
| T2DM | acinar_cell | *MLXIPL* | 0.535189476 | 0.00010249 | 0.011185597 | 0.402681487 |
| T2DM | acinar_cell | *CEP68* | 0.964270143 | 6.24E-06 | 0.001226297 | 0.024525945 |
| T2DM | acinar_cell | *LTF* | -0.914212248 | 0.000686743 | 0.049966895 | 1 |
| T2DM | acinar_cell | *BAK1* | 1.280540908 | 4.27E-06 | 0.000922451 | 0.016794411 |
| T2DM | acinar_cell | *ITIH4* | -0.7115692 | 9.74E-05 | 0.011185597 | 0.382739294 |
| T2DM | acinar_cell | *TCF3* | -1.425565827 | 2.98E-06 | 0.000700712 | 0.011697388 |
| T2DM | acinar_cell | *ST6GAL1* | 0.669957685 | 0.000389308 | 0.032544479 | 1 |
| T2DM | acinar_cell | *MARK3* | -1.620499992 | 4.46E-06 | 0.000922451 | 0.017526562 |
| T2DM | acinar_cell | *CWF19L1* | -0.496144474 | 7.08E-05 | 0.009592382 | 0.278179092 |
| T2DM | acinar_cell | *CCDC134* | 1.164240546 | 0.000204076 | 0.020045325 | 0.801813018 |
| T2DM | acinar_cell | *OIP5* | 2.012782462 | 0.000100362 | 0.011185597 | 0.394323735 |
| T2DM | acinar_cell | *HSD17B1* | -0.625785989 | 0.000339439 | 0.02899253 | 1 |
| T2DM | acinar_cell | *CNTNAP1* | 0.622289993 | 4.11E-05 | 0.006451962 | 0.161299052 |
| T2DM | acinar_cell | *PRR4* | 0.449767647 | 0.000539124 | 0.041533688 | 1 |
| T2DM | acinar_cell | *DNAH7* | -0.672412341 | 0.000178542 | 0.017986968 | 0.701491756 |
| T2DM | acinar_cell | *FLT3* | 0.775955002 | 0.000505201 | 0.039698698 | 1 |
| T2DM | acinar_cell | *OPRL1* | -0.856803103 | 0.000480377 | 0.038518359 | 1 |
| T2DM | acinar_cell | *PTPN12* | -0.610007861 | 0.000297198 | 0.026538452 | 1 |
| T2DM | acinar_cell | *MAU2* | -1.495475188 | 3.03E-06 | 0.000700712 | 0.011912098 |
| T2DM | acinar_cell | *NOTCH2* | -0.6194768 | 2.12E-09 | 1.67E-06 | 8.33E-06 |
| T2DM | acinar_cell | *MAN2C1* | -0.468478519 | 1.79E-05 | 0.003050403 | 0.070159278 |
| T2DM | acinar_cell | *NPC1* | -1.842344081 | 2.79E-05 | 0.004571552 | 0.109717244 |
| T2DM | acinar_cell | *GIGYF1* | -1.64654733 | 7.59E-05 | 0.00994128 | 0.298238411 |
| T2DM | acinar_cell | *HSD17B12* | -0.285134048 | 0.000105609 | 0.011214586 | 0.414939684 |
| T2DM | acinar_cell | *APIP* | -0.407448892 | 0.000553641 | 0.041831821 | 1 |
| T2DM | acinar_cell | *PTPN14* | -0.513777922 | 4.62E-05 | 0.006981613 | 0.18152193 |
| T2DM | acinar_cell | *JAZF1* | -1.600126564 | 3.25E-12 | 4.26E-09 | 1.28E-08 |
| T2DM | acinar_cell | *CLEC18A* | -2.220775946 | 3.04E-09 | 1.99E-06 | 1.19E-05 |
| T2DM | acinar_cell | *CILP2* | -0.723637829 | 9.55E-05 | 0.011185597 | 0.375337955 |
| T2DM | acinar_cell | *SGPL1* | -0.999672114 | 0.000668356 | 0.049546606 | 1 |
| T2DM | acinar_cell | *INO80E* | -0.744666203 | 0.000240927 | 0.022013989 | 0.946601511 |
| T2DM | acinar_cell | *IRX3* | 2.653787077 | 3.34E-23 | 1.31E-19 | 1.31E-19 |

| | | | | | | |
|---|---|---|---|---|---|---|
| T2DM | acinar_cell | *FARSA* | -1.332008435 | 5.62E-05 | 0.008181197 | 0.220892326 |
| T2DM | acinar_cell | *HLA-DQB1* | -0.42004321 | 1.13E-08 | 5.41E-06 | 4.46E-05 |
| T2DM | acinar_cell | *UBA7* | -0.593708176 | 1.41E-06 | 0.000462888 | 0.005554653 |
| T2DM | acinar_cell | *VMO1* | 0.584895189 | 0.000337597 | 0.02899253 | 1 |
| T2DM | acinar_cell | *CAMK1D* | 1.80842789 | 0.000227406 | 0.021273249 | 0.893476444 |
| T2DM | acinar_cell | *FAM43B* | 2.943995455 | 0.000165969 | 0.01716033 | 0.652092525 |
| T2DM | acinar_cell | *HLA-DRB1* | -0.329684647 | 1.49E-10 | 1.47E-07 | 5.87E-07 |
| T2DM | acinar_cell | *HLA-DQA1* | -0.208280171 | 0.000401532 | 0.032867099 | 1 |
| T2DM | acinar_cell | *PIWIL2* | -0.593794866 | 0.000218863 | 0.020973448 | 0.859911383 |
| T2DM | acinar_cell | *HLA-DRB5* | -0.229387518 | 1.49E-05 | 0.002711248 | 0.058700589 |
| T2DM | acinar_cell | *NOTCH4* | -0.5612611 | 1.24E-08 | 5.41E-06 | 4.87E-05 |
| T2DM | acinar_cell | *RNF5* | -0.55849212 | 2.20E-06 | 0.000575108 | 0.008626619 |
| T2DM | acinar_cell | *HCG27* | 0.514807343 | 1.54E-06 | 0.000464581 | 0.00603955 |
| T2DM | acinar_cell | *DNLZ* | 0.314544719 | 8.21E-05 | 0.010078575 | 0.322514391 |
| T2DM | acinar_cell | *ATF6B* | 0.537276606 | 7.07E-05 | 0.009592382 | 0.277749948 |
| T2DM | acinar_cell | *SLC12A8* | -2.535840194 | 1.43E-07 | 5.61E-05 | 0.000561213 |
| T2DM | acinar_cell | *PPT2* | 0.944873489 | 2.12E-06 | 0.000575108 | 0.008336986 |
| T2DM | acinar_cell | *HLA-DQB2* | 0.953737898 | 4.04E-09 | 2.27E-06 | 1.59E-05 |
| T2DM | acinar_cell | *HLA-B* | 0.376317742 | 1.52E-05 | 0.002711248 | 0.059647459 |
| T2DM | acinar_cell | *HLA-DQA2* | 0.595067619 | 2.11E-13 | 4.14E-10 | 8.28E-10 |
| T2DM | acinar_cell | *JMJD7* | 1.027246108 | 8.19E-05 | 0.010078575 | 0.321590816 |
| T2DM | alpha_cell | *RBM6* | -0.127977665 | 7.62E-08 | 2.72E-05 | 0.000299239 |
| T2DM | alpha_cell | *BAIAP3* | -0.185238573 | 0.000636709 | 0.041010293 | 1 |
| T2DM | alpha_cell | *MLXIPL* | 0.085565517 | 0.000273378 | 0.022377128 | 1 |
| T2DM | alpha_cell | *GIPR* | -0.150925617 | 0.000516749 | 0.035005266 | 1 |
| T2DM | alpha_cell | *CEP68* | 0.179209104 | 4.84E-07 | 0.000146417 | 0.00190342 |
| T2DM | alpha_cell | *BAK1* | 0.219087846 | 2.33E-06 | 0.00057226 | 0.009156166 |
| T2DM | alpha_cell | *ITIH4* | -0.170227305 | 0.000197303 | 0.018457222 | 0.775203311 |
| T2DM | alpha_cell | *TCF3* | -0.225101587 | 1.11E-05 | 0.001985953 | 0.04369096 |
| T2DM | alpha_cell | *MARK3* | -0.366412534 | 6.21E-08 | 2.44E-05 | 0.000244148 |
| T2DM | alpha_cell | *CWF19L1* | -0.083146528 | 4.32E-05 | 0.005853332 | 0.169746627 |
| T2DM | alpha_cell | *CCDC134* | 0.179308131 | 0.000616429 | 0.040365829 | 1 |
| T2DM | alpha_cell | *AP1B1* | -0.294874627 | 0.000228264 | 0.020382933 | 0.896849032 |
| T2DM | alpha_cell | *OIP5* | 0.318902279 | 0.000156486 | 0.015845631 | 0.61483363 |
| T2DM | alpha_cell | *GCDH* | -0.363032588 | 0.000121373 | 0.014025759 | 0.476875803 |
| T2DM | alpha_cell | *HSD17B1* | -0.103610128 | 0.000264989 | 0.022151985 | 1 |
| T2DM | alpha_cell | *CNTNAP1* | 0.095472266 | 0.00011457 | 0.013640744 | 0.450144552 |
| T2DM | alpha_cell | *NNT* | -0.155930566 | 0.000161884 | 0.015901092 | 0.636043678 |
| T2DM | alpha_cell | *DNAH7* | -0.124894315 | 2.88E-05 | 0.004192955 | 0.113209793 |
| T2DM | alpha_cell | *FLT3* | 0.127152456 | 0.000395166 | 0.028229227 | 1 |
| T2DM | alpha_cell | *OPRL1* | -0.142829009 | 0.000322327 | 0.02389476 | 1 |
| T2DM | alpha_cell | *PTPN12* | -0.093286377 | 0.000812371 | 0.047685791 | 1 |
| T2DM | alpha_cell | *MAU2* | -0.261600723 | 2.50E-06 | 0.00057883 | 0.009840118 |
| T2DM | alpha_cell | *PPARG* | -0.359325626 | 0.000157287 | 0.015845631 | 0.617979617 |
| T2DM | alpha_cell | *NOTCH2* | -0.096931937 | 1.21E-08 | 5.29E-06 | 4.76E-05 |
| T2DM | alpha_cell | *FXYD2* | -0.16098264 | 0.000320388 | 0.02389476 | 1 |
| T2DM | alpha_cell | *MAN2C1* | -0.080040997 | 6.86E-06 | 0.001347115 | 0.026942305 |
| T2DM | alpha_cell | *NPC1* | -0.364045405 | 1.03E-05 | 0.001919713 | 0.040313977 |
| T2DM | alpha_cell | *CNTN4* | -0.078155116 | 0.000457277 | 0.032082913 | 1 |
| T2DM | alpha_cell | *GIGYF1* | -0.308013921 | 0.000135135 | 0.015169824 | 0.530943843 |

| | | | | | | |
|---|---|---|---|---|---|---|
| T2DM | alpha_cell | *HSD17B12* | -0.047549048 | 7.55E-05 | 0.009569469 | 0.296653549 |
| T2DM | alpha_cell | *APIP* | -0.071289964 | 0.000226538 | 0.020382933 | 0.890066031 |
| T2DM | alpha_cell | *PTPN14* | -0.076779464 | 0.000234608 | 0.020483902 | 0.921775592 |
| T2DM | alpha_cell | *JAZF1* | -0.345268477 | 7.65E-17 | 1.50E-13 | 3.00E-13 |
| T2DM | alpha_cell | *NUS1* | -0.31179408 | 0.000861979 | 0.049740491 | 1 |
| T2DM | alpha_cell | *CC2D1B* | -0.086156546 | 0.000813171 | 0.047685791 | 1 |
| T2DM | alpha_cell | *CLEC18A* | -0.34917244 | 7.75E-09 | 5.08E-06 | 3.05E-05 |
| T2DM | alpha_cell | *AP3S2* | 0.288153372 | 0.000914793 | 0.049919746 | 1 |
| T2DM | alpha_cell | *EXTL1* | -0.207631412 | 0.000898848 | 0.049740491 | 1 |
| T2DM | alpha_cell | *CILP2* | -0.118780579 | 7.10E-05 | 0.00930407 | 0.279122105 |
| T2DM | alpha_cell | *ICA1L* | -0.067856921 | 0.000892756 | 0.049740491 | 1 |
| T2DM | alpha_cell | *CASP7* | 0.065229004 | 0.000305925 | 0.023568214 | 1 |
| T2DM | alpha_cell | *CKB* | -0.111437658 | 0.000255607 | 0.021832143 | 1 |
| T2DM | alpha_cell | *PCBD1* | -0.201531273 | 0.000394839 | 0.028229227 | 1 |
| T2DM | alpha_cell | *DNAJC21* | -0.292096367 | 0.000799849 | 0.047685791 | 1 |
| T2DM | alpha_cell | *INO80E* | -0.151417015 | 1.41E-05 | 0.002410525 | 0.055442065 |
| T2DM | alpha_cell | *PPP1R3B* | -0.151157566 | 0.000882254 | 0.049740491 | 1 |
| T2DM | alpha_cell | *IRX3* | 0.426096048 | 2.07E-23 | 8.13E-20 | 8.13E-20 |
| T2DM | alpha_cell | *FARSA* | -0.238267161 | 2.23E-05 | 0.003365322 | 0.087498378 |
| T2DM | alpha_cell | *HLA-DQB1* | -0.068420431 | 1.05E-08 | 5.16E-06 | 4.13E-05 |
| T2DM | alpha_cell | *UBA7* | -0.092152265 | 4.79E-06 | 0.000990221 | 0.018814201 |
| T2DM | alpha_cell | *VMO1* | 0.09171573 | 0.00046602 | 0.032122687 | 1 |
| T2DM | alpha_cell | *ZNF320* | 0.039540615 | 0.000702122 | 0.044494123 | 1 |
| T2DM | alpha_cell | *CAMK1D* | 0.340727812 | 1.70E-05 | 0.002775206 | 0.066604953 |
| T2DM | alpha_cell | *FAM43B* | 0.433126065 | 0.000537115 | 0.035768194 | 1 |
| T2DM | alpha_cell | *DMWD* | -0.158910531 | 0.000772949 | 0.047685791 | 1 |
| T2DM | alpha_cell | *NUTM2B* | -0.954190279 | 0.000151651 | 0.015845631 | 0.595838548 |
| T2DM | alpha_cell | *HLA-DRB1* | -0.05361681 | 2.58E-10 | 2.53E-07 | 1.01E-06 |
| T2DM | alpha_cell | *XRCC6* | -0.226310491 | 0.000297967 | 0.023414237 | 1 |
| T2DM | alpha_cell | *HLA-DQA1* | -0.034680007 | 0.00028432 | 0.02279782 | 1 |
| T2DM | alpha_cell | *PIWIL2* | -0.089785581 | 0.000792935 | 0.047685791 | 1 |
| T2DM | alpha_cell | *HLA-DRB5* | -0.036766336 | 3.33E-05 | 0.004674874 | 0.130896462 |
| T2DM | alpha_cell | *NOTCH4* | -0.092005969 | 7.35E-09 | 5.08E-06 | 2.89E-05 |
| T2DM | alpha_cell | *RNF5* | -0.091256173 | 1.97E-06 | 0.000551556 | 0.007721781 |
| T2DM | alpha_cell | *HCG27* | 0.082946359 | 2.27E-06 | 0.00057226 | 0.008899946 |
| T2DM | alpha_cell | *DNLZ* | 0.048814151 | 0.000176385 | 0.016902873 | 0.693017812 |
| T2DM | alpha_cell | *ATF6B* | 0.084600094 | 0.000147658 | 0.015845631 | 0.580149086 |
| T2DM | alpha_cell | *SLC12A8* | -0.522598855 | 2.31E-07 | 7.58E-05 | 0.00090948 |
| T2DM | alpha_cell | *PPT2* | 0.159753729 | 3.08E-06 | 0.000672542 | 0.012105754 |
| T2DM | alpha_cell | *HLA-DQB2* | 0.153646367 | 9.70E-09 | 5.16E-06 | 3.81E-05 |
| T2DM | alpha_cell | *HLA-B* | 0.062434979 | 1.89E-05 | 0.002972209 | 0.07430522 |
| T2DM | alpha_cell | *HLA-DQA2* | 0.095677146 | 5.34E-13 | 7.00E-10 | 2.10E-09 |
| T2DM | alpha_cell | *JMJD7* | 0.16592955 | 9.16E-05 | 0.011252725 | 0.360087211 |
| T2DM | beta_cell | *RBM6* | -0.073336338 | 5.02E-07 | 0.0001408 | 0.001971204 |
| T2DM | beta_cell | *MLXIPL* | 0.059033694 | 3.90E-05 | 0.004929304 | 0.153319364 |
| T2DM | beta_cell | *CEP68* | 0.126274583 | 1.11E-08 | 5.45E-06 | 4.36E-05 |
| T2DM | beta_cell | *BAK1* | 0.152283218 | 1.26E-07 | 3.80E-05 | 0.000494197 |
| T2DM | beta_cell | *ANAPC4* | 0.528951373 | 3.99E-05 | 0.004929304 | 0.156820791 |
| T2DM | beta_cell | *ITIH4* | -0.099962396 | 0.000388247 | 0.023884252 | 1 |
| T2DM | beta_cell | *TCF3* | -0.126373996 | 6.63E-05 | 0.007657555 | 0.260356855 |

| | | | | | | |
|---|---|---|---|---|---|---|
| T2DM | beta_cell | *ST6GAL1* | 0.065692652 | 0.000675566 | 0.033598695 | 1 |
| T2DM | beta_cell | *MARK3* | -0.241527256 | 7.12E-07 | 0.000186514 | 0.002797703 |
| T2DM | beta_cell | *CD59* | 0.173250668 | 0.000389449 | 0.023884252 | 1 |
| T2DM | beta_cell | *MGST2* | 0.187469391 | 0.000210526 | 0.01621875 | 0.827156231 |
| T2DM | beta_cell | *UIMC1* | 0.208035336 | 0.000769076 | 0.036611569 | 1 |
| T2DM | beta_cell | *CWF19L1* | -0.047557424 | 0.000117364 | 0.01152805 | 0.461122014 |
| T2DM | beta_cell | *CCDC134* | 0.114003257 | 0.000335673 | 0.023137856 | 1 |
| T2DM | beta_cell | *TOM1* | 0.31823681 | 5.32E-05 | 0.006338095 | 0.209157139 |
| T2DM | beta_cell | *RBL2* | 0.126759431 | 0.000363618 | 0.023420566 | 1 |
| T2DM | beta_cell | *NOMO1* | 0.114546712 | 0.000628535 | 0.032071586 | 1 |
| T2DM | beta_cell | *OIP5* | 0.200153358 | 8.69E-05 | 0.008981758 | 0.341306789 |
| T2DM | beta_cell | *ACBD5* | 0.385867982 | 0.000596166 | 0.031084775 | 1 |
| T2DM | beta_cell | *DHRS11* | 0.153928941 | 0.000238365 | 0.018010316 | 0.936536429 |
| T2DM | beta_cell | *RAI1* | 0.576488723 | 0.000687705 | 0.033774888 | 1 |
| T2DM | beta_cell | *HSD17B1* | -0.057466036 | 0.000849606 | 0.039406797 | 1 |
| T2DM | beta_cell | *CNTNAP1* | 0.059062163 | 8.35E-05 | 0.008868126 | 0.328120666 |
| T2DM | beta_cell | *PDHX* | 0.080199146 | 0.001061096 | 0.046843227 | 1 |
| T2DM | beta_cell | *PRR4* | 0.044526584 | 0.000913507 | 0.041616947 | 1 |
| T2DM | beta_cell | *CDK2AP1* | 0.080902947 | 4.01E-05 | 0.004929304 | 0.157737732 |
| T2DM | beta_cell | *SEMA5A* | 0.249219582 | 0.00017513 | 0.0146401 | 0.688084682 |
| T2DM | beta_cell | *NNT* | -0.085707213 | 0.000974102 | 0.043491454 | 1 |
| T2DM | beta_cell | *DNAH7* | -0.070473906 | 0.000111835 | 0.011266642 | 0.439399039 |
| T2DM | beta_cell | *BCL11A* | 0.268334069 | 0.000450811 | 0.025804744 | 1 |
| T2DM | beta_cell | *IAPP* | 0.125601849 | 6.27E-06 | 0.001071367 | 0.024641433 |
| T2DM | beta_cell | *FLT3* | 0.077767186 | 0.000347695 | 0.023420566 | 1 |
| T2DM | beta_cell | *BHLHE41* | 0.244659956 | 0.000245007 | 0.018162877 | 0.962632464 |
| T2DM | beta_cell | *OPRL1* | -0.085537254 | 0.000358434 | 0.023420566 | 1 |
| T2DM | beta_cell | *BECN1* | 0.60394207 | 1.94E-06 | 0.000362411 | 0.007610634 |
| T2DM | beta_cell | *PLEKHG3* | 0.569098148 | 0.000458816 | 0.025804744 | 1 |
| T2DM | beta_cell | *MAU2* | -0.147090622 | 3.80E-05 | 0.004929304 | 0.149321066 |
| T2DM | beta_cell | *SYT5* | 0.059729639 | 0.000400953 | 0.023884252 | 1 |
| T2DM | beta_cell | *TRAF3* | 0.119851979 | 0.001116036 | 0.048185758 | 1 |
| T2DM | beta_cell | *PPARG* | -0.202164404 | 0.000664537 | 0.033473921 | 1 |
| T2DM | beta_cell | *NOTCH2* | -0.057509769 | 2.65E-08 | 8.68E-06 | 0.000104114 |
| T2DM | beta_cell | *MAN2C1* | -0.045870579 | 2.17E-05 | 0.003417891 | 0.085447265 |
| T2DM | beta_cell | *NPC1* | -0.218059335 | 0.00015067 | 0.012869143 | 0.591980599 |
| T2DM | beta_cell | *POGZ* | 0.105791846 | 0.000192497 | 0.015126395 | 0.756319738 |
| T2DM | beta_cell | *CNTN4* | -0.045495729 | 0.000852527 | 0.039406797 | 1 |
| T2DM | beta_cell | *SEC16A* | 0.102123649 | 0.000407032 | 0.023884252 | 1 |
| T2DM | beta_cell | *HSD17B12* | -0.027255742 | 0.000190758 | 0.015126395 | 0.749488625 |
| T2DM | beta_cell | *APIP* | -0.040607943 | 0.000556977 | 0.030393921 | 1 |
| T2DM | beta_cell | *PTPN14* | -0.04531086 | 0.000383729 | 0.023884252 | 1 |
| T2DM | beta_cell | *ACMSD* | 0.722682214 | 0.000148217 | 0.012869143 | 0.582345149 |
| T2DM | beta_cell | *JAZF1* | -0.206359772 | 2.11E-15 | 4.15E-12 | 8.29E-12 |
| T2DM | beta_cell | *CLEC18A* | -0.205441803 | 2.16E-08 | 8.29E-06 | 8.47E-05 |
| T2DM | beta_cell | *AP3S2* | 0.429731809 | 5.26E-10 | 3.45E-07 | 2.07E-06 |
| T2DM | beta_cell | *EXTL1* | -0.123099829 | 0.001111691 | 0.048185758 | 1 |
| T2DM | beta_cell | *CILP2* | -0.071584956 | 7.40E-05 | 0.008311021 | 0.290885748 |
| T2DM | beta_cell | *CASP7* | 0.041769701 | 0.000135761 | 0.012122878 | 0.533406618 |
| T2DM | beta_cell | *POLR2J3* | 0.194452781 | 0.000584015 | 0.031008035 | 1 |

| | | | | | | |
|---|---|---|---|---|---|---|
| T2DM | beta_cell | *INO80E* | -0.085646542 | 7.65E-05 | 0.008347736 | 0.300518486 |
| T2DM | beta_cell | *PHYHD1* | 0.031071775 | 0.000704769 | 0.034185655 | 1 |
| T2DM | beta_cell | *IRX3* | 0.268091009 | 1.28E-24 | 5.05E-21 | 5.05E-21 |
| T2DM | beta_cell | *IMP3* | 0.290767858 | 1.70E-06 | 0.000334427 | 0.006688542 |
| T2DM | beta_cell | *FARSA* | -0.132118327 | 0.000190447 | 0.015126395 | 0.74826477 |
| T2DM | beta_cell | *HLA-DQB1* | -0.040459271 | 2.32E-08 | 8.29E-06 | 9.12E-05 |
| T2DM | beta_cell | *UBA7* | -0.055557383 | 4.98E-06 | 0.000889012 | 0.019558267 |
| T2DM | beta_cell | *ARL6IP4* | 0.119681523 | 0.000268719 | 0.019551793 | 1 |
| T2DM | beta_cell | *VMO1* | 0.057591949 | 0.000303409 | 0.02128739 | 1 |
| T2DM | beta_cell | *ZNF320* | 0.024276237 | 0.000568042 | 0.030573089 | 1 |
| T2DM | beta_cell | *CAMK1D* | 0.372391357 | 2.08E-10 | 2.04E-07 | 8.15E-07 |
| T2DM | beta_cell | *FAM43B* | 0.277842445 | 0.000298788 | 0.02128739 | 1 |
| T2DM | beta_cell | *ZBTB40* | 0.200950607 | 0.000121167 | 0.011611322 | 0.47606421 |
| T2DM | beta_cell | *NUTM2B* | -0.518862928 | 0.000601284 | 0.031084775 | 1 |
| T2DM | beta_cell | *ZNF567* | 0.189864267 | 0.000459743 | 0.025804744 | 1 |
| T2DM | beta_cell | *HLA-DRB1* | -0.032165101 | 3.60E-10 | 2.83E-07 | 1.41E-06 |
| T2DM | beta_cell | *HLA-DQA1* | -0.020614873 | 0.000362043 | 0.023420566 | 1 |
| T2DM | beta_cell | *DDX42* | 0.146854886 | 0.000407291 | 0.023884252 | 1 |
| T2DM | beta_cell | *HLA-DRB5* | -0.022358449 | 3.25E-05 | 0.004565161 | 0.127824511 |
| T2DM | beta_cell | *NOTCH4* | -0.054303094 | 1.66E-08 | 7.26E-06 | 6.54E-05 |
| T2DM | beta_cell | *RNF5* | -0.052157336 | 8.12E-06 | 0.001329303 | 0.03190328 |
| T2DM | beta_cell | *HCG27* | 0.051717086 | 1.10E-06 | 0.000260467 | 0.004332086 |
| T2DM | beta_cell | *TAS2R14* | 0.042655791 | 0.000921526 | 0.041616947 | 1 |
| T2DM | beta_cell | *DNLZ* | 0.030037997 | 0.000133023 | 0.012122878 | 0.522647812 |
| T2DM | beta_cell | *ATF6B* | 0.056804703 | 2.91E-05 | 0.004236584 | 0.114387778 |
| T2DM | beta_cell | *SLC12A8* | -0.307916471 | 1.31E-06 | 0.000285318 | 0.005135727 |
| T2DM | beta_cell | *PPT2* | 0.101443639 | 1.13E-06 | 0.000260467 | 0.004427935 |
| T2DM | beta_cell | *HLA-DQB2* | 0.09374773 | 7.75E-09 | 4.35E-06 | 3.04E-05 |
| T2DM | beta_cell | *HLA-B* | 0.043358 | 1.65E-06 | 0.000334427 | 0.006486625 |
| T2DM | beta_cell | *HLA-DQA2* | 0.059239238 | 2.29E-13 | 3.00E-10 | 8.99E-10 |
| T2DM | beta_cell | *JMJD7* | 0.108769846 | 2.44E-05 | 0.003683469 | 0.095770183 |
| T2DM | beta_cell | *NEAT1* | 0.181013908 | 0.000125359 | 0.011727034 | 0.492535408 |
| T2DM | beta_cell | *BCKDHA* | 0.184039446 | 0.000773418 | 0.036611569 | 1 |
| T2DM | beta_cell | *TAS2R31* | 0.939670117 | 0.000519751 | 0.028761995 | 1 |
| T2DM | delta_cell | *RBM6* | -0.589289967 | 3.28E-07 | 0.00014299 | 0.001286913 |
| T2DM | delta_cell | *CEP68* | 0.612190801 | 6.47E-05 | 0.012098448 | 0.254067404 |
| T2DM | delta_cell | *BAK1* | 0.868954619 | 7.60E-05 | 0.013569574 | 0.298530623 |
| T2DM | delta_cell | *TCF3* | -0.79741734 | 5.02E-05 | 0.010514442 | 0.197330373 |
| T2DM | delta_cell | *MARK3* | -0.739955226 | 0.000378489 | 0.046735952 | 1 |
| T2DM | delta_cell | *CWF19L1* | -0.360869172 | 0.000235152 | 0.032996855 | 0.923911944 |
| T2DM | delta_cell | *HSD17B1* | -0.53563126 | 0.000144275 | 0.022674288 | 0.566857199 |
| T2DM | delta_cell | *CNTNAP1* | 0.455305661 | 0.00041015 | 0.04883275 | 1 |
| T2DM | delta_cell | *MAU2* | -0.954947839 | 1.92E-05 | 0.005029363 | 0.075440439 |
| T2DM | delta_cell | *NOTCH2* | -0.451759965 | 1.34E-07 | 6.58E-05 | 0.000526797 |
| T2DM | delta_cell | *MAN2C1* | -0.410393508 | 6.23E-06 | 0.002038418 | 0.024461017 |
| T2DM | delta_cell | *HSD17B12* | -0.243635288 | 6.44E-05 | 0.012098448 | 0.253191129 |
| T2DM | delta_cell | *APIP* | -0.37375048 | 0.000133109 | 0.021791062 | 0.522985488 |
| T2DM | delta_cell | *JAZF1* | -1.34058846 | 1.26E-13 | 2.48E-10 | 4.97E-10 |
| T2DM | delta_cell | *CC2D1B* | -0.454690478 | 0.000266231 | 0.036069708 | 1 |
| T2DM | delta_cell | *CLEC18A* | -1.351171726 | 1.68E-06 | 0.00059883 | 0.006587126 |

| | | | | | | |
|---|---|---|---|---|---|---|
| T2DM | delta_cell | *CILP2* | -0.576932895 | 0.000193255 | 0.029203819 | 0.759299297 |
| T2DM | delta_cell | *INO80E* | -0.693548723 | 1.72E-05 | 0.004815893 | 0.067422498 |
| T2DM | delta_cell | *IRX3* | 1.436886734 | 2.43E-14 | 9.54E-11 | 9.54E-11 |
| T2DM | delta_cell | *FARSA* | -0.986281883 | 5.08E-05 | 0.010514442 | 0.199774403 |
| T2DM | delta_cell | *HLA-DQB1* | -0.369178488 | 4.22E-09 | 3.31E-06 | 1.66E-05 |
| T2DM | delta_cell | *UBA7* | -0.462951059 | 1.42E-05 | 0.004278471 | 0.055620128 |
| T2DM | delta_cell | *FAM43B* | 2.353355859 | 0.000380644 | 0.046735952 | 1 |
| T2DM | delta_cell | *HLA-DRB1* | -0.28851223 | 1.18E-10 | 1.16E-07 | 4.63E-07 |
| T2DM | delta_cell | *HLA-DQA1* | -0.185795562 | 0.000228339 | 0.032996855 | 0.897143811 |
| T2DM | delta_cell | *HLA-DRB5* | -0.181741181 | 4.63E-05 | 0.010514442 | 0.182102274 |
| T2DM | delta_cell | *NOTCH4* | -0.461254576 | 2.12E-08 | 1.19E-05 | 8.33E-05 |
| T2DM | delta_cell | *RNF5* | -0.47227765 | 6.86E-07 | 0.000269479 | 0.00269479 |
| T2DM | delta_cell | *HCG27* | 0.264525114 | 0.000118014 | 0.020159919 | 0.463678142 |
| T2DM | delta_cell | *PPT2* | 0.737696359 | 2.26E-05 | 0.005557908 | 0.088926522 |
| T2DM | delta_cell | *HLA-DQB2* | 0.774841324 | 8.73E-09 | 5.71E-06 | 3.43E-05 |
| T2DM | delta_cell | *HLA-B* | 0.22642686 | 0.000361743 | 0.046735952 | 1 |
| T2DM | delta_cell | *HLA-DQA2* | 0.466021723 | 1.55E-12 | 2.03E-09 | 6.09E-09 |
| T2DM | duct_epithelial_cell | *RBM6* | -0.396360465 | 5.41E-06 | 0.001516945 | 0.021237225 |
| T2DM | duct_epithelial_cell | *MLXIPL* | 0.315263694 | 5.88E-05 | 0.009569986 | 0.230889356 |
| T2DM | duct_epithelial_cell | *CEP68* | 0.689445793 | 1.65E-07 | 8.11E-05 | 0.000648466 |
| T2DM | duct_epithelial_cell | *BAK1* | 0.648257522 | 8.31E-05 | 0.011666462 | 0.326660924 |
| T2DM | duct_epithelial_cell | *TCF3* | -0.633451251 | 2.02E-05 | 0.004175602 | 0.079336445 |
| T2DM | duct_epithelial_cell | *CWF19L1* | -0.289998526 | 0.000168284 | 0.020036034 | 0.661189115 |
| T2DM | duct_epithelial_cell | *OIP5* | 1.150965603 | 5.13E-05 | 0.009170606 | 0.201753337 |
| T2DM | duct_epithelial_cell | *HSD17B1* | -0.402373928 | 0.000190602 | 0.021396452 | 0.748875821 |
| T2DM | duct_epithelial_cell | *CNTNAP1* | 0.379721894 | 2.40E-05 | 0.004716974 | 0.094339474 |
| T2DM | duct_epithelial_cell | *CDK2AP1* | 0.408049473 | 0.000324159 | 0.033516316 | 1 |
| T2DM | duct_epithelial_cell | *MAU2* | -0.796360824 | 6.38E-05 | 0.009644654 | 0.250761002 |
| T2DM | duct_epithelial_cell | *PPARG* | -1.228072302 | 0.00060309 | 0.049594302 | 1 |
| T2DM | duct_epithelial_cell | *NOTCH2* | -0.30598468 | 1.89E-08 | 1.06E-05 | 7.42E-05 |
| T2DM | duct_epithelial_cell | *MAN2C1* | -0.276497979 | 4.20E-05 | 0.007859605 | 0.165051704 |
| T2DM | duct_epithelial_cell | *NPC1* | -1.006442432 | 0.000349693 | 0.034348625 | 1 |
| T2DM | duct_epithelial_cell | *HSD17B12* | -0.155194742 | 0.000529534 | 0.047284978 | 1 |
| T2DM | duct_epithelial_cell | *APIP* | -0.261115217 | 0.000381595 | 0.036567956 | 1 |
| T2DM | duct_epithelial_cell | *PTPN14* | -0.294873672 | 7.10E-05 | 0.010332015 | 0.278964393 |
| T2DM | duct_epithelial_cell | *JAZF1* | -0.859560004 | 3.27E-10 | 4.29E-07 | 1.29E-06 |
| T2DM | duct_epithelial_cell | *CLEC18A* | -0.936561864 | 8.23E-06 | 0.002072492 | 0.032330103 |
| T2DM | duct_epithelial_cell | *AP3S2* | 0.942323587 | 0.000340759 | 0.034329243 | 1 |
| T2DM | duct_epithelial_cell | *EXTL1* | -0.773307442 | 0.00023386 | 0.024833409 | 0.918836119 |
| T2DM | duct_epithelial_cell | *CILP2* | -0.462547936 | 1.85E-05 | 0.004028615 | 0.072515074 |
| T2DM | duct_epithelial_cell | *TMC8* | -0.325015952 | 0.000442683 | 0.041411944 | 1 |
| T2DM | duct_epithelial_cell | *INO80E* | -0.474856647 | 0.000206493 | 0.022536401 | 0.811310454 |
| T2DM | duct_epithelial_cell | *RGS19* | -0.494641433 | 0.000605886 | 0.049594302 | 1 |
| T2DM | duct_epithelial_cell | *DSCAML1* | -0.580855511 | 0.000476747 | 0.043561353 | 1 |
| T2DM | duct_epithelial_cell | *FARSA* | -0.83738955 | 5.42E-05 | 0.009250605 | 0.212763919 |
| T2DM | duct_epithelial_cell | *HLA-DQB1* | -0.266646872 | 5.07E-09 | 3.32E-06 | 1.99E-05 |
| T2DM | duct_epithelial_cell | *UBA7* | -0.333804121 | 2.03E-06 | 0.000663067 | 0.007956801 |
| T2DM | duct_epithelial_cell | *ZNF320* | 0.154363537 | 0.000567373 | 0.049537962 | 1 |
| T2DM | duct_epithelial_cell | *CAMK1D* | 1.588267309 | 6.85E-07 | 0.000299006 | 0.002691052 |
| T2DM | duct_epithelial_cell | *FAM43B* | 1.716695352 | 9.65E-05 | 0.013078654 | 0.379280955 |

| | | | | | | |
|---|---|---|---|---|---|---|
| T2DM | duct_epithelial_cell | *HLA-DRB1* | -0.220196697 | 2.20E-11 | 4.33E-08 | 8.66E-08 |
| T2DM | duct_epithelial_cell | *HLA-DQA1* | -0.137572365 | 0.000189892 | 0.021396452 | 0.746086703 |
| T2DM | duct_epithelial_cell | *HLA-DRB5* | -0.143260641 | 8.44E-06 | 0.002072492 | 0.033159867 |
| T2DM | duct_epithelial_cell | *NOTCH4* | -0.3483444 | 4.68E-09 | 3.32E-06 | 1.84E-05 |
| T2DM | duct_epithelial_cell | *RNF5* | -0.329101887 | 2.63E-06 | 0.000796259 | 0.010351371 |
| T2DM | duct_epithelial_cell | *HCG27* | 0.255348226 | 6.09E-05 | 0.009569986 | 0.239249657 |
| T2DM | duct_epithelial_cell | *TAS2R14* | 0.278498928 | 0.000583378 | 0.049594302 | 1 |
| T2DM | duct_epithelial_cell | *DNLZ* | 0.1880497 | 0.000161511 | 0.01983057 | 0.634578248 |
| T2DM | duct_epithelial_cell | *ATF6B* | 0.329209106 | 0.000104229 | 0.013650573 | 0.4095172 |
| T2DM | duct_epithelial_cell | *SLC12A8* | -1.512075929 | 1.26E-06 | 0.00049485 | 0.0049485 |
| T2DM | duct_epithelial_cell | *PPT2* | 0.623624387 | 1.47E-06 | 0.000524197 | 0.005766172 |
| T2DM | duct_epithelial_cell | *HLA-DQB2* | 0.605560482 | 3.37E-09 | 3.31E-06 | 1.32E-05 |
| T2DM | duct_epithelial_cell | *HLA-B* | 0.23817545 | 1.13E-05 | 0.002606935 | 0.044317892 |
| T2DM | duct_epithelial_cell | *HLA-DQA2* | 0.389402711 | 5.20E-14 | 2.04E-10 | 2.04E-10 |
| T2DM | duct_epithelial_cell | *JMJD7* | 0.621408661 | 0.000121573 | 0.015408452 | 0.477662013 |
| T2DM | endothelial_cell | *RBM6* | -0.678296428 | 1.79E-08 | 1.41E-05 | 7.05E-05 |
| T2DM | endothelial_cell | *ZNF195* | -0.763597996 | 0.001207648 | 0.040554277 | 1 |
| T2DM | endothelial_cell | *GGNBP2* | -0.820058565 | 0.00045448 | 0.027055351 | 1 |
| T2DM | endothelial_cell | *BAIAP3* | -0.495360063 | 0.000427841 | 0.026682363 | 1 |
| T2DM | endothelial_cell | *GIPR* | -0.569743244 | 0.000126884 | 0.012085623 | 0.4985256 |
| T2DM | endothelial_cell | *CEP68* | 0.844275635 | 2.00E-06 | 0.000561312 | 0.007858364 |
| T2DM | endothelial_cell | *LTF* | -0.874550696 | 0.000445329 | 0.026918395 | 1 |
| T2DM | endothelial_cell | *SH2D2A* | -1.013734713 | 0.000444149 | 0.026918395 | 1 |
| T2DM | endothelial_cell | *BAK1* | 1.041957899 | 9.60E-06 | 0.001886494 | 0.037729887 |
| T2DM | endothelial_cell | *ARID4A* | -0.838090342 | 0.00179515 | 0.04551528 | 1 |
| T2DM | endothelial_cell | *GOPC* | -1.136448764 | 0.001481766 | 0.043247251 | 1 |
| T2DM | endothelial_cell | *HEBP2* | -0.563922837 | 0.001965166 | 0.047080096 | 1 |
| T2DM | endothelial_cell | *USP36* | -1.247026171 | 0.001031844 | 0.037888925 | 1 |
| T2DM | endothelial_cell | *ITIH4* | -1.001288235 | 1.85E-05 | 0.003155572 | 0.072578147 |
| T2DM | endothelial_cell | *HAGH* | -0.789619084 | 0.00030358 | 0.021299366 | 1 |
| T2DM | endothelial_cell | *SEC61A2* | -0.83795835 | 0.00144949 | 0.043247251 | 1 |
| T2DM | endothelial_cell | *ST6GAL1* | 0.412951834 | 0.00049378 | 0.027313218 | 1 |
| T2DM | endothelial_cell | *MARK3* | -1.567766718 | 2.85E-08 | 1.80E-05 | 0.00011184 |
| T2DM | endothelial_cell | *XAB2* | -1.393587676 | 0.001114765 | 0.038551247 | 1 |
| T2DM | endothelial_cell | *FDFT1* | -0.556197863 | 0.001486358 | 0.043247251 | 1 |
| T2DM | endothelial_cell | *POMGNT1* | -0.948738885 | 0.000580994 | 0.028754401 | 1 |
| T2DM | endothelial_cell | *KLHL42* | -0.490032923 | 0.001706068 | 0.045291503 | 1 |
| T2DM | endothelial_cell | *SUCO* | -0.585306732 | 0.001309611 | 0.042953994 | 1 |
| T2DM | endothelial_cell | *CWF19L1* | -0.443023717 | 2.52E-05 | 0.003937403 | 0.098937669 |
| T2DM | endothelial_cell | *IKZF5* | -1.146130644 | 0.00083447 | 0.034123028 | 1 |
| T2DM | endothelial_cell | *AP1B1* | -1.311736521 | 1.58E-05 | 0.002928447 | 0.06211393 |
| T2DM | endothelial_cell | *RBFOX2* | -0.981455883 | 0.001405291 | 0.043242585 | 1 |
| T2DM | endothelial_cell | *SYNGR1* | -0.465335764 | 0.001733041 | 0.045394121 | 1 |
| T2DM | endothelial_cell | *POLR3H* | -1.315319212 | 0.001052149 | 0.038019184 | 1 |
| T2DM | endothelial_cell | *RPAP1* | -2.0653448 | 0.000713829 | 0.032580379 | 1 |
| T2DM | endothelial_cell | *LEPROTL1* | -1.034850306 | 0.002027102 | 0.04797882 | 1 |
| T2DM | endothelial_cell | *TMEM248* | -0.713037551 | 0.00196345 | 0.047080096 | 1 |
| T2DM | endothelial_cell | *SLC1A1* | 0.441373635 | 0.001857496 | 0.046190526 | 1 |
| T2DM | endothelial_cell | *SMARCD2* | -1.139007104 | 0.001645038 | 0.044731778 | 1 |
| T2DM | endothelial_cell | *HSD17B1* | -0.575276589 | 2.61E-05 | 0.003937403 | 0.102372485 |

| | | | | | | |
|---|---|---|---|---|---|---|
| T2DM | endothelial_cell | *CNTNAP1* | 0.492789211 | 0.000141202 | 0.012112199 | 0.554782247 |
| T2DM | endothelial_cell | *SRSF9* | -0.715631559 | 0.001311906 | 0.042953994 | 1 |
| T2DM | endothelial_cell | *PEX7* | -1.03462421 | 0.0020112 | 0.047890929 | 1 |
| T2DM | endothelial_cell | *NNT* | -0.701015243 | 1.64E-05 | 0.002928447 | 0.064425842 |
| T2DM | endothelial_cell | *C3orf14* | -0.352109898 | 0.001812273 | 0.04551528 | 1 |
| T2DM | endothelial_cell | *NEK11* | -0.52615063 | 0.001491663 | 0.043247251 | 1 |
| T2DM | endothelial_cell | *CHMP3* | -0.677514135 | 0.000269511 | 0.020363641 | 1 |
| T2DM | endothelial_cell | *ANGPTL1* | -2.421681898 | 6.88E-05 | 0.00900829 | 0.270248704 |
| T2DM | endothelial_cell | *CCDC181* | -0.593307843 | 0.001363895 | 0.043242585 | 1 |
| T2DM | endothelial_cell | *DNAH7* | -0.574681073 | 5.27E-06 | 0.001380539 | 0.02070808 |
| T2DM | endothelial_cell | *SLC10A7* | -0.670810343 | 0.001137425 | 0.038860382 | 1 |
| T2DM | endothelial_cell | *DNAJC15* | -0.835666815 | 0.000844873 | 0.034123028 | 1 |
| T2DM | endothelial_cell | *FAM53C* | -0.913479004 | 0.000864049 | 0.034123028 | 1 |
| T2DM | endothelial_cell | *POLK* | -0.874059588 | 0.000425333 | 0.026682363 | 1 |
| T2DM | endothelial_cell | *PEPD* | -1.177789104 | 0.000239374 | 0.018809989 | 0.940499461 |
| T2DM | endothelial_cell | *PRMT1* | -0.842250404 | 0.000133896 | 0.012085623 | 0.526076359 |
| T2DM | endothelial_cell | *EIF2AK4* | -1.019689646 | 0.000141807 | 0.012112199 | 0.557161166 |
| T2DM | endothelial_cell | *MAU2* | -1.360412005 | 2.58E-07 | 0.000126905 | 0.001015237 |
| T2DM | endothelial_cell | *CDC16* | -1.030555668 | 0.000549184 | 0.028754401 | 1 |
| T2DM | endothelial_cell | *GTPBP3* | -0.459666922 | 0.001354569 | 0.043242585 | 1 |
| T2DM | endothelial_cell | *UCK1* | -1.212178165 | 0.001749248 | 0.045515201 | 1 |
| T2DM | endothelial_cell | *BHMT2* | -0.631466388 | 0.000625418 | 0.030336639 | 1 |
| T2DM | endothelial_cell | *HSD17B4* | -1.070193902 | 0.001650829 | 0.044731778 | 1 |
| T2DM | endothelial_cell | *THUMPD3* | -0.608367596 | 0.001818758 | 0.04551528 | 1 |
| T2DM | endothelial_cell | *NOTCH2* | -0.52519944 | 2.19E-09 | 2.15E-06 | 8.59E-06 |
| T2DM | endothelial_cell | *LRP4* | 0.376565228 | 0.000868491 | 0.034123028 | 1 |
| T2DM | endothelial_cell | *GCC2* | -0.665937393 | 0.001107964 | 0.038551247 | 1 |
| T2DM | endothelial_cell | *FXYD2* | -0.378299304 | 0.000328678 | 0.022655706 | 1 |
| T2DM | endothelial_cell | *TUBGCP4* | -0.97356872 | 0.000578812 | 0.028754401 | 1 |
| T2DM | endothelial_cell | *MYPN* | 1.699969761 | 0.00138856 | 0.043242585 | 1 |
| T2DM | endothelial_cell | *G3BP2* | -0.794657155 | 0.001019926 | 0.037804605 | 1 |
| T2DM | endothelial_cell | *USO1* | -0.65251791 | 0.001589967 | 0.044731778 | 1 |
| T2DM | endothelial_cell | *CERS5* | -0.579273434 | 0.001424752 | 0.043242585 | 1 |
| T2DM | endothelial_cell | *MAN2C1* | -0.407617552 | 9.26E-06 | 0.001886494 | 0.036372822 |
| T2DM | endothelial_cell | *OSGIN1* | -0.885890765 | 0.001677449 | 0.045141762 | 1 |
| T2DM | endothelial_cell | *VPS53* | -0.591885792 | 0.000507474 | 0.027313218 | 1 |
| T2DM | endothelial_cell | *NPC1* | -1.645400178 | 8.16E-06 | 0.001886494 | 0.032043317 |
| T2DM | endothelial_cell | *ZNF593* | -0.615236204 | 0.001639707 | 0.044731778 | 1 |
| T2DM | endothelial_cell | *NME7* | -0.562275133 | 0.00058548 | 0.028754401 | 1 |
| T2DM | endothelial_cell | *CIAO1* | -0.978169863 | 0.000758724 | 0.032882406 | 1 |
| T2DM | endothelial_cell | *ZNF514* | -0.592843332 | 0.00029552 | 0.021299366 | 1 |
| T2DM | endothelial_cell | *CNTN4* | -0.444949704 | 3.69E-05 | 0.005171633 | 0.144805734 |
| T2DM | endothelial_cell | *TBC1D7* | -1.928773453 | 0.000250179 | 0.019273585 | 0.982952844 |
| T2DM | endothelial_cell | *RAB19* | -1.783145754 | 0.000930445 | 0.036113394 | 1 |
| T2DM | endothelial_cell | *ADHFE1* | -0.698742184 | 0.00101686 | 0.037804605 | 1 |
| T2DM | endothelial_cell | *LRP12* | -1.497101513 | 0.000734209 | 0.032780767 | 1 |
| T2DM | endothelial_cell | *HSD17B12* | -0.266595156 | 8.69E-06 | 0.001886494 | 0.034149658 |
| T2DM | endothelial_cell | *APIP* | -0.380689437 | 0.000133459 | 0.012085623 | 0.524359446 |
| T2DM | endothelial_cell | *DOC2A* | -0.549272337 | 0.00181148 | 0.04551528 | 1 |
| T2DM | endothelial_cell | *DLG5* | -0.769613808 | 0.001885967 | 0.046603561 | 1 |

| | | | | | | |
|---|---|---|---|---|---|---|
| T2DM | endothelial_cell | *ARL14EP* | -1.143387574 | 0.001370842 | 0.043242585 | 1 |
| T2DM | endothelial_cell | *POC5* | -0.309748087 | 0.000167218 | 0.013687476 | 0.656998842 |
| T2DM | endothelial_cell | *JAZF1* | -1.047186529 | 6.21E-13 | 8.14E-10 | 2.44E-09 |
| T2DM | endothelial_cell | *NUS1* | -1.282717263 | 0.000133222 | 0.012085623 | 0.523428286 |
| T2DM | endothelial_cell | *CC2D1B* | -0.494962221 | 0.000146886 | 0.012279069 | 0.57711626 |
| T2DM | endothelial_cell | *DHRS4* | -1.396708293 | 0.000577566 | 0.028754401 | 1 |
| T2DM | endothelial_cell | *EXTL1* | -1.183738343 | 0.000209047 | 0.016762191 | 0.82134734 |
| T2DM | endothelial_cell | *FAIM* | -1.001047891 | 0.001629338 | 0.044731778 | 1 |
| T2DM | endothelial_cell | *CILP2* | -0.654900656 | 2.42E-05 | 0.003937403 | 0.095111087 |
| T2DM | endothelial_cell | *PDXK* | -0.568388564 | 0.000756027 | 0.032882406 | 1 |
| T2DM | endothelial_cell | *PAQR6* | -0.781780418 | 0.000288242 | 0.021299366 | 1 |
| T2DM | endothelial_cell | *ZNF75A* | -1.297106184 | 0.000767148 | 0.032882406 | 1 |
| T2DM | endothelial_cell | *DCAF16* | -1.047799596 | 0.000367648 | 0.0244048 | 1 |
| T2DM | endothelial_cell | *ICA1L* | -0.389793875 | 8.48E-05 | 0.010586276 | 0.333187321 |
| T2DM | endothelial_cell | *LMBRD2* | -0.668983455 | 0.001694287 | 0.045284719 | 1 |
| T2DM | endothelial_cell | *BRI3* | -0.67177911 | 0.001118565 | 0.038551247 | 1 |
| T2DM | endothelial_cell | *INPPL1* | -1.234384321 | 0.000705008 | 0.032580379 | 1 |
| T2DM | endothelial_cell | *VDAC2* | -0.673231507 | 0.000853178 | 0.034123028 | 1 |
| T2DM | endothelial_cell | *CASP7* | 0.281091409 | 0.000107316 | 0.012085623 | 0.4216434 |
| T2DM | endothelial_cell | *CEP57* | -0.923005722 | 0.000111024 | 0.012085623 | 0.436213554 |
| T2DM | endothelial_cell | *CKB* | -0.396060598 | 8.62E-05 | 0.010586276 | 0.338760836 |
| T2DM | endothelial_cell | *SGPL1* | -0.70122196 | 0.00010966 | 0.012085623 | 0.430852905 |
| T2DM | endothelial_cell | *PCBD1* | -0.434525043 | 0.000505719 | 0.027313218 | 1 |
| T2DM | endothelial_cell | *POP5* | -0.956966561 | 0.000658013 | 0.031528461 | 1 |
| T2DM | endothelial_cell | *ACAA2* | -0.634950482 | 0.0008245 | 0.034123028 | 1 |
| T2DM | endothelial_cell | *TP53I13* | -0.445200113 | 0.000847139 | 0.034123028 | 1 |
| T2DM | endothelial_cell | *TRANK1* | -1.025714141 | 0.000769962 | 0.032882406 | 1 |
| T2DM | endothelial_cell | *SAC3D1* | -1.714360834 | 0.00129798 | 0.042953994 | 1 |
| T2DM | endothelial_cell | *KIAA1586* | -0.730159087 | 0.000685035 | 0.032066829 | 1 |
| T2DM | endothelial_cell | *NDUFS5* | -0.576798043 | 0.001199363 | 0.040554277 | 1 |
| T2DM | endothelial_cell | *DNAJC21* | -1.094528042 | 0.00161708 | 0.044731778 | 1 |
| T2DM | endothelial_cell | *PPIP5K1* | -0.725439668 | 0.001794981 | 0.04551528 | 1 |
| T2DM | endothelial_cell | *TMEM129* | -0.454649596 | 0.001950753 | 0.047080096 | 1 |
| T2DM | endothelial_cell | *INO80E* | -0.825529505 | 1.24E-06 | 0.000444446 | 0.004888908 |
| T2DM | endothelial_cell | *NIPA1* | -1.101228315 | 0.000860929 | 0.034123028 | 1 |
| T2DM | endothelial_cell | *ZNF440* | -1.414143834 | 0.000937533 | 0.036113394 | 1 |
| T2DM | endothelial_cell | *CLSTN1* | -0.608395187 | 0.001484903 | 0.043247251 | 1 |
| T2DM | endothelial_cell | *CTSW* | -0.231812544 | 0.00150295 | 0.043247251 | 1 |
| T2DM | endothelial_cell | *LCLAT1* | -0.709260704 | 0.001507985 | 0.043247251 | 1 |
| T2DM | endothelial_cell | *YIF1A* | -0.494986967 | 0.000122243 | 0.012085623 | 0.480294439 |
| T2DM | endothelial_cell | *CCDC57* | -0.400671791 | 0.001073163 | 0.038019184 | 1 |
| T2DM | endothelial_cell | *COX8A* | -0.451589369 | 0.001604588 | 0.044731778 | 1 |
| T2DM | endothelial_cell | *DSCAML1* | -0.471742581 | 0.000503772 | 0.027313218 | 1 |
| T2DM | endothelial_cell | *ARIH2* | -0.576977038 | 0.00192149 | 0.047080096 | 1 |
| T2DM | endothelial_cell | *IRX3* | 1.426540934 | 1.07E-13 | 4.20E-10 | 4.20E-10 |
| T2DM | endothelial_cell | *TRIM73* | -0.720049302 | 0.001002393 | 0.037804605 | 1 |
| T2DM | endothelial_cell | *FARSA* | -1.200373149 | 1.62E-06 | 0.000488429 | 0.006349583 |
| T2DM | endothelial_cell | *HLA-DQB1* | -0.303729551 | 8.44E-07 | 0.000368647 | 0.003317825 |
| T2DM | endothelial_cell | *D2HGDH* | -1.104221046 | 0.000466473 | 0.027313218 | 1 |
| T2DM | endothelial_cell | *VMO1* | 0.502322966 | 9.69E-05 | 0.011541365 | 0.380865054 |

| | | | | | | |
|---|---|---|---|---|---|---|
| T2DM | endothelial_cell | *ZNF320* | 0.206563464 | 0.000685572 | 0.032066829 | 1 |
| T2DM | endothelial_cell | *FAM43B* | 2.309801041 | 0.000496952 | 0.027313218 | 1 |
| T2DM | endothelial_cell | *TRMT12* | -1.503835653 | 0.001019519 | 0.037804605 | 1 |
| T2DM | endothelial_cell | *MACC1* | -0.462137717 | 0.001381592 | 0.043242585 | 1 |
| T2DM | endothelial_cell | *PGP* | -0.835468015 | 0.000299756 | 0.021299366 | 1 |
| T2DM | endothelial_cell | *BRWD1* | -0.828077635 | 0.00211222 | 0.049694078 | 1 |
| T2DM | endothelial_cell | *DMWD* | -0.807232046 | 0.000502215 | 0.027313218 | 1 |
| T2DM | endothelial_cell | *NUTM2B* | -4.122132213 | 3.04E-05 | 0.004423456 | 0.1194333 |
| T2DM | endothelial_cell | *ASAH2* | -0.651951848 | 0.000558203 | 0.028754401 | 1 |
| T2DM | endothelial_cell | *HLA-DRB1* | -0.21373167 | 9.85E-07 | 0.000386982 | 0.003869822 |
| T2DM | endothelial_cell | *XPNPEP3* | -0.789762315 | 0.00143078 | 0.043242585 | 1 |
| T2DM | endothelial_cell | *SUPT3H* | -1.577804661 | 0.001074098 | 0.038019184 | 1 |
| T2DM | endothelial_cell | *XRCC6* | -0.843828692 | 4.65E-05 | 0.006300362 | 0.182710508 |
| T2DM | endothelial_cell | *HLA-DQA1* | -0.164031853 | 0.001336483 | 0.043242585 | 1 |
| T2DM | endothelial_cell | *PIWIL2* | -0.483050537 | 0.00053352 | 0.028327055 | 1 |
| T2DM | endothelial_cell | *KIAA0753* | -0.372273262 | 0.001546607 | 0.044033456 | 1 |
| T2DM | endothelial_cell | *RNF5* | -0.475844257 | 1.36E-06 | 0.000446462 | 0.005357548 |
| T2DM | endothelial_cell | *HMGN1* | -0.655627662 | 0.001790445 | 0.04551528 | 1 |
| T2DM | endothelial_cell | *HCG27* | 0.286918296 | 0.000721429 | 0.032580379 | 1 |
| T2DM | endothelial_cell | *DNLZ* | 0.257567697 | 0.000135344 | 0.012085623 | 0.531767416 |
| T2DM | endothelial_cell | *ATF6B* | 0.408338871 | 0.000372687 | 0.0244048 | 1 |
| T2DM | endothelial_cell | *IFRD2* | -1.164047888 | 0.001422404 | 0.043242585 | 1 |
| T2DM | endothelial_cell | *SLC12A8* | -0.606147151 | 0.000370283 | 0.0244048 | 1 |
| T2DM | endothelial_cell | *PPT2* | 0.333804544 | 0.000124781 | 0.012085623 | 0.49026372 |
| T2DM | endothelial_cell | *HLA-DQB2* | 0.780829438 | 3.21E-08 | 1.80E-05 | 0.000125992 |
| T2DM | endothelial_cell | *HLA-B* | 0.299083867 | 9.26E-06 | 0.001886494 | 0.036396578 |
| T2DM | endothelial_cell | *B3GALT4* | -0.372696383 | 0.000421652 | 0.026682363 | 1 |
| T2DM | endothelial_cell | *HLA-DQA2* | 0.515409689 | 4.72E-13 | 8.14E-10 | 1.86E-09 |
| T2DM | endothelial_cell | *JMJD7* | 0.840481171 | 0.000127324 | 0.012085623 | 0.50025792 |
| T2DM | endothelial_cell | *LINC01091* | -0.528013709 | 0.001949944 | 0.047080096 | 1 |
| T2DM | endothelial_cell | *TAS2R20* | 0.953515631 | 0.001729901 | 0.045394121 | 1 |
| T2DM | endothelial_cell | *ZNF432* | -1.2127724 | 0.001058698 | 0.038019184 | 1 |
| T2DM | epsilon_cell | *IRX3* | 14.2725765 | 1.07E-13 | 4.21E-10 | 4.21E-10 |
| T2DM | epsilon_cell | *UBA7* | -5.898332582 | 9.01E-06 | 0.007080607 | 0.035403033 |
| T2DM | epsilon_cell | *HLA-DRB1* | -2.484785461 | 3.92E-07 | 0.000513569 | 0.001540708 |
| T2DM | epsilon_cell | *HLA-DQB2* | 7.369618614 | 2.88E-06 | 0.002827766 | 0.011311064 |
| T2DM | epsilon_cell | *HLA-DQA2* | 6.105339943 | 4.16E-13 | 8.18E-10 | 1.64E-09 |
| T2DM | leukocyte | *GCLC* | -1.428864345 | 0.000663897 | 0.004990048 | 1 |
| T2DM | leukocyte | *NFYA* | -0.964070269 | 0.004191306 | 0.007244893 | 1 |
| T2DM | leukocyte | *STPG1* | -0.863510803 | 0.002186771 | 0.005356498 | 1 |
| T2DM | leukocyte | *LASP1* | -0.899031832 | 0.001194904 | 0.004990048 | 1 |
| T2DM | leukocyte | *SNX11* | -0.757163971 | 0.016818342 | 0.022627392 | 1 |
| T2DM | leukocyte | *TMEM176A* | -0.577384489 | 0.007669579 | 0.011409987 | 1 |
| T2DM | leukocyte | *M6PR* | -2.027633553 | 0.000544902 | 0.004990048 | 1 |
| T2DM | leukocyte | *DBNDD1* | -0.635397361 | 0.003914914 | 0.007085756 | 1 |
| T2DM | leukocyte | *CASP10* | -2.642188976 | 0.000485422 | 0.004990048 | 1 |
| T2DM | leukocyte | *NDUFAF7* | -2.066377509 | 0.000603261 | 0.004990048 | 1 |
| T2DM | leukocyte | *POLDIP2* | -0.690227958 | 0.004141384 | 0.007238288 | 1 |
| T2DM | leukocyte | *PLXND1* | -1.974513092 | 0.001070398 | 0.004990048 | 1 |
| T2DM | leukocyte | *KDM1A* | -0.906675734 | 0.002541208 | 0.00567203 | 1 |

| | | | | | | |
|---|---|---|---|---|---|---|
| T2DM | leukocyte | *RBM6* | -1.735769141 | 4.18E-08 | 3.28E-05 | 0.000164088 |
| T2DM | leukocyte | *RECQL* | -0.963409157 | 0.001364834 | 0.004990048 | 1 |
| T2DM | leukocyte | *SLC25A13* | -0.907358777 | 0.002124278 | 0.005299229 | 1 |
| T2DM | leukocyte | *MEOX1* | 1.986469378 | 0.01485814 | 0.020234881 | 1 |
| T2DM | leukocyte | *PON1* | -0.900627702 | 0.010006588 | 0.014296686 | 1 |
| T2DM | leukocyte | *KMT2E* | -1.002928621 | 0.000885577 | 0.004990048 | 1 |
| T2DM | leukocyte | *IBTK* | -0.981412908 | 0.000894939 | 0.004990048 | 1 |
| T2DM | leukocyte | *ZNF195* | -1.599118618 | 0.000175909 | 0.004990048 | 0.691147926 |
| T2DM | leukocyte | *GGNBP2* | -1.119045011 | 0.000280361 | 0.004990048 | 1 |
| T2DM | leukocyte | *TMEM98* | -0.57278085 | 0.013160217 | 0.018219343 | 1 |
| T2DM | leukocyte | *SYNRG* | -1.403074411 | 0.000787915 | 0.004990048 | 1 |
| T2DM | leukocyte | *SPATA20* | -0.897547164 | 0.004925232 | 0.007976602 | 1 |
| T2DM | leukocyte | *FARP2* | -2.937105307 | 0.000447871 | 0.004990048 | 1 |
| T2DM | leukocyte | *USH1C* | -1.028856737 | 0.000513416 | 0.004990048 | 1 |
| T2DM | leukocyte | *VPS41* | -1.291093741 | 0.000952718 | 0.004990048 | 1 |
| T2DM | leukocyte | *ELAC2* | -0.976412912 | 0.002150484 | 0.005327398 | 1 |
| T2DM | leukocyte | *CCDC124* | -0.827813723 | 0.001395912 | 0.004990048 | 1 |
| T2DM | leukocyte | *NOS2* | -13.7593998 | 0.004358629 | 0.007368783 | 1 |
| T2DM | leukocyte | *CD79B* | -5.294546636 | 0.000485669 | 0.004990048 | 1 |
| T2DM | leukocyte | *ST7L* | -1.517689988 | 0.000488531 | 0.004990048 | 1 |
| T2DM | leukocyte | *BAIAP3* | -1.033623076 | 0.000284037 | 0.004990048 | 1 |
| T2DM | leukocyte | *CYB561* | -0.842283897 | 0.001338136 | 0.004990048 | 1 |
| T2DM | leukocyte | *MGST1* | -0.581520965 | 0.005618136 | 0.008825931 | 1 |
| T2DM | leukocyte | *ST3GAL1* | -2.027859106 | 0.000986689 | 0.004990048 | 1 |
| T2DM | leukocyte | *IL32* | -0.392222555 | 0.004167721 | 0.007242343 | 1 |
| T2DM | leukocyte | *MED24* | -1.264584471 | 0.001327147 | 0.004990048 | 1 |
| T2DM | leukocyte | *RPS20* | -0.65462262 | 0.000870422 | 0.004990048 | 1 |
| T2DM | leukocyte | *REV3L* | -0.794515164 | 0.001565541 | 0.005009694 | 1 |
| T2DM | leukocyte | *ETV7* | -1.7246826 | 0.001537539 | 0.005006447 | 1 |
| T2DM | leukocyte | *NCAPD2* | -1.436717188 | 0.006211483 | 0.009563054 | 1 |
| T2DM | leukocyte | *GIPR* | -0.83944537 | 0.000584675 | 0.004990048 | 1 |
| T2DM | leukocyte | *FUZ* | -1.348331585 | 0.002508792 | 0.005648735 | 1 |
| T2DM | leukocyte | *LRRC23* | -0.630652651 | 0.004557734 | 0.007575016 | 1 |
| T2DM | leukocyte | *TSPAN9* | -0.725310539 | 0.004189302 | 0.007244617 | 1 |
| T2DM | leukocyte | *BTBD7* | -0.981798428 | 0.001946762 | 0.005192687 | 1 |
| T2DM | leukocyte | *APBA3* | -1.696335977 | 0.002030112 | 0.005251028 | 1 |
| T2DM | leukocyte | *TTC19* | -0.720075661 | 0.002680552 | 0.005854302 | 1 |
| T2DM | leukocyte | *RABGAP1* | -0.866435341 | 0.00276533 | 0.005956822 | 1 |
| T2DM | leukocyte | *QPCTL* | -3.575408647 | 0.000632971 | 0.004990048 | 1 |
| T2DM | leukocyte | *PPP5C* | -0.886272926 | 0.002090959 | 0.005283012 | 1 |
| T2DM | leukocyte | *ERCC1* | -1.180881082 | 0.000569107 | 0.004990048 | 1 |
| T2DM | leukocyte | *CD22* | 0.390616145 | 0.017115067 | 0.022950546 | 1 |
| T2DM | leukocyte | *LTF* | -2.888581529 | 0.000147724 | 0.004990048 | 0.580406627 |
| T2DM | leukocyte | *EXTL3* | -0.774945939 | 0.003143764 | 0.006311945 | 1 |
| T2DM | leukocyte | *CALCOCO1* | -2.811432088 | 0.002609605 | 0.005766669 | 1 |
| T2DM | leukocyte | *UBR7* | -0.800612329 | 0.008299371 | 0.012180885 | 1 |
| T2DM | leukocyte | *MAN2B2* | -1.341079903 | 0.001190253 | 0.004990048 | 1 |
| T2DM | leukocyte | *ANGEL1* | -0.906429592 | 0.026367296 | 0.033744986 | 1 |
| T2DM | leukocyte | *DDX11* | -0.908110519 | 0.004304863 | 0.007328649 | 1 |
| T2DM | leukocyte | *GPRC5A* | -0.897405994 | 0.017362675 | 0.023234996 | 1 |

| | | | | | | |
|---|---|---|---|---|---|---|
| T2DM | leukocyte | *UFL1* | -0.91299115 | 0.00108512 | 0.004990048 | 1 |
| T2DM | leukocyte | *POLA2* | -1.631148846 | 0.001556294 | 0.005009694 | 1 |
| T2DM | leukocyte | *ZC3H3* | -0.654098938 | 0.021345628 | 0.027937033 | 1 |
| T2DM | leukocyte | *MDH1* | -0.922675571 | 0.000928197 | 0.004990048 | 1 |
| T2DM | leukocyte | *MTMR11* | -3.277860881 | 0.001849217 | 0.005146173 | 1 |
| T2DM | leukocyte | *DPEP1* | -0.50049721 | 0.005080944 | 0.008170331 | 1 |
| T2DM | leukocyte | *NUDCD3* | -0.79984365 | 0.002138543 | 0.005317933 | 1 |
| T2DM | leukocyte | *TTC27* | -1.933217691 | 0.002321902 | 0.005502263 | 1 |
| T2DM | leukocyte | *SNAI2* | -2.060835242 | 0.010173454 | 0.014519251 | 1 |
| T2DM | leukocyte | *PLEKHB1* | -0.608203496 | 0.040160452 | 0.049776157 | 1 |
| T2DM | leukocyte | *NLRP2* | -1.629578988 | 0.007471603 | 0.011166196 | 1 |
| T2DM | leukocyte | *ZDHHC6* | -1.177976221 | 0.001205175 | 0.004990048 | 1 |
| T2DM | leukocyte | *RNH1* | -0.906211326 | 0.000911619 | 0.004990048 | 1 |
| T2DM | leukocyte | *NDUFS1* | -0.815597943 | 0.001539514 | 0.005006447 | 1 |
| T2DM | leukocyte | *TYMP* | 0.325878486 | 0.038967225 | 0.048496113 | 1 |
| T2DM | leukocyte | *CD44* | -0.477497987 | 0.001998713 | 0.005227473 | 1 |
| T2DM | leukocyte | *KCNG1* | -3.024046638 | 0.0039469 | 0.007116737 | 1 |
| T2DM | leukocyte | *AGPAT4* | -2.428707703 | 0.017389282 | 0.023249955 | 1 |
| T2DM | leukocyte | *SH2D2A* | -5.830663208 | 4.83E-05 | 0.004515102 | 0.189634264 |
| T2DM | leukocyte | *VEZT* | -0.914562293 | 0.002220076 | 0.005407737 | 1 |
| T2DM | leukocyte | *RABEP1* | -1.529581678 | 0.001659187 | 0.005041098 | 1 |
| T2DM | leukocyte | *ARID4A* | -1.342002685 | 0.000414299 | 0.004990048 | 1 |
| T2DM | leukocyte | *PNPLA6* | -1.110893246 | 0.002084413 | 0.005283012 | 1 |
| T2DM | leukocyte | *ABCF2* | -1.258873035 | 0.00146754 | 0.004990048 | 1 |
| T2DM | leukocyte | *UBA6* | -1.48229157 | 0.000809698 | 0.004990048 | 1 |
| T2DM | leukocyte | *APBA2* | -2.201225413 | 0.004157835 | 0.007242343 | 1 |
| T2DM | leukocyte | *ASTE1* | -0.908913894 | 0.018358311 | 0.024409047 | 1 |
| T2DM | leukocyte | *RNF19A* | -0.811799058 | 0.001044556 | 0.004990048 | 1 |
| T2DM | leukocyte | *SH3YL1* | -0.52717157 | 0.006287747 | 0.009657763 | 1 |
| T2DM | leukocyte | *FAM136A* | -0.872402786 | 0.001599255 | 0.005024491 | 1 |
| T2DM | leukocyte | *DAPK2* | -0.916625957 | 0.002687299 | 0.00586252 | 1 |
| T2DM | leukocyte | *TIMP2* | -1.979172603 | 0.000163873 | 0.004990048 | 0.643858015 |
| T2DM | leukocyte | *MYOM2* | 0.842142322 | 0.036301617 | 0.045437736 | 1 |
| T2DM | leukocyte | *RPL26L1* | -0.822486111 | 0.001334675 | 0.004990048 | 1 |
| T2DM | leukocyte | *NSUN2* | -1.405192125 | 0.000505035 | 0.004990048 | 1 |
| T2DM | leukocyte | *MRI1* | -1.023675552 | 0.002833601 | 0.006014705 | 1 |
| T2DM | leukocyte | *AGA* | -1.097216294 | 0.000790787 | 0.004990048 | 1 |
| T2DM | leukocyte | *BOD1L1* | -0.768082995 | 0.003481899 | 0.006621675 | 1 |
| T2DM | leukocyte | *DNAH5* | -1.381050145 | 0.001411192 | 0.004990048 | 1 |
| T2DM | leukocyte | *PNKP* | -1.609491749 | 0.009453093 | 0.013599854 | 1 |
| T2DM | leukocyte | *PHLPP2* | -0.786186158 | 0.003047568 | 0.006220403 | 1 |
| T2DM | leukocyte | *CTNS* | -1.045711424 | 0.004314018 | 0.007328649 | 1 |
| T2DM | leukocyte | *INPP4A* | -0.793301756 | 0.004039779 | 0.007166394 | 1 |
| T2DM | leukocyte | *PSMA4* | -0.855107094 | 0.001096181 | 0.004990048 | 1 |
| T2DM | leukocyte | *LSG1* | -0.591103462 | 0.009419611 | 0.013556649 | 1 |
| T2DM | leukocyte | *SPATA7* | -1.296864011 | 0.000763211 | 0.004990048 | 1 |
| T2DM | leukocyte | *TRIT1* | -1.939378471 | 0.002099339 | 0.005283012 | 1 |
| T2DM | leukocyte | *CNTLN* | -1.286391758 | 0.00150012 | 0.004993935 | 1 |
| T2DM | leukocyte | *PREX2* | -1.461421727 | 0.009525598 | 0.013691804 | 1 |
| T2DM | leukocyte | *WDR37* | -1.347846799 | 0.000771024 | 0.004990048 | 1 |

| | | | | | | |
|---|---|---|---|---|---|---|
| T2DM | leukocyte | *YTHDC2* | -1.486186564 | 0.000595861 | 0.004990048 | 1 |
| T2DM | leukocyte | *DTNBP1* | -1.015539499 | 0.001683376 | 0.005060368 | 1 |
| T2DM | leukocyte | *GOPC* | -1.071883033 | 0.000950009 | 0.004990048 | 1 |
| T2DM | leukocyte | *ZNF800* | -1.053739543 | 0.000988956 | 0.004990048 | 1 |
| T2DM | leukocyte | *MRPS10* | -1.308011858 | 0.000319688 | 0.004990048 | 1 |
| T2DM | leukocyte | *COL9A2* | -0.755239211 | 0.015078656 | 0.02047841 | 1 |
| T2DM | leukocyte | *ERCC8* | -2.591480017 | 0.014231571 | 0.019503259 | 1 |
| T2DM | leukocyte | *H6PD* | -2.574485568 | 0.000314915 | 0.004990048 | 1 |
| T2DM | leukocyte | *VAMP3* | -1.054202498 | 0.001402173 | 0.004990048 | 1 |
| T2DM | leukocyte | *RCN1* | -0.703674016 | 0.002380949 | 0.005560194 | 1 |
| T2DM | leukocyte | *RFC2* | -0.937235402 | 0.003581496 | 0.006727272 | 1 |
| T2DM | leukocyte | *JKAMP* | -0.827307045 | 0.002105315 | 0.005283012 | 1 |
| T2DM | leukocyte | *PTGER3* | -0.788448184 | 0.003699307 | 0.006868344 | 1 |
| T2DM | leukocyte | *BCAR1* | -0.688844764 | 0.003008944 | 0.006176667 | 1 |
| T2DM | leukocyte | *THOC3* | -0.67236824 | 0.003838189 | 0.007020598 | 1 |
| T2DM | leukocyte | *HEBP2* | -0.867835986 | 0.000588059 | 0.004990048 | 1 |
| T2DM | leukocyte | *RRP12* | -0.603402461 | 0.003889026 | 0.00705773 | 1 |
| T2DM | leukocyte | *FNIP2* | -2.07409232 | 0.000726881 | 0.004990048 | 1 |
| T2DM | leukocyte | *MRTO4* | -1.042568387 | 0.001824929 | 0.00513989 | 1 |
| T2DM | leukocyte | *MCF2L2* | -1.45995675 | 0.001649119 | 0.005035955 | 1 |
| T2DM | leukocyte | *LAMA3* | -1.37221642 | 0.003233692 | 0.006381841 | 1 |
| T2DM | leukocyte | *ANAPC4* | -3.101703136 | 0.014539148 | 0.01985302 | 1 |
| T2DM | leukocyte | *LY75* | -3.824525302 | 0.000425533 | 0.004990048 | 1 |
| T2DM | leukocyte | *ARID4B* | -0.721477862 | 0.003573448 | 0.006719453 | 1 |
| T2DM | leukocyte | *SDCCAG8* | -0.859408714 | 0.002261036 | 0.005456717 | 1 |
| T2DM | leukocyte | *KIF1B* | -0.879113167 | 0.001254139 | 0.004990048 | 1 |
| T2DM | leukocyte | *TBC1D22A* | -0.915593036 | 0.001979341 | 0.005222499 | 1 |
| T2DM | leukocyte | *SYNE2* | -0.844320674 | 0.001627093 | 0.005035955 | 1 |
| T2DM | leukocyte | *ATP9A* | -0.955157451 | 0.001160088 | 0.004990048 | 1 |
| T2DM | leukocyte | *CHRDL2* | -3.006135237 | 0.004698026 | 0.007719684 | 1 |
| T2DM | leukocyte | *GALC* | -1.250445961 | 0.001963946 | 0.005212073 | 1 |
| T2DM | leukocyte | *CYFIP2* | -0.722231983 | 0.002268219 | 0.005457772 | 1 |
| T2DM | leukocyte | *USP36* | -1.507700651 | 0.000361411 | 0.004990048 | 1 |
| T2DM | leukocyte | *MCOLN3* | -0.8098272 | 0.003450744 | 0.006578347 | 1 |
| T2DM | leukocyte | *ITIH4* | -2.659462704 | 6.12E-06 | 0.001302453 | 0.024052853 |
| T2DM | leukocyte | *PCGF2* | -0.849195549 | 0.002426379 | 0.005602682 | 1 |
| T2DM | leukocyte | *GDI2* | -0.923378838 | 0.001497309 | 0.004993935 | 1 |
| T2DM | leukocyte | *ATP11B* | -1.592267959 | 0.000571046 | 0.004990048 | 1 |
| T2DM | leukocyte | *LAMC2* | -0.563657716 | 0.007208645 | 0.01083503 | 1 |
| T2DM | leukocyte | *RASGRF1* | -0.61529224 | 0.005610721 | 0.008817809 | 1 |
| T2DM | leukocyte | *CROCC* | -1.305218734 | 0.003341635 | 0.006470236 | 1 |
| T2DM | leukocyte | *FLYWCH1* | -0.671761805 | 0.001465354 | 0.004990048 | 1 |
| T2DM | leukocyte | *OGFR* | -1.94469777 | 0.012489835 | 0.017401617 | 1 |
| T2DM | leukocyte | *GNA15* | 1.170007623 | 0.023310489 | 0.030176906 | 1 |
| T2DM | leukocyte | *RIMBP2* | -0.682471857 | 0.004147027 | 0.007238372 | 1 |
| T2DM | leukocyte | *MPC1* | -0.842231849 | 0.001221599 | 0.004990048 | 1 |
| T2DM | leukocyte | *BCAT1* | 0.261437359 | 0.03762152 | 0.046955195 | 1 |
| T2DM | leukocyte | *MRPS35* | -0.921280107 | 0.0018778 | 0.005146173 | 1 |
| T2DM | leukocyte | *ZNF112* | -1.058948305 | 0.008032106 | 0.011844895 | 1 |
| T2DM | leukocyte | *EIF4B* | -0.838832289 | 0.001063673 | 0.004990048 | 1 |

| | | | | | | |
|---|---|---|---|---|---|---|
| T2DM | leukocyte | *SLC6A16* | -2.294580416 | 0.003235589 | 0.006381841 | 1 |
| T2DM | leukocyte | *SPHK2* | -0.947441266 | 0.007130834 | 0.0107345 | 1 |
| T2DM | leukocyte | *HAGH* | -1.229489552 | 0.000167544 | 0.004990048 | 0.658280623 |
| T2DM | leukocyte | *RNF4* | -1.085039713 | 0.001057778 | 0.004990048 | 1 |
| T2DM | leukocyte | *TM7SF3* | -1.656627173 | 0.001067639 | 0.004990048 | 1 |
| T2DM | leukocyte | *HIPK2* | -0.727977565 | 0.002070719 | 0.005273744 | 1 |
| T2DM | leukocyte | *TMEM161A* | -0.999429138 | 0.002300463 | 0.005478715 | 1 |
| T2DM | leukocyte | *SNX24* | -2.180253528 | 0.000463166 | 0.004990048 | 1 |
| T2DM | leukocyte | *ABCA7* | -1.278163851 | 0.002262755 | 0.005456717 | 1 |
| T2DM | leukocyte | *PMS1* | -0.802627667 | 0.005021811 | 0.008092984 | 1 |
| T2DM | leukocyte | *IPO5* | -0.787793958 | 0.001512426 | 0.004993935 | 1 |
| T2DM | leukocyte | *OAT* | -0.809392091 | 0.00195994 | 0.00520663 | 1 |
| T2DM | leukocyte | *WDR18* | -1.054961702 | 0.001030636 | 0.004990048 | 1 |
| T2DM | leukocyte | *MCM10* | 1.425823234 | 0.023984173 | 0.030997966 | 1 |
| T2DM | leukocyte | *ERBB3* | -0.807094439 | 0.001338586 | 0.004990048 | 1 |
| T2DM | leukocyte | *ANKRD44* | -4.130194584 | 0.000641443 | 0.004990048 | 1 |
| T2DM | leukocyte | *ADAT1* | -0.832762068 | 0.004038082 | 0.007166394 | 1 |
| T2DM | leukocyte | *MYLK* | -0.579213538 | 0.027388899 | 0.034870702 | 1 |
| T2DM | leukocyte | *GSTO2* | -0.610169977 | 0.002018644 | 0.005244224 | 1 |
| T2DM | leukocyte | *SEC61A2* | -3.228544742 | 6.50E-05 | 0.004990048 | 0.255424531 |
| T2DM | leukocyte | *PRKCQ* | -2.068288013 | 0.00373057 | 0.006898954 | 1 |
| T2DM | leukocyte | *TBC1D1* | -0.911154064 | 0.002192234 | 0.005366534 | 1 |
| T2DM | leukocyte | *CTNNA2* | -0.93867699 | 0.011225196 | 0.015824828 | 1 |
| T2DM | leukocyte | *CD84* | 0.123392197 | 0.006872428 | 0.010409317 | 1 |
| T2DM | leukocyte | *ATXN3* | -1.373795761 | 0.002002379 | 0.005227473 | 1 |
| T2DM | leukocyte | *EML1* | -1.410668218 | 0.001291656 | 0.004990048 | 1 |
| T2DM | leukocyte | *STOML1* | -0.780414586 | 0.003008449 | 0.006176667 | 1 |
| T2DM | leukocyte | *DNTTIP2* | -0.808486394 | 0.00169768 | 0.005060368 | 1 |
| T2DM | leukocyte | *RRP15* | -0.837489855 | 0.004837896 | 0.007890449 | 1 |
| T2DM | leukocyte | *REEP1* | -0.647360586 | 0.016499807 | 0.022224114 | 1 |
| T2DM | leukocyte | *POLR1A* | -0.770414732 | 0.003234647 | 0.006381841 | 1 |
| T2DM | leukocyte | *LAPTM4A* | -0.801519632 | 0.001010268 | 0.004990048 | 1 |
| T2DM | leukocyte | *KIF2A* | -0.83194653 | 0.002058678 | 0.00526598 | 1 |
| T2DM | leukocyte | *MAST4* | -1.012979253 | 0.000903416 | 0.004990048 | 1 |
| T2DM | leukocyte | *DRD4* | -2.547539146 | 0.00800689 | 0.011817833 | 1 |
| T2DM | leukocyte | *PLA2G10* | -3.259409977 | 0.002503886 | 0.005644159 | 1 |
| T2DM | leukocyte | *GNB5* | -0.999810855 | 0.004663319 | 0.007686961 | 1 |
| T2DM | leukocyte | *RAB27A* | -0.837076688 | 0.001455844 | 0.004990048 | 1 |
| T2DM | leukocyte | *PTPN3* | -1.535327 | 0.000540717 | 0.004990048 | 1 |
| T2DM | leukocyte | *FGF10* | -2.309687671 | 0.009057444 | 0.013102613 | 1 |
| T2DM | leukocyte | *DGCR2* | -1.249814808 | 0.000730411 | 0.004990048 | 1 |
| T2DM | leukocyte | *ZXDC* | -2.355824837 | 0.000382836 | 0.004990048 | 1 |
| T2DM | leukocyte | *WIPI1* | -2.324799474 | 0.000339184 | 0.004990048 | 1 |
| T2DM | leukocyte | *GBA2* | -0.79693132 | 0.006162139 | 0.009503098 | 1 |
| T2DM | leukocyte | *ASNS* | -0.824540099 | 0.004243066 | 0.007289465 | 1 |
| T2DM | leukocyte | *AP3M2* | -0.978438268 | 0.033550533 | 0.042317832 | 1 |
| T2DM | leukocyte | *TESK2* | -2.534124281 | 0.034142903 | 0.042913457 | 1 |
| T2DM | leukocyte | *MAP4K4* | -0.821614138 | 0.000953654 | 0.004990048 | 1 |
| T2DM | leukocyte | *RPL31* | -0.716314947 | 0.0009325 | 0.004990048 | 1 |
| T2DM | leukocyte | *SEL1L* | -0.800918545 | 0.001171869 | 0.004990048 | 1 |

| | | | | | | |
|---|---|---|---|---|---|---|
| T2DM | leukocyte | *TCF3* | -0.985863748 | 0.000228119 | 0.004990048 | 0.896277925 |
| T2DM | leukocyte | *CPSF1* | -0.736122028 | 0.00470944 | 0.007722616 | 1 |
| T2DM | leukocyte | *LNX1* | -0.657083151 | 0.003518764 | 0.006649019 | 1 |
| T2DM | leukocyte | *ALDH3A2* | -0.584892532 | 0.002960675 | 0.006135605 | 1 |
| T2DM | leukocyte | *P4HA2* | -0.594831277 | 0.007535128 | 0.011246286 | 1 |
| T2DM | leukocyte | *TRNT1* | -2.975358518 | 0.000799578 | 0.004990048 | 1 |
| T2DM | leukocyte | *CRMP1* | -0.66756489 | 0.005490296 | 0.008659724 | 1 |
| T2DM | leukocyte | *DERL2* | -0.935326373 | 0.001118832 | 0.004990048 | 1 |
| T2DM | leukocyte | *XRCC1* | -1.205130269 | 0.002788538 | 0.005983705 | 1 |
| T2DM | leukocyte | *SCARB1* | -2.671600322 | 0.000112436 | 0.004990048 | 0.441759334 |
| T2DM | leukocyte | *NLE1* | -0.675992035 | 0.037893478 | 0.047264596 | 1 |
| T2DM | leukocyte | *SMARCE1* | -0.805967623 | 0.001251031 | 0.004990048 | 1 |
| T2DM | leukocyte | *ST6GAL1* | 2.253133459 | 0.002885684 | 0.00605886 | 1 |
| T2DM | leukocyte | *MRPS34* | -0.733441036 | 0.00175288 | 0.005084476 | 1 |
| T2DM | leukocyte | *TEAD2* | -0.98054012 | 0.015495981 | 0.02102338 | 1 |
| T2DM | leukocyte | *IPCEF1* | -1.737480256 | 0.001882517 | 0.005146173 | 1 |
| T2DM | leukocyte | *ZZEF1* | -1.342967564 | 0.000796946 | 0.004990048 | 1 |
| T2DM | leukocyte | *WSCD2* | -0.863923929 | 0.005965606 | 0.009262768 | 1 |
| T2DM | leukocyte | *TIPIN* | -0.80452621 | 0.039804351 | 0.049412732 | 1 |
| T2DM | leukocyte | *SEMA3A* | -2.347129754 | 0.004768588 | 0.007800075 | 1 |
| T2DM | leukocyte | *TTC38* | -1.93335306 | 0.000271357 | 0.004990048 | 1 |
| T2DM | leukocyte | *GRAMD4* | -1.26952199 | 0.002325499 | 0.005507466 | 1 |
| T2DM | leukocyte | *CELSR1* | -4.250503538 | 0.004405707 | 0.007420205 | 1 |
| T2DM | leukocyte | *TIMM21* | -1.320732039 | 0.001638521 | 0.005035955 | 1 |
| T2DM | leukocyte | *VPS9D1* | -0.73731257 | 0.011929128 | 0.016697379 | 1 |
| T2DM | leukocyte | *ZNF37A* | -0.902478244 | 0.001646766 | 0.005035955 | 1 |
| T2DM | leukocyte | *MARK3* | -1.17565035 | 6.87E-05 | 0.004990048 | 0.269803645 |
| T2DM | leukocyte | *CACNG5* | -24.06846005 | 0.005421995 | 0.008576094 | 1 |
| T2DM | leukocyte | *MOCOS* | -2.509446885 | 0.004673663 | 0.007686961 | 1 |
| T2DM | leukocyte | *DLG1* | -0.874575404 | 0.001522712 | 0.004993935 | 1 |
| T2DM | leukocyte | *SEC31B* | 1.339144188 | 0.013646007 | 0.018785971 | 1 |
| T2DM | leukocyte | *EXOSC7* | -0.707117989 | 0.003997637 | 0.007143456 | 1 |
| T2DM | leukocyte | *MKRN2* | -1.109779235 | 0.00154399 | 0.00500936 | 1 |
| T2DM | leukocyte | *MLH1* | -1.256127082 | 0.001285483 | 0.004990048 | 1 |
| T2DM | leukocyte | *TPD52* | -0.714739555 | 0.00157652 | 0.005009694 | 1 |
| T2DM | leukocyte | *ACACB* | -1.103228677 | 0.00139338 | 0.004990048 | 1 |
| T2DM | leukocyte | *GPATCH1* | -0.722664328 | 0.01237726 | 0.017257011 | 1 |
| T2DM | leukocyte | *ICAM3* | -0.817927666 | 0.005817506 | 0.009073831 | 1 |
| T2DM | leukocyte | *XAB2* | -0.939217425 | 0.001188586 | 0.004990048 | 1 |
| T2DM | leukocyte | *ACTL6B* | -0.78145194 | 0.002601088 | 0.005751212 | 1 |
| T2DM | leukocyte | *IL4R* | -0.751011122 | 0.005534512 | 0.008722462 | 1 |
| T2DM | leukocyte | *APBB1IP* | 0.327282912 | 0.012220785 | 0.017063064 | 1 |
| T2DM | leukocyte | *GPR137B* | -1.373578728 | 0.002488538 | 0.005622792 | 1 |
| T2DM | leukocyte | *NAALAD2* | -2.450580082 | 0.002680171 | 0.005854302 | 1 |
| T2DM | leukocyte | *MCCC1* | -0.920115552 | 0.002344725 | 0.005519727 | 1 |
| T2DM | leukocyte | *N4BP2* | -1.357439826 | 0.000796836 | 0.004990048 | 1 |
| T2DM | leukocyte | *MLLT10* | -1.184303823 | 0.001399123 | 0.004990048 | 1 |
| T2DM | leukocyte | *VDAC3* | -1.000366351 | 0.001127289 | 0.004990048 | 1 |
| T2DM | leukocyte | *PCM1* | -0.842120568 | 0.001253974 | 0.004990048 | 1 |
| T2DM | leukocyte | *TP73* | 2.10003024 | 0.022326102 | 0.029074993 | 1 |

| | | | | | | |
|---|---|---|---|---|---|---|
| T2DM | leukocyte | *TOLLIP* | -0.935875261 | 0.001599806 | 0.005024491 | 1 |
| T2DM | leukocyte | *CLUL1* | -2.277951571 | 0.002957007 | 0.006135605 | 1 |
| T2DM | leukocyte | *FKBP7* | -1.263604899 | 0.001014747 | 0.004990048 | 1 |
| T2DM | leukocyte | *LXN* | -0.611461741 | 0.007447677 | 0.011134674 | 1 |
| T2DM | leukocyte | *SAR1A* | -0.717903864 | 0.001685283 | 0.005060368 | 1 |
| T2DM | leukocyte | *CDC14A* | -1.329658085 | 0.002431457 | 0.005603046 | 1 |
| T2DM | leukocyte | *FDFT1* | -1.107362204 | 0.000326658 | 0.004990048 | 1 |
| T2DM | leukocyte | *SCGN* | -0.428558923 | 0.0022035 | 0.005384049 | 1 |
| T2DM | leukocyte | *DDX1* | -0.814521391 | 0.00169052 | 0.005060368 | 1 |
| T2DM | leukocyte | *RABL2B* | -1.317442782 | 0.002271178 | 0.005457772 | 1 |
| T2DM | leukocyte | *DCT* | -6.024250747 | 0.000691469 | 0.004990048 | 1 |
| T2DM | leukocyte | *SCTR* | -0.821125101 | 0.005469853 | 0.008631969 | 1 |
| T2DM | leukocyte | *RIF1* | -1.732123176 | 0.000460513 | 0.004990048 | 1 |
| T2DM | leukocyte | *COL5A3* | -4.359093491 | 0.008061957 | 0.011881256 | 1 |
| T2DM | leukocyte | *IGSF9B* | -1.601393195 | 0.004175593 | 0.007242343 | 1 |
| T2DM | leukocyte | *COL4A4* | -2.019336863 | 0.002112848 | 0.005283012 | 1 |
| T2DM | leukocyte | *ZNF506* | -0.8605112 | 0.002427835 | 0.005602682 | 1 |
| T2DM | leukocyte | *CADPS2* | -0.837108413 | 0.006412633 | 0.009838046 | 1 |
| T2DM | leukocyte | *ME2* | -2.167812806 | 0.00046042 | 0.004990048 | 1 |
| T2DM | leukocyte | *CCNT2* | -0.83660621 | 0.020200632 | 0.026544577 | 1 |
| T2DM | leukocyte | *SEMA5B* | 1.245841676 | 0.01191212 | 0.016679515 | 1 |
| T2DM | leukocyte | *BCKDHB* | -2.215515924 | 0.003788255 | 0.006955165 | 1 |
| T2DM | leukocyte | *DIS3* | -0.898143697 | 0.001483936 | 0.004990048 | 1 |
| T2DM | leukocyte | *PIBF1* | -1.334325207 | 0.002704389 | 0.005880214 | 1 |
| T2DM | leukocyte | *CHMP2B* | -0.807108602 | 0.001191797 | 0.004990048 | 1 |
| T2DM | leukocyte | *PPIE* | -1.014824571 | 0.001798923 | 0.005114303 | 1 |
| T2DM | leukocyte | *NCOA1* | -1.076397317 | 0.000988202 | 0.004990048 | 1 |
| T2DM | leukocyte | *AGBL5* | -1.43828591 | 0.00699554 | 0.010563211 | 1 |
| T2DM | leukocyte | *CAD* | -0.79388996 | 0.022533317 | 0.029296295 | 1 |
| T2DM | leukocyte | *CD59* | -0.532029465 | 0.003574359 | 0.006719453 | 1 |
| T2DM | leukocyte | *CD82* | -1.690263877 | 0.000738381 | 0.004990048 | 1 |
| T2DM | leukocyte | *MECOM* | -0.483524205 | 0.023575265 | 0.030489537 | 1 |
| T2DM | leukocyte | *SLC25A24* | -0.864467986 | 0.002532104 | 0.005660135 | 1 |
| T2DM | leukocyte | *PILRA* | 0.293835388 | 0.007075778 | 0.010655705 | 1 |
| T2DM | leukocyte | *ZNF213* | -1.007703976 | 0.001824291 | 0.00513989 | 1 |
| T2DM | leukocyte | *CPNE3* | -0.67828095 | 0.00211764 | 0.005286028 | 1 |
| T2DM | leukocyte | *RRN3* | -0.751115207 | 0.003090994 | 0.00625753 | 1 |
| T2DM | leukocyte | *WNT11* | -1.728972109 | 0.034521426 | 0.043375338 | 1 |
| T2DM | leukocyte | *DDHD2* | -0.81056631 | 0.017127507 | 0.022959391 | 1 |
| T2DM | leukocyte | *TTC39A* | -0.794503373 | 0.009527461 | 0.013691804 | 1 |
| T2DM | leukocyte | *EPS15* | -1.145499735 | 0.003257937 | 0.006405338 | 1 |
| T2DM | leukocyte | *ORC1* | -3.3018759 | 0.001395497 | 0.004990048 | 1 |
| T2DM | leukocyte | *MGST2* | -0.559290427 | 0.007872737 | 0.011663643 | 1 |
| T2DM | leukocyte | *USP40* | -1.45858206 | 0.000973614 | 0.004990048 | 1 |
| T2DM | leukocyte | *POMGNT1* | -1.286394022 | 0.00044349 | 0.004990048 | 1 |
| T2DM | leukocyte | *NFX1* | -1.418167157 | 0.001643622 | 0.005035955 | 1 |
| T2DM | leukocyte | *EIF2AK1* | -0.887321839 | 0.002563393 | 0.00569337 | 1 |
| T2DM | leukocyte | *ITPKC* | -1.079267221 | 0.002812805 | 0.006001505 | 1 |
| T2DM | leukocyte | *TMED2* | -0.686055135 | 0.001385412 | 0.004990048 | 1 |
| T2DM | leukocyte | *ACOX3* | -2.007729062 | 0.00133313 | 0.004990048 | 1 |

| | | | | | | |
|---|---|---|---|---|---|---|
| T2DM | leukocyte | *TRIP6* | -0.872984924 | 0.000263553 | 0.004990048 | 1 |
| T2DM | leukocyte | *PGS1* | -1.422810208 | 0.00130873 | 0.004990048 | 1 |
| T2DM | leukocyte | *PSMC5* | -0.781002202 | 0.000645362 | 0.004990048 | 1 |
| T2DM | leukocyte | *UIMC1* | -0.901849492 | 0.004270075 | 0.007310294 | 1 |
| T2DM | leukocyte | *SF3B2* | -0.7464054 | 0.001754 | 0.005084476 | 1 |
| T2DM | leukocyte | *KLHL42* | -0.854920577 | 0.000505813 | 0.004990048 | 1 |
| T2DM | leukocyte | *METTL2A* | -1.421627303 | 0.001126078 | 0.004990048 | 1 |
| T2DM | leukocyte | *GP6* | 1.115609996 | 0.026560182 | 0.033969712 | 1 |
| T2DM | leukocyte | *SLC15A1* | -1.488618627 | 0.013431038 | 0.018535492 | 1 |
| T2DM | leukocyte | *TGDS* | -0.853908664 | 0.004651116 | 0.007681478 | 1 |
| T2DM | leukocyte | *DOCK3* | -0.963926851 | 0.004231832 | 0.007276528 | 1 |
| T2DM | leukocyte | *ARHGAP28* | -2.178406119 | 0.00403488 | 0.007166394 | 1 |
| T2DM | leukocyte | *PPP1R13B* | -0.608925727 | 0.005853882 | 0.009116093 | 1 |
| T2DM | leukocyte | *ZNF343* | -1.018307115 | 0.011168098 | 0.015766962 | 1 |
| T2DM | leukocyte | *MAVS* | -1.750093288 | 0.001540684 | 0.005006447 | 1 |
| T2DM | leukocyte | *DYNLL1* | -0.805122577 | 0.000639536 | 0.004990048 | 1 |
| T2DM | leukocyte | *RBBP9* | -0.698488263 | 0.008417042 | 0.012330558 | 1 |
| T2DM | leukocyte | *SLC23A2* | -1.542464132 | 0.000880416 | 0.004990048 | 1 |
| T2DM | leukocyte | *SLC8B1* | 0.221868287 | 0.025347457 | 0.032577742 | 1 |
| T2DM | leukocyte | *TMEM230* | -0.841180015 | 0.001324296 | 0.004990048 | 1 |
| T2DM | leukocyte | *KIF16B* | -1.477410698 | 0.000484524 | 0.004990048 | 1 |
| T2DM | leukocyte | *ZNF302* | -0.89566518 | 0.000910515 | 0.004990048 | 1 |
| T2DM | leukocyte | *GRAMD1A* | -1.473779863 | 0.000825167 | 0.004990048 | 1 |
| T2DM | leukocyte | *CDIP1* | -0.89386001 | 0.003316455 | 0.006450669 | 1 |
| T2DM | leukocyte | *MLF2* | -0.951400261 | 0.001020114 | 0.004990048 | 1 |
| T2DM | leukocyte | *DDX24* | -0.769227876 | 0.00096598 | 0.004990048 | 1 |
| T2DM | leukocyte | *ANKRD24* | -6.103997059 | 0.010209665 | 0.014560353 | 1 |
| T2DM | leukocyte | *GPATCH2L* | -1.109621066 | 0.000834266 | 0.004990048 | 1 |
| T2DM | leukocyte | *SLC9A1* | -0.73744306 | 0.00508082 | 0.008170331 | 1 |
| T2DM | leukocyte | *SPTLC1* | -0.858314389 | 0.002114445 | 0.005283012 | 1 |
| T2DM | leukocyte | *NDUFB2* | -0.816682028 | 0.000208512 | 0.004990048 | 0.819242871 |
| T2DM | leukocyte | *MAEA* | -0.752236808 | 0.003842766 | 0.007022431 | 1 |
| T2DM | leukocyte | *STRN4* | -0.873035187 | 0.00164783 | 0.005035955 | 1 |
| T2DM | leukocyte | *LYZ* | 0.183052526 | 0.014319122 | 0.019616398 | 1 |
| T2DM | leukocyte | *MUL1* | -0.968334402 | 0.001235475 | 0.004990048 | 1 |
| T2DM | leukocyte | *SPG21* | -1.22953384 | 0.000842094 | 0.004990048 | 1 |
| T2DM | leukocyte | *USP48* | -0.805321708 | 0.005150195 | 0.008249407 | 1 |
| T2DM | leukocyte | *GLG1* | -0.881332168 | 0.001485487 | 0.004990048 | 1 |
| T2DM | leukocyte | *NRCAM* | -0.559561274 | 0.003432414 | 0.006559316 | 1 |
| T2DM | leukocyte | *TF* | -1.212887593 | 0.01108079 | 0.015660584 | 1 |
| T2DM | leukocyte | *MYO15A* | -1.940178764 | 0.007000542 | 0.010565863 | 1 |
| T2DM | leukocyte | *PITPNM3* | -2.367334508 | 0.00395964 | 0.007124893 | 1 |
| T2DM | leukocyte | *SPAG7* | -0.838760254 | 0.007425941 | 0.011106403 | 1 |
| T2DM | leukocyte | *TMEM101* | -1.176792055 | 0.000909852 | 0.004990048 | 1 |
| T2DM | leukocyte | *PPP2R3C* | -1.64909572 | 0.000189448 | 0.004990048 | 0.744340782 |
| T2DM | leukocyte | *OSGEP* | -1.595421824 | 0.000844803 | 0.004990048 | 1 |
| T2DM | leukocyte | *SCFD1* | -0.911386611 | 0.001591088 | 0.005021192 | 1 |
| T2DM | leukocyte | *RPGRIP1* | 0.920280361 | 0.027685941 | 0.035237467 | 1 |
| T2DM | leukocyte | *EZR* | -0.609380714 | 0.002007035 | 0.005232069 | 1 |
| T2DM | leukocyte | *NUP50* | -1.695391933 | 0.000395241 | 0.004990048 | 1 |

| | | | | | | |
|---|---|---|---|---|---|---|
| T2DM | leukocyte | *COMT* | -0.795921942 | 0.001541818 | 0.005006447 | 1 |
| T2DM | leukocyte | *ECHDC1* | -0.680667344 | 0.004208896 | 0.007263735 | 1 |
| T2DM | leukocyte | *CBX5* | -0.784322335 | 0.000897318 | 0.004990048 | 1 |
| T2DM | leukocyte | *SUCO* | -0.644923524 | 0.001237583 | 0.004990048 | 1 |
| T2DM | leukocyte | *MSH2* | -1.911124644 | 0.001269603 | 0.004990048 | 1 |
| T2DM | leukocyte | *TMEM38B* | -1.457310677 | 0.000228947 | 0.004990048 | 0.899531627 |
| T2DM | leukocyte | *PSMD5* | -0.810240428 | 0.002804573 | 0.006001505 | 1 |
| T2DM | leukocyte | *CRAT* | -1.507741632 | 0.001009443 | 0.004990048 | 1 |
| T2DM | leukocyte | *SH2D3C* | -1.166309154 | 0.005561602 | 0.008754621 | 1 |
| T2DM | leukocyte | *CWF19L1* | -1.305422578 | 3.69E-06 | 0.0010358 | 0.014501198 |
| T2DM | leukocyte | *SEMA4G* | -8.311742455 | 0.003367338 | 0.006494978 | 1 |
| T2DM | leukocyte | *IKZF5* | -1.521312302 | 0.000355785 | 0.004990048 | 1 |
| T2DM | leukocyte | *TLL2* | -3.427002804 | 0.004246876 | 0.007292822 | 1 |
| T2DM | leukocyte | *TPSD1* | -28.03606079 | 0.013460208 | 0.018569226 | 1 |
| T2DM | leukocyte | *SRPK1* | -1.106118874 | 0.000859466 | 0.004990048 | 1 |
| T2DM | leukocyte | *PGC* | -0.82129192 | 0.002176975 | 0.005349178 | 1 |
| T2DM | leukocyte | *EFHC1* | -0.735006668 | 0.004551083 | 0.007568852 | 1 |
| T2DM | leukocyte | *ITPR3* | -0.881294966 | 0.004476954 | 0.007504245 | 1 |
| T2DM | leukocyte | *IFT74* | -1.255052406 | 0.001184121 | 0.004990048 | 1 |
| T2DM | leukocyte | *ABL1* | -0.930310984 | 0.001708996 | 0.005068796 | 1 |
| T2DM | leukocyte | *SH3GLB1* | -0.936354294 | 0.001308215 | 0.004990048 | 1 |
| T2DM | leukocyte | *CDC7* | -1.008301261 | 0.013634382 | 0.018776546 | 1 |
| T2DM | leukocyte | *SYDE2* | -0.770986322 | 0.023435673 | 0.030318986 | 1 |
| T2DM | leukocyte | *ABLIM1* | -0.742553082 | 0.002269837 | 0.005457772 | 1 |
| T2DM | leukocyte | *ERMP1* | -0.832094529 | 0.003715448 | 0.006879357 | 1 |
| T2DM | leukocyte | *PALMD* | -0.829827341 | 0.000963684 | 0.004990048 | 1 |
| T2DM | leukocyte | *MZF1* | -0.815188332 | 0.009681744 | 0.013893197 | 1 |
| T2DM | leukocyte | *OCEL1* | -1.234973747 | 0.001148804 | 0.004990048 | 1 |
| T2DM | leukocyte | *STX1B* | 1.087933705 | 0.002766913 | 0.005956822 | 1 |
| T2DM | leukocyte | *HSD3B7* | -2.619124389 | 0.003515142 | 0.006649019 | 1 |
| T2DM | leukocyte | *HNRNPM* | -0.776272236 | 0.000715041 | 0.004990048 | 1 |
| T2DM | leukocyte | *TIMM13* | -0.920075878 | 0.00097808 | 0.004990048 | 1 |
| T2DM | leukocyte | *CEP170B* | -1.338101746 | 0.001469987 | 0.004990048 | 1 |
| T2DM | leukocyte | *POLR2E* | -1.251776141 | 0.000841893 | 0.004990048 | 1 |
| T2DM | leukocyte | *RASSF7* | -0.787740367 | 0.002062188 | 0.00526847 | 1 |
| T2DM | leukocyte | *ARVCF* | -1.505541902 | 0.005124583 | 0.008214804 | 1 |
| T2DM | leukocyte | *TRMT2A* | -0.781928245 | 0.004451098 | 0.007480054 | 1 |
| T2DM | leukocyte | *MED15* | -2.588917052 | 0.000313606 | 0.004990048 | 1 |
| T2DM | leukocyte | *SMARCB1* | -0.990768073 | 0.001009547 | 0.004990048 | 1 |
| T2DM | leukocyte | *DDT* | -1.029761866 | 0.000514737 | 0.004990048 | 1 |
| T2DM | leukocyte | *CABIN1* | -1.163573517 | 0.002012008 | 0.005238686 | 1 |
| T2DM | leukocyte | *SPECC1L* | -1.416315132 | 0.001104015 | 0.004990048 | 1 |
| T2DM | leukocyte | *PES1* | -0.887691843 | 0.002263794 | 0.005456717 | 1 |
| T2DM | leukocyte | *GGT1* | -5.648239162 | 0.024265207 | 0.031330265 | 1 |
| T2DM | leukocyte | *TOP3B* | -1.275302502 | 0.006126893 | 0.009458767 | 1 |
| T2DM | leukocyte | *CARD10* | -0.645628964 | 0.00313897 | 0.006311945 | 1 |
| T2DM | leukocyte | *SLC25A1* | -0.987447908 | 0.001909686 | 0.00515389 | 1 |
| T2DM | leukocyte | *HPS4* | -1.016862796 | 0.001822697 | 0.00513989 | 1 |
| T2DM | leukocyte | *GCAT* | -1.283583921 | 0.001696778 | 0.005060368 | 1 |
| T2DM | leukocyte | *CYP2D6* | 1.845368875 | 0.032049824 | 0.04052905 | 1 |

| | | | | | | |
|---|---|---|---|---|---|---|
| T2DM | leukocyte | *DDX17* | -0.977827296 | 0.000594533 | 0.004990048 | 1 |
| T2DM | leukocyte | *HSCB* | -1.563124227 | 0.00161946 | 0.005035955 | 1 |
| T2DM | leukocyte | *CBY1* | -1.000383837 | 0.001564068 | 0.005009694 | 1 |
| T2DM | leukocyte | *XBP1* | -0.661677334 | 0.001066792 | 0.004990048 | 1 |
| T2DM | leukocyte | *TIMP3* | -0.944070795 | 0.003891906 | 0.007059695 | 1 |
| T2DM | leukocyte | *CYB5R3* | -0.897784427 | 0.000982302 | 0.004990048 | 1 |
| T2DM | leukocyte | *MIOX* | -2.019339682 | 0.003868699 | 0.007040351 | 1 |
| T2DM | leukocyte | *LMF2* | -1.432956349 | 0.007914503 | 0.011703456 | 1 |
| T2DM | leukocyte | *RHBDD3* | -1.063084529 | 0.001385142 | 0.004990048 | 1 |
| T2DM | leukocyte | *TOM1* | -1.329489193 | 0.003395208 | 0.006510382 | 1 |
| T2DM | leukocyte | *CHKB* | -2.371893901 | 0.001120523 | 0.004990048 | 1 |
| T2DM | leukocyte | *TTLL12* | -1.327714846 | 0.002477926 | 0.005622792 | 1 |
| T2DM | leukocyte | *RBFOX2* | -0.931900436 | 0.000783894 | 0.004990048 | 1 |
| T2DM | leukocyte | *SYNGR1* | -3.06136173 | 3.83E-05 | 0.004122285 | 0.150356863 |
| T2DM | leukocyte | *MTMR3* | -1.199448758 | 0.001152168 | 0.004990048 | 1 |
| T2DM | leukocyte | *SAMM50* | -0.847071557 | 0.001809637 | 0.005122825 | 1 |
| T2DM | leukocyte | *FOXRED2* | -0.932878159 | 0.006028208 | 0.009335763 | 1 |
| T2DM | leukocyte | *SGSM3* | -1.350916927 | 0.001233325 | 0.004990048 | 1 |
| T2DM | leukocyte | *IFT27* | -1.19696262 | 0.000697676 | 0.004990048 | 1 |
| T2DM | leukocyte | *FAM118A* | -0.953811058 | 0.018533122 | 0.024600215 | 1 |
| T2DM | leukocyte | *ST13* | -0.807887552 | 0.000966174 | 0.004990048 | 1 |
| T2DM | leukocyte | *POLR3H* | -2.374418316 | 0.000127597 | 0.004990048 | 0.501330534 |
| T2DM | leukocyte | *PMM1* | -1.202020107 | 0.001443368 | 0.004990048 | 1 |
| T2DM | leukocyte | *DESI1* | -1.156733315 | 0.002458988 | 0.005622792 | 1 |
| T2DM | leukocyte | *KHNYN* | -1.325661705 | 0.000820625 | 0.004990048 | 1 |
| T2DM | leukocyte | *RBM23* | -1.008462902 | 0.000349558 | 0.004990048 | 1 |
| T2DM | leukocyte | *PRMT5* | -0.828868473 | 0.002982913 | 0.006152298 | 1 |
| T2DM | leukocyte | *VCPKMT* | -1.348729718 | 0.000995518 | 0.004990048 | 1 |
| T2DM | leukocyte | *SOS2* | -1.204871604 | 0.000960511 | 0.004990048 | 1 |
| T2DM | leukocyte | *CDKL1* | -1.486364166 | 0.006117025 | 0.00945096 | 1 |
| T2DM | leukocyte | *NIN* | -2.178566962 | 0.000526347 | 0.004990048 | 1 |
| T2DM | leukocyte | *TRIM9* | -0.650883898 | 0.004669876 | 0.007686961 | 1 |
| T2DM | leukocyte | *ATP6V1D* | -0.852266202 | 0.001060582 | 0.004990048 | 1 |
| T2DM | leukocyte | *PLEK2* | -1.055198599 | 0.021974983 | 0.028655728 | 1 |
| T2DM | leukocyte | *GSTZ1* | -1.252702994 | 0.003452844 | 0.006579158 | 1 |
| T2DM | leukocyte | *TMED8* | -1.100705876 | 0.000766617 | 0.004990048 | 1 |
| T2DM | leukocyte | *DAAM1* | -0.804173566 | 0.001809704 | 0.005122825 | 1 |
| T2DM | leukocyte | *DHRS7* | -1.009936421 | 0.00081463 | 0.004990048 | 1 |
| T2DM | leukocyte | *MTHFD1* | -3.821538098 | 0.000103131 | 0.004990048 | 0.405201463 |
| T2DM | leukocyte | *GSKIP* | -0.880610951 | 0.002219013 | 0.005407737 | 1 |
| T2DM | leukocyte | *TRIP11* | -0.768176929 | 0.001934848 | 0.005184924 | 1 |
| T2DM | leukocyte | *DCAF11* | -1.173244246 | 0.001649599 | 0.005035955 | 1 |
| T2DM | leukocyte | *EMC9* | -0.735235216 | 0.018552916 | 0.02460986 | 1 |
| T2DM | leukocyte | *NFATC4* | -1.188050934 | 0.003981018 | 0.007132431 | 1 |
| T2DM | leukocyte | *PYGB* | -1.149033212 | 0.00025647 | 0.004990048 | 1 |
| T2DM | leukocyte | *ABHD12* | -1.811240049 | 0.000445013 | 0.004990048 | 1 |
| T2DM | leukocyte | *PROCR* | -0.685447853 | 0.008458567 | 0.012372937 | 1 |
| T2DM | leukocyte | *CD40* | -0.991659145 | 0.002036196 | 0.005253355 | 1 |
| T2DM | leukocyte | *IFT52* | -0.885145072 | 0.00160976 | 0.005033031 | 1 |
| T2DM | leukocyte | *MYBL2* | -2.280296329 | 0.003078443 | 0.006253982 | 1 |

| | | | | | | |
|---|---|---|---|---|---|---|
| T2DM | leukocyte | *NELFCD* | -0.837769679 | 0.001831261 | 0.005146173 | 1 |
| T2DM | leukocyte | *MTG2* | -1.041518829 | 0.001407246 | 0.004990048 | 1 |
| T2DM | leukocyte | *DIDO1* | -0.936364311 | 0.007841821 | 0.011622224 | 1 |
| T2DM | leukocyte | *CDC25B* | -1.531088657 | 0.005440307 | 0.008601596 | 1 |
| T2DM | leukocyte | *ISM1* | 1.136723054 | 0.020179977 | 0.026526307 | 1 |
| T2DM | leukocyte | *ARFRP1* | -1.046028802 | 0.018974629 | 0.025084163 | 1 |
| T2DM | leukocyte | *SLC52A3* | -1.532614462 | 0.039357467 | 0.048919799 | 1 |
| T2DM | leukocyte | *SIRPB1* | 0.563584189 | 0.011627869 | 0.016328055 | 1 |
| T2DM | leukocyte | *PLCB4* | -0.742780749 | 0.004849549 | 0.00790211 | 1 |
| T2DM | leukocyte | *POFUT1* | -0.85826567 | 0.002408692 | 0.005586019 | 1 |
| T2DM | leukocyte | *NOP56* | -1.168510695 | 0.002761778 | 0.005955558 | 1 |
| T2DM | leukocyte | *MANBAL* | -0.822415792 | 0.001968347 | 0.005212073 | 1 |
| T2DM | leukocyte | *IDH3B* | -1.092230987 | 0.001587967 | 0.005020048 | 1 |
| T2DM | leukocyte | *MAPRE1* | -0.733782233 | 0.002033326 | 0.005252426 | 1 |
| T2DM | leukocyte | *OXT* | 0.678069564 | 0.022503191 | 0.029266811 | 1 |
| T2DM | leukocyte | *TTI1* | -1.389543498 | 0.001573379 | 0.005009694 | 1 |
| T2DM | leukocyte | *CHMP4B* | -0.759980447 | 0.001358605 | 0.004990048 | 1 |
| T2DM | leukocyte | *DHX35* | -1.577364077 | 0.001379777 | 0.004990048 | 1 |
| T2DM | leukocyte | *DNTTIP1* | -2.138792409 | 0.000363349 | 0.004990048 | 1 |
| T2DM | leukocyte | *MAP1LC3A* | -0.549608523 | 0.006467424 | 0.009910495 | 1 |
| T2DM | leukocyte | *APMAP* | -0.973762183 | 0.002740319 | 0.005930931 | 1 |
| T2DM | leukocyte | *VAPA* | -0.751651405 | 0.001095586 | 0.004990048 | 1 |
| T2DM | leukocyte | *MYOM1* | -1.99431643 | 0.005502975 | 0.008676239 | 1 |
| T2DM | leukocyte | *CEP192* | -0.72682958 | 0.005123138 | 0.008214804 | 1 |
| T2DM | leukocyte | *LIPG* | -0.926882076 | 0.00417942 | 0.007242343 | 1 |
| T2DM | leukocyte | *LAMA1* | -3.597953766 | 0.033634777 | 0.042410475 | 1 |
| T2DM | leukocyte | *POLI* | -1.326382944 | 0.009219333 | 0.013317191 | 1 |
| T2DM | leukocyte | *RIOK3* | -0.838465147 | 0.001034663 | 0.004990048 | 1 |
| T2DM | leukocyte | *NALCN* | -0.768392453 | 0.006059599 | 0.009380679 | 1 |
| T2DM | leukocyte | *FGF14* | -0.874561493 | 0.002488621 | 0.005622792 | 1 |
| T2DM | leukocyte | *FNDC3A* | -0.691821539 | 0.001784519 | 0.005100156 | 1 |
| T2DM | leukocyte | *KLF5* | -0.613112772 | 0.003972148 | 0.007124893 | 1 |
| T2DM | leukocyte | *ACP5* | 0.048580304 | 0.018464941 | 0.024526286 | 1 |
| T2DM | leukocyte | *DNAJC3* | -0.804873852 | 0.001136533 | 0.004990048 | 1 |
| T2DM | leukocyte | *UGGT2* | -0.840088947 | 0.001864415 | 0.005146173 | 1 |
| T2DM | leukocyte | *ARHGEF7* | -1.231573281 | 0.000944394 | 0.004990048 | 1 |
| T2DM | leukocyte | *PARP4* | -2.662817183 | 0.000296226 | 0.004990048 | 1 |
| T2DM | leukocyte | *MEDAG* | -1.443317845 | 0.002248678 | 0.005439981 | 1 |
| T2DM | leukocyte | *CLN5* | -1.159614357 | 0.000758285 | 0.004990048 | 1 |
| T2DM | leukocyte | *OLFM4* | -0.319149465 | 0.026626808 | 0.034038245 | 1 |
| T2DM | leukocyte | *MGRN1* | -1.16989571 | 0.001167074 | 0.004990048 | 1 |
| T2DM | leukocyte | *LONP2* | -0.854324292 | 0.002062333 | 0.00526847 | 1 |
| T2DM | leukocyte | *DHODH* | -2.663319001 | 0.000891827 | 0.004990048 | 1 |
| T2DM | leukocyte | *PARD6A* | -0.680701169 | 0.001680196 | 0.005060368 | 1 |
| T2DM | leukocyte | *MMP15* | -1.752532608 | 0.004423208 | 0.007442734 | 1 |
| T2DM | leukocyte | *USB1* | -1.28697494 | 0.001843279 | 0.005146173 | 1 |
| T2DM | leukocyte | *CYB5B* | -1.879024208 | 0.000256767 | 0.004990048 | 1 |
| T2DM | leukocyte | *CCDC113* | -1.204512358 | 0.002286386 | 0.005462378 | 1 |
| T2DM | leukocyte | *NME3* | -0.984715661 | 0.001379933 | 0.004990048 | 1 |
| T2DM | leukocyte | *VAC14* | -0.969824632 | 0.001847524 | 0.005146173 | 1 |

| | | | | | | |
|---|---|---|---|---|---|---|
| T2DM | leukocyte | *NPRL3* | -1.415555235 | 0.00125041 | 0.004990048 | 1 |
| T2DM | leukocyte | *NAGPA* | -4.843850528 | 0.000211205 | 0.004990048 | 0.829822961 |
| T2DM | leukocyte | *ZNF500* | -1.731083842 | 0.003217148 | 0.006372159 | 1 |
| T2DM | leukocyte | *NME4* | -0.971664431 | 0.001466394 | 0.004990048 | 1 |
| T2DM | leukocyte | *NOMO3* | -1.451540925 | 0.000821367 | 0.004990048 | 1 |
| T2DM | leukocyte | *LMF1* | -0.888241036 | 0.002917117 | 0.006099708 | 1 |
| T2DM | leukocyte | *MTHFSD* | -0.933025977 | 0.009301146 | 0.0134061 | 1 |
| T2DM | leukocyte | *CLCN7* | -2.005887427 | 0.008989989 | 0.013019413 | 1 |
| T2DM | leukocyte | *HAGHL* | -1.514489875 | 0.001349271 | 0.004990048 | 1 |
| T2DM | leukocyte | *METRN* | -0.773004761 | 0.000798073 | 0.004990048 | 1 |
| T2DM | leukocyte | *PIEZO1* | -1.106405545 | 0.001812368 | 0.005123568 | 1 |
| T2DM | leukocyte | *CLUAP1* | -0.805895302 | 0.019349111 | 0.025536667 | 1 |
| T2DM | leukocyte | *UBFD1* | -1.152935275 | 0.004093742 | 0.007206818 | 1 |
| T2DM | leukocyte | *AQP8* | -1.357640453 | 0.004859226 | 0.007912101 | 1 |
| T2DM | leukocyte | *USP31* | -0.657686882 | 0.003538686 | 0.006674746 | 1 |
| T2DM | leukocyte | *HMOX2* | -0.808623239 | 0.002369974 | 0.005551753 | 1 |
| T2DM | leukocyte | *RBL2* | -0.872177374 | 0.017833174 | 0.02375137 | 1 |
| T2DM | leukocyte | *KAT8* | -1.112933475 | 0.001112092 | 0.004990048 | 1 |
| T2DM | leukocyte | *NOMO1* | -0.972998349 | 0.039104503 | 0.048651549 | 1 |
| T2DM | leukocyte | *RNF40* | -1.751356857 | 0.000600558 | 0.004990048 | 1 |
| T2DM | leukocyte | *KNOP1* | -1.160861839 | 0.001292894 | 0.004990048 | 1 |
| T2DM | leukocyte | *TRIP4* | -0.926295945 | 0.003514315 | 0.006649019 | 1 |
| T2DM | leukocyte | *AP3B2* | -0.944357074 | 0.003480899 | 0.006621675 | 1 |
| T2DM | leukocyte | *IGDCC4* | 2.163151469 | 0.011348517 | 0.01597003 | 1 |
| T2DM | leukocyte | *RPAP1* | -1.417071741 | 0.001474379 | 0.004990048 | 1 |
| T2DM | leukocyte | *HOMER2* | -0.693018952 | 0.003177194 | 0.006330221 | 1 |
| T2DM | leukocyte | *ATP8B4* | 0.427771244 | 0.015641602 | 0.021198984 | 1 |
| T2DM | leukocyte | *DMXL2* | -2.244638205 | 0.026631093 | 0.034038245 | 1 |
| T2DM | leukocyte | *DNAJC17* | -0.815231554 | 0.005823452 | 0.009077881 | 1 |
| T2DM | leukocyte | *ZDHHC2* | -0.595082707 | 0.003073293 | 0.006249984 | 1 |
| T2DM | leukocyte | *TRIM35* | -2.25948884 | 0.000895372 | 0.004990048 | 1 |
| T2DM | leukocyte | *RIPK2* | -1.168931471 | 0.000846769 | 0.004990048 | 1 |
| T2DM | leukocyte | *NBN* | -1.316944618 | 0.00132638 | 0.004990048 | 1 |
| T2DM | leukocyte | *LAPTM4B* | -0.557320061 | 0.002839593 | 0.006020917 | 1 |
| T2DM | leukocyte | *RAB2A* | -0.64543304 | 0.001766551 | 0.00509228 | 1 |
| T2DM | leukocyte | *EIF3E* | -0.746330763 | 0.001217165 | 0.004990048 | 1 |
| T2DM | leukocyte | *ESRP1* | -0.711980386 | 0.003319861 | 0.006454098 | 1 |
| T2DM | leukocyte | *ARMC1* | -1.048053903 | 0.001231818 | 0.004990048 | 1 |
| T2DM | leukocyte | *SPAG1* | -0.822209255 | 0.002640106 | 0.005811191 | 1 |
| T2DM | leukocyte | *CHRAC1* | -0.715287367 | 0.002798108 | 0.005997385 | 1 |
| T2DM | leukocyte | *SNX16* | -2.377323377 | 0.003945538 | 0.007116737 | 1 |
| T2DM | leukocyte | *SH2D4A* | -0.986722598 | 0.002761634 | 0.005955558 | 1 |
| T2DM | leukocyte | *INTS10* | -1.245510633 | 0.000687899 | 0.004990048 | 1 |
| T2DM | leukocyte | *ERI1* | -2.639718052 | 0.000121452 | 0.004990048 | 0.47718579 |
| T2DM | leukocyte | *LEPROTL1* | -1.119639958 | 0.000644292 | 0.004990048 | 1 |
| T2DM | leukocyte | *ERICH1* | -1.28675167 | 0.001218018 | 0.004990048 | 1 |
| T2DM | leukocyte | *ARHGEF10* | -0.93470781 | 0.003299747 | 0.006432819 | 1 |
| T2DM | leukocyte | *KLHDC4* | -1.103957053 | 0.002873421 | 0.006055188 | 1 |
| T2DM | leukocyte | *MCM4* | -1.129700256 | 0.001915379 | 0.005158002 | 1 |
| T2DM | leukocyte | *FGL1* | -1.259245609 | 0.017772732 | 0.023686929 | 1 |

| | | | | | | |
|---|---|---|---|---|---|---|
| T2DM | leukocyte | *ECH1* | -0.814696664 | 0.001953924 | 0.005198138 | 1 |
| T2DM | leukocyte | *HNRNPL* | -0.924037187 | 0.00134024 | 0.004990048 | 1 |
| T2DM | leukocyte | *SARS2* | -1.150842262 | 0.000855283 | 0.004990048 | 1 |
| T2DM | leukocyte | *LIN7B* | -1.238645159 | 0.00097322 | 0.004990048 | 1 |
| T2DM | leukocyte | *PPP1R13L* | -0.868007503 | 0.009309012 | 0.013407297 | 1 |
| T2DM | leukocyte | *ERCC2* | -0.878432779 | 0.006294231 | 0.009663944 | 1 |
| T2DM | leukocyte | *KLC3* | -0.869928485 | 0.002136361 | 0.005317933 | 1 |
| T2DM | leukocyte | *AMH* | -1.008613128 | 0.002387666 | 0.005560842 | 1 |
| T2DM | leukocyte | *IL27RA* | -1.042331559 | 0.011883016 | 0.016650632 | 1 |
| T2DM | leukocyte | *PDCD5* | -0.673137756 | 0.003080361 | 0.006254644 | 1 |
| T2DM | leukocyte | *NUMBL* | -0.730333235 | 0.003599021 | 0.006745301 | 1 |
| T2DM | leukocyte | *SLC1A5* | -1.106708971 | 0.003393934 | 0.006510382 | 1 |
| T2DM | leukocyte | *PRKD2* | -1.159435189 | 0.000473446 | 0.004990048 | 1 |
| T2DM | leukocyte | *APLP1* | -0.390809449 | 0.007635994 | 0.011372941 | 1 |
| T2DM | leukocyte | *PLIN3* | -0.989206159 | 0.001005173 | 0.004990048 | 1 |
| T2DM | leukocyte | *MYH14* | -0.737324656 | 0.002245331 | 0.005438799 | 1 |
| T2DM | leukocyte | *ZNF175* | -0.826278276 | 0.00284597 | 0.006031185 | 1 |
| T2DM | leukocyte | *PLA2G4C* | -1.303546266 | 0.00071404 | 0.004990048 | 1 |
| T2DM | leukocyte | *CAPS* | -1.111885347 | 0.004212033 | 0.007263735 | 1 |
| T2DM | leukocyte | *PPP2R1A* | -0.752882343 | 0.002435569 | 0.005609232 | 1 |
| T2DM | leukocyte | *GCDH* | -1.148542946 | 0.000813417 | 0.004990048 | 1 |
| T2DM | leukocyte | *PIK3R2* | -2.883335097 | 0.00031284 | 0.004990048 | 1 |
| T2DM | leukocyte | *UPK1A* | -11.79217218 | 0.007644581 | 0.011381417 | 1 |
| T2DM | leukocyte | *COPE* | -0.805361992 | 0.000976309 | 0.004990048 | 1 |
| T2DM | leukocyte | *ETV2* | -0.999084487 | 0.01120143 | 0.015796991 | 1 |
| T2DM | leukocyte | *PBX4* | -3.321504638 | 0.005583095 | 0.008784934 | 1 |
| T2DM | leukocyte | *SIPA1L3* | -2.504933568 | 0.000346475 | 0.004990048 | 1 |
| T2DM | leukocyte | *ETHE1* | -1.251064428 | 0.004342757 | 0.007358292 | 1 |
| T2DM | leukocyte | *SMG9* | -0.73200505 | 0.00798289 | 0.011786837 | 1 |
| T2DM | leukocyte | *RUNDC3B* | -5.103683682 | 0.000372764 | 0.004990048 | 1 |
| T2DM | leukocyte | *PMPCB* | -1.080776613 | 0.001131443 | 0.004990048 | 1 |
| T2DM | leukocyte | *PON2* | -0.803891462 | 0.001381541 | 0.004990048 | 1 |
| T2DM | leukocyte | *ITGB8* | -0.809541994 | 0.002071109 | 0.005273744 | 1 |
| T2DM | leukocyte | *DNAH11* | -3.46069706 | 0.005969287 | 0.009262768 | 1 |
| T2DM | leukocyte | *CAV2* | -0.524476333 | 0.004347841 | 0.007358292 | 1 |
| T2DM | leukocyte | *CAV1* | -0.622282977 | 0.004150688 | 0.007238372 | 1 |
| T2DM | leukocyte | *ANKRD7* | -1.390313441 | 0.0086922 | 0.012653447 | 1 |
| T2DM | leukocyte | *TSPAN12* | -1.405591416 | 0.001160405 | 0.004990048 | 1 |
| T2DM | leukocyte | *CPVL* | 0.102469514 | 0.018763998 | 0.024856287 | 1 |
| T2DM | leukocyte | *ABHD11* | -1.340376918 | 0.000723128 | 0.004990048 | 1 |
| T2DM | leukocyte | *CHCHD2* | -0.627479515 | 0.00118224 | 0.004990048 | 1 |
| T2DM | leukocyte | *EIF3B* | -0.911392929 | 0.001431545 | 0.004990048 | 1 |
| T2DM | leukocyte | *WASL* | -0.700144733 | 0.003505857 | 0.006641519 | 1 |
| T2DM | leukocyte | *AGFG2* | -0.81619394 | 0.004171571 | 0.007242343 | 1 |
| T2DM | leukocyte | *C1GALT1* | -0.771098119 | 0.001900508 | 0.005146173 | 1 |
| T2DM | leukocyte | *RPA3* | -0.996796541 | 0.001126555 | 0.004990048 | 1 |
| T2DM | leukocyte | *ZNHIT1* | -0.746850053 | 0.002534023 | 0.005660135 | 1 |
| T2DM | leukocyte | *CLDN15* | -0.712096512 | 0.004546375 | 0.007568852 | 1 |
| T2DM | leukocyte | *TMEM106B* | -0.783578924 | 0.002020371 | 0.005244224 | 1 |
| T2DM | leukocyte | *ACTR3C* | -1.41468613 | 0.002865776 | 0.006053568 | 1 |

| | | | | | | |
|---|---|---|---|---|---|---|
| T2DM | leukocyte | *TSPAN13* | -0.592571061 | 0.002182467 | 0.005356498 | 1 |
| T2DM | leukocyte | *RARRES2* | -0.392961095 | 0.007575108 | 0.01129082 | 1 |
| T2DM | leukocyte | *AGR2* | -0.549001184 | 0.012789122 | 0.017761916 | 1 |
| T2DM | leukocyte | *TMEM176B* | -0.651848956 | 0.010184863 | 0.014530256 | 1 |
| T2DM | leukocyte | *COA1* | -1.183300943 | 0.000993881 | 0.004990048 | 1 |
| T2DM | leukocyte | *BLVRA* | -1.62724446 | 0.000692811 | 0.004990048 | 1 |
| T2DM | leukocyte | *TMEM248* | -1.231084166 | 0.000248086 | 0.004990048 | 0.974728953 |
| T2DM | leukocyte | *RHEB* | -0.795830285 | 0.000980608 | 0.004990048 | 1 |
| T2DM | leukocyte | *TBL2* | -0.599710545 | 0.037373799 | 0.04667567 | 1 |
| T2DM | leukocyte | *LIMK1* | -3.250791671 | 0.001261513 | 0.004990048 | 1 |
| T2DM | leukocyte | *SPIN1* | -1.382556247 | 0.000822662 | 0.004990048 | 1 |
| T2DM | leukocyte | *NMRK1* | -0.933822389 | 0.002452375 | 0.005621365 | 1 |
| T2DM | leukocyte | *C5* | -0.998018187 | 0.01394475 | 0.019143579 | 1 |
| T2DM | leukocyte | *OGN* | -2.030230147 | 0.001932457 | 0.005184924 | 1 |
| T2DM | leukocyte | *SUSD1* | -1.632709036 | 0.000520089 | 0.004990048 | 1 |
| T2DM | leukocyte | *DNM1* | -0.622175352 | 0.003356232 | 0.006483106 | 1 |
| T2DM | leukocyte | *AK1* | -0.706294981 | 0.001556677 | 0.005009694 | 1 |
| T2DM | leukocyte | *PLGRKT* | -0.726277552 | 0.004094074 | 0.007206818 | 1 |
| T2DM | leukocyte | *DOCK8* | 0.321740783 | 0.036966521 | 0.046211091 | 1 |
| T2DM | leukocyte | *MPDZ* | -0.686109972 | 0.00568568 | 0.008916678 | 1 |
| T2DM | leukocyte | *NPDC1* | -0.634682254 | 0.001678026 | 0.005059834 | 1 |
| T2DM | leukocyte | *HPS1* | -1.08494635 | 0.001213158 | 0.004990048 | 1 |
| T2DM | leukocyte | *CXCL12* | -0.906581388 | 0.0015945 | 0.005024491 | 1 |
| T2DM | leukocyte | *TRDMT1* | -2.001709351 | 0.001449036 | 0.004990048 | 1 |
| T2DM | leukocyte | *ATE1* | -1.510168703 | 0.000794129 | 0.004990048 | 1 |
| T2DM | leukocyte | *NSMCE4A* | -0.754608911 | 0.005214688 | 0.008330015 | 1 |
| T2DM | leukocyte | *PLEKHA1* | -0.679105723 | 0.002159233 | 0.005342335 | 1 |
| T2DM | leukocyte | *PALD1* | -6.174050286 | 0.000335809 | 0.004990048 | 1 |
| T2DM | leukocyte | *SPOCK2* | -1.2902233 | 0.004313562 | 0.007328649 | 1 |
| T2DM | leukocyte | *BMPR1A* | -2.064594499 | 0.000544659 | 0.004990048 | 1 |
| T2DM | leukocyte | *LIPA* | 0.234137531 | 0.032800875 | 0.041412158 | 1 |
| T2DM | leukocyte | *SFXN3* | -1.446403968 | 0.000819737 | 0.004990048 | 1 |
| T2DM | leukocyte | *NPM3* | -0.977596726 | 0.00271421 | 0.005895041 | 1 |
| T2DM | leukocyte | *TNKS2* | -1.016189559 | 0.001008124 | 0.004990048 | 1 |
| T2DM | leukocyte | *ACBD5* | -0.747992784 | 0.00336638 | 0.006494978 | 1 |
| T2DM | leukocyte | *LHPP* | -1.510993814 | 0.002916069 | 0.006099708 | 1 |
| T2DM | leukocyte | *GTPBP4* | -0.76064206 | 0.001563704 | 0.005009694 | 1 |
| T2DM | leukocyte | *NEURL1* | -0.899452818 | 0.007682424 | 0.011424771 | 1 |
| T2DM | leukocyte | *XPNPEP1* | -0.837816415 | 0.002416187 | 0.005594105 | 1 |
| T2DM | leukocyte | *SMC3* | -0.755052908 | 0.001266291 | 0.004990048 | 1 |
| T2DM | leukocyte | *RPL28* | -0.630827897 | 0.000551884 | 0.004990048 | 1 |
| T2DM | leukocyte | *PBLD* | -0.730385491 | 0.004124022 | 0.007233608 | 1 |
| T2DM | leukocyte | *KRT23* | -0.452358535 | 0.016136307 | 0.021794276 | 1 |
| T2DM | leukocyte | *DHRS11* | -0.646239203 | 0.013292343 | 0.018363438 | 1 |
| T2DM | leukocyte | *CWC25* | -1.095721975 | 0.001161716 | 0.004990048 | 1 |
| T2DM | leukocyte | *RAD51C* | -0.986355215 | 0.00140434 | 0.004990048 | 1 |
| T2DM | leukocyte | *TRIM37* | -0.852803569 | 0.002001952 | 0.005227473 | 1 |
| T2DM | leukocyte | *RPS6KB1* | -0.870836706 | 0.007525589 | 0.011238327 | 1 |
| T2DM | leukocyte | *TRIM16L* | -1.284495355 | 0.005414834 | 0.008574413 | 1 |
| T2DM | leukocyte | *CDK5RAP3* | -1.139410877 | 0.011012152 | 0.015569178 | 1 |

| | | | | | | |
|---|---|---|---|---|---|---|
| T2DM | leukocyte | *CAMTA2* | -1.141095156 | 0.010822618 | 0.015317747 | 1 |
| T2DM | leukocyte | *MED13* | -0.89776687 | 0.001055936 | 0.004990048 | 1 |
| T2DM | leukocyte | *RAI1* | -0.673441459 | 0.00466499 | 0.007686961 | 1 |
| T2DM | leukocyte | *CCDC47* | -0.857149549 | 0.000932454 | 0.004990048 | 1 |
| T2DM | leukocyte | *DRG2* | -0.897549129 | 0.003129915 | 0.006306376 | 1 |
| T2DM | leukocyte | *FTSJ3* | -1.004562575 | 0.00027474 | 0.004990048 | 1 |
| T2DM | leukocyte | *SMARCD2* | -2.197206397 | 0.000214415 | 0.004990048 | 0.84243803 |
| T2DM | leukocyte | *SYNGR2* | -0.77126975 | 0.002343644 | 0.005519727 | 1 |
| T2DM | leukocyte | *C17orf75* | -0.996923915 | 0.01254373 | 0.017458135 | 1 |
| T2DM | leukocyte | *DHX58* | -2.082019844 | 0.000279796 | 0.004990048 | 1 |
| T2DM | leukocyte | *NAGLU* | -0.866847586 | 0.004563039 | 0.007578894 | 1 |
| T2DM | leukocyte | *HSD17B1* | -1.432142816 | 2.02E-05 | 0.0030497 | 0.0792922 |
| T2DM | leukocyte | *RND2* | -2.311093896 | 0.000970824 | 0.004990048 | 1 |
| T2DM | leukocyte | *ABCC3* | -0.645245677 | 0.007002674 | 0.010565863 | 1 |
| T2DM | leukocyte | *MPP2* | -0.610599331 | 0.004219249 | 0.007267615 | 1 |
| T2DM | leukocyte | *HLF* | -0.519399701 | 0.006646919 | 0.010135575 | 1 |
| T2DM | leukocyte | *EFNB3* | -0.778147755 | 0.00405348 | 0.007180398 | 1 |
| T2DM | leukocyte | *RANGRF* | -1.176667098 | 0.000963712 | 0.004990048 | 1 |
| T2DM | leukocyte | *DPH1* | -1.611745784 | 0.013592994 | 0.01873268 | 1 |
| T2DM | leukocyte | *OCIAD1* | -0.779920545 | 0.001145778 | 0.004990048 | 1 |
| T2DM | leukocyte | *UBE2D3* | -0.721965831 | 0.002049674 | 0.00526208 | 1 |
| T2DM | leukocyte | *MAPK10* | -0.731914873 | 0.002200904 | 0.005384049 | 1 |
| T2DM | leukocyte | *ANXA10* | -0.7786026 | 0.025720346 | 0.032981475 | 1 |
| T2DM | leukocyte | *AADAT* | -2.53606174 | 0.002315938 | 0.005496836 | 1 |
| T2DM | leukocyte | *TRIM2* | -0.825917888 | 0.00227529 | 0.005460974 | 1 |
| T2DM | leukocyte | *SH3D19* | -1.832928053 | 0.001038413 | 0.004990048 | 1 |
| T2DM | leukocyte | *GLRB* | -0.846957621 | 0.002831418 | 0.006013319 | 1 |
| T2DM | leukocyte | *LRP2BP* | -0.9357478 | 0.003950458 | 0.007119885 | 1 |
| T2DM | leukocyte | *UFSP2* | -1.078475429 | 0.003047665 | 0.006220403 | 1 |
| T2DM | leukocyte | *KLHL5* | -1.124791563 | 0.001252398 | 0.004990048 | 1 |
| T2DM | leukocyte | *FAM149A* | -0.65599437 | 0.020236575 | 0.026582917 | 1 |
| T2DM | leukocyte | *DDX25* | -0.709936412 | 0.018599032 | 0.024662706 | 1 |
| T2DM | leukocyte | *HTATIP2* | -1.234129714 | 0.000942231 | 0.004990048 | 1 |
| T2DM | leukocyte | *B3GAT1* | -2.021826529 | 0.005539175 | 0.00872631 | 1 |
| T2DM | leukocyte | *P2RX3* | -16.43355389 | 0.004002769 | 0.007145551 | 1 |
| T2DM | leukocyte | *VWA5A* | -1.019029046 | 0.00228055 | 0.005462378 | 1 |
| T2DM | leukocyte | *DTX4* | -1.04890237 | 0.001581273 | 0.005009694 | 1 |
| T2DM | leukocyte | *FOXRED1* | -1.435888694 | 0.003859436 | 0.007033267 | 1 |
| T2DM | leukocyte | *ST3GAL4* | -1.709027579 | 0.000394789 | 0.004990048 | 1 |
| T2DM | leukocyte | *PRPF19* | -0.807035309 | 0.001704158 | 0.00506478 | 1 |
| T2DM | leukocyte | *CEP164* | -1.627101402 | 0.00249012 | 0.005622805 | 1 |
| T2DM | leukocyte | *PDHX* | -0.745663909 | 0.037231348 | 0.046512549 | 1 |
| T2DM | leukocyte | *SLC1A2* | -0.830109075 | 0.00790346 | 0.011693207 | 1 |
| T2DM | leukocyte | *CD5* | -3.516844017 | 0.008735701 | 0.012702654 | 1 |
| T2DM | leukocyte | *ACCS* | 0.44399644 | 0.008521753 | 0.012446828 | 1 |
| T2DM | leukocyte | *MDK* | -0.922701958 | 0.000830143 | 0.004990048 | 1 |
| T2DM | leukocyte | *MADD* | -1.291676607 | 0.000648825 | 0.004990048 | 1 |
| T2DM | leukocyte | *NAA40* | -1.080923308 | 0.002381727 | 0.005560194 | 1 |
| T2DM | leukocyte | *CD81* | -0.569421778 | 0.00330039 | 0.006432819 | 1 |
| T2DM | leukocyte | *SLC35F2* | -1.046449453 | 0.000753892 | 0.004990048 | 1 |

| | | | | | | |
|---|---|---|---|---|---|---|
| T2DM | leukocyte | *NUP98* | -0.941659985 | 0.001581764 | 0.005009694 | 1 |
| T2DM | leukocyte | *GTF2H1* | -1.655769513 | 0.000548363 | 0.004990048 | 1 |
| T2DM | leukocyte | *PTPN5* | 1.602081725 | 0.013609571 | 0.01874895 | 1 |
| T2DM | leukocyte | *PRDM4* | -0.717566536 | 0.002407395 | 0.005586019 | 1 |
| T2DM | leukocyte | *CORO1C* | -1.034788674 | 0.001968304 | 0.005212073 | 1 |
| T2DM | leukocyte | *LIN7A* | -1.972931545 | 0.003034164 | 0.006212209 | 1 |
| T2DM | leukocyte | *KRT18* | -0.297937015 | 0.004836763 | 0.007890449 | 1 |
| T2DM | leukocyte | *METAP2* | -0.861296138 | 0.001227902 | 0.004990048 | 1 |
| T2DM | leukocyte | *WNT5B* | -1.982947344 | 0.003853753 | 0.007029432 | 1 |
| T2DM | leukocyte | *ITFG2* | -4.188652684 | 0.00073354 | 0.004990048 | 1 |
| T2DM | leukocyte | *VPS29* | -0.833489826 | 0.000840502 | 0.004990048 | 1 |
| T2DM | leukocyte | *CREBL2* | -1.13469325 | 0.001424012 | 0.004990048 | 1 |
| T2DM | leukocyte | *GSG1* | -7.338813638 | 0.000560491 | 0.004990048 | 1 |
| T2DM | leukocyte | *LTBR* | -1.045131533 | 0.001028823 | 0.004990048 | 1 |
| T2DM | leukocyte | *DDX55* | -1.51665827 | 0.000448839 | 0.004990048 | 1 |
| T2DM | leukocyte | *STX2* | -1.11899745 | 0.00392177 | 0.007087688 | 1 |
| T2DM | leukocyte | *CAND1* | -1.067632478 | 0.000784804 | 0.004990048 | 1 |
| T2DM | leukocyte | *NUP107* | -1.179529768 | 0.001604631 | 0.005033031 | 1 |
| T2DM | leukocyte | *KRR1* | -0.922604249 | 0.001000259 | 0.004990048 | 1 |
| T2DM | leukocyte | *GNB3* | -1.019141462 | 0.004862399 | 0.007913988 | 1 |
| T2DM | leukocyte | *CDCA3* | -1.742801826 | 0.002739857 | 0.005930931 | 1 |
| T2DM | leukocyte | *CHPT1* | -1.578564733 | 0.000346543 | 0.004990048 | 1 |
| T2DM | leukocyte | *SPSB2* | -0.871624677 | 0.003759026 | 0.00692001 | 1 |
| T2DM | leukocyte | *ATN1* | -0.966515606 | 0.001347036 | 0.004990048 | 1 |
| T2DM | leukocyte | *NT5DC3* | -0.958372884 | 0.008256709 | 0.012133749 | 1 |
| T2DM | leukocyte | *GOLT1B* | -0.864454269 | 0.001473834 | 0.004990048 | 1 |
| T2DM | leukocyte | *PRKAB1* | -0.778373615 | 0.004012829 | 0.00714972 | 1 |
| T2DM | leukocyte | *C2CD5* | -0.811710026 | 0.003496398 | 0.006633196 | 1 |
| T2DM | leukocyte | *SRSF9* | -0.848976769 | 0.000467949 | 0.004990048 | 1 |
| T2DM | leukocyte | *FRK* | -0.911257458 | 0.002703108 | 0.005880214 | 1 |
| T2DM | leukocyte | *RSPH4A* | -0.661315635 | 0.04010597 | 0.049736615 | 1 |
| T2DM | leukocyte | *SMIM8* | -1.061769993 | 0.001780242 | 0.005098084 | 1 |
| T2DM | leukocyte | *NEDD9* | -0.486536804 | 0.003304003 | 0.006432819 | 1 |
| T2DM | leukocyte | *CEP85L* | -0.91681058 | 0.002809057 | 0.006001505 | 1 |
| T2DM | leukocyte | *HDDC2* | -0.724177086 | 0.001749249 | 0.005084476 | 1 |
| T2DM | leukocyte | *TPD52L1* | -0.738023286 | 0.003439566 | 0.006566174 | 1 |
| T2DM | leukocyte | *MTRF1L* | -0.866799342 | 0.009384808 | 0.01351151 | 1 |
| T2DM | leukocyte | *MRPL18* | -1.271877932 | 0.001037941 | 0.004990048 | 1 |
| T2DM | leukocyte | *RNF8* | -0.901076187 | 0.001892403 | 0.005146173 | 1 |
| T2DM | leukocyte | *MDGA1* | 1.17096093 | 0.025797579 | 0.033069719 | 1 |
| T2DM | leukocyte | *CD83* | 0.136086019 | 0.035637372 | 0.044677484 | 1 |
| T2DM | leukocyte | *GLP1R* | -0.833613595 | 0.004394328 | 0.007406827 | 1 |
| T2DM | leukocyte | *BACH2* | -0.785022087 | 0.002664499 | 0.005838039 | 1 |
| T2DM | leukocyte | *FBXL4* | -1.070077762 | 0.001894398 | 0.005146173 | 1 |
| T2DM | leukocyte | *ASCC3* | -0.729039031 | 0.002444699 | 0.005621365 | 1 |
| T2DM | leukocyte | *WASF1* | -0.763925161 | 0.0039214 | 0.007087688 | 1 |
| T2DM | leukocyte | *ALDH5A1* | -2.202861647 | 0.008182292 | 0.012040533 | 1 |
| T2DM | leukocyte | *C6orf62* | -1.067534508 | 0.000876046 | 0.004990048 | 1 |
| T2DM | leukocyte | *B3GAT2* | -1.327199492 | 0.001932464 | 0.005184924 | 1 |
| T2DM | leukocyte | *HBS1L* | -0.927027637 | 0.001382657 | 0.004990048 | 1 |

| | | | | | | |
|---|---|---|---|---|---|---|
| T2DM | leukocyte | *PEX7* | -0.840922653 | 0.002086737 | 0.005283012 | 1 |
| T2DM | leukocyte | *PHACTR2* | -0.698534007 | 0.001745108 | 0.005084476 | 1 |
| T2DM | leukocyte | *EPM2A* | -1.504844345 | 0.001330063 | 0.004990048 | 1 |
| T2DM | leukocyte | *TFEB* | -1.07878497 | 0.002097374 | 0.005283012 | 1 |
| T2DM | leukocyte | *PRPH2* | -1.168868063 | 0.003285945 | 0.006432725 | 1 |
| T2DM | leukocyte | *DNPH1* | -0.847029644 | 0.004910399 | 0.007970068 | 1 |
| T2DM | leukocyte | *DUSP22* | -1.138478052 | 0.001711853 | 0.005068796 | 1 |
| T2DM | leukocyte | *LAMA4* | -0.704320587 | 0.002815476 | 0.006001505 | 1 |
| T2DM | leukocyte | *FBRSL1* | -1.074035991 | 0.001795612 | 0.005114303 | 1 |
| T2DM | leukocyte | *ENPP5* | -0.672309966 | 0.009029068 | 0.013066375 | 1 |
| T2DM | leukocyte | *NUDT12* | -1.157991938 | 0.001864407 | 0.005146173 | 1 |
| T2DM | leukocyte | *CEP72* | -2.608847502 | 0.004762599 | 0.007793525 | 1 |
| T2DM | leukocyte | *SEMA5A* | -0.990927519 | 0.004756064 | 0.007788402 | 1 |
| T2DM | leukocyte | *GHR* | -2.257407784 | 0.004073993 | 0.007189 | 1 |
| T2DM | leukocyte | *DAP* | -0.687008885 | 0.003179167 | 0.006330941 | 1 |
| T2DM | leukocyte | *NME5* | -1.114920426 | 0.013934801 | 0.019136607 | 1 |
| T2DM | leukocyte | *BRD8* | -1.049182023 | 0.001824361 | 0.00513989 | 1 |
| T2DM | leukocyte | *NNT* | -1.871587808 | 8.17E-06 | 0.001605004 | 0.032100082 |
| T2DM | leukocyte | *TMCO6* | 1.610879378 | 0.008742766 | 0.012708224 | 1 |
| T2DM | leukocyte | *IK* | -0.910931217 | 0.001050855 | 0.004990048 | 1 |
| T2DM | leukocyte | *PDE8B* | -0.567481191 | 0.004172836 | 0.007242343 | 1 |
| T2DM | leukocyte | *MSH3* | -1.003351346 | 0.001345646 | 0.004990048 | 1 |
| T2DM | leukocyte | *CCNG1* | -1.753229959 | 0.000465753 | 0.004990048 | 1 |
| T2DM | leukocyte | *DROSHA* | -0.64144394 | 0.011452523 | 0.016104854 | 1 |
| T2DM | leukocyte | *GOLPH3* | -0.913993133 | 0.001203633 | 0.004990048 | 1 |
| T2DM | leukocyte | *NPR3* | -2.624992609 | 0.003002353 | 0.006172668 | 1 |
| T2DM | leukocyte | *RAD50* | -1.167960489 | 0.000894965 | 0.004990048 | 1 |
| T2DM | leukocyte | *WWC1* | -0.478427729 | 0.005687247 | 0.008916678 | 1 |
| T2DM | leukocyte | *SMAD5* | -0.80655399 | 0.001877098 | 0.005146173 | 1 |
| T2DM | leukocyte | *HMGXB3* | -1.105425812 | 0.001955421 | 0.005198138 | 1 |
| T2DM | leukocyte | *BNIP1* | -0.738044579 | 0.007331258 | 0.010998287 | 1 |
| T2DM | leukocyte | *STC2* | -0.64487368 | 0.004393914 | 0.007406827 | 1 |
| T2DM | leukocyte | *EHHADH* | -1.123335117 | 0.004613972 | 0.007636182 | 1 |
| T2DM | leukocyte | *ACTR8* | -0.843640323 | 0.002960832 | 0.006135605 | 1 |
| T2DM | leukocyte | *TIMMDC1* | -0.970706152 | 0.002314086 | 0.005496836 | 1 |
| T2DM | leukocyte | *CLDN16* | 3.276938594 | 0.037094069 | 0.046355787 | 1 |
| T2DM | leukocyte | *FAM162A* | -1.126067624 | 0.000481431 | 0.004990048 | 1 |
| T2DM | leukocyte | *RBP1* | -0.797175863 | 0.001031001 | 0.004990048 | 1 |
| T2DM | leukocyte | *RNF7* | -0.868655848 | 0.00093204 | 0.004990048 | 1 |
| T2DM | leukocyte | *SERPINI2* | -0.585890448 | 0.00301345 | 0.006182688 | 1 |
| T2DM | leukocyte | *TUSC2* | -1.201253543 | 0.000785245 | 0.004990048 | 1 |
| T2DM | leukocyte | *CYB561D2* | -0.846308109 | 0.001253719 | 0.004990048 | 1 |
| T2DM | leukocyte | *C3orf14* | -0.869213823 | 0.000287586 | 0.004990048 | 1 |
| T2DM | leukocyte | *CBLB* | -1.50840419 | 0.000839729 | 0.004990048 | 1 |
| T2DM | leukocyte | *IFT57* | -0.746376206 | 0.003600127 | 0.006745301 | 1 |
| T2DM | leukocyte | *NCBP2* | -0.82745132 | 0.001397066 | 0.004990048 | 1 |
| T2DM | leukocyte | *C3orf52* | -0.693997901 | 0.006484624 | 0.009925239 | 1 |
| T2DM | leukocyte | *SLC41A3* | -0.829307975 | 0.002870404 | 0.006055188 | 1 |
| T2DM | leukocyte | *ATP6V1A* | -1.057259765 | 0.000932952 | 0.004990048 | 1 |
| T2DM | leukocyte | *PODXL2* | -0.67307846 | 0.002482733 | 0.005622792 | 1 |

| | | | | | | |
|---|---|---|---|---|---|---|
| T2DM | leukocyte | *UPK1B* | -0.973521936 | 0.033413736 | 0.042158821 | 1 |
| T2DM | leukocyte | *NEK11* | -0.809661593 | 0.000866896 | 0.004990048 | 1 |
| T2DM | leukocyte | *PLSCR4* | -1.481048581 | 0.001308535 | 0.004990048 | 1 |
| T2DM | leukocyte | *COMMD2* | -0.864278953 | 0.001077225 | 0.004990048 | 1 |
| T2DM | leukocyte | *GORASP1* | -1.976914 | 0.001921118 | 0.005162839 | 1 |
| T2DM | leukocyte | *AADAC* | -0.876054103 | 0.000490129 | 0.004990048 | 1 |
| T2DM | leukocyte | *ARHGEF26* | -2.406945393 | 0.000658516 | 0.004990048 | 1 |
| T2DM | leukocyte | *PLCH1* | -4.196732888 | 0.000473442 | 0.004990048 | 1 |
| T2DM | leukocyte | *VIPR1* | -1.253984169 | 0.00873279 | 0.012702654 | 1 |
| T2DM | leukocyte | *NKTR* | -0.85753222 | 0.00146002 | 0.004990048 | 1 |
| T2DM | leukocyte | *NEK4* | -2.754307016 | 0.000227046 | 0.004990048 | 0.892064599 |
| T2DM | leukocyte | *KANSL3* | -1.987352679 | 0.000435573 | 0.004990048 | 1 |
| T2DM | leukocyte | *ACTR1B* | -1.565052313 | 0.000737354 | 0.004990048 | 1 |
| T2DM | leukocyte | *SLC35F5* | -0.828700315 | 0.002901074 | 0.006075864 | 1 |
| T2DM | leukocyte | *ACTR3* | -0.976944453 | 0.001182554 | 0.004990048 | 1 |
| T2DM | leukocyte | *POMC* | -1.516408885 | 0.000739904 | 0.004990048 | 1 |
| T2DM | leukocyte | *TANC1* | -0.619520301 | 0.014955657 | 0.020332448 | 1 |
| T2DM | leukocyte | *REEP6* | -1.048407312 | 0.003774706 | 0.006933529 | 1 |
| T2DM | leukocyte | *INO80B* | -2.887676554 | 0.001493022 | 0.004993935 | 1 |
| T2DM | leukocyte | *MOGS* | -1.500180137 | 0.001034635 | 0.004990048 | 1 |
| T2DM | leukocyte | *RTN4* | -0.753279846 | 0.000846671 | 0.004990048 | 1 |
| T2DM | leukocyte | *HTRA2* | -1.466010308 | 0.001092999 | 0.004990048 | 1 |
| T2DM | leukocyte | *GALNT3* | -0.974417278 | 0.002072829 | 0.005274707 | 1 |
| T2DM | leukocyte | *MRPL19* | -0.902719296 | 0.00247953 | 0.005622792 | 1 |
| T2DM | leukocyte | *WDR75* | -1.611380536 | 0.000961704 | 0.004990048 | 1 |
| T2DM | leukocyte | *FANCL* | -2.014975199 | 0.000622265 | 0.004990048 | 1 |
| T2DM | leukocyte | *GGCX* | -1.144228054 | 0.001300074 | 0.004990048 | 1 |
| T2DM | leukocyte | *SF3B1* | -1.024110795 | 0.000715886 | 0.004990048 | 1 |
| T2DM | leukocyte | *CHMP3* | -0.875364306 | 0.000348323 | 0.004990048 | 1 |
| T2DM | leukocyte | *IL1R2* | -0.850111275 | 0.004699792 | 0.007719684 | 1 |
| T2DM | leukocyte | *IL1R1* | -0.845027587 | 0.001494588 | 0.004993935 | 1 |
| T2DM | leukocyte | *IL1RL2* | 0.696973551 | 0.027076492 | 0.034528898 | 1 |
| T2DM | leukocyte | *IL18R1* | 0.744841791 | 0.010939137 | 0.015477086 | 1 |
| T2DM | leukocyte | *MLPH* | -1.185059039 | 0.009289744 | 0.013402558 | 1 |
| T2DM | leukocyte | *CNPPD1* | -0.696603954 | 0.00450429 | 0.007539852 | 1 |
| T2DM | leukocyte | *HDLBP* | -0.663921114 | 0.001662831 | 0.005041098 | 1 |
| T2DM | leukocyte | *PROC* | 1.026782579 | 0.013398735 | 0.01849741 | 1 |
| T2DM | leukocyte | *TAF1B* | -0.748408042 | 0.006565392 | 0.010030007 | 1 |
| T2DM | leukocyte | *STRN* | -1.326161539 | 0.00733119 | 0.010998287 | 1 |
| T2DM | leukocyte | *CEBPZ* | -0.853359866 | 0.001427745 | 0.004990048 | 1 |
| T2DM | leukocyte | *COX7A2L* | -0.738429924 | 0.001139888 | 0.004990048 | 1 |
| T2DM | leukocyte | *PNO1* | -0.951777508 | 0.001226047 | 0.004990048 | 1 |
| T2DM | leukocyte | *ORC4* | -0.984685929 | 0.000882593 | 0.004990048 | 1 |
| T2DM | leukocyte | *SUMO1* | -0.838552898 | 0.001060825 | 0.004990048 | 1 |
| T2DM | leukocyte | *ALMS1* | -1.091699212 | 0.002045556 | 0.00526208 | 1 |
| T2DM | leukocyte | *MARK1* | -1.556251007 | 0.000645406 | 0.004990048 | 1 |
| T2DM | leukocyte | *MORN1* | -1.373884879 | 0.01142549 | 0.016072593 | 1 |
| T2DM | leukocyte | *GPX7* | -1.029454696 | 0.000129536 | 0.004990048 | 0.50894764 |
| T2DM | leukocyte | *SCP2* | -0.890350897 | 0.000926339 | 0.004990048 | 1 |
| T2DM | leukocyte | *RALGPS2* | -0.834786808 | 0.002371046 | 0.005551753 | 1 |

| | | | | | | |
|---|---|---|---|---|---|---|
| T2DM | leukocyte | *ANGPTL1* | -1.650382203 | 0.001242094 | 0.004990048 | 1 |
| T2DM | leukocyte | *FAM20B* | -1.01535571 | 0.001688775 | 0.005060368 | 1 |
| T2DM | leukocyte | *WRAP73* | -3.533604644 | 0.000242611 | 0.004990048 | 0.953220402 |
| T2DM | leukocyte | *ICMT* | -1.209451046 | 0.000938061 | 0.004990048 | 1 |
| T2DM | leukocyte | *PARK7* | -0.73092898 | 0.001083626 | 0.004990048 | 1 |
| T2DM | leukocyte | *MECR* | -2.680948649 | 0.000396808 | 0.004990048 | 1 |
| T2DM | leukocyte | *EDEM3* | -1.046203159 | 0.0009762 | 0.004990048 | 1 |
| T2DM | leukocyte | *FBXO6* | -4.339072959 | 0.000490864 | 0.004990048 | 1 |
| T2DM | leukocyte | *SMG7* | -0.891951793 | 0.001623272 | 0.005035955 | 1 |
| T2DM | leukocyte | *PDC* | -8.959678821 | 0.003069158 | 0.006249984 | 1 |
| T2DM | leukocyte | *BCAS2* | -0.792603545 | 0.001662029 | 0.005041098 | 1 |
| T2DM | leukocyte | *AGMAT* | -1.365308538 | 0.002162346 | 0.005345956 | 1 |
| T2DM | leukocyte | *PLEKHM2* | -2.292911953 | 0.000540935 | 0.004990048 | 1 |
| T2DM | leukocyte | *CRYZ* | -0.60711091 | 0.00311239 | 0.006284318 | 1 |
| T2DM | leukocyte | *TMEM9* | -0.89793189 | 0.000892108 | 0.004990048 | 1 |
| T2DM | leukocyte | *WARS2* | -0.860916476 | 0.003770231 | 0.00692855 | 1 |
| T2DM | leukocyte | *C1orf109* | -0.651447594 | 0.004467511 | 0.007498014 | 1 |
| T2DM | leukocyte | *LGALS8* | -1.186041806 | 0.000653266 | 0.004990048 | 1 |
| T2DM | leukocyte | *HPCAL4* | -2.227629641 | 0.004563926 | 0.007578894 | 1 |
| T2DM | leukocyte | *BMP8B* | -0.769707258 | 0.003698137 | 0.006868344 | 1 |
| T2DM | leukocyte | *MYCL* | 0.415569513 | 0.011111624 | 0.015698515 | 1 |
| T2DM | leukocyte | *UAP1* | -0.639057774 | 0.002361332 | 0.005542217 | 1 |
| T2DM | leukocyte | *CTBS* | -1.145170821 | 0.000512605 | 0.004990048 | 1 |
| T2DM | leukocyte | *ZNHIT6* | -0.614051995 | 0.004577715 | 0.007595372 | 1 |
| T2DM | leukocyte | *CD46* | -0.712731187 | 0.001380579 | 0.004990048 | 1 |
| T2DM | leukocyte | *SLC2A1* | -1.200655775 | 0.002600525 | 0.005751212 | 1 |
| T2DM | leukocyte | *PRDX1* | -0.773853691 | 0.001045663 | 0.004990048 | 1 |
| T2DM | leukocyte | *CCDC181* | -0.726387925 | 0.001520184 | 0.004993935 | 1 |
| T2DM | leukocyte | *NSUN4* | -3.429375454 | 0.000103893 | 0.004990048 | 0.408197123 |
| T2DM | leukocyte | *TMED5* | -1.279690131 | 0.000625652 | 0.004990048 | 1 |
| T2DM | leukocyte | *DR1* | -1.158686626 | 0.001656334 | 0.005041098 | 1 |
| T2DM | leukocyte | *PTBP2* | -0.874613972 | 0.00366228 | 0.006825948 | 1 |
| T2DM | leukocyte | *NSL1* | -1.428785581 | 0.000909259 | 0.004990048 | 1 |
| T2DM | leukocyte | *RPA2* | -1.076506942 | 0.003272188 | 0.006415383 | 1 |
| T2DM | leukocyte | *ESYT2* | -1.026722293 | 0.001093365 | 0.004990048 | 1 |
| T2DM | leukocyte | *DDX59* | -0.7226067 | 0.004072711 | 0.007189 | 1 |
| T2DM | leukocyte | *TTR* | -0.34692105 | 0.001755445 | 0.005084476 | 1 |
| T2DM | leukocyte | *SPCS2* | -0.833878785 | 0.000902731 | 0.004990048 | 1 |
| T2DM | leukocyte | *ELOVL4* | -1.261566084 | 0.002176114 | 0.005349178 | 1 |
| T2DM | leukocyte | *FILIP1* | -0.847776018 | 0.007904596 | 0.011693207 | 1 |
| T2DM | leukocyte | *AKAP7* | -1.919868958 | 0.000747918 | 0.004990048 | 1 |
| T2DM | leukocyte | *RNF146* | -0.984718562 | 0.001899295 | 0.005146173 | 1 |
| T2DM | leukocyte | *MED28* | -1.412864746 | 0.000628253 | 0.004990048 | 1 |
| T2DM | leukocyte | *VAMP8* | -0.945889821 | 0.00107764 | 0.004990048 | 1 |
| T2DM | leukocyte | *ARMC2* | -0.82899864 | 0.001659286 | 0.005041098 | 1 |
| T2DM | leukocyte | *PKD2* | -0.710036634 | 0.002691383 | 0.005864439 | 1 |
| T2DM | leukocyte | *SPP1* | -0.441876644 | 0.000816109 | 0.004990048 | 1 |
| T2DM | leukocyte | *WDR35* | -0.759989532 | 0.004266699 | 0.007310294 | 1 |
| T2DM | leukocyte | *DNAH7* | -1.262699733 | 1.97E-05 | 0.0030497 | 0.077349156 |
| T2DM | leukocyte | *NDUFB3* | -0.818044079 | 0.001748308 | 0.005084476 | 1 |

| | | | | | | |
|---|---|---|---|---|---|---|
| T2DM | leukocyte | *GTF3C3* | -1.176224123 | 0.005372161 | 0.008521283 | 1 |
| T2DM | leukocyte | *CPSF3* | -1.247910774 | 0.00276556 | 0.005956822 | 1 |
| T2DM | leukocyte | *CCDC92* | -0.966432871 | 0.001218572 | 0.004990048 | 1 |
| T2DM | leukocyte | *CTNNAL1* | -0.583932805 | 0.012574682 | 0.017495017 | 1 |
| T2DM | leukocyte | *NEK6* | -0.797455508 | 0.020142633 | 0.026486122 | 1 |
| T2DM | leukocyte | *BSPRY* | -0.789235901 | 0.001485965 | 0.004990048 | 1 |
| T2DM | leukocyte | *RBM18* | -1.152019839 | 0.001442124 | 0.004990048 | 1 |
| T2DM | leukocyte | *INVS* | -0.574699626 | 0.037406129 | 0.046701202 | 1 |
| T2DM | leukocyte | *DCAF4* | -0.924598134 | 0.011268375 | 0.015873701 | 1 |
| T2DM | leukocyte | *IFI27L2* | -0.79136496 | 0.001291634 | 0.004990048 | 1 |
| T2DM | leukocyte | *ACOT2* | -1.033635609 | 0.02503435 | 0.032217478 | 1 |
| T2DM | leukocyte | *ZC2HC1C* | -2.70632553 | 0.002882742 | 0.006056841 | 1 |
| T2DM | leukocyte | *KLHL29* | -2.421204571 | 0.017081397 | 0.022921042 | 1 |
| T2DM | leukocyte | *AFTPH* | -0.865661324 | 0.00215522 | 0.005335765 | 1 |
| T2DM | leukocyte | *BCL11A* | -1.264367904 | 0.008947625 | 0.012962839 | 1 |
| T2DM | leukocyte | *CRIPT* | -1.089102983 | 0.000961858 | 0.004990048 | 1 |
| T2DM | leukocyte | *EPCAM* | -0.56357884 | 0.001551914 | 0.005009694 | 1 |
| T2DM | leukocyte | *SLC17A5* | -3.034610195 | 0.000420457 | 0.004990048 | 1 |
| T2DM | leukocyte | *CUTC* | -0.936213694 | 0.00642164 | 0.009848019 | 1 |
| T2DM | leukocyte | *PYROXD2* | -1.289813449 | 0.005175203 | 0.00827906 | 1 |
| T2DM | leukocyte | *HELLS* | -1.04033651 | 0.002429243 | 0.005602682 | 1 |
| T2DM | leukocyte | *PRLHR* | -1.156808491 | 0.004311172 | 0.007328649 | 1 |
| T2DM | leukocyte | *WDR11* | -1.556305086 | 0.000500258 | 0.004990048 | 1 |
| T2DM | leukocyte | *KANSL1* | -0.990045178 | 0.000854623 | 0.004990048 | 1 |
| T2DM | leukocyte | *HOXB3* | -1.033989148 | 0.010478804 | 0.014891758 | 1 |
| T2DM | leukocyte | *RCL1* | -1.637957472 | 0.000721894 | 0.004990048 | 1 |
| T2DM | leukocyte | *CD274* | -2.697980417 | 0.000919482 | 0.004990048 | 1 |
| T2DM | leukocyte | *PLEKHG1* | -0.80073283 | 0.006144448 | 0.009482144 | 1 |
| T2DM | leukocyte | *PCDHB10* | -1.53093968 | 0.00326102 | 0.006405338 | 1 |
| T2DM | leukocyte | *MRPS14* | -1.450548144 | 0.001905672 | 0.00515389 | 1 |
| T2DM | leukocyte | *GORAB* | -0.847963938 | 0.004009756 | 0.00714972 | 1 |
| T2DM | leukocyte | *SNX19* | -0.830698915 | 0.001538711 | 0.005006447 | 1 |
| T2DM | leukocyte | *KCNJ5* | 0.296473505 | 0.004227063 | 0.007272975 | 1 |
| T2DM | leukocyte | *SLC10A7* | -2.042170425 | 0.000136422 | 0.004990048 | 0.536001278 |
| T2DM | leukocyte | *MASTL* | -0.993572169 | 0.000557119 | 0.004990048 | 1 |
| T2DM | leukocyte | *MRC1* | 0.208506418 | 0.033924699 | 0.042693832 | 1 |
| T2DM | leukocyte | *IQSEC3* | -0.942332055 | 0.002529955 | 0.005660135 | 1 |
| T2DM | leukocyte | *DNAJC15* | -0.977503071 | 0.000468307 | 0.004990048 | 1 |
| T2DM | leukocyte | *PROSER1* | -1.208701955 | 0.001168108 | 0.004990048 | 1 |
| T2DM | leukocyte | *WBP4* | -0.893978193 | 0.001464603 | 0.004990048 | 1 |
| T2DM | leukocyte | *KBTBD7* | -2.004375322 | 0.000386433 | 0.004990048 | 1 |
| T2DM | leukocyte | *FAM53C* | -2.154167297 | 9.49E-05 | 0.004990048 | 0.373042938 |
| T2DM | leukocyte | *SIL1* | -0.940659034 | 0.000785516 | 0.004990048 | 1 |
| T2DM | leukocyte | *PLS1* | -0.691652117 | 0.004132712 | 0.007236049 | 1 |
| T2DM | leukocyte | *ZFP30* | -1.058651863 | 0.006202516 | 0.009552993 | 1 |
| T2DM | leukocyte | *UTP20* | -1.486452565 | 0.011582551 | 0.016276053 | 1 |
| T2DM | leukocyte | *TMPO* | -1.332560289 | 0.001221936 | 0.004990048 | 1 |
| T2DM | leukocyte | *PPP3CC* | -1.086139131 | 0.001776792 | 0.005098084 | 1 |
| T2DM | leukocyte | *EPHX2* | -1.06737385 | 0.001451368 | 0.004990048 | 1 |
| T2DM | leukocyte | *CRISPLD1* | -5.100608949 | 0.001910196 | 0.00515389 | 1 |

| | | | | | | |
|---|---|---|---|---|---|---|
| T2DM | leukocyte | *PYROXD1* | -0.777352521 | 0.00099709 | 0.004990048 | 1 |
| T2DM | leukocyte | *IAPP* | -0.265826773 | 0.009304768 | 0.0134061 | 1 |
| T2DM | leukocyte | *ZSCAN18* | -0.758922152 | 0.002023478 | 0.005244224 | 1 |
| T2DM | leukocyte | *LHX4* | -2.408571281 | 0.005360115 | 0.008509048 | 1 |
| T2DM | leukocyte | *TRMT1L* | -0.887683123 | 0.003423179 | 0.006544851 | 1 |
| T2DM | leukocyte | *CAT* | -2.190583055 | 0.000195861 | 0.004990048 | 0.769536992 |
| T2DM | leukocyte | *CSMD2* | 0.934624549 | 0.02602211 | 0.033324925 | 1 |
| T2DM | leukocyte | *ACVR2A* | -0.650387936 | 0.015156188 | 0.020576594 | 1 |
| T2DM | leukocyte | *POLK* | -1.442726322 | 0.000163167 | 0.004990048 | 0.641082067 |
| T2DM | leukocyte | *UBL3* | -0.788960724 | 0.001291827 | 0.004990048 | 1 |
| T2DM | leukocyte | *FMOD* | -1.129843569 | 0.008031848 | 0.011844895 | 1 |
| T2DM | leukocyte | *LDB3* | -1.306488411 | 0.005289143 | 0.008423609 | 1 |
| T2DM | leukocyte | *NAA60* | -1.209107528 | 0.000820878 | 0.004990048 | 1 |
| T2DM | leukocyte | *TRMT13* | -2.122958744 | 0.000767605 | 0.004990048 | 1 |
| T2DM | leukocyte | *RWDD3* | -0.899510598 | 0.001589447 | 0.005020048 | 1 |
| T2DM | leukocyte | *CCDC18* | -0.846493156 | 0.005048523 | 0.008126033 | 1 |
| T2DM | leukocyte | *NBPF14* | -2.24890083 | 0.00541875 | 0.008574413 | 1 |
| T2DM | leukocyte | *PMS2* | -1.686696829 | 0.003439983 | 0.006566174 | 1 |
| T2DM | leukocyte | *EEPD1* | 0.400009786 | 0.033846163 | 0.0426223 | 1 |
| T2DM | leukocyte | *KLHL7* | -1.264267097 | 0.001540418 | 0.005006447 | 1 |
| T2DM | leukocyte | *CBX3* | -0.978039582 | 0.001001635 | 0.004990048 | 1 |
| T2DM | leukocyte | *NPY* | -0.414439469 | 0.002531198 | 0.005660135 | 1 |
| T2DM | leukocyte | *INHBA* | -0.797520836 | 0.001872302 | 0.005146173 | 1 |
| T2DM | leukocyte | *CCZ1* | -1.265238774 | 0.002026753 | 0.00524925 | 1 |
| T2DM | leukocyte | *POLM* | -0.948792279 | 0.000857022 | 0.004990048 | 1 |
| T2DM | leukocyte | *SMU1* | -1.048839558 | 0.00089029 | 0.004990048 | 1 |
| T2DM | leukocyte | *SLC25A51* | -0.873494985 | 0.014366028 | 0.019667292 | 1 |
| T2DM | leukocyte | *ACO1* | -0.805991482 | 0.003627857 | 0.006781089 | 1 |
| T2DM | leukocyte | *RBM19* | -0.743449585 | 0.004365155 | 0.007376642 | 1 |
| T2DM | leukocyte | *BHLHE41* | -1.483682604 | 0.002338147 | 0.005515664 | 1 |
| T2DM | leukocyte | *ITPR2* | -2.146251146 | 0.004201666 | 0.007256417 | 1 |
| T2DM | leukocyte | *EBPL* | -0.748593909 | 0.002941595 | 0.006124816 | 1 |
| T2DM | leukocyte | *ATP7B* | -1.345465746 | 0.008261696 | 0.012134656 | 1 |
| T2DM | leukocyte | *TSFM* | -1.241350625 | 0.000911168 | 0.004990048 | 1 |
| T2DM | leukocyte | *CDK2* | -1.295681194 | 0.02558443 | 0.032828617 | 1 |
| T2DM | leukocyte | *IKZF4* | -1.1953966 | 0.00510499 | 0.00819008 | 1 |
| T2DM | leukocyte | *USP45* | -0.679118147 | 0.006080823 | 0.009402422 | 1 |
| T2DM | leukocyte | *C4BPB* | -1.966620721 | 0.01450015 | 0.019816031 | 1 |
| T2DM | leukocyte | *RAB38* | -2.033486542 | 0.021712615 | 0.028370092 | 1 |
| T2DM | leukocyte | *CKS2* | -0.783117235 | 0.004101137 | 0.00720956 | 1 |
| T2DM | leukocyte | *SNX21* | -0.841656004 | 0.007343511 | 0.011008262 | 1 |
| T2DM | leukocyte | *ARHGAP40* | -45.22893927 | 0.002976973 | 0.006152298 | 1 |
| T2DM | leukocyte | *NCOA3* | -0.892280669 | 0.003335795 | 0.00646588 | 1 |
| T2DM | leukocyte | *MATN4* | -6.229855173 | 0.002363236 | 0.005543376 | 1 |
| T2DM | leukocyte | *VAPB* | -0.856475562 | 0.000699098 | 0.004990048 | 1 |
| T2DM | leukocyte | *PARD6B* | -1.14516177 | 0.001684261 | 0.005060368 | 1 |
| T2DM | leukocyte | *ARFGEF2* | -1.146598651 | 0.00114194 | 0.004990048 | 1 |
| T2DM | leukocyte | *ZNFX1* | -3.222537919 | 0.000357426 | 0.004990048 | 1 |
| T2DM | leukocyte | *CSE1L* | -1.110427386 | 0.000981576 | 0.004990048 | 1 |
| T2DM | leukocyte | *BCAS4* | -0.617023099 | 0.004536403 | 0.007568852 | 1 |

| | | | | | | |
|---|---|---|---|---|---|---|
| T2DM | leukocyte | *KCNK15* | -0.60815035 | 0.035713862 | 0.044744823 | 1 |
| T2DM | leukocyte | *PEPD* | -1.621867208 | 0.000150642 | 0.004990048 | 0.591873793 |
| T2DM | leukocyte | *SNRNP27* | -0.86825024 | 0.001870566 | 0.005146173 | 1 |
| T2DM | leukocyte | *USP22* | -1.071050255 | 0.001331166 | 0.004990048 | 1 |
| T2DM | leukocyte | *BTN2A2* | -1.221388101 | 0.018969815 | 0.025084163 | 1 |
| T2DM | leukocyte | *MRS2* | -1.539724453 | 0.00082295 | 0.004990048 | 1 |
| T2DM | leukocyte | *SNRPC* | -0.99940405 | 0.000758611 | 0.004990048 | 1 |
| T2DM | leukocyte | *SERPINB6* | -0.860944243 | 0.000946095 | 0.004990048 | 1 |
| T2DM | leukocyte | *ABCC10* | -1.055061271 | 0.004523861 | 0.007557079 | 1 |
| T2DM | leukocyte | *PEX6* | -0.976882427 | 0.000938151 | 0.004990048 | 1 |
| T2DM | leukocyte | *NQO2* | -2.432109595 | 0.000132983 | 0.004990048 | 0.522489091 |
| T2DM | leukocyte | *UNC5CL* | -0.824756544 | 0.002667187 | 0.005838092 | 1 |
| T2DM | leukocyte | *ZNF391* | -0.615314774 | 0.027308493 | 0.0347796 | 1 |
| T2DM | leukocyte | *MED20* | -1.336386682 | 0.003003854 | 0.006172668 | 1 |
| T2DM | leukocyte | *GLO1* | -0.952531712 | 0.00142711 | 0.004990048 | 1 |
| T2DM | leukocyte | *KCNK17* | -0.579502191 | 0.012688018 | 0.017627731 | 1 |
| T2DM | leukocyte | *ATXN1* | -1.250136209 | 0.000800501 | 0.004990048 | 1 |
| T2DM | leukocyte | *LRRFIP1* | -0.746028632 | 0.001258937 | 0.004990048 | 1 |
| T2DM | leukocyte | *CXCL6* | -1.119661013 | 0.024407497 | 0.031503632 | 1 |
| T2DM | leukocyte | *SH3TC1* | -0.965412437 | 0.002316814 | 0.005496836 | 1 |
| T2DM | leukocyte | *MT1G* | -0.59465237 | 0.012158125 | 0.016987651 | 1 |
| T2DM | leukocyte | *DOK4* | -0.584527085 | 0.00687167 | 0.010409317 | 1 |
| T2DM | leukocyte | *CLYBL* | -2.266341215 | 0.001098395 | 0.004990048 | 1 |
| T2DM | leukocyte | *TM9SF2* | -0.799730691 | 0.001605125 | 0.005033031 | 1 |
| T2DM | leukocyte | *KIF25* | -12.68423923 | 0.001672962 | 0.005048438 | 1 |
| T2DM | leukocyte | *BMP4* | -2.226521724 | 0.000424868 | 0.004990048 | 1 |
| T2DM | leukocyte | *HS3ST3B1* | -1.963708204 | 0.00426155 | 0.007310294 | 1 |
| T2DM | leukocyte | *MRPS7* | -0.745186356 | 0.001955363 | 0.005198138 | 1 |
| T2DM | leukocyte | *GGA3* | -2.148854203 | 0.000899017 | 0.004990048 | 1 |
| T2DM | leukocyte | *DDX31* | -1.189914371 | 0.020674288 | 0.027094489 | 1 |
| T2DM | leukocyte | *PPDPF* | -0.691774601 | 0.001133692 | 0.004990048 | 1 |
| T2DM | leukocyte | *CHCHD5* | -0.818519396 | 0.001450118 | 0.004990048 | 1 |
| T2DM | leukocyte | *INSIG2* | -0.777463422 | 0.001730301 | 0.005070552 | 1 |
| T2DM | leukocyte | *POLR1B* | -0.693133205 | 0.039316172 | 0.048883937 | 1 |
| T2DM | leukocyte | *PSD4* | -1.007041247 | 0.012582047 | 0.017499066 | 1 |
| T2DM | leukocyte | *CLPP* | -0.900413638 | 0.001473385 | 0.004990048 | 1 |
| T2DM | leukocyte | *GPR108* | -1.217037268 | 0.002516231 | 0.005655212 | 1 |
| T2DM | leukocyte | *FOSB* | -1.448504369 | 0.001198396 | 0.004990048 | 1 |
| T2DM | leukocyte | *SYMPK* | -1.888559673 | 0.000633651 | 0.004990048 | 1 |
| T2DM | leukocyte | *NAPB* | -0.820153322 | 0.002527902 | 0.005660135 | 1 |
| T2DM | leukocyte | *PSMF1* | -1.081737118 | 0.000645153 | 0.004990048 | 1 |
| T2DM | leukocyte | *RBCK1* | -0.905505922 | 0.000854918 | 0.004990048 | 1 |
| T2DM | leukocyte | *FLRT3* | -0.734050793 | 0.003710576 | 0.006873574 | 1 |
| T2DM | leukocyte | *MGME1* | -3.710314628 | 0.000975892 | 0.004990048 | 1 |
| T2DM | leukocyte | *ITPA* | -1.254728547 | 0.000913313 | 0.004990048 | 1 |
| T2DM | leukocyte | *MCM8* | -0.898902588 | 0.012917338 | 0.017908335 | 1 |
| T2DM | leukocyte | *BANF2* | -11.65446282 | 0.004181065 | 0.007242343 | 1 |
| T2DM | leukocyte | *MRPS26* | -0.727724644 | 0.002869773 | 0.006055188 | 1 |
| T2DM | leukocyte | *ZNF436* | -2.250096463 | 0.000826037 | 0.004990048 | 1 |
| T2DM | leukocyte | *TUBGCP3* | -1.094031514 | 0.001881303 | 0.005146173 | 1 |

| | | | | | | |
|---|---|---|---|---|---|---|
| T2DM | leukocyte | *MCF2L* | -0.578315523 | 0.009292076 | 0.013402558 | 1 |
| T2DM | leukocyte | *IGFLR1* | 0.262185413 | 0.02311258 | 0.029979969 | 1 |
| T2DM | leukocyte | *NR1D1* | -0.782971473 | 0.001645607 | 0.005035955 | 1 |
| T2DM | leukocyte | *PRMT1* | -0.848327863 | 0.00031152 | 0.004990048 | 1 |
| T2DM | leukocyte | *PRR12* | -0.716975215 | 0.019627313 | 0.025877756 | 1 |
| T2DM | leukocyte | *FLRT1* | -1.980850408 | 0.014905758 | 0.020278644 | 1 |
| T2DM | leukocyte | *ASL* | -0.965246185 | 0.001762549 | 0.005084476 | 1 |
| T2DM | leukocyte | *WNK4* | -0.875712044 | 0.027139467 | 0.034597977 | 1 |
| T2DM | leukocyte | *BECN1* | -0.935306197 | 0.002268478 | 0.005457772 | 1 |
| T2DM | leukocyte | *TRAP1* | -0.681526921 | 0.005947416 | 0.009239777 | 1 |
| T2DM | leukocyte | *ZNF384* | -1.135812384 | 0.001870865 | 0.005146173 | 1 |
| T2DM | leukocyte | *RHOJ* | -1.473004702 | 0.003295613 | 0.006432819 | 1 |
| T2DM | leukocyte | *L3HYPDH* | -1.053127367 | 0.004667674 | 0.007686961 | 1 |
| T2DM | leukocyte | *HSPA2* | -1.537663813 | 0.001514806 | 0.004993935 | 1 |
| T2DM | leukocyte | *ZBTB1* | -1.304525483 | 0.000682057 | 0.004990048 | 1 |
| T2DM | leukocyte | *PLEKHG3* | -1.275057014 | 0.002052331 | 0.00526208 | 1 |
| T2DM | leukocyte | *FGD3* | 2.075541962 | 0.036040564 | 0.045139743 | 1 |
| T2DM | leukocyte | *BCL11B* | 1.330647702 | 0.023227893 | 0.030105511 | 1 |
| T2DM | leukocyte | *ABHD8* | -1.658620976 | 0.01209469 | 0.016905029 | 1 |
| T2DM | leukocyte | *HELB* | -3.906509739 | 0.033995775 | 0.042769367 | 1 |
| T2DM | leukocyte | *BEST3* | -2.291057948 | 0.002977793 | 0.006152298 | 1 |
| T2DM | leukocyte | *YEATS4* | -0.835943767 | 0.002144765 | 0.005323324 | 1 |
| T2DM | leukocyte | *LRRC61* | -1.32482192 | 0.002354548 | 0.005532907 | 1 |
| T2DM | leukocyte | *IDUA* | -0.874452106 | 0.003350186 | 0.006474609 | 1 |
| T2DM | leukocyte | *FGFRL1* | -0.973047533 | 0.000717867 | 0.004990048 | 1 |
| T2DM | leukocyte | *EMC1* | -0.991759325 | 0.003158747 | 0.006314657 | 1 |
| T2DM | leukocyte | *UBR4* | -1.012972077 | 0.001142279 | 0.004990048 | 1 |
| T2DM | leukocyte | *SYNGR3* | -0.962305975 | 0.007898016 | 0.011692277 | 1 |
| T2DM | leukocyte | *WFIKKN1* | -0.709916094 | 0.025982465 | 0.033285003 | 1 |
| T2DM | leukocyte | *KDM4B* | -2.319017504 | 0.000291106 | 0.004990048 | 1 |
| T2DM | leukocyte | *PNKD* | -1.223828948 | 0.000602943 | 0.004990048 | 1 |
| T2DM | leukocyte | *PTPN12* | -1.223507971 | 2.68E-05 | 0.003310187 | 0.105377589 |
| T2DM | leukocyte | *POR* | -0.984161892 | 0.001454273 | 0.004990048 | 1 |
| T2DM | leukocyte | *RBM48* | -1.771055964 | 0.000777522 | 0.004990048 | 1 |
| T2DM | leukocyte | *SRD5A3* | -1.14948588 | 0.002185945 | 0.005356498 | 1 |
| T2DM | leukocyte | *PPAT* | -1.170179192 | 0.000567724 | 0.004990048 | 1 |
| T2DM | leukocyte | *A4GALT* | -0.70650296 | 0.008661015 | 0.0126174 | 1 |
| T2DM | leukocyte | *RIBC2* | 2.659439609 | 0.034727086 | 0.043598559 | 1 |
| T2DM | leukocyte | *NAA38* | -0.752993581 | 0.001452697 | 0.004990048 | 1 |
| T2DM | leukocyte | *PRKRIP1* | -0.871733031 | 0.002000993 | 0.005227473 | 1 |
| T2DM | leukocyte | *VGF* | -0.465996176 | 0.001938867 | 0.00518571 | 1 |
| T2DM | leukocyte | *IRF5* | 0.335550847 | 0.01734104 | 0.023213951 | 1 |
| T2DM | leukocyte | *MTX2* | -1.343321612 | 0.000755784 | 0.004990048 | 1 |
| T2DM | leukocyte | *GAD1* | -3.428085776 | 0.001171242 | 0.004990048 | 1 |
| T2DM | leukocyte | *ORMDL1* | -0.868970527 | 0.001700593 | 0.005060368 | 1 |
| T2DM | leukocyte | *PSMG2* | -1.024881086 | 0.001389751 | 0.004990048 | 1 |
| T2DM | leukocyte | *TWSG1* | -1.430569749 | 0.000741055 | 0.004990048 | 1 |
| T2DM | leukocyte | *EIF2AK4* | -1.573692332 | 9.76E-05 | 0.004990048 | 0.383612134 |
| T2DM | leukocyte | *MYO5C* | -0.628550098 | 0.002821055 | 0.006006579 | 1 |
| T2DM | leukocyte | *CGNL1* | -1.095275434 | 0.000810496 | 0.004990048 | 1 |

| | | | | | | |
|---|---|---|---|---|---|---|
| T2DM | leukocyte | *TMOD2* | -1.015493414 | 0.0018238 | 0.00513989 | 1 |
| T2DM | leukocyte | *ELL3* | -2.81601822 | 0.001264379 | 0.004990048 | 1 |
| T2DM | leukocyte | *IVD* | -1.046808278 | 0.010716074 | 0.015172416 | 1 |
| T2DM | leukocyte | *CHAC1* | -1.113805837 | 0.001220085 | 0.004990048 | 1 |
| T2DM | leukocyte | *ACKR4* | -1.307945957 | 0.030804333 | 0.039016835 | 1 |
| T2DM | leukocyte | *ANAPC13* | -0.547320981 | 0.006772187 | 0.010281269 | 1 |
| T2DM | leukocyte | *SUMF2* | -1.069810028 | 0.001039941 | 0.004990048 | 1 |
| T2DM | leukocyte | *TPH1* | -2.343811566 | 0.025230044 | 0.032437448 | 1 |
| T2DM | leukocyte | *DCTD* | -1.101993809 | 0.001340066 | 0.004990048 | 1 |
| T2DM | leukocyte | *AP1M2* | -0.824401575 | 0.001671321 | 0.005048289 | 1 |
| T2DM | leukocyte | *CDKN2D* | -0.89028868 | 0.000891847 | 0.004990048 | 1 |
| T2DM | leukocyte | *MTUS1* | -0.716235428 | 0.003326555 | 0.006456709 | 1 |
| T2DM | leukocyte | *ADCY4* | -0.904755721 | 0.005737628 | 0.008970609 | 1 |
| T2DM | leukocyte | *DTD2* | -0.751874345 | 0.003968918 | 0.007124893 | 1 |
| T2DM | leukocyte | *HEATR5A* | -1.966354495 | 0.00053408 | 0.004990048 | 1 |
| T2DM | leukocyte | *MIS18BP1* | -1.553994303 | 0.006781737 | 0.010291791 | 1 |
| T2DM | leukocyte | *RNASE1* | 0.129733825 | 0.024768804 | 0.031917557 | 1 |
| T2DM | leukocyte | *EPB41L4A* | -0.983329166 | 0.004108547 | 0.007219356 | 1 |
| T2DM | leukocyte | *REEP5* | -0.765738578 | 0.001259869 | 0.004990048 | 1 |
| T2DM | leukocyte | *MAU2* | -3.027396751 | 5.44E-07 | 0.000266998 | 0.002135982 |
| T2DM | leukocyte | *SHC2* | -1.031123466 | 0.001341645 | 0.004990048 | 1 |
| T2DM | leukocyte | *CBFA2T3* | -0.838963967 | 0.003993181 | 0.007143456 | 1 |
| T2DM | leukocyte | *ERMARD* | -1.846198983 | 0.000943306 | 0.004990048 | 1 |
| T2DM | leukocyte | *PHF10* | -0.936832173 | 0.001462558 | 0.004990048 | 1 |
| T2DM | leukocyte | *SH3BP4* | -0.449167085 | 0.014696635 | 0.020042722 | 1 |
| T2DM | leukocyte | *DOCK6* | -0.653380058 | 0.004929245 | 0.007979813 | 1 |
| T2DM | leukocyte | *ECSIT* | -1.042456943 | 0.002097211 | 0.005283012 | 1 |
| T2DM | leukocyte | *CDC16* | -1.392999077 | 0.000312624 | 0.004990048 | 1 |
| T2DM | leukocyte | *DPP6* | -0.467088476 | 0.003755807 | 0.00692001 | 1 |
| T2DM | leukocyte | *GTPBP3* | -1.115431243 | 0.000252745 | 0.004990048 | 0.993033826 |
| T2DM | leukocyte | *PLVAP* | -0.428051888 | 0.005031832 | 0.008102486 | 1 |
| T2DM | leukocyte | *MRPL34* | -0.772557423 | 0.001144409 | 0.004990048 | 1 |
| T2DM | leukocyte | *LSM7* | -0.584959825 | 0.005750862 | 0.008987724 | 1 |
| T2DM | leukocyte | *TULP4* | -0.842454982 | 0.004533875 | 0.007568852 | 1 |
| T2DM | leukocyte | *QRSL1* | -0.977074485 | 0.00291991 | 0.006102301 | 1 |
| T2DM | leukocyte | *RSPH3* | -1.016353108 | 0.002571103 | 0.005704045 | 1 |
| T2DM | leukocyte | *STK33* | -0.790004127 | 0.001345613 | 0.004990048 | 1 |
| T2DM | leukocyte | *NDUFA10* | -0.887465792 | 0.001535089 | 0.005006447 | 1 |
| T2DM | leukocyte | *TRPM4* | -3.386663174 | 0.00647968 | 0.009921536 | 1 |
| T2DM | leukocyte | *UBAC1* | -0.922543249 | 0.001081721 | 0.004990048 | 1 |
| T2DM | leukocyte | *ATXN10* | -0.769132726 | 0.001497407 | 0.004993935 | 1 |
| T2DM | leukocyte | *PNPLA7* | -1.747112239 | 0.002673981 | 0.005846451 | 1 |
| T2DM | leukocyte | *PAK4* | -1.157499827 | 0.001419162 | 0.004990048 | 1 |
| T2DM | leukocyte | *LAMA5* | -0.601543333 | 0.003630087 | 0.006782032 | 1 |
| T2DM | leukocyte | *ADRM1* | -0.881125053 | 0.001122456 | 0.004990048 | 1 |
| T2DM | leukocyte | *UCK1* | -1.747940376 | 0.000334786 | 0.004990048 | 1 |
| T2DM | leukocyte | *MAP3K10* | -2.188931332 | 0.000699751 | 0.004990048 | 1 |
| T2DM | leukocyte | *SMPDL3B* | -0.725029338 | 0.00490524 | 0.007970068 | 1 |
| T2DM | leukocyte | *CLIP1* | -1.015893425 | 0.002244165 | 0.005438799 | 1 |
| T2DM | leukocyte | *ZNF317* | -1.173737904 | 0.002723893 | 0.005904436 | 1 |

| | | | | | | |
|---|---|---|---|---|---|---|
| T2DM | leukocyte | *DNMT1* | -1.510384722 | 0.000972553 | 0.004990048 | 1 |
| T2DM | leukocyte | *ZNF426* | -1.373758625 | 0.000643667 | 0.004990048 | 1 |
| T2DM | leukocyte | *LRP3* | -1.339976727 | 0.0037313 | 0.006898954 | 1 |
| T2DM | leukocyte | *HSD17B3* | -11.99808734 | 0.00110997 | 0.004990048 | 1 |
| T2DM | leukocyte | *SYNE1* | -0.804256832 | 0.001728099 | 0.005070552 | 1 |
| T2DM | leukocyte | *EPS8L1* | -2.782216789 | 0.002822399 | 0.006006579 | 1 |
| T2DM | leukocyte | *C1QL1* | -0.378338922 | 0.00579348 | 0.009041044 | 1 |
| T2DM | leukocyte | *F12* | -0.916714042 | 0.010590793 | 0.015022103 | 1 |
| T2DM | leukocyte | *TRAF3* | -0.628902095 | 0.011944409 | 0.016712814 | 1 |
| T2DM | leukocyte | *PDLIM4* | -1.027610012 | 0.001409141 | 0.004990048 | 1 |
| T2DM | leukocyte | *MGAT1* | -2.059578283 | 0.007920298 | 0.011707619 | 1 |
| T2DM | leukocyte | *DIAPH1* | -0.680986298 | 0.002691605 | 0.005864439 | 1 |
| T2DM | leukocyte | *EXOC4* | -1.791501352 | 0.001907032 | 0.00515389 | 1 |
| T2DM | leukocyte | *C1orf159* | -3.056494605 | 0.005889697 | 0.009160973 | 1 |
| T2DM | leukocyte | *ANO1* | -2.18536571 | 0.002384977 | 0.005560842 | 1 |
| T2DM | leukocyte | *PPFIA1* | -0.799900571 | 0.002166685 | 0.005345956 | 1 |
| T2DM | leukocyte | *TRAF7* | -1.208890208 | 0.001475579 | 0.004990048 | 1 |
| T2DM | leukocyte | *MAP1B* | -0.512050286 | 0.006557046 | 0.010024372 | 1 |
| T2DM | leukocyte | *CHD1L* | -2.11815861 | 0.001511325 | 0.004993935 | 1 |
| T2DM | leukocyte | *PIAS3* | -0.673943954 | 0.004649839 | 0.007681478 | 1 |
| T2DM | leukocyte | *RBM8A* | -0.71410442 | 0.000407956 | 0.004990048 | 1 |
| T2DM | leukocyte | *ZNF304* | -4.227554986 | 0.000358301 | 0.004990048 | 1 |
| T2DM | leukocyte | *ZNF132* | 1.013038418 | 0.021638323 | 0.028282425 | 1 |
| T2DM | leukocyte | *RHPN2* | -0.825114866 | 0.003997105 | 0.007143456 | 1 |
| T2DM | leukocyte | *GCH1* | -0.623415801 | 0.003372014 | 0.006497619 | 1 |
| T2DM | leukocyte | *CC2D1A* | -1.321419673 | 0.000946291 | 0.004990048 | 1 |
| T2DM | leukocyte | *CCT6B* | -1.459495126 | 0.001505307 | 0.004993935 | 1 |
| T2DM | leukocyte | *PPARG* | -1.672888635 | 0.000504141 | 0.004990048 | 1 |
| T2DM | leukocyte | *ENOSF1* | -2.449250356 | 0.000351962 | 0.004990048 | 1 |
| T2DM | leukocyte | *IMMT* | -0.984839075 | 0.00146531 | 0.004990048 | 1 |
| T2DM | leukocyte | *MRPL35* | -0.906918892 | 0.004508757 | 0.007541467 | 1 |
| T2DM | leukocyte | *CARD6* | -1.765061217 | 0.002251852 | 0.005441283 | 1 |
| T2DM | leukocyte | *INPP5K* | -0.814435384 | 0.013273188 | 0.018344791 | 1 |
| T2DM | leukocyte | *RPA1* | -1.050092607 | 0.003695203 | 0.006868344 | 1 |
| T2DM | leukocyte | *TMEM128* | -1.428920906 | 0.000852356 | 0.004990048 | 1 |
| T2DM | leukocyte | *SEC61G* | -0.711411651 | 0.000984117 | 0.004990048 | 1 |
| T2DM | leukocyte | *FIGNL1* | -2.774323139 | 0.003177194 | 0.006330221 | 1 |
| T2DM | leukocyte | *TRIM47* | -0.71189886 | 0.014542375 | 0.01985302 | 1 |
| T2DM | leukocyte | *EIF5A* | -0.789092685 | 0.001094465 | 0.004990048 | 1 |
| T2DM | leukocyte | *PRMT7* | -1.589310554 | 0.019920748 | 0.026220642 | 1 |
| T2DM | leukocyte | *ANKEF1* | -0.743934858 | 0.003973184 | 0.007124893 | 1 |
| T2DM | leukocyte | *BTBD3* | -0.759724872 | 0.001719721 | 0.005070552 | 1 |
| T2DM | leukocyte | *IGHMBP2* | -1.209758547 | 0.000863944 | 0.004990048 | 1 |
| T2DM | leukocyte | *LPIN3* | -0.998878311 | 0.029834786 | 0.03786204 | 1 |
| T2DM | leukocyte | *RBM38* | -1.589105724 | 0.000318796 | 0.004990048 | 1 |
| T2DM | leukocyte | *VSTM2L* | -0.684458925 | 0.00695867 | 0.010520217 | 1 |
| T2DM | leukocyte | *SERINC3* | -0.897198632 | 0.001504468 | 0.004993935 | 1 |
| T2DM | leukocyte | *BHMT2* | -1.29160758 | 0.000223793 | 0.004990048 | 0.879281151 |
| T2DM | leukocyte | *CASP9* | -1.589706182 | 0.011188432 | 0.015789997 | 1 |
| T2DM | leukocyte | *WASF3* | -0.934118777 | 0.002857656 | 0.006048134 | 1 |

| | | | | | | |
|---|---|---|---|---|---|---|
| T2DM | leukocyte | *SCO1* | -1.649567734 | 0.000991734 | 0.004990048 | 1 |
| T2DM | leukocyte | *CHI3L1* | -0.63671119 | 0.026755976 | 0.03416452 | 1 |
| T2DM | leukocyte | *SLC41A1* | -1.010113367 | 0.002600826 | 0.005751212 | 1 |
| T2DM | leukocyte | *TRPC4* | -2.602454065 | 0.002524154 | 0.005660135 | 1 |
| T2DM | leukocyte | *POSTN* | 0.734504327 | 0.021614705 | 0.028260957 | 1 |
| T2DM | leukocyte | *RFC3* | -0.718013495 | 0.004164904 | 0.007242343 | 1 |
| T2DM | leukocyte | *EPHB2* | -3.370558711 | 0.003302229 | 0.006432819 | 1 |
| T2DM | leukocyte | *BTBD2* | -2.229076483 | 0.000482028 | 0.004990048 | 1 |
| T2DM | leukocyte | *CNDP2* | -0.932737821 | 0.00162999 | 0.005035955 | 1 |
| T2DM | leukocyte | *WDR74* | -1.541436778 | 0.000414018 | 0.004990048 | 1 |
| T2DM | leukocyte | *GSTT2B* | -0.791792034 | 0.004889897 | 0.007948864 | 1 |
| T2DM | leukocyte | *SLC2A11* | -1.527510218 | 0.000829714 | 0.004990048 | 1 |
| T2DM | leukocyte | *C1QTNF6* | -1.405697128 | 0.004171706 | 0.007242343 | 1 |
| T2DM | leukocyte | *FAM83F* | -0.998346927 | 0.001700374 | 0.005060368 | 1 |
| T2DM | leukocyte | *GIMAP6* | -1.805320176 | 0.001143526 | 0.004990048 | 1 |
| T2DM | leukocyte | *AGAP3* | -3.171062937 | 0.000332234 | 0.004990048 | 1 |
| T2DM | leukocyte | *SFTPD* | -0.725344748 | 0.022666346 | 0.029430296 | 1 |
| T2DM | leukocyte | *IMPA1* | -0.917338521 | 0.000920869 | 0.004990048 | 1 |
| T2DM | leukocyte | *LRRCC1* | -1.404715895 | 0.002313982 | 0.005496836 | 1 |
| T2DM | leukocyte | *RRAS2* | -0.755546159 | 0.003029863 | 0.006206637 | 1 |
| T2DM | leukocyte | *HSD17B4* | -1.774697131 | 0.000304118 | 0.004990048 | 1 |
| T2DM | leukocyte | *COX16* | -0.911715604 | 0.000922951 | 0.004990048 | 1 |
| T2DM | leukocyte | *ELP3* | -1.956163395 | 0.000798895 | 0.004990048 | 1 |
| T2DM | leukocyte | *PEBP4* | -1.111314292 | 0.00825799 | 0.012133749 | 1 |
| T2DM | leukocyte | *MRO* | -2.477923163 | 0.002232339 | 0.005430873 | 1 |
| T2DM | leukocyte | *MRPS36* | -0.881140393 | 0.005304275 | 0.008434033 | 1 |
| T2DM | leukocyte | *THUMPD3* | -1.102785745 | 0.00047569 | 0.004990048 | 1 |
| T2DM | leukocyte | *CHL1* | -0.828646959 | 0.020074138 | 0.026413694 | 1 |
| T2DM | leukocyte | *REG4* | -0.963245662 | 0.004660367 | 0.007686961 | 1 |
| T2DM | leukocyte | *GSTM5* | -1.428407496 | 0.02562446 | 0.032869248 | 1 |
| T2DM | leukocyte | *GSTM3* | -0.593227703 | 0.006763593 | 0.010278358 | 1 |
| T2DM | leukocyte | *PTPN22* | -4.267293523 | 0.013569125 | 0.018706349 | 1 |
| T2DM | leukocyte | *SORT1* | -1.185799922 | 0.001001274 | 0.004990048 | 1 |
| T2DM | leukocyte | *NOTCH2* | -2.710476667 | 9.16E-12 | 3.60E-08 | 3.60E-08 |
| T2DM | leukocyte | *CEPT1* | -1.617346856 | 0.001226489 | 0.004990048 | 1 |
| T2DM | leukocyte | *NAPG* | -0.931443162 | 0.001599781 | 0.005024491 | 1 |
| T2DM | leukocyte | *SPIRE1* | -0.989911403 | 0.000999628 | 0.004990048 | 1 |
| T2DM | leukocyte | *FKBP11* | -0.546332889 | 0.002601146 | 0.005751212 | 1 |
| T2DM | leukocyte | *KIDINS220* | -1.010559837 | 0.00101178 | 0.004990048 | 1 |
| T2DM | leukocyte | *LPIN1* | -0.653818534 | 0.002997957 | 0.006172668 | 1 |
| T2DM | leukocyte | *IAH1* | -1.207787165 | 0.001070232 | 0.004990048 | 1 |
| T2DM | leukocyte | *ANO3* | 1.187725357 | 0.026116407 | 0.033434787 | 1 |
| T2DM | leukocyte | *IL2RA* | 0.247966468 | 0.019869571 | 0.026163627 | 1 |
| T2DM | leukocyte | *ECHDC3* | -0.853509333 | 0.003418695 | 0.006539461 | 1 |
| T2DM | leukocyte | *TMEM241* | -1.381755542 | 0.00803427 | 0.011844895 | 1 |
| T2DM | leukocyte | *AGO4* | -1.772710587 | 0.000578616 | 0.004990048 | 1 |
| T2DM | leukocyte | *PRPF38A* | -0.925608505 | 0.001536142 | 0.005006447 | 1 |
| T2DM | leukocyte | *DSC2* | -2.022227678 | 0.021230554 | 0.027795683 | 1 |
| T2DM | leukocyte | *TPGS2* | -0.985079357 | 0.005215535 | 0.008330015 | 1 |
| T2DM | leukocyte | *DAGLA* | -0.991581963 | 0.005294578 | 0.008425434 | 1 |

| | | | | | | |
|---|---|---|---|---|---|---|
| T2DM | leukocyte | *TIMM10* | -0.967744977 | 0.00111352 | 0.004990048 | 1 |
| T2DM | leukocyte | *FADS2* | -1.1187484 | 0.000990807 | 0.004990048 | 1 |
| T2DM | leukocyte | *TMEM165* | -0.754228989 | 0.001779103 | 0.005098084 | 1 |
| T2DM | leukocyte | *CLOCK* | -1.041416009 | 0.002273932 | 0.005460974 | 1 |
| T2DM | leukocyte | *GGACT* | -1.569794147 | 0.001227297 | 0.004990048 | 1 |
| T2DM | leukocyte | *CLDN10* | -0.371967617 | 0.008936713 | 0.012951806 | 1 |
| T2DM | leukocyte | *CARS2* | -0.979255087 | 0.000587232 | 0.004990048 | 1 |
| T2DM | leukocyte | *TMED7* | -0.84132643 | 0.001164116 | 0.004990048 | 1 |
| T2DM | leukocyte | *NREP* | -5.341076361 | 0.000449643 | 0.004990048 | 1 |
| T2DM | leukocyte | *WDR36* | -0.944908165 | 0.002542236 | 0.00567203 | 1 |
| T2DM | leukocyte | *OSTF1* | -1.232401548 | 0.002973934 | 0.006152298 | 1 |
| T2DM | leukocyte | *GOLM1* | -0.7170405 | 0.001515358 | 0.004993935 | 1 |
| T2DM | leukocyte | *SDS* | 0.053511191 | 0.017662348 | 0.023570678 | 1 |
| T2DM | leukocyte | *MSI1* | -1.61833251 | 0.001405623 | 0.004990048 | 1 |
| T2DM | leukocyte | *TBX3* | -1.460168199 | 0.039205779 | 0.04876211 | 1 |
| T2DM | leukocyte | *DTX1* | -2.276345209 | 0.002243494 | 0.005438799 | 1 |
| T2DM | leukocyte | *PNPLA8* | -0.739466483 | 0.001582344 | 0.005009694 | 1 |
| T2DM | leukocyte | *TES* | -0.594086705 | 0.002935031 | 0.006117633 | 1 |
| T2DM | leukocyte | *MDFIC* | -2.177790998 | 0.003147899 | 0.006311945 | 1 |
| T2DM | leukocyte | *MTO1* | -0.836939685 | 0.003160385 | 0.006314657 | 1 |
| T2DM | leukocyte | *LCA5* | -2.567617914 | 0.000511871 | 0.004990048 | 1 |
| T2DM | leukocyte | *LMO2* | -0.797758097 | 0.002533137 | 0.005660135 | 1 |
| T2DM | leukocyte | *RDH5* | -0.788380817 | 0.002824443 | 0.006006579 | 1 |
| T2DM | leukocyte | *TFCP2* | -3.002257977 | 0.000254102 | 0.004990048 | 0.998365013 |
| T2DM | leukocyte | *KRT7* | -0.409514011 | 0.004959246 | 0.00801517 | 1 |
| T2DM | leukocyte | *NHSL1* | -0.960719801 | 0.017764039 | 0.023686929 | 1 |
| T2DM | leukocyte | *AHI1* | -0.586367821 | 0.005152685 | 0.008249756 | 1 |
| T2DM | leukocyte | *HEY2* | -1.235253918 | 0.004371059 | 0.007383444 | 1 |
| T2DM | leukocyte | *PKIB* | -0.639767566 | 0.002171698 | 0.005349178 | 1 |
| T2DM | leukocyte | *SMPD2* | -2.229035862 | 0.001988573 | 0.005227473 | 1 |
| T2DM | leukocyte | *SEMA4F* | -1.098483353 | 0.008354601 | 0.012252792 | 1 |
| T2DM | leukocyte | *GNS* | -1.831341491 | 0.0003717 | 0.004990048 | 1 |
| T2DM | leukocyte | *MPHOSPH6* | -0.74998749 | 0.005025718 | 0.00809596 | 1 |
| T2DM | leukocyte | *AGT* | -0.576199827 | 0.002647241 | 0.005823633 | 1 |
| T2DM | leukocyte | *NTPCR* | -1.271041861 | 0.001106157 | 0.004990048 | 1 |
| T2DM | leukocyte | *USP37* | -1.057785027 | 0.005348156 | 0.008493495 | 1 |
| T2DM | leukocyte | *ARMC9* | -4.244227778 | 0.002144775 | 0.005323324 | 1 |
| T2DM | leukocyte | *COX5B* | -0.735357422 | 0.001189838 | 0.004990048 | 1 |
| T2DM | leukocyte | *REV1* | -0.863614497 | 0.001711098 | 0.005068796 | 1 |
| T2DM | leukocyte | *TSGA10* | -0.921754812 | 0.004801867 | 0.007847977 | 1 |
| T2DM | leukocyte | *TGFBRAP1* | -1.143072579 | 0.000704725 | 0.004990048 | 1 |
| T2DM | leukocyte | *GCC2* | -1.126216549 | 0.000305852 | 0.004990048 | 1 |
| T2DM | leukocyte | *MRPS9* | -0.763189653 | 0.003327777 | 0.006456709 | 1 |
| T2DM | leukocyte | *ANKRD36* | -1.191542424 | 0.002253378 | 0.005441623 | 1 |
| T2DM | leukocyte | *USP44* | -1.092577742 | 0.006918882 | 0.010467574 | 1 |
| T2DM | leukocyte | *DRAM1* | -1.837430328 | 0.000739617 | 0.004990048 | 1 |
| T2DM | leukocyte | *VILL* | 0.971094927 | 0.037747 | 0.047096845 | 1 |
| T2DM | leukocyte | *FLNB* | -0.684028107 | 0.00664647 | 0.010135575 | 1 |
| T2DM | leukocyte | *NEK3* | -0.54007957 | 0.015978862 | 0.021618784 | 1 |
| T2DM | leukocyte | *VPS36* | -1.46274435 | 0.000677815 | 0.004990048 | 1 |

| | | | | | | |
|---|---|---|---|---|---|---|
| T2DM | leukocyte | *RNASEH2B* | -1.858247699 | 0.000541972 | 0.004990048 | 1 |
| T2DM | leukocyte | *TBC1D4* | -0.944921279 | 0.002293916 | 0.005475575 | 1 |
| T2DM | leukocyte | *SUCLA2* | -1.124175981 | 0.000672471 | 0.004990048 | 1 |
| T2DM | leukocyte | *RCBTB1* | -1.056839353 | 0.022564493 | 0.029327123 | 1 |
| T2DM | leukocyte | *LMO7* | -0.506278274 | 0.003113969 | 0.006284318 | 1 |
| T2DM | leukocyte | *C7orf25* | -0.816632593 | 0.001726535 | 0.005070552 | 1 |
| T2DM | leukocyte | *RAPGEF5* | -0.609336852 | 0.004219122 | 0.007267615 | 1 |
| T2DM | leukocyte | *KDELR2* | -0.84563028 | 0.000990467 | 0.004990048 | 1 |
| T2DM | leukocyte | *TBRG4* | -2.108607999 | 0.000345928 | 0.004990048 | 1 |
| T2DM | leukocyte | *DBNL* | -2.788918408 | 0.000464129 | 0.004990048 | 1 |
| T2DM | leukocyte | *ALPK3* | -0.654655048 | 0.002510527 | 0.005649404 | 1 |
| T2DM | leukocyte | *CIB2* | -0.855905247 | 0.016006514 | 0.021640595 | 1 |
| T2DM | leukocyte | *NMT1* | -0.676316224 | 0.006673527 | 0.010162902 | 1 |
| T2DM | leukocyte | *CHAD* | -3.814557871 | 0.04011589 | 0.049736615 | 1 |
| T2DM | leukocyte | *TEX2* | -0.606998347 | 0.004913083 | 0.007970068 | 1 |
| T2DM | leukocyte | *LIMD2* | 0.687116169 | 0.008760919 | 0.0127299 | 1 |
| T2DM | leukocyte | *MRPL47* | -0.884538255 | 0.001091032 | 0.004990048 | 1 |
| T2DM | leukocyte | *SMPD4* | -2.303556546 | 0.000367973 | 0.004990048 | 1 |
| T2DM | leukocyte | *CCDC115* | -0.965893036 | 0.00246891 | 0.005622792 | 1 |
| T2DM | leukocyte | *BIN1* | -2.071793618 | 0.001624558 | 0.005035955 | 1 |
| T2DM | leukocyte | *HS6ST1* | -1.360128053 | 0.001035322 | 0.004990048 | 1 |
| T2DM | leukocyte | *GYPC* | -0.803361406 | 0.007748307 | 0.011505329 | 1 |
| T2DM | leukocyte | *NIPSNAP3A* | -1.206190941 | 0.003223727 | 0.006377655 | 1 |
| T2DM | leukocyte | *CDK9* | -1.078675544 | 0.001627877 | 0.005035955 | 1 |
| T2DM | leukocyte | *TXN* | -0.662740294 | 0.001285968 | 0.004990048 | 1 |
| T2DM | leukocyte | *TOR1B* | -1.096777685 | 0.019106901 | 0.025242439 | 1 |
| T2DM | leukocyte | *TOR1A* | -0.816664318 | 0.003137538 | 0.006311945 | 1 |
| T2DM | leukocyte | *SLC2A8* | -0.958638602 | 0.002301546 | 0.005478715 | 1 |
| T2DM | leukocyte | *ZFP37* | -0.625234876 | 0.024786215 | 0.031929522 | 1 |
| T2DM | leukocyte | *STX17* | -0.892823881 | 0.001344716 | 0.004990048 | 1 |
| T2DM | leukocyte | *PRPF4* | -0.867330893 | 0.026857781 | 0.034280595 | 1 |
| T2DM | leukocyte | *FPGS* | -1.155054258 | 0.001715357 | 0.005070552 | 1 |
| T2DM | leukocyte | *ATP6V1G1* | -0.676894416 | 0.001871604 | 0.005146173 | 1 |
| T2DM | leukocyte | *ARPC5L* | -0.698435761 | 0.001647279 | 0.005035955 | 1 |
| T2DM | leukocyte | *POLR1E* | -0.681973725 | 0.001365429 | 0.004990048 | 1 |
| T2DM | leukocyte | *APTX* | -0.779404644 | 0.00161061 | 0.005033031 | 1 |
| T2DM | leukocyte | *CD72* | -1.73992406 | 0.001884824 | 0.005146173 | 1 |
| T2DM | leukocyte | *TMEM8B* | -1.165879881 | 0.002107021 | 0.005283012 | 1 |
| T2DM | leukocyte | *GRHPR* | -0.890597769 | 0.001454914 | 0.004990048 | 1 |
| T2DM | leukocyte | *ALDH1B1* | -1.810152032 | 0.00152649 | 0.004997983 | 1 |
| T2DM | leukocyte | *ARHGEF39* | -3.200933671 | 0.003298828 | 0.006432819 | 1 |
| T2DM | leukocyte | *PPIL1* | -2.232730071 | 0.000553626 | 0.004990048 | 1 |
| T2DM | leukocyte | *GMPR* | -1.456185774 | 0.003001342 | 0.006172668 | 1 |
| T2DM | leukocyte | *KIAA0319* | -0.918845829 | 0.003519969 | 0.006649019 | 1 |
| T2DM | leukocyte | *LRRC1* | -0.822862055 | 0.002517426 | 0.005655212 | 1 |
| T2DM | leukocyte | *RIPK1* | -0.918797445 | 0.00210036 | 0.005283012 | 1 |
| T2DM | leukocyte | *TUBB2B* | -0.715860087 | 0.003753749 | 0.00692001 | 1 |
| T2DM | leukocyte | *UQCC2* | -0.84858173 | 0.002285542 | 0.005462378 | 1 |
| T2DM | leukocyte | *HMGA1* | -0.638663966 | 0.002163924 | 0.005345956 | 1 |
| T2DM | leukocyte | *CLPS* | -0.392786902 | 0.000986333 | 0.004990048 | 1 |

| | | | | | | |
|---|---|---|---|---|---|---|
| T2DM | leukocyte | *MTCH1* | -0.808826799 | 0.001287233 | 0.004990048 | 1 |
| T2DM | leukocyte | *ANKRD42* | -1.426701156 | 0.000517533 | 0.004990048 | 1 |
| T2DM | leukocyte | *CREBZF* | -1.213894718 | 0.001025649 | 0.004990048 | 1 |
| T2DM | leukocyte | *PRCP* | -1.003874379 | 0.011276024 | 0.015873701 | 1 |
| T2DM | leukocyte | *NARS2* | -1.038891667 | 0.00314737 | 0.006311945 | 1 |
| T2DM | leukocyte | *TTPA* | -5.418775892 | 0.023237858 | 0.030105511 | 1 |
| T2DM | leukocyte | *GGH* | -1.270156427 | 0.000563505 | 0.004990048 | 1 |
| T2DM | leukocyte | *SDCBP* | -1.443574974 | 0.001148447 | 0.004990048 | 1 |
| T2DM | leukocyte | *TRPC6* | -1.924232077 | 0.01882808 | 0.024919309 | 1 |
| T2DM | leukocyte | *MMP7* | -0.340146686 | 0.008240403 | 0.012116971 | 1 |
| T2DM | leukocyte | *FDX1* | -0.929154267 | 0.001124645 | 0.004990048 | 1 |
| T2DM | leukocyte | *C11orf1* | -0.844368439 | 0.002878873 | 0.006055188 | 1 |
| T2DM | leukocyte | *FXYD2* | -0.559969971 | 0.000428542 | 0.004990048 | 1 |
| T2DM | leukocyte | *CASP1* | 0.279956488 | 0.026892061 | 0.03430484 | 1 |
| T2DM | leukocyte | *ALKBH8* | -1.18616802 | 0.003733981 | 0.006900664 | 1 |
| T2DM | leukocyte | *TUBGCP4* | -1.643039384 | 0.000233239 | 0.004990048 | 0.916396285 |
| T2DM | leukocyte | *UACA* | -0.751670476 | 0.003841655 | 0.007022431 | 1 |
| T2DM | leukocyte | *PLCB2* | 0.479160051 | 0.017181718 | 0.023024205 | 1 |
| T2DM | leukocyte | *PAK6* | -0.5321167 | 0.002021956 | 0.005244224 | 1 |
| T2DM | leukocyte | *ADAM10* | -1.136251511 | 0.001186895 | 0.004990048 | 1 |
| T2DM | leukocyte | *DUOX1* | -0.820875929 | 0.028072811 | 0.035695169 | 1 |
| T2DM | leukocyte | *CYP19A1* | -6.005219016 | 0.020368026 | 0.026746649 | 1 |
| T2DM | leukocyte | *ZNF280D* | -1.135448806 | 0.001003102 | 0.004990048 | 1 |
| T2DM | leukocyte | *RSL24D1* | -0.899854216 | 0.000586022 | 0.004990048 | 1 |
| T2DM | leukocyte | *TTLL7* | -0.850177583 | 0.002297736 | 0.005478037 | 1 |
| T2DM | leukocyte | *SLC44A5* | -1.206666074 | 0.009208455 | 0.013306369 | 1 |
| T2DM | leukocyte | *RTCA* | -1.975335406 | 0.003162148 | 0.006314657 | 1 |
| T2DM | leukocyte | *KHK* | -4.30195504 | 4.29E-05 | 0.004212864 | 0.168514541 |
| T2DM | leukocyte | *DYNC2LI1* | -1.007764817 | 0.001708839 | 0.005068796 | 1 |
| T2DM | leukocyte | *ACTR2* | -0.854100069 | 0.000695749 | 0.004990048 | 1 |
| T2DM | leukocyte | *ATRAID* | -0.857796269 | 0.00138488 | 0.004990048 | 1 |
| T2DM | leukocyte | *LRPPRC* | -0.966612939 | 0.002055773 | 0.00526208 | 1 |
| T2DM | leukocyte | *DUSP5* | -1.703847167 | 0.000654789 | 0.004990048 | 1 |
| T2DM | leukocyte | *ARL3* | -0.849949957 | 0.000831833 | 0.004990048 | 1 |
| T2DM | leukocyte | *PLCE1* | -0.779004353 | 0.026752145 | 0.03416452 | 1 |
| T2DM | leukocyte | *DNAJC13* | -0.729646384 | 0.006016895 | 0.009321917 | 1 |
| T2DM | leukocyte | *FAM149B1* | -1.010070139 | 0.001144547 | 0.004990048 | 1 |
| T2DM | leukocyte | *NCOA4* | -1.93072748 | 0.000430795 | 0.004990048 | 1 |
| T2DM | leukocyte | *ASCC1* | -1.012227484 | 0.003160944 | 0.006314657 | 1 |
| T2DM | leukocyte | *RPS24* | -0.638709612 | 0.000833662 | 0.004990048 | 1 |
| T2DM | leukocyte | *DNA2* | -1.369646682 | 0.004552099 | 0.007568852 | 1 |
| T2DM | leukocyte | *MYPN* | 4.469546344 | 0.0013364 | 0.004990048 | 1 |
| T2DM | leukocyte | *ATIC* | -1.364886865 | 0.003146858 | 0.006311945 | 1 |
| T2DM | leukocyte | *SMARCAL1* | -1.13857457 | 0.004175878 | 0.007242343 | 1 |
| T2DM | leukocyte | *BARD1* | -0.992189794 | 0.002141902 | 0.005322917 | 1 |
| T2DM | leukocyte | *FASTKD1* | -1.059251973 | 0.002665676 | 0.005838039 | 1 |
| T2DM | leukocyte | *FAM117B* | -1.095547922 | 0.002110311 | 0.005283012 | 1 |
| T2DM | leukocyte | *WDR12* | -1.173139318 | 0.003672266 | 0.006838073 | 1 |
| T2DM | leukocyte | *SLC35A5* | -1.172170672 | 0.00334219 | 0.006470236 | 1 |
| T2DM | leukocyte | *USP8* | -1.139430094 | 0.000816848 | 0.004990048 | 1 |

| | | | | | | |
|---|---|---|---|---|---|---|
| T2DM | leukocyte | *SPPL2A* | -0.996064517 | 0.00089018 | 0.004990048 | 1 |
| T2DM | leukocyte | *APH1B* | -1.462187359 | 0.000912687 | 0.004990048 | 1 |
| T2DM | leukocyte | *HCN4* | -1.138478824 | 0.004348678 | 0.007358292 | 1 |
| T2DM | leukocyte | *SEMA7A* | -1.972573053 | 0.004322038 | 0.007335329 | 1 |
| T2DM | leukocyte | *AP1AR* | -0.760045876 | 0.002108818 | 0.005283012 | 1 |
| T2DM | leukocyte | *BBS7* | -1.178201981 | 0.000737534 | 0.004990048 | 1 |
| T2DM | leukocyte | *RAP1GDS1* | -1.082927469 | 0.001656864 | 0.005041098 | 1 |
| T2DM | leukocyte | *PRDM5* | -0.79245136 | 0.006838025 | 0.010369201 | 1 |
| T2DM | leukocyte | *G3BP2* | -0.765223659 | 0.000943288 | 0.004990048 | 1 |
| T2DM | leukocyte | *SCARB2* | -0.862315763 | 0.001020436 | 0.004990048 | 1 |
| T2DM | leukocyte | *CNOT6L* | -1.175537516 | 0.000666398 | 0.004990048 | 1 |
| T2DM | leukocyte | *USO1* | -0.754494891 | 0.001248093 | 0.004990048 | 1 |
| T2DM | leukocyte | *SHROOM3* | -1.686777976 | 0.000873548 | 0.004990048 | 1 |
| T2DM | leukocyte | *ANXA3* | -0.634422968 | 0.001738681 | 0.005075243 | 1 |
| T2DM | leukocyte | *PPA2* | -0.894361274 | 0.001408623 | 0.004990048 | 1 |
| T2DM | leukocyte | *MAPK8IP3* | -0.968534904 | 0.001670049 | 0.005048289 | 1 |
| T2DM | leukocyte | *RGS3* | -0.806351762 | 0.001968622 | 0.005212073 | 1 |
| T2DM | leukocyte | *ETV6* | -1.7140537 | 0.001573507 | 0.005009694 | 1 |
| T2DM | leukocyte | *ALG10* | -1.542369259 | 0.003263814 | 0.006405357 | 1 |
| T2DM | leukocyte | *SLCO1C1* | -13.76724257 | 0.00298259 | 0.006152298 | 1 |
| T2DM | leukocyte | *ZCRB1* | -0.823312164 | 0.001072709 | 0.004990048 | 1 |
| T2DM | leukocyte | *CLSTN3* | -1.520515455 | 0.000686025 | 0.004990048 | 1 |
| T2DM | leukocyte | *KLRG1* | -5.445018118 | 0.004165337 | 0.007242343 | 1 |
| T2DM | leukocyte | *TAPBPL* | -0.658268412 | 0.01730033 | 0.023167347 | 1 |
| T2DM | leukocyte | *PEX5* | -0.869588519 | 0.002562937 | 0.00569337 | 1 |
| T2DM | leukocyte | *SLC38A4* | -1.245590469 | 0.002897649 | 0.006071927 | 1 |
| T2DM | leukocyte | *LRIG3* | -0.876071543 | 0.002862937 | 0.006050822 | 1 |
| T2DM | leukocyte | *GLIPR1* | -1.041787783 | 0.001980535 | 0.005222499 | 1 |
| T2DM | leukocyte | *SNRPF* | -0.728228443 | 0.001451389 | 0.004990048 | 1 |
| T2DM | leukocyte | *AMDHD1* | -1.294098 | 0.018083375 | 0.024060135 | 1 |
| T2DM | leukocyte | *ASCL1* | -1.183105978 | 0.010544675 | 0.014967495 | 1 |
| T2DM | leukocyte | *RITA1* | -1.30717619 | 0.001696547 | 0.005060368 | 1 |
| T2DM | leukocyte | *GLTP* | -0.712897724 | 0.002149531 | 0.005327398 | 1 |
| T2DM | leukocyte | *MTMR6* | -1.21421201 | 0.000734291 | 0.004990048 | 1 |
| T2DM | leukocyte | *PDX1* | -1.237441824 | 0.00141349 | 0.004990048 | 1 |
| T2DM | leukocyte | *CCDC65* | -1.255133382 | 0.008172511 | 0.012030646 | 1 |
| T2DM | leukocyte | *KANSL2* | -2.239402189 | 0.000466573 | 0.004990048 | 1 |
| T2DM | leukocyte | *CERS5* | -1.382439286 | 0.000146657 | 0.004990048 | 0.57621634 |
| T2DM | leukocyte | *ZNF740* | -1.877781647 | 0.002001809 | 0.005227473 | 1 |
| T2DM | leukocyte | *WDFY2* | -1.031803084 | 0.001517262 | 0.004993935 | 1 |
| T2DM | leukocyte | *GRTP1* | -1.841729045 | 0.004988086 | 0.008055154 | 1 |
| T2DM | leukocyte | *TSSK4* | -6.087221262 | 0.031807513 | 0.040235582 | 1 |
| T2DM | leukocyte | *TMX1* | -1.334074983 | 0.000678144 | 0.004990048 | 1 |
| T2DM | leukocyte | *RAB15* | -1.212970354 | 0.003277362 | 0.006422321 | 1 |
| T2DM | leukocyte | *NIPA2* | -0.732662531 | 0.002338531 | 0.005515664 | 1 |
| T2DM | leukocyte | *SERF2* | -0.721970854 | 0.000547941 | 0.004990048 | 1 |
| T2DM | leukocyte | *BNIP2* | -1.539422107 | 0.000507537 | 0.004990048 | 1 |
| T2DM | leukocyte | *GTF2A2* | -0.825723089 | 0.001105674 | 0.004990048 | 1 |
| T2DM | leukocyte | *SRP14* | -0.667955548 | 0.001196941 | 0.004990048 | 1 |
| T2DM | leukocyte | *BAHD1* | -1.033879605 | 0.001879714 | 0.005146173 | 1 |

| | | | | | | |
|---|---|---|---|---|---|---|
| T2DM | leukocyte | *DISP2* | -0.555764793 | 0.022414807 | 0.029180839 | 1 |
| T2DM | leukocyte | *TLE3* | -2.251800618 | 0.000147076 | 0.004990048 | 0.577860509 |
| T2DM | leukocyte | *SCAPER* | -0.788482589 | 0.002392465 | 0.005565421 | 1 |
| T2DM | leukocyte | *TSPAN3* | -0.705982768 | 0.001450395 | 0.004990048 | 1 |
| T2DM | leukocyte | *MAN2C1* | -1.426190477 | 4.35E-07 | 0.000266998 | 0.001707834 |
| T2DM | leukocyte | *DNAJA4* | -0.802589262 | 0.00163242 | 0.005035955 | 1 |
| T2DM | leukocyte | *TPM1* | -0.494309567 | 0.002296295 | 0.005477925 | 1 |
| T2DM | leukocyte | *IGF1R* | -0.754629313 | 0.001633629 | 0.005035955 | 1 |
| T2DM | leukocyte | *ADAMTS17* | -1.552761068 | 0.029393271 | 0.037313785 | 1 |
| T2DM | leukocyte | *ULK3* | -0.852837726 | 0.003634748 | 0.006787511 | 1 |
| T2DM | leukocyte | *CELF6* | -1.046934241 | 0.002555128 | 0.005681968 | 1 |
| T2DM | leukocyte | *HAPLN3* | -5.349458718 | 0.014403674 | 0.019702973 | 1 |
| T2DM | leukocyte | *POLG* | -1.425621479 | 0.001361005 | 0.004990048 | 1 |
| T2DM | leukocyte | *WDR93* | -2.770882172 | 0.016438398 | 0.022152044 | 1 |
| T2DM | leukocyte | *UNC45A* | -0.98619815 | 0.001661583 | 0.005041098 | 1 |
| T2DM | leukocyte | *IQGAP1* | -0.935496147 | 0.001013865 | 0.004990048 | 1 |
| T2DM | leukocyte | *PMM2* | -0.888409208 | 0.001864925 | 0.005146173 | 1 |
| T2DM | leukocyte | *SLC5A2* | -3.061964399 | 0.007835788 | 0.01162205 | 1 |
| T2DM | leukocyte | *UQCRC2* | -0.996215272 | 0.000773466 | 0.004990048 | 1 |
| T2DM | leukocyte | *ARHGAP17* | -2.37579736 | 0.000388647 | 0.004990048 | 1 |
| T2DM | leukocyte | *TXNL4B* | -2.317075921 | 0.000307089 | 0.004990048 | 1 |
| T2DM | leukocyte | *MARVELD3* | -0.697111882 | 0.005702893 | 0.008935705 | 1 |
| T2DM | leukocyte | *KIFC3* | -1.043384224 | 0.001074925 | 0.004990048 | 1 |
| T2DM | leukocyte | *NUDT7* | -0.763573058 | 0.004076503 | 0.007189 | 1 |
| T2DM | leukocyte | *GCSH* | -0.854628837 | 0.001289991 | 0.004990048 | 1 |
| T2DM | leukocyte | *MAP1LC3B* | -0.750524154 | 0.001090405 | 0.004990048 | 1 |
| T2DM | leukocyte | *OSGIN1* | -0.681431267 | 0.002881691 | 0.006056841 | 1 |
| T2DM | leukocyte | *RPL3L* | -1.471998998 | 0.030333122 | 0.038457024 | 1 |
| T2DM | leukocyte | *RPS2* | -0.58838256 | 0.000816556 | 0.004990048 | 1 |
| T2DM | leukocyte | *PDPK1* | -1.172069039 | 0.001047532 | 0.004990048 | 1 |
| T2DM | leukocyte | *DEF8* | -1.046401037 | 0.000847008 | 0.004990048 | 1 |
| T2DM | leukocyte | *GAS8* | -1.736912505 | 0.000459988 | 0.004990048 | 1 |
| T2DM | leukocyte | *RANBP10* | -0.765457286 | 0.008424966 | 0.012337567 | 1 |
| T2DM | leukocyte | *PRPSAP2* | -0.994445752 | 0.005103685 | 0.00819008 | 1 |
| T2DM | leukocyte | *TOM1L1* | -0.828491217 | 0.002859829 | 0.006048134 | 1 |
| T2DM | leukocyte | *C17orf80* | -1.418560725 | 0.003226671 | 0.006380267 | 1 |
| T2DM | leukocyte | *VPS53* | -1.14048515 | 0.00012027 | 0.004990048 | 0.472541883 |
| T2DM | leukocyte | *NPEPPS* | -1.298291899 | 0.000596473 | 0.004990048 | 1 |
| T2DM | leukocyte | *SCRN2* | -2.148150358 | 4.21E-05 | 0.004212864 | 0.165304763 |
| T2DM | leukocyte | *RHBDL3* | -1.254712467 | 0.039959885 | 0.049590142 | 1 |
| T2DM | leukocyte | *SPACA3* | -0.957906609 | 0.00047332 | 0.004990048 | 1 |
| T2DM | leukocyte | *ARSG* | -2.727397664 | 0.009593467 | 0.013781621 | 1 |
| T2DM | leukocyte | *PTRH2* | -0.994561076 | 0.001888631 | 0.005146173 | 1 |
| T2DM | leukocyte | *GNAL* | -1.196901025 | 0.005346999 | 0.008493495 | 1 |
| T2DM | leukocyte | *RPRD1A* | -0.871424083 | 0.002425993 | 0.005602682 | 1 |
| T2DM | leukocyte | *NPC1* | -2.414904964 | 1.47E-05 | 0.00262752 | 0.057805445 |
| T2DM | leukocyte | *SLC14A1* | -1.28262906 | 0.005885695 | 0.009158375 | 1 |
| T2DM | leukocyte | *ZMYND15* | 0.174371915 | 0.019414795 | 0.025614751 | 1 |
| T2DM | leukocyte | *SAT2* | -1.125884968 | 0.000570278 | 0.004990048 | 1 |
| T2DM | leukocyte | *CCDC40* | -0.742166846 | 0.018240344 | 0.024260769 | 1 |

| | | | | | | |
|---|---|---|---|---|---|---|
| T2DM | leukocyte | *ARHGDIA* | -0.842196319 | 0.001404932 | 0.004990048 | 1 |
| T2DM | leukocyte | *TMC6* | -1.356730498 | 0.003983392 | 0.00713343 | 1 |
| T2DM | leukocyte | *TTYH2* | -1.496983794 | 0.004284001 | 0.007324561 | 1 |
| T2DM | leukocyte | *EIF4A3* | -0.965535846 | 0.002044525 | 0.00526208 | 1 |
| T2DM | leukocyte | *TBCD* | -1.516543627 | 0.001834774 | 0.005146173 | 1 |
| T2DM | leukocyte | *FN3KRP* | -0.945011036 | 0.013005221 | 0.018017459 | 1 |
| T2DM | leukocyte | *NARF* | -1.342749495 | 0.000531592 | 0.004990048 | 1 |
| T2DM | leukocyte | *TRIM65* | -1.433280828 | 0.001217105 | 0.004990048 | 1 |
| T2DM | leukocyte | *RNF157* | -0.995705956 | 0.003086135 | 0.00625753 | 1 |
| T2DM | leukocyte | *WDR45B* | -0.749896535 | 0.001785532 | 0.005100156 | 1 |
| T2DM | leukocyte | *TNFRSF11A* | -0.768294039 | 0.004239657 | 0.007286795 | 1 |
| T2DM | leukocyte | *ZCCHC2* | -1.258016922 | 0.001217796 | 0.004990048 | 1 |
| T2DM | leukocyte | *PMAIP1* | -0.808048112 | 0.002947794 | 0.006131224 | 1 |
| T2DM | leukocyte | *NT5C3B* | -0.887908077 | 0.003063531 | 0.006246297 | 1 |
| T2DM | leukocyte | *PIP4K2B* | -0.818679406 | 0.004577379 | 0.007595372 | 1 |
| T2DM | leukocyte | *TXNL4A* | -0.843461695 | 0.000714026 | 0.004990048 | 1 |
| T2DM | leukocyte | *PRDM15* | -0.944396723 | 0.024706665 | 0.031847929 | 1 |
| T2DM | leukocyte | *MVB12A* | -0.795020846 | 0.02467465 | 0.031817099 | 1 |
| T2DM | leukocyte | *SH3GL1* | -0.886206709 | 0.000909402 | 0.004990048 | 1 |
| T2DM | leukocyte | *SIRT3* | -1.21518775 | 0.00251948 | 0.005656594 | 1 |
| T2DM | leukocyte | *IFITM3* | -0.549721569 | 0.00317108 | 0.006327666 | 1 |
| T2DM | leukocyte | *COL6A1* | -1.379267586 | 0.000266417 | 0.004990048 | 1 |
| T2DM | leukocyte | *IFNAR1* | -0.930332214 | 0.000441628 | 0.004990048 | 1 |
| T2DM | leukocyte | *TMEM50B* | -0.656406803 | 0.022630461 | 0.029393415 | 1 |
| T2DM | leukocyte | *AKT1* | -1.21007756 | 0.003269208 | 0.00641274 | 1 |
| T2DM | leukocyte | *CBLC* | -0.926335015 | 0.001805178 | 0.005122825 | 1 |
| T2DM | leukocyte | *EVI5L* | -0.79531459 | 0.034032158 | 0.042787952 | 1 |
| T2DM | leukocyte | *CTU1* | -1.586681584 | 0.000548467 | 0.004990048 | 1 |
| T2DM | leukocyte | *RERE* | -0.757751322 | 0.001669466 | 0.005048289 | 1 |
| T2DM | leukocyte | *FHAD1* | -0.91770182 | 0.002795454 | 0.005995273 | 1 |
| T2DM | leukocyte | *ARHGEF19* | -1.545105159 | 0.007033761 | 0.010600555 | 1 |
| T2DM | leukocyte | *PGD* | -1.209033083 | 0.000674695 | 0.004990048 | 1 |
| T2DM | leukocyte | *CNKSR1* | -1.24409479 | 0.023292875 | 0.030164042 | 1 |
| T2DM | leukocyte | *ZNF593* | -0.90619315 | 0.000760243 | 0.004990048 | 1 |
| T2DM | leukocyte | *FCN3* | -0.505174115 | 0.002717448 | 0.005898814 | 1 |
| T2DM | leukocyte | *CELA3A* | -0.397489181 | 0.001920892 | 0.005162839 | 1 |
| T2DM | leukocyte | *NBPF3* | -1.136674833 | 0.001276507 | 0.004990048 | 1 |
| T2DM | leukocyte | *HSPG2* | -0.550198388 | 0.000777748 | 0.004990048 | 1 |
| T2DM | leukocyte | *ITGB3BP* | -0.787621787 | 0.007694534 | 0.011438451 | 1 |
| T2DM | leukocyte | *PRKACB* | -1.060824304 | 0.001075229 | 0.004990048 | 1 |
| T2DM | leukocyte | *PIGK* | -1.576796271 | 0.000681396 | 0.004990048 | 1 |
| T2DM | leukocyte | *RPS8* | -0.545337419 | 0.001330287 | 0.004990048 | 1 |
| T2DM | leukocyte | *SYPL2* | -2.603806348 | 0.003022693 | 0.006198413 | 1 |
| T2DM | leukocyte | *SLC44A3* | -0.817706378 | 0.004592102 | 0.007609603 | 1 |
| T2DM | leukocyte | *ZNF697* | -1.064675106 | 0.020895139 | 0.027374792 | 1 |
| T2DM | leukocyte | *CELSR2* | -0.942718615 | 0.007560095 | 0.011272719 | 1 |
| T2DM | leukocyte | *ALDH9A1* | -0.792071414 | 0.001728309 | 0.005070552 | 1 |
| T2DM | leukocyte | *ATP1B1* | -0.549150898 | 0.001482837 | 0.004990048 | 1 |
| T2DM | leukocyte | *TIPRL* | -0.783498861 | 0.001730916 | 0.005070552 | 1 |
| T2DM | leukocyte | *NME7* | -1.657684858 | 7.21E-05 | 0.004990048 | 0.28346605 |

| | | | | | | |
|---|---|---|---|---|---|---|
| T2DM | leukocyte | *MGST3* | -0.905964594 | 0.00076898 | 0.004990048 | 1 |
| T2DM | leukocyte | *LYPLAL1* | -1.089846502 | 0.001298726 | 0.004990048 | 1 |
| T2DM | leukocyte | *SNX27* | -1.394721884 | 0.001631488 | 0.005035955 | 1 |
| T2DM | leukocyte | *CTSK* | -3.219258878 | 0.0003559 | 0.004990048 | 1 |
| T2DM | leukocyte | *SELENBP1* | -0.989722676 | 0.004134701 | 0.007236049 | 1 |
| T2DM | leukocyte | *POGZ* | -0.70840513 | 0.024891902 | 0.032055156 | 1 |
| T2DM | leukocyte | *GABPB2* | -1.335479306 | 0.007160688 | 0.010771187 | 1 |
| T2DM | leukocyte | *SLC27A3* | -1.299766937 | 0.001180989 | 0.004990048 | 1 |
| T2DM | leukocyte | *KCNN3* | -1.509202552 | 0.00386553 | 0.007037843 | 1 |
| T2DM | leukocyte | *RIT1* | -1.534666646 | 0.00467085 | 0.007686961 | 1 |
| T2DM | leukocyte | *SRP9* | -0.891877916 | 0.000925113 | 0.004990048 | 1 |
| T2DM | leukocyte | *CNIH4* | -0.910263542 | 0.001208296 | 0.004990048 | 1 |
| T2DM | leukocyte | *PSEN2* | -1.753676851 | 0.001858231 | 0.005146173 | 1 |
| T2DM | leukocyte | *PYCR2* | -0.973628835 | 0.002212844 | 0.005400163 | 1 |
| T2DM | leukocyte | *LBR* | -1.122068468 | 0.00121062 | 0.004990048 | 1 |
| T2DM | leukocyte | *ARL8A* | -1.564033833 | 0.000297034 | 0.004990048 | 1 |
| T2DM | leukocyte | *EML4* | -1.523579062 | 0.000729394 | 0.004990048 | 1 |
| T2DM | leukocyte | *WDPCP* | -2.380285747 | 0.000951969 | 0.004990048 | 1 |
| T2DM | leukocyte | *REG3G* | -1.073734239 | 0.000396949 | 0.004990048 | 1 |
| T2DM | leukocyte | *CIAO1* | -1.385726126 | 0.000258864 | 0.004990048 | 1 |
| T2DM | leukocyte | *ZNF514* | -1.175210602 | 0.000145504 | 0.004990048 | 0.571683877 |
| T2DM | leukocyte | *TPRKB* | -0.793616232 | 0.001463243 | 0.004990048 | 1 |
| T2DM | leukocyte | *TEX261* | -1.031877823 | 0.001506851 | 0.004993935 | 1 |
| T2DM | leukocyte | *RALB* | -0.903470931 | 0.002555367 | 0.005681968 | 1 |
| T2DM | leukocyte | *RABL2A* | -1.092681957 | 0.013124248 | 0.018175949 | 1 |
| T2DM | leukocyte | *SLC20A1* | -1.408868034 | 0.001226435 | 0.004990048 | 1 |
| T2DM | leukocyte | *LYG1* | -4.029828205 | 0.015521245 | 0.021050387 | 1 |
| T2DM | leukocyte | *GULP1* | -0.841281203 | 0.003001481 | 0.006172668 | 1 |
| T2DM | leukocyte | *FAM171B* | -0.788424901 | 0.002234511 | 0.005432793 | 1 |
| T2DM | leukocyte | *METTL21A* | -0.901420654 | 0.002553282 | 0.005681968 | 1 |
| T2DM | leukocyte | *SUMF1* | -1.304557824 | 0.000905121 | 0.004990048 | 1 |
| T2DM | leukocyte | *HES6* | -2.396732778 | 0.000330607 | 0.004990048 | 1 |
| T2DM | leukocyte | *TAMM41* | -0.800657429 | 0.027772355 | 0.035327696 | 1 |
| T2DM | leukocyte | *RAB5A* | -0.845166056 | 0.001625514 | 0.005035955 | 1 |
| T2DM | leukocyte | *CNTN4* | -1.263988585 | 1.67E-05 | 0.002853237 | 0.065624451 |
| T2DM | leukocyte | *POMGNT2* | -1.720136249 | 0.000464141 | 0.004990048 | 1 |
| T2DM | leukocyte | *CTDSPL* | -0.823197167 | 0.002176637 | 0.005349178 | 1 |
| T2DM | leukocyte | *IL17RD* | -3.3046154 | 0.007031666 | 0.010600555 | 1 |
| T2DM | leukocyte | *UBA3* | -1.377551495 | 0.000975349 | 0.004990048 | 1 |
| T2DM | leukocyte | *ARL6IP5* | -0.843135471 | 0.001129177 | 0.004990048 | 1 |
| T2DM | leukocyte | *LIMD1* | -1.560390213 | 0.000944177 | 0.004990048 | 1 |
| T2DM | leukocyte | *NXPE3* | -0.791227933 | 0.004673983 | 0.007686961 | 1 |
| T2DM | leukocyte | *PHLDB2* | -0.644840148 | 0.002876416 | 0.006055188 | 1 |
| T2DM | leukocyte | *ABHD10* | -0.944087622 | 0.003848664 | 0.007027251 | 1 |
| T2DM | leukocyte | *TAGLN3* | -0.40338361 | 0.034006491 | 0.042769367 | 1 |
| T2DM | leukocyte | *EIF2A* | -0.762690333 | 0.001408741 | 0.004990048 | 1 |
| T2DM | leukocyte | *ALDH1L1* | -1.183939715 | 0.004040102 | 0.007166394 | 1 |
| T2DM | leukocyte | *TRPC1* | -1.055960787 | 0.00239187 | 0.005565421 | 1 |
| T2DM | leukocyte | *NCEH1* | -0.529028179 | 0.003817061 | 0.006991718 | 1 |
| T2DM | leukocyte | *AMT* | -1.602409667 | 0.001106268 | 0.004990048 | 1 |

| | | | | | | |
|---|---|---|---|---|---|---|
| T2DM | leukocyte | *DGKQ* | -0.882183178 | 0.006767631 | 0.010278358 | 1 |
| T2DM | leukocyte | *SCD5* | -0.738847567 | 0.002516666 | 0.005655212 | 1 |
| T2DM | leukocyte | *PYURF* | -0.736786418 | 0.00137726 | 0.004990048 | 1 |
| T2DM | leukocyte | *DDIT4L* | -2.499698304 | 0.000872363 | 0.004990048 | 1 |
| T2DM | leukocyte | *TIFA* | -1.128962477 | 0.001479176 | 0.004990048 | 1 |
| T2DM | leukocyte | *CYP4V2* | -1.599650439 | 0.008820729 | 0.012807333 | 1 |
| T2DM | leukocyte | *SSBP2* | -0.716495827 | 0.002652338 | 0.005824744 | 1 |
| T2DM | leukocyte | *GTF2H2* | -2.546809452 | 0.000384526 | 0.004990048 | 1 |
| T2DM | leukocyte | *BTF3* | -0.711440816 | 0.000754577 | 0.004990048 | 1 |
| T2DM | leukocyte | *COMMD10* | -1.031730163 | 0.002092974 | 0.005283012 | 1 |
| T2DM | leukocyte | *ATG12* | -0.88842369 | 0.00083294 | 0.004990048 | 1 |
| T2DM | leukocyte | *PCYOX1L* | -5.052444085 | 0.000356405 | 0.004990048 | 1 |
| T2DM | leukocyte | *TBC1D7* | -3.126644702 | 0.01438781 | 0.019689901 | 1 |
| T2DM | leukocyte | *ZMAT2* | -1.128557407 | 0.000940779 | 0.004990048 | 1 |
| T2DM | leukocyte | *KLHL3* | -3.90558463 | 0.000806026 | 0.004990048 | 1 |
| T2DM | leukocyte | *DCDC2* | -0.651026498 | 0.003342983 | 0.006470236 | 1 |
| T2DM | leukocyte | *TRIM7* | -3.243516302 | 0.000234126 | 0.004990048 | 0.919881559 |
| T2DM | leukocyte | *ANO7* | -2.472903418 | 0.002718957 | 0.005898831 | 1 |
| T2DM | leukocyte | *RPL7L1* | -0.835586685 | 0.001168068 | 0.004990048 | 1 |
| T2DM | leukocyte | *IRAK1BP1* | -0.7271178 | 0.003762062 | 0.00692001 | 1 |
| T2DM | leukocyte | *MMS22L* | -1.242061669 | 0.002969118 | 0.006149534 | 1 |
| T2DM | leukocyte | *FAXC* | -1.383257021 | 0.001686702 | 0.005060368 | 1 |
| T2DM | leukocyte | *SCML4* | -3.563017319 | 0.001057376 | 0.004990048 | 1 |
| T2DM | leukocyte | *ABRACL* | -0.866651642 | 0.003381645 | 0.006503419 | 1 |
| T2DM | leukocyte | *DYNLT1* | -0.769966651 | 0.000870207 | 0.004990048 | 1 |
| T2DM | leukocyte | *WTAP* | -0.789915565 | 0.000723976 | 0.004990048 | 1 |
| T2DM | leukocyte | *SLC22A3* | -4.417705238 | 0.030323351 | 0.038457024 | 1 |
| T2DM | leukocyte | *VWDE* | -0.879637952 | 0.002055628 | 0.00526208 | 1 |
| T2DM | leukocyte | *GNA12* | -1.350301607 | 0.002814787 | 0.006001505 | 1 |
| T2DM | leukocyte | *SDK1* | -0.964895534 | 0.006728394 | 0.010230596 | 1 |
| T2DM | leukocyte | *CCZ1B* | -1.311942237 | 0.00388011 | 0.007054583 | 1 |
| T2DM | leukocyte | *RBAK* | -1.795200806 | 0.003765042 | 0.00692225 | 1 |
| T2DM | leukocyte | *POMZP3* | -1.012892079 | 0.001812614 | 0.005123568 | 1 |
| T2DM | leukocyte | *GIGYF1* | -2.350878155 | 2.70E-05 | 0.003310187 | 0.105925972 |
| T2DM | leukocyte | *TRIM4* | -0.942661241 | 0.00016209 | 0.004990048 | 0.636852299 |
| T2DM | leukocyte | *TMEM209* | -1.444850695 | 0.000667218 | 0.004990048 | 1 |
| T2DM | leukocyte | *RAB19* | -8.873280475 | 8.65E-05 | 0.004990048 | 0.339700603 |
| T2DM | leukocyte | *CSGALNACT1* | -0.782310511 | 0.003491269 | 0.006628411 | 1 |
| T2DM | leukocyte | *CCDC25* | -0.751768737 | 0.001242625 | 0.004990048 | 1 |
| T2DM | leukocyte | *BIN3* | -2.584440495 | 0.000824746 | 0.004990048 | 1 |
| T2DM | leukocyte | *CHMP7* | -1.203086066 | 0.004518603 | 0.007554719 | 1 |
| T2DM | leukocyte | *STAR* | 0.541441768 | 0.021863505 | 0.028548259 | 1 |
| T2DM | leukocyte | *LACTB2* | -0.79529436 | 0.002099301 | 0.005283012 | 1 |
| T2DM | leukocyte | *TERF1* | -1.889498701 | 0.000343134 | 0.004990048 | 1 |
| T2DM | leukocyte | *LRP12* | -0.922881051 | 0.001409967 | 0.004990048 | 1 |
| T2DM | leukocyte | *ZNF7* | -0.752307689 | 0.003038546 | 0.006214704 | 1 |
| T2DM | leukocyte | *AK3* | -0.809659037 | 0.001327528 | 0.004990048 | 1 |
| T2DM | leukocyte | *CER1* | 2.880374493 | 0.016377537 | 0.022082136 | 1 |
| T2DM | leukocyte | *C9orf72* | -1.393477069 | 0.000567222 | 0.004990048 | 1 |
| T2DM | leukocyte | *FBXO10* | -1.203194681 | 0.005624773 | 0.008832827 | 1 |

| | | | | | | |
|---|---|---|---|---|---|---|
| T2DM | leukocyte | *SIGMAR1* | -1.172743868 | 0.000366016 | 0.004990048 | 1 |
| T2DM | leukocyte | *ZNF462* | -0.778879754 | 0.002445925 | 0.005621365 | 1 |
| T2DM | leukocyte | *UGCG* | -0.94813811 | 0.00089681 | 0.004990048 | 1 |
| T2DM | leukocyte | *ALAD* | -0.93353982 | 0.004866832 | 0.007914645 | 1 |
| T2DM | leukocyte | *SURF4* | -0.806888815 | 0.000640631 | 0.004990048 | 1 |
| T2DM | leukocyte | *SURF1* | -0.936589508 | 0.002372469 | 0.005551774 | 1 |
| T2DM | leukocyte | *SURF6* | -1.330926103 | 0.000996049 | 0.004990048 | 1 |
| T2DM | leukocyte | *SH3GLB2* | -1.461869036 | 0.000837461 | 0.004990048 | 1 |
| T2DM | leukocyte | *LCN2* | -0.360432103 | 0.003704245 | 0.006868344 | 1 |
| T2DM | leukocyte | *GPR107* | -1.167222199 | 0.00247985 | 0.005622792 | 1 |
| T2DM | leukocyte | *INPP5E* | -1.182369919 | 0.033901346 | 0.042678112 | 1 |
| T2DM | leukocyte | *SEC16A* | -0.853898289 | 0.02885791 | 0.03665785 | 1 |
| T2DM | leukocyte | *MSRB2* | -1.446712349 | 0.000529369 | 0.004990048 | 1 |
| T2DM | leukocyte | *RSU1* | -1.134782079 | 0.001712566 | 0.005068796 | 1 |
| T2DM | leukocyte | *GLUD1* | -1.070389926 | 0.001100103 | 0.004990048 | 1 |
| T2DM | leukocyte | *FRA10AC1* | -0.660584526 | 0.003200066 | 0.006353934 | 1 |
| T2DM | leukocyte | *DNAJB12* | -1.169007371 | 0.000955882 | 0.004990048 | 1 |
| T2DM | leukocyte | *LRRC27* | -1.892392806 | 0.038541074 | 0.048011376 | 1 |
| T2DM | leukocyte | *MTG1* | -0.769453972 | 0.002485025 | 0.005622792 | 1 |
| T2DM | leukocyte | *PAOX* | -5.372634293 | 7.53E-05 | 0.004990048 | 0.295792562 |
| T2DM | leukocyte | *GSTO1* | -1.117179882 | 0.001698944 | 0.005060368 | 1 |
| T2DM | leukocyte | *HSD17B12* | -0.880015286 | 1.65E-06 | 0.00053944 | 0.006473279 |
| T2DM | leukocyte | *APIP* | -1.241822897 | 6.30E-06 | 0.001302453 | 0.0247466 |
| T2DM | leukocyte | *PTPRJ* | -1.191776503 | 0.001873741 | 0.005146173 | 1 |
| T2DM | leukocyte | *CCDC82* | -0.9648036 | 0.000636617 | 0.004990048 | 1 |
| T2DM | leukocyte | *RPS3* | -0.596076518 | 0.000944203 | 0.004990048 | 1 |
| T2DM | leukocyte | *AASDHPPT* | -0.949857415 | 0.00135745 | 0.004990048 | 1 |
| T2DM | leukocyte | *GLB1L2* | -0.719089678 | 0.019269342 | 0.025439934 | 1 |
| T2DM | leukocyte | *TMC2* | -54.75723831 | 0.022064521 | 0.028762941 | 1 |
| T2DM | leukocyte | *EML3* | -4.043515278 | 0.000807338 | 0.004990048 | 1 |
| T2DM | leukocyte | *CCDC15* | -1.007571235 | 0.003390606 | 0.006510382 | 1 |
| T2DM | leukocyte | *CHEK1* | -1.102993945 | 0.002660561 | 0.00583507 | 1 |
| T2DM | leukocyte | *SIDT2* | -1.217430247 | 0.000666119 | 0.004990048 | 1 |
| T2DM | leukocyte | *TAGLN* | -0.872778267 | 0.000315791 | 0.004990048 | 1 |
| T2DM | leukocyte | *DUSP15* | -1.052205185 | 0.004730582 | 0.007750816 | 1 |
| T2DM | leukocyte | *DSN1* | -1.079967704 | 0.001873827 | 0.005146173 | 1 |
| T2DM | leukocyte | *YTHDF1* | -0.999975943 | 0.001400539 | 0.004990048 | 1 |
| T2DM | leukocyte | *CABLES2* | -1.130083362 | 0.003858247 | 0.007033267 | 1 |
| T2DM | leukocyte | *TRPT1* | -0.852957579 | 0.003301766 | 0.006432819 | 1 |
| T2DM | leukocyte | *MPP7* | -0.700200571 | 0.00370091 | 0.006868344 | 1 |
| T2DM | leukocyte | *DCUN1D2* | -1.021715528 | 0.005661404 | 0.008886798 | 1 |
| T2DM | leukocyte | *TMCO3* | -0.999393951 | 0.002423604 | 0.005602682 | 1 |
| T2DM | leukocyte | *TMEM218* | -1.413596789 | 0.001092705 | 0.004990048 | 1 |
| T2DM | leukocyte | *MIA2* | -0.898289579 | 0.000966553 | 0.004990048 | 1 |
| T2DM | leukocyte | *LYPD1* | -1.373967214 | 0.003798098 | 0.006966727 | 1 |
| T2DM | leukocyte | *LYPD6B* | -2.243744379 | 0.003812987 | 0.006987512 | 1 |
| T2DM | leukocyte | *WDR17* | -0.724679723 | 0.003888566 | 0.00705773 | 1 |
| T2DM | leukocyte | *CNDP1* | -3.249385655 | 0.027774816 | 0.035327696 | 1 |
| T2DM | leukocyte | *PRSS23* | -0.697770963 | 0.001833795 | 0.005146173 | 1 |
| T2DM | leukocyte | *MTMR12* | -0.754714678 | 0.001772148 | 0.005097196 | 1 |

| | | | | | | |
|---|---|---|---|---|---|---|
| T2DM | leukocyte | *DOCK1* | -0.93556329 | 0.001755779 | 0.005084476 | 1 |
| T2DM | leukocyte | *IL18* | 0.409843259 | 0.0322199 | 0.040731013 | 1 |
| T2DM | leukocyte | *TMEM86A* | -1.05318579 | 0.004149033 | 0.007238372 | 1 |
| T2DM | leukocyte | *PLBD2* | -2.168347339 | 0.008295045 | 0.012179085 | 1 |
| T2DM | leukocyte | *DLG5* | -0.898135253 | 0.001235026 | 0.004990048 | 1 |
| T2DM | leukocyte | *SLC2A13* | -0.736371047 | 0.001030178 | 0.004990048 | 1 |
| T2DM | leukocyte | *MAGI1* | -0.740701909 | 0.011254469 | 0.015860404 | 1 |
| T2DM | leukocyte | *CSNK1G3* | -0.946903382 | 0.004021403 | 0.00715584 | 1 |
| T2DM | leukocyte | *AKAP6* | -1.075517887 | 0.002456575 | 0.005621365 | 1 |
| T2DM | leukocyte | *MBIP* | -1.014165191 | 0.001606988 | 0.005033031 | 1 |
| T2DM | leukocyte | *MIPOL1* | -0.776550055 | 0.005082283 | 0.008170331 | 1 |
| T2DM | leukocyte | *TMEM18* | -0.96561974 | 0.001487295 | 0.004990249 | 1 |
| T2DM | leukocyte | *NDUFC2* | -0.740200384 | 0.000965435 | 0.004990048 | 1 |
| T2DM | leukocyte | *VIPAS39* | -1.659873532 | 0.000464176 | 0.004990048 | 1 |
| T2DM | leukocyte | *C4orf33* | -1.91961594 | 0.001669659 | 0.005048289 | 1 |
| T2DM | leukocyte | *QDPR* | -0.652275838 | 0.002686021 | 0.00586252 | 1 |
| T2DM | leukocyte | *TEX9* | -1.717461053 | 0.000635785 | 0.004990048 | 1 |
| T2DM | leukocyte | *ZNF827* | -0.81829939 | 0.002286997 | 0.005462378 | 1 |
| T2DM | leukocyte | *AKR1C2* | -2.619840941 | 0.000111701 | 0.004990048 | 0.438874434 |
| T2DM | leukocyte | *KIN* | -1.59008076 | 0.001093862 | 0.004990048 | 1 |
| T2DM | leukocyte | *TMEM45B* | -1.198750869 | 0.012054251 | 0.016860503 | 1 |
| T2DM | leukocyte | *BICD1* | -0.736773478 | 0.003980738 | 0.007132431 | 1 |
| T2DM | leukocyte | *GUF1* | -2.366468819 | 0.000437128 | 0.004990048 | 1 |
| T2DM | leukocyte | *CCL28* | -1.109334786 | 0.001917435 | 0.005160002 | 1 |
| T2DM | leukocyte | *CACUL1* | -1.346086059 | 0.000615783 | 0.004990048 | 1 |
| T2DM | leukocyte | *KCNE4* | -1.112068928 | 0.00307593 | 0.00625211 | 1 |
| T2DM | leukocyte | *RABGAP1L* | -1.036246187 | 0.001272821 | 0.004990048 | 1 |
| T2DM | leukocyte | *PTPN14* | -1.147239902 | 1.98E-05 | 0.0030497 | 0.077661022 |
| T2DM | leukocyte | *ARL14EP* | -1.298332702 | 0.00052144 | 0.004990048 | 1 |
| T2DM | leukocyte | *EPG5* | -1.530313673 | 0.000875404 | 0.004990048 | 1 |
| T2DM | leukocyte | *HAUS1* | -1.2609468 | 0.001050086 | 0.004990048 | 1 |
| T2DM | leukocyte | *SPC25* | -1.10275084 | 0.002096052 | 0.005283012 | 1 |
| T2DM | leukocyte | *PDK1* | -1.10054597 | 0.002147644 | 0.005327078 | 1 |
| T2DM | leukocyte | *PDE3B* | -0.53249322 | 0.003417578 | 0.006539461 | 1 |
| T2DM | leukocyte | *TGOLN2* | -0.941013385 | 0.000851156 | 0.004990048 | 1 |
| T2DM | leukocyte | *ATG10* | -0.703094281 | 0.02686433 | 0.034280595 | 1 |
| T2DM | leukocyte | *POC5* | -1.711392318 | 4.94E-06 | 0.001213482 | 0.019415718 |
| T2DM | leukocyte | *GUCY1A2* | -0.862683639 | 0.004085351 | 0.007197912 | 1 |
| T2DM | leukocyte | *ZNF773* | -0.659199775 | 0.019870772 | 0.026163627 | 1 |
| T2DM | leukocyte | *DCLRE1C* | -2.63974418 | 0.001761396 | 0.005084476 | 1 |
| T2DM | leukocyte | *ZSCAN1* | -0.990409847 | 0.00098127 | 0.004990048 | 1 |
| T2DM | leukocyte | *USP12* | -0.83632393 | 0.001880055 | 0.005146173 | 1 |
| T2DM | leukocyte | *CCDC50* | -0.625148441 | 0.006521126 | 0.009977222 | 1 |
| T2DM | leukocyte | *PLEKHH2* | -0.745902593 | 0.017374433 | 0.023242815 | 1 |
| T2DM | leukocyte | *GRIA4* | -1.216432932 | 0.001477159 | 0.004990048 | 1 |
| T2DM | leukocyte | *IGSF10* | -1.951943102 | 0.007412897 | 0.011095342 | 1 |
| T2DM | leukocyte | *SPEF2* | -0.885807228 | 0.013266205 | 0.018344791 | 1 |
| T2DM | leukocyte | *GPD1L* | -0.927526873 | 0.006646088 | 0.010135575 | 1 |
| T2DM | leukocyte | *CCNO* | -0.98058684 | 0.005411485 | 0.008573276 | 1 |
| T2DM | leukocyte | *RASGRP3* | -0.851618378 | 0.016011507 | 0.021640595 | 1 |

| | | | | | | |
|---|---|---|---|---|---|---|
| T2DM | leukocyte | *CATSPER3* | -4.751227344 | 0.004052228 | 0.007180398 | 1 |
| T2DM | leukocyte | *GPR180* | -0.847371852 | 0.001566306 | 0.005009694 | 1 |
| T2DM | leukocyte | *IFIT5* | -0.935678144 | 0.003596161 | 0.006744304 | 1 |
| T2DM | leukocyte | *MED21* | -1.554554185 | 0.000282974 | 0.004990048 | 1 |
| T2DM | leukocyte | *LGI2* | -1.203180185 | 0.01061915 | 0.015056889 | 1 |
| T2DM | leukocyte | *SRP19* | -0.779821318 | 0.001125607 | 0.004990048 | 1 |
| T2DM | leukocyte | *DAB2* | -1.23548529 | 0.001044733 | 0.004990048 | 1 |
| T2DM | leukocyte | *ACMSD* | -1.121232887 | 0.006665751 | 0.010154996 | 1 |
| T2DM | leukocyte | *CAST* | -0.818066281 | 0.001045649 | 0.004990048 | 1 |
| T2DM | leukocyte | *SYCP2L* | -2.135205088 | 0.017098061 | 0.022935569 | 1 |
| T2DM | leukocyte | *MERTK* | 0.457245146 | 0.036854538 | 0.046100439 | 1 |
| T2DM | leukocyte | *TMEM87B* | -0.965997047 | 0.002989958 | 0.006163456 | 1 |
| T2DM | leukocyte | *PLA2R1* | -2.975885562 | 0.003914054 | 0.007085756 | 1 |
| T2DM | leukocyte | *SLC25A27* | -1.399777192 | 0.000620196 | 0.004990048 | 1 |
| T2DM | leukocyte | *ASAP1* | -0.833816058 | 0.002053993 | 0.00526208 | 1 |
| T2DM | leukocyte | *NMRAL1* | -0.934259025 | 0.010333262 | 0.014709923 | 1 |
| T2DM | leukocyte | *CMTM7* | -0.798854794 | 0.002029984 | 0.005251028 | 1 |
| T2DM | leukocyte | *RPIA* | -0.958360986 | 0.001502434 | 0.004993935 | 1 |
| T2DM | leukocyte | *PTPRD* | -0.692030572 | 0.028631256 | 0.036393466 | 1 |
| T2DM | leukocyte | *GTF2E1* | -2.332693775 | 0.000625992 | 0.004990048 | 1 |
| T2DM | leukocyte | *CFDP1* | -0.7378749 | 0.003824536 | 0.007002145 | 1 |
| T2DM | leukocyte | *ZDHHC7* | -1.191314215 | 0.017803909 | 0.023720433 | 1 |
| T2DM | leukocyte | *JAZF1* | -1.976653238 | 1.46E-08 | 1.91E-05 | 5.74E-05 |
| T2DM | leukocyte | *CMIP* | -0.95771465 | 0.001340415 | 0.004990048 | 1 |
| T2DM | leukocyte | *PID1* | -1.659783995 | 0.009224111 | 0.013319197 | 1 |
| T2DM | leukocyte | *KCTD15* | -1.220925024 | 0.004100073 | 0.00720956 | 1 |
| T2DM | leukocyte | *LGI4* | 1.13749046 | 0.006647843 | 0.010135575 | 1 |
| T2DM | leukocyte | *DDAH1* | -2.145097492 | 0.000478761 | 0.004990048 | 1 |
| T2DM | leukocyte | *NUS1* | -1.127427492 | 0.000402033 | 0.004990048 | 1 |
| T2DM | leukocyte | *PPP2R5E* | -0.990913768 | 0.002303598 | 0.005478715 | 1 |
| T2DM | leukocyte | *ROBO3* | -2.562603018 | 0.00414361 | 0.007238335 | 1 |
| T2DM | leukocyte | *PITPNC1* | -1.034860508 | 0.001488698 | 0.004990695 | 1 |
| T2DM | leukocyte | *CC2D1B* | -2.243991756 | 0.000138459 | 0.004990048 | 0.544006304 |
| T2DM | leukocyte | *CEP112* | -0.738183732 | 0.003324367 | 0.006456709 | 1 |
| T2DM | leukocyte | *ABCA5* | -1.35912768 | 0.001185932 | 0.004990048 | 1 |
| T2DM | leukocyte | *SRSF12* | -1.466104343 | 0.008359669 | 0.01225565 | 1 |
| T2DM | leukocyte | *PDLIM3* | -0.572246802 | 0.005794184 | 0.009041044 | 1 |
| T2DM | leukocyte | *PDE1C* | -0.922566096 | 0.021497729 | 0.028126732 | 1 |
| T2DM | leukocyte | *RABGEF1* | -1.042776215 | 0.0007342 | 0.004990048 | 1 |
| T2DM | leukocyte | *MRPL39* | -0.983440696 | 0.00434609 | 0.007358292 | 1 |
| T2DM | leukocyte | *TSEN2* | -1.991582393 | 0.002914453 | 0.006099708 | 1 |
| T2DM | leukocyte | *FLCN* | -1.507357092 | 0.000825618 | 0.004990048 | 1 |
| T2DM | leukocyte | *MPPE1* | -1.103742451 | 0.002051425 | 0.00526208 | 1 |
| T2DM | leukocyte | *RAB6B* | -1.110918904 | 0.002387127 | 0.005560842 | 1 |
| T2DM | leukocyte | *ANKRD40* | -1.167458568 | 0.003200416 | 0.006353934 | 1 |
| T2DM | leukocyte | *ATP6V1C1* | -0.852452566 | 0.001864989 | 0.005146173 | 1 |
| T2DM | leukocyte | *MARVELD1* | -3.312087207 | 0.000694358 | 0.004990048 | 1 |
| T2DM | leukocyte | *TRMT44* | -0.7958612 | 0.005275088 | 0.00840804 | 1 |
| T2DM | leukocyte | *PPM1J* | -2.176716639 | 0.004541069 | 0.007568852 | 1 |
| T2DM | leukocyte | *DBI* | -0.936689713 | 0.00056956 | 0.004990048 | 1 |

| | | | | | | |
|---|---|---|---|---|---|---|
| T2DM | leukocyte | *LRGUK* | -3.13770258 | 0.001433896 | 0.004990048 | 1 |
| T2DM | leukocyte | *SETD9* | -1.161256733 | 0.000316666 | 0.004990048 | 1 |
| T2DM | leukocyte | *MIER3* | -2.106195334 | 0.000283205 | 0.004990048 | 1 |
| T2DM | leukocyte | *TTN* | -2.352383882 | 0.000795301 | 0.004990048 | 1 |
| T2DM | leukocyte | *DEPTOR* | -0.854788794 | 0.013911948 | 0.019114815 | 1 |
| T2DM | leukocyte | *RMND1* | -1.738436074 | 0.000450186 | 0.004990048 | 1 |
| T2DM | leukocyte | *TMBIM4* | -0.848409282 | 0.000964263 | 0.004990048 | 1 |
| T2DM | leukocyte | *PSD3* | -0.788888475 | 0.002583503 | 0.00572832 | 1 |
| T2DM | leukocyte | *MMP16* | -0.730042125 | 0.00493838 | 0.007991307 | 1 |
| T2DM | leukocyte | *ADK* | -0.970553546 | 0.002860126 | 0.006048134 | 1 |
| T2DM | leukocyte | *BATF* | -12.66145167 | 0.003963995 | 0.007124893 | 1 |
| T2DM | leukocyte | *DRAM2* | -1.328562029 | 0.005737341 | 0.008970609 | 1 |
| T2DM | leukocyte | *ART3* | -3.595878446 | 0.013914067 | 0.019114815 | 1 |
| T2DM | leukocyte | *CCT8* | -0.643752121 | 0.006266913 | 0.009633295 | 1 |
| T2DM | leukocyte | *CDK20* | -0.860710073 | 0.002814742 | 0.006001505 | 1 |
| T2DM | leukocyte | *TDRD9* | -0.644582269 | 0.001162778 | 0.004990048 | 1 |
| T2DM | leukocyte | *HKDC1* | -1.9475662 | 0.010256161 | 0.014621356 | 1 |
| T2DM | leukocyte | *ZDHHC5* | -1.103137618 | 0.001317447 | 0.004990048 | 1 |
| T2DM | leukocyte | *MED19* | -0.907293665 | 0.001264206 | 0.004990048 | 1 |
| T2DM | leukocyte | *MAPK13* | -0.90636517 | 0.008558595 | 0.012491352 | 1 |
| T2DM | leukocyte | *COX6A2* | -0.910984703 | 0.019842139 | 0.026143448 | 1 |
| T2DM | leukocyte | *MPV17L* | -0.847735259 | 0.003886712 | 0.00705773 | 1 |
| T2DM | leukocyte | *TATDN2* | -1.599882392 | 0.001391579 | 0.004990048 | 1 |
| T2DM | leukocyte | *EXOG* | -1.21448821 | 0.000871577 | 0.004990048 | 1 |
| T2DM | leukocyte | *ZFYVE9* | -0.988715594 | 0.002065792 | 0.005273744 | 1 |
| T2DM | leukocyte | *SLC6A1* | -5.032831268 | 0.004062338 | 0.007184034 | 1 |
| T2DM | leukocyte | *NECAP2* | -1.204455244 | 0.001213775 | 0.004990048 | 1 |
| T2DM | leukocyte | *STEAP2* | -1.121767354 | 0.012539299 | 0.017458135 | 1 |
| T2DM | leukocyte | *GATAD1* | -1.220404868 | 0.000948787 | 0.004990048 | 1 |
| T2DM | leukocyte | *CLEC18A* | -5.129525168 | 2.07E-08 | 2.03E-05 | 8.13E-05 |
| T2DM | leukocyte | *DHRS4* | -1.120878679 | 0.001070188 | 0.004990048 | 1 |
| T2DM | leukocyte | *DHRS1* | -1.017160373 | 0.007022112 | 0.010591123 | 1 |
| T2DM | leukocyte | *AASDH* | -0.665446764 | 0.007242914 | 0.010874058 | 1 |
| T2DM | leukocyte | *SLC38A10* | -1.083640923 | 0.001191495 | 0.004990048 | 1 |
| T2DM | leukocyte | *SLC37A3* | -0.709642384 | 0.002788176 | 0.005983705 | 1 |
| T2DM | leukocyte | *AP3S2* | -0.930391748 | 0.01051333 | 0.014928397 | 1 |
| T2DM | leukocyte | *SPPL3* | -1.090585659 | 0.00082517 | 0.004990048 | 1 |
| T2DM | leukocyte | *C12orf43* | -1.109460268 | 0.002480005 | 0.005622792 | 1 |
| T2DM | leukocyte | *PEX10* | -1.538053467 | 0.000985696 | 0.004990048 | 1 |
| T2DM | leukocyte | *LDLRAP1* | -0.756927214 | 0.001945242 | 0.005192687 | 1 |
| T2DM | leukocyte | *KRTCAP3* | -0.898779573 | 0.002330683 | 0.005513096 | 1 |
| T2DM | leukocyte | *PAFAH2* | -2.697328128 | 0.000640599 | 0.004990048 | 1 |
| T2DM | leukocyte | *EXTL1* | -6.151233395 | 3.88E-05 | 0.004122285 | 0.152524531 |
| T2DM | leukocyte | *TRIM63* | -14.28003871 | 0.000632682 | 0.004990048 | 1 |
| T2DM | leukocyte | *XKR8* | -1.740664029 | 0.00083304 | 0.004990048 | 1 |
| T2DM | leukocyte | *CNNM4* | -1.523492394 | 0.003491433 | 0.006628411 | 1 |
| T2DM | leukocyte | *ESYT3* | -1.746787929 | 0.00459748 | 0.007615304 | 1 |
| T2DM | leukocyte | *FAIM* | -0.973632464 | 0.001322913 | 0.004990048 | 1 |
| T2DM | leukocyte | *AHCYL2* | -0.996301166 | 0.007073861 | 0.010655705 | 1 |
| T2DM | leukocyte | *COPG2* | -0.915066436 | 0.002184763 | 0.005356498 | 1 |

| | | | | | | |
|---|---|---|---|---|---|---|
| T2DM | leukocyte | *RNF166* | -1.183964634 | 0.003089404 | 0.00625753 | 1 |
| T2DM | leukocyte | *NIT1* | -1.098430275 | 0.014133573 | 0.019389249 | 1 |
| T2DM | leukocyte | *NPM2* | -1.025776466 | 0.003584391 | 0.006728653 | 1 |
| T2DM | leukocyte | *VWA5B1* | 2.151589599 | 0.022317913 | 0.029073965 | 1 |
| T2DM | leukocyte | *CDA* | -10.47558898 | 0.005706297 | 0.008935847 | 1 |
| T2DM | leukocyte | *PINK1* | -1.207355754 | 0.00060505 | 0.004990048 | 1 |
| T2DM | leukocyte | *DMTN* | -0.853318667 | 0.013274177 | 0.018344791 | 1 |
| T2DM | leukocyte | *CCAR2* | -1.368456026 | 0.001036622 | 0.004990048 | 1 |
| T2DM | leukocyte | *EPB41* | -0.896401283 | 0.002412243 | 0.005588268 | 1 |
| T2DM | leukocyte | *MIS18A* | -1.276735999 | 0.000363949 | 0.004990048 | 1 |
| T2DM | leukocyte | *ALG8* | -0.844068754 | 0.003581943 | 0.006727272 | 1 |
| T2DM | leukocyte | *IFNAR2* | -1.752741924 | 0.002281568 | 0.005462378 | 1 |
| T2DM | leukocyte | *IFNGR2* | -0.964232459 | 0.001089632 | 0.004990048 | 1 |
| T2DM | leukocyte | *C1QC* | 0.047879092 | 0.01612283 | 0.021783562 | 1 |
| T2DM | leukocyte | *GJD2* | -0.602116492 | 0.001953954 | 0.005198138 | 1 |
| T2DM | leukocyte | *CLDN14* | -1.890399695 | 0.01183766 | 0.016598917 | 1 |
| T2DM | leukocyte | *HLCS* | -0.797894934 | 0.004804649 | 0.007849258 | 1 |
| T2DM | leukocyte | *THEM4* | -0.775893743 | 0.001855881 | 0.005146173 | 1 |
| T2DM | leukocyte | *UBR1* | -1.200380999 | 0.001549396 | 0.005009694 | 1 |
| T2DM | leukocyte | *TEPP* | 2.74706872 | 0.038520977 | 0.04800156 | 1 |
| T2DM | leukocyte | *ZDHHC1* | -0.985247514 | 0.001480231 | 0.004990048 | 1 |
| T2DM | leukocyte | *ZFYVE28* | -1.25696638 | 0.002548529 | 0.005679621 | 1 |
| T2DM | leukocyte | *LYPD5* | -9.337893481 | 0.022422233 | 0.029180839 | 1 |
| T2DM | leukocyte | *NPR2* | -0.798291537 | 0.038338723 | 0.047789607 | 1 |
| T2DM | leukocyte | *ATAD3B* | -2.37850639 | 0.000927511 | 0.004990048 | 1 |
| T2DM | leukocyte | *ANKLE1* | 0.750289931 | 0.000799038 | 0.004990048 | 1 |
| T2DM | leukocyte | *KALRN* | -1.166919992 | 0.007407499 | 0.011091488 | 1 |
| T2DM | leukocyte | *CILP2* | -3.410474918 | 4.94E-06 | 0.001213482 | 0.019394961 |
| T2DM | leukocyte | *TMPRSS3* | -2.376034051 | 0.01168416 | 0.016401238 | 1 |
| T2DM | leukocyte | *RSPH1* | -0.624588386 | 0.003250574 | 0.006400784 | 1 |
| T2DM | leukocyte | *SLC37A1* | -0.916496102 | 0.005337682 | 0.008483718 | 1 |
| T2DM | leukocyte | *NDUFV3* | -0.538649 | 0.018980712 | 0.025084163 | 1 |
| T2DM | leukocyte | *HSF2BP* | -3.124326463 | 0.000849859 | 0.004990048 | 1 |
| T2DM | leukocyte | *PDXK* | -1.27498898 | 0.000184241 | 0.004990048 | 0.723882111 |
| T2DM | leukocyte | *CSTB* | -1.116346326 | 0.000572296 | 0.004990048 | 1 |
| T2DM | leukocyte | *ICOSLG* | 0.356871198 | 0.038012188 | 0.047397615 | 1 |
| T2DM | leukocyte | *ZNF66* | -2.123637664 | 0.0112755 | 0.015873701 | 1 |
| T2DM | leukocyte | *FTCD* | -1.46745813 | 0.000461234 | 0.004990048 | 1 |
| T2DM | leukocyte | *SPATC1L* | -0.76277273 | 0.001564842 | 0.005009694 | 1 |
| T2DM | leukocyte | *LSS* | -0.945511562 | 0.006432806 | 0.009861293 | 1 |
| T2DM | leukocyte | *VAV2* | -1.40543938 | 0.002429399 | 0.005602682 | 1 |
| T2DM | leukocyte | *MCM3AP* | -1.228872994 | 0.011588159 | 0.016278112 | 1 |
| T2DM | leukocyte | *DIP2A* | -0.739488406 | 0.005470502 | 0.008631969 | 1 |
| T2DM | leukocyte | *ZNF208* | -0.941311289 | 0.00077862 | 0.004990048 | 1 |
| T2DM | leukocyte | *SLC2A6* | -1.711907136 | 0.001087114 | 0.004990048 | 1 |
| T2DM | leukocyte | *RDH13* | -1.088254597 | 0.00277077 | 0.005961859 | 1 |
| T2DM | leukocyte | *MPZL3* | -1.000640533 | 0.002775549 | 0.005965608 | 1 |
| T2DM | leukocyte | *S100A1* | -1.612139786 | 0.018364252 | 0.024409047 | 1 |
| T2DM | leukocyte | *VPS11* | -2.231250736 | 6.80E-05 | 0.004990048 | 0.267158568 |
| T2DM | leukocyte | *PAQR6* | -1.264047314 | 0.000475363 | 0.004990048 | 1 |

| | | | | | | |
|---|---|---|---|---|---|---|
| T2DM | leukocyte | *SLC25A44* | -1.556597483 | 0.001031227 | 0.004990048 | 1 |
| T2DM | leukocyte | *CCR5* | -108.9403803 | 0.008838241 | 0.012823283 | 1 |
| T2DM | leukocyte | *NBEAL2* | -0.653732898 | 0.005550278 | 0.008740298 | 1 |
| T2DM | leukocyte | *LY6E* | -1.111046757 | 0.000605008 | 0.004990048 | 1 |
| T2DM | leukocyte | *VPS28* | -0.745318804 | 0.004127745 | 0.00723368 | 1 |
| T2DM | leukocyte | *TONSL* | -4.7331673 | 0.00100142 | 0.004990048 | 1 |
| T2DM | leukocyte | *ZNF333* | -3.417668516 | 0.000884875 | 0.004990048 | 1 |
| T2DM | leukocyte | *RPL8* | -0.583391682 | 0.000902726 | 0.004990048 | 1 |
| T2DM | leukocyte | *MAML1* | -1.651152697 | 0.002478418 | 0.005622792 | 1 |
| T2DM | leukocyte | *LRWD1* | -1.266227308 | 0.004290068 | 0.00732537 | 1 |
| T2DM | leukocyte | *U2AF1L4* | -0.758062865 | 0.002492997 | 0.005623364 | 1 |
| T2DM | leukocyte | *ZNF382* | -0.828378842 | 0.008548665 | 0.012481496 | 1 |
| T2DM | leukocyte | *DUSP14* | -1.192484656 | 0.001599612 | 0.005024491 | 1 |
| T2DM | leukocyte | *PGAP3* | -0.955431924 | 0.003953822 | 0.00712268 | 1 |
| T2DM | leukocyte | *PRPSAP1* | -1.000339248 | 0.000583417 | 0.004990048 | 1 |
| T2DM | leukocyte | *SIGLEC11* | -12.59257378 | 0.018690336 | 0.024775415 | 1 |
| T2DM | leukocyte | *SPC24* | -2.539594273 | 0.00411682 | 0.007227428 | 1 |
| T2DM | leukocyte | *IP6K3* | 2.196481105 | 0.007729212 | 0.011485655 | 1 |
| T2DM | leukocyte | *LEMD2* | -1.829184097 | 0.002113782 | 0.005283012 | 1 |
| T2DM | leukocyte | *RPL26* | -0.625789956 | 0.000802861 | 0.004990048 | 1 |
| T2DM | leukocyte | *SNRNP25* | -0.819828595 | 0.001770435 | 0.005097196 | 1 |
| T2DM | leukocyte | *WDR90* | -1.436846809 | 0.002138249 | 0.005317933 | 1 |
| T2DM | leukocyte | *CCDC78* | -2.567996882 | 0.006298776 | 0.009667145 | 1 |
| T2DM | leukocyte | *ZNF75A* | -2.024259783 | 0.000169122 | 0.004990048 | 0.664481785 |
| T2DM | leukocyte | *ASRGL1* | -0.753391375 | 0.00236826 | 0.005551753 | 1 |
| T2DM | leukocyte | *TAF6L* | -1.044558049 | 0.012073688 | 0.01688168 | 1 |
| T2DM | leukocyte | *ZFPL1* | -1.025157016 | 0.002231945 | 0.005430873 | 1 |
| T2DM | leukocyte | *LRP5* | -1.816578183 | 0.00081092 | 0.004990048 | 1 |
| T2DM | leukocyte | *PDZK1IP1* | -0.671379358 | 0.001798597 | 0.005114303 | 1 |
| T2DM | leukocyte | *CTRC* | -0.467761302 | 0.002091084 | 0.005283012 | 1 |
| T2DM | leukocyte | *SDC3* | -1.746466573 | 0.00253143 | 0.005660135 | 1 |
| T2DM | leukocyte | *OMA1* | -1.187570129 | 0.007550307 | 0.011262397 | 1 |
| T2DM | leukocyte | *TM2D1* | -1.386490251 | 0.000694724 | 0.004990048 | 1 |
| T2DM | leukocyte | *FUBP1* | -0.785221387 | 0.001397041 | 0.004990048 | 1 |
| T2DM | leukocyte | *SNX7* | -0.722102423 | 0.00341469 | 0.006538167 | 1 |
| T2DM | leukocyte | *HFM1* | -1.739733908 | 0.003262174 | 0.006405338 | 1 |
| T2DM | leukocyte | *VCAM1* | -0.486720708 | 0.004551879 | 0.007568852 | 1 |
| T2DM | leukocyte | *ZNF496* | -0.903593596 | 0.002624958 | 0.005790825 | 1 |
| T2DM | leukocyte | *TRIM58* | -3.421142457 | 0.007959497 | 0.011761137 | 1 |
| T2DM | leukocyte | *PEA15* | -0.844893531 | 0.002098687 | 0.005283012 | 1 |
| T2DM | leukocyte | *FLVCR1* | -0.722712963 | 0.002448664 | 0.005621365 | 1 |
| T2DM | leukocyte | *TDRD5* | -2.735929873 | 0.009303471 | 0.0134061 | 1 |
| T2DM | leukocyte | *ACP6* | -1.279648192 | 0.001315027 | 0.004990048 | 1 |
| T2DM | leukocyte | *OXER1* | -2.969244011 | 0.014158311 | 0.019416406 | 1 |
| T2DM | leukocyte | *HAAO* | -1.019457339 | 0.003884999 | 0.00705773 | 1 |
| T2DM | leukocyte | *RFTN2* | -1.551451334 | 0.024216655 | 0.031277856 | 1 |
| T2DM | leukocyte | *KCNJ3* | -0.732769114 | 0.009096599 | 0.013154413 | 1 |
| T2DM | leukocyte | *DUSP19* | -1.434182084 | 0.001558346 | 0.005009694 | 1 |
| T2DM | leukocyte | *SMC6* | -1.658072028 | 0.000691871 | 0.004990048 | 1 |
| T2DM | leukocyte | *NOSTRIN* | -0.932815105 | 0.005200197 | 0.008312275 | 1 |

| | | | | | | |
|---|---|---|---|---|---|---|
| T2DM | leukocyte | *CTSS* | 0.176684482 | 0.030218638 | 0.038336786 | 1 |
| T2DM | leukocyte | *MSX1* | -0.988326092 | 0.003346871 | 0.006472193 | 1 |
| T2DM | leukocyte | *PACRGL* | -0.982335575 | 0.004195601 | 0.007249127 | 1 |
| T2DM | leukocyte | *ERCC3* | -1.142917186 | 0.004115426 | 0.007227428 | 1 |
| T2DM | leukocyte | *TDRD10* | -15.36832206 | 0.004035328 | 0.007166394 | 1 |
| T2DM | leukocyte | *CCNYL1* | -2.394711403 | 0.000484715 | 0.004990048 | 1 |
| T2DM | leukocyte | *DCAF16* | -1.32999112 | 0.000350925 | 0.004990048 | 1 |
| T2DM | leukocyte | *NPPC* | -10.25229789 | 0.000309087 | 0.004990048 | 1 |
| T2DM | leukocyte | *GNPDA2* | -0.727081657 | 0.006239131 | 0.009598099 | 1 |
| T2DM | leukocyte | *NIPAL1* | -0.698264979 | 0.012635689 | 0.017567454 | 1 |
| T2DM | leukocyte | *CGGBP1* | -0.793235969 | 0.004990163 | 0.008055197 | 1 |
| T2DM | leukocyte | *DAPL1* | -3.36453001 | 0.000121706 | 0.004990048 | 0.478184689 |
| T2DM | leukocyte | *EOGT* | -0.940741473 | 0.002084342 | 0.005283012 | 1 |
| T2DM | leukocyte | *POGLUT1* | -0.809901099 | 0.001306644 | 0.004990048 | 1 |
| T2DM | leukocyte | *ATP1A1* | -0.620402266 | 0.001689882 | 0.005060368 | 1 |
| T2DM | leukocyte | *ARPC2* | -0.977026803 | 0.000715118 | 0.004990048 | 1 |
| T2DM | leukocyte | *CCT3* | -0.632119156 | 0.00400471 | 0.007145551 | 1 |
| T2DM | leukocyte | *EOMES* | -3.787935813 | 0.034624333 | 0.04349073 | 1 |
| T2DM | leukocyte | *CWC22* | -1.002914776 | 0.001945836 | 0.005192687 | 1 |
| T2DM | leukocyte | *HDAC11* | -2.397552889 | 0.004133574 | 0.007236049 | 1 |
| T2DM | leukocyte | *SUCLG1* | -0.9229957 | 0.00108893 | 0.004990048 | 1 |
| T2DM | leukocyte | *ICA1L* | -1.249492095 | 1.01E-05 | 0.00188355 | 0.039554547 |
| T2DM | leukocyte | *RYBP* | -0.664636478 | 0.002934868 | 0.006117633 | 1 |
| T2DM | leukocyte | *GTPBP8* | -1.819736877 | 0.000142028 | 0.004990048 | 0.558027549 |
| T2DM | leukocyte | *CADPS* | -0.470678279 | 0.003234412 | 0.006381841 | 1 |
| T2DM | leukocyte | *ATXN7* | -1.099725226 | 0.000817462 | 0.004990048 | 1 |
| T2DM | leukocyte | *PSMD6* | -0.816347354 | 0.001345509 | 0.004990048 | 1 |
| T2DM | leukocyte | *PRICKLE2* | -0.888700014 | 0.005731819 | 0.008968665 | 1 |
| T2DM | leukocyte | *RPL9* | -0.658193072 | 0.000302985 | 0.004990048 | 1 |
| T2DM | leukocyte | *IL17RC* | -1.052284127 | 0.001146534 | 0.004990048 | 1 |
| T2DM | leukocyte | *CRELD1* | -0.611940538 | 0.009638017 | 0.013835502 | 1 |
| T2DM | leukocyte | *TTC14* | -1.569107604 | 0.000611825 | 0.004990048 | 1 |
| T2DM | leukocyte | *CXCL1* | -0.617622623 | 0.002544095 | 0.005672956 | 1 |
| T2DM | leukocyte | *CCDC158* | -2.008920993 | 0.01238746 | 0.017265105 | 1 |
| T2DM | leukocyte | *TM4SF18* | -0.80979119 | 0.010479947 | 0.014891758 | 1 |
| T2DM | leukocyte | *SNRK* | -0.662765603 | 0.002017787 | 0.005244224 | 1 |
| T2DM | leukocyte | *KIAA1143* | -0.69118531 | 0.00295314 | 0.006135605 | 1 |
| T2DM | leukocyte | *NMNAT3* | -0.707575885 | 0.009690188 | 0.013900237 | 1 |
| T2DM | leukocyte | *SMIM12* | -0.898930649 | 0.001386777 | 0.004990048 | 1 |
| T2DM | leukocyte | *TPRA1* | -1.717022647 | 0.000769184 | 0.004990048 | 1 |
| T2DM | leukocyte | *MEAF6* | -0.812802515 | 0.001327575 | 0.004990048 | 1 |
| T2DM | leukocyte | *DNALI1* | -0.939478016 | 0.002925031 | 0.006103547 | 1 |
| T2DM | leukocyte | *CAMK2N2* | -0.602340459 | 0.002008134 | 0.005232069 | 1 |
| T2DM | leukocyte | *RPN1* | -1.027985142 | 0.000724771 | 0.004990048 | 1 |
| T2DM | leukocyte | *IFT122* | -1.139365502 | 0.004016145 | 0.00714972 | 1 |
| T2DM | leukocyte | *RFC4* | -1.095212723 | 0.003240057 | 0.006387448 | 1 |
| T2DM | leukocyte | *TKT* | -0.995687853 | 0.000742366 | 0.004990048 | 1 |
| T2DM | leukocyte | *RFT1* | -1.753064676 | 0.000531944 | 0.004990048 | 1 |
| T2DM | leukocyte | *UVSSA* | -1.114070088 | 0.024557839 | 0.031676871 | 1 |
| T2DM | leukocyte | *ARHGEF3* | -0.864052959 | 0.003744284 | 0.006913201 | 1 |

| | | | | | | |
|---|---|---|---|---|---|---|
| T2DM | leukocyte | *LRPAP1* | -0.815145343 | 0.001004742 | 0.004990048 | 1 |
| T2DM | leukocyte | *PIGX* | -1.351806499 | 0.000496713 | 0.004990048 | 1 |
| T2DM | leukocyte | *EXO5* | -1.476760745 | 0.003555312 | 0.006699674 | 1 |
| T2DM | leukocyte | *ERMAP* | -3.289868963 | 0.000197665 | 0.004990048 | 0.776626095 |
| T2DM | leukocyte | *BDH2* | -1.040276259 | 0.000290675 | 0.004990048 | 1 |
| T2DM | leukocyte | *CDC25A* | -1.691258432 | 0.000768359 | 0.004990048 | 1 |
| T2DM | leukocyte | *TEX264* | -1.158829773 | 0.000696011 | 0.004990048 | 1 |
| T2DM | leukocyte | *PPM1M* | -1.536865809 | 0.000146929 | 0.004990048 | 0.57728302 |
| T2DM | leukocyte | *SAP30* | -0.870280723 | 0.002803912 | 0.006001505 | 1 |
| T2DM | leukocyte | *ANXA5* | -0.575800566 | 0.002318302 | 0.005497049 | 1 |
| T2DM | leukocyte | *CEP44* | -1.033443325 | 0.001697631 | 0.005060368 | 1 |
| T2DM | leukocyte | *HPGD* | -1.321237692 | 0.01505024 | 0.020446886 | 1 |
| T2DM | leukocyte | *ARFIP1* | -1.456814569 | 0.000471202 | 0.004990048 | 1 |
| T2DM | leukocyte | *ITGA2* | -0.597902199 | 0.005273148 | 0.00840804 | 1 |
| T2DM | leukocyte | *ELOVL7* | -0.939998663 | 0.001477123 | 0.004990048 | 1 |
| T2DM | leukocyte | *LMBRD2* | -1.143379761 | 0.000541593 | 0.004990048 | 1 |
| T2DM | leukocyte | *STARD4* | -1.272366881 | 0.000907668 | 0.004990048 | 1 |
| T2DM | leukocyte | *CMBL* | -0.984274313 | 0.00158071 | 0.005009694 | 1 |
| T2DM | leukocyte | *PRRC1* | -0.726042465 | 0.001935934 | 0.005184924 | 1 |
| T2DM | leukocyte | *AGGF1* | -1.281690447 | 0.000663804 | 0.004990048 | 1 |
| T2DM | leukocyte | *WDR41* | -1.518961263 | 0.000739852 | 0.004990048 | 1 |
| T2DM | leukocyte | *SERINC5* | -3.645017206 | 0.000773679 | 0.004990048 | 1 |
| T2DM | leukocyte | *RICTOR* | -1.356352724 | 0.000655658 | 0.004990048 | 1 |
| T2DM | leukocyte | *UTP15* | -0.80458784 | 0.003045956 | 0.006220403 | 1 |
| T2DM | leukocyte | *NSA2* | -0.57084823 | 0.004679217 | 0.00769232 | 1 |
| T2DM | leukocyte | *LEAP2* | -1.336050919 | 0.005417388 | 0.008574413 | 1 |
| T2DM | leukocyte | *DACT2* | -3.787985255 | 0.003704256 | 0.006868344 | 1 |
| T2DM | leukocyte | *PDSS2* | -0.929056159 | 0.001988352 | 0.005227473 | 1 |
| T2DM | leukocyte | *STK17A* | -0.798551455 | 0.002202992 | 0.005384049 | 1 |
| T2DM | leukocyte | *GALNT10* | -1.607683334 | 0.010984642 | 0.015535874 | 1 |
| T2DM | leukocyte | *SAP30L* | -1.27770722 | 0.001062853 | 0.004990048 | 1 |
| T2DM | leukocyte | *COG5* | -1.684647275 | 0.000540851 | 0.004990048 | 1 |
| T2DM | leukocyte | *SLU7* | -0.986920075 | 0.000956317 | 0.004990048 | 1 |
| T2DM | leukocyte | *CAMLG* | -0.763675203 | 0.001285527 | 0.004990048 | 1 |
| T2DM | leukocyte | *KIF6* | -1.642968787 | 0.002634482 | 0.005802062 | 1 |
| T2DM | leukocyte | *STEAP1* | -0.938425383 | 0.025066813 | 0.032248693 | 1 |
| T2DM | leukocyte | *FNDC1* | -4.330914903 | 0.039679886 | 0.049289368 | 1 |
| T2DM | leukocyte | *CHMP4C* | -0.890327877 | 0.001798609 | 0.005114303 | 1 |
| T2DM | leukocyte | *BRI3* | -1.237931406 | 0.000178434 | 0.004990048 | 0.701068301 |
| T2DM | leukocyte | *LMTK2* | -0.983778045 | 0.00104967 | 0.004990048 | 1 |
| T2DM | leukocyte | *PEX2* | -0.795842076 | 0.001761112 | 0.005084476 | 1 |
| T2DM | leukocyte | *MED30* | -0.939108521 | 0.001511626 | 0.004993935 | 1 |
| T2DM | leukocyte | *TNFRSF11B* | -2.809203028 | 0.003720809 | 0.006886038 | 1 |
| T2DM | leukocyte | *CSMD3* | -59.50096086 | 0.004301858 | 0.007328649 | 1 |
| T2DM | leukocyte | *OSGIN2* | -1.323287981 | 0.000742161 | 0.004990048 | 1 |
| T2DM | leukocyte | *DEFB1* | -0.409924668 | 0.003218576 | 0.006372159 | 1 |
| T2DM | leukocyte | *SUN1* | -0.73365236 | 0.001759714 | 0.005084476 | 1 |
| T2DM | leukocyte | *TMEM184A* | -1.007903516 | 0.00143947 | 0.004990048 | 1 |
| T2DM | leukocyte | *INTS1* | -0.956859144 | 0.00194218 | 0.005191039 | 1 |
| T2DM | leukocyte | *YWHAZ* | -0.832921544 | 0.000808463 | 0.004990048 | 1 |

| | | | | | | |
|---|---|---|---|---|---|---|
| T2DM | leukocyte | *DCAF13* | -0.933487823 | 0.002042933 | 0.00526208 | 1 |
| T2DM | leukocyte | *INTS8* | -1.157185747 | 0.00721561 | 0.010841351 | 1 |
| T2DM | leukocyte | *GEM* | -0.975402921 | 0.002348487 | 0.005521966 | 1 |
| T2DM | leukocyte | *RPP25L* | -1.548117303 | 0.000719816 | 0.004990048 | 1 |
| T2DM | leukocyte | *FAM219A* | -1.51139296 | 0.00081244 | 0.004990048 | 1 |
| T2DM | leukocyte | *NUDT2* | -1.076301198 | 0.000976088 | 0.004990048 | 1 |
| T2DM | leukocyte | *METTL2B* | -0.726064231 | 0.029344703 | 0.037264168 | 1 |
| T2DM | leukocyte | *FXN* | -1.803633372 | 0.002007521 | 0.005232069 | 1 |
| T2DM | leukocyte | *MAMDC2* | -2.108154744 | 0.002442916 | 0.005621365 | 1 |
| T2DM | leukocyte | *PRSS37* | -488.4734141 | 0.004352311 | 0.007361269 | 1 |
| T2DM | leukocyte | *C9orf64* | -1.276092682 | 0.001106838 | 0.004990048 | 1 |
| T2DM | leukocyte | *ANKS6* | -0.677910371 | 0.023972325 | 0.030992848 | 1 |
| T2DM | leukocyte | *CLDN3* | -0.989977434 | 0.000820193 | 0.004990048 | 1 |
| T2DM | leukocyte | *WNK2* | -1.538562703 | 0.00085634 | 0.004990048 | 1 |
| T2DM | leukocyte | *NOL6* | -1.83337211 | 0.001276297 | 0.004990048 | 1 |
| T2DM | leukocyte | *PIGO* | -2.186848831 | 8.05E-05 | 0.004990048 | 0.316381002 |
| T2DM | leukocyte | *ARHGAP12* | -1.016298104 | 0.001410139 | 0.004990048 | 1 |
| T2DM | leukocyte | *WRN* | -1.053011922 | 0.001612365 | 0.005033031 | 1 |
| T2DM | leukocyte | *CFL2* | -0.940221827 | 0.001565904 | 0.005009694 | 1 |
| T2DM | leukocyte | *PGM2L1* | -1.711984664 | 0.000541632 | 0.004990048 | 1 |
| T2DM | leukocyte | *PHYHIPL* | -0.918165446 | 0.002612801 | 0.005770487 | 1 |
| T2DM | leukocyte | *PKNOX2* | 3.201250027 | 0.021902887 | 0.028590181 | 1 |
| T2DM | leukocyte | *RPL36AL* | -0.710869845 | 0.000918301 | 0.004990048 | 1 |
| T2DM | leukocyte | *TMEM63C* | -0.812273896 | 0.002809746 | 0.006001505 | 1 |
| T2DM | leukocyte | *OXGR1* | -1.974735624 | 0.004505873 | 0.007539852 | 1 |
| T2DM | leukocyte | *VDAC2* | -0.872915457 | 0.000522061 | 0.004990048 | 1 |
| T2DM | leukocyte | *SNAPC4* | -0.912658998 | 0.003653109 | 0.006815321 | 1 |
| T2DM | leukocyte | *TSC1* | -1.333404783 | 0.001267956 | 0.004990048 | 1 |
| T2DM | leukocyte | *ZMYND19* | -0.89911052 | 0.002631911 | 0.005799651 | 1 |
| T2DM | leukocyte | *METTL17* | -1.326516733 | 0.00121593 | 0.004990048 | 1 |
| T2DM | leukocyte | *VWA2* | -2.473823449 | 0.001779024 | 0.005098084 | 1 |
| T2DM | leukocyte | *METTL3* | -0.940914285 | 0.004182954 | 0.007242343 | 1 |
| T2DM | leukocyte | *TRUB1* | -1.407986169 | 0.000577218 | 0.004990048 | 1 |
| T2DM | leukocyte | *ARHGAP42* | -1.366853391 | 0.004209721 | 0.007263735 | 1 |
| T2DM | leukocyte | *SLC39A13* | -0.903641063 | 0.001997654 | 0.005227473 | 1 |
| T2DM | leukocyte | *PSMC3* | -0.759394488 | 0.001380183 | 0.004990048 | 1 |
| T2DM | leukocyte | *CPSF2* | -0.789745512 | 0.002669158 | 0.005839155 | 1 |
| T2DM | leukocyte | *IFI27L1* | -1.005784671 | 0.00124447 | 0.004990048 | 1 |
| T2DM | leukocyte | *SMCO4* | -0.969864809 | 0.002283013 | 0.005462378 | 1 |
| T2DM | leukocyte | *ABTB2* | -1.236011644 | 0.006767654 | 0.010278358 | 1 |
| T2DM | leukocyte | *CEP57* | -0.903689932 | 0.000450011 | 0.004990048 | 1 |
| T2DM | leukocyte | *JAM3* | -0.712316176 | 0.004476914 | 0.007504245 | 1 |
| T2DM | leukocyte | *IKBIP* | -0.982282154 | 0.005682775 | 0.008916678 | 1 |
| T2DM | leukocyte | *ZFYVE19* | -0.731219261 | 0.016874261 | 0.022685009 | 1 |
| T2DM | leukocyte | *CKB* | -0.830102412 | 9.05E-05 | 0.004990048 | 0.35540689 |
| T2DM | leukocyte | *BTRC* | -1.647282273 | 0.002780672 | 0.005973352 | 1 |
| T2DM | leukocyte | *BAG5* | -0.914247685 | 0.002799492 | 0.005997385 | 1 |
| T2DM | leukocyte | *LARP6* | -0.82523617 | 0.001344363 | 0.004990048 | 1 |
| T2DM | leukocyte | *ZNF319* | -2.792984664 | 0.000626149 | 0.004990048 | 1 |
| T2DM | leukocyte | *SGPL1* | -1.955795675 | 2.81E-05 | 0.003340136 | 0.110224481 |

| | | | | | | |
|---|---|---|---|---|---|---|
| T2DM | leukocyte | *FRS2* | -1.112522672 | 0.00054824 | 0.004990048 | 1 |
| T2DM | leukocyte | *PCBD1* | -1.032508398 | 0.000186684 | 0.004990048 | 0.733481118 |
| T2DM | leukocyte | *COX11* | -1.113097137 | 0.000742702 | 0.004990048 | 1 |
| T2DM | leukocyte | *WBP1L* | -1.999895717 | 0.00123587 | 0.004990048 | 1 |
| T2DM | leukocyte | *CYB5R2* | -1.219119528 | 0.000176459 | 0.004990048 | 0.693308306 |
| T2DM | leukocyte | *TUB* | -1.351931353 | 0.000873117 | 0.004990048 | 1 |
| T2DM | leukocyte | *RIC3* | -0.726112857 | 0.001748635 | 0.005084476 | 1 |
| T2DM | leukocyte | *XRRA1* | -1.057135699 | 0.000904486 | 0.004990048 | 1 |
| T2DM | leukocyte | *CDYL2* | -0.853823715 | 0.007815845 | 0.011596849 | 1 |
| T2DM | leukocyte | *ZNF143* | -0.910379092 | 0.00234775 | 0.005521966 | 1 |
| T2DM | leukocyte | *CCDC68* | -0.852320408 | 0.007339284 | 0.011006125 | 1 |
| T2DM | leukocyte | *ZSCAN21* | -1.412478465 | 0.002429873 | 0.005602682 | 1 |
| T2DM | leukocyte | *RIMKLB* | -0.911048312 | 0.001738445 | 0.005075243 | 1 |
| T2DM | leukocyte | *GALR1* | 0.976284375 | 0.013267077 | 0.018344791 | 1 |
| T2DM | leukocyte | *ATF7IP2* | -1.355521819 | 0.000482346 | 0.004990048 | 1 |
| T2DM | leukocyte | *COG1* | -0.952702921 | 0.001053455 | 0.004990048 | 1 |
| T2DM | leukocyte | *B2M* | -0.61546798 | 0.00091579 | 0.004990048 | 1 |
| T2DM | leukocyte | *AP1G1* | -0.670156843 | 0.003105764 | 0.00627703 | 1 |
| T2DM | leukocyte | *SLFN5* | -0.745534751 | 0.002507225 | 0.005648444 | 1 |
| T2DM | leukocyte | *SAAL1* | -0.866666229 | 0.003261763 | 0.006405338 | 1 |
| T2DM | leukocyte | *SCNN1G* | -7.859895386 | 0.00338113 | 0.006503419 | 1 |
| T2DM | leukocyte | *DCTN5* | -1.074583005 | 0.00079179 | 0.004990048 | 1 |
| T2DM | leukocyte | *STAT6* | -1.67791014 | 0.000836455 | 0.004990048 | 1 |
| T2DM | leukocyte | *PATL1* | -0.862198362 | 0.000967732 | 0.004990048 | 1 |
| T2DM | leukocyte | *MRPL16* | -1.205704337 | 0.0163751 | 0.022082136 | 1 |
| T2DM | leukocyte | *GREM1* | -1.611931881 | 0.000586359 | 0.004990048 | 1 |
| T2DM | leukocyte | *TSC22D4* | -1.264917833 | 0.001297118 | 0.004990048 | 1 |
| T2DM | leukocyte | *MS4A14* | 0.179420661 | 0.020465312 | 0.02685645 | 1 |
| T2DM | leukocyte | *DIS3L* | -1.398748069 | 0.002598091 | 0.005751212 | 1 |
| T2DM | leukocyte | *RCCD1* | -2.970256587 | 2.98E-05 | 0.003343813 | 0.117033452 |
| T2DM | leukocyte | *MAPRE2* | -0.884859987 | 0.001864779 | 0.005146173 | 1 |
| T2DM | leukocyte | *NAT16* | -0.609572436 | 0.002742411 | 0.005930931 | 1 |
| T2DM | leukocyte | *GNGT2* | 0.362775212 | 0.015971717 | 0.02161656 | 1 |
| T2DM | leukocyte | *SLC27A4* | -0.879325819 | 0.010001799 | 0.014296244 | 1 |
| T2DM | leukocyte | *URM1* | -1.284891463 | 0.001088325 | 0.004990048 | 1 |
| T2DM | leukocyte | *CERCAM* | -0.762047499 | 0.006842593 | 0.010372125 | 1 |
| T2DM | leukocyte | *DOLPP1* | -1.127931173 | 0.001856891 | 0.005146173 | 1 |
| T2DM | leukocyte | *ENDOG* | -1.158664805 | 0.000840538 | 0.004990048 | 1 |
| T2DM | leukocyte | *RNF214* | -1.596415926 | 0.001456829 | 0.004990048 | 1 |
| T2DM | leukocyte | *POP5* | -1.08596641 | 0.000642636 | 0.004990048 | 1 |
| T2DM | leukocyte | *ACAA2* | -1.091732471 | 0.000422631 | 0.004990048 | 1 |
| T2DM | leukocyte | *FN3K* | -1.199474648 | 0.002745938 | 0.005931166 | 1 |
| T2DM | leukocyte | *ZNF23* | -0.847212877 | 0.003464956 | 0.006599036 | 1 |
| T2DM | leukocyte | *ZNF229* | -1.215738431 | 0.00281669 | 0.006001505 | 1 |
| T2DM | leukocyte | *ZNF180* | -0.917791783 | 0.004186401 | 0.007242787 | 1 |
| T2DM | leukocyte | *CA4* | -0.718099658 | 0.002528022 | 0.005660135 | 1 |
| T2DM | leukocyte | *GPX4* | -1.008395133 | 0.000413363 | 0.004990048 | 1 |
| T2DM | leukocyte | *ZNF641* | -1.452886146 | 0.001990325 | 0.005227473 | 1 |
| T2DM | leukocyte | *CACNB3* | -0.861325474 | 0.040056797 | 0.049694712 | 1 |
| T2DM | leukocyte | *TP53I13* | -1.113979874 | 0.000169555 | 0.004990048 | 0.666180856 |

| | | | | | | |
|---|---|---|---|---|---|---|
| T2DM | leukocyte | *TUBA1C* | -0.610821573 | 0.002001483 | 0.005227473 | 1 |
| T2DM | leukocyte | *ZNF528* | -3.839917834 | 0.00018687 | 0.004990048 | 0.734212755 |
| T2DM | leukocyte | *RAB4B* | -1.184025714 | 0.000841405 | 0.004990048 | 1 |
| T2DM | leukocyte | *TMC4* | -0.976455251 | 0.001216668 | 0.004990048 | 1 |
| T2DM | leukocyte | *C19orf33* | -0.759575132 | 0.00405982 | 0.007184034 | 1 |
| T2DM | leukocyte | *DNAAF3* | -3.091529722 | 0.003938597 | 0.007108734 | 1 |
| T2DM | leukocyte | *CHAF1A* | -1.493924839 | 0.00079042 | 0.004990048 | 1 |
| T2DM | leukocyte | *UBXN6* | -1.351553565 | 0.00036378 | 0.004990048 | 1 |
| T2DM | leukocyte | *NXN* | -0.703160834 | 0.001579054 | 0.005009694 | 1 |
| T2DM | leukocyte | *KIFC2* | -4.138293495 | 5.36E-05 | 0.004896114 | 0.21066247 |
| T2DM | leukocyte | *SLC43A2* | 0.746906879 | 0.015865051 | 0.021486999 | 1 |
| T2DM | leukocyte | *SERPINF2* | 0.391256044 | 0.020142665 | 0.026486122 | 1 |
| T2DM | leukocyte | *SRR* | -0.834201601 | 0.014756364 | 0.020103243 | 1 |
| T2DM | leukocyte | *TRPV3* | -4.540437638 | 0.001995066 | 0.005227473 | 1 |
| T2DM | leukocyte | *C19orf48* | -1.583657744 | 0.002486375 | 0.005622792 | 1 |
| T2DM | leukocyte | *ZNF83* | -1.220802688 | 0.001209996 | 0.004990048 | 1 |
| T2DM | leukocyte | *ZNF558* | -0.970904393 | 0.013813741 | 0.018990268 | 1 |
| T2DM | leukocyte | *NDUFV1* | -0.858358187 | 0.001481832 | 0.004990048 | 1 |
| T2DM | leukocyte | *ZNF232* | -0.791550158 | 0.004703931 | 0.007720459 | 1 |
| T2DM | leukocyte | *MIS12* | -1.150831874 | 0.001613307 | 0.005033031 | 1 |
| T2DM | leukocyte | *HID1* | -0.990838329 | 0.002249929 | 0.005439981 | 1 |
| T2DM | leukocyte | *SRP68* | -0.773231395 | 0.002023381 | 0.005244224 | 1 |
| T2DM | leukocyte | *TMEM68* | -1.233526164 | 0.001859308 | 0.005146173 | 1 |
| T2DM | leukocyte | *GSDMA* | -5.891767728 | 0.005874781 | 0.009145014 | 1 |
| T2DM | leukocyte | *MLST8* | -0.721880273 | 0.00346839 | 0.006602377 | 1 |
| T2DM | leukocyte | *ECI1* | -0.737660323 | 0.001896139 | 0.005146173 | 1 |
| T2DM | leukocyte | *ABCA3* | -0.978271876 | 0.00067897 | 0.004990048 | 1 |
| T2DM | leukocyte | *BSCL2* | -0.814322472 | 0.001161286 | 0.004990048 | 1 |
| T2DM | leukocyte | *C2CD3* | -1.703577565 | 0.001588703 | 0.005020048 | 1 |
| T2DM | leukocyte | *TRANK1* | -2.747800707 | 0.000290167 | 0.004990048 | 1 |
| T2DM | leukocyte | *RPSA* | -0.628417573 | 0.001007409 | 0.004990048 | 1 |
| T2DM | leukocyte | *ENTPD3* | -0.567803065 | 0.004948043 | 0.008003288 | 1 |
| T2DM | leukocyte | *ULK4* | -1.314636644 | 0.000709908 | 0.004990048 | 1 |
| T2DM | leukocyte | *LTBP3* | -0.704949293 | 0.004911531 | 0.007970068 | 1 |
| T2DM | leukocyte | *SAC3D1* | -1.091830511 | 0.001737653 | 0.005075243 | 1 |
| T2DM | leukocyte | *CCDC88B* | -2.316313041 | 0.009738092 | 0.01395367 | 1 |
| T2DM | leukocyte | *PBK* | -1.973902597 | 0.003587879 | 0.006731985 | 1 |
| T2DM | leukocyte | *KIAA1586* | -1.312382318 | 0.000420881 | 0.004990048 | 1 |
| T2DM | leukocyte | *FAM83B* | -0.781308482 | 0.021110353 | 0.027647525 | 1 |
| T2DM | leukocyte | *LMBRD1* | -1.17564691 | 0.000531474 | 0.004990048 | 1 |
| T2DM | leukocyte | *TTC39C* | -0.926749587 | 0.000909652 | 0.004990048 | 1 |
| T2DM | leukocyte | *POLR2J3* | -0.934527797 | 0.003669375 | 0.006835929 | 1 |
| T2DM | leukocyte | *COA6* | -1.016471014 | 0.001615335 | 0.005033031 | 1 |
| T2DM | leukocyte | *PXK* | -1.251533061 | 0.001319454 | 0.004990048 | 1 |
| T2DM | leukocyte | *ING5* | -1.721890804 | 0.000632201 | 0.004990048 | 1 |
| T2DM | leukocyte | *BMP1* | -0.795651907 | 0.023175792 | 0.030052042 | 1 |
| T2DM | leukocyte | *HEXIM2* | -1.080094517 | 0.004184301 | 0.007242343 | 1 |
| T2DM | leukocyte | *SERINC2* | -0.634174474 | 0.002055814 | 0.00526208 | 1 |
| T2DM | leukocyte | *TRAPPC11* | -0.738549807 | 0.003251709 | 0.006400784 | 1 |
| T2DM | leukocyte | *ING2* | -0.689482905 | 0.002208077 | 0.00539188 | 1 |

| | | | | | | |
|---|---|---|---|---|---|---|
| T2DM | leukocyte | *ADAM9* | -1.447951894 | 0.008818977 | 0.012807333 | 1 |
| T2DM | leukocyte | *NDUFS5* | -0.768257448 | 0.00051462 | 0.004990048 | 1 |
| T2DM | leukocyte | *DNAJC21* | -1.120974297 | 0.000358083 | 0.004990048 | 1 |
| T2DM | leukocyte | *CNNM3* | -0.918680296 | 0.001733988 | 0.00507285 | 1 |
| T2DM | leukocyte | *GSTM4* | -0.978433499 | 0.005100531 | 0.008189615 | 1 |
| T2DM | leukocyte | *SHOX2* | -6.812471475 | 0.014670683 | 0.020014276 | 1 |
| T2DM | leukocyte | *PPIP5K1* | -2.924976234 | 0.000103505 | 0.004990048 | 0.406669438 |
| T2DM | leukocyte | *ADAL* | -2.164157376 | 0.010034716 | 0.014326453 | 1 |
| T2DM | leukocyte | *LCMT2* | -1.250800258 | 0.006258406 | 0.009623983 | 1 |
| T2DM | leukocyte | *ZNF507* | -1.218795246 | 0.000846804 | 0.004990048 | 1 |
| T2DM | leukocyte | *NSG1* | -0.654071497 | 0.004269282 | 0.007310294 | 1 |
| T2DM | leukocyte | *ZBTB49* | -2.478368283 | 0.002104573 | 0.005283012 | 1 |
| T2DM | leukocyte | *GFM1* | -0.782912914 | 0.001726406 | 0.005070552 | 1 |
| T2DM | leukocyte | *FSTL5* | -0.557265061 | 0.007746658 | 0.011505329 | 1 |
| T2DM | leukocyte | *CTRB1* | -0.405400192 | 0.001935474 | 0.005184924 | 1 |
| T2DM | leukocyte | *CTRB2* | -0.373700599 | 0.00249323 | 0.005623364 | 1 |
| T2DM | leukocyte | *TMEM129* | -1.781773233 | 2.26E-05 | 0.003059063 | 0.088712829 |
| T2DM | leukocyte | *PPIC* | -0.443327504 | 0.009248729 | 0.013349836 | 1 |
| T2DM | leukocyte | *STXBP6* | -1.262149703 | 0.000167716 | 0.004990048 | 0.658956787 |
| T2DM | leukocyte | *MFF* | -0.923685092 | 0.001136151 | 0.004990048 | 1 |
| T2DM | leukocyte | *CPLX1* | -0.811547863 | 0.010307919 | 0.014679164 | 1 |
| T2DM | leukocyte | *ROR2* | -2.415549963 | 0.008139916 | 0.011987155 | 1 |
| T2DM | leukocyte | *FAM110B* | -0.92318018 | 0.003393466 | 0.006510382 | 1 |
| T2DM | leukocyte | *ATF5* | -1.147952167 | 0.001157301 | 0.004990048 | 1 |
| T2DM | leukocyte | *XPO6* | -1.773291024 | 0.000353409 | 0.004990048 | 1 |
| T2DM | leukocyte | *RGS14* | -1.280910982 | 0.003302703 | 0.006432819 | 1 |
| T2DM | leukocyte | *TBC1D10B* | -1.226468725 | 0.000970453 | 0.004990048 | 1 |
| T2DM | leukocyte | *PRELID1* | -0.70590814 | 0.001609136 | 0.005033031 | 1 |
| T2DM | leukocyte | *CXCL10* | -151.2856829 | 0.004227252 | 0.007272975 | 1 |
| T2DM | leukocyte | *NMD3* | -0.900167351 | 0.001596006 | 0.005024491 | 1 |
| T2DM | leukocyte | *STK32A* | -1.048871901 | 0.002925167 | 0.006103547 | 1 |
| T2DM | leukocyte | *SNUPN* | -2.817007253 | 0.000130039 | 0.004990048 | 0.510921471 |
| T2DM | leukocyte | *CRADD* | -0.950279779 | 0.003370986 | 0.006497619 | 1 |
| T2DM | leukocyte | *COL22A1* | -8.872209971 | 0.004409921 | 0.007423556 | 1 |
| T2DM | leukocyte | *SDC2* | -2.339104271 | 0.000486994 | 0.004990048 | 1 |
| T2DM | leukocyte | *CLIC4* | -0.514605602 | 0.004119538 | 0.007228971 | 1 |
| T2DM | leukocyte | *SLC38A11* | -7.51265852 | 0.001894773 | 0.005146173 | 1 |
| T2DM | leukocyte | *CCDC8* | -1.11378087 | 0.03138959 | 0.039719709 | 1 |
| T2DM | leukocyte | *HINT1* | -0.659693653 | 0.000817201 | 0.004990048 | 1 |
| T2DM | leukocyte | *DTWD2* | -2.393614844 | 0.015162527 | 0.020578089 | 1 |
| T2DM | leukocyte | *INO80E* | -1.759073169 | 2.92E-06 | 0.000882318 | 0.011470136 |
| T2DM | leukocyte | *C15orf40* | -0.930677341 | 0.001725302 | 0.005070552 | 1 |
| T2DM | leukocyte | *HIC2* | -2.914471466 | 0.016319557 | 0.022026637 | 1 |
| T2DM | leukocyte | *AGPAT2* | -1.006252125 | 0.000748625 | 0.004990048 | 1 |
| T2DM | leukocyte | *FASN* | -1.832256987 | 0.000154598 | 0.004990048 | 0.607417278 |
| T2DM | leukocyte | *MT1E* | -0.5452943 | 0.002115087 | 0.005283012 | 1 |
| T2DM | leukocyte | *LDB2* | -0.931912137 | 0.006959034 | 0.010520217 | 1 |
| T2DM | leukocyte | *RAC3* | -1.345209444 | 0.001051906 | 0.004990048 | 1 |
| T2DM | leukocyte | *LIMS1* | -1.189771932 | 0.000308825 | 0.004990048 | 1 |
| T2DM | leukocyte | *CSGALNACT2* | -2.48839572 | 0.001066774 | 0.004990048 | 1 |

| | | | | | | |
|---|---|---|---|---|---|---|
| T2DM | leukocyte | *AVEN* | -2.778287874 | 0.000420554 | 0.004990048 | 1 |
| T2DM | leukocyte | *CTNND2* | -1.055537944 | 0.00115483 | 0.004990048 | 1 |
| T2DM | leukocyte | *WNT10B* | -52.9403424 | 0.003805399 | 0.006976862 | 1 |
| T2DM | leukocyte | *CALML6* | 0.860964004 | 0.019673552 | 0.025930018 | 1 |
| T2DM | leukocyte | *TM4SF4* | -0.866361524 | 0.001422796 | 0.004990048 | 1 |
| T2DM | leukocyte | *OTUD3* | -0.696114094 | 0.001967748 | 0.005212073 | 1 |
| T2DM | leukocyte | *MAP3K2* | -1.281315797 | 0.000325004 | 0.004990048 | 1 |
| T2DM | leukocyte | *PUSL1* | -2.130968913 | 0.000505939 | 0.004990048 | 1 |
| T2DM | leukocyte | *IFFO2* | -0.831266966 | 0.003072943 | 0.006249984 | 1 |
| T2DM | leukocyte | *MYRIP* | -1.115106909 | 0.005923214 | 0.009205818 | 1 |
| T2DM | leukocyte | *FAM153A* | -1.612171711 | 0.016255507 | 0.021947728 | 1 |
| T2DM | leukocyte | *ZNF778* | -1.313979084 | 0.000768884 | 0.004990048 | 1 |
| T2DM | leukocyte | *NIPA1* | -1.230205202 | 0.000600431 | 0.004990048 | 1 |
| T2DM | leukocyte | *UBE2E1* | -1.656220988 | 0.000562565 | 0.004990048 | 1 |
| T2DM | leukocyte | *ANKK1* | -22.75887303 | 0.005285646 | 0.008421454 | 1 |
| T2DM | leukocyte | *USP47* | -1.13421842 | 0.00070269 | 0.004990048 | 1 |
| T2DM | leukocyte | *PDCD6IP* | -0.898533279 | 0.001568013 | 0.005009694 | 1 |
| T2DM | leukocyte | *CRTAP* | -1.76764773 | 0.000472911 | 0.004990048 | 1 |
| T2DM | leukocyte | *STX8* | -1.410924662 | 0.000308712 | 0.004990048 | 1 |
| T2DM | leukocyte | *NFRKB* | -1.680485202 | 0.000925771 | 0.004990048 | 1 |
| T2DM | leukocyte | *SETMAR* | -0.875261739 | 0.002724546 | 0.005904436 | 1 |
| T2DM | leukocyte | *CST2* | -2.058848875 | 0.001149827 | 0.004990048 | 1 |
| T2DM | leukocyte | *KRT8* | -0.373345541 | 0.003708402 | 0.006872788 | 1 |
| T2DM | leukocyte | *MGMT* | -1.217776344 | 0.001217118 | 0.004990048 | 1 |
| T2DM | leukocyte | *NFXL1* | -1.669807613 | 0.000416878 | 0.004990048 | 1 |
| T2DM | leukocyte | *KRT4* | -46.1237745 | 0.009020261 | 0.01305844 | 1 |
| T2DM | leukocyte | *SLC23A1* | -6.739266157 | 0.01111956 | 0.015704081 | 1 |
| T2DM | leukocyte | *NPAS2* | -0.707402623 | 0.004505215 | 0.007539852 | 1 |
| T2DM | leukocyte | *NUDT9* | -0.662188635 | 0.010002641 | 0.014296244 | 1 |
| T2DM | leukocyte | *IRX1* | -1.266769452 | 0.004313727 | 0.007328649 | 1 |
| T2DM | leukocyte | *CDH2* | -0.704123506 | 0.004296604 | 0.007328649 | 1 |
| T2DM | leukocyte | *HSPA4* | -1.006044331 | 0.000877627 | 0.004990048 | 1 |
| T2DM | leukocyte | *ARMC10* | -1.182300784 | 0.003492189 | 0.006628411 | 1 |
| T2DM | leukocyte | *ACYP2* | -0.851129931 | 0.001580744 | 0.005009694 | 1 |
| T2DM | leukocyte | *ATF7* | -0.96843867 | 0.002014617 | 0.005242007 | 1 |
| T2DM | leukocyte | *RASA4B* | -1.573140978 | 0.008659741 | 0.0126174 | 1 |
| T2DM | leukocyte | *KCNS3* | -1.170691489 | 0.004504208 | 0.007539852 | 1 |
| T2DM | leukocyte | *FOXN2* | -2.964932265 | 0.000311274 | 0.004990048 | 1 |
| T2DM | leukocyte | *GPR27* | -1.001978022 | 0.000904688 | 0.004990048 | 1 |
| T2DM | leukocyte | *MTSS1* | -1.215749294 | 0.002835292 | 0.006015044 | 1 |
| T2DM | leukocyte | *TMEM43* | -0.912517391 | 0.002381541 | 0.005560194 | 1 |
| T2DM | leukocyte | *RPS9* | -0.601048627 | 0.000974625 | 0.004990048 | 1 |
| T2DM | leukocyte | *NDUFA3* | -1.031950749 | 0.000782112 | 0.004990048 | 1 |
| T2DM | leukocyte | *ZNF160* | -0.95968865 | 0.001478085 | 0.004990048 | 1 |
| T2DM | leukocyte | *ZNF415* | -0.830342566 | 0.006475664 | 0.009919253 | 1 |
| T2DM | leukocyte | *TSNARE1* | 0.77092965 | 0.020539443 | 0.026935738 | 1 |
| T2DM | leukocyte | *SOX7* | -1.146413024 | 0.004310789 | 0.007328649 | 1 |
| T2DM | leukocyte | *TRMT61B* | -0.658268055 | 0.012887553 | 0.017882851 | 1 |
| T2DM | leukocyte | *INSR* | -0.652213096 | 0.002465422 | 0.005622792 | 1 |
| T2DM | leukocyte | *MFN1* | -0.872726339 | 0.006217827 | 0.009569073 | 1 |

| | | | | | | |
|---|---|---|---|---|---|---|
| T2DM | leukocyte | *KCNMB3* | -1.453535915 | 0.004545238 | 0.007568852 | 1 |
| T2DM | leukocyte | *ATP6V0E2* | -0.806723886 | 0.001382949 | 0.004990048 | 1 |
| T2DM | leukocyte | *JAGN1* | -1.036880469 | 0.001199988 | 0.004990048 | 1 |
| T2DM | leukocyte | *MORN4* | -0.614181095 | 0.005367637 | 0.008517547 | 1 |
| T2DM | leukocyte | *TRIM8* | -1.084412965 | 0.000951136 | 0.004990048 | 1 |
| T2DM | leukocyte | *CDC42BPG* | -0.646699281 | 0.003674923 | 0.006839778 | 1 |
| T2DM | leukocyte | *ZNF440* | -3.856935367 | 2.98E-05 | 0.003343813 | 0.11690834 |
| T2DM | leukocyte | *EXOSC1* | -1.292980566 | 0.000659169 | 0.004990048 | 1 |
| T2DM | leukocyte | *PGAM1* | -0.940360163 | 0.000297127 | 0.004990048 | 1 |
| T2DM | leukocyte | *ESCO2* | -2.46264608 | 0.038840071 | 0.048353181 | 1 |
| T2DM | leukocyte | *MRPL36* | -0.891293974 | 0.00239871 | 0.005576645 | 1 |
| T2DM | leukocyte | *MCC* | -1.155183677 | 0.00315035 | 0.006311945 | 1 |
| T2DM | leukocyte | *ZNF561* | -1.120272161 | 0.001317578 | 0.004990048 | 1 |
| T2DM | leukocyte | *PPID* | -1.166295511 | 0.008628556 | 0.012584111 | 1 |
| T2DM | leukocyte | *RXFP1* | -18.55370939 | 0.01229086 | 0.017154809 | 1 |
| T2DM | leukocyte | *MAP6* | -0.62889606 | 0.004071987 | 0.007189 | 1 |
| T2DM | leukocyte | *NMUR1* | -9.566563372 | 0.004004087 | 0.007145551 | 1 |
| T2DM | leukocyte | *CLSTN1* | -0.848860732 | 0.000579154 | 0.004990048 | 1 |
| T2DM | leukocyte | *PIK3CD* | 0.652869713 | 0.0403489 | 0.049993954 | 1 |
| T2DM | leukocyte | *ENC1* | -0.779299653 | 0.006716891 | 0.010221016 | 1 |
| T2DM | leukocyte | *RGS19* | -1.884059364 | 7.49E-05 | 0.004990048 | 0.294167597 |
| T2DM | leukocyte | *ANO5* | -1.070460769 | 0.004811812 | 0.007855834 | 1 |
| T2DM | leukocyte | *GPHN* | -1.673872633 | 0.000858472 | 0.004990048 | 1 |
| T2DM | leukocyte | *VAT1L* | -1.02416132 | 0.008130167 | 0.011977287 | 1 |
| T2DM | leukocyte | *LGALS4* | -0.479517328 | 0.005243219 | 0.008370827 | 1 |
| T2DM | leukocyte | *LRRC34* | -1.089844109 | 0.003895544 | 0.007059776 | 1 |
| T2DM | leukocyte | *SPATA5L1* | -2.072026045 | 0.000236253 | 0.004990048 | 0.928237022 |
| T2DM | leukocyte | *CTPS1* | -0.823882055 | 0.003998081 | 0.007143456 | 1 |
| T2DM | leukocyte | *METTL18* | -1.167431741 | 0.002939437 | 0.006123567 | 1 |
| T2DM | leukocyte | *PWWP2B* | -1.173310587 | 0.00136191 | 0.004990048 | 1 |
| T2DM | leukocyte | *PTEN* | -1.066006507 | 0.001373851 | 0.004990048 | 1 |
| T2DM | leukocyte | *CYP4F11* | -1.929601662 | 0.016876631 | 0.022685009 | 1 |
| T2DM | leukocyte | *TLN2* | -1.723271248 | 0.004100904 | 0.00720956 | 1 |
| T2DM | leukocyte | *TVP23B* | -1.645730519 | 0.000891893 | 0.004990048 | 1 |
| T2DM | leukocyte | *SRGAP2C* | -2.009794345 | 0.000357939 | 0.004990048 | 1 |
| T2DM | leukocyte | *ATPAF2* | -1.029461416 | 0.006988579 | 0.010556758 | 1 |
| T2DM | leukocyte | *ZNF556* | 0.702234848 | 0.020616989 | 0.027028412 | 1 |
| T2DM | leukocyte | *ZNF554* | -0.988790865 | 0.003612906 | 0.006763117 | 1 |
| T2DM | leukocyte | *GAP43* | -0.679881553 | 0.004812673 | 0.007855834 | 1 |
| T2DM | leukocyte | *REG1B* | -0.425822061 | 0.002108197 | 0.005283012 | 1 |
| T2DM | leukocyte | *NME6* | -1.593493769 | 0.002085199 | 0.005283012 | 1 |
| T2DM | leukocyte | *TEFM* | -1.202039766 | 0.001977631 | 0.005222499 | 1 |
| T2DM | leukocyte | *MBOAT1* | -1.951930053 | 0.001858722 | 0.005146173 | 1 |
| T2DM | leukocyte | *ZNF131* | -0.959118167 | 0.0017517 | 0.005084476 | 1 |
| T2DM | leukocyte | *BSG* | -0.822186516 | 0.000972268 | 0.004990048 | 1 |
| T2DM | leukocyte | *HINFP* | -2.85469019 | 0.000686573 | 0.004990048 | 1 |
| T2DM | leukocyte | *CERS6* | -0.916773649 | 0.001240775 | 0.004990048 | 1 |
| T2DM | leukocyte | *SPTLC3* | 0.961750613 | 0.01953957 | 0.025770718 | 1 |
| T2DM | leukocyte | *COPRS* | -0.655932605 | 0.00318631 | 0.006339351 | 1 |
| T2DM | leukocyte | *SUCLG2* | -1.764818565 | 0.000468597 | 0.004990048 | 1 |

| | | | | | | |
|---|---|---|---|---|---|---|
| T2DM | leukocyte | *CSDC2* | -2.582784505 | 0.003562158 | 0.006706143 | 1 |
| T2DM | leukocyte | *ARNT2* | -0.615592927 | 0.018942412 | 0.025058833 | 1 |
| T2DM | leukocyte | *FUT9* | -3.983409621 | 0.00500911 | 0.008075829 | 1 |
| T2DM | leukocyte | *ZNF24* | -1.067520402 | 0.000502474 | 0.004990048 | 1 |
| T2DM | leukocyte | *MANEA* | -1.078276715 | 0.001701386 | 0.005060368 | 1 |
| T2DM | leukocyte | *AFF1* | -1.045899366 | 0.001259109 | 0.004990048 | 1 |
| T2DM | leukocyte | *CTSW* | -1.297525671 | 4.04E-05 | 0.004175341 | 0.158662972 |
| T2DM | leukocyte | *SMPDL3A* | -2.403529781 | 0.002482924 | 0.005622792 | 1 |
| T2DM | leukocyte | *RAD9A* | -2.068342191 | 0.000130863 | 0.004990048 | 0.514158791 |
| T2DM | leukocyte | *EFEMP2* | -0.944317561 | 0.003347288 | 0.006472193 | 1 |
| T2DM | leukocyte | *FUT10* | -2.536994091 | 0.032872008 | 0.041488635 | 1 |
| T2DM | leukocyte | *NAA16* | -0.767507059 | 0.010647427 | 0.015091537 | 1 |
| T2DM | leukocyte | *OVOL1* | -3.039858746 | 0.000146505 | 0.004990048 | 0.575616975 |
| T2DM | leukocyte | *DMXL1* | -1.518704474 | 0.000971813 | 0.004990048 | 1 |
| T2DM | leukocyte | *NADSYN1* | -1.744571349 | 0.001562753 | 0.005009694 | 1 |
| T2DM | leukocyte | *RNASEH2C* | -1.160518068 | 0.000167405 | 0.004990048 | 0.657735293 |
| T2DM | leukocyte | *LCLAT1* | -0.992987978 | 0.001114044 | 0.004990048 | 1 |
| T2DM | leukocyte | *SH3RF3* | -1.379644903 | 0.003917094 | 0.007085756 | 1 |
| T2DM | leukocyte | *DCAKD* | -0.749434959 | 0.001842654 | 0.005146173 | 1 |
| T2DM | leukocyte | *TADA2B* | -1.125822224 | 0.001067856 | 0.004990048 | 1 |
| T2DM | leukocyte | *TRMT112* | -0.822925145 | 0.001460849 | 0.004990048 | 1 |
| T2DM | leukocyte | *NOC3L* | -1.031216665 | 0.000606939 | 0.004990048 | 1 |
| T2DM | leukocyte | *RHOD* | -0.61781187 | 0.002856967 | 0.006048134 | 1 |
| T2DM | leukocyte | *PARP14* | -1.040440931 | 0.000821415 | 0.004990048 | 1 |
| T2DM | leukocyte | *VANGL1* | -0.882735716 | 0.004136464 | 0.007236049 | 1 |
| T2DM | leukocyte | *IQCB1* | -2.59954037 | 0.000141837 | 0.004990048 | 0.557276859 |
| T2DM | leukocyte | *ZNF483* | -0.814692158 | 0.001276682 | 0.004990048 | 1 |
| T2DM | leukocyte | *SLC2A14* | -3.431139289 | 0.004851079 | 0.00790211 | 1 |
| T2DM | leukocyte | *MZT2A* | -0.705022468 | 0.003069339 | 0.006249984 | 1 |
| T2DM | leukocyte | *PPP1R3B* | -3.305220867 | 1.55E-06 | 0.00053944 | 0.006075666 |
| T2DM | leukocyte | *MAP3K11* | -0.912846754 | 0.001097722 | 0.004990048 | 1 |
| T2DM | leukocyte | *NDNF* | 0.832790883 | 0.018545683 | 0.024608575 | 1 |
| T2DM | leukocyte | *ARV1* | -0.852774382 | 0.001995184 | 0.005227473 | 1 |
| T2DM | leukocyte | *EHBP1L1* | -0.862104195 | 0.007841189 | 0.011622224 | 1 |
| T2DM | leukocyte | *PPP1R14B* | -0.787595219 | 0.001156535 | 0.004990048 | 1 |
| T2DM | leukocyte | *TNFRSF10D* | -1.021984799 | 0.006183334 | 0.009527184 | 1 |
| T2DM | leukocyte | *GMPPB* | -1.293042144 | 0.004583111 | 0.007597909 | 1 |
| T2DM | leukocyte | *NUDT18* | -0.906566628 | 0.003518478 | 0.006649019 | 1 |
| T2DM | leukocyte | *NUDT4* | -0.808911533 | 0.003932133 | 0.007103149 | 1 |
| T2DM | leukocyte | *PC* | -1.472002731 | 0.001900041 | 0.005146173 | 1 |
| T2DM | leukocyte | *UQCRH* | -0.829941122 | 0.00074452 | 0.004990048 | 1 |
| T2DM | leukocyte | *HEG1* | -0.884991961 | 0.003536242 | 0.006673341 | 1 |
| T2DM | leukocyte | *ENDOV* | -1.97433703 | 0.000373458 | 0.004990048 | 1 |
| T2DM | leukocyte | *DPY19L1* | -1.116013569 | 0.002044655 | 0.00526208 | 1 |
| T2DM | leukocyte | *GPR160* | -1.336498024 | 0.002877895 | 0.006055188 | 1 |
| T2DM | leukocyte | *XXYLT1* | -1.47289918 | 0.002302967 | 0.005478715 | 1 |
| T2DM | leukocyte | *GNG5* | -0.852485077 | 0.000682846 | 0.004990048 | 1 |
| T2DM | leukocyte | *C9orf131* | -1.488704562 | 0.025405496 | 0.032641659 | 1 |
| T2DM | leukocyte | *FAM53A* | -4.638204607 | 0.001109058 | 0.004990048 | 1 |
| T2DM | leukocyte | *SELP* | -5.283850847 | 0.004043181 | 0.007168617 | 1 |

| | | | | | | |
|---|---|---|---|---|---|---|
| T2DM | leukocyte | *CTU2* | -0.84446018 | 0.004343604 | 0.007358292 | 1 |
| T2DM | leukocyte | *ADCY6* | -0.955860791 | 0.004380564 | 0.007389967 | 1 |
| T2DM | leukocyte | *PODN* | -0.832177774 | 0.012684926 | 0.017627731 | 1 |
| T2DM | leukocyte | *RALGAPA1* | -0.914975478 | 0.000946322 | 0.004990048 | 1 |
| T2DM | leukocyte | *CNTNAP2* | 1.857300612 | 0.006080337 | 0.009402422 | 1 |
| T2DM | leukocyte | *GOLT1A* | -0.91090674 | 0.001771229 | 0.005097196 | 1 |
| T2DM | leukocyte | *NPAS4* | -2.622846402 | 2.61E-05 | 0.003310187 | 0.102412252 |
| T2DM | leukocyte | *ANGEL2* | -1.301788046 | 0.000977337 | 0.004990048 | 1 |
| T2DM | leukocyte | *ZNF266* | -0.815491597 | 0.000786814 | 0.004990048 | 1 |
| T2DM | leukocyte | *BRSK2* | -0.841741338 | 0.002924105 | 0.006103547 | 1 |
| T2DM | leukocyte | *BRMS1* | -0.848761625 | 0.001784384 | 0.005100156 | 1 |
| T2DM | leukocyte | *HRAS* | -0.850294402 | 0.001547305 | 0.005009694 | 1 |
| T2DM | leukocyte | *SRP72* | -0.705140829 | 0.001808421 | 0.005122825 | 1 |
| T2DM | leukocyte | *PCP2* | -3.539778913 | 0.002874248 | 0.006055188 | 1 |
| T2DM | leukocyte | *DENND6A* | -0.838593297 | 0.003192127 | 0.006345399 | 1 |
| T2DM | leukocyte | *YIF1A* | -1.030112933 | 6.99E-05 | 0.004990048 | 0.274636325 |
| T2DM | leukocyte | *NDUFA11* | -0.881275493 | 0.000776936 | 0.004990048 | 1 |
| T2DM | leukocyte | *RSRC1* | -1.966571373 | 0.000409593 | 0.004990048 | 1 |
| T2DM | leukocyte | *DHX36* | -0.971395911 | 0.000876301 | 0.004990048 | 1 |
| T2DM | leukocyte | *PSMD2* | -0.721594297 | 0.00312136 | 0.006295597 | 1 |
| T2DM | leukocyte | *PPM1E* | -0.582246269 | 0.014177854 | 0.019436423 | 1 |
| T2DM | leukocyte | *ZNF408* | -2.84662562 | 0.001404927 | 0.004990048 | 1 |
| T2DM | leukocyte | *ARHGAP1* | -0.839674326 | 0.003257194 | 0.006405338 | 1 |
| T2DM | leukocyte | *MED16* | -0.845220313 | 0.002182946 | 0.005356498 | 1 |
| T2DM | leukocyte | *CHST1* | -1.74250714 | 0.004302373 | 0.007328649 | 1 |
| T2DM | leukocyte | *DOLK* | -1.665701537 | 0.003229015 | 0.00638169 | 1 |
| T2DM | leukocyte | *CCNE2* | -1.076181217 | 0.0046347 | 0.007664031 | 1 |
| T2DM | leukocyte | *ZNF519* | -1.707286586 | 0.001725485 | 0.005070552 | 1 |
| T2DM | leukocyte | *TMEM9B* | -0.939356085 | 0.00094435 | 0.004990048 | 1 |
| T2DM | leukocyte | *EIF3F* | -0.974676361 | 0.00061698 | 0.004990048 | 1 |
| T2DM | leukocyte | *ZNF25* | -0.868305384 | 0.002246671 | 0.005438799 | 1 |
| T2DM | leukocyte | *PPP2R2D* | -0.877844924 | 0.001422397 | 0.004990048 | 1 |
| T2DM | leukocyte | *CLCF1* | -1.075587453 | 0.011199091 | 0.015796991 | 1 |
| T2DM | leukocyte | *MRPL48* | -1.000415258 | 0.006981015 | 0.010549388 | 1 |
| T2DM | leukocyte | *TMEM70* | -0.875408723 | 0.001671627 | 0.005048289 | 1 |
| T2DM | leukocyte | *RPS6KB2* | -1.394615527 | 0.000838237 | 0.004990048 | 1 |
| T2DM | leukocyte | *RMI2* | -2.429832314 | 0.017428645 | 0.023291546 | 1 |
| T2DM | leukocyte | *LINC01106* | -3.978473006 | 0.013808366 | 0.018989524 | 1 |
| T2DM | leukocyte | *SFN* | -1.531676464 | 0.022885878 | 0.029705522 | 1 |
| T2DM | leukocyte | *SWI5* | -1.195927645 | 0.005097276 | 0.008187734 | 1 |
| T2DM | leukocyte | *ZDHHC21* | -1.905826976 | 0.000284729 | 0.004990048 | 1 |
| T2DM | leukocyte | *TUBB6* | -0.433072572 | 0.007425023 | 0.011106403 | 1 |
| T2DM | leukocyte | *CHMP6* | -0.805481127 | 0.00882436 | 0.012807872 | 1 |
| T2DM | leukocyte | *DLEU1* | -1.132110683 | 0.001713251 | 0.005068796 | 1 |
| T2DM | leukocyte | *CCDC57* | -1.467162732 | 6.72E-05 | 0.004990048 | 0.264080744 |
| T2DM | leukocyte | *BNIP3* | -0.688164559 | 0.001497487 | 0.004993935 | 1 |
| T2DM | leukocyte | *SMIM19* | -0.771340486 | 0.001972587 | 0.005215543 | 1 |
| T2DM | leukocyte | *SLC35G1* | -1.270424497 | 0.00233845 | 0.005515664 | 1 |
| T2DM | leukocyte | *COX8A* | -0.711924501 | 0.000598243 | 0.004990048 | 1 |
| T2DM | leukocyte | *CDC26* | -0.976452424 | 0.001439577 | 0.004990048 | 1 |

| | | | | | | |
|---|---|---|---|---|---|---|
| T2DM | leukocyte | *RNPEP* | -1.654043387 | 0.001266403 | 0.004990048 | 1 |
| T2DM | leukocyte | *DMRTA1* | -0.774493344 | 0.005703932 | 0.008935705 | 1 |
| T2DM | leukocyte | *CLEC14A* | -0.517972886 | 0.006727842 | 0.010230596 | 1 |
| T2DM | leukocyte | *WDR25* | -2.554437946 | 0.000207905 | 0.004990048 | 0.816858929 |
| T2DM | leukocyte | *DIRAS1* | -1.108060772 | 0.004267683 | 0.007310294 | 1 |
| T2DM | leukocyte | *PHLDB3* | -0.748816191 | 0.005968456 | 0.009262768 | 1 |
| T2DM | leukocyte | *KBTBD11* | -0.769405783 | 0.003894271 | 0.007059776 | 1 |
| T2DM | leukocyte | *RMDN1* | -1.086197274 | 0.002047854 | 0.00526208 | 1 |
| T2DM | leukocyte | *LRRC37A* | -4.909541872 | 0.002377946 | 0.005560194 | 1 |
| T2DM | leukocyte | *ACSF3* | -1.242005563 | 0.001647806 | 0.005035955 | 1 |
| T2DM | leukocyte | *RUFY1* | -2.306702729 | 0.000600236 | 0.004990048 | 1 |
| T2DM | leukocyte | *VSIG10* | -1.506137984 | 0.000558809 | 0.004990048 | 1 |
| T2DM | leukocyte | *WSB2* | -0.795133974 | 0.002650966 | 0.005824744 | 1 |
| T2DM | leukocyte | *ANKLE2* | -0.708738968 | 0.00428685 | 0.00732537 | 1 |
| T2DM | leukocyte | *MUC20* | -0.892979624 | 0.00414514 | 0.007238335 | 1 |
| T2DM | leukocyte | *NFATC2IP* | -1.410383888 | 0.000844997 | 0.004990048 | 1 |
| T2DM | leukocyte | *DPP7* | -0.882538043 | 0.000944328 | 0.004990048 | 1 |
| T2DM | leukocyte | *MTHFR* | -1.030095374 | 0.001196488 | 0.004990048 | 1 |
| T2DM | leukocyte | *DEAF1* | -1.257717512 | 0.000952659 | 0.004990048 | 1 |
| T2DM | leukocyte | *TMEM80* | -0.703456414 | 0.004921036 | 0.007973094 | 1 |
| T2DM | leukocyte | *DSCAML1* | -1.457216242 | 0.000173295 | 0.004990048 | 0.680876638 |
| T2DM | leukocyte | *PUS1* | -1.845568318 | 0.00270243 | 0.005880214 | 1 |
| T2DM | leukocyte | *MAN1B1* | -1.631560654 | 0.000750498 | 0.004990048 | 1 |
| T2DM | leukocyte | *PPFIA3* | -0.719292051 | 0.003300307 | 0.006432819 | 1 |
| T2DM | leukocyte | *PAWR* | -0.825616772 | 0.003125779 | 0.006301276 | 1 |
| T2DM | leukocyte | *GPR4* | -0.71646584 | 0.002170583 | 0.005349178 | 1 |
| T2DM | leukocyte | *ACOT4* | -1.252986105 | 0.002067628 | 0.005273744 | 1 |
| T2DM | leukocyte | *ARIH2* | -0.992831098 | 0.000324166 | 0.004990048 | 1 |
| T2DM | leukocyte | *IRX3* | 2.479525057 | 0.000214266 | 0.004990048 | 0.841849721 |
| T2DM | leukocyte | *SAMD12* | -1.257305318 | 0.002454856 | 0.005621365 | 1 |
| T2DM | leukocyte | *CD163* | 0.076437943 | 0.0188306 | 0.024919309 | 1 |
| T2DM | leukocyte | *ACAD9* | -0.919927716 | 0.013717797 | 0.018871577 | 1 |
| T2DM | leukocyte | *AGTRAP* | -1.009416714 | 0.00163287 | 0.005035955 | 1 |
| T2DM | leukocyte | *THAP5* | -0.972571787 | 0.001237134 | 0.004990048 | 1 |
| T2DM | leukocyte | *CD151* | -0.75571375 | 0.001414358 | 0.004990048 | 1 |
| T2DM | leukocyte | *CHID1* | -0.841005062 | 0.00062838 | 0.004990048 | 1 |
| T2DM | leukocyte | *ZBTB41* | -1.236783043 | 0.000922013 | 0.004990048 | 1 |
| T2DM | leukocyte | *BET1L* | -1.434471291 | 0.000681959 | 0.004990048 | 1 |
| T2DM | leukocyte | *RIC8A* | -1.213586736 | 0.001307109 | 0.004990048 | 1 |
| T2DM | leukocyte | *IMP3* | -0.885365393 | 0.005984345 | 0.009278806 | 1 |
| T2DM | leukocyte | *GPR150* | -0.691208389 | 0.036459992 | 0.045621436 | 1 |
| T2DM | leukocyte | *ADAMTSL1* | -4.583646515 | 0.002960796 | 0.006135605 | 1 |
| T2DM | leukocyte | *ALS2CL* | -1.244688578 | 0.003362063 | 0.006491177 | 1 |
| T2DM | leukocyte | *STAP2* | -0.638720508 | 0.005468303 | 0.008631969 | 1 |
| T2DM | leukocyte | *DALRD3* | -0.94213439 | 0.005150368 | 0.008249407 | 1 |
| T2DM | leukocyte | *ZNF114* | -2.394914675 | 0.003613078 | 0.006763117 | 1 |
| T2DM | leukocyte | *PLEC* | -1.879929564 | 0.000200437 | 0.004990048 | 0.787515047 |
| T2DM | leukocyte | *WDR6* | -0.795045953 | 0.010276813 | 0.014645484 | 1 |
| T2DM | leukocyte | *AQP11* | -1.108022168 | 0.004276203 | 0.007317597 | 1 |
| T2DM | leukocyte | *ZFAND2A* | -1.234213165 | 0.000862116 | 0.004990048 | 1 |

| | | | | | | |
|---|---|---|---|---|---|---|
| T2DM | leukocyte | *DNAJC22* | -0.567435557 | 0.006162865 | 0.009503098 | 1 |
| T2DM | leukocyte | *NT5DC1* | -0.903959342 | 0.001620099 | 0.005035955 | 1 |
| T2DM | leukocyte | *COX14* | -0.880213417 | 0.003250661 | 0.006400784 | 1 |
| T2DM | leukocyte | *P4HTM* | -0.705810092 | 0.00105344 | 0.004990048 | 1 |
| T2DM | leukocyte | *ERBB4* | -8.10389048 | 0.00099931 | 0.004990048 | 1 |
| T2DM | leukocyte | *ERN1* | -1.287562731 | 0.00071811 | 0.004990048 | 1 |
| T2DM | leukocyte | *PARP10* | -1.147207877 | 0.002692657 | 0.005864439 | 1 |
| T2DM | leukocyte | *RPP25* | -1.580734262 | 0.000661243 | 0.004990048 | 1 |
| T2DM | leukocyte | *ZHX2* | -1.020715357 | 0.002166388 | 0.005345956 | 1 |
| T2DM | leukocyte | *HYI* | -1.109360803 | 0.001193911 | 0.004990048 | 1 |
| T2DM | leukocyte | *GAK* | -1.007598032 | 0.003826465 | 0.007002413 | 1 |
| T2DM | leukocyte | *TUFM* | -0.535873705 | 0.016440662 | 0.022152044 | 1 |
| T2DM | leukocyte | *RMI1* | 1.075494969 | 0.035363816 | 0.044348686 | 1 |
| T2DM | leukocyte | *C3orf38* | -1.153853862 | 0.010693997 | 0.015146617 | 1 |
| T2DM | leukocyte | *IGSF22* | -0.631232987 | 0.006661266 | 0.0101521 | 1 |
| T2DM | leukocyte | *HTR1F* | -2.804445841 | 0.000719889 | 0.004990048 | 1 |
| T2DM | leukocyte | *TMTC2* | -1.620334079 | 0.003114168 | 0.006284318 | 1 |
| T2DM | leukocyte | *FARSA* | -1.124243726 | 0.000199112 | 0.004990048 | 0.782310684 |
| T2DM | leukocyte | *SPTY2D1* | -0.988548427 | 0.001307717 | 0.004990048 | 1 |
| T2DM | leukocyte | *SAMD4B* | -0.793941729 | 0.002553381 | 0.005681968 | 1 |
| T2DM | leukocyte | *TCAIM* | -1.348474157 | 0.000956366 | 0.004990048 | 1 |
| T2DM | leukocyte | *CALR* | -0.67375109 | 0.002712146 | 0.005893818 | 1 |
| T2DM | leukocyte | *CDH4* | -1.71986147 | 0.033836145 | 0.0426223 | 1 |
| T2DM | leukocyte | *ELMOD2* | -1.713823487 | 0.000551374 | 0.004990048 | 1 |
| T2DM | leukocyte | *LINC00174* | -0.89985544 | 0.001992013 | 0.005227473 | 1 |
| T2DM | leukocyte | *GEMIN4* | -0.880987679 | 0.004537137 | 0.007568852 | 1 |
| T2DM | leukocyte | *LBX2* | 0.764693306 | 0.032436535 | 0.040978504 | 1 |
| T2DM | leukocyte | *CDC42EP4* | -0.803013 | 0.001561267 | 0.005009694 | 1 |
| T2DM | leukocyte | *ZBTB42* | -2.035176969 | 0.003140989 | 0.006311945 | 1 |
| T2DM | leukocyte | *SYCN* | -0.486766104 | 0.003390568 | 0.006510382 | 1 |
| T2DM | leukocyte | *CITED4* | -0.598955037 | 0.003964851 | 0.007124893 | 1 |
| T2DM | leukocyte | *B3GNT3* | -2.017155577 | 0.005261464 | 0.008396545 | 1 |
| T2DM | leukocyte | *PUF60* | -0.810545013 | 0.002829164 | 0.006011783 | 1 |
| T2DM | leukocyte | *TSHZ1* | -0.725131963 | 0.004646495 | 0.007680302 | 1 |
| T2DM | leukocyte | *EXOC3* | -1.579898515 | 0.001694982 | 0.005060368 | 1 |
| T2DM | leukocyte | *TH* | -5.582389465 | 0.000509783 | 0.004990048 | 1 |
| T2DM | leukocyte | *ZNF816* | -1.251922347 | 0.004157281 | 0.007242343 | 1 |
| T2DM | leukocyte | *CCDC66* | -2.300722206 | 0.000741253 | 0.004990048 | 1 |
| T2DM | leukocyte | *MCFD2* | -0.721247121 | 0.001301148 | 0.004990048 | 1 |
| T2DM | leukocyte | *SERTM1* | 0.580880493 | 0.03689519 | 0.046136602 | 1 |
| T2DM | leukocyte | *GLIPR1L2* | -1.394173687 | 0.011892024 | 0.016657313 | 1 |
| T2DM | leukocyte | *TSPYL5* | -0.732665846 | 0.001642437 | 0.005035955 | 1 |
| T2DM | leukocyte | *SHISA2* | -1.532976385 | 0.008505642 | 0.012427917 | 1 |
| T2DM | leukocyte | *CHST13* | -72.00732348 | 0.004314371 | 0.007328649 | 1 |
| T2DM | leukocyte | *PPA1* | -0.682190165 | 0.001273201 | 0.004990048 | 1 |
| T2DM | leukocyte | *PSMG4* | -0.765220616 | 0.001727307 | 0.005070552 | 1 |
| T2DM | leukocyte | *MAP6D1* | -1.8216795 | 0.003652414 | 0.006815321 | 1 |
| T2DM | leukocyte | *D2HGDH* | -4.171955643 | 2.20E-05 | 0.003059063 | 0.086305646 |
| T2DM | leukocyte | *FAM83H* | -1.340924571 | 0.003215505 | 0.006372159 | 1 |
| T2DM | leukocyte | *LRRC57* | -1.317781212 | 0.005849596 | 0.00911303 | 1 |

| | | | | | | |
|---|---|---|---|---|---|---|
| T2DM | leukocyte | *MRPL14* | -0.98545249 | 0.00075814 | 0.004990048 | 1 |
| T2DM | leukocyte | *ZFP82* | -0.844601262 | 0.003100472 | 0.00626956 | 1 |
| T2DM | leukocyte | *FKRP* | -0.823338434 | 0.003294337 | 0.006432819 | 1 |
| T2DM | leukocyte | *RPH3AL* | -0.639583452 | 0.004476207 | 0.007504245 | 1 |
| T2DM | leukocyte | *HIGD1A* | -0.734861199 | 0.00136029 | 0.004990048 | 1 |
| T2DM | leukocyte | *F2R* | -0.558241022 | 0.024037444 | 0.031056599 | 1 |
| T2DM | leukocyte | *ZNF707* | -1.095365507 | 0.004078463 | 0.007189 | 1 |
| T2DM | leukocyte | *OGFOD3* | -0.801123364 | 0.001459729 | 0.004990048 | 1 |
| T2DM | leukocyte | *ZNF467* | -1.165942347 | 0.00298244 | 0.006152298 | 1 |
| T2DM | leukocyte | *ZNF678* | -2.068100332 | 0.001412811 | 0.004990048 | 1 |
| T2DM | leukocyte | *TMEM45A* | -0.701153539 | 0.004227914 | 0.007272975 | 1 |
| T2DM | leukocyte | *SGSH* | -2.407388135 | 0.006912548 | 0.010462019 | 1 |
| T2DM | leukocyte | *GPR135* | -1.016006883 | 0.003851937 | 0.007029382 | 1 |
| T2DM | leukocyte | *ANKRD62* | -1.344221874 | 0.002386469 | 0.005560842 | 1 |
| T2DM | leukocyte | *GPR88* | -17.43621041 | 0.001281105 | 0.004990048 | 1 |
| T2DM | leukocyte | *RFX7* | -2.478893823 | 0.000173948 | 0.004990048 | 0.683442721 |
| T2DM | leukocyte | *ZNF329* | -1.177247887 | 0.003326377 | 0.006456709 | 1 |
| T2DM | leukocyte | *CCDC149* | -1.558394487 | 0.000612506 | 0.004990048 | 1 |
| T2DM | leukocyte | *IDH2* | -0.661457453 | 0.004522373 | 0.007557079 | 1 |
| T2DM | leukocyte | *TNRC18* | -1.147422751 | 0.001462705 | 0.004990048 | 1 |
| T2DM | leukocyte | *TDRKH* | -0.938673711 | 0.004704401 | 0.007720459 | 1 |
| T2DM | leukocyte | *ZNF708* | -0.704976481 | 0.002653676 | 0.005824744 | 1 |
| T2DM | leukocyte | *IST1* | -1.42915478 | 0.000558061 | 0.004990048 | 1 |
| T2DM | leukocyte | *ERCC6L2* | -2.815910316 | 0.000449268 | 0.004990048 | 1 |
| T2DM | leukocyte | *MRPL41* | -0.884678585 | 0.000948547 | 0.004990048 | 1 |
| T2DM | leukocyte | *TP53TG1* | -0.970123899 | 0.001236657 | 0.004990048 | 1 |
| T2DM | leukocyte | *UBA7* | -2.881967263 | 7.91E-07 | 0.000310956 | 0.003109565 |
| T2DM | leukocyte | *ARL6IP4* | -0.602685209 | 0.006008708 | 0.009312905 | 1 |
| T2DM | leukocyte | *FAM153B* | -1.494293524 | 0.000719489 | 0.004990048 | 1 |
| T2DM | leukocyte | *BACE2* | -0.521116682 | 0.003484507 | 0.006623429 | 1 |
| T2DM | leukocyte | *SYNM* | -0.712176105 | 0.018461231 | 0.024526286 | 1 |
| T2DM | leukocyte | *FIGN* | -0.859869469 | 0.003680487 | 0.006846891 | 1 |
| T2DM | leukocyte | *B4GALNT4* | -1.705567302 | 0.003864524 | 0.007037843 | 1 |
| T2DM | leukocyte | *ZNF804B* | -62.99075685 | 0.00405747 | 0.007184034 | 1 |
| T2DM | leukocyte | *YBEY* | -1.04694175 | 0.002460065 | 0.005622792 | 1 |
| T2DM | leukocyte | *NPLOC4* | -0.938806004 | 0.001626348 | 0.005035955 | 1 |
| T2DM | leukocyte | *CAPN12* | -2.027043666 | 0.000224696 | 0.004990048 | 0.882831304 |
| T2DM | leukocyte | *GLRX5* | -0.725478535 | 0.00560771 | 0.008817809 | 1 |
| T2DM | leukocyte | *MXRA7* | -0.819524984 | 0.000888024 | 0.004990048 | 1 |
| T2DM | leukocyte | *ADI1* | -2.013833381 | 0.000412691 | 0.004990048 | 1 |
| T2DM | leukocyte | *SATB1* | -1.446138687 | 0.000724927 | 0.004990048 | 1 |
| T2DM | leukocyte | *EPHB3* | -2.100978047 | 0.015948504 | 0.021592582 | 1 |
| T2DM | leukocyte | *SKA2* | -1.713666485 | 0.000235612 | 0.004990048 | 0.925718204 |
| T2DM | leukocyte | *TTC3* | -0.411872439 | 0.024427774 | 0.03151945 | 1 |
| T2DM | leukocyte | *BRICD5* | -1.614941184 | 0.021932498 | 0.028609822 | 1 |
| T2DM | leukocyte | *TSKU* | -0.942797093 | 0.00438019 | 0.007389967 | 1 |
| T2DM | leukocyte | *ANXA2* | -0.546065023 | 0.001480241 | 0.004990048 | 1 |
| T2DM | leukocyte | *SLC35D3* | -1.662307515 | 0.00191254 | 0.00515389 | 1 |
| T2DM | leukocyte | *GPIHBP1* | -7.728342202 | 0.00463315 | 0.007664031 | 1 |
| T2DM | leukocyte | *VMO1* | 0.096244748 | 0.010293573 | 0.014664049 | 1 |

| | | | | | | |
|---|---|---|---|---|---|---|
| T2DM | leukocyte | *COL18A1* | -0.450282639 | 0.002952213 | 0.006135605 | 1 |
| T2DM | leukocyte | *C11orf54* | -1.237165079 | 0.002615306 | 0.005772773 | 1 |
| T2DM | leukocyte | *CEP63* | -1.108931885 | 0.000959996 | 0.004990048 | 1 |
| T2DM | leukocyte | *HMGN4* | -1.0593016 | 0.005915453 | 0.009197394 | 1 |
| T2DM | leukocyte | *SPATA13* | -0.863023754 | 0.001892949 | 0.005146173 | 1 |
| T2DM | leukocyte | *ZNF320* | 1.07124065 | 4.40E-05 | 0.004219101 | 0.172983145 |
| T2DM | leukocyte | *SLC25A21* | -2.157171538 | 0.004213299 | 0.007263735 | 1 |
| T2DM | leukocyte | *LYSMD4* | -1.028737895 | 0.003398519 | 0.006511001 | 1 |
| T2DM | leukocyte | *BEGAIN* | -1.313845708 | 0.001888562 | 0.005146173 | 1 |
| T2DM | leukocyte | *GPC6* | -1.009829876 | 0.003334717 | 0.00646588 | 1 |
| T2DM | leukocyte | *FANCF* | -3.568264511 | 0.000137517 | 0.004990048 | 0.540304869 |
| T2DM | leukocyte | *SMDT1* | -0.706495554 | 0.000772156 | 0.004990048 | 1 |
| T2DM | leukocyte | *PTTG1IP* | -0.671332467 | 0.00862844 | 0.012584111 | 1 |
| T2DM | leukocyte | *ZNF623* | -0.713065231 | 0.00843111 | 0.012341964 | 1 |
| T2DM | leukocyte | *CCDC125* | -1.526050939 | 0.027237597 | 0.034700558 | 1 |
| T2DM | leukocyte | *BOLA2* | -0.91098447 | 0.001255185 | 0.004990048 | 1 |
| T2DM | leukocyte | *FHL3* | -0.748295361 | 0.016924399 | 0.022733662 | 1 |
| T2DM | leukocyte | *SF3A3* | -0.746161226 | 0.003795281 | 0.00696481 | 1 |
| T2DM | leukocyte | *PCBP3* | -1.592006069 | 0.00059242 | 0.004990048 | 1 |
| T2DM | leukocyte | *SFXN4* | -1.024614424 | 0.001294288 | 0.004990048 | 1 |
| T2DM | leukocyte | *MRPL54* | -0.89286298 | 0.000875046 | 0.004990048 | 1 |
| T2DM | leukocyte | *DGCR6* | -0.942907414 | 0.002773095 | 0.005963596 | 1 |
| T2DM | leukocyte | *NDUFB1* | -0.888964692 | 0.001130698 | 0.004990048 | 1 |
| T2DM | leukocyte | *TRMT12* | -1.052404413 | 0.001411711 | 0.004990048 | 1 |
| T2DM | leukocyte | *FIGLA* | -20.23652911 | 0.003162954 | 0.006314657 | 1 |
| T2DM | leukocyte | *CBX6* | -0.825556585 | 0.001033765 | 0.004990048 | 1 |
| T2DM | leukocyte | *MACC1* | -0.715144884 | 0.001025227 | 0.004990048 | 1 |
| T2DM | leukocyte | *KREMEN1* | -3.107581542 | 0.00069476 | 0.004990048 | 1 |
| T2DM | leukocyte | *NUDT14* | -0.70848157 | 0.009962727 | 0.014254754 | 1 |
| T2DM | leukocyte | *FAM3B* | -0.651355356 | 0.011690042 | 0.016403634 | 1 |
| T2DM | leukocyte | *TOB2* | -0.996203207 | 0.0021374 | 0.005317933 | 1 |
| T2DM | leukocyte | *NPIPA7* | -6.449713566 | 0.030976073 | 0.039221718 | 1 |
| T2DM | leukocyte | *TTC32* | -0.900168687 | 0.001509128 | 0.004993935 | 1 |
| T2DM | leukocyte | *TMEM236* | -0.618587133 | 0.00819304 | 0.012051837 | 1 |
| T2DM | leukocyte | *UQCR10* | -0.752902802 | 0.001355433 | 0.004990048 | 1 |
| T2DM | leukocyte | *NIPSNAP1* | -0.826896458 | 0.003377169 | 0.006503419 | 1 |
| T2DM | leukocyte | *CRELD2* | -0.639196654 | 0.005713361 | 0.008943345 | 1 |
| T2DM | leukocyte | *UBE2F* | -1.400144829 | 0.001410918 | 0.004990048 | 1 |
| T2DM | leukocyte | *GOLGA6L4* | -1.182007319 | 0.00113352 | 0.004990048 | 1 |
| T2DM | leukocyte | *PGP* | -1.554327387 | 5.48E-05 | 0.004896114 | 0.215429008 |
| T2DM | leukocyte | *ACOT1* | -5.458937874 | 0.003037228 | 0.006214704 | 1 |
| T2DM | leukocyte | *OAF* | -1.119392264 | 0.00161467 | 0.005033031 | 1 |
| T2DM | leukocyte | *TM2D3* | -0.883422428 | 0.001366362 | 0.004990048 | 1 |
| T2DM | leukocyte | *TOP1MT* | -2.300814502 | 0.000578558 | 0.004990048 | 1 |
| T2DM | leukocyte | *COPB2* | -1.058401361 | 0.000638646 | 0.004990048 | 1 |
| T2DM | leukocyte | *CCR10* | -1.653124753 | 0.01183482 | 0.016598917 | 1 |
| T2DM | leukocyte | *NCMAP* | -0.830135772 | 0.00454936 | 0.007568852 | 1 |
| T2DM | leukocyte | *TXNRD2* | -0.884847465 | 0.003147295 | 0.006311945 | 1 |
| T2DM | leukocyte | *ZFP1* | -1.177403457 | 0.001731912 | 0.005070552 | 1 |
| T2DM | leukocyte | *XPOT* | -0.856421377 | 0.00157985 | 0.005009694 | 1 |

| | | | | | | |
|---|---|---|---|---|---|---|
| T2DM | leukocyte | *C14orf180* | -89.7247995 | 0.004172038 | 0.007242343 | 1 |
| T2DM | leukocyte | *ZBTB40* | -1.115815589 | 0.018726689 | 0.024815232 | 1 |
| T2DM | leukocyte | *UBE2G2* | -0.920983289 | 0.001880186 | 0.005146173 | 1 |
| T2DM | leukocyte | *OSBP2* | -0.988412069 | 0.004177649 | 0.007242343 | 1 |
| T2DM | leukocyte | *TMED9* | -0.780891888 | 0.001226888 | 0.004990048 | 1 |
| T2DM | leukocyte | *BTBD6* | -1.198139929 | 0.00096713 | 0.004990048 | 1 |
| T2DM | leukocyte | *JAG2* | -0.668944852 | 0.014868916 | 0.02024254 | 1 |
| T2DM | leukocyte | *LCN12* | -3.301726179 | 0.000564782 | 0.004990048 | 1 |
| T2DM | leukocyte | *FAM227A* | -0.982753308 | 0.003088744 | 0.00625753 | 1 |
| T2DM | leukocyte | *MUC6* | -0.523793781 | 0.006269637 | 0.009633712 | 1 |
| T2DM | leukocyte | *NOC4L* | -1.524548903 | 0.000606222 | 0.004990048 | 1 |
| T2DM | leukocyte | *NDUFA6* | -0.538787563 | 0.014945668 | 0.020325902 | 1 |
| T2DM | leukocyte | *SIVA1* | -0.878938381 | 0.001449093 | 0.004990048 | 1 |
| T2DM | leukocyte | *BRI3BP* | -1.254840024 | 0.001443248 | 0.004990048 | 1 |
| T2DM | leukocyte | *UBOX5* | -0.861800427 | 0.003620322 | 0.00677345 | 1 |
| T2DM | leukocyte | *MAFF* | -0.528717463 | 0.006111407 | 0.009445995 | 1 |
| T2DM | leukocyte | *SEMA4B* | -0.818876275 | 0.005397465 | 0.008554513 | 1 |
| T2DM | leukocyte | *CIB1* | -0.871243451 | 0.000913201 | 0.004990048 | 1 |
| T2DM | leukocyte | *ANO9* | -1.236372237 | 0.007979378 | 0.011786082 | 1 |
| T2DM | leukocyte | *MFSD6L* | -2.363356557 | 0.008496678 | 0.012420662 | 1 |
| T2DM | leukocyte | *ZNF445* | -3.893548647 | 0.001031 | 0.004990048 | 1 |
| T2DM | leukocyte | *ZNF74* | -1.80463676 | 0.006892165 | 0.01043519 | 1 |
| T2DM | leukocyte | *RBM11* | -0.75665204 | 0.003025468 | 0.006200868 | 1 |
| T2DM | leukocyte | *RGPD2* | -2.578728554 | 0.004964624 | 0.008020562 | 1 |
| T2DM | leukocyte | *GAS2L1* | -2.074669025 | 0.00538798 | 0.008542928 | 1 |
| T2DM | leukocyte | *ATP6V0A2* | -1.454727861 | 0.001217265 | 0.004990048 | 1 |
| T2DM | leukocyte | *RAD51D* | -2.411118815 | 0.000627942 | 0.004990048 | 1 |
| T2DM | leukocyte | *SH3BGR* | -0.552111893 | 0.004604942 | 0.007624449 | 1 |
| T2DM | leukocyte | *MUC1* | -0.543729399 | 0.006552953 | 0.010022013 | 1 |
| T2DM | leukocyte | *IRF7* | -1.449058894 | 0.003558571 | 0.006702601 | 1 |
| T2DM | leukocyte | *FAM131C* | -0.646495023 | 0.002994181 | 0.006168923 | 1 |
| T2DM | leukocyte | *LSAMP* | -0.491055916 | 0.004335337 | 0.007353588 | 1 |
| T2DM | leukocyte | *OLFML2A* | -2.180874915 | 0.00992366 | 0.014209205 | 1 |
| T2DM | leukocyte | *PDIA2* | -0.39300887 | 0.012304611 | 0.017167904 | 1 |
| T2DM | leukocyte | *PCGF3* | -0.894860079 | 0.003214103 | 0.006372159 | 1 |
| T2DM | leukocyte | *P4HB* | -0.604353071 | 0.001856389 | 0.005146173 | 1 |
| T2DM | leukocyte | *PSMD13* | -1.199971464 | 5.69E-05 | 0.004970621 | 0.223677937 |
| T2DM | leukocyte | *BRWD1* | -1.018144936 | 0.000779239 | 0.004990048 | 1 |
| T2DM | leukocyte | *DRG1* | -0.841892081 | 0.001951757 | 0.005198138 | 1 |
| T2DM | leukocyte | *ADAMTSL5* | -5.773035253 | 0.017460565 | 0.023326269 | 1 |
| T2DM | leukocyte | *MORF4L1* | -0.69682556 | 0.00112431 | 0.004990048 | 1 |
| T2DM | leukocyte | *DMWD* | -2.004296925 | 2.19E-05 | 0.003059063 | 0.085914715 |
| T2DM | leukocyte | *PIGP* | -0.764218589 | 0.000851184 | 0.004990048 | 1 |
| T2DM | leukocyte | *NPAP1* | -21.94676388 | 0.03261288 | 0.041188044 | 1 |
| T2DM | leukocyte | *ARL17A* | -0.585626495 | 0.02244836 | 0.029205167 | 1 |
| T2DM | leukocyte | *TRIM69* | -0.845782191 | 0.006837875 | 0.010369201 | 1 |
| T2DM | leukocyte | *IFITM1* | -1.864900665 | 0.005787439 | 0.009037699 | 1 |
| T2DM | leukocyte | *SETD4* | -3.111031148 | 0.003192887 | 0.006345399 | 1 |
| T2DM | leukocyte | *ZNF529* | -1.621808803 | 0.001340277 | 0.004990048 | 1 |
| T2DM | leukocyte | *AIDA* | -0.850391 | 0.001257899 | 0.004990048 | 1 |

| | | | | | | |
|---|---|---|---|---|---|---|
| T2DM | leukocyte | *WWOX* | -0.907121591 | 0.001841007 | 0.005146173 | 1 |
| T2DM | leukocyte | *POLR1D* | -0.818261339 | 0.000942191 | 0.004990048 | 1 |
| T2DM | leukocyte | *EDARADD* | -0.934429486 | 0.002448677 | 0.005621365 | 1 |
| T2DM | leukocyte | *ZNF749* | -0.686454263 | 0.001860876 | 0.005146173 | 1 |
| T2DM | leukocyte | *TOR3A* | -0.991779491 | 0.001063246 | 0.004990048 | 1 |
| T2DM | leukocyte | *ZNF555* | -0.71199055 | 0.003759114 | 0.00692001 | 1 |
| T2DM | leukocyte | *NUDT17* | -1.37252592 | 0.001070023 | 0.004990048 | 1 |
| T2DM | leukocyte | *CYP4X1* | 1.060021271 | 0.023240014 | 0.030105511 | 1 |
| T2DM | leukocyte | *ZNF197* | -0.835042053 | 0.002114154 | 0.005283012 | 1 |
| T2DM | leukocyte | *RPS23* | -0.644083038 | 0.000664894 | 0.004990048 | 1 |
| T2DM | leukocyte | *BTN3A2* | -0.422781015 | 0.022572636 | 0.029328004 | 1 |
| T2DM | leukocyte | *ZNF396* | -2.394089777 | 0.002075284 | 0.005277533 | 1 |
| T2DM | leukocyte | *CEACAM19* | -0.955918552 | 0.000129441 | 0.004990048 | 0.508573382 |
| T2DM | leukocyte | *ZFP91* | -1.048569755 | 0.001524078 | 0.004994246 | 1 |
| T2DM | leukocyte | *BCR* | -0.861244991 | 0.001757875 | 0.005084476 | 1 |
| T2DM | leukocyte | *HYAL3* | -1.753851487 | 0.000720427 | 0.004990048 | 1 |
| T2DM | leukocyte | *RTN4RL2* | -0.884821303 | 0.004290022 | 0.00732537 | 1 |
| T2DM | leukocyte | *TMEM232* | -1.382058193 | 0.002478924 | 0.005622792 | 1 |
| T2DM | leukocyte | *FAM183A* | -0.719899733 | 0.004141428 | 0.007238288 | 1 |
| T2DM | leukocyte | *EMID1* | -1.118529054 | 0.003159932 | 0.006314657 | 1 |
| T2DM | leukocyte | *ESPN* | -19.10726172 | 0.000892658 | 0.004990048 | 1 |
| T2DM | leukocyte | *TMEM216* | -2.265994197 | 0.03949684 | 0.049077509 | 1 |
| T2DM | leukocyte | *LYPD6* | -3.071636627 | 0.025578735 | 0.032828617 | 1 |
| T2DM | leukocyte | *AKR1C1* | -0.620544223 | 0.006076084 | 0.009402422 | 1 |
| T2DM | leukocyte | *BCAM* | -0.697036513 | 0.001481963 | 0.004990048 | 1 |
| T2DM | leukocyte | *CHP1* | -0.758139691 | 0.001730815 | 0.005070552 | 1 |
| T2DM | leukocyte | *KCNJ11* | -2.918644184 | 0.001168114 | 0.004990048 | 1 |
| T2DM | leukocyte | *ZNF385C* | -3.624860173 | 0.000198431 | 0.004990048 | 0.779636481 |
| T2DM | leukocyte | *EXD3* | -0.929268494 | 0.00355 | 0.006692874 | 1 |
| T2DM | leukocyte | *RGPD1* | -95.8845659 | 0.004455179 | 0.007483711 | 1 |
| T2DM | leukocyte | *DHRS4L2* | -0.947159525 | 0.001364369 | 0.004990048 | 1 |
| T2DM | leukocyte | *PALM3* | -1.083179082 | 0.00391695 | 0.007085756 | 1 |
| T2DM | leukocyte | *KLHL17* | -1.439892246 | 0.004723182 | 0.00774192 | 1 |
| T2DM | leukocyte | *RINL* | -1.177011226 | 0.001460195 | 0.004990048 | 1 |
| T2DM | leukocyte | *ZNF490* | -1.51955664 | 0.003703315 | 0.006868344 | 1 |
| T2DM | leukocyte | *NRN1L* | -2.287168182 | 0.004336539 | 0.007353588 | 1 |
| T2DM | leukocyte | *RAB42* | 0.420149417 | 0.020496255 | 0.026888076 | 1 |
| T2DM | leukocyte | *MAPK12* | -0.871021854 | 0.01766751 | 0.023570678 | 1 |
| T2DM | leukocyte | *LINC00265* | -1.814810285 | 0.004069281 | 0.007189 | 1 |
| T2DM | leukocyte | *LAMTOR4* | -0.766086168 | 0.003627532 | 0.006781089 | 1 |
| T2DM | leukocyte | *PRKAR1B* | -1.063012397 | 0.00338979 | 0.006510382 | 1 |
| T2DM | leukocyte | *NUTM2B* | -28.44657379 | 6.79E-07 | 0.000296569 | 0.002669121 |
| T2DM | leukocyte | *AGAP4* | -1.95950769 | 0.005975289 | 0.009268422 | 1 |
| T2DM | leukocyte | *CENPP* | -0.733797515 | 0.004027728 | 0.007161461 | 1 |
| T2DM | leukocyte | *ZNF559* | -0.585295396 | 0.026643672 | 0.034043248 | 1 |
| T2DM | leukocyte | *FOCAD* | -0.96625666 | 0.002106908 | 0.005283012 | 1 |
| T2DM | leukocyte | *ZP3* | -1.201812319 | 0.004028208 | 0.007161461 | 1 |
| T2DM | leukocyte | *SRSF10* | -0.850344854 | 0.001798106 | 0.005114303 | 1 |
| T2DM | leukocyte | *RALGAPA2* | -0.965918359 | 0.001660899 | 0.005041098 | 1 |
| T2DM | leukocyte | *NDOR1* | -2.065289293 | 0.017080279 | 0.022921042 | 1 |

| | | | | | | |
|---|---|---|---|---|---|---|
| T2DM | leukocyte | *FBLL1* | -2.074422335 | 0.000690957 | 0.004990048 | 1 |
| T2DM | leukocyte | *ASAH2* | -2.728575462 | 6.64E-05 | 0.004990048 | 0.260896276 |
| T2DM | leukocyte | *S100A16* | -0.468857454 | 0.003748409 | 0.006917566 | 1 |
| T2DM | leukocyte | *PARVB* | -0.699452886 | 0.000841613 | 0.004990048 | 1 |
| T2DM | leukocyte | *UROS* | -1.139646853 | 0.00092461 | 0.004990048 | 1 |
| T2DM | leukocyte | *FAM221A* | -1.531107084 | 0.000118859 | 0.004990048 | 0.466998493 |
| T2DM | leukocyte | *NOXA1* | -1.591794211 | 0.003740306 | 0.006909103 | 1 |
| T2DM | leukocyte | *TMEM198* | -0.704443493 | 0.005765769 | 0.009007438 | 1 |
| T2DM | leukocyte | *ZNF548* | -0.926681448 | 0.013656631 | 0.018794011 | 1 |
| T2DM | leukocyte | *MTF1* | -0.894453101 | 0.002278362 | 0.005462378 | 1 |
| T2DM | leukocyte | *ENTPD8* | -5.486111681 | 0.004654622 | 0.007684038 | 1 |
| T2DM | leukocyte | *RPL14* | -0.591951808 | 0.001257755 | 0.004990048 | 1 |
| T2DM | leukocyte | *MSL1* | -0.908644251 | 0.003219327 | 0.006372159 | 1 |
| T2DM | leukocyte | *NOC2L* | -1.18516461 | 0.00069042 | 0.004990048 | 1 |
| T2DM | leukocyte | *ZNF567* | -0.780499824 | 0.008497516 | 0.012420662 | 1 |
| T2DM | leukocyte | *ARID2* | -1.227510754 | 0.001053487 | 0.004990048 | 1 |
| T2DM | leukocyte | *PLAC9* | -0.931639615 | 0.002081319 | 0.005283012 | 1 |
| T2DM | leukocyte | *CLDN4* | -0.503557171 | 0.002888329 | 0.00605886 | 1 |
| T2DM | leukocyte | *FAM47E* | -1.381187013 | 0.000396141 | 0.004990048 | 1 |
| T2DM | leukocyte | *ZNF527* | -1.039085417 | 0.01149312 | 0.01615616 | 1 |
| T2DM | leukocyte | *ZNF600* | -0.858700849 | 0.007619975 | 0.011353388 | 1 |
| T2DM | leukocyte | *PNRC2* | -0.776485673 | 0.001873534 | 0.005146173 | 1 |
| T2DM | leukocyte | *ZKSCAN3* | -1.749124658 | 0.004774591 | 0.007806645 | 1 |
| T2DM | leukocyte | *RRP7A* | -0.904563479 | 0.004674001 | 0.007686961 | 1 |
| T2DM | leukocyte | *S100A14* | -0.661953772 | 0.010448065 | 0.014857201 | 1 |
| T2DM | leukocyte | *CXCL17* | -0.680855687 | 0.0071798 | 0.010795804 | 1 |
| T2DM | leukocyte | *SPATA41* | -2.911091262 | 0.027217531 | 0.034686241 | 1 |
| T2DM | leukocyte | *BLOC1S2* | -0.959221271 | 0.001192914 | 0.004990048 | 1 |
| T2DM | leukocyte | *ZNF676* | -1.070293852 | 0.005297774 | 0.008427107 | 1 |
| T2DM | leukocyte | *WDSUB1* | -1.090641324 | 0.004406042 | 0.007420205 | 1 |
| T2DM | leukocyte | *ZNF681* | -5.277318764 | 0.000155258 | 0.004990048 | 0.61000834 |
| T2DM | leukocyte | *HRCT1* | 0.829605949 | 0.033694841 | 0.042472579 | 1 |
| T2DM | leukocyte | *MPHOSPH8* | -0.888008904 | 0.001478533 | 0.004990048 | 1 |
| T2DM | leukocyte | *SIRPB2* | 1.409416118 | 0.017391605 | 0.023249955 | 1 |
| T2DM | leukocyte | *LCOR* | -0.825270634 | 0.001520862 | 0.004993935 | 1 |
| T2DM | leukocyte | *SUPT5H* | -0.801425702 | 0.001393308 | 0.004990048 | 1 |
| T2DM | leukocyte | *XPNPEP3* | -1.493713065 | 0.000278549 | 0.004990048 | 1 |
| T2DM | leukocyte | *PPIA* | -0.725979565 | 0.001476077 | 0.004990048 | 1 |
| T2DM | leukocyte | *ZNF471* | -1.222633458 | 0.00432002 | 0.007335073 | 1 |
| T2DM | leukocyte | *ZNF493* | -1.231534615 | 0.001180752 | 0.004990048 | 1 |
| T2DM | leukocyte | *GTF2IRD2* | -1.30237722 | 0.000986601 | 0.004990048 | 1 |
| T2DM | leukocyte | *SUPT3H* | -0.775326446 | 0.003040316 | 0.006215089 | 1 |
| T2DM | leukocyte | *POM121* | -0.873061829 | 0.00444214 | 0.007470068 | 1 |
| T2DM | leukocyte | *LONP1* | -1.287546276 | 0.001520485 | 0.004993935 | 1 |
| T2DM | leukocyte | *ZNF34* | -1.197499345 | 0.004127339 | 0.00723368 | 1 |
| T2DM | leukocyte | *ZNF781* | -1.114679593 | 0.013254617 | 0.018343568 | 1 |
| T2DM | leukocyte | *EVL* | -1.65478403 | 0.008639226 | 0.012594998 | 1 |
| T2DM | leukocyte | *ZNF124* | -1.513468504 | 0.00156845 | 0.005009694 | 1 |
| T2DM | leukocyte | *XRCC6* | -0.970178489 | 0.00021594 | 0.004990048 | 0.848427746 |
| T2DM | leukocyte | *PIK3R4* | -1.522773826 | 0.001213932 | 0.004990048 | 1 |

| | | | | | | |
|---|---|---|---|---|---|---|
| T2DM | leukocyte | *C20orf96* | -1.768175097 | 0.001758926 | 0.005084476 | 1 |
| T2DM | leukocyte | *SULT1A1* | -2.007718901 | 0.000386199 | 0.004990048 | 1 |
| T2DM | leukocyte | *ARL9* | 0.917917038 | 0.02644921 | 0.033838797 | 1 |
| T2DM | leukocyte | *TPK1* | -1.8940338 | 0.012519174 | 0.017436312 | 1 |
| T2DM | leukocyte | *AFAP1* | -0.937177955 | 0.005292834 | 0.008425434 | 1 |
| T2DM | leukocyte | *MYO18A* | -0.813898468 | 0.001889502 | 0.005146173 | 1 |
| T2DM | leukocyte | *XRCC2* | -3.299565187 | 0.002825187 | 0.006006579 | 1 |
| T2DM | leukocyte | *MYO6* | -0.659163708 | 0.002239395 | 0.005438799 | 1 |
| T2DM | leukocyte | *ZNF502* | -3.987090878 | 0.000304896 | 0.004990048 | 1 |
| T2DM | leukocyte | *TRAPPC4* | -0.755923277 | 0.005090883 | 0.008180809 | 1 |
| T2DM | leukocyte | *ZNF33B* | -1.754526027 | 0.000411305 | 0.004990048 | 1 |
| T2DM | leukocyte | *AMZ2* | -1.121907863 | 0.001255722 | 0.004990048 | 1 |
| T2DM | leukocyte | *ZNF431* | -0.922187565 | 0.001756251 | 0.005084476 | 1 |
| T2DM | leukocyte | *DAPK1* | -1.567648393 | 0.001171866 | 0.004990048 | 1 |
| T2DM | leukocyte | *HLA-DQA1* | 0.060242445 | 0.039734421 | 0.049341511 | 1 |
| T2DM | leukocyte | *CLPSL2* | -0.855839702 | 0.024938975 | 0.032105254 | 1 |
| T2DM | leukocyte | *MVB12B* | -4.234584586 | 0.006689179 | 0.010182792 | 1 |
| T2DM | leukocyte | *ADA* | 0.407175881 | 0.016621141 | 0.022372204 | 1 |
| T2DM | leukocyte | *PPTC7* | -1.515158158 | 0.000313236 | 0.004990048 | 1 |
| T2DM | leukocyte | *ZFP28* | -1.293105065 | 0.001911562 | 0.00515389 | 1 |
| T2DM | leukocyte | *ANKRD36B* | -0.901331726 | 0.010510122 | 0.014928397 | 1 |
| T2DM | leukocyte | *SRGAP1* | -1.055408478 | 0.001857727 | 0.005146173 | 1 |
| T2DM | leukocyte | *FAM3C* | -0.746870765 | 0.001422877 | 0.004990048 | 1 |
| T2DM | leukocyte | *AP2A1* | -1.177249767 | 0.000942115 | 0.004990048 | 1 |
| T2DM | leukocyte | *WDR5B* | -2.681032311 | 0.002031765 | 0.005251844 | 1 |
| T2DM | leukocyte | *ZNF138* | -0.589198724 | 0.017659031 | 0.023570678 | 1 |
| T2DM | leukocyte | *ZNF429* | -0.854736644 | 0.002476874 | 0.005622792 | 1 |
| T2DM | leukocyte | *ZNF100* | -1.381870129 | 0.000348821 | 0.004990048 | 1 |
| T2DM | leukocyte | *ZNF763* | -2.183591177 | 0.00055343 | 0.004990048 | 1 |
| T2DM | leukocyte | *ARRDC1* | -0.793658939 | 0.004306391 | 0.007328649 | 1 |
| T2DM | leukocyte | *ZNF682* | -1.718034258 | 0.001256146 | 0.004990048 | 1 |
| T2DM | leukocyte | *ZNF257* | -1.458802554 | 0.00140436 | 0.004990048 | 1 |
| T2DM | leukocyte | *ADAM32* | 0.887186851 | 0.032403057 | 0.040949376 | 1 |
| T2DM | leukocyte | *ACSL5* | -1.405937635 | 0.004252727 | 0.007299678 | 1 |
| T2DM | leukocyte | *SND1* | -0.893797831 | 0.001518523 | 0.004993935 | 1 |
| T2DM | leukocyte | *TBC1D9B* | -1.903446289 | 0.000161616 | 0.004990048 | 0.634987616 |
| T2DM | leukocyte | *C6orf141* | -1.853175619 | 0.001168014 | 0.004990048 | 1 |
| T2DM | leukocyte | *GTF2E2* | -0.85683752 | 0.002175537 | 0.005349178 | 1 |
| T2DM | leukocyte | *ZNF165* | -1.007578118 | 0.001388452 | 0.004990048 | 1 |
| T2DM | leukocyte | *SVIL* | -0.817636939 | 0.002455796 | 0.005621365 | 1 |
| T2DM | leukocyte | *MRPL21* | -0.697110381 | 0.013318567 | 0.018393198 | 1 |
| T2DM | leukocyte | *ZNF98* | -1.207918835 | 0.004375225 | 0.007387305 | 1 |
| T2DM | leukocyte | *ZNF517* | -3.627894793 | 0.001832757 | 0.005146173 | 1 |
| T2DM | leukocyte | *SLC22A5* | -1.348298719 | 0.001907608 | 0.00515389 | 1 |
| T2DM | leukocyte | *C5AR1* | 0.102665797 | 0.016913469 | 0.022726751 | 1 |
| T2DM | leukocyte | *VEPH1* | -1.520596513 | 0.012841514 | 0.017828378 | 1 |
| T2DM | leukocyte | *SHPK* | -0.881785664 | 0.034935675 | 0.043839753 | 1 |
| T2DM | leukocyte | *PCDHB11* | -2.227241899 | 6.86E-05 | 0.004990048 | 0.269476384 |
| T2DM | leukocyte | *ATG7* | -1.809795524 | 0.000922697 | 0.004990048 | 1 |
| T2DM | leukocyte | *PIGN* | -0.769575861 | 0.004949857 | 0.008003288 | 1 |

| | | | | | | |
|---|---|---|---|---|---|---|
| T2DM | leukocyte | *HHLA3* | -0.796841339 | 0.004443255 | 0.007470068 | 1 |
| T2DM | leukocyte | *BCO2* | -0.789316236 | 0.016359556 | 0.022073041 | 1 |
| T2DM | leukocyte | *ENTPD6* | -1.119262678 | 0.000756197 | 0.004990048 | 1 |
| T2DM | leukocyte | *ENPP1* | -0.545517331 | 0.030802982 | 0.039016835 | 1 |
| T2DM | leukocyte | *CCDC154* | 3.778833164 | 0.007219191 | 0.010842585 | 1 |
| T2DM | leukocyte | *DPP4* | -0.650355355 | 0.001314124 | 0.004990048 | 1 |
| T2DM | leukocyte | *NMB* | -0.887206256 | 0.001872235 | 0.005146173 | 1 |
| T2DM | leukocyte | *RPS26* | -0.750638956 | 0.000245688 | 0.004990048 | 0.965309009 |
| T2DM | leukocyte | *LHFPL5* | -2.259058825 | 0.002467448 | 0.005622792 | 1 |
| T2DM | leukocyte | *ZNF780A* | -0.789268596 | 0.00280649 | 0.006001505 | 1 |
| T2DM | leukocyte | *ATAD3A* | -1.563314448 | 0.001251832 | 0.004990048 | 1 |
| T2DM | leukocyte | *FAM118B* | -2.069613207 | 0.00152093 | 0.004993935 | 1 |
| T2DM | leukocyte | *SLC9A8* | -3.225103823 | 0.00043222 | 0.004990048 | 1 |
| T2DM | leukocyte | *OCLN* | -0.475885312 | 0.020376001 | 0.026748181 | 1 |
| T2DM | leukocyte | *ZNF181* | -1.218822075 | 0.002982366 | 0.006152298 | 1 |
| T2DM | leukocyte | *TEAD4* | -0.722357006 | 0.004866125 | 0.007914645 | 1 |
| T2DM | leukocyte | *C2orf27A* | -1.552244243 | 0.000600819 | 0.004990048 | 1 |
| T2DM | leukocyte | *PLCG2* | -1.065024474 | 0.001027835 | 0.004990048 | 1 |
| T2DM | leukocyte | *RPL12* | -0.590559727 | 0.000843175 | 0.004990048 | 1 |
| T2DM | leukocyte | *VPS13A* | -0.826906416 | 0.003052657 | 0.006227356 | 1 |
| T2DM | leukocyte | *LEKR1* | -1.544801056 | 0.00266135 | 0.00583507 | 1 |
| T2DM | leukocyte | *ENTPD7* | -1.437156288 | 0.021761474 | 0.028424478 | 1 |
| T2DM | leukocyte | *AGAP9* | -1.813873022 | 0.002957593 | 0.006135605 | 1 |
| T2DM | leukocyte | *ZNF273* | -1.779739181 | 0.001257158 | 0.004990048 | 1 |
| T2DM | leukocyte | *ZNF84* | -1.937009714 | 0.001136568 | 0.004990048 | 1 |
| T2DM | leukocyte | *ZNF667* | -0.643486373 | 0.003761507 | 0.00692001 | 1 |
| T2DM | leukocyte | *CD2AP* | -0.905311151 | 0.001351135 | 0.004990048 | 1 |
| T2DM | leukocyte | *SFI1* | -1.574246888 | 0.009815346 | 0.01405924 | 1 |
| T2DM | leukocyte | *HIBCH* | -0.804056067 | 0.002749495 | 0.005935586 | 1 |
| T2DM | leukocyte | *TMEM229B* | -4.280835378 | 0.001497088 | 0.004993935 | 1 |
| T2DM | leukocyte | *SOWAHC* | -1.616310027 | 0.000918471 | 0.004990048 | 1 |
| T2DM | leukocyte | *SVIP* | -0.811978681 | 0.001000625 | 0.004990048 | 1 |
| T2DM | leukocyte | *DDRGK1* | -1.0055668 | 0.001022083 | 0.004990048 | 1 |
| T2DM | leukocyte | *ZNF607* | -0.924199593 | 0.012336937 | 0.017206895 | 1 |
| T2DM | leukocyte | *HSD17B11* | -1.24953762 | 0.003146298 | 0.006311945 | 1 |
| T2DM | leukocyte | *RPS6KL1* | -1.026642129 | 0.008441656 | 0.012352799 | 1 |
| T2DM | leukocyte | *QRICH1* | -1.454539834 | 0.001641379 | 0.005035955 | 1 |
| T2DM | leukocyte | *DDX42* | -0.673158489 | 0.007895467 | 0.011692277 | 1 |
| T2DM | leukocyte | *HELZ* | -0.986875346 | 0.001201476 | 0.004990048 | 1 |
| T2DM | leukocyte | *ZNF583* | -0.768843161 | 0.003848981 | 0.007027251 | 1 |
| T2DM | leukocyte | *ZNF568* | -0.873235196 | 0.010391614 | 0.014787632 | 1 |
| T2DM | leukocyte | *ZNF587* | -1.040322638 | 0.0019378 | 0.00518571 | 1 |
| T2DM | leukocyte | *TPM2* | -1.006406017 | 0.001851429 | 0.005146173 | 1 |
| T2DM | leukocyte | *ZNF789* | -2.25405706 | 0.002452266 | 0.005621365 | 1 |
| T2DM | leukocyte | *CCDC69* | -0.795040547 | 0.035363789 | 0.044348686 | 1 |
| T2DM | leukocyte | *NCOA6* | -0.825180242 | 0.001996106 | 0.005227473 | 1 |
| T2DM | leukocyte | *STK39* | -0.791497095 | 0.003291941 | 0.006432819 | 1 |
| T2DM | leukocyte | *PAPSS2* | -0.754781937 | 0.001898446 | 0.005146173 | 1 |
| T2DM | leukocyte | *CEP290* | -1.007450401 | 0.001876156 | 0.005146173 | 1 |
| T2DM | leukocyte | *ECI2* | -1.023576252 | 0.001148562 | 0.004990048 | 1 |

| | | | | | | |
|---|---|---|---|---|---|---|
| T2DM | leukocyte | *MSRB1* | -1.183750424 | 0.001780225 | 0.005098084 | 1 |
| T2DM | leukocyte | *COLGALT2* | -1.300942491 | 0.006565838 | 0.010030007 | 1 |
| T2DM | leukocyte | *PNP* | -0.478070405 | 0.017636202 | 0.023552902 | 1 |
| T2DM | leukocyte | *DENND4B* | -0.959907351 | 0.00226566 | 0.005457772 | 1 |
| T2DM | leukocyte | *CES1* | -1.619493644 | 0.014055065 | 0.019288281 | 1 |
| T2DM | leukocyte | *TSEN15* | -1.12719002 | 0.001298444 | 0.004990048 | 1 |
| T2DM | leukocyte | *LTN1* | -0.910675614 | 0.001888636 | 0.005146173 | 1 |
| T2DM | leukocyte | *TYW1* | -1.310020794 | 0.003574082 | 0.006719453 | 1 |
| T2DM | leukocyte | *MAP3K3* | -1.069677096 | 0.000687599 | 0.004990048 | 1 |
| T2DM | leukocyte | *C1orf174* | -1.151451946 | 0.001646885 | 0.005035955 | 1 |
| T2DM | leukocyte | *KIAA0753* | -1.995680574 | 0.000846398 | 0.004990048 | 1 |
| T2DM | leukocyte | *ATG9A* | -0.925715554 | 0.012894405 | 0.017882851 | 1 |
| T2DM | leukocyte | *TBKBP1* | -1.230156299 | 0.000152417 | 0.004990048 | 0.598845209 |
| T2DM | leukocyte | *CCDC167* | -0.701730103 | 0.003311258 | 0.00644375 | 1 |
| T2DM | leukocyte | *SOWAHA* | -0.816673774 | 0.002286778 | 0.005462378 | 1 |
| T2DM | leukocyte | *L3MBTL3* | -1.625741958 | 0.000675703 | 0.004990048 | 1 |
| T2DM | leukocyte | *MFAP3L* | -1.428859389 | 0.003174125 | 0.006330221 | 1 |
| T2DM | leukocyte | *PJA2* | -0.793952779 | 0.001465597 | 0.004990048 | 1 |
| T2DM | leukocyte | *INF2* | -1.356983108 | 0.001522629 | 0.004993935 | 1 |
| T2DM | leukocyte | *EFCAB2* | -0.947790945 | 0.002084734 | 0.005283012 | 1 |
| T2DM | leukocyte | *SAMD5* | -1.160822287 | 0.000927899 | 0.004990048 | 1 |
| T2DM | leukocyte | *SPRN* | -4.585796346 | 0.018048266 | 0.024021557 | 1 |
| T2DM | leukocyte | *C6orf163* | -1.526573889 | 0.00872343 | 0.012694206 | 1 |
| T2DM | leukocyte | *PCMTD2* | -0.875272968 | 0.001880653 | 0.005146173 | 1 |
| T2DM | leukocyte | *LIME1* | -1.617216303 | 0.00091412 | 0.004990048 | 1 |
| T2DM | leukocyte | *FOXO6* | -1.777186112 | 0.002401715 | 0.005580331 | 1 |
| T2DM | leukocyte | *SYS1* | -0.82076714 | 0.005263891 | 0.008397006 | 1 |
| T2DM | leukocyte | *INPP5B* | -1.725787139 | 0.000269811 | 0.004990048 | 1 |
| T2DM | leukocyte | *TRAF3IP1* | -2.841451179 | 0.000183983 | 0.004990048 | 0.722870511 |
| T2DM | leukocyte | *CLPSL1* | -0.465004828 | 0.003938843 | 0.007108734 | 1 |
| T2DM | leukocyte | *HSD17B8* | -0.766493887 | 0.019194823 | 0.02535007 | 1 |
| T2DM | leukocyte | *HLA-DOA* | 0.135485183 | 0.014428378 | 0.019724807 | 1 |
| T2DM | leukocyte | *HLA-DMA* | 0.173186217 | 0.009137408 | 0.013208564 | 1 |
| T2DM | leukocyte | *NOTCH4* | -1.657957596 | 1.39E-08 | 1.91E-05 | 5.45E-05 |
| T2DM | leukocyte | *RNF5* | -1.363808206 | 5.11E-07 | 0.000266998 | 0.002008765 |
| T2DM | leukocyte | *MRPL38* | -0.932708924 | 0.001567375 | 0.005009694 | 1 |
| T2DM | leukocyte | *SMIM5* | -0.687110155 | 0.005610194 | 0.008817809 | 1 |
| T2DM | leukocyte | *EHMT2* | -3.059663047 | 0.00177626 | 0.005098084 | 1 |
| T2DM | leukocyte | *LAYN* | -0.802710072 | 0.004757486 | 0.007788402 | 1 |
| T2DM | leukocyte | *SLC44A4* | -0.653560642 | 0.003655284 | 0.006816142 | 1 |
| T2DM | leukocyte | *MICA* | -0.493033314 | 0.009969826 | 0.014259718 | 1 |
| T2DM | leukocyte | *HLA-C* | -0.672251356 | 8.75E-05 | 0.004990048 | 0.343612634 |
| T2DM | leukocyte | *CCHCR1* | 1.104960484 | 0.017188048 | 0.023024835 | 1 |
| T2DM | leukocyte | *PSORS1C1* | -0.693009406 | 0.004706159 | 0.007720459 | 1 |
| T2DM | leukocyte | *DHX16* | -1.060696636 | 0.00224221 | 0.005438799 | 1 |
| T2DM | leukocyte | *ABCF1* | -0.879453573 | 0.003091338 | 0.00625753 | 1 |
| T2DM | leukocyte | *ZNF468* | -2.360136907 | 0.000538456 | 0.004990048 | 1 |
| T2DM | leukocyte | *HLA-G* | -0.640525432 | 0.033738212 | 0.04251361 | 1 |
| T2DM | leukocyte | *ASPDH* | -1.58928782 | 0.002743203 | 0.005930931 | 1 |
| T2DM | leukocyte | *ZNF783* | -4.15926394 | 0.008921677 | 0.012934786 | 1 |

| | | | | | | |
|---|---|---|---|---|---|---|
| T2DM | leukocyte | *ZNF425* | -2.82996556 | 0.0002709 | 0.004990048 | 1 |
| T2DM | leukocyte | *PCDHA5* | -1.131894833 | 0.025564993 | 0.032825117 | 1 |
| T2DM | leukocyte | *SPIRE2* | -0.721299971 | 0.007361738 | 0.011031377 | 1 |
| T2DM | leukocyte | *PABPN1L* | -123.7148303 | 0.002461812 | 0.005622792 | 1 |
| T2DM | leukocyte | *PKHD1L1* | -3.453667611 | 0.005004826 | 0.008072234 | 1 |
| T2DM | leukocyte | *SYCE1L* | -1.222358058 | 0.007536643 | 0.011246286 | 1 |
| T2DM | leukocyte | *TMEM231* | -0.882375444 | 0.00516683 | 0.008269033 | 1 |
| T2DM | leukocyte | *LINC00654* | -3.097117135 | 0.02509905 | 0.032279597 | 1 |
| T2DM | leukocyte | *SARNP* | -0.864766954 | 0.001725071 | 0.005070552 | 1 |
| T2DM | leukocyte | *MT1A* | -0.655141081 | 0.000743304 | 0.004990048 | 1 |
| T2DM | leukocyte | *ATP6AP1L* | 1.581890566 | 0.015596067 | 0.021144564 | 1 |
| T2DM | leukocyte | *CCDC85C* | -0.924701011 | 0.003149531 | 0.006311945 | 1 |
| T2DM | leukocyte | *RGL3* | -1.68226902 | 0.001693702 | 0.005060368 | 1 |
| T2DM | leukocyte | *CPT1B* | -0.736867713 | 0.038698483 | 0.048192183 | 1 |
| T2DM | leukocyte | *SMN2* | -0.926078785 | 0.014529949 | 0.01984985 | 1 |
| T2DM | leukocyte | *SERF1B* | -8.521975441 | 0.004457482 | 0.007484379 | 1 |
| T2DM | leukocyte | *HMGN1* | -0.747999037 | 0.000720246 | 0.004990048 | 1 |
| T2DM | leukocyte | *LCMT1* | -1.281438732 | 0.000951265 | 0.004990048 | 1 |
| T2DM | leukocyte | *LIN52* | -2.05318686 | 0.000931236 | 0.004990048 | 1 |
| T2DM | leukocyte | *DENND1C* | 0.312266086 | 0.016822248 | 0.022627392 | 1 |
| T2DM | leukocyte | *RNPS1* | -0.838467158 | 0.001898439 | 0.005146173 | 1 |
| T2DM | leukocyte | *ZDHHC11B* | -1.507850187 | 0.003282479 | 0.006429143 | 1 |
| T2DM | leukocyte | *HLA-A* | -0.562067715 | 0.002243945 | 0.005438799 | 1 |
| T2DM | leukocyte | *STK38L* | -1.590774832 | 0.002333954 | 0.005515664 | 1 |
| T2DM | leukocyte | *MAP10* | -0.912334038 | 0.009612523 | 0.013803948 | 1 |
| T2DM | leukocyte | *NUP62* | -0.966538628 | 0.00240985 | 0.005586019 | 1 |
| T2DM | leukocyte | *FAM106A* | -1.095370879 | 0.010435252 | 0.014844353 | 1 |
| T2DM | leukocyte | *ZNF254* | -1.209852877 | 0.000487455 | 0.004990048 | 1 |
| T2DM | leukocyte | *LINGO4* | -0.969858194 | 0.03117188 | 0.039456932 | 1 |
| T2DM | leukocyte | *ASIC3* | -0.91570891 | 0.034732372 | 0.043598559 | 1 |
| T2DM | leukocyte | *ARHGEF35* | -0.598720562 | 0.004579685 | 0.007595434 | 1 |
| T2DM | leukocyte | *DNLZ* | 0.913955604 | 0.005465788 | 0.008631969 | 1 |
| T2DM | leukocyte | *SUPT4H1* | -1.104563117 | 0.000988769 | 0.004990048 | 1 |
| T2DM | leukocyte | *MXD3* | -0.84964437 | 0.008304586 | 0.012183988 | 1 |
| T2DM | leukocyte | *COG8* | -1.228228176 | 0.003968716 | 0.007124893 | 1 |
| T2DM | leukocyte | *ERV3-1* | -1.160265333 | 0.017002764 | 0.022831121 | 1 |
| T2DM | leukocyte | *RBMXL1* | -0.993996714 | 0.001561114 | 0.005009694 | 1 |
| T2DM | leukocyte | *SRA1* | -0.654207886 | 0.004314317 | 0.007328649 | 1 |
| T2DM | leukocyte | *C8orf82* | -1.762631828 | 0.000935107 | 0.004990048 | 1 |
| T2DM | leukocyte | *LEPROT* | -0.918545098 | 0.00111231 | 0.004990048 | 1 |
| T2DM | leukocyte | *PPP1CB* | -0.739090099 | 0.00122175 | 0.004990048 | 1 |
| T2DM | leukocyte | *NCKIPSD* | -1.329542972 | 0.001807199 | 0.005122825 | 1 |
| T2DM | leukocyte | *ZNF888* | -1.28222639 | 0.003398845 | 0.006511001 | 1 |
| T2DM | leukocyte | *EMP2* | -1.004049098 | 0.001325264 | 0.004990048 | 1 |
| T2DM | leukocyte | *LTB4R* | -1.199453823 | 0.003511128 | 0.0066483 | 1 |
| T2DM | leukocyte | *DNASE1* | -0.723001594 | 0.010567569 | 0.014994575 | 1 |
| T2DM | leukocyte | *NUDT19* | -1.238724633 | 0.001573864 | 0.005009694 | 1 |
| T2DM | leukocyte | *AP1G2* | -3.810255428 | 0.000310161 | 0.004990048 | 1 |
| T2DM | leukocyte | *ZNF90* | -1.078964456 | 0.003987871 | 0.007138199 | 1 |
| T2DM | leukocyte | *REPIN1* | -1.015802629 | 0.001303688 | 0.004990048 | 1 |

| | | | | | | |
|---|---|---|---|---|---|---|
| T2DM | leukocyte | *TSPAN4* | -0.895127908 | 0.009698076 | 0.013904233 | 1 |
| T2DM | leukocyte | *CPNE1* | -0.924949951 | 0.023349512 | 0.030217467 | 1 |
| T2DM | leukocyte | *ARL16* | -0.89473237 | 0.000749374 | 0.004990048 | 1 |
| T2DM | leukocyte | *WEE2* | -4.532245946 | 0.014897743 | 0.020274761 | 1 |
| T2DM | leukocyte | *FIS1* | -0.688183804 | 0.00298298 | 0.006152298 | 1 |
| T2DM | leukocyte | *MBLAC1* | -0.957324521 | 0.035681502 | 0.04471854 | 1 |
| T2DM | leukocyte | *MEG3* | -0.573851413 | 0.001971741 | 0.005215543 | 1 |
| T2DM | leukocyte | *EML6* | -1.095932535 | 0.004011408 | 0.00714972 | 1 |
| T2DM | leukocyte | *C12orf71* | -15.07390486 | 0.002874686 | 0.006055188 | 1 |
| T2DM | leukocyte | *IFRD2* | -1.022283315 | 0.001056525 | 0.004990048 | 1 |
| T2DM | leukocyte | *ARHGEF28* | -1.258727395 | 0.001538969 | 0.005006447 | 1 |
| T2DM | leukocyte | *COL28A1* | -4.862041706 | 0.002070038 | 0.005273744 | 1 |
| T2DM | leukocyte | *UBXN2B* | -1.502190387 | 0.001394967 | 0.004990048 | 1 |
| T2DM | leukocyte | *C5orf49* | -0.752137285 | 0.003096775 | 0.006265309 | 1 |
| T2DM | leukocyte | *GOLGA8B* | -0.987444567 | 0.015993892 | 0.02163167 | 1 |
| T2DM | leukocyte | *VPS16* | -1.406776618 | 0.00484769 | 0.00790211 | 1 |
| T2DM | leukocyte | *LINC01139* | -2.813721705 | 0.001653825 | 0.005041098 | 1 |
| T2DM | leukocyte | *ATAD3C* | -0.844477123 | 0.002342559 | 0.005519727 | 1 |
| T2DM | leukocyte | *SYCE3* | -2.008810617 | 0.000754803 | 0.004990048 | 1 |
| T2DM | leukocyte | *PAM16* | -0.843553779 | 0.001569163 | 0.005009694 | 1 |
| T2DM | leukocyte | *LINC00339* | -1.006516357 | 0.003872597 | 0.007044182 | 1 |
| T2DM | leukocyte | *CELA3B* | -0.458415975 | 0.001786158 | 0.005100156 | 1 |
| T2DM | leukocyte | *VAMP2* | -0.686124656 | 0.0010718 | 0.004990048 | 1 |
| T2DM | leukocyte | *PPP3R1* | -1.466895892 | 0.000600557 | 0.004990048 | 1 |
| T2DM | leukocyte | *FANCG* | -1.772913414 | 0.000955863 | 0.004990048 | 1 |
| T2DM | leukocyte | *AP4M1* | -0.669751495 | 0.008365433 | 0.012259525 | 1 |
| T2DM | leukocyte | *CEBPD* | -1.808064488 | 0.005004431 | 0.008072234 | 1 |
| T2DM | leukocyte | *ZNF880* | -0.768076873 | 0.002479099 | 0.005622792 | 1 |
| T2DM | leukocyte | *TRIM16* | -1.215633028 | 0.003970676 | 0.007124893 | 1 |
| T2DM | leukocyte | *TIGD1* | -1.521031765 | 0.001437651 | 0.004990048 | 1 |
| T2DM | leukocyte | *SLC12A8* | -1.604749771 | 0.000287615 | 0.004990048 | 1 |
| T2DM | leukocyte | *UBA52* | -0.703142445 | 0.000950906 | 0.004990048 | 1 |
| T2DM | leukocyte | *EXOSC6* | -1.123275763 | 0.000387363 | 0.004990048 | 1 |
| T2DM | leukocyte | *CKMT1A* | -0.744090962 | 0.016547143 | 0.022280235 | 1 |
| T2DM | leukocyte | *HLA-DPB1* | 0.091693911 | 0.015007798 | 0.020396277 | 1 |
| T2DM | leukocyte | *LINC00115* | -2.456392039 | 0.002338787 | 0.005515664 | 1 |
| T2DM | leukocyte | *LINC00937* | -0.910334051 | 0.010651922 | 0.015092463 | 1 |
| T2DM | leukocyte | *TMEM185B* | -0.976572586 | 0.002565974 | 0.005695883 | 1 |
| T2DM | leukocyte | *CYP4F26P* | 3.972602344 | 0.018774468 | 0.024861774 | 1 |
| T2DM | leukocyte | *WDR46* | -0.723286695 | 0.011878383 | 0.016650077 | 1 |
| T2DM | leukocyte | *ZNF717* | -0.538742097 | 0.014407392 | 0.019702973 | 1 |
| T2DM | leukocyte | *PROB1* | -1.306233538 | 0.022983414 | 0.029822269 | 1 |
| T2DM | leukocyte | *ARL17B* | -3.097844847 | 6.88E-05 | 0.004990048 | 0.270507287 |
| T2DM | leukocyte | *DHFR* | -1.156010037 | 0.009700052 | 0.013904233 | 1 |
| T2DM | leukocyte | *PSMG3-AS1* | -0.887588404 | 0.018039728 | 0.02401833 | 1 |
| T2DM | leukocyte | *HSBP1* | -0.810407306 | 0.001218806 | 0.004990048 | 1 |
| T2DM | leukocyte | *RPS18* | -0.525052757 | 0.001979201 | 0.005222499 | 1 |
| T2DM | leukocyte | *LINC00865* | -2.083952393 | 0.005824724 | 0.009077881 | 1 |
| T2DM | leukocyte | *HLA-DQB2* | 1.629089729 | 5.86E-06 | 0.001302453 | 0.023005665 |
| T2DM | leukocyte | *LINC00665* | -0.65789508 | 0.004551841 | 0.007568852 | 1 |

| | | | | | | |
|---|---|---|---|---|---|---|
| T2DM | leukocyte | *LYRM9* | -1.038438861 | 0.002887639 | 0.00605886 | 1 |
| T2DM | leukocyte | *SNHG15* | -0.86421177 | 0.0021675 | 0.005345956 | 1 |
| T2DM | leukocyte | *RPS28* | -0.596345264 | 0.001012676 | 0.004990048 | 1 |
| T2DM | leukocyte | *TRIM26* | -1.281181754 | 0.000308598 | 0.004990048 | 1 |
| T2DM | leukocyte | *ZNF879* | -1.181381197 | 0.002891221 | 0.006061691 | 1 |
| T2DM | leukocyte | *PINLYP* | -2.045858022 | 0.001127855 | 0.004990048 | 1 |
| T2DM | leukocyte | *MCIDAS* | 32.60480758 | 0.012168262 | 0.01699577 | 1 |
| T2DM | leukocyte | *ANKRD65* | -0.834144893 | 0.00212908 | 0.005307839 | 1 |
| T2DM | leukocyte | *C12orf75* | -0.516021007 | 0.00203636 | 0.005253355 | 1 |
| T2DM | leukocyte | *B3GALT4* | -0.908980399 | 0.000223209 | 0.004990048 | 0.876987103 |
| T2DM | leukocyte | *ZNF737* | -1.341474264 | 0.001032343 | 0.004990048 | 1 |
| T2DM | leukocyte | *SHISA9* | -6.614470993 | 0.003502875 | 0.006639072 | 1 |
| T2DM | leukocyte | *HLA-DQA2* | 0.106096596 | 0.002630318 | 0.005799393 | 1 |
| T2DM | leukocyte | *LRRC37A2* | -0.941597064 | 0.028851778 | 0.03665785 | 1 |
| T2DM | leukocyte | *TXNDC5* | -0.947129092 | 0.001261997 | 0.004990048 | 1 |
| T2DM | leukocyte | *RBM14* | -1.250903121 | 0.001110784 | 0.004990048 | 1 |
| T2DM | leukocyte | *NME1* | -0.667157089 | 0.002329716 | 0.005513096 | 1 |
| T2DM | leukocyte | *CDRT4* | -1.893670782 | 0.003501191 | 0.006639072 | 1 |
| T2DM | leukocyte | *AMY2B* | -0.738178107 | 0.002651905 | 0.005824744 | 1 |
| T2DM | leukocyte | *LY6G5B* | -0.900077797 | 0.012963811 | 0.017966425 | 1 |
| T2DM | leukocyte | *PSMB9* | -0.523952756 | 0.02585622 | 0.033134079 | 1 |
| T2DM | leukocyte | *SMKR1* | -1.125863362 | 0.003087583 | 0.00625753 | 1 |
| T2DM | leukocyte | *LINC00886* | -1.214406425 | 0.00612136 | 0.00945394 | 1 |
| T2DM | leukocyte | *MIF* | -0.563358513 | 0.001379078 | 0.004990048 | 1 |
| T2DM | leukocyte | *NSUN6* | -0.873462901 | 0.002404422 | 0.005583319 | 1 |
| T2DM | leukocyte | *CRCP* | -1.130037308 | 0.000604271 | 0.004990048 | 1 |
| T2DM | leukocyte | *C8orf58* | -1.123880311 | 0.018480616 | 0.02453881 | 1 |
| T2DM | leukocyte | *PI4KA* | -1.52330134 | 0.001294005 | 0.004990048 | 1 |
| T2DM | leukocyte | *AMACR* | -0.672282643 | 0.008668135 | 0.012623092 | 1 |
| T2DM | leukocyte | *BGLAP* | -1.853468106 | 0.000265895 | 0.004990048 | 1 |
| T2DM | leukocyte | *MRPL20* | -0.724728392 | 0.003159245 | 0.006314657 | 1 |
| T2DM | leukocyte | *HLA-DMB* | 0.086745605 | 0.014704812 | 0.020046914 | 1 |
| T2DM | leukocyte | *DECR2* | -0.921673581 | 0.00274432 | 0.005930931 | 1 |
| T2DM | leukocyte | *GNG10* | -0.709423716 | 0.005185523 | 0.008292193 | 1 |
| T2DM | leukocyte | *CNTF* | 1.762028213 | 0.013512244 | 0.018634471 | 1 |
| T2DM | leukocyte | *EIF4EBP3* | -1.298784753 | 0.000676996 | 0.004990048 | 1 |
| T2DM | leukocyte | *MICAL3* | -0.862148341 | 0.002488683 | 0.005622792 | 1 |
| T2DM | leukocyte | *STON1* | -1.165418763 | 0.007154201 | 0.010765551 | 1 |
| T2DM | leukocyte | *VSIG8* | -2.238591961 | 0.004014351 | 0.00714972 | 1 |
| T2DM | leukocyte | *C4orf48* | -0.681563285 | 0.00127455 | 0.004990048 | 1 |
| T2DM | leukocyte | *IL10RB* | -2.394947145 | 0.000163305 | 0.004990048 | 0.641623898 |
| T2DM | leukocyte | *LEFTY1* | -6.86750909 | 0.007377549 | 0.011050853 | 1 |
| T2DM | leukocyte | *TTC4* | -0.899244728 | 0.003968606 | 0.007124893 | 1 |
| T2DM | leukocyte | *JMJD7* | 3.026241729 | 0.000297498 | 0.004990048 | 1 |
| T2DM | leukocyte | *APOBEC3D* | -3.599892946 | 0.003381032 | 0.006503419 | 1 |
| T2DM | leukocyte | *RN7SL832P* | -3.678375319 | 0.030342778 | 0.038457024 | 1 |
| T2DM | leukocyte | *OR2A7* | -3.077083834 | 0.021557533 | 0.028195588 | 1 |
| T2DM | leukocyte | *GSTA1* | -0.404312507 | 0.021911143 | 0.028591458 | 1 |
| T2DM | leukocyte | *GSTA2* | -0.643438344 | 0.004908057 | 0.007970068 | 1 |
| T2DM | leukocyte | *TMEM141* | -1.051579002 | 0.00294491 | 0.006128469 | 1 |

| | | | | | | |
|---|---|---|---|---|---|---|
| T2DM | leukocyte | *APOBEC3C* | -1.570228199 | 0.014590115 | 0.019911276 | 1 |
| T2DM | leukocyte | *C4A* | -7.488372484 | 0.003441016 | 0.006566174 | 1 |
| T2DM | leukocyte | *NEAT1* | -0.61506392 | 0.001794829 | 0.005114303 | 1 |
| T2DM | leukocyte | *LINC00920* | -6.521606844 | 0.014717594 | 0.02005738 | 1 |
| T2DM | leukocyte | *UBAP1L* | -0.893570024 | 0.005459927 | 0.008629144 | 1 |
| T2DM | leukocyte | *TWF2* | -1.151994253 | 0.003071092 | 0.006249984 | 1 |
| T2DM | leukocyte | *BCKDHA* | -0.816828188 | 0.007487522 | 0.011185732 | 1 |
| T2DM | leukocyte | *PCDHAC1* | -1.261017222 | 0.006606627 | 0.010088394 | 1 |
| T2DM | leukocyte | *FMN1* | -2.437940294 | 0.014366283 | 0.019667292 | 1 |
| T2DM | leukocyte | *TMEM150C* | -1.147359911 | 0.001265576 | 0.004990048 | 1 |
| T2DM | leukocyte | *LINC01091* | -0.675361649 | 0.001834364 | 0.005146173 | 1 |
| T2DM | leukocyte | *PDCD6* | -0.904142259 | 0.001135033 | 0.004990048 | 1 |
| T2DM | leukocyte | *YJEFN3* | -1.586877537 | 0.002453723 | 0.005621365 | 1 |
| T2DM | leukocyte | *PCDHA10* | -0.824736911 | 0.003442966 | 0.006566706 | 1 |
| T2DM | leukocyte | *ZNF718* | -1.04595972 | 0.009479433 | 0.013632757 | 1 |
| T2DM | leukocyte | *SNHG18* | -0.929659094 | 0.002219901 | 0.005407737 | 1 |
| T2DM | leukocyte | *ZNF550* | -1.073767802 | 0.001993623 | 0.005227473 | 1 |
| T2DM | leukocyte | *LINC01099* | -1.046671354 | 0.004266932 | 0.007310294 | 1 |
| T2DM | leukocyte | *PCDHA12* | -1.422625784 | 0.004917577 | 0.007973094 | 1 |
| T2DM | leukocyte | *ALG11* | -1.708805955 | 0.001052746 | 0.004990048 | 1 |
| T2DM | leukocyte | *ZNF260* | -1.553757152 | 0.004920056 | 0.007973094 | 1 |
| T2DM | leukocyte | *AP5B1* | -1.740230426 | 0.000908877 | 0.004990048 | 1 |
| T2DM | leukocyte | *SIGLEC12* | 1.61370917 | 0.015861815 | 0.021486999 | 1 |
| T2DM | leukocyte | *PABPC4L* | -2.448880663 | 0.004282986 | 0.007324561 | 1 |
| T2DM | leukocyte | *NPIPA2* | -9.458640847 | 0.003532643 | 0.006669753 | 1 |
| T2DM | leukocyte | *DPP3* | -0.789908468 | 0.007651919 | 0.011388027 | 1 |
| T2DM | leukocyte | *BRK1* | -0.845402762 | 0.000155094 | 0.004990048 | 0.609365836 |
| T2DM | leukocyte | *EID3* | -2.00382545 | 0.000789839 | 0.004990048 | 1 |
| T2DM | leukocyte | *SNHG9* | -0.842148829 | 0.001809743 | 0.005122825 | 1 |
| T2DM | leukocyte | *PCDHA3* | -1.201097577 | 0.017768219 | 0.023686929 | 1 |
| T2DM | leukocyte | *ZNF432* | -2.070774924 | 0.000218 | 0.004990048 | 0.856521355 |
| T2DM | leukocyte | *LINC01152* | -4.188406983 | 0.004062847 | 0.007184034 | 1 |
| T2DM | leukocyte | *SMLR1* | 2.794137908 | 0.025493777 | 0.032744377 | 1 |
| T2DM | leukocyte | *ZNF10* | -1.17285406 | 0.001520385 | 0.004993935 | 1 |
| T2DM | leukocyte | *TAS2R31* | -2.94653174 | 0.009943694 | 0.014232705 | 1 |
| T2DM | leukocyte | *GATC* | -1.289662895 | 0.000689278 | 0.004990048 | 1 |
| T2DM | leukocyte | *TEN1* | -2.465286945 | 0.001393359 | 0.004990048 | 1 |
| T2DM | leukocyte | *CHURC1* | -0.741855799 | 0.001648699 | 0.005035955 | 1 |
| T2DM | leukocyte | *LINC00638* | -6.620899375 | 0.002359166 | 0.005540445 | 1 |
| T2DM | leukocyte | *SLC25A21-AS1* | -0.872787887 | 0.003958076 | 0.007124893 | 1 |
| T2DM | leukocyte | *TMEM179* | -0.632997124 | 0.004077889 | 0.007189 | 1 |
| T2DM | leukocyte | *IQCH-AS1* | -1.115818679 | 0.036228238 | 0.045360341 | 1 |
| T2DM | leukocyte | *PWRN1* | -1.155614189 | 0.006176481 | 0.009520358 | 1 |
| T2DM | leukocyte | *TMEM178B* | -0.827312878 | 0.003186617 | 0.006339351 | 1 |
| T2DM | leukocyte | *LINC01003* | -1.093995761 | 0.001528844 | 0.005001523 | 1 |
| T2DM | leukocyte | *GOLGA8H* | -11.97788044 | 0.022264913 | 0.029014542 | 1 |
| T2DM | leukocyte | *MRPL12* | -1.179285139 | 0.001138119 | 0.004990048 | 1 |
| T2DM | leukocyte | *ZNF234* | -4.970248852 | 0.00124357 | 0.004990048 | 1 |
| T2DM | leukocyte | *LINC00667* | -1.005477284 | 0.001300671 | 0.004990048 | 1 |
| T2DM | leukocyte | *FSBP* | -1.243505938 | 0.004177894 | 0.007242343 | 1 |

| | | | | | | |
|---|---|---|---|---|---|---|
| T2DM | leukocyte | *MYO15B* | -1.728859762 | 0.008847026 | 0.012831291 | 1 |
| T2DM | leukocyte | *LINC00663* | -1.296100677 | 0.007790967 | 0.011564302 | 1 |
| T2DM | leukocyte | *ZNF285* | -2.105443343 | 0.003752385 | 0.00692001 | 1 |
| T2DM | leukocyte | *FBXO17* | -0.751493085 | 0.002485175 | 0.005622792 | 1 |
| T2DM | leukocyte | *LINC00528* | 0.493105847 | 0.012891246 | 0.017882851 | 1 |
| T2DM | leukocyte | *SNHG8* | -0.688593453 | 0.001151454 | 0.004990048 | 1 |
| T2DM | leukocyte | *MILR1* | 0.78206395 | 0.01001522 | 0.014303817 | 1 |
| T2DM | leukocyte | *GTF2H5* | -0.808830908 | 0.003217602 | 0.006372159 | 1 |
| T2DM | mesenchymal_cell | *RBM6* | -0.832993035 | 2.28E-07 | 9.23E-05 | 0.00089423 |
| T2DM | mesenchymal_cell | *CEP68* | 0.991739724 | 3.60E-05 | 0.005503398 | 0.141591933 |
| T2DM | mesenchymal_cell | *ERCC1* | -1.153465916 | 0.000886192 | 0.04145058 | 1 |
| T2DM | mesenchymal_cell | *BAK1* | 1.043658561 | 0.000806888 | 0.039440621 | 1 |
| T2DM | mesenchymal_cell | *ITIH4* | -1.186516391 | 0.000202922 | 0.019302292 | 0.797278978 |
| T2DM | mesenchymal_cell | *SEC61A2* | -2.103612755 | 0.000284997 | 0.024175174 | 1 |
| T2DM | mesenchymal_cell | *TCF3* | -1.378317002 | 1.69E-06 | 0.00044143 | 0.006621445 |
| T2DM | mesenchymal_cell | *SCARB1* | -1.55221091 | 0.000484728 | 0.032693637 | 1 |
| T2DM | mesenchymal_cell | *ST6GAL1* | 0.678213903 | 0.001099799 | 0.04910353 | 1 |
| T2DM | mesenchymal_cell | *TTC38* | -2.28800492 | 0.000140823 | 0.017224517 | 0.553294488 |
| T2DM | mesenchymal_cell | *MARK3* | -1.110702499 | 0.000254545 | 0.022224624 | 1 |
| T2DM | mesenchymal_cell | *CWF19L1* | -0.568030425 | 6.41E-05 | 0.008995669 | 0.251878739 |
| T2DM | mesenchymal_cell | *RBFOX2* | -1.381767956 | 0.000540163 | 0.034230682 | 1 |
| T2DM | mesenchymal_cell | *SAMM50* | -1.93889085 | 0.000595721 | 0.03493416 | 1 |
| T2DM | mesenchymal_cell | *VCPKMT* | -2.151750888 | 0.00084761 | 0.040123603 | 1 |
| T2DM | mesenchymal_cell | *NAGPA* | -2.194073586 | 0.000187438 | 0.019302292 | 0.736443443 |
| T2DM | mesenchymal_cell | *RPAP1* | -2.841836606 | 0.000726953 | 0.037577027 | 1 |
| T2DM | mesenchymal_cell | *ERI1* | -2.227055145 | 0.000193107 | 0.019302292 | 0.758715639 |
| T2DM | mesenchymal_cell | *GCDH* | -2.640186281 | 9.42E-06 | 0.001948128 | 0.03701444 |
| T2DM | mesenchymal_cell | *GTPBP4* | -1.378146135 | 0.001070366 | 0.04890078 | 1 |
| T2DM | mesenchymal_cell | *HSD17B1* | -0.834943316 | 3.26E-05 | 0.005331695 | 0.127960683 |
| T2DM | mesenchymal_cell | *RANGRF* | -1.676075749 | 0.000939078 | 0.043407488 | 1 |
| T2DM | mesenchymal_cell | *NNT* | -0.870618359 | 0.000564125 | 0.034810097 | 1 |
| T2DM | mesenchymal_cell | *FLT3* | 0.892234282 | 0.000206337 | 0.019302292 | 0.81069626 |
| T2DM | mesenchymal_cell | *PEPD* | -1.433613259 | 0.000704367 | 0.037398103 | 1 |
| T2DM | mesenchymal_cell | *PRMT1* | -1.095440188 | 0.000171485 | 0.018209846 | 0.67376429 |
| T2DM | mesenchymal_cell | *PTPN12* | -0.544488914 | 0.000146483 | 0.017224517 | 0.575530491 |
| T2DM | mesenchymal_cell | *MAU2* | -1.839981387 | 2.35E-07 | 9.23E-05 | 0.000923051 |
| T2DM | mesenchymal_cell | *NOTCH2* | -0.469993808 | 1.54E-07 | 7.57E-05 | 0.000605691 |
| T2DM | mesenchymal_cell | *RDH5* | -1.817409389 | 0.000813105 | 0.039440621 | 1 |
| T2DM | mesenchymal_cell | *BARD1* | -1.755912631 | 0.00053993 | 0.034230682 | 1 |
| T2DM | mesenchymal_cell | *TLE3* | -1.526438677 | 0.001098225 | 0.04910353 | 1 |
| T2DM | mesenchymal_cell | *MAN2C1* | -0.585036296 | 3.76E-06 | 0.000869995 | 0.014789907 |
| T2DM | mesenchymal_cell | *SCRN2* | -1.168868918 | 0.000205149 | 0.019302292 | 0.806031114 |
| T2DM | mesenchymal_cell | *NPC1* | -2.249177902 | 4.54E-07 | 0.000162039 | 0.001782432 |
| T2DM | mesenchymal_cell | *CNTN4* | -0.559274027 | 0.000412979 | 0.030615021 | 1 |
| T2DM | mesenchymal_cell | *TBC1D7* | -2.80847892 | 0.000139008 | 0.017224517 | 0.546164289 |
| T2DM | mesenchymal_cell | *GIGYF1* | -1.948111793 | 2.78E-05 | 0.004743979 | 0.109111517 |
| T2DM | mesenchymal_cell | *LRP12* | -1.391227627 | 0.000490945 | 0.032693637 | 1 |
| T2DM | mesenchymal_cell | *HSD17B12* | -0.29862748 | 0.000247506 | 0.022101199 | 0.972452734 |
| T2DM | mesenchymal_cell | *APIP* | -0.467343007 | 0.000436592 | 0.031766122 | 1 |
| T2DM | mesenchymal_cell | *DLG5* | -1.500810126 | 0.00069894 | 0.037398103 | 1 |

| | | | | | | |
|---|---|---|---|---|---|---|
| T2DM | mesenchymal_cell | *PTPN14* | -0.507679385 | 4.76E-05 | 0.006919878 | 0.1868367 |
| T2DM | mesenchymal_cell | *MED21* | -1.491956268 | 0.0007642 | 0.038178998 | 1 |
| T2DM | mesenchymal_cell | *JAZF1* | -0.728523206 | 2.26E-05 | 0.004029478 | 0.088648525 |
| T2DM | mesenchymal_cell | *NUS1* | -1.574690967 | 0.000375003 | 0.029467764 | 1 |
| T2DM | mesenchymal_cell | *CLEC18A* | -1.745018773 | 8.05E-08 | 4.52E-05 | 0.000316138 |
| T2DM | mesenchymal_cell | *CILP2* | -0.766779226 | 3.64E-05 | 0.005503398 | 0.143088338 |
| T2DM | mesenchymal_cell | *ZNF208* | -0.824008647 | 0.000649837 | 0.03596072 | 1 |
| T2DM | mesenchymal_cell | *COG5* | -2.006553946 | 0.000637915 | 0.035805262 | 1 |
| T2DM | mesenchymal_cell | *RPP25L* | -2.369124644 | 0.000504119 | 0.033011362 | 1 |
| T2DM | mesenchymal_cell | *INPPL1* | -1.681255807 | 0.000303121 | 0.024811746 | 1 |
| T2DM | mesenchymal_cell | *RCCD1* | -1.000728067 | 0.000575886 | 0.034810097 | 1 |
| T2DM | mesenchymal_cell | *POP5* | -1.530652014 | 0.000455067 | 0.0322272 | 1 |
| T2DM | mesenchymal_cell | *TMC8* | -0.603822429 | 0.00038285 | 0.029494452 | 1 |
| T2DM | mesenchymal_cell | *TRANK1* | -1.575310184 | 0.000833693 | 0.039946115 | 1 |
| T2DM | mesenchymal_cell | *DNAJC21* | -1.658127636 | 0.00010363 | 0.014040089 | 0.40716259 |
| T2DM | mesenchymal_cell | *TMEM129* | -0.678232886 | 0.000588093 | 0.03493416 | 1 |
| T2DM | mesenchymal_cell | *SNUPN* | -2.464324064 | 0.000289191 | 0.024175174 | 1 |
| T2DM | mesenchymal_cell | *INO80E* | -1.117707699 | 2.96E-06 | 0.000727342 | 0.011637476 |
| T2DM | mesenchymal_cell | *ZNF440* | -2.00850717 | 0.000153438 | 0.017224517 | 0.602858093 |
| T2DM | mesenchymal_cell | *RGS19* | -0.647135783 | 0.000700673 | 0.037398103 | 1 |
| T2DM | mesenchymal_cell | *RNASEH2C* | -0.781380437 | 0.000629677 | 0.035805262 | 1 |
| T2DM | mesenchymal_cell | *IRX3* | 2.559942734 | 1.64E-18 | 6.44E-15 | 6.44E-15 |
| T2DM | mesenchymal_cell | *FARSA* | -1.717183526 | 7.28E-07 | 0.000216 | 0.00286077 |
| T2DM | mesenchymal_cell | *HLA-DQB1* | -0.416255737 | 3.32E-08 | 2.61E-05 | 0.000130308 |
| T2DM | mesenchymal_cell | *D2HGDH* | -1.677046718 | 0.000459334 | 0.0322272 | 1 |
| T2DM | mesenchymal_cell | *UBA7* | -0.493104041 | 1.12E-05 | 0.002193575 | 0.043871498 |
| T2DM | mesenchymal_cell | *ZNF320* | 0.294540309 | 0.000471983 | 0.032533701 | 1 |
| T2DM | mesenchymal_cell | *TRMT12* | -1.587585472 | 0.000767661 | 0.038178998 | 1 |
| T2DM | mesenchymal_cell | *PGP* | -1.155218037 | 0.000150357 | 0.017224517 | 0.590753782 |
| T2DM | mesenchymal_cell | *DMWD* | -1.265854161 | 2.08E-05 | 0.00389804 | 0.081858834 |
| T2DM | mesenchymal_cell | *HLA-DRB1* | -0.294321389 | 4.37E-09 | 5.73E-06 | 1.72E-05 |
| T2DM | mesenchymal_cell | *HLA-DQA1* | -0.203574943 | 0.000410148 | 0.030615021 | 1 |
| T2DM | mesenchymal_cell | *PIWIL2* | -0.708460881 | 0.000143122 | 0.017224517 | 0.562325753 |
| T2DM | mesenchymal_cell | *HLA-DRB5* | -0.170683984 | 0.000164839 | 0.017990355 | 0.64765279 |
| T2DM | mesenchymal_cell | *TBKBP1* | -0.564162184 | 0.000727942 | 0.037577027 | 1 |
| T2DM | mesenchymal_cell | *NOTCH4* | -0.401109528 | 5.41E-07 | 0.000177081 | 0.002124967 |
| T2DM | mesenchymal_cell | *RNF5* | -0.675889309 | 7.70E-07 | 0.000216 | 0.003023994 |
| T2DM | mesenchymal_cell | *RNF39* | 0.410407318 | 0.000736429 | 0.037577027 | 1 |
| T2DM | mesenchymal_cell | *HCG27* | 0.541498994 | 4.17E-06 | 0.000911272 | 0.016402897 |
| T2DM | mesenchymal_cell | *ATF6B* | 0.539464648 | 0.00057024 | 0.034810097 | 1 |
| T2DM | mesenchymal_cell | *SLC12A8* | -3.719082151 | 4.46E-08 | 2.92E-05 | 0.000175174 |
| T2DM | mesenchymal_cell | *HLA-DQB2* | 1.063346586 | 1.10E-08 | 1.08E-05 | 4.34E-05 |
| T2DM | mesenchymal_cell | *HLA-B* | 0.357147446 | 0.000315463 | 0.025295016 | 1 |
| T2DM | mesenchymal_cell | *HLA-DQA2* | 0.679592645 | 3.50E-12 | 6.88E-09 | 1.38E-08 |
| T2DM | mesenchymal_cell | *JMJD7* | 1.089388615 | 0.000227778 | 0.020812572 | 0.894940598 |
| T2DM | mesenchymal_cell | *ZNF432* | -1.782769077 | 0.000632666 | 0.035805262 | 1 |
| T2DM | PP_cell | *RBM6* | -1.361582632 | 4.80E-07 | 0.000269598 | 0.001887187 |
| T2DM | PP_cell | *CEP68* | 1.467898161 | 0.000183382 | 0.036540786 | 0.720508395 |
| T2DM | PP_cell | *TCF3* | -1.625061494 | 0.000186006 | 0.036540786 | 0.730815714 |
| T2DM | PP_cell | *MARK3* | -1.772630664 | 0.000156907 | 0.034249235 | 0.616486224 |

| | | | | | | |
|---|---|---|---|---|---|---|
| T2DM | PP_cell | *HSD17B1* | -1.10320676 | 0.000297373 | 0.044937675 | 1 |
| T2DM | PP_cell | *NNT* | -1.540405998 | 0.000230743 | 0.040242285 | 0.906588954 |
| T2DM | PP_cell | *MAU2* | -1.542119812 | 0.000235575 | 0.040242285 | 0.925572548 |
| T2DM | PP_cell | *MAN2C1* | -0.940184891 | 9.48E-06 | 0.003385593 | 0.037241525 |
| T2DM | PP_cell | *HSD17B12* | -0.578912704 | 7.38E-05 | 0.020724605 | 0.290144464 |
| T2DM | PP_cell | *APIP* | -0.92296033 | 9.62E-05 | 0.025198778 | 0.377981672 |
| T2DM | PP_cell | *PTPN14* | -0.947736848 | 0.000296933 | 0.044937675 | 1 |
| T2DM | PP_cell | *JAZF1* | -1.307437947 | 8.11E-06 | 0.003186366 | 0.031863657 |
| T2DM | PP_cell | *CLEC18A* | -3.866515414 | 3.04E-08 | 2.39E-05 | 0.000119263 |
| T2DM | PP_cell | *CILP2* | -1.408555434 | 0.000146468 | 0.033851357 | 0.575473069 |
| T2DM | PP_cell | *INO80E* | -1.549179831 | 3.62E-05 | 0.011857222 | 0.142286666 |
| T2DM | PP_cell | *IRX3* | 1.862615791 | 2.83E-09 | 5.57E-06 | 1.11E-05 |
| T2DM | PP_cell | *HLA-DQB1* | -0.793797082 | 8.16E-08 | 5.34E-05 | 0.000320489 |
| T2DM | PP_cell | *ZNF320* | 0.590242902 | 0.000256329 | 0.041963114 | 1 |
| T2DM | PP_cell | *HLA-DRB1* | -0.609411662 | 6.07E-09 | 7.95E-06 | 2.38E-05 |
| T2DM | PP_cell | *NOTCH4* | -0.873298737 | 1.26E-06 | 0.00055176 | 0.004965841 |
| T2DM | PP_cell | *RNF5* | -1.138180221 | 1.07E-06 | 0.00052359 | 0.004188719 |
| T2DM | PP_cell | *HCG27* | 0.655231386 | 0.000134552 | 0.033040892 | 0.52865427 |
| T2DM | PP_cell | *DNLZ* | 0.591883554 | 0.000234483 | 0.040242285 | 0.921284405 |
| T2DM | PP_cell | *PPT2* | 1.669573276 | 6.45E-05 | 0.019487882 | 0.253342463 |
| T2DM | PP_cell | *HLA-DQB2* | 1.833401553 | 2.43E-08 | 2.39E-05 | 9.55E-05 |
| T2DM | PP_cell | *HLA-DQA2* | 1.133319958 | 5.71E-12 | 2.24E-08 | 2.24E-08 |
| T2DM_admissi | acinar_cell | *RBM6* | -0.917672389 | 3.41E-08 | 1.29E-05 | 0.000133958 |
| T2DM_admissi | acinar_cell | *MLXIPL* | 0.581660804 | 0.000240192 | 0.019660748 | 0.943715891 |
| T2DM_admissi | acinar_cell | *GIPR* | -0.988885771 | 0.00023392 | 0.019660748 | 0.919070657 |
| T2DM_admissi | acinar_cell | *CEP68* | 1.383347846 | 1.72E-08 | 8.44E-06 | 6.76E-05 |
| T2DM_admissi | acinar_cell | *BAK1* | 1.703537331 | 1.04E-07 | 3.39E-05 | 0.000406952 |
| T2DM_admissi | acinar_cell | *TCF3* | -1.613032022 | 4.25E-06 | 0.000695624 | 0.016694966 |
| T2DM_admissi | acinar_cell | *ST6GAL1* | 0.834487685 | 0.000121353 | 0.012225528 | 0.476795584 |
| T2DM_admissi | acinar_cell | *MARK3* | -1.837724277 | 6.00E-06 | 0.000942178 | 0.023554457 |
| T2DM_admissi | acinar_cell | *CWF19L1* | -0.495311941 | 0.000559218 | 0.036275307 | 1 |
| T2DM_admissi | acinar_cell | *CCDC134* | 1.316648143 | 0.000258682 | 0.020568623 | 1 |
| T2DM_admissi | acinar_cell | *OIP5* | 2.34197843 | 8.25E-05 | 0.008534212 | 0.324300047 |
| T2DM_admissi | acinar_cell | *GCDH* | -2.378889507 | 3.41E-05 | 0.004059448 | 0.133961794 |
| T2DM_admissi | acinar_cell | *HSD17B1* | -0.801494872 | 6.55E-05 | 0.006956183 | 0.257378789 |
| T2DM_admissi | acinar_cell | *CNTNAP1* | 0.778526985 | 8.06E-06 | 0.001218682 | 0.031685723 |
| T2DM_admissi | acinar_cell | *CDK2AP1* | 0.757848433 | 0.000359753 | 0.02666921 | 1 |
| T2DM_admissi | acinar_cell | *FAM53C* | -1.573862232 | 0.000234207 | 0.019660748 | 0.92019921 |
| T2DM_admissi | acinar_cell | *ZNF211* | 0.570030816 | 0.000404415 | 0.028374015 | 1 |
| T2DM_admissi | acinar_cell | *FLT3* | 0.959059462 | 0.000184549 | 0.017264156 | 0.725094549 |
| T2DM_admissi | acinar_cell | *OPRL1* | -1.138295147 | 5.46E-05 | 0.006131934 | 0.214617678 |
| T2DM_admissi | acinar_cell | *MAU2* | -1.772237155 | 1.49E-06 | 0.000307981 | 0.005851638 |
| T2DM_admissi | acinar_cell | *NOTCH2* | -0.818069013 | 6.03E-12 | 4.74E-09 | 2.37E-08 |
| T2DM_admissi | acinar_cell | *SPTBN5* | 0.888918408 | 0.000313897 | 0.024182417 | 1 |
| T2DM_admissi | acinar_cell | *MAN2C1* | -0.554363788 | 1.01E-05 | 0.001463258 | 0.039507953 |
| T2DM_admissi | acinar_cell | *NPC1* | -2.337687434 | 3.76E-06 | 0.000670899 | 0.014759771 |
| T2DM_admissi | acinar_cell | *GIGYF1* | -1.604713136 | 0.000794911 | 0.046615029 | 1 |
| T2DM_admissi | acinar_cell | *HSD17B12* | -0.419032516 | 7.17E-07 | 0.000165794 | 0.002818498 |
| T2DM_admissi | acinar_cell | *APIP* | -0.487161411 | 0.000328599 | 0.024828216 | 1 |
| T2DM_admissi | acinar_cell | *VIPAS39* | -1.521103227 | 0.000838423 | 0.04844358 | 1 |

| | | | | | | |
|---|---|---|---|---|---|---|
| T2DM_admissi | acinar_cell | *PTPN14* | -0.581332257 | 6.08E-05 | 0.006631241 | 0.238724675 |
| T2DM_admissi | acinar_cell | *POC5* | -0.496734478 | 2.42E-05 | 0.003062247 | 0.094929662 |
| T2DM_admissi | acinar_cell | *JAZF1* | -2.169299124 | 2.11E-16 | 4.15E-13 | 8.31E-13 |
| T2DM_admissi | acinar_cell | *NUS1* | -1.731001607 | 0.000489326 | 0.032790875 | 1 |
| T2DM_admissi | acinar_cell | *CLEC18A* | -3.071796998 | 9.71E-13 | 9.54E-10 | 3.82E-09 |
| T2DM_admissi | acinar_cell | *EXTL1* | -1.524619641 | 0.000631262 | 0.039368698 | 1 |
| T2DM_admissi | acinar_cell | *CILP2* | -0.896923928 | 2.59E-05 | 0.003182263 | 0.101832401 |
| T2DM_admissi | acinar_cell | *SRGAP2* | 1.207678053 | 0.000397462 | 0.028374015 | 1 |
| T2DM_admissi | acinar_cell | *ICA1L* | -0.530435873 | 0.000223007 | 0.019660748 | 0.876193496 |
| T2DM_admissi | acinar_cell | *BDH2* | -1.07991901 | 2.33E-05 | 0.003057615 | 0.091728454 |
| T2DM_admissi | acinar_cell | *SGPL1* | -1.259016583 | 0.000193568 | 0.017686746 | 0.760530065 |
| T2DM_admissi | acinar_cell | *PCBD1* | -1.314974227 | 0.00013913 | 0.01366607 | 0.546642801 |
| T2DM_admissi | acinar_cell | *TMEM129* | -0.736358925 | 0.000261754 | 0.020568623 | 1 |
| T2DM_admissi | acinar_cell | *INO80E* | -0.856284687 | 0.000240072 | 0.019660748 | 0.943243401 |
| T2DM_admissi | acinar_cell | *SNX32* | 0.877004217 | 0.00067808 | 0.041627725 | 1 |
| T2DM_admissi | acinar_cell | *PEAK1* | 0.766249697 | 0.000563195 | 0.036275307 | 1 |
| T2DM_admissi | acinar_cell | *YIF1A* | -0.69040049 | 0.00074901 | 0.045274764 | 1 |
| T2DM_admissi | acinar_cell | *IRX3* | 3.348519066 | 1.28E-27 | 5.03E-24 | 5.03E-24 |
| T2DM_admissi | acinar_cell | *FARSA* | -1.941585931 | 3.26E-07 | 8.55E-05 | 0.00128275 |
| T2DM_admissi | acinar_cell | *HLA-DQB1* | -0.465918295 | 3.61E-08 | 1.29E-05 | 0.000141977 |
| T2DM_admissi | acinar_cell | *UBA7* | -0.67488792 | 1.85E-06 | 0.000353372 | 0.00727256 |
| T2DM_admissi | acinar_cell | *VMO1* | 0.654322997 | 0.000486227 | 0.032790875 | 1 |
| T2DM_admissi | acinar_cell | *CAMK1D* | 1.994993552 | 0.000403902 | 0.028374015 | 1 |
| T2DM_admissi | acinar_cell | *HLA-DRB1* | -0.391837494 | 3.52E-11 | 2.31E-08 | 1.38E-07 |
| T2DM_admissi | acinar_cell | *HLA-DRB5* | -0.296784376 | 1.10E-06 | 0.000240316 | 0.004325685 |
| T2DM_admissi | acinar_cell | *LIME1* | -0.559481216 | 0.000160273 | 0.015358852 | 0.629712948 |
| T2DM_admissi | acinar_cell | *NOTCH4* | -0.677536694 | 2.24E-09 | 1.26E-06 | 8.81E-06 |
| T2DM_admissi | acinar_cell | *AGER* | 1.017394488 | 3.78E-05 | 0.004363289 | 0.14835181 |
| T2DM_admissi | acinar_cell | *RNF5* | -0.700274557 | 2.42E-07 | 6.79E-05 | 0.000950124 |
| T2DM_admissi | acinar_cell | *HLA-C* | -0.296841946 | 0.000868684 | 0.049464647 | 1 |
| T2DM_admissi | acinar_cell | *RNF39* | 0.41264696 | 0.000792894 | 0.046615029 | 1 |
| T2DM_admissi | acinar_cell | *HCG27* | 0.644736633 | 1.64E-07 | 4.96E-05 | 0.000644539 |
| T2DM_admissi | acinar_cell | *DNLZ* | 0.319975001 | 0.000492406 | 0.032790875 | 1 |
| T2DM_admissi | acinar_cell | *ATF6B* | 0.678103795 | 1.28E-05 | 0.001801329 | 0.050437218 |
| T2DM_admissi | acinar_cell | *SLC12A8* | -2.756983024 | 6.49E-07 | 0.000159432 | 0.002550905 |
| T2DM_admissi | acinar_cell | *PPT2* | 0.995200738 | 1.40E-05 | 0.00189481 | 0.054949485 |
| T2DM_admissi | acinar_cell | *HLA-DQB2* | 1.037743804 | 2.58E-08 | 1.13E-05 | 0.000101382 |
| T2DM_admissi | acinar_cell | *HLA-B* | 0.476511508 | 1.89E-06 | 0.000353372 | 0.007420812 |
| T2DM_admissi | acinar_cell | *HLA-DQA2* | 0.686929812 | 1.68E-13 | 2.20E-10 | 6.59E-10 |
| T2DM_admissi | acinar_cell | *JMJD7* | 1.380210135 | 4.15E-06 | 0.000695624 | 0.016313527 |
| T2DM_admissi | acinar_cell | *HMBS* | 0.472683596 | 0.000618503 | 0.039195121 | 1 |
| T2DM_admissi | alpha_cell | *RBM6* | -0.149497121 | 4.69E-08 | 1.54E-05 | 0.000184235 |
| T2DM_admissi | alpha_cell | *MLXIPL* | 0.099731408 | 0.000224435 | 0.016957799 | 0.881805564 |
| T2DM_admissi | alpha_cell | *GIPR* | -0.236088711 | 2.31E-06 | 0.000412433 | 0.009073524 |
| T2DM_admissi | alpha_cell | *CEP68* | 0.257404438 | 3.22E-10 | 1.81E-07 | 1.27E-06 |
| T2DM_admissi | alpha_cell | *BAK1* | 0.30734952 | 8.27E-09 | 3.61E-06 | 3.25E-05 |
| T2DM_admissi | alpha_cell | *TCF3* | -0.254591248 | 1.54E-05 | 0.001832074 | 0.060458432 |
| T2DM_admissi | alpha_cell | *ST6GAL1* | 0.132300359 | 0.000266377 | 0.018689229 | 1 |
| T2DM_admissi | alpha_cell | *MARK3* | -0.379411094 | 1.09E-06 | 0.000213571 | 0.004271414 |
| T2DM_admissi | alpha_cell | *CWF19L1* | -0.080432343 | 0.000578937 | 0.032037236 | 1 |

| | | | | | | |
|---|---|---|---|---|---|---|
| T2DM_admissi | alpha_cell | *CCDC134* | 0.215214016 | 0.000350063 | 0.022923293 | 1 |
| T2DM_admissi | alpha_cell | *OIP5* | 0.383048926 | 7.81E-05 | 0.006977228 | 0.306998051 |
| T2DM_admissi | alpha_cell | *GCDH* | -0.55419366 | 3.33E-07 | 8.73E-05 | 0.001309269 |
| T2DM_admissi | alpha_cell | *HSD17B1* | -0.130174058 | 6.72E-05 | 0.006291704 | 0.263910178 |
| T2DM_admissi | alpha_cell | *CNTNAP1* | 0.126151977 | 9.26E-06 | 0.001212892 | 0.036386767 |
| T2DM_admissi | alpha_cell | *CDK2AP1* | 0.13903664 | 0.000117437 | 0.010030681 | 0.46141131 |
| T2DM_admissi | alpha_cell | *DNAH7* | -0.118400909 | 0.000562342 | 0.031563453 | 1 |
| T2DM_admissi | alpha_cell | *FAM53C* | -0.274960102 | 0.00069799 | 0.035158991 | 1 |
| T2DM_admissi | alpha_cell | *ZNF211* | 0.095452181 | 0.000381416 | 0.024566916 | 1 |
| T2DM_admissi | alpha_cell | *POLK* | -0.268490104 | 0.000296626 | 0.020446376 | 1 |
| T2DM_admissi | alpha_cell | *FLT3* | 0.158052072 | 0.000127525 | 0.010660578 | 0.501047161 |
| T2DM_admissi | alpha_cell | *PEPD* | -0.31529553 | 0.001072009 | 0.049394903 | 1 |
| T2DM_admissi | alpha_cell | *OPRL1* | -0.186002403 | 4.62E-05 | 0.004771893 | 0.181331924 |
| T2DM_admissi | alpha_cell | *MAU2* | -0.300403202 | 2.57E-06 | 0.00043896 | 0.010096077 |
| T2DM_admissi | alpha_cell | *PPARG* | -0.426954029 | 9.38E-05 | 0.008186863 | 0.368408843 |
| T2DM_admissi | alpha_cell | *NOTCH2* | -0.129957182 | 3.02E-11 | 2.38E-08 | 1.19E-07 |
| T2DM_admissi | alpha_cell | *FXYD2* | -0.168490967 | 0.001053078 | 0.049256454 | 1 |
| T2DM_admissi | alpha_cell | *SPTBN5* | 0.13682184 | 0.00068474 | 0.035158991 | 1 |
| T2DM_admissi | alpha_cell | *MAN2C1* | -0.092479579 | 6.16E-06 | 0.000896698 | 0.024210836 |
| T2DM_admissi | alpha_cell | *NPC1* | -0.437954417 | 3.90E-06 | 0.000613072 | 0.015326804 |
| T2DM_admissi | alpha_cell | *POGZ* | 0.17835488 | 0.00047166 | 0.029413161 | 1 |
| T2DM_admissi | alpha_cell | *INPP5E* | 0.21361291 | 0.000654484 | 0.03428626 | 1 |
| T2DM_admissi | alpha_cell | *SEC16A* | 0.170509876 | 0.000980381 | 0.046408645 | 1 |
| T2DM_admissi | alpha_cell | *HSD17B12* | -0.067706944 | 9.45E-07 | 0.00019535 | 0.003711653 |
| T2DM_admissi | alpha_cell | *APIP* | -0.081259321 | 0.000256079 | 0.018632083 | 1 |
| T2DM_admissi | alpha_cell | *PTPN14* | -0.089288282 | 0.000198357 | 0.015586927 | 0.779346352 |
| T2DM_admissi | alpha_cell | *POC5* | -0.082896037 | 1.59E-05 | 0.001842081 | 0.062630766 |
| T2DM_admissi | alpha_cell | *JAZF1* | -0.446904569 | 6.21E-21 | 1.22E-17 | 2.44E-17 |
| T2DM_admissi | alpha_cell | *NUS1* | -0.35741459 | 0.000891992 | 0.042739447 | 1 |
| T2DM_admissi | alpha_cell | *CLEC18A* | -0.477032154 | 6.81E-12 | 6.69E-09 | 2.68E-08 |
| T2DM_admissi | alpha_cell | *AP3S2* | 0.447628921 | 7.46E-06 | 0.001046474 | 0.029301273 |
| T2DM_admissi | alpha_cell | *EXTL1* | -0.250666195 | 0.000488563 | 0.029413161 | 1 |
| T2DM_admissi | alpha_cell | *CILP2* | -0.146930627 | 1.91E-05 | 0.002147963 | 0.075178689 |
| T2DM_admissi | alpha_cell | *SRGAP2* | 0.215083835 | 0.000190566 | 0.015280298 | 0.748734597 |
| T2DM_admissi | alpha_cell | *ICA1L* | -0.084409289 | 0.00032451 | 0.021982728 | 1 |
| T2DM_admissi | alpha_cell | *BDH2* | -0.163632648 | 7.59E-05 | 0.00693296 | 0.298117292 |
| T2DM_admissi | alpha_cell | *CASP7* | 0.085962838 | 3.49E-05 | 0.003813143 | 0.137273134 |
| T2DM_admissi | alpha_cell | *CKB* | -0.125681949 | 0.000334135 | 0.022251122 | 1 |
| T2DM_admissi | alpha_cell | *SGPL1* | -0.203498448 | 0.000641113 | 0.034131535 | 1 |
| T2DM_admissi | alpha_cell | *PCBD1* | -0.246472313 | 0.000163423 | 0.013376821 | 0.642087409 |
| T2DM_admissi | alpha_cell | *TMEM129* | -0.121538711 | 0.000261922 | 0.018689229 | 1 |
| T2DM_admissi | alpha_cell | *INO80E* | -0.159785034 | 6.73E-05 | 0.006291704 | 0.264251573 |
| T2DM_admissi | alpha_cell | *RGS19* | -0.16105548 | 0.00087681 | 0.042530682 | 1 |
| T2DM_admissi | alpha_cell | *CTSW* | -0.055854212 | 0.000521981 | 0.029722681 | 1 |
| T2DM_admissi | alpha_cell | *SNX32* | 0.144599456 | 0.00050906 | 0.029413161 | 1 |
| T2DM_admissi | alpha_cell | *RNASEH2C* | -0.148594513 | 0.000489717 | 0.029413161 | 1 |
| T2DM_admissi | alpha_cell | *PEAK1* | 0.135169659 | 0.000220457 | 0.016957799 | 0.8661754 |
| T2DM_admissi | alpha_cell | *YIF1A* | -0.121024016 | 0.000634303 | 0.034131535 | 1 |
| T2DM_admissi | alpha_cell | *DSCAML1* | -0.177034314 | 0.000791004 | 0.038848205 | 1 |
| T2DM_admissi | alpha_cell | *IRX3* | 0.536645624 | 9.02E-28 | 3.54E-24 | 3.54E-24 |

| | | | | | | |
|---|---|---|---|---|---|---|
| T2DM_admissi | alpha_cell | *FARSA* | -0.332441966 | 2.65E-07 | 7.43E-05 | 0.001039905 |
| T2DM_admissi | alpha_cell | *HLA-DQB1* | -0.075349992 | 4.21E-08 | 1.54E-05 | 0.000165524 |
| T2DM_admissi | alpha_cell | *TH* | -1.042292572 | 1.00E-05 | 0.001268445 | 0.03932179 |
| T2DM_admissi | alpha_cell | *UBA7* | -0.10477671 | 6.08E-06 | 0.000896698 | 0.023879081 |
| T2DM_admissi | alpha_cell | *VMO1* | 0.104839533 | 0.000501913 | 0.029413161 | 1 |
| T2DM_admissi | alpha_cell | *CAMK1D* | 0.369614234 | 4.93E-05 | 0.004963234 | 0.193566125 |
| T2DM_admissi | alpha_cell | *PGP* | -0.192658065 | 0.000774329 | 0.038510624 | 1 |
| T2DM_admissi | alpha_cell | *DMWD* | -0.191704172 | 0.000418219 | 0.026502926 | 1 |
| T2DM_admissi | alpha_cell | *NUTM2B* | -1.170460409 | 5.29E-05 | 0.005200127 | 0.208005079 |
| T2DM_admissi | alpha_cell | *HLA-DRB1* | -0.064464684 | 3.79E-11 | 2.48E-08 | 1.49E-07 |
| T2DM_admissi | alpha_cell | *HLA-DRB5* | -0.049023679 | 1.49E-06 | 0.000278314 | 0.005844588 |
| T2DM_admissi | alpha_cell | *LIME1* | -0.080656509 | 0.000642844 | 0.034131535 | 1 |
| T2DM_admissi | alpha_cell | *NOTCH4* | -0.109749155 | 1.97E-09 | 9.68E-07 | 7.74E-06 |
| T2DM_admissi | alpha_cell | *AGER* | 0.173360815 | 3.62E-05 | 0.003844321 | 0.142239888 |
| T2DM_admissi | alpha_cell | *RNF5* | -0.111424488 | 4.36E-07 | 0.000107128 | 0.001714049 |
| T2DM_admissi | alpha_cell | *RNF39* | 0.074349178 | 0.000236913 | 0.017562828 | 0.930829869 |
| T2DM_admissi | alpha_cell | *HCG27* | 0.106766665 | 1.20E-07 | 3.62E-05 | 0.000470108 |
| T2DM_admissi | alpha_cell | *DNLZ* | 0.050773195 | 0.000690128 | 0.035158991 | 1 |
| T2DM_admissi | alpha_cell | *ATF6B* | 0.111944459 | 1.25E-05 | 0.001538842 | 0.049242949 |
| T2DM_admissi | alpha_cell | *SLC12A8* | -0.584921018 | 4.77E-07 | 0.000110128 | 0.001872172 |
| T2DM_admissi | alpha_cell | *PPT2* | 0.174663998 | 9.12E-06 | 0.001212892 | 0.035847154 |
| T2DM_admissi | alpha_cell | *EPPK1* | 0.184457966 | 0.001081181 | 0.049394903 | 1 |
| T2DM_admissi | alpha_cell | *HLA-DQB2* | 0.168518054 | 4.44E-08 | 1.54E-05 | 0.000174532 |
| T2DM_admissi | alpha_cell | *HLA-B* | 0.083202867 | 7.11E-07 | 0.000155169 | 0.002793047 |
| T2DM_admissi | alpha_cell | *HLA-DQA2* | 0.110993721 | 3.29E-13 | 4.30E-10 | 1.29E-09 |
| T2DM_admissi | alpha_cell | *JMJD7* | 0.22565654 | 3.70E-06 | 0.000606033 | 0.014544799 |
| T2DM_admissi | alpha_cell | *HMBS* | 0.078797719 | 0.000504554 | 0.029413161 | 1 |
| T2DM_admissi | beta_cell | *RBM6* | -0.089472409 | 9.63E-08 | 2.52E-05 | 0.000378454 |
| T2DM_admissi | beta_cell | *MLXIPL* | 0.065493865 | 7.20E-05 | 0.006022146 | 0.283040859 |
| T2DM_admissi | beta_cell | *GIPR* | -0.140781844 | 4.78E-06 | 0.000671393 | 0.018799009 |
| T2DM_admissi | beta_cell | *CEP68* | 0.17097216 | 1.71E-11 | 1.34E-08 | 6.72E-08 |
| T2DM_admissi | beta_cell | *BAK1* | 0.200817984 | 1.34E-09 | 5.86E-07 | 5.27E-06 |
| T2DM_admissi | beta_cell | *ANAPC4* | 0.5894558 | 6.83E-05 | 0.005935502 | 0.268363874 |
| T2DM_admissi | beta_cell | *SPTB* | 0.075127425 | 0.000972432 | 0.037828564 | 1 |
| T2DM_admissi | beta_cell | *GBA2* | 0.242060018 | 0.001448709 | 0.049068763 | 1 |
| T2DM_admissi | beta_cell | *TCF3* | -0.152854549 | 2.70E-05 | 0.002653239 | 0.10612956 |
| T2DM_admissi | beta_cell | *ST6GAL1* | 0.084556304 | 0.00014122 | 0.009905263 | 0.554853856 |
| T2DM_admissi | beta_cell | *MARK3* | -0.289370292 | 2.38E-07 | 5.49E-05 | 0.000933981 |
| T2DM_admissi | beta_cell | *MGST2* | 0.214497263 | 0.000225562 | 0.012851644 | 0.886232907 |
| T2DM_admissi | beta_cell | *CWF19L1* | -0.047184134 | 0.000887258 | 0.035571797 | 1 |
| T2DM_admissi | beta_cell | *CCDC134* | 0.133083445 | 0.000271066 | 0.014791934 | 1 |
| T2DM_admissi | beta_cell | *RBL2* | 0.147312338 | 0.000313303 | 0.015972513 | 1 |
| T2DM_admissi | beta_cell | *OIP5* | 0.236580253 | 5.46E-05 | 0.00487185 | 0.214361396 |
| T2DM_admissi | beta_cell | *GCDH* | -0.348198932 | 8.92E-07 | 0.000166842 | 0.003503673 |
| T2DM_admissi | beta_cell | *TMEM106B* | 0.322046765 | 0.000199984 | 0.012088287 | 0.785738668 |
| T2DM_admissi | beta_cell | *DHRS11* | 0.173553703 | 0.000313642 | 0.015972513 | 1 |
| T2DM_admissi | beta_cell | *HSD17B1* | -0.075791847 | 0.000129509 | 0.009251644 | 0.508840394 |
| T2DM_admissi | beta_cell | *CNTNAP1* | 0.076428915 | 9.49E-06 | 0.001165755 | 0.037304168 |
| T2DM_admissi | beta_cell | *PDHX* | 0.102276996 | 0.000281537 | 0.015152878 | 1 |
| T2DM_admissi | beta_cell | *CDK2AP1* | 0.09908492 | 1.21E-05 | 0.001403112 | 0.047705795 |

| | | | | | | |
|---|---|---|---|---|---|---|
| T2DM_admissi | beta_cell | *SEMA5A* | 0.256057274 | 0.000797535 | 0.032640766 | 1 |
| T2DM_admissi | beta_cell | *DNAH7* | -0.069282169 | 0.00095396 | 0.0374811 | 1 |
| T2DM_admissi | beta_cell | *BCL11A* | 0.291355041 | 0.000920874 | 0.036546613 | 1 |
| T2DM_admissi | beta_cell | *MTRF1* | 0.093708136 | 0.001354618 | 0.046686775 | 1 |
| T2DM_admissi | beta_cell | *FAM53C* | -0.18204409 | 0.001170592 | 0.043718899 | 1 |
| T2DM_admissi | beta_cell | *IAPP* | 0.120854455 | 0.000156412 | 0.010172146 | 0.614542121 |
| T2DM_admissi | beta_cell | *ZNF211* | 0.059781409 | 0.000244208 | 0.013707027 | 0.9594919 |
| T2DM_admissi | beta_cell | *POLK* | -0.187356743 | 0.000294191 | 0.015512286 | 1 |
| T2DM_admissi | beta_cell | *FLT3* | 0.096194449 | 0.000118757 | 0.008803714 | 0.466596851 |
| T2DM_admissi | beta_cell | *BHLHE41* | 0.303647006 | 7.52E-05 | 0.006152939 | 0.29534105 |
| T2DM_admissi | beta_cell | *OPRL1* | -0.112624089 | 4.36E-05 | 0.003982837 | 0.171262001 |
| T2DM_admissi | beta_cell | *BECN1* | 0.545672563 | 0.000183325 | 0.011433049 | 0.720282116 |
| T2DM_admissi | beta_cell | *ABHD8* | 0.144903167 | 0.000563252 | 0.025436984 | 1 |
| T2DM_admissi | beta_cell | *MAU2* | -0.188814793 | 4.23E-06 | 0.000639524 | 0.016627624 |
| T2DM_admissi | beta_cell | *TRAF3* | 0.152214014 | 0.000317092 | 0.015972513 | 1 |
| T2DM_admissi | beta_cell | *PPARG* | -0.257976455 | 0.000157928 | 0.010172146 | 0.620500916 |
| T2DM_admissi | beta_cell | *NOTCH2* | -0.078596618 | 3.75E-11 | 2.03E-08 | 1.47E-07 |
| T2DM_admissi | beta_cell | *OSTF1* | 0.303787997 | 0.000570955 | 0.025491847 | 1 |
| T2DM_admissi | beta_cell | *SPTBN5* | 0.083531269 | 0.000601802 | 0.025983316 | 1 |
| T2DM_admissi | beta_cell | *MAN2C1* | -0.054491869 | 1.15E-05 | 0.001363485 | 0.044995007 |
| T2DM_admissi | beta_cell | *NPC1* | -0.314073823 | 2.05E-06 | 0.000322273 | 0.008056832 |
| T2DM_admissi | beta_cell | *EVI5L* | 0.109395572 | 0.000854907 | 0.034628121 | 1 |
| T2DM_admissi | beta_cell | *POGZ* | 0.129752107 | 6.95E-05 | 0.005935502 | 0.273033113 |
| T2DM_admissi | beta_cell | *INPP5E* | 0.144423923 | 0.000196726 | 0.012077112 | 0.772935149 |
| T2DM_admissi | beta_cell | *SEC16A* | 0.127333067 | 0.000125792 | 0.009152551 | 0.494237775 |
| T2DM_admissi | beta_cell | *HSD17B12* | -0.040689741 | 1.27E-06 | 0.000221668 | 0.004972986 |
| T2DM_admissi | beta_cell | *APIP* | -0.048588944 | 0.00032732 | 0.015992755 | 1 |
| T2DM_admissi | beta_cell | *PTPN14* | -0.053574864 | 0.000259923 | 0.014383653 | 1 |
| T2DM_admissi | beta_cell | *POC5* | -0.049863539 | 1.89E-05 | 0.00199659 | 0.074189806 |
| T2DM_admissi | beta_cell | *JAZF1* | -0.279649462 | 8.48E-21 | 1.67E-17 | 3.33E-17 |
| T2DM_admissi | beta_cell | *NUS1* | -0.364154412 | 0.000153193 | 0.010172146 | 0.601896956 |
| T2DM_admissi | beta_cell | *CLEC18A* | -0.287486412 | 9.40E-12 | 9.24E-09 | 3.70E-08 |
| T2DM_admissi | beta_cell | *AP3S2* | 0.529091242 | 2.89E-11 | 1.89E-08 | 1.14E-07 |
| T2DM_admissi | beta_cell | *EXTL1* | -0.148429008 | 0.000626457 | 0.026466126 | 1 |
| T2DM_admissi | beta_cell | *CILP2* | -0.08918931 | 1.75E-05 | 0.001906949 | 0.068650149 |
| T2DM_admissi | beta_cell | *NIPAL1* | 0.190027915 | 0.001274794 | 0.044324475 | 1 |
| T2DM_admissi | beta_cell | *SRGAP2* | 0.147427752 | 3.68E-05 | 0.00343967 | 0.144466153 |
| T2DM_admissi | beta_cell | *ICA1L* | -0.050273851 | 0.000455578 | 0.021309112 | 1 |
| T2DM_admissi | beta_cell | *BDH2* | -0.098500052 | 0.000118675 | 0.008803714 | 0.466275681 |
| T2DM_admissi | beta_cell | *NOXRED1* | 0.182993344 | 0.001237502 | 0.043758779 | 1 |
| T2DM_admissi | beta_cell | *CASP7* | 0.053473924 | 2.14E-05 | 0.002160663 | 0.084265855 |
| T2DM_admissi | beta_cell | *CKB* | -0.074524332 | 0.000723814 | 0.029935413 | 1 |
| T2DM_admissi | beta_cell | *SGPL1* | -0.127274745 | 0.001048262 | 0.039986619 | 1 |
| T2DM_admissi | beta_cell | *PCBD1* | -0.172973032 | 0.00020885 | 0.012432913 | 0.820572242 |
| T2DM_admissi | beta_cell | *TMEM129* | -0.071668541 | 0.000444878 | 0.021059327 | 1 |
| T2DM_admissi | beta_cell | *INO80E* | -0.097146002 | 9.53E-05 | 0.007436229 | 0.374256362 |
| T2DM_admissi | beta_cell | *RGS19* | -0.09684472 | 0.00099728 | 0.038414816 | 1 |
| T2DM_admissi | beta_cell | *CTSW* | -0.033312158 | 0.000614171 | 0.026229112 | 1 |
| T2DM_admissi | beta_cell | *NAA16* | 0.140113658 | 0.000225697 | 0.012851644 | 0.886763456 |
| T2DM_admissi | beta_cell | *SNX32* | 0.08615544 | 0.000600844 | 0.025983316 | 1 |

| | | | | | | |
|---|---|---|---|---|---|---|
| T2DM_admissi | beta_cell | *RNASEH2C* | -0.087451046 | 0.001223398 | 0.043758779 | 1 |
| T2DM_admissi | beta_cell | *PEAK1* | 0.087363557 | 9.65E-05 | 0.007436229 | 0.37924768 |
| T2DM_admissi | beta_cell | *YIF1A* | -0.072033708 | 0.001247387 | 0.043758779 | 1 |
| T2DM_admissi | beta_cell | *FUT2* | 0.018003174 | 0.001215585 | 0.043758779 | 1 |
| T2DM_admissi | beta_cell | *DSCAML1* | -0.102738069 | 0.00138342 | 0.047264859 | 1 |
| T2DM_admissi | beta_cell | *IRX3* | 0.335582039 | 6.99E-29 | 2.75E-25 | 2.75E-25 |
| T2DM_admissi | beta_cell | *IMP3* | 0.311151925 | 8.39E-06 | 0.001063744 | 0.032976072 |
| T2DM_admissi | beta_cell | *COX14* | 0.214786618 | 1.93E-05 | 0.00199659 | 0.075870429 |
| T2DM_admissi | beta_cell | *FARSA* | -0.205706036 | 4.33E-07 | 8.96E-05 | 0.001703035 |
| T2DM_admissi | beta_cell | *HLA-DQB1* | -0.045033617 | 6.35E-08 | 1.87E-05 | 0.000249571 |
| T2DM_admissi | beta_cell | *TH* | -0.573273359 | 8.74E-05 | 0.007009383 | 0.343459777 |
| T2DM_admissi | beta_cell | *UBA7* | -0.063403913 | 5.83E-06 | 0.000763546 | 0.022906385 |
| T2DM_admissi | beta_cell | *ARL6IP4* | 0.163437347 | 1.50E-05 | 0.001686472 | 0.059026518 |
| T2DM_admissi | beta_cell | *ACTL10* | 0.588718588 | 0.000329706 | 0.015992755 | 1 |
| T2DM_admissi | beta_cell | *VMO1* | 0.064138833 | 0.000466182 | 0.021548592 | 1 |
| T2DM_admissi | beta_cell | *CAMK1D* | 0.388186819 | 8.26E-09 | 2.95E-06 | 3.25E-05 |
| T2DM_admissi | beta_cell | *EIF3C* | 0.089266644 | 0.001244805 | 0.043758779 | 1 |
| T2DM_admissi | beta_cell | *ZBTB40* | 0.194956304 | 0.001179487 | 0.043718899 | 1 |
| T2DM_admissi | beta_cell | *DMWD* | -0.117108282 | 0.000659064 | 0.027547469 | 1 |
| T2DM_admissi | beta_cell | *NUTM2B* | -0.659427124 | 0.000148797 | 0.01007971 | 0.584623173 |
| T2DM_admissi | beta_cell | *ZNF567* | 0.217070919 | 0.00049408 | 0.02257258 | 1 |
| T2DM_admissi | beta_cell | *HLA-DRB1* | -0.038914123 | 4.14E-11 | 2.03E-08 | 1.63E-07 |
| T2DM_admissi | beta_cell | *DPP4* | -0.092386595 | 0.001217937 | 0.043758779 | 1 |
| T2DM_admissi | beta_cell | *DDX42* | 0.181546603 | 0.000143701 | 0.009905263 | 0.564600006 |
| T2DM_admissi | beta_cell | *HLA-DRB5* | -0.029941797 | 1.30E-06 | 0.000221668 | 0.005098372 |
| T2DM_admissi | beta_cell | *LIME1* | -0.050855832 | 0.000436694 | 0.020924053 | 1 |
| T2DM_admissi | beta_cell | *NOTCH4* | -0.065366129 | 3.43E-09 | 1.35E-06 | 1.35E-05 |
| T2DM_admissi | beta_cell | *AGER* | 0.106240088 | 2.95E-05 | 0.002822459 | 0.115720809 |
| T2DM_admissi | beta_cell | *RNF5* | -0.066392615 | 7.79E-07 | 0.000152949 | 0.003058975 |
| T2DM_admissi | beta_cell | *RNF39* | 0.045907804 | 0.000182882 | 0.011433049 | 0.718544984 |
| T2DM_admissi | beta_cell | *HCG27* | 0.065889976 | 6.67E-08 | 1.87E-05 | 0.000262102 |
| T2DM_admissi | beta_cell | *DNLZ* | 0.031010656 | 0.000600957 | 0.025983316 | 1 |
| T2DM_admissi | beta_cell | *ATF6B* | 0.071512868 | 4.70E-06 | 0.000671393 | 0.018460966 |
| T2DM_admissi | beta_cell | *SLC12A8* | -0.372603521 | 3.52E-07 | 7.68E-05 | 0.00138325 |
| T2DM_admissi | beta_cell | *PPT2* | 0.108608215 | 5.80E-06 | 0.000763546 | 0.022782134 |
| T2DM_admissi | beta_cell | *EPPK1* | 0.111320142 | 0.001068923 | 0.040382672 | 1 |
| T2DM_admissi | beta_cell | *HLA-DQB2* | 0.102661529 | 3.80E-08 | 1.24E-05 | 0.000149188 |
| T2DM_admissi | beta_cell | *HLA-B* | 0.05496703 | 1.26E-07 | 3.10E-05 | 0.000495203 |
| T2DM_admissi | beta_cell | *HLA-DQA2* | 0.068739447 | 1.37E-13 | 1.79E-10 | 5.37E-10 |
| T2DM_admissi | beta_cell | *JMJD7* | 0.143087019 | 1.37E-06 | 0.000223998 | 0.005375952 |
| T2DM_admissi | beta_cell | *NEAT1* | 0.194897543 | 0.000327947 | 0.015992755 | 1 |
| T2DM_admissi | beta_cell | *HMBS* | 0.049813462 | 0.000296111 | 0.015512286 | 1 |
| T2DM_admissi | beta_cell | *TAS2R31* | 1.149306047 | 0.000222312 | 0.012851644 | 0.873462864 |
| T2DM_admissi | delta_cell | *RBM6* | -0.697546544 | 1.45E-07 | 5.19E-05 | 0.000571076 |
| T2DM_admissi | delta_cell | *GIPR* | -1.016910187 | 1.62E-06 | 0.000375439 | 0.006382465 |
| T2DM_admissi | delta_cell | *CEP68* | 0.893544316 | 3.93E-07 | 0.00012864 | 0.001543674 |
| T2DM_admissi | delta_cell | *BAK1* | 1.21828129 | 1.40E-06 | 0.00034298 | 0.005487678 |
| T2DM_admissi | delta_cell | *TCF3* | -0.97003699 | 1.78E-05 | 0.00291948 | 0.070067516 |
| T2DM_admissi | delta_cell | *CCDC134* | 1.089392269 | 0.000553806 | 0.049452358 | 1 |
| T2DM_admissi | delta_cell | *HSD17B1* | -0.684494651 | 2.39E-05 | 0.003755985 | 0.093899616 |

| | | | | | | |
|---|---|---|---|---|---|---|
| T2DM_admissi | delta_cell | *CNTNAP1* | 0.587059864 | 7.40E-05 | 0.010767235 | 0.29071535 |
| T2DM_admissi | delta_cell | *ZNF211* | 0.434321141 | 0.00045523 | 0.042585704 | 1 |
| T2DM_admissi | delta_cell | *FLT3* | 0.748179855 | 0.000493521 | 0.045094024 | 1 |
| T2DM_admissi | delta_cell | *MAU2* | -1.10271183 | 1.76E-05 | 0.00291948 | 0.06924163 |
| T2DM_admissi | delta_cell | *NOTCH2* | -0.597067685 | 1.34E-09 | 1.05E-06 | 5.27E-06 |
| T2DM_admissi | delta_cell | *SPTBN5* | 0.755357012 | 0.000229525 | 0.024373023 | 0.901801853 |
| T2DM_admissi | delta_cell | *MAN2C1* | -0.482073537 | 3.89E-06 | 0.000764624 | 0.01529248 |
| T2DM_admissi | delta_cell | *HSD17B12* | -0.342785955 | 1.01E-06 | 0.000282276 | 0.003951859 |
| T2DM_admissi | delta_cell | *APIP* | -0.433295364 | 0.000116723 | 0.014793741 | 0.458605974 |
| T2DM_admissi | delta_cell | *POC5* | -0.458654845 | 3.65E-06 | 0.000764624 | 0.014342599 |
| T2DM_admissi | delta_cell | *JAZF1* | -1.729441143 | 9.16E-17 | 3.31E-13 | 3.60E-13 |
| T2DM_admissi | delta_cell | *CLEC18A* | -1.825927381 | 1.81E-08 | 9.67E-06 | 7.10E-05 |
| T2DM_admissi | delta_cell | *EXTL1* | -1.249493135 | 0.000189991 | 0.02073537 | 0.746473323 |
| T2DM_admissi | delta_cell | *CILP2* | -0.686016295 | 0.00011542 | 0.014793741 | 0.453486596 |
| T2DM_admissi | delta_cell | *ICA1L* | -0.434319751 | 0.000149753 | 0.017169454 | 0.588378553 |
| T2DM_admissi | delta_cell | *CASP7* | 0.408223141 | 0.000145168 | 0.017169454 | 0.570364128 |
| T2DM_admissi | delta_cell | *TMEM129* | -0.610349057 | 9.42E-05 | 0.012755989 | 0.369923675 |
| T2DM_admissi | delta_cell | *INO80E* | -0.772968648 | 3.08E-05 | 0.004649185 | 0.12087882 |
| T2DM_admissi | delta_cell | *CTSW* | -0.299383266 | 0.000445746 | 0.042585704 | 1 |
| T2DM_admissi | delta_cell | *IRX3* | 1.785505209 | 1.68E-16 | 3.31E-13 | 6.62E-13 |
| T2DM_admissi | delta_cell | *FARSA* | -1.384289905 | 7.55E-07 | 0.000228121 | 0.002965573 |
| T2DM_admissi | delta_cell | *HLA-DQB1* | -0.405567506 | 1.97E-08 | 9.67E-06 | 7.74E-05 |
| T2DM_admissi | delta_cell | *UBA7* | -0.530855546 | 1.49E-05 | 0.002785186 | 0.058488899 |
| T2DM_admissi | delta_cell | *VMO1* | 0.488693456 | 0.000345278 | 0.034784577 | 1 |
| T2DM_admissi | delta_cell | *HLA-DRB1* | -0.345343194 | 1.98E-11 | 1.94E-08 | 7.77E-08 |
| T2DM_admissi | delta_cell | *HLA-DRB5* | -0.249901815 | 1.10E-06 | 0.000289213 | 0.004338202 |
| T2DM_admissi | delta_cell | *LIME1* | -0.443352062 | 0.000125976 | 0.015467485 | 0.494959534 |
| T2DM_admissi | delta_cell | *NOTCH4* | -0.54406471 | 9.05E-09 | 5.93E-06 | 3.56E-05 |
| T2DM_admissi | delta_cell | *RNF5* | -0.582259518 | 1.01E-07 | 3.97E-05 | 0.00039668 |
| T2DM_admissi | delta_cell | *RNF39* | 0.364687641 | 0.000152948 | 0.017169454 | 0.600930878 |
| T2DM_admissi | delta_cell | *HCG27* | 0.364711843 | 3.88E-06 | 0.000764624 | 0.015249476 |
| T2DM_admissi | delta_cell | *ATF6B* | 0.454474682 | 0.000377093 | 0.037039982 | 1 |
| T2DM_admissi | delta_cell | *PPT2* | 0.785923934 | 8.61E-05 | 0.01208281 | 0.338318676 |
| T2DM_admissi | delta_cell | *HLA-DQB2* | 0.861992704 | 2.58E-08 | 1.13E-05 | 0.000101274 |
| T2DM_admissi | delta_cell | *HLA-B* | 0.313166375 | 1.78E-05 | 0.00291948 | 0.069927771 |
| T2DM_admissi | delta_cell | *HLA-DQA2* | 0.547934947 | 4.80E-13 | 6.28E-10 | 1.89E-09 |
| T2DM_admissi | delta_cell | *JMJD7* | 0.873992207 | 0.000249297 | 0.025775954 | 0.979486255 |
| T2DM_admissi | duct_epithelial_cell | *RBM6* | -0.455215681 | 5.52E-06 | 0.000938953 | 0.021687736 |
| T2DM_admissi | duct_epithelial_cell | *MLXIPL* | 0.400919905 | 8.81E-06 | 0.00128249 | 0.034627235 |
| T2DM_admissi | duct_epithelial_cell | *QPCTL* | -0.599134472 | 0.00101381 | 0.040234944 | 1 |
| T2DM_admissi | duct_epithelial_cell | *CEP68* | 0.971470606 | 1.39E-10 | 1.10E-07 | 5.48E-07 |
| T2DM_admissi | duct_epithelial_cell | *BAK1* | 0.828457847 | 1.22E-05 | 0.001593379 | 0.047801356 |
| T2DM_admissi | duct_epithelial_cell | *SPTB* | 0.477256632 | 0.000205537 | 0.013929731 | 0.807556191 |
| T2DM_admissi | duct_epithelial_cell | *TCF3* | -0.849229647 | 6.66E-07 | 0.000153849 | 0.002615428 |
| T2DM_admissi | duct_epithelial_cell | *NLE1* | 0.660579568 | 0.000603365 | 0.028561705 | 1 |
| T2DM_admissi | duct_epithelial_cell | *POMGNT1* | -1.005103092 | 0.00022695 | 0.014861418 | 0.891685085 |
| T2DM_admissi | duct_epithelial_cell | *RBL2* | 0.7145644 | 0.000225706 | 0.014861418 | 0.886798004 |
| T2DM_admissi | duct_epithelial_cell | *ATP8B4* | 6.325634069 | 0.000443542 | 0.024895385 | 1 |
| T2DM_admissi | duct_epithelial_cell | *OIP5* | 1.51555746 | 3.51E-06 | 0.000690332 | 0.013806646 |
| T2DM_admissi | duct_epithelial_cell | *VIPR2* | 0.35387087 | 0.001384687 | 0.048145436 | 1 |

| | | | | | | |
|---|---|---|---|---|---|---|
| T2DM_admissi | duct_epithelial_cell | *HSD17B1* | -0.527428272 | 2.10E-05 | 0.00242323 | 0.082389816 |
| T2DM_admissi | duct_epithelial_cell | *CNTNAP1* | 0.518293382 | 5.30E-07 | 0.00013019 | 0.002083036 |
| T2DM_admissi | duct_epithelial_cell | *PDHX* | 0.582207783 | 0.000145242 | 0.010877533 | 0.570655733 |
| T2DM_admissi | duct_epithelial_cell | *PRR4* | 0.314470561 | 0.00106636 | 0.041668898 | 1 |
| T2DM_admissi | duct_epithelial_cell | *CDK2AP1* | 0.544495161 | 3.01E-05 | 0.003280029 | 0.118081053 |
| T2DM_admissi | duct_epithelial_cell | *ANGPTL1* | -1.292020529 | 0.000167205 | 0.012165687 | 0.656947117 |
| T2DM_admissi | duct_epithelial_cell | *SPP1* | -0.136591825 | 0.001176868 | 0.044037274 | 1 |
| T2DM_admissi | duct_epithelial_cell | *CCDC77* | -0.951320255 | 0.001237399 | 0.045124707 | 1 |
| T2DM_admissi | duct_epithelial_cell | *MTRF1* | 0.585008356 | 0.001071153 | 0.041668898 | 1 |
| T2DM_admissi | duct_epithelial_cell | *IAPP* | 0.185895078 | 0.001516532 | 0.049653787 | 1 |
| T2DM_admissi | duct_epithelial_cell | *ZNF211* | 0.38312796 | 0.000173968 | 0.012427671 | 0.683521903 |
| T2DM_admissi | duct_epithelial_cell | *MOCS1* | -0.478277235 | 0.000940181 | 0.038082164 | 1 |
| T2DM_admissi | duct_epithelial_cell | *OPRL1* | -0.590849243 | 0.000585491 | 0.028561705 | 1 |
| T2DM_admissi | duct_epithelial_cell | *ABHD8* | 0.724559408 | 0.000559596 | 0.02855392 | 1 |
| T2DM_admissi | duct_epithelial_cell | *WFIKKN1* | 0.542653258 | 0.00112449 | 0.042481929 | 1 |
| T2DM_admissi | duct_epithelial_cell | *CGNL1* | -0.333892151 | 0.000706544 | 0.031191127 | 1 |
| T2DM_admissi | duct_epithelial_cell | *MAU2* | -0.943573314 | 3.78E-05 | 0.003903605 | 0.148336996 |
| T2DM_admissi | duct_epithelial_cell | *SYT5* | 0.293694679 | 0.000335116 | 0.020256494 | 1 |
| T2DM_admissi | duct_epithelial_cell | *PPARG* | -1.484168775 | 0.000310885 | 0.019085423 | 1 |
| T2DM_admissi | duct_epithelial_cell | *NOTCH2* | -0.437455419 | 2.72E-12 | 3.49E-09 | 1.07E-08 |
| T2DM_admissi | duct_epithelial_cell | *FXYD2* | -0.545260372 | 9.92E-05 | 0.008856282 | 0.389676387 |
| T2DM_admissi | duct_epithelial_cell | *MYPN* | 1.66253831 | 0.0008421 | 0.034827494 | 1 |
| T2DM_admissi | duct_epithelial_cell | *BARD1* | -0.883475588 | 0.000498439 | 0.026826961 | 1 |
| T2DM_admissi | duct_epithelial_cell | *MAN2C1* | -0.314757374 | 4.99E-05 | 0.004781497 | 0.196041364 |
| T2DM_admissi | duct_epithelial_cell | *NPC1* | -1.316377076 | 4.74E-05 | 0.00466016 | 0.186406389 |
| T2DM_admissi | duct_epithelial_cell | *FN3KRP* | -0.234497612 | 0.001452347 | 0.048754417 | 1 |
| T2DM_admissi | duct_epithelial_cell | *CBLC* | -0.351369445 | 0.000614025 | 0.02872028 | 1 |
| T2DM_admissi | duct_epithelial_cell | *POGZ* | 0.661850791 | 0.000131211 | 0.010740163 | 0.51552781 |
| T2DM_admissi | duct_epithelial_cell | *CDCA7* | 0.85516248 | 0.000244167 | 0.015721334 | 0.959331978 |
| T2DM_admissi | duct_epithelial_cell | *IQGAP2* | -0.230896845 | 0.000371266 | 0.021451515 | 1 |
| T2DM_admissi | duct_epithelial_cell | *RAB19* | -1.080547489 | 0.000863335 | 0.035333774 | 1 |
| T2DM_admissi | duct_epithelial_cell | *ADHFE1* | -0.978211197 | 0.000142547 | 0.010877533 | 0.560066555 |
| T2DM_admissi | duct_epithelial_cell | *SEC16A* | 0.66389297 | 0.000137789 | 0.010877533 | 0.541372727 |
| T2DM_admissi | duct_epithelial_cell | *HSD17B12* | -0.220686316 | 1.82E-05 | 0.002300961 | 0.071329798 |
| T2DM_admissi | duct_epithelial_cell | *APIP* | -0.301149131 | 0.000365289 | 0.021421226 | 1 |
| T2DM_admissi | duct_epithelial_cell | *KLHL35* | 0.272404384 | 0.000724813 | 0.031198389 | 1 |
| T2DM_admissi | duct_epithelial_cell | *VIPAS39* | -0.911487495 | 0.000117543 | 0.010039689 | 0.461825686 |
| T2DM_admissi | duct_epithelial_cell | *PTPN14* | -0.37648567 | 1.02E-05 | 0.001434694 | 0.040171424 |
| T2DM_admissi | duct_epithelial_cell | *POC5* | -0.303037305 | 3.45E-05 | 0.003661289 | 0.135467677 |
| T2DM_admissi | duct_epithelial_cell | *JAZF1* | -1.093299348 | 3.55E-12 | 3.49E-09 | 1.39E-08 |
| T2DM_admissi | duct_epithelial_cell | *CLEC18A* | -1.24242675 | 2.67E-07 | 7.48E-05 | 0.001047565 |
| T2DM_admissi | duct_epithelial_cell | *AP3S2* | 1.344765318 | 8.72E-06 | 0.00128249 | 0.034249948 |
| T2DM_admissi | duct_epithelial_cell | *CIB4* | 5.079628327 | 0.000144303 | 0.010877533 | 0.56696724 |
| T2DM_admissi | duct_epithelial_cell | *EXTL1* | -1.033706446 | 1.88E-05 | 0.00230887 | 0.074016782 |
| T2DM_admissi | duct_epithelial_cell | *NPR2* | 0.499686046 | 0.001270341 | 0.045374278 | 1 |
| T2DM_admissi | duct_epithelial_cell | *CILP2* | -0.606908423 | 1.02E-06 | 0.00022217 | 0.003999058 |
| T2DM_admissi | duct_epithelial_cell | *ZNF208* | -0.55243716 | 0.001095196 | 0.042186525 | 1 |
| T2DM_admissi | duct_epithelial_cell | *LY6K* | 0.56567074 | 0.001232608 | 0.045124707 | 1 |
| T2DM_admissi | duct_epithelial_cell | *SRGAP2* | 0.67782838 | 0.001476655 | 0.048754417 | 1 |
| T2DM_admissi | duct_epithelial_cell | *BDH2* | -0.519519076 | 4.56E-06 | 0.000814206 | 0.017912535 |

| | | | | | | |
|---|---|---|---|---|---|---|
| T2DM_admissi | duct_epithelial_cell | *NOXRED1* | 1.306093369 | 0.000130405 | 0.010740163 | 0.512360815 |
| T2DM_admissi | duct_epithelial_cell | *SGPL1* | -0.795168893 | 8.28E-05 | 0.007569279 | 0.325479018 |
| T2DM_admissi | duct_epithelial_cell | *PROCA1* | 1.9049387 | 0.000971961 | 0.03896769 | 1 |
| T2DM_admissi | duct_epithelial_cell | *TMC4* | -0.470946782 | 0.001109883 | 0.042337199 | 1 |
| T2DM_admissi | duct_epithelial_cell | *TMC8* | -0.394778629 | 0.000205631 | 0.013929731 | 0.807924393 |
| T2DM_admissi | duct_epithelial_cell | *TMEM129* | -0.422408487 | 0.000768024 | 0.032101789 | 1 |
| T2DM_admissi | duct_epithelial_cell | *INO80E* | -0.496900947 | 0.00073053 | 0.031198389 | 1 |
| T2DM_admissi | duct_epithelial_cell | *SETMAR* | -1.110521327 | 0.000348731 | 0.02076009 | 1 |
| T2DM_admissi | duct_epithelial_cell | *RGS19* | -0.677246655 | 4.43E-05 | 0.004462198 | 0.174025709 |
| T2DM_admissi | duct_epithelial_cell | *ARNT2* | 0.640892573 | 0.00141205 | 0.048666184 | 1 |
| T2DM_admissi | duct_epithelial_cell | *CTSW* | -0.215087781 | 0.000505642 | 0.026846835 | 1 |
| T2DM_admissi | duct_epithelial_cell | *NAA16* | 0.679265879 | 0.000146732 | 0.010877533 | 0.576509248 |
| T2DM_admissi | duct_epithelial_cell | *OVOL1* | -0.298241019 | 0.000697058 | 0.031191127 | 1 |
| T2DM_admissi | duct_epithelial_cell | *PEAK1* | 0.529680392 | 0.00011557 | 0.010039689 | 0.454073705 |
| T2DM_admissi | duct_epithelial_cell | *FUT2* | 0.119891869 | 0.00068336 | 0.031191127 | 1 |
| T2DM_admissi | duct_epithelial_cell | *DSCAML1* | -0.760381345 | 6.95E-05 | 0.00649881 | 0.272950039 |
| T2DM_admissi | duct_epithelial_cell | *COX14* | 0.56408642 | 0.000702197 | 0.031191127 | 1 |
| T2DM_admissi | duct_epithelial_cell | *FARSA* | -1.243778387 | 1.83E-07 | 5.99E-05 | 0.000719087 |
| T2DM_admissi | duct_epithelial_cell | *HLA-DQB1* | -0.300419365 | 1.02E-08 | 4.99E-06 | 3.99E-05 |
| T2DM_admissi | duct_epithelial_cell | *UBA7* | -0.417785419 | 2.31E-07 | 6.99E-05 | 0.000908189 |
| T2DM_admissi | duct_epithelial_cell | *ARL6IP4* | 0.528641483 | 0.000290314 | 0.018105429 | 1 |
| T2DM_admissi | duct_epithelial_cell | *CAPN12* | -0.336526828 | 0.001240919 | 0.045124707 | 1 |
| T2DM_admissi | duct_epithelial_cell | *SKA2* | -0.678570855 | 0.000526556 | 0.027221563 | 1 |
| T2DM_admissi | duct_epithelial_cell | *TTC3* | 0.210857038 | 0.001381876 | 0.048145436 | 1 |
| T2DM_admissi | duct_epithelial_cell | *CAMK1D* | 1.732349981 | 2.46E-06 | 0.000509197 | 0.009674735 |
| T2DM_admissi | duct_epithelial_cell | *FAM43B* | 1.85462575 | 0.000248084 | 0.015721334 | 0.974722705 |
| T2DM_admissi | duct_epithelial_cell | *MACC1* | -0.342468461 | 0.001432362 | 0.048754417 | 1 |
| T2DM_admissi | duct_epithelial_cell | *EIF3C* | 0.575303362 | 0.000590231 | 0.028561705 | 1 |
| T2DM_admissi | duct_epithelial_cell | *ZBTB40* | 0.885897495 | 0.000576525 | 0.028561705 | 1 |
| T2DM_admissi | duct_epithelial_cell | *CYP4X1* | -1.105159374 | 0.000726651 | 0.031198389 | 1 |
| T2DM_admissi | duct_epithelial_cell | *HLA-DRB1* | -0.271411315 | 7.24E-13 | 1.42E-09 | 2.84E-09 |
| T2DM_admissi | duct_epithelial_cell | *SUPT3H* | -1.282318415 | 0.001470807 | 0.048754417 | 1 |
| T2DM_admissi | duct_epithelial_cell | *SULT1A1* | -1.283396342 | 0.000200722 | 0.013929731 | 0.788636253 |
| T2DM_admissi | duct_epithelial_cell | *ZNF257* | -0.692927908 | 0.000515928 | 0.027027768 | 1 |
| T2DM_admissi | duct_epithelial_cell | *ENPP1* | 0.343044762 | 0.000451595 | 0.024990389 | 1 |
| T2DM_admissi | duct_epithelial_cell | *DDX42* | 0.664541484 | 0.000592766 | 0.028561705 | 1 |
| T2DM_admissi | duct_epithelial_cell | *HLA-DRB5* | -0.201741163 | 4.88E-08 | 1.92E-05 | 0.00019176 |
| T2DM_admissi | duct_epithelial_cell | *LIME1* | -0.368262517 | 1.94E-05 | 0.00230887 | 0.076192715 |
| T2DM_admissi | duct_epithelial_cell | *NOTCH4* | -0.428274275 | 3.73E-10 | 2.44E-07 | 1.46E-06 |
| T2DM_admissi | duct_epithelial_cell | *AGER* | 0.689961647 | 5.74E-06 | 0.000938953 | 0.022534884 |
| T2DM_admissi | duct_epithelial_cell | *RNF5* | -0.423277698 | 1.48E-07 | 5.28E-05 | 0.000580277 |
| T2DM_admissi | duct_epithelial_cell | *HCG27* | 0.308420398 | 2.53E-05 | 0.002836075 | 0.099262618 |
| T2DM_admissi | duct_epithelial_cell | *TAS2R14* | 0.300388288 | 0.001251869 | 0.045124707 | 1 |
| T2DM_admissi | duct_epithelial_cell | *DNLZ* | 0.19656494 | 0.000602912 | 0.028561705 | 1 |
| T2DM_admissi | duct_epithelial_cell | *ATF6B* | 0.427973984 | 1.14E-05 | 0.001548334 | 0.044901683 |
| T2DM_admissi | duct_epithelial_cell | *SLC12A8* | -1.989841527 | 2.90E-08 | 1.27E-05 | 0.000113961 |
| T2DM_admissi | duct_epithelial_cell | *PPT2* | 0.686041571 | 4.06E-06 | 0.000760052 | 0.015961092 |
| T2DM_admissi | duct_epithelial_cell | *HLA-DQB2* | 0.67562156 | 9.58E-09 | 4.99E-06 | 3.76E-05 |
| T2DM_admissi | duct_epithelial_cell | *HLA-B* | 0.313222443 | 5.08E-07 | 0.00013019 | 0.001996931 |
| T2DM_admissi | duct_epithelial_cell | *LINC00957* | 0.660037204 | 0.000740011 | 0.031263462 | 1 |

| | | | | | | |
|---|---|---|---|---|---|---|
| T2DM_admissi | duct_epithelial_cell | *HLA-DQA2* | 0.456402136 | 1.67E-14 | 6.56E-11 | 6.56E-11 |
| T2DM_admissi | duct_epithelial_cell | *JMJD7* | 0.829807013 | 8.05E-06 | 0.001264843 | 0.031621076 |
| T2DM_admissi | duct_epithelial_cell | *OR2A7* | -0.710918959 | 0.000397925 | 0.022658667 | 1 |
| T2DM_admissi | duct_epithelial_cell | *P2RY11* | 0.356620863 | 0.000678696 | 0.031191127 | 1 |
| T2DM_admissi | duct_epithelial_cell | *HMBS* | 0.301635392 | 0.000478818 | 0.026128838 | 1 |
| T2DM_admissi | duct_epithelial_cell | *TAS2R31* | 5.076739661 | 0.001322073 | 0.046796633 | 1 |
| T2DM_admissi | duct_epithelial_cell | *LINC00638* | -1.203527521 | 0.001471201 | 0.048754417 | 1 |
| T2DM_admissi | endothelial_cell | *STPG1* | -0.947393964 | 0.000364913 | 0.020778905 | 1 |
| T2DM_admissi | endothelial_cell | *RBM6* | -0.811270483 | 4.68E-09 | 3.68E-06 | 1.84E-05 |
| T2DM_admissi | endothelial_cell | *ZNF195* | -1.019260226 | 0.000170881 | 0.012160846 | 0.671391076 |
| T2DM_admissi | endothelial_cell | *GGNBP2* | -0.96116196 | 0.000350574 | 0.020255957 | 1 |
| T2DM_admissi | endothelial_cell | *USH1C* | -0.510800703 | 0.000856727 | 0.036109444 | 1 |
| T2DM_admissi | endothelial_cell | *ST7L* | -0.493059249 | 0.00095333 | 0.037920116 | 1 |
| T2DM_admissi | endothelial_cell | *BAIAP3* | -0.538617934 | 0.000863906 | 0.036109444 | 1 |
| T2DM_admissi | endothelial_cell | *GIPR* | -0.776931599 | 5.47E-06 | 0.001022913 | 0.021481174 |
| T2DM_admissi | endothelial_cell | *CEP68* | 1.153696526 | 1.60E-08 | 8.99E-06 | 6.30E-05 |
| T2DM_admissi | endothelial_cell | *SH2D2A* | -1.152882244 | 0.000511534 | 0.027155979 | 1 |
| T2DM_admissi | endothelial_cell | *BAK1* | 1.400193703 | 2.30E-07 | 6.44E-05 | 0.000902101 |
| T2DM_admissi | endothelial_cell | *LY75* | -2.642110733 | 0.000764084 | 0.034068568 | 1 |
| T2DM_admissi | endothelial_cell | *USP36* | -1.415973698 | 0.001189558 | 0.043275676 | 1 |
| T2DM_admissi | endothelial_cell | *ITIH4* | -1.01871839 | 0.000150578 | 0.011134387 | 0.591622227 |
| T2DM_admissi | endothelial_cell | *HAGH* | -0.990081658 | 8.15E-05 | 0.007280483 | 0.320341263 |
| T2DM_admissi | endothelial_cell | *ST6GAL1* | 0.488340868 | 0.00033839 | 0.019843808 | 1 |
| T2DM_admissi | endothelial_cell | *MARK3* | -1.768967679 | 5.10E-08 | 2.22E-05 | 0.00020023 |
| T2DM_admissi | endothelial_cell | *XAB2* | -1.800909843 | 0.000247902 | 0.01521887 | 0.974007695 |
| T2DM_admissi | endothelial_cell | *POMGNT1* | -1.239574867 | 9.23E-05 | 0.007882797 | 0.36260864 |
| T2DM_admissi | endothelial_cell | *CWF19L1* | -0.443690577 | 0.000242183 | 0.015103786 | 0.95153852 |
| T2DM_admissi | endothelial_cell | *AP1B1* | -1.410928123 | 5.36E-05 | 0.005694071 | 0.210680612 |
| T2DM_admissi | endothelial_cell | *RPAP1* | -2.566365507 | 0.000254266 | 0.015369373 | 0.999009277 |
| T2DM_admissi | endothelial_cell | *HSD17B1* | -0.706761844 | 6.98E-06 | 0.001192418 | 0.027425624 |
| T2DM_admissi | endothelial_cell | *CNTNAP1* | 0.642616091 | 1.58E-05 | 0.00238634 | 0.062044836 |
| T2DM_admissi | endothelial_cell | *CDK2AP1* | 0.572409537 | 0.001317986 | 0.046505084 | 1 |
| T2DM_admissi | endothelial_cell | *NNT* | -0.701524966 | 0.000176146 | 0.012160846 | 0.692075835 |
| T2DM_admissi | endothelial_cell | *CHMP3* | -0.809475651 | 0.000153031 | 0.011134387 | 0.60125689 |
| T2DM_admissi | endothelial_cell | *ANGPTL1* | -2.661658956 | 0.000141483 | 0.011114854 | 0.555886464 |
| T2DM_admissi | endothelial_cell | *NSUN4* | -1.180872068 | 0.000233706 | 0.015052959 | 0.918230527 |
| T2DM_admissi | endothelial_cell | *DNAH7* | -0.56874061 | 8.85E-05 | 0.00772634 | 0.347685307 |
| T2DM_admissi | endothelial_cell | *FAM53C* | -1.310390386 | 3.22E-05 | 0.003858653 | 0.126674209 |
| T2DM_admissi | endothelial_cell | *ZNF211* | 0.464839267 | 0.000831032 | 0.035777083 | 1 |
| T2DM_admissi | endothelial_cell | *POLK* | -1.261893061 | 9.63E-06 | 0.001576051 | 0.037825214 |
| T2DM_admissi | endothelial_cell | *PEPD* | -1.531871668 | 3.24E-05 | 0.003858653 | 0.127271556 |
| T2DM_admissi | endothelial_cell | *PRMT1* | -0.831089675 | 0.001044954 | 0.039477155 | 1 |
| T2DM_admissi | endothelial_cell | *EIF2AK4* | -1.046985841 | 0.000677763 | 0.03287571 | 1 |
| T2DM_admissi | endothelial_cell | *MAU2* | -1.647081404 | 5.78E-08 | 2.27E-05 | 0.00022721 |
| T2DM_admissi | endothelial_cell | *GTPBP3* | -0.525748062 | 0.001433288 | 0.049835308 | 1 |
| T2DM_admissi | endothelial_cell | *UCK1* | -1.469528876 | 0.000965134 | 0.037920116 | 1 |
| T2DM_admissi | endothelial_cell | *BHMT2* | -0.807319103 | 0.0001425 | 0.011114854 | 0.559882318 |
| T2DM_admissi | endothelial_cell | *NOTCH2* | -0.701549428 | 3.59E-12 | 3.53E-09 | 1.41E-08 |
| T2DM_admissi | endothelial_cell | *FXYD2* | -0.409240797 | 0.000725621 | 0.033740124 | 1 |
| T2DM_admissi | endothelial_cell | *TUBGCP4* | -1.201084633 | 0.000221563 | 0.014508679 | 0.870520759 |

| | | | | | | |
|---|---|---|---|---|---|---|
| T2DM_admissi | endothelial_cell | *MAN2C1* | -0.482209945 | 5.05E-06 | 0.000992512 | 0.019850231 |
| T2DM_admissi | endothelial_cell | *NPC1* | -2.159062843 | 3.54E-07 | 9.28E-05 | 0.001392462 |
| T2DM_admissi | endothelial_cell | *FN3KRP* | -0.332813256 | 0.000764271 | 0.034068568 | 1 |
| T2DM_admissi | endothelial_cell | *TBC1D7* | -2.125393619 | 0.000447711 | 0.024677265 | 1 |
| T2DM_admissi | endothelial_cell | *SCML4* | -1.197437986 | 0.000944491 | 0.037920116 | 1 |
| T2DM_admissi | endothelial_cell | *LRP12* | -1.787899368 | 0.000452218 | 0.024677265 | 1 |
| T2DM_admissi | endothelial_cell | *HSD17B12* | -0.367295661 | 9.82E-08 | 3.22E-05 | 0.000386007 |
| T2DM_admissi | endothelial_cell | *APIP* | -0.452471488 | 7.83E-05 | 0.007158157 | 0.307800768 |
| T2DM_admissi | endothelial_cell | *SIDT2* | -0.833967003 | 0.000786411 | 0.034331211 | 1 |
| T2DM_admissi | endothelial_cell | *TAGLN* | -0.304003866 | 0.000837743 | 0.035777083 | 1 |
| T2DM_admissi | endothelial_cell | *VIPAS39* | -1.305832751 | 0.000142124 | 0.011114854 | 0.558405928 |
| T2DM_admissi | endothelial_cell | *POC5* | -0.452558566 | 1.72E-06 | 0.00039765 | 0.006760057 |
| T2DM_admissi | endothelial_cell | *JAZF1* | -1.30968796 | 4.94E-15 | 9.71E-12 | 1.94E-11 |
| T2DM_admissi | endothelial_cell | *NUS1* | -1.617075535 | 2.80E-05 | 0.003858653 | 0.109841156 |
| T2DM_admissi | endothelial_cell | *CC2D1B* | -0.512872069 | 0.000622255 | 0.031751159 | 1 |
| T2DM_admissi | endothelial_cell | *EXTL1* | -1.394711225 | 0.000144619 | 0.011114854 | 0.568206742 |
| T2DM_admissi | endothelial_cell | *CILP2* | -0.801690303 | 6.93E-06 | 0.001192418 | 0.027230803 |
| T2DM_admissi | endothelial_cell | *VPS11* | -0.763088693 | 0.001003555 | 0.038478225 | 1 |
| T2DM_admissi | endothelial_cell | *PAQR6* | -0.860425385 | 0.000518376 | 0.027155979 | 1 |
| T2DM_admissi | endothelial_cell | *DCAF16* | -1.153958598 | 0.000643364 | 0.031997201 | 1 |
| T2DM_admissi | endothelial_cell | *ICA1L* | -0.479835923 | 2.57E-05 | 0.003737767 | 0.100919719 |
| T2DM_admissi | endothelial_cell | *PIGX* | -1.261213886 | 0.000770478 | 0.034068568 | 1 |
| T2DM_admissi | endothelial_cell | *LMBRD2* | -0.787600357 | 0.001305288 | 0.046505084 | 1 |
| T2DM_admissi | endothelial_cell | *CASP7* | 0.334440789 | 6.10E-05 | 0.006147298 | 0.239744641 |
| T2DM_admissi | endothelial_cell | *TRUB1* | -1.123679328 | 0.001322266 | 0.046505084 | 1 |
| T2DM_admissi | endothelial_cell | *CEP57* | -1.095393916 | 6.60E-05 | 0.006487616 | 0.25950463 |
| T2DM_admissi | endothelial_cell | *CKB* | -0.440179938 | 0.000147104 | 0.011114854 | 0.577972388 |
| T2DM_admissi | endothelial_cell | *SGPL1* | -0.837266929 | 5.88E-05 | 0.006079628 | 0.231025873 |
| T2DM_admissi | endothelial_cell | *PCBD1* | -0.501434497 | 0.000481309 | 0.025904951 | 1 |
| T2DM_admissi | endothelial_cell | *POP5* | -1.211255282 | 0.000176424 | 0.012160846 | 0.693168247 |
| T2DM_admissi | endothelial_cell | *TP53I13* | -0.563042533 | 0.000241619 | 0.015103786 | 0.949321395 |
| T2DM_admissi | endothelial_cell | *RAB4B* | -0.903563806 | 0.001325673 | 0.046505084 | 1 |
| T2DM_admissi | endothelial_cell | *DNAJC21* | -1.312263765 | 0.001008719 | 0.038478225 | 1 |
| T2DM_admissi | endothelial_cell | *TMEM129* | -0.698594592 | 3.47E-05 | 0.004008419 | 0.136286236 |
| T2DM_admissi | endothelial_cell | *INO80E* | -0.919107318 | 2.66E-06 | 0.000549822 | 0.010446617 |
| T2DM_admissi | endothelial_cell | *ZNF561* | -1.302865432 | 0.001065408 | 0.039866566 | 1 |
| T2DM_admissi | endothelial_cell | *CTSW* | -0.2886368 | 0.000586327 | 0.03031154 | 1 |
| T2DM_admissi | endothelial_cell | *RNASEH2C* | -0.82963128 | 3.20E-05 | 0.003858653 | 0.125746115 |
| T2DM_admissi | endothelial_cell | *PEAK1* | 0.422822935 | 0.001176446 | 0.043198651 | 1 |
| T2DM_admissi | endothelial_cell | *YIF1A* | -0.619312828 | 2.90E-05 | 0.003858653 | 0.114041902 |
| T2DM_admissi | endothelial_cell | *DSCAML1* | -0.508623919 | 0.001103932 | 0.040918403 | 1 |
| T2DM_admissi | endothelial_cell | *IRX3* | 1.862193191 | 3.20E-17 | 1.26E-13 | 1.26E-13 |
| T2DM_admissi | endothelial_cell | *TRIM73* | -0.855138882 | 0.000677277 | 0.03287571 | 1 |
| T2DM_admissi | endothelial_cell | *FARSA* | -1.630786483 | 1.44E-08 | 8.99E-06 | 5.67E-05 |
| T2DM_admissi | endothelial_cell | *HLA-DQB1* | -0.34795308 | 9.22E-07 | 0.000226346 | 0.003621543 |
| T2DM_admissi | endothelial_cell | *HIGD1A* | -0.845340286 | 0.001007767 | 0.038478225 | 1 |
| T2DM_admissi | endothelial_cell | *VMO1* | 0.537899149 | 0.000282337 | 0.01680761 | 1 |
| T2DM_admissi | endothelial_cell | *PGP* | -1.093644219 | 3.84E-05 | 0.004311177 | 0.1508912 |
| T2DM_admissi | endothelial_cell | *DMWD* | -1.061027658 | 6.94E-05 | 0.006649354 | 0.272623496 |
| T2DM_admissi | endothelial_cell | *NUTM2B* | -4.722214114 | 3.24E-05 | 0.003858653 | 0.127335546 |

| | | | | | | |
|---|---|---|---|---|---|---|
| T2DM_admissi | endothelial_cell | *HLA-DRB1* | -0.273829592 | 4.90E-08 | 2.22E-05 | 0.000192575 |
| T2DM_admissi | endothelial_cell | *SUPT3H* | -1.882920213 | 0.000686564 | 0.032896455 | 1 |
| T2DM_admissi | endothelial_cell | *XRCC6* | -0.924098584 | 0.000104655 | 0.008748733 | 0.411190429 |
| T2DM_admissi | endothelial_cell | *SULT1A1* | -0.949156235 | 0.000641894 | 0.031997201 | 1 |
| T2DM_admissi | endothelial_cell | *DPP4* | -0.432257684 | 0.000926076 | 0.037920116 | 1 |
| T2DM_admissi | endothelial_cell | *RPS26* | -0.508230611 | 0.000771724 | 0.034068568 | 1 |
| T2DM_admissi | endothelial_cell | *LIME1* | -0.465950502 | 0.000191494 | 0.012802692 | 0.752381656 |
| T2DM_admissi | endothelial_cell | *AGER* | 0.537477713 | 0.000192252 | 0.012802692 | 0.755358834 |
| T2DM_admissi | endothelial_cell | *RNF5* | -0.603988788 | 9.65E-08 | 3.22E-05 | 0.000379185 |
| T2DM_admissi | endothelial_cell | *RNF39* | 0.335387062 | 0.000956764 | 0.037920116 | 1 |
| T2DM_admissi | endothelial_cell | *HCG27* | 0.396847607 | 4.74E-05 | 0.005170887 | 0.18615192 |
| T2DM_admissi | endothelial_cell | *DNLZ* | 0.262085657 | 0.000729934 | 0.033740124 | 1 |
| T2DM_admissi | endothelial_cell | *ATF6B* | 0.523983386 | 7.12E-05 | 0.006659332 | 0.279691925 |
| T2DM_admissi | endothelial_cell | *SLC12A8* | -0.646585786 | 0.000954247 | 0.037920116 | 1 |
| T2DM_admissi | endothelial_cell | *PPT2* | 0.338282209 | 0.000719541 | 0.033740124 | 1 |
| T2DM_admissi | endothelial_cell | *HLA-DQB2* | 0.862728881 | 1.07E-07 | 3.23E-05 | 0.000419863 |
| T2DM_admissi | endothelial_cell | *HLA-B* | 0.36963967 | 1.87E-06 | 0.000409116 | 0.007364094 |
| T2DM_admissi | endothelial_cell | *HLA-DQA2* | 0.59836796 | 2.78E-13 | 3.65E-10 | 1.09E-09 |
| T2DM_admissi | endothelial_cell | *JMJD7* | 1.11222595 | 1.03E-05 | 0.001620318 | 0.040507949 |
| T2DM_admissi | endothelial_cell | *GATC* | -1.271209676 | 0.000447138 | 0.024677265 | 1 |
| T2DM_admissi | epsilon_cell | *CLEC18A* | -13.12636886 | 1.43E-05 | 0.00802549 | 0.05617843 |
| T2DM_admissi | epsilon_cell | *CILP2* | -8.928464978 | 6.22E-05 | 0.024439789 | 0.244397894 |
| T2DM_admissi | epsilon_cell | *IRX3* | 16.63146655 | 4.96E-14 | 1.95E-10 | 1.95E-10 |
| T2DM_admissi | epsilon_cell | *HLA-DQB1* | -2.989706767 | 2.48E-05 | 0.012157563 | 0.097260502 |
| T2DM_admissi | epsilon_cell | *UBA7* | -6.863896022 | 6.99E-06 | 0.005490509 | 0.027452546 |
| T2DM_admissi | epsilon_cell | *HLA-DRB1* | -3.215039152 | 1.13E-08 | 1.48E-05 | 4.45E-05 |
| T2DM_admissi | epsilon_cell | *HLA-DRB5* | -3.242192178 | 3.88E-05 | 0.016942315 | 0.152480832 |
| T2DM_admissi | epsilon_cell | *NOTCH4* | -4.580842671 | 3.20E-06 | 0.00313839 | 0.012553561 |
| T2DM_admissi | epsilon_cell | *HLA-DQB2* | 7.958874207 | 1.10E-05 | 0.007227829 | 0.043366972 |
| T2DM_admissi | epsilon_cell | *HLA-DQA2* | 6.903375205 | 9.96E-13 | 1.96E-09 | 3.91E-09 |
| T2DM_admissi | leukocyte | *GCLC* | -1.777226517 | 0.000230506 | 0.001064322 | 0.905658417 |
| T2DM_admissi | leukocyte | *NFYA* | -1.295327638 | 0.000818101 | 0.001665451 | 1 |
| T2DM_admissi | leukocyte | *STPG1* | -1.194405255 | 0.000227691 | 0.001062643 | 0.894598986 |
| T2DM_admissi | leukocyte | *LASP1* | -1.156071019 | 0.000289798 | 0.001088708 | 1 |
| T2DM_admissi | leukocyte | *SNX11* | -0.996844458 | 0.006184704 | 0.008390781 | 1 |
| T2DM_admissi | leukocyte | *TMEM176A* | -0.704362991 | 0.00466412 | 0.006482252 | 1 |
| T2DM_admissi | leukocyte | *M6PR* | -2.662031075 | 7.86E-05 | 0.001062643 | 0.308697687 |
| T2DM_admissi | leukocyte | *DBNDD1* | -0.89719355 | 0.000394896 | 0.001181681 | 1 |
| T2DM_admissi | leukocyte | *CASP10* | -3.103899826 | 0.000363905 | 0.001146348 | 1 |
| T2DM_admissi | leukocyte | *NDUFAF7* | -2.684446827 | 0.000106089 | 0.001062643 | 0.416824957 |
| T2DM_admissi | leukocyte | *POLDIP2* | -0.928494899 | 0.000793606 | 0.001634654 | 1 |
| T2DM_admissi | leukocyte | *PLXND1* | -2.523457317 | 0.000276105 | 0.001088708 | 1 |
| T2DM_admissi | leukocyte | *KDM1A* | -1.238947564 | 0.000333481 | 0.00112758 | 1 |
| T2DM_admissi | leukocyte | *RBM6* | -2.183634435 | 1.97E-09 | 1.55E-06 | 7.74E-06 |
| T2DM_admissi | leukocyte | *RECQL* | -1.249751091 | 0.000302522 | 0.001093914 | 1 |
| T2DM_admissi | leukocyte | *SLC25A13* | -1.17739448 | 0.000524712 | 0.001319879 | 1 |
| T2DM_admissi | leukocyte | *MEOX1* | 2.739038574 | 0.003482997 | 0.004994414 | 1 |
| T2DM_admissi | leukocyte | *LIG3* | -2.520648822 | 0.010196484 | 0.013252394 | 1 |
| T2DM_admissi | leukocyte | *PON1* | -1.194037564 | 0.002974933 | 0.00434033 | 1 |
| T2DM_admissi | leukocyte | *KMT2E* | -1.308000306 | 0.000162215 | 0.001062643 | 0.63734135 |

| | | | | | | |
|---|---|---|---|---|---|---|
| T2DM_admissi | leukocyte | *IBTK* | -1.280650379 | 0.00016302 | 0.001062643 | 0.640507146 |
| T2DM_admissi | leukocyte | *ZNF195* | -2.149001534 | 1.16E-05 | 0.000784902 | 0.045524327 |
| T2DM_admissi | leukocyte | *GGNBP2* | -1.461434866 | 3.68E-05 | 0.001037577 | 0.144455994 |
| T2DM_admissi | leukocyte | *TMEM98* | -0.838791773 | 0.001586475 | 0.002539896 | 1 |
| T2DM_admissi | leukocyte | *SYNRG* | -1.844374593 | 0.000123741 | 0.001062643 | 0.48617915 |
| T2DM_admissi | leukocyte | *SPATA20* | -1.128727211 | 0.002098282 | 0.003195406 | 1 |
| T2DM_admissi | leukocyte | *FARP2* | -3.995350583 | 3.27E-05 | 0.001037577 | 0.128659701 |
| T2DM_admissi | leukocyte | *USH1C* | -1.391283292 | 4.39E-05 | 0.001037577 | 0.172425123 |
| T2DM_admissi | leukocyte | *VPS41* | -1.533464502 | 0.000640895 | 0.001440839 | 1 |
| T2DM_admissi | leukocyte | *ELAC2* | -1.223683473 | 0.000822047 | 0.001670341 | 1 |
| T2DM_admissi | leukocyte | *CCDC124* | -1.086942979 | 0.00026249 | 0.001083674 | 1 |
| T2DM_admissi | leukocyte | *NOS2* | -17.83406838 | 0.00130735 | 0.002206435 | 1 |
| T2DM_admissi | leukocyte | *CD79B* | -7.158616004 | 4.08E-05 | 0.001037577 | 0.160273336 |
| T2DM_admissi | leukocyte | *ST7L* | -2.316946458 | 3.65E-06 | 0.000477601 | 0.014328042 |
| T2DM_admissi | leukocyte | *BAIAP3* | -1.328055872 | 4.98E-05 | 0.00105496 | 0.195765772 |
| T2DM_admissi | leukocyte | *FMO3* | -2.910591801 | 0.03967202 | 0.047319784 | 1 |
| T2DM_admissi | leukocyte | *CYB561* | -1.110754946 | 0.000233594 | 0.001066706 | 0.917790791 |
| T2DM_admissi | leukocyte | *MGST1* | -0.758966088 | 0.001666895 | 0.002644017 | 1 |
| T2DM_admissi | leukocyte | *ST3GAL1* | -2.930973286 | 3.45E-05 | 0.001037577 | 0.135391413 |
| T2DM_admissi | leukocyte | *IL32* | -0.508145088 | 0.001242549 | 0.002152855 | 1 |
| T2DM_admissi | leukocyte | *MED24* | -1.589034104 | 0.000450283 | 0.001234587 | 1 |
| T2DM_admissi | leukocyte | *RPS20* | -0.846237704 | 0.000181242 | 0.001062643 | 0.712101733 |
| T2DM_admissi | leukocyte | *REV3L* | -0.997794937 | 0.000551288 | 0.001352066 | 1 |
| T2DM_admissi | leukocyte | *ETV7* | -2.260666013 | 0.000304351 | 0.00109505 | 1 |
| T2DM_admissi | leukocyte | *NCAPD2* | -2.015037791 | 0.000842499 | 0.001699271 | 1 |
| T2DM_admissi | leukocyte | *GIPR* | -1.146413114 | 4.41E-05 | 0.001037577 | 0.173223707 |
| T2DM_admissi | leukocyte | *FUZ* | -1.781768788 | 0.000512595 | 0.001308606 | 1 |
| T2DM_admissi | leukocyte | *LRRC23* | -0.832714164 | 0.001121265 | 0.002027359 | 1 |
| T2DM_admissi | leukocyte | *TSPAN9* | -0.940028546 | 0.001245334 | 0.002154446 | 1 |
| T2DM_admissi | leukocyte | *BTBD7* | -1.226192838 | 0.000762922 | 0.001593931 | 1 |
| T2DM_admissi | leukocyte | *APBA3* | -2.057867319 | 0.001128782 | 0.002034397 | 1 |
| T2DM_admissi | leukocyte | *TTC19* | -0.92954171 | 0.00074854 | 0.001571055 | 1 |
| T2DM_admissi | leukocyte | *DPF1* | -0.895518721 | 0.026494113 | 0.032167914 | 1 |
| T2DM_admissi | leukocyte | *RABGAP1* | -1.148093258 | 0.000561664 | 0.001362208 | 1 |
| T2DM_admissi | leukocyte | *QPCTL* | -4.644976554 | 0.000112659 | 0.001062643 | 0.442637755 |
| T2DM_admissi | leukocyte | *PPP5C* | -1.151662932 | 0.000505062 | 0.001300386 | 1 |
| T2DM_admissi | leukocyte | *ZBTB32* | -18.03485435 | 0.007378514 | 0.009853903 | 1 |
| T2DM_admissi | leukocyte | *ERCC1* | -1.533753213 | 9.89E-05 | 0.001062643 | 0.38869646 |
| T2DM_admissi | leukocyte | *CD22* | 0.551441641 | 0.003413181 | 0.004905043 | 1 |
| T2DM_admissi | leukocyte | *LTF* | -3.503531721 | 6.23E-05 | 0.001060479 | 0.24493909 |
| T2DM_admissi | leukocyte | *EXTL3* | -1.040078417 | 0.00056493 | 0.001362351 | 1 |
| T2DM_admissi | leukocyte | *CALCOCO1* | -3.895077848 | 0.000285739 | 0.001088708 | 1 |
| T2DM_admissi | leukocyte | *UBR7* | -1.06259107 | 0.002307723 | 0.003475294 | 1 |
| T2DM_admissi | leukocyte | *MAN2B2* | -1.746512749 | 0.000240878 | 0.001072513 | 0.94641157 |
| T2DM_admissi | leukocyte | *ANGEL1* | -1.245586312 | 0.007941201 | 0.010544434 | 1 |
| T2DM_admissi | leukocyte | *DDX11* | -1.210750865 | 0.000929601 | 0.001800431 | 1 |
| T2DM_admissi | leukocyte | *GPRC5A* | -1.191163327 | 0.006019213 | 0.008203083 | 1 |
| T2DM_admissi | leukocyte | *UFL1* | -1.187412637 | 0.000218534 | 0.001062643 | 0.85861972 |
| T2DM_admissi | leukocyte | *POLA2* | -2.051473124 | 0.000537295 | 0.001334229 | 1 |
| T2DM_admissi | leukocyte | *ZC3H3* | -0.905408698 | 0.005578986 | 0.007637573 | 1 |

| | | | | | | |
|---|---|---|---|---|---|---|
| T2DM_admissi | leukocyte | *MDH1* | -1.181492058 | 0.000225585 | 0.001062643 | 0.886324752 |
| T2DM_admissi | leukocyte | *MTMR11* | -4.141769386 | 0.0006215 | 0.001422226 | 1 |
| T2DM_admissi | leukocyte | *DPEP1* | -0.678594312 | 0.000951454 | 0.001823593 | 1 |
| T2DM_admissi | leukocyte | *NUDCD3* | -1.03233109 | 0.000566802 | 0.001364563 | 1 |
| T2DM_admissi | leukocyte | *TTC27* | -2.753760919 | 0.000160801 | 0.001062643 | 0.631785257 |
| T2DM_admissi | leukocyte | *SNAI2* | -2.86374324 | 0.001893925 | 0.002938875 | 1 |
| T2DM_admissi | leukocyte | *PLEKHB1* | -0.828175685 | 0.015071818 | 0.019071554 | 1 |
| T2DM_admissi | leukocyte | *NLRP2* | -1.692402026 | 0.01566447 | 0.019789615 | 1 |
| T2DM_admissi | leukocyte | *ZDHHC6* | -1.503465205 | 0.000325015 | 0.001118201 | 1 |
| T2DM_admissi | leukocyte | *RNH1* | -1.184590044 | 0.000162539 | 0.001062643 | 0.638614389 |
| T2DM_admissi | leukocyte | *NDUFS1* | -1.073984728 | 0.000285883 | 0.001088708 | 1 |
| T2DM_admissi | leukocyte | *TYMP* | 0.41040322 | 0.023718231 | 0.02906704 | 1 |
| T2DM_admissi | leukocyte | *CD44* | -0.606035453 | 0.000645127 | 0.001445924 | 1 |
| T2DM_admissi | leukocyte | *KCNG1* | -3.950360151 | 0.001055528 | 0.001948857 | 1 |
| T2DM_admissi | leukocyte | *AGPAT4* | -3.126085974 | 0.0077468 | 0.010314191 | 1 |
| T2DM_admissi | leukocyte | *SH2D2A* | -7.226293526 | 1.18E-05 | 0.000786127 | 0.046381497 |
| T2DM_admissi | leukocyte | *VEZT* | -1.173953634 | 0.000636145 | 0.001436444 | 1 |
| T2DM_admissi | leukocyte | *RABEP1* | -1.985737118 | 0.000382444 | 0.001165285 | 1 |
| T2DM_admissi | leukocyte | *ARID4A* | -1.708713912 | 9.21E-05 | 0.001062643 | 0.361763381 |
| T2DM_admissi | leukocyte | *PNPLA6* | -1.530915609 | 0.000224527 | 0.001062643 | 0.882166515 |
| T2DM_admissi | leukocyte | *ABCF2* | -1.63820331 | 0.000317182 | 0.001111111 | 1 |
| T2DM_admissi | leukocyte | *UBA6* | -1.91312135 | 0.000169637 | 0.001062643 | 0.666502898 |
| T2DM_admissi | leukocyte | *APBA2* | -2.847034854 | 0.001262481 | 0.002166065 | 1 |
| T2DM_admissi | leukocyte | *ASTE1* | -1.389096072 | 0.001717491 | 0.002713317 | 1 |
| T2DM_admissi | leukocyte | *RNF19A* | -1.042279028 | 0.000250999 | 0.001077213 | 0.986176822 |
| T2DM_admissi | leukocyte | *SH3YL1* | -0.701223748 | 0.001569397 | 0.002518856 | 1 |
| T2DM_admissi | leukocyte | *FAM136A* | -1.13998647 | 0.000334021 | 0.001128435 | 1 |
| T2DM_admissi | leukocyte | *DAPK2* | -1.217216984 | 0.000526313 | 0.001319879 | 1 |
| T2DM_admissi | leukocyte | *TIMP2* | -2.534760377 | 2.68E-05 | 0.000994138 | 0.105378619 |
| T2DM_admissi | leukocyte | *MYOM2* | 1.076891777 | 0.019873862 | 0.024725904 | 1 |
| T2DM_admissi | leukocyte | *RPL26L1* | -1.078205333 | 0.000253593 | 0.001077213 | 0.996368042 |
| T2DM_admissi | leukocyte | *NSUN2* | -1.824469792 | 8.56E-05 | 0.001062643 | 0.336291815 |
| T2DM_admissi | leukocyte | *MRI1* | -1.367050709 | 0.000524763 | 0.001319879 | 1 |
| T2DM_admissi | leukocyte | *AGA* | -1.340856698 | 0.000360412 | 0.001142338 | 1 |
| T2DM_admissi | leukocyte | *BOD1L1* | -0.994911379 | 0.00099491 | 0.001874821 | 1 |
| T2DM_admissi | leukocyte | *DNAH5* | -1.755302426 | 0.000416455 | 0.001199564 | 1 |
| T2DM_admissi | leukocyte | *PNKP* | -2.439925673 | 0.000621017 | 0.001422226 | 1 |
| T2DM_admissi | leukocyte | *PHLPP2* | -1.005648332 | 0.00097803 | 0.001856058 | 1 |
| T2DM_admissi | leukocyte | *CTNS* | -1.369831986 | 0.001144557 | 0.002052471 | 1 |
| T2DM_admissi | leukocyte | *INPP4A* | -1.026573218 | 0.001210971 | 0.002123116 | 1 |
| T2DM_admissi | leukocyte | *PSMA4* | -1.112121694 | 0.000221371 | 0.001062643 | 0.869764707 |
| T2DM_admissi | leukocyte | *LSG1* | -0.674324988 | 0.009979891 | 0.013000993 | 1 |
| T2DM_admissi | leukocyte | *SPATA7* | -1.683905222 | 0.00014378 | 0.001062643 | 0.564909784 |
| T2DM_admissi | leukocyte | *TRIT1* | -2.437147198 | 0.000773112 | 0.001608876 | 1 |
| T2DM_admissi | leukocyte | *CNTLN* | -1.661403181 | 0.000361687 | 0.001142338 | 1 |
| T2DM_admissi | leukocyte | *PREX2* | -1.975821467 | 0.002296037 | 0.003461677 | 1 |
| T2DM_admissi | leukocyte | *WDR37* | -1.758743163 | 0.000135001 | 0.001062643 | 0.530420329 |
| T2DM_admissi | leukocyte | *YTHDC2* | -1.901674973 | 0.000132564 | 0.001062643 | 0.520844053 |
| T2DM_admissi | leukocyte | *DTNBP1* | -1.324372344 | 0.000366352 | 0.001147001 | 1 |
| T2DM_admissi | leukocyte | *GOPC* | -1.394803609 | 0.000183343 | 0.001062643 | 0.720355442 |

| | | | | | | |
|---|---|---|---|---|---|---|
| T2DM_admissi | leukocyte | *ZNF800* | -1.373151948 | 0.000188849 | 0.001062643 | 0.741989002 |
| T2DM_admissi | leukocyte | *MRPS10* | -1.713380617 | 4.12E-05 | 0.001037577 | 0.161878263 |
| T2DM_admissi | leukocyte | *COL9A2* | -1.188866048 | 0.00087448 | 0.001741426 | 1 |
| T2DM_admissi | leukocyte | *ERCC8* | -3.151795815 | 0.009503279 | 0.012404779 | 1 |
| T2DM_admissi | leukocyte | *H6PD* | -3.3593954 | 4.33E-05 | 0.001037577 | 0.16997902 |
| T2DM_admissi | leukocyte | *VAMP3* | -1.414183308 | 0.000193532 | 0.001062643 | 0.760386358 |
| T2DM_admissi | leukocyte | *RCN1* | -0.942152666 | 0.000402648 | 0.001185781 | 1 |
| T2DM_admissi | leukocyte | *RFC2* | -1.156716251 | 0.001765172 | 0.002770339 | 1 |
| T2DM_admissi | leukocyte | *JKAMP* | -1.096092618 | 0.000394378 | 0.001181536 | 1 |
| T2DM_admissi | leukocyte | *PTGER3* | -1.02690803 | 0.001006743 | 0.001889818 | 1 |
| T2DM_admissi | leukocyte | *BCAR1* | -0.900466871 | 0.000742009 | 0.001563191 | 1 |
| T2DM_admissi | leukocyte | *THOC3* | -0.863215588 | 0.001243275 | 0.002152855 | 1 |
| T2DM_admissi | leukocyte | *HEBP2* | -1.132769012 | 9.52E-05 | 0.001062643 | 0.374059376 |
| T2DM_admissi | leukocyte | *RRP12* | -0.781649838 | 0.00114124 | 0.002049538 | 1 |
| T2DM_admissi | leukocyte | *FNIP2* | -2.662655796 | 0.000160904 | 0.001062643 | 0.632192491 |
| T2DM_admissi | leukocyte | *MRTO4* | -1.315779889 | 0.000621016 | 0.001422226 | 1 |
| T2DM_admissi | leukocyte | *MCF2L2* | -1.89796863 | 0.000372504 | 0.001155145 | 1 |
| T2DM_admissi | leukocyte | *LAMA3* | -1.799976149 | 0.000779795 | 0.001614233 | 1 |
| T2DM_admissi | leukocyte | *ANAPC4* | -4.202545254 | 0.003975773 | 0.005625067 | 1 |
| T2DM_admissi | leukocyte | *LY75* | -5.252901665 | 2.55E-05 | 0.000983968 | 0.100364704 |
| T2DM_admissi | leukocyte | *ARID4B* | -0.910239805 | 0.001386388 | 0.002298537 | 1 |
| T2DM_admissi | leukocyte | *SDCCAG8* | -1.077581536 | 0.000866754 | 0.001732185 | 1 |
| T2DM_admissi | leukocyte | *KIF1B* | -1.139966011 | 0.000273542 | 0.001088708 | 1 |
| T2DM_admissi | leukocyte | *TBC1D22A* | -1.190504502 | 0.000467569 | 0.001253701 | 1 |
| T2DM_admissi | leukocyte | *SYNE2* | -1.09668232 | 0.000370576 | 0.001151894 | 1 |
| T2DM_admissi | leukocyte | *ATP9A* | -1.225504142 | 0.000288326 | 0.001088708 | 1 |
| T2DM_admissi | leukocyte | *CHRDL2* | -3.914330824 | 0.001364379 | 0.002274982 | 1 |
| T2DM_admissi | leukocyte | *GALC* | -1.709738985 | 0.00023155 | 0.001066541 | 0.909759426 |
| T2DM_admissi | leukocyte | *CYFIP2* | -0.93210638 | 0.000610207 | 0.001411957 | 1 |
| T2DM_admissi | leukocyte | *USP36* | -1.962635313 | 5.37E-05 | 0.001060479 | 0.211098211 |
| T2DM_admissi | leukocyte | *MCOLN3* | -1.046225283 | 0.001014583 | 0.001900952 | 1 |
| T2DM_admissi | leukocyte | *ITIH4* | -3.145814509 | 3.27E-06 | 0.000443199 | 0.012852761 |
| T2DM_admissi | leukocyte | *PCGF2* | -1.121708506 | 0.000493558 | 0.001285934 | 1 |
| T2DM_admissi | leukocyte | *GDI2* | -1.21372164 | 0.000282909 | 0.001088708 | 1 |
| T2DM_admissi | leukocyte | *ATP11B* | -2.003055128 | 0.00016338 | 0.001062643 | 0.641918202 |
| T2DM_admissi | leukocyte | *LAMC2* | -0.799919165 | 0.000909588 | 0.001780653 | 1 |
| T2DM_admissi | leukocyte | *RASGRF1* | -0.806579714 | 0.001587031 | 0.002539896 | 1 |
| T2DM_admissi | leukocyte | *CROCC* | -1.496125495 | 0.003434652 | 0.004930489 | 1 |
| T2DM_admissi | leukocyte | *FLYWCH1* | -0.813263724 | 0.000807021 | 0.0016529 | 1 |
| T2DM_admissi | leukocyte | *OGFR* | -2.586776518 | 0.003848209 | 0.005466251 | 1 |
| T2DM_admissi | leukocyte | *GNA15* | 1.500361142 | 0.011398756 | 0.01468384 | 1 |
| T2DM_admissi | leukocyte | *RIMBP2* | -0.883125043 | 0.001251587 | 0.002156134 | 1 |
| T2DM_admissi | leukocyte | *MPC1* | -1.097015848 | 0.000248398 | 0.001077213 | 0.975954259 |
| T2DM_admissi | leukocyte | *BCAT1* | 0.332376064 | 0.02149917 | 0.026521268 | 1 |
| T2DM_admissi | leukocyte | *MRPS35* | -1.195924829 | 0.00044723 | 0.001232235 | 1 |
| T2DM_admissi | leukocyte | *ZNF112* | -1.451288524 | 0.001577422 | 0.00253068 | 1 |
| T2DM_admissi | leukocyte | *EIF4B* | -1.075082649 | 0.00026321 | 0.001083884 | 1 |
| T2DM_admissi | leukocyte | *SLC6A16* | -2.891145417 | 0.00125034 | 0.002155971 | 1 |
| T2DM_admissi | leukocyte | *SPHK2* | -1.268767015 | 0.001722598 | 0.002719068 | 1 |
| T2DM_admissi | leukocyte | *HAGH* | -1.629486475 | 1.43E-05 | 0.000815363 | 0.056284355 |

| | | | | | | |
|---|---|---|---|---|---|---|
| T2DM_admissi | leukocyte | *RNF4* | -1.406734698 | 0.000221557 | 0.001062643 | 0.870497955 |
| T2DM_admissi | leukocyte | *TM7SF3* | -2.241801656 | 0.000117326 | 0.001062643 | 0.460973836 |
| T2DM_admissi | leukocyte | *ST3GAL6* | -1.897151565 | 0.019828412 | 0.024684991 | 1 |
| T2DM_admissi | leukocyte | *HIPK2* | -0.946315304 | 0.000496541 | 0.001291992 | 1 |
| T2DM_admissi | leukocyte | *TMEM161A* | -1.269288332 | 0.000757952 | 0.001588262 | 1 |
| T2DM_admissi | leukocyte | *SNX24* | -2.734411782 | 0.000133633 | 0.001062643 | 0.525042836 |
| T2DM_admissi | leukocyte | *ABCA7* | -1.577062857 | 0.001048638 | 0.001940644 | 1 |
| T2DM_admissi | leukocyte | *PMS1* | -1.038019183 | 0.001598048 | 0.002554405 | 1 |
| T2DM_admissi | leukocyte | *IPO5* | -1.030735239 | 0.000305692 | 0.001096936 | 1 |
| T2DM_admissi | leukocyte | *OAT* | -1.065277695 | 0.000392938 | 0.001181536 | 1 |
| T2DM_admissi | leukocyte | *WDR18* | -1.320497151 | 0.000352236 | 0.001141109 | 1 |
| T2DM_admissi | leukocyte | *MCM10* | 1.778552636 | 0.014309991 | 0.01821904 | 1 |
| T2DM_admissi | leukocyte | *ERBB3* | -0.928371606 | 0.001330072 | 0.002233728 | 1 |
| T2DM_admissi | leukocyte | *ANKRD44* | -4.736159762 | 0.000661945 | 0.001464403 | 1 |
| T2DM_admissi | leukocyte | *ADAT1* | -1.091861739 | 0.001041187 | 0.001936841 | 1 |
| T2DM_admissi | leukocyte | *MYLK* | -0.72325961 | 0.016572229 | 0.02088271 | 1 |
| T2DM_admissi | leukocyte | *GSTO2* | -0.796039939 | 0.000458674 | 0.001240899 | 1 |
| T2DM_admissi | leukocyte | *SEC61A2* | -4.215026629 | 5.74E-06 | 0.000591121 | 0.022565957 |
| T2DM_admissi | leukocyte | *PRKCQ* | -2.600064422 | 0.001517649 | 0.002459919 | 1 |
| T2DM_admissi | leukocyte | *TBC1D1* | -1.154843115 | 0.000733218 | 0.001552225 | 1 |
| T2DM_admissi | leukocyte | *CTNNA2* | -1.218907627 | 0.004181961 | 0.005880789 | 1 |
| T2DM_admissi | leukocyte | *CD84* | 0.157568127 | 0.002678384 | 0.003956154 | 1 |
| T2DM_admissi | leukocyte | *ATXN3* | -1.682768003 | 0.000993841 | 0.001873994 | 1 |
| T2DM_admissi | leukocyte | *EML1* | -1.834378361 | 0.000272843 | 0.001088708 | 1 |
| T2DM_admissi | leukocyte | *STOML1* | -1.016535425 | 0.00077489 | 0.001611723 | 1 |
| T2DM_admissi | leukocyte | *DNTTIP2* | -1.025924841 | 0.000531424 | 0.001324223 | 1 |
| T2DM_admissi | leukocyte | *RRP15* | -1.171737709 | 0.000605464 | 0.00140595 | 1 |
| T2DM_admissi | leukocyte | *REEP1* | -0.738923736 | 0.017282712 | 0.02170143 | 1 |
| T2DM_admissi | leukocyte | *POLR1A* | -1.193839998 | 7.22E-05 | 0.001062643 | 0.283508196 |
| T2DM_admissi | leukocyte | *LAPTM4A* | -1.044843938 | 0.000193014 | 0.001062643 | 0.758352327 |
| T2DM_admissi | leukocyte | *KIF2A* | -1.102144572 | 0.000383472 | 0.001166145 | 1 |
| T2DM_admissi | leukocyte | *MAST4* | -1.283138129 | 0.000255134 | 0.001077348 | 1 |
| T2DM_admissi | leukocyte | *ADAM7* | -12.58453298 | 0.031105573 | 0.037488895 | 1 |
| T2DM_admissi | leukocyte | *DRD4* | -3.037591568 | 0.005952145 | 0.008117312 | 1 |
| T2DM_admissi | leukocyte | *PLA2G10* | -4.303997563 | 0.00051622 | 0.001313619 | 1 |
| T2DM_admissi | leukocyte | *PIGB* | -3.856460347 | 0.001835798 | 0.002861108 | 1 |
| T2DM_admissi | leukocyte | *GNB5* | -1.274452736 | 0.001705211 | 0.00269816 | 1 |
| T2DM_admissi | leukocyte | *RAB27A* | -1.059009384 | 0.00045964 | 0.001240899 | 1 |
| T2DM_admissi | leukocyte | *PTPN3* | -1.969133898 | 0.000113466 | 0.001062643 | 0.445809334 |
| T2DM_admissi | leukocyte | *SPTB* | -0.639069297 | 0.028695793 | 0.034733756 | 1 |
| T2DM_admissi | leukocyte | *FGF10* | -2.968624744 | 0.003523736 | 0.005041792 | 1 |
| T2DM_admissi | leukocyte | *DGCR2* | -1.593628919 | 0.000179231 | 0.001062643 | 0.704198541 |
| T2DM_admissi | leukocyte | *ZXDC* | -3.087626835 | 5.14E-05 | 0.00105496 | 0.201975993 |
| T2DM_admissi | leukocyte | *WIPI1* | -3.057868425 | 4.13E-05 | 0.001037577 | 0.162370677 |
| T2DM_admissi | leukocyte | *GBA2* | -1.02601368 | 0.002158819 | 0.003274904 | 1 |
| T2DM_admissi | leukocyte | *ASNS* | -1.067785054 | 0.001276678 | 0.002175489 | 1 |
| T2DM_admissi | leukocyte | *AP3M2* | -1.388100258 | 0.008717809 | 0.011474798 | 1 |
| T2DM_admissi | leukocyte | *TESK2* | -4.423708532 | 0.001296606 | 0.002195847 | 1 |
| T2DM_admissi | leukocyte | *MAP4K4* | -1.02274178 | 0.000346999 | 0.001140861 | 1 |
| T2DM_admissi | leukocyte | *RPL31* | -0.93362439 | 0.000174808 | 0.001062643 | 0.686819314 |

| | | | | | | |
|---|---|---|---|---|---|---|
| T2DM_admissi | leukocyte | *SEL1L* | -1.053656614 | 0.000203866 | 0.001062643 | 0.80098798 |
| T2DM_admissi | leukocyte | *TCF3* | -1.239911544 | 5.53E-05 | 0.001060479 | 0.217127421 |
| T2DM_admissi | leukocyte | *CPSF1* | -0.936072376 | 0.001770513 | 0.002776984 | 1 |
| T2DM_admissi | leukocyte | *CYBRD1* | 0.613252734 | 0.02542865 | 0.031027692 | 1 |
| T2DM_admissi | leukocyte | *LNX1* | -0.847562485 | 0.001058582 | 0.001952662 | 1 |
| T2DM_admissi | leukocyte | *ALDH3A2* | -0.720851117 | 0.001442794 | 0.002362958 | 1 |
| T2DM_admissi | leukocyte | *P4HA2* | -0.835148386 | 0.001100159 | 0.001999636 | 1 |
| T2DM_admissi | leukocyte | *TRNT1* | -3.800459015 | 0.000194998 | 0.001062643 | 0.766146907 |
| T2DM_admissi | leukocyte | *CRMP1* | -0.863380903 | 0.001784616 | 0.002793529 | 1 |
| T2DM_admissi | leukocyte | *DERL2* | -1.217344675 | 0.000224751 | 0.001062643 | 0.883047529 |
| T2DM_admissi | leukocyte | *XRCC1* | -1.593235744 | 0.000584615 | 0.001382042 | 1 |
| T2DM_admissi | leukocyte | *SCARB1* | -3.542337884 | 8.41E-06 | 0.000751223 | 0.033053797 |
| T2DM_admissi | leukocyte | *NLE1* | -0.925370102 | 0.013434864 | 0.017177215 | 1 |
| T2DM_admissi | leukocyte | *SMARCE1* | -1.045840222 | 0.000270036 | 0.001088175 | 1 |
| T2DM_admissi | leukocyte | *ST6GAL1* | 2.946475519 | 0.000700233 | 0.001515822 | 1 |
| T2DM_admissi | leukocyte | *MRPS34* | -0.937525273 | 0.000502584 | 0.001297407 | 1 |
| T2DM_admissi | leukocyte | *TEAD2* | -1.235331244 | 0.007985142 | 0.010595617 | 1 |
| T2DM_admissi | leukocyte | *IPCEF1* | -2.315414333 | 0.000314561 | 0.001109126 | 1 |
| T2DM_admissi | leukocyte | *ZZEF1* | -1.75193194 | 0.00014134 | 0.001062643 | 0.555323486 |
| T2DM_admissi | leukocyte | *WSCD2* | -1.144965995 | 0.001524805 | 0.002467446 | 1 |
| T2DM_admissi | leukocyte | *TIPIN* | -1.044012576 | 0.020310862 | 0.025197782 | 1 |
| T2DM_admissi | leukocyte | *SEMA3A* | -3.038998697 | 0.001479638 | 0.002406249 | 1 |
| T2DM_admissi | leukocyte | *TTC38* | -2.438334736 | 6.48E-05 | 0.001060479 | 0.254550592 |
| T2DM_admissi | leukocyte | *GRAMD4* | -1.712084781 | 0.000353859 | 0.001141109 | 1 |
| T2DM_admissi | leukocyte | *CELSR1* | -5.403626274 | 0.001638506 | 0.002606352 | 1 |
| T2DM_admissi | leukocyte | *TIMM21* | -1.714703199 | 0.000376178 | 0.001159218 | 1 |
| T2DM_admissi | leukocyte | *VPS9D1* | -0.94225174 | 0.005190968 | 0.00714622 | 1 |
| T2DM_admissi | leukocyte | *ZNF37A* | -1.214926943 | 0.000227999 | 0.001062643 | 0.895807654 |
| T2DM_admissi | leukocyte | *MARK3* | -1.489474946 | 1.15E-05 | 0.000784902 | 0.045123335 |
| T2DM_admissi | leukocyte | *CACNG5* | -31.37698597 | 0.001613412 | 0.002575821 | 1 |
| T2DM_admissi | leukocyte | *MOCOS* | -2.592837425 | 0.011010941 | 0.014230917 | 1 |
| T2DM_admissi | leukocyte | *DLG1* | -1.133027312 | 0.000353307 | 0.001141109 | 1 |
| T2DM_admissi | leukocyte | *SEC31B* | 1.387066119 | 0.026266526 | 0.031921181 | 1 |
| T2DM_admissi | leukocyte | *EXOSC7* | -0.915995257 | 0.001181262 | 0.002099858 | 1 |
| T2DM_admissi | leukocyte | *MKRN2* | -1.427905162 | 0.000394464 | 0.001181536 | 1 |
| T2DM_admissi | leukocyte | *MLH1* | -1.560055284 | 0.00050553 | 0.001300737 | 1 |
| T2DM_admissi | leukocyte | *TPD52* | -0.937155775 | 0.000312882 | 0.001108487 | 1 |
| T2DM_admissi | leukocyte | *ACACB* | -1.406391421 | 0.000394012 | 0.001181536 | 1 |
| T2DM_admissi | leukocyte | *GPATCH1* | -0.928803593 | 0.005168784 | 0.007118175 | 1 |
| T2DM_admissi | leukocyte | *ICAM3* | -1.177273125 | 0.000554469 | 0.001356582 | 1 |
| T2DM_admissi | leukocyte | *XAB2* | -1.226894216 | 0.000230115 | 0.001064322 | 0.904121667 |
| T2DM_admissi | leukocyte | *ACTL6B* | -0.997349485 | 0.000828367 | 0.00167593 | 1 |
| T2DM_admissi | leukocyte | *IL4R* | -0.999073317 | 0.001326385 | 0.002228985 | 1 |
| T2DM_admissi | leukocyte | *APBB1IP* | 0.419988889 | 0.005157647 | 0.007105328 | 1 |
| T2DM_admissi | leukocyte | *GPR137B* | -1.971586673 | 0.000158983 | 0.001062643 | 0.624643461 |
| T2DM_admissi | leukocyte | *NAALAD2* | -2.891152386 | 0.002062866 | 0.003151674 | 1 |
| T2DM_admissi | leukocyte | *MCCC1* | -1.181702045 | 0.000675868 | 0.00148591 | 1 |
| T2DM_admissi | leukocyte | *N4BP2* | -1.773769288 | 0.000137728 | 0.001062643 | 0.541134015 |
| T2DM_admissi | leukocyte | *MLLT10* | -1.540646778 | 0.000300047 | 0.001093914 | 1 |
| T2DM_admissi | leukocyte | *VDAC3* | -1.362936276 | 0.000113576 | 0.001062643 | 0.446239715 |

| | | | | | | |
|---|---|---|---|---|---|---|
| T2DM_admissi | leukocyte | *PCM1* | -1.105738399 | 0.000228716 | 0.00106363 | 0.898623815 |
| T2DM_admissi | leukocyte | *TP73* | 2.695310469 | 0.010747388 | 0.013917761 | 1 |
| T2DM_admissi | leukocyte | *TOLLIP* | -1.253002248 | 0.000237399 | 0.001070884 | 0.932739973 |
| T2DM_admissi | leukocyte | *CLUL1* | -2.851134351 | 0.001212494 | 0.00212484 | 1 |
| T2DM_admissi | leukocyte | *FKBP7* | -1.651692528 | 0.000186436 | 0.001062643 | 0.732506331 |
| T2DM_admissi | leukocyte | *LXN* | -0.766907094 | 0.003504115 | 0.005019201 | 1 |
| T2DM_admissi | leukocyte | *SAR1A* | -0.933328352 | 0.000382588 | 0.001165285 | 1 |
| T2DM_admissi | leukocyte | *CDC14A* | -1.788832671 | 0.000388286 | 0.001174424 | 1 |
| T2DM_admissi | leukocyte | *FDFT1* | -1.388024274 | 8.94E-05 | 0.001062643 | 0.351147435 |
| T2DM_admissi | leukocyte | *SCGN* | -0.531444846 | 0.00095912 | 0.001831992 | 1 |
| T2DM_admissi | leukocyte | *DDX1* | -1.060998994 | 0.000374125 | 0.001157429 | 1 |
| T2DM_admissi | leukocyte | *RABL2B* | -1.719171539 | 0.000530883 | 0.001324223 | 1 |
| T2DM_admissi | leukocyte | *DCT* | -7.868751649 | 0.000115704 | 0.001062643 | 0.454600304 |
| T2DM_admissi | leukocyte | *SCTR* | -1.068976512 | 0.001656018 | 0.002629949 | 1 |
| T2DM_admissi | leukocyte | *RIF1* | -2.324270335 | 4.34E-05 | 0.001037577 | 0.170568368 |
| T2DM_admissi | leukocyte | *COL5A3* | -5.82642821 | 0.002066808 | 0.003154814 | 1 |
| T2DM_admissi | leukocyte | *IGSF9B* | -2.092394167 | 0.001130718 | 0.002036018 | 1 |
| T2DM_admissi | leukocyte | *COL4A4* | -2.559079464 | 0.000702608 | 0.001519287 | 1 |
| T2DM_admissi | leukocyte | *ZNF506* | -1.113984933 | 0.000638919 | 0.001438574 | 1 |
| T2DM_admissi | leukocyte | *CADPS2* | -1.084273781 | 0.002131476 | 0.003238426 | 1 |
| T2DM_admissi | leukocyte | *ME2* | -2.811955282 | 7.74E-05 | 0.001062643 | 0.30409743 |
| T2DM_admissi | leukocyte | *CCNT2* | -1.167985236 | 0.004793328 | 0.006642945 | 1 |
| T2DM_admissi | leukocyte | *SEMA5B* | 1.560782599 | 0.006135351 | 0.008335337 | 1 |
| T2DM_admissi | leukocyte | *BCKDHB* | -3.001524529 | 0.00064489 | 0.001445924 | 1 |
| T2DM_admissi | leukocyte | *DIS3* | -1.173068686 | 0.000305713 | 0.001096936 | 1 |
| T2DM_admissi | leukocyte | *PIBF1* | -1.756353994 | 0.000593915 | 0.001393962 | 1 |
| T2DM_admissi | leukocyte | *CHMP2B* | -1.051443368 | 0.000240184 | 0.001072513 | 0.943681968 |
| T2DM_admissi | leukocyte | *PPIE* | -1.341332719 | 0.00033199 | 0.001127387 | 1 |
| T2DM_admissi | leukocyte | *NCOA1* | -1.401500078 | 0.00019103 | 0.001062643 | 0.750557985 |
| T2DM_admissi | leukocyte | *AGBL5* | -1.761618295 | 0.004059809 | 0.005725409 | 1 |
| T2DM_admissi | leukocyte | *CAD* | -0.987796467 | 0.013546734 | 0.017314613 | 1 |
| T2DM_admissi | leukocyte | *CD59* | -0.699551883 | 0.000861262 | 0.001725599 | 1 |
| T2DM_admissi | leukocyte | *CD82* | -1.840960193 | 0.001386493 | 0.002298537 | 1 |
| T2DM_admissi | leukocyte | *MECOM* | -0.64614616 | 0.008495847 | 0.011220229 | 1 |
| T2DM_admissi | leukocyte | *SLC25A24* | -1.127153678 | 0.000615567 | 0.001419345 | 1 |
| T2DM_admissi | leukocyte | *PILRA* | 0.365009562 | 0.003611498 | 0.005159846 | 1 |
| T2DM_admissi | leukocyte | *ZNF213* | -1.258755973 | 0.000705757 | 0.001522744 | 1 |
| T2DM_admissi | leukocyte | *CPNE3* | -0.885058063 | 0.000486203 | 0.001277166 | 1 |
| T2DM_admissi | leukocyte | *RRN3* | -0.991735372 | 0.000678903 | 0.001488511 | 1 |
| T2DM_admissi | leukocyte | *WNT11* | -2.466565449 | 0.008708526 | 0.01146642 | 1 |
| T2DM_admissi | leukocyte | *DDHD2* | -1.175866242 | 0.002627219 | 0.003884962 | 1 |
| T2DM_admissi | leukocyte | *TTC39A* | -1.028224696 | 0.0035167 | 0.005035392 | 1 |
| T2DM_admissi | leukocyte | *EPS15* | -1.506441558 | 0.000763091 | 0.001593931 | 1 |
| T2DM_admissi | leukocyte | *ORC1* | -4.401777083 | 0.000210819 | 0.001062643 | 0.828306001 |
| T2DM_admissi | leukocyte | *MGST2* | -0.732556231 | 0.002463545 | 0.00368033 | 1 |
| T2DM_admissi | leukocyte | *USP40* | -1.963100131 | 0.000112843 | 0.001062643 | 0.443358975 |
| T2DM_admissi | leukocyte | *POMGNT1* | -1.726002379 | 4.13E-05 | 0.001037577 | 0.162439754 |
| T2DM_admissi | leukocyte | *NFX1* | -1.781413728 | 0.000582188 | 0.001382042 | 1 |
| T2DM_admissi | leukocyte | *EIF2AK1* | -1.133408594 | 0.000805687 | 0.0016513 | 1 |
| T2DM_admissi | leukocyte | *ITPKC* | -1.386376071 | 0.000843296 | 0.001700004 | 1 |

| | | | | | | |
|---|---|---|---|---|---|---|
| T2DM_admissi | leukocyte | *TMED2* | -0.891294051 | 0.000301902 | 0.001093914 | 1 |
| T2DM_admissi | leukocyte | *ACOX3* | -2.641804118 | 0.00024002 | 0.001072513 | 0.943039422 |
| T2DM_admissi | leukocyte | *TRIP6* | -1.072680497 | 9.62E-05 | 0.001062643 | 0.378073479 |
| T2DM_admissi | leukocyte | *PGS1* | -1.846652125 | 0.000284923 | 0.001088708 | 1 |
| T2DM_admissi | leukocyte | *PSMC5* | -0.981568657 | 0.000191518 | 0.001062643 | 0.752474624 |
| T2DM_admissi | leukocyte | *UIMC1* | -1.196448771 | 0.0009752 | 0.001854014 | 1 |
| T2DM_admissi | leukocyte | *SF3B2* | -0.969237738 | 0.000408735 | 0.001188129 | 1 |
| T2DM_admissi | leukocyte | *KLHL42* | -1.07926389 | 0.000134038 | 0.001062643 | 0.526637187 |
| T2DM_admissi | leukocyte | *METTL2A* | -1.852201053 | 0.000223197 | 0.001062643 | 0.876939474 |
| T2DM_admissi | leukocyte | *GP6* | 1.331487197 | 0.021300401 | 0.026300841 | 1 |
| T2DM_admissi | leukocyte | *EDEM2* | -0.821957072 | 0.035729649 | 0.042825439 | 1 |
| T2DM_admissi | leukocyte | *SLC15A1* | -1.917761247 | 0.005599709 | 0.007663273 | 1 |
| T2DM_admissi | leukocyte | *TGDS* | -1.13436277 | 0.001073486 | 0.00196814 | 1 |
| T2DM_admissi | leukocyte | *DOCK3* | -1.250063629 | 0.001252303 | 0.002156134 | 1 |
| T2DM_admissi | leukocyte | *ARHGAP28* | -2.823584046 | 0.001186885 | 0.00210081 | 1 |
| T2DM_admissi | leukocyte | *PPP1R13B* | -0.801849675 | 0.001595528 | 0.002551417 | 1 |
| T2DM_admissi | leukocyte | *ZNF343* | -1.31843291 | 0.00426707 | 0.005987614 | 1 |
| T2DM_admissi | leukocyte | *MAVS* | -2.186909273 | 0.00057666 | 0.00137565 | 1 |
| T2DM_admissi | leukocyte | *DYNLL1* | -0.958185728 | 0.000408442 | 0.001188129 | 1 |
| T2DM_admissi | leukocyte | *RBBP9* | -0.931873921 | 0.002229739 | 0.003369478 | 1 |
| T2DM_admissi | leukocyte | *SLC23A2* | -2.029578124 | 0.00014063 | 0.001062643 | 0.552534844 |
| T2DM_admissi | leukocyte | *SLC8B1* | 0.294226537 | 0.009896577 | 0.012901013 | 1 |
| T2DM_admissi | leukocyte | *TMEM230* | -1.081926405 | 0.000327958 | 0.001122427 | 1 |
| T2DM_admissi | leukocyte | *KIF16B* | -2.007336844 | 3.73E-05 | 0.001037577 | 0.146451012 |
| T2DM_admissi | leukocyte | *ZNF302* | -1.128575858 | 0.000277506 | 0.001088708 | 1 |
| T2DM_admissi | leukocyte | *GRAMD1A* | -1.875746387 | 0.000213545 | 0.001062643 | 0.839020176 |
| T2DM_admissi | leukocyte | *CDIP1* | -1.210986483 | 0.000538327 | 0.001334229 | 1 |
| T2DM_admissi | leukocyte | *MLF2* | -1.284596759 | 0.000114245 | 0.001062643 | 0.448866708 |
| T2DM_admissi | leukocyte | *DDX24* | -1.005393766 | 0.000175326 | 0.001062643 | 0.688857688 |
| T2DM_admissi | leukocyte | *ANKRD24* | -7.270390555 | 0.007783273 | 0.010352227 | 1 |
| T2DM_admissi | leukocyte | *GPATCH2L* | -1.447429749 | 0.000149942 | 0.001062643 | 0.589122588 |
| T2DM_admissi | leukocyte | *SLC9A1* | -0.935315796 | 0.001993444 | 0.003065456 | 1 |
| T2DM_admissi | leukocyte | *SPTLC1* | -1.078213416 | 0.000783202 | 0.001618943 | 1 |
| T2DM_admissi | leukocyte | *YPEL3* | -0.855580465 | 0.022136152 | 0.027264245 | 1 |
| T2DM_admissi | leukocyte | *NDUFB2* | -1.025988656 | 5.06E-05 | 0.00105496 | 0.198989844 |
| T2DM_admissi | leukocyte | *MAEA* | -0.990684939 | 0.000927366 | 0.001800431 | 1 |
| T2DM_admissi | leukocyte | *STRN4* | -1.143026294 | 0.000337872 | 0.001134436 | 1 |
| T2DM_admissi | leukocyte | *LYZ* | 0.239279882 | 0.005355628 | 0.007354862 | 1 |
| T2DM_admissi | leukocyte | *MUL1* | -1.258386175 | 0.000260314 | 0.001083674 | 1 |
| T2DM_admissi | leukocyte | *SPG21* | -1.616141746 | 0.000134942 | 0.001062643 | 0.53018864 |
| T2DM_admissi | leukocyte | *USP48* | -1.132676532 | 0.000620252 | 0.001422226 | 1 |
| T2DM_admissi | leukocyte | *GLG1* | -1.146536041 | 0.000323498 | 0.001116892 | 1 |
| T2DM_admissi | leukocyte | *NRCAM* | -0.727189582 | 0.000940289 | 0.001810978 | 1 |
| T2DM_admissi | leukocyte | *TF* | -1.42871763 | 0.009247601 | 0.012099175 | 1 |
| T2DM_admissi | leukocyte | *MYO15A* | -2.367207131 | 0.004207349 | 0.005914143 | 1 |
| T2DM_admissi | leukocyte | *PITPNM3* | -3.071184511 | 0.001147637 | 0.002057055 | 1 |
| T2DM_admissi | leukocyte | *SPAG7* | -1.090895446 | 0.002455679 | 0.003671371 | 1 |
| T2DM_admissi | leukocyte | *TMEM101* | -1.54525538 | 0.000151389 | 0.001062643 | 0.594806942 |
| T2DM_admissi | leukocyte | *PPP2R3C* | -2.041845739 | 5.81E-05 | 0.001060479 | 0.228445543 |
| T2DM_admissi | leukocyte | *HAUS4* | -2.260254052 | 0.010797918 | 0.013978589 | 1 |

| | | | | | | |
|---|---|---|---|---|---|---|
| T2DM_admissi | leukocyte | *OSGEP* | -2.187161816 | 6.89E-05 | 0.001062643 | 0.270567973 |
| T2DM_admissi | leukocyte | *SCFD1* | -1.204706257 | 0.000282764 | 0.001088708 | 1 |
| T2DM_admissi | leukocyte | *RPGRIP1* | 1.073170251 | 0.0255251 | 0.031133606 | 1 |
| T2DM_admissi | leukocyte | *EZR* | -0.792401667 | 0.000475415 | 0.001261234 | 1 |
| T2DM_admissi | leukocyte | *NUP50* | -2.225846435 | 5.20E-05 | 0.00105496 | 0.20428185 |
| T2DM_admissi | leukocyte | *COMT* | -1.042041594 | 0.00031049 | 0.00110427 | 1 |
| T2DM_admissi | leukocyte | *ECHDC1* | -0.910342515 | 0.000869494 | 0.001735895 | 1 |
| T2DM_admissi | leukocyte | *CBX5* | -1.003780978 | 0.000218159 | 0.001062643 | 0.857145397 |
| T2DM_admissi | leukocyte | *SUCO* | -0.821086301 | 0.000347282 | 0.001140861 | 1 |
| T2DM_admissi | leukocyte | *MSH2* | -2.489090757 | 0.000261052 | 0.001083674 | 1 |
| T2DM_admissi | leukocyte | *TMEM38B* | -1.849744162 | 4.74E-05 | 0.00105496 | 0.186040769 |
| T2DM_admissi | leukocyte | *PSMD5* | -1.053221465 | 0.00072739 | 0.00154878 | 1 |
| T2DM_admissi | leukocyte | *CRAT* | -2.105776989 | 6.49E-05 | 0.001060479 | 0.25480805 |
| T2DM_admissi | leukocyte | *SH2D3C* | -1.437653629 | 0.002950434 | 0.004310096 | 1 |
| T2DM_admissi | leukocyte | *CWF19L1* | -1.523875677 | 2.61E-06 | 0.000379549 | 0.01024783 |
| T2DM_admissi | leukocyte | *SEMA4G* | -11.44057015 | 0.000447136 | 0.001232235 | 1 |
| T2DM_admissi | leukocyte | *IKZF5* | -1.898455397 | 0.000106091 | 0.001062643 | 0.416832586 |
| T2DM_admissi | leukocyte | *TLL2* | -4.439664429 | 0.001272706 | 0.002175059 | 1 |
| T2DM_admissi | leukocyte | *SRPK1* | -1.448458086 | 0.000146783 | 0.001062643 | 0.576710112 |
| T2DM_admissi | leukocyte | *PGC* | -0.90338697 | 0.003361423 | 0.004839513 | 1 |
| T2DM_admissi | leukocyte | *EFHC1* | -0.929240653 | 0.001807462 | 0.002824788 | 1 |
| T2DM_admissi | leukocyte | *ITPR3* | -1.158384608 | 0.001154325 | 0.00206595 | 1 |
| T2DM_admissi | leukocyte | *IFT74* | -1.648618961 | 0.000211209 | 0.001062643 | 0.829839706 |
| T2DM_admissi | leukocyte | *ABL1* | -1.204807509 | 0.000410101 | 0.001188264 | 1 |
| T2DM_admissi | leukocyte | *SH3GLB1* | -1.23175118 | 0.000235135 | 0.001067235 | 0.923845501 |
| T2DM_admissi | leukocyte | *CDC7* | -1.27115333 | 0.006828211 | 0.009178255 | 1 |
| T2DM_admissi | leukocyte | *SYDE2* | -1.118549273 | 0.004236143 | 0.005948466 | 1 |
| T2DM_admissi | leukocyte | *ABLIM1* | -0.925821119 | 0.000931159 | 0.001802228 | 1 |
| T2DM_admissi | leukocyte | *ERMP1* | -1.017938933 | 0.002018717 | 0.003094794 | 1 |
| T2DM_admissi | leukocyte | *PALMD* | -1.147407058 | 7.18E-05 | 0.001062643 | 0.28210407 |
| T2DM_admissi | leukocyte | *MZF1* | -1.028567735 | 0.004520324 | 0.006302467 | 1 |
| T2DM_admissi | leukocyte | *OCEL1* | -1.436969324 | 0.000999147 | 0.0018783 | 1 |
| T2DM_admissi | leukocyte | *CATSPERG* | -1.535981071 | 0.01735816 | 0.021768341 | 1 |
| T2DM_admissi | leukocyte | *STX1B* | 1.478049515 | 0.00040546 | 0.001187074 | 1 |
| T2DM_admissi | leukocyte | *HSD3B7* | -3.304388582 | 0.001359478 | 0.00226907 | 1 |
| T2DM_admissi | leukocyte | *HNRNPM* | -1.017321587 | 0.000114604 | 0.001062643 | 0.45027913 |
| T2DM_admissi | leukocyte | *TIMM13* | -1.193386244 | 0.000199541 | 0.001062643 | 0.783996927 |
| T2DM_admissi | leukocyte | *CEP170B* | -1.754533028 | 0.000286013 | 0.001088708 | 1 |
| T2DM_admissi | leukocyte | *POLR2E* | -1.632962351 | 0.000151541 | 0.001062643 | 0.595404507 |
| T2DM_admissi | leukocyte | *RASSF7* | -1.024178896 | 0.000492862 | 0.001284972 | 1 |
| T2DM_admissi | leukocyte | *ARVCF* | -1.9445141 | 0.001662235 | 0.002637968 | 1 |
| T2DM_admissi | leukocyte | *TRMT2A* | -1.051457096 | 0.000877943 | 0.001743901 | 1 |
| T2DM_admissi | leukocyte | *MED15* | -3.401443989 | 3.81E-05 | 0.001037577 | 0.149721787 |
| T2DM_admissi | leukocyte | *SMARCB1* | -1.276554038 | 0.000228752 | 0.00106363 | 0.898767356 |
| T2DM_admissi | leukocyte | *DDT* | -1.326872038 | 9.92E-05 | 0.001062643 | 0.389592333 |
| T2DM_admissi | leukocyte | *CABIN1* | -1.455249913 | 0.000779493 | 0.001614233 | 1 |
| T2DM_admissi | leukocyte | *SPECC1L* | -1.849859844 | 0.000209944 | 0.001062643 | 0.824870689 |
| T2DM_admissi | leukocyte | *PES1* | -1.064746602 | 0.00144433 | 0.002364489 | 1 |
| T2DM_admissi | leukocyte | *GGT1* | -7.268816778 | 0.011669093 | 0.015017316 | 1 |
| T2DM_admissi | leukocyte | *TOP3B* | -1.677372102 | 0.001713257 | 0.002707728 | 1 |

| | | | | | | |
|---|---|---|---|---|---|---|
| T2DM_admissi | leukocyte | *CARD10* | -0.873099865 | 0.000511503 | 0.001308382 | 1 |
| T2DM_admissi | leukocyte | *SLC25A1* | -1.305893498 | 0.000355988 | 0.001141109 | 1 |
| T2DM_admissi | leukocyte | *HPS4* | -1.336029015 | 0.000366375 | 0.001147001 | 1 |
| T2DM_admissi | leukocyte | *GCAT* | -1.695426657 | 0.000310601 | 0.00110427 | 1 |
| T2DM_admissi | leukocyte | *CYP2D6* | 2.359779955 | 0.017096211 | 0.02150849 | 1 |
| T2DM_admissi | leukocyte | *DDX17* | -1.250597428 | 0.000133186 | 0.001062643 | 0.523286278 |
| T2DM_admissi | leukocyte | *HSCB* | -2.13627546 | 0.00017853 | 0.001062643 | 0.701446125 |
| T2DM_admissi | leukocyte | *CBY1* | -1.291289348 | 0.000383869 | 0.00116645 | 1 |
| T2DM_admissi | leukocyte | *XBP1* | -0.861418859 | 0.000210797 | 0.001062643 | 0.828222477 |
| T2DM_admissi | leukocyte | *TIMP3* | -1.305847811 | 0.000513902 | 0.001310266 | 1 |
| T2DM_admissi | leukocyte | *CYB5R3* | -1.133816971 | 0.000294212 | 0.001089501 | 1 |
| T2DM_admissi | leukocyte | *MIOX* | -2.647424714 | 0.000986644 | 0.001867193 | 1 |
| T2DM_admissi | leukocyte | *LMF2* | -1.891402432 | 0.002294801 | 0.003461141 | 1 |
| T2DM_admissi | leukocyte | *RHBDD3* | -1.421945198 | 0.000198781 | 0.001062643 | 0.781009793 |
| T2DM_admissi | leukocyte | *TOM1* | -1.800848365 | 0.000557024 | 0.001357659 | 1 |
| T2DM_admissi | leukocyte | *CHKB* | -3.149890636 | 0.000167198 | 0.001062643 | 0.656920064 |
| T2DM_admissi | leukocyte | *TTLL12* | -1.74359837 | 0.000546772 | 0.001346876 | 1 |
| T2DM_admissi | leukocyte | *RBFOX2* | -1.166702083 | 0.00025475 | 0.001077348 | 1 |
| T2DM_admissi | leukocyte | *SYNGR1* | -3.442958371 | 5.62E-05 | 0.001060479 | 0.220738359 |
| T2DM_admissi | leukocyte | *MTMR3* | -1.611788659 | 0.000145003 | 0.001062643 | 0.569718461 |
| T2DM_admissi | leukocyte | *SAMM50* | -1.098984847 | 0.000430106 | 0.00121662 | 1 |
| T2DM_admissi | leukocyte | *FOXRED2* | -1.205355104 | 0.002024598 | 0.003102437 | 1 |
| T2DM_admissi | leukocyte | *SGSM3* | -1.750239801 | 0.000271187 | 0.001088708 | 1 |
| T2DM_admissi | leukocyte | *IFT27* | -1.553195103 | 0.00012971 | 0.001062643 | 0.509628963 |
| T2DM_admissi | leukocyte | *FAM118A* | -1.387805801 | 0.002878995 | 0.004219162 | 1 |
| T2DM_admissi | leukocyte | *ST13* | -1.054994398 | 0.000177694 | 0.001062643 | 0.698158242 |
| T2DM_admissi | leukocyte | *POLR3H* | -3.001417063 | 2.53E-05 | 0.000982543 | 0.099236821 |
| T2DM_admissi | leukocyte | *PMM1* | -1.562739589 | 0.000314712 | 0.001109126 | 1 |
| T2DM_admissi | leukocyte | *DESI1* | -1.490079763 | 0.000690354 | 0.001503881 | 1 |
| T2DM_admissi | leukocyte | *KHNYN* | -1.787262113 | 8.72E-05 | 0.001062643 | 0.342554084 |
| T2DM_admissi | leukocyte | *RBM23* | -1.183106384 | 0.000263454 | 0.001083884 | 1 |
| T2DM_admissi | leukocyte | *PRMT5* | -1.068038832 | 0.000873473 | 0.001740302 | 1 |
| T2DM_admissi | leukocyte | *VCPKMT* | -1.735003344 | 0.000230066 | 0.001064322 | 0.903929617 |
| T2DM_admissi | leukocyte | *SOS2* | -1.573739835 | 0.000175807 | 0.001062643 | 0.690746995 |
| T2DM_admissi | leukocyte | *CDKL1* | -1.941947615 | 0.00183567 | 0.002861108 | 1 |
| T2DM_admissi | leukocyte | *NIN* | -2.863731724 | 7.36E-05 | 0.001062643 | 0.289247624 |
| T2DM_admissi | leukocyte | *TRIM9* | -0.848571082 | 0.001335609 | 0.00223874 | 1 |
| T2DM_admissi | leukocyte | *ATP6V1D* | -1.118097467 | 0.000186842 | 0.001062643 | 0.734101701 |
| T2DM_admissi | leukocyte | *PLEK2* | -1.196602527 | 0.023838935 | 0.029205854 | 1 |
| T2DM_admissi | leukocyte | *GSTZ1* | -1.735266977 | 0.00042575 | 0.001211276 | 1 |
| T2DM_admissi | leukocyte | *TMED8* | -1.438257216 | 0.000131167 | 0.001062643 | 0.515355716 |
| T2DM_admissi | leukocyte | *DAAM1* | -1.049220876 | 0.000398981 | 0.001183984 | 1 |
| T2DM_admissi | leukocyte | *DHRS7* | -1.330464877 | 0.00012488 | 0.001062643 | 0.490655461 |
| T2DM_admissi | leukocyte | *MTHFD1* | -4.80012778 | 2.21E-05 | 0.000943039 | 0.08675955 |
| T2DM_admissi | leukocyte | *GSKIP* | -1.146537955 | 0.000530771 | 0.001324223 | 1 |
| T2DM_admissi | leukocyte | *TRIP11* | -0.998357873 | 0.000457192 | 0.001240899 | 1 |
| T2DM_admissi | leukocyte | *DCAF11* | -1.629243708 | 0.000143873 | 0.001062643 | 0.565278889 |
| T2DM_admissi | leukocyte | *EMC9* | -1.054093639 | 0.003322745 | 0.004789092 | 1 |
| T2DM_admissi | leukocyte | *NFATC4* | -1.509473569 | 0.001459549 | 0.002380476 | 1 |
| T2DM_admissi | leukocyte | *PYGB* | -1.30694931 | 0.000298034 | 0.001093914 | 1 |

| | | | | | | |
|---|---|---|---|---|---|---|
| T2DM_admissi | leukocyte | *ABHD12* | -2.35788063 | 6.99E-05 | 0.001062643 | 0.274451021 |
| T2DM_admissi | leukocyte | *PROCR* | -0.907175885 | 0.00243376 | 0.003641372 | 1 |
| T2DM_admissi | leukocyte | *CD40* | -1.289294913 | 0.000485099 | 0.001275739 | 1 |
| T2DM_admissi | leukocyte | *IFT52* | -1.14297458 | 0.000395819 | 0.001181845 | 1 |
| T2DM_admissi | leukocyte | *MYBL2* | -2.281668364 | 0.009990382 | 0.013010346 | 1 |
| T2DM_admissi | leukocyte | *PABPC1L* | -0.869347067 | 0.033532253 | 0.040240752 | 1 |
| T2DM_admissi | leukocyte | *SALL4* | 1.211765069 | 0.038588369 | 0.046055195 | 1 |
| T2DM_admissi | leukocyte | *NELFCD* | -1.09467205 | 0.000397059 | 0.001182749 | 1 |
| T2DM_admissi | leukocyte | *MTG2* | -1.337912443 | 0.000359627 | 0.001142338 | 1 |
| T2DM_admissi | leukocyte | *DIDO1* | -1.241059267 | 0.00217387 | 0.003295191 | 1 |
| T2DM_admissi | leukocyte | *SLC17A9* | 0.943544449 | 0.033294199 | 0.040005451 | 1 |
| T2DM_admissi | leukocyte | *CDC25B* | -1.951328269 | 0.002058493 | 0.003148236 | 1 |
| T2DM_admissi | leukocyte | *ISM1* | 1.373536883 | 0.014620414 | 0.018590163 | 1 |
| T2DM_admissi | leukocyte | *ARFRP1* | -1.402327115 | 0.006222167 | 0.008435756 | 1 |
| T2DM_admissi | leukocyte | *SLC52A3* | -2.618931339 | 0.002193676 | 0.00332137 | 1 |
| T2DM_admissi | leukocyte | *RSPO4* | -12.04942398 | 0.041404899 | 0.049207456 | 1 |
| T2DM_admissi | leukocyte | *SIRPB1* | 0.71231569 | 0.005535307 | 0.007588353 | 1 |
| T2DM_admissi | leukocyte | *PLCB4* | -1.026600087 | 0.000707769 | 0.001525411 | 1 |
| T2DM_admissi | leukocyte | *POFUT1* | -1.114498699 | 0.000608624 | 0.001409123 | 1 |
| T2DM_admissi | leukocyte | *NOP56* | -1.521869022 | 0.00069665 | 0.001511098 | 1 |
| T2DM_admissi | leukocyte | *MANBAL* | -1.034279431 | 0.000709863 | 0.001527411 | 1 |
| T2DM_admissi | leukocyte | *IDH3B* | -1.438304722 | 0.000297401 | 0.001093914 | 1 |
| T2DM_admissi | leukocyte | *MAPRE1* | -0.937752274 | 0.000603959 | 0.001404115 | 1 |
| T2DM_admissi | leukocyte | *OXT* | 0.837392365 | 0.014238522 | 0.018133924 | 1 |
| T2DM_admissi | leukocyte | *TTI1* | -1.808246501 | 0.000346307 | 0.001140861 | 1 |
| T2DM_admissi | leukocyte | *CHMP4B* | -0.98951019 | 0.000285623 | 0.001088708 | 1 |
| T2DM_admissi | leukocyte | *DHX35* | -1.914773801 | 0.000730757 | 0.001550629 | 1 |
| T2DM_admissi | leukocyte | *DNTTIP1* | -2.820829787 | 4.31E-05 | 0.001037577 | 0.169206096 |
| T2DM_admissi | leukocyte | *MAP1LC3A* | -0.712624221 | 0.002131373 | 0.003238426 | 1 |
| T2DM_admissi | leukocyte | *APMAP* | -1.223424317 | 0.001061493 | 0.001956194 | 1 |
| T2DM_admissi | leukocyte | *VAPA* | -0.979221662 | 0.000215853 | 0.001062643 | 0.848085919 |
| T2DM_admissi | leukocyte | *MYOM1* | -2.612206171 | 0.00156202 | 0.00251112 | 1 |
| T2DM_admissi | leukocyte | *CEP192* | -0.831446828 | 0.005346475 | 0.007344861 | 1 |
| T2DM_admissi | leukocyte | *LIPG* | -1.20015367 | 0.00125479 | 0.002158524 | 1 |
| T2DM_admissi | leukocyte | *LAMA1* | -5.340278402 | 0.006090991 | 0.008286531 | 1 |
| T2DM_admissi | leukocyte | *POLI* | -1.651358981 | 0.004803903 | 0.006652991 | 1 |
| T2DM_admissi | leukocyte | *RIOK3* | -1.09416542 | 0.000195848 | 0.001062643 | 0.769486466 |
| T2DM_admissi | leukocyte | *NALCN* | -1.03532759 | 0.001296574 | 0.002195847 | 1 |
| T2DM_admissi | leukocyte | *FGF14* | -1.197684024 | 0.000314317 | 0.001109126 | 1 |
| T2DM_admissi | leukocyte | *FNDC3A* | -0.894998087 | 0.000439009 | 0.001222199 | 1 |
| T2DM_admissi | leukocyte | *KLF5* | -0.791693598 | 0.001215115 | 0.002128483 | 1 |
| T2DM_admissi | leukocyte | *ACP5* | 0.061924463 | 0.008987999 | 0.011783065 | 1 |
| T2DM_admissi | leukocyte | *DNAJC3* | -1.046655313 | 0.000232125 | 0.001066588 | 0.912017395 |
| T2DM_admissi | leukocyte | *UGGT2* | -1.058260045 | 0.00065212 | 0.001450838 | 1 |
| T2DM_admissi | leukocyte | *ARHGEF7* | -1.605608118 | 0.000176958 | 0.001062643 | 0.695267655 |
| T2DM_admissi | leukocyte | *PARP4* | -3.188469956 | 0.000163881 | 0.001062643 | 0.643886519 |
| T2DM_admissi | leukocyte | *MEDAG* | -1.799120442 | 0.00092362 | 0.001797337 | 1 |
| T2DM_admissi | leukocyte | *CLN5* | -1.476995905 | 0.00019065 | 0.001062643 | 0.749063005 |
| T2DM_admissi | leukocyte | *OLFM4* | -0.393147603 | 0.017521939 | 0.02195271 | 1 |
| T2DM_admissi | leukocyte | *MGRN1* | -1.425700797 | 0.000577802 | 0.001376912 | 1 |

| | | | | | | |
|---|---|---|---|---|---|---|
| T2DM_admissi | leukocyte | *LONP2* | -1.123009051 | 0.000426613 | 0.001212854 | 1 |
| T2DM_admissi | leukocyte | *CBLN1* | -3.024895264 | 0.006174189 | 0.008379409 | 1 |
| T2DM_admissi | leukocyte | *DHODH* | -3.469955385 | 0.000166172 | 0.001062643 | 0.652889507 |
| T2DM_admissi | leukocyte | *PARD6A* | -0.836941149 | 0.000779357 | 0.001614233 | 1 |
| T2DM_admissi | leukocyte | *MMP15* | -2.418418988 | 0.000634049 | 0.001434874 | 1 |
| T2DM_admissi | leukocyte | *USB1* | -1.643704448 | 0.000540153 | 0.001334756 | 1 |
| T2DM_admissi | leukocyte | *CYB5B* | -2.434150093 | 3.80E-05 | 0.001037577 | 0.149436232 |
| T2DM_admissi | leukocyte | *CCDC113* | -1.54115815 | 0.000686575 | 0.001497995 | 1 |
| T2DM_admissi | leukocyte | *NME3* | -1.316767506 | 0.000198574 | 0.001062643 | 0.780196044 |
| T2DM_admissi | leukocyte | *VAC14* | -1.259032412 | 0.000437779 | 0.001222199 | 1 |
| T2DM_admissi | leukocyte | *NPRL3* | -1.968325591 | 9.49E-05 | 0.001062643 | 0.372741685 |
| T2DM_admissi | leukocyte | *NAGPA* | -5.896241893 | 8.73E-05 | 0.001062643 | 0.343193199 |
| T2DM_admissi | leukocyte | *ZNF500* | -2.358690542 | 0.000479548 | 0.001266209 | 1 |
| T2DM_admissi | leukocyte | *NME4* | -1.267850349 | 0.000305274 | 0.001096936 | 1 |
| T2DM_admissi | leukocyte | *NOMO3* | -1.995678031 | 6.31E-05 | 0.001060479 | 0.247779015 |
| T2DM_admissi | leukocyte | *LMF1* | -1.159613207 | 0.000724868 | 0.001544472 | 1 |
| T2DM_admissi | leukocyte | *MTHFSD* | -0.959758877 | 0.01995769 | 0.024814483 | 1 |
| T2DM_admissi | leukocyte | *CLCN7* | -2.766540007 | 0.001723207 | 0.002719068 | 1 |
| T2DM_admissi | leukocyte | *HAGHL* | -1.97298675 | 0.000280954 | 0.001088708 | 1 |
| T2DM_admissi | leukocyte | *METRN* | -1.015175504 | 0.000127661 | 0.001062643 | 0.501579748 |
| T2DM_admissi | leukocyte | *PIEZO1* | -1.445911501 | 0.000391038 | 0.00117953 | 1 |
| T2DM_admissi | leukocyte | *CLUAP1* | -1.046809396 | 0.008228709 | 0.01090408 | 1 |
| T2DM_admissi | leukocyte | *UBFD1* | -1.625907927 | 0.000428784 | 0.00121662 | 1 |
| T2DM_admissi | leukocyte | *AQP8* | -1.774550609 | 0.001364758 | 0.002274982 | 1 |
| T2DM_admissi | leukocyte | *USP31* | -0.892823253 | 0.000572979 | 0.001370771 | 1 |
| T2DM_admissi | leukocyte | *HMOX2* | -1.06032839 | 0.000526471 | 0.001319879 | 1 |
| T2DM_admissi | leukocyte | *RBL2* | -1.154488278 | 0.0063758 | 0.008617309 | 1 |
| T2DM_admissi | leukocyte | *KAT8* | -1.440987693 | 0.00024045 | 0.001072513 | 0.944726903 |
| T2DM_admissi | leukocyte | *NOMO1* | -1.620730822 | 0.002796275 | 0.004105592 | 1 |
| T2DM_admissi | leukocyte | *RNF40* | -2.300768506 | 8.81E-05 | 0.001062643 | 0.346213035 |
| T2DM_admissi | leukocyte | *KNOP1* | -1.51384267 | 0.000262374 | 0.001083674 | 1 |
| T2DM_admissi | leukocyte | *TRIP4* | -1.21687138 | 0.000851858 | 0.001707628 | 1 |
| T2DM_admissi | leukocyte | *AP3B2* | -1.222841564 | 0.000998375 | 0.001877747 | 1 |
| T2DM_admissi | leukocyte | *IGDCC4* | 2.574673866 | 0.008760215 | 0.011515184 | 1 |
| T2DM_admissi | leukocyte | *RPAP1* | -1.856272685 | 0.000291051 | 0.001088708 | 1 |
| T2DM_admissi | leukocyte | *HOMER2* | -0.900796859 | 0.000851414 | 0.001707609 | 1 |
| T2DM_admissi | leukocyte | *ATP8B4* | 0.551011462 | 0.006761475 | 0.009101006 | 1 |
| T2DM_admissi | leukocyte | *DMXL2* | -2.633031458 | 0.023693945 | 0.029046337 | 1 |
| T2DM_admissi | leukocyte | *DNAJC17* | -1.048261199 | 0.00203992 | 0.003122261 | 1 |
| T2DM_admissi | leukocyte | *SLC30A4* | -2.770880664 | 0.004000361 | 0.005655782 | 1 |
| T2DM_admissi | leukocyte | *ZDHHC2* | -0.771740872 | 0.000839262 | 0.001694482 | 1 |
| T2DM_admissi | leukocyte | *TRIM35* | -2.959037003 | 0.000154468 | 0.001062643 | 0.606906527 |
| T2DM_admissi | leukocyte | *RIPK2* | -1.573891349 | 9.29E-05 | 0.001062643 | 0.365049188 |
| T2DM_admissi | leukocyte | *NBN* | -1.544028597 | 0.001060548 | 0.001955371 | 1 |
| T2DM_admissi | leukocyte | *LAPTM4B* | -0.724455534 | 0.000738798 | 0.001559773 | 1 |
| T2DM_admissi | leukocyte | *RAB2A* | -0.806009184 | 0.000682076 | 0.001492137 | 1 |
| T2DM_admissi | leukocyte | *EIF3E* | -0.973755785 | 0.000241309 | 0.001072513 | 0.948101929 |
| T2DM_admissi | leukocyte | *ESRP1* | -0.926016305 | 0.000893114 | 0.001764226 | 1 |
| T2DM_admissi | leukocyte | *ARMC1* | -1.314530498 | 0.000422562 | 0.001206582 | 1 |
| T2DM_admissi | leukocyte | *SPAG1* | -1.103753325 | 0.000446215 | 0.001232235 | 1 |

| | | | | | | |
|---|---|---|---|---|---|---|
| T2DM_admissi | leukocyte | *CHRAC1* | -0.947212045 | 0.000574963 | 0.001373271 | 1 |
| T2DM_admissi | leukocyte | *SNX16* | -3.366527959 | 0.000384225 | 0.001166629 | 1 |
| T2DM_admissi | leukocyte | *GSDMD* | -0.653331091 | 0.017918344 | 0.022420756 | 1 |
| T2DM_admissi | leukocyte | *SH2D4A* | -1.28159625 | 0.00072053 | 0.001541079 | 1 |
| T2DM_admissi | leukocyte | *INTS10* | -1.626015584 | 0.000115889 | 0.001062643 | 0.455328819 |
| T2DM_admissi | leukocyte | *ERI1* | -3.371078226 | 1.96E-05 | 0.000917316 | 0.07705454 |
| T2DM_admissi | leukocyte | *LEPROTL1* | -1.417736888 | 0.000170966 | 0.001062643 | 0.67172406 |
| T2DM_admissi | leukocyte | *ERICH1* | -1.705504694 | 0.000191969 | 0.001062643 | 0.754246093 |
| T2DM_admissi | leukocyte | *ARHGEF10* | -1.171890118 | 0.001352833 | 0.002258938 | 1 |
| T2DM_admissi | leukocyte | *KLHDC4* | -1.458498642 | 0.000612989 | 0.001415201 | 1 |
| T2DM_admissi | leukocyte | *MCM4* | -1.468672506 | 0.000449543 | 0.001234067 | 1 |
| T2DM_admissi | leukocyte | *FGL1* | -1.649848127 | 0.006904477 | 0.009271254 | 1 |
| T2DM_admissi | leukocyte | *ECH1* | -1.053460013 | 0.000494612 | 0.001287826 | 1 |
| T2DM_admissi | leukocyte | *HNRNPL* | -1.204296839 | 0.000276899 | 0.001088708 | 1 |
| T2DM_admissi | leukocyte | *SARS2* | -1.419997357 | 0.000345346 | 0.001140861 | 1 |
| T2DM_admissi | leukocyte | *LIN7B* | -1.567434279 | 0.000282886 | 0.001088708 | 1 |
| T2DM_admissi | leukocyte | *PPP1R13L* | -1.114549015 | 0.00367753 | 0.005246556 | 1 |
| T2DM_admissi | leukocyte | *ERCC2* | -1.09157743 | 0.003145513 | 0.004560414 | 1 |
| T2DM_admissi | leukocyte | *KLC3* | -1.125919708 | 0.000546261 | 0.001346461 | 1 |
| T2DM_admissi | leukocyte | *AMH* | -1.301485756 | 0.00065171 | 0.001450838 | 1 |
| T2DM_admissi | leukocyte | *IL27RA* | -1.357585611 | 0.004371092 | 0.006122646 | 1 |
| T2DM_admissi | leukocyte | *PDCD5* | -0.86360704 | 0.000956865 | 0.001830343 | 1 |
| T2DM_admissi | leukocyte | *NUMBL* | -0.943425701 | 0.001070316 | 0.001964163 | 1 |
| T2DM_admissi | leukocyte | *SLC1A5* | -1.522350653 | 0.000455861 | 0.001240899 | 1 |
| T2DM_admissi | leukocyte | *PRKD2* | -1.359297768 | 0.000364428 | 0.001146348 | 1 |
| T2DM_admissi | leukocyte | *APLP1* | -0.59415927 | 0.00041862 | 0.001200552 | 1 |
| T2DM_admissi | leukocyte | *PLIN3* | -1.290478233 | 0.000189658 | 0.001062643 | 0.745165217 |
| T2DM_admissi | leukocyte | *MYH14* | -0.957621696 | 0.000556004 | 0.001357223 | 1 |
| T2DM_admissi | leukocyte | *CARD8* | -2.925615213 | 0.004102412 | 0.005783415 | 1 |
| T2DM_admissi | leukocyte | *ZNF175* | -1.056760303 | 0.000901455 | 0.001772681 | 1 |
| T2DM_admissi | leukocyte | *PLA2G4C* | -1.58282208 | 0.000351023 | 0.001141109 | 1 |
| T2DM_admissi | leukocyte | *CAPS* | -1.554546871 | 0.000500514 | 0.001295577 | 1 |
| T2DM_admissi | leukocyte | *PPP2R1A* | -0.973184658 | 0.000653669 | 0.001453462 | 1 |
| T2DM_admissi | leukocyte | *GCDH* | -1.54475702 | 8.96E-05 | 0.001062643 | 0.351988669 |
| T2DM_admissi | leukocyte | *PIK3R2* | -3.820458411 | 3.26E-05 | 0.001037577 | 0.128139788 |
| T2DM_admissi | leukocyte | *UPK1A* | -15.72725713 | 0.001971235 | 0.003037248 | 1 |
| T2DM_admissi | leukocyte | *COPE* | -1.049670009 | 0.000185313 | 0.001062643 | 0.728093799 |
| T2DM_admissi | leukocyte | *ETV2* | -1.248473228 | 0.005833348 | 0.007966363 | 1 |
| T2DM_admissi | leukocyte | *PBX4* | -4.463649335 | 0.001196954 | 0.002108893 | 1 |
| T2DM_admissi | leukocyte | *SIPA1L3* | -3.286399802 | 4.44E-05 | 0.001037577 | 0.174571559 |
| T2DM_admissi | leukocyte | *ETHE1* | -1.679514596 | 0.000866538 | 0.001732185 | 1 |
| T2DM_admissi | leukocyte | *SMG9* | -0.971557457 | 0.002193259 | 0.00332137 | 1 |
| T2DM_admissi | leukocyte | *RUNDC3B* | -6.699088217 | 4.84E-05 | 0.00105496 | 0.190174834 |
| T2DM_admissi | leukocyte | *PMPCB* | -1.411081767 | 0.00021778 | 0.001062643 | 0.855659014 |
| T2DM_admissi | leukocyte | *PON2* | -1.056741054 | 0.000254838 | 0.001077348 | 1 |
| T2DM_admissi | leukocyte | *ITGB8* | -1.042213449 | 0.000562603 | 0.001362351 | 1 |
| T2DM_admissi | leukocyte | *DNAH11* | -3.790504139 | 0.008803748 | 0.011560805 | 1 |
| T2DM_admissi | leukocyte | *CAV2* | -0.689281586 | 0.001113621 | 0.002017251 | 1 |
| T2DM_admissi | leukocyte | *CAV1* | -0.806446371 | 0.001231963 | 0.00214366 | 1 |
| T2DM_admissi | leukocyte | *ANKRD7* | -1.875145231 | 0.002081125 | 0.003172969 | 1 |

| | | | | | | |
|---|---|---|---|---|---|---|
| T2DM_admissi | leukocyte | *TSPAN12* | -1.827679302 | 0.000238563 | 0.00107244 | 0.937312655 |
| T2DM_admissi | leukocyte | *CPVL* | 0.131665233 | 0.008618747 | 0.011359631 | 1 |
| T2DM_admissi | leukocyte | *ABHD11* | -1.8276312 | 6.08E-05 | 0.001060479 | 0.238842064 |
| T2DM_admissi | leukocyte | *CRHR2* | -3.456432059 | 0.003804893 | 0.005411731 | 1 |
| T2DM_admissi | leukocyte | *CHCHD2* | -0.81341599 | 0.00025515 | 0.001077348 | 1 |
| T2DM_admissi | leukocyte | *EIF3B* | -1.202454527 | 0.000253204 | 0.001077213 | 0.994839733 |
| T2DM_admissi | leukocyte | *WASL* | -0.935938887 | 0.00068666 | 0.001497995 | 1 |
| T2DM_admissi | leukocyte | *AGFG2* | -1.096744403 | 0.000812031 | 0.001657387 | 1 |
| T2DM_admissi | leukocyte | *C1GALT1* | -1.003819095 | 0.000437305 | 0.001222199 | 1 |
| T2DM_admissi | leukocyte | *RPA3* | -1.276246841 | 0.000286456 | 0.001088708 | 1 |
| T2DM_admissi | leukocyte | *ZNHIT1* | -0.990100847 | 0.000498166 | 0.001293956 | 1 |
| T2DM_admissi | leukocyte | *CLDN15* | -0.857101239 | 0.002969479 | 0.004333983 | 1 |
| T2DM_admissi | leukocyte | *TMEM106B* | -0.996024385 | 0.000641223 | 0.001440839 | 1 |
| T2DM_admissi | leukocyte | *ACTR3C* | -1.853840418 | 0.000676556 | 0.00148591 | 1 |
| T2DM_admissi | leukocyte | *TSPAN13* | -0.766543767 | 0.000564771 | 0.001362351 | 1 |
| T2DM_admissi | leukocyte | *RARRES2* | -0.527936143 | 0.001803572 | 0.00281983 | 1 |
| T2DM_admissi | leukocyte | *AGR2* | -0.712859684 | 0.004924027 | 0.00680496 | 1 |
| T2DM_admissi | leukocyte | *TMEM176B* | -0.742186462 | 0.010932204 | 0.014143112 | 1 |
| T2DM_admissi | leukocyte | *COA1* | -1.526909865 | 0.00021953 | 0.001062643 | 0.862535208 |
| T2DM_admissi | leukocyte | *BLVRA* | -2.108009903 | 0.000131955 | 0.001062643 | 0.518449763 |
| T2DM_admissi | leukocyte | *TMEM248* | -1.501290407 | 0.000101505 | 0.001062643 | 0.398814075 |
| T2DM_admissi | leukocyte | *RHEB* | -1.03490321 | 0.000192701 | 0.001062643 | 0.757122312 |
| T2DM_admissi | leukocyte | *TBL2* | -0.809507233 | 0.014516761 | 0.018464342 | 1 |
| T2DM_admissi | leukocyte | *LIMK1* | -3.985246995 | 0.000584548 | 0.001382042 | 1 |
| T2DM_admissi | leukocyte | *SPIN1* | -1.825524546 | 0.000122025 | 0.001062643 | 0.479436847 |
| T2DM_admissi | leukocyte | *NMRK1* | -1.214806215 | 0.000608486 | 0.001409123 | 1 |
| T2DM_admissi | leukocyte | *C5* | -1.201027839 | 0.010059974 | 0.013087959 | 1 |
| T2DM_admissi | leukocyte | *OGN* | -2.335332716 | 0.001920639 | 0.002974454 | 1 |
| T2DM_admissi | leukocyte | *SUSD1* | -2.123714267 | 8.62E-05 | 0.001062643 | 0.338814241 |
| T2DM_admissi | leukocyte | *DNM1* | -0.807056303 | 0.000934548 | 0.001804129 | 1 |
| T2DM_admissi | leukocyte | *AK1* | -0.877048263 | 0.000631927 | 0.001433512 | 1 |
| T2DM_admissi | leukocyte | *PLGRKT* | -0.977462912 | 0.000776896 | 0.001613332 | 1 |
| T2DM_admissi | leukocyte | *DOCK8* | 0.426827069 | 0.016068788 | 0.020274331 | 1 |
| T2DM_admissi | leukocyte | *MPDZ* | -0.888843813 | 0.001831433 | 0.00285657 | 1 |
| T2DM_admissi | leukocyte | *RAPGEF1* | -1.49145861 | 0.013569589 | 0.017338184 | 1 |
| T2DM_admissi | leukocyte | *NPDC1* | -0.834890394 | 0.000324074 | 0.001116918 | 1 |
| T2DM_admissi | leukocyte | *HPS1* | -1.413786132 | 0.000244657 | 0.001077213 | 0.961255446 |
| T2DM_admissi | leukocyte | *CXCL12* | -1.192971016 | 0.000302005 | 0.001093914 | 1 |
| T2DM_admissi | leukocyte | *TRDMT1* | -2.922381013 | 5.25E-05 | 0.00105496 | 0.206083247 |
| T2DM_admissi | leukocyte | *ATE1* | -2.051988549 | 7.33E-05 | 0.001062643 | 0.288052387 |
| T2DM_admissi | leukocyte | *NSMCE4A* | -0.898672379 | 0.003805705 | 0.005411731 | 1 |
| T2DM_admissi | leukocyte | *PLEKHA1* | -0.889997402 | 0.000470822 | 0.001255055 | 1 |
| T2DM_admissi | leukocyte | *PALD1* | -8.137818544 | 3.93E-05 | 0.001037577 | 0.154462492 |
| T2DM_admissi | leukocyte | *SPOCK2* | -1.672349709 | 0.001290117 | 0.002190523 | 1 |
| T2DM_admissi | leukocyte | *BMPR1A* | -2.698333303 | 8.46E-05 | 0.001062643 | 0.332292679 |
| T2DM_admissi | leukocyte | *LIPA* | 0.30116464 | 0.016928086 | 0.021303795 | 1 |
| T2DM_admissi | leukocyte | *SFXN3* | -1.819124525 | 0.000251597 | 0.001077213 | 0.98852501 |
| T2DM_admissi | leukocyte | *NPM3* | -1.270694166 | 0.000698541 | 0.001512992 | 1 |
| T2DM_admissi | leukocyte | *TNKS2* | -1.321669321 | 0.000199078 | 0.001062643 | 0.782177451 |
| T2DM_admissi | leukocyte | *ACBD5* | -0.978211275 | 0.000851226 | 0.001707609 | 1 |

| | | | | | | |
|---|---|---|---|---|---|---|
| T2DM_admissi | leukocyte | *LHPP* | -1.922248683 | 0.000988441 | 0.001868906 | 1 |
| T2DM_admissi | leukocyte | *GTPBP4* | -0.984373897 | 0.00037057 | 0.001151894 | 1 |
| T2DM_admissi | leukocyte | *NEURL1* | -1.108037639 | 0.004282771 | 0.0060075 | 1 |
| T2DM_admissi | leukocyte | *XPNPEP1* | -1.096453993 | 0.00055339 | 0.001355918 | 1 |
| T2DM_admissi | leukocyte | *SMC3* | -0.984157457 | 0.000257346 | 0.001080629 | 1 |
| T2DM_admissi | leukocyte | *RPL28* | -0.797400352 | 0.000145806 | 0.001062643 | 0.572871619 |
| T2DM_admissi | leukocyte | *PBLD* | -0.803391323 | 0.006058011 | 0.008250235 | 1 |
| T2DM_admissi | leukocyte | *KRT23* | -0.569910104 | 0.008375175 | 0.011075753 | 1 |
| T2DM_admissi | leukocyte | *DHRS11* | -0.853626315 | 0.004444254 | 0.006207421 | 1 |
| T2DM_admissi | leukocyte | *CWC25* | -1.415395942 | 0.000262517 | 0.001083674 | 1 |
| T2DM_admissi | leukocyte | *RAD51C* | -1.32809728 | 0.000183525 | 0.001062643 | 0.72107126 |
| T2DM_admissi | leukocyte | *TRIM37* | -1.108096917 | 0.000478606 | 0.001265283 | 1 |
| T2DM_admissi | leukocyte | *RPS6KB1* | -1.19720188 | 0.001392879 | 0.002307176 | 1 |
| T2DM_admissi | leukocyte | *TRIM16L* | -1.573724749 | 0.003035961 | 0.0044228 | 1 |
| T2DM_admissi | leukocyte | *CDK5RAP3* | -1.795663648 | 0.000491798 | 0.001283904 | 1 |
| T2DM_admissi | leukocyte | *CAMTA2* | -1.628769943 | 0.001555533 | 0.002503881 | 1 |
| T2DM_admissi | leukocyte | *MED13* | -1.086276565 | 0.000566479 | 0.001364563 | 1 |
| T2DM_admissi | leukocyte | *CHRNE* | -1.750186516 | 0.011324599 | 0.014593096 | 1 |
| T2DM_admissi | leukocyte | *RAI1* | -0.872431975 | 0.001431013 | 0.00235249 | 1 |
| T2DM_admissi | leukocyte | *CCDC47* | -1.107212451 | 0.000199691 | 0.001062643 | 0.784584961 |
| T2DM_admissi | leukocyte | *DRG2* | -1.149054863 | 0.001000561 | 0.001880059 | 1 |
| T2DM_admissi | leukocyte | *FTSJ3* | -1.161542502 | 0.000253103 | 0.001077213 | 0.994440677 |
| T2DM_admissi | leukocyte | *SMARCD2* | -2.836191016 | 3.24E-05 | 0.001037577 | 0.127213381 |
| T2DM_admissi | leukocyte | *SYNGR2* | -1.02183286 | 0.000453642 | 0.001238727 | 1 |
| T2DM_admissi | leukocyte | *C17orf75* | -1.331117014 | 0.003736924 | 0.00532172 | 1 |
| T2DM_admissi | leukocyte | *DHX58* | -2.207822346 | 0.000803878 | 0.001649159 | 1 |
| T2DM_admissi | leukocyte | *NAGLU* | -1.120854008 | 0.001421682 | 0.002343313 | 1 |
| T2DM_admissi | leukocyte | *HSD17B1* | -1.845849378 | 1.76E-06 | 0.000300491 | 0.00691129 |
| T2DM_admissi | leukocyte | *RND2* | -3.009834275 | 0.000186129 | 0.001062643 | 0.731299531 |
| T2DM_admissi | leukocyte | *ABCC3* | -0.815159427 | 0.003041682 | 0.004429492 | 1 |
| T2DM_admissi | leukocyte | *MPP2* | -0.782034747 | 0.001433911 | 0.002354561 | 1 |
| T2DM_admissi | leukocyte | *HLF* | -0.696133411 | 0.001555344 | 0.002503881 | 1 |
| T2DM_admissi | leukocyte | *EFNB3* | -1.009223166 | 0.001185522 | 0.00210081 | 1 |
| T2DM_admissi | leukocyte | *RANGRF* | -1.533524262 | 0.000182444 | 0.001062643 | 0.716823804 |
| T2DM_admissi | leukocyte | *DPH1* | -2.208438844 | 0.00326706 | 0.004714021 | 1 |
| T2DM_admissi | leukocyte | *OCIAD1* | -1.008144288 | 0.000255558 | 0.001077348 | 1 |
| T2DM_admissi | leukocyte | *UBE2D3* | -0.926988064 | 0.00057468 | 0.001373271 | 1 |
| T2DM_admissi | leukocyte | *MAPK10* | -0.939288478 | 0.000631212 | 0.001433512 | 1 |
| T2DM_admissi | leukocyte | *ANXA10* | -0.926085907 | 0.021018377 | 0.025977101 | 1 |
| T2DM_admissi | leukocyte | *AADAT* | -3.454208563 | 0.000306971 | 0.001098442 | 1 |
| T2DM_admissi | leukocyte | *TRIM2* | -1.089591655 | 0.000461669 | 0.001244702 | 1 |
| T2DM_admissi | leukocyte | *SH3D19* | -2.353662999 | 0.000248664 | 0.001077213 | 0.977000516 |
| T2DM_admissi | leukocyte | *GLRB* | -1.101863651 | 0.000728689 | 0.001549254 | 1 |
| T2DM_admissi | leukocyte | *LRP2BP* | -1.209507447 | 0.001193305 | 0.002106243 | 1 |
| T2DM_admissi | leukocyte | *UFSP2* | -1.331268495 | 0.001465516 | 0.002388226 | 1 |
| T2DM_admissi | leukocyte | *KLHL5* | -1.462580774 | 0.000262575 | 0.001083674 | 1 |
| T2DM_admissi | leukocyte | *FAM149A* | -0.851413333 | 0.008755737 | 0.011515184 | 1 |
| T2DM_admissi | leukocyte | *DDX25* | -0.873614965 | 0.011762582 | 0.01512772 | 1 |
| T2DM_admissi | leukocyte | *HTATIP2* | -1.753897664 | 4.34E-05 | 0.001037577 | 0.1706127 |
| T2DM_admissi | leukocyte | *B3GAT1* | -2.759799707 | 0.000989068 | 0.001869192 | 1 |

| | | | | | | |
|---|---|---|---|---|---|---|
| T2DM_admissi | leukocyte | *P2RX3* | -21.25632707 | 0.001203284 | 0.002114239 | 1 |
| T2DM_admissi | leukocyte | *VWA5A* | -1.314392513 | 0.000618802 | 0.001422062 | 1 |
| T2DM_admissi | leukocyte | *DTX4* | -1.083602779 | 0.004524 | 0.00630499 | 1 |
| T2DM_admissi | leukocyte | *FOXRED1* | -1.841023219 | 0.001270485 | 0.002175059 | 1 |
| T2DM_admissi | leukocyte | *ST3GAL4* | -2.236603608 | 5.48E-05 | 0.001060479 | 0.215504129 |
| T2DM_admissi | leukocyte | *PRPF19* | -1.039669737 | 0.000438312 | 0.001222199 | 1 |
| T2DM_admissi | leukocyte | *CEP164* | -2.143917598 | 0.000527078 | 0.001319879 | 1 |
| T2DM_admissi | leukocyte | *PDHX* | -0.957208602 | 0.020007243 | 0.02486036 | 1 |
| T2DM_admissi | leukocyte | *SLC1A2* | -1.136139369 | 0.001565276 | 0.002514761 | 1 |
| T2DM_admissi | leukocyte | *CD5* | -5.423433126 | 0.000435505 | 0.001222199 | 1 |
| T2DM_admissi | leukocyte | *ACCS* | 0.556438337 | 0.004132877 | 0.005822185 | 1 |
| T2DM_admissi | leukocyte | *MDK* | -1.17991938 | 0.000200738 | 0.001062643 | 0.788699933 |
| T2DM_admissi | leukocyte | *MADD* | -1.706643986 | 8.87E-05 | 0.001062643 | 0.348533371 |
| T2DM_admissi | leukocyte | *NAA40* | -1.418332584 | 0.000525284 | 0.001319879 | 1 |
| T2DM_admissi | leukocyte | *CD81* | -0.72891622 | 0.001068616 | 0.001964163 | 1 |
| T2DM_admissi | leukocyte | *SLC35F2* | -1.228594976 | 0.000579868 | 0.001379559 | 1 |
| T2DM_admissi | leukocyte | *NUP98* | -1.224864554 | 0.000350744 | 0.001141109 | 1 |
| T2DM_admissi | leukocyte | *GTF2H1* | -2.189463155 | 7.03E-05 | 0.001062643 | 0.276313656 |
| T2DM_admissi | leukocyte | *PTPN5* | 2.033609424 | 0.006441369 | 0.008699944 | 1 |
| T2DM_admissi | leukocyte | *PRDM4* | -0.9115608 | 0.000797995 | 0.001641531 | 1 |
| T2DM_admissi | leukocyte | *CORO1C* | -1.341059905 | 0.000484774 | 0.001275738 | 1 |
| T2DM_admissi | leukocyte | *LIN7A* | -2.479228421 | 0.001194473 | 0.002107357 | 1 |
| T2DM_admissi | leukocyte | *KRT18* | -0.388379926 | 0.00139799 | 0.002313691 | 1 |
| T2DM_admissi | leukocyte | *METAP2* | -1.122039029 | 0.000249408 | 0.001077213 | 0.979925881 |
| T2DM_admissi | leukocyte | *SLC6A12* | -4.579694234 | 0.026754862 | 0.032464438 | 1 |
| T2DM_admissi | leukocyte | *WNT5B* | -2.568916303 | 0.001127338 | 0.00203366 | 1 |
| T2DM_admissi | leukocyte | *ITFG2* | -5.686198407 | 6.68E-05 | 0.001062643 | 0.262405946 |
| T2DM_admissi | leukocyte | *VPS29* | -1.055610373 | 0.000234505 | 0.001066706 | 0.921370438 |
| T2DM_admissi | leukocyte | *CREBL2* | -1.4793063 | 0.000297675 | 0.001093914 | 1 |
| T2DM_admissi | leukocyte | *GSG1* | -8.727928119 | 0.000358182 | 0.001141109 | 1 |
| T2DM_admissi | leukocyte | *LTBR* | -1.364026906 | 0.000194089 | 0.001062643 | 0.762576217 |
| T2DM_admissi | leukocyte | *CDK2AP1* | -0.685510709 | 0.030652637 | 0.037011128 | 1 |
| T2DM_admissi | leukocyte | *DDX55* | -1.998985225 | 5.73E-05 | 0.001060479 | 0.225084192 |
| T2DM_admissi | leukocyte | *STX2* | -1.348644987 | 0.002495066 | 0.003720348 | 1 |
| T2DM_admissi | leukocyte | *CAND1* | -1.386031936 | 0.000149291 | 0.001062643 | 0.586563491 |
| T2DM_admissi | leukocyte | *NUP107* | -1.523672139 | 0.000392298 | 0.001181536 | 1 |
| T2DM_admissi | leukocyte | *KRR1* | -1.234198794 | 0.000128703 | 0.001062643 | 0.505673346 |
| T2DM_admissi | leukocyte | *GNB3* | -1.319062775 | 0.001522192 | 0.002466266 | 1 |
| T2DM_admissi | leukocyte | *CDCA3* | -2.232253323 | 0.000845441 | 0.001700095 | 1 |
| T2DM_admissi | leukocyte | *CHPT1* | -2.225870489 | 1.14E-05 | 0.000784902 | 0.044873671 |
| T2DM_admissi | leukocyte | *SPSB2* | -1.135154579 | 0.00102818 | 0.001917284 | 1 |
| T2DM_admissi | leukocyte | *ATN1* | -1.264555989 | 0.000263792 | 0.00108414 | 1 |
| T2DM_admissi | leukocyte | *NT5DC3* | -1.235341015 | 0.003059814 | 0.00444981 | 1 |
| T2DM_admissi | leukocyte | *GOLT1B* | -1.127154927 | 0.000310304 | 0.00110427 | 1 |
| T2DM_admissi | leukocyte | *PRKAB1* | -1.030842256 | 0.000918065 | 0.001791892 | 1 |
| T2DM_admissi | leukocyte | *C2CD5* | -1.054965623 | 0.000961376 | 0.001831992 | 1 |
| T2DM_admissi | leukocyte | *SRSF9* | -1.086180388 | 9.88E-05 | 0.001062643 | 0.388293605 |
| T2DM_admissi | leukocyte | *FRK* | -1.184814984 | 0.000692418 | 0.001507191 | 1 |
| T2DM_admissi | leukocyte | *RSPH4A* | -0.91971161 | 0.01302109 | 0.016659024 | 1 |
| T2DM_admissi | leukocyte | *SMIM8* | -1.368804222 | 0.000458355 | 0.001240899 | 1 |

| | | | | | | |
|---|---|---|---|---|---|---|
| T2DM_admissi | leukocyte | *NEDD9* | -0.639877215 | 0.000776458 | 0.001613276 | 1 |
| T2DM_admissi | leukocyte | *CEP85L* | -1.206627771 | 0.000624682 | 0.001426963 | 1 |
| T2DM_admissi | leukocyte | *HDDC2* | -0.930487381 | 0.000468563 | 0.001253701 | 1 |
| T2DM_admissi | leukocyte | *TPD52L1* | -0.900408494 | 0.001905002 | 0.002952841 | 1 |
| T2DM_admissi | leukocyte | *MTRF1L* | -1.168648121 | 0.002314908 | 0.003484779 | 1 |
| T2DM_admissi | leukocyte | *MRPL18* | -1.610416114 | 0.000303116 | 0.001093914 | 1 |
| T2DM_admissi | leukocyte | *RNF8* | -1.179939007 | 0.00040218 | 0.001185421 | 1 |
| T2DM_admissi | leukocyte | *MDGA1* | 1.39677681 | 0.020717429 | 0.025637411 | 1 |
| T2DM_admissi | leukocyte | *CD83* | 0.173119755 | 0.020073179 | 0.024918648 | 1 |
| T2DM_admissi | leukocyte | *GLP1R* | -1.077409694 | 0.001363266 | 0.002274426 | 1 |
| T2DM_admissi | leukocyte | *BACH2* | -0.990991706 | 0.000971405 | 0.001848256 | 1 |
| T2DM_admissi | leukocyte | *FBXL4* | -1.390857876 | 0.000444629 | 0.001230246 | 1 |
| T2DM_admissi | leukocyte | *ASCC3* | -0.925342372 | 0.000821357 | 0.001670341 | 1 |
| T2DM_admissi | leukocyte | *WASF1* | -0.975816069 | 0.001350682 | 0.002257051 | 1 |
| T2DM_admissi | leukocyte | *ALDH5A1* | -2.619828145 | 0.006224295 | 0.008435756 | 1 |
| T2DM_admissi | leukocyte | *C6orf62* | -1.390647439 | 0.000162831 | 0.001062643 | 0.639761964 |
| T2DM_admissi | leukocyte | *B3GAT2* | -1.697068733 | 0.000563629 | 0.001362351 | 1 |
| T2DM_admissi | leukocyte | *HBS1L* | -1.204495514 | 0.000300659 | 0.001093914 | 1 |
| T2DM_admissi | leukocyte | *PEX7* | -1.060999137 | 0.000730915 | 0.001550629 | 1 |
| T2DM_admissi | leukocyte | *PHACTR2* | -0.906573833 | 0.000409147 | 0.001188129 | 1 |
| T2DM_admissi | leukocyte | *EPM2A* | -1.940422238 | 0.000318428 | 0.001113024 | 1 |
| T2DM_admissi | leukocyte | *TFEB* | -1.328619512 | 0.000982625 | 0.00186239 | 1 |
| T2DM_admissi | leukocyte | *PRPH2* | -1.56144284 | 0.000635697 | 0.001436259 | 1 |
| T2DM_admissi | leukocyte | *DNPH1* | -1.034982184 | 0.002792209 | 0.004101156 | 1 |
| T2DM_admissi | leukocyte | *DUSP22* | -1.466392773 | 0.0004418 | 0.001226736 | 1 |
| T2DM_admissi | leukocyte | *LAMA4* | -0.880880697 | 0.001154702 | 0.00206595 | 1 |
| T2DM_admissi | leukocyte | *FBRSL1* | -1.410995583 | 0.000359927 | 0.001142338 | 1 |
| T2DM_admissi | leukocyte | *ENPP5* | -0.954601414 | 0.0012606 | 0.002164729 | 1 |
| T2DM_admissi | leukocyte | *NUDT12* | -1.521118844 | 0.000378301 | 0.001163156 | 1 |
| T2DM_admissi | leukocyte | *CEP72* | -3.421569193 | 0.001280852 | 0.002180445 | 1 |
| T2DM_admissi | leukocyte | *SEMA5A* | -1.314309336 | 0.001125465 | 0.002031214 | 1 |
| T2DM_admissi | leukocyte | *GHR* | -2.925649706 | 0.001202829 | 0.002114239 | 1 |
| T2DM_admissi | leukocyte | *DAP* | -0.943840258 | 0.00042309 | 0.001206582 | 1 |
| T2DM_admissi | leukocyte | *NME5* | -1.179630449 | 0.023630528 | 0.028977635 | 1 |
| T2DM_admissi | leukocyte | *BRD8* | -1.363815679 | 0.000423565 | 0.001206806 | 1 |
| T2DM_admissi | leukocyte | *NNT* | -2.271988526 | 2.47E-06 | 0.000373794 | 0.009718639 |
| T2DM_admissi | leukocyte | *TMCO6* | 1.83285184 | 0.009458668 | 0.01235065 | 1 |
| T2DM_admissi | leukocyte | *IK* | -1.185951697 | 0.000206734 | 0.001062643 | 0.812258243 |
| T2DM_admissi | leukocyte | *PDE8B* | -0.744700075 | 0.001074713 | 0.001968161 | 1 |
| T2DM_admissi | leukocyte | *MSH3* | -1.383617976 | 0.000120158 | 0.001062643 | 0.472101847 |
| T2DM_admissi | leukocyte | *CCNG1* | -2.302766986 | 6.37E-05 | 0.001060479 | 0.250468893 |
| T2DM_admissi | leukocyte | *DROSHA* | -0.838087028 | 0.004055512 | 0.005721403 | 1 |
| T2DM_admissi | leukocyte | *GOLPH3* | -1.193698496 | 0.000234518 | 0.001066706 | 0.921422929 |
| T2DM_admissi | leukocyte | *NPR3* | -3.142788842 | 0.001998215 | 0.003071591 | 1 |
| T2DM_admissi | leukocyte | *RAD50* | -1.522959498 | 0.000164842 | 0.001062643 | 0.647665223 |
| T2DM_admissi | leukocyte | *WWC1* | -0.625159124 | 0.001671364 | 0.002648967 | 1 |
| T2DM_admissi | leukocyte | *SMAD5* | -1.033339142 | 0.000530532 | 0.001324223 | 1 |
| T2DM_admissi | leukocyte | *HMGXB3* | -1.424103961 | 0.000519466 | 0.001315067 | 1 |
| T2DM_admissi | leukocyte | *BNIP1* | -0.857197102 | 0.006748224 | 0.009086282 | 1 |
| T2DM_admissi | leukocyte | *STC2* | -0.830190958 | 0.001423975 | 0.002344844 | 1 |

| | | | | | | |
|---|---|---|---|---|---|---|
| T2DM_admissi | leukocyte | *EHHADH* | -1.651866226 | 0.00029057 | 0.001088708 | 1 |
| T2DM_admissi | leukocyte | *ACTR8* | -1.097741826 | 0.000769202 | 0.001602435 | 1 |
| T2DM_admissi | leukocyte | *TIMMDC1* | -1.281399182 | 0.000468045 | 0.001253701 | 1 |
| T2DM_admissi | leukocyte | *CLDN16* | 4.106104108 | 0.023069581 | 0.028333975 | 1 |
| T2DM_admissi | leukocyte | *NPHP3* | -0.844759627 | 0.040822402 | 0.048559255 | 1 |
| T2DM_admissi | leukocyte | *FAM162A* | -1.493198207 | 5.66E-05 | 0.001060479 | 0.222327818 |
| T2DM_admissi | leukocyte | *RBP1* | -1.03950343 | 0.000197142 | 0.001062643 | 0.774570943 |
| T2DM_admissi | leukocyte | *RNF7* | -1.042730367 | 0.000547191 | 0.001347063 | 1 |
| T2DM_admissi | leukocyte | *SERPINI2* | -0.765591945 | 0.000746897 | 0.001568868 | 1 |
| T2DM_admissi | leukocyte | *TUSC2* | -1.530932316 | 0.000197183 | 0.001062643 | 0.774732316 |
| T2DM_admissi | leukocyte | *CYB561D2* | -1.017823722 | 0.000737421 | 0.00155805 | 1 |
| T2DM_admissi | leukocyte | *C3orf14* | -1.012056348 | 0.000240007 | 0.001072513 | 0.942988506 |
| T2DM_admissi | leukocyte | *CBLB* | -1.961390988 | 0.000158653 | 0.001062643 | 0.623347203 |
| T2DM_admissi | leukocyte | *IFT57* | -1.017415778 | 0.000556488 | 0.001357223 | 1 |
| T2DM_admissi | leukocyte | *NCBP2* | -1.085952326 | 0.000264542 | 0.001084251 | 1 |
| T2DM_admissi | leukocyte | *C3orf52* | -0.90977287 | 0.001908005 | 0.002956054 | 1 |
| T2DM_admissi | leukocyte | *SLC41A3* | -1.168895449 | 0.000256946 | 0.001080629 | 1 |
| T2DM_admissi | leukocyte | *ATP6V1A* | -1.386836786 | 0.000158859 | 0.001062643 | 0.624156114 |
| T2DM_admissi | leukocyte | *PODXL2* | -0.873330576 | 0.000638522 | 0.001438504 | 1 |
| T2DM_admissi | leukocyte | *UPK1B* | -1.183457972 | 0.024495468 | 0.029954153 | 1 |
| T2DM_admissi | leukocyte | *NEK11* | -0.970254537 | 0.000517163 | 0.00131413 | 1 |
| T2DM_admissi | leukocyte | *PLSCR4* | -1.927935405 | 0.000273235 | 0.001088708 | 1 |
| T2DM_admissi | leukocyte | *COMMD2* | -1.115146212 | 0.000242952 | 0.001077213 | 0.954558961 |
| T2DM_admissi | leukocyte | *GORASP1* | -2.742669994 | 0.000181322 | 0.001062643 | 0.712415273 |
| T2DM_admissi | leukocyte | *AADAC* | -1.09262166 | 0.000155531 | 0.001062643 | 0.611081331 |
| T2DM_admissi | leukocyte | *ARHGEF26* | -3.109205832 | 0.000129445 | 0.001062643 | 0.508587559 |
| T2DM_admissi | leukocyte | *PLCH1* | -5.389787701 | 9.43E-05 | 0.001062643 | 0.370467839 |
| T2DM_admissi | leukocyte | *VIPR1* | -1.543022735 | 0.005002716 | 0.006908847 | 1 |
| T2DM_admissi | leukocyte | *NKTR* | -1.088344718 | 0.000442273 | 0.001227183 | 1 |
| T2DM_admissi | leukocyte | *NEK4* | -3.426629821 | 6.61E-05 | 0.001062643 | 0.259713669 |
| T2DM_admissi | leukocyte | *KANSL3* | -2.5452388 | 8.90E-05 | 0.001062643 | 0.349768662 |
| T2DM_admissi | leukocyte | *ACTR1B* | -2.019163103 | 0.000151884 | 0.001062643 | 0.596752628 |
| T2DM_admissi | leukocyte | *SLC35F5* | -1.037569495 | 0.001181139 | 0.002099858 | 1 |
| T2DM_admissi | leukocyte | *ACTR3* | -1.310398739 | 0.00015439 | 0.001062643 | 0.606599226 |
| T2DM_admissi | leukocyte | *TFCP2L1* | -2.435909075 | 0.031515647 | 0.037936573 | 1 |
| T2DM_admissi | leukocyte | *POMC* | -1.97770063 | 0.000129124 | 0.001062643 | 0.507329548 |
| T2DM_admissi | leukocyte | *TANC1* | -0.76089593 | 0.009327253 | 0.012187155 | 1 |
| T2DM_admissi | leukocyte | *REEP6* | -1.361806457 | 0.001065564 | 0.001960938 | 1 |
| T2DM_admissi | leukocyte | *INO80B* | -3.809404751 | 0.000267845 | 0.001087356 | 1 |
| T2DM_admissi | leukocyte | *MOGS* | -2.048948365 | 9.70E-05 | 0.001062643 | 0.38114229 |
| T2DM_admissi | leukocyte | *RTN4* | -0.983263889 | 0.000151224 | 0.001062643 | 0.594159347 |
| T2DM_admissi | leukocyte | *HTRA2* | -1.975023645 | 0.000129862 | 0.001062643 | 0.510226926 |
| T2DM_admissi | leukocyte | *GALNT3* | -1.266145887 | 0.00049985 | 0.001295455 | 1 |
| T2DM_admissi | leukocyte | *MRPL19* | -1.125899227 | 0.001027874 | 0.001917284 | 1 |
| T2DM_admissi | leukocyte | *WDR75* | -2.098193289 | 0.00018441 | 0.001062643 | 0.724547516 |
| T2DM_admissi | leukocyte | *FANCL* | -2.44703203 | 0.000300761 | 0.001093914 | 1 |
| T2DM_admissi | leukocyte | *GGCX* | -1.434332777 | 0.000453685 | 0.001238727 | 1 |
| T2DM_admissi | leukocyte | *SF3B1* | -1.336799014 | 0.00012217 | 0.001062643 | 0.480005816 |
| T2DM_admissi | leukocyte | *CHMP3* | -1.149163078 | 4.43E-05 | 0.001037577 | 0.173868056 |
| T2DM_admissi | leukocyte | *IL1R2* | -1.085212774 | 0.001694722 | 0.002682741 | 1 |

| | | | | | | |
|---|---|---|---|---|---|---|
| T2DM_admissi | leukocyte | *IL1R1* | -1.084701613 | 0.000391175 | 0.00117953 | 1 |
| T2DM_admissi | leukocyte | *IL1RL2* | 0.92176812 | 0.010993524 | 0.014213082 | 1 |
| T2DM_admissi | leukocyte | *IL18R1* | 0.872311251 | 0.009533837 | 0.012440533 | 1 |
| T2DM_admissi | leukocyte | *MLPH* | -1.243510844 | 0.017582558 | 0.022014617 | 1 |
| T2DM_admissi | leukocyte | *CNPPD1* | -0.958542526 | 0.000673995 | 0.001482714 | 1 |
| T2DM_admissi | leukocyte | *HDLBP* | -0.862707992 | 0.000378779 | 0.001163156 | 1 |
| T2DM_admissi | leukocyte | *PROC* | 1.307339246 | 0.006163637 | 0.008367978 | 1 |
| T2DM_admissi | leukocyte | *TAF1B* | -0.969811036 | 0.002185024 | 0.003310821 | 1 |
| T2DM_admissi | leukocyte | *STRN* | -1.983060023 | 0.00048696 | 0.001278066 | 1 |
| T2DM_admissi | leukocyte | *CEBPZ* | -1.104004592 | 0.000332277 | 0.001127387 | 1 |
| T2DM_admissi | leukocyte | *WIPF1* | 0.53536103 | 0.028660333 | 0.034701525 | 1 |
| T2DM_admissi | leukocyte | *COX7A2L* | -0.961781685 | 0.000227689 | 0.001062643 | 0.89458878 |
| T2DM_admissi | leukocyte | *PNO1* | -1.242649185 | 0.000241185 | 0.001072513 | 0.947615674 |
| T2DM_admissi | leukocyte | *ORC4* | -1.259132696 | 0.000216464 | 0.001062643 | 0.850487015 |
| T2DM_admissi | leukocyte | *SUMO1* | -1.091255855 | 0.000210525 | 0.001062643 | 0.827152733 |
| T2DM_admissi | leukocyte | *GRIN3B* | 1.655324108 | 0.018698396 | 0.023359618 | 1 |
| T2DM_admissi | leukocyte | *ALMS1* | -1.283039516 | 0.00161951 | 0.002582409 | 1 |
| T2DM_admissi | leukocyte | *MARK1* | -2.016444989 | 0.000120367 | 0.001062643 | 0.47292011 |
| T2DM_admissi | leukocyte | *MORN1* | -1.883199782 | 0.002562203 | 0.003798828 | 1 |
| T2DM_admissi | leukocyte | *GPX7* | -1.296257384 | 2.76E-05 | 0.000996218 | 0.108587745 |
| T2DM_admissi | leukocyte | *SCP2* | -1.166496651 | 0.000160307 | 0.001062643 | 0.629848064 |
| T2DM_admissi | leukocyte | *RALGPS2* | -1.084024377 | 0.000596612 | 0.001396953 | 1 |
| T2DM_admissi | leukocyte | *ANGPTL1* | -2.106936959 | 0.000335995 | 0.001131077 | 1 |
| T2DM_admissi | leukocyte | *FAM20B* | -1.356834822 | 0.00026212 | 0.001083674 | 1 |
| T2DM_admissi | leukocyte | *WRAP73* | -4.665931829 | 2.49E-05 | 0.000982543 | 0.097998175 |
| T2DM_admissi | leukocyte | *ICMT* | -1.702680899 | 5.09E-05 | 0.00105496 | 0.199877783 |
| T2DM_admissi | leukocyte | *PARK7* | -0.950643089 | 0.000218102 | 0.001062643 | 0.856921744 |
| T2DM_admissi | leukocyte | *MECR* | -3.405492484 | 9.08E-05 | 0.001062643 | 0.356741127 |
| T2DM_admissi | leukocyte | *EDEM3* | -1.352192302 | 0.000209662 | 0.001062643 | 0.823761382 |
| T2DM_admissi | leukocyte | *FBXO6* | -5.74109985 | 6.03E-05 | 0.001060479 | 0.236754076 |
| T2DM_admissi | leukocyte | *SMG7* | -1.162603518 | 0.000352333 | 0.001141109 | 1 |
| T2DM_admissi | leukocyte | *PDC* | -11.22957046 | 0.001247028 | 0.002154446 | 1 |
| T2DM_admissi | leukocyte | *BCAS2* | -1.036150788 | 0.000348745 | 0.001141096 | 1 |
| T2DM_admissi | leukocyte | *AGMAT* | -1.658953477 | 0.001188428 | 0.00210081 | 1 |
| T2DM_admissi | leukocyte | *TNNI3K* | -2.377200231 | 0.01012663 | 0.013165959 | 1 |
| T2DM_admissi | leukocyte | *PLEKHM2* | -2.982399248 | 9.07E-05 | 0.001062643 | 0.356169975 |
| T2DM_admissi | leukocyte | *CRYZ* | -0.7737903 | 0.001046493 | 0.001939468 | 1 |
| T2DM_admissi | leukocyte | *TMEM9* | -1.155482281 | 0.000199933 | 0.001062643 | 0.785538707 |
| T2DM_admissi | leukocyte | *WARS2* | -1.116215 | 0.001086838 | 0.001980605 | 1 |
| T2DM_admissi | leukocyte | *C1orf109* | -0.886041885 | 0.000768979 | 0.001602435 | 1 |
| T2DM_admissi | leukocyte | *LGALS8* | -1.510303482 | 0.000159678 | 0.001062643 | 0.627376664 |
| T2DM_admissi | leukocyte | *HPCAL4* | -2.596703333 | 0.004028017 | 0.005688742 | 1 |
| T2DM_admissi | leukocyte | *BMP8B* | -0.978297392 | 0.001330345 | 0.002233728 | 1 |
| T2DM_admissi | leukocyte | *MYCL* | 0.516033906 | 0.006093763 | 0.008287434 | 1 |
| T2DM_admissi | leukocyte | *UAP1* | -0.820570049 | 0.000683514 | 0.001493619 | 1 |
| T2DM_admissi | leukocyte | *CTBS* | -1.444627192 | 0.000137729 | 0.001062643 | 0.541138292 |
| T2DM_admissi | leukocyte | *ZNHIT6* | -0.891632421 | 0.000341923 | 0.001140092 | 1 |
| T2DM_admissi | leukocyte | *CD46* | -0.93195634 | 0.0002745 | 0.001088708 | 1 |
| T2DM_admissi | leukocyte | *SLC2A1* | -1.607361961 | 0.000453487 | 0.001238727 | 1 |
| T2DM_admissi | leukocyte | *PRDX1* | -0.998646996 | 0.000233558 | 0.001066706 | 0.917647447 |

| | | | | | | |
|---|---|---|---|---|---|---|
| T2DM_admissi | leukocyte | *CCDC181* | -0.916765635 | 0.000499453 | 0.001295455 | 1 |
| T2DM_admissi | leukocyte | *NSUN4* | -4.862028991 | 1.70E-06 | 0.000300491 | 0.006662293 |
| T2DM_admissi | leukocyte | *TMED5* | -1.670450046 | 0.000102911 | 0.001062643 | 0.404336322 |
| T2DM_admissi | leukocyte | *DR1* | -1.549263585 | 0.000253371 | 0.001077213 | 0.995495454 |
| T2DM_admissi | leukocyte | *PTBP2* | -1.179916941 | 0.000649459 | 0.001449765 | 1 |
| T2DM_admissi | leukocyte | *NSL1* | -1.863227227 | 0.000167962 | 0.001062643 | 0.659922099 |
| T2DM_admissi | leukocyte | *RPA2* | -1.629910305 | 0.000107356 | 0.001062643 | 0.421800419 |
| T2DM_admissi | leukocyte | *ESYT2* | -1.29616222 | 0.00033606 | 0.001131077 | 1 |
| T2DM_admissi | leukocyte | *DDX59* | -0.934764111 | 0.001228112 | 0.002139802 | 1 |
| T2DM_admissi | leukocyte | *TTR* | -0.450858609 | 0.000404717 | 0.001185781 | 1 |
| T2DM_admissi | leukocyte | *SPCS2* | -1.092172556 | 0.00015583 | 0.001062643 | 0.612256467 |
| T2DM_admissi | leukocyte | *ELOVL4* | -1.622213395 | 0.000607155 | 0.001408212 | 1 |
| T2DM_admissi | leukocyte | *FILIP1* | -1.166973644 | 0.001470828 | 0.002393904 | 1 |
| T2DM_admissi | leukocyte | *AKAP7* | -2.5829355 | 7.96E-05 | 0.001062643 | 0.312882749 |
| T2DM_admissi | leukocyte | *RNF146* | -1.285640034 | 0.000420414 | 0.001203063 | 1 |
| T2DM_admissi | leukocyte | *MED28* | -1.884402494 | 7.28E-05 | 0.001062643 | 0.286224319 |
| T2DM_admissi | leukocyte | *VAMP8* | -1.312717669 | 7.92E-05 | 0.001062643 | 0.31107182 |
| T2DM_admissi | leukocyte | *ARMC2* | -0.938316328 | 0.001956584 | 0.003020911 | 1 |
| T2DM_admissi | leukocyte | *PKD2* | -0.921506049 | 0.000704358 | 0.001521605 | 1 |
| T2DM_admissi | leukocyte | *SPP1* | -0.56576491 | 0.000192995 | 0.001062643 | 0.758277318 |
| T2DM_admissi | leukocyte | *WDR35* | -0.97837335 | 0.001373537 | 0.002283803 | 1 |
| T2DM_admissi | leukocyte | *DNAH7* | -1.503355227 | 9.84E-06 | 0.000784902 | 0.038675649 |
| T2DM_admissi | leukocyte | *NDUFB3* | -1.080453508 | 0.000323149 | 0.001116892 | 1 |
| T2DM_admissi | leukocyte | *GTF3C3* | -1.494006714 | 0.00209949 | 0.003196008 | 1 |
| T2DM_admissi | leukocyte | *CPSF3* | -1.790408362 | 0.000187795 | 0.001062643 | 0.737846422 |
| T2DM_admissi | leukocyte | *CCDC92* | -1.263133428 | 0.000235663 | 0.001067958 | 0.925919192 |
| T2DM_admissi | leukocyte | *CTNNAL1* | -0.859640056 | 0.00139445 | 0.002308804 | 1 |
| T2DM_admissi | leukocyte | *NEK6* | -1.050939467 | 0.007726739 | 0.010290968 | 1 |
| T2DM_admissi | leukocyte | *BSPRY* | -1.09604911 | 0.000123847 | 0.001062643 | 0.486594049 |
| T2DM_admissi | leukocyte | *RBM18* | -1.497903656 | 0.000313879 | 0.001109126 | 1 |
| T2DM_admissi | leukocyte | *INVS* | -0.744731292 | 0.018971373 | 0.023678058 | 1 |
| T2DM_admissi | leukocyte | *DCAF4* | -1.273805217 | 0.00238898 | 0.003585294 | 1 |
| T2DM_admissi | leukocyte | *IFI27L2* | -1.042227883 | 0.000227461 | 0.001062643 | 0.893694823 |
| T2DM_admissi | leukocyte | *ACOT2* | -1.61288263 | 0.002353129 | 0.003535543 | 1 |
| T2DM_admissi | leukocyte | *ZC2HC1C* | -3.54012098 | 0.000696898 | 0.001511098 | 1 |
| T2DM_admissi | leukocyte | *ESRRB* | -3.76291866 | 0.026716348 | 0.03242772 | 1 |
| T2DM_admissi | leukocyte | *KLHL29* | -2.484896429 | 0.033241611 | 0.039965205 | 1 |
| T2DM_admissi | leukocyte | *AFTPH* | -1.148303596 | 0.000400036 | 0.001184732 | 1 |
| T2DM_admissi | leukocyte | *BCL11A* | -1.673284279 | 0.002618754 | 0.003874551 | 1 |
| T2DM_admissi | leukocyte | *CRIPT* | -1.39060921 | 0.000245561 | 0.001077213 | 0.964810967 |
| T2DM_admissi | leukocyte | *EPCAM* | -0.733050003 | 0.000342586 | 0.001140694 | 1 |
| T2DM_admissi | leukocyte | *SLC17A5* | -4.17591212 | 2.42E-05 | 0.000982543 | 0.095239804 |
| T2DM_admissi | leukocyte | *CUTC* | -1.240813629 | 0.001677444 | 0.002656459 | 1 |
| T2DM_admissi | leukocyte | *PYROXD2* | -1.623882934 | 0.0021987 | 0.003327693 | 1 |
| T2DM_admissi | leukocyte | *HELLS* | -1.381811604 | 0.000459795 | 0.001240899 | 1 |
| T2DM_admissi | leukocyte | *PRLHR* | -1.497249043 | 0.001310342 | 0.002209586 | 1 |
| T2DM_admissi | leukocyte | *WDR11* | -2.058793728 | 6.20E-05 | 0.001060479 | 0.24344166 |
| T2DM_admissi | leukocyte | *KANSL1* | -1.324715674 | 0.000104018 | 0.001062643 | 0.4086878 |
| T2DM_admissi | leukocyte | *HOXB3* | -1.221591031 | 0.008526362 | 0.011249187 | 1 |
| T2DM_admissi | leukocyte | *RCL1* | -2.152997955 | 0.000110667 | 0.001062643 | 0.434809357 |

| | | | | | | |
|---|---|---|---|---|---|---|
| T2DM_admissi | leukocyte | *CD274* | -3.553891602 | 0.000146261 | 0.001062643 | 0.574658537 |
| T2DM_admissi | leukocyte | *PLEKHG1* | -1.115465237 | 0.000899484 | 0.001770578 | 1 |
| T2DM_admissi | leukocyte | *PCDHB10* | -1.976149224 | 0.00095535 | 0.001829225 | 1 |
| T2DM_admissi | leukocyte | *MRPS14* | -1.96304973 | 0.000257499 | 0.001080629 | 1 |
| T2DM_admissi | leukocyte | *SEC16B* | -6.852557318 | 0.025832603 | 0.031452215 | 1 |
| T2DM_admissi | leukocyte | *GORAB* | -1.098719337 | 0.001182379 | 0.002099858 | 1 |
| T2DM_admissi | leukocyte | *SNX19* | -1.054797096 | 0.000468145 | 0.001253701 | 1 |
| T2DM_admissi | leukocyte | *KCNJ5* | 0.338707598 | 0.004470448 | 0.006239571 | 1 |
| T2DM_admissi | leukocyte | *SLC10A7* | -2.335427234 | 0.000147859 | 0.001062643 | 0.580936275 |
| T2DM_admissi | leukocyte | *MASTL* | -1.299700743 | 8.58E-05 | 0.001062643 | 0.337122316 |
| T2DM_admissi | leukocyte | *MRC1* | 0.274803754 | 0.015033255 | 0.019028885 | 1 |
| T2DM_admissi | leukocyte | *IQSEC3* | -1.222404717 | 0.000655694 | 0.001454635 | 1 |
| T2DM_admissi | leukocyte | *CCDC77* | -4.447821626 | 0.006999018 | 0.009388577 | 1 |
| T2DM_admissi | leukocyte | *DNAJC15* | -1.201637 | 0.000183474 | 0.001062643 | 0.720870934 |
| T2DM_admissi | leukocyte | *PROSER1* | -1.57769204 | 0.000227639 | 0.001062643 | 0.894394762 |
| T2DM_admissi | leukocyte | *WBP4* | -1.163383692 | 0.000316279 | 0.001111111 | 1 |
| T2DM_admissi | leukocyte | *KBTBD7* | -2.555518337 | 8.28E-05 | 0.001062643 | 0.325222243 |
| T2DM_admissi | leukocyte | *FAM53C* | -3.010839386 | 2.08E-06 | 0.000340493 | 0.008174348 |
| T2DM_admissi | leukocyte | *SIL1* | -1.193157982 | 0.000211462 | 0.001062643 | 0.830832515 |
| T2DM_admissi | leukocyte | *PLS1* | -0.903157964 | 0.001124 | 0.002030435 | 1 |
| T2DM_admissi | leukocyte | *ZFP30* | -1.472955291 | 0.000924972 | 0.001797337 | 1 |
| T2DM_admissi | leukocyte | *UTP20* | -1.774318539 | 0.008758008 | 0.011515184 | 1 |
| T2DM_admissi | leukocyte | *TMPO* | -1.714802428 | 0.000294877 | 0.001090935 | 1 |
| T2DM_admissi | leukocyte | *APAF1* | -2.720094716 | 0.014740348 | 0.018718431 | 1 |
| T2DM_admissi | leukocyte | *PPP3CC* | -1.428409112 | 0.00034999 | 0.001141109 | 1 |
| T2DM_admissi | leukocyte | *EPHX2* | -1.378388626 | 0.000347639 | 0.00114108 | 1 |
| T2DM_admissi | leukocyte | *CRISPLD1* | -6.32255153 | 0.000817669 | 0.001665435 | 1 |
| T2DM_admissi | leukocyte | *PYROXD1* | -0.988440337 | 0.000272141 | 0.001088708 | 1 |
| T2DM_admissi | leukocyte | *IAPP* | -0.354839306 | 0.00253012 | 0.003761196 | 1 |
| T2DM_admissi | leukocyte | *ZSCAN18* | -0.981449627 | 0.000515889 | 0.001313619 | 1 |
| T2DM_admissi | leukocyte | *LHX4* | -2.561106414 | 0.01000729 | 0.013028046 | 1 |
| T2DM_admissi | leukocyte | *TRMT1L* | -1.148181845 | 0.000990556 | 0.001869863 | 1 |
| T2DM_admissi | leukocyte | *CAT* | -2.82114761 | 3.02E-05 | 0.001023384 | 0.118543044 |
| T2DM_admissi | leukocyte | *CSMD2* | 1.188666305 | 0.013797285 | 0.017611933 | 1 |
| T2DM_admissi | leukocyte | *ACVR2A* | -0.892246742 | 0.003753427 | 0.005343193 | 1 |
| T2DM_admissi | leukocyte | *POLK* | -1.990410665 | 6.06E-06 | 0.000591121 | 0.023790281 |
| T2DM_admissi | leukocyte | *UBL3* | -1.028154936 | 0.000264657 | 0.001084251 | 1 |
| T2DM_admissi | leukocyte | *FMOD* | -1.469660578 | 0.002706297 | 0.003991381 | 1 |
| T2DM_admissi | leukocyte | *LDB3* | -1.641819957 | 0.002299115 | 0.003463659 | 1 |
| T2DM_admissi | leukocyte | *NAA60* | -1.573025026 | 0.000152968 | 0.001062643 | 0.601010522 |
| T2DM_admissi | leukocyte | *TRMT13* | -2.867561606 | 7.72E-05 | 0.001062643 | 0.303455208 |
| T2DM_admissi | leukocyte | *RWDD3* | -1.180316695 | 0.000312871 | 0.001108487 | 1 |
| T2DM_admissi | leukocyte | *CCDC18* | -1.040243665 | 0.002723929 | 0.004011362 | 1 |
| T2DM_admissi | leukocyte | *NBPF14* | -2.905259199 | 0.001777146 | 0.002785164 | 1 |
| T2DM_admissi | leukocyte | *PMS2* | -2.279938028 | 0.000582285 | 0.001382042 | 1 |
| T2DM_admissi | leukocyte | *EEPD1* | 0.509704653 | 0.018676422 | 0.023339587 | 1 |
| T2DM_admissi | leukocyte | *KLHL7* | -1.664823773 | 0.00028603 | 0.001088708 | 1 |
| T2DM_admissi | leukocyte | *CBX3* | -1.274925145 | 0.000190986 | 0.001062643 | 0.750384386 |
| T2DM_admissi | leukocyte | *NPY* | -0.548489929 | 0.000508545 | 0.001305383 | 1 |
| T2DM_admissi | leukocyte | *INHBA* | -1.03594135 | 0.000441793 | 0.001226736 | 1 |

| | | | | | | |
|---|---|---|---|---|---|---|
| T2DM_admissi | leukocyte | *CCZ1* | -1.591348266 | 0.000733872 | 0.00155271 | 1 |
| T2DM_admissi | leukocyte | *POLM* | -1.130345551 | 0.000550809 | 0.001351735 | 1 |
| T2DM_admissi | leukocyte | *SMU1* | -1.32166197 | 0.000269967 | 0.001088175 | 1 |
| T2DM_admissi | leukocyte | *SLC25A51* | -1.074254969 | 0.008823461 | 0.01158282 | 1 |
| T2DM_admissi | leukocyte | *ACO1* | -1.059885845 | 0.000876769 | 0.001743468 | 1 |
| T2DM_admissi | leukocyte | *PLAU* | -1.352444354 | 0.005491125 | 0.007533042 | 1 |
| T2DM_admissi | leukocyte | *RBM19* | -0.968686036 | 0.001234676 | 0.002144697 | 1 |
| T2DM_admissi | leukocyte | *BHLHE41* | -1.916678644 | 0.000625991 | 0.001429122 | 1 |
| T2DM_admissi | leukocyte | *ITPR2* | -2.883491769 | 0.000821551 | 0.001670341 | 1 |
| T2DM_admissi | leukocyte | *EBPL* | -0.962901867 | 0.000876837 | 0.001743468 | 1 |
| T2DM_admissi | leukocyte | *ATP7B* | -1.199615242 | 0.040519269 | 0.048242487 | 1 |
| T2DM_admissi | leukocyte | *TSFM* | -1.52884364 | 0.00038064 | 0.001165285 | 1 |
| T2DM_admissi | leukocyte | *CDK2* | -1.898992123 | 0.004424872 | 0.006186948 | 1 |
| T2DM_admissi | leukocyte | *IKZF4* | -1.672939724 | 0.000651965 | 0.001450838 | 1 |
| T2DM_admissi | leukocyte | *USP45* | -0.887022722 | 0.001827387 | 0.002851392 | 1 |
| T2DM_admissi | leukocyte | *C4BPA* | -1.437022488 | 0.019826694 | 0.024684991 | 1 |
| T2DM_admissi | leukocyte | *C4BPB* | -2.601694522 | 0.004905193 | 0.006783704 | 1 |
| T2DM_admissi | leukocyte | *RAB38* | -2.639989793 | 0.009537417 | 0.012441073 | 1 |
| T2DM_admissi | leukocyte | *CKS2* | -1.036376727 | 0.000952488 | 0.001824634 | 1 |
| T2DM_admissi | leukocyte | *SNX21* | -1.197593145 | 0.000906081 | 0.001776443 | 1 |
| T2DM_admissi | leukocyte | *ARHGAP40* | -58.94706275 | 0.000759352 | 0.001590349 | 1 |
| T2DM_admissi | leukocyte | *NCOA3* | -1.199924368 | 0.000596353 | 0.001396953 | 1 |
| T2DM_admissi | leukocyte | *MATN4* | -7.947339205 | 0.000741472 | 0.001563191 | 1 |
| T2DM_admissi | leukocyte | *VAPB* | -1.090475316 | 0.00017374 | 0.001062643 | 0.682626023 |
| T2DM_admissi | leukocyte | *PARD6B* | -1.493059541 | 0.000367778 | 0.001148802 | 1 |
| T2DM_admissi | leukocyte | *ARFGEF2* | -1.480475998 | 0.000258553 | 0.001081954 | 1 |
| T2DM_admissi | leukocyte | *ZNFX1* | -4.252620508 | 4.17E-05 | 0.001037577 | 0.163956215 |
| T2DM_admissi | leukocyte | *CSE1L* | -1.467302125 | 0.000151727 | 0.001062643 | 0.59613369 |
| T2DM_admissi | leukocyte | *BCAS4* | -0.817634049 | 0.001069497 | 0.001964163 | 1 |
| T2DM_admissi | leukocyte | *KCNK15* | -0.937531277 | 0.004857828 | 0.006722933 | 1 |
| T2DM_admissi | leukocyte | *PEPD* | -2.17441162 | 9.87E-06 | 0.000784902 | 0.038786272 |
| T2DM_admissi | leukocyte | *PAIP2B* | -0.682523066 | 0.009199244 | 0.012039917 | 1 |
| T2DM_admissi | leukocyte | *SNRNP27* | -1.134451043 | 0.000408151 | 0.001188129 | 1 |
| T2DM_admissi | leukocyte | *USP22* | -1.352714598 | 0.000422422 | 0.001206582 | 1 |
| T2DM_admissi | leukocyte | *BTN2A2* | -1.704685839 | 0.00439469 | 0.006149122 | 1 |
| T2DM_admissi | leukocyte | *MRS2* | -2.013035618 | 0.000142327 | 0.001062643 | 0.559201945 |
| T2DM_admissi | leukocyte | *SNRPC* | -1.286046877 | 0.000163647 | 0.001062643 | 0.642970373 |
| T2DM_admissi | leukocyte | *SERPINB6* | -1.128785715 | 0.000162903 | 0.001062643 | 0.640047347 |
| T2DM_admissi | leukocyte | *ABCC10* | -1.318128962 | 0.002032632 | 0.003113533 | 1 |
| T2DM_admissi | leukocyte | *PEX6* | -1.31136354 | 0.000111873 | 0.001062643 | 0.43954816 |
| T2DM_admissi | leukocyte | *NQO2* | -3.2245601 | 1.05E-05 | 0.000784902 | 0.041261277 |
| T2DM_admissi | leukocyte | *UNC5CL* | -1.068329919 | 0.000713223 | 0.001532285 | 1 |
| T2DM_admissi | leukocyte | *ZNF391* | -0.721812392 | 0.024313259 | 0.029740596 | 1 |
| T2DM_admissi | leukocyte | *MED20* | -1.757438408 | 0.000687709 | 0.00149945 | 1 |
| T2DM_admissi | leukocyte | *TREM1* | 0.855141399 | 0.035208121 | 0.042213216 | 1 |
| T2DM_admissi | leukocyte | *GLO1* | -1.243288382 | 0.000293608 | 0.001088724 | 1 |
| T2DM_admissi | leukocyte | *KCNK17* | -0.75884301 | 0.004525343 | 0.00630499 | 1 |
| T2DM_admissi | leukocyte | *ATXN1* | -1.6252523 | 0.000149801 | 0.001062643 | 0.58856807 |
| T2DM_admissi | leukocyte | *LRRFIP1* | -0.948471833 | 0.000361419 | 0.001142338 | 1 |
| T2DM_admissi | leukocyte | *CXCL6* | -1.464722694 | 0.010433756 | 0.013529448 | 1 |

| | | | | | | |
|---|---|---|---|---|---|---|
| T2DM_admissi | leukocyte | *SH3TC1* | -1.26846405 | 0.000498284 | 0.001293956 | 1 |
| T2DM_admissi | leukocyte | *MT1G* | -0.794720148 | 0.003556237 | 0.00508459 | 1 |
| T2DM_admissi | leukocyte | *DOK4* | -0.779005779 | 0.001727187 | 0.002722068 | 1 |
| T2DM_admissi | leukocyte | *CLYBL* | -3.130328864 | 8.79E-05 | 0.001062643 | 0.345510206 |
| T2DM_admissi | leukocyte | *TM9SF2* | -1.039454013 | 0.000361028 | 0.001142338 | 1 |
| T2DM_admissi | leukocyte | *KIF25* | -17.1670307 | 0.000215541 | 0.001062643 | 0.846859759 |
| T2DM_admissi | leukocyte | *BMP4* | -2.728282973 | 0.000172409 | 0.001062643 | 0.67739325 |
| T2DM_admissi | leukocyte | *GRK4* | -1.036144209 | 0.020383963 | 0.02526454 | 1 |
| T2DM_admissi | leukocyte | *HS3ST3B1* | -2.544482696 | 0.001275324 | 0.002175489 | 1 |
| T2DM_admissi | leukocyte | *MRPS7* | -0.958969285 | 0.000526792 | 0.001319879 | 1 |
| T2DM_admissi | leukocyte | *GGA3* | -2.833563736 | 0.000139769 | 0.001062643 | 0.549152265 |
| T2DM_admissi | leukocyte | *DDX31* | -2.057317931 | 0.000501418 | 0.001296953 | 1 |
| T2DM_admissi | leukocyte | *PPDPF* | -0.901136667 | 0.000225682 | 0.001062643 | 0.88670341 |
| T2DM_admissi | leukocyte | *CHCHD5* | -1.058759837 | 0.000339538 | 0.001134896 | 1 |
| T2DM_admissi | leukocyte | *INSIG2* | -1.009712146 | 0.000401042 | 0.001184732 | 1 |
| T2DM_admissi | leukocyte | *PSD4* | -1.417687101 | 0.00224475 | 0.003390858 | 1 |
| T2DM_admissi | leukocyte | *CLPP* | -1.135056303 | 0.000488695 | 0.001280054 | 1 |
| T2DM_admissi | leukocyte | *SH2D3A* | -0.848557393 | 0.003519791 | 0.005037981 | 1 |
| T2DM_admissi | leukocyte | *GPR108* | -1.644588611 | 0.000383055 | 0.00116578 | 1 |
| T2DM_admissi | leukocyte | *FOSB* | -2.132914543 | 3.34E-05 | 0.001037577 | 0.131154462 |
| T2DM_admissi | leukocyte | *SYMPK* | -2.671307296 | 2.62E-05 | 0.000984894 | 0.103031996 |
| T2DM_admissi | leukocyte | *NAPB* | -1.061886653 | 0.00067078 | 0.001477964 | 1 |
| T2DM_admissi | leukocyte | *PSMF1* | -1.475105915 | 5.18E-05 | 0.00105496 | 0.203711379 |
| T2DM_admissi | leukocyte | *RBCK1* | -1.145650674 | 0.000242868 | 0.001077213 | 0.954227133 |
| T2DM_admissi | leukocyte | *FLRT3* | -0.939325223 | 0.001237836 | 0.002148171 | 1 |
| T2DM_admissi | leukocyte | *MGME1* | -5.36391051 | 3.37E-05 | 0.001037577 | 0.13257979 |
| T2DM_admissi | leukocyte | *ITPA* | -1.680628346 | 0.00011177 | 0.001062643 | 0.439142982 |
| T2DM_admissi | leukocyte | *MCM8* | -0.850570895 | 0.040732828 | 0.04848206 | 1 |
| T2DM_admissi | leukocyte | *BANF2* | -15.07019741 | 0.00127461 | 0.002175489 | 1 |
| T2DM_admissi | leukocyte | *MRPS26* | -1.033890296 | 0.000229196 | 0.001064322 | 0.900512547 |
| T2DM_admissi | leukocyte | *ZNF436* | -2.986750446 | 0.000112901 | 0.001062643 | 0.443589223 |
| T2DM_admissi | leukocyte | *TUBGCP3* | -1.424750916 | 0.000429587 | 0.00121662 | 1 |
| T2DM_admissi | leukocyte | *MCF2L* | -0.731536092 | 0.00420869 | 0.005914143 | 1 |
| T2DM_admissi | leukocyte | *IGFLR1* | 0.323274021 | 0.014836442 | 0.018816134 | 1 |
| T2DM_admissi | leukocyte | *NR1D1* | -0.998650702 | 0.000478869 | 0.001265283 | 1 |
| T2DM_admissi | leukocyte | *PRMT1* | -1.056206923 | 9.43E-05 | 0.001062643 | 0.370427589 |
| T2DM_admissi | leukocyte | *PRR12* | -1.02601996 | 0.003676795 | 0.005246556 | 1 |
| T2DM_admissi | leukocyte | *FLRT1* | -2.308406658 | 0.013585993 | 0.0173535 | 1 |
| T2DM_admissi | leukocyte | *ASL* | -1.272055412 | 0.000336673 | 0.001131556 | 1 |
| T2DM_admissi | leukocyte | *WNK4* | -1.052866515 | 0.020845151 | 0.025779225 | 1 |
| T2DM_admissi | leukocyte | *BECN1* | -1.233955639 | 0.00045962 | 0.001240899 | 1 |
| T2DM_admissi | leukocyte | *PRKCG* | 2.42891225 | 0.030761113 | 0.037119291 | 1 |
| T2DM_admissi | leukocyte | *TRAP1* | -0.916579895 | 0.001291189 | 0.002191396 | 1 |
| T2DM_admissi | leukocyte | *ZNF384* | -1.476334538 | 0.000437549 | 0.001222199 | 1 |
| T2DM_admissi | leukocyte | *RHOJ* | -1.926480232 | 0.000827975 | 0.00167593 | 1 |
| T2DM_admissi | leukocyte | *L3HYPDH* | -1.29938682 | 0.002394188 | 0.003590368 | 1 |
| T2DM_admissi | leukocyte | *HSPA2* | -1.972957408 | 0.000399946 | 0.001184732 | 1 |
| T2DM_admissi | leukocyte | *ZBTB1* | -1.705862985 | 0.000111699 | 0.001062643 | 0.438864665 |
| T2DM_admissi | leukocyte | *PLEKHG3* | -1.691839255 | 0.000373856 | 0.001157429 | 1 |
| T2DM_admissi | leukocyte | *FGD3* | 2.651625135 | 0.019816273 | 0.024684991 | 1 |

| | | | | | | |
|---|---|---|---|---|---|---|
| T2DM_admissi | leukocyte | *BCL11B* | 1.72842016 | 0.010333068 | 0.013412166 | 1 |
| T2DM_admissi | leukocyte | *ABHD8* | -1.827179569 | 0.016188562 | 0.020412343 | 1 |
| T2DM_admissi | leukocyte | *HELB* | -4.312242619 | 0.041774801 | 0.049602053 | 1 |
| T2DM_admissi | leukocyte | *BEST3* | -2.810124524 | 0.001530493 | 0.00247563 | 1 |
| T2DM_admissi | leukocyte | *YEATS4* | -1.07947024 | 0.00056519 | 0.001362351 | 1 |
| T2DM_admissi | leukocyte | *LRRC61* | -1.742866856 | 0.000500556 | 0.001295577 | 1 |
| T2DM_admissi | leukocyte | *IDUA* | -1.138574694 | 0.000891709 | 0.001763074 | 1 |
| T2DM_admissi | leukocyte | *FGFRL1* | -1.139263667 | 0.000570882 | 0.001370185 | 1 |
| T2DM_admissi | leukocyte | *EMC1* | -1.310123308 | 0.000693923 | 0.001507546 | 1 |
| T2DM_admissi | leukocyte | *UBR4* | -1.323099444 | 0.000219117 | 0.001062643 | 0.860909631 |
| T2DM_admissi | leukocyte | *SYNGR3* | -1.228042691 | 0.003190142 | 0.004611504 | 1 |
| T2DM_admissi | leukocyte | *WFIKKN1* | -0.966529926 | 0.008369221 | 0.011071606 | 1 |
| T2DM_admissi | leukocyte | *KDM4B* | -2.973868299 | 5.31E-05 | 0.001059005 | 0.208624049 |
| T2DM_admissi | leukocyte | *PNKD* | -1.578678598 | 0.000118571 | 0.001062643 | 0.465863777 |
| T2DM_admissi | leukocyte | *PTPN12* | -1.382109607 | 3.69E-05 | 0.001037577 | 0.145068517 |
| T2DM_admissi | leukocyte | *POR* | -1.292286402 | 0.00027649 | 0.001088708 | 1 |
| T2DM_admissi | leukocyte | *RBM48* | -2.291573668 | 0.000155257 | 0.001062643 | 0.610006318 |
| T2DM_admissi | leukocyte | *SRD5A3* | -1.448011213 | 0.000787513 | 0.001625926 | 1 |
| T2DM_admissi | leukocyte | *PPAT* | -1.458219192 | 0.000186948 | 0.001062643 | 0.734517318 |
| T2DM_admissi | leukocyte | *A4GALT* | -0.887717958 | 0.004114609 | 0.005798529 | 1 |
| T2DM_admissi | leukocyte | *NAA38* | -0.984954109 | 0.000291422 | 0.001088708 | 1 |
| T2DM_admissi | leukocyte | *PRKRIP1* | -1.111248592 | 0.000611436 | 0.001413969 | 1 |
| T2DM_admissi | leukocyte | *VGF* | -0.605521554 | 0.000459467 | 0.001240899 | 1 |
| T2DM_admissi | leukocyte | *IRF5* | 0.437239383 | 0.006997001 | 0.009388577 | 1 |
| T2DM_admissi | leukocyte | *MTX2* | -1.752758537 | 0.000131716 | 0.001062643 | 0.517511447 |
| T2DM_admissi | leukocyte | *GAD1* | -4.533065028 | 0.00018886 | 0.001062643 | 0.742030801 |
| T2DM_admissi | leukocyte | *ORMDL1* | -1.076037407 | 0.000724435 | 0.001544387 | 1 |
| T2DM_admissi | leukocyte | *PSMG2* | -1.331751081 | 0.000302241 | 0.001093914 | 1 |
| T2DM_admissi | leukocyte | *TWSG1* | -1.850449473 | 0.000146902 | 0.001062643 | 0.577177733 |
| T2DM_admissi | leukocyte | *EIF2AK4* | -1.978004253 | 2.04E-05 | 0.000922724 | 0.080277006 |
| T2DM_admissi | leukocyte | *MYO5C* | -0.800318612 | 0.000940037 | 0.001810978 | 1 |
| T2DM_admissi | leukocyte | *CGNL1* | -1.457328176 | 0.000106028 | 0.001062643 | 0.416583047 |
| T2DM_admissi | leukocyte | *TMOD2* | -1.320208255 | 0.000422617 | 0.001206582 | 1 |
| T2DM_admissi | leukocyte | *ELL3* | -3.933657146 | 8.95E-05 | 0.001062643 | 0.351646839 |
| T2DM_admissi | leukocyte | *IVD* | -1.547119577 | 0.001035702 | 0.001927651 | 1 |
| T2DM_admissi | leukocyte | *CHAC1* | -1.433968895 | 0.000292406 | 0.001088708 | 1 |
| T2DM_admissi | leukocyte | *ACKR4* | -1.958764946 | 0.004903628 | 0.006783704 | 1 |
| T2DM_admissi | leukocyte | *ANAPC13* | -0.655379513 | 0.004794959 | 0.006642945 | 1 |
| T2DM_admissi | leukocyte | *SUMF2* | -1.405398041 | 0.000178549 | 0.001062643 | 0.701519716 |
| T2DM_admissi | leukocyte | *TPH1* | -3.326853753 | 0.005725613 | 0.007827396 | 1 |
| T2DM_admissi | leukocyte | *DCTD* | -1.494527723 | 0.000154548 | 0.001062643 | 0.607217528 |
| T2DM_admissi | leukocyte | *KRI1* | -0.769196429 | 0.007792518 | 0.010361017 | 1 |
| T2DM_admissi | leukocyte | *AP1M2* | -1.059014268 | 0.000444384 | 0.001230246 | 1 |
| T2DM_admissi | leukocyte | *CDKN2D* | -1.096990179 | 0.000369118 | 0.00115009 | 1 |
| T2DM_admissi | leukocyte | *MTUS1* | -0.922010808 | 0.001010898 | 0.001894951 | 1 |
| T2DM_admissi | leukocyte | *ADCY4* | -1.113068848 | 0.003114812 | 0.004520908 | 1 |
| T2DM_admissi | leukocyte | *DTD2* | -0.973346757 | 0.001179286 | 0.002098466 | 1 |
| T2DM_admissi | leukocyte | *HEATR5A* | -2.572281707 | 8.13E-05 | 0.001062643 | 0.319276752 |
| T2DM_admissi | leukocyte | *MIS18BP1* | -1.589951983 | 0.015972755 | 0.020166116 | 1 |
| T2DM_admissi | leukocyte | *RNASE1* | 0.166433314 | 0.012236205 | 0.015721403 | 1 |

| | | | | | | |
|---|---|---|---|---|---|---|
| T2DM_admissi | leukocyte | *EPB41L4A* | -1.274955862 | 0.00120976 | 0.002121941 | 1 |
| T2DM_admissi | leukocyte | *REEP5* | -0.999977326 | 0.000248872 | 0.001077213 | 0.977816683 |
| T2DM_admissi | leukocyte | *MAU2* | -3.884143719 | 2.25E-08 | 9.82E-06 | 8.84E-05 |
| T2DM_admissi | leukocyte | *SHC2* | -1.346523336 | 0.000269623 | 0.001088175 | 1 |
| T2DM_admissi | leukocyte | *SYT5* | -0.581862198 | 0.012942436 | 0.016563788 | 1 |
| T2DM_admissi | leukocyte | *CBFA2T3* | -1.086459545 | 0.00118368 | 0.002100578 | 1 |
| T2DM_admissi | leukocyte | *ERMARD* | -2.565683585 | 6.40E-05 | 0.001060479 | 0.251410375 |
| T2DM_admissi | leukocyte | *PHF10* | -1.208411761 | 0.000356554 | 0.001141109 | 1 |
| T2DM_admissi | leukocyte | *SH3BP4* | -0.611022255 | 0.003889137 | 0.005520383 | 1 |
| T2DM_admissi | leukocyte | *DOCK6* | -0.866069908 | 0.001187489 | 0.00210081 | 1 |
| T2DM_admissi | leukocyte | *ECSIT* | -1.367169678 | 0.000449329 | 0.001234067 | 1 |
| T2DM_admissi | leukocyte | *CDC16* | -1.775659291 | 6.42E-05 | 0.001060479 | 0.252200512 |
| T2DM_admissi | leukocyte | *DPP6* | -0.6064402 | 0.001064443 | 0.001959791 | 1 |
| T2DM_admissi | leukocyte | *GTPBP3* | -1.419289432 | 5.11E-05 | 0.00105496 | 0.2008977 |
| T2DM_admissi | leukocyte | *PLVAP* | -0.555871679 | 0.001532012 | 0.002477069 | 1 |
| T2DM_admissi | leukocyte | *MRPL34* | -1.135540866 | 3.20E-05 | 0.001037577 | 0.125918259 |
| T2DM_admissi | leukocyte | *LSM7* | -0.779253753 | 0.001373424 | 0.002283803 | 1 |
| T2DM_admissi | leukocyte | *TULP4* | -1.06282395 | 0.001840206 | 0.002865703 | 1 |
| T2DM_admissi | leukocyte | *QRSL1* | -1.253389333 | 0.000897436 | 0.001768319 | 1 |
| T2DM_admissi | leukocyte | *RSPH3* | -1.287644316 | 0.000892082 | 0.001763074 | 1 |
| T2DM_admissi | leukocyte | *STK33* | -1.006870117 | 0.000378817 | 0.001163156 | 1 |
| T2DM_admissi | leukocyte | *NDUFA10* | -1.131054592 | 0.00044462 | 0.001230246 | 1 |
| T2DM_admissi | leukocyte | *TRPM4* | -4.51893535 | 0.001578052 | 0.00253068 | 1 |
| T2DM_admissi | leukocyte | *UBAC1* | -1.159639506 | 0.000351977 | 0.001141109 | 1 |
| T2DM_admissi | leukocyte | *ATXN10* | -1.001103399 | 0.000324398 | 0.001117056 | 1 |
| T2DM_admissi | leukocyte | *PNPLA7* | -2.259242687 | 0.000730598 | 0.001550629 | 1 |
| T2DM_admissi | leukocyte | *PAK4* | -1.500990042 | 0.000319281 | 0.001113024 | 1 |
| T2DM_admissi | leukocyte | *LAMA5* | -0.749036167 | 0.001630034 | 0.002594977 | 1 |
| T2DM_admissi | leukocyte | *ADRM1* | -1.147934358 | 0.000222459 | 0.001062643 | 0.874041516 |
| T2DM_admissi | leukocyte | *UCK1* | -2.311247511 | 3.70E-05 | 0.001037577 | 0.145346435 |
| T2DM_admissi | leukocyte | *MAP3K10* | -2.695001693 | 0.000283148 | 0.001088708 | 1 |
| T2DM_admissi | leukocyte | *SMPDL3B* | -0.932337049 | 0.001650726 | 0.002622605 | 1 |
| T2DM_admissi | leukocyte | *CLIP1* | -1.335733179 | 0.00047393 | 0.001259006 | 1 |
| T2DM_admissi | leukocyte | *ZNF317* | -1.615977533 | 0.000331197 | 0.001125668 | 1 |
| T2DM_admissi | leukocyte | *DNMT1* | -1.951672787 | 0.000209412 | 0.001062643 | 0.822779773 |
| T2DM_admissi | leukocyte | *ZNF426* | -1.800817883 | 9.97E-05 | 0.001062643 | 0.391890216 |
| T2DM_admissi | leukocyte | *LRP3* | -1.753950158 | 0.000960073 | 0.001831992 | 1 |
| T2DM_admissi | leukocyte | *HSD17B3* | -15.31944989 | 0.000292403 | 0.001088708 | 1 |
| T2DM_admissi | leukocyte | *SYNE1* | -1.048792543 | 0.000378936 | 0.001163156 | 1 |
| T2DM_admissi | leukocyte | *EPS8L1* | -3.308142505 | 0.002009223 | 0.003086098 | 1 |
| T2DM_admissi | leukocyte | *C1QL1* | -0.475821628 | 0.002539497 | 0.003772282 | 1 |
| T2DM_admissi | leukocyte | *F12* | -1.135210202 | 0.005901273 | 0.008050729 | 1 |
| T2DM_admissi | leukocyte | *TRAF3* | -0.805014264 | 0.005126471 | 0.007069815 | 1 |
| T2DM_admissi | leukocyte | *PDLIM4* | -1.353882779 | 0.00025312 | 0.001077213 | 0.99450767 |
| T2DM_admissi | leukocyte | *MGAT1* | -2.757371511 | 0.001984945 | 0.003054843 | 1 |
| T2DM_admissi | leukocyte | *DIAPH1* | -0.896100886 | 0.000592581 | 0.001392055 | 1 |
| T2DM_admissi | leukocyte | *EXOC4* | -2.19132229 | 0.000956534 | 0.001830343 | 1 |
| T2DM_admissi | leukocyte | *C1orf159* | -4.699250011 | 0.000230526 | 0.001064322 | 0.905738398 |
| T2DM_admissi | leukocyte | *ANO1* | -2.842668099 | 0.000588203 | 0.001384692 | 1 |
| T2DM_admissi | leukocyte | *PPFIA1* | -1.043883272 | 0.000499707 | 0.001295455 | 1 |

| | | | | | | |
|---|---|---|---|---|---|---|
| T2DM_admissi | leukocyte | *TRAF7* | -1.55748505 | 0.000366368 | 0.001147001 | 1 |
| T2DM_admissi | leukocyte | *MAP1B* | -0.706859186 | 0.001096833 | 0.001995936 | 1 |
| T2DM_admissi | leukocyte | *CHD1L* | -2.750361171 | 0.000339268 | 0.001134896 | 1 |
| T2DM_admissi | leukocyte | *PIAS3* | -0.894911408 | 0.001078377 | 0.001970875 | 1 |
| T2DM_admissi | leukocyte | *RBM8A* | -0.907216431 | 9.37E-05 | 0.001062643 | 0.368087993 |
| T2DM_admissi | leukocyte | *ZNF304* | -5.557267215 | 4.48E-05 | 0.001037577 | 0.176130471 |
| T2DM_admissi | leukocyte | *ZNF132* | 1.288392166 | 0.011060102 | 0.01427567 | 1 |
| T2DM_admissi | leukocyte | *RHPN2* | -1.073805902 | 0.001120187 | 0.002027274 | 1 |
| T2DM_admissi | leukocyte | *GCH1* | -0.7740785 | 0.001543386 | 0.002491358 | 1 |
| T2DM_admissi | leukocyte | *CC2D1A* | -1.709428089 | 0.000199054 | 0.001062643 | 0.782083905 |
| T2DM_admissi | leukocyte | *CCT6B* | -1.837033481 | 0.000511294 | 0.001308382 | 1 |
| T2DM_admissi | leukocyte | *PPARG* | -2.148497042 | 0.000101856 | 0.001062643 | 0.400192484 |
| T2DM_admissi | leukocyte | *ENOSF1* | -3.246643708 | 3.78E-05 | 0.001037577 | 0.148399938 |
| T2DM_admissi | leukocyte | *IMMT* | -1.116415638 | 0.001705836 | 0.00269816 | 1 |
| T2DM_admissi | leukocyte | *MRPL35* | -1.008056157 | 0.006031177 | 0.008216538 | 1 |
| T2DM_admissi | leukocyte | *SCLY* | -1.444074488 | 0.014817448 | 0.018801362 | 1 |
| T2DM_admissi | leukocyte | *CARD6* | -2.229047351 | 0.000791306 | 0.0016329 | 1 |
| T2DM_admissi | leukocyte | *INPP5K* | -1.11360495 | 0.003225771 | 0.004659579 | 1 |
| T2DM_admissi | leukocyte | *RPA1* | -1.352652026 | 0.001142425 | 0.00205052 | 1 |
| T2DM_admissi | leukocyte | *TMEM128* | -1.868569876 | 0.000148353 | 0.001062643 | 0.582877868 |
| T2DM_admissi | leukocyte | *SEC61G* | -0.920162629 | 0.000209505 | 0.001062643 | 0.823145577 |
| T2DM_admissi | leukocyte | *FIGNL1* | -3.506099831 | 0.00118274 | 0.002099858 | 1 |
| T2DM_admissi | leukocyte | *ITGB4* | -1.69873275 | 0.024298475 | 0.029731769 | 1 |
| T2DM_admissi | leukocyte | *TRIM47* | -1.07169433 | 0.001374948 | 0.002285182 | 1 |
| T2DM_admissi | leukocyte | *EIF5A* | -1.066540625 | 0.00012366 | 0.001062643 | 0.485859773 |
| T2DM_admissi | leukocyte | *PRMT7* | -2.968647785 | 0.000155389 | 0.001062643 | 0.610524729 |
| T2DM_admissi | leukocyte | *ANKEF1* | -0.963669932 | 0.001172498 | 0.002089227 | 1 |
| T2DM_admissi | leukocyte | *BTBD3* | -0.985525826 | 0.000404229 | 0.001185781 | 1 |
| T2DM_admissi | leukocyte | *IGHMBP2* | -1.615933028 | 0.000108443 | 0.001062643 | 0.42607118 |
| T2DM_admissi | leukocyte | *LPIN3* | -1.524999753 | 0.003915938 | 0.005546402 | 1 |
| T2DM_admissi | leukocyte | *RBM38* | -1.945485769 | 0.000126439 | 0.001062643 | 0.496780193 |
| T2DM_admissi | leukocyte | *VSTM2L* | -0.895186335 | 0.002137943 | 0.003246996 | 1 |
| T2DM_admissi | leukocyte | *SERINC3* | -1.221958192 | 0.000169831 | 0.001062643 | 0.667266435 |
| T2DM_admissi | leukocyte | *BHMT2* | -1.71936342 | 1.92E-05 | 0.000917316 | 0.075627446 |
| T2DM_admissi | leukocyte | *CASP9* | -2.320169778 | 0.00127933 | 0.002178798 | 1 |
| T2DM_admissi | leukocyte | *WASF3* | -1.151564869 | 0.001381684 | 0.002292499 | 1 |
| T2DM_admissi | leukocyte | *SCO1* | -2.117383026 | 0.000236302 | 0.001068391 | 0.928431734 |
| T2DM_admissi | leukocyte | *CHI3L1* | -0.859828313 | 0.009268405 | 0.012118325 | 1 |
| T2DM_admissi | leukocyte | *SLC41A1* | -1.319978049 | 0.000619279 | 0.001422062 | 1 |
| T2DM_admissi | leukocyte | *TRPC4* | -3.436202426 | 0.000522217 | 0.001319879 | 1 |
| T2DM_admissi | leukocyte | *POSTN* | 0.943266902 | 0.010286272 | 0.013355838 | 1 |
| T2DM_admissi | leukocyte | *RFC3* | -0.930907469 | 0.001231248 | 0.002143364 | 1 |
| T2DM_admissi | leukocyte | *EPHB2* | -4.670199203 | 0.000398282 | 0.001183526 | 1 |
| T2DM_admissi | leukocyte | *BTBD2* | -2.916611625 | 7.10E-05 | 0.001062643 | 0.279061979 |
| T2DM_admissi | leukocyte | *CNDP2* | -1.21714235 | 0.000348745 | 0.001141096 | 1 |
| T2DM_admissi | leukocyte | *WDR74* | -2.033735576 | 5.07E-05 | 0.00105496 | 0.199149664 |
| T2DM_admissi | leukocyte | *GSTT2B* | -1.012965485 | 0.001737572 | 0.002734049 | 1 |
| T2DM_admissi | leukocyte | *SLC2A11* | -1.98773599 | 0.000154486 | 0.001062643 | 0.606976195 |
| T2DM_admissi | leukocyte | *C1QTNF6* | -1.814308563 | 0.001297873 | 0.002196252 | 1 |
| T2DM_admissi | leukocyte | *FAM83F* | -1.297974513 | 0.000386707 | 0.001171451 | 1 |

| | | | | | | |
|---|---|---|---|---|---|---|
| T2DM_admissi | leukocyte | *GIMAP6* | -2.243865411 | 0.000437084 | 0.001222199 | 1 |
| T2DM_admissi | leukocyte | *AGAP3* | -4.225493327 | 3.19E-05 | 0.001037577 | 0.125158115 |
| T2DM_admissi | leukocyte | *SFTPD* | -0.817123714 | 0.0255313 | 0.031133606 | 1 |
| T2DM_admissi | leukocyte | *IMPA1* | -1.160789172 | 0.000264923 | 0.001084251 | 1 |
| T2DM_admissi | leukocyte | *LRRCC1* | -1.807027625 | 0.000651549 | 0.001450838 | 1 |
| T2DM_admissi | leukocyte | *RRAS2* | -0.968634583 | 0.000945687 | 0.0018187 | 1 |
| T2DM_admissi | leukocyte | *HSD17B4* | -2.278826677 | 5.48E-05 | 0.001060479 | 0.215257166 |
| T2DM_admissi | leukocyte | *RNF122* | -1.654311894 | 0.023195889 | 0.028480203 | 1 |
| T2DM_admissi | leukocyte | *COX16* | -1.188705985 | 0.00017167 | 0.001062643 | 0.674492986 |
| T2DM_admissi | leukocyte | *ELP3* | -2.552828158 | 0.000140954 | 0.001062643 | 0.553807443 |
| T2DM_admissi | leukocyte | *PEBP4* | -1.49003182 | 0.002065923 | 0.003154688 | 1 |
| T2DM_admissi | leukocyte | *MRO* | -3.146212774 | 0.000733248 | 0.001552225 | 1 |
| T2DM_admissi | leukocyte | *MRPS36* | -1.159259245 | 0.001419797 | 0.00234189 | 1 |
| T2DM_admissi | leukocyte | *THUMPD3* | -1.43473526 | 7.68E-05 | 0.001062643 | 0.301631291 |
| T2DM_admissi | leukocyte | *CHL1* | -1.174186612 | 0.004160564 | 0.005855516 | 1 |
| T2DM_admissi | leukocyte | *GSTM1* | 1.886652361 | 0.015026857 | 0.019026916 | 1 |
| T2DM_admissi | leukocyte | *REG4* | -1.266010709 | 0.001216263 | 0.00212877 | 1 |
| T2DM_admissi | leukocyte | *GSTM3* | -0.802086584 | 0.001446327 | 0.002366772 | 1 |
| T2DM_admissi | leukocyte | *PTPN22* | -5.380044828 | 0.006786309 | 0.009128178 | 1 |
| T2DM_admissi | leukocyte | *SORT1* | -1.549556467 | 0.000184058 | 0.001062643 | 0.723162563 |
| T2DM_admissi | leukocyte | *NOTCH2* | -3.595543145 | 3.59E-15 | 1.41E-11 | 1.41E-11 |
| T2DM_admissi | leukocyte | *CEPT1* | -2.105618369 | 0.000251344 | 0.001077213 | 0.987529581 |
| T2DM_admissi | leukocyte | *NAPG* | -1.218295627 | 0.000329808 | 0.001123865 | 1 |
| T2DM_admissi | leukocyte | *SPIRE1* | -1.308213192 | 0.000155076 | 0.001062643 | 0.609292331 |
| T2DM_admissi | leukocyte | *FKBP11* | -0.694365568 | 0.000870979 | 0.001737977 | 1 |
| T2DM_admissi | leukocyte | *KIDINS220* | -1.317181149 | 0.000193734 | 0.001062643 | 0.761182277 |
| T2DM_admissi | leukocyte | *LPIN1* | -0.928835273 | 0.000244777 | 0.001077213 | 0.961727012 |
| T2DM_admissi | leukocyte | *IAH1* | -1.546799661 | 0.000268014 | 0.001087356 | 1 |
| T2DM_admissi | leukocyte | *ANO3* | 1.537128967 | 0.01227202 | 0.015762265 | 1 |
| T2DM_admissi | leukocyte | *IL2RA* | 0.320234185 | 0.008893411 | 0.011670746 | 1 |
| T2DM_admissi | leukocyte | *ECHDC3* | -1.142901338 | 0.000650162 | 0.001449765 | 1 |
| T2DM_admissi | leukocyte | *TMEM241* | -1.873997697 | 0.001765571 | 0.002770339 | 1 |
| T2DM_admissi | leukocyte | *AGO4* | -2.142453465 | 0.000296917 | 0.001093914 | 1 |
| T2DM_admissi | leukocyte | *PRPF38A* | -1.197145723 | 0.000365397 | 0.001147001 | 1 |
| T2DM_admissi | leukocyte | *DSC2* | -2.662585988 | 0.008323846 | 0.011022321 | 1 |
| T2DM_admissi | leukocyte | *TPGS2* | -1.329248853 | 0.001042313 | 0.001936841 | 1 |
| T2DM_admissi | leukocyte | *DAGLA* | -1.280959405 | 0.00172711 | 0.002722068 | 1 |
| T2DM_admissi | leukocyte | *TIMM10* | -1.27149185 | 0.000194546 | 0.001062643 | 0.764372527 |
| T2DM_admissi | leukocyte | *FADS2* | -1.387887892 | 0.000379846 | 0.001164428 | 1 |
| T2DM_admissi | leukocyte | *TMEM165* | -0.98307595 | 0.000395725 | 0.001181845 | 1 |
| T2DM_admissi | leukocyte | *CLOCK* | -1.422228349 | 0.000288402 | 0.001088708 | 1 |
| T2DM_admissi | leukocyte | *GGACT* | -1.978415701 | 0.000394526 | 0.001181536 | 1 |
| T2DM_admissi | leukocyte | *CLDN10* | -0.470291776 | 0.00403532 | 0.005694962 | 1 |
| T2DM_admissi | leukocyte | *CARS2* | -1.262071864 | 0.000116357 | 0.001062643 | 0.457164898 |
| T2DM_admissi | leukocyte | *TMED7* | -1.095714157 | 0.000234091 | 0.001066706 | 0.919745256 |
| T2DM_admissi | leukocyte | *NREP* | -7.059101382 | 5.48E-05 | 0.001060479 | 0.215184128 |
| T2DM_admissi | leukocyte | *WDR36* | -1.22505921 | 0.000664312 | 0.001467987 | 1 |
| T2DM_admissi | leukocyte | *OSTF1* | -1.591051281 | 0.000850518 | 0.001707555 | 1 |
| T2DM_admissi | leukocyte | *GOLM1* | -0.905030727 | 0.00049712 | 0.001292643 | 1 |
| T2DM_admissi | leukocyte | *SDS* | 0.068840912 | 0.007928381 | 0.01053097 | 1 |

| | | | | | | |
|---|---|---|---|---|---|---|
| T2DM_admissi | leukocyte | *MSI1* | -2.104712703 | 0.000302921 | 0.001093914 | 1 |
| T2DM_admissi | leukocyte | *TBX3* | -1.85547274 | 0.022651031 | 0.027872189 | 1 |
| T2DM_admissi | leukocyte | *DTX1* | -2.968236399 | 0.000527803 | 0.001320853 | 1 |
| T2DM_admissi | leukocyte | *PNPLA8* | -0.94930456 | 0.000418982 | 0.001200714 | 1 |
| T2DM_admissi | leukocyte | *TES* | -0.772854273 | 0.000762808 | 0.001593931 | 1 |
| T2DM_admissi | leukocyte | *MDFIC* | -2.935459563 | 0.000535619 | 0.001332771 | 1 |
| T2DM_admissi | leukocyte | *MTO1* | -1.0831746 | 0.00089064 | 0.001763074 | 1 |
| T2DM_admissi | leukocyte | *LCA5* | -3.39723377 | 6.37E-05 | 0.001060479 | 0.250087316 |
| T2DM_admissi | leukocyte | *LMO2* | -1.054358091 | 0.000518093 | 0.00131413 | 1 |
| T2DM_admissi | leukocyte | *RDH5* | -1.005020976 | 0.00092827 | 0.001800431 | 1 |
| T2DM_admissi | leukocyte | *TFCP2* | -3.872271602 | 4.06E-05 | 0.001037577 | 0.159436432 |
| T2DM_admissi | leukocyte | *KRT7* | -0.509179057 | 0.002374896 | 0.00356552 | 1 |
| T2DM_admissi | leukocyte | *NHSL1* | -1.249402839 | 0.007326706 | 0.00979137 | 1 |
| T2DM_admissi | leukocyte | *AHI1* | -0.694981771 | 0.003926363 | 0.005559164 | 1 |
| T2DM_admissi | leukocyte | *HEY2* | -1.668168283 | 0.000813876 | 0.001659956 | 1 |
| T2DM_admissi | leukocyte | *PKIB* | -0.827274587 | 0.000564098 | 0.001362351 | 1 |
| T2DM_admissi | leukocyte | *SMPD2* | -3.532859914 | 2.02E-05 | 0.000922424 | 0.079328428 |
| T2DM_admissi | leukocyte | *SEMA4F* | -1.424014927 | 0.002939045 | 0.004295947 | 1 |
| T2DM_admissi | leukocyte | *GNS* | -2.400761455 | 4.93E-05 | 0.00105496 | 0.193716632 |
| T2DM_admissi | leukocyte | *MPHOSPH6* | -0.979579146 | 0.001435869 | 0.002354561 | 1 |
| T2DM_admissi | leukocyte | *AGT* | -0.717997197 | 0.00112093 | 0.002027359 | 1 |
| T2DM_admissi | leukocyte | *NTPCR* | -1.656395741 | 0.00021747 | 0.001062643 | 0.854438404 |
| T2DM_admissi | leukocyte | *USP37* | -1.350102105 | 0.001985756 | 0.003054843 | 1 |
| T2DM_admissi | leukocyte | *ARMC9* | -5.645997619 | 0.000382595 | 0.001165285 | 1 |
| T2DM_admissi | leukocyte | *COX5B* | -0.955574617 | 0.000248454 | 0.001077213 | 0.976174317 |
| T2DM_admissi | leukocyte | *REV1* | -1.090222981 | 0.000573219 | 0.001370771 | 1 |
| T2DM_admissi | leukocyte | *TSGA10* | -1.097311688 | 0.003497525 | 0.005013416 | 1 |
| T2DM_admissi | leukocyte | *TGFBRAP1* | -1.451305028 | 0.000182926 | 0.001062643 | 0.718715771 |
| T2DM_admissi | leukocyte | *GCC2* | -1.450160976 | 5.26E-05 | 0.00105496 | 0.2066448 |
| T2DM_admissi | leukocyte | *MRPS9* | -1.001699801 | 0.000803045 | 0.001648465 | 1 |
| T2DM_admissi | leukocyte | *ANKRD36* | -1.547168286 | 0.000560272 | 0.001360512 | 1 |
| T2DM_admissi | leukocyte | *USP44* | -1.506310493 | 0.001200032 | 0.002112301 | 1 |
| T2DM_admissi | leukocyte | *DRAM1* | -2.41796005 | 0.00011213 | 0.001062643 | 0.440557988 |
| T2DM_admissi | leukocyte | *VILL* | 1.254749787 | 0.019537855 | 0.024354135 | 1 |
| T2DM_admissi | leukocyte | *FLNB* | -0.849048521 | 0.00338513 | 0.004870075 | 1 |
| T2DM_admissi | leukocyte | *NEK3* | -0.643771953 | 0.012481241 | 0.016010663 | 1 |
| T2DM_admissi | leukocyte | *VPS36* | -1.912075863 | 0.000111441 | 0.001062643 | 0.437853386 |
| T2DM_admissi | leukocyte | *RNASEH2B* | -2.378468283 | 0.000117485 | 0.001062643 | 0.461598651 |
| T2DM_admissi | leukocyte | *CKAP2* | -0.842985948 | 0.015231496 | 0.019267401 | 1 |
| T2DM_admissi | leukocyte | *TBC1D4* | -1.25880329 | 0.000410023 | 0.001188264 | 1 |
| T2DM_admissi | leukocyte | *SUCLA2* | -1.484263248 | 9.40E-05 | 0.001062643 | 0.369412097 |
| T2DM_admissi | leukocyte | *RCBTB1* | -1.678425217 | 0.00162833 | 0.002593316 | 1 |
| T2DM_admissi | leukocyte | *LMO7* | -0.658064272 | 0.000830244 | 0.001678862 | 1 |
| T2DM_admissi | leukocyte | *C7orf25* | -1.054825067 | 0.00043008 | 0.00121662 | 1 |
| T2DM_admissi | leukocyte | *RAPGEF5* | -0.789971888 | 0.001251948 | 0.002156134 | 1 |
| T2DM_admissi | leukocyte | *KDELR2* | -1.101821505 | 0.000189568 | 0.001062643 | 0.744810935 |
| T2DM_admissi | leukocyte | *TBRG4* | -2.837089027 | 2.82E-05 | 0.001005591 | 0.11061503 |
| T2DM_admissi | leukocyte | *DBNL* | -3.894429377 | 2.12E-05 | 0.000929374 | 0.083187896 |
| T2DM_admissi | leukocyte | *ALPK3* | -0.850068339 | 0.000641391 | 0.001440839 | 1 |
| T2DM_admissi | leukocyte | *CIB2* | -1.196078254 | 0.003410828 | 0.004903456 | 1 |

| | | | | | | |
|---|---|---|---|---|---|---|
| T2DM_admissi | leukocyte | *NMT1* | -0.890646216 | 0.001886807 | 0.002928987 | 1 |
| T2DM_admissi | leukocyte | *TEX2* | -0.788257277 | 0.001486772 | 0.002415851 | 1 |
| T2DM_admissi | leukocyte | *LIMD2* | 0.819875412 | 0.006514919 | 0.008790219 | 1 |
| T2DM_admissi | leukocyte | *MRPL47* | -1.149415236 | 0.000222812 | 0.001062643 | 0.875429311 |
| T2DM_admissi | leukocyte | *SMPD4* | -3.019632001 | 4.87E-05 | 0.00105496 | 0.191417401 |
| T2DM_admissi | leukocyte | *CCDC115* | -1.261181669 | 0.000585605 | 0.001382291 | 1 |
| T2DM_admissi | leukocyte | *BIN1* | -2.768970682 | 0.000248471 | 0.001077213 | 0.976241107 |
| T2DM_admissi | leukocyte | *HS6ST1* | -1.773041907 | 0.000199083 | 0.001062643 | 0.782198214 |
| T2DM_admissi | leukocyte | *GYPC* | -1.082474025 | 0.001801937 | 0.002818981 | 1 |
| T2DM_admissi | leukocyte | *NIPSNAP3A* | -1.607497417 | 0.000638509 | 0.001438504 | 1 |
| T2DM_admissi | leukocyte | *CDK9* | -1.390788826 | 0.00040951 | 0.001188264 | 1 |
| T2DM_admissi | leukocyte | *TXN* | -0.867876425 | 0.000245505 | 0.001077213 | 0.9645884 |
| T2DM_admissi | leukocyte | *TOR1B* | -1.427439519 | 0.007977027 | 0.010588425 | 1 |
| T2DM_admissi | leukocyte | *TOR1A* | -1.04945802 | 0.000960081 | 0.001831992 | 1 |
| T2DM_admissi | leukocyte | *SLC2A8* | -1.245306063 | 0.000572093 | 0.001370771 | 1 |
| T2DM_admissi | leukocyte | *ZFP37* | -0.862923655 | 0.007041011 | 0.009435242 | 1 |
| T2DM_admissi | leukocyte | *STX17* | -1.126844414 | 0.00043147 | 0.001216752 | 1 |
| T2DM_admissi | leukocyte | *PRPF4* | -1.068908761 | 0.01764321 | 0.022083521 | 1 |
| T2DM_admissi | leukocyte | *FPGS* | -1.49959651 | 0.000398485 | 0.001183526 | 1 |
| T2DM_admissi | leukocyte | *ATP6V1G1* | -0.893341757 | 0.000356709 | 0.001141109 | 1 |
| T2DM_admissi | leukocyte | *ARPC5L* | -0.911946888 | 0.000350549 | 0.001141109 | 1 |
| T2DM_admissi | leukocyte | *POLR1E* | -0.844902513 | 0.000559375 | 0.001360165 | 1 |
| T2DM_admissi | leukocyte | *APTX* | -1.014417326 | 0.000355911 | 0.001141109 | 1 |
| T2DM_admissi | leukocyte | *CD72* | -2.135295753 | 0.00090773 | 0.001777903 | 1 |
| T2DM_admissi | leukocyte | *TMEM8B* | -1.507948796 | 0.000541563 | 0.001337398 | 1 |
| T2DM_admissi | leukocyte | *GRHPR* | -1.207650456 | 0.00017328 | 0.001062643 | 0.680817131 |
| T2DM_admissi | leukocyte | *ALDH1B1* | -2.325276179 | 0.000397514 | 0.001183205 | 1 |
| T2DM_admissi | leukocyte | *ARHGEF39* | -4.110511506 | 0.001029444 | 0.001918342 | 1 |
| T2DM_admissi | leukocyte | *PPIL1* | -2.90707901 | 9.18E-05 | 0.001062643 | 0.360759959 |
| T2DM_admissi | leukocyte | *GMPR* | -1.875099205 | 0.000887283 | 0.001758011 | 1 |
| T2DM_admissi | leukocyte | *KIAA0319* | -1.183606834 | 0.001075717 | 0.001968556 | 1 |
| T2DM_admissi | leukocyte | *LRRC1* | -1.07348821 | 0.000606881 | 0.001408212 | 1 |
| T2DM_admissi | leukocyte | *RIPK1* | -1.192116774 | 0.000517994 | 0.00131413 | 1 |
| T2DM_admissi | leukocyte | *TUBB2B* | -0.904731196 | 0.001441915 | 0.002362504 | 1 |
| T2DM_admissi | leukocyte | *UQCC2* | -1.10353636 | 0.000559287 | 0.001360165 | 1 |
| T2DM_admissi | leukocyte | *HMGA1* | -0.830682692 | 0.000521008 | 0.00131812 | 1 |
| T2DM_admissi | leukocyte | *TCF19* | 0.740616301 | 0.018904401 | 0.023601968 | 1 |
| T2DM_admissi | leukocyte | *CLPS* | -0.478683085 | 0.000478786 | 0.001265283 | 1 |
| T2DM_admissi | leukocyte | *MTCH1* | -1.060688859 | 0.000240844 | 0.001072513 | 0.946277674 |
| T2DM_admissi | leukocyte | *ANKRD42* | -1.821973125 | 0.000115081 | 0.001062643 | 0.452152297 |
| T2DM_admissi | leukocyte | *CREBZF* | -1.576281656 | 0.000208286 | 0.001062643 | 0.818355272 |
| T2DM_admissi | leukocyte | *PRCP* | -1.502482678 | 0.000970017 | 0.001846509 | 1 |
| T2DM_admissi | leukocyte | *NARS2* | -1.342960791 | 0.00089858 | 0.001769684 | 1 |
| T2DM_admissi | leukocyte | *TTPA* | -6.827610358 | 0.012867354 | 0.016478668 | 1 |
| T2DM_admissi | leukocyte | *GGH* | -1.450602252 | 0.000612591 | 0.001415201 | 1 |
| T2DM_admissi | leukocyte | *SDCBP* | -1.913648902 | 0.000177374 | 0.001062643 | 0.696902843 |
| T2DM_admissi | leukocyte | *TRPC6* | -2.572769304 | 0.00629731 | 0.008525889 | 1 |
| T2DM_admissi | leukocyte | *MMP7* | -0.425749426 | 0.004019699 | 0.005679035 | 1 |
| T2DM_admissi | leukocyte | *TRIM29* | -2.588815379 | 0.036296006 | 0.043438016 | 1 |
| T2DM_admissi | leukocyte | *FDX1* | -1.234049442 | 0.000167746 | 0.001062643 | 0.659073615 |

| | | | | | | |
|---|---|---|---|---|---|---|
| T2DM_admissi | leukocyte | *C11orf1* | -1.101146565 | 0.000722397 | 0.001544023 | 1 |
| T2DM_admissi | leukocyte | *FXYD2* | -0.719474536 | 8.28E-05 | 0.001062643 | 0.32529608 |
| T2DM_admissi | leukocyte | *CASP1* | 0.369207546 | 0.011123331 | 0.014352567 | 1 |
| T2DM_admissi | leukocyte | *ALKBH8* | -1.538406632 | 0.00106985 | 0.001964163 | 1 |
| T2DM_admissi | leukocyte | *NDUFAF1* | -0.910056521 | 0.024132572 | 0.029537967 | 1 |
| T2DM_admissi | leukocyte | *TUBGCP4* | -2.176158227 | 2.24E-05 | 0.000945312 | 0.087914044 |
| T2DM_admissi | leukocyte | *UACA* | -0.991901576 | 0.000905502 | 0.001776194 | 1 |
| T2DM_admissi | leukocyte | *PLCB2* | 0.623213361 | 0.007021127 | 0.009411808 | 1 |
| T2DM_admissi | leukocyte | *PAK6* | -0.744701687 | 0.000171244 | 0.001062643 | 0.672818401 |
| T2DM_admissi | leukocyte | *ADAM10* | -1.490861587 | 0.000215403 | 0.001062643 | 0.84631817 |
| T2DM_admissi | leukocyte | *DUOX1* | -1.046311539 | 0.014886983 | 0.01886805 | 1 |
| T2DM_admissi | leukocyte | *CYP19A1* | -6.060584827 | 0.041725615 | 0.049568731 | 1 |
| T2DM_admissi | leukocyte | *ZNF280D* | -1.479594995 | 0.00019234 | 0.001062643 | 0.755705576 |
| T2DM_admissi | leukocyte | *RSL24D1* | -1.158328125 | 0.000118304 | 0.001062643 | 0.464816961 |
| T2DM_admissi | leukocyte | *SPTBN5* | 2.238051489 | 0.023412327 | 0.028736967 | 1 |
| T2DM_admissi | leukocyte | *TTLL7* | -1.100078467 | 0.000600125 | 0.001399402 | 1 |
| T2DM_admissi | leukocyte | *SLC44A5* | -1.506439747 | 0.004682697 | 0.006501172 | 1 |
| T2DM_admissi | leukocyte | *RTCA* | -2.748943069 | 0.000353034 | 0.001141109 | 1 |
| T2DM_admissi | leukocyte | *KHK* | -4.658997144 | 0.000116076 | 0.001062643 | 0.456064063 |
| T2DM_admissi | leukocyte | *DYNC2LI1* | -1.323832114 | 0.000338108 | 0.001134436 | 1 |
| T2DM_admissi | leukocyte | *ACTR2* | -1.115878296 | 0.000116172 | 0.001062643 | 0.456438157 |
| T2DM_admissi | leukocyte | *ATRAID* | -1.111873607 | 0.000311489 | 0.001105548 | 1 |
| T2DM_admissi | leukocyte | *LRPPRC* | -1.265795529 | 0.000446806 | 0.001232235 | 1 |
| T2DM_admissi | leukocyte | *DUSP5* | -2.261191019 | 8.35E-05 | 0.001062643 | 0.328202187 |
| T2DM_admissi | leukocyte | *CALHM2* | 0.943767477 | 0.03038086 | 0.036705534 | 1 |
| T2DM_admissi | leukocyte | *ARL3* | -1.123928362 | 0.000120969 | 0.001062643 | 0.475289118 |
| T2DM_admissi | leukocyte | *PLCE1* | -1.063575097 | 0.008520223 | 0.011244863 | 1 |
| T2DM_admissi | leukocyte | *DNAJC13* | -0.940276108 | 0.002075931 | 0.003167503 | 1 |
| T2DM_admissi | leukocyte | *FAM149B1* | -1.181630839 | 0.000934216 | 0.001804129 | 1 |
| T2DM_admissi | leukocyte | *NCOA4* | -2.518350659 | 6.49E-05 | 0.001060479 | 0.254877796 |
| T2DM_admissi | leukocyte | *ASCC1* | -1.330226347 | 0.000740494 | 0.001562515 | 1 |
| T2DM_admissi | leukocyte | *RPS24* | -0.833078467 | 0.000150017 | 0.001062643 | 0.589416554 |
| T2DM_admissi | leukocyte | *DNA2* | -1.727388378 | 0.001854982 | 0.002885283 | 1 |
| T2DM_admissi | leukocyte | *MYPN* | 5.630758629 | 0.000438816 | 0.001222199 | 1 |
| T2DM_admissi | leukocyte | *ATIC* | -2.027053877 | 0.000136222 | 0.001062643 | 0.535214749 |
| T2DM_admissi | leukocyte | *SMARCAL1* | -1.475013669 | 0.001246252 | 0.002154446 | 1 |
| T2DM_admissi | leukocyte | *BARD1* | -1.286290619 | 0.000536477 | 0.001334061 | 1 |
| T2DM_admissi | leukocyte | *FASTKD1* | -1.495236989 | 0.000225624 | 0.001062643 | 0.886474765 |
| T2DM_admissi | leukocyte | *FAM117B* | -1.420298888 | 0.00052653 | 0.001319879 | 1 |
| T2DM_admissi | leukocyte | *WDR12* | -1.525126095 | 0.001019231 | 0.001906024 | 1 |
| T2DM_admissi | leukocyte | *SLC35A5* | -1.520474211 | 0.000929729 | 0.001800431 | 1 |
| T2DM_admissi | leukocyte | *USP8* | -1.491875732 | 0.000137836 | 0.001062643 | 0.541557876 |
| T2DM_admissi | leukocyte | *SPPL2A* | -1.301407532 | 0.000158862 | 0.001062643 | 0.624170074 |
| T2DM_admissi | leukocyte | *APH1B* | -1.903628823 | 0.000173017 | 0.001062643 | 0.679783598 |
| T2DM_admissi | leukocyte | *HCN4* | -1.283716773 | 0.00515737 | 0.007105328 | 1 |
| T2DM_admissi | leukocyte | *SEMA7A* | -2.56228247 | 0.001262475 | 0.002166065 | 1 |
| T2DM_admissi | leukocyte | *AP1AR* | -0.972301492 | 0.000623395 | 0.00142568 | 1 |
| T2DM_admissi | leukocyte | *BBS7* | -1.477146777 | 0.000232374 | 0.001066588 | 0.912999084 |
| T2DM_admissi | leukocyte | *RAP1GDS1* | -1.418355453 | 0.000338507 | 0.001134808 | 1 |
| T2DM_admissi | leukocyte | *PRDM5* | -1.073607959 | 0.001435724 | 0.002354561 | 1 |

| | | | | | | |
|---|---|---|---|---|---|---|
| T2DM_admissi | leukocyte | *NUP54* | -0.754425646 | 0.012867537 | 0.016478668 | 1 |
| T2DM_admissi | leukocyte | *G3BP2* | -0.982050502 | 0.000222806 | 0.001062643 | 0.875406107 |
| T2DM_admissi | leukocyte | *SCARB2* | -1.106321371 | 0.00024648 | 0.001077213 | 0.968419842 |
| T2DM_admissi | leukocyte | *CNOT6L* | -1.542352476 | 0.000102801 | 0.001062643 | 0.403905944 |
| T2DM_admissi | leukocyte | *USO1* | -0.976630713 | 0.00027881 | 0.001088708 | 1 |
| T2DM_admissi | leukocyte | *SHROOM3* | -2.201076836 | 0.000158099 | 0.001062643 | 0.621170776 |
| T2DM_admissi | leukocyte | *ANXA3* | -0.825102034 | 0.000395938 | 0.001181845 | 1 |
| T2DM_admissi | leukocyte | *PPA2* | -1.153703664 | 0.000340011 | 0.001135007 | 1 |
| T2DM_admissi | leukocyte | *MAPK8IP3* | -1.229710916 | 0.000517255 | 0.00131413 | 1 |
| T2DM_admissi | leukocyte | *RGS3* | -0.97246758 | 0.001167216 | 0.002082648 | 1 |
| T2DM_admissi | leukocyte | *ETV6* | -2.248326134 | 0.000310416 | 0.00110427 | 1 |
| T2DM_admissi | leukocyte | *ALG10* | -2.000498707 | 0.000903472 | 0.001773984 | 1 |
| T2DM_admissi | leukocyte | *SLCO1C1* | -17.787384 | 0.000845768 | 0.001700095 | 1 |
| T2DM_admissi | leukocyte | *ZCRB1* | -1.076330037 | 0.000199669 | 0.001062643 | 0.784499777 |
| T2DM_admissi | leukocyte | *C1RL* | -0.954128371 | 0.026116693 | 0.031756332 | 1 |
| T2DM_admissi | leukocyte | *CLSTN3* | -1.832690452 | 0.000371405 | 0.001152986 | 1 |
| T2DM_admissi | leukocyte | *KLRG1* | -6.887882011 | 0.001616298 | 0.002578334 | 1 |
| T2DM_admissi | leukocyte | *TAPBPL* | -0.783652113 | 0.013705866 | 0.017500925 | 1 |
| T2DM_admissi | leukocyte | *PEX5* | -1.127408759 | 0.000671083 | 0.001477964 | 1 |
| T2DM_admissi | leukocyte | *SLC38A4* | -1.479828505 | 0.002083931 | 0.003176014 | 1 |
| T2DM_admissi | leukocyte | *LRIG3* | -1.130988053 | 0.000811614 | 0.001657387 | 1 |
| T2DM_admissi | leukocyte | *GLIPR1* | -1.354315633 | 0.000469168 | 0.001253988 | 1 |
| T2DM_admissi | leukocyte | *SNRPF* | -0.927579791 | 0.00041857 | 0.001200552 | 1 |
| T2DM_admissi | leukocyte | *AMDHD1* | -1.344804387 | 0.032610662 | 0.039218638 | 1 |
| T2DM_admissi | leukocyte | *ASCL1* | -1.623816702 | 0.002263383 | 0.003416378 | 1 |
| T2DM_admissi | leukocyte | *RITA1* | -1.735045318 | 0.000290077 | 0.001088708 | 1 |
| T2DM_admissi | leukocyte | *GLTP* | -0.92624243 | 0.00052385 | 0.001319879 | 1 |
| T2DM_admissi | leukocyte | *MTMR6* | -1.484493775 | 0.000329628 | 0.001123865 | 1 |
| T2DM_admissi | leukocyte | *PDX1* | -1.610816008 | 0.000301206 | 0.001093914 | 1 |
| T2DM_admissi | leukocyte | *SUOX* | -1.026863164 | 0.010260724 | 0.013331476 | 1 |
| T2DM_admissi | leukocyte | *CCDC65* | -1.494268627 | 0.006162619 | 0.008367978 | 1 |
| T2DM_admissi | leukocyte | *SLC39A5* | -0.589463874 | 0.008770429 | 0.011524754 | 1 |
| T2DM_admissi | leukocyte | *KANSL2* | -2.948852334 | 6.12E-05 | 0.001060479 | 0.240409004 |
| T2DM_admissi | leukocyte | *CERS5* | -1.719815063 | 3.98E-05 | 0.001037577 | 0.15643039 |
| T2DM_admissi | leukocyte | *ZNF740* | -2.361802635 | 0.000723085 | 0.001544023 | 1 |
| T2DM_admissi | leukocyte | *WDFY2* | -1.359027915 | 0.000279569 | 0.001088708 | 1 |
| T2DM_admissi | leukocyte | *GRTP1* | -2.480013545 | 0.001005915 | 0.001889217 | 1 |
| T2DM_admissi | leukocyte | *TSSK4* | -9.407873864 | 0.003899775 | 0.005528505 | 1 |
| T2DM_admissi | leukocyte | *TMX1* | -1.742101815 | 0.000113325 | 0.001062643 | 0.445254978 |
| T2DM_admissi | leukocyte | *RAB15* | -1.648444242 | 0.000508805 | 0.001305383 | 1 |
| T2DM_admissi | leukocyte | *PTGR2* | -0.852743035 | 0.001735333 | 0.002732715 | 1 |
| T2DM_admissi | leukocyte | *NIPA2* | -0.951483969 | 0.000585778 | 0.001382291 | 1 |
| T2DM_admissi | leukocyte | *SERF2* | -0.960801483 | 6.31E-05 | 0.001060479 | 0.247915902 |
| T2DM_admissi | leukocyte | *BNIP2* | -2.015458774 | 7.51E-05 | 0.001062643 | 0.295088867 |
| T2DM_admissi | leukocyte | *GTF2A2* | -1.075957273 | 0.000217668 | 0.001062643 | 0.855217186 |
| T2DM_admissi | leukocyte | *SRP14* | -0.868669859 | 0.000247525 | 0.001077213 | 0.972525005 |
| T2DM_admissi | leukocyte | *BAHD1* | -1.344853681 | 0.000435788 | 0.001222199 | 1 |
| T2DM_admissi | leukocyte | *DISP2* | -0.759316893 | 0.006656438 | 0.008971919 | 1 |
| T2DM_admissi | leukocyte | *TLE3* | -2.898950157 | 2.13E-05 | 0.000929374 | 0.083643621 |
| T2DM_admissi | leukocyte | *SCAPER* | -1.033155065 | 0.000537953 | 0.001334229 | 1 |

| | | | | | | |
|---|---|---|---|---|---|---|
| T2DM_admissi | leukocyte | *TSPAN3* | -0.924194381 | 0.000287617 | 0.001088708 | 1 |
| T2DM_admissi | leukocyte | *MAN2C1* | -1.765243984 | 5.31E-08 | 2.09E-05 | 0.000208769 |
| T2DM_admissi | leukocyte | *DNAJA4* | -1.036262369 | 0.000403325 | 0.001185781 | 1 |
| T2DM_admissi | leukocyte | *TPM1* | -0.638141619 | 0.000617273 | 0.001419945 | 1 |
| T2DM_admissi | leukocyte | *IGF1R* | -0.998020375 | 0.000290486 | 0.001088708 | 1 |
| T2DM_admissi | leukocyte | *ADAMTS17* | -2.194978135 | 0.007398356 | 0.009873688 | 1 |
| T2DM_admissi | leukocyte | *ULK3* | -1.178723838 | 0.000471499 | 0.001255095 | 1 |
| T2DM_admissi | leukocyte | *CELF6* | -1.355372562 | 0.00068066 | 0.00149096 | 1 |
| T2DM_admissi | leukocyte | *HAPLN3* | -5.715834522 | 0.022943881 | 0.028197219 | 1 |
| T2DM_admissi | leukocyte | *POLG* | -1.856019572 | 0.000286627 | 0.001088708 | 1 |
| T2DM_admissi | leukocyte | *WDR93* | -3.643297849 | 0.006071877 | 0.008263389 | 1 |
| T2DM_admissi | leukocyte | *UNC45A* | -1.331082497 | 0.000222244 | 0.001062643 | 0.873197708 |
| T2DM_admissi | leukocyte | *IQGAP1* | -1.207944568 | 0.000222882 | 0.001062643 | 0.875703158 |
| T2DM_admissi | leukocyte | *PMM2* | -1.15046259 | 0.000457791 | 0.001240899 | 1 |
| T2DM_admissi | leukocyte | *SLC5A2* | -3.830548362 | 0.003808338 | 0.005413516 | 1 |
| T2DM_admissi | leukocyte | *UQCRC2* | -1.28650773 | 0.000158857 | 0.001062643 | 0.624148359 |
| T2DM_admissi | leukocyte | *ARHGAP17* | -3.113989739 | 5.24E-05 | 0.00105496 | 0.205701756 |
| T2DM_admissi | leukocyte | *TXNL4B* | -2.913567809 | 7.89E-05 | 0.001062643 | 0.309919203 |
| T2DM_admissi | leukocyte | *MARVELD3* | -0.872524087 | 0.002614905 | 0.003871124 | 1 |
| T2DM_admissi | leukocyte | *KIFC3* | -1.35536879 | 0.00021976 | 0.001062643 | 0.863436028 |
| T2DM_admissi | leukocyte | *NUDT7* | -0.975134668 | 0.00141961 | 0.00234189 | 1 |
| T2DM_admissi | leukocyte | *GCSH* | -1.12192738 | 0.000237896 | 0.001071897 | 0.934694307 |
| T2DM_admissi | leukocyte | *MAP1LC3B* | -0.974285937 | 0.000225936 | 0.001062643 | 0.887703987 |
| T2DM_admissi | leukocyte | *OSGIN1* | -0.86520937 | 0.000996946 | 0.001875957 | 1 |
| T2DM_admissi | leukocyte | *RPL3L* | -2.009349955 | 0.010123525 | 0.013165959 | 1 |
| T2DM_admissi | leukocyte | *RPS2* | -0.767631176 | 0.000145556 | 0.001062643 | 0.571888236 |
| T2DM_admissi | leukocyte | *PDPK1* | -1.46206059 | 0.000376118 | 0.001159218 | 1 |
| T2DM_admissi | leukocyte | *DEF8* | -1.359892724 | 0.000161735 | 0.001062643 | 0.635458179 |
| T2DM_admissi | leukocyte | *GAS8* | -2.230240651 | 9.13E-05 | 0.001062643 | 0.358689341 |
| T2DM_admissi | leukocyte | *RANBP10* | -0.931132959 | 0.005308192 | 0.007297371 | 1 |
| T2DM_admissi | leukocyte | *PRPSAP2* | -1.346159487 | 0.000975375 | 0.001854014 | 1 |
| T2DM_admissi | leukocyte | *TOM1L1* | -1.038024123 | 0.001152361 | 0.002063641 | 1 |
| T2DM_admissi | leukocyte | *C17orf80* | -1.658571488 | 0.002740338 | 0.004032504 | 1 |
| T2DM_admissi | leukocyte | *VPS53* | -1.387335499 | 4.72E-05 | 0.00105496 | 0.185384912 |
| T2DM_admissi | leukocyte | *NPEPPS* | -1.663574231 | 0.000129884 | 0.001062643 | 0.510312546 |
| T2DM_admissi | leukocyte | *SCRN2* | -2.38743929 | 7.51E-05 | 0.001062643 | 0.29491828 |
| T2DM_admissi | leukocyte | *RHBDL3* | -1.725274541 | 0.014011357 | 0.017861979 | 1 |
| T2DM_admissi | leukocyte | *SPACA3* | -1.26002807 | 6.35E-05 | 0.001060479 | 0.249378213 |
| T2DM_admissi | leukocyte | *ARSG* | -3.955638264 | 0.001084188 | 0.001978529 | 1 |
| T2DM_admissi | leukocyte | *PTRH2* | -1.299380959 | 0.00041362 | 0.001193182 | 1 |
| T2DM_admissi | leukocyte | *GNAL* | -1.532355483 | 0.001922619 | 0.002976348 | 1 |
| T2DM_admissi | leukocyte | *RPRD1A* | -1.155150813 | 0.000471165 | 0.001255055 | 1 |
| T2DM_admissi | leukocyte | *NPC1* | -3.17574767 | 7.17E-07 | 0.000156607 | 0.002818928 |
| T2DM_admissi | leukocyte | *SLC14A1* | -1.763733851 | 0.000986742 | 0.001867193 | 1 |
| T2DM_admissi | leukocyte | *ZMYND15* | 0.217951543 | 0.01103959 | 0.014258563 | 1 |
| T2DM_admissi | leukocyte | *SAT2* | -1.478350232 | 8.31E-05 | 0.001062643 | 0.326557984 |
| T2DM_admissi | leukocyte | *CCDC40* | -0.954026113 | 0.00829845 | 0.010992788 | 1 |
| T2DM_admissi | leukocyte | *ARHGDIA* | -1.044217743 | 0.000572315 | 0.001370771 | 1 |
| T2DM_admissi | leukocyte | *TMC6* | -1.762183859 | 0.001140789 | 0.002049538 | 1 |
| T2DM_admissi | leukocyte | *TTYH2* | -1.941464794 | 0.001270828 | 0.002175059 | 1 |

| | | | | | | |
|---|---|---|---|---|---|---|
| T2DM_admissi | leukocyte | *EIF4A3* | -1.267283501 | 0.000430434 | 0.001216672 | 1 |
| T2DM_admissi | leukocyte | *TBCD* | -1.731915111 | 0.001966969 | 0.003031864 | 1 |
| T2DM_admissi | leukocyte | *FN3KRP* | -1.994306512 | 5.13E-06 | 0.00058206 | 0.020168175 |
| T2DM_admissi | leukocyte | *NARF* | -1.684196952 | 0.00015697 | 0.001062643 | 0.616733414 |
| T2DM_admissi | leukocyte | *TRIM65* | -1.791914269 | 0.000434888 | 0.001222199 | 1 |
| T2DM_admissi | leukocyte | *RNF157* | -1.338657254 | 0.000539178 | 0.001334229 | 1 |
| T2DM_admissi | leukocyte | *WDR45B* | -0.993771488 | 0.000317077 | 0.001111111 | 1 |
| T2DM_admissi | leukocyte | *TNFRSF11A* | -1.004731991 | 0.001141357 | 0.002049538 | 1 |
| T2DM_admissi | leukocyte | *ZCCHC2* | -1.654812454 | 0.000214564 | 0.001062643 | 0.843023495 |
| T2DM_admissi | leukocyte | *PMAIP1* | -1.048230369 | 0.00079382 | 0.001634654 | 1 |
| T2DM_admissi | leukocyte | *NT5C3B* | -1.187725724 | 0.000569529 | 0.001367774 | 1 |
| T2DM_admissi | leukocyte | *PIP4K2B* | -1.056249781 | 0.001462159 | 0.002383744 | 1 |
| T2DM_admissi | leukocyte | *TXNL4A* | -1.08207845 | 0.00015903 | 0.001062643 | 0.624828441 |
| T2DM_admissi | leukocyte | *PRDM15* | -1.148983772 | 0.017458504 | 0.021887193 | 1 |
| T2DM_admissi | leukocyte | *MVB12A* | -1.024512353 | 0.011796018 | 0.015165757 | 1 |
| T2DM_admissi | leukocyte | *SH3GL1* | -1.162201159 | 0.000154263 | 0.001062643 | 0.606097776 |
| T2DM_admissi | leukocyte | *DUS3L* | -0.571253428 | 0.031361559 | 0.037774238 | 1 |
| T2DM_admissi | leukocyte | *SIRT3* | -1.569160481 | 0.000690507 | 0.001503881 | 1 |
| T2DM_admissi | leukocyte | *IFITM3* | -0.751621108 | 0.000449223 | 0.001234067 | 1 |
| T2DM_admissi | leukocyte | *COL6A1* | -1.715350837 | 8.00E-05 | 0.001062643 | 0.314508198 |
| T2DM_admissi | leukocyte | *IFNAR1* | -1.221086614 | 6.02E-05 | 0.001060479 | 0.23669113 |
| T2DM_admissi | leukocyte | *TMEM50B* | -0.834687084 | 0.011684702 | 0.01503248 | 1 |
| T2DM_admissi | leukocyte | *AKT1* | -1.574514128 | 0.000871616 | 0.001738366 | 1 |
| T2DM_admissi | leukocyte | *CBLC* | -1.296777581 | 0.000144717 | 0.001062643 | 0.568592915 |
| T2DM_admissi | leukocyte | *EVI5L* | -0.977524978 | 0.023434634 | 0.028755365 | 1 |
| T2DM_admissi | leukocyte | *CTU1* | -1.998753488 | 0.000152551 | 0.001062643 | 0.599373177 |
| T2DM_admissi | leukocyte | *RERE* | -0.981668501 | 0.000396455 | 0.001181845 | 1 |
| T2DM_admissi | leukocyte | *FHAD1* | -1.018344806 | 0.003907609 | 0.005536602 | 1 |
| T2DM_admissi | leukocyte | *ARHGEF19* | -2.064105641 | 0.001736146 | 0.002732899 | 1 |
| T2DM_admissi | leukocyte | *PGD* | -1.553169377 | 0.000145289 | 0.001062643 | 0.570841031 |
| T2DM_admissi | leukocyte | *CNKSR1* | -1.879556239 | 0.002869946 | 0.004209039 | 1 |
| T2DM_admissi | leukocyte | *ZNF593* | -1.163080371 | 0.00017062 | 0.001062643 | 0.670367715 |
| T2DM_admissi | leukocyte | *FCN3* | -0.554346945 | 0.004213681 | 0.005919039 | 1 |
| T2DM_admissi | leukocyte | *CELA3A* | -0.498332796 | 0.000716722 | 0.00153628 | 1 |
| T2DM_admissi | leukocyte | *NBPF3* | -1.206762638 | 0.002931971 | 0.004287203 | 1 |
| T2DM_admissi | leukocyte | *HSPG2* | -0.695554195 | 0.00021941 | 0.001062643 | 0.862063592 |
| T2DM_admissi | leukocyte | *ITGB3BP* | -0.999059486 | 0.003274579 | 0.004723135 | 1 |
| T2DM_admissi | leukocyte | *PRKACB* | -1.390004531 | 0.000193632 | 0.001062643 | 0.760781618 |
| T2DM_admissi | leukocyte | *PIGK* | -2.058041795 | 0.00011491 | 0.001062643 | 0.451479528 |
| T2DM_admissi | leukocyte | *RPS8* | -0.707771215 | 0.000290926 | 0.001088708 | 1 |
| T2DM_admissi | leukocyte | *SYPL2* | -3.403521086 | 0.000746877 | 0.001568868 | 1 |
| T2DM_admissi | leukocyte | *SLC44A3* | -1.067844931 | 0.001283311 | 0.00218274 | 1 |
| T2DM_admissi | leukocyte | *ZNF697* | -1.588666921 | 0.002716097 | 0.004001329 | 1 |
| T2DM_admissi | leukocyte | *CELSR2* | -1.318985404 | 0.001150475 | 0.002061203 | 1 |
| T2DM_admissi | leukocyte | *ALDH9A1* | -1.023222027 | 0.000429948 | 0.00121662 | 1 |
| T2DM_admissi | leukocyte | *ATP1B1* | -0.723241232 | 0.000271765 | 0.001088708 | 1 |
| T2DM_admissi | leukocyte | *TIPRL* | -1.034009434 | 0.000322493 | 0.001116501 | 1 |
| T2DM_admissi | leukocyte | *NME7* | -2.072911863 | 1.58E-05 | 0.000823546 | 0.062032466 |
| T2DM_admissi | leukocyte | *MGST3* | -1.210884339 | 9.20E-05 | 0.001062643 | 0.361458169 |
| T2DM_admissi | leukocyte | *LYPLAL1* | -1.40856458 | 0.000299327 | 0.001093914 | 1 |

| | | | | | | |
|---|---|---|---|---|---|---|
| T2DM_admissi | leukocyte | *SNX27* | -1.827986136 | 0.000328764 | 0.001123231 | 1 |
| T2DM_admissi | leukocyte | *CTSK* | -4.20183565 | 5.03E-05 | 0.00105496 | 0.197696363 |
| T2DM_admissi | leukocyte | *SELENBP1* | -1.282639974 | 0.001225924 | 0.002137886 | 1 |
| T2DM_admissi | leukocyte | *POGZ* | -0.929200113 | 0.01048804 | 0.013595351 | 1 |
| T2DM_admissi | leukocyte | *GABPB2* | -1.537660822 | 0.007071553 | 0.009472939 | 1 |
| T2DM_admissi | leukocyte | *SLC27A3* | -1.67170688 | 0.000284853 | 0.001088708 | 1 |
| T2DM_admissi | leukocyte | *KCNN3* | -1.964402961 | 0.001071932 | 0.001966209 | 1 |
| T2DM_admissi | leukocyte | *RIT1* | -2.301885864 | 0.00022341 | 0.001062643 | 0.87777902 |
| T2DM_admissi | leukocyte | *SRP9* | -1.156985432 | 0.000185664 | 0.001062643 | 0.729472241 |
| T2DM_admissi | leukocyte | *CNIH4* | -1.190461013 | 0.00023105 | 0.001065487 | 0.907794983 |
| T2DM_admissi | leukocyte | *PSEN2* | -2.372743932 | 0.00024964 | 0.001077213 | 0.980837293 |
| T2DM_admissi | leukocyte | *PYCR2* | -1.28164539 | 0.000458417 | 0.001240899 | 1 |
| T2DM_admissi | leukocyte | *LBR* | -1.403051129 | 0.000431278 | 0.001216752 | 1 |
| T2DM_admissi | leukocyte | *ARL8A* | -2.059921961 | 3.40E-05 | 0.001037577 | 0.133595061 |
| T2DM_admissi | leukocyte | *EML4* | -2.013844524 | 0.00010264 | 0.001062643 | 0.403271339 |
| T2DM_admissi | leukocyte | *WDPCP* | -3.012157918 | 0.000275449 | 0.001088708 | 1 |
| T2DM_admissi | leukocyte | *REG3G* | -1.396044586 | 6.17E-05 | 0.001060479 | 0.242595904 |
| T2DM_admissi | leukocyte | *CIAO1* | -1.721302756 | 7.90E-05 | 0.001062643 | 0.310306635 |
| T2DM_admissi | leukocyte | *ZNF514* | -1.39684746 | 8.59E-05 | 0.001062643 | 0.33731471 |
| T2DM_admissi | leukocyte | *TPRKB* | -1.052741496 | 0.000241111 | 0.001072513 | 0.947324343 |
| T2DM_admissi | leukocyte | *TEX261* | -1.342675458 | 0.000328386 | 0.001122913 | 1 |
| T2DM_admissi | leukocyte | *RALB* | -1.191639907 | 0.000537952 | 0.001334229 | 1 |
| T2DM_admissi | leukocyte | *RABL2A* | -1.532999515 | 0.002469624 | 0.003686608 | 1 |
| T2DM_admissi | leukocyte | *SLC20A1* | -1.803145526 | 0.000319545 | 0.001113024 | 1 |
| T2DM_admissi | leukocyte | *LYG1* | -5.086000046 | 0.007889432 | 0.010486326 | 1 |
| T2DM_admissi | leukocyte | *PHOSPHO2* | -1.640468732 | 0.026784241 | 0.032490053 | 1 |
| T2DM_admissi | leukocyte | *GULP1* | -1.105197846 | 0.000695868 | 0.001510534 | 1 |
| T2DM_admissi | leukocyte | *FAM171B* | -1.029190606 | 0.000517763 | 0.00131413 | 1 |
| T2DM_admissi | leukocyte | *METTL21A* | -1.184572136 | 0.000563312 | 0.001362351 | 1 |
| T2DM_admissi | leukocyte | *SUMF1* | -1.67622753 | 0.000208064 | 0.001062643 | 0.817484838 |
| T2DM_admissi | leukocyte | *HES6* | -3.144732111 | 4.18E-05 | 0.001037577 | 0.164102399 |
| T2DM_admissi | leukocyte | *ESPNL* | -10.22297641 | 0.021729763 | 0.026780502 | 1 |
| T2DM_admissi | leukocyte | *TAMM41* | -1.159958944 | 0.005552434 | 0.007609178 | 1 |
| T2DM_admissi | leukocyte | *RAB5A* | -1.11500926 | 0.000298731 | 0.001093914 | 1 |
| T2DM_admissi | leukocyte | *CNTN4* | -1.315850986 | 9.68E-05 | 0.001062643 | 0.380516251 |
| T2DM_admissi | leukocyte | *POMGNT2* | -2.199956163 | 9.84E-05 | 0.001062643 | 0.38664085 |
| T2DM_admissi | leukocyte | *CTDSPL* | -1.073241804 | 0.000508996 | 0.001305383 | 1 |
| T2DM_admissi | leukocyte | *IL17RD* | -4.263855649 | 0.00248512 | 0.003708332 | 1 |
| T2DM_admissi | leukocyte | *UBA3* | -1.816179111 | 0.000155729 | 0.001062643 | 0.611861149 |
| T2DM_admissi | leukocyte | *ARL6IP5* | -1.098313592 | 0.000224524 | 0.001062643 | 0.88215297 |
| T2DM_admissi | leukocyte | *LIMD1* | -1.887715891 | 0.000501955 | 0.001297006 | 1 |
| T2DM_admissi | leukocyte | *ZNF660* | -1.241162542 | 0.006375462 | 0.008617309 | 1 |
| T2DM_admissi | leukocyte | *NXPE3* | -1.065612793 | 0.000920186 | 0.001793131 | 1 |
| T2DM_admissi | leukocyte | *PHLDB2* | -0.836731232 | 0.000767361 | 0.001600298 | 1 |
| T2DM_admissi | leukocyte | *ABHD10* | -1.156948281 | 0.002062824 | 0.003151674 | 1 |
| T2DM_admissi | leukocyte | *TAGLN3* | -0.610153312 | 0.005280969 | 0.007267576 | 1 |
| T2DM_admissi | leukocyte | *EIF2A* | -0.979151071 | 0.000363032 | 0.001144746 | 1 |
| T2DM_admissi | leukocyte | *ALDH1L1* | -1.531094028 | 0.001219947 | 0.002133102 | 1 |
| T2DM_admissi | leukocyte | *TRPC1* | -1.371229193 | 0.000603216 | 0.001403431 | 1 |
| T2DM_admissi | leukocyte | *NCEH1* | -0.692310403 | 0.000990851 | 0.001869863 | 1 |

| | | | | | | |
|---|---|---|---|---|---|---|
| T2DM_admissi | leukocyte | *AMT* | -1.94844889 | 0.000560055 | 0.001360512 | 1 |
| T2DM_admissi | leukocyte | *DGKQ* | -1.131984065 | 0.002505467 | 0.003733022 | 1 |
| T2DM_admissi | leukocyte | *SCD5* | -0.959552072 | 0.000642019 | 0.001441424 | 1 |
| T2DM_admissi | leukocyte | *PYURF* | -0.955406642 | 0.000307613 | 0.001098736 | 1 |
| T2DM_admissi | leukocyte | *TBCK* | -1.081704545 | 0.029448788 | 0.035623241 | 1 |
| T2DM_admissi | leukocyte | *DDIT4L* | -3.053912935 | 0.000403871 | 0.001185781 | 1 |
| T2DM_admissi | leukocyte | *TIFA* | -1.454900506 | 0.000366135 | 0.001147001 | 1 |
| T2DM_admissi | leukocyte | *CYP4V2* | -2.729640126 | 0.000101335 | 0.001062643 | 0.398147047 |
| T2DM_admissi | leukocyte | *SSBP2* | -0.929715135 | 0.000693177 | 0.001507191 | 1 |
| T2DM_admissi | leukocyte | *GTF2H2* | -3.337861488 | 5.17E-05 | 0.00105496 | 0.203200223 |
| T2DM_admissi | leukocyte | *BTF3* | -0.906750989 | 0.000187693 | 0.001062643 | 0.73744639 |
| T2DM_admissi | leukocyte | *SPATA9* | -5.344456316 | 0.037824917 | 0.045185193 | 1 |
| T2DM_admissi | leukocyte | *COMMD10* | -1.340629541 | 0.000505904 | 0.00130085 | 1 |
| T2DM_admissi | leukocyte | *ATG12* | -1.137649812 | 0.000197521 | 0.001062643 | 0.776061393 |
| T2DM_admissi | leukocyte | *PCYOX1L* | -6.659780723 | 4.24E-05 | 0.001037577 | 0.166709596 |
| T2DM_admissi | leukocyte | *TBC1D7* | -4.025830225 | 0.006121747 | 0.008319732 | 1 |
| T2DM_admissi | leukocyte | *ZMAT2* | -1.472742309 | 0.000173545 | 0.001062643 | 0.681858303 |
| T2DM_admissi | leukocyte | *KLHL3* | -5.651439227 | 2.47E-05 | 0.000982543 | 0.097023021 |
| T2DM_admissi | leukocyte | *DCDC2* | -0.835255715 | 0.001057588 | 0.001951744 | 1 |
| T2DM_admissi | leukocyte | *TRIM7* | -4.047696277 | 6.50E-05 | 0.001060479 | 0.255575433 |
| T2DM_admissi | leukocyte | *ANO7* | -3.261386929 | 0.000583463 | 0.001382042 | 1 |
| T2DM_admissi | leukocyte | *RPL7L1* | -1.074749916 | 0.000280818 | 0.001088708 | 1 |
| T2DM_admissi | leukocyte | *IRAK1BP1* | -0.935606099 | 0.001182502 | 0.002099858 | 1 |
| T2DM_admissi | leukocyte | *MMS22L* | -1.620682348 | 0.000746052 | 0.001568868 | 1 |
| T2DM_admissi | leukocyte | *FAXC* | -1.747206274 | 0.000559437 | 0.001360165 | 1 |
| T2DM_admissi | leukocyte | *SCML4* | -5.397170559 | 1.60E-05 | 0.000823546 | 0.062695268 |
| T2DM_admissi | leukocyte | *TBC1D32* | -3.02095262 | 0.004670409 | 0.006486404 | 1 |
| T2DM_admissi | leukocyte | *ABRACL* | -1.102851927 | 0.001178045 | 0.002098083 | 1 |
| T2DM_admissi | leukocyte | *DYNLT1* | -1.005914377 | 0.00015454 | 0.001062643 | 0.607187259 |
| T2DM_admissi | leukocyte | *WTAP* | -0.993355158 | 0.000217538 | 0.001062643 | 0.854706217 |
| T2DM_admissi | leukocyte | *SLC22A3* | -5.579599767 | 0.017336605 | 0.021748251 | 1 |
| T2DM_admissi | leukocyte | *VWDE* | -1.077702926 | 0.001021081 | 0.001908576 | 1 |
| T2DM_admissi | leukocyte | *GNA12* | -1.848729188 | 0.00037412 | 0.001157429 | 1 |
| T2DM_admissi | leukocyte | *SDK1* | -1.317758375 | 0.001284566 | 0.002183928 | 1 |
| T2DM_admissi | leukocyte | *CCZ1B* | -1.943959068 | 0.000197602 | 0.001062643 | 0.776377221 |
| T2DM_admissi | leukocyte | *RBAK* | -2.282948416 | 0.001351129 | 0.002257051 | 1 |
| T2DM_admissi | leukocyte | *POMZP3* | -1.288791903 | 0.000555435 | 0.001357223 | 1 |
| T2DM_admissi | leukocyte | *GIGYF1* | -2.907118033 | 6.32E-06 | 0.000591121 | 0.024827101 |
| T2DM_admissi | leukocyte | *TRIM4* | -1.077949855 | 0.00017557 | 0.001062643 | 0.689815989 |
| T2DM_admissi | leukocyte | *TMEM209* | -1.88031952 | 0.000117163 | 0.001062643 | 0.460334237 |
| T2DM_admissi | leukocyte | *NCAPG2* | 0.853268087 | 0.02190513 | 0.026988164 | 1 |
| T2DM_admissi | leukocyte | *RAB19* | -9.349276857 | 0.000320616 | 0.001113794 | 1 |
| T2DM_admissi | leukocyte | *CSGALNACT1* | -1.00732677 | 0.001069833 | 0.001964163 | 1 |
| T2DM_admissi | leukocyte | *CCDC25* | -0.9612025 | 0.000329174 | 0.001123654 | 1 |
| T2DM_admissi | leukocyte | *BIN3* | -3.410826972 | 0.000123364 | 0.001062643 | 0.484696325 |
| T2DM_admissi | leukocyte | *CHMP7* | -1.630734713 | 0.000814135 | 0.001659956 | 1 |
| T2DM_admissi | leukocyte | *STAR* | 0.686441651 | 0.011456422 | 0.014753288 | 1 |
| T2DM_admissi | leukocyte | *LACTB2* | -1.039165054 | 0.000472339 | 0.001256287 | 1 |
| T2DM_admissi | leukocyte | *TERF1* | -2.474927629 | 4.52E-05 | 0.001037577 | 0.177425645 |
| T2DM_admissi | leukocyte | *LRP12* | -1.193745953 | 0.000327877 | 0.001122427 | 1 |

| | | | | | | |
|---|---|---|---|---|---|---|
| T2DM_admissi | leukocyte | *ZNF7* | -0.975871233 | 0.000825049 | 0.001673837 | 1 |
| T2DM_admissi | leukocyte | *AK3* | -1.054011622 | 0.00027807 | 0.001088708 | 1 |
| T2DM_admissi | leukocyte | *CER1* | 3.614150525 | 0.008794658 | 0.011552729 | 1 |
| T2DM_admissi | leukocyte | *C9orf72* | -1.801768118 | 0.000105854 | 0.001062643 | 0.415901882 |
| T2DM_admissi | leukocyte | *FBXO10* | -1.538131447 | 0.002076734 | 0.003167503 | 1 |
| T2DM_admissi | leukocyte | *SIGMAR1* | -1.445288982 | 0.000133405 | 0.001062643 | 0.524148991 |
| T2DM_admissi | leukocyte | *ZNF462* | -0.997434794 | 0.000737585 | 0.00155805 | 1 |
| T2DM_admissi | leukocyte | *UGCG* | -1.243977934 | 0.000150498 | 0.001062643 | 0.591306327 |
| T2DM_admissi | leukocyte | *ALAD* | -1.166562446 | 0.002208475 | 0.003341201 | 1 |
| T2DM_admissi | leukocyte | *SURF4* | -1.029152294 | 0.000152154 | 0.001062643 | 0.59781394 |
| T2DM_admissi | leukocyte | *SURF1* | -1.371770316 | 0.000107942 | 0.001062643 | 0.424104133 |
| T2DM_admissi | leukocyte | *SURF6* | -1.741282983 | 0.000179605 | 0.001062643 | 0.705668952 |
| T2DM_admissi | leukocyte | *SH3GLB2* | -1.976227268 | 8.58E-05 | 0.001062643 | 0.336929953 |
| T2DM_admissi | leukocyte | *LCN2* | -0.466037676 | 0.001097282 | 0.001995936 | 1 |
| T2DM_admissi | leukocyte | *GPR107* | -1.560968999 | 0.000431701 | 0.001216752 | 1 |
| T2DM_admissi | leukocyte | *SEC16A* | -1.104975242 | 0.013894192 | 0.017729874 | 1 |
| T2DM_admissi | leukocyte | *MSRB2* | -1.887815183 | 8.37E-05 | 0.001062643 | 0.328830178 |
| T2DM_admissi | leukocyte | *RSU1* | -1.472504436 | 0.000400481 | 0.001184732 | 1 |
| T2DM_admissi | leukocyte | *GLUD1* | -1.395666481 | 0.000214264 | 0.001062643 | 0.841843453 |
| T2DM_admissi | leukocyte | *FRA10AC1* | -0.863759443 | 0.000799728 | 0.001644234 | 1 |
| T2DM_admissi | leukocyte | *DNAJB12* | -1.522643006 | 0.000182165 | 0.001062643 | 0.715724594 |
| T2DM_admissi | leukocyte | *LRRC27* | -2.748338635 | 0.008952611 | 0.01174451 | 1 |
| T2DM_admissi | leukocyte | *MTG1* | -1.003521254 | 0.000599212 | 0.001399402 | 1 |
| T2DM_admissi | leukocyte | *PAOX* | -5.344629191 | 0.00061305 | 0.001415201 | 1 |
| T2DM_admissi | leukocyte | *GSTO1* | -1.449110941 | 0.000398525 | 0.001183526 | 1 |
| T2DM_admissi | leukocyte | *HSD17B12* | -1.214530076 | 8.74E-09 | 4.91E-06 | 3.44E-05 |
| T2DM_admissi | leukocyte | *APIP* | -1.560311378 | 7.98E-07 | 0.000164962 | 0.003134273 |
| T2DM_admissi | leukocyte | *PTPRJ* | -1.529329838 | 0.000518519 | 0.001314363 | 1 |
| T2DM_admissi | leukocyte | *CCDC82* | -1.330385958 | 4.19E-05 | 0.001037577 | 0.164572304 |
| T2DM_admissi | leukocyte | *RPS3* | -0.773802012 | 0.00018851 | 0.001062643 | 0.740654378 |
| T2DM_admissi | leukocyte | *AASDHPPT* | -1.236230589 | 0.000286953 | 0.001088708 | 1 |
| T2DM_admissi | leukocyte | *GLB1L2* | -1.031453847 | 0.003500084 | 0.005015255 | 1 |
| T2DM_admissi | leukocyte | *FADS1* | -0.677632152 | 0.017202149 | 0.021620999 | 1 |
| T2DM_admissi | leukocyte | *TMC2* | -67.09675862 | 0.014683741 | 0.018658608 | 1 |
| T2DM_admissi | leukocyte | *EML3* | -5.259789936 | 0.000150114 | 0.001062643 | 0.589796159 |
| T2DM_admissi | leukocyte | *CCDC15* | -1.443350337 | 0.000261304 | 0.001083674 | 1 |
| T2DM_admissi | leukocyte | *CHEK1* | -1.46351377 | 0.00052494 | 0.001319879 | 1 |
| T2DM_admissi | leukocyte | *SIDT2* | -1.662964211 | 5.26E-05 | 0.00105496 | 0.206772115 |
| T2DM_admissi | leukocyte | *TAGLN* | -1.268064892 | 5.31E-06 | 0.00058206 | 0.020846529 |
| T2DM_admissi | leukocyte | *DUSP15* | -1.404043681 | 0.001042246 | 0.001936841 | 1 |
| T2DM_admissi | leukocyte | *DSN1* | -1.343994225 | 0.000762259 | 0.001593931 | 1 |
| T2DM_admissi | leukocyte | *YTHDF1* | -1.30679638 | 0.00028185 | 0.001088708 | 1 |
| T2DM_admissi | leukocyte | *CABLES2* | -1.435668218 | 0.001407228 | 0.002326041 | 1 |
| T2DM_admissi | leukocyte | *TRPT1* | -1.094226811 | 0.001042764 | 0.001936841 | 1 |
| T2DM_admissi | leukocyte | *MPP7* | -0.905779834 | 0.001090174 | 0.001985764 | 1 |
| T2DM_admissi | leukocyte | *DCUN1D2* | -1.28378205 | 0.002494315 | 0.003720348 | 1 |
| T2DM_admissi | leukocyte | *TMCO3* | -1.337853675 | 0.000413253 | 0.001193 | 1 |
| T2DM_admissi | leukocyte | *TMEM218* | -1.847215946 | 0.000205728 | 0.001062643 | 0.808306477 |
| T2DM_admissi | leukocyte | *MIA2* | -1.162884873 | 0.000202255 | 0.001062643 | 0.794660443 |
| T2DM_admissi | leukocyte | *LYPD1* | -1.784198903 | 0.001077244 | 0.001970434 | 1 |

| | | | | | | |
|---|---|---|---|---|---|---|
| T2DM_admissi | leukocyte | *LYPD6B* | -2.912880154 | 0.001085931 | 0.00197991 | 1 |
| T2DM_admissi | leukocyte | *WDR17* | -0.888149297 | 0.002085011 | 0.003176428 | 1 |
| T2DM_admissi | leukocyte | *CNDP1* | -4.128689994 | 0.01501327 | 0.01901584 | 1 |
| T2DM_admissi | leukocyte | *PRSS23* | -0.909005241 | 0.000414095 | 0.001193676 | 1 |
| T2DM_admissi | leukocyte | *MTMR12* | -0.967329265 | 0.000491753 | 0.001283904 | 1 |
| T2DM_admissi | leukocyte | *DOCK1* | -1.221002989 | 0.000382471 | 0.001165285 | 1 |
| T2DM_admissi | leukocyte | *IL18* | 0.520542993 | 0.017974021 | 0.022483262 | 1 |
| T2DM_admissi | leukocyte | *TMEM86A* | -1.364488505 | 0.001235289 | 0.002144697 | 1 |
| T2DM_admissi | leukocyte | *PLBD2* | -2.50201669 | 0.008057845 | 0.010681267 | 1 |
| T2DM_admissi | leukocyte | *DLG5* | -1.161179773 | 0.000279821 | 0.001088708 | 1 |
| T2DM_admissi | leukocyte | *SLC2A13* | -0.905047208 | 0.00044978 | 0.001234067 | 1 |
| T2DM_admissi | leukocyte | *MAGI1* | -1.015300554 | 0.002509134 | 0.003735653 | 1 |
| T2DM_admissi | leukocyte | *CSNK1G3* | -1.252557605 | 0.000933556 | 0.001804129 | 1 |
| T2DM_admissi | leukocyte | *AKAP6* | -1.398174234 | 0.000615119 | 0.001419144 | 1 |
| T2DM_admissi | leukocyte | *MBIP* | -1.315824719 | 0.000370392 | 0.001151894 | 1 |
| T2DM_admissi | leukocyte | *MIPOL1* | -1.014348896 | 0.001455166 | 0.002374314 | 1 |
| T2DM_admissi | leukocyte | *TMEM18* | -1.256337418 | 0.000323458 | 0.001116892 | 1 |
| T2DM_admissi | leukocyte | *NDUFC2* | -0.97095267 | 0.000165953 | 0.001062643 | 0.652029189 |
| T2DM_admissi | leukocyte | *VIPAS39* | -2.28086421 | 2.86E-05 | 0.001008808 | 0.112393677 |
| T2DM_admissi | leukocyte | *C4orf33* | -2.494759427 | 0.000379943 | 0.001164428 | 1 |
| T2DM_admissi | leukocyte | *QDPR* | -0.843393347 | 0.000735593 | 0.001555514 | 1 |
| T2DM_admissi | leukocyte | *TEX9* | -2.159312593 | 0.000187134 | 0.001062643 | 0.735249325 |
| T2DM_admissi | leukocyte | *ZNF827* | -1.109550351 | 0.000321168 | 0.001114222 | 1 |
| T2DM_admissi | leukocyte | *AKR1C2* | -2.427607146 | 0.001844202 | 0.002870788 | 1 |
| T2DM_admissi | leukocyte | *KIN* | -2.120544726 | 0.000151972 | 0.001062643 | 0.597097274 |
| T2DM_admissi | leukocyte | *TMEM45B* | -1.626351386 | 0.003048292 | 0.004437473 | 1 |
| T2DM_admissi | leukocyte | *CENPU* | -1.97678006 | 0.031421426 | 0.037834748 | 1 |
| T2DM_admissi | leukocyte | *BICD1* | -0.94853568 | 0.00126018 | 0.002164729 | 1 |
| T2DM_admissi | leukocyte | *GUF1* | -3.355660512 | 1.44E-05 | 0.000815363 | 0.056591434 |
| T2DM_admissi | leukocyte | *CCL28* | -1.409987039 | 0.000602635 | 0.001403431 | 1 |
| T2DM_admissi | leukocyte | *CACUL1* | -1.758610809 | 9.95E-05 | 0.001062643 | 0.390897644 |
| T2DM_admissi | leukocyte | *KCNE4* | -1.415072403 | 0.001051401 | 0.001942151 | 1 |
| T2DM_admissi | leukocyte | *RABGAP1L* | -1.347470583 | 0.000267907 | 0.001087356 | 1 |
| T2DM_admissi | leukocyte | *PTPN14* | -1.426222139 | 3.93E-06 | 0.000498523 | 0.015454216 |
| T2DM_admissi | leukocyte | *ARL14EP* | -1.690241152 | 8.53E-05 | 0.001062643 | 0.335163675 |
| T2DM_admissi | leukocyte | *EPG5* | -2.031245559 | 0.000121899 | 0.001062643 | 0.478941375 |
| T2DM_admissi | leukocyte | *HAUS1* | -1.563480084 | 0.000409021 | 0.001188129 | 1 |
| T2DM_admissi | leukocyte | *SPC25* | -1.244057303 | 0.002537522 | 0.003770774 | 1 |
| T2DM_admissi | leukocyte | *PDK1* | -1.42432239 | 0.000550248 | 0.001351735 | 1 |
| T2DM_admissi | leukocyte | *PDE3B* | -0.748322417 | 0.000345415 | 0.001140861 | 1 |
| T2DM_admissi | leukocyte | *TGOLN2* | -1.272010232 | 8.78E-05 | 0.001062643 | 0.344865546 |
| T2DM_admissi | leukocyte | *ATG10* | -0.849483474 | 0.020001725 | 0.02486036 | 1 |
| T2DM_admissi | leukocyte | *POC5* | -2.508682321 | 5.75E-09 | 3.77E-06 | 2.26E-05 |
| T2DM_admissi | leukocyte | *GUCY1A2* | -1.118335662 | 0.001203608 | 0.002114239 | 1 |
| T2DM_admissi | leukocyte | *ZNF773* | -0.675518631 | 0.03790641 | 0.04526878 | 1 |
| T2DM_admissi | leukocyte | *DCLRE1C* | -3.302011758 | 0.000665729 | 0.001470292 | 1 |
| T2DM_admissi | leukocyte | *ZSCAN1* | -1.323024801 | 0.00012827 | 0.001062643 | 0.503973232 |
| T2DM_admissi | leukocyte | *USP12* | -1.10581564 | 0.000349765 | 0.001141109 | 1 |
| T2DM_admissi | leukocyte | *CCDC50* | -0.867042895 | 0.001030832 | 0.001919497 | 1 |
| T2DM_admissi | leukocyte | *PLEKHH2* | -0.93267329 | 0.009674979 | 0.012616326 | 1 |

| | | | | | | |
|---|---|---|---|---|---|---|
| T2DM_admissi | leukocyte | *GRIA4* | -1.478180535 | 0.000778117 | 0.001614162 | 1 |
| T2DM_admissi | leukocyte | *IGSF10* | -2.751854019 | 0.001024298 | 0.00191186 | 1 |
| T2DM_admissi | leukocyte | *SPEF2* | -1.04659789 | 0.010918708 | 0.014130304 | 1 |
| T2DM_admissi | leukocyte | *GPD1L* | -1.201541717 | 0.002225674 | 0.003365925 | 1 |
| T2DM_admissi | leukocyte | *CCNO* | -1.47706486 | 0.000267907 | 0.001087356 | 1 |
| T2DM_admissi | leukocyte | *RASGRP3* | -1.217692041 | 0.002736922 | 0.004028987 | 1 |
| T2DM_admissi | leukocyte | *CATSPER3* | -6.160649985 | 0.00118831 | 0.00210081 | 1 |
| T2DM_admissi | leukocyte | *GPR180* | -1.055977554 | 0.000608612 | 0.001409123 | 1 |
| T2DM_admissi | leukocyte | *IFIT5* | -1.194714341 | 0.001221451 | 0.002133102 | 1 |
| T2DM_admissi | leukocyte | *ZNF117* | -0.705311889 | 0.033295451 | 0.040005451 | 1 |
| T2DM_admissi | leukocyte | *MED21* | -2.026268275 | 3.85E-05 | 0.001037577 | 0.151322836 |
| T2DM_admissi | leukocyte | *LGI2* | -1.61202336 | 0.002904205 | 0.004252935 | 1 |
| T2DM_admissi | leukocyte | *SRP19* | -1.017467637 | 0.000218474 | 0.001062643 | 0.858385873 |
| T2DM_admissi | leukocyte | *DAB2* | -1.611732847 | 0.000199231 | 0.001062643 | 0.782777556 |
| T2DM_admissi | leukocyte | *ACMSD* | -1.472496421 | 0.001939246 | 0.002997363 | 1 |
| T2DM_admissi | leukocyte | *CAST* | -1.06249581 | 0.000212789 | 0.001062643 | 0.836049284 |
| T2DM_admissi | leukocyte | *SYCP2L* | -2.779538905 | 0.006929449 | 0.009301608 | 1 |
| T2DM_admissi | leukocyte | *MERTK* | 0.51247002 | 0.041848252 | 0.049674254 | 1 |
| T2DM_admissi | leukocyte | *TMEM87B* | -1.289881436 | 0.00056418 | 0.001362351 | 1 |
| T2DM_admissi | leukocyte | *PLA2R1* | -3.80528952 | 0.001331979 | 0.002235518 | 1 |
| T2DM_admissi | leukocyte | *SLC25A27* | -1.727301171 | 0.000238937 | 0.001072513 | 0.93878201 |
| T2DM_admissi | leukocyte | *ASAP1* | -1.067378127 | 0.000598646 | 0.001399402 | 1 |
| T2DM_admissi | leukocyte | *NMRAL1* | -1.353470804 | 0.001230549 | 0.002143098 | 1 |
| T2DM_admissi | leukocyte | *CMTM7* | -1.052119619 | 0.000407416 | 0.001188129 | 1 |
| T2DM_admissi | leukocyte | *RPIA* | -1.246043504 | 0.000330708 | 0.001124979 | 1 |
| T2DM_admissi | leukocyte | *PTPRD* | -0.763231286 | 0.035765346 | 0.042855153 | 1 |
| T2DM_admissi | leukocyte | *GTF2E1* | -3.045558897 | 0.000102677 | 0.001062643 | 0.403417638 |
| T2DM_admissi | leukocyte | *CFDP1* | -0.986098887 | 0.000773102 | 0.001608876 | 1 |
| T2DM_admissi | leukocyte | *ZDHHC7* | -1.420832697 | 0.01395508 | 0.01779601 | 1 |
| T2DM_admissi | leukocyte | *JAZF1* | -2.604488043 | 8.35E-11 | 1.09E-07 | 3.28E-07 |
| T2DM_admissi | leukocyte | *CMIP* | -1.24673524 | 0.000281529 | 0.001088708 | 1 |
| T2DM_admissi | leukocyte | *PID1* | -2.073288215 | 0.004668774 | 0.006486404 | 1 |
| T2DM_admissi | leukocyte | *KCTD15* | -1.576460448 | 0.001264578 | 0.002168716 | 1 |
| T2DM_admissi | leukocyte | *LGI4* | 1.197666815 | 0.012930074 | 0.01655336 | 1 |
| T2DM_admissi | leukocyte | *DDAH1* | -2.723884777 | 0.000114523 | 0.001062643 | 0.449962716 |
| T2DM_admissi | leukocyte | *NUS1* | -1.483571542 | 5.11E-05 | 0.00105496 | 0.200706188 |
| T2DM_admissi | leukocyte | *PPP2R5E* | -1.276879724 | 0.0006343 | 0.001434874 | 1 |
| T2DM_admissi | leukocyte | *ROBO3* | -3.32717089 | 0.001203755 | 0.002114239 | 1 |
| T2DM_admissi | leukocyte | *PITPNC1* | -1.360692402 | 0.000279557 | 0.001088708 | 1 |
| T2DM_admissi | leukocyte | *CC2D1B* | -2.489019686 | 0.000236055 | 0.001068391 | 0.927458548 |
| T2DM_admissi | leukocyte | *CEP112* | -0.956665531 | 0.000933461 | 0.001804129 | 1 |
| T2DM_admissi | leukocyte | *ABCA5* | -1.871051334 | 0.000103345 | 0.001062643 | 0.406042832 |
| T2DM_admissi | leukocyte | *SRSF12* | -1.961076702 | 0.002151571 | 0.003265736 | 1 |
| T2DM_admissi | leukocyte | *PDLIM3* | -0.749158135 | 0.00167717 | 0.002656459 | 1 |
| T2DM_admissi | leukocyte | *PDE1C* | -1.242195376 | 0.007085138 | 0.009487903 | 1 |
| T2DM_admissi | leukocyte | *RABGEF1* | -1.316895371 | 0.000207948 | 0.001062643 | 0.817028272 |
| T2DM_admissi | leukocyte | *MRPL39* | -1.364420709 | 0.00057789 | 0.001376912 | 1 |
| T2DM_admissi | leukocyte | *TSEN2* | -2.433564184 | 0.001556849 | 0.002504379 | 1 |
| T2DM_admissi | leukocyte | *FLCN* | -1.948246304 | 0.000170117 | 0.001062643 | 0.668387842 |
| T2DM_admissi | leukocyte | *MPPE1* | -1.468142051 | 0.000361342 | 0.001142338 | 1 |

| | | | | | | |
|---|---|---|---|---|---|---|
| T2DM_admissi | leukocyte | *RAB6B* | -1.43973676 | 0.000616973 | 0.001419945 | 1 |
| T2DM_admissi | leukocyte | *ACSS1* | -1.373461329 | 0.012609416 | 0.016169189 | 1 |
| T2DM_admissi | leukocyte | *ANKRD40* | -1.561285829 | 0.000605398 | 0.00140595 | 1 |
| T2DM_admissi | leukocyte | *ATP6V1C1* | -1.108000033 | 0.000435936 | 0.001222199 | 1 |
| T2DM_admissi | leukocyte | *MARVELD1* | -4.587680561 | 4.38E-05 | 0.001037577 | 0.171910543 |
| T2DM_admissi | leukocyte | *TRMT44* | -1.008193873 | 0.002111226 | 0.003212628 | 1 |
| T2DM_admissi | leukocyte | *PPM1J* | -2.754894717 | 0.00178231 | 0.00279103 | 1 |
| T2DM_admissi | leukocyte | *DBI* | -1.205349278 | 0.000114796 | 0.001062643 | 0.451034233 |
| T2DM_admissi | leukocyte | *HEATR3* | -0.733045361 | 0.020069623 | 0.024918648 | 1 |
| T2DM_admissi | leukocyte | *LRGUK* | -3.859319467 | 0.000648161 | 0.001447767 | 1 |
| T2DM_admissi | leukocyte | *SETD9* | -1.510326338 | 4.62E-05 | 0.001054402 | 0.181357219 |
| T2DM_admissi | leukocyte | *MIER3* | -2.81117627 | 2.50E-05 | 0.000982543 | 0.098247942 |
| T2DM_admissi | leukocyte | *TTN* | -3.148263733 | 9.42E-05 | 0.001062643 | 0.370050392 |
| T2DM_admissi | leukocyte | *DEPTOR* | -1.061043191 | 0.007914652 | 0.010516289 | 1 |
| T2DM_admissi | leukocyte | *RMND1* | -2.066343962 | 0.000285788 | 0.001088708 | 1 |
| T2DM_admissi | leukocyte | *TMBIM4* | -1.114161749 | 0.000162874 | 0.001062643 | 0.639931433 |
| T2DM_admissi | leukocyte | *PSD3* | -1.012805853 | 0.000764758 | 0.001596564 | 1 |
| T2DM_admissi | leukocyte | *FAM161B* | -1.541266533 | 0.007303299 | 0.00976341 | 1 |
| T2DM_admissi | leukocyte | *MMP16* | -0.92205008 | 0.002012898 | 0.003089327 | 1 |
| T2DM_admissi | leukocyte | *ADK* | -1.308858245 | 0.000467792 | 0.001253701 | 1 |
| T2DM_admissi | leukocyte | *BATF* | -16.43774177 | 0.001139796 | 0.002049538 | 1 |
| T2DM_admissi | leukocyte | *DRAM2* | -1.85687388 | 0.000783718 | 0.001618943 | 1 |
| T2DM_admissi | leukocyte | *CCT8* | -0.849012473 | 0.001713264 | 0.002707728 | 1 |
| T2DM_admissi | leukocyte | *CDK20* | -1.035098803 | 0.001777986 | 0.002785369 | 1 |
| T2DM_admissi | leukocyte | *TDRD9* | -0.788552894 | 0.000547822 | 0.001347773 | 1 |
| T2DM_admissi | leukocyte | *HKDC1* | -2.715928761 | 0.001845831 | 0.002872187 | 1 |
| T2DM_admissi | leukocyte | *ZDHHC5* | -1.42157641 | 0.000317351 | 0.001111111 | 1 |
| T2DM_admissi | leukocyte | *MED19* | -1.173396653 | 0.000286792 | 0.001088708 | 1 |
| T2DM_admissi | leukocyte | *MAPK13* | -1.31679312 | 0.000891541 | 0.001763074 | 1 |
| T2DM_admissi | leukocyte | *COX6A2* | -1.297672616 | 0.003898566 | 0.005528505 | 1 |
| T2DM_admissi | leukocyte | *MPV17L* | -1.141661049 | 0.000718915 | 0.0015393 | 1 |
| T2DM_admissi | leukocyte | *TATDN2* | -2.095556694 | 0.000270678 | 0.00108853 | 1 |
| T2DM_admissi | leukocyte | *EXOG* | -1.668753764 | 6.92E-05 | 0.001062643 | 0.272003371 |
| T2DM_admissi | leukocyte | *ZFYVE9* | -1.279521208 | 0.000524586 | 0.001319879 | 1 |
| T2DM_admissi | leukocyte | *SLC6A1* | -6.528151602 | 0.001187122 | 0.00210081 | 1 |
| T2DM_admissi | leukocyte | *NECAP2* | -1.582996266 | 0.000216346 | 0.001062643 | 0.850024067 |
| T2DM_admissi | leukocyte | *STEAP2* | -1.610437775 | 0.001822412 | 0.00284702 | 1 |
| T2DM_admissi | leukocyte | *GATAD1* | -1.581163918 | 0.000195265 | 0.001062643 | 0.767196767 |
| T2DM_admissi | leukocyte | *CLEC18A* | -6.985106048 | 3.13E-11 | 6.14E-08 | 1.23E-07 |
| T2DM_admissi | leukocyte | *DHRS4* | -1.43147627 | 0.000278836 | 0.001088708 | 1 |
| T2DM_admissi | leukocyte | *DHRS1* | -1.267799642 | 0.003469 | 0.004977977 | 1 |
| T2DM_admissi | leukocyte | *AASDH* | -0.877672846 | 0.002063147 | 0.003151674 | 1 |
| T2DM_admissi | leukocyte | *SLC38A10* | -1.417150712 | 0.000227097 | 0.001062643 | 0.892263005 |
| T2DM_admissi | leukocyte | *SLC37A3* | -0.91401549 | 0.000807729 | 0.0016529 | 1 |
| T2DM_admissi | leukocyte | *AP3S2* | -1.213408224 | 0.00370191 | 0.005275591 | 1 |
| T2DM_admissi | leukocyte | *SPPL3* | -1.410958972 | 0.000167506 | 0.001062643 | 0.658131783 |
| T2DM_admissi | leukocyte | *TNFRSF14* | 0.631283373 | 0.03498844 | 0.041962631 | 1 |
| T2DM_admissi | leukocyte | *C12orf43* | -1.465304978 | 0.000508588 | 0.001305383 | 1 |
| T2DM_admissi | leukocyte | *PEX10* | -2.055621085 | 0.000128032 | 0.001062643 | 0.503035919 |
| T2DM_admissi | leukocyte | *LDLRAP1* | -0.987399166 | 0.000438259 | 0.001222199 | 1 |

| | | | | | | |
|---|---|---|---|---|---|---|
| T2DM_admissi | leukocyte | *KRTCAP3* | -1.167097182 | 0.000584057 | 0.001382042 | 1 |
| T2DM_admissi | leukocyte | *PAFAH2* | -3.512677294 | 0.000110094 | 0.001062643 | 0.432559878 |
| T2DM_admissi | leukocyte | *EXTL1* | -7.476451428 | 1.36E-05 | 0.000815363 | 0.053486067 |
| T2DM_admissi | leukocyte | *TRIM63* | -16.23890482 | 0.000724199 | 0.001544387 | 1 |
| T2DM_admissi | leukocyte | *XKR8* | -2.241836613 | 0.000181387 | 0.001062643 | 0.712669125 |
| T2DM_admissi | leukocyte | *CNNM4* | -2.101934426 | 0.000456011 | 0.001240899 | 1 |
| T2DM_admissi | leukocyte | *ESYT3* | -2.446045641 | 0.000556459 | 0.001357223 | 1 |
| T2DM_admissi | leukocyte | *FAIM* | -1.272468676 | 0.000261847 | 0.001083674 | 1 |
| T2DM_admissi | leukocyte | *AHCYL2* | -1.407992445 | 0.000929772 | 0.001800431 | 1 |
| T2DM_admissi | leukocyte | *COPG2* | -1.375882413 | 6.14E-05 | 0.001060479 | 0.241279383 |
| T2DM_admissi | leukocyte | *RNF166* | -1.589406616 | 0.000550283 | 0.001351735 | 1 |
| T2DM_admissi | leukocyte | *NIT1* | -1.56019798 | 0.002430647 | 0.0036381 | 1 |
| T2DM_admissi | leukocyte | *NPM2* | -1.3748413 | 0.000684491 | 0.001494923 | 1 |
| T2DM_admissi | leukocyte | *FGF17* | -1.831402935 | 0.040298398 | 0.04799406 | 1 |
| T2DM_admissi | leukocyte | *VWA5B1* | 2.762222131 | 0.010721612 | 0.013888959 | 1 |
| T2DM_admissi | leukocyte | *CDA* | -13.25277571 | 0.002350694 | 0.003534587 | 1 |
| T2DM_admissi | leukocyte | *PINK1* | -1.664523749 | 3.91E-05 | 0.001037577 | 0.153708044 |
| T2DM_admissi | leukocyte | *DMTN* | -1.177611249 | 0.002951985 | 0.004310096 | 1 |
| T2DM_admissi | leukocyte | *CCAR2* | -1.783394478 | 0.00020023 | 0.001062643 | 0.786705405 |
| T2DM_admissi | leukocyte | *EPB41* | -1.16805185 | 0.000583625 | 0.001382042 | 1 |
| T2DM_admissi | leukocyte | *MIS18A* | -1.57399542 | 0.000131834 | 0.001062643 | 0.517977079 |
| T2DM_admissi | leukocyte | *ALG8* | -1.091983968 | 0.001045656 | 0.001938832 | 1 |
| T2DM_admissi | leukocyte | *IFNAR2* | -2.257395958 | 0.000630885 | 0.001433512 | 1 |
| T2DM_admissi | leukocyte | *IFNGR2* | -1.305847873 | 0.000119157 | 0.001062643 | 0.468169291 |
| T2DM_admissi | leukocyte | *C1QC* | 0.061458614 | 0.007217952 | 0.009655884 | 1 |
| T2DM_admissi | leukocyte | *GJD2* | -0.771499215 | 0.0005565 | 0.001357223 | 1 |
| T2DM_admissi | leukocyte | *CLDN14* | -2.379258711 | 0.005858792 | 0.007995552 | 1 |
| T2DM_admissi | leukocyte | *HLCS* | -1.059075656 | 0.001130716 | 0.002036018 | 1 |
| T2DM_admissi | leukocyte | *THEM4* | -0.979589181 | 0.000630594 | 0.001433512 | 1 |
| T2DM_admissi | leukocyte | *UBR1* | -1.559797479 | 0.000346551 | 0.001140861 | 1 |
| T2DM_admissi | leukocyte | *TEPP* | 3.505806874 | 0.021611165 | 0.026651057 | 1 |
| T2DM_admissi | leukocyte | *ZDHHC1* | -1.264985545 | 0.000385353 | 0.001168251 | 1 |
| T2DM_admissi | leukocyte | *ZFYVE28* | -1.537371617 | 0.001325599 | 0.002228618 | 1 |
| T2DM_admissi | leukocyte | *LYPD5* | -12.75124368 | 0.00668818 | 0.009008522 | 1 |
| T2DM_admissi | leukocyte | *NPR2* | -1.065551107 | 0.016178837 | 0.020406629 | 1 |
| T2DM_admissi | leukocyte | *ATAD3B* | -2.962663183 | 0.000333015 | 0.00112758 | 1 |
| T2DM_admissi | leukocyte | *ANKLE1* | 0.939725533 | 0.000258993 | 0.001081954 | 1 |
| T2DM_admissi | leukocyte | *KALRN* | -1.585488642 | 0.001550761 | 0.002501207 | 1 |
| T2DM_admissi | leukocyte | *CILP2* | -4.277925754 | 6.24E-07 | 0.000144219 | 0.002451725 |
| T2DM_admissi | leukocyte | *TMPRSS3* | -3.222606595 | 0.002930845 | 0.004287152 | 1 |
| T2DM_admissi | leukocyte | *RSPH1* | -0.797401527 | 0.001081912 | 0.001975294 | 1 |
| T2DM_admissi | leukocyte | *SLC37A1* | -1.192794318 | 0.001610792 | 0.002572683 | 1 |
| T2DM_admissi | leukocyte | *NDUFV3* | -0.821386892 | 0.001859808 | 0.002891644 | 1 |
| T2DM_admissi | leukocyte | *HSF2BP* | -3.527916446 | 0.001050136 | 0.001940726 | 1 |
| T2DM_admissi | leukocyte | *PDXK* | -1.599817091 | 4.47E-05 | 0.001037577 | 0.175578267 |
| T2DM_admissi | leukocyte | *CSTB* | -1.373379314 | 0.000227784 | 0.001062643 | 0.894963728 |
| T2DM_admissi | leukocyte | *ICOSLG* | 0.471226275 | 0.01717084 | 0.021595465 | 1 |
| T2DM_admissi | leukocyte | *ZNF66* | -3.31698346 | 0.000575437 | 0.001373567 | 1 |
| T2DM_admissi | leukocyte | *FTCD* | -1.812062988 | 0.000168588 | 0.001062643 | 0.66238095 |
| T2DM_admissi | leukocyte | *SPATC1L* | -0.956278311 | 0.000563304 | 0.001362351 | 1 |

| | | | | | | |
|---|---|---|---|---|---|---|
| T2DM_admissi | leukocyte | *LSS* | -1.28591662 | 0.001265865 | 0.002169029 | 1 |
| T2DM_admissi | leukocyte | *VAV2* | -1.781940255 | 0.000825909 | 0.001674406 | 1 |
| T2DM_admissi | leukocyte | *MCM3AP* | -1.651931859 | 0.003158215 | 0.004571187 | 1 |
| T2DM_admissi | leukocyte | *DIP2A* | -0.99178244 | 0.001191734 | 0.00210536 | 1 |
| T2DM_admissi | leukocyte | *ZNF208* | -1.162206736 | 0.000307472 | 0.001098736 | 1 |
| T2DM_admissi | leukocyte | *ADAMTS13* | -1.391231497 | 0.02009828 | 0.024941928 | 1 |
| T2DM_admissi | leukocyte | *SLC2A6* | -2.268560237 | 0.000165974 | 0.001062643 | 0.652110044 |
| T2DM_admissi | leukocyte | *RDH13* | -1.389990809 | 0.000886243 | 0.001757306 | 1 |
| T2DM_admissi | leukocyte | *MPZL3* | -1.397066856 | 0.000279933 | 0.001088708 | 1 |
| T2DM_admissi | leukocyte | *S100A1* | -2.128068139 | 0.006773537 | 0.009114118 | 1 |
| T2DM_admissi | leukocyte | *VPS11* | -3.05063363 | 2.17E-06 | 0.000340493 | 0.008512334 |
| T2DM_admissi | leukocyte | *PAQR6* | -1.602104869 | 0.000116949 | 0.001062643 | 0.459493111 |
| T2DM_admissi | leukocyte | *SLC25A44* | -1.983076932 | 0.000275902 | 0.001088708 | 1 |
| T2DM_admissi | leukocyte | *CCR5* | -139.2431028 | 0.003602379 | 0.00514869 | 1 |
| T2DM_admissi | leukocyte | *NBEAL2* | -0.844757055 | 0.001825595 | 0.002849726 | 1 |
| T2DM_admissi | leukocyte | *LY6E* | -1.394012364 | 0.000181968 | 0.001062643 | 0.714953713 |
| T2DM_admissi | leukocyte | *VPS28* | -0.967113013 | 0.00120611 | 0.002117283 | 1 |
| T2DM_admissi | leukocyte | *TONSL* | -6.0010195 | 0.000284979 | 0.001088708 | 1 |
| T2DM_admissi | leukocyte | *ZNF333* | -4.686519164 | 7.31E-05 | 0.001062643 | 0.287354317 |
| T2DM_admissi | leukocyte | *RPL8* | -0.759385973 | 0.000171089 | 0.001062643 | 0.672207467 |
| T2DM_admissi | leukocyte | *MAML1* | -2.170607284 | 0.000539646 | 0.001334341 | 1 |
| T2DM_admissi | leukocyte | *LRWD1* | -1.641762458 | 0.001276833 | 0.002175489 | 1 |
| T2DM_admissi | leukocyte | *CCDC116* | -9.420220801 | 0.007402757 | 0.009876208 | 1 |
| T2DM_admissi | leukocyte | *U2AF1L4* | -0.874173734 | 0.002416872 | 0.00362099 | 1 |
| T2DM_admissi | leukocyte | *ZNF382* | -1.042570627 | 0.003991494 | 0.005645278 | 1 |
| T2DM_admissi | leukocyte | *DUSP14* | -1.550946505 | 0.000356297 | 0.001141109 | 1 |
| T2DM_admissi | leukocyte | *PGAP3* | -1.241608917 | 0.00112341 | 0.002030303 | 1 |
| T2DM_admissi | leukocyte | *PRPSAP1* | -1.245422629 | 0.000195627 | 0.001062643 | 0.768619863 |
| T2DM_admissi | leukocyte | *SIGLEC11* | -15.01396169 | 0.014729643 | 0.018710885 | 1 |
| T2DM_admissi | leukocyte | *SPC24* | -3.08138128 | 0.0024608 | 0.003677627 | 1 |
| T2DM_admissi | leukocyte | *IP6K3* | 3.468228043 | 0.000253607 | 0.001077213 | 0.996421609 |
| T2DM_admissi | leukocyte | *LEMD2* | -2.161481418 | 0.001580523 | 0.002532576 | 1 |
| T2DM_admissi | leukocyte | *CXCL16* | 0.334616801 | 0.026396299 | 0.032068973 | 1 |
| T2DM_admissi | leukocyte | *RPL26* | -0.811795838 | 0.000155426 | 0.001062643 | 0.610666952 |
| T2DM_admissi | leukocyte | *SNRNP25* | -1.07611176 | 0.00035762 | 0.001141109 | 1 |
| T2DM_admissi | leukocyte | *WDR90* | -1.850662527 | 0.000581812 | 0.001382042 | 1 |
| T2DM_admissi | leukocyte | *CCDC78* | -3.158055414 | 0.003474434 | 0.004983954 | 1 |
| T2DM_admissi | leukocyte | *ZNF75A* | -2.588150178 | 2.88E-05 | 0.001008808 | 0.112986454 |
| T2DM_admissi | leukocyte | *ASRGL1* | -0.957333413 | 0.000779793 | 0.001614233 | 1 |
| T2DM_admissi | leukocyte | *TAF6L* | -1.496310189 | 0.001762386 | 0.002769765 | 1 |
| T2DM_admissi | leukocyte | *ZFPL1* | -1.382162344 | 0.000336045 | 0.001131077 | 1 |
| T2DM_admissi | leukocyte | *LRP5* | -2.451492336 | 8.44E-05 | 0.001062643 | 0.331797321 |
| T2DM_admissi | leukocyte | *TPCN2* | -1.181149749 | 0.014084501 | 0.017943581 | 1 |
| T2DM_admissi | leukocyte | *PDZK1IP1* | -0.845109303 | 0.000630759 | 0.001433512 | 1 |
| T2DM_admissi | leukocyte | *CTRC* | -0.60735686 | 0.000510227 | 0.001306833 | 1 |
| T2DM_admissi | leukocyte | *SDC3* | -2.262655119 | 0.000666674 | 0.001471551 | 1 |
| T2DM_admissi | leukocyte | *OMA1* | -1.648780042 | 0.001253605 | 0.002157431 | 1 |
| T2DM_admissi | leukocyte | *TM2D1* | -1.807331822 | 0.000120171 | 0.001062643 | 0.472152962 |
| T2DM_admissi | leukocyte | *FUBP1* | -1.071892691 | 0.000148135 | 0.001062643 | 0.582021926 |
| T2DM_admissi | leukocyte | *SNX7* | -0.938878629 | 0.000928852 | 0.001800431 | 1 |

| | | | | | | |
|---|---|---|---|---|---|---|
| T2DM_admissi | leukocyte | *HFM1* | -2.155049284 | 0.001524677 | 0.002467446 | 1 |
| T2DM_admissi | leukocyte | *VCAM1* | -0.633323819 | 0.001321422 | 0.002223498 | 1 |
| T2DM_admissi | leukocyte | *ZNF496* | -1.177312283 | 0.000650132 | 0.001449765 | 1 |
| T2DM_admissi | leukocyte | *TRIM58* | -3.970260769 | 0.007385401 | 0.009859749 | 1 |
| T2DM_admissi | leukocyte | *PEA15* | -1.141569885 | 0.000300193 | 0.001093914 | 1 |
| T2DM_admissi | leukocyte | *FLVCR1* | -0.942648991 | 0.000587451 | 0.001383749 | 1 |
| T2DM_admissi | leukocyte | *TDRD5* | -3.180418924 | 0.008543937 | 0.011268589 | 1 |
| T2DM_admissi | leukocyte | *ACP6* | -1.716402652 | 0.000177955 | 0.001062643 | 0.699185715 |
| T2DM_admissi | leukocyte | *OXER1* | -3.305103895 | 0.017532538 | 0.021958987 | 1 |
| T2DM_admissi | leukocyte | *HAAO* | -1.322966307 | 0.001116686 | 0.00202187 | 1 |
| T2DM_admissi | leukocyte | *RFTN2* | -2.329392329 | 0.003246427 | 0.004685971 | 1 |
| T2DM_admissi | leukocyte | *KCNJ3* | -0.953468497 | 0.003153848 | 0.004569126 | 1 |
| T2DM_admissi | leukocyte | *DUSP19* | -1.985667213 | 0.000138919 | 0.001062643 | 0.54581367 |
| T2DM_admissi | leukocyte | *SMC6* | -2.165306676 | 0.000116154 | 0.001062643 | 0.456370942 |
| T2DM_admissi | leukocyte | *NOSTRIN* | -1.296122481 | 0.000731693 | 0.001551442 | 1 |
| T2DM_admissi | leukocyte | *CTSS* | 0.223931536 | 0.01688082 | 0.021251118 | 1 |
| T2DM_admissi | leukocyte | *MSX1* | -1.241920695 | 0.001341362 | 0.002245911 | 1 |
| T2DM_admissi | leukocyte | *PACRGL* | -1.348041201 | 0.00063166 | 0.001433512 | 1 |
| T2DM_admissi | leukocyte | *ERCC3* | -1.556481392 | 0.000676583 | 0.00148591 | 1 |
| T2DM_admissi | leukocyte | *TDRD10* | -19.21544151 | 0.001764119 | 0.002770339 | 1 |
| T2DM_admissi | leukocyte | *CCNYL1* | -3.134343953 | 7.11E-05 | 0.001062643 | 0.27954506 |
| T2DM_admissi | leukocyte | *DCAF16* | -1.722270453 | 5.66E-05 | 0.001060479 | 0.222526642 |
| T2DM_admissi | leukocyte | *NPPC* | -13.44379912 | 3.87E-05 | 0.001037577 | 0.152144159 |
| T2DM_admissi | leukocyte | *GNPDA2* | -0.832918162 | 0.006423851 | 0.008679267 | 1 |
| T2DM_admissi | leukocyte | *NIPAL1* | -0.902699859 | 0.005040112 | 0.006955602 | 1 |
| T2DM_admissi | leukocyte | *CGGBP1* | -1.091766849 | 0.000775383 | 0.001611893 | 1 |
| T2DM_admissi | leukocyte | *DAPL1* | -4.432776455 | 1.06E-05 | 0.000784902 | 0.041777483 |
| T2DM_admissi | leukocyte | *EOGT* | -1.227689923 | 0.000475731 | 0.001261234 | 1 |
| T2DM_admissi | leukocyte | *POGLUT1* | -0.979910865 | 0.000716593 | 0.00153628 | 1 |
| T2DM_admissi | leukocyte | *IGFN1* | -1.112705409 | 0.012108257 | 0.0155621 | 1 |
| T2DM_admissi | leukocyte | *ATP1A1* | -0.809630357 | 0.000364707 | 0.001146348 | 1 |
| T2DM_admissi | leukocyte | *ARPC2* | -1.275678741 | 0.000121496 | 0.001062643 | 0.477355909 |
| T2DM_admissi | leukocyte | *CCT3* | -0.838654955 | 0.000896313 | 0.001768089 | 1 |
| T2DM_admissi | leukocyte | *EOMES* | -4.618098312 | 0.025055301 | 0.030610161 | 1 |
| T2DM_admissi | leukocyte | *CWC22* | -1.315476676 | 0.000407581 | 0.001188129 | 1 |
| T2DM_admissi | leukocyte | *HDAC11* | -3.2840892 | 0.000633156 | 0.001434874 | 1 |
| T2DM_admissi | leukocyte | *SUCLG1* | -1.205756119 | 0.000205742 | 0.001062643 | 0.808361382 |
| T2DM_admissi | leukocyte | *EFHB* | -0.592097665 | 0.021307755 | 0.026301656 | 1 |
| T2DM_admissi | leukocyte | *ICA1L* | -1.646545885 | 4.16E-07 | 0.000116632 | 0.001632847 |
| T2DM_admissi | leukocyte | *RYBP* | -0.869760861 | 0.000709374 | 0.001527196 | 1 |
| T2DM_admissi | leukocyte | *GTPBP8* | -2.339371497 | 2.09E-05 | 0.000929374 | 0.082305223 |
| T2DM_admissi | leukocyte | *CADPS* | -0.608737719 | 0.000924204 | 0.001797337 | 1 |
| T2DM_admissi | leukocyte | *ATXN7* | -1.427653836 | 0.000157213 | 0.001062643 | 0.617688845 |
| T2DM_admissi | leukocyte | *PSMD6* | -1.066985963 | 0.000267272 | 0.001087356 | 1 |
| T2DM_admissi | leukocyte | *PRICKLE2* | -1.178956293 | 0.00143203 | 0.002353177 | 1 |
| T2DM_admissi | leukocyte | *RPL9* | -0.866441362 | 3.52E-05 | 0.001037577 | 0.138320452 |
| T2DM_admissi | leukocyte | *IL17RC* | -1.330876146 | 0.00034681 | 0.001140861 | 1 |
| T2DM_admissi | leukocyte | *CRELD1* | -0.807019782 | 0.00298285 | 0.004348652 | 1 |
| T2DM_admissi | leukocyte | *TTC14* | -2.050700573 | 9.81E-05 | 0.001062643 | 0.385518976 |
| T2DM_admissi | leukocyte | *CXCL3* | 0.776451318 | 0.039013724 | 0.046548716 | 1 |

| | | | | | | |
|---|---|---|---|---|---|---|
| T2DM_admissi | leukocyte | *CXCL1* | -0.797304604 | 0.000701343 | 0.001517388 | 1 |
| T2DM_admissi | leukocyte | *CCDC158* | -2.947720244 | 0.001412542 | 0.002331881 | 1 |
| T2DM_admissi | leukocyte | *TM4SF18* | -1.077441152 | 0.00305275 | 0.004442316 | 1 |
| T2DM_admissi | leukocyte | *SNRK* | -0.863411923 | 0.000467192 | 0.001253701 | 1 |
| T2DM_admissi | leukocyte | *KIAA1143* | -0.886186343 | 0.000915655 | 0.001788965 | 1 |
| T2DM_admissi | leukocyte | *CCR1* | 0.823790035 | 0.020330622 | 0.025206379 | 1 |
| T2DM_admissi | leukocyte | *NMNAT3* | -0.943126301 | 0.002707653 | 0.003991883 | 1 |
| T2DM_admissi | leukocyte | *SMIM12* | -1.172263544 | 0.000286747 | 0.001088708 | 1 |
| T2DM_admissi | leukocyte | *TPRA1* | -2.244392185 | 0.000130987 | 0.001062643 | 0.514649526 |
| T2DM_admissi | leukocyte | *MEAF6* | -1.057489634 | 0.000280367 | 0.001088708 | 1 |
| T2DM_admissi | leukocyte | *DNALI1* | -1.214167976 | 0.000822629 | 0.001670341 | 1 |
| T2DM_admissi | leukocyte | *CAMK2N2* | -0.782277209 | 0.00048351 | 0.001273265 | 1 |
| T2DM_admissi | leukocyte | *RPN1* | -1.355511899 | 0.000105778 | 0.001062643 | 0.415602997 |
| T2DM_admissi | leukocyte | *IFT122* | -1.511462576 | 0.000900887 | 0.001772452 | 1 |
| T2DM_admissi | leukocyte | *RFC4* | -1.460514111 | 0.000637726 | 0.001438362 | 1 |
| T2DM_admissi | leukocyte | *TKT* | -1.299270013 | 0.000128597 | 0.001062643 | 0.505257585 |
| T2DM_admissi | leukocyte | *RFT1* | -2.281783405 | 8.78E-05 | 0.001062643 | 0.344838489 |
| T2DM_admissi | leukocyte | *UVSSA* | -1.502267941 | 0.0083601 | 0.011063265 | 1 |
| T2DM_admissi | leukocyte | *ARHGEF3* | -1.096239837 | 0.0013776 | 0.002288621 | 1 |
| T2DM_admissi | leukocyte | *LRPAP1* | -1.060980509 | 0.000196065 | 0.001062643 | 0.770339424 |
| T2DM_admissi | leukocyte | *PIGX* | -1.800960673 | 5.44E-05 | 0.001060479 | 0.213753512 |
| T2DM_admissi | leukocyte | *EXO5* | -2.06884535 | 0.000381652 | 0.001165285 | 1 |
| T2DM_admissi | leukocyte | *ERMAP* | -4.194032526 | 3.67E-05 | 0.001037577 | 0.144135583 |
| T2DM_admissi | leukocyte | *BDH2* | -1.409823235 | 1.94E-05 | 0.000917316 | 0.076244503 |
| T2DM_admissi | leukocyte | *CDC25A* | -1.907261646 | 0.000967203 | 0.001842046 | 1 |
| T2DM_admissi | leukocyte | *TEX264* | -1.533334026 | 9.49E-05 | 0.001062643 | 0.373053563 |
| T2DM_admissi | leukocyte | *PPM1M* | -1.764597161 | 0.000149746 | 0.001062643 | 0.588351348 |
| T2DM_admissi | leukocyte | *SAP30* | -1.125091975 | 0.000777515 | 0.001613765 | 1 |
| T2DM_admissi | leukocyte | *ANXA5* | -0.784367417 | 0.000306824 | 0.001098442 | 1 |
| T2DM_admissi | leukocyte | *CEP44* | -1.359913663 | 0.000327376 | 0.001122427 | 1 |
| T2DM_admissi | leukocyte | *HPGD* | -1.469042658 | 0.018704918 | 0.023360338 | 1 |
| T2DM_admissi | leukocyte | *NPY1R* | 2.41610017 | 0.019841025 | 0.024692869 | 1 |
| T2DM_admissi | leukocyte | *ARFIP1* | -1.923211392 | 5.94E-05 | 0.001060479 | 0.233226948 |
| T2DM_admissi | leukocyte | *ITGA2* | -0.812241592 | 0.000978042 | 0.001856058 | 1 |
| T2DM_admissi | leukocyte | *ELOVL7* | -1.172154786 | 0.0005639 | 0.001362351 | 1 |
| T2DM_admissi | leukocyte | *LMBRD2* | -1.489585801 | 8.85E-05 | 0.001062643 | 0.347543575 |
| T2DM_admissi | leukocyte | *STARD4* | -1.692451597 | 0.000123658 | 0.001062643 | 0.485853698 |
| T2DM_admissi | leukocyte | *CMBL* | -1.267346896 | 0.000402133 | 0.001185421 | 1 |
| T2DM_admissi | leukocyte | *PRRC1* | -0.935524685 | 0.000511831 | 0.001308382 | 1 |
| T2DM_admissi | leukocyte | *AGGF1* | -1.698005126 | 8.75E-05 | 0.001062643 | 0.343698281 |
| T2DM_admissi | leukocyte | *WDR41* | -2.037558416 | 8.24E-05 | 0.001062643 | 0.323651383 |
| T2DM_admissi | leukocyte | *SERINC5* | -4.834434479 | 0.000104959 | 0.001062643 | 0.412382787 |
| T2DM_admissi | leukocyte | *ERAP1* | -1.307398646 | 0.030850577 | 0.037204395 | 1 |
| T2DM_admissi | leukocyte | *RICTOR* | -1.771261863 | 0.000108502 | 0.001062643 | 0.426302798 |
| T2DM_admissi | leukocyte | *UTP15* | -1.043763457 | 0.000826843 | 0.001675216 | 1 |
| T2DM_admissi | leukocyte | *NSA2* | -0.76070312 | 0.001043597 | 0.001936841 | 1 |
| T2DM_admissi | leukocyte | *LEAP2* | -1.753469931 | 0.001498341 | 0.002433642 | 1 |
| T2DM_admissi | leukocyte | *DACT2* | -5.017711845 | 0.000825203 | 0.001673837 | 1 |
| T2DM_admissi | leukocyte | *PDSS2* | -1.195333633 | 0.000539261 | 0.001334229 | 1 |
| T2DM_admissi | leukocyte | *STK17A* | -1.020011603 | 0.000670091 | 0.001477434 | 1 |

| | | | | | | |
|---|---|---|---|---|---|---|
| T2DM_admissi | leukocyte | *GALNT10* | -2.243729434 | 0.002018824 | 0.003094794 | 1 |
| T2DM_admissi | leukocyte | *SAP30L* | -1.660689332 | 0.000214893 | 0.001062643 | 0.844316237 |
| T2DM_admissi | leukocyte | *HCN1* | -2.35428548 | 0.025864792 | 0.03148165 | 1 |
| T2DM_admissi | leukocyte | *COG5* | -2.197736319 | 8.64E-05 | 0.001062643 | 0.339325598 |
| T2DM_admissi | leukocyte | *SLU7* | -1.280033943 | 0.000194104 | 0.001062643 | 0.762635722 |
| T2DM_admissi | leukocyte | *CAMLG* | -0.990890728 | 0.000279692 | 0.001088708 | 1 |
| T2DM_admissi | leukocyte | *KIF6* | -1.89477151 | 0.002552798 | 0.003786313 | 1 |
| T2DM_admissi | leukocyte | *STEAP1* | -1.332056678 | 0.005670171 | 0.0077543 | 1 |
| T2DM_admissi | leukocyte | *FNDC1* | -7.272792598 | 0.002657186 | 0.003927797 | 1 |
| T2DM_admissi | leukocyte | *CHMP4C* | -1.157179264 | 0.000416748 | 0.001199564 | 1 |
| T2DM_admissi | leukocyte | *BRI3* | -1.604736423 | 2.38E-05 | 0.000982543 | 0.093468219 |
| T2DM_admissi | leukocyte | *LMTK2* | -1.262452765 | 0.000254324 | 0.001077348 | 0.999237943 |
| T2DM_admissi | leukocyte | *DLC1* | -0.860235866 | 0.026122796 | 0.031756332 | 1 |
| T2DM_admissi | leukocyte | *PEX2* | -1.034666441 | 0.000404315 | 0.001185781 | 1 |
| T2DM_admissi | leukocyte | *MED30* | -1.224918544 | 0.000318886 | 0.001113024 | 1 |
| T2DM_admissi | leukocyte | *TNFRSF11B* | -3.517407712 | 0.001580139 | 0.002532576 | 1 |
| T2DM_admissi | leukocyte | *CSMD3* | -76.95691984 | 0.001317259 | 0.002219344 | 1 |
| T2DM_admissi | leukocyte | *OSGIN2* | -1.727392108 | 0.000127815 | 0.001062643 | 0.502186632 |
| T2DM_admissi | leukocyte | *DEFB1* | -0.530694761 | 0.000907731 | 0.001777903 | 1 |
| T2DM_admissi | leukocyte | *SUN1* | -0.959750785 | 0.000371514 | 0.001152986 | 1 |
| T2DM_admissi | leukocyte | *TMEM184A* | -1.256221637 | 0.00055061 | 0.001351735 | 1 |
| T2DM_admissi | leukocyte | *INTS1* | -1.243744671 | 0.000458562 | 0.001240899 | 1 |
| T2DM_admissi | leukocyte | *YWHAZ* | -1.088496047 | 0.000140062 | 0.001062643 | 0.550302512 |
| T2DM_admissi | leukocyte | *DCAF13* | -1.208517927 | 0.000514767 | 0.001311621 | 1 |
| T2DM_admissi | leukocyte | *INTS8* | -1.485536019 | 0.002697746 | 0.003980265 | 1 |
| T2DM_admissi | leukocyte | *GEM* | -1.275889073 | 0.000537094 | 0.001334229 | 1 |
| T2DM_admissi | leukocyte | *RPP25L* | -2.008009748 | 0.000135786 | 0.001062643 | 0.533503315 |
| T2DM_admissi | leukocyte | *FAM219A* | -1.972303547 | 0.000144097 | 0.001062643 | 0.566158803 |
| T2DM_admissi | leukocyte | *NUDT2* | -1.410342659 | 0.000170993 | 0.001062643 | 0.671831158 |
| T2DM_admissi | leukocyte | *METTL2B* | -0.973882017 | 0.011016619 | 0.014233573 | 1 |
| T2DM_admissi | leukocyte | *FXN* | -2.391231695 | 0.000367288 | 0.001148802 | 1 |
| T2DM_admissi | leukocyte | *MAMDC2* | -2.657311368 | 0.000891773 | 0.001763074 | 1 |
| T2DM_admissi | leukocyte | *PRSS37* | -631.5623973 | 0.001341602 | 0.002245911 | 1 |
| T2DM_admissi | leukocyte | *C9orf64* | -1.663526597 | 0.000216596 | 0.001062643 | 0.851004 |
| T2DM_admissi | leukocyte | *ANKS6* | -0.910842348 | 0.008326355 | 0.011022321 | 1 |
| T2DM_admissi | leukocyte | *CLDN3* | -1.293197969 | 0.00014358 | 0.001062643 | 0.564125942 |
| T2DM_admissi | leukocyte | *WNK2* | -2.051011951 | 0.000110648 | 0.001062643 | 0.434734074 |
| T2DM_admissi | leukocyte | *NOL6* | -2.381356066 | 0.000273089 | 0.001088708 | 1 |
| T2DM_admissi | leukocyte | *PIGO* | -2.496044825 | 9.05E-05 | 0.001062643 | 0.355714904 |
| T2DM_admissi | leukocyte | *ARHGAP12* | -1.326465787 | 0.000289349 | 0.001088708 | 1 |
| T2DM_admissi | leukocyte | *WRN* | -1.369650154 | 0.000359514 | 0.001142338 | 1 |
| T2DM_admissi | leukocyte | *CFL2* | -1.220160275 | 0.000357463 | 0.001141109 | 1 |
| T2DM_admissi | leukocyte | *PGM2L1* | -2.426998197 | 1.99E-05 | 0.000920455 | 0.078238646 |
| T2DM_admissi | leukocyte | *PHYHIPL* | -1.196173797 | 0.000647175 | 0.00144721 | 1 |
| T2DM_admissi | leukocyte | *PKNOX2* | 3.754225265 | 0.019378653 | 0.024171024 | 1 |
| T2DM_admissi | leukocyte | *RPL36AL* | -0.96773142 | 8.68E-05 | 0.001062643 | 0.340943073 |
| T2DM_admissi | leukocyte | *TMEM63C* | -1.064996203 | 0.000655314 | 0.001454635 | 1 |
| T2DM_admissi | leukocyte | *OXGR1* | -2.564536738 | 0.001333371 | 0.002236898 | 1 |
| T2DM_admissi | leukocyte | *VDAC2* | -1.118475112 | 0.000110353 | 0.001062643 | 0.433577595 |
| T2DM_admissi | leukocyte | *SNAPC4* | -1.101283474 | 0.002281151 | 0.003441875 | 1 |

| | | | | | | |
|---|---|---|---|---|---|---|
| T2DM_admissi | leukocyte | *TSC1* | -1.761143576 | 0.000212999 | 0.001062643 | 0.836873432 |
| T2DM_admissi | leukocyte | *ZMYND19* | -1.167159965 | 0.000682954 | 0.001493225 | 1 |
| T2DM_admissi | leukocyte | *METTL17* | -1.703167181 | 0.0003025 | 0.001093914 | 1 |
| T2DM_admissi | leukocyte | *VWA2* | -3.230878352 | 0.000385199 | 0.001168251 | 1 |
| T2DM_admissi | leukocyte | *METTL3* | -1.203914947 | 0.001434226 | 0.002354561 | 1 |
| T2DM_admissi | leukocyte | *TRUB1* | -1.862438512 | 7.48E-05 | 0.001062643 | 0.293764887 |
| T2DM_admissi | leukocyte | *ARHGAP42* | -1.803726654 | 0.001018865 | 0.001906024 | 1 |
| T2DM_admissi | leukocyte | *SLC39A13* | -1.174458042 | 0.000475695 | 0.001261234 | 1 |
| T2DM_admissi | leukocyte | *PSMC3* | -1.003192765 | 0.000237115 | 0.001070831 | 0.93162334 |
| T2DM_admissi | leukocyte | *CPSF2* | -0.971269949 | 0.001312982 | 0.002213088 | 1 |
| T2DM_admissi | leukocyte | *IFI27L1* | -1.328651093 | 0.000207381 | 0.001062643 | 0.814798494 |
| T2DM_admissi | leukocyte | *SMCO4* | -1.282165428 | 0.000451137 | 0.001236066 | 1 |
| T2DM_admissi | leukocyte | *ABTB2* | -1.400668193 | 0.007597422 | 0.010125601 | 1 |
| T2DM_admissi | leukocyte | *LIPC* | -3.756794205 | 0.02694194 | 0.032665504 | 1 |
| T2DM_admissi | leukocyte | *CEP57* | -1.170101364 | 7.75E-05 | 0.001062643 | 0.304427756 |
| T2DM_admissi | leukocyte | *JAM3* | -0.913333932 | 0.001522832 | 0.002466285 | 1 |
| T2DM_admissi | leukocyte | *IKBIP* | -1.36524519 | 0.000827162 | 0.001675216 | 1 |
| T2DM_admissi | leukocyte | *ZFYVE19* | -0.969495627 | 0.005855195 | 0.007993419 | 1 |
| T2DM_admissi | leukocyte | *CKB* | -1.061306469 | 1.34E-05 | 0.000815363 | 0.052549056 |
| T2DM_admissi | leukocyte | *BTRC* | -2.310341855 | 0.000263243 | 0.001083884 | 1 |
| T2DM_admissi | leukocyte | *BAG5* | -1.226581178 | 0.000486292 | 0.001277166 | 1 |
| T2DM_admissi | leukocyte | *LARP6* | -1.05818875 | 0.000348365 | 0.001141096 | 1 |
| T2DM_admissi | leukocyte | *ZNF319* | -3.575353364 | 0.000139924 | 0.001062643 | 0.549761574 |
| T2DM_admissi | leukocyte | *SGPL1* | -2.590141115 | 1.40E-06 | 0.000261424 | 0.005489894 |
| T2DM_admissi | leukocyte | *FRS2* | -1.408183921 | 0.000141694 | 0.001062643 | 0.556716674 |
| T2DM_admissi | leukocyte | *PCBD1* | -1.368516117 | 1.65E-05 | 0.000823546 | 0.064771035 |
| T2DM_admissi | leukocyte | *COX11* | -1.45960564 | 0.000119192 | 0.001062643 | 0.468307316 |
| T2DM_admissi | leukocyte | *WBP1L* | -2.657657223 | 0.00018824 | 0.001062643 | 0.739594201 |
| T2DM_admissi | leukocyte | *CYB5R2* | -1.218896543 | 0.001106888 | 0.002007832 | 1 |
| T2DM_admissi | leukocyte | *TUB* | -1.660302698 | 0.000376872 | 0.001160448 | 1 |
| T2DM_admissi | leukocyte | *RIC3* | -0.944047993 | 0.000400509 | 0.001184732 | 1 |
| T2DM_admissi | leukocyte | *XRRA1* | -1.430292756 | 9.39E-05 | 0.001062643 | 0.368995534 |
| T2DM_admissi | leukocyte | *CDYL2* | -1.190854114 | 0.001250562 | 0.002155971 | 1 |
| T2DM_admissi | leukocyte | *ZNF143* | -1.180478807 | 0.000600151 | 0.001399402 | 1 |
| T2DM_admissi | leukocyte | *CCDC68* | -1.197648311 | 0.001049007 | 0.001940644 | 1 |
| T2DM_admissi | leukocyte | *ZSCAN21* | -1.739405836 | 0.001162961 | 0.002078832 | 1 |
| T2DM_admissi | leukocyte | *RIMKLB* | -1.159496383 | 0.000526386 | 0.001319879 | 1 |
| T2DM_admissi | leukocyte | *GALR1* | 1.254327345 | 0.005644425 | 0.007721777 | 1 |
| T2DM_admissi | leukocyte | *ATF7IP2* | -1.786307223 | 6.31E-05 | 0.001060479 | 0.247778172 |
| T2DM_admissi | leukocyte | *COG1* | -1.245204797 | 0.000195766 | 0.001062643 | 0.769163878 |
| T2DM_admissi | leukocyte | *B2M* | -0.804763919 | 0.00016278 | 0.001062643 | 0.639563174 |
| T2DM_admissi | leukocyte | *ACSM1* | -2.542457625 | 0.030746235 | 0.037112736 | 1 |
| T2DM_admissi | leukocyte | *AP1G1* | -0.890582855 | 0.000630118 | 0.001433512 | 1 |
| T2DM_admissi | leukocyte | *SLFN5* | -0.959603359 | 0.000714246 | 0.001532645 | 1 |
| T2DM_admissi | leukocyte | *SAAL1* | -1.124726735 | 0.000896719 | 0.001768089 | 1 |
| T2DM_admissi | leukocyte | *SCNN1G* | -10.2489531 | 0.00088648 | 0.001757306 | 1 |
| T2DM_admissi | leukocyte | *DCTN5* | -1.365347443 | 0.00020826 | 0.001062643 | 0.81825314 |
| T2DM_admissi | leukocyte | *STAT6* | -2.066876742 | 0.000344486 | 0.001140861 | 1 |
| T2DM_admissi | leukocyte | *PATL1* | -1.148487207 | 0.000131619 | 0.001062643 | 0.517130573 |
| T2DM_admissi | leukocyte | *MRPL16* | -1.719504917 | 0.002902567 | 0.00425212 | 1 |

| | | | | | | |
|---|---|---|---|---|---|---|
| T2DM_admissi | leukocyte | *GREM1* | -1.976147504 | 0.000246228 | 0.001077213 | 0.967430605 |
| T2DM_admissi | leukocyte | *TSC22D4* | -1.610269855 | 0.00036812 | 0.001148802 | 1 |
| T2DM_admissi | leukocyte | *MS4A14* | 0.22571605 | 0.011203302 | 0.014446266 | 1 |
| T2DM_admissi | leukocyte | *DIS3L* | -1.928827318 | 0.0003032 | 0.001093914 | 1 |
| T2DM_admissi | leukocyte | *RCCD1* | -3.528169267 | 1.60E-05 | 0.000823546 | 0.063014973 |
| T2DM_admissi | leukocyte | *MAPRE2* | -1.171618425 | 0.000339549 | 0.001134896 | 1 |
| T2DM_admissi | leukocyte | *NAT16* | -0.784982292 | 0.000793139 | 0.001634654 | 1 |
| T2DM_admissi | leukocyte | *GNGT2* | 0.450363958 | 0.00926693 | 0.012118325 | 1 |
| T2DM_admissi | leukocyte | *SLC27A4* | -1.099217771 | 0.00509664 | 0.007031144 | 1 |
| T2DM_admissi | leukocyte | *URM1* | -1.760816075 | 9.86E-05 | 0.001062643 | 0.387352738 |
| T2DM_admissi | leukocyte | *CERCAM* | -1.012993974 | 0.001765101 | 0.002770339 | 1 |
| T2DM_admissi | leukocyte | *DOLPP1* | -1.375568467 | 0.000961458 | 0.001831992 | 1 |
| T2DM_admissi | leukocyte | *ENDOG* | -1.51620786 | 0.000144202 | 0.001062643 | 0.566569864 |
| T2DM_admissi | leukocyte | *PRR15L* | -1.190907252 | 0.017243388 | 0.021665901 | 1 |
| T2DM_admissi | leukocyte | *RNF214* | -2.114135753 | 0.000245539 | 0.001077213 | 0.964724153 |
| T2DM_admissi | leukocyte | *POP5* | -1.441295159 | 8.14E-05 | 0.001062643 | 0.319874846 |
| T2DM_admissi | leukocyte | *ACAA2* | -1.355621413 | 0.000140088 | 0.001062643 | 0.550406884 |
| T2DM_admissi | leukocyte | *FN3K* | -1.425831895 | 0.001956787 | 0.003020911 | 1 |
| T2DM_admissi | leukocyte | *ZNF23* | -1.095346741 | 0.001010876 | 0.001894951 | 1 |
| T2DM_admissi | leukocyte | *ZNF229* | -1.566849498 | 0.000811757 | 0.001657387 | 1 |
| T2DM_admissi | leukocyte | *ZNF180* | -1.189015768 | 0.001249883 | 0.002155971 | 1 |
| T2DM_admissi | leukocyte | *CA4* | -0.859384494 | 0.001667648 | 0.002644145 | 1 |
| T2DM_admissi | leukocyte | *GPX4* | -1.298511093 | 7.63E-05 | 0.001062643 | 0.299936614 |
| T2DM_admissi | leukocyte | *ZNF641* | -1.877737305 | 0.000509434 | 0.001305652 | 1 |
| T2DM_admissi | leukocyte | *CACNB3* | -1.107402384 | 0.021663571 | 0.026707301 | 1 |
| T2DM_admissi | leukocyte | *TP53I13* | -1.497940381 | 1.09E-05 | 0.000784902 | 0.042799834 |
| T2DM_admissi | leukocyte | *TUBA1C* | -0.790744833 | 0.000502099 | 0.001297006 | 1 |
| T2DM_admissi | leukocyte | *ZNF528* | -4.966200857 | 2.63E-05 | 0.000984894 | 0.103413857 |
| T2DM_admissi | leukocyte | *RAB4B* | -1.583622578 | 0.000102549 | 0.001062643 | 0.402914732 |
| T2DM_admissi | leukocyte | *TMC4* | -1.276218712 | 0.000235232 | 0.001067235 | 0.924225727 |
| T2DM_admissi | leukocyte | *C19orf33* | -0.984565378 | 0.001195681 | 0.002107792 | 1 |
| T2DM_admissi | leukocyte | *DNAAF3* | -4.003524891 | 0.001163574 | 0.002078982 | 1 |
| T2DM_admissi | leukocyte | *CHAF1A* | -1.95956971 | 0.000128496 | 0.001062643 | 0.504860383 |
| T2DM_admissi | leukocyte | *UBXN6* | -1.745457028 | 6.20E-05 | 0.001060479 | 0.243750447 |
| T2DM_admissi | leukocyte | *NXN* | -0.87521019 | 0.000623762 | 0.00142569 | 1 |
| T2DM_admissi | leukocyte | *KIFC2* | -5.113235673 | 1.42E-05 | 0.000815363 | 0.055627794 |
| T2DM_admissi | leukocyte | *SLC43A2* | 0.822763903 | 0.02082174 | 0.02575838 | 1 |
| T2DM_admissi | leukocyte | *SERPINF2* | 0.483576516 | 0.012481715 | 0.016010663 | 1 |
| T2DM_admissi | leukocyte | *SRR* | -1.151448787 | 0.003415808 | 0.004907024 | 1 |
| T2DM_admissi | leukocyte | *TRPV3* | -5.830976545 | 0.00055451 | 0.001356582 | 1 |
| T2DM_admissi | leukocyte | *C19orf48* | -1.924179819 | 0.001388033 | 0.002300119 | 1 |
| T2DM_admissi | leukocyte | *ZNF83* | -1.589945713 | 0.000245779 | 0.001077213 | 0.965665826 |
| T2DM_admissi | leukocyte | *ZNF558* | -1.134194383 | 0.012354388 | 0.015862873 | 1 |
| T2DM_admissi | leukocyte | *NDUFV1* | -1.124701598 | 0.000291811 | 0.001088708 | 1 |
| T2DM_admissi | leukocyte | *ZNF232* | -0.955806768 | 0.002987516 | 0.004353839 | 1 |
| T2DM_admissi | leukocyte | *MIS12* | -1.47314116 | 0.000445821 | 0.001232235 | 1 |
| T2DM_admissi | leukocyte | *HID1* | -1.284757697 | 0.000569308 | 0.001367774 | 1 |
| T2DM_admissi | leukocyte | *SRP68* | -1.005531981 | 0.000479864 | 0.001266209 | 1 |
| T2DM_admissi | leukocyte | *TMEM68* | -1.635294084 | 0.000332563 | 0.001127387 | 1 |
| T2DM_admissi | leukocyte | *GSDMA* | -7.75289446 | 0.001615281 | 0.002577757 | 1 |

| | | | | | | |
|---|---|---|---|---|---|---|
| T2DM_admissi | leukocyte | *MLST8* | -0.89779186 | 0.001566624 | 0.002515434 | 1 |
| T2DM_admissi | leukocyte | *ECI1* | -0.951646797 | 0.00049104 | 0.001283629 | 1 |
| T2DM_admissi | leukocyte | *ABCA3* | -1.306255601 | 7.92E-05 | 0.001062643 | 0.311304753 |
| T2DM_admissi | leukocyte | *BSCL2* | -1.062717624 | 0.000226569 | 0.001062643 | 0.890188388 |
| T2DM_admissi | leukocyte | *C2CD3* | -2.239934548 | 0.000303985 | 0.001094736 | 1 |
| T2DM_admissi | leukocyte | *TRANK1* | -3.193616573 | 0.000248536 | 0.001077213 | 0.976496006 |
| T2DM_admissi | leukocyte | *RPSA* | -0.785113676 | 0.000351877 | 0.001141109 | 1 |
| T2DM_admissi | leukocyte | *ENTPD3* | -0.793507275 | 0.00063448 | 0.001434874 | 1 |
| T2DM_admissi | leukocyte | *ULK4* | -1.60785084 | 0.000315788 | 0.001111111 | 1 |
| T2DM_admissi | leukocyte | *LTBP3* | -0.91110141 | 0.001565565 | 0.002514761 | 1 |
| T2DM_admissi | leukocyte | *SAC3D1* | -1.409750383 | 0.000435739 | 0.001222199 | 1 |
| T2DM_admissi | leukocyte | *CCDC88B* | -3.070143536 | 0.002878876 | 0.004219162 | 1 |
| T2DM_admissi | leukocyte | *PBK* | -2.515840027 | 0.001243035 | 0.002152855 | 1 |
| T2DM_admissi | leukocyte | *KIAA1586* | -1.670731987 | 9.41E-05 | 0.001062643 | 0.369702719 |
| T2DM_admissi | leukocyte | *FAM83B* | -1.240063738 | 0.001454001 | 0.002373399 | 1 |
| T2DM_admissi | leukocyte | *LMBRD1* | -1.509322186 | 0.000109354 | 0.001062643 | 0.429651273 |
| T2DM_admissi | leukocyte | *TTC39C* | -1.165268292 | 0.000285599 | 0.001088708 | 1 |
| T2DM_admissi | leukocyte | *POLR2J3* | -1.259015351 | 0.000662408 | 0.001464605 | 1 |
| T2DM_admissi | leukocyte | *COA6* | -1.322649933 | 0.000358394 | 0.001141109 | 1 |
| T2DM_admissi | leukocyte | *PXK* | -1.566724148 | 0.000469914 | 0.001255055 | 1 |
| T2DM_admissi | leukocyte | *ING5* | -2.249471714 | 0.000102962 | 0.001062643 | 0.404539624 |
| T2DM_admissi | leukocyte | *BMP1* | -1.117915694 | 0.005520114 | 0.007570167 | 1 |
| T2DM_admissi | leukocyte | *HEXIM2* | -1.398381319 | 0.0012582 | 0.002162496 | 1 |
| T2DM_admissi | leukocyte | *SERINC2* | -0.82275479 | 0.000504823 | 0.001300386 | 1 |
| T2DM_admissi | leukocyte | *TRAPPC11* | -0.956692108 | 0.000913101 | 0.001784988 | 1 |
| T2DM_admissi | leukocyte | *ING2* | -0.872691212 | 0.00075207 | 0.001577074 | 1 |
| T2DM_admissi | leukocyte | *ADAM9* | -2.168300488 | 0.00064595 | 0.001446812 | 1 |
| T2DM_admissi | leukocyte | *NDUFS5* | -0.982474206 | 0.000111801 | 0.001062643 | 0.439265827 |
| T2DM_admissi | leukocyte | *DNAJC21* | -1.445287864 | 6.26E-05 | 0.001060479 | 0.245787879 |
| T2DM_admissi | leukocyte | *CNNM3* | -1.178663754 | 0.000472586 | 0.001256287 | 1 |
| T2DM_admissi | leukocyte | *GSTM4* | -1.193923714 | 0.002952015 | 0.004310096 | 1 |
| T2DM_admissi | leukocyte | *SHOX2* | -9.056925333 | 0.004769346 | 0.006614458 | 1 |
| T2DM_admissi | leukocyte | *PPIP5K1* | -3.781724437 | 1.26E-05 | 0.000801308 | 0.049681118 |
| T2DM_admissi | leukocyte | *ADAL* | -2.630816323 | 0.006479032 | 0.008744801 | 1 |
| T2DM_admissi | leukocyte | *LCMT2* | -1.587445469 | 0.002542761 | 0.003774544 | 1 |
| T2DM_admissi | leukocyte | *ZNF507* | -1.582531068 | 0.000163867 | 0.001062643 | 0.643833486 |
| T2DM_admissi | leukocyte | *NSG1* | -0.84698109 | 0.001287304 | 0.002187637 | 1 |
| T2DM_admissi | leukocyte | *ZBTB49* | -2.931646543 | 0.001555605 | 0.002503881 | 1 |
| T2DM_admissi | leukocyte | *GFM1* | -1.052713299 | 0.000247125 | 0.001077213 | 0.970953098 |
| T2DM_admissi | leukocyte | *FSTL5* | -0.716121291 | 0.002913785 | 0.004263785 | 1 |
| T2DM_admissi | leukocyte | *CTRB1* | -0.526342206 | 0.000463522 | 0.001247383 | 1 |
| T2DM_admissi | leukocyte | *CTRB2* | -0.483833702 | 0.000659523 | 0.001460691 | 1 |
| T2DM_admissi | leukocyte | *TMEM129* | -2.517988172 | 1.89E-07 | 6.20E-05 | 0.000744365 |
| T2DM_admissi | leukocyte | *PPIC* | -0.626773198 | 0.001370243 | 0.002282189 | 1 |
| T2DM_admissi | leukocyte | *STXBP6* | -1.535062433 | 6.85E-05 | 0.001062643 | 0.269238058 |
| T2DM_admissi | leukocyte | *MFF* | -1.213684997 | 0.000199444 | 0.001062643 | 0.783614881 |
| T2DM_admissi | leukocyte | *CPLX1* | -1.055615623 | 0.003700937 | 0.005275591 | 1 |
| T2DM_admissi | leukocyte | *ROR2* | -2.863495529 | 0.00635845 | 0.008599776 | 1 |
| T2DM_admissi | leukocyte | *FAM110B* | -1.206217531 | 0.000869376 | 0.001735895 | 1 |
| T2DM_admissi | leukocyte | *ATF5* | -1.501104798 | 0.000219146 | 0.001062643 | 0.861024893 |

| | | | | | | |
|---|---|---|---|---|---|---|
| T2DM_admissi | leukocyte | *XPO6* | -2.295435537 | 5.75E-05 | 0.001060479 | 0.225986801 |
| T2DM_admissi | leukocyte | *RGS14* | -1.692459799 | 0.000733207 | 0.001552225 | 1 |
| T2DM_admissi | leukocyte | *TBC1D10B* | -1.639533994 | 0.000124994 | 0.001062643 | 0.491100254 |
| T2DM_admissi | leukocyte | *PRELID1* | -0.925296229 | 0.000322532 | 0.001116501 | 1 |
| T2DM_admissi | leukocyte | *CXCL10* | -195.6300815 | 0.001291768 | 0.002191432 | 1 |
| T2DM_admissi | leukocyte | *NMD3* | -1.175479094 | 0.000336243 | 0.001131077 | 1 |
| T2DM_admissi | leukocyte | *STK32A* | -1.352606961 | 0.00084446 | 0.001700095 | 1 |
| T2DM_admissi | leukocyte | *P2RY12* | -54.83960411 | 0.02995557 | 0.03620284 | 1 |
| T2DM_admissi | leukocyte | *SNUPN* | -3.689059961 | 1.31E-05 | 0.000815363 | 0.051379311 |
| T2DM_admissi | leukocyte | *CRADD* | -1.152306782 | 0.001985765 | 0.003054843 | 1 |
| T2DM_admissi | leukocyte | *COL22A1* | -11.31033526 | 0.00159221 | 0.002547147 | 1 |
| T2DM_admissi | leukocyte | *SDC2* | -3.106508557 | 5.59E-05 | 0.001060479 | 0.219818442 |
| T2DM_admissi | leukocyte | *CLIC4* | -0.663469954 | 0.001293501 | 0.002192479 | 1 |
| T2DM_admissi | leukocyte | *SLC38A11* | -9.566700626 | 0.000580054 | 0.001379559 | 1 |
| T2DM_admissi | leukocyte | *CCDC8* | -1.26583248 | 0.033370471 | 0.040083333 | 1 |
| T2DM_admissi | leukocyte | *HINT1* | -0.867301173 | 0.000129419 | 0.001062643 | 0.508486459 |
| T2DM_admissi | leukocyte | *DTWD2* | -3.750684672 | 0.000932613 | 0.001804129 | 1 |
| T2DM_admissi | leukocyte | *INO80E* | -2.16849876 | 5.31E-07 | 0.000130474 | 0.002087587 |
| T2DM_admissi | leukocyte | *C15orf40* | -1.26772568 | 0.000204603 | 0.001062643 | 0.803886549 |
| T2DM_admissi | leukocyte | *HIC2* | -4.397351868 | 0.001621006 | 0.002583745 | 1 |
| T2DM_admissi | leukocyte | *AGPAT2* | -1.318005315 | 0.000122569 | 0.001062643 | 0.481571797 |
| T2DM_admissi | leukocyte | *FASN* | -2.445704264 | 1.12E-05 | 0.000784902 | 0.043905853 |
| T2DM_admissi | leukocyte | *MT1E* | -0.689142771 | 0.000727798 | 0.00154878 | 1 |
| T2DM_admissi | leukocyte | *LDB2* | -1.213847036 | 0.002229597 | 0.003369478 | 1 |
| T2DM_admissi | leukocyte | *RAC3* | -1.81161701 | 0.000124032 | 0.001062643 | 0.487320468 |
| T2DM_admissi | leukocyte | *LIMS1* | -1.541679281 | 4.77E-05 | 0.00105496 | 0.187498153 |
| T2DM_admissi | leukocyte | *CSGALNACT2* | -3.120716184 | 0.00035731 | 0.001141109 | 1 |
| T2DM_admissi | leukocyte | *AVEN* | -3.695255358 | 4.50E-05 | 0.001037577 | 0.176628351 |
| T2DM_admissi | leukocyte | *CTNND2* | -1.332756799 | 0.000357903 | 0.001141109 | 1 |
| T2DM_admissi | leukocyte | *WNT10B* | -68.77371524 | 0.001075 | 0.001968161 | 1 |
| T2DM_admissi | leukocyte | *CALML6* | 1.108703476 | 0.008980373 | 0.011776997 | 1 |
| T2DM_admissi | leukocyte | *TM4SF4* | -1.116901753 | 0.000347154 | 0.001140861 | 1 |
| T2DM_admissi | leukocyte | *OTUD3* | -0.880994092 | 0.000656048 | 0.001454635 | 1 |
| T2DM_admissi | leukocyte | *MAP3K2* | -1.60167061 | 9.29E-05 | 0.001062643 | 0.364919792 |
| T2DM_admissi | leukocyte | *PUSL1* | -2.903663989 | 3.76E-05 | 0.001037577 | 0.147621859 |
| T2DM_admissi | leukocyte | *IFFO2* | -1.070063746 | 0.000916949 | 0.001790602 | 1 |
| T2DM_admissi | leukocyte | *MYRIP* | -1.534760489 | 0.00098504 | 0.001866067 | 1 |
| T2DM_admissi | leukocyte | *FAM153A* | -1.885213293 | 0.014508272 | 0.018459521 | 1 |
| T2DM_admissi | leukocyte | *ZNF778* | -1.730083421 | 0.000116865 | 0.001062643 | 0.459161279 |
| T2DM_admissi | leukocyte | *NIPA1* | -1.548388929 | 0.000172043 | 0.001062643 | 0.675955544 |
| T2DM_admissi | leukocyte | *FOXD4* | -11.05726022 | 0.031142153 | 0.037521472 | 1 |
| T2DM_admissi | leukocyte | *UBE2E1* | -2.164931502 | 8.79E-05 | 0.001062643 | 0.345430993 |
| T2DM_admissi | leukocyte | *ANKK1* | -31.70882499 | 0.000724308 | 0.001544387 | 1 |
| T2DM_admissi | leukocyte | *USP47* | -1.481134078 | 0.000118548 | 0.001062643 | 0.465775241 |
| T2DM_admissi | leukocyte | *PDCD6IP* | -1.192718348 | 0.000261419 | 0.001083674 | 1 |
| T2DM_admissi | leukocyte | *CRTAP* | -2.312949096 | 6.93E-05 | 0.001062643 | 0.272162445 |
| T2DM_admissi | leukocyte | *STX8* | -1.719567348 | 0.000130833 | 0.001062643 | 0.51404391 |
| T2DM_admissi | leukocyte | *NFRKB* | -2.167056203 | 0.000202929 | 0.001062643 | 0.797306888 |
| T2DM_admissi | leukocyte | *SETMAR* | -1.156242966 | 0.000572873 | 0.001370771 | 1 |
| T2DM_admissi | leukocyte | *CST5* | -5.389778283 | 0.021155084 | 0.026137838 | 1 |

| | | | | | | |
|---|---|---|---|---|---|---|
| T2DM_admissi | leukocyte | *CST2* | -2.066268549 | 0.004537597 | 0.006319823 | 1 |
| T2DM_admissi | leukocyte | *KRT8* | -0.489235704 | 0.000940058 | 0.001810978 | 1 |
| T2DM_admissi | leukocyte | *MGMT* | -1.591985307 | 0.000234572 | 0.001066706 | 0.921634205 |
| T2DM_admissi | leukocyte | *NFXL1* | -2.045284314 | 0.000169783 | 0.001062643 | 0.667076808 |
| T2DM_admissi | leukocyte | *DNAJC18* | -1.087889488 | 0.015671919 | 0.019792661 | 1 |
| T2DM_admissi | leukocyte | *KRT4* | -51.43882419 | 0.011302308 | 0.014569149 | 1 |
| T2DM_admissi | leukocyte | *SLC23A1* | -9.163027958 | 0.002675067 | 0.003952741 | 1 |
| T2DM_admissi | leukocyte | *NPAS2* | -0.925053721 | 0.001233597 | 0.002144697 | 1 |
| T2DM_admissi | leukocyte | *NUDT9* | -0.916193796 | 0.001935341 | 0.002992504 | 1 |
| T2DM_admissi | leukocyte | *SMAGP* | -0.650918209 | 0.016266392 | 0.020503899 | 1 |
| T2DM_admissi | leukocyte | *IRX1* | -1.638732025 | 0.001318836 | 0.002220096 | 1 |
| T2DM_admissi | leukocyte | *CDH2* | -0.911317653 | 0.001305222 | 0.002204736 | 1 |
| T2DM_admissi | leukocyte | *HSPA4* | -1.314263107 | 0.000156349 | 0.001062643 | 0.614294538 |
| T2DM_admissi | leukocyte | *ARMC10* | -1.622866687 | 0.00048781 | 0.001279443 | 1 |
| T2DM_admissi | leukocyte | *ACYP2* | -1.108596645 | 0.000344073 | 0.001140861 | 1 |
| T2DM_admissi | leukocyte | *ATF7* | -1.271743531 | 0.000419518 | 0.001201375 | 1 |
| T2DM_admissi | leukocyte | *RASA4B* | -1.813044559 | 0.008493137 | 0.011220229 | 1 |
| T2DM_admissi | leukocyte | *KCNS3* | -1.500585191 | 0.001539544 | 0.002486242 | 1 |
| T2DM_admissi | leukocyte | *FOXN2* | -3.902908978 | 3.65E-05 | 0.001037577 | 0.14330485 |
| T2DM_admissi | leukocyte | *GPR27* | -1.257101682 | 0.000292613 | 0.001088708 | 1 |
| T2DM_admissi | leukocyte | *MTSS1* | -1.623651867 | 0.000524762 | 0.001319879 | 1 |
| T2DM_admissi | leukocyte | *TMEM43* | -1.208573657 | 0.000465213 | 0.001251077 | 1 |
| T2DM_admissi | leukocyte | *RPS9* | -0.776518161 | 0.000210516 | 0.001062643 | 0.827118445 |
| T2DM_admissi | leukocyte | *NDUFA3* | -1.325547825 | 0.000174576 | 0.001062643 | 0.685909955 |
| T2DM_admissi | leukocyte | *ZNF160* | -1.241081497 | 0.000348805 | 0.001141096 | 1 |
| T2DM_admissi | leukocyte | *ZNF415* | -1.032373402 | 0.003233768 | 0.004669414 | 1 |
| T2DM_admissi | leukocyte | *TSNARE1* | 0.893924759 | 0.019472332 | 0.024280163 | 1 |
| T2DM_admissi | leukocyte | *SOX7* | -1.487315205 | 0.001275825 | 0.002175489 | 1 |
| T2DM_admissi | leukocyte | *TRMT61B* | -0.948627321 | 0.00182438 | 0.002848962 | 1 |
| T2DM_admissi | leukocyte | *INSR* | -0.84001851 | 0.000694108 | 0.001507546 | 1 |
| T2DM_admissi | leukocyte | *MFN1* | -1.117941195 | 0.002298111 | 0.003463474 | 1 |
| T2DM_admissi | leukocyte | *KCNMB3* | -1.867750394 | 0.001515293 | 0.002457114 | 1 |
| T2DM_admissi | leukocyte | *ATP6V0E2* | -0.995704263 | 0.000595437 | 0.001396699 | 1 |
| T2DM_admissi | leukocyte | *JAGN1* | -1.347013937 | 0.00025214 | 0.001077213 | 0.990658174 |
| T2DM_admissi | leukocyte | *MORN4* | -0.766669808 | 0.002502255 | 0.003729651 | 1 |
| T2DM_admissi | leukocyte | *TRIM8* | -1.414258645 | 0.000177611 | 0.001062643 | 0.697833999 |
| T2DM_admissi | leukocyte | *CDC42BPG* | -0.842605584 | 0.000993994 | 0.001873994 | 1 |
| T2DM_admissi | leukocyte | *ZNF440* | -4.720424713 | 8.79E-06 | 0.000762289 | 0.034517466 |
| T2DM_admissi | leukocyte | *GAA* | -0.93547868 | 0.027219078 | 0.032986971 | 1 |
| T2DM_admissi | leukocyte | *EXOSC1* | -1.556182616 | 0.000362633 | 0.001144406 | 1 |
| T2DM_admissi | leukocyte | *PGAM1* | -0.941673888 | 0.001625039 | 0.002589124 | 1 |
| T2DM_admissi | leukocyte | *ESCO2* | -3.663441068 | 0.007511524 | 0.010017915 | 1 |
| T2DM_admissi | leukocyte | *MRPL36* | -1.168655191 | 0.0005351 | 0.001332323 | 1 |
| T2DM_admissi | leukocyte | *MCC* | -1.317196168 | 0.003404509 | 0.004896163 | 1 |
| T2DM_admissi | leukocyte | *ZNF561* | -1.478676232 | 0.000226005 | 0.001062643 | 0.887975257 |
| T2DM_admissi | leukocyte | *PPID* | -1.463496062 | 0.004146221 | 0.005838889 | 1 |
| T2DM_admissi | leukocyte | *RXFP1* | -23.0235108 | 0.00688134 | 0.009243345 | 1 |
| T2DM_admissi | leukocyte | *PTGER4* | 0.575402101 | 0.023607382 | 0.02895829 | 1 |
| T2DM_admissi | leukocyte | *MAP6* | -0.848786004 | 0.000745236 | 0.001568308 | 1 |
| T2DM_admissi | leukocyte | *NMUR1* | -12.35503457 | 0.001224969 | 0.002137168 | 1 |

| | | | | | | |
|---|---|---|---|---|---|---|
| T2DM_admissi | leukocyte | *CLSTN1* | -1.094537194 | 0.000113522 | 0.001062643 | 0.446026254 |
| T2DM_admissi | leukocyte | *PIK3CD* | 0.827074771 | 0.023870235 | 0.029235085 | 1 |
| T2DM_admissi | leukocyte | *ENC1* | -1.000608628 | 0.002466625 | 0.00368353 | 1 |
| T2DM_admissi | leukocyte | *RGS19* | -2.41159157 | 1.04E-05 | 0.000784902 | 0.040719109 |
| T2DM_admissi | leukocyte | *ANO5* | -1.430881243 | 0.00104469 | 0.001937955 | 1 |
| T2DM_admissi | leukocyte | *GPHN* | -2.23473752 | 0.000108403 | 0.001062643 | 0.425916779 |
| T2DM_admissi | leukocyte | *VAT1L* | -1.313223986 | 0.003157493 | 0.004571187 | 1 |
| T2DM_admissi | leukocyte | *LGALS4* | -0.624733371 | 0.00155719 | 0.002504379 | 1 |
| T2DM_admissi | leukocyte | *LRRC34* | -1.309927299 | 0.002545309 | 0.003776631 | 1 |
| T2DM_admissi | leukocyte | *SPATA5L1* | -2.298907482 | 0.000387521 | 0.001173013 | 1 |
| T2DM_admissi | leukocyte | *CTPS1* | -1.078294201 | 0.001049578 | 0.001940644 | 1 |
| T2DM_admissi | leukocyte | *METTL18* | -1.435350759 | 0.00146934 | 0.002392473 | 1 |
| T2DM_admissi | leukocyte | *PWWP2B* | -1.514260582 | 0.000323944 | 0.001116918 | 1 |
| T2DM_admissi | leukocyte | *FAM90A1* | -2.953888335 | 0.020536281 | 0.025437279 | 1 |
| T2DM_admissi | leukocyte | *PTEN* | -1.414251631 | 0.000221615 | 0.001062643 | 0.870725428 |
| T2DM_admissi | leukocyte | *CYP4F11* | -2.6416251 | 0.004435787 | 0.006197798 | 1 |
| T2DM_admissi | leukocyte | *TLN2* | -2.232812469 | 0.001216363 | 0.00212877 | 1 |
| T2DM_admissi | leukocyte | *TVP23B* | -2.123189404 | 0.000192474 | 0.001062643 | 0.756229851 |
| T2DM_admissi | leukocyte | *SRGAP2C* | -2.589407057 | 6.33E-05 | 0.001060479 | 0.248648755 |
| T2DM_admissi | leukocyte | *ATPAF2* | -1.385986474 | 0.001583346 | 0.002536064 | 1 |
| T2DM_admissi | leukocyte | *ZNF556* | 0.849534948 | 0.014848462 | 0.018825301 | 1 |
| T2DM_admissi | leukocyte | *ZNF554* | -1.321924375 | 0.000713688 | 0.001532285 | 1 |
| T2DM_admissi | leukocyte | *GAP43* | -0.885486936 | 0.001402859 | 0.002319795 | 1 |
| T2DM_admissi | leukocyte | *REG1B* | -0.543138087 | 0.000646628 | 0.001446812 | 1 |
| T2DM_admissi | leukocyte | *NME6* | -2.130193105 | 0.000344891 | 0.001140861 | 1 |
| T2DM_admissi | leukocyte | *SLFN12* | 0.949366891 | 0.017468491 | 0.021892728 | 1 |
| T2DM_admissi | leukocyte | *TEFM* | -1.595408364 | 0.000354786 | 0.001141109 | 1 |
| T2DM_admissi | leukocyte | *MBOAT1* | -2.548611782 | 0.000408649 | 0.001188129 | 1 |
| T2DM_admissi | leukocyte | *ZNF131* | -1.241616981 | 0.000425206 | 0.001210604 | 1 |
| T2DM_admissi | leukocyte | *BSG* | -1.07155044 | 0.000184465 | 0.001062643 | 0.72476164 |
| T2DM_admissi | leukocyte | *HINFP* | -3.740021595 | 0.00010931 | 0.001062643 | 0.429478663 |
| T2DM_admissi | leukocyte | *CERS6* | -1.180061426 | 0.000299393 | 0.001093914 | 1 |
| T2DM_admissi | leukocyte | *SPTLC3* | 1.242562244 | 0.008683657 | 0.011437509 | 1 |
| T2DM_admissi | leukocyte | *COPRS* | -0.892515946 | 0.000481825 | 0.001270529 | 1 |
| T2DM_admissi | leukocyte | *SUCLG2* | -2.311277193 | 6.75E-05 | 0.001062643 | 0.265060623 |
| T2DM_admissi | leukocyte | *CSDC2* | -3.477749986 | 0.000640759 | 0.001440839 | 1 |
| T2DM_admissi | leukocyte | *ARNT2* | -0.79340176 | 0.00851637 | 0.011243554 | 1 |
| T2DM_admissi | leukocyte | *FUT9* | -3.846298714 | 0.018413641 | 0.023025842 | 1 |
| T2DM_admissi | leukocyte | *ZNF24* | -1.395163294 | 7.64E-05 | 0.001062643 | 0.299988486 |
| T2DM_admissi | leukocyte | *MANEA* | -1.447657527 | 0.00024768 | 0.001077213 | 0.973134565 |
| T2DM_admissi | leukocyte | *AFF1* | -1.380616818 | 0.000212852 | 0.001062643 | 0.836294701 |
| T2DM_admissi | leukocyte | *CTSW* | -1.613310663 | 8.99E-06 | 0.000762289 | 0.035322435 |
| T2DM_admissi | leukocyte | *SMPDL3A* | -2.499379607 | 0.006206966 | 0.008418077 | 1 |
| T2DM_admissi | leukocyte | *RAD9A* | -2.225706262 | 0.000343324 | 0.001140861 | 1 |
| T2DM_admissi | leukocyte | *EFEMP2* | -1.228250785 | 0.00090195 | 0.001772767 | 1 |
| T2DM_admissi | leukocyte | *ZNF738* | -1.097930822 | 0.015350105 | 0.019411188 | 1 |
| T2DM_admissi | leukocyte | *FUT10* | -3.360521063 | 0.013954098 | 0.01779601 | 1 |
| T2DM_admissi | leukocyte | *NAA16* | -0.944571987 | 0.006252618 | 0.008471192 | 1 |
| T2DM_admissi | leukocyte | *SNX32* | 2.303848545 | 0.025358696 | 0.030951947 | 1 |
| T2DM_admissi | leukocyte | *OVOL1* | -3.590332715 | 9.58E-05 | 0.001062643 | 0.376552401 |

| | | | | | | |
|---|---|---|---|---|---|---|
| T2DM_admissi | leukocyte | *DMXL1* | -1.980416165 | 0.000182839 | 0.001062643 | 0.71837607 |
| T2DM_admissi | leukocyte | *NADSYN1* | -2.589837533 | 4.42E-05 | 0.001037577 | 0.17379775 |
| T2DM_admissi | leukocyte | *RNASEH2C* | -1.627705843 | 4.39E-06 | 0.000522919 | 0.017256316 |
| T2DM_admissi | leukocyte | *LCLAT1* | -1.266058666 | 0.000299642 | 0.001093914 | 1 |
| T2DM_admissi | leukocyte | *SH3RF3* | -1.689417445 | 0.002120783 | 0.003225922 | 1 |
| T2DM_admissi | leukocyte | *DCAKD* | -0.953269196 | 0.000568881 | 0.001367774 | 1 |
| T2DM_admissi | leukocyte | *TADA2B* | -1.467284721 | 0.000207701 | 0.001062643 | 0.816057272 |
| T2DM_admissi | leukocyte | *TRMT112* | -1.066580118 | 0.000333433 | 0.00112758 | 1 |
| T2DM_admissi | leukocyte | *NOC3L* | -1.277508906 | 0.000220224 | 0.001062643 | 0.865258392 |
| T2DM_admissi | leukocyte | *RHOD* | -0.769268084 | 0.001235231 | 0.002144697 | 1 |
| T2DM_admissi | leukocyte | *PARP14* | -1.333169013 | 0.000192341 | 0.001062643 | 0.755708282 |
| T2DM_admissi | leukocyte | *VANGL1* | -1.142805827 | 0.001241012 | 0.002152731 | 1 |
| T2DM_admissi | leukocyte | *IQCB1* | -3.30401219 | 2.59E-05 | 0.000984894 | 0.101796696 |
| T2DM_admissi | leukocyte | *ZNF483* | -1.064338787 | 0.000251631 | 0.001077213 | 0.988658231 |
| T2DM_admissi | leukocyte | *SLC2A14* | -4.498007751 | 0.001317889 | 0.002219454 | 1 |
| T2DM_admissi | leukocyte | *MZT2A* | -0.927192919 | 0.000706638 | 0.001523809 | 1 |
| T2DM_admissi | leukocyte | *PPP1R3B* | -4.008002979 | 4.01E-07 | 0.000116632 | 0.001574769 |
| T2DM_admissi | leukocyte | *MAP3K11* | -1.163031432 | 0.000297332 | 0.001093914 | 1 |
| T2DM_admissi | leukocyte | *NDNF* | 1.070652664 | 0.008460583 | 0.011181174 | 1 |
| T2DM_admissi | leukocyte | *ARV1* | -1.091432391 | 0.00057921 | 0.001379223 | 1 |
| T2DM_admissi | leukocyte | *EHBP1L1* | -1.143752579 | 0.002151944 | 0.003265736 | 1 |
| T2DM_admissi | leukocyte | *PPP1R14B* | -1.025034279 | 0.000234472 | 0.001066706 | 0.921241124 |
| T2DM_admissi | leukocyte | *TNFRSF10D* | -1.328345741 | 0.001964607 | 0.003029411 | 1 |
| T2DM_admissi | leukocyte | *GMPPB* | -1.782857158 | 0.000673398 | 0.001482231 | 1 |
| T2DM_admissi | leukocyte | *NUDT18* | -1.162929375 | 0.001128049 | 0.002034008 | 1 |
| T2DM_admissi | leukocyte | *NUDT4* | -1.055630721 | 0.001062857 | 0.00195779 | 1 |
| T2DM_admissi | leukocyte | *PC* | -1.856408852 | 0.000657462 | 0.001456947 | 1 |
| T2DM_admissi | leukocyte | *UQCRH* | -1.077384505 | 0.000139859 | 0.001062643 | 0.54950494 |
| T2DM_admissi | leukocyte | *HEG1* | -1.148768423 | 0.000989593 | 0.001869283 | 1 |
| T2DM_admissi | leukocyte | *ENDOV* | -2.618524279 | 4.04E-05 | 0.001037577 | 0.158857052 |
| T2DM_admissi | leukocyte | *DPY19L1* | -1.459187921 | 0.000452844 | 0.001238727 | 1 |
| T2DM_admissi | leukocyte | *GPR160* | -1.814609797 | 0.000431398 | 0.001216752 | 1 |
| T2DM_admissi | leukocyte | *XXYLT1* | -1.887699995 | 0.000678484 | 0.001488422 | 1 |
| T2DM_admissi | leukocyte | *GNG5* | -1.068940443 | 0.000211779 | 0.001062643 | 0.832077937 |
| T2DM_admissi | leukocyte | *C9orf131* | -2.189280042 | 0.004246013 | 0.005960195 | 1 |
| T2DM_admissi | leukocyte | *FAM53A* | -6.673183901 | 4.47E-05 | 0.001037577 | 0.175769511 |
| T2DM_admissi | leukocyte | *SELP* | -6.861387868 | 0.001165241 | 0.002080069 | 1 |
| T2DM_admissi | leukocyte | *CTU2* | -1.129625065 | 0.000904419 | 0.001774955 | 1 |
| T2DM_admissi | leukocyte | *ADCY6* | -1.224370076 | 0.001499318 | 0.002434224 | 1 |
| T2DM_admissi | leukocyte | *PODN* | -0.903703593 | 0.018548692 | 0.02318734 | 1 |
| T2DM_admissi | leukocyte | *RALGAPA1* | -1.160102459 | 0.000266121 | 0.001086558 | 1 |
| T2DM_admissi | leukocyte | *CNTNAP2* | 2.140173902 | 0.005962189 | 0.008128189 | 1 |
| T2DM_admissi | leukocyte | *GOLT1A* | -1.188390728 | 0.000388646 | 0.001174609 | 1 |
| T2DM_admissi | leukocyte | *NPAS4* | -3.090799315 | 1.63E-05 | 0.000823546 | 0.063924955 |
| T2DM_admissi | leukocyte | *ANGEL2* | -1.695024037 | 0.000188283 | 0.001062643 | 0.739764298 |
| T2DM_admissi | leukocyte | *ZNF266* | -1.201834523 | 1.68E-05 | 0.000823546 | 0.065883669 |
| T2DM_admissi | leukocyte | *BRSK2* | -1.090582435 | 0.000797779 | 0.001641531 | 1 |
| T2DM_admissi | leukocyte | *BRMS1* | -1.10434234 | 0.000406713 | 0.001188129 | 1 |
| T2DM_admissi | leukocyte | *HRAS* | -1.108637755 | 0.000330239 | 0.001124357 | 1 |
| T2DM_admissi | leukocyte | *SRP72* | -0.934912463 | 0.000320244 | 0.001113485 | 1 |

| | | | | | | |
|---|---|---|---|---|---|---|
| T2DM_admissi | leukocyte | *PCP2* | -4.538483928 | 0.000885684 | 0.001757306 | 1 |
| T2DM_admissi | leukocyte | *BTC* | -1.223298913 | 0.023936123 | 0.029306647 | 1 |
| T2DM_admissi | leukocyte | *DENND6A* | -1.135802748 | 0.000512918 | 0.001308606 | 1 |
| T2DM_admissi | leukocyte | *YIF1A* | -1.354090271 | 5.44E-06 | 0.00058206 | 0.021375196 |
| T2DM_admissi | leukocyte | *NDUFA11* | -1.138245836 | 0.000159382 | 0.001062643 | 0.626210511 |
| T2DM_admissi | leukocyte | *RSRC1* | -2.705583436 | 2.35E-05 | 0.000980682 | 0.092184077 |
| T2DM_admissi | leukocyte | *DHX36* | -1.267564478 | 0.000158751 | 0.001062643 | 0.623733252 |
| T2DM_admissi | leukocyte | *PSMD2* | -1.00292699 | 0.000352584 | 0.001141109 | 1 |
| T2DM_admissi | leukocyte | *PPM1E* | -0.787725332 | 0.003895789 | 0.005527828 | 1 |
| T2DM_admissi | leukocyte | *ZNF408* | -4.657685898 | 5.48E-06 | 0.00058206 | 0.021536203 |
| T2DM_admissi | leukocyte | *ARHGAP1* | -1.20927914 | 0.000227743 | 0.001062643 | 0.894802673 |
| T2DM_admissi | leukocyte | *MED16* | -1.030061692 | 0.00116105 | 0.002076362 | 1 |
| T2DM_admissi | leukocyte | *CHST1* | -2.261831542 | 0.001265353 | 0.002169029 | 1 |
| T2DM_admissi | leukocyte | *DOLK* | -2.396932581 | 0.000227442 | 0.001062643 | 0.893618444 |
| T2DM_admissi | leukocyte | *CCNE2* | -1.390021365 | 0.00146684 | 0.002389393 | 1 |
| T2DM_admissi | leukocyte | *ZNF519* | -2.312502586 | 0.000222204 | 0.001062643 | 0.873041316 |
| T2DM_admissi | leukocyte | *TMEM9B* | -1.244048487 | 0.000139383 | 0.001062643 | 0.547636976 |
| T2DM_admissi | leukocyte | *EIF3F* | -1.272882801 | 0.000100361 | 0.001062643 | 0.394317004 |
| T2DM_admissi | leukocyte | *ZNF25* | -1.114233535 | 0.000648013 | 0.001447767 | 1 |
| T2DM_admissi | leukocyte | *PPP2R2D* | -1.143314644 | 0.000301401 | 0.001093914 | 1 |
| T2DM_admissi | leukocyte | *CLCF1* | -1.473462864 | 0.00250693 | 0.003733786 | 1 |
| T2DM_admissi | leukocyte | *UCP2* | 0.698159211 | 0.029719973 | 0.035929161 | 1 |
| T2DM_admissi | leukocyte | *MRPL48* | -1.325122177 | 0.001881418 | 0.002921775 | 1 |
| T2DM_admissi | leukocyte | *TMEM70* | -1.109230973 | 0.000531509 | 0.001324223 | 1 |
| T2DM_admissi | leukocyte | *RPS6KB2* | -1.593263703 | 0.000903314 | 0.001773984 | 1 |
| T2DM_admissi | leukocyte | *RMI2* | -3.365733507 | 0.00417377 | 0.005871371 | 1 |
| T2DM_admissi | leukocyte | *LINC01106* | -5.549078687 | 0.002814391 | 0.004130646 | 1 |
| T2DM_admissi | leukocyte | *SFN* | -1.61567429 | 0.03681858 | 0.04402319 | 1 |
| T2DM_admissi | leukocyte | *SWI5* | -1.73374715 | 0.000412462 | 0.001192427 | 1 |
| T2DM_admissi | leukocyte | *ZDHHC21* | -2.522307322 | 2.95E-05 | 0.001015174 | 0.115729869 |
| T2DM_admissi | leukocyte | *LRRN1* | -1.099549218 | 0.040105684 | 0.04779352 | 1 |
| T2DM_admissi | leukocyte | *TUBB6* | -0.545794096 | 0.003336584 | 0.004805512 | 1 |
| T2DM_admissi | leukocyte | *CHMP6* | -1.07711738 | 0.002317512 | 0.003487363 | 1 |
| T2DM_admissi | leukocyte | *DLEU1* | -1.542335655 | 0.000202204 | 0.001062643 | 0.794459724 |
| T2DM_admissi | leukocyte | *CCDC57* | -1.707282229 | 5.47E-05 | 0.001060479 | 0.214827262 |
| T2DM_admissi | leukocyte | *BNIP3* | -0.911839746 | 0.000252516 | 0.001077213 | 0.9921368 |
| T2DM_admissi | leukocyte | *SMIM19* | -0.996845833 | 0.000504112 | 0.001299643 | 1 |
| T2DM_admissi | leukocyte | *SLC35G1* | -1.646849026 | 0.000599461 | 0.001399402 | 1 |
| T2DM_admissi | leukocyte | *COX8A* | -0.904942918 | 0.000147508 | 0.001062643 | 0.579559786 |
| T2DM_admissi | leukocyte | *CDC26* | -1.318514808 | 0.000181881 | 0.001062643 | 0.714611021 |
| T2DM_admissi | leukocyte | *CRLF3* | -3.500118841 | 0.030584613 | 0.036940346 | 1 |
| T2DM_admissi | leukocyte | *RNPEP* | -2.123212796 | 0.000319015 | 0.001113024 | 1 |
| T2DM_admissi | leukocyte | *DMRTA1* | -0.94529979 | 0.003336554 | 0.004805512 | 1 |
| T2DM_admissi | leukocyte | *CLEC14A* | -0.677241564 | 0.002054759 | 0.00314375 | 1 |
| T2DM_admissi | leukocyte | *WDR25* | -3.360620283 | 2.19E-05 | 0.000943039 | 0.085960713 |
| T2DM_admissi | leukocyte | *DIRAS1* | -1.521285809 | 0.000642999 | 0.001442799 | 1 |
| T2DM_admissi | leukocyte | *PHLDB3* | -1.01567001 | 0.001178536 | 0.002098083 | 1 |
| T2DM_admissi | leukocyte | *PRR15* | 1.280819452 | 0.02284815 | 0.028105943 | 1 |
| T2DM_admissi | leukocyte | *KBTBD11* | -0.990007471 | 0.001234086 | 0.002144697 | 1 |
| T2DM_admissi | leukocyte | *RMDN1* | -1.342980343 | 0.000913165 | 0.001784988 | 1 |

| | | | | | | |
|---|---|---|---|---|---|---|
| T2DM_admissi | leukocyte | *LRRC37A* | -6.377237393 | 0.000596559 | 0.001396953 | 1 |
| T2DM_admissi | leukocyte | *ACSF3* | -1.623656315 | 0.000344914 | 0.001140861 | 1 |
| T2DM_admissi | leukocyte | *RUFY1* | -3.029055823 | 8.87E-05 | 0.001062643 | 0.348349672 |
| T2DM_admissi | leukocyte | *VSIG10* | -2.010631667 | 6.14E-05 | 0.001060479 | 0.241357176 |
| T2DM_admissi | leukocyte | *WSB2* | -1.031735022 | 0.000692894 | 0.001507191 | 1 |
| T2DM_admissi | leukocyte | *ANKLE2* | -0.927855119 | 0.001143244 | 0.002051052 | 1 |
| T2DM_admissi | leukocyte | *FUT2* | 0.484416051 | 0.034574005 | 0.041478249 | 1 |
| T2DM_admissi | leukocyte | *MUC20* | -1.143141799 | 0.001410775 | 0.002330561 | 1 |
| T2DM_admissi | leukocyte | *NFATC2IP* | -1.81225401 | 0.000191082 | 0.001062643 | 0.75075993 |
| T2DM_admissi | leukocyte | *DPP7* | -1.150269409 | 0.000177633 | 0.001062643 | 0.697918634 |
| T2DM_admissi | leukocyte | *MTHFR* | -1.307946975 | 0.000345899 | 0.001140861 | 1 |
| T2DM_admissi | leukocyte | *DEAF1* | -1.640497966 | 0.000177614 | 0.001062643 | 0.697845796 |
| T2DM_admissi | leukocyte | *TMEM80* | -0.928533494 | 0.001242775 | 0.002152855 | 1 |
| T2DM_admissi | leukocyte | *DSCAML1* | -1.856832088 | 3.15E-05 | 0.001037577 | 0.123866891 |
| T2DM_admissi | leukocyte | *PUS1* | -2.506442467 | 0.000394547 | 0.001181536 | 1 |
| T2DM_admissi | leukocyte | *MAN1B1* | -2.13637584 | 0.000123564 | 0.001062643 | 0.485483188 |
| T2DM_admissi | leukocyte | *PPFIA3* | -0.929960626 | 0.00095148 | 0.001823593 | 1 |
| T2DM_admissi | leukocyte | *PAWR* | -1.080606126 | 0.000767029 | 0.001600298 | 1 |
| T2DM_admissi | leukocyte | *GPR4* | -1.012002184 | 0.000165267 | 0.001062643 | 0.64933435 |
| T2DM_admissi | leukocyte | *ACOT4* | -1.516211897 | 0.001185365 | 0.00210081 | 1 |
| T2DM_admissi | leukocyte | *ARIH2* | -1.248026773 | 8.45E-05 | 0.001062643 | 0.332145982 |
| T2DM_admissi | leukocyte | *IRX3* | 3.044270526 | 7.71E-05 | 0.001062643 | 0.302933631 |
| T2DM_admissi | leukocyte | *SAMD12* | -1.663458018 | 0.000490696 | 0.001283585 | 1 |
| T2DM_admissi | leukocyte | *CD163* | 0.099614432 | 0.00775154 | 0.010317005 | 1 |
| T2DM_admissi | leukocyte | *ACAD9* | -0.990432506 | 0.020991121 | 0.025951578 | 1 |
| T2DM_admissi | leukocyte | *IL17RA* | -2.084381882 | 0.001959754 | 0.003024302 | 1 |
| T2DM_admissi | leukocyte | *AGTRAP* | -1.321461545 | 0.000334392 | 0.001128716 | 1 |
| T2DM_admissi | leukocyte | *THAP5* | -1.266902697 | 0.000252063 | 0.001077213 | 0.990354673 |
| T2DM_admissi | leukocyte | *CD151* | -0.989975925 | 0.000275848 | 0.001088708 | 1 |
| T2DM_admissi | leukocyte | *CHID1* | -1.078075497 | 0.000137574 | 0.001062643 | 0.540527769 |
| T2DM_admissi | leukocyte | *ZBTB41* | -1.626005306 | 0.000151167 | 0.001062643 | 0.593936392 |
| T2DM_admissi | leukocyte | *BET1L* | -1.901150597 | 9.00E-05 | 0.001062643 | 0.353796905 |
| T2DM_admissi | leukocyte | *RIC8A* | -1.580200746 | 0.00027181 | 0.001088708 | 1 |
| T2DM_admissi | leukocyte | *IMP3* | -1.177390285 | 0.001474995 | 0.002399691 | 1 |
| T2DM_admissi | leukocyte | *DPY19L2* | -0.885028003 | 0.01601504 | 0.020213007 | 1 |
| T2DM_admissi | leukocyte | *GPR150* | -1.0685029 | 0.004912076 | 0.006790833 | 1 |
| T2DM_admissi | leukocyte | *ADAMTSL1* | -6.307461396 | 0.000374789 | 0.001157716 | 1 |
| T2DM_admissi | leukocyte | *ALS2CL* | -1.616311542 | 0.000924582 | 0.001797337 | 1 |
| T2DM_admissi | leukocyte | *STAP2* | -0.919502379 | 0.000503553 | 0.001299056 | 1 |
| T2DM_admissi | leukocyte | *DALRD3* | -1.211807432 | 0.001748321 | 0.002749861 | 1 |
| T2DM_admissi | leukocyte | *ZNF114* | -3.126763048 | 0.000949933 | 0.001823593 | 1 |
| T2DM_admissi | leukocyte | *PLEC* | -2.424678783 | 3.02E-05 | 0.001023384 | 0.118712585 |
| T2DM_admissi | leukocyte | *WDR6* | -1.135720559 | 0.001427796 | 0.002348599 | 1 |
| T2DM_admissi | leukocyte | *AQP11* | -1.497553873 | 0.000782449 | 0.001618875 | 1 |
| T2DM_admissi | leukocyte | *ZFAND2A* | -1.638569799 | 0.000119079 | 0.001062643 | 0.4678601 |
| T2DM_admissi | leukocyte | *DNAJC22* | -0.581443321 | 0.014632145 | 0.018599061 | 1 |
| T2DM_admissi | leukocyte | *NT5DC1* | -1.181190541 | 0.000339689 | 0.001134896 | 1 |
| T2DM_admissi | leukocyte | *COX14* | -1.09471094 | 0.001453383 | 0.002373399 | 1 |
| T2DM_admissi | leukocyte | *P4HTM* | -0.89187589 | 0.00031731 | 0.001111111 | 1 |
| T2DM_admissi | leukocyte | *ERBB4* | -10.56991689 | 0.000188808 | 0.001062643 | 0.741828503 |

| | | | | | | |
|---|---|---|---|---|---|---|
| T2DM_admissi | leukocyte | *ERN1* | -1.727323843 | 7.90E-05 | 0.001062643 | 0.310493651 |
| T2DM_admissi | leukocyte | *PARP10* | -1.25303375 | 0.004357112 | 0.006107418 | 1 |
| T2DM_admissi | leukocyte | *RPP25* | -2.087161602 | 9.19E-05 | 0.001062643 | 0.361145293 |
| T2DM_admissi | leukocyte | *ZHX2* | -1.395218816 | 0.000266316 | 0.001086558 | 1 |
| T2DM_admissi | leukocyte | *HYI* | -1.529071181 | 0.000102311 | 0.001062643 | 0.401978716 |
| T2DM_admissi | leukocyte | *GAK* | -1.268478825 | 0.001539584 | 0.002486242 | 1 |
| T2DM_admissi | leukocyte | *TUFM* | -0.700918415 | 0.006342999 | 0.008584789 | 1 |
| T2DM_admissi | leukocyte | *RMI1* | 1.505395216 | 0.010408068 | 0.013500594 | 1 |
| T2DM_admissi | leukocyte | *C3orf38* | -1.535880637 | 0.00312163 | 0.00452913 | 1 |
| T2DM_admissi | leukocyte | *IGSF22* | -0.830322713 | 0.00190518 | 0.002952841 | 1 |
| T2DM_admissi | leukocyte | *HTR1F* | -3.664832958 | 0.000120893 | 0.001062643 | 0.474987846 |
| T2DM_admissi | leukocyte | *TMTC2* | -2.041172685 | 0.001197776 | 0.002109396 | 1 |
| T2DM_admissi | leukocyte | *FARSA* | -1.504283262 | 1.49E-05 | 0.000823546 | 0.058604833 |
| T2DM_admissi | leukocyte | *SPTY2D1* | -1.272998576 | 0.000317582 | 0.001111111 | 1 |
| T2DM_admissi | leukocyte | *SAMD4B* | -1.035389452 | 0.000620658 | 0.001422226 | 1 |
| T2DM_admissi | leukocyte | *TCAIM* | -1.730274234 | 0.000227165 | 0.001062643 | 0.892529947 |
| T2DM_admissi | leukocyte | *FUCA1* | 0.464747681 | 0.039690366 | 0.047327299 | 1 |
| T2DM_admissi | leukocyte | *CALR* | -0.822370659 | 0.001453798 | 0.002373399 | 1 |
| T2DM_admissi | leukocyte | *CDH4* | -2.17499285 | 0.019577765 | 0.024396142 | 1 |
| T2DM_admissi | leukocyte | *NSUN7* | -1.552177597 | 0.019932018 | 0.024790408 | 1 |
| T2DM_admissi | leukocyte | *ELMOD2* | -2.181335944 | 0.000130968 | 0.001062643 | 0.514575223 |
| T2DM_admissi | leukocyte | *LINC00174* | -1.132501102 | 0.000713471 | 0.001532285 | 1 |
| T2DM_admissi | leukocyte | *GEMIN4* | -1.15171133 | 0.0012488 | 0.00215577 | 1 |
| T2DM_admissi | leukocyte | *LBX2* | 0.993621923 | 0.015621049 | 0.019747459 | 1 |
| T2DM_admissi | leukocyte | *CDC42EP4* | -1.029779077 | 0.000418069 | 0.001200552 | 1 |
| T2DM_admissi | leukocyte | *ZBTB42* | -2.631223179 | 0.000894783 | 0.001766634 | 1 |
| T2DM_admissi | leukocyte | *SYCN* | -0.630617031 | 0.000960647 | 0.001831992 | 1 |
| T2DM_admissi | leukocyte | *CITED4* | -0.768543637 | 0.001301771 | 0.002200323 | 1 |
| T2DM_admissi | leukocyte | *ZNF154* | -0.806209087 | 0.031078945 | 0.037468296 | 1 |
| T2DM_admissi | leukocyte | *B3GNT3* | -2.790898515 | 0.000783569 | 0.001618943 | 1 |
| T2DM_admissi | leukocyte | *PUF60* | -1.031618455 | 0.000947958 | 0.001821284 | 1 |
| T2DM_admissi | leukocyte | *TSHZ1* | -0.939521048 | 0.001421853 | 0.002343313 | 1 |
| T2DM_admissi | leukocyte | *ANKRD18A* | -0.618663299 | 0.026945507 | 0.032665504 | 1 |
| T2DM_admissi | leukocyte | *EXOC3* | -2.227441501 | 0.000118263 | 0.001062643 | 0.464653435 |
| T2DM_admissi | leukocyte | *TH* | -8.011435656 | 1.43E-05 | 0.000815363 | 0.056292998 |
| T2DM_admissi | leukocyte | *FAHD1* | -0.91652975 | 0.007364565 | 0.009838618 | 1 |
| T2DM_admissi | leukocyte | *ZNF816* | -1.62330718 | 0.001226853 | 0.002138556 | 1 |
| T2DM_admissi | leukocyte | *CCDC66* | -3.071433027 | 8.93E-05 | 0.001062643 | 0.350943157 |
| T2DM_admissi | leukocyte | *MCFD2* | -0.942864691 | 0.000255428 | 0.001077348 | 1 |
| T2DM_admissi | leukocyte | *SERTM1* | 0.727721588 | 0.022951747 | 0.028198065 | 1 |
| T2DM_admissi | leukocyte | *GLIPR1L2* | -1.871809288 | 0.003307723 | 0.00476919 | 1 |
| T2DM_admissi | leukocyte | *TSPYL5* | -0.935605896 | 0.000470326 | 0.001255055 | 1 |
| T2DM_admissi | leukocyte | *SHISA2* | -1.666029119 | 0.012863052 | 0.016478668 | 1 |
| T2DM_admissi | leukocyte | *CHST13* | -93.1325014 | 0.001321995 | 0.00222351 | 1 |
| T2DM_admissi | leukocyte | *PPA1* | -0.890773507 | 0.000252618 | 0.001077213 | 0.992537004 |
| T2DM_admissi | leukocyte | *PSMG4* | -1.002826599 | 0.000354155 | 0.001141109 | 1 |
| T2DM_admissi | leukocyte | *MAP6D1* | -2.189977696 | 0.002368633 | 0.003557476 | 1 |
| T2DM_admissi | leukocyte | *ZNF792* | -1.98986186 | 0.010966083 | 0.014182271 | 1 |
| T2DM_admissi | leukocyte | *D2HGDH* | -5.01441204 | 9.12E-06 | 0.000762289 | 0.035827577 |
| T2DM_admissi | leukocyte | *FAM83H* | -1.767356225 | 0.000730242 | 0.001550629 | 1 |

| | | | | | | |
|---|---|---|---|---|---|---|
| T2DM_admissi | leukocyte | *LRRC57* | -1.919883605 | 0.000477927 | 0.001265283 | 1 |
| T2DM_admissi | leukocyte | *MRPL14* | -1.268946232 | 0.00016187 | 0.001062643 | 0.635985676 |
| T2DM_admissi | leukocyte | *ZFP82* | -1.095473102 | 0.000847251 | 0.001701865 | 1 |
| T2DM_admissi | leukocyte | *FKRP* | -0.980119533 | 0.002340855 | 0.00352114 | 1 |
| T2DM_admissi | leukocyte | *RPH3AL* | -0.841891029 | 0.001135162 | 0.002043083 | 1 |
| T2DM_admissi | leukocyte | *HIGD1A* | -0.987642116 | 0.000180742 | 0.001062643 | 0.710134249 |
| T2DM_admissi | leukocyte | *F2R* | -0.58110218 | 0.041027208 | 0.048788106 | 1 |
| T2DM_admissi | leukocyte | *ZNF707* | -1.419400939 | 0.001206565 | 0.002117283 | 1 |
| T2DM_admissi | leukocyte | *OGFOD3* | -1.002901836 | 0.00052889 | 0.001321888 | 1 |
| T2DM_admissi | leukocyte | *ZNF467* | -1.525093844 | 0.000728073 | 0.00154878 | 1 |
| T2DM_admissi | leukocyte | *ZNF678* | -2.561065504 | 0.000584998 | 0.001382115 | 1 |
| T2DM_admissi | leukocyte | *TMEM45A* | -0.907675975 | 0.001276087 | 0.002175489 | 1 |
| T2DM_admissi | leukocyte | *SGSH* | -2.87520171 | 0.005014598 | 0.006922824 | 1 |
| T2DM_admissi | leukocyte | *GPR135* | -1.327847581 | 0.001017872 | 0.001906024 | 1 |
| T2DM_admissi | leukocyte | *ANKRD62* | -1.47520447 | 0.003736988 | 0.00532172 | 1 |
| T2DM_admissi | leukocyte | *RFX7* | -3.172938219 | 2.92E-05 | 0.001014102 | 0.11459356 |
| T2DM_admissi | leukocyte | *ZNF329* | -1.525579621 | 0.000934896 | 0.001804129 | 1 |
| T2DM_admissi | leukocyte | *CCDC149* | -2.018029329 | 0.000113785 | 0.001062643 | 0.447061732 |
| T2DM_admissi | leukocyte | *IDH2* | -0.881575509 | 0.000996157 | 0.001875957 | 1 |
| T2DM_admissi | leukocyte | *TNRC18* | -1.534425515 | 0.000214371 | 0.001062643 | 0.842265349 |
| T2DM_admissi | leukocyte | *TDRKH* | -1.183993505 | 0.001926379 | 0.002980994 | 1 |
| T2DM_admissi | leukocyte | *ZNF708* | -0.916147045 | 0.00068078 | 0.00149096 | 1 |
| T2DM_admissi | leukocyte | *IST1* | -1.86791749 | 8.72E-05 | 0.001062643 | 0.342519126 |
| T2DM_admissi | leukocyte | *ERCC6L2* | -3.655271071 | 7.42E-05 | 0.001062643 | 0.291353538 |
| T2DM_admissi | leukocyte | *MRPL41* | -1.153367941 | 0.000177926 | 0.001062643 | 0.699069693 |
| T2DM_admissi | leukocyte | *TP53TG1* | -1.29782835 | 0.000170531 | 0.001062643 | 0.670017084 |
| T2DM_admissi | leukocyte | *UBA7* | -3.381418837 | 4.67E-07 | 0.000122423 | 0.001836338 |
| T2DM_admissi | leukocyte | *ARL6IP4* | -0.765809608 | 0.002392577 | 0.003589322 | 1 |
| T2DM_admissi | leukocyte | *FAM153B* | -1.881380748 | 0.000212224 | 0.001062643 | 0.833829314 |
| T2DM_admissi | leukocyte | *BACE2* | -0.675997514 | 0.000978951 | 0.001856322 | 1 |
| T2DM_admissi | leukocyte | *SYNM* | -0.989862969 | 0.00438883 | 0.006143112 | 1 |
| T2DM_admissi | leukocyte | *FIGN* | -1.110064244 | 0.001108409 | 0.002009662 | 1 |
| T2DM_admissi | leukocyte | *B4GALNT4* | -2.245733975 | 0.000936459 | 0.001806258 | 1 |
| T2DM_admissi | leukocyte | *ZNF804B* | -80.25308503 | 0.001448937 | 0.002370055 | 1 |
| T2DM_admissi | leukocyte | *YBEY* | -1.352088513 | 0.000669219 | 0.001476341 | 1 |
| T2DM_admissi | leukocyte | *NPLOC4* | -1.233311448 | 0.000317067 | 0.001111111 | 1 |
| T2DM_admissi | leukocyte | *CAPN12* | -2.39031727 | 0.000154005 | 0.001062643 | 0.605087025 |
| T2DM_admissi | leukocyte | *GLRX5* | -0.890411556 | 0.003104643 | 0.004507814 | 1 |
| T2DM_admissi | leukocyte | *MXRA7* | -1.026724992 | 0.000292064 | 0.001088708 | 1 |
| T2DM_admissi | leukocyte | *ADI1* | -2.63798955 | 5.71E-05 | 0.001060479 | 0.224426415 |
| T2DM_admissi | leukocyte | *SATB1* | -1.885497607 | 0.000126399 | 0.001062643 | 0.496620196 |
| T2DM_admissi | leukocyte | *EPHB3* | -2.061069158 | 0.039722297 | 0.047351003 | 1 |
| T2DM_admissi | leukocyte | *SKA2* | -2.306397677 | 1.67E-05 | 0.000823546 | 0.065523347 |
| T2DM_admissi | leukocyte | *TTC3* | -0.501304712 | 0.017194497 | 0.021618297 | 1 |
| T2DM_admissi | leukocyte | *BRICD5* | -2.223139072 | 0.006069129 | 0.008262511 | 1 |
| T2DM_admissi | leukocyte | *TSKU* | -1.234533663 | 0.001172207 | 0.002089227 | 1 |
| T2DM_admissi | leukocyte | *ANXA2* | -0.707046304 | 0.000343614 | 0.001140861 | 1 |
| T2DM_admissi | leukocyte | *SLC35D3* | -1.901711156 | 0.002012349 | 0.003089327 | 1 |
| T2DM_admissi | leukocyte | *GPIHBP1* | -10.12804589 | 0.001247484 | 0.002154446 | 1 |
| T2DM_admissi | leukocyte | *VMO1* | 0.122276902 | 0.004574177 | 0.006361749 | 1 |

| | | | | | | |
|---|---|---|---|---|---|---|
| T2DM_admissi | leukocyte | *COL18A1* | -0.601360556 | 0.000553549 | 0.001355918 | 1 |
| T2DM_admissi | leukocyte | *C11orf54* | -1.487364036 | 0.001646977 | 0.002617707 | 1 |
| T2DM_admissi | leukocyte | *CEP63* | -1.420802931 | 0.000233164 | 0.001066706 | 0.916100579 |
| T2DM_admissi | leukocyte | *HMGN4* | -1.48192168 | 0.000809715 | 0.00165524 | 1 |
| T2DM_admissi | leukocyte | *SPATA13* | -1.095879136 | 0.000600546 | 0.001399493 | 1 |
| T2DM_admissi | leukocyte | *ZNF320* | 1.167157426 | 0.000108035 | 0.001062643 | 0.424471463 |
| T2DM_admissi | leukocyte | *SLC25A21* | -2.79546508 | 0.0012558 | 0.002159317 | 1 |
| T2DM_admissi | leukocyte | *CAMK1D* | -1.825586404 | 0.036757661 | 0.043963729 | 1 |
| T2DM_admissi | leukocyte | *LYSMD4* | -1.367630679 | 0.000705666 | 0.001522744 | 1 |
| T2DM_admissi | leukocyte | *BEGAIN* | -1.733829581 | 0.000361291 | 0.001142338 | 1 |
| T2DM_admissi | leukocyte | *GPC6* | -1.324814039 | 0.000809649 | 0.00165524 | 1 |
| T2DM_admissi | leukocyte | *FANCF* | -4.61198721 | 1.82E-05 | 0.000880427 | 0.071314623 |
| T2DM_admissi | leukocyte | *CALN1* | -3.786782836 | 0.032113142 | 0.038632129 | 1 |
| T2DM_admissi | leukocyte | *SMDT1* | -0.770500682 | 0.001422832 | 0.002343944 | 1 |
| T2DM_admissi | leukocyte | *PTTG1IP* | -0.870128362 | 0.003064707 | 0.004453119 | 1 |
| T2DM_admissi | leukocyte | *PLGLB1* | 1.841577787 | 0.030832648 | 0.037194189 | 1 |
| T2DM_admissi | leukocyte | *ZNF623* | -0.964654728 | 0.001935322 | 0.002992504 | 1 |
| T2DM_admissi | leukocyte | *CCDC125* | -2.155502785 | 0.006667828 | 0.008984189 | 1 |
| T2DM_admissi | leukocyte | *BOLA2* | -1.170928334 | 0.000309822 | 0.00110427 | 1 |
| T2DM_admissi | leukocyte | *FHL3* | -1.064259746 | 0.003129045 | 0.004538213 | 1 |
| T2DM_admissi | leukocyte | *SF3A3* | -0.965222655 | 0.001125039 | 0.002031214 | 1 |
| T2DM_admissi | leukocyte | *MX2* | 1.105026017 | 0.033403027 | 0.040110176 | 1 |
| T2DM_admissi | leukocyte | *PCBP3* | -1.662858192 | 0.001802311 | 0.002818981 | 1 |
| T2DM_admissi | leukocyte | *SFXN4* | -1.332038995 | 0.000274503 | 0.001088708 | 1 |
| T2DM_admissi | leukocyte | *MRPL54* | -1.17701341 | 0.000135586 | 0.001062643 | 0.532718844 |
| T2DM_admissi | leukocyte | *DGCR6* | -1.220915145 | 0.000752211 | 0.001577074 | 1 |
| T2DM_admissi | leukocyte | *NDUFB1* | -1.115170084 | 0.000381201 | 0.001165285 | 1 |
| T2DM_admissi | leukocyte | *TRMT12* | -1.350935277 | 0.000364539 | 0.001146348 | 1 |
| T2DM_admissi | leukocyte | *FIGLA* | -28.5588364 | 0.000290999 | 0.001088708 | 1 |
| T2DM_admissi | leukocyte | *CBX6* | -1.04055655 | 0.000321307 | 0.001114222 | 1 |
| T2DM_admissi | leukocyte | *MACC1* | -0.907341776 | 0.000290283 | 0.001088708 | 1 |
| T2DM_admissi | leukocyte | *KREMEN1* | -3.343142276 | 0.001504086 | 0.002439948 | 1 |
| T2DM_admissi | leukocyte | *NUDT14* | -1.007024463 | 0.001440644 | 0.002361407 | 1 |
| T2DM_admissi | leukocyte | *FAM3B* | -0.917787238 | 0.001999031 | 0.003071644 | 1 |
| T2DM_admissi | leukocyte | *TOB2* | -1.306122069 | 0.000462164 | 0.001244702 | 1 |
| T2DM_admissi | leukocyte | *NPIPA7* | -9.694237372 | 0.004792429 | 0.006642945 | 1 |
| T2DM_admissi | leukocyte | *TTC32* | -1.164209436 | 0.000357543 | 0.001141109 | 1 |
| T2DM_admissi | leukocyte | *TMEM236* | -0.804685422 | 0.002772944 | 0.004074381 | 1 |
| T2DM_admissi | leukocyte | *UQCR10* | -1.004138264 | 0.000201581 | 0.001062643 | 0.792013383 |
| T2DM_admissi | leukocyte | *EIF3C* | -0.723450041 | 0.02571067 | 0.03131346 | 1 |
| T2DM_admissi | leukocyte | *NIPSNAP1* | -1.073844898 | 0.000929134 | 0.001800431 | 1 |
| T2DM_admissi | leukocyte | *CRELD2* | -0.833132777 | 0.001727051 | 0.002722068 | 1 |
| T2DM_admissi | leukocyte | *UBE2F* | -1.885567312 | 0.000184225 | 0.001062643 | 0.723819126 |
| T2DM_admissi | leukocyte | *GOLGA6L4* | -1.362189066 | 0.001101942 | 0.002000708 | 1 |
| T2DM_admissi | leukocyte | *PGP* | -2.066880204 | 3.07E-06 | 0.000430544 | 0.012055222 |
| T2DM_admissi | leukocyte | *ACOT1* | -5.789016905 | 0.006254754 | 0.008471192 | 1 |
| T2DM_admissi | leukocyte | *OAF* | -1.515294394 | 0.000204824 | 0.001062643 | 0.80475315 |
| T2DM_admissi | leukocyte | *TM2D3* | -1.149715454 | 0.000289511 | 0.001088708 | 1 |
| T2DM_admissi | leukocyte | *TOP1MT* | -2.86817216 | 0.000190111 | 0.001062643 | 0.746946813 |
| T2DM_admissi | leukocyte | *COPB2* | -1.381792921 | 0.000105387 | 0.001062643 | 0.414066686 |

| | | | | | | |
|---|---|---|---|---|---|---|
| T2DM_admissi | leukocyte | *CCR10* | -2.043018205 | 0.006812645 | 0.009160466 | 1 |
| T2DM_admissi | leukocyte | *NCMAP* | -1.075943012 | 0.001379546 | 0.002289918 | 1 |
| T2DM_admissi | leukocyte | *WDR27* | -1.589284789 | 0.037333996 | 0.044625881 | 1 |
| T2DM_admissi | leukocyte | *TXNRD2* | -1.147732526 | 0.000862763 | 0.001726874 | 1 |
| T2DM_admissi | leukocyte | *ZFP1* | -1.525972782 | 0.000412752 | 0.001192427 | 1 |
| T2DM_admissi | leukocyte | *XPOT* | -1.102671867 | 0.000402118 | 0.001185421 | 1 |
| T2DM_admissi | leukocyte | *C14orf180* | -116.0093991 | 0.001272704 | 0.002175059 | 1 |
| T2DM_admissi | leukocyte | *ZBTB40* | -1.549569882 | 0.004512184 | 0.006293468 | 1 |
| T2DM_admissi | leukocyte | *UBE2G2* | -1.254686065 | 0.000229825 | 0.001064322 | 0.902981351 |
| T2DM_admissi | leukocyte | *OSBP2* | -1.280679381 | 0.001244716 | 0.0021544 | 1 |
| T2DM_admissi | leukocyte | *TMED9* | -1.01397052 | 0.000261051 | 0.001083674 | 1 |
| T2DM_admissi | leukocyte | *BTBD6* | -1.555971152 | 0.000193184 | 0.001062643 | 0.759020771 |
| T2DM_admissi | leukocyte | *JAG2* | -0.932371127 | 0.003148082 | 0.004562455 | 1 |
| T2DM_admissi | leukocyte | *LCN12* | -4.202507353 | 0.000134785 | 0.001062643 | 0.529569668 |
| T2DM_admissi | leukocyte | *FAM227A* | -1.234797872 | 0.001221552 | 0.002133102 | 1 |
| T2DM_admissi | leukocyte | *MUC6* | -0.676844748 | 0.002123689 | 0.003229092 | 1 |
| T2DM_admissi | leukocyte | *NOC4L* | -1.94718446 | 0.000139281 | 0.001062643 | 0.547236084 |
| T2DM_admissi | leukocyte | *NDUFA6* | -0.653164418 | 0.010276483 | 0.013347538 | 1 |
| T2DM_admissi | leukocyte | *SIVA1* | -1.146659831 | 0.000301445 | 0.001093914 | 1 |
| T2DM_admissi | leukocyte | *BRI3BP* | -1.644437812 | 0.000281585 | 0.001088708 | 1 |
| T2DM_admissi | leukocyte | *UBOX5* | -1.132940048 | 0.000877526 | 0.001743901 | 1 |
| T2DM_admissi | leukocyte | *MAFF* | -0.689395502 | 0.001872574 | 0.00290919 | 1 |
| T2DM_admissi | leukocyte | *SEMA4B* | -1.077331806 | 0.001452038 | 0.002373152 | 1 |
| T2DM_admissi | leukocyte | *CIB1* | -1.135254977 | 0.000170913 | 0.001062643 | 0.671518025 |
| T2DM_admissi | leukocyte | *ANO9* | -1.493856836 | 0.005298665 | 0.007289375 | 1 |
| T2DM_admissi | leukocyte | *INPP5J* | -5.600277778 | 0.019336302 | 0.024125859 | 1 |
| T2DM_admissi | leukocyte | *MFSD6L* | -3.140281072 | 0.002352167 | 0.003535449 | 1 |
| T2DM_admissi | leukocyte | *ZNF445* | -5.105975591 | 0.000181251 | 0.001062643 | 0.7121358 |
| T2DM_admissi | leukocyte | *ZNF74* | -2.474007279 | 0.001272196 | 0.002175059 | 1 |
| T2DM_admissi | leukocyte | *RBM11* | -0.977317559 | 0.000863888 | 0.001728216 | 1 |
| T2DM_admissi | leukocyte | *RGPD2* | -3.089009509 | 0.003419291 | 0.004910231 | 1 |
| T2DM_admissi | leukocyte | *CDK10* | -0.950564993 | 0.024768421 | 0.030278508 | 1 |
| T2DM_admissi | leukocyte | *GAS2L1* | -2.580003941 | 0.002609518 | 0.003864604 | 1 |
| T2DM_admissi | leukocyte | *ATP6V0A2* | -1.851408664 | 0.000342115 | 0.001140092 | 1 |
| T2DM_admissi | leukocyte | *RAD51D* | -3.149469752 | 0.000102292 | 0.001062643 | 0.401903466 |
| T2DM_admissi | leukocyte | *SH3BGR* | -0.589055473 | 0.008547841 | 0.011269955 | 1 |
| T2DM_admissi | leukocyte | *MUC1* | -0.711816531 | 0.001961288 | 0.003025481 | 1 |
| T2DM_admissi | leukocyte | *IRF7* | -1.90093109 | 0.000880389 | 0.001747877 | 1 |
| T2DM_admissi | leukocyte | *FAM131C* | -0.833114341 | 0.000876637 | 0.001743468 | 1 |
| T2DM_admissi | leukocyte | *LSAMP* | -0.619259146 | 0.001752349 | 0.002755094 | 1 |
| T2DM_admissi | leukocyte | *OLFML2A* | -2.950485112 | 0.002409884 | 0.003612528 | 1 |
| T2DM_admissi | leukocyte | *PDIA2* | -0.508488334 | 0.004842221 | 0.006703695 | 1 |
| T2DM_admissi | leukocyte | *PCGF3* | -1.171375668 | 0.000793564 | 0.001634654 | 1 |
| T2DM_admissi | leukocyte | *P4HB* | -0.791240466 | 0.000393419 | 0.001181536 | 1 |
| T2DM_admissi | leukocyte | *PSMD13* | -1.503095269 | 1.16E-05 | 0.000784902 | 0.045394405 |
| T2DM_admissi | leukocyte | *BRWD1* | -1.29100791 | 0.000210466 | 0.001062643 | 0.826920112 |
| T2DM_admissi | leukocyte | *DRG1* | -1.095136602 | 0.000456838 | 0.001240899 | 1 |
| T2DM_admissi | leukocyte | *ADAMTSL5* | -6.249663676 | 0.025205185 | 0.030783703 | 1 |
| T2DM_admissi | leukocyte | *MORF4L1* | -0.907836789 | 0.000222893 | 0.001062643 | 0.875745133 |
| T2DM_admissi | leukocyte | *DMWD* | -2.635494384 | 1.20E-06 | 0.000236063 | 0.004721267 |

| | | | | | | |
|---|---|---|---|---|---|---|
| T2DM_admissi | leukocyte | *PIGP* | -0.935612292 | 0.000381967 | 0.001165285 | 1 |
| T2DM_admissi | leukocyte | *NPAP1* | -34.04242575 | 0.003937051 | 0.005572289 | 1 |
| T2DM_admissi | leukocyte | *ARL17A* | -0.718674892 | 0.01482001 | 0.018801362 | 1 |
| T2DM_admissi | leukocyte | *TRIM69* | -1.084091021 | 0.002565069 | 0.003800209 | 1 |
| T2DM_admissi | leukocyte | *IFITM1* | -2.626470986 | 0.000722889 | 0.001544023 | 1 |
| T2DM_admissi | leukocyte | *SETD4* | -3.155018498 | 0.009293099 | 0.012146569 | 1 |
| T2DM_admissi | leukocyte | *ZNF529* | -2.070773622 | 0.000367831 | 0.001148802 | 1 |
| T2DM_admissi | leukocyte | *AIDA* | -1.065948817 | 0.00043669 | 0.001222199 | 1 |
| T2DM_admissi | leukocyte | *GSAP* | 1.01359245 | 0.036262779 | 0.043411474 | 1 |
| T2DM_admissi | leukocyte | *WWOX* | -1.184503018 | 0.000403325 | 0.001185781 | 1 |
| T2DM_admissi | leukocyte | *POLR1D* | -1.061198534 | 0.000190741 | 0.001062643 | 0.749420339 |
| T2DM_admissi | leukocyte | *EDARADD* | -1.196223484 | 0.000741698 | 0.001563191 | 1 |
| T2DM_admissi | leukocyte | *ZNF749* | -1.028917956 | 4.97E-05 | 0.00105496 | 0.195316419 |
| T2DM_admissi | leukocyte | *TOR3A* | -1.297218834 | 0.000195978 | 0.001062643 | 0.769996454 |
| T2DM_admissi | leukocyte | *ZNF555* | -0.931334722 | 0.000976779 | 0.001855786 | 1 |
| T2DM_admissi | leukocyte | *NUDT17* | -1.723077356 | 0.000353569 | 0.001141109 | 1 |
| T2DM_admissi | leukocyte | *CYP4X1* | 1.178114197 | 0.028231559 | 0.034192909 | 1 |
| T2DM_admissi | leukocyte | *ZNF197* | -1.083940707 | 0.000519192 | 0.001315067 | 1 |
| T2DM_admissi | leukocyte | *RPS23* | -0.836884324 | 0.000119577 | 0.001062643 | 0.469816405 |
| T2DM_admissi | leukocyte | *BTN3A2* | -0.540984918 | 0.011134532 | 0.014362303 | 1 |
| T2DM_admissi | leukocyte | *ZNF396* | -2.9556285 | 0.000943774 | 0.001816677 | 1 |
| T2DM_admissi | leukocyte | *SMYD4* | -0.83999768 | 0.038426465 | 0.045875899 | 1 |
| T2DM_admissi | leukocyte | *CEACAM19* | -1.104079968 | 0.000120328 | 0.001062643 | 0.472768327 |
| T2DM_admissi | leukocyte | *ZFP91* | -1.423100443 | 0.000182226 | 0.001062643 | 0.715967413 |
| T2DM_admissi | leukocyte | *BCR* | -1.083782441 | 0.00061619 | 0.001419945 | 1 |
| T2DM_admissi | leukocyte | *FSCN2* | -1.411749822 | 0.029671202 | 0.035881242 | 1 |
| T2DM_admissi | leukocyte | *HYAL3* | -1.690959384 | 0.00456662 | 0.006353488 | 1 |
| T2DM_admissi | leukocyte | *RTN4RL2* | -1.146229274 | 0.001289515 | 0.002190448 | 1 |
| T2DM_admissi | leukocyte | *TMEM232* | -1.753744715 | 0.000837789 | 0.001692377 | 1 |
| T2DM_admissi | leukocyte | *FAM183A* | -0.929439745 | 0.0012819 | 0.002181285 | 1 |
| T2DM_admissi | leukocyte | *EMID1* | -1.316523349 | 0.002510225 | 0.003735861 | 1 |
| T2DM_admissi | leukocyte | *ESPN* | -22.85668979 | 0.000546053 | 0.001346461 | 1 |
| T2DM_admissi | leukocyte | *TMEM216* | -2.57392235 | 0.041910459 | 0.04973307 | 1 |
| T2DM_admissi | leukocyte | *LYPD6* | -4.38488134 | 0.005566157 | 0.007625325 | 1 |
| T2DM_admissi | leukocyte | *AKR1C1* | -0.81493114 | 0.001723189 | 0.002719068 | 1 |
| T2DM_admissi | leukocyte | *BCAM* | -0.907318985 | 0.000319908 | 0.001113304 | 1 |
| T2DM_admissi | leukocyte | *CHP1* | -0.986171297 | 0.000392777 | 0.001181536 | 1 |
| T2DM_admissi | leukocyte | *KCNJ11* | -3.777764439 | 0.000256841 | 0.001080629 | 1 |
| T2DM_admissi | leukocyte | *ZNF385C* | -3.769828188 | 0.000762094 | 0.001593931 | 1 |
| T2DM_admissi | leukocyte | *EXD3* | -1.203719659 | 0.001019037 | 0.001906024 | 1 |
| T2DM_admissi | leukocyte | *C17orf97* | -0.710542864 | 0.02003966 | 0.024892768 | 1 |
| T2DM_admissi | leukocyte | *RGPD1* | -124.0718327 | 0.001368853 | 0.002280841 | 1 |
| T2DM_admissi | leukocyte | *DHRS4L2* | -1.234396017 | 0.000283338 | 0.001088708 | 1 |
| T2DM_admissi | leukocyte | *TRPV2* | -6.501321282 | 0.027388071 | 0.033181538 | 1 |
| T2DM_admissi | leukocyte | *MCRS1* | -1.232144052 | 0.026432292 | 0.032102775 | 1 |
| T2DM_admissi | leukocyte | *PALM3* | -1.404343039 | 0.001140371 | 0.002049538 | 1 |
| T2DM_admissi | leukocyte | *KLHL17* | -1.917047903 | 0.001067599 | 0.001963762 | 1 |
| T2DM_admissi | leukocyte | *RINL* | -1.534775514 | 0.00030645 | 0.001098442 | 1 |
| T2DM_admissi | leukocyte | *ZNF490* | -2.067432217 | 0.00059265 | 0.001392055 | 1 |
| T2DM_admissi | leukocyte | *NRN1L* | -2.962984086 | 0.001306466 | 0.002205889 | 1 |

| | | | | | | |
|---|---|---|---|---|---|---|
| T2DM_admissi | leukocyte | *RAB42* | 0.536621321 | 0.010042956 | 0.013070147 | 1 |
| T2DM_admissi | leukocyte | *MAPK12* | -1.174624278 | 0.005384099 | 0.007391378 | 1 |
| T2DM_admissi | leukocyte | *LINC00265* | -2.352949688 | 0.001195792 | 0.002107792 | 1 |
| T2DM_admissi | leukocyte | *LAMTOR4* | -1.035411936 | 0.000626514 | 0.001429485 | 1 |
| T2DM_admissi | leukocyte | *PRKAR1B* | -1.462169714 | 0.000455385 | 0.001240899 | 1 |
| T2DM_admissi | leukocyte | *NUTM2B* | -35.21701602 | 8.84E-08 | 3.16E-05 | 0.000347214 |
| T2DM_admissi | leukocyte | *AGAP4* | -2.650571357 | 0.001217516 | 0.00212984 | 1 |
| T2DM_admissi | leukocyte | *CENPP* | -0.868473856 | 0.003067831 | 0.00445601 | 1 |
| T2DM_admissi | leukocyte | *FOCAD* | -1.252064968 | 0.000528817 | 0.001321888 | 1 |
| T2DM_admissi | leukocyte | *ZP3* | -1.441700859 | 0.002690702 | 0.003971363 | 1 |
| T2DM_admissi | leukocyte | *SRSF10* | -1.095551642 | 0.000467907 | 0.001253701 | 1 |
| T2DM_admissi | leukocyte | *RALGAPA2* | -1.2446626 | 0.000423179 | 0.001206582 | 1 |
| T2DM_admissi | leukocyte | *NDOR1* | -2.764801708 | 0.005481748 | 0.007522804 | 1 |
| T2DM_admissi | leukocyte | *FBLL1* | -2.710995229 | 0.000114639 | 0.001062643 | 0.450415556 |
| T2DM_admissi | leukocyte | *ASAH2* | -3.109735936 | 7.67E-05 | 0.001062643 | 0.301435004 |
| T2DM_admissi | leukocyte | *S100A16* | -0.607917714 | 0.001078489 | 0.001970875 | 1 |
| T2DM_admissi | leukocyte | *PARVB* | -0.780470935 | 0.001192619 | 0.002105978 | 1 |
| T2DM_admissi | leukocyte | *UROS* | -1.485971944 | 0.000171911 | 0.001062643 | 0.675439891 |
| T2DM_admissi | leukocyte | *FAM221A* | -1.741622808 | 0.000140144 | 0.001062643 | 0.550627049 |
| T2DM_admissi | leukocyte | *NOXA1* | -2.063245968 | 0.001079586 | 0.001971964 | 1 |
| T2DM_admissi | leukocyte | *TMEM198* | -0.904827813 | 0.002037814 | 0.003120254 | 1 |
| T2DM_admissi | leukocyte | *ZNF548* | -1.203859086 | 0.005321688 | 0.007313365 | 1 |
| T2DM_admissi | leukocyte | *MTF1* | -1.124656163 | 0.00084582 | 0.001700095 | 1 |
| T2DM_admissi | leukocyte | *ZDHHC11* | -1.905047542 | 0.042045519 | 0.049878274 | 1 |
| T2DM_admissi | leukocyte | *ENTPD8* | -7.052387011 | 0.001553263 | 0.002503187 | 1 |
| T2DM_admissi | leukocyte | *RPL14* | -0.776668227 | 0.000232128 | 0.001066588 | 0.912030225 |
| T2DM_admissi | leukocyte | *FBF1* | 0.752503743 | 0.031949539 | 0.038447087 | 1 |
| T2DM_admissi | leukocyte | *MSL1* | -1.317669806 | 0.000202272 | 0.001062643 | 0.794728337 |
| T2DM_admissi | leukocyte | *NOC2L* | -1.462976828 | 0.000268775 | 0.001088175 | 1 |
| T2DM_admissi | leukocyte | *ZNF567* | -1.020931455 | 0.002750419 | 0.00404431 | 1 |
| T2DM_admissi | leukocyte | *ARID2* | -1.605700275 | 0.000193418 | 0.001062643 | 0.759938016 |
| T2DM_admissi | leukocyte | *PLAC9* | -1.297209781 | 0.000192612 | 0.001062643 | 0.756772468 |
| T2DM_admissi | leukocyte | *CLDN4* | -0.649104939 | 0.00083554 | 0.001688702 | 1 |
| T2DM_admissi | leukocyte | *FAM47E* | -1.81338515 | 5.21E-05 | 0.00105496 | 0.204778436 |
| T2DM_admissi | leukocyte | *ZNF527* | -1.368541751 | 0.003785739 | 0.005387239 | 1 |
| T2DM_admissi | leukocyte | *ZNF600* | -1.122207986 | 0.002417373 | 0.00362099 | 1 |
| T2DM_admissi | leukocyte | *PNRC2* | -1.015135217 | 0.000405841 | 0.001187305 | 1 |
| T2DM_admissi | leukocyte | *ZKSCAN3* | -2.578943427 | 0.000295553 | 0.001092406 | 1 |
| T2DM_admissi | leukocyte | *RRP7A* | -1.205292515 | 0.001043215 | 0.001936841 | 1 |
| T2DM_admissi | leukocyte | *S100A14* | -0.882552712 | 0.002980929 | 0.004347464 | 1 |
| T2DM_admissi | leukocyte | *CXCL17* | -0.846815711 | 0.003631067 | 0.005184034 | 1 |
| T2DM_admissi | leukocyte | *SPATA41* | -4.52602981 | 0.002820299 | 0.004137772 | 1 |
| T2DM_admissi | leukocyte | *BLOC1S2* | -1.345228416 | 7.71E-05 | 0.001062643 | 0.302956869 |
| T2DM_admissi | leukocyte | *ZNF676* | -1.516548278 | 0.000588672 | 0.001384966 | 1 |
| T2DM_admissi | leukocyte | *WDSUB1* | -1.379454948 | 0.001730967 | 0.002726932 | 1 |
| T2DM_admissi | leukocyte | *ZNF681* | -6.90602675 | 1.66E-05 | 0.000823546 | 0.06539431 |
| T2DM_admissi | leukocyte | *HRCT1* | 1.085653395 | 0.01562798 | 0.019749866 | 1 |
| T2DM_admissi | leukocyte | *MPHOSPH8* | -1.175454428 | 0.000251959 | 0.001077213 | 0.989945552 |
| T2DM_admissi | leukocyte | *SIRPB2* | 1.818155955 | 0.007612583 | 0.010142367 | 1 |
| T2DM_admissi | leukocyte | *LCOR* | -1.119789123 | 0.000182316 | 0.001062643 | 0.716318926 |

| | | | | | | |
|---|---|---|---|---|---|---|
| T2DM_admissi | leukocyte | *SUPT5H* | -1.103224624 | 0.000129689 | 0.001062643 | 0.509549855 |
| T2DM_admissi | leukocyte | *XPNPEP3* | -1.902099791 | 5.68E-05 | 0.001060479 | 0.223035205 |
| T2DM_admissi | leukocyte | *ZNF107* | -0.928099907 | 0.013206596 | 0.016890858 | 1 |
| T2DM_admissi | leukocyte | *PPIA* | -0.923210038 | 0.000436548 | 0.001222199 | 1 |
| T2DM_admissi | leukocyte | *ZNF471* | -1.579828263 | 0.00133821 | 0.002242144 | 1 |
| T2DM_admissi | leukocyte | *ZNF493* | -1.603981878 | 0.000238169 | 0.001071897 | 0.93576579 |
| T2DM_admissi | leukocyte | *GTF2IRD2* | -1.652034799 | 0.000278024 | 0.001088708 | 1 |
| T2DM_admissi | leukocyte | *SUPT3H* | -1.008315906 | 0.00080043 | 0.001644817 | 1 |
| T2DM_admissi | leukocyte | *POM121* | -1.10280023 | 0.001772647 | 0.002779222 | 1 |
| T2DM_admissi | leukocyte | *LONP1* | -1.669277955 | 0.0003491 | 0.001141108 | 1 |
| T2DM_admissi | leukocyte | *ZNF34* | -1.601774951 | 0.000845937 | 0.001700095 | 1 |
| T2DM_admissi | leukocyte | *ZNF781* | -1.666776259 | 0.001274512 | 0.002175489 | 1 |
| T2DM_admissi | leukocyte | *EVL* | -2.467119996 | 0.00066012 | 0.00146119 | 1 |
| T2DM_admissi | leukocyte | *ZNF124* | -1.969449232 | 0.000345092 | 0.001140861 | 1 |
| T2DM_admissi | leukocyte | *XRCC6* | -1.247954317 | 3.48E-05 | 0.001037577 | 0.136722944 |
| T2DM_admissi | leukocyte | *PIK3R4* | -2.081090121 | 0.000119841 | 0.001062643 | 0.470854157 |
| T2DM_admissi | leukocyte | *ZNF605* | -1.743702551 | 0.037753716 | 0.045113853 | 1 |
| T2DM_admissi | leukocyte | *C20orf96* | -2.529453 | 9.92E-05 | 0.001062643 | 0.389584622 |
| T2DM_admissi | leukocyte | *SULT1A1* | -2.840127228 | 1.26E-05 | 0.000801308 | 0.049374414 |
| T2DM_admissi | leukocyte | *ARL9* | 1.100118893 | 0.020670921 | 0.025595981 | 1 |
| T2DM_admissi | leukocyte | *TPK1* | -2.075558233 | 0.017295219 | 0.021704452 | 1 |
| T2DM_admissi | leukocyte | *AFAP1* | -1.241730576 | 0.001308836 | 0.002207994 | 1 |
| T2DM_admissi | leukocyte | *MYO18A* | -1.029724708 | 0.000627528 | 0.001430967 | 1 |
| T2DM_admissi | leukocyte | *XRCC2* | -4.283951736 | 0.000744929 | 0.001568308 | 1 |
| T2DM_admissi | leukocyte | *MYO6* | -0.857384073 | 0.000543678 | 0.001341777 | 1 |
| T2DM_admissi | leukocyte | *ZNF502* | -5.324954432 | 2.73E-05 | 0.000996218 | 0.107121511 |
| T2DM_admissi | leukocyte | *TRAPPC4* | -1.025475433 | 0.000947811 | 0.001821284 | 1 |
| T2DM_admissi | leukocyte | *ZNF33B* | -2.285785475 | 6.24E-05 | 0.001060479 | 0.245247423 |
| T2DM_admissi | leukocyte | *AMZ2* | -1.450151504 | 0.000286617 | 0.001088708 | 1 |
| T2DM_admissi | leukocyte | *ZNF431* | -1.21723087 | 0.000327913 | 0.001122427 | 1 |
| T2DM_admissi | leukocyte | *DAPK1* | -2.023508309 | 0.000268172 | 0.001087356 | 1 |
| T2DM_admissi | leukocyte | *HLA-DQA1* | 0.081961747 | 0.014940438 | 0.018929694 | 1 |
| T2DM_admissi | leukocyte | *CLPSL2* | -0.98177963 | 0.025249596 | 0.03082836 | 1 |
| T2DM_admissi | leukocyte | *MVB12B* | -6.449666797 | 0.000326887 | 0.001122427 | 1 |
| T2DM_admissi | leukocyte | *ADA* | 0.509160861 | 0.009184413 | 0.012024512 | 1 |
| T2DM_admissi | leukocyte | *PPTC7* | -1.905105834 | 8.08E-05 | 0.001062643 | 0.317569142 |
| T2DM_admissi | leukocyte | *ZFP28* | -1.682730698 | 0.000442736 | 0.001227599 | 1 |
| T2DM_admissi | leukocyte | *ANKRD36B* | -1.169845766 | 0.003869081 | 0.005493899 | 1 |
| T2DM_admissi | leukocyte | *SRGAP1* | -1.373399841 | 0.000427132 | 0.00121345 | 1 |
| T2DM_admissi | leukocyte | *FAM3C* | -0.972258093 | 0.000303587 | 0.001094307 | 1 |
| T2DM_admissi | leukocyte | *AP2A1* | -1.534755218 | 0.000176471 | 0.001062643 | 0.693352625 |
| T2DM_admissi | leukocyte | *WDR5B* | -3.616868823 | 0.000293596 | 0.001088724 | 1 |
| T2DM_admissi | leukocyte | *ZNF138* | -0.823911323 | 0.003900488 | 0.005528505 | 1 |
| T2DM_admissi | leukocyte | *ZNF429* | -1.109885897 | 0.000630536 | 0.001433512 | 1 |
| T2DM_admissi | leukocyte | *ZNF100* | -1.617957212 | 0.000270398 | 0.00108852 | 1 |
| T2DM_admissi | leukocyte | *ZNF763* | -2.558273687 | 0.000432348 | 0.001217703 | 1 |
| T2DM_admissi | leukocyte | *ARRDC1* | -1.020278816 | 0.001411149 | 0.002330561 | 1 |
| T2DM_admissi | leukocyte | *ZNF682* | -2.152312638 | 0.000439232 | 0.001222199 | 1 |
| T2DM_admissi | leukocyte | *ZNF257* | -1.967252043 | 0.000179376 | 0.001062643 | 0.704766546 |
| T2DM_admissi | leukocyte | *ADAM32* | 1.221966143 | 0.010370269 | 0.013456007 | 1 |

| | | | | | | |
|---|---|---|---|---|---|---|
| T2DM_admissi | leukocyte | *ACSL5* | -2.002812321 | 0.00039639 | 0.001181845 | 1 |
| T2DM_admissi | leukocyte | *SND1* | -1.180397834 | 0.00026953 | 0.001088175 | 1 |
| T2DM_admissi | leukocyte | *TBC1D9B* | -2.514910275 | 1.45E-05 | 0.000815363 | 0.057075439 |
| T2DM_admissi | leukocyte | *C6orf141* | -2.358437312 | 0.000325568 | 0.001119124 | 1 |
| T2DM_admissi | leukocyte | *GTF2E2* | -1.120576001 | 0.000488538 | 0.001280054 | 1 |
| T2DM_admissi | leukocyte | *ZNF165* | -1.214220671 | 0.000804222 | 0.001649159 | 1 |
| T2DM_admissi | leukocyte | *SVIL* | -1.064528834 | 0.000603308 | 0.001403431 | 1 |
| T2DM_admissi | leukocyte | *MRPL21* | -0.892696563 | 0.005831659 | 0.007966363 | 1 |
| T2DM_admissi | leukocyte | *ZNF98* | -1.56686078 | 0.001302049 | 0.002200323 | 1 |
| T2DM_admissi | leukocyte | *ZNF517* | -4.664270886 | 0.00049235 | 0.001284491 | 1 |
| T2DM_admissi | leukocyte | *SLC22A5* | -1.758282666 | 0.000429332 | 0.00121662 | 1 |
| T2DM_admissi | leukocyte | *C5AR1* | 0.131988615 | 0.007558634 | 0.010077324 | 1 |
| T2DM_admissi | leukocyte | *VEPH1* | -2.044367472 | 0.003616055 | 0.005164478 | 1 |
| T2DM_admissi | leukocyte | *SHPK* | -1.073244284 | 0.025550428 | 0.031147264 | 1 |
| T2DM_admissi | leukocyte | *PCDHB11* | -2.24222229 | 0.000489698 | 0.001281828 | 1 |
| T2DM_admissi | leukocyte | *MIB2* | -0.779218003 | 0.020325447 | 0.025206379 | 1 |
| T2DM_admissi | leukocyte | *ATG7* | -2.343631582 | 0.000189942 | 0.001062643 | 0.746283975 |
| T2DM_admissi | leukocyte | *PIGN* | -1.007581061 | 0.001371489 | 0.002283297 | 1 |
| T2DM_admissi | leukocyte | *HHLA3* | -1.042894178 | 0.001200501 | 0.002112301 | 1 |
| T2DM_admissi | leukocyte | *BCO2* | -1.131350507 | 0.00275964 | 0.004056351 | 1 |
| T2DM_admissi | leukocyte | *ENTPD6* | -1.364908047 | 0.000352841 | 0.001141109 | 1 |
| T2DM_admissi | leukocyte | *ENPP1* | -0.691231285 | 0.017296167 | 0.021704452 | 1 |
| T2DM_admissi | leukocyte | *CCDC154* | 3.971803916 | 0.014038508 | 0.017890788 | 1 |
| T2DM_admissi | leukocyte | *DPP4* | -0.872352031 | 0.000177716 | 0.001062643 | 0.698245185 |
| T2DM_admissi | leukocyte | *NMB* | -1.147265542 | 0.000468742 | 0.001253701 | 1 |
| T2DM_admissi | leukocyte | *RPS26* | -0.991744722 | 2.51E-05 | 0.000982543 | 0.098560903 |
| T2DM_admissi | leukocyte | *LHFPL5* | -3.010280316 | 0.000449743 | 0.001234067 | 1 |
| T2DM_admissi | leukocyte | *ZNF780A* | -1.015748561 | 0.000822309 | 0.001670341 | 1 |
| T2DM_admissi | leukocyte | *ATAD3A* | -2.119534608 | 0.000141487 | 0.001062643 | 0.555903068 |
| T2DM_admissi | leukocyte | *FAM118B* | -2.865160733 | 0.000134416 | 0.001062643 | 0.528121022 |
| T2DM_admissi | leukocyte | *SLC9A8* | -4.188347725 | 7.01E-05 | 0.001062643 | 0.27540394 |
| T2DM_admissi | leukocyte | *OCLN* | -0.633173702 | 0.007267206 | 0.009718466 | 1 |
| T2DM_admissi | leukocyte | *ZNF181* | -1.62289152 | 0.000582859 | 0.001382042 | 1 |
| T2DM_admissi | leukocyte | *TEAD4* | -0.940080207 | 0.0014347 | 0.002354561 | 1 |
| T2DM_admissi | leukocyte | *C2orf27A* | -1.923183519 | 0.000217329 | 0.001062643 | 0.853885941 |
| T2DM_admissi | leukocyte | *PLCG2* | -1.401947332 | 0.000170623 | 0.001062643 | 0.670376538 |
| T2DM_admissi | leukocyte | *RPL12* | -0.751822206 | 0.000218396 | 0.001062643 | 0.85807976 |
| T2DM_admissi | leukocyte | *VPS13A* | -1.053831934 | 0.001023321 | 0.001910945 | 1 |
| T2DM_admissi | leukocyte | *LEKR1* | -2.022804315 | 0.000621523 | 0.001422226 | 1 |
| T2DM_admissi | leukocyte | *ENTPD7* | -1.800835343 | 0.012385022 | 0.015897011 | 1 |
| T2DM_admissi | leukocyte | *AGAP9* | -2.634657559 | 0.000173195 | 0.001062643 | 0.680483761 |
| T2DM_admissi | leukocyte | *ZNF273* | -2.391428943 | 0.000163135 | 0.001062643 | 0.640956892 |
| T2DM_admissi | leukocyte | *ZNF84* | -2.509648706 | 0.00024473 | 0.001077213 | 0.961542927 |
| T2DM_admissi | leukocyte | *ZNF667* | -0.833130892 | 0.001101475 | 0.002000708 | 1 |
| T2DM_admissi | leukocyte | *CD2AP* | -1.177623262 | 0.000287423 | 0.001088708 | 1 |
| T2DM_admissi | leukocyte | *SFI1* | -1.981772842 | 0.004688365 | 0.006506741 | 1 |
| T2DM_admissi | leukocyte | *HIBCH* | -1.055006927 | 0.000631489 | 0.001433512 | 1 |
| T2DM_admissi | leukocyte | *TMEM229B* | -5.71691622 | 0.000225484 | 0.001062643 | 0.885928159 |
| T2DM_admissi | leukocyte | *SOWAHC* | -2.1167336 | 0.000159572 | 0.001062643 | 0.626956877 |
| T2DM_admissi | leukocyte | *SVIP* | -1.036473675 | 0.000258705 | 0.001081954 | 1 |

| | | | | | | |
|---|---|---|---|---|---|---|
| T2DM_admissi | leukocyte | *DDRGK1* | -1.314182978 | 0.000188623 | 0.001062643 | 0.741099511 |
| T2DM_admissi | leukocyte | *ZNF607* | -1.209103424 | 0.004403277 | 0.006158945 | 1 |
| T2DM_admissi | leukocyte | *HSD17B11* | -1.721694556 | 0.000401005 | 0.001184732 | 1 |
| T2DM_admissi | leukocyte | *RPS6KL1* | -1.352441081 | 0.002542942 | 0.003774544 | 1 |
| T2DM_admissi | leukocyte | *QRICH1* | -1.958486922 | 0.00022638 | 0.001062643 | 0.889445749 |
| T2DM_admissi | leukocyte | *DDX42* | -0.874424789 | 0.002684754 | 0.003964074 | 1 |
| T2DM_admissi | leukocyte | *HELZ* | -1.292914962 | 0.00022365 | 0.001062643 | 0.878722365 |
| T2DM_admissi | leukocyte | *ZNF583* | -0.999147513 | 0.001085952 | 0.00197991 | 1 |
| T2DM_admissi | leukocyte | *ZNF568* | -1.183655086 | 0.002515851 | 0.003742817 | 1 |
| T2DM_admissi | leukocyte | *ZNF587* | -1.351652314 | 0.000459849 | 0.001240899 | 1 |
| T2DM_admissi | leukocyte | *TPM2* | -1.311335302 | 0.000418014 | 0.001200552 | 1 |
| T2DM_admissi | leukocyte | *NBR2* | -1.49377426 | 0.021282535 | 0.026287042 | 1 |
| T2DM_admissi | leukocyte | *ZNF789* | -3.119015383 | 0.000266255 | 0.001086558 | 1 |
| T2DM_admissi | leukocyte | *NCOA6* | -1.066256224 | 0.000512657 | 0.001308606 | 1 |
| T2DM_admissi | leukocyte | *STK39* | -1.037825814 | 0.00080156 | 0.001646277 | 1 |
| T2DM_admissi | leukocyte | *PAPSS2* | -0.98188416 | 0.000440813 | 0.00122573 | 1 |
| T2DM_admissi | leukocyte | *CEP290* | -1.324863431 | 0.000375592 | 0.001159218 | 1 |
| T2DM_admissi | leukocyte | *ECI2* | -1.357403343 | 0.000176387 | 0.001062643 | 0.693026403 |
| T2DM_admissi | leukocyte | *MSRB1* | -1.56956626 | 0.000313494 | 0.001109126 | 1 |
| T2DM_admissi | leukocyte | *COLGALT2* | -1.582299618 | 0.004031055 | 0.005690987 | 1 |
| T2DM_admissi | leukocyte | *PNP* | -0.658495858 | 0.004463203 | 0.006231672 | 1 |
| T2DM_admissi | leukocyte | *DENND4B* | -1.277647225 | 0.000408145 | 0.001188129 | 1 |
| T2DM_admissi | leukocyte | *CES1* | -1.815604284 | 0.016622971 | 0.020933222 | 1 |
| T2DM_admissi | leukocyte | *TSEN15* | -1.458770959 | 0.000293725 | 0.001088724 | 1 |
| T2DM_admissi | leukocyte | *LTN1* | -1.165987238 | 0.000538967 | 0.001334229 | 1 |
| T2DM_admissi | leukocyte | *TYW1* | -1.82585336 | 0.000412012 | 0.001192043 | 1 |
| T2DM_admissi | leukocyte | *ITPRIPL1* | -1.387111191 | 0.009356954 | 0.012221899 | 1 |
| T2DM_admissi | leukocyte | *MAP3K3* | -1.392465817 | 0.000121164 | 0.001062643 | 0.476051442 |
| T2DM_admissi | leukocyte | *C1orf174* | -1.491613659 | 0.000390095 | 0.00117808 | 1 |
| T2DM_admissi | leukocyte | *KIAA0753* | -2.367907654 | 0.000572531 | 0.001370771 | 1 |
| T2DM_admissi | leukocyte | *ATG9A* | -1.323837955 | 0.001979182 | 0.003048297 | 1 |
| T2DM_admissi | leukocyte | *TBKBP1* | -1.483471158 | 7.11E-05 | 0.001062643 | 0.279303269 |
| T2DM_admissi | leukocyte | *CCDC167* | -0.928947126 | 0.000718761 | 0.0015393 | 1 |
| T2DM_admissi | leukocyte | *SOWAHA* | -1.058627014 | 0.000583116 | 0.001382042 | 1 |
| T2DM_admissi | leukocyte | *L3MBTL3* | -2.083789924 | 0.000150658 | 0.001062643 | 0.591934601 |
| T2DM_admissi | leukocyte | *MFAP3L* | -1.820766059 | 0.001074274 | 0.001968161 | 1 |
| T2DM_admissi | leukocyte | *PJA2* | -1.001390807 | 0.000482229 | 0.001270743 | 1 |
| T2DM_admissi | leukocyte | *INF2* | -1.706243919 | 0.000525219 | 0.001319879 | 1 |
| T2DM_admissi | leukocyte | *EFCAB2* | -1.169843818 | 0.00095111 | 0.001823593 | 1 |
| T2DM_admissi | leukocyte | *SAMD5* | -1.501130769 | 0.000195278 | 0.001062643 | 0.767245636 |
| T2DM_admissi | leukocyte | *DDO* | -1.819629079 | 0.035980531 | 0.043086713 | 1 |
| T2DM_admissi | leukocyte | *C6orf163* | -2.163496703 | 0.001223888 | 0.002136231 | 1 |
| T2DM_admissi | leukocyte | *PCMTD2* | -1.136515178 | 0.000446457 | 0.001232235 | 1 |
| T2DM_admissi | leukocyte | *LIME1* | -2.538334174 | 5.98E-06 | 0.000591121 | 0.023507009 |
| T2DM_admissi | leukocyte | *FOXO6* | -2.225596449 | 0.000944173 | 0.001816677 | 1 |
| T2DM_admissi | leukocyte | *SYS1* | -1.185789997 | 0.000453543 | 0.001238727 | 1 |
| T2DM_admissi | leukocyte | *INPP5B* | -2.448837417 | 6.92E-06 | 0.000632075 | 0.027179205 |
| T2DM_admissi | leukocyte | *TRAF3IP1* | -3.816928115 | 1.24E-05 | 0.000801308 | 0.048767472 |
| T2DM_admissi | leukocyte | *C2orf72* | -2.944760636 | 0.016581508 | 0.020887703 | 1 |
| T2DM_admissi | leukocyte | *CLPSL1* | -0.589231697 | 0.001483943 | 0.002412251 | 1 |

| | | | | | | |
|---|---|---|---|---|---|---|
| T2DM_admissi | leukocyte | *HSD17B8* | -1.134802918 | 0.002563207 | 0.003798883 | 1 |
| T2DM_admissi | leukocyte | *HLA-DOA* | 0.181542729 | 0.004351744 | 0.006102071 | 1 |
| T2DM_admissi | leukocyte | *HLA-DMA* | 0.225381317 | 0.003167051 | 0.004581496 | 1 |
| T2DM_admissi | leukocyte | *NOTCH4* | -2.053037299 | 9.78E-10 | 9.61E-07 | 3.84E-06 |
| T2DM_admissi | leukocyte | *RNF5* | -1.754115473 | 1.92E-08 | 9.45E-06 | 7.56E-05 |
| T2DM_admissi | leukocyte | *MRPL38* | -1.253838138 | 0.000217818 | 0.001062643 | 0.855805103 |
| T2DM_admissi | leukocyte | *SMIM5* | -0.914404772 | 0.001344621 | 0.002250006 | 1 |
| T2DM_admissi | leukocyte | *EHMT2* | -4.119046858 | 0.00025234 | 0.001077213 | 0.991445749 |
| T2DM_admissi | leukocyte | *LAYN* | -1.047804555 | 0.001348633 | 0.0022548 | 1 |
| T2DM_admissi | leukocyte | *SLC44A4* | -0.902994332 | 0.00047736 | 0.001264698 | 1 |
| T2DM_admissi | leukocyte | *MICA* | -0.694176129 | 0.001599441 | 0.002555593 | 1 |
| T2DM_admissi | leukocyte | *HLA-C* | -0.889884631 | 6.27E-06 | 0.000591121 | 0.024617451 |
| T2DM_admissi | leukocyte | *POU5F1* | 1.162174592 | 0.025624695 | 0.03122811 | 1 |
| T2DM_admissi | leukocyte | *CCHCR1* | 1.587022421 | 0.002912718 | 0.004263785 | 1 |
| T2DM_admissi | leukocyte | *PSORS1C1* | -0.853757464 | 0.002453971 | 0.003670214 | 1 |
| T2DM_admissi | leukocyte | *DHX16* | -1.359712847 | 0.000654818 | 0.00145437 | 1 |
| T2DM_admissi | leukocyte | *ABCF1* | -1.179434511 | 0.000557856 | 0.001358844 | 1 |
| T2DM_admissi | leukocyte | *LILRB3* | 1.0774421 | 0.006356045 | 0.008599484 | 1 |
| T2DM_admissi | leukocyte | *ZNF468* | -3.06663026 | 9.17E-05 | 0.001062643 | 0.360166742 |
| T2DM_admissi | leukocyte | *HLA-G* | -0.797913551 | 0.021408992 | 0.02641832 | 1 |
| T2DM_admissi | leukocyte | *HLA-F* | -0.387928407 | 0.041330829 | 0.049134289 | 1 |
| T2DM_admissi | leukocyte | *ASPDH* | -2.063059035 | 0.000719524 | 0.001539766 | 1 |
| T2DM_admissi | leukocyte | *ZNF783* | -5.645921916 | 0.002016482 | 0.003093619 | 1 |
| T2DM_admissi | leukocyte | *ZNF425* | -3.058128404 | 0.000619055 | 0.001422062 | 1 |
| T2DM_admissi | leukocyte | *PCDHA5* | -1.184475726 | 0.042111777 | 0.049941797 | 1 |
| T2DM_admissi | leukocyte | *SPIRE2* | -0.877877402 | 0.004549493 | 0.006334146 | 1 |
| T2DM_admissi | leukocyte | *PABPN1L* | -160.4110726 | 0.000637294 | 0.001438212 | 1 |
| T2DM_admissi | leukocyte | *PKHD1L1* | -4.534359442 | 0.001348607 | 0.0022548 | 1 |
| T2DM_admissi | leukocyte | *SYCE1L* | -1.717512783 | 0.001090835 | 0.001986047 | 1 |
| T2DM_admissi | leukocyte | *TMEM231* | -1.128337792 | 0.001867122 | 0.002901868 | 1 |
| T2DM_admissi | leukocyte | *LINC00654* | -4.319638988 | 0.006578289 | 0.008872673 | 1 |
| T2DM_admissi | leukocyte | *SAP25* | -0.897331859 | 0.005578238 | 0.007637573 | 1 |
| T2DM_admissi | leukocyte | *SARNP* | -1.111563708 | 0.000458243 | 0.001240899 | 1 |
| T2DM_admissi | leukocyte | *MT1A* | -0.58819192 | 0.008431635 | 0.011146666 | 1 |
| T2DM_admissi | leukocyte | *ATP6AP1L* | 1.911019546 | 0.011047445 | 0.01426402 | 1 |
| T2DM_admissi | leukocyte | *CCDC85C* | -1.193006965 | 0.000920526 | 0.001793131 | 1 |
| T2DM_admissi | leukocyte | *RGL3* | -2.093787655 | 0.000676991 | 0.001485976 | 1 |
| T2DM_admissi | leukocyte | *CPT1B* | -1.108240685 | 0.006836991 | 0.009186915 | 1 |
| T2DM_admissi | leukocyte | *SMN2* | -1.142501184 | 0.00872478 | 0.011480128 | 1 |
| T2DM_admissi | leukocyte | *SERF1B* | -9.641342252 | 0.00513136 | 0.007074074 | 1 |
| T2DM_admissi | leukocyte | *HMGN1* | -0.956223505 | 0.000169537 | 0.001062643 | 0.66611242 |
| T2DM_admissi | leukocyte | *LCMT1* | -1.664731779 | 0.000188225 | 0.001062643 | 0.739535909 |
| T2DM_admissi | leukocyte | *LIN52* | -2.67334193 | 0.000177187 | 0.001062643 | 0.696167054 |
| T2DM_admissi | leukocyte | *DENND1C* | 0.397977841 | 0.008043251 | 0.010665519 | 1 |
| T2DM_admissi | leukocyte | *DEFB134* | -126.8839692 | 0.009088221 | 0.01190254 | 1 |
| T2DM_admissi | leukocyte | *RNPS1* | -1.105369483 | 0.000368785 | 0.001149965 | 1 |
| T2DM_admissi | leukocyte | *ZDHHC11B* | -1.968397781 | 0.000842389 | 0.001699271 | 1 |
| T2DM_admissi | leukocyte | *HLA-A* | -0.675455802 | 0.001400948 | 0.00231761 | 1 |
| T2DM_admissi | leukocyte | *STK38L* | -1.906838653 | 0.001503349 | 0.002439759 | 1 |
| T2DM_admissi | leukocyte | *MAP10* | -1.273724397 | 0.00166241 | 0.002637968 | 1 |

| | | | | | | |
|---|---|---|---|---|---|---|
| T2DM_admissi | leukocyte | *NUP62* | -1.283461251 | 0.000456535 | 0.001240899 | 1 |
| T2DM_admissi | leukocyte | *FAM106A* | -1.557416912 | 0.001537599 | 0.002485079 | 1 |
| T2DM_admissi | leukocyte | *ZNF254* | -1.544648986 | 0.000107415 | 0.001062643 | 0.422034245 |
| T2DM_admissi | leukocyte | *ASIC3* | -1.461978257 | 0.003363832 | 0.004841208 | 1 |
| T2DM_admissi | leukocyte | *ARHGEF35* | -0.795517664 | 0.001049598 | 0.001940644 | 1 |
| T2DM_admissi | leukocyte | *DNLZ* | 0.843614386 | 0.025702941 | 0.03131346 | 1 |
| T2DM_admissi | leukocyte | *SUPT4H1* | -1.433306589 | 0.000200976 | 0.001062643 | 0.78963277 |
| T2DM_admissi | leukocyte | *MXD3* | -1.091943738 | 0.003170602 | 0.004584945 | 1 |
| T2DM_admissi | leukocyte | *COG8* | -1.68218127 | 0.00059938 | 0.001399402 | 1 |
| T2DM_admissi | leukocyte | *ERV3-1* | -1.518520597 | 0.006586003 | 0.008880029 | 1 |
| T2DM_admissi | leukocyte | *CCL14* | -1.463499289 | 0.009058573 | 0.011871625 | 1 |
| T2DM_admissi | leukocyte | *RBMXL1* | -1.177897618 | 0.001112188 | 0.002015584 | 1 |
| T2DM_admissi | leukocyte | *SRA1* | -0.829796622 | 0.001637436 | 0.002605704 | 1 |
| T2DM_admissi | leukocyte | *C8orf82* | -2.39953838 | 8.90E-05 | 0.001062643 | 0.349535996 |
| T2DM_admissi | leukocyte | *LEPROT* | -1.201028774 | 0.000208594 | 0.001062643 | 0.819565958 |
| T2DM_admissi | leukocyte | *PPP1CB* | -0.961524207 | 0.000252713 | 0.001077213 | 0.99290743 |
| T2DM_admissi | leukocyte | *NCKIPSD* | -1.651633988 | 0.000747099 | 0.001568868 | 1 |
| T2DM_admissi | leukocyte | *ZNF888* | -1.663123084 | 0.000950229 | 0.001823593 | 1 |
| T2DM_admissi | leukocyte | *EMP2* | -1.296503206 | 0.000310848 | 0.00110427 | 1 |
| T2DM_admissi | leukocyte | *LTB4R* | -1.519322012 | 0.001298522 | 0.002196252 | 1 |
| T2DM_admissi | leukocyte | *DNASE1* | -0.997022802 | 0.002163195 | 0.003280275 | 1 |
| T2DM_admissi | leukocyte | *NUDT19* | -1.577700341 | 0.000462209 | 0.001244702 | 1 |
| T2DM_admissi | leukocyte | *AP1G2* | -5.017113124 | 3.61E-05 | 0.001037577 | 0.141900553 |
| T2DM_admissi | leukocyte | *ZNF90* | -1.396545233 | 0.001188623 | 0.00210081 | 1 |
| T2DM_admissi | leukocyte | *REPIN1* | -1.321317564 | 0.000274851 | 0.001088708 | 1 |
| T2DM_admissi | leukocyte | *TSPAN4* | -1.059281027 | 0.007758345 | 0.010322566 | 1 |
| T2DM_admissi | leukocyte | *CPNE1* | -1.071078259 | 0.022360094 | 0.027531435 | 1 |
| T2DM_admissi | leukocyte | *ARL16* | -0.983284688 | 0.001271335 | 0.002175059 | 1 |
| T2DM_admissi | leukocyte | *WEE2* | -6.008691389 | 0.004985646 | 0.006887694 | 1 |
| T2DM_admissi | leukocyte | *FIS1* | -0.910187983 | 0.000634719 | 0.001434874 | 1 |
| T2DM_admissi | leukocyte | *MBLAC1* | -1.166208608 | 0.02602328 | 0.031654943 | 1 |
| T2DM_admissi | leukocyte | *MEG3* | -0.745451959 | 0.000471085 | 0.001255055 | 1 |
| T2DM_admissi | leukocyte | *EML6* | -1.401294014 | 0.00137335 | 0.002283803 | 1 |
| T2DM_admissi | leukocyte | *C12orf71* | -18.59740713 | 0.001379027 | 0.002289918 | 1 |
| T2DM_admissi | leukocyte | *IFRD2* | -1.310878662 | 0.000259129 | 0.001081954 | 1 |
| T2DM_admissi | leukocyte | *ARHGEF28* | -1.65078205 | 0.000302464 | 0.001093914 | 1 |
| T2DM_admissi | leukocyte | *COL28A1* | -5.356884138 | 0.003158761 | 0.004571187 | 1 |
| T2DM_admissi | leukocyte | *UBXN2B* | -2.001500338 | 0.000212397 | 0.001062643 | 0.834509526 |
| T2DM_admissi | leukocyte | *C5orf49* | -0.97562299 | 0.00084517 | 0.001700095 | 1 |
| T2DM_admissi | leukocyte | *GOLGA8B* | -1.564899211 | 0.00089687 | 0.001768089 | 1 |
| T2DM_admissi | leukocyte | *VPS16* | -1.719286995 | 0.002746813 | 0.004040519 | 1 |
| T2DM_admissi | leukocyte | *LINC01139* | -3.740431151 | 0.00027457 | 0.001088708 | 1 |
| T2DM_admissi | leukocyte | *ATAD3C* | -1.096658308 | 0.000587286 | 0.001383749 | 1 |
| T2DM_admissi | leukocyte | *SYCE3* | -2.447454202 | 0.000356519 | 0.001141109 | 1 |
| T2DM_admissi | leukocyte | *PAM16* | -1.094025213 | 0.000361466 | 0.001142338 | 1 |
| T2DM_admissi | leukocyte | *LINC00339* | -1.342114293 | 0.000807525 | 0.0016529 | 1 |
| T2DM_admissi | leukocyte | *CELA3B* | -0.595339461 | 0.000417534 | 0.001200552 | 1 |
| T2DM_admissi | leukocyte | *FAM228B* | -0.915860187 | 0.014781995 | 0.018765254 | 1 |
| T2DM_admissi | leukocyte | *VAMP2* | -0.874870838 | 0.000285613 | 0.001088708 | 1 |
| T2DM_admissi | leukocyte | *PPP3R1* | -1.917971454 | 9.51E-05 | 0.001062643 | 0.373805191 |

| | | | | | | |
|---|---|---|---|---|---|---|
| T2DM_admissi | leukocyte | *FANCG* | -1.85675868 | 0.002619192 | 0.003874551 | 1 |
| T2DM_admissi | leukocyte | *AP4M1* | -0.827680698 | 0.004587289 | 0.006377728 | 1 |
| T2DM_admissi | leukocyte | *CEBPD* | -2.395002754 | 0.001220472 | 0.002133102 | 1 |
| T2DM_admissi | leukocyte | *ZNF880* | -1.041696169 | 0.000357299 | 0.001141109 | 1 |
| T2DM_admissi | leukocyte | *TRIM16* | -1.57288474 | 0.001187005 | 0.00210081 | 1 |
| T2DM_admissi | leukocyte | *TIGD1* | -1.818262039 | 0.000919803 | 0.001793131 | 1 |
| T2DM_admissi | leukocyte | *SLC12A8* | -2.0365524 | 6.25E-05 | 0.001060479 | 0.245672064 |
| T2DM_admissi | leukocyte | *UBA52* | -0.924793909 | 0.000156343 | 0.001062643 | 0.61427195 |
| T2DM_admissi | leukocyte | *EXOSC6* | -1.572063801 | 1.56E-05 | 0.000823546 | 0.061296166 |
| T2DM_admissi | leukocyte | *CKMT1A* | -0.900440546 | 0.011643214 | 0.014988921 | 1 |
| T2DM_admissi | leukocyte | *HLA-DPB1* | 0.12353616 | 0.004366718 | 0.0061187 | 1 |
| T2DM_admissi | leukocyte | *C4B* | 9.838317641 | 0.028849496 | 0.034909045 | 1 |
| T2DM_admissi | leukocyte | *LINC00115* | -3.281844941 | 0.000404653 | 0.001185781 | 1 |
| T2DM_admissi | leukocyte | *LINC00937* | -1.174281962 | 0.004161008 | 0.005855516 | 1 |
| T2DM_admissi | leukocyte | *TMEM185B* | -1.271993517 | 0.000634268 | 0.001434874 | 1 |
| T2DM_admissi | leukocyte | *CYP4F26P* | 5.101007142 | 0.008670024 | 0.011423382 | 1 |
| T2DM_admissi | leukocyte | *WDR46* | -0.950941513 | 0.004013074 | 0.005671715 | 1 |
| T2DM_admissi | leukocyte | *ZNF717* | -0.57583955 | 0.022901668 | 0.028154147 | 1 |
| T2DM_admissi | leukocyte | *PROB1* | -1.751433683 | 0.00800372 | 0.010616683 | 1 |
| T2DM_admissi | leukocyte | *ARL17B* | -3.492727982 | 9.48E-05 | 0.001062643 | 0.372380393 |
| T2DM_admissi | leukocyte | *DHFR* | -1.451362075 | 0.004734509 | 0.006568462 | 1 |
| T2DM_admissi | leukocyte | *PSMG3-AS1* | -1.112366454 | 0.009937544 | 0.012950119 | 1 |
| T2DM_admissi | leukocyte | *HSBP1* | -1.055349601 | 0.000248401 | 0.001077213 | 0.975968932 |
| T2DM_admissi | leukocyte | *RPS18* | -0.689322909 | 0.000411446 | 0.001191282 | 1 |
| T2DM_admissi | leukocyte | *DIRC3* | -3.162012347 | 0.02478574 | 0.030290256 | 1 |
| T2DM_admissi | leukocyte | *LINC00865* | -2.697418603 | 0.001904447 | 0.002952841 | 1 |
| T2DM_admissi | leukocyte | *HLA-DQB2* | 1.901553831 | 4.20E-06 | 0.000516024 | 0.016512773 |
| T2DM_admissi | leukocyte | *GOLGA8N* | -0.896326801 | 0.035917485 | 0.043024329 | 1 |
| T2DM_admissi | leukocyte | *LINC00665* | -0.857710838 | 0.001293158 | 0.002192479 | 1 |
| T2DM_admissi | leukocyte | *LYRM9* | -1.318820729 | 0.00099658 | 0.001875957 | 1 |
| T2DM_admissi | leukocyte | *SNHG15* | -1.067351948 | 0.00098706 | 0.001867193 | 1 |
| T2DM_admissi | leukocyte | *RPS28* | -0.776832842 | 0.000195632 | 0.001062643 | 0.76864009 |
| T2DM_admissi | leukocyte | *TRIM26* | -1.667353336 | 4.42E-05 | 0.001037577 | 0.173600309 |
| T2DM_admissi | leukocyte | *ZNF879* | -1.518938127 | 0.000862778 | 0.001726874 | 1 |
| T2DM_admissi | leukocyte | *PINLYP* | -2.675762631 | 0.000211464 | 0.001062643 | 0.830842707 |
| T2DM_admissi | leukocyte | *MCIDAS* | 40.22496741 | 0.007131095 | 0.009546192 | 1 |
| T2DM_admissi | leukocyte | *ANKRD65* | -1.075855395 | 0.000568707 | 0.001367774 | 1 |
| T2DM_admissi | leukocyte | *C12orf75* | -0.666953245 | 0.000523752 | 0.001319879 | 1 |
| T2DM_admissi | leukocyte | *B3GALT4* | -1.025066448 | 0.0002936 | 0.001088724 | 1 |
| T2DM_admissi | leukocyte | *ARHGEF38* | -1.325691121 | 0.025954985 | 0.031581646 | 1 |
| T2DM_admissi | leukocyte | *ZNF737* | -1.708762809 | 0.000276838 | 0.001088708 | 1 |
| T2DM_admissi | leukocyte | *SHISA9* | -8.62926828 | 0.000921557 | 0.00179425 | 1 |
| T2DM_admissi | leukocyte | *HLA-DQA2* | 0.133093678 | 0.001029723 | 0.001918342 | 1 |
| T2DM_admissi | leukocyte | *LRRC37A2* | -1.013119767 | 0.040806102 | 0.048554566 | 1 |
| T2DM_admissi | leukocyte | *TXNDC5* | -1.224087515 | 0.000288896 | 0.001088708 | 1 |
| T2DM_admissi | leukocyte | *RBM14* | -1.60331009 | 0.000277333 | 0.001088708 | 1 |
| T2DM_admissi | leukocyte | *NME1* | -0.864483136 | 0.000599739 | 0.001399402 | 1 |
| T2DM_admissi | leukocyte | *CDRT4* | -2.461683886 | 0.000960674 | 0.001831992 | 1 |
| T2DM_admissi | leukocyte | *AMY2B* | -0.956584882 | 0.000704454 | 0.001521605 | 1 |
| T2DM_admissi | leukocyte | *LY6G5B* | -1.374908536 | 0.000961195 | 0.001831992 | 1 |

| | | | | | | |
|---|---|---|---|---|---|---|
| T2DM_admissi | leukocyte | *PSMB9* | -0.690788369 | 0.01060095 | 0.013737182 | 1 |
| T2DM_admissi | leukocyte | *PCDHGC3* | -1.032220271 | 0.014377331 | 0.018298844 | 1 |
| T2DM_admissi | leukocyte | *SMKR1* | -1.490724565 | 0.000654576 | 0.00145437 | 1 |
| T2DM_admissi | leukocyte | *LINC00886* | -1.419907469 | 0.00530385 | 0.007293954 | 1 |
| T2DM_admissi | leukocyte | *MIF* | -0.73746817 | 0.000269793 | 0.001088175 | 1 |
| T2DM_admissi | leukocyte | *NSUN6* | -1.137411409 | 0.000586128 | 0.001382291 | 1 |
| T2DM_admissi | leukocyte | *HLA-DOB* | 1.373041079 | 0.04012867 | 0.047806411 | 1 |
| T2DM_admissi | leukocyte | *CRCP* | -1.445470219 | 0.000135563 | 0.001062643 | 0.532627546 |
| T2DM_admissi | leukocyte | *C8orf58* | -1.493703042 | 0.006457602 | 0.008718872 | 1 |
| T2DM_admissi | leukocyte | *PI4KA* | -2.015883929 | 0.000212564 | 0.001062643 | 0.835164878 |
| T2DM_admissi | leukocyte | *AMACR* | -0.867708839 | 0.003208627 | 0.004636519 | 1 |
| T2DM_admissi | leukocyte | *ARHGDIG* | -0.956400518 | 0.036743413 | 0.04396007 | 1 |
| T2DM_admissi | leukocyte | *BGLAP* | -2.197752433 | 0.000169172 | 0.001062643 | 0.664677722 |
| T2DM_admissi | leukocyte | *MRPL20* | -0.928187212 | 0.001007197 | 0.001889818 | 1 |
| T2DM_admissi | leukocyte | *LINC00960* | -0.676222612 | 0.022452353 | 0.02763637 | 1 |
| T2DM_admissi | leukocyte | *HLA-DMB* | 0.112090143 | 0.006104672 | 0.008299397 | 1 |
| T2DM_admissi | leukocyte | *DECR2* | -1.199396062 | 0.000698208 | 0.001512992 | 1 |
| T2DM_admissi | leukocyte | *GNG10* | -0.923383754 | 0.001551644 | 0.002501604 | 1 |
| T2DM_admissi | leukocyte | *CNTF* | 2.428749645 | 0.003061297 | 0.00444981 | 1 |
| T2DM_admissi | leukocyte | *EIF4EBP3* | -1.743678953 | 7.21E-05 | 0.001062643 | 0.283393924 |
| T2DM_admissi | leukocyte | *MICAL3* | -1.117833746 | 0.000646334 | 0.001446812 | 1 |
| T2DM_admissi | leukocyte | *STON1* | -1.598139995 | 0.001335433 | 0.00223874 | 1 |
| T2DM_admissi | leukocyte | *VSIG8* | -2.931631651 | 0.001047648 | 0.001940644 | 1 |
| T2DM_admissi | leukocyte | *C4orf48* | -0.887153825 | 0.000264569 | 0.001084251 | 1 |
| T2DM_admissi | leukocyte | *IL10RB* | -2.60830257 | 0.000355018 | 0.001141109 | 1 |
| T2DM_admissi | leukocyte | *LEFTY1* | -8.836893835 | 0.002708851 | 0.003992152 | 1 |
| T2DM_admissi | leukocyte | *TTC4* | -1.183078835 | 0.00097834 | 0.001856058 | 1 |
| T2DM_admissi | leukocyte | *JMJD7* | 3.915993246 | 4.66E-05 | 0.00105496 | 0.183231631 |
| T2DM_admissi | leukocyte | *APOBEC3D* | -4.683153184 | 0.000911411 | 0.001783334 | 1 |
| T2DM_admissi | leukocyte | *RN7SL832P* | -3.97595982 | 0.041734121 | 0.049568731 | 1 |
| T2DM_admissi | leukocyte | *OR2A7* | -4.367177741 | 0.004552301 | 0.006335809 | 1 |
| T2DM_admissi | leukocyte | *GSTA1* | -0.575940482 | 0.004512267 | 0.006293468 | 1 |
| T2DM_admissi | leukocyte | *GSTA2* | -0.803233339 | 0.00225348 | 0.003402738 | 1 |
| T2DM_admissi | leukocyte | *TMEM141* | -1.376349329 | 0.000711028 | 0.00152908 | 1 |
| T2DM_admissi | leukocyte | *APOBEC3C* | -2.145408463 | 0.003698613 | 0.005274718 | 1 |
| T2DM_admissi | leukocyte | *C4A* | -9.795909819 | 0.000872342 | 0.001738931 | 1 |
| T2DM_admissi | leukocyte | *NEAT1* | -0.805588396 | 0.000374806 | 0.001157716 | 1 |
| T2DM_admissi | leukocyte | *LINC00920* | -7.121723127 | 0.020500159 | 0.025400544 | 1 |
| T2DM_admissi | leukocyte | *UBAP1L* | -1.145295136 | 0.001949274 | 0.003011678 | 1 |
| T2DM_admissi | leukocyte | *TWF2* | -1.713223903 | 0.000128204 | 0.001062643 | 0.503712707 |
| T2DM_admissi | leukocyte | *BCKDHA* | -1.093906146 | 0.001836756 | 0.002861464 | 1 |
| T2DM_admissi | leukocyte | *PCDHAC1* | -1.519056362 | 0.004426629 | 0.006187203 | 1 |
| T2DM_admissi | leukocyte | *FMN1* | -3.458587435 | 0.0025204 | 0.003748166 | 1 |
| T2DM_admissi | leukocyte | *TMEM150C* | -1.411466785 | 0.000561343 | 0.001362208 | 1 |
| T2DM_admissi | leukocyte | *LINC01091* | -0.843744187 | 0.000708981 | 0.001527186 | 1 |
| T2DM_admissi | leukocyte | *PDCD6* | -1.186853246 | 0.000202059 | 0.001062643 | 0.793889895 |
| T2DM_admissi | leukocyte | *YJEFN3* | -2.062197862 | 0.000617021 | 0.001419945 | 1 |
| T2DM_admissi | leukocyte | *PCDHA10* | -1.074194988 | 0.000919367 | 0.001793131 | 1 |
| T2DM_admissi | leukocyte | *ZNF718* | -1.245308625 | 0.00721516 | 0.009655437 | 1 |
| T2DM_admissi | leukocyte | *SNHG18* | -1.139573115 | 0.001106825 | 0.002007832 | 1 |

| | | | | | | |
|---|---|---|---|---|---|---|
| T2DM_admissi | leukocyte | *ZNF550* | -1.404622501 | 0.000435752 | 0.001222199 | 1 |
| T2DM_admissi | leukocyte | *LINC01099* | -1.35428029 | 0.001298112 | 0.002196252 | 1 |
| T2DM_admissi | leukocyte | *PCDHA12* | -1.345477128 | 0.020683168 | 0.025603077 | 1 |
| T2DM_admissi | leukocyte | *ALG11* | -2.250412161 | 0.000174784 | 0.001062643 | 0.686725808 |
| T2DM_admissi | leukocyte | *ZNF260* | -1.925898437 | 0.00242817 | 0.003635778 | 1 |
| T2DM_admissi | leukocyte | *AP5B1* | -2.281540775 | 0.000154824 | 0.001062643 | 0.608301941 |
| T2DM_admissi | leukocyte | *SIGLEC12* | 2.073687066 | 0.007012422 | 0.009403346 | 1 |
| T2DM_admissi | leukocyte | *PABPC4L* | -3.119492878 | 0.001549555 | 0.002500288 | 1 |
| T2DM_admissi | leukocyte | *NPIPA2* | -12.16490505 | 0.001100334 | 0.001999636 | 1 |
| T2DM_admissi | leukocyte | *BRK1* | -0.985399504 | 0.000125367 | 0.001062643 | 0.49256504 |
| T2DM_admissi | leukocyte | *EID3* | -2.612210552 | 0.000141253 | 0.001062643 | 0.554983004 |
| T2DM_admissi | leukocyte | *SNHG9* | -1.089269932 | 0.000447867 | 0.001233124 | 1 |
| T2DM_admissi | leukocyte | *TAS2R43* | 2.047477514 | 0.022858588 | 0.028109982 | 1 |
| T2DM_admissi | leukocyte | *TAS2R20* | 6.620813507 | 0.01727824 | 0.02170143 | 1 |
| T2DM_admissi | leukocyte | *ZNF432* | -2.648361038 | 3.90E-05 | 0.001037577 | 0.153391628 |
| T2DM_admissi | leukocyte | *LINC01152* | -5.439552091 | 0.001170724 | 0.002087959 | 1 |
| T2DM_admissi | leukocyte | *SMLR1* | 3.581537349 | 0.012748577 | 0.016342303 | 1 |
| T2DM_admissi | leukocyte | *ZNF10* | -1.532053215 | 0.000314756 | 0.001109126 | 1 |
| T2DM_admissi | leukocyte | *TAS2R31* | -3.798072186 | 0.003848218 | 0.005466251 | 1 |
| T2DM_admissi | leukocyte | *GATC* | -1.759726508 | 5.62E-05 | 0.001060479 | 0.220855921 |
| T2DM_admissi | leukocyte | *TEN1* | -3.718081662 | 2.75E-05 | 0.000996218 | 0.108211879 |
| T2DM_admissi | leukocyte | *CHURC1* | -0.993826069 | 0.000245015 | 0.001077213 | 0.962662807 |
| T2DM_admissi | leukocyte | *LINC00638* | -9.264564295 | 0.000214501 | 0.001062643 | 0.842772921 |
| T2DM_admissi | leukocyte | *SLC25A21-AS1* | -1.143609908 | 0.001022526 | 0.001910369 | 1 |
| T2DM_admissi | leukocyte | *TMEM179* | -0.822773481 | 0.001164965 | 0.002080069 | 1 |
| T2DM_admissi | leukocyte | *IQCH-AS1* | -1.688925576 | 0.005824122 | 0.007959296 | 1 |
| T2DM_admissi | leukocyte | *SH3RF3-AS1* | 2.056180122 | 0.033446595 | 0.040150221 | 1 |
| T2DM_admissi | leukocyte | *PWRN1* | -1.414462243 | 0.003551836 | 0.005080148 | 1 |
| T2DM_admissi | leukocyte | *TMEM178B* | -1.028412384 | 0.001428049 | 0.002348599 | 1 |
| T2DM_admissi | leukocyte | *LINC01003* | -1.435367977 | 0.000298098 | 0.001093914 | 1 |
| T2DM_admissi | leukocyte | *GOLGA8H* | -17.16109903 | 0.004388782 | 0.006143112 | 1 |
| T2DM_admissi | leukocyte | *MRPL12* | -1.53592783 | 0.000227357 | 0.001062643 | 0.893287558 |
| T2DM_admissi | leukocyte | *ZNF234* | -6.078769183 | 0.000592748 | 0.001392055 | 1 |
| T2DM_admissi | leukocyte | *LINC00667* | -1.313566531 | 0.000257712 | 0.001080629 | 1 |
| T2DM_admissi | leukocyte | *FSBP* | -1.610944854 | 0.001247059 | 0.002154446 | 1 |
| T2DM_admissi | leukocyte | *MYO15B* | -2.421083333 | 0.001427598 | 0.002348599 | 1 |
| T2DM_admissi | leukocyte | *LINC00663* | -1.762832754 | 0.001642552 | 0.00261173 | 1 |
| T2DM_admissi | leukocyte | *ZNF285* | -2.659241853 | 0.00145145 | 0.002373152 | 1 |
| T2DM_admissi | leukocyte | *FBXO17* | -0.97004186 | 0.000681935 | 0.001492137 | 1 |
| T2DM_admissi | leukocyte | *LINC00528* | 0.675104565 | 0.003061147 | 0.00444981 | 1 |
| T2DM_admissi | leukocyte | *SNHG8* | -0.907683019 | 0.000193558 | 0.001062643 | 0.760491319 |
| T2DM_admissi | leukocyte | *MILR1* | 0.911067779 | 0.009062625 | 0.011872975 | 1 |
| T2DM_admissi | leukocyte | *GTF2H5* | -1.056586612 | 0.000814703 | 0.001660254 | 1 |
| T2DM_admissi | mesenchymal_cell | *RBM6* | -1.008795298 | 4.98E-08 | 1.78E-05 | 0.000195838 |
| T2DM_admissi | mesenchymal_cell | *ST7L* | -0.689117811 | 0.001139084 | 0.042716695 | 1 |
| T2DM_admissi | mesenchymal_cell | *QPCTL* | -0.844205023 | 0.001333765 | 0.047893638 | 1 |
| T2DM_admissi | mesenchymal_cell | *CEP68* | 1.411373039 | 3.15E-07 | 7.72E-05 | 0.001235698 |
| T2DM_admissi | mesenchymal_cell | *ERCC1* | -1.311098627 | 0.00101286 | 0.039795261 | 1 |
| T2DM_admissi | mesenchymal_cell | *BAK1* | 1.460407929 | 4.54E-05 | 0.005098244 | 0.178330183 |
| T2DM_admissi | mesenchymal_cell | *ITIH4* | -1.214657596 | 0.000937995 | 0.037605957 | 1 |

| | | | | | | |
|---|---|---|---|---|---|---|
| T2DM_admissi | mesenchymal_cell | *SEC61A2* | -2.505298192 | 0.000170549 | 0.013173055 | 0.670085247 |
| T2DM_admissi | mesenchymal_cell | *TCF3* | -1.590072643 | 1.55E-06 | 0.000320518 | 0.006089847 |
| T2DM_admissi | mesenchymal_cell | *SCARB1* | -1.930855512 | 0.000159777 | 0.013173055 | 0.627762549 |
| T2DM_admissi | mesenchymal_cell | *ST6GAL1* | 0.863967727 | 0.000298616 | 0.020584892 | 1 |
| T2DM_admissi | mesenchymal_cell | *TTC38* | -2.388892258 | 0.00054556 | 0.028966309 | 1 |
| T2DM_admissi | mesenchymal_cell | *MARK3* | -1.312050078 | 0.000170992 | 0.013173055 | 0.671825824 |
| T2DM_admissi | mesenchymal_cell | *CWF19L1* | -0.577171169 | 0.00041099 | 0.025230903 | 1 |
| T2DM_admissi | mesenchymal_cell | *SAMM50* | -2.285792513 | 0.000429759 | 0.025274167 | 1 |
| T2DM_admissi | mesenchymal_cell | *VCPKMT* | -2.373582974 | 0.001365931 | 0.048349037 | 1 |
| T2DM_admissi | mesenchymal_cell | *NAGPA* | -2.317770218 | 0.000598168 | 0.030923723 | 1 |
| T2DM_admissi | mesenchymal_cell | *RPAP1* | -3.637474632 | 0.000168278 | 0.013173055 | 0.66116543 |
| T2DM_admissi | mesenchymal_cell | *ERI1* | -2.483738988 | 0.000298635 | 0.020584892 | 1 |
| T2DM_admissi | mesenchymal_cell | *GCDH* | -3.972603966 | 6.70E-09 | 3.76E-06 | 2.63E-05 |
| T2DM_admissi | mesenchymal_cell | *GTPBP4* | -1.553203469 | 0.001340876 | 0.047893638 | 1 |
| T2DM_admissi | mesenchymal_cell | *HSD17B1* | -1.066687834 | 3.89E-06 | 0.000694796 | 0.015285513 |
| T2DM_admissi | mesenchymal_cell | *RANGRF* | -1.987004976 | 0.000646088 | 0.032132659 | 1 |
| T2DM_admissi | mesenchymal_cell | *CHPT1* | -0.854228114 | 0.000902206 | 0.03663557 | 1 |
| T2DM_admissi | mesenchymal_cell | *PNO1* | -1.769137744 | 0.000851535 | 0.03663557 | 1 |
| T2DM_admissi | mesenchymal_cell | *NSUN4* | -1.55310097 | 0.000465454 | 0.026125274 | 1 |
| T2DM_admissi | mesenchymal_cell | *FAM53C* | -1.379782599 | 0.000904467 | 0.03663557 | 1 |
| T2DM_admissi | mesenchymal_cell | *FLT3* | 1.095587496 | 7.37E-05 | 0.007622994 | 0.289673777 |
| T2DM_admissi | mesenchymal_cell | *ARHGAP40* | -174.5105007 | 0.000706683 | 0.032874904 | 1 |
| T2DM_admissi | mesenchymal_cell | *PEPD* | -1.923956908 | 7.65E-05 | 0.00762496 | 0.300752721 |
| T2DM_admissi | mesenchymal_cell | *NQO2* | -1.409808506 | 0.000866784 | 0.03663557 | 1 |
| T2DM_admissi | mesenchymal_cell | *MOCS1* | -0.715443941 | 0.001183951 | 0.043474254 | 1 |
| T2DM_admissi | mesenchymal_cell | *OPRL1* | -1.128018206 | 0.00033848 | 0.021681357 | 1 |
| T2DM_admissi | mesenchymal_cell | *FOSB* | -0.507505117 | 0.001200012 | 0.043655975 | 1 |
| T2DM_admissi | mesenchymal_cell | *PRMT1* | -1.115317911 | 0.000874754 | 0.03663557 | 1 |
| T2DM_admissi | mesenchymal_cell | *MAU2* | -2.191713434 | 8.50E-08 | 2.52E-05 | 0.000333945 |
| T2DM_admissi | mesenchymal_cell | *UCK1* | -2.135998941 | 0.000493592 | 0.026790626 | 1 |
| T2DM_admissi | mesenchymal_cell | *ZNF426* | -1.820043046 | 0.001143608 | 0.042716695 | 1 |
| T2DM_admissi | mesenchymal_cell | *BHMT2* | -0.649636482 | 0.000833477 | 0.03663557 | 1 |
| T2DM_admissi | mesenchymal_cell | *NOTCH2* | -0.608528816 | 3.42E-09 | 2.32E-06 | 1.34E-05 |
| T2DM_admissi | mesenchymal_cell | *BARD1* | -2.082293504 | 0.000357953 | 0.022323789 | 1 |
| T2DM_admissi | mesenchymal_cell | *TLE3* | -1.719034397 | 0.001386054 | 0.048623257 | 1 |
| T2DM_admissi | mesenchymal_cell | *MAN2C1* | -0.690285984 | 2.08E-06 | 0.000388467 | 0.008157813 |
| T2DM_admissi | mesenchymal_cell | *NPC1* | -2.82101115 | 3.71E-08 | 1.62E-05 | 0.00014576 |
| T2DM_admissi | mesenchymal_cell | *FN3KRP* | -0.457124282 | 0.000896703 | 0.03663557 | 1 |
| T2DM_admissi | mesenchymal_cell | *SH3GL1* | -1.131238232 | 0.000655464 | 0.032191457 | 1 |
| T2DM_admissi | mesenchymal_cell | *IQGAP2* | -0.675302946 | 0.00084731 | 0.03663557 | 1 |
| T2DM_admissi | mesenchymal_cell | *TBC1D7* | -3.197388834 | 0.000161269 | 0.013173055 | 0.63362742 |
| T2DM_admissi | mesenchymal_cell | *GIGYF1* | -2.077441306 | 0.000101229 | 0.009469765 | 0.397730118 |
| T2DM_admissi | mesenchymal_cell | *LRP12* | -1.618228156 | 0.000420676 | 0.025274167 | 1 |
| T2DM_admissi | mesenchymal_cell | *HSD17B12* | -0.445768363 | 1.95E-06 | 0.000382824 | 0.007656471 |
| T2DM_admissi | mesenchymal_cell | *APIP* | -0.568257186 | 0.000199362 | 0.01477912 | 0.783293371 |
| T2DM_admissi | mesenchymal_cell | *DLG5* | -1.743000967 | 0.000615492 | 0.031406096 | 1 |
| T2DM_admissi | mesenchymal_cell | *PTPN14* | -0.584013607 | 4.70E-05 | 0.005130932 | 0.184713554 |
| T2DM_admissi | mesenchymal_cell | *POC5* | -0.54142268 | 4.07E-05 | 0.004993422 | 0.159789489 |
| T2DM_admissi | mesenchymal_cell | *MED21* | -1.743194581 | 0.000625175 | 0.031491163 | 1 |
| T2DM_admissi | mesenchymal_cell | *JAZF1* | -1.003695675 | 3.80E-07 | 8.29E-05 | 0.001492316 |

| | | | | | | |
|---|---|---|---|---|---|---|
| T2DM_admissi | mesenchymal_cell | *NUS1* | -2.052080203 | 5.53E-05 | 0.005867264 | 0.217088774 |
| T2DM_admissi | mesenchymal_cell | *CLEC18A* | -2.317742641 | 5.65E-10 | 7.41E-07 | 2.22E-06 |
| T2DM_admissi | mesenchymal_cell | *FAIM* | -2.057489394 | 0.000872893 | 0.03663557 | 1 |
| T2DM_admissi | mesenchymal_cell | *PINK1* | -1.385046949 | 0.000711247 | 0.032874904 | 1 |
| T2DM_admissi | mesenchymal_cell | *CILP2* | -0.908959207 | 2.06E-05 | 0.00270434 | 0.081130189 |
| T2DM_admissi | mesenchymal_cell | *VPS11* | -1.123211476 | 0.00032667 | 0.021391465 | 1 |
| T2DM_admissi | mesenchymal_cell | *PIGX* | -1.886835969 | 0.000497764 | 0.026790626 | 1 |
| T2DM_admissi | mesenchymal_cell | *BDH2* | -0.541377308 | 0.000594138 | 0.030923723 | 1 |
| T2DM_admissi | mesenchymal_cell | *WDR41* | -1.866888069 | 0.000485432 | 0.026790626 | 1 |
| T2DM_admissi | mesenchymal_cell | *COG5* | -2.371053993 | 0.000447834 | 0.025875581 | 1 |
| T2DM_admissi | mesenchymal_cell | *RPP25L* | -2.803855323 | 0.000342134 | 0.021681357 | 1 |
| T2DM_admissi | mesenchymal_cell | *INPPL1* | -1.738992166 | 0.001152448 | 0.042716695 | 1 |
| T2DM_admissi | mesenchymal_cell | *NOXRED1* | 1.974160866 | 0.001050685 | 0.0408727 | 1 |
| T2DM_admissi | mesenchymal_cell | *PATL1* | -1.424883953 | 0.000193107 | 0.014590691 | 0.758715931 |
| T2DM_admissi | mesenchymal_cell | *POP5* | -2.050475597 | 4.40E-05 | 0.005098244 | 0.172915693 |
| T2DM_admissi | mesenchymal_cell | *TMC8* | -0.635350995 | 0.001151033 | 0.042716695 | 1 |
| T2DM_admissi | mesenchymal_cell | *DNAJC21* | -1.940408167 | 7.76E-05 | 0.00762496 | 0.304998417 |
| T2DM_admissi | mesenchymal_cell | *TMEM129* | -1.006184883 | 9.19E-06 | 0.001388494 | 0.036100839 |
| T2DM_admissi | mesenchymal_cell | *SNUPN* | -3.016471702 | 0.000113599 | 0.010143837 | 0.446328813 |
| T2DM_admissi | mesenchymal_cell | *INO80E* | -1.260106438 | 4.58E-06 | 0.000778604 | 0.017993579 |
| T2DM_admissi | mesenchymal_cell | *SETMAR* | -1.545171202 | 0.000899826 | 0.03663557 | 1 |
| T2DM_admissi | mesenchymal_cell | *ZNF440* | -2.063461802 | 0.000717243 | 0.032874904 | 1 |
| T2DM_admissi | mesenchymal_cell | *RGS19* | -0.74680199 | 0.000668206 | 0.032412114 | 1 |
| T2DM_admissi | mesenchymal_cell | *CTSW* | -0.392843582 | 0.000719583 | 0.032874904 | 1 |
| T2DM_admissi | mesenchymal_cell | *RNASEH2C* | -1.199992217 | 4.95E-06 | 0.000778604 | 0.019465095 |
| T2DM_admissi | mesenchymal_cell | *YIF1A* | -0.811797212 | 0.000248022 | 0.017717767 | 0.974477174 |
| T2DM_admissi | mesenchymal_cell | *ZNF408* | -1.175637079 | 0.000116269 | 0.010151571 | 0.456820701 |
| T2DM_admissi | mesenchymal_cell | *IRX3* | 3.241867635 | 3.98E-22 | 1.56E-18 | 1.56E-18 |
| T2DM_admissi | mesenchymal_cell | *FARSA* | -2.352928212 | 3.54E-09 | 2.32E-06 | 1.39E-05 |
| T2DM_admissi | mesenchymal_cell | *HLA-DQB1* | -0.463173241 | 8.96E-08 | 2.52E-05 | 0.000352182 |
| T2DM_admissi | mesenchymal_cell | *UBA7* | -0.554616559 | 1.72E-05 | 0.002380813 | 0.067636404 |
| T2DM_admissi | mesenchymal_cell | *SKA2* | -1.485169919 | 0.000313577 | 0.02124212 | 1 |
| T2DM_admissi | mesenchymal_cell | *PGP* | -1.504349716 | 1.76E-05 | 0.002380813 | 0.069043565 |
| T2DM_admissi | mesenchymal_cell | *DMWD* | -1.564270928 | 4.77E-06 | 0.000778604 | 0.018743432 |
| T2DM_admissi | mesenchymal_cell | *HLA-DRB1* | -0.350155193 | 1.25E-09 | 1.22E-06 | 4.90E-06 |
| T2DM_admissi | mesenchymal_cell | *SULT1A1* | -2.439646225 | 8.14E-05 | 0.007796154 | 0.319642304 |
| T2DM_admissi | mesenchymal_cell | *DPP4* | -1.013606485 | 0.000692469 | 0.032874904 | 1 |
| T2DM_admissi | mesenchymal_cell | *HLA-DRB5* | -0.218868179 | 2.64E-05 | 0.003346348 | 0.103736796 |
| T2DM_admissi | mesenchymal_cell | *MAP3K3* | -1.008570719 | 0.001152186 | 0.042716695 | 1 |
| T2DM_admissi | mesenchymal_cell | *LIME1* | -0.64063141 | 0.000212419 | 0.015455469 | 0.834595348 |
| T2DM_admissi | mesenchymal_cell | *INPP5B* | -0.623404616 | 0.000995374 | 0.039503293 | 1 |
| T2DM_admissi | mesenchymal_cell | *NOTCH4* | -0.472180443 | 2.88E-07 | 7.53E-05 | 0.001129655 |
| T2DM_admissi | mesenchymal_cell | *AGER* | 1.058241946 | 0.000430992 | 0.025274167 | 1 |
| T2DM_admissi | mesenchymal_cell | *RNF5* | -0.847548481 | 6.98E-08 | 2.29E-05 | 0.000274267 |
| T2DM_admissi | mesenchymal_cell | *HLA-C* | -0.360993814 | 0.000462515 | 0.026125274 | 1 |
| T2DM_admissi | mesenchymal_cell | *RNF39* | 0.529068459 | 0.000153498 | 0.013110726 | 0.60309338 |
| T2DM_admissi | mesenchymal_cell | *HCG27* | 0.688441419 | 3.58E-07 | 8.28E-05 | 0.001407206 |
| T2DM_admissi | mesenchymal_cell | *ATF6B* | 0.695832921 | 0.000110757 | 0.010120089 | 0.435163814 |
| T2DM_admissi | mesenchymal_cell | *SLC12A8* | -4.340615803 | 2.78E-08 | 1.36E-05 | 0.000109063 |
| T2DM_admissi | mesenchymal_cell | *EXOSC6* | -0.99676793 | 0.00031978 | 0.0212952 | 1 |

| | | | | | | |
|---|---|---|---|---|---|---|
| T2DM_admissi | mesenchymal_cell | *HLA-DQB2* | 1.169254538 | 4.62E-08 | 1.78E-05 | 0.000181522 |
| T2DM_admissi | mesenchymal_cell | *HLA-B* | 0.464807414 | 4.54E-05 | 0.005098244 | 0.178438523 |
| T2DM_admissi | mesenchymal_cell | *HLA-DQA2* | 0.795977063 | 1.37E-12 | 2.70E-09 | 5.39E-09 |
| T2DM_admissi | mesenchymal_cell | *JMJD7* | 1.462676188 | 1.67E-05 | 0.002380813 | 0.065599331 |
| T2DM_admissi | mesenchymal_cell | *ZNF432* | -1.996733166 | 0.000870782 | 0.03663557 | 1 |
| T2DM_admissi | PP_cell | *RBM6* | -1.81883063 | 4.93E-09 | 3.88E-06 | 1.94E-05 |
| T2DM_admissi | PP_cell | *CEP68* | 1.893350509 | 2.70E-05 | 0.004616313 | 0.106175194 |
| T2DM_admissi | PP_cell | *HAGH* | -1.472834914 | 0.000365496 | 0.033396163 | 1 |
| T2DM_admissi | PP_cell | *TCF3* | -2.179196473 | 1.30E-05 | 0.00273703 | 0.051218292 |
| T2DM_admissi | PP_cell | *MARK3* | -2.451700438 | 5.43E-06 | 0.001523051 | 0.021322721 |
| T2DM_admissi | PP_cell | *XAB2* | -2.39607294 | 0.000661987 | 0.046561223 | 1 |
| T2DM_admissi | PP_cell | *POMGNT1* | -2.53178483 | 0.000132269 | 0.015284802 | 0.51968327 |
| T2DM_admissi | PP_cell | *AP1B1* | -1.969309206 | 0.000589187 | 0.045359675 | 1 |
| T2DM_admissi | PP_cell | *HSD17B1* | -1.523242402 | 1.39E-05 | 0.00273703 | 0.054740591 |
| T2DM_admissi | PP_cell | *NNT* | -1.870085171 | 0.00010062 | 0.013412311 | 0.395337315 |
| T2DM_admissi | PP_cell | *CHMP3* | -1.55574781 | 0.000546905 | 0.042975832 | 1 |
| T2DM_admissi | PP_cell | *NSUN4* | -2.049785755 | 0.000503321 | 0.040358146 | 1 |
| T2DM_admissi | PP_cell | *FAM53C* | -2.048141818 | 0.00018952 | 0.019595389 | 0.744624796 |
| T2DM_admissi | PP_cell | *POLK* | -2.224356017 | 7.18E-05 | 0.010840837 | 0.281913855 |
| T2DM_admissi | PP_cell | *FLT3* | 1.458972123 | 0.000674584 | 0.046561223 | 1 |
| T2DM_admissi | PP_cell | *PEPD* | -2.133657388 | 0.000463072 | 0.039552409 | 1 |
| T2DM_admissi | PP_cell | *OPRL1* | -2.418518409 | 4.18E-05 | 0.006850405 | 0.16440972 |
| T2DM_admissi | PP_cell | *MAU2* | -2.197272522 | 5.17E-06 | 0.001523051 | 0.020299609 |
| T2DM_admissi | PP_cell | *NOTCH2* | -0.684526113 | 5.24E-05 | 0.008241355 | 0.206033879 |
| T2DM_admissi | PP_cell | *FXYD2* | -1.318802502 | 0.000196573 | 0.019803442 | 0.772334251 |
| T2DM_admissi | PP_cell | *SPTBN5* | 1.782461617 | 0.000647098 | 0.046561223 | 1 |
| T2DM_admissi | PP_cell | *MAN2C1* | -1.171138855 | 1.60E-06 | 0.000522792 | 0.006273501 |
| T2DM_admissi | PP_cell | *NPC1* | -2.57407646 | 0.000131096 | 0.015284802 | 0.51507776 |
| T2DM_admissi | PP_cell | *GIGYF1* | -2.274522151 | 0.000435619 | 0.038034418 | 1 |
| T2DM_admissi | PP_cell | *HSD17B12* | -0.869213497 | 2.26E-07 | 9.86E-05 | 0.000887526 |
| T2DM_admissi | PP_cell | *APIP* | -1.184587531 | 1.34E-05 | 0.00273703 | 0.052499345 |
| T2DM_admissi | PP_cell | *TAGLN* | -0.689191748 | 0.000675487 | 0.046561223 | 1 |
| T2DM_admissi | PP_cell | *VIPAS39* | -2.336440006 | 0.000282381 | 0.027736872 | 1 |
| T2DM_admissi | PP_cell | *PTPN14* | -1.156551955 | 0.000122763 | 0.015072995 | 0.482335854 |
| T2DM_admissi | PP_cell | *JAZF1* | -1.771361636 | 1.45E-07 | 7.12E-05 | 0.000569428 |
| T2DM_admissi | PP_cell | *CLEC18A* | -5.518590188 | 6.10E-12 | 1.20E-08 | 2.39E-08 |
| T2DM_admissi | PP_cell | *CILP2* | -1.656738941 | 0.00010241 | 0.013412311 | 0.402369342 |
| T2DM_admissi | PP_cell | *PAQR6* | -2.228599221 | 0.00032287 | 0.030940393 | 1 |
| T2DM_admissi | PP_cell | *ICA1L* | -1.107101253 | 7.73E-05 | 0.010840837 | 0.303543428 |
| T2DM_admissi | PP_cell | *BDH2* | -1.970561528 | 1.09E-05 | 0.002516086 | 0.042773464 |
| T2DM_admissi | PP_cell | *PATL1* | -1.827613105 | 0.00043279 | 0.038034418 | 1 |
| T2DM_admissi | PP_cell | *TMEM129* | -1.48152618 | 2.41E-05 | 0.004517046 | 0.09485797 |
| T2DM_admissi | PP_cell | *INO80E* | -1.952126939 | 5.97E-06 | 0.001564084 | 0.02346126 |
| T2DM_admissi | PP_cell | *SNX32* | 1.726198841 | 0.000746807 | 0.047731588 | 1 |
| T2DM_admissi | PP_cell | *RNASEH2C* | -1.285030227 | 0.00015765 | 0.016740709 | 0.619406237 |
| T2DM_admissi | PP_cell | *FUT2* | 0.383651065 | 0.000753209 | 0.047731588 | 1 |
| T2DM_admissi | PP_cell | *IRX3* | 2.404052491 | 2.56E-11 | 3.35E-08 | 1.00E-07 |
| T2DM_admissi | PP_cell | *HLA-DQB1* | -0.87279556 | 2.90E-07 | 0.000113882 | 0.001138825 |
| T2DM_admissi | PP_cell | *UBA7* | -0.840128832 | 7.62E-05 | 0.010840837 | 0.299343966 |
| T2DM_admissi | PP_cell | *ZNF320* | 0.636280663 | 0.000607757 | 0.045359675 | 1 |

| | | | | | | |
|---|---|---|---|---|---|---|
| T2DM_admissi | PP_cell | *PGP* | -1.610840837 | 0.000141805 | 0.01564521 | 0.557150979 |
| T2DM_admissi | PP_cell | *DMWD* | -2.203318608 | 2.70E-05 | 0.004616313 | 0.105940259 |
| T2DM_admissi | PP_cell | *HLA-DRB1* | -0.725289247 | 1.74E-09 | 1.71E-06 | 6.86E-06 |
| T2DM_admissi | PP_cell | *XRCC6* | -1.465951481 | 0.000500782 | 0.040358146 | 1 |
| T2DM_admissi | PP_cell | *RPS26* | -1.012129253 | 0.000495987 | 0.040358146 | 1 |
| T2DM_admissi | PP_cell | *HLA-DRB5* | -0.598208043 | 0.000113004 | 0.014322283 | 0.44399076 |
| T2DM_admissi | PP_cell | *MAP3K3* | -2.078330593 | 0.000729982 | 0.047731588 | 1 |
| T2DM_admissi | PP_cell | *NOTCH4* | -1.028697056 | 6.88E-07 | 0.000245654 | 0.002702198 |
| T2DM_admissi | PP_cell | *AGER* | 1.467823985 | 0.000611876 | 0.045359675 | 1 |
| T2DM_admissi | PP_cell | *RNF5* | -1.49014384 | 2.75E-08 | 1.80E-05 | 0.000108129 |
| T2DM_admissi | PP_cell | *HLA-C* | -0.602623735 | 0.000143351 | 0.01564521 | 0.563227568 |
| T2DM_admissi | PP_cell | *RNF39* | 0.779320437 | 0.00034997 | 0.032738873 | 1 |
| T2DM_admissi | PP_cell | *HCG27* | 0.870171256 | 1.03E-05 | 0.002516086 | 0.040507031 |
| T2DM_admissi | PP_cell | *PPT2* | 1.621381669 | 0.000736949 | 0.047731588 | 1 |
| T2DM_admissi | PP_cell | *HLA-DQB2* | 2.024079354 | 8.46E-08 | 4.75E-05 | 0.000332538 |
| T2DM_admissi | PP_cell | *HLA-B* | 0.655651613 | 0.000701038 | 0.047489257 | 1 |
| T2DM_admissi | PP_cell | *HLA-DQA2* | 1.329526658 | 2.10E-12 | 8.25E-09 | 8.25E-09 |
| T2DM_insulin | acinar_cell | *CEP68* | 0.381914223 | 8.86E-05 | 0.03165813 | 0.348239435 |
| T2DM_insulin | acinar_cell | *CXCL11* | 0.160901485 | 2.24E-05 | 0.008797388 | 0.087973877 |
| T2DM_insulin | acinar_cell | *IRX3* | 0.755101666 | 6.24E-10 | 8.18E-07 | 2.45E-06 |
| T2DM_insulin | acinar_cell | *HLA-DQB1* | -0.171964483 | 3.05E-07 | 0.000239955 | 0.001199775 |
| T2DM_insulin | acinar_cell | *HLA-DRB1* | -0.10058692 | 1.86E-05 | 0.008797388 | 0.073019871 |
| T2DM_insulin | acinar_cell | *HLA-DRB5* | -0.117977391 | 1.07E-06 | 0.000702563 | 0.004215378 |
| T2DM_insulin | acinar_cell | *HLA-DMA* | 0.095423592 | 2.02E-05 | 0.008797388 | 0.079331571 |
| T2DM_insulin | acinar_cell | *NOTCH4* | -0.254798396 | 1.49E-08 | 1.47E-05 | 5.86E-05 |
| T2DM_insulin | acinar_cell | *HLA-DQB2* | 0.542625012 | 2.28E-13 | 8.94E-10 | 8.94E-10 |
| T2DM_insulin | acinar_cell | *HLA-DQA2* | 0.255276726 | 5.23E-12 | 1.03E-08 | 2.06E-08 |
| T2DM_insulin | acinar_cell | *HLA-DMB* | 0.471322054 | 1.43E-06 | 0.00080286 | 0.005620018 |
| T2DM_insulin | alpha_cell | *SEC61A2* | -0.187098915 | 5.36E-05 | 0.01913977 | 0.210537472 |
| T2DM_insulin | alpha_cell | *BTNL8* | -0.598152286 | 0.000141156 | 0.046216718 | 0.554600621 |
| T2DM_insulin | alpha_cell | *CXCL11* | 0.025145981 | 3.88E-05 | 0.015262061 | 0.152620611 |
| T2DM_insulin | alpha_cell | *IRX3* | 0.111838978 | 9.93E-09 | 9.76E-06 | 3.90E-05 |
| T2DM_insulin | alpha_cell | *HLA-DQB1* | -0.029615844 | 5.77E-08 | 4.54E-05 | 0.000226881 |
| T2DM_insulin | alpha_cell | *HLA-DRB1* | -0.017340854 | 7.50E-06 | 0.003684049 | 0.029472395 |
| T2DM_insulin | alpha_cell | *HLA-DRB5* | -0.020268158 | 5.42E-07 | 0.000354838 | 0.002129028 |
| T2DM_insulin | alpha_cell | *HLA-DMA* | 0.015113966 | 2.93E-05 | 0.012790155 | 0.115111395 |
| T2DM_insulin | alpha_cell | *NOTCH4* | -0.042507972 | 4.85E-09 | 6.35E-06 | 1.90E-05 |
| T2DM_insulin | alpha_cell | *HLA-DQB2* | 0.09129261 | 8.30E-14 | 3.26E-10 | 3.26E-10 |
| T2DM_insulin | alpha_cell | *HLA-DQA2* | 0.041771879 | 5.15E-12 | 1.01E-08 | 2.02E-08 |
| T2DM_insulin | alpha_cell | *HLA-DMB* | 0.078286206 | 1.54E-06 | 0.000865916 | 0.006061409 |
| T2DM_insulin | beta_cell | *CEP68* | 0.039071458 | 0.000107856 | 0.038524224 | 0.423766459 |
| T2DM_insulin | beta_cell | *SEC61A2* | -0.120962664 | 0.000158967 | 0.044612922 | 0.624580914 |
| T2DM_insulin | beta_cell | *BTNL8* | -0.354562033 | 0.000184842 | 0.048416252 | 0.726243778 |
| T2DM_insulin | beta_cell | *CXCL11* | 0.015726761 | 2.07E-05 | 0.00812182 | 0.081218203 |
| T2DM_insulin | beta_cell | *IRX3* | 0.073100721 | 9.52E-10 | 1.25E-06 | 3.74E-06 |
| T2DM_insulin | beta_cell | *HLA-DQB1* | -0.017843727 | 6.79E-08 | 5.33E-05 | 0.000266663 |
| T2DM_insulin | beta_cell | *ZBTB40* | 0.090516727 | 0.000149051 | 0.044612922 | 0.585620484 |
| T2DM_insulin | beta_cell | *HLA-DRB1* | -0.010548052 | 6.66E-06 | 0.003270615 | 0.026164918 |
| T2DM_insulin | beta_cell | *HLA-DRB5* | -0.01276389 | 2.04E-07 | 0.000133431 | 0.000800588 |
| T2DM_insulin | beta_cell | *HLA-DMA* | 0.00938426 | 1.86E-05 | 0.008111444 | 0.073002998 |

| | | | | | |
|---|---|---|---|---|---|
| T2DM_insulin beta_cell | *NOTCH4* | -0.025365906 | 7.70E-09 | 7.56E-06 | 3.03E-05 |
| T2DM_insulin beta_cell | *PPT2* | 0.036166097 | 0.000143706 | 0.044612922 | 0.564619317 |
| T2DM_insulin beta_cell | *HLA-DQB2* | 0.056215027 | 3.34E-14 | 1.31E-10 | 1.31E-10 |
| T2DM_insulin beta_cell | *HLA-DQA2* | 0.026276884 | 1.06E-12 | 2.07E-09 | 4.15E-09 |
| T2DM_insulin beta_cell | *HLA-DMB* | 0.049667965 | 5.22E-07 | 0.00029293 | 0.00205051 |
| T2DM_insulin delta_cell | *CXCL11* | 0.141559167 | 1.07E-05 | 0.005998557 | 0.041989899 |
| T2DM_insulin delta_cell | *IRX3* | 0.459690304 | 9.10E-08 | 8.93E-05 | 0.000357363 |
| T2DM_insulin delta_cell | *HLA-DQB1* | -0.149022494 | 2.04E-07 | 0.000160672 | 0.000803362 |
| T2DM_insulin delta_cell | *HLA-DRB1* | -0.088555554 | 1.48E-05 | 0.00727725 | 0.058218 |
| T2DM_insulin delta_cell | *HLA-DRB5* | -0.095340319 | 2.86E-06 | 0.001871247 | 0.011227481 |
| T2DM_insulin delta_cell | *HLA-DMA* | 0.080545045 | 1.73E-05 | 0.007530916 | 0.067778242 |
| T2DM_insulin delta_cell | *NOTCH4* | -0.20812461 | 3.08E-08 | 4.03E-05 | 0.000121029 |
| T2DM_insulin delta_cell | *PPT2* | 0.315168022 | 7.33E-05 | 0.026169415 | 0.287863566 |
| T2DM_insulin delta_cell | *HLA-DQB2* | 0.434498027 | 1.57E-12 | 6.19E-09 | 6.19E-09 |
| T2DM_insulin delta_cell | *HLA-DQA2* | 0.19381382 | 1.19E-10 | 2.33E-07 | 4.67E-07 |
| T2DM_insulin delta_cell | *HLA-DMB* | 0.290826069 | 5.22E-05 | 0.020521897 | 0.205218972 |
| T2DM_insulin duct_epithelial_cell | *CEP68* | 0.234199001 | 9.81E-05 | 0.038535138 | 0.385351383 |
| T2DM_insulin duct_epithelial_cell | *CXCL11* | 0.098927834 | 2.18E-05 | 0.009516526 | 0.085648735 |
| T2DM_insulin duct_epithelial_cell | *HLA-DQB1* | -0.110042694 | 1.27E-07 | 0.000124289 | 0.000497156 |
| T2DM_insulin duct_epithelial_cell | *HLA-DRB1* | -0.068210132 | 5.61E-06 | 0.003146666 | 0.022026661 |
| T2DM_insulin duct_epithelial_cell | *HLA-DRB5* | -0.07149655 | 1.12E-06 | 0.000734954 | 0.004409722 |
| T2DM_insulin duct_epithelial_cell | *HLA-DMA* | 0.060305274 | 1.29E-05 | 0.006346573 | 0.050772587 |
| T2DM_insulin duct_epithelial_cell | *NOTCH4* | -0.152321809 | 2.01E-08 | 2.63E-05 | 7.89E-05 |
| T2DM_insulin duct_epithelial_cell | *PPT2* | 0.228012512 | 0.000114864 | 0.041027155 | 0.451298709 |
| T2DM_insulin duct_epithelial_cell | *HLA-DQB2* | 0.351949515 | 5.18E-14 | 2.03E-10 | 2.03E-10 |
| T2DM_insulin duct_epithelial_cell | *HLA-DQA2* | 0.168431933 | 9.93E-13 | 1.95E-09 | 3.90E-09 |
| T2DM_insulin duct_epithelial_cell | *HLA-DMB* | 0.310217195 | 6.49E-07 | 0.000510102 | 0.00255051 |
| T2DM_insulin endothelial_cell | *CEP68* | 0.340240382 | 2.71E-05 | 0.017776184 | 0.106657104 |
| T2DM_insulin endothelial_cell | *ALOX5* | -0.275167177 | 0.000131013 | 0.039596298 | 0.514751877 |
| T2DM_insulin endothelial_cell | *EXTL1* | -0.602864096 | 3.53E-05 | 0.018233293 | 0.138565562 |
| T2DM_insulin endothelial_cell | *CXCL11* | 0.131191384 | 3.71E-05 | 0.018233293 | 0.145866343 |
| T2DM_insulin endothelial_cell | *IRX3* | 0.346737335 | 7.59E-05 | 0.027117923 | 0.298297152 |
| T2DM_insulin endothelial_cell | *HLA-DQB1* | -0.11929613 | 2.26E-05 | 0.017776184 | 0.088941071 |
| T2DM_insulin endothelial_cell | *ZBTB40* | 0.3601792 | 7.53E-05 | 0.027117923 | 0.295886741 |
| T2DM_insulin endothelial_cell | *AGAP4* | 0.669036674 | 0.000144909 | 0.04019126 | 0.569347519 |
| T2DM_insulin endothelial_cell | *HLA-DMA* | 0.089261146 | 1.78E-06 | 0.001750242 | 0.007000969 |
| T2DM_insulin endothelial_cell | *AGER* | 0.224427589 | 8.82E-05 | 0.028865377 | 0.34638453 |
| T2DM_insulin endothelial_cell | *PPT2* | 0.158199687 | 6.81E-05 | 0.027117923 | 0.267472909 |
| T2DM_insulin endothelial_cell | *HLA-DQB2* | 0.471006405 | 2.74E-13 | 1.07E-09 | 1.07E-09 |
| T2DM_insulin endothelial_cell | *B3GALT4* | -0.182651218 | 0.000153441 | 0.04019126 | 0.602868905 |
| T2DM_insulin endothelial_cell | *HLA-DQA2* | 0.232079786 | 9.73E-13 | 1.91E-09 | 3.82E-09 |
| T2DM_insulin endothelial_cell | *HLA-DMB* | 0.394982205 | 1.38E-08 | 1.81E-05 | 5.43E-05 |
| T2DM_insulin epsilon_cell | *HLA-DQB1* | -1.201763728 | 1.97E-05 | 0.019314483 | 0.077257934 |
| T2DM_insulin epsilon_cell | *HLA-DRB1* | -0.908622035 | 4.84E-05 | 0.037992038 | 0.19007735 |
| T2DM_insulin epsilon_cell | *HLA-DRB5* | -1.258157023 | 5.80E-05 | 0.037992038 | 0.227952228 |
| T2DM_insulin epsilon_cell | *NOTCH4* | -1.852077016 | 2.11E-06 | 0.002766217 | 0.008298652 |
| T2DM_insulin epsilon_cell | *HLA-DQB2* | 4.468038356 | 5.15E-10 | 1.01E-06 | 2.02E-06 |
| T2DM_insulin epsilon_cell | *HLA-DQA2* | 2.557526941 | 2.88E-11 | 1.13E-07 | 1.13E-07 |
| T2DM_insulin leukocyte | *GCLC* | -0.565541505 | 0.003164023 | 0.011330438 | 1 |
| T2DM_insulin leukocyte | *NFYA* | -0.434345602 | 0.004714976 | 0.011330438 | 1 |

| | | | | | |
|---|---|---|---|---|---|
| T2DM_insulin leukocyte | *STPG1* | -0.376850996 | 0.003403169 | 0.011330438 | 1 |
| T2DM_insulin leukocyte | *LASP1* | -0.361359054 | 0.004333866 | 0.011330438 | 1 |
| T2DM_insulin leukocyte | *SNX11* | -0.396423699 | 0.006110524 | 0.011570498 | 1 |
| T2DM_insulin leukocyte | *TMEM176A* | -0.286176937 | 0.003793655 | 0.011330438 | 1 |
| T2DM_insulin leukocyte | *M6PR* | -0.792072099 | 0.003089237 | 0.011330438 | 1 |
| T2DM_insulin leukocyte | *CASP10* | -0.914302735 | 0.008182764 | 0.013396068 | 1 |
| T2DM_insulin leukocyte | *NDUFAF7* | -0.825323439 | 0.002690484 | 0.011330438 | 1 |
| T2DM_insulin leukocyte | *POLDIP2* | -0.322509693 | 0.003339436 | 0.011330438 | 1 |
| T2DM_insulin leukocyte | *PLXND1* | -0.745386836 | 0.006827051 | 0.012014984 | 1 |
| T2DM_insulin leukocyte | *KDM1A* | -0.37889072 | 0.005726878 | 0.011570498 | 1 |
| T2DM_insulin leukocyte | *RBM6* | -0.526035123 | 0.000272626 | 0.011330438 | 1 |
| T2DM_insulin leukocyte | *RECQL* | -0.414611963 | 0.00253992 | 0.011330438 | 1 |
| T2DM_insulin leukocyte | *SLC25A13* | -0.385068578 | 0.004289336 | 0.011330438 | 1 |
| T2DM_insulin leukocyte | *MEOX1* | 0.857972498 | 0.021192548 | 0.028613581 | 1 |
| T2DM_insulin leukocyte | *PON1* | -0.461247917 | 0.003859138 | 0.011330438 | 1 |
| T2DM_insulin leukocyte | *KMT2E* | -0.409758839 | 0.002926449 | 0.011330438 | 1 |
| T2DM_insulin leukocyte | *IBTK* | -0.397196615 | 0.003231935 | 0.011330438 | 1 |
| T2DM_insulin leukocyte | *ZNF195* | -0.584019673 | 0.002690302 | 0.011330438 | 1 |
| T2DM_insulin leukocyte | *GGNBP2* | -0.396044914 | 0.004857462 | 0.011330438 | 1 |
| T2DM_insulin leukocyte | *TMEM98* | -0.250010414 | 0.017757714 | 0.024532369 | 1 |
| T2DM_insulin leukocyte | *SYNRG* | -0.550514809 | 0.003910735 | 0.011330438 | 1 |
| T2DM_insulin leukocyte | *SPATA20* | -0.314380853 | 0.030969845 | 0.039764876 | 1 |
| T2DM_insulin leukocyte | *FARP2* | -1.11190739 | 0.003604456 | 0.011330438 | 1 |
| T2DM_insulin leukocyte | *USH1C* | -0.400769519 | 0.003037007 | 0.011330438 | 1 |
| T2DM_insulin leukocyte | *VPS41* | -0.485304902 | 0.006516872 | 0.011788577 | 1 |
| T2DM_insulin leukocyte | *ELAC2* | -0.425635384 | 0.003387624 | 0.011330438 | 1 |
| T2DM_insulin leukocyte | *CCDC124* | -0.345255932 | 0.003507059 | 0.011330438 | 1 |
| T2DM_insulin leukocyte | *NOS2* | -6.065858118 | 0.005902199 | 0.011570498 | 1 |
| T2DM_insulin leukocyte | *CD79B* | -1.74382916 | 0.011839486 | 0.0176246 | 1 |
| T2DM_insulin leukocyte | *BAIAP3* | -0.371764103 | 0.004243011 | 0.011330438 | 1 |
| T2DM_insulin leukocyte | *CYB561* | -0.3441002 | 0.004097735 | 0.011330438 | 1 |
| T2DM_insulin leukocyte | *MGST1* | -0.223368061 | 0.019802598 | 0.026987307 | 1 |
| T2DM_insulin leukocyte | *ST3GAL1* | -0.907296126 | 0.001243759 | 0.011330438 | 1 |
| T2DM_insulin leukocyte | *IL32* | -0.171577065 | 0.006041374 | 0.011570498 | 1 |
| T2DM_insulin leukocyte | *MED24* | -0.542514635 | 0.002556039 | 0.011330438 | 1 |
| T2DM_insulin leukocyte | *RPS20* | -0.266174948 | 0.003022765 | 0.011330438 | 1 |
| T2DM_insulin leukocyte | *REV3L* | -0.310480558 | 0.006791066 | 0.012013551 | 1 |
| T2DM_insulin leukocyte | *MLXIPL* | -0.331994018 | 0.011089547 | 0.016732269 | 1 |
| T2DM_insulin leukocyte | *ETV7* | -0.709318058 | 0.004322448 | 0.011330438 | 1 |
| T2DM_insulin leukocyte | *NCAPD2* | -0.60054977 | 0.012225749 | 0.017949483 | 1 |
| T2DM_insulin leukocyte | *GIPR* | -0.319635333 | 0.004129497 | 0.011330438 | 1 |
| T2DM_insulin leukocyte | *FUZ* | -0.573565242 | 0.004859126 | 0.011330438 | 1 |
| T2DM_insulin leukocyte | *LRRC23* | -0.274271085 | 0.006883294 | 0.012078812 | 1 |
| T2DM_insulin leukocyte | *TSPAN9* | -0.31756978 | 0.006025039 | 0.011570498 | 1 |
| T2DM_insulin leukocyte | *BTBD7* | -0.372211147 | 0.010084117 | 0.015574095 | 1 |
| T2DM_insulin leukocyte | *APBA3* | -0.74055094 | 0.003168337 | 0.011330438 | 1 |
| T2DM_insulin leukocyte | *TTC19* | -0.318106906 | 0.003669982 | 0.011330438 | 1 |
| T2DM_insulin leukocyte | *DPF1* | -0.365723636 | 0.022490797 | 0.030128313 | 1 |
| T2DM_insulin leukocyte | *RABGAP1* | -0.374494497 | 0.004604647 | 0.011330438 | 1 |
| T2DM_insulin leukocyte | *PPP5C* | -0.376317404 | 0.004210505 | 0.011330438 | 1 |

| | | | | | |
|---|---|---|---|---|---|
| T2DM_insulin leukocyte | *ERCC1* | -0.465525391 | 0.002923165 | 0.011330438 | 1 |
| T2DM_insulin leukocyte | *CD22* | 0.175413819 | 0.019009025 | 0.026005146 | 1 |
| T2DM_insulin leukocyte | *LTF* | -1.070221822 | 0.002070942 | 0.011330438 | 1 |
| T2DM_insulin leukocyte | *EXTL3* | -0.312862549 | 0.00900676 | 0.014309095 | 1 |
| T2DM_insulin leukocyte | *CALCOCO1* | -1.020532469 | 0.016681697 | 0.023299818 | 1 |
| T2DM_insulin leukocyte | *UBR7* | -0.371351745 | 0.007318782 | 0.012496955 | 1 |
| T2DM_insulin leukocyte | *MAN2B2* | -0.55529429 | 0.003283936 | 0.011330438 | 1 |
| T2DM_insulin leukocyte | *ANGEL1* | -0.465720013 | 0.012439556 | 0.018194523 | 1 |
| T2DM_insulin leukocyte | *DDX11* | -0.389733309 | 0.007276555 | 0.012446489 | 1 |
| T2DM_insulin leukocyte | *GPRC5A* | -0.406012227 | 0.018390774 | 0.025300193 | 1 |
| T2DM_insulin leukocyte | *UFL1* | -0.370080091 | 0.003716544 | 0.011330438 | 1 |
| T2DM_insulin leukocyte | *POLA2* | -0.634877884 | 0.00698426 | 0.01217983 | 1 |
| T2DM_insulin leukocyte | *ZC3H3* | -0.31981356 | 0.013689534 | 0.019673073 | 1 |
| T2DM_insulin leukocyte | *MDH1* | -0.382241053 | 0.002655411 | 0.011330438 | 1 |
| T2DM_insulin leukocyte | *MTMR11* | -1.406200409 | 0.003435771 | 0.011330438 | 1 |
| T2DM_insulin leukocyte | *DPEP1* | -0.215938684 | 0.00809639 | 0.013304764 | 1 |
| T2DM_insulin leukocyte | *NUDCD3* | -0.318850073 | 0.007338873 | 0.012522047 | 1 |
| T2DM_insulin leukocyte | *TTC27* | -0.765637141 | 0.00823678 | 0.013448692 | 1 |
| T2DM_insulin leukocyte | *PLEKHB1* | -0.295347788 | 0.029044721 | 0.037637437 | 1 |
| T2DM_insulin leukocyte | *OSBPL5* | -1.16049184 | 0.000119308 | 0.011330438 | 0.468759837 |
| T2DM_insulin leukocyte | *NLRP2* | -0.9059345 | 0.001123434 | 0.011330438 | 1 |
| T2DM_insulin leukocyte | *ZDHHC6* | -0.58473328 | 0.000430801 | 0.011330438 | 1 |
| T2DM_insulin leukocyte | *RNH1* | -0.35649908 | 0.004264118 | 0.011330438 | 1 |
| T2DM_insulin leukocyte | *NDUFS1* | -0.323850858 | 0.005873766 | 0.011570498 | 1 |
| T2DM_insulin leukocyte | *TYMP* | 0.153270882 | 0.033417786 | 0.042546495 | 1 |
| T2DM_insulin leukocyte | *CD44* | -0.198636909 | 0.004860536 | 0.011330438 | 1 |
| T2DM_insulin leukocyte | *KCNG1* | -1.10782075 | 0.020721912 | 0.02807079 | 1 |
| T2DM_insulin leukocyte | *AGPAT4* | -1.265036237 | 0.006654477 | 0.01190541 | 1 |
| T2DM_insulin leukocyte | *VEZT* | -0.389037632 | 0.004365315 | 0.011330438 | 1 |
| T2DM_insulin leukocyte | *RABEP1* | -0.638305864 | 0.004038003 | 0.011330438 | 1 |
| T2DM_insulin leukocyte | *ARID4A* | -0.527802367 | 0.002351271 | 0.011330438 | 1 |
| T2DM_insulin leukocyte | *PNPLA6* | -0.49108763 | 0.002877526 | 0.011330438 | 1 |
| T2DM_insulin leukocyte | *ABCF2* | -0.527002217 | 0.003533519 | 0.011330438 | 1 |
| T2DM_insulin leukocyte | *UBA6* | -0.603807036 | 0.002801447 | 0.011330438 | 1 |
| T2DM_insulin leukocyte | *APBA2* | -0.964134218 | 0.005964901 | 0.011570498 | 1 |
| T2DM_insulin leukocyte | *ASTE1* | -0.455323953 | 0.009656486 | 0.015078655 | 1 |
| T2DM_insulin leukocyte | *RNF19A* | -0.329936647 | 0.003516425 | 0.011330438 | 1 |
| T2DM_insulin leukocyte | *SH3YL1* | -0.244608489 | 0.005482751 | 0.011427973 | 1 |
| T2DM_insulin leukocyte | *FAM136A* | -0.369799965 | 0.003384319 | 0.011330438 | 1 |
| T2DM_insulin leukocyte | *DAPK2* | -0.388250913 | 0.00535617 | 0.011421159 | 1 |
| T2DM_insulin leukocyte | *TIMP2* | -0.736351647 | 0.002128416 | 0.011330438 | 1 |
| T2DM_insulin leukocyte | *MYOM2* | 0.413552602 | 0.024320921 | 0.032293645 | 1 |
| T2DM_insulin leukocyte | *RPL26L1* | -0.339650369 | 0.003704413 | 0.011330438 | 1 |
| T2DM_insulin leukocyte | *NSUN2* | -0.569782392 | 0.002006361 | 0.011330438 | 1 |
| T2DM_insulin leukocyte | *MRI1* | -0.365599738 | 0.019520455 | 0.026658279 | 1 |
| T2DM_insulin leukocyte | *AGA* | -0.446437114 | 0.00277938 | 0.011330438 | 1 |
| T2DM_insulin leukocyte | *BOD1L1* | -0.294072151 | 0.014273622 | 0.020385699 | 1 |
| T2DM_insulin leukocyte | *DNAH5* | -0.576730161 | 0.003497438 | 0.011330438 | 1 |
| T2DM_insulin leukocyte | *PHLPP2* | -0.336745235 | 0.005436408 | 0.011421159 | 1 |
| T2DM_insulin leukocyte | *CTNS* | -0.461268403 | 0.005817315 | 0.011570498 | 1 |

| | | | | | |
|---|---|---|---|---|---|
| T2DM_insulin leukocyte | *INPP4A* | -0.348730257 | 0.005631766 | 0.01154792 | 1 |
| T2DM_insulin leukocyte | *PSMA4* | -0.36211056 | 0.002459396 | 0.011330438 | 1 |
| T2DM_insulin leukocyte | *SPATA7* | -0.440550864 | 0.012259358 | 0.017972767 | 1 |
| T2DM_insulin leukocyte | *TRIT1* | -0.783851926 | 0.006465558 | 0.011733569 | 1 |
| T2DM_insulin leukocyte | *CNTLN* | -0.519219029 | 0.005002628 | 0.011330438 | 1 |
| T2DM_insulin leukocyte | *PREX2* | -0.685375596 | 0.007735809 | 0.012941946 | 1 |
| T2DM_insulin leukocyte | *WDR37* | -0.545828979 | 0.002852106 | 0.011330438 | 1 |
| T2DM_insulin leukocyte | *YTHDC2* | -0.610991555 | 0.00198788 | 0.011330438 | 1 |
| T2DM_insulin leukocyte | *DTNBP1* | -0.427846932 | 0.003746212 | 0.011330438 | 1 |
| T2DM_insulin leukocyte | *ANO2* | -4.867917879 | 0.017964674 | 0.024774729 | 1 |
| T2DM_insulin leukocyte | *GOPC* | -0.445451195 | 0.002624715 | 0.011330438 | 1 |
| T2DM_insulin leukocyte | *ZNF800* | -0.432109543 | 0.003090495 | 0.011330438 | 1 |
| T2DM_insulin leukocyte | *MRPS10* | -0.536632052 | 0.001219757 | 0.011330438 | 1 |
| T2DM_insulin leukocyte | *COL9A2* | -0.417395122 | 0.003256655 | 0.011330438 | 1 |
| T2DM_insulin leukocyte | *ERCC8* | -1.223354224 | 0.011248158 | 0.016874384 | 1 |
| T2DM_insulin leukocyte | *H6PD* | -0.985549449 | 0.002518651 | 0.011330438 | 1 |
| T2DM_insulin leukocyte | *VAMP3* | -0.445625208 | 0.003099357 | 0.011330438 | 1 |
| T2DM_insulin leukocyte | *EPN3* | -0.7491647 | 0.027776318 | 0.036220762 | 1 |
| T2DM_insulin leukocyte | *RCN1* | -0.301213831 | 0.004389626 | 0.011330438 | 1 |
| T2DM_insulin leukocyte | *RFC2* | -0.426890974 | 0.003657863 | 0.011330438 | 1 |
| T2DM_insulin leukocyte | *JKAMP* | -0.372789394 | 0.002403979 | 0.011330438 | 1 |
| T2DM_insulin leukocyte | *PTGER3* | -0.265156172 | 0.032487459 | 0.041509993 | 1 |
| T2DM_insulin leukocyte | *BCAR1* | -0.307673952 | 0.003699131 | 0.011330438 | 1 |
| T2DM_insulin leukocyte | *THOC3* | -0.286583298 | 0.006946968 | 0.012141743 | 1 |
| T2DM_insulin leukocyte | *HEBP2* | -0.340630919 | 0.003125104 | 0.011330438 | 1 |
| T2DM_insulin leukocyte | *RRP12* | -0.263228275 | 0.005801061 | 0.011570498 | 1 |
| T2DM_insulin leukocyte | *FNIP2* | -0.683252357 | 0.014749587 | 0.020951239 | 1 |
| T2DM_insulin leukocyte | *MRTO4* | -0.420668392 | 0.005864173 | 0.011570498 | 1 |
| T2DM_insulin leukocyte | *MCF2L2* | -0.612887534 | 0.003803962 | 0.011330438 | 1 |
| T2DM_insulin leukocyte | *LAMA3* | -0.564312791 | 0.007987558 | 0.013208382 | 1 |
| T2DM_insulin leukocyte | *ANAPC4* | -1.244912853 | 0.031677574 | 0.040593995 | 1 |
| T2DM_insulin leukocyte | *LY75* | -1.591816297 | 0.001315154 | 0.011330438 | 1 |
| T2DM_insulin leukocyte | *ARID4B* | -0.343326814 | 0.002388908 | 0.011330438 | 1 |
| T2DM_insulin leukocyte | *SDCCAG8* | -0.361321372 | 0.004919529 | 0.011330438 | 1 |
| T2DM_insulin leukocyte | *KIF1B* | -0.362057415 | 0.003607504 | 0.011330438 | 1 |
| T2DM_insulin leukocyte | *TBC1D22A* | -0.386655301 | 0.004216516 | 0.011330438 | 1 |
| T2DM_insulin leukocyte | *SYNE2* | -0.354859357 | 0.003719953 | 0.011330438 | 1 |
| T2DM_insulin leukocyte | *ATP9A* | -0.381496919 | 0.004480967 | 0.011330438 | 1 |
| T2DM_insulin leukocyte | *CHRDL2* | -1.297402577 | 0.007524998 | 0.012715195 | 1 |
| T2DM_insulin leukocyte | *GALC* | -0.584664734 | 0.001521355 | 0.011330438 | 1 |
| T2DM_insulin leukocyte | *CYFIP2* | -0.308726378 | 0.004257862 | 0.011330438 | 1 |
| T2DM_insulin leukocyte | *USP36* | -0.602465533 | 0.001796019 | 0.011330438 | 1 |
| T2DM_insulin leukocyte | *MCOLN3* | -0.37024749 | 0.003402705 | 0.011330438 | 1 |
| T2DM_insulin leukocyte | *ITIH4* | -0.987894074 | 0.000233559 | 0.011330438 | 0.917654856 |
| T2DM_insulin leukocyte | *PCGF2* | -0.357606764 | 0.005154314 | 0.011330438 | 1 |
| T2DM_insulin leukocyte | *GDI2* | -0.397760676 | 0.002734295 | 0.011330438 | 1 |
| T2DM_insulin leukocyte | *ATP11B* | -0.601480369 | 0.004363187 | 0.011330438 | 1 |
| T2DM_insulin leukocyte | *LAMC2* | -0.273444115 | 0.004297445 | 0.011330438 | 1 |
| T2DM_insulin leukocyte | *RASGRF1* | -0.265999609 | 0.008718738 | 0.013987718 | 1 |
| T2DM_insulin leukocyte | *CROCC* | -0.521477218 | 0.010223956 | 0.0157344 | 1 |

| | | | | | |
|---|---|---|---|---|---|
| T2DM_insulin leukocyte | *FLYWCH1* | -0.285770156 | 0.003030177 | 0.011330438 | 1 |
| T2DM_insulin leukocyte | *OGFR* | -0.745576973 | 0.035916598 | 0.045360435 | 1 |
| T2DM_insulin leukocyte | *GNA15* | 0.55956578 | 0.017484518 | 0.024206016 | 1 |
| T2DM_insulin leukocyte | *RIMBP2* | -0.298711084 | 0.005985854 | 0.011570498 | 1 |
| T2DM_insulin leukocyte | *MPC1* | -0.349302197 | 0.003305072 | 0.011330438 | 1 |
| T2DM_insulin leukocyte | *MRPS35* | -0.388167476 | 0.004113265 | 0.011330438 | 1 |
| T2DM_insulin leukocyte | *ZNF112* | -0.413535096 | 0.023359502 | 0.03112224 | 1 |
| T2DM_insulin leukocyte | *EIF4B* | -0.310153268 | 0.008025396 | 0.01322829 | 1 |
| T2DM_insulin leukocyte | *SLC6A16* | -0.987475839 | 0.005508542 | 0.011434531 | 1 |
| T2DM_insulin leukocyte | *SPHK2* | -0.444225815 | 0.005718494 | 0.011563542 | 1 |
| T2DM_insulin leukocyte | *HAGH* | -0.454978063 | 0.002282787 | 0.011330438 | 1 |
| T2DM_insulin leukocyte | *RNF4* | -0.448922798 | 0.002998824 | 0.011330438 | 1 |
| T2DM_insulin leukocyte | *TM7SF3* | -0.639263637 | 0.005677957 | 0.011549238 | 1 |
| T2DM_insulin leukocyte | *HIPK2* | -0.305393369 | 0.00465116 | 0.011330438 | 1 |
| T2DM_insulin leukocyte | *TMEM161A* | -0.424596059 | 0.00455519 | 0.011330438 | 1 |
| T2DM_insulin leukocyte | *SNX24* | -0.78706839 | 0.005628821 | 0.01154792 | 1 |
| T2DM_insulin leukocyte | *PMS1* | -0.359229998 | 0.005947601 | 0.011570498 | 1 |
| T2DM_insulin leukocyte | *IPO5* | -0.328233724 | 0.003788234 | 0.011330438 | 1 |
| T2DM_insulin leukocyte | *OAT* | -0.340654793 | 0.004309874 | 0.011330438 | 1 |
| T2DM_insulin leukocyte | *WDR18* | -0.447150973 | 0.002309531 | 0.011330438 | 1 |
| T2DM_insulin leukocyte | *MCM10* | 0.656194117 | 0.022862666 | 0.030553543 | 1 |
| T2DM_insulin leukocyte | *ERBB3* | -0.297732898 | 0.00954347 | 0.014974558 | 1 |
| T2DM_insulin leukocyte | *ANKRD44* | -1.164609586 | 0.034997664 | 0.04437103 | 1 |
| T2DM_insulin leukocyte | *ADAT1* | -0.384101038 | 0.003673355 | 0.011330438 | 1 |
| T2DM_insulin leukocyte | *MYLK* | -0.330584942 | 0.005816777 | 0.011570498 | 1 |
| T2DM_insulin leukocyte | *GSTO2* | -0.253738239 | 0.004916493 | 0.011330438 | 1 |
| T2DM_insulin leukocyte | *SEC61A2* | -1.446070557 | 8.91E-05 | 0.011330438 | 0.350063187 |
| T2DM_insulin leukocyte | *PRKCQ* | -0.771572298 | 0.017794584 | 0.024566029 | 1 |
| T2DM_insulin leukocyte | *TBC1D1* | -0.368220527 | 0.00670018 | 0.011939628 | 1 |
| T2DM_insulin leukocyte | *CTNNA2* | -0.450894315 | 0.007630073 | 0.012827794 | 1 |
| T2DM_insulin leukocyte | *CD84* | 0.056434339 | 0.006769689 | 0.011991254 | 1 |
| T2DM_insulin leukocyte | *ATXN3* | -0.541817688 | 0.007597907 | 0.012795617 | 1 |
| T2DM_insulin leukocyte | *EML1* | -0.588503213 | 0.00327594 | 0.011330438 | 1 |
| T2DM_insulin leukocyte | *STOML1* | -0.337175812 | 0.004987021 | 0.011330438 | 1 |
| T2DM_insulin leukocyte | *DNTTIP2* | -0.37131139 | 0.001591168 | 0.011330438 | 1 |
| T2DM_insulin leukocyte | *RRP15* | -0.385567512 | 0.004489714 | 0.011330438 | 1 |
| T2DM_insulin leukocyte | *REEP1* | -0.328495017 | 0.007695741 | 0.012910575 | 1 |
| T2DM_insulin leukocyte | *LAPTM4A* | -0.32935271 | 0.00308402 | 0.011330438 | 1 |
| T2DM_insulin leukocyte | *KIF2A* | -0.356034361 | 0.003866565 | 0.011330438 | 1 |
| T2DM_insulin leukocyte | *MAST4* | -0.418760869 | 0.002651296 | 0.011330438 | 1 |
| T2DM_insulin leukocyte | *ADAM7* | -5.222734489 | 0.024262919 | 0.032238421 | 1 |
| T2DM_insulin leukocyte | *DRD4* | -1.177370247 | 0.007262027 | 0.012437884 | 1 |
| T2DM_insulin leukocyte | *PLA2G10* | -1.387181789 | 0.004830417 | 0.011330438 | 1 |
| T2DM_insulin leukocyte | *GNB5* | -0.401632515 | 0.012783538 | 0.018608509 | 1 |
| T2DM_insulin leukocyte | *RAB27A* | -0.36226585 | 0.002546028 | 0.011330438 | 1 |
| T2DM_insulin leukocyte | *PTPN3* | -0.611848429 | 0.002526301 | 0.011330438 | 1 |
| T2DM_insulin leukocyte | *FGF10* | -1.015595399 | 0.01194299 | 0.017706833 | 1 |
| T2DM_insulin leukocyte | *DGCR2* | -0.509439924 | 0.002560756 | 0.011330438 | 1 |
| T2DM_insulin leukocyte | *ZXDC* | -0.913397356 | 0.00255775 | 0.011330438 | 1 |
| T2DM_insulin leukocyte | *WIPI1* | -0.878030415 | 0.003030999 | 0.011330438 | 1 |

| | | | | | |
|---|---|---|---|---|---|
| T2DM_insulin leukocyte | *GBA2* | -0.352243108 | 0.008002877 | 0.013211473 | 1 |
| T2DM_insulin leukocyte | *ASNS* | -0.360546811 | 0.006163178 | 0.011570498 | 1 |
| T2DM_insulin leukocyte | *MAP4K4* | -0.33198617 | 0.003452479 | 0.011330438 | 1 |
| T2DM_insulin leukocyte | *RPL31* | -0.29341493 | 0.002977024 | 0.011330438 | 1 |
| T2DM_insulin leukocyte | *SEL1L* | -0.321025056 | 0.004376178 | 0.011330438 | 1 |
| T2DM_insulin leukocyte | *TCF3* | -0.377166134 | 0.002010309 | 0.011330438 | 1 |
| T2DM_insulin leukocyte | *CPSF1* | -0.30351695 | 0.010690394 | 0.016273754 | 1 |
| T2DM_insulin leukocyte | *CYBRD1* | 0.233802265 | 0.03190454 | 0.04084488 | 1 |
| T2DM_insulin leukocyte | *LNX1* | -0.288410352 | 0.005017169 | 0.011330438 | 1 |
| T2DM_insulin leukocyte | *ALDH3A2* | -0.257153708 | 0.004208665 | 0.011330438 | 1 |
| T2DM_insulin leukocyte | *P4HA2* | -0.264332667 | 0.009289389 | 0.014663725 | 1 |
| T2DM_insulin leukocyte | *TRNT1* | -1.201064552 | 0.003028507 | 0.011330438 | 1 |
| T2DM_insulin leukocyte | *CRMP1* | -0.310088491 | 0.004722294 | 0.011330438 | 1 |
| T2DM_insulin leukocyte | *DERL2* | -0.389817998 | 0.002928785 | 0.011330438 | 1 |
| T2DM_insulin leukocyte | *XRCC1* | -0.449712663 | 0.014516946 | 0.020652657 | 1 |
| T2DM_insulin leukocyte | *SCARB1* | -0.990169088 | 0.001715778 | 0.011330438 | 1 |
| T2DM_insulin leukocyte | *NLE1* | -0.326187909 | 0.028211626 | 0.036678848 | 1 |
| T2DM_insulin leukocyte | *SMARCE1* | -0.335665933 | 0.003240326 | 0.011330438 | 1 |
| T2DM_insulin leukocyte | *MRPS34* | -0.311762202 | 0.003572298 | 0.011330438 | 1 |
| T2DM_insulin leukocyte | *TEAD2* | -0.394719738 | 0.03280473 | 0.041847333 | 1 |
| T2DM_insulin leukocyte | *IPCEF1* | -0.541249248 | 0.033927041 | 0.043138946 | 1 |
| T2DM_insulin leukocyte | *ZZEF1* | -0.545623029 | 0.002837459 | 0.011330438 | 1 |
| T2DM_insulin leukocyte | *WSCD2* | -0.342053509 | 0.01709009 | 0.023777254 | 1 |
| T2DM_insulin leukocyte | *SEMA3A* | -1.02216679 | 0.007095311 | 0.01228624 | 1 |
| T2DM_insulin leukocyte | *TTC38* | -0.688943818 | 0.0044794 | 0.011330438 | 1 |
| T2DM_insulin leukocyte | *GRAMD4* | -0.492503438 | 0.009659704 | 0.015078655 | 1 |
| T2DM_insulin leukocyte | *CELSR1* | -1.564189888 | 0.021707528 | 0.029248586 | 1 |
| T2DM_insulin leukocyte | *TIMM21* | -0.553625554 | 0.003834688 | 0.011330438 | 1 |
| T2DM_insulin leukocyte | *VPS9D1* | -0.35556062 | 0.00790921 | 0.013134103 | 1 |
| T2DM_insulin leukocyte | *ZNF37A* | -0.338179163 | 0.009778132 | 0.01522119 | 1 |
| T2DM_insulin leukocyte | *MARK3* | -0.445912746 | 0.000941489 | 0.011330438 | 1 |
| T2DM_insulin leukocyte | *CACNG5* | -11.55243731 | 0.003457887 | 0.011330438 | 1 |
| T2DM_insulin leukocyte | *MOCOS* | -0.888595857 | 0.028226674 | 0.036686272 | 1 |
| T2DM_insulin leukocyte | *DLG1* | -0.367609914 | 0.003511113 | 0.011330438 | 1 |
| T2DM_insulin leukocyte | *EXOSC7* | -0.309798886 | 0.00573872 | 0.011570498 | 1 |
| T2DM_insulin leukocyte | *MKRN2* | -0.449149195 | 0.004999663 | 0.011330438 | 1 |
| T2DM_insulin leukocyte | *MLH1* | -0.504468673 | 0.004625569 | 0.011330438 | 1 |
| T2DM_insulin leukocyte | *TPD52* | -0.298078989 | 0.003889298 | 0.011330438 | 1 |
| T2DM_insulin leukocyte | *ACACB* | -0.505727361 | 0.001330679 | 0.011330438 | 1 |
| T2DM_insulin leukocyte | *GPATCH1* | -0.339826419 | 0.009982929 | 0.015458818 | 1 |
| T2DM_insulin leukocyte | *ICAM3* | -0.449843956 | 0.000891959 | 0.011330438 | 1 |
| T2DM_insulin leukocyte | *XAB2* | -0.380742281 | 0.003995237 | 0.011330438 | 1 |
| T2DM_insulin leukocyte | *ACTL6B* | -0.296918875 | 0.012198801 | 0.017937534 | 1 |
| T2DM_insulin leukocyte | *IL4R* | -0.328313565 | 0.007894112 | 0.013125674 | 1 |
| T2DM_insulin leukocyte | *APBB1IP* | 0.154934569 | 0.009366553 | 0.01477366 | 1 |
| T2DM_insulin leukocyte | *MCCC1* | -0.409341958 | 0.003025195 | 0.011330438 | 1 |
| T2DM_insulin leukocyte | *N4BP2* | -0.545566856 | 0.003149813 | 0.011330438 | 1 |
| T2DM_insulin leukocyte | *MLLT10* | -0.494446326 | 0.003480112 | 0.011330438 | 1 |
| T2DM_insulin leukocyte | *VDAC3* | -0.40218662 | 0.004129941 | 0.011330438 | 1 |
| T2DM_insulin leukocyte | *PCM1* | -0.342027895 | 0.004099719 | 0.011330438 | 1 |

| | | | | | |
|---|---|---|---|---|---|
| T2DM_insulin leukocyte | *TP73* | 0.999854502 | 0.017177872 | 0.023872903 | 1 |
| T2DM_insulin leukocyte | *TOLLIP* | -0.407644421 | 0.002602169 | 0.011330438 | 1 |
| T2DM_insulin leukocyte | *CLUL1* | -0.967545463 | 0.005686417 | 0.011549238 | 1 |
| T2DM_insulin leukocyte | *FKBP7* | -0.494335249 | 0.004858104 | 0.011330438 | 1 |
| T2DM_insulin leukocyte | *LXN* | -0.272973082 | 0.008870324 | 0.014148275 | 1 |
| T2DM_insulin leukocyte | *SAR1A* | -0.301339492 | 0.003879464 | 0.011330438 | 1 |
| T2DM_insulin leukocyte | *CDC14A* | -0.538141729 | 0.007192568 | 0.012373634 | 1 |
| T2DM_insulin leukocyte | *FDFT1* | -0.4207961 | 0.002781168 | 0.011330438 | 1 |
| T2DM_insulin leukocyte | *SCGN* | -0.173222476 | 0.006717872 | 0.011954039 | 1 |
| T2DM_insulin leukocyte | *DDX1* | -0.339142095 | 0.004186956 | 0.011330438 | 1 |
| T2DM_insulin leukocyte | *RABL2B* | -0.561983756 | 0.004343391 | 0.011330438 | 1 |
| T2DM_insulin leukocyte | *DCT* | -2.378300036 | 0.003344327 | 0.011330438 | 1 |
| T2DM_insulin leukocyte | *SCTR* | -0.378140095 | 0.005073487 | 0.011330438 | 1 |
| T2DM_insulin leukocyte | *RIF1* | -0.73358011 | 0.001155898 | 0.011330438 | 1 |
| T2DM_insulin leukocyte | *IGSF9B* | -0.654035219 | 0.01038376 | 0.015911777 | 1 |
| T2DM_insulin leukocyte | *COL4A4* | -0.998158544 | 0.000873662 | 0.011330438 | 1 |
| T2DM_insulin leukocyte | *ZNF506* | -0.360731669 | 0.005362683 | 0.011421159 | 1 |
| T2DM_insulin leukocyte | *CADPS2* | -0.3790714 | 0.006852324 | 0.012045987 | 1 |
| T2DM_insulin leukocyte | *ME2* | -0.82576655 | 0.003469261 | 0.011330438 | 1 |
| T2DM_insulin leukocyte | *CCNT2* | -0.408420868 | 0.013000509 | 0.018823927 | 1 |
| T2DM_insulin leukocyte | *SEMA5B* | 0.541401911 | 0.016672626 | 0.02329543 | 1 |
| T2DM_insulin leukocyte | *BCKDHB* | -0.827396218 | 0.017859878 | 0.024647509 | 1 |
| T2DM_insulin leukocyte | *DIS3* | -0.381406063 | 0.003115757 | 0.011330438 | 1 |
| T2DM_insulin leukocyte | *PIBF1* | -0.564037872 | 0.005479412 | 0.011427973 | 1 |
| T2DM_insulin leukocyte | *CHMP2B* | -0.336276712 | 0.003098431 | 0.011330438 | 1 |
| T2DM_insulin leukocyte | *PPIE* | -0.472130794 | 0.001466655 | 0.011330438 | 1 |
| T2DM_insulin leukocyte | *NCOA1* | -0.441963593 | 0.00305097 | 0.011330438 | 1 |
| T2DM_insulin leukocyte | *AGBL5* | -0.591816644 | 0.015058598 | 0.021336182 | 1 |
| T2DM_insulin leukocyte | *CD59* | -0.237188925 | 0.004438014 | 0.011330438 | 1 |
| T2DM_insulin leukocyte | *CD82* | -0.659741138 | 0.003907293 | 0.011330438 | 1 |
| T2DM_insulin leukocyte | *MECOM* | -0.252955093 | 0.009474171 | 0.014889607 | 1 |
| T2DM_insulin leukocyte | *SLC25A24* | -0.36863357 | 0.004795 | 0.011330438 | 1 |
| T2DM_insulin leukocyte | *PILRA* | 0.114968145 | 0.020980744 | 0.028366601 | 1 |
| T2DM_insulin leukocyte | *ZNF213* | -0.415288491 | 0.004889193 | 0.011330438 | 1 |
| T2DM_insulin leukocyte | *CPNE3* | -0.289279783 | 0.004090418 | 0.011330438 | 1 |
| T2DM_insulin leukocyte | *RRN3* | -0.329593582 | 0.004458098 | 0.011330438 | 1 |
| T2DM_insulin leukocyte | *WNT11* | -0.87941585 | 0.018508352 | 0.025435228 | 1 |
| T2DM_insulin leukocyte | *DDHD2* | -0.449413889 | 0.003787071 | 0.011330438 | 1 |
| T2DM_insulin leukocyte | *TTC39A* | -0.322128739 | 0.021305848 | 0.028756673 | 1 |
| T2DM_insulin leukocyte | *EPS15* | -0.417044901 | 0.018950806 | 0.025970602 | 1 |
| T2DM_insulin leukocyte | *ORC1* | -1.371513276 | 0.003642034 | 0.011330438 | 1 |
| T2DM_insulin leukocyte | *MGST2* | -0.258429149 | 0.007149298 | 0.012352504 | 1 |
| T2DM_insulin leukocyte | *USP40* | -0.594310005 | 0.003243732 | 0.011330438 | 1 |
| T2DM_insulin leukocyte | *POMGNT1* | -0.516240759 | 0.002015314 | 0.011330438 | 1 |
| T2DM_insulin leukocyte | *NFX1* | -0.517932376 | 0.011787111 | 0.017562214 | 1 |
| T2DM_insulin leukocyte | *EIF2AK1* | -0.343580668 | 0.010529395 | 0.016078505 | 1 |
| T2DM_insulin leukocyte | *ITPKC* | -0.474166455 | 0.004038809 | 0.011330438 | 1 |
| T2DM_insulin leukocyte | *TMED2* | -0.2870407 | 0.003382424 | 0.011330438 | 1 |
| T2DM_insulin leukocyte | *ACOX3* | -0.741635364 | 0.009421248 | 0.014830162 | 1 |
| T2DM_insulin leukocyte | *TRIP6* | -0.314886729 | 0.003939952 | 0.011330438 | 1 |

| | | | | | |
|---|---|---|---|---|---|
| T2DM_insulin leukocyte | *PGS1* | -0.59621858 | 0.003175137 | 0.011330438 | 1 |
| T2DM_insulin leukocyte | *PSMC5* | -0.29178857 | 0.005235188 | 0.011389288 | 1 |
| T2DM_insulin leukocyte | *UIMC1* | -0.42939284 | 0.002880127 | 0.011330438 | 1 |
| T2DM_insulin leukocyte | *SH3BP2* | 0.428448654 | 0.030111886 | 0.038853727 | 1 |
| T2DM_insulin leukocyte | *SF3B2* | -0.290759688 | 0.007584987 | 0.01278254 | 1 |
| T2DM_insulin leukocyte | *KLHL42* | -0.315062145 | 0.004994363 | 0.011330438 | 1 |
| T2DM_insulin leukocyte | *METTL2A* | -0.587381844 | 0.00320061 | 0.011330438 | 1 |
| T2DM_insulin leukocyte | *GP6* | 0.489585412 | 0.032997854 | 0.04205273 | 1 |
| T2DM_insulin leukocyte | *SLC15A1* | -0.688437246 | 0.012266726 | 0.017976861 | 1 |
| T2DM_insulin leukocyte | *TGDS* | -0.304669748 | 0.026965004 | 0.035279887 | 1 |
| T2DM_insulin leukocyte | *DOCK3* | -0.421348298 | 0.006166137 | 0.011570498 | 1 |
| T2DM_insulin leukocyte | *ARHGAP28* | -0.950990743 | 0.005964885 | 0.011570498 | 1 |
| T2DM_insulin leukocyte | *PPP1R13B* | -0.270033395 | 0.007427306 | 0.012600123 | 1 |
| T2DM_insulin leukocyte | *ZNF343* | -0.48684168 | 0.007876379 | 0.013101733 | 1 |
| T2DM_insulin leukocyte | *MAVS* | -0.6661146 | 0.00828078 | 0.013483293 | 1 |
| T2DM_insulin leukocyte | *DYNLL1* | -0.302080968 | 0.005014278 | 0.011330438 | 1 |
| T2DM_insulin leukocyte | *RBBP9* | -0.331925785 | 0.006092507 | 0.011570498 | 1 |
| T2DM_insulin leukocyte | *SLC23A2* | -0.65276168 | 0.002046311 | 0.011330438 | 1 |
| T2DM_insulin leukocyte | *SLC8B1* | 0.101438846 | 0.025126781 | 0.033251304 | 1 |
| T2DM_insulin leukocyte | *TMEM230* | -0.340816075 | 0.004378451 | 0.011330438 | 1 |
| T2DM_insulin leukocyte | *DZANK1* | 0.829987872 | 0.039205179 | 0.049166022 | 1 |
| T2DM_insulin leukocyte | *KIF16B* | -0.53514013 | 0.005632591 | 0.01154792 | 1 |
| T2DM_insulin leukocyte | *ZNF302* | -0.349740634 | 0.004553185 | 0.011330438 | 1 |
| T2DM_insulin leukocyte | *GRAMD1A* | -0.591564405 | 0.003277635 | 0.011330438 | 1 |
| T2DM_insulin leukocyte | *CDIP1* | -0.412438375 | 0.00299454 | 0.011330438 | 1 |
| T2DM_insulin leukocyte | *MLF2* | -0.403170759 | 0.002293895 | 0.011330438 | 1 |
| T2DM_insulin leukocyte | *DDX24* | -0.31722076 | 0.002870602 | 0.011330438 | 1 |
| T2DM_insulin leukocyte | *ANKRD24* | -3.392234056 | 0.001766176 | 0.011330438 | 1 |
| T2DM_insulin leukocyte | *GPATCH2L* | -0.451999682 | 0.002870087 | 0.011330438 | 1 |
| T2DM_insulin leukocyte | *SLC9A1* | -0.338323529 | 0.00486595 | 0.011330438 | 1 |
| T2DM_insulin leukocyte | *SPTLC1* | -0.371578499 | 0.003559204 | 0.011330438 | 1 |
| T2DM_insulin leukocyte | *NDUFB2* | -0.299818573 | 0.00286119 | 0.011330438 | 1 |
| T2DM_insulin leukocyte | *MAEA* | -0.349604132 | 0.003250809 | 0.011330438 | 1 |
| T2DM_insulin leukocyte | *STRN4* | -0.358898127 | 0.004594557 | 0.011330438 | 1 |
| T2DM_insulin leukocyte | *LYZ* | 0.082626442 | 0.015451226 | 0.021810191 | 1 |
| T2DM_insulin leukocyte | *MUL1* | -0.378461136 | 0.005678355 | 0.011549238 | 1 |
| T2DM_insulin leukocyte | *SPG21* | -0.492792464 | 0.003377584 | 0.011330438 | 1 |
| T2DM_insulin leukocyte | *USP48* | -0.377280095 | 0.004093751 | 0.011330438 | 1 |
| T2DM_insulin leukocyte | *GLG1* | -0.372831423 | 0.003235166 | 0.011330438 | 1 |
| T2DM_insulin leukocyte | *NRCAM* | -0.242327124 | 0.005506349 | 0.011434531 | 1 |
| T2DM_insulin leukocyte | *TF* | -0.786117147 | 0.000310481 | 0.011330438 | 1 |
| T2DM_insulin leukocyte | *MYO15A* | -0.8659475 | 0.008373473 | 0.013599474 | 1 |
| T2DM_insulin leukocyte | *PITPNM3* | -1.032927834 | 0.005888205 | 0.011570498 | 1 |
| T2DM_insulin leukocyte | *SPAG7* | -0.363306054 | 0.011082454 | 0.016732269 | 1 |
| T2DM_insulin leukocyte | *TMEM101* | -0.498662468 | 0.002077387 | 0.011330438 | 1 |
| T2DM_insulin leukocyte | *PPP2R3C* | -0.5652262 | 0.00507099 | 0.011330438 | 1 |
| T2DM_insulin leukocyte | *OSGEP* | -0.658175636 | 0.002559763 | 0.011330438 | 1 |
| T2DM_insulin leukocyte | *SCFD1* | -0.38168289 | 0.003771763 | 0.011330438 | 1 |
| T2DM_insulin leukocyte | *RPGRIP1* | 0.414057357 | 0.03001153 | 0.038749688 | 1 |
| T2DM_insulin leukocyte | *EZR* | -0.258259637 | 0.00413246 | 0.011330438 | 1 |

| | | | | | |
|---|---|---|---|---|---|
| T2DM_insulin leukocyte | *NUP50* | -0.63472853 | 0.003663209 | 0.011330438 | 1 |
| T2DM_insulin leukocyte | *COMT* | -0.332778309 | 0.003728466 | 0.011330438 | 1 |
| T2DM_insulin leukocyte | *ECHDC1* | -0.294282162 | 0.006717382 | 0.011954039 | 1 |
| T2DM_insulin leukocyte | *CBX5* | -0.283432254 | 0.008569027 | 0.01382216 | 1 |
| T2DM_insulin leukocyte | *SUCO* | -0.263283483 | 0.003870652 | 0.011330438 | 1 |
| T2DM_insulin leukocyte | *MSH2* | -0.79317213 | 0.003390638 | 0.011330438 | 1 |
| T2DM_insulin leukocyte | *TMEM38B* | -0.562776848 | 0.001827051 | 0.011330438 | 1 |
| T2DM_insulin leukocyte | *PSMD5* | -0.345546367 | 0.005242456 | 0.011392572 | 1 |
| T2DM_insulin leukocyte | *CRAT* | -0.622212737 | 0.00295694 | 0.011330438 | 1 |
| T2DM_insulin leukocyte | *SH2D3C* | -0.527450231 | 0.006021447 | 0.011570498 | 1 |
| T2DM_insulin leukocyte | *CWF19L1* | -0.384482141 | 0.00282779 | 0.011330438 | 1 |
| T2DM_insulin leukocyte | *IKZF5* | -0.595127583 | 0.002213786 | 0.011330438 | 1 |
| T2DM_insulin leukocyte | *TLL2* | -1.478850211 | 0.006877082 | 0.012075281 | 1 |
| T2DM_insulin leukocyte | *SRPK1* | -0.451534134 | 0.002879817 | 0.011330438 | 1 |
| T2DM_insulin leukocyte | *PGC* | -0.268709195 | 0.028047069 | 0.036525334 | 1 |
| T2DM_insulin leukocyte | *EFHC1* | -0.311230367 | 0.008497156 | 0.013744474 | 1 |
| T2DM_insulin leukocyte | *ITPR3* | -0.384107357 | 0.006652623 | 0.01190541 | 1 |
| T2DM_insulin leukocyte | *IFT74* | -0.499145795 | 0.004728288 | 0.011330438 | 1 |
| T2DM_insulin leukocyte | *ABL1* | -0.38229773 | 0.004750584 | 0.011330438 | 1 |
| T2DM_insulin leukocyte | *SH3GLB1* | -0.384878197 | 0.003803641 | 0.011330438 | 1 |
| T2DM_insulin leukocyte | *CDC7* | -0.431776515 | 0.020674028 | 0.028029074 | 1 |
| T2DM_insulin leukocyte | *SYDE2* | -0.423013481 | 0.006454876 | 0.01173015 | 1 |
| T2DM_insulin leukocyte | *ABLIM1* | -0.281746146 | 0.011179792 | 0.016816769 | 1 |
| T2DM_insulin leukocyte | *ERMP1* | -0.340816605 | 0.009237524 | 0.01459945 | 1 |
| T2DM_insulin leukocyte | *PALMD* | -0.238850462 | 0.037413253 | 0.047138726 | 1 |
| T2DM_insulin leukocyte | *MZF1* | -0.42624986 | 0.003045367 | 0.011330438 | 1 |
| T2DM_insulin leukocyte | *OCEL1* | -0.609521027 | 0.000439623 | 0.011330438 | 1 |
| T2DM_insulin leukocyte | *CATSPERG* | -0.530818271 | 0.03841391 | 0.048297041 | 1 |
| T2DM_insulin leukocyte | *HSD3B7* | -1.272088491 | 0.001901227 | 0.011330438 | 1 |
| T2DM_insulin leukocyte | *HNRNPM* | -0.313229745 | 0.002782093 | 0.011330438 | 1 |
| T2DM_insulin leukocyte | *TIMM13* | -0.376770909 | 0.003103998 | 0.011330438 | 1 |
| T2DM_insulin leukocyte | *CEP170B* | -0.573154414 | 0.002843013 | 0.011330438 | 1 |
| T2DM_insulin leukocyte | *POLR2E* | -0.510272444 | 0.002871156 | 0.011330438 | 1 |
| T2DM_insulin leukocyte | *RASSF7* | -0.325480266 | 0.005292398 | 0.011421159 | 1 |
| T2DM_insulin leukocyte | *ARVCF* | -0.720225117 | 0.003354943 | 0.011330438 | 1 |
| T2DM_insulin leukocyte | *TRMT2A* | -0.329449652 | 0.008662858 | 0.013926501 | 1 |
| T2DM_insulin leukocyte | *MED15* | -0.977374415 | 0.002880363 | 0.011330438 | 1 |
| T2DM_insulin leukocyte | *LZTR1* | -0.733528692 | 0.001065262 | 0.011330438 | 1 |
| T2DM_insulin leukocyte | *SMARCB1* | -0.39291042 | 0.004288162 | 0.011330438 | 1 |
| T2DM_insulin leukocyte | *DDT* | -0.418049637 | 0.002012132 | 0.011330438 | 1 |
| T2DM_insulin leukocyte | *CABIN1* | -0.500184783 | 0.00363511 | 0.011330438 | 1 |
| T2DM_insulin leukocyte | *SPECC1L* | -0.570797083 | 0.003973589 | 0.011330438 | 1 |
| T2DM_insulin leukocyte | *PES1* | -0.447349049 | 0.000750326 | 0.011330438 | 1 |
| T2DM_insulin leukocyte | *GGT1* | -2.561983811 | 0.02518975 | 0.03330179 | 1 |
| T2DM_insulin leukocyte | *TOP3B* | -0.508564895 | 0.016652973 | 0.023284531 | 1 |
| T2DM_insulin leukocyte | *CARD10* | -0.243231909 | 0.014784782 | 0.020989512 | 1 |
| T2DM_insulin leukocyte | *SLC25A1* | -0.430481928 | 0.003035231 | 0.011330438 | 1 |
| T2DM_insulin leukocyte | *HPS4* | -0.463245125 | 0.001863203 | 0.011330438 | 1 |
| T2DM_insulin leukocyte | *GCAT* | -0.516185106 | 0.005693638 | 0.011549238 | 1 |
| T2DM_insulin leukocyte | *CYP2D6* | 0.898544489 | 0.022220528 | 0.029827281 | 1 |

| | | | | | |
|---|---|---|---|---|---|
| T2DM_insulin leukocyte | *DDX17* | -0.382981169 | 0.003216187 | 0.011330438 | 1 |
| T2DM_insulin leukocyte | *HSCB* | -0.701899572 | 0.001930294 | 0.011330438 | 1 |
| T2DM_insulin leukocyte | *CBY1* | -0.41567525 | 0.00399552 | 0.011330438 | 1 |
| T2DM_insulin leukocyte | *XBP1* | -0.271104019 | 0.003315147 | 0.011330438 | 1 |
| T2DM_insulin leukocyte | *TIMP3* | -0.423085548 | 0.004598189 | 0.011330438 | 1 |
| T2DM_insulin leukocyte | *CYB5R3* | -0.342822883 | 0.005840791 | 0.011570498 | 1 |
| T2DM_insulin leukocyte | *MIOX* | -0.90614853 | 0.004518991 | 0.011330438 | 1 |
| T2DM_insulin leukocyte | *LMF2* | -0.757618488 | 0.002100102 | 0.011330438 | 1 |
| T2DM_insulin leukocyte | *RHBDD3* | -0.459675233 | 0.002455187 | 0.011330438 | 1 |
| T2DM_insulin leukocyte | *TOM1* | -0.576127543 | 0.005422199 | 0.011421159 | 1 |
| T2DM_insulin leukocyte | *CHKB* | -0.845653562 | 0.010935682 | 0.016550349 | 1 |
| T2DM_insulin leukocyte | *BIK* | -0.752054001 | 0.003885368 | 0.011330438 | 1 |
| T2DM_insulin leukocyte | *ARSA* | -0.327582379 | 0.023970402 | 0.031871306 | 1 |
| T2DM_insulin leukocyte | *TTLL12* | -0.575288334 | 0.00407748 | 0.011330438 | 1 |
| T2DM_insulin leukocyte | *RBFOX2* | -0.374271999 | 0.003130684 | 0.011330438 | 1 |
| T2DM_insulin leukocyte | *SYNGR1* | -1.03660307 | 0.002256621 | 0.011330438 | 1 |
| T2DM_insulin leukocyte | *MTMR3* | -0.451198602 | 0.007397133 | 0.012565211 | 1 |
| T2DM_insulin leukocyte | *APOL4* | -1.068913195 | 0.023322073 | 0.031082912 | 1 |
| T2DM_insulin leukocyte | *SAMM50* | -0.358505864 | 0.003823604 | 0.011330438 | 1 |
| T2DM_insulin leukocyte | *FOXRED2* | -0.404555847 | 0.009086208 | 0.014418301 | 1 |
| T2DM_insulin leukocyte | *SGSM3* | -0.574446908 | 0.00261566 | 0.011330438 | 1 |
| T2DM_insulin leukocyte | *IFT27* | -0.470225843 | 0.003526654 | 0.011330438 | 1 |
| T2DM_insulin leukocyte | *ST13* | -0.331411305 | 0.003021577 | 0.011330438 | 1 |
| T2DM_insulin leukocyte | *POLR3H* | -0.788138561 | 0.005343911 | 0.011421159 | 1 |
| T2DM_insulin leukocyte | *PMM1* | -0.507283918 | 0.003227823 | 0.011330438 | 1 |
| T2DM_insulin leukocyte | *DESI1* | -0.500817462 | 0.004077637 | 0.011330438 | 1 |
| T2DM_insulin leukocyte | *KHNYN* | -0.551791062 | 0.002284054 | 0.011330438 | 1 |
| T2DM_insulin leukocyte | *RBM23* | -0.377308835 | 0.003385743 | 0.011330438 | 1 |
| T2DM_insulin leukocyte | *PRMT5* | -0.346045316 | 0.006614218 | 0.011871102 | 1 |
| T2DM_insulin leukocyte | *VCPKMT* | -0.573839473 | 0.002155512 | 0.011330438 | 1 |
| T2DM_insulin leukocyte | *SOS2* | -0.492174983 | 0.00313188 | 0.011330438 | 1 |
| T2DM_insulin leukocyte | *CDKL1* | -0.633232808 | 0.01051625 | 0.016070925 | 1 |
| T2DM_insulin leukocyte | *NIN* | -0.858506491 | 0.002764773 | 0.011330438 | 1 |
| T2DM_insulin leukocyte | *TRIM9* | -0.284861483 | 0.006685931 | 0.011938173 | 1 |
| T2DM_insulin leukocyte | *ATP6V1D* | -0.339290662 | 0.004302941 | 0.011330438 | 1 |
| T2DM_insulin leukocyte | *PLEK2* | -0.518123156 | 0.013741184 | 0.019732863 | 1 |
| T2DM_insulin leukocyte | *GSTZ1* | -0.560262593 | 0.004171742 | 0.011330438 | 1 |
| T2DM_insulin leukocyte | *TMED8* | -0.435244929 | 0.003564484 | 0.011330438 | 1 |
| T2DM_insulin leukocyte | *DAAM1* | -0.347049478 | 0.003185137 | 0.011330438 | 1 |
| T2DM_insulin leukocyte | *DHRS7* | -0.406598489 | 0.00315317 | 0.011330438 | 1 |
| T2DM_insulin leukocyte | *MTHFD1* | -1.444546752 | 0.001303173 | 0.011330438 | 1 |
| T2DM_insulin leukocyte | *TELO2* | -0.375320606 | 0.033551041 | 0.04269447 | 1 |
| T2DM_insulin leukocyte | *GSKIP* | -0.375629486 | 0.004256702 | 0.011330438 | 1 |
| T2DM_insulin leukocyte | *TRIP11* | -0.325117274 | 0.004052375 | 0.011330438 | 1 |
| T2DM_insulin leukocyte | *DCAF11* | -0.457072415 | 0.007240305 | 0.012418942 | 1 |
| T2DM_insulin leukocyte | *EMC9* | -0.326020025 | 0.022202503 | 0.029813272 | 1 |
| T2DM_insulin leukocyte | *NFATC4* | -0.503400733 | 0.007521986 | 0.012715195 | 1 |
| T2DM_insulin leukocyte | *PYGB* | -0.300262626 | 0.036385021 | 0.045922502 | 1 |
| T2DM_insulin leukocyte | *ABHD12* | -0.742740717 | 0.001607238 | 0.011330438 | 1 |
| T2DM_insulin leukocyte | *PROCR* | -0.29768486 | 0.01224417 | 0.017963907 | 1 |

| | | | | | |
|---|---|---|---|---|---|
| T2DM_insulin leukocyte | *CD40* | -0.420728879 | 0.004143784 | 0.011330438 | 1 |
| T2DM_insulin leukocyte | *IFT52* | -0.366728647 | 0.004202105 | 0.011330438 | 1 |
| T2DM_insulin leukocyte | *MYBL2* | -1.288397491 | 0.000249019 | 0.011330438 | 0.978396877 |
| T2DM_insulin leukocyte | *NELFCD* | -0.356850861 | 0.003640897 | 0.011330438 | 1 |
| T2DM_insulin leukocyte | *MTG2* | -0.418441209 | 0.004951303 | 0.011330438 | 1 |
| T2DM_insulin leukocyte | *DIDO1* | -0.389300593 | 0.015459124 | 0.021810191 | 1 |
| T2DM_insulin leukocyte | *CDC25B* | -0.809665977 | 0.001281867 | 0.011330438 | 1 |
| T2DM_insulin leukocyte | *ISM1* | 0.523936992 | 0.019006359 | 0.026005146 | 1 |
| T2DM_insulin leukocyte | *ARFRP1* | -0.558916202 | 0.006034574 | 0.011570498 | 1 |
| T2DM_insulin leukocyte | *SLC52A3* | -0.85171196 | 0.012134148 | 0.017862521 | 1 |
| T2DM_insulin leukocyte | *SIRPB1* | 0.249406983 | 0.014445255 | 0.020587417 | 1 |
| T2DM_insulin leukocyte | *PLCB4* | -0.273349509 | 0.023161966 | 0.030911469 | 1 |
| T2DM_insulin leukocyte | *POFUT1* | -0.366580245 | 0.004522933 | 0.011330438 | 1 |
| T2DM_insulin leukocyte | *NOP56* | -0.529716821 | 0.002956834 | 0.011330438 | 1 |
| T2DM_insulin leukocyte | *MANBAL* | -0.342919879 | 0.004700695 | 0.011330438 | 1 |
| T2DM_insulin leukocyte | *IDH3B* | -0.483813559 | 0.002181469 | 0.011330438 | 1 |
| T2DM_insulin leukocyte | *MAPRE1* | -0.299940076 | 0.005734687 | 0.011570498 | 1 |
| T2DM_insulin leukocyte | *OXT* | 0.321099867 | 0.017936624 | 0.024744731 | 1 |
| T2DM_insulin leukocyte | *TTI1* | -0.583982611 | 0.00361596 | 0.011330438 | 1 |
| T2DM_insulin leukocyte | *CHMP4B* | -0.317947401 | 0.003328954 | 0.011330438 | 1 |
| T2DM_insulin leukocyte | *DHX35* | -0.529512267 | 0.018660082 | 0.025625817 | 1 |
| T2DM_insulin leukocyte | *DNTTIP1* | -0.913622962 | 0.000849382 | 0.011330438 | 1 |
| T2DM_insulin leukocyte | *APMAP* | -0.32469175 | 0.028676636 | 0.037197261 | 1 |
| T2DM_insulin leukocyte | *VAPA* | -0.310180972 | 0.003165587 | 0.011330438 | 1 |
| T2DM_insulin leukocyte | *MYOM1* | -0.821429252 | 0.012257705 | 0.017972767 | 1 |
| T2DM_insulin leukocyte | *CEP192* | -0.301747031 | 0.010909147 | 0.016529903 | 1 |
| T2DM_insulin leukocyte | *LIPG* | -0.406186971 | 0.005967207 | 0.011570498 | 1 |
| T2DM_insulin leukocyte | *RIOK3* | -0.338792663 | 0.003684611 | 0.011330438 | 1 |
| T2DM_insulin leukocyte | *NALCN* | -0.357138074 | 0.005201553 | 0.011378137 | 1 |
| T2DM_insulin leukocyte | *FGF14* | -0.358277186 | 0.006641187 | 0.011903843 | 1 |
| T2DM_insulin leukocyte | *FNDC3A* | -0.283009861 | 0.005120693 | 0.011330438 | 1 |
| T2DM_insulin leukocyte | *KLF5* | -0.262271981 | 0.00695439 | 0.01214931 | 1 |
| T2DM_insulin leukocyte | *ACP5* | 0.023219064 | 0.013632035 | 0.019604782 | 1 |
| T2DM_insulin leukocyte | *DNAJC3* | -0.33487729 | 0.00301717 | 0.011330438 | 1 |
| T2DM_insulin leukocyte | *UGGT2* | -0.36284928 | 0.003246161 | 0.011330438 | 1 |
| T2DM_insulin leukocyte | *ARHGEF7* | -0.504899513 | 0.002983619 | 0.011330438 | 1 |
| T2DM_insulin leukocyte | *PARP4* | -0.980167824 | 0.003526026 | 0.011330438 | 1 |
| T2DM_insulin leukocyte | *MEDAG* | -0.56216776 | 0.009140759 | 0.014469799 | 1 |
| T2DM_insulin leukocyte | *CLN5* | -0.468882852 | 0.002856726 | 0.011330438 | 1 |
| T2DM_insulin leukocyte | *OLFM4* | -0.185166659 | 0.004839151 | 0.011330438 | 1 |
| T2DM_insulin leukocyte | *MGRN1* | -0.490485568 | 0.002865333 | 0.011330438 | 1 |
| T2DM_insulin leukocyte | *LONP2* | -0.36878124 | 0.003575189 | 0.011330438 | 1 |
| T2DM_insulin leukocyte | *CBLN1* | -1.220835331 | 0.00538328 | 0.011421159 | 1 |
| T2DM_insulin leukocyte | *DHODH* | -1.034804621 | 0.004685842 | 0.011330438 | 1 |
| T2DM_insulin leukocyte | *PARD6A* | -0.250953614 | 0.011177743 | 0.016816769 | 1 |
| T2DM_insulin leukocyte | *MMP15* | -0.793961846 | 0.004733698 | 0.011330438 | 1 |
| T2DM_insulin leukocyte | *USB1* | -0.501089317 | 0.007902171 | 0.013129703 | 1 |
| T2DM_insulin leukocyte | *CYB5B* | -0.736887476 | 0.001688551 | 0.011330438 | 1 |
| T2DM_insulin leukocyte | *CCDC113* | -0.471972155 | 0.008841559 | 0.014115597 | 1 |
| T2DM_insulin leukocyte | *NME3* | -0.389554504 | 0.005571288 | 0.011495826 | 1 |

| | | | | | |
|---|---|---|---|---|---|
| T2DM_insulin leukocyte | *VAC14* | -0.396000952 | 0.005352502 | 0.011421159 | 1 |
| T2DM_insulin leukocyte | *NPRL3* | -0.549499806 | 0.006067331 | 0.011570498 | 1 |
| T2DM_insulin leukocyte | *ZNF500* | -0.697658333 | 0.00929655 | 0.014669134 | 1 |
| T2DM_insulin leukocyte | *NME4* | -0.410377325 | 0.003248917 | 0.011330438 | 1 |
| T2DM_insulin leukocyte | *NOMO3* | -0.627940359 | 0.001522976 | 0.011330438 | 1 |
| T2DM_insulin leukocyte | *LMF1* | -0.386525084 | 0.004552365 | 0.011330438 | 1 |
| T2DM_insulin leukocyte | *CLCN7* | -0.845400496 | 0.015872253 | 0.022312015 | 1 |
| T2DM_insulin leukocyte | *HAGHL* | -0.632027648 | 0.003388431 | 0.011330438 | 1 |
| T2DM_insulin leukocyte | *METRN* | -0.316322207 | 0.002647164 | 0.011330438 | 1 |
| T2DM_insulin leukocyte | *PIEZO1* | -0.435367963 | 0.007171258 | 0.012363261 | 1 |
| T2DM_insulin leukocyte | *UBFD1* | -0.486972976 | 0.007903219 | 0.013129703 | 1 |
| T2DM_insulin leukocyte | *AQP8* | -0.62979078 | 0.004213624 | 0.011330438 | 1 |
| T2DM_insulin leukocyte | *USP31* | -0.273661454 | 0.007851538 | 0.013077022 | 1 |
| T2DM_insulin leukocyte | *HMOX2* | -0.34097445 | 0.004992742 | 0.011330438 | 1 |
| T2DM_insulin leukocyte | *RBL2* | -0.45129475 | 0.007248203 | 0.01242504 | 1 |
| T2DM_insulin leukocyte | *KAT8* | -0.456171068 | 0.003417362 | 0.011330438 | 1 |
| T2DM_insulin leukocyte | *NOMO1* | -0.557703655 | 0.009588166 | 0.015025219 | 1 |
| T2DM_insulin leukocyte | *RNF40* | -0.703600578 | 0.002530191 | 0.011330438 | 1 |
| T2DM_insulin leukocyte | *KNOP1* | -0.483700633 | 0.003316591 | 0.011330438 | 1 |
| T2DM_insulin leukocyte | *TRIP4* | -0.436313259 | 0.002598953 | 0.011330438 | 1 |
| T2DM_insulin leukocyte | *AP3B2* | -0.409386402 | 0.005528654 | 0.011449692 | 1 |
| T2DM_insulin leukocyte | *IGDCC4* | 0.93605991 | 0.016400367 | 0.0229804 | 1 |
| T2DM_insulin leukocyte | *RPAP1* | -0.599477226 | 0.003213587 | 0.011330438 | 1 |
| T2DM_insulin leukocyte | *HOMER2* | -0.29017935 | 0.006813683 | 0.012014984 | 1 |
| T2DM_insulin leukocyte | *ATP8B4* | 0.198765334 | 0.013882969 | 0.019921908 | 1 |
| T2DM_insulin leukocyte | *DNAJC17* | -0.346756394 | 0.010190289 | 0.015699813 | 1 |
| T2DM_insulin leukocyte | *ZDHHC2* | -0.256848114 | 0.005128109 | 0.011330438 | 1 |
| T2DM_insulin leukocyte | *TRIM35* | -0.928329508 | 0.002796007 | 0.011330438 | 1 |
| T2DM_insulin leukocyte | *RIPK2* | -0.459233943 | 0.004081553 | 0.011330438 | 1 |
| T2DM_insulin leukocyte | *NBN* | -0.606578761 | 0.001199823 | 0.011330438 | 1 |
| T2DM_insulin leukocyte | *LAPTM4B* | -0.239436987 | 0.004972911 | 0.011330438 | 1 |
| T2DM_insulin leukocyte | *RAB2A* | -0.283026057 | 0.002668337 | 0.011330438 | 1 |
| T2DM_insulin leukocyte | *EIF3E* | -0.308615297 | 0.003388411 | 0.011330438 | 1 |
| T2DM_insulin leukocyte | *ESRP1* | -0.307056926 | 0.005535537 | 0.011449692 | 1 |
| T2DM_insulin leukocyte | *ARMC1* | -0.393935347 | 0.007800134 | 0.01301113 | 1 |
| T2DM_insulin leukocyte | *SPAG1* | -0.360758585 | 0.003853345 | 0.011330438 | 1 |
| T2DM_insulin leukocyte | *CHRAC1* | -0.312315186 | 0.004251168 | 0.011330438 | 1 |
| T2DM_insulin leukocyte | *SNX16* | -1.184185446 | 0.001659828 | 0.011330438 | 1 |
| T2DM_insulin leukocyte | *GSDMD* | -0.285535427 | 0.009176301 | 0.014520212 | 1 |
| T2DM_insulin leukocyte | *SH2D4A* | -0.455675886 | 0.002463314 | 0.011330438 | 1 |
| T2DM_insulin leukocyte | *INTS10* | -0.5023353 | 0.002710264 | 0.011330438 | 1 |
| T2DM_insulin leukocyte | *ERI1* | -0.991783708 | 0.001561649 | 0.011330438 | 1 |
| T2DM_insulin leukocyte | *LEPROTL1* | -0.460183212 | 0.002125354 | 0.011330438 | 1 |
| T2DM_insulin leukocyte | *ERICH1* | -0.565694123 | 0.00183944 | 0.011330438 | 1 |
| T2DM_insulin leukocyte | *ARHGEF10* | -0.429840945 | 0.003077097 | 0.011330438 | 1 |
| T2DM_insulin leukocyte | *KLHDC4* | -0.475587949 | 0.004906897 | 0.011330438 | 1 |
| T2DM_insulin leukocyte | *MCM4* | -0.477988089 | 0.004027445 | 0.011330438 | 1 |
| T2DM_insulin leukocyte | *FGL1* | -0.714927812 | 0.003199838 | 0.011330438 | 1 |
| T2DM_insulin leukocyte | *ECH1* | -0.347181222 | 0.003838459 | 0.011330438 | 1 |
| T2DM_insulin leukocyte | *HNRNPL* | -0.390867833 | 0.002960807 | 0.011330438 | 1 |

| | | | | | |
|---|---|---|---|---|---|
| T2DM_insulin leukocyte | *SARS2* | -0.49384191 | 0.001723605 | 0.011330438 | 1 |
| T2DM_insulin leukocyte | *LIN7B* | -0.444087352 | 0.009591054 | 0.015025219 | 1 |
| T2DM_insulin leukocyte | *PPP1R13L* | -0.322650398 | 0.034222526 | 0.043466792 | 1 |
| T2DM_insulin leukocyte | *ERCC2* | -0.424878993 | 0.003796119 | 0.011330438 | 1 |
| T2DM_insulin leukocyte | *KLC3* | -0.366065987 | 0.00464928 | 0.011330438 | 1 |
| T2DM_insulin leukocyte | *AMH* | -0.390141939 | 0.01006797 | 0.015561391 | 1 |
| T2DM_insulin leukocyte | *IL27RA* | -0.45018901 | 0.017312459 | 0.024010113 | 1 |
| T2DM_insulin leukocyte | *PDCD5* | -0.283019015 | 0.006414816 | 0.01170553 | 1 |
| T2DM_insulin leukocyte | *NUMBL* | -0.317193004 | 0.005609994 | 0.011546185 | 1 |
| T2DM_insulin leukocyte | *SLC1A5* | -0.433432785 | 0.011960788 | 0.017706833 | 1 |
| T2DM_insulin leukocyte | *PRKD2* | -0.530286293 | 0.000461984 | 0.011330438 | 1 |
| T2DM_insulin leukocyte | *APLP1* | -0.194028323 | 0.003716102 | 0.011330438 | 1 |
| T2DM_insulin leukocyte | *PLIN3* | -0.479409329 | 0.000479898 | 0.011330438 | 1 |
| T2DM_insulin leukocyte | *MYH14* | -0.297566452 | 0.006905548 | 0.012100405 | 1 |
| T2DM_insulin leukocyte | *ZNF175* | -0.354567416 | 0.005034542 | 0.011330438 | 1 |
| T2DM_insulin leukocyte | *PLA2G4C* | -0.581254921 | 0.000948139 | 0.011330438 | 1 |
| T2DM_insulin leukocyte | *CAPS* | -0.502252539 | 0.004629566 | 0.011330438 | 1 |
| T2DM_insulin leukocyte | *PPP2R1A* | -0.322360535 | 0.004468708 | 0.011330438 | 1 |
| T2DM_insulin leukocyte | *GCDH* | -0.484263738 | 0.001985648 | 0.011330438 | 1 |
| T2DM_insulin leukocyte | *PIK3R2* | -1.070636558 | 0.003370375 | 0.011330438 | 1 |
| T2DM_insulin leukocyte | *UPK1A* | -5.077792385 | 0.011869113 | 0.017638109 | 1 |
| T2DM_insulin leukocyte | *COPE* | -0.330554937 | 0.003031622 | 0.011330438 | 1 |
| T2DM_insulin leukocyte | *ETV2* | -0.447870476 | 0.012752161 | 0.018577398 | 1 |
| T2DM_insulin leukocyte | *PBX4* | -1.696481594 | 0.001931212 | 0.011330438 | 1 |
| T2DM_insulin leukocyte | *SIPA1L3* | -0.957434276 | 0.002739451 | 0.011330438 | 1 |
| T2DM_insulin leukocyte | *SMG9* | -0.338748903 | 0.007163494 | 0.012363261 | 1 |
| T2DM_insulin leukocyte | *RUNDC3B* | -1.97289297 | 0.002583682 | 0.011330438 | 1 |
| T2DM_insulin leukocyte | *PMPCB* | -0.447244004 | 0.00316606 | 0.011330438 | 1 |
| T2DM_insulin leukocyte | *PON2* | -0.314408099 | 0.006139663 | 0.011570498 | 1 |
| T2DM_insulin leukocyte | *ITGB8* | -0.336755519 | 0.005009738 | 0.011330438 | 1 |
| T2DM_insulin leukocyte | *CAV2* | -0.232806436 | 0.005556517 | 0.011472179 | 1 |
| T2DM_insulin leukocyte | *CAV1* | -0.272803579 | 0.005910735 | 0.011570498 | 1 |
| T2DM_insulin leukocyte | *ANKRD7* | -0.630736506 | 0.009118969 | 0.014446947 | 1 |
| T2DM_insulin leukocyte | *TSPAN12* | -0.587897238 | 0.002918331 | 0.011330438 | 1 |
| T2DM_insulin leukocyte | *CPVL* | 0.048595012 | 0.014628652 | 0.020794491 | 1 |
| T2DM_insulin leukocyte | *ABHD11* | -0.487736287 | 0.007045837 | 0.012237751 | 1 |
| T2DM_insulin leukocyte | *CHCHD2* | -0.259837127 | 0.003263117 | 0.011330438 | 1 |
| T2DM_insulin leukocyte | *EIF3B* | -0.38219711 | 0.003403732 | 0.011330438 | 1 |
| T2DM_insulin leukocyte | *WASL* | -0.308149004 | 0.004882524 | 0.011330438 | 1 |
| T2DM_insulin leukocyte | *AGFG2* | -0.354304589 | 0.006445847 | 0.01173015 | 1 |
| T2DM_insulin leukocyte | *C1GALT1* | -0.325114214 | 0.004130289 | 0.011330438 | 1 |
| T2DM_insulin leukocyte | *RPA3* | -0.408457736 | 0.003462529 | 0.011330438 | 1 |
| T2DM_insulin leukocyte | *ZNHIT1* | -0.356434983 | 0.001597245 | 0.011330438 | 1 |
| T2DM_insulin leukocyte | *TMEM106B* | -0.316807949 | 0.006253483 | 0.011570498 | 1 |
| T2DM_insulin leukocyte | *ACTR3C* | -0.627027151 | 0.003791544 | 0.011330438 | 1 |
| T2DM_insulin leukocyte | *TSPAN13* | -0.249573148 | 0.004698924 | 0.011330438 | 1 |
| T2DM_insulin leukocyte | *RARRES2* | -0.194625381 | 0.003764899 | 0.011330438 | 1 |
| T2DM_insulin leukocyte | *AGR2* | -0.240073693 | 0.017089931 | 0.023777254 | 1 |
| T2DM_insulin leukocyte | *TMEM176B* | -0.289234245 | 0.012509024 | 0.018258541 | 1 |
| T2DM_insulin leukocyte | *COA1* | -0.465953056 | 0.004514265 | 0.011330438 | 1 |

| | | | | | |
|---|---|---|---|---|---|
| T2DM_insulin leukocyte | *BLVRA* | -0.639198668 | 0.00351128 | 0.011330438 | 1 |
| T2DM_insulin leukocyte | *TMEM248* | -0.482302931 | 0.001663315 | 0.011330438 | 1 |
| T2DM_insulin leukocyte | *RHEB* | -0.329571192 | 0.00279003 | 0.011330438 | 1 |
| T2DM_insulin leukocyte | *TBL2* | -0.286628111 | 0.029303174 | 0.037934817 | 1 |
| T2DM_insulin leukocyte | *LIMK1* | -1.265413112 | 0.00596733 | 0.011570498 | 1 |
| T2DM_insulin leukocyte | *SPIN1* | -0.564485199 | 0.002774174 | 0.011330438 | 1 |
| T2DM_insulin leukocyte | *NMRK1* | -0.401221556 | 0.004358955 | 0.011330438 | 1 |
| T2DM_insulin leukocyte | *OGN* | -0.768908703 | 0.010106815 | 0.015598378 | 1 |
| T2DM_insulin leukocyte | *SUSD1* | -0.658698991 | 0.002164003 | 0.011330438 | 1 |
| T2DM_insulin leukocyte | *DNM1* | -0.269619197 | 0.005365164 | 0.011421159 | 1 |
| T2DM_insulin leukocyte | *AK1* | -0.308629693 | 0.00245802 | 0.011330438 | 1 |
| T2DM_insulin leukocyte | *PLGRKT* | -0.302487693 | 0.008815257 | 0.014103329 | 1 |
| T2DM_insulin leukocyte | *DOCK8* | 0.160544787 | 0.022593718 | 0.03024556 | 1 |
| T2DM_insulin leukocyte | *MPDZ* | -0.306389427 | 0.006828572 | 0.012014984 | 1 |
| T2DM_insulin leukocyte | *NPDC1* | -0.261407754 | 0.004586218 | 0.011330438 | 1 |
| T2DM_insulin leukocyte | *HPS1* | -0.436317631 | 0.004365264 | 0.011330438 | 1 |
| T2DM_insulin leukocyte | *CXCL12* | -0.387336409 | 0.003131284 | 0.011330438 | 1 |
| T2DM_insulin leukocyte | *TRDMT1* | -0.867370971 | 0.002503889 | 0.011330438 | 1 |
| T2DM_insulin leukocyte | *ATE1* | -0.61887747 | 0.002598768 | 0.011330438 | 1 |
| T2DM_insulin leukocyte | *NSMCE4A* | -0.334474662 | 0.006682587 | 0.011938173 | 1 |
| T2DM_insulin leukocyte | *PLEKHA1* | -0.286443021 | 0.004595086 | 0.011330438 | 1 |
| T2DM_insulin leukocyte | *PALD1* | -2.379624257 | 0.002465348 | 0.011330438 | 1 |
| T2DM_insulin leukocyte | *SPOCK2* | -0.564108074 | 0.006264017 | 0.011570498 | 1 |
| T2DM_insulin leukocyte | *BMPR1A* | -0.822659823 | 0.002543935 | 0.011330438 | 1 |
| T2DM_insulin leukocyte | *LIPA* | 0.114779164 | 0.021898479 | 0.029455366 | 1 |
| T2DM_insulin leukocyte | *SFXN3* | -0.636895386 | 0.001249065 | 0.011330438 | 1 |
| T2DM_insulin leukocyte | *NPM3* | -0.410257536 | 0.005845515 | 0.011570498 | 1 |
| T2DM_insulin leukocyte | *TNKS2* | -0.413529924 | 0.003376674 | 0.011330438 | 1 |
| T2DM_insulin leukocyte | *ACBD5* | -0.324035284 | 0.005395131 | 0.011421159 | 1 |
| T2DM_insulin leukocyte | *LHPP* | -0.52614122 | 0.023188934 | 0.030936952 | 1 |
| T2DM_insulin leukocyte | *GTPBP4* | -0.319456891 | 0.003619769 | 0.011330438 | 1 |
| T2DM_insulin leukocyte | *XPNPEP1* | -0.365559884 | 0.003737708 | 0.011330438 | 1 |
| T2DM_insulin leukocyte | *SMC3* | -0.312506623 | 0.003472074 | 0.011330438 | 1 |
| T2DM_insulin leukocyte | *RPL28* | -0.236783966 | 0.004509535 | 0.011330438 | 1 |
| T2DM_insulin leukocyte | *PBLD* | -0.321454403 | 0.005683776 | 0.011549238 | 1 |
| T2DM_insulin leukocyte | *DHRS11* | -0.324537331 | 0.006457703 | 0.01173015 | 1 |
| T2DM_insulin leukocyte | *CWC25* | -0.467802507 | 0.002384136 | 0.011330438 | 1 |
| T2DM_insulin leukocyte | *RAD51C* | -0.388196339 | 0.005898268 | 0.011570498 | 1 |
| T2DM_insulin leukocyte | *TRIM37* | -0.361336054 | 0.00413243 | 0.011330438 | 1 |
| T2DM_insulin leukocyte | *RPS6KB1* | -0.438688086 | 0.003185712 | 0.011330438 | 1 |
| T2DM_insulin leukocyte | *TRIM16L* | -0.525121925 | 0.012750249 | 0.018577398 | 1 |
| T2DM_insulin leukocyte | *CDK5RAP3* | -0.452883292 | 0.026862354 | 0.03518073 | 1 |
| T2DM_insulin leukocyte | *CAMTA2* | -0.612408751 | 0.002736154 | 0.011330438 | 1 |
| T2DM_insulin leukocyte | *MED13* | -0.396282717 | 0.001541122 | 0.011330438 | 1 |
| T2DM_insulin leukocyte | *RAI1* | -0.29553625 | 0.00653212 | 0.011799861 | 1 |
| T2DM_insulin leukocyte | *CCDC47* | -0.347831244 | 0.003256434 | 0.011330438 | 1 |
| T2DM_insulin leukocyte | *DRG2* | -0.406327892 | 0.003388971 | 0.011330438 | 1 |
| T2DM_insulin leukocyte | *FTSJ3* | -0.26603219 | 0.034822643 | 0.044188081 | 1 |
| T2DM_insulin leukocyte | *SMARCD2* | -0.808923579 | 0.002835511 | 0.011330438 | 1 |
| T2DM_insulin leukocyte | *SYNGR2* | -0.334823473 | 0.003808855 | 0.011330438 | 1 |

| | | | | | |
|---|---|---|---|---|---|
| T2DM_insulin leukocyte | *C17orf75* | -0.542103853 | 0.002942518 | 0.011330438 | 1 |
| T2DM_insulin leukocyte | *DHX58* | -0.808854439 | 0.001988301 | 0.011330438 | 1 |
| T2DM_insulin leukocyte | *NAGLU* | -0.371193154 | 0.007802042 | 0.01301113 | 1 |
| T2DM_insulin leukocyte | *HSD17B1* | -0.453709079 | 0.003093626 | 0.011330438 | 1 |
| T2DM_insulin leukocyte | *RND2* | -0.947926447 | 0.00303749 | 0.011330438 | 1 |
| T2DM_insulin leukocyte | *MPP2* | -0.263149157 | 0.006909306 | 0.012100405 | 1 |
| T2DM_insulin leukocyte | *HLF* | -0.234924297 | 0.007166891 | 0.012363261 | 1 |
| T2DM_insulin leukocyte | *EFNB3* | -0.342354577 | 0.00560962 | 0.011546185 | 1 |
| T2DM_insulin leukocyte | *RANGRF* | -0.445016244 | 0.006244546 | 0.011570498 | 1 |
| T2DM_insulin leukocyte | *DPH1* | -0.761590748 | 0.010636892 | 0.016217442 | 1 |
| T2DM_insulin leukocyte | *OCIAD1* | -0.327259345 | 0.002797704 | 0.011330438 | 1 |
| T2DM_insulin leukocyte | *UBE2D3* | -0.286974204 | 0.007267265 | 0.012441431 | 1 |
| T2DM_insulin leukocyte | *MAPK10* | -0.356826267 | 0.001076692 | 0.011330438 | 1 |
| T2DM_insulin leukocyte | *AADAT* | -1.065122042 | 0.005067889 | 0.011330438 | 1 |
| T2DM_insulin leukocyte | *TRIM2* | -0.357814794 | 0.003778268 | 0.011330438 | 1 |
| T2DM_insulin leukocyte | *SH3D19* | -0.73705854 | 0.003863272 | 0.011330438 | 1 |
| T2DM_insulin leukocyte | *GLRB* | -0.355396705 | 0.006066341 | 0.011570498 | 1 |
| T2DM_insulin leukocyte | *LRP2BP* | -0.427511793 | 0.003922531 | 0.011330438 | 1 |
| T2DM_insulin leukocyte | *UFSP2* | -0.445006467 | 0.007405254 | 0.012573571 | 1 |
| T2DM_insulin leukocyte | *KLHL5* | -0.469207423 | 0.00319368 | 0.011330438 | 1 |
| T2DM_insulin leukocyte | *FAM149A* | -0.402580343 | 0.001799895 | 0.011330438 | 1 |
| T2DM_insulin leukocyte | *DDX25* | -0.337768108 | 0.014176376 | 0.020276295 | 1 |
| T2DM_insulin leukocyte | *HTATIP2* | -0.417896825 | 0.01416225 | 0.020263467 | 1 |
| T2DM_insulin leukocyte | *B3GAT1* | -0.797999212 | 0.016473865 | 0.023066934 | 1 |
| T2DM_insulin leukocyte | *P2RX3* | -7.271840225 | 0.005277282 | 0.011419801 | 1 |
| T2DM_insulin leukocyte | *VWA5A* | -0.42196077 | 0.005649707 | 0.011549238 | 1 |
| T2DM_insulin leukocyte | *DTX4* | -0.401812235 | 0.008022295 | 0.01322829 | 1 |
| T2DM_insulin leukocyte | *FOXRED1* | -0.509517765 | 0.024708238 | 0.032752586 | 1 |
| T2DM_insulin leukocyte | *ST3GAL4* | -0.644150279 | 0.003437384 | 0.011330438 | 1 |
| T2DM_insulin leukocyte | *PRPF19* | -0.334305545 | 0.004414305 | 0.011330438 | 1 |
| T2DM_insulin leukocyte | *CEP164* | -0.730362777 | 0.002940136 | 0.011330438 | 1 |
| T2DM_insulin leukocyte | *PDHX* | -0.358692429 | 0.02815391 | 0.03662805 | 1 |
| T2DM_insulin leukocyte | *SLC1A2* | -0.429091756 | 0.002633316 | 0.011330438 | 1 |
| T2DM_insulin leukocyte | *ACCS* | 0.212196981 | 0.00588614 | 0.011570498 | 1 |
| T2DM_insulin leukocyte | *MDK* | -0.369404693 | 0.003375135 | 0.011330438 | 1 |
| T2DM_insulin leukocyte | *MADD* | -0.516587095 | 0.002811053 | 0.011330438 | 1 |
| T2DM_insulin leukocyte | *NAA40* | -0.453139851 | 0.005274513 | 0.011419801 | 1 |
| T2DM_insulin leukocyte | *CD81* | -0.2285803 | 0.009774226 | 0.015221139 | 1 |
| T2DM_insulin leukocyte | *SLC35F2* | -0.468819725 | 0.000944901 | 0.011330438 | 1 |
| T2DM_insulin leukocyte | *NUP98* | -0.395108384 | 0.003687721 | 0.011330438 | 1 |
| T2DM_insulin leukocyte | *GTF2H1* | -0.630959097 | 0.003916689 | 0.011330438 | 1 |
| T2DM_insulin leukocyte | *PTPN5* | 0.737218575 | 0.012877588 | 0.018704637 | 1 |
| T2DM_insulin leukocyte | *PRDM4* | -0.311732589 | 0.003877646 | 0.011330438 | 1 |
| T2DM_insulin leukocyte | *CORO1C* | -0.442472565 | 0.003744308 | 0.011330438 | 1 |
| T2DM_insulin leukocyte | *LIN7A* | -0.854896938 | 0.004898804 | 0.011330438 | 1 |
| T2DM_insulin leukocyte | *KRT18* | -0.131258702 | 0.006543282 | 0.011804448 | 1 |
| T2DM_insulin leukocyte | *METAP2* | -0.353623225 | 0.0036488 | 0.011330438 | 1 |
| T2DM_insulin leukocyte | *SLC6A12* | -2.453276506 | 0.002807123 | 0.011330438 | 1 |
| T2DM_insulin leukocyte | *WNT5B* | -0.866298807 | 0.005682284 | 0.011549238 | 1 |
| T2DM_insulin leukocyte | *ITFG2* | -1.294567137 | 0.022249391 | 0.029855825 | 1 |

| | | | | | |
|---|---|---|---|---|---|
| T2DM_insulin leukocyte | *VPS29* | -0.361497835 | 0.001512074 | 0.011330438 | 1 |
| T2DM_insulin leukocyte | *CREBL2* | -0.474929991 | 0.003449979 | 0.011330438 | 1 |
| T2DM_insulin leukocyte | *GSG1* | -3.15523743 | 0.001157251 | 0.011330438 | 1 |
| T2DM_insulin leukocyte | *LTBR* | -0.466103773 | 0.001342379 | 0.011330438 | 1 |
| T2DM_insulin leukocyte | *CDK2AP1* | -0.269803627 | 0.032163152 | 0.041122365 | 1 |
| T2DM_insulin leukocyte | *DDX55* | -0.596788642 | 0.002485907 | 0.011330438 | 1 |
| T2DM_insulin leukocyte | *STX2* | -0.524384571 | 0.003067554 | 0.011330438 | 1 |
| T2DM_insulin leukocyte | *CAND1* | -0.432188188 | 0.00290352 | 0.011330438 | 1 |
| T2DM_insulin leukocyte | *NUP107* | -0.506960328 | 0.002973769 | 0.011330438 | 1 |
| T2DM_insulin leukocyte | *KRR1* | -0.355910443 | 0.005427012 | 0.011421159 | 1 |
| T2DM_insulin leukocyte | *GNB3* | -0.451610818 | 0.006267428 | 0.011570498 | 1 |
| T2DM_insulin leukocyte | *CDCA3* | -0.725989768 | 0.006268755 | 0.011570498 | 1 |
| T2DM_insulin leukocyte | *CHPT1* | -0.704217992 | 0.000471827 | 0.011330438 | 1 |
| T2DM_insulin leukocyte | *SPSB2* | -0.379569121 | 0.005706991 | 0.011549238 | 1 |
| T2DM_insulin leukocyte | *ATN1* | -0.416590893 | 0.002471801 | 0.011330438 | 1 |
| T2DM_insulin leukocyte | *NT5DC3* | -0.416316662 | 0.011955986 | 0.017706833 | 1 |
| T2DM_insulin leukocyte | *GOLT1B* | -0.36078003 | 0.003649102 | 0.011330438 | 1 |
| T2DM_insulin leukocyte | *PRKAB1* | -0.339648784 | 0.005957321 | 0.011570498 | 1 |
| T2DM_insulin leukocyte | *C2CD5* | -0.354503738 | 0.005208598 | 0.011381858 | 1 |
| T2DM_insulin leukocyte | *SRSF9* | -0.342617078 | 0.001983476 | 0.011330438 | 1 |
| T2DM_insulin leukocyte | *FRK* | -0.394079035 | 0.004489367 | 0.011330438 | 1 |
| T2DM_insulin leukocyte | *SMIM8* | -0.446642972 | 0.003986446 | 0.011330438 | 1 |
| T2DM_insulin leukocyte | *NEDD9* | -0.21981801 | 0.003641889 | 0.011330438 | 1 |
| T2DM_insulin leukocyte | *CEP85L* | -0.376495621 | 0.007193021 | 0.012373634 | 1 |
| T2DM_insulin leukocyte | *HDDC2* | -0.301902462 | 0.004261145 | 0.011330438 | 1 |
| T2DM_insulin leukocyte | *TPD52L1* | -0.316349862 | 0.006016755 | 0.011570498 | 1 |
| T2DM_insulin leukocyte | *MTRF1L* | -0.413666405 | 0.006615107 | 0.011871102 | 1 |
| T2DM_insulin leukocyte | *MRPL18* | -0.506577208 | 0.004213634 | 0.011330438 | 1 |
| T2DM_insulin leukocyte | *RNF8* | -0.391856096 | 0.003082626 | 0.011330438 | 1 |
| T2DM_insulin leukocyte | *MDGA1* | 0.579194949 | 0.015718251 | 0.022135129 | 1 |
| T2DM_insulin leukocyte | *CD83* | 0.071919961 | 0.015003052 | 0.021272822 | 1 |
| T2DM_insulin leukocyte | *GLP1R* | -0.37223282 | 0.005334916 | 0.011421159 | 1 |
| T2DM_insulin leukocyte | *BACH2* | -0.364842607 | 0.002226395 | 0.011330438 | 1 |
| T2DM_insulin leukocyte | *FBXL4* | -0.452496412 | 0.004010429 | 0.011330438 | 1 |
| T2DM_insulin leukocyte | *ASCC3* | -0.310404647 | 0.004712389 | 0.011330438 | 1 |
| T2DM_insulin leukocyte | *WASF1* | -0.331064814 | 0.006177398 | 0.011570498 | 1 |
| T2DM_insulin leukocyte | *ALDH5A1* | -0.909773973 | 0.016739724 | 0.023364254 | 1 |
| T2DM_insulin leukocyte | *C6orf62* | -0.432779083 | 0.003126565 | 0.011330438 | 1 |
| T2DM_insulin leukocyte | *B3GAT2* | -0.567219551 | 0.003701155 | 0.011330438 | 1 |
| T2DM_insulin leukocyte | *HBS1L* | -0.386478167 | 0.003492135 | 0.011330438 | 1 |
| T2DM_insulin leukocyte | *PEX7* | -0.351798568 | 0.004797674 | 0.011330438 | 1 |
| T2DM_insulin leukocyte | *PHACTR2* | -0.294508675 | 0.003838333 | 0.011330438 | 1 |
| T2DM_insulin leukocyte | *EPM2A* | -0.637894905 | 0.002881586 | 0.011330438 | 1 |
| T2DM_insulin leukocyte | *TFEB* | -0.472977938 | 0.003133999 | 0.011330438 | 1 |
| T2DM_insulin leukocyte | *DNPH1* | -0.30459741 | 0.02670841 | 0.03503751 | 1 |
| T2DM_insulin leukocyte | *DUSP22* | -0.491119459 | 0.003041582 | 0.011330438 | 1 |
| T2DM_insulin leukocyte | *LAMA4* | -0.316810748 | 0.003246894 | 0.011330438 | 1 |
| T2DM_insulin leukocyte | *FBRSL1* | -0.464714328 | 0.003085215 | 0.011330438 | 1 |
| T2DM_insulin leukocyte | *ENPP5* | -0.364172667 | 0.001948167 | 0.011330438 | 1 |
| T2DM_insulin leukocyte | *NUDT12* | -0.448864248 | 0.008252684 | 0.013448692 | 1 |

| | | | | | |
|---|---|---|---|---|---|
| T2DM_insulin leukocyte | *CEP72* | -0.976936616 | 0.020591616 | 0.027936622 | 1 |
| T2DM_insulin leukocyte | *SEMA5A* | -0.407005972 | 0.011085331 | 0.016732269 | 1 |
| T2DM_insulin leukocyte | *GHR* | -0.99167914 | 0.005708481 | 0.011549238 | 1 |
| T2DM_insulin leukocyte | *DAP* | -0.299282131 | 0.004878829 | 0.011330438 | 1 |
| T2DM_insulin leukocyte | *NME5* | -0.708080336 | 0.000624238 | 0.011330438 | 1 |
| T2DM_insulin leukocyte | *BRD8* | -0.442842708 | 0.003947587 | 0.011330438 | 1 |
| T2DM_insulin leukocyte | *NNT* | -0.581736209 | 0.002388512 | 0.011330438 | 1 |
| T2DM_insulin leukocyte | *IK* | -0.373893065 | 0.003219683 | 0.011330438 | 1 |
| T2DM_insulin leukocyte | *PDE8B* | -0.253333743 | 0.005087455 | 0.011330438 | 1 |
| T2DM_insulin leukocyte | *MSH3* | -0.462111429 | 0.001218372 | 0.011330438 | 1 |
| T2DM_insulin leukocyte | *CCNG1* | -0.666101872 | 0.003584313 | 0.011330438 | 1 |
| T2DM_insulin leukocyte | *DROSHA* | -0.294361287 | 0.011027492 | 0.016664237 | 1 |
| T2DM_insulin leukocyte | *GOLPH3* | -0.373693304 | 0.003731589 | 0.011330438 | 1 |
| T2DM_insulin leukocyte | *NPR3* | -0.973674473 | 0.015902696 | 0.022346814 | 1 |
| T2DM_insulin leukocyte | *RAD50* | -0.477329334 | 0.002943009 | 0.011330438 | 1 |
| T2DM_insulin leukocyte | *WWC1* | -0.218848027 | 0.005590903 | 0.011524093 | 1 |
| T2DM_insulin leukocyte | *SMAD5* | -0.360071455 | 0.002362911 | 0.011330438 | 1 |
| T2DM_insulin leukocyte | *HMGXB3* | -0.522731144 | 0.00133689 | 0.011330438 | 1 |
| T2DM_insulin leukocyte | *BNIP1* | -0.275838046 | 0.028146526 | 0.03662805 | 1 |
| T2DM_insulin leukocyte | *STC2* | -0.287448584 | 0.005415614 | 0.011421159 | 1 |
| T2DM_insulin leukocyte | *EHHADH* | -0.453666562 | 0.012208192 | 0.017937915 | 1 |
| T2DM_insulin leukocyte | *ACTR8* | -0.364174978 | 0.004953199 | 0.011330438 | 1 |
| T2DM_insulin leukocyte | *TIMMDC1* | -0.413455259 | 0.004475242 | 0.011330438 | 1 |
| T2DM_insulin leukocyte | *NPHP3* | -0.436829585 | 0.007735136 | 0.012941946 | 1 |
| T2DM_insulin leukocyte | *FAM162A* | -0.489389995 | 0.000889515 | 0.011330438 | 1 |
| T2DM_insulin leukocyte | *RBP1* | -0.328248189 | 0.003074189 | 0.011330438 | 1 |
| T2DM_insulin leukocyte | *RNF7* | -0.353596472 | 0.003160369 | 0.011330438 | 1 |
| T2DM_insulin leukocyte | *SERPINI2* | -0.24271276 | 0.007106147 | 0.012294167 | 1 |
| T2DM_insulin leukocyte | *TUSC2* | -0.472909484 | 0.003782343 | 0.011330438 | 1 |
| T2DM_insulin leukocyte | *CYB561D2* | -0.276497559 | 0.020943875 | 0.0283265 | 1 |
| T2DM_insulin leukocyte | *C3orf14* | -0.368733969 | 0.000752545 | 0.011330438 | 1 |
| T2DM_insulin leukocyte | *CBLB* | -0.585053553 | 0.004551365 | 0.011330438 | 1 |
| T2DM_insulin leukocyte | *IFT57* | -0.325360834 | 0.005437404 | 0.011421159 | 1 |
| T2DM_insulin leukocyte | *NCBP2* | -0.354333661 | 0.002724287 | 0.011330438 | 1 |
| T2DM_insulin leukocyte | *C3orf52* | -0.308831057 | 0.007964284 | 0.013183733 | 1 |
| T2DM_insulin leukocyte | *SLC41A3* | -0.323851088 | 0.010765074 | 0.016355753 | 1 |
| T2DM_insulin leukocyte | *ATP6V1A* | -0.428986077 | 0.003261433 | 0.011330438 | 1 |
| T2DM_insulin leukocyte | *PODXL2* | -0.287796529 | 0.004601127 | 0.011330438 | 1 |
| T2DM_insulin leukocyte | *UPK1B* | -0.619182613 | 0.003041659 | 0.011330438 | 1 |
| T2DM_insulin leukocyte | *NEK11* | -0.328628147 | 0.00306511 | 0.011330438 | 1 |
| T2DM_insulin leukocyte | *PLSCR4* | -0.618419193 | 0.003284071 | 0.011330438 | 1 |
| T2DM_insulin leukocyte | *COMMD2* | -0.346998251 | 0.004034438 | 0.011330438 | 1 |
| T2DM_insulin leukocyte | *GORASP1* | -0.821998088 | 0.004720749 | 0.011330438 | 1 |
| T2DM_insulin leukocyte | *ARHGEF26* | -0.967369173 | 0.002710012 | 0.011330438 | 1 |
| T2DM_insulin leukocyte | *PLCH1* | -1.786826065 | 0.001114391 | 0.011330438 | 1 |
| T2DM_insulin leukocyte | *VIPR1* | -0.575932919 | 0.008333442 | 0.013546584 | 1 |
| T2DM_insulin leukocyte | *NKTR* | -0.369245445 | 0.002683856 | 0.011330438 | 1 |
| T2DM_insulin leukocyte | *NEK4* | -1.064432526 | 0.001801762 | 0.011330438 | 1 |
| T2DM_insulin leukocyte | *KANSL3* | -0.792571631 | 0.002119876 | 0.011330438 | 1 |
| T2DM_insulin leukocyte | *ACTR1B* | -0.638565528 | 0.002555215 | 0.011330438 | 1 |

| | | | | | |
|---|---|---|---|---|---|
| T2DM_insulin leukocyte | *SLC35F5* | -0.374960501 | 0.003160842 | 0.011330438 | 1 |
| T2DM_insulin leukocyte | *ACTR3* | -0.398157991 | 0.003788516 | 0.011330438 | 1 |
| T2DM_insulin leukocyte | *POMC* | -0.611284473 | 0.002885869 | 0.011330438 | 1 |
| T2DM_insulin leukocyte | *TANC1* | -0.337107878 | 0.003724807 | 0.011330438 | 1 |
| T2DM_insulin leukocyte | *REEP6* | -0.42416665 | 0.01026069 | 0.015772399 | 1 |
| T2DM_insulin leukocyte | *INO80B* | -1.409980431 | 0.000681249 | 0.011330438 | 1 |
| T2DM_insulin leukocyte | *MOGS* | -0.628386502 | 0.002608811 | 0.011330438 | 1 |
| T2DM_insulin leukocyte | *RTN4* | -0.308250654 | 0.002777948 | 0.011330438 | 1 |
| T2DM_insulin leukocyte | *HTRA2* | -0.600246278 | 0.00340352 | 0.011330438 | 1 |
| T2DM_insulin leukocyte | *GALNT3* | -0.413342633 | 0.004215366 | 0.011330438 | 1 |
| T2DM_insulin leukocyte | *MRPL19* | -0.381244702 | 0.005122072 | 0.011330438 | 1 |
| T2DM_insulin leukocyte | *WDR75* | -0.665273422 | 0.002828431 | 0.011330438 | 1 |
| T2DM_insulin leukocyte | *FANCL* | -0.797927754 | 0.002996041 | 0.011330438 | 1 |
| T2DM_insulin leukocyte | *GGCX* | -0.460266606 | 0.004601164 | 0.011330438 | 1 |
| T2DM_insulin leukocyte | *SF3B1* | -0.413934797 | 0.002738803 | 0.011330438 | 1 |
| T2DM_insulin leukocyte | *CHMP3* | -0.330328726 | 0.003113216 | 0.011330438 | 1 |
| T2DM_insulin leukocyte | *IL1R2* | -0.376024743 | 0.006161515 | 0.011570498 | 1 |
| T2DM_insulin leukocyte | *IL1R1* | -0.346260022 | 0.004364949 | 0.011330438 | 1 |
| T2DM_insulin leukocyte | *IL1RL2* | 0.333946697 | 0.020342674 | 0.027646616 | 1 |
| T2DM_insulin leukocyte | *IL18R1* | 0.363944297 | 0.006457352 | 0.01173015 | 1 |
| T2DM_insulin leukocyte | *MLPH* | -0.569617079 | 0.006165863 | 0.011570498 | 1 |
| T2DM_insulin leukocyte | *CNPPD1* | -0.308762114 | 0.005817226 | 0.011570498 | 1 |
| T2DM_insulin leukocyte | *HDLBP* | -0.280890731 | 0.003564365 | 0.011330438 | 1 |
| T2DM_insulin leukocyte | *TAF1B* | -0.323078309 | 0.010160807 | 0.015674052 | 1 |
| T2DM_insulin leukocyte | *STRN* | -0.674863734 | 0.002798592 | 0.011330438 | 1 |
| T2DM_insulin leukocyte | *CEBPZ* | -0.368593594 | 0.002550589 | 0.011330438 | 1 |
| T2DM_insulin leukocyte | *WIPF1* | 0.210456607 | 0.030301132 | 0.039046621 | 1 |
| T2DM_insulin leukocyte | *COX7A2L* | -0.305243057 | 0.003218929 | 0.011330438 | 1 |
| T2DM_insulin leukocyte | *PNO1* | -0.394656171 | 0.003321377 | 0.011330438 | 1 |
| T2DM_insulin leukocyte | *ORC4* | -0.432950865 | 0.001360445 | 0.011330438 | 1 |
| T2DM_insulin leukocyte | *SUMO1* | -0.300465759 | 0.010180232 | 0.015697854 | 1 |
| T2DM_insulin leukocyte | *GRIN3B* | 0.674744095 | 0.015788467 | 0.022226043 | 1 |
| T2DM_insulin leukocyte | *ALMS1* | -0.468524174 | 0.003745378 | 0.011330438 | 1 |
| T2DM_insulin leukocyte | *MARK1* | -0.640166464 | 0.002110239 | 0.011330438 | 1 |
| T2DM_insulin leukocyte | *MORN1* | -0.609514944 | 0.013967271 | 0.020020945 | 1 |
| T2DM_insulin leukocyte | *GPX7* | -0.416839348 | 0.000686968 | 0.011330438 | 1 |
| T2DM_insulin leukocyte | *SCP2* | -0.374551955 | 0.002273078 | 0.011330438 | 1 |
| T2DM_insulin leukocyte | *RALGPS2* | -0.356299743 | 0.004488596 | 0.011330438 | 1 |
| T2DM_insulin leukocyte | *ANGPTL1* | -0.676616255 | 0.003733272 | 0.011330438 | 1 |
| T2DM_insulin leukocyte | *FAM20B* | -0.425569882 | 0.003938948 | 0.011330438 | 1 |
| T2DM_insulin leukocyte | *WRAP73* | -1.272405824 | 0.003794504 | 0.011330438 | 1 |
| T2DM_insulin leukocyte | *ICMT* | -0.416779232 | 0.012510051 | 0.018258541 | 1 |
| T2DM_insulin leukocyte | *PARK7* | -0.302986543 | 0.003004443 | 0.011330438 | 1 |
| T2DM_insulin leukocyte | *MECR* | -1.028877964 | 0.002903308 | 0.011330438 | 1 |
| T2DM_insulin leukocyte | *EDEM3* | -0.425100086 | 0.003337509 | 0.011330438 | 1 |
| T2DM_insulin leukocyte | *FBXO6* | -1.502975024 | 0.008174317 | 0.0133932 | 1 |
| T2DM_insulin leukocyte | *SMG7* | -0.37348163 | 0.003842577 | 0.011330438 | 1 |
| T2DM_insulin leukocyte | *PDC* | -4.893622002 | 0.000396668 | 0.011330438 | 1 |
| T2DM_insulin leukocyte | *RGS2* | 0.190214401 | 0.030438997 | 0.039211416 | 1 |
| T2DM_insulin leukocyte | *BCAS2* | -0.334785438 | 0.003617622 | 0.011330438 | 1 |

| | | | | | |
|---|---|---|---|---|---|
| T2DM_insulin leukocyte | *AGMAT* | -0.636225176 | 0.001744044 | 0.011330438 | 1 |
| T2DM_insulin leukocyte | *TNNI3K* | -1.018633166 | 0.005523632 | 0.011446386 | 1 |
| T2DM_insulin leukocyte | *PLEKHM2* | -0.81062677 | 0.00737938 | 0.012556771 | 1 |
| T2DM_insulin leukocyte | *CRYZ* | -0.273694341 | 0.003505796 | 0.011330438 | 1 |
| T2DM_insulin leukocyte | *TMEM9* | -0.274678155 | 0.025990816 | 0.0341989 | 1 |
| T2DM_insulin leukocyte | *WARS2* | -0.382951302 | 0.004764099 | 0.011330438 | 1 |
| T2DM_insulin leukocyte | *C1orf109* | -0.286698773 | 0.006127751 | 0.011570498 | 1 |
| T2DM_insulin leukocyte | *LGALS8* | -0.468286233 | 0.003199008 | 0.011330438 | 1 |
| T2DM_insulin leukocyte | *HPCAL4* | -0.891084817 | 0.012944934 | 0.018774693 | 1 |
| T2DM_insulin leukocyte | *BMP8B* | -0.355555546 | 0.003310712 | 0.011330438 | 1 |
| T2DM_insulin leukocyte | *MYCL* | 0.196936006 | 0.008392722 | 0.013614784 | 1 |
| T2DM_insulin leukocyte | *MFAP2* | -0.306443585 | 0.007872068 | 0.013100109 | 1 |
| T2DM_insulin leukocyte | *UAP1* | -0.276427872 | 0.003964145 | 0.011330438 | 1 |
| T2DM_insulin leukocyte | *CTBS* | -0.465126739 | 0.001995372 | 0.011330438 | 1 |
| T2DM_insulin leukocyte | *ZNHIT6* | -0.280215789 | 0.004593194 | 0.011330438 | 1 |
| T2DM_insulin leukocyte | *CD46* | -0.295845116 | 0.003632651 | 0.011330438 | 1 |
| T2DM_insulin leukocyte | *SLC2A1* | -0.545557713 | 0.002723417 | 0.011330438 | 1 |
| T2DM_insulin leukocyte | *PRDX1* | -0.31246056 | 0.003740648 | 0.011330438 | 1 |
| T2DM_insulin leukocyte | *CCDC181* | -0.298396 | 0.004327397 | 0.011330438 | 1 |
| T2DM_insulin leukocyte | *NSUN4* | -1.34982018 | 0.000818493 | 0.011330438 | 1 |
| T2DM_insulin leukocyte | *TMED5* | -0.51384095 | 0.00262675 | 0.011330438 | 1 |
| T2DM_insulin leukocyte | *DR1* | -0.459917965 | 0.006233864 | 0.011570498 | 1 |
| T2DM_insulin leukocyte | *PTBP2* | -0.353998358 | 0.009984304 | 0.015458818 | 1 |
| T2DM_insulin leukocyte | *NSL1* | -0.582159913 | 0.003069494 | 0.011330438 | 1 |
| T2DM_insulin leukocyte | *RPA2* | -0.476728418 | 0.004332145 | 0.011330438 | 1 |
| T2DM_insulin leukocyte | *ESYT2* | -0.448543779 | 0.001779092 | 0.011330438 | 1 |
| T2DM_insulin leukocyte | *DDX59* | -0.316448223 | 0.005859842 | 0.011570498 | 1 |
| T2DM_insulin leukocyte | *TTR* | -0.146073919 | 0.003904589 | 0.011330438 | 1 |
| T2DM_insulin leukocyte | *SPCS2* | -0.347721534 | 0.002430468 | 0.011330438 | 1 |
| T2DM_insulin leukocyte | *ELOVL4* | -0.537066511 | 0.004259247 | 0.011330438 | 1 |
| T2DM_insulin leukocyte | *FILIP1* | -0.370775056 | 0.01093949 | 0.016550349 | 1 |
| T2DM_insulin leukocyte | *AKAP7* | -0.759583661 | 0.003479523 | 0.011330438 | 1 |
| T2DM_insulin leukocyte | *RNF146* | -0.416049851 | 0.004050446 | 0.011330438 | 1 |
| T2DM_insulin leukocyte | *MED28* | -0.547950497 | 0.003676076 | 0.011330438 | 1 |
| T2DM_insulin leukocyte | *VAMP8* | -0.370499777 | 0.005028291 | 0.011330438 | 1 |
| T2DM_insulin leukocyte | *ARMC2* | -0.354049166 | 0.003255776 | 0.011330438 | 1 |
| T2DM_insulin leukocyte | *PKD2* | -0.304938675 | 0.004755074 | 0.011330438 | 1 |
| T2DM_insulin leukocyte | *SPP1* | -0.151233882 | 0.012090587 | 0.017818423 | 1 |
| T2DM_insulin leukocyte | *WDR35* | -0.326122744 | 0.00722632 | 0.012409183 | 1 |
| T2DM_insulin leukocyte | *DNAH7* | -0.448547763 | 0.000895678 | 0.011330438 | 1 |
| T2DM_insulin leukocyte | *NDUFB3* | -0.339571797 | 0.004426711 | 0.011330438 | 1 |
| T2DM_insulin leukocyte | *GTF3C3* | -0.484719457 | 0.011971485 | 0.017715828 | 1 |
| T2DM_insulin leukocyte | *CPSF3* | -0.399770364 | 0.035723856 | 0.045204196 | 1 |
| T2DM_insulin leukocyte | *CCDC92* | -0.404907794 | 0.002992497 | 0.011330438 | 1 |
| T2DM_insulin leukocyte | *CTNNAL1* | -0.275000662 | 0.010038661 | 0.015522195 | 1 |
| T2DM_insulin leukocyte | *NEK6* | -0.415227333 | 0.008040798 | 0.013235147 | 1 |
| T2DM_insulin leukocyte | *BSPRY* | -0.289011688 | 0.010810761 | 0.016411836 | 1 |
| T2DM_insulin leukocyte | *RBM18* | -0.481315466 | 0.003546527 | 0.011330438 | 1 |
| T2DM_insulin leukocyte | *DCAF4* | -0.455531418 | 0.006237141 | 0.011570498 | 1 |
| T2DM_insulin leukocyte | *IFI27L2* | -0.313334673 | 0.005256432 | 0.011402654 | 1 |

| | | | | | |
|---|---|---|---|---|---|
| T2DM_insulin leukocyte | *ACOT2* | -0.502229054 | 0.01707643 | 0.023775087 | 1 |
| T2DM_insulin leukocyte | *ZC2HC1C* | -0.967066616 | 0.019670587 | 0.026832745 | 1 |
| T2DM_insulin leukocyte | *KLHL29* | -1.038358011 | 0.025059381 | 0.033184465 | 1 |
| T2DM_insulin leukocyte | *AFTPH* | -0.39729848 | 0.002039831 | 0.011330438 | 1 |
| T2DM_insulin leukocyte | *BCL11A* | -0.58684687 | 0.00786667 | 0.013096672 | 1 |
| T2DM_insulin leukocyte | *CRIPT* | -0.418638416 | 0.005438361 | 0.011421159 | 1 |
| T2DM_insulin leukocyte | *EPCAM* | -0.236089609 | 0.003682837 | 0.011330438 | 1 |
| T2DM_insulin leukocyte | *SLC17A5* | -0.985605259 | 0.012099738 | 0.017825223 | 1 |
| T2DM_insulin leukocyte | *CUTC* | -0.435538605 | 0.005479916 | 0.011427973 | 1 |
| T2DM_insulin leukocyte | *PYROXD2* | -0.526089794 | 0.012487102 | 0.018238596 | 1 |
| T2DM_insulin leukocyte | *HELLS* | -0.457949725 | 0.003459714 | 0.011330438 | 1 |
| T2DM_insulin leukocyte | *PRLHR* | -0.505662091 | 0.006272609 | 0.011570498 | 1 |
| T2DM_insulin leukocyte | *WDR11* | -0.715739353 | 0.000454178 | 0.011330438 | 1 |
| T2DM_insulin leukocyte | *KANSL1* | -0.436891348 | 0.001267503 | 0.011330438 | 1 |
| T2DM_insulin leukocyte | *HOXB3* | -0.383792754 | 0.037420638 | 0.047138726 | 1 |
| T2DM_insulin leukocyte | *RCL1* | -0.661210874 | 0.002791391 | 0.011330438 | 1 |
| T2DM_insulin leukocyte | *CD274* | -1.022404786 | 0.005941154 | 0.011570498 | 1 |
| T2DM_insulin leukocyte | *PLEKHG1* | -0.4114543 | 0.002041721 | 0.011330438 | 1 |
| T2DM_insulin leukocyte | *PCDHB10* | -0.708116194 | 0.002874748 | 0.011330438 | 1 |
| T2DM_insulin leukocyte | *MRPS14* | -0.691822695 | 0.001180855 | 0.011330438 | 1 |
| T2DM_insulin leukocyte | *GORAB* | -0.370431705 | 0.005897083 | 0.011570498 | 1 |
| T2DM_insulin leukocyte | *SNX19* | -0.369647094 | 0.002019897 | 0.011330438 | 1 |
| T2DM_insulin leukocyte | *KCNJ5* | 0.120072287 | 0.011151669 | 0.016813088 | 1 |
| T2DM_insulin leukocyte | *SLC10A7* | -0.813411496 | 0.000874074 | 0.011330438 | 1 |
| T2DM_insulin leukocyte | *MASTL* | -0.369759752 | 0.004895644 | 0.011330438 | 1 |
| T2DM_insulin leukocyte | *MRC1* | 0.09701072 | 0.030646738 | 0.039440233 | 1 |
| T2DM_insulin leukocyte | *IQSEC3* | -0.402649884 | 0.004706873 | 0.011330438 | 1 |
| T2DM_insulin leukocyte | *MTRF1* | -0.735887312 | 0.017410567 | 0.024120634 | 1 |
| T2DM_insulin leukocyte | *DNAJC15* | -0.3642826 | 0.004293672 | 0.011330438 | 1 |
| T2DM_insulin leukocyte | *PROSER1* | -0.52205623 | 0.002128983 | 0.011330438 | 1 |
| T2DM_insulin leukocyte | *WBP4* | -0.373813547 | 0.00356584 | 0.011330438 | 1 |
| T2DM_insulin leukocyte | *KBTBD7* | -0.770246243 | 0.002811609 | 0.011330438 | 1 |
| T2DM_insulin leukocyte | *FAM53C* | -0.704090287 | 0.005197331 | 0.011378137 | 1 |
| T2DM_insulin leukocyte | *SIL1* | -0.39902703 | 0.001807469 | 0.011330438 | 1 |
| T2DM_insulin leukocyte | *PLS1* | -0.276717171 | 0.011959303 | 0.017706833 | 1 |
| T2DM_insulin leukocyte | *ZFP30* | -0.573125406 | 0.001171879 | 0.011330438 | 1 |
| T2DM_insulin leukocyte | *UTP20* | -0.824222564 | 0.002166145 | 0.011330438 | 1 |
| T2DM_insulin leukocyte | *TMPO* | -0.594264428 | 0.001583451 | 0.011330438 | 1 |
| T2DM_insulin leukocyte | *PPP3CC* | -0.448741557 | 0.004678522 | 0.011330438 | 1 |
| T2DM_insulin leukocyte | *EPHX2* | -0.445617606 | 0.003590721 | 0.011330438 | 1 |
| T2DM_insulin leukocyte | *CRISPLD1* | -2.091618184 | 0.005301842 | 0.011421159 | 1 |
| T2DM_insulin leukocyte | *TRIM6* | 0.427546555 | 0.029382847 | 0.038012909 | 1 |
| T2DM_insulin leukocyte | *PYROXD1* | -0.305352843 | 0.004624747 | 0.011330438 | 1 |
| T2DM_insulin leukocyte | *IAPP* | -0.115970631 | 0.012944363 | 0.018774693 | 1 |
| T2DM_insulin leukocyte | *ZSCAN18* | -0.35919134 | 0.001373235 | 0.011330438 | 1 |
| T2DM_insulin leukocyte | *LHX4* | -0.954824137 | 0.015603605 | 0.021989442 | 1 |
| T2DM_insulin leukocyte | *TRMT1L* | -0.367028156 | 0.008026532 | 0.01322829 | 1 |
| T2DM_insulin leukocyte | *CAT* | -0.793249258 | 0.003134776 | 0.011330438 | 1 |
| T2DM_insulin leukocyte | *CSMD2* | 0.461216787 | 0.016122916 | 0.022599692 | 1 |
| T2DM_insulin leukocyte | *ACVR2A* | -0.315282214 | 0.009911464 | 0.01538607 | 1 |

| | | | | | |
|---|---|---|---|---|---|
| T2DM_insulin leukocyte | *POLK* | -0.581888522 | 0.000865742 | 0.011330438 | 1 |
| T2DM_insulin leukocyte | *UBL3* | -0.329674497 | 0.003226065 | 0.011330438 | 1 |
| T2DM_insulin leukocyte | *LDB3* | -0.649938516 | 0.002372946 | 0.011330438 | 1 |
| T2DM_insulin leukocyte | *NAA60* | -0.507191551 | 0.002110512 | 0.011330438 | 1 |
| T2DM_insulin leukocyte | *TRMT13* | -0.793510862 | 0.005878264 | 0.011570498 | 1 |
| T2DM_insulin leukocyte | *RWDD3* | -0.401128383 | 0.002037526 | 0.011330438 | 1 |
| T2DM_insulin leukocyte | *CCDC18* | -0.441663989 | 0.001352213 | 0.011330438 | 1 |
| T2DM_insulin leukocyte | *NBPF14* | -1.004265412 | 0.006520592 | 0.011788627 | 1 |
| T2DM_insulin leukocyte | *PMS2* | -0.751375096 | 0.004308299 | 0.011330438 | 1 |
| T2DM_insulin leukocyte | *EEPD1* | 0.182293674 | 0.034155581 | 0.04341549 | 1 |
| T2DM_insulin leukocyte | *KLHL7* | -0.527466814 | 0.003799871 | 0.011330438 | 1 |
| T2DM_insulin leukocyte | *CBX3* | -0.402113322 | 0.003045755 | 0.011330438 | 1 |
| T2DM_insulin leukocyte | *NPY* | -0.180579403 | 0.00395095 | 0.011330438 | 1 |
| T2DM_insulin leukocyte | *INHBA* | -0.337296389 | 0.003963964 | 0.011330438 | 1 |
| T2DM_insulin leukocyte | *CCZ1* | -0.512228371 | 0.006200341 | 0.011570498 | 1 |
| T2DM_insulin leukocyte | *POLM* | -0.321747635 | 0.013272584 | 0.01915093 | 1 |
| T2DM_insulin leukocyte | *SMU1* | -0.449281751 | 0.001819871 | 0.011330438 | 1 |
| T2DM_insulin leukocyte | *SLC25A51* | -0.453995179 | 0.005318884 | 0.011421159 | 1 |
| T2DM_insulin leukocyte | *ACO1* | -0.356217634 | 0.004860835 | 0.011330438 | 1 |
| T2DM_insulin leukocyte | *PLAU* | -0.430974253 | 0.025865484 | 0.034056798 | 1 |
| T2DM_insulin leukocyte | *RBM19* | -0.331326889 | 0.00539023 | 0.011421159 | 1 |
| T2DM_insulin leukocyte | *BHLHE41* | -0.602731999 | 0.006761369 | 0.011982597 | 1 |
| T2DM_insulin leukocyte | *ITPR2* | -0.923368484 | 0.0069807 | 0.012179027 | 1 |
| T2DM_insulin leukocyte | *EBPL* | -0.327078143 | 0.004424872 | 0.011330438 | 1 |
| T2DM_insulin leukocyte | *TSFM* | -0.514709226 | 0.002592139 | 0.011330438 | 1 |
| T2DM_insulin leukocyte | *CDK2* | -0.574338713 | 0.030180711 | 0.038904203 | 1 |
| T2DM_insulin leukocyte | *IKZF4* | -0.518242423 | 0.007828833 | 0.013044735 | 1 |
| T2DM_insulin leukocyte | *USP45* | -0.307679014 | 0.006476588 | 0.011737322 | 1 |
| T2DM_insulin leukocyte | *C4BPA* | -0.563868367 | 0.021338523 | 0.028790885 | 1 |
| T2DM_insulin leukocyte | *C4BPB* | -0.947651275 | 0.009868388 | 0.015333268 | 1 |
| T2DM_insulin leukocyte | *RAB38* | -1.074038824 | 0.007914713 | 0.013137687 | 1 |
| T2DM_insulin leukocyte | *CKS2* | -0.322504863 | 0.009617503 | 0.015048653 | 1 |
| T2DM_insulin leukocyte | *SNX21* | -0.369467769 | 0.009940992 | 0.015425812 | 1 |
| T2DM_insulin leukocyte | *ARHGAP40* | -19.40751693 | 0.005242797 | 0.011392572 | 1 |
| T2DM_insulin leukocyte | *NCOA3* | -0.352120444 | 0.011176993 | 0.016816769 | 1 |
| T2DM_insulin leukocyte | *MATN4* | -2.459016 | 0.00856981 | 0.01382216 | 1 |
| T2DM_insulin leukocyte | *VAPB* | -0.33937835 | 0.00325642 | 0.011330438 | 1 |
| T2DM_insulin leukocyte | *PARD6B* | -0.479375217 | 0.003975234 | 0.011330438 | 1 |
| T2DM_insulin leukocyte | *ARFGEF2* | -0.474831549 | 0.003168186 | 0.011330438 | 1 |
| T2DM_insulin leukocyte | *ZNFX1* | -1.165719555 | 0.004675276 | 0.011330438 | 1 |
| T2DM_insulin leukocyte | *CSE1L* | -0.48787321 | 0.001514475 | 0.011330438 | 1 |
| T2DM_insulin leukocyte | *BCAS4* | -0.281909711 | 0.004503436 | 0.011330438 | 1 |
| T2DM_insulin leukocyte | *KCNK15* | -0.333954554 | 0.011528292 | 0.017228854 | 1 |
| T2DM_insulin leukocyte | *PEPD* | -0.671067556 | 0.000592362 | 0.011330438 | 1 |
| T2DM_insulin leukocyte | *SNRNP27* | -0.353514202 | 0.005540481 | 0.011451105 | 1 |
| T2DM_insulin leukocyte | *USP22* | -0.447938179 | 0.003282037 | 0.011330438 | 1 |
| T2DM_insulin leukocyte | *BTN2A2* | -0.81287246 | 0.000625447 | 0.011330438 | 1 |
| T2DM_insulin leukocyte | *MRS2* | -0.622564826 | 0.00305012 | 0.011330438 | 1 |
| T2DM_insulin leukocyte | *SNRPC* | -0.405074226 | 0.002791477 | 0.011330438 | 1 |
| T2DM_insulin leukocyte | *SERPINB6* | -0.369966884 | 0.001857828 | 0.011330438 | 1 |

| | | | | | |
|---|---|---|---|---|---|
| T2DM_insulin leukocyte | *ABCC10* | -0.46405767 | 0.006229991 | 0.011570498 | 1 |
| T2DM_insulin leukocyte | *PEX6* | -0.353041439 | 0.008815927 | 0.014103329 | 1 |
| T2DM_insulin leukocyte | *NQO2* | -0.853726501 | 0.003303956 | 0.011330438 | 1 |
| T2DM_insulin leukocyte | *UNC5CL* | -0.357747384 | 0.004317224 | 0.011330438 | 1 |
| T2DM_insulin leukocyte | *MED20* | -0.551037578 | 0.007358277 | 0.01254259 | 1 |
| T2DM_insulin leukocyte | *GLO1* | -0.375863301 | 0.005841356 | 0.011570498 | 1 |
| T2DM_insulin leukocyte | *KCNK17* | -0.258492875 | 0.014877736 | 0.021110374 | 1 |
| T2DM_insulin leukocyte | *ATXN1* | -0.498466282 | 0.003408645 | 0.011330438 | 1 |
| T2DM_insulin leukocyte | *LRRFIP1* | -0.327422863 | 0.00193088 | 0.011330438 | 1 |
| T2DM_insulin leukocyte | *CXCL6* | -0.587494964 | 0.009685331 | 0.015106656 | 1 |
| T2DM_insulin leukocyte | *SH3TC1* | -0.399546304 | 0.005750525 | 0.011570498 | 1 |
| T2DM_insulin leukocyte | *DOK4* | -0.277734417 | 0.004903336 | 0.011330438 | 1 |
| T2DM_insulin leukocyte | *CLYBL* | -1.197767802 | 0.00015761 | 0.011330438 | 0.619248013 |
| T2DM_insulin leukocyte | *TM9SF2* | -0.338824071 | 0.003411338 | 0.011330438 | 1 |
| T2DM_insulin leukocyte | *KIF25* | -5.272867074 | 0.004210899 | 0.011330438 | 1 |
| T2DM_insulin leukocyte | *BMP4* | -0.885837979 | 0.002131073 | 0.011330438 | 1 |
| T2DM_insulin leukocyte | *GRK4* | -0.392857378 | 0.026803376 | 0.035115194 | 1 |
| T2DM_insulin leukocyte | *HS3ST3B1* | -0.858406489 | 0.006203871 | 0.011570498 | 1 |
| T2DM_insulin leukocyte | *MRPS7* | -0.318714237 | 0.003714411 | 0.011330438 | 1 |
| T2DM_insulin leukocyte | *GGA3* | -0.784447034 | 0.007927444 | 0.013147711 | 1 |
| T2DM_insulin leukocyte | *DDX31* | -0.697660692 | 0.00296073 | 0.011330438 | 1 |
| T2DM_insulin leukocyte | *PPDPF* | -0.286013842 | 0.00319833 | 0.011330438 | 1 |
| T2DM_insulin leukocyte | *CHCHD5* | -0.345398756 | 0.003244247 | 0.011330438 | 1 |
| T2DM_insulin leukocyte | *INSIG2* | -0.334334302 | 0.003164748 | 0.011330438 | 1 |
| T2DM_insulin leukocyte | *PSD4* | -0.538642515 | 0.003458532 | 0.011330438 | 1 |
| T2DM_insulin leukocyte | *CLPP* | -0.381408472 | 0.003171922 | 0.011330438 | 1 |
| T2DM_insulin leukocyte | *GPR108* | -0.547626658 | 0.002900775 | 0.011330438 | 1 |
| T2DM_insulin leukocyte | *SYMPK* | -0.627860011 | 0.01283694 | 0.018659392 | 1 |
| T2DM_insulin leukocyte | *NAPB* | -0.363476647 | 0.00337014 | 0.011330438 | 1 |
| T2DM_insulin leukocyte | *PSMF1* | -0.416483371 | 0.004008266 | 0.011330438 | 1 |
| T2DM_insulin leukocyte | *RBCK1* | -0.327938251 | 0.008163838 | 0.013387195 | 1 |
| T2DM_insulin leukocyte | *FLRT3* | -0.314730066 | 0.006423294 | 0.01170553 | 1 |
| T2DM_insulin leukocyte | *MGME1* | -1.38033372 | 0.007206758 | 0.012381002 | 1 |
| T2DM_insulin leukocyte | *ITPA* | -0.519091641 | 0.002655507 | 0.011330438 | 1 |
| T2DM_insulin leukocyte | *BANF2* | -5.098721679 | 0.006054316 | 0.011570498 | 1 |
| T2DM_insulin leukocyte | *MRPS26* | -0.319160447 | 0.004180701 | 0.011330438 | 1 |
| T2DM_insulin leukocyte | *ZNF436* | -0.982559711 | 0.001380938 | 0.011330438 | 1 |
| T2DM_insulin leukocyte | *TUBGCP3* | -0.474933467 | 0.003118181 | 0.011330438 | 1 |
| T2DM_insulin leukocyte | *MCF2L* | -0.288365353 | 0.004495895 | 0.011330438 | 1 |
| T2DM_insulin leukocyte | *IGFLR1* | 0.119541563 | 0.023284509 | 0.031043378 | 1 |
| T2DM_insulin leukocyte | *NR1D1* | -0.35160186 | 0.001959391 | 0.011330438 | 1 |
| T2DM_insulin leukocyte | *PRMT1* | -0.328937777 | 0.002195526 | 0.011330438 | 1 |
| T2DM_insulin leukocyte | *PRR12* | -0.359349352 | 0.010412468 | 0.01593712 | 1 |
| T2DM_insulin leukocyte | *FLRT1* | -0.85678716 | 0.021066873 | 0.028463461 | 1 |
| T2DM_insulin leukocyte | *ASL* | -0.397342354 | 0.004799229 | 0.011330438 | 1 |
| T2DM_insulin leukocyte | *WNK4* | -0.417818348 | 0.020930352 | 0.028317959 | 1 |
| T2DM_insulin leukocyte | *BECN1* | -0.398998228 | 0.00433658 | 0.011330438 | 1 |
| T2DM_insulin leukocyte | *PRKCG* | 0.950701884 | 0.033239223 | 0.042332871 | 1 |
| T2DM_insulin leukocyte | *TRAP1* | -0.319134384 | 0.004780363 | 0.011330438 | 1 |
| T2DM_insulin leukocyte | *ZNF384* | -0.479884117 | 0.003998137 | 0.011330438 | 1 |

| | | | | | |
|---|---|---|---|---|---|
| T2DM_insulin leukocyte | *RHOJ* | -0.630068225 | 0.005895611 | 0.011570498 | 1 |
| T2DM_insulin leukocyte | *L3HYPDH* | -0.366339994 | 0.031102258 | 0.039908809 | 1 |
| T2DM_insulin leukocyte | *HSPA2* | -0.606311968 | 0.006150867 | 0.011570498 | 1 |
| T2DM_insulin leukocyte | *ZBTB1* | -0.550525579 | 0.001689545 | 0.011330438 | 1 |
| T2DM_insulin leukocyte | *PLEKHG3* | -0.530594679 | 0.004955695 | 0.011330438 | 1 |
| T2DM_insulin leukocyte | *FGD3* | 1.021883191 | 0.023757581 | 0.031609731 | 1 |
| T2DM_insulin leukocyte | *BCL11B* | 0.631671749 | 0.018268081 | 0.025166652 | 1 |
| T2DM_insulin leukocyte | *ABHD8* | -0.925703557 | 0.002155881 | 0.011330438 | 1 |
| T2DM_insulin leukocyte | *BEST3* | -0.760517777 | 0.030801373 | 0.039574426 | 1 |
| T2DM_insulin leukocyte | *YEATS4* | -0.349337792 | 0.004957622 | 0.011330438 | 1 |
| T2DM_insulin leukocyte | *LRRC61* | -0.568218002 | 0.004270228 | 0.011330438 | 1 |
| T2DM_insulin leukocyte | *IDUA* | -0.373064325 | 0.006114467 | 0.011570498 | 1 |
| T2DM_insulin leukocyte | *FGFRL1* | -0.342963138 | 0.009010104 | 0.014309095 | 1 |
| T2DM_insulin leukocyte | *EMC1* | -0.440724866 | 0.004060208 | 0.011330438 | 1 |
| T2DM_insulin leukocyte | *UBR4* | -0.405708391 | 0.004318864 | 0.011330438 | 1 |
| T2DM_insulin leukocyte | *SYNGR3* | -0.449693221 | 0.006543672 | 0.011804448 | 1 |
| T2DM_insulin leukocyte | *WFIKKN1* | -0.369304345 | 0.011176563 | 0.016816769 | 1 |
| T2DM_insulin leukocyte | *KDM4B* | -0.850898529 | 0.00359064 | 0.011330438 | 1 |
| T2DM_insulin leukocyte | *PNKD* | -0.486590895 | 0.002811901 | 0.011330438 | 1 |
| T2DM_insulin leukocyte | *PTPN12* | -0.484782356 | 0.000268045 | 0.011330438 | 1 |
| T2DM_insulin leukocyte | *POR* | -0.399903185 | 0.004600543 | 0.011330438 | 1 |
| T2DM_insulin leukocyte | *RBM48* | -0.720133033 | 0.002759561 | 0.011330438 | 1 |
| T2DM_insulin leukocyte | *SRD5A3* | -0.456030836 | 0.007756461 | 0.012968143 | 1 |
| T2DM_insulin leukocyte | *PPAT* | -0.438748381 | 0.004644202 | 0.011330438 | 1 |
| T2DM_insulin leukocyte | *A4GALT* | -0.33821976 | 0.005907386 | 0.011570498 | 1 |
| T2DM_insulin leukocyte | *NAA38* | -0.315144601 | 0.003511521 | 0.011330438 | 1 |
| T2DM_insulin leukocyte | *PRKRIP1* | -0.366345019 | 0.00444645 | 0.011330438 | 1 |
| T2DM_insulin leukocyte | *VGF* | -0.203311552 | 0.003054782 | 0.011330438 | 1 |
| T2DM_insulin leukocyte | *IRF5* | 0.143334133 | 0.025986728 | 0.0341989 | 1 |
| T2DM_insulin leukocyte | *MTX2* | -0.544656551 | 0.002773786 | 0.011330438 | 1 |
| T2DM_insulin leukocyte | *GAD1* | -1.356648176 | 0.004896973 | 0.011330438 | 1 |
| T2DM_insulin leukocyte | *ORMDL1* | -0.335273803 | 0.007997089 | 0.013211473 | 1 |
| T2DM_insulin leukocyte | *PSMG2* | -0.443020841 | 0.002470116 | 0.011330438 | 1 |
| T2DM_insulin leukocyte | *TWSG1* | -0.587870167 | 0.002388933 | 0.011330438 | 1 |
| T2DM_insulin leukocyte | *EIF2AK4* | -0.540638185 | 0.003365563 | 0.011330438 | 1 |
| T2DM_insulin leukocyte | *MYO5C* | -0.264037461 | 0.00599231 | 0.011570498 | 1 |
| T2DM_insulin leukocyte | *CGNL1* | -0.483822533 | 0.001192046 | 0.011330438 | 1 |
| T2DM_insulin leukocyte | *TMOD2* | -0.428651094 | 0.003944196 | 0.011330438 | 1 |
| T2DM_insulin leukocyte | *IVD* | -0.548399866 | 0.003407245 | 0.011330438 | 1 |
| T2DM_insulin leukocyte | *CHAC1* | -0.467771354 | 0.002927055 | 0.011330438 | 1 |
| T2DM_insulin leukocyte | *ANAPC13* | -0.248066463 | 0.007179136 | 0.012365991 | 1 |
| T2DM_insulin leukocyte | *SUMF2* | -0.446521145 | 0.002714053 | 0.011330438 | 1 |
| T2DM_insulin leukocyte | *TPH1* | -1.444369607 | 0.002520195 | 0.011330438 | 1 |
| T2DM_insulin leukocyte | *DCTD* | -0.457009947 | 0.003573072 | 0.011330438 | 1 |
| T2DM_insulin leukocyte | *KRI1* | -0.261573814 | 0.022685113 | 0.030347228 | 1 |
| T2DM_insulin leukocyte | *AP1M2* | -0.340618937 | 0.004444643 | 0.011330438 | 1 |
| T2DM_insulin leukocyte | *CDKN2D* | -0.367765362 | 0.002642567 | 0.011330438 | 1 |
| T2DM_insulin leukocyte | *MTUS1* | -0.317171277 | 0.004401639 | 0.011330438 | 1 |
| T2DM_insulin leukocyte | *ADCY4* | -0.400538554 | 0.007387544 | 0.012565211 | 1 |
| T2DM_insulin leukocyte | *DTD2* | -0.32106972 | 0.007048225 | 0.012237751 | 1 |

| | | | | | |
|---|---|---|---|---|---|
| T2DM_insulin leukocyte | *HEATR5A* | -0.771170033 | 0.002929405 | 0.011330438 | 1 |
| T2DM_insulin leukocyte | *RNASE1* | 0.062154081 | 0.018472137 | 0.025394341 | 1 |
| T2DM_insulin leukocyte | *EPB41L4A* | -0.429992576 | 0.005981001 | 0.011570498 | 1 |
| T2DM_insulin leukocyte | *REEP5* | -0.315659916 | 0.003589701 | 0.011330438 | 1 |
| T2DM_insulin leukocyte | *MAU2* | -1.077235628 | 9.41E-05 | 0.011330438 | 0.369891368 |
| T2DM_insulin leukocyte | *SHC2* | -0.428345819 | 0.003520295 | 0.011330438 | 1 |
| T2DM_insulin leukocyte | *SYT5* | -0.223847386 | 0.016050115 | 0.02251371 | 1 |
| T2DM_insulin leukocyte | *CBFA2T3* | -0.366589701 | 0.005862513 | 0.011570498 | 1 |
| T2DM_insulin leukocyte | *ERMARD* | -0.54999317 | 0.030929426 | 0.039725961 | 1 |
| T2DM_insulin leukocyte | *PHF10* | -0.398723843 | 0.003011108 | 0.011330438 | 1 |
| T2DM_insulin leukocyte | *SH3BP4* | -0.221804119 | 0.008312321 | 0.01352344 | 1 |
| T2DM_insulin leukocyte | *DOCK6* | -0.282881454 | 0.007664417 | 0.012869015 | 1 |
| T2DM_insulin leukocyte | *ECSIT* | -0.443679075 | 0.004132441 | 0.011330438 | 1 |
| T2DM_insulin leukocyte | *CDC16* | -0.567254805 | 0.001302723 | 0.011330438 | 1 |
| T2DM_insulin leukocyte | *DPP6* | -0.203667962 | 0.005640229 | 0.01154792 | 1 |
| T2DM_insulin leukocyte | *GTPBP3* | -0.460070193 | 0.000945288 | 0.011330438 | 1 |
| T2DM_insulin leukocyte | *PLVAP* | -0.202163067 | 0.003708947 | 0.011330438 | 1 |
| T2DM_insulin leukocyte | *MRPL34* | -0.227611962 | 0.035817005 | 0.045274858 | 1 |
| T2DM_insulin leukocyte | *LSM7* | -0.283732008 | 0.003343354 | 0.011330438 | 1 |
| T2DM_insulin leukocyte | *TULP4* | -0.407568552 | 0.002629794 | 0.011330438 | 1 |
| T2DM_insulin leukocyte | *QRSL1* | -0.423071105 | 0.004761364 | 0.011330438 | 1 |
| T2DM_insulin leukocyte | *RSPH3* | -0.416396678 | 0.006816704 | 0.012014984 | 1 |
| T2DM_insulin leukocyte | *STK33* | -0.352455982 | 0.001728792 | 0.011330438 | 1 |
| T2DM_insulin leukocyte | *NDUFA10* | -0.404452917 | 0.001563668 | 0.011330438 | 1 |
| T2DM_insulin leukocyte | *TRPM4* | -1.519167353 | 0.007470252 | 0.012656584 | 1 |
| T2DM_insulin leukocyte | *UBAC1* | -0.363622482 | 0.004774087 | 0.011330438 | 1 |
| T2DM_insulin leukocyte | *ATXN10* | -0.327098546 | 0.003096744 | 0.011330438 | 1 |
| T2DM_insulin leukocyte | *PNPLA7* | -0.748160347 | 0.004849545 | 0.011330438 | 1 |
| T2DM_insulin leukocyte | *PAK4* | -0.479206241 | 0.003808582 | 0.011330438 | 1 |
| T2DM_insulin leukocyte | *CEP85* | 0.545672407 | 0.017469477 | 0.024193717 | 1 |
| T2DM_insulin leukocyte | *LAMA5* | -0.267459201 | 0.004613154 | 0.011330438 | 1 |
| T2DM_insulin leukocyte | *ADRM1* | -0.364085043 | 0.003189914 | 0.011330438 | 1 |
| T2DM_insulin leukocyte | *UCK1* | -0.691716263 | 0.001875758 | 0.011330438 | 1 |
| T2DM_insulin leukocyte | *MAP3K10* | -0.807444544 | 0.006163438 | 0.011570498 | 1 |
| T2DM_insulin leukocyte | *SMPDL3B* | -0.313097247 | 0.007787316 | 0.013008659 | 1 |
| T2DM_insulin leukocyte | *CLIP1* | -0.409995895 | 0.006901894 | 0.012100405 | 1 |
| T2DM_insulin leukocyte | *ZNF317* | -0.511434518 | 0.004225063 | 0.011330438 | 1 |
| T2DM_insulin leukocyte | *DNMT1* | -0.600002564 | 0.004102409 | 0.011330438 | 1 |
| T2DM_insulin leukocyte | *ZNF426* | -0.588792613 | 0.001355992 | 0.011330438 | 1 |
| T2DM_insulin leukocyte | *LRP3* | -0.597516 | 0.00461557 | 0.011330438 | 1 |
| T2DM_insulin leukocyte | *HSD17B3* | -4.534844037 | 0.006935014 | 0.012128132 | 1 |
| T2DM_insulin leukocyte | *SYNE1* | -0.34044098 | 0.003667824 | 0.011330438 | 1 |
| T2DM_insulin leukocyte | *EPS8L1* | -1.269865257 | 0.002827863 | 0.011330438 | 1 |
| T2DM_insulin leukocyte | *C1QL1* | -0.169651198 | 0.006722703 | 0.011957221 | 1 |
| T2DM_insulin leukocyte | *F12* | -0.355426797 | 0.029949931 | 0.038682866 | 1 |
| T2DM_insulin leukocyte | *TRAF3* | -0.266252171 | 0.01974129 | 0.026913091 | 1 |
| T2DM_insulin leukocyte | *PDLIM4* | -0.428832153 | 0.003516124 | 0.011330438 | 1 |
| T2DM_insulin leukocyte | *DIAPH1* | -0.292952095 | 0.00468578 | 0.011330438 | 1 |
| T2DM_insulin leukocyte | *EXOC4* | -0.809857728 | 0.00211187 | 0.011330438 | 1 |
| T2DM_insulin leukocyte | *C1orf159* | -1.086820815 | 0.031954617 | 0.040882347 | 1 |

| | | | | | |
|---|---|---|---|---|---|
| T2DM_insulin leukocyte | *ANO1* | -0.933759052 | 0.004468508 | 0.011330438 | 1 |
| T2DM_insulin leukocyte | *PPFIA1* | -0.339652999 | 0.004342465 | 0.011330438 | 1 |
| T2DM_insulin leukocyte | *TRAF7* | -0.516572582 | 0.002919711 | 0.011330438 | 1 |
| T2DM_insulin leukocyte | *MAP1B* | -0.263598087 | 0.002172447 | 0.011330438 | 1 |
| T2DM_insulin leukocyte | *CHD1L* | -0.869362457 | 0.004340794 | 0.011330438 | 1 |
| T2DM_insulin leukocyte | *PIAS3* | -0.269650296 | 0.013114871 | 0.018958178 | 1 |
| T2DM_insulin leukocyte | *RBM8A* | -0.256438367 | 0.005425419 | 0.011421159 | 1 |
| T2DM_insulin leukocyte | *ZNF304* | -1.650069347 | 0.002277186 | 0.011330438 | 1 |
| T2DM_insulin leukocyte | *RHPN2* | -0.403728365 | 0.002034698 | 0.011330438 | 1 |
| T2DM_insulin leukocyte | *GCH1* | -0.26819332 | 0.005733562 | 0.011570498 | 1 |
| T2DM_insulin leukocyte | *CC2D1A* | -0.511001003 | 0.005102516 | 0.011330438 | 1 |
| T2DM_insulin leukocyte | *CCT6B* | -0.573575026 | 0.006293454 | 0.011592584 | 1 |
| T2DM_insulin leukocyte | *PPARG* | -0.658276665 | 0.002714241 | 0.011330438 | 1 |
| T2DM_insulin leukocyte | *ENOSF1* | -0.962308309 | 0.002099959 | 0.011330438 | 1 |
| T2DM_insulin leukocyte | *IMMT* | -0.440949359 | 0.001806589 | 0.011330438 | 1 |
| T2DM_insulin leukocyte | *MRPL35* | -0.435602041 | 0.002805897 | 0.011330438 | 1 |
| T2DM_insulin leukocyte | *SCLY* | -0.641221303 | 0.00643377 | 0.011719185 | 1 |
| T2DM_insulin leukocyte | *CARD6* | -0.746216405 | 0.004668734 | 0.011330438 | 1 |
| T2DM_insulin leukocyte | *INPP5K* | -0.372965895 | 0.012987208 | 0.01881517 | 1 |
| T2DM_insulin leukocyte | *RPA1* | -0.434355233 | 0.008528351 | 0.013783584 | 1 |
| T2DM_insulin leukocyte | *TMEM128* | -0.582476438 | 0.002900371 | 0.011330438 | 1 |
| T2DM_insulin leukocyte | *SEC61G* | -0.300120457 | 0.002327158 | 0.011330438 | 1 |
| T2DM_insulin leukocyte | *FIGNL1* | -0.960158249 | 0.025321335 | 0.033452429 | 1 |
| T2DM_insulin leukocyte | *ITGB4* | -0.754416607 | 0.01176968 | 0.017542896 | 1 |
| T2DM_insulin leukocyte | *TRIM47* | -0.407693342 | 0.002173653 | 0.011330438 | 1 |
| T2DM_insulin leukocyte | *EIF5A* | -0.332492101 | 0.00258145 | 0.011330438 | 1 |
| T2DM_insulin leukocyte | *PRMT7* | -0.744146098 | 0.016961867 | 0.023640715 | 1 |
| T2DM_insulin leukocyte | *ANKEF1* | -0.324582005 | 0.005909121 | 0.011570498 | 1 |
| T2DM_insulin leukocyte | *BTBD3* | -0.315124106 | 0.004395696 | 0.011330438 | 1 |
| T2DM_insulin leukocyte | *LPIN3* | -0.469895722 | 0.025190359 | 0.03330179 | 1 |
| T2DM_insulin leukocyte | *RBM38* | -0.674710704 | 0.000814609 | 0.011330438 | 1 |
| T2DM_insulin leukocyte | *VSTM2L* | -0.332854922 | 0.004041079 | 0.011330438 | 1 |
| T2DM_insulin leukocyte | *SERINC3* | -0.338895006 | 0.008637826 | 0.013903325 | 1 |
| T2DM_insulin leukocyte | *BHMT2* | -0.525155982 | 0.001012806 | 0.011330438 | 1 |
| T2DM_insulin leukocyte | *WASF3* | -0.385122884 | 0.007066757 | 0.012258405 | 1 |
| T2DM_insulin leukocyte | *SCO1* | -0.625331706 | 0.006249616 | 0.011570498 | 1 |
| T2DM_insulin leukocyte | *CHI3L1* | -0.281779882 | 0.03176648 | 0.040694652 | 1 |
| T2DM_insulin leukocyte | *SLC41A1* | -0.440309069 | 0.004035272 | 0.011330438 | 1 |
| T2DM_insulin leukocyte | *TRPC4* | -1.111157428 | 0.00472902 | 0.011330438 | 1 |
| T2DM_insulin leukocyte | *POSTN* | 0.347994626 | 0.017127185 | 0.023820429 | 1 |
| T2DM_insulin leukocyte | *RFC3* | -0.313325346 | 0.00616235 | 0.011570498 | 1 |
| T2DM_insulin leukocyte | *EPHB2* | -1.327843221 | 0.011229491 | 0.016859255 | 1 |
| T2DM_insulin leukocyte | *BTBD2* | -0.879816418 | 0.002545173 | 0.011330438 | 1 |
| T2DM_insulin leukocyte | *CNDP2* | -0.396045892 | 0.003386465 | 0.011330438 | 1 |
| T2DM_insulin leukocyte | *WDR74* | -0.606977654 | 0.002321278 | 0.011330438 | 1 |
| T2DM_insulin leukocyte | *GSTT2B* | -0.349491762 | 0.006508568 | 0.011778978 | 1 |
| T2DM_insulin leukocyte | *SLC2A11* | -0.599059618 | 0.004083623 | 0.011330438 | 1 |
| T2DM_insulin leukocyte | *C1QTNF6* | -0.543559644 | 0.015238676 | 0.021555956 | 1 |
| T2DM_insulin leukocyte | *FAM83F* | -0.419867442 | 0.003840291 | 0.011330438 | 1 |
| T2DM_insulin leukocyte | *GIMAP6* | -0.767142854 | 0.002464942 | 0.011330438 | 1 |

| | | | | | |
|---|---|---|---|---|---|
| T2DM_insulin leukocyte | *AGAP3* | -1.196193841 | 0.003022483 | 0.011330438 | 1 |
| T2DM_insulin leukocyte | *SFTPD* | -0.353802367 | 0.014889157 | 0.021118953 | 1 |
| T2DM_insulin leukocyte | *IMPA1* | -0.365586117 | 0.003818653 | 0.011330438 | 1 |
| T2DM_insulin leukocyte | *LRRCC1* | -0.569551189 | 0.006811495 | 0.012014984 | 1 |
| T2DM_insulin leukocyte | *RRAS2* | -0.336719728 | 0.003800791 | 0.011330438 | 1 |
| T2DM_insulin leukocyte | *HSD17B4* | -0.706709728 | 0.001629765 | 0.011330438 | 1 |
| T2DM_insulin leukocyte | *COX16* | -0.374288031 | 0.002888887 | 0.011330438 | 1 |
| T2DM_insulin leukocyte | *ELP3* | -0.794547381 | 0.002850236 | 0.011330438 | 1 |
| T2DM_insulin leukocyte | *PEBP4* | -0.563381385 | 0.00335523 | 0.011330438 | 1 |
| T2DM_insulin leukocyte | *MRO* | -1.079605091 | 0.003523045 | 0.011330438 | 1 |
| T2DM_insulin leukocyte | *MRPS36* | -0.373887019 | 0.009559814 | 0.014994215 | 1 |
| T2DM_insulin leukocyte | *THUMPD3* | -0.444920676 | 0.002015358 | 0.011330438 | 1 |
| T2DM_insulin leukocyte | *CHL1* | -0.37423793 | 0.0214443 | 0.028923671 | 1 |
| T2DM_insulin leukocyte | *REG4* | -0.433082732 | 0.005322814 | 0.011421159 | 1 |
| T2DM_insulin leukocyte | *GSTM3* | -0.285062341 | 0.004361806 | 0.011330438 | 1 |
| T2DM_insulin leukocyte | *SORT1* | -0.487596438 | 0.003041003 | 0.011330438 | 1 |
| T2DM_insulin leukocyte | *NOTCH2* | -0.766084709 | 2.42E-05 | 0.011330438 | 0.095227714 |
| T2DM_insulin leukocyte | *CEPT1* | -0.690500975 | 0.002501193 | 0.011330438 | 1 |
| T2DM_insulin leukocyte | *NAPG* | -0.387882753 | 0.00399114 | 0.011330438 | 1 |
| T2DM_insulin leukocyte | *SPIRE1* | -0.406532479 | 0.003074123 | 0.011330438 | 1 |
| T2DM_insulin leukocyte | *FKBP11* | -0.219772988 | 0.007965941 | 0.013183733 | 1 |
| T2DM_insulin leukocyte | *KIDINS220* | -0.415401421 | 0.003076985 | 0.011330438 | 1 |
| T2DM_insulin leukocyte | *LPIN1* | -0.291712591 | 0.003723416 | 0.011330438 | 1 |
| T2DM_insulin leukocyte | *IAH1* | -0.505318873 | 0.002716242 | 0.011330438 | 1 |
| T2DM_insulin leukocyte | *ANO3* | 0.592814456 | 0.015012335 | 0.021278306 | 1 |
| T2DM_insulin leukocyte | *IL2RA* | 0.118918448 | 0.014427105 | 0.020582461 | 1 |
| T2DM_insulin leukocyte | *ECHDC3* | -0.393037512 | 0.003148162 | 0.011330438 | 1 |
| T2DM_insulin leukocyte | *TMEM241* | -0.694565364 | 0.003515808 | 0.011330438 | 1 |
| T2DM_insulin leukocyte | *AGO4* | -0.766439522 | 0.001116769 | 0.011330438 | 1 |
| T2DM_insulin leukocyte | *PRPF38A* | -0.383107991 | 0.004078004 | 0.011330438 | 1 |
| T2DM_insulin leukocyte | *DSC2* | -0.878225244 | 0.028401949 | 0.036901871 | 1 |
| T2DM_insulin leukocyte | *TPGS2* | -0.43656694 | 0.006691385 | 0.011938173 | 1 |
| T2DM_insulin leukocyte | *DAGLA* | -0.408026623 | 0.011952944 | 0.017706833 | 1 |
| T2DM_insulin leukocyte | *TIMM10* | -0.387563225 | 0.004236374 | 0.011330438 | 1 |
| T2DM_insulin leukocyte | *FADS2* | -0.450499275 | 0.003674852 | 0.011330438 | 1 |
| T2DM_insulin leukocyte | *TMEM165* | -0.323311856 | 0.003343971 | 0.011330438 | 1 |
| T2DM_insulin leukocyte | *CLOCK* | -0.425902358 | 0.006256363 | 0.011570498 | 1 |
| T2DM_insulin leukocyte | *GGACT* | -0.621475297 | 0.005059132 | 0.011330438 | 1 |
| T2DM_insulin leukocyte | *CLDN10* | -0.144558423 | 0.02603377 | 0.034243952 | 1 |
| T2DM_insulin leukocyte | *CARS2* | -0.3683072 | 0.004625477 | 0.011330438 | 1 |
| T2DM_insulin leukocyte | *TMED7* | -0.34812527 | 0.003244743 | 0.011330438 | 1 |
| T2DM_insulin leukocyte | *NREP* | -1.897873876 | 0.006307309 | 0.011612661 | 1 |
| T2DM_insulin leukocyte | *WDR36* | -0.406576318 | 0.004443411 | 0.011330438 | 1 |
| T2DM_insulin leukocyte | *OSTF1* | -0.535750744 | 0.004674836 | 0.011330438 | 1 |
| T2DM_insulin leukocyte | *GOLM1* | -0.326857238 | 0.001539842 | 0.011330438 | 1 |
| T2DM_insulin leukocyte | *SDS* | 0.025387089 | 0.013673725 | 0.019657544 | 1 |
| T2DM_insulin leukocyte | *MSI1* | -0.674591565 | 0.003545608 | 0.011330438 | 1 |
| T2DM_insulin leukocyte | *DTX1* | -0.974732692 | 0.004151366 | 0.011330438 | 1 |
| T2DM_insulin leukocyte | *PNPLA8* | -0.319043537 | 0.002829205 | 0.011330438 | 1 |
| T2DM_insulin leukocyte | *TES* | -0.263718194 | 0.003824123 | 0.011330438 | 1 |

| | | | | | |
|---|---|---|---|---|---|
| T2DM_insulin leukocyte | *MDFIC* | -0.71540406 | 0.03359747 | 0.04273372 | 1 |
| T2DM_insulin leukocyte | *MTO1* | -0.362603838 | 0.005090053 | 0.011330438 | 1 |
| T2DM_insulin leukocyte | *LCA5* | -1.037010293 | 0.002112914 | 0.011330438 | 1 |
| T2DM_insulin leukocyte | *LMO2* | -0.340416279 | 0.004768332 | 0.011330438 | 1 |
| T2DM_insulin leukocyte | *RDH5* | -0.32751783 | 0.00657785 | 0.011838925 | 1 |
| T2DM_insulin leukocyte | *TFCP2* | -1.142994346 | 0.002283225 | 0.011330438 | 1 |
| T2DM_insulin leukocyte | *KRT7* | -0.175195634 | 0.008462539 | 0.013705406 | 1 |
| T2DM_insulin leukocyte | *NHSL1* | -0.470368547 | 0.011012152 | 0.016647459 | 1 |
| T2DM_insulin leukocyte | *AHI1* | -0.297325335 | 0.001889416 | 0.011330438 | 1 |
| T2DM_insulin leukocyte | *HEY2* | -0.646977484 | 0.001075833 | 0.011330438 | 1 |
| T2DM_insulin leukocyte | *PKIB* | -0.272094654 | 0.004288856 | 0.011330438 | 1 |
| T2DM_insulin leukocyte | *SEMA4F* | -0.542124082 | 0.004355462 | 0.011330438 | 1 |
| T2DM_insulin leukocyte | *GNS* | -0.707179048 | 0.002605503 | 0.011330438 | 1 |
| T2DM_insulin leukocyte | *MPHOSPH6* | -0.329121292 | 0.007001948 | 0.012199846 | 1 |
| T2DM_insulin leukocyte | *AGT* | -0.257102949 | 0.003302228 | 0.011330438 | 1 |
| T2DM_insulin leukocyte | *NTPCR* | -0.523162607 | 0.003270314 | 0.011330438 | 1 |
| T2DM_insulin leukocyte | *USP37* | -0.479721434 | 0.005657624 | 0.011549238 | 1 |
| T2DM_insulin leukocyte | *ARMC9* | -1.365597458 | 0.030514547 | 0.039295856 | 1 |
| T2DM_insulin leukocyte | *COX5B* | -0.292675514 | 0.004715011 | 0.011330438 | 1 |
| T2DM_insulin leukocyte | *REV1* | -0.32598141 | 0.009509178 | 0.014932678 | 1 |
| T2DM_insulin leukocyte | *TSGA10* | -0.328360302 | 0.027766156 | 0.036219531 | 1 |
| T2DM_insulin leukocyte | *TGFBRAP1* | -0.459983515 | 0.002824319 | 0.011330438 | 1 |
| T2DM_insulin leukocyte | *GCC2* | -0.432383327 | 0.0023955 | 0.011330438 | 1 |
| T2DM_insulin leukocyte | *MRPS9* | -0.344589972 | 0.003689397 | 0.011330438 | 1 |
| T2DM_insulin leukocyte | *ANKRD36* | -0.507349542 | 0.004384686 | 0.011330438 | 1 |
| T2DM_insulin leukocyte | *USP44* | -0.483297672 | 0.00887283 | 0.014148275 | 1 |
| T2DM_insulin leukocyte | *PLXNC1* | 0.635493624 | 0.032715616 | 0.041747209 | 1 |
| T2DM_insulin leukocyte | *DRAM1* | -0.750225035 | 0.002543788 | 0.011330438 | 1 |
| T2DM_insulin leukocyte | *VILL* | 0.493857891 | 0.020642988 | 0.027996651 | 1 |
| T2DM_insulin leukocyte | *FLNB* | -0.301386456 | 0.008806255 | 0.014103329 | 1 |
| T2DM_insulin leukocyte | *VPS36* | -0.598350726 | 0.002325649 | 0.011330438 | 1 |
| T2DM_insulin leukocyte | *RNASEH2B* | -0.733557393 | 0.002779114 | 0.011330438 | 1 |
| T2DM_insulin leukocyte | *CKAP2* | -0.362436995 | 0.008600709 | 0.013849257 | 1 |
| T2DM_insulin leukocyte | *TBC1D4* | -0.413277697 | 0.003484653 | 0.011330438 | 1 |
| T2DM_insulin leukocyte | *SUCLA2* | -0.449080959 | 0.002923286 | 0.011330438 | 1 |
| T2DM_insulin leukocyte | *RCBTB1* | -0.49847322 | 0.018451713 | 0.025375143 | 1 |
| T2DM_insulin leukocyte | *LMO7* | -0.218385796 | 0.005216612 | 0.011389288 | 1 |
| T2DM_insulin leukocyte | *C7orf25* | -0.377391508 | 0.001512846 | 0.011330438 | 1 |
| T2DM_insulin leukocyte | *RAPGEF5* | -0.266683198 | 0.006085441 | 0.011570498 | 1 |
| T2DM_insulin leukocyte | *KDELR2* | -0.347685328 | 0.003016845 | 0.011330438 | 1 |
| T2DM_insulin leukocyte | *TBRG4* | -0.802345626 | 0.002859048 | 0.011330438 | 1 |
| T2DM_insulin leukocyte | *DBNL* | -1.095741113 | 0.00258868 | 0.011330438 | 1 |
| T2DM_insulin leukocyte | *ALPK3* | -0.280066325 | 0.004625155 | 0.011330438 | 1 |
| T2DM_insulin leukocyte | *NMT1* | -0.30917423 | 0.006598289 | 0.011859412 | 1 |
| T2DM_insulin leukocyte | *MYCBPAP* | -0.575550068 | 0.039628049 | 0.049648789 | 1 |
| T2DM_insulin leukocyte | *TEX2* | -0.268655234 | 0.006392535 | 0.011698309 | 1 |
| T2DM_insulin leukocyte | *LIMD2* | 0.265091449 | 0.026743834 | 0.035059586 | 1 |
| T2DM_insulin leukocyte | *MRPL47* | -0.370592648 | 0.002723549 | 0.011330438 | 1 |
| T2DM_insulin leukocyte | *SMPD4* | -0.889909515 | 0.002575331 | 0.011330438 | 1 |
| T2DM_insulin leukocyte | *CCDC115* | -0.394118162 | 0.006816675 | 0.012014984 | 1 |

| | | | | | | |
|---|---|---|---|---|---|---|
| T2DM_insulin | leukocyte | *BIN1* | -0.785506398 | 0.008861539 | 0.014141749 | 1 |
| T2DM_insulin | leukocyte | *HS6ST1* | -0.559616083 | 0.003107936 | 0.011330438 | 1 |
| T2DM_insulin | leukocyte | *NIPSNAP3A* | -0.555406729 | 0.002967832 | 0.011330438 | 1 |
| T2DM_insulin | leukocyte | *CDK9* | -0.455354587 | 0.003572433 | 0.011330438 | 1 |
| T2DM_insulin | leukocyte | *TXN* | -0.275863325 | 0.003333812 | 0.011330438 | 1 |
| T2DM_insulin | leukocyte | *TOR1B* | -0.602072565 | 0.004833289 | 0.011330438 | 1 |
| T2DM_insulin | leukocyte | *TOR1A* | -0.357055208 | 0.004668762 | 0.011330438 | 1 |
| T2DM_insulin | leukocyte | *SLC2A8* | -0.408741844 | 0.00441254 | 0.011330438 | 1 |
| T2DM_insulin | leukocyte | *ZFP37* | -0.333804867 | 0.00866032 | 0.013926501 | 1 |
| T2DM_insulin | leukocyte | *STX17* | -0.3567617 | 0.00500898 | 0.011330438 | 1 |
| T2DM_insulin | leukocyte | *PRPF4* | -0.48139405 | 0.007117869 | 0.012309025 | 1 |
| T2DM_insulin | leukocyte | *FPGS* | -0.485935046 | 0.003857486 | 0.011330438 | 1 |
| T2DM_insulin | leukocyte | *ATP6V1G1* | -0.285256648 | 0.004094072 | 0.011330438 | 1 |
| T2DM_insulin | leukocyte | *ARPC5L* | -0.292173419 | 0.003925076 | 0.011330438 | 1 |
| T2DM_insulin | leukocyte | *POLR1E* | -0.240808026 | 0.013264829 | 0.019146772 | 1 |
| T2DM_insulin | leukocyte | *APTX* | -0.333030693 | 0.003156488 | 0.011330438 | 1 |
| T2DM_insulin | leukocyte | *CD72* | -0.619999849 | 0.015273151 | 0.021585687 | 1 |
| T2DM_insulin | leukocyte | *TMEM8B* | -0.510157466 | 0.003207138 | 0.011330438 | 1 |
| T2DM_insulin | leukocyte | *GRHPR* | -0.445081857 | 0.000491958 | 0.011330438 | 1 |
| T2DM_insulin | leukocyte | *ALDH1B1* | -0.735668343 | 0.004776262 | 0.011330438 | 1 |
| T2DM_insulin | leukocyte | *ARHGEF39* | -1.139369099 | 0.021956068 | 0.029522721 | 1 |
| T2DM_insulin | leukocyte | *PPIL1* | -0.891844959 | 0.002514068 | 0.011330438 | 1 |
| T2DM_insulin | leukocyte | *GMPR* | -0.624206987 | 0.005327502 | 0.011421159 | 1 |
| T2DM_insulin | leukocyte | *KIAA0319* | -0.395210578 | 0.005968343 | 0.011570498 | 1 |
| T2DM_insulin | leukocyte | *LRRC1* | -0.356427604 | 0.004148258 | 0.011330438 | 1 |
| T2DM_insulin | leukocyte | *RIPK1* | -0.385587187 | 0.004692661 | 0.011330438 | 1 |
| T2DM_insulin | leukocyte | *TUBB2B* | -0.298414965 | 0.008134719 | 0.013356168 | 1 |
| T2DM_insulin | leukocyte | *UQCC2* | -0.315162148 | 0.013077567 | 0.018918174 | 1 |
| T2DM_insulin | leukocyte | *HMGA1* | -0.275820064 | 0.003717152 | 0.011330438 | 1 |
| T2DM_insulin | leukocyte | *TCF19* | 0.317902931 | 0.011168282 | 0.016816769 | 1 |
| T2DM_insulin | leukocyte | *CLPS* | -0.121904259 | 0.025112447 | 0.033243532 | 1 |
| T2DM_insulin | leukocyte | *MTCH1* | -0.344050931 | 0.002706634 | 0.011330438 | 1 |
| T2DM_insulin | leukocyte | *FCHSD2* | 0.570944103 | 0.022523923 | 0.030162404 | 1 |
| T2DM_insulin | leukocyte | *ANKRD42* | -0.547880032 | 0.003497671 | 0.011330438 | 1 |
| T2DM_insulin | leukocyte | *CREBZF* | -0.498348257 | 0.003149901 | 0.011330438 | 1 |
| T2DM_insulin | leukocyte | *PRCP* | -0.405829832 | 0.02483244 | 0.032906123 | 1 |
| T2DM_insulin | leukocyte | *NARS2* | -0.446197612 | 0.005466208 | 0.011427973 | 1 |
| T2DM_insulin | leukocyte | *GGH* | -0.592158012 | 0.000428553 | 0.011330438 | 1 |
| T2DM_insulin | leukocyte | *SDCBP* | -0.515815086 | 0.010931157 | 0.016550349 | 1 |
| T2DM_insulin | leukocyte | *NEK1* | -0.594917839 | 0.022333153 | 0.02993755 | 1 |
| T2DM_insulin | leukocyte | *TRPC6* | -0.961907664 | 0.01010778 | 0.015598378 | 1 |
| T2DM_insulin | leukocyte | *MMP7* | -0.150094737 | 0.010655114 | 0.016227482 | 1 |
| T2DM_insulin | leukocyte | *FDX1* | -0.388497415 | 0.002850786 | 0.011330438 | 1 |
| T2DM_insulin | leukocyte | *C11orf1* | -0.360914658 | 0.005261647 | 0.011402654 | 1 |
| T2DM_insulin | leukocyte | *FXYD2* | -0.219543804 | 0.00248926 | 0.011330438 | 1 |
| T2DM_insulin | leukocyte | *CASP1* | 0.137783213 | 0.017037806 | 0.023738134 | 1 |
| T2DM_insulin | leukocyte | *ALKBH8* | -0.515806042 | 0.00574126 | 0.011570498 | 1 |
| T2DM_insulin | leukocyte | *TUBGCP4* | -0.625409348 | 0.002152186 | 0.011330438 | 1 |
| T2DM_insulin | leukocyte | *UACA* | -0.301423575 | 0.011105119 | 0.016749333 | 1 |
| T2DM_insulin | leukocyte | *PLCB2* | 0.212691315 | 0.020510355 | 0.027845607 | 1 |

| | | | | | |
|---|---|---|---|---|---|
| T2DM_insulin leukocyte | *PAK6* | -0.194204064 | 0.013589664 | 0.019553021 | 1 |
| T2DM_insulin leukocyte | *ADAM10* | -0.503980141 | 0.001633107 | 0.011330438 | 1 |
| T2DM_insulin leukocyte | *DUOX1* | -0.47545423 | 0.005327499 | 0.011421159 | 1 |
| T2DM_insulin leukocyte | *CYP19A1* | -3.133711061 | 0.008016375 | 0.013228197 | 1 |
| T2DM_insulin leukocyte | *ZNF280D* | -0.46652118 | 0.003068955 | 0.011330438 | 1 |
| T2DM_insulin leukocyte | *RSL24D1* | -0.369623444 | 0.001978568 | 0.011330438 | 1 |
| T2DM_insulin leukocyte | *TTLL7* | -0.366665638 | 0.003973316 | 0.011330438 | 1 |
| T2DM_insulin leukocyte | *SLC44A5* | -0.57571944 | 0.00649371 | 0.011762927 | 1 |
| T2DM_insulin leukocyte | *RTCA* | -0.832722531 | 0.006419692 | 0.01170553 | 1 |
| T2DM_insulin leukocyte | *DYNC2LI1* | -0.485639499 | 0.000929361 | 0.011330438 | 1 |
| T2DM_insulin leukocyte | *ACTR2* | -0.34982288 | 0.002345381 | 0.011330438 | 1 |
| T2DM_insulin leukocyte | *ATRAID* | -0.343201821 | 0.005069227 | 0.011330438 | 1 |
| T2DM_insulin leukocyte | *LRPPRC* | -0.395904902 | 0.00568891 | 0.011549238 | 1 |
| T2DM_insulin leukocyte | *MYOF* | -0.228404235 | 0.011207015 | 0.01684018 | 1 |
| T2DM_insulin leukocyte | *DUSP5* | -0.709090151 | 0.001891986 | 0.011330438 | 1 |
| T2DM_insulin leukocyte | *CALHM2* | 0.434426892 | 0.012082884 | 0.017813752 | 1 |
| T2DM_insulin leukocyte | *ARL3* | -0.335797003 | 0.003825091 | 0.011330438 | 1 |
| T2DM_insulin leukocyte | *PLCE1* | -0.472214236 | 0.003268066 | 0.011330438 | 1 |
| T2DM_insulin leukocyte | *DNAJC13* | -0.332670185 | 0.006081121 | 0.011570498 | 1 |
| T2DM_insulin leukocyte | *FAM149B1* | -0.470016116 | 0.000915869 | 0.011330438 | 1 |
| T2DM_insulin leukocyte | *NCOA4* | -0.757830343 | 0.002470802 | 0.011330438 | 1 |
| T2DM_insulin leukocyte | *ASCC1* | -0.42095989 | 0.007169074 | 0.012363261 | 1 |
| T2DM_insulin leukocyte | *RPS24* | -0.260213765 | 0.00286433 | 0.011330438 | 1 |
| T2DM_insulin leukocyte | *DNA2* | -0.517990088 | 0.018754009 | 0.025727828 | 1 |
| T2DM_insulin leukocyte | *MYPN* | 1.90015633 | 0.002812337 | 0.011330438 | 1 |
| T2DM_insulin leukocyte | *ATIC* | -0.687857878 | 0.001114182 | 0.011330438 | 1 |
| T2DM_insulin leukocyte | *SMARCAL1* | -0.497938489 | 0.006065427 | 0.011570498 | 1 |
| T2DM_insulin leukocyte | *BARD1* | -0.405238591 | 0.006023979 | 0.011570498 | 1 |
| T2DM_insulin leukocyte | *FASTKD1* | -0.408266976 | 0.011208214 | 0.01684018 | 1 |
| T2DM_insulin leukocyte | *FAM117B* | -0.494107566 | 0.002387677 | 0.011330438 | 1 |
| T2DM_insulin leukocyte | *WDR12* | -0.532980984 | 0.0038393 | 0.011330438 | 1 |
| T2DM_insulin leukocyte | *SLC35A5* | -0.502641844 | 0.005845463 | 0.011570498 | 1 |
| T2DM_insulin leukocyte | *USP8* | -0.462484062 | 0.002921556 | 0.011330438 | 1 |
| T2DM_insulin leukocyte | *SPPL2A* | -0.410223961 | 0.00271793 | 0.011330438 | 1 |
| T2DM_insulin leukocyte | *APH1B* | -0.580333548 | 0.003938231 | 0.011330438 | 1 |
| T2DM_insulin leukocyte | *HCN4* | -0.545577741 | 0.002757786 | 0.011330438 | 1 |
| T2DM_insulin leukocyte | *SEMA7A* | -0.863777609 | 0.006195285 | 0.011570498 | 1 |
| T2DM_insulin leukocyte | *AP1AR* | -0.335824823 | 0.002923186 | 0.011330438 | 1 |
| T2DM_insulin leukocyte | *BBS7* | -0.4370972 | 0.006090107 | 0.011570498 | 1 |
| T2DM_insulin leukocyte | *RAP1GDS1* | -0.46117313 | 0.003341169 | 0.011330438 | 1 |
| T2DM_insulin leukocyte | *PRDM5* | -0.362192551 | 0.006772358 | 0.011991254 | 1 |
| T2DM_insulin leukocyte | *NUP54* | -0.288349889 | 0.016659805 | 0.023285796 | 1 |
| T2DM_insulin leukocyte | *G3BP2* | -0.316355166 | 0.002746937 | 0.011330438 | 1 |
| T2DM_insulin leukocyte | *SCARB2* | -0.394042997 | 0.001008861 | 0.011330438 | 1 |
| T2DM_insulin leukocyte | *CNOT6L* | -0.462806155 | 0.003337758 | 0.011330438 | 1 |
| T2DM_insulin leukocyte | *USO1* | -0.316732581 | 0.002997002 | 0.011330438 | 1 |
| T2DM_insulin leukocyte | *SHROOM3* | -0.684709558 | 0.003080778 | 0.011330438 | 1 |
| T2DM_insulin leukocyte | *ANXA3* | -0.262529855 | 0.004530624 | 0.011330438 | 1 |
| T2DM_insulin leukocyte | *PPA2* | -0.368485003 | 0.003956416 | 0.011330438 | 1 |
| T2DM_insulin leukocyte | *EGF* | -1.042037141 | 0.003933695 | 0.011330438 | 1 |

| | | | | | |
|---|---|---|---|---|---|
| T2DM_insulin leukocyte | *MAPK8IP3* | -0.407702204 | 0.003749578 | 0.011330438 | 1 |
| T2DM_insulin leukocyte | *RGS3* | -0.372289274 | 0.001747667 | 0.011330438 | 1 |
| T2DM_insulin leukocyte | *ETV6* | -0.677828977 | 0.00618112 | 0.011570498 | 1 |
| T2DM_insulin leukocyte | *ALG10* | -0.66759482 | 0.005284336 | 0.011419801 | 1 |
| T2DM_insulin leukocyte | *SLCO1C1* | -6.163891783 | 0.003587287 | 0.011330438 | 1 |
| T2DM_insulin leukocyte | *ZCRB1* | -0.338301545 | 0.003240498 | 0.011330438 | 1 |
| T2DM_insulin leukocyte | *C1RL* | -0.423956346 | 0.01280758 | 0.018623605 | 1 |
| T2DM_insulin leukocyte | *CLSTN3* | -0.463085595 | 0.023512287 | 0.031315178 | 1 |
| T2DM_insulin leukocyte | *KLRG1* | -1.825404623 | 0.035361567 | 0.044774605 | 1 |
| T2DM_insulin leukocyte | *VAMP1* | 0.304506649 | 0.010536303 | 0.016082803 | 1 |
| T2DM_insulin leukocyte | *TAPBPL* | -0.327935797 | 0.009390089 | 0.014787038 | 1 |
| T2DM_insulin leukocyte | *PEX5* | -0.361983643 | 0.005960283 | 0.011570498 | 1 |
| T2DM_insulin leukocyte | *SLC38A4* | -0.534017567 | 0.005156229 | 0.011330438 | 1 |
| T2DM_insulin leukocyte | *LRIG3* | -0.394543558 | 0.00326244 | 0.011330438 | 1 |
| T2DM_insulin leukocyte | *GLIPR1* | -0.441702411 | 0.004069273 | 0.011330438 | 1 |
| T2DM_insulin leukocyte | *SNRPF* | -0.285596386 | 0.006228594 | 0.011570498 | 1 |
| T2DM_insulin leukocyte | *ASCL1* | -0.559301226 | 0.008088711 | 0.013302865 | 1 |
| T2DM_insulin leukocyte | *GLTP* | -0.303174333 | 0.004253295 | 0.011330438 | 1 |
| T2DM_insulin leukocyte | *MTMR6* | -0.476415731 | 0.00370785 | 0.011330438 | 1 |
| T2DM_insulin leukocyte | *PDX1* | -0.516516772 | 0.003517674 | 0.011330438 | 1 |
| T2DM_insulin leukocyte | *SUOX* | -0.489569672 | 0.002057217 | 0.011330438 | 1 |
| T2DM_insulin leukocyte | *CCDC65* | -0.529888108 | 0.014447953 | 0.020587417 | 1 |
| T2DM_insulin leukocyte | *SLC39A5* | -0.200706255 | 0.024625779 | 0.032654299 | 1 |
| T2DM_insulin leukocyte | *KANSL2* | -0.901176322 | 0.002038252 | 0.011330438 | 1 |
| T2DM_insulin leukocyte | *CERS5* | -0.43465262 | 0.008927125 | 0.014211781 | 1 |
| T2DM_insulin leukocyte | *ZNF740* | -0.740944222 | 0.007567178 | 0.012760276 | 1 |
| T2DM_insulin leukocyte | *WDFY2* | -0.443429491 | 0.002830929 | 0.011330438 | 1 |
| T2DM_insulin leukocyte | *GRTP1* | -0.768752285 | 0.01024981 | 0.015761841 | 1 |
| T2DM_insulin leukocyte | *TSSK4* | -2.717042255 | 0.03581348 | 0.045274858 | 1 |
| T2DM_insulin leukocyte | *TMX1* | -0.53781707 | 0.002691404 | 0.011330438 | 1 |
| T2DM_insulin leukocyte | *RAB15* | -0.611212646 | 0.001171768 | 0.011330438 | 1 |
| T2DM_insulin leukocyte | *PTGR2* | -0.270510459 | 0.012346115 | 0.01808646 | 1 |
| T2DM_insulin leukocyte | *NIPA2* | -0.312660179 | 0.004440694 | 0.011330438 | 1 |
| T2DM_insulin leukocyte | *SERF2* | -0.279682917 | 0.00335894 | 0.011330438 | 1 |
| T2DM_insulin leukocyte | *BNIP2* | -0.610952881 | 0.002506766 | 0.011330438 | 1 |
| T2DM_insulin leukocyte | *GTF2A2* | -0.3406286 | 0.003200401 | 0.011330438 | 1 |
| T2DM_insulin leukocyte | *SRP14* | -0.272306528 | 0.003815703 | 0.011330438 | 1 |
| T2DM_insulin leukocyte | *BAHD1* | -0.462175165 | 0.002334275 | 0.011330438 | 1 |
| T2DM_insulin leukocyte | *DISP2* | -0.320730551 | 0.003897015 | 0.011330438 | 1 |
| T2DM_insulin leukocyte | *TLE3* | -0.834580049 | 0.002057482 | 0.011330438 | 1 |
| T2DM_insulin leukocyte | *SCAPER* | -0.345422488 | 0.003567792 | 0.011330438 | 1 |
| T2DM_insulin leukocyte | *TSPAN3* | -0.287701694 | 0.0044737 | 0.011330438 | 1 |
| T2DM_insulin leukocyte | *MAN2C1* | -0.398117299 | 0.002002643 | 0.011330438 | 1 |
| T2DM_insulin leukocyte | *DNAJA4* | -0.326831803 | 0.004955237 | 0.011330438 | 1 |
| T2DM_insulin leukocyte | *TPM1* | -0.208321646 | 0.004881979 | 0.011330438 | 1 |
| T2DM_insulin leukocyte | *IGF1R* | -0.311763621 | 0.00436457 | 0.011330438 | 1 |
| T2DM_insulin leukocyte | *ULK3* | -0.373606802 | 0.005258175 | 0.011402654 | 1 |
| T2DM_insulin leukocyte | *CELF6* | -0.448228055 | 0.004666427 | 0.011330438 | 1 |
| T2DM_insulin leukocyte | *POLG* | -0.586166019 | 0.003918539 | 0.011330438 | 1 |
| T2DM_insulin leukocyte | *WDR93* | -1.215286476 | 0.021174523 | 0.028599073 | 1 |

| | | | | | |
|---|---|---|---|---|---|
| T2DM_insulin leukocyte | *UNC45A* | -0.448583188 | 0.001727362 | 0.011330438 | 1 |
| T2DM_insulin leukocyte | *IQGAP1* | -0.36381092 | 0.005112719 | 0.011330438 | 1 |
| T2DM_insulin leukocyte | *PMM2* | -0.376990708 | 0.003831385 | 0.011330438 | 1 |
| T2DM_insulin leukocyte | *SLC5A2* | -1.445472142 | 0.005964912 | 0.011570498 | 1 |
| T2DM_insulin leukocyte | *UQCRC2* | -0.369055828 | 0.006365395 | 0.011665654 | 1 |
| T2DM_insulin leukocyte | *ARHGAP17* | -0.923233851 | 0.002528572 | 0.011330438 | 1 |
| T2DM_insulin leukocyte | *TXNL4B* | -0.872233072 | 0.002918902 | 0.011330438 | 1 |
| T2DM_insulin leukocyte | *MARVELD3* | -0.287071945 | 0.012643828 | 0.018433247 | 1 |
| T2DM_insulin leukocyte | *KIFC3* | -0.420732352 | 0.003870591 | 0.011330438 | 1 |
| T2DM_insulin leukocyte | *NUDT7* | -0.332567495 | 0.006139498 | 0.011570498 | 1 |
| T2DM_insulin leukocyte | *GCSH* | -0.355981073 | 0.003320912 | 0.011330438 | 1 |
| T2DM_insulin leukocyte | *MAP1LC3B* | -0.309315037 | 0.003192407 | 0.011330438 | 1 |
| T2DM_insulin leukocyte | *ADAD2* | -4.616328537 | 0.031338017 | 0.040198194 | 1 |
| T2DM_insulin leukocyte | *OSGIN1* | -0.290659234 | 0.005362043 | 0.011421159 | 1 |
| T2DM_insulin leukocyte | *RPS2* | -0.23765556 | 0.003061809 | 0.011330438 | 1 |
| T2DM_insulin leukocyte | *PDPK1* | -0.444504392 | 0.006474553 | 0.011737322 | 1 |
| T2DM_insulin leukocyte | *DEF8* | -0.433960844 | 0.002433845 | 0.011330438 | 1 |
| T2DM_insulin leukocyte | *GAS8* | -0.612470201 | 0.006813743 | 0.012014984 | 1 |
| T2DM_insulin leukocyte | *RANBP10* | -0.346575591 | 0.008976579 | 0.014278939 | 1 |
| T2DM_insulin leukocyte | *PRPSAP2* | -0.428789785 | 0.008168022 | 0.013388469 | 1 |
| T2DM_insulin leukocyte | *TOM1L1* | -0.356535541 | 0.004931885 | 0.011330438 | 1 |
| T2DM_insulin leukocyte | *C17orf80* | -0.4989399 | 0.023256166 | 0.031016115 | 1 |
| T2DM_insulin leukocyte | *VPS53* | -0.370746313 | 0.006174219 | 0.011570498 | 1 |
| T2DM_insulin leukocyte | *NPEPPS* | -0.505448626 | 0.00341291 | 0.011330438 | 1 |
| T2DM_insulin leukocyte | *ARSG* | -1.159688955 | 0.015844156 | 0.02228049 | 1 |
| T2DM_insulin leukocyte | *PTRH2* | -0.421401265 | 0.003927702 | 0.011330438 | 1 |
| T2DM_insulin leukocyte | *GNAL* | -0.54814979 | 0.00520173 | 0.011378137 | 1 |
| T2DM_insulin leukocyte | *RPRD1A* | -0.375843122 | 0.004171026 | 0.011330438 | 1 |
| T2DM_insulin leukocyte | *NPC1* | -0.788049583 | 0.001952964 | 0.011330438 | 1 |
| T2DM_insulin leukocyte | *SLC14A1* | -0.538056878 | 0.011381545 | 0.017041955 | 1 |
| T2DM_insulin leukocyte | *ZMYND15* | 0.082696382 | 0.015169563 | 0.021477915 | 1 |
| T2DM_insulin leukocyte | *SAT2* | -0.455902113 | 0.002242751 | 0.011330438 | 1 |
| T2DM_insulin leukocyte | *CCDC40* | -0.316774201 | 0.027301851 | 0.035684954 | 1 |
| T2DM_insulin leukocyte | *ARHGDIA* | -0.33637781 | 0.005203999 | 0.011378137 | 1 |
| T2DM_insulin leukocyte | *TMC6* | -0.591292312 | 0.005978406 | 0.011570498 | 1 |
| T2DM_insulin leukocyte | *TTYH2* | -0.65521257 | 0.006168821 | 0.011570498 | 1 |
| T2DM_insulin leukocyte | *EIF4A3* | -0.454589225 | 0.001471211 | 0.011330438 | 1 |
| T2DM_insulin leukocyte | *TBCD* | -0.634746742 | 0.004281538 | 0.011330438 | 1 |
| T2DM_insulin leukocyte | *FN3KRP* | -0.53622622 | 0.002021448 | 0.011330438 | 1 |
| T2DM_insulin leukocyte | *NARF* | -0.536615551 | 0.002423634 | 0.011330438 | 1 |
| T2DM_insulin leukocyte | *TRIM65* | -0.470339972 | 0.020055713 | 0.027303845 | 1 |
| T2DM_insulin leukocyte | *RNF157* | -0.451132164 | 0.003316589 | 0.011330438 | 1 |
| T2DM_insulin leukocyte | *WDR45B* | -0.309174788 | 0.004785012 | 0.011330438 | 1 |
| T2DM_insulin leukocyte | *TNFRSF11A* | -0.343461747 | 0.005103196 | 0.011330438 | 1 |
| T2DM_insulin leukocyte | *ZCCHC2* | -0.505356875 | 0.004422233 | 0.011330438 | 1 |
| T2DM_insulin leukocyte | *PMAIP1* | -0.34812995 | 0.005018329 | 0.011330438 | 1 |
| T2DM_insulin leukocyte | *NT5C3B* | -0.417742388 | 0.00227405 | 0.011330438 | 1 |
| T2DM_insulin leukocyte | *PIP4K2B* | -0.355181615 | 0.007047586 | 0.012237751 | 1 |
| T2DM_insulin leukocyte | *TXNL4A* | -0.332992193 | 0.003426907 | 0.011330438 | 1 |
| T2DM_insulin leukocyte | *MVB12A* | -0.340656726 | 0.034983381 | 0.044367238 | 1 |

| | | | | | |
|---|---|---|---|---|---|
| T2DM_insulin leukocyte | *SH3GL1* | -0.36623676 | 0.00267431 | 0.011330438 | 1 |
| T2DM_insulin leukocyte | *DUS3L* | -0.253978736 | 0.015956828 | 0.022398848 | 1 |
| T2DM_insulin leukocyte | *SIRT3* | -0.523113936 | 0.004389567 | 0.011330438 | 1 |
| T2DM_insulin leukocyte | *IFITM3* | -0.233107337 | 0.006128947 | 0.011570498 | 1 |
| T2DM_insulin leukocyte | *COL6A1* | -0.549626211 | 0.001459855 | 0.011330438 | 1 |
| T2DM_insulin leukocyte | *IFNAR1* | -0.388070067 | 0.001323952 | 0.011330438 | 1 |
| T2DM_insulin leukocyte | *TMEM50B* | -0.371653116 | 0.004693995 | 0.011330438 | 1 |
| T2DM_insulin leukocyte | *AKT1* | -0.521030953 | 0.005535138 | 0.011449692 | 1 |
| T2DM_insulin leukocyte | *CBLC* | -0.385350527 | 0.004461019 | 0.011330438 | 1 |
| T2DM_insulin leukocyte | *CTU1* | -0.661017034 | 0.001611746 | 0.011330438 | 1 |
| T2DM_insulin leukocyte | *RERE* | -0.31814621 | 0.003839294 | 0.011330438 | 1 |
| T2DM_insulin leukocyte | *FHAD1* | -0.352160966 | 0.011975882 | 0.017715828 | 1 |
| T2DM_insulin leukocyte | *PGD* | -0.523085152 | 0.001273277 | 0.011330438 | 1 |
| T2DM_insulin leukocyte | *CNKSR1* | -0.684314544 | 0.006267912 | 0.011570498 | 1 |
| T2DM_insulin leukocyte | *ZNF593* | -0.364038543 | 0.003048654 | 0.011330438 | 1 |
| T2DM_insulin leukocyte | *FCN3* | -0.170502251 | 0.026656782 | 0.034981461 | 1 |
| T2DM_insulin leukocyte | *CELA3A* | -0.173635269 | 0.002993084 | 0.011330438 | 1 |
| T2DM_insulin leukocyte | *HSPG2* | -0.267597514 | 0.000343122 | 0.011330438 | 1 |
| T2DM_insulin leukocyte | *ITGB3BP* | -0.281636931 | 0.036834317 | 0.046444812 | 1 |
| T2DM_insulin leukocyte | *PRKACB* | -0.432084845 | 0.003527139 | 0.011330438 | 1 |
| T2DM_insulin leukocyte | *PIGK* | -0.635408405 | 0.002712496 | 0.011330438 | 1 |
| T2DM_insulin leukocyte | *RPS8* | -0.222505796 | 0.004125345 | 0.011330438 | 1 |
| T2DM_insulin leukocyte | *SYPL2* | -1.108549531 | 0.00568288 | 0.011549238 | 1 |
| T2DM_insulin leukocyte | *SLC44A3* | -0.360243567 | 0.00623325 | 0.011570498 | 1 |
| T2DM_insulin leukocyte | *ZNF697* | -0.499907053 | 0.017511487 | 0.024234812 | 1 |
| T2DM_insulin leukocyte | *CELSR2* | -0.437962375 | 0.006562276 | 0.011821724 | 1 |
| T2DM_insulin leukocyte | *ALDH9A1* | -0.333912613 | 0.003809817 | 0.011330438 | 1 |
| T2DM_insulin leukocyte | *ATP1B1* | -0.231337691 | 0.00336095 | 0.011330438 | 1 |
| T2DM_insulin leukocyte | *TIPRL* | -0.33960932 | 0.002933346 | 0.011330438 | 1 |
| T2DM_insulin leukocyte | *NME7* | -0.559428802 | 0.003344815 | 0.011330438 | 1 |
| T2DM_insulin leukocyte | *MGST3* | -0.34343253 | 0.005219437 | 0.011389288 | 1 |
| T2DM_insulin leukocyte | *LYPLAL1* | -0.446475212 | 0.003898932 | 0.011330438 | 1 |
| T2DM_insulin leukocyte | *SNX27* | -0.622974872 | 0.002054282 | 0.011330438 | 1 |
| T2DM_insulin leukocyte | *CTSK* | -1.104437086 | 0.007286352 | 0.012457822 | 1 |
| T2DM_insulin leukocyte | *SELENBP1* | -0.43315574 | 0.005973539 | 0.011570498 | 1 |
| T2DM_insulin leukocyte | *POGZ* | -0.33125534 | 0.021585119 | 0.029103614 | 1 |
| T2DM_insulin leukocyte | *GABPB2* | -0.583304597 | 0.010081907 | 0.015574095 | 1 |
| T2DM_insulin leukocyte | *SLC27A3* | -0.455552045 | 0.012769292 | 0.018595459 | 1 |
| T2DM_insulin leukocyte | *KCNN3* | -0.690379164 | 0.003793503 | 0.011330438 | 1 |
| T2DM_insulin leukocyte | *RIT1* | -0.560142433 | 0.023711609 | 0.031559739 | 1 |
| T2DM_insulin leukocyte | *SRP9* | -0.365633915 | 0.002933885 | 0.011330438 | 1 |
| T2DM_insulin leukocyte | *CNIH4* | -0.369856204 | 0.00396411 | 0.011330438 | 1 |
| T2DM_insulin leukocyte | *PSEN2* | -0.705628689 | 0.006089523 | 0.011570498 | 1 |
| T2DM_insulin leukocyte | *PYCR2* | -0.416226896 | 0.00416222 | 0.011330438 | 1 |
| T2DM_insulin leukocyte | *LBR* | -0.457533777 | 0.003843419 | 0.011330438 | 1 |
| T2DM_insulin leukocyte | *ARL8A* | -0.598263413 | 0.002434064 | 0.011330438 | 1 |
| T2DM_insulin leukocyte | *EML4* | -0.593501924 | 0.003942776 | 0.011330438 | 1 |
| T2DM_insulin leukocyte | *WDPCP* | -0.979957395 | 0.002882823 | 0.011330438 | 1 |
| T2DM_insulin leukocyte | *REG3G* | -0.433433218 | 0.001735703 | 0.011330438 | 1 |
| T2DM_insulin leukocyte | *CIAO1* | -0.551225466 | 0.001455717 | 0.011330438 | 1 |

| | | | | | |
|---|---|---|---|---|---|
| T2DM_insulin leukocyte | *ZNF514* | -0.436291074 | 0.0020076 | 0.011330438 | 1 |
| T2DM_insulin leukocyte | *TPRKB* | -0.329892926 | 0.00376396 | 0.011330438 | 1 |
| T2DM_insulin leukocyte | *TEX261* | -0.432082811 | 0.003605607 | 0.011330438 | 1 |
| T2DM_insulin leukocyte | *RALB* | -0.412937117 | 0.002525238 | 0.011330438 | 1 |
| T2DM_insulin leukocyte | *RABL2A* | -0.518822308 | 0.009884997 | 0.015351048 | 1 |
| T2DM_insulin leukocyte | *SLC20A1* | -0.569600995 | 0.004197736 | 0.011330438 | 1 |
| T2DM_insulin leukocyte | *LYG1* | -1.623504592 | 0.032709476 | 0.041747209 | 1 |
| T2DM_insulin leukocyte | *GULP1* | -0.369492473 | 0.004302897 | 0.011330438 | 1 |
| T2DM_insulin leukocyte | *FAM171B* | -0.327405235 | 0.005420429 | 0.011421159 | 1 |
| T2DM_insulin leukocyte | *METTL21A* | -0.382401445 | 0.00505439 | 0.011330438 | 1 |
| T2DM_insulin leukocyte | *SUMF1* | -0.535704949 | 0.002835609 | 0.011330438 | 1 |
| T2DM_insulin leukocyte | *HES6* | -0.913639519 | 0.002719343 | 0.011330438 | 1 |
| T2DM_insulin leukocyte | *TAMM41* | -0.385898975 | 0.020166528 | 0.027426199 | 1 |
| T2DM_insulin leukocyte | *RAB5A* | -0.35367034 | 0.003869248 | 0.011330438 | 1 |
| T2DM_insulin leukocyte | *CNTN4* | -0.469134922 | 0.000465254 | 0.011330438 | 1 |
| T2DM_insulin leukocyte | *POMGNT2* | -0.694388094 | 0.001964678 | 0.011330438 | 1 |
| T2DM_insulin leukocyte | *CTDSPL* | -0.34928384 | 0.004389224 | 0.011330438 | 1 |
| T2DM_insulin leukocyte | *IL17RD* | -1.408312198 | 0.01186402 | 0.017638109 | 1 |
| T2DM_insulin leukocyte | *UBA3* | -0.52890423 | 0.005548074 | 0.011460768 | 1 |
| T2DM_insulin leukocyte | *ARL6IP5* | -0.348489114 | 0.003196885 | 0.011330438 | 1 |
| T2DM_insulin leukocyte | *LIMD1* | -0.654986272 | 0.002362431 | 0.011330438 | 1 |
| T2DM_insulin leukocyte | *NXPE3* | -0.29849482 | 0.019409079 | 0.026525186 | 1 |
| T2DM_insulin leukocyte | *PHLDB2* | -0.277562443 | 0.004948203 | 0.011330438 | 1 |
| T2DM_insulin leukocyte | *ABHD10* | -0.468103702 | 0.001694 | 0.011330438 | 1 |
| T2DM_insulin leukocyte | *TAGLN3* | -0.217708819 | 0.012198769 | 0.017937534 | 1 |
| T2DM_insulin leukocyte | *EIF2A* | -0.317651446 | 0.003580659 | 0.011330438 | 1 |
| T2DM_insulin leukocyte | *ALDH1L1* | -0.517906636 | 0.005870655 | 0.011570498 | 1 |
| T2DM_insulin leukocyte | *TRPC1* | -0.449940132 | 0.004591737 | 0.011330438 | 1 |
| T2DM_insulin leukocyte | *NCEH1* | -0.238341174 | 0.004304179 | 0.011330438 | 1 |
| T2DM_insulin leukocyte | *AMT* | -0.641950349 | 0.004201916 | 0.011330438 | 1 |
| T2DM_insulin leukocyte | *DGKQ* | -0.359269173 | 0.015699092 | 0.022116074 | 1 |
| T2DM_insulin leukocyte | *SCD5* | -0.316125598 | 0.004629857 | 0.011330438 | 1 |
| T2DM_insulin leukocyte | *PYURF* | -0.304515852 | 0.003772962 | 0.011330438 | 1 |
| T2DM_insulin leukocyte | *DDIT4L* | -0.873066718 | 0.010872922 | 0.016483075 | 1 |
| T2DM_insulin leukocyte | *TIFA* | -0.478391552 | 0.003171893 | 0.011330438 | 1 |
| T2DM_insulin leukocyte | *CYP4V2* | -0.736003638 | 0.0083023 | 0.013512733 | 1 |
| T2DM_insulin leukocyte | *SSBP2* | -0.30739021 | 0.004737 | 0.011330438 | 1 |
| T2DM_insulin leukocyte | *IQGAP2* | 0.220452038 | 0.016901014 | 0.023572625 | 1 |
| T2DM_insulin leukocyte | *GTF2H2* | -0.986241063 | 0.002597369 | 0.011330438 | 1 |
| T2DM_insulin leukocyte | *BTF3* | -0.286686252 | 0.0029416 | 0.011330438 | 1 |
| T2DM_insulin leukocyte | *SPATA9* | -2.522025812 | 0.013591067 | 0.019553021 | 1 |
| T2DM_insulin leukocyte | *COMMD10* | -0.438012982 | 0.004219835 | 0.011330438 | 1 |
| T2DM_insulin leukocyte | *ATG12* | -0.357295311 | 0.003241993 | 0.011330438 | 1 |
| T2DM_insulin leukocyte | *PCYOX1L* | -1.833838425 | 0.004529217 | 0.011330438 | 1 |
| T2DM_insulin leukocyte | *ZMAT2* | -0.458653177 | 0.003233544 | 0.011330438 | 1 |
| T2DM_insulin leukocyte | *KLHL3* | -1.271159493 | 0.016898931 | 0.023572625 | 1 |
| T2DM_insulin leukocyte | *DCDC2* | -0.283904266 | 0.005062746 | 0.011330438 | 1 |
| T2DM_insulin leukocyte | *TRIM7* | -1.448836108 | 0.000318412 | 0.011330438 | 1 |
| T2DM_insulin leukocyte | *ANO7* | -0.957640616 | 0.010989601 | 0.016619762 | 1 |
| T2DM_insulin leukocyte | *RPL7L1* | -0.345641996 | 0.003263617 | 0.011330438 | 1 |

| | | | | | |
|---|---|---|---|---|---|
| T2DM_insulin leukocyte | *IRAK1BP1* | -0.322390522 | 0.004891247 | 0.011330438 | 1 |
| T2DM_insulin leukocyte | *MMS22L* | -0.572632237 | 0.00269678 | 0.011330438 | 1 |
| T2DM_insulin leukocyte | *FAXC* | -0.582879048 | 0.003745892 | 0.011330438 | 1 |
| T2DM_insulin leukocyte | *TBC1D32* | -1.357199145 | 0.001371865 | 0.011330438 | 1 |
| T2DM_insulin leukocyte | *ABRACL* | -0.416697815 | 0.002023635 | 0.011330438 | 1 |
| T2DM_insulin leukocyte | *DYNLT1* | -0.323836723 | 0.002159197 | 0.011330438 | 1 |
| T2DM_insulin leukocyte | *WTAP* | -0.333713068 | 0.001758601 | 0.011330438 | 1 |
| T2DM_insulin leukocyte | *SLC22A3* | -2.232886056 | 0.016486172 | 0.023075942 | 1 |
| T2DM_insulin leukocyte | *VWDE* | -0.357589252 | 0.006059453 | 0.011570498 | 1 |
| T2DM_insulin leukocyte | *GNA12* | -0.583029289 | 0.004722009 | 0.011330438 | 1 |
| T2DM_insulin leukocyte | *SDK1* | -0.407964298 | 0.01207629 | 0.017813386 | 1 |
| T2DM_insulin leukocyte | *CCZ1B* | -0.543042573 | 0.008835973 | 0.014115597 | 1 |
| T2DM_insulin leukocyte | *RBAK* | -0.819941238 | 0.003747698 | 0.011330438 | 1 |
| T2DM_insulin leukocyte | *POMZP3* | -0.373672749 | 0.011709209 | 0.017459386 | 1 |
| T2DM_insulin leukocyte | *GIGYF1* | -0.881796562 | 0.000562569 | 0.011330438 | 1 |
| T2DM_insulin leukocyte | *TRIM4* | -0.345228058 | 0.002479636 | 0.011330438 | 1 |
| T2DM_insulin leukocyte | *TMEM209* | -0.589445664 | 0.002359268 | 0.011330438 | 1 |
| T2DM_insulin leukocyte | *RAB19* | -3.69904795 | 0.000337227 | 0.011330438 | 1 |
| T2DM_insulin leukocyte | *CSGALNACT1* | -0.331047338 | 0.006780192 | 0.011999719 | 1 |
| T2DM_insulin leukocyte | *CCDC25* | -0.314933371 | 0.003046172 | 0.011330438 | 1 |
| T2DM_insulin leukocyte | *BIN3* | -0.963958472 | 0.006286735 | 0.011585639 | 1 |
| T2DM_insulin leukocyte | *CHMP7* | -0.600044697 | 0.00192066 | 0.011330438 | 1 |
| T2DM_insulin leukocyte | *STAR* | 0.247238967 | 0.021829315 | 0.029382453 | 1 |
| T2DM_insulin leukocyte | *LACTB2* | -0.340244954 | 0.003945221 | 0.011330438 | 1 |
| T2DM_insulin leukocyte | *TERF1* | -0.733938049 | 0.00231611 | 0.011330438 | 1 |
| T2DM_insulin leukocyte | *LRP12* | -0.38168704 | 0.003823501 | 0.011330438 | 1 |
| T2DM_insulin leukocyte | *ZNF7* | -0.324681958 | 0.005079482 | 0.011330438 | 1 |
| T2DM_insulin leukocyte | *AK3* | -0.337032032 | 0.003423005 | 0.011330438 | 1 |
| T2DM_insulin leukocyte | *CER1* | 1.300058342 | 0.017632378 | 0.024384939 | 1 |
| T2DM_insulin leukocyte | *C9orf72* | -0.569451182 | 0.002032258 | 0.011330438 | 1 |
| T2DM_insulin leukocyte | *FBXO10* | -0.498194894 | 0.012025316 | 0.017759823 | 1 |
| T2DM_insulin leukocyte | *SIGMAR1* | -0.429120106 | 0.004287119 | 0.011330438 | 1 |
| T2DM_insulin leukocyte | *ZNF462* | -0.337227652 | 0.004057433 | 0.011330438 | 1 |
| T2DM_insulin leukocyte | *UGCG* | -0.364597823 | 0.005150618 | 0.011330438 | 1 |
| T2DM_insulin leukocyte | *ALAD* | -0.39201923 | 0.009595762 | 0.015026604 | 1 |
| T2DM_insulin leukocyte | *SURF4* | -0.327381663 | 0.002412776 | 0.011330438 | 1 |
| T2DM_insulin leukocyte | *SURF1* | -0.354701825 | 0.011694451 | 0.017450626 | 1 |
| T2DM_insulin leukocyte | *SURF6* | -0.546704591 | 0.00305831 | 0.011330438 | 1 |
| T2DM_insulin leukocyte | *SH3GLB2* | -0.612594695 | 0.002169436 | 0.011330438 | 1 |
| T2DM_insulin leukocyte | *LCN2* | -0.156437907 | 0.005791384 | 0.011570498 | 1 |
| T2DM_insulin leukocyte | *GPR107* | -0.47995759 | 0.00642179 | 0.01170553 | 1 |
| T2DM_insulin leukocyte | *INPP5E* | -0.52932459 | 0.037506605 | 0.047231876 | 1 |
| T2DM_insulin leukocyte | *SEC16A* | -0.420633031 | 0.018363951 | 0.025280997 | 1 |
| T2DM_insulin leukocyte | *MSRB2* | -0.616272887 | 0.001222098 | 0.011330438 | 1 |
| T2DM_insulin leukocyte | *RSU1* | -0.489544364 | 0.003041568 | 0.011330438 | 1 |
| T2DM_insulin leukocyte | *GLUD1* | -0.441813198 | 0.003169566 | 0.011330438 | 1 |
| T2DM_insulin leukocyte | *FRA10AC1* | -0.280016678 | 0.006196536 | 0.011570498 | 1 |
| T2DM_insulin leukocyte | *DNAJB12* | -0.466595113 | 0.003876396 | 0.011330438 | 1 |
| T2DM_insulin leukocyte | *MTG1* | -0.300394955 | 0.009680131 | 0.015104542 | 1 |
| T2DM_insulin leukocyte | *GSTO1* | -0.466922055 | 0.004062897 | 0.011330438 | 1 |

| | | | | | |
|---|---|---|---|---|---|
| T2DM_insulin leukocyte | *HSD17B12* | -0.286040659 | 0.000644306 | 0.011330438 | 1 |
| T2DM_insulin leukocyte | *APIP* | -0.418677371 | 0.000852188 | 0.011330438 | 1 |
| T2DM_insulin leukocyte | *PTPRJ* | -0.511493919 | 0.003462188 | 0.011330438 | 1 |
| T2DM_insulin leukocyte | *CCDC82* | -0.408122795 | 0.001551558 | 0.011330438 | 1 |
| T2DM_insulin leukocyte | *RPS3* | -0.246337929 | 0.00275894 | 0.011330438 | 1 |
| T2DM_insulin leukocyte | *AASDHPPT* | -0.395782652 | 0.003455703 | 0.011330438 | 1 |
| T2DM_insulin leukocyte | *GLB1L2* | -0.306422782 | 0.028925257 | 0.037494997 | 1 |
| T2DM_insulin leukocyte | *FADS1* | -0.254121412 | 0.024459418 | 0.032455608 | 1 |
| T2DM_insulin leukocyte | *TMC2* | -23.12039674 | 0.034229131 | 0.043466792 | 1 |
| T2DM_insulin leukocyte | *EML3* | -1.334214472 | 0.015454808 | 0.021810191 | 1 |
| T2DM_insulin leukocyte | *CHEK1* | -0.471389747 | 0.00491309 | 0.011330438 | 1 |
| T2DM_insulin leukocyte | *SIDT2* | -0.539552116 | 0.000953716 | 0.011330438 | 1 |
| T2DM_insulin leukocyte | *TAGLN* | -0.424764986 | 0.000123018 | 0.011330438 | 0.483337044 |
| T2DM_insulin leukocyte | *DUSP15* | -0.46355847 | 0.006408551 | 0.01170553 | 1 |
| T2DM_insulin leukocyte | *DSN1* | -0.458712772 | 0.003813873 | 0.011330438 | 1 |
| T2DM_insulin leukocyte | *YTHDF1* | -0.41805711 | 0.00343894 | 0.011330438 | 1 |
| T2DM_insulin leukocyte | *CABLES2* | -0.619611946 | 0.000519617 | 0.011330438 | 1 |
| T2DM_insulin leukocyte | *TRPT1* | -0.360660068 | 0.006500686 | 0.011770137 | 1 |
| T2DM_insulin leukocyte | *MPP7* | -0.307171541 | 0.005282616 | 0.011419801 | 1 |
| T2DM_insulin leukocyte | *DCUN1D2* | -0.413378119 | 0.014202539 | 0.020306323 | 1 |
| T2DM_insulin leukocyte | *TMCO3* | -0.417880628 | 0.005473492 | 0.011427973 | 1 |
| T2DM_insulin leukocyte | *TMEM218* | -0.587860868 | 0.002932581 | 0.011330438 | 1 |
| T2DM_insulin leukocyte | *MIA2* | -0.357275903 | 0.004038456 | 0.011330438 | 1 |
| T2DM_insulin leukocyte | *LYPD1* | -0.598119703 | 0.005778164 | 0.011570498 | 1 |
| T2DM_insulin leukocyte | *LYPD6B* | -0.977676872 | 0.005752819 | 0.011570498 | 1 |
| T2DM_insulin leukocyte | *WDR17* | -0.313424072 | 0.006234177 | 0.011570498 | 1 |
| T2DM_insulin leukocyte | *CNDP1* | -1.469002732 | 0.029322954 | 0.037947921 | 1 |
| T2DM_insulin leukocyte | *PRSS23* | -0.310645559 | 0.002376219 | 0.011330438 | 1 |
| T2DM_insulin leukocyte | *MTMR12* | -0.320251473 | 0.00366507 | 0.011330438 | 1 |
| T2DM_insulin leukocyte | *DOCK1* | -0.397461318 | 0.003595691 | 0.011330438 | 1 |
| T2DM_insulin leukocyte | *IL18* | 0.200304253 | 0.021857519 | 0.02941034 | 1 |
| T2DM_insulin leukocyte | *TMEM86A* | -0.461290576 | 0.005953587 | 0.011570498 | 1 |
| T2DM_insulin leukocyte | *PLBD2* | -0.89744972 | 0.016696268 | 0.023311883 | 1 |
| T2DM_insulin leukocyte | *DLG5* | -0.376949624 | 0.002976407 | 0.011330438 | 1 |
| T2DM_insulin leukocyte | *SLC2A13* | -0.296838403 | 0.003753065 | 0.011330438 | 1 |
| T2DM_insulin leukocyte | *MAGI1* | -0.354947947 | 0.007798014 | 0.01301113 | 1 |
| T2DM_insulin leukocyte | *CSNK1G3* | -0.42829251 | 0.004372078 | 0.011330438 | 1 |
| T2DM_insulin leukocyte | *AKAP6* | -0.476973754 | 0.003258932 | 0.011330438 | 1 |
| T2DM_insulin leukocyte | *MBIP* | -0.412186496 | 0.004976622 | 0.011330438 | 1 |
| T2DM_insulin leukocyte | *MIPOL1* | -0.348049131 | 0.005945903 | 0.011570498 | 1 |
| T2DM_insulin leukocyte | *TMEM18* | -0.403443878 | 0.003640021 | 0.011330438 | 1 |
| T2DM_insulin leukocyte | *NDUFC2* | -0.309766108 | 0.00248209 | 0.011330438 | 1 |
| T2DM_insulin leukocyte | *VIPAS39* | -0.655276379 | 0.0024683 | 0.011330438 | 1 |
| T2DM_insulin leukocyte | *C4orf33* | -0.653947524 | 0.018988712 | 0.026005146 | 1 |
| T2DM_insulin leukocyte | *QDPR* | -0.260311131 | 0.008691742 | 0.013955805 | 1 |
| T2DM_insulin leukocyte | *TEX9* | -0.691307838 | 0.002596648 | 0.011330438 | 1 |
| T2DM_insulin leukocyte | *ZNF827* | -0.292523741 | 0.0169182 | 0.023588221 | 1 |
| T2DM_insulin leukocyte | *AKR1C2* | -1.169492231 | 0.000158058 | 0.011330438 | 0.621008552 |
| T2DM_insulin leukocyte | *DPYSL4* | -0.401648757 | 0.030762229 | 0.039537062 | 1 |
| T2DM_insulin leukocyte | *KIN* | -0.657689699 | 0.00309228 | 0.011330438 | 1 |

| | | | | | |
|---|---|---|---|---|---|
| T2DM_insulin leukocyte | *TMEM45B* | -0.547928033 | 0.011947651 | 0.017706833 | 1 |
| T2DM_insulin leukocyte | *CENPU* | -0.753909802 | 0.038780115 | 0.048695133 | 1 |
| T2DM_insulin leukocyte | *BICD1* | -0.32507456 | 0.00538292 | 0.011421159 | 1 |
| T2DM_insulin leukocyte | *GUF1* | -0.892851859 | 0.003656805 | 0.011330438 | 1 |
| T2DM_insulin leukocyte | *CCL28* | -0.474868457 | 0.003621348 | 0.011330438 | 1 |
| T2DM_insulin leukocyte | *CACUL1* | -0.539451301 | 0.00264452 | 0.011330438 | 1 |
| T2DM_insulin leukocyte | *RABGAP1L* | -0.42896996 | 0.00348082 | 0.011330438 | 1 |
| T2DM_insulin leukocyte | *PTPN14* | -0.323029887 | 0.008484965 | 0.013736063 | 1 |
| T2DM_insulin leukocyte | *ARL14EP* | -0.521824839 | 0.002253755 | 0.011330438 | 1 |
| T2DM_insulin leukocyte | *EPG5* | -0.595461907 | 0.004561825 | 0.011330438 | 1 |
| T2DM_insulin leukocyte | *HAUS1* | -0.556765385 | 0.001527957 | 0.011330438 | 1 |
| T2DM_insulin leukocyte | *SPC25* | -0.538705101 | 0.000995138 | 0.011330438 | 1 |
| T2DM_insulin leukocyte | *PDK1* | -0.475077126 | 0.003707497 | 0.011330438 | 1 |
| T2DM_insulin leukocyte | *PDE3B* | -0.194865767 | 0.018940634 | 0.02596572 | 1 |
| T2DM_insulin leukocyte | *TGOLN2* | -0.397790974 | 0.002010137 | 0.011330438 | 1 |
| T2DM_insulin leukocyte | *POC5* | -0.535496108 | 0.001745404 | 0.011330438 | 1 |
| T2DM_insulin leukocyte | *GUCY1A2* | -0.377481922 | 0.005917362 | 0.011570498 | 1 |
| T2DM_insulin leukocyte | *ZNF773* | -0.285118923 | 0.027355924 | 0.03574374 | 1 |
| T2DM_insulin leukocyte | *DCLRE1C* | -0.841717283 | 0.028913349 | 0.03749193 | 1 |
| T2DM_insulin leukocyte | *ZSCAN1* | -0.411974269 | 0.002672526 | 0.011330438 | 1 |
| T2DM_insulin leukocyte | *USP12* | -0.328244009 | 0.007527499 | 0.012715195 | 1 |
| T2DM_insulin leukocyte | *CCDC50* | -0.30024461 | 0.004210119 | 0.011330438 | 1 |
| T2DM_insulin leukocyte | *PLEKHH2* | -0.351754203 | 0.014002178 | 0.02006366 | 1 |
| T2DM_insulin leukocyte | *GRIA4* | -0.481693426 | 0.005821967 | 0.011570498 | 1 |
| T2DM_insulin leukocyte | *IGSF10* | -0.843614584 | 0.011243144 | 0.016873305 | 1 |
| T2DM_insulin leukocyte | *GPD1L* | -0.426092977 | 0.00631117 | 0.011614327 | 1 |
| T2DM_insulin leukocyte | *CCNO* | -0.345882436 | 0.031624355 | 0.040539018 | 1 |
| T2DM_insulin leukocyte | *RASGRP3* | -0.431422186 | 0.007520333 | 0.012715195 | 1 |
| T2DM_insulin leukocyte | *CATSPER3* | -2.066498621 | 0.006176679 | 0.011570498 | 1 |
| T2DM_insulin leukocyte | *GPR180* | -0.360056155 | 0.003248359 | 0.011330438 | 1 |
| T2DM_insulin leukocyte | *IFIT5* | -0.390864721 | 0.007716102 | 0.012933688 | 1 |
| T2DM_insulin leukocyte | *PRDM8* | 0.438129831 | 0.014465998 | 0.020593083 | 1 |
| T2DM_insulin leukocyte | *ZNF117* | -0.279669529 | 0.033555746 | 0.04269447 | 1 |
| T2DM_insulin leukocyte | *MED21* | -0.602997993 | 0.002037626 | 0.011330438 | 1 |
| T2DM_insulin leukocyte | *LGI2* | -0.521665902 | 0.015241142 | 0.021555956 | 1 |
| T2DM_insulin leukocyte | *SRP19* | -0.32124025 | 0.003291874 | 0.011330438 | 1 |
| T2DM_insulin leukocyte | *DAB2* | -0.50944838 | 0.003066024 | 0.011330438 | 1 |
| T2DM_insulin leukocyte | *ACMSD* | -0.50446431 | 0.007497189 | 0.012691278 | 1 |
| T2DM_insulin leukocyte | *CAST* | -0.336202359 | 0.003168035 | 0.011330438 | 1 |
| T2DM_insulin leukocyte | *SYCP2L* | -0.883160875 | 0.030731225 | 0.039510139 | 1 |
| T2DM_insulin leukocyte | *TMEM87B* | -0.413843861 | 0.005335969 | 0.011421159 | 1 |
| T2DM_insulin leukocyte | *PLA2R1* | -1.301776435 | 0.005701281 | 0.011549238 | 1 |
| T2DM_insulin leukocyte | *SLC25A27* | -0.638184157 | 0.000630436 | 0.011330438 | 1 |
| T2DM_insulin leukocyte | *ASAP1* | -0.353048599 | 0.004252105 | 0.011330438 | 1 |
| T2DM_insulin leukocyte | *NMRAL1* | -0.414007003 | 0.012797735 | 0.01861618 | 1 |
| T2DM_insulin leukocyte | *CMTM7* | -0.302633083 | 0.010445139 | 0.015974679 | 1 |
| T2DM_insulin leukocyte | *RPIA* | -0.392333359 | 0.004420467 | 0.011330438 | 1 |
| T2DM_insulin leukocyte | *PTPRD* | -0.322373477 | 0.025533853 | 0.033699197 | 1 |
| T2DM_insulin leukocyte | *GTF2E1* | -0.936728512 | 0.002625953 | 0.011330438 | 1 |
| T2DM_insulin leukocyte | *CFDP1* | -0.333549718 | 0.004183903 | 0.011330438 | 1 |

| | | | | | |
|---|---|---|---|---|---|
| T2DM_insulin leukocyte | *JAZF1* | -0.578341542 | 0.00028188 | 0.011330438 | 1 |
| T2DM_insulin leukocyte | *CMIP* | -0.398616024 | 0.003454751 | 0.011330438 | 1 |
| T2DM_insulin leukocyte | *KCTD15* | -0.517351376 | 0.007715348 | 0.012933688 | 1 |
| T2DM_insulin leukocyte | *LGI4* | 0.528884913 | 0.005706326 | 0.011549238 | 1 |
| T2DM_insulin leukocyte | *DDAH1* | -0.863863558 | 0.002064397 | 0.011330438 | 1 |
| T2DM_insulin leukocyte | *NUS1* | -0.452896705 | 0.001846235 | 0.011330438 | 1 |
| T2DM_insulin leukocyte | *PPP2R5E* | -0.457439281 | 0.002054719 | 0.011330438 | 1 |
| T2DM_insulin leukocyte | *THY1* | 0.395461013 | 0.020726231 | 0.02807079 | 1 |
| T2DM_insulin leukocyte | *ROBO3* | -1.122611444 | 0.005937349 | 0.011570498 | 1 |
| T2DM_insulin leukocyte | *PITPNC1* | -0.442862354 | 0.002900724 | 0.011330438 | 1 |
| T2DM_insulin leukocyte | *CEP112* | -0.308581172 | 0.007176963 | 0.012365991 | 1 |
| T2DM_insulin leukocyte | *ABCA5* | -0.565260679 | 0.003138887 | 0.011330438 | 1 |
| T2DM_insulin leukocyte | *SRSF12* | -0.604743531 | 0.017179799 | 0.023872903 | 1 |
| T2DM_insulin leukocyte | *PDLIM3* | -0.268208941 | 0.004613694 | 0.011330438 | 1 |
| T2DM_insulin leukocyte | *PDE1C* | -0.484107117 | 0.008223787 | 0.013443435 | 1 |
| T2DM_insulin leukocyte | *RABGEF1* | -0.427074606 | 0.002452276 | 0.011330438 | 1 |
| T2DM_insulin leukocyte | *MRPL39* | -0.487051396 | 0.001975445 | 0.011330438 | 1 |
| T2DM_insulin leukocyte | *TSEN2* | -0.946003995 | 0.001953755 | 0.011330438 | 1 |
| T2DM_insulin leukocyte | *FLCN* | -0.618839923 | 0.002635413 | 0.011330438 | 1 |
| T2DM_insulin leukocyte | *MPPE1* | -0.590154734 | 0.000305529 | 0.011330438 | 1 |
| T2DM_insulin leukocyte | *RAB6B* | -0.473583892 | 0.004564305 | 0.011330438 | 1 |
| T2DM_insulin leukocyte | *ANKRD40* | -0.46199588 | 0.010609645 | 0.016188464 | 1 |
| T2DM_insulin leukocyte | *ATP6V1C1* | -0.360216091 | 0.003981966 | 0.011330438 | 1 |
| T2DM_insulin leukocyte | *MARVELD1* | -1.287145066 | 0.003884788 | 0.011330438 | 1 |
| T2DM_insulin leukocyte | *TRMT44* | -0.342324104 | 0.008575847 | 0.013826222 | 1 |
| T2DM_insulin leukocyte | *PPM1J* | -0.93487919 | 0.007586888 | 0.01278254 | 1 |
| T2DM_insulin leukocyte | *DBI* | -0.351151529 | 0.004660716 | 0.011330438 | 1 |
| T2DM_insulin leukocyte | *HEATR3* | -0.357000045 | 0.004352045 | 0.011330438 | 1 |
| T2DM_insulin leukocyte | *LRGUK* | -1.204118229 | 0.007368727 | 0.012552105 | 1 |
| T2DM_insulin leukocyte | *SETD9* | -0.545652125 | 0.000209953 | 0.011330438 | 0.824906532 |
| T2DM_insulin leukocyte | *MIER3* | -0.866245229 | 0.001073432 | 0.011330438 | 1 |
| T2DM_insulin leukocyte | *TTN* | -0.950813499 | 0.002978291 | 0.011330438 | 1 |
| T2DM_insulin leukocyte | *DEPTOR* | -0.399308955 | 0.011842439 | 0.0176246 | 1 |
| T2DM_insulin leukocyte | *RMND1* | -0.645026773 | 0.004347143 | 0.011330438 | 1 |
| T2DM_insulin leukocyte | *TMBIM4* | -0.358751835 | 0.002232859 | 0.011330438 | 1 |
| T2DM_insulin leukocyte | *PSD3* | -0.337481157 | 0.004745627 | 0.011330438 | 1 |
| T2DM_insulin leukocyte | *FAM161B* | -0.480233189 | 0.035296194 | 0.044706237 | 1 |
| T2DM_insulin leukocyte | *MMP16* | -0.302775395 | 0.010655868 | 0.016227482 | 1 |
| T2DM_insulin leukocyte | *ADK* | -0.419000433 | 0.004796867 | 0.011330438 | 1 |
| T2DM_insulin leukocyte | *BATF* | -5.549494021 | 0.005673737 | 0.011549238 | 1 |
| T2DM_insulin leukocyte | *DRAM2* | -0.595294648 | 0.006700656 | 0.011939628 | 1 |
| T2DM_insulin leukocyte | *ART3* | -2.418196674 | 0.000291117 | 0.011330438 | 1 |
| T2DM_insulin leukocyte | *CCT8* | -0.31408483 | 0.00348458 | 0.011330438 | 1 |
| T2DM_insulin leukocyte | *CDK20* | -0.347598136 | 0.008225507 | 0.013443435 | 1 |
| T2DM_insulin leukocyte | *SFXN2* | -0.278923491 | 0.038732577 | 0.048654903 | 1 |
| T2DM_insulin leukocyte | *TDRD9* | -0.246008645 | 0.006621867 | 0.01187463 | 1 |
| T2DM_insulin leukocyte | *HKDC1* | -0.901977145 | 0.009206611 | 0.014556448 | 1 |
| T2DM_insulin leukocyte | *ZDHHC5* | -0.478196771 | 0.002287329 | 0.011330438 | 1 |
| T2DM_insulin leukocyte | *MED19* | -0.376039948 | 0.003422256 | 0.011330438 | 1 |
| T2DM_insulin leukocyte | *MAPK13* | -0.419135739 | 0.007737498 | 0.012941946 | 1 |

| | | | | | |
|---|---|---|---|---|---|
| T2DM_insulin leukocyte | *COX6A2* | -0.483134425 | 0.006809146 | 0.012014984 | 1 |
| T2DM_insulin leukocyte | *MPV17L* | -0.389489044 | 0.003663513 | 0.011330438 | 1 |
| T2DM_insulin leukocyte | *TATDN2* | -0.660820173 | 0.003827663 | 0.011330438 | 1 |
| T2DM_insulin leukocyte | *EXOG* | -0.487109301 | 0.00344694 | 0.011330438 | 1 |
| T2DM_insulin leukocyte | *ZFYVE9* | -0.427789594 | 0.003503384 | 0.011330438 | 1 |
| T2DM_insulin leukocyte | *SLC6A1* | -2.210740022 | 0.005697247 | 0.011549238 | 1 |
| T2DM_insulin leukocyte | *NECAP2* | -0.502784574 | 0.003089724 | 0.011330438 | 1 |
| T2DM_insulin leukocyte | *STEAP2* | -0.598315918 | 0.003535162 | 0.011330438 | 1 |
| T2DM_insulin leukocyte | *GATAD1* | -0.508267616 | 0.002566415 | 0.011330438 | 1 |
| T2DM_insulin leukocyte | *CLEC18A* | -1.83091237 | 1.17E-05 | 0.011330438 | 0.045954047 |
| T2DM_insulin leukocyte | *DHRS4* | -0.453733596 | 0.003721783 | 0.011330438 | 1 |
| T2DM_insulin leukocyte | *DHRS1* | -0.443799638 | 0.009980412 | 0.015458818 | 1 |
| T2DM_insulin leukocyte | *AASDH* | -0.279398233 | 0.013515641 | 0.019473031 | 1 |
| T2DM_insulin leukocyte | *SLC38A10* | -0.433937311 | 0.004471104 | 0.011330438 | 1 |
| T2DM_insulin leukocyte | *SLC37A3* | -0.310910537 | 0.004108342 | 0.011330438 | 1 |
| T2DM_insulin leukocyte | *AP3S2* | -0.449802996 | 0.006739042 | 0.011963834 | 1 |
| T2DM_insulin leukocyte | *SPPL3* | -0.437509172 | 0.003295018 | 0.011330438 | 1 |
| T2DM_insulin leukocyte | *TNFRSF14* | 0.253286936 | 0.033140316 | 0.042220591 | 1 |
| T2DM_insulin leukocyte | *C12orf43* | -0.462926245 | 0.005682523 | 0.011549238 | 1 |
| T2DM_insulin leukocyte | *PEX10* | -0.620606785 | 0.003591204 | 0.011330438 | 1 |
| T2DM_insulin leukocyte | *LDLRAP1* | -0.312498749 | 0.005076665 | 0.011330438 | 1 |
| T2DM_insulin leukocyte | *KRTCAP3* | -0.382860303 | 0.004499189 | 0.011330438 | 1 |
| T2DM_insulin leukocyte | *PAFAH2* | -1.091355922 | 0.002480948 | 0.011330438 | 1 |
| T2DM_insulin leukocyte | *EXTL1* | -3.064453867 | 7.12E-06 | 0.009328675 | 0.027986024 |
| T2DM_insulin leukocyte | *TRIM63* | -5.836546447 | 0.002217401 | 0.011330438 | 1 |
| T2DM_insulin leukocyte | *XKR8* | -0.69643567 | 0.003404615 | 0.011330438 | 1 |
| T2DM_insulin leukocyte | *ESYT3* | -0.663718368 | 0.018335926 | 0.025251264 | 1 |
| T2DM_insulin leukocyte | *FAIM* | -0.408186856 | 0.003190046 | 0.011330438 | 1 |
| T2DM_insulin leukocyte | *AHCYL2* | -0.522484342 | 0.001974658 | 0.011330438 | 1 |
| T2DM_insulin leukocyte | *COPG2* | -0.36009087 | 0.008264452 | 0.013462285 | 1 |
| T2DM_insulin leukocyte | *RNF166* | -0.516491164 | 0.00469421 | 0.011330438 | 1 |
| T2DM_insulin leukocyte | *NIT1* | -0.51795305 | 0.011256834 | 0.016880954 | 1 |
| T2DM_insulin leukocyte | *NPM2* | -0.442558467 | 0.005912175 | 0.011570498 | 1 |
| T2DM_insulin leukocyte | *VWA5B1* | 1.022524956 | 0.017375888 | 0.024081081 | 1 |
| T2DM_insulin leukocyte | *CDA* | -4.722123183 | 0.006343997 | 0.011637991 | 1 |
| T2DM_insulin leukocyte | *PINK1* | -0.478022874 | 0.002937502 | 0.011330438 | 1 |
| T2DM_insulin leukocyte | *DMTN* | -0.432214832 | 0.006004406 | 0.011570498 | 1 |
| T2DM_insulin leukocyte | *CCAR2* | -0.563250643 | 0.003100146 | 0.011330438 | 1 |
| T2DM_insulin leukocyte | *EPB41* | -0.39310644 | 0.003560592 | 0.011330438 | 1 |
| T2DM_insulin leukocyte | *MIS18A* | -0.465996889 | 0.004368789 | 0.011330438 | 1 |
| T2DM_insulin leukocyte | *ALG8* | -0.367802119 | 0.005430462 | 0.011421159 | 1 |
| T2DM_insulin leukocyte | *IFNAR2* | -0.540949744 | 0.039151526 | 0.049114415 | 1 |
| T2DM_insulin leukocyte | *IFNGR2* | -0.399197707 | 0.003054851 | 0.011330438 | 1 |
| T2DM_insulin leukocyte | *C1QC* | 0.022194789 | 0.014555101 | 0.020697427 | 1 |
| T2DM_insulin leukocyte | *GJD2* | -0.237101409 | 0.007550408 | 0.012742935 | 1 |
| T2DM_insulin leukocyte | *CLDN14* | -0.868600063 | 0.011299228 | 0.016931605 | 1 |
| T2DM_insulin leukocyte | *HLCS* | -0.346528756 | 0.007303826 | 0.012482268 | 1 |
| T2DM_insulin leukocyte | *THEM4* | -0.329634073 | 0.003773879 | 0.011330438 | 1 |
| T2DM_insulin leukocyte | *UBR1* | -0.501982654 | 0.003737258 | 0.011330438 | 1 |
| T2DM_insulin leukocyte | *TEPP* | 1.351496883 | 0.025745806 | 0.033956116 | 1 |

| | | | | | |
|---|---|---|---|---|---|
| T2DM_insulin leukocyte | *ZDHHC1* | -0.425670929 | 0.002628268 | 0.011330438 | 1 |
| T2DM_insulin leukocyte | *ZFYVE28* | -0.552547041 | 0.00366495 | 0.011330438 | 1 |
| T2DM_insulin leukocyte | *NPR2* | -0.365674088 | 0.037683103 | 0.047423738 | 1 |
| T2DM_insulin leukocyte | *ATAD3B* | -0.907636997 | 0.005636686 | 0.01154792 | 1 |
| T2DM_insulin leukocyte | *ANKLE1* | 0.225351963 | 0.027375915 | 0.035757969 | 1 |
| T2DM_insulin leukocyte | *KALRN* | -0.50059701 | 0.011852826 | 0.017633379 | 1 |
| T2DM_insulin leukocyte | *CILP2* | -0.903447851 | 0.008040065 | 0.013235147 | 1 |
| T2DM_insulin leukocyte | *TMPRSS3* | -1.091059708 | 0.011201419 | 0.01684018 | 1 |
| T2DM_insulin leukocyte | *RSPH1* | -0.289986568 | 0.002761647 | 0.011330438 | 1 |
| T2DM_insulin leukocyte | *SLC37A1* | -0.46048703 | 0.002167886 | 0.011330438 | 1 |
| T2DM_insulin leukocyte | *NDUFV3* | -0.303846239 | 0.003746816 | 0.011330438 | 1 |
| T2DM_insulin leukocyte | *HSF2BP* | -1.075279632 | 0.011902769 | 0.01766754 | 1 |
| T2DM_insulin leukocyte | *PDXK* | -0.496676605 | 0.001416963 | 0.011330438 | 1 |
| T2DM_insulin leukocyte | *CSTB* | -0.423341935 | 0.004219872 | 0.011330438 | 1 |
| T2DM_insulin leukocyte | *ICOSLG* | 0.168408026 | 0.031979884 | 0.040901356 | 1 |
| T2DM_insulin leukocyte | *ZNF66* | -1.048665273 | 0.006124202 | 0.011570498 | 1 |
| T2DM_insulin leukocyte | *FTCD* | -0.528304637 | 0.005745235 | 0.011570498 | 1 |
| T2DM_insulin leukocyte | *LSS* | -0.480062775 | 0.002440963 | 0.011330438 | 1 |
| T2DM_insulin leukocyte | *VAV2* | -0.59288871 | 0.005081835 | 0.011330438 | 1 |
| T2DM_insulin leukocyte | *DIP2A* | -0.289282703 | 0.017291128 | 0.023988997 | 1 |
| T2DM_insulin leukocyte | *ZNF208* | -0.37459832 | 0.003397884 | 0.011330438 | 1 |
| T2DM_insulin leukocyte | *SLC2A6* | -0.674328009 | 0.004818753 | 0.011330438 | 1 |
| T2DM_insulin leukocyte | *RDH13* | -0.43742184 | 0.008427202 | 0.013659437 | 1 |
| T2DM_insulin leukocyte | *MPZL3* | -0.418060716 | 0.006184642 | 0.011570498 | 1 |
| T2DM_insulin leukocyte | *S100A1* | -0.72507826 | 0.020162147 | 0.027426199 | 1 |
| T2DM_insulin leukocyte | *VPS11* | -0.750161213 | 0.003351872 | 0.011330438 | 1 |
| T2DM_insulin leukocyte | *PAQR6* | -0.444621746 | 0.007095266 | 0.01228624 | 1 |
| T2DM_insulin leukocyte | *SLC25A44* | -0.629188354 | 0.003662667 | 0.011330438 | 1 |
| T2DM_insulin leukocyte | *NBEAL2* | -0.243122638 | 0.023869945 | 0.031748481 | 1 |
| T2DM_insulin leukocyte | *LY6K* | -0.710907274 | 0.006794557 | 0.012014318 | 1 |
| T2DM_insulin leukocyte | *LY6E* | -0.387662726 | 0.008765064 | 0.014050565 | 1 |
| T2DM_insulin leukocyte | *VPS28* | -0.310615397 | 0.008833383 | 0.014115597 | 1 |
| T2DM_insulin leukocyte | *TONSL* | -1.658740058 | 0.011546086 | 0.017248887 | 1 |
| T2DM_insulin leukocyte | *ZNF333* | -1.213350747 | 0.009720181 | 0.015148984 | 1 |
| T2DM_insulin leukocyte | *RPL8* | -0.239674356 | 0.002816812 | 0.011330438 | 1 |
| T2DM_insulin leukocyte | *MAML1* | -0.708992459 | 0.004424457 | 0.011330438 | 1 |
| T2DM_insulin leukocyte | *LRWD1* | -0.566493522 | 0.005127468 | 0.011330438 | 1 |
| T2DM_insulin leukocyte | *U2AF1L4* | -0.299748319 | 0.008807585 | 0.014103329 | 1 |
| T2DM_insulin leukocyte | *ZNF382* | -0.403856081 | 0.004981329 | 0.011330438 | 1 |
| T2DM_insulin leukocyte | *DUSP14* | -0.501357385 | 0.003654565 | 0.011330438 | 1 |
| T2DM_insulin leukocyte | *PGAP3* | -0.411329223 | 0.006573409 | 0.011836354 | 1 |
| T2DM_insulin leukocyte | *PRPSAP1* | -0.430422272 | 0.001188807 | 0.011330438 | 1 |
| T2DM_insulin leukocyte | *SIGLEC11* | -6.970993462 | 0.004348883 | 0.011330438 | 1 |
| T2DM_insulin leukocyte | *DBF4B* | -0.61446535 | 0.002733766 | 0.011330438 | 1 |
| T2DM_insulin leukocyte | *LEMD2* | -0.796922865 | 0.003352358 | 0.011330438 | 1 |
| T2DM_insulin leukocyte | *CXCL16* | 0.142592095 | 0.017188056 | 0.023872903 | 1 |
| T2DM_insulin leukocyte | *RPL26* | -0.253888615 | 0.00289386 | 0.011330438 | 1 |
| T2DM_insulin leukocyte | *SNRNP25* | -0.354184418 | 0.003090902 | 0.011330438 | 1 |
| T2DM_insulin leukocyte | *WDR90* | -0.554014916 | 0.009507715 | 0.014932678 | 1 |
| T2DM_insulin leukocyte | *CCDC78* | -1.329084303 | 0.0019542 | 0.011330438 | 1 |

| | | | | | |
|---|---|---|---|---|---|
| T2DM_insulin leukocyte | *AMDHD2* | 0.338864378 | 0.035786064 | 0.045268334 | 1 |
| T2DM_insulin leukocyte | *ZNF75A* | -0.74881333 | 0.002305786 | 0.011330438 | 1 |
| T2DM_insulin leukocyte | *ASRGL1* | -0.310471276 | 0.006070473 | 0.011570498 | 1 |
| T2DM_insulin leukocyte | *TAF6L* | -0.457165713 | 0.016112744 | 0.022593495 | 1 |
| T2DM_insulin leukocyte | *ZFPL1* | -0.428230351 | 0.005146911 | 0.011330438 | 1 |
| T2DM_insulin leukocyte | *LRP5* | -0.701507544 | 0.004611466 | 0.011330438 | 1 |
| T2DM_insulin leukocyte | *PDZK1IP1* | -0.282710085 | 0.00398448 | 0.011330438 | 1 |
| T2DM_insulin leukocyte | *CTRC* | -0.193199799 | 0.0053709 | 0.011421159 | 1 |
| T2DM_insulin leukocyte | *SDC3* | -0.757293704 | 0.004129414 | 0.011330438 | 1 |
| T2DM_insulin leukocyte | *OMA1* | -0.518475585 | 0.010620982 | 0.016199471 | 1 |
| T2DM_insulin leukocyte | *TM2D1* | -0.559613538 | 0.002711556 | 0.011330438 | 1 |
| T2DM_insulin leukocyte | *FUBP1* | -0.319654367 | 0.004381225 | 0.011330438 | 1 |
| T2DM_insulin leukocyte | *SNX7* | -0.310647614 | 0.005798689 | 0.011570498 | 1 |
| T2DM_insulin leukocyte | *HFM1* | -0.691729429 | 0.010398763 | 0.015922346 | 1 |
| T2DM_insulin leukocyte | *VCAM1* | -0.215194843 | 0.006000145 | 0.011570498 | 1 |
| T2DM_insulin leukocyte | *ZNF496* | -0.385387615 | 0.004941923 | 0.011330438 | 1 |
| T2DM_insulin leukocyte | *TRIM58* | -1.497755759 | 0.010928751 | 0.016550349 | 1 |
| T2DM_insulin leukocyte | *PEA15* | -0.354985986 | 0.004641362 | 0.011330438 | 1 |
| T2DM_insulin leukocyte | *FLVCR1* | -0.312544641 | 0.004104826 | 0.011330438 | 1 |
| T2DM_insulin leukocyte | *TDRD5* | -1.233444 | 0.010218876 | 0.015732745 | 1 |
| T2DM_insulin leukocyte | *C1orf115* | -0.270924882 | 0.01192899 | 0.017699775 | 1 |
| T2DM_insulin leukocyte | *ACP6* | -0.474025609 | 0.009136466 | 0.014468834 | 1 |
| T2DM_insulin leukocyte | *HAAO* | -0.438209432 | 0.006554132 | 0.011815361 | 1 |
| T2DM_insulin leukocyte | *RFTN2* | -0.926778276 | 0.003187884 | 0.011330438 | 1 |
| T2DM_insulin leukocyte | *KCNJ3* | -0.339825065 | 0.008054367 | 0.01325193 | 1 |
| T2DM_insulin leukocyte | *DUSP19* | -0.557673345 | 0.00704619 | 0.012237751 | 1 |
| T2DM_insulin leukocyte | *SMC6* | -0.669443651 | 0.002694174 | 0.011330438 | 1 |
| T2DM_insulin leukocyte | *NOSTRIN* | -0.331881293 | 0.029421391 | 0.038050246 | 1 |
| T2DM_insulin leukocyte | *CTSS* | 0.094867565 | 0.010803454 | 0.016407719 | 1 |
| T2DM_insulin leukocyte | *MSX1* | -0.417264062 | 0.006660242 | 0.01190541 | 1 |
| T2DM_insulin leukocyte | *PACRGL* | -0.460582736 | 0.00327642 | 0.011330438 | 1 |
| T2DM_insulin leukocyte | *ERCC3* | -0.510958298 | 0.004957256 | 0.011330438 | 1 |
| T2DM_insulin leukocyte | *TDRD10* | -5.703590787 | 0.019409496 | 0.026525186 | 1 |
| T2DM_insulin leukocyte | *CCNYL1* | -0.947236697 | 0.002501513 | 0.011330438 | 1 |
| T2DM_insulin leukocyte | *DCAF16* | -0.5362442 | 0.00159406 | 0.011330438 | 1 |
| T2DM_insulin leukocyte | *NPPC* | -3.842139012 | 0.003061418 | 0.011330438 | 1 |
| T2DM_insulin leukocyte | *GNPDA2* | -0.262238147 | 0.030713528 | 0.039500311 | 1 |
| T2DM_insulin leukocyte | *NIPAL1* | -0.293428675 | 0.021696364 | 0.029243572 | 1 |
| T2DM_insulin leukocyte | *CGGBP1* | -0.3870249 | 0.002693154 | 0.011330438 | 1 |
| T2DM_insulin leukocyte | *DAPL1* | -1.28795251 | 0.00127205 | 0.011330438 | 1 |
| T2DM_insulin leukocyte | *EOGT* | -0.396517854 | 0.00448535 | 0.011330438 | 1 |
| T2DM_insulin leukocyte | *POGLUT1* | -0.302528801 | 0.008532223 | 0.013784171 | 1 |
| T2DM_insulin leukocyte | *ATP1A1* | -0.265936112 | 0.003195317 | 0.011330438 | 1 |
| T2DM_insulin leukocyte | *ARPC2* | -0.395002785 | 0.002729737 | 0.011330438 | 1 |
| T2DM_insulin leukocyte | *CCT3* | -0.267563307 | 0.007623729 | 0.01282334 | 1 |
| T2DM_insulin leukocyte | *EOMES* | -2.258760509 | 0.005785824 | 0.011570498 | 1 |
| T2DM_insulin leukocyte | *CWC22* | -0.41597801 | 0.00487682 | 0.011330438 | 1 |
| T2DM_insulin leukocyte | *HDAC11* | -0.995966707 | 0.009065429 | 0.014391139 | 1 |
| T2DM_insulin leukocyte | *SUCLG1* | -0.373625447 | 0.00377429 | 0.011330438 | 1 |
| T2DM_insulin leukocyte | *ICA1L* | -0.338972896 | 0.008688239 | 0.013955805 | 1 |

| | | | | | |
|---|---|---|---|---|---|
| T2DM_insulin leukocyte | *RYBP* | -0.304724342 | 0.002814385 | 0.011330438 | 1 |
| T2DM_insulin leukocyte | *GTPBP8* | -0.629638667 | 0.003931259 | 0.011330438 | 1 |
| T2DM_insulin leukocyte | *CADPS* | -0.197830273 | 0.006707722 | 0.0119468 | 1 |
| T2DM_insulin leukocyte | *ATXN7* | -0.444116834 | 0.003069617 | 0.011330438 | 1 |
| T2DM_insulin leukocyte | *PSMD6* | -0.344743313 | 0.003018834 | 0.011330438 | 1 |
| T2DM_insulin leukocyte | *PRICKLE2* | -0.425563567 | 0.00375562 | 0.011330438 | 1 |
| T2DM_insulin leukocyte | *RPL9* | -0.243371301 | 0.003431722 | 0.011330438 | 1 |
| T2DM_insulin leukocyte | *IL17RC* | -0.443210259 | 0.002697713 | 0.011330438 | 1 |
| T2DM_insulin leukocyte | *CRELD1* | -0.239446139 | 0.026505757 | 0.034818161 | 1 |
| T2DM_insulin leukocyte | *TTC14* | -0.629319784 | 0.002610898 | 0.011330438 | 1 |
| T2DM_insulin leukocyte | *CXCL3* | 0.319215589 | 0.032605824 | 0.041634151 | 1 |
| T2DM_insulin leukocyte | *CXCL1* | -0.252259119 | 0.006929696 | 0.01212774 | 1 |
| T2DM_insulin leukocyte | *CCDC158* | -1.018807153 | 0.005464875 | 0.011427973 | 1 |
| T2DM_insulin leukocyte | *TM4SF18* | -0.402330797 | 0.005342098 | 0.011421159 | 1 |
| T2DM_insulin leukocyte | *SNRK* | -0.274337769 | 0.005116814 | 0.011330438 | 1 |
| T2DM_insulin leukocyte | *KIAA1143* | -0.295609055 | 0.005355124 | 0.011421159 | 1 |
| T2DM_insulin leukocyte | *CCR1* | 0.328862419 | 0.019675525 | 0.026832745 | 1 |
| T2DM_insulin leukocyte | *NMNAT3* | -0.354256752 | 0.004555984 | 0.011330438 | 1 |
| T2DM_insulin leukocyte | *SMIM12* | -0.370648803 | 0.003878506 | 0.011330438 | 1 |
| T2DM_insulin leukocyte | *TPRA1* | -0.698360303 | 0.00272807 | 0.011330438 | 1 |
| T2DM_insulin leukocyte | *MEAF6* | -0.332337145 | 0.004040166 | 0.011330438 | 1 |
| T2DM_insulin leukocyte | *DNALI1* | -0.354487984 | 0.013918344 | 0.01996538 | 1 |
| T2DM_insulin leukocyte | *CAMK2N2* | -0.258548778 | 0.003678575 | 0.011330438 | 1 |
| T2DM_insulin leukocyte | *RPN1* | -0.393824711 | 0.004561017 | 0.011330438 | 1 |
| T2DM_insulin leukocyte | *IFT122* | -0.508234257 | 0.004937721 | 0.011330438 | 1 |
| T2DM_insulin leukocyte | *RFC4* | -0.489057982 | 0.003980633 | 0.011330438 | 1 |
| T2DM_insulin leukocyte | *TKT* | -0.403443006 | 0.002751823 | 0.011330438 | 1 |
| T2DM_insulin leukocyte | *RFT1* | -0.733270709 | 0.001503502 | 0.011330438 | 1 |
| T2DM_insulin leukocyte | *UVSSA* | -0.56692256 | 0.012204651 | 0.017937915 | 1 |
| T2DM_insulin leukocyte | *ARHGEF3* | -0.35223382 | 0.009634749 | 0.01506364 | 1 |
| T2DM_insulin leukocyte | *LRPAP1* | -0.286666579 | 0.01128224 | 0.016912599 | 1 |
| T2DM_insulin leukocyte | *PIGX* | -0.537204736 | 0.002432949 | 0.011330438 | 1 |
| T2DM_insulin leukocyte | *ERMAP* | -1.260737254 | 0.001781679 | 0.011330438 | 1 |
| T2DM_insulin leukocyte | *BDH2* | -0.393608978 | 0.002671363 | 0.011330438 | 1 |
| T2DM_insulin leukocyte | *CDC25A* | -0.673132863 | 0.00335888 | 0.011330438 | 1 |
| T2DM_insulin leukocyte | *TEX264* | -0.464220527 | 0.002922636 | 0.011330438 | 1 |
| T2DM_insulin leukocyte | *PPM1M* | -0.530856109 | 0.004074402 | 0.011330438 | 1 |
| T2DM_insulin leukocyte | *SAP30* | -0.362376402 | 0.006414542 | 0.01170553 | 1 |
| T2DM_insulin leukocyte | *ANXA5* | -0.242297808 | 0.004987871 | 0.011330438 | 1 |
| T2DM_insulin leukocyte | *CEP44* | -0.393237866 | 0.008894759 | 0.014171739 | 1 |
| T2DM_insulin leukocyte | *HPGD* | -0.58804244 | 0.017777174 | 0.024550621 | 1 |
| T2DM_insulin leukocyte | *NPY1R* | 0.87241736 | 0.03416983 | 0.043419555 | 1 |
| T2DM_insulin leukocyte | *ARFIP1* | -0.562639086 | 0.003094933 | 0.011330438 | 1 |
| T2DM_insulin leukocyte | *ITGA2* | -0.277964342 | 0.004495674 | 0.011330438 | 1 |
| T2DM_insulin leukocyte | *ELOVL7* | -0.422487185 | 0.001748107 | 0.011330438 | 1 |
| T2DM_insulin leukocyte | *LMBRD2* | -0.437360998 | 0.003748705 | 0.011330438 | 1 |
| T2DM_insulin leukocyte | *STARD4* | -0.510806887 | 0.003527325 | 0.011330438 | 1 |
| T2DM_insulin leukocyte | *CMBL* | -0.404329414 | 0.00446913 | 0.011330438 | 1 |
| T2DM_insulin leukocyte | *PRRC1* | -0.310630957 | 0.003670198 | 0.011330438 | 1 |
| T2DM_insulin leukocyte | *AGGF1* | -0.520550038 | 0.002457601 | 0.011330438 | 1 |

| | | | | | |
|---|---|---|---|---|---|
| T2DM_insulin leukocyte | *WDR41* | -0.600335236 | 0.003484972 | 0.011330438 | 1 |
| T2DM_insulin leukocyte | *SERINC5* | -1.237008545 | 0.012443035 | 0.018194523 | 1 |
| T2DM_insulin leukocyte | *RICTOR* | -0.545709167 | 0.002672519 | 0.011330438 | 1 |
| T2DM_insulin leukocyte | *UTP15* | -0.347258761 | 0.005088912 | 0.011330438 | 1 |
| T2DM_insulin leukocyte | *KLKB1* | -0.895488398 | 0.000706106 | 0.011330438 | 1 |
| T2DM_insulin leukocyte | *NSA2* | -0.246795441 | 0.007395085 | 0.012565211 | 1 |
| T2DM_insulin leukocyte | *LEAP2* | -0.610668572 | 0.005360994 | 0.011421159 | 1 |
| T2DM_insulin leukocyte | *DACT2* | -1.630995857 | 0.006193081 | 0.011570498 | 1 |
| T2DM_insulin leukocyte | *PDSS2* | -0.391328118 | 0.004332732 | 0.011330438 | 1 |
| T2DM_insulin leukocyte | *STK17A* | -0.333093726 | 0.005153927 | 0.011330438 | 1 |
| T2DM_insulin leukocyte | *GALNT10* | -0.63457573 | 0.027884054 | 0.036349186 | 1 |
| T2DM_insulin leukocyte | *SAP30L* | -0.508331899 | 0.004335175 | 0.011330438 | 1 |
| T2DM_insulin leukocyte | *COG5* | -0.67443415 | 0.002413593 | 0.011330438 | 1 |
| T2DM_insulin leukocyte | *SLU7* | -0.400688257 | 0.003307859 | 0.011330438 | 1 |
| T2DM_insulin leukocyte | *CAMLG* | -0.301263053 | 0.005405322 | 0.011421159 | 1 |
| T2DM_insulin leukocyte | *KIF6* | -0.723656254 | 0.00371173 | 0.011330438 | 1 |
| T2DM_insulin leukocyte | *STEAP1* | -0.547924804 | 0.004164478 | 0.011330438 | 1 |
| T2DM_insulin leukocyte | *FNDC1* | -2.148555803 | 0.025388589 | 0.033530006 | 1 |
| T2DM_insulin leukocyte | *CHMP4C* | -0.37526769 | 0.003949616 | 0.011330438 | 1 |
| T2DM_insulin leukocyte | *BRI3* | -0.456590766 | 0.002462321 | 0.011330438 | 1 |
| T2DM_insulin leukocyte | *LMTK2* | -0.404587685 | 0.003156922 | 0.011330438 | 1 |
| T2DM_insulin leukocyte | *DLC1* | -0.391812489 | 0.010731237 | 0.016316962 | 1 |
| T2DM_insulin leukocyte | *PEX2* | -0.33548661 | 0.003873795 | 0.011330438 | 1 |
| T2DM_insulin leukocyte | *MED30* | -0.373194281 | 0.005752009 | 0.011570498 | 1 |
| T2DM_insulin leukocyte | *TNFRSF11B* | -1.049593972 | 0.017586545 | 0.024330118 | 1 |
| T2DM_insulin leukocyte | *CSMD3* | -26.01378869 | 0.006250441 | 0.011570498 | 1 |
| T2DM_insulin leukocyte | *OSGIN2* | -0.536088182 | 0.002756014 | 0.011330438 | 1 |
| T2DM_insulin leukocyte | *DEFB1* | -0.174803713 | 0.005924885 | 0.011570498 | 1 |
| T2DM_insulin leukocyte | *SUN1* | -0.307110376 | 0.004126842 | 0.011330438 | 1 |
| T2DM_insulin leukocyte | *TMEM184A* | -0.419100316 | 0.003701682 | 0.011330438 | 1 |
| T2DM_insulin leukocyte | *INTS1* | -0.405038982 | 0.004059765 | 0.011330438 | 1 |
| T2DM_insulin leukocyte | *YWHAZ* | -0.336182368 | 0.003059057 | 0.011330438 | 1 |
| T2DM_insulin leukocyte | *DCAF13* | -0.402538154 | 0.003579135 | 0.011330438 | 1 |
| T2DM_insulin leukocyte | *INTS8* | -0.43068713 | 0.028493211 | 0.036983755 | 1 |
| T2DM_insulin leukocyte | *GEM* | -0.402297495 | 0.005986455 | 0.011570498 | 1 |
| T2DM_insulin leukocyte | *RPP25L* | -0.614017754 | 0.003300845 | 0.011330438 | 1 |
| T2DM_insulin leukocyte | *FAM219A* | -0.613524941 | 0.002905907 | 0.011330438 | 1 |
| T2DM_insulin leukocyte | *NUDT2* | -0.456156917 | 0.002204617 | 0.011330438 | 1 |
| T2DM_insulin leukocyte | *NIPSNAP3B* | 0.689142634 | 0.031780008 | 0.040698713 | 1 |
| T2DM_insulin leukocyte | *FXN* | -0.774801812 | 0.003650786 | 0.011330438 | 1 |
| T2DM_insulin leukocyte | *MAMDC2* | -0.88630404 | 0.005259091 | 0.011402654 | 1 |
| T2DM_insulin leukocyte | *PRSS37* | -214.900987 | 0.005997088 | 0.011570498 | 1 |
| T2DM_insulin leukocyte | *C9orf64* | -0.541199589 | 0.002443 | 0.011330438 | 1 |
| T2DM_insulin leukocyte | *ANKS6* | -0.38620189 | 0.004843056 | 0.011330438 | 1 |
| T2DM_insulin leukocyte | *CLDN3* | -0.394568295 | 0.003487324 | 0.011330438 | 1 |
| T2DM_insulin leukocyte | *WNK2* | -0.635616513 | 0.002552572 | 0.011330438 | 1 |
| T2DM_insulin leukocyte | *NOL6* | -0.762769838 | 0.003327706 | 0.011330438 | 1 |
| T2DM_insulin leukocyte | *PIGO* | -0.857800213 | 0.000704467 | 0.011330438 | 1 |
| T2DM_insulin leukocyte | *ARHGAP12* | -0.419360165 | 0.003905286 | 0.011330438 | 1 |
| T2DM_insulin leukocyte | *WRN* | -0.442209108 | 0.003719059 | 0.011330438 | 1 |

| | | | | | |
|---|---|---|---|---|---|
| T2DM_insulin leukocyte | *CFL2* | -0.394395991 | 0.003665459 | 0.011330438 | 1 |
| T2DM_insulin leukocyte | *PGM2L1* | -0.69319338 | 0.002153388 | 0.011330438 | 1 |
| T2DM_insulin leukocyte | *PHYHIPL* | -0.396197484 | 0.004439621 | 0.011330438 | 1 |
| T2DM_insulin leukocyte | *RPL36AL* | -0.279814087 | 0.004265608 | 0.011330438 | 1 |
| T2DM_insulin leukocyte | *TMEM63C* | -0.34952437 | 0.00485826 | 0.011330438 | 1 |
| T2DM_insulin leukocyte | *AKR1E2* | -1.560388847 | 0.015442927 | 0.021810191 | 1 |
| T2DM_insulin leukocyte | *OXGR1* | -0.865874525 | 0.006368752 | 0.011665654 | 1 |
| T2DM_insulin leukocyte | *VDAC2* | -0.350479671 | 0.002279672 | 0.011330438 | 1 |
| T2DM_insulin leukocyte | *SNAPC4* | -0.396869855 | 0.0056275 | 0.01154792 | 1 |
| T2DM_insulin leukocyte | *TSC1* | -0.560317122 | 0.003008556 | 0.011330438 | 1 |
| T2DM_insulin leukocyte | *ZMYND19* | -0.38568912 | 0.004709429 | 0.011330438 | 1 |
| T2DM_insulin leukocyte | *METTL17* | -0.536309177 | 0.00416882 | 0.011330438 | 1 |
| T2DM_insulin leukocyte | *VWA2* | -0.952425783 | 0.008405285 | 0.013629535 | 1 |
| T2DM_insulin leukocyte | *METTL3* | -0.421676421 | 0.004929603 | 0.011330438 | 1 |
| T2DM_insulin leukocyte | *TRUB1* | -0.613676225 | 0.001015015 | 0.011330438 | 1 |
| T2DM_insulin leukocyte | *ARHGAP42* | -0.564883821 | 0.009571913 | 0.015007201 | 1 |
| T2DM_insulin leukocyte | *SLC39A13* | -0.383186654 | 0.004094464 | 0.011330438 | 1 |
| T2DM_insulin leukocyte | *PSMC3* | -0.296744414 | 0.00618016 | 0.011570498 | 1 |
| T2DM_insulin leukocyte | *CPSF2* | -0.33509177 | 0.005245396 | 0.011392572 | 1 |
| T2DM_insulin leukocyte | *IFI27L1* | -0.409061699 | 0.004024051 | 0.011330438 | 1 |
| T2DM_insulin leukocyte | *SMCO4* | -0.419313505 | 0.003862396 | 0.011330438 | 1 |
| T2DM_insulin leukocyte | *ABTB2* | -0.522955469 | 0.012078101 | 0.017813386 | 1 |
| T2DM_insulin leukocyte | *LIPC* | -1.772988074 | 0.008555995 | 0.013816894 | 1 |
| T2DM_insulin leukocyte | *CEP57* | -0.348170839 | 0.003063118 | 0.011330438 | 1 |
| T2DM_insulin leukocyte | *JAM3* | -0.319099017 | 0.005282573 | 0.011419801 | 1 |
| T2DM_insulin leukocyte | *IKBIP* | -0.442128347 | 0.006396781 | 0.01170063 | 1 |
| T2DM_insulin leukocyte | *ZFYVE19* | -0.382225582 | 0.006219203 | 0.011570498 | 1 |
| T2DM_insulin leukocyte | *CKB* | -0.318330342 | 0.001006933 | 0.011330438 | 1 |
| T2DM_insulin leukocyte | *BTRC* | -0.696241864 | 0.005619308 | 0.011547208 | 1 |
| T2DM_insulin leukocyte | *BAG5* | -0.406639603 | 0.0035898 | 0.011330438 | 1 |
| T2DM_insulin leukocyte | *LARP6* | -0.350596709 | 0.002845899 | 0.011330438 | 1 |
| T2DM_insulin leukocyte | *ZNF319* | -1.119392955 | 0.002677278 | 0.011330438 | 1 |
| T2DM_insulin leukocyte | *SGPL1* | -0.610628846 | 0.004173998 | 0.011330438 | 1 |
| T2DM_insulin leukocyte | *FRS2* | -0.438992464 | 0.002815126 | 0.011330438 | 1 |
| T2DM_insulin leukocyte | *PCBD1* | -0.349375363 | 0.005614792 | 0.011547208 | 1 |
| T2DM_insulin leukocyte | *COX11* | -0.448860689 | 0.002884726 | 0.011330438 | 1 |
| T2DM_insulin leukocyte | *WBP1L* | -0.847273005 | 0.002717757 | 0.011330438 | 1 |
| T2DM_insulin leukocyte | *CYB5R2* | -0.432891322 | 0.003531479 | 0.011330438 | 1 |
| T2DM_insulin leukocyte | *TUB* | -0.473127314 | 0.010721684 | 0.016308748 | 1 |
| T2DM_insulin leukocyte | *RIC3* | -0.305750179 | 0.003889852 | 0.011330438 | 1 |
| T2DM_insulin leukocyte | *XRRA1* | -0.44047128 | 0.00245376 | 0.011330438 | 1 |
| T2DM_insulin leukocyte | *TRIM66* | -0.409733892 | 0.03483087 | 0.044188081 | 1 |
| T2DM_insulin leukocyte | *CDYL2* | -0.425430196 | 0.003693717 | 0.011330438 | 1 |
| T2DM_insulin leukocyte | *ZNF143* | -0.387815047 | 0.00452676 | 0.011330438 | 1 |
| T2DM_insulin leukocyte | *CCDC68* | -0.372170251 | 0.010334708 | 0.015871143 | 1 |
| T2DM_insulin leukocyte | *ZSCAN21* | -0.649878938 | 0.002244941 | 0.011330438 | 1 |
| T2DM_insulin leukocyte | *RIMKLB* | -0.389237133 | 0.003380696 | 0.011330438 | 1 |
| T2DM_insulin leukocyte | *GALR1* | 0.45634572 | 0.011220651 | 0.016852423 | 1 |
| T2DM_insulin leukocyte | *ATF7IP2* | -0.578481588 | 0.00110284 | 0.011330438 | 1 |
| T2DM_insulin leukocyte | *COG1* | -0.405902819 | 0.002233554 | 0.011330438 | 1 |

| | | | | | |
|---|---|---|---|---|---|
| T2DM_insulin leukocyte | *B2M* | -0.258643731 | 0.00227492 | 0.011330438 | 1 |
| T2DM_insulin leukocyte | *AP1G1* | -0.288273365 | 0.005330394 | 0.011421159 | 1 |
| T2DM_insulin leukocyte | *SLFN5* | -0.313948803 | 0.005302727 | 0.011421159 | 1 |
| T2DM_insulin leukocyte | *SAAL1* | -0.373847908 | 0.005438316 | 0.011421159 | 1 |
| T2DM_insulin leukocyte | *SCNN1G* | -3.400900227 | 0.005472142 | 0.011427973 | 1 |
| T2DM_insulin leukocyte | *DCTN5* | -0.489581162 | 0.000811187 | 0.011330438 | 1 |
| T2DM_insulin leukocyte | *STAT6* | -0.64832844 | 0.004693768 | 0.011330438 | 1 |
| T2DM_insulin leukocyte | *PATL1* | -0.341070056 | 0.004245456 | 0.011330438 | 1 |
| T2DM_insulin leukocyte | *MRPL16* | -0.566883292 | 0.013426599 | 0.019358939 | 1 |
| T2DM_insulin leukocyte | *GREM1* | -0.603120831 | 0.004837303 | 0.011330438 | 1 |
| T2DM_insulin leukocyte | *TSC22D4* | -0.468250517 | 0.009098639 | 0.014422203 | 1 |
| T2DM_insulin leukocyte | *MS4A14* | 0.082140011 | 0.020110635 | 0.027369132 | 1 |
| T2DM_insulin leukocyte | *DIS3L* | -0.623434539 | 0.00327785 | 0.011330438 | 1 |
| T2DM_insulin leukocyte | *MS4A8* | -1.226604317 | 0.003568434 | 0.011330438 | 1 |
| T2DM_insulin leukocyte | *RCCD1* | -1.05024455 | 0.001221718 | 0.011330438 | 1 |
| T2DM_insulin leukocyte | *MAPRE2* | -0.322612004 | 0.012971593 | 0.018806416 | 1 |
| T2DM_insulin leukocyte | *NAT16* | -0.278836722 | 0.00268877 | 0.011330438 | 1 |
| T2DM_insulin leukocyte | *GNGT2* | 0.169820767 | 0.013480916 | 0.019430124 | 1 |
| T2DM_insulin leukocyte | *SLC27A4* | -0.373441554 | 0.016565185 | 0.02317828 | 1 |
| T2DM_insulin leukocyte | *URM1* | -0.490283327 | 0.0063255 | 0.011629804 | 1 |
| T2DM_insulin leukocyte | *CERCAM* | -0.313556586 | 0.014787215 | 0.020989512 | 1 |
| T2DM_insulin leukocyte | *DOLPP1* | -0.444169493 | 0.007261272 | 0.012437884 | 1 |
| T2DM_insulin leukocyte | *ENDOG* | -0.50146401 | 0.001547599 | 0.011330438 | 1 |
| T2DM_insulin leukocyte | *PRR15L* | -0.508970593 | 0.010375595 | 0.01590871 | 1 |
| T2DM_insulin leukocyte | *RNF214* | -0.636925935 | 0.005403632 | 0.011421159 | 1 |
| T2DM_insulin leukocyte | *POP5* | -0.464917386 | 0.001371243 | 0.011330438 | 1 |
| T2DM_insulin leukocyte | *ACAA2* | -0.423888821 | 0.002712932 | 0.011330438 | 1 |
| T2DM_insulin leukocyte | *FN3K* | -0.492483117 | 0.007070213 | 0.012258988 | 1 |
| T2DM_insulin leukocyte | *ZNF23* | -0.364270149 | 0.005902344 | 0.011570498 | 1 |
| T2DM_insulin leukocyte | *ZNF229* | -0.523681898 | 0.004824736 | 0.011330438 | 1 |
| T2DM_insulin leukocyte | *ZNF180* | -0.401684968 | 0.006041373 | 0.011570498 | 1 |
| T2DM_insulin leukocyte | *CA4* | -0.23208988 | 0.032511834 | 0.041527632 | 1 |
| T2DM_insulin leukocyte | *GPX4* | -0.39931714 | 0.002189546 | 0.011330438 | 1 |
| T2DM_insulin leukocyte | *ZNF641* | -0.596251344 | 0.005447329 | 0.011427973 | 1 |
| T2DM_insulin leukocyte | *CACNB3* | -0.518868135 | 0.006742027 | 0.011963834 | 1 |
| T2DM_insulin leukocyte | *TP53I13* | -0.417545482 | 0.002017771 | 0.011330438 | 1 |
| T2DM_insulin leukocyte | *TUBA1C* | -0.245764355 | 0.006461362 | 0.011731372 | 1 |
| T2DM_insulin leukocyte | *ZNF528* | -1.273386879 | 0.006647988 | 0.011905364 | 1 |
| T2DM_insulin leukocyte | *RAB4B* | -0.496656693 | 0.002156118 | 0.011330438 | 1 |
| T2DM_insulin leukocyte | *TMC4* | -0.384316004 | 0.005286999 | 0.011419801 | 1 |
| T2DM_insulin leukocyte | *C19orf33* | -0.331774012 | 0.005971782 | 0.011570498 | 1 |
| T2DM_insulin leukocyte | *DNAAF3* | -1.338566582 | 0.006249165 | 0.011570498 | 1 |
| T2DM_insulin leukocyte | *CHAF1A* | -0.491689807 | 0.015536755 | 0.021910951 | 1 |
| T2DM_insulin leukocyte | *UBXN6* | -0.548414889 | 0.001530924 | 0.011330438 | 1 |
| T2DM_insulin leukocyte | *NXN* | -0.253824953 | 0.012472159 | 0.018223545 | 1 |
| T2DM_insulin leukocyte | *KIFC2* | -1.603203147 | 0.000608472 | 0.011330438 | 1 |
| T2DM_insulin leukocyte | *SERPINF2* | 0.18079335 | 0.018670684 | 0.025631418 | 1 |
| T2DM_insulin leukocyte | *TRPV3* | -2.013613347 | 0.002675485 | 0.011330438 | 1 |
| T2DM_insulin leukocyte | *ZNF83* | -0.506003361 | 0.003297576 | 0.011330438 | 1 |
| T2DM_insulin leukocyte | *NDUFV1* | -0.353815111 | 0.004108447 | 0.011330438 | 1 |

| | | | | | |
|---|---|---|---|---|---|
| T2DM_insulin leukocyte | *ZNF232* | -0.357879901 | 0.005118586 | 0.011330438 | 1 |
| T2DM_insulin leukocyte | *MIS12* | -0.423294481 | 0.011060241 | 0.0167073 | 1 |
| T2DM_insulin leukocyte | *HID1* | -0.429881154 | 0.003690569 | 0.011330438 | 1 |
| T2DM_insulin leukocyte | *SRP68* | -0.330762526 | 0.003823025 | 0.011330438 | 1 |
| T2DM_insulin leukocyte | *TMEM68* | -0.521648066 | 0.003943423 | 0.011330438 | 1 |
| T2DM_insulin leukocyte | *MLST8* | -0.291467122 | 0.009734893 | 0.015165898 | 1 |
| T2DM_insulin leukocyte | *ECI1* | -0.312224759 | 0.003979104 | 0.011330438 | 1 |
| T2DM_insulin leukocyte | *ABCA3* | -0.377699104 | 0.0040564 | 0.011330438 | 1 |
| T2DM_insulin leukocyte | *BSCL2* | -0.293444163 | 0.010346792 | 0.015879901 | 1 |
| T2DM_insulin leukocyte | *C2CD3* | -0.721369485 | 0.003398192 | 0.011330438 | 1 |
| T2DM_insulin leukocyte | *TRANK1* | -0.983329303 | 0.004500204 | 0.011330438 | 1 |
| T2DM_insulin leukocyte | *RPSA* | -0.261376767 | 0.002734542 | 0.011330438 | 1 |
| T2DM_insulin leukocyte | *ENTPD3* | -0.276270812 | 0.002740996 | 0.011330438 | 1 |
| T2DM_insulin leukocyte | *LTBP3* | -0.342448835 | 0.002761603 | 0.011330438 | 1 |
| T2DM_insulin leukocyte | *SAC3D1* | -0.461838238 | 0.003710106 | 0.011330438 | 1 |
| T2DM_insulin leukocyte | *PBK* | -0.881527516 | 0.004385711 | 0.011330438 | 1 |
| T2DM_insulin leukocyte | *KIAA1586* | -0.567663735 | 0.000833351 | 0.011330438 | 1 |
| T2DM_insulin leukocyte | *FAM83B* | -0.408805507 | 0.008216769 | 0.013442029 | 1 |
| T2DM_insulin leukocyte | *LMBRD1* | -0.452287749 | 0.003505909 | 0.011330438 | 1 |
| T2DM_insulin leukocyte | *TTC39C* | -0.354904841 | 0.005392458 | 0.011421159 | 1 |
| T2DM_insulin leukocyte | *POLR2J3* | -0.412002293 | 0.005020768 | 0.011330438 | 1 |
| T2DM_insulin leukocyte | *COA6* | -0.427280956 | 0.003691379 | 0.011330438 | 1 |
| T2DM_insulin leukocyte | *PXK* | -0.525778115 | 0.003120724 | 0.011330438 | 1 |
| T2DM_insulin leukocyte | *ING5* | -0.691084276 | 0.002660361 | 0.011330438 | 1 |
| T2DM_insulin leukocyte | *BMP1* | -0.383058499 | 0.016642856 | 0.023278669 | 1 |
| T2DM_insulin leukocyte | *HEXIM2* | -0.472463728 | 0.006065926 | 0.011570498 | 1 |
| T2DM_insulin leukocyte | *SERINC2* | -0.260755972 | 0.005504041 | 0.011434531 | 1 |
| T2DM_insulin leukocyte | *TRAPPC11* | -0.30865433 | 0.00705804 | 0.012248692 | 1 |
| T2DM_insulin leukocyte | *ING2* | -0.279099773 | 0.006648096 | 0.011905364 | 1 |
| T2DM_insulin leukocyte | *ADAM9* | -0.559074485 | 0.026752008 | 0.035059586 | 1 |
| T2DM_insulin leukocyte | *NDUFS5* | -0.318959517 | 0.001585707 | 0.011330438 | 1 |
| T2DM_insulin leukocyte | *DNAJC21* | -0.44089136 | 0.002104658 | 0.011330438 | 1 |
| T2DM_insulin leukocyte | *CNNM3* | -0.38539215 | 0.003996315 | 0.011330438 | 1 |
| T2DM_insulin leukocyte | *GSTM4* | -0.420494444 | 0.008376362 | 0.013599474 | 1 |
| T2DM_insulin leukocyte | *PPIP5K1* | -1.118636481 | 0.001144655 | 0.011330438 | 1 |
| T2DM_insulin leukocyte | *ADAL* | -1.086508599 | 0.004633936 | 0.011330438 | 1 |
| T2DM_insulin leukocyte | *LCMT2* | -0.642776998 | 0.002087009 | 0.011330438 | 1 |
| T2DM_insulin leukocyte | *ZNF507* | -0.492142039 | 0.003160737 | 0.011330438 | 1 |
| T2DM_insulin leukocyte | *NSG1* | -0.286136645 | 0.006174833 | 0.011570498 | 1 |
| T2DM_insulin leukocyte | *ZBTB49* | -1.130873226 | 0.002114816 | 0.011330438 | 1 |
| T2DM_insulin leukocyte | *GFM1* | -0.336292592 | 0.003193083 | 0.011330438 | 1 |
| T2DM_insulin leukocyte | *FSTL5* | -0.231018323 | 0.015598211 | 0.021989442 | 1 |
| T2DM_insulin leukocyte | *CTRB1* | -0.171545473 | 0.004060609 | 0.011330438 | 1 |
| T2DM_insulin leukocyte | *CTRB2* | -0.15630148 | 0.005593394 | 0.011524093 | 1 |
| T2DM_insulin leukocyte | *TMEM129* | -0.521509432 | 0.006587852 | 0.011847079 | 1 |
| T2DM_insulin leukocyte | *PPIC* | -0.191958234 | 0.013562968 | 0.019526897 | 1 |
| T2DM_insulin leukocyte | *STXBP6* | -0.44370613 | 0.003755839 | 0.011330438 | 1 |
| T2DM_insulin leukocyte | *MFF* | -0.39023505 | 0.002597048 | 0.011330438 | 1 |
| T2DM_insulin leukocyte | *CPLX1* | -0.372069366 | 0.009985869 | 0.015458818 | 1 |
| T2DM_insulin leukocyte | *ROR2* | -1.046575002 | 0.012023369 | 0.017759823 | 1 |

| | | | | | |
|---|---|---|---|---|---|
| T2DM_insulin leukocyte | *FAM110B* | -0.410256516 | 0.004346117 | 0.011330438 | 1 |
| T2DM_insulin leukocyte | *ATF5* | -0.469307138 | 0.003617104 | 0.011330438 | 1 |
| T2DM_insulin leukocyte | *XPO6* | -0.699777946 | 0.00201325 | 0.011330438 | 1 |
| T2DM_insulin leukocyte | *RGS14* | -0.545991175 | 0.006082335 | 0.011570498 | 1 |
| T2DM_insulin leukocyte | *TBC1D10B* | -0.503594271 | 0.003005201 | 0.011330438 | 1 |
| T2DM_insulin leukocyte | *PRELID1* | -0.303287545 | 0.002991778 | 0.011330438 | 1 |
| T2DM_insulin leukocyte | *CXCL10* | -66.23312583 | 0.006079991 | 0.011570498 | 1 |
| T2DM_insulin leukocyte | *CXCL11* | 0.505924112 | 0.000837989 | 0.011330438 | 1 |
| T2DM_insulin leukocyte | *NMD3* | -0.383279139 | 0.003238183 | 0.011330438 | 1 |
| T2DM_insulin leukocyte | *STK32A* | -0.478872735 | 0.002922229 | 0.011330438 | 1 |
| T2DM_insulin leukocyte | *P2RY12* | -22.24136849 | 0.026625431 | 0.034951994 | 1 |
| T2DM_insulin leukocyte | *SNUPN* | -1.085430348 | 0.001239623 | 0.011330438 | 1 |
| T2DM_insulin leukocyte | *CRADD* | -0.397386506 | 0.007241485 | 0.012418942 | 1 |
| T2DM_insulin leukocyte | *COL22A1* | -3.77316749 | 0.007991111 | 0.013208698 | 1 |
| T2DM_insulin leukocyte | *SDC2* | -0.774095796 | 0.011458711 | 0.017150962 | 1 |
| T2DM_insulin leukocyte | *CLIC4* | -0.22190576 | 0.006730691 | 0.011961858 | 1 |
| T2DM_insulin leukocyte | *SLC38A11* | -2.801342717 | 0.011174177 | 0.016816769 | 1 |
| T2DM_insulin leukocyte | *HINT1* | -0.278175699 | 0.001991596 | 0.011330438 | 1 |
| T2DM_insulin leukocyte | *INO80E* | -0.652204731 | 0.000145849 | 0.011330438 | 0.573042456 |
| T2DM_insulin leukocyte | *C15orf40* | -0.403335229 | 0.002929553 | 0.011330438 | 1 |
| T2DM_insulin leukocyte | *HIC2* | -1.429474088 | 0.009869645 | 0.015333268 | 1 |
| T2DM_insulin leukocyte | *AGPAT2* | -0.405140343 | 0.002947395 | 0.011330438 | 1 |
| T2DM_insulin leukocyte | *FASN* | -0.704599481 | 0.001436789 | 0.011330438 | 1 |
| T2DM_insulin leukocyte | *MT1E* | -0.221750356 | 0.006182477 | 0.011570498 | 1 |
| T2DM_insulin leukocyte | *LDB2* | -0.380063402 | 0.015908535 | 0.022347027 | 1 |
| T2DM_insulin leukocyte | *RAC3* | -0.565997707 | 0.002531027 | 0.011330438 | 1 |
| T2DM_insulin leukocyte | *LIMS1* | -0.440586005 | 0.003427615 | 0.011330438 | 1 |
| T2DM_insulin leukocyte | *CSGALNACT2* | -0.8683514 | 0.012371949 | 0.0181108 | 1 |
| T2DM_insulin leukocyte | *AVEN* | -1.07139535 | 0.002889704 | 0.011330438 | 1 |
| T2DM_insulin leukocyte | *CTNND2* | -0.390847185 | 0.008390186 | 0.013614784 | 1 |
| T2DM_insulin leukocyte | *WNT10B* | -22.43087438 | 0.007234287 | 0.012417437 | 1 |
| T2DM_insulin leukocyte | *CALML6* | 0.392235096 | 0.019926208 | 0.02713694 | 1 |
| T2DM_insulin leukocyte | *TM4SF4* | -0.367713913 | 0.003018642 | 0.011330438 | 1 |
| T2DM_insulin leukocyte | *OTUD3* | -0.302630805 | 0.003203919 | 0.011330438 | 1 |
| T2DM_insulin leukocyte | *MAP3K2* | -0.472579099 | 0.003683687 | 0.011330438 | 1 |
| T2DM_insulin leukocyte | *PUSL1* | -0.803890414 | 0.004056652 | 0.011330438 | 1 |
| T2DM_insulin leukocyte | *IFFO2* | -0.339212288 | 0.008140504 | 0.013360083 | 1 |
| T2DM_insulin leukocyte | *MYRIP* | -0.505578356 | 0.00627263 | 0.011570498 | 1 |
| T2DM_insulin leukocyte | *FAM153A* | -0.806008112 | 0.008493435 | 0.013744113 | 1 |
| T2DM_insulin leukocyte | *ZNF778* | -0.550872262 | 0.002007522 | 0.011330438 | 1 |
| T2DM_insulin leukocyte | *NIPA1* | -0.465922901 | 0.00441612 | 0.011330438 | 1 |
| T2DM_insulin leukocyte | *FOXD4* | -4.719343257 | 0.020532051 | 0.027865433 | 1 |
| T2DM_insulin leukocyte | *UBE2E1* | -0.660899391 | 0.002571218 | 0.011330438 | 1 |
| T2DM_insulin leukocyte | *ANKK1* | -10.26234136 | 0.005871055 | 0.011570498 | 1 |
| T2DM_insulin leukocyte | *USP47* | -0.458009188 | 0.002723515 | 0.011330438 | 1 |
| T2DM_insulin leukocyte | *PDCD6IP* | -0.375534371 | 0.003795834 | 0.011330438 | 1 |
| T2DM_insulin leukocyte | *CRTAP* | -0.698585042 | 0.002476464 | 0.011330438 | 1 |
| T2DM_insulin leukocyte | *STX8* | -0.553624943 | 0.001928292 | 0.011330438 | 1 |
| T2DM_insulin leukocyte | *NFRKB* | -0.687542077 | 0.002993486 | 0.011330438 | 1 |
| T2DM_insulin leukocyte | *SETMAR* | -0.381800987 | 0.004183998 | 0.011330438 | 1 |

| | | | | | |
|---|---|---|---|---|---|
| T2DM_insulin leukocyte | *CST2* | -0.914131437 | 0.001567204 | 0.011330438 | 1 |
| T2DM_insulin leukocyte | *KRT8* | -0.163504727 | 0.00537056 | 0.011421159 | 1 |
| T2DM_insulin leukocyte | *MGMT* | -0.508473964 | 0.003089417 | 0.011330438 | 1 |
| T2DM_insulin leukocyte | *NFXL1* | -0.728889653 | 0.000739404 | 0.011330438 | 1 |
| T2DM_insulin leukocyte | *DNAJC18* | -0.370962045 | 0.037984892 | 0.047788229 | 1 |
| T2DM_insulin leukocyte | *KRT4* | -23.45540345 | 0.003627706 | 0.011330438 | 1 |
| T2DM_insulin leukocyte | *SLC23A1* | -3.136630693 | 0.009639694 | 0.015065376 | 1 |
| T2DM_insulin leukocyte | *NPAS2* | -0.30171064 | 0.007960885 | 0.013183733 | 1 |
| T2DM_insulin leukocyte | *NUDT9* | -0.31296437 | 0.007661664 | 0.012869015 | 1 |
| T2DM_insulin leukocyte | *SMAGP* | -0.246005569 | 0.022201564 | 0.029813272 | 1 |
| T2DM_insulin leukocyte | *IRX1* | -0.553811322 | 0.006268213 | 0.011570498 | 1 |
| T2DM_insulin leukocyte | *CDH2* | -0.30822971 | 0.006178444 | 0.011570498 | 1 |
| T2DM_insulin leukocyte | *HSPA4* | -0.417030227 | 0.002518881 | 0.011330438 | 1 |
| T2DM_insulin leukocyte | *ARMC10* | -0.495009009 | 0.007391048 | 0.012565211 | 1 |
| T2DM_insulin leukocyte | *ACYP2* | -0.353763255 | 0.004020356 | 0.011330438 | 1 |
| T2DM_insulin leukocyte | *ATF7* | -0.412607876 | 0.003951144 | 0.011330438 | 1 |
| T2DM_insulin leukocyte | *KCNS3* | -0.509024524 | 0.006821251 | 0.012014984 | 1 |
| T2DM_insulin leukocyte | *FOXN2* | -1.065141233 | 0.004547183 | 0.011330438 | 1 |
| T2DM_insulin leukocyte | *GPR27* | -0.409873375 | 0.002942552 | 0.011330438 | 1 |
| T2DM_insulin leukocyte | *MTSS1* | -0.570029877 | 0.002170411 | 0.011330438 | 1 |
| T2DM_insulin leukocyte | *TMEM43* | -0.383677432 | 0.005140766 | 0.011330438 | 1 |
| T2DM_insulin leukocyte | *RPS9* | -0.250260597 | 0.002631542 | 0.011330438 | 1 |
| T2DM_insulin leukocyte | *NDUFA3* | -0.40892478 | 0.00354856 | 0.011330438 | 1 |
| T2DM_insulin leukocyte | *ZNF160* | -0.400507314 | 0.003659608 | 0.011330438 | 1 |
| T2DM_insulin leukocyte | *ZNF415* | -0.372622503 | 0.007441356 | 0.01261851 | 1 |
| T2DM_insulin leukocyte | *TSNARE1* | 0.377729142 | 0.012915607 | 0.018752927 | 1 |
| T2DM_insulin leukocyte | *SOX7* | -0.502103131 | 0.00617052 | 0.011570498 | 1 |
| T2DM_insulin leukocyte | *TRMT61B* | -0.339662557 | 0.004940432 | 0.011330438 | 1 |
| T2DM_insulin leukocyte | *INSR* | -0.272971934 | 0.005508869 | 0.011434531 | 1 |
| T2DM_insulin leukocyte | *MFN1* | -0.421618572 | 0.003786161 | 0.011330438 | 1 |
| T2DM_insulin leukocyte | *KCNMB3* | -0.73855048 | 0.00158685 | 0.011330438 | 1 |
| T2DM_insulin leukocyte | *ATP6V0E2* | -0.256138196 | 0.026126213 | 0.034342552 | 1 |
| T2DM_insulin leukocyte | *JAGN1* | -0.431695334 | 0.003138198 | 0.011330438 | 1 |
| T2DM_insulin leukocyte | *MORN4* | -0.313150598 | 0.001874092 | 0.011330438 | 1 |
| T2DM_insulin leukocyte | *TRIM8* | -0.445410962 | 0.002946648 | 0.011330438 | 1 |
| T2DM_insulin leukocyte | *CDC42BPG* | -0.27395463 | 0.007028353 | 0.012235001 | 1 |
| T2DM_insulin leukocyte | *ZNF440* | -1.490590022 | 0.000408232 | 0.011330438 | 1 |
| T2DM_insulin leukocyte | *GAA* | -0.535221889 | 0.001463817 | 0.011330438 | 1 |
| T2DM_insulin leukocyte | *EXOSC1* | -0.463775202 | 0.007447489 | 0.012623462 | 1 |
| T2DM_insulin leukocyte | *PGAM1* | -0.258285634 | 0.029504274 | 0.038132332 | 1 |
| T2DM_insulin leukocyte | *ESCO2* | -1.377568917 | 0.01136028 | 0.017016599 | 1 |
| T2DM_insulin leukocyte | *MRPL36* | -0.401429604 | 0.002743396 | 0.011330438 | 1 |
| T2DM_insulin leukocyte | *MCC* | -0.501421712 | 0.004993281 | 0.011330438 | 1 |
| T2DM_insulin leukocyte | *ZNF561* | -0.466272268 | 0.003405283 | 0.011330438 | 1 |
| T2DM_insulin leukocyte | *PPID* | -0.497850687 | 0.014054796 | 0.020131715 | 1 |
| T2DM_insulin leukocyte | *PTGER4* | 0.223306443 | 0.026961799 | 0.035279887 | 1 |
| T2DM_insulin leukocyte | *MAP6* | -0.255274079 | 0.010645991 | 0.016225019 | 1 |
| T2DM_insulin leukocyte | *ZNF584* | -0.555314022 | 0.012638746 | 0.018432677 | 1 |
| T2DM_insulin leukocyte | *NMUR1* | -4.222725203 | 0.005392539 | 0.011421159 | 1 |
| T2DM_insulin leukocyte | *CLSTN1* | -0.363582826 | 0.001243694 | 0.011330438 | 1 |

| | | | | | |
|---|---|---|---|---|---|
| T2DM_insulin leukocyte | *PIK3CD* | 0.31521382 | 0.030138006 | 0.038874664 | 1 |
| T2DM_insulin leukocyte | *RGS19* | -0.681144014 | 0.001711128 | 0.011330438 | 1 |
| T2DM_insulin leukocyte | *ANO5* | -0.457957741 | 0.008238711 | 0.013448692 | 1 |
| T2DM_insulin leukocyte | *GPHN* | -0.693300925 | 0.002492935 | 0.011330438 | 1 |
| T2DM_insulin leukocyte | *VAT1L* | -0.430802212 | 0.014742683 | 0.020949006 | 1 |
| T2DM_insulin leukocyte | *LGALS4* | -0.20498604 | 0.008944173 | 0.014233153 | 1 |
| T2DM_insulin leukocyte | *LRRC34* | -0.422976865 | 0.014128623 | 0.020230087 | 1 |
| T2DM_insulin leukocyte | *SPATA5L1* | -0.84738071 | 0.000988606 | 0.011330438 | 1 |
| T2DM_insulin leukocyte | *CTPS1* | -0.342954864 | 0.008676029 | 0.013941971 | 1 |
| T2DM_insulin leukocyte | *METTL18* | -0.50497683 | 0.004833625 | 0.011330438 | 1 |
| T2DM_insulin leukocyte | *PWWP2B* | -0.486895935 | 0.003600333 | 0.011330438 | 1 |
| T2DM_insulin leukocyte | *PTEN* | -0.511495145 | 0.000769611 | 0.011330438 | 1 |
| T2DM_insulin leukocyte | *CYP4F11* | -0.957103912 | 0.009428273 | 0.014830386 | 1 |
| T2DM_insulin leukocyte | *TLN2* | -0.756178618 | 0.005798688 | 0.011570498 | 1 |
| T2DM_insulin leukocyte | *TVP23B* | -0.68590019 | 0.00241844 | 0.011330438 | 1 |
| T2DM_insulin leukocyte | *SRGAP2C* | -0.802817726 | 0.001789179 | 0.011330438 | 1 |
| T2DM_insulin leukocyte | *ATPAF2* | -0.519686802 | 0.002856337 | 0.011330438 | 1 |
| T2DM_insulin leukocyte | *ZNF556* | 0.364769474 | 0.008437781 | 0.013670944 | 1 |
| T2DM_insulin leukocyte | *ZNF554* | -0.428970623 | 0.00568274 | 0.011549238 | 1 |
| T2DM_insulin leukocyte | *GAP43* | -0.303513313 | 0.005833652 | 0.011570498 | 1 |
| T2DM_insulin leukocyte | *REG1B* | -0.181080686 | 0.004183306 | 0.011330438 | 1 |
| T2DM_insulin leukocyte | *NME6* | -0.678219411 | 0.004110865 | 0.011330438 | 1 |
| T2DM_insulin leukocyte | *TEFM* | -0.52495125 | 0.003082134 | 0.011330438 | 1 |
| T2DM_insulin leukocyte | *MBOAT1* | -0.786628361 | 0.006011699 | 0.011570498 | 1 |
| T2DM_insulin leukocyte | *ZNF131* | -0.408751402 | 0.003484413 | 0.011330438 | 1 |
| T2DM_insulin leukocyte | *BSG* | -0.337292762 | 0.003035827 | 0.011330438 | 1 |
| T2DM_insulin leukocyte | *HINFP* | -1.155906451 | 0.00260241 | 0.011330438 | 1 |
| T2DM_insulin leukocyte | *CERS6* | -0.396469312 | 0.002219286 | 0.011330438 | 1 |
| T2DM_insulin leukocyte | *SPTLC3* | 0.4702501 | 0.012384151 | 0.018121911 | 1 |
| T2DM_insulin leukocyte | *COPRS* | -0.286131917 | 0.004831139 | 0.011330438 | 1 |
| T2DM_insulin leukocyte | *SUCLG2* | -0.698376633 | 0.00242723 | 0.011330438 | 1 |
| T2DM_insulin leukocyte | *CSDC2* | -1.298379813 | 0.001329879 | 0.011330438 | 1 |
| T2DM_insulin leukocyte | *ARNT2* | -0.302434473 | 0.011555593 | 0.017256528 | 1 |
| T2DM_insulin leukocyte | *ZNF24* | -0.435679175 | 0.001866995 | 0.011330438 | 1 |
| T2DM_insulin leukocyte | *MANEA* | -0.483003283 | 0.002076853 | 0.011330438 | 1 |
| T2DM_insulin leukocyte | *AFF1* | -0.455296512 | 0.002102277 | 0.011330438 | 1 |
| T2DM_insulin leukocyte | *CTSW* | -0.304718103 | 0.034696574 | 0.044046152 | 1 |
| T2DM_insulin leukocyte | *PDE3A* | -0.607523448 | 0.003214231 | 0.011330438 | 1 |
| T2DM_insulin leukocyte | *RAD9A* | -0.722276389 | 0.00343613 | 0.011330438 | 1 |
| T2DM_insulin leukocyte | *EFEMP2* | -0.415968375 | 0.004640716 | 0.011330438 | 1 |
| T2DM_insulin leukocyte | *ZNF738* | -0.39410355 | 0.028443045 | 0.03694305 | 1 |
| T2DM_insulin leukocyte | *FUT10* | -1.292894237 | 0.017226304 | 0.023915953 | 1 |
| T2DM_insulin leukocyte | *NAA16* | -0.369647542 | 0.007048621 | 0.012237751 | 1 |
| T2DM_insulin leukocyte | *OVOL1* | -0.937244567 | 0.010337046 | 0.015871143 | 1 |
| T2DM_insulin leukocyte | *DMXL1* | -0.574884316 | 0.006235508 | 0.011570498 | 1 |
| T2DM_insulin leukocyte | *NADSYN1* | -0.6940019 | 0.005850482 | 0.011570498 | 1 |
| T2DM_insulin leukocyte | *RNASEH2C* | -0.393416741 | 0.005191815 | 0.011376821 | 1 |
| T2DM_insulin leukocyte | *LCLAT1* | -0.417805985 | 0.002658624 | 0.011330438 | 1 |
| T2DM_insulin leukocyte | *SH3RF3* | -0.602511968 | 0.005786159 | 0.011570498 | 1 |
| T2DM_insulin leukocyte | *DCAKD* | -0.296716584 | 0.006910998 | 0.012100405 | 1 |

| | | | | | |
|---|---|---|---|---|---|
| T2DM_insulin leukocyte | *TADA2B* | -0.463168259 | 0.00319114 | 0.011330438 | 1 |
| T2DM_insulin leukocyte | *TRMT112* | -0.350535158 | 0.002983979 | 0.011330438 | 1 |
| T2DM_insulin leukocyte | *NOC3L* | -0.364312512 | 0.007972699 | 0.013189362 | 1 |
| T2DM_insulin leukocyte | *RHOD* | -0.260170521 | 0.005933194 | 0.011570498 | 1 |
| T2DM_insulin leukocyte | *PARP14* | -0.386695036 | 0.006455969 | 0.01173015 | 1 |
| T2DM_insulin leukocyte | *VANGL1* | -0.401426768 | 0.004283251 | 0.011330438 | 1 |
| T2DM_insulin leukocyte | *IQCB1* | -0.959133235 | 0.002103446 | 0.011330438 | 1 |
| T2DM_insulin leukocyte | *ZNF483* | -0.328765047 | 0.004407417 | 0.011330438 | 1 |
| T2DM_insulin leukocyte | *SLC2A14* | -1.514148344 | 0.006471729 | 0.011737322 | 1 |
| T2DM_insulin leukocyte | *MZT2A* | -0.2967081 | 0.006344764 | 0.011637991 | 1 |
| T2DM_insulin leukocyte | *PPP1R3B* | -0.872110158 | 0.005481386 | 0.011427973 | 1 |
| T2DM_insulin leukocyte | *MAP3K11* | -0.366653342 | 0.004079055 | 0.011330438 | 1 |
| T2DM_insulin leukocyte | *NDNF* | 0.392141326 | 0.01515631 | 0.021466886 | 1 |
| T2DM_insulin leukocyte | *ARV1* | -0.367731128 | 0.003503923 | 0.011330438 | 1 |
| T2DM_insulin leukocyte | *EHBP1L1* | -0.339073548 | 0.021979593 | 0.029544243 | 1 |
| T2DM_insulin leukocyte | *PPP1R14B* | -0.335139516 | 0.002455239 | 0.011330438 | 1 |
| T2DM_insulin leukocyte | *PEAK1* | -0.248051825 | 0.039106414 | 0.049073491 | 1 |
| T2DM_insulin leukocyte | *TNFRSF10D* | -0.481059842 | 0.004756847 | 0.011330438 | 1 |
| T2DM_insulin leukocyte | *GMPPB* | -0.533979375 | 0.010331867 | 0.015871143 | 1 |
| T2DM_insulin leukocyte | *NUDT18* | -0.395982839 | 0.005233009 | 0.011389288 | 1 |
| T2DM_insulin leukocyte | *NUDT4* | -0.321103467 | 0.012163186 | 0.01789856 | 1 |
| T2DM_insulin leukocyte | *PC* | -0.630829351 | 0.003554348 | 0.011330438 | 1 |
| T2DM_insulin leukocyte | *UQCRH* | -0.338955189 | 0.002550785 | 0.011330438 | 1 |
| T2DM_insulin leukocyte | *HEG1* | -0.384790682 | 0.005468475 | 0.011427973 | 1 |
| T2DM_insulin leukocyte | *HAP1* | -0.464531946 | 0.019512291 | 0.026656394 | 1 |
| T2DM_insulin leukocyte | *ENDOV* | -0.682060198 | 0.007089531 | 0.01228624 | 1 |
| T2DM_insulin leukocyte | *DPY19L1* | -0.483068036 | 0.003457426 | 0.011330438 | 1 |
| T2DM_insulin leukocyte | *TUBB8* | -11.11014481 | 0.028589115 | 0.037095981 | 1 |
| T2DM_insulin leukocyte | *GPR160* | -0.558806391 | 0.006337688 | 0.011637991 | 1 |
| T2DM_insulin leukocyte | *XXYLT1* | -0.539871716 | 0.01439186 | 0.020539636 | 1 |
| T2DM_insulin leukocyte | *GNG5* | -0.346042594 | 0.002527846 | 0.011330438 | 1 |
| T2DM_insulin leukocyte | *FAM53A* | -1.771314737 | 0.00636738 | 0.011665654 | 1 |
| T2DM_insulin leukocyte | *SELP* | -2.305806808 | 0.005995265 | 0.011570498 | 1 |
| T2DM_insulin leukocyte | *CTU2* | -0.385855411 | 0.004308474 | 0.011330438 | 1 |
| T2DM_insulin leukocyte | *SNX31* | -2.918590283 | 0.017713548 | 0.024479962 | 1 |
| T2DM_insulin leukocyte | *ADCY6* | -0.445995667 | 0.003588054 | 0.011330438 | 1 |
| T2DM_insulin leukocyte | *PODN* | -0.362873686 | 0.017275635 | 0.023975969 | 1 |
| T2DM_insulin leukocyte | *RALGAPA1* | -0.391215724 | 0.001958767 | 0.011330438 | 1 |
| T2DM_insulin leukocyte | *CNTNAP2* | 0.71645744 | 0.020439616 | 0.027759161 | 1 |
| T2DM_insulin leukocyte | *GOLT1A* | -0.387705408 | 0.003561207 | 0.011330438 | 1 |
| T2DM_insulin leukocyte | *NPAS4* | -0.858475851 | 0.002568772 | 0.011330438 | 1 |
| T2DM_insulin leukocyte | *ANGEL2* | -0.538749476 | 0.002800016 | 0.011330438 | 1 |
| T2DM_insulin leukocyte | *ZNF266* | -0.241186494 | 0.029621794 | 0.038271631 | 1 |
| T2DM_insulin leukocyte | *BRSK2* | -0.362389844 | 0.005012834 | 0.011330438 | 1 |
| T2DM_insulin leukocyte | *BRMS1* | -0.359020169 | 0.003796728 | 0.011330438 | 1 |
| T2DM_insulin leukocyte | *HRAS* | -0.358991279 | 0.003414828 | 0.011330438 | 1 |
| T2DM_insulin leukocyte | *SRP72* | -0.30311158 | 0.003304915 | 0.011330438 | 1 |
| T2DM_insulin leukocyte | *BTC* | -0.631057733 | 0.003351562 | 0.011330438 | 1 |
| T2DM_insulin leukocyte | *DENND6A* | -0.350270935 | 0.0069791 | 0.012179027 | 1 |
| T2DM_insulin leukocyte | *YIF1A* | -0.333605519 | 0.00478687 | 0.011330438 | 1 |

| | | | | | |
|---|---|---|---|---|---|
| T2DM_insulin leukocyte | *NDUFA11* | -0.35342381 | 0.003152509 | 0.011330438 | 1 |
| T2DM_insulin leukocyte | *RSRC1* | -0.771494222 | 0.002391131 | 0.011330438 | 1 |
| T2DM_insulin leukocyte | *DHX36* | -0.395904426 | 0.002971239 | 0.011330438 | 1 |
| T2DM_insulin leukocyte | *PSMD2* | -0.36447357 | 0.001075252 | 0.011330438 | 1 |
| T2DM_insulin leukocyte | *PPM1E* | -0.317724339 | 0.00336958 | 0.011330438 | 1 |
| T2DM_insulin leukocyte | *ARHGAP1* | -0.379725776 | 0.003559644 | 0.011330438 | 1 |
| T2DM_insulin leukocyte | *MED16* | -0.370404053 | 0.00326676 | 0.011330438 | 1 |
| T2DM_insulin leukocyte | *CHST1* | -0.763438426 | 0.006141929 | 0.011570498 | 1 |
| T2DM_insulin leukocyte | *DOLK* | -0.721072963 | 0.005227256 | 0.011389288 | 1 |
| T2DM_insulin leukocyte | *CCNE2* | -0.45432017 | 0.008836427 | 0.014115597 | 1 |
| T2DM_insulin leukocyte | *ZNF519* | -0.798390606 | 0.001326497 | 0.011330438 | 1 |
| T2DM_insulin leukocyte | *TMEM9B* | -0.387195517 | 0.002830402 | 0.011330438 | 1 |
| T2DM_insulin leukocyte | *SCUBE2* | -0.225423603 | 0.031916595 | 0.040847004 | 1 |
| T2DM_insulin leukocyte | *EIF3F* | -0.360699877 | 0.005508516 | 0.011434531 | 1 |
| T2DM_insulin leukocyte | *ZNF25* | -0.377088176 | 0.003652389 | 0.011330438 | 1 |
| T2DM_insulin leukocyte | *PPP2R2D* | -0.367873165 | 0.003407357 | 0.011330438 | 1 |
| T2DM_insulin leukocyte | *CLCF1* | -0.564173706 | 0.003564675 | 0.011330438 | 1 |
| T2DM_insulin leukocyte | *UCP2* | 0.276587426 | 0.030107355 | 0.038853727 | 1 |
| T2DM_insulin leukocyte | *MRPL48* | -0.447268011 | 0.008241297 | 0.013448692 | 1 |
| T2DM_insulin leukocyte | *TMEM70* | -0.372967846 | 0.003353014 | 0.011330438 | 1 |
| T2DM_insulin leukocyte | *RPS6KB2* | -0.533559327 | 0.005126234 | 0.011330438 | 1 |
| T2DM_insulin leukocyte | *RMI2* | -1.061371942 | 0.022910774 | 0.03059702 | 1 |
| T2DM_insulin leukocyte | *LINC01106* | -1.533901077 | 0.037574015 | 0.047301604 | 1 |
| T2DM_insulin leukocyte | *SWI5* | -0.520289161 | 0.007604428 | 0.012801112 | 1 |
| T2DM_insulin leukocyte | *ZDHHC21* | -0.700064362 | 0.003501649 | 0.011330438 | 1 |
| T2DM_insulin leukocyte | *TUBB6* | -0.210464178 | 0.004372489 | 0.011330438 | 1 |
| T2DM_insulin leukocyte | *CHMP6* | -0.39465118 | 0.00494429 | 0.011330438 | 1 |
| T2DM_insulin leukocyte | *DLEU1* | -0.483501072 | 0.003349281 | 0.011330438 | 1 |
| T2DM_insulin leukocyte | *CCDC57* | -0.586568505 | 0.00048168 | 0.011330438 | 1 |
| T2DM_insulin leukocyte | *BNIP3* | -0.296019692 | 0.002772765 | 0.011330438 | 1 |
| T2DM_insulin leukocyte | *SMIM19* | -0.331568341 | 0.003572065 | 0.011330438 | 1 |
| T2DM_insulin leukocyte | *SLC35G1* | -0.542348792 | 0.004425853 | 0.011330438 | 1 |
| T2DM_insulin leukocyte | *COX8A* | -0.28311788 | 0.002789273 | 0.011330438 | 1 |
| T2DM_insulin leukocyte | *CDC26* | -0.396205643 | 0.004621055 | 0.011330438 | 1 |
| T2DM_insulin leukocyte | *RNPEP* | -0.694361419 | 0.003034441 | 0.011330438 | 1 |
| T2DM_insulin leukocyte | *DMRTA1* | -0.330373416 | 0.009794458 | 0.015234531 | 1 |
| T2DM_insulin leukocyte | *CLEC14A* | -0.206989596 | 0.017679851 | 0.024441989 | 1 |
| T2DM_insulin leukocyte | *ZNF575* | 0.43815385 | 0.025245581 | 0.033363569 | 1 |
| T2DM_insulin leukocyte | *WDR25* | -0.975288309 | 0.001921069 | 0.011330438 | 1 |
| T2DM_insulin leukocyte | *DIRAS1* | -0.477282755 | 0.007011102 | 0.012210382 | 1 |
| T2DM_insulin leukocyte | *PHLDB3* | -0.320404547 | 0.009964255 | 0.015455806 | 1 |
| T2DM_insulin leukocyte | *PRR15* | 0.49133965 | 0.027902954 | 0.03636176 | 1 |
| T2DM_insulin leukocyte | *KBTBD11* | -0.338134464 | 0.005454858 | 0.011427973 | 1 |
| T2DM_insulin leukocyte | *RMDN1* | -0.427933897 | 0.007794866 | 0.01301113 | 1 |
| T2DM_insulin leukocyte | *LRRC37A* | -2.097470603 | 0.004461958 | 0.011330438 | 1 |
| T2DM_insulin leukocyte | *ACSF3* | -0.518387763 | 0.00400821 | 0.011330438 | 1 |
| T2DM_insulin leukocyte | *RUFY1* | -0.922656931 | 0.002641592 | 0.011330438 | 1 |
| T2DM_insulin leukocyte | *VSIG10* | -0.618869846 | 0.001896051 | 0.011330438 | 1 |
| T2DM_insulin leukocyte | *WSB2* | -0.362251095 | 0.002704689 | 0.011330438 | 1 |
| T2DM_insulin leukocyte | *ANKLE2* | -0.274293368 | 0.015459756 | 0.021810191 | 1 |

| | | | | | |
|---|---|---|---|---|---|
| T2DM_insulin leukocyte | *MUC20* | -0.393781462 | 0.005617309 | 0.011547208 | 1 |
| T2DM_insulin leukocyte | *NFATC2IP* | -0.590645697 | 0.002200286 | 0.011330438 | 1 |
| T2DM_insulin leukocyte | *DPP7* | -0.361409759 | 0.003015336 | 0.011330438 | 1 |
| T2DM_insulin leukocyte | *MTHFR* | -0.423551167 | 0.003523072 | 0.011330438 | 1 |
| T2DM_insulin leukocyte | *DEAF1* | -0.515736083 | 0.002998328 | 0.011330438 | 1 |
| T2DM_insulin leukocyte | *TMEM80* | -0.318280248 | 0.005317479 | 0.011421159 | 1 |
| T2DM_insulin leukocyte | *DSCAML1* | -0.524301961 | 0.003081186 | 0.011330438 | 1 |
| T2DM_insulin leukocyte | *PUS1* | -0.822145466 | 0.00342091 | 0.011330438 | 1 |
| T2DM_insulin leukocyte | *MAN1B1* | -0.666964284 | 0.002543742 | 0.011330438 | 1 |
| T2DM_insulin leukocyte | *PPFIA3* | -0.307885694 | 0.005868755 | 0.011570498 | 1 |
| T2DM_insulin leukocyte | *UBE2U* | -1.751350449 | 0.01222977 | 0.017949483 | 1 |
| T2DM_insulin leukocyte | *PAWR* | -0.359789424 | 0.004788909 | 0.011330438 | 1 |
| T2DM_insulin leukocyte | *ACOT4* | -0.482082669 | 0.00942894 | 0.014830386 | 1 |
| T2DM_insulin leukocyte | *ARIH2* | -0.413616448 | 0.001034846 | 0.011330438 | 1 |
| T2DM_insulin leukocyte | *SAMD12* | -0.397560118 | 0.035913357 | 0.045360435 | 1 |
| T2DM_insulin leukocyte | *CD163* | 0.036451809 | 0.014140686 | 0.020239984 | 1 |
| T2DM_insulin leukocyte | *ACAD9* | -0.39315852 | 0.021038653 | 0.028435111 | 1 |
| T2DM_insulin leukocyte | *AGTRAP* | -0.422970929 | 0.003836492 | 0.011330438 | 1 |
| T2DM_insulin leukocyte | *THAP5* | -0.403388339 | 0.003337886 | 0.011330438 | 1 |
| T2DM_insulin leukocyte | *CD151* | -0.319008166 | 0.003164337 | 0.011330438 | 1 |
| T2DM_insulin leukocyte | *CHID1* | -0.370234368 | 0.000976893 | 0.011330438 | 1 |
| T2DM_insulin leukocyte | *ZBTB41* | -0.509277439 | 0.002802521 | 0.011330438 | 1 |
| T2DM_insulin leukocyte | *BET1L* | -0.555036615 | 0.003989728 | 0.011330438 | 1 |
| T2DM_insulin leukocyte | *RIC8A* | -0.518190793 | 0.002642347 | 0.011330438 | 1 |
| T2DM_insulin leukocyte | *IMP3* | -0.412856224 | 0.004990619 | 0.011330438 | 1 |
| T2DM_insulin leukocyte | *ADAMTSL1* | -1.477882219 | 0.035825791 | 0.045274858 | 1 |
| T2DM_insulin leukocyte | *ALS2CL* | -0.547692716 | 0.004704276 | 0.011330438 | 1 |
| T2DM_insulin leukocyte | *STAP2* | -0.306548789 | 0.00349304 | 0.011330438 | 1 |
| T2DM_insulin leukocyte | *DALRD3* | -0.408100358 | 0.007944789 | 0.013170918 | 1 |
| T2DM_insulin leukocyte | *ZNF114* | -1.027425282 | 0.006242973 | 0.011570498 | 1 |
| T2DM_insulin leukocyte | *PLEC* | -0.829289548 | 0.000326703 | 0.011330438 | 1 |
| T2DM_insulin leukocyte | *WDR6* | -0.305091384 | 0.030984656 | 0.039770897 | 1 |
| T2DM_insulin leukocyte | *AQP11* | -0.397042146 | 0.024923174 | 0.033015223 | 1 |
| T2DM_insulin leukocyte | *ZFAND2A* | -0.485247792 | 0.004109137 | 0.011330438 | 1 |
| T2DM_insulin leukocyte | *ZNF223* | -0.474287827 | 0.015955534 | 0.022398848 | 1 |
| T2DM_insulin leukocyte | *NT5DC1* | -0.381218586 | 0.003590969 | 0.011330438 | 1 |
| T2DM_insulin leukocyte | *COX14* | -0.36690426 | 0.00720544 | 0.012381002 | 1 |
| T2DM_insulin leukocyte | *P4HTM* | -0.304523666 | 0.00196134 | 0.011330438 | 1 |
| T2DM_insulin leukocyte | *ERBB4* | -3.46078935 | 0.00208142 | 0.011330438 | 1 |
| T2DM_insulin leukocyte | *ERN1* | -0.492196467 | 0.004619167 | 0.011330438 | 1 |
| T2DM_insulin leukocyte | *PARP10* | -0.447371973 | 0.010366132 | 0.01590337 | 1 |
| T2DM_insulin leukocyte | *RPP25* | -0.634611127 | 0.002748375 | 0.011330438 | 1 |
| T2DM_insulin leukocyte | *ZHX2* | -0.419039971 | 0.005822957 | 0.011570498 | 1 |
| T2DM_insulin leukocyte | *HYI* | -0.476677802 | 0.002289019 | 0.011330438 | 1 |
| T2DM_insulin leukocyte | *GAK* | -0.408620922 | 0.01019349 | 0.015699813 | 1 |
| T2DM_insulin leukocyte | *TUFM* | -0.224527964 | 0.027677836 | 0.036128312 | 1 |
| T2DM_insulin leukocyte | *RMI1* | 0.71299633 | 0.002245518 | 0.011330438 | 1 |
| T2DM_insulin leukocyte | *C3orf38* | -0.519460992 | 0.01182827 | 0.017616858 | 1 |
| T2DM_insulin leukocyte | *IGSF22* | -0.284547396 | 0.007378721 | 0.012556771 | 1 |
| T2DM_insulin leukocyte | *HTR1F* | -1.129733171 | 0.002842751 | 0.011330438 | 1 |

| | | | | | |
|---|---|---|---|---|---|
| T2DM_insulin leukocyte | *TMTC2* | -0.711054974 | 0.004486731 | 0.011330438 | 1 |
| T2DM_insulin leukocyte | *FARSA* | -0.46115118 | 0.00082987 | 0.011330438 | 1 |
| T2DM_insulin leukocyte | *SPTY2D1* | -0.400354737 | 0.004350963 | 0.011330438 | 1 |
| T2DM_insulin leukocyte | *SAMD4B* | -0.294462936 | 0.01424323 | 0.020357093 | 1 |
| T2DM_insulin leukocyte | *TCAIM* | -0.556890582 | 0.002807481 | 0.011330438 | 1 |
| T2DM_insulin leukocyte | *CALR* | -0.269074635 | 0.008709477 | 0.013978569 | 1 |
| T2DM_insulin leukocyte | *NSUN7* | -0.648808423 | 0.014281454 | 0.020389474 | 1 |
| T2DM_insulin leukocyte | *ELMOD2* | -0.644409725 | 0.004437332 | 0.011330438 | 1 |
| T2DM_insulin leukocyte | *LINC00174* | -0.387058856 | 0.003582136 | 0.011330438 | 1 |
| T2DM_insulin leukocyte | *GEMIN4* | -0.382915771 | 0.006887812 | 0.012081345 | 1 |
| T2DM_insulin leukocyte | *LBX2* | 0.418005848 | 0.010429722 | 0.015957312 | 1 |
| T2DM_insulin leukocyte | *CDC42EP4* | -0.326502651 | 0.004844525 | 0.011330438 | 1 |
| T2DM_insulin leukocyte | *ZBTB42* | -0.977153236 | 0.001893751 | 0.011330438 | 1 |
| T2DM_insulin leukocyte | *SYCN* | -0.205474833 | 0.006744691 | 0.011963834 | 1 |
| T2DM_insulin leukocyte | *CITED4* | -0.239249405 | 0.011708825 | 0.017459386 | 1 |
| T2DM_insulin leukocyte | *ABCA13* | -7.613769403 | 0.013184544 | 0.019044879 | 1 |
| T2DM_insulin leukocyte | *B3GNT3* | -0.785172184 | 0.017341243 | 0.024041546 | 1 |
| T2DM_insulin leukocyte | *PUF60* | -0.315719125 | 0.010848938 | 0.016457713 | 1 |
| T2DM_insulin leukocyte | *TSHZ1* | -0.328813695 | 0.004929506 | 0.011330438 | 1 |
| T2DM_insulin leukocyte | *EXOC3* | -0.698833266 | 0.002353939 | 0.011330438 | 1 |
| T2DM_insulin leukocyte | *TH* | -1.963095063 | 0.007422432 | 0.012597294 | 1 |
| T2DM_insulin leukocyte | *FAHD1* | -0.284332831 | 0.036295596 | 0.045824356 | 1 |
| T2DM_insulin leukocyte | *ZNF816* | -0.570297651 | 0.004238369 | 0.011330438 | 1 |
| T2DM_insulin leukocyte | *CCDC66* | -0.92812874 | 0.002869591 | 0.011330438 | 1 |
| T2DM_insulin leukocyte | *MCFD2* | -0.304645428 | 0.002926637 | 0.011330438 | 1 |
| T2DM_insulin leukocyte | *SERTM1* | 0.290340036 | 0.022316639 | 0.029925623 | 1 |
| T2DM_insulin leukocyte | *TSPYL5* | -0.297256826 | 0.005142414 | 0.011330438 | 1 |
| T2DM_insulin leukocyte | *SHISA2* | -0.739322181 | 0.005438786 | 0.011421159 | 1 |
| T2DM_insulin leukocyte | *CHST13* | -31.45259824 | 0.006315263 | 0.011616417 | 1 |
| T2DM_insulin leukocyte | *PPA1* | -0.284970465 | 0.003196051 | 0.011330438 | 1 |
| T2DM_insulin leukocyte | *PSMG4* | -0.319837221 | 0.004118625 | 0.011330438 | 1 |
| T2DM_insulin leukocyte | *MAP6D1* | -0.719789885 | 0.011874789 | 0.017638109 | 1 |
| T2DM_insulin leukocyte | *ZNF792* | -0.687456752 | 0.026891909 | 0.035207701 | 1 |
| T2DM_insulin leukocyte | *D2HGDH* | -1.444056986 | 0.001291882 | 0.011330438 | 1 |
| T2DM_insulin leukocyte | *FAM83H* | -0.561524362 | 0.006878208 | 0.012075281 | 1 |
| T2DM_insulin leukocyte | *LRRC57* | -0.64904548 | 0.00294387 | 0.011330438 | 1 |
| T2DM_insulin leukocyte | *MRPL14* | -0.409754968 | 0.00215976 | 0.011330438 | 1 |
| T2DM_insulin leukocyte | *ZFP82* | -0.359561186 | 0.005814166 | 0.011570498 | 1 |
| T2DM_insulin leukocyte | *LSMEM1* | -0.497795825 | 0.031605006 | 0.040527438 | 1 |
| T2DM_insulin leukocyte | *FKRP* | -0.437325559 | 0.000627561 | 0.011330438 | 1 |
| T2DM_insulin leukocyte | *RPH3AL* | -0.286822196 | 0.005233452 | 0.011389288 | 1 |
| T2DM_insulin leukocyte | *HIGD1A* | -0.303588304 | 0.003749279 | 0.011330438 | 1 |
| T2DM_insulin leukocyte | *F2R* | -0.302897021 | 0.007317884 | 0.012496955 | 1 |
| T2DM_insulin leukocyte | *ZNF707* | -0.479150408 | 0.005923221 | 0.011570498 | 1 |
| T2DM_insulin leukocyte | *SLC25A41* | -2.16713561 | 0.006540054 | 0.011804448 | 1 |
| T2DM_insulin leukocyte | *OGFOD3* | -0.350240407 | 0.002304918 | 0.011330438 | 1 |
| T2DM_insulin leukocyte | *ZNF467* | -0.470016172 | 0.008735443 | 0.014008798 | 1 |
| T2DM_insulin leukocyte | *TMEM45A* | -0.307217674 | 0.006039377 | 0.011570498 | 1 |
| T2DM_insulin leukocyte | *GPR135* | -0.434901378 | 0.0067314 | 0.011961858 | 1 |
| T2DM_insulin leukocyte | *ANKRD62* | -0.610868344 | 0.002497963 | 0.011330438 | 1 |

| | | | | | |
|---|---|---|---|---|---|
| T2DM_insulin leukocyte | *RFX7* | -0.899583253 | 0.002842547 | 0.011330438 | 1 |
| T2DM_insulin leukocyte | *ZNF329* | -0.53601877 | 0.003409886 | 0.011330438 | 1 |
| T2DM_insulin leukocyte | *CCDC149* | -0.641853332 | 0.00199505 | 0.011330438 | 1 |
| T2DM_insulin leukocyte | *IDH2* | -0.289895072 | 0.006415634 | 0.01170553 | 1 |
| T2DM_insulin leukocyte | *TNRC18* | -0.471001506 | 0.004220817 | 0.011330438 | 1 |
| T2DM_insulin leukocyte | *TDRKH* | -0.455084011 | 0.002683387 | 0.011330438 | 1 |
| T2DM_insulin leukocyte | *ZNF708* | -0.308047833 | 0.004020608 | 0.011330438 | 1 |
| T2DM_insulin leukocyte | *IST1* | -0.570102611 | 0.002563584 | 0.011330438 | 1 |
| T2DM_insulin leukocyte | *ERCC6L2* | -1.08629148 | 0.003022039 | 0.011330438 | 1 |
| T2DM_insulin leukocyte | *MRPL41* | -0.36271875 | 0.002991709 | 0.011330438 | 1 |
| T2DM_insulin leukocyte | *TP53TG1* | -0.429016554 | 0.001752747 | 0.011330438 | 1 |
| T2DM_insulin leukocyte | *UBA7* | -0.607278084 | 0.02266661 | 0.030332803 | 1 |
| T2DM_insulin leukocyte | *ARL6IP4* | -0.269591471 | 0.007103179 | 0.012294167 | 1 |
| T2DM_insulin leukocyte | *FAM153B* | -0.516123162 | 0.010502269 | 0.016055804 | 1 |
| T2DM_insulin leukocyte | *BACE2* | -0.229937733 | 0.004747622 | 0.011330438 | 1 |
| T2DM_insulin leukocyte | *SYNM* | -0.379445821 | 0.00595998 | 0.011570498 | 1 |
| T2DM_insulin leukocyte | *FIGN* | -0.355177417 | 0.008592016 | 0.013846609 | 1 |
| T2DM_insulin leukocyte | *ZNF804B* | -27.79333892 | 0.005478722 | 0.011427973 | 1 |
| T2DM_insulin leukocyte | *YBEY* | -0.43471954 | 0.005880819 | 0.011570498 | 1 |
| T2DM_insulin leukocyte | *NPLOC4* | -0.392656067 | 0.003888992 | 0.011330438 | 1 |
| T2DM_insulin leukocyte | *CAPN12* | -0.69597781 | 0.005522049 | 0.011446386 | 1 |
| T2DM_insulin leukocyte | *GLRX5* | -0.276382395 | 0.020805392 | 0.028168292 | 1 |
| T2DM_insulin leukocyte | *MXRA7* | -0.330230828 | 0.003347997 | 0.011330438 | 1 |
| T2DM_insulin leukocyte | *ADI1* | -0.788570428 | 0.002450006 | 0.011330438 | 1 |
| T2DM_insulin leukocyte | *SATB1* | -0.602064639 | 0.002053171 | 0.011330438 | 1 |
| T2DM_insulin leukocyte | *SKA2* | -0.688336584 | 0.001213597 | 0.011330438 | 1 |
| T2DM_insulin leukocyte | *TSKU* | -0.419546931 | 0.005476941 | 0.011427973 | 1 |
| T2DM_insulin leukocyte | *ANXA2* | -0.217769334 | 0.005491767 | 0.011434531 | 1 |
| T2DM_insulin leukocyte | *SLC35D3* | -0.742181816 | 0.002403266 | 0.011330438 | 1 |
| T2DM_insulin leukocyte | *GPIHBP1* | -3.397099897 | 0.006403567 | 0.01170553 | 1 |
| T2DM_insulin leukocyte | *VMO1* | 0.042759594 | 0.012522983 | 0.018270627 | 1 |
| T2DM_insulin leukocyte | *ALG12* | -0.548850736 | 0.024288174 | 0.032261067 | 1 |
| T2DM_insulin leukocyte | *COL18A1* | -0.154319945 | 0.025636747 | 0.033823634 | 1 |
| T2DM_insulin leukocyte | *C11orf54* | -0.485637878 | 0.009655755 | 0.015078655 | 1 |
| T2DM_insulin leukocyte | *CEP63* | -0.466784585 | 0.002329638 | 0.011330438 | 1 |
| T2DM_insulin leukocyte | *HMGN4* | -0.511872599 | 0.003574689 | 0.011330438 | 1 |
| T2DM_insulin leukocyte | *SPATA13* | -0.364559812 | 0.004046104 | 0.011330438 | 1 |
| T2DM_insulin leukocyte | *ZNF320* | 0.400707154 | 0.000815771 | 0.011330438 | 1 |
| T2DM_insulin leukocyte | *SLC25A21* | -0.948918873 | 0.005823913 | 0.011570498 | 1 |
| T2DM_insulin leukocyte | *LYSMD4* | -0.433740873 | 0.006823756 | 0.012014984 | 1 |
| T2DM_insulin leukocyte | *BEGAIN* | -0.542844185 | 0.004920048 | 0.011330438 | 1 |
| T2DM_insulin leukocyte | *GPC6* | -0.422445612 | 0.007153708 | 0.01235469 | 1 |
| T2DM_insulin leukocyte | *FANCF* | -1.310354623 | 0.00216298 | 0.011330438 | 1 |
| T2DM_insulin leukocyte | *SMDT1* | -0.214706359 | 0.025189167 | 0.03330179 | 1 |
| T2DM_insulin leukocyte | *PTTG1IP* | -0.318264799 | 0.006381585 | 0.011683713 | 1 |
| T2DM_insulin leukocyte | *ZNF623* | -0.324695272 | 0.008598972 | 0.013849257 | 1 |
| T2DM_insulin leukocyte | *CCDC125* | -0.692507142 | 0.028168705 | 0.036635168 | 1 |
| T2DM_insulin leukocyte | *BOLA2* | -0.383023783 | 0.002966467 | 0.011330438 | 1 |
| T2DM_insulin leukocyte | *FHL3* | -0.318886126 | 0.025780827 | 0.033972625 | 1 |
| T2DM_insulin leukocyte | *SF3A3* | -0.337940023 | 0.004081178 | 0.011330438 | 1 |

| | | | | | |
|---|---|---|---|---|---|
| T2DM_insulin leukocyte | *MX2* | 0.468675642 | 0.023087276 | 0.030822259 | 1 |
| T2DM_insulin leukocyte | *PCBP3* | -0.451303076 | 0.032919538 | 0.041980157 | 1 |
| T2DM_insulin leukocyte | *SFXN4* | -0.427613779 | 0.003269612 | 0.011330438 | 1 |
| T2DM_insulin leukocyte | *MRPL54* | -0.343665356 | 0.005018123 | 0.011330438 | 1 |
| T2DM_insulin leukocyte | *DGCR6* | -0.44044187 | 0.002203877 | 0.011330438 | 1 |
| T2DM_insulin leukocyte | *NDUFB1* | -0.344997386 | 0.005643368 | 0.01154833 | 1 |
| T2DM_insulin leukocyte | *TRMT12* | -0.454489673 | 0.002529034 | 0.011330438 | 1 |
| T2DM_insulin leukocyte | *FIGLA* | -9.367362161 | 0.002765244 | 0.011330438 | 1 |
| T2DM_insulin leukocyte | *CBX6* | -0.307162521 | 0.007490357 | 0.012685177 | 1 |
| T2DM_insulin leukocyte | *MACC1* | -0.276492076 | 0.005422085 | 0.011421159 | 1 |
| T2DM_insulin leukocyte | *KREMEN1* | -1.31871176 | 0.00161763 | 0.011330438 | 1 |
| T2DM_insulin leukocyte | *NUDT14* | -0.330926458 | 0.008368044 | 0.013597207 | 1 |
| T2DM_insulin leukocyte | *FAM3B* | -0.296166641 | 0.012028223 | 0.017759823 | 1 |
| T2DM_insulin leukocyte | *TOB2* | -0.421864699 | 0.004396446 | 0.011330438 | 1 |
| T2DM_insulin leukocyte | *TTC32* | -0.394406812 | 0.002324645 | 0.011330438 | 1 |
| T2DM_insulin leukocyte | *LRRC55* | -4.226014083 | 0.010874047 | 0.016483075 | 1 |
| T2DM_insulin leukocyte | *TMEM236* | -0.24972324 | 0.019380678 | 0.026504241 | 1 |
| T2DM_insulin leukocyte | *UQCR10* | -0.331942648 | 0.00197286 | 0.011330438 | 1 |
| T2DM_insulin leukocyte | *NIPSNAP1* | -0.369775832 | 0.004088462 | 0.011330438 | 1 |
| T2DM_insulin leukocyte | *CRELD2* | -0.288653576 | 0.006257378 | 0.011570498 | 1 |
| T2DM_insulin leukocyte | *UBE2F* | -0.620194176 | 0.001951181 | 0.011330438 | 1 |
| T2DM_insulin leukocyte | *GOLGA6L4* | -0.54392796 | 0.001033654 | 0.011330438 | 1 |
| T2DM_insulin leukocyte | *PGP* | -0.594555501 | 0.000724498 | 0.011330438 | 1 |
| T2DM_insulin leukocyte | *ACOT1* | -1.913000408 | 0.022892694 | 0.03058327 | 1 |
| T2DM_insulin leukocyte | *OAF* | -0.476851811 | 0.003256507 | 0.011330438 | 1 |
| T2DM_insulin leukocyte | *TM2D3* | -0.368106125 | 0.003474376 | 0.011330438 | 1 |
| T2DM_insulin leukocyte | *MROH7* | -0.693702738 | 0.005069518 | 0.011330438 | 1 |
| T2DM_insulin leukocyte | *TOP1MT* | -0.810472433 | 0.007918974 | 0.013139209 | 1 |
| T2DM_insulin leukocyte | *COPB2* | -0.421207876 | 0.002913454 | 0.011330438 | 1 |
| T2DM_insulin leukocyte | *LRRC19* | -1.420270692 | 0.000463591 | 0.011330438 | 1 |
| T2DM_insulin leukocyte | *CCR10* | -0.883051748 | 0.003227301 | 0.011330438 | 1 |
| T2DM_insulin leukocyte | *NCMAP* | -0.360358238 | 0.006975839 | 0.012179027 | 1 |
| T2DM_insulin leukocyte | *WDR27* | -0.885195094 | 0.003496396 | 0.011330438 | 1 |
| T2DM_insulin leukocyte | *TXNRD2* | -0.382417649 | 0.005180237 | 0.011364127 | 1 |
| T2DM_insulin leukocyte | *ZFP1* | -0.515834805 | 0.002643123 | 0.011330438 | 1 |
| T2DM_insulin leukocyte | *XPOT* | -0.346590197 | 0.005094979 | 0.011330438 | 1 |
| T2DM_insulin leukocyte | *C14orf180* | -39.27993671 | 0.006008628 | 0.011570498 | 1 |
| T2DM_insulin leukocyte | *UBE2G2* | -0.378528235 | 0.005131638 | 0.011330438 | 1 |
| T2DM_insulin leukocyte | *OSBP2* | -0.432097596 | 0.006087732 | 0.011570498 | 1 |
| T2DM_insulin leukocyte | *TMED9* | -0.325012148 | 0.003207231 | 0.011330438 | 1 |
| T2DM_insulin leukocyte | *BTBD6* | -0.490562833 | 0.003080107 | 0.011330438 | 1 |
| T2DM_insulin leukocyte | *JAG2* | -0.335985246 | 0.007370248 | 0.012552105 | 1 |
| T2DM_insulin leukocyte | *LCN12* | -1.236458084 | 0.004677223 | 0.011330438 | 1 |
| T2DM_insulin leukocyte | *FAM227A* | -0.403392231 | 0.007805711 | 0.013011726 | 1 |
| T2DM_insulin leukocyte | *MUC6* | -0.221094108 | 0.011494306 | 0.017184601 | 1 |
| T2DM_insulin leukocyte | *NOC4L* | -0.568304968 | 0.00511306 | 0.011330438 | 1 |
| T2DM_insulin leukocyte | *SIVA1* | -0.364011069 | 0.003862839 | 0.011330438 | 1 |
| T2DM_insulin leukocyte | *BRI3BP* | -0.524736726 | 0.003519705 | 0.011330438 | 1 |
| T2DM_insulin leukocyte | *UBOX5* | -0.37599118 | 0.005426416 | 0.011421159 | 1 |
| T2DM_insulin leukocyte | *MAFF* | -0.232256954 | 0.008333047 | 0.013546584 | 1 |

| | | | | | |
|---|---|---|---|---|---|
| T2DM_insulin leukocyte | *SEMA4B* | -0.367704088 | 0.006205334 | 0.011570498 | 1 |
| T2DM_insulin leukocyte | *CIB1* | -0.356884175 | 0.002925933 | 0.011330438 | 1 |
| T2DM_insulin leukocyte | *MFSD6L* | -0.994750163 | 0.015249311 | 0.021559749 | 1 |
| T2DM_insulin leukocyte | *AQP12B* | -2.361272454 | 0.009789489 | 0.015232833 | 1 |
| T2DM_insulin leukocyte | *ZNF445* | -1.425170523 | 0.008501895 | 0.01374648 | 1 |
| T2DM_insulin leukocyte | *RBM11* | -0.347778571 | 0.002832307 | 0.011330438 | 1 |
| T2DM_insulin leukocyte | *RGPD2* | -1.210370701 | 0.003871736 | 0.011330438 | 1 |
| T2DM_insulin leukocyte | *ATP6V0A2* | -0.573295052 | 0.005230469 | 0.011389288 | 1 |
| T2DM_insulin leukocyte | *RAD51D* | -0.965820896 | 0.002697702 | 0.011330438 | 1 |
| T2DM_insulin leukocyte | *SH3BGR* | -0.288339665 | 0.00118922 | 0.011330438 | 1 |
| T2DM_insulin leukocyte | *MUC1* | -0.247973847 | 0.006606399 | 0.011868561 | 1 |
| T2DM_insulin leukocyte | *IRF7* | -0.562450881 | 0.013199319 | 0.019059215 | 1 |
| T2DM_insulin leukocyte | *FAM131C* | -0.276600058 | 0.005403581 | 0.011421159 | 1 |
| T2DM_insulin leukocyte | *LSAMP* | -0.194952791 | 0.013107648 | 0.018954711 | 1 |
| T2DM_insulin leukocyte | *OLFML2A* | -0.968621206 | 0.012120574 | 0.017849227 | 1 |
| T2DM_insulin leukocyte | *PDIA2* | -0.184519304 | 0.010037751 | 0.015522195 | 1 |
| T2DM_insulin leukocyte | *PCGF3* | -0.410187578 | 0.003090921 | 0.011330438 | 1 |
| T2DM_insulin leukocyte | *P4HB* | -0.247139038 | 0.005304979 | 0.011421159 | 1 |
| T2DM_insulin leukocyte | *PSMD13* | -0.380722041 | 0.005150126 | 0.011330438 | 1 |
| T2DM_insulin leukocyte | *BRWD1* | -0.424554276 | 0.002146679 | 0.011330438 | 1 |
| T2DM_insulin leukocyte | *DRG1* | -0.359721299 | 0.003742224 | 0.011330438 | 1 |
| T2DM_insulin leukocyte | *ZNF696* | -1.072648625 | 0.005121459 | 0.011330438 | 1 |
| T2DM_insulin leukocyte | *MORF4L1* | -0.288169278 | 0.003169234 | 0.011330438 | 1 |
| T2DM_insulin leukocyte | *DMWD* | -0.767392145 | 0.000370609 | 0.011330438 | 1 |
| T2DM_insulin leukocyte | *PIGP* | -0.341266042 | 0.001103028 | 0.011330438 | 1 |
| T2DM_insulin leukocyte | *NPAP1* | -12.21267383 | 0.009195844 | 0.014545279 | 1 |
| T2DM_insulin leukocyte | *ARL17A* | -0.246858467 | 0.035051283 | 0.044424674 | 1 |
| T2DM_insulin leukocyte | *TRIM69* | -0.406685307 | 0.004388924 | 0.011330438 | 1 |
| T2DM_insulin leukocyte | *IFITM1* | -0.998013997 | 0.001216467 | 0.011330438 | 1 |
| T2DM_insulin leukocyte | *SETD4* | -1.122246569 | 0.019811763 | 0.026990436 | 1 |
| T2DM_insulin leukocyte | *ZNF529* | -0.704284576 | 0.002281809 | 0.011330438 | 1 |
| T2DM_insulin leukocyte | *AIDA* | -0.35683787 | 0.003029502 | 0.011330438 | 1 |
| T2DM_insulin leukocyte | *WWOX* | -0.382603689 | 0.004005154 | 0.011330438 | 1 |
| T2DM_insulin leukocyte | *POLR1D* | -0.339376214 | 0.002658352 | 0.011330438 | 1 |
| T2DM_insulin leukocyte | *EDARADD* | -0.391297904 | 0.005451952 | 0.011427973 | 1 |
| T2DM_insulin leukocyte | *ZNF749* | -0.291659399 | 0.003779995 | 0.011330438 | 1 |
| T2DM_insulin leukocyte | *TOR3A* | -0.393687081 | 0.004424954 | 0.011330438 | 1 |
| T2DM_insulin leukocyte | *ZNF555* | -0.299630041 | 0.007557367 | 0.012749204 | 1 |
| T2DM_insulin leukocyte | *NUDT17* | -0.514850262 | 0.007185974 | 0.012372346 | 1 |
| T2DM_insulin leukocyte | *CYP4X1* | 0.529742388 | 0.012977034 | 0.018807365 | 1 |
| T2DM_insulin leukocyte | *ZNF197* | -0.355301397 | 0.004172373 | 0.011330438 | 1 |
| T2DM_insulin leukocyte | *RPS23* | -0.262373407 | 0.002388004 | 0.011330438 | 1 |
| T2DM_insulin leukocyte | *ZNF396* | -0.887822912 | 0.01237172 | 0.0181108 | 1 |
| T2DM_insulin leukocyte | *CEACAM19* | -0.234761186 | 0.03948901 | 0.049506165 | 1 |
| T2DM_insulin leukocyte | *ZFP91* | -0.469967432 | 0.001856194 | 0.011330438 | 1 |
| T2DM_insulin leukocyte | *BCR* | -0.371199902 | 0.003141419 | 0.011330438 | 1 |
| T2DM_insulin leukocyte | *HYAL3* | -0.900307662 | 0.000143192 | 0.011330438 | 0.562602546 |
| T2DM_insulin leukocyte | *RTN4RL2* | -0.387187443 | 0.00618863 | 0.011570498 | 1 |
| T2DM_insulin leukocyte | *TMEM232* | -0.610694351 | 0.003401999 | 0.011330438 | 1 |
| T2DM_insulin leukocyte | *FAM183A* | -0.310822351 | 0.006693763 | 0.011938173 | 1 |

| | | | | | |
|---|---|---|---|---|---|
| T2DM_insulin leukocyte | *EMID1* | -0.515967953 | 0.002857774 | 0.011330438 | 1 |
| T2DM_insulin leukocyte | *ESPN* | -9.513394721 | 0.000290637 | 0.011330438 | 1 |
| T2DM_insulin leukocyte | *AKR1C1* | -0.289446671 | 0.00505807 | 0.011330438 | 1 |
| T2DM_insulin leukocyte | *BCAM* | -0.264855573 | 0.008161045 | 0.013387195 | 1 |
| T2DM_insulin leukocyte | *CHP1* | -0.321719033 | 0.003588813 | 0.011330438 | 1 |
| T2DM_insulin leukocyte | *KCNJ11* | -1.222408147 | 0.002896176 | 0.011330438 | 1 |
| T2DM_insulin leukocyte | *ZNF385C* | -1.257369253 | 0.004693662 | 0.011330438 | 1 |
| T2DM_insulin leukocyte | *EXD3* | -0.405463035 | 0.005325236 | 0.011421159 | 1 |
| T2DM_insulin leukocyte | *RGPD1* | -41.84265848 | 0.006555736 | 0.011815361 | 1 |
| T2DM_insulin leukocyte | *DHRS4L2* | -0.397527708 | 0.003240977 | 0.011330438 | 1 |
| T2DM_insulin leukocyte | *TRPV2* | -2.426590169 | 0.038134128 | 0.047960624 | 1 |
| T2DM_insulin leukocyte | *FANCA* | -0.690416315 | 0.001990282 | 0.011330438 | 1 |
| T2DM_insulin leukocyte | *PALM3* | -0.472112284 | 0.005882613 | 0.011570498 | 1 |
| T2DM_insulin leukocyte | *KLHL17* | -0.645988848 | 0.00549326 | 0.011434531 | 1 |
| T2DM_insulin leukocyte | *RINL* | -0.486848357 | 0.003932348 | 0.011330438 | 1 |
| T2DM_insulin leukocyte | *ZNF490* | -0.701677969 | 0.0033271 | 0.011330438 | 1 |
| T2DM_insulin leukocyte | *NRN1L* | -1.000419772 | 0.006272425 | 0.011570498 | 1 |
| T2DM_insulin leukocyte | *NWD1* | -0.960640745 | 0.030710953 | 0.039500311 | 1 |
| T2DM_insulin leukocyte | *RAB42* | 0.170336011 | 0.039610076 | 0.049642102 | 1 |
| T2DM_insulin leukocyte | *MAPK12* | -0.491658757 | 0.003351247 | 0.011330438 | 1 |
| T2DM_insulin leukocyte | *LINC00265* | -0.793635308 | 0.005924983 | 0.011570498 | 1 |
| T2DM_insulin leukocyte | *LAMTOR4* | -0.322317971 | 0.007343038 | 0.012522047 | 1 |
| T2DM_insulin leukocyte | *PRKAR1B* | -0.433133859 | 0.008918916 | 0.014204468 | 1 |
| T2DM_insulin leukocyte | *NUTM2B* | -5.82873248 | 0.025784263 | 0.033972625 | 1 |
| T2DM_insulin leukocyte | *CENPP* | -0.274924919 | 0.018259219 | 0.025163265 | 1 |
| T2DM_insulin leukocyte | *ZNF559* | -0.315357697 | 0.008889588 | 0.014169246 | 1 |
| T2DM_insulin leukocyte | *FOCAD* | -0.406811966 | 0.004572967 | 0.011330438 | 1 |
| T2DM_insulin leukocyte | *ZP3* | -0.473171118 | 0.013126205 | 0.018967583 | 1 |
| T2DM_insulin leukocyte | *SERPINA5* | -0.235624607 | 0.026595122 | 0.034923875 | 1 |
| T2DM_insulin leukocyte | *SRSF10* | -0.340208282 | 0.00622249 | 0.011570498 | 1 |
| T2DM_insulin leukocyte | *RALGAPA2* | -0.409434277 | 0.003498212 | 0.011330438 | 1 |
| T2DM_insulin leukocyte | *NDOR1* | -1.100472486 | 0.005373477 | 0.011421159 | 1 |
| T2DM_insulin leukocyte | *FBLL1* | -0.846044951 | 0.002434118 | 0.011330438 | 1 |
| T2DM_insulin leukocyte | *ASAH2* | -1.182125383 | 0.000153423 | 0.011330438 | 0.602797695 |
| T2DM_insulin leukocyte | *S100A16* | -0.20429716 | 0.005663364 | 0.011549238 | 1 |
| T2DM_insulin leukocyte | *PARVB* | -0.213647032 | 0.025488856 | 0.033651114 | 1 |
| T2DM_insulin leukocyte | *UROS* | -0.466559593 | 0.002972357 | 0.011330438 | 1 |
| T2DM_insulin leukocyte | *FAM221A* | -0.606436802 | 0.000840856 | 0.011330438 | 1 |
| T2DM_insulin leukocyte | *NOXA1* | -0.704037211 | 0.004970256 | 0.011330438 | 1 |
| T2DM_insulin leukocyte | *TMEM198* | -0.302575506 | 0.009387871 | 0.014787038 | 1 |
| T2DM_insulin leukocyte | *ZNF548* | -0.419268727 | 0.01451836 | 0.020652657 | 1 |
| T2DM_insulin leukocyte | *MTF1* | -0.388008713 | 0.003737907 | 0.011330438 | 1 |
| T2DM_insulin leukocyte | *RPL14* | -0.253517041 | 0.00247875 | 0.011330438 | 1 |
| T2DM_insulin leukocyte | *FBF1* | 0.395945093 | 0.004479916 | 0.011330438 | 1 |
| T2DM_insulin leukocyte | *MSL1* | -0.365519834 | 0.009433391 | 0.01483145 | 1 |
| T2DM_insulin leukocyte | *NOC2L* | -0.473403126 | 0.00298649 | 0.011330438 | 1 |
| T2DM_insulin leukocyte | *ZNF567* | -0.378746554 | 0.005152142 | 0.011330438 | 1 |
| T2DM_insulin leukocyte | *ARID2* | -0.501870033 | 0.003348049 | 0.011330438 | 1 |
| T2DM_insulin leukocyte | *PLAC9* | -0.347582608 | 0.011875805 | 0.017638109 | 1 |
| T2DM_insulin leukocyte | *CLDN4* | -0.220213153 | 0.004316051 | 0.011330438 | 1 |

| | | | | | |
|---|---|---|---|---|---|
| T2DM_insulin leukocyte | *FAM47E* | -0.742157502 | 3.05E-05 | 0.011330438 | 0.119762241 |
| T2DM_insulin leukocyte | *ZNF527* | -0.547897269 | 0.003509691 | 0.011330438 | 1 |
| T2DM_insulin leukocyte | *ZNF600* | -0.335519257 | 0.022375512 | 0.029984102 | 1 |
| T2DM_insulin leukocyte | *PNRC2* | -0.312086343 | 0.006186569 | 0.011570498 | 1 |
| T2DM_insulin leukocyte | *ZKSCAN3* | -1.123973507 | 7.12E-05 | 0.011330438 | 0.279911498 |
| T2DM_insulin leukocyte | *RRP7A* | -0.362819878 | 0.012943323 | 0.018774693 | 1 |
| T2DM_insulin leukocyte | *S100A14* | -0.268816124 | 0.022736312 | 0.030405368 | 1 |
| T2DM_insulin leukocyte | *CXCL17* | -0.2903567 | 0.012025357 | 0.017759823 | 1 |
| T2DM_insulin leukocyte | *BLOC1S2* | -0.377897686 | 0.005165054 | 0.011343487 | 1 |
| T2DM_insulin leukocyte | *ZNF676* | -0.593674909 | 0.000704098 | 0.011330438 | 1 |
| T2DM_insulin leukocyte | *WDSUB1* | -0.489857659 | 0.005085306 | 0.011330438 | 1 |
| T2DM_insulin leukocyte | *ZNF681* | -1.70261127 | 0.0075137 | 0.01271375 | 1 |
| T2DM_insulin leukocyte | *HRCT1* | 0.416385363 | 0.019549495 | 0.026687509 | 1 |
| T2DM_insulin leukocyte | *MPHOSPH8* | -0.377530551 | 0.003072169 | 0.011330438 | 1 |
| T2DM_insulin leukocyte | *SIRPB2* | 0.668146961 | 0.013521193 | 0.019473889 | 1 |
| T2DM_insulin leukocyte | *LCOR* | -0.350236971 | 0.003202826 | 0.011330438 | 1 |
| T2DM_insulin leukocyte | *SUPT5H* | -0.328551297 | 0.004103324 | 0.011330438 | 1 |
| T2DM_insulin leukocyte | *XPNPEP3* | -0.577477189 | 0.002084592 | 0.011330438 | 1 |
| T2DM_insulin leukocyte | *ZNF107* | -0.433659939 | 0.003546155 | 0.011330438 | 1 |
| T2DM_insulin leukocyte | *SFTA2* | -0.517330401 | 0.001226094 | 0.011330438 | 1 |
| T2DM_insulin leukocyte | *PPIA* | -0.319795301 | 0.002156123 | 0.011330438 | 1 |
| T2DM_insulin leukocyte | *ZNF471* | -0.539652948 | 0.00579371 | 0.011570498 | 1 |
| T2DM_insulin leukocyte | *ZNF493* | -0.509784833 | 0.003271213 | 0.011330438 | 1 |
| T2DM_insulin leukocyte | *GTF2IRD2* | -0.54168307 | 0.002687578 | 0.011330438 | 1 |
| T2DM_insulin leukocyte | *SUPT3H* | -0.330426749 | 0.005662698 | 0.011549238 | 1 |
| T2DM_insulin leukocyte | *POM121* | -0.36093775 | 0.009984817 | 0.015458818 | 1 |
| T2DM_insulin leukocyte | *LONP1* | -0.546569657 | 0.003193912 | 0.011330438 | 1 |
| T2DM_insulin leukocyte | *ZNF34* | -0.52326592 | 0.006042847 | 0.011570498 | 1 |
| T2DM_insulin leukocyte | *ZNF781* | -0.688335414 | 0.000806899 | 0.011330438 | 1 |
| T2DM_insulin leukocyte | *EVL* | -0.716196247 | 0.012787726 | 0.018608509 | 1 |
| T2DM_insulin leukocyte | *ZNF124* | -0.634711137 | 0.003678361 | 0.011330438 | 1 |
| T2DM_insulin leukocyte | *XRCC6* | -0.351288938 | 0.003342337 | 0.011330438 | 1 |
| T2DM_insulin leukocyte | *PIK3R4* | -0.625638221 | 0.003591506 | 0.011330438 | 1 |
| T2DM_insulin leukocyte | *C20orf96* | -0.85959903 | 0.000864504 | 0.011330438 | 1 |
| T2DM_insulin leukocyte | *SULT1A1* | -0.836079749 | 0.001204895 | 0.011330438 | 1 |
| T2DM_insulin leukocyte | *ARL9* | 0.413636237 | 0.028457146 | 0.036949149 | 1 |
| T2DM_insulin leukocyte | *TPK1* | -0.961016617 | 0.00551209 | 0.011434531 | 1 |
| T2DM_insulin leukocyte | *AFAP1* | -0.465150963 | 0.00243028 | 0.011330438 | 1 |
| T2DM_insulin leukocyte | *MYO18A* | -0.338477348 | 0.00464805 | 0.011330438 | 1 |
| T2DM_insulin leukocyte | *XRCC2* | -1.417950988 | 0.004938616 | 0.011330438 | 1 |
| T2DM_insulin leukocyte | *MYO6* | -0.280577351 | 0.004373228 | 0.011330438 | 1 |
| T2DM_insulin leukocyte | *ZNF502* | -1.425937896 | 0.004669587 | 0.011330438 | 1 |
| T2DM_insulin leukocyte | *TRAPPC4* | -0.293521732 | 0.01718922 | 0.023872903 | 1 |
| T2DM_insulin leukocyte | *ZNF33B* | -0.739572597 | 0.001106486 | 0.011330438 | 1 |
| T2DM_insulin leukocyte | *AMZ2* | -0.463066436 | 0.003538038 | 0.011330438 | 1 |
| T2DM_insulin leukocyte | *ZNF431* | -0.394935029 | 0.00333554 | 0.011330438 | 1 |
| T2DM_insulin leukocyte | *DAPK1* | -0.598306887 | 0.006658589 | 0.01190541 | 1 |
| T2DM_insulin leukocyte | *HLA-DQA1* | 0.031017801 | 0.020370785 | 0.027675247 | 1 |
| T2DM_insulin leukocyte | *CLPSL2* | -0.386777618 | 0.026430573 | 0.034731011 | 1 |
| T2DM_insulin leukocyte | *ADA* | 0.191154451 | 0.013780659 | 0.019782319 | 1 |

| | | | | | |
|---|---|---|---|---|---|
| T2DM_insulin leukocyte | *PPTC7* | -0.647836868 | 0.000736538 | 0.011330438 | 1 |
| T2DM_insulin leukocyte | *KRT39* | -2.617445017 | 0.029087598 | 0.037680572 | 1 |
| T2DM_insulin leukocyte | *ZFP28* | -0.542855245 | 0.004315959 | 0.011330438 | 1 |
| T2DM_insulin leukocyte | *ANKRD36B* | -0.363736024 | 0.023711975 | 0.031559739 | 1 |
| T2DM_insulin leukocyte | *SRGAP1* | -0.448181814 | 0.00379294 | 0.011330438 | 1 |
| T2DM_insulin leukocyte | *FAM3C* | -0.312104055 | 0.003499966 | 0.011330438 | 1 |
| T2DM_insulin leukocyte | *AP2A1* | -0.482552963 | 0.002982254 | 0.011330438 | 1 |
| T2DM_insulin leukocyte | *WDR5B* | -1.050371471 | 0.008096638 | 0.013304764 | 1 |
| T2DM_insulin leukocyte | *ZNF138* | -0.294902358 | 0.009284727 | 0.014662255 | 1 |
| T2DM_insulin leukocyte | *ZNF429* | -0.362618257 | 0.0049195 | 0.011330438 | 1 |
| T2DM_insulin leukocyte | *ZNF100* | -0.559173761 | 0.001525504 | 0.011330438 | 1 |
| T2DM_insulin leukocyte | *ZNF763* | -0.688561892 | 0.017057582 | 0.023757263 | 1 |
| T2DM_insulin leukocyte | *ARRDC1* | -0.335259241 | 0.008251165 | 0.013448692 | 1 |
| T2DM_insulin leukocyte | *ZNF682* | -0.801537949 | 0.000978287 | 0.011330438 | 1 |
| T2DM_insulin leukocyte | *ZNF257* | -0.643211648 | 0.002037917 | 0.011330438 | 1 |
| T2DM_insulin leukocyte | *ACSL5* | -0.657638585 | 0.003398323 | 0.011330438 | 1 |
| T2DM_insulin leukocyte | *SND1* | -0.370457775 | 0.003987927 | 0.011330438 | 1 |
| T2DM_insulin leukocyte | *SULT1A2* | -1.307756356 | 0.008645183 | 0.013909469 | 1 |
| T2DM_insulin leukocyte | *TBC1D9B* | -0.681115216 | 0.003106155 | 0.011330438 | 1 |
| T2DM_insulin leukocyte | *C6orf141* | -0.682511434 | 0.008814089 | 0.014103329 | 1 |
| T2DM_insulin leukocyte | *GTF2E2* | -0.35181142 | 0.005836297 | 0.011570498 | 1 |
| T2DM_insulin leukocyte | *ZNF165* | -0.316057875 | 0.028037736 | 0.036525287 | 1 |
| T2DM_insulin leukocyte | *SVIL* | -0.357950851 | 0.003678401 | 0.011330438 | 1 |
| T2DM_insulin leukocyte | *MRPL21* | -0.392805947 | 0.002249957 | 0.011330438 | 1 |
| T2DM_insulin leukocyte | *ZNF98* | -0.521883528 | 0.006995385 | 0.01219382 | 1 |
| T2DM_insulin leukocyte | *ZNF517* | -1.598885584 | 0.002627339 | 0.011330438 | 1 |
| T2DM_insulin leukocyte | *SLC22A5* | -0.579268391 | 0.003484115 | 0.011330438 | 1 |
| T2DM_insulin leukocyte | *C5AR1* | 0.047349833 | 0.01581625 | 0.022249211 | 1 |
| T2DM_insulin leukocyte | *VEPH1* | -0.629379949 | 0.024076894 | 0.03200207 | 1 |
| T2DM_insulin leukocyte | *OGDHL* | -0.33717463 | 0.037250685 | 0.046954745 | 1 |
| T2DM_insulin leukocyte | *PCDHB11* | -0.820283717 | 0.001319015 | 0.011330438 | 1 |
| T2DM_insulin leukocyte | *ATG7* | -0.750961332 | 0.002600439 | 0.011330438 | 1 |
| T2DM_insulin leukocyte | *PIGN* | -0.295717131 | 0.018006185 | 0.024823263 | 1 |
| T2DM_insulin leukocyte | *HHLA3* | -0.281737744 | 0.027571723 | 0.036001761 | 1 |
| T2DM_insulin leukocyte | *BCO2* | -0.386549577 | 0.010010391 | 0.015490676 | 1 |
| T2DM_insulin leukocyte | *ENTPD6* | -0.382728034 | 0.011635004 | 0.017368514 | 1 |
| T2DM_insulin leukocyte | *ENPP1* | -0.286217648 | 0.013062888 | 0.0189039 | 1 |
| T2DM_insulin leukocyte | *DPP4* | -0.258299512 | 0.005186672 | 0.011371894 | 1 |
| T2DM_insulin leukocyte | *NMB* | -0.365008887 | 0.005069755 | 0.011330438 | 1 |
| T2DM_insulin leukocyte | *RPS26* | -0.244103088 | 0.009002729 | 0.014308949 | 1 |
| T2DM_insulin leukocyte | *LHFPL5* | -0.955705633 | 0.005024911 | 0.011330438 | 1 |
| T2DM_insulin leukocyte | *ZNF780A* | -0.301344512 | 0.012449393 | 0.018197048 | 1 |
| T2DM_insulin leukocyte | *ATAD3A* | -0.679585279 | 0.002121491 | 0.011330438 | 1 |
| T2DM_insulin leukocyte | *FAM118B* | -0.815604387 | 0.006198804 | 0.011570498 | 1 |
| T2DM_insulin leukocyte | *CCDC180* | -1.030556352 | 0.001698503 | 0.011330438 | 1 |
| T2DM_insulin leukocyte | *SLC9A8* | -1.175820672 | 0.004941312 | 0.011330438 | 1 |
| T2DM_insulin leukocyte | *ZNF181* | -0.541957702 | 0.003822741 | 0.011330438 | 1 |
| T2DM_insulin leukocyte | *TEAD4* | -0.320773695 | 0.006162821 | 0.011570498 | 1 |
| T2DM_insulin leukocyte | *C2orf27A* | -0.770557068 | 0.000190598 | 0.011330438 | 0.748859662 |
| T2DM_insulin leukocyte | *PLCG2* | -0.426202 | 0.004005785 | 0.011330438 | 1 |

| | | | | | |
|---|---|---|---|---|---|
| T2DM_insulin leukocyte | *RPL12* | -0.258510761 | 0.001369832 | 0.011330438 | 1 |
| T2DM_insulin leukocyte | *VPS13A* | -0.340015444 | 0.007624159 | 0.01282334 | 1 |
| T2DM_insulin leukocyte | *LEKR1* | -0.68518361 | 0.003511536 | 0.011330438 | 1 |
| T2DM_insulin leukocyte | *ENTPD7* | -0.63606405 | 0.026114544 | 0.034338702 | 1 |
| T2DM_insulin leukocyte | *AGAP9* | -0.699352468 | 0.012067713 | 0.017811436 | 1 |
| T2DM_insulin leukocyte | *ZNF273* | -0.836214773 | 0.000900687 | 0.011330438 | 1 |
| T2DM_insulin leukocyte | *ZNF84* | -0.718545831 | 0.008182887 | 0.013396068 | 1 |
| T2DM_insulin leukocyte | *ZNF667* | -0.279648292 | 0.005810083 | 0.011570498 | 1 |
| T2DM_insulin leukocyte | *CD2AP* | -0.37434493 | 0.003697072 | 0.011330438 | 1 |
| T2DM_insulin leukocyte | *SFI1* | -0.821886126 | 0.003145331 | 0.011330438 | 1 |
| T2DM_insulin leukocyte | *HIBCH* | -0.330956595 | 0.006936094 | 0.012128132 | 1 |
| T2DM_insulin leukocyte | *TMEM229B* | -1.782062334 | 0.003785929 | 0.011330438 | 1 |
| T2DM_insulin leukocyte | *SOWAHC* | -0.676239656 | 0.002385013 | 0.011330438 | 1 |
| T2DM_insulin leukocyte | *SVIP* | -0.312510677 | 0.005536883 | 0.011449692 | 1 |
| T2DM_insulin leukocyte | *DDRGK1* | -0.406929526 | 0.003597657 | 0.011330438 | 1 |
| T2DM_insulin leukocyte | *ZNF607* | -0.459293548 | 0.006447892 | 0.01173015 | 1 |
| T2DM_insulin leukocyte | *HSD17B11* | -0.522308401 | 0.006852076 | 0.012045987 | 1 |
| T2DM_insulin leukocyte | *RPS6KL1* | -0.470065201 | 0.008249404 | 0.013448692 | 1 |
| T2DM_insulin leukocyte | *QRICH1* | -0.637283544 | 0.002513601 | 0.011330438 | 1 |
| T2DM_insulin leukocyte | *DDX42* | -0.310139081 | 0.007340148 | 0.012522047 | 1 |
| T2DM_insulin leukocyte | *HELZ* | -0.404942066 | 0.003604046 | 0.011330438 | 1 |
| T2DM_insulin leukocyte | *ZNF583* | -0.335431529 | 0.005741534 | 0.011570498 | 1 |
| T2DM_insulin leukocyte | *ZNF568* | -0.357035432 | 0.021729677 | 0.029268393 | 1 |
| T2DM_insulin leukocyte | *ZNF587* | -0.44011988 | 0.004072719 | 0.011330438 | 1 |
| T2DM_insulin leukocyte | *TPM2* | -0.444341056 | 0.002605638 | 0.011330438 | 1 |
| T2DM_insulin leukocyte | *CNGA1* | -0.538646993 | 0.019555443 | 0.026687509 | 1 |
| T2DM_insulin leukocyte | *ZNF789* | -0.843708224 | 0.013002835 | 0.018823927 | 1 |
| T2DM_insulin leukocyte | *CCDC69* | -0.384554944 | 0.025784235 | 0.033972625 | 1 |
| T2DM_insulin leukocyte | *NCOA6* | -0.350429717 | 0.004037417 | 0.011330438 | 1 |
| T2DM_insulin leukocyte | *STK39* | -0.336633739 | 0.006177557 | 0.011570498 | 1 |
| T2DM_insulin leukocyte | *PAPSS2* | -0.321193519 | 0.003791594 | 0.011330438 | 1 |
| T2DM_insulin leukocyte | *CEP290* | -0.43400103 | 0.003346379 | 0.011330438 | 1 |
| T2DM_insulin leukocyte | *ECI2* | -0.410749791 | 0.004262661 | 0.011330438 | 1 |
| T2DM_insulin leukocyte | *MSRB1* | -0.501094713 | 0.003762835 | 0.011330438 | 1 |
| T2DM_insulin leukocyte | *OXCT2* | -0.408378873 | 0.011878452 | 0.017638109 | 1 |
| T2DM_insulin leukocyte | *PNP* | -0.206830295 | 0.024509644 | 0.032511274 | 1 |
| T2DM_insulin leukocyte | *PAX9* | 9.2144022 | 0.018773371 | 0.025745402 | 1 |
| T2DM_insulin leukocyte | *DENND4B* | -0.375795149 | 0.008840655 | 0.014115597 | 1 |
| T2DM_insulin leukocyte | *CES1* | -0.760843896 | 0.01149256 | 0.017184601 | 1 |
| T2DM_insulin leukocyte | *TSEN15* | -0.466093409 | 0.00357652 | 0.011330438 | 1 |
| T2DM_insulin leukocyte | *LTN1* | -0.369029439 | 0.005813337 | 0.011570498 | 1 |
| T2DM_insulin leukocyte | *TYW1* | -0.560918052 | 0.006282885 | 0.011583977 | 1 |
| T2DM_insulin leukocyte | *MAP3K3* | -0.430490888 | 0.002767233 | 0.011330438 | 1 |
| T2DM_insulin leukocyte | *C1orf174* | -0.467815108 | 0.005091994 | 0.011330438 | 1 |
| T2DM_insulin leukocyte | *ATG9A* | -0.440086494 | 0.009609781 | 0.015042561 | 1 |
| T2DM_insulin leukocyte | *TBKBP1* | -0.486520764 | 0.001034731 | 0.011330438 | 1 |
| T2DM_insulin leukocyte | *CCDC167* | -0.310808936 | 0.004374792 | 0.011330438 | 1 |
| T2DM_insulin leukocyte | *SOWAHA* | -0.347485439 | 0.004470154 | 0.011330438 | 1 |
| T2DM_insulin leukocyte | *L3MBTL3* | -0.783338136 | 0.000333475 | 0.011330438 | 1 |
| T2DM_insulin leukocyte | *MFAP3L* | -0.639141094 | 0.003841795 | 0.011330438 | 1 |

| | | | | | |
|---|---|---|---|---|---|
| T2DM_insulin leukocyte | *PJA2* | -0.314237749 | 0.005811565 | 0.011570498 | 1 |
| T2DM_insulin leukocyte | *INF2* | -0.552113725 | 0.004719644 | 0.011330438 | 1 |
| T2DM_insulin leukocyte | *EFCAB2* | -0.36350752 | 0.009714423 | 0.015146019 | 1 |
| T2DM_insulin leukocyte | *SAMD5* | -0.472818107 | 0.00313046 | 0.011330438 | 1 |
| T2DM_insulin leukocyte | *PCMTD2* | -0.369843421 | 0.0040125 | 0.011330438 | 1 |
| T2DM_insulin leukocyte | *LIME1* | -0.508926311 | 0.022275027 | 0.029880021 | 1 |
| T2DM_insulin leukocyte | *FOXO6* | -0.729703044 | 0.006330824 | 0.011634148 | 1 |
| T2DM_insulin leukocyte | *SYS1* | -0.428397614 | 0.001420755 | 0.011330438 | 1 |
| T2DM_insulin leukocyte | *TRAF3IP1* | -1.120430582 | 0.001235771 | 0.011330438 | 1 |
| T2DM_insulin leukocyte | *HLA-DOA* | 0.060652508 | 0.016438781 | 0.023026015 | 1 |
| T2DM_insulin leukocyte | *HLA-DMA* | 0.111033216 | 0.000251167 | 0.011330438 | 0.986833637 |
| T2DM_insulin leukocyte | *NOTCH4* | -0.767082432 | 8.84E-09 | 3.47E-05 | 3.47E-05 |
| T2DM_insulin leukocyte | *RNF5* | -0.53270214 | 1.73E-05 | 0.011330438 | 0.068046959 |
| T2DM_insulin leukocyte | *MRPL38* | -0.408727452 | 0.002403922 | 0.011330438 | 1 |
| T2DM_insulin leukocyte | *SMIM5* | -0.355905597 | 0.001674324 | 0.011330438 | 1 |
| T2DM_insulin leukocyte | *EHMT2* | -1.093109944 | 0.014451539 | 0.020587417 | 1 |
| T2DM_insulin leukocyte | *LAYN* | -0.357895834 | 0.005831052 | 0.011570498 | 1 |
| T2DM_insulin leukocyte | *SLC44A4* | -0.323303968 | 0.001635604 | 0.011330438 | 1 |
| T2DM_insulin leukocyte | *HLA-C* | -0.205662859 | 0.008564152 | 0.01382216 | 1 |
| T2DM_insulin leukocyte | *POU5F1* | 0.427890717 | 0.038517994 | 0.048412411 | 1 |
| T2DM_insulin leukocyte | *CCHCR1* | 0.565686713 | 0.007539713 | 0.012730353 | 1 |
| T2DM_insulin leukocyte | *PSORS1C1* | -0.461964562 | 3.67E-05 | 0.011330438 | 0.144213616 |
| T2DM_insulin leukocyte | *DHX16* | -0.43858722 | 0.005638366 | 0.01154792 | 1 |
| T2DM_insulin leukocyte | *ABCF1* | -0.358664669 | 0.008217805 | 0.013442029 | 1 |
| T2DM_insulin leukocyte | *LILRB3* | 0.474608558 | 0.002469842 | 0.011330438 | 1 |
| T2DM_insulin leukocyte | *ZNF468* | -0.829221755 | 0.007731708 | 0.012941946 | 1 |
| T2DM_insulin leukocyte | *HLA-F* | -0.196955787 | 0.009094763 | 0.014422203 | 1 |
| T2DM_insulin leukocyte | *ASPDH* | -0.682054049 | 0.004867715 | 0.011330438 | 1 |
| T2DM_insulin leukocyte | *ZNF425* | -1.015976152 | 0.004185605 | 0.011330438 | 1 |
| T2DM_insulin leukocyte | *PCDHA5* | -0.625486943 | 0.00687742 | 0.012075281 | 1 |
| T2DM_insulin leukocyte | *SPIRE2* | -0.421763052 | 0.000597543 | 0.011330438 | 1 |
| T2DM_insulin leukocyte | *PABPN1L* | -52.46063309 | 0.004913665 | 0.011330438 | 1 |
| T2DM_insulin leukocyte | *PKHD1L1* | -1.420167652 | 0.011466008 | 0.017155348 | 1 |
| T2DM_insulin leukocyte | *SYCE1L* | -0.565621415 | 0.006757905 | 0.011981863 | 1 |
| T2DM_insulin leukocyte | *TMEM231* | -0.31811524 | 0.027214216 | 0.035594093 | 1 |
| T2DM_insulin leukocyte | *SARNP* | -0.365427409 | 0.003720954 | 0.011330438 | 1 |
| T2DM_insulin leukocyte | *ATP6AP1L* | 0.763897868 | 0.010526892 | 0.016078505 | 1 |
| T2DM_insulin leukocyte | *CCDC85C* | -0.414382581 | 0.003749417 | 0.011330438 | 1 |
| T2DM_insulin leukocyte | *RGL3* | -0.760587101 | 0.001876829 | 0.011330438 | 1 |
| T2DM_insulin leukocyte | *CPT1B* | -0.391780954 | 0.016048708 | 0.02251371 | 1 |
| T2DM_insulin leukocyte | *SMN2* | -0.401623294 | 0.020251028 | 0.027531588 | 1 |
| T2DM_insulin leukocyte | *HMGN1* | -0.307894511 | 0.00229431 | 0.011330438 | 1 |
| T2DM_insulin leukocyte | *LCMT1* | -0.530714455 | 0.002718093 | 0.011330438 | 1 |
| T2DM_insulin leukocyte | *LIN52* | -0.841231856 | 0.002966512 | 0.011330438 | 1 |
| T2DM_insulin leukocyte | *DENND1C* | 0.143919653 | 0.015800908 | 0.02223559 | 1 |
| T2DM_insulin leukocyte | *RNPS1* | -0.347866056 | 0.004765537 | 0.011330438 | 1 |
| T2DM_insulin leukocyte | *ZDHHC11B* | -0.630974367 | 0.007040946 | 0.012237751 | 1 |
| T2DM_insulin leukocyte | *GOLGA8O* | -0.926363234 | 0.029454057 | 0.038079957 | 1 |
| T2DM_insulin leukocyte | *HLA-A* | -0.18540097 | 0.027241386 | 0.035617773 | 1 |
| T2DM_insulin leukocyte | *STK38L* | -0.646945311 | 0.006691425 | 0.011938173 | 1 |

| | | | | | |
|---|---|---|---|---|---|
| T2DM_insulin leukocyte | *MAP10* | -0.400078897 | 0.012867366 | 0.018696702 | 1 |
| T2DM_insulin leukocyte | *NUP62* | -0.409691194 | 0.004840043 | 0.011330438 | 1 |
| T2DM_insulin leukocyte | *FAM106A* | -0.477512039 | 0.01445998 | 0.020591976 | 1 |
| T2DM_insulin leukocyte | *ZNF254* | -0.491622209 | 0.001907146 | 0.011330438 | 1 |
| T2DM_insulin leukocyte | *ASIC3* | -0.558929182 | 0.004755687 | 0.011330438 | 1 |
| T2DM_insulin leukocyte | *ARHGEF35* | -0.26352774 | 0.006264819 | 0.011570498 | 1 |
| T2DM_insulin leukocyte | *SUPT4H1* | -0.449291087 | 0.003338352 | 0.011330438 | 1 |
| T2DM_insulin leukocyte | *MXD3* | -0.325520495 | 0.026751582 | 0.035059586 | 1 |
| T2DM_insulin leukocyte | *COG8* | -0.57794153 | 0.002985665 | 0.011330438 | 1 |
| T2DM_insulin leukocyte | *CCL14* | -0.490252866 | 0.027694148 | 0.036137598 | 1 |
| T2DM_insulin leukocyte | *RBMXL1* | -0.417357315 | 0.003623072 | 0.011330438 | 1 |
| T2DM_insulin leukocyte | *SRA1* | -0.288660253 | 0.005803089 | 0.011570498 | 1 |
| T2DM_insulin leukocyte | *C8orf82* | -0.78630796 | 0.001221274 | 0.011330438 | 1 |
| T2DM_insulin leukocyte | *LEPROT* | -0.377109027 | 0.003365785 | 0.011330438 | 1 |
| T2DM_insulin leukocyte | *PPP1CB* | -0.303362437 | 0.003643427 | 0.011330438 | 1 |
| T2DM_insulin leukocyte | *NCKIPSD* | -0.606992416 | 0.001805956 | 0.011330438 | 1 |
| T2DM_insulin leukocyte | *ZNF888* | -0.555699487 | 0.005423041 | 0.011421159 | 1 |
| T2DM_insulin leukocyte | *EMP2* | -0.457019798 | 0.00136924 | 0.011330438 | 1 |
| T2DM_insulin leukocyte | *LTB4R* | -0.497469548 | 0.008002195 | 0.013211473 | 1 |
| T2DM_insulin leukocyte | *DNASE1* | -0.270123314 | 0.036402017 | 0.045929199 | 1 |
| T2DM_insulin leukocyte | *NUDT19* | -0.48022597 | 0.007272107 | 0.012444298 | 1 |
| T2DM_insulin leukocyte | *AP1G2* | -1.394297832 | 0.003839769 | 0.011330438 | 1 |
| T2DM_insulin leukocyte | *ZNF90* | -0.471548767 | 0.005845952 | 0.011570498 | 1 |
| T2DM_insulin leukocyte | *REPIN1* | -0.424433699 | 0.00325328 | 0.011330438 | 1 |
| T2DM_insulin leukocyte | *TSPAN4* | -0.443326949 | 0.005016395 | 0.011330438 | 1 |
| T2DM_insulin leukocyte | *CPNE1* | -0.475029873 | 0.01073797 | 0.016320884 | 1 |
| T2DM_insulin leukocyte | *ARL16* | -0.339273563 | 0.005111851 | 0.011330438 | 1 |
| T2DM_insulin leukocyte | *FIS1* | -0.324136951 | 0.002185838 | 0.011330438 | 1 |
| T2DM_insulin leukocyte | *VSTM5* | -1.309638953 | 0.008993536 | 0.014300123 | 1 |
| T2DM_insulin leukocyte | *AS3MT* | 0.445814483 | 0.035526359 | 0.04496877 | 1 |
| T2DM_insulin leukocyte | *MEG3* | -0.243124575 | 0.004080852 | 0.011330438 | 1 |
| T2DM_insulin leukocyte | *EML6* | -0.463692017 | 0.007660241 | 0.012869015 | 1 |
| T2DM_insulin leukocyte | *IFRD2* | -0.420834933 | 0.003144344 | 0.011330438 | 1 |
| T2DM_insulin leukocyte | *ARHGEF28* | -0.493533492 | 0.0065229 | 0.011788627 | 1 |
| T2DM_insulin leukocyte | *COL28A1* | -1.582428358 | 0.028103024 | 0.036586077 | 1 |
| T2DM_insulin leukocyte | *UBXN2B* | -0.610198218 | 0.00446171 | 0.011330438 | 1 |
| T2DM_insulin leukocyte | *C5orf49* | -0.324582957 | 0.005170691 | 0.011349522 | 1 |
| T2DM_insulin leukocyte | *GOLGA8B* | -0.470145032 | 0.011986499 | 0.01772486 | 1 |
| T2DM_insulin leukocyte | *VPS16* | -0.687365875 | 0.002569351 | 0.011330438 | 1 |
| T2DM_insulin leukocyte | *LINC01139* | -0.873774834 | 0.032337636 | 0.041332002 | 1 |
| T2DM_insulin leukocyte | *ATAD3C* | -0.362797424 | 0.004187811 | 0.011330438 | 1 |
| T2DM_insulin leukocyte | *SYCE3* | -0.708425691 | 0.009255718 | 0.014622322 | 1 |
| T2DM_insulin leukocyte | *PAM16* | -0.351189111 | 0.003936204 | 0.011330438 | 1 |
| T2DM_insulin leukocyte | *LINC00339* | -0.43759451 | 0.005946134 | 0.011570498 | 1 |
| T2DM_insulin leukocyte | *CELA3B* | -0.193289085 | 0.003913085 | 0.011330438 | 1 |
| T2DM_insulin leukocyte | *FAM228B* | -0.415700725 | 0.005331701 | 0.011421159 | 1 |
| T2DM_insulin leukocyte | *VAMP2* | -0.274599157 | 0.004135924 | 0.011330438 | 1 |
| T2DM_insulin leukocyte | *PPP3R1* | -0.58653996 | 0.002651754 | 0.011330438 | 1 |
| T2DM_insulin leukocyte | *FANCG* | -0.723395841 | 0.00315344 | 0.011330438 | 1 |
| T2DM_insulin leukocyte | *AP4M1* | -0.345600552 | 0.002876377 | 0.011330438 | 1 |

| | | | | | |
|---|---|---|---|---|---|
| T2DM_insulin leukocyte | *ZNF880* | -0.299189459 | 0.009825673 | 0.015277036 | 1 |
| T2DM_insulin leukocyte | *TRIM16* | -0.539388521 | 0.005115002 | 0.011330438 | 1 |
| T2DM_insulin leukocyte | *TIGD1* | -0.512098217 | 0.018753447 | 0.025727828 | 1 |
| T2DM_insulin leukocyte | *SLC12A8* | -0.572653467 | 0.004589103 | 0.011330438 | 1 |
| T2DM_insulin leukocyte | *UBA52* | -0.292513227 | 0.002599994 | 0.011330438 | 1 |
| T2DM_insulin leukocyte | *EXOSC6* | -0.453304905 | 0.001707418 | 0.011330438 | 1 |
| T2DM_insulin leukocyte | *CKMT1A* | -0.380900872 | 0.007201043 | 0.012381002 | 1 |
| T2DM_insulin leukocyte | *HLA-DPB1* | 0.037553481 | 0.029109656 | 0.037696717 | 1 |
| T2DM_insulin leukocyte | *C4B* | 4.1925103 | 0.019009107 | 0.026005146 | 1 |
| T2DM_insulin leukocyte | *LINC00115* | -1.118074238 | 0.002409351 | 0.011330438 | 1 |
| T2DM_insulin leukocyte | *LINC00937* | -0.422733054 | 0.009380101 | 0.014783159 | 1 |
| T2DM_insulin leukocyte | *TMEM185B* | -0.397774597 | 0.007134677 | 0.012332664 | 1 |
| T2DM_insulin leukocyte | *CYP4F26P* | 1.872775862 | 0.01523544 | 0.021555956 | 1 |
| T2DM_insulin leukocyte | *WDR46* | -0.328880113 | 0.012218816 | 0.017946814 | 1 |
| T2DM_insulin leukocyte | *PROB1* | -0.541810364 | 0.038838184 | 0.048752468 | 1 |
| T2DM_insulin leukocyte | *ARL17B* | -1.005463166 | 0.004657294 | 0.011330438 | 1 |
| T2DM_insulin leukocyte | *PSMG3-AS1* | -0.488562338 | 0.004353362 | 0.011330438 | 1 |
| T2DM_insulin leukocyte | *HSBP1* | -0.340063522 | 0.002948509 | 0.011330438 | 1 |
| T2DM_insulin leukocyte | *RPS18* | -0.230868585 | 0.002887662 | 0.011330438 | 1 |
| T2DM_insulin leukocyte | *DIRC3* | -1.282970241 | 0.021814954 | 0.029373186 | 1 |
| T2DM_insulin leukocyte | *LINC00865* | -0.956306035 | 0.005573823 | 0.011495826 | 1 |
| T2DM_insulin leukocyte | *HLA-DQB2* | 0.923627024 | 1.82E-08 | 3.58E-05 | 7.17E-05 |
| T2DM_insulin leukocyte | *GOLGA8N* | -0.43928232 | 0.009622369 | 0.015050274 | 1 |
| T2DM_insulin leukocyte | *LINC00665* | -0.295119643 | 0.00530576 | 0.011421159 | 1 |
| T2DM_insulin leukocyte | *LYRM9* | -0.475123583 | 0.002825278 | 0.011330438 | 1 |
| T2DM_insulin leukocyte | *SNHG15* | -0.375445591 | 0.003522952 | 0.011330438 | 1 |
| T2DM_insulin leukocyte | *RPS28* | -0.247473055 | 0.002808497 | 0.011330438 | 1 |
| T2DM_insulin leukocyte | *TRIM26* | -0.540227584 | 0.00086055 | 0.011330438 | 1 |
| T2DM_insulin leukocyte | *ZNF879* | -0.489946374 | 0.006802398 | 0.012014984 | 1 |
| T2DM_insulin leukocyte | *PINLYP* | -0.934757375 | 0.001116856 | 0.011330438 | 1 |
| T2DM_insulin leukocyte | *MCIDAS* | 13.9724884 | 0.018592545 | 0.025541996 | 1 |
| T2DM_insulin leukocyte | *ANKRD65* | -0.349417476 | 0.004825047 | 0.011330438 | 1 |
| T2DM_insulin leukocyte | *C12orf75* | -0.219070074 | 0.004120292 | 0.011330438 | 1 |
| T2DM_insulin leukocyte | *B3GALT4* | -0.430354274 | 0.000129127 | 0.011330438 | 0.507338592 |
| T2DM_insulin leukocyte | *ZNF737* | -0.562125629 | 0.002593935 | 0.011330438 | 1 |
| T2DM_insulin leukocyte | *SHISA9* | -2.858655668 | 0.005706998 | 0.011549238 | 1 |
| T2DM_insulin leukocyte | *HLA-DQA2* | 0.049393661 | 0.002158542 | 0.011330438 | 1 |
| T2DM_insulin leukocyte | *TXNDC5* | -0.410464912 | 0.002205425 | 0.011330438 | 1 |
| T2DM_insulin leukocyte | *RBM14* | -0.494399809 | 0.004755591 | 0.011330438 | 1 |
| T2DM_insulin leukocyte | *NME1* | -0.271645803 | 0.006616878 | 0.011871102 | 1 |
| T2DM_insulin leukocyte | *CDRT4* | -0.818750653 | 0.005684462 | 0.011549238 | 1 |
| T2DM_insulin leukocyte | *AMY2B* | -0.316518865 | 0.004759224 | 0.011330438 | 1 |
| T2DM_insulin leukocyte | *LY6G5B* | -0.46830038 | 0.004627321 | 0.011330438 | 1 |
| T2DM_insulin leukocyte | *PCDHGC3* | -0.412663642 | 0.013731586 | 0.019726289 | 1 |
| T2DM_insulin leukocyte | *SMKR1* | -0.483159177 | 0.005410826 | 0.011421159 | 1 |
| T2DM_insulin leukocyte | *C1QTNF9* | -6.569878705 | 0.005509345 | 0.011434531 | 1 |
| T2DM_insulin leukocyte | *LINC00886* | -0.538344923 | 0.007770921 | 0.012986793 | 1 |
| T2DM_insulin leukocyte | *MIF* | -0.229114712 | 0.004373402 | 0.011330438 | 1 |
| T2DM_insulin leukocyte | *NSUN6* | -0.372880993 | 0.004536381 | 0.011330438 | 1 |
| T2DM_insulin leukocyte | *HLA-DOB* | 1.051727575 | 7.53E-05 | 0.011330438 | 0.2957142 |

| | | | | | |
|---|---|---|---|---|---|
| T2DM_insulin leukocyte | *CRCP* | -0.478426315 | 0.001469144 | 0.011330438 | 1 |
| T2DM_insulin leukocyte | *ARHGAP8* | -0.378175263 | 0.03295012 | 0.042005522 | 1 |
| T2DM_insulin leukocyte | *C8orf58* | -0.450156287 | 0.038735693 | 0.048654903 | 1 |
| T2DM_insulin leukocyte | *PI4KA* | -0.555344402 | 0.010190313 | 0.015699813 | 1 |
| T2DM_insulin leukocyte | *AMACR* | -0.322296846 | 0.005842147 | 0.011570498 | 1 |
| T2DM_insulin leukocyte | *BGLAP* | -0.536761777 | 0.020712155 | 0.02807079 | 1 |
| T2DM_insulin leukocyte | *MRPL20* | -0.291203132 | 0.00937426 | 0.014779882 | 1 |
| T2DM_insulin leukocyte | *LINC00960* | -0.26934178 | 0.022048848 | 0.029627197 | 1 |
| T2DM_insulin leukocyte | *HLA-DMB* | 0.042961937 | 0.00812928 | 0.013352818 | 1 |
| T2DM_insulin leukocyte | *DECR2* | -0.403386704 | 0.004086854 | 0.011330438 | 1 |
| T2DM_insulin leukocyte | *GNG10* | -0.35734606 | 0.002040353 | 0.011330438 | 1 |
| T2DM_insulin leukocyte | *EIF4EBP3* | -0.54263243 | 0.001867623 | 0.011330438 | 1 |
| T2DM_insulin leukocyte | *PSG4* | 1.171562885 | 0.024404605 | 0.032393815 | 1 |
| T2DM_insulin leukocyte | *MICAL3* | -0.3697521 | 0.004489043 | 0.011330438 | 1 |
| T2DM_insulin leukocyte | *STON1* | -0.413768793 | 0.03646214 | 0.045990288 | 1 |
| T2DM_insulin leukocyte | *VSIG8* | -0.98042627 | 0.005777578 | 0.011570498 | 1 |
| T2DM_insulin leukocyte | *C4orf48* | -0.283041961 | 0.003382105 | 0.011330438 | 1 |
| T2DM_insulin leukocyte | *IL10RB* | -1.051503705 | 0.000288167 | 0.011330438 | 1 |
| T2DM_insulin leukocyte | *PLA2G4B* | -0.547849311 | 0.010814519 | 0.016411836 | 1 |
| T2DM_insulin leukocyte | *LEFTY1* | -3.312597981 | 0.004640363 | 0.011330438 | 1 |
| T2DM_insulin leukocyte | *TTC4* | -0.392479993 | 0.005885718 | 0.011570498 | 1 |
| T2DM_insulin leukocyte | *APOBEC3D* | -1.436796555 | 0.010396906 | 0.015922346 | 1 |
| T2DM_insulin leukocyte | *OR2A7* | -1.513413683 | 0.013290739 | 0.019170086 | 1 |
| T2DM_insulin leukocyte | *GSTA1* | -0.214533079 | 0.007723566 | 0.012940678 | 1 |
| T2DM_insulin leukocyte | *GSTA2* | -0.225763337 | 0.030615025 | 0.039412331 | 1 |
| T2DM_insulin leukocyte | *TMEM141* | -0.441952788 | 0.006192631 | 0.011570498 | 1 |
| T2DM_insulin leukocyte | *APOBEC3C* | -0.795157797 | 0.006742885 | 0.011963834 | 1 |
| T2DM_insulin leukocyte | *C4A* | -3.545617072 | 0.002412801 | 0.011330438 | 1 |
| T2DM_insulin leukocyte | *NEAT1* | -0.264683523 | 0.003248406 | 0.011330438 | 1 |
| T2DM_insulin leukocyte | *LINC00920* | -2.877695001 | 0.018386941 | 0.025300193 | 1 |
| T2DM_insulin leukocyte | *UBAP1L* | -0.389151902 | 0.008032804 | 0.013233077 | 1 |
| T2DM_insulin leukocyte | *TWF2* | -0.469240763 | 0.008252299 | 0.013448692 | 1 |
| T2DM_insulin leukocyte | *BCKDHA* | -0.37166958 | 0.007685644 | 0.012899144 | 1 |
| T2DM_insulin leukocyte | *PCDHAC1* | -0.594819068 | 0.005009925 | 0.011330438 | 1 |
| T2DM_insulin leukocyte | *FMN1* | -1.160384626 | 0.010701243 | 0.016283959 | 1 |
| T2DM_insulin leukocyte | *TMEM150C* | -0.467430959 | 0.004016792 | 0.011330438 | 1 |
| T2DM_insulin leukocyte | *LINC01091* | -0.256544372 | 0.009523845 | 0.014949735 | 1 |
| T2DM_insulin leukocyte | *PDCD6* | -0.372920405 | 0.003275469 | 0.011330438 | 1 |
| T2DM_insulin leukocyte | *YJEFN3* | -0.693340196 | 0.003743563 | 0.011330438 | 1 |
| T2DM_insulin leukocyte | *PCDHA10* | -0.359477746 | 0.005224238 | 0.011389288 | 1 |
| T2DM_insulin leukocyte | *ZNF718* | -0.514863321 | 0.005153241 | 0.011330438 | 1 |
| T2DM_insulin leukocyte | *SNHG18* | -0.42575361 | 0.002148721 | 0.011330438 | 1 |
| T2DM_insulin leukocyte | *ZNF550* | -0.443592861 | 0.005152505 | 0.011330438 | 1 |
| T2DM_insulin leukocyte | *LINC01099* | -0.458828804 | 0.006066812 | 0.011570498 | 1 |
| T2DM_insulin leukocyte | *PCDHA12* | -0.533551846 | 0.020860099 | 0.028232631 | 1 |
| T2DM_insulin leukocyte | *ALG11* | -0.715540503 | 0.002656244 | 0.011330438 | 1 |
| T2DM_insulin leukocyte | *ZNF260* | -0.657872546 | 0.00909968 | 0.014422203 | 1 |
| T2DM_insulin leukocyte | *AP5B1* | -0.676295516 | 0.004743013 | 0.011330438 | 1 |
| T2DM_insulin leukocyte | *SIGLEC12* | 0.750997584 | 0.013933833 | 0.019980303 | 1 |
| T2DM_insulin leukocyte | *PABPC4L* | -1.038637158 | 0.007957079 | 0.013183733 | 1 |

| | | | | | | |
|---|---|---|---|---|---|---|
| T2DM_insulin | leukocyte | *NPIPA2* | -4.197011854 | 0.004576772 | 0.011330438 | 1 |
| T2DM_insulin | leukocyte | *DPP3* | -0.437181807 | 0.001223074 | 0.011330438 | 1 |
| T2DM_insulin | leukocyte | *BRK1* | -0.345043681 | 0.000719742 | 0.011330438 | 1 |
| T2DM_insulin | leukocyte | *EID3* | -0.817568395 | 0.002702579 | 0.011330438 | 1 |
| T2DM_insulin | leukocyte | *SNHG9* | -0.359543451 | 0.003526569 | 0.011330438 | 1 |
| T2DM_insulin | leukocyte | *PCDHA3* | -0.566801301 | 0.014272664 | 0.020385699 | 1 |
| T2DM_insulin | leukocyte | *ZNF432* | -0.81558778 | 0.001423981 | 0.011330438 | 1 |
| T2DM_insulin | leukocyte | *LINC01152* | -1.816432384 | 0.006342273 | 0.011637991 | 1 |
| T2DM_insulin | leukocyte | *SMLR1* | 1.339436849 | 0.018991134 | 0.026005146 | 1 |
| T2DM_insulin | leukocyte | *ZNF10* | -0.498285801 | 0.003169122 | 0.011330438 | 1 |
| T2DM_insulin | leukocyte | *TAS2R31* | -1.337406762 | 0.010377711 | 0.01590871 | 1 |
| T2DM_insulin | leukocyte | *GATC* | -0.565792069 | 0.001108311 | 0.011330438 | 1 |
| T2DM_insulin | leukocyte | *TEN1* | -1.267859522 | 0.000317619 | 0.011330438 | 1 |
| T2DM_insulin | leukocyte | *CHURC1* | -0.314503749 | 0.003471107 | 0.011330438 | 1 |
| T2DM_insulin | leukocyte | *LINC00638* | -2.552447193 | 0.010231243 | 0.015739449 | 1 |
| T2DM_insulin | leukocyte | *SLC25A21-AS1* | -0.388256683 | 0.004987372 | 0.011330438 | 1 |
| T2DM_insulin | leukocyte | *TMEM179* | -0.275057153 | 0.006261158 | 0.011570498 | 1 |
| T2DM_insulin | leukocyte | *IQCH-AS1* | -0.726085978 | 0.002832662 | 0.011330438 | 1 |
| T2DM_insulin | leukocyte | *PWRN1* | -0.523448731 | 0.006588411 | 0.011847079 | 1 |
| T2DM_insulin | leukocyte | *TMEM178B* | -0.359689963 | 0.004974657 | 0.011330438 | 1 |
| T2DM_insulin | leukocyte | *LINC01003* | -0.468000233 | 0.002980379 | 0.011330438 | 1 |
| T2DM_insulin | leukocyte | *GOLGA8H* | -5.851381256 | 0.014442598 | 0.020587417 | 1 |
| T2DM_insulin | leukocyte | *MRPL12* | -0.488581669 | 0.003146133 | 0.011330438 | 1 |
| T2DM_insulin | leukocyte | *ZNF234* | -1.480198237 | 0.035180684 | 0.044574302 | 1 |
| T2DM_insulin | leukocyte | *LINC00667* | -0.416509144 | 0.003522296 | 0.011330438 | 1 |
| T2DM_insulin | leukocyte | *FSBP* | -0.543430247 | 0.006105397 | 0.011570498 | 1 |
| T2DM_insulin | leukocyte | *MYO15B* | -0.671975565 | 0.025818969 | 0.034006949 | 1 |
| T2DM_insulin | leukocyte | *LINC00663* | -0.50643063 | 0.022751971 | 0.030415956 | 1 |
| T2DM_insulin | leukocyte | *ZNF285* | -0.923740002 | 0.005345829 | 0.011421159 | 1 |
| T2DM_insulin | leukocyte | *FBXO17* | -0.310002953 | 0.006264802 | 0.011570498 | 1 |
| T2DM_insulin | leukocyte | *LINC00528* | 0.25440588 | 0.00494796 | 0.011330438 | 1 |
| T2DM_insulin | leukocyte | *SNHG8* | -0.286956147 | 0.003003806 | 0.011330438 | 1 |
| T2DM_insulin | leukocyte | *MILR1* | 0.300556946 | 0.030158494 | 0.038888324 | 1 |
| T2DM_insulin | leukocyte | *GTF2H5* | -0.360603902 | 0.004011862 | 0.011330438 | 1 |
| T2DM_insulin | mesenchymal_cell | *CEP68* | 0.436056151 | 6.91E-05 | 0.027162253 | 0.271622528 |
| T2DM_insulin | mesenchymal_cell | *ART3* | -0.534017296 | 0.00012061 | 0.036452112 | 0.473877459 |
| T2DM_insulin | mesenchymal_cell | *CLEC18A* | -0.574388243 | 0.000109248 | 0.036452112 | 0.429234216 |
| T2DM_insulin | mesenchymal_cell | *CXCL11* | 0.192913695 | 9.37E-06 | 0.005258949 | 0.036812643 |
| T2DM_insulin | mesenchymal_cell | *IRX3* | 0.680752573 | 3.15E-07 | 0.00031844 | 0.001238045 |
| T2DM_insulin | mesenchymal_cell | *HLA-DQB1* | -0.173538259 | 4.55E-07 | 0.000357208 | 0.001786042 |
| T2DM_insulin | mesenchymal_cell | *HLA-DRB1* | -0.097067272 | 2.23E-05 | 0.010964775 | 0.0877182 |
| T2DM_insulin | mesenchymal_cell | *HLA-DRB5* | -0.095556022 | 3.83E-06 | 0.00250842 | 0.015050521 |
| T2DM_insulin | mesenchymal_cell | *HLA-DMA* | 0.107214345 | 3.68E-05 | 0.016059429 | 0.144534863 |
| T2DM_insulin | mesenchymal_cell | *NOTCH4* | -0.18667715 | 3.24E-07 | 0.00031844 | 0.001273758 |
| T2DM_insulin | mesenchymal_cell | *PSORS1C1* | -0.123193155 | 0.000111982 | 0.036452112 | 0.439977742 |
| T2DM_insulin | mesenchymal_cell | *HLA-DQB2* | 0.635383115 | 7.49E-14 | 2.94E-10 | 2.94E-10 |
| T2DM_insulin | mesenchymal_cell | *HLA-DQA2* | 0.295092303 | 3.69E-11 | 7.24E-08 | 1.45E-07 |
| T2DM_insulin | PP_cell | *CEP68* | 0.726964944 | 4.94E-05 | 0.021584462 | 0.194260157 |
| T2DM_insulin | PP_cell | *CXCL11* | 0.351345787 | 2.99E-06 | 0.00348512 | 0.011756182 |
| T2DM_insulin | PP_cell | *IRX3* | 0.663567975 | 3.55E-06 | 0.00348512 | 0.013940481 |

| | | | | | | |
|---|---|---|---|---|---|---|
| T2DM_insulin | PP_cell | *HLA-DQB1* | -0.299322458 | 9.41E-06 | 0.006162119 | 0.036972712 |
| T2DM_insulin | PP_cell | *HLA-DRB5* | -0.240279488 | 9.42E-05 | 0.036993087 | 0.369930867 |
| T2DM_insulin | PP_cell | *HLA-DMA* | 0.207347426 | 6.52E-06 | 0.005126934 | 0.02563467 |
| T2DM_insulin | PP_cell | *NOTCH4* | -0.346599657 | 2.53E-05 | 0.014177966 | 0.099245764 |
| T2DM_insulin | PP_cell | *HLA-DQB2* | 1.132702893 | 4.37E-14 | 1.72E-10 | 1.72E-10 |
| T2DM_insulin | PP_cell | *HLA-DQA2* | 0.53818504 | 7.84E-13 | 1.54E-09 | 3.08E-09 |
| T2DM_insulin | PP_cell | *PSMB9* | 0.445144878 | 0.000121512 | 0.043402021 | 0.47742223 |
| T2DM_insulin | PP_cell | *HLA-DMB* | 0.523790308 | 4.32E-05 | 0.021236049 | 0.169888393 |

c. Identified trait-associated genes from CTS TWAS analysis in subcutaneous adipose tissue

| disorder | celltype | gene | Estimates | Pvalues | Pvalues_BH | Pvalue_Bonferroni |
|---|---|---|---|---|---|---|
| CAD | adipocyte | *SEMA3F* | -0.221590916 | 0.000214112 | 0.040791534 | 1 |
| CAD | adipocyte | *UHRF1BP1* | 0.298009851 | 5.89E-06 | 0.004627148 | 0.037017185 |
| CAD | adipocyte | *ATP2A3* | 0.776529784 | 6.04E-05 | 0.019972965 | 0.379486343 |
| CAD | adipocyte | *TIPIN* | 0.183881257 | 0.000172384 | 0.037604821 | 1 |
| CAD | adipocyte | *KCNH4* | 0.381232879 | 0.000323666 | 0.048449646 | 1 |
| CAD | adipocyte | *ABHD12* | 0.67395607 | 7.58E-05 | 0.022684977 | 0.476384519 |
| CAD | adipocyte | *MAP1LC3A* | 0.467660301 | 4.09E-05 | 0.016877267 | 0.257029754 |
| CAD | adipocyte | *PIGU* | 0.894812704 | 0.000242112 | 0.043490239 | 1 |
| CAD | adipocyte | *ICAM2* | 0.669562833 | 9.07E-05 | 0.025854379 | 0.570353888 |
| CAD | adipocyte | *BCHE* | -0.393518576 | 0.000298047 | 0.047841246 | 1 |
| CAD | adipocyte | *UMPS* | 0.946003793 | 0.000110083 | 0.026618961 | 0.692092993 |
| CAD | adipocyte | *PLXNA1* | 0.480526332 | 9.65E-05 | 0.025854379 | 0.6067602 |
| CAD | adipocyte | *GGCX* | 0.496745831 | 8.00E-06 | 0.005589586 | 0.050306278 |
| CAD | adipocyte | *EIF2B2* | 0.411332606 | 0.000263931 | 0.046092571 | 1 |
| CAD | adipocyte | *HIF3A* | 0.433052568 | 0.000176049 | 0.037604821 | 1 |
| CAD | adipocyte | *SNRPC* | -0.474334587 | 1.41E-05 | 0.008893002 | 0.088930016 |
| CAD | adipocyte | *GRK4* | 0.141948429 | 0.000283195 | 0.04685383 | 1 |
| CAD | adipocyte | *CARF* | -0.646418916 | 1.87E-07 | 0.000426176 | 0.00117815 |
| CAD | adipocyte | *TMEM91* | 0.63717179 | 4.01E-05 | 0.016877267 | 0.252224167 |
| CAD | adipocyte | *NBEAL1* | -0.444578326 | 2.71E-07 | 0.000426176 | 0.001704705 |
| CAD | adipocyte | *AMT* | -0.204062848 | 0.000132447 | 0.030840525 | 0.832694187 |
| CAD | adipocyte | *PSD2* | 2.17767934 | 2.96E-05 | 0.014308366 | 0.18600876 |
| CAD | adipocyte | *DISP1* | 0.450123067 | 4.30E-05 | 0.016877267 | 0.270036268 |
| CAD | adipocyte | *LMOD1* | -0.377136493 | 0.000235932 | 0.043490239 | 1 |
| CAD | adipocyte | *ICA1L* | 0.951207371 | 2.86E-08 | 0.000180026 | 0.000180026 |
| CAD | adipocyte | *APEH* | -0.555952414 | 4.69E-06 | 0.004215517 | 0.029508622 |
| CAD | adipocyte | *RNF123* | 0.313329824 | 0.000179441 | 0.037604821 | 1 |
| CAD | adipocyte | *IRS1* | -0.19917963 | 2.62E-07 | 0.000426176 | 0.001649937 |
| CAD | adipocyte | *PCSK9* | 2.689171795 | 1.68E-05 | 0.009473447 | 0.105762348 |
| CAD | adipocyte | *LPL* | -0.268188618 | 5.10E-07 | 0.000640972 | 0.003204859 |
| CAD | adipocyte | *CRLF3* | 0.943074885 | 5.72E-05 | 0.019972965 | 0.359459753 |
| CAD | adipocyte | *TMEM136* | 0.507744052 | 0.000100568 | 0.025854379 | 0.632271404 |
| CAD | adipocyte | *HLA-DRB1* | -0.307984642 | 0.000278516 | 0.04685383 | 1 |
| CAD | adipocyte | *C6orf106* | 0.419851132 | 2.07E-06 | 0.00217275 | 0.013036501 |
| CAD | adipocyte | *WDR5B* | 0.650672199 | 0.000310899 | 0.047841246 | 1 |
| CAD | adipocyte | *TMEM116* | -0.240956522 | 0.000311992 | 0.047841246 | 1 |
| CAD | adipocyte | *CENPW* | 0.955158191 | 1.81E-05 | 0.009473447 | 0.113681364 |
| CAD | adipocyte | *SLC2A1-AS1* | -0.762757322 | 0.000197736 | 0.039838503 | 1 |
| CAD | adipocyte | *HLA-B* | 0.338224767 | 0.000102809 | 0.025854379 | 0.646359484 |
| CAD | adipocyte | *LY6G5B* | -0.132040605 | 0.000202773 | 0.039838503 | 1 |
| CAD | adipocyte | *BCKDHA* | -1.129618427 | 5.77E-05 | 0.019972965 | 0.362990495 |
| CAD | adipocyte | *HMBS* | 0.315926285 | 7.04E-05 | 0.022136592 | 0.442731837 |
| CAD | ASPC | *SEMA3F* | -0.103732254 | 4.86E-05 | 0.015457113 | 0.305567695 |
| CAD | ASPC | *UHRF1BP1* | 0.124191885 | 9.58E-06 | 0.004631262 | 0.060206403 |
| CAD | ASPC | *TIE1* | -0.177831755 | 5.34E-05 | 0.015534196 | 0.335549187 |
| CAD | ASPC | *TIPIN* | 0.078025641 | 0.000170755 | 0.029820393 | 1 |

| | | | | | | |
|---|---|---|---|---|---|---|
| CAD | ASPC | *TXNDC16* | -0.33173762 | 0.000197394 | 0.033230556 | 1 |
| CAD | ASPC | *ABHD12* | 0.28063681 | 0.000316538 | 0.041459946 | 1 |
| CAD | ASPC | *ACTR5* | -1.033868473 | 5.51E-05 | 0.015534196 | 0.346311173 |
| CAD | ASPC | *MAP1LC3A* | 0.226139623 | 7.43E-06 | 0.00389067 | 0.046688039 |
| CAD | ASPC | *TRPV4* | -0.343788772 | 0.000345196 | 0.044290788 | 1 |
| CAD | ASPC | *FAM162A* | -0.375655998 | 0.000384067 | 0.04829254 | 1 |
| CAD | ASPC | *BCHE* | -0.184491979 | 7.31E-05 | 0.016420961 | 0.459786899 |
| CAD | ASPC | *UMPS* | 0.427901908 | 0.000149645 | 0.027671181 | 0.940820153 |
| CAD | ASPC | *PLXNA1* | 0.195331479 | 0.000207577 | 0.033230556 | 1 |
| CAD | ASPC | *GGCX* | 0.207094791 | 1.50E-05 | 0.006300934 | 0.094514008 |
| CAD | ASPC | *SNRPC* | -0.213002031 | 5.08E-06 | 0.002903781 | 0.031941586 |
| CAD | ASPC | *GRK4* | 0.061333311 | 0.000211837 | 0.033230556 | 1 |
| CAD | ASPC | *KLHDC10* | -0.823757593 | 6.63E-07 | 0.000595882 | 0.004171176 |
| CAD | ASPC | *CARF* | -0.306024165 | 1.99E-08 | 6.25E-05 | 0.000124963 |
| CAD | ASPC | *TMEM91* | 0.255929869 | 0.000147902 | 0.027671181 | 0.929860174 |
| CAD | ASPC | *NME7* | -0.725379606 | 1.14E-05 | 0.005125916 | 0.071762817 |
| CAD | ASPC | *NBEAL1* | -0.209114158 | 3.02E-08 | 6.33E-05 | 0.000189878 |
| CAD | ASPC | *AMT* | -0.089613284 | 7.17E-05 | 0.016420961 | 0.450533169 |
| CAD | ASPC | *PSD2* | 0.87523765 | 6.78E-05 | 0.01639443 | 0.426255178 |
| CAD | ASPC | *DNAJB12* | -0.78380191 | 0.000259089 | 0.035410716 | 1 |
| CAD | ASPC | *HSD17B12* | -0.096571969 | 0.000397366 | 0.048985109 | 1 |
| CAD | ASPC | *MIA3* | -1.065284976 | 1.25E-06 | 0.000953986 | 0.00782848 |
| CAD | ASPC | *DISP1* | 0.180375159 | 0.000146245 | 0.027671181 | 0.919441536 |
| CAD | ASPC | *LMOD1* | -0.176370233 | 6.50E-05 | 0.01639443 | 0.408586436 |
| CAD | ASPC | *ICA1L* | 0.417197428 | 8.62E-08 | 0.000135474 | 0.000541897 |
| CAD | ASPC | *APEH* | -0.249414271 | 1.37E-06 | 0.000953986 | 0.008585874 |
| CAD | ASPC | *RNF123* | 0.130583737 | 0.00024005 | 0.034297727 | 1 |
| CAD | ASPC | *OXGR1* | -0.702937453 | 0.000207575 | 0.033230556 | 1 |
| CAD | ASPC | *SP2* | -0.736479984 | 0.000132346 | 0.027028612 | 0.832060354 |
| CAD | ASPC | *IRS1* | -0.083623143 | 4.92E-07 | 0.000595882 | 0.003091012 |
| CAD | ASPC | *PCSK9* | 1.130722607 | 1.79E-05 | 0.006638179 | 0.112849039 |
| CAD | ASPC | *C3AR1* | -0.313060551 | 5.68E-05 | 0.015534196 | 0.357286508 |
| CAD | ASPC | *DHX36* | -0.954814542 | 0.000168117 | 0.029820393 | 1 |
| CAD | ASPC | *LPL* | -0.114242134 | 6.03E-07 | 0.000595882 | 0.003793195 |
| CAD | ASPC | *CRLF3* | 0.480703962 | 6.77E-05 | 0.01639443 | 0.425724706 |
| CAD | ASPC | *TMEM136* | 0.20562618 | 0.000229205 | 0.033511954 | 1 |
| CAD | ASPC | *FES* | -2.641727231 | 1.69E-10 | 1.06E-06 | 1.06E-06 |
| CAD | ASPC | *MSL1* | -0.146200964 | 0.000221995 | 0.033230556 | 1 |
| CAD | ASPC | *HLA-DRB1* | -0.144947497 | 3.89E-05 | 0.013589761 | 0.244615706 |
| CAD | ASPC | *MYO6* | -0.350440827 | 0.000286279 | 0.038294408 | 1 |
| CAD | ASPC | *C6orf106* | 0.182447629 | 1.99E-06 | 0.001248183 | 0.012481826 |
| CAD | ASPC | *TMEM116* | -0.105032181 | 0.000218257 | 0.033230556 | 1 |
| CAD | ASPC | *CENPW* | 0.407364533 | 1.61E-05 | 0.006307341 | 0.100917452 |
| CAD | ASPC | *SLC2A1-AS1* | -0.403038027 | 0.000133273 | 0.027028612 | 0.837886985 |
| CAD | ASPC | *LY6G5B* | -0.054745207 | 0.00024549 | 0.034297727 | 1 |
| CAD | ASPC | *BCKDHA* | -0.497624099 | 0.000102667 | 0.022257458 | 0.645466277 |
| CAD | ASPC | *HMBS* | 0.136985474 | 4.92E-05 | 0.015457113 | 0.309142265 |
| CAD | dendritic_cell | *SEMA3F* | -1.391341494 | 2.79E-05 | 0.023097142 | 0.175119318 |

| | | | | | | |
|---|---|---|---|---|---|---|
| CAD | dendritic_cell | *SNRPC* | -3.689527095 | 7.72E-06 | 0.008089947 | 0.048539681 |
| CAD | dendritic_cell | *CARF* | -3.367792099 | 1.64E-06 | 0.003646194 | 0.010284978 |
| CAD | dendritic_cell | *NBEAL1* | -1.862827742 | 5.96E-06 | 0.007500221 | 0.037501107 |
| CAD | dendritic_cell | *AMT* | -1.25360808 | 2.94E-05 | 0.023097142 | 0.18477714 |
| CAD | dendritic_cell | *APEH* | -3.590223364 | 1.74E-06 | 0.003646194 | 0.010938581 |
| CAD | dendritic_cell | *IRS1* | -1.257473217 | 5.34E-07 | 0.003355722 | 0.003355722 |
| CAD | dendritic_cell | *LPL* | -1.282476941 | 5.05E-06 | 0.007500221 | 0.031772337 |
| CAD | dendritic_cell | *TMEM136* | 3.648367324 | 5.58E-05 | 0.035109913 | 0.351099134 |
| CAD | dendritic_cell | *TMEM116* | -1.926988759 | 3.72E-05 | 0.02599999 | 0.233999913 |
| CAD | endothelial | *IRS1* | -0.274259811 | 5.15E-06 | 0.032401592 | 0.032401592 |
| CAD | macrophage | *SEMA3F* | -0.263144251 | 0.000144074 | 0.032349767 | 0.905793488 |
| CAD | macrophage | *MDH1* | 0.78203347 | 0.000121008 | 0.029252727 | 0.760779337 |
| CAD | macrophage | *UHRF1BP1* | 0.354523512 | 2.64E-06 | 0.003080364 | 0.016591875 |
| CAD | macrophage | *ATP2A3* | 0.749544586 | 0.000220828 | 0.041674027 | 1 |
| CAD | macrophage | *TIPIN* | 0.223714337 | 7.67E-05 | 0.026786429 | 0.482155727 |
| CAD | macrophage | *MAP1LC3A* | 0.599096317 | 2.80E-06 | 0.003080364 | 0.017595837 |
| CAD | macrophage | *PIGU* | 0.979480105 | 0.000179371 | 0.036377522 | 1 |
| CAD | macrophage | *ICAM2* | 0.786207857 | 0.000119309 | 0.029252727 | 0.750093104 |
| CAD | macrophage | *UMPS* | 1.113638595 | 2.85E-05 | 0.012820952 | 0.17949333 |
| CAD | macrophage | *PLXNA1* | 0.540850653 | 0.000106247 | 0.029252727 | 0.667972403 |
| CAD | macrophage | *GGCX* | 0.59278735 | 2.94E-06 | 0.003080364 | 0.018482183 |
| CAD | macrophage | *EIF2B2* | 0.490354702 | 0.000125628 | 0.029252727 | 0.789823635 |
| CAD | macrophage | *SNRPC* | -0.513523004 | 4.17E-05 | 0.015436555 | 0.262421439 |
| CAD | macrophage | *GRK4* | 0.17360833 | 0.000121463 | 0.029252727 | 0.763640727 |
| CAD | macrophage | *POLR1E* | 0.396736085 | 0.000279306 | 0.04856531 | 1 |
| CAD | macrophage | *CARF* | -0.635769084 | 4.94E-06 | 0.004435187 | 0.031046311 |
| CAD | macrophage | *TMEM91* | 0.665303042 | 0.000123444 | 0.029252727 | 0.776090143 |
| CAD | macrophage | *NBEAL1* | -0.437086757 | 5.82E-06 | 0.004570773 | 0.036566187 |
| CAD | macrophage | *AMT* | -0.229171098 | 0.000190142 | 0.037356887 | 1 |
| CAD | macrophage | *PSD2* | 2.40181989 | 3.36E-05 | 0.014097326 | 0.211459892 |
| CAD | macrophage | *DISP1* | 0.436238582 | 0.000273893 | 0.04856531 | 1 |
| CAD | macrophage | *ICA1L* | 0.99225737 | 5.63E-08 | 0.000354242 | 0.000354242 |
| CAD | macrophage | *APEH* | -0.621867313 | 8.56E-06 | 0.005380989 | 0.053809885 |
| CAD | macrophage | *RNF123* | 0.37806941 | 8.69E-05 | 0.028754713 | 0.546339543 |
| CAD | macrophage | *OXGR1* | -1.882530782 | 0.000285815 | 0.04856531 | 1 |
| CAD | macrophage | *IRS1* | -0.220309976 | 1.17E-06 | 0.002455802 | 0.007367405 |
| CAD | macrophage | *PCSK9* | 3.087734753 | 1.33E-05 | 0.007623321 | 0.083856532 |
| CAD | macrophage | *LPL* | -0.27562718 | 6.82E-06 | 0.004767267 | 0.042905401 |
| CAD | macrophage | *CRLF3* | 0.998024257 | 3.97E-05 | 0.015436555 | 0.249595005 |
| CAD | macrophage | *TMEM136* | 0.560939249 | 0.000108357 | 0.029252727 | 0.681241564 |
| CAD | macrophage | *C6orf106* | 0.508731156 | 4.56E-07 | 0.001434038 | 0.002868076 |
| CAD | macrophage | *WDR5B* | 0.78221233 | 0.000151789 | 0.032906705 | 0.954294459 |
| CAD | macrophage | *CENPW* | 1.017399875 | 1.76E-05 | 0.009236475 | 0.110837703 |
| CAD | macrophage | *FAM216A* | 1.298266167 | 0.000172868 | 0.036227388 | 1 |
| CAD | macrophage | *HLA-B* | 0.404780832 | 0.000101934 | 0.029252727 | 0.640856696 |
| CAD | macrophage | *LY6G5B* | -0.152006742 | 0.000225373 | 0.041674027 | 1 |
| CAD | macrophage | *HMBS* | 0.388676293 | 2.38E-05 | 0.011497335 | 0.14946536 |
| CAD | mast_cell | *UHRF1BP1* | 2.04877728 | 8.46E-06 | 0.013302876 | 0.053211506 |
| CAD | mast_cell | *NBEAL1* | -2.379430572 | 2.61E-05 | 0.027390538 | 0.164343229 |
| CAD | mast_cell | *ICA1L* | 4.616567135 | 4.31E-06 | 0.009781714 | 0.027106953 |

| | | | | | | |
|---|---|---|---|---|---|---|
| CAD | mast_cell | *IRS1* | -1.411102537 | 1.43E-06 | 0.009003131 | 0.009003131 |
| CAD | mast_cell | *C6orf106* | 2.758628743 | 4.67E-06 | 0.009781714 | 0.029345141 |
| CAD | mast_cell | *CENPW* | 6.793961742 | 2.51E-05 | 0.027390538 | 0.157900886 |
| CAD | monocyte | *UHRF1BP1* | 2.46799926 | 4.18E-06 | 0.024892658 | 0.026282733 |
| CAD | monocyte | *GGCX* | 3.97377307 | 7.92E-06 | 0.024892658 | 0.049785316 |
| CAD | monocyte | *PSD2* | 19.22397939 | 2.29E-05 | 0.036005012 | 0.14402005 |
| CAD | monocyte | *C6orf106* | 2.635714297 | 1.88E-05 | 0.036005012 | 0.11816803 |
| CAD | neutrophil | *APEH* | -7.136494515 | 4.90E-06 | 0.030794764 | 0.030794764 |
| CAD | nk_cell | *UHRF1BP1* | 1.180005646 | 3.12E-06 | 0.011458714 | 0.019630595 |
| CAD | nk_cell | *ICA1L* | 2.115676742 | 1.44E-05 | 0.030089982 | 0.090269947 |
| CAD | nk_cell | *C6orf106* | 1.493949951 | 3.65E-06 | 0.011458714 | 0.022917428 |
| CAD | nk_cell | *HMBS* | 1.420051413 | 2.15E-05 | 0.033769955 | 0.135079821 |
| CAD | pericyte | *SCYL3* | 2.404764574 | 0.000691333 | 0.034565119 | 1 |
| CAD | pericyte | *C1orf112* | 4.195217761 | 0.002666086 | 0.04058519 | 1 |
| CAD | pericyte | *SEMA3F* | -0.840001029 | 0.003161089 | 0.042947791 | 1 |
| CAD | pericyte | *MAD1L1* | 3.691983979 | 0.004510872 | 0.048175643 | 1 |
| CAD | pericyte | *GCFC2* | 3.796914094 | 0.001195597 | 0.034565119 | 1 |
| CAD | pericyte | *SYNRG* | 3.674134832 | 0.001671349 | 0.036359072 | 1 |
| CAD | pericyte | *FARP2* | 2.68234475 | 0.002302145 | 0.0387427 | 1 |
| CAD | pericyte | *TRAPPC6A* | 4.701123488 | 0.002812169 | 0.041045659 | 1 |
| CAD | pericyte | *SEC62* | 3.600776409 | 0.002706132 | 0.040799638 | 1 |
| CAD | pericyte | *CEP68* | 1.800167874 | 0.00240101 | 0.039224575 | 1 |
| CAD | pericyte | *ZBTB32* | 8.249767128 | 0.001967337 | 0.037807709 | 1 |
| CAD | pericyte | *MDH1* | 3.795762311 | 6.44E-06 | 0.006066092 | 0.040481254 |
| CAD | pericyte | *WWTR1* | 2.953378199 | 0.002317549 | 0.0387427 | 1 |
| CAD | pericyte | *OSBPL5* | 4.540502771 | 0.001841116 | 0.036727251 | 1 |
| CAD | pericyte | *UBR2* | 2.89255571 | 0.003654215 | 0.045313356 | 1 |
| CAD | pericyte | *TYMP* | 2.480346308 | 0.004371098 | 0.047876468 | 1 |
| CAD | pericyte | *VRK2* | 3.008917209 | 0.002408253 | 0.039224575 | 1 |
| CAD | pericyte | *ARID4A* | 3.697075733 | 0.001706429 | 0.036710139 | 1 |
| CAD | pericyte | *FUT8* | 3.077198524 | 0.003729778 | 0.045499723 | 1 |
| CAD | pericyte | *CUL3* | 3.059680566 | 0.004504653 | 0.048175643 | 1 |
| CAD | pericyte | *CNTLN* | 3.010213113 | 0.00453137 | 0.048175643 | 1 |
| CAD | pericyte | *POLR2B* | 4.037871857 | 0.001508311 | 0.035381167 | 1 |
| CAD | pericyte | *CP* | 6.049222457 | 0.001033014 | 0.034565119 | 1 |
| CAD | pericyte | *ANO2* | 1.401616476 | 0.001542189 | 0.035381167 | 1 |
| CAD | pericyte | *NFE2L3* | 5.837800876 | 0.001442871 | 0.035044707 | 1 |
| CAD | pericyte | *FOXN3* | 2.699326958 | 0.002594985 | 0.040259323 | 1 |
| CAD | pericyte | *ANAPC4* | 4.071455837 | 0.00076225 | 0.034565119 | 1 |
| CAD | pericyte | *GINM1* | 3.993290461 | 0.001225319 | 0.034565119 | 1 |
| CAD | pericyte | *DCBLD2* | 3.746807166 | 0.001326446 | 0.034565119 | 1 |
| CAD | pericyte | *GDI2* | 3.206624442 | 0.000840795 | 0.034565119 | 1 |
| CAD | pericyte | *ATP11B* | 1.543743004 | 0.0039128 | 0.046327258 | 1 |
| CAD | pericyte | *CDK14* | 2.749751761 | 0.002233093 | 0.0387427 | 1 |
| CAD | pericyte | *QSER1* | 3.65541932 | 0.00400081 | 0.046493697 | 1 |
| CAD | pericyte | *VMP1* | 3.250988474 | 0.00225513 | 0.0387427 | 1 |
| CAD | pericyte | *EPN1* | 5.133347195 | 0.000758704 | 0.034565119 | 1 |
| CAD | pericyte | *UHRF1BP1* | 1.430436014 | 5.95E-06 | 0.006066092 | 0.037409797 |
| CAD | pericyte | *IPO5* | 4.204587674 | 0.003064059 | 0.042566384 | 1 |
| CAD | pericyte | *TIE1* | -1.548642603 | 0.003619156 | 0.045269918 | 1 |

| | | | | | | |
|---|---|---|---|---|---|---|
| CAD | pericyte | *GOLGA5* | 4.308104802 | 0.002587681 | 0.040259323 | 1 |
| CAD | pericyte | *ZFAT* | 3.559244673 | 0.001860157 | 0.036727251 | 1 |
| CAD | pericyte | *SP100* | 3.484208914 | 0.001148585 | 0.034565119 | 1 |
| CAD | pericyte | *RASSF1* | 3.430331184 | 0.000554357 | 0.03183793 | 1 |
| CAD | pericyte | *IFI35* | 1.726455197 | 0.004081215 | 0.046721986 | 1 |
| CAD | pericyte | *CYFIP1* | 3.515955182 | 0.002885572 | 0.041307015 | 1 |
| CAD | pericyte | *DNAJA2* | 5.164818912 | 0.00136537 | 0.034565119 | 1 |
| CAD | pericyte | *PDE8A* | 2.566185777 | 0.000544939 | 0.031722516 | 1 |
| CAD | pericyte | *LMAN1* | 4.393925693 | 0.001068614 | 0.034565119 | 1 |
| CAD | pericyte | *TIPIN* | 1.019859543 | 5.59E-05 | 0.01463766 | 0.351303829 |
| CAD | pericyte | *SRI* | 4.934207568 | 0.000930888 | 0.034565119 | 1 |
| CAD | pericyte | *EIF4G3* | 2.832928012 | 0.002618989 | 0.04035683 | 1 |
| CAD | pericyte | *VPS9D1* | 5.036455231 | 0.003661394 | 0.045313356 | 1 |
| CAD | pericyte | *SLC25A3* | 3.894230231 | 0.000636717 | 0.033638992 | 1 |
| CAD | pericyte | *PLD1* | 3.803857057 | 0.000801745 | 0.034565119 | 1 |
| CAD | pericyte | *RBMS2* | 3.840440959 | 0.000974969 | 0.034565119 | 1 |
| CAD | pericyte | *GPATCH1* | 3.385502205 | 0.001348387 | 0.034565119 | 1 |
| CAD | pericyte | *DYNC1I2* | 3.57259653 | 0.001191409 | 0.034565119 | 1 |
| CAD | pericyte | *SIRT6* | 8.445021878 | 0.001066758 | 0.034565119 | 1 |
| CAD | pericyte | *PIK3C3* | 3.68475275 | 0.001391933 | 0.034565119 | 1 |
| CAD | pericyte | *SYNJ2* | 3.259704286 | 0.0008359 | 0.034565119 | 1 |
| CAD | pericyte | *PCM1* | 2.929488247 | 0.001881052 | 0.036727251 | 1 |
| CAD | pericyte | *OSBPL6* | 5.266993336 | 0.001217616 | 0.034565119 | 1 |
| CAD | pericyte | *XRCC5* | 3.364299597 | 0.002148529 | 0.038369728 | 1 |
| CAD | pericyte | *REXO1* | 5.702562963 | 0.001270341 | 0.034565119 | 1 |
| CAD | pericyte | *DDX1* | 4.111486557 | 0.002040657 | 0.037841263 | 1 |
| CAD | pericyte | *DNM2* | 3.278581816 | 0.003731554 | 0.045499723 | 1 |
| CAD | pericyte | *PSEN1* | 2.850044375 | 0.001751286 | 0.036710139 | 1 |
| CAD | pericyte | *ZNF506* | 3.853885075 | 0.001800908 | 0.036710139 | 1 |
| CAD | pericyte | *ERC1* | 2.781947333 | 0.002989156 | 0.042197406 | 1 |
| CAD | pericyte | *PDS5B* | 3.626441193 | 0.000874403 | 0.034565119 | 1 |
| CAD | pericyte | *GSTP1* | 4.689993827 | 0.001824293 | 0.036727251 | 1 |
| CAD | pericyte | *WBP11* | 4.976813369 | 0.002263436 | 0.0387427 | 1 |
| CAD | pericyte | *NCOA1* | 2.828170316 | 0.002720757 | 0.040835321 | 1 |
| CAD | pericyte | *TAF9* | 7.345843502 | 0.001150591 | 0.034565119 | 1 |
| CAD | pericyte | *USP40* | 2.933215626 | 0.001092664 | 0.034565119 | 1 |
| CAD | pericyte | *GMCL1* | 4.552259288 | 0.000242308 | 0.024799812 | 1 |
| CAD | pericyte | *ERGIC2* | 3.863485844 | 0.0028637 | 0.041199267 | 1 |
| CAD | pericyte | *AAMDC* | 1.020439608 | 0.003647829 | 0.045313356 | 1 |
| CAD | pericyte | *ANAPC5* | 3.497721369 | 0.001871093 | 0.036727251 | 1 |
| CAD | pericyte | *SLC23A2* | 4.373012875 | 0.000611756 | 0.033530993 | 1 |
| CAD | pericyte | *OAS1* | 5.190702367 | 0.004615517 | 0.048380004 | 1 |
| CAD | pericyte | *FUS* | 3.548043758 | 0.001195498 | 0.034565119 | 1 |
| CAD | pericyte | *ZNF302* | 1.417183543 | 0.001610433 | 0.035525597 | 1 |
| CAD | pericyte | *KCNH4* | 2.033516584 | 0.000276123 | 0.024799812 | 1 |
| CAD | pericyte | *PAPOLA* | 3.492393349 | 0.001802942 | 0.036710139 | 1 |
| CAD | pericyte | *USP48* | 2.310012955 | 0.002015796 | 0.03783077 | 1 |
| CAD | pericyte | *TXNL1* | 4.217870654 | 0.000937342 | 0.034565119 | 1 |
| CAD | pericyte | *PPP2R3C* | 3.344782097 | 0.003306185 | 0.043629875 | 1 |
| CAD | pericyte | *WAC* | 2.558871747 | 0.004711952 | 0.048688525 | 1 |

| | | | | | | |
|---|---|---|---|---|---|---|
| CAD | pericyte | *PCSK5* | 3.175746228 | 0.001122483 | 0.034565119 | 1 |
| CAD | pericyte | *NRP1* | 3.040647372 | 0.000943238 | 0.034565119 | 1 |
| CAD | pericyte | *CDC34* | 4.670760258 | 0.002817569 | 0.041045659 | 1 |
| CAD | pericyte | *MTAP* | 5.930281529 | 0.000728695 | 0.034565119 | 1 |
| CAD | pericyte | *TIMP3* | 2.739839051 | 0.003097889 | 0.042805339 | 1 |
| CAD | pericyte | *TTLL1* | 5.901324077 | 0.001796617 | 0.036710139 | 1 |
| CAD | pericyte | *AP1B1* | 3.565266524 | 0.00132675 | 0.034565119 | 1 |
| CAD | pericyte | *RBFOX2* | 3.360857466 | 0.001383213 | 0.034565119 | 1 |
| CAD | pericyte | *RBM23* | 4.355239287 | 0.000312898 | 0.026229157 | 1 |
| CAD | pericyte | *SOS2* | 3.075931611 | 0.003315514 | 0.043629875 | 1 |
| CAD | pericyte | *CNIH1* | 4.864110871 | 0.000102384 | 0.019164322 | 0.643690792 |
| CAD | pericyte | *ABHD12* | 2.54308014 | 0.000168857 | 0.023591244 | 1 |
| CAD | pericyte | *NDRG3* | 5.043301573 | 0.000275639 | 0.024799812 | 1 |
| CAD | pericyte | *SLC17A9* | 4.532982307 | 0.002701184 | 0.040799638 | 1 |
| CAD | pericyte | *RNF24* | 2.984916104 | 0.001415442 | 0.034895087 | 1 |
| CAD | pericyte | *MYL9* | 3.133467718 | 0.000220697 | 0.024799812 | 1 |
| CAD | pericyte | *MAP1LC3A* | 2.357713297 | 7.72E-06 | 0.006066092 | 0.04852874 |
| CAD | pericyte | *SMCHD1* | 2.713185508 | 0.004467859 | 0.048175643 | 1 |
| CAD | pericyte | *STK24* | 3.428107551 | 0.00295084 | 0.041877951 | 1 |
| CAD | pericyte | *ARHGEF7* | 3.11697757 | 0.001810621 | 0.036720571 | 1 |
| CAD | pericyte | *VWA8* | 2.540636448 | 0.002031749 | 0.037841263 | 1 |
| CAD | pericyte | *TSC22D1* | 3.963397298 | 0.000180824 | 0.024188147 | 1 |
| CAD | pericyte | *MGRN1* | 2.144440914 | 0.001002788 | 0.034565119 | 1 |
| CAD | pericyte | *NFAT5* | 2.714121643 | 0.002206691 | 0.0387427 | 1 |
| CAD | pericyte | *SLC7A5* | 16.93761836 | 0.001301763 | 0.034565119 | 1 |
| CAD | pericyte | *KNOP1* | 3.233316422 | 0.000347469 | 0.02668961 | 1 |
| CAD | pericyte | *CSK* | 5.686466947 | 0.002830764 | 0.041045659 | 1 |
| CAD | pericyte | *HERC1* | 3.316922177 | 0.000820416 | 0.034565119 | 1 |
| CAD | pericyte | *DNAJC17* | 4.427413679 | 0.001468671 | 0.035377516 | 1 |
| CAD | pericyte | *RIPK2* | 3.21990022 | 0.004666904 | 0.048577522 | 1 |
| CAD | pericyte | *IKBKB* | 3.705364279 | 0.002458476 | 0.039429683 | 1 |
| CAD | pericyte | *GDAP1* | 3.71487096 | 0.004138385 | 0.046816261 | 1 |
| CAD | pericyte | *ERICH1* | 3.573615736 | 0.001579865 | 0.035381167 | 1 |
| CAD | pericyte | *FCGRT* | 1.965607948 | 0.003616842 | 0.045269918 | 1 |
| CAD | pericyte | *AES* | 4.351227853 | 0.002721489 | 0.040835321 | 1 |
| CAD | pericyte | *RPS16* | 3.609493574 | 0.000469948 | 0.029253131 | 1 |
| CAD | pericyte | *PRKD2* | 2.482379388 | 0.00137544 | 0.034565119 | 1 |
| CAD | pericyte | *FZR1* | 5.541650955 | 0.00496253 | 0.049919086 | 1 |
| CAD | pericyte | *CYTH2* | 5.40018269 | 0.000119173 | 0.020812171 | 0.749238147 |
| CAD | pericyte | *TFPT* | 3.767571397 | 0.003030023 | 0.04236944 | 1 |
| CAD | pericyte | *ZNF14* | 4.95682183 | 0.002254938 | 0.0387427 | 1 |
| CAD | pericyte | *EPHB6* | 2.54849866 | 0.004243112 | 0.047403738 | 1 |
| CAD | pericyte | *USP42* | 3.963756349 | 0.003331675 | 0.043637999 | 1 |
| CAD | pericyte | *AP1S1* | 10.0467973 | 0.001280214 | 0.034565119 | 1 |
| CAD | pericyte | *PHF14* | 2.755171989 | 0.004930161 | 0.04975268 | 1 |
| CAD | pericyte | *LIMK1* | 4.789382296 | 0.003310079 | 0.043629875 | 1 |
| CAD | pericyte | *PDLIM1* | 1.775273258 | 0.003504641 | 0.04451248 | 1 |
| CAD | pericyte | *CCNJ* | 3.548703756 | 0.003446096 | 0.044125465 | 1 |
| CAD | pericyte | *DNMBP* | 3.525342879 | 0.004307933 | 0.04750524 | 1 |
| CAD | pericyte | *DDX50* | 4.39362338 | 0.000929892 | 0.034565119 | 1 |

| | | | | | | |
|---|---|---|---|---|---|---|
| CAD | pericyte | *MAPK8* | 2.934278606 | 0.003155595 | 0.042947791 | 1 |
| CAD | pericyte | *CCSER2* | 3.217179942 | 0.002915369 | 0.041562188 | 1 |
| CAD | pericyte | *LIPA* | 2.183278497 | 0.001852848 | 0.036727251 | 1 |
| CAD | pericyte | *MLLT6* | 4.648598126 | 0.000725689 | 0.034565119 | 1 |
| CAD | pericyte | *RPS6KB1* | 4.425609713 | 0.000938312 | 0.034565119 | 1 |
| CAD | pericyte | *NUP88* | 3.691075763 | 0.000618155 | 0.033530993 | 1 |
| CAD | pericyte | *ICAM2* | 3.773813364 | 1.91E-05 | 0.008013521 | 0.120202812 |
| CAD | pericyte | *UTP6* | 4.061802187 | 0.002237323 | 0.0387427 | 1 |
| CAD | pericyte | *CYTH1* | 2.153337772 | 0.003629082 | 0.045269918 | 1 |
| CAD | pericyte | *RAB5C* | 2.531031766 | 0.004476245 | 0.048175643 | 1 |
| CAD | pericyte | *WFS1* | 5.134169596 | 0.003931524 | 0.046378327 | 1 |
| CAD | pericyte | *SH3D19* | 1.652067099 | 0.004956657 | 0.049919086 | 1 |
| CAD | pericyte | *RAPGEF2* | 3.129770025 | 0.0003532 | 0.02668961 | 1 |
| CAD | pericyte | *UFSP2* | 3.946964647 | 0.004087364 | 0.046721986 | 1 |
| CAD | pericyte | *KLF3* | 3.396408771 | 0.004398726 | 0.047928574 | 1 |
| CAD | pericyte | *KLHL5* | 2.434609927 | 0.000258268 | 0.024799812 | 1 |
| CAD | pericyte | *NDUFS8* | 5.679724576 | 0.001977392 | 0.037807709 | 1 |
| CAD | pericyte | *CAPRIN2* | 4.171559637 | 0.000369817 | 0.02668961 | 1 |
| CAD | pericyte | *LTA4H* | 4.59916246 | 0.001450604 | 0.035076726 | 1 |
| CAD | pericyte | *VPS29* | 3.856802249 | 0.002439553 | 0.039413454 | 1 |
| CAD | pericyte | *CNOT2* | 2.997514074 | 0.004252426 | 0.047403738 | 1 |
| CAD | pericyte | *GNPTAB* | 3.322095745 | 0.003401378 | 0.04373101 | 1 |
| CAD | pericyte | *PHC1* | 6.972203899 | 0.001741883 | 0.036710139 | 1 |
| CAD | pericyte | *COL12A1* | 3.147418103 | 0.001804252 | 0.036710139 | 1 |
| CAD | pericyte | *SMIM8* | 3.414275129 | 0.000251703 | 0.024799812 | 1 |
| CAD | pericyte | *MDN1* | 3.713876383 | 2.82E-05 | 0.009733533 | 0.177208387 |
| CAD | pericyte | *ASCC3* | 2.930084401 | 0.004140264 | 0.046816261 | 1 |
| CAD | pericyte | *PACRG* | 5.459163417 | 0.002089184 | 0.037961558 | 1 |
| CAD | pericyte | *TFEB* | 4.383481269 | 0.003836891 | 0.045686612 | 1 |
| CAD | pericyte | *SENP6* | 3.370569789 | 0.00097229 | 0.034565119 | 1 |
| CAD | pericyte | *DAP* | 3.865739654 | 0.003032646 | 0.04236944 | 1 |
| CAD | pericyte | *SMAD5* | 3.236212799 | 0.004697781 | 0.048657243 | 1 |
| CAD | pericyte | *BNIP1* | 1.159325533 | 0.000539065 | 0.031722516 | 1 |
| CAD | pericyte | *BCHE* | -1.755679842 | 0.000952712 | 0.034565119 | 1 |
| CAD | pericyte | *PRKAR2A* | 1.96163462 | 0.000696822 | 0.034565119 | 1 |
| CAD | pericyte | *UMPS* | 2.428337985 | 0.00204645 | 0.037841263 | 1 |
| CAD | pericyte | *PLXNA1* | 2.733729442 | 1.32E-05 | 0.007247662 | 0.082706997 |
| CAD | pericyte | *ABTB1* | 4.039819271 | 0.003768322 | 0.045499723 | 1 |
| CAD | pericyte | *COMMD2* | 3.072830064 | 0.003392661 | 0.04373101 | 1 |
| CAD | pericyte | *ABCC5* | 2.962910953 | 0.004906039 | 0.049710001 | 1 |
| CAD | pericyte | *MOB1A* | 4.824648085 | 0.001506997 | 0.035381167 | 1 |
| CAD | pericyte | *STAM2* | 4.037041714 | 0.001345612 | 0.034565119 | 1 |
| CAD | pericyte | *PPM1G* | 3.57118261 | 0.00282777 | 0.041045659 | 1 |
| CAD | pericyte | *GGCX* | 2.886440983 | 3.52E-07 | 0.000630892 | 0.002216029 |
| CAD | pericyte | *SF3B1* | 2.99683344 | 0.002817236 | 0.041045659 | 1 |
| CAD | pericyte | *STRN* | 3.69161126 | 0.002301201 | 0.0387427 | 1 |
| CAD | pericyte | *CEBPZ* | 2.832741396 | 0.00084954 | 0.034565119 | 1 |
| CAD | pericyte | *PRKD3* | 3.953454577 | 0.000223257 | 0.024799812 | 1 |
| CAD | pericyte | *RMDN2* | 4.281700567 | 0.003348499 | 0.043652553 | 1 |
| CAD | pericyte | *BCL9* | 3.620276981 | 0.001540266 | 0.035381167 | 1 |

| | | | | | | |
|---|---|---|---|---|---|---|
| CAD | pericyte | *TMEM59* | 3.195770435 | 0.002451195 | 0.039413454 | 1 |
| CAD | pericyte | *DOCK7* | 3.611866859 | 0.001355326 | 0.034565119 | 1 |
| CAD | pericyte | *TMED5* | 3.711086359 | 0.003129063 | 0.042947791 | 1 |
| CAD | pericyte | *PRRC2C* | 3.768798157 | 0.00010364 | 0.019164322 | 0.651586934 |
| CAD | pericyte | *RCOR3* | 3.728377314 | 0.001310196 | 0.034565119 | 1 |
| CAD | pericyte | *MTFR1L* | 4.16001967 | 0.003165282 | 0.042947791 | 1 |
| CAD | pericyte | *KMT2A* | 3.256681657 | 0.003159688 | 0.042947791 | 1 |
| CAD | pericyte | *SENP5* | 3.460121535 | 0.002144069 | 0.038369728 | 1 |
| CAD | pericyte | *PTBP3* | 3.438015323 | 0.003468474 | 0.044321742 | 1 |
| CAD | pericyte | *CNTRL* | 3.585048376 | 0.002647132 | 0.040572748 | 1 |
| CAD | pericyte | *EIF2B2* | 2.448029269 | 1.23E-05 | 0.007247662 | 0.077214998 |
| CAD | pericyte | *CNRIP1* | 3.231902415 | 0.001595456 | 0.035443925 | 1 |
| CAD | pericyte | *CAAP1* | 3.280540886 | 0.002504235 | 0.039558105 | 1 |
| CAD | pericyte | *DNAJC15* | 4.483060294 | 0.001067563 | 0.034565119 | 1 |
| CAD | pericyte | *SMAD9* | 5.137230669 | 0.001581376 | 0.035381167 | 1 |
| CAD | pericyte | *SIL1* | 2.667869854 | 0.004042147 | 0.046642937 | 1 |
| CAD | pericyte | *LIAS* | 3.907281532 | 0.001880837 | 0.036727251 | 1 |
| CAD | pericyte | *ACVR2A* | 3.224030048 | 0.00353772 | 0.044751802 | 1 |
| CAD | pericyte | *POLK* | 2.96620409 | 0.004483075 | 0.048175643 | 1 |
| CAD | pericyte | *SPATA1* | 3.81622916 | 0.002365266 | 0.039029998 | 1 |
| CAD | pericyte | *PMS2* | 4.450256 | 0.001822924 | 0.036727251 | 1 |
| CAD | pericyte | *KLHL7* | 4.273744335 | 0.001248573 | 0.034565119 | 1 |
| CAD | pericyte | *FAM126A* | 3.045214971 | 0.001311659 | 0.034565119 | 1 |
| CAD | pericyte | *FKBP9* | 2.395829026 | 0.004310887 | 0.04750524 | 1 |
| CAD | pericyte | *PKN1* | 3.713590471 | 0.00483306 | 0.049407237 | 1 |
| CAD | pericyte | *SPATS2* | 4.356807576 | 0.000186222 | 0.02439126 | 1 |
| CAD | pericyte | *TTC21B* | 3.345600731 | 0.003260157 | 0.043609808 | 1 |
| CAD | pericyte | *PLA2G12A* | 5.035488941 | 0.000385709 | 0.027075465 | 1 |
| CAD | pericyte | *AGO2* | 4.003009683 | 0.002087928 | 0.037961558 | 1 |
| CAD | pericyte | *RNF114* | 4.216647936 | 9.57E-05 | 0.019164322 | 0.601424773 |
| CAD | pericyte | *HIF3A* | 2.67661951 | 1.48E-05 | 0.007247662 | 0.092772625 |
| CAD | pericyte | *SNRPC* | -1.95404542 | 0.000390443 | 0.027075465 | 1 |
| CAD | pericyte | *NUP153* | 3.719042932 | 0.001272805 | 0.034565119 | 1 |
| CAD | pericyte | *CNOT1* | 3.000800584 | 0.003346505 | 0.043652553 | 1 |
| CAD | pericyte | *GRK4* | 0.718504444 | 0.000264292 | 0.024799812 | 1 |
| CAD | pericyte | *MIF4GD* | 7.111473896 | 0.002007319 | 0.03783077 | 1 |
| CAD | pericyte | *SYMPK* | 4.781206362 | 0.000100082 | 0.019164322 | 0.629212563 |
| CAD | pericyte | *PANK2* | 4.542451716 | 0.00081655 | 0.034565119 | 1 |
| CAD | pericyte | *PSMF1* | 2.753160324 | 0.004385239 | 0.047915501 | 1 |
| CAD | pericyte | *DSTN* | 3.376728596 | 0.003710298 | 0.045499723 | 1 |
| CAD | pericyte | *MMP24* | 3.139574244 | 0.000511458 | 0.030624163 | 1 |
| CAD | pericyte | *KLC1* | 3.658202346 | 0.00122408 | 0.034565119 | 1 |
| CAD | pericyte | *SBDS* | 4.501009353 | 0.000801157 | 0.034565119 | 1 |
| CAD | pericyte | *BECN1* | 4.321553903 | 0.000704235 | 0.034565119 | 1 |
| CAD | pericyte | *RAP1B* | 2.905937027 | 0.003277247 | 0.043629875 | 1 |
| CAD | pericyte | *HIP1* | 3.143105329 | 0.001940643 | 0.037540992 | 1 |
| CAD | pericyte | *PTPN12* | 3.048039231 | 0.00316968 | 0.042947791 | 1 |
| CAD | pericyte | *SUMF2* | 3.610861493 | 0.002705766 | 0.040799638 | 1 |
| CAD | pericyte | *SPCS3* | 4.53156904 | 0.001199236 | 0.034565119 | 1 |
| CAD | pericyte | *SERGEF* | 4.55584959 | 0.00036759 | 0.02668961 | 1 |

| | | | | | | |
|---|---|---|---|---|---|---|
| CAD | pericyte | *SLC44A2* | 4.145688195 | 0.002993485 | 0.042197406 | 1 |
| CAD | pericyte | *RPL36* | 4.872481976 | 7.59E-05 | 0.017044725 | 0.4771149 |
| CAD | pericyte | *BST2* | 5.26484452 | 0.001848781 | 0.036727251 | 1 |
| CAD | pericyte | *TUBGCP2* | 3.380731399 | 0.001272522 | 0.034565119 | 1 |
| CAD | pericyte | *PAK4* | 5.379457115 | 0.002765539 | 0.040951088 | 1 |
| CAD | pericyte | *POMT1* | 3.197509904 | 0.004430709 | 0.048110312 | 1 |
| CAD | pericyte | *TBC1D5* | 3.004262577 | 0.000968262 | 0.034565119 | 1 |
| CAD | pericyte | *RFTN1* | 2.683854827 | 0.002658815 | 0.040572748 | 1 |
| CAD | pericyte | *EXOC4* | 2.240351284 | 0.003080816 | 0.042663195 | 1 |
| CAD | pericyte | *ACAP3* | 3.162912595 | 0.001105115 | 0.034565119 | 1 |
| CAD | pericyte | *NINJ1* | 6.054795201 | 0.000348836 | 0.02668961 | 1 |
| CAD | pericyte | *NPHP4* | 5.846101908 | 0.001026575 | 0.034565119 | 1 |
| CAD | pericyte | *MAP1B* | 0.829582041 | 0.004880433 | 0.049649327 | 1 |
| CAD | pericyte | *LGALS3* | 3.165126372 | 0.003867295 | 0.045886261 | 1 |
| CAD | pericyte | *TERF2* | 3.70798481 | 0.003931867 | 0.046378327 | 1 |
| CAD | pericyte | *PTPRA* | 3.424787121 | 0.001192258 | 0.034565119 | 1 |
| CAD | pericyte | *MUTYH* | 5.031306702 | 0.000808747 | 0.034565119 | 1 |
| CAD | pericyte | *POMP* | 4.142657365 | 0.003363162 | 0.043686357 | 1 |
| CAD | pericyte | *FAM83F* | 14.60253017 | 0.002133601 | 0.038369728 | 1 |
| CAD | pericyte | *SWAP70* | 4.006896955 | 0.001167534 | 0.034565119 | 1 |
| CAD | pericyte | *SBF2* | 2.607812576 | 0.003106103 | 0.042824708 | 1 |
| CAD | pericyte | *AP4B1* | 4.295968362 | 0.000734679 | 0.034565119 | 1 |
| CAD | pericyte | *NARS* | 5.11439336 | 0.000623246 | 0.033530993 | 1 |
| CAD | pericyte | *FBXO18* | 4.556672659 | 0.000624006 | 0.033530993 | 1 |
| CAD | pericyte | *ZCCHC11* | 2.856926287 | 0.002760293 | 0.040951088 | 1 |
| CAD | pericyte | *B4GALNT1* | 5.524660666 | 0.000391899 | 0.027075465 | 1 |
| CAD | pericyte | *SP110* | 3.386616997 | 0.002319299 | 0.0387427 | 1 |
| CAD | pericyte | *USP37* | 3.32305998 | 0.002559324 | 0.04022617 | 1 |
| CAD | pericyte | *GCC2* | 3.12738949 | 0.000222414 | 0.024799812 | 1 |
| CAD | pericyte | *SLC41A2* | 3.696464144 | 0.002258815 | 0.0387427 | 1 |
| CAD | pericyte | *LRCH1* | 3.636385442 | 0.001260111 | 0.034565119 | 1 |
| CAD | pericyte | *NUPL2* | 3.442438126 | 0.002496347 | 0.039558105 | 1 |
| CAD | pericyte | *UGGT1* | 4.32934789 | 0.001303159 | 0.034565119 | 1 |
| CAD | pericyte | *STX17* | 4.328440272 | 0.00114673 | 0.034565119 | 1 |
| CAD | pericyte | *GARNL3* | 4.447571015 | 0.000849967 | 0.034565119 | 1 |
| CAD | pericyte | *POLR1E* | 1.520237938 | 0.0003326 | 0.02668961 | 1 |
| CAD | pericyte | *KIF13A* | 3.153543798 | 0.001552697 | 0.035381167 | 1 |
| CAD | pericyte | *CCDC90B* | 3.308421828 | 0.00317788 | 0.042966298 | 1 |
| CAD | pericyte | *NARS2* | 4.214122074 | 0.001373231 | 0.034565119 | 1 |
| CAD | pericyte | *TGS1* | 2.310075679 | 0.0014407 | 0.035044707 | 1 |
| CAD | pericyte | *DDX60* | 4.879464436 | 0.001572799 | 0.035381167 | 1 |
| CAD | pericyte | *FDX1* | 3.364202183 | 0.004260198 | 0.047403738 | 1 |
| CAD | pericyte | *RMDN3* | 4.137442004 | 0.000153828 | 0.023345686 | 0.967113914 |
| CAD | pericyte | *UACA* | 2.814770892 | 0.004486999 | 0.048175643 | 1 |
| CAD | pericyte | *RTCA* | 2.128005169 | 0.003242378 | 0.043464458 | 1 |
| CAD | pericyte | *IFT172* | 4.192255227 | 0.000287491 | 0.02510354 | 1 |
| CAD | pericyte | *DYNC2LI1* | 1.678222111 | 0.004494161 | 0.048175643 | 1 |
| CAD | pericyte | *MYOF* | 1.74698411 | 0.000921189 | 0.034565119 | 1 |
| CAD | pericyte | *ARL3* | 4.165583606 | 6.96E-05 | 0.017044725 | 0.43763544 |
| CAD | pericyte | *RBP4* | 3.530692465 | 0.002498219 | 0.039558105 | 1 |

| | | | | | | |
|---|---|---|---|---|---|---|
| CAD | pericyte | *DNAJC13* | 3.719460461 | 0.002075726 | 0.037961558 | 1 |
| CAD | pericyte | *RPS24* | 3.636815578 | 0.00394327 | 0.046425731 | 1 |
| CAD | pericyte | *CARF* | -2.236187403 | 0.000273785 | 0.024799812 | 1 |
| CAD | pericyte | *NAB1* | 4.476168076 | 0.001112811 | 0.034565119 | 1 |
| CAD | pericyte | *COX17* | 4.350826067 | 0.003769344 | 0.045499723 | 1 |
| CAD | pericyte | *USP8* | 2.672618544 | 0.004278378 | 0.04743944 | 1 |
| CAD | pericyte | *ARHGAP24* | 2.873769406 | 0.001300198 | 0.034565119 | 1 |
| CAD | pericyte | *FAM13A* | 2.189972261 | 0.002329369 | 0.0387427 | 1 |
| CAD | pericyte | *HERC3* | 3.586617781 | 0.002293256 | 0.0387427 | 1 |
| CAD | pericyte | *BBS7* | 4.89830935 | 0.001804268 | 0.036710139 | 1 |
| CAD | pericyte | *KIAA1109* | 3.079702443 | 0.002322515 | 0.0387427 | 1 |
| CAD | pericyte | *NUP54* | 3.531837738 | 4.16E-05 | 0.011764439 | 0.261701757 |
| CAD | pericyte | *SCARB2* | 3.331922826 | 0.00364002 | 0.045313356 | 1 |
| CAD | pericyte | *GAS2L3* | 5.595436093 | 0.001514734 | 0.035381167 | 1 |
| CAD | pericyte | *LPAR6* | 3.259539316 | 0.002472896 | 0.039525186 | 1 |
| CAD | pericyte | *TMX1* | 4.560906624 | 0.00229943 | 0.0387427 | 1 |
| CAD | pericyte | *C14orf79* | 6.531394012 | 0.0015749 | 0.035381167 | 1 |
| CAD | pericyte | *TCF12* | 2.832696453 | 0.002002524 | 0.03783077 | 1 |
| CAD | pericyte | *SRP14* | 3.810800023 | 0.000234057 | 0.024799812 | 1 |
| CAD | pericyte | *IGF1R* | 3.26219708 | 0.002547339 | 0.040138142 | 1 |
| CAD | pericyte | *CRTC3* | 3.712593612 | 0.001327138 | 0.034565119 | 1 |
| CAD | pericyte | *ZFHX3* | 2.749070033 | 0.004832366 | 0.049407237 | 1 |
| CAD | pericyte | *CMTM3* | 4.948967545 | 0.003484451 | 0.044401111 | 1 |
| CAD | pericyte | *MYO19* | 4.31673453 | 9.03E-05 | 0.018940359 | 0.567683036 |
| CAD | pericyte | *ASXL3* | 3.951623917 | 0.003126806 | 0.042947791 | 1 |
| CAD | pericyte | *NARF* | 2.868612566 | 0.003952732 | 0.046447144 | 1 |
| CAD | pericyte | *SIRT3* | 2.196636073 | 0.004314537 | 0.04750524 | 1 |
| CAD | pericyte | *IFITM3* | 4.765690638 | 0.001091014 | 0.034565119 | 1 |
| CAD | pericyte | *RCN3* | 4.371571947 | 0.004071619 | 0.046721986 | 1 |
| CAD | pericyte | *GPN2* | 6.090005453 | 0.000269721 | 0.024799812 | 1 |
| CAD | pericyte | *MOB3C* | 4.421824269 | 0.002862034 | 0.041199267 | 1 |
| CAD | pericyte | *GPR161* | 3.556351995 | 0.00380667 | 0.045499723 | 1 |
| CAD | pericyte | *TIPRL* | 4.690862192 | 0.001276686 | 0.034565119 | 1 |
| CAD | pericyte | *TMCO1* | 3.688361678 | 0.004532253 | 0.048175643 | 1 |
| CAD | pericyte | *MGST3* | 3.469895402 | 0.002267714 | 0.0387427 | 1 |
| CAD | pericyte | *SEMA6C* | 4.409712812 | 0.001258013 | 0.034565119 | 1 |
| CAD | pericyte | *TP53BP2* | 4.570292395 | 0.001783878 | 0.036710139 | 1 |
| CAD | pericyte | *NVL* | 3.738117475 | 0.003191505 | 0.04305792 | 1 |
| CAD | pericyte | *GUK1* | 4.578691831 | 0.001969665 | 0.037807709 | 1 |
| CAD | pericyte | *WDPCP* | 0.908228056 | 0.001519591 | 0.035381167 | 1 |
| CAD | pericyte | *METTL21A* | 4.49767137 | 0.001379445 | 0.034565119 | 1 |
| CAD | pericyte | *NBEAL1* | -1.432755448 | 0.000561128 | 0.03183793 | 1 |
| CAD | pericyte | *FANCD2* | 8.225913785 | 3.68E-05 | 0.011577693 | 0.231553869 |
| CAD | pericyte | *TAMM41* | 4.947286774 | 2.94E-05 | 0.009733533 | 0.18493712 |
| CAD | pericyte | *CTDSPL* | 2.997417223 | 0.004083241 | 0.046721986 | 1 |
| CAD | pericyte | *LIMD1* | 2.769471922 | 0.002244623 | 0.0387427 | 1 |
| CAD | pericyte | *AMT* | -1.038125222 | 0.00022673 | 0.024799812 | 1 |
| CAD | pericyte | *NDUFS6* | 4.530473502 | 0.003353616 | 0.043652553 | 1 |
| CAD | pericyte | *LHFPL2* | 3.834336123 | 0.003289019 | 0.043629875 | 1 |
| CAD | pericyte | *SSBP2* | 3.052396807 | 0.001092001 | 0.034565119 | 1 |

| | | | | | | |
|---|---|---|---|---|---|---|
| CAD | pericyte | *PPIP5K2* | 2.629181347 | 0.003770401 | 0.045499723 | 1 |
| CAD | pericyte | *PSD2* | 6.366845172 | 0.002567888 | 0.040259323 | 1 |
| CAD | pericyte | *FAM193B* | 3.796985675 | 0.002791336 | 0.041045659 | 1 |
| CAD | pericyte | *IRAK1BP1* | 4.023954539 | 0.001283455 | 0.034565119 | 1 |
| CAD | pericyte | *PHIP* | 2.629051789 | 0.003818257 | 0.045551014 | 1 |
| CAD | pericyte | *TACC1* | 2.917098954 | 0.001733612 | 0.036710139 | 1 |
| CAD | pericyte | *AK3* | 3.266846624 | 0.001895285 | 0.036871353 | 1 |
| CAD | pericyte | *ZNF462* | 3.95217111 | 0.002160471 | 0.038369728 | 1 |
| CAD | pericyte | *ASTN2* | 3.164435489 | 0.003578692 | 0.045088657 | 1 |
| CAD | pericyte | *WDR31* | 6.113602844 | 9.04E-05 | 0.018940359 | 0.568210755 |
| CAD | pericyte | *ADIRF* | 4.202985337 | 0.000306383 | 0.026030132 | 1 |
| CAD | pericyte | *PDCD11* | 3.954712176 | 0.000167542 | 0.023591244 | 1 |
| CAD | pericyte | *TTC12* | 1.05104256 | 0.001111543 | 0.034565119 | 1 |
| CAD | pericyte | *COMMD7* | 5.132706378 | 0.001866658 | 0.036727251 | 1 |
| CAD | pericyte | *ARID5B* | 2.231262545 | 0.003397302 | 0.04373101 | 1 |
| CAD | pericyte | *SAP18* | 3.66828496 | 0.002154865 | 0.038369728 | 1 |
| CAD | pericyte | *DOCK1* | 2.671084915 | 0.004341295 | 0.047633025 | 1 |
| CAD | pericyte | *PIP4K2A* | 2.62288063 | 0.004919361 | 0.049723513 | 1 |
| CAD | pericyte | *BTBD11* | 5.492264959 | 0.001594264 | 0.035443925 | 1 |
| CAD | pericyte | *UBE3B* | 1.607811848 | 0.004614695 | 0.048380004 | 1 |
| CAD | pericyte | *DLG5* | 1.722073118 | 0.003960805 | 0.046447144 | 1 |
| CAD | pericyte | *FAM177A1* | 4.570446205 | 0.000358804 | 0.02668961 | 1 |
| CAD | pericyte | *MIPOL1* | 2.514529209 | 0.000958083 | 0.034565119 | 1 |
| CAD | pericyte | *C4orf33* | 3.783398771 | 0.001347713 | 0.034565119 | 1 |
| CAD | pericyte | *VPS26B* | 4.538053034 | 0.001309506 | 0.034565119 | 1 |
| CAD | pericyte | *ASAP2* | 3.357674781 | 0.000693334 | 0.034565119 | 1 |
| CAD | pericyte | *BICD1* | 3.444159652 | 0.002830325 | 0.041045659 | 1 |
| CAD | pericyte | *RNF219* | 4.622785372 | 0.002039455 | 0.037841263 | 1 |
| CAD | pericyte | *TGOLN2* | 4.183189095 | 0.000881975 | 0.034565119 | 1 |
| CAD | pericyte | *CWF19L2* | 3.657490214 | 0.000361841 | 0.02668961 | 1 |
| CAD | pericyte | *MBNL1* | 2.361716282 | 0.002748895 | 0.040951088 | 1 |
| CAD | pericyte | *ZNF117* | 0.532316627 | 0.00362587 | 0.045269918 | 1 |
| CAD | pericyte | *CAST* | 1.682488583 | 0.004910118 | 0.049710001 | 1 |
| CAD | pericyte | *TUBGCP5* | 3.427655571 | 0.003317175 | 0.043629875 | 1 |
| CAD | pericyte | *CFDP1* | 3.121525519 | 0.004389904 | 0.047915501 | 1 |
| CAD | pericyte | *ROBO4* | 2.574286029 | 0.004508324 | 0.048175643 | 1 |
| CAD | pericyte | *CEP112* | 1.909034034 | 0.004627327 | 0.048406 | 1 |
| CAD | pericyte | *DISP1* | 2.516570645 | 7.01E-06 | 0.006066092 | 0.044063478 |
| CAD | pericyte | *TSEN2* | 2.183563441 | 0.001619291 | 0.035596085 | 1 |
| CAD | pericyte | *TTC39B* | 3.044763772 | 0.001659459 | 0.036225767 | 1 |
| CAD | pericyte | *DEPTOR* | 4.390581024 | 0.001396459 | 0.034565119 | 1 |
| CAD | pericyte | *DCK* | 3.456647247 | 0.00234551 | 0.038878835 | 1 |
| CAD | pericyte | *DRAM2* | 2.524377985 | 0.004648746 | 0.048468767 | 1 |
| CAD | pericyte | *MED19* | 1.468612102 | 0.001283055 | 0.034565119 | 1 |
| CAD | pericyte | *VPS8* | 2.371974915 | 0.002115206 | 0.03832363 | 1 |
| CAD | pericyte | *RPUSD3* | 6.45472002 | 0.001012058 | 0.034565119 | 1 |
| CAD | pericyte | *FCHO2* | 3.191090996 | 0.002603228 | 0.040259323 | 1 |
| CAD | pericyte | *DYRK1A* | 3.22168275 | 0.001281253 | 0.034565119 | 1 |
| CAD | pericyte | *AUTS2* | 2.597851626 | 0.002398997 | 0.039224575 | 1 |
| CAD | pericyte | *MITD1* | 2.629006749 | 0.004191708 | 0.047143595 | 1 |

| | | | | | | |
|---|---|---|---|---|---|---|
| CAD | pericyte | *DYNC1I1* | 4.852560715 | 0.001248224 | 0.034565119 | 1 |
| CAD | pericyte | *ELK4* | 5.166087504 | 0.000223333 | 0.024799812 | 1 |
| CAD | pericyte | *NIT1* | 1.963739034 | 0.000689459 | 0.034565119 | 1 |
| CAD | pericyte | *RCAN1* | 3.23629042 | 0.001392516 | 0.034565119 | 1 |
| CAD | pericyte | *PSMD4* | 4.939954011 | 0.000869191 | 0.034565119 | 1 |
| CAD | pericyte | *CCDC107* | 2.865391125 | 0.000965899 | 0.034565119 | 1 |
| CAD | pericyte | *ORAI2* | 3.542122937 | 0.00348881 | 0.044401111 | 1 |
| CAD | pericyte | *DVL3* | 4.719185762 | 0.00026382 | 0.024799812 | 1 |
| CAD | pericyte | *PGAP3* | 7.307081413 | 0.001420891 | 0.034895087 | 1 |
| CAD | pericyte | *ALDH16A1* | 6.17315118 | 7.18E-05 | 0.017044725 | 0.451351658 |
| CAD | pericyte | *LARP4* | 1.909409901 | 0.002483291 | 0.039525186 | 1 |
| CAD | pericyte | *JMJD8* | 2.040324181 | 0.000876619 | 0.034565119 | 1 |
| CAD | pericyte | *RBP7* | 5.37624543 | 0.00265384 | 0.040572748 | 1 |
| CAD | pericyte | *NFIA* | 2.447366609 | 0.003233348 | 0.043436019 | 1 |
| CAD | pericyte | *ZNF496* | 5.202218944 | 0.001864239 | 0.036727251 | 1 |
| CAD | pericyte | *HAAO* | 4.331462091 | 0.000847494 | 0.034565119 | 1 |
| CAD | pericyte | *SMC6* | 3.011246322 | 0.004892074 | 0.049687353 | 1 |
| CAD | pericyte | *PDLIM5* | 3.314496056 | 0.001162704 | 0.034565119 | 1 |
| CAD | pericyte | *DCAF16* | 4.663366424 | 0.001482205 | 0.035381167 | 1 |
| CAD | pericyte | *PBXIP1* | 3.582026732 | 0.003020791 | 0.04236944 | 1 |
| CAD | pericyte | *LRRC58* | 4.260607164 | 0.004687923 | 0.048657243 | 1 |
| CAD | pericyte | *ICA1L* | 5.205607431 | 1.43E-10 | 9.02E-07 | 9.02E-07 |
| CAD | pericyte | *GMPS* | 3.921825358 | 0.004267618 | 0.047403738 | 1 |
| CAD | pericyte | *FAM208A* | 2.488056698 | 0.004414376 | 0.048015883 | 1 |
| CAD | pericyte | *ABLIM2* | 4.389299737 | 0.004105537 | 0.046759984 | 1 |
| CAD | pericyte | *RNF123* | 1.89376635 | 1.50E-05 | 0.007247662 | 0.094219612 |
| CAD | pericyte | *ANXA5* | 2.583722139 | 0.003801288 | 0.045499723 | 1 |
| CAD | pericyte | *CEP44* | 3.913936931 | 0.004644921 | 0.048468767 | 1 |
| CAD | pericyte | *PRRC1* | 4.759746456 | 0.000584995 | 0.03283807 | 1 |
| CAD | pericyte | *UBLCP1* | 2.136551619 | 0.002349922 | 0.038878835 | 1 |
| CAD | pericyte | *SAP30L* | 4.25204543 | 0.002213158 | 0.0387427 | 1 |
| CAD | pericyte | *RPS14* | 3.309296935 | 0.002860511 | 0.041199267 | 1 |
| CAD | pericyte | *BRI3* | 2.579891237 | 0.004259717 | 0.047403738 | 1 |
| CAD | pericyte | *SLC4A2* | 6.357169276 | 0.001774032 | 0.036710139 | 1 |
| CAD | pericyte | *FAM219A* | 4.462591411 | 0.001092026 | 0.034565119 | 1 |
| CAD | pericyte | *HGSNAT* | 3.205750646 | 0.000893034 | 0.034565119 | 1 |
| CAD | pericyte | *HNRNPK* | 4.260110329 | 0.000897632 | 0.034565119 | 1 |
| CAD | pericyte | *TRMT10B* | 4.098016419 | 0.002087684 | 0.037961558 | 1 |
| CAD | pericyte | *VCP* | 2.920182606 | 0.000702762 | 0.034565119 | 1 |
| CAD | pericyte | *PIGO* | 5.35602936 | 0.000886781 | 0.034565119 | 1 |
| CAD | pericyte | *CFL2* | 4.53061333 | 0.000250297 | 0.024799812 | 1 |
| CAD | pericyte | *ZCCHC24* | 4.74493911 | 0.000255589 | 0.024799812 | 1 |
| CAD | pericyte | *PGM2L1* | 3.733817168 | 0.000750694 | 0.034565119 | 1 |
| CAD | pericyte | *OXGR1* | -8.580191245 | 0.000274715 | 0.024799812 | 1 |
| CAD | pericyte | *QSOX2* | 7.880868466 | 0.001219607 | 0.034565119 | 1 |
| CAD | pericyte | *TSC1* | 3.689631196 | 0.000846561 | 0.034565119 | 1 |
| CAD | pericyte | *JAM3* | 2.543445403 | 0.004137472 | 0.046816261 | 1 |
| CAD | pericyte | *COX11* | 2.173081871 | 0.004772654 | 0.048973259 | 1 |
| CAD | pericyte | *STXBP4* | 2.512519949 | 0.004517567 | 0.048175643 | 1 |
| CAD | pericyte | *ILK* | 3.703393302 | 0.002171598 | 0.038458701 | 1 |

| | | | | | | |
|---|---|---|---|---|---|---|
| CAD | pericyte | *TMEM41B* | 4.47306786 | 0.001204535 | 0.034565119 | 1 |
| CAD | pericyte | *C16orf45* | 3.076320001 | 0.003385983 | 0.04373101 | 1 |
| CAD | pericyte | *RPL13* | 3.165097033 | 0.004520319 | 0.048175643 | 1 |
| CAD | pericyte | *ZNF641* | 1.547706478 | 0.00074729 | 0.034565119 | 1 |
| CAD | pericyte | *TMC4* | 6.400121385 | 0.000505431 | 0.030554299 | 1 |
| CAD | pericyte | *LENG8* | 3.47849893 | 0.000303537 | 0.026030132 | 1 |
| CAD | pericyte | *KIFC2* | 1.407585209 | 0.001087866 | 0.034565119 | 1 |
| CAD | pericyte | *CYB5D2* | 4.829639065 | 0.000882558 | 0.034565119 | 1 |
| CAD | pericyte | *CD320* | 6.367250175 | 0.002078383 | 0.037961558 | 1 |
| CAD | pericyte | *SPRYD3* | 6.818258775 | 0.001181923 | 0.034565119 | 1 |
| CAD | pericyte | *NDUFV1* | 4.294883634 | 0.000403435 | 0.027401174 | 1 |
| CAD | pericyte | *DDB1* | 4.029068447 | 0.004542106 | 0.048175643 | 1 |
| CAD | pericyte | *UBTD2* | 2.80747068 | 0.002444124 | 0.039413454 | 1 |
| CAD | pericyte | *MLKL* | 1.170000313 | 0.001038053 | 0.034565119 | 1 |
| CAD | pericyte | *STX18* | 3.985171954 | 0.001776575 | 0.036710139 | 1 |
| CAD | pericyte | *IRS1* | -0.769770763 | 2.54E-05 | 0.009404623 | 0.159878585 |
| CAD | pericyte | *SLC38A11* | 4.817348977 | 0.003734253 | 0.045499723 | 1 |
| CAD | pericyte | *MT1E* | 4.8579075 | 0.004567754 | 0.048296703 | 1 |
| CAD | pericyte | *LIMS1* | 1.941256035 | 0.00177014 | 0.036710139 | 1 |
| CAD | pericyte | *MAP3K2* | 3.270818469 | 0.002066709 | 0.037961558 | 1 |
| CAD | pericyte | *NFXL1* | 5.314073113 | 0.000159107 | 0.023345686 | 1 |
| CAD | pericyte | *TSNARE1* | 1.795758602 | 0.001057294 | 0.034565119 | 1 |
| CAD | pericyte | *MFN1* | 3.162051545 | 0.001020591 | 0.034565119 | 1 |
| CAD | pericyte | *KCNMB3* | 2.016339482 | 0.001527235 | 0.035381167 | 1 |
| CAD | pericyte | *ATP6V0E2* | 7.018598501 | 0.001323853 | 0.034565119 | 1 |
| CAD | pericyte | *LRRC8C* | 5.311808206 | 0.003697865 | 0.045496047 | 1 |
| CAD | pericyte | *TBCA* | 3.201602502 | 0.000481918 | 0.029498045 | 1 |
| CAD | pericyte | *BCL2L1* | 3.769659866 | 0.000418796 | 0.027715494 | 1 |
| CAD | pericyte | *GPHN* | 1.741425049 | 0.004043328 | 0.046642937 | 1 |
| CAD | pericyte | *MTBP* | 4.396134538 | 0.00367724 | 0.045338952 | 1 |
| CAD | pericyte | *SPTLC3* | 4.357486631 | 0.002327238 | 0.0387427 | 1 |
| CAD | pericyte | *ZMAT3* | 4.858261622 | 0.00380272 | 0.045499723 | 1 |
| CAD | pericyte | *ZNF738* | 1.535812418 | 0.002478189 | 0.039525186 | 1 |
| CAD | pericyte | *LCLAT1* | 3.185480555 | 0.004716289 | 0.048688525 | 1 |
| CAD | pericyte | *ABLIM3* | 3.14774859 | 0.003776256 | 0.045499723 | 1 |
| CAD | pericyte | *GLRX* | 3.670569502 | 0.001542134 | 0.035381167 | 1 |
| CAD | pericyte | *MAP3K11* | 5.19680289 | 0.002310491 | 0.0387427 | 1 |
| CAD | pericyte | *OLR1* | 11.394545 | 0.002989048 | 0.042197406 | 1 |
| CAD | pericyte | *RNF213* | 2.933704632 | 0.004221856 | 0.047313387 | 1 |
| CAD | pericyte | *GOLIM4* | 3.67694795 | 0.000107773 | 0.019359081 | 0.677567833 |
| CAD | pericyte | *USMG5* | 3.966200661 | 0.001579948 | 0.035381167 | 1 |
| CAD | pericyte | *MGA* | 2.266363506 | 0.00460391 | 0.048380004 | 1 |
| CAD | pericyte | *ANKRD36C* | 3.013946714 | 0.004000776 | 0.046493697 | 1 |
| CAD | pericyte | *LARP7* | 3.717767429 | 0.004190701 | 0.047143595 | 1 |
| CAD | pericyte | *FGFBP3* | 6.605565741 | 0.002746413 | 0.040951088 | 1 |
| CAD | pericyte | *RSRC1* | 1.912780807 | 0.003806721 | 0.045499723 | 1 |
| CAD | pericyte | *TP53I11* | 4.20375773 | 0.001226967 | 0.034565119 | 1 |
| CAD | pericyte | *LPL* | -0.970330091 | 0.000132461 | 0.021915375 | 0.832784233 |
| CAD | pericyte | *IP6K1* | 4.042201555 | 0.000197649 | 0.024799812 | 1 |
| CAD | pericyte | *TCP11L1* | 2.425898278 | 0.002593555 | 0.040259323 | 1 |

| | | | | | | |
|---|---|---|---|---|---|---|
| CAD | pericyte | *CRLF3* | 4.881265311 | 4.01E-07 | 0.000630892 | 0.002523569 |
| CAD | pericyte | *ZNF843* | 8.853645608 | 0.003971798 | 0.046447144 | 1 |
| CAD | pericyte | *ANKLE2* | 4.583680175 | 0.000838087 | 0.034565119 | 1 |
| CAD | pericyte | *DPP7* | 4.370121227 | 0.0028909 | 0.041307015 | 1 |
| CAD | pericyte | *TBL1XR1* | 2.494373098 | 0.003518516 | 0.044598608 | 1 |
| CAD | pericyte | *RPLP2* | 3.325944979 | 0.003868255 | 0.045886261 | 1 |
| CAD | pericyte | *ZNF354C* | 4.359602467 | 0.002158885 | 0.038369728 | 1 |
| CAD | pericyte | *IMP3* | 5.197800643 | 0.002201547 | 0.0387427 | 1 |
| CAD | pericyte | *MLF1* | 4.8230641 | 0.000251198 | 0.024799812 | 1 |
| CAD | pericyte | *LCORL* | 2.945021425 | 0.004875315 | 0.049649327 | 1 |
| CAD | pericyte | *GAK* | 3.012425185 | 0.004854059 | 0.049541343 | 1 |
| CAD | pericyte | *HLA-DQB1* | -0.694463762 | 0.004739529 | 0.048768277 | 1 |
| CAD | pericyte | *TIGD5* | 3.504630002 | 0.000159673 | 0.023345686 | 1 |
| CAD | pericyte | *PDXDC1* | 3.024274546 | 0.002145919 | 0.038369728 | 1 |
| CAD | pericyte | *ZNF609* | 2.909497172 | 0.000734346 | 0.034565119 | 1 |
| CAD | pericyte | *ZNF594* | 4.49837042 | 0.001550229 | 0.035381167 | 1 |
| CAD | pericyte | *D2HGDH* | 2.801083356 | 0.001649909 | 0.036142776 | 1 |
| CAD | pericyte | *MRPL14* | 4.125239637 | 4.30E-05 | 0.011764439 | 0.2705821 |
| CAD | pericyte | *ZFP82* | 4.895864489 | 0.000441179 | 0.028579527 | 1 |
| CAD | pericyte | *ZBTB20* | 2.289460932 | 0.002737571 | 0.040951088 | 1 |
| CAD | pericyte | *TNRC18* | 3.490482467 | 0.002023802 | 0.037841263 | 1 |
| CAD | pericyte | *IST1* | 2.952757778 | 0.004544004 | 0.048175643 | 1 |
| CAD | pericyte | *TRAK1* | 3.550469286 | 0.000149662 | 0.023345686 | 0.94092692 |
| CAD | pericyte | *ACBD3* | 3.644057134 | 0.002833437 | 0.041045659 | 1 |
| CAD | pericyte | *PLCXD3* | 2.169902144 | 7.59E-05 | 0.017044725 | 0.477252294 |
| CAD | pericyte | *ZNF721* | 2.08278047 | 0.00398938 | 0.046493697 | 1 |
| CAD | pericyte | *WBP2NL* | 5.448575769 | 0.001130628 | 0.034565119 | 1 |
| CAD | pericyte | *BMP8A* | 3.738499192 | 0.001281931 | 0.034565119 | 1 |
| CAD | pericyte | *CBX6* | 5.48465981 | 0.002606258 | 0.040259323 | 1 |
| CAD | pericyte | *PP2D1* | 2.271926299 | 0.0046975 | 0.048657243 | 1 |
| CAD | pericyte | *CSF1* | 2.056088561 | 0.00040533 | 0.027401174 | 1 |
| CAD | pericyte | *WDR27* | 3.228987893 | 0.004159408 | 0.046948296 | 1 |
| CAD | pericyte | *ZNF93* | 7.47474192 | 0.00099913 | 0.034565119 | 1 |
| CAD | pericyte | *ZFP90* | 4.605398575 | 0.001443706 | 0.035044707 | 1 |
| CAD | pericyte | *KIAA0825* | 3.602732079 | 0.001371893 | 0.034565119 | 1 |
| CAD | pericyte | *SP1* | 2.282234066 | 0.001784315 | 0.036710139 | 1 |
| CAD | pericyte | *C16orf52* | 4.047427873 | 0.001091844 | 0.034565119 | 1 |
| CAD | pericyte | *ZNF529* | 2.60066073 | 0.004570787 | 0.048296703 | 1 |
| CAD | pericyte | *TOR3A* | 6.962793684 | 0.000738184 | 0.034565119 | 1 |
| CAD | pericyte | *KANK3* | 4.77801562 | 0.004130867 | 0.046816261 | 1 |
| CAD | pericyte | *DNAH17* | 6.612273004 | 0.000562114 | 0.03183793 | 1 |
| CAD | pericyte | *TMEM220* | 4.050754365 | 0.001073547 | 0.034565119 | 1 |
| CAD | pericyte | *SRSF10* | 3.296896947 | 0.003742515 | 0.045499723 | 1 |
| CAD | pericyte | *DPYD* | 2.67671768 | 0.002768286 | 0.040951088 | 1 |
| CAD | pericyte | *FAM196A* | 3.551557026 | 0.002635485 | 0.040511729 | 1 |
| CAD | pericyte | *NELFB* | 5.077245121 | 0.001774939 | 0.036710139 | 1 |
| CAD | pericyte | *ADAT2* | 3.230468773 | 0.000483267 | 0.029498045 | 1 |
| CAD | pericyte | *ZNF527* | 4.335257906 | 0.001732782 | 0.036710139 | 1 |
| CAD | pericyte | *ZKSCAN3* | 3.612903674 | 0.004617147 | 0.048380004 | 1 |
| CAD | pericyte | *HLA-DRB1* | -1.289824696 | 0.002602991 | 0.040259323 | 1 |

| | | | | | | |
|---|---|---|---|---|---|---|
| CAD | pericyte | *MPHOSPH8* | 3.462505422 | 0.00093578 | 0.034565119 | 1 |
| CAD | pericyte | *SUPT5H* | 4.722758103 | 0.000427361 | 0.027987662 | 1 |
| CAD | pericyte | *SUPT3H* | 3.470581501 | 0.001900162 | 0.036871353 | 1 |
| CAD | pericyte | *ZNF565* | 2.307038224 | 0.003045297 | 0.042451851 | 1 |
| CAD | pericyte | *ASB13* | 4.908318374 | 0.002888159 | 0.041307015 | 1 |
| CAD | pericyte | *HDAC2* | 4.068177824 | 0.00320024 | 0.043083312 | 1 |
| CAD | pericyte | *HRH1* | 3.270751559 | 0.004775029 | 0.048973259 | 1 |
| CAD | pericyte | *TOMM7* | 4.581025293 | 0.001702013 | 0.036710139 | 1 |
| CAD | pericyte | *ZNF431* | 3.576047794 | 0.000737881 | 0.034565119 | 1 |
| CAD | pericyte | *STRN3* | 3.220815693 | 0.001517584 | 0.035381167 | 1 |
| CAD | pericyte | *C6orf106* | 2.152385103 | 3.59E-07 | 0.000630892 | 0.00225739 |
| CAD | pericyte | *NHLRC2* | 3.202288745 | 0.003325636 | 0.043637999 | 1 |
| CAD | pericyte | *KPNA5* | 3.457953298 | 0.002689469 | 0.040799638 | 1 |
| CAD | pericyte | *ANKRD36B* | 2.266576107 | 0.004037464 | 0.046642937 | 1 |
| CAD | pericyte | *SRGAP1* | 3.854319287 | 0.000209295 | 0.024799812 | 1 |
| CAD | pericyte | *PCDHB16* | 0.998989102 | 0.003797193 | 0.045499723 | 1 |
| CAD | pericyte | *WDR5B* | 3.825592757 | 1.72E-05 | 0.007744296 | 0.10842014 |
| CAD | pericyte | *ZMYM1* | 4.400470382 | 0.002004403 | 0.03783077 | 1 |
| CAD | pericyte | *IGF2R* | 2.864486511 | 0.003755802 | 0.045499723 | 1 |
| CAD | pericyte | *ZNF682* | 4.56621341 | 0.001978485 | 0.037807709 | 1 |
| CAD | pericyte | *KANK2* | 3.806203596 | 0.004200169 | 0.047154399 | 1 |
| CAD | pericyte | *GTF2E2* | 3.999741379 | 0.000373578 | 0.02668961 | 1 |
| CAD | pericyte | *RAMP2-AS1* | 3.22909061 | 0.000671252 | 0.034565119 | 1 |
| CAD | pericyte | *ZNF517* | 8.691643751 | 0.002407879 | 0.039224575 | 1 |
| CAD | pericyte | *ZNF665* | 1.708177087 | 0.002015005 | 0.03783077 | 1 |
| CAD | pericyte | *ZNF624* | 6.821127258 | 0.000455112 | 0.028612907 | 1 |
| CAD | pericyte | *ZNF71* | 2.437020037 | 0.0004116 | 0.027529057 | 1 |
| CAD | pericyte | *NOL8* | 4.138965355 | 0.002178041 | 0.038464447 | 1 |
| CAD | pericyte | *MRPL42* | 2.450027811 | 0.000286026 | 0.02510354 | 1 |
| CAD | pericyte | *HIBCH* | 2.701356111 | 0.000240298 | 0.024799812 | 1 |
| CAD | pericyte | *MIER1* | 3.827007101 | 0.000612679 | 0.033530993 | 1 |
| CAD | pericyte | *DDRGK1* | 5.648809134 | 0.001540181 | 0.035381167 | 1 |
| CAD | pericyte | *QRICH1* | 1.801601747 | 0.000445615 | 0.028579527 | 1 |
| CAD | pericyte | *TMEM116* | -0.931292619 | 0.004028805 | 0.046642937 | 1 |
| CAD | pericyte | *GFPT1* | 3.268438067 | 0.000362535 | 0.02668961 | 1 |
| CAD | pericyte | *MGEA5* | 2.151236093 | 0.004733765 | 0.048768277 | 1 |
| CAD | pericyte | *ZNF808* | 2.907952974 | 0.000914681 | 0.034565119 | 1 |
| CAD | pericyte | *LRBA* | 3.20191687 | 0.00109497 | 0.034565119 | 1 |
| CAD | pericyte | *MDM4* | 3.340283357 | 0.002448805 | 0.039413454 | 1 |
| CAD | pericyte | *PAPSS2* | 2.708535894 | 0.00175745 | 0.036710139 | 1 |
| CAD | pericyte | *FAN1* | 4.938337323 | 0.001056788 | 0.034565119 | 1 |
| CAD | pericyte | *ZNF652* | 3.65770237 | 0.002407831 | 0.039224575 | 1 |
| CAD | pericyte | *SFT2D1* | 4.907739119 | 0.000543662 | 0.031722516 | 1 |
| CAD | pericyte | *APRT* | 5.346361213 | 0.002431577 | 0.039413454 | 1 |
| CAD | pericyte | *GIGYF2* | 3.134016675 | 0.003067055 | 0.042566384 | 1 |
| CAD | pericyte | *PRRT1* | 1.48079951 | 0.00397464 | 0.046447144 | 1 |
| CAD | pericyte | *GPANK1* | 1.963770721 | 0.003301698 | 0.043629875 | 1 |
| CAD | pericyte | *BAG6* | 2.722636176 | 4.21E-05 | 0.011764439 | 0.264445996 |
| CAD | pericyte | *HLA-F* | 1.674338485 | 0.002274939 | 0.0387427 | 1 |
| CAD | pericyte | *TRIM13* | 3.042914182 | 0.003586566 | 0.045097485 | 1 |

| | | | | | | |
|---|---|---|---|---|---|---|
| CAD | pericyte | *C1QTNF9B* | 103.5436118 | 0.001245677 | 0.034565119 | 1 |
| CAD | pericyte | *NYNRIN* | 7.058282963 | 0.000122576 | 0.020828029 | 0.770637071 |
| CAD | pericyte | *ZDHHC11B* | 2.766246091 | 0.001606312 | 0.035525597 | 1 |
| CAD | pericyte | *HCG27* | 0.896686186 | 0.003009462 | 0.042327712 | 1 |
| CAD | pericyte | *TAS2R14* | 1.23168468 | 0.004094769 | 0.046721986 | 1 |
| CAD | pericyte | *SIPA1* | 3.355260063 | 0.000138213 | 0.022280634 | 0.868944715 |
| CAD | pericyte | *ERV3-1* | 0.546794345 | 0.00329795 | 0.043629875 | 1 |
| CAD | pericyte | *HEXA* | 4.815038311 | 0.001342803 | 0.034565119 | 1 |
| CAD | pericyte | *ATF6B* | 1.395555111 | 0.004341198 | 0.047633025 | 1 |
| CAD | pericyte | *CLIC1* | 4.338273689 | 0.000321382 | 0.026240618 | 1 |
| CAD | pericyte | *LYRM4* | 3.919191255 | 0.001094517 | 0.034565119 | 1 |
| CAD | pericyte | *NPEPL1* | 4.718929779 | 0.001482203 | 0.035381167 | 1 |
| CAD | pericyte | *VPS52* | 4.726416383 | 0.000339524 | 0.02668961 | 1 |
| CAD | pericyte | *ERCC6* | 4.832539015 | 0.000450035 | 0.028579527 | 1 |
| CAD | pericyte | *NOL7* | 4.288877401 | 0.00315438 | 0.042947791 | 1 |
| CAD | pericyte | *TAS2R46* | -2.123746346 | 0.004052953 | 0.046668347 | 1 |
| CAD | pericyte | *RPL41* | 7.289181563 | 0.000633183 | 0.033638992 | 1 |
| CAD | pericyte | *PATL2* | 4.412844325 | 0.00357491 | 0.045088657 | 1 |
| CAD | pericyte | *RPS18* | 1.874068814 | 0.004592177 | 0.048380004 | 1 |
| CAD | pericyte | *GOLGA8N* | 1.18408029 | 0.003677885 | 0.045338952 | 1 |
| CAD | pericyte | *HLA-B* | 1.990746341 | 1.04E-05 | 0.007247662 | 0.065381515 |
| CAD | pericyte | *HLA-DQA2* | 0.790246572 | 0.002922552 | 0.041570331 | 1 |
| CAD | pericyte | *LY6G5B* | -0.487358664 | 0.00344317 | 0.044125465 | 1 |
| CAD | pericyte | *LINC00886* | 3.646827178 | 0.000770118 | 0.034565119 | 1 |
| CAD | pericyte | *ATP5J2* | 3.875994622 | 0.001226934 | 0.034565119 | 1 |
| CAD | pericyte | *ARFGAP3* | 3.727519562 | 0.002009334 | 0.03783077 | 1 |
| CAD | pericyte | *AP5Z1* | 4.243302583 | 0.001569204 | 0.035381167 | 1 |
| CAD | pericyte | *FGF14-IT1* | 20.30203132 | 0.001749884 | 0.036710139 | 1 |
| CAD | pericyte | *APOBEC3C* | 9.836927512 | 0.000851356 | 0.034565119 | 1 |
| CAD | pericyte | *UBE2V1* | 5.880509351 | 0.003384064 | 0.04373101 | 1 |
| CAD | pericyte | *SHANK3* | 1.618754259 | 0.000320212 | 0.026240618 | 1 |
| CAD | pericyte | *ZNF260* | 5.236772316 | 0.000180541 | 0.024188147 | 1 |
| CAD | pericyte | *FPGT* | 5.227068267 | 0.002803991 | 0.041045659 | 1 |
| CAD | pericyte | *HMBS* | 1.731297513 | 2.52E-05 | 0.009404623 | 0.158595615 |
| CAD | pericyte | *ZNF253* | 3.880334046 | 0.004288976 | 0.047473231 | 1 |
| CAD | pericyte | *IQCH-AS1* | 1.690887274 | 0.001874232 | 0.036727251 | 1 |
| CAD | pericyte | *HCG17* | 2.962373842 | 0.001726688 | 0.036710139 | 1 |
| CAD | SMC | *SEMA3F* | -0.874892695 | 6.25E-05 | 0.026695616 | 0.392878817 |
| CAD | SMC | *UHRF1BP1* | 0.968415398 | 3.02E-05 | 0.023734972 | 0.189879774 |
| CAD | SMC | *TIE1* | -1.355144352 | 0.000141175 | 0.049309179 | 0.887565229 |
| CAD | SMC | *MAP1LC3A* | 1.545607634 | 8.52E-05 | 0.031506883 | 0.535617018 |
| CAD | SMC | *BCHE* | -1.588307505 | 4.94E-05 | 0.026695616 | 0.310339763 |
| CAD | SMC | *GGCX* | 1.581107544 | 4.98E-05 | 0.026695616 | 0.313217273 |
| CAD | SMC | *SNRPC* | -1.66935696 | 1.20E-05 | 0.015148454 | 0.075742272 |
| CAD | SMC | *CARF* | -2.12352287 | 3.45E-07 | 0.001646819 | 0.00216688 |
| CAD | SMC | *NBEAL1* | -1.324082782 | 2.35E-06 | 0.003698719 | 0.014794877 |
| CAD | SMC | *AMT* | -0.768482982 | 5.96E-05 | 0.026695616 | 0.374903169 |
| CAD | SMC | *PSD2* | 7.329073662 | 5.39E-05 | 0.026695616 | 0.338785099 |
| CAD | SMC | *ICA1L* | 2.128041663 | 4.99E-05 | 0.026695616 | 0.31400899 |
| CAD | SMC | *APEH* | -1.624248794 | 1.60E-05 | 0.016742249 | 0.100453491 |

| | | | | | | |
|---|---|---|---|---|---|---|
| CAD | SMC | *IRS1* | -0.708415481 | 5.24E-07 | 0.001646819 | 0.003293639 |
| CAD | SMC | *LPL* | -0.873115565 | 1.67E-06 | 0.003502718 | 0.010508153 |
| CAD | SMC | *HLA-DRB1* | -1.165749013 | 6.44E-05 | 0.026695616 | 0.405120765 |
| CAD | SMC | *C6orf106* | 1.26103989 | 2.92E-05 | 0.023734972 | 0.183853564 |
| CAD | SMC | *HMBS* | 1.141207005 | 6.79E-05 | 0.026695616 | 0.427129864 |
| CAD | t_cell | *UHRF1BP1* | 1.977884802 | 4.34E-06 | 0.013386897 | 0.027297398 |
| CAD | t_cell | *TIPIN* | 1.354168686 | 5.68E-05 | 0.035679599 | 0.356795986 |
| CAD | t_cell | *MAP1LC3A* | 1.803879275 | 0.000101043 | 0.048866166 | 0.635260163 |
| CAD | t_cell | *PLXNA1* | 2.998193277 | 8.63E-05 | 0.046372005 | 0.542740491 |
| CAD | t_cell | *GGCX* | 3.229618247 | 6.84E-06 | 0.013386897 | 0.043027462 |
| CAD | t_cell | *SNRPC* | -3.036346637 | 5.52E-05 | 0.035679599 | 0.347224131 |
| CAD | t_cell | *GRK4* | 1.105180925 | 4.69E-05 | 0.035679599 | 0.294765933 |
| CAD | t_cell | *DISP1* | 2.571158793 | 5.19E-05 | 0.035679599 | 0.326350902 |
| CAD | t_cell | *ICA1L* | 3.527256797 | 1.06E-05 | 0.013386897 | 0.066934487 |
| CAD | t_cell | *APEH* | -3.662138562 | 3.23E-06 | 0.013386897 | 0.02030928 |
| CAD | t_cell | *TMEM136* | 3.18648354 | 8.85E-05 | 0.046372005 | 0.556464058 |
| CAD | t_cell | *C6orf106* | 2.582831971 | 8.94E-06 | 0.013386897 | 0.056212832 |
| CAD | t_cell | *HMBS* | 2.361056464 | 2.28E-05 | 0.023891527 | 0.143349162 |
| T2DM | adipocyte | *CFH* | 0.764379222 | 9.89E-07 | 0.000141342 | 0.006219028 |
| T2DM | adipocyte | *SEMA3F* | -0.190854127 | 3.12E-05 | 0.001056656 | 0.196089012 |
| T2DM | adipocyte | *TMEM176A* | 0.219908802 | 0.001354056 | 0.012355519 | 1 |
| T2DM | adipocyte | *M6PR* | 0.419785141 | 0.010413941 | 0.04844569 | 1 |
| T2DM | adipocyte | *RBM6* | -0.17523276 | 2.68E-07 | 6.01E-05 | 0.001683482 |
| T2DM | adipocyte | *RECQL* | 1.175220336 | 0.000281424 | 0.00440128 | 1 |
| T2DM | adipocyte | *WDR54* | 0.70627487 | 0.00270842 | 0.020040713 | 1 |
| T2DM | adipocyte | *ABCB4* | 1.001079099 | 0.001327592 | 0.012202593 | 1 |
| T2DM | adipocyte | *GGNBP2* | -0.424286223 | 0.000120741 | 0.002547302 | 0.759095875 |
| T2DM | adipocyte | *TMEM132A* | 1.60654627 | 8.38E-05 | 0.001941003 | 0.527046922 |
| T2DM | adipocyte | *RALA* | 0.820748405 | 0.005177898 | 0.030234016 | 1 |
| T2DM | adipocyte | *NOS2* | 3.880728858 | 0.005261284 | 0.030505142 | 1 |
| T2DM | adipocyte | *TSR3* | 1.541420111 | 0.00019984 | 0.003539133 | 1 |
| T2DM | adipocyte | *STARD3NL* | 0.839936845 | 0.001254863 | 0.01176218 | 1 |
| T2DM | adipocyte | *CD4* | 0.980551654 | 0.000119054 | 0.002528693 | 0.748493043 |
| T2DM | adipocyte | *SYT7* | 0.55652033 | 0.003183525 | 0.022463322 | 1 |
| T2DM | adipocyte | *LARS2* | 0.551611562 | 0.004555499 | 0.028028472 | 1 |
| T2DM | adipocyte | *PPP5C* | 0.518226947 | 0.008359992 | 0.041846552 | 1 |
| T2DM | adipocyte | *CEP68* | 0.616962994 | 5.39E-11 | 3.39E-08 | 3.39E-07 |
| T2DM | adipocyte | *ZBTB32* | -0.942418472 | 0.003261523 | 0.022707856 | 1 |
| T2DM | adipocyte | *ALOX5* | 0.646167739 | 0.001585896 | 0.013733509 | 1 |
| T2DM | adipocyte | *PSMC4* | 0.33698729 | 0.010647862 | 0.049259093 | 1 |
| T2DM | adipocyte | *ANGEL1* | 1.447897452 | 1.39E-05 | 0.00073252 | 0.0873849 |
| T2DM | adipocyte | *NPC1L1* | -0.471021975 | 0.010113087 | 0.047519414 | 1 |
| T2DM | adipocyte | *IL20RA* | 4.104859967 | 6.79E-05 | 0.001674964 | 0.427115719 |
| T2DM | adipocyte | *WWTR1* | 0.633493032 | 0.003699172 | 0.024415616 | 1 |
| T2DM | adipocyte | *OSBPL5* | 1.080298787 | 0.000637107 | 0.007514999 | 1 |
| T2DM | adipocyte | *AQR* | 0.66982432 | 0.003788725 | 0.024853488 | 1 |
| T2DM | adipocyte | *AKAP11* | 0.818175845 | 0.007214815 | 0.038407743 | 1 |
| T2DM | adipocyte | *NR1H3* | 0.526236778 | 0.002056225 | 0.01648866 | 1 |
| T2DM | adipocyte | *KPNA6* | 0.550717486 | 0.005367302 | 0.030981545 | 1 |
| T2DM | adipocyte | *VIM* | 0.614120093 | 0.001878237 | 0.01547857 | 1 |

| | | | | | | |
|---|---|---|---|---|---|---|
| T2DM | adipocyte | *CD44* | 0.526024245 | 0.000654659 | 0.007678812 | 1 |
| T2DM | adipocyte | *ANK1* | 1.423116226 | 1.79E-05 | 0.000809781 | 0.112497183 |
| T2DM | adipocyte | *BAK1* | 0.190650893 | 0.003936908 | 0.02530812 | 1 |
| T2DM | adipocyte | *GRN* | 0.905312668 | 0.000175866 | 0.003310394 | 1 |
| T2DM | adipocyte | *ARID4A* | 0.819788873 | 0.002392262 | 0.018520378 | 1 |
| T2DM | adipocyte | *FUT8* | 0.743157927 | 0.003354501 | 0.02302506 | 1 |
| T2DM | adipocyte | *PIAS1* | 0.689994919 | 0.002530596 | 0.019099469 | 1 |
| T2DM | adipocyte | *SLC4A7* | 0.718593705 | 0.000776328 | 0.008653858 | 1 |
| T2DM | adipocyte | *ADSS* | 1.052472473 | 0.000228959 | 0.003808107 | 1 |
| T2DM | adipocyte | *CUL3* | 0.779974163 | 0.000922927 | 0.009622628 | 1 |
| T2DM | adipocyte | *TUBG2* | -0.26452018 | 3.39E-09 | 1.71E-06 | 2.13E-05 |
| T2DM | adipocyte | *MAT2B* | 1.010371431 | 0.000890194 | 0.009413933 | 1 |
| T2DM | adipocyte | *C6* | -0.546502407 | 0.000159428 | 0.003103179 | 1 |
| T2DM | adipocyte | *RAB27B* | 0.278389779 | 0.000254202 | 0.0040489 | 1 |
| T2DM | adipocyte | *THAP3* | -0.890113302 | 0.00579028 | 0.032892943 | 1 |
| T2DM | adipocyte | *MED17* | 0.739541107 | 0.000574488 | 0.00702686 | 1 |
| T2DM | adipocyte | *LCP2* | 0.484788301 | 0.002197241 | 0.017354337 | 1 |
| T2DM | adipocyte | *CNTLN* | 0.811551415 | 7.33E-05 | 0.001763187 | 0.461054154 |
| T2DM | adipocyte | *EPHA3* | 0.867238019 | 1.76E-05 | 0.000808689 | 0.110790407 |
| T2DM | adipocyte | *DSG2* | 0.755850368 | 0.002727662 | 0.020127714 | 1 |
| T2DM | adipocyte | *YTHDC2* | 0.788873807 | 0.002213593 | 0.017461556 | 1 |
| T2DM | adipocyte | *POLR2B* | 0.837200965 | 0.00423846 | 0.026727379 | 1 |
| T2DM | adipocyte | *CP* | 1.344824929 | 2.74E-05 | 0.000990203 | 0.172295278 |
| T2DM | adipocyte | *DTNBP1* | 0.975384042 | 0.000358553 | 0.005239142 | 1 |
| T2DM | adipocyte | *TSPAN17* | 0.298367294 | 0.002280457 | 0.017854583 | 1 |
| T2DM | adipocyte | *SNX29* | 0.66116833 | 0.000779621 | 0.008675178 | 1 |
| T2DM | adipocyte | *RFC2* | 0.733499593 | 0.006349311 | 0.034985203 | 1 |
| T2DM | adipocyte | *DKK3* | 0.790824293 | 4.89E-05 | 0.001371325 | 0.30717688 |
| T2DM | adipocyte | *NFE2L3* | 1.54164017 | 5.52E-06 | 0.000480968 | 0.034733837 |
| T2DM | adipocyte | *MAPK9* | 0.539318011 | 0.003840951 | 0.024997991 | 1 |
| T2DM | adipocyte | *CYBA* | 0.89991572 | 4.93E-05 | 0.001378479 | 0.31015769 |
| T2DM | adipocyte | *ALX4* | 2.577028198 | 0.001115823 | 0.010825898 | 1 |
| T2DM | adipocyte | *ANAPC4* | 0.860883176 | 0.000246189 | 0.003980202 | 1 |
| T2DM | adipocyte | *OPN3* | 0.190354052 | 0.006797818 | 0.036779587 | 1 |
| T2DM | adipocyte | *TBC1D22A* | 0.724654501 | 0.000788248 | 0.008709522 | 1 |
| T2DM | adipocyte | *ITIH4* | 0.152838918 | 4.95E-06 | 0.000450593 | 0.031090909 |
| T2DM | adipocyte | *IL17RB* | -1.353714564 | 0.006273714 | 0.03462936 | 1 |
| T2DM | adipocyte | *GDI2* | 0.441787055 | 0.008914577 | 0.043649492 | 1 |
| T2DM | adipocyte | *CDK14* | 0.674537563 | 0.006246445 | 0.034509139 | 1 |
| T2DM | adipocyte | *YBX3* | 0.842289866 | 0.000316771 | 0.004833828 | 1 |
| T2DM | adipocyte | *PIGV* | -0.417823727 | 0.001365861 | 0.012445176 | 1 |
| T2DM | adipocyte | *QSER1* | 0.940243491 | 0.001473338 | 0.013046308 | 1 |
| T2DM | adipocyte | *VMP1* | 0.701413668 | 0.00334571 | 0.02302506 | 1 |
| T2DM | adipocyte | *EPN1* | 1.136931632 | 0.00092171 | 0.009622628 | 1 |
| T2DM | adipocyte | *CASP8* | 0.545284989 | 0.006331166 | 0.034915828 | 1 |
| T2DM | adipocyte | *LPAR2* | 0.576507786 | 0.000663823 | 0.007725842 | 1 |
| T2DM | adipocyte | *SUGP2* | 0.40964013 | 0.004031716 | 0.025812014 | 1 |
| T2DM | adipocyte | *BTBD1* | 1.139223691 | 2.74E-05 | 0.000990203 | 0.172148999 |
| T2DM | adipocyte | *ANKRD44* | 0.574626748 | 0.001349826 | 0.012351141 | 1 |
| T2DM | adipocyte | *NDUFB4* | 0.98445736 | 0.002421655 | 0.018680915 | 1 |

| | | | | | | |
|---|---|---|---|---|---|---|
| T2DM | adipocyte | *MAP2K4* | 0.705229424 | 0.008589326 | 0.042824024 | 1 |
| T2DM | adipocyte | *TBC1D1* | 0.934350217 | 5.66E-05 | 0.001496027 | 0.356054526 |
| T2DM | adipocyte | *NGEF* | 3.376260606 | 8.60E-05 | 0.001965411 | 0.540488082 |
| T2DM | adipocyte | *CD84* | 0.312654823 | 0.002907422 | 0.021024181 | 1 |
| T2DM | adipocyte | *SP100* | 0.822949264 | 0.000184469 | 0.003395587 | 1 |
| T2DM | adipocyte | *TP53BP1* | 0.514800611 | 0.006438641 | 0.035384382 | 1 |
| T2DM | adipocyte | *HDAC4* | 0.756187092 | 0.005314677 | 0.030739072 | 1 |
| T2DM | adipocyte | *RASSF1* | 0.456071698 | 0.00585076 | 0.03304917 | 1 |
| T2DM | adipocyte | *IFI35* | 0.281330371 | 0.007811263 | 0.040586289 | 1 |
| T2DM | adipocyte | *CYFIP1* | 0.825653207 | 0.001756928 | 0.014708134 | 1 |
| T2DM | adipocyte | *PITX1* | -1.692220578 | 0.000809597 | 0.008852065 | 1 |
| T2DM | adipocyte | *NUCKS1* | 0.666278828 | 0.005082777 | 0.029948844 | 1 |
| T2DM | adipocyte | *GAL* | 2.16886325 | 6.18E-05 | 0.001599534 | 0.388686728 |
| T2DM | adipocyte | *NEDD4* | 0.436746677 | 0.000518157 | 0.006450799 | 1 |
| T2DM | adipocyte | *GNB5* | 0.312437273 | 0.0076886 | 0.040114714 | 1 |
| T2DM | adipocyte | *ASNS* | 0.790365161 | 0.000767918 | 0.008576883 | 1 |
| T2DM | adipocyte | *TESK2* | 0.675530933 | 0.007722798 | 0.040259728 | 1 |
| T2DM | adipocyte | *CDC42* | 0.896228628 | 0.002625271 | 0.019596375 | 1 |
| T2DM | adipocyte | *OSBPL3* | 0.481152252 | 0.004578058 | 0.028052873 | 1 |
| T2DM | adipocyte | *SNX13* | 0.774845634 | 0.003217307 | 0.022589029 | 1 |
| T2DM | adipocyte | *VASH1* | 0.732362097 | 0.004566608 | 0.028037365 | 1 |
| T2DM | adipocyte | *TRIP13* | -2.005608658 | 0.008027182 | 0.041298601 | 1 |
| T2DM | adipocyte | *CYBRD1* | 0.372224028 | 0.002712684 | 0.020040713 | 1 |
| T2DM | adipocyte | *EPN2* | 0.594862383 | 0.009162562 | 0.044546498 | 1 |
| T2DM | adipocyte | *LIMS2* | 0.385313162 | 0.004394274 | 0.027428067 | 1 |
| T2DM | adipocyte | *TRHDE* | -0.332164558 | 0.005901356 | 0.033072047 | 1 |
| T2DM | adipocyte | *NFATC3* | 0.435217387 | 0.004242869 | 0.026728373 | 1 |
| T2DM | adipocyte | *ALPK1* | 0.771948225 | 0.001571757 | 0.013648668 | 1 |
| T2DM | adipocyte | *SDHA* | 0.424472736 | 0.000599499 | 0.007210362 | 1 |
| T2DM | adipocyte | *NTN4* | 0.694217484 | 0.000187367 | 0.003398243 | 1 |
| T2DM | adipocyte | *SRI* | 1.030843077 | 0.003622072 | 0.024071846 | 1 |
| T2DM | adipocyte | *EIF4G3* | 0.524750291 | 0.008161683 | 0.041544539 | 1 |
| T2DM | adipocyte | *CELSR1* | 1.431501887 | 9.02E-05 | 0.002025613 | 0.567171542 |
| T2DM | adipocyte | *ADD2* | 1.380294361 | 5.62E-05 | 0.001489992 | 0.353128009 |
| T2DM | adipocyte | *SLC25A3* | 0.759484579 | 0.000392846 | 0.005550157 | 1 |
| T2DM | adipocyte | *PLD1* | 0.830097306 | 0.00052476 | 0.006520091 | 1 |
| T2DM | adipocyte | *MCM6* | 0.542705765 | 0.01026032 | 0.047996007 | 1 |
| T2DM | adipocyte | *NT5C2* | 0.746235806 | 0.000215349 | 0.00364933 | 1 |
| T2DM | adipocyte | *XAB2* | -0.48547129 | 0.009518505 | 0.045716457 | 1 |
| T2DM | adipocyte | *STXBP2* | 0.530917414 | 2.81E-05 | 0.00099459 | 0.176736318 |
| T2DM | adipocyte | *CTTNBP2* | -0.434086241 | 0.004311195 | 0.027050381 | 1 |
| T2DM | adipocyte | *DNAJC10* | 1.018560533 | 0.000791894 | 0.008730615 | 1 |
| T2DM | adipocyte | *DYNC1I2* | 0.647079526 | 0.00375519 | 0.024695481 | 1 |
| T2DM | adipocyte | *LRCH4* | -0.29646909 | 0.002442471 | 0.018795368 | 1 |
| T2DM | adipocyte | *FBLN1* | 0.254986415 | 0.00258478 | 0.019415187 | 1 |
| T2DM | adipocyte | *SYNJ2* | 0.466604419 | 0.003536994 | 0.023782977 | 1 |
| T2DM | adipocyte | *FGF20* | 6.040854257 | 1.79E-05 | 0.000809781 | 0.112559625 |
| T2DM | adipocyte | *XRCC5* | 0.809099199 | 0.001504447 | 0.013241122 | 1 |
| T2DM | adipocyte | *DNM2* | 0.805017403 | 0.003875434 | 0.025066724 | 1 |
| T2DM | adipocyte | *DCT* | -0.608937308 | 0.000495701 | 0.006341444 | 1 |

| | | | | | | |
|---|---|---|---|---|---|---|
| T2DM | adipocyte | *MAGI3* | 0.745646169 | 0.004178907 | 0.026484666 | 1 |
| T2DM | adipocyte | *STK17B* | 0.935480056 | 0.000187879 | 0.003398243 | 1 |
| T2DM | adipocyte | *HSPB11* | 0.815089593 | 0.004437814 | 0.027624292 | 1 |
| T2DM | adipocyte | *COBLL1* | -0.392292339 | 1.00E-07 | 2.74E-05 | 0.000631291 |
| T2DM | adipocyte | *GSK3B* | 0.687534539 | 0.002503212 | 0.0190529 | 1 |
| T2DM | adipocyte | *ERC1* | 0.624853406 | 0.005228882 | 0.030410711 | 1 |
| T2DM | adipocyte | *PDS5B* | 0.720265589 | 0.001957609 | 0.015997616 | 1 |
| T2DM | adipocyte | *RPS5* | 0.914464519 | 0.001650366 | 0.014136036 | 1 |
| T2DM | adipocyte | *GSTP1* | 1.086662925 | 0.000267929 | 0.004221729 | 1 |
| T2DM | adipocyte | *APOB* | -0.472356908 | 0.000417312 | 0.005670594 | 1 |
| T2DM | adipocyte | *NCOA1* | 0.677165604 | 0.001705566 | 0.014412493 | 1 |
| T2DM | adipocyte | *CD59* | 0.667750992 | 0.000169408 | 0.003217726 | 1 |
| T2DM | adipocyte | *CD82* | 0.37047083 | 0.002573876 | 0.019356409 | 1 |
| T2DM | adipocyte | *SCAMP1* | 0.758073932 | 0.000321544 | 0.0048855 | 1 |
| T2DM | adipocyte | *CPNE3* | 0.361956193 | 0.001768754 | 0.014787441 | 1 |
| T2DM | adipocyte | *EPS15* | 0.27662111 | 0.009279367 | 0.044980246 | 1 |
| T2DM | adipocyte | *ATG16L1* | 1.071698881 | 0.000758308 | 0.00851336 | 1 |
| T2DM | adipocyte | *B4GALT1* | 0.869701109 | 0.000666042 | 0.007725842 | 1 |
| T2DM | adipocyte | *SNX10* | -0.066533515 | 0.00356262 | 0.023853237 | 1 |
| T2DM | adipocyte | *LPCAT2* | 0.370471582 | 0.009498041 | 0.045678294 | 1 |
| T2DM | adipocyte | *KLHL42* | 0.506807019 | 0.008728394 | 0.043068751 | 1 |
| T2DM | adipocyte | *C3orf18* | 0.7174714 | 1.06E-06 | 0.000145821 | 0.006675688 |
| T2DM | adipocyte | *FKBP1A* | 1.060387943 | 0.000416952 | 0.005670594 | 1 |
| T2DM | adipocyte | *XRN2* | 0.813792406 | 0.002731427 | 0.020131868 | 1 |
| T2DM | adipocyte | *TESC* | 1.517006739 | 1.08E-05 | 0.000656895 | 0.067660206 |
| T2DM | adipocyte | *SLC23A2* | 0.857380962 | 0.002238588 | 0.017614524 | 1 |
| T2DM | adipocyte | *OAS1* | 1.235067132 | 0.000162424 | 0.00313738 | 1 |
| T2DM | adipocyte | *FUS* | 0.793764919 | 0.001272858 | 0.011908421 | 1 |
| T2DM | adipocyte | *FXYD5* | 0.960672319 | 2.85E-05 | 0.001002409 | 0.179431168 |
| T2DM | adipocyte | *GMIP* | 1.396835781 | 4.25E-05 | 0.001220134 | 0.26720942 |
| T2DM | adipocyte | *PAPOLA* | 0.713195312 | 0.003349764 | 0.02302506 | 1 |
| T2DM | adipocyte | *RGS1* | 1.637143415 | 9.06E-06 | 0.000586394 | 0.056934528 |
| T2DM | adipocyte | *YPEL3* | 0.170050086 | 0.003865621 | 0.025034168 | 1 |
| T2DM | adipocyte | *LYZ* | 0.47339008 | 0.000599812 | 0.007210362 | 1 |
| T2DM | adipocyte | *PABPC4* | -0.544410526 | 7.15E-07 | 0.000115199 | 0.004492766 |
| T2DM | adipocyte | *PDPR* | 0.841234029 | 0.000662755 | 0.007725842 | 1 |
| T2DM | adipocyte | *EXOC1* | 0.741068517 | 0.005400241 | 0.031090947 | 1 |
| T2DM | adipocyte | *TXNL1* | 0.811639033 | 0.006643138 | 0.036170411 | 1 |
| T2DM | adipocyte | *IL5RA* | 3.446024549 | 0.001165317 | 0.011138941 | 1 |
| T2DM | adipocyte | *SEL1L3* | 1.052914472 | 3.03E-06 | 0.000307456 | 0.019062268 |
| T2DM | adipocyte | *MYO15A* | 0.603381346 | 0.006688796 | 0.036377559 | 1 |
| T2DM | adipocyte | *NLRP1* | 0.629521203 | 0.003510858 | 0.023694794 | 1 |
| T2DM | adipocyte | *RGS17* | -0.358517217 | 0.008959638 | 0.043835988 | 1 |
| T2DM | adipocyte | *PPP2R3C* | 0.727715421 | 0.001789167 | 0.014898665 | 1 |
| T2DM | adipocyte | *OSGEP* | 0.965173375 | 0.003973687 | 0.025492416 | 1 |
| T2DM | adipocyte | *DPYSL2* | 0.684478201 | 0.000186024 | 0.003395587 | 1 |
| T2DM | adipocyte | *COMT* | 0.896829204 | 0.005780324 | 0.032892943 | 1 |
| T2DM | adipocyte | *VNN3* | 1.842480246 | 0.000485084 | 0.006236657 | 1 |
| T2DM | adipocyte | *SUCO* | 0.776361091 | 0.008621256 | 0.042949155 | 1 |
| T2DM | adipocyte | *MSH2* | 1.197510096 | 0.000579698 | 0.007060862 | 1 |

| | | | | | | |
|---|---|---|---|---|---|---|
| T2DM | adipocyte | *TMEM38B* | 1.385424986 | 0.001592145 | 0.013763932 | 1 |
| T2DM | adipocyte | *CWF19L1* | -0.219023162 | 0.000689935 | 0.00792984 | 1 |
| T2DM | adipocyte | *CREM* | 0.933986768 | 0.001678244 | 0.014340975 | 1 |
| T2DM | adipocyte | *TPSD1* | 4.618711421 | 8.90E-07 | 0.000133232 | 0.005595729 |
| T2DM | adipocyte | *ITPR3* | 0.963868155 | 1.40E-05 | 0.00073252 | 0.088044726 |
| T2DM | adipocyte | *NRP1* | 0.67754133 | 0.000818938 | 0.008923161 | 1 |
| T2DM | adipocyte | *STX1B* | -0.378202731 | 0.003922893 | 0.02524384 | 1 |
| T2DM | adipocyte | *MARCH2* | 0.655916617 | 0.002009011 | 0.016276616 | 1 |
| T2DM | adipocyte | *P2RX6* | -0.532809909 | 0.000212255 | 0.00360662 | 1 |
| T2DM | adipocyte | *GGT5* | 1.144281104 | 2.80E-06 | 0.000294676 | 0.017619489 |
| T2DM | adipocyte | *PPM1F* | 0.739679172 | 0.003230084 | 0.022589029 | 1 |
| T2DM | adipocyte | *CYTH4* | 1.048193259 | 1.89E-05 | 0.000811251 | 0.118583918 |
| T2DM | adipocyte | *CARD10* | 0.43846241 | 0.004814827 | 0.028912435 | 1 |
| T2DM | adipocyte | *SH3BP1* | 1.316999528 | 0.000176999 | 0.003311883 | 1 |
| T2DM | adipocyte | *TIMP3* | 0.675128371 | 0.002007054 | 0.016276616 | 1 |
| T2DM | adipocyte | *RBFOX2* | 0.647579076 | 0.008656931 | 0.042990619 | 1 |
| T2DM | adipocyte | *KIAA0930* | 0.391161992 | 0.007400835 | 0.039028017 | 1 |
| T2DM | adipocyte | *KCTD17* | 2.115888198 | 2.22E-05 | 0.000837403 | 0.139846262 |
| T2DM | adipocyte | *DESI1* | 0.218738712 | 0.00368312 | 0.024400185 | 1 |
| T2DM | adipocyte | *ATP6V1D* | -0.237885876 | 0.008226822 | 0.041544539 | 1 |
| T2DM | adipocyte | *PLTP* | 0.229999721 | 0.000410487 | 0.005646146 | 1 |
| T2DM | adipocyte | *CD40* | 0.656988348 | 0.007351583 | 0.038905221 | 1 |
| T2DM | adipocyte | *NDRG3* | 0.879694589 | 0.001530309 | 0.013362569 | 1 |
| T2DM | adipocyte | *BMP7* | 1.40148894 | 1.47E-05 | 0.000753745 | 0.092713048 |
| T2DM | adipocyte | *DIDO1* | 0.560285318 | 0.000921055 | 0.009622628 | 1 |
| T2DM | adipocyte | *SLC17A9* | 0.827625597 | 0.000211149 | 0.00360662 | 1 |
| T2DM | adipocyte | *CCM2L* | 1.252679923 | 5.57E-06 | 0.000480968 | 0.035039513 |
| T2DM | adipocyte | *METTL4* | 1.173531699 | 0.001301862 | 0.012083315 | 1 |
| T2DM | adipocyte | *LPIN2* | 0.915705159 | 0.002058788 | 0.01648866 | 1 |
| T2DM | adipocyte | *SMCHD1* | 0.724734368 | 0.001106765 | 0.010771256 | 1 |
| T2DM | adipocyte | *MYL12A* | 0.86622856 | 0.002793565 | 0.020448966 | 1 |
| T2DM | adipocyte | *HTR2A* | 2.666267786 | 1.57E-05 | 0.000762794 | 0.098400458 |
| T2DM | adipocyte | *UGGT2* | 0.497491732 | 0.002523791 | 0.019099469 | 1 |
| T2DM | adipocyte | *SGCG* | -0.308387529 | 0.010739493 | 0.049500875 | 1 |
| T2DM | adipocyte | *PARP4* | 0.858361675 | 0.000922772 | 0.009622628 | 1 |
| T2DM | adipocyte | *MEDAG* | 1.126795294 | 1.38E-05 | 0.00073252 | 0.087022641 |
| T2DM | adipocyte | *TSC22D1* | 0.817516052 | 1.98E-05 | 0.00081257 | 0.124611157 |
| T2DM | adipocyte | *CTCF* | 0.612030164 | 0.00510579 | 0.029999308 | 1 |
| T2DM | adipocyte | *COG4* | -0.349994898 | 0.009627281 | 0.045962686 | 1 |
| T2DM | adipocyte | *HCFC1R1* | 1.658250251 | 9.11E-05 | 0.002031326 | 0.572833803 |
| T2DM | adipocyte | *LMF1* | 0.786001341 | 0.004133895 | 0.026252322 | 1 |
| T2DM | adipocyte | *SLC7A5* | 3.794377007 | 0.000151891 | 0.003021954 | 0.954937616 |
| T2DM | adipocyte | *NUBP1* | 1.020484999 | 0.010157678 | 0.047657704 | 1 |
| T2DM | adipocyte | *UBE2I* | 0.642222544 | 0.005991369 | 0.033423016 | 1 |
| T2DM | adipocyte | *RBL2* | 0.434718079 | 9.51E-06 | 0.000597863 | 0.059786299 |
| T2DM | adipocyte | *KAT8* | 0.672799301 | 0.007269238 | 0.038599407 | 1 |
| T2DM | adipocyte | *SYT17* | -0.298796919 | 0.009887741 | 0.046765514 | 1 |
| T2DM | adipocyte | *TMC5* | -0.687403667 | 0.000367401 | 0.005285702 | 1 |
| T2DM | adipocyte | *CSK* | 1.382143911 | 5.20E-05 | 0.001416342 | 0.327174962 |
| T2DM | adipocyte | *HERC1* | 0.773147105 | 0.000649606 | 0.007633777 | 1 |

| | | | | | | |
|---|---|---|---|---|---|---|
| T2DM | adipocyte | *RASL12* | 1.357354471 | 1.12E-05 | 0.000673225 | 0.070688618 |
| T2DM | adipocyte | *FAH* | -0.485420726 | 0.002511024 | 0.019089246 | 1 |
| T2DM | adipocyte | *EHD4* | 0.681245114 | 0.002174737 | 0.01721986 | 1 |
| T2DM | adipocyte | *DTWD1* | 0.513574469 | 0.00198019 | 0.01612624 | 1 |
| T2DM | adipocyte | *PDGFRL* | 0.590157678 | 6.35E-05 | 0.001613043 | 0.398922286 |
| T2DM | adipocyte | *EYA1* | 0.188187795 | 0.001020307 | 0.010362093 | 1 |
| T2DM | adipocyte | *CPQ* | 0.27527137 | 0.007398758 | 0.039028017 | 1 |
| T2DM | adipocyte | *NIPAL2* | 0.634344275 | 3.27E-05 | 0.00106579 | 0.205718058 |
| T2DM | adipocyte | *IKBKB* | 1.100782326 | 2.02E-05 | 0.000815077 | 0.127121943 |
| T2DM | adipocyte | *PLAT* | 1.099421187 | 1.99E-06 | 0.000232105 | 0.012533667 |
| T2DM | adipocyte | *GDAP1* | 0.561125515 | 0.003390039 | 0.023141342 | 1 |
| T2DM | adipocyte | *ZC2HC1A* | 0.559802735 | 0.003919029 | 0.02524384 | 1 |
| T2DM | adipocyte | *CHRAC1* | 0.813296474 | 0.00480571 | 0.028912435 | 1 |
| T2DM | adipocyte | *MTMR9* | 0.963292594 | 0.003479172 | 0.02351995 | 1 |
| T2DM | adipocyte | *LEPROTL1* | 1.202418843 | 0.000184977 | 0.003395587 | 1 |
| T2DM | adipocyte | *ERICH1* | 0.820863426 | 0.001710054 | 0.014420117 | 1 |
| T2DM | adipocyte | *TUSC3* | 0.913826001 | 7.67E-06 | 0.000560411 | 0.048195372 |
| T2DM | adipocyte | *DMPK* | -0.633729011 | 0.001008486 | 0.0102761 | 1 |
| T2DM | adipocyte | *AES* | 1.127924545 | 0.000209299 | 0.003599172 | 1 |
| T2DM | adipocyte | *CCDC61* | -0.393706857 | 0.00654905 | 0.035865745 | 1 |
| T2DM | adipocyte | *IL27RA* | 0.623349306 | 0.000734851 | 0.008338307 | 1 |
| T2DM | adipocyte | *CCNE1* | -1.031300076 | 0.002627604 | 0.019596375 | 1 |
| T2DM | adipocyte | *CCDC9* | 0.87557837 | 0.003297773 | 0.022858988 | 1 |
| T2DM | adipocyte | *FZR1* | 1.363492573 | 0.003211271 | 0.022583065 | 1 |
| T2DM | adipocyte | *TGFB1* | 0.685904041 | 0.001948181 | 0.015948197 | 1 |
| T2DM | adipocyte | *CEACAM4* | 3.867447631 | 4.38E-05 | 0.001246899 | 0.27556471 |
| T2DM | adipocyte | *BABAM1* | 1.386788022 | 0.009738066 | 0.046311059 | 1 |
| T2DM | adipocyte | *TYK2* | 0.628629227 | 0.00328245 | 0.022786903 | 1 |
| T2DM | adipocyte | *GRIN2D* | 1.843591306 | 0.000345553 | 0.005139885 | 1 |
| T2DM | adipocyte | *PPP2R1A* | 0.956618108 | 0.009258536 | 0.044913899 | 1 |
| T2DM | adipocyte | *GCDH* | -0.814793763 | 1.60E-06 | 0.00020495 | 0.010042538 |
| T2DM | adipocyte | *LILRB5* | 1.043640293 | 3.37E-05 | 0.001075499 | 0.212118229 |
| T2DM | adipocyte | *ELL* | 0.660309572 | 0.000930504 | 0.009685559 | 1 |
| T2DM | adipocyte | *COPE* | 0.85076177 | 0.008755044 | 0.043103336 | 1 |
| T2DM | adipocyte | *ATP13A1* | 0.626073601 | 2.11E-08 | 7.36E-06 | 0.000132473 |
| T2DM | adipocyte | *SIPA1L3* | 0.659517115 | 0.004077407 | 0.026030006 | 1 |
| T2DM | adipocyte | *ZNF85* | 0.550982683 | 0.007447249 | 0.039180632 | 1 |
| T2DM | adipocyte | *PON3* | 1.434003395 | 0.000598767 | 0.007210362 | 1 |
| T2DM | adipocyte | *OGDH* | 0.853498091 | 0.00150587 | 0.013241122 | 1 |
| T2DM | adipocyte | *ADAP1* | 2.935422501 | 2.16E-05 | 0.000829507 | 0.135652422 |
| T2DM | adipocyte | *MET* | 0.596700903 | 0.002220618 | 0.017495019 | 1 |
| T2DM | adipocyte | *CPVL* | 0.884052789 | 3.54E-09 | 1.71E-06 | 2.22E-05 |
| T2DM | adipocyte | *NOD1* | 0.87843836 | 0.000869102 | 0.009308421 | 1 |
| T2DM | adipocyte | *SNX8* | 0.919718244 | 0.00954735 | 0.045746239 | 1 |
| T2DM | adipocyte | *USP42* | 0.967372248 | 0.001649967 | 0.014136036 | 1 |
| T2DM | adipocyte | *C1GALT1* | 0.673472347 | 0.010716402 | 0.049466976 | 1 |
| T2DM | adipocyte | *PHF14* | 0.711288099 | 0.00256435 | 0.019307868 | 1 |
| T2DM | adipocyte | *MEST* | -0.155737546 | 0.004895119 | 0.029226604 | 1 |
| T2DM | adipocyte | *POU6F2* | -0.499549368 | 0.001392089 | 0.012592899 | 1 |
| T2DM | adipocyte | *TSPAN13* | 1.261935624 | 0.007048994 | 0.037630362 | 1 |

| | | | | | | |
|---|---|---|---|---|---|---|
| T2DM | adipocyte | *TMEM176B* | 0.169417322 | 0.001142359 | 0.010981664 | 1 |
| T2DM | adipocyte | *POLD2* | -0.89755853 | 1.38E-05 | 0.00073252 | 0.086688834 |
| T2DM | adipocyte | *GCK* | -8.371810539 | 0.00106838 | 0.010577802 | 1 |
| T2DM | adipocyte | *CLIP2* | 1.193949724 | 0.000126474 | 0.002615595 | 0.795140953 |
| T2DM | adipocyte | *LIMK1* | 1.520733399 | 9.07E-05 | 0.002030351 | 0.570528668 |
| T2DM | adipocyte | *SLC1A1* | 0.131986568 | 0.00788161 | 0.040841088 | 1 |
| T2DM | adipocyte | *TNFSF8* | 0.802986079 | 0.001593788 | 0.013763932 | 1 |
| T2DM | adipocyte | *KDM4C* | 0.66596964 | 0.007819808 | 0.040597139 | 1 |
| T2DM | adipocyte | *PTGDS* | 2.246511405 | 0.000124015 | 0.002586077 | 0.779682675 |
| T2DM | adipocyte | *DNMBP* | 0.937996929 | 0.000794477 | 0.008732301 | 1 |
| T2DM | adipocyte | *MAPK8* | 0.662292363 | 0.005430961 | 0.031125299 | 1 |
| T2DM | adipocyte | *PLEKHA1* | 1.028647866 | 0.000135134 | 0.002758392 | 0.849584688 |
| T2DM | adipocyte | *MICU1* | 0.855149992 | 0.000417605 | 0.005670594 | 1 |
| T2DM | adipocyte | *CCSER2* | 0.77416386 | 0.001786488 | 0.014898665 | 1 |
| T2DM | adipocyte | *SFXN3* | 0.787579871 | 0.001402236 | 0.012666465 | 1 |
| T2DM | adipocyte | *ACBD5* | 0.596322728 | 0.0096915 | 0.046159439 | 1 |
| T2DM | adipocyte | *SHOC2* | 0.701470644 | 0.004461108 | 0.027714414 | 1 |
| T2DM | adipocyte | *DHRS11* | -0.220256912 | 0.000104531 | 0.002281892 | 0.657184821 |
| T2DM | adipocyte | *MLLT6* | 1.050115026 | 0.000250208 | 0.004012908 | 1 |
| T2DM | adipocyte | *PSMB3* | 0.843422682 | 0.000296604 | 0.004604319 | 1 |
| T2DM | adipocyte | *RGS9* | 2.076251829 | 0.000206857 | 0.003597496 | 1 |
| T2DM | adipocyte | *WNT3* | -0.657698936 | 0.001711056 | 0.014420117 | 1 |
| T2DM | adipocyte | *TRIM37* | 0.852432454 | 0.0019914 | 0.016196549 | 1 |
| T2DM | adipocyte | *GALK1* | 2.097169553 | 0.0001894 | 0.00340696 | 1 |
| T2DM | adipocyte | *RNF167* | 0.731998878 | 0.002930904 | 0.021107208 | 1 |
| T2DM | adipocyte | *C1QBP* | 1.148463733 | 0.002160888 | 0.017153409 | 1 |
| T2DM | adipocyte | *ICAM2* | 0.56103263 | 1.84E-05 | 0.000811251 | 0.115397001 |
| T2DM | adipocyte | *CYTH1* | 0.685765043 | 9.65E-07 | 0.000141146 | 0.006069257 |
| T2DM | adipocyte | *HSD17B1* | -0.184124619 | 9.38E-08 | 2.68E-05 | 0.000589879 |
| T2DM | adipocyte | *DLX4* | -3.960192685 | 0.000153333 | 0.003025687 | 0.964007162 |
| T2DM | adipocyte | *VAT1* | 0.765386333 | 1.69E-06 | 0.000210988 | 0.010630286 |
| T2DM | adipocyte | *MPP2* | 1.746431803 | 6.55E-06 | 0.000515083 | 0.041206604 |
| T2DM | adipocyte | *EFTUD2* | 0.579679096 | 0.008632635 | 0.042959913 | 1 |
| T2DM | adipocyte | *FAM20A* | 0.82677242 | 2.10E-05 | 0.000819911 | 0.132005746 |
| T2DM | adipocyte | *UBE2D3* | 0.805770098 | 0.00102648 | 0.010375372 | 1 |
| T2DM | adipocyte | *NDUFC1* | 1.009751646 | 0.009417381 | 0.045543902 | 1 |
| T2DM | adipocyte | *CPE* | 0.769568268 | 0.000444926 | 0.005901368 | 1 |
| T2DM | adipocyte | *WFS1* | 1.01169357 | 0.000395929 | 0.005568695 | 1 |
| T2DM | adipocyte | *CPZ* | 15.63312756 | 3.33E-05 | 0.00107474 | 0.209574232 |
| T2DM | adipocyte | *SLC2A9* | 0.92330994 | 3.19E-07 | 6.46E-05 | 0.00200306 |
| T2DM | adipocyte | *STIM2* | 0.69519742 | 0.001378728 | 0.012508027 | 1 |
| T2DM | adipocyte | *RAPGEF2* | 0.716120982 | 0.000253796 | 0.0040489 | 1 |
| T2DM | adipocyte | *KLF3* | 0.915196911 | 0.000692889 | 0.007949253 | 1 |
| T2DM | adipocyte | *UGDH* | 0.879599022 | 0.001960665 | 0.015997616 | 1 |
| T2DM | adipocyte | *NRXN2* | 2.33109861 | 3.20E-05 | 0.00106579 | 0.201275181 |
| T2DM | adipocyte | *EIF4G2* | 0.869008347 | 0.003377874 | 0.023102217 | 1 |
| T2DM | adipocyte | *MADD* | 0.887418004 | 0.000220303 | 0.003723247 | 1 |
| T2DM | adipocyte | *CARS* | 0.49879453 | 0.003981077 | 0.025513793 | 1 |
| T2DM | adipocyte | *RPS13* | 0.476250767 | 0.003021886 | 0.021564811 | 1 |
| T2DM | adipocyte | *TCIRG1* | 0.851892952 | 2.16E-05 | 0.000829507 | 0.136039172 |

| | | | | | | |
|---|---|---|---|---|---|---|
| T2DM | adipocyte | *PPFIBP1* | 0.497262648 | 0.000663963 | 0.007725842 | 1 |
| T2DM | adipocyte | *COQ5* | 0.07234904 | 0.005893757 | 0.033072047 | 1 |
| T2DM | adipocyte | *GLI1* | 1.921890507 | 1.40E-05 | 0.00073252 | 0.08813191 |
| T2DM | adipocyte | *LTA4H* | 1.026242544 | 0.000850992 | 0.009161281 | 1 |
| T2DM | adipocyte | *WNT5B* | 0.945398661 | 0.000453831 | 0.005967377 | 1 |
| T2DM | adipocyte | *PRR4* | 0.275086819 | 0.001570089 | 0.013648668 | 1 |
| T2DM | adipocyte | *PARP11* | 0.958170076 | 0.001505179 | 0.013241122 | 1 |
| T2DM | adipocyte | *CREBL2* | 0.780273017 | 7.64E-05 | 0.001825352 | 0.480067492 |
| T2DM | adipocyte | *CDK2AP1* | 0.174198826 | 0.000121527 | 0.002555327 | 0.764042895 |
| T2DM | adipocyte | *MGP* | 0.325767265 | 0.000185657 | 0.003395587 | 1 |
| T2DM | adipocyte | *ENDOU* | -0.313010383 | 0.001204705 | 0.011392753 | 1 |
| T2DM | adipocyte | *GNB3* | -0.073961703 | 0.008669948 | 0.043009506 | 1 |
| T2DM | adipocyte | *GNPTAB* | 0.859509296 | 0.000480017 | 0.006196857 | 1 |
| T2DM | adipocyte | *ST8SIA1* | 0.54236406 | 0.001090066 | 0.010645331 | 1 |
| T2DM | adipocyte | *C2CD5* | 0.863780173 | 0.000246196 | 0.003980202 | 1 |
| T2DM | adipocyte | *PHC1* | 1.85674003 | 3.13E-05 | 0.001056656 | 0.196538082 |
| T2DM | adipocyte | *RNGTT* | 0.777139745 | 0.003219344 | 0.022589029 | 1 |
| T2DM | adipocyte | *UST* | 0.403078725 | 0.001076875 | 0.010621557 | 1 |
| T2DM | adipocyte | *PHACTR1* | 0.753646532 | 1.73E-05 | 0.000808689 | 0.108674985 |
| T2DM | adipocyte | *CAP2* | -0.275429925 | 0.004615713 | 0.028228589 | 1 |
| T2DM | adipocyte | *ASCC3* | 0.660605435 | 0.004558154 | 0.028028472 | 1 |
| T2DM | adipocyte | *BVES* | -0.176448178 | 0.004183426 | 0.026486608 | 1 |
| T2DM | adipocyte | *VNN2* | 0.479537313 | 0.002043513 | 0.016459063 | 1 |
| T2DM | adipocyte | *PHACTR2* | 0.723003338 | 0.002016815 | 0.016318811 | 1 |
| T2DM | adipocyte | *TFEB* | 1.068375586 | 0.000496261 | 0.006341444 | 1 |
| T2DM | adipocyte | *FAM120B* | 0.38156168 | 0.002530404 | 0.019099469 | 1 |
| T2DM | adipocyte | *TMEM30A* | 0.616413414 | 0.005956173 | 0.033285742 | 1 |
| T2DM | adipocyte | *SENP6* | 0.697663815 | 0.00310228 | 0.022013581 | 1 |
| T2DM | adipocyte | *FBRSL1* | 0.662871483 | 0.008260009 | 0.041544539 | 1 |
| T2DM | adipocyte | *LY86* | 1.028509732 | 0.002517407 | 0.019099469 | 1 |
| T2DM | adipocyte | *DAP* | 0.634438771 | 0.005837915 | 0.033035977 | 1 |
| T2DM | adipocyte | *LOX* | 1.113430365 | 0.002296339 | 0.017956569 | 1 |
| T2DM | adipocyte | *RNF130* | 0.656120365 | 0.008681667 | 0.043011538 | 1 |
| T2DM | adipocyte | *TXNDC15* | 0.7144802 | 0.010737058 | 0.049500875 | 1 |
| T2DM | adipocyte | *SMAD5* | 0.770651254 | 0.002631266 | 0.019600438 | 1 |
| T2DM | adipocyte | *HGD* | -1.320979504 | 0.007573462 | 0.039657458 | 1 |
| T2DM | adipocyte | *NPHP3* | 0.107808574 | 0.005000643 | 0.02954797 | 1 |
| T2DM | adipocyte | *IFT57* | 1.030840522 | 0.008172419 | 0.041544539 | 1 |
| T2DM | adipocyte | *NKTR* | 0.756883814 | 4.96E-05 | 0.001380058 | 0.311893204 |
| T2DM | adipocyte | *MOB1A* | 1.126577681 | 0.000265271 | 0.004190353 | 1 |
| T2DM | adipocyte | *KCNIP3* | 0.848582684 | 0.001279126 | 0.011931548 | 1 |
| T2DM | adipocyte | *SLC35F5* | 0.484347175 | 0.003438628 | 0.023295965 | 1 |
| T2DM | adipocyte | *ACTR3* | 0.89744095 | 0.000168363 | 0.003217315 | 1 |
| T2DM | adipocyte | *DNAJC27* | -0.502851118 | 0.004800039 | 0.028912435 | 1 |
| T2DM | adipocyte | *STAM2* | 0.774731675 | 0.008357332 | 0.041846552 | 1 |
| T2DM | adipocyte | *NRBP1* | -0.578067683 | 0.000232277 | 0.003853101 | 1 |
| T2DM | adipocyte | *ITGA4* | 0.579071796 | 0.000291766 | 0.004540422 | 1 |
| T2DM | adipocyte | *PPM1G* | -0.593110357 | 0.007948496 | 0.040960814 | 1 |
| T2DM | adipocyte | *POLE4* | 1.488679429 | 0.000348008 | 0.005151093 | 1 |
| T2DM | adipocyte | *GLS* | 0.685697292 | 0.002793966 | 0.020448966 | 1 |

| | | | | | | |
|---|---|---|---|---|---|---|
| T2DM | adipocyte | *GNLY* | 0.690811035 | 0.001819419 | 0.015070735 | 1 |
| T2DM | adipocyte | *SF3B1* | 0.610823683 | 0.008250096 | 0.041544539 | 1 |
| T2DM | adipocyte | *ST3GAL5* | 0.56364018 | 0.003425143 | 0.023240953 | 1 |
| T2DM | adipocyte | *PLCD4* | -0.775197862 | 0.010527706 | 0.048849149 | 1 |
| T2DM | adipocyte | *CHMP3* | -0.380248432 | 0.002363577 | 0.018345446 | 1 |
| T2DM | adipocyte | *MLPH* | 1.117931276 | 8.05E-06 | 0.000574639 | 0.050592825 |
| T2DM | adipocyte | *STRN* | 0.832764151 | 0.002937113 | 0.021127722 | 1 |
| T2DM | adipocyte | *SDC1* | 0.632385185 | 0.002868686 | 0.020898528 | 1 |
| T2DM | adipocyte | *WIPF1* | 0.542276035 | 0.003373119 | 0.023101091 | 1 |
| T2DM | adipocyte | *THADA* | 0.66518505 | 0.008891201 | 0.043578441 | 1 |
| T2DM | adipocyte | *CD207* | 10.86255209 | 2.33E-06 | 0.000261239 | 0.014629362 |
| T2DM | adipocyte | *GPX7* | -0.160542639 | 0.000750496 | 0.008440726 | 1 |
| T2DM | adipocyte | *RALGPS2* | 0.607115106 | 0.000502354 | 0.006370945 | 1 |
| T2DM | adipocyte | *TMEM59* | 0.775426282 | 0.002349983 | 0.018271006 | 1 |
| T2DM | adipocyte | *MRPL37* | 0.519530518 | 0.003856688 | 0.025028633 | 1 |
| T2DM | adipocyte | *QSOX1* | 0.778094028 | 0.001145861 | 0.010998516 | 1 |
| T2DM | adipocyte | *RAP1A* | 0.771611773 | 0.000392096 | 0.005550157 | 1 |
| T2DM | adipocyte | *LEPR* | 0.444714963 | 0.009651803 | 0.046040127 | 1 |
| T2DM | adipocyte | *SMG7* | 0.742583318 | 0.004078186 | 0.026030006 | 1 |
| T2DM | adipocyte | *NCF2* | 1.133497521 | 4.22E-05 | 0.001215782 | 0.265040475 |
| T2DM | adipocyte | *RGS2* | 1.20677052 | 0.001351612 | 0.012351141 | 1 |
| T2DM | adipocyte | *OLFML3* | 0.626549858 | 0.004602447 | 0.028174864 | 1 |
| T2DM | adipocyte | *PLA2G2D* | -1.901428539 | 0.000736084 | 0.008338307 | 1 |
| T2DM | adipocyte | *DR1* | 0.943373092 | 0.004788141 | 0.028912435 | 1 |
| T2DM | adipocyte | *PRRC2C* | 0.63517815 | 0.0021692 | 0.017197683 | 1 |
| T2DM | adipocyte | *TNFSF4* | -0.523472503 | 0.000262631 | 0.004159091 | 1 |
| T2DM | adipocyte | *CD3EAP* | -0.478245255 | 6.39E-05 | 0.001613043 | 0.401647662 |
| T2DM | adipocyte | *KMT2A* | 0.817625442 | 0.001038438 | 0.010445535 | 1 |
| T2DM | adipocyte | *ATF6* | 0.695232144 | 0.003144761 | 0.022214732 | 1 |
| T2DM | adipocyte | *ELOVL4* | 0.206042391 | 0.000404379 | 0.00563134 | 1 |
| T2DM | adipocyte | *SGIP1* | 0.54975099 | 0.001300303 | 0.012083315 | 1 |
| T2DM | adipocyte | *FOXO3* | 0.661244695 | 0.005195516 | 0.030297489 | 1 |
| T2DM | adipocyte | *ABCG2* | 1.236278879 | 0.000118017 | 0.002518058 | 0.741973759 |
| T2DM | adipocyte | *SPP1* | 0.583013732 | 0.007468857 | 0.039228656 | 1 |
| T2DM | adipocyte | *MFSD1* | 0.852838238 | 0.000864594 | 0.009291798 | 1 |
| T2DM | adipocyte | *DNAH7* | -0.203154649 | 1.23E-05 | 0.000707371 | 0.077313341 |
| T2DM | adipocyte | *SENP5* | 0.722725923 | 0.005000249 | 0.02954797 | 1 |
| T2DM | adipocyte | *PTBP3* | 0.913911923 | 3.64E-05 | 0.001121192 | 0.228723166 |
| T2DM | adipocyte | *FKBP15* | 0.79537558 | 0.000339965 | 0.005088951 | 1 |
| T2DM | adipocyte | *CNTRL* | 0.753415218 | 0.003595876 | 0.023999231 | 1 |
| T2DM | adipocyte | *BSPRY* | 6.071620623 | 0.00205408 | 0.01648866 | 1 |
| T2DM | adipocyte | *HDHD3* | -0.066646452 | 0.007886286 | 0.040841088 | 1 |
| T2DM | adipocyte | *TTLL5* | 0.837862338 | 0.002755386 | 0.020260946 | 1 |
| T2DM | adipocyte | *ATAD2B* | 0.722177739 | 0.004470427 | 0.027717528 | 1 |
| T2DM | adipocyte | *HOXB3* | -0.341458288 | 0.001419412 | 0.012784877 | 1 |
| T2DM | adipocyte | *GORAB* | 0.727427844 | 0.008230563 | 0.041544539 | 1 |
| T2DM | adipocyte | *MRC1* | 0.41501117 | 0.001905733 | 0.015641442 | 1 |
| T2DM | adipocyte | *IQSEC3* | -0.793795321 | 0.009406835 | 0.045527921 | 1 |
| T2DM | adipocyte | *DNAJC15* | 0.841462783 | 0.008087307 | 0.041430225 | 1 |
| T2DM | adipocyte | *SMAD9* | 1.161151873 | 1.89E-05 | 0.000811251 | 0.118855822 |

| | | | | | | |
|---|---|---|---|---|---|---|
| T2DM | adipocyte | *APAF1* | 0.444069844 | 0.009099593 | 0.044377023 | 1 |
| T2DM | adipocyte | *ADRA1A* | 0.058121312 | 0.001136188 | 0.010939068 | 1 |
| T2DM | adipocyte | *CRISPLD1* | 1.352290637 | 7.15E-06 | 0.000534934 | 0.04493448 |
| T2DM | adipocyte | *TEX14* | 0.600179752 | 8.71E-05 | 0.001970146 | 0.547700578 |
| T2DM | adipocyte | *ECHDC2* | -0.143086218 | 0.004342014 | 0.02716243 | 1 |
| T2DM | adipocyte | *A1BG* | 1.109928622 | 0.000234735 | 0.003883633 | 1 |
| T2DM | adipocyte | *SEC22A* | 0.420200402 | 0.000242343 | 0.003967742 | 1 |
| T2DM | adipocyte | *POPDC2* | 0.921445903 | 0.008643914 | 0.042959913 | 1 |
| T2DM | adipocyte | *RNF115* | 0.840287616 | 0.000615093 | 0.00733793 | 1 |
| T2DM | adipocyte | *TNFSF10* | 0.64785499 | 0.00392214 | 0.02524384 | 1 |
| T2DM | adipocyte | *GPSM2* | 0.54971554 | 0.001502767 | 0.013241122 | 1 |
| T2DM | adipocyte | *ACVR2A* | 0.876413839 | 0.000458125 | 0.006000481 | 1 |
| T2DM | adipocyte | *FLT3* | 2.183967441 | 3.75E-07 | 7.37E-05 | 0.002358153 |
| T2DM | adipocyte | *ODF2L* | 0.355329756 | 0.008258315 | 0.041544539 | 1 |
| T2DM | adipocyte | *PMS2* | 0.707442328 | 0.003608708 | 0.024033845 | 1 |
| T2DM | adipocyte | *SMU1* | 0.810590182 | 0.008768643 | 0.043128627 | 1 |
| T2DM | adipocyte | *SSPN* | 0.266867213 | 0.003546426 | 0.023800752 | 1 |
| T2DM | adipocyte | *PKN1* | 0.613973643 | 0.009503294 | 0.045678294 | 1 |
| T2DM | adipocyte | *EBPL* | 0.211910655 | 0.00825341 | 0.041544539 | 1 |
| T2DM | adipocyte | *NLN* | 0.256203225 | 0.006509391 | 0.03571077 | 1 |
| T2DM | adipocyte | *ATF1* | 0.632181746 | 0.004768237 | 0.0289083 | 1 |
| T2DM | adipocyte | *ARHGAP9* | 0.618237316 | 0.008112178 | 0.041444425 | 1 |
| T2DM | adipocyte | *SPATS2* | 0.597203904 | 0.009483103 | 0.045678294 | 1 |
| T2DM | adipocyte | *IKZF4* | -0.603113614 | 0.009133045 | 0.044442304 | 1 |
| T2DM | adipocyte | *SARDH* | 3.628396033 | 3.28E-05 | 0.00106579 | 0.206327659 |
| T2DM | adipocyte | *NMI* | 0.654669894 | 0.008114854 | 0.041444425 | 1 |
| T2DM | adipocyte | *RAB38* | 1.183407398 | 0.007918186 | 0.040871623 | 1 |
| T2DM | adipocyte | *AGO2* | 0.849838776 | 0.004500805 | 0.027823562 | 1 |
| T2DM | adipocyte | *MOGAT1* | -0.278551903 | 0.008131253 | 0.041460817 | 1 |
| T2DM | adipocyte | *PEPD* | 0.376602151 | 0.010184763 | 0.047713563 | 1 |
| T2DM | adipocyte | *ATP8A1* | 0.297328833 | 0.001493067 | 0.013202408 | 1 |
| T2DM | adipocyte | *HIF3A* | 0.262376207 | 0.002991146 | 0.021369695 | 1 |
| T2DM | adipocyte | *SERPINB6* | 0.882890409 | 0.005371383 | 0.030981545 | 1 |
| T2DM | adipocyte | *GLO1* | 0.971871514 | 0.000112857 | 0.002446652 | 0.709529196 |
| T2DM | adipocyte | *EREG* | 4.13997679 | 0.000987361 | 0.010109996 | 1 |
| T2DM | adipocyte | *CNOT1* | 0.604734678 | 0.003912098 | 0.02524384 | 1 |
| T2DM | adipocyte | *GOT2* | 0.934959598 | 0.000367185 | 0.005285702 | 1 |
| T2DM | adipocyte | *KIF25* | 0.704615511 | 0.010535939 | 0.048849149 | 1 |
| T2DM | adipocyte | *GRK4* | 0.098315549 | 0.001022503 | 0.010362093 | 1 |
| T2DM | adipocyte | *GGA3* | 0.98251968 | 0.00554258 | 0.031707189 | 1 |
| T2DM | adipocyte | *OPRL1* | -0.418742811 | 6.00E-06 | 0.000483445 | 0.037717875 |
| T2DM | adipocyte | *PPDPF* | 1.225300117 | 0.00032492 | 0.004910514 | 1 |
| T2DM | adipocyte | *INSIG2* | 1.124793193 | 0.000905164 | 0.009519534 | 1 |
| T2DM | adipocyte | *GPCPD1* | 0.777295187 | 0.002899595 | 0.021024181 | 1 |
| T2DM | adipocyte | *PANK2* | 0.90090037 | 0.004560701 | 0.028028472 | 1 |
| T2DM | adipocyte | *DSTN* | 0.860448261 | 0.000581809 | 0.007061457 | 1 |
| T2DM | adipocyte | *ITPA* | 1.018302034 | 0.008256892 | 0.041544539 | 1 |
| T2DM | adipocyte | *FAM110A* | 1.564570044 | 0.00494167 | 0.029476547 | 1 |
| T2DM | adipocyte | *ZNF436* | 0.407753671 | 0.000549389 | 0.006785877 | 1 |
| T2DM | adipocyte | *KLC1* | 0.631933376 | 0.005423931 | 0.031125299 | 1 |

| | | | | | | |
|---|---|---|---|---|---|---|
| T2DM | adipocyte | *MCF2L* | 0.360426531 | 0.000225547 | 0.003776376 | 1 |
| T2DM | adipocyte | *F10* | 1.097500609 | 2.79E-05 | 0.00099459 | 0.175114341 |
| T2DM | adipocyte | *CAPNS1* | 0.539251922 | 0.006124787 | 0.034016372 | 1 |
| T2DM | adipocyte | *COX6B1* | 1.030038626 | 0.010341288 | 0.048231217 | 1 |
| T2DM | adipocyte | *PRR12* | 0.397632483 | 0.000189914 | 0.00340696 | 1 |
| T2DM | adipocyte | *BECN1* | 1.085849189 | 0.001298695 | 0.012083315 | 1 |
| T2DM | adipocyte | *RHOJ* | 0.839152956 | 3.79E-05 | 0.001141194 | 0.238473742 |
| T2DM | adipocyte | *PZP* | 0.780000146 | 0.001756605 | 0.014708134 | 1 |
| T2DM | adipocyte | *AIF1L* | 1.067660929 | 7.59E-06 | 0.000560411 | 0.047690493 |
| T2DM | adipocyte | *CANX* | 0.844961384 | 0.001176336 | 0.011188536 | 1 |
| T2DM | adipocyte | *FGD3* | 1.133544362 | 2.04E-05 | 0.000815077 | 0.128505435 |
| T2DM | adipocyte | *RAP1B* | 0.684772549 | 0.002389112 | 0.018520378 | 1 |
| T2DM | adipocyte | *PTPN12* | 0.695260378 | 0.001897194 | 0.015591714 | 1 |
| T2DM | adipocyte | *FGL2* | 0.394612055 | 0.000784899 | 0.008687779 | 1 |
| T2DM | adipocyte | *STEAP4* | 0.456891704 | 0.004532133 | 0.027934825 | 1 |
| T2DM | adipocyte | *MGAT3* | -0.650617663 | 0.000905469 | 0.009519534 | 1 |
| T2DM | adipocyte | *BAIAP2L2* | 0.509808503 | 0.003856235 | 0.025028633 | 1 |
| T2DM | adipocyte | *RIBC2* | 0.233952283 | 0.001532644 | 0.013364399 | 1 |
| T2DM | adipocyte | *EMC4* | 0.603082085 | 0.000768059 | 0.008576883 | 1 |
| T2DM | adipocyte | *OPN1SW* | -0.160429842 | 0.007334765 | 0.038848919 | 1 |
| T2DM | adipocyte | *PDE11A* | -0.161246721 | 0.005179264 | 0.030234016 | 1 |
| T2DM | adipocyte | *VPS13C* | 0.510340372 | 0.005872664 | 0.033072047 | 1 |
| T2DM | adipocyte | *SERGEF* | 1.038827253 | 0.000412918 | 0.005655801 | 1 |
| T2DM | adipocyte | *PLD2* | 0.558363022 | 0.000435679 | 0.005815531 | 1 |
| T2DM | adipocyte | *SLC44A2* | 0.952090445 | 0.000726986 | 0.008280007 | 1 |
| T2DM | adipocyte | *EPB41L4A* | 0.765835795 | 0.0029121 | 0.021024181 | 1 |
| T2DM | adipocyte | *CDO1* | -0.40359913 | 0.005110428 | 0.029999308 | 1 |
| T2DM | adipocyte | *SHC2* | 0.297148466 | 0.003252883 | 0.022672808 | 1 |
| T2DM | adipocyte | *RPL36* | 0.781329372 | 0.000420526 | 0.005685695 | 1 |
| T2DM | adipocyte | *GTPBP3* | -0.462592469 | 0.002624401 | 0.019596375 | 1 |
| T2DM | adipocyte | *BST2* | 1.048125462 | 3.79E-05 | 0.001141194 | 0.238509597 |
| T2DM | adipocyte | *PAK4* | 1.043171752 | 0.010861781 | 0.049988844 | 1 |
| T2DM | adipocyte | *TMEM160* | 1.937026736 | 0.000211878 | 0.00360662 | 1 |
| T2DM | adipocyte | *CLIP1* | 0.574440429 | 0.010128656 | 0.047557029 | 1 |
| T2DM | adipocyte | *C12orf65* | 0.678791037 | 0.00024627 | 0.003980202 | 1 |
| T2DM | adipocyte | *PKDREJ* | -0.249177658 | 0.004478121 | 0.027737878 | 1 |
| T2DM | adipocyte | *LILRB2* | 1.408989614 | 3.43E-05 | 0.001075499 | 0.215877464 |
| T2DM | adipocyte | *COX4I2* | 1.376794788 | 0.004792403 | 0.028912435 | 1 |
| T2DM | adipocyte | *PPT1* | 0.610538964 | 0.010547135 | 0.048865025 | 1 |
| T2DM | adipocyte | *TBC1D5* | 0.553180826 | 0.007432807 | 0.0391374 | 1 |
| T2DM | adipocyte | *AOC2* | -0.763259584 | 0.001455074 | 0.012939249 | 1 |
| T2DM | adipocyte | *EXOC4* | 0.658793218 | 0.000123694 | 0.002586077 | 0.777663355 |
| T2DM | adipocyte | *NPHP4* | 1.197406636 | 0.004641039 | 0.028355893 | 1 |
| T2DM | adipocyte | *GCH1* | 0.521856907 | 0.000634652 | 0.007500112 | 1 |
| T2DM | adipocyte | *MATN3* | -1.591784376 | 0.000716352 | 0.008203473 | 1 |
| T2DM | adipocyte | *EMILIN2* | 0.249336372 | 0.008408904 | 0.042024469 | 1 |
| T2DM | adipocyte | *TRIM5* | 0.675029965 | 0.003857602 | 0.025028633 | 1 |
| T2DM | adipocyte | *TRIM22* | 0.592382583 | 0.002898197 | 0.021024181 | 1 |
| T2DM | adipocyte | *CARD6* | 0.705657401 | 0.008082889 | 0.041430225 | 1 |
| T2DM | adipocyte | *XAF1* | 0.388507107 | 0.000341717 | 0.005103031 | 1 |

| | | | | | | |
|---|---|---|---|---|---|---|
| T2DM | adipocyte | *TERF2* | 0.729236749 | 0.010059221 | 0.04740804 | 1 |
| T2DM | adipocyte | *PTPRA* | 0.673529154 | 0.003584843 | 0.02395102 | 1 |
| T2DM | adipocyte | *CTNNBL1* | 0.734522821 | 0.009480767 | 0.045678294 | 1 |
| T2DM | adipocyte | *DMGDH* | -0.67194466 | 0.001737461 | 0.014603498 | 1 |
| T2DM | adipocyte | *TRPC4* | -0.759377869 | 0.001063107 | 0.010575556 | 1 |
| T2DM | adipocyte | *CNDP2* | 1.358591353 | 0.000349155 | 0.005151093 | 1 |
| T2DM | adipocyte | *FAM83F* | -1.283697185 | 0.000398325 | 0.005589884 | 1 |
| T2DM | adipocyte | *TMTC1* | 0.649846888 | 0.005128847 | 0.030051316 | 1 |
| T2DM | adipocyte | *IPO8* | 0.893384724 | 0.003547209 | 0.023800752 | 1 |
| T2DM | adipocyte | *SWAP70* | 0.860759671 | 0.000168882 | 0.003217451 | 1 |
| T2DM | adipocyte | *SBF2* | 0.594211795 | 0.005220051 | 0.030387463 | 1 |
| T2DM | adipocyte | *MICAL2* | 0.268351631 | 0.009073723 | 0.044290759 | 1 |
| T2DM | adipocyte | *EIF2S1* | 0.801483423 | 0.009613637 | 0.045962686 | 1 |
| T2DM | adipocyte | *EDEM1* | 0.946486543 | 0.005495095 | 0.031464171 | 1 |
| T2DM | adipocyte | *SYT6* | 1.752143904 | 0.000972241 | 0.010020462 | 1 |
| T2DM | adipocyte | *VAV3* | 0.557335153 | 0.000465393 | 0.006057822 | 1 |
| T2DM | adipocyte | *AP4B1* | 0.638956533 | 0.003969031 | 0.025488558 | 1 |
| T2DM | adipocyte | *PPHLN1* | 0.442721579 | 0.006878716 | 0.037121449 | 1 |
| T2DM | adipocyte | *YWHAQ* | 0.849034948 | 0.000379469 | 0.005434446 | 1 |
| T2DM | adipocyte | *KIAA1468* | 0.817873095 | 0.000560221 | 0.006892587 | 1 |
| T2DM | adipocyte | *ECHDC3* | -0.260312995 | 0.006976399 | 0.037359985 | 1 |
| T2DM | adipocyte | *IL15RA* | 1.206360335 | 0.000176681 | 0.003311883 | 1 |
| T2DM | adipocyte | *KLRD1* | 0.765582541 | 0.00830355 | 0.04166354 | 1 |
| T2DM | adipocyte | *PRH2* | -0.562464026 | 0.001578295 | 0.013686543 | 1 |
| T2DM | adipocyte | *DDB2* | 0.804660762 | 6.37E-05 | 0.001613043 | 0.400562279 |
| T2DM | adipocyte | *ACP2* | 0.370871459 | 0.008245188 | 0.041544539 | 1 |
| T2DM | adipocyte | *APLNR* | 0.785415136 | 0.000359857 | 0.005239142 | 1 |
| T2DM | adipocyte | *TMEM165* | 0.410898204 | 0.009492052 | 0.045678294 | 1 |
| T2DM | adipocyte | *WDR36* | 0.88618615 | 0.004274647 | 0.026874704 | 1 |
| T2DM | adipocyte | *OSTF1* | 1.089476395 | 8.65E-05 | 0.001970098 | 0.543746972 |
| T2DM | adipocyte | *P2RX4* | 0.453859961 | 1.17E-05 | 0.000693307 | 0.07349055 |
| T2DM | adipocyte | *SRPK2* | 0.680627156 | 0.003077591 | 0.021863065 | 1 |
| T2DM | adipocyte | *ANKRD6* | 0.718622608 | 0.004758197 | 0.028875276 | 1 |
| T2DM | adipocyte | *NT5E* | 0.499932231 | 2.11E-05 | 0.000820178 | 0.132868758 |
| T2DM | adipocyte | *ITGA7* | -0.104717847 | 0.009628258 | 0.045962686 | 1 |
| T2DM | adipocyte | *FAIM2* | 1.446440877 | 0.00562669 | 0.032129882 | 1 |
| T2DM | adipocyte | *MICAL1* | 0.870820028 | 4.86E-06 | 0.000449002 | 0.030532108 |
| T2DM | adipocyte | *SMYD5* | -0.508064275 | 0.009529926 | 0.045736368 | 1 |
| T2DM | adipocyte | *KCNMB4* | 0.441537428 | 0.004854535 | 0.029094817 | 1 |
| T2DM | adipocyte | *USP15* | 0.609289161 | 0.003341737 | 0.02302506 | 1 |
| T2DM | adipocyte | *MPHOSPH6* | 1.001510418 | 0.000452964 | 0.005967377 | 1 |
| T2DM | adipocyte | *SP110* | 0.599646638 | 0.001347475 | 0.012351141 | 1 |
| T2DM | adipocyte | *USP37* | 0.583732651 | 0.006696731 | 0.036389239 | 1 |
| T2DM | adipocyte | *TMBIM1* | -0.329310004 | 0.003799593 | 0.024853488 | 1 |
| T2DM | adipocyte | *ARMC9* | 1.008604993 | 0.000726094 | 0.008280007 | 1 |
| T2DM | adipocyte | *ANKRD36* | 0.720039266 | 0.001678854 | 0.014340975 | 1 |
| T2DM | adipocyte | *APPL2* | 0.671058719 | 0.009553811 | 0.045746239 | 1 |
| T2DM | adipocyte | *SLC41A2* | 0.759587865 | 0.006393332 | 0.035166124 | 1 |
| T2DM | adipocyte | *RNASEH2B* | 0.437165499 | 0.008217411 | 0.041544539 | 1 |
| T2DM | adipocyte | *LRCH1* | 0.862564406 | 1.37E-05 | 0.00073252 | 0.085982155 |

| | | | | | | |
|---|---|---|---|---|---|---|
| T2DM | adipocyte | *RCBTB2* | 0.734564137 | 0.007132519 | 0.03800182 | 1 |
| T2DM | adipocyte | *KDELR2* | 1.165659101 | 0.001684773 | 0.014365585 | 1 |
| T2DM | adipocyte | *NACAD* | -1.136271143 | 0.001255356 | 0.01176218 | 1 |
| T2DM | adipocyte | *ALPK3* | -0.407247126 | 0.002480623 | 0.018972842 | 1 |
| T2DM | adipocyte | *TM6SF1* | 0.737156122 | 2.51E-06 | 0.000272372 | 0.015797588 |
| T2DM | adipocyte | *EPRS* | 0.883153268 | 0.008640647 | 0.042959913 | 1 |
| T2DM | adipocyte | *IL1RN* | 1.333343051 | 3.23E-05 | 0.00106579 | 0.20310753 |
| T2DM | adipocyte | *PRPF4* | 0.303764808 | 0.001349073 | 0.012351141 | 1 |
| T2DM | adipocyte | *GARNL3* | 0.98159536 | 0.000186333 | 0.003395587 | 1 |
| T2DM | adipocyte | *LMX1B* | 2.072385723 | 0.003031484 | 0.021584304 | 1 |
| T2DM | adipocyte | *NOV* | 0.442510907 | 0.005298346 | 0.030672835 | 1 |
| T2DM | adipocyte | *TLN1* | 0.619275051 | 0.002653101 | 0.01973607 | 1 |
| T2DM | adipocyte | *DENND4C* | 0.73176609 | 0.000892181 | 0.009413933 | 1 |
| T2DM | adipocyte | *PPIL1* | 2.636399447 | 0.001878503 | 0.01547857 | 1 |
| T2DM | adipocyte | *TCF19* | 0.307686947 | 0.008069553 | 0.041430225 | 1 |
| T2DM | adipocyte | *FLOT1* | 0.648525548 | 9.90E-05 | 0.002191322 | 0.622335484 |
| T2DM | adipocyte | *MTCH1* | 1.035741836 | 0.001126925 | 0.010905597 | 1 |
| T2DM | adipocyte | *FHDC1* | 1.78308491 | 4.09E-05 | 0.001200631 | 0.256934995 |
| T2DM | adipocyte | *ARRB1* | 0.804232577 | 0.0004395 | 0.005854102 | 1 |
| T2DM | adipocyte | *PRCP* | 0.731365655 | 6.67E-05 | 0.001671925 | 0.419653295 |
| T2DM | adipocyte | *SULF1* | 0.400842079 | 0.001051044 | 0.010522157 | 1 |
| T2DM | adipocyte | *NEK1* | 0.546322052 | 0.010404338 | 0.04844569 | 1 |
| T2DM | adipocyte | *DDX60* | 0.991386099 | 1.81E-05 | 0.000811251 | 0.113729208 |
| T2DM | adipocyte | *CASP1* | 1.14623416 | 0.000671285 | 0.007766803 | 1 |
| T2DM | adipocyte | *MAP2K5* | 0.681537338 | 0.003866419 | 0.025034168 | 1 |
| T2DM | adipocyte | *NDUFAF1* | -0.291752875 | 0.009861135 | 0.046754867 | 1 |
| T2DM | adipocyte | *UACA* | 0.654316253 | 0.006943269 | 0.037214266 | 1 |
| T2DM | adipocyte | *SPTBN5* | 0.194785005 | 6.86E-05 | 0.001683725 | 0.431033646 |
| T2DM | adipocyte | *DNASE2B* | 2.657413221 | 0.000749371 | 0.008440726 | 1 |
| T2DM | adipocyte | *IFT172* | 0.706498571 | 0.002136226 | 0.017000572 | 1 |
| T2DM | adipocyte | *ADCY3* | 0.736702629 | 0.006896604 | 0.037133048 | 1 |
| T2DM | adipocyte | *LHCGR* | -0.428163129 | 0.000468471 | 0.006072732 | 1 |
| T2DM | adipocyte | *CENPO* | 1.847249263 | 0.001248107 | 0.011729218 | 1 |
| T2DM | adipocyte | *MYOF* | 0.310240051 | 0.000140222 | 0.002831013 | 0.881575368 |
| T2DM | adipocyte | *BTBD16* | -0.416541805 | 0.001516538 | 0.013316309 | 1 |
| T2DM | adipocyte | *DNAJC13* | 0.866155967 | 0.001306337 | 0.012095643 | 1 |
| T2DM | adipocyte | *RPS24* | 0.810214443 | 0.001452027 | 0.012939249 | 1 |
| T2DM | adipocyte | *BARD1* | 0.834899484 | 0.005264527 | 0.030505142 | 1 |
| T2DM | adipocyte | *CARF* | 0.332538407 | 0.000462058 | 0.006039415 | 1 |
| T2DM | adipocyte | *FAM117B* | 1.058595135 | 6.89E-05 | 0.001685133 | 0.433079084 |
| T2DM | adipocyte | *SENP7* | 0.781045944 | 0.000304682 | 0.004674872 | 1 |
| T2DM | adipocyte | *COX17* | 1.165384951 | 0.000587069 | 0.007111569 | 1 |
| T2DM | adipocyte | *HERC3* | 0.779862665 | 0.004705123 | 0.028691668 | 1 |
| T2DM | adipocyte | *PRKG2* | 0.550382208 | 0.00423088 | 0.026706366 | 1 |
| T2DM | adipocyte | *BBS7* | 1.054639996 | 0.003380633 | 0.023102217 | 1 |
| T2DM | adipocyte | *KIAA1109* | 0.684294447 | 0.00204261 | 0.016459063 | 1 |
| T2DM | adipocyte | *PRDM5* | 0.768402701 | 0.000149915 | 0.002992119 | 0.942517403 |
| T2DM | adipocyte | *NAAA* | 0.433146056 | 0.002602092 | 0.019521902 | 1 |
| T2DM | adipocyte | *NUP54* | 0.398713172 | 0.008191483 | 0.041544539 | 1 |
| T2DM | adipocyte | *ANXA3* | 1.216698876 | 6.81E-06 | 0.000524448 | 0.042826518 |

| | | | | | | |
|---|---|---|---|---|---|---|
| T2DM | adipocyte | *PPA2* | 0.565121159 | 0.000982695 | 0.010086671 | 1 |
| T2DM | adipocyte | *ETV6* | 0.814435169 | 0.000195635 | 0.003484289 | 1 |
| T2DM | adipocyte | *PRICKLE1* | 0.700574794 | 6.79E-05 | 0.001674964 | 0.426635906 |
| T2DM | adipocyte | *C1RL* | 0.356901821 | 0.003835483 | 0.024988274 | 1 |
| T2DM | adipocyte | *CLSTN3* | 0.38545978 | 0.006621746 | 0.036169344 | 1 |
| T2DM | adipocyte | *POC1B* | 0.9308265 | 0.006389436 | 0.035166124 | 1 |
| T2DM | adipocyte | *GAS2L3* | 1.257514668 | 6.76E-05 | 0.001674964 | 0.425309236 |
| T2DM | adipocyte | *ACVRL1* | 0.31651146 | 0.008512664 | 0.042475493 | 1 |
| T2DM | adipocyte | *LPAR6* | 0.7772991 | 0.001077806 | 0.010621557 | 1 |
| T2DM | adipocyte | *RHOF* | -0.379467331 | 0.005881161 | 0.033072047 | 1 |
| T2DM | adipocyte | *TMX1* | 0.958625931 | 0.006136646 | 0.034031853 | 1 |
| T2DM | adipocyte | *STON2* | 0.661449096 | 0.000224782 | 0.003776376 | 1 |
| T2DM | adipocyte | *PTGR2* | -0.107727273 | 0.009117781 | 0.044402393 | 1 |
| T2DM | adipocyte | *AK7* | 2.940897851 | 0.000202395 | 0.003564307 | 1 |
| T2DM | adipocyte | *FBLN5* | 0.899606498 | 0.000304363 | 0.004674872 | 1 |
| T2DM | adipocyte | *TCF12* | 0.649315222 | 0.001618013 | 0.013915793 | 1 |
| T2DM | adipocyte | *SRP14* | 0.679047423 | 8.38E-05 | 0.001941003 | 0.527019318 |
| T2DM | adipocyte | *UBE2Q2* | 0.912089745 | 0.000361578 | 0.005239142 | 1 |
| T2DM | adipocyte | *HMG20A* | 1.020282125 | 2.84E-06 | 0.000294676 | 0.017850812 |
| T2DM | adipocyte | *MAN2C1* | -0.113730888 | 5.27E-05 | 0.00142143 | 0.331193085 |
| T2DM | adipocyte | *TPM1* | 0.791972636 | 2.82E-05 | 0.00099459 | 0.177037022 |
| T2DM | adipocyte | *IGF1R* | 0.76177836 | 1.85E-05 | 0.000811251 | 0.116030552 |
| T2DM | adipocyte | *ZFHX3* | 0.67836978 | 0.004339959 | 0.02716243 | 1 |
| T2DM | adipocyte | *NUDT7* | -0.268510534 | 0.006785617 | 0.036776874 | 1 |
| T2DM | adipocyte | *CMTM3* | 1.148711714 | 0.002020306 | 0.016326047 | 1 |
| T2DM | adipocyte | *MYO19* | 1.1178276 | 1.37E-06 | 0.000179833 | 0.00863198 |
| T2DM | adipocyte | *RASL10B* | -0.395535951 | 0.002416832 | 0.018666613 | 1 |
| T2DM | adipocyte | *LRRC46* | -0.54402862 | 6.62E-07 | 0.00011251 | 0.004162887 |
| T2DM | adipocyte | *ASXL3* | 0.989336668 | 0.002394953 | 0.018520378 | 1 |
| T2DM | adipocyte | *C18orf8* | -0.419876368 | 0.007761436 | 0.040394163 | 1 |
| T2DM | adipocyte | *TMC6* | 0.197214851 | 0.005199776 | 0.030297489 | 1 |
| T2DM | adipocyte | *TBCD* | 0.512749427 | 0.005907414 | 0.033072047 | 1 |
| T2DM | adipocyte | *FN3KRP* | -0.129665377 | 0.008273855 | 0.041575074 | 1 |
| T2DM | adipocyte | *SECTM1* | 0.653730029 | 0.006493449 | 0.035654421 | 1 |
| T2DM | adipocyte | *WDR45B* | 0.566734529 | 0.002132114 | 0.017000572 | 1 |
| T2DM | adipocyte | *RNF165* | 0.607092928 | 0.005791972 | 0.032892943 | 1 |
| T2DM | adipocyte | *DUS3L* | 0.246383802 | 0.003693318 | 0.024415616 | 1 |
| T2DM | adipocyte | *CCDC97* | 1.124771898 | 0.00146974 | 0.013032798 | 1 |
| T2DM | adipocyte | *SIRT3* | 0.440611585 | 0.000939595 | 0.009764021 | 1 |
| T2DM | adipocyte | *IFITM3* | 1.141323712 | 0.000188101 | 0.003398243 | 1 |
| T2DM | adipocyte | *GEMIN7* | -0.630234768 | 0.008483049 | 0.042361343 | 1 |
| T2DM | adipocyte | *MYO1F* | 0.609039875 | 9.89E-05 | 0.002191322 | 0.621551 |
| T2DM | adipocyte | *CACNG8* | 3.41823286 | 4.43E-06 | 0.000428187 | 0.027832156 |
| T2DM | adipocyte | *RCN3* | 1.14882009 | 6.29E-05 | 0.001608673 | 0.395604388 |
| T2DM | adipocyte | *SH3BGRL3* | 0.879748078 | 0.0005888 | 0.007118824 | 1 |
| T2DM | adipocyte | *SERBP1* | 0.808330824 | 0.007237723 | 0.038464552 | 1 |
| T2DM | adipocyte | *GPR161* | 0.711687758 | 0.002913011 | 0.021024181 | 1 |
| T2DM | adipocyte | *ADCY10* | -0.732998793 | 0.00411214 | 0.026167029 | 1 |
| T2DM | adipocyte | *RGL1* | 0.73171285 | 0.000507114 | 0.006389226 | 1 |
| T2DM | adipocyte | *TARS2* | 0.489176846 | 0.007955799 | 0.040964871 | 1 |

| | | | | | | |
|---|---|---|---|---|---|---|
| T2DM | adipocyte | *SNX27* | 0.829107696 | 0.004795995 | 0.028912435 | 1 |
| T2DM | adipocyte | *IKBKE* | 0.877549185 | 0.005159797 | 0.030176414 | 1 |
| T2DM | adipocyte | *TP53BP2* | 1.094271449 | 0.000386186 | 0.005518078 | 1 |
| T2DM | adipocyte | *KCNN3* | 0.819117951 | 0.000165391 | 0.003170158 | 1 |
| T2DM | adipocyte | *RIT1* | 0.915153976 | 0.001961852 | 0.015997616 | 1 |
| T2DM | adipocyte | *CEP170* | 0.456935913 | 0.009168629 | 0.044546498 | 1 |
| T2DM | adipocyte | *GUK1* | 1.025093359 | 0.001134355 | 0.010938173 | 1 |
| T2DM | adipocyte | *CDC42BPA* | 0.508447581 | 0.003251363 | 0.022672808 | 1 |
| T2DM | adipocyte | *CNIH3* | 0.519923548 | 0.008796636 | 0.043206604 | 1 |
| T2DM | adipocyte | *WNT9A* | 0.609776518 | 1.73E-06 | 0.000210988 | 0.010851002 |
| T2DM | adipocyte | *PLEKHA6* | 0.607310075 | 0.000242158 | 0.003967742 | 1 |
| T2DM | adipocyte | *EML4* | 0.657256367 | 0.010079482 | 0.047432414 | 1 |
| T2DM | adipocyte | *EXOC6B* | 0.556256467 | 0.010530439 | 0.048849149 | 1 |
| T2DM | adipocyte | *TEX261* | 1.506282879 | 0.008875518 | 0.043560015 | 1 |
| T2DM | adipocyte | *SLC20A1* | 0.965085602 | 0.000129511 | 0.002660892 | 0.814232849 |
| T2DM | adipocyte | *ZNF385B* | 0.437622444 | 0.000471018 | 0.006093191 | 1 |
| T2DM | adipocyte | *GULP1* | 0.540285036 | 0.000245287 | 0.003980202 | 1 |
| T2DM | adipocyte | *METTL21A* | 1.000656225 | 0.001788103 | 0.014898665 | 1 |
| T2DM | adipocyte | *NBEAL1* | 0.246547929 | 0.000195241 | 0.003484289 | 1 |
| T2DM | adipocyte | *FANCD2* | 1.337625442 | 0.000256402 | 0.00407071 | 1 |
| T2DM | adipocyte | *TAMM41* | 0.743986374 | 0.001817708 | 0.015070735 | 1 |
| T2DM | adipocyte | *OSBPL10* | 0.611020099 | 9.02E-06 | 0.000586394 | 0.056719715 |
| T2DM | adipocyte | *STAC* | 1.337692689 | 0.000510905 | 0.006411295 | 1 |
| T2DM | adipocyte | *ARL6IP5* | 0.815485138 | 0.005324015 | 0.030764784 | 1 |
| T2DM | adipocyte | *LIMD1* | 0.721025791 | 0.000981723 | 0.010086671 | 1 |
| T2DM | adipocyte | *PLA1A* | -0.19324565 | 0.008078923 | 0.041430225 | 1 |
| T2DM | adipocyte | *LYAR* | 1.402324803 | 0.003318577 | 0.02297786 | 1 |
| T2DM | adipocyte | *ATP10D* | 0.403886085 | 0.000510858 | 0.006411295 | 1 |
| T2DM | adipocyte | *CISD2* | 0.655709912 | 0.002704142 | 0.020040713 | 1 |
| T2DM | adipocyte | *USP53* | 0.729158593 | 0.001703746 | 0.014412493 | 1 |
| T2DM | adipocyte | *SSBP2* | 0.665651213 | 0.001607709 | 0.013865111 | 1 |
| T2DM | adipocyte | *BHMT* | -0.149650564 | 3.67E-05 | 0.001125274 | 0.23068112 |
| T2DM | adipocyte | *YIPF5* | 1.041556095 | 0.006644954 | 0.036170411 | 1 |
| T2DM | adipocyte | *PSD2* | 1.035864073 | 0.009457434 | 0.045667347 | 1 |
| T2DM | adipocyte | *FGD2* | 0.583922411 | 0.002858739 | 0.020850222 | 1 |
| T2DM | adipocyte | *TPBG* | 1.177504268 | 2.06E-05 | 0.000815077 | 0.129750605 |
| T2DM | adipocyte | *PHIP* | 0.805904672 | 0.000410222 | 0.005646146 | 1 |
| T2DM | adipocyte | *CCDC25* | 0.895451347 | 2.19E-05 | 0.000830044 | 0.13771129 |
| T2DM | adipocyte | *DOCK5* | 0.910104674 | 0.000128092 | 0.002640372 | 0.80531354 |
| T2DM | adipocyte | *ADHFE1* | -0.100549272 | 0.002346416 | 0.018271006 | 1 |
| T2DM | adipocyte | *MTDH* | 0.702854709 | 0.00705083 | 0.037630362 | 1 |
| T2DM | adipocyte | *MAL2* | -0.258812623 | 0.002625128 | 0.019596375 | 1 |
| T2DM | adipocyte | *FAM135B* | 1.53732109 | 0.002855575 | 0.020850222 | 1 |
| T2DM | adipocyte | *ZNF7* | 0.669299841 | 0.000740302 | 0.008371002 | 1 |
| T2DM | adipocyte | *SLC39A4* | -0.514129359 | 0.003358355 | 0.02302506 | 1 |
| T2DM | adipocyte | *ZNF462* | 0.829990131 | 0.006585376 | 0.036001965 | 1 |
| T2DM | adipocyte | *SEC16A* | 0.372538297 | 0.005150028 | 0.030147322 | 1 |
| T2DM | adipocyte | *RSU1* | 0.735810951 | 0.001625295 | 0.01395933 | 1 |
| T2DM | adipocyte | *GLUD1* | 0.874357775 | 0.000124224 | 0.002586077 | 0.780995261 |
| T2DM | adipocyte | *HTR7* | 1.677230592 | 5.89E-06 | 0.000480968 | 0.037034185 |

| | | | | | | |
|---|---|---|---|---|---|---|
| T2DM | adipocyte | *HSD17B12* | -0.255688562 | 1.92E-07 | 4.65E-05 | 0.001208565 |
| T2DM | adipocyte | *APIP* | -0.091999262 | 0.000185045 | 0.003395587 | 1 |
| T2DM | adipocyte | *PTPRJ* | 0.663039157 | 7.20E-05 | 0.001748913 | 0.452968523 |
| T2DM | adipocyte | *C11orf49* | 0.574555771 | 0.003802935 | 0.024853488 | 1 |
| T2DM | adipocyte | *ENDOD1* | 1.232872896 | 1.65E-05 | 0.000780926 | 0.103714757 |
| T2DM | adipocyte | *CAPN5* | 0.67824038 | 0.000227879 | 0.003800196 | 1 |
| T2DM | adipocyte | *ZC3H12C* | 0.97131676 | 0.000389873 | 0.00554555 | 1 |
| T2DM | adipocyte | *C11orf52* | -2.552265357 | 0.000207141 | 0.003597496 | 1 |
| T2DM | adipocyte | *TMC2* | -1.157335125 | 0.004666382 | 0.028483051 | 1 |
| T2DM | adipocyte | *MS4A3* | -19.30758114 | 0.004819506 | 0.028912435 | 1 |
| T2DM | adipocyte | *PLCH2* | 6.316497239 | 8.41E-05 | 0.001941003 | 0.528909605 |
| T2DM | adipocyte | *FEZ1* | 1.43974977 | 5.89E-06 | 0.000480968 | 0.037034511 |
| T2DM | adipocyte | *DUSP15* | -0.347916772 | 0.010869214 | 0.049988844 | 1 |
| T2DM | adipocyte | *DOC2A* | -0.504302092 | 0.000605694 | 0.007267173 | 1 |
| T2DM | adipocyte | *TMCO3* | 1.071840931 | 0.000395081 | 0.005568695 | 1 |
| T2DM | adipocyte | *SAP18* | 1.016675254 | 0.002787268 | 0.020447552 | 1 |
| T2DM | adipocyte | *LYPD6B* | -0.857145976 | 0.000402499 | 0.00563134 | 1 |
| T2DM | adipocyte | *PDCD4* | 0.889178639 | 0.00034582 | 0.005139885 | 1 |
| T2DM | adipocyte | *CCDC102B* | 0.86061998 | 0.000876744 | 0.009344489 | 1 |
| T2DM | adipocyte | *RGS18* | 1.223820944 | 5.45E-05 | 0.001458864 | 0.342833054 |
| T2DM | adipocyte | *C11orf53* | -2.414625293 | 0.009784821 | 0.046465231 | 1 |
| T2DM | adipocyte | *DOCK1* | 0.487441472 | 0.009540318 | 0.045746239 | 1 |
| T2DM | adipocyte | *TEX12* | -5.3479217 | 0.004045928 | 0.025876652 | 1 |
| T2DM | adipocyte | *PIP4K2A* | 0.534444983 | 0.002485481 | 0.0189869 | 1 |
| T2DM | adipocyte | *PRSS53* | -0.347911058 | 0.006796022 | 0.036779587 | 1 |
| T2DM | adipocyte | *IPMK* | 1.025272771 | 0.000156726 | 0.003069586 | 0.985337166 |
| T2DM | adipocyte | *GXYLT1* | 0.848371123 | 0.004574178 | 0.028052873 | 1 |
| T2DM | adipocyte | *FAM177A1* | 1.08906613 | 7.98E-05 | 0.00187981 | 0.501909399 |
| T2DM | adipocyte | *ANKRD50* | 0.954152208 | 0.004088439 | 0.026055765 | 1 |
| T2DM | adipocyte | *PTPRO* | 0.642552893 | 0.001799942 | 0.014968563 | 1 |
| T2DM | adipocyte | *VTI1A* | 0.600162142 | 0.000947462 | 0.009829531 | 1 |
| T2DM | adipocyte | *AKR1C2* | -0.494163244 | 0.001088052 | 0.010645331 | 1 |
| T2DM | adipocyte | *INPP1* | 0.811100112 | 7.26E-05 | 0.001755852 | 0.456521498 |
| T2DM | adipocyte | *MFSD6* | 0.762173772 | 0.007857667 | 0.040760025 | 1 |
| T2DM | adipocyte | *ASAP2* | 0.756268701 | 0.000847216 | 0.009161281 | 1 |
| T2DM | adipocyte | *BICD1* | 0.756158255 | 0.001133186 | 0.010938173 | 1 |
| T2DM | adipocyte | *RABGAP1L* | 0.62035515 | 8.57E-05 | 0.001965411 | 0.539062506 |
| T2DM | adipocyte | *ARL14EP* | -0.644559217 | 0.000153522 | 0.003025687 | 0.965194031 |
| T2DM | adipocyte | *G6PC2* | 29.7675433 | 1.52E-05 | 0.000758566 | 0.095579353 |
| T2DM | adipocyte | *POC5* | -0.315334081 | 0.005424765 | 0.031125299 | 1 |
| T2DM | adipocyte | *CWF19L2* | 0.636158133 | 0.004790059 | 0.028912435 | 1 |
| T2DM | adipocyte | *ZFP36L2* | 1.096258459 | 6.95E-07 | 0.000115022 | 0.004370837 |
| T2DM | adipocyte | *TMEM123* | 0.805062834 | 0.002497385 | 0.0190529 | 1 |
| T2DM | adipocyte | *CLEC4F* | 2.120937655 | 1.98E-05 | 0.00081257 | 0.124355278 |
| T2DM | adipocyte | *SAR1B* | 0.625702834 | 0.000205822 | 0.003597496 | 1 |
| T2DM | adipocyte | *TMEM87B* | 0.439035202 | 0.006003735 | 0.033462306 | 1 |
| T2DM | adipocyte | *PTPRR* | 0.80050968 | 0.002091497 | 0.016708059 | 1 |
| T2DM | adipocyte | *PLA2R1* | 0.595974087 | 0.000531234 | 0.006574546 | 1 |
| T2DM | adipocyte | *FRMD1* | -0.537788999 | 0.000419737 | 0.005685695 | 1 |
| T2DM | adipocyte | *ASAP1* | 0.476136703 | 0.010176803 | 0.047711827 | 1 |

| | | | | | | |
|---|---|---|---|---|---|---|
| T2DM | adipocyte | *LPCAT1* | 1.102209875 | 0.000101976 | 0.002241686 | 0.641122203 |
| T2DM | adipocyte | *ZDHHC7* | 0.824175101 | 0.000662476 | 0.007725842 | 1 |
| T2DM | adipocyte | *FAM92B* | -7.971437932 | 0.000627426 | 0.007442698 | 1 |
| T2DM | adipocyte | *JAZF1* | -0.687593177 | 8.43E-05 | 0.001941003 | 0.529893923 |
| T2DM | adipocyte | *MCOLN2* | 0.298601313 | 0.006014725 | 0.033476156 | 1 |
| T2DM | adipocyte | *PPP2R5E* | 0.448186375 | 0.001615986 | 0.013915793 | 1 |
| T2DM | adipocyte | *ANKRD29* | 0.635800463 | 1.63E-05 | 0.000780895 | 0.102297184 |
| T2DM | adipocyte | *THY1* | 0.874920889 | 2.37E-05 | 0.000879907 | 0.148704348 |
| T2DM | adipocyte | *UBASH3B* | 0.645231737 | 0.008210358 | 0.041544539 | 1 |
| T2DM | adipocyte | *ROBO4* | 0.398039711 | 0.002623483 | 0.019596375 | 1 |
| T2DM | adipocyte | *PRKCA* | 0.707253467 | 0.000571195 | 0.007000196 | 1 |
| T2DM | adipocyte | *LRRK1* | 0.921454054 | 3.82E-05 | 0.001144351 | 0.240313649 |
| T2DM | adipocyte | *UCHL1* | -0.620795896 | 0.002432522 | 0.018741746 | 1 |
| T2DM | adipocyte | *GBP5* | 0.466328341 | 0.006542274 | 0.035859878 | 1 |
| T2DM | adipocyte | *PDLIM3* | 0.319294432 | 0.004967843 | 0.029508585 | 1 |
| T2DM | adipocyte | *L3MBTL4* | 0.901687691 | 0.000254385 | 0.0040489 | 1 |
| T2DM | adipocyte | *PDE1C* | 0.645970095 | 0.003420819 | 0.023240953 | 1 |
| T2DM | adipocyte | *TSEN2* | 0.471576751 | 1.49E-05 | 0.000753745 | 0.093464409 |
| T2DM | adipocyte | *PPP4R1* | 0.884541067 | 0.001827511 | 0.015117845 | 1 |
| T2DM | adipocyte | *CDK19* | 0.679582199 | 0.004962888 | 0.029508585 | 1 |
| T2DM | adipocyte | *TTC39B* | 0.495653445 | 0.001090439 | 0.010645331 | 1 |
| T2DM | adipocyte | *PDIA4* | 1.318608462 | 5.43E-05 | 0.001458864 | 0.34139159 |
| T2DM | adipocyte | *DEPTOR* | 0.830002631 | 7.35E-05 | 0.001763187 | 0.461954897 |
| T2DM | adipocyte | *UGT2B4* | -0.565452863 | 0.000236192 | 0.003897484 | 1 |
| T2DM | adipocyte | *BATF* | 1.677742192 | 3.25E-05 | 0.00106579 | 0.204382481 |
| T2DM | adipocyte | *NDUFAF6* | -0.491914766 | 0.001053909 | 0.010534064 | 1 |
| T2DM | adipocyte | *ANKRD9* | -0.696056893 | 0.001084953 | 0.010641344 | 1 |
| T2DM | adipocyte | *GDF6* | 1.374229753 | 3.96E-05 | 0.00117329 | 0.248737547 |
| T2DM | adipocyte | *NODAL* | -0.977897297 | 0.00471417 | 0.028718982 | 1 |
| T2DM | adipocyte | *VPS8* | 0.586561007 | 4.20E-05 | 0.001215782 | 0.264128323 |
| T2DM | adipocyte | *NTAN1* | 0.550792948 | 0.009900572 | 0.046765514 | 1 |
| T2DM | adipocyte | *FCHO2* | 0.818108346 | 0.000184991 | 0.003395587 | 1 |
| T2DM | adipocyte | *TIMP4* | 0.47206275 | 0.006091647 | 0.03386223 | 1 |
| T2DM | adipocyte | *TMED6* | -2.295466403 | 0.000372668 | 0.005349235 | 1 |
| T2DM | adipocyte | *CLEC18A* | -0.583201732 | 1.48E-08 | 5.47E-06 | 9.31E-05 |
| T2DM | adipocyte | *IL34* | 1.354643133 | 0.001081247 | 0.010621557 | 1 |
| T2DM | adipocyte | *DYRK1A* | 0.595312142 | 0.005101858 | 0.029999308 | 1 |
| T2DM | adipocyte | *KCNJ15* | -0.271561055 | 0.004971767 | 0.029508585 | 1 |
| T2DM | adipocyte | *CREB3L1* | 0.312686005 | 0.009894447 | 0.046765514 | 1 |
| T2DM | adipocyte | *SLC37A3* | 0.851852563 | 0.005121844 | 0.030038276 | 1 |
| T2DM | adipocyte | *AP3S2* | 0.342180425 | 2.25E-09 | 1.29E-06 | 1.41E-05 |
| T2DM | adipocyte | *C12orf43* | 0.378483751 | 0.001815122 | 0.015070735 | 1 |
| T2DM | adipocyte | *TRIM63* | 0.273161379 | 0.000275625 | 0.00432134 | 1 |
| T2DM | adipocyte | *GALNT14* | 2.070954725 | 4.29E-05 | 0.001226752 | 0.269885417 |
| T2DM | adipocyte | *LRRC43* | -1.49108461 | 0.000672044 | 0.007766803 | 1 |
| T2DM | adipocyte | *MRAS* | -0.172376841 | 0.006580497 | 0.036001965 | 1 |
| T2DM | adipocyte | *CLSTN2* | -0.308781836 | 0.008129879 | 0.041460817 | 1 |
| T2DM | adipocyte | *COLEC12* | 0.73384098 | 3.72E-05 | 0.001129902 | 0.233889771 |
| T2DM | adipocyte | *AUTS2* | 0.599232275 | 0.001127507 | 0.010905597 | 1 |
| T2DM | adipocyte | *CD1E* | 0.846022462 | 2.31E-05 | 0.000864716 | 0.145272239 |

| | | | | | | |
|---|---|---|---|---|---|---|
| T2DM | adipocyte | *NCF1* | 1.711637034 | 1.99E-05 | 0.00081257 | 0.12513576 |
| T2DM | adipocyte | *ELK4* | 0.943977741 | 0.001016771 | 0.010343757 | 1 |
| T2DM | adipocyte | *RNF166* | 0.697724627 | 0.002454044 | 0.01883831 | 1 |
| T2DM | adipocyte | *CDC42SE2* | 0.81515593 | 0.000392345 | 0.005550157 | 1 |
| T2DM | adipocyte | *IFNAR2* | 0.225237393 | 0.005413524 | 0.031125299 | 1 |
| T2DM | adipocyte | *ARHGAP27* | 1.252384942 | 0.000387482 | 0.005524036 | 1 |
| T2DM | adipocyte | *STARD9* | 0.509456394 | 0.000878415 | 0.009344489 | 1 |
| T2DM | adipocyte | *ACE* | 0.758565397 | 0.000349852 | 0.005151093 | 1 |
| T2DM | adipocyte | *EFCAB14* | 0.992421369 | 0.00174404 | 0.014639223 | 1 |
| T2DM | adipocyte | *CALM3* | 0.702303402 | 0.00232867 | 0.018186764 | 1 |
| T2DM | adipocyte | *SLC37A1* | 0.96796504 | 8.94E-05 | 0.002015343 | 0.562280806 |
| T2DM | adipocyte | *C21orf33* | -0.464048343 | 0.009833842 | 0.046660651 | 1 |
| T2DM | adipocyte | *ZNF66* | 0.520648933 | 0.005394243 | 0.031084883 | 1 |
| T2DM | adipocyte | *VAV2* | 0.614502689 | 0.010831468 | 0.049888235 | 1 |
| T2DM | adipocyte | *MPZL3* | 1.456612847 | 8.13E-06 | 0.000574639 | 0.051142869 |
| T2DM | adipocyte | *FDPS* | 0.647206018 | 7.92E-05 | 0.001875094 | 0.498196445 |
| T2DM | adipocyte | *VPS28* | 0.389379533 | 0.0002733 | 0.004295599 | 1 |
| T2DM | adipocyte | *RPL8* | 0.839247034 | 0.000949191 | 0.009831245 | 1 |
| T2DM | adipocyte | *MFSD12* | 1.072374407 | 0.001367976 | 0.012446407 | 1 |
| T2DM | adipocyte | *YDJC* | 3.345103903 | 9.12E-06 | 0.000586394 | 0.057333373 |
| T2DM | adipocyte | *AP2M1* | 0.798398536 | 0.000848521 | 0.009161281 | 1 |
| T2DM | adipocyte | *IKZF3* | 0.633615561 | 0.001527028 | 0.013352465 | 1 |
| T2DM | adipocyte | *ASGR2* | 3.454988775 | 5.72E-06 | 0.000480968 | 0.035950989 |
| T2DM | adipocyte | *JMJD8* | 0.258676182 | 0.008092304 | 0.041430225 | 1 |
| T2DM | adipocyte | *ZG16B* | 21.34553997 | 0.000207732 | 0.003597825 | 1 |
| T2DM | adipocyte | *RBP7* | 1.333645457 | 1.97E-05 | 0.00081257 | 0.123934031 |
| T2DM | adipocyte | *LAPTM5* | 0.622265265 | 5.82E-05 | 0.001523337 | 0.365600973 |
| T2DM | adipocyte | *PEF1* | 1.516198987 | 0.002274619 | 0.017853345 | 1 |
| T2DM | adipocyte | *CAMK2N1* | 1.52649623 | 8.81E-06 | 0.000586394 | 0.055393238 |
| T2DM | adipocyte | *NFIA* | 0.559664339 | 0.004750963 | 0.02885923 | 1 |
| T2DM | adipocyte | *KCNT2* | 0.725083155 | 6.84E-06 | 0.000524448 | 0.043004767 |
| T2DM | adipocyte | *VCAM1* | 0.422801952 | 0.005662398 | 0.032304441 | 1 |
| T2DM | adipocyte | *EXTL2* | -0.233622473 | 0.009992411 | 0.047128496 | 1 |
| T2DM | adipocyte | *ZNF496* | 0.991969947 | 0.000115616 | 0.002489307 | 0.726877698 |
| T2DM | adipocyte | *LMX1A* | 0.207099458 | 0.001434404 | 0.012846296 | 1 |
| T2DM | adipocyte | *DISC1* | 0.518529586 | 0.004090511 | 0.026055765 | 1 |
| T2DM | adipocyte | *FAM84A* | -1.009402764 | 0.000407858 | 0.005639191 | 1 |
| T2DM | adipocyte | *CCDC74A* | 4.552711132 | 6.29E-05 | 0.001608673 | 0.39573355 |
| T2DM | adipocyte | *PDLIM5* | 0.749725326 | 0.000570374 | 0.007000196 | 1 |
| T2DM | adipocyte | *RNF149* | 0.823338958 | 0.000850809 | 0.009161281 | 1 |
| T2DM | adipocyte | *TDRD10* | 0.947723329 | 1.94E-05 | 0.00081257 | 0.12226285 |
| T2DM | adipocyte | *PBXIP1* | 1.083702139 | 0.006933592 | 0.037194107 | 1 |
| T2DM | adipocyte | *LMOD3* | -0.946359077 | 0.010375097 | 0.048353027 | 1 |
| T2DM | adipocyte | *SLC22A15* | 0.595184364 | 0.000610637 | 0.007298626 | 1 |
| T2DM | adipocyte | *LMOD1* | -0.202531392 | 0.009898886 | 0.046765514 | 1 |
| T2DM | adipocyte | *ARPC2* | 0.74152321 | 0.002814256 | 0.020573521 | 1 |
| T2DM | adipocyte | *SPTA1* | -0.101018681 | 0.000551795 | 0.006802222 | 1 |
| T2DM | adipocyte | *AIM2* | 1.178674646 | 4.62E-06 | 0.000440302 | 0.029059958 |
| T2DM | adipocyte | *CADPS* | 1.612863307 | 6.29E-05 | 0.001608673 | 0.395671678 |
| T2DM | adipocyte | *GMPS* | 0.985579515 | 0.003227171 | 0.022589029 | 1 |

| | | | | | | |
|---|---|---|---|---|---|---|
| T2DM | adipocyte | *TIPARP* | 0.553265267 | 0.0061384 | 0.034031853 | 1 |
| T2DM | adipocyte | *DNASE1L3* | 0.794755251 | 3.42E-05 | 0.001075499 | 0.215287667 |
| T2DM | adipocyte | *GYG1* | 0.679400875 | 0.006927667 | 0.037194059 | 1 |
| T2DM | adipocyte | *HPS3* | 0.531042605 | 0.009682545 | 0.04615175 | 1 |
| T2DM | adipocyte | *PLB1* | 1.460982445 | 1.90E-05 | 0.000811251 | 0.119253961 |
| T2DM | adipocyte | *CDCP1* | 1.362816944 | 1.34E-05 | 0.00073252 | 0.084515187 |
| T2DM | adipocyte | *DTX3L* | 0.612824696 | 0.000311964 | 0.004772055 | 1 |
| T2DM | adipocyte | *YEATS2* | 1.00133127 | 0.000289944 | 0.004523265 | 1 |
| T2DM | adipocyte | *MEAF6* | 1.153183021 | 0.001000838 | 0.010214718 | 1 |
| T2DM | adipocyte | *RPN1* | 0.972862306 | 0.001693371 | 0.014406257 | 1 |
| T2DM | adipocyte | *RFT1* | -0.63346933 | 2.07E-05 | 0.000815077 | 0.130412398 |
| T2DM | adipocyte | *DNAJB14* | 0.598200369 | 0.001169586 | 0.011158103 | 1 |
| T2DM | adipocyte | *BDH2* | -0.203240409 | 0.002916031 | 0.021024181 | 1 |
| T2DM | adipocyte | *ATRIP* | -2.833899613 | 0.000190209 | 0.00340696 | 1 |
| T2DM | adipocyte | *SHISA5* | 0.790833282 | 0.008335753 | 0.041791768 | 1 |
| T2DM | adipocyte | *RNF123* | 0.32241205 | 4.77E-07 | 8.82E-05 | 0.002998102 |
| T2DM | adipocyte | *GRM2* | -6.724349312 | 0.004979896 | 0.029508585 | 1 |
| T2DM | adipocyte | *NPY1R* | -0.398811617 | 0.009785266 | 0.046465231 | 1 |
| T2DM | adipocyte | *IL15* | 0.72767915 | 0.00068043 | 0.00783958 | 1 |
| T2DM | adipocyte | *ITGA2* | 1.00884688 | 0.000463999 | 0.006052207 | 1 |
| T2DM | adipocyte | *PRRC1* | 0.900958549 | 0.001713572 | 0.014421988 | 1 |
| T2DM | adipocyte | *RICTOR* | 0.720287621 | 0.004201324 | 0.026573166 | 1 |
| T2DM | adipocyte | *KLKB1* | 0.159077342 | 0.004315691 | 0.027051592 | 1 |
| T2DM | adipocyte | *MB21D1* | 1.232617948 | 4.13E-05 | 0.001202961 | 0.259839665 |
| T2DM | adipocyte | *DCBLD1* | 0.57425 | 0.001413879 | 0.012753312 | 1 |
| T2DM | adipocyte | *TMEM200A* | 0.9118899 | 5.09E-05 | 0.001392051 | 0.320171735 |
| T2DM | adipocyte | *PDSS2* | 0.688102657 | 0.007932894 | 0.040913952 | 1 |
| T2DM | adipocyte | *STXBP5* | 0.690996857 | 0.003739099 | 0.024615411 | 1 |
| T2DM | adipocyte | *PI16* | 0.865858014 | 1.82E-05 | 0.000811251 | 0.114402364 |
| T2DM | adipocyte | *RPS14* | 0.592491347 | 0.003341938 | 0.02302506 | 1 |
| T2DM | adipocyte | *CAMLG* | 0.462689541 | 0.000767242 | 0.008576883 | 1 |
| T2DM | adipocyte | *SUN1* | 0.513272448 | 0.00051439 | 0.006418731 | 1 |
| T2DM | adipocyte | *OXR1* | 0.518713693 | 0.004883135 | 0.02918324 | 1 |
| T2DM | adipocyte | *COX6C* | 0.981210616 | 0.0048604 | 0.029102223 | 1 |
| T2DM | adipocyte | *DCAF13* | 1.25073111 | 0.0008804 | 0.009349786 | 1 |
| T2DM | adipocyte | *INTS8* | 0.528337831 | 0.001696653 | 0.014410427 | 1 |
| T2DM | adipocyte | *TMEM65* | 0.679624591 | 0.003353831 | 0.02302506 | 1 |
| T2DM | adipocyte | *SYK* | 0.602976206 | 0.001062483 | 0.010575556 | 1 |
| T2DM | adipocyte | *HNRNPK* | 0.724852434 | 0.009734395 | 0.046311059 | 1 |
| T2DM | adipocyte | *MARCH8* | 0.980402223 | 0.001453024 | 0.012939249 | 1 |
| T2DM | adipocyte | *PGM2L1* | 0.742851123 | 4.11E-05 | 0.001202864 | 0.258615685 |
| T2DM | adipocyte | *TAF3* | 0.875659114 | 0.000142107 | 0.002845314 | 0.893428674 |
| T2DM | adipocyte | *SLC18A2* | 0.666578298 | 0.003700983 | 0.024415616 | 1 |
| T2DM | adipocyte | *PDZD8* | 0.373810982 | 0.002134958 | 0.017000572 | 1 |
| T2DM | adipocyte | *QSOX2* | 1.707068339 | 0.000348577 | 0.005151093 | 1 |
| T2DM | adipocyte | *SLC39A13* | 1.728447244 | 8.70E-05 | 0.001970146 | 0.547017198 |
| T2DM | adipocyte | *LIPC* | 0.382530717 | 0.007405825 | 0.039028017 | 1 |
| T2DM | adipocyte | *JAM3* | 0.659704788 | 0.000205779 | 0.003597496 | 1 |
| T2DM | adipocyte | *TRMT61A* | -0.595228678 | 2.12E-07 | 4.95E-05 | 0.001335348 |
| T2DM | adipocyte | *ASPG* | -0.988695728 | 0.009969636 | 0.047056383 | 1 |

| | | | | | | |
|---|---|---|---|---|---|---|
| T2DM | adipocyte | *PCBD1* | -0.197592139 | 7.93E-05 | 0.001875094 | 0.498774903 |
| T2DM | adipocyte | *ZNF202* | 1.139821371 | 0.002447059 | 0.018807652 | 1 |
| T2DM | adipocyte | *C2* | 1.356936051 | 1.70E-14 | 2.13E-11 | 1.07E-10 |
| T2DM | adipocyte | *ILK* | 0.877976456 | 0.001102246 | 0.010743911 | 1 |
| T2DM | adipocyte | *DCHS1* | 0.926094849 | 5.95E-05 | 0.001546933 | 0.374357706 |
| T2DM | adipocyte | *NETO1* | 0.472284336 | 0.00328375 | 0.022786903 | 1 |
| T2DM | adipocyte | *LMO1* | 0.686484898 | 0.007049181 | 0.037630362 | 1 |
| T2DM | adipocyte | *RPL27A* | 0.715202499 | 0.00204462 | 0.016459063 | 1 |
| T2DM | adipocyte | *AKIP1* | 1.385954465 | 0.005816759 | 0.032945911 | 1 |
| T2DM | adipocyte | *C16orf46* | -4.325466012 | 0.000965127 | 0.009963466 | 1 |
| T2DM | adipocyte | *CLEC4E* | 0.193806214 | 0.009881113 | 0.046765514 | 1 |
| T2DM | adipocyte | *SEC11C* | 1.656934143 | 2.87E-05 | 0.00100347 | 0.180624615 |
| T2DM | adipocyte | *SAAL1* | -1.143359087 | 0.001424751 | 0.012814606 | 1 |
| T2DM | adipocyte | *TSC22D4* | 1.218491553 | 0.000608824 | 0.007290808 | 1 |
| T2DM | adipocyte | *AKTIP* | 1.228173057 | 0.002951245 | 0.021180912 | 1 |
| T2DM | adipocyte | *EVA1C* | 0.282044057 | 0.001222572 | 0.01150645 | 1 |
| T2DM | adipocyte | *SNRPD1* | 1.061352999 | 0.01009615 | 0.047475315 | 1 |
| T2DM | adipocyte | *GATAD2A* | 1.153292684 | 1.79E-06 | 0.00021176 | 0.011223265 |
| T2DM | adipocyte | *RPL13* | 0.62893005 | 0.005670783 | 0.032322949 | 1 |
| T2DM | adipocyte | *ZNF641* | 0.241751299 | 0.000919236 | 0.009622628 | 1 |
| T2DM | adipocyte | *CD320* | 1.493991849 | 1.43E-05 | 0.000736344 | 0.089833997 |
| T2DM | adipocyte | *SPRYD3* | 1.441309212 | 0.003707865 | 0.024435377 | 1 |
| T2DM | adipocyte | *PRDX2* | 1.007686971 | 0.00043405 | 0.005815531 | 1 |
| T2DM | adipocyte | *TMC8* | 0.643675423 | 7.06E-06 | 0.00053468 | 0.044378444 |
| T2DM | adipocyte | *KCTD5* | 1.376818755 | 0.000805607 | 0.008823777 | 1 |
| T2DM | adipocyte | *BSCL2* | -1.140561591 | 0.000426581 | 0.005742857 | 1 |
| T2DM | adipocyte | *TTC21A* | 0.930610372 | 0.004818414 | 0.028912435 | 1 |
| T2DM | adipocyte | *LTBP3* | 0.576039944 | 1.39E-07 | 3.49E-05 | 0.00087268 |
| T2DM | adipocyte | *UBTD2* | 0.602864235 | 0.001159099 | 0.011108623 | 1 |
| T2DM | adipocyte | *PXK* | 0.605674101 | 2.76E-05 | 0.000991922 | 0.173586403 |
| T2DM | adipocyte | *RFWD3* | 0.338899414 | 0.005904888 | 0.033072047 | 1 |
| T2DM | adipocyte | *RHOH* | 0.495173603 | 0.001203427 | 0.011392753 | 1 |
| T2DM | adipocyte | *FNTA* | 0.678725396 | 0.008370575 | 0.041866194 | 1 |
| T2DM | adipocyte | *WFDC13* | 1.720045629 | 0.004283067 | 0.026900743 | 1 |
| T2DM | adipocyte | *NPNT* | 0.603992159 | 0.007522385 | 0.039476824 | 1 |
| T2DM | adipocyte | *INPP5D* | 0.868359934 | 1.76E-05 | 0.000808689 | 0.110582397 |
| T2DM | adipocyte | *STXBP6* | 0.648330724 | 0.000153032 | 0.003025687 | 0.962112518 |
| T2DM | adipocyte | *FEM1B* | 0.658575253 | 0.000170238 | 0.003223745 | 1 |
| T2DM | adipocyte | *IRS1* | -0.328047295 | 1.62E-28 | 1.02E-24 | 1.02E-24 |
| T2DM | adipocyte | *FAM110B* | 0.893178887 | 0.003558713 | 0.02385248 | 1 |
| T2DM | adipocyte | *XPO6* | 1.085224831 | 0.000162683 | 0.00313738 | 1 |
| T2DM | adipocyte | *PGM2* | 1.133916743 | 0.00063916 | 0.007525092 | 1 |
| T2DM | adipocyte | *PTK2* | 0.5386819 | 0.005796943 | 0.032892943 | 1 |
| T2DM | adipocyte | *RNASE6* | 2.103075752 | 8.80E-06 | 0.000586394 | 0.055335973 |
| T2DM | adipocyte | *INO80E* | -0.393220926 | 0.002473584 | 0.018950029 | 1 |
| T2DM | adipocyte | *CSGALNACT2* | 0.924553483 | 0.000330701 | 0.004973968 | 1 |
| T2DM | adipocyte | *GUSB* | 0.955661344 | 0.004224866 | 0.026695207 | 1 |
| T2DM | adipocyte | *SEMA3E* | 0.973314737 | 4.98E-07 | 8.95E-05 | 0.003132297 |
| T2DM | adipocyte | *METTL7B* | -1.061263747 | 0.000249662 | 0.004012908 | 1 |
| T2DM | adipocyte | *DLGAP1* | 0.883849423 | 3.29E-05 | 0.00106579 | 0.206763313 |

| | | | | | | |
|---|---|---|---|---|---|---|
| T2DM | adipocyte | *HSPA4* | 0.947131599 | 0.0076448 | 0.039952498 | 1 |
| T2DM | adipocyte | *GTSF1* | 2.628623377 | 0.000983479 | 0.010086671 | 1 |
| T2DM | adipocyte | *TMEM133* | -1.588163696 | 0.004159336 | 0.026387231 | 1 |
| T2DM | adipocyte | *TTLL6* | -4.003436965 | 0.005239568 | 0.030444698 | 1 |
| T2DM | adipocyte | *KCNS3* | 0.14911544 | 0.003222904 | 0.022589029 | 1 |
| T2DM | adipocyte | *AKAP13* | 0.631998552 | 3.21E-05 | 0.00106579 | 0.201748085 |
| T2DM | adipocyte | *MTSS1* | 0.616549926 | 0.000443447 | 0.005894191 | 1 |
| T2DM | adipocyte | *SOX7* | 0.773530485 | 0.005924079 | 0.033135839 | 1 |
| T2DM | adipocyte | *ALK* | -0.315866881 | 0.008721476 | 0.043068751 | 1 |
| T2DM | adipocyte | *MFN1* | 0.527389817 | 0.002474626 | 0.018950029 | 1 |
| T2DM | adipocyte | *MCC* | 0.637054516 | 0.002766633 | 0.020319888 | 1 |
| T2DM | adipocyte | *HOPX* | 0.937878308 | 0.006892188 | 0.037133048 | 1 |
| T2DM | adipocyte | *LRRC8C* | 1.423193812 | 0.000118153 | 0.002518058 | 0.742827234 |
| T2DM | adipocyte | *RSL1D1* | 0.822047205 | 0.009210927 | 0.044717451 | 1 |
| T2DM | adipocyte | *BCL2L1* | 0.577301706 | 0.002526009 | 0.019099469 | 1 |
| T2DM | adipocyte | *NMUR1* | -0.591328069 | 0.010072257 | 0.047432414 | 1 |
| T2DM | adipocyte | *P2RY6* | 1.058400233 | 0.000493994 | 0.006338247 | 1 |
| T2DM | adipocyte | *RGS19* | -0.300975515 | 1.26E-05 | 0.000715288 | 0.079396947 |
| T2DM | adipocyte | *TCEA2* | 1.542145457 | 0.002087381 | 0.01669639 | 1 |
| T2DM | adipocyte | *CAMTA1* | 0.728845234 | 0.003635871 | 0.024138034 | 1 |
| T2DM | adipocyte | *FAM90A1* | 4.473581946 | 0.009332141 | 0.045201211 | 1 |
| T2DM | adipocyte | *AQP4* | -0.541617074 | 0.000360308 | 0.005239142 | 1 |
| T2DM | adipocyte | *SRGAP2C* | 0.621303189 | 0.001686305 | 0.014365585 | 1 |
| T2DM | adipocyte | *ANKRD20A4* | -1.758564063 | 0.006145269 | 0.034039917 | 1 |
| T2DM | adipocyte | *FRMD3* | 0.799311503 | 0.000126426 | 0.002615595 | 0.794839576 |
| T2DM | adipocyte | *MALT1* | 0.738384346 | 0.003399212 | 0.023153678 | 1 |
| T2DM | adipocyte | *MBOAT1* | 0.737172638 | 0.004750949 | 0.02885923 | 1 |
| T2DM | adipocyte | *C1QTNF4* | -0.087690876 | 0.000206375 | 0.003597496 | 1 |
| T2DM | adipocyte | *SPTLC3* | 0.945913832 | 0.003796126 | 0.024853488 | 1 |
| T2DM | adipocyte | *SUCLG2* | 0.530149811 | 0.002709799 | 0.020040713 | 1 |
| T2DM | adipocyte | *RCAN2* | 0.82308841 | 0.000119568 | 0.002531055 | 0.751723382 |
| T2DM | adipocyte | *PRSS27* | 1.407412261 | 0.001040068 | 0.010445535 | 1 |
| T2DM | adipocyte | *FGGY* | 0.479155345 | 0.010293991 | 0.04811771 | 1 |
| T2DM | adipocyte | *FIBP* | 1.829169315 | 1.07E-07 | 2.81E-05 | 0.000674133 |
| T2DM | adipocyte | *CTSW* | -0.059942727 | 0.001521365 | 0.013340055 | 1 |
| T2DM | adipocyte | *PDE3A* | 0.526138074 | 0.000454648 | 0.005967377 | 1 |
| T2DM | adipocyte | *AGAP5* | -22.08900287 | 0.001303084 | 0.012083315 | 1 |
| T2DM | adipocyte | *CFL1* | 0.950387865 | 0.001205055 | 0.011392753 | 1 |
| T2DM | adipocyte | *RAB37* | 0.838153084 | 0.001211859 | 0.011439876 | 1 |
| T2DM | adipocyte | *DCP2* | 0.789755246 | 0.003354941 | 0.02302506 | 1 |
| T2DM | adipocyte | *SNX32* | 0.388836355 | 6.74E-05 | 0.001674964 | 0.424025182 |
| T2DM | adipocyte | *RNASEH2C* | -0.34933528 | 0.002889538 | 0.021024181 | 1 |
| T2DM | adipocyte | *MYEOV* | -0.096652008 | 0.000875886 | 0.009344489 | 1 |
| T2DM | adipocyte | *ANKRD13D* | 0.570013439 | 0.007669757 | 0.040049637 | 1 |
| T2DM | adipocyte | *LCLAT1* | 0.613614643 | 0.006804206 | 0.036782496 | 1 |
| T2DM | adipocyte | *HPSE2* | 0.25317182 | 0.004397565 | 0.027428067 | 1 |
| T2DM | adipocyte | *ABLIM3* | 0.784356173 | 3.45E-05 | 0.001075499 | 0.216774434 |
| T2DM | adipocyte | *MAP3K11* | 1.169583767 | 0.000622001 | 0.007406292 | 1 |
| T2DM | adipocyte | *C1QA* | 1.085445581 | 5.50E-05 | 0.001465574 | 0.345875434 |
| T2DM | adipocyte | *OLR1* | 1.960011507 | 7.20E-05 | 0.001748913 | 0.452411491 |

| | | | | | | |
|---|---|---|---|---|---|---|
| T2DM | adipocyte | *SMARCC1* | 0.809208443 | 0.002153219 | 0.017114141 | 1 |
| T2DM | adipocyte | *VEGFB* | -0.343145057 | 6.07E-06 | 0.000483445 | 0.038192146 |
| T2DM | adipocyte | *PEAK1* | 0.627878506 | 0.004410112 | 0.027479064 | 1 |
| T2DM | adipocyte | *C1orf100* | 2.038951947 | 0.005846527 | 0.03304917 | 1 |
| T2DM | adipocyte | *RNF213* | 0.707358722 | 0.001834272 | 0.015153835 | 1 |
| T2DM | adipocyte | *PLK3* | 1.098552802 | 3.43E-05 | 0.001075499 | 0.215856446 |
| T2DM | adipocyte | *GPR160* | 0.974242617 | 0.000432573 | 0.005811082 | 1 |
| T2DM | adipocyte | *USMG5* | 0.638846501 | 0.010696184 | 0.049409926 | 1 |
| T2DM | adipocyte | *HOXB2* | -0.41194558 | 0.000211654 | 0.00360662 | 1 |
| T2DM | adipocyte | *SLC25A30* | 1.040767131 | 0.003130974 | 0.022160689 | 1 |
| T2DM | adipocyte | *LEMD3* | 0.737978147 | 0.006779517 | 0.036775516 | 1 |
| T2DM | adipocyte | *FAM53A* | -0.779784815 | 0.000792935 | 0.008730615 | 1 |
| T2DM | adipocyte | *LINGO2* | 1.019061709 | 0.000113427 | 0.002450574 | 0.713116961 |
| T2DM | adipocyte | *ANKRD36C* | 0.659525382 | 0.000504393 | 0.006370945 | 1 |
| T2DM | adipocyte | *IL20RB* | 0.256516478 | 0.003801043 | 0.024853488 | 1 |
| T2DM | adipocyte | *KLF14* | -4.144368103 | 1.22E-11 | 9.61E-09 | 7.68E-08 |
| T2DM | adipocyte | *SH3PXD2B* | 0.26581017 | 0.000184859 | 0.003395587 | 1 |
| T2DM | adipocyte | *C4orf32* | 0.652723597 | 0.003027507 | 0.021580426 | 1 |
| T2DM | adipocyte | *NLRP6* | -1.18558318 | 0.006775718 | 0.036775516 | 1 |
| T2DM | adipocyte | *CHST2* | 2.479023329 | 2.19E-06 | 0.000250757 | 0.013791616 |
| T2DM | adipocyte | *TP53I11* | 0.836347139 | 2.47E-05 | 0.000914004 | 0.155380665 |
| T2DM | adipocyte | *TMEM9B* | 1.255912575 | 1.19E-05 | 0.00069764 | 0.074647431 |
| T2DM | adipocyte | *LPL* | -0.108594705 | 0.007895865 | 0.040844634 | 1 |
| T2DM | adipocyte | *A2M* | 0.78179535 | 3.48E-05 | 0.00107846 | 0.218927389 |
| T2DM | adipocyte | *B3GALT6* | 0.84336391 | 0.00543029 | 0.031125299 | 1 |
| T2DM | adipocyte | *TCP11L1* | 0.409984402 | 0.001461597 | 0.012978897 | 1 |
| T2DM | adipocyte | *COX8A* | 1.56364293 | 0.000633701 | 0.007500112 | 1 |
| T2DM | adipocyte | *B3GNT4* | -4.277927175 | 0.000528743 | 0.006556624 | 1 |
| T2DM | adipocyte | *CDC26* | -0.211909475 | 0.008893135 | 0.043578441 | 1 |
| T2DM | adipocyte | *SLCO3A1* | 0.793720353 | 1.74E-05 | 0.000808689 | 0.109337141 |
| T2DM | adipocyte | *RNF152* | 0.730239805 | 0.007575729 | 0.039657458 | 1 |
| T2DM | adipocyte | *ZNF843* | -1.359888779 | 0.008042997 | 0.041346132 | 1 |
| T2DM | adipocyte | *ANKLE2* | 1.052052596 | 0.000886688 | 0.009400688 | 1 |
| T2DM | adipocyte | *GCNT4* | 3.722898889 | 8.35E-06 | 0.00058364 | 0.052527596 |
| T2DM | adipocyte | *DSCAML1* | 0.890115753 | 7.65E-07 | 0.000120195 | 0.004807798 |
| T2DM | adipocyte | *B3GNT8* | 3.518552654 | 1.06E-05 | 0.000654035 | 0.066711538 |
| T2DM | adipocyte | *MAN1B1* | 1.103276009 | 0.008279305 | 0.041575074 | 1 |
| T2DM | adipocyte | *C8orf31* | 2.095438606 | 0.000802543 | 0.008805568 | 1 |
| T2DM | adipocyte | *TIMM22* | 1.051019158 | 0.003512573 | 0.023694794 | 1 |
| T2DM | adipocyte | *PAWR* | 0.90695264 | 0.007899964 | 0.040844634 | 1 |
| T2DM | adipocyte | *RBM44* | 0.666748865 | 0.003533736 | 0.023782977 | 1 |
| T2DM | adipocyte | *ATOX1* | 1.044967196 | 0.000103152 | 0.002259646 | 0.648518533 |
| T2DM | adipocyte | *RPLP2* | 0.726758339 | 0.010426293 | 0.048447972 | 1 |
| T2DM | adipocyte | *C12orf54* | -1.220315466 | 0.000177723 | 0.003315555 | 1 |
| T2DM | adipocyte | *FAM87B* | -0.66042153 | 0.001926052 | 0.015787597 | 1 |
| T2DM | adipocyte | *RIC8A* | -0.640988779 | 0.007276517 | 0.038605453 | 1 |
| T2DM | adipocyte | *IMP3* | 1.439465427 | 5.01E-05 | 0.001382515 | 0.315251067 |
| T2DM | adipocyte | *C2orf73* | -0.810372662 | 0.001174204 | 0.011185187 | 1 |
| T2DM | adipocyte | *GRAMD1C* | 1.273161531 | 3.03E-05 | 0.001045532 | 0.1902868 |
| T2DM | adipocyte | *LCORL* | 0.789784818 | 0.001064891 | 0.010576279 | 1 |

| | | | | | | |
|---|---|---|---|---|---|---|
| T2DM | adipocyte | *SH2B1* | -0.168994256 | 0.004807134 | 0.028912435 | 1 |
| T2DM | adipocyte | *ZC3H12D* | 1.834672502 | 4.60E-05 | 0.00130246 | 0.289146215 |
| T2DM | adipocyte | *PLEC* | 0.703530256 | 0.001378611 | 0.012508027 | 1 |
| T2DM | adipocyte | *SH2D4B* | 3.934799115 | 0.000164304 | 0.003158959 | 1 |
| T2DM | adipocyte | *PRSS36* | 1.86333488 | 1.24E-05 | 0.000707371 | 0.077681096 |
| T2DM | adipocyte | *CSRNP3* | 0.634026458 | 0.003412822 | 0.023221226 | 1 |
| T2DM | adipocyte | *STX19* | 0.793239662 | 3.20E-06 | 0.000319674 | 0.020139452 |
| T2DM | adipocyte | *GDPD4* | 1.95710504 | 0.004469026 | 0.027717528 | 1 |
| T2DM | adipocyte | *TRIM73* | -0.413378367 | 0.005897379 | 0.033072047 | 1 |
| T2DM | adipocyte | *SNX18* | 0.626891154 | 0.005617811 | 0.032108344 | 1 |
| T2DM | adipocyte | *FARSA* | -0.768161049 | 3.10E-05 | 0.001056656 | 0.195035001 |
| T2DM | adipocyte | *CALR* | 1.095036139 | 8.00E-09 | 3.35E-06 | 5.03E-05 |
| T2DM | adipocyte | *HLA-DQB1* | -0.278126995 | 2.31E-14 | 2.42E-11 | 1.45E-10 |
| T2DM | adipocyte | *EGR3* | 1.544769408 | 1.24E-05 | 0.000707371 | 0.077810771 |
| T2DM | adipocyte | *CIITA* | 0.512456488 | 0.000449166 | 0.005932583 | 1 |
| T2DM | adipocyte | *SERTAD2* | 0.942401585 | 0.000407803 | 0.005639191 | 1 |
| T2DM | adipocyte | *B3GNT3* | 0.836203141 | 0.000361772 | 0.005239142 | 1 |
| T2DM | adipocyte | *HCLS1* | 1.105307163 | 1.98E-05 | 0.00081257 | 0.124402675 |
| T2DM | adipocyte | *RNF182* | -0.264910014 | 0.001324646 | 0.012193337 | 1 |
| T2DM | adipocyte | *SSTR2* | -0.873074047 | 0.000504649 | 0.006370945 | 1 |
| T2DM | adipocyte | *SHISA2* | 16.18860592 | 4.75E-06 | 0.000445914 | 0.029876251 |
| T2DM | adipocyte | *SRSF8* | 1.428092523 | 0.002948299 | 0.021180912 | 1 |
| T2DM | adipocyte | *ZDHHC20* | 0.839646425 | 0.001538672 | 0.013398383 | 1 |
| T2DM | adipocyte | *ANKRD34A* | -0.507232716 | 0.004525394 | 0.027920657 | 1 |
| T2DM | adipocyte | *SLC26A11* | 0.968335242 | 0.000748523 | 0.008440726 | 1 |
| T2DM | adipocyte | *SLC25A41* | -2.802729414 | 0.000826912 | 0.008978922 | 1 |
| T2DM | adipocyte | *DDX60L* | 0.635981382 | 0.000680835 | 0.00783958 | 1 |
| T2DM | adipocyte | *SGSH* | 1.28785795 | 0.005977016 | 0.033372555 | 1 |
| T2DM | adipocyte | *SETD2* | 0.499550205 | 0.006898584 | 0.037133048 | 1 |
| T2DM | adipocyte | *ATG9B* | 1.792505125 | 4.80E-05 | 0.001353931 | 0.30192658 |
| T2DM | adipocyte | *ZBTB20* | 0.517679724 | 0.00412989 | 0.026252322 | 1 |
| T2DM | adipocyte | *RFX7* | 0.729246173 | 0.003187754 | 0.022467949 | 1 |
| T2DM | adipocyte | *CHST15* | 0.561373795 | 0.001311 | 0.012116168 | 1 |
| T2DM | adipocyte | *TNRC18* | 0.787209783 | 0.00225429 | 0.017715898 | 1 |
| T2DM | adipocyte | *SYNM* | -0.638896134 | 0.010418097 | 0.04844569 | 1 |
| T2DM | adipocyte | *CAPN12* | 1.048067136 | 4.07E-05 | 0.001200631 | 0.255952316 |
| T2DM | adipocyte | *FES* | 0.973279841 | 0.003395732 | 0.023153678 | 1 |
| T2DM | adipocyte | *CLEC4G* | 1.20980179 | 0.004977429 | 0.029508585 | 1 |
| T2DM | adipocyte | *TTC3* | 0.110542788 | 0.002004762 | 0.016276616 | 1 |
| T2DM | adipocyte | *ZNF320* | 0.10905931 | 0.001165806 | 0.011138941 | 1 |
| T2DM | adipocyte | *BEGAIN* | -1.92598974 | 0.00119412 | 0.011340529 | 1 |
| T2DM | adipocyte | *GPR19* | 1.801685374 | 9.44E-06 | 0.000597863 | 0.059366531 |
| T2DM | adipocyte | *URAD* | -1.100499544 | 0.000208751 | 0.003599172 | 1 |
| T2DM | adipocyte | *GPR132* | 0.62261515 | 0.009439018 | 0.045613457 | 1 |
| T2DM | adipocyte | *RP1L1* | 0.417553178 | 0.000817087 | 0.008918444 | 1 |
| T2DM | adipocyte | *BMP8A* | 1.117345023 | 6.01E-07 | 0.000105007 | 0.003780258 |
| T2DM | adipocyte | *CBX6* | 1.37028163 | 8.35E-05 | 0.001941003 | 0.524727468 |
| T2DM | adipocyte | *KCTD16* | 0.468541623 | 0.002961827 | 0.021232615 | 1 |
| T2DM | adipocyte | *ZNF703* | -0.438673504 | 0.000408117 | 0.005639191 | 1 |
| T2DM | adipocyte | *TBX1* | 1.279941101 | 3.04E-05 | 0.001045558 | 0.191337077 |

| | | | | | | |
|---|---|---|---|---|---|---|
| T2DM | adipocyte | *EIF3C* | 0.207605752 | 0.001079593 | 0.010621557 | 1 |
| T2DM | adipocyte | *CNTN2* | -0.81596607 | 0.00445839 | 0.027714414 | 1 |
| T2DM | adipocyte | *PCDH9* | -0.163765126 | 0.008734832 | 0.043068751 | 1 |
| T2DM | adipocyte | *MROH7* | 0.564526514 | 0.004720045 | 0.028726937 | 1 |
| T2DM | adipocyte | *CSF1* | 0.238451586 | 0.006016869 | 0.033476156 | 1 |
| T2DM | adipocyte | *WDR27* | 0.612319876 | 0.007913199 | 0.040871623 | 1 |
| T2DM | adipocyte | *PROS1* | 0.481060884 | 0.000877935 | 0.009344489 | 1 |
| T2DM | adipocyte | *TMEM173* | 0.612356294 | 0.000414158 | 0.005660454 | 1 |
| T2DM | adipocyte | *ZNF93* | 1.541737566 | 0.002185708 | 0.017284962 | 1 |
| T2DM | adipocyte | *SEPT5* | 2.59471611 | 1.65E-05 | 0.000780926 | 0.103863217 |
| T2DM | adipocyte | *OSBP2* | 0.743552346 | 0.003567863 | 0.023862929 | 1 |
| T2DM | adipocyte | *TMED9* | 0.838771318 | 0.000304867 | 0.004674872 | 1 |
| T2DM | adipocyte | *ZFP90* | 1.088623147 | 0.000411315 | 0.005646146 | 1 |
| T2DM | adipocyte | *NDUFA6* | 0.091868924 | 0.008741169 | 0.043068751 | 1 |
| T2DM | adipocyte | *TMEM106A* | 0.914911505 | 9.02E-06 | 0.000586394 | 0.056692166 |
| T2DM | adipocyte | *TNFAIP8L1* | 0.916799617 | 0.000100433 | 0.00221551 | 0.631420448 |
| T2DM | adipocyte | *SV2B* | 0.454948862 | 0.009614555 | 0.045962686 | 1 |
| T2DM | adipocyte | *LSAMP* | 0.349959433 | 0.005809995 | 0.032937275 | 1 |
| T2DM | adipocyte | *AHNAK2* | 0.808922939 | 0.000221672 | 0.003736334 | 1 |
| T2DM | adipocyte | *C16orf52* | 0.833010369 | 0.00298186 | 0.021351882 | 1 |
| T2DM | adipocyte | *ADARB2* | 0.747931715 | 0.003527746 | 0.02377164 | 1 |
| T2DM | adipocyte | *PIGP* | -0.073395017 | 0.00524789 | 0.030464898 | 1 |
| T2DM | adipocyte | *POMK* | 0.846539624 | 0.008773901 | 0.043128627 | 1 |
| T2DM | adipocyte | *LINC00839* | 0.47795353 | 8.23E-05 | 0.001930952 | 0.517495226 |
| T2DM | adipocyte | *RTN4RL1* | -0.566014263 | 0.00588638 | 0.033072047 | 1 |
| T2DM | adipocyte | *RASA3* | -0.420031756 | 0.008207096 | 0.041544539 | 1 |
| T2DM | adipocyte | *GSAP* | 0.561280611 | 0.000513388 | 0.006418731 | 1 |
| T2DM | adipocyte | *BPIFB4* | 11.57705254 | 0.002552861 | 0.019244405 | 1 |
| T2DM | adipocyte | *TOR3A* | 1.322290961 | 0.005891982 | 0.033072047 | 1 |
| T2DM | adipocyte | *CYP4X1* | 0.428234372 | 0.007542743 | 0.039550645 | 1 |
| T2DM | adipocyte | *KPNA4* | 0.769440287 | 0.008189514 | 0.041544539 | 1 |
| T2DM | adipocyte | *RGS7BP* | -0.476156107 | 0.000729797 | 0.008296988 | 1 |
| T2DM | adipocyte | *ZNF396* | -0.303331117 | 0.007370663 | 0.038973391 | 1 |
| T2DM | adipocyte | *TNFRSF18* | 2.615952489 | 5.88E-06 | 0.000480968 | 0.036936708 |
| T2DM | adipocyte | *C3orf70* | 0.238294431 | 0.00361928 | 0.024071846 | 1 |
| T2DM | adipocyte | *ANGPTL5* | 1.459609997 | 0.002503149 | 0.0190529 | 1 |
| T2DM | adipocyte | *EPOR* | 0.554063078 | 0.005054329 | 0.029809163 | 1 |
| T2DM | adipocyte | *GJA4* | 0.535892448 | 0.001524602 | 0.013349825 | 1 |
| T2DM | adipocyte | *IFT140* | 1.13799096 | 0.00022585 | 0.003776376 | 1 |
| T2DM | adipocyte | *TET3* | 0.989974879 | 5.92E-05 | 0.001545232 | 0.372400866 |
| T2DM | adipocyte | *TRPV2* | 1.219979816 | 1.56E-05 | 0.000762794 | 0.097900691 |
| T2DM | adipocyte | *ADH1A* | -0.294064835 | 0.000483992 | 0.006235371 | 1 |
| T2DM | adipocyte | *SEMA4D* | 0.894404031 | 0.000580637 | 0.007060862 | 1 |
| T2DM | adipocyte | *MCRS1* | 0.582417773 | 0.000446794 | 0.005913671 | 1 |
| T2DM | adipocyte | *GFRAL* | 0.286125553 | 0.00810307 | 0.041444425 | 1 |
| T2DM | adipocyte | *FAM166A* | -1.427467166 | 0.00015628 | 0.003069586 | 0.982531971 |
| T2DM | adipocyte | *NUTM2B* | -4.29072942 | 0.001058984 | 0.010567984 | 1 |
| T2DM | adipocyte | *NCR3LG1* | 2.626047421 | 5.76E-06 | 0.000480968 | 0.036214359 |
| T2DM | adipocyte | *RUFY4* | -2.101814439 | 0.004266982 | 0.026853368 | 1 |
| T2DM | adipocyte | *PLSCR1* | 0.94445442 | 9.14E-06 | 0.000586394 | 0.05746665 |

| | | | | | | |
|---|---|---|---|---|---|---|
| T2DM | adipocyte | *CERKL* | 0.709652594 | 0.00619427 | 0.034250988 | 1 |
| T2DM | adipocyte | *SRSF10* | 0.697683675 | 0.003269384 | 0.022737411 | 1 |
| T2DM | adipocyte | *FAM72B* | 3.367369414 | 9.74E-06 | 0.00060642 | 0.061248469 |
| T2DM | adipocyte | *SUMO2* | 1.272837086 | 0.000162662 | 0.00313738 | 1 |
| T2DM | adipocyte | *DPYD* | 0.657709361 | 0.000302179 | 0.004667817 | 1 |
| T2DM | adipocyte | *TMEM201* | 0.741925288 | 0.001066544 | 0.010576279 | 1 |
| T2DM | adipocyte | *BEND4* | 1.797202197 | 0.000339005 | 0.005086696 | 1 |
| T2DM | adipocyte | *LRRK2* | 0.475004763 | 0.000134049 | 0.002745173 | 0.842768051 |
| T2DM | adipocyte | *C9orf152* | -1.098978411 | 0.001048747 | 0.010515902 | 1 |
| T2DM | adipocyte | *ALKBH2* | 0.422730281 | 0.001889217 | 0.015546476 | 1 |
| T2DM | adipocyte | *SP6* | -3.610627343 | 0.007790635 | 0.040512589 | 1 |
| T2DM | adipocyte | *S100A13* | 0.893473229 | 0.000245391 | 0.003980202 | 1 |
| T2DM | adipocyte | *LINC00994* | -0.343449302 | 0.003110988 | 0.022050487 | 1 |
| T2DM | adipocyte | *HLA-DRB1* | -0.502807016 | 9.14E-15 | 1.91E-11 | 5.74E-11 |
| T2DM | adipocyte | *WDSUB1* | 0.38191761 | 0.005905496 | 0.033072047 | 1 |
| T2DM | adipocyte | *SIRPB2* | 1.025772399 | 0.00010676 | 0.002322483 | 0.67119759 |
| T2DM | adipocyte | *GTF2IRD2* | 0.254824154 | 0.004515556 | 0.027887328 | 1 |
| T2DM | adipocyte | *SUPT3H* | 0.730695756 | 0.003775605 | 0.024803791 | 1 |
| T2DM | adipocyte | *GIMAP5* | 0.388644393 | 0.002908494 | 0.021024181 | 1 |
| T2DM | adipocyte | *NTNG2* | 1.372858608 | 0.000324482 | 0.004910514 | 1 |
| T2DM | adipocyte | *ASB13* | 1.220925097 | 0.009470525 | 0.045678294 | 1 |
| T2DM | adipocyte | *ZNF124* | 0.883399408 | 0.010691133 | 0.049409926 | 1 |
| T2DM | adipocyte | *RFX8* | 0.391359864 | 0.002350219 | 0.018271006 | 1 |
| T2DM | adipocyte | *SULT1A1* | -0.182331866 | 0.003351075 | 0.02302506 | 1 |
| T2DM | adipocyte | *TPK1* | 0.491638427 | 0.000201679 | 0.00356168 | 1 |
| T2DM | adipocyte | *SULF2* | 0.351290111 | 0.003828468 | 0.024968444 | 1 |
| T2DM | adipocyte | *HRH1* | 0.770856311 | 1.88E-05 | 0.000811251 | 0.118394548 |
| T2DM | adipocyte | *FAM180B* | -0.229117488 | 0.00774328 | 0.040333057 | 1 |
| T2DM | adipocyte | *TOMM7* | 1.148598791 | 0.000404862 | 0.00563134 | 1 |
| T2DM | adipocyte | *NF1* | 0.465058056 | 0.007234515 | 0.038464552 | 1 |
| T2DM | adipocyte | *HLA-DQA1* | -0.271327555 | 2.86E-06 | 0.000294676 | 0.017975214 |
| T2DM | adipocyte | *MVB12B* | 0.853404216 | 0.002047451 | 0.016460777 | 1 |
| T2DM | adipocyte | *SRGAP1* | 0.764812672 | 0.000504542 | 0.006370945 | 1 |
| T2DM | adipocyte | *FUT11* | 0.894453394 | 7.82E-06 | 0.00056523 | 0.049175051 |
| T2DM | adipocyte | *ANXA4* | 0.81371009 | 0.000503954 | 0.006370945 | 1 |
| T2DM | adipocyte | *GAL3ST4* | 1.049560038 | 1.51E-05 | 0.00075802 | 0.094752491 |
| T2DM | adipocyte | *SLC25A29* | 0.466301618 | 0.003052549 | 0.021709701 | 1 |
| T2DM | adipocyte | *FAM110D* | 0.888543349 | 0.001039245 | 0.010445535 | 1 |
| T2DM | adipocyte | *C5AR1* | 0.875908347 | 3.91E-05 | 0.001165288 | 0.245875773 |
| T2DM | adipocyte | *SLC2A10* | 0.386753973 | 0.008674416 | 0.043009506 | 1 |
| T2DM | adipocyte | *TTC30A* | -0.347302231 | 0.002519432 | 0.019099469 | 1 |
| T2DM | adipocyte | *HOXA4* | -0.70493413 | 0.008739149 | 0.043068751 | 1 |
| T2DM | adipocyte | *CCDC154* | 0.633386346 | 0.008969509 | 0.043850159 | 1 |
| T2DM | adipocyte | *MFAP5* | 0.771258881 | 0.000404732 | 0.00563134 | 1 |
| T2DM | adipocyte | *PSAP* | 0.722661284 | 0.000868865 | 0.009308421 | 1 |
| T2DM | adipocyte | *AC122129.1* | -1.520997823 | 0.000823497 | 0.008957307 | 1 |
| T2DM | adipocyte | *SGTB* | 0.779630279 | 0.000892429 | 0.009413933 | 1 |
| T2DM | adipocyte | *PRIM1* | -0.473658544 | 0.009073436 | 0.044290759 | 1 |
| T2DM | adipocyte | *SFI1* | 1.18351339 | 0.00035854 | 0.005239142 | 1 |
| T2DM | adipocyte | *MIER1* | 0.950159608 | 0.000117748 | 0.002518058 | 0.740279664 |

| | | | | | | |
|---|---|---|---|---|---|---|
| T2DM | adipocyte | *ZNF251* | 0.667401615 | 0.00325026 | 0.022672808 | 1 |
| T2DM | adipocyte | *HSD17B11* | 1.089528713 | 1.93E-05 | 0.00081257 | 0.121080489 |
| T2DM | adipocyte | *ANTXRL* | 49.05056692 | 0.000577508 | 0.007050089 | 1 |
| T2DM | adipocyte | *TMEM116* | -0.151914443 | 0.002988643 | 0.021369695 | 1 |
| T2DM | adipocyte | *ZNF568* | 0.809304801 | 2.59E-05 | 0.000951173 | 0.162824078 |
| T2DM | adipocyte | *HLA-DRB5* | -0.146844218 | 2.91E-12 | 2.61E-09 | 1.83E-08 |
| T2DM | adipocyte | *MAFK* | 1.417604824 | 0.000362498 | 0.005239142 | 1 |
| T2DM | adipocyte | *ARC* | 3.961974817 | 0.002655752 | 0.01973607 | 1 |
| T2DM | adipocyte | *LRBA* | 0.626660561 | 0.005785799 | 0.032892943 | 1 |
| T2DM | adipocyte | *ZNF534* | -0.419219839 | 0.006919307 | 0.037180926 | 1 |
| T2DM | adipocyte | *APCDD1L* | 1.450452342 | 0.007290411 | 0.038646554 | 1 |
| T2DM | adipocyte | *CAPZA2* | 0.894335203 | 0.000781054 | 0.008675772 | 1 |
| T2DM | adipocyte | *RASGEF1A* | 4.631596808 | 3.45E-05 | 0.001075499 | 0.216674704 |
| T2DM | adipocyte | *APRT* | 1.51376987 | 0.001114095 | 0.010825837 | 1 |
| T2DM | adipocyte | *AP001007.1* | -3.38839742 | 0.008740581 | 0.043068751 | 1 |
| T2DM | adipocyte | *CHML* | 0.267620331 | 0.001278961 | 0.011931548 | 1 |
| T2DM | adipocyte | *CENPW* | -0.804076636 | 2.42E-06 | 0.000267384 | 0.01524086 |
| T2DM | adipocyte | *FAM229B* | -0.188462982 | 0.001319407 | 0.012162916 | 1 |
| T2DM | adipocyte | *HIST2H2BF* | -2.608753817 | 0.006179019 | 0.034196737 | 1 |
| T2DM | adipocyte | *PCMTD2* | 0.87476091 | 0.000783987 | 0.008687779 | 1 |
| T2DM | adipocyte | *SYS1* | 1.818272839 | 0.00013989 | 0.002831013 | 0.879490359 |
| T2DM | adipocyte | *SYT15* | 0.487306406 | 0.004812494 | 0.028912435 | 1 |
| T2DM | adipocyte | *OXLD1* | 0.351114616 | 0.00819164 | 0.041544539 | 1 |
| T2DM | adipocyte | *HLA-DRA* | 0.54641325 | 1.44E-14 | 2.13E-11 | 9.08E-11 |
| T2DM | adipocyte | *NOTCH4* | -0.048997122 | 7.28E-08 | 2.18E-05 | 0.000457512 |
| T2DM | adipocyte | *AGER* | 0.363928118 | 0.000173491 | 0.003275486 | 1 |
| T2DM | adipocyte | *PRRT1* | 0.293665451 | 0.000209527 | 0.003599172 | 1 |
| T2DM | adipocyte | *DXO* | -0.232100148 | 0.002914245 | 0.021024181 | 1 |
| T2DM | adipocyte | *NELFE* | 0.367855265 | 5.25E-05 | 0.00142143 | 0.330366371 |
| T2DM | adipocyte | *BAG6* | 0.368017155 | 0.000247283 | 0.003986336 | 1 |
| T2DM | adipocyte | *NFKBIL1* | 0.347997864 | 0.004362491 | 0.027263402 | 1 |
| T2DM | adipocyte | *PSORS1C1* | 0.036261917 | 0.007386071 | 0.039022039 | 1 |
| T2DM | adipocyte | *LINC01123* | -4.321343682 | 0.006992317 | 0.037413357 | 1 |
| T2DM | adipocyte | *HLA-E* | 0.239039498 | 0.003608419 | 0.024033845 | 1 |
| T2DM | adipocyte | *TBC1D8* | 0.643347496 | 0.001442026 | 0.012896188 | 1 |
| T2DM | adipocyte | *GABBR1* | 0.912357782 | 0.000498387 | 0.006355699 | 1 |
| T2DM | adipocyte | *FAM71F2* | -0.610359779 | 0.001079258 | 0.010621557 | 1 |
| T2DM | adipocyte | *AC016586.1* | 1.87905792 | 0.008152069 | 0.041533273 | 1 |
| T2DM | adipocyte | *MT1M* | 1.968696954 | 2.19E-05 | 0.000830044 | 0.137787289 |
| T2DM | adipocyte | *SAMD9* | 0.680965183 | 0.001430805 | 0.012832337 | 1 |
| T2DM | adipocyte | *SERF1B* | -0.425481635 | 0.004998023 | 0.02954797 | 1 |
| T2DM | adipocyte | *DENND6B* | 1.847001735 | 0.000135873 | 0.002764505 | 0.854232184 |
| T2DM | adipocyte | *EIF3CL* | 0.928507677 | 0.004974449 | 0.029508585 | 1 |
| T2DM | adipocyte | *LIN52* | 0.436809431 | 0.000158918 | 0.003102843 | 0.999115573 |
| T2DM | adipocyte | *NYNRIN* | 1.04348915 | 8.62E-06 | 0.000586394 | 0.054219238 |
| T2DM | adipocyte | *HCG27* | 0.298011338 | 4.12E-11 | 2.88E-08 | 2.59E-07 |
| T2DM | adipocyte | *COLQ* | -0.752882762 | 0.0066446 | 0.036170411 | 1 |
| T2DM | adipocyte | *RNF208* | -1.958249612 | 0.001389922 | 0.012591415 | 1 |
| T2DM | adipocyte | *FGFR1OP* | 0.584031652 | 0.001703486 | 0.014412493 | 1 |
| T2DM | adipocyte | *CCDC183* | -1.273508416 | 0.001698445 | 0.014410427 | 1 |

| | | | | | | |
|---|---|---|---|---|---|---|
| T2DM | adipocyte | *DNLZ* | 0.170490289 | 0.000140492 | 0.002831013 | 0.883275902 |
| T2DM | adipocyte | *SUPT4H1* | 0.546793635 | 0.005102332 | 0.029999308 | 1 |
| T2DM | adipocyte | *ARHGAP19* | 0.893379115 | 0.001446042 | 0.012913729 | 1 |
| T2DM | adipocyte | *SIPA1* | 0.585200173 | 5.87E-06 | 0.000480968 | 0.03688265 |
| T2DM | adipocyte | *HEXA* | 1.002333791 | 0.001312408 | 0.012116168 | 1 |
| T2DM | adipocyte | *LBH* | 1.255990692 | 3.46E-05 | 0.001075499 | 0.217250747 |
| T2DM | adipocyte | *ATF6B* | 0.383303086 | 1.31E-05 | 0.00073252 | 0.082364985 |
| T2DM | adipocyte | *SLC35F6* | 1.123356018 | 0.001023518 | 0.010362093 | 1 |
| T2DM | adipocyte | *PPM1N* | 1.13417759 | 1.41E-05 | 0.00073252 | 0.088634939 |
| T2DM | adipocyte | *TAX1BP3* | 0.835779078 | 0.003469046 | 0.023476742 | 1 |
| T2DM | adipocyte | *AP1G2* | 1.44283898 | 1.32E-05 | 0.00073252 | 0.083260242 |
| T2DM | adipocyte | *TM6SF2* | 7.793426366 | 5.04E-05 | 0.001382515 | 0.316595862 |
| T2DM | adipocyte | *NEURL1B* | 1.062067614 | 1.09E-05 | 0.000658243 | 0.068457236 |
| T2DM | adipocyte | *STARD10* | 0.205254919 | 4.61E-07 | 8.78E-05 | 0.002897928 |
| T2DM | adipocyte | *UBE2QL1* | -0.72027187 | 0.000321711 | 0.0048855 | 1 |
| T2DM | adipocyte | *KCNU1* | -0.575383117 | 6.67E-09 | 2.99E-06 | 4.19E-05 |
| T2DM | adipocyte | *MYL5* | -1.145183156 | 0.010305725 | 0.048136772 | 1 |
| T2DM | adipocyte | *AC016773.1* | -0.126425555 | 0.004484752 | 0.027751613 | 1 |
| T2DM | adipocyte | *RNU6ATAC35I* | -1.63956826 | 0.000198178 | 0.00351961 | 1 |
| T2DM | adipocyte | *OR2A25* | -4.879314571 | 0.006916954 | 0.037180926 | 1 |
| T2DM | adipocyte | *SLC12A8* | -0.296450436 | 0.006628662 | 0.036170411 | 1 |
| T2DM | adipocyte | *UBA52* | 0.544507094 | 0.000435267 | 0.005815531 | 1 |
| T2DM | adipocyte | *AC062017.1* | -0.43499878 | 4.98E-05 | 0.00138011 | 0.313284889 |
| T2DM | adipocyte | *ADRA2B* | 3.683808588 | 1.55E-05 | 0.000762794 | 0.097408865 |
| T2DM | adipocyte | *LINC00205* | -0.315635917 | 0.009105513 | 0.044377023 | 1 |
| T2DM | adipocyte | *LINC01133* | 1.068094997 | 2.06E-05 | 0.000815077 | 0.129606343 |
| T2DM | adipocyte | *LINC01117* | 0.593624599 | 0.000141662 | 0.002845314 | 0.890631344 |
| T2DM | adipocyte | *LINC00184* | -0.731623967 | 2.04E-05 | 0.000815077 | 0.128165778 |
| T2DM | adipocyte | *LINC00237* | 3.868591965 | 2.60E-05 | 0.000951173 | 0.163601693 |
| T2DM | adipocyte | *NOL7* | 1.096669996 | 0.001038228 | 0.010445535 | 1 |
| T2DM | adipocyte | *ELFN1* | 2.469362438 | 3.65E-06 | 0.000358488 | 0.022943226 |
| T2DM | adipocyte | *ZNF503-AS1* | -0.727986228 | 0.006826789 | 0.036872873 | 1 |
| T2DM | adipocyte | *C14orf132* | 0.531057602 | 0.001429689 | 0.012832337 | 1 |
| T2DM | adipocyte | *AC108463.2* | 0.533912224 | 0.010622448 | 0.049177709 | 1 |
| T2DM | adipocyte | *LINC01013* | 1.978791086 | 1.07E-06 | 0.000145821 | 0.006707778 |
| T2DM | adipocyte | *LINC00954* | 2.380491555 | 2.95E-05 | 0.001026284 | 0.185757444 |
| T2DM | adipocyte | *PATL2* | 0.995209793 | 0.000665206 | 0.007725842 | 1 |
| T2DM | adipocyte | *U91324.1* | -1.454020211 | 0.002351081 | 0.018271006 | 1 |
| T2DM | adipocyte | *CISD3* | 1.95834008 | 0.000359065 | 0.005239142 | 1 |
| T2DM | adipocyte | *KLHL7-AS1* | -1.254075414 | 0.00051456 | 0.006418731 | 1 |
| T2DM | adipocyte | *TAPBP* | 0.458038068 | 0.000297797 | 0.004611451 | 1 |
| T2DM | adipocyte | *HLA-DQB2* | 0.121208435 | 1.18E-08 | 4.63E-06 | 7.41E-05 |
| T2DM | adipocyte | *TNF* | 0.384831809 | 0.000624122 | 0.007417495 | 1 |
| T2DM | adipocyte | *HLA-B* | 0.341252922 | 3.08E-07 | 6.46E-05 | 0.001935017 |
| T2DM | adipocyte | *HLA-DQA2* | 0.348931095 | 6.50E-20 | 2.04E-16 | 4.08E-16 |
| T2DM | adipocyte | *TXNDC5* | 1.407660784 | 0.008704479 | 0.043068751 | 1 |
| T2DM | adipocyte | *ASB14* | 0.399320632 | 0.005008862 | 0.02956875 | 1 |
| T2DM | adipocyte | *LY6G5B* | -0.087987012 | 0.001220469 | 0.01150388 | 1 |
| T2DM | adipocyte | *AQP1* | 0.60624312 | 0.003133586 | 0.022160689 | 1 |
| T2DM | adipocyte | *ARHGEF25* | 0.428670073 | 0.007637621 | 0.03994819 | 1 |

| | | | | | | |
|---|---|---|---|---|---|---|
| T2DM | adipocyte | *ARFGAP3* | 0.84166245 | 0.010220351 | 0.047844634 | 1 |
| T2DM | adipocyte | *C22orf39* | 1.284009584 | 0.003880551 | 0.025074024 | 1 |
| T2DM | adipocyte | *MRPL20* | 0.753455254 | 0.004883214 | 0.02918324 | 1 |
| T2DM | adipocyte | *DECR2* | -0.758914556 | 0.007465371 | 0.039228656 | 1 |
| T2DM | adipocyte | *FGF14-IT1* | 2.976190043 | 1.61E-05 | 0.000777737 | 0.101105782 |
| T2DM | adipocyte | *NAT6* | 0.570802532 | 0.0007228 | 0.008262261 | 1 |
| T2DM | adipocyte | *IL10RB* | 0.491910824 | 0.002279214 | 0.017854583 | 1 |
| T2DM | adipocyte | *PLA2G4B* | 0.361756813 | 4.76E-08 | 1.57E-05 | 0.000299179 |
| T2DM | adipocyte | *JMJD7* | 0.389662243 | 5.66E-08 | 1.78E-05 | 0.000355575 |
| T2DM | adipocyte | *APOBEC3C* | 1.691517613 | 6.57E-05 | 0.001651245 | 0.41281126 |
| T2DM | adipocyte | *SRP14-AS1* | -0.090384607 | 0.002755356 | 0.020260946 | 1 |
| T2DM | adipocyte | *FMN1* | 0.212776739 | 0.003699321 | 0.024415616 | 1 |
| T2DM | adipocyte | *DNAH10OS* | -3.545076407 | 1.75E-06 | 0.000210988 | 0.010971375 |
| T2DM | adipocyte | *LINC00989* | 1.262272662 | 0.000845497 | 0.009161281 | 1 |
| T2DM | adipocyte | *LINC01021* | -0.85663683 | 0.000237303 | 0.003905567 | 1 |
| T2DM | adipocyte | *FAM218A* | -0.956086447 | 0.000326677 | 0.004925224 | 1 |
| T2DM | adipocyte | *SNHG18* | -0.387157483 | 0.00096151 | 0.009942461 | 1 |
| T2DM | adipocyte | *TRNP1* | 4.394937353 | 3.21E-05 | 0.00106579 | 0.202001671 |
| T2DM | adipocyte | *PRKDC* | 0.721544836 | 0.0070668 | 0.037683607 | 1 |
| T2DM | adipocyte | *TAS2R20* | 0.171306383 | 0.00495688 | 0.029508585 | 1 |
| T2DM | adipocyte | *CTSO* | 1.208350575 | 7.72E-05 | 0.001839518 | 0.485632843 |
| T2DM | adipocyte | *HMBS* | 0.251004888 | 3.71E-05 | 0.001129902 | 0.232985147 |
| T2DM | adipocyte | *KIAA1147* | 0.501300782 | 0.010317158 | 0.048154397 | 1 |
| T2DM | adipocyte | *ZBED6* | -1.545672783 | 0.00099243 | 0.010145383 | 1 |
| T2DM | adipocyte | *FNTB* | -0.97784501 | 0.003822201 | 0.024953455 | 1 |
| T2DM | adipocyte | *LINC00638* | -0.975924438 | 0.003196497 | 0.022504342 | 1 |
| T2DM | adipocyte | *LINC00639* | 0.388641127 | 1.27E-06 | 0.000170304 | 0.00800429 |
| T2DM | adipocyte | *LINC00924* | 0.593163606 | 0.003426811 | 0.023240953 | 1 |
| T2DM | adipocyte | *UBE2Q2L* | -0.869992386 | 0.002679385 | 0.019888182 | 1 |
| T2DM | adipocyte | *AC083843.1* | -5.068093842 | 3.10E-07 | 6.46E-05 | 0.001947622 |
| T2DM | adipocyte | *AC009120.5* | -0.766769313 | 0.000467578 | 0.006072732 | 1 |
| T2DM | adipocyte | *AC139100.2* | -3.246647442 | 0.003675859 | 0.02437777 | 1 |
| T2DM | adipocyte | *LINC00906* | -4.487878605 | 0.000423263 | 0.005710415 | 1 |
| T2DM | adipocyte | *AC009060.1* | -1.344609235 | 8.40E-07 | 0.000128759 | 0.005279123 |
| T2DM | adipocyte | *LINC01050* | 3.679485564 | 5.74E-05 | 0.001511181 | 0.361172254 |
| T2DM | ASPC | *SEMA3F* | -0.096741292 | 7.48E-07 | 0.000120508 | 0.004699814 |
| T2DM | ASPC | *RBM6* | -0.079597947 | 3.97E-08 | 1.08E-05 | 0.000249439 |
| T2DM | ASPC | *ZNF195* | -0.153620091 | 0.001442074 | 0.047717469 | 1 |
| T2DM | ASPC | *GGNBP2* | -0.25000736 | 5.91E-07 | 0.000103162 | 0.003713832 |
| T2DM | ASPC | *CEP68* | 0.245456724 | 2.88E-09 | 1.21E-06 | 1.81E-05 |
| T2DM | ASPC | *TUBG2* | -0.110336912 | 6.95E-09 | 2.57E-06 | 4.37E-05 |
| T2DM | ASPC | *ITIH4* | 0.062848257 | 8.50E-06 | 0.000937955 | 0.053463458 |
| T2DM | ASPC | *MARK3* | -0.749038275 | 1.34E-08 | 4.20E-06 | 8.41E-05 |
| T2DM | ASPC | *CTTNBP2* | -0.317636302 | 7.86E-06 | 0.000898377 | 0.049410749 |
| T2DM | ASPC | *LRCH4* | -0.14215588 | 0.000604895 | 0.02535315 | 1 |
| T2DM | ASPC | *KCNN2* | -1.047576559 | 0.000195433 | 0.011591383 | 1 |
| T2DM | ASPC | *COBLL1* | -0.185926792 | 3.28E-08 | 9.37E-06 | 0.000206172 |
| T2DM | ASPC | *C3orf18* | 0.312414855 | 7.18E-07 | 0.000118807 | 0.004514649 |
| T2DM | ASPC | *PABPC4* | -0.282447305 | 3.69E-09 | 1.45E-06 | 2.32E-05 |
| T2DM | ASPC | *SLC26A4* | -2.626610255 | 1.30E-05 | 0.001338862 | 0.081670593 |

| | | | | | | |
|---|---|---|---|---|---|---|
| T2DM | ASPC | *ALKBH5* | -0.172363312 | 0.001388297 | 0.046181083 | 1 |
| T2DM | ASPC | *MAP3K1* | -0.457906806 | 0.000186854 | 0.011188076 | 1 |
| T2DM | ASPC | *CWF19L1* | -0.109131886 | 7.91E-05 | 0.006092493 | 0.49758588 |
| T2DM | ASPC | *PPP6R2* | -0.582986119 | 1.72E-05 | 0.001693128 | 0.108360169 |
| T2DM | ASPC | *RBL2* | 0.16452262 | 0.000172124 | 0.01071429 | 1 |
| T2DM | ASPC | *EYA1* | 0.097524976 | 8.27E-05 | 0.006186681 | 0.519681226 |
| T2DM | ASPC | *FCER2* | -0.844183701 | 0.001344165 | 0.045191249 | 1 |
| T2DM | ASPC | *DMPK* | -0.350039494 | 0.000131028 | 0.008684047 | 0.823772703 |
| T2DM | ASPC | *GCDH* | -0.406548977 | 3.68E-07 | 7.47E-05 | 0.002315465 |
| T2DM | ASPC | *ATP13A1* | 0.241990386 | 5.01E-07 | 9.27E-05 | 0.003151374 |
| T2DM | ASPC | *CPVL* | 0.336552916 | 0.000205491 | 0.011847527 | 1 |
| T2DM | ASPC | *POLD2* | -0.456998034 | 7.54E-06 | 0.000877953 | 0.047409467 |
| T2DM | ASPC | *DHRS11* | -0.080503842 | 0.001141685 | 0.040099288 | 1 |
| T2DM | ASPC | *HSD17B1* | -0.073961377 | 4.37E-07 | 8.33E-05 | 0.002749998 |
| T2DM | ASPC | *VAT1* | 0.246105557 | 0.000981559 | 0.036088085 | 1 |
| T2DM | ASPC | *OCIAD1* | -0.824101951 | 9.11E-05 | 0.00665014 | 0.57297608 |
| T2DM | ASPC | *WFS1* | 1.686905901 | 9.50E-16 | 1.19E-12 | 5.97E-12 |
| T2DM | ASPC | *CDK2AP1* | 0.073023402 | 0.000133714 | 0.008756845 | 0.840657101 |
| T2DM | ASPC | *ENDOU* | -0.134817063 | 0.000964786 | 0.035891167 | 1 |
| T2DM | ASPC | *MAPK14* | -0.460714536 | 0.000474577 | 0.021691757 | 1 |
| T2DM | ASPC | *BVES* | -0.086748399 | 0.000890731 | 0.033533095 | 1 |
| T2DM | ASPC | *DNAJC27* | -0.30780762 | 0.000207289 | 0.011847527 | 1 |
| T2DM | ASPC | *NRBP1* | -0.361965393 | 5.54E-07 | 9.96E-05 | 0.003484782 |
| T2DM | ASPC | *PPM1G* | -0.584331956 | 2.60E-07 | 5.44E-05 | 0.001631963 |
| T2DM | ASPC | *CHMP3* | -0.252924967 | 5.22E-06 | 0.000643271 | 0.032792114 |
| T2DM | ASPC | *GPX7* | -0.070032292 | 0.000498356 | 0.022334612 | 1 |
| T2DM | ASPC | *CD3EAP* | -0.218131156 | 2.66E-05 | 0.002535376 | 0.167334825 |
| T2DM | ASPC | *ELOVL4* | 0.079166003 | 0.001260062 | 0.043054409 | 1 |
| T2DM | ASPC | *DNAH7* | -0.085607986 | 1.42E-05 | 0.001443717 | 0.089510469 |
| T2DM | ASPC | *CCDC92* | -0.173238351 | 0.000778477 | 0.030976472 | 1 |
| T2DM | ASPC | *HOXB3* | -0.196695 | 3.46E-05 | 0.003192162 | 0.217726579 |
| T2DM | ASPC | *ADRA1A* | 0.028195977 | 0.00020533 | 0.011847527 | 1 |
| T2DM | ASPC | *TEX14* | 0.237116055 | 0.000363997 | 0.018019309 | 1 |
| T2DM | ASPC | *RASL11A* | -0.480398725 | 0.000810268 | 0.031838487 | 1 |
| T2DM | ASPC | *PMS2* | 0.380866758 | 0.001458645 | 0.047893484 | 1 |
| T2DM | ASPC | *CBX3* | -0.35222834 | 0.001382543 | 0.046181083 | 1 |
| T2DM | ASPC | *GRK4* | 0.043976745 | 0.000521425 | 0.022765292 | 1 |
| T2DM | ASPC | *OPRL1* | -0.177965771 | 5.22E-06 | 0.000643271 | 0.032806816 |
| T2DM | ASPC | *ZNF436* | 0.191511649 | 0.000180319 | 0.010900652 | 1 |
| T2DM | ASPC | *PRR12* | 0.154633602 | 0.000864612 | 0.03274588 | 1 |
| T2DM | ASPC | *DPP6* | -0.517957771 | 7.50E-06 | 0.000877953 | 0.047138105 |
| T2DM | ASPC | *GTPBP3* | -0.23137619 | 0.000580262 | 0.02473877 | 1 |
| T2DM | ASPC | *C12orf65* | 0.295382239 | 0.000516883 | 0.022724794 | 1 |
| T2DM | ASPC | *DMGDH* | -0.631110652 | 6.34E-05 | 0.005313786 | 0.398533929 |
| T2DM | ASPC | *DSTYK* | -0.373550831 | 0.000336908 | 0.017055971 | 1 |
| T2DM | ASPC | *TMCC2* | -0.152706805 | 0.001272106 | 0.043230992 | 1 |
| T2DM | ASPC | *TMBIM1* | -0.169384473 | 0.000771763 | 0.030904937 | 1 |
| T2DM | ASPC | *AOAH* | -0.184587414 | 8.50E-05 | 0.006286584 | 0.534359649 |
| T2DM | ASPC | *SYTL2* | -0.44852495 | 0.000111133 | 0.00784068 | 0.698695832 |
| T2DM | ASPC | *DCUN1D5* | -0.171995794 | 0.000995465 | 0.036386559 | 1 |

| | | | | | | |
|---|---|---|---|---|---|---|
| T2DM | ASPC | *SPTBN5* | 0.083403987 | 5.45E-05 | 0.004692591 | 0.34255913 |
| T2DM | ASPC | *CARF* | 0.137213106 | 0.001011208 | 0.036537162 | 1 |
| T2DM | ASPC | *HMG20A* | 0.530863258 | 8.33E-06 | 0.000935353 | 0.052379763 |
| T2DM | ASPC | *MAN2C1* | -0.050077148 | 2.55E-05 | 0.002465384 | 0.160249978 |
| T2DM | ASPC | *MYO19* | 0.558207061 | 1.13E-05 | 0.001184543 | 0.071072584 |
| T2DM | ASPC | *VPS53* | -0.232949511 | 0.000815381 | 0.03184039 | 1 |
| T2DM | ASPC | *LRRC46* | -0.196642538 | 6.76E-05 | 0.005518794 | 0.424947108 |
| T2DM | ASPC | *C18orf8* | -0.286005601 | 7.37E-05 | 0.00594361 | 0.463601572 |
| T2DM | ASPC | *NPC1* | -0.468582225 | 2.92E-06 | 0.000374823 | 0.01836632 |
| T2DM | ASPC | *SIRT3* | 0.191940441 | 0.001249311 | 0.042920317 | 1 |
| T2DM | ASPC | *WNT9A* | 0.190400532 | 0.001118194 | 0.039494863 | 1 |
| T2DM | ASPC | *NBEAL1* | 0.097861943 | 0.000707487 | 0.028882929 | 1 |
| T2DM | ASPC | *PDGFC* | -0.547543016 | 0.000106867 | 0.007634933 | 0.671874086 |
| T2DM | ASPC | *BHMT* | -0.058974412 | 0.000130695 | 0.008684047 | 0.821679856 |
| T2DM | ASPC | *PPIP5K2* | -0.373742703 | 7.77E-05 | 0.006092493 | 0.488374861 |
| T2DM | ASPC | *CCDC25* | 0.337373839 | 0.000976051 | 0.036088085 | 1 |
| T2DM | ASPC | *HSD17B12* | -0.118720526 | 1.29E-08 | 4.20E-06 | 8.10E-05 |
| T2DM | ASPC | *APIP* | -0.039787202 | 0.000131221 | 0.008684047 | 0.824984453 |
| T2DM | ASPC | *EIF3M* | -0.317052308 | 0.000382375 | 0.018645994 | 1 |
| T2DM | ASPC | *MS4A3* | -23.83923076 | 0.000752483 | 0.030326033 | 1 |
| T2DM | ASPC | *DOC2A* | -0.227282965 | 0.000254849 | 0.013464191 | 1 |
| T2DM | ASPC | *ARL14EP* | -0.419477803 | 1.40E-07 | 3.03E-05 | 0.000879158 |
| T2DM | ASPC | *POC5* | -0.18697486 | 0.0001768 | 0.010875031 | 1 |
| T2DM | ASPC | *ZFP36L2* | 0.699977937 | 0.000638253 | 0.026399305 | 1 |
| T2DM | ASPC | *SAR1B* | 0.280721637 | 0.000500892 | 0.022334612 | 1 |
| T2DM | ASPC | *JAZF1* | -0.94577446 | 1.03E-19 | 3.25E-16 | 6.50E-16 |
| T2DM | ASPC | *LONRF1* | -0.391282685 | 4.59E-05 | 0.004007116 | 0.288512359 |
| T2DM | ASPC | *TSEN2* | 0.231567756 | 2.59E-06 | 0.000347135 | 0.016287986 |
| T2DM | ASPC | *NDUFAF6* | -0.35428124 | 8.25E-07 | 0.000129675 | 0.005186994 |
| T2DM | ASPC | *TIMP4* | 0.350323437 | 3.50E-05 | 0.003192162 | 0.220259182 |
| T2DM | ASPC | *CLEC18A* | -0.233539788 | 8.32E-08 | 1.94E-05 | 0.000523226 |
| T2DM | ASPC | *KCNJ15* | -0.147562737 | 0.000359647 | 0.017945254 | 1 |
| T2DM | ASPC | *AP3S2* | 0.155672466 | 2.10E-10 | 1.20E-07 | 1.32E-06 |
| T2DM | ASPC | *CD1E* | 0.290247583 | 0.000738419 | 0.029951242 | 1 |
| T2DM | ASPC | *ZNF276* | -0.403333172 | 0.000476135 | 0.021691757 | 1 |
| T2DM | ASPC | *DIP2A* | -0.213117459 | 0.000633287 | 0.026367398 | 1 |
| T2DM | ASPC | *SPTA1* | -0.03980573 | 0.001324195 | 0.044759217 | 1 |
| T2DM | ASPC | *ICA1L* | -0.205210514 | 0.000582366 | 0.02473877 | 1 |
| T2DM | ASPC | *RFT1* | -0.311220657 | 2.36E-06 | 0.000329077 | 0.014808448 |
| T2DM | ASPC | *BDH2* | -0.104622491 | 0.000339112 | 0.017055971 | 1 |
| T2DM | ASPC | *ATRIP* | -2.073539478 | 0.000404757 | 0.019574666 | 1 |
| T2DM | ASPC | *RNF123* | 0.132246554 | 1.18E-06 | 0.00017043 | 0.007440407 |
| T2DM | ASPC | *CCDC171* | -0.669134995 | 0.001002207 | 0.036421235 | 1 |
| T2DM | ASPC | *REEP3* | -0.270809409 | 0.000675714 | 0.027766099 | 1 |
| T2DM | ASPC | *TRMT61A* | -0.242105443 | 1.19E-06 | 0.00017043 | 0.007498926 |
| T2DM | ASPC | *PCBD1* | -0.082948836 | 9.20E-05 | 0.00665014 | 0.578562153 |
| T2DM | ASPC | *C2* | 0.569757758 | 9.57E-12 | 7.52E-09 | 6.02E-08 |
| T2DM | ASPC | *SAAL1* | -1.388346099 | 4.86E-11 | 3.06E-08 | 3.06E-07 |
| T2DM | ASPC | *VKORC1* | -0.139161516 | 0.001216508 | 0.042022988 | 1 |
| T2DM | ASPC | *GATAD2A* | 0.523224161 | 0.00108684 | 0.038604294 | 1 |

| | | | | | | |
|---|---|---|---|---|---|---|
| T2DM | ASPC | *PRDX2* | 0.540195284 | 0.000298362 | 0.015250408 | 1 |
| T2DM | ASPC | *LTBP3* | 0.212134933 | 1.10E-05 | 0.001167508 | 0.068882965 |
| T2DM | ASPC | *KIF5C* | -0.447183754 | 0.000832691 | 0.032117356 | 1 |
| T2DM | ASPC | *PXK* | 0.245461256 | 0.000244671 | 0.013035983 | 1 |
| T2DM | ASPC | *TAP1* | -0.41895812 | 6.11E-07 | 0.000103819 | 0.003841287 |
| T2DM | ASPC | *IRS1* | -0.136490968 | 7.89E-27 | 4.96E-23 | 4.96E-23 |
| T2DM | ASPC | *HINT1* | -0.468028148 | 0.000858678 | 0.032718236 | 1 |
| T2DM | ASPC | *INO80E* | -0.201037743 | 0.000449833 | 0.021263909 | 1 |
| T2DM | ASPC | *EMB* | -0.102186082 | 0.000153486 | 0.009747156 | 0.96496846 |
| T2DM | ASPC | *TTLL6* | -3.192859463 | 1.08E-05 | 0.001167508 | 0.068169643 |
| T2DM | ASPC | *RGS19* | -0.156173857 | 1.21E-07 | 2.71E-05 | 0.000760006 |
| T2DM | ASPC | *GPHN* | -0.157305904 | 0.00102897 | 0.036756448 | 1 |
| T2DM | ASPC | *LAMB2* | -0.224872252 | 3.00E-05 | 0.002815123 | 0.188613252 |
| T2DM | ASPC | *C1QTNF4* | -0.036808658 | 0.000219036 | 0.012406138 | 1 |
| T2DM | ASPC | *FIBP* | 0.56002451 | 0.000577571 | 0.02473877 | 1 |
| T2DM | ASPC | *CTSW* | -0.030025635 | 0.000234254 | 0.012806576 | 1 |
| T2DM | ASPC | *SNX32* | 0.134265567 | 0.000948163 | 0.035482749 | 1 |
| T2DM | ASPC | *RNASEH2C* | -0.180398813 | 0.000382588 | 0.018645994 | 1 |
| T2DM | ASPC | *VEGFB* | -0.147195656 | 7.13E-06 | 0.000862185 | 0.044833627 |
| T2DM | ASPC | *HOXB2* | -0.197602128 | 3.78E-05 | 0.003347352 | 0.237662018 |
| T2DM | ASPC | *TMEM81* | 0.899130936 | 0.001505259 | 0.048781267 | 1 |
| T2DM | ASPC | *KLF14* | -2.083697308 | 1.17E-09 | 5.24E-07 | 7.33E-06 |
| T2DM | ASPC | *NLRP6* | -0.706375427 | 0.000178166 | 0.010875031 | 1 |
| T2DM | ASPC | *DHX36* | -0.675761309 | 0.000504352 | 0.022334612 | 1 |
| T2DM | ASPC | *PACS1* | -0.741476592 | 0.000206936 | 0.011847527 | 1 |
| T2DM | ASPC | *C12orf54* | -0.451445786 | 0.001474048 | 0.048017301 | 1 |
| T2DM | ASPC | *RIC8A* | -0.378162587 | 0.000530866 | 0.023017606 | 1 |
| T2DM | ASPC | *IMP3* | 0.775609035 | 6.46E-05 | 0.005345211 | 0.406236063 |
| T2DM | ASPC | *STX19* | 0.305841237 | 5.73E-05 | 0.004868127 | 0.360241431 |
| T2DM | ASPC | *FARSA* | -0.405925595 | 8.79E-07 | 0.000131546 | 0.005524923 |
| T2DM | ASPC | *CALR* | 0.440759087 | 8.12E-05 | 0.006147697 | 0.510258832 |
| T2DM | ASPC | *HLA-DQB1* | -0.118991923 | 1.63E-15 | 1.70E-12 | 1.02E-11 |
| T2DM | ASPC | *B3GNT3* | 0.3303422 | 0.000831996 | 0.032117356 | 1 |
| T2DM | ASPC | *ZNF320* | 0.047357455 | 0.000852887 | 0.032695734 | 1 |
| T2DM | ASPC | *RP1L1* | 0.193375194 | 0.000260628 | 0.013654734 | 1 |
| T2DM | ASPC | *BMP8A* | 0.673547317 | 3.87E-11 | 2.70E-08 | 2.43E-07 |
| T2DM | ASPC | *ZNF703* | -0.208699789 | 7.83E-05 | 0.006092493 | 0.492174465 |
| T2DM | ASPC | *EIF3C* | 0.093357971 | 0.000504456 | 0.022334612 | 1 |
| T2DM | ASPC | *LINC00839* | 0.172278781 | 0.000801426 | 0.031689102 | 1 |
| T2DM | ASPC | *RASA3* | -0.32193868 | 3.60E-05 | 0.003235473 | 0.226483132 |
| T2DM | ASPC | *RGS7BP* | -0.222756493 | 0.000227461 | 0.012655278 | 1 |
| T2DM | ASPC | *HLA-DRB1* | -0.238742615 | 8.66E-19 | 1.82E-15 | 5.45E-15 |
| T2DM | ASPC | *MYO6* | -0.291850842 | 7.95E-05 | 0.006092493 | 0.499584436 |
| T2DM | ASPC | *FAM180B* | -0.12815577 | 0.000465897 | 0.021691757 | 1 |
| T2DM | ASPC | *ZNF33B* | -0.148576799 | 0.000227356 | 0.012655278 | 1 |
| T2DM | ASPC | *HLA-DQA1* | -0.138715743 | 1.16E-08 | 4.06E-06 | 7.32E-05 |
| T2DM | ASPC | *SLC39A10* | -0.793589939 | 0.000410171 | 0.019685073 | 1 |
| T2DM | ASPC | *PRIM1* | -0.312002109 | 0.000128334 | 0.008684047 | 0.806837869 |
| T2DM | ASPC | *ZNF568* | 0.301340994 | 0.001212077 | 0.042022988 | 1 |
| T2DM | ASPC | *HLA-DRB5* | -0.062002939 | 1.85E-12 | 1.66E-09 | 1.16E-08 |

| | | | | | | |
|---|---|---|---|---|---|---|
| T2DM | ASPC | *MAP3K3* | -0.869466303 | 0.000244486 | 0.013035983 | 1 |
| T2DM | ASPC | *CENPW* | -0.339110382 | 2.72E-06 | 0.00035681 | 0.017126884 |
| T2DM | ASPC | *HLA-DMA* | -0.332674388 | 0.001213993 | 0.042022988 | 1 |
| T2DM | ASPC | *HLA-DRA* | 0.19011728 | 6.14E-10 | 2.97E-07 | 3.86E-06 |
| T2DM | ASPC | *NOTCH4* | -0.020647335 | 7.74E-08 | 1.94E-05 | 0.000486327 |
| T2DM | ASPC | *PRRT1* | 0.117580679 | 0.000417274 | 0.019874264 | 1 |
| T2DM | ASPC | *NELFE* | 0.143158185 | 0.000234067 | 0.012806576 | 1 |
| T2DM | ASPC | *LINC01123* | -2.328555628 | 0.000604284 | 0.02535315 | 1 |
| T2DM | ASPC | *LIN52* | 0.190494094 | 0.00026619 | 0.013830896 | 1 |
| T2DM | ASPC | *HCG27* | 0.120987579 | 2.64E-10 | 1.38E-07 | 1.66E-06 |
| T2DM | ASPC | *COLQ* | -0.862679598 | 0.000112241 | 0.00784068 | 0.705661221 |
| T2DM | ASPC | *FGFR1OP* | 0.298580269 | 0.00045562 | 0.02137676 | 1 |
| T2DM | ASPC | *DNLZ* | 0.07159585 | 0.000149358 | 0.009581745 | 0.939010991 |
| T2DM | ASPC | *ATF6B* | 0.145022118 | 0.000158677 | 0.009976045 | 0.997604454 |
| T2DM | ASPC | *CLIC1* | -0.393610867 | 0.000469645 | 0.021691757 | 1 |
| T2DM | ASPC | *STARD10* | 0.084608787 | 8.49E-07 | 0.000130182 | 0.005337448 |
| T2DM | ASPC | *KCNU1* | -0.27096725 | 2.60E-06 | 0.000347135 | 0.016315343 |
| T2DM | ASPC | *HLA-DQB2* | 0.050391484 | 4.47E-08 | 1.17E-05 | 0.000281054 |
| T2DM | ASPC | *HLA-B* | 0.117846814 | 0.00013626 | 0.008831647 | 0.856669738 |
| T2DM | ASPC | *HLA-DQA2* | 0.140298349 | 4.61E-18 | 7.24E-15 | 2.90E-14 |
| T2DM | ASPC | *LY6G5B* | -0.0363689 | 0.001462629 | 0.047893484 | 1 |
| T2DM | ASPC | *NAT6* | 0.238077074 | 0.001028377 | 0.036756448 | 1 |
| T2DM | ASPC | *PLA2G4B* | 0.149892606 | 8.10E-08 | 1.94E-05 | 0.000509264 |
| T2DM | ASPC | *JMJD7* | 0.170357159 | 1.93E-08 | 5.77E-06 | 0.000121084 |
| T2DM | ASPC | *ZNF286B* | -0.471647557 | 0.000289228 | 0.014904718 | 1 |
| T2DM | ASPC | *DNAH10OS* | -1.424994459 | 0.000237501 | 0.012872126 | 1 |
| T2DM | ASPC | *HMBS* | 0.111089802 | 1.71E-05 | 0.001693128 | 0.107272389 |
| T2DM | ASPC | *AC083843.1* | -1.926762604 | 0.000128697 | 0.008684047 | 0.80912068 |
| T2DM | ASPC | *AC009060.1* | -0.639968192 | 3.97E-07 | 7.80E-05 | 0.002495959 |
| T2DM | b_cell | *SCYL3* | -3.375263656 | 0.010003819 | 0.01514059 | 1 |
| T2DM | b_cell | *CFH* | -1.1806019 | 0.00104436 | 0.00415349 | 1 |
| T2DM | b_cell | *NFYA* | -4.039083573 | 0.018639561 | 0.025420156 | 1 |
| T2DM | b_cell | *NIPAL3* | -3.266449655 | 0.002880379 | 0.0061721 | 1 |
| T2DM | b_cell | *SEMA3F* | -3.973851516 | 2.40E-08 | 1.88E-05 | 0.000150607 |
| T2DM | b_cell | *KRIT1* | -2.710701776 | 0.019041882 | 0.025867829 | 1 |
| T2DM | b_cell | *RAD52* | -2.238958496 | 0.001223614 | 0.004253251 | 1 |
| T2DM | b_cell | *BAD* | -3.677071297 | 0.001210941 | 0.00424133 | 1 |
| T2DM | b_cell | *HS3ST1* | 2.578205087 | 0.001104201 | 0.004159248 | 1 |
| T2DM | b_cell | *LASP1* | -2.824958158 | 0.005398709 | 0.009426623 | 1 |
| T2DM | b_cell | *CYP26B1* | -3.045661746 | 0.001389574 | 0.004333459 | 1 |
| T2DM | b_cell | *ICA1* | -1.487035948 | 0.000851116 | 0.00415349 | 1 |
| T2DM | b_cell | *DBNDD1* | 2.181357913 | 0.00444587 | 0.008235471 | 1 |
| T2DM | b_cell | *ALS2* | -2.274568891 | 0.001710345 | 0.004728644 | 1 |
| T2DM | b_cell | *POLDIP2* | -2.349978097 | 0.00108122 | 0.00415349 | 1 |
| T2DM | b_cell | *CD38* | 3.248700956 | 0.01941137 | 0.026290238 | 1 |
| T2DM | b_cell | *RBM6* | -3.42204252 | 1.48E-08 | 1.55E-05 | 9.31E-05 |
| T2DM | b_cell | *ARHGAP33* | -4.455217316 | 0.000208002 | 0.00415349 | 1 |
| T2DM | b_cell | *PDK4* | -2.081572066 | 0.002146238 | 0.005293604 | 1 |
| T2DM | b_cell | *SLC22A16* | -4.386290566 | 0.007662136 | 0.012198493 | 1 |
| T2DM | b_cell | *CDC27* | -2.144328387 | 0.015250718 | 0.02144515 | 1 |

| | | | | | | |
|---|---|---|---|---|---|---|
| T2DM | b_cell | *MEOX1* | -2.187270768 | 0.001922643 | 0.005009388 | 1 |
| T2DM | b_cell | *LIG3* | -2.514069238 | 0.008021819 | 0.012633561 | 1 |
| T2DM | b_cell | *CIAPIN1* | -3.635111282 | 0.003100894 | 0.006438348 | 1 |
| T2DM | b_cell | *COPZ2* | -1.588428947 | 0.006325575 | 0.010529227 | 1 |
| T2DM | b_cell | *GCFC2* | -2.218846763 | 0.002142824 | 0.005287258 | 1 |
| T2DM | b_cell | *WDR54* | -3.781275549 | 0.001065054 | 0.00415349 | 1 |
| T2DM | b_cell | *ABCB4* | 1.922821413 | 0.000742859 | 0.00415349 | 1 |
| T2DM | b_cell | *ZNF195* | -3.270109455 | 0.000119501 | 0.00415349 | 0.751305386 |
| T2DM | b_cell | *PDK2* | -2.103741776 | 0.000925732 | 0.00415349 | 1 |
| T2DM | b_cell | *GGNBP2* | -3.189891986 | 4.27E-05 | 0.003541347 | 0.268475418 |
| T2DM | b_cell | *TMEM98* | -2.24372607 | 0.002343299 | 0.005521859 | 1 |
| T2DM | b_cell | *ABCC8* | -6.048137671 | 0.000964859 | 0.00415349 | 1 |
| T2DM | b_cell | *CCL18* | 3.972838018 | 0.001834341 | 0.004898539 | 1 |
| T2DM | b_cell | *SYNRG* | -2.468006121 | 0.003747306 | 0.007300686 | 1 |
| T2DM | b_cell | *TMEM132A* | -6.495409801 | 0.001395315 | 0.00434199 | 1 |
| T2DM | b_cell | *SPATA20* | -2.525180943 | 0.000878632 | 0.00415349 | 1 |
| T2DM | b_cell | *TNFRSF12A* | 4.822493343 | 0.009839563 | 0.014920726 | 1 |
| T2DM | b_cell | *MAP3K9* | 1.082665317 | 0.000360191 | 0.00415349 | 1 |
| T2DM | b_cell | *RALA* | -2.843955707 | 0.007117334 | 0.011508915 | 1 |
| T2DM | b_cell | *BAIAP2L1* | -2.047313338 | 0.000998419 | 0.00415349 | 1 |
| T2DM | b_cell | *FARP2* | -2.556586218 | 0.008364551 | 0.013052353 | 1 |
| T2DM | b_cell | *IFRD1* | -1.645502067 | 0.000897586 | 0.00415349 | 1 |
| T2DM | b_cell | *GTF2IRD1* | -1.891849131 | 0.001013956 | 0.00415349 | 1 |
| T2DM | b_cell | *VPS41* | -2.686919567 | 0.020970588 | 0.028207549 | 1 |
| T2DM | b_cell | *ELAC2* | -2.391471404 | 0.002111624 | 0.00524942 | 1 |
| T2DM | b_cell | *CCDC124* | -3.85119685 | 0.015004414 | 0.021165077 | 1 |
| T2DM | b_cell | *NOS2* | -15.98288291 | 0.001775796 | 0.004814329 | 1 |
| T2DM | b_cell | *TRAPPC6A* | -4.554402357 | 0.02130711 | 0.028592914 | 1 |
| T2DM | b_cell | *CD79B* | 0.232925934 | 0.000701233 | 0.00415349 | 1 |
| T2DM | b_cell | *ST7L* | -2.677675129 | 0.000488072 | 0.00415349 | 1 |
| T2DM | b_cell | *PAX6* | -3.913913266 | 0.01796005 | 0.024653894 | 1 |
| T2DM | b_cell | *RPUSD1* | -5.044526256 | 0.000779644 | 0.00415349 | 1 |
| T2DM | b_cell | *CDK11A* | -3.868048133 | 0.001297076 | 0.004268027 | 1 |
| T2DM | b_cell | *DLEC1* | -2.315228832 | 0.000927561 | 0.00415349 | 1 |
| T2DM | b_cell | *SS18L2* | -2.677206982 | 0.000507615 | 0.00415349 | 1 |
| T2DM | b_cell | *MPND* | 2.94859826 | 0.00042565 | 0.00415349 | 1 |
| T2DM | b_cell | *MGST1* | -1.042667076 | 0.000946275 | 0.00415349 | 1 |
| T2DM | b_cell | *NFIX* | -1.53218653 | 0.001330317 | 0.004274973 | 1 |
| T2DM | b_cell | *MMP25* | -2.899163257 | 0.002628143 | 0.005859268 | 1 |
| T2DM | b_cell | *IL32* | -2.945921372 | 0.001341134 | 0.004286583 | 1 |
| T2DM | b_cell | *PKD1* | -2.088304133 | 0.00371661 | 0.00726565 | 1 |
| T2DM | b_cell | *MAPK8IP2* | 1.596017048 | 0.000853241 | 0.00415349 | 1 |
| T2DM | b_cell | *MED24* | -2.939715125 | 0.00084004 | 0.00415349 | 1 |
| T2DM | b_cell | *RHOBTB2* | 3.911629621 | 0.011022478 | 0.016348443 | 1 |
| T2DM | b_cell | *RPS20* | -3.081203518 | 0.032823921 | 0.041825979 | 1 |
| T2DM | b_cell | *POMT2* | -4.078848147 | 0.002230419 | 0.005401538 | 1 |
| T2DM | b_cell | *MLXIPL* | -0.718446051 | 0.037450066 | 0.04702388 | 1 |
| T2DM | b_cell | *METTL13* | -2.345193347 | 0.001079567 | 0.00415349 | 1 |
| T2DM | b_cell | *DYRK4* | -2.446932595 | 0.013366462 | 0.019212266 | 1 |
| T2DM | b_cell | *STARD3NL* | -2.342626144 | 0.001416019 | 0.004364365 | 1 |

| | | | | | | |
|---|---|---|---|---|---|---|
| T2DM | b_cell | *ZNF200* | -3.462914396 | 0.014451067 | 0.02052267 | 1 |
| T2DM | b_cell | *CD4* | -1.901323826 | 0.000745872 | 0.00415349 | 1 |
| T2DM | b_cell | *LRRC23* | 3.516713244 | 0.015691275 | 0.021981071 | 1 |
| T2DM | b_cell | *HFE* | -3.375158171 | 0.000956007 | 0.00415349 | 1 |
| T2DM | b_cell | *SCMH1* | -2.236646391 | 0.008962375 | 0.013820567 | 1 |
| T2DM | b_cell | *MRC2* | -1.49232407 | 0.00098002 | 0.00415349 | 1 |
| T2DM | b_cell | *TSPAN9* | -1.764373503 | 0.000230135 | 0.00415349 | 1 |
| T2DM | b_cell | *MKS1* | -4.737729966 | 0.006855629 | 0.011183534 | 1 |
| T2DM | b_cell | *UTP18* | -2.432326072 | 0.001069542 | 0.00415349 | 1 |
| T2DM | b_cell | *TTC19* | -1.933632222 | 0.004075104 | 0.007751945 | 1 |
| T2DM | b_cell | *DPF1* | -10.20033597 | 0.006872962 | 0.011205995 | 1 |
| T2DM | b_cell | *SYT7* | -4.648227963 | 0.0103724 | 0.015574702 | 1 |
| T2DM | b_cell | *LARS2* | -2.591390435 | 0.016038971 | 0.022393296 | 1 |
| T2DM | b_cell | *PPP5C* | -2.786891216 | 0.008152092 | 0.012781098 | 1 |
| T2DM | b_cell | *ZBTB32* | -3.130699379 | 0.000688848 | 0.00415349 | 1 |
| T2DM | b_cell | *TMEM159* | -2.177566502 | 0.000619447 | 0.00415349 | 1 |
| T2DM | b_cell | *ERCC1* | -1.838305293 | 0.000945041 | 0.00415349 | 1 |
| T2DM | b_cell | *LTF* | -4.638232474 | 0.013132177 | 0.018936238 | 1 |
| T2DM | b_cell | *ALOX5* | 1.740307136 | 0.000891398 | 0.00415349 | 1 |
| T2DM | b_cell | *MAP4K5* | -2.381295893 | 0.002123183 | 0.005267208 | 1 |
| T2DM | b_cell | *PSMC4* | -1.797899757 | 0.003777338 | 0.007340997 | 1 |
| T2DM | b_cell | *MAN2B2* | -4.020697113 | 0.007545642 | 0.012052706 | 1 |
| T2DM | b_cell | *CLDN11* | -1.590566017 | 0.000824354 | 0.00415349 | 1 |
| T2DM | b_cell | *ANGEL1* | -3.08866868 | 0.005363945 | 0.009387268 | 1 |
| T2DM | b_cell | *RNF14* | -2.210952155 | 0.001342844 | 0.004286796 | 1 |
| T2DM | b_cell | *DDX11* | -3.053783316 | 0.001209975 | 0.00424031 | 1 |
| T2DM | b_cell | *HEBP1* | -4.801202375 | 0.002814648 | 0.006087269 | 1 |
| T2DM | b_cell | *TACC3* | -2.404970915 | 0.000501719 | 0.00415349 | 1 |
| T2DM | b_cell | *MDH1* | -1.685775095 | 0.000763601 | 0.00415349 | 1 |
| T2DM | b_cell | *COX15* | -3.459713139 | 0.022995191 | 0.030609944 | 1 |
| T2DM | b_cell | *CCDC88C* | -2.989973396 | 0.02563645 | 0.033543467 | 1 |
| T2DM | b_cell | *NPC1L1* | 4.387833726 | 0.003086627 | 0.006415083 | 1 |
| T2DM | b_cell | *NUDCD3* | -2.632038839 | 0.004161301 | 0.007865091 | 1 |
| T2DM | b_cell | *CHDH* | -6.433667218 | 0.001603796 | 0.004609717 | 1 |
| T2DM | b_cell | *IL20RA* | -6.943303926 | 0.000751221 | 0.00415349 | 1 |
| T2DM | b_cell | *GLT8D1* | -1.887087914 | 0.000779985 | 0.00415349 | 1 |
| T2DM | b_cell | *RUFY3* | -2.339029469 | 0.00353152 | 0.007021717 | 1 |
| T2DM | b_cell | *WWTR1* | -1.845877254 | 0.001613681 | 0.004616359 | 1 |
| T2DM | b_cell | *TTC27* | -2.812811002 | 0.007265561 | 0.01168549 | 1 |
| T2DM | b_cell | *VSIG2* | -6.330866366 | 0.002904939 | 0.006192483 | 1 |
| T2DM | b_cell | *PHLDB1* | -1.298161017 | 0.000258962 | 0.00415349 | 1 |
| T2DM | b_cell | *MARCO* | -1.388063793 | 0.010332213 | 0.015536623 | 1 |
| T2DM | b_cell | *MNAT1* | -1.887251552 | 0.001570278 | 0.004559973 | 1 |
| T2DM | b_cell | *OSBPL5* | -2.433953906 | 0.001424752 | 0.004380342 | 1 |
| T2DM | b_cell | *AQR* | -2.125181851 | 0.002176438 | 0.005332528 | 1 |
| T2DM | b_cell | *CPS1* | -2.17602579 | 0.000490567 | 0.00415349 | 1 |
| T2DM | b_cell | *NLRP2* | -3.700348126 | 0.003230197 | 0.006613998 | 1 |
| T2DM | b_cell | *ZDHHC6* | -2.593363832 | 0.001141689 | 0.00418468 | 1 |
| T2DM | b_cell | *RNH1* | -2.634630075 | 0.00537885 | 0.009404013 | 1 |
| T2DM | b_cell | *NDUFS1* | -2.709285765 | 0.0049986 | 0.008943142 | 1 |

| | | | | | | |
|---|---|---|---|---|---|---|
| T2DM | b_cell | *BIRC3* | 1.051560382 | 0.000778761 | 0.00415349 | 1 |
| T2DM | b_cell | *GLRX2* | -3.740968543 | 0.011534011 | 0.016962609 | 1 |
| T2DM | b_cell | *SNAPC1* | -2.726717109 | 0.001078022 | 0.00415349 | 1 |
| T2DM | b_cell | *GCLM* | -1.764947048 | 0.001286502 | 0.004268027 | 1 |
| T2DM | b_cell | *UBR2* | -2.384996486 | 0.010049134 | 0.015187237 | 1 |
| T2DM | b_cell | *NR1H3* | -1.588607937 | 0.00280211 | 0.006072688 | 1 |
| T2DM | b_cell | *TYMP* | -2.577251298 | 0.005022125 | 0.008977566 | 1 |
| T2DM | b_cell | *TOMM34* | 3.545594833 | 0.001410649 | 0.004364365 | 1 |
| T2DM | b_cell | *KPNA6* | -2.582369575 | 0.011669075 | 0.017109019 | 1 |
| T2DM | b_cell | *VIM* | -2.110101171 | 0.008071794 | 0.012693189 | 1 |
| T2DM | b_cell | *FAS* | -1.947716263 | 0.003020431 | 0.006329816 | 1 |
| T2DM | b_cell | *CD44* | -1.908609151 | 0.001318865 | 0.004274973 | 1 |
| T2DM | b_cell | *BTN3A1* | -2.880136388 | 0.00427266 | 0.008004235 | 1 |
| T2DM | b_cell | *MIPEP* | -2.471340039 | 0.001529085 | 0.004502743 | 1 |
| T2DM | b_cell | *INSRR* | -6.676747217 | 0.01015616 | 0.015315849 | 1 |
| T2DM | b_cell | *VRK2* | -2.576439501 | 0.013350987 | 0.019194524 | 1 |
| T2DM | b_cell | *VEZT* | -2.526134791 | 0.003632061 | 0.007178486 | 1 |
| T2DM | b_cell | *SNX1* | -2.741909463 | 0.005981568 | 0.010125504 | 1 |
| T2DM | b_cell | *ARNTL2* | -3.967726799 | 0.007871521 | 0.012459278 | 1 |
| T2DM | b_cell | *SLC39A9* | -3.082592711 | 0.003850067 | 0.007434083 | 1 |
| T2DM | b_cell | *ANK1* | -2.892260708 | 0.001228252 | 0.004253251 | 1 |
| T2DM | b_cell | *RABEP1* | -2.220522349 | 0.020281496 | 0.02737436 | 1 |
| T2DM | b_cell | *CENPQ* | -3.205128247 | 0.00018568 | 0.00415349 | 1 |
| T2DM | b_cell | *IFT88* | -2.562380418 | 0.002816217 | 0.006088569 | 1 |
| T2DM | b_cell | *ZCCHC8* | -3.050103272 | 0.008789046 | 0.013606681 | 1 |
| T2DM | b_cell | *FUT8* | -2.435062639 | 0.032860224 | 0.04186266 | 1 |
| T2DM | b_cell | *UBA6* | -2.954010317 | 0.000777081 | 0.00415349 | 1 |
| T2DM | b_cell | *PIAS1* | -2.326265063 | 0.010213889 | 0.015384456 | 1 |
| T2DM | b_cell | *SLC4A7* | -2.641597464 | 0.012276602 | 0.017841654 | 1 |
| T2DM | b_cell | *ASTE1* | -2.607703962 | 0.003900729 | 0.007513983 | 1 |
| T2DM | b_cell | *MYOC* | -1.463884977 | 0.006961459 | 0.011309223 | 1 |
| T2DM | b_cell | *SH3YL1* | -2.738724483 | 0.000187687 | 0.00415349 | 1 |
| T2DM | b_cell | *VCL* | -2.68387919 | 0.001130924 | 0.004182741 | 1 |
| T2DM | b_cell | *ADSS* | -3.197886346 | 0.026373101 | 0.0343928 | 1 |
| T2DM | b_cell | *STAP1* | 0.25188342 | 0.000605455 | 0.00415349 | 1 |
| T2DM | b_cell | *TBC1D23* | -2.491596873 | 0.002745658 | 0.006006247 | 1 |
| T2DM | b_cell | *CUL3* | -2.356236859 | 0.037467096 | 0.047035869 | 1 |
| T2DM | b_cell | *MYOM2* | -3.756971728 | 0.015610917 | 0.021878251 | 1 |
| T2DM | b_cell | *TUBG2* | -3.024972643 | 2.14E-08 | 1.88E-05 | 0.000134363 |
| T2DM | b_cell | *NSUN2* | -3.758345923 | 0.0044698 | 0.008272485 | 1 |
| T2DM | b_cell | *MRI1* | -2.473534778 | 0.00060246 | 0.00415349 | 1 |
| T2DM | b_cell | *AGA* | -2.97126949 | 0.001015977 | 0.00415349 | 1 |
| T2DM | b_cell | *MAT2B* | -3.733670385 | 0.01035767 | 0.015560018 | 1 |
| T2DM | b_cell | *CLEC16A* | -2.704596156 | 0.037539342 | 0.047107754 | 1 |
| T2DM | b_cell | *SKIV2L2* | -2.420676626 | 0.005740307 | 0.009822893 | 1 |
| T2DM | b_cell | *C6* | -1.714968331 | 0.000460429 | 0.00415349 | 1 |
| T2DM | b_cell | *RAI14* | -1.999815837 | 0.001407669 | 0.004357467 | 1 |
| T2DM | b_cell | *PNKP* | -2.274916129 | 0.000249704 | 0.00415349 | 1 |
| T2DM | b_cell | *SPDL1* | -2.962968416 | 0.003159562 | 0.006514977 | 1 |
| T2DM | b_cell | *PQLC2* | -2.99513993 | 0.001085135 | 0.00415349 | 1 |

| | | | | | | |
|---|---|---|---|---|---|---|
| T2DM | b_cell | *PSMA4* | -2.891097762 | 0.001576205 | 0.004562749 | 1 |
| T2DM | b_cell | *LSG1* | -2.402239773 | 0.014507729 | 0.020589185 | 1 |
| T2DM | b_cell | *THAP3* | -3.717361098 | 0.000702111 | 0.00415349 | 1 |
| T2DM | b_cell | *SPATA7* | -2.695217624 | 0.00213692 | 0.005285137 | 1 |
| T2DM | b_cell | *MED17* | -3.035306507 | 0.012020882 | 0.017522672 | 1 |
| T2DM | b_cell | *RETSAT* | -1.363716572 | 0.000671938 | 0.00415349 | 1 |
| T2DM | b_cell | *CAPG* | 2.784638865 | 0.026154122 | 0.034135554 | 1 |
| T2DM | b_cell | *LCP2* | -1.660103299 | 0.00084096 | 0.00415349 | 1 |
| T2DM | b_cell | *CUL7* | -3.138082833 | 0.007272638 | 0.011687903 | 1 |
| T2DM | b_cell | *CTNNA1* | -2.256383438 | 0.00498091 | 0.00892419 | 1 |
| T2DM | b_cell | *CNTLN* | -2.309619979 | 0.003258064 | 0.006651595 | 1 |
| T2DM | b_cell | *EPHA3* | -1.549378681 | 0.000792268 | 0.00415349 | 1 |
| T2DM | b_cell | *DSG2* | -2.42168621 | 0.001070355 | 0.00415349 | 1 |
| T2DM | b_cell | *WDR37* | -2.594907393 | 0.00337911 | 0.006811307 | 1 |
| T2DM | b_cell | *YTHDC2* | -2.521710381 | 0.021064776 | 0.028316068 | 1 |
| T2DM | b_cell | *ATP6V1H* | -2.451364308 | 0.00222806 | 0.005400083 | 1 |
| T2DM | b_cell | *POLR2B* | -2.731568646 | 0.004161181 | 0.007865091 | 1 |
| T2DM | b_cell | *CP* | -4.200895852 | 0.006782402 | 0.011092862 | 1 |
| T2DM | b_cell | *ANO2* | -1.344218187 | 0.001073884 | 0.00415349 | 1 |
| T2DM | b_cell | *GOPC* | -2.945710036 | 0.00392822 | 0.007554824 | 1 |
| T2DM | b_cell | *TSPAN17* | -2.012135803 | 0.025945804 | 0.033919998 | 1 |
| T2DM | b_cell | *CC2D2A* | -2.039254526 | 0.001556921 | 0.004533747 | 1 |
| T2DM | b_cell | *RRM2B* | -3.607131523 | 0.006089447 | 0.010252906 | 1 |
| T2DM | b_cell | *SNX29* | -2.236204533 | 0.021951678 | 0.029320204 | 1 |
| T2DM | b_cell | *LMO3* | -1.271904307 | 0.000951189 | 0.00415349 | 1 |
| T2DM | b_cell | *MRPS10* | -2.186893457 | 0.000676945 | 0.00415349 | 1 |
| T2DM | b_cell | *CELF2* | -1.831900146 | 0.003668273 | 0.007211517 | 1 |
| T2DM | b_cell | *ERCC8* | -2.296253939 | 0.004912292 | 0.008839032 | 1 |
| T2DM | b_cell | *ADAMTS6* | -2.743270112 | 0.01670006 | 0.023192684 | 1 |
| T2DM | b_cell | *H6PD* | -2.717205323 | 0.001054771 | 0.00415349 | 1 |
| T2DM | b_cell | *UTS2* | -3.603877751 | 0.001037905 | 0.00415349 | 1 |
| T2DM | b_cell | *RFC2* | 4.865347958 | 0.003052258 | 0.006376763 | 1 |
| T2DM | b_cell | *JKAMP* | -2.182103419 | 0.001929866 | 0.005023674 | 1 |
| T2DM | b_cell | *DKK3* | -2.919592969 | 0.002028635 | 0.005163442 | 1 |
| T2DM | b_cell | *ARHGEF5* | 3.525003147 | 0.000796448 | 0.00415349 | 1 |
| T2DM | b_cell | *MCUR1* | -3.595902536 | 0.002934235 | 0.006222119 | 1 |
| T2DM | b_cell | *LIMA1* | -0.966068128 | 0.00064196 | 0.00415349 | 1 |
| T2DM | b_cell | *PTGER3* | -1.148257139 | 0.000786975 | 0.00415349 | 1 |
| T2DM | b_cell | *PIK3CB* | -2.208883387 | 0.001120383 | 0.004182741 | 1 |
| T2DM | b_cell | *THOC3* | -6.389288991 | 0.010654997 | 0.015915412 | 1 |
| T2DM | b_cell | *RRP12* | -2.742986272 | 0.004870605 | 0.0087917 | 1 |
| T2DM | b_cell | *ALX4* | 4.273299062 | 0.00086429 | 0.00415349 | 1 |
| T2DM | b_cell | *FOXN3* | -2.150765954 | 0.003655534 | 0.007196818 | 1 |
| T2DM | b_cell | *AKR7A2* | -3.276509929 | 0.001848652 | 0.004914366 | 1 |
| T2DM | b_cell | *MRTO4* | -4.897283562 | 0.003675271 | 0.007220759 | 1 |
| T2DM | b_cell | *MCF2L2* | -3.22533897 | 0.001315804 | 0.004274973 | 1 |
| T2DM | b_cell | *ANAPC4* | -2.628172964 | 0.007622746 | 0.012151167 | 1 |
| T2DM | b_cell | *SDCCAG8* | -1.846394094 | 0.005782021 | 0.009883514 | 1 |
| T2DM | b_cell | *TBC1D22A* | -2.225614381 | 0.029561211 | 0.038060892 | 1 |
| T2DM | b_cell | *PLEKHH1* | -2.730045123 | 0.000539268 | 0.00415349 | 1 |

| | | | | | | |
|---|---|---|---|---|---|---|
| T2DM | b_cell | *ATP9A* | -1.381158965 | 0.000654912 | 0.00415349 | 1 |
| T2DM | b_cell | *GALC* | -3.603693337 | 0.019018909 | 0.025842205 | 1 |
| T2DM | b_cell | *FAM114A2* | -2.969820068 | 0.002021705 | 0.005157371 | 1 |
| T2DM | b_cell | *MRPL43* | -3.685158132 | 0.004177205 | 0.00787705 | 1 |
| T2DM | b_cell | *ITIH4* | 2.396217227 | 0.003433041 | 0.006885994 | 1 |
| T2DM | b_cell | *TRAF1* | -4.49079585 | 0.007487931 | 0.011975737 | 1 |
| T2DM | b_cell | *PCGF2* | -1.691742803 | 0.011818237 | 0.017267315 | 1 |
| T2DM | b_cell | *IL17RB* | -5.456494484 | 0.000363204 | 0.00415349 | 1 |
| T2DM | b_cell | *TRAF3IP2* | -2.363155854 | 0.001500766 | 0.004478079 | 1 |
| T2DM | b_cell | *DCBLD2* | -1.862267133 | 0.001035146 | 0.00415349 | 1 |
| T2DM | b_cell | *TMCC3* | -3.830165129 | 0.006075381 | 0.010240193 | 1 |
| T2DM | b_cell | *PITHD1* | -2.694386737 | 0.001508251 | 0.004480341 | 1 |
| T2DM | b_cell | *ATP11B* | -1.844838414 | 0.036902263 | 0.046438056 | 1 |
| T2DM | b_cell | *LAMC2* | -1.540460168 | 0.029552054 | 0.038056895 | 1 |
| T2DM | b_cell | *ZC3H11A* | -5.272275561 | 0.001306404 | 0.004274973 | 1 |
| T2DM | b_cell | *RIOK2* | -3.782501173 | 0.013231037 | 0.019051081 | 1 |
| T2DM | b_cell | *FLYWCH1* | -2.549381675 | 0.001162783 | 0.004195495 | 1 |
| T2DM | b_cell | *DNAJC25* | -2.629482888 | 0.00107537 | 0.00415349 | 1 |
| T2DM | b_cell | *YBX3* | -2.680536903 | 0.004678166 | 0.008562338 | 1 |
| T2DM | b_cell | *OGFR* | -2.846783235 | 0.004665251 | 0.008548654 | 1 |
| T2DM | b_cell | *PIGV* | 2.374084755 | 0.025607131 | 0.033519057 | 1 |
| T2DM | b_cell | *QSER1* | -2.567191581 | 0.00233441 | 0.005509725 | 1 |
| T2DM | b_cell | *NCKAP1* | -1.632910346 | 0.001399408 | 0.004344868 | 1 |
| T2DM | b_cell | *SFSWAP* | -2.508139186 | 0.003696584 | 0.00724956 | 1 |
| T2DM | b_cell | *TNK2* | -2.809461644 | 0.031779416 | 0.040592684 | 1 |
| T2DM | b_cell | *VMP1* | -2.274865245 | 0.003549335 | 0.007050448 | 1 |
| T2DM | b_cell | *EIF4B* | -2.956108414 | 0.022027188 | 0.029408565 | 1 |
| T2DM | b_cell | *SLC6A16* | -2.788624195 | 0.000676226 | 0.00415349 | 1 |
| T2DM | b_cell | *ISOC2* | -6.098943102 | 0.005814914 | 0.009915476 | 1 |
| T2DM | b_cell | *EPN1* | -3.87823696 | 0.026274523 | 0.034271354 | 1 |
| T2DM | b_cell | *ADCK1* | -2.205988491 | 0.000994795 | 0.00415349 | 1 |
| T2DM | b_cell | *HAGH* | -2.372669898 | 0.00083513 | 0.00415349 | 1 |
| T2DM | b_cell | *CASP8* | -2.836585927 | 0.023702895 | 0.031369643 | 1 |
| T2DM | b_cell | *DMRT3* | -3.78824253 | 0.001265548 | 0.004257394 | 1 |
| T2DM | b_cell | *ST3GAL6* | -1.698973648 | 0.000288232 | 0.00415349 | 1 |
| T2DM | b_cell | *ATP2C2* | -4.167179453 | 0.002170872 | 0.005323038 | 1 |
| T2DM | b_cell | *CDON* | -1.29631533 | 0.000760911 | 0.00415349 | 1 |
| T2DM | b_cell | *TAF2* | -2.442655287 | 0.028472723 | 0.036832924 | 1 |
| T2DM | b_cell | *TMEM161A* | -2.873231705 | 0.001362996 | 0.004298348 | 1 |
| T2DM | b_cell | *LPAR2* | -2.635730514 | 0.003028629 | 0.006342768 | 1 |
| T2DM | b_cell | *ABCA7* | 3.501944263 | 0.005451016 | 0.009490594 | 1 |
| T2DM | b_cell | *DDX20* | -3.604981632 | 0.001207912 | 0.004235439 | 1 |
| T2DM | b_cell | *BTBD1* | -2.649210406 | 0.006154166 | 0.010342486 | 1 |
| T2DM | b_cell | *CHI3L2* | 3.758746356 | 0.000569804 | 0.00415349 | 1 |
| T2DM | b_cell | *PMS1* | -2.795981728 | 0.005580449 | 0.009641728 | 1 |
| T2DM | b_cell | *TAF11* | -3.281789951 | 0.001462538 | 0.004424917 | 1 |
| T2DM | b_cell | *ANKS1A* | -2.376742567 | 0.003230728 | 0.006613998 | 1 |
| T2DM | b_cell | *UHRF1BP1* | -1.596795655 | 0.005365064 | 0.009387268 | 1 |
| T2DM | b_cell | *IPO5* | -3.234921069 | 0.017888009 | 0.024587212 | 1 |
| T2DM | b_cell | *WDR18* | -3.195481403 | 0.000151732 | 0.00415349 | 0.953941209 |

| | | | | | | |
|---|---|---|---|---|---|---|
| T2DM | b_cell | *MCM10* | 1.340029832 | 0.000536589 | 0.00415349 | 1 |
| T2DM | b_cell | *ZC3H15* | -3.212298057 | 0.006373444 | 0.010586483 | 1 |
| T2DM | b_cell | *TMEM206* | -2.953101848 | 0.001303941 | 0.004274973 | 1 |
| T2DM | b_cell | *SEC61A2* | -2.150404601 | 0.000889126 | 0.00415349 | 1 |
| T2DM | b_cell | *ASB1* | -3.59392663 | 0.00052815 | 0.00415349 | 1 |
| T2DM | b_cell | *PPP2R5A* | -2.099609452 | 0.002093683 | 0.005240042 | 1 |
| T2DM | b_cell | *TIE1* | -1.885157245 | 0.001455721 | 0.004416626 | 1 |
| T2DM | b_cell | *NFYC* | -2.961784898 | 0.013779049 | 0.019692858 | 1 |
| T2DM | b_cell | *ZMYND12* | -2.212060983 | 0.031157302 | 0.039911564 | 1 |
| T2DM | b_cell | *SLC9A3* | -3.27860902 | 0.023615237 | 0.031282975 | 1 |
| T2DM | b_cell | *NGEF* | -10.39759385 | 0.017292079 | 0.023882974 | 1 |
| T2DM | b_cell | *CD84* | -1.787192959 | 0.0029587 | 0.006264583 | 1 |
| T2DM | b_cell | *ZNRD1* | -3.855274972 | 0.002875551 | 0.006165959 | 1 |
| T2DM | b_cell | *ATXN3* | -1.910225376 | 0.014278862 | 0.020305634 | 1 |
| T2DM | b_cell | *GOLGA5* | -2.200037842 | 0.00088787 | 0.00415349 | 1 |
| T2DM | b_cell | *EML1* | -1.685196725 | 0.000516996 | 0.00415349 | 1 |
| T2DM | b_cell | *ZFAT* | 2.165447189 | 0.001689828 | 0.004704627 | 1 |
| T2DM | b_cell | *MTFR1* | -3.121017984 | 0.002912824 | 0.006193075 | 1 |
| T2DM | b_cell | *STAG3* | -4.369091274 | 0.023806783 | 0.031477023 | 1 |
| T2DM | b_cell | *MYO9A* | -2.134637339 | 0.00455116 | 0.00839095 | 1 |
| T2DM | b_cell | *STOML1* | 3.382351063 | 0.001010011 | 0.00415349 | 1 |
| T2DM | b_cell | *METTL22* | -4.146247478 | 0.016753931 | 0.023252089 | 1 |
| T2DM | b_cell | *TP53BP1* | -2.487992581 | 0.01132362 | 0.016699882 | 1 |
| T2DM | b_cell | *RRP15* | -1.826119588 | 0.006583053 | 0.01084014 | 1 |
| T2DM | b_cell | *DHX8* | -2.546280448 | 0.001762446 | 0.004791201 | 1 |
| T2DM | b_cell | *SYT1* | -2.452742656 | 0.000360627 | 0.00415349 | 1 |
| T2DM | b_cell | *ROGDI* | 3.778977459 | 0.00092501 | 0.00415349 | 1 |
| T2DM | b_cell | *HDAC4* | -2.399610102 | 0.002983927 | 0.006295285 | 1 |
| T2DM | b_cell | *RASSF1* | -3.079005349 | 0.023706292 | 0.031369643 | 1 |
| T2DM | b_cell | *IFI35* | -2.296577802 | 0.010980516 | 0.016304795 | 1 |
| T2DM | b_cell | *HEATR6* | -3.535050067 | 0.003267316 | 0.006662866 | 1 |
| T2DM | b_cell | *POLR1A* | -2.660826519 | 0.000124011 | 0.00415349 | 0.779659076 |
| T2DM | b_cell | *STON1-GTF2A* | -1.142081436 | 0.001485967 | 0.004462053 | 1 |
| T2DM | b_cell | *SRBD1* | -2.703650054 | 0.016198246 | 0.022590589 | 1 |
| T2DM | b_cell | *CYFIP1* | -2.360147262 | 0.002265043 | 0.005437315 | 1 |
| T2DM | b_cell | *PSME4* | -2.595561417 | 0.015706535 | 0.02198775 | 1 |
| T2DM | b_cell | *PITX1* | -7.165558835 | 0.000741905 | 0.00415349 | 1 |
| T2DM | b_cell | *SDK2* | 1.621458273 | 0.000623138 | 0.00415349 | 1 |
| T2DM | b_cell | *NUCKS1* | -2.496921854 | 0.007673105 | 0.012209772 | 1 |
| T2DM | b_cell | *DNAJA2* | -2.649121908 | 0.001094327 | 0.00415349 | 1 |
| T2DM | b_cell | *KCNAB2* | -2.492443684 | 0.000523804 | 0.00415349 | 1 |
| T2DM | b_cell | *GAL* | 2.167610245 | 0.000669577 | 0.00415349 | 1 |
| T2DM | b_cell | *NEDD4* | -1.86553069 | 0.003007773 | 0.006311705 | 1 |
| T2DM | b_cell | *PIGB* | -3.228820499 | 0.036660254 | 0.04619824 | 1 |
| T2DM | b_cell | *PFN2* | -3.425283654 | 0.000573776 | 0.00415349 | 1 |
| T2DM | b_cell | *DAPP1* | 3.074260299 | 0.00751724 | 0.012013444 | 1 |
| T2DM | b_cell | *FGF10* | -1.441495766 | 0.003652474 | 0.007196562 | 1 |
| T2DM | b_cell | *TMEM260* | -2.875449618 | 0.0073503 | 0.011794624 | 1 |
| T2DM | b_cell | *CLTCL1* | -1.880027446 | 0.001836733 | 0.004898539 | 1 |
| T2DM | b_cell | *DGCR2* | -2.966135098 | 0.002748956 | 0.006007705 | 1 |

| | | | | | | |
|---|---|---|---|---|---|---|
| T2DM | b_cell | *ZXDC* | -2.617388622 | 0.003483209 | 0.006956459 | 1 |
| T2DM | b_cell | *WIPI1* | -2.141766371 | 0.001095351 | 0.00415349 | 1 |
| T2DM | b_cell | *NDST1* | -2.795811508 | 0.002520702 | 0.005742541 | 1 |
| T2DM | b_cell | *ASNS* | -3.975327475 | 0.00897753 | 0.013835751 | 1 |
| T2DM | b_cell | *AP3M2* | -3.728964721 | 0.016367825 | 0.022801798 | 1 |
| T2DM | b_cell | *ST6GALNAC2* | -2.68584872 | 0.00129586 | 0.004268027 | 1 |
| T2DM | b_cell | *EIF2B3* | -1.747048527 | 0.001148981 | 0.004187618 | 1 |
| T2DM | b_cell | *CAMK2A* | 4.557039322 | 0.000829285 | 0.00415349 | 1 |
| T2DM | b_cell | *CDC42* | -2.900678973 | 0.030190841 | 0.038800044 | 1 |
| T2DM | b_cell | *OSBPL3* | -2.470308005 | 0.00124716 | 0.004253251 | 1 |
| T2DM | b_cell | *RAD18* | -2.982058541 | 0.002574439 | 0.005806222 | 1 |
| T2DM | b_cell | *SNX13* | -2.550780046 | 0.006050885 | 0.010210172 | 1 |
| T2DM | b_cell | *VASH1* | -3.504445154 | 0.001785746 | 0.004835048 | 1 |
| T2DM | b_cell | *TRIP13* | -12.67843471 | 0.003218644 | 0.006598061 | 1 |
| T2DM | b_cell | *MBD3* | -4.539487558 | 0.000821791 | 0.00415349 | 1 |
| T2DM | b_cell | *HLTF* | -3.109251255 | 0.000796574 | 0.00415349 | 1 |
| T2DM | b_cell | *CPSF1* | -2.322519286 | 0.000897467 | 0.00415349 | 1 |
| T2DM | b_cell | *CYBRD1* | -1.815934763 | 0.006211962 | 0.010403464 | 1 |
| T2DM | b_cell | *EPN2* | -2.39969645 | 0.002311145 | 0.005492478 | 1 |
| T2DM | b_cell | *LIMS2* | -1.614655213 | 0.000973881 | 0.00415349 | 1 |
| T2DM | b_cell | *AFF4* | -2.478440846 | 0.006215872 | 0.010405595 | 1 |
| T2DM | b_cell | *TRHDE* | -1.146611411 | 0.000770688 | 0.00415349 | 1 |
| T2DM | b_cell | *TRNT1* | -2.440879166 | 0.001630391 | 0.004644435 | 1 |
| T2DM | b_cell | *ACADVL* | -3.110623584 | 0.006891717 | 0.011227838 | 1 |
| T2DM | b_cell | *CRMP1* | -1.966584968 | 0.000489627 | 0.00415349 | 1 |
| T2DM | b_cell | *AP1M1* | -2.83570507 | 0.023412211 | 0.031033643 | 1 |
| T2DM | b_cell | *PVR* | -2.950051717 | 0.002909259 | 0.006192483 | 1 |
| T2DM | b_cell | *XRCC1* | -4.591490419 | 0.026998987 | 0.035121587 | 1 |
| T2DM | b_cell | *MOV10L1* | -2.899217545 | 0.000720181 | 0.00415349 | 1 |
| T2DM | b_cell | *TP63* | -1.782083486 | 0.001034028 | 0.00415349 | 1 |
| T2DM | b_cell | *ALPK1* | -2.550521985 | 0.007153937 | 0.01153843 | 1 |
| T2DM | b_cell | *LLGL2* | -2.700146086 | 0.009522326 | 0.014506146 | 1 |
| T2DM | b_cell | *PDE8A* | -1.868320312 | 0.004486785 | 0.008299035 | 1 |
| T2DM | b_cell | *NLE1* | 2.961892927 | 0.000398595 | 0.00415349 | 1 |
| T2DM | b_cell | *SMARCE1* | -2.325413101 | 0.001903949 | 0.004987601 | 1 |
| T2DM | b_cell | *GSDMB* | -1.599790725 | 0.000749424 | 0.00415349 | 1 |
| T2DM | b_cell | *PICALM* | -2.02164284 | 0.004604411 | 0.008469259 | 1 |
| T2DM | b_cell | *GLI2* | -1.815633987 | 0.001125342 | 0.004182741 | 1 |
| T2DM | b_cell | *CLNS1A* | -1.771723279 | 0.034247022 | 0.043400731 | 1 |
| T2DM | b_cell | *ATP2A3* | 3.499140209 | 0.006575487 | 0.010833356 | 1 |
| T2DM | b_cell | *CA12* | -2.254236633 | 0.000921372 | 0.00415349 | 1 |
| T2DM | b_cell | *NTN4* | -1.467246206 | 0.000779032 | 0.00415349 | 1 |
| T2DM | b_cell | *LMAN1* | -3.090945676 | 0.010574688 | 0.015806719 | 1 |
| T2DM | b_cell | *ENO1* | -3.357731407 | 0.003375013 | 0.006805229 | 1 |
| T2DM | b_cell | *SRI* | -2.669245887 | 0.001536115 | 0.004508014 | 1 |
| T2DM | b_cell | *EIF4G3* | -2.295832861 | 0.00982756 | 0.014909718 | 1 |
| T2DM | b_cell | *TTC38* | -1.685140649 | 0.000727429 | 0.00415349 | 1 |
| T2DM | b_cell | *GRAMD4* | -3.568624215 | 0.028143574 | 0.036437119 | 1 |
| T2DM | b_cell | *CELSR1* | -5.307758293 | 0.011806534 | 0.017254226 | 1 |
| T2DM | b_cell | *ADD2* | 6.417120915 | 0.004164608 | 0.007865091 | 1 |

| | | | | | | |
|---|---|---|---|---|---|---|
| T2DM | b_cell | *VPS9D1* | -4.182321698 | 0.00778116 | 0.012350455 | 1 |
| T2DM | b_cell | *MARK3* | -2.643960891 | 0.000819829 | 0.00415349 | 1 |
| T2DM | b_cell | *SLC25A3* | -2.814945611 | 0.024941912 | 0.032709596 | 1 |
| T2DM | b_cell | *FSCN1* | -3.039245636 | 0.000947764 | 0.00415349 | 1 |
| T2DM | b_cell | *MOCOS* | 2.385659013 | 0.000956093 | 0.00415349 | 1 |
| T2DM | b_cell | *PLD1* | -2.029662855 | 0.001709943 | 0.004728644 | 1 |
| T2DM | b_cell | *BCAP29* | -2.636940291 | 0.004994722 | 0.008941292 | 1 |
| T2DM | b_cell | *TUBA3D* | -8.888597431 | 0.001195449 | 0.004217141 | 1 |
| T2DM | b_cell | *MKRN2* | -2.319769855 | 0.000545435 | 0.00415349 | 1 |
| T2DM | b_cell | *MCM6* | 4.041597342 | 0.001011184 | 0.00415349 | 1 |
| T2DM | b_cell | *RBMS2* | -1.719347166 | 0.001036544 | 0.00415349 | 1 |
| T2DM | b_cell | *MLH1* | -2.693600859 | 0.001913564 | 0.004996087 | 1 |
| T2DM | b_cell | *FMO4* | -2.564372869 | 0.001288659 | 0.004268027 | 1 |
| T2DM | b_cell | *RGS11* | -1.976755087 | 0.000682361 | 0.00415349 | 1 |
| T2DM | b_cell | *TPD52* | 1.020310663 | 0.000547649 | 0.00415349 | 1 |
| T2DM | b_cell | *GPATCH1* | -2.310300961 | 0.00222303 | 0.005396212 | 1 |
| T2DM | b_cell | *ICAM3* | 3.816748555 | 0.00056781 | 0.00415349 | 1 |
| T2DM | b_cell | *NT5C2* | -2.222842859 | 0.028873959 | 0.037275249 | 1 |
| T2DM | b_cell | *CAMSAP3* | -17.56000254 | 0.000705411 | 0.00415349 | 1 |
| T2DM | b_cell | *XAB2* | -3.491634615 | 0.000119179 | 0.00415349 | 0.749280959 |
| T2DM | b_cell | *STXBP2* | -2.50908899 | 0.024524711 | 0.032263414 | 1 |
| T2DM | b_cell | *MAP2K7* | -2.524789306 | 0.001087787 | 0.00415349 | 1 |
| T2DM | b_cell | *CTTNBP2* | -2.560505123 | 0.000147947 | 0.00415349 | 0.930144415 |
| T2DM | b_cell | *TM9SF3* | -2.643319646 | 0.006557527 | 0.010809431 | 1 |
| T2DM | b_cell | *DYNC1I2* | -2.652262492 | 0.019643439 | 0.026593089 | 1 |
| T2DM | b_cell | *APBB1IP* | -1.826103006 | 0.002265004 | 0.005437315 | 1 |
| T2DM | b_cell | *LRCH4* | -8.729537457 | 2.15E-05 | 0.002553516 | 0.135336327 |
| T2DM | b_cell | *CAPZB* | -2.500969339 | 0.004038786 | 0.007696831 | 1 |
| T2DM | b_cell | *GPR137B* | -2.244697737 | 0.001627065 | 0.004639164 | 1 |
| T2DM | b_cell | *NAALAD2* | -1.903749743 | 0.006836064 | 0.011163204 | 1 |
| T2DM | b_cell | *FGFR1* | -1.892910833 | 0.000988058 | 0.00415349 | 1 |
| T2DM | b_cell | *FBLN1* | -1.13247148 | 0.001505108 | 0.004478191 | 1 |
| T2DM | b_cell | *PIAS2* | -2.308841913 | 0.001721244 | 0.004747873 | 1 |
| T2DM | b_cell | *AMPH* | -1.525412752 | 0.003007023 | 0.006311705 | 1 |
| T2DM | b_cell | *MCCC1* | -1.949228478 | 0.003700068 | 0.00724956 | 1 |
| T2DM | b_cell | *FAP* | -1.580855575 | 0.001322819 | 0.004274973 | 1 |
| T2DM | b_cell | *ACER3* | -1.68682931 | 0.000470368 | 0.00415349 | 1 |
| T2DM | b_cell | *PIK3C3* | -2.821647838 | 0.015568516 | 0.02182856 | 1 |
| T2DM | b_cell | *N4BP2* | -3.157735712 | 0.010950873 | 0.016268464 | 1 |
| T2DM | b_cell | *TULP3* | -2.314024444 | 0.001808392 | 0.004858702 | 1 |
| T2DM | b_cell | *SYNJ2* | -1.865725934 | 0.000875011 | 0.00415349 | 1 |
| T2DM | b_cell | *ZCWPW1* | -2.484752723 | 0.004752984 | 0.00865764 | 1 |
| T2DM | b_cell | *FGF20* | -23.86789365 | 0.00073649 | 0.00415349 | 1 |
| T2DM | b_cell | *PCM1* | -2.36615375 | 0.009431719 | 0.014389036 | 1 |
| T2DM | b_cell | *MYH7B* | -3.792308087 | 0.014193958 | 0.020216904 | 1 |
| T2DM | b_cell | *FKBP7* | -1.943734748 | 0.004825135 | 0.008742254 | 1 |
| T2DM | b_cell | *OSBPL6* | -2.885306677 | 0.001837875 | 0.004898539 | 1 |
| T2DM | b_cell | *XRCC5* | -2.592685065 | 0.014784276 | 0.02090615 | 1 |
| T2DM | b_cell | *LXN* | -2.47524569 | 0.001498463 | 0.004475552 | 1 |
| T2DM | b_cell | *REXO1* | -4.004429831 | 0.004419444 | 0.008201016 | 1 |

| | | | | | | |
|---|---|---|---|---|---|---|
| T2DM | b_cell | *SAR1A* | -2.803594211 | 0.003848469 | 0.00743328 | 1 |
| T2DM | b_cell | *RAPGEF3* | -1.101073567 | 0.031232294 | 0.039999477 | 1 |
| T2DM | b_cell | *FDFT1* | -2.733899273 | 0.002850746 | 0.00613369 | 1 |
| T2DM | b_cell | *SCGN* | -12.04419182 | 0.004044766 | 0.007705893 | 1 |
| T2DM | b_cell | *DDX1* | -2.720068722 | 0.002797384 | 0.006066627 | 1 |
| T2DM | b_cell | *DNM2* | -2.860401328 | 0.01205136 | 0.017563027 | 1 |
| T2DM | b_cell | *RABL2B* | -2.023346441 | 0.000838038 | 0.00415349 | 1 |
| T2DM | b_cell | *DDX43* | -3.323762909 | 0.000164913 | 0.00415349 | 1 |
| T2DM | b_cell | *DCT* | -3.985777737 | 0.000853124 | 0.00415349 | 1 |
| T2DM | b_cell | *SLC35C2* | -2.724242452 | 0.001561909 | 0.004541962 | 1 |
| T2DM | b_cell | *SCTR* | -1.926483723 | 0.000836334 | 0.00415349 | 1 |
| T2DM | b_cell | *RIF1* | -2.575204541 | 0.003180732 | 0.006549103 | 1 |
| T2DM | b_cell | *KCNN2* | -5.77417447 | 0.000274179 | 0.00415349 | 1 |
| T2DM | b_cell | *PSEN1* | -2.656777566 | 0.008386388 | 0.013079936 | 1 |
| T2DM | b_cell | *CPOX* | -2.817023804 | 0.000781846 | 0.00415349 | 1 |
| T2DM | b_cell | *MOK* | -2.367866918 | 0.001144846 | 0.00418468 | 1 |
| T2DM | b_cell | *RSBN1* | -3.241801636 | 0.003762111 | 0.007320455 | 1 |
| T2DM | b_cell | *MAGI3* | -2.144299985 | 0.002141696 | 0.005287258 | 1 |
| T2DM | b_cell | *COL4A4* | 0.822508531 | 0.000811902 | 0.00415349 | 1 |
| T2DM | b_cell | *PKP1* | -3.784700613 | 0.011215438 | 0.016575331 | 1 |
| T2DM | b_cell | *ZNF506* | -3.088900647 | 0.000836359 | 0.00415349 | 1 |
| T2DM | b_cell | *KIAA0141* | -2.525152266 | 0.000999544 | 0.00415349 | 1 |
| T2DM | b_cell | *PCDHA6* | -40.96342839 | 0.000320069 | 0.00415349 | 1 |
| T2DM | b_cell | *PCDHGA2* | 4.285410689 | 0.008246815 | 0.012910291 | 1 |
| T2DM | b_cell | *HSPB11* | -2.355051387 | 0.001908151 | 0.004988168 | 1 |
| T2DM | b_cell | *WDR70* | -2.192059278 | 0.003801907 | 0.007368246 | 1 |
| T2DM | b_cell | *PGR* | -1.849642434 | 0.00037183 | 0.00415349 | 1 |
| T2DM | b_cell | *COBLL1* | -1.446402746 | 3.16E-05 | 0.003200692 | 0.198442887 |
| T2DM | b_cell | *GEMIN5* | -3.222810791 | 0.010695232 | 0.015967923 | 1 |
| T2DM | b_cell | *GSK3B* | -2.224421813 | 0.006230697 | 0.010416354 | 1 |
| T2DM | b_cell | *ERC1* | -2.193010429 | 0.016853774 | 0.023364741 | 1 |
| T2DM | b_cell | *LYRM2* | -2.096328934 | 0.003011497 | 0.006317412 | 1 |
| T2DM | b_cell | *ULK2* | -2.084443082 | 0.001377874 | 0.004323793 | 1 |
| T2DM | b_cell | *PLOD1* | -3.083633601 | 0.001505664 | 0.004478191 | 1 |
| T2DM | b_cell | *PIBF1* | -2.39503615 | 0.005789816 | 0.00989146 | 1 |
| T2DM | b_cell | *TDRD3* | -2.472593635 | 0.018938116 | 0.025749121 | 1 |
| T2DM | b_cell | *PDS5B* | -2.566416795 | 0.021044678 | 0.0282951 | 1 |
| T2DM | b_cell | *OXCT1* | -2.132761399 | 0.002428501 | 0.005628918 | 1 |
| T2DM | b_cell | *ZNF324* | -2.787206076 | 0.000879212 | 0.00415349 | 1 |
| T2DM | b_cell | *ZNF586* | -2.372895594 | 0.014544902 | 0.020623319 | 1 |
| T2DM | b_cell | *ZNF446* | 5.52041346 | 0.039855283 | 0.049765673 | 1 |
| T2DM | b_cell | *CHMP2B* | -2.880881751 | 0.001231482 | 0.004253251 | 1 |
| T2DM | b_cell | *PPIE* | -3.359071099 | 0.002749192 | 0.006007705 | 1 |
| T2DM | b_cell | *ZMPSTE24* | -2.395432027 | 0.001123239 | 0.004182741 | 1 |
| T2DM | b_cell | *NOA1* | 2.117757224 | 0.001921176 | 0.005007644 | 1 |
| T2DM | b_cell | *GSTP1* | -2.350578203 | 0.001455717 | 0.004416626 | 1 |
| T2DM | b_cell | *WBP11* | -3.197491258 | 0.002358334 | 0.005540753 | 1 |
| T2DM | b_cell | *COL16A1* | -1.863269003 | 0.00026702 | 0.00415349 | 1 |
| T2DM | b_cell | *APOB* | -3.186748825 | 0.000513309 | 0.00415349 | 1 |
| T2DM | b_cell | *NCOA1* | -2.199654566 | 0.010783176 | 0.016064888 | 1 |

| | | | | | | |
|---|---|---|---|---|---|---|
| T2DM | b_cell | *CD59* | -2.776110975 | 0.014465345 | 0.020538307 | 1 |
| T2DM | b_cell | *TAF9* | -5.232103916 | 0.017847394 | 0.024536804 | 1 |
| T2DM | b_cell | *FCN1* | -2.36281841 | 0.000439142 | 0.00415349 | 1 |
| T2DM | b_cell | *SCAMP1* | -2.763894925 | 0.018350729 | 0.025102488 | 1 |
| T2DM | b_cell | *OVGP1* | 3.060131608 | 0.021918027 | 0.02929131 | 1 |
| T2DM | b_cell | *ABCB1* | -1.778598993 | 0.001726815 | 0.004752113 | 1 |
| T2DM | b_cell | *ZNF213* | -1.914111284 | 0.002274933 | 0.005445221 | 1 |
| T2DM | b_cell | *CPNE3* | -1.821880473 | 0.014090839 | 0.020083678 | 1 |
| T2DM | b_cell | *RRN3* | -3.064266946 | 0.002814288 | 0.006087269 | 1 |
| T2DM | b_cell | *CTTN* | -1.64355228 | 0.00091826 | 0.00415349 | 1 |
| T2DM | b_cell | *EPS15* | -1.792388197 | 0.016612443 | 0.0230812 | 1 |
| T2DM | b_cell | *USP40* | -2.253994111 | 0.002276225 | 0.005445221 | 1 |
| T2DM | b_cell | *MAST2* | -2.732574525 | 0.002560361 | 0.005798627 | 1 |
| T2DM | b_cell | *B4GALT1* | -2.808983655 | 0.017619707 | 0.024271494 | 1 |
| T2DM | b_cell | *EIF2AK1* | -2.543430506 | 0.006191893 | 0.010383684 | 1 |
| T2DM | b_cell | *SNX10* | -1.947534493 | 0.000487552 | 0.00415349 | 1 |
| T2DM | b_cell | *SEPHS1* | -2.378754269 | 0.001684719 | 0.004704627 | 1 |
| T2DM | b_cell | *ITPKC* | -1.764065779 | 0.000907704 | 0.00415349 | 1 |
| T2DM | b_cell | *CEACAM6* | 19.19164661 | 0.004132034 | 0.007836531 | 1 |
| T2DM | b_cell | *TMED2* | -3.750220723 | 0.010987715 | 0.016307783 | 1 |
| T2DM | b_cell | *ACOX3* | -2.953573252 | 0.019864859 | 0.026852369 | 1 |
| T2DM | b_cell | *HSD17B14* | -3.40985085 | 0.000211722 | 0.00415349 | 1 |
| T2DM | b_cell | *TRIP6* | -3.625112552 | 0.001177491 | 0.004202203 | 1 |
| T2DM | b_cell | *ACHE* | -3.326927427 | 0.007381339 | 0.01183235 | 1 |
| T2DM | b_cell | *SRRT* | -3.186275394 | 0.00226736 | 0.005440798 | 1 |
| T2DM | b_cell | *PIGS* | -2.091528376 | 0.005422023 | 0.009463702 | 1 |
| T2DM | b_cell | *ATXN7L3* | 2.823194272 | 0.000721295 | 0.00415349 | 1 |
| T2DM | b_cell | *PGS1* | -3.494663639 | 0.00391995 | 0.007541225 | 1 |
| T2DM | b_cell | *PSMC5* | -2.304313328 | 0.00044889 | 0.00415349 | 1 |
| T2DM | b_cell | *LPCAT2* | -2.20451751 | 0.000841343 | 0.00415349 | 1 |
| T2DM | b_cell | *OGFOD1* | -3.87595762 | 0.00128836 | 0.004268027 | 1 |
| T2DM | b_cell | *TXNDC16* | -2.930519989 | 0.030572225 | 0.039234043 | 1 |
| T2DM | b_cell | *GMCL1* | -2.979930331 | 0.008835664 | 0.013662031 | 1 |
| T2DM | b_cell | *PTHLH* | -3.026742399 | 0.001237728 | 0.004253251 | 1 |
| T2DM | b_cell | *PHACTR3* | -1.776973108 | 0.00324785 | 0.006636086 | 1 |
| T2DM | b_cell | *ERGIC2* | -2.911358797 | 0.002664602 | 0.005916635 | 1 |
| T2DM | b_cell | *AAMDC* | -1.979453837 | 0.000203484 | 0.00415349 | 1 |
| T2DM | b_cell | *METTL2A* | -4.339864195 | 0.001347304 | 0.004288861 | 1 |
| T2DM | b_cell | *DDX18* | -2.246363881 | 0.002894895 | 0.006192483 | 1 |
| T2DM | b_cell | *ASAP3* | -2.092504327 | 0.000837651 | 0.00415349 | 1 |
| T2DM | b_cell | *EDEM2* | -4.470580018 | 0.001476018 | 0.004446444 | 1 |
| T2DM | b_cell | *DOCK3* | -1.637675367 | 0.000630441 | 0.00415349 | 1 |
| T2DM | b_cell | *C3orf18* | 2.945655293 | 2.52E-05 | 0.002754251 | 0.158145663 |
| T2DM | b_cell | *FKBP1A* | -4.311739379 | 0.009638394 | 0.014665194 | 1 |
| T2DM | b_cell | *NSFL1C* | -3.53001098 | 0.016388517 | 0.022820511 | 1 |
| T2DM | b_cell | *ZNF343* | -2.656815786 | 0.002445342 | 0.005640408 | 1 |
| T2DM | b_cell | *CPXM1* | -2.673899587 | 0.000736746 | 0.00415349 | 1 |
| T2DM | b_cell | *MAVS* | -2.6313504 | 0.004690865 | 0.008580584 | 1 |
| T2DM | b_cell | *LZTS3* | -8.82034779 | 0.001671925 | 0.004700981 | 1 |
| T2DM | b_cell | *XRN2* | -2.685864174 | 0.004653459 | 0.008534509 | 1 |

| | | | | | | |
|---|---|---|---|---|---|---|
| T2DM | b_cell | *TESC* | -3.018078231 | 0.000794543 | 0.00415349 | 1 |
| T2DM | b_cell | *ANAPC5* | -2.809220353 | 0.022651298 | 0.030184127 | 1 |
| T2DM | b_cell | *SLC23A2* | -2.92207721 | 0.006323609 | 0.010529227 | 1 |
| T2DM | b_cell | *TMEM230* | -2.620971639 | 0.035473089 | 0.044818993 | 1 |
| T2DM | b_cell | *DZANK1* | -3.008270784 | 0.00087049 | 0.00415349 | 1 |
| T2DM | b_cell | *TASP1* | -2.418990235 | 0.002276047 | 0.005445221 | 1 |
| T2DM | b_cell | *KIF16B* | -2.307985937 | 0.001743921 | 0.004783999 | 1 |
| T2DM | b_cell | *FUS* | -2.451511404 | 0.007850695 | 0.012432574 | 1 |
| T2DM | b_cell | *FXYD5* | -3.829498498 | 0.006054049 | 0.010212451 | 1 |
| T2DM | b_cell | *ZNF302* | -2.555866246 | 0.001605059 | 0.00460987 | 1 |
| T2DM | b_cell | *FXYD3* | -10.48123361 | 0.003125631 | 0.006466219 | 1 |
| T2DM | b_cell | *GANAB* | -2.994980449 | 0.003206122 | 0.006581388 | 1 |
| T2DM | b_cell | *GMIP* | -4.329441426 | 0.001693335 | 0.004705788 | 1 |
| T2DM | b_cell | *ANKRD24* | -2.89810452 | 0.0023322 | 0.005509725 | 1 |
| T2DM | b_cell | *GPATCH2L* | -2.403383734 | 0.005439243 | 0.009478105 | 1 |
| T2DM | b_cell | *SLC9A1* | -2.005013072 | 0.000961402 | 0.00415349 | 1 |
| T2DM | b_cell | *SPTLC1* | -2.934642416 | 0.003552879 | 0.007055259 | 1 |
| T2DM | b_cell | *PAPOLA* | -2.527949654 | 0.037362434 | 0.046951354 | 1 |
| T2DM | b_cell | *RGS1* | -3.279919591 | 0.000785618 | 0.00415349 | 1 |
| T2DM | b_cell | *MUL1* | -4.141756851 | 0.000572285 | 0.00415349 | 1 |
| T2DM | b_cell | *SPG21* | -3.571581874 | 0.032706048 | 0.041691591 | 1 |
| T2DM | b_cell | *GNPTG* | -3.196561887 | 0.003023275 | 0.006333666 | 1 |
| T2DM | b_cell | *ZNF268* | -2.95951786 | 0.00048593 | 0.00415349 | 1 |
| T2DM | b_cell | *PABPC4* | -5.954913462 | 0.000886336 | 0.00415349 | 1 |
| T2DM | b_cell | *CERS4* | -3.656888064 | 0.01483073 | 0.020957698 | 1 |
| T2DM | b_cell | *USP48* | -2.657333502 | 0.004158858 | 0.007865091 | 1 |
| T2DM | b_cell | *EXOC1* | -2.445399259 | 0.004054729 | 0.007722533 | 1 |
| T2DM | b_cell | *NLRC4* | -4.485171803 | 0.007199755 | 0.011606375 | 1 |
| T2DM | b_cell | *LAMB1* | -1.658092564 | 0.000379509 | 0.00415349 | 1 |
| T2DM | b_cell | *SLC26A4* | -4.463649678 | 0.000496259 | 0.00415349 | 1 |
| T2DM | b_cell | *DLD* | -2.652119617 | 0.0028379 | 0.006114421 | 1 |
| T2DM | b_cell | *TXNL1* | -2.770182685 | 0.005432211 | 0.009475664 | 1 |
| T2DM | b_cell | *IL5RA* | -5.836255743 | 0.00064688 | 0.00415349 | 1 |
| T2DM | b_cell | *ITGA6* | -1.537133664 | 0.00065836 | 0.00415349 | 1 |
| T2DM | b_cell | *RAPGEF4* | -1.36463148 | 0.00085731 | 0.00415349 | 1 |
| T2DM | b_cell | *SEL1L3* | 0.741786954 | 0.000587248 | 0.00415349 | 1 |
| T2DM | b_cell | *TF* | -2.527424472 | 0.000792931 | 0.00415349 | 1 |
| T2DM | b_cell | *MYO15A* | -2.975227988 | 0.00341914 | 0.006872165 | 1 |
| T2DM | b_cell | *ALKBH5* | -3.580572913 | 0.000115285 | 0.00415349 | 0.724797561 |
| T2DM | b_cell | *NLRP1* | -2.55300229 | 0.004175448 | 0.00787705 | 1 |
| T2DM | b_cell | *SPAG7* | -2.718123081 | 0.008251624 | 0.012911389 | 1 |
| T2DM | b_cell | *ZFHX4* | -1.290898879 | 0.000712636 | 0.00415349 | 1 |
| T2DM | b_cell | *RGS17* | -3.165775833 | 0.000507757 | 0.00415349 | 1 |
| T2DM | b_cell | *TMEM101* | -3.083233416 | 0.00059441 | 0.00415349 | 1 |
| T2DM | b_cell | *CMA1* | -7.734924999 | 0.003344625 | 0.006768457 | 1 |
| T2DM | b_cell | *PPP2R3C* | -2.557419076 | 0.0083468 | 0.013034961 | 1 |
| T2DM | b_cell | *OSGEP* | -3.575151378 | 0.023629668 | 0.031295496 | 1 |
| T2DM | b_cell | *SLC22A17* | -4.187575298 | 0.000211767 | 0.00415349 | 1 |
| T2DM | b_cell | *SCFD1* | -2.082583587 | 0.014865234 | 0.020997018 | 1 |
| T2DM | b_cell | *RPGRIP1* | -3.000056861 | 0.006358275 | 0.010566872 | 1 |

| | | | | | | |
|---|---|---|---|---|---|---|
| T2DM | b_cell | *TOX4* | -3.105534597 | 0.005737082 | 0.009822722 | 1 |
| T2DM | b_cell | *RFFL* | -3.493663368 | 0.001264471 | 0.004257394 | 1 |
| T2DM | b_cell | *MFSD11* | -2.461600023 | 0.007993276 | 0.012601236 | 1 |
| T2DM | b_cell | *DPYSL2* | -1.891802993 | 0.001345973 | 0.004286796 | 1 |
| T2DM | b_cell | *COMT* | -2.859224403 | 0.002820106 | 0.00609273 | 1 |
| T2DM | b_cell | *VNN3* | 2.155433581 | 0.000456095 | 0.00415349 | 1 |
| T2DM | b_cell | *SEC22C* | -3.129650106 | 0.026834312 | 0.034943521 | 1 |
| T2DM | b_cell | *FMO2* | -1.338245281 | 0.002059848 | 0.005205091 | 1 |
| T2DM | b_cell | *PDE6C* | 4.245244076 | 0.004253757 | 0.007980713 | 1 |
| T2DM | b_cell | *CWF19L1* | -4.403292261 | 0.000320068 | 0.00415349 | 1 |
| T2DM | b_cell | *IKZF5* | -3.181055364 | 0.003066428 | 0.006387091 | 1 |
| T2DM | b_cell | *BLNK* | 2.822264498 | 0.004956587 | 0.008893283 | 1 |
| T2DM | b_cell | *WAC* | -2.464293489 | 0.005717965 | 0.009800666 | 1 |
| T2DM | b_cell | *CREM* | -2.3849626 | 0.002260803 | 0.005435438 | 1 |
| T2DM | b_cell | *TPSD1* | -17.29627764 | 0.002663718 | 0.005916635 | 1 |
| T2DM | b_cell | *ITPR3* | -1.79387546 | 0.000807996 | 0.00415349 | 1 |
| T2DM | b_cell | *IFT74* | -2.258174909 | 0.003990601 | 0.007635091 | 1 |
| T2DM | b_cell | *IL12RB1* | 4.34811806 | 0.000628177 | 0.00415349 | 1 |
| T2DM | b_cell | *ACOT7* | -3.049031406 | 0.000533232 | 0.00415349 | 1 |
| T2DM | b_cell | *CDC7* | -3.254095299 | 0.008882677 | 0.013721226 | 1 |
| T2DM | b_cell | *PCSK5* | -1.299541134 | 0.000935792 | 0.00415349 | 1 |
| T2DM | b_cell | *RAB18* | -2.079692789 | 0.00772619 | 0.01228181 | 1 |
| T2DM | b_cell | *NRP1* | -1.890492816 | 0.002845038 | 0.006127699 | 1 |
| T2DM | b_cell | *PRTFDC1* | -2.020372622 | 0.00073579 | 0.00415349 | 1 |
| T2DM | b_cell | *PALMD* | -1.08316833 | 0.000998089 | 0.00415349 | 1 |
| T2DM | b_cell | *TSPAN15* | -1.528080626 | 0.000732688 | 0.00415349 | 1 |
| T2DM | b_cell | *CATSPERG* | -3.319787403 | 0.035851209 | 0.045242182 | 1 |
| T2DM | b_cell | *STX1B* | -4.890370015 | 0.000115939 | 0.00415349 | 0.728905463 |
| T2DM | b_cell | *HSD3B7* | -2.443497043 | 0.00160046 | 0.004605077 | 1 |
| T2DM | b_cell | *MARCH2* | -2.410422548 | 0.005093773 | 0.009038822 | 1 |
| T2DM | b_cell | *CDC34* | -2.46380286 | 0.000665842 | 0.00415349 | 1 |
| T2DM | b_cell | *MTAP* | -3.291157584 | 0.001884376 | 0.004965245 | 1 |
| T2DM | b_cell | *POLR2E* | -1.965749834 | 0.000615794 | 0.00415349 | 1 |
| T2DM | b_cell | *MADCAM1* | -4.569969387 | 0.006722513 | 0.011026465 | 1 |
| T2DM | b_cell | *TRMT2A* | -3.084138788 | 0.000829007 | 0.00415349 | 1 |
| T2DM | b_cell | *ZDHHC8* | -2.198621692 | 0.001461094 | 0.0044241 | 1 |
| T2DM | b_cell | *MED15* | -2.468291204 | 0.002900221 | 0.006192483 | 1 |
| T2DM | b_cell | *LZTR1* | -3.40738675 | 0.002117312 | 0.005257323 | 1 |
| T2DM | b_cell | *CECR2* | -1.780722316 | 0.000644988 | 0.00415349 | 1 |
| T2DM | b_cell | *SMARCB1* | -2.792791533 | 0.012776153 | 0.018479173 | 1 |
| T2DM | b_cell | *P2RX6* | -3.352308958 | 0.000545863 | 0.00415349 | 1 |
| T2DM | b_cell | *SLC7A4* | -6.931451597 | 0.018173526 | 0.024892584 | 1 |
| T2DM | b_cell | *DDT* | -5.208945248 | 0.020756624 | 0.027955633 | 1 |
| T2DM | b_cell | *SUSD2* | -1.971267861 | 0.039014613 | 0.048783785 | 1 |
| T2DM | b_cell | *GGT5* | -2.287361563 | 0.0011173 | 0.004182741 | 1 |
| T2DM | b_cell | *SPECC1L* | -3.452793186 | 0.019131241 | 0.025972385 | 1 |
| T2DM | b_cell | *YPEL1* | -3.841970089 | 0.004946484 | 0.008877689 | 1 |
| T2DM | b_cell | *PRODH* | -23.01017197 | 0.006324216 | 0.010529227 | 1 |
| T2DM | b_cell | *TOP3B* | -2.483422138 | 0.000943123 | 0.00415349 | 1 |
| T2DM | b_cell | *CRYBB3* | -6.530549943 | 0.002514388 | 0.005735834 | 1 |

| | | | | | | |
|---|---|---|---|---|---|---|
| T2DM | b_cell | *CYTH4* | -2.897381551 | 0.00159912 | 0.004603327 | 1 |
| T2DM | b_cell | *CARD10* | -2.972883806 | 0.00190003 | 0.004985596 | 1 |
| T2DM | b_cell | *LRP5L* | -4.805292787 | 0.002187508 | 0.005349464 | 1 |
| T2DM | b_cell | *SLC25A1* | -2.452801195 | 0.001163087 | 0.004195495 | 1 |
| T2DM | b_cell | *LGALS2* | 3.547738772 | 0.001773594 | 0.004810434 | 1 |
| T2DM | b_cell | *GGA1* | -2.394884114 | 0.000374741 | 0.00415349 | 1 |
| T2DM | b_cell | *SH3BP1* | -3.047354232 | 0.000711855 | 0.00415349 | 1 |
| T2DM | b_cell | *HPS4* | -2.801639615 | 0.002549342 | 0.00579036 | 1 |
| T2DM | b_cell | *PIK3IP1* | -3.548578573 | 0.017209644 | 0.023784794 | 1 |
| T2DM | b_cell | *TRIOBP* | -2.271247697 | 0.007008268 | 0.011379386 | 1 |
| T2DM | b_cell | *GCAT* | -3.75076969 | 0.001787418 | 0.004835411 | 1 |
| T2DM | b_cell | *EIF3L* | -3.898586865 | 0.038613878 | 0.048304545 | 1 |
| T2DM | b_cell | *CCDC134* | 5.268238947 | 0.006168537 | 0.010360188 | 1 |
| T2DM | b_cell | *PICK1* | -2.954430652 | 0.002687727 | 0.005933194 | 1 |
| T2DM | b_cell | *SLC16A8* | -5.679312412 | 0.00063548 | 0.00415349 | 1 |
| T2DM | b_cell | *KDELR3* | -2.354010398 | 0.002102627 | 0.005241681 | 1 |
| T2DM | b_cell | *CYP2D6* | 2.788404475 | 0.021277433 | 0.028565283 | 1 |
| T2DM | b_cell | *DDX17* | -2.023460723 | 0.008271416 | 0.012939138 | 1 |
| T2DM | b_cell | *FBXO7* | -2.984027694 | 0.021301925 | 0.028592059 | 1 |
| T2DM | b_cell | *POLDIP3* | -2.362470292 | 0.000956474 | 0.00415349 | 1 |
| T2DM | b_cell | *TIMP3* | -1.509317601 | 0.001358767 | 0.004298348 | 1 |
| T2DM | b_cell | *PPP6R2* | -2.941186835 | 0.002373455 | 0.00555544 | 1 |
| T2DM | b_cell | *CYB5R3* | -2.627492333 | 0.001909899 | 0.004990664 | 1 |
| T2DM | b_cell | *RHBDD3* | -2.175941225 | 0.000988139 | 0.00415349 | 1 |
| T2DM | b_cell | *TTLL1* | -4.789640176 | 0.002988448 | 0.006296833 | 1 |
| T2DM | b_cell | *RASL10A* | -3.587222137 | 0.002568986 | 0.005799671 | 1 |
| T2DM | b_cell | *HMGXB4* | -3.167232583 | 0.005558292 | 0.009618899 | 1 |
| T2DM | b_cell | *TOM1* | -1.622338116 | 0.003003042 | 0.006308941 | 1 |
| T2DM | b_cell | *NEFH* | 4.285176179 | 0.001218004 | 0.004251853 | 1 |
| T2DM | b_cell | *BIK* | -11.52453267 | 0.001285851 | 0.004268027 | 1 |
| T2DM | b_cell | *MCAT* | -3.63204921 | 0.000585571 | 0.00415349 | 1 |
| T2DM | b_cell | *ARSA* | -1.246022982 | 0.032131844 | 0.040992878 | 1 |
| T2DM | b_cell | *TSPO* | -3.252967519 | 0.011050494 | 0.016370042 | 1 |
| T2DM | b_cell | *TTLL12* | -3.156250838 | 0.000682159 | 0.00415349 | 1 |
| T2DM | b_cell | *RBFOX2* | -1.77063323 | 0.001475248 | 0.004446254 | 1 |
| T2DM | b_cell | *MTMR3* | -2.20014695 | 0.002666042 | 0.005916635 | 1 |
| T2DM | b_cell | *APOL4* | -3.000467888 | 0.00691238 | 0.011255668 | 1 |
| T2DM | b_cell | *SAMM50* | -1.893101802 | 0.0008909 | 0.00415349 | 1 |
| T2DM | b_cell | *IFT27* | -2.620044121 | 0.000980687 | 0.00415349 | 1 |
| T2DM | b_cell | *KIAA0930* | -2.695832794 | 0.006232913 | 0.010416354 | 1 |
| T2DM | b_cell | *NCF4* | -2.145222266 | 0.002219868 | 0.005393569 | 1 |
| T2DM | b_cell | *KCTD17* | -7.586352817 | 0.001163154 | 0.004195495 | 1 |
| T2DM | b_cell | *EP300* | -2.552092304 | 0.01860552 | 0.025386012 | 1 |
| T2DM | b_cell | *ACO2* | -2.416515475 | 0.004135671 | 0.007838698 | 1 |
| T2DM | b_cell | *POLR3H* | -2.713253322 | 0.000944161 | 0.00415349 | 1 |
| T2DM | b_cell | *ZBED4* | -3.320553868 | 0.003001088 | 0.006308941 | 1 |
| T2DM | b_cell | *HDAC10* | -4.144000442 | 0.001532328 | 0.004505963 | 1 |
| T2DM | b_cell | *SDR39U1* | -3.504155294 | 0.001811104 | 0.004861831 | 1 |
| T2DM | b_cell | *RBM23* | -2.842852928 | 0.002607602 | 0.005849897 | 1 |
| T2DM | b_cell | *VCPKMT* | -2.935033291 | 0.005522565 | 0.009579216 | 1 |

| | | | | | | |
|---|---|---|---|---|---|---|
| T2DM | b_cell | *SOS2* | -2.46830877 | 0.015508528 | 0.021774108 | 1 |
| T2DM | b_cell | *PYGL* | -1.083262673 | 0.00284829 | 0.006132603 | 1 |
| T2DM | b_cell | *CNIH1* | -3.477658762 | 0.00262015 | 0.005854674 | 1 |
| T2DM | b_cell | *ATP6V1D* | -2.086422347 | 0.000110228 | 0.00415349 | 0.693002723 |
| T2DM | b_cell | *GSTZ1* | -2.519983972 | 0.001094962 | 0.00415349 | 1 |
| T2DM | b_cell | *TMED8* | -4.316405287 | 0.009728285 | 0.014780504 | 1 |
| T2DM | b_cell | *DAAM1* | -1.617342317 | 0.001281207 | 0.004266508 | 1 |
| T2DM | b_cell | *ISM2* | 8.070929796 | 3.06E-05 | 0.003163865 | 0.192425653 |
| T2DM | b_cell | *PPM1A* | -2.316559774 | 0.016550838 | 0.023010862 | 1 |
| T2DM | b_cell | *SLC10A1* | -2.104883543 | 0.000718933 | 0.00415349 | 1 |
| T2DM | b_cell | *DICER1* | -2.319166011 | 0.004676331 | 0.008561472 | 1 |
| T2DM | b_cell | *ZFYVE21* | -3.439576718 | 0.014961165 | 0.021123029 | 1 |
| T2DM | b_cell | *MTHFD1* | -1.574350559 | 0.000756741 | 0.00415349 | 1 |
| T2DM | b_cell | *VRK1* | -3.507565145 | 0.00712769 | 0.011510862 | 1 |
| T2DM | b_cell | *CCNB1IP1* | -2.988382345 | 0.001959249 | 0.005073227 | 1 |
| T2DM | b_cell | *TRIP11* | -2.343986434 | 0.002408778 | 0.00560266 | 1 |
| T2DM | b_cell | *PCK2* | -4.260609334 | 0.002203446 | 0.005373572 | 1 |
| T2DM | b_cell | *KIAA0391* | 7.363580634 | 0.014979237 | 0.02113871 | 1 |
| T2DM | b_cell | *PLTP* | -1.326846786 | 0.018610924 | 0.025386609 | 1 |
| T2DM | b_cell | *GSS* | 4.205003358 | 0.027524716 | 0.035724172 | 1 |
| T2DM | b_cell | *PYGB* | -2.065057043 | 0.000938136 | 0.00415349 | 1 |
| T2DM | b_cell | *ABHD12* | -2.316014417 | 0.005692338 | 0.009767394 | 1 |
| T2DM | b_cell | *PROCR* | -1.953334453 | 0.000284532 | 0.00415349 | 1 |
| T2DM | b_cell | *NINL* | -1.867965228 | 0.002293725 | 0.005477692 | 1 |
| T2DM | b_cell | *CD40* | 4.197494407 | 0.016516584 | 0.022973399 | 1 |
| T2DM | b_cell | *UQCC1* | -3.186081643 | 0.001151038 | 0.004188961 | 1 |
| T2DM | b_cell | *IFT52* | -2.666803059 | 0.010509003 | 0.015721688 | 1 |
| T2DM | b_cell | *DOK5* | -2.27781799 | 0.000984231 | 0.00415349 | 1 |
| T2DM | b_cell | *CSTF1* | -4.141089299 | 0.002834337 | 0.006110932 | 1 |
| T2DM | b_cell | *BMP7* | -2.89032556 | 0.00080467 | 0.00415349 | 1 |
| T2DM | b_cell | *TPD52L2* | -2.24883263 | 0.001095539 | 0.00415349 | 1 |
| T2DM | b_cell | *NELFCD* | -3.246371479 | 0.003314657 | 0.006731024 | 1 |
| T2DM | b_cell | *CTSZ* | -3.333973768 | 0.00824312 | 0.01290772 | 1 |
| T2DM | b_cell | *PRPF6* | -2.703742468 | 0.001394068 | 0.00434199 | 1 |
| T2DM | b_cell | *MTG2* | -2.755487199 | 0.003067063 | 0.006387091 | 1 |
| T2DM | b_cell | *TCFL5* | -3.756750324 | 0.036791979 | 0.046336372 | 1 |
| T2DM | b_cell | *DIDO1* | -2.280779227 | 0.018005695 | 0.024711156 | 1 |
| T2DM | b_cell | *SLC17A9* | 7.365269293 | 0.009561755 | 0.014558413 | 1 |
| T2DM | b_cell | *BIRC7* | 2.048612739 | 0.006051074 | 0.010210172 | 1 |
| T2DM | b_cell | *NKAIN4* | -7.735347706 | 0.000252422 | 0.00415349 | 1 |
| T2DM | b_cell | *PTK6* | 1.435285451 | 0.000607601 | 0.00415349 | 1 |
| T2DM | b_cell | *GMEB2* | -4.603109931 | 0.009440742 | 0.014399307 | 1 |
| T2DM | b_cell | *RNF24* | -2.889239231 | 0.002255765 | 0.005429181 | 1 |
| T2DM | b_cell | *NDUFAF5* | -1.169470137 | 0.031718193 | 0.040539191 | 1 |
| T2DM | b_cell | *TRIB3* | -4.149476652 | 0.000429021 | 0.00415349 | 1 |
| T2DM | b_cell | *SLC52A3* | -6.168425173 | 0.020531043 | 0.027681464 | 1 |
| T2DM | b_cell | *ANGPT4* | -3.439620738 | 0.0005927 | 0.00415349 | 1 |
| T2DM | b_cell | *RSPO4* | 3.483520492 | 0.001684971 | 0.004704627 | 1 |
| T2DM | b_cell | *SIRPB1* | 1.173668143 | 0.000556243 | 0.00415349 | 1 |
| T2DM | b_cell | *SEC23B* | -2.293469655 | 0.002312869 | 0.005492478 | 1 |

| | | | | | | |
|---|---|---|---|---|---|---|
| T2DM | b_cell | *CCM2L* | -2.710879946 | 0.000869925 | 0.00415349 | 1 |
| T2DM | b_cell | *MYL9* | -2.535144337 | 0.006004335 | 0.010153108 | 1 |
| T2DM | b_cell | *MROH8* | -4.068050022 | 0.002366332 | 0.005545309 | 1 |
| T2DM | b_cell | *TTI1* | -3.820767227 | 0.016252711 | 0.022661521 | 1 |
| T2DM | b_cell | *RPRD1B* | -2.458179996 | 0.001196749 | 0.004217466 | 1 |
| T2DM | b_cell | *PXMP4* | -4.793144551 | 0.000618686 | 0.00415349 | 1 |
| T2DM | b_cell | *ACTR5* | -2.862744847 | 0.000705848 | 0.00415349 | 1 |
| T2DM | b_cell | *WFDC2* | 6.238655467 | 0.001244622 | 0.004253251 | 1 |
| T2DM | b_cell | *DHX35* | -3.498299306 | 0.00370593 | 0.007256052 | 1 |
| T2DM | b_cell | *DNTTIP1* | -3.290409598 | 0.00401921 | 0.007671152 | 1 |
| T2DM | b_cell | *PIGU* | -2.972429425 | 0.023276823 | 0.030891682 | 1 |
| T2DM | b_cell | *APMAP* | -1.822801302 | 0.001042821 | 0.00415349 | 1 |
| T2DM | b_cell | *CELF4* | -2.586804958 | 0.000646694 | 0.00415349 | 1 |
| T2DM | b_cell | *METTL4* | -3.703767385 | 0.014260737 | 0.020293629 | 1 |
| T2DM | b_cell | *LPIN2* | -2.779565007 | 0.00479125 | 0.008705951 | 1 |
| T2DM | b_cell | *MYL12A* | -3.348225409 | 0.006682032 | 0.010976978 | 1 |
| T2DM | b_cell | *LIPG* | -8.529539768 | 0.001385718 | 0.004327874 | 1 |
| T2DM | b_cell | *NALCN* | -2.406503193 | 0.000970361 | 0.00415349 | 1 |
| T2DM | b_cell | *HTR2A* | -6.204314653 | 0.000795384 | 0.00415349 | 1 |
| T2DM | b_cell | *KLF5* | -3.845685986 | 0.001247395 | 0.004253251 | 1 |
| T2DM | b_cell | *STK24* | -2.938601033 | 0.004866208 | 0.008790778 | 1 |
| T2DM | b_cell | *DNAJC3* | -3.024550565 | 0.012837654 | 0.018558366 | 1 |
| T2DM | b_cell | *UGGT2* | -1.856889092 | 0.004164012 | 0.007865091 | 1 |
| T2DM | b_cell | *ARHGEF7* | -2.197970524 | 0.002579408 | 0.005814536 | 1 |
| T2DM | b_cell | *SGCG* | -1.553494436 | 0.000771683 | 0.00415349 | 1 |
| T2DM | b_cell | *PARP4* | -2.509583259 | 0.003842848 | 0.007424704 | 1 |
| T2DM | b_cell | *RGCC* | -1.039358779 | 0.007651937 | 0.012191439 | 1 |
| T2DM | b_cell | *VWA8* | -1.958009921 | 0.005369454 | 0.009390197 | 1 |
| T2DM | b_cell | *KATNAL1* | -2.452576625 | 0.001241319 | 0.004253251 | 1 |
| T2DM | b_cell | *DHRS12* | -1.832201842 | 0.000932102 | 0.00415349 | 1 |
| T2DM | b_cell | *MEDAG* | -1.707948507 | 0.000914992 | 0.00415349 | 1 |
| T2DM | b_cell | *TSC22D1* | -2.222883379 | 0.004306192 | 0.008050261 | 1 |
| T2DM | b_cell | *MGRN1* | -2.049715481 | 0.002985615 | 0.006296733 | 1 |
| T2DM | b_cell | *CORO1A* | 2.96120032 | 0.019278384 | 0.026143917 | 1 |
| T2DM | b_cell | *GDPD3* | -2.607928959 | 0.001472922 | 0.004442975 | 1 |
| T2DM | b_cell | *NFAT5* | -1.967914595 | 0.002672434 | 0.005916798 | 1 |
| T2DM | b_cell | *LONP2* | -2.625788565 | 0.004795869 | 0.008709882 | 1 |
| T2DM | b_cell | *PLLP* | -4.187960293 | 0.001760496 | 0.004789372 | 1 |
| T2DM | b_cell | *ACD* | -3.978716542 | 0.000791621 | 0.00415349 | 1 |
| T2DM | b_cell | *USB1* | -5.37574589 | 0.004929092 | 0.008859119 | 1 |
| T2DM | b_cell | *SETD6* | -2.33284321 | 0.001099317 | 0.00415349 | 1 |
| T2DM | b_cell | *COG4* | -2.976526751 | 0.000195994 | 0.00415349 | 1 |
| T2DM | b_cell | *SLC7A6OS* | -1.264345027 | 0.000702014 | 0.00415349 | 1 |
| T2DM | b_cell | *HCFC1R1* | -2.51556755 | 0.000784844 | 0.00415349 | 1 |
| T2DM | b_cell | *NPRL3* | -1.962989744 | 0.007402421 | 0.011857075 | 1 |
| T2DM | b_cell | *NAGPA* | -3.52024079 | 0.002550471 | 0.00579083 | 1 |
| T2DM | b_cell | *CRISPLD2* | -1.707310485 | 0.002766562 | 0.006028396 | 1 |
| T2DM | b_cell | *NME4* | -2.684794517 | 0.000664975 | 0.00415349 | 1 |
| T2DM | b_cell | *NOMO3* | -3.69788154 | 0.00029631 | 0.00415349 | 1 |
| T2DM | b_cell | *LMF1* | -2.743244286 | 0.008276307 | 0.012943568 | 1 |

| | | | | | | |
|---|---|---|---|---|---|---|
| T2DM | b_cell | *NARFL* | -4.126152844 | 0.026510764 | 0.034543663 | 1 |
| T2DM | b_cell | *MTHFSD* | -2.224550025 | 0.002639316 | 0.005882091 | 1 |
| T2DM | b_cell | *CLCN7* | -4.23762877 | 0.004107975 | 0.007801017 | 1 |
| T2DM | b_cell | *HAGHL* | 5.700646563 | 0.018536551 | 0.025307705 | 1 |
| T2DM | b_cell | *SLC7A5* | -10.19872452 | 0.00173731 | 0.004775894 | 1 |
| T2DM | b_cell | *FBXO31* | -2.396686373 | 0.000778953 | 0.00415349 | 1 |
| T2DM | b_cell | *STUB1* | -4.416544107 | 0.001160957 | 0.004195495 | 1 |
| T2DM | b_cell | *RHBDL1* | -5.387922955 | 0.009401935 | 0.014357534 | 1 |
| T2DM | b_cell | *UBE2I* | -2.42509653 | 0.002247668 | 0.005418361 | 1 |
| T2DM | b_cell | *EARS2* | -2.230011578 | 0.000589078 | 0.00415349 | 1 |
| T2DM | b_cell | *BFAR* | -2.878428049 | 0.001591733 | 0.004593791 | 1 |
| T2DM | b_cell | *RRN3P2* | -3.96462591 | 0.001683289 | 0.004704627 | 1 |
| T2DM | b_cell | *STX4* | 3.328473522 | 0.03703728 | 0.046598634 | 1 |
| T2DM | b_cell | *KAT8* | -3.159954015 | 0.023323823 | 0.030930259 | 1 |
| T2DM | b_cell | *NOMO1* | -2.711693195 | 0.000533092 | 0.00415349 | 1 |
| T2DM | b_cell | *SYT17* | -2.334816745 | 0.000228955 | 0.00415349 | 1 |
| T2DM | b_cell | *TMC5* | -3.395833851 | 0.00034721 | 0.00415349 | 1 |
| T2DM | b_cell | *KNOP1* | -3.411339837 | 0.018541004 | 0.025307705 | 1 |
| T2DM | b_cell | *IQCH* | -2.355517384 | 0.001174234 | 0.004202203 | 1 |
| T2DM | b_cell | *LACTB* | -2.866284126 | 0.001327678 | 0.004274973 | 1 |
| T2DM | b_cell | *HERC1* | -2.197963872 | 0.009016851 | 0.01388074 | 1 |
| T2DM | b_cell | *TRIP4* | -3.100789119 | 0.017844894 | 0.024536804 | 1 |
| T2DM | b_cell | *RASL12* | -3.423587248 | 0.000883762 | 0.00415349 | 1 |
| T2DM | b_cell | *AP3B2* | 1.304166441 | 0.018135572 | 0.024859113 | 1 |
| T2DM | b_cell | *TTC23* | -2.523075612 | 0.002817691 | 0.00608966 | 1 |
| T2DM | b_cell | *FAH* | -1.806215268 | 0.000766762 | 0.00415349 | 1 |
| T2DM | b_cell | *RPAP1* | -2.243183526 | 0.000538327 | 0.00415349 | 1 |
| T2DM | b_cell | *EHD4* | -2.597879328 | 0.002671479 | 0.005916798 | 1 |
| T2DM | b_cell | *ATP8B4* | -1.365073955 | 0.000768642 | 0.00415349 | 1 |
| T2DM | b_cell | *DTWD1* | -1.760066603 | 0.004294511 | 0.008039295 | 1 |
| T2DM | b_cell | *DNAJC17* | -2.849131826 | 0.002007811 | 0.005131344 | 1 |
| T2DM | b_cell | *PDGFRL* | -1.63888045 | 0.001755248 | 0.004789372 | 1 |
| T2DM | b_cell | *ZFAND1* | -2.10586449 | 0.001507133 | 0.00448007 | 1 |
| T2DM | b_cell | *RIPK2* | -3.27673178 | 0.003478289 | 0.006953391 | 1 |
| T2DM | b_cell | *NBN* | -2.782971996 | 0.002884646 | 0.006179137 | 1 |
| T2DM | b_cell | *CPQ* | -1.691481565 | 0.004647675 | 0.008526388 | 1 |
| T2DM | b_cell | *NIPAL2* | -1.832086341 | 0.00349882 | 0.006971457 | 1 |
| T2DM | b_cell | *IKBKB* | -2.751697017 | 0.018123865 | 0.024857055 | 1 |
| T2DM | b_cell | *PLAT* | -1.817724361 | 0.000961922 | 0.00415349 | 1 |
| T2DM | b_cell | *RAB2A* | -1.621843057 | 0.01699049 | 0.023523279 | 1 |
| T2DM | b_cell | *ZC2HC1A* | -2.781424971 | 0.008309921 | 0.012989675 | 1 |
| T2DM | b_cell | *ARMC1* | -2.168534299 | 0.000309199 | 0.00415349 | 1 |
| T2DM | b_cell | *SPAG1* | -2.491054008 | 0.005063209 | 0.009007468 | 1 |
| T2DM | b_cell | *CHRAC1* | -2.905684984 | 0.001378964 | 0.004323793 | 1 |
| T2DM | b_cell | *SNX16* | -2.714099115 | 0.001804607 | 0.00485268 | 1 |
| T2DM | b_cell | *TSTA3* | -5.4952586 | 0.001297544 | 0.004268027 | 1 |
| T2DM | b_cell | *MTMR9* | -2.352536437 | 0.001464867 | 0.004429831 | 1 |
| T2DM | b_cell | *LEPROTL1* | -3.198438692 | 0.003952919 | 0.007588398 | 1 |
| T2DM | b_cell | *TNFRSF10A* | -1.657658321 | 0.009701808 | 0.014750972 | 1 |
| T2DM | b_cell | *ERICH1* | -2.458305651 | 0.003950256 | 0.007585602 | 1 |

| | | | | | | |
|---|---|---|---|---|---|---|
| T2DM | b_cell | *TUSC3* | -1.69512199 | 0.000853972 | 0.00415349 | 1 |
| T2DM | b_cell | *ARHGEF10* | -1.682147501 | 0.002073484 | 0.005226942 | 1 |
| T2DM | b_cell | *MCM4* | -3.854965445 | 0.0034366 | 0.006887461 | 1 |
| T2DM | b_cell | *MAN2B1* | -2.169556229 | 0.004168907 | 0.007870846 | 1 |
| T2DM | b_cell | *TULP2* | 4.309709561 | 0.000326074 | 0.00415349 | 1 |
| T2DM | b_cell | *NUCB1* | -2.773573932 | 0.003309746 | 0.00672757 | 1 |
| T2DM | b_cell | *SARS2* | -4.91459233 | 0.004250461 | 0.007980713 | 1 |
| T2DM | b_cell | *SNRNP70* | -2.982160211 | 0.005740113 | 0.009822893 | 1 |
| T2DM | b_cell | *FCGRT* | -2.968809251 | 0.002905755 | 0.006192483 | 1 |
| T2DM | b_cell | *PEX11G* | -4.884140415 | 0.006729338 | 0.011034781 | 1 |
| T2DM | b_cell | *ERCC2* | -4.82881336 | 0.001489448 | 0.004462053 | 1 |
| T2DM | b_cell | *PLEKHJ1* | -3.535265735 | 0.001328987 | 0.004274973 | 1 |
| T2DM | b_cell | *CD37* | 0.69673569 | 0.000618435 | 0.00415349 | 1 |
| T2DM | b_cell | *STX10* | 4.770372633 | 0.013410625 | 0.019249452 | 1 |
| T2DM | b_cell | *FCER2* | 0.757720419 | 0.000664872 | 0.00415349 | 1 |
| T2DM | b_cell | *DMPK* | -5.838222598 | 0.002031573 | 0.005166868 | 1 |
| T2DM | b_cell | *IL4I1* | -2.761835883 | 0.005433398 | 0.009475664 | 1 |
| T2DM | b_cell | *AES* | -3.06649751 | 0.004431496 | 0.008216105 | 1 |
| T2DM | b_cell | *C19orf53* | -4.832549877 | 0.021424755 | 0.028715467 | 1 |
| T2DM | b_cell | *TIMM44* | -2.991244994 | 0.007075589 | 0.011462054 | 1 |
| T2DM | b_cell | *OLFM2* | -1.719532364 | 0.001043909 | 0.00415349 | 1 |
| T2DM | b_cell | *RASAL3* | -2.829973031 | 0.00108989 | 0.00415349 | 1 |
| T2DM | b_cell | *SYDE1* | -3.305183375 | 0.000650346 | 0.00415349 | 1 |
| T2DM | b_cell | *AURKC* | 1.637588099 | 0.000333013 | 0.00415349 | 1 |
| T2DM | b_cell | *CCNE1* | -5.441570269 | 0.000695626 | 0.00415349 | 1 |
| T2DM | b_cell | *PDCD5* | -3.271050408 | 0.025975298 | 0.033951496 | 1 |
| T2DM | b_cell | *ANKRD27* | -2.038727502 | 0.000834919 | 0.00415349 | 1 |
| T2DM | b_cell | *PRX* | 2.90036599 | 0.001355589 | 0.004296951 | 1 |
| T2DM | b_cell | *CCDC94* | -4.526817486 | 0.002799774 | 0.006069716 | 1 |
| T2DM | b_cell | *POLR2I* | -3.726606425 | 0.01042743 | 0.015642389 | 1 |
| T2DM | b_cell | *CLIP3* | -2.601563677 | 0.000424863 | 0.00415349 | 1 |
| T2DM | b_cell | *SLC1A5* | -2.033765251 | 0.001254338 | 0.004255815 | 1 |
| T2DM | b_cell | *PRKD2* | -3.499168365 | 0.034610474 | 0.043808346 | 1 |
| T2DM | b_cell | *CACTIN* | -3.176510662 | 0.000779752 | 0.00415349 | 1 |
| T2DM | b_cell | *CCDC9* | 3.594703856 | 0.000638093 | 0.00415349 | 1 |
| T2DM | b_cell | *FZR1* | 4.993671446 | 0.026613789 | 0.03467072 | 1 |
| T2DM | b_cell | *BBC3* | -7.294402098 | 0.016430556 | 0.022868919 | 1 |
| T2DM | b_cell | *TGFB1* | -2.888309365 | 0.005111926 | 0.009060806 | 1 |
| T2DM | b_cell | *PLIN3* | -2.314088019 | 0.00096329 | 0.00415349 | 1 |
| T2DM | b_cell | *MRPL4* | -3.289454256 | 0.003182432 | 0.006549248 | 1 |
| T2DM | b_cell | *ICAM5* | -24.14618897 | 1.95E-05 | 0.002553516 | 0.122475857 |
| T2DM | b_cell | *CD33* | -2.937779931 | 0.001672855 | 0.004701493 | 1 |
| T2DM | b_cell | *TYK2* | -2.960378516 | 0.015168863 | 0.021353928 | 1 |
| T2DM | b_cell | *RABAC1* | -2.49197783 | 0.000678823 | 0.00415349 | 1 |
| T2DM | b_cell | *PTPRS* | -1.346113863 | 0.00184301 | 0.004901439 | 1 |
| T2DM | b_cell | *GRIN2D* | -3.081597883 | 0.000760627 | 0.00415349 | 1 |
| T2DM | b_cell | *CARD8* | -2.739330649 | 0.002188332 | 0.005349464 | 1 |
| T2DM | b_cell | *LIG1* | 1.942751446 | 0.000861856 | 0.00415349 | 1 |
| T2DM | b_cell | *ZNF175* | -4.866636538 | 0.000960193 | 0.00415349 | 1 |
| T2DM | b_cell | *PLA2G4C* | -2.592148089 | 0.003198426 | 0.006575704 | 1 |

| | | | | | | |
|---|---|---|---|---|---|---|
| T2DM | b_cell | *CAPS* | -2.951817396 | 0.017309084 | 0.023901211 | 1 |
| T2DM | b_cell | *MIER2* | -4.952147018 | 0.003804082 | 0.007370189 | 1 |
| T2DM | b_cell | *PPP2R1A* | -3.47461645 | 0.009430759 | 0.014389036 | 1 |
| T2DM | b_cell | *TNPO2* | -2.945076913 | 0.029304378 | 0.037768886 | 1 |
| T2DM | b_cell | *GCDH* | -2.364779193 | 8.73E-05 | 0.00415349 | 0.548926274 |
| T2DM | b_cell | *LILRB5* | -1.702928171 | 0.000781406 | 0.00415349 | 1 |
| T2DM | b_cell | *MAST1* | -6.4073774 | 0.000778133 | 0.00415349 | 1 |
| T2DM | b_cell | *PRPF31* | -3.629411881 | 0.001433699 | 0.004395857 | 1 |
| T2DM | b_cell | *TFPT* | 4.912634511 | 0.010900137 | 0.016208411 | 1 |
| T2DM | b_cell | *ELL* | -2.086528205 | 0.002849775 | 0.00613369 | 1 |
| T2DM | b_cell | *UPK1A* | -9.563144271 | 0.005934915 | 0.010062787 | 1 |
| T2DM | b_cell | *COPE* | 3.941713664 | 0.007094116 | 0.01148319 | 1 |
| T2DM | b_cell | *LSR* | -2.388676212 | 0.000800522 | 0.00415349 | 1 |
| T2DM | b_cell | *KXD1* | -2.648898794 | 0.001057707 | 0.00415349 | 1 |
| T2DM | b_cell | *HPN* | -3.508040605 | 0.000196382 | 0.00415349 | 1 |
| T2DM | b_cell | *SIPA1L3* | 2.626765924 | 0.018966458 | 0.02578208 | 1 |
| T2DM | b_cell | *SMG9* | -2.64593443 | 0.001377547 | 0.004323793 | 1 |
| T2DM | b_cell | *GTPBP10* | -2.446876664 | 0.005316507 | 0.009320937 | 1 |
| T2DM | b_cell | *PON3* | -3.00471603 | 0.000819334 | 0.00415349 | 1 |
| T2DM | b_cell | *PON2* | -2.683382198 | 0.002448905 | 0.005641723 | 1 |
| T2DM | b_cell | *HBP1* | -2.642424537 | 0.002909907 | 0.006192483 | 1 |
| T2DM | b_cell | *WDR91* | -3.238573003 | 0.001919632 | 0.005005693 | 1 |
| T2DM | b_cell | *DLX5* | 29.21440982 | 0.001639404 | 0.004657671 | 1 |
| T2DM | b_cell | *STEAP1B* | 5.097095729 | 0.003298889 | 0.00670977 | 1 |
| T2DM | b_cell | *PTN* | -2.844380329 | 0.001109303 | 0.004166182 | 1 |
| T2DM | b_cell | *MPP6* | -2.018884942 | 0.001063104 | 0.00415349 | 1 |
| T2DM | b_cell | *DFNA5* | -2.135892911 | 0.00148589 | 0.004462053 | 1 |
| T2DM | b_cell | *OGDH* | -2.746199691 | 0.008112475 | 0.012728507 | 1 |
| T2DM | b_cell | *ADAP1* | -8.647902349 | 0.001611467 | 0.004615623 | 1 |
| T2DM | b_cell | *H2AFV* | -3.137119249 | 0.008840873 | 0.013666725 | 1 |
| T2DM | b_cell | *CAV1* | -1.284595869 | 0.000932087 | 0.00415349 | 1 |
| T2DM | b_cell | *MET* | -1.722042478 | 0.000738492 | 0.00415349 | 1 |
| T2DM | b_cell | *RNF32* | -2.279818101 | 0.000481139 | 0.00415349 | 1 |
| T2DM | b_cell | *LMBR1* | -2.842393325 | 0.007906077 | 0.01250453 | 1 |
| T2DM | b_cell | *HOXA2* | -7.580133757 | 0.001505789 | 0.004478191 | 1 |
| T2DM | b_cell | *HOXA5* | -2.013278282 | 0.000572819 | 0.00415349 | 1 |
| T2DM | b_cell | *HOXA6* | -2.189534707 | 0.0005229 | 0.00415349 | 1 |
| T2DM | b_cell | *CPED1* | -2.168980228 | 0.003490508 | 0.00696378 | 1 |
| T2DM | b_cell | *HIBADH* | -2.203302346 | 0.002317293 | 0.005497668 | 1 |
| T2DM | b_cell | *CPVL* | -1.604608769 | 0.003960298 | 0.007597923 | 1 |
| T2DM | b_cell | *ABHD11* | -3.491238852 | 0.020440112 | 0.027576605 | 1 |
| T2DM | b_cell | *PLEKHA8* | -2.129604778 | 0.00081538 | 0.00415349 | 1 |
| T2DM | b_cell | *NOD1* | -2.718118699 | 0.003728331 | 0.007274989 | 1 |
| T2DM | b_cell | *CHCHD2* | -3.120342242 | 0.027287386 | 0.03543809 | 1 |
| T2DM | b_cell | *CCL24* | -4.81138075 | 0.039499999 | 0.049361259 | 1 |
| T2DM | b_cell | *NPTX2* | -90.55829759 | 0.002402724 | 0.005596247 | 1 |
| T2DM | b_cell | *CYP3A5* | -2.866649402 | 0.001541217 | 0.004509332 | 1 |
| T2DM | b_cell | *ZKSCAN1* | -2.482179933 | 0.003491305 | 0.00696378 | 1 |
| T2DM | b_cell | *EIF3B* | -2.847447929 | 0.001546525 | 0.004516025 | 1 |
| T2DM | b_cell | *SNX8* | -3.100911743 | 0.003124687 | 0.006466219 | 1 |

| | | | | | | |
|---|---|---|---|---|---|---|
| T2DM | b_cell | *TAF6* | -2.771579686 | 0.002170823 | 0.005323038 | 1 |
| T2DM | b_cell | *AIMP2* | -2.468536458 | 0.00100756 | 0.00415349 | 1 |
| T2DM | b_cell | *RBM28* | -1.525661398 | 0.000438288 | 0.00415349 | 1 |
| T2DM | b_cell | *USP42* | -2.508949075 | 0.002585532 | 0.005820315 | 1 |
| T2DM | b_cell | *IMPDH1* | -3.752832947 | 0.00141682 | 0.004364365 | 1 |
| T2DM | b_cell | *LSM5* | -3.209085436 | 0.001560594 | 0.004540237 | 1 |
| T2DM | b_cell | *AP1S1* | -6.087466593 | 0.002003822 | 0.005129117 | 1 |
| T2DM | b_cell | *PLOD3* | -3.761047029 | 0.002697639 | 0.005946726 | 1 |
| T2DM | b_cell | *RPA3* | -3.028108041 | 0.005027018 | 0.008978654 | 1 |
| T2DM | b_cell | *ZNHIT1* | -3.479807236 | 0.005869287 | 0.009978422 | 1 |
| T2DM | b_cell | *PHF14* | -2.351131093 | 0.008592043 | 0.013360914 | 1 |
| T2DM | b_cell | *ZNF862* | -2.859539737 | 0.009709316 | 0.014758818 | 1 |
| T2DM | b_cell | *MEST* | -1.534375668 | 0.000161587 | 0.00415349 | 1 |
| T2DM | b_cell | *ACTR3C* | -3.411256815 | 0.019239627 | 0.026096986 | 1 |
| T2DM | b_cell | *POU6F2* | -2.022807241 | 0.000764724 | 0.00415349 | 1 |
| T2DM | b_cell | *TSPAN13* | 2.304097606 | 0.000900895 | 0.00415349 | 1 |
| T2DM | b_cell | *RARRES2* | -1.701407995 | 0.00116156 | 0.004195495 | 1 |
| T2DM | b_cell | *COA1* | -2.093026593 | 0.015088496 | 0.021274137 | 1 |
| T2DM | b_cell | *BLVRA* | -2.866028803 | 0.001798291 | 0.004852128 | 1 |
| T2DM | b_cell | *URGCP* | -2.361467774 | 0.001323393 | 0.004274973 | 1 |
| T2DM | b_cell | *POLD2* | -5.716579706 | 0.000142915 | 0.00415349 | 0.898507226 |
| T2DM | b_cell | *GCK* | -29.96085443 | 0.000674614 | 0.00415349 | 1 |
| T2DM | b_cell | *YKT6* | -2.59011713 | 0.00106616 | 0.00415349 | 1 |
| T2DM | b_cell | *TBL2* | -1.991376743 | 0.023904486 | 0.031588461 | 1 |
| T2DM | b_cell | *LIMK1* | -4.576190723 | 0.006839766 | 0.011166349 | 1 |
| T2DM | b_cell | *SPATA6L* | -2.066019077 | 0.000678202 | 0.00415349 | 1 |
| T2DM | b_cell | *SLC1A1* | -1.143651516 | 0.018152304 | 0.024868933 | 1 |
| T2DM | b_cell | *FKTN* | -3.069438371 | 0.003701461 | 0.00724956 | 1 |
| T2DM | b_cell | *NMRK1* | -1.892603823 | 0.003338976 | 0.006765112 | 1 |
| T2DM | b_cell | *TMEM245* | -2.544996555 | 0.003815655 | 0.007388059 | 1 |
| T2DM | b_cell | *MEGF9* | -2.882447962 | 0.008853493 | 0.013682868 | 1 |
| T2DM | b_cell | *TRIM14* | -1.874639333 | 0.000389554 | 0.00415349 | 1 |
| T2DM | b_cell | *CORO2A* | -2.613608532 | 0.00048421 | 0.00415349 | 1 |
| T2DM | b_cell | *C5* | -2.007046626 | 0.001189752 | 0.004210557 | 1 |
| T2DM | b_cell | *OGN* | -2.632929699 | 0.00171359 | 0.004735535 | 1 |
| T2DM | b_cell | *ECM2* | -1.361193253 | 0.000908176 | 0.00415349 | 1 |
| T2DM | b_cell | *PTGR1* | -1.607448747 | 0.001907736 | 0.004988168 | 1 |
| T2DM | b_cell | *SUSD1* | -2.321967828 | 0.013466792 | 0.019316843 | 1 |
| T2DM | b_cell | *TNFSF8* | -2.405900887 | 0.00061375 | 0.00415349 | 1 |
| T2DM | b_cell | *PLGRKT* | -2.410278224 | 0.000869055 | 0.00415349 | 1 |
| T2DM | b_cell | *DOCK8* | -1.741688439 | 0.036428241 | 0.045915066 | 1 |
| T2DM | b_cell | *ELAVL2* | -5.596532804 | 0.000109473 | 0.00415349 | 0.688258843 |
| T2DM | b_cell | *FUBP3* | -2.681245469 | 0.00246212 | 0.005658298 | 1 |
| T2DM | b_cell | *MPDZ* | -1.526295165 | 0.00094022 | 0.00415349 | 1 |
| T2DM | b_cell | *PTGDS* | -5.222324725 | 0.001838493 | 0.004898539 | 1 |
| T2DM | b_cell | *SHB* | -11.6498139 | 0.024882974 | 0.032652736 | 1 |
| T2DM | b_cell | *DVL1* | -3.006506891 | 0.001054232 | 0.00415349 | 1 |
| T2DM | b_cell | *PDLIM1* | -2.215239951 | 0.001257685 | 0.004257394 | 1 |
| T2DM | b_cell | *CCNJ* | -3.613433861 | 0.019894166 | 0.026886204 | 1 |
| T2DM | b_cell | *ATRNL1* | -2.054827458 | 0.00133606 | 0.004282824 | 1 |

| | | | | | | |
|---|---|---|---|---|---|---|
| T2DM | b_cell | *HPS1* | -3.697362726 | 0.010144019 | 0.015301212 | 1 |
| T2DM | b_cell | *PHYH* | -2.11687967 | 0.000893365 | 0.00415349 | 1 |
| T2DM | b_cell | *DNMBP* | -2.043279198 | 0.001245509 | 0.004253251 | 1 |
| T2DM | b_cell | *TRDMT1* | -3.644730036 | 0.007021689 | 0.011392351 | 1 |
| T2DM | b_cell | *DDX50* | -2.898105134 | 0.004659412 | 0.008540445 | 1 |
| T2DM | b_cell | *MAPK8* | -2.323061949 | 0.005680492 | 0.009761317 | 1 |
| T2DM | b_cell | *ATE1* | -2.203241625 | 0.007126567 | 0.011510862 | 1 |
| T2DM | b_cell | *NSMCE4A* | -2.42383207 | 0.001076014 | 0.00415349 | 1 |
| T2DM | b_cell | *PLEKHA1* | -2.614833873 | 0.005682587 | 0.009761317 | 1 |
| T2DM | b_cell | *SPOCK2* | -2.688452508 | 0.000945186 | 0.00415349 | 1 |
| T2DM | b_cell | *MICU1* | -2.282086915 | 0.007794424 | 0.012363109 | 1 |
| T2DM | b_cell | *CCSER2* | -2.365043401 | 0.004753646 | 0.00865764 | 1 |
| T2DM | b_cell | *BMPR1A* | -1.315362064 | 0.001789192 | 0.00483813 | 1 |
| T2DM | b_cell | *SFXN3* | -3.429580439 | 0.002033283 | 0.00516834 | 1 |
| T2DM | b_cell | *KAZALD1* | -6.882134042 | 0.000243208 | 0.00415349 | 1 |
| T2DM | b_cell | *ANKRD26* | -2.147777862 | 0.012768807 | 0.018475832 | 1 |
| T2DM | b_cell | *ACBD5* | -2.861078028 | 0.009031848 | 0.013898681 | 1 |
| T2DM | b_cell | *MTPAP* | -3.634049171 | 0.008780641 | 0.013603719 | 1 |
| T2DM | b_cell | *PITRM1* | -3.657981665 | 0.005209774 | 0.00917989 | 1 |
| T2DM | b_cell | *MAP3K8* | -2.732974852 | 0.001721714 | 0.004747873 | 1 |
| T2DM | b_cell | *SHOC2* | -2.437495259 | 0.011576287 | 0.017008049 | 1 |
| T2DM | b_cell | *TFAM* | -3.262544584 | 0.000628561 | 0.00415349 | 1 |
| T2DM | b_cell | *PBLD* | -1.501398197 | 0.010610052 | 0.015852042 | 1 |
| T2DM | b_cell | *DHRS11* | -3.350133776 | 8.63E-06 | 0.001459962 | 0.054250397 |
| T2DM | b_cell | *MLLT6* | -3.08758265 | 0.004639224 | 0.008518341 | 1 |
| T2DM | b_cell | *CASC3* | -2.899099431 | 0.002641695 | 0.005883223 | 1 |
| T2DM | b_cell | *RAPGEFL1* | -2.767893098 | 0.000776033 | 0.00415349 | 1 |
| T2DM | b_cell | *RGS9* | -2.939818973 | 0.00079474 | 0.00415349 | 1 |
| T2DM | b_cell | *WNT3* | -5.396627944 | 0.000244867 | 0.00415349 | 1 |
| T2DM | b_cell | *ASPA* | -1.506586956 | 0.000776133 | 0.00415349 | 1 |
| T2DM | b_cell | *RPS6KB1* | -2.763276629 | 0.00295925 | 0.006264583 | 1 |
| T2DM | b_cell | *TRIM16L* | -2.626938769 | 0.000389977 | 0.00415349 | 1 |
| T2DM | b_cell | *CDK5RAP3* | -2.89310652 | 0.001084707 | 0.00415349 | 1 |
| T2DM | b_cell | *CBX1* | -2.41420642 | 0.002684359 | 0.005929925 | 1 |
| T2DM | b_cell | *RECQL5* | -3.366530874 | 0.003063878 | 0.006386805 | 1 |
| T2DM | b_cell | *PIGL* | -2.440495671 | 0.002640386 | 0.005882391 | 1 |
| T2DM | b_cell | *GALK1* | -5.168404161 | 0.001835795 | 0.004898539 | 1 |
| T2DM | b_cell | *MED13* | -2.466588374 | 0.011305125 | 0.016680431 | 1 |
| T2DM | b_cell | *HOXB6* | -2.51405328 | 0.000217998 | 0.00415349 | 1 |
| T2DM | b_cell | *C1QBP* | -2.684710253 | 0.001268118 | 0.004257394 | 1 |
| T2DM | b_cell | *GOSR1* | -2.379551001 | 0.001983393 | 0.005095868 | 1 |
| T2DM | b_cell | *DRG2* | -2.254919243 | 0.000772983 | 0.00415349 | 1 |
| T2DM | b_cell | *FTSJ3* | -2.425156622 | 0.007024562 | 0.011394071 | 1 |
| T2DM | b_cell | *AKAP10* | -2.617471194 | 0.012872605 | 0.018600337 | 1 |
| T2DM | b_cell | *ALDH3A1* | 4.464634225 | 0.004701809 | 0.008595602 | 1 |
| T2DM | b_cell | *SMARCD2* | -2.443893666 | 0.000715132 | 0.00415349 | 1 |
| T2DM | b_cell | *ICAM2* | -1.803274466 | 0.002959932 | 0.006264583 | 1 |
| T2DM | b_cell | *SYNGR2* | -3.671353672 | 0.000135658 | 0.00415349 | 0.85288485 |
| T2DM | b_cell | *DDX5* | -1.645715607 | 0.006463983 | 0.010691676 | 1 |
| T2DM | b_cell | *PEX12* | -5.324333995 | 0.000355719 | 0.00415349 | 1 |

| | | | | | | |
|---|---|---|---|---|---|---|
| T2DM | b_cell | *DHX58* | -4.04489077 | 0.002257846 | 0.005430406 | 1 |
| T2DM | b_cell | *RAB5C* | -2.723993245 | 0.001950471 | 0.005052579 | 1 |
| T2DM | b_cell | *NAGLU* | -2.155232565 | 0.000702495 | 0.00415349 | 1 |
| T2DM | b_cell | *HSD17B1* | -3.602720923 | 3.55E-05 | 0.003315867 | 0.223109368 |
| T2DM | b_cell | *EZH1* | -2.220399884 | 0.008604784 | 0.013373053 | 1 |
| T2DM | b_cell | *DLX4* | -27.89909689 | 0.000690794 | 0.00415349 | 1 |
| T2DM | b_cell | *VAT1* | -1.894632968 | 0.00400394 | 0.007651297 | 1 |
| T2DM | b_cell | *LRRC59* | -5.064682428 | 0.004671802 | 0.008555671 | 1 |
| T2DM | b_cell | *ALOX12* | -5.127907124 | 0.001699813 | 0.004717848 | 1 |
| T2DM | b_cell | *MPP2* | -4.173974865 | 0.000858961 | 0.00415349 | 1 |
| T2DM | b_cell | *EFNB3* | 12.35120387 | 0.000488194 | 0.00415349 | 1 |
| T2DM | b_cell | *FAM20A* | -2.092329562 | 0.001415836 | 0.004364365 | 1 |
| T2DM | b_cell | *YWHAE* | -2.509999492 | 0.00382751 | 0.007404171 | 1 |
| T2DM | b_cell | *DPH1* | -3.779522437 | 0.001184613 | 0.004202203 | 1 |
| T2DM | b_cell | *DHRS7B* | -2.371109537 | 0.001814027 | 0.004867153 | 1 |
| T2DM | b_cell | *RCVRN* | -9.777939495 | 0.009179487 | 0.014086267 | 1 |
| T2DM | b_cell | *SLC9A3R1* | -3.902248674 | 0.016785285 | 0.023285324 | 1 |
| T2DM | b_cell | *MYH3* | -2.905352773 | 0.002123807 | 0.005267208 | 1 |
| T2DM | b_cell | *TMEM104* | -3.370726996 | 0.003911052 | 0.007531021 | 1 |
| T2DM | b_cell | *TMEM97* | 3.043279804 | 0.000234131 | 0.00415349 | 1 |
| T2DM | b_cell | *RAB34* | -2.709902872 | 0.000726469 | 0.00415349 | 1 |
| T2DM | b_cell | *SLAIN2* | -2.277619701 | 0.005131484 | 0.009087785 | 1 |
| T2DM | b_cell | *OCIAD1* | -3.138169606 | 0.010348213 | 0.015549526 | 1 |
| T2DM | b_cell | *CWH43* | -21.13677122 | 0.006070631 | 0.010234931 | 1 |
| T2DM | b_cell | *KIAA1211* | 2.717587058 | 0.005803692 | 0.009904519 | 1 |
| T2DM | b_cell | *AREG* | 2.208403215 | 0.000619817 | 0.00415349 | 1 |
| T2DM | b_cell | *UBE2D3* | -2.282703061 | 0.035038402 | 0.044305397 | 1 |
| T2DM | b_cell | *TBC1D9* | -2.584514578 | 0.004523262 | 0.008354216 | 1 |
| T2DM | b_cell | *CPE* | -2.01955216 | 0.001802813 | 0.004852128 | 1 |
| T2DM | b_cell | *WFS1* | -1.92361648 | 0.003516031 | 0.006997559 | 1 |
| T2DM | b_cell | *GRPEL1* | -3.413557172 | 0.001245704 | 0.004253251 | 1 |
| T2DM | b_cell | *AADAT* | -2.792458707 | 0.000971607 | 0.00415349 | 1 |
| T2DM | b_cell | *CPZ* | -55.57194883 | 0.000703053 | 0.00415349 | 1 |
| T2DM | b_cell | *TRIM2* | -1.811754752 | 0.000972976 | 0.00415349 | 1 |
| T2DM | b_cell | *SLC2A9* | -2.056772441 | 0.003092175 | 0.006423584 | 1 |
| T2DM | b_cell | *SH3D19* | -1.026633746 | 0.001161322 | 0.004195495 | 1 |
| T2DM | b_cell | *RAPGEF2* | -2.032788096 | 0.006586255 | 0.010842572 | 1 |
| T2DM | b_cell | *LRP2BP* | -3.013104597 | 0.023718954 | 0.031374093 | 1 |
| T2DM | b_cell | *UFSP2* | -3.455745833 | 0.008995004 | 0.013857288 | 1 |
| T2DM | b_cell | *KLF3* | -2.861572757 | 0.01248997 | 0.018108734 | 1 |
| T2DM | b_cell | *KLHL5* | -2.095167336 | 0.001759483 | 0.004789372 | 1 |
| T2DM | b_cell | *FAM149A* | -1.143002599 | 0.030363716 | 0.039006269 | 1 |
| T2DM | b_cell | *UGDH* | -2.287998492 | 0.001159728 | 0.004195495 | 1 |
| T2DM | b_cell | *HTATIP2* | -2.598342017 | 0.013048202 | 0.018819465 | 1 |
| T2DM | b_cell | *CTSC* | -1.840443715 | 0.000824453 | 0.00415349 | 1 |
| T2DM | b_cell | *ELP4* | -2.316185379 | 0.003031363 | 0.006344326 | 1 |
| T2DM | b_cell | *C11orf63* | -2.045943483 | 0.000974923 | 0.00415349 | 1 |
| T2DM | b_cell | *SNX15* | -3.523803048 | 0.005506712 | 0.009566611 | 1 |
| T2DM | b_cell | *DTX4* | -4.046114362 | 0.030825557 | 0.039518817 | 1 |
| T2DM | b_cell | *PUS3* | -2.796003819 | 0.002522531 | 0.005743989 | 1 |

| | | | | | | |
|---|---|---|---|---|---|---|
| T2DM | b_cell | *DCPS* | 4.433319336 | 0.026973426 | 0.035095598 | 1 |
| T2DM | b_cell | *FOXRED1* | -2.086241222 | 0.006923075 | 0.011270163 | 1 |
| T2DM | b_cell | *NRXN2* | -8.49252357 | 0.027405232 | 0.035576439 | 1 |
| T2DM | b_cell | *MS4A4A* | -1.55382395 | 0.002149293 | 0.00529906 | 1 |
| T2DM | b_cell | *CCDC86* | -6.70433606 | 0.000741921 | 0.00415349 | 1 |
| T2DM | b_cell | *PRPF19* | -2.880853331 | 0.001822631 | 0.004880273 | 1 |
| T2DM | b_cell | *HPX* | -3.466101275 | 0.025858735 | 0.033813201 | 1 |
| T2DM | b_cell | *CHORDC1* | -3.152425191 | 0.005156494 | 0.00911667 | 1 |
| T2DM | b_cell | *PANX1* | -2.798325112 | 0.000986956 | 0.00415349 | 1 |
| T2DM | b_cell | *EIF4G2* | -2.962966413 | 0.014519345 | 0.020591726 | 1 |
| T2DM | b_cell | *GALNT18* | -1.602827274 | 0.000769066 | 0.00415349 | 1 |
| T2DM | b_cell | *CBL* | -1.971912384 | 0.004442243 | 0.008231177 | 1 |
| T2DM | b_cell | *KIAA1549L* | -3.734572772 | 0.002565695 | 0.005799421 | 1 |
| T2DM | b_cell | *MADD* | -3.202978835 | 0.00620625 | 0.010399439 | 1 |
| T2DM | b_cell | *NAA40* | -2.348392405 | 0.000874605 | 0.00415349 | 1 |
| T2DM | b_cell | *CARS* | -2.185898461 | 0.012273332 | 0.017841026 | 1 |
| T2DM | b_cell | *SLC22A18* | -1.753410911 | 0.013313217 | 0.019162133 | 1 |
| T2DM | b_cell | *C11orf21* | -6.161266677 | 0.011676624 | 0.017116095 | 1 |
| T2DM | b_cell | *ELMOD1* | -7.231971099 | 0.000287042 | 0.00415349 | 1 |
| T2DM | b_cell | *NDUFS8* | -4.211002766 | 0.017429338 | 0.024044445 | 1 |
| T2DM | b_cell | *TCIRG1* | -3.005085774 | 0.005188977 | 0.009153507 | 1 |
| T2DM | b_cell | *GTF2H1* | -3.043441145 | 0.007237109 | 0.011651653 | 1 |
| T2DM | b_cell | *PPFIBP1* | -2.048928203 | 0.003238886 | 0.006622074 | 1 |
| T2DM | b_cell | *PRDM4* | -3.210894458 | 0.003057955 | 0.006378687 | 1 |
| T2DM | b_cell | *ASIC1* | -4.898730399 | 0.000147743 | 0.00415349 | 0.928859746 |
| T2DM | b_cell | *CAPRIN2* | -3.560490511 | 0.006642384 | 0.010920677 | 1 |
| T2DM | b_cell | *KCTD10* | -2.393176117 | 0.001035337 | 0.00415349 | 1 |
| T2DM | b_cell | *CYP27B1* | 2.862520396 | 0.000560853 | 0.00415349 | 1 |
| T2DM | b_cell | *ACSS3* | -1.005052951 | 0.003640341 | 0.007185816 | 1 |
| T2DM | b_cell | *GLI1* | -4.838202763 | 0.000783139 | 0.00415349 | 1 |
| T2DM | b_cell | *METAP2* | -3.086712002 | 0.022515569 | 0.030009621 | 1 |
| T2DM | b_cell | *LTA4H* | -3.225879053 | 0.004545328 | 0.008382656 | 1 |
| T2DM | b_cell | *TRPV4* | -6.206534964 | 0.010283803 | 0.015478638 | 1 |
| T2DM | b_cell | *ITFG2* | -2.891737008 | 0.005059219 | 0.009005468 | 1 |
| T2DM | b_cell | *GPN3* | -4.491610592 | 0.023593294 | 0.031260492 | 1 |
| T2DM | b_cell | *VPS29* | -2.099444595 | 0.001296006 | 0.004268027 | 1 |
| T2DM | b_cell | *KCNA1* | -6.993348112 | 0.000778274 | 0.00415349 | 1 |
| T2DM | b_cell | *CREBL2* | -2.060398848 | 0.003442575 | 0.006894407 | 1 |
| T2DM | b_cell | *LTBR* | -2.742645442 | 0.002662256 | 0.005916438 | 1 |
| T2DM | b_cell | *MGP* | -1.433561142 | 0.006700724 | 0.011002208 | 1 |
| T2DM | b_cell | *RASAL1* | 2.69705815 | 0.000908371 | 0.00415349 | 1 |
| T2DM | b_cell | *EIF2B1* | -3.353156203 | 0.003610812 | 0.007151285 | 1 |
| T2DM | b_cell | *DDX55* | -2.803023278 | 0.00026941 | 0.00415349 | 1 |
| T2DM | b_cell | *ENDOU* | -7.154481256 | 3.06E-06 | 0.000686434 | 0.019220161 |
| T2DM | b_cell | *STX2* | -2.31787418 | 0.01048307 | 0.015699228 | 1 |
| T2DM | b_cell | *CAND1* | -2.384048047 | 0.000982954 | 0.00415349 | 1 |
| T2DM | b_cell | *NUP107* | -2.125114341 | 0.002078068 | 0.005230315 | 1 |
| T2DM | b_cell | *CNOT2* | -2.588079492 | 0.006236149 | 0.010418993 | 1 |
| T2DM | b_cell | *KRR1* | -3.543745311 | 0.003047142 | 0.006373048 | 1 |
| T2DM | b_cell | *COPS7A* | -3.54504864 | 0.00079857 | 0.00415349 | 1 |

| | | | | | | |
|---|---|---|---|---|---|---|
| T2DM | b_cell | *GNB3* | -2.381377925 | 0.000958674 | 0.00415349 | 1 |
| T2DM | b_cell | *CDCA3* | -4.963642342 | 0.00362033 | 0.007162057 | 1 |
| T2DM | b_cell | *CHPT1* | -1.833403334 | 0.001178963 | 0.004202203 | 1 |
| T2DM | b_cell | *GNPTAB* | -2.415026956 | 0.004355776 | 0.008109199 | 1 |
| T2DM | b_cell | *SPSB2* | -3.100219863 | 0.00107955 | 0.00415349 | 1 |
| T2DM | b_cell | *ATN1* | -2.220882847 | 0.001322162 | 0.004274973 | 1 |
| T2DM | b_cell | *NT5DC3* | -1.968622212 | 0.001201064 | 0.004223203 | 1 |
| T2DM | b_cell | *PRKAB1* | 2.964847831 | 0.004631391 | 0.008508929 | 1 |
| T2DM | b_cell | *ST8SIA1* | -1.697904618 | 0.000756756 | 0.00415349 | 1 |
| T2DM | b_cell | *C2CD5* | -2.58715319 | 0.030030475 | 0.038601839 | 1 |
| T2DM | b_cell | *COL12A1* | -1.22020005 | 0.000748097 | 0.00415349 | 1 |
| T2DM | b_cell | *TDP2* | -3.217048583 | 0.002350156 | 0.005533109 | 1 |
| T2DM | b_cell | *RWDD1* | -2.654090652 | 0.001290514 | 0.004268027 | 1 |
| T2DM | b_cell | *RSPH4A* | -2.321364091 | 0.039614619 | 0.049494656 | 1 |
| T2DM | b_cell | *SMIM8* | -1.842099141 | 0.001224327 | 0.004253251 | 1 |
| T2DM | b_cell | *MCM9* | -2.637791045 | 0.004898435 | 0.00882673 | 1 |
| T2DM | b_cell | *FAM184A* | -1.509049883 | 0.001603314 | 0.004609717 | 1 |
| T2DM | b_cell | *GABRR2* | -3.629228799 | 0.001242707 | 0.004253251 | 1 |
| T2DM | b_cell | *HDDC2* | -2.703349671 | 0.001876777 | 0.004957688 | 1 |
| T2DM | b_cell | *TPD52L1* | -1.77785774 | 0.001662633 | 0.004679659 | 1 |
| T2DM | b_cell | *UST* | -1.222682284 | 0.000939926 | 0.00415349 | 1 |
| T2DM | b_cell | *TULP1* | -21.87127735 | 0.001261901 | 0.004257394 | 1 |
| T2DM | b_cell | *SLC26A8* | -1.937289356 | 0.01738441 | 0.023989417 | 1 |
| T2DM | b_cell | *MAPK14* | -2.596733012 | 0.001171943 | 0.004202203 | 1 |
| T2DM | b_cell | *RNF8* | -1.880550106 | 0.000714409 | 0.00415349 | 1 |
| T2DM | b_cell | *PHACTR1* | -2.547220933 | 0.006128846 | 0.010302689 | 1 |
| T2DM | b_cell | *MDGA1* | 3.096544505 | 0.002300903 | 0.005481934 | 1 |
| T2DM | b_cell | *CD83* | -3.901040989 | 0.000382373 | 0.00415349 | 1 |
| T2DM | b_cell | *MDN1* | -2.714699752 | 0.003666133 | 0.007210886 | 1 |
| T2DM | b_cell | *CAP2* | -1.527290858 | 0.000390426 | 0.00415349 | 1 |
| T2DM | b_cell | *GPR63* | -3.436918537 | 0.005791993 | 0.009892492 | 1 |
| T2DM | b_cell | *ASCC3* | -2.528369688 | 0.011539357 | 0.016962609 | 1 |
| T2DM | b_cell | *BVES* | -5.948613494 | 4.12E-05 | 0.003541347 | 0.25906912 |
| T2DM | b_cell | *WASF1* | -1.604138271 | 0.001077182 | 0.00415349 | 1 |
| T2DM | b_cell | *GPLD1* | -1.315344358 | 0.008366653 | 0.013052394 | 1 |
| T2DM | b_cell | *ALDH5A1* | -2.408619717 | 0.015105423 | 0.021293228 | 1 |
| T2DM | b_cell | *VNN1* | -2.220079209 | 0.00393953 | 0.007571944 | 1 |
| T2DM | b_cell | *VNN2* | 3.661233567 | 0.000214484 | 0.00415349 | 1 |
| T2DM | b_cell | *ACOT13* | -2.60692866 | 0.00155919 | 0.004538253 | 1 |
| T2DM | b_cell | *C6orf62* | -2.261433452 | 0.005366286 | 0.009387268 | 1 |
| T2DM | b_cell | *B3GAT2* | -5.453929399 | 0.014163876 | 0.020178629 | 1 |
| T2DM | b_cell | *HBS1L* | -2.817188521 | 0.009280917 | 0.014213206 | 1 |
| T2DM | b_cell | *PEX7* | -1.97417118 | 0.034206553 | 0.043366928 | 1 |
| T2DM | b_cell | *SLC16A10* | -1.874115853 | 0.003141661 | 0.006486859 | 1 |
| T2DM | b_cell | *PHACTR2* | -2.373787145 | 0.004427801 | 0.008213295 | 1 |
| T2DM | b_cell | *PACRG* | -2.773279738 | 0.000654985 | 0.00415349 | 1 |
| T2DM | b_cell | *TFEB* | -3.462868674 | 0.011394238 | 0.016792212 | 1 |
| T2DM | b_cell | *TBP* | -3.510049822 | 0.026222679 | 0.034210829 | 1 |
| T2DM | b_cell | *PRPH2* | -3.870121085 | 0.000191884 | 0.00415349 | 1 |
| T2DM | b_cell | *DUSP22* | -2.711891369 | 0.001883499 | 0.004965013 | 1 |

| | | | | | | |
|---|---|---|---|---|---|---|
| T2DM | b_cell | *EXOC2* | -2.82357638 | 0.023032224 | 0.030639779 | 1 |
| T2DM | b_cell | *TMEM30A* | -2.714321183 | 0.003390038 | 0.006828956 | 1 |
| T2DM | b_cell | *SENP6* | -2.282999821 | 0.007144169 | 0.01153155 | 1 |
| T2DM | b_cell | *TTK* | 1.46099654 | 0.001148942 | 0.004187618 | 1 |
| T2DM | b_cell | *LAMA4* | -0.96293489 | 0.00067751 | 0.00415349 | 1 |
| T2DM | b_cell | *CLIC5* | -1.905795369 | 0.0011901 | 0.004210557 | 1 |
| T2DM | b_cell | *LY86* | 1.90066135 | 0.000866449 | 0.00415349 | 1 |
| T2DM | b_cell | *TBX18* | -1.729728922 | 0.001035116 | 0.00415349 | 1 |
| T2DM | b_cell | *PCDHB2* | -2.502324023 | 0.031398272 | 0.040179308 | 1 |
| T2DM | b_cell | *NUDT12* | -3.720427996 | 0.001644639 | 0.004657671 | 1 |
| T2DM | b_cell | *C7* | -1.969279601 | 0.00062602 | 0.00415349 | 1 |
| T2DM | b_cell | *PAPD7* | -2.866243266 | 0.005278677 | 0.009275306 | 1 |
| T2DM | b_cell | *GHR* | -0.803556098 | 0.003839122 | 0.007422066 | 1 |
| T2DM | b_cell | *NME5* | -1.982475754 | 0.0031119 | 0.006452677 | 1 |
| T2DM | b_cell | *NNT* | -2.571135035 | 0.001606246 | 0.004610639 | 1 |
| T2DM | b_cell | *PFDN1* | -2.121919927 | 0.00407306 | 0.007750402 | 1 |
| T2DM | b_cell | *LOX* | -1.7849671 | 0.000781312 | 0.00415349 | 1 |
| T2DM | b_cell | *APBB3* | 3.189533837 | 0.023168542 | 0.030775539 | 1 |
| T2DM | b_cell | *TMCO6* | 1.950994122 | 0.001177611 | 0.004202203 | 1 |
| T2DM | b_cell | *SPARC* | -1.539381537 | 0.001429392 | 0.004387981 | 1 |
| T2DM | b_cell | *IK* | -3.623429262 | 0.002974151 | 0.006280982 | 1 |
| T2DM | b_cell | *PDE8B* | -1.441795765 | 0.001155748 | 0.004195495 | 1 |
| T2DM | b_cell | *CLK4* | -2.609722504 | 0.002725402 | 0.005972326 | 1 |
| T2DM | b_cell | *PCDHB15* | -2.89788564 | 0.002134725 | 0.005283865 | 1 |
| T2DM | b_cell | *HAVCR1* | 2.949947956 | 0.001683798 | 0.004704627 | 1 |
| T2DM | b_cell | *RNF130* | -1.868412888 | 0.001613927 | 0.004616359 | 1 |
| T2DM | b_cell | *ARSB* | -2.684380919 | 0.014600267 | 0.020683163 | 1 |
| T2DM | b_cell | *CLINT1* | -2.186752029 | 0.004023303 | 0.007676633 | 1 |
| T2DM | b_cell | *BTNL8* | -2.443795442 | 0.002510936 | 0.005734251 | 1 |
| T2DM | b_cell | *TTC1* | -2.938834044 | 0.010186761 | 0.015354632 | 1 |
| T2DM | b_cell | *MSH3* | -2.446581497 | 0.000568925 | 0.00415349 | 1 |
| T2DM | b_cell | *RASGRF2* | -1.276676749 | 0.000753486 | 0.00415349 | 1 |
| T2DM | b_cell | *POLR3G* | -2.088298667 | 0.000590728 | 0.00415349 | 1 |
| T2DM | b_cell | *DROSHA* | -2.391064766 | 0.011539538 | 0.016962609 | 1 |
| T2DM | b_cell | *CDH6* | -1.926609846 | 0.005034218 | 0.008986408 | 1 |
| T2DM | b_cell | *SLC27A6* | -4.755554572 | 0.000648915 | 0.00415349 | 1 |
| T2DM | b_cell | *TARS* | -2.493141004 | 0.001384417 | 0.00432604 | 1 |
| T2DM | b_cell | *LNPEP* | -2.052136709 | 0.014101915 | 0.02009491 | 1 |
| T2DM | b_cell | *SLC12A7* | -3.679402465 | 0.000943402 | 0.00415349 | 1 |
| T2DM | b_cell | *IL4* | 3.87452784 | 0.002036174 | 0.005172292 | 1 |
| T2DM | b_cell | *GNPDA1* | -3.550874431 | 0.003650906 | 0.007196562 | 1 |
| T2DM | b_cell | *PPWD1* | -2.686572394 | 0.008691162 | 0.013481702 | 1 |
| T2DM | b_cell | *TRIM23* | -3.331312882 | 0.004816322 | 0.008733839 | 1 |
| T2DM | b_cell | *TRAPPC13* | -2.732189517 | 0.017316022 | 0.02390554 | 1 |
| T2DM | b_cell | *TXNDC15* | -2.713388853 | 0.001094596 | 0.00415349 | 1 |
| T2DM | b_cell | *SMAD5* | -2.598037786 | 0.006088228 | 0.010252906 | 1 |
| T2DM | b_cell | *HMGXB3* | -3.094312583 | 0.009295132 | 0.014228998 | 1 |
| T2DM | b_cell | *ATP6V0E1* | -2.93240931 | 0.002899974 | 0.006192483 | 1 |
| T2DM | b_cell | *CNTN3* | -1.593735306 | 0.000780145 | 0.00415349 | 1 |
| T2DM | b_cell | *ACTR8* | 4.399238711 | 0.033638268 | 0.042749907 | 1 |

| | | | | | | |
|---|---|---|---|---|---|---|
| T2DM | b_cell | *HGD* | 7.209160087 | 0.001889022 | 0.004970573 | 1 |
| T2DM | b_cell | *NIT2* | -2.494621441 | 0.001040441 | 0.00415349 | 1 |
| T2DM | b_cell | *FAM162A* | -2.539437565 | 0.00034879 | 0.00415349 | 1 |
| T2DM | b_cell | *PCCB* | -2.605642331 | 0.001388576 | 0.004333459 | 1 |
| T2DM | b_cell | *RBP1* | -5.636014522 | 0.000748587 | 0.00415349 | 1 |
| T2DM | b_cell | *RNF7* | -3.427208338 | 0.009717833 | 0.014768193 | 1 |
| T2DM | b_cell | *BCHE* | -2.972750837 | 0.001007798 | 0.00415349 | 1 |
| T2DM | b_cell | *PDCD10* | -3.080633987 | 0.005057426 | 0.009004825 | 1 |
| T2DM | b_cell | *WNT5A* | -5.911079934 | 0.000852234 | 0.00415349 | 1 |
| T2DM | b_cell | *PRKAR2A* | -2.623673301 | 0.000951228 | 0.00415349 | 1 |
| T2DM | b_cell | *HYAL1* | -2.023332119 | 0.005399279 | 0.009426623 | 1 |
| T2DM | b_cell | *CYB561D2* | -3.928483387 | 0.000562308 | 0.00415349 | 1 |
| T2DM | b_cell | *IFT57* | 3.535205348 | 0.009060753 | 0.013931268 | 1 |
| T2DM | b_cell | *IQCG* | -3.048727731 | 0.003433687 | 0.006885994 | 1 |
| T2DM | b_cell | *GBE1* | -1.228046818 | 0.000917108 | 0.00415349 | 1 |
| T2DM | b_cell | *UMPS* | -3.202083183 | 0.000325404 | 0.00415349 | 1 |
| T2DM | b_cell | *NCBP2* | -3.440638288 | 0.007957149 | 0.012564143 | 1 |
| T2DM | b_cell | *SLC41A3* | -2.282530036 | 0.000669043 | 0.00415349 | 1 |
| T2DM | b_cell | *PLXNA1* | -2.532979189 | 0.002433225 | 0.005628918 | 1 |
| T2DM | b_cell | *ABTB1* | -3.862407843 | 0.004752872 | 0.00865764 | 1 |
| T2DM | b_cell | *EFCC1* | -5.032147658 | 5.68E-05 | 0.00415349 | 0.357051481 |
| T2DM | b_cell | *NEK11* | -2.572658258 | 0.002221942 | 0.005395656 | 1 |
| T2DM | b_cell | *PLSCR4* | -1.879456996 | 0.00217617 | 0.005332528 | 1 |
| T2DM | b_cell | *WDR48* | -2.659330361 | 0.034065016 | 0.043213631 | 1 |
| T2DM | b_cell | *COMMD2* | -2.442775146 | 0.001105544 | 0.004159248 | 1 |
| T2DM | b_cell | *ABCC5* | -2.228431652 | 0.000691313 | 0.00415349 | 1 |
| T2DM | b_cell | *ARHGEF26* | -1.819687381 | 0.001236965 | 0.004253251 | 1 |
| T2DM | b_cell | *KLHL24* | -2.035440256 | 0.003545344 | 0.007044747 | 1 |
| T2DM | b_cell | *PLCH1* | 6.653995392 | 0.000652977 | 0.00415349 | 1 |
| T2DM | b_cell | *NKTR* | -2.042638633 | 0.023097997 | 0.030704248 | 1 |
| T2DM | b_cell | *CLCN2* | -6.719941987 | 0.007394315 | 0.01185013 | 1 |
| T2DM | b_cell | *MOB1A* | -3.322893037 | 0.004860715 | 0.008786462 | 1 |
| T2DM | b_cell | *LMAN2L* | -2.551108514 | 0.020207098 | 0.027285658 | 1 |
| T2DM | b_cell | *PIKFYVE* | -2.671064325 | 0.033390998 | 0.042461408 | 1 |
| T2DM | b_cell | *KCNIP3* | -3.725016799 | 0.001604273 | 0.004609717 | 1 |
| T2DM | b_cell | *FAHD2A* | 4.73688396 | 0.004227632 | 0.007948302 | 1 |
| T2DM | b_cell | *NCL* | -3.29879143 | 0.018229102 | 0.024952398 | 1 |
| T2DM | b_cell | *ACTR1B* | -3.657413795 | 0.004917936 | 0.008844125 | 1 |
| T2DM | b_cell | *SLC35F5* | -2.363664006 | 0.023698062 | 0.031369643 | 1 |
| T2DM | b_cell | *ACTR3* | -2.677746377 | 0.0100317 | 0.015168181 | 1 |
| T2DM | b_cell | *STEAP3* | -1.926140699 | 0.014515218 | 0.020590518 | 1 |
| T2DM | b_cell | *TFCP2L1* | -3.36954788 | 0.00334381 | 0.006768457 | 1 |
| T2DM | b_cell | *TP53I3* | -2.110467369 | 0.001077159 | 0.00415349 | 1 |
| T2DM | b_cell | *DNAJC27* | -5.184777975 | 0.002163017 | 0.005312843 | 1 |
| T2DM | b_cell | *STAM2* | -2.815437648 | 0.005732359 | 0.009817309 | 1 |
| T2DM | b_cell | *GPD2* | -2.590055779 | 0.003634705 | 0.007180327 | 1 |
| T2DM | b_cell | *CENPA* | -2.831929393 | 0.000828 | 0.00415349 | 1 |
| T2DM | b_cell | *TANC1* | -1.398969168 | 0.000441953 | 0.00415349 | 1 |
| T2DM | b_cell | *NRBP1* | -5.502505186 | 0.000378761 | 0.00415349 | 1 |
| T2DM | b_cell | *ITGB6* | -5.385867188 | 0.0001827 | 0.00415349 | 1 |

| | | | | | | |
|---|---|---|---|---|---|---|
| T2DM | b_cell | *PPM1G* | -5.118786276 | 0.000933119 | 0.00415349 | 1 |
| T2DM | b_cell | *PCSK4* | -4.091980257 | 0.002767518 | 0.006028396 | 1 |
| T2DM | b_cell | *INO80B* | -2.847154606 | 0.001683346 | 0.004704627 | 1 |
| T2DM | b_cell | *MOGS* | -1.737234696 | 0.000231945 | 0.00415349 | 1 |
| T2DM | b_cell | *GRB14* | -2.141265465 | 0.000699267 | 0.00415349 | 1 |
| T2DM | b_cell | *CLIP4* | -1.769346191 | 0.001068231 | 0.00415349 | 1 |
| T2DM | b_cell | *SPTBN1* | -1.210782594 | 0.000730498 | 0.00415349 | 1 |
| T2DM | b_cell | *RTN4* | -2.153328179 | 0.000466719 | 0.00415349 | 1 |
| T2DM | b_cell | *GALNT3* | 1.443192712 | 0.000901951 | 0.00415349 | 1 |
| T2DM | b_cell | *POLE4* | -3.602033313 | 0.001931883 | 0.005024672 | 1 |
| T2DM | b_cell | *MRPL19* | -2.202160869 | 0.000824712 | 0.00415349 | 1 |
| T2DM | b_cell | *LANCL1* | -3.352652647 | 0.013226546 | 0.019050469 | 1 |
| T2DM | b_cell | *WDR75* | -3.269201325 | 0.005608608 | 0.009663282 | 1 |
| T2DM | b_cell | *EFEMP1* | -1.206470215 | 0.00067803 | 0.00415349 | 1 |
| T2DM | b_cell | *GLS* | -2.277038844 | 0.009341686 | 0.014282874 | 1 |
| T2DM | b_cell | *GGCX* | -2.996063169 | 0.018429567 | 0.025188411 | 1 |
| T2DM | b_cell | *GNLY* | -1.749568378 | 0.000735448 | 0.00415349 | 1 |
| T2DM | b_cell | *SF3B1* | -2.432125421 | 0.013390461 | 0.019233683 | 1 |
| T2DM | b_cell | *ST3GAL5* | -1.541678373 | 0.000668345 | 0.00415349 | 1 |
| T2DM | b_cell | *PLCD4* | -2.670367668 | 0.000697013 | 0.00415349 | 1 |
| T2DM | b_cell | *CHMP3* | -6.584488908 | 7.15E-05 | 0.00415349 | 0.449400994 |
| T2DM | b_cell | *ZNF142* | -3.497639875 | 0.011273838 | 0.016638174 | 1 |
| T2DM | b_cell | *MLPH* | -2.467910791 | 0.000814993 | 0.00415349 | 1 |
| T2DM | b_cell | *HDLBP* | -2.52381883 | 0.009093564 | 0.01397488 | 1 |
| T2DM | b_cell | *STK25* | -2.228199597 | 0.000795542 | 0.00415349 | 1 |
| T2DM | b_cell | *TPO* | -1.675407776 | 0.000906472 | 0.00415349 | 1 |
| T2DM | b_cell | *TAF1B* | -3.066120009 | 0.005016541 | 0.008972687 | 1 |
| T2DM | b_cell | *STRN* | -2.694500985 | 0.00522868 | 0.009202887 | 1 |
| T2DM | b_cell | *CEBPZ* | -2.646479755 | 0.001608003 | 0.004610639 | 1 |
| T2DM | b_cell | *PRKD3* | -2.454388649 | 0.003626529 | 0.007172063 | 1 |
| T2DM | b_cell | *SLC25A12* | -2.340354218 | 0.001016686 | 0.00415349 | 1 |
| T2DM | b_cell | *SDC1* | 11.46841039 | 0.036675737 | 0.046208488 | 1 |
| T2DM | b_cell | *SOS1* | -1.914760294 | 0.00241656 | 0.005613599 | 1 |
| T2DM | b_cell | *WIPF1* | -2.223360459 | 0.001704171 | 0.004725848 | 1 |
| T2DM | b_cell | *PNO1* | -2.247733863 | 0.000784446 | 0.00415349 | 1 |
| T2DM | b_cell | *ORC4* | -2.731905545 | 0.034484616 | 0.043663005 | 1 |
| T2DM | b_cell | *THADA* | -2.44668957 | 0.009694475 | 0.014743387 | 1 |
| T2DM | b_cell | *ARID3A* | -2.981175177 | 0.002442479 | 0.0056392 | 1 |
| T2DM | b_cell | *CD207* | -48.84425168 | 0.003236832 | 0.006620026 | 1 |
| T2DM | b_cell | *VAX2* | -4.767930397 | 0.000998332 | 0.00415349 | 1 |
| T2DM | b_cell | *MSH6* | -3.57836944 | 0.005575752 | 0.009640408 | 1 |
| T2DM | b_cell | *ALMS1* | -2.259045853 | 0.001643978 | 0.004657671 | 1 |
| T2DM | b_cell | *DNAJC16* | -3.109280047 | 0.033135057 | 0.042187142 | 1 |
| T2DM | b_cell | *MORN1* | -2.171483354 | 0.001524702 | 0.004498266 | 1 |
| T2DM | b_cell | *SCP2* | -2.101717737 | 0.002567407 | 0.005799671 | 1 |
| T2DM | b_cell | *RALGPS2* | 0.415190076 | 0.000573988 | 0.00415349 | 1 |
| T2DM | b_cell | *CEP104* | -1.995839705 | 0.001615118 | 0.004617667 | 1 |
| T2DM | b_cell | *TMEM59* | -2.551371718 | 0.003797036 | 0.007363346 | 1 |
| T2DM | b_cell | *WRAP73* | -3.799945771 | 0.001264154 | 0.004257394 | 1 |
| T2DM | b_cell | *MRPL37* | -2.065831042 | 0.002472179 | 0.005676621 | 1 |

| | | | | | | |
|---|---|---|---|---|---|---|
| T2DM | b_cell | *ICMT* | -4.297744841 | 0.00078716 | 0.00415349 | 1 |
| T2DM | b_cell | *QSOX1* | -3.816961402 | 0.011065312 | 0.016384271 | 1 |
| T2DM | b_cell | *KCNC4* | -3.252316044 | 0.000378118 | 0.00415349 | 1 |
| T2DM | b_cell | *EDEM3* | -2.893317613 | 0.006230718 | 0.010416354 | 1 |
| T2DM | b_cell | *RAP1A* | -2.434043513 | 0.026056892 | 0.034022779 | 1 |
| T2DM | b_cell | *CAPZA1* | -2.967559198 | 0.006750627 | 0.011063918 | 1 |
| T2DM | b_cell | *DOCK7* | -2.274362495 | 0.002621051 | 0.005854674 | 1 |
| T2DM | b_cell | *FBXO6* | -4.054411172 | 0.002585675 | 0.005820315 | 1 |
| T2DM | b_cell | *C1orf21* | -1.463010109 | 0.000359853 | 0.00415349 | 1 |
| T2DM | b_cell | *LEPR* | -2.633953673 | 0.017529602 | 0.024168554 | 1 |
| T2DM | b_cell | *MFN2* | -2.645219471 | 0.000994056 | 0.00415349 | 1 |
| T2DM | b_cell | *MIIP* | -4.580106132 | 0.000823754 | 0.00415349 | 1 |
| T2DM | b_cell | *SMG7* | -2.75221234 | 0.005203451 | 0.009176464 | 1 |
| T2DM | b_cell | *NCF2* | -1.94525508 | 0.000767442 | 0.00415349 | 1 |
| T2DM | b_cell | *RGS2* | -4.921110911 | 0.003056585 | 0.006378687 | 1 |
| T2DM | b_cell | *OLFML3* | -2.043752997 | 0.000764747 | 0.00415349 | 1 |
| T2DM | b_cell | *TNNI3K* | -3.914975175 | 0.032999866 | 0.042032042 | 1 |
| T2DM | b_cell | *CFHR3* | -3.230928039 | 0.031540822 | 0.0403453 | 1 |
| T2DM | b_cell | *CRYZ* | -3.47599271 | 0.008014588 | 0.012625336 | 1 |
| T2DM | b_cell | *TTF2* | -3.147193233 | 0.002671872 | 0.005916798 | 1 |
| T2DM | b_cell | *TMEM9* | -2.345404416 | 0.000498629 | 0.00415349 | 1 |
| T2DM | b_cell | *WARS2* | -2.303513026 | 0.000968232 | 0.00415349 | 1 |
| T2DM | b_cell | *OSCP1* | -2.044975071 | 0.001296277 | 0.004268027 | 1 |
| T2DM | b_cell | *GNPAT* | -3.630675381 | 0.011111417 | 0.016433188 | 1 |
| T2DM | b_cell | *C1orf109* | -2.942400937 | 0.000497264 | 0.00415349 | 1 |
| T2DM | b_cell | *TBCE* | -3.615931156 | 0.001537852 | 0.004509332 | 1 |
| T2DM | b_cell | *LGALS8* | -2.424060959 | 0.000798222 | 0.00415349 | 1 |
| T2DM | b_cell | *MTR* | -2.44979179 | 0.004247702 | 0.007978877 | 1 |
| T2DM | b_cell | *KCNQ4* | -3.99357694 | 0.00013129 | 0.00415349 | 0.825419198 |
| T2DM | b_cell | *ACADM* | -2.558956326 | 0.006949396 | 0.011300363 | 1 |
| T2DM | b_cell | *PADI2* | 3.508580771 | 0.001045716 | 0.00415349 | 1 |
| T2DM | b_cell | *MFAP2* | 2.999633556 | 0.027032554 | 0.035150707 | 1 |
| T2DM | b_cell | *CTBS* | -3.297591432 | 0.033222159 | 0.042272356 | 1 |
| T2DM | b_cell | *ZNHIT6* | -2.996725897 | 0.00415296 | 0.007865091 | 1 |
| T2DM | b_cell | *PLA2G2D* | 3.82041253 | 0.001322831 | 0.004274973 | 1 |
| T2DM | b_cell | *RBBP5* | -2.285318593 | 0.020426782 | 0.027564538 | 1 |
| T2DM | b_cell | *KIF17* | -5.153590182 | 0.001487521 | 0.004462053 | 1 |
| T2DM | b_cell | *CD46* | -1.939511899 | 0.002620805 | 0.005854674 | 1 |
| T2DM | b_cell | *PRDX1* | -3.363446423 | 0.025192933 | 0.033025014 | 1 |
| T2DM | b_cell | *NSUN4* | -4.318894314 | 0.018804093 | 0.025607386 | 1 |
| T2DM | b_cell | *TMED5* | -2.45361015 | 0.00209753 | 0.005241324 | 1 |
| T2DM | b_cell | *DR1* | -2.962966441 | 0.002623289 | 0.005854674 | 1 |
| T2DM | b_cell | *CNN3* | -2.054260931 | 0.001094953 | 0.00415349 | 1 |
| T2DM | b_cell | *PRRC2C* | -2.282303923 | 0.021829063 | 0.029187435 | 1 |
| T2DM | b_cell | *ABCD3* | -2.991622959 | 0.012328776 | 0.017909199 | 1 |
| T2DM | b_cell | *VAMP4* | -2.549062727 | 0.007234228 | 0.011651653 | 1 |
| T2DM | b_cell | *PTBP2* | -1.766444058 | 0.034050847 | 0.043213631 | 1 |
| T2DM | b_cell | *TNFSF4* | -4.36670182 | 4.30E-05 | 0.003541347 | 0.270371555 |
| T2DM | b_cell | *DARS2* | -3.820347871 | 0.000982082 | 0.00415349 | 1 |
| T2DM | b_cell | *DIEXF* | -4.34590255 | 0.000911291 | 0.00415349 | 1 |

| | | | | | | |
|---|---|---|---|---|---|---|
| T2DM | b_cell | *RCAN3* | -2.850049064 | 0.001340003 | 0.004286583 | 1 |
| T2DM | b_cell | *RCOR3* | -2.533163464 | 0.004968445 | 0.008909473 | 1 |
| T2DM | b_cell | *NSL1* | -2.83353828 | 0.004628075 | 0.008505322 | 1 |
| T2DM | b_cell | *RPA2* | -3.727763829 | 0.009545661 | 0.014538172 | 1 |
| T2DM | b_cell | *SLC5A9* | 5.405636794 | 0.002495688 | 0.005713909 | 1 |
| T2DM | b_cell | *OSBPL9* | -2.519162588 | 0.010821131 | 0.016109981 | 1 |
| T2DM | b_cell | *TXNDC12* | -3.37144205 | 0.016411049 | 0.022846826 | 1 |
| T2DM | b_cell | *ESYT2* | -2.23155513 | 0.002425087 | 0.005623947 | 1 |
| T2DM | b_cell | *CD3EAP* | -2.861670901 | 3.64E-05 | 0.003315867 | 0.228794843 |
| T2DM | b_cell | *A4GNT* | -4.022141334 | 0.003065769 | 0.006387091 | 1 |
| T2DM | b_cell | *KMT2A* | -2.5665764 | 0.008012746 | 0.012625336 | 1 |
| T2DM | b_cell | *IFT46* | -3.856828079 | 0.001646611 | 0.004658975 | 1 |
| T2DM | b_cell | *CAMSAP2* | -2.544024064 | 0.010949265 | 0.016268464 | 1 |
| T2DM | b_cell | *ATF6* | -2.505122985 | 0.006512338 | 0.01075468 | 1 |
| T2DM | b_cell | *LRMP* | 1.39338076 | 0.00070634 | 0.00415349 | 1 |
| T2DM | b_cell | *ATP10B* | -4.75328095 | 0.000844185 | 0.00415349 | 1 |
| T2DM | b_cell | *ELOVL4* | 4.422870266 | 0.0031193 | 0.006465696 | 1 |
| T2DM | b_cell | *FILIP1* | -1.65012217 | 0.000695115 | 0.00415349 | 1 |
| T2DM | b_cell | *SGIP1* | -1.907047984 | 0.00100945 | 0.00415349 | 1 |
| T2DM | b_cell | *AKAP7* | -2.368703812 | 0.000669839 | 0.00415349 | 1 |
| T2DM | b_cell | *ALDH8A1* | -1.623825059 | 0.032153808 | 0.041001711 | 1 |
| T2DM | b_cell | *MED28* | -3.44363845 | 0.003263489 | 0.006657222 | 1 |
| T2DM | b_cell | *SLC16A7* | -2.166080994 | 0.003243325 | 0.006628995 | 1 |
| T2DM | b_cell | *VAMP8* | -2.4089548 | 0.00105595 | 0.00415349 | 1 |
| T2DM | b_cell | *FOXO3* | -2.243276564 | 0.005251658 | 0.009235574 | 1 |
| T2DM | b_cell | *ABCG2* | -2.652849222 | 0.000716803 | 0.00415349 | 1 |
| T2DM | b_cell | *SPP1* | 2.553759481 | 0.035282633 | 0.044596283 | 1 |
| T2DM | b_cell | *FAM47E-STBL* | -2.39758851 | 0.001704823 | 0.004725848 | 1 |
| T2DM | b_cell | *MFSD1* | -2.040308238 | 0.001345252 | 0.004286796 | 1 |
| T2DM | b_cell | *RAB3GAP2* | -2.612939245 | 0.016717734 | 0.023212101 | 1 |
| T2DM | b_cell | *UBN1* | -2.581609828 | 0.003499589 | 0.006971457 | 1 |
| T2DM | b_cell | *UCHL3* | -3.15357996 | 0.000878716 | 0.00415349 | 1 |
| T2DM | b_cell | *DNAH7* | -2.700036802 | 8.75E-07 | 0.000261873 | 0.005499342 |
| T2DM | b_cell | *NDUFB3* | -4.290008958 | 0.013140263 | 0.018943552 | 1 |
| T2DM | b_cell | *TRPM6* | -2.603076903 | 0.001714463 | 0.004735864 | 1 |
| T2DM | b_cell | *GDA* | 5.666563429 | 0.006239175 | 0.010421279 | 1 |
| T2DM | b_cell | *C2orf40* | -2.7868853 | 0.001389278 | 0.004333459 | 1 |
| T2DM | b_cell | *CPSF3* | -2.536497488 | 0.001626118 | 0.004638568 | 1 |
| T2DM | b_cell | *PIGZ* | -1.644348664 | 0.00238953 | 0.00557776 | 1 |
| T2DM | b_cell | *SENP5* | -2.416724844 | 0.006896909 | 0.011233385 | 1 |
| T2DM | b_cell | *CCDC92* | -2.848558285 | 0.000121522 | 0.00415349 | 0.764005755 |
| T2DM | b_cell | *FKBP15* | -2.661245405 | 0.008666318 | 0.013453122 | 1 |
| T2DM | b_cell | *CTNNAL1* | -1.136547365 | 0.013578872 | 0.019450984 | 1 |
| T2DM | b_cell | *WDR34* | 2.599624689 | 0.017462559 | 0.024081401 | 1 |
| T2DM | b_cell | *TRIM32* | -4.424807795 | 0.003055497 | 0.006378687 | 1 |
| T2DM | b_cell | *PHF19* | -2.775164855 | 0.000664202 | 0.00415349 | 1 |
| T2DM | b_cell | *BSPRY* | 3.592264276 | 0.000683124 | 0.00415349 | 1 |
| T2DM | b_cell | *PPP6C* | -2.300421817 | 0.001228208 | 0.004253251 | 1 |
| T2DM | b_cell | *HDHD3* | -1.72814791 | 5.59E-05 | 0.00415349 | 0.351655286 |
| T2DM | b_cell | *HSDL2* | -1.332128811 | 0.001400914 | 0.004345113 | 1 |

| | | | | | | |
|---|---|---|---|---|---|---|
| T2DM | b_cell | *INVS* | -2.310146509 | 0.006764132 | 0.011080276 | 1 |
| T2DM | b_cell | *KDSR* | -2.167692865 | 0.001194547 | 0.004217141 | 1 |
| T2DM | b_cell | *DNAL1* | -2.424376653 | 0.001975556 | 0.005092436 | 1 |
| T2DM | b_cell | *ACOT2* | -1.062733775 | 0.023272917 | 0.030891682 | 1 |
| T2DM | b_cell | *TTLL5* | -2.776507417 | 0.021676978 | 0.029008762 | 1 |
| T2DM | b_cell | *FLVCR2* | -2.068800456 | 0.000750772 | 0.00415349 | 1 |
| T2DM | b_cell | *PPP4R4* | 2.368834269 | 0.002969165 | 0.00627678 | 1 |
| T2DM | b_cell | *ZC2HC1C* | -2.566068123 | 0.000875494 | 0.00415349 | 1 |
| T2DM | b_cell | *ALDH6A1* | -2.500858228 | 0.000958497 | 0.00415349 | 1 |
| T2DM | b_cell | *ESRRB* | -2.451917832 | 0.000766291 | 0.00415349 | 1 |
| T2DM | b_cell | *COQ6* | 3.847653299 | 0.003742631 | 0.0072961 | 1 |
| T2DM | b_cell | *GPR75* | -12.38847224 | 0.003393339 | 0.006833415 | 1 |
| T2DM | b_cell | *KLHL29* | -2.017396067 | 0.001756635 | 0.004789372 | 1 |
| T2DM | b_cell | *ATAD2B* | -2.420797277 | 0.005539872 | 0.009597457 | 1 |
| T2DM | b_cell | *ATL2* | -2.649807 | 0.005095891 | 0.00904003 | 1 |
| T2DM | b_cell | *CNRIP1* | -3.483058799 | 0.000821625 | 0.00415349 | 1 |
| T2DM | b_cell | *CRIPT* | -4.311108572 | 0.003901008 | 0.007513983 | 1 |
| T2DM | b_cell | *EPCAM* | -6.565237157 | 8.54E-05 | 0.00415349 | 0.536721219 |
| T2DM | b_cell | *SLC17A5* | -2.863008059 | 0.010802958 | 0.016090546 | 1 |
| T2DM | b_cell | *OGFRL1* | -2.450354606 | 0.006775447 | 0.011087255 | 1 |
| T2DM | b_cell | *CUTC* | -2.672071691 | 0.004858731 | 0.008785402 | 1 |
| T2DM | b_cell | *PYROXD2* | -1.130666119 | 0.023837326 | 0.031510779 | 1 |
| T2DM | b_cell | *MXI1* | -3.292205124 | 0.005239665 | 0.00921964 | 1 |
| T2DM | b_cell | *C10orf88* | -2.55166707 | 0.001071504 | 0.00415349 | 1 |
| T2DM | b_cell | *HELLS* | 2.588883146 | 0.000961049 | 0.00415349 | 1 |
| T2DM | b_cell | *AVPI1* | -4.065183163 | 0.00650947 | 0.010752769 | 1 |
| T2DM | b_cell | *WDR11* | -2.71596649 | 0.012540485 | 0.018170553 | 1 |
| T2DM | b_cell | *C10orf76* | -2.304585184 | 0.00336317 | 0.006790062 | 1 |
| T2DM | b_cell | *KCNIP2* | -1.286301139 | 0.005861979 | 0.009968694 | 1 |
| T2DM | b_cell | *KANSL1* | -1.93228895 | 0.007948015 | 0.012555067 | 1 |
| T2DM | b_cell | *HOXB5* | -3.25377919 | 0.000179608 | 0.00415349 | 1 |
| T2DM | b_cell | *HOXB3* | -3.166965642 | 4.33E-05 | 0.003541347 | 0.272190538 |
| T2DM | b_cell | *CD274* | -2.821369509 | 0.0008521 | 0.00415349 | 1 |
| T2DM | b_cell | *NUP43* | -2.825330587 | 0.005977397 | 0.010123894 | 1 |
| T2DM | b_cell | *LRP11* | -1.997253235 | 0.000363345 | 0.00415349 | 1 |
| T2DM | b_cell | *CCDC170* | -1.438787796 | 0.001219062 | 0.004252746 | 1 |
| T2DM | b_cell | *CYSTM1* | -2.36123335 | 0.003487135 | 0.006959878 | 1 |
| T2DM | b_cell | *PCDHB10* | 1.515253367 | 0.000604554 | 0.00415349 | 1 |
| T2DM | b_cell | *PCDHB14* | -6.364385849 | 0.003780152 | 0.007342551 | 1 |
| T2DM | b_cell | *SEC16B* | -5.298623517 | 0.002771125 | 0.006028396 | 1 |
| T2DM | b_cell | *ACAT2* | -4.036727159 | 0.025530177 | 0.033425286 | 1 |
| T2DM | b_cell | *SNX19* | -2.74139202 | 0.012560328 | 0.018195112 | 1 |
| T2DM | b_cell | *MASTL* | -3.243424834 | 5.76E-05 | 0.00415349 | 0.36207734 |
| T2DM | b_cell | *MRC1* | -1.26514285 | 0.000880874 | 0.00415349 | 1 |
| T2DM | b_cell | *IQSEC3* | -5.767730268 | 0.002441065 | 0.005638124 | 1 |
| T2DM | b_cell | *CCDC77* | -5.080854859 | 0.002233815 | 0.005401538 | 1 |
| T2DM | b_cell | *ENOX1* | -2.135477985 | 0.001008078 | 0.00415349 | 1 |
| T2DM | b_cell | *WBP4* | -2.371218511 | 0.002507555 | 0.005732727 | 1 |
| T2DM | b_cell | *SMAD9* | -1.975198499 | 0.000793945 | 0.00415349 | 1 |
| T2DM | b_cell | *SIL1* | -2.506858716 | 0.012137753 | 0.017668454 | 1 |

| | | | | | | |
|---|---|---|---|---|---|---|
| T2DM | b_cell | *ARL1* | -2.427046424 | 0.001948064 | 0.005048426 | 1 |
| T2DM | b_cell | *APAF1* | -2.691490937 | 0.003754141 | 0.007309385 | 1 |
| T2DM | b_cell | *EPHX2* | -1.967407745 | 0.000266952 | 0.00415349 | 1 |
| T2DM | b_cell | *ZNF706* | -2.881410727 | 0.01615135 | 0.022535183 | 1 |
| T2DM | b_cell | *CRISPLD1* | -2.965840186 | 0.00085648 | 0.00415349 | 1 |
| T2DM | b_cell | *RDH10* | -1.597314745 | 0.006218199 | 0.010405595 | 1 |
| T2DM | b_cell | *SCPEP1* | -1.674072635 | 0.028066073 | 0.036344263 | 1 |
| T2DM | b_cell | *TRIM6* | -3.575233124 | 0.000451844 | 0.00415349 | 1 |
| T2DM | b_cell | *CEP89* | -2.170475488 | 0.018425214 | 0.025187937 | 1 |
| T2DM | b_cell | *ECHDC2* | -1.60575332 | 7.78E-05 | 0.00415349 | 0.4893596 |
| T2DM | b_cell | *PRB2* | -2.656539515 | 0.001992251 | 0.005104028 | 1 |
| T2DM | b_cell | *PYROXD1* | -3.257545324 | 0.037217684 | 0.046797516 | 1 |
| T2DM | b_cell | *ZSCAN18* | -3.319406899 | 0.0083 | 0.012977394 | 1 |
| T2DM | b_cell | *PDZRN3* | -1.039086857 | 0.000780357 | 0.00415349 | 1 |
| T2DM | b_cell | *CSTA* | -2.48134839 | 0.001132587 | 0.00418468 | 1 |
| T2DM | b_cell | *POPDC2* | 3.836382643 | 0.000949717 | 0.00415349 | 1 |
| T2DM | b_cell | *B4GALT4* | 3.888274705 | 0.00748252 | 0.011970128 | 1 |
| T2DM | b_cell | *CD80* | 2.619353772 | 0.000770999 | 0.00415349 | 1 |
| T2DM | b_cell | *CAT* | -1.502003248 | 0.000938839 | 0.00415349 | 1 |
| T2DM | b_cell | *GJB6* | -101.253067 | 0.000816978 | 0.00415349 | 1 |
| T2DM | b_cell | *TMEM39B* | -2.785994651 | 0.000818015 | 0.00415349 | 1 |
| T2DM | b_cell | *RNF115* | -2.657903744 | 0.011107511 | 0.016431276 | 1 |
| T2DM | b_cell | *POLR3GL* | 4.947444528 | 0.004710112 | 0.008602378 | 1 |
| T2DM | b_cell | *TNFSF10* | -2.559126298 | 0.002310778 | 0.005492478 | 1 |
| T2DM | b_cell | *LIAS* | -1.793660858 | 0.000719097 | 0.00415349 | 1 |
| T2DM | b_cell | *CLCC1* | -3.959679108 | 0.004456779 | 0.008250815 | 1 |
| T2DM | b_cell | *GPSM2* | -2.083061422 | 0.003981449 | 0.007624542 | 1 |
| T2DM | b_cell | *CXCR4* | 1.063092046 | 0.000834358 | 0.00415349 | 1 |
| T2DM | b_cell | *ACVR2A* | -2.535963971 | 0.005221939 | 0.009196171 | 1 |
| T2DM | b_cell | *POLK* | -2.552854201 | 0.008123868 | 0.012743202 | 1 |
| T2DM | b_cell | *FLT3* | 2.67866812 | 0.00071352 | 0.00415349 | 1 |
| T2DM | b_cell | *MTIF3* | -2.209487949 | 0.005813514 | 0.009915476 | 1 |
| T2DM | b_cell | *RASL11A* | -5.043479395 | 0.000151944 | 0.00415349 | 0.955274923 |
| T2DM | b_cell | *FMOD* | -2.109617825 | 0.000619541 | 0.00415349 | 1 |
| T2DM | b_cell | *SERAC1* | -2.153544636 | 0.000589099 | 0.00415349 | 1 |
| T2DM | b_cell | *LDB3* | -1.895197882 | 0.005595265 | 0.009653521 | 1 |
| T2DM | b_cell | *FAM35A* | -2.451256569 | 0.028709332 | 0.037100837 | 1 |
| T2DM | b_cell | *FAM213A* | -1.410146971 | 0.000558586 | 0.00415349 | 1 |
| T2DM | b_cell | *NAA60* | -2.666982509 | 0.000827061 | 0.00415349 | 1 |
| T2DM | b_cell | *ODF2L* | -2.066427787 | 0.037977559 | 0.047591172 | 1 |
| T2DM | b_cell | *SPATA1* | -2.879313571 | 0.006315427 | 0.010520692 | 1 |
| T2DM | b_cell | *LRRC39* | -1.960301086 | 0.005219643 | 0.009194703 | 1 |
| T2DM | b_cell | *RWDD3* | 6.506375737 | 0.03170764 | 0.040533943 | 1 |
| T2DM | b_cell | *CCDC18* | -2.452127482 | 0.0025595 | 0.005798627 | 1 |
| T2DM | b_cell | *RPAP2* | -2.381223141 | 0.003060212 | 0.006381278 | 1 |
| T2DM | b_cell | *BBS9* | -2.549037339 | 0.002605577 | 0.005849897 | 1 |
| T2DM | b_cell | *KIAA0087* | -8.870259009 | 0.000163374 | 0.00415349 | 1 |
| T2DM | b_cell | *KLHL7* | -3.102300989 | 0.020768058 | 0.027959054 | 1 |
| T2DM | b_cell | *HERPUD2* | -2.881119296 | 0.00293732 | 0.006226208 | 1 |
| T2DM | b_cell | *CBX3* | -3.677818523 | 0.000988382 | 0.00415349 | 1 |

| | | | | | | |
|---|---|---|---|---|---|---|
| T2DM | b_cell | *FAM126A* | -2.486520256 | 0.014275331 | 0.020305206 | 1 |
| T2DM | b_cell | *FKBP9* | -1.837345622 | 0.001175376 | 0.004202203 | 1 |
| T2DM | b_cell | *CCZ1* | -2.846606319 | 0.027031062 | 0.035150707 | 1 |
| T2DM | b_cell | *POLM* | -2.204929701 | 0.000417201 | 0.00415349 | 1 |
| T2DM | b_cell | *SMU1* | -3.182965397 | 0.025356029 | 0.033224959 | 1 |
| T2DM | b_cell | *GLIPR2* | -3.472456416 | 0.004622091 | 0.008496808 | 1 |
| T2DM | b_cell | *DCAF10* | -2.370734917 | 0.003563875 | 0.007074861 | 1 |
| T2DM | b_cell | *BICC1* | -1.217594364 | 0.001037626 | 0.00415349 | 1 |
| T2DM | b_cell | *CISD1* | -2.419488829 | 0.000499812 | 0.00415349 | 1 |
| T2DM | b_cell | *SLC25A16* | -1.34170596 | 0.003430559 | 0.006885994 | 1 |
| T2DM | b_cell | *RBM19* | -3.358706096 | 0.030747033 | 0.039442276 | 1 |
| T2DM | b_cell | *ACADS* | -1.985297409 | 0.000679137 | 0.00415349 | 1 |
| T2DM | b_cell | *SSPN* | -1.680183655 | 0.018277293 | 0.025012918 | 1 |
| T2DM | b_cell | *CCDC91* | -2.204998685 | 0.022098175 | 0.029490814 | 1 |
| T2DM | b_cell | *PKN1* | -3.124263975 | 0.002623015 | 0.005854674 | 1 |
| T2DM | b_cell | *ATP7B* | -1.415991943 | 0.004581843 | 0.008437623 | 1 |
| T2DM | b_cell | *NLN* | -1.430676179 | 0.0036449 | 0.007192558 | 1 |
| T2DM | b_cell | *CENPK* | 1.619807751 | 0.003283106 | 0.006688769 | 1 |
| T2DM | b_cell | *OPTN* | -2.33066638 | 0.00051231 | 0.00415349 | 1 |
| T2DM | b_cell | *ATF1* | -2.767526746 | 0.037728053 | 0.047325672 | 1 |
| T2DM | b_cell | *TSFM* | -2.764006455 | 0.002931424 | 0.006221257 | 1 |
| T2DM | b_cell | *ARHGAP9* | -4.518982965 | 0.003138873 | 0.006486603 | 1 |
| T2DM | b_cell | *SPATS2* | -2.776103925 | 0.004037952 | 0.007696831 | 1 |
| T2DM | b_cell | *PDE1B* | -4.486950437 | 0.001431985 | 0.004393796 | 1 |
| T2DM | b_cell | *SMUG1* | -1.347017462 | 0.033153057 | 0.042201512 | 1 |
| T2DM | b_cell | *SARDH* | -6.228489623 | 0.000774276 | 0.00415349 | 1 |
| T2DM | b_cell | *DBH* | -15.40306296 | 0.01029352 | 0.015489555 | 1 |
| T2DM | b_cell | *STIL* | -4.159773394 | 0.001768781 | 0.004801523 | 1 |
| T2DM | b_cell | *NDUFAF4* | -1.983769469 | 0.009957394 | 0.015073955 | 1 |
| T2DM | b_cell | *TTC21B* | -2.714961785 | 0.012115873 | 0.017652303 | 1 |
| T2DM | b_cell | *NMI* | -3.533042986 | 0.010410473 | 0.015624407 | 1 |
| T2DM | b_cell | *ACVR1C* | -1.252371807 | 0.001276473 | 0.004257394 | 1 |
| T2DM | b_cell | *SLC36A1* | -1.932743408 | 0.017919382 | 0.024603441 | 1 |
| T2DM | b_cell | *PLA2G12A* | -3.396359831 | 0.02804748 | 0.03632767 | 1 |
| T2DM | b_cell | *C4BPB* | -3.858930555 | 0.001141692 | 0.00418468 | 1 |
| T2DM | b_cell | *RAB38* | -3.40334134 | 0.000562443 | 0.00415349 | 1 |
| T2DM | b_cell | *AGO2* | -2.864661605 | 0.005154735 | 0.00911667 | 1 |
| T2DM | b_cell | *CHPF* | -2.910721345 | 0.001250393 | 0.004253251 | 1 |
| T2DM | b_cell | *DNPEP* | -2.864491263 | 0.001622931 | 0.004634377 | 1 |
| T2DM | b_cell | *MOGAT1* | -2.004087611 | 0.000952656 | 0.00415349 | 1 |
| T2DM | b_cell | *FAM124B* | -3.086517267 | 0.021234156 | 0.028522663 | 1 |
| T2DM | b_cell | *FAM210B* | -2.436345926 | 0.004933496 | 0.008861968 | 1 |
| T2DM | b_cell | *SNX21* | -1.545771519 | 0.000782193 | 0.00415349 | 1 |
| T2DM | b_cell | *WFDC3* | -1.926507062 | 0.001456285 | 0.004416626 | 1 |
| T2DM | b_cell | *SLC12A5* | -6.746398934 | 0.001106325 | 0.004159248 | 1 |
| T2DM | b_cell | *SDC4* | -2.850803115 | 0.000366199 | 0.00415349 | 1 |
| T2DM | b_cell | *VAPB* | -3.170707507 | 0.00090174 | 0.00415349 | 1 |
| T2DM | b_cell | *TOX2* | -1.641191595 | 0.002582499 | 0.005819416 | 1 |
| T2DM | b_cell | *ARFGEF2* | -2.560095275 | 0.006197482 | 0.010387515 | 1 |
| T2DM | b_cell | *CDH26* | -2.578689958 | 0.000800821 | 0.00415349 | 1 |

| | | | | | | |
|---|---|---|---|---|---|---|
| T2DM | b_cell | *STX16* | -2.940383109 | 0.004008878 | 0.007653756 | 1 |
| T2DM | b_cell | *RNF114* | -3.1886052 | 0.002659178 | 0.005911687 | 1 |
| T2DM | b_cell | *KCNK15* | 2.383181579 | 0.000661471 | 0.00415349 | 1 |
| T2DM | b_cell | *PEPD* | -1.566173374 | 0.002902158 | 0.006192483 | 1 |
| T2DM | b_cell | *CHST8* | -7.884668299 | 0.004782039 | 0.00869424 | 1 |
| T2DM | b_cell | *STAMBP* | -3.232362199 | 0.015572246 | 0.021828921 | 1 |
| T2DM | b_cell | *USP22* | -2.884028419 | 0.019327932 | 0.026193118 | 1 |
| T2DM | b_cell | *HIF3A* | -1.486091871 | 0.002366481 | 0.005545309 | 1 |
| T2DM | b_cell | *BTN2A2* | 1.82054532 | 0.000623458 | 0.00415349 | 1 |
| T2DM | b_cell | *MRS2* | -2.783655464 | 0.007552109 | 0.012059971 | 1 |
| T2DM | b_cell | *SERPINB6* | -2.583959779 | 0.002113683 | 0.005250385 | 1 |
| T2DM | b_cell | *HIST1H1D* | -4.232709092 | 0.016306413 | 0.022721281 | 1 |
| T2DM | b_cell | *PEX6* | -1.87389138 | 0.000887522 | 0.00415349 | 1 |
| T2DM | b_cell | *NQO2* | -2.437523453 | 0.001576315 | 0.004562749 | 1 |
| T2DM | b_cell | *RPS10* | 3.161495308 | 0.000637496 | 0.00415349 | 1 |
| T2DM | b_cell | *HIST1H2BJ* | 3.543831124 | 0.000100393 | 0.00415349 | 0.631172505 |
| T2DM | b_cell | *MED20* | -2.962186573 | 0.000401221 | 0.00415349 | 1 |
| T2DM | b_cell | *KLHDC3* | -2.313007071 | 0.000637283 | 0.00415349 | 1 |
| T2DM | b_cell | *MEA1* | -3.581344926 | 0.001126725 | 0.004182741 | 1 |
| T2DM | b_cell | *KLHL31* | -1.455800908 | 0.002304557 | 0.005484718 | 1 |
| T2DM | b_cell | *COL21A1* | -1.850715916 | 0.001035005 | 0.00415349 | 1 |
| T2DM | b_cell | *RPP40* | -2.958935497 | 0.021349314 | 0.028637324 | 1 |
| T2DM | b_cell | *EEF1E1* | -3.556983974 | 0.000789724 | 0.00415349 | 1 |
| T2DM | b_cell | *EREG* | 3.035003836 | 0.000673398 | 0.00415349 | 1 |
| T2DM | b_cell | *MYRF* | -4.362561785 | 0.022424094 | 0.029894037 | 1 |
| T2DM | b_cell | *CNOT1* | -2.469560536 | 0.031299117 | 0.040068734 | 1 |
| T2DM | b_cell | *MT1G* | -19.40280996 | 0.000358648 | 0.00415349 | 1 |
| T2DM | b_cell | *CLYBL* | -2.994208531 | 0.0058962 | 0.010013347 | 1 |
| T2DM | b_cell | *TMTC4* | -2.096506157 | 0.000859434 | 0.00415349 | 1 |
| T2DM | b_cell | *KIF25* | -5.341675287 | 0.005396642 | 0.009426623 | 1 |
| T2DM | b_cell | *TEKT3* | -2.48995554 | 0.009309822 | 0.014244549 | 1 |
| T2DM | b_cell | *MRPS7* | -4.351957357 | 0.011035422 | 0.016358965 | 1 |
| T2DM | b_cell | *GGA3* | -2.873015915 | 0.001890351 | 0.004970573 | 1 |
| T2DM | b_cell | *NUP85* | -2.796055149 | 0.002701366 | 0.005946726 | 1 |
| T2DM | b_cell | *NT5C* | -3.887356951 | 0.029425287 | 0.037909175 | 1 |
| T2DM | b_cell | *PPP1R12C* | -3.967989403 | 0.015269654 | 0.021466975 | 1 |
| T2DM | b_cell | *OPRL1* | -8.372908773 | 7.03E-08 | 4.02E-05 | 0.000441671 |
| T2DM | b_cell | *PLGLB2* | -30.95093466 | 0.032158333 | 0.041001711 | 1 |
| T2DM | b_cell | *CHCHD5* | -4.927615682 | 0.038278883 | 0.047911474 | 1 |
| T2DM | b_cell | *PAX8* | -2.68616972 | 0.008978823 | 0.013835751 | 1 |
| T2DM | b_cell | *POLR1B* | -2.435227776 | 0.000816746 | 0.00415349 | 1 |
| T2DM | b_cell | *SLC25A23* | -2.414781466 | 0.024524283 | 0.032263414 | 1 |
| T2DM | b_cell | *GTF2F1* | -3.7039432 | 0.007633206 | 0.012164757 | 1 |
| T2DM | b_cell | *CLPP* | -2.26238209 | 0.00232628 | 0.005506523 | 1 |
| T2DM | b_cell | *SH2D3A* | -3.32352947 | 0.000630116 | 0.00415349 | 1 |
| T2DM | b_cell | *TRIP10* | -1.808493267 | 0.001269521 | 0.004257394 | 1 |
| T2DM | b_cell | *GPR108* | -3.214954972 | 0.003470519 | 0.006942141 | 1 |
| T2DM | b_cell | *RTN2* | -3.162776637 | 0.00058851 | 0.00415349 | 1 |
| T2DM | b_cell | *SDCBP2* | -6.127888269 | 0.018384659 | 0.02514343 | 1 |
| T2DM | b_cell | *PANK2* | -2.772263107 | 0.00328321 | 0.006688769 | 1 |

| | | | | | | |
|---|---|---|---|---|---|---|
| T2DM | b_cell | *PSMF1* | -2.858612602 | 0.000835718 | 0.00415349 | 1 |
| T2DM | b_cell | *DTD1* | -3.228925789 | 0.024190118 | 0.031896659 | 1 |
| T2DM | b_cell | *RRBP1* | -2.596499525 | 0.003323342 | 0.006745057 | 1 |
| T2DM | b_cell | *BMP2* | -3.655852044 | 0.000790375 | 0.00415349 | 1 |
| T2DM | b_cell | *DSTN* | -2.444477585 | 0.002566246 | 0.005799421 | 1 |
| T2DM | b_cell | *MGME1* | -1.887290772 | 0.0007855 | 0.00415349 | 1 |
| T2DM | b_cell | *MCM8* | -1.903672306 | 0.02267889 | 0.030208089 | 1 |
| T2DM | b_cell | *TMEM74B* | -5.090612994 | 0.03037273 | 0.039009878 | 1 |
| T2DM | b_cell | *FAM110A* | -3.045311459 | 0.000797902 | 0.00415349 | 1 |
| T2DM | b_cell | *MRPS26* | -1.94187464 | 0.009096471 | 0.013975932 | 1 |
| T2DM | b_cell | *NCLN* | -4.14595086 | 0.001252847 | 0.004255347 | 1 |
| T2DM | b_cell | *ZNF436* | -1.72640595 | 0.013498047 | 0.019352844 | 1 |
| T2DM | b_cell | *MMP24* | -2.391819705 | 0.005062227 | 0.009007468 | 1 |
| T2DM | b_cell | *UROD* | -3.560568293 | 0.021887659 | 0.02925956 | 1 |
| T2DM | b_cell | *ST3GAL3* | -2.410511341 | 0.005104202 | 0.009050883 | 1 |
| T2DM | b_cell | *KLC1* | -2.739462238 | 0.015129707 | 0.021313123 | 1 |
| T2DM | b_cell | *XRCC3* | -2.40323865 | 0.001860375 | 0.004935095 | 1 |
| T2DM | b_cell | *MCF2L* | -1.659919261 | 0.003895962 | 0.007508864 | 1 |
| T2DM | b_cell | *F10* | -1.855754058 | 0.00079427 | 0.00415349 | 1 |
| T2DM | b_cell | *PCID2* | -2.67873717 | 0.001269493 | 0.004257394 | 1 |
| T2DM | b_cell | *PROZ* | 3.810884155 | 0.000605906 | 0.00415349 | 1 |
| T2DM | b_cell | *IGFLR1* | 8.578908151 | 0.001142231 | 0.00418468 | 1 |
| T2DM | b_cell | *TSKS* | -3.015111054 | 0.001029 | 0.00415349 | 1 |
| T2DM | b_cell | *FLRT1* | -2.772802508 | 0.001651558 | 0.004668493 | 1 |
| T2DM | b_cell | *DNAJC8* | -3.248342471 | 0.000917015 | 0.00415349 | 1 |
| T2DM | b_cell | *ZNF384* | -3.930583337 | 0.019661189 | 0.026611388 | 1 |
| T2DM | b_cell | *EMG1* | -4.134218746 | 0.011782406 | 0.017226973 | 1 |
| T2DM | b_cell | *ATG14* | -2.223992224 | 0.001905549 | 0.004988168 | 1 |
| T2DM | b_cell | *RHOJ* | -1.605225842 | 0.00073256 | 0.00415349 | 1 |
| T2DM | b_cell | *L3HYPDH* | -1.491929434 | 0.002436329 | 0.005631324 | 1 |
| T2DM | b_cell | *ZBTB1* | -2.374218115 | 0.003004449 | 0.006308941 | 1 |
| T2DM | b_cell | *PZP* | -1.712672971 | 0.001036108 | 0.00415349 | 1 |
| T2DM | b_cell | *RHOT1* | -2.159792586 | 0.004901238 | 0.00882925 | 1 |
| T2DM | b_cell | *WDR60* | -2.098903282 | 0.00205292 | 0.005193845 | 1 |
| T2DM | b_cell | *AIF1L* | -2.249131933 | 0.00085753 | 0.00415349 | 1 |
| T2DM | b_cell | *MAP2K2* | -3.086765482 | 0.001087177 | 0.00415349 | 1 |
| T2DM | b_cell | *CANX* | -2.948302999 | 0.015528618 | 0.021796924 | 1 |
| T2DM | b_cell | *IPPK* | -2.974966793 | 0.000593601 | 0.00415349 | 1 |
| T2DM | b_cell | *FGD3* | -2.608842482 | 0.001094285 | 0.00415349 | 1 |
| T2DM | b_cell | *ABHD8* | -3.377307249 | 0.005803761 | 0.009904519 | 1 |
| T2DM | b_cell | *MASP1* | -3.013168892 | 0.000453339 | 0.00415349 | 1 |
| T2DM | b_cell | *ATP13A4* | -3.080259019 | 0.003718232 | 0.007266559 | 1 |
| T2DM | b_cell | *HRASLS* | -4.473414163 | 0.002554697 | 0.005795069 | 1 |
| T2DM | b_cell | *HELB* | -3.537856665 | 0.00121511 | 0.004245409 | 1 |
| T2DM | b_cell | *RAP1B* | -2.312464787 | 0.013369439 | 0.019212266 | 1 |
| T2DM | b_cell | *TAS2R4* | -3.976267295 | 0.038232033 | 0.047871897 | 1 |
| T2DM | b_cell | *LRRC61* | -1.936529152 | 0.016018585 | 0.022374772 | 1 |
| T2DM | b_cell | *IDUA* | 3.787385772 | 0.002249901 | 0.005421667 | 1 |
| T2DM | b_cell | *EMC1* | -3.515483632 | 0.00692513 | 0.011270591 | 1 |
| T2DM | b_cell | *PLA2G5* | -2.310522164 | 0.010044381 | 0.015183703 | 1 |

| | | | | | | |
|---|---|---|---|---|---|---|
| T2DM | b_cell | *UBR4* | -2.738490477 | 0.000694207 | 0.00415349 | 1 |
| T2DM | b_cell | *SIN3B* | -3.24301504 | 0.011921475 | 0.017406018 | 1 |
| T2DM | b_cell | *WFIKKN1* | -29.40014337 | 0.000194111 | 0.00415349 | 1 |
| T2DM | b_cell | *WDR24* | -6.080284202 | 0.000410548 | 0.00415349 | 1 |
| T2DM | b_cell | *CHTF18* | 6.026345571 | 0.021920728 | 0.02929131 | 1 |
| T2DM | b_cell | *SMARCA4* | -3.146817712 | 0.001594843 | 0.004599438 | 1 |
| T2DM | b_cell | *TICAM1* | -4.309910007 | 0.003384924 | 0.006820839 | 1 |
| T2DM | b_cell | *AAMP* | -2.914157088 | 0.000525137 | 0.00415349 | 1 |
| T2DM | b_cell | *PNKD* | -3.368277586 | 0.001882109 | 0.004963761 | 1 |
| T2DM | b_cell | *ZNF835* | -4.026663422 | 0.000937959 | 0.00415349 | 1 |
| T2DM | b_cell | *HIP1* | -2.057022748 | 0.002509311 | 0.005734251 | 1 |
| T2DM | b_cell | *PTPN12* | -2.28571625 | 0.003423405 | 0.006878539 | 1 |
| T2DM | b_cell | *POR* | -1.883253847 | 0.001032833 | 0.00415349 | 1 |
| T2DM | b_cell | *FGL2* | -2.245346086 | 0.008105744 | 0.012724298 | 1 |
| T2DM | b_cell | *STEAP4* | -2.979562668 | 0.003399225 | 0.006843076 | 1 |
| T2DM | b_cell | *GNAI1* | -1.337127279 | 0.000579118 | 0.00415349 | 1 |
| T2DM | b_cell | *SGCE* | -1.867293565 | 0.00114452 | 0.00418468 | 1 |
| T2DM | b_cell | *RBM48* | -1.951325499 | 0.000691607 | 0.00415349 | 1 |
| T2DM | b_cell | *SRD5A3* | -2.872705083 | 0.001709558 | 0.004728644 | 1 |
| T2DM | b_cell | *DGCR6L* | 3.479844857 | 0.001638092 | 0.004657671 | 1 |
| T2DM | b_cell | *VPREB3* | -29.44487946 | 0.007986523 | 0.012595482 | 1 |
| T2DM | b_cell | *MGAT3* | -2.636145346 | 0.000730522 | 0.00415349 | 1 |
| T2DM | b_cell | *ATF4* | -3.695960984 | 0.018518341 | 0.025293246 | 1 |
| T2DM | b_cell | *A4GALT* | -1.962683015 | 0.002903354 | 0.006192483 | 1 |
| T2DM | b_cell | *CDC42EP1* | -2.510980837 | 0.00099712 | 0.00415349 | 1 |
| T2DM | b_cell | *TPST2* | -1.818110307 | 0.000969628 | 0.00415349 | 1 |
| T2DM | b_cell | *TST* | -2.169334634 | 0.002142048 | 0.005287258 | 1 |
| T2DM | b_cell | *APOL5* | -13.90452253 | 0.000836223 | 0.00415349 | 1 |
| T2DM | b_cell | *RIBC2* | 3.22195524 | 2.07E-05 | 0.002553516 | 0.130421625 |
| T2DM | b_cell | *EMC4* | -2.643210233 | 0.010073606 | 0.015216906 | 1 |
| T2DM | b_cell | *SPECC1* | -1.743740692 | 0.001084987 | 0.00415349 | 1 |
| T2DM | b_cell | *POT1* | -2.966672376 | 0.027761633 | 0.036009364 | 1 |
| T2DM | b_cell | *CDHR3* | -1.54971589 | 0.001758265 | 0.004789372 | 1 |
| T2DM | b_cell | *PRKRIP1* | -2.833704059 | 0.001466257 | 0.004431902 | 1 |
| T2DM | b_cell | *FOXP2* | -1.221182755 | 0.000327926 | 0.00415349 | 1 |
| T2DM | b_cell | *STRIP2* | 4.713785465 | 0.024578965 | 0.032314503 | 1 |
| T2DM | b_cell | *FLNC* | -2.67041952 | 0.001978817 | 0.005092436 | 1 |
| T2DM | b_cell | *IRF5* | -1.75803375 | 0.020951533 | 0.028187949 | 1 |
| T2DM | b_cell | *KLHDC10* | -2.14857325 | 0.002574797 | 0.005806222 | 1 |
| T2DM | b_cell | *OPN1SW* | -4.204634985 | 0.003447123 | 0.006899733 | 1 |
| T2DM | b_cell | *HOXD3* | -1.895698873 | 0.001038585 | 0.00415349 | 1 |
| T2DM | b_cell | *PDE11A* | -3.801876678 | 0.000425489 | 0.00415349 | 1 |
| T2DM | b_cell | *CHN1* | -1.542553839 | 0.00086598 | 0.00415349 | 1 |
| T2DM | b_cell | *GAD1* | 3.054293693 | 0.000704688 | 0.00415349 | 1 |
| T2DM | b_cell | *ORMDL1* | -1.647643116 | 0.009833823 | 0.01491562 | 1 |
| T2DM | b_cell | *HOXD9* | -4.458143566 | 0.014711191 | 0.020812164 | 1 |
| T2DM | b_cell | *PSMG2* | -2.406060699 | 0.002487495 | 0.005702562 | 1 |
| T2DM | b_cell | *TWSG1* | -3.512080066 | 0.012485609 | 0.018108734 | 1 |
| T2DM | b_cell | *WDFY4* | 1.341069212 | 0.001311366 | 0.004274973 | 1 |
| T2DM | b_cell | *EIF2AK4* | -3.450911061 | 0.000661949 | 0.00415349 | 1 |

| | | | | | | |
|---|---|---|---|---|---|---|
| T2DM | b_cell | *TMOD2* | -2.782432024 | 0.001126097 | 0.004182741 | 1 |
| T2DM | b_cell | *IVD* | -3.293868161 | 0.011516512 | 0.016952543 | 1 |
| T2DM | b_cell | *ARPP19* | -3.346416007 | 0.004509893 | 0.008336871 | 1 |
| T2DM | b_cell | *VPS13C* | -2.338133727 | 0.012458407 | 0.018076622 | 1 |
| T2DM | b_cell | *CALML4* | -9.504612481 | 0.002445642 | 0.005640408 | 1 |
| T2DM | b_cell | *ACKR4* | -2.966082345 | 0.004319419 | 0.008068011 | 1 |
| T2DM | b_cell | *ANAPC13* | -2.058311069 | 0.001365795 | 0.004299827 | 1 |
| T2DM | b_cell | *SUMF2* | -2.88653654 | 0.003575379 | 0.007093217 | 1 |
| T2DM | b_cell | *SPCS3* | -3.578107609 | 0.017602514 | 0.024253124 | 1 |
| T2DM | b_cell | *RPAIN* | -2.88923673 | 0.000534918 | 0.00415349 | 1 |
| T2DM | b_cell | *PLD2* | 3.696803858 | 0.000469896 | 0.00415349 | 1 |
| T2DM | b_cell | *ATP1B2* | -6.253034289 | 0.007511082 | 0.012006654 | 1 |
| T2DM | b_cell | *CCL4* | -2.137866203 | 0.001273186 | 0.004257394 | 1 |
| T2DM | b_cell | *PHF20L1* | -2.570596153 | 0.004003449 | 0.007651297 | 1 |
| T2DM | b_cell | *PUS7L* | -2.969193202 | 0.034244147 | 0.043400731 | 1 |
| T2DM | b_cell | *CDKN2D* | 1.995579961 | 0.000950346 | 0.00415349 | 1 |
| T2DM | b_cell | *MTUS1* | -2.398164576 | 0.001247954 | 0.004253251 | 1 |
| T2DM | b_cell | *NGDN* | -2.717083927 | 0.001437947 | 0.004399209 | 1 |
| T2DM | b_cell | *RIPK3* | -3.325593266 | 0.002089379 | 0.005231352 | 1 |
| T2DM | b_cell | *ADCY4* | -2.354657069 | 0.00168529 | 0.004704627 | 1 |
| T2DM | b_cell | *RAB2B* | -2.499870903 | 0.001340626 | 0.004286583 | 1 |
| T2DM | b_cell | *MIS18BP1* | -2.026215697 | 0.021145398 | 0.028418366 | 1 |
| T2DM | b_cell | *RNASE1* | -3.072875881 | 0.007980896 | 0.012594351 | 1 |
| T2DM | b_cell | *TEP1* | -4.034180991 | 0.013182544 | 0.018998056 | 1 |
| T2DM | b_cell | *EPB41L4A* | -1.33225142 | 0.000861657 | 0.00415349 | 1 |
| T2DM | b_cell | *CDO1* | -1.90391342 | 0.000808237 | 0.00415349 | 1 |
| T2DM | b_cell | *REEP5* | -2.704689738 | 0.005651127 | 0.009724561 | 1 |
| T2DM | b_cell | *QRICH2* | -2.463901407 | 0.001686877 | 0.004704627 | 1 |
| T2DM | b_cell | *SEC14L1* | -2.152059531 | 0.001505744 | 0.004478191 | 1 |
| T2DM | b_cell | *RHBDF2* | 2.627498781 | 0.00043941 | 0.00415349 | 1 |
| T2DM | b_cell | *AANAT* | 4.384476811 | 0.000314425 | 0.00415349 | 1 |
| T2DM | b_cell | *TMEM8A* | -3.73725296 | 0.001684068 | 0.004704627 | 1 |
| T2DM | b_cell | *SHC2* | -1.88908054 | 0.005522419 | 0.009579216 | 1 |
| T2DM | b_cell | *ERMARD* | -3.407672087 | 0.001592154 | 0.004593791 | 1 |
| T2DM | b_cell | *PHF10* | -3.534296567 | 0.009405741 | 0.014359857 | 1 |
| T2DM | b_cell | *GALNT8* | -3.407383162 | 0.000136575 | 0.00415349 | 0.858647105 |
| T2DM | b_cell | *DOCK6* | -1.573253217 | 0.001361654 | 0.004298348 | 1 |
| T2DM | b_cell | *ECSIT* | -3.942967643 | 0.011514921 | 0.016952543 | 1 |
| T2DM | b_cell | *ELOF1* | -3.763534975 | 0.000804031 | 0.00415349 | 1 |
| T2DM | b_cell | *CDC16* | -2.53424995 | 0.005065762 | 0.009009461 | 1 |
| T2DM | b_cell | *THEM6* | -3.035722353 | 0.000387436 | 0.00415349 | 1 |
| T2DM | b_cell | *TOMM40* | -4.192427636 | 0.00383874 | 0.007422066 | 1 |
| T2DM | b_cell | *DPP6* | -3.324911122 | 0.000197695 | 0.00415349 | 1 |
| T2DM | b_cell | *FAM98C* | 3.567761705 | 0.011627471 | 0.017055842 | 1 |
| T2DM | b_cell | *ATP8B3* | -3.037532837 | 0.002646808 | 0.005892522 | 1 |
| T2DM | b_cell | *GTPBP3* | -6.137383896 | 0.011707868 | 0.017144273 | 1 |
| T2DM | b_cell | *PLVAP* | -1.356953446 | 0.003646866 | 0.007194179 | 1 |
| T2DM | b_cell | *BST2* | -2.766301464 | 0.00123678 | 0.004253251 | 1 |
| T2DM | b_cell | *DDA1* | -2.510620557 | 0.000754581 | 0.00415349 | 1 |
| T2DM | b_cell | *TULP4* | -2.542778425 | 0.003512031 | 0.00699181 | 1 |

| | | | | | | |
|---|---|---|---|---|---|---|
| T2DM | b_cell | *RTN4IP1* | -3.082341449 | 0.004705661 | 0.008599361 | 1 |
| T2DM | b_cell | *RSPH3* | -2.340248424 | 0.012015824 | 0.017521682 | 1 |
| T2DM | b_cell | *ACTN4* | -2.755884666 | 0.003788212 | 0.007350767 | 1 |
| T2DM | b_cell | *NDUFA10* | -1.788772357 | 0.018320089 | 0.025066028 | 1 |
| T2DM | b_cell | *PXDN* | -1.29423725 | 0.00077909 | 0.00415349 | 1 |
| T2DM | b_cell | *PGPEP1* | 4.15305438 | 0.005685226 | 0.009763184 | 1 |
| T2DM | b_cell | *LSM4* | -2.908996009 | 0.013766317 | 0.019683611 | 1 |
| T2DM | b_cell | *CAMSAP1* | -2.888923841 | 0.001862829 | 0.0049373 | 1 |
| T2DM | b_cell | *UBAC1* | -3.489999613 | 0.010139607 | 0.015298227 | 1 |
| T2DM | b_cell | *TNNT3* | -3.32915027 | 0.000274594 | 0.00415349 | 1 |
| T2DM | b_cell | *ATXN10* | -2.511421412 | 0.031488236 | 0.040286231 | 1 |
| T2DM | b_cell | *TUBGCP2* | -2.311476556 | 0.002220225 | 0.005393569 | 1 |
| T2DM | b_cell | *CYP2E1* | 5.529261419 | 0.007091902 | 0.011482562 | 1 |
| T2DM | b_cell | *PAK4* | -2.662463562 | 0.000994102 | 0.00415349 | 1 |
| T2DM | b_cell | *ZNF337* | -2.425769025 | 0.001802637 | 0.004852128 | 1 |
| T2DM | b_cell | *POMT1* | -3.214339861 | 0.00305163 | 0.006376763 | 1 |
| T2DM | b_cell | *UCK1* | 2.633472572 | 0.011120228 | 0.016442351 | 1 |
| T2DM | b_cell | *MAP3K10* | -2.663197806 | 0.00076077 | 0.00415349 | 1 |
| T2DM | b_cell | *SMPDL3B* | 4.917875167 | 0.000595175 | 0.00415349 | 1 |
| T2DM | b_cell | *ATPIF1* | -3.905038526 | 0.001654133 | 0.00466975 | 1 |
| T2DM | b_cell | *THEMIS2* | -1.836611384 | 0.002868234 | 0.006154645 | 1 |
| T2DM | b_cell | *CLIP1* | -2.457251999 | 0.010503711 | 0.015719312 | 1 |
| T2DM | b_cell | *ZNF317* | -2.164724447 | 0.000905939 | 0.00415349 | 1 |
| T2DM | b_cell | *PPAN* | 5.92268815 | 0.002086364 | 0.005230315 | 1 |
| T2DM | b_cell | *C12orf65* | -2.596395302 | 0.037936419 | 0.047549096 | 1 |
| T2DM | b_cell | *PKDREJ* | -7.684208752 | 0.000780036 | 0.00415349 | 1 |
| T2DM | b_cell | *HSD17B3* | -2.772834844 | 0.00072583 | 0.00415349 | 1 |
| T2DM | b_cell | *HABP4* | -1.959321108 | 0.000785261 | 0.00415349 | 1 |
| T2DM | b_cell | *EPS8L1* | 4.071358507 | 0.000742531 | 0.00415349 | 1 |
| T2DM | b_cell | *LILRB2* | -2.799481389 | 0.000741477 | 0.00415349 | 1 |
| T2DM | b_cell | *COX4I2* | 2.384390916 | 0.000714448 | 0.00415349 | 1 |
| T2DM | b_cell | *ATP6V1E1* | -2.713206084 | 0.000511428 | 0.00415349 | 1 |
| T2DM | b_cell | *PPT1* | -2.161875852 | 0.000885905 | 0.00415349 | 1 |
| T2DM | b_cell | *HAUS8* | -3.222609324 | 0.000366753 | 0.00415349 | 1 |
| T2DM | b_cell | *TBC1D5* | -2.108522306 | 0.009249905 | 0.014177023 | 1 |
| T2DM | b_cell | *RFTN1* | -1.63776523 | 0.001961497 | 0.005076957 | 1 |
| T2DM | b_cell | *KCNC3* | -3.864980448 | 0.003577153 | 0.007094499 | 1 |
| T2DM | b_cell | *KIF3A* | -2.827062523 | 0.003172583 | 0.006535397 | 1 |
| T2DM | b_cell | *AOC3* | -1.734644881 | 0.000617526 | 0.00415349 | 1 |
| T2DM | b_cell | *ACLY* | -3.313928253 | 0.006941204 | 0.011290906 | 1 |
| T2DM | b_cell | *AOC2* | -4.83385892 | 0.000850949 | 0.00415349 | 1 |
| T2DM | b_cell | *DIAPH1* | -3.121680134 | 0.004198444 | 0.00790288 | 1 |
| T2DM | b_cell | *EXOC4* | -1.788409393 | 0.014247453 | 0.020283907 | 1 |
| T2DM | b_cell | *ACAP3* | -2.185715678 | 0.00171893 | 0.004746118 | 1 |
| T2DM | b_cell | *C1orf159* | -3.113320056 | 0.002275116 | 0.005445221 | 1 |
| T2DM | b_cell | *PPFIA1* | -2.423525594 | 0.002276665 | 0.005445221 | 1 |
| T2DM | b_cell | *NINJ1* | -2.766508356 | 0.000715088 | 0.00415349 | 1 |
| T2DM | b_cell | *NPHP4* | -2.399297035 | 0.000945983 | 0.00415349 | 1 |
| T2DM | b_cell | *MAP1B* | -1.03692284 | 0.00645822 | 0.010687768 | 1 |
| T2DM | b_cell | *CKMT2* | -2.004082513 | 0.000743414 | 0.00415349 | 1 |

| | | | | | | |
|---|---|---|---|---|---|---|
| T2DM | b_cell | *ZCCHC9* | -3.421644218 | 0.010905006 | 0.016211816 | 1 |
| T2DM | b_cell | *KHDRBS3* | -1.73253915 | 0.000937738 | 0.00415349 | 1 |
| T2DM | b_cell | *CHD1L* | -3.283747255 | 0.007614829 | 0.012141625 | 1 |
| T2DM | b_cell | *RBM8A* | -2.074897008 | 0.000350903 | 0.00415349 | 1 |
| T2DM | b_cell | *ZSCAN5A* | -3.355842738 | 0.007066929 | 0.011450974 | 1 |
| T2DM | b_cell | *ZNF132* | -2.581472917 | 0.024270593 | 0.031982648 | 1 |
| T2DM | b_cell | *RHPN2* | -4.073722896 | 0.000110755 | 0.00415349 | 0.696318957 |
| T2DM | b_cell | *C19orf12* | -1.370201995 | 0.001110091 | 0.004166654 | 1 |
| T2DM | b_cell | *ACTR10* | -2.981476041 | 0.018753048 | 0.025547219 | 1 |
| T2DM | b_cell | *GCH1* | -2.224897625 | 0.002418277 | 0.005614368 | 1 |
| T2DM | b_cell | *LGALS3* | -2.567890011 | 0.003691969 | 0.007246772 | 1 |
| T2DM | b_cell | *DNAJB1* | -2.370389612 | 0.001010966 | 0.00415349 | 1 |
| T2DM | b_cell | *FBXW9* | -2.985542803 | 0.006125873 | 0.010300445 | 1 |
| T2DM | b_cell | *C19orf57* | -5.341525869 | 0.001296772 | 0.004268027 | 1 |
| T2DM | b_cell | *MATN3* | -11.14742095 | 0.000907669 | 0.00415349 | 1 |
| T2DM | b_cell | *CCT6B* | -3.596108583 | 0.001878185 | 0.004959322 | 1 |
| T2DM | b_cell | *RAF1* | -2.807388936 | 0.015242445 | 0.021438312 | 1 |
| T2DM | b_cell | *ENOSF1* | -1.799803582 | 0.001364586 | 0.004299827 | 1 |
| T2DM | b_cell | *EMILIN2* | -1.847365297 | 0.002553221 | 0.005794981 | 1 |
| T2DM | b_cell | *TRIM5* | -3.093104555 | 0.009806571 | 0.014881466 | 1 |
| T2DM | b_cell | *CNGA4* | -4.729602058 | 0.000716237 | 0.00415349 | 1 |
| T2DM | b_cell | *RRP8* | -4.153044097 | 0.034054371 | 0.043213631 | 1 |
| T2DM | b_cell | *IMMT* | -2.174991848 | 0.005682395 | 0.009761317 | 1 |
| T2DM | b_cell | *CARD6* | -4.024203102 | 0.007269634 | 0.011686062 | 1 |
| T2DM | b_cell | *INPP5K* | -2.260583293 | 0.000652643 | 0.00415349 | 1 |
| T2DM | b_cell | *MYBBP1A* | -2.459056214 | 0.001029053 | 0.00415349 | 1 |
| T2DM | b_cell | *RPA1* | -2.54805234 | 0.001329487 | 0.004274973 | 1 |
| T2DM | b_cell | *SERPINF1* | -1.912763701 | 0.001294809 | 0.004268027 | 1 |
| T2DM | b_cell | *TMEM128* | -4.855758037 | 0.011230038 | 0.016585212 | 1 |
| T2DM | b_cell | *SEC61G* | -2.476650701 | 0.002328266 | 0.005509149 | 1 |
| T2DM | b_cell | *FIGNL1* | -4.102347372 | 0.003653024 | 0.007196562 | 1 |
| T2DM | b_cell | *WBP2* | -3.068400892 | 0.001689652 | 0.004704627 | 1 |
| T2DM | b_cell | *EIF5A* | -2.857071031 | 0.00047959 | 0.00415349 | 1 |
| T2DM | b_cell | *KDM6B* | -4.400902933 | 0.008487929 | 0.013218631 | 1 |
| T2DM | b_cell | *XAF1* | -1.562780602 | 0.001937987 | 0.005028529 | 1 |
| T2DM | b_cell | *REEP2* | -3.439798404 | 0.005916272 | 0.01004201 | 1 |
| T2DM | b_cell | *PRMT7* | -3.173221177 | 0.008080505 | 0.012700534 | 1 |
| T2DM | b_cell | *BTBD3* | -1.922446865 | 0.000789813 | 0.00415349 | 1 |
| T2DM | b_cell | *PCNA* | 5.496606086 | 0.017631052 | 0.024281801 | 1 |
| T2DM | b_cell | *PTPRA* | -2.465540774 | 0.0105103 | 0.015721688 | 1 |
| T2DM | b_cell | *IGHMBP2* | -3.692685143 | 0.001363198 | 0.004298348 | 1 |
| T2DM | b_cell | *MMACHC* | -3.471652512 | 0.00045131 | 0.00415349 | 1 |
| T2DM | b_cell | *CTNNBL1* | -3.111649527 | 0.009503153 | 0.014483958 | 1 |
| T2DM | b_cell | *LPIN3* | -2.390085583 | 0.000964397 | 0.00415349 | 1 |
| T2DM | b_cell | *SERINC3* | -2.916135516 | 0.000716398 | 0.00415349 | 1 |
| T2DM | b_cell | *DMGDH* | -1.70519456 | 0.00043228 | 0.00415349 | 1 |
| T2DM | b_cell | *BHMT2* | -1.147448228 | 0.001284114 | 0.004268027 | 1 |
| T2DM | b_cell | *KANK4* | -1.572325736 | 0.00166006 | 0.004679659 | 1 |
| T2DM | b_cell | *ATP8A2* | -3.001406284 | 0.000772093 | 0.00415349 | 1 |
| T2DM | b_cell | *USPL1* | -3.053015809 | 0.018866496 | 0.025664149 | 1 |

| | | | | | | |
|---|---|---|---|---|---|---|
| T2DM | b_cell | *POMP* | -3.543449435 | 0.019690944 | 0.026645924 | 1 |
| T2DM | b_cell | *WASF3* | -1.441393918 | 0.00075239 | 0.00415349 | 1 |
| T2DM | b_cell | *RNF17* | 4.284521797 | 0.006229212 | 0.010416354 | 1 |
| T2DM | b_cell | *PEMT* | -2.072228413 | 0.000852121 | 0.00415349 | 1 |
| T2DM | b_cell | *SCO1* | -3.674657085 | 0.001817238 | 0.004869982 | 1 |
| T2DM | b_cell | *TMCC2* | -6.252197041 | 8.37E-06 | 0.001459962 | 0.052614512 |
| T2DM | b_cell | *COG6* | -2.504729257 | 0.027818287 | 0.036075406 | 1 |
| T2DM | b_cell | *TRPC4* | -3.525716307 | 0.000809574 | 0.00415349 | 1 |
| T2DM | b_cell | *RFC3* | -2.446251893 | 0.00202522 | 0.00516315 | 1 |
| T2DM | b_cell | *PRAM1* | -3.04536007 | 0.000166793 | 0.00415349 | 1 |
| T2DM | b_cell | *ZNF414* | -2.68063959 | 0.000781971 | 0.00415349 | 1 |
| T2DM | b_cell | *CNDP2* | -2.941908743 | 0.001590603 | 0.004593531 | 1 |
| T2DM | b_cell | *RTN3* | -1.87358227 | 0.001723631 | 0.004750754 | 1 |
| T2DM | b_cell | *C1QTNF6* | -2.492625542 | 0.000893266 | 0.00415349 | 1 |
| T2DM | b_cell | *FAM83F* | -5.616160089 | 0.000655027 | 0.00415349 | 1 |
| T2DM | b_cell | *GIMAP4* | -2.244766951 | 0.000353173 | 0.00415349 | 1 |
| T2DM | b_cell | *ADCK2* | -3.381276858 | 0.001379039 | 0.004323793 | 1 |
| T2DM | b_cell | *AGAP3* | -2.601123755 | 0.001328542 | 0.004274973 | 1 |
| T2DM | b_cell | *ACTR3B* | -3.924902211 | 0.004556419 | 0.00839637 | 1 |
| T2DM | b_cell | *DYDC2* | -13.08560353 | 0.000916549 | 0.00415349 | 1 |
| T2DM | b_cell | *TMTC1* | -1.267328734 | 0.000912737 | 0.00415349 | 1 |
| T2DM | b_cell | *IPO8* | -3.202775443 | 0.010407571 | 0.015623782 | 1 |
| T2DM | b_cell | *LARS* | -2.123851736 | 0.008329819 | 0.013014308 | 1 |
| T2DM | b_cell | *SPINK5* | -4.74757619 | 0.007013997 | 0.011382808 | 1 |
| T2DM | b_cell | *IMPA1* | -3.12737456 | 0.018826476 | 0.025625038 | 1 |
| T2DM | b_cell | *LRRCC1* | -2.542845107 | 0.006346334 | 0.010555397 | 1 |
| T2DM | b_cell | *AMPD3* | -2.227072367 | 0.000616958 | 0.00415349 | 1 |
| T2DM | b_cell | *SBF2* | -1.976721577 | 0.004368072 | 0.008127276 | 1 |
| T2DM | b_cell | *MICAL2* | -2.527641928 | 0.033736035 | 0.042865492 | 1 |
| T2DM | b_cell | *HSD17B4* | -2.504786522 | 0.002157047 | 0.00530985 | 1 |
| T2DM | b_cell | *DPF2* | -2.635807091 | 0.001457078 | 0.004416899 | 1 |
| T2DM | b_cell | *GSC* | -6.999022296 | 0.00076662 | 0.00415349 | 1 |
| T2DM | b_cell | *CATSPERB* | -1.834320842 | 0.000337355 | 0.00415349 | 1 |
| T2DM | b_cell | *COX16* | -3.714940853 | 0.012576464 | 0.018214289 | 1 |
| T2DM | b_cell | *MED6* | -1.663379313 | 0.001828836 | 0.004890949 | 1 |
| T2DM | b_cell | *EIF2S1* | -3.316588513 | 0.028207857 | 0.036512826 | 1 |
| T2DM | b_cell | *ELP3* | -2.695037802 | 0.002548413 | 0.005790341 | 1 |
| T2DM | b_cell | *ADAMDEC1* | -9.664414298 | 0.007774655 | 0.012350455 | 1 |
| T2DM | b_cell | *MRO* | -3.963871893 | 0.001316183 | 0.004274973 | 1 |
| T2DM | b_cell | *THUMPD3* | -3.226699168 | 0.000863548 | 0.00415349 | 1 |
| T2DM | b_cell | *CNTN6* | -2.211966795 | 0.000660622 | 0.00415349 | 1 |
| T2DM | b_cell | *KATNBL1* | -2.251053081 | 0.003372135 | 0.00680379 | 1 |
| T2DM | b_cell | *GSTM1* | 2.987440095 | 0.027095726 | 0.035225564 | 1 |
| T2DM | b_cell | *GSTM5* | -2.311545534 | 0.006385406 | 0.010597953 | 1 |
| T2DM | b_cell | *SYT6* | -7.778472568 | 0.000953655 | 0.00415349 | 1 |
| T2DM | b_cell | *VAV3* | 2.763880967 | 0.008438676 | 0.013145182 | 1 |
| T2DM | b_cell | *PSRC1* | 2.870600959 | 0.00208456 | 0.005230315 | 1 |
| T2DM | b_cell | *WNT2B* | -2.453083048 | 0.001274684 | 0.004257394 | 1 |
| T2DM | b_cell | *CEPT1* | -2.196084415 | 0.002179389 | 0.005337677 | 1 |
| T2DM | b_cell | *AP4B1* | -3.05893396 | 0.005425554 | 0.009467238 | 1 |

| | | | | | | |
|---|---|---|---|---|---|---|
| T2DM | b_cell | *NAPG* | -2.894049176 | 0.00348177 | 0.006955795 | 1 |
| T2DM | b_cell | *SPIRE1* | -2.374596012 | 0.000610173 | 0.00415349 | 1 |
| T2DM | b_cell | *PPHLN1* | -2.206246309 | 0.039253876 | 0.049073199 | 1 |
| T2DM | b_cell | *FKBP11* | -3.958132334 | 0.03535704 | 0.044681349 | 1 |
| T2DM | b_cell | *YWHAQ* | -2.770193326 | 0.03208475 | 0.040941104 | 1 |
| T2DM | b_cell | *LPIN1* | -2.231975011 | 0.000582745 | 0.00415349 | 1 |
| T2DM | b_cell | *LDHA* | -2.566113711 | 0.001753046 | 0.004789372 | 1 |
| T2DM | b_cell | *ANO3* | -1.38230312 | 0.000897697 | 0.00415349 | 1 |
| T2DM | b_cell | *FST* | -2.891566663 | 0.000527142 | 0.00415349 | 1 |
| T2DM | b_cell | *NARS* | -2.742996928 | 0.001826503 | 0.004888559 | 1 |
| T2DM | b_cell | *FBXO18* | -3.041984091 | 0.020227332 | 0.027307115 | 1 |
| T2DM | b_cell | *ECHDC3* | -1.935002472 | 0.000731779 | 0.00415349 | 1 |
| T2DM | b_cell | *IL15RA* | -2.107454435 | 0.000953886 | 0.00415349 | 1 |
| T2DM | b_cell | *TMEM241* | -1.93502678 | 0.00135478 | 0.004296951 | 1 |
| T2DM | b_cell | *EMP1* | -1.963816046 | 0.001795459 | 0.004852128 | 1 |
| T2DM | b_cell | *KLRD1* | -1.841008791 | 0.001171049 | 0.004202203 | 1 |
| T2DM | b_cell | *PRH2* | -8.380945723 | 3.36E-05 | 0.003301266 | 0.211281053 |
| T2DM | b_cell | *DDB2* | -2.561321735 | 0.031628406 | 0.040449102 | 1 |
| T2DM | b_cell | *ACP2* | 4.737673813 | 0.031119902 | 0.03987178 | 1 |
| T2DM | b_cell | *HOOK1* | 2.503928497 | 0.000645627 | 0.00415349 | 1 |
| T2DM | b_cell | *ZCCHC11* | -2.462513584 | 0.017901659 | 0.024589847 | 1 |
| T2DM | b_cell | *PRPF38A* | -4.073080497 | 0.008224216 | 0.012881327 | 1 |
| T2DM | b_cell | *FHOD3* | -1.542642938 | 0.000437475 | 0.00415349 | 1 |
| T2DM | b_cell | *TPGS2* | -2.289092739 | 0.00286411 | 0.006149816 | 1 |
| T2DM | b_cell | *SLC43A3* | -4.60108621 | 0.019288356 | 0.0261518 | 1 |
| T2DM | b_cell | *TIMM10* | -3.829803224 | 0.00016575 | 0.00415349 | 1 |
| T2DM | b_cell | *DHX34* | -3.379330551 | 0.000517389 | 0.00415349 | 1 |
| T2DM | b_cell | *APLNR* | -3.121007176 | 0.001452123 | 0.004416445 | 1 |
| T2DM | b_cell | *FADS2* | -2.30987193 | 0.001248419 | 0.004253251 | 1 |
| T2DM | b_cell | *TMEM165* | -1.931199343 | 0.005728033 | 0.009813953 | 1 |
| T2DM | b_cell | *CLOCK* | -2.686838125 | 0.015214834 | 0.021404266 | 1 |
| T2DM | b_cell | *CLDN10* | 1.9830185 | 0.000724359 | 0.00415349 | 1 |
| T2DM | b_cell | *DZIP1* | -2.10533887 | 0.001008346 | 0.00415349 | 1 |
| T2DM | b_cell | *BIVM* | -4.545071601 | 0.007268487 | 0.011686062 | 1 |
| T2DM | b_cell | *ERCC5* | -6.556128562 | 0.003344934 | 0.006768457 | 1 |
| T2DM | b_cell | *CARS2* | -2.386764896 | 0.0007558 | 0.00415349 | 1 |
| T2DM | b_cell | *STT3A* | -2.618467432 | 0.001876471 | 0.004957688 | 1 |
| T2DM | b_cell | *KLB* | -1.262694001 | 0.002588902 | 0.005823408 | 1 |
| T2DM | b_cell | *WDR36* | -2.816778133 | 0.003200629 | 0.006578083 | 1 |
| T2DM | b_cell | *NAA35* | -2.848352617 | 0.018106213 | 0.024838264 | 1 |
| T2DM | b_cell | *TMEM2* | -1.894505638 | 0.001281242 | 0.004266508 | 1 |
| T2DM | b_cell | *AGTPBP1* | -2.373704371 | 0.00234573 | 0.005525518 | 1 |
| T2DM | b_cell | *GOLM1* | -3.230157597 | 0.001165756 | 0.004200062 | 1 |
| T2DM | b_cell | *ADAM19* | 0.924390378 | 0.000206512 | 0.00415349 | 1 |
| T2DM | b_cell | *USP30* | -2.372185397 | 0.01706689 | 0.023618652 | 1 |
| T2DM | b_cell | *HRK* | 2.759343825 | 0.000735654 | 0.00415349 | 1 |
| T2DM | b_cell | *P2RX4* | -1.777786245 | 0.018134625 | 0.024859113 | 1 |
| T2DM | b_cell | *DTX1* | -1.934525976 | 0.000246648 | 0.00415349 | 1 |
| T2DM | b_cell | *DMTF1* | -2.187689876 | 0.029220958 | 0.037676818 | 1 |
| T2DM | b_cell | *CCDC146* | -2.094402741 | 0.003337746 | 0.006765112 | 1 |

| | | | | | | |
|---|---|---|---|---|---|---|
| T2DM | b_cell | *TMEM60* | 4.377980428 | 0.027133422 | 0.035252702 | 1 |
| T2DM | b_cell | *POM121C* | -3.147763023 | 0.000287394 | 0.00415349 | 1 |
| T2DM | b_cell | *SRPK2* | -2.277522369 | 0.013970697 | 0.01994409 | 1 |
| T2DM | b_cell | *TES* | -3.189200895 | 0.004917059 | 0.008844125 | 1 |
| T2DM | b_cell | *MDFIC* | -1.748463935 | 0.001380285 | 0.004323793 | 1 |
| T2DM | b_cell | *ANKRD6* | -3.096153186 | 0.03175449 | 0.040577334 | 1 |
| T2DM | b_cell | *KHDC1* | -2.081270638 | 0.004103332 | 0.007798563 | 1 |
| T2DM | b_cell | *SNX14* | -2.760116931 | 0.010673996 | 0.015940004 | 1 |
| T2DM | b_cell | *NT5E* | -1.633560358 | 0.003230243 | 0.006613998 | 1 |
| T2DM | b_cell | *PRR5L* | -1.93848078 | 0.001061693 | 0.00415349 | 1 |
| T2DM | b_cell | *NAT10* | -2.868736472 | 0.002608283 | 0.005849897 | 1 |
| T2DM | b_cell | *ITGA7* | -1.368140334 | 0.000210575 | 0.00415349 | 1 |
| T2DM | b_cell | *RDH5* | -1.353064974 | 0.001074471 | 0.00415349 | 1 |
| T2DM | b_cell | *BLOC1S1* | -3.807857337 | 0.002417053 | 0.005613599 | 1 |
| T2DM | b_cell | *PPP1R1A* | -2.836681567 | 0.001575433 | 0.004562749 | 1 |
| T2DM | b_cell | *B4GALNT1* | 3.831126234 | 0.001764332 | 0.004791515 | 1 |
| T2DM | b_cell | *FAIM2* | -5.56623375 | 0.000564108 | 0.00415349 | 1 |
| T2DM | b_cell | *KRT7* | 2.19688025 | 0.000262479 | 0.00415349 | 1 |
| T2DM | b_cell | *SLC26A10* | -8.936858455 | 0.000349064 | 0.00415349 | 1 |
| T2DM | b_cell | *MAP7* | -1.632714629 | 0.001151348 | 0.004188961 | 1 |
| T2DM | b_cell | *AHI1* | -1.408974818 | 0.008109743 | 0.012727398 | 1 |
| T2DM | b_cell | *PKIB* | -2.715581654 | 0.000748564 | 0.00415349 | 1 |
| T2DM | b_cell | *SMPD2* | 3.133588362 | 0.008033256 | 0.012648405 | 1 |
| T2DM | b_cell | *MICAL1* | -3.237533605 | 0.009422731 | 0.014382304 | 1 |
| T2DM | b_cell | *PRADC1* | 4.10490284 | 0.00305754 | 0.006378687 | 1 |
| T2DM | b_cell | *SMYD5* | -3.303244171 | 0.000188732 | 0.00415349 | 1 |
| T2DM | b_cell | *DYSF* | -2.207133254 | 0.00055407 | 0.00415349 | 1 |
| T2DM | b_cell | *KCNMB4* | 2.4613805 | 0.001092844 | 0.00415349 | 1 |
| T2DM | b_cell | *USP15* | -2.350425637 | 0.017246861 | 0.023825757 | 1 |
| T2DM | b_cell | *KLHL36* | -2.448333409 | 0.002322241 | 0.005505253 | 1 |
| T2DM | b_cell | *MPHOSPH6* | 4.486635415 | 0.013562755 | 0.019432325 | 1 |
| T2DM | b_cell | *FHOD1* | -2.112857425 | 0.000622461 | 0.00415349 | 1 |
| T2DM | b_cell | *CCDC102A* | -3.802915527 | 0.000594152 | 0.00415349 | 1 |
| T2DM | b_cell | *ZNF670* | 2.516337312 | 0.006956809 | 0.01130459 | 1 |
| T2DM | b_cell | *KCNK1* | -1.130231855 | 0.038262543 | 0.047900559 | 1 |
| T2DM | b_cell | *NTPCR* | -2.716122392 | 0.002774341 | 0.006030781 | 1 |
| T2DM | b_cell | *RNASEL* | -2.240374819 | 0.004700923 | 0.008595602 | 1 |
| T2DM | b_cell | *PIGC* | -2.600836197 | 0.004113105 | 0.007807696 | 1 |
| T2DM | b_cell | *SP110* | 2.88814323 | 0.017847432 | 0.024536804 | 1 |
| T2DM | b_cell | *SLC19A3* | -1.107010541 | 0.001756088 | 0.004789372 | 1 |
| T2DM | b_cell | *TMBIM1* | -2.503185179 | 0.000104931 | 0.00415349 | 0.65970417 |
| T2DM | b_cell | *ARMC9* | -3.147993759 | 0.002808976 | 0.006081279 | 1 |
| T2DM | b_cell | *TMEM127* | -3.177932619 | 0.001802481 | 0.004852128 | 1 |
| T2DM | b_cell | *GCC2* | -2.731676442 | 0.011696619 | 0.017137415 | 1 |
| T2DM | b_cell | *ANKRD36* | -2.585549552 | 0.004587289 | 0.008445179 | 1 |
| T2DM | b_cell | *USP44* | -3.397143359 | 0.000661223 | 0.00415349 | 1 |
| T2DM | b_cell | *APPL2* | -2.449756921 | 0.00307692 | 0.006405496 | 1 |
| T2DM | b_cell | *DRAM1* | -2.522628975 | 0.00222733 | 0.005400083 | 1 |
| T2DM | b_cell | *SLC41A2* | -2.563345685 | 0.002325085 | 0.005505767 | 1 |
| T2DM | b_cell | *NEK3* | -2.2937332 | 0.019061182 | 0.025888454 | 1 |

| | | | | | | |
|---|---|---|---|---|---|---|
| T2DM | b_cell | *VPS36* | -2.607611173 | 0.007986727 | 0.012595482 | 1 |
| T2DM | b_cell | *TBC1D4* | -2.148144906 | 0.002563862 | 0.005799421 | 1 |
| T2DM | b_cell | *SUCLA2* | -2.276241752 | 0.001355424 | 0.004296951 | 1 |
| T2DM | b_cell | *RCBTB1* | -3.502115663 | 0.000226669 | 0.00415349 | 1 |
| T2DM | b_cell | *MED4* | -3.366587883 | 0.006335539 | 0.010543022 | 1 |
| T2DM | b_cell | *RCBTB2* | -2.208510555 | 0.001371779 | 0.004316504 | 1 |
| T2DM | b_cell | *SETDB2* | -2.87645904 | 0.006891709 | 0.011227838 | 1 |
| T2DM | b_cell | *SPDYE1* | -2.399280266 | 0.00168432 | 0.004704627 | 1 |
| T2DM | b_cell | *CHST12* | -3.051082076 | 0.002167881 | 0.005319854 | 1 |
| T2DM | b_cell | *GPNMB* | -1.666070264 | 0.000935809 | 0.00415349 | 1 |
| T2DM | b_cell | *KDELR2* | -3.751306412 | 0.008917913 | 0.01376551 | 1 |
| T2DM | b_cell | *NUPL2* | -2.814263322 | 0.002142035 | 0.005287258 | 1 |
| T2DM | b_cell | *ZDHHC4* | -3.337246616 | 0.002006188 | 0.005130808 | 1 |
| T2DM | b_cell | *AOAH* | -1.727337772 | 0.000114157 | 0.00415349 | 0.717706652 |
| T2DM | b_cell | *TBRG4* | 3.476110907 | 0.003404901 | 0.006850117 | 1 |
| T2DM | b_cell | *DDX56* | -4.415362281 | 0.008102003 | 0.012721602 | 1 |
| T2DM | b_cell | *HUS1* | -2.13395844 | 0.001839672 | 0.004898539 | 1 |
| T2DM | b_cell | *NACAD* | -13.6801777 | 1.64E-05 | 0.002349192 | 0.103364449 |
| T2DM | b_cell | *TTC5* | -3.417642804 | 0.000883865 | 0.00415349 | 1 |
| T2DM | b_cell | *MTHFS* | -1.854764604 | 0.020600026 | 0.027762564 | 1 |
| T2DM | b_cell | *ALPK3* | -1.966516526 | 0.000461898 | 0.00415349 | 1 |
| T2DM | b_cell | *TM6SF1* | -1.879753825 | 0.002129049 | 0.005276048 | 1 |
| T2DM | b_cell | *RSAD1* | -3.28481083 | 0.001378319 | 0.004323793 | 1 |
| T2DM | b_cell | *NMT1* | -1.606660951 | 0.023100222 | 0.030704248 | 1 |
| T2DM | b_cell | *MYCBPAP* | -2.170423671 | 0.023324377 | 0.030930259 | 1 |
| T2DM | b_cell | *BRIP1* | 2.514392382 | 0.000795272 | 0.00415349 | 1 |
| T2DM | b_cell | *RTP4* | -6.374417257 | 0.02473917 | 0.03248437 | 1 |
| T2DM | b_cell | *MRPL47* | -4.663031074 | 0.006505706 | 0.010749375 | 1 |
| T2DM | b_cell | *BLK* | 0.107248443 | 0.000596316 | 0.00415349 | 1 |
| T2DM | b_cell | *EPRS* | -3.169293269 | 0.007298665 | 0.011723738 | 1 |
| T2DM | b_cell | *RPS6KC1* | -2.359056903 | 0.003721022 | 0.007268006 | 1 |
| T2DM | b_cell | *CBWD2* | -3.883037518 | 0.007653827 | 0.012191439 | 1 |
| T2DM | b_cell | *IL1RN* | 2.646739442 | 0.001200433 | 0.004223203 | 1 |
| T2DM | b_cell | *SAP130* | -3.655084448 | 0.005938266 | 0.010065752 | 1 |
| T2DM | b_cell | *GYPC* | -2.199204032 | 0.00179918 | 0.004852128 | 1 |
| T2DM | b_cell | *LRRC8A* | -2.420730657 | 0.001140016 | 0.00418468 | 1 |
| T2DM | b_cell | *TXN* | -2.791636707 | 0.001293664 | 0.004268027 | 1 |
| T2DM | b_cell | *ODF2* | -3.816136155 | 0.016881393 | 0.023392841 | 1 |
| T2DM | b_cell | *TOR1B* | -2.513623081 | 0.001176227 | 0.004202203 | 1 |
| T2DM | b_cell | *TOR1A* | -3.092045967 | 0.000215792 | 0.00415349 | 1 |
| T2DM | b_cell | *SLC2A8* | -3.415277204 | 0.001780387 | 0.004822616 | 1 |
| T2DM | b_cell | *ZFP37* | -2.428509586 | 0.002103554 | 0.005241681 | 1 |
| T2DM | b_cell | *ZNF189* | -3.37399592 | 0.003656217 | 0.007196818 | 1 |
| T2DM | b_cell | *ALDOB* | -4.392585582 | 0.002724304 | 0.005972002 | 1 |
| T2DM | b_cell | *STX17* | -3.161990659 | 0.010768187 | 0.01604636 | 1 |
| T2DM | b_cell | *PRPF4* | 5.027421728 | 0.001514732 | 0.004480664 | 1 |
| T2DM | b_cell | *FPGS* | -2.618800577 | 0.000795122 | 0.00415349 | 1 |
| T2DM | b_cell | *TEX10* | -2.379783617 | 0.001318945 | 0.004274973 | 1 |
| T2DM | b_cell | *GARNL3* | -2.788528372 | 0.002904305 | 0.006192483 | 1 |
| T2DM | b_cell | *GABBR2* | -5.207256931 | 0.000819611 | 0.00415349 | 1 |

| | | | | | | |
|---|---|---|---|---|---|---|
| T2DM | b_cell | *PSMB7* | -2.91660185 | 0.00598859 | 0.010134661 | 1 |
| T2DM | b_cell | *RABEPK* | -2.267827488 | 0.00100898 | 0.00415349 | 1 |
| T2DM | b_cell | *GOLGA1* | -1.920079397 | 0.001035108 | 0.00415349 | 1 |
| T2DM | b_cell | *LMX1B* | -6.635856855 | 0.000670082 | 0.00415349 | 1 |
| T2DM | b_cell | *DSCC1* | -1.485439268 | 0.002649663 | 0.005895393 | 1 |
| T2DM | b_cell | *DERL1* | -3.739518899 | 0.029907271 | 0.038459196 | 1 |
| T2DM | b_cell | *NOV* | -3.613207582 | 0.003137882 | 0.006486603 | 1 |
| T2DM | b_cell | *POLR1E* | -2.335637617 | 0.012851254 | 0.018573755 | 1 |
| T2DM | b_cell | *IL11RA* | -2.743349131 | 0.001982519 | 0.005095706 | 1 |
| T2DM | b_cell | *APTX* | -2.64909815 | 0.003430253 | 0.006885994 | 1 |
| T2DM | b_cell | *TLN1* | -1.882961872 | 0.002151504 | 0.005302433 | 1 |
| T2DM | b_cell | *SPAG8* | 6.357293505 | 0.001893287 | 0.00497413 | 1 |
| T2DM | b_cell | *CD72* | -3.813823212 | 0.009349055 | 0.014290666 | 1 |
| T2DM | b_cell | *TMEM8B* | 4.282589129 | 0.034460154 | 0.043644438 | 1 |
| T2DM | b_cell | *GRHPR* | -3.466474626 | 0.000670422 | 0.00415349 | 1 |
| T2DM | b_cell | *ARHGEF39* | 2.563600051 | 0.012285288 | 0.017850151 | 1 |
| T2DM | b_cell | *DENND4C* | -2.163870253 | 0.006439817 | 0.010668545 | 1 |
| T2DM | b_cell | *PPIL1* | 4.582634952 | 0.001051978 | 0.00415349 | 1 |
| T2DM | b_cell | *KIF13A* | -1.840940777 | 0.001906654 | 0.004988168 | 1 |
| T2DM | b_cell | *TMEM63B* | -2.090936602 | 0.002618282 | 0.005854674 | 1 |
| T2DM | b_cell | *FRS3* | 5.354708101 | 0.010337889 | 0.015541442 | 1 |
| T2DM | b_cell | *SLC22A23* | -1.820740842 | 0.001161632 | 0.004195495 | 1 |
| T2DM | b_cell | *LRRC1* | -1.876788135 | 0.000566673 | 0.00415349 | 1 |
| T2DM | b_cell | *FOXF2* | -6.26417658 | 0.038000038 | 0.047593002 | 1 |
| T2DM | b_cell | *BPHL* | -1.356840855 | 0.002434322 | 0.005628918 | 1 |
| T2DM | b_cell | *RIPK1* | -2.396862324 | 0.001986853 | 0.005098507 | 1 |
| T2DM | b_cell | *TCF19* | -3.409479342 | 0.018876564 | 0.025670984 | 1 |
| T2DM | b_cell | *FLOT1* | -2.190187406 | 0.00724707 | 0.011664702 | 1 |
| T2DM | b_cell | *TPMT* | -4.889284757 | 0.013964477 | 0.019939737 | 1 |
| T2DM | b_cell | *MTCH1* | -3.298062981 | 0.026848619 | 0.034954912 | 1 |
| T2DM | b_cell | *FHDC1* | -2.819327169 | 0.000795021 | 0.00415349 | 1 |
| T2DM | b_cell | *MYO7A* | -2.335180332 | 0.001459605 | 0.004422426 | 1 |
| T2DM | b_cell | *ARRB1* | -2.283086717 | 0.00361094 | 0.007151285 | 1 |
| T2DM | b_cell | *CCDC90B* | -2.57005949 | 0.001353106 | 0.004296454 | 1 |
| T2DM | b_cell | *SYTL2* | -1.853473531 | 0.000336674 | 0.00415349 | 1 |
| T2DM | b_cell | *PI15* | 3.716220006 | 0.000860475 | 0.00415349 | 1 |
| T2DM | b_cell | *TTPA* | -1.750889185 | 0.000109299 | 0.00415349 | 0.68716342 |
| T2DM | b_cell | *GGH* | -2.338607496 | 0.001072658 | 0.00415349 | 1 |
| T2DM | b_cell | *SULF1* | -1.198573309 | 0.001036981 | 0.00415349 | 1 |
| T2DM | b_cell | *TGS1* | -2.666462548 | 0.023592587 | 0.031260492 | 1 |
| T2DM | b_cell | *NEK1* | -2.281002966 | 0.004799347 | 0.008710593 | 1 |
| T2DM | b_cell | *DDX60* | -3.391275512 | 0.005688802 | 0.009764542 | 1 |
| T2DM | b_cell | *DCUN1D5* | -1.847916374 | 0.00019955 | 0.00415349 | 1 |
| T2DM | b_cell | *TRIM29* | -3.922371448 | 0.000990863 | 0.00415349 | 1 |
| T2DM | b_cell | *POU2F3* | 2.444668768 | 0.001321923 | 0.004274973 | 1 |
| T2DM | b_cell | *PPP2R1B* | -1.176261094 | 0.000766213 | 0.00415349 | 1 |
| T2DM | b_cell | *FDX1* | -2.798743809 | 0.002186115 | 0.005349464 | 1 |
| T2DM | b_cell | *FXYD6* | -5.580562733 | 0.028879869 | 0.037275249 | 1 |
| T2DM | b_cell | *TMPRSS13* | -7.710194496 | 0.013519835 | 0.019379663 | 1 |
| T2DM | b_cell | *MAP2K5* | -2.311481032 | 0.019990017 | 0.027004134 | 1 |

| | | | | | | |
|---|---|---|---|---|---|---|
| T2DM | b_cell | *NDUFAF1* | -4.473947778 | 9.80E-05 | 0.00415349 | 0.616373777 |
| T2DM | b_cell | *ITGA11* | -1.545300234 | 0.000528881 | 0.00415349 | 1 |
| T2DM | b_cell | *PAQR5* | -3.521350134 | 0.000971397 | 0.00415349 | 1 |
| T2DM | b_cell | *ITPKA* | 4.195093288 | 0.000893254 | 0.00415349 | 1 |
| T2DM | b_cell | *UACA* | -1.926294902 | 0.001882234 | 0.004963761 | 1 |
| T2DM | b_cell | *PLCB2* | -1.477520484 | 0.011536694 | 0.016962609 | 1 |
| T2DM | b_cell | *RSL24D1* | -3.686918355 | 0.014555025 | 0.02063302 | 1 |
| T2DM | b_cell | *RABGGTB* | -3.947783382 | 0.008036127 | 0.012649758 | 1 |
| T2DM | b_cell | *DNASE2B* | -18.03079914 | 0.001807127 | 0.004857377 | 1 |
| T2DM | b_cell | *DBT* | -3.126063636 | 0.007979942 | 0.012594351 | 1 |
| T2DM | b_cell | *RTCA* | -2.202906312 | 0.032824779 | 0.041825979 | 1 |
| T2DM | b_cell | *IFT172* | -2.495197067 | 0.007578502 | 0.012092904 | 1 |
| T2DM | b_cell | *KHK* | -2.224380738 | 0.00232204 | 0.005505253 | 1 |
| T2DM | b_cell | *ADCY3* | -2.858168758 | 0.001145663 | 0.004185233 | 1 |
| T2DM | b_cell | *DYNC2LI1* | -2.64718591 | 0.001050017 | 0.00415349 | 1 |
| T2DM | b_cell | *LHCGR* | -2.641038131 | 0.000567292 | 0.00415349 | 1 |
| T2DM | b_cell | *THUMPD2* | -2.74785646 | 0.002741899 | 0.006000111 | 1 |
| T2DM | b_cell | *PREB* | -2.364452856 | 0.000563291 | 0.00415349 | 1 |
| T2DM | b_cell | *ABCG5* | -3.030497015 | 0.005583425 | 0.009641728 | 1 |
| T2DM | b_cell | *ATRAID* | -3.607673058 | 0.013218788 | 0.019043657 | 1 |
| T2DM | b_cell | *LRPPRC* | -2.656844484 | 0.00505611 | 0.009004825 | 1 |
| T2DM | b_cell | *DTNB* | -2.172633604 | 0.002384625 | 0.005571214 | 1 |
| T2DM | b_cell | *CYP2C9* | 2.260552838 | 0.00876558 | 0.013583732 | 1 |
| T2DM | b_cell | *CH25H* | -2.311041337 | 0.012348374 | 0.017925243 | 1 |
| T2DM | b_cell | *ATAD1* | -3.029511264 | 0.005964094 | 0.010106808 | 1 |
| T2DM | b_cell | *BTBD16* | -6.444061394 | 0.000141387 | 0.00415349 | 0.888899315 |
| T2DM | b_cell | *TACC2* | -1.339820994 | 0.0013179 | 0.004274973 | 1 |
| T2DM | b_cell | *CALHM2* | -2.237662085 | 0.00076256 | 0.00415349 | 1 |
| T2DM | b_cell | *ARL3* | -2.605636762 | 0.002154675 | 0.005308166 | 1 |
| T2DM | b_cell | *PLCE1* | -1.756853322 | 0.000565038 | 0.00415349 | 1 |
| T2DM | b_cell | *RBP4* | -2.016601971 | 0.00134232 | 0.004286796 | 1 |
| T2DM | b_cell | *DNAJC13* | -2.73835387 | 0.006998162 | 0.011365912 | 1 |
| T2DM | b_cell | *FAM149B1* | -2.885046795 | 0.002301507 | 0.005481934 | 1 |
| T2DM | b_cell | *ATIC* | -3.778256543 | 0.000723584 | 0.00415349 | 1 |
| T2DM | b_cell | *SMARCAL1* | -3.265717732 | 0.029999401 | 0.038569781 | 1 |
| T2DM | b_cell | *BARD1* | -3.238608682 | 0.001528728 | 0.004502743 | 1 |
| T2DM | b_cell | *STAT4* | -1.810752708 | 0.000388302 | 0.00415349 | 1 |
| T2DM | b_cell | *ASNSD1* | -2.292189206 | 0.000363602 | 0.00415349 | 1 |
| T2DM | b_cell | *METTL5* | -3.778297523 | 0.007060679 | 0.011446748 | 1 |
| T2DM | b_cell | *NAB1* | -3.16078966 | 0.008493436 | 0.013223931 | 1 |
| T2DM | b_cell | *MDH1B* | 2.272270516 | 0.000762522 | 0.00415349 | 1 |
| T2DM | b_cell | *ITGAV* | -2.271433271 | 0.005587851 | 0.009643377 | 1 |
| T2DM | b_cell | *COX17* | 3.969861845 | 0.016763149 | 0.023259748 | 1 |
| T2DM | b_cell | *USP8* | -2.137655281 | 0.002350713 | 0.005533109 | 1 |
| T2DM | b_cell | *SPPL2A* | -2.891672165 | 0.012877482 | 0.018601696 | 1 |
| T2DM | b_cell | *UBL7* | -3.373253106 | 0.001581782 | 0.004576467 | 1 |
| T2DM | b_cell | *ARHGAP24* | -1.985327886 | 0.005918314 | 0.010042765 | 1 |
| T2DM | b_cell | *FAM13A* | -1.504500201 | 0.003353536 | 0.006777924 | 1 |
| T2DM | b_cell | *HERC3* | -2.703340258 | 0.008540945 | 0.013291317 | 1 |
| T2DM | b_cell | *AP1AR* | -3.621613436 | 0.005836636 | 0.009939039 | 1 |

| | | | | | | |
|---|---|---|---|---|---|---|
| T2DM | b_cell | *PRKG2* | -2.01954635 | 0.000626333 | 0.00415349 | 1 |
| T2DM | b_cell | *BBS7* | -2.480485291 | 0.001250768 | 0.004253251 | 1 |
| T2DM | b_cell | *KIAA1109* | -2.343054143 | 0.02222628 | 0.029649188 | 1 |
| T2DM | b_cell | *MMRN1* | -2.305801004 | 0.001440924 | 0.004405146 | 1 |
| T2DM | b_cell | *PRDM5* | -1.672981631 | 0.001376596 | 0.004323793 | 1 |
| T2DM | b_cell | *TRPC3* | -4.365160313 | 0.001320001 | 0.004274973 | 1 |
| T2DM | b_cell | *NAAA* | -2.465915029 | 0.006676823 | 0.010971559 | 1 |
| T2DM | b_cell | *NUP54* | -2.147617057 | 0.007278497 | 0.01169433 | 1 |
| T2DM | b_cell | *BMP2K* | -2.361404561 | 0.001348678 | 0.004290463 | 1 |
| T2DM | b_cell | *G3BP2* | -3.443670032 | 0.001362329 | 0.004298348 | 1 |
| T2DM | b_cell | *SCARB2* | -2.71386275 | 0.005994924 | 0.010139921 | 1 |
| T2DM | b_cell | *CNOT6L* | -3.021885946 | 0.010825186 | 0.016112202 | 1 |
| T2DM | b_cell | *ANXA3* | -2.280233488 | 0.000868304 | 0.00415349 | 1 |
| T2DM | b_cell | *ENPEP* | -2.251986429 | 0.000641087 | 0.00415349 | 1 |
| T2DM | b_cell | *PAPSS1* | -2.100411973 | 0.007307467 | 0.011730073 | 1 |
| T2DM | b_cell | *SLC39A8* | -3.07504758 | 0.005316292 | 0.009320937 | 1 |
| T2DM | b_cell | *RNF185* | -3.150081778 | 0.002135878 | 0.005284637 | 1 |
| T2DM | b_cell | *B4GALNT3* | -6.576249432 | 0.000565277 | 0.00415349 | 1 |
| T2DM | b_cell | *ERP27* | 2.943133306 | 0.001725948 | 0.004752113 | 1 |
| T2DM | b_cell | *ETV6* | -2.300517022 | 0.005606427 | 0.009662174 | 1 |
| T2DM | b_cell | *KIF21A* | -1.311255092 | 0.002606411 | 0.005849897 | 1 |
| T2DM | b_cell | *CPNE8* | -2.287009018 | 0.000992652 | 0.00415349 | 1 |
| T2DM | b_cell | *ALG10* | -3.956681079 | 0.00109163 | 0.00415349 | 1 |
| T2DM | b_cell | *SLCO1C1* | -2.853605043 | 0.002488011 | 0.005702562 | 1 |
| T2DM | b_cell | *PRICKLE1* | -1.26092767 | 0.001175111 | 0.004202203 | 1 |
| T2DM | b_cell | *C1RL* | -1.778153981 | 0.006432023 | 0.010661251 | 1 |
| T2DM | b_cell | *CLSTN3* | -3.447888921 | 0.024259641 | 0.031974919 | 1 |
| T2DM | b_cell | *KLRG1* | -2.587821003 | 0.000393747 | 0.00415349 | 1 |
| T2DM | b_cell | *VAMP1* | -2.346807944 | 0.019331329 | 0.026193118 | 1 |
| T2DM | b_cell | *TAPBPL* | -3.069828007 | 0.020850232 | 0.028063671 | 1 |
| T2DM | b_cell | *CD27* | -3.526645496 | 0.00088007 | 0.00415349 | 1 |
| T2DM | b_cell | *PEX5* | 3.027463231 | 0.005104888 | 0.009050883 | 1 |
| T2DM | b_cell | *PPFIA2* | -1.36634773 | 0.000960383 | 0.00415349 | 1 |
| T2DM | b_cell | *LGR5* | -3.475395036 | 0.000704018 | 0.00415349 | 1 |
| T2DM | b_cell | *POC1B* | -3.363144688 | 0.007195271 | 0.01160212 | 1 |
| T2DM | b_cell | *SNRPF* | -4.192410528 | 0.016121649 | 0.022498736 | 1 |
| T2DM | b_cell | *AMDHD1* | -2.983058912 | 0.007698436 | 0.012243883 | 1 |
| T2DM | b_cell | *GAS2L3* | -1.930505522 | 0.000775402 | 0.00415349 | 1 |
| T2DM | b_cell | *TMEM132B* | -4.130220751 | 0.002358367 | 0.005540753 | 1 |
| T2DM | b_cell | *TDG* | -2.723564243 | 0.002393328 | 0.005579107 | 1 |
| T2DM | b_cell | *MMAB* | -2.686051869 | 0.000421042 | 0.00415349 | 1 |
| T2DM | b_cell | *GLTP* | -2.521555279 | 0.001234374 | 0.004253251 | 1 |
| T2DM | b_cell | *MTMR6* | -2.202519936 | 0.000923709 | 0.00415349 | 1 |
| T2DM | b_cell | *SLC46A3* | -1.877174802 | 0.006530975 | 0.010776966 | 1 |
| T2DM | b_cell | *CCDC65* | -2.660751231 | 0.000539556 | 0.00415349 | 1 |
| T2DM | b_cell | *ACVRL1* | -2.827240645 | 0.002706049 | 0.005952834 | 1 |
| T2DM | b_cell | *GPR84* | -41.20241497 | 2.55E-05 | 0.002754251 | 0.160621577 |
| T2DM | b_cell | *KANSL2* | -1.704017622 | 0.00109177 | 0.00415349 | 1 |
| T2DM | b_cell | *CERS5* | -3.115028057 | 0.002433424 | 0.005628918 | 1 |
| T2DM | b_cell | *LMBR1L* | -3.956673399 | 0.010862003 | 0.01615935 | 1 |

| | | | | | | |
|---|---|---|---|---|---|---|
| T2DM | b_cell | *ZNF740* | -2.495050102 | 0.001746365 | 0.004788224 | 1 |
| T2DM | b_cell | *WDFY2* | -2.574349748 | 0.026440308 | 0.034466144 | 1 |
| T2DM | b_cell | *HNRNPA1L2* | 6.648396955 | 0.002555103 | 0.005795069 | 1 |
| T2DM | b_cell | *LPAR6* | -2.390800323 | 0.001988807 | 0.005101414 | 1 |
| T2DM | b_cell | *VPS33A* | -3.720472404 | 0.001178565 | 0.004202203 | 1 |
| T2DM | b_cell | *METTL21C* | 9.94631631 | 0.007125797 | 0.011510862 | 1 |
| T2DM | b_cell | *GRTP1* | -1.908624837 | 0.002098741 | 0.005241681 | 1 |
| T2DM | b_cell | *CBLN3* | 1.205482453 | 0.011912525 | 0.01739699 | 1 |
| T2DM | b_cell | *TSSK4* | -11.35702262 | 0.003013504 | 0.006319514 | 1 |
| T2DM | b_cell | *TMX1* | -3.11668851 | 0.00201096 | 0.005137305 | 1 |
| T2DM | b_cell | *RTN1* | -1.336057926 | 0.000902054 | 0.00415349 | 1 |
| T2DM | b_cell | *C14orf37* | -2.501482927 | 0.002617453 | 0.005854674 | 1 |
| T2DM | b_cell | *RAB15* | -2.488824544 | 0.000416802 | 0.00415349 | 1 |
| T2DM | b_cell | *STON2* | -1.799108744 | 0.000904439 | 0.00415349 | 1 |
| T2DM | b_cell | *PTGR2* | -2.665839965 | 0.000151425 | 0.00415349 | 0.952007968 |
| T2DM | b_cell | *AK7* | -4.283486956 | 0.000835628 | 0.00415349 | 1 |
| T2DM | b_cell | *FBLN5* | -1.449058493 | 0.000880857 | 0.00415349 | 1 |
| T2DM | b_cell | *C14orf79* | -2.73008636 | 0.000932714 | 0.00415349 | 1 |
| T2DM | b_cell | *DUOXA1* | -3.460516375 | 0.01443742 | 0.020507921 | 1 |
| T2DM | b_cell | *MFAP1* | -3.400222439 | 0.001437194 | 0.004399046 | 1 |
| T2DM | b_cell | *TCF12* | -2.035340546 | 0.033327062 | 0.042388679 | 1 |
| T2DM | b_cell | *SORD* | 5.776604338 | 0.010816368 | 0.016106705 | 1 |
| T2DM | b_cell | *ZSCAN29* | -3.51335868 | 0.001272674 | 0.004257394 | 1 |
| T2DM | b_cell | *LYSMD2* | -4.82673012 | 0.004261992 | 0.007989011 | 1 |
| T2DM | b_cell | *BAHD1* | -3.071293677 | 0.001056931 | 0.00415349 | 1 |
| T2DM | b_cell | *DISP2* | 1.78077475 | 0.029832564 | 0.038378827 | 1 |
| T2DM | b_cell | *ANP32A* | -2.365880222 | 0.001289724 | 0.004268027 | 1 |
| T2DM | b_cell | *COMMD4* | -5.122395834 | 0.004153775 | 0.007865091 | 1 |
| T2DM | b_cell | *UBE2Q2* | -2.70182006 | 0.003312357 | 0.006729773 | 1 |
| T2DM | b_cell | *PSTPIP1* | -2.840963105 | 0.002456884 | 0.005651273 | 1 |
| T2DM | b_cell | *HMG20A* | -2.051450543 | 0.010474782 | 0.01569776 | 1 |
| T2DM | b_cell | *SCAPER* | -2.314156349 | 0.007117727 | 0.011508915 | 1 |
| T2DM | b_cell | *NEIL1* | -2.026478583 | 0.000294205 | 0.00415349 | 1 |
| T2DM | b_cell | *MAN2C1* | -2.587038128 | 1.57E-07 | 7.05E-05 | 0.000986842 |
| T2DM | b_cell | *DNAJA4* | -2.617719514 | 0.000887185 | 0.00415349 | 1 |
| T2DM | b_cell | *TPM1* | -2.310600647 | 0.002859051 | 0.006143148 | 1 |
| T2DM | b_cell | *IGF1R* | -1.833641258 | 0.001598278 | 0.004603327 | 1 |
| T2DM | b_cell | *CYP11A1* | -2.260624284 | 0.015163154 | 0.021350672 | 1 |
| T2DM | b_cell | *BBS4* | -2.807590933 | 0.005556287 | 0.009618899 | 1 |
| T2DM | b_cell | *ADAMTS17* | -1.728507144 | 0.000628444 | 0.00415349 | 1 |
| T2DM | b_cell | *ULK3* | -4.682732494 | 0.009204933 | 0.014111538 | 1 |
| T2DM | b_cell | *SCAMP2* | -3.720519709 | 0.000280617 | 0.00415349 | 1 |
| T2DM | b_cell | *HAPLN3* | -2.961430995 | 0.000724905 | 0.00415349 | 1 |
| T2DM | b_cell | *POLG* | -3.408926714 | 0.002423837 | 0.005623123 | 1 |
| T2DM | b_cell | *WDR93* | -1.836482707 | 0.000325348 | 0.00415349 | 1 |
| T2DM | b_cell | *TICRR* | -4.961413346 | 0.000624757 | 0.00415349 | 1 |
| T2DM | b_cell | *MFGE8* | -1.782833621 | 0.000765952 | 0.00415349 | 1 |
| T2DM | b_cell | *UNC45A* | -2.741573863 | 0.009496071 | 0.014476673 | 1 |
| T2DM | b_cell | *ST8SIA2* | -17.87136709 | 0.000548192 | 0.00415349 | 1 |
| T2DM | b_cell | *IQGAP1* | -2.198408854 | 0.008736599 | 0.013545498 | 1 |

| | | | | | | |
|---|---|---|---|---|---|---|
| T2DM | b_cell | *CRTC3* | -2.471879605 | 0.004068491 | 0.007746397 | 1 |
| T2DM | b_cell | *SH3GL3* | 2.077203579 | 0.007905883 | 0.01250453 | 1 |
| T2DM | b_cell | *SEC11A* | -2.50658806 | 0.002232002 | 0.005401538 | 1 |
| T2DM | b_cell | *TGFB1I1* | -2.826878844 | 0.001237399 | 0.004253251 | 1 |
| T2DM | b_cell | *PARN* | -2.365928716 | 0.003983802 | 0.007626725 | 1 |
| T2DM | b_cell | *FTO* | -2.217019747 | 0.003864006 | 0.007456417 | 1 |
| T2DM | b_cell | *TXNL4B* | -3.742004944 | 0.001700436 | 0.004717848 | 1 |
| T2DM | b_cell | *MARVELD3* | 2.07518945 | 0.000495653 | 0.00415349 | 1 |
| T2DM | b_cell | *ZFHX3* | -1.937913044 | 0.002131689 | 0.005278429 | 1 |
| T2DM | b_cell | *KIFC3* | -3.28303727 | 0.002656011 | 0.005906736 | 1 |
| T2DM | b_cell | *ADAMTS18* | -1.867898698 | 0.001019609 | 0.00415349 | 1 |
| T2DM | b_cell | *NUDT7* | -4.820310411 | 0.000441058 | 0.00415349 | 1 |
| T2DM | b_cell | *GCSH* | -2.339180918 | 0.007865096 | 0.012452243 | 1 |
| T2DM | b_cell | *CMTM3* | -2.070741498 | 0.00083707 | 0.00415349 | 1 |
| T2DM | b_cell | *MAP1LC3B* | -2.911812621 | 0.004710989 | 0.008602378 | 1 |
| T2DM | b_cell | *TLDC1* | -4.061118115 | 0.021332441 | 0.028620797 | 1 |
| T2DM | b_cell | *RPS2* | -3.461653108 | 0.003971847 | 0.007613111 | 1 |
| T2DM | b_cell | *NDUFB10* | -4.506489514 | 0.005165412 | 0.009127303 | 1 |
| T2DM | b_cell | *TIGD7* | -1.891737211 | 0.009520024 | 0.014506146 | 1 |
| T2DM | b_cell | *DEF8* | -2.267121347 | 0.00032291 | 0.00415349 | 1 |
| T2DM | b_cell | *GALNS* | -3.516678656 | 0.000978233 | 0.00415349 | 1 |
| T2DM | b_cell | *GAS8* | -1.783179932 | 0.026063287 | 0.034024063 | 1 |
| T2DM | b_cell | *COPS3* | -2.490025358 | 0.001053331 | 0.00415349 | 1 |
| T2DM | b_cell | *PRPSAP2* | 3.086942773 | 0.03799251 | 0.047593002 | 1 |
| T2DM | b_cell | *DDX52* | -3.123320992 | 0.008077564 | 0.012699085 | 1 |
| T2DM | b_cell | *RASL10B* | -8.784215917 | 0.00031851 | 0.00415349 | 1 |
| T2DM | b_cell | *UNC45B* | -12.17135217 | 0.002971566 | 0.006279742 | 1 |
| T2DM | b_cell | *TOM1L1* | -1.973683396 | 0.001451698 | 0.004416445 | 1 |
| T2DM | b_cell | *C17orf80* | -2.81717983 | 0.002227295 | 0.005400083 | 1 |
| T2DM | b_cell | *VPS53* | -2.882526982 | 0.000452213 | 0.00415349 | 1 |
| T2DM | b_cell | *SPATA22* | -1.292703322 | 0.008927694 | 0.013777224 | 1 |
| T2DM | b_cell | *NPEPPS* | -2.449750389 | 0.003052975 | 0.006376763 | 1 |
| T2DM | b_cell | *LRRC46* | -10.18784906 | 3.03E-07 | 0.000118892 | 0.001902273 |
| T2DM | b_cell | *SCRN2* | -1.239499644 | 0.005651908 | 0.009724561 | 1 |
| T2DM | b_cell | *RHBDL3* | -5.668837294 | 0.00370007 | 0.00724956 | 1 |
| T2DM | b_cell | *ARSG* | -3.31593192 | 0.000815446 | 0.00415349 | 1 |
| T2DM | b_cell | *ABCA8* | -1.138001377 | 0.001036839 | 0.00415349 | 1 |
| T2DM | b_cell | *G6PC3* | -6.02585033 | 0.004716058 | 0.008609133 | 1 |
| T2DM | b_cell | *IMPA2* | -3.124276744 | 0.002080289 | 0.005230315 | 1 |
| T2DM | b_cell | *GNAL* | -2.498804433 | 0.001496568 | 0.004474047 | 1 |
| T2DM | b_cell | *RPRD1A* | -3.05947999 | 0.003438521 | 0.006889095 | 1 |
| T2DM | b_cell | *C18orf21* | -3.194096297 | 0.003611654 | 0.007151285 | 1 |
| T2DM | b_cell | *GALNT1* | -1.98099541 | 0.001038329 | 0.00415349 | 1 |
| T2DM | b_cell | *ASXL3* | -2.626681826 | 0.00112697 | 0.004182741 | 1 |
| T2DM | b_cell | *ADCYAP1* | 11.36745673 | 0.003666805 | 0.007210886 | 1 |
| T2DM | b_cell | *ESCO1* | -2.928938683 | 0.031039798 | 0.039785364 | 1 |
| T2DM | b_cell | *GREB1L* | -1.337078728 | 0.000400435 | 0.00415349 | 1 |
| T2DM | b_cell | *C18orf8* | -4.92130791 | 0.000616005 | 0.00415349 | 1 |
| T2DM | b_cell | *NPC1* | -3.544592192 | 0.000679492 | 0.00415349 | 1 |
| T2DM | b_cell | *SLC14A1* | 4.796848855 | 0.003411959 | 0.006859926 | 1 |

| | | | | | | |
|---|---|---|---|---|---|---|
| T2DM | b_cell | *SLC13A5* | -19.44969786 | 0.001517397 | 0.004483023 | 1 |
| T2DM | b_cell | *CCDC40* | -1.778349165 | 0.002366466 | 0.005545309 | 1 |
| T2DM | b_cell | *CARD14* | -2.491382442 | 0.00075891 | 0.00415349 | 1 |
| T2DM | b_cell | *EIF4A3* | 4.145292751 | 0.032020632 | 0.040883501 | 1 |
| T2DM | b_cell | *TBCD* | -2.444145738 | 0.003730752 | 0.007277456 | 1 |
| T2DM | b_cell | *FN3KRP* | -4.065331941 | 0.00015539 | 0.00415349 | 0.976933957 |
| T2DM | b_cell | *NARF* | -3.522077335 | 0.005800608 | 0.009904515 | 1 |
| T2DM | b_cell | *CBX8* | 1.132781629 | 0.000750317 | 0.00415349 | 1 |
| T2DM | b_cell | *SECTM1* | -4.51277725 | 0.001721831 | 0.004747873 | 1 |
| T2DM | b_cell | *RNF157* | -1.631420503 | 0.001585955 | 0.004584321 | 1 |
| T2DM | b_cell | *RNF165* | -2.21042233 | 0.000830271 | 0.00415349 | 1 |
| T2DM | b_cell | *SMAD4* | -3.260058709 | 0.00936217 | 0.014303758 | 1 |
| T2DM | b_cell | *PIP4K2B* | -2.612377629 | 0.000985558 | 0.00415349 | 1 |
| T2DM | b_cell | *GRB7* | 2.730495212 | 0.001083148 | 0.00415349 | 1 |
| T2DM | b_cell | *FKBP10* | -2.38415863 | 0.000613128 | 0.00415349 | 1 |
| T2DM | b_cell | *TXNL4A* | -2.863177367 | 0.00071686 | 0.00415349 | 1 |
| T2DM | b_cell | *NFIC* | -1.653221368 | 0.000816968 | 0.00415349 | 1 |
| T2DM | b_cell | *TPGS1* | -2.515166072 | 0.001412145 | 0.004364365 | 1 |
| T2DM | b_cell | *PRDM15* | -1.979522739 | 0.028698727 | 0.037094757 | 1 |
| T2DM | b_cell | *PFKL* | -4.07333039 | 0.003132886 | 0.006479096 | 1 |
| T2DM | b_cell | *MVB12A* | -3.082181693 | 0.003798804 | 0.007364502 | 1 |
| T2DM | b_cell | *DUS3L* | 5.687405448 | 0.000126796 | 0.00415349 | 0.797167409 |
| T2DM | b_cell | *TMEM91* | -6.004163001 | 0.002403351 | 0.005596247 | 1 |
| T2DM | b_cell | *SIRT3* | -1.930072334 | 0.015787996 | 0.02209195 | 1 |
| T2DM | b_cell | *IFITM3* | -2.268675774 | 0.001150961 | 0.004188961 | 1 |
| T2DM | b_cell | *COL6A1* | -1.344195171 | 0.000395938 | 0.00415349 | 1 |
| T2DM | b_cell | *IFNAR1* | -3.227447548 | 0.000772329 | 0.00415349 | 1 |
| T2DM | b_cell | *AKT1* | -2.831158996 | 0.00100038 | 0.00415349 | 1 |
| T2DM | b_cell | *SAE1* | -2.365684006 | 0.004353835 | 0.008107986 | 1 |
| T2DM | b_cell | *GEMIN7* | 3.110322609 | 0.006056625 | 0.010214055 | 1 |
| T2DM | b_cell | *WTIP* | -1.825267932 | 0.000725611 | 0.00415349 | 1 |
| T2DM | b_cell | *CAPN10* | -2.235834351 | 0.002453643 | 0.005646432 | 1 |
| T2DM | b_cell | *MYO1F* | -1.494893994 | 0.001309387 | 0.004274973 | 1 |
| T2DM | b_cell | *CACNG8* | -6.40561757 | 0.000912255 | 0.00415349 | 1 |
| T2DM | b_cell | *FBN3* | -3.855335468 | 0.004886855 | 0.008810915 | 1 |
| T2DM | b_cell | *EVI5L* | 4.430735338 | 0.019968104 | 0.026980328 | 1 |
| T2DM | b_cell | *CTU1* | -3.381292986 | 0.000736728 | 0.00415349 | 1 |
| T2DM | b_cell | *RCN3* | -1.955497746 | 0.000826321 | 0.00415349 | 1 |
| T2DM | b_cell | *RERE* | -2.008763839 | 0.016005683 | 0.022361718 | 1 |
| T2DM | b_cell | *ARHGEF19* | -1.801570219 | 0.00343661 | 0.006887461 | 1 |
| T2DM | b_cell | *PEX14* | -1.854137324 | 0.000891724 | 0.00415349 | 1 |
| T2DM | b_cell | *IL22RA1* | -5.387029887 | 0.005167799 | 0.009128955 | 1 |
| T2DM | b_cell | *ZNF593* | -5.821334541 | 0.000845848 | 0.00415349 | 1 |
| T2DM | b_cell | *C1orf216* | -2.870105775 | 0.001292744 | 0.004268027 | 1 |
| T2DM | b_cell | *GPN2* | -2.15988537 | 0.00057269 | 0.00415349 | 1 |
| T2DM | b_cell | *NBPF3* | -2.693771591 | 0.000273027 | 0.00415349 | 1 |
| T2DM | b_cell | *HSPG2* | -1.716031433 | 0.00100613 | 0.00415349 | 1 |
| T2DM | b_cell | *ITGB3BP* | -2.201167053 | 0.011630093 | 0.017055842 | 1 |
| T2DM | b_cell | *SERBP1* | -3.186363008 | 0.018856781 | 0.025660733 | 1 |
| T2DM | b_cell | *PIGK* | -3.386402194 | 0.008630409 | 0.013403998 | 1 |

| | | | | | | |
|---|---|---|---|---|---|---|
| T2DM | b_cell | *RPS8* | -2.869491001 | 0.026221075 | 0.034210829 | 1 |
| T2DM | b_cell | *MOB3C* | 2.698925504 | 0.000792641 | 0.00415349 | 1 |
| T2DM | b_cell | *CYP4B1* | -1.405802617 | 0.010023468 | 0.01515938 | 1 |
| T2DM | b_cell | *SYPL2* | -1.961678998 | 0.015556521 | 0.021821474 | 1 |
| T2DM | b_cell | *ZNF697* | -2.531448768 | 0.000904288 | 0.00415349 | 1 |
| T2DM | b_cell | *CELSR2* | -6.950922277 | 0.003139611 | 0.006486603 | 1 |
| T2DM | b_cell | *ATP1B1* | -2.025734998 | 0.000557388 | 0.00415349 | 1 |
| T2DM | b_cell | *TIPRL* | -3.420512058 | 0.003348031 | 0.006770366 | 1 |
| T2DM | b_cell | *NME7* | -3.298993746 | 0.003722941 | 0.007268984 | 1 |
| T2DM | b_cell | *CREG1* | -3.20958665 | 0.002649973 | 0.005895393 | 1 |
| T2DM | b_cell | *GPA33* | -7.262696879 | 0.00074398 | 0.00415349 | 1 |
| T2DM | b_cell | *RXRG* | 14.64213081 | 0.005052723 | 0.00900155 | 1 |
| T2DM | b_cell | *TMCO1* | -3.210705391 | 0.01732051 | 0.023906487 | 1 |
| T2DM | b_cell | *MGST3* | -2.040376413 | 0.001357934 | 0.004298348 | 1 |
| T2DM | b_cell | *ADCY10* | -2.824822889 | 0.000517667 | 0.00415349 | 1 |
| T2DM | b_cell | *RFWD2* | -2.391546339 | 0.009000794 | 0.013862811 | 1 |
| T2DM | b_cell | *PPOX* | -2.687786847 | 0.006080733 | 0.010246468 | 1 |
| T2DM | b_cell | *RRNAD1* | -4.074514489 | 0.000435723 | 0.00415349 | 1 |
| T2DM | b_cell | *PIGM* | -2.641159646 | 0.001319598 | 0.004274973 | 1 |
| T2DM | b_cell | *RGL1* | -1.442532953 | 0.00100044 | 0.00415349 | 1 |
| T2DM | b_cell | *TUFT1* | -2.793182392 | 0.000273207 | 0.00415349 | 1 |
| T2DM | b_cell | *TARS2* | -2.265220149 | 0.004904304 | 0.008832242 | 1 |
| T2DM | b_cell | *SNX27* | -3.218610412 | 0.014626853 | 0.020706828 | 1 |
| T2DM | b_cell | *SELENBP1* | -1.704683155 | 0.000967567 | 0.00415349 | 1 |
| T2DM | b_cell | *SEMA6C* | -5.257311573 | 0.001978289 | 0.005092436 | 1 |
| T2DM | b_cell | *ARNT* | -2.777261899 | 0.005044357 | 0.008996844 | 1 |
| T2DM | b_cell | *POGZ* | -2.34782919 | 0.005833774 | 0.009937291 | 1 |
| T2DM | b_cell | *GABPB2* | -1.8777424 | 0.013681111 | 0.019570681 | 1 |
| T2DM | b_cell | *DYRK3* | -3.413142752 | 0.001744063 | 0.004783999 | 1 |
| T2DM | b_cell | *EIF2D* | -3.40007494 | 0.001753165 | 0.004789372 | 1 |
| T2DM | b_cell | *HHIPL2* | -6.978922182 | 0.009132025 | 0.014023704 | 1 |
| T2DM | b_cell | *TP53BP2* | -3.105503313 | 0.002337297 | 0.005511847 | 1 |
| T2DM | b_cell | *RAB13* | -2.583297307 | 0.000496272 | 0.00415349 | 1 |
| T2DM | b_cell | *S100A8* | 2.259582486 | 0.001488793 | 0.004462053 | 1 |
| T2DM | b_cell | *HAX1* | -3.647040097 | 0.011705136 | 0.017144273 | 1 |
| T2DM | b_cell | *CREB3L4* | -3.611698394 | 0.000660644 | 0.00415349 | 1 |
| T2DM | b_cell | *KCNN3* | -2.211395318 | 0.001096934 | 0.00415349 | 1 |
| T2DM | b_cell | *GATAD2B* | -2.432223055 | 0.029879856 | 0.038431804 | 1 |
| T2DM | b_cell | *RIT1* | -3.075080469 | 0.0018955 | 0.004975787 | 1 |
| T2DM | b_cell | *FLG* | -2.408043536 | 0.00329466 | 0.006703407 | 1 |
| T2DM | b_cell | *GALNT2* | -2.49240753 | 0.002708436 | 0.00595502 | 1 |
| T2DM | b_cell | *SCCPDH* | -3.01253715 | 0.002515454 | 0.005736184 | 1 |
| T2DM | b_cell | *CEP170* | -2.52805778 | 0.008673002 | 0.013456852 | 1 |
| T2DM | b_cell | *NVL* | -2.70366571 | 0.003251334 | 0.006641045 | 1 |
| T2DM | b_cell | *SDE2* | -5.538589286 | 0.009686022 | 0.014734096 | 1 |
| T2DM | b_cell | *FBXO28* | -2.961330117 | 0.020766914 | 0.027959054 | 1 |
| T2DM | b_cell | *GUK1* | -2.160995831 | 0.001025868 | 0.00415349 | 1 |
| T2DM | b_cell | *CDC42BPA* | -1.663495259 | 0.002210204 | 0.005381337 | 1 |
| T2DM | b_cell | *CNIH3* | -2.409463514 | 0.003707645 | 0.007257149 | 1 |
| T2DM | b_cell | *PYCR2* | 4.767466441 | 0.004158358 | 0.007865091 | 1 |

| | | | | | | |
|---|---|---|---|---|---|---|
| T2DM | b_cell | *WNT9A* | -1.988683165 | 0.00726139 | 0.011681771 | 1 |
| T2DM | b_cell | *PLEKHA6* | -1.82902939 | 0.002095493 | 0.005240399 | 1 |
| T2DM | b_cell | *ABCG8* | -6.796602836 | 0.039983083 | 0.049905428 | 1 |
| T2DM | b_cell | *EML4* | -2.502532543 | 0.011789736 | 0.017233683 | 1 |
| T2DM | b_cell | *CHAC2* | -4.986766257 | 0.005775971 | 0.009875857 | 1 |
| T2DM | b_cell | *WDPCP* | -0.901591253 | 0.000763196 | 0.00415349 | 1 |
| T2DM | b_cell | *ASXL2* | -2.693290752 | 0.011179032 | 0.016525411 | 1 |
| T2DM | b_cell | *CIAO1* | -3.409299236 | 0.00148831 | 0.004462053 | 1 |
| T2DM | b_cell | *ZNF514* | -2.147510012 | 0.000464575 | 0.00415349 | 1 |
| T2DM | b_cell | *EXOC6B* | -2.104689091 | 0.010331633 | 0.015536623 | 1 |
| T2DM | b_cell | *TEX261* | -5.509796319 | 0.02142584 | 0.028715467 | 1 |
| T2DM | b_cell | *THNSL2* | -1.11289579 | 0.008419604 | 0.01312523 | 1 |
| T2DM | b_cell | *TMEM177* | -3.674428653 | 0.000796266 | 0.00415349 | 1 |
| T2DM | b_cell | *RABL2A* | -4.058828411 | 0.002314602 | 0.005493356 | 1 |
| T2DM | b_cell | *FBLN7* | -1.879292251 | 0.007131531 | 0.011514107 | 1 |
| T2DM | b_cell | *ZC3H8* | -2.899508847 | 0.007779525 | 0.012350455 | 1 |
| T2DM | b_cell | *FAHD2B* | -3.918946068 | 0.000941691 | 0.00415349 | 1 |
| T2DM | b_cell | *LYG1* | -2.45087312 | 0.000401739 | 0.00415349 | 1 |
| T2DM | b_cell | *AFF3* | 1.62829122 | 0.018137228 | 0.024859113 | 1 |
| T2DM | b_cell | *UBXN4* | -2.61649168 | 0.005364084 | 0.009387268 | 1 |
| T2DM | b_cell | *THSD7B* | -1.203656626 | 0.004133992 | 0.007837879 | 1 |
| T2DM | b_cell | *POLR2D* | -3.713888959 | 0.00175654 | 0.004789372 | 1 |
| T2DM | b_cell | *ZNF385B* | -1.4992655 | 0.001455583 | 0.004416626 | 1 |
| T2DM | b_cell | *TMEFF2* | -1.785501539 | 0.001446854 | 0.004415713 | 1 |
| T2DM | b_cell | *GULP1* | -1.404988394 | 0.001234293 | 0.004253251 | 1 |
| T2DM | b_cell | *FAM171B* | -2.458435283 | 0.001243932 | 0.004253251 | 1 |
| T2DM | b_cell | *HSPD1* | -2.867588756 | 0.006312534 | 0.010520692 | 1 |
| T2DM | b_cell | *PTH2R* | -1.735137781 | 0.002224693 | 0.005398166 | 1 |
| T2DM | b_cell | *SUMF1* | -2.145744804 | 0.006846208 | 0.011171064 | 1 |
| T2DM | b_cell | *ESPNL* | -20.97842319 | 0.015076308 | 0.02126172 | 1 |
| T2DM | b_cell | *FANCD2* | 1.117384309 | 0.000558719 | 0.00415349 | 1 |
| T2DM | b_cell | *OSBPL10* | 2.304481624 | 0.000968677 | 0.00415349 | 1 |
| T2DM | b_cell | *SLC25A38* | 2.132088492 | 0.00123874 | 0.004253251 | 1 |
| T2DM | b_cell | *CTDSPL* | -3.297356445 | 0.005682262 | 0.009761317 | 1 |
| T2DM | b_cell | *STAC* | -2.042078781 | 0.000769299 | 0.00415349 | 1 |
| T2DM | b_cell | *ARL6IP5* | -2.60151402 | 0.003628642 | 0.007173984 | 1 |
| T2DM | b_cell | *TMF1* | -2.803702534 | 0.002574221 | 0.005806222 | 1 |
| T2DM | b_cell | *COL8A1* | -1.225673879 | 0.00070237 | 0.00415349 | 1 |
| T2DM | b_cell | *NXPE3* | -4.264973467 | 0.002924491 | 0.006211579 | 1 |
| T2DM | b_cell | *PLA1A* | -6.925889166 | 8.63E-06 | 0.001459962 | 0.054273343 |
| T2DM | b_cell | *RABL3* | -3.381481189 | 0.004937932 | 0.008867403 | 1 |
| T2DM | b_cell | *ADPRH* | -2.702684282 | 0.00402715 | 0.007681642 | 1 |
| T2DM | b_cell | *IGSF11* | -3.946187399 | 0.014318148 | 0.020352295 | 1 |
| T2DM | b_cell | *BOC* | -1.566419779 | 0.000720254 | 0.00415349 | 1 |
| T2DM | b_cell | *AGTR1* | -1.661864667 | 0.000713945 | 0.00415349 | 1 |
| T2DM | b_cell | *EIF2A* | -2.167813759 | 0.004187429 | 0.007886869 | 1 |
| T2DM | b_cell | *TCTA* | -2.530430474 | 0.00318132 | 0.006549103 | 1 |
| T2DM | b_cell | *EAF2* | 1.458866437 | 0.000945068 | 0.00415349 | 1 |
| T2DM | b_cell | *ILDR1* | 2.36066766 | 0.005585563 | 0.009642074 | 1 |
| T2DM | b_cell | *MUC4* | -2.316388224 | 0.002542522 | 0.005783227 | 1 |

| | | | | | | |
|---|---|---|---|---|---|---|
| T2DM | b_cell | *DGKQ* | -2.767417118 | 0.000978115 | 0.00415349 | 1 |
| T2DM | b_cell | *LYAR* | -2.381659939 | 0.000811432 | 0.00415349 | 1 |
| T2DM | b_cell | *CENPC* | -2.474369839 | 0.018675437 | 0.025463559 | 1 |
| T2DM | b_cell | *ATP10D* | -2.291112911 | 0.014270958 | 0.020303579 | 1 |
| T2DM | b_cell | *PYURF* | -5.387013404 | 0.005038084 | 0.008990003 | 1 |
| T2DM | b_cell | *CISD2* | -3.279997339 | 0.015281374 | 0.021478649 | 1 |
| T2DM | b_cell | *CCNA2* | 3.640025524 | 0.005141118 | 0.009099722 | 1 |
| T2DM | b_cell | *USP53* | -1.75830297 | 0.001972283 | 0.005091111 | 1 |
| T2DM | b_cell | *MARCH1* | 2.091028916 | 0.027903328 | 0.036170767 | 1 |
| T2DM | b_cell | *PDGFC* | -1.648689334 | 0.000615618 | 0.00415349 | 1 |
| T2DM | b_cell | *CBR4* | -2.392770619 | 0.007778721 | 0.012350455 | 1 |
| T2DM | b_cell | *NDUFS6* | -4.420749156 | 0.008350519 | 0.013036929 | 1 |
| T2DM | b_cell | *FAM105A* | -2.585821597 | 0.000359752 | 0.00415349 | 1 |
| T2DM | b_cell | *LHFPL2* | -3.120713431 | 0.037420592 | 0.046996257 | 1 |
| T2DM | b_cell | *SSBP2* | -2.153828671 | 0.01511588 | 0.021299033 | 1 |
| T2DM | b_cell | *BHMT* | -3.244199097 | 3.58E-07 | 0.000124872 | 0.00224769 |
| T2DM | b_cell | *CRHBP* | -3.028139574 | 0.000285458 | 0.00415349 | 1 |
| T2DM | b_cell | *RASA1* | -1.955340578 | 0.006500479 | 0.010743562 | 1 |
| T2DM | b_cell | *PPIP5K2* | -3.158879866 | 0.000871639 | 0.00415349 | 1 |
| T2DM | b_cell | *SPATA9* | -1.519350227 | 0.001855731 | 0.004929017 | 1 |
| T2DM | b_cell | *TSLP* | -3.502919249 | 0.002016004 | 0.005148098 | 1 |
| T2DM | b_cell | *FEM1C* | -3.619156624 | 0.00780636 | 0.012371713 | 1 |
| T2DM | b_cell | *COMMD10* | -2.374320787 | 0.008400039 | 0.013097978 | 1 |
| T2DM | b_cell | *YIPF5* | -2.896467848 | 0.001515012 | 0.004480664 | 1 |
| T2DM | b_cell | *SLC25A48* | -3.381295087 | 0.018438169 | 0.02519469 | 1 |
| T2DM | b_cell | *GABRB2* | -2.267598993 | 0.00033327 | 0.00415349 | 1 |
| T2DM | b_cell | *MYLK4* | -4.761565992 | 0.001262379 | 0.004257394 | 1 |
| T2DM | b_cell | *TBC1D7* | -3.937026208 | 0.00816355 | 0.012795871 | 1 |
| T2DM | b_cell | *GFOD1* | -1.233282957 | 0.001879436 | 0.004960544 | 1 |
| T2DM | b_cell | *PSD2* | 5.156820308 | 0.000205275 | 0.00415349 | 1 |
| T2DM | b_cell | *ZMAT2* | -2.129073588 | 0.000919631 | 0.00415349 | 1 |
| T2DM | b_cell | *KLHL3* | -1.654761972 | 0.001133617 | 0.00418468 | 1 |
| T2DM | b_cell | *TRIM7* | 3.030571314 | 0.001547917 | 0.004517818 | 1 |
| T2DM | b_cell | *HIGD2A* | -4.14486112 | 0.000871178 | 0.00415349 | 1 |
| T2DM | b_cell | *FAM193B* | -2.716285304 | 0.003204946 | 0.006581388 | 1 |
| T2DM | b_cell | *MUT* | -3.052915364 | 0.001863562 | 0.0049373 | 1 |
| T2DM | b_cell | *DOK3* | -2.495436312 | 0.005115298 | 0.009064228 | 1 |
| T2DM | b_cell | *ABT1* | -2.643444084 | 0.002298224 | 0.005481387 | 1 |
| T2DM | b_cell | *MLIP* | -3.732476373 | 0.008060071 | 0.012684272 | 1 |
| T2DM | b_cell | *TPBG* | -2.143689315 | 0.000795897 | 0.00415349 | 1 |
| T2DM | b_cell | *IRAK1BP1* | -2.518147945 | 0.002680888 | 0.005924338 | 1 |
| T2DM | b_cell | *PHIP* | -2.29193495 | 0.009279647 | 0.014213206 | 1 |
| T2DM | b_cell | *PM20D2* | -4.494557721 | 0.022955957 | 0.03056419 | 1 |
| T2DM | b_cell | *RNF217* | -2.104155556 | 0.00169061 | 0.004704627 | 1 |
| T2DM | b_cell | *SLC18B1* | -1.551317845 | 0.028374667 | 0.036713631 | 1 |
| T2DM | b_cell | *AIG1* | -1.623753231 | 0.001222093 | 0.004252746 | 1 |
| T2DM | b_cell | *DYNLT1* | -2.151267623 | 0.000784088 | 0.00415349 | 1 |
| T2DM | b_cell | *PNLDC1* | -4.77830098 | 0.028879 | 0.037275249 | 1 |
| T2DM | b_cell | *SLC22A3* | -1.619845175 | 0.000769197 | 0.00415349 | 1 |
| T2DM | b_cell | *GNA12* | -2.874813863 | 0.011356641 | 0.016740727 | 1 |

| | | | | | | |
|---|---|---|---|---|---|---|
| T2DM | b_cell | *C7orf50* | -2.231994455 | 0.002229006 | 0.005400293 | 1 |
| T2DM | b_cell | *SDK1* | -1.249932521 | 0.000668461 | 0.00415349 | 1 |
| T2DM | b_cell | *RBAK* | -5.4563899 | 0.006232095 | 0.010416354 | 1 |
| T2DM | b_cell | *EGFR* | -1.21965986 | 0.000349598 | 0.00415349 | 1 |
| T2DM | b_cell | *POMZP3* | -3.60499258 | 0.002591326 | 0.00582516 | 1 |
| T2DM | b_cell | *CCT6A* | -3.33442415 | 0.007797517 | 0.012363932 | 1 |
| T2DM | b_cell | *PSPH* | -1.873139309 | 0.011578567 | 0.017008049 | 1 |
| T2DM | b_cell | *GIGYF1* | -4.358264084 | 0.010095436 | 0.015238897 | 1 |
| T2DM | b_cell | *TRIM4* | -2.483405825 | 0.002324611 | 0.005505767 | 1 |
| T2DM | b_cell | *TMEM209* | -1.898231944 | 0.001613546 | 0.004616359 | 1 |
| T2DM | b_cell | *AGBL3* | -1.87940193 | 0.001014846 | 0.00415349 | 1 |
| T2DM | b_cell | *EPHA1* | -4.674970144 | 0.005174041 | 0.009136916 | 1 |
| T2DM | b_cell | *NCAPG2* | -2.74273134 | 0.006380225 | 0.010594949 | 1 |
| T2DM | b_cell | *FBXO25* | -2.564531142 | 0.001489717 | 0.004462053 | 1 |
| T2DM | b_cell | *CSGALNACT1* | -2.905369945 | 0.007679867 | 0.01221744 | 1 |
| T2DM | b_cell | *SLC25A37* | -2.441285193 | 0.001929225 | 0.005023674 | 1 |
| T2DM | b_cell | *DOCK5* | -2.982756789 | 0.006040826 | 0.010198355 | 1 |
| T2DM | b_cell | *ST18* | -2.269690115 | 0.000568711 | 0.00415349 | 1 |
| T2DM | b_cell | *RGS20* | -2.187930747 | 0.000948587 | 0.00415349 | 1 |
| T2DM | b_cell | *TACC1* | -2.019281079 | 0.001985685 | 0.005097591 | 1 |
| T2DM | b_cell | *ADHFE1* | -1.589857377 | 2.58E-05 | 0.002754251 | 0.162500803 |
| T2DM | b_cell | *SLC26A7* | -1.872528206 | 0.00143453 | 0.004395857 | 1 |
| T2DM | b_cell | *ATP6V0D2* | -3.400546318 | 0.000769838 | 0.00415349 | 1 |
| T2DM | b_cell | *MTDH* | -2.549540018 | 0.015550867 | 0.021818412 | 1 |
| T2DM | b_cell | *MAL2* | -2.450910572 | 0.00037746 | 0.00415349 | 1 |
| T2DM | b_cell | *FAM135B* | 3.242738254 | 0.000855412 | 0.00415349 | 1 |
| T2DM | b_cell | *ZNF7* | -2.452974166 | 0.01703017 | 0.023573024 | 1 |
| T2DM | b_cell | *SLC39A4* | -11.73746627 | 0.005118262 | 0.009066925 | 1 |
| T2DM | b_cell | *AK3* | -2.490373239 | 0.001748266 | 0.004789372 | 1 |
| T2DM | b_cell | *CER1* | -5.998825586 | 0.013544082 | 0.019409993 | 1 |
| T2DM | b_cell | *PLIN2* | -2.833351376 | 0.00369184 | 0.007246772 | 1 |
| T2DM | b_cell | *HAUS6* | -3.520201706 | 0.003949805 | 0.007585602 | 1 |
| T2DM | b_cell | *C9orf72* | -2.582897537 | 0.011096984 | 0.016423432 | 1 |
| T2DM | b_cell | *SIGMAR1* | -3.323990196 | 0.00074131 | 0.00415349 | 1 |
| T2DM | b_cell | *CBWD5* | -2.329169517 | 0.000876585 | 0.00415349 | 1 |
| T2DM | b_cell | *ZNF462* | -2.117754124 | 0.001365474 | 0.004299827 | 1 |
| T2DM | b_cell | *ALAD* | -1.868100315 | 0.013596095 | 0.019466784 | 1 |
| T2DM | b_cell | *ASTN2* | -3.003601408 | 0.009032927 | 0.013898681 | 1 |
| T2DM | b_cell | *WDR31* | -3.481382976 | 0.004144957 | 0.007853931 | 1 |
| T2DM | b_cell | *POLE3* | 4.145351651 | 0.007587237 | 0.01210377 | 1 |
| T2DM | b_cell | *SURF1* | -3.884389072 | 0.003970977 | 0.007613111 | 1 |
| T2DM | b_cell | *SURF6* | 3.521826567 | 0.003231791 | 0.006614021 | 1 |
| T2DM | b_cell | *ASB6* | -3.188967242 | 0.001011951 | 0.00415349 | 1 |
| T2DM | b_cell | *NTMT1* | -3.681625263 | 0.008329642 | 0.013014308 | 1 |
| T2DM | b_cell | *INPP5E* | -4.6056781 | 0.001641937 | 0.004657671 | 1 |
| T2DM | b_cell | *DPH7* | -3.461722531 | 0.001679468 | 0.004704627 | 1 |
| T2DM | b_cell | *SLC39A12* | -5.616842503 | 0.010209098 | 0.015380925 | 1 |
| T2DM | b_cell | *RSU1* | -2.509964667 | 0.00520573 | 0.00917791 | 1 |
| T2DM | b_cell | *PARD3* | -1.218863192 | 0.000816818 | 0.00415349 | 1 |
| T2DM | b_cell | *NRBF2* | -2.517525303 | 0.000227303 | 0.00415349 | 1 |

| | | | | | | |
|---|---|---|---|---|---|---|
| T2DM | b_cell | *ADIRF* | -2.236249719 | 0.002104321 | 0.005241681 | 1 |
| T2DM | b_cell | *HTR7* | -2.97456167 | 0.000827465 | 0.00415349 | 1 |
| T2DM | b_cell | *RPP30* | -3.232018047 | 0.005601239 | 0.009655878 | 1 |
| T2DM | b_cell | *DNAJB12* | -3.20119436 | 0.003034254 | 0.006348205 | 1 |
| T2DM | b_cell | *PLEKHS1* | 4.821546397 | 0.004338833 | 0.008089632 | 1 |
| T2DM | b_cell | *MTG1* | -2.264314316 | 0.002728484 | 0.005976996 | 1 |
| T2DM | b_cell | *PAOX* | 2.334122424 | 0.000874361 | 0.00415349 | 1 |
| T2DM | b_cell | *GSTO1* | -2.539762154 | 0.000503462 | 0.00415349 | 1 |
| T2DM | b_cell | *PDCD11* | -2.372098015 | 0.001311551 | 0.004274973 | 1 |
| T2DM | b_cell | *RGS10* | -2.198268754 | 0.000509283 | 0.00415349 | 1 |
| T2DM | b_cell | *GAS2* | -3.743717632 | 0.002332301 | 0.005509725 | 1 |
| T2DM | b_cell | *LIN7C* | -4.142201704 | 0.010066058 | 0.015209158 | 1 |
| T2DM | b_cell | *ZNF215* | 1.81735271 | 0.001549668 | 0.004517818 | 1 |
| T2DM | b_cell | *HSD17B12* | -5.87741537 | 3.17E-08 | 1.99E-05 | 0.000199456 |
| T2DM | b_cell | *APIP* | -2.559157027 | 1.46E-06 | 0.000397014 | 0.009156278 |
| T2DM | b_cell | *GLYAT* | -1.275453442 | 0.004335344 | 0.008085527 | 1 |
| T2DM | b_cell | *PTPRJ* | -1.965577604 | 0.005293862 | 0.009291769 | 1 |
| T2DM | b_cell | *CCDC82* | -2.59097601 | 0.000498397 | 0.00415349 | 1 |
| T2DM | b_cell | *SERPINH1* | -1.963641488 | 0.001885273 | 0.004965526 | 1 |
| T2DM | b_cell | *CAPN5* | -2.594080762 | 0.001977815 | 0.005092436 | 1 |
| T2DM | b_cell | *ZC3H12C* | -2.796442805 | 0.004822464 | 0.008739934 | 1 |
| T2DM | b_cell | *DRD2* | -11.73897648 | 0.001501744 | 0.004478191 | 1 |
| T2DM | b_cell | *C11orf52* | -20.42499785 | 0.000763737 | 0.00415349 | 1 |
| T2DM | b_cell | *NPAT* | -3.025364288 | 0.010491645 | 0.015704993 | 1 |
| T2DM | b_cell | *GRIK4* | -2.214150878 | 0.010266943 | 0.015456961 | 1 |
| T2DM | b_cell | *ADAM33* | -1.860634083 | 0.000598253 | 0.00415349 | 1 |
| T2DM | b_cell | *MTA2* | -4.68239045 | 0.029214805 | 0.037676611 | 1 |
| T2DM | b_cell | *TMC2* | -8.443923635 | 0.021486722 | 0.02878479 | 1 |
| T2DM | b_cell | *EML3* | -2.785592844 | 0.00184114 | 0.004898539 | 1 |
| T2DM | b_cell | *INCENP* | -3.082576481 | 0.000841612 | 0.00415349 | 1 |
| T2DM | b_cell | *MS4A3* | -53.31500836 | 0.000626499 | 0.00415349 | 1 |
| T2DM | b_cell | *PLCH2* | -9.13954135 | 0.000815644 | 0.00415349 | 1 |
| T2DM | b_cell | *CPSF7* | -3.14222021 | 0.009106701 | 0.013988231 | 1 |
| T2DM | b_cell | *FEZ1* | -2.678519181 | 0.001144629 | 0.00418468 | 1 |
| T2DM | b_cell | *COMMD7* | -2.873345292 | 0.001568931 | 0.004558164 | 1 |
| T2DM | b_cell | *KIAA1755* | -2.255162304 | 0.003735151 | 0.007283776 | 1 |
| T2DM | b_cell | *CABLES2* | 2.986994337 | 0.001382608 | 0.00432604 | 1 |
| T2DM | b_cell | *TRPT1* | -2.867948406 | 0.000937662 | 0.00415349 | 1 |
| T2DM | b_cell | *DOC2A* | -9.23606195 | 2.17E-06 | 0.000505081 | 0.013637198 |
| T2DM | b_cell | *TAOK2* | -3.982011911 | 0.00175277 | 0.004789372 | 1 |
| T2DM | b_cell | *MPP7* | -2.84180318 | 0.00016581 | 0.00415349 | 1 |
| T2DM | b_cell | *CWC15* | -3.347371494 | 0.003824547 | 0.007400716 | 1 |
| T2DM | b_cell | *TMCO3* | -3.301188435 | 0.013403518 | 0.019243644 | 1 |
| T2DM | b_cell | *SAP18* | -3.645806523 | 0.004689892 | 0.008580584 | 1 |
| T2DM | b_cell | *LYPD6B* | -4.147080759 | 0.000516268 | 0.00415349 | 1 |
| T2DM | b_cell | *WDR17* | -4.423525895 | 0.021978495 | 0.029349787 | 1 |
| T2DM | b_cell | *CCDC102B* | -1.732039907 | 0.000944498 | 0.00415349 | 1 |
| T2DM | b_cell | *RGS18* | -2.829005341 | 0.00079404 | 0.00415349 | 1 |
| T2DM | b_cell | *PRSS23* | -1.242425199 | 0.000810419 | 0.00415349 | 1 |
| T2DM | b_cell | *MTMR12* | -2.380443827 | 0.002477906 | 0.005683544 | 1 |

| | | | | | | |
|---|---|---|---|---|---|---|
| T2DM | b_cell | *PPP1R1C* | 7.864047083 | 0.000438378 | 0.00415349 | 1 |
| T2DM | b_cell | *C11orf53* | 20.40444414 | 0.002490504 | 0.005706196 | 1 |
| T2DM | b_cell | *CCT5* | -2.447425869 | 0.001684817 | 0.004704627 | 1 |
| T2DM | b_cell | *FAM173B* | -1.878927082 | 0.000552759 | 0.00415349 | 1 |
| T2DM | b_cell | *DOCK1* | -1.987038493 | 0.001962395 | 0.005077193 | 1 |
| T2DM | b_cell | *TEX12* | -18.12237358 | 0.000743588 | 0.00415349 | 1 |
| T2DM | b_cell | *PIP4K2A* | -2.588165714 | 0.008754342 | 0.013569661 | 1 |
| T2DM | b_cell | *CRIM1* | -1.14843401 | 0.000622211 | 0.00415349 | 1 |
| T2DM | b_cell | *ABCB9* | -6.133236019 | 0.000160648 | 0.00415349 | 1 |
| T2DM | b_cell | *RILPL2* | -2.781038438 | 0.002037207 | 0.005172827 | 1 |
| T2DM | b_cell | *PRSS53* | -11.4155732 | 0.000107481 | 0.00415349 | 0.675733104 |
| T2DM | b_cell | *GPR158* | -2.708769501 | 0.002334644 | 0.005509725 | 1 |
| T2DM | b_cell | *THRB* | -1.63039062 | 0.001530756 | 0.004503445 | 1 |
| T2DM | b_cell | *TMEM86A* | -3.691267162 | 0.002247495 | 0.005418361 | 1 |
| T2DM | b_cell | *BTBD11* | -2.229780329 | 0.000764222 | 0.00415349 | 1 |
| T2DM | b_cell | *UBE3B* | -3.045591738 | 0.012134756 | 0.017668183 | 1 |
| T2DM | b_cell | *ANK3* | -1.852087806 | 0.00608655 | 0.010252906 | 1 |
| T2DM | b_cell | *IPMK* | -2.635400615 | 0.002085581 | 0.005230315 | 1 |
| T2DM | b_cell | *DLG5* | -1.960915363 | 0.000461923 | 0.00415349 | 1 |
| T2DM | b_cell | *GXYLT1* | -3.296973024 | 0.033809265 | 0.042949858 | 1 |
| T2DM | b_cell | *MIPOL1* | -1.557179659 | 0.001453416 | 0.004416445 | 1 |
| T2DM | b_cell | *EXT2* | -2.56551698 | 0.014966212 | 0.02112541 | 1 |
| T2DM | b_cell | *TMEM18* | -4.040101192 | 0.002928077 | 0.006217095 | 1 |
| T2DM | b_cell | *KCTD14* | 8.478584624 | 0.004177198 | 0.00787705 | 1 |
| T2DM | b_cell | *THRSP* | -2.012249602 | 0.000576632 | 0.00415349 | 1 |
| T2DM | b_cell | *NDUFC2* | -4.675778456 | 0.012228277 | 0.017783757 | 1 |
| T2DM | b_cell | *ME3* | -2.108772214 | 0.003772326 | 0.007333523 | 1 |
| T2DM | b_cell | *ADAMTS12* | -1.15725487 | 0.000895027 | 0.00415349 | 1 |
| T2DM | b_cell | *PTPRO* | -3.422819538 | 0.001815409 | 0.004867153 | 1 |
| T2DM | b_cell | *VPS26B* | -3.859413059 | 0.006578006 | 0.010834667 | 1 |
| T2DM | b_cell | *NCAPD3* | -2.932010581 | 0.000837949 | 0.00415349 | 1 |
| T2DM | b_cell | *QDPR* | -2.001557462 | 0.00035814 | 0.00415349 | 1 |
| T2DM | b_cell | *FAM160B1* | -2.13276358 | 0.001297143 | 0.004268027 | 1 |
| T2DM | b_cell | *ANO4* | -1.990766462 | 0.000981513 | 0.00415349 | 1 |
| T2DM | b_cell | *AKR1C2* | -1.539833162 | 0.000571431 | 0.00415349 | 1 |
| T2DM | b_cell | *DPYSL4* | 2.327079079 | 0.015699297 | 0.021987409 | 1 |
| T2DM | b_cell | *ADAM8* | -3.070650309 | 0.000860837 | 0.00415349 | 1 |
| T2DM | b_cell | *ITIH2* | 1.309823599 | 0.000647013 | 0.00415349 | 1 |
| T2DM | b_cell | *PIGF* | -3.068112667 | 0.003204129 | 0.006581388 | 1 |
| T2DM | b_cell | *INPP1* | -2.334322547 | 0.001571217 | 0.004560592 | 1 |
| T2DM | b_cell | *MFSD6* | -3.082238526 | 0.006023983 | 0.010177114 | 1 |
| T2DM | b_cell | *RNF144A* | -1.376001839 | 0.000862097 | 0.00415349 | 1 |
| T2DM | b_cell | *ASAP2* | -1.990119267 | 0.00210601 | 0.005241681 | 1 |
| T2DM | b_cell | *ADAM17* | -2.68596385 | 0.012798051 | 0.018505369 | 1 |
| T2DM | b_cell | *TMEM45B* | -7.62661839 | 0.000948411 | 0.00415349 | 1 |
| T2DM | b_cell | *ACSL1* | -1.065998161 | 0.001930523 | 0.005023674 | 1 |
| T2DM | b_cell | *CCDC122* | -1.957950488 | 0.004234049 | 0.007955609 | 1 |
| T2DM | b_cell | *GABRA2* | -3.234050605 | 0.000741699 | 0.00415349 | 1 |
| T2DM | b_cell | *CENPJ* | -1.782405341 | 0.001075537 | 0.00415349 | 1 |
| T2DM | b_cell | *PARP8* | -1.716447411 | 0.003290134 | 0.006696366 | 1 |

| | | | | | | |
|---|---|---|---|---|---|---|
| T2DM | b_cell | *CACUL1* | -2.768570054 | 0.006878606 | 0.011212288 | 1 |
| T2DM | b_cell | *BEND6* | -2.205228987 | 0.001122957 | 0.004182741 | 1 |
| T2DM | b_cell | *PTPN14* | -1.357371058 | 0.000711401 | 0.00415349 | 1 |
| T2DM | b_cell | *ARL14EP* | -4.116970227 | 7.99E-05 | 0.00415349 | 0.502061196 |
| T2DM | b_cell | *EPG5* | -3.258842288 | 0.007439445 | 0.011907279 | 1 |
| T2DM | b_cell | *SPC25* | -1.487842098 | 0.008902611 | 0.013745264 | 1 |
| T2DM | b_cell | *G6PC2* | -129.8404747 | 0.000735525 | 0.00415349 | 1 |
| T2DM | b_cell | *TGOLN2* | -3.19252448 | 0.02430088 | 0.032009156 | 1 |
| T2DM | b_cell | *POC5* | -3.857219895 | 6.82E-05 | 0.00415349 | 0.428817201 |
| T2DM | b_cell | *TADA1* | -4.026917831 | 0.003462809 | 0.006928923 | 1 |
| T2DM | b_cell | *GUCY1A2* | -1.592181535 | 0.000770926 | 0.00415349 | 1 |
| T2DM | b_cell | *CWF19L2* | -2.254304056 | 0.005479982 | 0.009527833 | 1 |
| T2DM | b_cell | *ZNF547* | 2.526872744 | 0.00081339 | 0.00415349 | 1 |
| T2DM | b_cell | *ZNF773* | -5.325607811 | 0.000279864 | 0.00415349 | 1 |
| T2DM | b_cell | *NMT2* | -2.070080115 | 0.000582844 | 0.00415349 | 1 |
| T2DM | b_cell | *ZNF837* | 2.386076655 | 0.000864859 | 0.00415349 | 1 |
| T2DM | b_cell | *ZFP36L2* | -2.253058155 | 0.010446588 | 0.015663654 | 1 |
| T2DM | b_cell | *PLEKHH2* | -0.837033418 | 0.022397663 | 0.029865134 | 1 |
| T2DM | b_cell | *TMEM123* | -3.186290274 | 0.004930984 | 0.008859987 | 1 |
| T2DM | b_cell | *IGSF10* | -2.053164938 | 0.00084623 | 0.00415349 | 1 |
| T2DM | b_cell | *SPEF2* | -2.271790112 | 0.001416839 | 0.004364365 | 1 |
| T2DM | b_cell | *SPARCL1* | -1.182505352 | 0.00082201 | 0.00415349 | 1 |
| T2DM | b_cell | *CCNO* | 16.95256394 | 0.004771306 | 0.008682258 | 1 |
| T2DM | b_cell | *CLEC4F* | -7.452449587 | 0.000745704 | 0.00415349 | 1 |
| T2DM | b_cell | *PELO* | -2.811066805 | 0.000803136 | 0.00415349 | 1 |
| T2DM | b_cell | *GPR180* | -2.04965028 | 0.001136568 | 0.00418468 | 1 |
| T2DM | b_cell | *WDR78* | -2.594799926 | 0.000964805 | 0.00415349 | 1 |
| T2DM | b_cell | *IFIT5* | -3.501969844 | 0.007119162 | 0.011508915 | 1 |
| T2DM | b_cell | *PRDM8* | 1.898051487 | 0.002357417 | 0.005540753 | 1 |
| T2DM | b_cell | *HHEX* | -2.90261709 | 0.000765529 | 0.00415349 | 1 |
| T2DM | b_cell | *ZNF117* | -0.83984852 | 0.01219759 | 0.017747339 | 1 |
| T2DM | b_cell | *RAB3C* | -1.999682021 | 0.000963757 | 0.00415349 | 1 |
| T2DM | b_cell | *RAD17* | -2.92734376 | 0.001617102 | 0.004621237 | 1 |
| T2DM | b_cell | *MED21* | -3.035019048 | 0.002714917 | 0.005958936 | 1 |
| T2DM | b_cell | *SRP19* | -3.347377049 | 0.00193617 | 0.005026068 | 1 |
| T2DM | b_cell | *CENPH* | -2.192407461 | 0.015549607 | 0.021818412 | 1 |
| T2DM | b_cell | *CARHSP1* | -2.494992795 | 0.001183774 | 0.004202203 | 1 |
| T2DM | b_cell | *TEKT5* | -6.035760901 | 0.00231 | 0.005492478 | 1 |
| T2DM | b_cell | *BANK1* | 0.285783067 | 0.000589609 | 0.00415349 | 1 |
| T2DM | b_cell | *CAST* | -2.101544469 | 0.010955748 | 0.016271862 | 1 |
| T2DM | b_cell | *SCOC* | -2.250199731 | 0.001483499 | 0.004462053 | 1 |
| T2DM | b_cell | *CLGN* | -17.53940553 | 0.00164431 | 0.004657671 | 1 |
| T2DM | b_cell | *SYCP2L* | -2.327478841 | 0.000896494 | 0.00415349 | 1 |
| T2DM | b_cell | *AHCTF1* | -2.437345282 | 0.003158545 | 0.006514977 | 1 |
| T2DM | b_cell | *MERTK* | -0.958294745 | 0.007458182 | 0.011934231 | 1 |
| T2DM | b_cell | *TMEM87B* | -2.708128996 | 0.004867447 | 0.008790778 | 1 |
| T2DM | b_cell | *PTPRR* | -3.277151045 | 0.001068254 | 0.00415349 | 1 |
| T2DM | b_cell | *PLA2R1* | -1.375308006 | 0.001973224 | 0.005091111 | 1 |
| T2DM | b_cell | *SLC25A27* | -1.606586365 | 0.000532785 | 0.00415349 | 1 |
| T2DM | b_cell | *FRMD1* | -3.02441433 | 0.000777164 | 0.00415349 | 1 |

| | | | | | | |
|---|---|---|---|---|---|---|
| T2DM | b_cell | *ASAP1* | -1.917515819 | 0.00193623 | 0.005026068 | 1 |
| T2DM | b_cell | *INO80C* | -3.1413363 | 0.000793032 | 0.00415349 | 1 |
| T2DM | b_cell | *LPCAT1* | -3.809689703 | 0.003169544 | 0.006531275 | 1 |
| T2DM | b_cell | *C16orf89* | -2.46149781 | 0.002923217 | 0.006210972 | 1 |
| T2DM | b_cell | *TEX29* | -3.203695998 | 0.00149586 | 0.004474047 | 1 |
| T2DM | b_cell | *ADPRHL1* | -3.945390928 | 0.000971556 | 0.00415349 | 1 |
| T2DM | b_cell | *TUBGCP5* | -3.329322312 | 0.01404933 | 0.020029056 | 1 |
| T2DM | b_cell | *LURAP1L* | -1.22022194 | 0.002855974 | 0.006140735 | 1 |
| T2DM | b_cell | *CFDP1* | -2.475214877 | 0.004108158 | 0.007801017 | 1 |
| T2DM | b_cell | *ZDHHC7* | -2.297820755 | 0.002232532 | 0.005401538 | 1 |
| T2DM | b_cell | *FAM92B* | 9.391818755 | 0.000626578 | 0.00415349 | 1 |
| T2DM | b_cell | *JAZF1* | -3.805325815 | 0.001360872 | 0.004298348 | 1 |
| T2DM | b_cell | *MCOLN2* | 1.748532598 | 0.000379874 | 0.00415349 | 1 |
| T2DM | b_cell | *CACNA2D1* | -1.134951523 | 0.000716507 | 0.00415349 | 1 |
| T2DM | b_cell | *ZUFSP* | -2.928067184 | 0.002021135 | 0.005157371 | 1 |
| T2DM | b_cell | *HS3ST3A1* | -3.314567428 | 0.004518996 | 0.008348788 | 1 |
| T2DM | b_cell | *PPP2R5E* | -2.02307191 | 0.036187027 | 0.045638291 | 1 |
| T2DM | b_cell | *ANKRD29* | -1.697866691 | 0.001680489 | 0.004704627 | 1 |
| T2DM | b_cell | *THY1* | -1.784549663 | 0.000904696 | 0.00415349 | 1 |
| T2DM | b_cell | *TBCEL* | -2.035101357 | 0.001054618 | 0.00415349 | 1 |
| T2DM | b_cell | *UBASH3B* | -2.37268643 | 0.000946112 | 0.00415349 | 1 |
| T2DM | b_cell | *ROBO4* | -1.887325374 | 0.001227516 | 0.004253251 | 1 |
| T2DM | b_cell | *ROBO3* | -2.14414072 | 0.000557519 | 0.00415349 | 1 |
| T2DM | b_cell | *CC2D1B* | -3.134422657 | 0.024231171 | 0.031944091 | 1 |
| T2DM | b_cell | *PRKCA* | -2.667390966 | 0.006122324 | 0.010297514 | 1 |
| T2DM | b_cell | *LRRK1* | -2.903930022 | 0.005689123 | 0.009764542 | 1 |
| T2DM | b_cell | *CEP112* | -1.439163047 | 0.001327077 | 0.004274973 | 1 |
| T2DM | b_cell | *UCHL1* | -2.654160258 | 0.000715993 | 0.00415349 | 1 |
| T2DM | b_cell | *MIA3* | -3.078110647 | 0.005523247 | 0.009579216 | 1 |
| T2DM | b_cell | *DISP1* | 3.769616498 | 0.01685075 | 0.023364741 | 1 |
| T2DM | b_cell | *FAM167A* | 2.84204038 | 0.000544755 | 0.00415349 | 1 |
| T2DM | b_cell | *NEIL2* | 4.324465369 | 0.002033795 | 0.00516834 | 1 |
| T2DM | b_cell | *LONRF1* | -2.311394063 | 0.000258717 | 0.00415349 | 1 |
| T2DM | b_cell | *CCSAP* | -4.187002425 | 0.002748024 | 0.006007705 | 1 |
| T2DM | b_cell | *GBP5* | -2.750068452 | 0.001106795 | 0.004159248 | 1 |
| T2DM | b_cell | *FAM69A* | -2.118998731 | 0.000980861 | 0.00415349 | 1 |
| T2DM | b_cell | *PDLIM3* | -1.994465816 | 0.002105641 | 0.005241681 | 1 |
| T2DM | b_cell | *NCAM2* | -1.624533211 | 0.000965656 | 0.00415349 | 1 |
| T2DM | b_cell | *L3MBTL4* | -2.262926168 | 0.004191363 | 0.007891914 | 1 |
| T2DM | b_cell | *PDE1C* | -1.728289969 | 0.000824662 | 0.00415349 | 1 |
| T2DM | b_cell | *RABGEF1* | -2.443375837 | 0.001840589 | 0.004898539 | 1 |
| T2DM | b_cell | *FLCN* | -2.637372436 | 0.00251187 | 0.005734251 | 1 |
| T2DM | b_cell | *CXXC1* | -3.51045755 | 0.001646209 | 0.004658975 | 1 |
| T2DM | b_cell | *PPP4R1* | -2.564783508 | 0.0026758 | 0.00592142 | 1 |
| T2DM | b_cell | *APCDD1* | -1.342260442 | 0.001828952 | 0.004890949 | 1 |
| T2DM | b_cell | *EPHB1* | -3.715147338 | 0.004130663 | 0.007836293 | 1 |
| T2DM | b_cell | *ANKRD40* | -2.547134212 | 0.00060489 | 0.00415349 | 1 |
| T2DM | b_cell | *CYP2U1* | -3.525907118 | 0.002110599 | 0.005248946 | 1 |
| T2DM | b_cell | *FBXL18* | -2.938859451 | 0.001867202 | 0.004940699 | 1 |
| T2DM | b_cell | *PROM2* | -11.79959471 | 0.000925185 | 0.00415349 | 1 |

| | | | | | | |
|---|---|---|---|---|---|---|
| T2DM | b_cell | *AK9* | -2.223660564 | 0.001652202 | 0.004668493 | 1 |
| T2DM | b_cell | *CDK19* | -2.637026842 | 0.017109165 | 0.023661531 | 1 |
| T2DM | b_cell | *GTF3C6* | 3.410248043 | 0.003653798 | 0.007196562 | 1 |
| T2DM | b_cell | *TTC39B* | -2.758568641 | 0.029124693 | 0.037568106 | 1 |
| T2DM | b_cell | *AGPAT5* | 3.280696503 | 0.028839738 | 0.037246392 | 1 |
| T2DM | b_cell | *MMS19* | -2.873495796 | 0.001515633 | 0.004480664 | 1 |
| T2DM | b_cell | *GOLGA7B* | 2.389452872 | 0.002212437 | 0.005382969 | 1 |
| T2DM | b_cell | *TRMT44* | -2.315676585 | 0.001105067 | 0.004159248 | 1 |
| T2DM | b_cell | *C16orf87* | 3.734318402 | 0.002236593 | 0.005404098 | 1 |
| T2DM | b_cell | *HEATR3* | -2.116058653 | 0.014206592 | 0.020230316 | 1 |
| T2DM | b_cell | *CNOT8* | -3.145519674 | 0.015335374 | 0.021544916 | 1 |
| T2DM | b_cell | *LRGUK* | -2.494204199 | 0.019092842 | 0.025925853 | 1 |
| T2DM | b_cell | *SETD9* | -2.144904482 | 7.51E-05 | 0.00415349 | 0.47184207 |
| T2DM | b_cell | *FAM126B* | -2.390434924 | 0.002671029 | 0.005916798 | 1 |
| T2DM | b_cell | *ALS2CR12* | -2.235879557 | 0.000380854 | 0.00415349 | 1 |
| T2DM | b_cell | *SPAG17* | -1.340177536 | 0.000858964 | 0.00415349 | 1 |
| T2DM | b_cell | *DEPTOR* | -2.524325356 | 0.001640634 | 0.004657671 | 1 |
| T2DM | b_cell | *RRAGA* | -5.469018808 | 0.004126781 | 0.007831293 | 1 |
| T2DM | b_cell | *TMBIM4* | -2.552165061 | 0.011954906 | 0.017446118 | 1 |
| T2DM | b_cell | *FAM161B* | -2.984210764 | 0.002711873 | 0.005955133 | 1 |
| T2DM | b_cell | *WIF1* | 6.523682805 | 0.000318477 | 0.00415349 | 1 |
| T2DM | b_cell | *UGT2B4* | -3.944489934 | 0.000773565 | 0.00415349 | 1 |
| T2DM | b_cell | *MMP16* | -1.529461466 | 0.000328938 | 0.00415349 | 1 |
| T2DM | b_cell | *KCNMA1* | -1.339255559 | 0.000727168 | 0.00415349 | 1 |
| T2DM | b_cell | *BATF* | 6.172341521 | 0.021758041 | 0.02909866 | 1 |
| T2DM | b_cell | *DCK* | 2.932168199 | 0.002104619 | 0.005241681 | 1 |
| T2DM | b_cell | *DPY19L4* | -2.750761678 | 0.001643211 | 0.004657671 | 1 |
| T2DM | b_cell | *NDUFAF6* | -3.320631627 | 0.000157637 | 0.00415349 | 0.991063433 |
| T2DM | b_cell | *DRAM2* | -2.400920213 | 0.006208963 | 0.010401212 | 1 |
| T2DM | b_cell | *ADAMTSL3* | -1.254349516 | 0.001521384 | 0.004492692 | 1 |
| T2DM | b_cell | *WHAMM* | -3.654174686 | 0.000112083 | 0.00415349 | 0.704664708 |
| T2DM | b_cell | *CXCL13* | -4.436461081 | 0.000840811 | 0.00415349 | 1 |
| T2DM | b_cell | *N6AMT1* | -2.3437714 | 0.001263134 | 0.004257394 | 1 |
| T2DM | b_cell | *RWDD2B* | -2.057619724 | 0.002889227 | 0.006184116 | 1 |
| T2DM | b_cell | *ANKRD9* | -3.309285109 | 0.000308926 | 0.00415349 | 1 |
| T2DM | b_cell | *SFXN2* | -2.171558063 | 0.001708365 | 0.004728644 | 1 |
| T2DM | b_cell | *C14orf2* | -4.028909452 | 0.002448369 | 0.005641723 | 1 |
| T2DM | b_cell | *SH3RF2* | -1.502840627 | 0.003750646 | 0.007304929 | 1 |
| T2DM | b_cell | *GDF6* | -5.769948101 | 0.004270659 | 0.008002871 | 1 |
| T2DM | b_cell | *NODAL* | -3.452651263 | 0.000784616 | 0.00415349 | 1 |
| T2DM | b_cell | *MED19* | -2.252699766 | 0.002081902 | 0.005230315 | 1 |
| T2DM | b_cell | *WDYHV1* | -3.764811231 | 0.001095467 | 0.00415349 | 1 |
| T2DM | b_cell | *ATAD2* | -3.613636466 | 0.013480678 | 0.01933235 | 1 |
| T2DM | b_cell | *SASS6* | -3.138357432 | 0.000703287 | 0.00415349 | 1 |
| T2DM | b_cell | *ITGAD* | -2.631710301 | 0.004925083 | 0.008854445 | 1 |
| T2DM | b_cell | *VPS8* | -1.977809702 | 0.023073359 | 0.030688007 | 1 |
| T2DM | b_cell | *MPV17L* | -2.287518524 | 0.029436779 | 0.037916212 | 1 |
| T2DM | b_cell | *RPUSD3* | -3.561373499 | 0.001980891 | 0.005095689 | 1 |
| T2DM | b_cell | *GHRL* | -2.854946838 | 0.001032023 | 0.00415349 | 1 |
| T2DM | b_cell | *EXOG* | -3.493546807 | 0.004184832 | 0.007886524 | 1 |

| | | | | | | |
|---|---|---|---|---|---|---|
| T2DM | b_cell | *SLC6A1* | -5.869322975 | 0.00039679 | 0.00415349 | 1 |
| T2DM | b_cell | *FCHO2* | -2.242541766 | 0.003338223 | 0.006765112 | 1 |
| T2DM | b_cell | *TIMP4* | -1.846186095 | 0.005534098 | 0.009595387 | 1 |
| T2DM | b_cell | *CPT2* | 3.772475992 | 0.001759719 | 0.004789372 | 1 |
| T2DM | b_cell | *GATAD1* | -1.86738726 | 0.005485073 | 0.009534048 | 1 |
| T2DM | b_cell | *TMED6* | -11.49925373 | 0.00072031 | 0.00415349 | 1 |
| T2DM | b_cell | *CLEC18A* | -15.58335124 | 1.07E-07 | 5.60E-05 | 0.000672084 |
| T2DM | b_cell | *DHRS1* | -2.507799642 | 0.005344666 | 0.009365082 | 1 |
| T2DM | b_cell | *AASDH* | -2.97500771 | 0.009941124 | 0.015058607 | 1 |
| T2DM | b_cell | *RNF111* | -1.86864765 | 0.001933304 | 0.005024672 | 1 |
| T2DM | b_cell | *DYRK1A* | -2.316331538 | 0.029771716 | 0.038308387 | 1 |
| T2DM | b_cell | *KCNJ15* | -6.497013324 | 0.004161339 | 0.007865091 | 1 |
| T2DM | b_cell | *LCA5L* | -2.068275324 | 0.031766161 | 0.040583997 | 1 |
| T2DM | b_cell | *CREB3L1* | -2.606824369 | 0.002910575 | 0.006192483 | 1 |
| T2DM | b_cell | *C2CD2* | -1.428898395 | 0.002871394 | 0.006159146 | 1 |
| T2DM | b_cell | *SLC38A10* | -3.244661502 | 0.010161151 | 0.015319701 | 1 |
| T2DM | b_cell | *C9orf43* | -4.55724165 | 0.000409933 | 0.00415349 | 1 |
| T2DM | b_cell | *WDR19* | -2.674655737 | 0.002379383 | 0.005562581 | 1 |
| T2DM | b_cell | *SLC37A3* | -2.590844679 | 0.002047708 | 0.005191106 | 1 |
| T2DM | b_cell | *SPPL3* | -2.618600894 | 0.005834029 | 0.009937291 | 1 |
| T2DM | b_cell | *RADIL* | -2.055738441 | 0.000576956 | 0.00415349 | 1 |
| T2DM | b_cell | *LDLRAP1* | -4.806603431 | 0.014597038 | 0.020683163 | 1 |
| T2DM | b_cell | *PAFAH2* | -4.42380213 | 0.001176983 | 0.004202203 | 1 |
| T2DM | b_cell | *TRIM63* | 4.246996513 | 8.82E-06 | 0.001459962 | 0.055478563 |
| T2DM | b_cell | *WDR66* | -2.501608273 | 0.000474103 | 0.00415349 | 1 |
| T2DM | b_cell | *UBXN11* | -3.151942782 | 0.001892558 | 0.00497413 | 1 |
| T2DM | b_cell | *GALNT14* | 6.970275852 | 0.004349813 | 0.008105298 | 1 |
| T2DM | b_cell | *RHPN1* | -3.673614771 | 0.003780473 | 0.007342551 | 1 |
| T2DM | b_cell | *TPRG1L* | -4.487768162 | 0.000722209 | 0.00415349 | 1 |
| T2DM | b_cell | *LRRC43* | -6.980479379 | 0.00074285 | 0.00415349 | 1 |
| T2DM | b_cell | *XKR8* | -4.078630244 | 0.000619466 | 0.00415349 | 1 |
| T2DM | b_cell | *EYA3* | -2.757844698 | 0.01107784 | 0.01639896 | 1 |
| T2DM | b_cell | *MRAS* | -1.552633386 | 0.000199448 | 0.00415349 | 1 |
| T2DM | b_cell | *ABHD3* | -1.843093803 | 0.006009749 | 0.01015953 | 1 |
| T2DM | b_cell | *CLSTN2* | -1.238795119 | 0.000698732 | 0.00415349 | 1 |
| T2DM | b_cell | *COLEC12* | -1.321193817 | 0.000812435 | 0.00415349 | 1 |
| T2DM | b_cell | *RNF207* | -5.415984905 | 0.003084176 | 0.006414229 | 1 |
| T2DM | b_cell | *AUTS2* | -1.683377148 | 0.002125615 | 0.005269615 | 1 |
| T2DM | b_cell | *MITD1* | -3.257701951 | 0.017789178 | 0.024478127 | 1 |
| T2DM | b_cell | *TSPAN33* | -2.819882607 | 0.03205197 | 0.04090758 | 1 |
| T2DM | b_cell | *AHCYL2* | -2.447917722 | 0.011768191 | 0.017218203 | 1 |
| T2DM | b_cell | *B4GALT5* | -1.953563114 | 0.00112418 | 0.004182741 | 1 |
| T2DM | b_cell | *CD1D* | -9.289950222 | 0.00029189 | 0.00415349 | 1 |
| T2DM | b_cell | *FAM86C1* | 9.551293411 | 0.014256012 | 0.020291498 | 1 |
| T2DM | b_cell | *NCF1* | 2.014491469 | 0.000692732 | 0.00415349 | 1 |
| T2DM | b_cell | *PPP1R9A* | -1.267017367 | 0.000669214 | 0.00415349 | 1 |
| T2DM | b_cell | *ZFAND2B* | -3.984580373 | 0.004261072 | 0.007989011 | 1 |
| T2DM | b_cell | *GDPD5* | -1.655837088 | 0.000689846 | 0.00415349 | 1 |
| T2DM | b_cell | *DYNC1I1* | -2.945899309 | 0.002240106 | 0.005409545 | 1 |
| T2DM | b_cell | *TMED4* | -2.910569616 | 0.001798069 | 0.004852128 | 1 |

| | | | | | | |
|---|---|---|---|---|---|---|
| T2DM | b_cell | *COPG2* | -1.558521802 | 0.00039266 | 0.00415349 | 1 |
| T2DM | b_cell | *ELK4* | -3.432802741 | 0.009282555 | 0.014213206 | 1 |
| T2DM | b_cell | *DUSP23* | -3.273892044 | 0.000925075 | 0.00415349 | 1 |
| T2DM | b_cell | *RNF166* | -4.121640867 | 0.030224595 | 0.038835486 | 1 |
| T2DM | b_cell | *F11R* | -2.166676504 | 0.001545731 | 0.004515804 | 1 |
| T2DM | b_cell | *NIT1* | -1.781191273 | 0.004779321 | 0.00869424 | 1 |
| T2DM | b_cell | *ZNF276* | -3.225301132 | 0.000354974 | 0.00415349 | 1 |
| T2DM | b_cell | *CDA* | -3.796824411 | 0.012776995 | 0.018479173 | 1 |
| T2DM | b_cell | *B4GALT3* | -2.938516593 | 0.004315283 | 0.008064858 | 1 |
| T2DM | b_cell | *DMTN* | -1.69226696 | 0.012217586 | 0.017772319 | 1 |
| T2DM | b_cell | *FAM160B2* | -2.692492519 | 0.001183936 | 0.004202203 | 1 |
| T2DM | b_cell | *NDUFS2* | -3.019549028 | 0.003755259 | 0.007309385 | 1 |
| T2DM | b_cell | *SLC5A11* | -1.739049985 | 0.004980206 | 0.00892419 | 1 |
| T2DM | b_cell | *CCAR2* | -3.44067186 | 0.003355002 | 0.006777924 | 1 |
| T2DM | b_cell | *EPB41* | -1.671809456 | 0.005384615 | 0.009411475 | 1 |
| T2DM | b_cell | *ALG8* | -3.544895335 | 0.007560371 | 0.0120701 | 1 |
| T2DM | b_cell | *MRPL10* | -2.831862432 | 0.000842941 | 0.00415349 | 1 |
| T2DM | b_cell | *SV2A* | -10.8591521 | 0.002672709 | 0.005916798 | 1 |
| T2DM | b_cell | *RCAN1* | -2.385316506 | 0.002467903 | 0.005668874 | 1 |
| T2DM | b_cell | *UBE2Z* | -3.858575849 | 0.003403763 | 0.006850019 | 1 |
| T2DM | b_cell | *CLIC6* | -2.760652857 | 0.024540175 | 0.032270253 | 1 |
| T2DM | b_cell | *CBR3* | -3.054836335 | 0.023259707 | 0.030890109 | 1 |
| T2DM | b_cell | *SIM2* | 4.982189217 | 0.001662691 | 0.004679659 | 1 |
| T2DM | b_cell | *HLCS* | -1.991788742 | 0.005770454 | 0.009869109 | 1 |
| T2DM | b_cell | *ARHGAP27* | -2.509670005 | 0.00108273 | 0.00415349 | 1 |
| T2DM | b_cell | *PADI4* | -6.877818543 | 0.001286341 | 0.004268027 | 1 |
| T2DM | b_cell | *PSMD4* | -3.600924575 | 0.010715021 | 0.015989873 | 1 |
| T2DM | b_cell | *STARD9* | -1.457914096 | 0.001079965 | 0.00415349 | 1 |
| T2DM | b_cell | *THEM4* | -2.1694898 | 0.002276998 | 0.005445221 | 1 |
| T2DM | b_cell | *TCHH* | 2.587653784 | 0.002357633 | 0.005540753 | 1 |
| T2DM | b_cell | *MED8* | -2.305832031 | 0.004215941 | 0.007929499 | 1 |
| T2DM | b_cell | *RSPRY1* | -3.449079541 | 0.009305301 | 0.014241097 | 1 |
| T2DM | b_cell | *CCDC17* | -5.72040005 | 0.000678947 | 0.00415349 | 1 |
| T2DM | b_cell | *ACE* | -2.147735979 | 0.000725459 | 0.00415349 | 1 |
| T2DM | b_cell | *TEPP* | 15.88903369 | 0.027957529 | 0.036233557 | 1 |
| T2DM | b_cell | *EFCAB14* | -3.19965421 | 0.008785519 | 0.013604571 | 1 |
| T2DM | b_cell | *LYPD5* | -5.455213819 | 0.008091267 | 0.012711094 | 1 |
| T2DM | b_cell | *CCDC117* | -4.332365846 | 0.03475101 | 0.043959678 | 1 |
| T2DM | b_cell | *CCDC107* | -2.359455588 | 0.000394149 | 0.00415349 | 1 |
| T2DM | b_cell | *NPR2* | -2.497019263 | 0.001349172 | 0.004290463 | 1 |
| T2DM | b_cell | *ZNF235* | -2.744364976 | 0.001406359 | 0.004357467 | 1 |
| T2DM | b_cell | *DFFA* | -2.608035501 | 0.007099156 | 0.01148839 | 1 |
| T2DM | b_cell | *ATAD3B* | -4.129361376 | 0.017908735 | 0.024594193 | 1 |
| T2DM | b_cell | *SSU72* | -2.411177446 | 0.001157779 | 0.004195495 | 1 |
| T2DM | b_cell | *NR2F6* | -2.738155638 | 0.003700626 | 0.00724956 | 1 |
| T2DM | b_cell | *ANKLE1* | -5.666936747 | 0.033234514 | 0.04227952 | 1 |
| T2DM | b_cell | *CILP2* | -21.59349666 | 0.000906488 | 0.00415349 | 1 |
| T2DM | b_cell | *RSPH1* | -6.534232369 | 0.002462399 | 0.005658298 | 1 |
| T2DM | b_cell | *U2AF1* | -5.851754798 | 0.010523244 | 0.015737306 | 1 |
| T2DM | b_cell | *HSF2BP* | -2.857098137 | 0.016598812 | 0.023072458 | 1 |

| | | | | | | |
|---|---|---|---|---|---|---|
| T2DM | b_cell | *CSTB* | -2.287768564 | 0.000418267 | 0.00415349 | 1 |
| T2DM | b_cell | *C21orf33* | -10.20757509 | 0.000239707 | 0.00415349 | 1 |
| T2DM | b_cell | *ICOSLG* | 4.932735736 | 0.004738392 | 0.008639869 | 1 |
| T2DM | b_cell | *LRRC3* | -4.551785391 | 0.003877076 | 0.007477048 | 1 |
| T2DM | b_cell | *ITGB2* | -2.577421252 | 0.005034108 | 0.008986408 | 1 |
| T2DM | b_cell | *LSS* | -2.050031196 | 0.000743532 | 0.00415349 | 1 |
| T2DM | b_cell | *VAV2* | 3.236683813 | 0.033604216 | 0.042715266 | 1 |
| T2DM | b_cell | *MCM3AP* | -2.071165947 | 0.025465787 | 0.033354876 | 1 |
| T2DM | b_cell | *PCNT* | -2.079307044 | 0.012678063 | 0.018348752 | 1 |
| T2DM | b_cell | *DIP2A* | -3.807927279 | 0.001180447 | 0.004202203 | 1 |
| T2DM | b_cell | *PRMT2* | -2.735410477 | 0.000643462 | 0.00415349 | 1 |
| T2DM | b_cell | *ZNF208* | -1.753387739 | 0.000963083 | 0.00415349 | 1 |
| T2DM | b_cell | *ADAMTS13* | -2.624230036 | 0.001215288 | 0.004245409 | 1 |
| T2DM | b_cell | *ZNF714* | -3.113598841 | 0.014829102 | 0.020957698 | 1 |
| T2DM | b_cell | *GPSM1* | -3.794711156 | 0.000105494 | 0.00415349 | 0.663238559 |
| T2DM | b_cell | *C19orf47* | -3.253453225 | 0.000994078 | 0.00415349 | 1 |
| T2DM | b_cell | *HIPK4* | 5.103047495 | 0.011519379 | 0.016952793 | 1 |
| T2DM | b_cell | *RDH13* | -4.949105909 | 0.005090281 | 0.009037019 | 1 |
| T2DM | b_cell | *MPZL3* | -3.127227353 | 0.000829362 | 0.00415349 | 1 |
| T2DM | b_cell | *PCSK7* | -3.556861275 | 0.009369391 | 0.014311312 | 1 |
| T2DM | b_cell | *S100A1* | 2.185683552 | 0.000354984 | 0.00415349 | 1 |
| T2DM | b_cell | *ZBTB7B* | -4.229066703 | 0.010013665 | 0.015150372 | 1 |
| T2DM | b_cell | *VPS11* | -3.635829208 | 0.000768804 | 0.00415349 | 1 |
| T2DM | b_cell | *FDPS* | -2.053832744 | 0.009628739 | 0.014654051 | 1 |
| T2DM | b_cell | *PAQR6* | 1.457417143 | 0.001508992 | 0.004480341 | 1 |
| T2DM | b_cell | *SLC25A44* | -3.235336653 | 0.000719719 | 0.00415349 | 1 |
| T2DM | b_cell | *NBEAL2* | -4.018207818 | 0.017124521 | 0.02367756 | 1 |
| T2DM | b_cell | *FCRL3* | 0.247268661 | 0.000688334 | 0.00415349 | 1 |
| T2DM | b_cell | *FGFR4* | 6.366103897 | 0.000568109 | 0.00415349 | 1 |
| T2DM | b_cell | *NACC1* | 2.974182518 | 0.000832857 | 0.00415349 | 1 |
| T2DM | b_cell | *LY6K* | 9.491267365 | 0.001762693 | 0.004791201 | 1 |
| T2DM | b_cell | *TONSL* | -4.139060411 | 0.000946635 | 0.00415349 | 1 |
| T2DM | b_cell | *MUM1* | -1.878727017 | 0.002378849 | 0.005562581 | 1 |
| T2DM | b_cell | *ZNF333* | -3.532671858 | 0.01880536 | 0.025607386 | 1 |
| T2DM | b_cell | *PPP1R16A* | -1.945292152 | 0.002507162 | 0.005732727 | 1 |
| T2DM | b_cell | *ORAI2* | 1.914868517 | 0.000902701 | 0.00415349 | 1 |
| T2DM | b_cell | *LRWD1* | 3.367872937 | 0.00299911 | 0.006308941 | 1 |
| T2DM | b_cell | *FBXL13* | -2.591323437 | 0.001098816 | 0.00415349 | 1 |
| T2DM | b_cell | *MFSD12* | -4.03522387 | 0.005571013 | 0.009636761 | 1 |
| T2DM | b_cell | *YDJC* | 11.64466786 | 0.027230072 | 0.035370963 | 1 |
| T2DM | b_cell | *CCDC116* | 3.622106885 | 0.003913439 | 0.007533311 | 1 |
| T2DM | b_cell | *DVL3* | -3.195798458 | 0.018091046 | 0.024822873 | 1 |
| T2DM | b_cell | *FBXO27* | -1.819731655 | 0.002770426 | 0.006028396 | 1 |
| T2DM | b_cell | *DMKN* | 3.592561015 | 0.006194319 | 0.010384982 | 1 |
| T2DM | b_cell | *U2AF1L4* | 1.554351401 | 0.009337062 | 0.014279278 | 1 |
| T2DM | b_cell | *BDH1* | -2.864548354 | 0.000791049 | 0.00415349 | 1 |
| T2DM | b_cell | *COX7A1* | -2.995063401 | 0.000423889 | 0.00415349 | 1 |
| T2DM | b_cell | *DUSP14* | -1.965578691 | 0.001582713 | 0.004577055 | 1 |
| T2DM | b_cell | *LRRC56* | 2.479629843 | 0.007499225 | 0.01199075 | 1 |
| T2DM | b_cell | *PGAP3* | -4.534820317 | 0.001809396 | 0.004859322 | 1 |

| | | | | | | |
|---|---|---|---|---|---|---|
| T2DM | b_cell | *IKZF3* | 0.524756296 | 0.000593644 | 0.00415349 | 1 |
| T2DM | b_cell | *ACOX1* | -2.134420737 | 0.000752231 | 0.00415349 | 1 |
| T2DM | b_cell | *PRPSAP1* | -2.898378417 | 0.001091198 | 0.00415349 | 1 |
| T2DM | b_cell | *CYGB* | -3.123412089 | 0.000454953 | 0.00415349 | 1 |
| T2DM | b_cell | *ZNF577* | -2.341454375 | 0.007786587 | 0.01235595 | 1 |
| T2DM | b_cell | *ALDH16A1* | -3.907483114 | 0.003498938 | 0.006971457 | 1 |
| T2DM | b_cell | *SIGLEC11* | -5.658964094 | 0.000537031 | 0.00415349 | 1 |
| T2DM | b_cell | *MPP3* | -2.287855697 | 0.000636546 | 0.00415349 | 1 |
| T2DM | b_cell | *ASB16* | 3.84180257 | 0.003974329 | 0.007615547 | 1 |
| T2DM | b_cell | *PLCD3* | -2.143806383 | 0.002537462 | 0.005775896 | 1 |
| T2DM | b_cell | *LARP4* | -2.40860372 | 0.011042421 | 0.016361937 | 1 |
| T2DM | b_cell | *SPC24* | -11.1343531 | 0.024896891 | 0.03266418 | 1 |
| T2DM | b_cell | *IP6K3* | -14.48957876 | 0.000430164 | 0.00415349 | 1 |
| T2DM | b_cell | *SCIMP* | 2.452500333 | 0.010879048 | 0.016180879 | 1 |
| T2DM | b_cell | *ASGR2* | -13.70802094 | 0.002983567 | 0.006295285 | 1 |
| T2DM | b_cell | *TNFSF13* | 6.527957867 | 0.000432997 | 0.00415349 | 1 |
| T2DM | b_cell | *SENP3* | 2.754040988 | 0.001395172 | 0.00434199 | 1 |
| T2DM | b_cell | *SNRNP25* | -5.091749174 | 0.000870354 | 0.00415349 | 1 |
| T2DM | b_cell | *WDR90* | -2.912619828 | 0.001312368 | 0.004274973 | 1 |
| T2DM | b_cell | *JMJD8* | 5.472444295 | 0.000309581 | 0.00415349 | 1 |
| T2DM | b_cell | *MEIOB* | -4.914093327 | 0.00214029 | 0.005287258 | 1 |
| T2DM | b_cell | *FLYWCH2* | -4.490837644 | 0.023018455 | 0.030627942 | 1 |
| T2DM | b_cell | *ZG16B* | -39.67287119 | 0.000685917 | 0.00415349 | 1 |
| T2DM | b_cell | *NXF1* | -2.480144382 | 0.002475412 | 0.005679896 | 1 |
| T2DM | b_cell | *TPCN2* | -1.773141906 | 0.001142941 | 0.00418468 | 1 |
| T2DM | b_cell | *CYP4A22* | 13.85686537 | 0.000164089 | 0.00415349 | 1 |
| T2DM | b_cell | *PDZK1IP1* | 50.0294544 | 0.005261659 | 0.009250573 | 1 |
| T2DM | b_cell | *COA7* | 3.319526397 | 0.000711564 | 0.00415349 | 1 |
| T2DM | b_cell | *C1orf123* | -3.903861328 | 0.000160237 | 0.00415349 | 1 |
| T2DM | b_cell | *ACOT11* | -2.96955266 | 0.00028419 | 0.00415349 | 1 |
| T2DM | b_cell | *USP24* | -2.262944625 | 0.005539703 | 0.009597457 | 1 |
| T2DM | b_cell | *ZSWIM5* | -2.771276853 | 0.024042397 | 0.031746467 | 1 |
| T2DM | b_cell | *RBP7* | -2.28418746 | 0.000952701 | 0.00415349 | 1 |
| T2DM | b_cell | *FBLIM1* | -2.441557966 | 0.001425385 | 0.004380342 | 1 |
| T2DM | b_cell | *SLC25A34* | 6.510478124 | 0.03678094 | 0.046331751 | 1 |
| T2DM | b_cell | *PDPN* | -2.512638168 | 0.000987781 | 0.00415349 | 1 |
| T2DM | b_cell | *LRRC38* | -13.85302421 | 0.003186431 | 0.006555332 | 1 |
| T2DM | b_cell | *LAPTM5* | 3.661378134 | 0.016989302 | 0.023523279 | 1 |
| T2DM | b_cell | *PEF1* | -2.406564506 | 0.001015127 | 0.00415349 | 1 |
| T2DM | b_cell | *SYNC* | -2.546584315 | 0.002507278 | 0.005732727 | 1 |
| T2DM | b_cell | *RBBP4* | -3.048800822 | 0.033869444 | 0.043008926 | 1 |
| T2DM | b_cell | *TSSK3* | -2.710240312 | 0.011625167 | 0.017055842 | 1 |
| T2DM | b_cell | *CAMK2N1* | -3.388937261 | 0.000886169 | 0.00415349 | 1 |
| T2DM | b_cell | *TTLL10* | -6.865654613 | 0.001821167 | 0.00487843 | 1 |
| T2DM | b_cell | *NFIA* | -1.81320088 | 0.004394389 | 0.008166575 | 1 |
| T2DM | b_cell | *FUBP1* | -2.591940564 | 0.029694131 | 0.038216377 | 1 |
| T2DM | b_cell | *NEXN* | -1.903188242 | 0.001031711 | 0.00415349 | 1 |
| T2DM | b_cell | *B3GALT2* | -3.591853436 | 0.000641107 | 0.00415349 | 1 |
| T2DM | b_cell | *NTNG1* | -1.700839003 | 0.008515063 | 0.013254321 | 1 |
| T2DM | b_cell | *HFM1* | -1.89749287 | 0.000762199 | 0.00415349 | 1 |

| | | | | | | |
|---|---|---|---|---|---|---|
| T2DM | b_cell | *KCNT2* | -1.397063097 | 0.001003615 | 0.00415349 | 1 |
| T2DM | b_cell | *VCAM1* | -2.378101645 | 0.000917022 | 0.00415349 | 1 |
| T2DM | b_cell | *EXTL2* | -3.426499345 | 0.000135037 | 0.00415349 | 0.848980689 |
| T2DM | b_cell | *ZNF496* | -4.172011702 | 0.011005533 | 0.016330373 | 1 |
| T2DM | b_cell | *TRIM58* | 1.565429874 | 0.000750414 | 0.00415349 | 1 |
| T2DM | b_cell | *IGSF8* | -2.442678606 | 0.000777949 | 0.00415349 | 1 |
| T2DM | b_cell | *VANGL2* | -5.449040586 | 0.004321138 | 0.008068011 | 1 |
| T2DM | b_cell | *FCRLB* | -6.731204671 | 0.006109893 | 0.010281826 | 1 |
| T2DM | b_cell | *KLHDC9* | 2.637182803 | 0.007089299 | 0.011481305 | 1 |
| T2DM | b_cell | *FLVCR1* | -2.246098376 | 0.000776495 | 0.00415349 | 1 |
| T2DM | b_cell | *TDRD5* | -3.758746429 | 0.004335106 | 0.008085527 | 1 |
| T2DM | b_cell | *SNED1* | -1.962151161 | 0.0022631 | 0.005437295 | 1 |
| T2DM | b_cell | *C1orf115* | -2.238220439 | 0.009163635 | 0.014068809 | 1 |
| T2DM | b_cell | *CNST* | -2.785169888 | 0.005084681 | 0.009030336 | 1 |
| T2DM | b_cell | *PM20D1* | -4.019811256 | 0.000653793 | 0.00415349 | 1 |
| T2DM | b_cell | *HAAO* | -3.495073974 | 0.039778051 | 0.049679103 | 1 |
| T2DM | b_cell | *B3GALNT2* | -2.990450332 | 0.024980091 | 0.032752832 | 1 |
| T2DM | b_cell | *PUS10* | -2.376357645 | 0.001512564 | 0.004480664 | 1 |
| T2DM | b_cell | *RFTN2* | -2.136115489 | 0.001167257 | 0.004202203 | 1 |
| T2DM | b_cell | *DISC1* | -2.892603288 | 0.002405944 | 0.005600211 | 1 |
| T2DM | b_cell | *FAM84A* | -5.354689141 | 0.000735964 | 0.00415349 | 1 |
| T2DM | b_cell | *NUP35* | -3.783130161 | 0.002437745 | 0.005632525 | 1 |
| T2DM | b_cell | *FBXO41* | 5.844560531 | 0.025493334 | 0.033384002 | 1 |
| T2DM | b_cell | *CCDC74A* | -11.77841635 | 0.000732703 | 0.00415349 | 1 |
| T2DM | b_cell | *NOSTRIN* | -1.73258055 | 0.000679255 | 0.00415349 | 1 |
| T2DM | b_cell | *SGPP2* | 7.111813532 | 0.008362011 | 0.01305163 | 1 |
| T2DM | b_cell | *BBS5* | -2.893298526 | 0.001982236 | 0.005095706 | 1 |
| T2DM | b_cell | *HPGDS* | -2.841876969 | 0.003652468 | 0.007196562 | 1 |
| T2DM | b_cell | *PDLIM5* | -2.251269794 | 0.005712164 | 0.009793394 | 1 |
| T2DM | b_cell | *RPRD2* | -2.693060544 | 0.010449475 | 0.015664246 | 1 |
| T2DM | b_cell | *PACRGL* | -2.628995276 | 0.019724944 | 0.026686189 | 1 |
| T2DM | b_cell | *C1QTNF7* | -1.316385576 | 0.000840174 | 0.00415349 | 1 |
| T2DM | b_cell | *LYSMD1* | -3.135994665 | 0.000602588 | 0.00415349 | 1 |
| T2DM | b_cell | *RNF149* | -2.576748517 | 0.007415932 | 0.011872667 | 1 |
| T2DM | b_cell | *DHX57* | -2.494207306 | 0.001650417 | 0.004667644 | 1 |
| T2DM | b_cell | *S100A9* | 3.569102349 | 0.002122004 | 0.005266891 | 1 |
| T2DM | b_cell | *S100A12* | -23.71319269 | 0.001530052 | 0.004503445 | 1 |
| T2DM | b_cell | *TDRD10* | -2.57564385 | 0.001220227 | 0.004252746 | 1 |
| T2DM | b_cell | *DCAF16* | -2.593353548 | 0.001181207 | 0.004202203 | 1 |
| T2DM | b_cell | *NIPAL1* | -2.753908439 | 0.000783484 | 0.00415349 | 1 |
| T2DM | b_cell | *ANTXR2* | -1.915861798 | 0.003484657 | 0.006957141 | 1 |
| T2DM | b_cell | *CGGBP1* | -1.2451399 | 0.020071308 | 0.027108123 | 1 |
| T2DM | b_cell | *DCST2* | -4.059286689 | 0.001014237 | 0.00415349 | 1 |
| T2DM | b_cell | *LMOD3* | -3.194602312 | 0.0007689 | 0.00415349 | 1 |
| T2DM | b_cell | *NBPF10* | 2.191623925 | 0.001399452 | 0.004344868 | 1 |
| T2DM | b_cell | *SLC22A15* | -1.781161277 | 0.001313826 | 0.004274973 | 1 |
| T2DM | b_cell | *SLC15A2* | 1.897214677 | 0.000754768 | 0.00415349 | 1 |
| T2DM | b_cell | *LRRC58* | -3.234927995 | 0.002934401 | 0.006222119 | 1 |
| T2DM | b_cell | *FSTL1* | -1.375422863 | 0.001206541 | 0.004235305 | 1 |
| T2DM | b_cell | *LMOD1* | -1.906568657 | 7.45E-05 | 0.00415349 | 0.468584183 |

| | | | | | | |
|---|---|---|---|---|---|---|
| T2DM | b_cell | *TMEM183A* | -3.399905733 | 0.0012072 | 0.004235305 | 1 |
| T2DM | b_cell | *IGFBP7* | -1.883961302 | 0.001277684 | 0.004259172 | 1 |
| T2DM | b_cell | *ARPC2* | -2.770623491 | 0.011022912 | 0.016348443 | 1 |
| T2DM | b_cell | *GLB1L* | 3.460743273 | 0.005075396 | 0.009021491 | 1 |
| T2DM | b_cell | *NUAK2* | 1.007148654 | 0.000537553 | 0.00415349 | 1 |
| T2DM | b_cell | *SPTA1* | -2.72375554 | 9.67E-06 | 0.001520543 | 0.060821708 |
| T2DM | b_cell | *PYHIN1* | -2.11309112 | 0.00020172 | 0.00415349 | 1 |
| T2DM | b_cell | *AIM2* | 2.259900685 | 0.000719332 | 0.00415349 | 1 |
| T2DM | b_cell | *EFHB* | -1.025024174 | 0.016826764 | 0.023337715 | 1 |
| T2DM | b_cell | *ICA1L* | -2.920860187 | 0.000141636 | 0.00415349 | 0.890465267 |
| T2DM | b_cell | *CD200R1* | -2.008518584 | 0.000563676 | 0.00415349 | 1 |
| T2DM | b_cell | *GTPBP8* | -2.510758402 | 0.001144349 | 0.00418468 | 1 |
| T2DM | b_cell | *SPICE1* | -2.664072524 | 0.004353743 | 0.008107986 | 1 |
| T2DM | b_cell | *CADPS* | 3.705738002 | 0.001756031 | 0.004789372 | 1 |
| T2DM | b_cell | *GMPS* | -3.217200516 | 0.007987623 | 0.012595482 | 1 |
| T2DM | b_cell | *DCLK3* | 4.244141066 | 0.000323872 | 0.00415349 | 1 |
| T2DM | b_cell | *DNASE1L3* | -2.950979415 | 0.002520982 | 0.005742541 | 1 |
| T2DM | b_cell | *CRELD1* | -2.22506811 | 0.004173246 | 0.007876672 | 1 |
| T2DM | b_cell | *PCOLCE2* | -1.199785169 | 0.000847194 | 0.00415349 | 1 |
| T2DM | b_cell | *TTC14* | -2.305309768 | 0.008347185 | 0.013034961 | 1 |
| T2DM | b_cell | *RCHY1* | -3.456846119 | 0.030822047 | 0.039518817 | 1 |
| T2DM | b_cell | *CCDC158* | -2.556563937 | 0.002233582 | 0.005401538 | 1 |
| T2DM | b_cell | *GYG1* | -3.346548651 | 0.003959972 | 0.007597923 | 1 |
| T2DM | b_cell | *RYK* | -2.5000412 | 0.00490877 | 0.008837754 | 1 |
| T2DM | b_cell | *PLB1* | -2.549462822 | 0.00077854 | 0.00415349 | 1 |
| T2DM | b_cell | *CDCP1* | -2.601469265 | 0.000773297 | 0.00415349 | 1 |
| T2DM | b_cell | *LZTFL1* | -2.088392653 | 0.000777402 | 0.00415349 | 1 |
| T2DM | b_cell | *FYCO1* | -3.488070695 | 0.005177374 | 0.009138167 | 1 |
| T2DM | b_cell | *LRRC2* | -1.77326771 | 0.001760086 | 0.004789372 | 1 |
| T2DM | b_cell | *DTX3L* | -2.945853979 | 0.016028412 | 0.022383524 | 1 |
| T2DM | b_cell | *NMNAT3* | -2.597184999 | 0.011352317 | 0.016738277 | 1 |
| T2DM | b_cell | *SMIM12* | -3.021864568 | 0.002070217 | 0.005222896 | 1 |
| T2DM | b_cell | *YEATS2* | -2.999820333 | 0.010368051 | 0.015571891 | 1 |
| T2DM | b_cell | *MEAF6* | -3.237976499 | 0.002932016 | 0.006221257 | 1 |
| T2DM | b_cell | *DNALI1* | -3.689811156 | 0.003004152 | 0.006308941 | 1 |
| T2DM | b_cell | *RPN1* | -3.591876745 | 0.016857399 | 0.023364741 | 1 |
| T2DM | b_cell | *RFC4* | 2.006990758 | 0.000555692 | 0.00415349 | 1 |
| T2DM | b_cell | *RFT1* | -4.921638407 | 2.12E-05 | 0.002553516 | 0.133380651 |
| T2DM | b_cell | *GNL3* | -2.870121453 | 0.001657898 | 0.004678277 | 1 |
| T2DM | b_cell | *FAM208A* | -2.797250515 | 0.022672093 | 0.030205435 | 1 |
| T2DM | b_cell | *LRPAP1* | -3.656716989 | 0.011313075 | 0.016688246 | 1 |
| T2DM | b_cell | *SLC51A* | -2.146178503 | 0.000819258 | 0.00415349 | 1 |
| T2DM | b_cell | *PIGX* | -3.170227873 | 0.001227113 | 0.004253251 | 1 |
| T2DM | b_cell | *ABLIM2* | -2.146998425 | 0.000824886 | 0.00415349 | 1 |
| T2DM | b_cell | *EXO5* | 2.755354151 | 0.030570739 | 0.039234043 | 1 |
| T2DM | b_cell | *ERMAP* | -2.575746355 | 0.001222164 | 0.004252746 | 1 |
| T2DM | b_cell | *METAP1* | -3.71957509 | 0.005599561 | 0.009655634 | 1 |
| T2DM | b_cell | *SLC9B2* | -2.707869426 | 0.000678038 | 0.00415349 | 1 |
| T2DM | b_cell | *BDH2* | -2.139842978 | 2.13E-05 | 0.002553516 | 0.134178794 |
| T2DM | b_cell | *ZNF589* | -3.520468317 | 0.001234435 | 0.004253251 | 1 |

| | | | | | | |
|---|---|---|---|---|---|---|
| T2DM | b_cell | *ATRIP* | -7.433397301 | 0.00040168 | 0.00415349 | 1 |
| T2DM | b_cell | *SHISA5* | -3.013563685 | 0.001453341 | 0.004416445 | 1 |
| T2DM | b_cell | *RNF123* | 2.866237907 | 0.026702554 | 0.03477915 | 1 |
| T2DM | b_cell | *RAD54L2* | -2.912685577 | 0.005828553 | 0.009935761 | 1 |
| T2DM | b_cell | *GRM2* | 4.121973072 | 0.000576181 | 0.00415349 | 1 |
| T2DM | b_cell | *POC1A* | 4.83035528 | 0.001021429 | 0.00415349 | 1 |
| T2DM | b_cell | *PPM1M* | -3.389440043 | 0.000965907 | 0.00415349 | 1 |
| T2DM | b_cell | *C4orf3* | -3.346885604 | 0.002773716 | 0.006030781 | 1 |
| T2DM | b_cell | *ANXA5* | -2.192132412 | 0.003479743 | 0.006953955 | 1 |
| T2DM | b_cell | *MAP9* | -2.480783977 | 0.000766969 | 0.00415349 | 1 |
| T2DM | b_cell | *CEP44* | -2.837120021 | 0.002380045 | 0.005562581 | 1 |
| T2DM | b_cell | *C4orf45* | -1.995600795 | 0.000499439 | 0.00415349 | 1 |
| T2DM | b_cell | *TMEM144* | -1.799267204 | 0.000901271 | 0.00415349 | 1 |
| T2DM | b_cell | *NPY1R* | -1.350808901 | 0.00076854 | 0.00415349 | 1 |
| T2DM | b_cell | *IL15* | -2.446743384 | 0.001344837 | 0.004286796 | 1 |
| T2DM | b_cell | *ARFIP1* | -2.536413612 | 0.007780077 | 0.012350455 | 1 |
| T2DM | b_cell | *OTUD4* | -3.189547513 | 0.010347527 | 0.015549526 | 1 |
| T2DM | b_cell | *ITGA2* | -2.540607821 | 0.000893056 | 0.00415349 | 1 |
| T2DM | b_cell | *NDUFAF2* | -2.392651921 | 0.000786062 | 0.00415349 | 1 |
| T2DM | b_cell | *LMBRD2* | -3.252194422 | 0.003122704 | 0.006466219 | 1 |
| T2DM | b_cell | *PGGT1B* | -3.169955677 | 0.035086008 | 0.044356673 | 1 |
| T2DM | b_cell | *CMBL* | -2.345820088 | 0.000783093 | 0.00415349 | 1 |
| T2DM | b_cell | *C5orf63* | -2.347816582 | 0.000653531 | 0.00415349 | 1 |
| T2DM | b_cell | *PRRC1* | -2.443111921 | 0.002511424 | 0.005734251 | 1 |
| T2DM | b_cell | *F2RL1* | -5.779156681 | 0.03414589 | 0.043298752 | 1 |
| T2DM | b_cell | *WDR41* | -2.272679068 | 0.000512185 | 0.00415349 | 1 |
| T2DM | b_cell | *NDUFS4* | -2.579677453 | 0.003635276 | 0.007180327 | 1 |
| T2DM | b_cell | *RHOBTB3* | -1.054529397 | 0.002244464 | 0.005416869 | 1 |
| T2DM | b_cell | *TIGD6* | -3.795057536 | 0.000823477 | 0.00415349 | 1 |
| T2DM | b_cell | *SERINC5* | -2.424228717 | 0.004158923 | 0.007865091 | 1 |
| T2DM | b_cell | *ENPP6* | -7.705723322 | 0.002006785 | 0.005130808 | 1 |
| T2DM | b_cell | *PRIMPOL* | -3.067774984 | 0.001048055 | 0.00415349 | 1 |
| T2DM | b_cell | *ERAP1* | -1.846325352 | 0.002290315 | 0.005472904 | 1 |
| T2DM | b_cell | *RICTOR* | -2.510606059 | 0.012122359 | 0.01765422 | 1 |
| T2DM | b_cell | *ANKRA2* | -2.448032537 | 0.001231509 | 0.004253251 | 1 |
| T2DM | b_cell | *UBLCP1* | -2.576056874 | 0.008249352 | 0.012911047 | 1 |
| T2DM | b_cell | *UTP15* | -3.271224769 | 0.000799577 | 0.00415349 | 1 |
| T2DM | b_cell | *TLR3* | -2.431320791 | 0.000912937 | 0.00415349 | 1 |
| T2DM | b_cell | *CCDC127* | -3.26174108 | 0.00112476 | 0.004182741 | 1 |
| T2DM | b_cell | *GRIK2* | -1.953816674 | 0.000419027 | 0.00415349 | 1 |
| T2DM | b_cell | *MB21D1* | 3.778320011 | 0.011267455 | 0.016632658 | 1 |
| T2DM | b_cell | *TXLNB* | -6.095416346 | 0.004407258 | 0.008188071 | 1 |
| T2DM | b_cell | *DCBLD1* | -1.838173009 | 0.001707286 | 0.004728644 | 1 |
| T2DM | b_cell | *TMEM200A* | -2.999512917 | 0.001259763 | 0.004257394 | 1 |
| T2DM | b_cell | *PDSS2* | -2.480820298 | 0.026045045 | 0.034020664 | 1 |
| T2DM | b_cell | *STXBP5* | -2.466501249 | 0.020611253 | 0.027771742 | 1 |
| T2DM | b_cell | *PI16* | -1.400657356 | 0.000866259 | 0.00415349 | 1 |
| T2DM | b_cell | *STK17A* | -3.047714246 | 0.003121954 | 0.006466219 | 1 |
| T2DM | b_cell | *GALNT10* | -3.017711851 | 0.023081083 | 0.030691788 | 1 |
| T2DM | b_cell | *SAP30L* | -3.791789199 | 0.002779951 | 0.006035067 | 1 |

| | | | | | | |
|---|---|---|---|---|---|---|
| T2DM | b_cell | *HCN1* | -3.207414741 | 0.001833909 | 0.004898539 | 1 |
| T2DM | b_cell | *RP9* | -4.504492301 | 0.003354028 | 0.006777924 | 1 |
| T2DM | b_cell | *KCNK5* | -4.214872308 | 0.000786096 | 0.00415349 | 1 |
| T2DM | b_cell | *KIAA1324L* | -1.844101855 | 0.000449069 | 0.00415349 | 1 |
| T2DM | b_cell | *USP49* | -2.548095777 | 0.002048621 | 0.00519116 | 1 |
| T2DM | b_cell | *ZNF704* | -1.542340084 | 0.001323809 | 0.004274973 | 1 |
| T2DM | b_cell | *FNDC1* | -1.842462357 | 0.001607452 | 0.004610639 | 1 |
| T2DM | b_cell | *BRI3* | -2.713560286 | 0.019796821 | 0.02677767 | 1 |
| T2DM | b_cell | *LMTK2* | -2.79585164 | 0.005049711 | 0.009000686 | 1 |
| T2DM | b_cell | *CTSB* | -2.051827706 | 0.002889921 | 0.006184116 | 1 |
| T2DM | b_cell | *PEX2* | -2.652755053 | 0.001237387 | 0.004253251 | 1 |
| T2DM | b_cell | *TNFRSF11B* | -2.75024945 | 0.001031673 | 0.00415349 | 1 |
| T2DM | b_cell | *DEFB1* | -4.217548622 | 0.000543858 | 0.00415349 | 1 |
| T2DM | b_cell | *SUN1* | -1.905892029 | 0.008797654 | 0.013615691 | 1 |
| T2DM | b_cell | *OXR1* | -2.178993531 | 0.010601265 | 0.015842679 | 1 |
| T2DM | b_cell | *TMEM184A* | 1.698552115 | 0.001121097 | 0.004182741 | 1 |
| T2DM | b_cell | *SLC4A2* | -4.574966016 | 0.007308207 | 0.011730073 | 1 |
| T2DM | b_cell | *FASTK* | -2.257749839 | 0.000841612 | 0.00415349 | 1 |
| T2DM | b_cell | *ALDH7A1* | -2.296368578 | 0.008000943 | 0.012610159 | 1 |
| T2DM | b_cell | *COX6C* | -2.82686743 | 0.00208212 | 0.005230315 | 1 |
| T2DM | b_cell | *YWHAZ* | -2.245854936 | 0.02175749 | 0.02909866 | 1 |
| T2DM | b_cell | *SLC25A32* | -2.968790525 | 0.001215053 | 0.004245409 | 1 |
| T2DM | b_cell | *DCAF13* | -3.892963743 | 0.008633724 | 0.013405834 | 1 |
| T2DM | b_cell | *INTS8* | -2.295534143 | 0.006535255 | 0.010781199 | 1 |
| T2DM | b_cell | *FREM1* | -1.272757416 | 0.000591027 | 0.00415349 | 1 |
| T2DM | b_cell | *RPP25L* | 5.569348884 | 0.035607032 | 0.044961118 | 1 |
| T2DM | b_cell | *FAM219A* | -3.144339013 | 0.014625529 | 0.020706828 | 1 |
| T2DM | b_cell | *TMEM65* | -3.044931398 | 0.015135458 | 0.021316448 | 1 |
| T2DM | b_cell | *CCDC171* | -2.529262579 | 0.00113101 | 0.004182741 | 1 |
| T2DM | b_cell | *NIPSNAP3B* | -1.831774639 | 0.000424439 | 0.00415349 | 1 |
| T2DM | b_cell | *TMC1* | -2.629606823 | 0.00130479 | 0.004274973 | 1 |
| T2DM | b_cell | *HGSNAT* | -2.600803 | 0.028590828 | 0.036978098 | 1 |
| T2DM | b_cell | *C9orf64* | -2.227099126 | 0.002693561 | 0.005942501 | 1 |
| T2DM | b_cell | *SVEP1* | -1.125578516 | 0.000874529 | 0.00415349 | 1 |
| T2DM | b_cell | *ZNF367* | 4.431814718 | 0.000497619 | 0.00415349 | 1 |
| T2DM | b_cell | *NOL6* | -3.686759344 | 0.001572548 | 0.004562116 | 1 |
| T2DM | b_cell | *AQP3* | -4.867149376 | 0.004108346 | 0.007801017 | 1 |
| T2DM | b_cell | *TRMT10B* | -2.940868735 | 0.005048917 | 0.009000686 | 1 |
| T2DM | b_cell | *VCP* | -2.777145047 | 0.010725263 | 0.015993768 | 1 |
| T2DM | b_cell | *PIGO* | -4.246827092 | 0.000884044 | 0.00415349 | 1 |
| T2DM | b_cell | *LRFN5* | -1.453359344 | 0.004081215 | 0.007761223 | 1 |
| T2DM | b_cell | *WRN* | -2.405709336 | 0.003815588 | 0.007388059 | 1 |
| T2DM | b_cell | *CFL2* | 3.935455943 | 0.018741294 | 0.025536739 | 1 |
| T2DM | b_cell | *ZCCHC24* | -2.010147724 | 0.001255966 | 0.004257394 | 1 |
| T2DM | b_cell | *PGM2L1* | 3.865984847 | 0.001323971 | 0.004274973 | 1 |
| T2DM | b_cell | *GJB2* | 5.572377096 | 0.006565229 | 0.010819291 | 1 |
| T2DM | b_cell | *CRYL1* | -3.192847759 | 0.004593753 | 0.008454603 | 1 |
| T2DM | b_cell | *REEP3* | -2.601216446 | 0.000408795 | 0.00415349 | 1 |
| T2DM | b_cell | *LRR1* | -2.745401115 | 0.001222318 | 0.004252746 | 1 |
| T2DM | b_cell | *RPL36AL* | -2.776170023 | 0.028924735 | 0.037325494 | 1 |

| | | | | | | |
|---|---|---|---|---|---|---|
| T2DM | b_cell | *KLHDC2* | -2.438334042 | 0.00421434 | 0.007929499 | 1 |
| T2DM | b_cell | *AKR1E2* | -2.310217063 | 0.000192899 | 0.00415349 | 1 |
| T2DM | b_cell | *ATP5C1* | -3.332728149 | 0.007659466 | 0.012197332 | 1 |
| T2DM | b_cell | *TAF3* | -2.490771772 | 0.014840372 | 0.02096661 | 1 |
| T2DM | b_cell | *VDAC2* | -2.23516037 | 0.00142502 | 0.004380342 | 1 |
| T2DM | b_cell | *SLC18A2* | -2.591810591 | 0.001861942 | 0.004937169 | 1 |
| T2DM | b_cell | *PDZD8* | -2.036308838 | 0.017737795 | 0.024412767 | 1 |
| T2DM | b_cell | *QSOX2* | 2.204167545 | 0.000837813 | 0.00415349 | 1 |
| T2DM | b_cell | *CLEC1B* | 1.345831876 | 0.016000533 | 0.022359491 | 1 |
| T2DM | b_cell | *SNAPC4* | -2.626873309 | 0.015312201 | 0.021517168 | 1 |
| T2DM | b_cell | *SDCCAG3* | -3.508633574 | 0.003873656 | 0.007472745 | 1 |
| T2DM | b_cell | *AK8* | -3.999094696 | 0.001136398 | 0.00418468 | 1 |
| T2DM | b_cell | *TSC1* | -2.936996873 | 0.011899085 | 0.017381401 | 1 |
| T2DM | b_cell | *ZMYND19* | -6.522962419 | 0.001864638 | 0.004938071 | 1 |
| T2DM | b_cell | *STOX1* | -1.392801944 | 0.002050552 | 0.005192035 | 1 |
| T2DM | b_cell | *RET* | 1.979515268 | 0.000768399 | 0.00415349 | 1 |
| T2DM | b_cell | *STK32C* | -2.632518892 | 0.00091926 | 0.00415349 | 1 |
| T2DM | b_cell | *NSMF* | -2.77107269 | 0.000836352 | 0.00415349 | 1 |
| T2DM | b_cell | *METTL3* | -3.120446443 | 0.002784247 | 0.006042306 | 1 |
| T2DM | b_cell | *TRUB1* | -3.430381187 | 0.00234045 | 0.005517214 | 1 |
| T2DM | b_cell | *HSPA12A* | -1.286653014 | 0.003152001 | 0.006503654 | 1 |
| T2DM | b_cell | *UBTD1* | -2.700221997 | 0.001434754 | 0.004395857 | 1 |
| T2DM | b_cell | *ARHGAP42* | -1.646316269 | 0.001001437 | 0.00415349 | 1 |
| T2DM | b_cell | *OTOGL* | -8.497172763 | 0.0155089 | 0.021774108 | 1 |
| T2DM | b_cell | *TTC7B* | -2.208529533 | 0.001078247 | 0.00415349 | 1 |
| T2DM | b_cell | *SLC39A13* | -3.427430024 | 0.00156479 | 0.004548235 | 1 |
| T2DM | b_cell | *PSMC3* | -2.505263935 | 0.002825023 | 0.006097224 | 1 |
| T2DM | b_cell | *TC2N* | -1.465891003 | 0.000765772 | 0.00415349 | 1 |
| T2DM | b_cell | *CLMN* | -1.50701031 | 0.000836408 | 0.00415349 | 1 |
| T2DM | b_cell | *PTER* | -2.68246924 | 0.001315702 | 0.004274973 | 1 |
| T2DM | b_cell | *ABTB2* | -2.454845941 | 0.001087602 | 0.00415349 | 1 |
| T2DM | b_cell | *LIPC* | -2.404468431 | 0.001544764 | 0.004515077 | 1 |
| T2DM | b_cell | *CEP57* | -3.523061214 | 0.00118213 | 0.004202203 | 1 |
| T2DM | b_cell | *TCP11L2* | -2.988055253 | 0.003787627 | 0.007350767 | 1 |
| T2DM | b_cell | *JAM3* | -2.777092669 | 0.004815829 | 0.008733839 | 1 |
| T2DM | b_cell | *GPT2* | -1.703801376 | 0.001118523 | 0.004182741 | 1 |
| T2DM | b_cell | *IKBIP* | -3.294910048 | 0.006820265 | 0.01114609 | 1 |
| T2DM | b_cell | *HIF1AN* | -2.790707523 | 0.001203847 | 0.004229472 | 1 |
| T2DM | b_cell | *NDUFB8* | -4.198128827 | 0.001886516 | 0.004966719 | 1 |
| T2DM | b_cell | *AVPR1A* | -2.386347968 | 0.009064151 | 0.013933085 | 1 |
| T2DM | b_cell | *TRMT61A* | -8.550116694 | 3.55E-07 | 0.000124872 | 0.002231469 |
| T2DM | b_cell | *BTRC* | -2.156920319 | 0.002356569 | 0.005540753 | 1 |
| T2DM | b_cell | *LARP6* | -1.996875869 | 0.001213924 | 0.004245409 | 1 |
| T2DM | b_cell | *ASPG* | -9.034077269 | 0.001536611 | 0.004508014 | 1 |
| T2DM | b_cell | *GABRB3* | 4.326163425 | 0.004097785 | 0.007790375 | 1 |
| T2DM | b_cell | *TBATA* | -13.56933477 | 0.038986494 | 0.048758323 | 1 |
| T2DM | b_cell | *PCBD1* | -3.788448162 | 6.05E-07 | 0.000190271 | 0.003805422 |
| T2DM | b_cell | *COX11* | -2.09902807 | 0.001096807 | 0.00415349 | 1 |
| T2DM | b_cell | *ZNF202* | -4.288029109 | 0.021928606 | 0.029295611 | 1 |
| T2DM | b_cell | *FAM227B* | -1.774502554 | 0.002943343 | 0.006236871 | 1 |

| | | | | | | |
|---|---|---|---|---|---|---|
| T2DM | b_cell | *ILK* | -2.325682492 | 0.002301225 | 0.005481934 | 1 |
| T2DM | b_cell | *TAF10* | -2.847645603 | 0.000749219 | 0.00415349 | 1 |
| T2DM | b_cell | *DCHS1* | -2.612448438 | 0.000965165 | 0.00415349 | 1 |
| T2DM | b_cell | *NETO1* | 0.641539089 | 0.000512436 | 0.00415349 | 1 |
| T2DM | b_cell | *USP54* | -2.165997368 | 0.002766871 | 0.006028396 | 1 |
| T2DM | b_cell | *C11orf74* | -1.977205577 | 0.000558391 | 0.00415349 | 1 |
| T2DM | b_cell | *CYB5R2* | 2.56911625 | 0.000958303 | 0.00415349 | 1 |
| T2DM | b_cell | *TUB* | -2.752316769 | 0.000232335 | 0.00415349 | 1 |
| T2DM | b_cell | *RIC3* | -3.352876182 | 0.001296872 | 0.004268027 | 1 |
| T2DM | b_cell | *LMO1* | -5.656328835 | 0.001693844 | 0.004705788 | 1 |
| T2DM | b_cell | *XRRA1* | -2.281747068 | 0.000540991 | 0.00415349 | 1 |
| T2DM | b_cell | *PRTG* | -3.323481797 | 0.001441289 | 0.004405146 | 1 |
| T2DM | b_cell | *AKIP1* | -4.847108576 | 0.00660203 | 0.010862853 | 1 |
| T2DM | b_cell | *C16orf46* | -35.38745058 | 0.000463193 | 0.00415349 | 1 |
| T2DM | b_cell | *TMEM41B* | -2.828289722 | 0.002963502 | 0.006266914 | 1 |
| T2DM | b_cell | *TMX3* | -2.930247931 | 0.004229105 | 0.007948694 | 1 |
| T2DM | b_cell | *CCDC68* | -2.569607989 | 0.002088819 | 0.005231352 | 1 |
| T2DM | b_cell | *ZNF3* | -3.048817594 | 0.002056553 | 0.005198853 | 1 |
| T2DM | b_cell | *CLEC4D* | -7.960683487 | 0.011037813 | 0.016358965 | 1 |
| T2DM | b_cell | *ZSCAN21* | -1.803699899 | 0.024498653 | 0.032242628 | 1 |
| T2DM | b_cell | *RIMKLB* | -2.140170682 | 0.001022636 | 0.00415349 | 1 |
| T2DM | b_cell | *BEAN1* | -2.767302277 | 0.001687926 | 0.004704627 | 1 |
| T2DM | b_cell | *TK2* | -2.2856759 | 0.000216071 | 0.00415349 | 1 |
| T2DM | b_cell | *SEC11C* | 3.383482165 | 0.001435945 | 0.004397363 | 1 |
| T2DM | b_cell | *GALR1* | -3.843978373 | 0.000600384 | 0.00415349 | 1 |
| T2DM | b_cell | *CENPV* | -2.285077786 | 0.000731652 | 0.00415349 | 1 |
| T2DM | b_cell | *SPDYE6* | -4.616367671 | 0.005846771 | 0.009953602 | 1 |
| T2DM | b_cell | *ATF7IP2* | -2.865688839 | 0.005266788 | 0.009257002 | 1 |
| T2DM | b_cell | *ZNF592* | -2.418921866 | 0.001118424 | 0.004182741 | 1 |
| T2DM | b_cell | *ACSM1* | 1.378690801 | 0.000494287 | 0.00415349 | 1 |
| T2DM | b_cell | *AP1G1* | -2.780561479 | 0.004297016 | 0.008039295 | 1 |
| T2DM | b_cell | *SLFN5* | -2.115691352 | 0.001055094 | 0.00415349 | 1 |
| T2DM | b_cell | *CATSPER2* | -2.540643836 | 0.002029404 | 0.005163442 | 1 |
| T2DM | b_cell | *C16orf45* | -2.687632747 | 0.00473145 | 0.008629715 | 1 |
| T2DM | b_cell | *SAAL1* | -5.924701085 | 0.000642557 | 0.00415349 | 1 |
| T2DM | b_cell | *YPEL4* | -6.995007687 | 0.006383132 | 0.010596976 | 1 |
| T2DM | b_cell | *FAM111A* | -2.698746042 | 0.00735995 | 0.011804082 | 1 |
| T2DM | b_cell | *SCNN1G* | -14.66349402 | 0.00178714 | 0.004835411 | 1 |
| T2DM | b_cell | *ANKDD1A* | -2.913043676 | 0.002081777 | 0.005230315 | 1 |
| T2DM | b_cell | *DCTN5* | -2.147333656 | 0.009060135 | 0.013931268 | 1 |
| T2DM | b_cell | *STX3* | -2.514440675 | 0.000973322 | 0.00415349 | 1 |
| T2DM | b_cell | *NYAP1* | -3.972106022 | 0.003085724 | 0.006415083 | 1 |
| T2DM | b_cell | *TSC22D4* | -2.045595228 | 0.001040041 | 0.00415349 | 1 |
| T2DM | b_cell | *MS4A6E* | -2.938392456 | 0.001333679 | 0.004280164 | 1 |
| T2DM | b_cell | *MS4A7* | -1.705713436 | 0.000306033 | 0.00415349 | 1 |
| T2DM | b_cell | *MS4A14* | -2.429595711 | 0.00033497 | 0.00415349 | 1 |
| T2DM | b_cell | *DIS3L* | -1.810511495 | 0.00103111 | 0.00415349 | 1 |
| T2DM | b_cell | *CCNDBP1* | -2.818539488 | 0.00160854 | 0.004610639 | 1 |
| T2DM | b_cell | *SMAD3* | -2.015814308 | 0.0026281 | 0.005859268 | 1 |
| T2DM | b_cell | *RCCD1* | -2.648750603 | 0.001021223 | 0.00415349 | 1 |

| | | | | | | |
|---|---|---|---|---|---|---|
| T2DM | b_cell | *AKTIP* | -3.396049106 | 0.003544883 | 0.007044747 | 1 |
| T2DM | b_cell | *EVA1C* | -1.488273268 | 0.005855005 | 0.009962224 | 1 |
| T2DM | b_cell | *CNPY4* | -1.901063628 | 0.036823804 | 0.046367165 | 1 |
| T2DM | b_cell | *NKX3-1* | -33.34760944 | 0.002878545 | 0.006170274 | 1 |
| T2DM | b_cell | *DUSP18* | -4.022616064 | 0.003216926 | 0.006598061 | 1 |
| T2DM | b_cell | *TEF* | -1.338191808 | 0.012651449 | 0.018318668 | 1 |
| T2DM | b_cell | *MEI1* | 3.170972575 | 0.001249068 | 0.004253251 | 1 |
| T2DM | b_cell | *PBX3* | -2.465518621 | 0.008820939 | 0.013645975 | 1 |
| T2DM | b_cell | *SNRPD1* | -2.387878533 | 0.000966304 | 0.00415349 | 1 |
| T2DM | b_cell | *URM1* | -3.936852231 | 0.001550822 | 0.004518081 | 1 |
| T2DM | b_cell | *CERCAM* | -1.837445966 | 0.004162739 | 0.007865091 | 1 |
| T2DM | b_cell | *ENDOG* | 2.12418002 | 0.006603811 | 0.010862941 | 1 |
| T2DM | b_cell | *ISLR2* | -3.680409448 | 0.002078338 | 0.005230315 | 1 |
| T2DM | b_cell | *SP2* | -3.775719899 | 0.001043838 | 0.00415349 | 1 |
| T2DM | b_cell | *GPRC5B* | -2.795059356 | 0.001200221 | 0.004223203 | 1 |
| T2DM | b_cell | *KATNAL2* | -1.354151022 | 0.003517476 | 0.006998218 | 1 |
| T2DM | b_cell | *RNF214* | -2.421693962 | 0.001574878 | 0.004562749 | 1 |
| T2DM | b_cell | *ENGASE* | -2.047105791 | 0.002821049 | 0.00609273 | 1 |
| T2DM | b_cell | *RBFOX3* | -3.834984253 | 0.001127387 | 0.004182741 | 1 |
| T2DM | b_cell | *MYO5B* | -1.524331543 | 0.011774766 | 0.017219808 | 1 |
| T2DM | b_cell | *RRM1* | -3.172374527 | 0.002410381 | 0.005604315 | 1 |
| T2DM | b_cell | *TRIM68* | -4.163539403 | 0.00055 | 0.00415349 | 1 |
| T2DM | b_cell | *FN3K* | -2.168015407 | 0.001067435 | 0.00415349 | 1 |
| T2DM | b_cell | *ZNF229* | -6.945447857 | 0.034097168 | 0.043245692 | 1 |
| T2DM | b_cell | *ZNF646* | -4.008907535 | 0.025630227 | 0.033542306 | 1 |
| T2DM | b_cell | *VKORC1* | -2.789114454 | 4.34E-05 | 0.003541347 | 0.27268372 |
| T2DM | b_cell | *SMG8* | -7.287317017 | 0.000873216 | 0.00415349 | 1 |
| T2DM | b_cell | *RAB8A* | -4.267612415 | 0.011485341 | 0.016917561 | 1 |
| T2DM | b_cell | *PROCA1* | -4.161139905 | 0.000732582 | 0.00415349 | 1 |
| T2DM | b_cell | *CACNB3* | 1.749311892 | 0.000942134 | 0.00415349 | 1 |
| T2DM | b_cell | *TP53I13* | -1.97422078 | 0.003428716 | 0.006885994 | 1 |
| T2DM | b_cell | *ZNF528* | -2.34022893 | 0.001027653 | 0.00415349 | 1 |
| T2DM | b_cell | *ZNF701* | -3.942312301 | 0.00670394 | 0.011004614 | 1 |
| T2DM | b_cell | *SERTAD3* | -5.849055589 | 0.002711665 | 0.005955133 | 1 |
| T2DM | b_cell | *TMC4* | -3.67925255 | 0.000942856 | 0.00415349 | 1 |
| T2DM | b_cell | *TTYH1* | -2.740147547 | 0.001473454 | 0.004442975 | 1 |
| T2DM | b_cell | *LENG8* | -2.320993244 | 0.002805346 | 0.006076587 | 1 |
| T2DM | b_cell | *CDC42EP5* | -2.924332291 | 0.000593952 | 0.00415349 | 1 |
| T2DM | b_cell | *ZNF283* | 3.501323872 | 0.014492226 | 0.020571828 | 1 |
| T2DM | b_cell | *PPP1R14A* | -5.359387124 | 0.011926991 | 0.017410028 | 1 |
| T2DM | b_cell | *SPINT2* | -2.351373407 | 0.000909341 | 0.00415349 | 1 |
| T2DM | b_cell | *UBXN6* | -3.144246838 | 0.000459418 | 0.00415349 | 1 |
| T2DM | b_cell | *ZNF444* | -2.625055473 | 0.000899207 | 0.00415349 | 1 |
| T2DM | b_cell | *GLOD4* | -3.718901827 | 0.011584862 | 0.017013322 | 1 |
| T2DM | b_cell | *KIFC2* | -2.789110931 | 9.20E-05 | 0.00415349 | 0.57822136 |
| T2DM | b_cell | *SERPINF2* | 5.100325797 | 0.008951194 | 0.0138101 | 1 |
| T2DM | b_cell | *CYB5D2* | -2.894339082 | 0.005082354 | 0.009028754 | 1 |
| T2DM | b_cell | *ZNF83* | -2.198919629 | 0.001643935 | 0.004657671 | 1 |
| T2DM | b_cell | *CD320* | -4.521274675 | 0.005910152 | 0.010034331 | 1 |
| T2DM | b_cell | *SPRYD3* | -2.780672648 | 0.000904163 | 0.00415349 | 1 |

| | | | | | | |
|---|---|---|---|---|---|---|
| T2DM | b_cell | *SOAT2* | -3.306504673 | 0.010723205 | 0.015993768 | 1 |
| T2DM | b_cell | *ZNF558* | -2.487943984 | 0.014919139 | 0.021068425 | 1 |
| T2DM | b_cell | *NDUFV1* | -2.793005036 | 0.003285421 | 0.006688938 | 1 |
| T2DM | b_cell | *CDK2AP2* | -3.812246461 | 0.001696409 | 0.004710831 | 1 |
| T2DM | b_cell | *NUDT8* | -3.322741153 | 0.007118573 | 0.011508915 | 1 |
| T2DM | b_cell | *MIS12* | -3.155435258 | 0.001328859 | 0.004274973 | 1 |
| T2DM | b_cell | *HID1* | -3.310637187 | 0.002429357 | 0.005628918 | 1 |
| T2DM | b_cell | *ATP5H* | -4.148624276 | 0.006754972 | 0.011068154 | 1 |
| T2DM | b_cell | *MGAT5B* | -7.944749639 | 0.013044908 | 0.018819031 | 1 |
| T2DM | b_cell | *TMC8* | -2.058301992 | 0.004783746 | 0.008694829 | 1 |
| T2DM | b_cell | *TMEM68* | -3.8829746 | 0.005161028 | 0.009122121 | 1 |
| T2DM | b_cell | *GSDMA* | -5.545198703 | 0.001324909 | 0.004274973 | 1 |
| T2DM | b_cell | *KRT24* | -7.355358998 | 0.000358361 | 0.00415349 | 1 |
| T2DM | b_cell | *ZNF598* | -2.995764444 | 0.000630196 | 0.00415349 | 1 |
| T2DM | b_cell | *MLST8* | -3.584912 | 0.000656686 | 0.00415349 | 1 |
| T2DM | b_cell | *ECI1* | -3.776986961 | 0.00582997 | 0.009935761 | 1 |
| T2DM | b_cell | *ABCA3* | -2.738885137 | 0.002189117 | 0.005349464 | 1 |
| T2DM | b_cell | *KCTD5* | -3.365059912 | 0.001680549 | 0.004704627 | 1 |
| T2DM | b_cell | *SDHAF2* | -5.092388171 | 0.002029168 | 0.005163442 | 1 |
| T2DM | b_cell | *DDB1* | -3.052233274 | 0.003710093 | 0.007257422 | 1 |
| T2DM | b_cell | *VPS37C* | -3.266622177 | 0.002365395 | 0.005545309 | 1 |
| T2DM | b_cell | *TRANK1* | -2.926346035 | 0.003494609 | 0.006968159 | 1 |
| T2DM | b_cell | *TTC21A* | -4.166944229 | 0.039342313 | 0.049173981 | 1 |
| T2DM | b_cell | *ULK4* | -1.639107024 | 0.000686705 | 0.00415349 | 1 |
| T2DM | b_cell | *LTBP3* | -1.457508225 | 0.026883641 | 0.034986017 | 1 |
| T2DM | b_cell | *SCARA5* | -1.501323706 | 0.000492836 | 0.00415349 | 1 |
| T2DM | b_cell | *COPS6* | -2.513517928 | 0.001036923 | 0.00415349 | 1 |
| T2DM | b_cell | *NUDT16L1* | 2.733383751 | 0.001015407 | 0.00415349 | 1 |
| T2DM | b_cell | *KIAA1586* | -2.251109034 | 0.00054008 | 0.00415349 | 1 |
| T2DM | b_cell | *RNF187* | 4.496429952 | 0.003478355 | 0.006953391 | 1 |
| T2DM | b_cell | *PTGDR* | -2.095506429 | 0.000855097 | 0.00415349 | 1 |
| T2DM | b_cell | *GLYCTK* | -2.345465986 | 0.00282507 | 0.006097224 | 1 |
| T2DM | b_cell | *UBTD2* | -2.11895697 | 0.002960407 | 0.006264583 | 1 |
| T2DM | b_cell | *NT5DC2* | -3.938637078 | 0.001480308 | 0.004457229 | 1 |
| T2DM | b_cell | *COA6* | 5.250817399 | 0.007236482 | 0.011651653 | 1 |
| T2DM | b_cell | *KIF5C* | -3.723376746 | 0.000212495 | 0.00415349 | 1 |
| T2DM | b_cell | *ACOX2* | -1.997396908 | 0.000704388 | 0.00415349 | 1 |
| T2DM | b_cell | *FAM107A* | -3.075648639 | 0.000606957 | 0.00415349 | 1 |
| T2DM | b_cell | *CX3CR1* | -2.523316889 | 0.000870942 | 0.00415349 | 1 |
| T2DM | b_cell | *SCN11A* | -1.238738905 | 0.006731862 | 0.01103604 | 1 |
| T2DM | b_cell | *SEPT2* | -1.621646868 | 0.013462555 | 0.019315172 | 1 |
| T2DM | b_cell | *DTYMK* | 4.828026847 | 0.010129726 | 0.015286987 | 1 |
| T2DM | b_cell | *TAP1* | -4.96687974 | 3.49E-05 | 0.003315867 | 0.219296058 |
| T2DM | b_cell | *ING5* | -2.526194456 | 0.001644668 | 0.004657671 | 1 |
| T2DM | b_cell | *MLKL* | -1.525663752 | 0.012167104 | 0.01770708 | 1 |
| T2DM | b_cell | *RHOH* | 1.263991466 | 0.000549116 | 0.00415349 | 1 |
| T2DM | b_cell | *KLHL30* | -4.213663181 | 0.001088562 | 0.00415349 | 1 |
| T2DM | b_cell | *REEP4* | 2.462199516 | 0.000796796 | 0.00415349 | 1 |
| T2DM | b_cell | *BMP1* | -2.396032388 | 0.001292711 | 0.004268027 | 1 |
| T2DM | b_cell | *ATXN2L* | -2.537963143 | 0.001090564 | 0.00415349 | 1 |

| | | | | | | |
|---|---|---|---|---|---|---|
| T2DM | b_cell | *SERINC2* | -3.885371573 | 0.002296784 | 0.005480032 | 1 |
| T2DM | b_cell | *TRAPPC11* | -3.058989805 | 0.011772169 | 0.017219808 | 1 |
| T2DM | b_cell | *TMEM223* | -3.392867571 | 0.000634388 | 0.00415349 | 1 |
| T2DM | b_cell | *SLC20A2* | -2.363201483 | 0.003566055 | 0.007076954 | 1 |
| T2DM | b_cell | *ZSWIM1* | 6.088752808 | 0.000190481 | 0.00415349 | 1 |
| T2DM | b_cell | *NBPF9* | -3.506286987 | 0.001276247 | 0.004257394 | 1 |
| T2DM | b_cell | *NDUFS5* | -4.053962008 | 0.001944974 | 0.005044575 | 1 |
| T2DM | b_cell | *ZNF30* | -2.524674514 | 0.004636397 | 0.008515638 | 1 |
| T2DM | b_cell | *FAM84B* | -2.316701197 | 0.037311678 | 0.046896946 | 1 |
| T2DM | b_cell | *SLC16A4* | -1.864213529 | 0.017163583 | 0.023726351 | 1 |
| T2DM | b_cell | *TMEM208* | -2.206627665 | 0.000458678 | 0.00415349 | 1 |
| T2DM | b_cell | *LRP1B* | -1.157151796 | 0.000480767 | 0.00415349 | 1 |
| T2DM | b_cell | *NPNT* | -3.224831023 | 0.001098807 | 0.00415349 | 1 |
| T2DM | b_cell | *GSTM4* | -1.750126474 | 0.017575932 | 0.024221807 | 1 |
| T2DM | b_cell | *TCTN2* | 1.973363045 | 0.02418809 | 0.031896659 | 1 |
| T2DM | b_cell | *ABHD15* | -1.993087712 | 0.006474648 | 0.0107065 | 1 |
| T2DM | b_cell | *SNTB2* | -2.348448725 | 0.002366113 | 0.005545309 | 1 |
| T2DM | b_cell | *ZNF507* | -3.351081746 | 0.000355352 | 0.00415349 | 1 |
| T2DM | b_cell | *STX18* | -2.926440964 | 0.008062967 | 0.012685655 | 1 |
| T2DM | b_cell | *ZBTB49* | -3.977304625 | 0.002903999 | 0.006192483 | 1 |
| T2DM | b_cell | *GFM1* | -2.890782663 | 0.02300233 | 0.030612966 | 1 |
| T2DM | b_cell | *TNIP2* | -3.897457699 | 0.000776515 | 0.00415349 | 1 |
| T2DM | b_cell | *ZNF608* | -1.546261814 | 0.00079049 | 0.00415349 | 1 |
| T2DM | b_cell | *LETM1* | -1.996950847 | 0.00078125 | 0.00415349 | 1 |
| T2DM | b_cell | *TMEM129* | -4.163590292 | 0.012339318 | 0.017916233 | 1 |
| T2DM | b_cell | *CEP120* | -2.987685905 | 0.002918703 | 0.006203477 | 1 |
| T2DM | b_cell | *STXBP6* | -1.532524683 | 0.000840979 | 0.00415349 | 1 |
| T2DM | b_cell | *MFF* | -3.143541712 | 0.006184674 | 0.010374344 | 1 |
| T2DM | b_cell | *LGALS9* | -4.121759117 | 0.035524358 | 0.044874752 | 1 |
| T2DM | b_cell | *CPLX1* | -2.85551265 | 0.002994585 | 0.006307188 | 1 |
| T2DM | b_cell | *FEM1B* | -2.742832377 | 0.021540639 | 0.028850873 | 1 |
| T2DM | b_cell | *MFSD7* | 3.546352843 | 0.002235572 | 0.005403707 | 1 |
| T2DM | b_cell | *COL4A3* | 2.860341047 | 0.006718916 | 0.011026318 | 1 |
| T2DM | b_cell | *IRS1* | -3.368038833 | 1.08E-16 | 6.80E-13 | 6.80E-13 |
| T2DM | b_cell | *UPF3A* | -2.01866161 | 0.001996421 | 0.005112626 | 1 |
| T2DM | b_cell | *HSPBAP1* | -1.875635013 | 0.005437863 | 0.009478105 | 1 |
| T2DM | b_cell | *PARM1* | -1.712313274 | 0.001509363 | 0.004480341 | 1 |
| T2DM | b_cell | *FAM110B* | -1.654005471 | 0.000857132 | 0.00415349 | 1 |
| T2DM | b_cell | *ATF5* | -2.690388418 | 0.000696379 | 0.00415349 | 1 |
| T2DM | b_cell | *CPT1C* | -4.100526893 | 0.002621733 | 0.005854674 | 1 |
| T2DM | b_cell | *PCSK9* | 5.422854789 | 0.000962224 | 0.00415349 | 1 |
| T2DM | b_cell | *RGS14* | -2.429967377 | 0.005889895 | 0.010005408 | 1 |
| T2DM | b_cell | *RAB24* | -4.677744855 | 0.00018973 | 0.00415349 | 1 |
| T2DM | b_cell | *CXCL10* | -2.413281643 | 0.031827394 | 0.040645709 | 1 |
| T2DM | b_cell | *SH3TC2* | -3.298129155 | 0.000985488 | 0.00415349 | 1 |
| T2DM | b_cell | *STK32A* | -1.965870733 | 0.000242198 | 0.00415349 | 1 |
| T2DM | b_cell | *P2RY12* | -1.97853172 | 0.000689466 | 0.00415349 | 1 |
| T2DM | b_cell | *SLC33A1* | -3.13583929 | 0.00384231 | 0.007424704 | 1 |
| T2DM | b_cell | *SNUPN* | -3.553910122 | 0.000599462 | 0.00415349 | 1 |
| T2DM | b_cell | *PTK2* | -2.146407776 | 0.003862778 | 0.007456335 | 1 |

| | | | | | | |
|---|---|---|---|---|---|---|
| T2DM | b_cell | *PTAFR* | -2.14117482 | 0.000910368 | 0.00415349 | 1 |
| T2DM | b_cell | *RNASE6* | 1.470463166 | 0.000620016 | 0.00415349 | 1 |
| T2DM | b_cell | *NPR1* | -1.84031771 | 0.000807284 | 0.00415349 | 1 |
| T2DM | b_cell | *CLIC4* | -2.435347735 | 0.002164179 | 0.005312843 | 1 |
| T2DM | b_cell | *SLC38A11* | 1.065156294 | 0.000780331 | 0.00415349 | 1 |
| T2DM | b_cell | *CCDC8* | -6.160424919 | 0.000691183 | 0.00415349 | 1 |
| T2DM | b_cell | *METTL15* | -2.528472261 | 0.003110516 | 0.006451934 | 1 |
| T2DM | b_cell | *HINT1* | -4.012235618 | 0.002421252 | 0.0056192 | 1 |
| T2DM | b_cell | *DTWD2* | -1.744314019 | 0.001076652 | 0.00415349 | 1 |
| T2DM | b_cell | *CLIC3* | 5.952970398 | 0.002610913 | 0.005849897 | 1 |
| T2DM | b_cell | *INO80E* | -5.458778195 | 0.000165813 | 0.00415349 | 1 |
| T2DM | b_cell | *DFFB* | -4.874492126 | 0.014512104 | 0.020590518 | 1 |
| T2DM | b_cell | *C15orf40* | -2.793918885 | 0.016160589 | 0.022543071 | 1 |
| T2DM | b_cell | *FAM103A1* | 1.303353977 | 0.004706601 | 0.008599361 | 1 |
| T2DM | b_cell | *RGPD8* | -1.220959464 | 0.030789555 | 0.039488766 | 1 |
| T2DM | b_cell | *LUZP1* | -2.909592738 | 0.002706092 | 0.005952834 | 1 |
| T2DM | b_cell | *HEXDC* | -2.795569868 | 0.003125033 | 0.006466219 | 1 |
| T2DM | b_cell | *LRRC45* | -3.726196792 | 3.95E-05 | 0.003541347 | 0.248645331 |
| T2DM | b_cell | *CHRNA5* | 3.779574329 | 0.009194141 | 0.01410187 | 1 |
| T2DM | b_cell | *FASN* | -1.431444065 | 0.003603985 | 0.007143208 | 1 |
| T2DM | b_cell | *MT1E* | -3.82986335 | 0.00054614 | 0.00415349 | 1 |
| T2DM | b_cell | *DCXR* | -4.425155379 | 0.001932797 | 0.005024672 | 1 |
| T2DM | b_cell | *LIMS1* | -1.679516242 | 0.001077267 | 0.00415349 | 1 |
| T2DM | b_cell | *UGP2* | -1.525126973 | 0.002113552 | 0.005250385 | 1 |
| T2DM | b_cell | *CSGALNACT2* | -2.552914247 | 0.002288964 | 0.005471754 | 1 |
| T2DM | b_cell | *AVEN* | -2.611134528 | 0.002162016 | 0.005312843 | 1 |
| T2DM | b_cell | *CALML6* | -7.176188695 | 0.013233032 | 0.019051081 | 1 |
| T2DM | b_cell | *TPST1* | -1.449364735 | 0.000463452 | 0.00415349 | 1 |
| T2DM | b_cell | *OTUD3* | -2.692851847 | 0.000253286 | 0.00415349 | 1 |
| T2DM | b_cell | *BRD3* | -4.016405697 | 0.004543279 | 0.008382656 | 1 |
| T2DM | b_cell | *ZFPM2* | -1.321468019 | 0.001067021 | 0.00415349 | 1 |
| T2DM | b_cell | *ZNF747* | -4.277521071 | 0.001320962 | 0.004274973 | 1 |
| T2DM | b_cell | *MAP3K2* | -2.561997363 | 0.01581583 | 0.022121051 | 1 |
| T2DM | b_cell | *PUSL1* | -5.482124315 | 0.000624447 | 0.00415349 | 1 |
| T2DM | b_cell | *MYO7B* | -3.017771833 | 0.000738407 | 0.00415349 | 1 |
| T2DM | b_cell | *CNTROB* | -3.134264337 | 0.000848592 | 0.00415349 | 1 |
| T2DM | b_cell | *KCNAB3* | -5.562463472 | 0.002022096 | 0.005157371 | 1 |
| T2DM | b_cell | *NIPA1* | -3.955468967 | 0.005397334 | 0.009426623 | 1 |
| T2DM | b_cell | *RNF150* | -1.093309929 | 0.001351883 | 0.00429474 | 1 |
| T2DM | b_cell | *HOXD4* | -7.820013601 | 0.000307827 | 0.00415349 | 1 |
| T2DM | b_cell | *CHRNB1* | 2.384008225 | 0.000932917 | 0.00415349 | 1 |
| T2DM | b_cell | *ANKK1* | -4.115082292 | 0.006245653 | 0.010429328 | 1 |
| T2DM | b_cell | *PDCD6IP* | -2.381501623 | 0.012335046 | 0.017914168 | 1 |
| T2DM | b_cell | *ZNF212* | -4.220965403 | 0.0241707 | 0.03189112 | 1 |
| T2DM | b_cell | *GLB1* | -2.882317429 | 0.001779519 | 0.004822342 | 1 |
| T2DM | b_cell | *FAXDC2* | -2.243780579 | 0.002332987 | 0.005509725 | 1 |
| T2DM | b_cell | *CRTAP* | -3.380132814 | 0.005081625 | 0.009028754 | 1 |
| T2DM | b_cell | *ELP5* | -4.923165983 | 0.001643201 | 0.004657671 | 1 |
| T2DM | b_cell | *STX8* | -2.476207186 | 0.007957757 | 0.012564143 | 1 |
| T2DM | b_cell | *NFRKB* | -3.034885116 | 0.01476652 | 0.020885739 | 1 |

| | | | | | | |
|---|---|---|---|---|---|---|
| T2DM | b_cell | *SEMA3E* | -1.700173492 | 0.001075501 | 0.00415349 | 1 |
| T2DM | b_cell | *DCLK2* | -2.270963715 | 0.000604414 | 0.00415349 | 1 |
| T2DM | b_cell | *GPRC5C* | -2.889950327 | 0.013938788 | 0.019907579 | 1 |
| T2DM | b_cell | *ADORA2B* | -3.296011195 | 0.001198097 | 0.004219852 | 1 |
| T2DM | b_cell | *MGMT* | -2.35989825 | 0.005474718 | 0.009521315 | 1 |
| T2DM | b_cell | *METTL7B* | -8.326542675 | 0.000765698 | 0.00415349 | 1 |
| T2DM | b_cell | *CD14* | -2.169773079 | 0.001232776 | 0.004253251 | 1 |
| T2DM | b_cell | *KRT72* | 5.638053088 | 0.006284364 | 0.010480053 | 1 |
| T2DM | b_cell | *HSD17B13* | -3.042683649 | 0.002434393 | 0.005628918 | 1 |
| T2DM | b_cell | *TMC7* | -2.839432059 | 0.000192501 | 0.00415349 | 1 |
| T2DM | b_cell | *SERPINB9* | -3.679444367 | 0.024164202 | 0.03188924 | 1 |
| T2DM | b_cell | *SMAGP* | -4.043444023 | 0.010479322 | 0.01569776 | 1 |
| T2DM | b_cell | *IRX2* | -2.29210819 | 0.005155636 | 0.00911667 | 1 |
| T2DM | b_cell | *EMB* | -1.931325218 | 2.36E-05 | 0.002703191 | 0.148675482 |
| T2DM | b_cell | *DLGAP1* | -1.656825976 | 0.00115883 | 0.004195495 | 1 |
| T2DM | b_cell | *NUDCD2* | -4.212551553 | 0.011487366 | 0.016917561 | 1 |
| T2DM | b_cell | *HSPA4* | -2.901339818 | 0.00221091 | 0.005381337 | 1 |
| T2DM | b_cell | *SLC26A5* | -3.213451923 | 0.029264903 | 0.037725743 | 1 |
| T2DM | b_cell | *COMMD5* | -4.957390299 | 0.001117814 | 0.004182741 | 1 |
| T2DM | b_cell | *GTSF1* | 6.305780694 | 0.00065822 | 0.00415349 | 1 |
| T2DM | b_cell | *ARMC10* | -3.058816575 | 0.017533904 | 0.024169186 | 1 |
| T2DM | b_cell | *RNF34* | -3.541888147 | 0.007795032 | 0.012363109 | 1 |
| T2DM | b_cell | *ACYP2* | -2.063696601 | 0.003998327 | 0.007647546 | 1 |
| T2DM | b_cell | *TRABD* | -2.718625873 | 0.030538842 | 0.039207208 | 1 |
| T2DM | b_cell | *TMEM133* | -6.352668511 | 0.000830663 | 0.00415349 | 1 |
| T2DM | b_cell | *TTLL6* | 5.541634661 | 0.001158502 | 0.004195495 | 1 |
| T2DM | b_cell | *HTRA3* | -1.475452601 | 0.002359769 | 0.005541976 | 1 |
| T2DM | b_cell | *FOXN2* | -2.3068902 | 0.002433261 | 0.005628918 | 1 |
| T2DM | b_cell | *KBTBD2* | -3.098365068 | 0.001339382 | 0.004286583 | 1 |
| T2DM | b_cell | *MTSS1* | -1.91099381 | 0.002103236 | 0.005241681 | 1 |
| T2DM | b_cell | *TSEN34* | 4.272901138 | 0.004820431 | 0.008738769 | 1 |
| T2DM | b_cell | *NDUFA3* | -2.594194528 | 0.000883932 | 0.00415349 | 1 |
| T2DM | b_cell | *NUDT6* | -1.793757308 | 0.001549574 | 0.004517818 | 1 |
| T2DM | b_cell | *ZNF160* | -2.85594945 | 0.005448898 | 0.009489535 | 1 |
| T2DM | b_cell | *ZNF415* | 2.068128543 | 0.026010256 | 0.033983059 | 1 |
| T2DM | b_cell | *FPR1* | -1.954467509 | 0.000772333 | 0.00415349 | 1 |
| T2DM | b_cell | *FEZ2* | -2.297417888 | 0.003299922 | 0.00670977 | 1 |
| T2DM | b_cell | *SOX7* | -3.955911498 | 0.001018789 | 0.00415349 | 1 |
| T2DM | b_cell | *C11orf24* | -2.817131439 | 0.000793626 | 0.00415349 | 1 |
| T2DM | b_cell | *ALK* | -1.920323556 | 0.000500875 | 0.00415349 | 1 |
| T2DM | b_cell | *GIMAP8* | -1.911180319 | 0.000316989 | 0.00415349 | 1 |
| T2DM | b_cell | *KCNMB3* | -1.537546476 | 0.004983931 | 0.008927058 | 1 |
| T2DM | b_cell | *MORN4* | -1.400565493 | 0.034719067 | 0.043936951 | 1 |
| T2DM | b_cell | *GRIK1* | -1.850624324 | 0.001062079 | 0.00415349 | 1 |
| T2DM | b_cell | *CLDN20* | -2.70590727 | 0.002324918 | 0.005505767 | 1 |
| T2DM | b_cell | *CANT1* | -3.340342407 | 0.001052402 | 0.00415349 | 1 |
| T2DM | b_cell | *CHST11* | -2.782038836 | 0.002026826 | 0.00516315 | 1 |
| T2DM | b_cell | *PGAM1* | -2.100313544 | 0.023156095 | 0.030765505 | 1 |
| T2DM | b_cell | *LURAP1* | -2.333057029 | 0.000576938 | 0.00415349 | 1 |
| T2DM | b_cell | *KCND3* | -2.383633489 | 0.002613837 | 0.005854361 | 1 |

| | | | | | | |
|---|---|---|---|---|---|---|
| T2DM | b_cell | *MRPL36* | -3.25331529 | 0.013994705 | 0.019969294 | 1 |
| T2DM | b_cell | *ZNF581* | -8.466770678 | 0.001023413 | 0.00415349 | 1 |
| T2DM | b_cell | *MCC* | -2.424226786 | 0.016900144 | 0.023413663 | 1 |
| T2DM | b_cell | *KRT27* | 3.694864004 | 0.000662622 | 0.00415349 | 1 |
| T2DM | b_cell | *DLK2* | -9.83558497 | 0.000494217 | 0.00415349 | 1 |
| T2DM | b_cell | *ZNF561* | -4.478226275 | 0.000168308 | 0.00415349 | 1 |
| T2DM | b_cell | *HOPX* | 8.552568498 | 0.019820346 | 0.026803725 | 1 |
| T2DM | b_cell | *RSL1D1* | -3.788788732 | 0.015187297 | 0.021375092 | 1 |
| T2DM | b_cell | *COL24A1* | -1.498475351 | 0.000293288 | 0.00415349 | 1 |
| T2DM | b_cell | *ETFDH* | -1.28463112 | 0.00695241 | 0.011300363 | 1 |
| T2DM | b_cell | *TBCA* | -2.679357644 | 0.014433173 | 0.020506522 | 1 |
| T2DM | b_cell | *MAP6* | -3.516798539 | 0.000694012 | 0.00415349 | 1 |
| T2DM | b_cell | *BCL2L1* | -2.362319191 | 0.003671678 | 0.007215955 | 1 |
| T2DM | b_cell | *PLRG1* | -3.847252913 | 0.004541664 | 0.008382656 | 1 |
| T2DM | b_cell | *NMUR1* | -3.107306577 | 0.000993378 | 0.00415349 | 1 |
| T2DM | b_cell | *P2RY6* | -2.718428866 | 0.000650733 | 0.00415349 | 1 |
| T2DM | b_cell | *TCEA2* | -2.421615437 | 0.00080789 | 0.00415349 | 1 |
| T2DM | b_cell | *ANO5* | -2.323262434 | 0.000761802 | 0.00415349 | 1 |
| T2DM | b_cell | *GPHN* | -1.936299425 | 0.000244817 | 0.00415349 | 1 |
| T2DM | b_cell | *VAT1L* | -3.111400796 | 0.001445466 | 0.004415364 | 1 |
| T2DM | b_cell | *CAMTA1* | -2.486689381 | 0.002605015 | 0.005849897 | 1 |
| T2DM | b_cell | *LGALS4* | 1.936083949 | 0.000988611 | 0.00415349 | 1 |
| T2DM | b_cell | *LRRC34* | -1.42941197 | 0.007928583 | 0.012530669 | 1 |
| T2DM | b_cell | *SPATA5L1* | -2.793506498 | 0.000380554 | 0.00415349 | 1 |
| T2DM | b_cell | *SYCE1* | 2.186376857 | 0.000937896 | 0.00415349 | 1 |
| T2DM | b_cell | *NXNL1* | -5.162806938 | 0.011990772 | 0.017490948 | 1 |
| T2DM | b_cell | *BCL2* | -1.188850372 | 0.001733203 | 0.004766687 | 1 |
| T2DM | b_cell | *RHNO1* | 1.503101969 | 0.000811829 | 0.00415349 | 1 |
| T2DM | b_cell | *METTL18* | -5.258700318 | 0.000521338 | 0.00415349 | 1 |
| T2DM | b_cell | *PWWP2B* | -3.652340708 | 0.00095668 | 0.00415349 | 1 |
| T2DM | b_cell | *ZNF570* | -6.02356882 | 0.004415285 | 0.00820056 | 1 |
| T2DM | b_cell | *NINJ2* | -3.050613999 | 0.003218745 | 0.006598061 | 1 |
| T2DM | b_cell | *FAM90A1* | -8.038741751 | 0.00078291 | 0.00415349 | 1 |
| T2DM | b_cell | *TRAPPC12* | -2.963825354 | 0.003270266 | 0.00666672 | 1 |
| T2DM | b_cell | *C3AR1* | -2.32496777 | 0.000342029 | 0.00415349 | 1 |
| T2DM | b_cell | *AQP4* | -3.106190298 | 0.000764357 | 0.00415349 | 1 |
| T2DM | b_cell | *TLN2* | -1.042961682 | 0.001676417 | 0.004704627 | 1 |
| T2DM | b_cell | *TVP23B* | -2.847567943 | 0.001310775 | 0.004274973 | 1 |
| T2DM | b_cell | *ATPAF2* | -2.512071692 | 0.000962246 | 0.00415349 | 1 |
| T2DM | b_cell | *CYP4F22* | -7.64966402 | 0.000113501 | 0.00415349 | 0.713579293 |
| T2DM | b_cell | *ZNF554* | -2.97338364 | 0.000130562 | 0.00415349 | 0.820845203 |
| T2DM | b_cell | *ANKRD20A4* | 2.598918476 | 0.000652692 | 0.00415349 | 1 |
| T2DM | b_cell | *LAMB2* | -2.22549115 | 7.63E-05 | 0.00415349 | 0.479852511 |
| T2DM | b_cell | *ORMDL3* | -1.831901776 | 0.001195986 | 0.004217141 | 1 |
| T2DM | b_cell | *SERF1A* | -13.53151723 | 0.000722249 | 0.00415349 | 1 |
| T2DM | b_cell | *LRRC15* | 2.098379034 | 0.000824906 | 0.00415349 | 1 |
| T2DM | b_cell | *MOB3A* | -2.612649335 | 0.000525045 | 0.00415349 | 1 |
| T2DM | b_cell | *NME6* | -1.862283154 | 0.02934067 | 0.03780791 | 1 |
| T2DM | b_cell | *SLFN12* | -2.426849647 | 0.009563583 | 0.014558413 | 1 |
| T2DM | b_cell | *FRMD3* | -1.62536412 | 0.000932139 | 0.00415349 | 1 |

| | | | | | | |
|---|---|---|---|---|---|---|
| T2DM | b_cell | *MTBP* | -3.248895372 | 0.002192705 | 0.005351554 | 1 |
| T2DM | b_cell | *ISG20* | 1.589025331 | 0.000703376 | 0.00415349 | 1 |
| T2DM | b_cell | *MBOAT1* | -2.390681455 | 0.001124062 | 0.004182741 | 1 |
| T2DM | b_cell | *C5orf34* | -2.507294857 | 0.000881471 | 0.00415349 | 1 |
| T2DM | b_cell | *C1QTNF4* | -2.676516417 | 4.31E-06 | 0.000935333 | 0.027124645 |
| T2DM | b_cell | *ZNF131* | -2.587097423 | 0.03739434 | 0.046977395 | 1 |
| T2DM | b_cell | *HINFP* | -3.059459205 | 0.001318046 | 0.004274973 | 1 |
| T2DM | b_cell | *SPTLC3* | -3.230141128 | 0.007354101 | 0.011797713 | 1 |
| T2DM | b_cell | *COPRS* | -4.294242744 | 0.011957265 | 0.017446118 | 1 |
| T2DM | b_cell | *CLEC12A* | -0.795779929 | 0.037205597 | 0.046791676 | 1 |
| T2DM | b_cell | *SUCLG2* | -2.260301835 | 0.028730961 | 0.037121157 | 1 |
| T2DM | b_cell | *CSDC2* | -5.59100548 | 0.003191345 | 0.006563292 | 1 |
| T2DM | b_cell | *RCAN2* | -1.462138142 | 0.000754679 | 0.00415349 | 1 |
| T2DM | b_cell | *IL16* | -1.80200288 | 0.000811995 | 0.00415349 | 1 |
| T2DM | b_cell | *PRSS27* | -9.806522803 | 0.018820092 | 0.025621897 | 1 |
| T2DM | b_cell | *FGGY* | -2.1664022 | 0.006180056 | 0.010371415 | 1 |
| T2DM | b_cell | *ZNF24* | -2.680955903 | 0.002045542 | 0.005187706 | 1 |
| T2DM | b_cell | *CARNS1* | -7.364324986 | 0.001158693 | 0.004195495 | 1 |
| T2DM | b_cell | *CTSW* | -2.067628321 | 5.95E-07 | 0.000190271 | 0.003742868 |
| T2DM | b_cell | *SNTG2* | -1.310054383 | 0.000548197 | 0.00415349 | 1 |
| T2DM | b_cell | *PDE3A* | -1.411726363 | 0.002560203 | 0.005798627 | 1 |
| T2DM | b_cell | *RAD9A* | -1.566049219 | 0.037475758 | 0.047037351 | 1 |
| T2DM | b_cell | *EFEMP2* | -2.07922349 | 0.001978489 | 0.005092436 | 1 |
| T2DM | b_cell | *AGAP5* | -79.57737126 | 0.000693314 | 0.00415349 | 1 |
| T2DM | b_cell | *ZMAT3* | -4.009156664 | 0.013184113 | 0.018998056 | 1 |
| T2DM | b_cell | *ZFAND4* | -2.663057569 | 0.002390976 | 0.00557776 | 1 |
| T2DM | b_cell | *SLFN11* | -2.679336581 | 0.002669768 | 0.005916798 | 1 |
| T2DM | b_cell | *CCL19* | -13.30335815 | 0.000516171 | 0.00415349 | 1 |
| T2DM | b_cell | *FUT10* | -2.779544201 | 0.011590521 | 0.017017657 | 1 |
| T2DM | b_cell | *EFCAB12* | -2.859364382 | 0.01162961 | 0.017055842 | 1 |
| T2DM | b_cell | *FAM192A* | -2.522527389 | 0.01249206 | 0.018108734 | 1 |
| T2DM | b_cell | *CBWD1* | -3.370935123 | 0.004472019 | 0.008274157 | 1 |
| T2DM | b_cell | *HOXC5* | 4.383294514 | 0.004417934 | 0.008201016 | 1 |
| T2DM | b_cell | *DCP2* | -2.969960511 | 0.012988651 | 0.018746476 | 1 |
| T2DM | b_cell | *SNX32* | 5.890521572 | 9.18E-06 | 0.00148026 | 0.057730126 |
| T2DM | b_cell | *CYP7B1* | -1.517644331 | 0.000665016 | 0.00415349 | 1 |
| T2DM | b_cell | *METAP1D* | -1.767131638 | 0.00087045 | 0.00415349 | 1 |
| T2DM | b_cell | *NADSYN1* | -2.895103025 | 0.001608994 | 0.004610639 | 1 |
| T2DM | b_cell | *MYEOV* | -2.122976137 | 3.63E-05 | 0.003315867 | 0.228051724 |
| T2DM | b_cell | *ANKRD13D* | -3.293586615 | 0.016907162 | 0.023418225 | 1 |
| T2DM | b_cell | *MRGPRF* | -4.412734251 | 0.038568858 | 0.048264811 | 1 |
| T2DM | b_cell | *ADH6* | -2.507460569 | 0.000624474 | 0.00415349 | 1 |
| T2DM | b_cell | *GXYLT2* | -1.686472538 | 0.000535297 | 0.00415349 | 1 |
| T2DM | b_cell | *HPSE2* | -1.49913753 | 0.013980745 | 0.019953903 | 1 |
| T2DM | b_cell | *DCAKD* | -2.165548958 | 0.013324859 | 0.019170112 | 1 |
| T2DM | b_cell | *HECTD4* | -2.387098877 | 0.006527552 | 0.010774145 | 1 |
| T2DM | b_cell | *BNC2* | -1.454721246 | 0.001536031 | 0.004508014 | 1 |
| T2DM | b_cell | *TRMT112* | -2.692859761 | 0.003433525 | 0.006885994 | 1 |
| T2DM | b_cell | *RHOD* | -1.844277087 | 0.027119038 | 0.035241297 | 1 |
| T2DM | b_cell | *AHSA2* | -2.763466275 | 0.001395763 | 0.00434199 | 1 |

| | | | | | | |
|---|---|---|---|---|---|---|
| T2DM | b_cell | *ABLIM3* | -1.449374925 | 0.000807139 | 0.00415349 | 1 |
| T2DM | b_cell | *VANGL1* | -2.274888835 | 0.001363274 | 0.004298348 | 1 |
| T2DM | b_cell | *GLRX* | -1.322441984 | 0.000793694 | 0.00415349 | 1 |
| T2DM | b_cell | *IQCB1* | -2.483109141 | 0.009757241 | 0.014813758 | 1 |
| T2DM | b_cell | *GOLGB1* | -2.382055642 | 0.006611723 | 0.010873111 | 1 |
| T2DM | b_cell | *DMRT2* | -2.136502189 | 0.00105446 | 0.00415349 | 1 |
| T2DM | b_cell | *ZNF483* | -1.992462545 | 0.002071965 | 0.005225209 | 1 |
| T2DM | b_cell | *SLC2A14* | -3.74175292 | 0.001262024 | 0.004257394 | 1 |
| T2DM | b_cell | *MZT2A* | 2.822140497 | 0.000658693 | 0.00415349 | 1 |
| T2DM | b_cell | *MAP3K11* | -3.731927164 | 0.003768765 | 0.007328867 | 1 |
| T2DM | b_cell | *C1QA* | -2.994995499 | 0.002547603 | 0.005790341 | 1 |
| T2DM | b_cell | *NDNF* | -2.966915477 | 0.000529361 | 0.00415349 | 1 |
| T2DM | b_cell | *OLR1* | 4.829195796 | 0.000540519 | 0.00415349 | 1 |
| T2DM | b_cell | *GLIPR1L1* | -1.738176191 | 0.00169113 | 0.004704627 | 1 |
| T2DM | b_cell | *ARV1* | 3.527261869 | 0.01674477 | 0.023244506 | 1 |
| T2DM | b_cell | *CCDC36* | -1.511663562 | 0.001230143 | 0.004253251 | 1 |
| T2DM | b_cell | *THAP2* | 3.151964749 | 0.005558368 | 0.009618899 | 1 |
| T2DM | b_cell | *PPP1R14B* | -6.078588003 | 0.015427458 | 0.021669444 | 1 |
| T2DM | b_cell | *SMARCC1* | -2.509717679 | 0.036872341 | 0.046409692 | 1 |
| T2DM | b_cell | *ZNF417* | -3.609441481 | 0.000512403 | 0.00415349 | 1 |
| T2DM | b_cell | *VEGFB* | -4.804863577 | 1.52E-06 | 0.000397014 | 0.009563996 |
| T2DM | b_cell | *PEAK1* | -2.231568086 | 0.019182876 | 0.026030082 | 1 |
| T2DM | b_cell | *TNFRSF10D* | -2.201146483 | 0.000539863 | 0.00415349 | 1 |
| T2DM | b_cell | *TNFRSF10C* | 2.851942428 | 0.004572703 | 0.008423258 | 1 |
| T2DM | b_cell | *GMPPB* | -3.134348779 | 0.002562116 | 0.005799421 | 1 |
| T2DM | b_cell | *CSPG4* | -2.193623073 | 0.001468551 | 0.004434572 | 1 |
| T2DM | b_cell | *SNX33* | -3.118198857 | 0.000831177 | 0.00415349 | 1 |
| T2DM | b_cell | *NUDT18* | -5.527737396 | 0.000769048 | 0.00415349 | 1 |
| T2DM | b_cell | *SULT1B1* | -3.274602188 | 0.005538546 | 0.009597457 | 1 |
| T2DM | b_cell | *NUDT4* | -2.357505633 | 0.000823504 | 0.00415349 | 1 |
| T2DM | b_cell | *SCAI* | -2.770869984 | 0.009007069 | 0.013869077 | 1 |
| T2DM | b_cell | *C1orf100* | -5.493302673 | 0.00104627 | 0.00415349 | 1 |
| T2DM | b_cell | *AGFG1* | -1.816815492 | 0.002716093 | 0.005958936 | 1 |
| T2DM | b_cell | *RNF213* | -2.390051547 | 0.00550685 | 0.009566611 | 1 |
| T2DM | b_cell | *PLK3* | -2.452715791 | 0.000965931 | 0.00415349 | 1 |
| T2DM | b_cell | *TUBB8* | -8.695605593 | 0.000209892 | 0.00415349 | 1 |
| T2DM | b_cell | *GOLIM4* | -3.06192802 | 0.007402345 | 0.011857075 | 1 |
| T2DM | b_cell | *USMG5* | -3.059902005 | 0.015964428 | 0.022318959 | 1 |
| T2DM | b_cell | *HOXB2* | -5.14770554 | 8.19E-06 | 0.001459962 | 0.051485995 |
| T2DM | b_cell | *XXYLT1* | -2.374239392 | 0.001345537 | 0.004286796 | 1 |
| T2DM | b_cell | *UBXN2A* | -2.987718984 | 0.006018232 | 0.010171136 | 1 |
| T2DM | b_cell | *CCS* | -3.063894763 | 0.015797155 | 0.022099848 | 1 |
| T2DM | b_cell | *NRROS* | -1.988622731 | 0.007151775 | 0.01153843 | 1 |
| T2DM | b_cell | *CEP19* | -2.754082878 | 0.002868317 | 0.006154645 | 1 |
| T2DM | b_cell | *SLC25A30* | -4.344020213 | 0.003980813 | 0.007624542 | 1 |
| T2DM | b_cell | *CTSF* | -2.025243719 | 0.001041813 | 0.00415349 | 1 |
| T2DM | b_cell | *SOCS7* | -2.274643437 | 0.001093019 | 0.00415349 | 1 |
| T2DM | b_cell | *TLR6* | -3.136146119 | 0.002598389 | 0.005838481 | 1 |
| T2DM | b_cell | *RGMB* | -4.408421982 | 0.0248154 | 0.032577661 | 1 |
| T2DM | b_cell | *FAM53A* | -3.13845621 | 0.000383485 | 0.00415349 | 1 |

| | | | | | | |
|---|---|---|---|---|---|---|
| T2DM | b_cell | *CTU2* | -3.660801601 | 0.001037095 | 0.00415349 | 1 |
| T2DM | b_cell | *MGA* | -2.404341469 | 0.00635017 | 0.010558984 | 1 |
| T2DM | b_cell | *SNX31* | -3.179981104 | 0.017101118 | 0.023655606 | 1 |
| T2DM | b_cell | *DDX23* | -4.056547981 | 0.009193195 | 0.01410187 | 1 |
| T2DM | b_cell | *GLIS1* | -2.014484962 | 0.002330355 | 0.005509725 | 1 |
| T2DM | b_cell | *RALGAPA1* | -2.374471648 | 0.011759124 | 0.017208942 | 1 |
| T2DM | b_cell | *LIG4* | -2.340151674 | 0.005583829 | 0.009641728 | 1 |
| T2DM | b_cell | *LINGO2* | -2.997564496 | 0.002492088 | 0.005707744 | 1 |
| T2DM | b_cell | *BBS1* | 4.090632353 | 0.001550013 | 0.004517818 | 1 |
| T2DM | b_cell | *ANKRD36C* | -2.357961802 | 0.004392223 | 0.008165287 | 1 |
| T2DM | b_cell | *IL20RB* | 3.863370752 | 0.000112781 | 0.00415349 | 0.709057091 |
| T2DM | b_cell | *ZNF497* | 2.06196391 | 0.000578987 | 0.00415349 | 1 |
| T2DM | b_cell | *KLF14* | 3.161417969 | 0.001483752 | 0.004462053 | 1 |
| T2DM | b_cell | *ANGEL2* | -2.468669886 | 0.001803964 | 0.00485268 | 1 |
| T2DM | b_cell | *ZNF266* | -3.260298828 | 0.000193598 | 0.00415349 | 1 |
| T2DM | b_cell | *BRSK2* | -4.764520784 | 0.007819082 | 0.012385631 | 1 |
| T2DM | b_cell | *SH3PXD2B* | -1.140154041 | 0.004986056 | 0.00892832 | 1 |
| T2DM | b_cell | *LARP7* | -2.3535576 | 0.001504312 | 0.004478191 | 1 |
| T2DM | b_cell | *FGFBP3* | -4.044656454 | 0.000775262 | 0.00415349 | 1 |
| T2DM | b_cell | *C4orf32* | -1.454225473 | 0.000764962 | 0.00415349 | 1 |
| T2DM | b_cell | *HRAS* | -4.075639274 | 0.001154478 | 0.004195495 | 1 |
| T2DM | b_cell | *C4orf26* | 5.964263424 | 0.002999792 | 0.006308941 | 1 |
| T2DM | b_cell | *BTC* | -2.579434388 | 0.001595611 | 0.004599544 | 1 |
| T2DM | b_cell | *RSRC1* | -2.539587166 | 0.00531289 | 0.009319794 | 1 |
| T2DM | b_cell | *PTDSS2* | -3.296620664 | 0.001099082 | 0.00415349 | 1 |
| T2DM | b_cell | *FUT1* | -5.826629294 | 0.023358821 | 0.030969403 | 1 |
| T2DM | b_cell | *DHX36* | -2.906319314 | 0.003323713 | 0.006745057 | 1 |
| T2DM | b_cell | *FBXW8* | -2.636283602 | 0.002390834 | 0.00557776 | 1 |
| T2DM | b_cell | *CHST2* | -5.40108234 | 0.001143132 | 0.00418468 | 1 |
| T2DM | b_cell | *ATR* | -1.982440214 | 0.002367985 | 0.005546767 | 1 |
| T2DM | b_cell | *PACS1* | -2.422747006 | 0.039714227 | 0.049609248 | 1 |
| T2DM | b_cell | *SH3BP5L* | -3.180016993 | 0.003684634 | 0.007236894 | 1 |
| T2DM | b_cell | *CADM2* | -1.456209915 | 0.000696633 | 0.00415349 | 1 |
| T2DM | b_cell | *PSMD2* | -1.758739082 | 0.002735562 | 0.005990413 | 1 |
| T2DM | b_cell | *ARHGAP1* | -3.299039043 | 0.004759789 | 0.008666318 | 1 |
| T2DM | b_cell | *TP53I11* | -3.55516967 | 0.013585384 | 0.01945588 | 1 |
| T2DM | b_cell | *PHYHD1* | -1.98224552 | 0.001660844 | 0.004679659 | 1 |
| T2DM | b_cell | *ZNF519* | -3.139775362 | 0.004645113 | 0.008524175 | 1 |
| T2DM | b_cell | *BANF1* | 3.29744767 | 9.01E-05 | 0.00415349 | 0.566339812 |
| T2DM | b_cell | *NRIP3* | -4.091039226 | 9.27E-05 | 0.00415349 | 0.58267855 |
| T2DM | b_cell | *SCUBE2* | -2.081152067 | 0.000469978 | 0.00415349 | 1 |
| T2DM | b_cell | *SMAD2* | -2.117300578 | 0.0047532 | 0.00865764 | 1 |
| T2DM | b_cell | *EIF3F* | -4.067758395 | 0.019186162 | 0.026030082 | 1 |
| T2DM | b_cell | *ZNF25* | -2.960859651 | 0.000398584 | 0.00415349 | 1 |
| T2DM | b_cell | *CLTB* | -2.726801488 | 0.005459035 | 0.009501286 | 1 |
| T2DM | b_cell | *PCSK1* | -2.972293948 | 0.001251553 | 0.004253251 | 1 |
| T2DM | b_cell | *LPL* | -1.20148785 | 0.000108595 | 0.00415349 | 0.682734157 |
| T2DM | b_cell | *TBC1D10C* | 2.217684986 | 0.001534926 | 0.004508014 | 1 |
| T2DM | b_cell | *PPP2R2D* | -2.23962309 | 0.005599445 | 0.009655634 | 1 |
| T2DM | b_cell | *MCTP1* | -1.127198079 | 0.000703788 | 0.00415349 | 1 |

| | | | | | | |
|---|---|---|---|---|---|---|
| T2DM | b_cell | *UBQLNL* | 1.730861363 | 0.000698806 | 0.00415349 | 1 |
| T2DM | b_cell | *LIPT2* | -9.036101027 | 0.001378099 | 0.004323793 | 1 |
| T2DM | b_cell | *KCNE3* | -2.27571268 | 0.000891026 | 0.00415349 | 1 |
| T2DM | b_cell | *UCP3* | 1.758245182 | 0.001204192 | 0.004229472 | 1 |
| T2DM | b_cell | *MRPL48* | -3.301044032 | 0.011099895 | 0.016423874 | 1 |
| T2DM | b_cell | *RAB6A* | -2.245085159 | 0.002272615 | 0.005445221 | 1 |
| T2DM | b_cell | *RPS6KB2* | -3.251884023 | 0.001984381 | 0.005096324 | 1 |
| T2DM | b_cell | *RMI2* | -3.901202958 | 0.005630793 | 0.009698848 | 1 |
| T2DM | b_cell | *TOM1L2* | -2.25435518 | 0.002066724 | 0.005216175 | 1 |
| T2DM | b_cell | *TEX26* | -4.964138662 | 0.019138067 | 0.025976043 | 1 |
| T2DM | b_cell | *GPR156* | -4.270644875 | 0.00361531 | 0.007154376 | 1 |
| T2DM | b_cell | *TTLL11* | -2.110559683 | 0.005680193 | 0.009761317 | 1 |
| T2DM | b_cell | *LINC01106* | -31.36037923 | 0.003411175 | 0.006859926 | 1 |
| T2DM | b_cell | *MSRA* | -2.952972949 | 0.019314197 | 0.026181189 | 1 |
| T2DM | b_cell | *CTDNEP1* | -4.047920768 | 0.005091319 | 0.009037019 | 1 |
| T2DM | b_cell | *ETV4* | 5.308385353 | 0.000462102 | 0.00415349 | 1 |
| T2DM | b_cell | *SWI5* | -2.781994723 | 0.000598046 | 0.00415349 | 1 |
| T2DM | b_cell | *ZDHHC21* | -2.731698757 | 0.004796184 | 0.008709882 | 1 |
| T2DM | b_cell | *A2M* | -1.71143368 | 0.001234602 | 0.004253251 | 1 |
| T2DM | b_cell | *LRRN1* | -2.673001295 | 0.000855408 | 0.00415349 | 1 |
| T2DM | b_cell | *B3GALT6* | -4.465034502 | 0.001270329 | 0.004257394 | 1 |
| T2DM | b_cell | *ZNF613* | -2.308596051 | 0.004880838 | 0.008805116 | 1 |
| T2DM | b_cell | *IP6K1* | -2.896576717 | 0.011218873 | 0.016576512 | 1 |
| T2DM | b_cell | *SSNA1* | -2.464491821 | 0.000736426 | 0.00415349 | 1 |
| T2DM | b_cell | *CHMP6* | -4.487999909 | 0.017796824 | 0.024483289 | 1 |
| T2DM | b_cell | *UFSP1* | 2.732934046 | 0.00100457 | 0.00415349 | 1 |
| T2DM | b_cell | *TMEM39A* | -2.617495495 | 0.006683612 | 0.010976978 | 1 |
| T2DM | b_cell | *TCP11L1* | -2.346052213 | 0.023121962 | 0.030726649 | 1 |
| T2DM | b_cell | *GPX2* | -4.341192671 | 0.001494134 | 0.004473152 | 1 |
| T2DM | b_cell | *CCDC57* | -2.267788662 | 0.001080349 | 0.00415349 | 1 |
| T2DM | b_cell | *CIDEA* | -1.020754167 | 0.004997238 | 0.008943142 | 1 |
| T2DM | b_cell | *SMIM19* | -1.426641717 | 0.004863991 | 0.008789857 | 1 |
| T2DM | b_cell | *ZNF404* | -1.997299461 | 0.00645618 | 0.010687204 | 1 |
| T2DM | b_cell | *SLC35G1* | -3.077614242 | 0.004252037 | 0.007980713 | 1 |
| T2DM | b_cell | *ZNF135* | -3.039727758 | 0.000453166 | 0.00415349 | 1 |
| T2DM | b_cell | *COX8A* | -4.67398397 | 0.03444624 | 0.043635606 | 1 |
| T2DM | b_cell | *B3GNT4* | 2.230947841 | 0.000583053 | 0.00415349 | 1 |
| T2DM | b_cell | *CDC26* | -4.273464298 | 6.65E-05 | 0.00415349 | 0.418105885 |
| T2DM | b_cell | *RNPEP* | -2.376779848 | 0.001316219 | 0.004274973 | 1 |
| T2DM | b_cell | *SPRYD4* | -2.756062406 | 0.002710796 | 0.005955133 | 1 |
| T2DM | b_cell | *VPS37D* | -2.743546437 | 0.029650276 | 0.038167748 | 1 |
| T2DM | b_cell | *SLCO3A1* | -2.064157941 | 0.001875666 | 0.004957688 | 1 |
| T2DM | b_cell | *WDR25* | -3.444777384 | 0.007925693 | 0.012529757 | 1 |
| T2DM | b_cell | *PLA2G16* | -0.954773572 | 0.005151736 | 0.009115948 | 1 |
| T2DM | b_cell | *PRR15* | -10.78243907 | 0.010417812 | 0.015631691 | 1 |
| T2DM | b_cell | *RMDN1* | -2.259249464 | 0.005635807 | 0.009702168 | 1 |
| T2DM | b_cell | *RNF152* | -2.500557438 | 0.000781399 | 0.00415349 | 1 |
| T2DM | b_cell | *ACSF3* | -3.64661457 | 0.004365801 | 0.008125456 | 1 |
| T2DM | b_cell | *BOK* | -1.811447996 | 0.001140857 | 0.00418468 | 1 |
| T2DM | b_cell | *ZNF843* | -4.604982184 | 0.000408919 | 0.00415349 | 1 |

| | | | | | | |
|---|---|---|---|---|---|---|
| T2DM | b_cell | *C8orf59* | -2.228792064 | 0.002209747 | 0.005381337 | 1 |
| T2DM | b_cell | *PFN4* | -1.761837076 | 0.024685716 | 0.032431982 | 1 |
| T2DM | b_cell | *RUFY1* | -2.554561483 | 0.024141841 | 0.031866419 | 1 |
| T2DM | b_cell | *BASP1* | -1.580293401 | 0.001008665 | 0.00415349 | 1 |
| T2DM | b_cell | *VSIG10* | -1.442127088 | 0.00366013 | 0.007202266 | 1 |
| T2DM | b_cell | *IRX5* | -2.831222189 | 0.002083663 | 0.005230315 | 1 |
| T2DM | b_cell | *PNMA1* | -6.666966453 | 0.006217851 | 0.010405595 | 1 |
| T2DM | b_cell | *ANKLE2* | -2.683581293 | 0.003263093 | 0.006657222 | 1 |
| T2DM | b_cell | *GCNT4* | 1.942211153 | 0.000591279 | 0.00415349 | 1 |
| T2DM | b_cell | *MUC20* | 2.906054936 | 0.000828041 | 0.00415349 | 1 |
| T2DM | b_cell | *SHMT1* | -1.41369607 | 0.004072988 | 0.007750402 | 1 |
| T2DM | b_cell | *SEC24C* | -4.090227865 | 0.023761899 | 0.031424287 | 1 |
| T2DM | b_cell | *MTHFR* | -3.382374922 | 0.001912502 | 0.004995388 | 1 |
| T2DM | b_cell | *C19orf18* | -2.159743516 | 0.000662165 | 0.00415349 | 1 |
| T2DM | b_cell | *TMEM80* | -2.092257628 | 0.001181798 | 0.004202203 | 1 |
| T2DM | b_cell | *WDR73* | -2.250200474 | 0.037133301 | 0.046710097 | 1 |
| T2DM | b_cell | *POLE* | -3.440753743 | 0.004544598 | 0.008382656 | 1 |
| T2DM | b_cell | *DSCAML1* | -2.05967857 | 0.00208668 | 0.005230315 | 1 |
| T2DM | b_cell | *EPS8L2* | -3.548463835 | 0.000176861 | 0.00415349 | 1 |
| T2DM | b_cell | *TALDO1* | -2.141341853 | 0.003443365 | 0.006894407 | 1 |
| T2DM | b_cell | *RIMKLA* | -3.14055934 | 0.028685462 | 0.037085235 | 1 |
| T2DM | b_cell | *B3GNT8* | 1.191678397 | 0.000585367 | 0.00415349 | 1 |
| T2DM | b_cell | *AP006621.1* | -1.930436538 | 0.004855763 | 0.008782561 | 1 |
| T2DM | b_cell | *MAN1B1* | -3.964980345 | 0.006254772 | 0.01043901 | 1 |
| T2DM | b_cell | *TOP3A* | -3.327174758 | 0.00578748 | 0.009890156 | 1 |
| T2DM | b_cell | *ZBTB38* | -3.034118377 | 0.00806822 | 0.012690742 | 1 |
| T2DM | b_cell | *C8orf31* | -3.181442535 | 0.00079737 | 0.00415349 | 1 |
| T2DM | b_cell | *AC024940.1* | 0.721604092 | 0.000557887 | 0.00415349 | 1 |
| T2DM | b_cell | *TIMM22* | -2.905248694 | 0.001587179 | 0.004585751 | 1 |
| T2DM | b_cell | *PPFIA3* | -3.230747372 | 0.004370339 | 0.008129088 | 1 |
| T2DM | b_cell | *MAGEF1* | -2.945555204 | 0.000759264 | 0.00415349 | 1 |
| T2DM | b_cell | *PAWR* | -3.224905341 | 0.006792549 | 0.011106569 | 1 |
| T2DM | b_cell | *NAP1L5* | -4.106456624 | 0.000899121 | 0.00415349 | 1 |
| T2DM | b_cell | *NIM1K* | -3.566461542 | 0.002627931 | 0.005859268 | 1 |
| T2DM | b_cell | *ACOT4* | -3.421010062 | 0.002094965 | 0.005240399 | 1 |
| T2DM | b_cell | *ARIH2* | -2.077203831 | 0.005990463 | 0.010135103 | 1 |
| T2DM | b_cell | *RBM44* | -3.294332569 | 0.001195972 | 0.004217141 | 1 |
| T2DM | b_cell | *IRX3* | -2.437018359 | 0.017702526 | 0.024369561 | 1 |
| T2DM | b_cell | *ATOX1* | 4.092198403 | 0.021236613 | 0.028522663 | 1 |
| T2DM | b_cell | *TBL1XR1* | -2.278237398 | 0.004813544 | 0.008733839 | 1 |
| T2DM | b_cell | *C18orf32* | 3.857839221 | 0.000723663 | 0.00415349 | 1 |
| T2DM | b_cell | *C12orf54* | -14.58898047 | 1.78E-05 | 0.00243787 | 0.112142015 |
| T2DM | b_cell | *GBA* | -3.259883804 | 0.000266344 | 0.00415349 | 1 |
| T2DM | b_cell | *ACAD9* | -1.300939279 | 0.008145037 | 0.012773221 | 1 |
| T2DM | b_cell | *IL17RA* | -2.182366215 | 0.007568325 | 0.012079731 | 1 |
| T2DM | b_cell | *CD163L1* | -1.431407625 | 0.001185065 | 0.004202203 | 1 |
| T2DM | b_cell | *THAP5* | -4.553950907 | 0.018191424 | 0.024906246 | 1 |
| T2DM | b_cell | *CD151* | -2.865680324 | 0.004322103 | 0.008068011 | 1 |
| T2DM | b_cell | *POLR2L* | -2.788154325 | 0.002301942 | 0.005481934 | 1 |
| T2DM | b_cell | *FAM87B* | -11.96725444 | 0.000767058 | 0.00415349 | 1 |

| | | | | | | |
|---|---|---|---|---|---|---|
| T2DM | b_cell | *ZBTB41* | -2.591593034 | 0.002065215 | 0.005214462 | 1 |
| T2DM | b_cell | *ZNF354C* | -3.96463231 | 0.007809639 | 0.012373791 | 1 |
| T2DM | b_cell | *CENPBD1* | -6.191745357 | 0.001243687 | 0.004253251 | 1 |
| T2DM | b_cell | *RIC8A* | -4.771121462 | 0.00017318 | 0.00415349 | 1 |
| T2DM | b_cell | *IMP3* | -2.216657796 | 0.001902292 | 0.004987368 | 1 |
| T2DM | b_cell | *ODF3B* | -3.593354019 | 0.003530339 | 0.007021588 | 1 |
| T2DM | b_cell | *DPY19L2* | -3.011443258 | 0.00897605 | 0.013835751 | 1 |
| T2DM | b_cell | *C2orf73* | -3.401350836 | 0.000771783 | 0.00415349 | 1 |
| T2DM | b_cell | *FAM26E* | -4.454470954 | 0.020587662 | 0.02775185 | 1 |
| T2DM | b_cell | *ALS2CL* | -1.767061788 | 0.007379198 | 0.011831934 | 1 |
| T2DM | b_cell | *MLF1* | -1.802922449 | 0.000811255 | 0.00415349 | 1 |
| T2DM | b_cell | *PRSS42* | 8.031992183 | 0.01391631 | 0.019879991 | 1 |
| T2DM | b_cell | *C2orf69* | -3.125362022 | 0.001903967 | 0.004987601 | 1 |
| T2DM | b_cell | *GRAMD1C* | -2.611313698 | 0.000881328 | 0.00415349 | 1 |
| T2DM | b_cell | *TSSK6* | -9.605284493 | 0.000264252 | 0.00415349 | 1 |
| T2DM | b_cell | *GOLGA8Q* | 55.59921652 | 8.45E-05 | 0.00415349 | 0.531433074 |
| T2DM | b_cell | *ZNF114* | -5.685720356 | 0.000378039 | 0.00415349 | 1 |
| T2DM | b_cell | *ZNF518B* | -3.794029698 | 0.000730802 | 0.00415349 | 1 |
| T2DM | b_cell | *LCORL* | -2.623124235 | 0.012673838 | 0.018346861 | 1 |
| T2DM | b_cell | *ZNF454* | -2.588052764 | 0.004428673 | 0.008213295 | 1 |
| T2DM | b_cell | *SH2B1* | -1.971077985 | 6.12E-05 | 0.00415349 | 0.38466138 |
| T2DM | b_cell | *ZC3H12D* | -5.657108118 | 0.008188296 | 0.012831459 | 1 |
| T2DM | b_cell | *KDELC2* | -2.232123102 | 0.000773918 | 0.00415349 | 1 |
| T2DM | b_cell | *PLEC* | -3.437318992 | 0.013439523 | 0.019286528 | 1 |
| T2DM | b_cell | *SH2D4B* | 5.244609013 | 0.000869817 | 0.00415349 | 1 |
| T2DM | b_cell | *RNF212* | -1.3907771 | 0.021478384 | 0.028779753 | 1 |
| T2DM | b_cell | *PRSS36* | -3.828748843 | 0.000785753 | 0.00415349 | 1 |
| T2DM | b_cell | *WDR6* | -2.048248162 | 0.035829212 | 0.045223501 | 1 |
| T2DM | b_cell | *TMPRSS9* | -2.18850399 | 0.001400308 | 0.004345113 | 1 |
| T2DM | b_cell | *AQP11* | -2.804837042 | 0.001403238 | 0.004350178 | 1 |
| T2DM | b_cell | *ZFAND2A* | -5.334853185 | 0.005461691 | 0.009501286 | 1 |
| T2DM | b_cell | *ZNF223* | -2.276127179 | 0.013806267 | 0.019727273 | 1 |
| T2DM | b_cell | *FAM220A* | -3.794129283 | 0.003988418 | 0.007633238 | 1 |
| T2DM | b_cell | *BEND3* | -4.359654258 | 0.00086418 | 0.00415349 | 1 |
| T2DM | b_cell | *NT5DC1* | -2.649702535 | 0.006950987 | 0.011300363 | 1 |
| T2DM | b_cell | *AC006111.1* | -11.92756937 | 0.002083555 | 0.005230315 | 1 |
| T2DM | b_cell | *COX14* | -1.664734626 | 0.001021943 | 0.00415349 | 1 |
| T2DM | b_cell | *CA8* | -2.324326882 | 0.001890033 | 0.004970573 | 1 |
| T2DM | b_cell | *ERBB4* | -1.074269936 | 0.000779029 | 0.00415349 | 1 |
| T2DM | b_cell | *GPR35* | -2.939271553 | 0.001693318 | 0.004705788 | 1 |
| T2DM | b_cell | *CSRNP3* | -3.267196952 | 0.002375164 | 0.005557371 | 1 |
| T2DM | b_cell | *ZNF713* | -3.543076921 | 0.010247489 | 0.015431369 | 1 |
| T2DM | b_cell | *KCTD12* | -1.977688327 | 0.000597349 | 0.00415349 | 1 |
| T2DM | b_cell | *FAM219B* | -1.666285097 | 0.000906368 | 0.00415349 | 1 |
| T2DM | b_cell | *CPNE7* | 3.242933904 | 0.001496035 | 0.004474047 | 1 |
| T2DM | b_cell | *GDPD4* | 2.820200213 | 0.000685668 | 0.00415349 | 1 |
| T2DM | b_cell | *MPI* | -4.038431259 | 0.009176161 | 0.0140846 | 1 |
| T2DM | b_cell | *TRIM73* | -14.64993544 | 3.46E-05 | 0.003315867 | 0.217572195 |
| T2DM | b_cell | *EFCAB13* | -2.44168286 | 0.010764037 | 0.016043978 | 1 |
| T2DM | b_cell | *HYI* | -1.904676689 | 0.001527544 | 0.004502743 | 1 |

| | | | | | | |
|---|---|---|---|---|---|---|
| T2DM | b_cell | *GAK* | -3.360741321 | 0.015920816 | 0.022262938 | 1 |
| T2DM | b_cell | *TUFM* | -2.222373875 | 0.035828033 | 0.045223501 | 1 |
| T2DM | b_cell | *CTC1* | 4.130955233 | 0.00267277 | 0.005916798 | 1 |
| T2DM | b_cell | *SNX18* | -2.88073723 | 0.003082964 | 0.006413831 | 1 |
| T2DM | b_cell | *C14orf39* | -1.85240714 | 0.001267838 | 0.004257394 | 1 |
| T2DM | b_cell | *TRIML2* | 6.994462034 | 0.000680425 | 0.00415349 | 1 |
| T2DM | b_cell | *FARSA* | -7.261001037 | 0.002811118 | 0.00608382 | 1 |
| T2DM | b_cell | *SPTY2D1* | -3.767364639 | 0.022181319 | 0.029595491 | 1 |
| T2DM | b_cell | *TCAIM* | -2.262677854 | 0.001126562 | 0.004182741 | 1 |
| T2DM | b_cell | *FUCA1* | -2.325414281 | 0.00290438 | 0.006192483 | 1 |
| T2DM | b_cell | *ZNF664* | -3.742268181 | 0.001667248 | 0.004689926 | 1 |
| T2DM | b_cell | *CALR* | -2.360361831 | 0.028341534 | 0.036678309 | 1 |
| T2DM | b_cell | *NSUN7* | -2.12013204 | 0.005281664 | 0.00927796 | 1 |
| T2DM | b_cell | *ARID3B* | -4.776748979 | 0.002620207 | 0.005854674 | 1 |
| T2DM | b_cell | *EGR3* | -5.831063194 | 0.00215875 | 0.005311961 | 1 |
| T2DM | b_cell | *LINC00174* | -2.340329524 | 0.005509762 | 0.009569025 | 1 |
| T2DM | b_cell | *GEMIN4* | 2.728202623 | 0.025994646 | 0.033969724 | 1 |
| T2DM | b_cell | *FJX1* | -6.227682734 | 0.004728095 | 0.008626098 | 1 |
| T2DM | b_cell | *CIITA* | 2.559415907 | 0.001137048 | 0.00418468 | 1 |
| T2DM | b_cell | *ZBTB42* | -9.088951685 | 0.001876257 | 0.004957688 | 1 |
| T2DM | b_cell | *LACC1* | -4.779250475 | 0.005861432 | 0.009968694 | 1 |
| T2DM | b_cell | *MAF1* | -3.284897469 | 0.001130029 | 0.004182741 | 1 |
| T2DM | b_cell | *FCER1A* | 4.824925177 | 0.006600795 | 0.010862853 | 1 |
| T2DM | b_cell | *PCED1B* | -2.017546829 | 0.001384443 | 0.00432604 | 1 |
| T2DM | b_cell | *PIPOX* | -6.454716909 | 0.001061915 | 0.00415349 | 1 |
| T2DM | b_cell | *CDH5* | -1.54231513 | 0.003223623 | 0.006605906 | 1 |
| T2DM | b_cell | *GIPC3* | 2.414514929 | 0.001798947 | 0.004852128 | 1 |
| T2DM | b_cell | *CITED4* | 4.354758112 | 0.005582775 | 0.009641728 | 1 |
| T2DM | b_cell | *TIGD5* | -3.780187918 | 0.001662853 | 0.004679659 | 1 |
| T2DM | b_cell | *PDXDC1* | -2.508079854 | 0.008671747 | 0.013456852 | 1 |
| T2DM | b_cell | *ZNF154* | -2.271649516 | 0.002443325 | 0.0056392 | 1 |
| T2DM | b_cell | *R3HDM2* | -2.19573356 | 0.004006573 | 0.007653388 | 1 |
| T2DM | b_cell | *SSC5D* | -1.552995869 | 0.010093863 | 0.015238897 | 1 |
| T2DM | b_cell | *ZADH2* | -3.531178462 | 0.00672129 | 0.011026465 | 1 |
| T2DM | b_cell | *SEPT1* | -2.608719207 | 0.000685941 | 0.00415349 | 1 |
| T2DM | b_cell | *EXOC3* | -3.148169244 | 0.00443911 | 0.008227796 | 1 |
| T2DM | b_cell | *LYNX1* | -4.365727939 | 0.000200693 | 0.00415349 | 1 |
| T2DM | b_cell | *FAHD1* | -1.634942192 | 0.018710467 | 0.025505791 | 1 |
| T2DM | b_cell | *TDRP* | -2.104620004 | 0.001103766 | 0.004159248 | 1 |
| T2DM | b_cell | *ZNF816* | 3.048704528 | 0.001853236 | 0.004924469 | 1 |
| T2DM | b_cell | *FGD6* | -2.406709828 | 0.001515015 | 0.004480664 | 1 |
| T2DM | b_cell | *HCLS1* | -2.348014337 | 0.00127641 | 0.004257394 | 1 |
| T2DM | b_cell | *ZNF609* | -2.27046068 | 0.007305588 | 0.011730073 | 1 |
| T2DM | b_cell | *CCDC66* | -1.989917825 | 0.024539857 | 0.032270253 | 1 |
| T2DM | b_cell | *DEFB124* | -12.12256525 | 0.000409677 | 0.00415349 | 1 |
| T2DM | b_cell | *MCFD2* | -2.025333283 | 0.001130029 | 0.004182741 | 1 |
| T2DM | b_cell | *CYP8B1* | -2.952107444 | 0.001267338 | 0.004257394 | 1 |
| T2DM | b_cell | *GLIPR1L2* | -2.290566049 | 0.000583449 | 0.00415349 | 1 |
| T2DM | b_cell | *RNF182* | -5.221387678 | 3.07E-05 | 0.003163865 | 0.19299578 |
| T2DM | b_cell | *MB21D2* | -2.84140794 | 0.002004492 | 0.005129117 | 1 |

| | | | | | | |
|---|---|---|---|---|---|---|
| T2DM | b_cell | *SSTR2* | 2.965367189 | 0.000713229 | 0.00415349 | 1 |
| T2DM | b_cell | *ZNF594* | -2.244126661 | 0.000825932 | 0.00415349 | 1 |
| T2DM | b_cell | *PCGF5* | -2.757212322 | 0.021264595 | 0.028554145 | 1 |
| T2DM | b_cell | *SHISA2* | -65.34565203 | 0.000882667 | 0.00415349 | 1 |
| T2DM | b_cell | *CHST13* | -14.05803651 | 0.00013667 | 0.00415349 | 0.859243212 |
| T2DM | b_cell | *SRSF8* | -3.828681992 | 0.001382645 | 0.00432604 | 1 |
| T2DM | b_cell | *SLC36A4* | -3.898113641 | 0.008608358 | 0.013373053 | 1 |
| T2DM | b_cell | *ZDHHC20* | -2.801734431 | 0.014982359 | 0.02113871 | 1 |
| T2DM | b_cell | *PSMG4* | -1.717853973 | 0.000712977 | 0.00415349 | 1 |
| T2DM | b_cell | *CAPS2* | -2.518648228 | 0.003716282 | 0.00726565 | 1 |
| T2DM | b_cell | *ZNF792* | -5.046131474 | 0.000324823 | 0.00415349 | 1 |
| T2DM | b_cell | *KCTD2* | -3.117476031 | 0.001451747 | 0.004416445 | 1 |
| T2DM | b_cell | *D2HGDH* | -2.057095684 | 0.002701424 | 0.005946726 | 1 |
| T2DM | b_cell | *CMTR2* | -3.487740044 | 0.00455677 | 0.00839637 | 1 |
| T2DM | b_cell | *LRRC57* | -3.648318238 | 0.019621694 | 0.026569371 | 1 |
| T2DM | b_cell | *MRPL14* | -2.617225741 | 0.000590556 | 0.00415349 | 1 |
| T2DM | b_cell | *BBS12* | -3.833935377 | 0.001091164 | 0.00415349 | 1 |
| T2DM | b_cell | *ZFP82* | -3.570646405 | 0.010759358 | 0.016040806 | 1 |
| T2DM | b_cell | *LSMEM1* | -3.239555771 | 0.03684847 | 0.046388933 | 1 |
| T2DM | b_cell | *AEN* | -4.315059015 | 0.000612983 | 0.00415349 | 1 |
| T2DM | b_cell | *FKRP* | -2.03409794 | 0.001082217 | 0.00415349 | 1 |
| T2DM | b_cell | *TRAPPC5* | -7.122398001 | 0.019841013 | 0.026825903 | 1 |
| T2DM | b_cell | *RPH3AL* | -1.787266608 | 0.001448005 | 0.004416445 | 1 |
| T2DM | b_cell | *ANKRD34A* | -5.849385119 | 0.000171674 | 0.00415349 | 1 |
| T2DM | b_cell | *SLC26A11* | -2.672718063 | 0.001679303 | 0.004704627 | 1 |
| T2DM | b_cell | *HIGD1A* | -4.670714585 | 0.00174309 | 0.004783999 | 1 |
| T2DM | b_cell | *ADIPOQ* | -1.183900086 | 0.000356772 | 0.00415349 | 1 |
| T2DM | b_cell | *F2R* | -2.14474071 | 0.002562868 | 0.005799421 | 1 |
| T2DM | b_cell | *HIST3H2A* | 5.100444676 | 0.022034434 | 0.029411993 | 1 |
| T2DM | b_cell | *POLR2A* | -2.854857249 | 0.001485408 | 0.004462053 | 1 |
| T2DM | b_cell | *TMEM132C* | -1.086382309 | 0.001399203 | 0.004344868 | 1 |
| T2DM | b_cell | *SLC25A41* | -14.67112113 | 0.000540011 | 0.00415349 | 1 |
| T2DM | b_cell | *TMEM136* | -2.389249435 | 0.005223465 | 0.009196282 | 1 |
| T2DM | b_cell | *TMEM132E* | -17.09258131 | 0.006277329 | 0.010471098 | 1 |
| T2DM | b_cell | *ZNF322* | -2.447300385 | 0.000752944 | 0.00415349 | 1 |
| T2DM | b_cell | *CCL13* | -4.000356955 | 0.000552708 | 0.00415349 | 1 |
| T2DM | b_cell | *DDX60L* | -2.909957669 | 0.013019124 | 0.018786145 | 1 |
| T2DM | b_cell | *ACBD4* | -3.508524594 | 0.003145119 | 0.006491583 | 1 |
| T2DM | b_cell | *SGSH* | -5.05633082 | 0.005441983 | 0.009480118 | 1 |
| T2DM | b_cell | *SETD2* | -2.065633175 | 0.00808491 | 0.012704282 | 1 |
| T2DM | b_cell | *MRPS23* | -2.753236764 | 0.001092962 | 0.00415349 | 1 |
| T2DM | b_cell | *GPR135* | -1.763746694 | 0.001291844 | 0.004268027 | 1 |
| T2DM | b_cell | *ZFP41* | -3.364470017 | 0.005069629 | 0.009013789 | 1 |
| T2DM | b_cell | *ATG9B* | -5.823265753 | 0.000772825 | 0.00415349 | 1 |
| T2DM | b_cell | *HKR1* | -3.214625299 | 0.015978714 | 0.022333965 | 1 |
| T2DM | b_cell | *ZBTB20* | -1.719399888 | 0.005888558 | 0.010005408 | 1 |
| T2DM | b_cell | *C3orf58* | -2.944308863 | 0.002736741 | 0.005990909 | 1 |
| T2DM | b_cell | *AMIGO1* | -3.322794566 | 0.00551373 | 0.009573273 | 1 |
| T2DM | b_cell | *RELL1* | -2.230536758 | 0.001739753 | 0.004780517 | 1 |
| T2DM | b_cell | *CLDN7* | -3.246783797 | 0.000796805 | 0.00415349 | 1 |

| | | | | | | |
|---|---|---|---|---|---|---|
| T2DM | b_cell | *ADO* | -3.500597827 | 0.001010547 | 0.00415349 | 1 |
| T2DM | b_cell | *CHST15* | -3.360441724 | 0.00543932 | 0.009478105 | 1 |
| T2DM | b_cell | *PTCHD3* | 2.594388875 | 0.001242459 | 0.004253251 | 1 |
| T2DM | b_cell | *WRB* | -2.58403273 | 0.006170903 | 0.010360188 | 1 |
| T2DM | b_cell | *TNRC18* | -2.322773916 | 0.002776207 | 0.006031103 | 1 |
| T2DM | b_cell | *FAM181B* | 1.820954798 | 0.000581901 | 0.00415349 | 1 |
| T2DM | b_cell | *NOP10* | -3.04444445 | 0.000238835 | 0.00415349 | 1 |
| T2DM | b_cell | *FAM89A* | -1.844685381 | 0.000330372 | 0.00415349 | 1 |
| T2DM | b_cell | *TDRKH* | -5.066146714 | 0.003510783 | 0.006991541 | 1 |
| T2DM | b_cell | *ZNF708* | -2.806175912 | 0.002700897 | 0.005946726 | 1 |
| T2DM | b_cell | *IST1* | -2.845945502 | 0.001989608 | 0.005101414 | 1 |
| T2DM | b_cell | *ERCC6L2* | -2.398896466 | 0.00483432 | 0.008753851 | 1 |
| T2DM | b_cell | *MRPL41* | -3.057855659 | 0.000901169 | 0.00415349 | 1 |
| T2DM | b_cell | *HHIPL1* | -7.31714412 | 0.006122492 | 0.010297514 | 1 |
| T2DM | b_cell | *FAM153B* | -5.793367082 | 0.005889934 | 0.010005408 | 1 |
| T2DM | b_cell | *BACE2* | -1.962689158 | 0.002722638 | 0.005970431 | 1 |
| T2DM | b_cell | *SYNM* | -2.179886472 | 0.000764423 | 0.00415349 | 1 |
| T2DM | b_cell | *IZUMO1* | 2.895429552 | 0.000201034 | 0.00415349 | 1 |
| T2DM | b_cell | *C8orf33* | -2.671745158 | 0.001176508 | 0.004202203 | 1 |
| T2DM | b_cell | *FBXL6* | -5.980244518 | 0.014697991 | 0.020802852 | 1 |
| T2DM | b_cell | *KBTBD3* | 3.909921571 | 0.002852646 | 0.006135678 | 1 |
| T2DM | b_cell | *CLN8* | 3.204749634 | 0.024688956 | 0.032431982 | 1 |
| T2DM | b_cell | *CACNB4* | -1.437201333 | 0.001047025 | 0.00415349 | 1 |
| T2DM | b_cell | *TRAPPC6B* | -2.903841961 | 0.002698017 | 0.005946726 | 1 |
| T2DM | b_cell | *NPLOC4* | -1.902253849 | 0.000665477 | 0.00415349 | 1 |
| T2DM | b_cell | *TSHZ2* | -1.214773871 | 0.000872096 | 0.00415349 | 1 |
| T2DM | b_cell | *CAPN12* | 3.389991089 | 0.000303652 | 0.00415349 | 1 |
| T2DM | b_cell | *FES* | -4.722470276 | 0.005750946 | 0.009838421 | 1 |
| T2DM | b_cell | *CAV3* | 1.88245946 | 0.00926982 | 0.014204084 | 1 |
| T2DM | b_cell | *MXRA7* | -1.86649911 | 0.037885861 | 0.047495196 | 1 |
| T2DM | b_cell | *MFSD5* | -2.916838531 | 0.001446037 | 0.004415364 | 1 |
| T2DM | b_cell | *ADI1* | -2.988413414 | 0.001297991 | 0.004268027 | 1 |
| T2DM | b_cell | *SPNS3* | 1.196597489 | 0.000634847 | 0.00415349 | 1 |
| T2DM | b_cell | *CLEC4G* | -7.466829933 | 0.000938915 | 0.00415349 | 1 |
| T2DM | b_cell | *SATB1* | -3.36540154 | 0.001296718 | 0.004268027 | 1 |
| T2DM | b_cell | *ACTL10* | -8.23913151 | 0.006823091 | 0.01114781 | 1 |
| T2DM | b_cell | *TRAK1* | -2.395882579 | 0.018537507 | 0.025307705 | 1 |
| T2DM | b_cell | *SKA2* | -3.488481668 | 0.005050806 | 0.009000686 | 1 |
| T2DM | b_cell | *KCNB2* | -1.63525743 | 0.000549505 | 0.00415349 | 1 |
| T2DM | b_cell | *BRICD5* | -4.932604914 | 0.001542081 | 0.004509332 | 1 |
| T2DM | b_cell | *TSKU* | -1.873113126 | 0.000855078 | 0.00415349 | 1 |
| T2DM | b_cell | *GRID1* | -2.35417706 | 0.000174037 | 0.00415349 | 1 |
| T2DM | b_cell | *ACBD3* | -2.448159155 | 0.002096963 | 0.005241324 | 1 |
| T2DM | b_cell | *PLCXD3* | -2.052615456 | 0.00196426 | 0.005077837 | 1 |
| T2DM | b_cell | *GPIHBP1* | -2.784681934 | 0.000279487 | 0.00415349 | 1 |
| T2DM | b_cell | *COL18A1* | -2.830636561 | 0.003333258 | 0.006760063 | 1 |
| T2DM | b_cell | *SLC25A18* | -2.433316579 | 0.000736306 | 0.00415349 | 1 |
| T2DM | b_cell | *ZNF721* | -2.126098718 | 0.008194448 | 0.0128379 | 1 |
| T2DM | b_cell | *C11orf54* | -2.089448226 | 0.00449152 | 0.008305349 | 1 |
| T2DM | b_cell | *CEP63* | -2.001157492 | 0.001390486 | 0.004334153 | 1 |

| | | | | | | |
|---|---|---|---|---|---|---|
| T2DM | b_cell | *HMGN4* | -4.517189267 | 0.003341633 | 0.006768314 | 1 |
| T2DM | b_cell | *C12orf60* | -4.241080578 | 0.006548758 | 0.010797808 | 1 |
| T2DM | b_cell | *LYSMD4* | -1.912188957 | 0.002908809 | 0.006192483 | 1 |
| T2DM | b_cell | *WBP2NL* | -3.840759632 | 0.009746603 | 0.014804758 | 1 |
| T2DM | b_cell | *GAS6* | -3.622396502 | 0.001030521 | 0.00415349 | 1 |
| T2DM | b_cell | *BEGAIN* | -7.508694473 | 0.000710123 | 0.00415349 | 1 |
| T2DM | b_cell | *ARHGEF37* | -2.770441012 | 0.00174355 | 0.004783999 | 1 |
| T2DM | b_cell | *CSMD1* | -1.534375873 | 0.000650913 | 0.00415349 | 1 |
| T2DM | b_cell | *GPR19* | -6.396969508 | 0.003578396 | 0.007094727 | 1 |
| T2DM | b_cell | *FANCF* | 3.621836454 | 0.000895589 | 0.00415349 | 1 |
| T2DM | b_cell | *C2CD4C* | -4.727121645 | 0.02369336 | 0.031369643 | 1 |
| T2DM | b_cell | *RUVBL2* | -2.485723752 | 0.002716448 | 0.005958936 | 1 |
| T2DM | b_cell | *GDPGP1* | 2.818889771 | 0.000249544 | 0.00415349 | 1 |
| T2DM | b_cell | *CTNNA3* | -2.02227647 | 0.001193412 | 0.004217141 | 1 |
| T2DM | b_cell | *PTTG1IP* | -2.880789275 | 0.012442371 | 0.018057522 | 1 |
| T2DM | b_cell | *CCBE1* | -1.449063264 | 0.002451129 | 0.00564478 | 1 |
| T2DM | b_cell | *ZNF623* | -3.558303297 | 0.001894136 | 0.004974283 | 1 |
| T2DM | b_cell | *CCDC125* | -1.635800789 | 0.000927995 | 0.00415349 | 1 |
| T2DM | b_cell | *FHL3* | -5.117080725 | 0.000248827 | 0.00415349 | 1 |
| T2DM | b_cell | *C19orf71* | -5.058477881 | 0.000628728 | 0.00415349 | 1 |
| T2DM | b_cell | *LRIT3* | -6.280431928 | 0.001859059 | 0.004935095 | 1 |
| T2DM | b_cell | *SF3A3* | -2.892488483 | 0.006108872 | 0.010281826 | 1 |
| T2DM | b_cell | *GRIN2A* | -2.749169359 | 0.001514537 | 0.004480664 | 1 |
| T2DM | b_cell | *URAD* | -14.29963012 | 0.000205508 | 0.00415349 | 1 |
| T2DM | b_cell | *MX2* | -3.499544638 | 0.024026269 | 0.031733855 | 1 |
| T2DM | b_cell | *KLRC4* | -5.285025768 | 0.006340751 | 0.010548901 | 1 |
| T2DM | b_cell | *ACSM5* | -1.575576172 | 0.004296212 | 0.008039295 | 1 |
| T2DM | b_cell | *AC131971.1* | -11.33432006 | 0.002912747 | 0.006193075 | 1 |
| T2DM | b_cell | *SERHL2* | 3.650004313 | 0.00117495 | 0.004202203 | 1 |
| T2DM | b_cell | *PCBP3* | -1.038584974 | 0.000360005 | 0.00415349 | 1 |
| T2DM | b_cell | *TANGO2* | -3.187758433 | 0.01335003 | 0.019194524 | 1 |
| T2DM | b_cell | *SFXN4* | -3.218466715 | 0.000721537 | 0.00415349 | 1 |
| T2DM | b_cell | *FAM167B* | 4.841140083 | 0.001139251 | 0.00418468 | 1 |
| T2DM | b_cell | *DGCR6* | -6.553169404 | 0.006548072 | 0.010797808 | 1 |
| T2DM | b_cell | *NDUFB1* | -2.628041786 | 0.021616963 | 0.028934607 | 1 |
| T2DM | b_cell | *FAM19A1* | -1.703441262 | 0.000809326 | 0.00415349 | 1 |
| T2DM | b_cell | *TRMT12* | 2.850790444 | 0.002758317 | 0.00602346 | 1 |
| T2DM | b_cell | *CMTM4* | -3.507298238 | 0.002457544 | 0.005651273 | 1 |
| T2DM | b_cell | *KREMEN1* | -1.669558878 | 0.000387814 | 0.00415349 | 1 |
| T2DM | b_cell | *AIFM3* | 8.273845114 | 0.011464809 | 0.016892256 | 1 |
| T2DM | b_cell | *KCTD16* | -2.145240363 | 0.002366152 | 0.005545309 | 1 |
| T2DM | b_cell | *B3GALT5* | 2.042132976 | 0.000616052 | 0.00415349 | 1 |
| T2DM | b_cell | *ZNF703* | -5.927359936 | 4.61E-06 | 0.000965814 | 0.02897442 |
| T2DM | b_cell | *KCTD8* | -1.415020042 | 0.002686214 | 0.005931938 | 1 |
| T2DM | b_cell | *C9orf66* | -7.646805906 | 0.000995237 | 0.00415349 | 1 |
| T2DM | b_cell | *TUBA8* | -2.348777267 | 0.003795532 | 0.007362699 | 1 |
| T2DM | b_cell | *FAM162B* | -2.802406548 | 0.003746126 | 0.00730065 | 1 |
| T2DM | b_cell | *BTBD9* | -2.167572084 | 0.018606449 | 0.025386012 | 1 |
| T2DM | b_cell | *NUDT14* | -3.846349784 | 0.010911728 | 0.016217974 | 1 |
| T2DM | b_cell | *ANKRD45* | 14.18652827 | 0.011752205 | 0.01720282 | 1 |

| | | | | | | |
|---|---|---|---|---|---|---|
| T2DM | b_cell | *MAATS1* | -2.614821862 | 0.014021342 | 0.019998225 | 1 |
| T2DM | b_cell | *GPR39* | -3.722818267 | 0.001452273 | 0.004416445 | 1 |
| T2DM | b_cell | *FAM3B* | -1.491860287 | 0.015685246 | 0.021977522 | 1 |
| T2DM | b_cell | *NPIPA7* | -12.21754111 | 0.001625135 | 0.004637868 | 1 |
| T2DM | b_cell | *TTC32* | -2.45856292 | 0.000962362 | 0.00415349 | 1 |
| T2DM | b_cell | *PP2D1* | -2.567205343 | 0.005971928 | 0.010117357 | 1 |
| T2DM | b_cell | *DENND5A* | -2.800218333 | 0.00644394 | 0.010672564 | 1 |
| T2DM | b_cell | *TMEM236* | -3.624378439 | 0.000759233 | 0.00415349 | 1 |
| T2DM | b_cell | *TBX1* | -3.226145705 | 0.000860791 | 0.00415349 | 1 |
| T2DM | b_cell | *CNTN2* | -4.801289657 | 0.001015419 | 0.00415349 | 1 |
| T2DM | b_cell | *UBE2F* | -4.758080136 | 0.000480552 | 0.00415349 | 1 |
| T2DM | b_cell | *GOLGA6L4* | 6.391932014 | 0.020512204 | 0.027661997 | 1 |
| T2DM | b_cell | *PCDH9* | -1.023389062 | 0.00034476 | 0.00415349 | 1 |
| T2DM | b_cell | *OAF* | -1.660769491 | 0.000862827 | 0.00415349 | 1 |
| T2DM | b_cell | *TM2D3* | -3.437970985 | 0.021605624 | 0.028925588 | 1 |
| T2DM | b_cell | *CLECL1* | 1.91202211 | 0.000236109 | 0.00415349 | 1 |
| T2DM | b_cell | *CSF1* | -1.621970138 | 0.006404371 | 0.010621019 | 1 |
| T2DM | b_cell | *PLA2G6* | -1.425016127 | 0.006181277 | 0.010371415 | 1 |
| T2DM | b_cell | *A3GALT2* | -3.479591123 | 0.013350826 | 0.019194524 | 1 |
| T2DM | b_cell | *SS18L1* | -3.306016737 | 0.011028145 | 0.016352346 | 1 |
| T2DM | b_cell | *THAP7* | -4.267730432 | 0.001296943 | 0.004268027 | 1 |
| T2DM | b_cell | *CCR10* | -4.523951275 | 0.001536406 | 0.004508014 | 1 |
| T2DM | b_cell | *WDR27* | -2.673000593 | 0.009942465 | 0.015058607 | 1 |
| T2DM | b_cell | *TXNRD2* | -3.160235398 | 0.002827968 | 0.006101385 | 1 |
| T2DM | b_cell | *PROS1* | -2.016879156 | 0.001221407 | 0.004252746 | 1 |
| T2DM | b_cell | *HDDC3* | -2.809015998 | 0.000924097 | 0.00415349 | 1 |
| T2DM | b_cell | *ZFP1* | -3.004989191 | 0.00029755 | 0.00415349 | 1 |
| T2DM | b_cell | *TMEM173* | -3.699454924 | 0.034761432 | 0.043964016 | 1 |
| T2DM | b_cell | *SNN* | -7.036807538 | 0.007404653 | 0.01185763 | 1 |
| T2DM | b_cell | *NELL2* | 2.628064721 | 0.004349803 | 0.008105298 | 1 |
| T2DM | b_cell | *KRBA2* | -5.764444632 | 0.000499275 | 0.00415349 | 1 |
| T2DM | b_cell | *ZNF93* | -3.044808631 | 0.000981281 | 0.00415349 | 1 |
| T2DM | b_cell | *SEPT5* | -8.81171918 | 0.000931242 | 0.00415349 | 1 |
| T2DM | b_cell | *ATL3* | -2.473046951 | 0.001135143 | 0.00418468 | 1 |
| T2DM | b_cell | *NDUFA12* | -3.376578143 | 0.002294061 | 0.005477692 | 1 |
| T2DM | b_cell | *UBE2G2* | -2.42124096 | 0.001397303 | 0.004344631 | 1 |
| T2DM | b_cell | *OSBP2* | -2.526525457 | 0.001502847 | 0.004478191 | 1 |
| T2DM | b_cell | *TUSC5* | -1.425837183 | 0.005026602 | 0.008978654 | 1 |
| T2DM | b_cell | *TMED9* | -2.75167264 | 0.002777212 | 0.006031202 | 1 |
| T2DM | b_cell | *BTBD6* | -2.877751359 | 0.000125984 | 0.00415349 | 0.792063379 |
| T2DM | b_cell | *H1FX* | -3.659644165 | 0.001691181 | 0.004704627 | 1 |
| T2DM | b_cell | *RBM43* | -3.608886673 | 0.001360928 | 0.004298348 | 1 |
| T2DM | b_cell | *FAM227A* | -2.815209323 | 0.000558562 | 0.00415349 | 1 |
| T2DM | b_cell | *NOC4L* | -3.099397558 | 0.002197869 | 0.005362051 | 1 |
| T2DM | b_cell | *SORCS2* | -2.225050856 | 0.001304112 | 0.004274973 | 1 |
| T2DM | b_cell | *TMEM106A* | -3.263390927 | 0.034021795 | 0.043193664 | 1 |
| T2DM | b_cell | *SIVA1* | -3.281765686 | 0.001310793 | 0.004274973 | 1 |
| T2DM | b_cell | *UBOX5* | -2.220216065 | 0.000536656 | 0.00415349 | 1 |
| T2DM | b_cell | *MAFF* | -2.8894883 | 0.001064881 | 0.00415349 | 1 |
| T2DM | b_cell | *BRF1* | -3.329723592 | 0.005187011 | 0.009152608 | 1 |

| | | | | | | |
|---|---|---|---|---|---|---|
| T2DM | b_cell | *FLRT2* | -1.204371884 | 0.000485896 | 0.00415349 | 1 |
| T2DM | b_cell | *ADSSL1* | -4.000222507 | 0.001238245 | 0.004253251 | 1 |
| T2DM | b_cell | *C6orf120* | -5.525566254 | 0.001973443 | 0.005091111 | 1 |
| T2DM | b_cell | *DDX51* | -1.918642564 | 0.002981376 | 0.006294128 | 1 |
| T2DM | b_cell | *SIGIRR* | -3.52932228 | 0.018180227 | 0.024896338 | 1 |
| T2DM | b_cell | *IFITM2* | -3.371541445 | 0.03349585 | 0.042586129 | 1 |
| T2DM | b_cell | *KIAA0825* | -2.375981308 | 0.001914507 | 0.004996474 | 1 |
| T2DM | b_cell | *TMEM105* | -36.76324549 | 0.021563163 | 0.028874889 | 1 |
| T2DM | b_cell | *TCN2* | -3.197173018 | 0.016266947 | 0.022671314 | 1 |
| T2DM | b_cell | *GAS2L1* | -2.672729204 | 0.000372716 | 0.00415349 | 1 |
| T2DM | b_cell | *ATP6V0A2* | -2.705959086 | 0.001541731 | 0.004509332 | 1 |
| T2DM | b_cell | *HS6ST3* | 1.892513845 | 0.000891085 | 0.00415349 | 1 |
| T2DM | b_cell | *TNFAIP8L1* | -2.683858385 | 0.002055021 | 0.005197071 | 1 |
| T2DM | b_cell | *MAPK11* | -3.490784967 | 0.000456484 | 0.00415349 | 1 |
| T2DM | b_cell | *SH3BGR* | -1.872087304 | 0.000326376 | 0.00415349 | 1 |
| T2DM | b_cell | *FAM174B* | -2.975315173 | 0.004896849 | 0.008826403 | 1 |
| T2DM | b_cell | *MUC1* | 6.217562596 | 0.000877685 | 0.00415349 | 1 |
| T2DM | b_cell | *SV2B* | -3.702018867 | 0.001215482 | 0.004245409 | 1 |
| T2DM | b_cell | *PDE6G* | 2.310684837 | 0.000281083 | 0.00415349 | 1 |
| T2DM | b_cell | *TLCD2* | -3.029749231 | 0.00152809 | 0.004502743 | 1 |
| T2DM | b_cell | *LSAMP* | -1.406852452 | 0.00096022 | 0.00415349 | 1 |
| T2DM | b_cell | *AHNAK2* | -2.08998048 | 0.000740049 | 0.00415349 | 1 |
| T2DM | b_cell | *SP1* | -2.761525884 | 0.010485296 | 0.015699228 | 1 |
| T2DM | b_cell | *PCGF3* | -2.809242424 | 0.005025412 | 0.008978654 | 1 |
| T2DM | b_cell | *PBX1* | -1.22632872 | 0.00094843 | 0.00415349 | 1 |
| T2DM | b_cell | *PMEL* | 2.436681775 | 0.004392397 | 0.008165287 | 1 |
| T2DM | b_cell | *SNAI3* | -2.648191737 | 0.00142551 | 0.004380342 | 1 |
| T2DM | b_cell | *C6orf201* | -2.894010352 | 0.026382017 | 0.034397292 | 1 |
| T2DM | b_cell | *MYBL1* | -2.277916594 | 6.12E-05 | 0.00415349 | 0.384682556 |
| T2DM | b_cell | *C16orf52* | -2.204507199 | 0.001752728 | 0.004789372 | 1 |
| T2DM | b_cell | *ADARB2* | -2.530621008 | 0.000794912 | 0.00415349 | 1 |
| T2DM | b_cell | *SRL* | 2.171924614 | 0.037241046 | 0.046817527 | 1 |
| T2DM | b_cell | *KCNIP4* | -1.276992096 | 0.000487902 | 0.00415349 | 1 |
| T2DM | b_cell | *SLC52A2* | -2.657197472 | 0.000788923 | 0.00415349 | 1 |
| T2DM | b_cell | *PIGP* | -2.285277657 | 0.000315782 | 0.00415349 | 1 |
| T2DM | b_cell | *NPIPB4* | -2.579242606 | 0.001515885 | 0.004480664 | 1 |
| T2DM | b_cell | *ZNF829* | -3.56946376 | 0.014048102 | 0.020029056 | 1 |
| T2DM | b_cell | *TRIM69* | 3.648250836 | 0.00476316 | 0.008669945 | 1 |
| T2DM | b_cell | *PTCH1* | -3.687016832 | 0.016485154 | 0.022939833 | 1 |
| T2DM | b_cell | *RTN4RL1* | -1.976770772 | 0.000774333 | 0.00415349 | 1 |
| T2DM | b_cell | *PLK5* | -6.655288564 | 0.003931944 | 0.007559673 | 1 |
| T2DM | b_cell | *RASA3* | -2.986154074 | 0.000356835 | 0.00415349 | 1 |
| T2DM | b_cell | *ZNF566* | -3.804545987 | 0.002973304 | 0.006280982 | 1 |
| T2DM | b_cell | *ZNF284* | -4.926438668 | 0.000791699 | 0.00415349 | 1 |
| T2DM | b_cell | *GSAP* | -1.964191642 | 0.00403819 | 0.007696831 | 1 |
| T2DM | b_cell | *C2orf76* | -3.674456936 | 0.002693833 | 0.005942501 | 1 |
| T2DM | b_cell | *WWOX* | -1.765944173 | 0.00935702 | 0.014299364 | 1 |
| T2DM | b_cell | *CYP4Z1* | -2.438655406 | 0.001512184 | 0.004480664 | 1 |
| T2DM | b_cell | *POLR1D* | -3.210197625 | 0.014802398 | 0.020927069 | 1 |
| T2DM | b_cell | *FFAR4* | -1.882224725 | 0.00077263 | 0.00415349 | 1 |

| | | | | | | |
|---|---|---|---|---|---|---|
| T2DM | b_cell | *BPIFB4* | -106.5077561 | 0.03215045 | 0.041001711 | 1 |
| T2DM | b_cell | *CYP4F12* | -1.986958944 | 0.002474499 | 0.005679874 | 1 |
| T2DM | b_cell | *ZNF749* | -4.183532142 | 0.000614763 | 0.00415349 | 1 |
| T2DM | b_cell | *NBPF12* | -2.74015684 | 0.001241389 | 0.004253251 | 1 |
| T2DM | b_cell | *GPAT2* | 3.781731701 | 0.000947326 | 0.00415349 | 1 |
| T2DM | b_cell | *TOR3A* | 4.593990421 | 0.008833808 | 0.013662031 | 1 |
| T2DM | b_cell | *CYP4X1* | -1.747797934 | 0.002161746 | 0.005312843 | 1 |
| T2DM | b_cell | *CCDC30* | -3.072108137 | 0.002903274 | 0.006192483 | 1 |
| T2DM | b_cell | *GLDN* | -1.657047279 | 0.000750253 | 0.00415349 | 1 |
| T2DM | b_cell | *FCAR* | 2.647240101 | 0.000707224 | 0.00415349 | 1 |
| T2DM | b_cell | *KPNA4* | -2.74147558 | 0.005670526 | 0.009753925 | 1 |
| T2DM | b_cell | *ZNF501* | -2.999032424 | 0.000349991 | 0.00415349 | 1 |
| T2DM | b_cell | *ZNF197* | -2.671903102 | 0.00114448 | 0.00418468 | 1 |
| T2DM | b_cell | *FAM228A* | -2.082332588 | 0.01372956 | 0.019635519 | 1 |
| T2DM | b_cell | *PCLO* | -1.230945189 | 0.001541466 | 0.004509332 | 1 |
| T2DM | b_cell | *RGS7BP* | -7.940169131 | 1.13E-05 | 0.001732428 | 0.071029568 |
| T2DM | b_cell | *ZNF396* | -4.609994422 | 9.41E-05 | 0.00415349 | 0.591323452 |
| T2DM | b_cell | *TMEM222* | -2.259361878 | 0.00132142 | 0.004274973 | 1 |
| T2DM | b_cell | *CLCNKA* | -7.0656199 | 0.023995158 | 0.031699424 | 1 |
| T2DM | b_cell | *SEPT10* | -2.174352802 | 0.00238928 | 0.00557776 | 1 |
| T2DM | b_cell | *SMYD4* | -2.35265467 | 0.001486939 | 0.004462053 | 1 |
| T2DM | b_cell | *CEACAM19* | -2.916353535 | 7.95E-05 | 0.00415349 | 0.499818054 |
| T2DM | b_cell | *NF2* | -2.958623031 | 0.011692797 | 0.017135808 | 1 |
| T2DM | b_cell | *MIR22HG* | -1.579922996 | 0.000697602 | 0.00415349 | 1 |
| T2DM | b_cell | *PDE2A* | -2.027594397 | 0.001267271 | 0.004257394 | 1 |
| T2DM | b_cell | *C17orf58* | -1.641901943 | 0.00396782 | 0.007610032 | 1 |
| T2DM | b_cell | *CYP27C1* | -1.744387157 | 0.012879463 | 0.018601696 | 1 |
| T2DM | b_cell | *BCR* | -2.318740124 | 0.001050823 | 0.00415349 | 1 |
| T2DM | b_cell | *HYAL3* | 1.22365861 | 0.011573554 | 0.017008049 | 1 |
| T2DM | b_cell | *ZSCAN30* | -3.392859016 | 0.019352847 | 0.026216624 | 1 |
| T2DM | b_cell | *LILRB4* | -1.922717987 | 0.006297687 | 0.010499485 | 1 |
| T2DM | b_cell | *KRT16* | -3.297342208 | 0.034741018 | 0.043955882 | 1 |
| T2DM | b_cell | *TRABD2A* | -1.99539973 | 0.036096422 | 0.045533348 | 1 |
| T2DM | b_cell | *TMEM17* | -3.407998507 | 0.002253647 | 0.005426533 | 1 |
| T2DM | b_cell | *TNFRSF18* | -7.464942528 | 0.000813102 | 0.00415349 | 1 |
| T2DM | b_cell | *RTN4RL2* | -7.259870879 | 0.001236012 | 0.004253251 | 1 |
| T2DM | b_cell | *TMEM232* | -2.358007642 | 0.002336863 | 0.005511847 | 1 |
| T2DM | b_cell | *EFCAB6* | -2.276291999 | 0.003895363 | 0.007508864 | 1 |
| T2DM | b_cell | *KANK3* | -1.993184034 | 0.000672311 | 0.00415349 | 1 |
| T2DM | b_cell | *EMID1* | -5.292147054 | 0.000836046 | 0.00415349 | 1 |
| T2DM | b_cell | *TMEM216* | -2.299187304 | 0.007604923 | 0.012128907 | 1 |
| T2DM | b_cell | *RPS19BP1* | -2.415348536 | 0.000895172 | 0.00415349 | 1 |
| T2DM | b_cell | *PLCD1* | -3.317116349 | 0.039878483 | 0.049784754 | 1 |
| T2DM | b_cell | *ENTPD5* | -2.257643818 | 0.001413463 | 0.004364365 | 1 |
| T2DM | b_cell | *HEATR4* | -1.502846951 | 0.024045916 | 0.031746467 | 1 |
| T2DM | b_cell | *MRPS21* | -2.966354306 | 0.005460748 | 0.009501286 | 1 |
| T2DM | b_cell | *ANGPTL5* | -2.402647224 | 0.000827497 | 0.00415349 | 1 |
| T2DM | b_cell | *TSPYL4* | -3.501607798 | 0.006813549 | 0.011138009 | 1 |
| T2DM | b_cell | *MT1X* | -2.262070217 | 0.001407496 | 0.004357467 | 1 |
| T2DM | b_cell | *WDR86* | -2.285922525 | 0.000609467 | 0.00415349 | 1 |

| | | | | | | |
|---|---|---|---|---|---|---|
| T2DM | b_cell | *EPOR* | -2.611310541 | 0.001978781 | 0.005092436 | 1 |
| T2DM | b_cell | *FPR3* | -1.589183366 | 0.000770876 | 0.00415349 | 1 |
| T2DM | b_cell | *GJA4* | -3.219909458 | 0.002863504 | 0.006149816 | 1 |
| T2DM | b_cell | *TLR5* | -2.06982225 | 0.000817137 | 0.00415349 | 1 |
| T2DM | b_cell | *PLEKHN1* | -4.763933701 | 0.009325318 | 0.014264787 | 1 |
| T2DM | b_cell | *ZNF385C* | 4.597137106 | 0.001271967 | 0.004257394 | 1 |
| T2DM | b_cell | *TET3* | -1.560016958 | 0.000797021 | 0.00415349 | 1 |
| T2DM | b_cell | *ISG15* | -4.048435223 | 0.002101324 | 0.005241681 | 1 |
| T2DM | b_cell | *DHRS4L2* | -3.862556625 | 0.002268656 | 0.005441832 | 1 |
| T2DM | b_cell | *TRPV2* | -4.483989812 | 0.034488704 | 0.043663005 | 1 |
| T2DM | b_cell | *C2orf88* | -1.703009016 | 0.001318228 | 0.004274973 | 1 |
| T2DM | b_cell | *ADH1A* | -4.396496723 | 7.05E-05 | 0.00415349 | 0.44322801 |
| T2DM | b_cell | *SEMA4D* | -2.495236562 | 0.000895102 | 0.00415349 | 1 |
| T2DM | b_cell | *DNAH17* | -2.893587147 | 0.000543865 | 0.00415349 | 1 |
| T2DM | b_cell | *ZFP69* | -3.063867499 | 0.001054995 | 0.00415349 | 1 |
| T2DM | b_cell | *TMEM220* | -2.6930003 | 0.002987178 | 0.006296833 | 1 |
| T2DM | b_cell | *CCDC157* | 1.529281534 | 0.000658614 | 0.00415349 | 1 |
| T2DM | b_cell | *GFRAL* | 1.081319551 | 0.000340518 | 0.00415349 | 1 |
| T2DM | b_cell | *CPSF4L* | -7.769889732 | 0.001271061 | 0.004257394 | 1 |
| T2DM | b_cell | *S100A3* | -5.549526366 | 0.003639239 | 0.007185816 | 1 |
| T2DM | b_cell | *C6orf132* | -2.336869301 | 0.000894425 | 0.00415349 | 1 |
| T2DM | b_cell | *MAPK12* | -5.248805249 | 0.001012541 | 0.00415349 | 1 |
| T2DM | b_cell | *AGRN* | -3.949166824 | 0.002762851 | 0.00602499 | 1 |
| T2DM | b_cell | *FAM166A* | -12.54578729 | 0.000726047 | 0.00415349 | 1 |
| T2DM | b_cell | *ZNF626* | -2.601114813 | 0.000914647 | 0.00415349 | 1 |
| T2DM | b_cell | *SMTNL2* | -18.10919853 | 0.000708188 | 0.00415349 | 1 |
| T2DM | b_cell | *ZC3H6* | -2.59586987 | 0.005020612 | 0.008977414 | 1 |
| T2DM | b_cell | *LINC00265* | -3.892191445 | 0.026881475 | 0.034986017 | 1 |
| T2DM | b_cell | *PRKAR1B* | -2.069632453 | 0.000656015 | 0.00415349 | 1 |
| T2DM | b_cell | *NUTM2B* | -112.1635725 | 5.90E-06 | 0.001158498 | 0.037071924 |
| T2DM | b_cell | *NCR3LG1* | -5.420622242 | 0.000812477 | 0.00415349 | 1 |
| T2DM | b_cell | *AGAP4* | -2.844423843 | 0.000622811 | 0.00415349 | 1 |
| T2DM | b_cell | *HYKK* | -2.839617748 | 0.003109624 | 0.006451934 | 1 |
| T2DM | b_cell | *RUFY4* | -6.484638015 | 0.000688018 | 0.00415349 | 1 |
| T2DM | b_cell | *ZNF669* | -7.272886864 | 0.018405125 | 0.025165946 | 1 |
| T2DM | b_cell | *CENPP* | -1.59531241 | 0.003016005 | 0.006322649 | 1 |
| T2DM | b_cell | *PLSCR1* | -2.951326667 | 0.013638977 | 0.019523736 | 1 |
| T2DM | b_cell | *ZNF559* | -1.665272303 | 0.02645203 | 0.034474278 | 1 |
| T2DM | b_cell | *JAKMIP3* | -1.943388159 | 0.013648714 | 0.019533227 | 1 |
| T2DM | b_cell | *PDCD1* | 9.896631152 | 0.000426788 | 0.00415349 | 1 |
| T2DM | b_cell | *CERKL* | -5.753052912 | 0.006401083 | 0.010618367 | 1 |
| T2DM | b_cell | *COL25A1* | -1.447750194 | 0.000568796 | 0.00415349 | 1 |
| T2DM | b_cell | *NBR1* | -2.633928915 | 0.001916099 | 0.004998553 | 1 |
| T2DM | b_cell | *NDOR1* | -4.913100908 | 0.007532742 | 0.012035158 | 1 |
| T2DM | b_cell | *SUMO2* | -3.026996221 | 0.003696869 | 0.00724956 | 1 |
| T2DM | b_cell | *GOLGA8M* | -2.360079051 | 0.001331551 | 0.004275516 | 1 |
| T2DM | b_cell | *DPYD* | -2.045143886 | 0.009887343 | 0.014985951 | 1 |
| T2DM | b_cell | *S100A16* | -2.730740427 | 0.001254045 | 0.004255815 | 1 |
| T2DM | b_cell | *PARVB* | -2.146060665 | 0.000772837 | 0.00415349 | 1 |
| T2DM | b_cell | *SLC4A5* | 1.384691634 | 0.000658286 | 0.00415349 | 1 |

| | | | | | | |
|---|---|---|---|---|---|---|
| T2DM | b_cell | *UROS* | -2.845115758 | 0.004671784 | 0.008555671 | 1 |
| T2DM | b_cell | *FAM221A* | -2.096111461 | 0.001357874 | 0.004298348 | 1 |
| T2DM | b_cell | *FSIP2* | -1.299297814 | 0.004640883 | 0.0085189 | 1 |
| T2DM | b_cell | *NOXA1* | -2.55560802 | 0.000857486 | 0.00415349 | 1 |
| T2DM | b_cell | *SKOR1* | 1.245125638 | 0.000528989 | 0.00415349 | 1 |
| T2DM | b_cell | *TMEM201* | -3.060882014 | 0.005933054 | 0.010062344 | 1 |
| T2DM | b_cell | *ZDHHC11* | -3.315957353 | 0.000584356 | 0.00415349 | 1 |
| T2DM | b_cell | *FAM26F* | 1.081287018 | 0.000456821 | 0.00415349 | 1 |
| T2DM | b_cell | *BEND4* | 1.23264612 | 0.000511857 | 0.00415349 | 1 |
| T2DM | b_cell | *ZNF563* | -2.641271985 | 0.004286982 | 0.008028673 | 1 |
| T2DM | b_cell | *FBF1* | -1.651960424 | 0.002083878 | 0.005230315 | 1 |
| T2DM | b_cell | *MSL1* | -2.252966362 | 0.037582805 | 0.047152883 | 1 |
| T2DM | b_cell | *LRRK2* | -2.039276001 | 0.013318499 | 0.019165348 | 1 |
| T2DM | b_cell | *FAM196A* | -1.733338146 | 0.000728423 | 0.00415349 | 1 |
| T2DM | b_cell | *UTS2B* | -2.573737839 | 0.002805842 | 0.006076587 | 1 |
| T2DM | b_cell | *C9orf152* | -6.816983025 | 0.000784832 | 0.00415349 | 1 |
| T2DM | b_cell | *SLC15A5* | -1.631603892 | 0.018459968 | 0.025218996 | 1 |
| T2DM | b_cell | *LIPI* | -2.153056918 | 0.004185244 | 0.007886524 | 1 |
| T2DM | b_cell | *ADAT2* | 4.039205479 | 0.016380381 | 0.022814235 | 1 |
| T2DM | b_cell | *ZNF567* | -2.799945458 | 0.001573193 | 0.004562116 | 1 |
| T2DM | b_cell | *ANKDD1B* | -3.17018129 | 0.000620235 | 0.00415349 | 1 |
| T2DM | b_cell | *ALKBH2* | 5.855149318 | 0.02100871 | 0.028252783 | 1 |
| T2DM | b_cell | *LITAF* | -2.036606722 | 0.001267694 | 0.004257394 | 1 |
| T2DM | b_cell | *SF3B3* | -3.17901423 | 0.005711663 | 0.009793394 | 1 |
| T2DM | b_cell | *PLAC9* | -1.947771188 | 0.000351503 | 0.00415349 | 1 |
| T2DM | b_cell | *CLDN4* | 3.47865006 | 0.021225711 | 0.028520206 | 1 |
| T2DM | b_cell | *FAM47E* | -1.9100111 | 0.008799197 | 0.013615691 | 1 |
| T2DM | b_cell | *ZNF527* | -3.96347063 | 0.000619972 | 0.00415349 | 1 |
| T2DM | b_cell | *S100A13* | -1.787728714 | 0.001144004 | 0.00418468 | 1 |
| T2DM | b_cell | *ZNF33A* | -2.635219981 | 0.007939239 | 0.012544357 | 1 |
| T2DM | b_cell | *ZNF600* | -2.811984318 | 0.006932129 | 0.011279062 | 1 |
| T2DM | b_cell | *LINC00994* | -8.9425626 | 0.001251526 | 0.004253251 | 1 |
| T2DM | b_cell | *TSPYL1* | -3.829301821 | 0.010724058 | 0.015993768 | 1 |
| T2DM | b_cell | *PNRC2* | -3.736278368 | 0.002761099 | 0.006023719 | 1 |
| T2DM | b_cell | *ZKSCAN3* | -3.249875555 | 0.000747875 | 0.00415349 | 1 |
| T2DM | b_cell | *BLOC1S2* | -3.711883181 | 0.000179666 | 0.00415349 | 1 |
| T2DM | b_cell | *AKR1C3* | -1.690104382 | 0.0008159 | 0.00415349 | 1 |
| T2DM | b_cell | *WDSUB1* | -2.272934811 | 0.013775826 | 0.019692728 | 1 |
| T2DM | b_cell | *S100A4* | -2.060422026 | 0.00107968 | 0.00415349 | 1 |
| T2DM | b_cell | *COLCA1* | -5.840174367 | 0.000192485 | 0.00415349 | 1 |
| T2DM | b_cell | *ACADSB* | -2.667443928 | 0.000298551 | 0.00415349 | 1 |
| T2DM | b_cell | *HRCT1* | -3.435178378 | 0.000496526 | 0.00415349 | 1 |
| T2DM | b_cell | *MPHOSPH8* | -2.393132899 | 0.006026626 | 0.010177114 | 1 |
| T2DM | b_cell | *SIRPB2* | -2.48702857 | 0.000788332 | 0.00415349 | 1 |
| T2DM | b_cell | *SRGAP3* | -1.599562542 | 0.033863313 | 0.043008926 | 1 |
| T2DM | b_cell | *XPNPEP3* | -2.250637352 | 0.000280829 | 0.00415349 | 1 |
| T2DM | b_cell | *PPIA* | -2.789494192 | 0.001184668 | 0.004202203 | 1 |
| T2DM | b_cell | *ZNF471* | -2.552072198 | 0.002400233 | 0.00559313 | 1 |
| T2DM | b_cell | *GRM7* | -1.404466777 | 0.001682096 | 0.004704627 | 1 |
| T2DM | b_cell | *SUPT3H* | -2.628743683 | 0.009945037 | 0.015058875 | 1 |

| | | | | | | |
|---|---|---|---|---|---|---|
| T2DM | b_cell | *POM121* | -3.300338152 | 0.001814998 | 0.004867153 | 1 |
| T2DM | b_cell | *GIMAP5* | -2.422463815 | 0.001801302 | 0.004852128 | 1 |
| T2DM | b_cell | *STK31* | -3.2237099 | 0.0008017 | 0.00415349 | 1 |
| T2DM | b_cell | *ZKSCAN7* | 4.192726043 | 0.002903872 | 0.006192483 | 1 |
| T2DM | b_cell | *ZNF565* | -1.713573573 | 0.000986045 | 0.00415349 | 1 |
| T2DM | b_cell | *WDR5* | -2.873187486 | 0.002448307 | 0.005641723 | 1 |
| T2DM | b_cell | *LONP1* | -2.93850172 | 0.007063811 | 0.011448873 | 1 |
| T2DM | b_cell | *SLC35F1* | -2.754732119 | 0.001076564 | 0.00415349 | 1 |
| T2DM | b_cell | *ZNF34* | -4.043499388 | 0.00332678 | 0.006749102 | 1 |
| T2DM | b_cell | *ZNF781* | -2.77374112 | 0.003766654 | 0.007327028 | 1 |
| T2DM | b_cell | *INCA1* | -5.683815047 | 0.016507248 | 0.022965494 | 1 |
| T2DM | b_cell | *ZNF774* | -6.352359468 | 0.004297769 | 0.008039295 | 1 |
| T2DM | b_cell | *EVL* | -2.498657915 | 0.005920675 | 0.01004406 | 1 |
| T2DM | b_cell | *ZNF124* | -2.962551338 | 0.002788809 | 0.006050117 | 1 |
| T2DM | b_cell | *ZNF569* | -1.985328804 | 0.000766041 | 0.00415349 | 1 |
| T2DM | b_cell | *PIK3R4* | -2.808276512 | 0.005293959 | 0.009291769 | 1 |
| T2DM | b_cell | *ZNF775* | -6.428019097 | 2.42E-05 | 0.002713223 | 0.151940467 |
| T2DM | b_cell | *ZNF605* | -2.309427571 | 0.00093971 | 0.00415349 | 1 |
| T2DM | b_cell | *RFX8* | -2.232730312 | 0.001905781 | 0.004988168 | 1 |
| T2DM | b_cell | *MYL6B* | -2.583283354 | 0.00078597 | 0.00415349 | 1 |
| T2DM | b_cell | *ZNF799* | -4.173387759 | 0.001338637 | 0.004286583 | 1 |
| T2DM | b_cell | *C20orf96* | -2.624742928 | 0.000812024 | 0.00415349 | 1 |
| T2DM | b_cell | *SULT1A1* | -2.239685635 | 6.03E-05 | 0.00415349 | 0.379262452 |
| T2DM | b_cell | *ANAPC7* | -2.469349069 | 0.001412913 | 0.004364365 | 1 |
| T2DM | b_cell | *C1orf186* | 0.321856767 | 0.000568814 | 0.00415349 | 1 |
| T2DM | b_cell | *SPTSSB* | -7.897075766 | 0.000982958 | 0.00415349 | 1 |
| T2DM | b_cell | *SULF2* | -2.238925726 | 0.002164138 | 0.005312843 | 1 |
| T2DM | b_cell | *XRCC2* | -5.074999293 | 0.002063333 | 0.005211801 | 1 |
| T2DM | b_cell | *MYO6* | -2.156005531 | 0.000234949 | 0.00415349 | 1 |
| T2DM | b_cell | *HDAC2* | -2.720339464 | 0.001512263 | 0.004480664 | 1 |
| T2DM | b_cell | *ADH1B* | -1.091147299 | 0.000848218 | 0.00415349 | 1 |
| T2DM | b_cell | *ZNF502* | -2.336634589 | 0.001763914 | 0.004791515 | 1 |
| T2DM | b_cell | *TECPR2* | -2.589954214 | 0.002038998 | 0.005175284 | 1 |
| T2DM | b_cell | *FAM180B* | -4.118369582 | 5.57E-06 | 0.001129145 | 0.035003506 |
| T2DM | b_cell | *ZFP62* | -3.95390027 | 0.002245582 | 0.005417488 | 1 |
| T2DM | b_cell | *ERI2* | -1.540269701 | 0.001052634 | 0.00415349 | 1 |
| T2DM | b_cell | *TRPV1* | -2.426745764 | 0.024725343 | 0.032472996 | 1 |
| T2DM | b_cell | *ZNF33B* | -4.258273904 | 4.27E-05 | 0.003541347 | 0.268766274 |
| T2DM | b_cell | *AMZ2* | -2.619700467 | 0.000650672 | 0.00415349 | 1 |
| T2DM | b_cell | *ZNF431* | -2.503397893 | 0.002104754 | 0.005241681 | 1 |
| T2DM | b_cell | *NF1* | -2.110038731 | 0.0066521 | 0.010933791 | 1 |
| T2DM | b_cell | *DAPK1* | -1.677671281 | 0.001275178 | 0.004257394 | 1 |
| T2DM | b_cell | *TLE1* | -1.474595343 | 0.000528085 | 0.00415349 | 1 |
| T2DM | b_cell | *STRN3* | -2.312340041 | 0.00421637 | 0.007929499 | 1 |
| T2DM | b_cell | *ZNF239* | -2.375519933 | 0.006400725 | 0.010618367 | 1 |
| T2DM | b_cell | *MVB12B* | -2.767573176 | 0.004330324 | 0.008080958 | 1 |
| T2DM | b_cell | *PPTC7* | -3.250142865 | 0.011242472 | 0.016599676 | 1 |
| T2DM | b_cell | *NHLRC2* | -2.467403677 | 0.002610563 | 0.005849897 | 1 |
| T2DM | b_cell | *ZFP28* | -2.821175355 | 0.000823782 | 0.00415349 | 1 |
| T2DM | b_cell | *KIAA1211L* | -1.79510514 | 0.014615532 | 0.020700123 | 1 |

| | | | | | | |
|---|---|---|---|---|---|---|
| T2DM | b_cell | *LAMB3* | -2.992232962 | 0.001264867 | 0.004257394 | 1 |
| T2DM | b_cell | *KPNA5* | -2.956416482 | 0.010477476 | 0.01569776 | 1 |
| T2DM | b_cell | *ANKRD36B* | -2.965021548 | 0.021358761 | 0.028643885 | 1 |
| T2DM | b_cell | *HCAR1* | -2.76320177 | 0.007749853 | 0.012316311 | 1 |
| T2DM | b_cell | *SRGAP1* | -1.984267705 | 0.002207655 | 0.005381337 | 1 |
| T2DM | b_cell | *AP2A1* | -3.281898052 | 0.036243611 | 0.045691515 | 1 |
| T2DM | b_cell | *PCDHB16* | -2.22527663 | 0.015198069 | 0.021385465 | 1 |
| T2DM | b_cell | *HIST1H3E* | 2.212947334 | 0.000212031 | 0.00415349 | 1 |
| T2DM | b_cell | *ZNF585A* | -1.985999662 | 0.00977604 | 0.014838717 | 1 |
| T2DM | b_cell | *FUT11* | -1.929883608 | 0.002219946 | 0.005393569 | 1 |
| T2DM | b_cell | *ANXA4* | -2.51380546 | 0.008424423 | 0.013129486 | 1 |
| T2DM | b_cell | *WDR5B* | 5.137711188 | 0.0002365 | 0.00415349 | 1 |
| T2DM | b_cell | *ZNF138* | -2.689120992 | 0.013399957 | 0.019242926 | 1 |
| T2DM | b_cell | *ZNF429* | -2.90784073 | 0.002253037 | 0.005426533 | 1 |
| T2DM | b_cell | *ZNF470* | -4.052435379 | 0.000575904 | 0.00415349 | 1 |
| T2DM | b_cell | *ZNF100* | -2.954333065 | 0.020505406 | 0.027658762 | 1 |
| T2DM | b_cell | *GMFB* | -3.404270515 | 0.00361261 | 0.007151285 | 1 |
| T2DM | b_cell | *ZNF763* | -2.491695691 | 0.00975606 | 0.014813758 | 1 |
| T2DM | b_cell | *ARRDC1* | -2.812357307 | 0.000502691 | 0.00415349 | 1 |
| T2DM | b_cell | *IGF2R* | -2.59354175 | 0.001726235 | 0.004752113 | 1 |
| T2DM | b_cell | *GAL3ST4* | -3.902182657 | 0.004844586 | 0.008769913 | 1 |
| T2DM | b_cell | *SLC25A29* | -2.468210647 | 0.004945366 | 0.008877689 | 1 |
| T2DM | b_cell | *ZNF682* | -1.900575121 | 0.000703452 | 0.00415349 | 1 |
| T2DM | b_cell | *ADAM32* | -1.959243229 | 0.000687881 | 0.00415349 | 1 |
| T2DM | b_cell | *LRRC8B* | -1.59927482 | 0.000776044 | 0.00415349 | 1 |
| T2DM | b_cell | *ABCB8* | -3.266610785 | 0.000259809 | 0.00415349 | 1 |
| T2DM | b_cell | *SND1* | -2.535668723 | 0.003785002 | 0.007349075 | 1 |
| T2DM | b_cell | *NEK5* | -2.690592081 | 0.000646057 | 0.00415349 | 1 |
| T2DM | b_cell | *PIWIL2* | -2.092858123 | 0.002586822 | 0.005820813 | 1 |
| T2DM | b_cell | *C1D* | -3.266781331 | 0.000547992 | 0.00415349 | 1 |
| T2DM | b_cell | *TBC1D9B* | -3.45177848 | 0.000148413 | 0.00415349 | 0.933074415 |
| T2DM | b_cell | *FAM110D* | -4.291682327 | 0.001419644 | 0.004370862 | 1 |
| T2DM | b_cell | *KANK2* | -1.869040488 | 0.000964861 | 0.00415349 | 1 |
| T2DM | b_cell | *C6orf141* | -3.33262889 | 0.000469161 | 0.00415349 | 1 |
| T2DM | b_cell | *CCL4L2* | -3.588224166 | 0.001972025 | 0.005091111 | 1 |
| T2DM | b_cell | *GTF2E2* | -2.294604135 | 0.003919158 | 0.007541225 | 1 |
| T2DM | b_cell | *RAMP2-AS1* | -1.781857535 | 0.002836491 | 0.00611348 | 1 |
| T2DM | b_cell | *ZNF720* | -2.920953487 | 0.000504395 | 0.00415349 | 1 |
| T2DM | b_cell | *DDI2* | -2.63770505 | 0.002518539 | 0.005741137 | 1 |
| T2DM | b_cell | *MRPL21* | 2.481887017 | 0.011709481 | 0.017144273 | 1 |
| T2DM | b_cell | *ZNF786* | -3.95846371 | 0.001160636 | 0.004195495 | 1 |
| T2DM | b_cell | *ZNF517* | -3.177798221 | 0.000768152 | 0.00415349 | 1 |
| T2DM | b_cell | *SLC22A5* | -1.224083998 | 0.036194133 | 0.045638291 | 1 |
| T2DM | b_cell | *C5AR1* | -2.343006372 | 0.00276131 | 0.006023719 | 1 |
| T2DM | b_cell | *DCHS2* | -2.175506518 | 0.000393625 | 0.00415349 | 1 |
| T2DM | b_cell | *SHPK* | -2.171629172 | 0.010712954 | 0.015989873 | 1 |
| T2DM | b_cell | *MAP3K5* | -2.317813798 | 0.014300101 | 0.020331238 | 1 |
| T2DM | b_cell | *STMN3* | -4.42574149 | 0.024921736 | 0.032689955 | 1 |
| T2DM | b_cell | *GYPE* | 4.223858782 | 0.000326856 | 0.00415349 | 1 |
| T2DM | b_cell | *PCDHB11* | -6.070609884 | 0.000130163 | 0.00415349 | 0.818337432 |

| | | | | | | |
|---|---|---|---|---|---|---|
| T2DM | b_cell | *SLC2A10* | -1.885404656 | 0.003141798 | 0.006486859 | 1 |
| T2DM | b_cell | *ZNF665* | -3.008255636 | 0.004780728 | 0.00869424 | 1 |
| T2DM | b_cell | *RPF2* | -2.715669752 | 0.002569124 | 0.005799671 | 1 |
| T2DM | b_cell | *MYO5A* | -1.816396552 | 0.003347526 | 0.006770366 | 1 |
| T2DM | b_cell | *ATG7* | -1.839233753 | 0.002052826 | 0.005193845 | 1 |
| T2DM | b_cell | *TTC30A* | -5.361177582 | 5.90E-05 | 0.00415349 | 0.37071456 |
| T2DM | b_cell | *RAB40C* | -2.627421065 | 0.00118288 | 0.004202203 | 1 |
| T2DM | b_cell | *HHLA3* | -6.267094621 | 0.000730418 | 0.00415349 | 1 |
| T2DM | b_cell | *HOXA4* | -4.748679537 | 0.000560784 | 0.00415349 | 1 |
| T2DM | b_cell | *ENTPD6* | -2.668157505 | 0.000436816 | 0.00415349 | 1 |
| T2DM | b_cell | *C5orf42* | -2.407415592 | 0.005139254 | 0.009098983 | 1 |
| T2DM | b_cell | *MFAP5* | -1.154630246 | 0.000784724 | 0.00415349 | 1 |
| T2DM | b_cell | *CDC42SE1* | -3.551031459 | 0.02390614 | 0.031588461 | 1 |
| T2DM | b_cell | *MPEG1* | -1.932810548 | 0.024878571 | 0.032652736 | 1 |
| T2DM | b_cell | *PDCD1LG2* | -1.707139484 | 0.001797412 | 0.004852128 | 1 |
| T2DM | b_cell | *KLHL14* | 0.536498365 | 0.000551538 | 0.00415349 | 1 |
| T2DM | b_cell | *FAM114A1* | -2.3196151 | 0.002775099 | 0.006030781 | 1 |
| T2DM | b_cell | *PHF2* | -3.970715057 | 0.009885343 | 0.014985951 | 1 |
| T2DM | b_cell | *PSAP* | -2.530679783 | 0.01400788 | 0.019983559 | 1 |
| T2DM | b_cell | *S100A10* | -2.755395103 | 0.000872434 | 0.00415349 | 1 |
| T2DM | b_cell | *MAP1LC3C* | -3.601031557 | 0.003373623 | 0.00680461 | 1 |
| T2DM | b_cell | *TAF13* | -1.906756205 | 0.002049381 | 0.00519116 | 1 |
| T2DM | b_cell | *AC122129.1* | 8.607518991 | 0.001344108 | 0.004286796 | 1 |
| T2DM | b_cell | *OCLN* | -2.499327667 | 0.004540892 | 0.008382656 | 1 |
| T2DM | b_cell | *ZNF181* | 4.000665923 | 0.001629075 | 0.00464279 | 1 |
| T2DM | b_cell | *SGTB* | -2.310098831 | 0.001184376 | 0.004202203 | 1 |
| T2DM | b_cell | *ADH5* | -2.200146558 | 0.000875753 | 0.00415349 | 1 |
| T2DM | b_cell | *ZNF677* | -2.230460195 | 0.00077994 | 0.00415349 | 1 |
| T2DM | b_cell | *ZNF311* | -3.220514323 | 0.000621786 | 0.00415349 | 1 |
| T2DM | b_cell | *ZNF71* | -2.602920804 | 0.000326471 | 0.00415349 | 1 |
| T2DM | b_cell | *VPS13A* | -1.944153681 | 0.018867456 | 0.025664149 | 1 |
| T2DM | b_cell | *LEKR1* | -2.217046562 | 0.000411549 | 0.00415349 | 1 |
| T2DM | b_cell | *MRPL42* | -2.840177714 | 0.002274464 | 0.005445221 | 1 |
| T2DM | b_cell | *ZNF335* | -3.163461659 | 0.004980874 | 0.00892419 | 1 |
| T2DM | b_cell | *AGAP9* | 3.882506019 | 0.000790049 | 0.00415349 | 1 |
| T2DM | b_cell | *ZNF84* | -2.849069195 | 0.002044499 | 0.005187152 | 1 |
| T2DM | b_cell | *MAK16* | -2.347244049 | 0.000674711 | 0.00415349 | 1 |
| T2DM | b_cell | *ZNF667* | -3.315977567 | 0.000223988 | 0.00415349 | 1 |
| T2DM | b_cell | *PRIM1* | -4.879350087 | 0.000103461 | 0.00415349 | 0.650459526 |
| T2DM | b_cell | *SULT1C4* | -2.806765788 | 0.002829024 | 0.00610157 | 1 |
| T2DM | b_cell | *SFI1* | -3.463300977 | 0.02418745 | 0.031896659 | 1 |
| T2DM | b_cell | *ZNF649* | -4.891626779 | 0.015777967 | 0.022082832 | 1 |
| T2DM | b_cell | *ADH4* | -2.091798105 | 0.000259395 | 0.00415349 | 1 |
| T2DM | b_cell | *ZNF248* | -2.297929236 | 0.000399819 | 0.00415349 | 1 |
| T2DM | b_cell | *HIBCH* | -2.067247523 | 0.00132808 | 0.004274973 | 1 |
| T2DM | b_cell | *ZNF544* | -2.462567122 | 0.006771125 | 0.011083068 | 1 |
| T2DM | b_cell | *SOWAHC* | -4.06057328 | 0.000521527 | 0.00415349 | 1 |
| T2DM | b_cell | *MIER1* | -2.523739192 | 0.025443278 | 0.033332338 | 1 |
| T2DM | b_cell | *SVIP* | -2.602732138 | 0.00112045 | 0.004182741 | 1 |
| T2DM | b_cell | *ZNF251* | -2.720914848 | 0.009452785 | 0.01441418 | 1 |

| | | | | | | |
|---|---|---|---|---|---|---|
| T2DM | b_cell | *DDRGK1* | -3.617852026 | 0.002393228 | 0.005579107 | 1 |
| T2DM | b_cell | *ZNF334* | -2.029192516 | 0.003284425 | 0.006688938 | 1 |
| T2DM | b_cell | *HSD17B11* | -3.116623316 | 0.007228176 | 0.011646218 | 1 |
| T2DM | b_cell | *RPS6KL1* | -2.124913231 | 0.007672651 | 0.012209772 | 1 |
| T2DM | b_cell | *QRICH1* | -3.065042637 | 0.001187348 | 0.004205554 | 1 |
| T2DM | b_cell | *DDX42* | -2.426059732 | 0.037748131 | 0.047331971 | 1 |
| T2DM | b_cell | *SLC29A3* | -2.622509582 | 0.00060693 | 0.00415349 | 1 |
| T2DM | b_cell | *ANTXRL* | -99.74881711 | 0.000638503 | 0.00415349 | 1 |
| T2DM | b_cell | *STYX* | -3.57091811 | 0.006766928 | 0.01108168 | 1 |
| T2DM | b_cell | *TMEM116* | -2.649733298 | 1.99E-05 | 0.002553516 | 0.124859896 |
| T2DM | b_cell | *GFPT1* | -2.34797275 | 0.005283669 | 0.009278889 | 1 |
| T2DM | b_cell | *UVRAG* | -1.722001362 | 0.008887104 | 0.013724692 | 1 |
| T2DM | b_cell | *MGEA5* | -1.863032768 | 0.001192486 | 0.004216626 | 1 |
| T2DM | b_cell | *MT1F* | -12.28503663 | 0.000747774 | 0.00415349 | 1 |
| T2DM | b_cell | *ZNF587* | -3.083719303 | 0.002026313 | 0.00516315 | 1 |
| T2DM | b_cell | *FLVCR1-AS1* | -1.571566282 | 0.018973024 | 0.02578543 | 1 |
| T2DM | b_cell | *SH3BGRL2* | -1.842565391 | 0.00051976 | 0.00415349 | 1 |
| T2DM | b_cell | *TMA16* | -3.208383553 | 0.010296484 | 0.015490307 | 1 |
| T2DM | b_cell | *HLA-DRB5* | -3.304232537 | 6.51E-10 | 1.37E-06 | 4.10E-06 |
| T2DM | b_cell | *CNGA1* | -1.996824062 | 0.004449718 | 0.00824017 | 1 |
| T2DM | b_cell | *MAFK* | -3.952304269 | 0.003001089 | 0.006308941 | 1 |
| T2DM | b_cell | *HIST1H4E* | 7.83495451 | 0.006835331 | 0.011163204 | 1 |
| T2DM | b_cell | *C20orf203* | 3.92182252 | 0.002666109 | 0.005916635 | 1 |
| T2DM | b_cell | *ZNF627* | -2.419972953 | 0.001270508 | 0.004257394 | 1 |
| T2DM | b_cell | *ARC* | -15.11191252 | 0.000817826 | 0.00415349 | 1 |
| T2DM | b_cell | *MMP17* | -4.743117982 | 0.002907374 | 0.006192483 | 1 |
| T2DM | b_cell | *CCDC69* | -2.04707828 | 0.001355997 | 0.004296951 | 1 |
| T2DM | b_cell | *MDM4* | -2.560093763 | 0.013376223 | 0.019217622 | 1 |
| T2DM | b_cell | *ZNF534* | -9.71724259 | 0.000737746 | 0.00415349 | 1 |
| T2DM | b_cell | *NCOA6* | -2.374454339 | 0.00277077 | 0.006028396 | 1 |
| T2DM | b_cell | *FAM19A2* | -2.08938242 | 0.000958674 | 0.00415349 | 1 |
| T2DM | b_cell | *TTC37* | -2.349241946 | 0.004799317 | 0.008710593 | 1 |
| T2DM | b_cell | *PAPSS2* | -1.653757298 | 0.001229086 | 0.004253251 | 1 |
| T2DM | b_cell | *FAN1* | -2.307998961 | 0.001186047 | 0.004203312 | 1 |
| T2DM | b_cell | *CEP290* | -3.09309254 | 0.012118235 | 0.017652303 | 1 |
| T2DM | b_cell | *ANKRD13B* | -2.849804059 | 0.004655828 | 0.008536363 | 1 |
| T2DM | b_cell | *ECI2* | -2.422929003 | 0.00126519 | 0.004257394 | 1 |
| T2DM | b_cell | *ZNF652* | -2.731643701 | 0.027894248 | 0.036166454 | 1 |
| T2DM | b_cell | *APCDD1L* | 4.74929762 | 0.005812182 | 0.009915476 | 1 |
| T2DM | b_cell | *RCSD1* | 2.717110866 | 0.011223851 | 0.01657997 | 1 |
| T2DM | b_cell | *RASSF9* | -2.022170505 | 0.000805097 | 0.00415349 | 1 |
| T2DM | b_cell | *FAM169A* | 4.478679177 | 0.031085039 | 0.03983523 | 1 |
| T2DM | b_cell | *TMEM184B* | -2.776888085 | 0.002583572 | 0.005819748 | 1 |
| T2DM | b_cell | *SCAMP5* | -2.462790919 | 0.025657193 | 0.033563623 | 1 |
| T2DM | b_cell | *ALPK2* | -2.76443354 | 0.003120235 | 0.006465696 | 1 |
| T2DM | b_cell | *SFT2D1* | -2.804378513 | 0.002560066 | 0.005798627 | 1 |
| T2DM | b_cell | *ARHGAP11A* | -3.706103948 | 0.003003477 | 0.006308941 | 1 |
| T2DM | b_cell | *HMGN2* | -1.897708325 | 0.002623097 | 0.005854674 | 1 |
| T2DM | b_cell | *DENND4B* | -3.793962115 | 0.001330702 | 0.004274973 | 1 |
| T2DM | b_cell | *CD3E* | -2.942804452 | 0.001833393 | 0.004898539 | 1 |

| | | | | | | |
|---|---|---|---|---|---|---|
| T2DM | b_cell | *TSEN15* | -2.544006855 | 0.001335209 | 0.004282824 | 1 |
| T2DM | b_cell | *TYW1* | -1.762586377 | 0.025247605 | 0.033089784 | 1 |
| T2DM | b_cell | *ITPRIPL1* | 6.094194836 | 0.000874521 | 0.00415349 | 1 |
| T2DM | b_cell | *PRMT6* | 3.863782827 | 0.001308229 | 0.004274973 | 1 |
| T2DM | b_cell | *SHISA4* | -6.244737754 | 0.000918839 | 0.00415349 | 1 |
| T2DM | b_cell | *CAPZA2* | -2.509862561 | 0.003941878 | 0.00757414 | 1 |
| T2DM | b_cell | *MAP3K3* | -2.72845042 | 0.001971576 | 0.005091111 | 1 |
| T2DM | b_cell | *C1orf174* | -3.075653924 | 0.00186557 | 0.004938459 | 1 |
| T2DM | b_cell | *RASGEF1A* | -6.441703652 | 0.00079683 | 0.00415349 | 1 |
| T2DM | b_cell | *KIAA0753* | -3.388929919 | 0.002680288 | 0.005924338 | 1 |
| T2DM | b_cell | *DCLRE1A* | 6.024948797 | 0.031298344 | 0.040068734 | 1 |
| T2DM | b_cell | *APRT* | 4.489391795 | 0.023271461 | 0.030891682 | 1 |
| T2DM | b_cell | *TBKBP1* | -3.602326111 | 0.000193149 | 0.00415349 | 1 |
| T2DM | b_cell | *ZFP2* | -3.015008869 | 0.002313358 | 0.005492478 | 1 |
| T2DM | b_cell | *L3MBTL3* | 3.132041349 | 0.010435381 | 0.015650582 | 1 |
| T2DM | b_cell | *MFAP3L* | -4.288923864 | 0.001642056 | 0.004657671 | 1 |
| T2DM | b_cell | *NAGA* | -2.634358854 | 0.000746297 | 0.00415349 | 1 |
| T2DM | b_cell | *SMG5* | -3.243797239 | 0.002208515 | 0.005381337 | 1 |
| T2DM | b_cell | *TGM2* | -1.43876793 | 0.017895531 | 0.024589847 | 1 |
| T2DM | b_cell | *PJA2* | -2.596958653 | 0.003362257 | 0.006790062 | 1 |
| T2DM | b_cell | *AL590822.1* | -2.432208072 | 0.002498636 | 0.005718574 | 1 |
| T2DM | b_cell | *AP001007.1* | 2.316977172 | 0.000574616 | 0.00415349 | 1 |
| T2DM | b_cell | *ZNF525* | -4.731892541 | 0.020661443 | 0.027833403 | 1 |
| T2DM | b_cell | *AC105020.1* | 4.637474563 | 0.001538939 | 0.004509332 | 1 |
| T2DM | b_cell | *EFCAB2* | -2.419547433 | 0.002750573 | 0.006008635 | 1 |
| T2DM | b_cell | *CHML* | 3.493103892 | 0.00011984 | 0.00415349 | 0.753431988 |
| T2DM | b_cell | *CAPN8* | -9.622755267 | 0.000517543 | 0.00415349 | 1 |
| T2DM | b_cell | *CENPW* | -7.209136652 | 1.58E-06 | 0.000397014 | 0.009925341 |
| T2DM | b_cell | *FAM229B* | -3.471455544 | 1.47E-05 | 0.002158387 | 0.092489541 |
| T2DM | b_cell | *DDO* | -3.658243541 | 0.001336552 | 0.004282824 | 1 |
| T2DM | b_cell | *HIST2H2BF* | -9.662892845 | 0.00077543 | 0.00415349 | 1 |
| T2DM | b_cell | *RBM20* | -2.963789699 | 0.000561631 | 0.00415349 | 1 |
| T2DM | b_cell | *C6orf163* | -3.287234038 | 0.000706771 | 0.00415349 | 1 |
| T2DM | b_cell | *PCMTD2* | -2.06369875 | 0.002217553 | 0.005393328 | 1 |
| T2DM | b_cell | *OOEP* | 1.767887411 | 0.002858243 | 0.006143148 | 1 |
| T2DM | b_cell | *SYS1* | -5.46496135 | 0.037746903 | 0.047331971 | 1 |
| T2DM | b_cell | *INPP5B* | -1.849249111 | 0.000491878 | 0.00415349 | 1 |
| T2DM | b_cell | *TRAF3IP1* | -2.448063313 | 0.000937027 | 0.00415349 | 1 |
| T2DM | b_cell | *GIGYF2* | -2.38941434 | 0.005245929 | 0.00922808 | 1 |
| T2DM | b_cell | *RUFY2* | -2.711249637 | 0.006520542 | 0.010765402 | 1 |
| T2DM | b_cell | *PHACTR4* | -2.873435081 | 0.002263305 | 0.005437295 | 1 |
| T2DM | b_cell | *C10orf128* | -1.699949421 | 0.002085363 | 0.005230315 | 1 |
| T2DM | b_cell | *ZDBF2* | -2.961554407 | 0.00369557 | 0.00724956 | 1 |
| T2DM | b_cell | *RING1* | -2.843957316 | 0.001179406 | 0.004202203 | 1 |
| T2DM | b_cell | *HSD17B8* | -4.527466055 | 0.003233376 | 0.006615111 | 1 |
| T2DM | b_cell | *HLA-DOA* | 0.777485983 | 0.000893959 | 0.00415349 | 1 |
| T2DM | b_cell | *HLA-DRA* | 4.797787017 | 2.53E-07 | 0.000106041 | 0.001590611 |
| T2DM | b_cell | *COL15A1* | -1.869246488 | 0.000798757 | 0.00415349 | 1 |
| T2DM | b_cell | *NOTCH4* | -1.043344402 | 1.47E-07 | 7.05E-05 | 0.000921051 |
| T2DM | b_cell | *MRPL38* | 3.050179928 | 0.000777001 | 0.00415349 | 1 |

| | | | | | | |
|---|---|---|---|---|---|---|
| T2DM | b_cell | *DXO* | -3.692109475 | 4.16E-05 | 0.003541347 | 0.261722181 |
| T2DM | b_cell | *SKIV2L* | -1.840202688 | 0.000346296 | 0.00415349 | 1 |
| T2DM | b_cell | *LAYN* | -4.010030285 | 0.000794899 | 0.00415349 | 1 |
| T2DM | b_cell | *CARD16* | 4.005556652 | 0.00640723 | 0.010622957 | 1 |
| T2DM | b_cell | *ABHD16A* | 1.884264007 | 0.000848728 | 0.00415349 | 1 |
| T2DM | b_cell | *ZNF814* | -2.233066319 | 0.001350355 | 0.004292052 | 1 |
| T2DM | b_cell | *MICA* | -2.227089345 | 0.004613409 | 0.008483329 | 1 |
| T2DM | b_cell | *HLA-C* | -2.195150795 | 0.017701293 | 0.024369561 | 1 |
| T2DM | b_cell | *CCHCR1* | -1.43175024 | 0.031390285 | 0.040177264 | 1 |
| T2DM | b_cell | *ABCF1* | -2.826299644 | 0.006373124 | 0.010586483 | 1 |
| T2DM | b_cell | *LILRB3* | -2.337439299 | 0.002565968 | 0.005799421 | 1 |
| T2DM | b_cell | *LINC01123* | -142.894731 | 2.08E-06 | 0.000503113 | 0.013080929 |
| T2DM | b_cell | *HLA-E* | -1.919674084 | 0.010984158 | 0.016306352 | 1 |
| T2DM | b_cell | *ZNF468* | -3.781609411 | 0.000875716 | 0.00415349 | 1 |
| T2DM | b_cell | *HCG9* | -2.371654539 | 0.015116317 | 0.021299033 | 1 |
| T2DM | b_cell | *TBC1D8* | -2.684926548 | 0.005853309 | 0.009962034 | 1 |
| T2DM | b_cell | *HLA-F* | -2.77008319 | 0.013660723 | 0.019545964 | 1 |
| T2DM | b_cell | *ZFP57* | 2.098759503 | 0.002087301 | 0.005230315 | 1 |
| T2DM | b_cell | *FAM153C* | 2.166524379 | 0.000624313 | 0.00415349 | 1 |
| T2DM | b_cell | *TRIM27* | -2.575534204 | 0.000873521 | 0.00415349 | 1 |
| T2DM | b_cell | *MALRD1* | -1.492621716 | 0.001759815 | 0.004789372 | 1 |
| T2DM | b_cell | *MROH6* | -3.42872942 | 0.008702128 | 0.013495383 | 1 |
| T2DM | b_cell | *ATXN2* | -2.3127805 | 0.00704264 | 0.011420448 | 1 |
| T2DM | b_cell | *ZNF155* | -3.031647566 | 0.003206425 | 0.006581388 | 1 |
| T2DM | b_cell | *ZNF425* | -3.481811079 | 0.0237106 | 0.031369643 | 1 |
| T2DM | b_cell | *LRRC10B* | -20.20671041 | 0.024495329 | 0.032242628 | 1 |
| T2DM | b_cell | *C12orf73* | -4.703232817 | 0.001470914 | 0.004439577 | 1 |
| T2DM | b_cell | *PCDHGA1* | -5.819889246 | 0.000954764 | 0.00415349 | 1 |
| T2DM | b_cell | *PCDHA5* | -23.72212863 | 0.008607294 | 0.013373053 | 1 |
| T2DM | b_cell | *PCDHA4* | -13.88354595 | 0.000962335 | 0.00415349 | 1 |
| T2DM | b_cell | *TRIM13* | -2.653348632 | 0.006777327 | 0.011087445 | 1 |
| T2DM | b_cell | *SLFN12L* | -1.737593328 | 0.00099112 | 0.00415349 | 1 |
| T2DM | b_cell | *SYCE1L* | 4.841616054 | 0.000261736 | 0.00415349 | 1 |
| T2DM | b_cell | *TMEM231* | -3.022869975 | 0.002304855 | 0.005484718 | 1 |
| T2DM | b_cell | *FAM71F2* | -2.663081137 | 0.000787591 | 0.00415349 | 1 |
| T2DM | b_cell | *LINC00654* | -5.105679205 | 0.003817421 | 0.007389201 | 1 |
| T2DM | b_cell | *TMEM170B* | -1.307216811 | 0.004833985 | 0.008753851 | 1 |
| T2DM | b_cell | *NT5M* | -2.582293224 | 0.003727075 | 0.007274797 | 1 |
| T2DM | b_cell | *PRR13* | -4.935527694 | 0.001298778 | 0.004268382 | 1 |
| T2DM | b_cell | *MT1A* | -7.127917317 | 0.000757272 | 0.00415349 | 1 |
| T2DM | b_cell | *MT1M* | -3.353979339 | 0.000786021 | 0.00415349 | 1 |
| T2DM | b_cell | *CFI* | -2.099445643 | 0.000642258 | 0.00415349 | 1 |
| T2DM | b_cell | *EXOC3L4* | -5.628350132 | 0.000417467 | 0.00415349 | 1 |
| T2DM | b_cell | *TP53TG3D* | 3.730198145 | 0.001623174 | 0.004634377 | 1 |
| T2DM | b_cell | *ATP6AP1L* | -1.954719382 | 0.010202014 | 0.015373936 | 1 |
| T2DM | b_cell | *NAP1L4* | -2.35117147 | 0.001839246 | 0.004898539 | 1 |
| T2DM | b_cell | *CPT1B* | -3.35852535 | 0.026049861 | 0.034020664 | 1 |
| T2DM | b_cell | *SERF1B* | -11.54604795 | 1.72E-05 | 0.002401215 | 0.108054694 |
| T2DM | b_cell | *DENND6B* | -5.571693285 | 0.010853184 | 0.016150051 | 1 |
| T2DM | b_cell | *EIF3CL* | -6.181603035 | 0.015704988 | 0.02198775 | 1 |

| | | | | | | |
|---|---|---|---|---|---|---|
| T2DM | b_cell | *LCMT1* | -2.704345873 | 0.001963316 | 0.005077485 | 1 |
| T2DM | b_cell | *CDPF1* | -4.052554778 | 0.027103051 | 0.035227802 | 1 |
| T2DM | b_cell | *LIN52* | -1.591879919 | 0.020900763 | 0.028125663 | 1 |
| T2DM | b_cell | *C17orf107* | 2.969913157 | 0.000123042 | 0.00415349 | 0.773563201 |
| T2DM | b_cell | *LINC00487* | -8.103591091 | 0.027606154 | 0.035822474 | 1 |
| T2DM | b_cell | *RFPL3S* | -4.283554779 | 0.011058192 | 0.016377586 | 1 |
| T2DM | b_cell | *C1QTNF9B* | -35.78163448 | 0.000778685 | 0.00415349 | 1 |
| T2DM | b_cell | *RNPS1* | -2.754250058 | 0.02172341 | 0.029064711 | 1 |
| T2DM | b_cell | *NYNRIN* | -4.177405603 | 0.003708863 | 0.007257274 | 1 |
| T2DM | b_cell | *ZDHHC11B* | -2.766876325 | 0.001523964 | 0.004498197 | 1 |
| T2DM | b_cell | *GOLGA8O* | 10.72392606 | 0.003369349 | 0.006800352 | 1 |
| T2DM | b_cell | *HCG27* | 3.827581758 | 0.000104754 | 0.00415349 | 0.658586001 |
| T2DM | b_cell | *LNP1* | -3.091642338 | 0.000707775 | 0.00415349 | 1 |
| T2DM | b_cell | *VGLL3* | -1.367084734 | 0.000421545 | 0.00415349 | 1 |
| T2DM | b_cell | *COLQ* | -2.589243184 | 0.000588287 | 0.00415349 | 1 |
| T2DM | b_cell | *METTL6* | -2.598240849 | 0.008877619 | 0.013716783 | 1 |
| T2DM | b_cell | *SACM1L* | -2.884403043 | 0.028759454 | 0.037150336 | 1 |
| T2DM | b_cell | *SLC48A1* | -2.150336644 | 0.000676979 | 0.00415349 | 1 |
| T2DM | b_cell | *TAS2R14* | 5.228706712 | 4.62E-05 | 0.00372663 | 0.29067713 |
| T2DM | b_cell | *ZRSR1* | -12.91323532 | 0.003031391 | 0.006344326 | 1 |
| T2DM | b_cell | *AC084018.1* | -3.589912295 | 0.000194256 | 0.00415349 | 1 |
| T2DM | b_cell | *C17orf51* | -3.572379172 | 0.000553908 | 0.00415349 | 1 |
| T2DM | b_cell | *OR2A42* | -4.209193765 | 0.001328055 | 0.004274973 | 1 |
| T2DM | b_cell | *RNF208* | -7.617167744 | 0.000747814 | 0.00415349 | 1 |
| T2DM | b_cell | *MAP10* | 5.819270537 | 0.033109551 | 0.042163206 | 1 |
| T2DM | b_cell | *NUP62* | -3.62555075 | 0.001498495 | 0.004475552 | 1 |
| T2DM | b_cell | *FGFR1OP* | -2.608462538 | 0.015562412 | 0.021824868 | 1 |
| T2DM | b_cell | *FAM106A* | -3.062931791 | 0.033169799 | 0.042214276 | 1 |
| T2DM | b_cell | *ZNF254* | -2.5644718 | 0.003164487 | 0.006522993 | 1 |
| T2DM | b_cell | *LINGO4* | -28.19990474 | 0.038615977 | 0.048304545 | 1 |
| T2DM | b_cell | *ARHGEF35* | -8.135187911 | 0.004419401 | 0.008201016 | 1 |
| T2DM | b_cell | *DNLZ* | 4.141783976 | 0.000432783 | 0.00415349 | 1 |
| T2DM | b_cell | *NOTCH2NL* | 4.167598155 | 0.021393308 | 0.028684096 | 1 |
| T2DM | b_cell | *GSTM2* | -2.070983472 | 0.003258615 | 0.006651595 | 1 |
| T2DM | b_cell | *ARHGAP19* | 8.208579267 | 0.004911172 | 0.008839032 | 1 |
| T2DM | b_cell | *LCAT* | -2.020795637 | 0.002411875 | 0.005605714 | 1 |
| T2DM | b_cell | *SIPA1* | 6.077781143 | 0.000143012 | 0.00415349 | 0.899118423 |
| T2DM | b_cell | *ERV3-1* | -0.815069555 | 0.034058465 | 0.043213631 | 1 |
| T2DM | b_cell | *GBP7* | -2.162775575 | 0.025730792 | 0.033652899 | 1 |
| T2DM | b_cell | *RBMXL1* | -3.139346516 | 0.000701532 | 0.00415349 | 1 |
| T2DM | b_cell | *SRA1* | -2.674290386 | 0.00110408 | 0.004159248 | 1 |
| T2DM | b_cell | *TMEM110* | -6.387440901 | 0.006447231 | 0.010675201 | 1 |
| T2DM | b_cell | *HEXA* | -3.534620117 | 0.01455969 | 0.02063498 | 1 |
| T2DM | b_cell | *LEPROT* | -2.791299878 | 0.002189584 | 0.005349464 | 1 |
| T2DM | b_cell | *LBH* | 2.539340656 | 0.001003594 | 0.00415349 | 1 |
| T2DM | b_cell | *PPP1CB* | -2.915757221 | 0.012535306 | 0.018167235 | 1 |
| T2DM | b_cell | *NCKIPSD* | -2.852436884 | 0.002190158 | 0.005349464 | 1 |
| T2DM | b_cell | *SLC35F6* | -3.998418841 | 0.002406993 | 0.005600579 | 1 |
| T2DM | b_cell | *CLIC1* | -4.313183533 | 0.001467968 | 0.004434572 | 1 |
| T2DM | b_cell | *ATP6V1G2* | 2.31403171 | 0.00013469 | 0.00415349 | 0.846795207 |

| | | | | | | |
|---|---|---|---|---|---|---|
| T2DM | b_cell | *EMP2* | -2.104036745 | 0.000882181 | 0.00415349 | 1 |
| T2DM | b_cell | *LTB4R* | -3.183223046 | 0.028682153 | 0.037085235 | 1 |
| T2DM | b_cell | *LTB4R2* | -6.698066758 | 0.000590948 | 0.00415349 | 1 |
| T2DM | b_cell | *GALT* | -2.807651152 | 0.00270898 | 0.00595502 | 1 |
| T2DM | b_cell | *ITGA1* | -1.265649576 | 0.000884076 | 0.00415349 | 1 |
| T2DM | b_cell | *NUDT19* | -2.865426433 | 0.000483785 | 0.00415349 | 1 |
| T2DM | b_cell | *ZNF726* | -3.118750228 | 0.000595541 | 0.00415349 | 1 |
| T2DM | b_cell | *TAX1BP3* | -3.121662453 | 0.001262571 | 0.004257394 | 1 |
| T2DM | b_cell | *AP1G2* | 4.054433394 | 0.001270783 | 0.004257394 | 1 |
| T2DM | b_cell | *ZNF90* | 3.818002305 | 0.007799564 | 0.01236406 | 1 |
| T2DM | b_cell | *TM6SF2* | -40.4165487 | 0.001103408 | 0.004159248 | 1 |
| T2DM | b_cell | *REPIN1* | -2.342850737 | 0.000952957 | 0.00415349 | 1 |
| T2DM | b_cell | *SMIM7* | -2.708401116 | 0.00460108 | 0.008465609 | 1 |
| T2DM | b_cell | *TSPAN4* | -2.30855337 | 0.000986288 | 0.00415349 | 1 |
| T2DM | b_cell | *CPNE1* | -2.001741618 | 0.034330337 | 0.043497547 | 1 |
| T2DM | b_cell | *ARL16* | -1.486649898 | 0.002546781 | 0.005790341 | 1 |
| T2DM | b_cell | *WEE2* | -3.673071741 | 0.000513394 | 0.00415349 | 1 |
| T2DM | b_cell | *PAXIP1-AS2* | -3.281381968 | 0.001802859 | 0.004852128 | 1 |
| T2DM | b_cell | *LYRM4* | -3.16149855 | 0.016261982 | 0.022669419 | 1 |
| T2DM | b_cell | *PRCD* | -2.650328107 | 0.002332011 | 0.005509725 | 1 |
| T2DM | b_cell | *C17orf67* | -2.112903378 | 0.000764635 | 0.00415349 | 1 |
| T2DM | b_cell | *FIS1* | -3.184513985 | 0.005728838 | 0.009813953 | 1 |
| T2DM | b_cell | *COLCA2* | -3.978248687 | 0.000353491 | 0.00415349 | 1 |
| T2DM | b_cell | *NEURL1B* | -2.267365063 | 0.00091157 | 0.00415349 | 1 |
| T2DM | b_cell | *VSTM5* | 2.662243197 | 0.001414558 | 0.004364365 | 1 |
| T2DM | b_cell | *PPME1* | -3.214400779 | 0.00165308 | 0.004668875 | 1 |
| T2DM | b_cell | *STARD10* | 5.467965613 | 3.03E-08 | 1.99E-05 | 0.000190693 |
| T2DM | b_cell | *CPEB1* | -2.125626088 | 0.000573288 | 0.00415349 | 1 |
| T2DM | b_cell | *C10orf105* | -5.414749874 | 0.00097714 | 0.00415349 | 1 |
| T2DM | b_cell | *AC005082.1* | 8.86424131 | 0.000819444 | 0.00415349 | 1 |
| T2DM | b_cell | *ACSM4* | -1.971424698 | 0.024327998 | 0.032038149 | 1 |
| T2DM | b_cell | *COL28A1* | -3.323881556 | 0.001039728 | 0.00415349 | 1 |
| T2DM | b_cell | *GRID2IP* | -5.374760383 | 0.006495929 | 0.010738866 | 1 |
| T2DM | b_cell | *UBXN2B* | -2.707425044 | 0.024617066 | 0.032357829 | 1 |
| T2DM | b_cell | *UBE2QL1* | -4.160795181 | 0.000748146 | 0.00415349 | 1 |
| T2DM | b_cell | *GOLGA8B* | -2.513951329 | 0.0016608 | 0.004679659 | 1 |
| T2DM | b_cell | *KCNU1* | -4.638746075 | 6.70E-06 | 0.001276926 | 0.042138574 |
| T2DM | b_cell | *MYL5* | -4.165020799 | 0.000815776 | 0.00415349 | 1 |
| T2DM | b_cell | *NPEPL1* | -3.442289342 | 0.00231273 | 0.005492478 | 1 |
| T2DM | b_cell | *LINC01139* | -4.631852411 | 0.004253535 | 0.007980713 | 1 |
| T2DM | b_cell | *TSTD1* | -2.885709844 | 0.001046678 | 0.00415349 | 1 |
| T2DM | b_cell | *LINC01138* | -2.444871784 | 0.001451892 | 0.004416445 | 1 |
| T2DM | b_cell | *CYB5RL* | 3.132119444 | 0.00113 | 0.004182741 | 1 |
| T2DM | b_cell | *ATAD3C* | 1.328083907 | 0.000389744 | 0.00415349 | 1 |
| T2DM | b_cell | *CCDC7* | -2.256885305 | 0.00177045 | 0.004803979 | 1 |
| T2DM | b_cell | *RNASEK* | -3.573203827 | 0.002107975 | 0.005244494 | 1 |
| T2DM | b_cell | *RNU6ATAC35I* | -14.33418847 | 0.000768238 | 0.00415349 | 1 |
| T2DM | b_cell | *PPP3R1* | 3.415291518 | 0.002159648 | 0.005312091 | 1 |
| T2DM | b_cell | *FANCG* | 1.120824934 | 0.002155794 | 0.005308842 | 1 |
| T2DM | b_cell | *C19orf73* | -18.47846744 | 0.002610281 | 0.005849897 | 1 |

| | | | | | | |
|---|---|---|---|---|---|---|
| T2DM | b_cell | *OR2A25* | -96.59267549 | 0.004185992 | 0.007886524 | 1 |
| T2DM | b_cell | *XKR9* | -3.156378029 | 0.001249892 | 0.004253251 | 1 |
| T2DM | b_cell | *C1orf229* | -6.034149185 | 0.003312971 | 0.006729773 | 1 |
| T2DM | b_cell | *SLC12A8* | -2.935234713 | 0.000147874 | 0.00415349 | 0.929686336 |
| T2DM | b_cell | *APOL6* | -1.380803959 | 0.000778051 | 0.00415349 | 1 |
| T2DM | b_cell | *BTBD19* | -2.275647824 | 0.001379995 | 0.004323793 | 1 |
| T2DM | b_cell | *FAM185A* | -3.138523697 | 0.02732651 | 0.035481571 | 1 |
| T2DM | b_cell | *AC062017.1* | -2.740084698 | 8.32E-05 | 0.00415349 | 0.523250171 |
| T2DM | b_cell | *ADRA2B* | -22.76529689 | 0.006437064 | 0.010666796 | 1 |
| T2DM | b_cell | *EXOSC6* | -3.062789013 | 0.000256857 | 0.00415349 | 1 |
| T2DM | b_cell | *TINCR* | 2.857127595 | 0.000616068 | 0.00415349 | 1 |
| T2DM | b_cell | *LINC00205* | -4.24738275 | 0.00019647 | 0.00415349 | 1 |
| T2DM | b_cell | *AC147651.3* | -7.636611249 | 0.000556856 | 0.00415349 | 1 |
| T2DM | b_cell | *HLA-DPB1* | 2.488147903 | 0.006862388 | 0.011191656 | 1 |
| T2DM | b_cell | *OSER1-AS1* | -2.392899571 | 0.000861292 | 0.00415349 | 1 |
| T2DM | b_cell | *LINC01133* | -1.968159996 | 0.000804343 | 0.00415349 | 1 |
| T2DM | b_cell | *LINC01117* | -2.118789175 | 0.000975746 | 0.00415349 | 1 |
| T2DM | b_cell | *RASAL2-AS1* | 1.670221881 | 0.000706804 | 0.00415349 | 1 |
| T2DM | b_cell | *AC069213.1* | -13.44571427 | 0.002760616 | 0.006023719 | 1 |
| T2DM | b_cell | *LINC00240* | -1.45855501 | 0.006768514 | 0.01108168 | 1 |
| T2DM | b_cell | *LINC00863* | -2.987033963 | 0.008556513 | 0.013312249 | 1 |
| T2DM | b_cell | *LINC00184* | -7.700596799 | 0.0001655 | 0.00415349 | 1 |
| T2DM | b_cell | *TMEM233* | -1.96270788 | 0.001748647 | 0.004789372 | 1 |
| T2DM | b_cell | *AL049840.1* | -2.888766269 | 5.04E-05 | 0.004007391 | 0.316583896 |
| T2DM | b_cell | *LINC00237* | -13.89579231 | 0.000705731 | 0.00415349 | 1 |
| T2DM | b_cell | *PLEKHM1* | -2.509031239 | 0.005308402 | 0.009314519 | 1 |
| T2DM | b_cell | *AP000350.6* | 1.965337056 | 0.016607029 | 0.023078778 | 1 |
| T2DM | b_cell | *SFTA1P* | -10.44409513 | 0.00058195 | 0.00415349 | 1 |
| T2DM | b_cell | *FAM225B* | -4.318338032 | 0.008600462 | 0.013370698 | 1 |
| T2DM | b_cell | *SLC26A6* | 6.128960329 | 0.000369211 | 0.00415349 | 1 |
| T2DM | b_cell | *LINC00626* | 90.78425927 | 0.001383132 | 0.00432604 | 1 |
| T2DM | b_cell | *ERCC6* | -2.838046438 | 0.002988667 | 0.006296833 | 1 |
| T2DM | b_cell | *NOL7* | -3.256561493 | 0.01654959 | 0.023010862 | 1 |
| T2DM | b_cell | *ELFN1* | -9.869034231 | 0.001451517 | 0.004416445 | 1 |
| T2DM | b_cell | *ZNF503-AS1* | 4.232537284 | 0.002538699 | 0.00577662 | 1 |
| T2DM | b_cell | *LINC00937* | -6.563417755 | 0.012981023 | 0.018739769 | 1 |
| T2DM | b_cell | *FTCDNL1* | -1.430787588 | 0.002996981 | 0.006308941 | 1 |
| T2DM | b_cell | *AC064836.3* | 3.750804023 | 0.000682249 | 0.00415349 | 1 |
| T2DM | b_cell | *TMEM185B* | -5.104814496 | 0.000707175 | 0.00415349 | 1 |
| T2DM | b_cell | *HSBP1L1* | -5.067117393 | 0.002188306 | 0.005349464 | 1 |
| T2DM | b_cell | *TAS2R46* | 2.46725562 | 0.006314531 | 0.010520692 | 1 |
| T2DM | b_cell | *AC093901.1* | 1.118885658 | 0.000521111 | 0.00415349 | 1 |
| T2DM | b_cell | *C14orf132* | -2.112004969 | 0.001119591 | 0.004182741 | 1 |
| T2DM | b_cell | *AC009299.3* | -3.611928834 | 0.001127756 | 0.004182741 | 1 |
| T2DM | b_cell | *LINC01068* | -4.652214046 | 0.024300891 | 0.032009156 | 1 |
| T2DM | b_cell | *AC108463.2* | -5.007331234 | 0.031665118 | 0.040487817 | 1 |
| T2DM | b_cell | *HCG20* | -9.374595608 | 0.00072269 | 0.00415349 | 1 |
| T2DM | b_cell | *C19orf24* | -4.005468974 | 0.000323717 | 0.00415349 | 1 |
| T2DM | b_cell | *LINC01013* | 2.6702539 | 0.000560164 | 0.00415349 | 1 |
| T2DM | b_cell | *AC073133.2* | -36.27610161 | 0.001055161 | 0.00415349 | 1 |

| | | | | | | |
|---|---|---|---|---|---|---|
| T2DM | b_cell | *PROB1* | -15.10074403 | 0.005979336 | 0.01012445 | 1 |
| T2DM | b_cell | *ARL17B* | -2.226784707 | 0.000412709 | 0.00415349 | 1 |
| T2DM | b_cell | *DHFR* | -2.50577943 | 0.001147053 | 0.004185446 | 1 |
| T2DM | b_cell | *LINC00954* | -7.810349293 | 0.012910866 | 0.018642769 | 1 |
| T2DM | b_cell | *AC067945.3* | -4.992605911 | 0.006460012 | 0.01068792 | 1 |
| T2DM | b_cell | *RPL41* | 2.428238103 | 0.000863845 | 0.00415349 | 1 |
| T2DM | b_cell | *PATL2* | 3.411153165 | 0.002591534 | 0.00582516 | 1 |
| T2DM | b_cell | *ZNF492* | -4.210058944 | 0.002240569 | 0.005409545 | 1 |
| T2DM | b_cell | *U91324.1* | -6.853446194 | 0.000838171 | 0.00415349 | 1 |
| T2DM | b_cell | *ZNF688* | -5.265702406 | 0.032026601 | 0.040883501 | 1 |
| T2DM | b_cell | *CISD3* | -3.223763072 | 0.001072381 | 0.00415349 | 1 |
| T2DM | b_cell | *PSMG3-AS1* | -3.54606779 | 0.003048426 | 0.006373613 | 1 |
| T2DM | b_cell | *AC112229.4* | 2.084734576 | 0.000860291 | 0.00415349 | 1 |
| T2DM | b_cell | *KLHL7-AS1* | 4.51175836 | 0.000709739 | 0.00415349 | 1 |
| T2DM | b_cell | *AC092171.4* | -5.125408832 | 0.010567628 | 0.015799924 | 1 |
| T2DM | b_cell | *DIRC3* | -1.056479866 | 0.000632304 | 0.00415349 | 1 |
| T2DM | b_cell | *AC107021.1* | -1.467209274 | 0.000736681 | 0.00415349 | 1 |
| T2DM | b_cell | *LINC00865* | -5.702098882 | 0.00712124 | 0.011509314 | 1 |
| T2DM | b_cell | *AC009495.3* | 6.662204021 | 0.003721284 | 0.007268006 | 1 |
| T2DM | b_cell | *HLA-DQB2* | 2.779721298 | 1.71E-09 | 2.69E-06 | 1.07E-05 |
| T2DM | b_cell | *LYRM9* | -5.518647266 | 0.028040448 | 0.036326046 | 1 |
| T2DM | b_cell | *LINC00342* | -2.339307575 | 0.00073687 | 0.00415349 | 1 |
| T2DM | b_cell | *LINC00702* | -2.875354911 | 0.00058778 | 0.00415349 | 1 |
| T2DM | b_cell | *LINC00472* | -1.902150895 | 0.001707761 | 0.004728644 | 1 |
| T2DM | b_cell | *AC073133.1* | -5.614461227 | 0.00080336 | 0.00415349 | 1 |
| T2DM | b_cell | *RPS28* | -2.34068346 | 0.00077551 | 0.00415349 | 1 |
| T2DM | b_cell | *AC123768.3* | -3.103690112 | 0.000804857 | 0.00415349 | 1 |
| T2DM | b_cell | *TRIM26* | -3.056031514 | 0.009929409 | 0.015046083 | 1 |
| T2DM | b_cell | *ZNF736* | -2.377993277 | 0.014357988 | 0.02040431 | 1 |
| T2DM | b_cell | *PINLYP* | -2.319923312 | 0.003092761 | 0.006423584 | 1 |
| T2DM | b_cell | *ANKRD65* | -4.753374883 | 0.010077247 | 0.015218749 | 1 |
| T2DM | b_cell | *LINC00957* | -6.194304129 | 0.004511512 | 0.008337412 | 1 |
| T2DM | b_cell | *LINC00484* | -1.728685094 | 0.00616361 | 0.010355589 | 1 |
| T2DM | b_cell | *KIAA0040* | 3.436432886 | 0.009049041 | 0.013920068 | 1 |
| T2DM | b_cell | *B3GALT4* | 4.357793868 | 0.038001729 | 0.047593002 | 1 |
| T2DM | b_cell | *LINC00941* | 1.772468306 | 0.000601772 | 0.00415349 | 1 |
| T2DM | b_cell | *ZBTB22* | 1.431124659 | 0.00059587 | 0.00415349 | 1 |
| T2DM | b_cell | *B3GNT9* | -5.69742876 | 0.001414277 | 0.004364365 | 1 |
| T2DM | b_cell | *CDKN2AIPNL* | -2.853553085 | 0.000345862 | 0.00415349 | 1 |
| T2DM | b_cell | *GS1-124K5.4* | 2.271485914 | 2.35E-05 | 0.002703191 | 0.147985686 |
| T2DM | b_cell | *LINC00959* | -3.541773507 | 0.011529478 | 0.016962609 | 1 |
| T2DM | b_cell | *HLA-DQA2* | 2.893716478 | 2.43E-11 | 7.64E-08 | 1.53E-07 |
| T2DM | b_cell | *AC073257.2* | -18.73284982 | 0.030608899 | 0.039273092 | 1 |
| T2DM | b_cell | *IRGM* | 1.197316309 | 0.000521014 | 0.00415349 | 1 |
| T2DM | b_cell | *LRRC37A2* | -2.939076832 | 0.007254622 | 0.01167387 | 1 |
| T2DM | b_cell | *TXNDC5* | -7.873335588 | 0.002514346 | 0.005735834 | 1 |
| T2DM | b_cell | *ASB14* | -2.519102081 | 0.035536122 | 0.044880594 | 1 |
| T2DM | b_cell | *NME1* | -2.03093543 | 0.012017416 | 0.017521682 | 1 |
| T2DM | b_cell | *TNFSF12* | -3.821990371 | 0.004300205 | 0.00804146 | 1 |
| T2DM | b_cell | *CDRT4* | -6.443851096 | 0.000581559 | 0.00415349 | 1 |

| | | | | | | |
|---|---|---|---|---|---|---|
| T2DM | b_cell | *LILRA4* | 16.08723344 | 0.000453032 | 0.00415349 | 1 |
| T2DM | b_cell | *LY6G5B* | -2.604576728 | 0.000275994 | 0.00415349 | 1 |
| T2DM | b_cell | *COX19* | -3.455104004 | 0.005039092 | 0.008990003 | 1 |
| T2DM | b_cell | *AQP1* | -2.321818478 | 0.001239283 | 0.004253251 | 1 |
| T2DM | b_cell | *ARHGEF25* | 6.807860578 | 0.001500337 | 0.004478079 | 1 |
| T2DM | b_cell | *MIF* | -1.671317171 | 0.012242502 | 0.017800325 | 1 |
| T2DM | b_cell | *NSUN6* | -2.760720079 | 0.005545381 | 0.009604355 | 1 |
| T2DM | b_cell | *HLA-DOB* | 0.565738917 | 0.000187814 | 0.00415349 | 1 |
| T2DM | b_cell | *TDGF1* | -4.670417303 | 0.023280546 | 0.030891682 | 1 |
| T2DM | b_cell | *CD302* | -3.767099384 | 0.00602566 | 0.010177114 | 1 |
| T2DM | b_cell | *ATP5J2* | -2.823218924 | 0.000953103 | 0.00415349 | 1 |
| T2DM | b_cell | *ARHGAP8* | -5.368887169 | 0.000452597 | 0.00415349 | 1 |
| T2DM | b_cell | *ATP5O* | 3.076615855 | 0.001539911 | 0.004509332 | 1 |
| T2DM | b_cell | *PLEKHO2* | -4.444887817 | 0.002961663 | 0.006265134 | 1 |
| T2DM | b_cell | *PWP2* | -14.91024184 | 0.000146021 | 0.00415349 | 0.918034822 |
| T2DM | b_cell | *PI4KA* | -2.751675277 | 0.003107359 | 0.006449641 | 1 |
| T2DM | b_cell | *AKAP2* | -9.388645796 | 0.000785735 | 0.00415349 | 1 |
| T2DM | b_cell | *LINC00969* | -2.385038843 | 0.002768285 | 0.006028396 | 1 |
| T2DM | b_cell | *ARHGDIG* | -4.234533406 | 0.002256474 | 0.005429181 | 1 |
| T2DM | b_cell | *TCP10L* | -16.79382758 | 0.023318835 | 0.030930259 | 1 |
| T2DM | b_cell | *ARFGAP3* | -3.05706301 | 0.005339743 | 0.009359064 | 1 |
| T2DM | b_cell | *EIF6* | -2.62301232 | 0.000849546 | 0.00415349 | 1 |
| T2DM | b_cell | *MRPL20* | -2.978766831 | 0.008433212 | 0.013139926 | 1 |
| T2DM | b_cell | *AP5Z1* | -2.215377161 | 0.0011653 | 0.004200062 | 1 |
| T2DM | b_cell | *PSG4* | -4.014225601 | 0.005209724 | 0.00917989 | 1 |
| T2DM | b_cell | *STON1* | -2.499075206 | 0.000503115 | 0.00415349 | 1 |
| T2DM | b_cell | *VSIG8* | -7.96180063 | 0.005571755 | 0.009636761 | 1 |
| T2DM | b_cell | *C7orf73* | -3.135553648 | 0.009204458 | 0.014111538 | 1 |
| T2DM | b_cell | *FGF14-IT1* | -8.063418213 | 0.00078717 | 0.00415349 | 1 |
| T2DM | b_cell | *TICAM2* | -8.795882951 | 0.001837119 | 0.004898539 | 1 |
| T2DM | b_cell | *PALM2* | -1.459818345 | 0.007902965 | 0.01250453 | 1 |
| T2DM | b_cell | *NAT6* | -2.777803815 | 0.017223709 | 0.023799002 | 1 |
| T2DM | b_cell | *WDR92* | -3.148794983 | 0.017092685 | 0.023649144 | 1 |
| T2DM | b_cell | *PLA2G4B* | 7.162455158 | 2.04E-05 | 0.002553516 | 0.12798183 |
| T2DM | b_cell | *LEFTY1* | -25.50565222 | 0.002452889 | 0.005646432 | 1 |
| T2DM | b_cell | *JMJD7* | 5.814184237 | 5.82E-09 | 7.32E-06 | 3.66E-05 |
| T2DM | b_cell | *APOBEC3D* | 2.991574773 | 0.000510872 | 0.00415349 | 1 |
| T2DM | b_cell | *RN7SL832P* | -3.985866142 | 0.005358719 | 0.009384476 | 1 |
| T2DM | b_cell | *LINC01011* | -4.284988 | 0.001727148 | 0.004752113 | 1 |
| T2DM | b_cell | *TMEM199* | -2.977507204 | 0.001426379 | 0.004380872 | 1 |
| T2DM | b_cell | *P2RY11* | 3.260465171 | 0.00243205 | 0.005628918 | 1 |
| T2DM | b_cell | *LILRA6* | -1.938373258 | 0.016634876 | 0.023107261 | 1 |
| T2DM | b_cell | *APOBEC3C* | 4.863482113 | 0.001901566 | 0.004987368 | 1 |
| T2DM | b_cell | *CCDC13* | -3.503692138 | 6.38E-05 | 0.00415349 | 0.401010015 |
| T2DM | b_cell | *ASPRV1* | -9.767198051 | 0.000821803 | 0.00415349 | 1 |
| T2DM | b_cell | *OR2AE1* | 3.778210095 | 0.000312751 | 0.00415349 | 1 |
| T2DM | b_cell | *CRYBB2* | -9.815783301 | 0.000606171 | 0.00415349 | 1 |
| T2DM | b_cell | *N4BP2L2* | -2.048053678 | 0.008783332 | 0.013604536 | 1 |
| T2DM | b_cell | *LINC00847* | 2.941190969 | 0.000947576 | 0.00415349 | 1 |
| T2DM | b_cell | *NEAT1* | -1.379614076 | 0.002192715 | 0.005351554 | 1 |

| | | | | | | |
|---|---|---|---|---|---|---|
| T2DM | b_cell | *RAB30-AS1* | -2.938842304 | 0.002889519 | 0.006184116 | 1 |
| T2DM | b_cell | *LINC00920* | 6.999198104 | 0.010015102 | 0.015150372 | 1 |
| T2DM | b_cell | *UBAP1L* | -4.352471158 | 0.005576931 | 0.009640408 | 1 |
| T2DM | b_cell | *PGAM5* | 6.601501624 | 0.006171291 | 0.010360188 | 1 |
| T2DM | b_cell | *DNAJC3-AS1* | -3.406041104 | 0.001947361 | 0.005048426 | 1 |
| T2DM | b_cell | *LINC00926* | 0.31188538 | 0.000571465 | 0.00415349 | 1 |
| T2DM | b_cell | *BCKDHA* | -6.904943644 | 0.004868695 | 0.008790778 | 1 |
| T2DM | b_cell | *CDK11B* | -3.616381092 | 0.014702093 | 0.020803974 | 1 |
| T2DM | b_cell | *PCDHAC1* | 3.436251214 | 0.000885971 | 0.00415349 | 1 |
| T2DM | b_cell | *ABHD14A* | -2.320237012 | 0.001305397 | 0.004274973 | 1 |
| T2DM | b_cell | *SRP14-AS1* | -2.586566804 | 1.48E-05 | 0.002158387 | 0.092810658 |
| T2DM | b_cell | *CCDC153* | -2.862996731 | 0.017900901 | 0.024589847 | 1 |
| T2DM | b_cell | *C5orf17* | -2.227433062 | 0.000890476 | 0.00415349 | 1 |
| T2DM | b_cell | *FMN1* | -1.240576939 | 0.003213217 | 0.006593178 | 1 |
| T2DM | b_cell | *LINC01060* | -3.09547813 | 0.004001135 | 0.007650588 | 1 |
| T2DM | b_cell | *NAIP* | -3.432448193 | 0.000293292 | 0.00415349 | 1 |
| T2DM | b_cell | *PDCD6* | -3.124438451 | 0.024436511 | 0.032174313 | 1 |
| T2DM | b_cell | *LINC00603* | -3.731348292 | 0.000773522 | 0.00415349 | 1 |
| T2DM | b_cell | *YJEFN3* | -2.887629981 | 0.001754368 | 0.004789372 | 1 |
| T2DM | b_cell | *DNAH10OS* | -24.73889696 | 3.23E-05 | 0.003222973 | 0.203047301 |
| T2DM | b_cell | *PCDHA10* | -5.91841976 | 0.001860187 | 0.004935095 | 1 |
| T2DM | b_cell | *ZNF718* | -3.122701595 | 0.007926013 | 0.012529757 | 1 |
| T2DM | b_cell | *LINC00989* | -5.212492429 | 0.005355439 | 0.009381345 | 1 |
| T2DM | b_cell | *LINC01021* | -6.02240624 | 0.000765566 | 0.00415349 | 1 |
| T2DM | b_cell | *FAM218A* | -5.61292944 | 0.000746393 | 0.00415349 | 1 |
| T2DM | b_cell | *GLI4* | -2.482114464 | 0.00114699 | 0.004185446 | 1 |
| T2DM | b_cell | *SNHG18* | -3.49248081 | 0.000124397 | 0.00415349 | 0.782086169 |
| T2DM | b_cell | *ZNF345* | -3.140407843 | 0.001174184 | 0.004202203 | 1 |
| T2DM | b_cell | *SHANK3* | -1.414744167 | 0.002700132 | 0.005946726 | 1 |
| T2DM | b_cell | *CCDC71L* | -2.149680788 | 0.000861935 | 0.00415349 | 1 |
| T2DM | b_cell | *TMEM200B* | 5.256175945 | 0.000139969 | 0.00415349 | 0.879982297 |
| T2DM | b_cell | *PCDHGB6* | -2.503312244 | 0.006844115 | 0.011170547 | 1 |
| T2DM | b_cell | *TRNP1* | 2.142958287 | 0.000602773 | 0.00415349 | 1 |
| T2DM | b_cell | *PCDHGA5* | -3.232856507 | 0.000398345 | 0.00415349 | 1 |
| T2DM | b_cell | *ATXN7L3B* | -2.869582504 | 0.001461565 | 0.0044241 | 1 |
| T2DM | b_cell | *PRKDC* | -2.643400653 | 0.00624961 | 0.010433165 | 1 |
| T2DM | b_cell | *PCDHGA6* | -2.137234662 | 0.017431936 | 0.024044445 | 1 |
| T2DM | b_cell | *PCDHGA11* | -4.574169168 | 0.004718528 | 0.008611143 | 1 |
| T2DM | b_cell | *CLDN23* | -5.406132123 | 0.002679554 | 0.005924338 | 1 |
| T2DM | b_cell | *ZNF260* | -3.283328393 | 0.030433063 | 0.039079384 | 1 |
| T2DM | b_cell | *ARMS2* | -6.976457166 | 0.022724563 | 0.030262514 | 1 |
| T2DM | b_cell | *FPGT* | -4.622366181 | 0.004854303 | 0.008782447 | 1 |
| T2DM | b_cell | *MPV17L2* | -3.9107709 | 0.018146507 | 0.02486641 | 1 |
| T2DM | b_cell | *DPP3* | -2.240907431 | 0.007207482 | 0.011615853 | 1 |
| T2DM | b_cell | *EID3* | -2.200466238 | 0.004880185 | 0.008805116 | 1 |
| T2DM | b_cell | *EBLN2* | -3.507924161 | 0.004958691 | 0.00889452 | 1 |
| T2DM | b_cell | *POLR2M* | -2.504825188 | 0.000839483 | 0.00415349 | 1 |
| T2DM | b_cell | *TAS2R20* | 4.912451307 | 0.002138583 | 0.00528717 | 1 |
| T2DM | b_cell | *LINC00346* | 5.347433474 | 0.00625864 | 0.010442694 | 1 |
| T2DM | b_cell | *CTSO* | -3.786758229 | 0.016054908 | 0.02241057 | 1 |

| | | | | | | |
|---|---|---|---|---|---|---|
| T2DM | b_cell | *LINC00944* | -3.647071149 | 0.004255555 | 0.007981704 | 1 |
| T2DM | b_cell | *ZNF10* | -4.311645387 | 0.007013128 | 0.011382808 | 1 |
| T2DM | b_cell | *AC093668.2* | 3.13217992 | 0.000166119 | 0.00415349 | 1 |
| T2DM | b_cell | *PGA5* | -36.11125732 | 0.00488473 | 0.008809609 | 1 |
| T2DM | b_cell | *HP* | -9.336714664 | 0.001169223 | 0.004202203 | 1 |
| T2DM | b_cell | *KIAA1147* | -2.211468256 | 0.002369767 | 0.005548873 | 1 |
| T2DM | b_cell | *ZBED6* | -6.389777016 | 0.000744424 | 0.00415349 | 1 |
| T2DM | b_cell | *FNTB* | -3.447810471 | 0.000728224 | 0.00415349 | 1 |
| T2DM | b_cell | *OVCH1-AS1* | -2.714913278 | 0.001106615 | 0.004159248 | 1 |
| T2DM | b_cell | *CUX1* | -2.26704558 | 0.008563146 | 0.013319272 | 1 |
| T2DM | b_cell | *TEN1* | 4.55132804 | 0.011616014 | 0.017051105 | 1 |
| T2DM | b_cell | *CHURC1* | -2.167304232 | 0.006354295 | 0.010563049 | 1 |
| T2DM | b_cell | *C17orf49* | -10.69691173 | 0.001991831 | 0.005104028 | 1 |
| T2DM | b_cell | *ZNF578* | -3.05204769 | 0.005175212 | 0.009136916 | 1 |
| T2DM | b_cell | *LINC00640* | -2.236683187 | 0.008097946 | 0.012718407 | 1 |
| T2DM | b_cell | *LINC00638* | -12.0046324 | 0.0003305 | 0.00415349 | 1 |
| T2DM | b_cell | *CEP95* | -2.500231108 | 0.003082773 | 0.006413831 | 1 |
| T2DM | b_cell | *LINC00639* | -1.275665136 | 0.007153484 | 0.01153843 | 1 |
| T2DM | b_cell | *LINC00648* | -2.244842318 | 0.000622689 | 0.00415349 | 1 |
| T2DM | b_cell | *LINC00924* | 2.01477873 | 0.000643324 | 0.00415349 | 1 |
| T2DM | b_cell | *UBE2Q2L* | 9.715207214 | 0.024646498 | 0.032389743 | 1 |
| T2DM | b_cell | *IQCH-AS1* | -2.22355772 | 0.027971477 | 0.036244162 | 1 |
| T2DM | b_cell | *AC083843.1* | -82.3179028 | 1.07E-06 | 0.000307053 | 0.006755169 |
| T2DM | b_cell | *SH3RF3-AS1* | -6.433696099 | 0.000387111 | 0.00415349 | 1 |
| T2DM | b_cell | *PWRN1* | -2.937271852 | 0.000699218 | 0.00415349 | 1 |
| T2DM | b_cell | *HOXB7* | -3.382459907 | 0.000396534 | 0.00415349 | 1 |
| T2DM | b_cell | *FRRS1L* | -6.566331083 | 0.001598982 | 0.004603327 | 1 |
| T2DM | b_cell | *LINC00562* | -1.78016897 | 0.01872635 | 0.025521908 | 1 |
| T2DM | b_cell | *AC009120.5* | -4.579159048 | 0.000865332 | 0.00415349 | 1 |
| T2DM | b_cell | *LINC00565* | -25.8229175 | 0.007722949 | 0.012279763 | 1 |
| T2DM | b_cell | *TMEM178B* | -2.043318285 | 0.000643094 | 0.00415349 | 1 |
| T2DM | b_cell | *LINC01003* | 3.540877657 | 0.037398101 | 0.046977395 | 1 |
| T2DM | b_cell | *C15orf65* | -3.782978336 | 0.000350043 | 0.00415349 | 1 |
| T2DM | b_cell | *HPR* | -9.126713918 | 0.012489684 | 0.018108734 | 1 |
| T2DM | b_cell | *AC123768.4* | 14.74861635 | 0.00229566 | 0.005479428 | 1 |
| T2DM | b_cell | *LINC00667* | -2.018306697 | 0.005517012 | 0.009576326 | 1 |
| T2DM | b_cell | *LINC00672* | -3.202790426 | 0.000554449 | 0.00415349 | 1 |
| T2DM | b_cell | *LINC00909* | -2.62249661 | 0.004320844 | 0.008068011 | 1 |
| T2DM | b_cell | *LINC00526* | -4.437133374 | 0.000756616 | 0.00415349 | 1 |
| T2DM | b_cell | *LINC00668* | 26.96616856 | 0.000340465 | 0.00415349 | 1 |
| T2DM | b_cell | *STRADA* | -2.578295309 | 0.000257648 | 0.00415349 | 1 |
| T2DM | b_cell | *MYO15B* | -2.917574565 | 0.004846606 | 0.008771045 | 1 |
| T2DM | b_cell | *ILF3-AS1* | -2.425827695 | 0.022382806 | 0.029851654 | 1 |
| T2DM | b_cell | *AC139100.2* | 7.420224654 | 0.000741702 | 0.00415349 | 1 |
| T2DM | b_cell | *LINC01140* | -3.863128772 | 0.027710148 | 0.03595 | 1 |
| T2DM | b_cell | *LINC00906* | -22.26918142 | 0.000740014 | 0.00415349 | 1 |
| T2DM | b_cell | *ZNF582-AS1* | -3.927321236 | 0.001083443 | 0.00415349 | 1 |
| T2DM | b_cell | *ZNF285* | 2.666562018 | 0.004007469 | 0.007653388 | 1 |
| T2DM | b_cell | *AC137932.1* | -11.40011967 | 0.001936239 | 0.005026068 | 1 |
| T2DM | b_cell | *AC003005.2* | -4.481354193 | 0.036089547 | 0.045533348 | 1 |

| | | | | | | |
|---|---|---|---|---|---|---|
| T2DM | b_cell | *AL132989.1* | 11.04332495 | 0.002679809 | 0.005924338 | 1 |
| T2DM | b_cell | *LINC00664* | -9.141385578 | 0.005633273 | 0.009700462 | 1 |
| T2DM | b_cell | *FBXO17* | -1.680510972 | 0.000887615 | 0.00415349 | 1 |
| T2DM | b_cell | *SNHG8* | -3.477805982 | 0.008953519 | 0.013810298 | 1 |
| T2DM | b_cell | *HCG17* | -3.3925443 | 0.001086671 | 0.00415349 | 1 |
| T2DM | b_cell | *YTHDF3-AS1* | 4.225645696 | 0.000555294 | 0.00415349 | 1 |
| T2DM | b_cell | *LINC01050* | 3.36723417 | 0.000470605 | 0.00415349 | 1 |
| T2DM | b_cell | *MILR1* | -1.587356545 | 0.028969811 | 0.037375991 | 1 |
| T2DM | b_cell | *GTF2H5* | -2.038826299 | 0.00163917 | 0.004657671 | 1 |
| T2DM | b_cell | *FGF14-AS2* | -3.84452316 | 0.001686282 | 0.004704627 | 1 |
| T2DM | b_cell | *CASC15* | -1.757599237 | 0.000970607 | 0.00415349 | 1 |
| T2DM | b_cell | *LINC01023* | -3.380023898 | 0.00213065 | 0.005277935 | 1 |
| T2DM | b_cell | *AP000347.2* | -3.789144799 | 0.000913661 | 0.00415349 | 1 |
| T2DM | b_cell | *DOC2B* | -2.803496331 | 0.000130921 | 0.00415349 | 0.823103115 |
| T2DM | b_cell | *POLR2J4* | -3.163324751 | 0.001033777 | 0.00415349 | 1 |
| T2DM | b_cell | *PAXIP1-AS1* | -4.433409256 | 0.005702768 | 0.00978262 | 1 |
| T2DM | dendritic_cell | *SCYL3* | -2.193668409 | 2.11E-05 | 0.000132591 | 0.1328079 |
| T2DM | dendritic_cell | *C1orf112* | -3.058118146 | 0.000122629 | 0.000316231 | 0.770969986 |
| T2DM | dendritic_cell | *CFH* | -0.774813401 | 2.88E-05 | 0.000143316 | 0.180935704 |
| T2DM | dendritic_cell | *NFYA* | -3.693804157 | 7.84E-05 | 0.000233255 | 0.493100715 |
| T2DM | dendritic_cell | *STPG1* | -1.743973135 | 0.000307568 | 0.000637547 | 1 |
| T2DM | dendritic_cell | *NIPAL3* | -2.69493883 | 0.00731835 | 0.010430847 | 1 |
| T2DM | dendritic_cell | *ENPP4* | -1.350113303 | 0.005818053 | 0.008439801 | 1 |
| T2DM | dendritic_cell | *SEMA3F* | -1.653328867 | 7.81E-11 | 9.82E-08 | 4.91E-07 |
| T2DM | dendritic_cell | *KRIT1* | -2.299044823 | 4.64E-05 | 0.000170668 | 0.291707101 |
| T2DM | dendritic_cell | *RAD52* | -2.576230854 | 2.96E-05 | 0.000144362 | 0.186394227 |
| T2DM | dendritic_cell | *BAD* | -4.855675111 | 6.08E-05 | 0.000198694 | 0.382013136 |
| T2DM | dendritic_cell | *WNT16* | -3.251542418 | 1.53E-05 | 0.000130129 | 0.095958192 |
| T2DM | dendritic_cell | *HECW1* | -0.815825147 | 0.000310905 | 0.000643193 | 1 |
| T2DM | dendritic_cell | *MAD1L1* | -2.981368186 | 3.27E-05 | 0.000148375 | 0.205499903 |
| T2DM | dendritic_cell | *LASP1* | -2.719609833 | 0.000163756 | 0.000391609 | 1 |
| T2DM | dendritic_cell | *TMEM176A* | 1.578229467 | 0.005759382 | 0.008370143 | 1 |
| T2DM | dendritic_cell | *CYP26B1* | -3.345006402 | 0.000218139 | 0.000483583 | 1 |
| T2DM | dendritic_cell | *ICA1* | -1.661212701 | 3.23E-05 | 0.000148254 | 0.203082823 |
| T2DM | dendritic_cell | *ALS2* | -2.008043165 | 2.82E-05 | 0.000142583 | 0.177582799 |
| T2DM | dendritic_cell | *POLDIP2* | -2.107169808 | 3.48E-05 | 0.000152319 | 0.218729695 |
| T2DM | dendritic_cell | *CD38* | 1.565290853 | 0.002779518 | 0.00432974 | 1 |
| T2DM | dendritic_cell | *RBM6* | -1.726166856 | 1.27E-11 | 2.65E-08 | 7.96E-08 |
| T2DM | dendritic_cell | *RECQL* | -3.151733029 | 0.000769947 | 0.001383835 | 1 |
| T2DM | dendritic_cell | *PDK4* | -2.513208644 | 0.000228935 | 0.000502553 | 1 |
| T2DM | dendritic_cell | *SLC22A16* | 1.103134458 | 0.000274884 | 0.000581492 | 1 |
| T2DM | dendritic_cell | *CDC27* | -1.587718492 | 0.006097153 | 0.00880202 | 1 |
| T2DM | dendritic_cell | *MEOX1* | -1.016435218 | 0.000101497 | 0.000275404 | 0.638111817 |
| T2DM | dendritic_cell | *FAM214B* | -3.078536088 | 0.000168782 | 0.000401185 | 1 |
| T2DM | dendritic_cell | *COPZ2* | -0.936727465 | 0.005816379 | 0.008439321 | 1 |
| T2DM | dendritic_cell | *GCFC2* | -2.292742155 | 5.63E-05 | 0.000190114 | 0.353909541 |
| T2DM | dendritic_cell | *WDR54* | -4.298492675 | 0.009663272 | 0.013437955 | 1 |
| T2DM | dendritic_cell | *ABCB4* | 1.279888038 | 6.58E-05 | 0.00020828 | 0.413851803 |
| T2DM | dendritic_cell | *ZNF195* | -2.565714285 | 1.85E-07 | 2.17E-05 | 0.001165393 |
| T2DM | dendritic_cell | *PDK2* | -1.98237575 | 3.58E-05 | 0.000154392 | 0.224794535 |

| | | | | | | |
|---|---|---|---|---|---|---|
| T2DM | dendritic_cell | *GGNBP2* | -3.231843503 | 2.60E-09 | 1.26E-06 | 1.64E-05 |
| T2DM | dendritic_cell | *TMEM98* | -1.128841061 | 0.000166239 | 0.000396042 | 1 |
| T2DM | dendritic_cell | *ABCC8* | 3.766502637 | 0.0001494 | 0.000365052 | 0.93927989 |
| T2DM | dendritic_cell | *CCL18* | -0.845067263 | 0.023999009 | 0.031303272 | 1 |
| T2DM | dendritic_cell | *SYNRG* | -2.174960711 | 3.32E-05 | 0.000149182 | 0.208884504 |
| T2DM | dendritic_cell | *TMEM132A* | -3.125914756 | 4.91E-05 | 0.000175728 | 0.308929784 |
| T2DM | dendritic_cell | *ZNF263* | -2.745788325 | 2.51E-05 | 0.000138591 | 0.157690725 |
| T2DM | dendritic_cell | *SPATA20* | -2.786329483 | 0.010619165 | 0.014657012 | 1 |
| T2DM | dendritic_cell | *RALA* | -2.955309222 | 0.000342514 | 0.000693298 | 1 |
| T2DM | dendritic_cell | *BAIAP2L1* | -2.058527546 | 0.000202043 | 0.000456665 | 1 |
| T2DM | dendritic_cell | *IFRD1* | -1.805600812 | 0.000196145 | 0.000446312 | 1 |
| T2DM | dendritic_cell | *GTF2IRD1* | -1.387751113 | 0.000276905 | 0.000584196 | 1 |
| T2DM | dendritic_cell | *VPS41* | -2.365127688 | 0.000355539 | 0.000717353 | 1 |
| T2DM | dendritic_cell | *ARHGAP44* | -3.583743228 | 1.96E-05 | 0.000131266 | 0.123374623 |
| T2DM | dendritic_cell | *ELAC2* | -2.174031678 | 0.01618793 | 0.02169086 | 1 |
| T2DM | dendritic_cell | *CCDC124* | -2.020475415 | 0.00059487 | 0.001104533 | 1 |
| T2DM | dendritic_cell | *CEACAM21* | -2.152539569 | 1.19E-05 | 0.000130129 | 0.074789371 |
| T2DM | dendritic_cell | *NOS2* | -7.871905059 | 5.55E-05 | 0.000188762 | 0.349194179 |
| T2DM | dendritic_cell | *TRAPPC6A* | -3.51541572 | 6.97E-05 | 0.000215325 | 0.437971485 |
| T2DM | dendritic_cell | *CD79B* | -2.271510656 | 8.11E-05 | 0.000238485 | 0.509641976 |
| T2DM | dendritic_cell | *ST7L* | -1.507529007 | 3.28E-05 | 0.000148688 | 0.206527929 |
| T2DM | dendritic_cell | *PAX6* | -1.583669006 | 0.002680451 | 0.004194125 | 1 |
| T2DM | dendritic_cell | *RPUSD1* | 3.449426247 | 0.000122213 | 0.000315287 | 0.768353754 |
| T2DM | dendritic_cell | *BAIAP3* | 4.326448976 | 0.000104114 | 0.000280929 | 0.654563766 |
| T2DM | dendritic_cell | *TSR3* | -3.779174686 | 0.000878674 | 0.001547811 | 1 |
| T2DM | dendritic_cell | *PIGQ* | -2.527053681 | 4.96E-05 | 0.000176714 | 0.312077675 |
| T2DM | dendritic_cell | *PSMB1* | -2.971915854 | 4.14E-05 | 0.000162632 | 0.260374271 |
| T2DM | dendritic_cell | *CDK11A* | -2.167010208 | 7.01E-05 | 0.000216062 | 0.440550202 |
| T2DM | dendritic_cell | *DLEC1* | -2.317631045 | 6.19E-05 | 0.000200487 | 0.389429305 |
| T2DM | dendritic_cell | *SS18L2* | -2.853021162 | 7.66E-06 | 0.000130129 | 0.048140599 |
| T2DM | dendritic_cell | *MPND* | -2.340080778 | 0.021124195 | 0.027766635 | 1 |
| T2DM | dendritic_cell | *MGST1* | -0.925410991 | 2.44E-05 | 0.000137267 | 0.153190047 |
| T2DM | dendritic_cell | *NFIX* | -1.293729148 | 7.26E-05 | 0.000220421 | 0.456712535 |
| T2DM | dendritic_cell | *MMP25* | 2.948606952 | 0.001000475 | 0.001733257 | 1 |
| T2DM | dendritic_cell | *IL32* | -2.758982619 | 0.000521761 | 0.000985374 | 1 |
| T2DM | dendritic_cell | *PKD1* | -1.615821715 | 0.001115942 | 0.001905513 | 1 |
| T2DM | dendritic_cell | *MED24* | -2.156383944 | 0.021202351 | 0.027851897 | 1 |
| T2DM | dendritic_cell | *RHOBTB2* | -3.467482549 | 0.000119355 | 0.00030982 | 0.750384967 |
| T2DM | dendritic_cell | *SEC62* | -2.508266867 | 4.51E-05 | 0.000168939 | 0.28356706 |
| T2DM | dendritic_cell | *RPS20* | -3.155702439 | 0.001008117 | 0.001745533 | 1 |
| T2DM | dendritic_cell | *POMT2* | -2.831920898 | 2.54E-05 | 0.000138965 | 0.159408034 |
| T2DM | dendritic_cell | *MLXIPL* | -0.363459929 | 0.02291587 | 0.029965074 | 1 |
| T2DM | dendritic_cell | *SPRTN* | -3.950438652 | 0.000163363 | 0.000391034 | 1 |
| T2DM | dendritic_cell | *METTL13* | 2.891277856 | 0.010964412 | 0.015090468 | 1 |
| T2DM | dendritic_cell | *DYRK4* | 1.995019786 | 2.91E-05 | 0.000143677 | 0.183044906 |
| T2DM | dendritic_cell | *ZNF200* | -2.010634213 | 0.000317374 | 0.000653992 | 1 |
| T2DM | dendritic_cell | *CD4* | 0.678508706 | 5.52E-05 | 0.000188075 | 0.346999169 |
| T2DM | dendritic_cell | *SCMH1* | -1.89995489 | 0.000389887 | 0.000774477 | 1 |
| T2DM | dendritic_cell | *CLCN6* | -3.25353678 | 4.60E-05 | 0.000170199 | 0.28891741 |
| T2DM | dendritic_cell | *MRC2* | -1.169826543 | 4.57E-05 | 0.000169903 | 0.287315178 |

| | | | | | | |
|---|---|---|---|---|---|---|
| T2DM | dendritic_cell | *TSPAN9* | -1.653417535 | 1.28E-06 | 6.80E-05 | 0.008068719 |
| T2DM | dendritic_cell | *MKS1* | -3.931524728 | 0.000131106 | 0.000331963 | 0.82426336 |
| T2DM | dendritic_cell | *UTP18* | -3.847718139 | 0.000195418 | 0.00044482 | 1 |
| T2DM | dendritic_cell | *TTC19* | -1.794507045 | 0.000818095 | 0.001457456 | 1 |
| T2DM | dendritic_cell | *DPF1* | 2.186504086 | 0.008235663 | 0.011606728 | 1 |
| T2DM | dendritic_cell | *SYT7* | 3.269383553 | 1.58E-05 | 0.000130129 | 0.099596006 |
| T2DM | dendritic_cell | *LARS2* | -2.032390779 | 0.008714728 | 0.012227069 | 1 |
| T2DM | dendritic_cell | *PPP5C* | -2.030491912 | 0.000466022 | 0.000895989 | 1 |
| T2DM | dendritic_cell | *ZBTB32* | -2.283194879 | 1.16E-05 | 0.000130129 | 0.072757506 |
| T2DM | dendritic_cell | *TMEM159* | -2.178255178 | 2.81E-05 | 0.000142583 | 0.176739822 |
| T2DM | dendritic_cell | *ERCC1* | -2.407225493 | 2.75E-05 | 0.000141637 | 0.172797224 |
| T2DM | dendritic_cell | *LTF* | -2.144287304 | 0.001683294 | 0.002757391 | 1 |
| T2DM | dendritic_cell | *ALOX5* | 0.996338663 | 8.01E-05 | 0.000236464 | 0.503668506 |
| T2DM | dendritic_cell | *MAP4K5* | -2.756213983 | 0.00011592 | 0.000302403 | 0.728792078 |
| T2DM | dendritic_cell | *MAN2B2* | -3.169747554 | 2.35E-05 | 0.00013573 | 0.147444388 |
| T2DM | dendritic_cell | *CLDN11* | -1.454640263 | 2.73E-05 | 0.000141269 | 0.171501151 |
| T2DM | dendritic_cell | *ANGEL1* | -1.939357754 | 7.53E-05 | 0.000225984 | 0.473435841 |
| T2DM | dendritic_cell | *RNF14* | -2.890883478 | 0.00282977 | 0.004399299 | 1 |
| T2DM | dendritic_cell | *HEBP1* | -2.616818289 | 3.05E-06 | 0.000100866 | 0.019164578 |
| T2DM | dendritic_cell | *TACC3* | -2.764641386 | 0.008727341 | 0.012236573 | 1 |
| T2DM | dendritic_cell | *ZC3H3* | -1.866025629 | 0.001290789 | 0.002172161 | 1 |
| T2DM | dendritic_cell | *MDH1* | -1.642275086 | 1.62E-05 | 0.000130129 | 0.102085146 |
| T2DM | dendritic_cell | *SLC30A9* | -2.025120783 | 6.11E-05 | 0.000199045 | 0.384356232 |
| T2DM | dendritic_cell | *MTMR11* | -0.539152094 | 0.032393564 | 0.041410805 | 1 |
| T2DM | dendritic_cell | *COX15* | -2.879032762 | 3.97E-05 | 0.000159657 | 0.249703129 |
| T2DM | dendritic_cell | *CCDC88C* | -2.046987592 | 0.00040762 | 0.000803608 | 1 |
| T2DM | dendritic_cell | *NPC1L1* | 2.559027737 | 0.004936511 | 0.007261545 | 1 |
| T2DM | dendritic_cell | *NUDCD3* | -1.718633618 | 0.000117794 | 0.00030678 | 0.740567776 |
| T2DM | dendritic_cell | *CHDH* | -3.108621284 | 6.08E-05 | 0.000198694 | 0.382092714 |
| T2DM | dendritic_cell | *IL20RA* | -3.421687277 | 1.47E-05 | 0.000130129 | 0.092213595 |
| T2DM | dendritic_cell | *GLT8D1* | -2.062570839 | 0.000109772 | 0.000291319 | 0.690134975 |
| T2DM | dendritic_cell | *RUFY3* | -2.101882375 | 0.002254032 | 0.003596726 | 1 |
| T2DM | dendritic_cell | *WWTR1* | -2.184956069 | 0.00013991 | 0.000347949 | 0.879615797 |
| T2DM | dendritic_cell | *TTC27* | -2.129894497 | 6.16E-05 | 0.000200088 | 0.387262562 |
| T2DM | dendritic_cell | *VSIG2* | -2.965771295 | 0.000163342 | 0.000391034 | 1 |
| T2DM | dendritic_cell | *PHLDB1* | -1.005743782 | 1.97E-05 | 0.000131266 | 0.123856119 |
| T2DM | dendritic_cell | *MNAT1* | -1.72160428 | 2.52E-05 | 0.000138904 | 0.158488967 |
| T2DM | dendritic_cell | *OSBPL5* | -1.806171963 | 1.61E-05 | 0.000130129 | 0.101186782 |
| T2DM | dendritic_cell | *AQR* | -2.477034659 | 0.000489962 | 0.00093406 | 1 |
| T2DM | dendritic_cell | *CPS1* | -1.466827927 | 3.70E-05 | 0.000156008 | 0.232882473 |
| T2DM | dendritic_cell | *ZDHHC6* | -3.316985279 | 0.00066355 | 0.001210955 | 1 |
| T2DM | dendritic_cell | *RNH1* | -1.701056793 | 0.017657302 | 0.023504437 | 1 |
| T2DM | dendritic_cell | *NDUFS1* | -2.49192029 | 0.000146387 | 0.000359449 | 0.920332529 |
| T2DM | dendritic_cell | *BIRC3* | 0.460125147 | 4.96E-05 | 0.000176609 | 0.311542809 |
| T2DM | dendritic_cell | *AKAP11* | -3.012540104 | 0.000793125 | 0.001419407 | 1 |
| T2DM | dendritic_cell | *GLRX2* | -2.089245869 | 0.000244416 | 0.000528818 | 1 |
| T2DM | dendritic_cell | *SNAPC1* | -3.317758011 | 3.65E-05 | 0.000155217 | 0.229377576 |
| T2DM | dendritic_cell | *GCLM* | -1.389452131 | 0.000106049 | 0.0002842 | 0.66673253 |
| T2DM | dendritic_cell | *UBR2* | -1.858369845 | 3.15E-05 | 0.000147669 | 0.197876566 |
| T2DM | dendritic_cell | *HSF2* | -2.361183092 | 4.52E-05 | 0.000168939 | 0.284155591 |

| | | | | | | |
|---|---|---|---|---|---|---|
| T2DM | dendritic_cell | *TOMM34* | -4.249765421 | 0.025145825 | 0.032677098 | 1 |
| T2DM | dendritic_cell | *KPNA6* | -2.170049938 | 0.001999048 | 0.003223395 | 1 |
| T2DM | dendritic_cell | *VIM* | -2.150933538 | 0.000792847 | 0.001419314 | 1 |
| T2DM | dendritic_cell | *FAS* | -1.195060472 | 0.000918889 | 0.001607864 | 1 |
| T2DM | dendritic_cell | *RNASET2* | 0.95025421 | 0.000145396 | 0.000357912 | 0.914106644 |
| T2DM | dendritic_cell | *BTN3A1* | -1.39619802 | 0.003288095 | 0.005032193 | 1 |
| T2DM | dendritic_cell | *MIPEP* | -3.370265049 | 0.000502081 | 0.000953074 | 1 |
| T2DM | dendritic_cell | *INSRR* | -2.588953543 | 0.003727541 | 0.00564292 | 1 |
| T2DM | dendritic_cell | *B4GALT7* | -2.375435129 | 0.030265842 | 0.0389203 | 1 |
| T2DM | dendritic_cell | *VRK2* | -2.40152523 | 0.002068241 | 0.00332427 | 1 |
| T2DM | dendritic_cell | *VEZT* | -2.134578422 | 2.99E-05 | 0.000145022 | 0.1881478 |
| T2DM | dendritic_cell | *SNX1* | -2.299235629 | 0.000209308 | 0.000468799 | 1 |
| T2DM | dendritic_cell | *ARNTL2* | -3.549483058 | 0.012690999 | 0.017271301 | 1 |
| T2DM | dendritic_cell | *ANK1* | -2.996963925 | 8.27E-05 | 0.000241985 | 0.519625129 |
| T2DM | dendritic_cell | *RABEP1* | -1.743097297 | 0.00073749 | 0.001330826 | 1 |
| T2DM | dendritic_cell | *BAK1* | 2.057078747 | 0.000175448 | 0.000411275 | 1 |
| T2DM | dendritic_cell | *GRN* | 1.927154638 | 0.008607032 | 0.012086757 | 1 |
| T2DM | dendritic_cell | *CENPQ* | -1.910888863 | 6.60E-06 | 0.000130129 | 0.041511758 |
| T2DM | dendritic_cell | *ARID4A* | -2.770255765 | 0.000380838 | 0.000759812 | 1 |
| T2DM | dendritic_cell | *IFT88* | -1.863286731 | 2.13E-05 | 0.000132591 | 0.134057118 |
| T2DM | dendritic_cell | *ZCCHC8* | -2.430275077 | 3.43E-05 | 0.00015116 | 0.215889475 |
| T2DM | dendritic_cell | *FUT8* | -2.117430236 | 4.26E-05 | 0.000164317 | 0.267834056 |
| T2DM | dendritic_cell | *UBA6* | -3.248227455 | 4.27E-05 | 0.000164485 | 0.268562746 |
| T2DM | dendritic_cell | *PIAS1* | -2.392232657 | 0.000833513 | 0.001480728 | 1 |
| T2DM | dendritic_cell | *EFCAB1* | 2.508538083 | 0.002069568 | 0.003325166 | 1 |
| T2DM | dendritic_cell | *ASTE1* | -1.518654554 | 0.003903804 | 0.005882842 | 1 |
| T2DM | dendritic_cell | *SH3YL1* | -2.114639858 | 8.04E-07 | 5.27E-05 | 0.005051836 |
| T2DM | dendritic_cell | *VCL* | -2.436307389 | 5.40E-06 | 0.000125356 | 0.033972248 |
| T2DM | dendritic_cell | *ADSS* | -2.992221271 | 0.000100121 | 0.000272936 | 0.629463847 |
| T2DM | dendritic_cell | *STAP1* | -3.099355417 | 1.53E-05 | 0.000130129 | 0.096227843 |
| T2DM | dendritic_cell | *TBC1D23* | -2.345652434 | 3.95E-05 | 0.000159553 | 0.248615947 |
| T2DM | dendritic_cell | *CUL3* | -2.233479236 | 0.000225589 | 0.000496344 | 1 |
| T2DM | dendritic_cell | *MYOM2* | -1.394333085 | 0.000134553 | 0.000338374 | 0.845934657 |
| T2DM | dendritic_cell | *TUBG2* | -2.215875161 | 9.02E-13 | 2.84E-09 | 5.67E-09 |
| T2DM | dendritic_cell | *NSUN2* | -3.747480758 | 0.000139393 | 0.000346801 | 0.876365834 |
| T2DM | dendritic_cell | *MRI1* | -1.973919354 | 2.66E-05 | 0.000140322 | 0.167513997 |
| T2DM | dendritic_cell | *AGA* | -4.892147117 | 0.000238469 | 0.000518774 | 1 |
| T2DM | dendritic_cell | *BOD1L1* | -2.207756604 | 0.008421897 | 0.011847945 | 1 |
| T2DM | dendritic_cell | *CLEC16A* | -2.848983177 | 0.000169274 | 0.000401831 | 1 |
| T2DM | dendritic_cell | *SKIV2L2* | -1.853007655 | 4.57E-05 | 0.000169903 | 0.287192478 |
| T2DM | dendritic_cell | *DNAH5* | -1.397558325 | 1.14E-05 | 0.000130129 | 0.071574772 |
| T2DM | dendritic_cell | *C6* | -1.382093459 | 4.20E-06 | 0.000116241 | 0.026397212 |
| T2DM | dendritic_cell | *RAI14* | -1.958276442 | 0.000798319 | 0.001427483 | 1 |
| T2DM | dendritic_cell | *PNKP* | -1.654725263 | 0.010616668 | 0.014656783 | 1 |
| T2DM | dendritic_cell | *PHLPP2* | -2.308370297 | 0.003942468 | 0.005933541 | 1 |
| T2DM | dendritic_cell | *CTNS* | -1.003999185 | 0.003916039 | 0.005899865 | 1 |
| T2DM | dendritic_cell | *THAP3* | 2.453223232 | 0.02149451 | 0.028212105 | 1 |
| T2DM | dendritic_cell | *SPATA7* | -2.918529594 | 3.99E-05 | 0.000159733 | 0.25062154 |
| T2DM | dendritic_cell | *RETSAT* | -1.073408011 | 2.27E-05 | 0.000133629 | 0.142644016 |
| T2DM | dendritic_cell | *CAPG* | -2.413726117 | 1.15E-05 | 0.000130129 | 0.072413659 |

| | | | | | | |
|---|---|---|---|---|---|---|
| T2DM | dendritic_cell | *AP2S1* | 2.431270068 | 0.01198517 | 0.016362815 | 1 |
| T2DM | dendritic_cell | *LCP2* | 1.52053169 | 0.000720708 | 0.001303054 | 1 |
| T2DM | dendritic_cell | *CTNNA1* | -2.170363269 | 0.00017476 | 0.000410213 | 1 |
| T2DM | dendritic_cell | *CNTLN* | -2.388492986 | 0.006454241 | 0.00927917 | 1 |
| T2DM | dendritic_cell | *EPHA3* | -1.137480703 | 1.56E-05 | 0.000130129 | 0.097805616 |
| T2DM | dendritic_cell | *DSG2* | -1.542835451 | 4.24E-05 | 0.000164065 | 0.266702975 |
| T2DM | dendritic_cell | *WDR37* | -2.741471153 | 0.001060214 | 0.00182169 | 1 |
| T2DM | dendritic_cell | *YTHDC2* | -2.132942243 | 9.43E-05 | 0.000264058 | 0.592976912 |
| T2DM | dendritic_cell | *ATP6V1H* | -2.971831993 | 0.00071937 | 0.001301864 | 1 |
| T2DM | dendritic_cell | *POLR2B* | -2.81666641 | 0.000124101 | 0.000319108 | 0.780220161 |
| T2DM | dendritic_cell | *ANO2* | -0.766304756 | 4.84E-05 | 0.000173951 | 0.304240434 |
| T2DM | dendritic_cell | *GOPC* | -2.824805458 | 3.92E-05 | 0.000159069 | 0.246398001 |
| T2DM | dendritic_cell | *TSPAN17* | -1.047056446 | 0.006728832 | 0.009651874 | 1 |
| T2DM | dendritic_cell | *CC2D2A* | -1.932808732 | 2.27E-05 | 0.000133654 | 0.143009879 |
| T2DM | dendritic_cell | *SNX29* | -1.930122986 | 0.014639886 | 0.019730111 | 1 |
| T2DM | dendritic_cell | *LMO3* | -0.826084841 | 4.35E-05 | 0.000165783 | 0.273388198 |
| T2DM | dendritic_cell | *MRPS10* | -1.044829881 | 0.000606727 | 0.001123231 | 1 |
| T2DM | dendritic_cell | *CELF2* | -1.710267951 | 4.24E-05 | 0.000164065 | 0.266392437 |
| T2DM | dendritic_cell | *ERCC8* | -2.048648856 | 0.008470745 | 0.011911334 | 1 |
| T2DM | dendritic_cell | *ADAMTS6* | -1.477451827 | 2.05E-05 | 0.00013202 | 0.128577286 |
| T2DM | dendritic_cell | *H6PD* | -1.866669289 | 0.004304576 | 0.006422134 | 1 |
| T2DM | dendritic_cell | *UTS2* | -1.969069929 | 0.000222196 | 0.000490387 | 1 |
| T2DM | dendritic_cell | *RCN1* | -3.707259332 | 7.77E-06 | 0.000130129 | 0.048832391 |
| T2DM | dendritic_cell | *RFC2* | 3.184069368 | 0.010095525 | 0.013992628 | 1 |
| T2DM | dendritic_cell | *JKAMP* | -1.951541426 | 0.00033667 | 0.000684776 | 1 |
| T2DM | dendritic_cell | *DKK3* | -1.214362749 | 3.30E-05 | 0.00014892 | 0.207743437 |
| T2DM | dendritic_cell | *ARHGEF5* | -1.777147223 | 2.27E-05 | 0.000133629 | 0.142443649 |
| T2DM | dendritic_cell | *NFE2L3* | 2.190403483 | 0.000133116 | 0.000335836 | 0.836903049 |
| T2DM | dendritic_cell | *LIMA1* | -0.842492543 | 5.03E-05 | 0.000178067 | 0.316413142 |
| T2DM | dendritic_cell | *PTGER3* | -0.912105271 | 1.55E-05 | 0.000130129 | 0.097392735 |
| T2DM | dendritic_cell | *FAM160A2* | -1.473752806 | 0.006666728 | 0.009564974 | 1 |
| T2DM | dendritic_cell | *CYBA* | 1.853315205 | 0.00216172 | 0.003462607 | 1 |
| T2DM | dendritic_cell | *THOC3* | 2.540354323 | 0.000206374 | 0.000464046 | 1 |
| T2DM | dendritic_cell | *RRP12* | 1.968056598 | 0.000399248 | 0.000789828 | 1 |
| T2DM | dendritic_cell | *ALX4* | -2.298083397 | 1.73E-05 | 0.000130129 | 0.108459936 |
| T2DM | dendritic_cell | *FOXN3* | -2.337587575 | 0.000341549 | 0.000692013 | 1 |
| T2DM | dendritic_cell | *AKR7A2* | -3.049103321 | 0.000123719 | 0.000318518 | 0.777820839 |
| T2DM | dendritic_cell | *MRTO4* | -1.95723396 | 7.23E-06 | 0.000130129 | 0.045435465 |
| T2DM | dendritic_cell | *MCF2L2* | 1.778640512 | 0.001299748 | 0.002185481 | 1 |
| T2DM | dendritic_cell | *ANAPC4* | -2.274622783 | 0.000438838 | 0.000852588 | 1 |
| T2DM | dendritic_cell | *LY75* | 0.673222464 | 4.36E-05 | 0.000165912 | 0.27392003 |
| T2DM | dendritic_cell | *OPN3* | 1.081552618 | 7.69E-06 | 0.000130129 | 0.048333069 |
| T2DM | dendritic_cell | *SDCCAG8* | -1.395821273 | 0.0060654 | 0.008764231 | 1 |
| T2DM | dendritic_cell | *PLEKHH1* | -2.217287435 | 6.06E-05 | 0.000198694 | 0.380829477 |
| T2DM | dendritic_cell | *ATP9A* | -1.572405282 | 8.92E-06 | 0.000130129 | 0.056070046 |
| T2DM | dendritic_cell | *GALC* | -3.03791809 | 1.55E-05 | 0.000130129 | 0.097264734 |
| T2DM | dendritic_cell | *FAM114A2* | -2.426274219 | 0.000101062 | 0.00027458 | 0.635378574 |
| T2DM | dendritic_cell | *GINM1* | -2.504755956 | 3.98E-05 | 0.000159733 | 0.250502046 |
| T2DM | dendritic_cell | *MRPL43* | -2.111638891 | 0.000110212 | 0.00029224 | 0.692901701 |
| T2DM | dendritic_cell | *ITIH4* | 1.173659891 | 3.50E-05 | 0.000152633 | 0.219740725 |

| | | | | | | |
|---|---|---|---|---|---|---|
| T2DM | dendritic_cell | *TRAF1* | 0.872519559 | 0.000105773 | 0.0002837 | 0.664992172 |
| T2DM | dendritic_cell | *IL17RB* | -2.662206986 | 5.20E-06 | 0.000124852 | 0.032665839 |
| T2DM | dendritic_cell | *TRAF3IP2* | -3.020191148 | 0.000165026 | 0.000393914 | 1 |
| T2DM | dendritic_cell | *DCBLD2* | -1.892614043 | 1.54E-05 | 0.000130129 | 0.096541994 |
| T2DM | dendritic_cell | *MSH4* | -1.396907242 | 0.000217379 | 0.00048236 | 1 |
| T2DM | dendritic_cell | *F7* | 8.671003172 | 0.000954337 | 0.001662488 | 1 |
| T2DM | dendritic_cell | *TMCC3* | 1.664922333 | 0.001008947 | 0.00174649 | 1 |
| T2DM | dendritic_cell | *PITHD1* | -2.802771971 | 0.019548081 | 0.025884327 | 1 |
| T2DM | dendritic_cell | *ATP11B* | -1.205641501 | 0.010342471 | 0.014309665 | 1 |
| T2DM | dendritic_cell | *LAMC2* | -0.649879313 | 0.007805524 | 0.011057532 | 1 |
| T2DM | dendritic_cell | *CDK14* | -1.930697615 | 3.23E-05 | 0.000148254 | 0.203358287 |
| T2DM | dendritic_cell | *CROCC* | -1.43227539 | 0.002857028 | 0.004430718 | 1 |
| T2DM | dendritic_cell | *ZC3H11A* | 2.943705789 | 9.44E-05 | 0.000264058 | 0.593776606 |
| T2DM | dendritic_cell | *RIOK2* | -2.284974979 | 2.42E-05 | 0.000136859 | 0.152324267 |
| T2DM | dendritic_cell | *FLYWCH1* | -1.268405694 | 0.002362143 | 0.003749254 | 1 |
| T2DM | dendritic_cell | *DNAJC25* | -2.941649497 | 0.004205131 | 0.006288692 | 1 |
| T2DM | dendritic_cell | *YBX3* | -2.564080741 | 0.007852418 | 0.011111446 | 1 |
| T2DM | dendritic_cell | *PIGV* | -2.930493118 | 1.84E-07 | 2.17E-05 | 0.001156284 |
| T2DM | dendritic_cell | *NCKAP1* | -1.78336351 | 6.91E-05 | 0.000214095 | 0.434185034 |
| T2DM | dendritic_cell | *SFSWAP* | -2.664863756 | 0.000431133 | 0.00084178 | 1 |
| T2DM | dendritic_cell | *VMP1* | -2.526567271 | 0.000968707 | 0.001684253 | 1 |
| T2DM | dendritic_cell | *EIF4B* | -1.655813541 | 0.016313701 | 0.021840766 | 1 |
| T2DM | dendritic_cell | *SLC6A16* | -2.08490832 | 2.16E-05 | 0.000132663 | 0.13569244 |
| T2DM | dendritic_cell | *ISOC2* | -1.617729522 | 1.57E-05 | 0.000130129 | 0.098512694 |
| T2DM | dendritic_cell | *ADCK1* | -3.127067963 | 0.000328798 | 0.000671661 | 1 |
| T2DM | dendritic_cell | *TM7SF3* | -2.569486416 | 0.000504189 | 0.000956787 | 1 |
| T2DM | dendritic_cell | *DMRT3* | -1.84852502 | 3.76E-05 | 0.000156608 | 0.236164384 |
| T2DM | dendritic_cell | *ST3GAL6* | -1.853666204 | 0.00136647 | 0.002286664 | 1 |
| T2DM | dendritic_cell | *ATP2C2* | -1.971702872 | 7.53E-05 | 0.000225984 | 0.47339257 |
| T2DM | dendritic_cell | *CDON* | -0.964898683 | 3.86E-05 | 0.000158005 | 0.242686012 |
| T2DM | dendritic_cell | *TAF2* | -2.007105066 | 0.006740261 | 0.009662469 | 1 |
| T2DM | dendritic_cell | *TMEM161A* | -2.305102085 | 3.69E-05 | 0.000155835 | 0.232276103 |
| T2DM | dendritic_cell | *LPAR2* | 2.887760918 | 0.018463791 | 0.024520882 | 1 |
| T2DM | dendritic_cell | *CHI3L2* | -1.292186664 | 0.003034593 | 0.004682987 | 1 |
| T2DM | dendritic_cell | *PMS1* | -2.524899504 | 5.62E-05 | 0.000189964 | 0.353142661 |
| T2DM | dendritic_cell | *TAF11* | -2.206484029 | 0.001326771 | 0.002224376 | 1 |
| T2DM | dendritic_cell | *ANKS1A* | -2.353940948 | 0.007445104 | 0.010592298 | 1 |
| T2DM | dendritic_cell | *IPO5* | -2.196855958 | 5.36E-05 | 0.000184297 | 0.336696229 |
| T2DM | dendritic_cell | *WDR18* | -1.964638418 | 0.000552399 | 0.001036077 | 1 |
| T2DM | dendritic_cell | *MCM10* | 0.963176839 | 0.007249612 | 0.010344601 | 1 |
| T2DM | dendritic_cell | *ANKRD44* | 1.037449749 | 0.00021515 | 0.000478136 | 1 |
| T2DM | dendritic_cell | *NDUFB4* | -2.903211923 | 0.000173099 | 0.000407754 | 1 |
| T2DM | dendritic_cell | *ZC3H15* | -2.868519373 | 2.49E-05 | 0.000138038 | 0.156396498 |
| T2DM | dendritic_cell | *MAP2K4* | -2.610029319 | 0.000229655 | 0.000503783 | 1 |
| T2DM | dendritic_cell | *TMEM206* | -2.410719657 | 1.64E-05 | 0.000130129 | 0.1032013 |
| T2DM | dendritic_cell | *SEC61A2* | -3.030019197 | 2.81E-05 | 0.000142583 | 0.176961893 |
| T2DM | dendritic_cell | *ASB1* | -4.626792202 | 3.55E-05 | 0.000154001 | 0.223282296 |
| T2DM | dendritic_cell | *TBC1D1* | -2.786857758 | 0.007036146 | 0.010051408 | 1 |
| T2DM | dendritic_cell | *PPP2R5A* | -1.984026479 | 2.84E-05 | 0.000142583 | 0.178647033 |
| T2DM | dendritic_cell | *TIE1* | -1.143027455 | 0.000111416 | 0.000294439 | 0.700470674 |

| | | | | | | |
|---|---|---|---|---|---|---|
| T2DM | dendritic_cell | *SLC9A3* | -1.223114908 | 0.011552846 | 0.015837176 | 1 |
| T2DM | dendritic_cell | *NGEF* | -2.45898306 | 1.89E-05 | 0.000130129 | 0.118558977 |
| T2DM | dendritic_cell | *CD84* | 1.172384803 | 3.20E-05 | 0.000148254 | 0.201306213 |
| T2DM | dendritic_cell | *ATXN3* | -1.306175751 | 0.010565194 | 0.014598544 | 1 |
| T2DM | dendritic_cell | *GOLGA5* | -3.316729033 | 3.42E-05 | 0.000151054 | 0.21479908 |
| T2DM | dendritic_cell | *EML1* | -1.936539937 | 3.30E-05 | 0.00014892 | 0.207704793 |
| T2DM | dendritic_cell | *ZFAT* | -1.99317894 | 2.39E-05 | 0.000136538 | 0.150269051 |
| T2DM | dendritic_cell | *MTFR1* | -2.725888279 | 9.51E-06 | 0.000130129 | 0.059807639 |
| T2DM | dendritic_cell | *STAG3* | -2.707884843 | 0.000471117 | 0.000904125 | 1 |
| T2DM | dendritic_cell | *MYO9A* | -1.891427343 | 8.76E-05 | 0.000251946 | 0.550501911 |
| T2DM | dendritic_cell | *SP100* | -2.410228361 | 0.000310429 | 0.000642418 | 1 |
| T2DM | dendritic_cell | *STOML1* | -6.392875053 | 6.77E-05 | 0.000211932 | 0.425669091 |
| T2DM | dendritic_cell | *METTL22* | -3.049962701 | 2.32E-05 | 0.000135087 | 0.146164558 |
| T2DM | dendritic_cell | *TP53BP1* | -1.567013883 | 0.000103882 | 0.000280899 | 0.653105835 |
| T2DM | dendritic_cell | *RRP15* | -0.893340867 | 0.016839251 | 0.022510817 | 1 |
| T2DM | dendritic_cell | *DHX8* | -2.389757967 | 0.000157381 | 0.00038002 | 0.989452937 |
| T2DM | dendritic_cell | *SYT1* | -1.89516508 | 2.96E-06 | 9.90E-05 | 0.018624023 |
| T2DM | dendritic_cell | *ROGDI* | 2.692575033 | 0.000109166 | 0.000290323 | 0.686324481 |
| T2DM | dendritic_cell | *HDAC4* | -2.242621593 | 3.58E-05 | 0.000154397 | 0.225013307 |
| T2DM | dendritic_cell | *RASSF1* | -2.436684963 | 0.00566055 | 0.008237509 | 1 |
| T2DM | dendritic_cell | *IFI35* | -1.358674168 | 0.009315006 | 0.012993886 | 1 |
| T2DM | dendritic_cell | *HEATR6* | -2.492065792 | 3.30E-05 | 0.000148822 | 0.207159843 |
| T2DM | dendritic_cell | *POLR1A* | -1.424181217 | 7.10E-06 | 0.000130129 | 0.044643375 |
| T2DM | dendritic_cell | *STON1-GTF2A* | -0.439453977 | 0.001123721 | 0.001917707 | 1 |
| T2DM | dendritic_cell | *SRBD1* | -2.700886031 | 0.000179051 | 0.000417853 | 1 |
| T2DM | dendritic_cell | *CYFIP1* | -2.824613753 | 0.000287182 | 0.000601837 | 1 |
| T2DM | dendritic_cell | *PSME4* | -2.41770244 | 0.000344791 | 0.000697235 | 1 |
| T2DM | dendritic_cell | *PITX1* | -3.519712688 | 1.48E-05 | 0.000130129 | 0.093168388 |
| T2DM | dendritic_cell | *SDK2* | 2.819501882 | 0.00032795 | 0.000670729 | 1 |
| T2DM | dendritic_cell | *NUCKS1* | -2.446660335 | 0.000769155 | 0.001382807 | 1 |
| T2DM | dendritic_cell | *DNAJA2* | -2.802240574 | 1.89E-05 | 0.000130129 | 0.118570345 |
| T2DM | dendritic_cell | *KCNAB2* | 1.357969027 | 0.011883653 | 0.016234795 | 1 |
| T2DM | dendritic_cell | *GAL* | 2.980364424 | 0.000133664 | 0.000336812 | 0.840345279 |
| T2DM | dendritic_cell | *NEDD4* | -1.673466096 | 0.001302532 | 0.002189578 | 1 |
| T2DM | dendritic_cell | *PIGB* | -2.784628576 | 1.32E-05 | 0.000130129 | 0.082848485 |
| T2DM | dendritic_cell | *PFN2* | -1.694428643 | 6.15E-06 | 0.000130129 | 0.038637814 |
| T2DM | dendritic_cell | *DAPP1* | 0.217728487 | 3.49E-05 | 0.000152633 | 0.219700009 |
| T2DM | dendritic_cell | *FGF10* | -0.815570541 | 0.000409495 | 0.000805938 | 1 |
| T2DM | dendritic_cell | *TMEM260* | -2.795789967 | 0.000107593 | 0.00028699 | 0.676436115 |
| T2DM | dendritic_cell | *CLTCL1* | -1.828952372 | 0.005491818 | 0.008009061 | 1 |
| T2DM | dendritic_cell | *DGCR2* | -2.709393706 | 2.17E-05 | 0.000132663 | 0.136318553 |
| T2DM | dendritic_cell | *MNT* | -2.229867077 | 0.002201454 | 0.003521767 | 1 |
| T2DM | dendritic_cell | *ZXDC* | -2.761494902 | 0.000258505 | 0.00055355 | 1 |
| T2DM | dendritic_cell | *POLB* | -2.240532449 | 2.51E-05 | 0.000138591 | 0.157575509 |
| T2DM | dendritic_cell | *WIPI1* | -2.698396495 | 3.24E-05 | 0.000148254 | 0.20364904 |
| T2DM | dendritic_cell | *NDST1* | -2.174142808 | 3.87E-05 | 0.000158078 | 0.243281689 |
| T2DM | dendritic_cell | *ASNS* | -1.958467431 | 0.00065088 | 0.001190252 | 1 |
| T2DM | dendritic_cell | *AP3M2* | -2.448049747 | 1.76E-05 | 0.000130129 | 0.110688314 |
| T2DM | dendritic_cell | *ST6GALNAC2* | -1.832239136 | 8.15E-05 | 0.000239417 | 0.512243638 |
| T2DM | dendritic_cell | *EIF2B3* | -1.949195136 | 2.37E-05 | 0.000136299 | 0.148838991 |

| | | | | | | |
|---|---|---|---|---|---|---|
| T2DM | dendritic_cell | *CAMK2A* | -7.460759105 | 4.32E-06 | 0.000116241 | 0.027166043 |
| T2DM | dendritic_cell | *OSBPL3* | 1.50669945 | 0.036812976 | 0.046765646 | 1 |
| T2DM | dendritic_cell | *RAD18* | -3.149062569 | 0.000175566 | 0.000411399 | 1 |
| T2DM | dendritic_cell | *SNX13* | -2.531268244 | 0.004284783 | 0.006395639 | 1 |
| T2DM | dendritic_cell | *TRIP13* | -3.240565702 | 1.54E-05 | 0.000130129 | 0.096828904 |
| T2DM | dendritic_cell | *TCF3* | -3.640521391 | 1.98E-05 | 0.000131266 | 0.12424933 |
| T2DM | dendritic_cell | *MBD3* | -2.489836932 | 1.07E-05 | 0.000130129 | 0.067340297 |
| T2DM | dendritic_cell | *HLTF* | -2.636928035 | 1.05E-05 | 0.000130129 | 0.066242069 |
| T2DM | dendritic_cell | *CPSF1* | -3.84441457 | 0.000145609 | 0.000358007 | 0.915440758 |
| T2DM | dendritic_cell | *CYBRD1* | -1.225974523 | 0.00030929 | 0.000640482 | 1 |
| T2DM | dendritic_cell | *EPN2* | -2.323902833 | 4.16E-05 | 0.000162756 | 0.261270636 |
| T2DM | dendritic_cell | *LIMS2* | -1.450377932 | 4.37E-05 | 0.000166228 | 0.274940738 |
| T2DM | dendritic_cell | *AFF4* | -2.337458277 | 0.000316968 | 0.000653511 | 1 |
| T2DM | dendritic_cell | *TRHDE* | -0.85227413 | 1.44E-05 | 0.000130129 | 0.090235733 |
| T2DM | dendritic_cell | *NFATC3* | -1.630147403 | 0.000639112 | 0.001171799 | 1 |
| T2DM | dendritic_cell | *TRNT1* | -3.243667294 | 0.000492826 | 0.000938055 | 1 |
| T2DM | dendritic_cell | *ACADVL* | -2.923272988 | 6.09E-05 | 0.000198694 | 0.38267495 |
| T2DM | dendritic_cell | *CRMP1* | -2.432224418 | 7.23E-06 | 0.000130129 | 0.045447938 |
| T2DM | dendritic_cell | *DERL2* | -3.185659768 | 2.96E-05 | 0.000144362 | 0.185980099 |
| T2DM | dendritic_cell | *MOV10L1* | -1.266354415 | 4.62E-05 | 0.00017055 | 0.290515312 |
| T2DM | dendritic_cell | *TP63* | -2.80022982 | 0.00025891 | 0.00055404 | 1 |
| T2DM | dendritic_cell | *PDE8A* | -1.652503746 | 0.000794489 | 0.001421443 | 1 |
| T2DM | dendritic_cell | *NLE1* | -2.63716558 | 0.00043211 | 0.000843165 | 1 |
| T2DM | dendritic_cell | *GSDMB* | -1.331556126 | 1.61E-05 | 0.000130129 | 0.101368931 |
| T2DM | dendritic_cell | *PICALM* | -2.180023389 | 0.000913816 | 0.00160077 | 1 |
| T2DM | dendritic_cell | *GLI2* | -0.833373559 | 5.79E-05 | 0.00019326 | 0.364101989 |
| T2DM | dendritic_cell | *ATP2A3* | -1.318760913 | 4.21E-05 | 0.000163644 | 0.264939358 |
| T2DM | dendritic_cell | *CA12* | -1.066274162 | 2.88E-05 | 0.000143316 | 0.181367431 |
| T2DM | dendritic_cell | *NTN4* | -1.25206538 | 1.62E-05 | 0.000130129 | 0.102075498 |
| T2DM | dendritic_cell | *LMAN1* | -1.960903667 | 2.36E-05 | 0.000136166 | 0.148556688 |
| T2DM | dendritic_cell | *ENO1* | -3.269390935 | 4.19E-05 | 0.000163153 | 0.263492207 |
| T2DM | dendritic_cell | *SRI* | -3.343536913 | 0.000103969 | 0.000280899 | 0.653652337 |
| T2DM | dendritic_cell | *EIF4G3* | -2.331859967 | 0.000386896 | 0.000769509 | 1 |
| T2DM | dendritic_cell | *TTC38* | -2.165739836 | 2.18E-05 | 0.000132858 | 0.136878652 |
| T2DM | dendritic_cell | *ADD2* | 4.492135656 | 0.002931627 | 0.004538572 | 1 |
| T2DM | dendritic_cell | *VPS9D1* | 2.542660975 | 0.007765305 | 0.011005517 | 1 |
| T2DM | dendritic_cell | *MARK3* | -2.875183846 | 6.36E-07 | 4.82E-05 | 0.004001322 |
| T2DM | dendritic_cell | *SLC25A3* | -2.021844793 | 0.000254307 | 0.00054642 | 1 |
| T2DM | dendritic_cell | *FSCN1* | 1.396040147 | 8.57E-05 | 0.000248152 | 0.538738793 |
| T2DM | dendritic_cell | *MOCOS* | -1.06768231 | 0.004217786 | 0.006306116 | 1 |
| T2DM | dendritic_cell | *PLD1* | -2.254044623 | 4.87E-05 | 0.000174599 | 0.305894707 |
| T2DM | dendritic_cell | *BCAP29* | -2.561120736 | 0.002445728 | 0.003866304 | 1 |
| T2DM | dendritic_cell | *TUBA3D* | -4.036524031 | 8.03E-05 | 0.000236857 | 0.504897191 |
| T2DM | dendritic_cell | *MKRN2* | -2.807636511 | 4.40E-06 | 0.000117102 | 0.02763616 |
| T2DM | dendritic_cell | *MCM6* | -2.466328237 | 0.01685816 | 0.022523052 | 1 |
| T2DM | dendritic_cell | *RBMS2* | -2.383321941 | 6.73E-05 | 0.00021117 | 0.422973975 |
| T2DM | dendritic_cell | *MLH1* | -2.305708346 | 0.000183411 | 0.000425343 | 1 |
| T2DM | dendritic_cell | *FMO4* | -2.12027869 | 1.62E-05 | 0.000130129 | 0.101603052 |
| T2DM | dendritic_cell | *RGS11* | -1.218420627 | 0.001185127 | 0.002010495 | 1 |
| T2DM | dendritic_cell | *TPD52* | 1.512355231 | 0.030852446 | 0.039569426 | 1 |

| | | | | | | |
|---|---|---|---|---|---|---|
| T2DM | dendritic_cell | *NT5C2* | -1.838330485 | 0.019661809 | 0.026012017 | 1 |
| T2DM | dendritic_cell | *CAMSAP3* | -8.688586872 | 1.28E-05 | 0.000130129 | 0.080214259 |
| T2DM | dendritic_cell | *XAB2* | -3.037727374 | 2.30E-07 | 2.41E-05 | 0.001446 |
| T2DM | dendritic_cell | *STXBP2* | 2.331011189 | 1.26E-06 | 6.80E-05 | 0.007923074 |
| T2DM | dendritic_cell | *MAP2K7* | -3.705729157 | 7.92E-05 | 0.000234792 | 0.497919339 |
| T2DM | dendritic_cell | *NMRK2* | -3.411678765 | 0.023960655 | 0.03125973 | 1 |
| T2DM | dendritic_cell | *CTTNBP2* | -3.955825888 | 1.61E-07 | 2.11E-05 | 0.001010447 |
| T2DM | dendritic_cell | *TM9SF3* | -2.539039017 | 0.000317036 | 0.000653511 | 1 |
| T2DM | dendritic_cell | *DYNC1I2* | -1.98973752 | 6.11E-05 | 0.000198972 | 0.384015622 |
| T2DM | dendritic_cell | *APBB1IP* | -1.966802221 | 0.000164032 | 0.000391917 | 1 |
| T2DM | dendritic_cell | *SIRT6* | -5.757078935 | 7.53E-05 | 0.000225984 | 0.473176209 |
| T2DM | dendritic_cell | *CAPZB* | -2.514365349 | 8.77E-05 | 0.000252107 | 0.55160953 |
| T2DM | dendritic_cell | *NAALAD2* | -1.130993747 | 0.000849982 | 0.001507003 | 1 |
| T2DM | dendritic_cell | *FGFR1* | -1.560613681 | 9.97E-06 | 0.000130129 | 0.062650386 |
| T2DM | dendritic_cell | *FBLN1* | -0.829093328 | 9.16E-05 | 0.000259066 | 0.575645116 |
| T2DM | dendritic_cell | *PIAS2* | -2.745462312 | 0.000815803 | 0.001453785 | 1 |
| T2DM | dendritic_cell | *AMPH* | -0.715226935 | 0.000194989 | 0.000444164 | 1 |
| T2DM | dendritic_cell | *MCCC1* | -1.48109674 | 0.000482831 | 0.000922102 | 1 |
| T2DM | dendritic_cell | *FAP* | -0.745145354 | 5.93E-05 | 0.000196192 | 0.372568376 |
| T2DM | dendritic_cell | *ACER3* | -2.580261641 | 6.93E-05 | 0.000214503 | 0.435539369 |
| T2DM | dendritic_cell | *PIK3C3* | -2.647781011 | 5.62E-05 | 0.000189964 | 0.353050075 |
| T2DM | dendritic_cell | *TULP3* | -1.410007894 | 2.00E-05 | 0.000131266 | 0.125959949 |
| T2DM | dendritic_cell | *SYNJ2* | -1.213088545 | 2.43E-05 | 0.00013716 | 0.152933027 |
| T2DM | dendritic_cell | *FGF20* | 6.542447252 | 4.71E-05 | 0.000171433 | 0.295952857 |
| T2DM | dendritic_cell | *PCM1* | -1.942398753 | 3.86E-05 | 0.000158005 | 0.242985942 |
| T2DM | dendritic_cell | *FKBP7* | -0.956761464 | 0.001905988 | 0.003089184 | 1 |
| T2DM | dendritic_cell | *OSBPL6* | -2.870103913 | 4.09E-05 | 0.000161687 | 0.257337685 |
| T2DM | dendritic_cell | *XRCC5* | -2.613232995 | 0.001393662 | 0.002326594 | 1 |
| T2DM | dendritic_cell | *REXO1* | -2.348544095 | 1.57E-05 | 0.000130129 | 0.098427965 |
| T2DM | dendritic_cell | *SAR1A* | -2.185346337 | 2.71E-05 | 0.000141017 | 0.170304414 |
| T2DM | dendritic_cell | *RAPGEF3* | -0.448193748 | 0.030442517 | 0.0391075 | 1 |
| T2DM | dendritic_cell | *FDFT1* | -2.347780953 | 4.33E-05 | 0.000165469 | 0.272265586 |
| T2DM | dendritic_cell | *SCGN* | 4.869645387 | 3.33E-05 | 0.0001492 | 0.209178674 |
| T2DM | dendritic_cell | *DDX1* | -1.954068175 | 1.83E-05 | 0.000130129 | 0.114890919 |
| T2DM | dendritic_cell | *DNM2* | -2.888901755 | 0.000352841 | 0.000712595 | 1 |
| T2DM | dendritic_cell | *RIMS1* | -2.437232055 | 2.06E-05 | 0.000132198 | 0.129289163 |
| T2DM | dendritic_cell | *RABL2B* | -2.873115637 | 3.52E-05 | 0.000153194 | 0.221211599 |
| T2DM | dendritic_cell | *DDX43* | 1.332291432 | 0.004841507 | 0.007140172 | 1 |
| T2DM | dendritic_cell | *DCT* | -1.95647152 | 1.83E-05 | 0.000130129 | 0.115295737 |
| T2DM | dendritic_cell | *SLC35C2* | -2.994932394 | 0.020594119 | 0.027137964 | 1 |
| T2DM | dendritic_cell | *SCTR* | -2.024457736 | 1.89E-05 | 0.000130129 | 0.119068431 |
| T2DM | dendritic_cell | *RIF1* | -2.187402104 | 0.000882261 | 0.00155198 | 1 |
| T2DM | dendritic_cell | *KCNN2* | -2.766797415 | 4.34E-06 | 0.000116241 | 0.027316657 |
| T2DM | dendritic_cell | *PSEN1* | -2.37727837 | 0.002470915 | 0.003901216 | 1 |
| T2DM | dendritic_cell | *CPOX* | -1.992457334 | 8.00E-06 | 0.000130129 | 0.050307307 |
| T2DM | dendritic_cell | *MOK* | -3.18533285 | 5.33E-05 | 0.000183801 | 0.33511121 |
| T2DM | dendritic_cell | *RSBN1* | -3.414090773 | 0.000248145 | 0.000535377 | 1 |
| T2DM | dendritic_cell | *MAGI3* | -1.606370578 | 1.87E-05 | 0.000130129 | 0.117744637 |
| T2DM | dendritic_cell | *COL4A4* | -1.826151809 | 1.46E-05 | 0.000130129 | 0.091646907 |
| T2DM | dendritic_cell | *STK17B* | 1.173566513 | 0.000202165 | 0.000456705 | 1 |

| | | | | | | |
|---|---|---|---|---|---|---|
| T2DM | dendritic_cell | *ZNF506* | -3.845006313 | 1.31E-05 | 0.000130129 | 0.082477561 |
| T2DM | dendritic_cell | *PCDHA6* | -14.24733449 | 0.000178419 | 0.000416533 | 1 |
| T2DM | dendritic_cell | *PCDHGA2* | -4.668473767 | 1.42E-06 | 7.19E-05 | 0.008917093 |
| T2DM | dendritic_cell | *HSPB11* | -2.863838437 | 0.004656286 | 0.006885368 | 1 |
| T2DM | dendritic_cell | *WDR70* | -2.264649338 | 0.000170675 | 0.000404308 | 1 |
| T2DM | dendritic_cell | *PGR* | -1.295090543 | 6.17E-06 | 0.000130129 | 0.038767838 |
| T2DM | dendritic_cell | *C5orf22* | -3.510715336 | 0.004611721 | 0.006828518 | 1 |
| T2DM | dendritic_cell | *COBLL1* | -1.505220214 | 1.85E-09 | 1.06E-06 | 1.16E-05 |
| T2DM | dendritic_cell | *GEMIN5* | -2.250521057 | 0.002154905 | 0.003454332 | 1 |
| T2DM | dendritic_cell | *GSK3B* | -2.234613895 | 0.00032639 | 0.000667974 | 1 |
| T2DM | dendritic_cell | *ERC1* | -2.122815895 | 0.000124836 | 0.000320348 | 0.784846297 |
| T2DM | dendritic_cell | *LYRM2* | -1.398999681 | 0.0028341 | 0.004403853 | 1 |
| T2DM | dendritic_cell | *ULK2* | -1.87265359 | 4.64E-05 | 0.000170668 | 0.292013296 |
| T2DM | dendritic_cell | *PLOD1* | -3.972755508 | 0.000205112 | 0.000462034 | 1 |
| T2DM | dendritic_cell | *PIBF1* | -2.032514556 | 3.44E-05 | 0.00015116 | 0.216007158 |
| T2DM | dendritic_cell | *TDRD3* | -2.086903453 | 3.81E-05 | 0.000157482 | 0.239511042 |
| T2DM | dendritic_cell | *PDS5B* | -2.583634431 | 0.000113086 | 0.000297331 | 0.710970342 |
| T2DM | dendritic_cell | *OXCT1* | -1.234633879 | 5.73E-05 | 0.000191853 | 0.360491399 |
| T2DM | dendritic_cell | *EPYC* | 3.883454704 | 0.000276789 | 0.000584146 | 1 |
| T2DM | dendritic_cell | *ZNF324* | -1.569396293 | 1.47E-05 | 0.000130129 | 0.092629938 |
| T2DM | dendritic_cell | *ZNF446* | -2.9441948 | 1.72E-05 | 0.000130129 | 0.108082832 |
| T2DM | dendritic_cell | *ZNF264* | -2.68973036 | 6.24E-05 | 0.000201036 | 0.392019806 |
| T2DM | dendritic_cell | *RPS5* | -3.60265309 | 0.000270865 | 0.000574537 | 1 |
| T2DM | dendritic_cell | *CHMP2B* | -2.093366194 | 2.18E-05 | 0.000132858 | 0.137268493 |
| T2DM | dendritic_cell | *PPIE* | -2.465861834 | 0.003478805 | 0.005295701 | 1 |
| T2DM | dendritic_cell | *ZMPSTE24* | -3.580271475 | 0.000133484 | 0.000336494 | 0.839215715 |
| T2DM | dendritic_cell | *NOA1* | -2.281831408 | 3.15E-05 | 0.000147792 | 0.198336941 |
| T2DM | dendritic_cell | *COL16A1* | -1.313176687 | 2.25E-06 | 8.72E-05 | 0.014168366 |
| T2DM | dendritic_cell | *APOB* | -1.545312491 | 9.81E-06 | 0.000130129 | 0.061694695 |
| T2DM | dendritic_cell | *NCOA1* | -2.175072725 | 0.000153253 | 0.000372727 | 0.963498579 |
| T2DM | dendritic_cell | *CD59* | -1.838727745 | 8.36E-05 | 0.000244014 | 0.525362809 |
| T2DM | dendritic_cell | *CD82* | -1.329959518 | 0.011642945 | 0.015937121 | 1 |
| T2DM | dendritic_cell | *TAF9* | -4.35723173 | 0.02144803 | 0.028156977 | 1 |
| T2DM | dendritic_cell | *FCN1* | 0.301708097 | 4.53E-05 | 0.0001691 | 0.28493287 |
| T2DM | dendritic_cell | *SCAMP1* | -2.254557789 | 0.000506599 | 0.000961071 | 1 |
| T2DM | dendritic_cell | *HACE1* | -2.145047841 | 1.25E-05 | 0.000130129 | 0.078773617 |
| T2DM | dendritic_cell | *SEH1L* | -2.443011546 | 0.023038266 | 0.030106335 | 1 |
| T2DM | dendritic_cell | *OVGP1* | -4.527986581 | 0.000100123 | 0.000272936 | 0.629472297 |
| T2DM | dendritic_cell | *SLC25A24* | -2.203953431 | 0.000372445 | 0.000744587 | 1 |
| T2DM | dendritic_cell | *PILRA* | 0.508035768 | 0.001554879 | 0.002567102 | 1 |
| T2DM | dendritic_cell | *ABCB1* | -1.019773852 | 0.000146619 | 0.000359587 | 0.921793195 |
| T2DM | dendritic_cell | *ZNF213* | -1.610733022 | 0.012799005 | 0.017409637 | 1 |
| T2DM | dendritic_cell | *RRN3* | -2.481774087 | 0.000135445 | 0.000339529 | 0.851539751 |
| T2DM | dendritic_cell | *CTTN* | -1.980994267 | 2.14E-05 | 0.000132591 | 0.134474244 |
| T2DM | dendritic_cell | *MTIF2* | -2.972271004 | 3.15E-06 | 0.000102388 | 0.019780889 |
| T2DM | dendritic_cell | *EPS15* | -1.363011343 | 0.005945793 | 0.008603379 | 1 |
| T2DM | dendritic_cell | *ATG16L1* | -2.458082235 | 0.000158852 | 0.000382645 | 0.998703272 |
| T2DM | dendritic_cell | *USP40* | -1.707336582 | 2.31E-05 | 0.00013467 | 0.145226448 |
| T2DM | dendritic_cell | *EIF2AK1* | -2.079694943 | 0.000483106 | 0.000922346 | 1 |
| T2DM | dendritic_cell | *SNX10* | -0.58461076 | 0.002258471 | 0.003602894 | 1 |

| | | | | | | |
|---|---|---|---|---|---|---|
| T2DM | dendritic_cell | *SEPHS1* | -2.729186653 | 0.005453058 | 0.007958072 | 1 |
| T2DM | dendritic_cell | *ITPKC* | -2.810754828 | 0.014253753 | 0.019259262 | 1 |
| T2DM | dendritic_cell | *TMED2* | -3.665030521 | 0.000180001 | 0.000419758 | 1 |
| T2DM | dendritic_cell | *ACOX3* | -1.863415848 | 0.000160879 | 0.000386342 | 1 |
| T2DM | dendritic_cell | *HSD17B14* | 1.311335593 | 0.000916611 | 0.00160477 | 1 |
| T2DM | dendritic_cell | *TRIP6* | -1.996416986 | 9.41E-06 | 0.000130129 | 0.059187523 |
| T2DM | dendritic_cell | *ACHE* | -1.309823361 | 0.000730864 | 0.001319628 | 1 |
| T2DM | dendritic_cell | *SRRT* | -2.288915159 | 0.001313726 | 0.002206625 | 1 |
| T2DM | dendritic_cell | *PIGS* | -1.051965496 | 0.02253716 | 0.029500547 | 1 |
| T2DM | dendritic_cell | *PGS1* | -3.651161304 | 0.000548458 | 0.001030839 | 1 |
| T2DM | dendritic_cell | *PSMC5* | -2.008904374 | 9.32E-06 | 0.000130129 | 0.058577274 |
| T2DM | dendritic_cell | *LPCAT2* | 1.499415318 | 0.008090979 | 0.011428439 | 1 |
| T2DM | dendritic_cell | *OGFOD1* | -2.527476308 | 0.012539266 | 0.017074802 | 1 |
| T2DM | dendritic_cell | *GMCL1* | -2.088848041 | 6.37E-05 | 0.000203711 | 0.400546829 |
| T2DM | dendritic_cell | *KLHL42* | -2.350431032 | 0.003784228 | 0.005719097 | 1 |
| T2DM | dendritic_cell | *PTHLH* | -1.462820296 | 3.89E-05 | 0.000158595 | 0.244711384 |
| T2DM | dendritic_cell | *PHACTR3* | -1.13296712 | 0.000943997 | 0.001646757 | 1 |
| T2DM | dendritic_cell | *ERGIC2* | -2.647880525 | 2.82E-05 | 0.000142583 | 0.177096731 |
| T2DM | dendritic_cell | *AAMDC* | -1.395161721 | 1.68E-05 | 0.000130129 | 0.105930045 |
| T2DM | dendritic_cell | *METTL2A* | -2.84113774 | 1.71E-05 | 0.000130129 | 0.107341338 |
| T2DM | dendritic_cell | *ALG6* | -4.082539024 | 0.000174151 | 0.000409479 | 1 |
| T2DM | dendritic_cell | *GP6* | 1.566754919 | 0.001149626 | 0.001957043 | 1 |
| T2DM | dendritic_cell | *DDX18* | -2.277238314 | 0.008135534 | 0.011488189 | 1 |
| T2DM | dendritic_cell | *ASAP3* | -1.616582316 | 1.15E-05 | 0.000130129 | 0.072231198 |
| T2DM | dendritic_cell | *EDEM2* | -2.941740072 | 3.73E-06 | 0.000110889 | 0.023474111 |
| T2DM | dendritic_cell | *TPX2* | -1.14446761 | 0.000930054 | 0.001624236 | 1 |
| T2DM | dendritic_cell | *DOCK3* | -1.263221379 | 0.0219814 | 0.028815067 | 1 |
| T2DM | dendritic_cell | *C3orf18* | 5.813505379 | 7.48E-08 | 1.34E-05 | 0.000470112 |
| T2DM | dendritic_cell | *FKBP1A* | 2.432623314 | 0.035633085 | 0.045376789 | 1 |
| T2DM | dendritic_cell | *NSFL1C* | -2.634696516 | 4.00E-05 | 0.000159994 | 0.251788267 |
| T2DM | dendritic_cell | *ZNF343* | -1.266837418 | 6.63E-05 | 0.000209164 | 0.416654956 |
| T2DM | dendritic_cell | *CPXM1* | -1.347477687 | 0.000328165 | 0.000670951 | 1 |
| T2DM | dendritic_cell | *MAVS* | -2.19575845 | 0.015625887 | 0.020995799 | 1 |
| T2DM | dendritic_cell | *LZTS3* | -2.249492311 | 5.53E-06 | 0.000126533 | 0.034796519 |
| T2DM | dendritic_cell | *XRN2* | -2.662583139 | 0.005722477 | 0.008320354 | 1 |
| T2DM | dendritic_cell | *TESC* | -5.173796612 | 5.16E-05 | 0.000180471 | 0.324125445 |
| T2DM | dendritic_cell | *RBBP9* | -3.856675053 | 0.000462426 | 0.000890709 | 1 |
| T2DM | dendritic_cell | *ANAPC5* | -2.590323443 | 3.97E-05 | 0.000159642 | 0.249361346 |
| T2DM | dendritic_cell | *TMEM230* | -2.008879502 | 0.001135192 | 0.001935707 | 1 |
| T2DM | dendritic_cell | *DZANK1* | -4.011961358 | 0.000911443 | 0.001597947 | 1 |
| T2DM | dendritic_cell | *TASP1* | -2.260671969 | 4.97E-05 | 0.000176792 | 0.312699197 |
| T2DM | dendritic_cell | *OAS1* | 2.189876011 | 0.000514405 | 0.000973237 | 1 |
| T2DM | dendritic_cell | *FUS* | -2.509521959 | 0.001328028 | 0.002225889 | 1 |
| T2DM | dendritic_cell | *ZNF302* | -1.473145124 | 0.000104089 | 0.000280929 | 0.65440599 |
| T2DM | dendritic_cell | *FXYD3* | -4.7520426 | 0.000265939 | 0.000565999 | 1 |
| T2DM | dendritic_cell | *KCNH4* | 1.323521207 | 2.31E-05 | 0.00013467 | 0.145287726 |
| T2DM | dendritic_cell | *GANAB* | -2.872090631 | 0.000110362 | 0.000292515 | 0.693845035 |
| T2DM | dendritic_cell | *GMIP* | 2.210671604 | 0.000243721 | 0.000528045 | 1 |
| T2DM | dendritic_cell | *MLF2* | -4.295428337 | 0.000688208 | 0.001252319 | 1 |
| T2DM | dendritic_cell | *GPATCH2L* | -1.990471206 | 0.006077211 | 0.00877928 | 1 |

| | | | | | | |
|---|---|---|---|---|---|---|
| T2DM | dendritic_cell | *SLC9A1* | -3.647865803 | 0.00034677 | 0.000700785 | 1 |
| T2DM | dendritic_cell | *SPTLC1* | -2.418591553 | 0.000140419 | 0.000348663 | 0.88281477 |
| T2DM | dendritic_cell | *PAPOLA* | -2.553324843 | 0.002955858 | 0.004572707 | 1 |
| T2DM | dendritic_cell | *RGS1* | 0.406643501 | 3.31E-05 | 0.000149067 | 0.208117187 |
| T2DM | dendritic_cell | *MRPS33* | -4.074890696 | 0.002848628 | 0.004420964 | 1 |
| T2DM | dendritic_cell | *MAEA* | -2.365459772 | 9.88E-05 | 0.000271395 | 0.621277742 |
| T2DM | dendritic_cell | *STRN4* | -2.723437922 | 3.33E-05 | 0.000149262 | 0.209414535 |
| T2DM | dendritic_cell | *LYZ* | 0.559833268 | 2.73E-05 | 0.000141272 | 0.171787169 |
| T2DM | dendritic_cell | *GNPTG* | -3.088026723 | 0.016810606 | 0.022477303 | 1 |
| T2DM | dendritic_cell | *ZNF268* | -2.75716459 | 5.31E-06 | 0.000124995 | 0.0333737 |
| T2DM | dendritic_cell | *GOLGA3* | -1.865174495 | 0.00394311 | 0.005933541 | 1 |
| T2DM | dendritic_cell | *PABPC4* | -1.750620569 | 0.011993162 | 0.016370172 | 1 |
| T2DM | dendritic_cell | *CERS4* | -1.649410858 | 1.17E-05 | 0.000130129 | 0.073459786 |
| T2DM | dendritic_cell | *USP48* | -2.345977819 | 0.000125124 | 0.000320559 | 0.786652614 |
| T2DM | dendritic_cell | *PDPR* | -2.441061705 | 0.000987763 | 0.001714067 | 1 |
| T2DM | dendritic_cell | *EXOC1* | -2.675099695 | 0.000481935 | 0.000920949 | 1 |
| T2DM | dendritic_cell | *NLRC4* | 1.772933863 | 0.00020179 | 0.000456516 | 1 |
| T2DM | dendritic_cell | *LAMB1* | -1.949790555 | 3.82E-05 | 0.000157656 | 0.240268276 |
| T2DM | dendritic_cell | *SLC26A4* | 3.95757656 | 0.00579412 | 0.008412848 | 1 |
| T2DM | dendritic_cell | *DLD* | -2.206279365 | 5.22E-05 | 0.000181742 | 0.328226954 |
| T2DM | dendritic_cell | *TXNL1* | -2.780767959 | 0.000954243 | 0.001662488 | 1 |
| T2DM | dendritic_cell | *ITGA6* | -1.697776901 | 1.15E-05 | 0.000130129 | 0.072464296 |
| T2DM | dendritic_cell | *RAPGEF4* | -1.57395521 | 2.13E-05 | 0.000132591 | 0.133922146 |
| T2DM | dendritic_cell | *SEL1L3* | 1.663116161 | 0.00013885 | 0.000345585 | 0.872947394 |
| T2DM | dendritic_cell | *TF* | -1.234153122 | 1.77E-05 | 0.000130129 | 0.111371025 |
| T2DM | dendritic_cell | *ALKBH5* | -3.384734712 | 4.90E-07 | 3.82E-05 | 0.003080466 |
| T2DM | dendritic_cell | *NLRP1* | -2.532624175 | 8.90E-05 | 0.000254619 | 0.559566531 |
| T2DM | dendritic_cell | *ZFHX4* | -0.881536895 | 1.32E-05 | 0.000130129 | 0.08269833 |
| T2DM | dendritic_cell | *RGS17* | -1.223549855 | 3.52E-06 | 0.000107473 | 0.022139448 |
| T2DM | dendritic_cell | *PPP2R3C* | -2.409089569 | 0.001399474 | 0.002335676 | 1 |
| T2DM | dendritic_cell | *HAUS4* | -1.096906402 | 0.004143884 | 0.006208914 | 1 |
| T2DM | dendritic_cell | *SLC22A17* | -3.821305354 | 4.46E-06 | 0.000117421 | 0.028070048 |
| T2DM | dendritic_cell | *SCFD1* | -1.716196089 | 0.002745024 | 0.004285565 | 1 |
| T2DM | dendritic_cell | *RPGRIP1* | 1.202397276 | 1.32E-05 | 0.000130129 | 0.083006045 |
| T2DM | dendritic_cell | *TOX4* | -3.010462144 | 0.000178097 | 0.000416129 | 1 |
| T2DM | dendritic_cell | *TGM1* | -1.025014341 | 0.035756742 | 0.045497397 | 1 |
| T2DM | dendritic_cell | *AGO1* | -3.553542188 | 0.000111738 | 0.000294672 | 0.702498104 |
| T2DM | dendritic_cell | *MFSD11* | -1.908483545 | 0.001975968 | 0.00319027 | 1 |
| T2DM | dendritic_cell | *VNN3* | 0.951498929 | 2.18E-05 | 0.000132858 | 0.137260644 |
| T2DM | dendritic_cell | *SEC22C* | -2.362062079 | 0.000126313 | 0.000322425 | 0.794132265 |
| T2DM | dendritic_cell | *FMO2* | -1.220125984 | 0.002673952 | 0.004186039 | 1 |
| T2DM | dendritic_cell | *SUCO* | -2.888435923 | 4.14E-05 | 0.000162565 | 0.259984464 |
| T2DM | dendritic_cell | *TMEM38B* | 2.452383323 | 0.000823691 | 0.001464935 | 1 |
| T2DM | dendritic_cell | *PDE6C* | -1.700067749 | 3.03E-05 | 0.000145705 | 0.190644207 |
| T2DM | dendritic_cell | *CWF19L1* | -2.132464328 | 7.72E-09 | 2.70E-06 | 4.85E-05 |
| T2DM | dendritic_cell | *IKZF5* | -3.247949939 | 0.000272952 | 0.00057799 | 1 |
| T2DM | dendritic_cell | *BLNK* | 1.04403656 | 0.0001828 | 0.000424082 | 1 |
| T2DM | dendritic_cell | *WAC* | -2.583729253 | 0.000198969 | 0.000451596 | 1 |
| T2DM | dendritic_cell | *TPSD1* | 22.03336547 | 0.000104521 | 0.000281545 | 0.657126516 |
| T2DM | dendritic_cell | *ITPR3* | -1.025849892 | 1.73E-05 | 0.000130129 | 0.108561774 |

| | | | | | | |
|---|---|---|---|---|---|---|
| T2DM | dendritic_cell | *IFT74* | -1.535676197 | 0.000605375 | 0.001121058 | 1 |
| T2DM | dendritic_cell | *IL12RB1* | 2.602429611 | 0.001024244 | 0.001769073 | 1 |
| T2DM | dendritic_cell | *ACOT7* | -2.91345364 | 1.76E-05 | 0.000130129 | 0.110803762 |
| T2DM | dendritic_cell | *PCSK5* | -1.234060311 | 1.86E-05 | 0.000130129 | 0.117118829 |
| T2DM | dendritic_cell | *NRP1* | -2.027232442 | 9.76E-05 | 0.000268845 | 0.613731092 |
| T2DM | dendritic_cell | *PRTFDC1* | -1.678023581 | 1.36E-05 | 0.000130129 | 0.085202321 |
| T2DM | dendritic_cell | *PALMD* | -0.898221069 | 3.21E-05 | 0.000148254 | 0.201583462 |
| T2DM | dendritic_cell | *TSPAN15* | -1.622557967 | 1.35E-05 | 0.000130129 | 0.085076823 |
| T2DM | dendritic_cell | *OCEL1* | -1.080270382 | 0.000252309 | 0.000542869 | 1 |
| T2DM | dendritic_cell | *CATSPERG* | -1.977054636 | 0.000409652 | 0.000805938 | 1 |
| T2DM | dendritic_cell | *STX1B* | -3.252817773 | 2.82E-06 | 9.76E-05 | 0.017756652 |
| T2DM | dendritic_cell | *HSD3B7* | -2.09492535 | 0.000673949 | 0.001227794 | 1 |
| T2DM | dendritic_cell | *ATP5D* | -2.665283383 | 5.27E-06 | 0.00012486 | 0.033103234 |
| T2DM | dendritic_cell | *MARCH2* | -1.967684884 | 0.024657236 | 0.032081962 | 1 |
| T2DM | dendritic_cell | *CDC34* | -4.139125106 | 1.43E-05 | 0.000130129 | 0.089615996 |
| T2DM | dendritic_cell | *MTAP* | -3.524864328 | 3.99E-05 | 0.000159733 | 0.250588092 |
| T2DM | dendritic_cell | *CEP170B* | -1.171985836 | 0.00026813 | 0.000569696 | 1 |
| T2DM | dendritic_cell | *POLR2E* | -2.166162731 | 7.03E-06 | 0.000130129 | 0.04418977 |
| T2DM | dendritic_cell | *MADCAM1* | -3.265311808 | 4.68E-05 | 0.000171433 | 0.294297087 |
| T2DM | dendritic_cell | *TRMT2A* | -4.191850715 | 0.000276242 | 0.000583385 | 1 |
| T2DM | dendritic_cell | *ZDHHC8* | -2.172209237 | 0.000449781 | 0.000870245 | 1 |
| T2DM | dendritic_cell | *MED15* | -2.376098884 | 8.84E-05 | 0.00025332 | 0.555531253 |
| T2DM | dendritic_cell | *SERPIND1* | -1.231319961 | 0.029101286 | 0.03749176 | 1 |
| T2DM | dendritic_cell | *LZTR1* | -3.587623756 | 5.08E-05 | 0.000178852 | 0.319157961 |
| T2DM | dendritic_cell | *CECR2* | -1.609019635 | 0.000285897 | 0.000599745 | 1 |
| T2DM | dendritic_cell | *SMARCB1* | -1.670866454 | 0.001227382 | 0.002074899 | 1 |
| T2DM | dendritic_cell | *P2RX6* | -1.629633188 | 1.06E-05 | 0.000130129 | 0.066698377 |
| T2DM | dendritic_cell | *SLC7A4* | 1.772422345 | 5.36E-05 | 0.000184412 | 0.337105151 |
| T2DM | dendritic_cell | *DDT* | -2.962349688 | 7.82E-06 | 0.000130129 | 0.049139781 |
| T2DM | dendritic_cell | *GGT5* | -2.119788817 | 4.18E-05 | 0.000162947 | 0.262996311 |
| T2DM | dendritic_cell | *SPECC1L* | -1.753594314 | 0.002528744 | 0.003984514 | 1 |
| T2DM | dendritic_cell | *UPB1* | 1.134118823 | 0.017525663 | 0.023344035 | 1 |
| T2DM | dendritic_cell | *TOP3B* | -2.656743865 | 0.000211601 | 0.000471919 | 1 |
| T2DM | dendritic_cell | *CYTH4* | 0.999260628 | 8.15E-05 | 0.000239417 | 0.512351651 |
| T2DM | dendritic_cell | *CARD10* | -1.437947935 | 6.98E-05 | 0.000215635 | 0.438817612 |
| T2DM | dendritic_cell | *LRP5L* | -3.116829626 | 4.96E-06 | 0.000123581 | 0.031158336 |
| T2DM | dendritic_cell | *SLC25A1* | -2.767918121 | 0.000109097 | 0.000290306 | 0.685892958 |
| T2DM | dendritic_cell | *LGALS2* | 0.409761706 | 3.21E-05 | 0.000148254 | 0.201679314 |
| T2DM | dendritic_cell | *GGA1* | -2.540333255 | 0.002367587 | 0.003756946 | 1 |
| T2DM | dendritic_cell | *SH3BP1* | 1.296718424 | 6.85E-05 | 0.00021325 | 0.430949189 |
| T2DM | dendritic_cell | *HPS4* | -3.112321628 | 0.004547136 | 0.006747189 | 1 |
| T2DM | dendritic_cell | *PIK3IP1* | -1.928463997 | 1.11E-05 | 0.000130129 | 0.06994956 |
| T2DM | dendritic_cell | *TRIOBP* | -1.615019291 | 0.000973261 | 0.001691236 | 1 |
| T2DM | dendritic_cell | *TFIP11* | 1.947406686 | 0.0029252 | 0.004529737 | 1 |
| T2DM | dendritic_cell | *GCAT* | -1.975789274 | 0.004858892 | 0.007164131 | 1 |
| T2DM | dendritic_cell | *ANKRD54* | -2.145117771 | 8.93E-05 | 0.000255186 | 0.561410083 |
| T2DM | dendritic_cell | *EIF3L* | -3.608566855 | 3.94E-06 | 0.000112647 | 0.024782301 |
| T2DM | dendritic_cell | *CCDC134* | -4.37135277 | 0.00059938 | 0.001111593 | 1 |
| T2DM | dendritic_cell | *SLC16A8* | 1.799726911 | 0.00020006 | 0.000453579 | 1 |
| T2DM | dendritic_cell | *KDELR3* | -1.539027623 | 0.000492394 | 0.000937802 | 1 |

| | | | | | | |
|---|---|---|---|---|---|---|
| T2DM | dendritic_cell | *CYP2D6* | -1.761798884 | 0.00321223 | 0.004930491 | 1 |
| T2DM | dendritic_cell | *DDX17* | -1.767169627 | 0.000161236 | 0.000387053 | 1 |
| T2DM | dendritic_cell | *FBXO7* | -2.598878421 | 0.002823825 | 0.004393315 | 1 |
| T2DM | dendritic_cell | *POLDIP3* | -3.49803044 | 0.014324305 | 0.019333814 | 1 |
| T2DM | dendritic_cell | *TIMP3* | -1.860811277 | 5.91E-05 | 0.000195724 | 0.371288157 |
| T2DM | dendritic_cell | *PPP6R2* | -2.90017983 | 2.85E-06 | 9.78E-05 | 0.017915787 |
| T2DM | dendritic_cell | *CYB5R3* | -3.143806635 | 0.000818533 | 0.001457824 | 1 |
| T2DM | dendritic_cell | *RHBDD3* | -1.801494165 | 2.62E-05 | 0.000139569 | 0.165008661 |
| T2DM | dendritic_cell | *TTLL1* | -2.20247609 | 9.99E-06 | 0.000130129 | 0.062826234 |
| T2DM | dendritic_cell | *RASL10A* | -1.777432384 | 0.000134976 | 0.000339031 | 0.84859412 |
| T2DM | dendritic_cell | *AP1B1* | -2.918127435 | 0.004865229 | 0.007170111 | 1 |
| T2DM | dendritic_cell | *HMGXB4* | -2.043525702 | 2.14E-05 | 0.000132591 | 0.134684066 |
| T2DM | dendritic_cell | *TOM1* | -1.230269837 | 0.0048676 | 0.007171925 | 1 |
| T2DM | dendritic_cell | *NEFH* | 1.696826992 | 8.52E-05 | 0.000247534 | 0.535515389 |
| T2DM | dendritic_cell | *BIK* | -5.824620599 | 2.74E-05 | 0.000141298 | 0.171960218 |
| T2DM | dendritic_cell | *THOC5* | -2.969010434 | 0.001273153 | 0.00214708 | 1 |
| T2DM | dendritic_cell | *TSPO* | -2.581798043 | 0.001111998 | 0.001900796 | 1 |
| T2DM | dendritic_cell | *TTLL12* | 2.608159821 | 0.000384714 | 0.000765895 | 1 |
| T2DM | dendritic_cell | *RBFOX2* | -1.642024462 | 2.20E-05 | 0.000132957 | 0.138455531 |
| T2DM | dendritic_cell | *MTMR3* | -1.785434202 | 0.000243821 | 0.000528045 | 1 |
| T2DM | dendritic_cell | *SAMM50* | -2.758751811 | 4.42E-05 | 0.000167565 | 0.277870784 |
| T2DM | dendritic_cell | *IFT27* | -4.004633411 | 0.009638878 | 0.013409964 | 1 |
| T2DM | dendritic_cell | *NCF4* | 1.390954001 | 0.000285661 | 0.00059945 | 1 |
| T2DM | dendritic_cell | *SLC25A17* | -2.109877711 | 2.36E-05 | 0.000136158 | 0.148276135 |
| T2DM | dendritic_cell | *KCTD17* | 5.111170371 | 0.00054043 | 0.001016967 | 1 |
| T2DM | dendritic_cell | *EP300* | -2.456568334 | 0.001041236 | 0.001793984 | 1 |
| T2DM | dendritic_cell | *ACO2* | -1.999772649 | 0.000332905 | 0.00067861 | 1 |
| T2DM | dendritic_cell | *POLR3H* | -1.515605193 | 0.000577709 | 0.001075871 | 1 |
| T2DM | dendritic_cell | *ZBED4* | -3.755704095 | 0.001614676 | 0.002655367 | 1 |
| T2DM | dendritic_cell | *HDAC10* | 1.884415712 | 0.007613174 | 0.01080696 | 1 |
| T2DM | dendritic_cell | *KHNYN* | -1.237309479 | 0.00365557 | 0.005545987 | 1 |
| T2DM | dendritic_cell | *SDR39U1* | -1.563880526 | 8.05E-06 | 0.000130129 | 0.050586964 |
| T2DM | dendritic_cell | *RBM23* | -3.13766802 | 0.000450003 | 0.000870245 | 1 |
| T2DM | dendritic_cell | *SOS2* | -1.913229505 | 2.79E-05 | 0.000142583 | 0.17521545 |
| T2DM | dendritic_cell | *PYGL* | -0.670526493 | 0.00158894 | 0.002616466 | 1 |
| T2DM | dendritic_cell | *TRIM9* | -1.063325389 | 0.003129173 | 0.004819478 | 1 |
| T2DM | dendritic_cell | *DDHD1* | -2.375175526 | 7.11E-05 | 0.000218276 | 0.446877515 |
| T2DM | dendritic_cell | *CNIH1* | -2.670196214 | 5.90E-05 | 0.000195554 | 0.370771014 |
| T2DM | dendritic_cell | *ATP6V1D* | -2.679988626 | 2.30E-06 | 8.72E-05 | 0.014457695 |
| T2DM | dendritic_cell | *PIGH* | -1.718944397 | 0.00343057 | 0.005232411 | 1 |
| T2DM | dendritic_cell | *VTI1B* | -2.637567286 | 3.52E-05 | 0.000153235 | 0.221424979 |
| T2DM | dendritic_cell | *TMED8* | -3.861307104 | 7.26E-05 | 0.000220421 | 0.456706562 |
| T2DM | dendritic_cell | *DAAM1* | -1.585181727 | 3.72E-05 | 0.000156097 | 0.234146131 |
| T2DM | dendritic_cell | *PPM1A* | -1.839842756 | 0.004271296 | 0.006381568 | 1 |
| T2DM | dendritic_cell | *SLC10A1* | -2.138559379 | 1.79E-05 | 0.000130129 | 0.112537803 |
| T2DM | dendritic_cell | *DICER1* | -2.094235905 | 0.000252077 | 0.000542556 | 1 |
| T2DM | dendritic_cell | *ZFYVE21* | -2.378398093 | 0.000189756 | 0.000435401 | 1 |
| T2DM | dendritic_cell | *MTHFD1* | -1.891273491 | 1.59E-05 | 0.000130129 | 0.099659737 |
| T2DM | dendritic_cell | *TELO2* | -3.009440148 | 2.30E-05 | 0.00013467 | 0.144805544 |
| T2DM | dendritic_cell | *GSKIP* | -3.845444662 | 0.000134134 | 0.00033759 | 0.843299569 |

| | | | | | | |
|---|---|---|---|---|---|---|
| T2DM | dendritic_cell | *VRK1* | -2.897674391 | 0.024431972 | 0.031826394 | 1 |
| T2DM | dendritic_cell | *CCNB1IP1* | -2.698097024 | 0.000194307 | 0.000443267 | 1 |
| T2DM | dendritic_cell | *TRIP11* | -2.156028169 | 2.90E-05 | 0.000143473 | 0.182210142 |
| T2DM | dendritic_cell | *KIAA0391* | -1.868738492 | 1.83E-05 | 0.000130129 | 0.114760572 |
| T2DM | dendritic_cell | *EMC9* | -1.351681362 | 0.000112892 | 0.000297092 | 0.709751771 |
| T2DM | dendritic_cell | *GSS* | -1.907259189 | 0.019579925 | 0.02591811 | 1 |
| T2DM | dendritic_cell | *PYGB* | -1.847093286 | 0.000100398 | 0.000273484 | 0.631200688 |
| T2DM | dendritic_cell | *PROCR* | -1.813066313 | 1.21E-05 | 0.000130129 | 0.076192989 |
| T2DM | dendritic_cell | *NINL* | -1.321211334 | 0.000112255 | 0.000295787 | 0.705747665 |
| T2DM | dendritic_cell | *CD40* | 2.576988165 | 0.003924166 | 0.005910693 | 1 |
| T2DM | dendritic_cell | *UQCC1* | -2.671798668 | 4.02E-06 | 0.000113708 | 0.025243284 |
| T2DM | dendritic_cell | *IFT52* | -1.895105837 | 0.001140248 | 0.001942222 | 1 |
| T2DM | dendritic_cell | *NDRG3* | -3.217720702 | 0.000453449 | 0.000876101 | 1 |
| T2DM | dendritic_cell | *DOK5* | -1.544619435 | 1.58E-05 | 0.000130129 | 0.099335529 |
| T2DM | dendritic_cell | *CSTF1* | 2.317937346 | 0.020826848 | 0.027415912 | 1 |
| T2DM | dendritic_cell | *BMP7* | -1.414167488 | 1.74E-05 | 0.000130129 | 0.109629715 |
| T2DM | dendritic_cell | *TPD52L2* | -2.298613823 | 0.037773015 | 0.047849879 | 1 |
| T2DM | dendritic_cell | *NELFCD* | -1.897185704 | 1.62E-05 | 0.000130129 | 0.101978024 |
| T2DM | dendritic_cell | *CTSZ* | 1.391080482 | 0.002226217 | 0.003558068 | 1 |
| T2DM | dendritic_cell | *PRPF6* | -2.706195175 | 2.05E-05 | 0.000132198 | 0.128922059 |
| T2DM | dendritic_cell | *MTG2* | -2.060257853 | 0.009453006 | 0.013168856 | 1 |
| T2DM | dendritic_cell | *TCFL5* | -2.097692689 | 0.000188049 | 0.000432589 | 1 |
| T2DM | dendritic_cell | *DIDO1* | -1.745247984 | 0.017147387 | 0.022874097 | 1 |
| T2DM | dendritic_cell | *GID8* | -2.540768158 | 0.018305237 | 0.02431545 | 1 |
| T2DM | dendritic_cell | *SLC17A9* | 0.687064028 | 2.63E-05 | 0.000139569 | 0.165268449 |
| T2DM | dendritic_cell | *BIRC7* | -2.89624834 | 0.001030113 | 0.001778232 | 1 |
| T2DM | dendritic_cell | *NKAIN4* | -3.538367578 | 6.57E-06 | 0.000130129 | 0.041323487 |
| T2DM | dendritic_cell | *PTK6* | 1.453183372 | 4.32E-05 | 0.000165275 | 0.271366949 |
| T2DM | dendritic_cell | *CDC25B* | -2.112306241 | 1.80E-05 | 0.000130129 | 0.112983492 |
| T2DM | dendritic_cell | *RNF24* | -2.727769411 | 4.84E-05 | 0.000173951 | 0.304209095 |
| T2DM | dendritic_cell | *TRIB3* | -2.721879276 | 7.44E-06 | 0.000130129 | 0.046769849 |
| T2DM | dendritic_cell | *SLC52A3* | 2.243600689 | 0.000705156 | 0.001279087 | 1 |
| T2DM | dendritic_cell | *ANGPT4* | -3.464095059 | 7.04E-06 | 0.000130129 | 0.044257886 |
| T2DM | dendritic_cell | *RSPO4* | -2.86382647 | 0.000219134 | 0.000485447 | 1 |
| T2DM | dendritic_cell | *SIRPB1* | 0.990303745 | 0.000464228 | 0.000893359 | 1 |
| T2DM | dendritic_cell | *SEC23B* | -1.652170296 | 0.008705983 | 0.012217526 | 1 |
| T2DM | dendritic_cell | *CCM2L* | -1.313771722 | 2.15E-05 | 0.000132663 | 0.135218904 |
| T2DM | dendritic_cell | *MYL9* | -1.962757031 | 0.007345267 | 0.010457359 | 1 |
| T2DM | dendritic_cell | *MROH8* | -2.453383968 | 0.000160134 | 0.000384848 | 1 |
| T2DM | dendritic_cell | *MANBAL* | -0.811014389 | 0.032828471 | 0.041958243 | 1 |
| T2DM | dendritic_cell | *IDH3B* | -4.286166043 | 0.004123513 | 0.006182811 | 1 |
| T2DM | dendritic_cell | *TTI1* | -3.910519196 | 0.000785706 | 0.001408134 | 1 |
| T2DM | dendritic_cell | *RPRD1B* | -3.248114281 | 5.45E-05 | 0.000186487 | 0.342389687 |
| T2DM | dendritic_cell | *BPI* | -2.352331389 | 0.000167206 | 0.000398191 | 1 |
| T2DM | dendritic_cell | *DHX35* | -3.635520613 | 5.94E-05 | 0.000196476 | 0.373528319 |
| T2DM | dendritic_cell | *DNTTIP1* | -1.852670586 | 0.001478757 | 0.002455612 | 1 |
| T2DM | dendritic_cell | *PIGU* | -2.012137875 | 0.00015025 | 0.000366843 | 0.944620116 |
| T2DM | dendritic_cell | *APMAP* | -2.786280456 | 0.000323278 | 0.000663116 | 1 |
| T2DM | dendritic_cell | *CELF4* | -1.637788877 | 1.04E-05 | 0.000130129 | 0.065483289 |
| T2DM | dendritic_cell | *RBFA* | -2.670778661 | 0.008332818 | 0.011738388 | 1 |

| | | | | | | |
|---|---|---|---|---|---|---|
| T2DM | dendritic_cell | *METTL4* | -3.745259321 | 0.000509791 | 0.00096567 | 1 |
| T2DM | dendritic_cell | *LPIN2* | -2.883781069 | 0.000312787 | 0.000645811 | 1 |
| T2DM | dendritic_cell | *SMCHD1* | -2.336874223 | 0.002778575 | 0.004329344 | 1 |
| T2DM | dendritic_cell | *MYL12A* | -3.373358136 | 0.001490639 | 0.002472589 | 1 |
| T2DM | dendritic_cell | *CEP76* | -2.718981077 | 9.47E-05 | 0.000264178 | 0.595437268 |
| T2DM | dendritic_cell | *CEP192* | -1.126765903 | 0.037864492 | 0.047956096 | 1 |
| T2DM | dendritic_cell | *LIPG* | 4.710998924 | 0.00431495 | 0.006436083 | 1 |
| T2DM | dendritic_cell | *NALCN* | -1.933909713 | 2.08E-05 | 0.000132272 | 0.131054049 |
| T2DM | dendritic_cell | *HTR2A* | -3.040136241 | 1.69E-05 | 0.000130129 | 0.106210177 |
| T2DM | dendritic_cell | *STK24* | -2.993979939 | 6.70E-05 | 0.00021043 | 0.421071276 |
| T2DM | dendritic_cell | *DNAJC3* | -3.163605886 | 0.000460366 | 0.000887284 | 1 |
| T2DM | dendritic_cell | *UGGT2* | -1.547160035 | 0.000206231 | 0.000464046 | 1 |
| T2DM | dendritic_cell | *ARHGEF7* | -2.414058827 | 0.000128331 | 0.000326514 | 0.806817113 |
| T2DM | dendritic_cell | *SGCG* | -0.760662745 | 1.59E-05 | 0.000130129 | 0.100015621 |
| T2DM | dendritic_cell | *PARP4* | -2.70760046 | 0.000219237 | 0.000485504 | 1 |
| T2DM | dendritic_cell | *VWA8* | -1.792442774 | 0.000995034 | 0.001724305 | 1 |
| T2DM | dendritic_cell | *KATNAL1* | -2.409778841 | 1.18E-05 | 0.000130129 | 0.074075459 |
| T2DM | dendritic_cell | *DHRS12* | -1.992232103 | 2.48E-05 | 0.000138038 | 0.155984943 |
| T2DM | dendritic_cell | *MEDAG* | -0.836912409 | 2.09E-05 | 0.000132352 | 0.131690012 |
| T2DM | dendritic_cell | *TSC22D1* | -1.894723621 | 9.32E-05 | 0.000262227 | 0.585816156 |
| T2DM | dendritic_cell | *CLN5* | -1.996492804 | 0.000256642 | 0.00055031 | 1 |
| T2DM | dendritic_cell | *MGRN1* | -1.391061358 | 0.029055569 | 0.037440533 | 1 |
| T2DM | dendritic_cell | *CORO1A* | 1.114795377 | 0.00086491 | 0.001529532 | 1 |
| T2DM | dendritic_cell | *NFAT5* | -1.889269581 | 2.99E-05 | 0.000145022 | 0.188237969 |
| T2DM | dendritic_cell | *LONP2* | -2.322429391 | 1.48E-05 | 0.000130129 | 0.093053848 |
| T2DM | dendritic_cell | *PLLP* | -3.587413597 | 0.000224696 | 0.000494803 | 1 |
| T2DM | dendritic_cell | *CTCF* | -2.464481595 | 0.001805387 | 0.002940536 | 1 |
| T2DM | dendritic_cell | *ACD* | -2.315593797 | 0.00061907 | 0.001141376 | 1 |
| T2DM | dendritic_cell | *USB1* | -4.216855813 | 3.23E-05 | 0.000148254 | 0.203362816 |
| T2DM | dendritic_cell | *SETD6* | -3.459135592 | 0.001123123 | 0.001917207 | 1 |
| T2DM | dendritic_cell | *COG4* | -2.79621663 | 3.76E-07 | 3.28E-05 | 0.002360988 |
| T2DM | dendritic_cell | *SLC7A6OS* | -1.077564361 | 0.000260997 | 0.000557555 | 1 |
| T2DM | dendritic_cell | *MON1B* | -3.170822224 | 0.000382317 | 0.000761596 | 1 |
| T2DM | dendritic_cell | *HCFC1R1* | -3.29488076 | 1.87E-05 | 0.000130129 | 0.117720455 |
| T2DM | dendritic_cell | *NPRL3* | -1.377066736 | 0.005461661 | 0.007966928 | 1 |
| T2DM | dendritic_cell | *TAF1C* | -1.306519628 | 8.09E-06 | 0.000130129 | 0.050891381 |
| T2DM | dendritic_cell | *NAGPA* | -3.108094042 | 0.000526933 | 0.000993352 | 1 |
| T2DM | dendritic_cell | *COTL1* | 1.307564669 | 0.000494674 | 0.000941004 | 1 |
| T2DM | dendritic_cell | *CRISPLD2* | -1.148143883 | 0.000447831 | 0.00086738 | 1 |
| T2DM | dendritic_cell | *NME4* | -2.756352019 | 1.30E-05 | 0.000130129 | 0.081847177 |
| T2DM | dendritic_cell | *NOMO3* | -3.68767325 | 2.17E-05 | 0.000132663 | 0.136435805 |
| T2DM | dendritic_cell | *LMF1* | -2.412134155 | 0.000198423 | 0.000450519 | 1 |
| T2DM | dendritic_cell | *NARFL* | -2.961615462 | 0.001073831 | 0.00184257 | 1 |
| T2DM | dendritic_cell | *MTHFSD* | -1.100499131 | 0.000631844 | 0.001161181 | 1 |
| T2DM | dendritic_cell | *CLCN7* | -4.290605355 | 0.00030002 | 0.000624683 | 1 |
| T2DM | dendritic_cell | *HAGHL* | -5.576149373 | 0.000205778 | 0.000463202 | 1 |
| T2DM | dendritic_cell | *SLC7A5* | -2.934072524 | 1.33E-05 | 0.000130129 | 0.083704871 |
| T2DM | dendritic_cell | *FBXO31* | -2.120350742 | 9.51E-06 | 0.000130129 | 0.05981592 |
| T2DM | dendritic_cell | *STUB1* | -3.561003182 | 4.12E-06 | 0.000114478 | 0.02587197 |
| T2DM | dendritic_cell | *UBE2I* | -2.294185063 | 0.000112965 | 0.00029716 | 0.710211288 |

| | | | | | | |
|---|---|---|---|---|---|---|
| T2DM | dendritic_cell | *EARS2* | -1.443103082 | 0.000714948 | 0.001294234 | 1 |
| T2DM | dendritic_cell | *CPPED1* | 1.814018902 | 0.012877411 | 0.017508712 | 1 |
| T2DM | dendritic_cell | *HMOX2* | -1.751333453 | 0.001438261 | 0.002393422 | 1 |
| T2DM | dendritic_cell | *BFAR* | -3.624305676 | 0.000143388 | 0.000354537 | 0.901482281 |
| T2DM | dendritic_cell | *KAT8* | -2.386516724 | 0.001104343 | 0.001889766 | 1 |
| T2DM | dendritic_cell | *SYT17* | -2.593288293 | 0.00273762 | 0.004275066 | 1 |
| T2DM | dendritic_cell | *RNF40* | -3.685522686 | 2.36E-05 | 0.000136166 | 0.148515691 |
| T2DM | dendritic_cell | *KNOP1* | -2.34758317 | 5.01E-05 | 0.000177579 | 0.315165433 |
| T2DM | dendritic_cell | *IQCH* | -3.252497356 | 0.000114558 | 0.000300218 | 0.720224081 |
| T2DM | dendritic_cell | *CSK* | 2.650686852 | 0.003024454 | 0.004671926 | 1 |
| T2DM | dendritic_cell | *HERC1* | -2.203328402 | 0.000779432 | 0.001398692 | 1 |
| T2DM | dendritic_cell | *TRIP4* | -2.230317607 | 0.001839487 | 0.002991426 | 1 |
| T2DM | dendritic_cell | *RASL12* | -5.8234156 | 6.36E-05 | 0.00020369 | 0.399970964 |
| T2DM | dendritic_cell | *AP3B2* | -0.988162384 | 0.001629346 | 0.002675992 | 1 |
| T2DM | dendritic_cell | *IGDCC4* | 0.56772706 | 0.000813761 | 0.001450969 | 1 |
| T2DM | dendritic_cell | *CTSH* | 0.954044112 | 3.31E-05 | 0.000149067 | 0.208246607 |
| T2DM | dendritic_cell | *TTC23* | -2.112496742 | 9.83E-05 | 0.000270222 | 0.617726699 |
| T2DM | dendritic_cell | *FAH* | -1.05344053 | 1.49E-05 | 0.000130129 | 0.093401581 |
| T2DM | dendritic_cell | *RPAP1* | -2.51768849 | 5.77E-06 | 0.000129602 | 0.036288538 |
| T2DM | dendritic_cell | *HOMER2* | 1.418610318 | 0.000240411 | 0.000522275 | 1 |
| T2DM | dendritic_cell | *ATP8B4* | 1.173894788 | 0.000272434 | 0.000577113 | 1 |
| T2DM | dendritic_cell | *DTWD1* | -1.489205871 | 0.000620709 | 0.001143728 | 1 |
| T2DM | dendritic_cell | *DNAJC17* | -3.05689447 | 5.26E-05 | 0.000182418 | 0.330906608 |
| T2DM | dendritic_cell | *PDGFRL* | -0.924207975 | 7.14E-05 | 0.000218403 | 0.448818655 |
| T2DM | dendritic_cell | *ZDHHC2* | -2.540650797 | 5.18E-06 | 0.000124852 | 0.032553496 |
| T2DM | dendritic_cell | *ZFAND1* | -1.740347139 | 0.015632983 | 0.020995799 | 1 |
| T2DM | dendritic_cell | *RIPK2* | 1.844802363 | 0.00376195 | 0.005688163 | 1 |
| T2DM | dendritic_cell | *EYA1* | -0.473282873 | 0.032916927 | 0.042062749 | 1 |
| T2DM | dendritic_cell | *NBN* | -3.119911496 | 0.000558351 | 0.001045059 | 1 |
| T2DM | dendritic_cell | *CPQ* | -1.460494756 | 0.001185633 | 0.00201081 | 1 |
| T2DM | dendritic_cell | *NIPAL2* | -1.617496556 | 0.000710663 | 0.001288333 | 1 |
| T2DM | dendritic_cell | *PLAT* | -1.331683533 | 2.63E-05 | 0.00013962 | 0.165589541 |
| T2DM | dendritic_cell | *RAB2A* | -1.129965859 | 0.005660595 | 0.008237509 | 1 |
| T2DM | dendritic_cell | *ZC2HC1A* | -1.765162819 | 2.63E-05 | 0.000139569 | 0.165150482 |
| T2DM | dendritic_cell | *ARMC1* | -1.739605919 | 0.000172219 | 0.000406891 | 1 |
| T2DM | dendritic_cell | *SPAG1* | -1.513651048 | 0.000308958 | 0.000640045 | 1 |
| T2DM | dendritic_cell | *CHRAC1* | 3.443441238 | 0.026332115 | 0.034085353 | 1 |
| T2DM | dendritic_cell | *SNX16* | -2.553485009 | 6.36E-05 | 0.00020369 | 0.399832725 |
| T2DM | dendritic_cell | *ERI1* | -3.505956118 | 0.001540545 | 0.002546781 | 1 |
| T2DM | dendritic_cell | *LEPROTL1* | -3.226740267 | 0.000147727 | 0.000361526 | 0.928759639 |
| T2DM | dendritic_cell | *ERICH1* | -2.492469439 | 0.000103864 | 0.000280899 | 0.652992614 |
| T2DM | dendritic_cell | *TUSC3* | -1.83243826 | 2.55E-05 | 0.000139164 | 0.160545497 |
| T2DM | dendritic_cell | *ARHGEF10* | -1.083793683 | 0.000276428 | 0.000583581 | 1 |
| T2DM | dendritic_cell | *KLHDC4* | -1.918218753 | 0.003468428 | 0.005282463 | 1 |
| T2DM | dendritic_cell | *MAN2B1* | 1.357520114 | 7.79E-05 | 0.000231975 | 0.489466575 |
| T2DM | dendritic_cell | *TULP2* | -1.740152055 | 0.003657655 | 0.005547811 | 1 |
| T2DM | dendritic_cell | *NUCB1* | -2.40345138 | 0.000512858 | 0.000971187 | 1 |
| T2DM | dendritic_cell | *NFKBIB* | 2.834801493 | 0.004559834 | 0.006764436 | 1 |
| T2DM | dendritic_cell | *TUBB4A* | 6.457156272 | 0.00249256 | 0.003931441 | 1 |
| T2DM | dendritic_cell | *SARS2* | -1.802512119 | 1.32E-05 | 0.000130129 | 0.082675477 |

| | | | | | | |
|---|---|---|---|---|---|---|
| T2DM | dendritic_cell | *SNRNP70* | -2.569624323 | 1.91E-05 | 0.000130716 | 0.119918312 |
| T2DM | dendritic_cell | *FCGRT* | -1.804645758 | 0.011640161 | 0.01593678 | 1 |
| T2DM | dendritic_cell | *PEX11G* | -2.17296874 | 0.000160385 | 0.000385304 | 1 |
| T2DM | dendritic_cell | *ERCC2* | -5.703819303 | 6.39E-05 | 0.000203993 | 0.401459144 |
| T2DM | dendritic_cell | *PLEKHJ1* | 2.533958543 | 0.013353033 | 0.018127946 | 1 |
| T2DM | dendritic_cell | *CD37* | 1.567105932 | 0.000266666 | 0.000567161 | 1 |
| T2DM | dendritic_cell | *AMH* | 1.520471432 | 9.41E-05 | 0.00026388 | 0.591881912 |
| T2DM | dendritic_cell | *FCER2* | -6.309525435 | 2.13E-06 | 8.62E-05 | 0.013414247 |
| T2DM | dendritic_cell | *DMPK* | -2.243257339 | 3.64E-07 | 3.22E-05 | 0.002286357 |
| T2DM | dendritic_cell | *IL4I1* | 0.556025718 | 2.01E-05 | 0.000131266 | 0.126097471 |
| T2DM | dendritic_cell | *AES* | -3.18464963 | 0.002587746 | 0.004065006 | 1 |
| T2DM | dendritic_cell | *C19orf53* | -2.779624424 | 4.64E-05 | 0.000170668 | 0.291433117 |
| T2DM | dendritic_cell | *TIMM44* | -2.037592687 | 0.000218345 | 0.000483869 | 1 |
| T2DM | dendritic_cell | *CCDC61* | -4.102043839 | 0.000159213 | 0.000383074 | 1 |
| T2DM | dendritic_cell | *IL27RA* | 2.448198732 | 9.49E-05 | 0.000264262 | 0.596438771 |
| T2DM | dendritic_cell | *C19orf44* | -1.003101734 | 0.002246274 | 0.003587078 | 1 |
| T2DM | dendritic_cell | *OLFM2* | -0.973967229 | 0.000320573 | 0.000658641 | 1 |
| T2DM | dendritic_cell | *RASAL3* | -2.453836472 | 0.007541973 | 0.010713146 | 1 |
| T2DM | dendritic_cell | *SYDE1* | -1.63067352 | 1.40E-05 | 0.000130129 | 0.088204123 |
| T2DM | dendritic_cell | *CCNE1* | -3.573095367 | 1.32E-05 | 0.000130129 | 0.083070603 |
| T2DM | dendritic_cell | *PDCD5* | -1.986139559 | 0.00031993 | 0.000658181 | 1 |
| T2DM | dendritic_cell | *ANKRD27* | -2.993801647 | 0.00428764 | 0.006398384 | 1 |
| T2DM | dendritic_cell | *RPS16* | -2.992165846 | 1.98E-05 | 0.000131266 | 0.12441207 |
| T2DM | dendritic_cell | *CCDC94* | 2.513301541 | 0.038237555 | 0.048389595 | 1 |
| T2DM | dendritic_cell | *TBCB* | -2.534670167 | 6.86E-05 | 0.00021325 | 0.431009049 |
| T2DM | dendritic_cell | *POLR2I* | 3.14197113 | 0.000870203 | 0.001535494 | 1 |
| T2DM | dendritic_cell | *CLIP3* | -1.602536585 | 0.000300635 | 0.000625443 | 1 |
| T2DM | dendritic_cell | *SLC1A5* | -2.313092654 | 0.010033138 | 0.013921505 | 1 |
| T2DM | dendritic_cell | *PRKD2* | -2.8897037 | 2.56E-05 | 0.0001392 | 0.160888823 |
| T2DM | dendritic_cell | *CACTIN* | 3.432621441 | 0.000740415 | 0.001335722 | 1 |
| T2DM | dendritic_cell | *FZR1* | -3.799890429 | 6.09E-05 | 0.000198694 | 0.38275368 |
| T2DM | dendritic_cell | *BBC3* | -7.221708875 | 0.000155335 | 0.000376191 | 0.976591907 |
| T2DM | dendritic_cell | *CEACAM4* | 0.555882354 | 3.06E-05 | 0.00014573 | 0.192072428 |
| T2DM | dendritic_cell | *MRPL4* | -2.642783558 | 0.035468033 | 0.045184909 | 1 |
| T2DM | dendritic_cell | *ICAM5* | -17.87758622 | 6.54E-07 | 4.90E-05 | 0.004113657 |
| T2DM | dendritic_cell | *CD33* | 0.786979496 | 4.51E-05 | 0.000168939 | 0.28326329 |
| T2DM | dendritic_cell | *BABAM1* | 3.595134367 | 0.003717803 | 0.005629534 | 1 |
| T2DM | dendritic_cell | *TYK2* | 2.51482837 | 0.008737688 | 0.012248349 | 1 |
| T2DM | dendritic_cell | *RABAC1* | -3.10484284 | 6.63E-06 | 0.000130129 | 0.041703156 |
| T2DM | dendritic_cell | *PTPRS* | -0.794240925 | 6.68E-05 | 0.0002101 | 0.419990108 |
| T2DM | dendritic_cell | *GRWD1* | -1.570210804 | 1.74E-05 | 0.000130129 | 0.109199352 |
| T2DM | dendritic_cell | *GRIN2D* | -2.562944607 | 1.37E-05 | 0.000130129 | 0.086181305 |
| T2DM | dendritic_cell | *CCDC114* | 1.367092986 | 0.000323694 | 0.000663752 | 1 |
| T2DM | dendritic_cell | *CARD8* | -2.784649052 | 0.002003421 | 0.003228789 | 1 |
| T2DM | dendritic_cell | *ZNF175* | -2.816414672 | 6.89E-06 | 0.000130129 | 0.043328886 |
| T2DM | dendritic_cell | *CAPS* | -1.316421041 | 0.000276031 | 0.000583385 | 1 |
| T2DM | dendritic_cell | *BCAT2* | -1.922663145 | 1.50E-05 | 0.000130129 | 0.094030982 |
| T2DM | dendritic_cell | *PPP2R1A* | -2.77297733 | 6.32E-05 | 0.000202959 | 0.397394247 |
| T2DM | dendritic_cell | *TNPO2* | -2.267508184 | 0.000320341 | 0.00065838 | 1 |
| T2DM | dendritic_cell | *GCDH* | -4.065383513 | 3.43E-09 | 1.54E-06 | 2.16E-05 |

| | | | | | | |
|---|---|---|---|---|---|---|
| T2DM | dendritic_cell | *MAST1* | 6.682821425 | 0.000515822 | 0.00097533 | 1 |
| T2DM | dendritic_cell | *PRPF31* | -2.891510775 | 0.000163757 | 0.000391609 | 1 |
| T2DM | dendritic_cell | *TFPT* | -2.555453306 | 3.55E-05 | 0.000154001 | 0.223301737 |
| T2DM | dendritic_cell | *PDE4C* | -2.41440915 | 0.001167891 | 0.001983932 | 1 |
| T2DM | dendritic_cell | *UPK1A* | 5.514511465 | 0.000153381 | 0.000372894 | 0.964304864 |
| T2DM | dendritic_cell | *LSR* | -1.557679118 | 0.017050708 | 0.022759618 | 1 |
| T2DM | dendritic_cell | *KXD1* | -3.692947852 | 0.01799621 | 0.02392518 | 1 |
| T2DM | dendritic_cell | *HPN* | -1.678080787 | 2.68E-06 | 9.51E-05 | 0.016870871 |
| T2DM | dendritic_cell | *ZNF14* | -2.46142173 | 1.49E-05 | 0.000130129 | 0.093837384 |
| T2DM | dendritic_cell | *ATP13A1* | 2.296652424 | 0.006110382 | 0.00881707 | 1 |
| T2DM | dendritic_cell | *ZNF85* | 2.933765032 | 0.002197753 | 0.003516741 | 1 |
| T2DM | dendritic_cell | *ETHE1* | 1.796831297 | 0.000301701 | 0.000627454 | 1 |
| T2DM | dendritic_cell | *GTPBP10* | -1.551404864 | 0.003030071 | 0.004679454 | 1 |
| T2DM | dendritic_cell | *RASA4* | 1.843095337 | 0.000841566 | 0.001493768 | 1 |
| T2DM | dendritic_cell | *PON3* | 3.448227251 | 0.001023068 | 0.001767526 | 1 |
| T2DM | dendritic_cell | *HBP1* | -2.663969208 | 4.01E-05 | 0.000160039 | 0.252220757 |
| T2DM | dendritic_cell | *WDR91* | -3.129154759 | 3.05E-05 | 0.000145705 | 0.191747657 |
| T2DM | dendritic_cell | *PTN* | -1.39274105 | 2.85E-05 | 0.000142609 | 0.17940268 |
| T2DM | dendritic_cell | *MPP6* | -1.278734441 | 1.50E-05 | 0.000130129 | 0.094052637 |
| T2DM | dendritic_cell | *OGDH* | -2.839822231 | 0.000403112 | 0.000796438 | 1 |
| T2DM | dendritic_cell | *ADAP1* | 3.715434248 | 0.000157262 | 0.00038002 | 0.988706624 |
| T2DM | dendritic_cell | *H2AFV* | -2.434344304 | 0.026332521 | 0.034085353 | 1 |
| T2DM | dendritic_cell | *CAV1* | -1.307490808 | 2.85E-05 | 0.000142609 | 0.179281611 |
| T2DM | dendritic_cell | *MET* | -1.161359921 | 1.59E-05 | 0.000130129 | 0.099836715 |
| T2DM | dendritic_cell | *RNF32* | -1.723468405 | 0.011659998 | 0.015953516 | 1 |
| T2DM | dendritic_cell | *LMBR1* | -2.72317701 | 0.00013389 | 0.00033711 | 0.841764209 |
| T2DM | dendritic_cell | *HOXA2* | -3.400556713 | 4.51E-05 | 0.000168939 | 0.283840233 |
| T2DM | dendritic_cell | *HOXA5* | -1.516261689 | 1.24E-05 | 0.000130129 | 0.077662694 |
| T2DM | dendritic_cell | *HOXA6* | -1.409282455 | 8.38E-06 | 0.000130129 | 0.052665197 |
| T2DM | dendritic_cell | *BRAT1* | -1.797739676 | 0.020976026 | 0.027589179 | 1 |
| T2DM | dendritic_cell | *CPED1* | -1.647603892 | 1.91E-05 | 0.000130738 | 0.120148624 |
| T2DM | dendritic_cell | *HIBADH* | -1.980493913 | 2.66E-05 | 0.000140322 | 0.167521927 |
| T2DM | dendritic_cell | *CPVL* | 0.36939185 | 1.84E-05 | 0.000130129 | 0.115392137 |
| T2DM | dendritic_cell | *PLEKHA8* | -3.217894887 | 0.005679899 | 0.008262269 | 1 |
| T2DM | dendritic_cell | *NOD1* | -2.869119466 | 0.000264185 | 0.000563027 | 1 |
| T2DM | dendritic_cell | *CRHR2* | -4.264889275 | 0.008364924 | 0.011775701 | 1 |
| T2DM | dendritic_cell | *EPHB6* | 2.278076171 | 0.000916291 | 0.001604658 | 1 |
| T2DM | dendritic_cell | *CHCHD2* | -2.072681816 | 0.004598908 | 0.006814361 | 1 |
| T2DM | dendritic_cell | *CCL24* | -1.850600498 | 0.017386144 | 0.023168013 | 1 |
| T2DM | dendritic_cell | *NPTX2* | -34.95127532 | 0.000641183 | 0.001175252 | 1 |
| T2DM | dendritic_cell | *CYP3A5* | -2.652338457 | 2.19E-05 | 0.000132858 | 0.137392658 |
| T2DM | dendritic_cell | *ZKSCAN1* | -2.53343698 | 0.000172387 | 0.000406985 | 1 |
| T2DM | dendritic_cell | *TAF6* | -2.077621091 | 0.000958472 | 0.001667842 | 1 |
| T2DM | dendritic_cell | *AIMP2* | -2.675568117 | 1.81E-05 | 0.000130129 | 0.113814385 |
| T2DM | dendritic_cell | *TFR2* | -2.032579834 | 0.000190752 | 0.000437207 | 1 |
| T2DM | dendritic_cell | *RBM28* | -1.706120893 | 4.93E-06 | 0.000123581 | 0.030991576 |
| T2DM | dendritic_cell | *USP42* | -2.420972762 | 0.000125799 | 0.000321635 | 0.790900733 |
| T2DM | dendritic_cell | *LSM5* | -2.696023595 | 1.75E-05 | 0.000130129 | 0.109856117 |
| T2DM | dendritic_cell | *C1GALT1* | -2.248275786 | 6.70E-05 | 0.000210469 | 0.421359114 |
| T2DM | dendritic_cell | *RPA3* | -1.956251205 | 1.75E-05 | 0.000130129 | 0.109736897 |

| | | | | | | |
|---|---|---|---|---|---|---|
| T2DM | dendritic_cell | *ZNHIT1* | -1.899875395 | 0.001280919 | 0.00215844 | 1 |
| T2DM | dendritic_cell | *PHF14* | -2.080997491 | 5.04E-05 | 0.000178067 | 0.316809256 |
| T2DM | dendritic_cell | *CEP41* | -1.504355148 | 0.000201379 | 0.000456076 | 1 |
| T2DM | dendritic_cell | *ZNF862* | -1.916272341 | 8.98E-05 | 0.000255768 | 0.564480534 |
| T2DM | dendritic_cell | *MEST* | -1.00228762 | 1.77E-06 | 8.21E-05 | 0.011135414 |
| T2DM | dendritic_cell | *ACTR3C* | -1.716115641 | 0.000459583 | 0.000886047 | 1 |
| T2DM | dendritic_cell | *POU6F2* | -1.433531751 | 1.38E-05 | 0.000130129 | 0.086576 |
| T2DM | dendritic_cell | *TSPAN13* | -3.897395552 | 2.00E-05 | 0.000131266 | 0.125507583 |
| T2DM | dendritic_cell | *RARRES2* | -1.982554115 | 9.45E-05 | 0.000264058 | 0.59389366 |
| T2DM | dendritic_cell | *TMEM176B* | 1.432408335 | 7.30E-06 | 0.000130129 | 0.045893895 |
| T2DM | dendritic_cell | *MRPL32* | -1.153167965 | 0.020553991 | 0.027102127 | 1 |
| T2DM | dendritic_cell | *COA1* | -1.460016122 | 0.003193942 | 0.004906797 | 1 |
| T2DM | dendritic_cell | *BLVRA* | -1.909106706 | 0.00180588 | 0.002940577 | 1 |
| T2DM | dendritic_cell | *URGCP* | -2.555577955 | 3.77E-05 | 0.000156929 | 0.236962076 |
| T2DM | dendritic_cell | *POLD2* | -5.142937179 | 3.09E-08 | 7.19E-06 | 0.000194143 |
| T2DM | dendritic_cell | *GCK* | -14.78293148 | 1.23E-05 | 0.000130129 | 0.077077428 |
| T2DM | dendritic_cell | *YKT6* | 3.169196502 | 0.003006806 | 0.004645807 | 1 |
| T2DM | dendritic_cell | *TBL2* | -0.855707629 | 0.00745481 | 0.010601309 | 1 |
| T2DM | dendritic_cell | *LIMK1* | 1.241615648 | 8.24E-05 | 0.000241604 | 0.517756563 |
| T2DM | dendritic_cell | *SPATA6L* | -1.828153181 | 1.74E-05 | 0.000130129 | 0.109616069 |
| T2DM | dendritic_cell | *FKTN* | -2.686099706 | 1.88E-05 | 0.000130129 | 0.118281471 |
| T2DM | dendritic_cell | *NMRK1* | -1.485457522 | 0.034085411 | 0.04351167 | 1 |
| T2DM | dendritic_cell | *TMEM245* | -2.313938423 | 7.50E-05 | 0.000225784 | 0.47166284 |
| T2DM | dendritic_cell | *MEGF9* | -1.604666439 | 0.031331204 | 0.040142507 | 1 |
| T2DM | dendritic_cell | *CORO2A* | -2.057696807 | 7.10E-06 | 0.000130129 | 0.044666093 |
| T2DM | dendritic_cell | *C5* | -1.598559545 | 9.35E-05 | 0.000262424 | 0.587566712 |
| T2DM | dendritic_cell | *OGN* | -1.5041021 | 1.95E-05 | 0.000131266 | 0.122676237 |
| T2DM | dendritic_cell | *ECM2* | -1.253295992 | 2.24E-05 | 0.000133629 | 0.14098685 |
| T2DM | dendritic_cell | *PTGR1* | -1.107162231 | 0.00050834 | 0.000963502 | 1 |
| T2DM | dendritic_cell | *TNFSF8* | 0.999878934 | 8.63E-05 | 0.000249123 | 0.542340314 |
| T2DM | dendritic_cell | *PLGRKT* | -1.554524749 | 0.004656686 | 0.006885368 | 1 |
| T2DM | dendritic_cell | *KDM4C* | -2.494242418 | 0.000831847 | 0.001478185 | 1 |
| T2DM | dendritic_cell | *DOCK8* | 1.374038365 | 0.003682331 | 0.00557851 | 1 |
| T2DM | dendritic_cell | *ELAVL2* | -2.476068041 | 3.39E-06 | 0.000105474 | 0.02130581 |
| T2DM | dendritic_cell | *FUBP3* | -3.07913542 | 0.000147721 | 0.000361526 | 0.928720597 |
| T2DM | dendritic_cell | *RGP1* | -2.984230797 | 0.000191533 | 0.000438272 | 1 |
| T2DM | dendritic_cell | *MPDZ* | -2.001993158 | 2.69E-05 | 0.000140623 | 0.168888645 |
| T2DM | dendritic_cell | *PTGDS* | -6.03992673 | 7.11E-05 | 0.000218276 | 0.447296425 |
| T2DM | dendritic_cell | *SHB* | -3.316955073 | 4.43E-05 | 0.000167569 | 0.278719717 |
| T2DM | dendritic_cell | *DVL1* | -4.073181521 | 0.003234637 | 0.004957621 | 1 |
| T2DM | dendritic_cell | *PDLIM1* | -2.119657092 | 0.001217298 | 0.002059513 | 1 |
| T2DM | dendritic_cell | *ATRNL1* | -1.658651021 | 0.008862286 | 0.012398129 | 1 |
| T2DM | dendritic_cell | *HPS1* | -3.504363226 | 0.000369754 | 0.000740804 | 1 |
| T2DM | dendritic_cell | *PHYH* | -1.042610271 | 9.45E-05 | 0.000264058 | 0.594294055 |
| T2DM | dendritic_cell | *DNMBP* | -3.041029569 | 0.000629234 | 0.0011574 | 1 |
| T2DM | dendritic_cell | *TRDMT1* | -1.853921188 | 1.59E-05 | 0.000130129 | 0.099854427 |
| T2DM | dendritic_cell | *DDX50* | -2.667632176 | 5.30E-05 | 0.000183002 | 0.333246745 |
| T2DM | dendritic_cell | *MAPK8* | -2.405799368 | 0.001021706 | 0.001766399 | 1 |
| T2DM | dendritic_cell | *ATE1* | -1.823198311 | 0.000386265 | 0.000768738 | 1 |
| T2DM | dendritic_cell | *NSMCE4A* | -2.501058588 | 1.32E-05 | 0.000130129 | 0.08272605 |

| | | | | | | |
|---|---|---|---|---|---|---|
| T2DM | dendritic_cell | *PLEKHA1* | -2.693548235 | 0.001381722 | 0.002309728 | 1 |
| T2DM | dendritic_cell | *SPOCK2* | -1.808414063 | 0.000131016 | 0.000331963 | 0.823697181 |
| T2DM | dendritic_cell | *MICU1* | -2.165872269 | 0.00055176 | 0.001035476 | 1 |
| T2DM | dendritic_cell | *CCSER2* | -2.39907865 | 0.000190463 | 0.000436703 | 1 |
| T2DM | dendritic_cell | *BMPR1A* | -1.025702181 | 0.000866006 | 0.001530667 | 1 |
| T2DM | dendritic_cell | *KAZALD1* | -3.242198586 | 4.31E-06 | 0.000116241 | 0.027069522 |
| T2DM | dendritic_cell | *TNKS2* | -3.021289721 | 0.000244073 | 0.000528405 | 1 |
| T2DM | dendritic_cell | *ANKRD26* | -1.485723714 | 0.00412574 | 0.006184675 | 1 |
| T2DM | dendritic_cell | *PITRM1* | -3.210668438 | 4.61E-05 | 0.00017047 | 0.290139411 |
| T2DM | dendritic_cell | *MAP3K8* | -3.123599132 | 0.001011441 | 0.001750326 | 1 |
| T2DM | dendritic_cell | *SHOC2* | -2.309968675 | 0.000363657 | 0.000731854 | 1 |
| T2DM | dendritic_cell | *TFAM* | -2.606250201 | 3.10E-06 | 0.000101654 | 0.01946434 |
| T2DM | dendritic_cell | *PBLD* | -0.551334218 | 0.03724733 | 0.047269674 | 1 |
| T2DM | dendritic_cell | *DHRS11* | -1.466724582 | 9.69E-08 | 1.56E-05 | 0.00060896 |
| T2DM | dendritic_cell | *MLLT6* | -2.812570094 | 6.88E-05 | 0.000213638 | 0.432789783 |
| T2DM | dendritic_cell | *PSMB3* | 2.520311522 | 5.20E-05 | 0.000181132 | 0.326943688 |
| T2DM | dendritic_cell | *RAPGEFL1* | -1.829558587 | 4.60E-05 | 0.000170331 | 0.289392571 |
| T2DM | dendritic_cell | *RGS9* | -5.087855408 | 3.49E-05 | 0.000152633 | 0.219570297 |
| T2DM | dendritic_cell | *WNT3* | 2.160230089 | 0.001468652 | 0.002440766 | 1 |
| T2DM | dendritic_cell | *ASPA* | -1.418005504 | 1.42E-05 | 0.000130129 | 0.089446643 |
| T2DM | dendritic_cell | *RAD51C* | -1.575678561 | 0.001003482 | 0.001737987 | 1 |
| T2DM | dendritic_cell | *SEPT4* | -1.833438271 | 9.42E-07 | 5.75E-05 | 0.005919726 |
| T2DM | dendritic_cell | *TRIM37* | -2.754733198 | 0.000713929 | 0.001292762 | 1 |
| T2DM | dendritic_cell | *PNPO* | 1.544487869 | 4.85E-05 | 0.000174265 | 0.304964441 |
| T2DM | dendritic_cell | *RPS6KB1* | -2.142843307 | 2.11E-05 | 0.000132591 | 0.13259023 |
| T2DM | dendritic_cell | *TRIM16L* | -1.581283232 | 0.031619059 | 0.040486562 | 1 |
| T2DM | dendritic_cell | *CDK5RAP3* | -1.382965156 | 3.64E-05 | 0.000155217 | 0.229068902 |
| T2DM | dendritic_cell | *RECQL5* | -2.098632033 | 0.002095278 | 0.003363036 | 1 |
| T2DM | dendritic_cell | *PIGL* | -2.325293243 | 3.73E-05 | 0.000156278 | 0.23457315 |
| T2DM | dendritic_cell | *GALK1* | -5.211129871 | 3.35E-05 | 0.00015 | 0.210750312 |
| T2DM | dendritic_cell | *CAMTA2* | -1.776601312 | 0.000361278 | 0.000727532 | 1 |
| T2DM | dendritic_cell | *MED13* | -2.307408708 | 0.000180989 | 0.000421377 | 1 |
| T2DM | dendritic_cell | *HOXB6* | -1.526349891 | 2.48E-06 | 9.04E-05 | 0.015565054 |
| T2DM | dendritic_cell | *NUP88* | -2.541338646 | 4.25E-05 | 0.000164104 | 0.267161873 |
| T2DM | dendritic_cell | *C1QBP* | 3.453108352 | 0.002301123 | 0.003662572 | 1 |
| T2DM | dendritic_cell | *GOSR1* | -2.971529989 | 0.000337819 | 0.000685765 | 1 |
| T2DM | dendritic_cell | *DRG2* | -4.042914877 | 0.016538865 | 0.022123371 | 1 |
| T2DM | dendritic_cell | *FTSJ3* | -1.055681093 | 0.000695854 | 0.001264402 | 1 |
| T2DM | dendritic_cell | *AKAP10* | -2.465302029 | 0.003096106 | 0.004773227 | 1 |
| T2DM | dendritic_cell | *ALDH3A1* | 2.029320002 | 0.001917161 | 0.003103293 | 1 |
| T2DM | dendritic_cell | *SMARCD2* | -4.361833384 | 0.000211218 | 0.000471732 | 1 |
| T2DM | dendritic_cell | *ICAM2* | -1.466879785 | 0.032348202 | 0.041369639 | 1 |
| T2DM | dendritic_cell | *UTP6* | -3.057193919 | 8.09E-05 | 0.000238113 | 0.508486728 |
| T2DM | dendritic_cell | *DDX5* | -1.272596492 | 0.001344207 | 0.002252407 | 1 |
| T2DM | dendritic_cell | *C17orf75* | -1.784681577 | 1.84E-05 | 0.000130129 | 0.115372171 |
| T2DM | dendritic_cell | *PEX12* | 1.294951737 | 0.005997319 | 0.008671837 | 1 |
| T2DM | dendritic_cell | *NAGLU* | -2.010828284 | 9.51E-06 | 0.000130129 | 0.059776893 |
| T2DM | dendritic_cell | *HSD17B1* | -1.333641262 | 4.73E-06 | 0.000121447 | 0.029754592 |
| T2DM | dendritic_cell | *EZH1* | -1.646875451 | 0.000192146 | 0.000439441 | 1 |
| T2DM | dendritic_cell | *DLX4* | -13.71297921 | 1.32E-05 | 0.000130129 | 0.083009169 |

| | | | | | | |
|---|---|---|---|---|---|---|
| T2DM | dendritic_cell | *VAT1* | -1.356952775 | 0.000904434 | 0.001587875 | 1 |
| T2DM | dendritic_cell | *LRRC59* | -1.728611734 | 1.57E-05 | 0.000130129 | 0.098686873 |
| T2DM | dendritic_cell | *ALOX12* | -5.0896165 | 3.25E-05 | 0.000148254 | 0.204272784 |
| T2DM | dendritic_cell | *MPP2* | -7.011119907 | 0.021747839 | 0.028526739 | 1 |
| T2DM | dendritic_cell | *EFNB3* | -13.47242257 | 0.000711239 | 0.001289006 | 1 |
| T2DM | dendritic_cell | *YWHAE* | -2.74544216 | 0.000282214 | 0.000594199 | 1 |
| T2DM | dendritic_cell | *DPH1* | -1.846141934 | 1.77E-05 | 0.000130129 | 0.11115905 |
| T2DM | dendritic_cell | *DHRS7B* | -2.369486715 | 2.91E-05 | 0.000143677 | 0.182977061 |
| T2DM | dendritic_cell | *RCVRN* | -4.091014947 | 0.002359565 | 0.003746107 | 1 |
| T2DM | dendritic_cell | *MYH3* | -2.243179124 | 0.000576106 | 0.001073496 | 1 |
| T2DM | dendritic_cell | *TMEM104* | -2.978428907 | 0.005579681 | 0.008125887 | 1 |
| T2DM | dendritic_cell | *TMEM97* | 1.315156981 | 1.44E-05 | 0.000130129 | 0.090328755 |
| T2DM | dendritic_cell | *RAB34* | -2.545812546 | 0.007948232 | 0.011236909 | 1 |
| T2DM | dendritic_cell | *SLAIN2* | -2.09112696 | 0.000977993 | 0.00169899 | 1 |
| T2DM | dendritic_cell | *OCIAD1* | -3.053742946 | 1.13E-05 | 0.000130129 | 0.070773646 |
| T2DM | dendritic_cell | *CWH43* | 2.45941183 | 2.04E-05 | 0.00013202 | 0.128287001 |
| T2DM | dendritic_cell | *KIAA1211* | 1.029321496 | 0.000263879 | 0.000562566 | 1 |
| T2DM | dendritic_cell | *AREG* | 0.49851903 | 2.71E-05 | 0.000141035 | 0.170652039 |
| T2DM | dendritic_cell | *UBE2D3* | -2.324405189 | 0.000542121 | 0.001019843 | 1 |
| T2DM | dendritic_cell | *NDUFC1* | -3.392254279 | 4.34E-05 | 0.000165564 | 0.272683276 |
| T2DM | dendritic_cell | *CPE* | -1.754324855 | 5.10E-05 | 0.000179321 | 0.320625122 |
| T2DM | dendritic_cell | *WFS1* | -1.640091974 | 0.001043155 | 0.001795814 | 1 |
| T2DM | dendritic_cell | *GRPEL1* | -3.175266666 | 2.67E-05 | 0.000140322 | 0.168090382 |
| T2DM | dendritic_cell | *AADAT* | -1.362320572 | 2.45E-05 | 0.000137669 | 0.153806948 |
| T2DM | dendritic_cell | *CPZ* | -27.18580184 | 1.42E-05 | 0.000130129 | 0.089500924 |
| T2DM | dendritic_cell | *TRIM2* | -1.506049562 | 6.31E-05 | 0.00020269 | 0.396664235 |
| T2DM | dendritic_cell | *SLC2A9* | 0.844629778 | 2.09E-05 | 0.000132272 | 0.131171126 |
| T2DM | dendritic_cell | *SH3D19* | -1.089544636 | 7.92E-05 | 0.000234792 | 0.497994684 |
| T2DM | dendritic_cell | *RAPGEF2* | -1.920192789 | 0.001352862 | 0.002265702 | 1 |
| T2DM | dendritic_cell | *LRP2BP* | -1.714108176 | 0.000992803 | 0.001720913 | 1 |
| T2DM | dendritic_cell | *UFSP2* | -2.148262183 | 1.65E-05 | 0.000130129 | 0.103652964 |
| T2DM | dendritic_cell | *KLF3* | -2.904762208 | 0.000396539 | 0.000785456 | 1 |
| T2DM | dendritic_cell | *UGDH* | -1.758163873 | 2.00E-05 | 0.000131266 | 0.125995024 |
| T2DM | dendritic_cell | *HTATIP2* | -1.063294207 | 0.000224119 | 0.000493707 | 1 |
| T2DM | dendritic_cell | *CTSC* | -2.3601463 | 0.000583026 | 0.001084785 | 1 |
| T2DM | dendritic_cell | *ELP4* | -2.319859424 | 9.33E-05 | 0.000262424 | 0.586805021 |
| T2DM | dendritic_cell | *SC5D* | -2.476475543 | 1.92E-05 | 0.000131132 | 0.121005476 |
| T2DM | dendritic_cell | *C11orf63* | -2.121933118 | 1.84E-05 | 0.000130129 | 0.115680663 |
| T2DM | dendritic_cell | *DTX4* | -3.13552386 | 5.63E-05 | 0.000190114 | 0.353993104 |
| T2DM | dendritic_cell | *DCPS* | -2.647304961 | 0.030988292 | 0.039727446 | 1 |
| T2DM | dendritic_cell | *FOXRED1* | -1.350246531 | 0.008970542 | 0.012543995 | 1 |
| T2DM | dendritic_cell | *NRXN2* | 3.929101941 | 0.000397667 | 0.000787196 | 1 |
| T2DM | dendritic_cell | *MS4A4A* | 1.014930246 | 0.004397999 | 0.00654598 | 1 |
| T2DM | dendritic_cell | *CCDC86* | -3.291490996 | 1.49E-05 | 0.000130129 | 0.093748471 |
| T2DM | dendritic_cell | *PRPF19* | -3.053130247 | 0.004865124 | 0.007170111 | 1 |
| T2DM | dendritic_cell | *HPX* | -1.458079428 | 0.008146005 | 0.011493253 | 1 |
| T2DM | dendritic_cell | *FOLR3* | 0.90381921 | 0.000661156 | 0.001207385 | 1 |
| T2DM | dendritic_cell | *PANX1* | -3.030605211 | 0.001438953 | 0.00239394 | 1 |
| T2DM | dendritic_cell | *EIF4G2* | -2.840687607 | 0.000100452 | 0.000273513 | 0.631542447 |
| T2DM | dendritic_cell | *GALNT18* | -1.058486349 | 3.96E-05 | 0.000159553 | 0.249055425 |

| | | | | | | |
|---|---|---|---|---|---|---|
| T2DM | dendritic_cell | *MADD* | 2.187472765 | 0.003950007 | 0.005938233 | 1 |
| T2DM | dendritic_cell | *NAA40* | -2.105320926 | 3.40E-05 | 0.000150835 | 0.21373269 |
| T2DM | dendritic_cell | *CARS* | -1.533161313 | 0.000754592 | 0.001358179 | 1 |
| T2DM | dendritic_cell | *SLC22A18* | 0.80397183 | 0.01017346 | 0.014088226 | 1 |
| T2DM | dendritic_cell | *C11orf21* | 0.909428597 | 0.023149399 | 0.030238992 | 1 |
| T2DM | dendritic_cell | *ELMOD1* | -3.35515439 | 7.39E-06 | 0.000130129 | 0.046436465 |
| T2DM | dendritic_cell | *RPS13* | -2.027123203 | 0.015327609 | 0.020612765 | 1 |
| T2DM | dendritic_cell | *AIP* | -2.056682002 | 3.71E-05 | 0.000156008 | 0.233243037 |
| T2DM | dendritic_cell | *TCIRG1* | 2.320040173 | 0.034369381 | 0.043847464 | 1 |
| T2DM | dendritic_cell | *GTF2H1* | -2.540555085 | 5.45E-05 | 0.000186495 | 0.342590688 |
| T2DM | dendritic_cell | *PPFIBP1* | -1.319444268 | 4.63E-05 | 0.000170668 | 0.291197156 |
| T2DM | dendritic_cell | *PRDM4* | -2.943053123 | 4.94E-05 | 0.000176169 | 0.310409751 |
| T2DM | dendritic_cell | *ASIC1* | -2.167808165 | 5.27E-06 | 0.00012486 | 0.033152725 |
| T2DM | dendritic_cell | *CAPRIN2* | -2.59633336 | 2.08E-05 | 0.000132272 | 0.13106817 |
| T2DM | dendritic_cell | *KCTD10* | -1.56854481 | 0.000132586 | 0.000334831 | 0.83356955 |
| T2DM | dendritic_cell | *CYP27B1* | -10.44567329 | 3.78E-05 | 0.00015701 | 0.237802482 |
| T2DM | dendritic_cell | *ACSS3* | -0.583166448 | 0.001675947 | 0.002746788 | 1 |
| T2DM | dendritic_cell | *GLI1* | 3.735523162 | 0.000163392 | 0.000391034 | 1 |
| T2DM | dendritic_cell | *METAP2* | -2.880811237 | 0.000230567 | 0.000505431 | 1 |
| T2DM | dendritic_cell | *WNT5B* | 1.24890567 | 6.36E-05 | 0.00020369 | 0.400047156 |
| T2DM | dendritic_cell | *TRPV4* | -3.638517569 | 0.001169491 | 0.001986113 | 1 |
| T2DM | dendritic_cell | *ITFG2* | -2.631188999 | 7.61E-05 | 0.000227931 | 0.478654062 |
| T2DM | dendritic_cell | *PARP11* | -2.342209597 | 0.034365181 | 0.043847464 | 1 |
| T2DM | dendritic_cell | *ARPC3* | 2.126438463 | 0.005859801 | 0.008490567 | 1 |
| T2DM | dendritic_cell | *GPN3* | -3.010581292 | 3.54E-05 | 0.000153735 | 0.222454765 |
| T2DM | dendritic_cell | *VPS29* | -2.466847799 | 0.020264676 | 0.026737465 | 1 |
| T2DM | dendritic_cell | *AKAP3* | -1.643622767 | 0.000132923 | 0.000335484 | 0.835689843 |
| T2DM | dendritic_cell | *KCNA1* | -3.364510782 | 1.86E-05 | 0.000130129 | 0.116768533 |
| T2DM | dendritic_cell | *CREBL2* | 2.508540156 | 0.004718173 | 0.006969726 | 1 |
| T2DM | dendritic_cell | *NAA25* | -2.60441201 | 0.000106007 | 0.0002842 | 0.666467503 |
| T2DM | dendritic_cell | *SCNN1A* | -1.938063488 | 5.18E-05 | 0.000180889 | 0.325419808 |
| T2DM | dendritic_cell | *CDK2AP1* | 1.743781933 | 5.33E-08 | 1.06E-05 | 0.00033521 |
| T2DM | dendritic_cell | *MGP* | -0.983644251 | 0.001294073 | 0.002176521 | 1 |
| T2DM | dendritic_cell | *RASAL1* | 1.160747099 | 5.39E-05 | 0.000185203 | 0.339065465 |
| T2DM | dendritic_cell | *ARHGDIB* | 1.344145621 | 0.000279402 | 0.000589069 | 1 |
| T2DM | dendritic_cell | *EIF2B1* | -2.500559372 | 2.16E-05 | 0.000132663 | 0.135756299 |
| T2DM | dendritic_cell | *DDX55* | -2.097961015 | 4.17E-05 | 0.000162756 | 0.2622353 |
| T2DM | dendritic_cell | *ENDOU* | -3.037334506 | 1.07E-07 | 1.57E-05 | 0.000672842 |
| T2DM | dendritic_cell | *STX2* | -1.371586216 | 0.032142474 | 0.041131637 | 1 |
| T2DM | dendritic_cell | *CAND1* | -2.47661915 | 7.03E-05 | 0.000216443 | 0.441976621 |
| T2DM | dendritic_cell | *MDM1* | -1.800851063 | 0.016507501 | 0.022086116 | 1 |
| T2DM | dendritic_cell | *NUP107* | -2.366332947 | 0.000245126 | 0.000530137 | 1 |
| T2DM | dendritic_cell | *CNOT2* | -2.708145036 | 0.00011865 | 0.000308883 | 0.745951576 |
| T2DM | dendritic_cell | *KRR1* | -3.252825311 | 0.003285991 | 0.005030198 | 1 |
| T2DM | dendritic_cell | *COPS7A* | -3.289614147 | 0.000337746 | 0.000685765 | 1 |
| T2DM | dendritic_cell | *CDCA3* | -1.832851281 | 0.000878907 | 0.001547811 | 1 |
| T2DM | dendritic_cell | *CHPT1* | -1.03861259 | 0.000428066 | 0.000837093 | 1 |
| T2DM | dendritic_cell | *GNPTAB* | -2.504936297 | 0.002253551 | 0.003596726 | 1 |
| T2DM | dendritic_cell | *SPSB2* | 2.354296763 | 0.000355417 | 0.000717337 | 1 |
| T2DM | dendritic_cell | *ATN1* | -2.054824276 | 5.47E-05 | 0.000187017 | 0.344111176 |

| | | | | | | |
|---|---|---|---|---|---|---|
| T2DM | dendritic_cell | *NT5DC3* | -1.56410334 | 0.023229239 | 0.03033068 | 1 |
| T2DM | dendritic_cell | *PRKAB1* | -1.795493365 | 0.000879282 | 0.001548039 | 1 |
| T2DM | dendritic_cell | *ST8SIA1* | -2.662280656 | 4.44E-05 | 0.000167569 | 0.279169928 |
| T2DM | dendritic_cell | *C2CD5* | -2.658505767 | 0.000200945 | 0.000455259 | 1 |
| T2DM | dendritic_cell | *PHC1* | -3.597812881 | 0.000103556 | 0.000280508 | 0.651058221 |
| T2DM | dendritic_cell | *COL12A1* | -0.99011898 | 1.31E-05 | 0.000130129 | 0.082055056 |
| T2DM | dendritic_cell | *TDP2* | -2.759182315 | 2.51E-05 | 0.000138674 | 0.158088101 |
| T2DM | dendritic_cell | *RWDD1* | -2.326146482 | 8.52E-05 | 0.000247534 | 0.535663068 |
| T2DM | dendritic_cell | *SMIM8* | -2.358939815 | 0.000232508 | 0.000508445 | 1 |
| T2DM | dendritic_cell | *MCM9* | -2.793108749 | 0.000432517 | 0.000843283 | 1 |
| T2DM | dendritic_cell | *FAM184A* | -1.478091905 | 0.000410559 | 0.000807377 | 1 |
| T2DM | dendritic_cell | *RNGTT* | -2.616456241 | 0.004374428 | 0.006517069 | 1 |
| T2DM | dendritic_cell | *GABRR2* | -1.614525909 | 3.63E-05 | 0.000155082 | 0.227965408 |
| T2DM | dendritic_cell | *HDDC2* | -1.955694197 | 9.04E-05 | 0.000257005 | 0.568494281 |
| T2DM | dendritic_cell | *TPD52L1* | -1.130035355 | 0.000141201 | 0.000350328 | 0.887730603 |
| T2DM | dendritic_cell | *UST* | -1.626048336 | 6.86E-05 | 0.00021325 | 0.43119157 |
| T2DM | dendritic_cell | *TULP1* | 5.299963828 | 4.25E-05 | 0.000164104 | 0.267008546 |
| T2DM | dendritic_cell | *MAPK14* | -2.967960481 | 6.19E-06 | 0.000130129 | 0.03891543 |
| T2DM | dendritic_cell | *MRPL18* | -2.076357009 | 0.004085189 | 0.006128271 | 1 |
| T2DM | dendritic_cell | *RNF8* | -2.520906022 | 0.00028065 | 0.000591104 | 1 |
| T2DM | dendritic_cell | *PHACTR1* | -2.504876259 | 0.00129032 | 0.002171952 | 1 |
| T2DM | dendritic_cell | *CD83* | 0.33414034 | 0.000107011 | 0.000285924 | 0.672778477 |
| T2DM | dendritic_cell | *MDN1* | -2.143541403 | 0.000136661 | 0.000341761 | 0.859188125 |
| T2DM | dendritic_cell | *CAP2* | -1.908693767 | 5.45E-06 | 0.000125532 | 0.034270102 |
| T2DM | dendritic_cell | *GPR63* | -1.518701353 | 0.00083129 | 0.001477614 | 1 |
| T2DM | dendritic_cell | *ASCC3* | -2.333895394 | 9.29E-05 | 0.000261514 | 0.583959816 |
| T2DM | dendritic_cell | *BVES* | -1.887007069 | 0.000267884 | 0.000569367 | 1 |
| T2DM | dendritic_cell | *WASF1* | -0.86829423 | 9.53E-05 | 0.000265129 | 0.599192196 |
| T2DM | dendritic_cell | *ALDH5A1* | -1.301883182 | 0.000617362 | 0.001138895 | 1 |
| T2DM | dendritic_cell | *VNN1* | 0.940406486 | 0.008390799 | 0.011806838 | 1 |
| T2DM | dendritic_cell | *VNN2* | 3.580376418 | 0.000219577 | 0.000486085 | 1 |
| T2DM | dendritic_cell | *ACOT13* | -3.251286778 | 8.43E-05 | 0.000245628 | 0.529818907 |
| T2DM | dendritic_cell | *C6orf62* | -2.046772261 | 0.002280747 | 0.003635664 | 1 |
| T2DM | dendritic_cell | *B3GAT2* | -2.628789206 | 1.92E-05 | 0.000130865 | 0.120396124 |
| T2DM | dendritic_cell | *HBS1L* | -1.818098441 | 0.000106423 | 0.00028472 | 0.669083016 |
| T2DM | dendritic_cell | *PERP* | -1.254848783 | 0.000169136 | 0.000401721 | 1 |
| T2DM | dendritic_cell | *SLC16A10* | 1.058940859 | 3.19E-05 | 0.000148219 | 0.200709321 |
| T2DM | dendritic_cell | *PACRG* | -2.348315801 | 8.03E-06 | 0.000130129 | 0.050497184 |
| T2DM | dendritic_cell | *TFEB* | 2.584468469 | 0.008355217 | 0.01176467 | 1 |
| T2DM | dendritic_cell | *FAM120B* | -1.45449116 | 0.003147048 | 0.004844636 | 1 |
| T2DM | dendritic_cell | *GUCA1B* | -2.476206083 | 3.11E-05 | 0.000146698 | 0.195611294 |
| T2DM | dendritic_cell | *PRPH2* | -2.614734151 | 3.20E-05 | 0.000148254 | 0.201441753 |
| T2DM | dendritic_cell | *DUSP22* | -3.143750061 | 0.005661567 | 0.008237509 | 1 |
| T2DM | dendritic_cell | *EXOC2* | -2.693591028 | 6.07E-05 | 0.000198694 | 0.381749064 |
| T2DM | dendritic_cell | *TMEM30A* | -2.207992686 | 6.09E-05 | 0.000198694 | 0.38275682 |
| T2DM | dendritic_cell | *SENP6* | -2.194797915 | 9.85E-05 | 0.000270827 | 0.61938224 |
| T2DM | dendritic_cell | *TTK* | -3.005807789 | 7.23E-05 | 0.000219971 | 0.45436527 |
| T2DM | dendritic_cell | *LAMA4* | -0.941909238 | 2.27E-05 | 0.000133654 | 0.14296667 |
| T2DM | dendritic_cell | *CLIC5* | -0.905759755 | 4.27E-05 | 0.000164485 | 0.268305845 |
| T2DM | dendritic_cell | *LY86* | 0.635443844 | 4.47E-05 | 0.000168467 | 0.281338222 |

| | | | | | | |
|---|---|---|---|---|---|---|
| T2DM | dendritic_cell | *TBX18* | -1.151117294 | 3.86E-05 | 0.000158005 | 0.242813764 |
| T2DM | dendritic_cell | *NUDT12* | -2.506214096 | 6.17E-06 | 0.000130129 | 0.038795913 |
| T2DM | dendritic_cell | *CEP72* | 1.553847591 | 0.020053403 | 0.026492066 | 1 |
| T2DM | dendritic_cell | *C7* | -1.135732362 | 1.08E-05 | 0.000130129 | 0.068175018 |
| T2DM | dendritic_cell | *PAPD7* | -2.009193767 | 4.17E-05 | 0.000162756 | 0.262206976 |
| T2DM | dendritic_cell | *GHR* | -0.574377158 | 0.003378205 | 0.005156294 | 1 |
| T2DM | dendritic_cell | *NME5* | -1.285531631 | 0.000906216 | 0.001590114 | 1 |
| T2DM | dendritic_cell | *NNT* | -1.56744237 | 3.00E-05 | 0.000145028 | 0.188390959 |
| T2DM | dendritic_cell | *PFDN1* | -1.770427077 | 0.001537974 | 0.002543199 | 1 |
| T2DM | dendritic_cell | *LOX* | -1.376685086 | 1.46E-05 | 0.000130129 | 0.092096342 |
| T2DM | dendritic_cell | *TMCO6* | -1.484541715 | 7.98E-06 | 0.000130129 | 0.050185504 |
| T2DM | dendritic_cell | *SPARC* | -1.30492992 | 2.14E-05 | 0.000132591 | 0.134302718 |
| T2DM | dendritic_cell | *IK* | -2.72477187 | 8.53E-05 | 0.000247826 | 0.536542756 |
| T2DM | dendritic_cell | *PDE8B* | -0.99931902 | 4.05E-05 | 0.000160731 | 0.254315471 |
| T2DM | dendritic_cell | *CLK4* | -2.132021056 | 1.85E-05 | 0.000130129 | 0.116466164 |
| T2DM | dendritic_cell | *PCDHB15* | -1.063090987 | 0.000869599 | 0.00153547 | 1 |
| T2DM | dendritic_cell | *HAVCR1* | 0.807281514 | 5.76E-05 | 0.000192599 | 0.362368546 |
| T2DM | dendritic_cell | *RNF130* | -2.424015793 | 0.002374106 | 0.00376621 | 1 |
| T2DM | dendritic_cell | *ARSB* | 1.230472281 | 0.000337072 | 0.000685287 | 1 |
| T2DM | dendritic_cell | *CLINT1* | -2.155579304 | 0.000395188 | 0.000783275 | 1 |
| T2DM | dendritic_cell | *TTC1* | -2.978794405 | 0.001137334 | 0.001938835 | 1 |
| T2DM | dendritic_cell | *MSH3* | -1.776166893 | 6.89E-06 | 0.000130129 | 0.043311007 |
| T2DM | dendritic_cell | *RASGRF2* | -1.089300134 | 1.43E-05 | 0.000130129 | 0.08973391 |
| T2DM | dendritic_cell | *POLR3G* | -2.46722072 | 2.33E-05 | 0.000135352 | 0.146721152 |
| T2DM | dendritic_cell | *DROSHA* | -1.876520565 | 0.002783411 | 0.00433473 | 1 |
| T2DM | dendritic_cell | *CDH6* | -0.593100762 | 0.01747904 | 0.023286867 | 1 |
| T2DM | dendritic_cell | *TARS* | -3.33468619 | 0.009222988 | 0.012871237 | 1 |
| T2DM | dendritic_cell | *LNPEP* | -1.724884308 | 0.003803082 | 0.005744829 | 1 |
| T2DM | dendritic_cell | *SLC12A7* | 1.864631614 | 0.020182966 | 0.026640838 | 1 |
| T2DM | dendritic_cell | *GNPDA1* | -3.84560279 | 0.000197228 | 0.000447967 | 1 |
| T2DM | dendritic_cell | *SKP1* | -2.702001632 | 7.24E-05 | 0.000220017 | 0.455088465 |
| T2DM | dendritic_cell | *PPWD1* | -1.451607378 | 0.00564049 | 0.008212543 | 1 |
| T2DM | dendritic_cell | *TRIM23* | -2.568958599 | 2.39E-05 | 0.000136538 | 0.150099988 |
| T2DM | dendritic_cell | *TRAPPC13* | -1.847860574 | 0.000266964 | 0.000567604 | 1 |
| T2DM | dendritic_cell | *TXNDC15* | -3.061508518 | 2.13E-05 | 0.000132591 | 0.133745235 |
| T2DM | dendritic_cell | *SMAD5* | -2.232615479 | 9.92E-05 | 0.000271818 | 0.623452194 |
| T2DM | dendritic_cell | *HMGXB3* | -2.340121327 | 4.17E-05 | 0.000162756 | 0.262091842 |
| T2DM | dendritic_cell | *ATP6V0E1* | -3.213106259 | 0.000184955 | 0.000427598 | 1 |
| T2DM | dendritic_cell | *CNTN3* | -1.075950268 | 1.50E-05 | 0.000130129 | 0.094479774 |
| T2DM | dendritic_cell | *ACTR8* | -5.098055027 | 0.000333803 | 0.000679605 | 1 |
| T2DM | dendritic_cell | *HGD* | -7.680753502 | 2.40E-05 | 0.000136538 | 0.150601572 |
| T2DM | dendritic_cell | *NIT2* | -3.047864995 | 5.29E-05 | 0.00018288 | 0.332652997 |
| T2DM | dendritic_cell | *FAM162A* | -3.870343121 | 1.99E-05 | 0.000131266 | 0.1251956 |
| T2DM | dendritic_cell | *PCCB* | -2.47543742 | 3.97E-05 | 0.000159657 | 0.249652376 |
| T2DM | dendritic_cell | *RBP1* | -2.674871636 | 1.98E-05 | 0.000131266 | 0.124281223 |
| T2DM | dendritic_cell | *BCHE* | -1.385882235 | 3.60E-05 | 0.000154842 | 0.226378662 |
| T2DM | dendritic_cell | *PDCD10* | -2.60722126 | 2.72E-05 | 0.000141161 | 0.170966844 |
| T2DM | dendritic_cell | *WNT5A* | -3.446929157 | 0.014499463 | 0.019553438 | 1 |
| T2DM | dendritic_cell | *PRKAR2A* | -1.885291381 | 9.28E-06 | 0.000130129 | 0.058351106 |
| T2DM | dendritic_cell | *C3orf14* | -1.454539577 | 6.59E-05 | 0.000208427 | 0.414352861 |

| | | | | | | |
|---|---|---|---|---|---|---|
| T2DM | dendritic_cell | *IFT57* | -2.936754529 | 0.037194136 | 0.047211696 | 1 |
| T2DM | dendritic_cell | *IQCG* | -3.262380942 | 0.000125004 | 0.000320514 | 0.785899919 |
| T2DM | dendritic_cell | *GBE1* | -1.358401451 | 1.77E-05 | 0.000130129 | 0.110999198 |
| T2DM | dendritic_cell | *UMPS* | -2.773045461 | 7.82E-07 | 5.27E-05 | 0.00491526 |
| T2DM | dendritic_cell | *SLC41A3* | -3.221103585 | 9.65E-05 | 0.000267366 | 0.606653051 |
| T2DM | dendritic_cell | *PLXNA1* | -1.513083554 | 0.012930103 | 0.017576553 | 1 |
| T2DM | dendritic_cell | *ABTB1* | -2.975983523 | 8.90E-05 | 0.000254619 | 0.559652775 |
| T2DM | dendritic_cell | *EFCC1* | -1.735862969 | 7.09E-07 | 5.24E-05 | 0.004459284 |
| T2DM | dendritic_cell | *NEK11* | -3.085321215 | 0.000765369 | 0.001376394 | 1 |
| T2DM | dendritic_cell | *PLSCR4* | -1.423312017 | 4.49E-05 | 0.000168807 | 0.282274387 |
| T2DM | dendritic_cell | *WDR48* | -2.144416618 | 0.002337666 | 0.003715535 | 1 |
| T2DM | dendritic_cell | *COMMD2* | -2.200413856 | 2.00E-05 | 0.000131266 | 0.125667968 |
| T2DM | dendritic_cell | *GORASP1* | 2.846993376 | 0.002407744 | 0.003813929 | 1 |
| T2DM | dendritic_cell | *ABCC5* | -2.982062095 | 1.60E-05 | 0.000130129 | 0.100758493 |
| T2DM | dendritic_cell | *ARHGEF26* | -1.298641994 | 3.85E-05 | 0.000158005 | 0.242028684 |
| T2DM | dendritic_cell | *KLHL24* | -1.700312485 | 0.000488209 | 0.000931241 | 1 |
| T2DM | dendritic_cell | *PLCH1* | -2.881794342 | 0.001102755 | 0.001887564 | 1 |
| T2DM | dendritic_cell | *ZBTB47* | -1.882549821 | 0.001702314 | 0.002784196 | 1 |
| T2DM | dendritic_cell | *NKTR* | -1.864091553 | 0.001290056 | 0.002171952 | 1 |
| T2DM | dendritic_cell | *CLCN2* | -2.292870271 | 3.25E-05 | 0.000148254 | 0.204316232 |
| T2DM | dendritic_cell | *NEK4* | -3.338658295 | 6.83E-05 | 0.000212807 | 0.429230795 |
| T2DM | dendritic_cell | *LMAN2L* | -1.598386256 | 0.000932271 | 0.001627656 | 1 |
| T2DM | dendritic_cell | *PIKFYVE* | -2.611841219 | 0.000172092 | 0.000406746 | 1 |
| T2DM | dendritic_cell | *KCNIP3* | -1.790673886 | 6.64E-05 | 0.000209348 | 0.417351448 |
| T2DM | dendritic_cell | *FAHD2A* | 2.315808433 | 0.033381767 | 0.042639408 | 1 |
| T2DM | dendritic_cell | *NCL* | -2.859340553 | 1.29E-05 | 0.000130129 | 0.081263739 |
| T2DM | dendritic_cell | *ACTR1B* | -3.211175629 | 0.007547406 | 0.010718442 | 1 |
| T2DM | dendritic_cell | *SLC35F5* | -1.888702238 | 0.001065744 | 0.001830192 | 1 |
| T2DM | dendritic_cell | *ACTR3* | 1.670768495 | 0.014843164 | 0.019991211 | 1 |
| T2DM | dendritic_cell | *TFCP2L1* | -2.154746343 | 0.000593711 | 0.001103031 | 1 |
| T2DM | dendritic_cell | *TP53I3* | -2.888683401 | 0.000398742 | 0.000789076 | 1 |
| T2DM | dendritic_cell | *DNAJC27* | -4.612894077 | 7.74E-07 | 5.27E-05 | 0.004863863 |
| T2DM | dendritic_cell | *STAM2* | -2.658772908 | 5.82E-05 | 0.000193956 | 0.365801201 |
| T2DM | dendritic_cell | *GPD2* | -2.542689381 | 4.20E-05 | 0.000163316 | 0.264081713 |
| T2DM | dendritic_cell | *CENPA* | 2.613017496 | 0.037468325 | 0.047511771 | 1 |
| T2DM | dendritic_cell | *TANC1* | -1.632550132 | 6.50E-06 | 0.000130129 | 0.04088667 |
| T2DM | dendritic_cell | *NRBP1* | -3.40908557 | 7.45E-09 | 2.70E-06 | 4.68E-05 |
| T2DM | dendritic_cell | *ITGB6* | -2.270903578 | 2.05E-06 | 8.62E-05 | 0.012912259 |
| T2DM | dendritic_cell | *ITGA4* | 0.965043565 | 8.96E-05 | 0.000255437 | 0.563494458 |
| T2DM | dendritic_cell | *PPM1G* | -2.917723575 | 1.07E-07 | 1.57E-05 | 0.000673463 |
| T2DM | dendritic_cell | *PCSK4* | -1.822026769 | 0.000266526 | 0.000567056 | 1 |
| T2DM | dendritic_cell | *INO80B* | -2.058135528 | 0.001043724 | 0.001796302 | 1 |
| T2DM | dendritic_cell | *MOGS* | -1.736043452 | 3.62E-05 | 0.00015508 | 0.227400497 |
| T2DM | dendritic_cell | *GRB14* | -2.147983165 | 1.75E-05 | 0.000130129 | 0.11012589 |
| T2DM | dendritic_cell | *CLIP4* | -1.794208068 | 1.58E-05 | 0.000130129 | 0.099579159 |
| T2DM | dendritic_cell | *SPTBN1* | -1.341996717 | 1.05E-05 | 0.000130129 | 0.066007711 |
| T2DM | dendritic_cell | *RTN4* | -2.292098224 | 2.57E-06 | 9.16E-05 | 0.016130242 |
| T2DM | dendritic_cell | *POLE4* | -2.620728453 | 2.01E-05 | 0.000131405 | 0.126411233 |
| T2DM | dendritic_cell | *MRPL19* | -2.188912569 | 9.07E-06 | 0.000130129 | 0.056992473 |
| T2DM | dendritic_cell | *LANCL1* | -2.820341547 | 2.94E-05 | 0.000143842 | 0.184634521 |

| | | | | | | |
|---|---|---|---|---|---|---|
| T2DM | dendritic_cell | *WDR75* | -3.096503147 | 5.94E-05 | 0.000196476 | 0.373696958 |
| T2DM | dendritic_cell | *EFEMP1* | -1.086542557 | 1.38E-05 | 0.000130129 | 0.086808578 |
| T2DM | dendritic_cell | *GLS* | -2.319634063 | 0.00030007 | 0.000624683 | 1 |
| T2DM | dendritic_cell | *GGCX* | -1.337714826 | 0.035048883 | 0.044662149 | 1 |
| T2DM | dendritic_cell | *GNLY* | -1.741396956 | 1.35E-05 | 0.000130129 | 0.085108646 |
| T2DM | dendritic_cell | *SF3B1* | -2.193016277 | 8.55E-05 | 0.000248016 | 0.537402399 |
| T2DM | dendritic_cell | *ST3GAL5* | -2.625226979 | 1.49E-05 | 0.000130129 | 0.093505182 |
| T2DM | dendritic_cell | *PLCD4* | -5.573797221 | 0.000101977 | 0.000276468 | 0.641128973 |
| T2DM | dendritic_cell | *CHMP3* | -4.665259148 | 1.03E-08 | 3.07E-06 | 6.45E-05 |
| T2DM | dendritic_cell | *HDLBP* | -1.957218031 | 0.000223276 | 0.000492021 | 1 |
| T2DM | dendritic_cell | *PASK* | -3.638444364 | 0.001926486 | 0.003116783 | 1 |
| T2DM | dendritic_cell | *STK25* | -3.690480834 | 0.000562118 | 0.001050545 | 1 |
| T2DM | dendritic_cell | *TPO* | -0.758817892 | 4.59E-05 | 0.000170199 | 0.288511731 |
| T2DM | dendritic_cell | *TAF1B* | -2.053607498 | 1.21E-05 | 0.000130129 | 0.076143866 |
| T2DM | dendritic_cell | *STRN* | -2.255244518 | 3.18E-05 | 0.000148219 | 0.200021596 |
| T2DM | dendritic_cell | *CEBPZ* | -2.233483491 | 0.014923462 | 0.02009075 | 1 |
| T2DM | dendritic_cell | *PRKD3* | -2.535456732 | 0.000380933 | 0.000759812 | 1 |
| T2DM | dendritic_cell | *SLC25A12* | -2.465451501 | 1.22E-05 | 0.000130129 | 0.076647224 |
| T2DM | dendritic_cell | *RMDN2* | -3.095492701 | 7.13E-05 | 0.000218327 | 0.448443015 |
| T2DM | dendritic_cell | *SDC1* | 3.48516549 | 1.25E-06 | 6.80E-05 | 0.007847386 |
| T2DM | dendritic_cell | *SOS1* | -1.607567827 | 2.67E-05 | 0.000140322 | 0.168105465 |
| T2DM | dendritic_cell | *COX7A2L* | -2.881879563 | 2.60E-05 | 0.0001395 | 0.163443348 |
| T2DM | dendritic_cell | *PNO1* | 2.831344172 | 0.003515763 | 0.005345491 | 1 |
| T2DM | dendritic_cell | *ORC4* | -1.561628671 | 2.20E-05 | 0.000132957 | 0.138231166 |
| T2DM | dendritic_cell | *THADA* | -2.170132708 | 5.38E-05 | 0.000185023 | 0.338406724 |
| T2DM | dendritic_cell | *ARID3A* | -2.409932824 | 1.89E-05 | 0.000130129 | 0.118985701 |
| T2DM | dendritic_cell | *CD207* | -23.13576349 | 0.000188945 | 0.000434015 | 1 |
| T2DM | dendritic_cell | *GRIN3B* | 1.436790793 | 6.24E-06 | 0.000130129 | 0.039228426 |
| T2DM | dendritic_cell | *VAX2* | -1.871800677 | 1.79E-05 | 0.000130129 | 0.112808741 |
| T2DM | dendritic_cell | *MSH6* | -3.62010005 | 0.001546022 | 0.002553821 | 1 |
| T2DM | dendritic_cell | *PLEKHA3* | -3.154048851 | 5.46E-05 | 0.000186642 | 0.343233723 |
| T2DM | dendritic_cell | *ALMS1* | -2.280576442 | 5.65E-05 | 0.000190479 | 0.355243399 |
| T2DM | dendritic_cell | *BCL9* | -1.43434722 | 8.86E-05 | 0.000253766 | 0.55701542 |
| T2DM | dendritic_cell | *DHCR24* | -2.810748044 | 3.57E-05 | 0.000154392 | 0.224699127 |
| T2DM | dendritic_cell | *DNAJC16* | -2.640077431 | 0.000430569 | 0.000841568 | 1 |
| T2DM | dendritic_cell | *MORN1* | -1.958129049 | 0.002444963 | 0.003866068 | 1 |
| T2DM | dendritic_cell | *GPX7* | -1.47560658 | 0.00022579 | 0.000496344 | 1 |
| T2DM | dendritic_cell | *SCP2* | -1.918493314 | 8.35E-05 | 0.000243975 | 0.525033318 |
| T2DM | dendritic_cell | *RALGPS2* | -1.561681334 | 2.99E-05 | 0.000145022 | 0.188025067 |
| T2DM | dendritic_cell | *CEP104* | -2.023621288 | 0.001782585 | 0.002905655 | 1 |
| T2DM | dendritic_cell | *FAM20B* | -3.487034881 | 0.000608665 | 0.001126156 | 1 |
| T2DM | dendritic_cell | *TMEM59* | -2.238853142 | 5.59E-05 | 0.000189353 | 0.351439082 |
| T2DM | dendritic_cell | *WRAP73* | -4.3294061 | 0.011585214 | 0.015871919 | 1 |
| T2DM | dendritic_cell | *ICMT* | -3.074598598 | 6.14E-06 | 0.000130129 | 0.038609262 |
| T2DM | dendritic_cell | *PARK7* | -2.487656475 | 0.027346152 | 0.035353744 | 1 |
| T2DM | dendritic_cell | *KCNC4* | -1.481232768 | 5.41E-06 | 0.000125356 | 0.033993974 |
| T2DM | dendritic_cell | *EDEM3* | -2.525857129 | 2.60E-05 | 0.0001395 | 0.16342904 |
| T2DM | dendritic_cell | *CAPZA1* | -2.951896182 | 0.000311376 | 0.000643955 | 1 |
| T2DM | dendritic_cell | *DOCK7* | -2.240569398 | 3.72E-05 | 0.000156008 | 0.233700212 |
| T2DM | dendritic_cell | *FBXO6* | 2.217547037 | 0.008544527 | 0.012009712 | 1 |

| | | | | | | |
|---|---|---|---|---|---|---|
| T2DM | dendritic_cell | *C1orf21* | -1.619353691 | 8.56E-06 | 0.000130129 | 0.053808891 |
| T2DM | dendritic_cell | *DNAJC6* | 1.961611854 | 0.002140283 | 0.003432642 | 1 |
| T2DM | dendritic_cell | *LEPR* | -1.903618288 | 7.52E-05 | 0.000225984 | 0.472695088 |
| T2DM | dendritic_cell | *MFN2* | 1.925337765 | 0.000373111 | 0.000745629 | 1 |
| T2DM | dendritic_cell | *MIIP* | -2.294555163 | 6.93E-06 | 0.000130129 | 0.043592446 |
| T2DM | dendritic_cell | *SMG7* | -2.92063751 | 0.00040781 | 0.000803731 | 1 |
| T2DM | dendritic_cell | *NCF2* | 1.293440888 | 0.000126558 | 0.000322918 | 0.79567011 |
| T2DM | dendritic_cell | *RGS2* | 0.465695003 | 3.66E-05 | 0.000155321 | 0.230340717 |
| T2DM | dendritic_cell | *OLFML3* | -1.095516972 | 1.65E-05 | 0.000130129 | 0.103820431 |
| T2DM | dendritic_cell | *TNNI3K* | -1.798471665 | 0.012659782 | 0.017235177 | 1 |
| T2DM | dendritic_cell | *CFHR3* | -1.255771282 | 0.015010771 | 0.02019964 | 1 |
| T2DM | dendritic_cell | *CRYZ* | -1.52665408 | 3.68E-05 | 0.000155415 | 0.23125792 |
| T2DM | dendritic_cell | *TMEM9* | -1.711610413 | 3.95E-05 | 0.000159553 | 0.248040202 |
| T2DM | dendritic_cell | *WARS2* | -1.898326965 | 0.000132525 | 0.000334831 | 0.833182093 |
| T2DM | dendritic_cell | *OSCP1* | -2.090135184 | 0.000103927 | 0.000280899 | 0.653391434 |
| T2DM | dendritic_cell | *GNPAT* | -3.085118083 | 0.000284168 | 0.000596914 | 1 |
| T2DM | dendritic_cell | *C1orf109* | -1.999222203 | 4.62E-06 | 0.000119532 | 0.029046261 |
| T2DM | dendritic_cell | *TBCE* | -3.099925161 | 3.21E-06 | 0.000102388 | 0.020170372 |
| T2DM | dendritic_cell | *LGALS8* | -1.623533078 | 2.78E-05 | 0.000142506 | 0.174712826 |
| T2DM | dendritic_cell | *MTR* | -2.14360752 | 5.61E-05 | 0.000189964 | 0.352786559 |
| T2DM | dendritic_cell | *KCNQ4* | -1.538556716 | 0.007350893 | 0.010462999 | 1 |
| T2DM | dendritic_cell | *ACADM* | -2.35691456 | 0.001192578 | 0.002020954 | 1 |
| T2DM | dendritic_cell | *PADI2* | 0.844526421 | 4.47E-05 | 0.000168467 | 0.281340033 |
| T2DM | dendritic_cell | *KLHL12* | -2.970782583 | 0.006114322 | 0.008820731 | 1 |
| T2DM | dendritic_cell | *ZNHIT6* | -2.235583633 | 3.40E-05 | 0.000150765 | 0.213483003 |
| T2DM | dendritic_cell | *PLA2G2D* | 4.003330553 | 0.000750572 | 0.001352493 | 1 |
| T2DM | dendritic_cell | *RBBP5* | -1.47627353 | 0.005744986 | 0.008351151 | 1 |
| T2DM | dendritic_cell | *KIF17* | -2.973840925 | 0.000992772 | 0.001720913 | 1 |
| T2DM | dendritic_cell | *CD46* | -1.840803845 | 5.64E-05 | 0.00019044 | 0.354871642 |
| T2DM | dendritic_cell | *NSUN4* | -3.334501203 | 0.001695526 | 0.002773815 | 1 |
| T2DM | dendritic_cell | *TMED5* | -2.2521702 | 2.49E-05 | 0.000138038 | 0.156370933 |
| T2DM | dendritic_cell | *CNN3* | -2.448610511 | 2.71E-05 | 0.000141017 | 0.170257289 |
| T2DM | dendritic_cell | *PRRC2C* | -2.189946602 | 0.002251521 | 0.003594544 | 1 |
| T2DM | dendritic_cell | *ABCD3* | -2.132391154 | 2.32E-05 | 0.000134676 | 0.145577221 |
| T2DM | dendritic_cell | *PTBP2* | -1.273963256 | 0.002111427 | 0.003388091 | 1 |
| T2DM | dendritic_cell | *TNFSF4* | -2.026893557 | 4.43E-07 | 3.61E-05 | 0.002782745 |
| T2DM | dendritic_cell | *DARS2* | -3.892384181 | 7.76E-06 | 0.000130129 | 0.048784089 |
| T2DM | dendritic_cell | *IRF6* | 0.97260404 | 0.021132315 | 0.027771502 | 1 |
| T2DM | dendritic_cell | *SERPINC1* | 1.847129852 | 0.001182151 | 0.002005988 | 1 |
| T2DM | dendritic_cell | *RCAN3* | 1.7382438 | 0.00051775 | 0.000978387 | 1 |
| T2DM | dendritic_cell | *RCOR3* | -2.29866903 | 4.53E-05 | 0.000169094 | 0.28475398 |
| T2DM | dendritic_cell | *MTFR1L* | -2.933897287 | 0.00468543 | 0.006924612 | 1 |
| T2DM | dendritic_cell | *NSL1* | -2.679048088 | 0.000210132 | 0.00047032 | 1 |
| T2DM | dendritic_cell | *RPA2* | -3.740154452 | 0.00048582 | 0.000926965 | 1 |
| T2DM | dendritic_cell | *SLC5A9* | -4.826154773 | 0.000501781 | 0.000952793 | 1 |
| T2DM | dendritic_cell | *OSBPL9* | -2.093011819 | 2.94E-05 | 0.000143842 | 0.184980505 |
| T2DM | dendritic_cell | *TXNDC12* | -2.957606787 | 9.40E-05 | 0.000263609 | 0.591011219 |
| T2DM | dendritic_cell | *ESYT2* | -2.273080872 | 0.000131599 | 0.000333078 | 0.827364652 |
| T2DM | dendritic_cell | *COLEC11* | -2.514865458 | 0.001225838 | 0.002072847 | 1 |
| T2DM | dendritic_cell | *KMT2A* | -2.089451551 | 3.26E-05 | 0.000148254 | 0.205035781 |

| | | | | | | |
|---|---|---|---|---|---|---|
| T2DM | dendritic_cell | *IFT46* | -2.746679506 | 2.12E-05 | 0.000132591 | 0.133566575 |
| T2DM | dendritic_cell | *DDX59* | -2.796374109 | 7.80E-05 | 0.000232445 | 0.490690474 |
| T2DM | dendritic_cell | *CAMSAP2* | -2.353548307 | 0.000450589 | 0.000871111 | 1 |
| T2DM | dendritic_cell | *ATF6* | -2.334571662 | 0.007650427 | 0.010857389 | 1 |
| T2DM | dendritic_cell | *LRMP* | 1.361395069 | 0.003192699 | 0.004906502 | 1 |
| T2DM | dendritic_cell | *ATP10B* | -6.534993973 | 0.000202852 | 0.000457763 | 1 |
| T2DM | dendritic_cell | *SPCS2* | -2.737801186 | 3.78E-05 | 0.00015701 | 0.23786948 |
| T2DM | dendritic_cell | *FILIP1* | -1.737521965 | 4.70E-05 | 0.000171433 | 0.295219596 |
| T2DM | dendritic_cell | *SGIP1* | -1.934796918 | 3.22E-05 | 0.000148254 | 0.202556614 |
| T2DM | dendritic_cell | *AKAP7* | -1.515224239 | 1.29E-05 | 0.000130129 | 0.080872256 |
| T2DM | dendritic_cell | *TCF21* | 8.237706565 | 0.00026921 | 0.000571605 | 1 |
| T2DM | dendritic_cell | *SLC16A7* | -1.272135193 | 1.54E-05 | 0.000130129 | 0.096965753 |
| T2DM | dendritic_cell | *VAMP8* | 1.662400041 | 0.013007336 | 0.017670078 | 1 |
| T2DM | dendritic_cell | *FOXO3* | -2.373774802 | 0.001308218 | 0.00219796 | 1 |
| T2DM | dendritic_cell | *TGIF2* | -3.71370901 | 0.011455054 | 0.015714656 | 1 |
| T2DM | dendritic_cell | *ABCG2* | -1.743514461 | 1.33E-05 | 0.000130129 | 0.083643248 |
| T2DM | dendritic_cell | *SPP1* | -3.314218759 | 7.46E-05 | 0.000224758 | 0.468846203 |
| T2DM | dendritic_cell | *FAM47E-STBL* | -1.106395913 | 9.35E-05 | 0.00026248 | 0.587955184 |
| T2DM | dendritic_cell | *MFSD1* | 1.789235348 | 0.001619104 | 0.002661953 | 1 |
| T2DM | dendritic_cell | *RAB3GAP2* | -2.41174259 | 8.77E-05 | 0.000252107 | 0.551132099 |
| T2DM | dendritic_cell | *UBN1* | -2.461650239 | 0.00047479 | 0.000910062 | 1 |
| T2DM | dendritic_cell | *UCHL3* | 3.668565658 | 0.001166902 | 0.001982787 | 1 |
| T2DM | dendritic_cell | *WDR35* | -1.715657149 | 1.05E-05 | 0.000130129 | 0.065925601 |
| T2DM | dendritic_cell | *DNAH7* | -1.758394469 | 9.78E-10 | 6.15E-07 | 6.15E-06 |
| T2DM | dendritic_cell | *NDUFB3* | -2.045052978 | 4.54E-05 | 0.000169108 | 0.285116416 |
| T2DM | dendritic_cell | *GDA* | -1.601806752 | 5.26E-05 | 0.000182418 | 0.330747593 |
| T2DM | dendritic_cell | *C2orf40* | -1.675773534 | 0.000190419 | 0.000436703 | 1 |
| T2DM | dendritic_cell | *CPSF3* | -3.346201882 | 0.000333192 | 0.0006788 | 1 |
| T2DM | dendritic_cell | *PIGZ* | -0.837861218 | 0.008522119 | 0.011980895 | 1 |
| T2DM | dendritic_cell | *SENP5* | -1.942468461 | 9.08E-05 | 0.000257652 | 0.570843869 |
| T2DM | dendritic_cell | *CCDC92* | -2.422133299 | 1.02E-07 | 1.57E-05 | 0.00064403 |
| T2DM | dendritic_cell | *PTBP3* | 1.751643082 | 0.007797292 | 0.01104836 | 1 |
| T2DM | dendritic_cell | *WDR34* | -2.999084435 | 0.000262979 | 0.000561217 | 1 |
| T2DM | dendritic_cell | *TRIM32* | -3.94647322 | 0.000157124 | 0.000379937 | 0.987837473 |
| T2DM | dendritic_cell | *PHF19* | 2.790364573 | 0.005992086 | 0.008666262 | 1 |
| T2DM | dendritic_cell | *BSPRY* | -6.047930844 | 8.09E-06 | 0.000130129 | 0.050867175 |
| T2DM | dendritic_cell | *PPP6C* | -2.592472909 | 2.76E-05 | 0.000141906 | 0.173267349 |
| T2DM | dendritic_cell | *HDHD3* | -0.767953192 | 4.03E-05 | 0.000160416 | 0.253121859 |
| T2DM | dendritic_cell | *HSDL2* | -1.034585523 | 0.000138625 | 0.000345162 | 0.871532999 |
| T2DM | dendritic_cell | *INVS* | -1.966977955 | 8.71E-05 | 0.000251049 | 0.547316547 |
| T2DM | dendritic_cell | *KDSR* | -2.826771306 | 5.20E-05 | 0.000181132 | 0.326624992 |
| T2DM | dendritic_cell | *DNAL1* | -2.212236592 | 0.000865121 | 0.001529532 | 1 |
| T2DM | dendritic_cell | *IRF2BPL* | -1.842235005 | 5.98E-05 | 0.000197204 | 0.375925173 |
| T2DM | dendritic_cell | *TTLL5* | -2.718341518 | 0.000126196 | 0.000322256 | 0.793393478 |
| T2DM | dendritic_cell | *FLVCR2* | 1.646668063 | 0.000263312 | 0.000561548 | 1 |
| T2DM | dendritic_cell | *ZC2HC1C* | 3.112283166 | 0.020586473 | 0.027133575 | 1 |
| T2DM | dendritic_cell | *ALDH6A1* | -1.321486329 | 2.48E-05 | 0.000138038 | 0.15569034 |
| T2DM | dendritic_cell | *ESRRB* | -1.253957974 | 9.14E-05 | 0.000258721 | 0.574360975 |
| T2DM | dendritic_cell | *COQ6* | -3.084908591 | 0.032346805 | 0.041369639 | 1 |
| T2DM | dendritic_cell | *GPR75* | -5.990738845 | 0.00018163 | 0.000422044 | 1 |

| | | | | | | |
|---|---|---|---|---|---|---|
| T2DM | dendritic_cell | *KLHL29* | -1.721534108 | 4.10E-05 | 0.000161722 | 0.257784453 |
| T2DM | dendritic_cell | *ATAD2B* | -2.154585549 | 0.016187621 | 0.02169086 | 1 |
| T2DM | dendritic_cell | *ATL2* | -1.912819679 | 2.82E-05 | 0.000142583 | 0.177591747 |
| T2DM | dendritic_cell | *CNRIP1* | -3.009603295 | 2.18E-06 | 8.66E-05 | 0.01368641 |
| T2DM | dendritic_cell | *CRIPT* | -2.6388355 | 9.10E-06 | 0.000130129 | 0.057212084 |
| T2DM | dendritic_cell | *EPCAM* | 1.41938507 | 0.005079773 | 0.00745658 | 1 |
| T2DM | dendritic_cell | *SLC17A5* | -1.944701291 | 0.004006976 | 0.006018122 | 1 |
| T2DM | dendritic_cell | *OGFRL1* | 1.398327889 | 0.000170328 | 0.000403638 | 1 |
| T2DM | dendritic_cell | *CUTC* | -2.303124192 | 9.94E-05 | 0.000271934 | 0.625176505 |
| T2DM | dendritic_cell | *MXI1* | -2.573220456 | 0.030799729 | 0.039509874 | 1 |
| T2DM | dendritic_cell | *C10orf88* | -4.041414077 | 0.000208915 | 0.000468087 | 1 |
| T2DM | dendritic_cell | *TCTN3* | -3.100206029 | 0.000302751 | 0.000629221 | 1 |
| T2DM | dendritic_cell | *AVPI1* | -2.397293315 | 0.000252443 | 0.000542972 | 1 |
| T2DM | dendritic_cell | *WDR11* | -2.600999613 | 0.00229532 | 0.003654262 | 1 |
| T2DM | dendritic_cell | *C10orf76* | -2.397884609 | 0.00451802 | 0.006707153 | 1 |
| T2DM | dendritic_cell | *KCNIP2* | -0.608907786 | 0.000479136 | 0.000916437 | 1 |
| T2DM | dendritic_cell | *C10orf95* | -1.691657231 | 0.001911486 | 0.003096498 | 1 |
| T2DM | dendritic_cell | *KANSL1* | -1.737900427 | 0.001042912 | 0.001795814 | 1 |
| T2DM | dendritic_cell | *HOXB5* | -2.109477093 | 1.88E-06 | 8.36E-05 | 0.011815003 |
| T2DM | dendritic_cell | *HOXB3* | -1.927393173 | 3.73E-08 | 8.09E-06 | 0.000234625 |
| T2DM | dendritic_cell | *CAAP1* | -2.130207343 | 7.06E-05 | 0.000217126 | 0.443804702 |
| T2DM | dendritic_cell | *CD274* | 1.609404439 | 0.000184368 | 0.000427247 | 1 |
| T2DM | dendritic_cell | *NUP43* | -1.881104452 | 0.000119326 | 0.00030982 | 0.7502012 |
| T2DM | dendritic_cell | *LRP11* | -2.265906479 | 2.03E-06 | 8.62E-05 | 0.012786229 |
| T2DM | dendritic_cell | *CCDC170* | -1.072628634 | 0.013677253 | 0.018517618 | 1 |
| T2DM | dendritic_cell | *CYSTM1* | -1.962874867 | 2.97E-05 | 0.000144362 | 0.186450048 |
| T2DM | dendritic_cell | *WDR55* | -2.693480189 | 1.98E-05 | 0.000131266 | 0.124427532 |
| T2DM | dendritic_cell | *PCDHB14* | -1.637329822 | 1.82E-05 | 0.000130129 | 0.114185256 |
| T2DM | dendritic_cell | *SEC16B* | -2.982904836 | 0.000445273 | 0.000863755 | 1 |
| T2DM | dendritic_cell | *GORAB* | -3.725698521 | 0.001037486 | 0.001788504 | 1 |
| T2DM | dendritic_cell | *ACAT2* | -2.741098558 | 0.000185281 | 0.000428101 | 1 |
| T2DM | dendritic_cell | *SNX19* | -2.003173763 | 0.002997992 | 0.004633327 | 1 |
| T2DM | dendritic_cell | *MASTL* | -1.492086055 | 0.000427246 | 0.000835748 | 1 |
| T2DM | dendritic_cell | *MRC1* | 0.575316002 | 5.53E-05 | 0.000188394 | 0.347775155 |
| T2DM | dendritic_cell | *IQSEC3* | -2.605051074 | 1.17E-05 | 0.000130129 | 0.073740732 |
| T2DM | dendritic_cell | *CCDC77* | -1.944201885 | 2.13E-06 | 8.62E-05 | 0.013365352 |
| T2DM | dendritic_cell | *DNAJC15* | -3.11151632 | 0.003194452 | 0.004906797 | 1 |
| T2DM | dendritic_cell | *WBP4* | -1.85026923 | 5.65E-05 | 0.00019044 | 0.354979772 |
| T2DM | dendritic_cell | *SMAD9* | -2.205961025 | 1.68E-05 | 0.000130129 | 0.105412203 |
| T2DM | dendritic_cell | *SIL1* | -1.976204765 | 5.77E-05 | 0.000192599 | 0.362471356 |
| T2DM | dendritic_cell | *ZFP30* | -1.456218794 | 0.004777801 | 0.007054494 | 1 |
| T2DM | dendritic_cell | *APAF1* | 1.834296638 | 0.001882109 | 0.003052843 | 1 |
| T2DM | dendritic_cell | *ADRA1A* | 0.383431172 | 0.009698152 | 0.013474537 | 1 |
| T2DM | dendritic_cell | *EPHX2* | -1.753648422 | 0.00902127 | 0.012612124 | 1 |
| T2DM | dendritic_cell | *ZNF706* | -2.020567996 | 2.83E-05 | 0.000142583 | 0.17811635 |
| T2DM | dendritic_cell | *CRISPLD1* | -5.231816115 | 0.000110678 | 0.000293091 | 0.695835635 |
| T2DM | dendritic_cell | *RDH10* | -0.924333169 | 0.009990606 | 0.013868611 | 1 |
| T2DM | dendritic_cell | *SCPEP1* | -0.768028739 | 0.01368298 | 0.018519892 | 1 |
| T2DM | dendritic_cell | *TRIM6* | -1.846007419 | 1.70E-05 | 0.000130129 | 0.106997266 |
| T2DM | dendritic_cell | *CEP89* | -1.077873901 | 0.006839752 | 0.009793104 | 1 |

| | | | | | | |
|---|---|---|---|---|---|---|
| T2DM | dendritic_cell | *ECHDC2* | -1.250498804 | 3.58E-07 | 3.22E-05 | 0.002251777 |
| T2DM | dendritic_cell | *PRB2* | -1.512413444 | 8.31E-05 | 0.000243001 | 0.522452699 |
| T2DM | dendritic_cell | *PYROXD1* | -2.360991705 | 5.83E-06 | 0.000129745 | 0.036661569 |
| T2DM | dendritic_cell | *A1BG* | 1.742186526 | 1.65E-05 | 0.000130129 | 0.103886611 |
| T2DM | dendritic_cell | *ZSCAN18* | -2.77158733 | 0.004976245 | 0.007316513 | 1 |
| T2DM | dendritic_cell | *PDZRN3* | -1.190241778 | 1.57E-05 | 0.000130129 | 0.098664623 |
| T2DM | dendritic_cell | *LHX4* | -3.098132881 | 6.39E-06 | 0.000130129 | 0.040174386 |
| T2DM | dendritic_cell | *POPDC2* | -2.360075783 | 2.89E-05 | 0.000143461 | 0.181908168 |
| T2DM | dendritic_cell | *B4GALT4* | -3.899619063 | 6.77E-05 | 0.000211932 | 0.425849439 |
| T2DM | dendritic_cell | *CD80* | 0.504823433 | 3.45E-05 | 0.00015138 | 0.21677679 |
| T2DM | dendritic_cell | *CAT* | -0.980472311 | 0.000203295 | 0.000458598 | 1 |
| T2DM | dendritic_cell | *GJB6* | -33.48911075 | 0.000765138 | 0.001376373 | 1 |
| T2DM | dendritic_cell | *RNF115* | -2.715605458 | 0.000819425 | 0.001458998 | 1 |
| T2DM | dendritic_cell | *POLR3GL* | -3.09956726 | 2.84E-05 | 0.000142583 | 0.178375536 |
| T2DM | dendritic_cell | *TNFSF10* | -1.736738595 | 2.67E-05 | 0.000140322 | 0.167844732 |
| T2DM | dendritic_cell | *ZNF639* | -1.740133057 | 4.38E-05 | 0.000166492 | 0.275595895 |
| T2DM | dendritic_cell | *LIAS* | -2.155861005 | 1.39E-05 | 0.000130129 | 0.08722452 |
| T2DM | dendritic_cell | *CLCC1* | -2.565157106 | 1.82E-05 | 0.000130129 | 0.114137316 |
| T2DM | dendritic_cell | *GPSM2* | -1.771165787 | 0.000692646 | 0.001259302 | 1 |
| T2DM | dendritic_cell | *CXCR4* | 0.605948309 | 6.09E-05 | 0.000198694 | 0.382961866 |
| T2DM | dendritic_cell | *ACVR2A* | -2.63501795 | 0.001999786 | 0.003223758 | 1 |
| T2DM | dendritic_cell | *POLK* | -2.287711688 | 7.24E-05 | 0.000220017 | 0.455136514 |
| T2DM | dendritic_cell | *FLT3* | 0.09018112 | 2.80E-05 | 0.000142583 | 0.175993954 |
| T2DM | dendritic_cell | *MTIF3* | -1.426526014 | 0.000145303 | 0.000357891 | 0.913519676 |
| T2DM | dendritic_cell | *RASL11A* | -5.062226414 | 1.98E-07 | 2.17E-05 | 0.001244276 |
| T2DM | dendritic_cell | *FMOD* | -3.178168142 | 1.59E-05 | 0.000130129 | 0.099880629 |
| T2DM | dendritic_cell | *SERAC1* | -3.668174237 | 9.33E-06 | 0.000130129 | 0.058660151 |
| T2DM | dendritic_cell | *LDB3* | 1.28673379 | 0.016945764 | 0.022633954 | 1 |
| T2DM | dendritic_cell | *OPN4* | 2.014172868 | 7.96E-05 | 0.00023544 | 0.500308939 |
| T2DM | dendritic_cell | *FAM35A* | -1.978627435 | 0.001706022 | 0.00278881 | 1 |
| T2DM | dendritic_cell | *FAM213A* | -1.449022413 | 1.21E-05 | 0.000130129 | 0.075828346 |
| T2DM | dendritic_cell | *NAA60* | -3.954221201 | 3.67E-05 | 0.000155353 | 0.230679311 |
| T2DM | dendritic_cell | *ODF2L* | -1.41693615 | 0.000548723 | 0.001031029 | 1 |
| T2DM | dendritic_cell | *SPATA1* | -2.821047076 | 0.000332991 | 0.00067861 | 1 |
| T2DM | dendritic_cell | *LRRC39* | -0.980203548 | 0.002839287 | 0.004409733 | 1 |
| T2DM | dendritic_cell | *RWDD3* | 3.318991179 | 0.000567299 | 0.001059284 | 1 |
| T2DM | dendritic_cell | *CCDC18* | -2.410503673 | 4.62E-05 | 0.00017055 | 0.290617341 |
| T2DM | dendritic_cell | *RPAP2* | -1.996015378 | 2.53E-05 | 0.000138949 | 0.158885139 |
| T2DM | dendritic_cell | *BBS9* | -2.517321503 | 1.71E-05 | 0.000130129 | 0.107600437 |
| T2DM | dendritic_cell | *PMS2* | -2.064881747 | 0.00120188 | 0.002035071 | 1 |
| T2DM | dendritic_cell | *KIAA0087* | 2.107351418 | 0.000492809 | 0.000938055 | 1 |
| T2DM | dendritic_cell | *KLHL7* | -2.565848392 | 3.45E-05 | 0.00015138 | 0.216723826 |
| T2DM | dendritic_cell | *HERPUD2* | -3.08508154 | 8.56E-05 | 0.000248016 | 0.537957274 |
| T2DM | dendritic_cell | *CBX3* | -3.578793271 | 2.30E-06 | 8.72E-05 | 0.014481974 |
| T2DM | dendritic_cell | *FAM126A* | -1.881324922 | 4.96E-05 | 0.000176609 | 0.311714498 |
| T2DM | dendritic_cell | *FKBP9* | -2.086480141 | 0.000124316 | 0.000319402 | 0.781575562 |
| T2DM | dendritic_cell | *CCZ1* | -1.585696447 | 0.000991221 | 0.001719119 | 1 |
| T2DM | dendritic_cell | *POLM* | -0.938516846 | 0.000331934 | 0.000677114 | 1 |
| T2DM | dendritic_cell | *DCAF10* | -2.19150895 | 4.11E-05 | 0.000161841 | 0.258136434 |
| T2DM | dendritic_cell | *TRIM24* | -3.060291003 | 0.001545348 | 0.002553377 | 1 |

| | | | | | | |
|---|---|---|---|---|---|---|
| T2DM | dendritic_cell | *BICC1* | -0.768852458 | 5.66E-05 | 0.000190501 | 0.355855553 |
| T2DM | dendritic_cell | *SLC25A16* | -0.96535142 | 0.003871737 | 0.005835917 | 1 |
| T2DM | dendritic_cell | *ZWINT* | -1.277466709 | 0.005843871 | 0.008469437 | 1 |
| T2DM | dendritic_cell | *RBM19* | -2.74574881 | 5.99E-05 | 0.000197214 | 0.376283572 |
| T2DM | dendritic_cell | *ACADS* | -1.876918102 | 1.14E-05 | 0.000130129 | 0.071840192 |
| T2DM | dendritic_cell | *SSPN* | -0.981552257 | 0.004172976 | 0.006248035 | 1 |
| T2DM | dendritic_cell | *CCDC91* | -2.098547036 | 0.00103188 | 0.001780793 | 1 |
| T2DM | dendritic_cell | *PKN1* | -2.672835844 | 0.024936732 | 0.032432196 | 1 |
| T2DM | dendritic_cell | *ATP7B* | -0.554562834 | 0.006927397 | 0.009907313 | 1 |
| T2DM | dendritic_cell | *NLN* | 1.790250647 | 0.005018192 | 0.007367906 | 1 |
| T2DM | dendritic_cell | *OPTN* | -1.510790531 | 1.81E-06 | 8.21E-05 | 0.011357651 |
| T2DM | dendritic_cell | *ATF1* | -2.323766796 | 0.000598695 | 0.00111065 | 1 |
| T2DM | dendritic_cell | *TSFM* | 2.486037206 | 0.010769235 | 0.014847847 | 1 |
| T2DM | dendritic_cell | *ARHGAP9* | 1.449981451 | 0.00014013 | 0.00034822 | 0.880997222 |
| T2DM | dendritic_cell | *SPATS2* | -2.888095244 | 0.000921388 | 0.001611085 | 1 |
| T2DM | dendritic_cell | *IKZF4* | -2.62550939 | 1.73E-06 | 8.21E-05 | 0.010881772 |
| T2DM | dendritic_cell | *SARDH* | -3.052800919 | 1.62E-05 | 0.000130129 | 0.101621016 |
| T2DM | dendritic_cell | *DBH* | -5.391746385 | 0.006232832 | 0.008971111 | 1 |
| T2DM | dendritic_cell | *STIL* | -3.80186116 | 0.000477418 | 0.000913986 | 1 |
| T2DM | dendritic_cell | *TTC21B* | -2.427077276 | 9.28E-05 | 0.000261455 | 0.58349465 |
| T2DM | dendritic_cell | *NMI* | -2.868229451 | 0.000145644 | 0.000358007 | 0.915665358 |
| T2DM | dendritic_cell | *ACVR1C* | -0.804912418 | 4.01E-05 | 0.000159994 | 0.251831031 |
| T2DM | dendritic_cell | *SLC36A1* | -0.802380938 | 0.034550972 | 0.044061249 | 1 |
| T2DM | dendritic_cell | *PLA2G12A* | -3.056981756 | 5.91E-05 | 0.000195748 | 0.371530011 |
| T2DM | dendritic_cell | *C4BPB* | -1.889273613 | 0.002481475 | 0.003915206 | 1 |
| T2DM | dendritic_cell | *RAB38* | -5.681056735 | 6.38E-06 | 0.000130129 | 0.040085607 |
| T2DM | dendritic_cell | *CKS2* | 1.959985676 | 0.000168943 | 0.000401416 | 1 |
| T2DM | dendritic_cell | *CHPF* | -1.404549817 | 0.009593248 | 0.013349436 | 1 |
| T2DM | dendritic_cell | *DNPEP* | -3.531881436 | 0.009053349 | 0.012648534 | 1 |
| T2DM | dendritic_cell | *MOGAT1* | -0.980159042 | 2.26E-05 | 0.000133629 | 0.142000792 |
| T2DM | dendritic_cell | *FAM124B* | -1.311915126 | 0.006034466 | 0.008721537 | 1 |
| T2DM | dendritic_cell | *FAM210B* | -1.360999582 | 0.000240741 | 0.000522631 | 1 |
| T2DM | dendritic_cell | *SNX21* | -1.490520885 | 1.49E-05 | 0.000130129 | 0.093525262 |
| T2DM | dendritic_cell | *WFDC3* | -0.599256094 | 0.009049199 | 0.012645546 | 1 |
| T2DM | dendritic_cell | *SLC12A5* | -1.732499268 | 0.010877642 | 0.014977602 | 1 |
| T2DM | dendritic_cell | *VAPB* | -1.760101907 | 0.000129287 | 0.000328414 | 0.812825028 |
| T2DM | dendritic_cell | *TOX2* | -0.661740285 | 0.004044338 | 0.006072785 | 1 |
| T2DM | dendritic_cell | *ARFGEF2* | -2.612743263 | 0.002068481 | 0.00332427 | 1 |
| T2DM | dendritic_cell | *CSE1L* | -3.297103086 | 0.000411151 | 0.000808288 | 1 |
| T2DM | dendritic_cell | *CDH26* | -2.514705814 | 1.68E-05 | 0.000130129 | 0.105892427 |
| T2DM | dendritic_cell | *STX16* | -2.847419379 | 5.85E-05 | 0.000194469 | 0.36770091 |
| T2DM | dendritic_cell | *NEURL2* | 1.397251047 | 8.65E-06 | 0.000130129 | 0.054370504 |
| T2DM | dendritic_cell | *FASTKD3* | -2.109304704 | 9.08E-05 | 0.000257652 | 0.570611181 |
| T2DM | dendritic_cell | *PEPD* | -1.480653944 | 0.002945038 | 0.004558211 | 1 |
| T2DM | dendritic_cell | *CHST8* | 4.038933924 | 0.005119097 | 0.007510796 | 1 |
| T2DM | dendritic_cell | *STAMBP* | -2.436545679 | 3.64E-05 | 0.000155217 | 0.2285862 |
| T2DM | dendritic_cell | *ATP8A1* | -1.28350772 | 0.011123608 | 0.015299524 | 1 |
| T2DM | dendritic_cell | *USP22* | -2.683299267 | 0.000335857 | 0.000683344 | 1 |
| T2DM | dendritic_cell | *HIF3A* | -1.017121469 | 0.000312355 | 0.000645335 | 1 |
| T2DM | dendritic_cell | *BTN2A2* | 0.792018885 | 6.05E-05 | 0.000198694 | 0.380633552 |

| | | | | | | |
|---|---|---|---|---|---|---|
| T2DM | dendritic_cell | *MRS2* | -1.921691887 | 0.002039502 | 0.003281052 | 1 |
| T2DM | dendritic_cell | *SERPINB6* | -2.81705021 | 6.23E-05 | 0.000200944 | 0.391640156 |
| T2DM | dendritic_cell | *HIST1H1D* | 1.406239906 | 0.025909157 | 0.033571902 | 1 |
| T2DM | dendritic_cell | *PEX6* | -0.826140838 | 0.030492148 | 0.039152747 | 1 |
| T2DM | dendritic_cell | *NQO2* | -2.485702235 | 3.38E-05 | 0.000150628 | 0.212284371 |
| T2DM | dendritic_cell | *RPS10* | -2.756075728 | 2.14E-05 | 0.000132591 | 0.134238792 |
| T2DM | dendritic_cell | *HIST1H2BJ* | 3.073264431 | 5.29E-05 | 0.00018288 | 0.332789951 |
| T2DM | dendritic_cell | *MED20* | -2.63712074 | 0.00047719 | 0.000913827 | 1 |
| T2DM | dendritic_cell | *TBCC* | 1.198345788 | 0.001583485 | 0.002609533 | 1 |
| T2DM | dendritic_cell | *TCP11* | -2.043939115 | 0.004635262 | 0.006860144 | 1 |
| T2DM | dendritic_cell | *KLHDC3* | -3.176179927 | 0.018800849 | 0.024963239 | 1 |
| T2DM | dendritic_cell | *MEA1* | -2.181442856 | 0.001418606 | 0.002365096 | 1 |
| T2DM | dendritic_cell | *KLHL31* | -0.72602311 | 0.00017381 | 0.000409115 | 1 |
| T2DM | dendritic_cell | *COL21A1* | -1.247099902 | 5.96E-05 | 0.000196766 | 0.37444658 |
| T2DM | dendritic_cell | *GLO1* | -2.304455088 | 0.008829448 | 0.01236044 | 1 |
| T2DM | dendritic_cell | *CPNE5* | -2.578050519 | 0.000899547 | 0.001579736 | 1 |
| T2DM | dendritic_cell | *RIOK1* | 2.206039584 | 0.002266604 | 0.003614035 | 1 |
| T2DM | dendritic_cell | *RPP40* | -1.374879908 | 0.019695369 | 0.026046443 | 1 |
| T2DM | dendritic_cell | *NUP153* | -2.267187988 | 9.97E-05 | 0.000272455 | 0.626919111 |
| T2DM | dendritic_cell | *EEF1E1* | -3.032592808 | 1.52E-05 | 0.000130129 | 0.095394777 |
| T2DM | dendritic_cell | *EREG* | 3.507027646 | 7.31E-05 | 0.000221298 | 0.459579525 |
| T2DM | dendritic_cell | *CNOT1* | -2.287169048 | 0.001060804 | 0.001822207 | 1 |
| T2DM | dendritic_cell | *BBS2* | -1.028679165 | 0.037410307 | 0.047466923 | 1 |
| T2DM | dendritic_cell | *MT1G* | -9.396862952 | 5.28E-06 | 0.00012486 | 0.033212695 |
| T2DM | dendritic_cell | *GOT2* | -3.378412372 | 0.000134195 | 0.000337609 | 0.843684599 |
| T2DM | dendritic_cell | *CLYBL* | -2.911603073 | 7.03E-05 | 0.000216424 | 0.441720841 |
| T2DM | dendritic_cell | *KIF25* | -3.270648166 | 0.021788674 | 0.028568383 | 1 |
| T2DM | dendritic_cell | *TEKT3* | 1.040115384 | 0.005814459 | 0.008438482 | 1 |
| T2DM | dendritic_cell | *MRPS7* | -2.886720615 | 4.78E-05 | 0.000172826 | 0.300409056 |
| T2DM | dendritic_cell | *NUP85* | -2.314812135 | 0.000102434 | 0.000277588 | 0.644003908 |
| T2DM | dendritic_cell | *MIF4GD* | 1.896059906 | 0.000143485 | 0.000354537 | 0.902091375 |
| T2DM | dendritic_cell | *NT5C* | -3.810188784 | 0.000917296 | 0.001605523 | 1 |
| T2DM | dendritic_cell | *DDX31* | -2.341260308 | 3.78E-05 | 0.00015701 | 0.237817536 |
| T2DM | dendritic_cell | *PPDPF* | 2.735249953 | 0.000369481 | 0.000740493 | 1 |
| T2DM | dendritic_cell | *PLGLB2* | -11.75377414 | 0.011615032 | 0.01590584 | 1 |
| T2DM | dendritic_cell | *CHCHD5* | -2.686593138 | 0.000239126 | 0.000519843 | 1 |
| T2DM | dendritic_cell | *INSIG2* | -2.804352685 | 2.75E-05 | 0.000141613 | 0.172598966 |
| T2DM | dendritic_cell | *POLR1B* | -1.506338821 | 0.001940153 | 0.003136473 | 1 |
| T2DM | dendritic_cell | *SLC25A23* | -1.127195907 | 0.012046317 | 0.01643559 | 1 |
| T2DM | dendritic_cell | *GTF2F1* | -2.038823304 | 9.56E-05 | 0.000265796 | 0.601229456 |
| T2DM | dendritic_cell | *CLPP* | -1.983697872 | 0.008758576 | 0.012274893 | 1 |
| T2DM | dendritic_cell | *SH2D3A* | -5.870413715 | 0.000392905 | 0.000779242 | 1 |
| T2DM | dendritic_cell | *TRIP10* | -2.399413085 | 0.00011537 | 0.000301467 | 0.725330112 |
| T2DM | dendritic_cell | *GPR108* | -2.085575201 | 0.00043089 | 0.000841568 | 1 |
| T2DM | dendritic_cell | *RTN2* | -3.997639126 | 9.24E-06 | 0.000130129 | 0.058092403 |
| T2DM | dendritic_cell | *SYMPK* | -3.479883616 | 5.24E-05 | 0.000181969 | 0.329257775 |
| T2DM | dendritic_cell | *SDCBP2* | -3.267932621 | 0.000955601 | 0.001663768 | 1 |
| T2DM | dendritic_cell | *PANK2* | -3.033735381 | 0.002700437 | 0.004224347 | 1 |
| T2DM | dendritic_cell | *DTD1* | -2.733841131 | 3.99E-05 | 0.000159738 | 0.25094829 |
| T2DM | dendritic_cell | *RBCK1* | -2.031563553 | 0.006031419 | 0.008719137 | 1 |

| | | | | | | |
|---|---|---|---|---|---|---|
| T2DM | dendritic_cell | *RRBP1* | -2.409326822 | 0.000171148 | 0.000405275 | 1 |
| T2DM | dendritic_cell | *BMP2* | -1.794237347 | 1.67E-05 | 0.000130129 | 0.105055451 |
| T2DM | dendritic_cell | *PCSK2* | -3.74825744 | 4.14E-05 | 0.000162565 | 0.260103875 |
| T2DM | dendritic_cell | *MKKS* | -2.663472088 | 2.26E-05 | 0.000133629 | 0.142257968 |
| T2DM | dendritic_cell | *DSTN* | -2.343314758 | 3.39E-05 | 0.000150715 | 0.213047805 |
| T2DM | dendritic_cell | *ITPA* | -3.708741264 | 0.000851755 | 0.001509722 | 1 |
| T2DM | dendritic_cell | *TMEM74B* | 3.125314643 | 0.000227223 | 0.000499144 | 1 |
| T2DM | dendritic_cell | *FAM110A* | 3.179776102 | 0.00054563 | 0.00102583 | 1 |
| T2DM | dendritic_cell | *NCLN* | -3.684570466 | 0.000174799 | 0.000410213 | 1 |
| T2DM | dendritic_cell | *GDF5* | 1.538618013 | 0.00028855 | 0.000604101 | 1 |
| T2DM | dendritic_cell | *MMP24* | -1.879275035 | 0.015756846 | 0.02114478 | 1 |
| T2DM | dendritic_cell | *UROD* | -1.64660557 | 0.000196619 | 0.000446908 | 1 |
| T2DM | dendritic_cell | *ST3GAL3* | -2.026311924 | 0.000125551 | 0.000321262 | 0.789341417 |
| T2DM | dendritic_cell | *KLC1* | -2.182574897 | 0.000259104 | 0.000554266 | 1 |
| T2DM | dendritic_cell | *XRCC3* | -2.711306881 | 0.0020057 | 0.003231634 | 1 |
| T2DM | dendritic_cell | *MCF2L* | -1.197001975 | 0.003549369 | 0.005393088 | 1 |
| T2DM | dendritic_cell | *F10* | -1.560217084 | 1.56E-05 | 0.000130129 | 0.098062682 |
| T2DM | dendritic_cell | *PCID2* | -2.505848457 | 1.20E-05 | 0.000130129 | 0.075482939 |
| T2DM | dendritic_cell | *PROZ* | 1.933618165 | 3.59E-05 | 0.000154525 | 0.225761434 |
| T2DM | dendritic_cell | *IGFLR1* | 6.916979295 | 0.000337569 | 0.000685718 | 1 |
| T2DM | dendritic_cell | *CAPNS1* | -2.376142271 | 0.000557765 | 0.001044273 | 1 |
| T2DM | dendritic_cell | *COX6B1* | -2.593839072 | 1.67E-05 | 0.000130129 | 0.105153283 |
| T2DM | dendritic_cell | *PRDX5* | -3.440943571 | 2.39E-06 | 8.93E-05 | 0.015056025 |
| T2DM | dendritic_cell | *SCAF1* | -2.97667823 | 1.78E-05 | 0.000130129 | 0.11195823 |
| T2DM | dendritic_cell | *TSKS* | 3.45045114 | 0.000322645 | 0.000662225 | 1 |
| T2DM | dendritic_cell | *FLRT1* | -1.617273497 | 0.001815976 | 0.002955543 | 1 |
| T2DM | dendritic_cell | *SBDS* | -2.121060672 | 3.08E-05 | 0.000146146 | 0.193936176 |
| T2DM | dendritic_cell | *BECN1* | -2.570279996 | 0.000125191 | 0.000320601 | 0.787076439 |
| T2DM | dendritic_cell | *DNAJC8* | -3.575587534 | 9.08E-05 | 0.000257652 | 0.570687789 |
| T2DM | dendritic_cell | *ZNF384* | -3.368313139 | 0.002213128 | 0.003539541 | 1 |
| T2DM | dendritic_cell | *EMG1* | -3.038164279 | 3.93E-05 | 0.000159487 | 0.247363833 |
| T2DM | dendritic_cell | *ATG14* | -2.672213772 | 0.000161474 | 0.000387476 | 1 |
| T2DM | dendritic_cell | *RHOJ* | -2.033099074 | 1.26E-05 | 0.000130129 | 0.078996164 |
| T2DM | dendritic_cell | *L3HYPDH* | -0.852727978 | 0.001099308 | 0.001882177 | 1 |
| T2DM | dendritic_cell | *ZBTB1* | -2.468825422 | 0.00028335 | 0.00059571 | 1 |
| T2DM | dendritic_cell | *PZP* | -1.928002106 | 4.75E-05 | 0.000172258 | 0.298350505 |
| T2DM | dendritic_cell | *RHOT1* | -1.75428562 | 0.004913177 | 0.007230604 | 1 |
| T2DM | dendritic_cell | *WDR60* | -1.550924372 | 6.48E-05 | 0.000205702 | 0.407085172 |
| T2DM | dendritic_cell | *AIF1L* | -1.389980292 | 2.08E-05 | 0.000132272 | 0.131016434 |
| T2DM | dendritic_cell | *CANX* | -2.877105713 | 0.000574644 | 0.00107109 | 1 |
| T2DM | dendritic_cell | *IPPK* | -3.555282136 | 3.56E-06 | 0.000108108 | 0.022378395 |
| T2DM | dendritic_cell | *FGD3* | 1.23177055 | 0.000110867 | 0.000293113 | 0.697022421 |
| T2DM | dendritic_cell | *MASP1* | -1.400441617 | 1.42E-05 | 0.000130129 | 0.089222232 |
| T2DM | dendritic_cell | *HRASLS* | -2.172066296 | 0.000106253 | 0.000284504 | 0.668015379 |
| T2DM | dendritic_cell | *HELB* | -3.958071146 | 0.000343658 | 0.00069539 | 1 |
| T2DM | dendritic_cell | *RAP1B* | -2.35203302 | 0.000655684 | 0.001198688 | 1 |
| T2DM | dendritic_cell | *RAB3IP* | -3.304298324 | 4.93E-06 | 0.000123581 | 0.030965659 |
| T2DM | dendritic_cell | *YEATS4* | -1.046345747 | 0.012871947 | 0.017505069 | 1 |
| T2DM | dendritic_cell | *LRRC61* | -0.853620996 | 0.003375732 | 0.00515377 | 1 |
| T2DM | dendritic_cell | *IDUA* | -1.711859907 | 1.48E-05 | 0.000130129 | 0.093064492 |

| | | | | | | |
|---|---|---|---|---|---|---|
| T2DM | dendritic_cell | *TMEM175* | -2.007160282 | 0.004278451 | 0.006390739 | 1 |
| T2DM | dendritic_cell | *EMC1* | -2.697679984 | 0.000243254 | 0.000527659 | 1 |
| T2DM | dendritic_cell | *UBR4* | -2.209555768 | 1.87E-05 | 0.000130129 | 0.117683123 |
| T2DM | dendritic_cell | *SIN3B* | -3.100155247 | 0.000101109 | 0.000274588 | 0.635671143 |
| T2DM | dendritic_cell | *WFIKKN1* | -10.88706902 | 5.06E-05 | 0.000178475 | 0.318041688 |
| T2DM | dendritic_cell | *WDR24* | 2.56967766 | 0.000783131 | 0.001403918 | 1 |
| T2DM | dendritic_cell | *CHTF18* | -3.082695583 | 0.030428791 | 0.039097856 | 1 |
| T2DM | dendritic_cell | *SMARCA4* | -3.463844135 | 0.000113125 | 0.000297331 | 0.711216045 |
| T2DM | dendritic_cell | *TICAM1* | 2.943784417 | 0.005511185 | 0.008031715 | 1 |
| T2DM | dendritic_cell | *AAMP* | -2.683148022 | 2.76E-06 | 9.65E-05 | 0.017373817 |
| T2DM | dendritic_cell | *PNKD* | -2.518662136 | 5.59E-06 | 0.000127198 | 0.035116141 |
| T2DM | dendritic_cell | *HIP1* | -2.403813917 | 0.001473349 | 0.002447278 | 1 |
| T2DM | dendritic_cell | *PTPN12* | -2.538483659 | 0.000821785 | 0.001462786 | 1 |
| T2DM | dendritic_cell | *POR* | -1.613816383 | 1.51E-05 | 0.000130129 | 0.094711641 |
| T2DM | dendritic_cell | *FGL2* | 0.86183092 | 2.77E-05 | 0.000142478 | 0.174393052 |
| T2DM | dendritic_cell | *STEAP4* | -1.693982888 | 5.05E-05 | 0.000178302 | 0.317556613 |
| T2DM | dendritic_cell | *GNAI1* | -1.515091637 | 7.10E-06 | 0.000130129 | 0.044653297 |
| T2DM | dendritic_cell | *SGCE* | -1.682138099 | 2.47E-05 | 0.000138038 | 0.155602141 |
| T2DM | dendritic_cell | *RBM48* | -3.410170123 | 2.84E-05 | 0.000142583 | 0.178324752 |
| T2DM | dendritic_cell | *CASD1* | -2.60376241 | 4.80E-05 | 0.000173305 | 0.301551404 |
| T2DM | dendritic_cell | *SRD5A3* | -3.018697935 | 0.00533682 | 0.007801113 | 1 |
| T2DM | dendritic_cell | *VPREB3* | -13.226426 | 0.001049968 | 0.00180606 | 1 |
| T2DM | dendritic_cell | *MGAT3* | -1.289720411 | 1.50E-05 | 0.000130129 | 0.094222412 |
| T2DM | dendritic_cell | *ATF4* | -3.355092981 | 0.000201684 | 0.00045644 | 1 |
| T2DM | dendritic_cell | *A4GALT* | -1.130162642 | 0.010366662 | 0.014339978 | 1 |
| T2DM | dendritic_cell | *CDC42EP1* | -1.743904241 | 3.88E-05 | 0.000158329 | 0.243928703 |
| T2DM | dendritic_cell | *TPST2* | -2.184945857 | 2.99E-05 | 0.000145022 | 0.188036184 |
| T2DM | dendritic_cell | *BAIAP2L2* | -1.57202961 | 0.000908835 | 0.001593821 | 1 |
| T2DM | dendritic_cell | *TST* | -1.174330381 | 0.000355322 | 0.000717337 | 1 |
| T2DM | dendritic_cell | *APOL5* | 2.367140246 | 0.002639863 | 0.00414299 | 1 |
| T2DM | dendritic_cell | *RIBC2* | 1.850140213 | 9.51E-07 | 5.75E-05 | 0.005981455 |
| T2DM | dendritic_cell | *EMC4* | -1.842107915 | 0.024435502 | 0.031826394 | 1 |
| T2DM | dendritic_cell | *POT1* | -2.347553275 | 5.35E-05 | 0.000184213 | 0.336189117 |
| T2DM | dendritic_cell | *CDHR3* | -1.177907537 | 8.73E-05 | 0.000251173 | 0.548561599 |
| T2DM | dendritic_cell | *PRKRIP1* | -2.238382865 | 7.23E-06 | 0.000130129 | 0.045479122 |
| T2DM | dendritic_cell | *FOXP2* | -1.008866252 | 2.01E-06 | 8.62E-05 | 0.012615283 |
| T2DM | dendritic_cell | *STRIP2* | -1.328124661 | 0.002572668 | 0.004045614 | 1 |
| T2DM | dendritic_cell | *FLNC* | -1.564242265 | 0.000422347 | 0.000827711 | 1 |
| T2DM | dendritic_cell | *IRF5* | 0.881354642 | 5.67E-05 | 0.000190803 | 0.35661016 |
| T2DM | dendritic_cell | *KLHDC10* | -2.187330886 | 0.000100995 | 0.000274517 | 0.634957004 |
| T2DM | dendritic_cell | *OPN1SW* | -1.497117717 | 2.76E-07 | 2.75E-05 | 0.001735614 |
| T2DM | dendritic_cell | *HOXD1* | -6.732083493 | 0.001429069 | 0.002380015 | 1 |
| T2DM | dendritic_cell | *HOXD3* | -2.539201404 | 0.000513794 | 0.000972373 | 1 |
| T2DM | dendritic_cell | *PDE11A* | -1.588264151 | 2.83E-05 | 0.000142583 | 0.177980738 |
| T2DM | dendritic_cell | *CHN1* | -1.737758806 | 6.17E-05 | 0.00020013 | 0.388011983 |
| T2DM | dendritic_cell | *GAD1* | -3.909573988 | 8.80E-06 | 0.000130129 | 0.055315094 |
| T2DM | dendritic_cell | *ORMDL1* | -0.895593123 | 0.034755399 | 0.044312958 | 1 |
| T2DM | dendritic_cell | *PSMG2* | -1.823335166 | 0.00253498 | 0.00399234 | 1 |
| T2DM | dendritic_cell | *WDFY4* | 0.199905773 | 3.72E-05 | 0.000156097 | 0.234002748 |
| T2DM | dendritic_cell | *EIF2AK4* | -2.744061394 | 4.54E-05 | 0.000169373 | 0.285731443 |

| | | | | | | |
|---|---|---|---|---|---|---|
| T2DM | dendritic_cell | *TMOD2* | -2.221214536 | 1.67E-05 | 0.000130129 | 0.105161277 |
| T2DM | dendritic_cell | *IVD* | -1.94556179 | 0.0037532 | 0.005676298 | 1 |
| T2DM | dendritic_cell | *KNSTRN* | -0.456446199 | 0.025318691 | 0.032874558 | 1 |
| T2DM | dendritic_cell | *ARPP19* | -2.973654681 | 3.39E-05 | 0.000150715 | 0.213082529 |
| T2DM | dendritic_cell | *VPS13C* | -2.343178709 | 0.000371796 | 0.00074371 | 1 |
| T2DM | dendritic_cell | *CALML4* | -2.98612076 | 2.27E-05 | 0.000133629 | 0.142702273 |
| T2DM | dendritic_cell | *ACKR4* | -1.43107481 | 5.12E-05 | 0.000179577 | 0.322032017 |
| T2DM | dendritic_cell | *SUMF2* | -2.943410443 | 0.000612269 | 0.00113116 | 1 |
| T2DM | dendritic_cell | *SERGEF* | -2.442429355 | 0.022954663 | 0.030009559 | 1 |
| T2DM | dendritic_cell | *DCTD* | -2.211112617 | 0.000478937 | 0.000916334 | 1 |
| T2DM | dendritic_cell | *RPAIN* | -2.434536313 | 3.29E-06 | 0.000103864 | 0.020669007 |
| T2DM | dendritic_cell | *USP6* | -0.753795948 | 0.017662013 | 0.023505731 | 1 |
| T2DM | dendritic_cell | *ATP1B2* | -2.771114762 | 0.0011085 | 0.001896364 | 1 |
| T2DM | dendritic_cell | *CCL4* | -1.781060782 | 0.002481644 | 0.003915206 | 1 |
| T2DM | dendritic_cell | *PHF20L1* | -2.632772315 | 9.13E-05 | 0.000258601 | 0.57383492 |
| T2DM | dendritic_cell | *PUS7L* | -2.470934373 | 0.000966349 | 0.001681084 | 1 |
| T2DM | dendritic_cell | *SLC44A2* | -2.600702775 | 4.71E-05 | 0.000171433 | 0.296068607 |
| T2DM | dendritic_cell | *CDKN2D* | -2.967513914 | 4.24E-05 | 0.000164065 | 0.266705082 |
| T2DM | dendritic_cell | *MTUS1* | -1.589449528 | 8.91E-06 | 0.000130129 | 0.055996077 |
| T2DM | dendritic_cell | *NGDN* | -3.231938182 | 6.07E-05 | 0.000198694 | 0.381413407 |
| T2DM | dendritic_cell | *ADCY4* | -1.207934555 | 7.43E-05 | 0.000224335 | 0.46734229 |
| T2DM | dendritic_cell | *RAB2B* | -2.314846155 | 2.86E-05 | 0.000142839 | 0.179834802 |
| T2DM | dendritic_cell | *BCL2L2* | -3.105376188 | 3.12E-05 | 0.00014679 | 0.196111452 |
| T2DM | dendritic_cell | *PARP2* | -1.953342282 | 7.56E-05 | 0.000226754 | 0.475504076 |
| T2DM | dendritic_cell | *RNASE1* | -1.917527648 | 0.018127483 | 0.024089513 | 1 |
| T2DM | dendritic_cell | *DAD1* | -2.506985468 | 9.01E-05 | 0.000256231 | 0.566218939 |
| T2DM | dendritic_cell | *TEP1* | -3.822894154 | 0.000526037 | 0.000991959 | 1 |
| T2DM | dendritic_cell | *EPB41L4A* | -1.982152004 | 3.57E-05 | 0.000154296 | 0.224191957 |
| T2DM | dendritic_cell | *CDO1* | -0.932178117 | 1.75E-05 | 0.000130129 | 0.110024681 |
| T2DM | dendritic_cell | *REEP5* | -2.810168508 | 0.000219816 | 0.000486271 | 1 |
| T2DM | dendritic_cell | *QRICH2* | -1.981240126 | 0.000204202 | 0.000460314 | 1 |
| T2DM | dendritic_cell | *SEC14L1* | -1.886462589 | 0.004933885 | 0.007259381 | 1 |
| T2DM | dendritic_cell | *ERMARD* | -2.840936889 | 7.61E-06 | 0.000130129 | 0.04785638 |
| T2DM | dendritic_cell | *PHF10* | -3.508041305 | 0.000147297 | 0.000361035 | 0.926055224 |
| T2DM | dendritic_cell | *GALNT8* | -2.534049412 | 0.000403997 | 0.000797716 | 1 |
| T2DM | dendritic_cell | *DOCK6* | -1.744343146 | 0.000458553 | 0.000884876 | 1 |
| T2DM | dendritic_cell | *ECSIT* | -3.309286879 | 0.001954313 | 0.003157741 | 1 |
| T2DM | dendritic_cell | *ELOF1* | -2.963815175 | 1.07E-05 | 0.000130129 | 0.067490154 |
| T2DM | dendritic_cell | *CDC16* | -1.89339414 | 0.000282535 | 0.000594495 | 1 |
| T2DM | dendritic_cell | *THEM6* | -1.85336176 | 1.53E-05 | 0.000130129 | 0.096495679 |
| T2DM | dendritic_cell | *TOMM40* | -3.324433927 | 0.000207203 | 0.000465078 | 1 |
| T2DM | dendritic_cell | *DPP6* | -3.109088318 | 1.85E-07 | 2.17E-05 | 0.001161248 |
| T2DM | dendritic_cell | *ATP8B3* | -1.214616661 | 0.000530676 | 0.001000108 | 1 |
| T2DM | dendritic_cell | *GTPBP3* | -2.127978937 | 3.32E-07 | 3.11E-05 | 0.0020848 |
| T2DM | dendritic_cell | *PLVAP* | -0.583953994 | 0.000570108 | 0.001064213 | 1 |
| T2DM | dendritic_cell | *BST2* | -2.76800039 | 2.38E-05 | 0.000136538 | 0.149385638 |
| T2DM | dendritic_cell | *DDA1* | -3.516396383 | 0.000370347 | 0.00074152 | 1 |
| T2DM | dendritic_cell | *TULP4* | -1.889341514 | 4.44E-05 | 0.000167569 | 0.279062469 |
| T2DM | dendritic_cell | *ACTN4* | -2.347802291 | 2.60E-05 | 0.0001395 | 0.163442277 |
| T2DM | dendritic_cell | *STK33* | -1.672535832 | 3.82E-06 | 0.000111457 | 0.024019463 |

| | | | | | | |
|---|---|---|---|---|---|---|
| T2DM | dendritic_cell | *CACNG6* | 3.509641711 | 0.019113452 | 0.025346187 | 1 |
| T2DM | dendritic_cell | *PXDN* | -1.267141559 | 1.46E-05 | 0.000130129 | 0.091759739 |
| T2DM | dendritic_cell | *PGPEP1* | -1.513530533 | 0.00500545 | 0.007350915 | 1 |
| T2DM | dendritic_cell | *HRC* | -1.189491344 | 0.030325572 | 0.038989135 | 1 |
| T2DM | dendritic_cell | *CAMSAP1* | -3.685480191 | 0.000462774 | 0.000890833 | 1 |
| T2DM | dendritic_cell | *UBAC1* | -2.879734773 | 0.000143896 | 0.000355053 | 0.904674004 |
| T2DM | dendritic_cell | *TNNT3* | -1.617931002 | 6.42E-06 | 0.000130129 | 0.040381903 |
| T2DM | dendritic_cell | *ATXN10* | -2.52223748 | 0.000172354 | 0.000406985 | 1 |
| T2DM | dendritic_cell | *TUBGCP2* | -2.2743961 | 0.011554816 | 0.015837176 | 1 |
| T2DM | dendritic_cell | *CYP2E1* | -2.772422085 | 0.000320096 | 0.000658308 | 1 |
| T2DM | dendritic_cell | *HBZ* | 8.867427792 | 4.93E-05 | 0.000176105 | 0.310121665 |
| T2DM | dendritic_cell | *PAK4* | -3.510533419 | 2.78E-05 | 0.000142561 | 0.174921798 |
| T2DM | dendritic_cell | *ZNF337* | -1.867551972 | 0.000244432 | 0.000528818 | 1 |
| T2DM | dendritic_cell | *CEP85* | -1.589847071 | 5.00E-05 | 0.000177555 | 0.31436107 |
| T2DM | dendritic_cell | *UCK1* | -2.086354236 | 7.97E-05 | 0.000235557 | 0.501029584 |
| T2DM | dendritic_cell | *YIPF2* | -3.072123061 | 8.25E-06 | 0.000130129 | 0.051877206 |
| T2DM | dendritic_cell | *TMEM160* | -5.821838281 | 0.00204088 | 0.003282429 | 1 |
| T2DM | dendritic_cell | *MAP3K10* | -3.391365675 | 0.00043648 | 0.000848831 | 1 |
| T2DM | dendritic_cell | *SMPDL3B* | 3.758003312 | 3.64E-05 | 0.000155217 | 0.228955552 |
| T2DM | dendritic_cell | *ATPIF1* | -2.36542385 | 0.010059632 | 0.013949031 | 1 |
| T2DM | dendritic_cell | *THEMIS2* | 0.65294706 | 2.13E-05 | 0.000132591 | 0.134023759 |
| T2DM | dendritic_cell | *CLIP1* | -2.192725308 | 0.000125706 | 0.000321527 | 0.790313034 |
| T2DM | dendritic_cell | *ZNF317* | -3.161448006 | 0.001280724 | 0.00215844 | 1 |
| T2DM | dendritic_cell | *PPAN* | 4.83909408 | 0.019393275 | 0.025691456 | 1 |
| T2DM | dendritic_cell | *ZNF236* | -2.699399839 | 9.04E-06 | 0.000130129 | 0.056847779 |
| T2DM | dendritic_cell | *C12orf65* | -1.668045708 | 0.022283305 | 0.029179706 | 1 |
| T2DM | dendritic_cell | *PKDREJ* | -2.487301265 | 0.000839217 | 0.00149044 | 1 |
| T2DM | dendritic_cell | *HABP4* | -2.973512696 | 3.03E-05 | 0.000145705 | 0.19047883 |
| T2DM | dendritic_cell | *EPS8L1* | -8.611875122 | 6.83E-06 | 0.000130129 | 0.042939224 |
| T2DM | dendritic_cell | *LILRB2* | 0.550347833 | 3.82E-05 | 0.000157656 | 0.240250224 |
| T2DM | dendritic_cell | *COX4I2* | -2.844353217 | 1.13E-05 | 0.000130129 | 0.071190957 |
| T2DM | dendritic_cell | *ATP6V1E1* | -3.259356406 | 4.12E-06 | 0.000114478 | 0.025871446 |
| T2DM | dendritic_cell | *PPT1* | 1.689126861 | 0.000454336 | 0.000877546 | 1 |
| T2DM | dendritic_cell | *HAUS8* | 1.721985532 | 0.003179786 | 0.004887852 | 1 |
| T2DM | dendritic_cell | *TBC1D5* | -2.180826177 | 0.000404213 | 0.000797853 | 1 |
| T2DM | dendritic_cell | *RFTN1* | -1.87444791 | 0.000317985 | 0.000654823 | 1 |
| T2DM | dendritic_cell | *KCNC3* | 0.677549232 | 7.42E-05 | 0.000224195 | 0.466773683 |
| T2DM | dendritic_cell | *KIF3A* | -1.351578249 | 2.46E-05 | 0.000137915 | 0.154879105 |
| T2DM | dendritic_cell | *AOC3* | -1.701483357 | 1.42E-05 | 0.000130129 | 0.088986573 |
| T2DM | dendritic_cell | *ACLY* | -2.711608231 | 0.000432462 | 0.000843283 | 1 |
| T2DM | dendritic_cell | *AOC2* | -2.371222873 | 1.84E-05 | 0.000130129 | 0.115626989 |
| T2DM | dendritic_cell | *DIAPH1* | 1.417497768 | 0.030526288 | 0.039183089 | 1 |
| T2DM | dendritic_cell | *EXOC4* | -1.595523967 | 0.001378516 | 0.002305595 | 1 |
| T2DM | dendritic_cell | *ACAP3* | -1.025445169 | 4.10E-05 | 0.000161687 | 0.257567872 |
| T2DM | dendritic_cell | *C1orf159* | -2.94595708 | 0.024516461 | 0.031918118 | 1 |
| T2DM | dendritic_cell | *ANO1* | -3.581140943 | 0.00018726 | 0.000431238 | 1 |
| T2DM | dendritic_cell | *PPFIA1* | -2.025234442 | 2.37E-05 | 0.000136431 | 0.149118832 |
| T2DM | dendritic_cell | *NINJ1* | -4.480719584 | 0.003971558 | 0.005967778 | 1 |
| T2DM | dendritic_cell | *MAP1B* | -0.580691623 | 0.020178495 | 0.026640838 | 1 |
| T2DM | dendritic_cell | *CKMT2* | -1.85106502 | 5.20E-05 | 0.000181132 | 0.326872693 |

| | | | | | | |
|---|---|---|---|---|---|---|
| T2DM | dendritic_cell | *ZCCHC9* | -2.230439941 | 6.93E-05 | 0.000214503 | 0.435655951 |
| T2DM | dendritic_cell | *KHDRBS3* | -2.550242535 | 7.75E-05 | 0.000231138 | 0.487237902 |
| T2DM | dendritic_cell | *RBM8A* | -2.684856228 | 6.17E-05 | 0.00020013 | 0.388031954 |
| T2DM | dendritic_cell | *ZNF304* | -2.118512265 | 2.22E-05 | 0.000133527 | 0.139857982 |
| T2DM | dendritic_cell | *ZSCAN5A* | -3.266112488 | 8.45E-05 | 0.000245949 | 0.531004495 |
| T2DM | dendritic_cell | *RHPN2* | -2.862374514 | 7.59E-07 | 5.26E-05 | 0.004774762 |
| T2DM | dendritic_cell | *C19orf12* | -1.080388239 | 0.000147677 | 0.000361526 | 0.928444326 |
| T2DM | dendritic_cell | *ACTR10* | -2.848470472 | 8.72E-05 | 0.00025117 | 0.548084604 |
| T2DM | dendritic_cell | *GCH1* | 2.585430559 | 0.003214754 | 0.00493316 | 1 |
| T2DM | dendritic_cell | *LGALS3* | -2.3567048 | 3.95E-05 | 0.000159553 | 0.248239526 |
| T2DM | dendritic_cell | *DNAJB1* | -2.16226956 | 0.008223838 | 0.011595261 | 1 |
| T2DM | dendritic_cell | *FBXW9* | 2.460732771 | 3.65E-05 | 0.000155217 | 0.229565252 |
| T2DM | dendritic_cell | *C19orf57* | -5.671347692 | 0.000237539 | 0.000517466 | 1 |
| T2DM | dendritic_cell | *MATN3* | -4.167075382 | 1.30E-05 | 0.000130129 | 0.081976081 |
| T2DM | dendritic_cell | *CCT6B* | -1.979365684 | 2.80E-05 | 0.000142583 | 0.176097602 |
| T2DM | dendritic_cell | *RAF1* | -2.657009973 | 6.68E-05 | 0.0002101 | 0.419885496 |
| T2DM | dendritic_cell | *ENOSF1* | -1.641532191 | 5.43E-05 | 0.000186187 | 0.341279905 |
| T2DM | dendritic_cell | *EMILIN2* | 1.214466834 | 0.000247516 | 0.000534571 | 1 |
| T2DM | dendritic_cell | *TRIM5* | -2.63347646 | 0.01107935 | 0.015241986 | 1 |
| T2DM | dendritic_cell | *CNGA4* | 3.102497774 | 0.001093651 | 0.00187351 | 1 |
| T2DM | dendritic_cell | *TRIM22* | -2.927985631 | 6.56E-05 | 0.000207869 | 0.412620702 |
| T2DM | dendritic_cell | *CARD6* | -3.162922087 | 3.41E-05 | 0.000150922 | 0.214420493 |
| T2DM | dendritic_cell | *INPP5K* | -1.744417974 | 1.47E-05 | 0.000130129 | 0.092251662 |
| T2DM | dendritic_cell | *RPA1* | -2.379841041 | 4.18E-05 | 0.000162813 | 0.262617649 |
| T2DM | dendritic_cell | *SERPINF1* | -1.244309944 | 2.75E-05 | 0.000141613 | 0.172626493 |
| T2DM | dendritic_cell | *TMEM128* | -2.491805855 | 6.20E-06 | 0.000130129 | 0.038951277 |
| T2DM | dendritic_cell | *FIGNL1* | -2.979595257 | 4.71E-05 | 0.000171433 | 0.295852304 |
| T2DM | dendritic_cell | *WBP2* | -3.831822472 | 0.000124837 | 0.000320348 | 0.784851686 |
| T2DM | dendritic_cell | *EIF5A* | -1.185198706 | 0.035732173 | 0.045475338 | 1 |
| T2DM | dendritic_cell | *XAF1* | -1.603842918 | 0.00228391 | 0.003639783 | 1 |
| T2DM | dendritic_cell | *REEP2* | -1.246277843 | 0.002666924 | 0.004176078 | 1 |
| T2DM | dendritic_cell | *PRMT7* | -1.853251853 | 0.000157561 | 0.000380263 | 0.990584709 |
| T2DM | dendritic_cell | *TERF2* | -2.654747546 | 0.021779547 | 0.028562372 | 1 |
| T2DM | dendritic_cell | *BTBD3* | -3.232949408 | 0.009591584 | 0.013349436 | 1 |
| T2DM | dendritic_cell | *PCNA* | -2.478082557 | 0.000174711 | 0.000410213 | 1 |
| T2DM | dendritic_cell | *PTPRA* | -2.133491516 | 3.89E-05 | 0.000158595 | 0.244646053 |
| T2DM | dendritic_cell | *IGHMBP2* | -1.274324182 | 0.020326301 | 0.026807522 | 1 |
| T2DM | dendritic_cell | *MMACHC* | -1.630671129 | 1.05E-05 | 0.000130129 | 0.065902455 |
| T2DM | dendritic_cell | *MUTYH* | -2.978952519 | 0.000121893 | 0.00031472 | 0.766343918 |
| T2DM | dendritic_cell | *LPIN3* | -1.343823191 | 1.51E-05 | 0.000130129 | 0.095012724 |
| T2DM | dendritic_cell | *SERINC3* | -2.07818084 | 0.000436499 | 0.000848831 | 1 |
| T2DM | dendritic_cell | *DMGDH* | -1.563898196 | 2.71E-06 | 9.51E-05 | 0.017024811 |
| T2DM | dendritic_cell | *BHMT2* | -0.558200874 | 0.000823193 | 0.001464463 | 1 |
| T2DM | dendritic_cell | *KANK4* | -0.721894599 | 0.000104161 | 0.000280934 | 0.654858129 |
| T2DM | dendritic_cell | *ATP8A2* | -2.300360966 | 1.43E-05 | 0.000130129 | 0.090156281 |
| T2DM | dendritic_cell | *USPL1* | -2.971836636 | 9.68E-05 | 0.000267843 | 0.60880667 |
| T2DM | dendritic_cell | *POMP* | -3.162616977 | 6.01E-05 | 0.000197705 | 0.377615697 |
| T2DM | dendritic_cell | *WASF3* | -1.158649296 | 1.32E-05 | 0.000130129 | 0.083137263 |
| T2DM | dendritic_cell | *RNF17* | 1.228611033 | 0.000178408 | 0.000416533 | 1 |
| T2DM | dendritic_cell | *PEMT* | -2.096322506 | 6.17E-05 | 0.00020013 | 0.388051699 |

| | | | | | | |
|---|---|---|---|---|---|---|
| T2DM | dendritic_cell | *SCO1* | -3.244697734 | 9.58E-06 | 0.000130129 | 0.060233769 |
| T2DM | dendritic_cell | *DSTYK* | -3.205260681 | 8.70E-07 | 5.49E-05 | 0.005467606 |
| T2DM | dendritic_cell | *COG6* | -1.817757914 | 0.000120258 | 0.000311632 | 0.756059505 |
| T2DM | dendritic_cell | *TRPC4* | -4.721970018 | 1.34E-05 | 0.000130129 | 0.084019629 |
| T2DM | dendritic_cell | *RFC3* | -2.31433256 | 9.97E-05 | 0.000272455 | 0.626825277 |
| T2DM | dendritic_cell | *BTBD2* | -3.249143457 | 0.000111648 | 0.000294558 | 0.701931275 |
| T2DM | dendritic_cell | *PRAM1* | 0.798037245 | 0.000468726 | 0.000900085 | 1 |
| T2DM | dendritic_cell | *ZNF414* | -5.699812176 | 0.000425362 | 0.000832582 | 1 |
| T2DM | dendritic_cell | *CNDP2* | 2.568317748 | 0.025350371 | 0.032905662 | 1 |
| T2DM | dendritic_cell | *RTN3* | -1.416721022 | 2.01E-05 | 0.000131266 | 0.126146353 |
| T2DM | dendritic_cell | *GSTT2B* | -0.768026262 | 0.037495076 | 0.047536105 | 1 |
| T2DM | dendritic_cell | *SLC2A11* | -5.561238498 | 0.000144267 | 0.00035569 | 0.907008658 |
| T2DM | dendritic_cell | *C1QTNF6* | -1.666790419 | 1.94E-05 | 0.000131266 | 0.122024098 |
| T2DM | dendritic_cell | *FAM83F* | -2.766208967 | 1.17E-05 | 0.000130129 | 0.073700036 |
| T2DM | dendritic_cell | *ADCK2* | -1.201043557 | 0.00535562 | 0.007823137 | 1 |
| T2DM | dendritic_cell | *ACTR3B* | -2.410269357 | 1.23E-05 | 0.000130129 | 0.077527952 |
| T2DM | dendritic_cell | *DYDC2* | -6.124227691 | 3.41E-05 | 0.000150922 | 0.21446 |
| T2DM | dendritic_cell | *TMTC1* | -1.888064304 | 4.20E-05 | 0.000163316 | 0.264068669 |
| T2DM | dendritic_cell | *IPO8* | -2.324220585 | 2.41E-05 | 0.000136859 | 0.151475522 |
| T2DM | dendritic_cell | *IMPA1* | -1.76141713 | 0.000507937 | 0.000963319 | 1 |
| T2DM | dendritic_cell | *LRRCC1* | 2.055848708 | 0.001655049 | 0.002713953 | 1 |
| T2DM | dendritic_cell | *SWAP70* | -2.613618826 | 5.56E-05 | 0.000188762 | 0.349775903 |
| T2DM | dendritic_cell | *AMPD3* | 1.144928534 | 0.001508123 | 0.002499096 | 1 |
| T2DM | dendritic_cell | *SBF2* | -1.905642178 | 6.20E-05 | 0.000200487 | 0.389714063 |
| T2DM | dendritic_cell | *MICAL2* | 1.21608644 | 7.68E-05 | 0.000229715 | 0.483090798 |
| T2DM | dendritic_cell | *RRAS2* | -0.809740925 | 0.003160835 | 0.004864669 | 1 |
| T2DM | dendritic_cell | *HSD17B4* | -2.425541793 | 3.74E-05 | 0.00015637 | 0.234868368 |
| T2DM | dendritic_cell | *DPF2* | -2.845227778 | 0.001157485 | 0.001968382 | 1 |
| T2DM | dendritic_cell | *GSC* | -5.526984426 | 1.38E-05 | 0.000130129 | 0.086822877 |
| T2DM | dendritic_cell | *CATSPERB* | -1.197438353 | 1.53E-05 | 0.000130129 | 0.096335615 |
| T2DM | dendritic_cell | *COX16* | 1.209016064 | 0.000256944 | 0.00055077 | 1 |
| T2DM | dendritic_cell | *MED6* | -1.576207412 | 0.003337445 | 0.005099032 | 1 |
| T2DM | dendritic_cell | *EIF2S1* | -2.619707835 | 0.031115922 | 0.039882936 | 1 |
| T2DM | dendritic_cell | *ELP3* | -3.06890145 | 0.000148453 | 0.000363021 | 0.933326498 |
| T2DM | dendritic_cell | *MRO* | -2.108728013 | 0.000120077 | 0.00031131 | 0.754926228 |
| T2DM | dendritic_cell | *THUMPD3* | -2.985334541 | 0.000647628 | 0.001184993 | 1 |
| T2DM | dendritic_cell | *EDEM1* | 1.692422253 | 0.000206636 | 0.000464267 | 1 |
| T2DM | dendritic_cell | *CNTN6* | -1.349943129 | 1.65E-05 | 0.000130129 | 0.103774719 |
| T2DM | dendritic_cell | *KATNBL1* | -2.364232188 | 0.000236912 | 0.000516279 | 1 |
| T2DM | dendritic_cell | *GSTM5* | -1.04865109 | 0.00070321 | 0.001276662 | 1 |
| T2DM | dendritic_cell | *SYT6* | -3.701244469 | 2.84E-05 | 0.000142583 | 0.17868227 |
| T2DM | dendritic_cell | *VAV3* | 1.001268505 | 0.000118953 | 0.000309415 | 0.747856013 |
| T2DM | dendritic_cell | *PSRC1* | -1.860835831 | 0.004594844 | 0.006809945 | 1 |
| T2DM | dendritic_cell | *CEPT1* | -2.170338735 | 4.64E-05 | 0.000170668 | 0.291928778 |
| T2DM | dendritic_cell | *AP4B1* | -2.290175363 | 0.000707488 | 0.001282947 | 1 |
| T2DM | dendritic_cell | *NAPG* | -2.562270043 | 7.41E-05 | 0.000223904 | 0.465945082 |
| T2DM | dendritic_cell | *SPIRE1* | -1.737805624 | 2.44E-06 | 9.02E-05 | 0.015326018 |
| T2DM | dendritic_cell | *PPHLN1* | -1.8625751 | 0.004380062 | 0.006522372 | 1 |
| T2DM | dendritic_cell | *YWHAQ* | -2.618402799 | 0.003791713 | 0.005729031 | 1 |
| T2DM | dendritic_cell | *LPIN1* | -1.817583545 | 2.24E-05 | 0.000133629 | 0.1411271 |

| | | | | | | |
|---|---|---|---|---|---|---|
| T2DM | dendritic_cell | *LDHA* | -3.297900742 | 0.000312131 | 0.000645303 | 1 |
| T2DM | dendritic_cell | *ANO3* | -0.673245354 | 2.58E-05 | 0.0001395 | 0.162336217 |
| T2DM | dendritic_cell | *FST* | -1.543866275 | 1.15E-05 | 0.000130129 | 0.072071211 |
| T2DM | dendritic_cell | *TIMM17A* | -2.736967595 | 0.001640317 | 0.002691198 | 1 |
| T2DM | dendritic_cell | *NARS* | -3.453678925 | 0.000328833 | 0.000671661 | 1 |
| T2DM | dendritic_cell | *KIAA1468* | -2.203692761 | 0.012422555 | 0.016926876 | 1 |
| T2DM | dendritic_cell | *FBXO18* | -2.262372705 | 3.39E-05 | 0.000150765 | 0.213441916 |
| T2DM | dendritic_cell | *ECHDC3* | -1.048813334 | 1.50E-05 | 0.000130129 | 0.094533476 |
| T2DM | dendritic_cell | *IL15RA* | -3.059091955 | 4.61E-05 | 0.00017047 | 0.290124641 |
| T2DM | dendritic_cell | *TMEM241* | -2.182541207 | 3.16E-05 | 0.000147838 | 0.198694563 |
| T2DM | dendritic_cell | *EMP1* | -1.550450275 | 2.21E-05 | 0.000132957 | 0.13885564 |
| T2DM | dendritic_cell | *KLRD1* | -1.712551975 | 2.60E-05 | 0.0001395 | 0.163456684 |
| T2DM | dendritic_cell | *DDB2* | -2.05229801 | 0.002431702 | 0.003848002 | 1 |
| T2DM | dendritic_cell | *ACP2* | 2.071857001 | 0.039425501 | 0.049822738 | 1 |
| T2DM | dendritic_cell | *HOOK1* | -3.133637586 | 4.76E-05 | 0.000172368 | 0.299075816 |
| T2DM | dendritic_cell | *ZCCHC11* | -1.917277883 | 2.61E-05 | 0.0001395 | 0.164028378 |
| T2DM | dendritic_cell | *PRPF38A* | -2.939200834 | 2.22E-05 | 0.000133234 | 0.139362332 |
| T2DM | dendritic_cell | *FHOD3* | -0.997186618 | 7.59E-06 | 0.000130129 | 0.047719721 |
| T2DM | dendritic_cell | *TPGS2* | -2.152576095 | 0.01786522 | 0.023761083 | 1 |
| T2DM | dendritic_cell | *SLC43A3* | -3.671762765 | 0.000121721 | 0.000314533 | 0.765259424 |
| T2DM | dendritic_cell | *APLNR* | -2.033932151 | 8.25E-05 | 0.000241877 | 0.518825704 |
| T2DM | dendritic_cell | *FADS2* | -2.835454001 | 0.000109756 | 0.000291319 | 0.690035001 |
| T2DM | dendritic_cell | *TMEM165* | -1.731448882 | 0.004192693 | 0.006273075 | 1 |
| T2DM | dendritic_cell | *CLOCK* | -2.414533049 | 5.93E-05 | 0.000196287 | 0.372944942 |
| T2DM | dendritic_cell | *DZIP1* | -1.176884319 | 6.41E-05 | 0.000204385 | 0.402843251 |
| T2DM | dendritic_cell | *BIVM* | -1.927075457 | 1.47E-05 | 0.000130129 | 0.092228514 |
| T2DM | dendritic_cell | *CARS2* | -2.083548559 | 6.62E-06 | 0.000130129 | 0.041602881 |
| T2DM | dendritic_cell | *KLB* | -0.733512311 | 0.000219727 | 0.000486246 | 1 |
| T2DM | dendritic_cell | *WDR36* | -3.116203811 | 0.001913883 | 0.003099583 | 1 |
| T2DM | dendritic_cell | *OSTF1* | -2.472964244 | 0.025255517 | 0.032799305 | 1 |
| T2DM | dendritic_cell | *UBQLN1* | -2.509253097 | 6.68E-05 | 0.00021013 | 0.42026003 |
| T2DM | dendritic_cell | *NAA35* | -2.624651906 | 6.75E-05 | 0.000211759 | 0.424365302 |
| T2DM | dendritic_cell | *CTSL* | -2.57106441 | 7.13E-05 | 0.000218327 | 0.448370766 |
| T2DM | dendritic_cell | *TMEM2* | -1.754930392 | 1.72E-05 | 0.000130129 | 0.107944638 |
| T2DM | dendritic_cell | *GOLM1* | -2.352246006 | 1.76E-05 | 0.000130129 | 0.110947971 |
| T2DM | dendritic_cell | *ADAM19* | 0.867576336 | 0.000637876 | 0.001170556 | 1 |
| T2DM | dendritic_cell | *USP30* | -1.361344891 | 0.003494027 | 0.005315013 | 1 |
| T2DM | dendritic_cell | *HRK* | -4.864823082 | 0.000378957 | 0.000756591 | 1 |
| T2DM | dendritic_cell | *P2RX4* | 2.27576138 | 5.18E-06 | 0.000124852 | 0.032569835 |
| T2DM | dendritic_cell | *DTX1* | -0.993624317 | 4.86E-06 | 0.000123437 | 0.030575821 |
| T2DM | dendritic_cell | *DMTF1* | -1.701014554 | 0.003361784 | 0.005133723 | 1 |
| T2DM | dendritic_cell | *CCDC146* | -1.75454451 | 9.76E-05 | 0.000268845 | 0.613683424 |
| T2DM | dendritic_cell | *TMEM60* | -2.290232747 | 0.005945165 | 0.008603379 | 1 |
| T2DM | dendritic_cell | *POM121C* | -3.578037687 | 1.13E-06 | 6.47E-05 | 0.00712059 |
| T2DM | dendritic_cell | *SRPK2* | -2.215734651 | 0.000143814 | 0.00035499 | 0.904159834 |
| T2DM | dendritic_cell | *TES* | -2.175704466 | 0.006798063 | 0.00973785 | 1 |
| T2DM | dendritic_cell | *MDFIC* | -1.692916988 | 1.95E-05 | 0.000131266 | 0.122573279 |
| T2DM | dendritic_cell | *MTO1* | -1.939146074 | 4.61E-05 | 0.00017047 | 0.290048535 |
| T2DM | dendritic_cell | *ANKRD6* | -2.733383289 | 3.50E-05 | 0.0001528 | 0.220337097 |
| T2DM | dendritic_cell | *KHDC1* | -1.000257838 | 0.00345355 | 0.005262353 | 1 |

| | | | | | | |
|---|---|---|---|---|---|---|
| T2DM | dendritic_cell | *SNX14* | -2.755363353 | 0.000733364 | 0.001323761 | 1 |
| T2DM | dendritic_cell | *NT5E* | -1.05642946 | 0.000482686 | 0.000922102 | 1 |
| T2DM | dendritic_cell | *PRR5L* | -1.941234154 | 5.41E-05 | 0.000185738 | 0.340271376 |
| T2DM | dendritic_cell | *NAT10* | -2.311203747 | 9.45E-05 | 0.000264058 | 0.594394772 |
| T2DM | dendritic_cell | *ATP5G2* | -3.294614784 | 0.000163819 | 0.000391609 | 1 |
| T2DM | dendritic_cell | *ITGA7* | -0.878628321 | 9.03E-06 | 0.000130129 | 0.056784239 |
| T2DM | dendritic_cell | *RDH5* | -0.746928079 | 2.64E-05 | 0.000139727 | 0.165855965 |
| T2DM | dendritic_cell | *BLOC1S1* | -2.73108398 | 0.000133469 | 0.000336494 | 0.839119044 |
| T2DM | dendritic_cell | *PPP1R1A* | -1.871039055 | 2.84E-05 | 0.000142583 | 0.178798956 |
| T2DM | dendritic_cell | *B4GALNT1* | 2.555797266 | 0.000937498 | 0.001635872 | 1 |
| T2DM | dendritic_cell | *FAIM2* | -2.701842277 | 1.14E-05 | 0.000130129 | 0.07188983 |
| T2DM | dendritic_cell | *SLC26A10* | -6.240879724 | 0.001089105 | 0.00186674 | 1 |
| T2DM | dendritic_cell | *MAP7* | -1.69062519 | 9.60E-05 | 0.000266479 | 0.603841111 |
| T2DM | dendritic_cell | *AHI1* | -0.749995784 | 0.009328967 | 0.013009053 | 1 |
| T2DM | dendritic_cell | *PKIB* | 1.045256541 | 0.00013255 | 0.000334831 | 0.833338812 |
| T2DM | dendritic_cell | *SMPD2* | 1.389544377 | 0.005458767 | 0.007964555 | 1 |
| T2DM | dendritic_cell | *MICAL1* | 2.819084026 | 6.09E-05 | 0.000198694 | 0.383122082 |
| T2DM | dendritic_cell | *SMYD5* | -3.147158195 | 7.35E-07 | 5.26E-05 | 0.004621501 |
| T2DM | dendritic_cell | *USP15* | -2.063537442 | 0.01160339 | 0.015893358 | 1 |
| T2DM | dendritic_cell | *KLHL36* | -1.899078136 | 9.47E-05 | 0.000264178 | 0.5954561 |
| T2DM | dendritic_cell | *MPHOSPH6* | 2.875741479 | 0.000271482 | 0.000575652 | 1 |
| T2DM | dendritic_cell | *FHOD1* | -3.71175155 | 0.001626286 | 0.002672128 | 1 |
| T2DM | dendritic_cell | *CCDC102A* | -3.038137331 | 6.40E-05 | 0.00020421 | 0.402174543 |
| T2DM | dendritic_cell | *ZNF670* | -1.32128097 | 0.00222641 | 0.003558068 | 1 |
| T2DM | dendritic_cell | *KCNK1* | -0.404419715 | 0.023672065 | 0.03089605 | 1 |
| T2DM | dendritic_cell | *NTPCR* | -2.478460878 | 0.0004241 | 0.00083037 | 1 |
| T2DM | dendritic_cell | *RNASEL* | 2.303472594 | 0.004279497 | 0.006390783 | 1 |
| T2DM | dendritic_cell | *PIGC* | -1.778896851 | 6.78E-05 | 0.000211937 | 0.426205281 |
| T2DM | dendritic_cell | *USP37* | -2.564390736 | 0.003285833 | 0.005030198 | 1 |
| T2DM | dendritic_cell | *SLC19A3* | -0.653865451 | 0.000448002 | 0.000867444 | 1 |
| T2DM | dendritic_cell | *TMBIM1* | -2.972611341 | 1.66E-07 | 2.12E-05 | 0.001041141 |
| T2DM | dendritic_cell | *ARMC9* | -3.163142101 | 6.28E-05 | 0.000202109 | 0.394770889 |
| T2DM | dendritic_cell | *GCC2* | -2.134744605 | 3.08E-05 | 0.000146146 | 0.193814663 |
| T2DM | dendritic_cell | *ANKRD36* | -2.463599328 | 7.67E-05 | 0.000229371 | 0.481908466 |
| T2DM | dendritic_cell | *USP44* | -1.875248587 | 9.15E-05 | 0.000258875 | 0.574962153 |
| T2DM | dendritic_cell | *APPL2* | -2.557389338 | 7.52E-05 | 0.000225984 | 0.472489294 |
| T2DM | dendritic_cell | *DRAM1* | -2.040993412 | 0.004475483 | 0.006648715 | 1 |
| T2DM | dendritic_cell | *SLC41A2* | 1.617657424 | 0.001087291 | 0.00186414 | 1 |
| T2DM | dendritic_cell | *NEK3* | -1.330150719 | 0.002355936 | 0.003741291 | 1 |
| T2DM | dendritic_cell | *VPS36* | -2.050361078 | 7.39E-05 | 0.000223611 | 0.464887953 |
| T2DM | dendritic_cell | *TBC1D4* | -1.657746404 | 1.52E-05 | 0.000130129 | 0.095439407 |
| T2DM | dendritic_cell | *LRCH1* | -2.443696293 | 0.00023346 | 0.000510172 | 1 |
| T2DM | dendritic_cell | *SUCLA2* | -1.591318408 | 0.000138515 | 0.000345162 | 0.870846846 |
| T2DM | dendritic_cell | *RCBTB1* | -2.877908917 | 0.000119145 | 0.000309531 | 0.749064265 |
| T2DM | dendritic_cell | *MED4* | -2.260607252 | 7.33E-06 | 0.000130129 | 0.046080148 |
| T2DM | dendritic_cell | *SPDYE1* | -1.900670983 | 0.000137893 | 0.000343885 | 0.866933128 |
| T2DM | dendritic_cell | *CHST12* | -2.680000934 | 4.84E-05 | 0.000173951 | 0.304185485 |
| T2DM | dendritic_cell | *GPNMB* | -2.092728643 | 3.65E-05 | 0.000155245 | 0.229762574 |
| T2DM | dendritic_cell | *NUPL2* | -2.989547761 | 9.36E-05 | 0.000262599 | 0.588483778 |
| T2DM | dendritic_cell | *ZDHHC4* | -2.032400775 | 0.000193122 | 0.000441031 | 1 |

| | | | | | | |
|---|---|---|---|---|---|---|
| T2DM | dendritic_cell | *AOAH* | 0.52128504 | 0.000907881 | 0.001592592 | 1 |
| T2DM | dendritic_cell | *TBRG4* | -4.247338117 | 0.000326084 | 0.000667565 | 1 |
| T2DM | dendritic_cell | *DDX56* | -3.718588791 | 0.019904325 | 0.02631171 | 1 |
| T2DM | dendritic_cell | *HUS1* | -2.16635677 | 0.020606069 | 0.027142332 | 1 |
| T2DM | dendritic_cell | *NACAD* | -6.332362012 | 9.57E-08 | 1.56E-05 | 0.000601546 |
| T2DM | dendritic_cell | *DBNL* | -2.631308031 | 0.008044171 | 0.011364877 | 1 |
| T2DM | dendritic_cell | *MTHFS* | -0.725847395 | 0.00868491 | 0.012190674 | 1 |
| T2DM | dendritic_cell | *TM6SF1* | 2.151910194 | 7.40E-05 | 0.00022367 | 0.465233048 |
| T2DM | dendritic_cell | *RSAD1* | -3.539342397 | 4.07E-05 | 0.00016095 | 0.255602516 |
| T2DM | dendritic_cell | *RTP4* | -2.711448939 | 1.29E-05 | 0.000130129 | 0.081374081 |
| T2DM | dendritic_cell | *MRPL47* | -4.068271797 | 0.002332794 | 0.003710164 | 1 |
| T2DM | dendritic_cell | *BLK* | -3.136222239 | 3.95E-07 | 3.40E-05 | 0.002483714 |
| T2DM | dendritic_cell | *EPRS* | -3.234131526 | 0.000289845 | 0.00060661 | 1 |
| T2DM | dendritic_cell | *RPS6KC1* | -2.359905537 | 0.00628955 | 0.009046535 | 1 |
| T2DM | dendritic_cell | *CBWD2* | -3.533805613 | 0.000367876 | 0.000737983 | 1 |
| T2DM | dendritic_cell | *IL1RN* | -4.731531007 | 0.000217434 | 0.00048236 | 1 |
| T2DM | dendritic_cell | *SAP130* | -2.523527331 | 9.56E-06 | 0.000130129 | 0.060087929 |
| T2DM | dendritic_cell | *UGGT1* | -2.502558702 | 2.58E-05 | 0.0001395 | 0.162146348 |
| T2DM | dendritic_cell | *GYPC* | -1.74523105 | 6.33E-05 | 0.000203099 | 0.398277601 |
| T2DM | dendritic_cell | *NIPSNAP3A* | -3.475355367 | 0.000272446 | 0.000577113 | 1 |
| T2DM | dendritic_cell | *LRRC8A* | -2.242474117 | 0.000109471 | 0.000290796 | 0.688244538 |
| T2DM | dendritic_cell | *TXN* | 2.074012308 | 0.010433468 | 0.014426042 | 1 |
| T2DM | dendritic_cell | *ODF2* | -3.689837436 | 0.000191566 | 0.000438272 | 1 |
| T2DM | dendritic_cell | *TOR1B* | -1.811245187 | 0.000258859 | 0.00055404 | 1 |
| T2DM | dendritic_cell | *TOR1A* | -2.412000933 | 4.11E-06 | 0.000114478 | 0.025835534 |
| T2DM | dendritic_cell | *SLC2A8* | -2.296082796 | 0.000656248 | 0.00119937 | 1 |
| T2DM | dendritic_cell | *ZFP37* | -1.461585935 | 0.000246924 | 0.000533475 | 1 |
| T2DM | dendritic_cell | *ZNF189* | -2.260283194 | 3.09E-05 | 0.000146243 | 0.194356852 |
| T2DM | dendritic_cell | *ALDOB* | -2.063057612 | 0.000157392 | 0.00038002 | 0.989526007 |
| T2DM | dendritic_cell | *STX17* | -2.390756038 | 2.05E-05 | 0.000132198 | 0.129090772 |
| T2DM | dendritic_cell | *FPGS* | -2.226881351 | 8.07E-06 | 0.000130129 | 0.050706946 |
| T2DM | dendritic_cell | *TEX10* | -2.8042273 | 3.68E-05 | 0.000155453 | 0.231469507 |
| T2DM | dendritic_cell | *GARNL3* | -2.662879095 | 4.03E-05 | 0.000160482 | 0.253560771 |
| T2DM | dendritic_cell | *GABBR2* | 1.428494544 | 4.12E-05 | 0.000162249 | 0.25927341 |
| T2DM | dendritic_cell | *PSMB7* | -2.801154784 | 0.000107433 | 0.000286804 | 0.675429422 |
| T2DM | dendritic_cell | *RABEPK* | -2.99844839 | 3.23E-05 | 0.000148254 | 0.202971178 |
| T2DM | dendritic_cell | *GOLGA1* | -2.297326518 | 2.54E-05 | 0.000139104 | 0.159969425 |
| T2DM | dendritic_cell | *LMX1B* | -3.259036692 | 1.25E-05 | 0.000130129 | 0.078791147 |
| T2DM | dendritic_cell | *DERL1* | -3.798965723 | 0.001852089 | 0.003011141 | 1 |
| T2DM | dendritic_cell | *NOV* | -1.326467428 | 5.56E-05 | 0.000188762 | 0.349703258 |
| T2DM | dendritic_cell | *POLR1E* | -1.010247379 | 0.020578223 | 0.027128389 | 1 |
| T2DM | dendritic_cell | *IL11RA* | -2.346059864 | 0.000551749 | 0.001035476 | 1 |
| T2DM | dendritic_cell | *APTX* | -2.306384009 | 0.000120301 | 0.000311632 | 0.756331388 |
| T2DM | dendritic_cell | *TLN1* | -2.163616233 | 0.001207993 | 0.002044871 | 1 |
| T2DM | dendritic_cell | *SPAG8* | -1.992549076 | 0.000391747 | 0.000777435 | 1 |
| T2DM | dendritic_cell | *CD72* | -3.031926091 | 0.000159205 | 0.000383074 | 1 |
| T2DM | dendritic_cell | *TMEM8B* | -4.083490081 | 0.006982825 | 0.009984313 | 1 |
| T2DM | dendritic_cell | *GRHPR* | -2.198955276 | 0.00325397 | 0.004984822 | 1 |
| T2DM | dendritic_cell | *ARHGEF39* | -1.62415624 | 0.000235951 | 0.000514721 | 1 |
| T2DM | dendritic_cell | *DENND4C* | -2.057131886 | 0.00015188 | 0.000369904 | 0.954871354 |

| | | | | | | |
|---|---|---|---|---|---|---|
| T2DM | dendritic_cell | *PPIL1* | 3.083241152 | 0.00014278 | 0.000353408 | 0.897656133 |
| T2DM | dendritic_cell | *KIF13A* | -1.701625306 | 2.51E-05 | 0.000138674 | 0.158064126 |
| T2DM | dendritic_cell | *FRS3* | 2.555206038 | 0.000213112 | 0.000474614 | 1 |
| T2DM | dendritic_cell | *SLC22A23* | -2.436225798 | 4.89E-05 | 0.00017503 | 0.307178511 |
| T2DM | dendritic_cell | *LRRC1* | -2.458931476 | 0.000236464 | 0.000515661 | 1 |
| T2DM | dendritic_cell | *FOXF2* | -2.426162167 | 0.019105318 | 0.025340745 | 1 |
| T2DM | dendritic_cell | *BPHL* | -0.76170962 | 0.000644486 | 0.001180252 | 1 |
| T2DM | dendritic_cell | *RIPK1* | -2.178095675 | 2.39E-05 | 0.000136538 | 0.150412276 |
| T2DM | dendritic_cell | *TUBB2B* | -1.076824331 | 0.025062874 | 0.032576036 | 1 |
| T2DM | dendritic_cell | *UQCC2* | -3.072292492 | 0.03604183 | 0.04582305 | 1 |
| T2DM | dendritic_cell | *TPMT* | 1.321121514 | 8.55E-05 | 0.000248016 | 0.537820065 |
| T2DM | dendritic_cell | *NRM* | 1.426127086 | 0.000409699 | 0.000805938 | 1 |
| T2DM | dendritic_cell | *FAM8A1* | -1.676738626 | 1.50E-05 | 0.000130129 | 0.094243247 |
| T2DM | dendritic_cell | *FGFBP2* | -1.612694709 | 3.82E-05 | 0.000157656 | 0.240137508 |
| T2DM | dendritic_cell | *FHDC1* | -5.85790367 | 0.001903542 | 0.003086015 | 1 |
| T2DM | dendritic_cell | *CCDC90B* | -2.63145903 | 3.05E-05 | 0.000145705 | 0.191492644 |
| T2DM | dendritic_cell | *SYTL2* | -2.068461828 | 8.37E-07 | 5.43E-05 | 0.005262485 |
| T2DM | dendritic_cell | *RAB30* | -1.954177358 | 1.74E-05 | 0.000130129 | 0.109649406 |
| T2DM | dendritic_cell | *PRCP* | 2.465526734 | 0.001631911 | 0.002679506 | 1 |
| T2DM | dendritic_cell | *NARS2* | -2.057621345 | 2.13E-05 | 0.000132591 | 0.133615653 |
| T2DM | dendritic_cell | *PI15* | -10.94016903 | 7.22E-05 | 0.000219971 | 0.453879054 |
| T2DM | dendritic_cell | *TTPA* | -0.692259509 | 0.000370686 | 0.000741726 | 1 |
| T2DM | dendritic_cell | *GGH* | -0.894161502 | 0.000476045 | 0.000911911 | 1 |
| T2DM | dendritic_cell | *SULF1* | -1.017581564 | 4.56E-05 | 0.000169574 | 0.286410148 |
| T2DM | dendritic_cell | *TGS1* | -1.682096228 | 0.000305758 | 0.000634422 | 1 |
| T2DM | dendritic_cell | *NEK1* | -2.124079445 | 0.000217397 | 0.00048236 | 1 |
| T2DM | dendritic_cell | *DDX60* | -3.405868052 | 0.000181058 | 0.000421377 | 1 |
| T2DM | dendritic_cell | *DCUN1D5* | -2.266806479 | 1.98E-07 | 2.17E-05 | 0.001246556 |
| T2DM | dendritic_cell | *TRIM29* | -1.75343063 | 5.72E-05 | 0.000191518 | 0.359478447 |
| T2DM | dendritic_cell | *SLC37A4* | -3.165098045 | 1.51E-05 | 0.000130129 | 0.095191086 |
| T2DM | dendritic_cell | *POU2F3* | -1.314859264 | 0.013633613 | 0.018468978 | 1 |
| T2DM | dendritic_cell | *PPP2R1B* | -0.957006562 | 1.43E-05 | 0.000130129 | 0.089881605 |
| T2DM | dendritic_cell | *FXYD6* | -2.682892393 | 7.95E-05 | 0.00023544 | 0.500108586 |
| T2DM | dendritic_cell | *TMPRSS13* | 0.67937278 | 2.47E-05 | 0.000137939 | 0.155043821 |
| T2DM | dendritic_cell | *CASP1* | 3.065103986 | 0.004078578 | 0.006119814 | 1 |
| T2DM | dendritic_cell | *MAP2K5* | -2.075612897 | 0.000172463 | 0.00040701 | 1 |
| T2DM | dendritic_cell | *NDUFAF1* | -3.245727405 | 4.23E-07 | 3.57E-05 | 0.002656491 |
| T2DM | dendritic_cell | *ITGA11* | -0.785773711 | 9.85E-06 | 0.000130129 | 0.06194791 |
| T2DM | dendritic_cell | *RMDN3* | -1.528612859 | 0.000260028 | 0.000556053 | 1 |
| T2DM | dendritic_cell | *ITPKA* | -7.46190411 | 0.002227626 | 0.003559106 | 1 |
| T2DM | dendritic_cell | *UACA* | -1.852315143 | 2.93E-05 | 0.000143842 | 0.18405832 |
| T2DM | dendritic_cell | *PLCB2* | 0.696016729 | 0.000124187 | 0.000319199 | 0.780761926 |
| T2DM | dendritic_cell | *SPTBN5* | 1.843618417 | 5.91E-06 | 0.000129745 | 0.03712732 |
| T2DM | dendritic_cell | *SLC44A5* | -2.145668617 | 0.000124903 | 0.000320386 | 0.785265729 |
| T2DM | dendritic_cell | *DNASE2B* | 3.330432217 | 3.42E-05 | 0.000151156 | 0.215264551 |
| T2DM | dendritic_cell | *DBT* | -2.225508708 | 0.000113532 | 0.000297904 | 0.713778236 |
| T2DM | dendritic_cell | *IFT172* | -2.201226196 | 0.003068214 | 0.004733708 | 1 |
| T2DM | dendritic_cell | *ADCY3* | -2.53286659 | 1.04E-05 | 0.000130129 | 0.065083801 |
| T2DM | dendritic_cell | *DYNC2LI1* | -1.589421503 | 1.18E-05 | 0.000130129 | 0.073972159 |
| T2DM | dendritic_cell | *LHCGR* | -1.285366191 | 1.07E-05 | 0.000130129 | 0.067255958 |

| | | | | | | |
|---|---|---|---|---|---|---|
| T2DM | dendritic_cell | *THUMPD2* | -2.562359494 | 3.32E-05 | 0.000149182 | 0.209004674 |
| T2DM | dendritic_cell | *PREB* | -1.572990057 | 2.20E-05 | 0.000132957 | 0.138582715 |
| T2DM | dendritic_cell | *CENPO* | 3.702366601 | 0.009792721 | 0.013602925 | 1 |
| T2DM | dendritic_cell | *LRPPRC* | -2.626317966 | 7.94E-05 | 0.000235099 | 0.498880686 |
| T2DM | dendritic_cell | *DTNB* | -1.865945385 | 0.006848449 | 0.009801093 | 1 |
| T2DM | dendritic_cell | *STAMBPL1* | -1.672548729 | 6.13E-05 | 0.000199479 | 0.385394096 |
| T2DM | dendritic_cell | *ATAD1* | -2.341497501 | 2.54E-05 | 0.000138965 | 0.159671176 |
| T2DM | dendritic_cell | *BTBD16* | -1.668261779 | 1.26E-06 | 6.80E-05 | 0.007946719 |
| T2DM | dendritic_cell | *TACC2* | -1.472596827 | 0.000230775 | 0.00050571 | 1 |
| T2DM | dendritic_cell | *CALHM2* | -2.21152418 | 0.008776811 | 0.012297707 | 1 |
| T2DM | dendritic_cell | *ARL3* | -2.404496598 | 0.000970419 | 0.001686763 | 1 |
| T2DM | dendritic_cell | *PLCE1* | -2.402426289 | 6.77E-06 | 0.000130129 | 0.042555799 |
| T2DM | dendritic_cell | *RBP4* | -2.258294826 | 5.77E-05 | 0.000192649 | 0.362757548 |
| T2DM | dendritic_cell | *DNAJC13* | -2.842169613 | 0.001160133 | 0.001972351 | 1 |
| T2DM | dendritic_cell | *FAM149B1* | -2.305698338 | 2.54E-05 | 0.000138965 | 0.159662894 |
| T2DM | dendritic_cell | *ASCC1* | -1.990113951 | 1.53E-05 | 0.000130129 | 0.096388083 |
| T2DM | dendritic_cell | *DNA2* | -0.68198214 | 0.001186167 | 0.002011174 | 1 |
| T2DM | dendritic_cell | *ATIC* | -1.97210621 | 5.05E-06 | 0.000124087 | 0.031766323 |
| T2DM | dendritic_cell | *SMARCAL1* | -1.94334093 | 1.56E-05 | 0.000130129 | 0.098380761 |
| T2DM | dendritic_cell | *STAT4* | -1.076548489 | 4.11E-06 | 0.000114478 | 0.025844662 |
| T2DM | dendritic_cell | *ASNSD1* | -2.984838831 | 2.61E-05 | 0.0001395 | 0.163812164 |
| T2DM | dendritic_cell | *METTL5* | -3.954568431 | 0.000751365 | 0.001353534 | 1 |
| T2DM | dendritic_cell | *NAB1* | -2.958677521 | 6.77E-05 | 0.000211932 | 0.425936134 |
| T2DM | dendritic_cell | *FASTKD1* | -2.407162478 | 0.010875869 | 0.014977602 | 1 |
| T2DM | dendritic_cell | *MDH1B* | -3.743523866 | 1.21E-05 | 0.000130129 | 0.076158149 |
| T2DM | dendritic_cell | *FAM117B* | 1.930154016 | 0.002540156 | 0.003998488 | 1 |
| T2DM | dendritic_cell | *ITGAV* | -2.013433941 | 0.002746611 | 0.004286977 | 1 |
| T2DM | dendritic_cell | *SENP7* | -2.173670162 | 7.18E-05 | 0.000219346 | 0.451340125 |
| T2DM | dendritic_cell | *USP8* | -2.289020001 | 0.000824118 | 0.00146528 | 1 |
| T2DM | dendritic_cell | *SPPL2A* | -2.935082224 | 0.000105019 | 0.000282159 | 0.660252808 |
| T2DM | dendritic_cell | *PPCDC* | -1.425288148 | 0.003179133 | 0.004887852 | 1 |
| T2DM | dendritic_cell | *UBL7* | 1.988812346 | 9.19E-05 | 0.000259612 | 0.577635812 |
| T2DM | dendritic_cell | *ARHGAP24* | -1.686395574 | 3.38E-05 | 0.000150628 | 0.212385118 |
| T2DM | dendritic_cell | *FAM13A* | -1.469830139 | 0.000366604 | 0.000735901 | 1 |
| T2DM | dendritic_cell | *HERC3* | -2.782629793 | 0.000393163 | 0.000779507 | 1 |
| T2DM | dendritic_cell | *AP1AR* | -3.117502744 | 4.36E-05 | 0.000166069 | 0.274345688 |
| T2DM | dendritic_cell | *PRKG2* | -1.332905976 | 1.08E-05 | 0.000130129 | 0.068199455 |
| T2DM | dendritic_cell | *BBS7* | -3.422787297 | 0.000100638 | 0.000273902 | 0.63271281 |
| T2DM | dendritic_cell | *KIAA1109* | -2.223153959 | 9.18E-05 | 0.000259486 | 0.577096166 |
| T2DM | dendritic_cell | *MMRN1* | -0.77766981 | 6.78E-06 | 0.000130129 | 0.042622456 |
| T2DM | dendritic_cell | *PRDM5* | -1.747265391 | 8.79E-05 | 0.000252457 | 0.552629122 |
| T2DM | dendritic_cell | *TRPC3* | -3.38712504 | 0.000432867 | 0.000843595 | 1 |
| T2DM | dendritic_cell | *NAAA* | 2.115475573 | 0.004198774 | 0.006280679 | 1 |
| T2DM | dendritic_cell | *NUP54* | -1.460956233 | 0.000290271 | 0.000607299 | 1 |
| T2DM | dendritic_cell | *G3BP2* | -3.634177595 | 5.55E-05 | 0.000188762 | 0.349073776 |
| T2DM | dendritic_cell | *SCARB2* | -2.36952862 | 3.67E-05 | 0.000155353 | 0.230753042 |
| T2DM | dendritic_cell | *CNOT6L* | -2.489358183 | 0.000143427 | 0.000354537 | 0.901725242 |
| T2DM | dendritic_cell | *ANXA3* | -1.26095651 | 2.17E-05 | 0.000132663 | 0.136133301 |
| T2DM | dendritic_cell | *GSTCD* | -2.594671411 | 0.00275736 | 0.004301618 | 1 |
| T2DM | dendritic_cell | *ENPEP* | -1.342017893 | 1.04E-05 | 0.000130129 | 0.065145175 |

| | | | | | | |
|---|---|---|---|---|---|---|
| T2DM | dendritic_cell | *PAPSS1* | -1.627538192 | 0.003297486 | 0.005045338 | 1 |
| T2DM | dendritic_cell | *FBN2* | 0.460430885 | 0.010889005 | 0.014989965 | 1 |
| T2DM | dendritic_cell | *RNF185* | -2.353407268 | 1.09E-05 | 0.000130129 | 0.068298818 |
| T2DM | dendritic_cell | *PARVG* | 0.698405349 | 7.97E-05 | 0.000235555 | 0.500789845 |
| T2DM | dendritic_cell | *B4GALNT3* | -2.172429377 | 6.65E-06 | 0.000130129 | 0.041800086 |
| T2DM | dendritic_cell | *ERP27* | 1.569639092 | 0.000215291 | 0.000478281 | 1 |
| T2DM | dendritic_cell | *ETV6* | 1.274347293 | 0.000638719 | 0.001171419 | 1 |
| T2DM | dendritic_cell | *KIF21A* | -0.916639309 | 0.000462665 | 0.000890833 | 1 |
| T2DM | dendritic_cell | *CPNE8* | -1.488998574 | 0.030422019 | 0.039097146 | 1 |
| T2DM | dendritic_cell | *ALG10* | -6.043295581 | 0.00409206 | 0.006137114 | 1 |
| T2DM | dendritic_cell | *PRICKLE1* | -0.956403863 | 4.47E-05 | 0.00016845 | 0.280974379 |
| T2DM | dendritic_cell | *C1RL* | -1.262452112 | 0.002337944 | 0.003715535 | 1 |
| T2DM | dendritic_cell | *CLSTN3* | -1.650241802 | 0.000227107 | 0.000499065 | 1 |
| T2DM | dendritic_cell | *KLRG1* | -1.783089038 | 0.000196549 | 0.000446908 | 1 |
| T2DM | dendritic_cell | *TAPBPL* | -1.617215278 | 0.000415462 | 0.000815488 | 1 |
| T2DM | dendritic_cell | *CD27* | -1.916484257 | 2.60E-05 | 0.0001395 | 0.163322929 |
| T2DM | dendritic_cell | *PEX5* | -1.763607531 | 2.42E-05 | 0.000136859 | 0.151876179 |
| T2DM | dendritic_cell | *PIANP* | 4.644530729 | 0.000368995 | 0.000739755 | 1 |
| T2DM | dendritic_cell | *PPFIA2* | -1.241261738 | 0.000208905 | 0.000468087 | 1 |
| T2DM | dendritic_cell | *GLIPR1* | 1.022296286 | 0.000143967 | 0.000355088 | 0.90511983 |
| T2DM | dendritic_cell | *POC1B* | -3.281333089 | 8.33E-05 | 0.000243529 | 0.523830489 |
| T2DM | dendritic_cell | *SNRPF* | -2.722093661 | 0.000239707 | 0.000520925 | 1 |
| T2DM | dendritic_cell | *GAS2L3* | 2.303141944 | 0.036734063 | 0.046674829 | 1 |
| T2DM | dendritic_cell | *TMEM132B* | -1.578938366 | 5.66E-05 | 0.000190501 | 0.355740738 |
| T2DM | dendritic_cell | *TDG* | -2.84781463 | 0.000432062 | 0.000843165 | 1 |
| T2DM | dendritic_cell | *MMAB* | -3.065878942 | 1.60E-05 | 0.000130129 | 0.100334444 |
| T2DM | dendritic_cell | *GLTP* | 2.326242812 | 0.036926369 | 0.046890746 | 1 |
| T2DM | dendritic_cell | *MTMR6* | -2.23749777 | 0.020657818 | 0.027204797 | 1 |
| T2DM | dendritic_cell | *CCDC65* | -2.407969105 | 0.004419497 | 0.006574865 | 1 |
| T2DM | dendritic_cell | *ACVRL1* | -1.202150457 | 5.19E-05 | 0.000181102 | 0.325984194 |
| T2DM | dendritic_cell | *GPR84* | 2.87781046 | 0.000412231 | 0.000809652 | 1 |
| T2DM | dendritic_cell | *BRCA2* | 1.217650813 | 0.000115226 | 0.000301342 | 0.724425085 |
| T2DM | dendritic_cell | *KANSL2* | -1.689445113 | 0.003547162 | 0.005391926 | 1 |
| T2DM | dendritic_cell | *CERS5* | -3.166817921 | 6.93E-05 | 0.000214503 | 0.435428393 |
| T2DM | dendritic_cell | *LMBR1L* | -3.789407209 | 9.66E-05 | 0.000267513 | 0.607254735 |
| T2DM | dendritic_cell | *ZNF740* | -2.280725857 | 0.000189184 | 0.000434245 | 1 |
| T2DM | dendritic_cell | *HNRNPA1L2* | 2.367213222 | 0.000171723 | 0.000406332 | 1 |
| T2DM | dendritic_cell | *LPAR6* | -2.374200996 | 0.019386288 | 0.025691456 | 1 |
| T2DM | dendritic_cell | *RHOF* | -1.469132058 | 3.74E-06 | 0.000110889 | 0.023528997 |
| T2DM | dendritic_cell | *METTL21C* | -4.343662337 | 0.03034628 | 0.039007782 | 1 |
| T2DM | dendritic_cell | *GRTP1* | -0.968760343 | 0.001765108 | 0.002880154 | 1 |
| T2DM | dendritic_cell | *TSSK4* | -5.261681434 | 0.000206354 | 0.000464046 | 1 |
| T2DM | dendritic_cell | *TMX1* | -2.770768503 | 1.82E-05 | 0.000130129 | 0.114718676 |
| T2DM | dendritic_cell | *RTN1* | 0.429813719 | 4.35E-05 | 0.000165783 | 0.273334704 |
| T2DM | dendritic_cell | *C14orf37* | -2.012204284 | 1.94E-05 | 0.000131266 | 0.122204826 |
| T2DM | dendritic_cell | *RAB15* | -2.114223318 | 0.000291685 | 0.000609649 | 1 |
| T2DM | dendritic_cell | *STON2* | -3.07637858 | 0.000292718 | 0.000611199 | 1 |
| T2DM | dendritic_cell | *AK7* | -2.077936832 | 2.00E-05 | 0.000131266 | 0.126008155 |
| T2DM | dendritic_cell | *SLC24A4* | 0.326842172 | 5.04E-05 | 0.000178067 | 0.316567585 |
| T2DM | dendritic_cell | *FBLN5* | -1.296594481 | 2.11E-05 | 0.000132591 | 0.132474211 |

| | | | | | | |
|---|---|---|---|---|---|---|
| T2DM | dendritic_cell | *C14orf79* | 3.659930717 | 0.00335376 | 0.005122713 | 1 |
| T2DM | dendritic_cell | *NIPA2* | -3.743673595 | 0.000291276 | 0.000609092 | 1 |
| T2DM | dendritic_cell | *DUOXA1* | -1.626542299 | 0.001865569 | 0.003030707 | 1 |
| T2DM | dendritic_cell | *MFAP1* | -2.989464107 | 0.000111544 | 0.000294532 | 0.701279787 |
| T2DM | dendritic_cell | *TCF12* | -2.106683123 | 0.000192899 | 0.000440682 | 1 |
| T2DM | dendritic_cell | *SORD* | 2.960583503 | 0.001388323 | 0.002319529 | 1 |
| T2DM | dendritic_cell | *BAHD1* | -3.770577954 | 0.000188925 | 0.000434015 | 1 |
| T2DM | dendritic_cell | *ANP32A* | -2.870591663 | 3.86E-05 | 0.000158005 | 0.242465747 |
| T2DM | dendritic_cell | *COMMD4* | -5.173708177 | 2.15E-05 | 0.000132663 | 0.135447676 |
| T2DM | dendritic_cell | *UBE2Q2* | -2.83326836 | 0.000201901 | 0.000456601 | 1 |
| T2DM | dendritic_cell | *PSTPIP1* | 1.536373014 | 9.75E-05 | 0.000268845 | 0.612995065 |
| T2DM | dendritic_cell | *HMG20A* | -1.626635777 | 0.029342126 | 0.037763347 | 1 |
| T2DM | dendritic_cell | *SCAPER* | -1.892701173 | 0.000136653 | 0.000341761 | 0.859137216 |
| T2DM | dendritic_cell | *NEIL1* | -2.260664992 | 1.29E-06 | 6.80E-05 | 0.008092565 |
| T2DM | dendritic_cell | *MAN2C1* | -1.184629124 | 2.02E-07 | 2.17E-05 | 0.001271197 |
| T2DM | dendritic_cell | *TPM1* | -1.731714404 | 2.31E-05 | 0.00013467 | 0.145028457 |
| T2DM | dendritic_cell | *IGF1R* | -1.87585535 | 3.94E-05 | 0.000159553 | 0.247823598 |
| T2DM | dendritic_cell | *CYP11A1* | -1.026318359 | 0.002649152 | 0.004152386 | 1 |
| T2DM | dendritic_cell | *BBS4* | -2.220791642 | 0.000135273 | 0.000339235 | 0.850463226 |
| T2DM | dendritic_cell | *ADAMTS17* | -1.281678434 | 9.76E-06 | 0.000130129 | 0.061339698 |
| T2DM | dendritic_cell | *ULK3* | -3.546763185 | 2.00E-05 | 0.000131266 | 0.125908768 |
| T2DM | dendritic_cell | *SCAMP2* | -3.496541988 | 0.000447688 | 0.00086738 | 1 |
| T2DM | dendritic_cell | *HAPLN3* | -2.925253071 | 6.51E-05 | 0.000206523 | 0.409327623 |
| T2DM | dendritic_cell | *POLG* | -3.682122774 | 9.67E-05 | 0.000267581 | 0.607677017 |
| T2DM | dendritic_cell | *WDR93* | -0.979065801 | 6.87E-05 | 0.000213479 | 0.431867434 |
| T2DM | dendritic_cell | *MFGE8* | -2.685707581 | 3.01E-05 | 0.000145251 | 0.18955253 |
| T2DM | dendritic_cell | *ST8SIA2* | -8.230533708 | 1.99E-05 | 0.000131266 | 0.12501241 |
| T2DM | dendritic_cell | *IQGAP1* | -1.618283221 | 0.017782859 | 0.023656545 | 1 |
| T2DM | dendritic_cell | *CRTC3* | -2.696801375 | 0.00031945 | 0.000657408 | 1 |
| T2DM | dendritic_cell | *SH3GL3* | -1.870831334 | 0.000181407 | 0.000421784 | 1 |
| T2DM | dendritic_cell | *SEC11A* | -2.753985503 | 5.98E-05 | 0.000197204 | 0.376067492 |
| T2DM | dendritic_cell | *PARN* | -1.950546546 | 8.00E-05 | 0.00023621 | 0.502890884 |
| T2DM | dendritic_cell | *FTO* | -1.926805009 | 2.40E-05 | 0.000136783 | 0.151145758 |
| T2DM | dendritic_cell | *TXNL4B* | -3.356083607 | 1.05E-05 | 0.000130129 | 0.066162919 |
| T2DM | dendritic_cell | *MARVELD3* | -2.616599944 | 0.000153516 | 0.000372935 | 0.965156235 |
| T2DM | dendritic_cell | *KIFC3* | -2.10767426 | 1.73E-05 | 0.000130129 | 0.108706666 |
| T2DM | dendritic_cell | *ADAMTS18* | -1.277325541 | 4.83E-05 | 0.000173951 | 0.303920584 |
| T2DM | dendritic_cell | *NUDT7* | -1.601595798 | 2.07E-06 | 8.62E-05 | 0.013001057 |
| T2DM | dendritic_cell | *GCSH* | -1.201317678 | 0.000839735 | 0.001490938 | 1 |
| T2DM | dendritic_cell | *MAP1LC3B* | -2.589887908 | 0.006150551 | 0.008864859 | 1 |
| T2DM | dendritic_cell | *TLDC1* | -3.509345357 | 6.24E-05 | 0.000201138 | 0.392420322 |
| T2DM | dendritic_cell | *NDUFB10* | -3.040689007 | 1.54E-05 | 0.000130129 | 0.097033539 |
| T2DM | dendritic_cell | *TIGD7* | -1.200068992 | 0.008584099 | 0.01205994 | 1 |
| T2DM | dendritic_cell | *DEF8* | 1.201214436 | 0.025795697 | 0.033441175 | 1 |
| T2DM | dendritic_cell | *GALNS* | -3.420581403 | 1.37E-05 | 0.000130129 | 0.086150925 |
| T2DM | dendritic_cell | *RANBP10* | -2.898774214 | 0.000413858 | 0.000812593 | 1 |
| T2DM | dendritic_cell | *PRPSAP2* | -2.23776889 | 0.000274267 | 0.000580383 | 1 |
| T2DM | dendritic_cell | *DDX52* | -2.659386248 | 9.03E-05 | 0.000256854 | 0.56790431 |
| T2DM | dendritic_cell | *RASL10B* | -3.62233396 | 2.70E-05 | 0.000140817 | 0.169684085 |
| T2DM | dendritic_cell | *UNC45B* | 3.723094877 | 0.001074324 | 0.001842913 | 1 |

| | | | | | | |
|---|---|---|---|---|---|---|
| T2DM | dendritic_cell | *TOM1L1* | -1.889800268 | 4.30E-05 | 0.000165019 | 0.270301267 |
| T2DM | dendritic_cell | *VPS53* | -3.130759571 | 1.38E-06 | 7.13E-05 | 0.008703449 |
| T2DM | dendritic_cell | *SPATA22* | -0.781721639 | 0.004155809 | 0.006223813 | 1 |
| T2DM | dendritic_cell | *SGSM2* | -3.240703013 | 3.19E-05 | 0.000148219 | 0.200746031 |
| T2DM | dendritic_cell | *NPEPPS* | -2.642015444 | 7.95E-05 | 0.000235328 | 0.499601415 |
| T2DM | dendritic_cell | *LRRC46* | -4.056618547 | 8.27E-09 | 2.74E-06 | 5.20E-05 |
| T2DM | dendritic_cell | *RHBDL3* | -2.471882917 | 4.07E-05 | 0.00016095 | 0.255749921 |
| T2DM | dendritic_cell | *ABCA8* | -0.825432009 | 3.32E-05 | 0.000149087 | 0.208448485 |
| T2DM | dendritic_cell | *IMPA2* | 2.559934336 | 0.004453254 | 0.00661882 | 1 |
| T2DM | dendritic_cell | *GNAL* | -2.009113904 | 2.31E-05 | 0.00013467 | 0.145308601 |
| T2DM | dendritic_cell | *RPRD1A* | -2.917945043 | 4.03E-05 | 0.000160482 | 0.253433169 |
| T2DM | dendritic_cell | *C18orf21* | 1.620958422 | 2.54E-05 | 0.000138965 | 0.159606839 |
| T2DM | dendritic_cell | *GALNT1* | -2.821717358 | 0.000207156 | 0.000465078 | 1 |
| T2DM | dendritic_cell | *ASXL3* | -2.039840973 | 3.12E-05 | 0.000146698 | 0.195842297 |
| T2DM | dendritic_cell | *ESCO1* | -3.210940056 | 0.000303651 | 0.000630468 | 1 |
| T2DM | dendritic_cell | *GREB1L* | -0.905166728 | 5.77E-06 | 0.000129602 | 0.03625956 |
| T2DM | dendritic_cell | *C18orf8* | -4.438224144 | 2.21E-06 | 8.70E-05 | 0.013919268 |
| T2DM | dendritic_cell | *NPC1* | -3.063030382 | 1.70E-07 | 2.14E-05 | 0.001071552 |
| T2DM | dendritic_cell | *SLC13A5* | -7.484276639 | 0.000430789 | 0.000841568 | 1 |
| T2DM | dendritic_cell | *CCDC40* | -1.391955039 | 0.001115969 | 0.001905513 | 1 |
| T2DM | dendritic_cell | *EIF4A3* | 1.922297926 | 0.000214505 | 0.000477022 | 1 |
| T2DM | dendritic_cell | *TBCD* | -2.132714284 | 0.000194508 | 0.000443521 | 1 |
| T2DM | dendritic_cell | *FN3KRP* | -1.439403588 | 0.000370208 | 0.000741478 | 1 |
| T2DM | dendritic_cell | *NARF* | -2.71287443 | 3.21E-05 | 0.000148254 | 0.201980977 |
| T2DM | dendritic_cell | *TRIM65* | -2.073287566 | 0.011259156 | 0.015465657 | 1 |
| T2DM | dendritic_cell | *CBX8* | -2.753085177 | 1.44E-05 | 0.000130129 | 0.090651102 |
| T2DM | dendritic_cell | *SECTM1* | 2.144273641 | 5.50E-05 | 0.000187663 | 0.34548698 |
| T2DM | dendritic_cell | *RNF157* | -0.820019083 | 0.002588081 | 0.004065006 | 1 |
| T2DM | dendritic_cell | *RNF165* | -2.095356855 | 2.06E-05 | 0.000132272 | 0.129749038 |
| T2DM | dendritic_cell | *MAPK4* | 1.816275435 | 6.00E-05 | 0.000197604 | 0.377226341 |
| T2DM | dendritic_cell | *SMAD4* | -2.42095429 | 1.51E-05 | 0.000130129 | 0.094723461 |
| T2DM | dendritic_cell | *GRB7* | -2.556926325 | 0.00038654 | 0.000769044 | 1 |
| T2DM | dendritic_cell | *FKBP10* | -1.101761923 | 3.28E-05 | 0.000148653 | 0.206033586 |
| T2DM | dendritic_cell | *NFIC* | -1.876022818 | 4.28E-05 | 0.0001645 | 0.268793543 |
| T2DM | dendritic_cell | *TPGS1* | 2.553016122 | 0.000457838 | 0.000883766 | 1 |
| T2DM | dendritic_cell | *PFKL* | -2.538682525 | 2.53E-05 | 0.000138949 | 0.158957943 |
| T2DM | dendritic_cell | *MVB12A* | -2.184257796 | 0.003447207 | 0.005253961 | 1 |
| T2DM | dendritic_cell | *DPP9* | -2.752996034 | 0.024306646 | 0.031678251 | 1 |
| T2DM | dendritic_cell | *TMEM91* | -3.469002134 | 1.77E-05 | 0.000130129 | 0.111183997 |
| T2DM | dendritic_cell | *SIRT3* | -1.233851824 | 0.009671915 | 0.013444026 | 1 |
| T2DM | dendritic_cell | *IFITM3* | -2.979335923 | 5.23E-05 | 0.00018182 | 0.32854905 |
| T2DM | dendritic_cell | *COL6A1* | -1.238655106 | 3.20E-06 | 0.000102388 | 0.020087949 |
| T2DM | dendritic_cell | *IFNAR1* | -2.439993769 | 4.82E-06 | 0.000123143 | 0.030293218 |
| T2DM | dendritic_cell | *TRPM2* | 1.014347605 | 0.02579763 | 0.033441175 | 1 |
| T2DM | dendritic_cell | *URB1* | -1.336953939 | 0.005934682 | 0.008593124 | 1 |
| T2DM | dendritic_cell | *SAE1* | -1.853404178 | 0.012755054 | 0.017353609 | 1 |
| T2DM | dendritic_cell | *NTN5* | -2.274012411 | 0.000153465 | 0.000372935 | 0.964831863 |
| T2DM | dendritic_cell | *GEMIN7* | -7.061268538 | 1.35E-06 | 7.03E-05 | 0.008502415 |
| T2DM | dendritic_cell | *WTIP* | -3.050469736 | 0.000100039 | 0.000272936 | 0.628946451 |
| T2DM | dendritic_cell | *MYO1F* | 1.276480372 | 9.74E-05 | 0.000268845 | 0.612588902 |

| | | | | | | |
|---|---|---|---|---|---|---|
| T2DM | dendritic_cell | *CACNG8* | -10.07656306 | 0.000149167 | 0.000364623 | 0.937810332 |
| T2DM | dendritic_cell | *FBN3* | 1.579846198 | 0.030496421 | 0.039152747 | 1 |
| T2DM | dendritic_cell | *EVI5L* | -1.316549006 | 1.75E-05 | 0.000130129 | 0.110286613 |
| T2DM | dendritic_cell | *CTU1* | 2.463802034 | 0.000496974 | 0.000944807 | 1 |
| T2DM | dendritic_cell | *RCN3* | 2.250326518 | 0.002375132 | 0.00376621 | 1 |
| T2DM | dendritic_cell | *RERE* | -1.731213281 | 0.000210531 | 0.000470868 | 1 |
| T2DM | dendritic_cell | *ARHGEF19* | -0.563683323 | 0.004770979 | 0.007046076 | 1 |
| T2DM | dendritic_cell | *PEX14* | -2.587691186 | 2.48E-05 | 0.000138038 | 0.15572715 |
| T2DM | dendritic_cell | *SH3BGRL3* | -3.567846364 | 0.003198335 | 0.004911561 | 1 |
| T2DM | dendritic_cell | *IL22RA1* | -2.024942509 | 0.002194465 | 0.003512373 | 1 |
| T2DM | dendritic_cell | *ZNF593* | -2.653645996 | 0.000184148 | 0.000426895 | 1 |
| T2DM | dendritic_cell | *MAP3K6* | -2.728516688 | 2.79E-06 | 9.71E-05 | 0.017567326 |
| T2DM | dendritic_cell | *GPN2* | -4.731398924 | 0.001307499 | 0.002197339 | 1 |
| T2DM | dendritic_cell | *NBPF3* | -1.158017679 | 1.84E-05 | 0.000130129 | 0.115378991 |
| T2DM | dendritic_cell | *HSPG2* | -1.125399325 | 0.000110766 | 0.000293091 | 0.696385273 |
| T2DM | dendritic_cell | *ITGB3BP* | -1.243026926 | 0.019659156 | 0.026012017 | 1 |
| T2DM | dendritic_cell | *SERBP1* | -2.878302902 | 0.008618122 | 0.012099628 | 1 |
| T2DM | dendritic_cell | *PIGK* | -3.111695649 | 7.02E-05 | 0.000216424 | 0.441655714 |
| T2DM | dendritic_cell | *RPS8* | -1.980007479 | 0.024856175 | 0.032334113 | 1 |
| T2DM | dendritic_cell | *MOB3C* | -4.617692427 | 0.003330245 | 0.005090506 | 1 |
| T2DM | dendritic_cell | *CYP4B1* | -0.735408255 | 0.030666882 | 0.039355519 | 1 |
| T2DM | dendritic_cell | *SYPL2* | -0.936719943 | 0.001363563 | 0.002282408 | 1 |
| T2DM | dendritic_cell | *ZNF697* | -2.876542972 | 2.61E-05 | 0.0001395 | 0.164206606 |
| T2DM | dendritic_cell | *CELSR2* | 1.641916793 | 8.55E-05 | 0.000248016 | 0.537759216 |
| T2DM | dendritic_cell | *GPR161* | -4.032969838 | 0.00039062 | 0.000775688 | 1 |
| T2DM | dendritic_cell | *ATP1B1* | 0.975901559 | 0.000140095 | 0.00034822 | 0.880778502 |
| T2DM | dendritic_cell | *TIPRL* | -3.664216943 | 0.000222404 | 0.000490443 | 1 |
| T2DM | dendritic_cell | *NME7* | -2.858367073 | 6.56E-06 | 0.000130129 | 0.041239949 |
| T2DM | dendritic_cell | *CREG1* | -2.294738767 | 0.025710284 | 0.033341698 | 1 |
| T2DM | dendritic_cell | *GPA33* | 1.639190912 | 0.000104076 | 0.000280929 | 0.654324434 |
| T2DM | dendritic_cell | *RXRG* | -3.475866696 | 3.46E-05 | 0.000151535 | 0.217300611 |
| T2DM | dendritic_cell | *TMCO1* | -2.159270324 | 1.34E-05 | 0.000130129 | 0.084003865 |
| T2DM | dendritic_cell | *XCL2* | 1.438243889 | 0.000808058 | 0.001442846 | 1 |
| T2DM | dendritic_cell | *MGST3* | -2.142328346 | 2.16E-05 | 0.000132663 | 0.13553815 |
| T2DM | dendritic_cell | *ADCY10* | -3.050773119 | 5.87E-06 | 0.000129745 | 0.036901661 |
| T2DM | dendritic_cell | *RFWD2* | -2.438389236 | 0.000777372 | 0.001395585 | 1 |
| T2DM | dendritic_cell | *PPOX* | -1.843277415 | 0.001152102 | 0.001960288 | 1 |
| T2DM | dendritic_cell | *RGS5* | -2.459472146 | 0.003031396 | 0.00468035 | 1 |
| T2DM | dendritic_cell | *RRNAD1* | -2.092372391 | 0.012037567 | 0.016427216 | 1 |
| T2DM | dendritic_cell | *PIGM* | -2.277902591 | 4.57E-05 | 0.000169903 | 0.28747552 |
| T2DM | dendritic_cell | *SELENBP1* | -1.419366562 | 4.42E-05 | 0.000167565 | 0.277991156 |
| T2DM | dendritic_cell | *CERS2* | -1.835542212 | 3.32E-05 | 0.000149087 | 0.208572545 |
| T2DM | dendritic_cell | *ENSA* | -2.996382689 | 0.000638131 | 0.001170683 | 1 |
| T2DM | dendritic_cell | *SEMA6C* | -4.474181815 | 0.000228663 | 0.000502133 | 1 |
| T2DM | dendritic_cell | *ARNT* | -2.112603222 | 1.98E-05 | 0.000131266 | 0.124374738 |
| T2DM | dendritic_cell | *POGZ* | -2.031992296 | 0.000162572 | 0.000389963 | 1 |
| T2DM | dendritic_cell | *GABPB2* | -1.160725411 | 0.005502659 | 0.008023009 | 1 |
| T2DM | dendritic_cell | *IKBKE* | -4.226905617 | 0.000115032 | 0.000301209 | 0.72320338 |
| T2DM | dendritic_cell | *DYRK3* | -3.730348099 | 0.000892949 | 0.001569463 | 1 |
| T2DM | dendritic_cell | *EIF2D* | -2.319567574 | 9.99E-05 | 0.00027268 | 0.627981135 |

| | | | | | | |
|---|---|---|---|---|---|---|
| T2DM | dendritic_cell | *HHIPL2* | -2.886632831 | 0.002334613 | 0.003712118 | 1 |
| T2DM | dendritic_cell | *TP53BP2* | 2.068933017 | 0.002144982 | 0.003439302 | 1 |
| T2DM | dendritic_cell | *RAB13* | -5.047881951 | 1.36E-05 | 0.000130129 | 0.085423223 |
| T2DM | dendritic_cell | *CREB3L4* | -1.603790881 | 3.74E-05 | 0.000156414 | 0.235245968 |
| T2DM | dendritic_cell | *KCNN3* | -1.084272504 | 2.70E-05 | 0.000140757 | 0.169471889 |
| T2DM | dendritic_cell | *GATAD2B* | -2.344317741 | 0.000772138 | 0.00138698 | 1 |
| T2DM | dendritic_cell | *RIT1* | 2.602420789 | 0.002031483 | 0.003270661 | 1 |
| T2DM | dendritic_cell | *GALNT2* | -2.46689486 | 4.55E-05 | 0.000169568 | 0.286230166 |
| T2DM | dendritic_cell | *SCCPDH* | -1.498039159 | 1.47E-05 | 0.000130129 | 0.092416014 |
| T2DM | dendritic_cell | *SNAP47* | -1.323860158 | 0.000499057 | 0.000947928 | 1 |
| T2DM | dendritic_cell | *NVL* | -2.505923216 | 3.74E-05 | 0.000156414 | 0.235134464 |
| T2DM | dendritic_cell | *SDE2* | -4.295689467 | 4.53E-06 | 0.000118089 | 0.028459413 |
| T2DM | dendritic_cell | *FBXO28* | -3.035535313 | 0.000285373 | 0.000599046 | 1 |
| T2DM | dendritic_cell | *CNIH4* | -3.308867373 | 2.04E-05 | 0.00013202 | 0.128449402 |
| T2DM | dendritic_cell | *GUK1* | -2.611818486 | 3.05E-05 | 0.000145705 | 0.191624875 |
| T2DM | dendritic_cell | *CDC42BPA* | -1.581327376 | 8.66E-05 | 0.000250092 | 0.544700774 |
| T2DM | dendritic_cell | *CNIH3* | -2.125762274 | 0.000187325 | 0.000431238 | 1 |
| T2DM | dendritic_cell | *PSEN2* | -3.488259098 | 8.77E-05 | 0.000252107 | 0.551524281 |
| T2DM | dendritic_cell | *WNT9A* | -1.150545796 | 0.001247654 | 0.002105772 | 1 |
| T2DM | dendritic_cell | *PLEKHA6* | -1.155267654 | 0.000138055 | 0.000344151 | 0.867949484 |
| T2DM | dendritic_cell | *ABCG8* | 4.541937511 | 1.03E-05 | 0.000130129 | 0.064881213 |
| T2DM | dendritic_cell | *EML4* | -2.499311198 | 0.000237909 | 0.000517805 | 1 |
| T2DM | dendritic_cell | *CHAC2* | -4.051335561 | 0.000573318 | 0.001069423 | 1 |
| T2DM | dendritic_cell | *WDPCP* | -0.749048047 | 2.94E-05 | 0.000143842 | 0.184818285 |
| T2DM | dendritic_cell | *ASXL2* | -2.65758897 | 0.000237942 | 0.000517805 | 1 |
| T2DM | dendritic_cell | *CIAO1* | -3.151653007 | 7.58E-05 | 0.000227248 | 0.476767091 |
| T2DM | dendritic_cell | *ZNF514* | -0.971105477 | 0.001870108 | 0.003037294 | 1 |
| T2DM | dendritic_cell | *EXOC6B* | -1.799954908 | 4.48E-05 | 0.000168727 | 0.281942855 |
| T2DM | dendritic_cell | *TEX261* | -5.565769213 | 0.000322739 | 0.000662225 | 1 |
| T2DM | dendritic_cell | *THNSL2* | -0.458548016 | 0.002645333 | 0.004147605 | 1 |
| T2DM | dendritic_cell | *RALB* | -2.981491186 | 4.30E-05 | 0.000165006 | 0.270093366 |
| T2DM | dendritic_cell | *TMEM177* | 1.688371759 | 9.19E-05 | 0.000259669 | 0.578022133 |
| T2DM | dendritic_cell | *RABL2A* | -2.075478038 | 1.10E-05 | 0.000130129 | 0.068939577 |
| T2DM | dendritic_cell | *FBLN7* | -1.037633552 | 0.036907843 | 0.046876688 | 1 |
| T2DM | dendritic_cell | *ZC3H8* | -2.295323991 | 0.001569004 | 0.002588382 | 1 |
| T2DM | dendritic_cell | *LIPT1* | -1.298471743 | 0.019923929 | 0.026332088 | 1 |
| T2DM | dendritic_cell | *FAHD2B* | -2.325198373 | 9.44E-05 | 0.000264058 | 0.593333614 |
| T2DM | dendritic_cell | *LYG1* | -1.148050014 | 1.32E-05 | 0.000130129 | 0.083189106 |
| T2DM | dendritic_cell | *AFF3* | -1.381119631 | 0.006275661 | 0.009028623 | 1 |
| T2DM | dendritic_cell | *UBXN4* | -2.565434996 | 0.000112843 | 0.000297087 | 0.709443633 |
| T2DM | dendritic_cell | *THSD7B* | -0.658884215 | 0.000306326 | 0.000635392 | 1 |
| T2DM | dendritic_cell | *ZNF385B* | -0.733661995 | 4.16E-05 | 0.000162756 | 0.261567329 |
| T2DM | dendritic_cell | *TMEFF2* | -2.407090583 | 0.016271169 | 0.021788464 | 1 |
| T2DM | dendritic_cell | *PHOSPHO2* | -0.934605084 | 0.005945869 | 0.008603379 | 1 |
| T2DM | dendritic_cell | *GULP1* | -1.643388417 | 0.000115907 | 0.000302403 | 0.728706761 |
| T2DM | dendritic_cell | *FAM171B* | -1.662657165 | 0.031858832 | 0.040785273 | 1 |
| T2DM | dendritic_cell | *HSPD1* | -2.744115612 | 0.000644661 | 0.001180252 | 1 |
| T2DM | dendritic_cell | *PTH2R* | -1.185966363 | 0.021030601 | 0.027655175 | 1 |
| T2DM | dendritic_cell | *SUMF1* | -2.1437284 | 0.000578911 | 0.001077765 | 1 |
| T2DM | dendritic_cell | *ESPNL* | -10.29375441 | 0.001584435 | 0.002610415 | 1 |

| | | | | | | |
|---|---|---|---|---|---|---|
| T2DM | dendritic_cell | *FANCD2* | -3.839586735 | 0.000158093 | 0.000381209 | 0.993927803 |
| T2DM | dendritic_cell | *TAMM41* | -2.146934264 | 0.000318775 | 0.000656233 | 1 |
| T2DM | dendritic_cell | *OSBPL10* | -1.763621282 | 0.00066805 | 0.001218517 | 1 |
| T2DM | dendritic_cell | *FAM198A* | -1.431198103 | 0.028826428 | 0.037160499 | 1 |
| T2DM | dendritic_cell | *CSRNP1* | -2.573549116 | 0.000211531 | 0.000471919 | 1 |
| T2DM | dendritic_cell | *CTDSPL* | -2.604103563 | 3.76E-06 | 0.000110889 | 0.023608589 |
| T2DM | dendritic_cell | *STAC* | -1.555980961 | 1.40E-05 | 0.000130129 | 0.087759456 |
| T2DM | dendritic_cell | *ARL6IP5* | -2.123739809 | 3.14E-05 | 0.000147278 | 0.197204722 |
| T2DM | dendritic_cell | *TMF1* | -3.202361332 | 0.000234641 | 0.000512219 | 1 |
| T2DM | dendritic_cell | *LIMD1* | -2.674634269 | 0.003635631 | 0.005517067 | 1 |
| T2DM | dendritic_cell | *ZNF660* | -1.077829542 | 0.013519424 | 0.018330087 | 1 |
| T2DM | dendritic_cell | *COL8A1* | -1.011562506 | 1.64E-05 | 0.000130129 | 0.102974647 |
| T2DM | dendritic_cell | *NXPE3* | -4.06044805 | 6.10E-05 | 0.000198694 | 0.383197256 |
| T2DM | dendritic_cell | *PLA1A* | 0.638877312 | 0.029473078 | 0.037916358 | 1 |
| T2DM | dendritic_cell | *RABL3* | -3.346777051 | 0.000181203 | 0.000421466 | 1 |
| T2DM | dendritic_cell | *IGSF11* | 2.265050406 | 0.001034745 | 0.001784757 | 1 |
| T2DM | dendritic_cell | *BOC* | -1.142521893 | 1.72E-05 | 0.000130129 | 0.108214122 |
| T2DM | dendritic_cell | *AGTR1* | -0.952174083 | 1.39E-05 | 0.000130129 | 0.087544976 |
| T2DM | dendritic_cell | *EIF2A* | -1.248877755 | 0.00079995 | 0.001429586 | 1 |
| T2DM | dendritic_cell | *AMT* | -0.621181583 | 0.006845128 | 0.00979857 | 1 |
| T2DM | dendritic_cell | *TCTA* | -2.035637196 | 0.000302415 | 0.000628732 | 1 |
| T2DM | dendritic_cell | *MUC4* | -0.953046685 | 0.000337139 | 0.000685287 | 1 |
| T2DM | dendritic_cell | *DGKQ* | -1.329530478 | 0.035771333 | 0.045506753 | 1 |
| T2DM | dendritic_cell | *LYAR* | -4.874992876 | 0.000403223 | 0.000796438 | 1 |
| T2DM | dendritic_cell | *CENPC* | -1.822934413 | 0.000165565 | 0.000394732 | 1 |
| T2DM | dendritic_cell | *PYURF* | -3.44668603 | 8.47E-06 | 0.000130129 | 0.053245719 |
| T2DM | dendritic_cell | *CISD2* | 2.778023545 | 0.001660081 | 0.002721493 | 1 |
| T2DM | dendritic_cell | *TIFA* | 1.714302067 | 9.89E-05 | 0.000271395 | 0.621765554 |
| T2DM | dendritic_cell | *CCNA2* | -2.207184429 | 8.63E-06 | 0.000130129 | 0.054263115 |
| T2DM | dendritic_cell | *USP53* | -1.464316212 | 4.15E-05 | 0.000162756 | 0.261137582 |
| T2DM | dendritic_cell | *MARCH1* | 0.737738428 | 0.000214572 | 0.000477022 | 1 |
| T2DM | dendritic_cell | *PDGFC* | -1.814325148 | 2.97E-06 | 9.90E-05 | 0.018702164 |
| T2DM | dendritic_cell | *CBR4* | -2.239126619 | 0.00010089 | 0.000274348 | 0.634293501 |
| T2DM | dendritic_cell | *CYP4V2* | -1.011197177 | 0.003459545 | 0.00527021 | 1 |
| T2DM | dendritic_cell | *NDUFS6* | -3.831582644 | 1.47E-05 | 0.000130129 | 0.092516385 |
| T2DM | dendritic_cell | *NKD2* | 4.623733164 | 0.005528088 | 0.008054482 | 1 |
| T2DM | dendritic_cell | *FAM105A* | 0.635367808 | 0.000147581 | 0.000361526 | 0.927843907 |
| T2DM | dendritic_cell | *LHFPL2* | -3.465559888 | 0.000466747 | 0.000897107 | 1 |
| T2DM | dendritic_cell | *SSBP2* | -2.113660991 | 0.00015606 | 0.000377655 | 0.981147606 |
| T2DM | dendritic_cell | *BHMT* | -1.287816238 | 9.85E-09 | 3.07E-06 | 6.19E-05 |
| T2DM | dendritic_cell | *CRHBP* | -2.755404687 | 1.63E-05 | 0.000130129 | 0.102305632 |
| T2DM | dendritic_cell | *RASA1* | -1.656266196 | 0.000719686 | 0.001302062 | 1 |
| T2DM | dendritic_cell | *PPIP5K2* | -3.297877255 | 1.40E-06 | 7.15E-05 | 0.008797375 |
| T2DM | dendritic_cell | *GTF2H2* | -2.193885617 | 0.000581019 | 0.00108137 | 1 |
| T2DM | dendritic_cell | *SPATA9* | -0.829212137 | 6.78E-05 | 0.000211932 | 0.425984168 |
| T2DM | dendritic_cell | *TSLP* | -1.418807205 | 0.000440869 | 0.000856269 | 1 |
| T2DM | dendritic_cell | *COMMD10* | -2.150772456 | 0.000141382 | 0.00035064 | 0.888871439 |
| T2DM | dendritic_cell | *SLC25A48* | -1.176419364 | 0.013541714 | 0.018356351 | 1 |
| T2DM | dendritic_cell | *GABRB2* | -1.824912914 | 0.000701548 | 0.001274381 | 1 |
| T2DM | dendritic_cell | *PCYOX1L* | 1.159061008 | 0.001879388 | 0.003049216 | 1 |

| | | | | | | |
|---|---|---|---|---|---|---|
| T2DM | dendritic_cell | *MYLK4* | -2.480429743 | 1.53E-05 | 0.000130129 | 0.096119517 |
| T2DM | dendritic_cell | *TBC1D7* | -1.676938792 | 2.69E-05 | 0.000140726 | 0.169221707 |
| T2DM | dendritic_cell | *GFOD1* | -1.013645 | 0.001191044 | 0.002018898 | 1 |
| T2DM | dendritic_cell | *ZMAT2* | -1.957348758 | 0.033136712 | 0.042334994 | 1 |
| T2DM | dendritic_cell | *KLHL3* | -1.761830659 | 0.000154063 | 0.00037383 | 0.968593842 |
| T2DM | dendritic_cell | *HIGD2A* | -2.593783258 | 0.035876047 | 0.045630732 | 1 |
| T2DM | dendritic_cell | *FAM193B* | -3.007435061 | 0.000271933 | 0.000576413 | 1 |
| T2DM | dendritic_cell | *MUT* | -3.11135745 | 3.44E-05 | 0.000151319 | 0.216386182 |
| T2DM | dendritic_cell | *ABT1* | 1.76730275 | 0.017353437 | 0.023134237 | 1 |
| T2DM | dendritic_cell | *MLIP* | 2.446421096 | 0.005784431 | 0.00840072 | 1 |
| T2DM | dendritic_cell | *FGD2* | 0.368586128 | 3.95E-05 | 0.000159553 | 0.24861899 |
| T2DM | dendritic_cell | *ANO7* | -1.284348215 | 0.000552917 | 0.00103643 | 1 |
| T2DM | dendritic_cell | *RPL7L1* | -1.711712168 | 0.002243566 | 0.003583663 | 1 |
| T2DM | dendritic_cell | *TPBG* | -2.038230129 | 1.65E-05 | 0.000130129 | 0.103690548 |
| T2DM | dendritic_cell | *IRAK1BP1* | -2.053476366 | 4.59E-05 | 0.000170199 | 0.288613723 |
| T2DM | dendritic_cell | *PHIP* | -2.291662917 | 0.001278968 | 0.002156307 | 1 |
| T2DM | dendritic_cell | *MMS22L* | -2.047985467 | 8.83E-06 | 0.000130129 | 0.055522229 |
| T2DM | dendritic_cell | *PM20D2* | -4.405251059 | 5.86E-05 | 0.000194679 | 0.368528224 |
| T2DM | dendritic_cell | *RNF217* | -2.683077385 | 0.00017506 | 0.000410673 | 1 |
| T2DM | dendritic_cell | *ABRACL* | 1.640877387 | 0.038983749 | 0.049294214 | 1 |
| T2DM | dendritic_cell | *AIG1* | -1.779397162 | 5.98E-05 | 0.000197204 | 0.375847163 |
| T2DM | dendritic_cell | *DYNLT1* | -3.148403822 | 5.01E-05 | 0.000177579 | 0.315202561 |
| T2DM | dendritic_cell | *PNLDC1* | -1.887354167 | 0.015512619 | 0.020852648 | 1 |
| T2DM | dendritic_cell | *SLC22A3* | -2.382097681 | 1.85E-05 | 0.000130129 | 0.116424792 |
| T2DM | dendritic_cell | *C7orf50* | -2.478961424 | 0.000713384 | 0.001292148 | 1 |
| T2DM | dendritic_cell | *SDK1* | -0.956221112 | 0.001626568 | 0.002672128 | 1 |
| T2DM | dendritic_cell | *CCZ1B* | -1.580009564 | 0.033684588 | 0.04301747 | 1 |
| T2DM | dendritic_cell | *RBAK* | -3.709781471 | 1.36E-05 | 0.000130129 | 0.085311022 |
| T2DM | dendritic_cell | *EGFR* | -1.253993778 | 2.15E-06 | 8.62E-05 | 0.013536988 |
| T2DM | dendritic_cell | *POMZP3* | -2.165814337 | 0.000592789 | 0.001101645 | 1 |
| T2DM | dendritic_cell | *CCT6A* | -3.077944207 | 0.001292591 | 0.002174611 | 1 |
| T2DM | dendritic_cell | *GIGYF1* | -3.266009933 | 3.83E-06 | 0.000111457 | 0.024074693 |
| T2DM | dendritic_cell | *TRIM4* | -0.998611503 | 0.001960162 | 0.003166377 | 1 |
| T2DM | dendritic_cell | *AGBL3* | -2.544295651 | 4.82E-05 | 0.000173736 | 0.302783087 |
| T2DM | dendritic_cell | *EPHA1* | 2.072549365 | 9.76E-05 | 0.000268845 | 0.61346201 |
| T2DM | dendritic_cell | *FBXO25* | -3.256406201 | 0.000119096 | 0.000309531 | 0.748758418 |
| T2DM | dendritic_cell | *CSGALNACT1* | -2.210081446 | 1.21E-05 | 0.000130129 | 0.07638351 |
| T2DM | dendritic_cell | *CCDC25* | -1.908525018 | 0.0039467 | 0.005936101 | 1 |
| T2DM | dendritic_cell | *BIN3* | -2.26305097 | 9.70E-05 | 0.000268182 | 0.61011435 |
| T2DM | dendritic_cell | *SLC25A37* | -2.516327503 | 4.24E-05 | 0.000164065 | 0.266442982 |
| T2DM | dendritic_cell | *CHMP7* | -1.55251719 | 0.000152037 | 0.000370057 | 0.955856096 |
| T2DM | dendritic_cell | *DOCK5* | 1.592268648 | 0.001281929 | 0.002159562 | 1 |
| T2DM | dendritic_cell | *ST18* | 0.261377704 | 3.90E-05 | 0.000158736 | 0.245247631 |
| T2DM | dendritic_cell | *TACC1* | -2.03651964 | 2.63E-05 | 0.000139569 | 0.165239341 |
| T2DM | dendritic_cell | *ADHFE1* | -0.932972324 | 1.16E-06 | 6.49E-05 | 0.007267925 |
| T2DM | dendritic_cell | *SLC26A7* | -1.259241914 | 0.001389634 | 0.002321101 | 1 |
| T2DM | dendritic_cell | *MTDH* | -2.586743125 | 0.000270826 | 0.000574537 | 1 |
| T2DM | dendritic_cell | *MAL2* | -1.705384168 | 7.92E-06 | 0.000130129 | 0.049780604 |
| T2DM | dendritic_cell | *UTP23* | -1.85724885 | 2.42E-05 | 0.000136859 | 0.152255298 |
| T2DM | dendritic_cell | *FAM135B* | 4.576214179 | 0.014862921 | 0.020013533 | 1 |

| | | | | | | |
|---|---|---|---|---|---|---|
| T2DM | dendritic_cell | *ZNF7* | -1.828069714 | 0.00314352 | 0.004840389 | 1 |
| T2DM | dendritic_cell | *SLC39A4* | -4.17008124 | 2.69E-07 | 2.73E-05 | 0.001690929 |
| T2DM | dendritic_cell | *AK3* | -2.478540299 | 5.56E-05 | 0.000188762 | 0.349617708 |
| T2DM | dendritic_cell | *CER1* | -2.531647761 | 0.003221665 | 0.004941353 | 1 |
| T2DM | dendritic_cell | *PLIN2* | -2.369388836 | 2.46E-05 | 0.000137905 | 0.15472938 |
| T2DM | dendritic_cell | *HAUS6* | -3.867680358 | 0.00071328 | 0.001292148 | 1 |
| T2DM | dendritic_cell | *C9orf72* | 1.665947954 | 0.002886968 | 0.00447384 | 1 |
| T2DM | dendritic_cell | *SIGMAR1* | -1.782380787 | 3.63E-05 | 0.000155082 | 0.227969962 |
| T2DM | dendritic_cell | *CBWD5* | -3.237582282 | 2.67E-05 | 0.000140322 | 0.168025903 |
| T2DM | dendritic_cell | *ZNF462* | -2.778071593 | 0.006497126 | 0.009334422 | 1 |
| T2DM | dendritic_cell | *ASTN2* | -1.974384103 | 2.19E-05 | 0.000132858 | 0.137507839 |
| T2DM | dendritic_cell | *WDR31* | -2.187182498 | 5.56E-05 | 0.000188762 | 0.349624214 |
| T2DM | dendritic_cell | *SURF6* | -2.64189421 | 1.47E-05 | 0.000130129 | 0.092675522 |
| T2DM | dendritic_cell | *NTMT1* | 1.898331581 | 0.016496263 | 0.022075778 | 1 |
| T2DM | dendritic_cell | *CIZ1* | -2.396138029 | 0.000137227 | 0.00034273 | 0.862744893 |
| T2DM | dendritic_cell | *SH3GLB2* | -2.617460292 | 0.000114913 | 0.000301024 | 0.722457227 |
| T2DM | dendritic_cell | *LRSAM1* | -3.33145594 | 3.84E-05 | 0.000158005 | 0.241482034 |
| T2DM | dendritic_cell | *INPP5E* | -3.127595129 | 1.48E-05 | 0.000130129 | 0.093219791 |
| T2DM | dendritic_cell | *DPH7* | -2.958768375 | 1.35E-05 | 0.000130129 | 0.085100535 |
| T2DM | dendritic_cell | *SLC39A12* | -2.235175826 | 0.000172545 | 0.00040705 | 1 |
| T2DM | dendritic_cell | *RSU1* | -2.329871939 | 7.28E-05 | 0.000220759 | 0.45785401 |
| T2DM | dendritic_cell | *PARD3* | -1.329162573 | 1.96E-05 | 0.000131266 | 0.123158895 |
| T2DM | dendritic_cell | *NRBF2* | -1.933913543 | 2.58E-05 | 0.0001395 | 0.161922545 |
| T2DM | dendritic_cell | *ADIRF* | -1.972026283 | 3.86E-05 | 0.000158005 | 0.242453508 |
| T2DM | dendritic_cell | *HTR7* | 0.287862686 | 3.10E-05 | 0.000146367 | 0.194699444 |
| T2DM | dendritic_cell | *RPP30* | -2.762032318 | 0.011911953 | 0.016269922 | 1 |
| T2DM | dendritic_cell | *FRA10AC1* | -1.607154453 | 0.003098304 | 0.004775445 | 1 |
| T2DM | dendritic_cell | *DNAJB12* | -3.170079326 | 0.000184995 | 0.000427598 | 1 |
| T2DM | dendritic_cell | *PLEKHS1* | -5.65890567 | 0.000280328 | 0.000590624 | 1 |
| T2DM | dendritic_cell | *FUOM* | 2.458670949 | 0.038459905 | 0.048661184 | 1 |
| T2DM | dendritic_cell | *MTG1* | -1.174432277 | 0.000573841 | 0.00106991 | 1 |
| T2DM | dendritic_cell | *PAOX* | -6.153405053 | 0.000250198 | 0.000539101 | 1 |
| T2DM | dendritic_cell | *PDCD11* | -2.87866153 | 0.001481609 | 0.002459698 | 1 |
| T2DM | dendritic_cell | *RGS10* | 1.2085119 | 0.000225758 | 0.000496344 | 1 |
| T2DM | dendritic_cell | *GAS2* | -1.433939428 | 1.49E-05 | 0.000130129 | 0.093632555 |
| T2DM | dendritic_cell | *LIN7C* | -3.106739133 | 2.67E-05 | 0.000140322 | 0.167786553 |
| T2DM | dendritic_cell | *HSD17B12* | -2.049171837 | 2.81E-07 | 2.76E-05 | 0.00176478 |
| T2DM | dendritic_cell | *APIP* | -0.973872447 | 1.45E-07 | 1.98E-05 | 0.000910796 |
| T2DM | dendritic_cell | *EIF3M* | -4.159208494 | 1.81E-05 | 0.000130129 | 0.113518761 |
| T2DM | dendritic_cell | *GLYAT* | -0.637108916 | 0.000312382 | 0.000645335 | 1 |
| T2DM | dendritic_cell | *PTPRJ* | 1.395875564 | 0.000137291 | 0.00034273 | 0.863148744 |
| T2DM | dendritic_cell | *C11orf49* | -1.815542414 | 9.81E-05 | 0.000269984 | 0.616913982 |
| T2DM | dendritic_cell | *ENDOD1* | -4.400240729 | 0.000184961 | 0.000427598 | 1 |
| T2DM | dendritic_cell | *CCDC82* | -1.761214519 | 5.50E-05 | 0.000187755 | 0.345845204 |
| T2DM | dendritic_cell | *SERPINH1* | -1.916174125 | 0.000672247 | 0.001225334 | 1 |
| T2DM | dendritic_cell | *CAPN5* | -2.288277929 | 0.000372471 | 0.000744587 | 1 |
| T2DM | dendritic_cell | *INTS4* | -1.832437796 | 0.000751823 | 0.001353971 | 1 |
| T2DM | dendritic_cell | *RPS3* | -2.020425266 | 0.003811899 | 0.005756764 | 1 |
| T2DM | dendritic_cell | *ZC3H12C* | -2.331937556 | 0.031417758 | 0.040245201 | 1 |
| T2DM | dendritic_cell | *DRD2* | 1.698406539 | 3.09E-05 | 0.000146197 | 0.194149121 |

| | | | | | | |
|---|---|---|---|---|---|---|
| T2DM | dendritic_cell | *C11orf52* | -10.09199042 | 1.46E-05 | 0.000130129 | 0.091978414 |
| T2DM | dendritic_cell | *P4HA3* | -1.590480303 | 0.006093086 | 0.008798171 | 1 |
| T2DM | dendritic_cell | *ADAM33* | -0.996662552 | 1.12E-05 | 0.000130129 | 0.07048345 |
| T2DM | dendritic_cell | *MTA2* | -2.151029065 | 0.000103698 | 0.00028065 | 0.65194998 |
| T2DM | dendritic_cell | *TMC2* | -2.190936875 | 1.50E-05 | 0.000130129 | 0.09442719 |
| T2DM | dendritic_cell | *ROM1* | -0.95551781 | 0.007280318 | 0.010383702 | 1 |
| T2DM | dendritic_cell | *EML3* | -2.468971036 | 9.89E-05 | 0.000271395 | 0.621741418 |
| T2DM | dendritic_cell | *INCENP* | 2.381229131 | 0.000184548 | 0.000427506 | 1 |
| T2DM | dendritic_cell | *MS4A3* | -26.27469128 | 1.11E-05 | 0.000130129 | 0.069571371 |
| T2DM | dendritic_cell | *PLCH2* | -4.533630875 | 1.56E-05 | 0.000130129 | 0.097803274 |
| T2DM | dendritic_cell | *CPSF7* | -3.089542822 | 0.000120564 | 0.000311929 | 0.757988588 |
| T2DM | dendritic_cell | *CCDC15* | -2.187648167 | 0.000283405 | 0.00059571 | 1 |
| T2DM | dendritic_cell | *FEZ1* | -3.976057543 | 0.000333386 | 0.000678976 | 1 |
| T2DM | dendritic_cell | *DUSP15* | -4.434571947 | 1.15E-06 | 6.49E-05 | 0.007248421 |
| T2DM | dendritic_cell | *COMMD7* | -3.261074426 | 4.32E-05 | 0.000165275 | 0.271381354 |
| T2DM | dendritic_cell | *KIAA1755* | -1.220856781 | 0.001741797 | 0.002843593 | 1 |
| T2DM | dendritic_cell | *CNBD2* | -5.5631528 | 0.000175881 | 0.000411983 | 1 |
| T2DM | dendritic_cell | *TRPT1* | -2.830949263 | 0.000111642 | 0.000294558 | 0.701893995 |
| T2DM | dendritic_cell | *TAOK2* | -2.428681389 | 1.00E-05 | 0.000130129 | 0.062950625 |
| T2DM | dendritic_cell | *MPP7* | -1.641502974 | 0.000291322 | 0.000609092 | 1 |
| T2DM | dendritic_cell | *CWC15* | -2.386287218 | 0.00489319 | 0.007206251 | 1 |
| T2DM | dendritic_cell | *ARID5B* | -1.567997618 | 0.000116115 | 0.000302786 | 0.730017292 |
| T2DM | dendritic_cell | *TMCO3* | -3.132363809 | 9.93E-05 | 0.000271912 | 0.624579958 |
| T2DM | dendritic_cell | *SAP18* | -2.962029259 | 5.00E-05 | 0.000177555 | 0.314628247 |
| T2DM | dendritic_cell | *LYPD6B* | -7.903201733 | 0.00040432 | 0.000797853 | 1 |
| T2DM | dendritic_cell | *PDCD4* | -2.305254698 | 9.94E-05 | 0.000271912 | 0.624841371 |
| T2DM | dendritic_cell | *CCDC102B* | -1.545734612 | 1.32E-05 | 0.000130129 | 0.082936987 |
| T2DM | dendritic_cell | *CNDP1* | -1.309456612 | 0.030949569 | 0.039685895 | 1 |
| T2DM | dendritic_cell | *RGS18* | 0.29488578 | 3.18E-05 | 0.000148215 | 0.19979362 |
| T2DM | dendritic_cell | *PRSS23* | -1.30407401 | 1.69E-05 | 0.000130129 | 0.106377383 |
| T2DM | dendritic_cell | *MTMR12* | -2.418653156 | 0.000132174 | 0.000334264 | 0.830980287 |
| T2DM | dendritic_cell | *C11orf53* | -10.27039635 | 1.11E-05 | 0.000130129 | 0.069829399 |
| T2DM | dendritic_cell | *FAM173B* | -2.995559607 | 1.55E-05 | 0.000130129 | 0.097441755 |
| T2DM | dendritic_cell | *DOCK1* | -1.790751999 | 2.24E-05 | 0.000133629 | 0.140798418 |
| T2DM | dendritic_cell | *TEX12* | -8.894706136 | 1.50E-05 | 0.000130129 | 0.094153588 |
| T2DM | dendritic_cell | *CRIM1* | -2.012081345 | 1.13E-05 | 0.000130129 | 0.071105341 |
| T2DM | dendritic_cell | *ABCB9* | -4.510584471 | 4.26E-07 | 3.57E-05 | 0.002676552 |
| T2DM | dendritic_cell | *RILPL2* | 1.269316341 | 0.022219517 | 0.029109002 | 1 |
| T2DM | dendritic_cell | *PRSS53* | -4.177248363 | 2.39E-05 | 0.000136538 | 0.150070878 |
| T2DM | dendritic_cell | *GPR158* | -1.288102907 | 0.000113181 | 0.000297355 | 0.711571397 |
| T2DM | dendritic_cell | *THRB* | -1.469328204 | 1.97E-05 | 0.000131266 | 0.124011964 |
| T2DM | dendritic_cell | *TMEM86A* | 0.850899695 | 0.025016744 | 0.032529528 | 1 |
| T2DM | dendritic_cell | *BTBD11* | 0.801834345 | 5.36E-05 | 0.000184297 | 0.336711074 |
| T2DM | dendritic_cell | *UBE3B* | -2.021009557 | 0.000145331 | 0.000357891 | 0.913695978 |
| T2DM | dendritic_cell | *PLBD2* | -1.762165292 | 0.000483529 | 0.000922873 | 1 |
| T2DM | dendritic_cell | *DLG5* | -1.636873158 | 1.28E-05 | 0.000130129 | 0.08063366 |
| T2DM | dendritic_cell | *FAM177A1* | -2.515668211 | 0.000855059 | 0.001514724 | 1 |
| T2DM | dendritic_cell | *MIPOL1* | -1.792962736 | 0.000321832 | 0.000661013 | 1 |
| T2DM | dendritic_cell | *EXT2* | -1.678891578 | 0.0010383 | 0.001789417 | 1 |
| T2DM | dendritic_cell | *TMEM18* | -3.559645332 | 0.029016789 | 0.037398228 | 1 |

| | | | | | | |
|---|---|---|---|---|---|---|
| T2DM | dendritic_cell | *THRSP* | -0.954796696 | 1.27E-05 | 0.000130129 | 0.079881652 |
| T2DM | dendritic_cell | *NDUFC2* | -3.032904395 | 4.52E-05 | 0.000168987 | 0.284405468 |
| T2DM | dendritic_cell | *ME3* | -1.597119609 | 0.000137621 | 0.000343343 | 0.865223513 |
| T2DM | dendritic_cell | *ADAMTS12* | -1.042479998 | 2.17E-05 | 0.000132663 | 0.136169281 |
| T2DM | dendritic_cell | *VIPAS39* | -3.865057333 | 1.41E-05 | 0.000130129 | 0.088936427 |
| T2DM | dendritic_cell | *ANKRD50* | -3.246885256 | 0.004443688 | 0.006606164 | 1 |
| T2DM | dendritic_cell | *C4orf33* | -2.452733595 | 3.15E-05 | 0.00014771 | 0.198079635 |
| T2DM | dendritic_cell | *PTPRO* | 0.892070325 | 6.53E-05 | 0.000206892 | 0.410473292 |
| T2DM | dendritic_cell | *VPS26B* | -3.317351733 | 3.71E-05 | 0.000156008 | 0.233375607 |
| T2DM | dendritic_cell | *VTI1A* | -1.858088538 | 0.000601273 | 0.001114447 | 1 |
| T2DM | dendritic_cell | *QDPR* | -0.871291296 | 0.000856545 | 0.001516159 | 1 |
| T2DM | dendritic_cell | *FAM160B1* | -2.529875569 | 3.72E-05 | 0.000156008 | 0.233613745 |
| T2DM | dendritic_cell | *ANO4* | -0.974126751 | 2.30E-05 | 0.000134478 | 0.14442977 |
| T2DM | dendritic_cell | *TEX9* | -3.240736308 | 0.001288387 | 0.00216986 | 1 |
| T2DM | dendritic_cell | *AKR1C2* | -1.059395465 | 7.60E-06 | 0.000130129 | 0.047794685 |
| T2DM | dendritic_cell | *ADAM8* | 0.721779502 | 4.52E-05 | 0.000168939 | 0.283986439 |
| T2DM | dendritic_cell | *ITIH2* | 4.310750271 | 0.005121367 | 0.007512373 | 1 |
| T2DM | dendritic_cell | *KIN* | -1.902352362 | 0.005225092 | 0.007644904 | 1 |
| T2DM | dendritic_cell | *PIGF* | -3.160952407 | 6.28E-05 | 0.000202109 | 0.394921028 |
| T2DM | dendritic_cell | *MFSD6* | -2.959854126 | 0.004961242 | 0.007296217 | 1 |
| T2DM | dendritic_cell | *RNF144A* | -1.805451578 | 3.07E-05 | 0.00014601 | 0.19327357 |
| T2DM | dendritic_cell | *ASAP2* | -1.724835917 | 3.66E-05 | 0.000155321 | 0.230320399 |
| T2DM | dendritic_cell | *ADAM17* | -2.697633976 | 0.001875436 | 0.00304359 | 1 |
| T2DM | dendritic_cell | *TMEM45B* | -3.614209249 | 3.39E-05 | 0.000150715 | 0.213111485 |
| T2DM | dendritic_cell | *ACSL1* | -0.945502437 | 0.000627033 | 0.001154027 | 1 |
| T2DM | dendritic_cell | *BICD1* | -2.801545877 | 0.000243309 | 0.000527659 | 1 |
| T2DM | dendritic_cell | *CCDC122* | -0.905067491 | 0.000496535 | 0.000944258 | 1 |
| T2DM | dendritic_cell | *GUF1* | -3.271019658 | 0.000516316 | 0.000975971 | 1 |
| T2DM | dendritic_cell | *GABRA2* | -1.566752975 | 3.84E-05 | 0.000158005 | 0.241662399 |
| T2DM | dendritic_cell | *CENPJ* | -2.381762505 | 0.001048582 | 0.001804169 | 1 |
| T2DM | dendritic_cell | *PARP8* | 1.380766066 | 0.006747963 | 0.009670491 | 1 |
| T2DM | dendritic_cell | *RABGAP1L* | -1.85745206 | 0.002641585 | 0.004144471 | 1 |
| T2DM | dendritic_cell | *PTPN14* | -2.014266747 | 5.09E-05 | 0.000179042 | 0.319829827 |
| T2DM | dendritic_cell | *RNF219* | -3.670683338 | 0.002651071 | 0.004153174 | 1 |
| T2DM | dendritic_cell | *ARL14EP* | -4.256865821 | 3.57E-05 | 0.000154392 | 0.224602392 |
| T2DM | dendritic_cell | *EPG5* | -2.499135616 | 0.001710019 | 0.00279389 | 1 |
| T2DM | dendritic_cell | *G6PC2* | -63.57787203 | 1.52E-05 | 0.000130129 | 0.095589966 |
| T2DM | dendritic_cell | *TGOLN2* | -3.266579444 | 0.000611373 | 0.001130169 | 1 |
| T2DM | dendritic_cell | *POC5* | -3.297059252 | 4.40E-08 | 9.21E-06 | 0.000276352 |
| T2DM | dendritic_cell | *GUCY1A2* | -0.84305366 | 2.12E-05 | 0.000132591 | 0.133337138 |
| T2DM | dendritic_cell | *CWF19L2* | -2.023479131 | 0.000127699 | 0.000325169 | 0.802842019 |
| T2DM | dendritic_cell | *ZNF547* | -3.022066716 | 0.037724482 | 0.047798029 | 1 |
| T2DM | dendritic_cell | *ZNF773* | -1.73447946 | 0.029172197 | 0.037565159 | 1 |
| T2DM | dendritic_cell | *NMT2* | -1.759377377 | 1.88E-05 | 0.000130129 | 0.118026062 |
| T2DM | dendritic_cell | *ZNF837* | -1.328134206 | 0.007902234 | 0.011179421 | 1 |
| T2DM | dendritic_cell | *CCDC50* | -1.269300827 | 0.012130484 | 0.016543244 | 1 |
| T2DM | dendritic_cell | *IGSF10* | -1.866459471 | 0.030798908 | 0.039509874 | 1 |
| T2DM | dendritic_cell | *SPEF2* | -2.177641614 | 0.000357274 | 0.000720161 | 1 |
| T2DM | dendritic_cell | *SPARCL1* | -1.105044814 | 3.96E-05 | 0.000159553 | 0.248975401 |
| T2DM | dendritic_cell | *MBNL1* | -1.873896223 | 0.002716713 | 0.004245581 | 1 |

| | | | | | | |
|---|---|---|---|---|---|---|
| T2DM | dendritic_cell | *CCNO* | 5.214008521 | 0.000113402 | 0.000297811 | 0.712959529 |
| T2DM | dendritic_cell | *CLEC4F* | 0.641349101 | 3.05E-05 | 0.000145705 | 0.191702546 |
| T2DM | dendritic_cell | *SAR1B* | -1.486027747 | 0.004997652 | 0.007341176 | 1 |
| T2DM | dendritic_cell | *GPR180* | -1.542410894 | 8.95E-05 | 0.000255275 | 0.5625897 |
| T2DM | dendritic_cell | *WDR78* | -1.947999193 | 1.25E-05 | 0.000130129 | 0.078404227 |
| T2DM | dendritic_cell | *IFIT5* | -1.874798026 | 0.004177237 | 0.006251437 | 1 |
| T2DM | dendritic_cell | *PRDM8* | -1.407066213 | 6.82E-05 | 0.000212634 | 0.428611475 |
| T2DM | dendritic_cell | *HHEX* | 1.823581973 | 0.010155504 | 0.014066458 | 1 |
| T2DM | dendritic_cell | *RAB3C* | -1.450112328 | 6.20E-05 | 0.000200487 | 0.389962138 |
| T2DM | dendritic_cell | *RAD17* | -1.801530159 | 0.000106493 | 0.000284729 | 0.669519946 |
| T2DM | dendritic_cell | *CENPH* | -1.122640039 | 0.010665957 | 0.014715135 | 1 |
| T2DM | dendritic_cell | *CARHSP1* | -2.069639371 | 1.13E-05 | 0.000130129 | 0.071187263 |
| T2DM | dendritic_cell | *TEKT5* | -3.003475651 | 9.49E-05 | 0.000264262 | 0.596421801 |
| T2DM | dendritic_cell | *BANK1* | -2.156739962 | 2.25E-05 | 0.000133629 | 0.141598896 |
| T2DM | dendritic_cell | *CAST* | -1.643442862 | 0.000649574 | 0.001188209 | 1 |
| T2DM | dendritic_cell | *SCOC* | -2.500309089 | 0.002533019 | 0.003990251 | 1 |
| T2DM | dendritic_cell | *CLGN* | 2.481262707 | 2.85E-05 | 0.000142609 | 0.179130018 |
| T2DM | dendritic_cell | *SYCP2L* | -2.767701904 | 0.016050447 | 0.021529583 | 1 |
| T2DM | dendritic_cell | *AHCTF1* | -2.484821066 | 9.58E-05 | 0.000266132 | 0.602256269 |
| T2DM | dendritic_cell | *PTPRR* | -1.657948952 | 3.30E-05 | 0.00014892 | 0.20757356 |
| T2DM | dendritic_cell | *PLA2R1* | -1.283706767 | 0.000251561 | 0.000541632 | 1 |
| T2DM | dendritic_cell | *SLC25A27* | -1.161399188 | 0.001241924 | 0.002097227 | 1 |
| T2DM | dendritic_cell | *FRMD1* | -4.125694132 | 1.75E-05 | 0.000130129 | 0.110113255 |
| T2DM | dendritic_cell | *ASAP1* | -1.95484644 | 0.011806716 | 0.01614372 | 1 |
| T2DM | dendritic_cell | *INO80C* | -2.98884381 | 2.48E-05 | 0.000138038 | 0.156085077 |
| T2DM | dendritic_cell | *LPCAT1* | -2.420705595 | 1.82E-05 | 0.000130129 | 0.114637318 |
| T2DM | dendritic_cell | *C16orf89* | -1.14101204 | 0.000210689 | 0.000471053 | 1 |
| T2DM | dendritic_cell | *TUBGCP5* | -2.736785522 | 3.43E-05 | 0.000151156 | 0.215699683 |
| T2DM | dendritic_cell | *LURAP1L* | -0.833786683 | 0.013760585 | 0.018616914 | 1 |
| T2DM | dendritic_cell | *CFDP1* | -2.477053248 | 7.98E-05 | 0.000235864 | 0.501918739 |
| T2DM | dendritic_cell | *FAM92B* | -16.78615815 | 1.31E-05 | 0.000130129 | 0.082182275 |
| T2DM | dendritic_cell | *C7orf31* | -1.975624067 | 3.99E-05 | 0.000159738 | 0.25091245 |
| T2DM | dendritic_cell | *JAZF1* | -3.554047468 | 7.22E-10 | 5.04E-07 | 4.54E-06 |
| T2DM | dendritic_cell | *FBXO36* | -2.446966625 | 4.77E-05 | 0.000172691 | 0.299964857 |
| T2DM | dendritic_cell | *MCOLN2* | 0.322104155 | 2.33E-05 | 0.00013522 | 0.146442798 |
| T2DM | dendritic_cell | *CACNA2D1* | -1.126983125 | 9.93E-06 | 0.000130129 | 0.062453691 |
| T2DM | dendritic_cell | *ZUFSP* | -2.884305472 | 0.009971687 | 0.013845405 | 1 |
| T2DM | dendritic_cell | *HS3ST3A1* | -1.45153365 | 0.00068161 | 0.001241031 | 1 |
| T2DM | dendritic_cell | *ANKRD29* | -1.142346287 | 9.28E-05 | 0.000261455 | 0.583567217 |
| T2DM | dendritic_cell | *THY1* | -1.824950426 | 2.49E-05 | 0.000138038 | 0.156343321 |
| T2DM | dendritic_cell | *TBCEL* | -3.259052311 | 0.000558812 | 0.00104547 | 1 |
| T2DM | dendritic_cell | *UBASH3B* | 1.399986658 | 0.00029713 | 0.000619035 | 1 |
| T2DM | dendritic_cell | *ROBO4* | -2.041418948 | 0.000302921 | 0.000629366 | 1 |
| T2DM | dendritic_cell | *ROBO3* | -2.770433889 | 4.43E-05 | 0.000167569 | 0.278709202 |
| T2DM | dendritic_cell | *CC2D1B* | -1.96228644 | 0.000181653 | 0.000422044 | 1 |
| T2DM | dendritic_cell | *LRRK1* | 1.207927631 | 0.000201462 | 0.000456102 | 1 |
| T2DM | dendritic_cell | *CEP112* | -1.579850284 | 0.000235276 | 0.000513426 | 1 |
| T2DM | dendritic_cell | *GAL3ST2* | 5.985696782 | 0.028121605 | 0.036289107 | 1 |
| T2DM | dendritic_cell | *ENPP3* | -1.260292465 | 1.40E-05 | 0.000130129 | 0.088060379 |
| T2DM | dendritic_cell | *MIA3* | -2.717272953 | 1.79E-05 | 0.000130129 | 0.112488057 |

| | | | | | | |
|---|---|---|---|---|---|---|
| T2DM | dendritic_cell | *FAM167A* | 1.755651602 | 2.93E-05 | 0.000143842 | 0.184102425 |
| T2DM | dendritic_cell | *CCSAP* | 2.055104405 | 0.00122547 | 0.002072781 | 1 |
| T2DM | dendritic_cell | *GBP5* | 2.63332235 | 0.003216666 | 0.004934889 | 1 |
| T2DM | dendritic_cell | *FAM69A* | -2.444673152 | 0.000175381 | 0.000411272 | 1 |
| T2DM | dendritic_cell | *NCAM2* | -1.017469531 | 4.92E-05 | 0.000175768 | 0.309176476 |
| T2DM | dendritic_cell | *L3MBTL4* | -2.174610872 | 0.000104908 | 0.000281983 | 0.659558021 |
| T2DM | dendritic_cell | *PDE1C* | -3.109130433 | 0.003204442 | 0.004919738 | 1 |
| T2DM | dendritic_cell | *RABGEF1* | -0.803969129 | 0.027487996 | 0.035529817 | 1 |
| T2DM | dendritic_cell | *MRPL39* | -0.986825168 | 0.000442695 | 0.00085955 | 1 |
| T2DM | dendritic_cell | *TSEN2* | -0.808462984 | 0.037940144 | 0.048042233 | 1 |
| T2DM | dendritic_cell | *FLCN* | -1.714868052 | 5.87E-05 | 0.000194716 | 0.368792169 |
| T2DM | dendritic_cell | *DPH3* | 3.198596498 | 0.013546349 | 0.018358676 | 1 |
| T2DM | dendritic_cell | *CXXC1* | -3.719288834 | 6.16E-05 | 0.000200088 | 0.38700298 |
| T2DM | dendritic_cell | *APCDD1* | -0.882200931 | 0.000473981 | 0.000908824 | 1 |
| T2DM | dendritic_cell | *EPHB1* | -3.485282388 | 1.54E-05 | 0.000130129 | 0.096814854 |
| T2DM | dendritic_cell | *ANKRD40* | -3.201001956 | 1.22E-05 | 0.000130129 | 0.0769839 |
| T2DM | dendritic_cell | *CYP2U1* | -4.172370403 | 0.000165034 | 0.000393914 | 1 |
| T2DM | dendritic_cell | *FBXL18* | -3.281055553 | 8.58E-05 | 0.000248187 | 0.539309679 |
| T2DM | dendritic_cell | *PROM2* | -5.602469881 | 2.68E-05 | 0.000140354 | 0.168287603 |
| T2DM | dendritic_cell | *AK9* | -2.445389368 | 0.000672404 | 0.001225334 | 1 |
| T2DM | dendritic_cell | *CDK19* | -2.4415996 | 0.006538632 | 0.009391908 | 1 |
| T2DM | dendritic_cell | *GTF3C6* | -2.880145433 | 0.001859626 | 0.003022614 | 1 |
| T2DM | dendritic_cell | *TTC39B* | -2.214999639 | 0.002460847 | 0.003887272 | 1 |
| T2DM | dendritic_cell | *MMS19* | -2.166091161 | 6.74E-06 | 0.000130129 | 0.042367171 |
| T2DM | dendritic_cell | *GOLGA7B* | 1.895023966 | 0.022988877 | 0.03004804 | 1 |
| T2DM | dendritic_cell | *TRMT44* | -2.070720213 | 2.23E-05 | 0.000133527 | 0.139936419 |
| T2DM | dendritic_cell | *C16orf87* | -2.363219556 | 0.009148459 | 0.012775735 | 1 |
| T2DM | dendritic_cell | *CNOT8* | -2.781974873 | 0.000310075 | 0.000641898 | 1 |
| T2DM | dendritic_cell | *LRGUK* | -1.220838169 | 0.00367175 | 0.005566504 | 1 |
| T2DM | dendritic_cell | *SETD9* | -1.417376834 | 2.40E-06 | 8.93E-05 | 0.015097868 |
| T2DM | dendritic_cell | *PDIA4* | -4.130106205 | 0.000142459 | 0.000352896 | 0.895639301 |
| T2DM | dendritic_cell | *KDM8* | -4.530751016 | 1.52E-05 | 0.000130129 | 0.095250575 |
| T2DM | dendritic_cell | *FAM126B* | -1.881874975 | 1.72E-05 | 0.000130129 | 0.107912256 |
| T2DM | dendritic_cell | *ALS2CR12* | -3.040710351 | 0.000514661 | 0.000973427 | 1 |
| T2DM | dendritic_cell | *SPAG17* | -0.825283392 | 1.98E-05 | 0.000131266 | 0.124448988 |
| T2DM | dendritic_cell | *RRAGA* | -3.489912344 | 1.88E-05 | 0.000130129 | 0.11821709 |
| T2DM | dendritic_cell | *TMBIM4* | -1.282910982 | 0.035493192 | 0.045207799 | 1 |
| T2DM | dendritic_cell | *FAM161B* | 3.96355495 | 0.010826917 | 0.014917561 | 1 |
| T2DM | dendritic_cell | *WIF1* | 4.139235563 | 5.45E-05 | 0.000186487 | 0.342333553 |
| T2DM | dendritic_cell | *UGT2B4* | -1.933730094 | 1.61E-05 | 0.000130129 | 0.101137431 |
| T2DM | dendritic_cell | *MMP16* | -1.147063006 | 5.53E-06 | 0.000126533 | 0.03474882 |
| T2DM | dendritic_cell | *KCNMA1* | -1.621898286 | 2.51E-05 | 0.000138591 | 0.157716398 |
| T2DM | dendritic_cell | *BATF* | -7.002700401 | 9.79E-05 | 0.000269514 | 0.615570459 |
| T2DM | dendritic_cell | *DCK* | 1.752397051 | 0.000981729 | 0.001704538 | 1 |
| T2DM | dendritic_cell | *DPY19L4* | -3.020168692 | 3.16E-05 | 0.000147838 | 0.198579387 |
| T2DM | dendritic_cell | *NDUFAF6* | -3.23676683 | 2.13E-08 | 5.54E-06 | 0.000134033 |
| T2DM | dendritic_cell | *C8orf37* | -2.25087797 | 8.03E-05 | 0.000236857 | 0.504978609 |
| T2DM | dendritic_cell | *ADAMTSL3* | -0.80723635 | 0.000145663 | 0.000358007 | 0.915783089 |
| T2DM | dendritic_cell | *WHAMM* | -2.368984793 | 3.41E-06 | 0.000105619 | 0.021440671 |
| T2DM | dendritic_cell | *CXCL13* | -6.428405312 | 8.20E-05 | 0.000240846 | 0.515650625 |

| | | | | | | |
|---|---|---|---|---|---|---|
| T2DM | dendritic_cell | *N6AMT1* | -1.045112844 | 0.012979682 | 0.017638812 | 1 |
| T2DM | dendritic_cell | *RWDD2B* | -1.188079658 | 0.037651826 | 0.047715587 | 1 |
| T2DM | dendritic_cell | *ANKRD9* | -5.909065132 | 2.41E-05 | 0.000136859 | 0.151725009 |
| T2DM | dendritic_cell | *SFXN2* | -1.764428679 | 0.026175242 | 0.033902709 | 1 |
| T2DM | dendritic_cell | *C14orf2* | -2.004528424 | 4.26E-06 | 0.000116241 | 0.026783359 |
| T2DM | dendritic_cell | *SH3RF2* | -0.890340202 | 0.031185691 | 0.039964215 | 1 |
| T2DM | dendritic_cell | *GDF6* | -2.82409885 | 0.000225713 | 0.000496344 | 1 |
| T2DM | dendritic_cell | *NODAL* | -1.686031469 | 1.71E-05 | 0.000130129 | 0.10744077 |
| T2DM | dendritic_cell | *MED19* | -1.247738814 | 0.004229931 | 0.006322772 | 1 |
| T2DM | dendritic_cell | *WDYHV1* | -2.65408013 | 0.000105487 | 0.000283088 | 0.663197746 |
| T2DM | dendritic_cell | *SASS6* | -2.45125033 | 5.97E-06 | 0.000129884 | 0.037536384 |
| T2DM | dendritic_cell | *ITGAD* | 1.25001335 | 0.000691433 | 0.001257671 | 1 |
| T2DM | dendritic_cell | *RPUSD3* | -3.2938754 | 3.27E-05 | 0.000148375 | 0.205396032 |
| T2DM | dendritic_cell | *GHRL* | -4.722024311 | 0.001317625 | 0.002211991 | 1 |
| T2DM | dendritic_cell | *NTAN1* | -3.057969091 | 0.000109482 | 0.000290796 | 0.688315303 |
| T2DM | dendritic_cell | *FCHO2* | -1.888348892 | 0.037541606 | 0.0475855 | 1 |
| T2DM | dendritic_cell | *CPT2* | -1.532359603 | 2.24E-05 | 0.000133629 | 0.140879628 |
| T2DM | dendritic_cell | *NECAP2* | -3.340904858 | 4.05E-05 | 0.000160731 | 0.25459741 |
| T2DM | dendritic_cell | *STEAP2* | -0.426959257 | 0.02228741 | 0.029179706 | 1 |
| T2DM | dendritic_cell | *GATAD1* | -1.35309281 | 0.015447365 | 0.020769372 | 1 |
| T2DM | dendritic_cell | *SUSD3* | 0.916647084 | 6.01E-05 | 0.00019776 | 0.37791967 |
| T2DM | dendritic_cell | *TMED6* | -5.634123545 | 1.45E-05 | 0.000130129 | 0.091320916 |
| T2DM | dendritic_cell | *IL34* | -1.70110914 | 2.45E-05 | 0.000137669 | 0.153993604 |
| T2DM | dendritic_cell | *DHRS1* | -1.186868734 | 0.002589251 | 0.00406559 | 1 |
| T2DM | dendritic_cell | *AASDH* | -2.294238114 | 0.000411792 | 0.000809044 | 1 |
| T2DM | dendritic_cell | *RNF111* | -2.038622271 | 4.93E-05 | 0.000176105 | 0.30998688 |
| T2DM | dendritic_cell | *DYRK1A* | -2.310778158 | 0.000204909 | 0.000461743 | 1 |
| T2DM | dendritic_cell | *KCNJ15* | -2.920103104 | 0.000295145 | 0.000615447 | 1 |
| T2DM | dendritic_cell | *LCA5L* | -0.93285515 | 0.00255953 | 0.004027764 | 1 |
| T2DM | dendritic_cell | *CREB3L1* | -1.247381302 | 0.000155322 | 0.000376191 | 0.976511755 |
| T2DM | dendritic_cell | *C2CD2* | -0.845113865 | 0.011847834 | 0.016192899 | 1 |
| T2DM | dendritic_cell | *SLC38A10* | -2.625132367 | 3.08E-05 | 0.000146146 | 0.193721762 |
| T2DM | dendritic_cell | *C9orf43* | 1.43734231 | 0.036970303 | 0.046937055 | 1 |
| T2DM | dendritic_cell | *WDR19* | -2.243003104 | 2.19E-05 | 0.000132858 | 0.137404836 |
| T2DM | dendritic_cell | *SLC37A3* | -1.401780192 | 1.53E-05 | 0.000130129 | 0.095959984 |
| T2DM | dendritic_cell | *AP3S2* | 1.005597557 | 0.011309634 | 0.015531601 | 1 |
| T2DM | dendritic_cell | *FAM213B* | 2.005434706 | 0.000347765 | 0.00070257 | 1 |
| T2DM | dendritic_cell | *C12orf43* | -1.156129697 | 0.007022917 | 0.010037072 | 1 |
| T2DM | dendritic_cell | *PEX10* | -2.479999808 | 2.53E-06 | 9.14E-05 | 0.015902407 |
| T2DM | dendritic_cell | *RADIL* | -1.547230518 | 6.68E-06 | 0.000130129 | 0.042010444 |
| T2DM | dendritic_cell | *LDLRAP1* | -2.653212938 | 1.34E-05 | 0.000130129 | 0.084361964 |
| T2DM | dendritic_cell | *WDR66* | -1.290401631 | 0.010800366 | 0.01488424 | 1 |
| T2DM | dendritic_cell | *UBXN11* | 1.190642797 | 0.001635943 | 0.002685424 | 1 |
| T2DM | dendritic_cell | *GALNT14* | 1.604148328 | 6.46E-05 | 0.00020528 | 0.406043875 |
| T2DM | dendritic_cell | *HPD* | -1.032279694 | 0.010835409 | 0.01492599 | 1 |
| T2DM | dendritic_cell | *RHPN1* | 1.713529898 | 0.000459486 | 0.000886047 | 1 |
| T2DM | dendritic_cell | *TPRG1L* | -5.383774762 | 1.06E-05 | 0.000130129 | 0.066777798 |
| T2DM | dendritic_cell | *LRRC43* | -3.450997965 | 1.39E-05 | 0.000130129 | 0.087694015 |
| T2DM | dendritic_cell | *XKR8* | -3.395303534 | 0.003702727 | 0.005608057 | 1 |
| T2DM | dendritic_cell | *EYA3* | -2.374889201 | 0.004068457 | 0.006106085 | 1 |

| | | | | | | |
|---|---|---|---|---|---|---|
| T2DM | dendritic_cell | *MRAS* | -1.317606602 | 1.89E-06 | 8.36E-05 | 0.011877938 |
| T2DM | dendritic_cell | *CLSTN2* | -1.018730389 | 1.20E-05 | 0.000130129 | 0.075177306 |
| T2DM | dendritic_cell | *COLEC12* | -2.358129659 | 4.76E-05 | 0.000172368 | 0.299230197 |
| T2DM | dendritic_cell | *RNF207* | 3.447827814 | 0.001401951 | 0.002339189 | 1 |
| T2DM | dendritic_cell | *AUTS2* | -1.809623552 | 5.07E-05 | 0.000178852 | 0.318969644 |
| T2DM | dendritic_cell | *MITD1* | -2.569484724 | 0.000493845 | 0.00093971 | 1 |
| T2DM | dendritic_cell | *TSPAN33* | 0.347133067 | 0.000284608 | 0.00059764 | 1 |
| T2DM | dendritic_cell | *B4GALT5* | -2.221139071 | 2.29E-05 | 0.000134433 | 0.144246523 |
| T2DM | dendritic_cell | *CD1D* | 0.282426516 | 4.60E-05 | 0.000170199 | 0.288909204 |
| T2DM | dendritic_cell | *FAM86C1* | -2.834401307 | 1.15E-05 | 0.000130129 | 0.07214258 |
| T2DM | dendritic_cell | *CD1E* | 0.195532904 | 1.89E-05 | 0.000130129 | 0.118910914 |
| T2DM | dendritic_cell | *NCF1* | 2.154006706 | 9.46E-05 | 0.000264178 | 0.594972119 |
| T2DM | dendritic_cell | *PPP1R9A* | -1.303182385 | 1.02E-05 | 0.000130129 | 0.063841382 |
| T2DM | dendritic_cell | *ZFAND2B* | -2.85839941 | 0.000174161 | 0.000409479 | 1 |
| T2DM | dendritic_cell | *GDPD5* | -1.441014101 | 1.10E-05 | 0.000130129 | 0.069270215 |
| T2DM | dendritic_cell | *DYNC1I1* | -3.443659808 | 9.25E-05 | 0.000261081 | 0.581427753 |
| T2DM | dendritic_cell | *TMED4* | -3.206719153 | 4.71E-05 | 0.000171433 | 0.296405813 |
| T2DM | dendritic_cell | *COPG2* | -0.745897398 | 6.90E-05 | 0.000213886 | 0.43354647 |
| T2DM | dendritic_cell | *ELK4* | -3.394292514 | 0.000848402 | 0.001505051 | 1 |
| T2DM | dendritic_cell | *DUSP23* | 1.945720883 | 8.09E-05 | 0.000238113 | 0.508608926 |
| T2DM | dendritic_cell | *RNF166* | 2.478869526 | 0.000551913 | 0.001035476 | 1 |
| T2DM | dendritic_cell | *NIT1* | -1.234391018 | 0.003849751 | 0.005808347 | 1 |
| T2DM | dendritic_cell | *ZNF276* | -4.230914547 | 0.000608498 | 0.001126156 | 1 |
| T2DM | dendritic_cell | *FGF17* | -3.541591727 | 0.021708325 | 0.028480851 | 1 |
| T2DM | dendritic_cell | *CDA* | -1.467565098 | 0.000142851 | 0.000353444 | 0.898101696 |
| T2DM | dendritic_cell | *DMTN* | -0.840569868 | 0.002288707 | 0.003645313 | 1 |
| T2DM | dendritic_cell | *NDUFS2* | -2.086147104 | 0.001379731 | 0.002307012 | 1 |
| T2DM | dendritic_cell | *SLC5A11* | -0.658856682 | 0.009126101 | 0.012747344 | 1 |
| T2DM | dendritic_cell | *FCER1G* | 0.878891992 | 2.16E-05 | 0.000132663 | 0.135999362 |
| T2DM | dendritic_cell | *CCAR2* | -3.281695964 | 8.96E-05 | 0.000255437 | 0.56324219 |
| T2DM | dendritic_cell | *CDC42SE2* | 1.562576017 | 0.01770955 | 0.023564009 | 1 |
| T2DM | dendritic_cell | *RAPGEF6* | -2.619639008 | 1.11E-05 | 0.000130129 | 0.069633175 |
| T2DM | dendritic_cell | *EPB41* | -1.323927018 | 0.001069188 | 0.001835605 | 1 |
| T2DM | dendritic_cell | *ALG8* | -3.266375112 | 2.88E-05 | 0.000143316 | 0.181171459 |
| T2DM | dendritic_cell | *MRPL10* | -2.011197393 | 0.000243823 | 0.000528045 | 1 |
| T2DM | dendritic_cell | *SV2A* | -5.703101903 | 0.000116543 | 0.000303775 | 0.732705779 |
| T2DM | dendritic_cell | *UBE2Z* | -3.465953195 | 0.005149357 | 0.007549909 | 1 |
| T2DM | dendritic_cell | *CLIC6* | -1.060895847 | 0.000123056 | 0.00031707 | 0.773650713 |
| T2DM | dendritic_cell | *CBR3* | -1.815097095 | 0.000179426 | 0.000418571 | 1 |
| T2DM | dendritic_cell | *SIM2* | -1.947247657 | 0.00055657 | 0.001042656 | 1 |
| T2DM | dendritic_cell | *HLCS* | -1.093374779 | 0.000880688 | 0.00155008 | 1 |
| T2DM | dendritic_cell | *PADI4* | 5.993426829 | 0.000405179 | 0.000799297 | 1 |
| T2DM | dendritic_cell | *PSMD4* | -3.686292704 | 0.000433529 | 0.000844402 | 1 |
| T2DM | dendritic_cell | *STARD9* | -1.880355135 | 8.62E-05 | 0.000249123 | 0.542239362 |
| T2DM | dendritic_cell | *TCHH* | 0.767148705 | 9.18E-05 | 0.000259486 | 0.576978186 |
| T2DM | dendritic_cell | *RSPRY1* | -3.281344971 | 0.000286352 | 0.000600436 | 1 |
| T2DM | dendritic_cell | *CCDC17* | 1.263231907 | 0.000432573 | 0.000843283 | 1 |
| T2DM | dendritic_cell | *ACE* | -2.291623562 | 1.85E-05 | 0.000130129 | 0.116218457 |
| T2DM | dendritic_cell | *TEPP* | -12.28572541 | 9.94E-06 | 0.000130129 | 0.062476172 |
| T2DM | dendritic_cell | *EFCAB14* | -3.203429379 | 0.000187471 | 0.000431416 | 1 |

| | | | | | | |
|---|---|---|---|---|---|---|
| T2DM | dendritic_cell | *ATP6V0D1* | -2.574187521 | 0.00218064 | 0.003491134 | 1 |
| T2DM | dendritic_cell | *ZFYVE28* | -2.1334006 | 5.25E-06 | 0.00012486 | 0.033004567 |
| T2DM | dendritic_cell | *C16orf86* | -1.975635074 | 0.020285553 | 0.026759394 | 1 |
| T2DM | dendritic_cell | *FAM131B* | 4.779350911 | 0.000874933 | 0.001541678 | 1 |
| T2DM | dendritic_cell | *LYPD5* | 1.014359585 | 0.005200054 | 0.007613587 | 1 |
| T2DM | dendritic_cell | *CCDC117* | -4.165593264 | 0.006120216 | 0.008825183 | 1 |
| T2DM | dendritic_cell | *CCDC107* | -2.32126498 | 2.91E-06 | 9.83E-05 | 0.018285302 |
| T2DM | dendritic_cell | *NPR2* | -1.91123897 | 6.99E-05 | 0.000215653 | 0.439499975 |
| T2DM | dendritic_cell | *ZNF235* | -2.974739446 | 6.79E-05 | 0.000212178 | 0.427055625 |
| T2DM | dendritic_cell | *DFFA* | -1.866602789 | 0.001501844 | 0.002490003 | 1 |
| T2DM | dendritic_cell | *ATAD3B* | 2.14988792 | 0.003075355 | 0.004743562 | 1 |
| T2DM | dendritic_cell | *SSU72* | -3.182831219 | 0.017375039 | 0.023158123 | 1 |
| T2DM | dendritic_cell | *NR2F6* | -1.956326203 | 0.000797804 | 0.001426969 | 1 |
| T2DM | dendritic_cell | *ANKLE1* | -2.265775856 | 0.012981515 | 0.017638812 | 1 |
| T2DM | dendritic_cell | *CILP2* | 4.262230256 | 3.75E-05 | 0.000156594 | 0.235830561 |
| T2DM | dendritic_cell | *RSPH1* | -2.478032712 | 0.000572326 | 0.001068036 | 1 |
| T2DM | dendritic_cell | *U2AF1* | -2.373916669 | 0.008269242 | 0.01165144 | 1 |
| T2DM | dendritic_cell | *C21orf33* | -4.384520056 | 1.16E-05 | 0.000130129 | 0.072622066 |
| T2DM | dendritic_cell | *ZNF66* | -2.271564903 | 0.002432527 | 0.003848339 | 1 |
| T2DM | dendritic_cell | *LRRC3* | -2.00948753 | 0.000465231 | 0.000894741 | 1 |
| T2DM | dendritic_cell | *ITGB2* | 0.905406642 | 0.0088576 | 0.012394332 | 1 |
| T2DM | dendritic_cell | *FAM207A* | 1.717291078 | 0.000100883 | 0.000274348 | 0.63424966 |
| T2DM | dendritic_cell | *LSS* | -1.745090964 | 1.86E-05 | 0.000130129 | 0.116677058 |
| T2DM | dendritic_cell | *MCM3AP* | -1.046463553 | 0.01389535 | 0.018791152 | 1 |
| T2DM | dendritic_cell | *PCNT* | -1.055133696 | 0.022170732 | 0.029051144 | 1 |
| T2DM | dendritic_cell | *DIP2A* | -2.906539924 | 0.0003974 | 0.000786914 | 1 |
| T2DM | dendritic_cell | *S100B* | -0.461556838 | 0.026596316 | 0.034412645 | 1 |
| T2DM | dendritic_cell | *PRMT2* | -2.25525333 | 0.001413479 | 0.002357172 | 1 |
| T2DM | dendritic_cell | *ZNF208* | -1.530777357 | 3.70E-05 | 0.000155835 | 0.232350694 |
| T2DM | dendritic_cell | *FCN2* | 1.108967082 | 0.0255271 | 0.033117804 | 1 |
| T2DM | dendritic_cell | *GPSM1* | -2.391950741 | 7.62E-07 | 5.26E-05 | 0.004789753 |
| T2DM | dendritic_cell | *C19orf47* | 2.877263241 | 0.002287836 | 0.003645116 | 1 |
| T2DM | dendritic_cell | *HIPK4* | -2.392315174 | 1.16E-05 | 0.000130129 | 0.072658995 |
| T2DM | dendritic_cell | *RDH13* | -3.804157176 | 1.58E-05 | 0.000130129 | 0.099184411 |
| T2DM | dendritic_cell | *MPZL3* | 2.129395834 | 9.26E-05 | 0.000261233 | 0.582027577 |
| T2DM | dendritic_cell | *PCSK7* | -3.475679776 | 1.16E-05 | 0.000130129 | 0.073141373 |
| T2DM | dendritic_cell | *S100A1* | 3.234298039 | 1.67E-05 | 0.000130129 | 0.105214739 |
| T2DM | dendritic_cell | *ZBTB7B* | -2.881197794 | 0.00018469 | 0.00042752 | 1 |
| T2DM | dendritic_cell | *FDPS* | -1.506857593 | 0.034440115 | 0.04392879 | 1 |
| T2DM | dendritic_cell | *PAQR6* | -4.414092192 | 3.18E-06 | 0.000102388 | 0.019987097 |
| T2DM | dendritic_cell | *PMF1* | -2.137030012 | 0.000620519 | 0.001143713 | 1 |
| T2DM | dendritic_cell | *FCRL3* | -1.438378744 | 0.000721063 | 0.001303054 | 1 |
| T2DM | dendritic_cell | *FGFR4* | 6.628596635 | 0.000189053 | 0.000434104 | 1 |
| T2DM | dendritic_cell | *NACC1* | 2.367549658 | 0.000147933 | 0.00036189 | 0.930056787 |
| T2DM | dendritic_cell | *LY6K* | 3.164886639 | 6.79E-05 | 0.000212178 | 0.42711415 |
| T2DM | dendritic_cell | *LY6E* | -1.925642392 | 0.023178802 | 0.030271111 | 1 |
| T2DM | dendritic_cell | *TONSL* | -2.068732211 | 2.08E-05 | 0.000132272 | 0.130689322 |
| T2DM | dendritic_cell | *MUM1* | -1.534598112 | 0.000238668 | 0.000519027 | 1 |
| T2DM | dendritic_cell | *ZNF333* | -3.219708579 | 0.000887064 | 0.001559556 | 1 |
| T2DM | dendritic_cell | *PPP1R16A* | -1.134257141 | 0.000720937 | 0.001303054 | 1 |

| | | | | | | |
|---|---|---|---|---|---|---|
| T2DM | dendritic_cell | *ORAI2* | 2.378445166 | 0.001825319 | 0.002969922 | 1 |
| T2DM | dendritic_cell | *RPL8* | -3.362797542 | 0.001553861 | 0.002566095 | 1 |
| T2DM | dendritic_cell | *LRWD1* | -0.992385403 | 0.035050588 | 0.044662149 | 1 |
| T2DM | dendritic_cell | *FBXL13* | -3.014296994 | 1.23E-05 | 0.000130129 | 0.077501324 |
| T2DM | dendritic_cell | *MFSD12* | -3.795576385 | 0.007027854 | 0.010041846 | 1 |
| T2DM | dendritic_cell | *CCDC116* | 1.949101411 | 0.035689291 | 0.04542996 | 1 |
| T2DM | dendritic_cell | *DVL3* | -2.677141691 | 0.002826434 | 0.0043952 | 1 |
| T2DM | dendritic_cell | *AP2M1* | -2.4712517 | 0.000599969 | 0.001112357 | 1 |
| T2DM | dendritic_cell | *DMKN* | 1.335978314 | 0.008838735 | 0.012370688 | 1 |
| T2DM | dendritic_cell | *BDH1* | 3.615399975 | 0.02018125 | 0.026640838 | 1 |
| T2DM | dendritic_cell | *COX7A1* | -2.084020746 | 1.64E-05 | 0.000130129 | 0.103225612 |
| T2DM | dendritic_cell | *DUSP14* | -1.8974736 | 0.000387046 | 0.000769562 | 1 |
| T2DM | dendritic_cell | *PGAP3* | -3.201117757 | 1.55E-05 | 0.000130129 | 0.097497504 |
| T2DM | dendritic_cell | *IKZF3* | -1.472172229 | 1.87E-05 | 0.000130129 | 0.117652068 |
| T2DM | dendritic_cell | *ACOX1* | -1.859352325 | 5.20E-06 | 0.000124852 | 0.032711128 |
| T2DM | dendritic_cell | *PRPSAP1* | -3.292750444 | 1.73E-05 | 0.000130129 | 0.108842278 |
| T2DM | dendritic_cell | *CYGB* | -2.424576419 | 6.11E-06 | 0.000130129 | 0.038410516 |
| T2DM | dendritic_cell | *ZNF577* | 1.347485727 | 0.000990041 | 0.001717546 | 1 |
| T2DM | dendritic_cell | *TMEM143* | -1.500092275 | 0.003324483 | 0.005082934 | 1 |
| T2DM | dendritic_cell | *ALDH16A1* | -3.290456445 | 0.000365445 | 0.000734042 | 1 |
| T2DM | dendritic_cell | *SIGLEC11* | 1.293334895 | 0.000274158 | 0.000580347 | 1 |
| T2DM | dendritic_cell | *MPP3* | -3.791216552 | 0.002560036 | 0.004027764 | 1 |
| T2DM | dendritic_cell | *NAGS* | 5.166860026 | 0.00011947 | 0.000309992 | 0.751110105 |
| T2DM | dendritic_cell | *ASB16* | -2.239513371 | 0.00019993 | 0.00045345 | 1 |
| T2DM | dendritic_cell | *PLCD3* | -1.235595843 | 0.000417719 | 0.000819152 | 1 |
| T2DM | dendritic_cell | *RACGAP1* | 2.078996243 | 8.85E-05 | 0.000253593 | 0.556382949 |
| T2DM | dendritic_cell | *LARP4* | -1.650794077 | 0.000172014 | 0.000406714 | 1 |
| T2DM | dendritic_cell | *SPC24* | 2.681838864 | 0.000360146 | 0.000725485 | 1 |
| T2DM | dendritic_cell | *IP6K3* | -5.338565975 | 0.000108734 | 0.000289666 | 0.683610839 |
| T2DM | dendritic_cell | *LEMD2* | -2.710755704 | 0.00031342 | 0.000646904 | 1 |
| T2DM | dendritic_cell | *ASGR2* | 0.839209643 | 2.80E-05 | 0.000142583 | 0.175866714 |
| T2DM | dendritic_cell | *SENP3* | -1.869224951 | 8.58E-06 | 0.000130129 | 0.053933638 |
| T2DM | dendritic_cell | *SNRNP25* | 1.695302276 | 0.000186525 | 0.000430185 | 1 |
| T2DM | dendritic_cell | *WDR90* | -1.525130016 | 0.000186668 | 0.000430357 | 1 |
| T2DM | dendritic_cell | *MEIOB* | 1.991769961 | 6.84E-05 | 0.000212936 | 0.429806955 |
| T2DM | dendritic_cell | *FLYWCH2* | -3.422291843 | 7.09E-05 | 0.000217986 | 0.445999661 |
| T2DM | dendritic_cell | *ZG16B* | -19.52754691 | 1.29E-05 | 0.000130129 | 0.08100574 |
| T2DM | dendritic_cell | *TTC9C* | -2.042331962 | 1.31E-05 | 0.000130129 | 0.082443705 |
| T2DM | dendritic_cell | *NXF1* | -2.509560266 | 0.000663286 | 0.001210824 | 1 |
| T2DM | dendritic_cell | *RPL29* | -2.380308457 | 6.64E-05 | 0.000209348 | 0.417440613 |
| T2DM | dendritic_cell | *TPCN2* | -2.088267044 | 0.018876139 | 0.025047338 | 1 |
| T2DM | dendritic_cell | *CYP4A22* | 2.748338872 | 1.11E-05 | 0.000130129 | 0.069872299 |
| T2DM | dendritic_cell | *PDZK1IP1* | -28.96802219 | 0.00028297 | 0.000595194 | 1 |
| T2DM | dendritic_cell | *COA7* | 4.165036673 | 0.010058088 | 0.013949031 | 1 |
| T2DM | dendritic_cell | *SLC1A7* | -0.564061452 | 0.017240939 | 0.022994014 | 1 |
| T2DM | dendritic_cell | *C1orf123* | -4.155107702 | 0.000164073 | 0.000391917 | 1 |
| T2DM | dendritic_cell | *ACOT11* | -3.00262538 | 0.000677246 | 0.001233444 | 1 |
| T2DM | dendritic_cell | *USP24* | -1.642840392 | 5.85E-05 | 0.000194469 | 0.367741133 |
| T2DM | dendritic_cell | *NOL9* | -2.414562804 | 9.28E-05 | 0.000261455 | 0.5831944 |
| T2DM | dendritic_cell | *ZSWIM5* | -1.984548776 | 0.000329603 | 0.000673015 | 1 |

| | | | | | | |
|---|---|---|---|---|---|---|
| T2DM | dendritic_cell | *RBP7* | -4.010461175 | 0.001981101 | 0.003197736 | 1 |
| T2DM | dendritic_cell | *FBLIM1* | -2.031196401 | 0.000192711 | 0.000440572 | 1 |
| T2DM | dendritic_cell | *SLC25A34* | -3.454189488 | 8.92E-06 | 0.000130129 | 0.056065863 |
| T2DM | dendritic_cell | *PDPN* | -1.2028275 | 3.02E-05 | 0.000145545 | 0.190081519 |
| T2DM | dendritic_cell | *LRRC38* | 4.019730192 | 5.04E-05 | 0.000178067 | 0.316799576 |
| T2DM | dendritic_cell | *LAPTM5* | 2.328328248 | 0.003843382 | 0.005800131 | 1 |
| T2DM | dendritic_cell | *PEF1* | 3.378699732 | 0.024052738 | 0.031366846 | 1 |
| T2DM | dendritic_cell | *SYNC* | -2.247630735 | 0.000811699 | 0.001448114 | 1 |
| T2DM | dendritic_cell | *RBBP4* | -3.090937601 | 0.000535251 | 0.00100834 | 1 |
| T2DM | dendritic_cell | *TSSK3* | -1.378474136 | 0.027697023 | 0.035785282 | 1 |
| T2DM | dendritic_cell | *CAMK2N1* | -2.30129917 | 2.04E-05 | 0.00013202 | 0.127991347 |
| T2DM | dendritic_cell | *TTLL10* | -3.203368486 | 9.98E-05 | 0.000272512 | 0.62732305 |
| T2DM | dendritic_cell | *NFIA* | -1.480559135 | 4.66E-05 | 0.00017094 | 0.292819451 |
| T2DM | dendritic_cell | *OMA1* | -1.420479492 | 0.001785107 | 0.002909011 | 1 |
| T2DM | dendritic_cell | *TM2D1* | -2.584350026 | 7.21E-05 | 0.00021994 | 0.453516916 |
| T2DM | dendritic_cell | *FUBP1* | -2.553482988 | 0.001951132 | 0.00315341 | 1 |
| T2DM | dendritic_cell | *NEXN* | -2.178762553 | 2.07E-05 | 0.000132272 | 0.130365124 |
| T2DM | dendritic_cell | *B3GALT2* | -3.965501101 | 7.71E-06 | 0.000130129 | 0.048477854 |
| T2DM | dendritic_cell | *NTNG1* | -0.795461992 | 0.009374424 | 0.013065175 | 1 |
| T2DM | dendritic_cell | *HFM1* | -2.136606153 | 1.44E-05 | 0.000130129 | 0.090630351 |
| T2DM | dendritic_cell | *KCNT2* | -0.81892777 | 2.50E-05 | 0.000138369 | 0.156910949 |
| T2DM | dendritic_cell | *VCAM1* | -2.548145018 | 0.000105255 | 0.000282673 | 0.661737383 |
| T2DM | dendritic_cell | *EXTL2* | -2.154452955 | 6.15E-07 | 4.72E-05 | 0.003867998 |
| T2DM | dendritic_cell | *ZNF496* | -3.066744066 | 6.33E-05 | 0.000203087 | 0.397870123 |
| T2DM | dendritic_cell | *TRIM58* | -1.962225012 | 0.000693014 | 0.001259606 | 1 |
| T2DM | dendritic_cell | *IGSF8* | -2.147324307 | 1.42E-05 | 0.000130129 | 0.089285625 |
| T2DM | dendritic_cell | *VANGL2* | -3.190126939 | 0.00734386 | 0.010457359 | 1 |
| T2DM | dendritic_cell | *FCRLB* | 2.5291572 | 4.17E-05 | 0.000162756 | 0.262362997 |
| T2DM | dendritic_cell | *LMX1A* | -0.612173989 | 0.007343958 | 0.010457359 | 1 |
| T2DM | dendritic_cell | *TDRD5* | -1.71246602 | 0.002714737 | 0.004243549 | 1 |
| T2DM | dendritic_cell | *SNED1* | -1.141217074 | 1.52E-05 | 0.000130129 | 0.095732162 |
| T2DM | dendritic_cell | *C1orf115* | -1.244816984 | 0.018846183 | 0.025018149 | 1 |
| T2DM | dendritic_cell | *ACP6* | -1.836922689 | 0.000142582 | 0.000353058 | 0.896413397 |
| T2DM | dendritic_cell | *CNST* | -2.523338432 | 3.85E-05 | 0.000158005 | 0.242118687 |
| T2DM | dendritic_cell | *PM20D1* | -3.294354724 | 8.76E-06 | 0.000130129 | 0.05509453 |
| T2DM | dendritic_cell | *B3GALNT2* | -2.243492926 | 0.001263872 | 0.002131999 | 1 |
| T2DM | dendritic_cell | *PUS10* | -3.16643121 | 0.000212499 | 0.000473584 | 1 |
| T2DM | dendritic_cell | *RFTN2* | -1.732784292 | 1.65E-05 | 0.000130129 | 0.103759354 |
| T2DM | dendritic_cell | *DISC1* | 1.255550905 | 0.001112437 | 0.00190103 | 1 |
| T2DM | dendritic_cell | *FAM84A* | -2.648674239 | 1.37E-05 | 0.000130129 | 0.086004159 |
| T2DM | dendritic_cell | *FRZB* | -3.006803401 | 0.008804051 | 0.012330378 | 1 |
| T2DM | dendritic_cell | *NUP35* | -3.320988304 | 0.016859151 | 0.022523052 | 1 |
| T2DM | dendritic_cell | *FBXO41* | 0.628872679 | 5.26E-05 | 0.000182337 | 0.330394353 |
| T2DM | dendritic_cell | *SMC6* | -3.00029531 | 8.93E-05 | 0.000255201 | 0.561696359 |
| T2DM | dendritic_cell | *CCDC74A* | -5.74777301 | 1.56E-05 | 0.000130129 | 0.097969993 |
| T2DM | dendritic_cell | *H3F3A* | 2.210434689 | 0.001932664 | 0.003125973 | 1 |
| T2DM | dendritic_cell | *NOSTRIN* | -1.338711979 | 2.16E-05 | 0.000132663 | 0.135981567 |
| T2DM | dendritic_cell | *SGPP2* | -3.173652205 | 0.00015031 | 0.000366846 | 0.944996459 |
| T2DM | dendritic_cell | *BBS5* | 2.19281313 | 0.000276175 | 0.000583385 | 1 |
| T2DM | dendritic_cell | *HPGDS* | -1.927775905 | 0.000129959 | 0.000329781 | 0.817051724 |

| | | | | | | |
|---|---|---|---|---|---|---|
| T2DM | dendritic_cell | *PDLIM5* | -2.178439847 | 9.09E-05 | 0.000257652 | 0.571214587 |
| T2DM | dendritic_cell | *RPRD2* | -2.298081151 | 5.69E-05 | 0.000191255 | 0.357982245 |
| T2DM | dendritic_cell | *CTSS* | 0.844270418 | 0.00033792 | 0.000685765 | 1 |
| T2DM | dendritic_cell | *PACRGL* | -1.671035129 | 0.000211173 | 0.000471732 | 1 |
| T2DM | dendritic_cell | *C1QTNF7* | -1.20835426 | 2.07E-05 | 0.000132272 | 0.129907188 |
| T2DM | dendritic_cell | *LYSMD1* | -1.519663816 | 1.22E-05 | 0.000130129 | 0.076878526 |
| T2DM | dendritic_cell | *DHX57* | -2.392952532 | 3.38E-05 | 0.00015067 | 0.212595974 |
| T2DM | dendritic_cell | *S100A9* | -3.744959584 | 0.001095038 | 0.001875376 | 1 |
| T2DM | dendritic_cell | *S100A12* | -10.54006469 | 0.000133789 | 0.000336992 | 0.841130988 |
| T2DM | dendritic_cell | *TDRD10* | -2.766134853 | 2.58E-05 | 0.0001395 | 0.162387936 |
| T2DM | dendritic_cell | *DCAF16* | -3.78950651 | 0.000252968 | 0.00054373 | 1 |
| T2DM | dendritic_cell | *GNPDA2* | -1.131298091 | 0.010138124 | 0.01404548 | 1 |
| T2DM | dendritic_cell | *NIPAL1* | -2.008995104 | 1.40E-05 | 0.000130129 | 0.087740684 |
| T2DM | dendritic_cell | *DCST2* | -2.981096345 | 2.78E-05 | 0.000142506 | 0.174656624 |
| T2DM | dendritic_cell | *LMOD3* | -5.559815849 | 3.22E-05 | 0.000148254 | 0.202613985 |
| T2DM | dendritic_cell | *SLC22A15* | 2.337519526 | 0.009173904 | 0.012805581 | 1 |
| T2DM | dendritic_cell | *LRRC58* | -3.336729937 | 0.008436641 | 0.011866032 | 1 |
| T2DM | dendritic_cell | *FSTL1* | -1.032121996 | 2.82E-05 | 0.000142583 | 0.177127374 |
| T2DM | dendritic_cell | *LMOD1* | -2.089652303 | 0.000557057 | 0.001043258 | 1 |
| T2DM | dendritic_cell | *TMEM183A* | -4.463506765 | 5.11E-05 | 0.000179577 | 0.32150512 |
| T2DM | dendritic_cell | *IGFBP7* | -1.426942844 | 2.27E-05 | 0.000133629 | 0.142519909 |
| T2DM | dendritic_cell | *GLB1L* | -1.318723003 | 5.17E-05 | 0.000180883 | 0.325228076 |
| T2DM | dendritic_cell | *SERPINI1* | -2.798459611 | 1.57E-05 | 0.000130129 | 0.098459965 |
| T2DM | dendritic_cell | *SPTA1* | -1.021328788 | 1.26E-06 | 6.80E-05 | 0.007908573 |
| T2DM | dendritic_cell | *PYHIN1* | -1.16514864 | 1.77E-06 | 8.21E-05 | 0.011141934 |
| T2DM | dendritic_cell | *AIM2* | 1.526427485 | 5.57E-05 | 0.000188849 | 0.350125192 |
| T2DM | dendritic_cell | *EFHB* | -0.421242454 | 0.005784124 | 0.00840072 | 1 |
| T2DM | dendritic_cell | *ICA1L* | -3.230025744 | 2.03E-07 | 2.17E-05 | 0.001278496 |
| T2DM | dendritic_cell | *CD200R1* | 0.653654596 | 0.000131065 | 0.000331963 | 0.824007099 |
| T2DM | dendritic_cell | *GTPBP8* | -3.838876003 | 0.000773243 | 0.001388569 | 1 |
| T2DM | dendritic_cell | *SPICE1* | -2.262698574 | 0.000556061 | 0.001042014 | 1 |
| T2DM | dendritic_cell | *CADPS* | -1.293298108 | 1.57E-05 | 0.000130129 | 0.098537294 |
| T2DM | dendritic_cell | *PPM1K* | -3.622494553 | 0.000151311 | 0.000368718 | 0.951293191 |
| T2DM | dendritic_cell | *GMPS* | -2.798261677 | 5.70E-05 | 0.000191255 | 0.358137412 |
| T2DM | dendritic_cell | *TIPARP* | -2.187458969 | 0.001537531 | 0.002543136 | 1 |
| T2DM | dendritic_cell | *RPP14* | -4.032420854 | 0.001911468 | 0.003096498 | 1 |
| T2DM | dendritic_cell | *DNASE1L3* | 0.566383001 | 2.32E-05 | 0.000134676 | 0.145584271 |
| T2DM | dendritic_cell | *CRELD1* | -1.44348628 | 0.000596716 | 0.001107632 | 1 |
| T2DM | dendritic_cell | *PCOLCE2* | -0.880729486 | 1.54E-05 | 0.000130129 | 0.096953546 |
| T2DM | dendritic_cell | *TTC14* | -1.593982736 | 7.20E-05 | 0.000219612 | 0.45241623 |
| T2DM | dendritic_cell | *RCHY1* | -2.951395004 | 1.11E-05 | 0.000130129 | 0.069519162 |
| T2DM | dendritic_cell | *CCDC158* | -1.247062542 | 8.69E-05 | 0.000250673 | 0.546217167 |
| T2DM | dendritic_cell | *GYG1* | -3.088887195 | 0.000509358 | 0.00096514 | 1 |
| T2DM | dendritic_cell | *RYK* | -2.548788904 | 0.000250214 | 0.000539101 | 1 |
| T2DM | dendritic_cell | *PLB1* | 0.602352828 | 4.03E-05 | 0.000160416 | 0.253135748 |
| T2DM | dendritic_cell | *CDCP1* | 0.489832965 | 3.75E-05 | 0.000156475 | 0.235494198 |
| T2DM | dendritic_cell | *LZTFL1* | -3.055334344 | 1.99E-05 | 0.000131266 | 0.125397867 |
| T2DM | dendritic_cell | *FYCO1* | -3.391988425 | 0.000115826 | 0.000302403 | 0.728198969 |
| T2DM | dendritic_cell | *LRRC2* | -0.849658605 | 7.20E-05 | 0.000219612 | 0.452619978 |
| T2DM | dendritic_cell | *SMIM12* | -2.866044853 | 2.94E-05 | 0.000143842 | 0.184533631 |

| | | | | | | |
|---|---|---|---|---|---|---|
| T2DM | dendritic_cell | *TPRA1* | -3.248049444 | 5.83E-05 | 0.000194287 | 0.366619155 |
| T2DM | dendritic_cell | *MEAF6* | -3.079947246 | 5.39E-05 | 0.000185203 | 0.339106275 |
| T2DM | dendritic_cell | *DNALI1* | -2.256224079 | 0.000324157 | 0.000664484 | 1 |
| T2DM | dendritic_cell | *RPN1* | -3.639010104 | 0.00077956 | 0.001398692 | 1 |
| T2DM | dendritic_cell | *RFC4* | -2.404094324 | 0.000230544 | 0.000505431 | 1 |
| T2DM | dendritic_cell | *RFT1* | -5.460933371 | 6.62E-08 | 1.22E-05 | 0.000416153 |
| T2DM | dendritic_cell | *SFMBT1* | -3.449620176 | 0.000104624 | 0.000281579 | 0.657769445 |
| T2DM | dendritic_cell | *GNL3* | -2.720633738 | 0.019372865 | 0.025679359 | 1 |
| T2DM | dendritic_cell | *FAM208A* | -2.525727778 | 0.000234261 | 0.000511565 | 1 |
| T2DM | dendritic_cell | *LRPAP1* | -2.66467545 | 0.026798011 | 0.034652221 | 1 |
| T2DM | dendritic_cell | *SLC51A* | -1.677222619 | 1.61E-05 | 0.000130129 | 0.101343123 |
| T2DM | dendritic_cell | *PIGX* | -3.566275231 | 1.85E-05 | 0.000130129 | 0.116550016 |
| T2DM | dendritic_cell | *ABLIM2* | -3.161947676 | 3.22E-05 | 0.000148254 | 0.202583706 |
| T2DM | dendritic_cell | *EXO5* | -2.593585403 | 4.94E-06 | 0.000123581 | 0.031085566 |
| T2DM | dendritic_cell | *C1orf50* | 2.806707088 | 0.002832668 | 0.004402715 | 1 |
| T2DM | dendritic_cell | *ERMAP* | -1.903519433 | 0.001344591 | 0.00225245 | 1 |
| T2DM | dendritic_cell | *METAP1* | -3.154724732 | 0.004403946 | 0.00655328 | 1 |
| T2DM | dendritic_cell | *DNAJB14* | -1.95861266 | 0.001089718 | 0.001867281 | 1 |
| T2DM | dendritic_cell | *SLC9B2* | -2.344933075 | 1.21E-05 | 0.000130129 | 0.076324664 |
| T2DM | dendritic_cell | *BDH2* | -1.593210991 | 3.34E-08 | 7.49E-06 | 0.000209689 |
| T2DM | dendritic_cell | *CDC25A* | -1.984634831 | 8.83E-05 | 0.00025332 | 0.555301695 |
| T2DM | dendritic_cell | *ZNF589* | -1.692047565 | 1.77E-05 | 0.000130129 | 0.111494631 |
| T2DM | dendritic_cell | *ATRIP* | 1.971543499 | 7.88E-05 | 0.000234049 | 0.495431538 |
| T2DM | dendritic_cell | *SHISA5* | -3.606552958 | 7.16E-05 | 0.000218897 | 0.450052178 |
| T2DM | dendritic_cell | *APEH* | -2.30309945 | 6.16E-05 | 0.000200088 | 0.387370177 |
| T2DM | dendritic_cell | *RNF123* | 1.546460962 | 0.00204468 | 0.0032877 | 1 |
| T2DM | dendritic_cell | *MST1R* | 1.491713869 | 0.022754808 | 0.029760657 | 1 |
| T2DM | dendritic_cell | *RAD54L2* | -2.542039967 | 0.000364114 | 0.000732538 | 1 |
| T2DM | dendritic_cell | *GRM2* | 1.728245491 | 2.81E-05 | 0.000142583 | 0.176564121 |
| T2DM | dendritic_cell | *POC1A* | 1.572037989 | 4.65E-05 | 0.000170752 | 0.29232794 |
| T2DM | dendritic_cell | *PPM1M* | 0.916157316 | 0.000535365 | 0.00100834 | 1 |
| T2DM | dendritic_cell | *C4orf3* | -3.289134794 | 0.000221733 | 0.000489824 | 1 |
| T2DM | dendritic_cell | *ANXA5* | -1.989113945 | 0.000171723 | 0.000406332 | 1 |
| T2DM | dendritic_cell | *MAP9* | -2.236419331 | 1.39E-05 | 0.000130129 | 0.087400064 |
| T2DM | dendritic_cell | *CEP44* | -3.188951411 | 0.000104577 | 0.000281573 | 0.657473725 |
| T2DM | dendritic_cell | *C4orf45* | -2.502156705 | 6.03E-06 | 0.000130129 | 0.037941869 |
| T2DM | dendritic_cell | *TMEM144* | -2.96333454 | 6.43E-05 | 0.000204784 | 0.404038428 |
| T2DM | dendritic_cell | *NPY1R* | -1.202353575 | 1.36E-05 | 0.000130129 | 0.085522451 |
| T2DM | dendritic_cell | *IL15* | 1.158558489 | 0.000135133 | 0.000339235 | 0.849579481 |
| T2DM | dendritic_cell | *ARFIP1* | -1.957104861 | 4.52E-05 | 0.000168939 | 0.284141784 |
| T2DM | dendritic_cell | *OTUD4* | -3.018074498 | 8.93E-05 | 0.000255186 | 0.56132951 |
| T2DM | dendritic_cell | *ITGA2* | 3.160683066 | 0.007155811 | 0.010217712 | 1 |
| T2DM | dendritic_cell | *NDUFAF2* | -2.658799895 | 6.49E-05 | 0.000205851 | 0.407791046 |
| T2DM | dendritic_cell | *LMBRD2* | -2.795311607 | 2.50E-05 | 0.000138591 | 0.157444445 |
| T2DM | dendritic_cell | *PGGT1B* | -3.420189234 | 0.000745596 | 0.001344296 | 1 |
| T2DM | dendritic_cell | *CMBL* | -1.103905774 | 1.54E-05 | 0.000130129 | 0.096596454 |
| T2DM | dendritic_cell | *C5orf63* | -2.457227323 | 8.78E-06 | 0.000130129 | 0.055218706 |
| T2DM | dendritic_cell | *F2RL1* | 1.751970224 | 0.012446672 | 0.016956063 | 1 |
| T2DM | dendritic_cell | *WDR41* | -2.542476893 | 3.67E-06 | 0.000110808 | 0.023047988 |
| T2DM | dendritic_cell | *NDUFS4* | -2.555032292 | 7.52E-05 | 0.000225984 | 0.472849179 |

| | | | | | | |
|---|---|---|---|---|---|---|
| T2DM | dendritic_cell | *RHOBTB3* | -1.007178028 | 0.001019796 | 0.001763813 | 1 |
| T2DM | dendritic_cell | *TIGD6* | -6.288130774 | 6.22E-05 | 0.000200868 | 0.391181564 |
| T2DM | dendritic_cell | *SERINC5* | -2.474754396 | 0.004611588 | 0.006828518 | 1 |
| T2DM | dendritic_cell | *CASP3* | -2.933093341 | 0.034895569 | 0.044482653 | 1 |
| T2DM | dendritic_cell | *PRIMPOL* | -2.613337136 | 4.47E-06 | 0.000117421 | 0.028101705 |
| T2DM | dendritic_cell | *ERAP1* | -1.277895959 | 0.019993106 | 0.026417961 | 1 |
| T2DM | dendritic_cell | *RICTOR* | -2.315455227 | 6.65E-05 | 0.000209613 | 0.418387266 |
| T2DM | dendritic_cell | *ANKRA2* | -2.885622342 | 4.07E-05 | 0.00016095 | 0.255622809 |
| T2DM | dendritic_cell | *UBLCP1* | -1.698261322 | 0.002799688 | 0.004358999 | 1 |
| T2DM | dendritic_cell | *UTP15* | -2.8168622 | 2.21E-05 | 0.000132957 | 0.138784476 |
| T2DM | dendritic_cell | *TLR3* | -3.430477368 | 7.70E-05 | 0.00023001 | 0.484170576 |
| T2DM | dendritic_cell | *CCDC127* | 1.71975747 | 0.000107477 | 0.000286804 | 0.675709083 |
| T2DM | dendritic_cell | *GDF9* | -2.379176719 | 0.014569729 | 0.019643981 | 1 |
| T2DM | dendritic_cell | *GRIK2* | -0.946952101 | 9.78E-06 | 0.000130129 | 0.061472351 |
| T2DM | dendritic_cell | *MB21D1* | -2.874187066 | 2.43E-05 | 0.000137123 | 0.152754531 |
| T2DM | dendritic_cell | *TXLNB* | 2.186110025 | 0.0001574 | 0.00038002 | 0.989571324 |
| T2DM | dendritic_cell | *DCBLD1* | -2.232400289 | 0.007328438 | 0.010440491 | 1 |
| T2DM | dendritic_cell | *TMEM200A* | -1.686265255 | 4.21E-05 | 0.000163644 | 0.264868736 |
| T2DM | dendritic_cell | *PDSS2* | -2.387729002 | 8.47E-05 | 0.000246303 | 0.532377167 |
| T2DM | dendritic_cell | *STXBP5* | -2.215128024 | 6.43E-05 | 0.000204784 | 0.404027976 |
| T2DM | dendritic_cell | *PI16* | -1.280242864 | 2.01E-05 | 0.000131406 | 0.126543782 |
| T2DM | dendritic_cell | *DAGLB* | -1.883536819 | 0.010540857 | 0.014568118 | 1 |
| T2DM | dendritic_cell | *STK17A* | -2.544098157 | 1.27E-05 | 0.000130129 | 0.079705993 |
| T2DM | dendritic_cell | *GALNT10* | -2.660906607 | 2.94E-05 | 0.000143842 | 0.184776099 |
| T2DM | dendritic_cell | *SAP30L* | -2.87178945 | 2.41E-05 | 0.000136859 | 0.151720541 |
| T2DM | dendritic_cell | *RPS14* | -2.740920423 | 0.000331799 | 0.00067706 | 1 |
| T2DM | dendritic_cell | *HCN1* | -1.575925596 | 5.65E-05 | 0.000190501 | 0.355500581 |
| T2DM | dendritic_cell | *SLU7* | -2.757479425 | 0.011922454 | 0.016280727 | 1 |
| T2DM | dendritic_cell | *RP9* | -2.77918335 | 4.29E-06 | 0.000116241 | 0.02694674 |
| T2DM | dendritic_cell | *CAMLG* | -1.383787866 | 0.004788073 | 0.007068001 | 1 |
| T2DM | dendritic_cell | *KCNK5* | -6.409815814 | 2.82E-05 | 0.000142583 | 0.177041451 |
| T2DM | dendritic_cell | *KIAA1324L* | -1.522359709 | 0.011366667 | 0.015606516 | 1 |
| T2DM | dendritic_cell | *USP49* | -2.184012015 | 3.51E-05 | 0.00015289 | 0.220620665 |
| T2DM | dendritic_cell | *ZNF704* | -1.8957741 | 8.62E-05 | 0.000249123 | 0.542093941 |
| T2DM | dendritic_cell | *FNDC1* | -1.085580789 | 8.56E-05 | 0.000248016 | 0.538194939 |
| T2DM | dendritic_cell | *BRI3* | -2.203178506 | 0.004589993 | 0.006805964 | 1 |
| T2DM | dendritic_cell | *LMTK2* | -2.294993724 | 0.000130372 | 0.000330638 | 0.819651819 |
| T2DM | dendritic_cell | *PEX2* | -2.918527462 | 2.24E-05 | 0.000133629 | 0.140930517 |
| T2DM | dendritic_cell | *TNFRSF11B* | -2.0902302 | 3.24E-05 | 0.000148254 | 0.203806431 |
| T2DM | dendritic_cell | *DEFB1* | -1.793778824 | 3.95E-05 | 0.000159553 | 0.248430655 |
| T2DM | dendritic_cell | *SUN1* | -1.387705372 | 0.000642752 | 0.001177786 | 1 |
| T2DM | dendritic_cell | *OXR1* | -1.922610547 | 0.000337362 | 0.000685519 | 1 |
| T2DM | dendritic_cell | *GPER1* | -1.041059366 | 0.000508212 | 0.000963502 | 1 |
| T2DM | dendritic_cell | *TMEM184A* | 1.354992993 | 0.000342357 | 0.000693204 | 1 |
| T2DM | dendritic_cell | *INTS1* | -3.332224721 | 2.42E-05 | 0.000136859 | 0.151951526 |
| T2DM | dendritic_cell | *FASTK* | -3.081736921 | 3.63E-05 | 0.000155209 | 0.228312928 |
| T2DM | dendritic_cell | *ALDH7A1* | -1.546963399 | 0.002854485 | 0.004427867 | 1 |
| T2DM | dendritic_cell | *COX6C* | -3.338837993 | 0.000121179 | 0.000313261 | 0.76185129 |
| T2DM | dendritic_cell | *YWHAZ* | -1.528598944 | 0.001719604 | 0.00280882 | 1 |
| T2DM | dendritic_cell | *SLC25A32* | -3.601913811 | 6.26E-05 | 0.000201484 | 0.393295935 |

| | | | | | | |
|---|---|---|---|---|---|---|
| T2DM | dendritic_cell | *DCAF13* | -3.717738518 | 0.000408788 | 0.000805153 | 1 |
| T2DM | dendritic_cell | *INTS8* | -1.879732061 | 0.002308724 | 0.003672811 | 1 |
| T2DM | dendritic_cell | *FREM1* | -1.093624121 | 7.65E-06 | 0.000130129 | 0.048071151 |
| T2DM | dendritic_cell | *FAM219A* | -2.744841186 | 5.28E-05 | 0.000182762 | 0.332079343 |
| T2DM | dendritic_cell | *CCDC171* | -2.585926498 | 5.88E-06 | 0.000129745 | 0.036975643 |
| T2DM | dendritic_cell | *SYK* | 0.49298794 | 5.68E-05 | 0.000191039 | 0.357243373 |
| T2DM | dendritic_cell | *NIPSNAP3B* | -1.679028415 | 2.02E-05 | 0.000131766 | 0.127286223 |
| T2DM | dendritic_cell | *TMC1* | -1.770449986 | 2.89E-05 | 0.000143316 | 0.181437449 |
| T2DM | dendritic_cell | *HGSNAT* | -2.521699344 | 0.000194647 | 0.000443547 | 1 |
| T2DM | dendritic_cell | *C9orf64* | -1.607281757 | 0.001243149 | 0.002098732 | 1 |
| T2DM | dendritic_cell | *HNRNPK* | -2.991288294 | 0.000929571 | 0.001623843 | 1 |
| T2DM | dendritic_cell | *SVEP1* | -0.915209209 | 1.96E-05 | 0.000131266 | 0.123002875 |
| T2DM | dendritic_cell | *NOL6* | 3.029513312 | 0.023947918 | 0.031249597 | 1 |
| T2DM | dendritic_cell | *AQP3* | 1.668240966 | 5.20E-05 | 0.000181132 | 0.326780336 |
| T2DM | dendritic_cell | *TRMT10B* | -2.132483765 | 2.10E-05 | 0.000132395 | 0.13199752 |
| T2DM | dendritic_cell | *VCP* | -2.039522899 | 0.000107285 | 0.000286533 | 0.674499156 |
| T2DM | dendritic_cell | *PIGO* | 3.022633367 | 0.00773712 | 0.010972992 | 1 |
| T2DM | dendritic_cell | *LRFN5* | -0.821740096 | 0.000921499 | 0.001611085 | 1 |
| T2DM | dendritic_cell | *WRN* | -2.156561034 | 8.28E-05 | 0.000242223 | 0.520536513 |
| T2DM | dendritic_cell | *CFL2* | -2.774424473 | 0.011431823 | 0.015689123 | 1 |
| T2DM | dendritic_cell | *ZCCHC24* | -1.971763095 | 2.26E-05 | 0.000133629 | 0.142123816 |
| T2DM | dendritic_cell | *PGM2L1* | -2.238605715 | 0.000590871 | 0.001098406 | 1 |
| T2DM | dendritic_cell | *GJB2* | 2.038451731 | 0.000328452 | 0.000671318 | 1 |
| T2DM | dendritic_cell | *REEP3* | -3.278209685 | 2.28E-06 | 8.72E-05 | 0.01432789 |
| T2DM | dendritic_cell | *C10orf25* | 1.212641261 | 4.01E-05 | 0.000160039 | 0.252167632 |
| T2DM | dendritic_cell | *KLHDC2* | -2.057907229 | 0.000261604 | 0.000558664 | 1 |
| T2DM | dendritic_cell | *OXGR1* | 2.062596148 | 6.21E-05 | 0.000200487 | 0.390147284 |
| T2DM | dendritic_cell | *ATP5C1* | -2.818364948 | 0.002305909 | 0.003669261 | 1 |
| T2DM | dendritic_cell | *TAF3* | -2.156324233 | 0.000217011 | 0.000481931 | 1 |
| T2DM | dendritic_cell | *VDAC2* | -3.108295717 | 0.004337497 | 0.006466645 | 1 |
| T2DM | dendritic_cell | *SLC18A2* | 2.845127304 | 0.008151777 | 0.011498815 | 1 |
| T2DM | dendritic_cell | *QSOX2* | 3.426102614 | 0.009469871 | 0.013189428 | 1 |
| T2DM | dendritic_cell | *CLEC1B* | 0.474490557 | 0.019665201 | 0.026012017 | 1 |
| T2DM | dendritic_cell | *SNAPC4* | -1.213173355 | 0.020202992 | 0.026661673 | 1 |
| T2DM | dendritic_cell | *SDCCAG3* | -3.194817372 | 6.88E-05 | 0.000213638 | 0.432829994 |
| T2DM | dendritic_cell | *AK8* | -3.393416005 | 0.021207689 | 0.02785309 | 1 |
| T2DM | dendritic_cell | *TSC1* | -2.561784347 | 0.000563954 | 0.001053663 | 1 |
| T2DM | dendritic_cell | *ZMYND19* | -6.62953808 | 3.58E-05 | 0.000154397 | 0.225110767 |
| T2DM | dendritic_cell | *STOX1* | -0.626223409 | 0.001610631 | 0.002649408 | 1 |
| T2DM | dendritic_cell | *STK32C* | 2.187022217 | 0.024538154 | 0.031933632 | 1 |
| T2DM | dendritic_cell | *CASP7* | -1.055701336 | 0.035943452 | 0.045707217 | 1 |
| T2DM | dendritic_cell | *VWA2* | 2.288914537 | 0.002967509 | 0.004588473 | 1 |
| T2DM | dendritic_cell | *METTL3* | -2.681239994 | 6.22E-05 | 0.000200868 | 0.391290891 |
| T2DM | dendritic_cell | *TRUB1* | -2.044738184 | 0.002085749 | 0.003349044 | 1 |
| T2DM | dendritic_cell | *HSPA12A* | -0.81115803 | 0.000447826 | 0.00086738 | 1 |
| T2DM | dendritic_cell | *UBTD1* | -3.787793121 | 0.000657269 | 0.001200887 | 1 |
| T2DM | dendritic_cell | *ARHGAP42* | -1.146863877 | 1.37E-05 | 0.000130129 | 0.08586774 |
| T2DM | dendritic_cell | *OTOGL* | 2.324177811 | 0.003256683 | 0.004987763 | 1 |
| T2DM | dendritic_cell | *TTC7B* | -1.690775166 | 1.09E-05 | 0.000130129 | 0.068818476 |
| T2DM | dendritic_cell | *SLC39A13* | -2.833431103 | 3.64E-05 | 0.000155217 | 0.229132895 |

| | | | | | | |
|---|---|---|---|---|---|---|
| T2DM | dendritic_cell | *TC2N* | -0.913409315 | 0.000409572 | 0.000805938 | 1 |
| T2DM | dendritic_cell | *IFI27L1* | -3.091758861 | 0.000104686 | 0.000281627 | 0.658162354 |
| T2DM | dendritic_cell | *CLMN* | -1.897163715 | 2.82E-05 | 0.000142583 | 0.177345652 |
| T2DM | dendritic_cell | *PTER* | -3.355987494 | 7.23E-05 | 0.000219971 | 0.454459215 |
| T2DM | dendritic_cell | *ABTB2* | -3.368494398 | 5.11E-05 | 0.000179549 | 0.321213089 |
| T2DM | dendritic_cell | *LIPC* | -1.171432415 | 5.04E-05 | 0.000178067 | 0.316960031 |
| T2DM | dendritic_cell | *CEP57* | -2.887226423 | 1.32E-06 | 6.92E-05 | 0.008302271 |
| T2DM | dendritic_cell | *TCP11L2* | -2.964723455 | 7.27E-05 | 0.000220547 | 0.457194339 |
| T2DM | dendritic_cell | *JAM3* | -1.837007874 | 9.34E-05 | 0.000262424 | 0.587323598 |
| T2DM | dendritic_cell | *GPT2* | -0.98900269 | 6.95E-05 | 0.000214823 | 0.436681491 |
| T2DM | dendritic_cell | *IKBIP* | -2.529591382 | 0.001522543 | 0.00251967 | 1 |
| T2DM | dendritic_cell | *HIF1AN* | -4.031983597 | 0.000787213 | 0.001410432 | 1 |
| T2DM | dendritic_cell | *NDUFB8* | -2.311569836 | 1.58E-05 | 0.000130129 | 0.099626214 |
| T2DM | dendritic_cell | *SPINT1* | 0.578608494 | 4.68E-05 | 0.000171433 | 0.294057107 |
| T2DM | dendritic_cell | *TRMT61A* | -4.581219468 | 9.41E-07 | 5.75E-05 | 0.005913032 |
| T2DM | dendritic_cell | *BTRC* | -2.374851629 | 0.000141177 | 0.000350328 | 0.887582146 |
| T2DM | dendritic_cell | *LARP6* | -2.281683894 | 0.000622811 | 0.001147264 | 1 |
| T2DM | dendritic_cell | *ASPG* | 1.581208204 | 2.68E-05 | 0.000140354 | 0.168424563 |
| T2DM | dendritic_cell | *TBATA* | -5.010526329 | 0.022081537 | 0.028940301 | 1 |
| T2DM | dendritic_cell | *PCBD1* | -1.914662469 | 7.26E-09 | 2.70E-06 | 4.56E-05 |
| T2DM | dendritic_cell | *COX11* | -2.429021298 | 0.001441935 | 0.002398266 | 1 |
| T2DM | dendritic_cell | *ZNF202* | -3.521644711 | 0.008382978 | 0.011798474 | 1 |
| T2DM | dendritic_cell | *FAM227B* | -1.5775226 | 0.001056093 | 0.001815602 | 1 |
| T2DM | dendritic_cell | *STXBP4* | -2.066299392 | 1.04E-05 | 0.000130129 | 0.065565561 |
| T2DM | dendritic_cell | *C2* | -1.263290554 | 0.010007734 | 0.013889321 | 1 |
| T2DM | dendritic_cell | *APBB1* | -3.346294082 | 0.000341172 | 0.000691473 | 1 |
| T2DM | dendritic_cell | *ILK* | -2.5122368 | 0.000191209 | 0.000437776 | 1 |
| T2DM | dendritic_cell | *TAF10* | -4.201659068 | 9.92E-05 | 0.000271818 | 0.623550041 |
| T2DM | dendritic_cell | *DCHS1* | -2.853050987 | 4.68E-05 | 0.000171433 | 0.294121508 |
| T2DM | dendritic_cell | *NETO1* | -1.818692872 | 0.000177906 | 0.000415951 | 1 |
| T2DM | dendritic_cell | *USP54* | -1.948918098 | 0.000594651 | 0.001104452 | 1 |
| T2DM | dendritic_cell | *C11orf74* | -2.128396983 | 7.02E-06 | 0.000130129 | 0.044156033 |
| T2DM | dendritic_cell | *WDR88* | -1.990346614 | 9.05E-06 | 0.000130129 | 0.056913291 |
| T2DM | dendritic_cell | *TUB* | -1.605798461 | 1.77E-06 | 8.21E-05 | 0.011157175 |
| T2DM | dendritic_cell | *RIC3* | -2.809376451 | 2.71E-05 | 0.000141017 | 0.170349108 |
| T2DM | dendritic_cell | *LMO1* | -4.582291657 | 0.001375462 | 0.002301099 | 1 |
| T2DM | dendritic_cell | *XRRA1* | -1.485954966 | 2.65E-05 | 0.000139973 | 0.166427394 |
| T2DM | dendritic_cell | *RPL27A* | -3.073688306 | 0.00087233 | 0.001538385 | 1 |
| T2DM | dendritic_cell | *AKIP1* | -3.518132368 | 2.05E-05 | 0.000132198 | 0.129185581 |
| T2DM | dendritic_cell | *C16orf46* | 7.168726258 | 6.37E-05 | 0.000203708 | 0.400286844 |
| T2DM | dendritic_cell | *TMEM41B* | -2.665369931 | 3.78E-05 | 0.00015701 | 0.237557924 |
| T2DM | dendritic_cell | *ZNF143* | -3.06373426 | 0.000288099 | 0.000603357 | 1 |
| T2DM | dendritic_cell | *TMX3* | -2.26240286 | 1.68E-05 | 0.000130129 | 0.105738928 |
| T2DM | dendritic_cell | *CCDC68* | -1.163772122 | 0.000165325 | 0.000394397 | 1 |
| T2DM | dendritic_cell | *CLEC4E* | 0.550461189 | 1.08E-05 | 0.000130129 | 0.067758125 |
| T2DM | dendritic_cell | *ZNF3* | -2.777592693 | 0.000459401 | 0.000886047 | 1 |
| T2DM | dendritic_cell | *CLEC4D* | -3.731548891 | 0.001356749 | 0.002271607 | 1 |
| T2DM | dendritic_cell | *RIMKLB* | -1.317783839 | 7.12E-05 | 0.000218276 | 0.447519911 |
| T2DM | dendritic_cell | *BEAN1* | -2.927680807 | 6.29E-05 | 0.000202356 | 0.395605628 |
| T2DM | dendritic_cell | *TK2* | -2.196075647 | 1.64E-06 | 8.10E-05 | 0.010281905 |

| | | | | | | |
|---|---|---|---|---|---|---|
| T2DM | dendritic_cell | *SEC11C* | -3.66587089 | 2.94E-05 | 0.000143842 | 0.184592795 |
| T2DM | dendritic_cell | *GALR1* | -1.71585414 | 2.62E-05 | 0.0001395 | 0.164424321 |
| T2DM | dendritic_cell | *CENPV* | -2.543925619 | 0.034288643 | 0.043762221 | 1 |
| T2DM | dendritic_cell | *SPDYE6* | 2.964647655 | 0.004231789 | 0.006324045 | 1 |
| T2DM | dendritic_cell | *ATF7IP2* | -2.301987922 | 0.00661444 | 0.009496456 | 1 |
| T2DM | dendritic_cell | *ZNF592* | -2.810684008 | 0.001506799 | 0.00249756 | 1 |
| T2DM | dendritic_cell | *ACSM1* | 1.078778565 | 3.36E-05 | 0.000150196 | 0.211325786 |
| T2DM | dendritic_cell | *AP1G1* | -2.304938038 | 1.79E-05 | 0.000130129 | 0.11249304 |
| T2DM | dendritic_cell | *SLFN5* | -1.927066676 | 7.96E-06 | 0.000130129 | 0.050020868 |
| T2DM | dendritic_cell | *CATSPER2* | -2.40591823 | 0.000805186 | 0.001438535 | 1 |
| T2DM | dendritic_cell | *C16orf45* | -2.65748442 | 0.000218038 | 0.000483529 | 1 |
| T2DM | dendritic_cell | *SAAL1* | -7.136608845 | 9.06E-07 | 5.64E-05 | 0.005694152 |
| T2DM | dendritic_cell | *YPEL4* | -2.343786637 | 4.32E-05 | 0.000165428 | 0.271798385 |
| T2DM | dendritic_cell | *SCNN1G* | 3.129962806 | 0.000123936 | 0.000318817 | 0.779187827 |
| T2DM | dendritic_cell | *ANKDD1A* | -2.590407765 | 0.000115185 | 0.000301342 | 0.724170501 |
| T2DM | dendritic_cell | *MTMR10* | -1.760556316 | 0.011240081 | 0.01544283 | 1 |
| T2DM | dendritic_cell | *YWHAB* | -2.820362998 | 0.000331116 | 0.000675885 | 1 |
| T2DM | dendritic_cell | *NYAP1* | -1.36406629 | 0.001744778 | 0.00284772 | 1 |
| T2DM | dendritic_cell | *MS4A6E* | -1.332656574 | 0.000170314 | 0.000403638 | 1 |
| T2DM | dendritic_cell | *MS4A7* | 0.788449074 | 0.001619928 | 0.002662611 | 1 |
| T2DM | dendritic_cell | *MS4A14* | 0.501139473 | 0.000400982 | 0.000792759 | 1 |
| T2DM | dendritic_cell | *DIS3L* | -2.569362676 | 0.00275981 | 0.004304372 | 1 |
| T2DM | dendritic_cell | *CCNDBP1* | -2.455007283 | 2.14E-05 | 0.000132591 | 0.134712881 |
| T2DM | dendritic_cell | *SMAD3* | -1.723562864 | 6.61E-05 | 0.000208955 | 0.41577984 |
| T2DM | dendritic_cell | *RCCD1* | -3.483601761 | 0.001686655 | 0.002760739 | 1 |
| T2DM | dendritic_cell | *EVA1C* | -1.070098268 | 0.004151094 | 0.006218234 | 1 |
| T2DM | dendritic_cell | *NUDT21* | -3.346657443 | 0.000211361 | 0.000471884 | 1 |
| T2DM | dendritic_cell | *NKX3-1* | -10.73268895 | 7.10E-06 | 0.000130129 | 0.04463667 |
| T2DM | dendritic_cell | *TEF* | -0.784471044 | 0.009813053 | 0.013628157 | 1 |
| T2DM | dendritic_cell | *MEI1* | 1.255699488 | 0.006792944 | 0.009732734 | 1 |
| T2DM | dendritic_cell | *PBX3* | -2.352640608 | 0.001541575 | 0.002547814 | 1 |
| T2DM | dendritic_cell | *SNRPD1* | -4.17316774 | 0.000958294 | 0.001667842 | 1 |
| T2DM | dendritic_cell | *SAMD14* | 2.603245713 | 0.000146649 | 0.000359587 | 0.921980467 |
| T2DM | dendritic_cell | *URM1* | -3.099071802 | 0.001863325 | 0.003027843 | 1 |
| T2DM | dendritic_cell | *CERCAM* | -1.053418434 | 0.001724949 | 0.00281682 | 1 |
| T2DM | dendritic_cell | *ENDOG* | -2.232586673 | 0.002260836 | 0.003605753 | 1 |
| T2DM | dendritic_cell | *ISLR2* | -1.718305589 | 0.00017268 | 0.000407216 | 1 |
| T2DM | dendritic_cell | *SP2* | -4.507288087 | 0.000255015 | 0.000547568 | 1 |
| T2DM | dendritic_cell | *COQ7* | -1.708199133 | 1.55E-05 | 0.000130129 | 0.097139387 |
| T2DM | dendritic_cell | *GPRC5B* | -1.644285246 | 1.78E-05 | 0.000130129 | 0.111640771 |
| T2DM | dendritic_cell | *KATNAL2* | -0.808889638 | 0.000982336 | 0.00170512 | 1 |
| T2DM | dendritic_cell | *RNF214* | -3.237727916 | 0.000297004 | 0.000619035 | 1 |
| T2DM | dendritic_cell | *ENGASE* | -1.610228738 | 0.005183517 | 0.007592911 | 1 |
| T2DM | dendritic_cell | *RBFOX3* | -6.049376555 | 0.000275513 | 0.000582628 | 1 |
| T2DM | dendritic_cell | *MYO5B* | -0.711461672 | 0.001424723 | 0.002374035 | 1 |
| T2DM | dendritic_cell | *RRM1* | -1.581719052 | 0.000106144 | 0.000284331 | 0.667324972 |
| T2DM | dendritic_cell | *OR51E2* | -1.279319995 | 0.032389945 | 0.041410805 | 1 |
| T2DM | dendritic_cell | *TRIM68* | 1.561945083 | 0.003680735 | 0.00557851 | 1 |
| T2DM | dendritic_cell | *FN3K* | -1.950213933 | 4.83E-05 | 0.000173951 | 0.303604205 |
| T2DM | dendritic_cell | *ZNF23* | -1.883973912 | 3.20E-05 | 0.000148254 | 0.201082962 |

| | | | | | | |
|---|---|---|---|---|---|---|
| T2DM | dendritic_cell | *ZNF229* | -3.138896333 | 0.008592272 | 0.012068725 | 1 |
| T2DM | dendritic_cell | *VKORC1* | -3.457860463 | 1.98E-07 | 2.17E-05 | 0.001243768 |
| T2DM | dendritic_cell | *SMG8* | -3.554046913 | 2.04E-05 | 0.00013202 | 0.128162882 |
| T2DM | dendritic_cell | *RAB8A* | 2.815356182 | 0.004904984 | 0.007220238 | 1 |
| T2DM | dendritic_cell | *GATAD2A* | -2.065373156 | 0.010588231 | 0.014627161 | 1 |
| T2DM | dendritic_cell | *RPL13* | -2.836932942 | 0.00317766 | 0.004886973 | 1 |
| T2DM | dendritic_cell | *ZNF641* | 1.732810147 | 1.88E-06 | 8.36E-05 | 0.011821181 |
| T2DM | dendritic_cell | *CACNB3* | -3.250589901 | 0.022552527 | 0.029514517 | 1 |
| T2DM | dendritic_cell | *TP53I13* | -1.547588832 | 0.00872171 | 0.012231405 | 1 |
| T2DM | dendritic_cell | *ZNF528* | -2.422634631 | 2.08E-05 | 0.000132272 | 0.130554366 |
| T2DM | dendritic_cell | *ZNF701* | -2.723212406 | 7.13E-06 | 0.000130129 | 0.044795442 |
| T2DM | dendritic_cell | *TMC4* | -5.192410095 | 0.00032263 | 0.000662225 | 1 |
| T2DM | dendritic_cell | *TTYH1* | -1.340880868 | 4.52E-05 | 0.000168939 | 0.284042017 |
| T2DM | dendritic_cell | *LENG8* | -2.326801773 | 0.000434138 | 0.000845024 | 1 |
| T2DM | dendritic_cell | *CDC42EP5* | -3.399196013 | 4.09E-05 | 0.000161687 | 0.257421295 |
| T2DM | dendritic_cell | *PPP1R14A* | -4.599320579 | 0.014179779 | 0.019163428 | 1 |
| T2DM | dendritic_cell | *SPINT2* | 1.004058265 | 0.007279456 | 0.010383702 | 1 |
| T2DM | dendritic_cell | *PSCA* | 3.732105427 | 2.87E-06 | 9.78E-05 | 0.018016915 |
| T2DM | dendritic_cell | *UBXN6* | -2.051327356 | 5.04E-06 | 0.000124087 | 0.031669922 |
| T2DM | dendritic_cell | *ZNF444* | -2.136074804 | 1.41E-05 | 0.000130129 | 0.088382722 |
| T2DM | dendritic_cell | *FAM57A* | -1.749244361 | 0.000861212 | 0.001523907 | 1 |
| T2DM | dendritic_cell | *GLOD4* | -3.186012589 | 0.003240527 | 0.004965438 | 1 |
| T2DM | dendritic_cell | *KIFC2* | -1.228666976 | 2.96E-06 | 9.90E-05 | 0.01862996 |
| T2DM | dendritic_cell | *SERPINF2* | -1.719174338 | 0.006350148 | 0.009131606 | 1 |
| T2DM | dendritic_cell | *CYB5D2* | -2.202046253 | 9.43E-05 | 0.000264058 | 0.592894969 |
| T2DM | dendritic_cell | *ZNF83* | -2.476747001 | 0.000134769 | 0.000338782 | 0.847292697 |
| T2DM | dendritic_cell | *CD320* | -2.094625605 | 2.14E-05 | 0.000132591 | 0.134468815 |
| T2DM | dendritic_cell | *SPRYD3* | -1.472648179 | 1.54E-05 | 0.000130129 | 0.096619034 |
| T2DM | dendritic_cell | *SOAT2* | -1.452500897 | 0.001873525 | 0.003042059 | 1 |
| T2DM | dendritic_cell | *ZNF558* | -1.224824918 | 0.007515383 | 0.010680202 | 1 |
| T2DM | dendritic_cell | *NDUFV1* | -2.658569062 | 0.000388641 | 0.000772246 | 1 |
| T2DM | dendritic_cell | *CDK2AP2* | 1.901511839 | 5.50E-05 | 0.000187773 | 0.34606491 |
| T2DM | dendritic_cell | *PRDX2* | -1.856619194 | 0.000303247 | 0.000629836 | 1 |
| T2DM | dendritic_cell | *ZNF232* | -0.918047649 | 0.019537628 | 0.025875936 | 1 |
| T2DM | dendritic_cell | *CD300C* | -1.879293196 | 0.014322833 | 0.019333814 | 1 |
| T2DM | dendritic_cell | *HID1* | -1.549602034 | 0.000127609 | 0.000325072 | 0.802277463 |
| T2DM | dendritic_cell | *ATP5H* | -2.936131648 | 4.70E-05 | 0.000171433 | 0.295302646 |
| T2DM | dendritic_cell | *SRP68* | -3.088685642 | 0.011744162 | 0.01606168 | 1 |
| T2DM | dendritic_cell | *MGAT5B* | -2.723911751 | 0.000115307 | 0.000301429 | 0.724936409 |
| T2DM | dendritic_cell | *CYP7A1* | 2.980736133 | 1.60E-05 | 0.000130129 | 0.100769806 |
| T2DM | dendritic_cell | *GSDMA* | -2.676871424 | 4.49E-05 | 0.000168807 | 0.28241433 |
| T2DM | dendritic_cell | *KRT24* | -2.725816952 | 0.000129982 | 0.000329781 | 0.817196891 |
| T2DM | dendritic_cell | *TMEM99* | -1.95328714 | 7.22E-05 | 0.000219971 | 0.454036614 |
| T2DM | dendritic_cell | *ZNF598* | -1.572199581 | 1.26E-05 | 0.000130129 | 0.079052839 |
| T2DM | dendritic_cell | *MLST8* | -3.184686683 | 1.11E-05 | 0.000130129 | 0.069817165 |
| T2DM | dendritic_cell | *ECI1* | -2.359552679 | 0.000190983 | 0.000437418 | 1 |
| T2DM | dendritic_cell | *ABCA3* | -2.358050796 | 0.000194565 | 0.000443521 | 1 |
| T2DM | dendritic_cell | *KCTD5* | -3.877393682 | 0.026767863 | 0.034620357 | 1 |
| T2DM | dendritic_cell | *SDHAF2* | -3.334229117 | 1.53E-05 | 0.000130129 | 0.096390425 |
| T2DM | dendritic_cell | *DDB1* | -2.271247014 | 2.26E-05 | 0.000133629 | 0.142161746 |

| | | | | | | |
|---|---|---|---|---|---|---|
| T2DM | dendritic_cell | *VWCE* | -1.812803633 | 1.89E-05 | 0.000130129 | 0.118719702 |
| T2DM | dendritic_cell | *BSCL2* | 1.147067856 | 5.70E-05 | 0.000191255 | 0.358492615 |
| T2DM | dendritic_cell | *TRANK1* | -3.020693414 | 0.000156381 | 0.000378287 | 0.983168102 |
| T2DM | dendritic_cell | *TTC21A* | -4.037549003 | 0.000966621 | 0.001681092 | 1 |
| T2DM | dendritic_cell | *ULK4* | -0.908898477 | 0.000255421 | 0.000548252 | 1 |
| T2DM | dendritic_cell | *NAALADL1* | -2.202459534 | 1.97E-05 | 0.000131266 | 0.123617702 |
| T2DM | dendritic_cell | *SCARA5* | -0.916327811 | 7.10E-06 | 0.000130129 | 0.04465681 |
| T2DM | dendritic_cell | *COPS6* | -2.884109858 | 0.001142685 | 0.001945845 | 1 |
| T2DM | dendritic_cell | *NUDT16L1* | 2.390785939 | 0.000408218 | 0.000804284 | 1 |
| T2DM | dendritic_cell | *KIAA1586* | -3.144904019 | 4.32E-05 | 0.000165275 | 0.271366814 |
| T2DM | dendritic_cell | *RNF187* | -2.212447378 | 0.001968197 | 0.00317854 | 1 |
| T2DM | dendritic_cell | *ZCCHC4* | -1.714047097 | 4.37E-05 | 0.000166135 | 0.274620988 |
| T2DM | dendritic_cell | *PTGDR* | -1.197051952 | 2.29E-05 | 0.000134411 | 0.14408819 |
| T2DM | dendritic_cell | *GLYCTK* | -1.631738634 | 0.000368209 | 0.000738414 | 1 |
| T2DM | dendritic_cell | *UBTD2* | -2.109273487 | 0.007449201 | 0.01059573 | 1 |
| T2DM | dendritic_cell | *POLR2J3* | 1.529049485 | 0.010601591 | 0.014642399 | 1 |
| T2DM | dendritic_cell | *NT5DC2* | -1.747471282 | 6.45E-05 | 0.00020528 | 0.405684358 |
| T2DM | dendritic_cell | *COA6* | 2.566366013 | 0.000283996 | 0.000596751 | 1 |
| T2DM | dendritic_cell | *KIF5C* | -3.222282861 | 7.99E-07 | 5.27E-05 | 0.005024867 |
| T2DM | dendritic_cell | *PDHB* | -4.412883334 | 1.23E-05 | 0.000130129 | 0.077051799 |
| T2DM | dendritic_cell | *PXK* | -1.247051086 | 0.024469538 | 0.031864123 | 1 |
| T2DM | dendritic_cell | *ACOX2* | -0.973488536 | 4.97E-05 | 0.000176792 | 0.312745146 |
| T2DM | dendritic_cell | *FAM107A* | -1.422701361 | 1.94E-05 | 0.000131266 | 0.121701097 |
| T2DM | dendritic_cell | *CX3CR1* | 0.814347039 | 0.000221176 | 0.000488765 | 1 |
| T2DM | dendritic_cell | *TAP1* | -4.027457818 | 0.001149883 | 0.001957043 | 1 |
| T2DM | dendritic_cell | *ING5* | -2.210722529 | 1.93E-05 | 0.000131132 | 0.121053462 |
| T2DM | dendritic_cell | *RFWD3* | -1.372494326 | 0.007816744 | 0.011070932 | 1 |
| T2DM | dendritic_cell | *RHOH* | -2.536335802 | 0.001472492 | 0.0024465 | 1 |
| T2DM | dendritic_cell | *KLHL30* | 1.422915918 | 0.00247679 | 0.00390951 | 1 |
| T2DM | dendritic_cell | *REEP4* | 1.624684953 | 9.59E-05 | 0.000266301 | 0.602906173 |
| T2DM | dendritic_cell | *BMP1* | -2.379891438 | 0.000222943 | 0.000491459 | 1 |
| T2DM | dendritic_cell | *ATXN2L* | -2.97077839 | 2.02E-05 | 0.000131411 | 0.126758681 |
| T2DM | dendritic_cell | *SERINC2* | -2.259833855 | 0.006172326 | 0.008894205 | 1 |
| T2DM | dendritic_cell | *TRAPPC11* | -3.125800976 | 0.0004917 | 0.00093696 | 1 |
| T2DM | dendritic_cell | *TMEM223* | -2.953388346 | 6.79E-06 | 0.000130129 | 0.042669649 |
| T2DM | dendritic_cell | *SLC20A2* | -2.47106681 | 0.000207881 | 0.000466268 | 1 |
| T2DM | dendritic_cell | *ZSWIM1* | -2.242513695 | 0.001324653 | 0.002222603 | 1 |
| T2DM | dendritic_cell | *NBPF9* | -2.279597633 | 1.49E-05 | 0.000130129 | 0.093568944 |
| T2DM | dendritic_cell | *WFDC13* | 11.46225287 | 1.33E-05 | 0.000130129 | 0.083601051 |
| T2DM | dendritic_cell | *NDUFS5* | -3.351576935 | 1.46E-05 | 0.000130129 | 0.091568395 |
| T2DM | dendritic_cell | *ZNF30* | -1.587817092 | 0.000912603 | 0.00159909 | 1 |
| T2DM | dendritic_cell | *FAM84B* | -0.991278357 | 0.011657622 | 0.015953516 | 1 |
| T2DM | dendritic_cell | *SLC16A4* | -1.070265198 | 0.015577787 | 0.020935773 | 1 |
| T2DM | dendritic_cell | *TMEM208* | -3.581884207 | 0.000590035 | 0.001097175 | 1 |
| T2DM | dendritic_cell | *LRP1B* | -1.111257449 | 9.61E-05 | 0.000266566 | 0.604304301 |
| T2DM | dendritic_cell | *DNAJC21* | -2.902917797 | 3.91E-05 | 0.00015902 | 0.246059929 |
| T2DM | dendritic_cell | *NPNT* | -2.485028936 | 2.88E-05 | 0.000143316 | 0.181190139 |
| T2DM | dendritic_cell | *CNNM3* | -4.369476271 | 0.000335248 | 0.000682326 | 1 |
| T2DM | dendritic_cell | *SNTB2* | -2.669418648 | 0.000117049 | 0.000304968 | 0.735888012 |
| T2DM | dendritic_cell | *ZNF507* | -2.614589712 | 7.54E-07 | 5.26E-05 | 0.004739055 |

| | | | | | | |
|---|---|---|---|---|---|---|
| T2DM | dendritic_cell | *STX18* | -2.667372077 | 5.85E-05 | 0.000194469 | 0.36750825 |
| T2DM | dendritic_cell | *GFM1* | -2.042491011 | 0.001392776 | 0.002325733 | 1 |
| T2DM | dendritic_cell | *ZNF608* | -1.494754978 | 1.68E-05 | 0.000130129 | 0.105799972 |
| T2DM | dendritic_cell | *INPP5D* | 1.437091779 | 0.000355991 | 0.000717804 | 1 |
| T2DM | dendritic_cell | *LETM1* | -3.518634907 | 3.84E-05 | 0.000158005 | 0.241181904 |
| T2DM | dendritic_cell | *TMEM129* | -1.998572807 | 0.001113053 | 0.001901567 | 1 |
| T2DM | dendritic_cell | *CEP120* | -3.052052571 | 0.000862661 | 0.001526042 | 1 |
| T2DM | dendritic_cell | *STXBP6* | -1.199742235 | 1.89E-05 | 0.000130129 | 0.11853736 |
| T2DM | dendritic_cell | *MFF* | -2.922532667 | 2.77E-05 | 0.000142478 | 0.174364994 |
| T2DM | dendritic_cell | *CPLX1* | -1.244554637 | 0.000367359 | 0.000737181 | 1 |
| T2DM | dendritic_cell | *SIGLEC7* | 0.59327489 | 5.29E-05 | 0.00018288 | 0.332841037 |
| T2DM | dendritic_cell | *FEM1B* | 2.700011041 | 0.00257492 | 0.004048142 | 1 |
| T2DM | dendritic_cell | *MFSD7* | -3.994609245 | 0.002519698 | 0.003971257 | 1 |
| T2DM | dendritic_cell | *COL4A3* | -1.513612898 | 8.22E-06 | 0.000130129 | 0.051702888 |
| T2DM | dendritic_cell | *IRS1* | -2.067308676 | 4.91E-27 | 3.09E-23 | 3.09E-23 |
| T2DM | dendritic_cell | *UPF3A* | -1.559658162 | 0.000136165 | 0.000340927 | 0.856068204 |
| T2DM | dendritic_cell | *FAM110B* | -1.971889551 | 1.72E-05 | 0.000130129 | 0.107884067 |
| T2DM | dendritic_cell | *CPT1C* | -2.311313169 | 0.000325462 | 0.000666509 | 1 |
| T2DM | dendritic_cell | *PCSK9* | -10.54719935 | 0.000182637 | 0.000423861 | 1 |
| T2DM | dendritic_cell | *RGS14* | 0.933073876 | 0.001109862 | 0.001897661 | 1 |
| T2DM | dendritic_cell | *RAB24* | -2.932951864 | 2.70E-06 | 9.51E-05 | 0.016994485 |
| T2DM | dendritic_cell | *CXCL10* | -0.987890554 | 0.011186763 | 0.015376953 | 1 |
| T2DM | dendritic_cell | *SH3TC2* | -1.658542291 | 6.14E-05 | 0.000199709 | 0.386036749 |
| T2DM | dendritic_cell | *GPRIN1* | -5.887395948 | 0.01498151 | 0.020164579 | 1 |
| T2DM | dendritic_cell | *STK32A* | -0.910700319 | 5.39E-06 | 0.000125356 | 0.033891316 |
| T2DM | dendritic_cell | *SLC33A1* | -2.638776932 | 2.76E-05 | 0.000142011 | 0.173537403 |
| T2DM | dendritic_cell | *SNUPN* | -2.153019824 | 2.46E-06 | 9.04E-05 | 0.015453926 |
| T2DM | dendritic_cell | *PTK2* | -2.218512067 | 0.000109056 | 0.000290306 | 0.685635395 |
| T2DM | dendritic_cell | *PTAFR* | 1.044107549 | 8.88E-05 | 0.000254328 | 0.558505152 |
| T2DM | dendritic_cell | *RNASE6* | 0.332753213 | 3.06E-05 | 0.000145994 | 0.192658111 |
| T2DM | dendritic_cell | *NPR1* | -1.834428177 | 1.78E-05 | 0.000130129 | 0.111999532 |
| T2DM | dendritic_cell | *CD52* | 1.923781163 | 0.002816505 | 0.004383012 | 1 |
| T2DM | dendritic_cell | *CLIC4* | -2.715726715 | 0.000135174 | 0.000339235 | 0.849839984 |
| T2DM | dendritic_cell | *SLC38A11* | -2.982466344 | 3.52E-06 | 0.000107473 | 0.022106604 |
| T2DM | dendritic_cell | *CCDC8* | 2.751077711 | 0.000106425 | 0.00028472 | 0.669092299 |
| T2DM | dendritic_cell | *METTL15* | -2.102006911 | 3.58E-05 | 0.000154407 | 0.225293975 |
| T2DM | dendritic_cell | *HINT1* | -3.746742315 | 0.001597429 | 0.002629755 | 1 |
| T2DM | dendritic_cell | *DTWD2* | -2.565723567 | 0.003864143 | 0.005825867 | 1 |
| T2DM | dendritic_cell | *CLIC3* | -3.789062304 | 3.96E-05 | 0.000159553 | 0.249062083 |
| T2DM | dendritic_cell | *INO80E* | -3.662215228 | 9.90E-08 | 1.56E-05 | 0.000622597 |
| T2DM | dendritic_cell | *DFFB* | -4.785325558 | 0.000169374 | 0.000401831 | 1 |
| T2DM | dendritic_cell | *C15orf40* | -1.616141181 | 0.005685896 | 0.008269078 | 1 |
| T2DM | dendritic_cell | *FAM103A1* | -1.622741642 | 0.000637199 | 0.001169656 | 1 |
| T2DM | dendritic_cell | *LUZP1* | -2.819096906 | 1.62E-05 | 0.000130129 | 0.101584648 |
| T2DM | dendritic_cell | *HEXDC* | -2.719078732 | 0.000473534 | 0.000908486 | 1 |
| T2DM | dendritic_cell | *LRRC45* | -2.392106236 | 3.88E-06 | 0.000111792 | 0.02437066 |
| T2DM | dendritic_cell | *DCXR* | -2.16336764 | 1.16E-05 | 0.000130129 | 0.072946431 |
| T2DM | dendritic_cell | *LIMS1* | -1.812325351 | 9.12E-05 | 0.00025857 | 0.573509008 |
| T2DM | dendritic_cell | *UGP2* | -1.209910754 | 7.33E-05 | 0.000221641 | 0.460569852 |
| T2DM | dendritic_cell | *CSGALNACT2* | 1.916316133 | 0.005179673 | 0.007589047 | 1 |

| | | | | | | |
|---|---|---|---|---|---|---|
| T2DM | dendritic_cell | *AVEN* | -3.192232705 | 0.00063133 | 0.001160576 | 1 |
| T2DM | dendritic_cell | *CTNND2* | -0.957637608 | 3.07E-05 | 0.00014601 | 0.193317644 |
| T2DM | dendritic_cell | *CALML6* | -2.661941716 | 0.006462432 | 0.009288823 | 1 |
| T2DM | dendritic_cell | *TPST1* | -1.330363298 | 2.16E-05 | 0.000132663 | 0.135707951 |
| T2DM | dendritic_cell | *OTUD3* | -1.959064223 | 0.000153217 | 0.000372727 | 0.963272557 |
| T2DM | dendritic_cell | *BRD3* | -3.867785893 | 0.003336721 | 0.005099032 | 1 |
| T2DM | dendritic_cell | *ZFPM2* | -1.327802195 | 1.70E-05 | 0.000130129 | 0.1070349 |
| T2DM | dendritic_cell | *ZNF747* | -4.282477648 | 0.000112481 | 0.000296259 | 0.707169776 |
| T2DM | dendritic_cell | *MAP3K2* | -2.446812404 | 8.37E-05 | 0.000244394 | 0.526423644 |
| T2DM | dendritic_cell | *PUSL1* | 2.526075887 | 0.00019677 | 0.000447088 | 1 |
| T2DM | dendritic_cell | *MYO7B* | -1.574391701 | 2.65E-05 | 0.00014009 | 0.1667067 |
| T2DM | dendritic_cell | *CNTROB* | -5.089801229 | 6.44E-05 | 0.000204952 | 0.40457442 |
| T2DM | dendritic_cell | *KCNAB3* | 0.787953085 | 0.000246178 | 0.000532046 | 1 |
| T2DM | dendritic_cell | *RNF150* | -0.887236297 | 8.44E-05 | 0.00024577 | 0.530372047 |
| T2DM | dendritic_cell | *HOXD4* | -2.978325751 | 0.000136028 | 0.000340721 | 0.855209455 |
| T2DM | dendritic_cell | *PDCD6IP* | -1.782987576 | 7.88E-05 | 0.000234051 | 0.495720698 |
| T2DM | dendritic_cell | *ZNF212* | -2.3811525 | 1.78E-05 | 0.000130129 | 0.11199033 |
| T2DM | dendritic_cell | *FAM161A* | 1.846441129 | 0.000524752 | 0.000989834 | 1 |
| T2DM | dendritic_cell | *GLB1* | -3.260352335 | 6.76E-05 | 0.000211802 | 0.424875549 |
| T2DM | dendritic_cell | *FAXDC2* | -1.841281551 | 4.40E-05 | 0.000166942 | 0.276622953 |
| T2DM | dendritic_cell | *CRTAP* | -3.326231154 | 0.004527855 | 0.006720166 | 1 |
| T2DM | dendritic_cell | *ELP5* | -4.176875433 | 3.92E-05 | 0.000159095 | 0.24659723 |
| T2DM | dendritic_cell | *STX8* | -2.27441199 | 7.54E-05 | 0.00022622 | 0.474157511 |
| T2DM | dendritic_cell | *NFRKB* | -2.169754197 | 0.013451969 | 0.018250439 | 1 |
| T2DM | dendritic_cell | *SP7* | -7.85875112 | 6.76E-05 | 0.000211802 | 0.42468981 |
| T2DM | dendritic_cell | *SEMA3E* | -0.995641504 | 2.94E-05 | 0.000143842 | 0.184646293 |
| T2DM | dendritic_cell | *GPRC5C* | -1.27227069 | 0.000628449 | 0.001156294 | 1 |
| T2DM | dendritic_cell | *ADORA2B* | 0.578701606 | 0.000691549 | 0.001257671 | 1 |
| T2DM | dendritic_cell | *MGMT* | -1.876436872 | 0.000186078 | 0.000429628 | 1 |
| T2DM | dendritic_cell | *METTL7B* | -4.108560099 | 1.47E-05 | 0.000130129 | 0.092550966 |
| T2DM | dendritic_cell | *NFXL1* | -2.820822216 | 5.24E-05 | 0.000181969 | 0.329363989 |
| T2DM | dendritic_cell | *CD14* | 1.359510868 | 0.001201658 | 0.002035071 | 1 |
| T2DM | dendritic_cell | *KRT72* | 2.072544963 | 0.000480245 | 0.000918 | 1 |
| T2DM | dendritic_cell | *TMC7* | -1.305795845 | 4.33E-06 | 0.000116241 | 0.027199005 |
| T2DM | dendritic_cell | *ARL6IP1* | -3.29065633 | 3.34E-05 | 0.000149646 | 0.210102487 |
| T2DM | dendritic_cell | *SMAGP* | -1.906791907 | 0.00039076 | 0.000775722 | 1 |
| T2DM | dendritic_cell | *IRX2* | -1.087504618 | 0.000364523 | 0.000733127 | 1 |
| T2DM | dendritic_cell | *DLGAP1* | -1.570177496 | 4.64E-05 | 0.000170668 | 0.291487042 |
| T2DM | dendritic_cell | *STAT2* | -1.924869258 | 0.000489984 | 0.00093406 | 1 |
| T2DM | dendritic_cell | *HSPA4* | -2.982953001 | 0.015639148 | 0.020995799 | 1 |
| T2DM | dendritic_cell | *SLC26A5* | -2.667464742 | 0.00032021 | 0.000658326 | 1 |
| T2DM | dendritic_cell | *COMMD5* | -4.509048599 | 4.34E-06 | 0.000116241 | 0.027310582 |
| T2DM | dendritic_cell | *GTSF1* | -13.38742699 | 0.000405648 | 0.000799972 | 1 |
| T2DM | dendritic_cell | *ARMC10* | -2.122880017 | 0.000609457 | 0.001126958 | 1 |
| T2DM | dendritic_cell | *RNF34* | -2.740774799 | 1.67E-05 | 0.000130129 | 0.105168428 |
| T2DM | dendritic_cell | *ACYP2* | -1.762305643 | 3.77E-05 | 0.000156929 | 0.236818337 |
| T2DM | dendritic_cell | *TRABD* | 2.117553217 | 0.01253005 | 0.017065949 | 1 |
| T2DM | dendritic_cell | *TMEM133* | -11.48701999 | 0.013644697 | 0.018480011 | 1 |
| T2DM | dendritic_cell | *TTLL6* | -16.23038174 | 7.53E-07 | 5.26E-05 | 0.004733311 |
| T2DM | dendritic_cell | *KCNS3* | -0.573155171 | 0.025159571 | 0.032681451 | 1 |

| | | | | | | |
|---|---|---|---|---|---|---|
| T2DM | dendritic_cell | *GPR37* | -1.365603463 | 0.030053992 | 0.038655779 | 1 |
| T2DM | dendritic_cell | *HTRA3* | -1.098055816 | 0.000758314 | 0.001364489 | 1 |
| T2DM | dendritic_cell | *KBTBD2* | -3.588862201 | 0.000491803 | 0.00093696 | 1 |
| T2DM | dendritic_cell | *MTSS1* | -2.165473614 | 0.004426628 | 0.00658374 | 1 |
| T2DM | dendritic_cell | *OSCAR* | 0.673712426 | 2.67E-05 | 0.000140322 | 0.167843234 |
| T2DM | dendritic_cell | *PAQR8* | 1.581395129 | 0.000214029 | 0.000476319 | 1 |
| T2DM | dendritic_cell | *NUDT6* | -1.854810022 | 0.000182061 | 0.000422835 | 1 |
| T2DM | dendritic_cell | *ZNF160* | -2.582075589 | 0.000615648 | 0.001137068 | 1 |
| T2DM | dendritic_cell | *FPR1* | 0.50390791 | 7.11E-05 | 0.000218276 | 0.447222099 |
| T2DM | dendritic_cell | *FEZ2* | -1.934229659 | 2.63E-05 | 0.000139569 | 0.165170403 |
| T2DM | dendritic_cell | *SOX7* | -2.244737813 | 3.04E-05 | 0.000145705 | 0.191125962 |
| T2DM | dendritic_cell | *C11orf24* | -4.045614583 | 2.62E-05 | 0.0001395 | 0.16446993 |
| T2DM | dendritic_cell | *ALK* | -2.098724642 | 1.14E-05 | 0.000130129 | 0.071456783 |
| T2DM | dendritic_cell | *TRMT61B* | -0.686022175 | 0.021655145 | 0.02841701 | 1 |
| T2DM | dendritic_cell | *MFN1* | -1.890928428 | 0.003682231 | 0.00557851 | 1 |
| T2DM | dendritic_cell | *GIMAP8* | -2.278975275 | 0.000132612 | 0.000334831 | 0.833729025 |
| T2DM | dendritic_cell | *ATP6V0E2* | -3.245364945 | 2.92E-05 | 0.000143842 | 0.183720011 |
| T2DM | dendritic_cell | *GRIK1* | 2.166443089 | 0.000844234 | 0.00149808 | 1 |
| T2DM | dendritic_cell | *CLDN20* | -1.121702415 | 5.09E-05 | 0.000179042 | 0.319948564 |
| T2DM | dendritic_cell | *CANT1* | -3.095035012 | 0.001603927 | 0.002639762 | 1 |
| T2DM | dendritic_cell | *CHST11* | 0.914080309 | 0.008201898 | 0.011566921 | 1 |
| T2DM | dendritic_cell | *LURAP1* | -1.179006569 | 0.001573612 | 0.002595304 | 1 |
| T2DM | dendritic_cell | *KCND3* | 2.11790329 | 0.00012181 | 0.000314634 | 0.765819483 |
| T2DM | dendritic_cell | *ZNF581* | -8.07491864 | 4.74E-05 | 0.00017213 | 0.297785592 |
| T2DM | dendritic_cell | *MCC* | -1.564737716 | 4.23E-05 | 0.000163993 | 0.265668015 |
| T2DM | dendritic_cell | *KRT27* | 2.719677336 | 0.001771954 | 0.002889075 | 1 |
| T2DM | dendritic_cell | *DLK2* | -4.96126724 | 4.69E-05 | 0.000171433 | 0.294966302 |
| T2DM | dendritic_cell | *ZNF561* | -2.458579477 | 0.000364898 | 0.000733647 | 1 |
| T2DM | dendritic_cell | *HOPX* | -2.216605208 | 3.59E-05 | 0.000154407 | 0.22543439 |
| T2DM | dendritic_cell | *LRRC8C* | -4.349799835 | 0.001161439 | 0.001974039 | 1 |
| T2DM | dendritic_cell | *RSL1D1* | -3.106317446 | 0.000248725 | 0.000536444 | 1 |
| T2DM | dendritic_cell | *PPID* | -3.292353705 | 8.81E-05 | 0.000252837 | 0.553966414 |
| T2DM | dendritic_cell | *COL24A1* | -0.659085348 | 0.000315107 | 0.000650173 | 1 |
| T2DM | dendritic_cell | *ETFDH* | -0.566305543 | 0.020929692 | 0.027533997 | 1 |
| T2DM | dendritic_cell | *TBCA* | -2.485617597 | 0.000686491 | 0.001249557 | 1 |
| T2DM | dendritic_cell | *MAP6* | -1.716340405 | 1.27E-05 | 0.000130129 | 0.079956489 |
| T2DM | dendritic_cell | *BCL2L1* | -2.307346406 | 0.003664128 | 0.005556288 | 1 |
| T2DM | dendritic_cell | *PLRG1* | -3.437813965 | 2.97E-05 | 0.000144362 | 0.186574293 |
| T2DM | dendritic_cell | *NMUR1* | 3.526140639 | 0.00045203 | 0.000873628 | 1 |
| T2DM | dendritic_cell | *P2RY6* | 0.453756086 | 3.98E-05 | 0.000159733 | 0.250229322 |
| T2DM | dendritic_cell | *ZIK1* | -3.969629296 | 2.34E-06 | 8.80E-05 | 0.014700281 |
| T2DM | dendritic_cell | *ANO5* | 2.509688273 | 0.000854982 | 0.001514724 | 1 |
| T2DM | dendritic_cell | *GPHN* | -1.974824231 | 4.36E-07 | 3.60E-05 | 0.002738552 |
| T2DM | dendritic_cell | *CAMTA1* | -2.621685175 | 0.005830239 | 0.008453578 | 1 |
| T2DM | dendritic_cell | *LGALS4* | -1.723451978 | 0.000869945 | 0.00153547 | 1 |
| T2DM | dendritic_cell | *LRRC34* | -0.706758614 | 0.020919763 | 0.027526695 | 1 |
| T2DM | dendritic_cell | *SPATA5L1* | -2.435316618 | 2.55E-05 | 0.000139164 | 0.160594608 |
| T2DM | dendritic_cell | *SYCE1* | -1.602034714 | 0.002075834 | 0.003334382 | 1 |
| T2DM | dendritic_cell | *NXNL1* | -2.07683792 | 0.003425715 | 0.005226272 | 1 |
| T2DM | dendritic_cell | *BCL2* | -0.766371388 | 0.003597834 | 0.005462348 | 1 |

| | | | | | | |
|---|---|---|---|---|---|---|
| T2DM | dendritic_cell | *RHNO1* | -2.321106738 | 0.024396348 | 0.031788568 | 1 |
| T2DM | dendritic_cell | *KNDC1* | -2.030698064 | 0.000205642 | 0.000463063 | 1 |
| T2DM | dendritic_cell | *METTL18* | 2.393126069 | 0.008798348 | 0.012325137 | 1 |
| T2DM | dendritic_cell | *PWWP2B* | -1.664978153 | 1.35E-05 | 0.000130129 | 0.084666035 |
| T2DM | dendritic_cell | *NINJ2* | -1.513343948 | 1.72E-05 | 0.000130129 | 0.108267551 |
| T2DM | dendritic_cell | *FAM90A1* | -3.945065868 | 1.62E-05 | 0.000130129 | 0.102129176 |
| T2DM | dendritic_cell | *TRAPPC12* | -2.34996202 | 9.20E-06 | 0.000130129 | 0.057856974 |
| T2DM | dendritic_cell | *C3AR1* | 0.906309212 | 0.000332648 | 0.000678351 | 1 |
| T2DM | dendritic_cell | *AQP4* | -1.524515557 | 1.56E-05 | 0.000130129 | 0.098183425 |
| T2DM | dendritic_cell | *TLN2* | -0.877032058 | 0.000286417 | 0.000600436 | 1 |
| T2DM | dendritic_cell | *TVP23B* | -3.460041652 | 0.000211469 | 0.000471919 | 1 |
| T2DM | dendritic_cell | *CYP4F22* | -4.153848964 | 1.16E-05 | 0.000130129 | 0.072838991 |
| T2DM | dendritic_cell | *C20orf196* | -2.613233676 | 2.01E-05 | 0.000131266 | 0.126120401 |
| T2DM | dendritic_cell | *ZNF554* | -3.099088866 | 0.0001699 | 0.000402927 | 1 |
| T2DM | dendritic_cell | *ANKRD20A4* | 1.731133418 | 4.70E-05 | 0.000171433 | 0.295785962 |
| T2DM | dendritic_cell | *LAMB2* | -2.146205592 | 2.69E-08 | 6.51E-06 | 0.00016932 |
| T2DM | dendritic_cell | *ORMDL3* | -1.083902645 | 0.001579503 | 0.002603653 | 1 |
| T2DM | dendritic_cell | *SERF1A* | -6.624154888 | 1.47E-05 | 0.000130129 | 0.09231215 |
| T2DM | dendritic_cell | *LRRC15* | -3.617312376 | 0.002086028 | 0.003349044 | 1 |
| T2DM | dendritic_cell | *SLFN12* | -1.178256996 | 0.013678355 | 0.018517618 | 1 |
| T2DM | dendritic_cell | *FRMD3* | -1.833744159 | 2.28E-05 | 0.000133984 | 0.143496477 |
| T2DM | dendritic_cell | *MTBP* | -3.761410315 | 0.000375432 | 0.000750028 | 1 |
| T2DM | dendritic_cell | *C5orf34* | -2.324546427 | 3.67E-05 | 0.000155353 | 0.230862408 |
| T2DM | dendritic_cell | *ZNF131* | -2.705775142 | 0.000180276 | 0.000420244 | 1 |
| T2DM | dendritic_cell | *HINFP* | -2.849533681 | 1.93E-05 | 0.000131132 | 0.121296934 |
| T2DM | dendritic_cell | *SPTLC3* | -2.33405113 | 2.59E-05 | 0.0001395 | 0.162678347 |
| T2DM | dendritic_cell | *COPRS* | -1.948656941 | 5.33E-05 | 0.000183801 | 0.335253702 |
| T2DM | dendritic_cell | *CLEC12A* | 0.323236857 | 0.002339567 | 0.003717174 | 1 |
| T2DM | dendritic_cell | *SUCLG2* | -1.859147921 | 0.000312454 | 0.000645335 | 1 |
| T2DM | dendritic_cell | *CSDC2* | 1.364025796 | 4.06E-05 | 0.00016095 | 0.255240847 |
| T2DM | dendritic_cell | *RCAN2* | -2.45663753 | 2.72E-05 | 0.000141161 | 0.171086994 |
| T2DM | dendritic_cell | *IL16* | -1.188290414 | 1.68E-05 | 0.000130129 | 0.105510295 |
| T2DM | dendritic_cell | *GNB2* | -4.07109722 | 1.67E-05 | 0.000130129 | 0.104756069 |
| T2DM | dendritic_cell | *PRSS27* | -4.90818514 | 0.005199107 | 0.007613587 | 1 |
| T2DM | dendritic_cell | *FGGY* | -1.836696744 | 0.001109109 | 0.00189689 | 1 |
| T2DM | dendritic_cell | *MANEA* | -2.919939966 | 0.000150786 | 0.000367725 | 0.947994149 |
| T2DM | dendritic_cell | *FIBP* | -2.428720292 | 0.000292157 | 0.000610432 | 1 |
| T2DM | dendritic_cell | *CARNS1* | -3.526697605 | 3.80E-05 | 0.000157482 | 0.239090969 |
| T2DM | dendritic_cell | *CTSW* | -0.715879852 | 2.14E-06 | 8.62E-05 | 0.013461822 |
| T2DM | dendritic_cell | *SNTG2* | -1.095021233 | 7.53E-06 | 0.000130129 | 0.047343108 |
| T2DM | dendritic_cell | *PDE3A* | -1.508546459 | 0.003600915 | 0.005465705 | 1 |
| T2DM | dendritic_cell | *SMPDL3A* | -2.974417897 | 0.000125919 | 0.000321811 | 0.791654324 |
| T2DM | dendritic_cell | *EFEMP2* | -1.830217904 | 0.000631096 | 0.001160485 | 1 |
| T2DM | dendritic_cell | *AGAP5* | -39.3522295 | 1.25E-05 | 0.000130129 | 0.078741101 |
| T2DM | dendritic_cell | *ZMAT3* | -3.607737617 | 0.010611777 | 0.014653249 | 1 |
| T2DM | dendritic_cell | *ZFAND4* | -2.098221631 | 6.05E-05 | 0.000198694 | 0.380305516 |
| T2DM | dendritic_cell | *ZNF738* | 1.621053477 | 0.002916978 | 0.004518117 | 1 |
| T2DM | dendritic_cell | *SLFN11* | -1.927088658 | 0.000233948 | 0.000511061 | 1 |
| T2DM | dendritic_cell | *CCL19* | -6.513535916 | 8.74E-06 | 0.000130129 | 0.054953381 |
| T2DM | dendritic_cell | *FUT10* | -1.5024471 | 8.23E-05 | 0.000241604 | 0.517550609 |

| | | | | | | |
|---|---|---|---|---|---|---|
| T2DM | dendritic_cell | *EFCAB12* | -1.227497677 | 0.002411852 | 0.003819474 | 1 |
| T2DM | dendritic_cell | *FAM192A* | -1.533376936 | 0.010127216 | 0.014033459 | 1 |
| T2DM | dendritic_cell | *RAB43* | 1.55585294 | 2.09E-05 | 0.000132336 | 0.131497224 |
| T2DM | dendritic_cell | *RAB37* | 3.416783583 | 0.000771104 | 0.001385519 | 1 |
| T2DM | dendritic_cell | *CYP7B1* | -1.460622717 | 1.62E-05 | 0.000130129 | 0.101890151 |
| T2DM | dendritic_cell | *METAP1D* | -1.781809256 | 1.66E-05 | 0.000130129 | 0.104539764 |
| T2DM | dendritic_cell | *RNASEH2C* | -3.871428334 | 2.26E-06 | 8.72E-05 | 0.014221885 |
| T2DM | dendritic_cell | *MYEOV* | -0.861696528 | 2.15E-06 | 8.62E-05 | 0.013539245 |
| T2DM | dendritic_cell | *MRGPRF* | 2.751787124 | 0.014292128 | 0.019303887 | 1 |
| T2DM | dendritic_cell | *LCLAT1* | -3.119248877 | 0.000300515 | 0.000625401 | 1 |
| T2DM | dendritic_cell | *ADH6* | -0.96183892 | 0.000171919 | 0.000406642 | 1 |
| T2DM | dendritic_cell | *GXYLT2* | -1.063662192 | 7.88E-06 | 0.000130129 | 0.049554938 |
| T2DM | dendritic_cell | *HPSE2* | -0.960682876 | 0.009669039 | 0.013443001 | 1 |
| T2DM | dendritic_cell | *DCAKD* | -0.928876757 | 0.013607424 | 0.018437473 | 1 |
| T2DM | dendritic_cell | *HECTD4* | -2.125857749 | 9.01E-05 | 0.000256231 | 0.56627151 |
| T2DM | dendritic_cell | *BNC2* | -1.260701523 | 3.67E-05 | 0.000155353 | 0.231009362 |
| T2DM | dendritic_cell | *TRMT112* | 1.933924494 | 0.000236865 | 0.000516279 | 1 |
| T2DM | dendritic_cell | *NOC3L* | -3.425766445 | 0.000809833 | 0.001445195 | 1 |
| T2DM | dendritic_cell | *RHOD* | -0.702060094 | 0.013878016 | 0.018771748 | 1 |
| T2DM | dendritic_cell | *AHSA2* | -2.198254348 | 9.61E-06 | 0.000130129 | 0.060442337 |
| T2DM | dendritic_cell | *ABLIM3* | -1.024243694 | 1.77E-05 | 0.000130129 | 0.111524072 |
| T2DM | dendritic_cell | *VANGL1* | -2.285722124 | 0.000127446 | 0.000324921 | 0.801254372 |
| T2DM | dendritic_cell | *GLRX* | -1.279545991 | 1.52E-05 | 0.000130129 | 0.09534772 |
| T2DM | dendritic_cell | *IQCB1* | -1.625025078 | 0.003577863 | 0.005434652 | 1 |
| T2DM | dendritic_cell | *GOLGB1* | -2.397293907 | 0.000233053 | 0.00050946 | 1 |
| T2DM | dendritic_cell | *DMRT2* | -1.006557446 | 4.00E-05 | 0.000159945 | 0.251433618 |
| T2DM | dendritic_cell | *ZNF483* | -1.109561876 | 0.000752428 | 0.001354672 | 1 |
| T2DM | dendritic_cell | *MZT2A* | 1.80233932 | 0.020603855 | 0.027142332 | 1 |
| T2DM | dendritic_cell | *MAP3K11* | 2.610261629 | 0.001577323 | 0.002600741 | 1 |
| T2DM | dendritic_cell | *C1QA* | 2.115043201 | 0.004139071 | 0.00620318 | 1 |
| T2DM | dendritic_cell | *NDNF* | -1.999788818 | 8.34E-06 | 0.000130129 | 0.052417535 |
| T2DM | dendritic_cell | *OLR1* | 0.388803664 | 2.69E-05 | 0.000140726 | 0.169293969 |
| T2DM | dendritic_cell | *GLIPR1L1* | -1.926878867 | 0.001995558 | 0.003218593 | 1 |
| T2DM | dendritic_cell | *ARV1* | -2.516576537 | 1.62E-05 | 0.000130129 | 0.101821171 |
| T2DM | dendritic_cell | *CCDC36* | -1.747507431 | 0.000126151 | 0.000322256 | 0.793110119 |
| T2DM | dendritic_cell | *THAP2* | -2.470971017 | 3.56E-05 | 0.000154222 | 0.223930651 |
| T2DM | dendritic_cell | *PPP1R14B* | -5.570081921 | 0.004984865 | 0.007325818 | 1 |
| T2DM | dendritic_cell | *SMARCC1* | -2.632059521 | 0.00081439 | 0.001451679 | 1 |
| T2DM | dendritic_cell | *VEGFB* | -2.801653379 | 5.75E-10 | 4.52E-07 | 3.61E-06 |
| T2DM | dendritic_cell | *PEAK1* | -2.28440333 | 0.000231258 | 0.000506415 | 1 |
| T2DM | dendritic_cell | *TNFRSF10D* | -1.313047628 | 2.84E-05 | 0.000142583 | 0.17878869 |
| T2DM | dendritic_cell | *GMPPB* | -1.470961262 | 0.000186749 | 0.000430385 | 1 |
| T2DM | dendritic_cell | *CSPG4* | -1.571894831 | 9.34E-05 | 0.000262424 | 0.587373441 |
| T2DM | dendritic_cell | *SNX33* | -1.463684643 | 1.18E-05 | 0.000130129 | 0.074255992 |
| T2DM | dendritic_cell | *NUDT18* | -3.461130709 | 1.19E-05 | 0.000130129 | 0.074735881 |
| T2DM | dendritic_cell | *SULT1B1* | 2.308697234 | 0.007827266 | 0.011078348 | 1 |
| T2DM | dendritic_cell | *NUDT4* | -1.79838179 | 1.09E-05 | 0.000130129 | 0.068612448 |
| T2DM | dendritic_cell | *SCAI* | -2.02363725 | 2.07E-05 | 0.000132272 | 0.130025948 |
| T2DM | dendritic_cell | *C1orf100* | -2.131949243 | 1.83E-05 | 0.000130129 | 0.115324076 |
| T2DM | dendritic_cell | *AGFG1* | -1.648884727 | 0.000257269 | 0.000551279 | 1 |

| | | | | | | |
|---|---|---|---|---|---|---|
| T2DM | dendritic_cell | *HAP1* | 1.564753697 | 0.000392013 | 0.000777717 | 1 |
| T2DM | dendritic_cell | *RNF213* | -2.539213298 | 0.000443508 | 0.000860598 | 1 |
| T2DM | dendritic_cell | *PLK3* | -4.16959181 | 0.000325334 | 0.000666509 | 1 |
| T2DM | dendritic_cell | *DPY19L1* | -3.101861232 | 5.70E-05 | 0.000191255 | 0.358603824 |
| T2DM | dendritic_cell | *TUBB8* | -5.123572697 | 3.90E-06 | 0.000111912 | 0.0245087 |
| T2DM | dendritic_cell | *GPR160* | -4.098053179 | 0.000498149 | 0.000946754 | 1 |
| T2DM | dendritic_cell | *XXYLT1* | -2.901887206 | 9.09E-05 | 0.000257652 | 0.57118971 |
| T2DM | dendritic_cell | *UBXN2A* | -2.498081037 | 2.85E-05 | 0.000142609 | 0.179349485 |
| T2DM | dendritic_cell | *CCS* | -2.604649255 | 0.008898809 | 0.012446454 | 1 |
| T2DM | dendritic_cell | *NRROS* | 1.221635406 | 0.000869272 | 0.00153547 | 1 |
| T2DM | dendritic_cell | *CEP19* | 1.578303257 | 0.005138423 | 0.007535635 | 1 |
| T2DM | dendritic_cell | *SLC25A30* | -4.235030178 | 0.004655691 | 0.006885368 | 1 |
| T2DM | dendritic_cell | *CTSF* | -1.480202925 | 0.000401919 | 0.000794362 | 1 |
| T2DM | dendritic_cell | *LEMD3* | -2.340491613 | 6.87E-05 | 0.000213481 | 0.432085107 |
| T2DM | dendritic_cell | *C16orf91* | 4.315141128 | 5.12E-05 | 0.000179577 | 0.321886076 |
| T2DM | dendritic_cell | *SOCS7* | -3.268143457 | 6.62E-05 | 0.000209064 | 0.416247304 |
| T2DM | dendritic_cell | *TLR6* | 0.958530488 | 3.79E-05 | 0.000157227 | 0.238355645 |
| T2DM | dendritic_cell | *RGMB* | -1.323249349 | 1.90E-05 | 0.000130241 | 0.119300808 |
| T2DM | dendritic_cell | *FAM53A* | -6.300504984 | 4.71E-05 | 0.000171433 | 0.296407006 |
| T2DM | dendritic_cell | *CTU2* | 1.818351652 | 0.000499068 | 0.000947928 | 1 |
| T2DM | dendritic_cell | *MGA* | -1.898308098 | 0.000222222 | 0.000490387 | 1 |
| T2DM | dendritic_cell | *DDX23* | -3.709405544 | 0.00010486 | 0.000281973 | 0.659252581 |
| T2DM | dendritic_cell | *GLIS1* | -1.014764811 | 0.00240567 | 0.003811605 | 1 |
| T2DM | dendritic_cell | *RALGAPA1* | -1.801201426 | 4.44E-05 | 0.000167569 | 0.278829103 |
| T2DM | dendritic_cell | *LIG4* | -1.579084224 | 0.00176977 | 0.002886264 | 1 |
| T2DM | dendritic_cell | *SNAPC5* | -2.758514026 | 0.000173064 | 0.000407754 | 1 |
| T2DM | dendritic_cell | *LINGO2* | -3.121368527 | 0.010376572 | 0.01435053 | 1 |
| T2DM | dendritic_cell | *ANKRD36C* | -1.608611288 | 4.04E-05 | 0.000160731 | 0.254205291 |
| T2DM | dendritic_cell | *TMEM81* | 4.349020574 | 5.60E-06 | 0.000127198 | 0.035233738 |
| T2DM | dendritic_cell | *ZNF497* | -5.775903374 | 0.001029129 | 0.001777021 | 1 |
| T2DM | dendritic_cell | *KLF14* | -18.44192976 | 1.54E-10 | 1.39E-07 | 9.69E-07 |
| T2DM | dendritic_cell | *ANGEL2* | -3.182352653 | 0.000461982 | 0.000890125 | 1 |
| T2DM | dendritic_cell | *ZNF266* | -1.816338472 | 2.58E-05 | 0.0001395 | 0.162388504 |
| T2DM | dendritic_cell | *BRSK2* | 3.345795034 | 0.00039603 | 0.000784696 | 1 |
| T2DM | dendritic_cell | *SH3PXD2B* | -0.765367837 | 0.001987476 | 0.003206379 | 1 |
| T2DM | dendritic_cell | *LARP7* | -2.864272555 | 5.96E-05 | 0.0001969 | 0.37489735 |
| T2DM | dendritic_cell | *FGFBP3* | 3.942918221 | 0.009649005 | 0.013421083 | 1 |
| T2DM | dendritic_cell | *C4orf32* | -1.988597244 | 2.25E-05 | 0.000133629 | 0.141552552 |
| T2DM | dendritic_cell | *HRAS* | 2.305929073 | 0.008015636 | 0.011327108 | 1 |
| T2DM | dendritic_cell | *BTC* | -1.268447368 | 5.27E-05 | 0.000182528 | 0.331287912 |
| T2DM | dendritic_cell | *NLRP6* | 3.230517778 | 0.005447532 | 0.007951854 | 1 |
| T2DM | dendritic_cell | *NDUFA11* | -2.058557211 | 2.97E-05 | 0.000144362 | 0.186660551 |
| T2DM | dendritic_cell | *RSRC1* | -2.0919942 | 0.000327203 | 0.00066942 | 1 |
| T2DM | dendritic_cell | *FUT1* | -2.584358473 | 0.005173102 | 0.007581187 | 1 |
| T2DM | dendritic_cell | *DHX36* | -3.248770565 | 0.000158355 | 0.000381594 | 0.995577532 |
| T2DM | dendritic_cell | *FBXW8* | -2.508559255 | 3.46E-05 | 0.000151535 | 0.217233455 |
| T2DM | dendritic_cell | *SLC22A1* | -2.006896597 | 1.41E-05 | 0.000130129 | 0.088666931 |
| T2DM | dendritic_cell | *CHST2* | 1.963406565 | 4.09E-05 | 0.000161687 | 0.257139543 |
| T2DM | dendritic_cell | *ATR* | -2.27004289 | 0.000231902 | 0.000507472 | 1 |
| T2DM | dendritic_cell | *TVP23C* | -3.362434979 | 0.000137114 | 0.000342676 | 0.862032622 |

| | | | | | | |
|---|---|---|---|---|---|---|
| T2DM | dendritic_cell | *PACS1* | -2.716078144 | 4.17E-05 | 0.000162756 | 0.261861974 |
| T2DM | dendritic_cell | *CADM2* | -1.076299874 | 3.24E-05 | 0.000148254 | 0.203674657 |
| T2DM | dendritic_cell | *ARHGAP1* | -2.536986084 | 0.005214958 | 0.007633629 | 1 |
| T2DM | dendritic_cell | *CHST1* | 2.350814385 | 0.000115096 | 0.000301252 | 0.72360842 |
| T2DM | dendritic_cell | *GOLGA8A* | -1.915666405 | 7.90E-05 | 0.000234395 | 0.496682981 |
| T2DM | dendritic_cell | *TP53I11* | -2.549506428 | 0.001427111 | 0.002377384 | 1 |
| T2DM | dendritic_cell | *PHYHD1* | -2.019557348 | 0.001015133 | 0.00175623 | 1 |
| T2DM | dendritic_cell | *ZNF519* | -2.638475471 | 3.62E-05 | 0.00015508 | 0.227304375 |
| T2DM | dendritic_cell | *BANF1* | 1.449835019 | 3.10E-06 | 0.000101654 | 0.019517653 |
| T2DM | dendritic_cell | *CHRNA7* | -1.515624419 | 0.001138677 | 0.001940597 | 1 |
| T2DM | dendritic_cell | *NRIP3* | -2.683459846 | 4.46E-06 | 0.000117421 | 0.028044701 |
| T2DM | dendritic_cell | *SCUBE2* | -3.141474817 | 1.59E-05 | 0.000130129 | 0.099958163 |
| T2DM | dendritic_cell | *SMAD2* | -1.739156422 | 0.00043605 | 0.000848482 | 1 |
| T2DM | dendritic_cell | *EIF3F* | -3.184954214 | 0.007595131 | 0.010783782 | 1 |
| T2DM | dendritic_cell | *ZNF25* | -1.921015225 | 3.05E-05 | 0.00014573 | 0.191974548 |
| T2DM | dendritic_cell | *CLTB* | -1.836074706 | 0.01610277 | 0.021595161 | 1 |
| T2DM | dendritic_cell | *PCSK1* | -1.457308362 | 3.43E-05 | 0.000151156 | 0.215330911 |
| T2DM | dendritic_cell | *LPL* | -1.049318326 | 1.08E-06 | 6.35E-05 | 0.006799521 |
| T2DM | dendritic_cell | *TBC1D10C* | -1.66222135 | 0.007312811 | 0.010425316 | 1 |
| T2DM | dendritic_cell | *PPP2R2D* | -1.678194381 | 0.000381502 | 0.000760463 | 1 |
| T2DM | dendritic_cell | *MCTP1* | -1.61607746 | 0.002642125 | 0.004144471 | 1 |
| T2DM | dendritic_cell | *LRRC25* | 0.782443091 | 1.43E-05 | 0.000130129 | 0.089740534 |
| T2DM | dendritic_cell | *UBQLNL* | -3.325801451 | 0.000110035 | 0.000291895 | 0.691791445 |
| T2DM | dendritic_cell | *LIPT2* | 2.861782301 | 0.000200427 | 0.000454249 | 1 |
| T2DM | dendritic_cell | *KCNE3* | -4.280604034 | 0.000812651 | 0.001449401 | 1 |
| T2DM | dendritic_cell | *UCP3* | 1.978824837 | 0.007014045 | 0.010026671 | 1 |
| T2DM | dendritic_cell | *UCP2* | 1.604774037 | 0.001490947 | 0.002472589 | 1 |
| T2DM | dendritic_cell | *RAB6A* | -2.52094779 | 0.000251561 | 0.000541632 | 1 |
| T2DM | dendritic_cell | *RPS6KB2* | -2.018095603 | 4.29E-05 | 0.000165006 | 0.269904742 |
| T2DM | dendritic_cell | *RMI2* | -1.503553579 | 0.000167963 | 0.000399582 | 1 |
| T2DM | dendritic_cell | *TOM1L2* | -2.550133884 | 9.72E-05 | 0.000268519 | 0.611149662 |
| T2DM | dendritic_cell | *GPR156* | -1.498520144 | 5.20E-05 | 0.000181132 | 0.326679471 |
| T2DM | dendritic_cell | *B3GNTL1* | 2.236502956 | 0.001684119 | 0.002757845 | 1 |
| T2DM | dendritic_cell | *TTLL11* | -1.338852292 | 0.002753521 | 0.004296696 | 1 |
| T2DM | dendritic_cell | *LINC01106* | -11.8933798 | 0.001036965 | 0.001788096 | 1 |
| T2DM | dendritic_cell | *MSRA* | -3.098894453 | 2.12E-05 | 0.000132591 | 0.133180954 |
| T2DM | dendritic_cell | *CTDNEP1* | -3.931352423 | 0.0015215 | 0.002518883 | 1 |
| T2DM | dendritic_cell | *ETV4* | -1.405345571 | 0.009371324 | 0.01306375 | 1 |
| T2DM | dendritic_cell | *SWI5* | -1.692258148 | 1.62E-05 | 0.000130129 | 0.102070192 |
| T2DM | dendritic_cell | *ZDHHC21* | -2.474922685 | 1.71E-05 | 0.000130129 | 0.107603676 |
| T2DM | dendritic_cell | *A2M* | -1.646068641 | 2.20E-05 | 0.000132957 | 0.138068859 |
| T2DM | dendritic_cell | *ARL4D* | -2.205295672 | 9.00E-05 | 0.000256231 | 0.565867434 |
| T2DM | dendritic_cell | *LRRN1* | -3.722458577 | 2.36E-05 | 0.000136158 | 0.148219775 |
| T2DM | dendritic_cell | *B3GALT6* | -2.455807662 | 4.12E-05 | 0.000162249 | 0.259042126 |
| T2DM | dendritic_cell | *ZNF613* | -1.497936631 | 0.007086344 | 0.010120819 | 1 |
| T2DM | dendritic_cell | *IP6K1* | -2.529070182 | 0.001217016 | 0.002059513 | 1 |
| T2DM | dendritic_cell | *UFSP1* | 1.133380993 | 0.027617466 | 0.035689827 | 1 |
| T2DM | dendritic_cell | *TMEM39A* | -2.061557135 | 0.000325378 | 0.000666509 | 1 |
| T2DM | dendritic_cell | *GPX2* | -2.651740781 | 1.99E-05 | 0.000131266 | 0.124951776 |
| T2DM | dendritic_cell | *CCDC57* | -2.120057572 | 6.82E-05 | 0.000212634 | 0.428670162 |

| | | | | | | |
|---|---|---|---|---|---|---|
| T2DM | dendritic_cell | *CIDEA* | -0.457613671 | 0.002842378 | 0.004413443 | 1 |
| T2DM | dendritic_cell | *SLC35G1* | -1.414163271 | 5.45E-05 | 0.000186501 | 0.342788525 |
| T2DM | dendritic_cell | *ZNF135* | -2.146927917 | 4.28E-06 | 0.000116241 | 0.026900023 |
| T2DM | dendritic_cell | *COX8A* | -4.80158706 | 0.000167981 | 0.000399582 | 1 |
| T2DM | dendritic_cell | *B3GNT4* | -10.4986249 | 1.49E-05 | 0.000130129 | 0.093432704 |
| T2DM | dendritic_cell | *CDC26* | -2.162186743 | 0.000946695 | 0.001650584 | 1 |
| T2DM | dendritic_cell | *RNPEP* | -2.877070836 | 0.000874234 | 0.001540877 | 1 |
| T2DM | dendritic_cell | *SPRYD4* | -1.462907068 | 0.002598867 | 0.004079669 | 1 |
| T2DM | dendritic_cell | *WDR25* | -2.999278177 | 0.000143518 | 0.000354537 | 0.902295882 |
| T2DM | dendritic_cell | *PLA2G16* | -0.562744625 | 0.002847858 | 0.004420861 | 1 |
| T2DM | dendritic_cell | *PRR15* | -4.217747103 | 0.003589178 | 0.005450522 | 1 |
| T2DM | dendritic_cell | *RMDN1* | -1.885895264 | 0.00043355 | 0.000844402 | 1 |
| T2DM | dendritic_cell | *RNF152* | -3.91756518 | 0.002679189 | 0.004193195 | 1 |
| T2DM | dendritic_cell | *LRRC37A* | -2.072445627 | 0.000598299 | 0.001110244 | 1 |
| T2DM | dendritic_cell | *ACSF3* | -2.669036105 | 3.62E-05 | 0.000155082 | 0.227889875 |
| T2DM | dendritic_cell | *BOK* | -1.433431601 | 5.13E-05 | 0.000179577 | 0.32228298 |
| T2DM | dendritic_cell | *ZNF843* | 3.712461674 | 0.00747301 | 0.010624788 | 1 |
| T2DM | dendritic_cell | *C8orf59* | -2.190211258 | 0.000157713 | 0.000380484 | 0.991540049 |
| T2DM | dendritic_cell | *RUFY1* | -1.941018751 | 0.003419983 | 0.005218795 | 1 |
| T2DM | dendritic_cell | *BASP1* | 0.886273726 | 0.000113514 | 0.000297904 | 0.713664653 |
| T2DM | dendritic_cell | *LRRC37A3* | -2.520235091 | 2.89E-05 | 0.000143353 | 0.181628236 |
| T2DM | dendritic_cell | *VSIG10* | -0.846784113 | 0.001412296 | 0.002355825 | 1 |
| T2DM | dendritic_cell | *IRX5* | -1.356410218 | 9.26E-05 | 0.000261234 | 0.582291186 |
| T2DM | dendritic_cell | *PNMA1* | -1.83282819 | 9.64E-06 | 0.000130129 | 0.060626181 |
| T2DM | dendritic_cell | *ANKLE2* | -2.401115071 | 6.84E-05 | 0.000212936 | 0.429917302 |
| T2DM | dendritic_cell | *GCNT4* | 5.025271435 | 7.18E-05 | 0.000219346 | 0.451413593 |
| T2DM | dendritic_cell | *THAP4* | -2.255797457 | 0.00045 | 0.000870245 | 1 |
| T2DM | dendritic_cell | *SHMT1* | -0.96848696 | 0.002414106 | 0.003822081 | 1 |
| T2DM | dendritic_cell | *DPP7* | -3.167777805 | 0.007993931 | 0.011298975 | 1 |
| T2DM | dendritic_cell | *SEC24C* | -2.079718696 | 4.48E-06 | 0.000117421 | 0.028181049 |
| T2DM | dendritic_cell | *MTHFR* | -3.151123274 | 0.008567159 | 0.01203883 | 1 |
| T2DM | dendritic_cell | *C19orf18* | -1.975564025 | 1.81E-05 | 0.000130129 | 0.114020298 |
| T2DM | dendritic_cell | *TMEM80* | -1.488449648 | 0.003446969 | 0.005253961 | 1 |
| T2DM | dendritic_cell | *WDR73* | -1.560286009 | 0.002422653 | 0.003834648 | 1 |
| T2DM | dendritic_cell | *POLE* | -2.337270388 | 3.54E-05 | 0.00015375 | 0.222629321 |
| T2DM | dendritic_cell | *DSCAML1* | -1.664989416 | 6.09E-05 | 0.000198694 | 0.382980801 |
| T2DM | dendritic_cell | *EPS8L2* | -1.683334441 | 2.49E-06 | 9.04E-05 | 0.015639386 |
| T2DM | dendritic_cell | *TALDO1* | -1.669719867 | 0.003491145 | 0.005311914 | 1 |
| T2DM | dendritic_cell | *RIMKLA* | -1.292366431 | 0.010688993 | 0.014743682 | 1 |
| T2DM | dendritic_cell | *B3GNT8* | 4.636096042 | 0.000104314 | 0.000281228 | 0.655824122 |
| T2DM | dendritic_cell | *AP006621.1* | -0.980715455 | 0.000365288 | 0.000734042 | 1 |
| T2DM | dendritic_cell | *MAN1B1* | -3.950614455 | 0.000119889 | 0.00031095 | 0.753743959 |
| T2DM | dendritic_cell | *FBXO39* | 0.915542255 | 0.007492782 | 0.01065049 | 1 |
| T2DM | dendritic_cell | *ZBTB38* | -2.059310253 | 5.01E-06 | 0.000124087 | 0.031529039 |
| T2DM | dendritic_cell | *C8orf31* | -2.026263342 | 1.60E-05 | 0.000130129 | 0.1008431 |
| T2DM | dendritic_cell | *AC024940.1* | -2.584712632 | 0.003168906 | 0.004875897 | 1 |
| T2DM | dendritic_cell | *PPFIA3* | 1.313665841 | 9.56E-05 | 0.000265792 | 0.600956637 |
| T2DM | dendritic_cell | *MAGEF1* | 3.631401944 | 0.016239392 | 0.021750545 | 1 |
| T2DM | dendritic_cell | *SAMD9L* | -1.587367901 | 0.006261249 | 0.00900995 | 1 |
| T2DM | dendritic_cell | *PAWR* | -2.763131661 | 4.27E-05 | 0.000164485 | 0.268604735 |

| | | | | | | |
|---|---|---|---|---|---|---|
| T2DM | dendritic_cell | *NAP1L5* | 3.341862523 | 0.007322443 | 0.010434316 | 1 |
| T2DM | dendritic_cell | *ACOT4* | -1.604544103 | 8.26E-05 | 0.000241985 | 0.519515452 |
| T2DM | dendritic_cell | *ARIH2* | -1.433366566 | 0.000224885 | 0.000495046 | 1 |
| T2DM | dendritic_cell | *RBM44* | -3.757496868 | 0.004282254 | 0.006393382 | 1 |
| T2DM | dendritic_cell | *IRX3* | -1.177878295 | 0.002118098 | 0.003397929 | 1 |
| T2DM | dendritic_cell | *TBL1XR1* | -2.200056337 | 6.19E-05 | 0.000200394 | 0.388965425 |
| T2DM | dendritic_cell | *C18orf32* | -2.564187962 | 0.000704451 | 0.001278545 | 1 |
| T2DM | dendritic_cell | *RPLP2* | -2.73761491 | 0.002159687 | 0.003460233 | 1 |
| T2DM | dendritic_cell | *C12orf54* | -9.032849939 | 9.01E-08 | 1.56E-05 | 0.000566392 |
| T2DM | dendritic_cell | *THAP5* | -1.83766046 | 1.68E-05 | 0.000130129 | 0.105540007 |
| T2DM | dendritic_cell | *CD151* | -2.501922446 | 0.000132025 | 0.000334022 | 0.830043621 |
| T2DM | dendritic_cell | *POLR2L* | -2.021231396 | 7.30E-05 | 0.000221211 | 0.459012662 |
| T2DM | dendritic_cell | *FAM87B* | -4.187483523 | 8.69E-06 | 0.000130129 | 0.054616044 |
| T2DM | dendritic_cell | *ZCCHC3* | 2.543456305 | 0.00550972 | 0.008031442 | 1 |
| T2DM | dendritic_cell | *PCDHB9* | 1.716101396 | 0.004346691 | 0.00647728 | 1 |
| T2DM | dendritic_cell | *ZNF619* | -3.098607978 | 6.95E-05 | 0.000214823 | 0.436735906 |
| T2DM | dendritic_cell | *ZBTB41* | -1.844641079 | 3.65E-05 | 0.000155217 | 0.229377383 |
| T2DM | dendritic_cell | *ZNF354C* | -2.746119766 | 2.99E-05 | 0.000145022 | 0.187694066 |
| T2DM | dendritic_cell | *CENPBD1* | -3.173916885 | 0.000524737 | 0.000989834 | 1 |
| T2DM | dendritic_cell | *BET1L* | -1.993039262 | 0.010229613 | 0.014162867 | 1 |
| T2DM | dendritic_cell | *IMP3* | -2.877863539 | 0.002580949 | 0.004056607 | 1 |
| T2DM | dendritic_cell | *DPY19L2* | -1.045172634 | 2.53E-05 | 0.000138949 | 0.158783988 |
| T2DM | dendritic_cell | *C2orf73* | -1.668082059 | 1.60E-05 | 0.000130129 | 0.100716294 |
| T2DM | dendritic_cell | *FAM26E* | -1.613188059 | 0.000128729 | 0.000327178 | 0.809318246 |
| T2DM | dendritic_cell | *MLF1* | -3.227478306 | 0.000365361 | 0.000734042 | 1 |
| T2DM | dendritic_cell | *C2orf69* | -3.016577651 | 0.037418599 | 0.047467864 | 1 |
| T2DM | dendritic_cell | *GRAMD1C* | -3.510405814 | 5.28E-05 | 0.000182762 | 0.331912377 |
| T2DM | dendritic_cell | *STAP2* | 2.144244929 | 0.010534649 | 0.014562739 | 1 |
| T2DM | dendritic_cell | *TSSK6* | -4.702975223 | 0.001518455 | 0.002515554 | 1 |
| T2DM | dendritic_cell | *NDUFV2* | 3.103247994 | 0.002178499 | 0.003488595 | 1 |
| T2DM | dendritic_cell | *DALRD3* | -1.592515882 | 0.000566245 | 0.001057631 | 1 |
| T2DM | dendritic_cell | *ZNF114* | -2.726196251 | 6.45E-06 | 0.000130129 | 0.040552874 |
| T2DM | dendritic_cell | *ZNF518B* | -4.419116301 | 0.000105499 | 0.000283088 | 0.66327472 |
| T2DM | dendritic_cell | *SH2B1* | -1.874601244 | 2.02E-06 | 8.62E-05 | 0.012676407 |
| T2DM | dendritic_cell | *ZC3H12D* | 0.753557968 | 4.31E-05 | 0.000165275 | 0.270944709 |
| T2DM | dendritic_cell | *KDELC2* | -2.875301878 | 0.000146261 | 0.000359336 | 0.919540634 |
| T2DM | dendritic_cell | *PLEC* | -3.158097137 | 0.000423948 | 0.000830331 | 1 |
| T2DM | dendritic_cell | *SH2D4B* | -3.091514709 | 1.51E-05 | 0.000130129 | 0.095011595 |
| T2DM | dendritic_cell | *RNF212* | -0.620135819 | 0.023494615 | 0.030670815 | 1 |
| T2DM | dendritic_cell | *PRSS36* | 1.244035653 | 4.82E-05 | 0.000173736 | 0.302822334 |
| T2DM | dendritic_cell | *TMPRSS9* | -1.394219393 | 7.26E-05 | 0.000220401 | 0.456229364 |
| T2DM | dendritic_cell | *AQP11* | -2.669786888 | 4.70E-05 | 0.000171433 | 0.295729516 |
| T2DM | dendritic_cell | *TMEM11* | -1.380717697 | 1.55E-05 | 0.000130129 | 0.097630151 |
| T2DM | dendritic_cell | *ZFAND2A* | -2.786692954 | 4.71E-05 | 0.000171433 | 0.296019735 |
| T2DM | dendritic_cell | *FAM220A* | -1.874865604 | 0.000468005 | 0.000898975 | 1 |
| T2DM | dendritic_cell | *BEND3* | 4.348588966 | 0.000443036 | 0.000859947 | 1 |
| T2DM | dendritic_cell | *NT5DC1* | -2.326375576 | 5.80E-05 | 0.000193363 | 0.364490064 |
| T2DM | dendritic_cell | *AC006111.1* | -5.716916323 | 9.94E-05 | 0.000271912 | 0.624748108 |
| T2DM | dendritic_cell | *LINC00843* | -4.083937162 | 2.09E-05 | 0.000132336 | 0.131542087 |
| T2DM | dendritic_cell | *COX14* | -1.16680698 | 2.81E-05 | 0.000142583 | 0.176950737 |

| | | | | | | |
|---|---|---|---|---|---|---|
| T2DM | dendritic_cell | *P4HTM* | -2.293739748 | 0.000202074 | 0.000456665 | 1 |
| T2DM | dendritic_cell | *CA8* | -1.051638195 | 0.000153848 | 0.000373452 | 0.96724013 |
| T2DM | dendritic_cell | *ERBB4* | -1.061835058 | 1.47E-05 | 0.000130129 | 0.092311933 |
| T2DM | dendritic_cell | *GPR35* | -1.129219994 | 0.000363007 | 0.00073078 | 1 |
| T2DM | dendritic_cell | *CSRNP3* | -3.043711539 | 0.013512464 | 0.018324604 | 1 |
| T2DM | dendritic_cell | *KCTD12* | -1.509836777 | 0.014367768 | 0.019388314 | 1 |
| T2DM | dendritic_cell | *FAM219B* | -2.231184706 | 4.05E-05 | 0.000160731 | 0.254468952 |
| T2DM | dendritic_cell | *CPNE7* | -3.888644278 | 0.002036321 | 0.003276772 | 1 |
| T2DM | dendritic_cell | *GDPD4* | 6.005000466 | 0.00228912 | 0.003645313 | 1 |
| T2DM | dendritic_cell | *MPI* | -1.643461561 | 5.85E-05 | 0.000194493 | 0.367980164 |
| T2DM | dendritic_cell | *TRIM73* | -5.220441155 | 9.17E-06 | 0.000130129 | 0.057665892 |
| T2DM | dendritic_cell | *EFCAB13* | -1.665403953 | 0.000153644 | 0.000373102 | 0.965960478 |
| T2DM | dendritic_cell | *GAK* | -3.057691109 | 0.00489278 | 0.007206251 | 1 |
| T2DM | dendritic_cell | *CTC1* | 2.036630479 | 0.005152062 | 0.007552113 | 1 |
| T2DM | dendritic_cell | *C14orf39* | -0.838306493 | 8.27E-05 | 0.000241985 | 0.519783006 |
| T2DM | dendritic_cell | *TRIML2* | -29.47334871 | 6.57E-06 | 0.000130129 | 0.041299995 |
| T2DM | dendritic_cell | *FARSA* | -5.936830067 | 2.02E-09 | 1.06E-06 | 1.27E-05 |
| T2DM | dendritic_cell | *SPTY2D1* | -3.172933376 | 7.18E-05 | 0.000219379 | 0.45170175 |
| T2DM | dendritic_cell | *TCAIM* | -1.849783615 | 3.16E-05 | 0.000147848 | 0.198856232 |
| T2DM | dendritic_cell | *ZNF664* | -3.810177301 | 1.70E-05 | 0.000130129 | 0.107167618 |
| T2DM | dendritic_cell | *CDH4* | -1.928094642 | 0.00038221 | 0.000761596 | 1 |
| T2DM | dendritic_cell | *HLA-DQB1* | 0.180768239 | 0.002708046 | 0.004235196 | 1 |
| T2DM | dendritic_cell | *ARID3B* | -4.450424048 | 2.04E-05 | 0.00013202 | 0.128543928 |
| T2DM | dendritic_cell | *EGR3* | 0.986426551 | 4.75E-05 | 0.00017227 | 0.298717034 |
| T2DM | dendritic_cell | *LINC00174* | -1.119807854 | 0.001155194 | 0.001965017 | 1 |
| T2DM | dendritic_cell | *GEMIN4* | -1.289598654 | 3.13E-05 | 0.000147135 | 0.196719215 |
| T2DM | dendritic_cell | *CIITA* | 0.288245017 | 2.91E-05 | 0.000143677 | 0.182939553 |
| T2DM | dendritic_cell | *LACC1* | 1.723221799 | 0.00012751 | 0.000324952 | 0.801657413 |
| T2DM | dendritic_cell | *MAF1* | 2.118772965 | 0.000159748 | 0.000384214 | 1 |
| T2DM | dendritic_cell | *FCER1A* | 0.115623507 | 2.82E-05 | 0.000142583 | 0.177575997 |
| T2DM | dendritic_cell | *PCED1B* | -1.823407119 | 1.09E-05 | 0.000130129 | 0.068525168 |
| T2DM | dendritic_cell | *PIPOX* | -2.36187115 | 1.27E-06 | 6.80E-05 | 0.007962661 |
| T2DM | dendritic_cell | *FOXS1* | 1.944369052 | 8.95E-05 | 0.000255275 | 0.562390428 |
| T2DM | dendritic_cell | *ATOH7* | 8.434049206 | 0.002460085 | 0.003887045 | 1 |
| T2DM | dendritic_cell | *CDH5* | -0.787429358 | 0.000370673 | 0.000741726 | 1 |
| T2DM | dendritic_cell | *GIPC3* | -1.969326028 | 0.006140261 | 0.008852058 | 1 |
| T2DM | dendritic_cell | *CITED4* | -2.798082999 | 1.82E-05 | 0.000130129 | 0.11466022 |
| T2DM | dendritic_cell | *TIGD5* | -1.717987283 | 5.01E-05 | 0.000177579 | 0.315130366 |
| T2DM | dendritic_cell | *PDXDC1* | -1.953961093 | 5.24E-05 | 0.000182013 | 0.32962563 |
| T2DM | dendritic_cell | *R3HDM2* | -2.216048846 | 0.000101302 | 0.000274993 | 0.636883952 |
| T2DM | dendritic_cell | *B3GNT3* | 3.488915822 | 3.38E-07 | 3.12E-05 | 0.002123773 |
| T2DM | dendritic_cell | *SSC5D* | -0.673818897 | 0.005770992 | 0.008385077 | 1 |
| T2DM | dendritic_cell | *ZADH2* | -3.280586645 | 6.06E-05 | 0.000198694 | 0.381202881 |
| T2DM | dendritic_cell | *SEPT1* | 2.902701273 | 0.002537814 | 0.003995803 | 1 |
| T2DM | dendritic_cell | *EXOC3* | -2.177033025 | 0.005839449 | 0.00846498 | 1 |
| T2DM | dendritic_cell | *TDRD6* | -0.723701666 | 0.007373152 | 0.010492305 | 1 |
| T2DM | dendritic_cell | *LYNX1* | -2.165100422 | 9.08E-06 | 0.000130129 | 0.05707005 |
| T2DM | dendritic_cell | *TDRP* | -1.624839541 | 3.88E-05 | 0.000158329 | 0.243984679 |
| T2DM | dendritic_cell | *FGD6* | -3.064448489 | 8.39E-05 | 0.00024479 | 0.52752322 |
| T2DM | dendritic_cell | *HCLS1* | 1.383159573 | 0.000167923 | 0.000399582 | 1 |

| | | | | | | |
|---|---|---|---|---|---|---|
| T2DM | dendritic_cell | *ZNF609* | -2.224089866 | 0.001021854 | 0.001766399 | 1 |
| T2DM | dendritic_cell | *CCDC66* | -1.442655921 | 0.004391305 | 0.006537564 | 1 |
| T2DM | dendritic_cell | *DEFB124* | -5.438612127 | 1.50E-05 | 0.000130129 | 0.094316619 |
| T2DM | dendritic_cell | *MCFD2* | -2.955181708 | 0.003229865 | 0.004951514 | 1 |
| T2DM | dendritic_cell | *CYP8B1* | -1.434735544 | 3.62E-05 | 0.00015508 | 0.227502504 |
| T2DM | dendritic_cell | *MB21D2* | -2.487391792 | 0.016468595 | 0.022043444 | 1 |
| T2DM | dendritic_cell | *SSTR2* | -10.02979641 | 0.000522768 | 0.000986683 | 1 |
| T2DM | dendritic_cell | *ZNF594* | -3.582903329 | 0.002881251 | 0.004466081 | 1 |
| T2DM | dendritic_cell | *PCGF5* | -2.733375943 | 0.000162807 | 0.000390227 | 1 |
| T2DM | dendritic_cell | *SHISA2* | 3.313053545 | 2.88E-05 | 0.000143316 | 0.181203375 |
| T2DM | dendritic_cell | *CHST13* | 4.056003072 | 0.00285282 | 0.004426377 | 1 |
| T2DM | dendritic_cell | *SLC36A4* | -3.832534163 | 4.78E-05 | 0.000172826 | 0.300544433 |
| T2DM | dendritic_cell | *ZDHHC20* | -2.402622721 | 0.015648411 | 0.021003749 | 1 |
| T2DM | dendritic_cell | *PSMG4* | -2.831435546 | 0.000822648 | 0.001463909 | 1 |
| T2DM | dendritic_cell | *ZNF443* | -2.051997771 | 0.009474087 | 0.013192378 | 1 |
| T2DM | dendritic_cell | *CAPS2* | -2.002053502 | 3.41E-05 | 0.000150922 | 0.214216074 |
| T2DM | dendritic_cell | *ZNF792* | -2.600704204 | 3.17E-05 | 0.000147987 | 0.199282485 |
| T2DM | dendritic_cell | *D2HGDH* | -1.998366328 | 0.010304915 | 0.014260841 | 1 |
| T2DM | dendritic_cell | *CMTR2* | -3.68698955 | 0.000196267 | 0.000446429 | 1 |
| T2DM | dendritic_cell | *MRPL14* | -3.100670848 | 5.16E-05 | 0.000180477 | 0.324317312 |
| T2DM | dendritic_cell | *BBS12* | 1.964890316 | 0.000109447 | 0.000290796 | 0.68809307 |
| T2DM | dendritic_cell | *ZFP82* | -2.388112756 | 9.94E-05 | 0.000271912 | 0.624853171 |
| T2DM | dendritic_cell | *LSMEM1* | -1.560655213 | 0.000154757 | 0.000375257 | 0.972955247 |
| T2DM | dendritic_cell | *AEN* | 3.425776899 | 0.002994313 | 0.004628779 | 1 |
| T2DM | dendritic_cell | *FKRP* | -1.168848954 | 0.002504146 | 0.003947734 | 1 |
| T2DM | dendritic_cell | *TRAPPC5* | 1.301676991 | 0.000184684 | 0.00042752 | 1 |
| T2DM | dendritic_cell | *RPH3AL* | -2.009636834 | 0.000668082 | 0.001218517 | 1 |
| T2DM | dendritic_cell | *ANKRD34A* | -2.775198886 | 2.15E-06 | 8.62E-05 | 0.013533062 |
| T2DM | dendritic_cell | *SLC26A11* | -3.253115391 | 0.000411328 | 0.000808383 | 1 |
| T2DM | dendritic_cell | *HIGD1A* | -2.106507291 | 0.000120952 | 0.000312803 | 0.760423613 |
| T2DM | dendritic_cell | *ADIPOQ* | -0.706272551 | 0.000112162 | 0.000295666 | 0.70516254 |
| T2DM | dendritic_cell | *F2R* | -1.138700216 | 0.003819383 | 0.005766682 | 1 |
| T2DM | dendritic_cell | *HIST3H2A* | -1.956011829 | 0.003487539 | 0.005307713 | 1 |
| T2DM | dendritic_cell | *POLR2A* | -3.217301837 | 3.25E-05 | 0.000148254 | 0.204431868 |
| T2DM | dendritic_cell | *TMEM132C* | -0.839892973 | 8.58E-05 | 0.000248187 | 0.539167722 |
| T2DM | dendritic_cell | *SLC25A41* | -7.207858568 | 9.41E-06 | 0.000130129 | 0.059164399 |
| T2DM | dendritic_cell | *TMEM136* | 1.873221423 | 0.006880463 | 0.009844668 | 1 |
| T2DM | dendritic_cell | *TMEM132E* | 4.403958952 | 0.001058423 | 0.00181911 | 1 |
| T2DM | dendritic_cell | *ZNF322* | -2.332974944 | 0.000306734 | 0.000636029 | 1 |
| T2DM | dendritic_cell | *DDX60L* | -2.752136303 | 0.000107811 | 0.000287452 | 0.677810805 |
| T2DM | dendritic_cell | *ACBD4* | -2.881056911 | 6.61E-05 | 0.000208955 | 0.415820571 |
| T2DM | dendritic_cell | *SETD2* | -1.829550033 | 0.000344118 | 0.000696097 | 1 |
| T2DM | dendritic_cell | *MRPS23* | -4.444116999 | 0.00295945 | 0.004577137 | 1 |
| T2DM | dendritic_cell | *GPR135* | -1.456568063 | 0.000245566 | 0.000530906 | 1 |
| T2DM | dendritic_cell | *ZFP41* | -1.299345847 | 0.009033872 | 0.012626934 | 1 |
| T2DM | dendritic_cell | *ATG9B* | -2.790110505 | 2.02E-05 | 0.000131411 | 0.12681187 |
| T2DM | dendritic_cell | *HKR1* | -2.099174205 | 1.85E-05 | 0.000130129 | 0.116105396 |
| T2DM | dendritic_cell | *ZBTB20* | -1.471368798 | 4.33E-05 | 0.000165469 | 0.272362718 |
| T2DM | dendritic_cell | *C3orf58* | 1.936026464 | 0.006117553 | 0.008823367 | 1 |
| T2DM | dendritic_cell | *RELL1* | -2.844467277 | 0.00191633 | 0.003102746 | 1 |

| | | | | | | |
|---|---|---|---|---|---|---|
| T2DM | dendritic_cell | *RFX7* | -2.404691302 | 0.007306202 | 0.010418256 | 1 |
| T2DM | dendritic_cell | *ADO* | -5.555563834 | 0.003857102 | 0.005818042 | 1 |
| T2DM | dendritic_cell | *SNRPE* | 2.189893688 | 0.000788245 | 0.001411477 | 1 |
| T2DM | dendritic_cell | *CHST15* | -3.258617478 | 0.000317756 | 0.000654565 | 1 |
| T2DM | dendritic_cell | *IDH2* | -2.591709749 | 1.24E-05 | 0.000130129 | 0.078144391 |
| T2DM | dendritic_cell | *PTCHD3* | 0.918308169 | 0.000558903 | 0.00104547 | 1 |
| T2DM | dendritic_cell | *TNRC18* | -2.362973492 | 0.011372279 | 0.015610812 | 1 |
| T2DM | dendritic_cell | *FAM181B* | -4.011256629 | 0.003841717 | 0.005799009 | 1 |
| T2DM | dendritic_cell | *NOP10* | -4.015421275 | 1.43E-05 | 0.000130129 | 0.089802657 |
| T2DM | dendritic_cell | *FAM89A* | -1.229941584 | 1.13E-05 | 0.000130129 | 0.071007633 |
| T2DM | dendritic_cell | *TDRKH* | -2.518082278 | 0.000178114 | 0.000416129 | 1 |
| T2DM | dendritic_cell | *ZNF708* | -2.450535207 | 3.29E-05 | 0.000148822 | 0.206877797 |
| T2DM | dendritic_cell | *ERCC6L2* | -1.940415535 | 0.000256305 | 0.000549775 | 1 |
| T2DM | dendritic_cell | *MRPL41* | -2.948767051 | 0.000905969 | 0.001590114 | 1 |
| T2DM | dendritic_cell | *HHIPL1* | -1.694442952 | 1.05E-05 | 0.000130129 | 0.066073964 |
| T2DM | dendritic_cell | *FAM153B* | -2.193768257 | 7.50E-05 | 0.000225758 | 0.471383479 |
| T2DM | dendritic_cell | *BACE2* | -1.314336784 | 0.006495696 | 0.009334422 | 1 |
| T2DM | dendritic_cell | *SYNM* | -4.342663271 | 5.84E-05 | 0.000194469 | 0.367420627 |
| T2DM | dendritic_cell | *C8orf33* | -2.821915354 | 6.18E-05 | 0.000200324 | 0.38862855 |
| T2DM | dendritic_cell | *FBXL6* | -4.256469494 | 0.001022212 | 0.001766533 | 1 |
| T2DM | dendritic_cell | *KBTBD3* | -3.146259366 | 3.01E-05 | 0.000145247 | 0.189220949 |
| T2DM | dendritic_cell | *CACNB4* | -1.016965845 | 0.00015483 | 0.000375257 | 0.973417415 |
| T2DM | dendritic_cell | *TRAPPC6B* | -2.346187647 | 4.12E-05 | 0.000162249 | 0.259255755 |
| T2DM | dendritic_cell | *NPLOC4* | -2.017714294 | 0.000135796 | 0.000340275 | 0.853751072 |
| T2DM | dendritic_cell | *TSHZ2* | -1.594819242 | 3.01E-05 | 0.000145247 | 0.189024111 |
| T2DM | dendritic_cell | *CAPN12* | -2.424072963 | 0.000868188 | 0.001534092 | 1 |
| T2DM | dendritic_cell | *FES* | -3.564741385 | 0.030469319 | 0.039133935 | 1 |
| T2DM | dendritic_cell | *GLRX5* | -4.548975387 | 0.000100153 | 0.000272936 | 0.629663754 |
| T2DM | dendritic_cell | *CAV3* | -0.868780842 | 0.001461374 | 0.002429313 | 1 |
| T2DM | dendritic_cell | *MXRA7* | -0.90499762 | 0.003471681 | 0.005286136 | 1 |
| T2DM | dendritic_cell | *MFSD5* | -1.834562985 | 6.37E-05 | 0.000203711 | 0.400698687 |
| T2DM | dendritic_cell | *ADI1* | -1.77638292 | 8.72E-06 | 0.000130129 | 0.054800553 |
| T2DM | dendritic_cell | *SPNS3* | 1.329556366 | 5.12E-05 | 0.000179577 | 0.32194742 |
| T2DM | dendritic_cell | *CLEC4G* | -3.659258521 | 2.17E-05 | 0.000132663 | 0.136510506 |
| T2DM | dendritic_cell | *ACTL10* | -3.335138866 | 0.001693403 | 0.002771064 | 1 |
| T2DM | dendritic_cell | *TRAK1* | -2.027701565 | 0.008143183 | 0.011493253 | 1 |
| T2DM | dendritic_cell | *SKA2* | -2.639564068 | 2.35E-05 | 0.00013573 | 0.147538178 |
| T2DM | dendritic_cell | *KCNB2* | -0.735445146 | 2.81E-05 | 0.000142583 | 0.176634724 |
| T2DM | dendritic_cell | *TSKU* | -1.761083793 | 1.82E-05 | 0.000130129 | 0.114121264 |
| T2DM | dendritic_cell | *GRID1* | -2.302741535 | 4.93E-07 | 3.82E-05 | 0.003097741 |
| T2DM | dendritic_cell | *ACBD3* | -2.278756652 | 3.04E-05 | 0.000145705 | 0.19095668 |
| T2DM | dendritic_cell | *PLCXD3* | -1.044967204 | 0.000136232 | 0.00034096 | 0.856490904 |
| T2DM | dendritic_cell | *GPIHBP1* | -1.473089725 | 0.000268641 | 0.000570589 | 1 |
| T2DM | dendritic_cell | *VMO1* | 0.563538358 | 1.47E-05 | 0.000130129 | 0.092464466 |
| T2DM | dendritic_cell | *COL18A1* | -2.071270264 | 0.000276236 | 0.000583385 | 1 |
| T2DM | dendritic_cell | *SLC25A18* | -1.125713118 | 2.55E-05 | 0.000139164 | 0.160444026 |
| T2DM | dendritic_cell | *ZNF721* | -1.718814898 | 0.000704872 | 0.001278942 | 1 |
| T2DM | dendritic_cell | *LENG9* | -1.705874747 | 0.000270574 | 0.000574307 | 1 |
| T2DM | dendritic_cell | *C11orf54* | -0.889205762 | 0.005321589 | 0.007780659 | 1 |
| T2DM | dendritic_cell | *CEP63* | -1.264695162 | 0.000294237 | 0.000613961 | 1 |

| | | | | | | |
|---|---|---|---|---|---|---|
| T2DM | dendritic_cell | *HMGN4* | 1.689415813 | 0.017082267 | 0.022796903 | 1 |
| T2DM | dendritic_cell | *ZNF320* | 0.968944402 | 0.000617289 | 0.001138895 | 1 |
| T2DM | dendritic_cell | *C12orf60* | -2.578852189 | 2.45E-05 | 0.000137669 | 0.154051573 |
| T2DM | dendritic_cell | *GAS6* | -2.302967428 | 0.025156214 | 0.032681451 | 1 |
| T2DM | dendritic_cell | *BEGAIN* | 8.271119367 | 0.001642454 | 0.002694001 | 1 |
| T2DM | dendritic_cell | *ARHGEF37* | -2.312286824 | 3.66E-05 | 0.000155321 | 0.230252985 |
| T2DM | dendritic_cell | *FAM43B* | 3.313116743 | 9.68E-05 | 0.000267843 | 0.608776258 |
| T2DM | dendritic_cell | *CSMD1* | -1.01151803 | 1.11E-05 | 0.000130129 | 0.069667788 |
| T2DM | dendritic_cell | *GPR19* | -3.134604939 | 0.000169327 | 0.000401831 | 1 |
| T2DM | dendritic_cell | *FANCF* | -7.017681612 | 0.000142145 | 0.000352391 | 0.893663494 |
| T2DM | dendritic_cell | *C2CD4C* | -2.087627259 | 0.005341601 | 0.007804473 | 1 |
| T2DM | dendritic_cell | *CCBE1* | -0.7099805 | 9.59E-05 | 0.00026632 | 0.603214244 |
| T2DM | dendritic_cell | *ZNF623* | -3.192851536 | 0.000119114 | 0.000309531 | 0.748872341 |
| T2DM | dendritic_cell | *CCDC125* | -2.669847076 | 0.000166241 | 0.000396042 | 1 |
| T2DM | dendritic_cell | *FHL3* | -4.426492671 | 0.003745721 | 0.005667714 | 1 |
| T2DM | dendritic_cell | *C19orf71* | -2.614007597 | 5.92E-06 | 0.000129745 | 0.037236752 |
| T2DM | dendritic_cell | *LRIT3* | -3.620722485 | 0.001325832 | 0.002223395 | 1 |
| T2DM | dendritic_cell | *GRIN2A* | -1.720465053 | 0.000181097 | 0.000421377 | 1 |
| T2DM | dendritic_cell | *URAD* | -6.631812149 | 3.99E-06 | 0.000113496 | 0.025082598 |
| T2DM | dendritic_cell | *GPR132* | 0.635025774 | 5.13E-05 | 0.000179577 | 0.322340026 |
| T2DM | dendritic_cell | *MX2* | 1.399808306 | 0.005439837 | 0.007942465 | 1 |
| T2DM | dendritic_cell | *ACSM5* | -0.934813205 | 0.001241374 | 0.002096861 | 1 |
| T2DM | dendritic_cell | *AC131971.1* | -5.304883264 | 0.000177878 | 0.000415951 | 1 |
| T2DM | dendritic_cell | *PCBP3* | -0.43790115 | 0.000212899 | 0.000474307 | 1 |
| T2DM | dendritic_cell | *SFXN4* | -3.968360731 | 1.79E-05 | 0.000130129 | 0.112792026 |
| T2DM | dendritic_cell | *FAM167B* | -5.342750795 | 0.00775737 | 0.01099675 | 1 |
| T2DM | dendritic_cell | *DGCR6* | -2.247311917 | 0.00406296 | 0.006099291 | 1 |
| T2DM | dendritic_cell | *FAM19A1* | -2.481076987 | 2.38E-05 | 0.000136538 | 0.14954127 |
| T2DM | dendritic_cell | *TRMT12* | -3.485446411 | 0.006105333 | 0.008811806 | 1 |
| T2DM | dendritic_cell | *BMP8A* | 2.136727655 | 9.74E-08 | 1.56E-05 | 0.000612652 |
| T2DM | dendritic_cell | *FIGLA* | -6.219305865 | 0.008818007 | 0.012347173 | 1 |
| T2DM | dendritic_cell | *KREMEN1* | -0.804153943 | 0.000539071 | 0.001014713 | 1 |
| T2DM | dendritic_cell | *AIFM3* | 1.41996396 | 3.07E-05 | 0.00014601 | 0.192990572 |
| T2DM | dendritic_cell | *KCTD16* | -0.967587156 | 7.44E-05 | 0.000224335 | 0.467739209 |
| T2DM | dendritic_cell | *ZNF703* | -2.034341947 | 5.40E-08 | 1.06E-05 | 0.000339245 |
| T2DM | dendritic_cell | *KCTD8* | -0.810436312 | 0.00021048 | 0.000470868 | 1 |
| T2DM | dendritic_cell | *C9orf66* | 7.545073445 | 0.000294809 | 0.000614953 | 1 |
| T2DM | dendritic_cell | *TUBA8* | -1.58431814 | 0.002946196 | 0.004558881 | 1 |
| T2DM | dendritic_cell | *NPIPA5* | -0.695487664 | 0.001678377 | 0.002750053 | 1 |
| T2DM | dendritic_cell | *FAM162B* | -1.381345059 | 0.021041908 | 0.027664257 | 1 |
| T2DM | dendritic_cell | *BTBD9* | -1.996720662 | 0.000430658 | 0.000841568 | 1 |
| T2DM | dendritic_cell | *ANKRD45* | -3.609298664 | 2.20E-05 | 0.000132957 | 0.13841848 |
| T2DM | dendritic_cell | *MAATS1* | -1.1969916 | 0.02660206 | 0.034412994 | 1 |
| T2DM | dendritic_cell | *GPR39* | 1.503220838 | 0.000202822 | 0.000457763 | 1 |
| T2DM | dendritic_cell | *FAM3B* | -0.558052893 | 0.007931996 | 0.011219001 | 1 |
| T2DM | dendritic_cell | *NPIPA7* | -5.028979794 | 0.000173375 | 0.000408242 | 1 |
| T2DM | dendritic_cell | *TTC32* | -2.874288552 | 0.004318827 | 0.006440338 | 1 |
| T2DM | dendritic_cell | *PP2D1* | -1.826708098 | 0.006540885 | 0.009392997 | 1 |
| T2DM | dendritic_cell | *DENND5A* | -2.92559578 | 0.000123388 | 0.000317796 | 0.775740456 |
| T2DM | dendritic_cell | *TMEM236* | 4.014340533 | 0.001134044 | 0.001934273 | 1 |

| | | | | | | |
|---|---|---|---|---|---|---|
| T2DM | dendritic_cell | *VPS33B* | -2.983838955 | 0.004611444 | 0.006828518 | 1 |
| T2DM | dendritic_cell | *TBX1* | -1.572093358 | 1.09E-05 | 0.000130129 | 0.068706165 |
| T2DM | dendritic_cell | *UQCR10* | -3.510177179 | 7.05E-06 | 0.000130129 | 0.044305858 |
| T2DM | dendritic_cell | *EIF3C* | 1.737630407 | 0.001262744 | 0.002130669 | 1 |
| T2DM | dendritic_cell | *NIPSNAP1* | -1.845210142 | 8.05E-05 | 0.000237314 | 0.506191118 |
| T2DM | dendritic_cell | *CNTN2* | -2.35737002 | 2.39E-05 | 0.000136538 | 0.150513259 |
| T2DM | dendritic_cell | *CRELD2* | -3.748041737 | 0.000232208 | 0.000507965 | 1 |
| T2DM | dendritic_cell | *UBE2F* | -3.587008624 | 8.25E-06 | 0.000130129 | 0.051841807 |
| T2DM | dendritic_cell | *PCDH9* | -0.765701332 | 3.28E-06 | 0.000103864 | 0.020652012 |
| T2DM | dendritic_cell | *ACOT1* | -0.772140968 | 0.023133467 | 0.030224461 | 1 |
| T2DM | dendritic_cell | *OAF* | -1.521662999 | 0.000115698 | 0.0003022 | 0.727395426 |
| T2DM | dendritic_cell | *TACSTD2* | 0.895760424 | 0.010777494 | 0.014855975 | 1 |
| T2DM | dendritic_cell | *CLECL1* | 0.270635262 | 1.36E-05 | 0.000130129 | 0.085754255 |
| T2DM | dendritic_cell | *MROH7* | 4.386372185 | 0.008144851 | 0.011493253 | 1 |
| T2DM | dendritic_cell | *GDF3* | 2.236106722 | 0.000912222 | 0.001598868 | 1 |
| T2DM | dendritic_cell | *PLA2G6* | -0.862990923 | 0.008716691 | 0.012227094 | 1 |
| T2DM | dendritic_cell | *SS18L1* | -2.727602726 | 0.004496911 | 0.006677392 | 1 |
| T2DM | dendritic_cell | *TOP1MT* | -1.179299481 | 0.005092532 | 0.007473564 | 1 |
| T2DM | dendritic_cell | *THAP7* | -2.273311181 | 1.20E-05 | 0.000130129 | 0.075481372 |
| T2DM | dendritic_cell | *CCR10* | -5.044358433 | 0.004378902 | 0.006522188 | 1 |
| T2DM | dendritic_cell | *WDR27* | -2.493371115 | 0.000872789 | 0.001538762 | 1 |
| T2DM | dendritic_cell | *TXNRD2* | -2.514110707 | 4.70E-05 | 0.000171433 | 0.295632539 |
| T2DM | dendritic_cell | *PROS1* | -2.227521986 | 6.09E-05 | 0.000198694 | 0.382871458 |
| T2DM | dendritic_cell | *HDDC3* | -2.492389211 | 0.004482074 | 0.006656933 | 1 |
| T2DM | dendritic_cell | *ZFP1* | -4.038005007 | 1.68E-06 | 8.15E-05 | 0.010592214 |
| T2DM | dendritic_cell | *LPAR5* | 0.79122484 | 0.011471246 | 0.015732924 | 1 |
| T2DM | dendritic_cell | *TMEM173* | -2.433549192 | 9.64E-05 | 0.000267256 | 0.606136079 |
| T2DM | dendritic_cell | *NELL2* | 1.212708791 | 0.011455427 | 0.015714656 | 1 |
| T2DM | dendritic_cell | *KRBA2* | -2.548216911 | 2.71E-05 | 0.000141035 | 0.170522715 |
| T2DM | dendritic_cell | *ZNF93* | 2.800238928 | 0.000625184 | 0.001150961 | 1 |
| T2DM | dendritic_cell | *SEPT5* | -4.300172741 | 2.25E-05 | 0.000133629 | 0.141542592 |
| T2DM | dendritic_cell | *ATL3* | -2.230742581 | 1.86E-05 | 0.000130129 | 0.117045605 |
| T2DM | dendritic_cell | *NDUFA12* | -3.363997317 | 4.26E-05 | 0.000164317 | 0.267835983 |
| T2DM | dendritic_cell | *UBE2G2* | -2.198593259 | 1.35E-05 | 0.000130129 | 0.084960374 |
| T2DM | dendritic_cell | *OSBP2* | -3.101793287 | 0.000162746 | 0.000390227 | 1 |
| T2DM | dendritic_cell | *TUSC5* | -0.788763006 | 0.000612001 | 0.001130996 | 1 |
| T2DM | dendritic_cell | *BTBD6* | -1.691327438 | 1.80E-06 | 8.21E-05 | 0.011336657 |
| T2DM | dendritic_cell | *RBM43* | 2.311010998 | 0.021435869 | 0.02814689 | 1 |
| T2DM | dendritic_cell | *ZFP90* | -3.115195997 | 0.00078764 | 0.001410795 | 1 |
| T2DM | dendritic_cell | *FAM227A* | -1.850534062 | 0.000946717 | 0.001650584 | 1 |
| T2DM | dendritic_cell | *SORCS2* | -1.182314972 | 3.71E-05 | 0.000156008 | 0.233540838 |
| T2DM | dendritic_cell | *TMEM106A* | 1.939210599 | 6.67E-05 | 0.000209934 | 0.419238266 |
| T2DM | dendritic_cell | *SIVA1* | -2.424068061 | 2.81E-05 | 0.000142583 | 0.176894451 |
| T2DM | dendritic_cell | *UBOX5* | -2.723978707 | 2.06E-05 | 0.000132272 | 0.129539456 |
| T2DM | dendritic_cell | *MAFF* | -2.573757285 | 8.47E-05 | 0.000246303 | 0.532506743 |
| T2DM | dendritic_cell | *BRF1* | -1.830346068 | 1.59E-05 | 0.000130129 | 0.099869379 |
| T2DM | dendritic_cell | *CIB1* | -1.555066066 | 0.038054134 | 0.048167171 | 1 |
| T2DM | dendritic_cell | *FLRT2* | -0.928514556 | 6.70E-06 | 0.000130129 | 0.042096951 |
| T2DM | dendritic_cell | *ADSSL1* | -1.965430867 | 3.28E-05 | 0.000148688 | 0.206422041 |
| T2DM | dendritic_cell | *C6orf120* | -3.457267025 | 9.49E-06 | 0.000130129 | 0.059633743 |

| | | | | | | |
|---|---|---|---|---|---|---|
| T2DM | dendritic_cell | *DDX51* | -1.104131429 | 0.002375222 | 0.00376621 | 1 |
| T2DM | dendritic_cell | *SIGIRR* | -2.068873383 | 0.003124262 | 0.004814273 | 1 |
| T2DM | dendritic_cell | *IFITM2* | -2.536512533 | 0.00019912 | 0.000451773 | 1 |
| T2DM | dendritic_cell | *GP1BA* | -1.139879666 | 0.027937116 | 0.036065842 | 1 |
| T2DM | dendritic_cell | *KIAA0825* | -2.473748882 | 2.46E-05 | 0.000137735 | 0.154401092 |
| T2DM | dendritic_cell | *KLHL33* | -1.010215725 | 0.036512886 | 0.046403177 | 1 |
| T2DM | dendritic_cell | *CDK10* | -1.402759282 | 0.011845189 | 0.016192803 | 1 |
| T2DM | dendritic_cell | *TMEM105* | -15.41961267 | 0.006913946 | 0.009890324 | 1 |
| T2DM | dendritic_cell | *TCN2* | -1.43519667 | 0.002767634 | 0.004314435 | 1 |
| T2DM | dendritic_cell | *GAS2L1* | -3.277931147 | 2.07E-06 | 8.62E-05 | 0.013012855 |
| T2DM | dendritic_cell | *ATP6V0A2* | -2.852960337 | 0.001684448 | 0.002757845 | 1 |
| T2DM | dendritic_cell | *TNFAIP8L1* | 1.69671041 | 1.93E-05 | 0.000131132 | 0.12119059 |
| T2DM | dendritic_cell | *MAPK11* | -2.066580954 | 7.50E-06 | 0.000130129 | 0.047128955 |
| T2DM | dendritic_cell | *FAM174B* | -2.064103357 | 4.90E-05 | 0.000175258 | 0.30775259 |
| T2DM | dendritic_cell | *SV2B* | -3.497325785 | 0.007161859 | 0.010224026 | 1 |
| T2DM | dendritic_cell | *TLCD2* | -1.438888587 | 2.45E-05 | 0.00013771 | 0.154235716 |
| T2DM | dendritic_cell | *LSAMP* | -1.597234806 | 8.95E-05 | 0.000255275 | 0.562627143 |
| T2DM | dendritic_cell | *AHNAK2* | -1.68037781 | 1.35E-05 | 0.000130129 | 0.085131326 |
| T2DM | dendritic_cell | *SP1* | -1.97163208 | 0.000176971 | 0.000414229 | 1 |
| T2DM | dendritic_cell | *PDIA2* | -1.582952782 | 0.016986039 | 0.022678226 | 1 |
| T2DM | dendritic_cell | *PCGF3* | -2.290422184 | 0.016550573 | 0.022134323 | 1 |
| T2DM | dendritic_cell | *PBX1* | -1.162930868 | 1.71E-05 | 0.000130129 | 0.10759032 |
| T2DM | dendritic_cell | *UBE2L3* | -2.825798189 | 0.002454254 | 0.003878807 | 1 |
| T2DM | dendritic_cell | *PMEL* | -1.590999793 | 0.00041643 | 0.000817133 | 1 |
| T2DM | dendritic_cell | *SNAI3* | -1.838374355 | 0.00013497 | 0.000339031 | 0.848559188 |
| T2DM | dendritic_cell | *C6orf201* | -1.325884296 | 0.001325432 | 0.002223317 | 1 |
| T2DM | dendritic_cell | *MYBL1* | -1.357049581 | 5.89E-06 | 0.000129745 | 0.037061735 |
| T2DM | dendritic_cell | *C16orf52* | -2.848083466 | 0.000339957 | 0.000689455 | 1 |
| T2DM | dendritic_cell | *ADARB2* | -1.214362152 | 2.14E-05 | 0.000132591 | 0.134517796 |
| T2DM | dendritic_cell | *KCNIP4* | -0.83200426 | 1.17E-05 | 0.000130129 | 0.073655771 |
| T2DM | dendritic_cell | *PIGP* | -0.768536296 | 0.00044832 | 0.000867793 | 1 |
| T2DM | dendritic_cell | *NPIPB4* | -2.690646508 | 0.002444184 | 0.003865808 | 1 |
| T2DM | dendritic_cell | *ZNF829* | -1.883840935 | 0.01796182 | 0.02388451 | 1 |
| T2DM | dendritic_cell | *IFITM1* | -2.985044887 | 0.028381369 | 0.036601778 | 1 |
| T2DM | dendritic_cell | *POMK* | 2.92689575 | 0.00046463 | 0.000893857 | 1 |
| T2DM | dendritic_cell | *PTCH1* | -1.682329109 | 1.20E-05 | 0.000130129 | 0.075281784 |
| T2DM | dendritic_cell | *RTN4RL1* | -1.080838924 | 1.55E-05 | 0.000130129 | 0.097255594 |
| T2DM | dendritic_cell | *PLK5* | -2.526527821 | 0.00394851 | 0.005937403 | 1 |
| T2DM | dendritic_cell | *RASA3* | -2.105074048 | 2.51E-07 | 2.59E-05 | 0.00157917 |
| T2DM | dendritic_cell | *ZNF566* | -1.880443146 | 3.70E-06 | 0.000110889 | 0.023281838 |
| T2DM | dendritic_cell | *ZNF529* | -2.100457557 | 0.002159355 | 0.003460233 | 1 |
| T2DM | dendritic_cell | *C2orf76* | -2.900326073 | 3.26E-05 | 0.000148254 | 0.205013022 |
| T2DM | dendritic_cell | *WWOX* | -1.405719624 | 0.000206694 | 0.000464267 | 1 |
| T2DM | dendritic_cell | *CYP4Z1* | -1.193953189 | 4.60E-05 | 0.000170199 | 0.288998604 |
| T2DM | dendritic_cell | *CCDC84* | -2.316859831 | 3.65E-05 | 0.000155217 | 0.229560812 |
| T2DM | dendritic_cell | *POLR1D* | -3.074962736 | 0.000265454 | 0.000565157 | 1 |
| T2DM | dendritic_cell | *FFAR4* | -0.973150441 | 6.31E-05 | 0.00020269 | 0.396562904 |
| T2DM | dendritic_cell | *BPIFB4* | -49.86473718 | 0.005904374 | 0.00855318 | 1 |
| T2DM | dendritic_cell | *CYP4F12* | -1.151524396 | 0.00471813 | 0.006969726 | 1 |
| T2DM | dendritic_cell | *ZNF749* | -1.3554712 | 0.016010956 | 0.021481196 | 1 |

| | | | | | | |
|---|---|---|---|---|---|---|
| T2DM | dendritic_cell | *KLHL32* | -1.775626555 | 0.005222571 | 0.007642994 | 1 |
| T2DM | dendritic_cell | *NBPF12* | -3.112132288 | 3.71E-05 | 0.000156008 | 0.232973308 |
| T2DM | dendritic_cell | *GPAT2* | 2.050743342 | 0.000202644 | 0.000457623 | 1 |
| T2DM | dendritic_cell | *TOR3A* | 3.01761621 | 0.002465622 | 0.003893837 | 1 |
| T2DM | dendritic_cell | *CYP4X1* | -0.860354999 | 7.87E-05 | 0.000233981 | 0.494869694 |
| T2DM | dendritic_cell | *CCDC30* | -2.948699087 | 7.24E-05 | 0.000220017 | 0.455215052 |
| T2DM | dendritic_cell | *GLDN* | -1.834804579 | 0.005338566 | 0.007801851 | 1 |
| T2DM | dendritic_cell | *FCAR* | 1.942082343 | 0.005280932 | 0.00772301 | 1 |
| T2DM | dendritic_cell | *ZNF501* | -1.819407786 | 5.38E-06 | 0.000125356 | 0.033817928 |
| T2DM | dendritic_cell | *ZNF197* | -2.028841804 | 2.40E-05 | 0.000136746 | 0.15096764 |
| T2DM | dendritic_cell | *PCLO* | -0.809366292 | 6.80E-05 | 0.000212384 | 0.427741908 |
| T2DM | dendritic_cell | *RGS7BP* | -5.185828476 | 8.73E-07 | 5.49E-05 | 0.005487467 |
| T2DM | dendritic_cell | *ZNF396* | -2.079717233 | 2.13E-06 | 8.62E-05 | 0.013408304 |
| T2DM | dendritic_cell | *TMEM222* | -1.703749735 | 0.000220617 | 0.000487699 | 1 |
| T2DM | dendritic_cell | *CLCNKA* | -3.124242564 | 0.004427557 | 0.00658374 | 1 |
| T2DM | dendritic_cell | *SEPT10* | -1.767946983 | 9.70E-05 | 0.000268128 | 0.609722654 |
| T2DM | dendritic_cell | *SMYD4* | -2.137280938 | 0.001874988 | 0.00304359 | 1 |
| T2DM | dendritic_cell | *CEACAM19* | -1.629194912 | 0.005964595 | 0.008628489 | 1 |
| T2DM | dendritic_cell | *NF2* | -2.123714134 | 0.000248917 | 0.000536674 | 1 |
| T2DM | dendritic_cell | *MIR22HG* | -1.854829784 | 2.73E-05 | 0.000141272 | 0.171753175 |
| T2DM | dendritic_cell | *PDE2A* | -1.023879361 | 4.95E-05 | 0.000176378 | 0.310954404 |
| T2DM | dendritic_cell | *C17orf58* | -0.937133804 | 0.000742591 | 0.001339264 | 1 |
| T2DM | dendritic_cell | *CYP27C1* | -0.777565853 | 0.009697668 | 0.013474537 | 1 |
| T2DM | dendritic_cell | *BCR* | -2.161126061 | 2.13E-05 | 0.000132591 | 0.133863358 |
| T2DM | dendritic_cell | *ZSCAN30* | -2.821568045 | 3.81E-05 | 0.000157482 | 0.239530511 |
| T2DM | dendritic_cell | *LILRB4* | 1.330804173 | 0.000180442 | 0.000420345 | 1 |
| T2DM | dendritic_cell | *QRFPR* | 2.199840361 | 3.26E-05 | 0.000148254 | 0.204981764 |
| T2DM | dendritic_cell | *TMEM17* | -1.793917169 | 0.000174414 | 0.000409856 | 1 |
| T2DM | dendritic_cell | *TNFRSF18* | -3.672380369 | 1.68E-05 | 0.000130129 | 0.105696188 |
| T2DM | dendritic_cell | *RTN4RL2* | -6.380410131 | 0.000475338 | 0.000910836 | 1 |
| T2DM | dendritic_cell | *TMEM232* | -1.451552434 | 4.24E-05 | 0.000164065 | 0.266770334 |
| T2DM | dendritic_cell | *EFCAB6* | -1.378177183 | 0.000110815 | 0.000293098 | 0.696693705 |
| T2DM | dendritic_cell | *KANK3* | -1.299255543 | 1.23E-05 | 0.000130129 | 0.077505386 |
| T2DM | dendritic_cell | *EMID1* | -4.294180155 | 1.01E-05 | 0.000130129 | 0.06372367 |
| T2DM | dendritic_cell | *CYP4A11* | -1.524795795 | 0.012100284 | 0.016505638 | 1 |
| T2DM | dendritic_cell | *C3orf70* | -0.961307795 | 0.004344575 | 0.006475662 | 1 |
| T2DM | dendritic_cell | *PLCD1* | -2.014603308 | 0.001769167 | 0.002886029 | 1 |
| T2DM | dendritic_cell | *ENTPD5* | -3.110292321 | 0.00026019 | 0.00055621 | 1 |
| T2DM | dendritic_cell | *ANGPTL5* | -2.810147716 | 2.07E-05 | 0.000132272 | 0.130424848 |
| T2DM | dendritic_cell | *FNBP1* | 1.350612155 | 0.001816011 | 0.002955543 | 1 |
| T2DM | dendritic_cell | *WDR86* | -1.59726052 | 1.52E-05 | 0.000130129 | 0.095628224 |
| T2DM | dendritic_cell | *EPOR* | -2.143933863 | 0.000277679 | 0.000585633 | 1 |
| T2DM | dendritic_cell | *FPR3* | 0.413478916 | 5.00E-05 | 0.000177555 | 0.314524723 |
| T2DM | dendritic_cell | *GJA4* | -2.330320029 | 0.001073044 | 0.001841722 | 1 |
| T2DM | dendritic_cell | *IFT140* | -2.715395998 | 0.000193805 | 0.00044243 | 1 |
| T2DM | dendritic_cell | *TLR5* | 1.740390367 | 0.00021234 | 0.000473397 | 1 |
| T2DM | dendritic_cell | *PLEKHN1* | 1.868699962 | 0.014317226 | 0.019332559 | 1 |
| T2DM | dendritic_cell | *ZNF385C* | -3.525762507 | 1.88E-05 | 0.000130129 | 0.11833726 |
| T2DM | dendritic_cell | *TET3* | 1.303629222 | 0.000231373 | 0.00050649 | 1 |
| T2DM | dendritic_cell | *ISG15* | -2.101805121 | 3.13E-05 | 0.000147228 | 0.19699153 |

| | | | | | | |
|---|---|---|---|---|---|---|
| T2DM | dendritic_cell | *TRPV2* | 2.361893162 | 0.000602431 | 0.001116264 | 1 |
| T2DM | dendritic_cell | *C2orf88* | -1.281663712 | 3.86E-05 | 0.000158005 | 0.242583792 |
| T2DM | dendritic_cell | *ADH1A* | -1.951887426 | 1.50E-06 | 7.56E-05 | 0.009445273 |
| T2DM | dendritic_cell | *SEMA4D* | -4.054266618 | 0.001384154 | 0.002313179 | 1 |
| T2DM | dendritic_cell | *FAM69C* | -2.840198644 | 0.012375062 | 0.016865903 | 1 |
| T2DM | dendritic_cell | *DNAH17* | -4.132070183 | 7.87E-06 | 0.000130129 | 0.049505612 |
| T2DM | dendritic_cell | *ZFP69* | -1.23696385 | 2.09E-05 | 0.000132272 | 0.131214167 |
| T2DM | dendritic_cell | *TMEM220* | -2.215161523 | 3.26E-05 | 0.000148254 | 0.204961616 |
| T2DM | dendritic_cell | *CCDC157* | -2.480938869 | 1.68E-05 | 0.000130129 | 0.105656447 |
| T2DM | dendritic_cell | *GFRAL* | -1.460980946 | 0.005576761 | 0.008123517 | 1 |
| T2DM | dendritic_cell | *LDLRAD2* | -1.361292633 | 7.34E-06 | 0.000130129 | 0.046137113 |
| T2DM | dendritic_cell | *CPSF4L* | -3.286638614 | 0.000146561 | 0.000359587 | 0.921431546 |
| T2DM | dendritic_cell | *KLHL17* | -3.754551578 | 9.73E-05 | 0.000268565 | 0.611523271 |
| T2DM | dendritic_cell | *S100A3* | -1.932473862 | 0.003315303 | 0.005070131 | 1 |
| T2DM | dendritic_cell | *TREML4* | 2.932702366 | 3.56E-05 | 0.000154118 | 0.223625466 |
| T2DM | dendritic_cell | *C6orf132* | -1.272850046 | 2.04E-05 | 0.00013202 | 0.128239516 |
| T2DM | dendritic_cell | *MAPK12* | -2.439933203 | 4.17E-05 | 0.000162756 | 0.262049335 |
| T2DM | dendritic_cell | *AGRN* | -2.026176226 | 0.000151916 | 0.000369904 | 0.955093244 |
| T2DM | dendritic_cell | *FAM166A* | -6.188789247 | 1.37E-05 | 0.000130129 | 0.086334818 |
| T2DM | dendritic_cell | *ZNF626* | -2.519147535 | 2.60E-05 | 0.0001395 | 0.16358762 |
| T2DM | dendritic_cell | *SMTNL2* | -8.891438616 | 1.35E-05 | 0.000130129 | 0.084881652 |
| T2DM | dendritic_cell | *ZC3H6* | -1.948885755 | 2.39E-05 | 0.000136538 | 0.150546766 |
| T2DM | dendritic_cell | *PRKAR1B* | -2.715549999 | 0.000809725 | 0.001445195 | 1 |
| T2DM | dendritic_cell | *NUTM2B* | -44.07455886 | 3.50E-07 | 3.19E-05 | 0.002199121 |
| T2DM | dendritic_cell | *NCR3LG1* | 0.948510318 | 3.53E-05 | 0.000153469 | 0.221915606 |
| T2DM | dendritic_cell | *ZNF793* | -0.570194608 | 0.02815918 | 0.036330138 | 1 |
| T2DM | dendritic_cell | *HYKK* | -1.691014325 | 0.000165362 | 0.000394397 | 1 |
| T2DM | dendritic_cell | *RUFY4* | 3.368536445 | 9.48E-05 | 0.000264262 | 0.596310232 |
| T2DM | dendritic_cell | *ZNF669* | -3.79584603 | 0.01939425 | 0.025691456 | 1 |
| T2DM | dendritic_cell | *CENPP* | -0.841518054 | 0.025440801 | 0.033012656 | 1 |
| T2DM | dendritic_cell | *ZP3* | -1.442963759 | 0.028770213 | 0.037095638 | 1 |
| T2DM | dendritic_cell | *PDCD1* | 5.395867842 | 0.000608986 | 0.001126419 | 1 |
| T2DM | dendritic_cell | *CERKL* | 1.778033325 | 1.55E-05 | 0.000130129 | 0.097374816 |
| T2DM | dendritic_cell | *COL25A1* | -0.840251353 | 1.06E-05 | 0.000130129 | 0.066677773 |
| T2DM | dendritic_cell | *SRSF10* | -2.391389792 | 0.000114197 | 0.000299522 | 0.717955257 |
| T2DM | dendritic_cell | *NBR1* | -3.311591373 | 0.000339537 | 0.000688825 | 1 |
| T2DM | dendritic_cell | *NDOR1* | 3.032076567 | 0.004614176 | 0.006830545 | 1 |
| T2DM | dendritic_cell | *FAM72B* | 2.468260828 | 4.35E-05 | 0.000165783 | 0.273541661 |
| T2DM | dendritic_cell | *SUMO2* | -3.072990164 | 0.000292579 | 0.00061111 | 1 |
| T2DM | dendritic_cell | *GOLGA8M* | -1.53882678 | 5.70E-05 | 0.000191255 | 0.358254619 |
| T2DM | dendritic_cell | *S100A16* | -2.951426055 | 0.017643267 | 0.023490729 | 1 |
| T2DM | dendritic_cell | *RHCE* | -1.466508801 | 0.000210137 | 0.00047032 | 1 |
| T2DM | dendritic_cell | *PARVB* | -1.352646724 | 0.032232754 | 0.041238772 | 1 |
| T2DM | dendritic_cell | *UROS* | -2.30501938 | 0.017273681 | 0.023032796 | 1 |
| T2DM | dendritic_cell | *FAM221A* | -1.948484613 | 0.000434061 | 0.000845024 | 1 |
| T2DM | dendritic_cell | *FSIP2* | -0.809203103 | 0.002572305 | 0.004045614 | 1 |
| T2DM | dendritic_cell | *NOXA1* | 2.876965913 | 0.003442785 | 0.005249767 | 1 |
| T2DM | dendritic_cell | *SKOR1* | 7.230426505 | 5.45E-05 | 0.000186487 | 0.342330955 |
| T2DM | dendritic_cell | *TMEM201* | -2.119768547 | 0.00276124 | 0.004305535 | 1 |
| T2DM | dendritic_cell | *ZDHHC11* | -2.230542279 | 3.62E-05 | 0.00015508 | 0.227432495 |

| | | | | | | |
|---|---|---|---|---|---|---|
| T2DM | dendritic_cell | *FAM26F* | 0.928992849 | 0.002225523 | 0.003558068 | 1 |
| T2DM | dendritic_cell | *LINC00910* | -2.439022919 | 4.55E-06 | 0.000118249 | 0.028616302 |
| T2DM | dendritic_cell | *BEND4* | -6.128950164 | 0.000586604 | 0.001091118 | 1 |
| T2DM | dendritic_cell | *FBF1* | -0.691985835 | 0.028098018 | 0.036266113 | 1 |
| T2DM | dendritic_cell | *MSL1* | -1.263983954 | 0.006663402 | 0.009562384 | 1 |
| T2DM | dendritic_cell | *FAM196A* | -2.631200486 | 3.26E-05 | 0.000148254 | 0.204832421 |
| T2DM | dendritic_cell | *UTS2B* | -1.243160356 | 0.000513528 | 0.000972162 | 1 |
| T2DM | dendritic_cell | *C9orf152* | -5.825483007 | 1.80E-05 | 0.000130129 | 0.113079822 |
| T2DM | dendritic_cell | *NOC2L* | -0.956359884 | 0.029176184 | 0.037565159 | 1 |
| T2DM | dendritic_cell | *SLC15A5* | -0.724372591 | 0.00417723 | 0.006251437 | 1 |
| T2DM | dendritic_cell | *LIPI* | -1.024546864 | 0.000467208 | 0.000897719 | 1 |
| T2DM | dendritic_cell | *ZNF567* | -3.78115652 | 0.00061717 | 0.001138895 | 1 |
| T2DM | dendritic_cell | *ANKDD1B* | -1.564011907 | 0.000260675 | 0.000557057 | 1 |
| T2DM | dendritic_cell | *LITAF* | -2.315025792 | 0.011706159 | 0.016013191 | 1 |
| T2DM | dendritic_cell | *VSTM1* | 2.56116666 | 0.001562649 | 0.002579252 | 1 |
| T2DM | dendritic_cell | *SF3B3* | -2.900950539 | 4.88E-05 | 0.000174774 | 0.306553831 |
| T2DM | dendritic_cell | *PLAC9* | -1.704628843 | 1.74E-06 | 8.21E-05 | 0.010947428 |
| T2DM | dendritic_cell | *CLDN4* | 1.390040305 | 0.008334798 | 0.011738547 | 1 |
| T2DM | dendritic_cell | *ZNF573* | -3.575562126 | 0.0006043 | 0.001119398 | 1 |
| T2DM | dendritic_cell | *FAM47E* | -0.862738136 | 0.00198674 | 0.003206014 | 1 |
| T2DM | dendritic_cell | *ZNF527* | -4.543207722 | 0.002802568 | 0.004362403 | 1 |
| T2DM | dendritic_cell | *S100A13* | -1.415660907 | 3.47E-05 | 0.000152237 | 0.218460027 |
| T2DM | dendritic_cell | *ZNF33A* | -2.424373438 | 0.002651007 | 0.004153174 | 1 |
| T2DM | dendritic_cell | *C15orf61* | -4.175824444 | 0.012366315 | 0.016861206 | 1 |
| T2DM | dendritic_cell | *PNRC2* | -4.047390355 | 0.000106519 | 0.000284729 | 0.669683717 |
| T2DM | dendritic_cell | *ZKSCAN3* | -4.297123622 | 3.19E-05 | 0.000148219 | 0.200805743 |
| T2DM | dendritic_cell | *RRP7A* | -3.181259321 | 0.00497737 | 0.007316513 | 1 |
| T2DM | dendritic_cell | *SLC35E2B* | -1.047814073 | 0.012375125 | 0.016865903 | 1 |
| T2DM | dendritic_cell | *ALG1L* | 3.125370302 | 4.44E-05 | 0.000167621 | 0.2794248 |
| T2DM | dendritic_cell | *BLOC1S2* | -1.054628815 | 0.01820556 | 0.024188157 | 1 |
| T2DM | dendritic_cell | *HLA-DRB1* | 0.258091429 | 0.002348561 | 0.003730522 | 1 |
| T2DM | dendritic_cell | *AKR1C3* | -1.434890734 | 1.46E-05 | 0.000130129 | 0.091927766 |
| T2DM | dendritic_cell | *WDSUB1* | -1.396027183 | 0.000340299 | 0.000689925 | 1 |
| T2DM | dendritic_cell | *ZNF79* | -2.092695949 | 0.000155879 | 0.000377363 | 0.980010808 |
| T2DM | dendritic_cell | *S100A4* | -1.295377917 | 0.000280005 | 0.000590141 | 1 |
| T2DM | dendritic_cell | *COLCA1* | -2.698030766 | 3.87E-06 | 0.000111792 | 0.024306707 |
| T2DM | dendritic_cell | *ACADSB* | -3.008955107 | 1.95E-05 | 0.000131266 | 0.122413045 |
| T2DM | dendritic_cell | *SEMA4A* | 0.866598765 | 4.74E-05 | 0.000172258 | 0.298209649 |
| T2DM | dendritic_cell | *HRCT1* | -2.270769401 | 2.39E-05 | 0.000136538 | 0.150015983 |
| T2DM | dendritic_cell | *MPHOSPH8* | -2.048886609 | 5.58E-05 | 0.000188986 | 0.350569696 |
| T2DM | dendritic_cell | *SIRPB2* | 0.627557068 | 4.16E-05 | 0.000162756 | 0.261240089 |
| T2DM | dendritic_cell | *SUPT5H* | -3.432693152 | 0.000701804 | 0.001274478 | 1 |
| T2DM | dendritic_cell | *XPNPEP3* | -1.555843107 | 6.99E-05 | 0.000215653 | 0.439469192 |
| T2DM | dendritic_cell | *ZNF107* | -1.04692306 | 0.039099385 | 0.049430491 | 1 |
| T2DM | dendritic_cell | *PPIA* | -3.816769669 | 0.000222348 | 0.000490443 | 1 |
| T2DM | dendritic_cell | *ZNF471* | -1.833301822 | 0.000263072 | 0.000561225 | 1 |
| T2DM | dendritic_cell | *GRM7* | -0.811069237 | 0.00054911 | 0.001031448 | 1 |
| T2DM | dendritic_cell | *SUPT3H* | -2.59682401 | 0.000123825 | 0.00031866 | 0.778486271 |
| T2DM | dendritic_cell | *POM121* | -3.050563748 | 9.34E-05 | 0.000262424 | 0.587389672 |
| T2DM | dendritic_cell | *GIMAP5* | -1.722743617 | 0.000213527 | 0.00047537 | 1 |

| | | | | | | |
|---|---|---|---|---|---|---|
| T2DM | dendritic_cell | *STK31* | -1.57357765 | 1.77E-05 | 0.000130129 | 0.111428213 |
| T2DM | dendritic_cell | *ZKSCAN7* | -2.504320589 | 1.66E-05 | 0.000130129 | 0.104251992 |
| T2DM | dendritic_cell | *ZNF565* | -2.110442846 | 7.88E-05 | 0.000234049 | 0.495481952 |
| T2DM | dendritic_cell | *NTNG2* | 3.953625095 | 0.000345687 | 0.000698822 | 1 |
| T2DM | dendritic_cell | *WDR5* | -2.154449802 | 5.31E-05 | 0.000183169 | 0.333734193 |
| T2DM | dendritic_cell | *LONP1* | -1.890329355 | 0.010257147 | 0.014197861 | 1 |
| T2DM | dendritic_cell | *ASB13* | -2.554336876 | 1.58E-05 | 0.000130129 | 0.099308962 |
| T2DM | dendritic_cell | *SLC35F1* | -2.035851233 | 1.80E-05 | 0.000130129 | 0.11313304 |
| T2DM | dendritic_cell | *ZNF34* | -3.689083739 | 0.026192908 | 0.033918601 | 1 |
| T2DM | dendritic_cell | *ZNF781* | -1.134053524 | 0.01708985 | 0.022802183 | 1 |
| T2DM | dendritic_cell | *INCA1* | 3.327566069 | 1.68E-05 | 0.000130129 | 0.10537042 |
| T2DM | dendritic_cell | *ZNF774* | 1.335984596 | 3.18E-05 | 0.000148219 | 0.200143591 |
| T2DM | dendritic_cell | *EVL* | -1.75170706 | 0.009330018 | 0.013009053 | 1 |
| T2DM | dendritic_cell | *ZNF124* | 2.042099732 | 0.015635451 | 0.020995799 | 1 |
| T2DM | dendritic_cell | *ZNF569* | -2.746514109 | 0.000140195 | 0.000348245 | 0.881409003 |
| T2DM | dendritic_cell | *PIK3R4* | -1.967202292 | 0.000561057 | 0.001048875 | 1 |
| T2DM | dendritic_cell | *ZNF775* | -2.623143695 | 5.63E-05 | 0.000190114 | 0.353675327 |
| T2DM | dendritic_cell | *ZNF605* | -4.115843244 | 0.000949261 | 0.001654562 | 1 |
| T2DM | dendritic_cell | *RFX8* | -1.133969022 | 7.82E-05 | 0.000232754 | 0.4915754 |
| T2DM | dendritic_cell | *MYL6B* | -4.729049018 | 0.000725261 | 0.001309886 | 1 |
| T2DM | dendritic_cell | *ZNF799* | -3.023414149 | 0.000577722 | 0.001075871 | 1 |
| T2DM | dendritic_cell | *C20orf96* | -5.039970695 | 0.000135239 | 0.000339235 | 0.850245049 |
| T2DM | dendritic_cell | *SULT1A1* | -1.96047656 | 2.56E-05 | 0.0001392 | 0.160915668 |
| T2DM | dendritic_cell | *ANAPC7* | -2.705627019 | 8.80E-05 | 0.000252633 | 0.553266286 |
| T2DM | dendritic_cell | *TPK1* | 1.827995076 | 0.011474965 | 0.015734592 | 1 |
| T2DM | dendritic_cell | *C1orf186* | 0.533961578 | 3.36E-05 | 0.000150196 | 0.211196847 |
| T2DM | dendritic_cell | *SPTSSB* | -7.142578421 | 0.000871174 | 0.001536777 | 1 |
| T2DM | dendritic_cell | *SULF2* | 1.437047921 | 0.000237805 | 0.000517805 | 1 |
| T2DM | dendritic_cell | *XRCC2* | -2.468602603 | 3.01E-05 | 0.000145247 | 0.189358466 |
| T2DM | dendritic_cell | *MYO6* | -1.885471241 | 3.04E-07 | 2.90E-05 | 0.001910852 |
| T2DM | dendritic_cell | *HDAC2* | -3.459240652 | 0.000240543 | 0.000522382 | 1 |
| T2DM | dendritic_cell | *ADH1B* | -0.831957385 | 1.80E-05 | 0.000130129 | 0.113084499 |
| T2DM | dendritic_cell | *HRH1* | -2.130975616 | 3.07E-05 | 0.000145994 | 0.192711895 |
| T2DM | dendritic_cell | *ZNF502* | -1.514187774 | 0.020883314 | 0.027484487 | 1 |
| T2DM | dendritic_cell | *TECPR2* | -3.131274299 | 0.000182389 | 0.00042344 | 1 |
| T2DM | dendritic_cell | *FAM180B* | -2.227530823 | 2.01E-08 | 5.54E-06 | 0.000126186 |
| T2DM | dendritic_cell | *ZFP62* | -3.739425966 | 3.92E-05 | 0.00015902 | 0.246163489 |
| T2DM | dendritic_cell | *ERI2* | -1.561427791 | 5.52E-05 | 0.000188057 | 0.346777451 |
| T2DM | dendritic_cell | *TOMM7* | -3.184064451 | 4.82E-05 | 0.000173757 | 0.303032964 |
| T2DM | dendritic_cell | *TRPV1* | -1.259957053 | 0.001939517 | 0.003136251 | 1 |
| T2DM | dendritic_cell | *ZNF33B* | -3.106637712 | 8.04E-07 | 5.27E-05 | 0.005056782 |
| T2DM | dendritic_cell | *AMZ2* | 1.647413892 | 0.027824769 | 0.035930801 | 1 |
| T2DM | dendritic_cell | *ZNF431* | -2.545354838 | 4.33E-05 | 0.000165469 | 0.272115269 |
| T2DM | dendritic_cell | *NF1* | -2.014205331 | 0.000881461 | 0.001551007 | 1 |
| T2DM | dendritic_cell | *HLA-DQA1* | 0.170515217 | 8.71E-05 | 0.000251049 | 0.547536947 |
| T2DM | dendritic_cell | *TLE1* | -1.665310843 | 1.16E-05 | 0.000130129 | 0.073207477 |
| T2DM | dendritic_cell | *STRN3* | -2.061791291 | 2.90E-05 | 0.000143483 | 0.182366795 |
| T2DM | dendritic_cell | *ZNF239* | -1.173465013 | 0.004986105 | 0.007325927 | 1 |
| T2DM | dendritic_cell | *MVB12B* | -2.992509488 | 0.000438467 | 0.000852131 | 1 |
| T2DM | dendritic_cell | *NHLRC2* | -2.693060966 | 0.000210823 | 0.000471186 | 1 |

| | | | | | | |
|---|---|---|---|---|---|---|
| T2DM | dendritic_cell | *ZFP28* | -3.696048317 | 2.63E-05 | 0.000139569 | 0.165389419 |
| T2DM | dendritic_cell | *LAMB3* | -1.474866693 | 3.19E-05 | 0.000148219 | 0.200247282 |
| T2DM | dendritic_cell | *KPNA5* | -2.764281371 | 6.46E-05 | 0.00020528 | 0.405998311 |
| T2DM | dendritic_cell | *ANKRD36B* | -1.747864851 | 5.73E-05 | 0.000191733 | 0.360074991 |
| T2DM | dendritic_cell | *HCAR1* | -1.314665312 | 0.000618923 | 0.001141376 | 1 |
| T2DM | dendritic_cell | *SRGAP1* | -2.35200659 | 0.000315309 | 0.000650377 | 1 |
| T2DM | dendritic_cell | *FAM3C* | -3.408652364 | 3.01E-05 | 0.000145247 | 0.189402014 |
| T2DM | dendritic_cell | *AP2A1* | -2.531876496 | 0.003498758 | 0.005320922 | 1 |
| T2DM | dendritic_cell | *ZNF585A* | 1.991447143 | 3.90E-05 | 0.000158801 | 0.245505831 |
| T2DM | dendritic_cell | *FUT11* | 3.222057408 | 0.000287291 | 0.000601865 | 1 |
| T2DM | dendritic_cell | *ANXA4* | -2.205142066 | 4.51E-05 | 0.000168939 | 0.283735237 |
| T2DM | dendritic_cell | *WDR5B* | -1.615728181 | 0.002651637 | 0.004153174 | 1 |
| T2DM | dendritic_cell | *ZNF138* | -2.060804163 | 0.000305182 | 0.000633437 | 1 |
| T2DM | dendritic_cell | *ZNF429* | -1.858256424 | 8.35E-06 | 0.000130129 | 0.05252366 |
| T2DM | dendritic_cell | *ZNF470* | 1.911372456 | 0.039287522 | 0.049658354 | 1 |
| T2DM | dendritic_cell | *ZNF100* | -1.804649105 | 0.001587046 | 0.002614031 | 1 |
| T2DM | dendritic_cell | *GMFB* | -3.118224289 | 9.87E-05 | 0.000271152 | 0.620395666 |
| T2DM | dendritic_cell | *ZMYM1* | -2.342535958 | 2.27E-05 | 0.000133629 | 0.142697806 |
| T2DM | dendritic_cell | *ARRDC1* | 1.904297947 | 0.001485517 | 0.002465534 | 1 |
| T2DM | dendritic_cell | *IGF2R* | -2.809014297 | 3.29E-05 | 0.000148822 | 0.207052372 |
| T2DM | dendritic_cell | *GAL3ST4* | -1.96589416 | 0.000229572 | 0.000503777 | 1 |
| T2DM | dendritic_cell | *ZNF682* | -2.520067339 | 1.50E-05 | 0.000130129 | 0.094542685 |
| T2DM | dendritic_cell | *ZNF772* | -1.205371219 | 0.003088626 | 0.004762863 | 1 |
| T2DM | dendritic_cell | *ADAM32* | -1.951740833 | 1.21E-05 | 0.000130129 | 0.076190838 |
| T2DM | dendritic_cell | *LRRC8B* | -2.243629039 | 7.73E-05 | 0.000230621 | 0.485687798 |
| T2DM | dendritic_cell | *ABCB8* | -1.944676246 | 2.97E-05 | 0.000144362 | 0.186523034 |
| T2DM | dendritic_cell | *SND1* | -2.453407154 | 0.001239347 | 0.002094 | 1 |
| T2DM | dendritic_cell | *NEK5* | -2.407710013 | 0.000192794 | 0.000440603 | 1 |
| T2DM | dendritic_cell | *ENTPD4* | -2.691414684 | 0.000869738 | 0.00153547 | 1 |
| T2DM | dendritic_cell | *C1D* | -3.038572085 | 2.30E-06 | 8.72E-05 | 0.014462076 |
| T2DM | dendritic_cell | *TBC1D9B* | -2.699792878 | 0.000162872 | 0.000390235 | 1 |
| T2DM | dendritic_cell | *FAM110D* | -3.732091146 | 0.000264977 | 0.000564333 | 1 |
| T2DM | dendritic_cell | *KANK2* | -1.744020054 | 1.52E-05 | 0.000130129 | 0.095605601 |
| T2DM | dendritic_cell | *C6orf141* | -1.479107866 | 2.25E-05 | 0.000133629 | 0.141411117 |
| T2DM | dendritic_cell | *CCL4L2* | -1.185316637 | 1.04E-05 | 0.000130129 | 0.065278073 |
| T2DM | dendritic_cell | *GTF2E2* | -1.923774758 | 0.000177053 | 0.000414265 | 1 |
| T2DM | dendritic_cell | *RAMP2-AS1* | -1.685879284 | 0.006226821 | 0.008966565 | 1 |
| T2DM | dendritic_cell | *ZNF720* | -2.332448316 | 2.54E-06 | 9.14E-05 | 0.015987354 |
| T2DM | dendritic_cell | *DDI2* | -2.076814302 | 1.66E-05 | 0.000130129 | 0.104292136 |
| T2DM | dendritic_cell | *ZNF786* | -2.747423967 | 0.039564909 | 0.049988863 | 1 |
| T2DM | dendritic_cell | *ZNF517* | -1.556082597 | 1.61E-05 | 0.000130129 | 0.101322027 |
| T2DM | dendritic_cell | *C5AR1* | 2.072996035 | 0.000114284 | 0.000299626 | 0.718502364 |
| T2DM | dendritic_cell | *DCHS2* | -1.109001126 | 0.00022006 | 0.00048664 | 1 |
| T2DM | dendritic_cell | *SHPK* | -0.984061347 | 0.001637277 | 0.002686912 | 1 |
| T2DM | dendritic_cell | *STMN3* | -1.876936955 | 4.44E-05 | 0.000167569 | 0.279084847 |
| T2DM | dendritic_cell | *GYPE* | -1.049274892 | 0.010048815 | 0.013940181 | 1 |
| T2DM | dendritic_cell | *PCDHB11* | -5.757104002 | 2.27E-05 | 0.000133629 | 0.142716197 |
| T2DM | dendritic_cell | *SLC2A10* | -1.08029502 | 0.000174001 | 0.000409411 | 1 |
| T2DM | dendritic_cell | *RPF2* | -2.553438926 | 0.001201637 | 0.002035071 | 1 |
| T2DM | dendritic_cell | *TTC30A* | -2.642867151 | 1.01E-06 | 6.03E-05 | 0.00632648 |

| | | | | | | |
|---|---|---|---|---|---|---|
| T2DM | dendritic_cell | *RAB40C* | -2.129607699 | 1.47E-05 | 0.000130129 | 0.092370584 |
| T2DM | dendritic_cell | *PIGN* | -1.032095227 | 0.029391201 | 0.037818764 | 1 |
| T2DM | dendritic_cell | *HHLA3* | -2.877623221 | 1.98E-06 | 8.62E-05 | 0.012462784 |
| T2DM | dendritic_cell | *HOXA4* | -2.285411812 | 1.28E-05 | 0.000130129 | 0.080495119 |
| T2DM | dendritic_cell | *BCO2* | -1.78762224 | 1.43E-05 | 0.000130129 | 0.08987535 |
| T2DM | dendritic_cell | *ENTPD6* | -3.56962447 | 0.000104391 | 0.000281314 | 0.65630532 |
| T2DM | dendritic_cell | *CCDC154* | 2.007286359 | 1.13E-05 | 0.000130129 | 0.070974961 |
| T2DM | dendritic_cell | *C5orf42* | -1.967381609 | 0.000150392 | 0.000366904 | 0.945511845 |
| T2DM | dendritic_cell | *ZNF841* | 2.072840677 | 0.016141928 | 0.021643059 | 1 |
| T2DM | dendritic_cell | *MFAP5* | -1.049986718 | 1.50E-05 | 0.000130129 | 0.094194635 |
| T2DM | dendritic_cell | *MPEG1* | 0.632302269 | 0.000241882 | 0.000524927 | 1 |
| T2DM | dendritic_cell | *PDCD1LG2* | -1.233222334 | 0.015637468 | 0.020995799 | 1 |
| T2DM | dendritic_cell | *KLHL14* | -1.787133182 | 0.000128457 | 0.000326703 | 0.807609011 |
| T2DM | dendritic_cell | *FAM114A1* | -1.614600671 | 3.10E-05 | 0.000146367 | 0.194961221 |
| T2DM | dendritic_cell | *PHF2* | -4.117241074 | 0.000264503 | 0.000563513 | 1 |
| T2DM | dendritic_cell | *PSAP* | -2.407489796 | 0.004801272 | 0.007085821 | 1 |
| T2DM | dendritic_cell | *S100A10* | -2.835901894 | 0.001034272 | 0.001784431 | 1 |
| T2DM | dendritic_cell | *MAP1LC3C* | -1.614614181 | 0.000355693 | 0.000717435 | 1 |
| T2DM | dendritic_cell | *AC122129.1* | -3.367022719 | 1.61E-05 | 0.000130129 | 0.101185601 |
| T2DM | dendritic_cell | *CCDC180* | -1.471762779 | 0.000473999 | 0.000908824 | 1 |
| T2DM | dendritic_cell | *ADH5* | -1.77077462 | 9.52E-06 | 0.000130129 | 0.059826229 |
| T2DM | dendritic_cell | *ZNF677* | -2.256554654 | 1.59E-05 | 0.000130129 | 0.100172306 |
| T2DM | dendritic_cell | *ZNF311* | -1.450739106 | 0.01269181 | 0.017271301 | 1 |
| T2DM | dendritic_cell | *FCHSD1* | -5.770392862 | 2.34E-05 | 0.000135538 | 0.147059176 |
| T2DM | dendritic_cell | *ZNF71* | -1.946507082 | 6.89E-06 | 0.000130129 | 0.043345541 |
| T2DM | dendritic_cell | *RPL12* | -1.247690466 | 0.025830371 | 0.033476715 | 1 |
| T2DM | dendritic_cell | *VPS13A* | -1.320013153 | 0.004594445 | 0.006809945 | 1 |
| T2DM | dendritic_cell | *LEKR1* | -2.210105353 | 8.63E-06 | 0.000130129 | 0.054251868 |
| T2DM | dendritic_cell | *NOL8* | -2.295664196 | 1.97E-05 | 0.000131266 | 0.124075979 |
| T2DM | dendritic_cell | *MRPL42* | -1.95998327 | 0.003549642 | 0.005393088 | 1 |
| T2DM | dendritic_cell | *ZNF335* | -2.305995693 | 0.005828205 | 0.008452578 | 1 |
| T2DM | dendritic_cell | *AGAP9* | 3.726476871 | 0.016849771 | 0.022520092 | 1 |
| T2DM | dendritic_cell | *ZNF273* | -2.540166347 | 0.000366394 | 0.000735713 | 1 |
| T2DM | dendritic_cell | *ZNF84* | -3.02132913 | 7.49E-05 | 0.000225673 | 0.470980316 |
| T2DM | dendritic_cell | *MAK16* | -3.635398747 | 1.79E-05 | 0.000130129 | 0.112530848 |
| T2DM | dendritic_cell | *ZNF667* | -1.657763847 | 1.97E-06 | 8.62E-05 | 0.012391841 |
| T2DM | dendritic_cell | *PRIM1* | -3.434121959 | 0.00464621 | 0.006874729 | 1 |
| T2DM | dendritic_cell | *SFI1* | -2.623628303 | 3.26E-05 | 0.000148254 | 0.204670288 |
| T2DM | dendritic_cell | *ADH4* | -0.850759534 | 3.50E-05 | 0.000152692 | 0.220029215 |
| T2DM | dendritic_cell | *ZNF248* | -1.858615696 | 1.78E-06 | 8.21E-05 | 0.011169893 |
| T2DM | dendritic_cell | *HIBCH* | -1.682485634 | 8.19E-06 | 0.000130129 | 0.051477277 |
| T2DM | dendritic_cell | *ZNF544* | -1.423684621 | 0.003176942 | 0.004886973 | 1 |
| T2DM | dendritic_cell | *SOWAHC* | -2.33115688 | 7.78E-06 | 0.000130129 | 0.048934916 |
| T2DM | dendritic_cell | *AC135178.1* | -5.05732314 | 0.014292921 | 0.019303887 | 1 |
| T2DM | dendritic_cell | *MIER1* | -2.395296535 | 0.003771896 | 0.00570183 | 1 |
| T2DM | dendritic_cell | *SVIP* | -2.359248423 | 1.84E-05 | 0.000130129 | 0.115471247 |
| T2DM | dendritic_cell | *ZNF251* | -2.347819792 | 0.000222017 | 0.000490278 | 1 |
| T2DM | dendritic_cell | *DDRGK1* | -1.993314901 | 1.46E-05 | 0.000130129 | 0.09178201 |
| T2DM | dendritic_cell | *ZNF334* | -1.276955982 | 0.002869295 | 0.004448645 | 1 |
| T2DM | dendritic_cell | *HSD17B11* | -3.04306163 | 0.003742532 | 0.005664251 | 1 |

| | | | | | | |
|---|---|---|---|---|---|---|
| T2DM | dendritic_cell | *RPS6KL1* | -0.967257469 | 0.001053271 | 0.001811245 | 1 |
| T2DM | dendritic_cell | *QRICH1* | -1.863142706 | 0.00055257 | 0.00103609 | 1 |
| T2DM | dendritic_cell | *DDX42* | -1.927917633 | 0.004569634 | 0.006777374 | 1 |
| T2DM | dendritic_cell | *SLC29A3* | 1.225856159 | 0.000184906 | 0.000427598 | 1 |
| T2DM | dendritic_cell | *ANTXRL* | -49.32070661 | 1.10E-05 | 0.000130129 | 0.069190861 |
| T2DM | dendritic_cell | *STYX* | -3.275819478 | 0.011224641 | 0.015424988 | 1 |
| T2DM | dendritic_cell | *TMEM116* | -1.76045582 | 8.59E-07 | 5.49E-05 | 0.005401408 |
| T2DM | dendritic_cell | *ZKSCAN8* | -2.429559396 | 0.001786689 | 0.002910835 | 1 |
| T2DM | dendritic_cell | *MYL4* | 1.615953713 | 3.86E-05 | 0.000158005 | 0.242397416 |
| T2DM | dendritic_cell | *ZNF813* | -2.960579889 | 5.72E-06 | 0.000129346 | 0.035958199 |
| T2DM | dendritic_cell | *GFPT1* | -1.572941429 | 4.71E-05 | 0.000171433 | 0.296062942 |
| T2DM | dendritic_cell | *MGEA5* | -1.581672793 | 7.42E-06 | 0.000130129 | 0.046680267 |
| T2DM | dendritic_cell | *MT1F* | -5.577119409 | 3.10E-05 | 0.000146367 | 0.194920879 |
| T2DM | dendritic_cell | *SH3BGRL2* | -1.675449509 | 2.96E-05 | 0.000144362 | 0.185820961 |
| T2DM | dendritic_cell | *ZNF808* | -2.489044811 | 0.003033317 | 0.004682166 | 1 |
| T2DM | dendritic_cell | *TMA16* | -2.621894873 | 0.024305088 | 0.031678251 | 1 |
| T2DM | dendritic_cell | *MAFK* | -2.488563868 | 1.57E-05 | 0.000130129 | 0.098899489 |
| T2DM | dendritic_cell | *HIST1H4E* | 1.93916679 | 6.45E-05 | 0.00020528 | 0.405809072 |
| T2DM | dendritic_cell | *C20orf203* | -3.786578346 | 0.002723871 | 0.004254653 | 1 |
| T2DM | dendritic_cell | *ARC* | 2.994871233 | 3.96E-05 | 0.000159553 | 0.248798891 |
| T2DM | dendritic_cell | *TLK1* | -2.219189922 | 4.81E-05 | 0.000173596 | 0.302230844 |
| T2DM | dendritic_cell | *LRBA* | -2.23299909 | 0.000151268 | 0.000368718 | 0.95101989 |
| T2DM | dendritic_cell | *MMP17* | -1.238398891 | 1.56E-05 | 0.000130129 | 0.098272687 |
| T2DM | dendritic_cell | *CCDC69* | -2.398779831 | 0.001419594 | 0.002366116 | 1 |
| T2DM | dendritic_cell | *MDM4* | -2.619059968 | 0.000551104 | 0.001034884 | 1 |
| T2DM | dendritic_cell | *NCOA6* | -2.374461974 | 0.000342066 | 0.000692838 | 1 |
| T2DM | dendritic_cell | *FAM19A2* | -2.473525035 | 0.014769027 | 0.019895623 | 1 |
| T2DM | dendritic_cell | *TTC37* | -1.958444534 | 4.30E-05 | 0.000165006 | 0.270114152 |
| T2DM | dendritic_cell | *TUSC1* | 2.517315399 | 0.000130585 | 0.000331043 | 0.820986234 |
| T2DM | dendritic_cell | *FAN1* | -3.200650836 | 8.24E-05 | 0.000241664 | 0.518128356 |
| T2DM | dendritic_cell | *CEP290* | -2.565498221 | 2.42E-05 | 0.000136859 | 0.152194893 |
| T2DM | dendritic_cell | *ANKRD13B* | -1.274665424 | 0.000478255 | 0.000915309 | 1 |
| T2DM | dendritic_cell | *ECI2* | -2.939957612 | 4.75E-05 | 0.00017227 | 0.298716802 |
| T2DM | dendritic_cell | *ZNF652* | -2.711824716 | 0.000118929 | 0.000309415 | 0.747706857 |
| T2DM | dendritic_cell | *RPL10A* | -2.883069203 | 0.000254665 | 0.000547003 | 1 |
| T2DM | dendritic_cell | *APCDD1L* | -2.021085891 | 1.54E-05 | 0.000130129 | 0.096979649 |
| T2DM | dendritic_cell | *RCSD1* | 1.182862684 | 0.001521667 | 0.002518883 | 1 |
| T2DM | dendritic_cell | *RASSF9* | -3.818851196 | 7.44E-05 | 0.000224335 | 0.467721031 |
| T2DM | dendritic_cell | *TMEM184B* | -2.392397794 | 4.07E-05 | 0.00016095 | 0.255649619 |
| T2DM | dendritic_cell | *ALPK2* | -1.19521476 | 0.000143707 | 0.000354864 | 0.90348424 |
| T2DM | dendritic_cell | *SFT2D1* | -2.975896256 | 0.000108668 | 0.000289612 | 0.683194628 |
| T2DM | dendritic_cell | *ARHGAP11A* | 3.204390229 | 0.019265254 | 0.025542103 | 1 |
| T2DM | dendritic_cell | *HMGN2* | -1.845163373 | 0.003748487 | 0.005670534 | 1 |
| T2DM | dendritic_cell | *DENND4B* | -2.340557367 | 0.031974959 | 0.040925604 | 1 |
| T2DM | dendritic_cell | *CD3E* | -1.62315434 | 0.006820974 | 0.009768442 | 1 |
| T2DM | dendritic_cell | *TSEN15* | -2.81249431 | 2.90E-05 | 0.000143473 | 0.18212307 |
| T2DM | dendritic_cell | *TYW1* | -1.056530062 | 0.002902912 | 0.00449744 | 1 |
| T2DM | dendritic_cell | *ITPRIPL1* | -2.17850531 | 0.000929492 | 0.001623843 | 1 |
| T2DM | dendritic_cell | *PRMT6* | 1.893708141 | 0.0001205 | 0.00031189 | 0.757580598 |
| T2DM | dendritic_cell | *SHISA4* | -3.245495566 | 2.61E-05 | 0.0001395 | 0.164181469 |

| | | | | | | |
|---|---|---|---|---|---|---|
| T2DM | dendritic_cell | *CAPZA2* | -2.177260966 | 0.027826796 | 0.035930801 | 1 |
| T2DM | dendritic_cell | *MAP3K3* | -3.27722099 | 0.000243638 | 0.000528045 | 1 |
| T2DM | dendritic_cell | *C1orf174* | 1.854921534 | 0.000423171 | 0.000829066 | 1 |
| T2DM | dendritic_cell | *KIAA0753* | -2.774497693 | 1.79E-05 | 0.000130129 | 0.112737872 |
| T2DM | dendritic_cell | *DCLRE1A* | -2.150303615 | 0.000445731 | 0.000864377 | 1 |
| T2DM | dendritic_cell | *APRT* | -4.812889785 | 0.000252787 | 0.000543527 | 1 |
| T2DM | dendritic_cell | *TBKBP1* | -1.9269865 | 8.54E-06 | 0.000130129 | 0.053688015 |
| T2DM | dendritic_cell | *ZFP2* | -2.432976211 | 0.000122105 | 0.000315137 | 0.76767272 |
| T2DM | dendritic_cell | *MFAP3L* | 4.446264759 | 0.00665007 | 0.009545431 | 1 |
| T2DM | dendritic_cell | *SMG5* | -2.717148857 | 0.014700013 | 0.019806897 | 1 |
| T2DM | dendritic_cell | *PJA2* | -2.603061732 | 8.61E-05 | 0.000249121 | 0.541590085 |
| T2DM | dendritic_cell | *AL590822.1* | -1.205762899 | 0.000388147 | 0.000771508 | 1 |
| T2DM | dendritic_cell | *AP001007.1* | 12.09530817 | 0.000146421 | 0.000359449 | 0.920548088 |
| T2DM | dendritic_cell | *ZNF525* | -3.233373349 | 3.78E-05 | 0.00015701 | 0.237714605 |
| T2DM | dendritic_cell | *AC105020.1* | -3.868727306 | 5.88E-06 | 0.000129745 | 0.036990041 |
| T2DM | dendritic_cell | *EFCAB2* | -2.494374891 | 0.000110753 | 0.000293091 | 0.69630371 |
| T2DM | dendritic_cell | *CHML* | 1.635791364 | 4.87E-06 | 0.000123437 | 0.030612305 |
| T2DM | dendritic_cell | *ECT2L* | -2.346842391 | 3.24E-05 | 0.000148254 | 0.203949067 |
| T2DM | dendritic_cell | *CENPW* | -3.29152932 | 4.94E-09 | 2.07E-06 | 3.11E-05 |
| T2DM | dendritic_cell | *FAM229B* | -1.626718161 | 1.29E-07 | 1.80E-05 | 0.000809527 |
| T2DM | dendritic_cell | *DDO* | -3.853795933 | 0.00052072 | 0.000983703 | 1 |
| T2DM | dendritic_cell | *HIST2H2BF* | 2.253439528 | 4.43E-05 | 0.000167569 | 0.278217865 |
| T2DM | dendritic_cell | *RBM20* | -1.635549661 | 1.18E-05 | 0.000130129 | 0.074140965 |
| T2DM | dendritic_cell | *C6orf163* | -1.772649572 | 1.39E-05 | 0.000130129 | 0.087124473 |
| T2DM | dendritic_cell | *PCMTD2* | -2.464822132 | 0.005233207 | 0.007654996 | 1 |
| T2DM | dendritic_cell | *OOEP* | -1.801130322 | 7.12E-05 | 0.000218276 | 0.447683796 |
| T2DM | dendritic_cell | *FOXO6* | -2.857685554 | 1.96E-05 | 0.000131266 | 0.123185486 |
| T2DM | dendritic_cell | *INPP5B* | -0.749852578 | 0.001487912 | 0.002468857 | 1 |
| T2DM | dendritic_cell | *TRAF3IP1* | -2.773593189 | 1.88E-05 | 0.000130129 | 0.118430074 |
| T2DM | dendritic_cell | *GIGYF2* | -2.321867785 | 7.68E-05 | 0.000229665 | 0.4827549 |
| T2DM | dendritic_cell | *C2orf72* | -2.304636354 | 0.024521157 | 0.031918118 | 1 |
| T2DM | dendritic_cell | *RUFY2* | -2.176775606 | 2.59E-05 | 0.0001395 | 0.162951449 |
| T2DM | dendritic_cell | *PHACTR4* | -2.565643957 | 1.54E-05 | 0.000130129 | 0.096926292 |
| T2DM | dendritic_cell | *C10orf128* | -1.465341059 | 0.003228205 | 0.004950177 | 1 |
| T2DM | dendritic_cell | *SYT15* | -2.03648259 | 0.007524303 | 0.010690462 | 1 |
| T2DM | dendritic_cell | *ZDBF2* | -2.926756452 | 0.001324248 | 0.002222516 | 1 |
| T2DM | dendritic_cell | *RING1* | -2.55205973 | 0.000535878 | 0.001009003 | 1 |
| T2DM | dendritic_cell | *HSD17B8* | -1.864731288 | 0.003862211 | 0.005824351 | 1 |
| T2DM | dendritic_cell | *OXLD1* | 3.311065652 | 0.000521919 | 0.000985376 | 1 |
| T2DM | dendritic_cell | *HLA-DOA* | 0.308952644 | 3.76E-05 | 0.000156608 | 0.236095842 |
| T2DM | dendritic_cell | *HLA-DMA* | 0.855852832 | 0.000262852 | 0.000561137 | 1 |
| T2DM | dendritic_cell | *TAP2* | -1.112370384 | 0.001437378 | 0.002392585 | 1 |
| T2DM | dendritic_cell | *HLA-DRA* | 0.372220895 | 2.88E-06 | 9.78E-05 | 0.018094041 |
| T2DM | dendritic_cell | *COL15A1* | -1.958196044 | 2.53E-05 | 0.000138965 | 0.159266116 |
| T2DM | dendritic_cell | *NOTCH4* | -0.492908669 | 1.55E-10 | 1.39E-07 | 9.72E-07 |
| T2DM | dendritic_cell | *AGER* | 3.357864943 | 1.35E-05 | 0.000130129 | 0.084714993 |
| T2DM | dendritic_cell | *PRRT1* | 2.25935391 | 0.000400713 | 0.000792476 | 1 |
| T2DM | dendritic_cell | *DXO* | -2.935795494 | 4.81E-07 | 3.82E-05 | 0.003024204 |
| T2DM | dendritic_cell | *LAYN* | -1.977488734 | 0.038929298 | 0.049235263 | 1 |
| T2DM | dendritic_cell | *C6orf48* | -2.803396285 | 0.001081577 | 0.001854848 | 1 |

| | | | | | | |
|---|---|---|---|---|---|---|
| T2DM | dendritic_cell | *CARD16* | -2.776679074 | 0.000137136 | 0.000342676 | 0.862173503 |
| T2DM | dendritic_cell | *MSH5* | -1.076812485 | 0.020089329 | 0.026533952 | 1 |
| T2DM | dendritic_cell | *ABHD16A* | 1.168635751 | 0.00271936 | 0.004248662 | 1 |
| T2DM | dendritic_cell | *ZNF814* | -2.711483065 | 7.76E-05 | 0.000231367 | 0.487951979 |
| T2DM | dendritic_cell | *MICB* | 0.501743823 | 0.025981113 | 0.033658203 | 1 |
| T2DM | dendritic_cell | *MICA* | -0.854164506 | 0.004896738 | 0.007209787 | 1 |
| T2DM | dendritic_cell | *ABCF1* | -1.719859448 | 0.013508982 | 0.018323834 | 1 |
| T2DM | dendritic_cell | *LILRB3* | 0.694246956 | 3.90E-05 | 0.000158736 | 0.245138662 |
| T2DM | dendritic_cell | *LINC01123* | -54.84200216 | 1.54E-07 | 2.06E-05 | 0.000968846 |
| T2DM | dendritic_cell | *HLA-E* | -1.062835952 | 0.014475068 | 0.019524727 | 1 |
| T2DM | dendritic_cell | *ZNF468* | -1.834068123 | 2.21E-05 | 0.000132957 | 0.138940306 |
| T2DM | dendritic_cell | *HCG9* | -0.88918704 | 0.005418492 | 0.007913138 | 1 |
| T2DM | dendritic_cell | *HLA-G* | 1.115482256 | 0.03626752 | 0.046100667 | 1 |
| T2DM | dendritic_cell | *FAM153C* | 4.453326534 | 0.002588233 | 0.004065006 | 1 |
| T2DM | dendritic_cell | *TRIM27* | -2.490532155 | 0.000671618 | 0.001224612 | 1 |
| T2DM | dendritic_cell | *MALRD1* | -0.87405942 | 0.000779993 | 0.001398692 | 1 |
| T2DM | dendritic_cell | *MROH6* | 0.812948586 | 0.000122794 | 0.000316526 | 0.772007484 |
| T2DM | dendritic_cell | *ATXN2* | -2.037725202 | 8.13E-05 | 0.000238955 | 0.510886018 |
| T2DM | dendritic_cell | *FAM216A* | -2.594384318 | 7.04E-05 | 0.000216691 | 0.442700593 |
| T2DM | dendritic_cell | *ZNF155* | -2.253896195 | 0.000929044 | 0.001623825 | 1 |
| T2DM | dendritic_cell | *ZNF783* | -1.975001266 | 0.033776223 | 0.043125734 | 1 |
| T2DM | dendritic_cell | *ZNF425* | -1.733167686 | 0.004804756 | 0.007089298 | 1 |
| T2DM | dendritic_cell | *LRRC10B* | -8.270015469 | 0.008136934 | 0.011488189 | 1 |
| T2DM | dendritic_cell | *C12orf73* | -2.794663794 | 9.03E-06 | 0.000130129 | 0.056748244 |
| T2DM | dendritic_cell | *PCDHGA1* | 1.079650866 | 0.001175746 | 0.001995658 | 1 |
| T2DM | dendritic_cell | *PCDHA4* | -6.312224387 | 5.54E-05 | 0.000188509 | 0.348176042 |
| T2DM | dendritic_cell | *TRIM13* | -2.256976949 | 6.57E-05 | 0.00020797 | 0.41302912 |
| T2DM | dendritic_cell | *SLFN12L* | -1.403065513 | 3.98E-05 | 0.000159733 | 0.250197685 |
| T2DM | dendritic_cell | *CLLU1OS* | -1.992957208 | 0.025353113 | 0.032905662 | 1 |
| T2DM | dendritic_cell | *SYCE1L* | -1.185126574 | 0.01501782 | 0.020204801 | 1 |
| T2DM | dendritic_cell | *TMEM231* | -1.565150855 | 0.000856592 | 0.001516159 | 1 |
| T2DM | dendritic_cell | *FAM71F2* | -2.283375452 | 1.16E-05 | 0.000130129 | 0.073232398 |
| T2DM | dendritic_cell | *AC016586.1* | 11.20089419 | 4.73E-06 | 0.000121447 | 0.029707778 |
| T2DM | dendritic_cell | *LINC00654* | -2.434511982 | 0.000231078 | 0.000506197 | 1 |
| T2DM | dendritic_cell | *SAP25* | -1.52275587 | 0.00930704 | 0.012985655 | 1 |
| T2DM | dendritic_cell | *PRR13* | -3.699777976 | 0.002017969 | 0.003250568 | 1 |
| T2DM | dendritic_cell | *MT1A* | 2.810839521 | 6.40E-05 | 0.00020421 | 0.40229298 |
| T2DM | dendritic_cell | *CFI* | -1.295671871 | 1.91E-05 | 0.000130716 | 0.119997658 |
| T2DM | dendritic_cell | *SAMD9* | 2.110347846 | 0.00066121 | 0.001207385 | 1 |
| T2DM | dendritic_cell | *EXOC3L4* | -2.724474231 | 7.19E-06 | 0.000130129 | 0.045173105 |
| T2DM | dendritic_cell | *ATP6AP1L* | -1.2345319 | 0.022644565 | 0.0296288 | 1 |
| T2DM | dendritic_cell | *NAP1L4* | -2.510800512 | 0.000247769 | 0.000534748 | 1 |
| T2DM | dendritic_cell | *SMN2* | -1.154247591 | 0.004665498 | 0.006896776 | 1 |
| T2DM | dendritic_cell | *SERF1B* | -4.610902924 | 1.12E-06 | 6.47E-05 | 0.007057887 |
| T2DM | dendritic_cell | *DENND6B* | -5.467356806 | 0.000634267 | 0.001164613 | 1 |
| T2DM | dendritic_cell | *EIF3CL* | -3.035191796 | 0.001605584 | 0.002641797 | 1 |
| T2DM | dendritic_cell | *LCMT1* | -3.173900341 | 0.000159153 | 0.000383074 | 1 |
| T2DM | dendritic_cell | *LIN52* | -1.007612558 | 0.019805508 | 0.026186589 | 1 |
| T2DM | dendritic_cell | *C17orf107* | 1.702383468 | 3.40E-05 | 0.00015089 | 0.213962438 |
| T2DM | dendritic_cell | *LINC00487* | -3.255890526 | 0.009477127 | 0.013193689 | 1 |

| | | | | | | |
|---|---|---|---|---|---|---|
| T2DM | dendritic_cell | *RFPL3S* | 0.847901832 | 0.024146403 | 0.031482463 | 1 |
| T2DM | dendritic_cell | *C1QTNF9B* | -17.47381475 | 1.69E-05 | 0.000130129 | 0.106065917 |
| T2DM | dendritic_cell | *RNPS1* | -2.056084089 | 0.00089595 | 0.001574297 | 1 |
| T2DM | dendritic_cell | *NYNRIN* | 3.960269162 | 0.001124852 | 0.001919117 | 1 |
| T2DM | dendritic_cell | *ZDHHC11B* | -2.12644643 | 0.000187119 | 0.000431079 | 1 |
| T2DM | dendritic_cell | *HCG27* | 1.107887573 | 0.001174923 | 0.001994799 | 1 |
| T2DM | dendritic_cell | *HLA-A* | -1.455041745 | 0.006080006 | 0.0087813 | 1 |
| T2DM | dendritic_cell | *LNP1* | -1.73091569 | 1.54E-05 | 0.000130129 | 0.0969135 |
| T2DM | dendritic_cell | *VGLL3* | -0.849993893 | 1.57E-05 | 0.000130129 | 0.098698077 |
| T2DM | dendritic_cell | *COLQ* | -1.977665078 | 3.52E-06 | 0.000107473 | 0.022124889 |
| T2DM | dendritic_cell | *METTL6* | -1.789122133 | 0.037431476 | 0.047474619 | 1 |
| T2DM | dendritic_cell | *SACM1L* | -2.583236765 | 0.000127356 | 0.000324823 | 0.800688025 |
| T2DM | dendritic_cell | *SLC48A1* | -4.049040726 | 5.08E-05 | 0.000178852 | 0.319250231 |
| T2DM | dendritic_cell | *ZRSR1* | -6.297622115 | 0.000138611 | 0.000345162 | 0.871445974 |
| T2DM | dendritic_cell | *AC084018.1* | -1.327234971 | 6.33E-05 | 0.000203087 | 0.398051235 |
| T2DM | dendritic_cell | *C17orf51* | -1.722037244 | 1.09E-05 | 0.000130129 | 0.068645007 |
| T2DM | dendritic_cell | *OR2A42* | -1.724047225 | 0.000173103 | 0.000407754 | 1 |
| T2DM | dendritic_cell | *RNF208* | -7.40707516 | 1.20E-05 | 0.000130129 | 0.075466512 |
| T2DM | dendritic_cell | *MAP10* | 2.711142649 | 0.001707965 | 0.00279126 | 1 |
| T2DM | dendritic_cell | *ZNF580* | -1.753162639 | 0.001526251 | 0.002525142 | 1 |
| T2DM | dendritic_cell | *FAM106A* | -1.03336274 | 0.029297086 | 0.0377131 | 1 |
| T2DM | dendritic_cell | *ZNF254* | -2.891094615 | 0.000447599 | 0.00086738 | 1 |
| T2DM | dendritic_cell | *TCTEX1D2* | 2.50589079 | 0.000282544 | 0.000594495 | 1 |
| T2DM | dendritic_cell | *LINGO4* | -10.19882263 | 0.026569503 | 0.034385027 | 1 |
| T2DM | dendritic_cell | *CCDC183* | 4.36672638 | 0.00037584 | 0.000750606 | 1 |
| T2DM | dendritic_cell | *ARHGEF35* | -3.35705735 | 0.000806288 | 0.001440094 | 1 |
| T2DM | dendritic_cell | *DNLZ* | 1.472714414 | 0.000101897 | 0.000276371 | 0.640628839 |
| T2DM | dendritic_cell | *SUPT4H1* | 2.495114423 | 8.72E-05 | 0.00025117 | 0.548304701 |
| T2DM | dendritic_cell | *MXD3* | -1.24342611 | 0.019019342 | 0.025232032 | 1 |
| T2DM | dendritic_cell | *GSTM2* | -1.228036718 | 0.000882682 | 0.001552285 | 1 |
| T2DM | dendritic_cell | *ARHGAP19* | 4.528895963 | 0.007826003 | 0.011078348 | 1 |
| T2DM | dendritic_cell | *LCAT* | -1.142378496 | 0.035670318 | 0.045415004 | 1 |
| T2DM | dendritic_cell | *SIPA1* | 3.050726001 | 6.84E-06 | 0.000130129 | 0.042993794 |
| T2DM | dendritic_cell | *GBP7* | -0.788610585 | 0.013984985 | 0.018904235 | 1 |
| T2DM | dendritic_cell | *SRA1* | -2.206973697 | 0.000188692 | 0.00043375 | 1 |
| T2DM | dendritic_cell | *TMEM110* | 3.167564941 | 6.52E-05 | 0.000206799 | 0.410082336 |
| T2DM | dendritic_cell | *HEXA* | -3.355714144 | 0.000208652 | 0.000467831 | 1 |
| T2DM | dendritic_cell | *LEPROT* | -2.907986404 | 6.04E-05 | 0.000198694 | 0.379998328 |
| T2DM | dendritic_cell | *LBH* | -1.382156756 | 1.66E-05 | 0.000130129 | 0.104213178 |
| T2DM | dendritic_cell | *PPP1CB* | 1.504191366 | 0.004819742 | 0.007109741 | 1 |
| T2DM | dendritic_cell | *NCKIPSD* | -2.345905695 | 0.000186147 | 0.000429628 | 1 |
| T2DM | dendritic_cell | *SLC35F6* | -4.004858739 | 6.65E-05 | 0.000209593 | 0.418138953 |
| T2DM | dendritic_cell | *CLIC1* | -4.543890839 | 1.18E-07 | 1.68E-05 | 0.000740711 |
| T2DM | dendritic_cell | *EMP2* | -2.10345082 | 3.05E-05 | 0.000145705 | 0.191677961 |
| T2DM | dendritic_cell | *PPM1N* | 2.285728252 | 1.15E-05 | 0.000130129 | 0.072082599 |
| T2DM | dendritic_cell | *LTB4R2* | -2.765839697 | 7.09E-05 | 0.000217986 | 0.445805981 |
| T2DM | dendritic_cell | *GALT* | -2.4497528 | 0.016158247 | 0.02166032 | 1 |
| T2DM | dendritic_cell | *ITGA1* | -1.566993559 | 4.16E-05 | 0.000162756 | 0.261253937 |
| T2DM | dendritic_cell | *NUDT19* | -2.615888322 | 6.48E-05 | 0.000205719 | 0.40732303 |
| T2DM | dendritic_cell | *ZNF726* | -1.855501874 | 1.03E-05 | 0.000130129 | 0.064560748 |

| | | | | | | |
|---|---|---|---|---|---|---|
| T2DM | dendritic_cell | *TAX1BP3* | -3.436726083 | 0.000186372 | 0.000429988 | 1 |
| T2DM | dendritic_cell | *AP1G2* | 3.253432431 | 0.000559341 | 0.001045979 | 1 |
| T2DM | dendritic_cell | *ZNF90* | -2.685511663 | 2.57E-05 | 0.0001395 | 0.161649952 |
| T2DM | dendritic_cell | *TM6SF2* | -19.49458068 | 3.38E-05 | 0.000150628 | 0.212233345 |
| T2DM | dendritic_cell | *TSPAN4* | -2.537612415 | 1.01E-05 | 0.000130129 | 0.063668064 |
| T2DM | dendritic_cell | *CPNE1* | -1.011186513 | 0.002028709 | 0.003267031 | 1 |
| T2DM | dendritic_cell | *ARL16* | -0.500209055 | 0.025593358 | 0.033196914 | 1 |
| T2DM | dendritic_cell | *AC079776.2* | -5.401436017 | 0.028220565 | 0.036401866 | 1 |
| T2DM | dendritic_cell | *WEE2* | -2.174862429 | 0.000426996 | 0.00083552 | 1 |
| T2DM | dendritic_cell | *PAXIP1-AS2* | -2.568000653 | 1.73E-05 | 0.000130129 | 0.108835947 |
| T2DM | dendritic_cell | *LYRM4* | -2.479094006 | 3.19E-05 | 0.000148219 | 0.200837398 |
| T2DM | dendritic_cell | *PRCD* | -1.29426585 | 0.000124477 | 0.000319684 | 0.782586932 |
| T2DM | dendritic_cell | *FIS1* | -1.706482573 | 0.002824617 | 0.004393461 | 1 |
| T2DM | dendritic_cell | *COLCA2* | -2.225952449 | 1.40E-05 | 0.000130129 | 0.088303584 |
| T2DM | dendritic_cell | *MBLAC1* | 1.055880238 | 7.31E-05 | 0.000221298 | 0.459636522 |
| T2DM | dendritic_cell | *NEURL1B* | -2.076557979 | 2.79E-05 | 0.000142583 | 0.175357461 |
| T2DM | dendritic_cell | *PPME1* | -2.072968728 | 2.05E-05 | 0.00013202 | 0.128587958 |
| T2DM | dendritic_cell | *STARD10* | 1.699637783 | 7.16E-07 | 5.24E-05 | 0.004502917 |
| T2DM | dendritic_cell | *NUTM2D* | 2.443473512 | 0.002034747 | 0.003275078 | 1 |
| T2DM | dendritic_cell | *CPEB1* | -1.0386422 | 1.02E-05 | 0.000130129 | 0.064165706 |
| T2DM | dendritic_cell | *C10orf105* | 1.29071659 | 8.41E-05 | 0.000245308 | 0.528883907 |
| T2DM | dendritic_cell | *AC005082.1* | -5.606919793 | 3.17E-05 | 0.000147987 | 0.199338507 |
| T2DM | dendritic_cell | *SMTNL1* | -0.683855626 | 0.002836161 | 0.004405966 | 1 |
| T2DM | dendritic_cell | *TBC1D26* | -18.55296059 | 0.013048897 | 0.017722709 | 1 |
| T2DM | dendritic_cell | *ACSM4* | -0.84697563 | 0.00623148 | 0.008971111 | 1 |
| T2DM | dendritic_cell | *COL28A1* | -1.336091753 | 1.86E-05 | 0.000130129 | 0.116721379 |
| T2DM | dendritic_cell | *PHB2* | -4.821325123 | 5.42E-06 | 0.000125356 | 0.03409685 |
| T2DM | dendritic_cell | *GRID2IP* | -1.666575778 | 3.87E-05 | 0.000158005 | 0.243011756 |
| T2DM | dendritic_cell | *UBE2QL1* | -2.050265309 | 1.46E-05 | 0.000130129 | 0.091508303 |
| T2DM | dendritic_cell | *GOLGA8B* | -2.062792666 | 2.49E-05 | 0.000138038 | 0.156239992 |
| T2DM | dendritic_cell | *KCNU1* | -2.170214156 | 2.20E-08 | 5.54E-06 | 0.000138598 |
| T2DM | dendritic_cell | *MYL5* | -2.294632384 | 1.48E-05 | 0.000130129 | 0.093222497 |
| T2DM | dendritic_cell | *NPEPL1* | -2.954561481 | 4.24E-05 | 0.000164065 | 0.266298319 |
| T2DM | dendritic_cell | *TSTD1* | 3.108167071 | 0.001520486 | 0.002518256 | 1 |
| T2DM | dendritic_cell | *LINC01138* | -3.373865715 | 0.000247721 | 0.000534748 | 1 |
| T2DM | dendritic_cell | *ATAD3C* | 4.240048286 | 9.52E-05 | 0.000264948 | 0.598518135 |
| T2DM | dendritic_cell | *CCDC7* | -2.101169036 | 1.92E-05 | 0.000131024 | 0.120672931 |
| T2DM | dendritic_cell | *AC016773.1* | -1.447290775 | 3.76E-06 | 0.000110889 | 0.023619441 |
| T2DM | dendritic_cell | *AP000350.4* | -0.539152885 | 0.01395719 | 0.018870721 | 1 |
| T2DM | dendritic_cell | *NBPF1* | -2.265733399 | 0.000137321 | 0.00034273 | 0.863336432 |
| T2DM | dendritic_cell | *RNU6ATAC35I* | 5.331937703 | 5.71E-05 | 0.000191408 | 0.359080889 |
| T2DM | dendritic_cell | *C6orf226* | -3.144383395 | 0.000207432 | 0.000465426 | 1 |
| T2DM | dendritic_cell | *PPP3R1* | -2.575795137 | 0.001832216 | 0.002980373 | 1 |
| T2DM | dendritic_cell | *C19orf73* | -8.641557111 | 0.00017628 | 0.000412764 | 1 |
| T2DM | dendritic_cell | *ZNF880* | -0.944480421 | 0.004461494 | 0.0066295 | 1 |
| T2DM | dendritic_cell | *OR2A25* | -44.86766204 | 0.000292852 | 0.000611274 | 1 |
| T2DM | dendritic_cell | *TIGD1* | 0.973261816 | 0.001565491 | 0.002583266 | 1 |
| T2DM | dendritic_cell | *XKR9* | -1.378888074 | 0.000111478 | 0.000294481 | 0.700864404 |
| T2DM | dendritic_cell | *C1orf229* | -2.541377637 | 9.51E-05 | 0.000264869 | 0.59807355 |
| T2DM | dendritic_cell | *SLC12A8* | -3.189911126 | 3.33E-06 | 0.000104276 | 0.020959488 |

| | | | | | | |
|---|---|---|---|---|---|---|
| T2DM | dendritic_cell | *APOL6* | -1.490686671 | 1.49E-05 | 0.000130129 | 0.09365191 |
| T2DM | dendritic_cell | *BTBD19* | -2.363511607 | 0.013409482 | 0.018196722 | 1 |
| T2DM | dendritic_cell | *FAM185A* | -2.77386346 | 0.000308977 | 0.000640045 | 1 |
| T2DM | dendritic_cell | *AC062017.1* | -1.517994637 | 4.85E-07 | 3.82E-05 | 0.003051148 |
| T2DM | dendritic_cell | *ADRA2B* | 2.600953077 | 4.51E-05 | 0.000168939 | 0.283510626 |
| T2DM | dendritic_cell | *EXOSC6* | -1.984188613 | 1.67E-06 | 8.15E-05 | 0.010486152 |
| T2DM | dendritic_cell | *VPS52* | -3.370455506 | 3.07E-05 | 0.00014601 | 0.193299808 |
| T2DM | dendritic_cell | *TINCR* | 2.80214262 | 0.00014975 | 0.000365765 | 0.941479776 |
| T2DM | dendritic_cell | *LINC00205* | -2.293694986 | 4.97E-06 | 0.000123581 | 0.031265927 |
| T2DM | dendritic_cell | *AC147651.3* | -4.824521003 | 0.00774885 | 0.010987149 | 1 |
| T2DM | dendritic_cell | *HLA-DPB1* | 0.311327534 | 2.24E-05 | 0.000133629 | 0.140541652 |
| T2DM | dendritic_cell | *OSER1-AS1* | -3.843300921 | 0.000749003 | 0.001350053 | 1 |
| T2DM | dendritic_cell | *LINC01133* | -0.961436874 | 1.77E-05 | 0.000130129 | 0.11122059 |
| T2DM | dendritic_cell | *LINC01117* | -1.133569292 | 2.59E-05 | 0.0001395 | 0.162820002 |
| T2DM | dendritic_cell | *RASAL2-AS1* | -5.811362099 | 1.04E-05 | 0.000130129 | 0.065206 |
| T2DM | dendritic_cell | *AC069213.1* | -4.006374024 | 0.038051266 | 0.048167171 | 1 |
| T2DM | dendritic_cell | *LINC00240* | -0.798186861 | 0.00396919 | 0.005965647 | 1 |
| T2DM | dendritic_cell | *LINC00184* | -3.657384789 | 2.20E-06 | 8.70E-05 | 0.013851064 |
| T2DM | dendritic_cell | *TMEM233* | -0.926988475 | 7.84E-05 | 0.000233163 | 0.492672688 |
| T2DM | dendritic_cell | *AL049840.1* | -1.957931495 | 0.001235767 | 0.002088513 | 1 |
| T2DM | dendritic_cell | *LINC00237* | -6.848620336 | 1.33E-05 | 0.000130129 | 0.083525851 |
| T2DM | dendritic_cell | *PLEKHM1* | -1.898802059 | 0.00035874 | 0.000722883 | 1 |
| T2DM | dendritic_cell | *SFTA1P* | -4.964018291 | 1.46E-05 | 0.000130129 | 0.091822033 |
| T2DM | dendritic_cell | *SLC26A6* | -1.805009167 | 0.000436886 | 0.000849321 | 1 |
| T2DM | dendritic_cell | *MIAT* | -2.393698896 | 0.001446783 | 0.002405693 | 1 |
| T2DM | dendritic_cell | *LINC00626* | -23.24062449 | 0.00057341 | 0.001069423 | 1 |
| T2DM | dendritic_cell | *ERCC6* | -2.617681423 | 5.76E-05 | 0.000192599 | 0.362214003 |
| T2DM | dendritic_cell | *LINC00115* | -1.501304038 | 0.00063216 | 0.001161423 | 1 |
| T2DM | dendritic_cell | *NOL7* | -3.071026377 | 0.000721004 | 0.001303054 | 1 |
| T2DM | dendritic_cell | *ELFN1* | 11.14691671 | 0.000935177 | 0.001632275 | 1 |
| T2DM | dendritic_cell | *ZNF503-AS1* | -4.420950165 | 3.81E-05 | 0.000157482 | 0.239505029 |
| T2DM | dendritic_cell | *LINC00937* | 2.149460319 | 0.000955383 | 0.001663768 | 1 |
| T2DM | dendritic_cell | *AC064836.3* | -2.877618174 | 1.98E-05 | 0.000131266 | 0.124687149 |
| T2DM | dendritic_cell | *HSBP1L1* | -3.571611045 | 1.26E-05 | 0.000130129 | 0.079063775 |
| T2DM | dendritic_cell | *TAS2R46* | 1.329128803 | 0.011023187 | 0.015168041 | 1 |
| T2DM | dendritic_cell | *AC093901.1* | 2.020997275 | 5.89E-05 | 0.000195504 | 0.370479492 |
| T2DM | dendritic_cell | *C14orf132* | -2.322198859 | 0.000159848 | 0.000384308 | 1 |
| T2DM | dendritic_cell | *AC009299.3* | -4.343670888 | 0.000479987 | 0.000917785 | 1 |
| T2DM | dendritic_cell | *SLC2A1-AS1* | -1.788897612 | 2.64E-05 | 0.000139936 | 0.166243865 |
| T2DM | dendritic_cell | *LINC01068* | -1.960441513 | 0.007946508 | 0.011236909 | 1 |
| T2DM | dendritic_cell | *AC108463.2* | 2.720232434 | 5.96E-06 | 0.000129884 | 0.037498334 |
| T2DM | dendritic_cell | *HCG20* | -5.229948 | 3.82E-06 | 0.000111457 | 0.024017061 |
| T2DM | dendritic_cell | *C19orf24* | -1.971651074 | 3.18E-06 | 0.000102388 | 0.019969657 |
| T2DM | dendritic_cell | *AC073133.2* | 9.479784394 | 0.000779839 | 0.001398692 | 1 |
| T2DM | dendritic_cell | *PROB1* | -6.256661525 | 0.001139307 | 0.001941145 | 1 |
| T2DM | dendritic_cell | *ARL17B* | -2.187476812 | 8.47E-06 | 0.000130129 | 0.053261917 |
| T2DM | dendritic_cell | *DHFR* | -0.801369322 | 0.016208014 | 0.021713144 | 1 |
| T2DM | dendritic_cell | *LINC00954* | 2.821250083 | 0.000168745 | 0.000401185 | 1 |
| T2DM | dendritic_cell | *AC067945.3* | -2.161912642 | 0.006196526 | 0.00892703 | 1 |
| T2DM | dendritic_cell | *RPL41* | -4.657942143 | 0.010621723 | 0.014657325 | 1 |

| | | | | | | |
|---|---|---|---|---|---|---|
| T2DM | dendritic_cell | *PATL2* | -3.796236444 | 0.002402278 | 0.003808149 | 1 |
| T2DM | dendritic_cell | *U91324.1* | -6.265477329 | 1.50E-05 | 0.000130129 | 0.094228294 |
| T2DM | dendritic_cell | *ZNF688* | 3.603513075 | 0.000616203 | 0.001137759 | 1 |
| T2DM | dendritic_cell | *CISD3* | -4.63833809 | 0.029134569 | 0.037526948 | 1 |
| T2DM | dendritic_cell | *PSMG3-AS1* | -1.210738943 | 2.55E-05 | 0.000139164 | 0.160595162 |
| T2DM | dendritic_cell | *PPP5D1* | -2.063266397 | 0.00010368 | 0.00028065 | 0.65183884 |
| T2DM | dendritic_cell | *AC092171.4* | -2.479166783 | 0.000129715 | 0.00032937 | 0.815520772 |
| T2DM | dendritic_cell | *HLA-DPA1* | 0.325898825 | 4.39E-05 | 0.000166492 | 0.275711354 |
| T2DM | dendritic_cell | *RPS18* | -2.684537237 | 0.000186112 | 0.000429628 | 1 |
| T2DM | dendritic_cell | *DIRC3* | -1.11639209 | 9.09E-06 | 0.000130129 | 0.057156524 |
| T2DM | dendritic_cell | *AC107021.1* | -0.922793498 | 1.85E-05 | 0.000130129 | 0.116010909 |
| T2DM | dendritic_cell | *LINC00865* | -2.640524467 | 0.000849395 | 0.001506388 | 1 |
| T2DM | dendritic_cell | *AC009495.3* | -2.079384535 | 2.07E-05 | 0.000132272 | 0.129981911 |
| T2DM | dendritic_cell | *HLA-DQB2* | 0.264856691 | 2.93E-07 | 2.83E-05 | 0.001839182 |
| T2DM | dendritic_cell | *TNF* | 1.001318466 | 3.11E-05 | 0.000146698 | 0.195782522 |
| T2DM | dendritic_cell | *LYRM9* | -2.370511126 | 1.00E-05 | 0.000130129 | 0.062920276 |
| T2DM | dendritic_cell | *LINC00342* | -1.621809252 | 1.41E-05 | 0.000130129 | 0.088392324 |
| T2DM | dendritic_cell | *LINC00702* | -1.466895226 | 1.38E-05 | 0.000130129 | 0.086492357 |
| T2DM | dendritic_cell | *LINC00472* | -1.65699133 | 0.000724843 | 0.001309508 | 1 |
| T2DM | dendritic_cell | *AC073133.1* | 0.697685139 | 0.006212075 | 0.00894738 | 1 |
| T2DM | dendritic_cell | *RPS28* | -3.435913052 | 2.10E-05 | 0.000132395 | 0.13188359 |
| T2DM | dendritic_cell | *AC123768.3* | -1.761751935 | 1.99E-05 | 0.000131266 | 0.125173904 |
| T2DM | dendritic_cell | *C19orf81* | 4.229260529 | 0.000128748 | 0.000327178 | 0.809438629 |
| T2DM | dendritic_cell | *LINC00957* | 2.410426818 | 3.84E-05 | 0.000158005 | 0.241220365 |
| T2DM | dendritic_cell | *LINC00484* | -0.610605932 | 0.011851053 | 0.016193777 | 1 |
| T2DM | dendritic_cell | *KIAA0040* | -2.366268033 | 3.28E-05 | 0.000148688 | 0.20649058 |
| T2DM | dendritic_cell | *B3GALT4* | -3.338556991 | 0.005926132 | 0.008582721 | 1 |
| T2DM | dendritic_cell | *LINC00941* | -11.94635734 | 0.020791155 | 0.027374657 | 1 |
| T2DM | dendritic_cell | *LINC00545* | 5.61209939 | 6.20E-05 | 0.000200487 | 0.389903739 |
| T2DM | dendritic_cell | *B3GNT9* | -3.301996275 | 9.44E-05 | 0.000264058 | 0.593613482 |
| T2DM | dendritic_cell | *CDKN2AIPNL* | -1.32159677 | 1.26E-05 | 0.000130129 | 0.079015497 |
| T2DM | dendritic_cell | *ZNF737* | -3.165276638 | 3.80E-05 | 0.000157482 | 0.239199513 |
| T2DM | dendritic_cell | *HLA-DQA2* | 0.395600533 | 5.84E-08 | 1.11E-05 | 0.000367327 |
| T2DM | dendritic_cell | *AC073257.2* | -8.580022662 | 0.006740829 | 0.009662469 | 1 |
| T2DM | dendritic_cell | *IRGM* | -1.226845874 | 0.004001523 | 0.006011368 | 1 |
| T2DM | dendritic_cell | *LRRC37A2* | -2.23885487 | 0.011187241 | 0.015376953 | 1 |
| T2DM | dendritic_cell | *TXNDC5* | -3.856662853 | 9.76E-05 | 0.000268845 | 0.613772449 |
| T2DM | dendritic_cell | *ASB14* | -1.531802673 | 0.03144519 | 0.040272134 | 1 |
| T2DM | dendritic_cell | *NME1* | -0.848912072 | 0.007683381 | 0.010899236 | 1 |
| T2DM | dendritic_cell | *TNFSF12* | -1.672273406 | 0.009390223 | 0.013084293 | 1 |
| T2DM | dendritic_cell | *CDRT4* | 2.346418875 | 0.018115993 | 0.024079334 | 1 |
| T2DM | dendritic_cell | *APOBEC3G* | -0.747700966 | 0.021146186 | 0.027783923 | 1 |
| T2DM | dendritic_cell | *ADSL* | -1.710446102 | 1.15E-05 | 0.000130129 | 0.072077059 |
| T2DM | dendritic_cell | *COX19* | -3.636162944 | 0.001703504 | 0.002785417 | 1 |
| T2DM | dendritic_cell | *AQP1* | -1.267863704 | 1.79E-05 | 0.000130129 | 0.112341251 |
| T2DM | dendritic_cell | *C1QTNF9* | -3.248665359 | 0.019581839 | 0.02591811 | 1 |
| T2DM | dendritic_cell | *ARHGEF25* | -2.49468991 | 0.011169902 | 0.015359837 | 1 |
| T2DM | dendritic_cell | *LINC00886* | -3.089593164 | 4.64E-05 | 0.000170668 | 0.291591244 |
| T2DM | dendritic_cell | *NSUN6* | -2.076883787 | 2.20E-05 | 0.000132957 | 0.138195166 |
| T2DM | dendritic_cell | *HLA-DOB* | 0.487684548 | 9.55E-06 | 0.000130129 | 0.060010398 |

| | | | | | | |
|---|---|---|---|---|---|---|
| T2DM | dendritic_cell | *TDGF1* | -1.662165608 | 0.01462184 | 0.019710015 | 1 |
| T2DM | dendritic_cell | *ARHGAP8* | -2.614680465 | 7.65E-06 | 0.000130129 | 0.048076509 |
| T2DM | dendritic_cell | *ATP5O* | -2.238034229 | 4.97E-05 | 0.000176745 | 0.312308544 |
| T2DM | dendritic_cell | *PI4KA* | -2.780872773 | 8.47E-05 | 0.000246303 | 0.532240843 |
| T2DM | dendritic_cell | *AKAP2* | 6.766119428 | 0.000142461 | 0.000352896 | 0.895650832 |
| T2DM | dendritic_cell | *LINC00969* | -1.755623815 | 0.000145466 | 0.000357942 | 0.91454165 |
| T2DM | dendritic_cell | *ARHGDIG* | -1.91475412 | 0.000174451 | 0.000409856 | 1 |
| T2DM | dendritic_cell | *TCP10L* | 1.966132784 | 2.61E-05 | 0.0001395 | 0.163846177 |
| T2DM | dendritic_cell | *ARFGAP3* | -2.798646356 | 4.87E-05 | 0.000174599 | 0.30602207 |
| T2DM | dendritic_cell | *C22orf39* | -4.160513978 | 0.007226157 | 0.010313473 | 1 |
| T2DM | dendritic_cell | *DECR2* | 4.506045758 | 0.00249985 | 0.003941951 | 1 |
| T2DM | dendritic_cell | *CCDC169* | 1.449888435 | 0.006740016 | 0.009662469 | 1 |
| T2DM | dendritic_cell | *AP5Z1* | -3.029871162 | 0.000125256 | 0.000320637 | 0.78748475 |
| T2DM | dendritic_cell | *PSG4* | -1.873996562 | 0.000382433 | 0.000761596 | 1 |
| T2DM | dendritic_cell | *STON1* | -1.458452478 | 1.47E-05 | 0.000130129 | 0.092625688 |
| T2DM | dendritic_cell | *C7orf73* | -2.469116198 | 2.92E-05 | 0.000143842 | 0.183847508 |
| T2DM | dendritic_cell | *FGF14-IT1* | -3.955588748 | 1.64E-05 | 0.000130129 | 0.103399763 |
| T2DM | dendritic_cell | *TICAM2* | 2.306233215 | 0.000144982 | 0.000357311 | 0.911499767 |
| T2DM | dendritic_cell | *PALM2* | -0.708375773 | 0.000646952 | 0.0011841 | 1 |
| T2DM | dendritic_cell | *NAT6* | 4.918125604 | 6.10E-05 | 0.000198694 | 0.38328145 |
| T2DM | dendritic_cell | *WDR92* | -1.609605219 | 0.000257419 | 0.000551411 | 1 |
| T2DM | dendritic_cell | *PLA2G4B* | 3.344958174 | 2.61E-11 | 4.11E-08 | 1.64E-07 |
| T2DM | dendritic_cell | *TTC4* | 0.877208801 | 0.001436605 | 0.002391933 | 1 |
| T2DM | dendritic_cell | *JMJD7* | 2.194893035 | 0.000128305 | 0.000326514 | 0.806654711 |
| T2DM | dendritic_cell | *APOBEC3D* | -3.790466606 | 0.014388351 | 0.019411923 | 1 |
| T2DM | dendritic_cell | *RN7SL832P* | -1.771416301 | 0.01338818 | 0.018171738 | 1 |
| T2DM | dendritic_cell | *LINC01011* | 2.065967428 | 0.003929837 | 0.005917816 | 1 |
| T2DM | dendritic_cell | *TMEM199* | 3.047143017 | 0.001891959 | 0.003068029 | 1 |
| T2DM | dendritic_cell | *P2RY11* | 2.015180274 | 0.002645442 | 0.004147605 | 1 |
| T2DM | dendritic_cell | *LILRA6* | 0.938872275 | 0.000120406 | 0.000311776 | 0.756992594 |
| T2DM | dendritic_cell | *APOBEC3C* | 1.541668148 | 7.12E-05 | 0.000218297 | 0.447945841 |
| T2DM | dendritic_cell | *CCDC13* | -1.514458583 | 1.60E-06 | 8.00E-05 | 0.010080103 |
| T2DM | dendritic_cell | *ASPRV1* | -4.532071225 | 3.22E-05 | 0.000148254 | 0.202129671 |
| T2DM | dendritic_cell | *N4BP2L2* | -1.769460436 | 4.87E-05 | 0.000174599 | 0.306072096 |
| T2DM | dendritic_cell | *LINC00847* | -4.856007993 | 0.000109113 | 0.000290306 | 0.685993801 |
| T2DM | dendritic_cell | *NEAT1* | -1.301004194 | 2.88E-05 | 0.000143316 | 0.181210081 |
| T2DM | dendritic_cell | *RAB30-AS1* | -2.374005757 | 9.10E-06 | 0.000130129 | 0.057220311 |
| T2DM | dendritic_cell | *UBAP1L* | -3.583819498 | 8.03E-06 | 0.000130129 | 0.050488984 |
| T2DM | dendritic_cell | *PGAM5* | 3.883824005 | 0.000799107 | 0.001428487 | 1 |
| T2DM | dendritic_cell | *DNAJC3-AS1* | -1.991960889 | 2.84E-05 | 0.000142583 | 0.178297738 |
| T2DM | dendritic_cell | *LINC00926* | -2.230694966 | 3.24E-05 | 0.000148254 | 0.203624115 |
| T2DM | dendritic_cell | *BCKDHA* | -3.114879354 | 0.000455143 | 0.000878834 | 1 |
| T2DM | dendritic_cell | *CDK11B* | -3.182040089 | 0.013206486 | 0.017932867 | 1 |
| T2DM | dendritic_cell | *ABHD14A* | -2.531181535 | 0.000110436 | 0.000292588 | 0.69431121 |
| T2DM | dendritic_cell | *SRP14-AS1* | -1.03268298 | 1.79E-05 | 0.000130129 | 0.112601279 |
| T2DM | dendritic_cell | *CCDC153* | 1.572631817 | 7.74E-05 | 0.000230873 | 0.486450351 |
| T2DM | dendritic_cell | *C5orf17* | -2.327047215 | 1.55E-05 | 0.000130129 | 0.097570348 |
| T2DM | dendritic_cell | *FMN1* | 0.851131835 | 2.58E-05 | 0.0001395 | 0.162423771 |
| T2DM | dendritic_cell | *LINC01060* | -1.63654117 | 4.90E-05 | 0.000175324 | 0.308044762 |
| T2DM | dendritic_cell | *ZNF286B* | -2.332456681 | 1.09E-06 | 6.36E-05 | 0.006868387 |

| | | | | | | |
|---|---|---|---|---|---|---|
| T2DM | dendritic_cell | *PDCD6* | -2.898502594 | 0.000855676 | 0.001515391 | 1 |
| T2DM | dendritic_cell | *LINC00603* | -1.770095713 | 2.21E-05 | 0.000132957 | 0.13892514 |
| T2DM | dendritic_cell | *YJEFN3* | -2.657999159 | 0.025057747 | 0.032576036 | 1 |
| T2DM | dendritic_cell | *DNAH10OS* | -11.75559916 | 1.93E-07 | 2.17E-05 | 0.001215398 |
| T2DM | dendritic_cell | *PCDHA10* | 4.379433745 | 0.022226911 | 0.029112623 | 1 |
| T2DM | dendritic_cell | *LINC00989* | -2.304349429 | 7.52E-06 | 0.000130129 | 0.0473029 |
| T2DM | dendritic_cell | *LINC01021* | -4.460883185 | 1.37E-05 | 0.000130129 | 0.086018998 |
| T2DM | dendritic_cell | *FAM218A* | -4.340677354 | 1.24E-05 | 0.000130129 | 0.077837379 |
| T2DM | dendritic_cell | *ATP6V1E2* | -0.721137253 | 0.003127996 | 0.004818846 | 1 |
| T2DM | dendritic_cell | *SNHG18* | -1.639458908 | 1.82E-06 | 8.21E-05 | 0.011414813 |
| T2DM | dendritic_cell | *ZNF345* | -2.046102725 | 3.30E-06 | 0.000103892 | 0.020778301 |
| T2DM | dendritic_cell | *SHANK3* | -1.001787899 | 0.000542809 | 0.001020832 | 1 |
| T2DM | dendritic_cell | *ZNF550* | -1.974887593 | 3.04E-05 | 0.000145705 | 0.190853908 |
| T2DM | dendritic_cell | *CCDC71L* | -2.535835697 | 3.50E-05 | 0.000152633 | 0.219790988 |
| T2DM | dendritic_cell | *TRNP1* | -6.513555078 | 1.23E-05 | 0.000130129 | 0.077409621 |
| T2DM | dendritic_cell | *PCDHGA5* | -2.208763782 | 3.61E-05 | 0.00015508 | 0.227238928 |
| T2DM | dendritic_cell | *ALG11* | -1.287690598 | 0.038547606 | 0.048762334 | 1 |
| T2DM | dendritic_cell | *ATXN7L3B* | -3.783434835 | 0.000125093 | 0.000320559 | 0.786460163 |
| T2DM | dendritic_cell | *PRKDC* | -2.781412071 | 0.000470875 | 0.000903937 | 1 |
| T2DM | dendritic_cell | *PCDHGA6* | -1.050618312 | 0.010073823 | 0.013965629 | 1 |
| T2DM | dendritic_cell | *CLDN23* | 2.596515385 | 2.73E-05 | 0.000141269 | 0.171403738 |
| T2DM | dendritic_cell | *ARMS2* | -2.985108572 | 0.005810568 | 0.008434781 | 1 |
| T2DM | dendritic_cell | *FPGT* | -4.964733822 | 0.000632774 | 0.001162212 | 1 |
| T2DM | dendritic_cell | *EID3* | -1.010087041 | 0.008235421 | 0.011606728 | 1 |
| T2DM | dendritic_cell | *SNHG9* | -0.848351321 | 0.007656094 | 0.010862979 | 1 |
| T2DM | dendritic_cell | *TAS2R43* | 0.976275979 | 0.000418691 | 0.000820801 | 1 |
| T2DM | dendritic_cell | *EBLN2* | -2.080917042 | 0.000190855 | 0.000437284 | 1 |
| T2DM | dendritic_cell | *POLR2M* | 2.895733181 | 0.027714207 | 0.035800128 | 1 |
| T2DM | dendritic_cell | *TAS2R20* | 2.097021733 | 1.68E-06 | 8.15E-05 | 0.010533523 |
| T2DM | dendritic_cell | *LINC00346* | -3.240835536 | 0.000154827 | 0.000375257 | 0.973400114 |
| T2DM | dendritic_cell | *CTSO* | -3.840119282 | 0.001042874 | 0.001795814 | 1 |
| T2DM | dendritic_cell | *ZNF432* | -3.231108884 | 0.003941015 | 0.005933228 | 1 |
| T2DM | dendritic_cell | *LINC00944* | -1.73736216 | 0.000214113 | 0.000476338 | 1 |
| T2DM | dendritic_cell | *AC093668.2* | -1.312901593 | 0.018864332 | 0.025036955 | 1 |
| T2DM | dendritic_cell | *CLEC12B* | 0.432155754 | 0.002773043 | 0.004321795 | 1 |
| T2DM | dendritic_cell | *PGA5* | -15.22912999 | 0.000921185 | 0.001611085 | 1 |
| T2DM | dendritic_cell | *ZNF253* | -3.263925549 | 0.000380372 | 0.000759175 | 1 |
| T2DM | dendritic_cell | *HP* | -3.736328804 | 0.000195117 | 0.000444296 | 1 |
| T2DM | dendritic_cell | *KIAA1147* | -2.055464321 | 0.00018847 | 0.000433398 | 1 |
| T2DM | dendritic_cell | *GATC* | -2.610747 | 0.013740439 | 0.018593659 | 1 |
| T2DM | dendritic_cell | *ZBED6* | -3.140589496 | 1.48E-05 | 0.000130129 | 0.093104937 |
| T2DM | dendritic_cell | *OVCH1-AS1* | 2.153933375 | 0.000980682 | 0.001703189 | 1 |
| T2DM | dendritic_cell | *CUX1* | -1.800999267 | 0.016986125 | 0.022678226 | 1 |
| T2DM | dendritic_cell | *TEN1* | -3.699709357 | 0.002403702 | 0.003809447 | 1 |
| T2DM | dendritic_cell | *C17orf49* | -3.75925821 | 0.001314462 | 0.002207271 | 1 |
| T2DM | dendritic_cell | *ZNF578* | -1.506937176 | 0.01072529 | 0.014790502 | 1 |
| T2DM | dendritic_cell | *LINC00640* | -0.92564234 | 0.001923145 | 0.003112178 | 1 |
| T2DM | dendritic_cell | *LINC00638* | -5.733854002 | 5.08E-06 | 0.000124224 | 0.031925665 |
| T2DM | dendritic_cell | *CEP95* | -2.1967739 | 4.59E-05 | 0.000170199 | 0.288505473 |
| T2DM | dendritic_cell | *LINC00639* | -0.609802834 | 0.000644067 | 0.00117985 | 1 |

| | | | | | | |
|---|---|---|---|---|---|---|
| T2DM | dendritic_cell | *LINC00648* | -1.09641821 | 1.20E-05 | 0.000130129 | 0.075289614 |
| T2DM | dendritic_cell | *LINC00924* | -3.713581149 | 0.00029716 | 0.000619035 | 1 |
| T2DM | dendritic_cell | *ITGB3* | -0.956991178 | 0.015273495 | 0.020544387 | 1 |
| T2DM | dendritic_cell | *UBE2Q2L* | -4.085912346 | 9.14E-06 | 0.000130129 | 0.057447616 |
| T2DM | dendritic_cell | *IQCH-AS1* | -1.231353007 | 0.017603459 | 0.023442692 | 1 |
| T2DM | dendritic_cell | *AC083843.1* | 6.684915562 | 0.000897338 | 0.001576296 | 1 |
| T2DM | dendritic_cell | *PWRN1* | -2.645928793 | 0.003944806 | 0.005934672 | 1 |
| T2DM | dendritic_cell | *HOXB7* | -4.013139148 | 9.65E-06 | 0.000130129 | 0.060641578 |
| T2DM | dendritic_cell | *FRRS1L* | -3.201344722 | 5.23E-05 | 0.000181896 | 0.328867582 |
| T2DM | dendritic_cell | *LINC00562* | -0.804216118 | 0.00330578 | 0.005056798 | 1 |
| T2DM | dendritic_cell | *AC009120.5* | -2.510029717 | 1.23E-05 | 0.000130129 | 0.077350498 |
| T2DM | dendritic_cell | *TMEM178B* | -3.708506609 | 3.00E-05 | 0.000145189 | 0.188745497 |
| T2DM | dendritic_cell | *LINC01003* | -1.854740456 | 0.000255855 | 0.000548998 | 1 |
| T2DM | dendritic_cell | *HPR* | 3.060869973 | 0.000194313 | 0.000443267 | 1 |
| T2DM | dendritic_cell | *LINC00662* | -3.051545914 | 0.000158135 | 0.000381209 | 0.99419178 |
| T2DM | dendritic_cell | *LINC00667* | -1.116744328 | 0.002712308 | 0.004240806 | 1 |
| T2DM | dendritic_cell | *LINC00672* | 1.286176491 | 0.007821148 | 0.011074675 | 1 |
| T2DM | dendritic_cell | *LINC00909* | -1.561603377 | 7.60E-05 | 0.000227569 | 0.477667604 |
| T2DM | dendritic_cell | *DYNLL2* | -1.665353696 | 5.14E-06 | 0.000124852 | 0.032336555 |
| T2DM | dendritic_cell | *STRADA* | -3.764946596 | 8.31E-06 | 0.000130129 | 0.052275625 |
| T2DM | dendritic_cell | *ILF3-AS1* | -1.584696964 | 0.011565055 | 0.015847756 | 1 |
| T2DM | dendritic_cell | *AC139100.2* | 1.347563524 | 3.43E-05 | 0.000151156 | 0.215574912 |
| T2DM | dendritic_cell | *LINC01140* | -1.588475627 | 0.000416932 | 0.000817863 | 1 |
| T2DM | dendritic_cell | *LINC00906* | -10.9380733 | 1.48E-05 | 0.000130129 | 0.092806295 |
| T2DM | dendritic_cell | *ZNF582-AS1* | -2.233991414 | 3.43E-05 | 0.000151156 | 0.215409502 |
| T2DM | dendritic_cell | *AC137932.1* | -5.233273163 | 9.91E-05 | 0.000271741 | 0.622830128 |
| T2DM | dendritic_cell | *AC003005.2* | -1.978601967 | 0.009153508 | 0.012779948 | 1 |
| T2DM | dendritic_cell | *AL132989.1* | 5.699669461 | 0.000206622 | 0.000464267 | 1 |
| T2DM | dendritic_cell | *LINC00664* | -4.121772216 | 0.000623934 | 0.001148996 | 1 |
| T2DM | dendritic_cell | *FBXO17* | -1.246141286 | 1.96E-05 | 0.000131266 | 0.123109278 |
| T2DM | dendritic_cell | *AC009060.1* | -4.258736306 | 2.11E-08 | 5.54E-06 | 0.00013272 |
| T2DM | dendritic_cell | *SNHG8* | -1.836065202 | 7.69E-05 | 0.000229746 | 0.483386216 |
| T2DM | dendritic_cell | *HCG17* | -2.02274734 | 1.08E-06 | 6.35E-05 | 0.006784887 |
| T2DM | dendritic_cell | *YTHDF3-AS1* | -2.802495281 | 0.000203557 | 0.000459025 | 1 |
| T2DM | dendritic_cell | *LINC01050* | -9.951884954 | 0.000381222 | 0.000760147 | 1 |
| T2DM | dendritic_cell | *MILR1* | 0.70197471 | 8.07E-05 | 0.000237677 | 0.507202876 |
| T2DM | dendritic_cell | *GTF2H5* | -1.721153545 | 0.022652485 | 0.029632995 | 1 |
| T2DM | dendritic_cell | *FGF14-AS2* | -1.833435322 | 6.99E-05 | 0.000215653 | 0.43930155 |
| T2DM | dendritic_cell | *LINC01023* | -2.335538858 | 0.000180454 | 0.000420345 | 1 |
| T2DM | dendritic_cell | *DOC2B* | -1.155730453 | 8.12E-06 | 0.000130129 | 0.05104376 |
| T2DM | dendritic_cell | *AC144652.1* | 1.53777269 | 7.86E-06 | 0.000130129 | 0.049395156 |
| T2DM | dendritic_cell | *PAXIP1-AS1* | -2.397924705 | 2.94E-05 | 0.000143842 | 0.184840984 |
| T2DM | endothelial | *SCYL3* | -0.431063574 | 0.00017101 | 0.000602318 | 1 |
| T2DM | endothelial | *C1orf112* | -0.51057461 | 0.000192989 | 0.000666662 | 1 |
| T2DM | endothelial | *CFH* | 0.219201368 | 0.019177782 | 0.032446371 | 1 |
| T2DM | endothelial | *NFYA* | -0.592647121 | 0.000115038 | 0.000432561 | 0.723242488 |
| T2DM | endothelial | *NIPAL3* | 0.341372171 | 0.001196119 | 0.003084303 | 1 |
| T2DM | endothelial | *ENPP4* | 0.289313065 | 0.002808298 | 0.006316912 | 1 |
| T2DM | endothelial | *SEMA3F* | 0.084162971 | 0.002657916 | 0.006039146 | 1 |
| T2DM | endothelial | *KRIT1* | -0.294367975 | 0.019842499 | 0.033391272 | 1 |

| | | | | | | |
|---|---|---|---|---|---|---|
| T2DM | endothelial | *RAD52* | -0.44936793 | 0.002156825 | 0.005071038 | 1 |
| T2DM | endothelial | *BAD* | -0.87158875 | 2.72E-05 | 0.000131431 | 0.170728743 |
| T2DM | endothelial | *HS3ST1* | -0.638044949 | 0.003335704 | 0.007330154 | 1 |
| T2DM | endothelial | *WNT16* | -0.788931257 | 7.68E-06 | 4.83E-05 | 0.048293472 |
| T2DM | endothelial | *HECW1* | -0.236020394 | 0.00014112 | 0.000514762 | 0.887223574 |
| T2DM | endothelial | *MAD1L1* | -0.46242686 | 0.000568952 | 0.001646112 | 1 |
| T2DM | endothelial | *LASP1* | -0.454806758 | 0.000253035 | 0.000833334 | 1 |
| T2DM | endothelial | *CYP26B1* | -0.652415286 | 3.02E-06 | 2.45E-05 | 0.018994646 |
| T2DM | endothelial | *ICA1* | 0.194495643 | 8.42E-06 | 5.21E-05 | 0.052941579 |
| T2DM | endothelial | *CD38* | -0.455297678 | 2.36E-07 | 5.29E-06 | 0.001486005 |
| T2DM | endothelial | *RBM6* | -0.283376695 | 6.38E-08 | 4.46E-06 | 0.000400905 |
| T2DM | endothelial | *RECQL* | -0.399765585 | 0.00884623 | 0.016827911 | 1 |
| T2DM | endothelial | *ARHGAP33* | -0.541165497 | 0.001806077 | 0.004365554 | 1 |
| T2DM | endothelial | *PDK4* | -0.323149148 | 0.004378851 | 0.009201699 | 1 |
| T2DM | endothelial | *SLC22A16* | -0.33556269 | 0.019199691 | 0.032470876 | 1 |
| T2DM | endothelial | *MEOX1* | 0.108708099 | 1.03E-06 | 1.21E-05 | 0.006487585 |
| T2DM | endothelial | *CIAPIN1* | -0.579575992 | 0.002669674 | 0.006055556 | 1 |
| T2DM | endothelial | *COPZ2* | -0.225075834 | 0.008367456 | 0.016033586 | 1 |
| T2DM | endothelial | *WDR54* | 0.426022397 | 4.10E-06 | 3.05E-05 | 0.025794451 |
| T2DM | endothelial | *ABCB4* | -0.840934328 | 2.07E-06 | 1.91E-05 | 0.013004389 |
| T2DM | endothelial | *ZNF195* | -0.572299129 | 2.41E-06 | 2.12E-05 | 0.015176336 |
| T2DM | endothelial | *PDK2* | -0.598491827 | 4.37E-05 | 0.000195005 | 0.27476197 |
| T2DM | endothelial | *GGNBP2* | -0.532160029 | 2.75E-07 | 5.77E-06 | 0.001726631 |
| T2DM | endothelial | *TMEM98* | -0.330763098 | 0.000212937 | 0.000725603 | 1 |
| T2DM | endothelial | *ABCC8* | -0.709750495 | 2.28E-07 | 5.21E-06 | 0.001433272 |
| T2DM | endothelial | *SYNRG* | -0.349593736 | 0.003793458 | 0.008181974 | 1 |
| T2DM | endothelial | *TMEM132A* | 0.704765388 | 2.18E-06 | 1.99E-05 | 0.013721333 |
| T2DM | endothelial | *TAC1* | 1.092351332 | 0.00024699 | 0.000818569 | 1 |
| T2DM | endothelial | *ZNF263* | -0.377296868 | 0.028363886 | 0.045665493 | 1 |
| T2DM | endothelial | *SPATA20* | -0.591432666 | 0.000729593 | 0.002030324 | 1 |
| T2DM | endothelial | *TNFRSF12A* | -0.959524918 | 3.33E-05 | 0.00015534 | 0.209492131 |
| T2DM | endothelial | *MAP3K9* | 0.581014112 | 0.000753359 | 0.002083752 | 1 |
| T2DM | endothelial | *BAIAP2L1* | 0.203313702 | 3.09E-06 | 2.47E-05 | 0.019408907 |
| T2DM | endothelial | *IFRD1* | -0.369689424 | 6.52E-05 | 0.000268847 | 0.410135276 |
| T2DM | endothelial | *GTF2IRD1* | -0.293184513 | 0.003388645 | 0.007414722 | 1 |
| T2DM | endothelial | *VPS41* | -0.266243976 | 0.023110994 | 0.038136173 | 1 |
| T2DM | endothelial | *ARHGAP44* | -0.6249543 | 7.34E-05 | 0.000295797 | 0.461739745 |
| T2DM | endothelial | *ELAC2* | -0.378451878 | 0.011972419 | 0.021771311 | 1 |
| T2DM | endothelial | *CCDC124* | -0.462533989 | 0.000647171 | 0.001828658 | 1 |
| T2DM | endothelial | *CEACAM21* | -0.516260112 | 0.004020058 | 0.008599026 | 1 |
| T2DM | endothelial | *NOS2* | -3.810749868 | 4.50E-05 | 0.000199196 | 0.282858838 |
| T2DM | endothelial | *TRAPPC6A* | -0.548337838 | 0.007902206 | 0.015248977 | 1 |
| T2DM | endothelial | *CD79B* | 0.223107825 | 0.026534001 | 0.043050132 | 1 |
| T2DM | endothelial | *ST7L* | -0.251419565 | 0.001194904 | 0.003082626 | 1 |
| T2DM | endothelial | *RPUSD1* | -1.66197501 | 8.48E-06 | 5.23E-05 | 0.053336337 |
| T2DM | endothelial | *BAIAP3* | 0.671762554 | 7.08E-07 | 9.70E-06 | 0.004452003 |
| T2DM | endothelial | *TSR3* | -0.73220614 | 0.001175695 | 0.003040556 | 1 |
| T2DM | endothelial | *PIGQ* | -0.557735421 | 0.001510332 | 0.003753145 | 1 |
| T2DM | endothelial | *PSMB1* | -0.483325221 | 0.001620492 | 0.003979701 | 1 |
| T2DM | endothelial | *CDK11A* | -0.678916947 | 2.76E-05 | 0.000132992 | 0.17337978 |

| | | | | | | |
|---|---|---|---|---|---|---|
| T2DM | endothelial | *DLEC1* | -0.502739874 | 8.27E-07 | 1.06E-05 | 0.005197173 |
| T2DM | endothelial | *MGST1* | -0.302026489 | 5.95E-06 | 3.99E-05 | 0.037393155 |
| T2DM | endothelial | *NFIX* | -0.335687968 | 0.000365184 | 0.001132105 | 1 |
| T2DM | endothelial | *IL32* | 0.397721212 | 0.001494376 | 0.003723797 | 1 |
| T2DM | endothelial | *PKD1* | -0.299531636 | 0.015889341 | 0.027672101 | 1 |
| T2DM | endothelial | *MAPK8IP2* | -1.188580238 | 0.0106665 | 0.019712019 | 1 |
| T2DM | endothelial | *MED24* | -0.482151886 | 0.000622176 | 0.001773977 | 1 |
| T2DM | endothelial | *POMT2* | -0.596775744 | 1.39E-05 | 7.67E-05 | 0.087521103 |
| T2DM | endothelial | *MLXIPL* | -0.088701131 | 0.019746739 | 0.033259418 | 1 |
| T2DM | endothelial | *METTL13* | -0.463993862 | 0.027804691 | 0.044914721 | 1 |
| T2DM | endothelial | *STARD3NL* | 0.315942156 | 0.00769455 | 0.014898564 | 1 |
| T2DM | endothelial | *NCAPD2* | -0.239628376 | 0.003885817 | 0.008349328 | 1 |
| T2DM | endothelial | *CD4* | -0.581782276 | 1.42E-06 | 1.52E-05 | 0.00889831 |
| T2DM | endothelial | *HFE* | -0.395482215 | 0.000308552 | 0.000981713 | 1 |
| T2DM | endothelial | *CLCN6* | -0.531227747 | 0.000356235 | 0.001110388 | 1 |
| T2DM | endothelial | *MRC2* | -0.30442084 | 0.004543747 | 0.009490545 | 1 |
| T2DM | endothelial | *TSPAN9* | 0.164332039 | 0.00033389 | 0.00104906 | 1 |
| T2DM | endothelial | *MKS1* | -0.59139077 | 0.012954821 | 0.02325727 | 1 |
| T2DM | endothelial | *UTP18* | -0.570351936 | 0.00053752 | 0.001562361 | 1 |
| T2DM | endothelial | *SYT7* | 0.21711409 | 2.11E-07 | 4.94E-06 | 0.001329494 |
| T2DM | endothelial | *CEP68* | 0.427199789 | 1.55E-08 | 2.38E-06 | 9.75E-05 |
| T2DM | endothelial | *ZBTB32* | -0.942598836 | 0.000950316 | 0.002531624 | 1 |
| T2DM | endothelial | *TMEM159* | -0.534143219 | 2.50E-06 | 2.16E-05 | 0.015698617 |
| T2DM | endothelial | *ERCC1* | -0.459271934 | 1.13E-05 | 6.50E-05 | 0.071352158 |
| T2DM | endothelial | *LTF* | 0.346724873 | 5.67E-07 | 8.58E-06 | 0.003564805 |
| T2DM | endothelial | *ALOX5* | -0.522288041 | 0.00226157 | 0.005280086 | 1 |
| T2DM | endothelial | *MAN2B2* | -0.578156576 | 0.000352221 | 0.001098419 | 1 |
| T2DM | endothelial | *CLDN11* | 0.273973041 | 0.002072303 | 0.004901644 | 1 |
| T2DM | endothelial | *ANGEL1* | 0.381112796 | 0.007227084 | 0.014119538 | 1 |
| T2DM | endothelial | *HEBP1* | -0.470014731 | 0.009532817 | 0.017920669 | 1 |
| T2DM | endothelial | *MDH1* | -0.447831095 | 1.40E-05 | 7.69E-05 | 0.087847055 |
| T2DM | endothelial | *COX15* | -0.464115601 | 0.001433939 | 0.003594567 | 1 |
| T2DM | endothelial | *CCDC88C* | 0.279917425 | 0.000830935 | 0.002265433 | 1 |
| T2DM | endothelial | *NPC1L1* | -0.79171236 | 2.05E-07 | 4.92E-06 | 0.001289508 |
| T2DM | endothelial | *NUDCD3* | -0.267096026 | 0.008787736 | 0.016726763 | 1 |
| T2DM | endothelial | *IL20RA* | -2.220890972 | 3.36E-06 | 2.63E-05 | 0.021118736 |
| T2DM | endothelial | *GLT8D1* | -0.387584558 | 0.004857433 | 0.010019252 | 1 |
| T2DM | endothelial | *RUFY3* | -0.25829531 | 0.015842264 | 0.027597759 | 1 |
| T2DM | endothelial | *WWTR1* | 0.243986234 | 0.004357531 | 0.009165539 | 1 |
| T2DM | endothelial | *TTC27* | -0.312186017 | 0.009938306 | 0.018535192 | 1 |
| T2DM | endothelial | *VSIG2* | 0.228734493 | 5.09E-07 | 8.20E-06 | 0.003200394 |
| T2DM | endothelial | *PHLDB1* | -0.254782259 | 3.13E-05 | 0.000147921 | 0.19658685 |
| T2DM | endothelial | *MNAT1* | -0.328617051 | 0.005115253 | 0.010492528 | 1 |
| T2DM | endothelial | *CPS1* | -0.368605948 | 9.71E-05 | 0.00037652 | 0.610715802 |
| T2DM | endothelial | *NLRP2* | -0.450388264 | 0.000296653 | 0.00095059 | 1 |
| T2DM | endothelial | *RNH1* | -0.345434928 | 0.002921771 | 0.006532423 | 1 |
| T2DM | endothelial | *NDUFS1* | -0.32911223 | 0.009874553 | 0.018443647 | 1 |
| T2DM | endothelial | *GLRX2* | -0.328183276 | 0.019871657 | 0.033431391 | 1 |
| T2DM | endothelial | *SNAPC1* | -0.600435832 | 2.62E-05 | 0.000128492 | 0.164726345 |
| T2DM | endothelial | *GCLM* | -0.304407094 | 0.008884626 | 0.016876874 | 1 |

| | | | | | | |
|---|---|---|---|---|---|---|
| T2DM | endothelial | *HSF2* | -0.535505642 | 0.002111247 | 0.004975041 | 1 |
| T2DM | endothelial | *TYMP* | -0.393285794 | 0.001010867 | 0.00266694 | 1 |
| T2DM | endothelial | *FAS* | -0.322942079 | 0.000593826 | 0.001710208 | 1 |
| T2DM | endothelial | *RNASET2* | -0.476098494 | 1.62E-05 | 8.73E-05 | 0.10171696 |
| T2DM | endothelial | *CD44* | -0.390567981 | 6.18E-06 | 4.09E-05 | 0.038850527 |
| T2DM | endothelial | *MIPEP* | -0.496353158 | 0.000389809 | 0.001192569 | 1 |
| T2DM | endothelial | *INSRR* | -0.650116672 | 2.55E-05 | 0.000126075 | 0.16024165 |
| T2DM | endothelial | *SNX1* | -0.318419456 | 0.005373814 | 0.010962092 | 1 |
| T2DM | endothelial | *SLC39A9* | -0.356956928 | 0.007602948 | 0.014753004 | 1 |
| T2DM | endothelial | *ANK1* | 0.354363945 | 3.38E-06 | 2.63E-05 | 0.021238091 |
| T2DM | endothelial | *RABEP1* | -0.23953574 | 0.025815201 | 0.042003149 | 1 |
| T2DM | endothelial | *BAK1* | 0.352497584 | 0.001613558 | 0.003967321 | 1 |
| T2DM | endothelial | *GRN* | -0.433782655 | 0.002085628 | 0.004929453 | 1 |
| T2DM | endothelial | *CENPQ* | -0.313929288 | 0.00666784 | 0.013198687 | 1 |
| T2DM | endothelial | *ZCCHC8* | -0.363551892 | 0.010390452 | 0.019275531 | 1 |
| T2DM | endothelial | *UBA6* | -0.285047018 | 0.02066604 | 0.034564351 | 1 |
| T2DM | endothelial | *PIAS1* | -0.258609983 | 0.016715007 | 0.028822614 | 1 |
| T2DM | endothelial | *SLC4A7* | -0.41190955 | 5.12E-06 | 3.59E-05 | 0.032193292 |
| T2DM | endothelial | *MYOC* | -0.220306096 | 0.002310961 | 0.005381115 | 1 |
| T2DM | endothelial | *SH3YL1* | -0.447402318 | 1.79E-05 | 9.40E-05 | 0.112484998 |
| T2DM | endothelial | *VCL* | -0.352899157 | 0.002380406 | 0.00551626 | 1 |
| T2DM | endothelial | *STAP1* | -0.87801265 | 0.000646242 | 0.001826853 | 1 |
| T2DM | endothelial | *TBC1D23* | -0.408201152 | 0.000857489 | 0.002325726 | 1 |
| T2DM | endothelial | *TUBG2* | -0.493449641 | 1.11E-11 | 1.75E-08 | 6.98E-08 |
| T2DM | endothelial | *MFAP3* | 0.243029495 | 0.004202949 | 0.00891496 | 1 |
| T2DM | endothelial | *MRI1* | -0.395991381 | 0.000783989 | 0.002155199 | 1 |
| T2DM | endothelial | *DNAH5* | -0.571575507 | 1.05E-07 | 4.46E-06 | 0.000658366 |
| T2DM | endothelial | *C6* | -0.439122217 | 4.66E-08 | 3.81E-06 | 0.000293085 |
| T2DM | endothelial | *RAI14* | 0.240055053 | 6.17E-05 | 0.000256549 | 0.387602265 |
| T2DM | endothelial | *PNKP* | -0.27713452 | 0.014101208 | 0.024980078 | 1 |
| T2DM | endothelial | *PHLPP2* | -0.384687592 | 0.004428179 | 0.009279987 | 1 |
| T2DM | endothelial | *PQLC2* | -0.823011398 | 4.11E-06 | 3.05E-05 | 0.02584259 |
| T2DM | endothelial | *INPP4A* | -0.264350095 | 0.015578632 | 0.027183696 | 1 |
| T2DM | endothelial | *PSMA4* | -0.45263017 | 0.006054708 | 0.012149999 | 1 |
| T2DM | endothelial | *THAP3* | -0.799870414 | 3.34E-07 | 6.48E-06 | 0.002101024 |
| T2DM | endothelial | *TDP1* | -0.545147506 | 0.00234293 | 0.005441447 | 1 |
| T2DM | endothelial | *SPATA7* | -0.46014696 | 0.000490668 | 0.001449087 | 1 |
| T2DM | endothelial | *RETSAT* | -0.329305627 | 4.86E-06 | 3.48E-05 | 0.030576517 |
| T2DM | endothelial | *CAPG* | -0.54516918 | 3.18E-07 | 6.29E-06 | 0.00199941 |
| T2DM | endothelial | *AP2S1* | -0.566854432 | 0.003124216 | 0.006925932 | 1 |
| T2DM | endothelial | *LCP2* | -0.413883344 | 0.002416063 | 0.005584481 | 1 |
| T2DM | endothelial | *DSG2* | 0.149063088 | 9.24E-07 | 1.12E-05 | 0.005811978 |
| T2DM | endothelial | *ATP6V1H* | -0.411427959 | 0.001193637 | 0.003081888 | 1 |
| T2DM | endothelial | *CP* | 0.399516779 | 0.002633617 | 0.005999112 | 1 |
| T2DM | endothelial | *ANO2* | 0.088579085 | 1.40E-06 | 1.50E-05 | 0.008782862 |
| T2DM | endothelial | *GOPC* | -0.325067332 | 0.016624263 | 0.028689746 | 1 |
| T2DM | endothelial | *CC2D2A* | -0.2774046 | 0.028081071 | 0.045314603 | 1 |
| T2DM | endothelial | *RRM2B* | -0.539630845 | 0.000263101 | 0.000859282 | 1 |
| T2DM | endothelial | *SNX29* | -0.343026176 | 0.000308889 | 0.00098229 | 1 |
| T2DM | endothelial | *LMO3* | -0.268628564 | 1.69E-06 | 1.68E-05 | 0.010652486 |

| | | | | | | |
|---|---|---|---|---|---|---|
| T2DM | endothelial | *MRPS10* | -0.191745018 | 0.004558737 | 0.009512373 | 1 |
| T2DM | endothelial | *CELF2* | -0.315600953 | 4.38E-05 | 0.000195348 | 0.275635447 |
| T2DM | endothelial | *ADAMTS6* | 0.190981831 | 0.016319506 | 0.028218023 | 1 |
| T2DM | endothelial | *H6PD* | -0.307335613 | 0.008107949 | 0.015598125 | 1 |
| T2DM | endothelial | *RCN1* | 0.26838322 | 0.012158633 | 0.022048263 | 1 |
| T2DM | endothelial | *RFC2* | -0.468296022 | 0.025710705 | 0.041868178 | 1 |
| T2DM | endothelial | *JKAMP* | -0.35556569 | 0.001959005 | 0.004674104 | 1 |
| T2DM | endothelial | *ARHGEF5* | -0.996191122 | 7.51E-06 | 4.75E-05 | 0.04723477 |
| T2DM | endothelial | *NFE2L3* | 0.404966631 | 0.01499944 | 0.026326487 | 1 |
| T2DM | endothelial | *LIMA1* | -0.179580146 | 0.001729618 | 0.004206617 | 1 |
| T2DM | endothelial | *PTGER3* | -0.315390246 | 5.34E-07 | 8.34E-06 | 0.003355676 |
| T2DM | endothelial | *CYBA* | -0.469303247 | 7.18E-05 | 0.000290472 | 0.451684518 |
| T2DM | endothelial | *THOC3* | -0.954794831 | 8.54E-06 | 5.25E-05 | 0.053686221 |
| T2DM | endothelial | *RRP12* | -0.374151278 | 0.012118589 | 0.021994678 | 1 |
| T2DM | endothelial | *ALX4* | -1.026764216 | 1.82E-07 | 4.62E-06 | 0.001142583 |
| T2DM | endothelial | *FOXN3* | -0.312404385 | 0.001988011 | 0.004728953 | 1 |
| T2DM | endothelial | *AKR7A2* | -0.648356426 | 3.33E-05 | 0.00015534 | 0.209395462 |
| T2DM | endothelial | *MRTO4* | -0.682946206 | 0.000712722 | 0.001990636 | 1 |
| T2DM | endothelial | *SDCCAG8* | -0.217872741 | 0.015994275 | 0.027808631 | 1 |
| T2DM | endothelial | *PLEKHH1* | -0.448448749 | 4.50E-07 | 7.56E-06 | 0.002828192 |
| T2DM | endothelial | *ATP9A* | -0.468365042 | 6.44E-06 | 4.22E-05 | 0.040504894 |
| T2DM | endothelial | *CHRDL2* | -0.349926796 | 0.026228394 | 0.042576276 | 1 |
| T2DM | endothelial | *GALC* | -0.505413796 | 0.000368189 | 0.001140298 | 1 |
| T2DM | endothelial | *MRPL43* | -0.47115293 | 0.000748382 | 0.002073634 | 1 |
| T2DM | endothelial | *ITIH4* | 0.153851333 | 0.006205069 | 0.012400277 | 1 |
| T2DM | endothelial | *TRAF1* | -0.521286427 | 2.56E-07 | 5.50E-06 | 0.001606469 |
| T2DM | endothelial | *PCGF2* | 0.189764982 | 0.000216816 | 0.000736426 | 1 |
| T2DM | endothelial | *IL17RB* | -1.800663386 | 6.01E-06 | 4.01E-05 | 0.037766002 |
| T2DM | endothelial | *TRAF3IP2* | -0.36777115 | 0.020041275 | 0.033644726 | 1 |
| T2DM | endothelial | *DCBLD2* | -0.453525124 | 4.71E-06 | 3.40E-05 | 0.029605686 |
| T2DM | endothelial | *F7* | -2.675520135 | 0.003231248 | 0.007120524 | 1 |
| T2DM | endothelial | *TMCC3* | 0.268775706 | 3.73E-05 | 0.000170819 | 0.234193092 |
| T2DM | endothelial | *PITHD1* | -0.5012835 | 0.00432203 | 0.009124447 | 1 |
| T2DM | endothelial | *ATP11B* | -0.20862937 | 0.019627302 | 0.033117779 | 1 |
| T2DM | endothelial | *CDK14* | -0.284086145 | 0.011172342 | 0.020497497 | 1 |
| T2DM | endothelial | *CROCC* | -0.274356897 | 0.003321048 | 0.007303991 | 1 |
| T2DM | endothelial | *ZC3H11A* | -1.040561696 | 2.06E-06 | 1.91E-05 | 0.012965362 |
| T2DM | endothelial | *RIOK2* | -0.530353878 | 0.001512762 | 0.003757699 | 1 |
| T2DM | endothelial | *FLYWCH1* | -0.242139116 | 0.003347221 | 0.007342631 | 1 |
| T2DM | endothelial | *DNAJC25* | -0.588140545 | 5.35E-06 | 3.69E-05 | 0.033640067 |
| T2DM | endothelial | *PIGV* | -0.744621867 | 1.74E-08 | 2.54E-06 | 0.000109201 |
| T2DM | endothelial | *TNK2* | -0.350035979 | 0.002813427 | 0.006323924 | 1 |
| T2DM | endothelial | *VMP1* | -0.343403156 | 0.00150194 | 0.003736723 | 1 |
| T2DM | endothelial | *SPHK2* | -0.575718074 | 2.69E-05 | 0.000130835 | 0.169169922 |
| T2DM | endothelial | *ISOC2* | -0.782702725 | 3.00E-07 | 6.14E-06 | 0.001886468 |
| T2DM | endothelial | *ADCK1* | -0.559368123 | 4.78E-06 | 3.44E-05 | 0.030050923 |
| T2DM | endothelial | *CASP8* | -0.440052335 | 0.000225379 | 0.000759821 | 1 |
| T2DM | endothelial | *DMRT3* | -0.544529013 | 4.18E-07 | 7.31E-06 | 0.002625533 |
| T2DM | endothelial | *ST3GAL6* | -0.37523067 | 1.12E-05 | 6.45E-05 | 0.070408133 |
| T2DM | endothelial | *ATP2C2* | 0.729008655 | 0.01390186 | 0.024682573 | 1 |

| | | | | | | |
|---|---|---|---|---|---|---|
| T2DM | endothelial | *TMEM161A* | -0.642195808 | 0.000256294 | 0.00084098 | 1 |
| T2DM | endothelial | *LPAR2* | -0.533740791 | 7.71E-05 | 0.000308262 | 0.484587721 |
| T2DM | endothelial | *ABCA7* | -0.537775044 | 0.011173043 | 0.020497497 | 1 |
| T2DM | endothelial | *DDX20* | -0.605443369 | 0.012184288 | 0.022082046 | 1 |
| T2DM | endothelial | *CHI3L2* | -0.318450425 | 0.006909072 | 0.013601733 | 1 |
| T2DM | endothelial | *TAF11* | -0.278065742 | 0.017924863 | 0.030631588 | 1 |
| T2DM | endothelial | *NTHL1* | 0.423214304 | 6.80E-06 | 4.40E-05 | 0.04273833 |
| T2DM | endothelial | *IPO5* | -0.432769748 | 0.004774997 | 0.009884889 | 1 |
| T2DM | endothelial | *WDR18* | -0.418932308 | 0.000221273 | 0.000748731 | 1 |
| T2DM | endothelial | *ANKRD44* | -0.345952077 | 0.001297475 | 0.003302856 | 1 |
| T2DM | endothelial | *TMEM206* | -0.49270412 | 0.004996897 | 0.010276575 | 1 |
| T2DM | endothelial | *SEC61A2* | -0.525083256 | 0.002805794 | 0.006313539 | 1 |
| T2DM | endothelial | *ASB1* | -0.551083534 | 5.67E-08 | 4.25E-06 | 0.000356782 |
| T2DM | endothelial | *TBC1D1* | 0.24463192 | 0.022365308 | 0.037090659 | 1 |
| T2DM | endothelial | *TIE1* | 0.119139572 | 1.45E-06 | 1.54E-05 | 0.009106465 |
| T2DM | endothelial | *NFYC* | -0.339153967 | 0.013627622 | 0.024269832 | 1 |
| T2DM | endothelial | *SLC9A3* | -0.293090797 | 0.002252714 | 0.005266943 | 1 |
| T2DM | endothelial | *NGEF* | 0.89140738 | 0.000101541 | 0.00039045 | 0.638386014 |
| T2DM | endothelial | *CD84* | -0.318477891 | 0.005426204 | 0.01104388 | 1 |
| T2DM | endothelial | *GOLGA5* | -0.399089552 | 0.010665544 | 0.019712019 | 1 |
| T2DM | endothelial | *EML1* | 0.230343512 | 0.002678386 | 0.006068112 | 1 |
| T2DM | endothelial | *ZFAT* | -0.512497429 | 9.26E-05 | 0.000361095 | 0.582151813 |
| T2DM | endothelial | *MTFR1* | -0.440395605 | 0.000512686 | 0.001502579 | 1 |
| T2DM | endothelial | *STAG3* | -0.629797532 | 7.97E-06 | 4.98E-05 | 0.050078822 |
| T2DM | endothelial | *STOML1* | -0.982565978 | 0.001395132 | 0.00351833 | 1 |
| T2DM | endothelial | *METTL22* | -0.640236733 | 0.000145763 | 0.000527886 | 0.916410874 |
| T2DM | endothelial | *TP53BP1* | -0.334393807 | 0.00689936 | 0.013589059 | 1 |
| T2DM | endothelial | *SYT1* | 0.18348757 | 0.007026931 | 0.013775589 | 1 |
| T2DM | endothelial | *ROGDI* | -1.252789997 | 7.85E-05 | 0.000312789 | 0.493268124 |
| T2DM | endothelial | *HDAC4* | -0.350421814 | 0.003876235 | 0.008331587 | 1 |
| T2DM | endothelial | *FGFR3* | 0.177478723 | 4.87E-06 | 3.48E-05 | 0.030620863 |
| T2DM | endothelial | *IFI35* | 0.425998293 | 6.98E-05 | 0.000284285 | 0.438936456 |
| T2DM | endothelial | *HEATR6* | -0.6016317 | 0.000509349 | 0.0014936 | 1 |
| T2DM | endothelial | *POLR1A* | -0.327007553 | 7.21E-07 | 9.76E-06 | 0.004534887 |
| T2DM | endothelial | *STON1-GTF2A* | -0.085365174 | 0.004658454 | 0.009685085 | 1 |
| T2DM | endothelial | *SRBD1* | -0.331965339 | 0.007437159 | 0.014480464 | 1 |
| T2DM | endothelial | *PSME4* | -0.361607053 | 0.001238273 | 0.003164643 | 1 |
| T2DM | endothelial | *PITX1* | -0.962754642 | 1.09E-07 | 4.46E-06 | 0.000684129 |
| T2DM | endothelial | *DNAJA2* | -0.537949956 | 0.003109461 | 0.006905398 | 1 |
| T2DM | endothelial | *KCNAB2* | -0.554985447 | 2.21E-06 | 2.00E-05 | 0.013870226 |
| T2DM | endothelial | *GAL* | -0.77191696 | 1.29E-07 | 4.46E-06 | 0.000809275 |
| T2DM | endothelial | *NEDD4* | 0.269128261 | 5.91E-05 | 0.000247902 | 0.371357928 |
| T2DM | endothelial | *PIGB* | -0.475875303 | 0.000552827 | 0.001603888 | 1 |
| T2DM | endothelial | *DAPP1* | -0.45250086 | 2.46E-05 | 0.000122388 | 0.154802217 |
| T2DM | endothelial | *FGF10* | -0.241950035 | 0.000136097 | 0.000500376 | 0.855642227 |
| T2DM | endothelial | *CLTCL1* | -0.363707546 | 0.002097348 | 0.004949709 | 1 |
| T2DM | endothelial | *DGCR2* | -0.403565713 | 0.005153849 | 0.010554479 | 1 |
| T2DM | endothelial | *MNT* | -0.476598115 | 0.000559343 | 0.001621295 | 1 |
| T2DM | endothelial | *WIPI1* | -0.500569654 | 1.88E-06 | 1.81E-05 | 0.011848936 |
| T2DM | endothelial | *ASNS* | -0.604742632 | 5.86E-05 | 0.000246461 | 0.368213347 |

| | | | | | | |
|---|---|---|---|---|---|---|
| T2DM | endothelial | *ST6GALNAC2* | 0.453111273 | 0.000386096 | 0.001184089 | 1 |
| T2DM | endothelial | *TESK2* | -0.42962501 | 0.004680122 | 0.009714073 | 1 |
| T2DM | endothelial | *EIF2B3* | -0.419634355 | 1.33E-05 | 7.41E-05 | 0.083536379 |
| T2DM | endothelial | *CAMK2A* | -1.487797091 | 3.77E-08 | 3.45E-06 | 0.000236768 |
| T2DM | endothelial | *OSBPL3* | -0.492750416 | 1.20E-05 | 6.81E-05 | 0.0755147 |
| T2DM | endothelial | *RAD18* | -0.504864324 | 0.00066968 | 0.001882951 | 1 |
| T2DM | endothelial | *VASH1* | 0.337320492 | 0.00269759 | 0.006105021 | 1 |
| T2DM | endothelial | *TRIP13* | -1.173588664 | 1.15E-07 | 4.46E-06 | 0.000722777 |
| T2DM | endothelial | *MBD3* | -0.71716433 | 5.13E-06 | 3.59E-05 | 0.032254166 |
| T2DM | endothelial | *HLTF* | -0.457072987 | 6.55E-05 | 0.00026952 | 0.411825971 |
| T2DM | endothelial | *CPSF1* | -0.655432302 | 0.000926164 | 0.002478839 | 1 |
| T2DM | endothelial | *LIMS2* | 0.184761551 | 5.26E-06 | 3.65E-05 | 0.033045247 |
| T2DM | endothelial | *TRHDE* | -0.311494823 | 3.78E-07 | 6.87E-06 | 0.002377518 |
| T2DM | endothelial | *ACADVL* | -0.497368318 | 3.77E-05 | 0.00017297 | 0.237314518 |
| T2DM | endothelial | *CRMP1* | -0.44234145 | 1.04E-07 | 4.46E-06 | 0.000655031 |
| T2DM | endothelial | *DERL2* | -0.482098071 | 0.009140033 | 0.017289448 | 1 |
| T2DM | endothelial | *AP1M1* | -0.319829649 | 0.007583005 | 0.01471885 | 1 |
| T2DM | endothelial | *PVR* | 0.384145084 | 0.002562688 | 0.005856641 | 1 |
| T2DM | endothelial | *MOV10L1* | -0.26055485 | 0.007843411 | 0.01514947 | 1 |
| T2DM | endothelial | *TP63* | -0.464511302 | 1.58E-06 | 1.61E-05 | 0.009911078 |
| T2DM | endothelial | *LLGL2* | 0.344758634 | 0.000930381 | 0.002489067 | 1 |
| T2DM | endothelial | *NLE1* | -0.708295549 | 0.000380104 | 0.001168563 | 1 |
| T2DM | endothelial | *GSDMB* | -0.40991466 | 3.61E-06 | 2.76E-05 | 0.022681997 |
| T2DM | endothelial | *PICALM* | -0.25064253 | 0.010879855 | 0.020029765 | 1 |
| T2DM | endothelial | *GLI2* | -0.270759732 | 9.36E-06 | 5.64E-05 | 0.058852359 |
| T2DM | endothelial | *ATP2A3* | 0.294714812 | 2.68E-05 | 0.00013071 | 0.16874713 |
| T2DM | endothelial | *CA12* | -0.358353875 | 1.63E-06 | 1.65E-05 | 0.010252397 |
| T2DM | endothelial | *NTN4* | 0.269107256 | 0.004971034 | 0.010230078 | 1 |
| T2DM | endothelial | *ENO1* | -0.527543344 | 0.000490301 | 0.001449087 | 1 |
| T2DM | endothelial | *TTC38* | -0.498886365 | 3.11E-06 | 2.48E-05 | 0.019551887 |
| T2DM | endothelial | *GRAMD4* | -0.37231183 | 0.028550229 | 0.045894985 | 1 |
| T2DM | endothelial | *CELSR1* | 0.30766299 | 6.74E-06 | 4.37E-05 | 0.042390949 |
| T2DM | endothelial | *ADD2* | 0.277758413 | 1.58E-06 | 1.61E-05 | 0.009925817 |
| T2DM | endothelial | *VPS9D1* | -0.556754439 | 0.003328427 | 0.00731672 | 1 |
| T2DM | endothelial | *MARK3* | -0.345766049 | 0.001857896 | 0.004477037 | 1 |
| T2DM | endothelial | *SLC25A3* | -0.317310109 | 0.029120042 | 0.046664632 | 1 |
| T2DM | endothelial | *TUBA3D* | -1.229821879 | 4.84E-06 | 3.47E-05 | 0.030452652 |
| T2DM | endothelial | *MKRN2* | -0.466525442 | 0.01344462 | 0.02400634 | 1 |
| T2DM | endothelial | *MCM6* | -0.538412134 | 0.001025071 | 0.002699882 | 1 |
| T2DM | endothelial | *RBMS2* | 0.255323332 | 0.004324371 | 0.009126325 | 1 |
| T2DM | endothelial | *MLH1* | -0.359679211 | 0.006912285 | 0.013601733 | 1 |
| T2DM | endothelial | *FMO4* | -0.581721105 | 3.47E-05 | 0.00016082 | 0.218393295 |
| T2DM | endothelial | *RGS11* | -0.258898556 | 0.001565534 | 0.003862839 | 1 |
| T2DM | endothelial | *TPD52* | 0.270646219 | 1.24E-05 | 7.00E-05 | 0.07822162 |
| T2DM | endothelial | *GPATCH1* | -0.453554524 | 0.001213024 | 0.003116584 | 1 |
| T2DM | endothelial | *ICAM3* | -0.327965003 | 0.00809521 | 0.015578385 | 1 |
| T2DM | endothelial | *XAB2* | -0.988925727 | 4.75E-06 | 3.42E-05 | 0.029855845 |
| T2DM | endothelial | *STXBP2* | -0.357999958 | 0.000972502 | 0.002580886 | 1 |
| T2DM | endothelial | *MAP2K7* | -0.581119344 | 0.000703222 | 0.001968459 | 1 |
| T2DM | endothelial | *CTTNBP2* | -0.545531858 | 1.76E-05 | 9.32E-05 | 0.110597935 |

| | | | | | | |
|---|---|---|---|---|---|---|
| T2DM | endothelial | *APBB1IP* | -0.335928919 | 3.58E-06 | 2.75E-05 | 0.022484402 |
| T2DM | endothelial | *LRCH4* | -0.748655862 | 3.26E-09 | 7.89E-07 | 2.05E-05 |
| T2DM | endothelial | *SIRT6* | -0.916430244 | 2.70E-06 | 2.27E-05 | 0.016968106 |
| T2DM | endothelial | *CAPZB* | -0.324088888 | 0.005978301 | 0.012015851 | 1 |
| T2DM | endothelial | *GPR137B* | -0.259223352 | 0.009980558 | 0.018597442 | 1 |
| T2DM | endothelial | *NAALAD2* | -0.305982616 | 0.00017752 | 0.000623501 | 1 |
| T2DM | endothelial | *FGFR1* | -0.418052348 | 1.91E-05 | 9.94E-05 | 0.119803355 |
| T2DM | endothelial | *FBLN1* | -0.28080265 | 7.94E-05 | 0.000315556 | 0.498986836 |
| T2DM | endothelial | *PIAS2* | -0.310668842 | 0.012646142 | 0.022787703 | 1 |
| T2DM | endothelial | *AMPH* | -0.230947286 | 0.000102596 | 0.00039364 | 0.645020041 |
| T2DM | endothelial | *MCCC1* | -0.323152581 | 0.000377461 | 0.001162713 | 1 |
| T2DM | endothelial | *FAP* | -0.257293537 | 0.008326483 | 0.015979425 | 1 |
| T2DM | endothelial | *N4BP2* | -0.47947117 | 0.000133114 | 0.000491419 | 0.836886115 |
| T2DM | endothelial | *TULP3* | -0.473898515 | 9.82E-05 | 0.000379887 | 0.617316773 |
| T2DM | endothelial | *SYNJ2* | 0.196676746 | 3.92E-06 | 2.95E-05 | 0.024656181 |
| T2DM | endothelial | *ZCWPW1* | -0.375872523 | 0.002584624 | 0.005898196 | 1 |
| T2DM | endothelial | *FGF20* | -6.628472593 | 5.42E-07 | 8.39E-06 | 0.003407501 |
| T2DM | endothelial | *OSBPL6* | -0.58802671 | 6.13E-05 | 0.000255545 | 0.385105672 |
| T2DM | endothelial | *REXO1* | -0.475822455 | 0.015452038 | 0.027007774 | 1 |
| T2DM | endothelial | *SAR1A* | -0.476597184 | 4.60E-05 | 0.000202822 | 0.28922356 |
| T2DM | endothelial | *RAPGEF3* | 0.108278012 | 2.63E-06 | 2.23E-05 | 0.016506656 |
| T2DM | endothelial | *FDFT1* | -0.472021058 | 0.000618919 | 0.001767095 | 1 |
| T2DM | endothelial | *SCGN* | -1.8257099 | 0.000172176 | 0.000606088 | 1 |
| T2DM | endothelial | *DDX1* | -0.357176519 | 0.012318715 | 0.022274306 | 1 |
| T2DM | endothelial | *RIMS1* | -0.671641729 | 5.31E-07 | 8.34E-06 | 0.003336019 |
| T2DM | endothelial | *RABL2B* | -0.638931782 | 1.45E-05 | 7.93E-05 | 0.090933708 |
| T2DM | endothelial | *DCT* | -0.54508517 | 1.47E-07 | 4.46E-06 | 0.00092344 |
| T2DM | endothelial | *SLC35C2* | -0.444005681 | 0.007032945 | 0.013783081 | 1 |
| T2DM | endothelial | *SCTR* | -0.401242311 | 1.95E-07 | 4.75E-06 | 0.00122639 |
| T2DM | endothelial | *KCNN2* | 0.235109404 | 1.53E-06 | 1.60E-05 | 0.009644293 |
| T2DM | endothelial | *PSEN1* | -0.307186071 | 0.009476424 | 0.0178218 | 1 |
| T2DM | endothelial | *CPOX* | -0.628796106 | 7.10E-05 | 0.000288112 | 0.446286113 |
| T2DM | endothelial | *MOK* | -0.540860659 | 0.000495193 | 0.001458893 | 1 |
| T2DM | endothelial | *RSBN1* | -0.351580104 | 0.016279313 | 0.028191566 | 1 |
| T2DM | endothelial | *COL4A4* | -0.581544187 | 2.67E-05 | 0.00013023 | 0.167865959 |
| T2DM | endothelial | *PKP1* | 0.201859838 | 1.63E-06 | 1.65E-05 | 0.010263194 |
| T2DM | endothelial | *STK17B* | -0.434249505 | 3.95E-05 | 0.000179093 | 0.248043715 |
| T2DM | endothelial | *ZNF506* | -0.50826834 | 0.001307799 | 0.003323417 | 1 |
| T2DM | endothelial | *PCDHA6* | -2.82277665 | 0.000693834 | 0.001945643 | 1 |
| T2DM | endothelial | *PCDHGA2* | -1.224785562 | 1.35E-08 | 2.24E-06 | 8.51E-05 |
| T2DM | endothelial | *HSPB11* | -0.368162502 | 0.012689906 | 0.022853463 | 1 |
| T2DM | endothelial | *WDR70* | -0.257697336 | 0.017408917 | 0.029888002 | 1 |
| T2DM | endothelial | *PGR* | -0.349985847 | 7.98E-08 | 4.46E-06 | 0.000501816 |
| T2DM | endothelial | *COBLL1* | -0.335460034 | 3.88E-05 | 0.000176958 | 0.243691535 |
| T2DM | endothelial | *GEMIN5* | -0.433473709 | 0.000246726 | 0.000818124 | 1 |
| T2DM | endothelial | *ULK2* | -0.411556337 | 9.01E-05 | 0.000352411 | 0.566677419 |
| T2DM | endothelial | *PLOD1* | -0.682187932 | 3.20E-05 | 0.000150476 | 0.201314137 |
| T2DM | endothelial | *OXCT1* | -0.279789125 | 0.004660773 | 0.009686704 | 1 |
| T2DM | endothelial | *EPYC* | -1.483235821 | 0.002326762 | 0.005407893 | 1 |
| T2DM | endothelial | *ZNF324* | -0.68431896 | 0.002262579 | 0.005280086 | 1 |

| | | | | | | |
|---|---|---|---|---|---|---|
| T2DM | endothelial | *ZNF446* | -0.93666229 | 1.18E-05 | 6.71E-05 | 0.074159865 |
| T2DM | endothelial | *CHMP2B* | -0.406791223 | 0.007319592 | 0.014291389 | 1 |
| T2DM | endothelial | *PPIE* | -0.484924198 | 0.000723955 | 0.002018082 | 1 |
| T2DM | endothelial | *ZMPSTE24* | -0.542336071 | 0.0002916 | 0.000937743 | 1 |
| T2DM | endothelial | *NOA1* | -0.466594732 | 0.004339318 | 0.009149009 | 1 |
| T2DM | endothelial | *GSTP1* | -0.412588659 | 0.01368043 | 0.02435132 | 1 |
| T2DM | endothelial | *COL16A1* | -0.390251704 | 3.36E-08 | 3.30E-06 | 0.000211293 |
| T2DM | endothelial | *APOB* | -0.50131976 | 7.28E-08 | 4.46E-06 | 0.00045781 |
| T2DM | endothelial | *CD59* | 0.275863542 | 0.000110626 | 0.00041898 | 0.695507557 |
| T2DM | endothelial | *TAF9* | -0.63505072 | 0.008482735 | 0.016210016 | 1 |
| T2DM | endothelial | *FCN1* | -0.492863156 | 4.73E-07 | 7.85E-06 | 0.002975259 |
| T2DM | endothelial | *SCAMP1* | 0.29127413 | 0.006073917 | 0.01218077 | 1 |
| T2DM | endothelial | *SEH1L* | -0.423453846 | 0.009325239 | 0.017605938 | 1 |
| T2DM | endothelial | *SLC25A24* | -0.316725597 | 0.010814237 | 0.019944003 | 1 |
| T2DM | endothelial | *ABCB1* | 0.140173665 | 1.50E-06 | 1.58E-05 | 0.009415019 |
| T2DM | endothelial | *RRN3* | -0.415059371 | 0.00838891 | 0.016060043 | 1 |
| T2DM | endothelial | *MTIF2* | -0.597243659 | 1.47E-05 | 8.05E-05 | 0.092549409 |
| T2DM | endothelial | *USP40* | -0.372281453 | 0.00093178 | 0.002491749 | 1 |
| T2DM | endothelial | *MAST2* | -0.340216607 | 0.009816888 | 0.018368684 | 1 |
| T2DM | endothelial | *EIF2AK1* | -0.333174371 | 0.018889397 | 0.032036051 | 1 |
| T2DM | endothelial | *SNX10* | -0.181962164 | 3.15E-07 | 6.28E-06 | 0.001979677 |
| T2DM | endothelial | *ITPKC* | 0.329857796 | 0.005894949 | 0.011859694 | 1 |
| T2DM | endothelial | *TMED2* | -0.503325929 | 0.002832454 | 0.0063576 | 1 |
| T2DM | endothelial | *HSD17B14* | -0.843938449 | 4.20E-08 | 3.59E-06 | 0.000263746 |
| T2DM | endothelial | *TRIP6* | -0.601131418 | 1.10E-05 | 6.38E-05 | 0.069199674 |
| T2DM | endothelial | *ACHE* | -0.28365219 | 0.000488777 | 0.001446107 | 1 |
| T2DM | endothelial | *SRRT* | -0.300978951 | 0.028987364 | 0.046502567 | 1 |
| T2DM | endothelial | *LPCAT2* | 0.254001464 | 0.000743648 | 0.002061426 | 1 |
| T2DM | endothelial | *TXNDC16* | -0.357893299 | 0.00652799 | 0.012942753 | 1 |
| T2DM | endothelial | *GMCL1* | -0.368585188 | 0.011358303 | 0.020807009 | 1 |
| T2DM | endothelial | *PTHLH* | -0.534155698 | 8.81E-07 | 1.09E-05 | 0.005541802 |
| T2DM | endothelial | *PHACTR3* | -0.29192622 | 0.001505648 | 0.003744466 | 1 |
| T2DM | endothelial | *AAMDC* | -0.177895514 | 0.020122321 | 0.033753745 | 1 |
| T2DM | endothelial | *GP6* | -0.885988973 | 1.65E-05 | 8.85E-05 | 0.103900539 |
| T2DM | endothelial | *EDEM2* | -0.500110883 | 0.000149444 | 0.000537839 | 0.939557311 |
| T2DM | endothelial | *TPX2* | -0.275584772 | 0.001719473 | 0.004185252 | 1 |
| T2DM | endothelial | *DOCK3* | -0.337760035 | 1.11E-05 | 6.40E-05 | 0.069747484 |
| T2DM | endothelial | *C3orf18* | -0.279546843 | 0.028877449 | 0.04634989 | 1 |
| T2DM | endothelial | *FKBP1A* | 0.388961594 | 0.000596431 | 0.001714569 | 1 |
| T2DM | endothelial | *NSFL1C* | -0.515981889 | 0.000501833 | 0.001474998 | 1 |
| T2DM | endothelial | *CPXM1* | -0.328340845 | 3.46E-05 | 0.00016052 | 0.217664618 |
| T2DM | endothelial | *MAVS* | -0.399189415 | 0.002365795 | 0.005490495 | 1 |
| T2DM | endothelial | *LZTS3* | -0.634362265 | 3.25E-08 | 3.24E-06 | 0.000204257 |
| T2DM | endothelial | *TESC* | 0.251882504 | 1.80E-06 | 1.74E-05 | 0.011300952 |
| T2DM | endothelial | *TMEM230* | -0.348799966 | 0.00571286 | 0.011537664 | 1 |
| T2DM | endothelial | *DZANK1* | -0.656884441 | 0.000170442 | 0.000600993 | 1 |
| T2DM | endothelial | *TASP1* | -0.351395117 | 0.002515517 | 0.005766004 | 1 |
| T2DM | endothelial | *KIF16B* | -0.296217722 | 0.025613162 | 0.041750051 | 1 |
| T2DM | endothelial | *FXYD5* | 0.397832465 | 0.000223196 | 0.00075402 | 1 |
| T2DM | endothelial | *ZNF302* | -0.212754387 | 0.019149309 | 0.032413736 | 1 |

| | | | | | | |
|---|---|---|---|---|---|---|
| T2DM | endothelial | *FXYD3* | -1.080458893 | 2.00E-06 | 1.88E-05 | 0.012604182 |
| T2DM | endothelial | *KCNH4* | 0.375621555 | 3.44E-06 | 2.67E-05 | 0.021606541 |
| T2DM | endothelial | *GANAB* | -0.424871471 | 0.005119631 | 0.010498082 | 1 |
| T2DM | endothelial | *GMIP* | -0.843031295 | 3.98E-06 | 2.98E-05 | 0.025009566 |
| T2DM | endothelial | *MLF2* | -0.608113497 | 0.00049279 | 0.001453858 | 1 |
| T2DM | endothelial | *NECAP1* | -0.371998369 | 0.029266111 | 0.046842169 | 1 |
| T2DM | endothelial | *ANKRD24* | -0.494501353 | 1.70E-05 | 9.06E-05 | 0.106861702 |
| T2DM | endothelial | *GPATCH2L* | -0.308754493 | 0.006890054 | 0.013574982 | 1 |
| T2DM | endothelial | *SLC9A1* | -0.626408095 | 3.87E-05 | 0.000176958 | 0.243568924 |
| T2DM | endothelial | *SPTLC1* | -0.340539368 | 0.009829131 | 0.018376196 | 1 |
| T2DM | endothelial | *RGS1* | -0.549106791 | 1.36E-07 | 4.46E-06 | 0.000856582 |
| T2DM | endothelial | *MRPS33* | -0.783675976 | 1.05E-06 | 1.22E-05 | 0.006594444 |
| T2DM | endothelial | *MAEA* | -0.389545029 | 0.001763723 | 0.004276331 | 1 |
| T2DM | endothelial | *STRN4* | -0.520204318 | 0.006938493 | 0.013644763 | 1 |
| T2DM | endothelial | *LYZ* | -0.337911832 | 0.006324865 | 0.012599628 | 1 |
| T2DM | endothelial | *MUL1* | -0.730205107 | 2.97E-05 | 0.000141657 | 0.186845231 |
| T2DM | endothelial | *SPG21* | -0.5130874 | 0.000299805 | 0.000958247 | 1 |
| T2DM | endothelial | *GNPTG* | -0.399413046 | 0.018629978 | 0.031655858 | 1 |
| T2DM | endothelial | *ZNF268* | -0.591958763 | 0.014938934 | 0.026234937 | 1 |
| T2DM | endothelial | *GOLGA3* | -0.361862409 | 0.002059887 | 0.004877782 | 1 |
| T2DM | endothelial | *PABPC4* | -0.839083875 | 7.92E-11 | 4.68E-08 | 4.98E-07 |
| T2DM | endothelial | *CERS4* | -0.467274571 | 3.52E-07 | 6.65E-06 | 0.002215918 |
| T2DM | endothelial | *PDPR* | -0.324848113 | 0.029121795 | 0.046664632 | 1 |
| T2DM | endothelial | *NLRC4* | -0.653353516 | 1.03E-06 | 1.20E-05 | 0.006457018 |
| T2DM | endothelial | *SLC26A4* | -1.147247057 | 1.29E-07 | 4.46E-06 | 0.000813887 |
| T2DM | endothelial | *DLD* | -0.359797922 | 0.015591604 | 0.027198783 | 1 |
| T2DM | endothelial | *IL5RA* | -1.278848728 | 1.60E-07 | 4.49E-06 | 0.001002976 |
| T2DM | endothelial | *ITGA6* | 0.143803795 | 3.69E-06 | 2.80E-05 | 0.023200046 |
| T2DM | endothelial | *RAPGEF4* | 0.262782967 | 0.004346261 | 0.009149009 | 1 |
| T2DM | endothelial | *SEL1L3* | 0.308458569 | 2.78E-05 | 0.000133901 | 0.175008546 |
| T2DM | endothelial | *TF* | -0.626792766 | 8.04E-07 | 1.04E-05 | 0.005053715 |
| T2DM | endothelial | *MYO15A* | -0.566995093 | 0.003970164 | 0.008510201 | 1 |
| T2DM | endothelial | *ALKBH5* | -0.590638569 | 1.21E-05 | 6.86E-05 | 0.076320947 |
| T2DM | endothelial | *NLRP1* | 0.207427687 | 0.028572755 | 0.045919456 | 1 |
| T2DM | endothelial | *ZFHX4* | -0.360096329 | 3.82E-07 | 6.92E-06 | 0.002402035 |
| T2DM | endothelial | *RGS17* | -0.492117079 | 5.48E-08 | 4.20E-06 | 0.000344453 |
| T2DM | endothelial | *TMEM101* | -0.937368851 | 3.50E-05 | 0.000161593 | 0.219928647 |
| T2DM | endothelial | *SLC22A17* | -0.821063105 | 0.000180964 | 0.000632495 | 1 |
| T2DM | endothelial | *RPGRIP1* | -0.45010557 | 0.000104251 | 0.000398434 | 0.655424238 |
| T2DM | endothelial | *TGM1* | -0.24104729 | 0.023514279 | 0.038750793 | 1 |
| T2DM | endothelial | *AGO1* | -0.52414472 | 0.000619532 | 0.001768043 | 1 |
| T2DM | endothelial | *RFFL* | -0.712864851 | 0.001626649 | 0.003991702 | 1 |
| T2DM | endothelial | *MFSD11* | -0.32574449 | 0.002778263 | 0.006258309 | 1 |
| T2DM | endothelial | *COMT* | -0.504631172 | 0.000149377 | 0.000537839 | 0.939132676 |
| T2DM | endothelial | *TMEM38B* | -0.692816996 | 2.80E-06 | 2.32E-05 | 0.017627183 |
| T2DM | endothelial | *CWF19L1* | -0.426891807 | 3.00E-06 | 2.43E-05 | 0.018844071 |
| T2DM | endothelial | *IKZF5* | -0.459555467 | 0.005552258 | 0.011263971 | 1 |
| T2DM | endothelial | *BLNK* | -0.459809083 | 4.23E-06 | 3.12E-05 | 0.026624508 |
| T2DM | endothelial | *CREM* | -0.423877337 | 0.000967695 | 0.002571386 | 1 |
| T2DM | endothelial | *TPSD1* | -3.503457643 | 0.001550503 | 0.003833272 | 1 |

| | | | | | | |
|---|---|---|---|---|---|---|
| T2DM | endothelial | *ITPR3* | 0.344844776 | 0.002176047 | 0.00510859 | 1 |
| T2DM | endothelial | *DSP* | 0.26883727 | 3.58E-07 | 6.68E-06 | 0.002249485 |
| T2DM | endothelial | *IL12RB1* | -0.497397986 | 0.002503389 | 0.005749743 | 1 |
| T2DM | endothelial | *ACOT7* | -0.691699556 | 2.00E-05 | 0.000103315 | 0.125734403 |
| T2DM | endothelial | *PCSK5* | 0.191238572 | 6.86E-05 | 0.000280275 | 0.431329307 |
| T2DM | endothelial | *PRTFDC1* | -0.43771845 | 1.77E-07 | 4.60E-06 | 0.001112141 |
| T2DM | endothelial | *PALMD* | 0.215336716 | 0.000228818 | 0.000768471 | 1 |
| T2DM | endothelial | *OCEL1* | -0.373369045 | 0.000228251 | 0.000767952 | 1 |
| T2DM | endothelial | *CATSPERG* | -0.322657784 | 0.014925043 | 0.026217866 | 1 |
| T2DM | endothelial | *STX1B* | -0.725699998 | 7.35E-08 | 4.46E-06 | 0.000461898 |
| T2DM | endothelial | *HSD3B7* | -0.497912006 | 0.000149802 | 0.000538789 | 0.941802706 |
| T2DM | endothelial | *ATP5D* | -0.690626516 | 0.000219395 | 0.000743979 | 1 |
| T2DM | endothelial | *CDC34* | -0.752287362 | 0.000546587 | 0.001586516 | 1 |
| T2DM | endothelial | *MTAP* | -0.641933527 | 6.03E-05 | 0.000252274 | 0.379419401 |
| T2DM | endothelial | *POLR2E* | -0.670802515 | 2.19E-05 | 0.000111182 | 0.13753178 |
| T2DM | endothelial | *MADCAM1* | -0.608455806 | 6.52E-07 | 9.23E-06 | 0.004099877 |
| T2DM | endothelial | *TRMT2A* | -0.692175827 | 1.12E-06 | 1.28E-05 | 0.007067237 |
| T2DM | endothelial | *MED15* | -0.362129773 | 0.004043725 | 0.008632563 | 1 |
| T2DM | endothelial | *LZTR1* | -0.501431705 | 0.007383115 | 0.014393067 | 1 |
| T2DM | endothelial | *CECR2* | -0.374082789 | 4.82E-05 | 0.000210606 | 0.303272326 |
| T2DM | endothelial | *P2RX6* | -0.485752538 | 6.60E-08 | 4.46E-06 | 0.00041522 |
| T2DM | endothelial | *SLC7A4* | -0.718647591 | 0.001551801 | 0.003834973 | 1 |
| T2DM | endothelial | *DDT* | -0.759086008 | 0.000249641 | 0.000825616 | 1 |
| T2DM | endothelial | *GGT5* | -0.686051488 | 3.05E-05 | 0.000144957 | 0.191976597 |
| T2DM | endothelial | *SPECC1L* | -0.387783962 | 0.001822195 | 0.004401131 | 1 |
| T2DM | endothelial | *PPIL2* | -0.214660999 | 0.014618233 | 0.025758081 | 1 |
| T2DM | endothelial | *UPB1* | 0.303696826 | 4.96E-05 | 0.00021471 | 0.311548951 |
| T2DM | endothelial | *YPEL1* | -0.447389568 | 0.016046392 | 0.027883821 | 1 |
| T2DM | endothelial | *PPM1F* | 0.332596982 | 0.001634461 | 0.004009304 | 1 |
| T2DM | endothelial | *CRYBB3* | 0.351993041 | 2.77E-06 | 2.32E-05 | 0.01742135 |
| T2DM | endothelial | *CYTH4* | -0.561364969 | 1.67E-06 | 1.68E-05 | 0.010511653 |
| T2DM | endothelial | *CARD10* | 0.138823857 | 5.11E-07 | 8.20E-06 | 0.003212457 |
| T2DM | endothelial | *SLC25A1* | -0.651273965 | 1.03E-05 | 6.02E-05 | 0.064487292 |
| T2DM | endothelial | *GGA1* | -0.530760857 | 9.41E-06 | 5.66E-05 | 0.059139068 |
| T2DM | endothelial | *SH3BP1* | -0.72808404 | 1.86E-07 | 4.62E-06 | 0.001169803 |
| T2DM | endothelial | *HPS4* | -0.474961694 | 0.001161157 | 0.003005433 | 1 |
| T2DM | endothelial | *GCAT* | -0.394022946 | 0.015546798 | 0.027135681 | 1 |
| T2DM | endothelial | *ANKRD54* | -0.467330069 | 1.16E-05 | 6.63E-05 | 0.073116809 |
| T2DM | endothelial | *EIF3L* | -0.542401903 | 0.000509019 | 0.001493328 | 1 |
| T2DM | endothelial | *CCDC134* | -0.762187413 | 4.69E-05 | 0.000206146 | 0.294582699 |
| T2DM | endothelial | *SLC16A8* | -0.9846124 | 0.000396545 | 0.001209058 | 1 |
| T2DM | endothelial | *SLC5A4* | 0.058677602 | 9.67E-05 | 0.000375061 | 0.607973974 |
| T2DM | endothelial | *CYP2D6* | -0.35248522 | 0.008110715 | 0.015598674 | 1 |
| T2DM | endothelial | *POLDIP3* | -0.619104514 | 0.000181741 | 0.000634075 | 1 |
| T2DM | endothelial | *PPP6R2* | -0.395330256 | 0.00062694 | 0.001784324 | 1 |
| T2DM | endothelial | *RHBDD3* | -0.621661405 | 3.91E-05 | 0.00017766 | 0.245526658 |
| T2DM | endothelial | *TTLL1* | -0.567539694 | 0.012509778 | 0.022593787 | 1 |
| T2DM | endothelial | *RASL10A* | 0.478879612 | 0.000330666 | 0.00104153 | 1 |
| T2DM | endothelial | *AP1B1* | -0.458781202 | 0.001452847 | 0.003630385 | 1 |
| T2DM | endothelial | *TOM1* | -0.278736899 | 0.001064608 | 0.00279232 | 1 |

| | | | | | | |
|---|---|---|---|---|---|---|
| T2DM | endothelial | *NEFH* | -0.862468492 | 0.003257714 | 0.007173817 | 1 |
| T2DM | endothelial | *CHKB* | -0.324885032 | 0.002452368 | 0.005649702 | 1 |
| T2DM | endothelial | *BIK* | -2.375910744 | 3.02E-06 | 2.45E-05 | 0.018976595 |
| T2DM | endothelial | *MCAT* | -0.802067507 | 9.74E-06 | 5.80E-05 | 0.061209273 |
| T2DM | endothelial | *THOC5* | -0.60381187 | 3.19E-05 | 0.000150409 | 0.20079573 |
| T2DM | endothelial | *TSPO* | -0.42485199 | 0.002703518 | 0.006109639 | 1 |
| T2DM | endothelial | *TTLL12* | -0.984634521 | 2.01E-06 | 1.88E-05 | 0.012640386 |
| T2DM | endothelial | *MTMR3* | -0.251950426 | 0.009799427 | 0.018351804 | 1 |
| T2DM | endothelial | *FOXRED2* | -0.266585996 | 0.003913875 | 0.008398133 | 1 |
| T2DM | endothelial | *IFT27* | -0.645128876 | 0.000819271 | 0.002239459 | 1 |
| T2DM | endothelial | *KIAA0930* | -0.432013557 | 4.27E-05 | 0.000191908 | 0.268356403 |
| T2DM | endothelial | *NCF4* | -0.38163191 | 0.000222706 | 0.000753174 | 1 |
| T2DM | endothelial | *SLC25A17* | -0.453276393 | 0.000946307 | 0.002523084 | 1 |
| T2DM | endothelial | *KCTD17* | -1.717146623 | 8.98E-06 | 5.46E-05 | 0.056433417 |
| T2DM | endothelial | *EP300* | -0.336341886 | 0.003543414 | 0.007703127 | 1 |
| T2DM | endothelial | *ACO2* | -0.396550804 | 0.001078898 | 0.002818728 | 1 |
| T2DM | endothelial | *POLR3H* | -0.326207431 | 0.00082031 | 0.00224035 | 1 |
| T2DM | endothelial | *ZBED4* | -0.430510506 | 0.011952685 | 0.021743787 | 1 |
| T2DM | endothelial | *SDR39U1* | -0.380965039 | 6.38E-06 | 4.19E-05 | 0.040121907 |
| T2DM | endothelial | *RBM23* | -0.39748635 | 0.007219244 | 0.014108607 | 1 |
| T2DM | endothelial | *SOS2* | -0.306927629 | 0.006643864 | 0.013160041 | 1 |
| T2DM | endothelial | *PYGL* | -0.164555032 | 0.001882961 | 0.004525296 | 1 |
| T2DM | endothelial | *TRIM9* | -0.243326617 | 0.015505252 | 0.027076072 | 1 |
| T2DM | endothelial | *DDHD1* | -0.419531587 | 0.000133695 | 0.000492695 | 0.840537733 |
| T2DM | endothelial | *ATP6V1D* | -0.398457109 | 0.000183432 | 0.000638914 | 1 |
| T2DM | endothelial | *PIGH* | -0.376357731 | 0.00773658 | 0.014966116 | 1 |
| T2DM | endothelial | *VTI1B* | -0.622099153 | 9.84E-05 | 0.000380432 | 0.618582699 |
| T2DM | endothelial | *GSTZ1* | -0.455749044 | 0.000651016 | 0.001837869 | 1 |
| T2DM | endothelial | *TMED8* | -0.597083677 | 3.00E-05 | 0.000143075 | 0.188858378 |
| T2DM | endothelial | *ISM2* | 0.944588507 | 0.001231305 | 0.003149396 | 1 |
| T2DM | endothelial | *PPM1A* | -0.273721018 | 0.023753912 | 0.039053568 | 1 |
| T2DM | endothelial | *ZFYVE21* | 0.324687621 | 0.024112383 | 0.039560164 | 1 |
| T2DM | endothelial | *MTHFD1* | -0.518722136 | 9.61E-06 | 5.75E-05 | 0.060392159 |
| T2DM | endothelial | *TELO2* | -0.697926959 | 0.001391231 | 0.0035099 | 1 |
| T2DM | endothelial | *PAPLN* | -0.447286568 | 2.14E-06 | 1.96E-05 | 0.013482115 |
| T2DM | endothelial | *CCNB1IP1* | -0.511448239 | 0.000352069 | 0.001098419 | 1 |
| T2DM | endothelial | *PCK2* | -0.626807412 | 4.95E-07 | 8.08E-06 | 0.003113046 |
| T2DM | endothelial | *KIAA0391* | -0.900629782 | 4.38E-07 | 7.49E-06 | 0.002754872 |
| T2DM | endothelial | *PLTP* | -0.185116976 | 0.008878867 | 0.016874678 | 1 |
| T2DM | endothelial | *GSS* | -0.461589754 | 9.13E-05 | 0.000356869 | 0.57420295 |
| T2DM | endothelial | *PYGB* | -0.402142343 | 0.000215587 | 0.000733041 | 1 |
| T2DM | endothelial | *ABHD12* | -0.340959664 | 0.002389997 | 0.005536444 | 1 |
| T2DM | endothelial | *PROCR* | -0.315623183 | 0.000708054 | 0.001979341 | 1 |
| T2DM | endothelial | *NINL* | -0.360172855 | 7.04E-06 | 4.50E-05 | 0.044263848 |
| T2DM | endothelial | *CD40* | 0.396520479 | 0.000372983 | 0.001152308 | 1 |
| T2DM | endothelial | *UQCC1* | -0.417993195 | 0.00024652 | 0.000818124 | 1 |
| T2DM | endothelial | *DOK5* | -0.582612286 | 0.000423365 | 0.001278432 | 1 |
| T2DM | endothelial | *BMP7* | -0.531028144 | 1.26E-07 | 4.46E-06 | 0.000791554 |
| T2DM | endothelial | *TPD52L2* | -0.471616275 | 0.001641496 | 0.004020291 | 1 |
| T2DM | endothelial | *CTSZ* | -0.446568364 | 0.000520589 | 0.001519472 | 1 |

| | | | | | | |
|---|---|---|---|---|---|---|
| T2DM | endothelial | *PRPF6* | -0.345944676 | 0.022534385 | 0.037291807 | 1 |
| T2DM | endothelial | *MTG2* | -0.341137728 | 0.008871933 | 0.016869739 | 1 |
| T2DM | endothelial | *GID8* | -0.430535309 | 0.012576161 | 0.022681102 | 1 |
| T2DM | endothelial | *SLC17A9* | -0.686416712 | 8.44E-06 | 5.21E-05 | 0.053070626 |
| T2DM | endothelial | *BIRC7* | -0.750450432 | 1.57E-05 | 8.51E-05 | 0.098634844 |
| T2DM | endothelial | *NKAIN4* | -0.975624568 | 1.46E-07 | 4.46E-06 | 0.000918214 |
| T2DM | endothelial | *PTK6* | -1.273389751 | 1.41E-07 | 4.46E-06 | 0.000886575 |
| T2DM | endothelial | *GMEB2* | -0.516758403 | 0.009634294 | 0.018075441 | 1 |
| T2DM | endothelial | *CDC25B* | 0.331700469 | 0.004205491 | 0.008917342 | 1 |
| T2DM | endothelial | *RNF24* | -0.447865824 | 7.13E-07 | 9.74E-06 | 0.004480485 |
| T2DM | endothelial | *SLC52A3* | 0.339889292 | 1.61E-06 | 1.64E-05 | 0.010127367 |
| T2DM | endothelial | *ANGPT4* | -0.686148361 | 1.46E-07 | 4.46E-06 | 0.000919017 |
| T2DM | endothelial | *RSPO4* | -0.659990336 | 2.92E-06 | 2.38E-05 | 0.018381447 |
| T2DM | endothelial | *CCM2L* | 0.117120187 | 6.87E-07 | 9.54E-06 | 0.00431844 |
| T2DM | endothelial | *MYL9* | -0.354878202 | 0.00405672 | 0.008651492 | 1 |
| T2DM | endothelial | *MROH8* | -0.489864372 | 7.78E-07 | 1.03E-05 | 0.004894063 |
| T2DM | endothelial | *MANBAL* | -0.182541499 | 0.029169539 | 0.046711383 | 1 |
| T2DM | endothelial | *IDH3B* | -0.527041195 | 0.016283213 | 0.028191566 | 1 |
| T2DM | endothelial | *MAPRE1* | -0.432472768 | 0.025222947 | 0.041178049 | 1 |
| T2DM | endothelial | *TTI1* | -0.402105528 | 0.022404521 | 0.037126654 | 1 |
| T2DM | endothelial | *RPRD1B* | -0.44793376 | 0.00129828 | 0.003302856 | 1 |
| T2DM | endothelial | *PXMP4* | -0.681119586 | 0.000141111 | 0.000514762 | 0.887165339 |
| T2DM | endothelial | *WFDC2* | -0.981919489 | 0.017507487 | 0.029999884 | 1 |
| T2DM | endothelial | *DHX35* | -0.585148014 | 6.14E-05 | 0.000255953 | 0.386233612 |
| T2DM | endothelial | *DNTTIP1* | -0.377025863 | 0.000542426 | 0.001575166 | 1 |
| T2DM | endothelial | *APMAP* | -0.474870735 | 4.83E-06 | 3.47E-05 | 0.030371037 |
| T2DM | endothelial | *CELF4* | -0.754235942 | 5.93E-07 | 8.81E-06 | 0.003729757 |
| T2DM | endothelial | *RBFA* | -0.503625925 | 0.002931717 | 0.006552329 | 1 |
| T2DM | endothelial | *LPIN2* | -0.47112718 | 5.63E-05 | 0.00023904 | 0.354018369 |
| T2DM | endothelial | *CEP76* | -0.478126371 | 0.00189895 | 0.004555017 | 1 |
| T2DM | endothelial | *LIPG* | -1.445543699 | 0.000256184 | 0.00084098 | 1 |
| T2DM | endothelial | *NALCN* | -0.656641827 | 1.21E-05 | 6.86E-05 | 0.076265008 |
| T2DM | endothelial | *HTR2A* | -0.679202454 | 1.20E-07 | 4.46E-06 | 0.000752266 |
| T2DM | endothelial | *KLF5* | -0.472830628 | 0.009405306 | 0.017725167 | 1 |
| T2DM | endothelial | *STK24* | -0.357549703 | 0.00576302 | 0.011616579 | 1 |
| T2DM | endothelial | *DNAJC3* | -0.328631635 | 0.01472422 | 0.025932524 | 1 |
| T2DM | endothelial | *ARHGEF7* | -0.259121984 | 0.024379462 | 0.039967061 | 1 |
| T2DM | endothelial | *SGCG* | -0.369290317 | 3.30E-07 | 6.45E-06 | 0.002077059 |
| T2DM | endothelial | *KATNAL1* | -0.399431237 | 0.000685384 | 0.001922807 | 1 |
| T2DM | endothelial | *DHRS12* | -0.460617652 | 0.004277745 | 0.009049187 | 1 |
| T2DM | endothelial | *MEDAG* | -0.546579564 | 0.000130687 | 0.000485025 | 0.821631964 |
| T2DM | endothelial | *MGRN1* | -0.318131594 | 0.001910134 | 0.004576604 | 1 |
| T2DM | endothelial | *CORO1A* | -0.706857912 | 3.87E-07 | 6.99E-06 | 0.002432471 |
| T2DM | endothelial | *GDPD3* | 0.476485366 | 0.000143205 | 0.000520862 | 0.900329817 |
| T2DM | endothelial | *PLLP* | 0.220485691 | 1.69E-06 | 1.68E-05 | 0.010636965 |
| T2DM | endothelial | *ACD* | -0.474904208 | 0.002320545 | 0.005395439 | 1 |
| T2DM | endothelial | *USB1* | -0.840233204 | 3.88E-05 | 0.000176958 | 0.244202094 |
| T2DM | endothelial | *COG4* | -0.419640408 | 0.002853809 | 0.006400963 | 1 |
| T2DM | endothelial | *SLC7A6OS* | -0.241806548 | 0.000817055 | 0.002235346 | 1 |
| T2DM | endothelial | *HCFC1R1* | -0.781305439 | 0.000528514 | 0.001540459 | 1 |

| | | | | | | |
|---|---|---|---|---|---|---|
| T2DM | endothelial | *NAGPA* | -0.599697615 | 0.000286334 | 0.000923644 | 1 |
| T2DM | endothelial | *COTL1* | -0.537791397 | 5.68E-07 | 8.58E-06 | 0.00357103 |
| T2DM | endothelial | *CRISPLD2* | -0.299195775 | 0.000847057 | 0.002303395 | 1 |
| T2DM | endothelial | *NME4* | -0.71443428 | 3.50E-07 | 6.64E-06 | 0.002199921 |
| T2DM | endothelial | *NOMO3* | -0.765317828 | 0.000112571 | 0.000425188 | 0.70773689 |
| T2DM | endothelial | *NARFL* | -0.581632534 | 0.001006527 | 0.002657723 | 1 |
| T2DM | endothelial | *CLCN7* | -0.657304742 | 1.21E-05 | 6.86E-05 | 0.076263054 |
| T2DM | endothelial | *HAGHL* | -0.728059143 | 6.26E-08 | 4.46E-06 | 0.000393574 |
| T2DM | endothelial | *SLC7A5* | -1.538308046 | 3.54E-07 | 6.65E-06 | 0.002223044 |
| T2DM | endothelial | *FBXO31* | 0.254993725 | 0.014135522 | 0.025026761 | 1 |
| T2DM | endothelial | *STUB1* | -0.687446522 | 2.19E-05 | 0.000111454 | 0.137980161 |
| T2DM | endothelial | *RHBDL1* | -0.485532397 | 1.02E-06 | 1.20E-05 | 0.006382832 |
| T2DM | endothelial | *NUBP1* | -0.469913557 | 0.017338772 | 0.029775706 | 1 |
| T2DM | endothelial | *EARS2* | -0.336214936 | 6.54E-05 | 0.000269165 | 0.411015478 |
| T2DM | endothelial | *CPPED1* | -0.453446309 | 0.000965304 | 0.002566116 | 1 |
| T2DM | endothelial | *BFAR* | -0.405093525 | 0.014813934 | 0.026059094 | 1 |
| T2DM | endothelial | *STX4* | -0.511835619 | 0.000778719 | 0.002143523 | 1 |
| T2DM | endothelial | *KAT8* | -0.458276064 | 0.003724464 | 0.008049401 | 1 |
| T2DM | endothelial | *NOMO1* | -0.752350091 | 0.001160939 | 0.003005433 | 1 |
| T2DM | endothelial | *SYT17* | -0.464667672 | 2.64E-08 | 3.16E-06 | 0.000165924 |
| T2DM | endothelial | *TMC5* | -0.738733171 | 3.02E-08 | 3.24E-06 | 0.000190059 |
| T2DM | endothelial | *RNF40* | -0.450278668 | 0.015217354 | 0.026671732 | 1 |
| T2DM | endothelial | *IQCH* | -0.391942316 | 0.016155531 | 0.028034729 | 1 |
| T2DM | endothelial | *LACTB* | -0.566978287 | 7.12E-05 | 0.000288609 | 0.44734406 |
| T2DM | endothelial | *CSK* | -0.613567274 | 0.0019699 | 0.004692976 | 1 |
| T2DM | endothelial | *TRIP4* | -0.347191363 | 0.00260296 | 0.00593573 | 1 |
| T2DM | endothelial | *RASL12* | -0.614821257 | 2.11E-07 | 4.94E-06 | 0.001328136 |
| T2DM | endothelial | *AP3B2* | -0.207223398 | 0.002339317 | 0.005435065 | 1 |
| T2DM | endothelial | *IGDCC4* | 0.094035058 | 0.010157531 | 0.018899201 | 1 |
| T2DM | endothelial | *CTSH* | 0.210677691 | 0.000677051 | 0.001902825 | 1 |
| T2DM | endothelial | *FAH* | -0.449109255 | 2.49E-07 | 5.43E-06 | 0.001565276 |
| T2DM | endothelial | *RPAP1* | -0.701911263 | 6.97E-05 | 0.000284107 | 0.438376745 |
| T2DM | endothelial | *HOMER2* | -0.418660697 | 0.00540503 | 0.011011479 | 1 |
| T2DM | endothelial | *EHD4* | 0.300214861 | 0.002069851 | 0.004897687 | 1 |
| T2DM | endothelial | *ATP8B4* | -0.41503094 | 1.73E-06 | 1.70E-05 | 0.010845325 |
| T2DM | endothelial | *DTWD1* | -0.291190473 | 0.007369876 | 0.014376175 | 1 |
| T2DM | endothelial | *PDGFRL* | -0.397547319 | 1.70E-05 | 9.08E-05 | 0.10714532 |
| T2DM | endothelial | *RIPK2* | -0.459323281 | 0.002238155 | 0.005234851 | 1 |
| T2DM | endothelial | *NBN* | -0.421522005 | 0.001474523 | 0.003677241 | 1 |
| T2DM | endothelial | *CPQ* | -0.257468531 | 0.001985417 | 0.004725322 | 1 |
| T2DM | endothelial | *IKBKB* | 0.256657238 | 0.016307066 | 0.028212032 | 1 |
| T2DM | endothelial | *PLAT* | 0.183883383 | 2.26E-06 | 2.02E-05 | 0.014230444 |
| T2DM | endothelial | *GDAP1* | 0.339019817 | 0.000571724 | 0.001652612 | 1 |
| T2DM | endothelial | *ARMC1* | -0.326488081 | 0.000596778 | 0.001714782 | 1 |
| T2DM | endothelial | *CHRAC1* | -0.617564869 | 0.012884348 | 0.023150585 | 1 |
| T2DM | endothelial | *SNX16* | -0.478647792 | 0.001215739 | 0.00312101 | 1 |
| T2DM | endothelial | *GSDMD* | 0.297535483 | 0.017992473 | 0.030697063 | 1 |
| T2DM | endothelial | *ERI1* | -0.485084107 | 0.001695233 | 0.004136324 | 1 |
| T2DM | endothelial | *TNFRSF10A* | 0.226845169 | 9.39E-07 | 1.13E-05 | 0.005903392 |
| T2DM | endothelial | *TUSC3* | 0.144652149 | 1.83E-06 | 1.77E-05 | 0.011506059 |

| | | | | | | |
|---|---|---|---|---|---|---|
| T2DM | endothelial | *KLHDC4* | -0.310048143 | 0.014364604 | 0.025368052 | 1 |
| T2DM | endothelial | *MCM4* | -0.468375017 | 0.008198975 | 0.015758775 | 1 |
| T2DM | endothelial | *MAN2B1* | -0.3239622 | 0.005679883 | 0.01147844 | 1 |
| T2DM | endothelial | *TULP2* | 0.464352294 | 0.017579303 | 0.030090139 | 1 |
| T2DM | endothelial | *NFKBIB* | -0.952784199 | 1.02E-05 | 6.01E-05 | 0.064143222 |
| T2DM | endothelial | *TUBB4A* | -1.710559299 | 0.001985732 | 0.004725322 | 1 |
| T2DM | endothelial | *SARS2* | -0.470261411 | 4.08E-05 | 0.000184409 | 0.256513593 |
| T2DM | endothelial | *SNRNP70* | -0.412054911 | 0.00025434 | 0.00083645 | 1 |
| T2DM | endothelial | *PPP1R37* | -0.774997307 | 5.84E-06 | 3.93E-05 | 0.036702736 |
| T2DM | endothelial | *FCGRT* | -0.317804475 | 0.004189231 | 0.008891862 | 1 |
| T2DM | endothelial | *PEX11G* | -0.641743592 | 4.69E-05 | 0.00020632 | 0.295057833 |
| T2DM | endothelial | *ERCC2* | -0.881917256 | 8.68E-07 | 1.08E-05 | 0.00545977 |
| T2DM | endothelial | *PLEKHJ1* | -0.735391491 | 1.55E-05 | 8.40E-05 | 0.097166735 |
| T2DM | endothelial | *CD37* | -0.558685558 | 0.000406358 | 0.001232998 | 1 |
| T2DM | endothelial | *AMH* | -0.652631572 | 0.000345957 | 0.001082644 | 1 |
| T2DM | endothelial | *DKKL1* | 0.458806216 | 0.009393834 | 0.017714167 | 1 |
| T2DM | endothelial | *STX10* | -0.648813708 | 1.52E-05 | 8.31E-05 | 0.095852637 |
| T2DM | endothelial | *FCER2* | -0.897103081 | 2.88E-08 | 3.21E-06 | 0.00018086 |
| T2DM | endothelial | *DMPK* | -0.691127061 | 6.51E-05 | 0.00026881 | 0.409397203 |
| T2DM | endothelial | *IL4I1* | -0.375593987 | 0.004024471 | 0.008603145 | 1 |
| T2DM | endothelial | *C19orf53* | -0.612835053 | 0.006379656 | 0.012680651 | 1 |
| T2DM | endothelial | *TIMM44* | -0.417800491 | 0.004310948 | 0.009104108 | 1 |
| T2DM | endothelial | *CCDC61* | -0.774432035 | 4.47E-05 | 0.000198297 | 0.281184517 |
| T2DM | endothelial | *IL27RA* | 0.480827906 | 4.71E-05 | 0.000206538 | 0.295968796 |
| T2DM | endothelial | *C19orf44* | -0.220274602 | 0.017157564 | 0.029512888 | 1 |
| T2DM | endothelial | *OLFM2* | -0.2272799 | 0.000322906 | 0.001021183 | 1 |
| T2DM | endothelial | *RASAL3* | -0.563758505 | 0.000485507 | 0.001439659 | 1 |
| T2DM | endothelial | *ZNF419* | 0.242874315 | 0.004036139 | 0.008622224 | 1 |
| T2DM | endothelial | *CCNE1* | -0.868480479 | 1.24E-07 | 4.46E-06 | 0.000781286 |
| T2DM | endothelial | *ANKRD27* | -0.365985376 | 0.012209749 | 0.02211544 | 1 |
| T2DM | endothelial | *PRX* | -0.579921578 | 0.017984804 | 0.030697063 | 1 |
| T2DM | endothelial | *CCDC94* | -0.571924691 | 0.009829672 | 0.018376196 | 1 |
| T2DM | endothelial | *CLIP3* | -0.336276513 | 0.000959161 | 0.002551944 | 1 |
| T2DM | endothelial | *SLC1A5* | -0.496926743 | 0.000283998 | 0.00091752 | 1 |
| T2DM | endothelial | *PRKD2* | 0.218764214 | 0.013066105 | 0.023423611 | 1 |
| T2DM | endothelial | *CACTIN* | -1.017390644 | 0.000196163 | 0.000675746 | 1 |
| T2DM | endothelial | *FZR1* | -0.726114596 | 0.000944406 | 0.002519087 | 1 |
| T2DM | endothelial | *BBC3* | -1.043183143 | 0.000624707 | 0.001780388 | 1 |
| T2DM | endothelial | *CEACAM4* | -1.286506753 | 1.21E-07 | 4.46E-06 | 0.000758936 |
| T2DM | endothelial | *PLIN3* | -0.614802071 | 8.32E-05 | 0.000328994 | 0.522770776 |
| T2DM | endothelial | *ICAM5* | 0.831780056 | 3.32E-05 | 0.000155218 | 0.208923644 |
| T2DM | endothelial | *CD33* | -0.498308782 | 4.07E-06 | 3.03E-05 | 0.025601951 |
| T2DM | endothelial | *BABAM1* | -0.696180831 | 0.003762896 | 0.008121292 | 1 |
| T2DM | endothelial | *TYK2* | -0.351806547 | 0.022812744 | 0.037723231 | 1 |
| T2DM | endothelial | *PTPRS* | -0.287009498 | 2.73E-05 | 0.000131899 | 0.171468118 |
| T2DM | endothelial | *GRWD1* | -0.464134617 | 2.53E-06 | 2.17E-05 | 0.015896907 |
| T2DM | endothelial | *GRIN2D* | -0.69482675 | 1.70E-07 | 4.55E-06 | 0.001068503 |
| T2DM | endothelial | *CARD8* | 0.217383 | 0.019572056 | 0.033033428 | 1 |
| T2DM | endothelial | *ZNF175* | -0.310708281 | 0.028234113 | 0.045514837 | 1 |
| T2DM | endothelial | *CAPS* | -0.429302022 | 0.000857112 | 0.002325726 | 1 |

| | | | | | | |
|---|---|---|---|---|---|---|
| T2DM | endothelial | *PPP2R1A* | -0.411908763 | 0.015383589 | 0.026903095 | 1 |
| T2DM | endothelial | *TNPO2* | -0.427011654 | 0.002313316 | 0.005384605 | 1 |
| T2DM | endothelial | *GCDH* | -0.838952423 | 1.95E-09 | 6.14E-07 | 1.23E-05 |
| T2DM | endothelial | *LILRB5* | -0.513850634 | 1.11E-06 | 1.27E-05 | 0.006995428 |
| T2DM | endothelial | *MAST1* | -1.997236121 | 0.000481578 | 0.001429844 | 1 |
| T2DM | endothelial | *PRPF31* | -0.528729514 | 0.000359277 | 0.001117653 | 1 |
| T2DM | endothelial | *PDE4C* | -0.507874125 | 0.004225664 | 0.008948047 | 1 |
| T2DM | endothelial | *ELL* | -0.345413588 | 0.014803614 | 0.026048229 | 1 |
| T2DM | endothelial | *UPK1A* | -1.089117487 | 2.36E-05 | 0.000118517 | 0.148620854 |
| T2DM | endothelial | *COPE* | -0.6377566 | 0.000655794 | 0.001848038 | 1 |
| T2DM | endothelial | *LSR* | 0.230744752 | 3.55E-05 | 0.000163614 | 0.223006249 |
| T2DM | endothelial | *KXD1* | -0.581879844 | 0.003175736 | 0.007022811 | 1 |
| T2DM | endothelial | *HPN* | -0.997530211 | 1.52E-06 | 1.59E-05 | 0.009551738 |
| T2DM | endothelial | *ZNF14* | -0.487681898 | 0.012825872 | 0.023065291 | 1 |
| T2DM | endothelial | *SIPA1L3* | -0.316622907 | 0.01272874 | 0.022903717 | 1 |
| T2DM | endothelial | *GTPBP10* | -0.253975506 | 0.016465231 | 0.028435152 | 1 |
| T2DM | endothelial | *RASA4* | 0.250210515 | 1.36E-07 | 4.46E-06 | 0.000856882 |
| T2DM | endothelial | *PON3* | -0.702496397 | 0.013850434 | 0.024633007 | 1 |
| T2DM | endothelial | *HBP1* | -0.327347003 | 0.012725046 | 0.022903626 | 1 |
| T2DM | endothelial | *WDR91* | -0.626313462 | 1.63E-05 | 8.74E-05 | 0.102303819 |
| T2DM | endothelial | *STEAP1B* | 0.252620256 | 6.36E-07 | 9.11E-06 | 0.00400021 |
| T2DM | endothelial | *PTN* | -0.705920627 | 1.57E-06 | 1.61E-05 | 0.009865173 |
| T2DM | endothelial | *MPP6* | -0.409696097 | 3.54E-07 | 6.65E-06 | 0.00222704 |
| T2DM | endothelial | *DFNA5* | -0.392571848 | 9.61E-06 | 5.75E-05 | 0.060431603 |
| T2DM | endothelial | *OGDH* | -0.39054176 | 0.00141435 | 0.003556808 | 1 |
| T2DM | endothelial | *ADAP1* | -0.82258382 | 1.24E-07 | 4.46E-06 | 0.000780779 |
| T2DM | endothelial | *MET* | 0.106105448 | 1.23E-06 | 1.37E-05 | 0.007746747 |
| T2DM | endothelial | *RNF32* | 0.245848747 | 0.000473657 | 0.001409982 | 1 |
| T2DM | endothelial | *HOXA2* | -1.014478296 | 8.06E-07 | 1.04E-05 | 0.005064551 |
| T2DM | endothelial | *HOXA5* | -0.485514689 | 3.12E-06 | 2.48E-05 | 0.01963757 |
| T2DM | endothelial | *HOXA6* | -0.578808557 | 2.24E-06 | 2.01E-05 | 0.014080993 |
| T2DM | endothelial | *BRAT1* | -0.433613537 | 0.000264924 | 0.000864335 | 1 |
| T2DM | endothelial | *CPED1* | -0.364676281 | 3.84E-06 | 2.90E-05 | 0.024126322 |
| T2DM | endothelial | *HIBADH* | -0.388560106 | 0.00032094 | 0.001015986 | 1 |
| T2DM | endothelial | *CPVL* | -0.285347791 | 0.013189055 | 0.023617085 | 1 |
| T2DM | endothelial | *ABHD11* | -0.330999958 | 0.001663908 | 0.004065679 | 1 |
| T2DM | endothelial | *PLEKHA8* | -0.488858526 | 0.001507793 | 0.003748318 | 1 |
| T2DM | endothelial | *NOD1* | 0.272466875 | 0.004129897 | 0.008789661 | 1 |
| T2DM | endothelial | *CRHR2* | 2.081200943 | 2.35E-06 | 2.08E-05 | 0.014778149 |
| T2DM | endothelial | *EPHB6* | -0.374974932 | 2.17E-06 | 1.98E-05 | 0.013649917 |
| T2DM | endothelial | *CYP3A5* | -0.608644385 | 1.81E-05 | 9.48E-05 | 0.113849917 |
| T2DM | endothelial | *SNX8* | -0.531058422 | 0.000190314 | 0.000659533 | 1 |
| T2DM | endothelial | *AIMP2* | -0.603781449 | 0.00035892 | 0.001117249 | 1 |
| T2DM | endothelial | *TFR2* | -0.629641751 | 0.01023977 | 0.019040945 | 1 |
| T2DM | endothelial | *RBM28* | -0.415758414 | 9.87E-05 | 0.00038115 | 0.620512851 |
| T2DM | endothelial | *LSM5* | -0.545487158 | 0.004391436 | 0.009215273 | 1 |
| T2DM | endothelial | *AP1S1* | -0.904877373 | 0.010463891 | 0.019400319 | 1 |
| T2DM | endothelial | *C1GALT1* | -0.408870892 | 0.000939318 | 0.002509773 | 1 |
| T2DM | endothelial | *ZNF862* | -0.347818723 | 0.00905358 | 0.017164112 | 1 |
| T2DM | endothelial | *MEST* | -0.321609611 | 1.44E-07 | 4.46E-06 | 0.000907247 |

| | | | | | | |
|---|---|---|---|---|---|---|
| T2DM | endothelial | *ACTR3C* | -0.455346114 | 2.61E-05 | 0.000128256 | 0.164167771 |
| T2DM | endothelial | *POU6F2* | -0.425393366 | 1.54E-07 | 4.49E-06 | 0.000970675 |
| T2DM | endothelial | *TSPAN13* | 0.191812799 | 1.54E-06 | 1.60E-05 | 0.009697543 |
| T2DM | endothelial | *RARRES2* | -0.445723424 | 2.41E-05 | 0.000119929 | 0.151230486 |
| T2DM | endothelial | *MRPL32* | -0.257353618 | 0.016289722 | 0.028191566 | 1 |
| T2DM | endothelial | *BLVRA* | -0.404632707 | 0.000166261 | 0.000588893 | 1 |
| T2DM | endothelial | *URGCP* | -0.476046804 | 0.00095463 | 0.002540966 | 1 |
| T2DM | endothelial | *POLD2* | -0.861750506 | 2.67E-09 | 7.82E-07 | 1.68E-05 |
| T2DM | endothelial | *GCK* | -3.84532531 | 1.07E-07 | 4.46E-06 | 0.000670667 |
| T2DM | endothelial | *TBL2* | -0.172265887 | 0.022529339 | 0.037291807 | 1 |
| T2DM | endothelial | *CLIP2* | -0.37802612 | 0.030479865 | 0.048648619 | 1 |
| T2DM | endothelial | *LIMK1* | -0.689976817 | 1.65E-06 | 1.66E-05 | 0.010380471 |
| T2DM | endothelial | *SPATA6L* | 0.197297346 | 5.74E-06 | 3.90E-05 | 0.03610517 |
| T2DM | endothelial | *SLC1A1* | 0.251303528 | 1.00E-05 | 5.94E-05 | 0.063163168 |
| T2DM | endothelial | *MEGF9* | -0.40426556 | 2.03E-05 | 0.000104458 | 0.127647821 |
| T2DM | endothelial | *CORO2A* | 0.241552001 | 7.55E-06 | 4.76E-05 | 0.047439229 |
| T2DM | endothelial | *C5* | -0.397262923 | 5.81E-06 | 3.92E-05 | 0.036555068 |
| T2DM | endothelial | *OGN* | -0.483733776 | 2.70E-05 | 0.000131067 | 0.169994293 |
| T2DM | endothelial | *ECM2* | -0.344607214 | 2.89E-06 | 2.37E-05 | 0.018178567 |
| T2DM | endothelial | *SUSD1* | -0.24421983 | 0.017114845 | 0.029447463 | 1 |
| T2DM | endothelial | *TNFSF8* | -0.671460348 | 2.25E-07 | 5.17E-06 | 0.001417344 |
| T2DM | endothelial | *AK1* | -0.52739646 | 0.004026954 | 0.008605526 | 1 |
| T2DM | endothelial | *PLGRKT* | -0.287538746 | 0.006066848 | 0.012170477 | 1 |
| T2DM | endothelial | *DOCK8* | -0.259397489 | 0.000884346 | 0.002389563 | 1 |
| T2DM | endothelial | *ELAVL2* | 0.353454158 | 0.001381774 | 0.003487441 | 1 |
| T2DM | endothelial | *FUBP3* | -0.33884312 | 0.019752273 | 0.033259418 | 1 |
| T2DM | endothelial | *MPDZ* | 0.224122366 | 0.016242042 | 0.028155557 | 1 |
| T2DM | endothelial | *PTGDS* | -0.723007255 | 2.07E-07 | 4.93E-06 | 0.001301052 |
| T2DM | endothelial | *SHB* | 0.759408662 | 2.22E-06 | 2.01E-05 | 0.013984821 |
| T2DM | endothelial | *DVL1* | -0.611117854 | 0.004524149 | 0.009455893 | 1 |
| T2DM | endothelial | *PDLIM1* | 0.254071109 | 8.91E-05 | 0.000348725 | 0.560052822 |
| T2DM | endothelial | *ATRNL1* | -0.402964877 | 6.39E-05 | 0.000264548 | 0.401848489 |
| T2DM | endothelial | *HPS1* | -0.575821378 | 2.46E-05 | 0.000122388 | 0.154820586 |
| T2DM | endothelial | *PHYH* | -0.219217764 | 0.0035127 | 0.007641643 | 1 |
| T2DM | endothelial | *DNMBP* | -0.379971048 | 0.00478991 | 0.009909235 | 1 |
| T2DM | endothelial | *DDX50* | -0.353523795 | 0.012860997 | 0.023115233 | 1 |
| T2DM | endothelial | *SPOCK2* | -0.357980354 | 0.003793614 | 0.008181974 | 1 |
| T2DM | endothelial | *BMPR1A* | -0.247886968 | 0.000132781 | 0.000491305 | 0.834793626 |
| T2DM | endothelial | *KAZALD1* | 0.63977556 | 0.000762816 | 0.002105277 | 1 |
| T2DM | endothelial | *ACBD5* | -0.414817326 | 0.003716763 | 0.00803552 | 1 |
| T2DM | endothelial | *MTPAP* | -0.504289692 | 0.003616275 | 0.007853376 | 1 |
| T2DM | endothelial | *MAP3K8* | -0.357190096 | 0.006789049 | 0.013413811 | 1 |
| T2DM | endothelial | *TFAM* | -0.684366284 | 0.00025707 | 0.000843087 | 1 |
| T2DM | endothelial | *PBLD* | -0.130935442 | 0.019725861 | 0.033259418 | 1 |
| T2DM | endothelial | *DHRS11* | -0.457362548 | 3.38E-07 | 6.50E-06 | 0.002126118 |
| T2DM | endothelial | *PSMB3* | -0.40299124 | 0.014075955 | 0.024956439 | 1 |
| T2DM | endothelial | *RAPGEFL1* | -0.398432802 | 0.006798956 | 0.013429166 | 1 |
| T2DM | endothelial | *RGS9* | -0.797932995 | 5.32E-07 | 8.34E-06 | 0.003345498 |
| T2DM | endothelial | *WNT3* | -1.192512236 | 1.24E-07 | 4.46E-06 | 0.000778668 |
| T2DM | endothelial | *ASPA* | -0.373257047 | 2.49E-07 | 5.43E-06 | 0.001568311 |

| | | | | | | |
|---|---|---|---|---|---|---|
| T2DM | endothelial | *RAD51C* | -0.346386439 | 0.000107019 | 0.000407529 | 0.672830434 |
| T2DM | endothelial | *SEPT4* | -0.795964039 | 3.70E-06 | 2.81E-05 | 0.023278629 |
| T2DM | endothelial | *TRIM37* | -0.346067356 | 0.016245033 | 0.028155557 | 1 |
| T2DM | endothelial | *RPS6KB1* | -0.397006455 | 0.00485713 | 0.010019252 | 1 |
| T2DM | endothelial | *TRIM16L* | -0.48015531 | 1.27E-06 | 1.41E-05 | 0.008013955 |
| T2DM | endothelial | *CDK5RAP3* | -0.455321951 | 0.017240455 | 0.029639251 | 1 |
| T2DM | endothelial | *RECQL5* | -0.377958784 | 0.007217657 | 0.014108607 | 1 |
| T2DM | endothelial | *PIGL* | -0.330652714 | 0.010629341 | 0.019656694 | 1 |
| T2DM | endothelial | *GALK1* | 0.513477918 | 0.030709881 | 0.048966022 | 1 |
| T2DM | endothelial | *CAMTA2* | -0.330013468 | 0.031432927 | 0.049975103 | 1 |
| T2DM | endothelial | *MED13* | -0.281263379 | 0.008926228 | 0.016949319 | 1 |
| T2DM | endothelial | *GOSR1* | -0.304427794 | 0.024800272 | 0.040593416 | 1 |
| T2DM | endothelial | *DRG2* | -0.621746098 | 0.003343672 | 0.007337405 | 1 |
| T2DM | endothelial | *FTSJ3* | -0.233086074 | 0.000104657 | 0.000399577 | 0.657979461 |
| T2DM | endothelial | *AKAP10* | -0.331584963 | 0.004756342 | 0.009859256 | 1 |
| T2DM | endothelial | *SMARCD2* | -0.638549689 | 2.15E-06 | 1.96E-05 | 0.013503689 |
| T2DM | endothelial | *ICAM2* | 0.177558692 | 4.65E-07 | 7.74E-06 | 0.002923861 |
| T2DM | endothelial | *UTP6* | -0.453717216 | 0.001712098 | 0.004172078 | 1 |
| T2DM | endothelial | *NAGLU* | -0.636482813 | 1.93E-06 | 1.83E-05 | 0.012114588 |
| T2DM | endothelial | *HSD17B1* | -0.330185137 | 3.52E-08 | 3.41E-06 | 0.00022157 |
| T2DM | endothelial | *DLX4* | -2.264192517 | 1.29E-07 | 4.46E-06 | 0.000808573 |
| T2DM | endothelial | *LRRC59* | -0.845265775 | 6.52E-05 | 0.000268847 | 0.410132252 |
| T2DM | endothelial | *MPP2* | 0.78343134 | 0.000228694 | 0.000768467 | 1 |
| T2DM | endothelial | *EFNB3* | -3.258165161 | 2.76E-05 | 0.000133046 | 0.173625594 |
| T2DM | endothelial | *FAM20A* | -0.348387588 | 0.009333237 | 0.01761575 | 1 |
| T2DM | endothelial | *DPH1* | -0.929031138 | 2.40E-05 | 0.000119809 | 0.150668421 |
| T2DM | endothelial | *DHRS7B* | -0.311803461 | 0.023853557 | 0.03918665 | 1 |
| T2DM | endothelial | *RCVRN* | -1.022436788 | 3.72E-05 | 0.000170735 | 0.233736009 |
| T2DM | endothelial | *SLC9A3R1* | -0.475811162 | 0.011547354 | 0.021097999 | 1 |
| T2DM | endothelial | *MYH3* | -0.594268185 | 0.000781985 | 0.002150629 | 1 |
| T2DM | endothelial | *TMEM104* | -0.554922758 | 7.01E-06 | 4.49E-05 | 0.044094776 |
| T2DM | endothelial | *RAB34* | -0.568968442 | 0.000611278 | 0.001747661 | 1 |
| T2DM | endothelial | *CWH43* | -1.931885114 | 2.27E-06 | 2.03E-05 | 0.014256729 |
| T2DM | endothelial | *AREG* | 0.259450081 | 8.25E-06 | 5.13E-05 | 0.051871385 |
| T2DM | endothelial | *CPE* | -0.35012298 | 0.007874644 | 0.015200457 | 1 |
| T2DM | endothelial | *GRPEL1* | -0.670307458 | 0.006889197 | 0.013574982 | 1 |
| T2DM | endothelial | *AADAT* | -0.482799871 | 3.18E-07 | 6.29E-06 | 0.001996713 |
| T2DM | endothelial | *CPZ* | -4.515660531 | 1.28E-07 | 4.46E-06 | 0.000805829 |
| T2DM | endothelial | *SLC2A9* | -0.428522329 | 0.001545671 | 0.003824334 | 1 |
| T2DM | endothelial | *SH3D19* | -0.245778349 | 0.00186037 | 0.004479565 | 1 |
| T2DM | endothelial | *RAPGEF2* | 0.237803505 | 0.002755872 | 0.006214551 | 1 |
| T2DM | endothelial | *LRP2BP* | 0.293261035 | 0.004960356 | 0.010211447 | 1 |
| T2DM | endothelial | *KLHL5* | 0.210697697 | 0.008679408 | 0.016535588 | 1 |
| T2DM | endothelial | *FAM149A* | 0.262902771 | 0.000913684 | 0.002452745 | 1 |
| T2DM | endothelial | *UGDH* | -0.552086427 | 6.78E-06 | 4.39E-05 | 0.042596019 |
| T2DM | endothelial | *DDX25* | 0.570501898 | 1.44E-06 | 1.53E-05 | 0.00907271 |
| T2DM | endothelial | *SC5D* | -0.689228335 | 1.02E-05 | 6.01E-05 | 0.064286794 |
| T2DM | endothelial | *C11orf63* | -0.555887669 | 4.14E-05 | 0.000186941 | 0.260221843 |
| T2DM | endothelial | *SNX15* | -0.255850307 | 0.007125932 | 0.013943584 | 1 |
| T2DM | endothelial | *DCPS* | -0.459869431 | 0.016151463 | 0.028034729 | 1 |

| | | | | | | |
|---|---|---|---|---|---|---|
| T2DM | endothelial | *FOXRED1* | -0.304582269 | 0.002293589 | 0.005344624 | 1 |
| T2DM | endothelial | *NRXN2* | -0.691746952 | 1.25E-07 | 4.46E-06 | 0.000788811 |
| T2DM | endothelial | *MS4A4A* | -0.234194375 | 0.003852988 | 0.008290121 | 1 |
| T2DM | endothelial | *PRPF19* | -0.624076117 | 0.00050697 | 0.001488415 | 1 |
| T2DM | endothelial | *HPX* | 0.344154063 | 0.016163286 | 0.028040447 | 1 |
| T2DM | endothelial | *CHORDC1* | -0.347777804 | 0.026235727 | 0.042577185 | 1 |
| T2DM | endothelial | *PANX1* | -0.624883326 | 1.94E-05 | 0.000100825 | 0.122199664 |
| T2DM | endothelial | *GALNT18* | 0.119126778 | 3.82E-06 | 2.89E-05 | 0.023999755 |
| T2DM | endothelial | *MADD* | -0.447062304 | 0.00349532 | 0.007609099 | 1 |
| T2DM | endothelial | *NAA40* | -0.530730966 | 0.005749554 | 0.011593151 | 1 |
| T2DM | endothelial | *C11orf21* | -0.61098879 | 0.000182574 | 0.00063663 | 1 |
| T2DM | endothelial | *AIP* | -0.534589125 | 0.00903364 | 0.01714292 | 1 |
| T2DM | endothelial | *NDUFS8* | -0.612513681 | 0.000868471 | 0.002353482 | 1 |
| T2DM | endothelial | *GTF2H1* | -0.347817019 | 0.007703053 | 0.014905845 | 1 |
| T2DM | endothelial | *PTPN5* | 0.230588037 | 1.53E-06 | 1.60E-05 | 0.009607069 |
| T2DM | endothelial | *PPFIBP1* | 0.249313143 | 0.003086802 | 0.006857499 | 1 |
| T2DM | endothelial | *ASIC1* | -0.505311939 | 2.56E-08 | 3.16E-06 | 0.000161065 |
| T2DM | endothelial | *CAPRIN2* | -0.485252966 | 0.001517057 | 0.003763905 | 1 |
| T2DM | endothelial | *CYP27B1* | -1.808938511 | 2.41E-07 | 5.33E-06 | 0.001517018 |
| T2DM | endothelial | *ACSS3* | -0.14521981 | 0.002254978 | 0.005270279 | 1 |
| T2DM | endothelial | *GLI1* | -0.657778584 | 1.17E-07 | 4.46E-06 | 0.000733796 |
| T2DM | endothelial | *METAP2* | -0.350420472 | 0.009802029 | 0.018351804 | 1 |
| T2DM | endothelial | *TRPV4* | -0.796898409 | 1.34E-05 | 7.43E-05 | 0.083944564 |
| T2DM | endothelial | *ITFG2* | -0.462302722 | 0.000111716 | 0.000422597 | 0.702356668 |
| T2DM | endothelial | *PARP11* | -0.374293054 | 0.008782223 | 0.016721332 | 1 |
| T2DM | endothelial | *GPN3* | -0.712809232 | 2.62E-05 | 0.000128359 | 0.164427407 |
| T2DM | endothelial | *VPS29* | -0.35177669 | 0.010122678 | 0.018839929 | 1 |
| T2DM | endothelial | *AKAP3* | -0.402649688 | 9.10E-06 | 5.52E-05 | 0.057208971 |
| T2DM | endothelial | *KCNA1* | -0.898837813 | 1.34E-07 | 4.46E-06 | 0.000840137 |
| T2DM | endothelial | *CREBL2* | -0.360050712 | 0.006749554 | 0.013344165 | 1 |
| T2DM | endothelial | *NAA25* | -0.363330618 | 0.005196418 | 0.010634726 | 1 |
| T2DM | endothelial | *CDK2AP1* | 0.325046167 | 3.58E-05 | 0.000164702 | 0.224817615 |
| T2DM | endothelial | *RASAL1* | -1.274812321 | 3.20E-05 | 0.000150476 | 0.201170532 |
| T2DM | endothelial | *EIF2B1* | -0.459027835 | 0.006018025 | 0.012084101 | 1 |
| T2DM | endothelial | *DDX55* | -0.370113664 | 0.000461686 | 0.001378261 | 1 |
| T2DM | endothelial | *CAND1* | -0.343561827 | 0.004346761 | 0.009149009 | 1 |
| T2DM | endothelial | *MDM1* | -0.302087278 | 0.025588908 | 0.041721333 | 1 |
| T2DM | endothelial | *KRR1* | -0.447580058 | 0.002506965 | 0.005754395 | 1 |
| T2DM | endothelial | *COPS7A* | -0.593275347 | 0.000161948 | 0.000575888 | 1 |
| T2DM | endothelial | *GNB3* | -0.122159086 | 0.013393171 | 0.023934868 | 1 |
| T2DM | endothelial | *CDCA3* | -0.403837825 | 0.000274806 | 0.00089103 | 1 |
| T2DM | endothelial | *CHPT1* | -0.239054467 | 3.21E-05 | 0.000150789 | 0.202056937 |
| T2DM | endothelial | *GNPTAB* | -0.342306696 | 0.00625107 | 0.012479765 | 1 |
| T2DM | endothelial | *NT5DC3* | -0.402481232 | 6.02E-05 | 0.000252274 | 0.378664158 |
| T2DM | endothelial | *ST8SIA1* | -0.457525443 | 8.72E-07 | 1.08E-05 | 0.005479528 |
| T2DM | endothelial | *PHC1* | 0.5272557 | 0.002653619 | 0.006031562 | 1 |
| T2DM | endothelial | *COL12A1* | -0.344385874 | 4.42E-07 | 7.50E-06 | 0.002776518 |
| T2DM | endothelial | *TDP2* | -0.400730417 | 0.01747966 | 0.029968537 | 1 |
| T2DM | endothelial | *SMIM8* | -0.337450377 | 0.009534697 | 0.017920669 | 1 |
| T2DM | endothelial | *FAM184A* | 0.293647274 | 0.004230922 | 0.008956164 | 1 |

| | | | | | | |
|---|---|---|---|---|---|---|
| T2DM | endothelial | *GABRR2* | -0.539097214 | 0.00535551 | 0.010928299 | 1 |
| T2DM | endothelial | *HDDC2* | 0.284081394 | 0.013479427 | 0.024048002 | 1 |
| T2DM | endothelial | *TPD52L1* | 0.165356539 | 2.23E-06 | 2.01E-05 | 0.014014386 |
| T2DM | endothelial | *UST* | -0.31961088 | 1.57E-06 | 1.61E-05 | 0.009850838 |
| T2DM | endothelial | *TULP1* | 1.922178119 | 4.38E-06 | 3.21E-05 | 0.027567236 |
| T2DM | endothelial | *SLC26A8* | 0.273923252 | 3.20E-05 | 0.000150476 | 0.201337353 |
| T2DM | endothelial | *PHACTR1* | 0.272370637 | 0.000968843 | 0.002573348 | 1 |
| T2DM | endothelial | *MDGA1* | -0.804496961 | 3.95E-07 | 7.06E-06 | 0.002486081 |
| T2DM | endothelial | *CD83* | -0.534750889 | 8.43E-08 | 4.46E-06 | 0.000530172 |
| T2DM | endothelial | *CAP2* | -0.448513899 | 1.65E-06 | 1.66E-05 | 0.010359918 |
| T2DM | endothelial | *BVES* | -0.511345557 | 1.04E-08 | 1.89E-06 | 6.53E-05 |
| T2DM | endothelial | *WASF1* | -0.22369341 | 0.000189289 | 0.000657855 | 1 |
| T2DM | endothelial | *GPLD1* | -0.166913448 | 0.013410053 | 0.023951421 | 1 |
| T2DM | endothelial | *ALDH5A1* | -0.324090575 | 0.010878588 | 0.020029765 | 1 |
| T2DM | endothelial | *VNN1* | -0.263804518 | 0.000495668 | 0.001459608 | 1 |
| T2DM | endothelial | *VNN2* | -0.438274573 | 0.015258501 | 0.026728948 | 1 |
| T2DM | endothelial | *ACOT13* | -0.368961497 | 0.022038914 | 0.036597636 | 1 |
| T2DM | endothelial | *C6orf62* | -0.364179876 | 0.000601042 | 0.001723327 | 1 |
| T2DM | endothelial | *PERP* | 0.292908446 | 0.000100337 | 0.000386767 | 0.630816875 |
| T2DM | endothelial | *SLC16A10* | -0.338482203 | 0.000190401 | 0.000659533 | 1 |
| T2DM | endothelial | *PRPH2* | -0.605830964 | 4.30E-06 | 3.16E-05 | 0.027024115 |
| T2DM | endothelial | *DNPH1* | -0.219583253 | 0.02206311 | 0.036628141 | 1 |
| T2DM | endothelial | *LAMA4* | -0.253435982 | 0.000760232 | 0.002099901 | 1 |
| T2DM | endothelial | *CLIC5* | 0.125859718 | 1.48E-06 | 1.57E-05 | 0.009322578 |
| T2DM | endothelial | *FBRSL1* | -0.386669919 | 0.019895069 | 0.033447758 | 1 |
| T2DM | endothelial | *LY86* | -0.560599388 | 3.32E-07 | 6.46E-06 | 0.00208552 |
| T2DM | endothelial | *TBX18* | -0.354452301 | 8.53E-07 | 1.08E-05 | 0.005362045 |
| T2DM | endothelial | *NUDT12* | -0.636129829 | 0.000231436 | 0.000774782 | 1 |
| T2DM | endothelial | *CEP72* | -0.407641035 | 0.023599024 | 0.038849715 | 1 |
| T2DM | endothelial | *C7* | -0.635904097 | 8.67E-05 | 0.000340357 | 0.544911304 |
| T2DM | endothelial | *PAPD7* | -0.459600348 | 0.000180986 | 0.000632495 | 1 |
| T2DM | endothelial | *NNT* | -0.333578158 | 0.00011445 | 0.000430865 | 0.719544557 |
| T2DM | endothelial | *LOX* | -0.578492767 | 4.06E-06 | 3.03E-05 | 0.025534957 |
| T2DM | endothelial | *APBB3* | -0.332775891 | 0.016247552 | 0.028155557 | 1 |
| T2DM | endothelial | *TMCO6* | -0.777109461 | 0.000488965 | 0.001446107 | 1 |
| T2DM | endothelial | *SPARC* | -0.33341731 | 0.000188112 | 0.000654488 | 1 |
| T2DM | endothelial | *PCDHB6* | 0.63109252 | 0.012435673 | 0.022466401 | 1 |
| T2DM | endothelial | *PDE8B* | -0.340916685 | 2.89E-06 | 2.37E-05 | 0.018192383 |
| T2DM | endothelial | *CLK4* | -0.39178391 | 0.002640922 | 0.006011397 | 1 |
| T2DM | endothelial | *HAVCR1* | -0.320548439 | 8.46E-05 | 0.00033386 | 0.531839692 |
| T2DM | endothelial | *RNF130* | -0.355251576 | 0.000740942 | 0.002055738 | 1 |
| T2DM | endothelial | *ARSB* | -0.333192177 | 0.002873967 | 0.006439285 | 1 |
| T2DM | endothelial | *CLINT1* | -0.293267082 | 0.014964001 | 0.02627162 | 1 |
| T2DM | endothelial | *BTNL8* | 0.11843206 | 5.76E-06 | 3.91E-05 | 0.036228914 |
| T2DM | endothelial | *TTC1* | -0.382596892 | 0.004794474 | 0.009912154 | 1 |
| T2DM | endothelial | *MSH3* | -0.299207587 | 0.001495468 | 0.003725043 | 1 |
| T2DM | endothelial | *RASGRF2* | 0.211847846 | 6.86E-05 | 0.000280275 | 0.431343657 |
| T2DM | endothelial | *POLR3G* | -0.551264711 | 7.36E-05 | 0.000296135 | 0.462563216 |
| T2DM | endothelial | *CDH6* | -0.165063022 | 0.000381022 | 0.001170813 | 1 |
| T2DM | endothelial | *SLC27A6* | 0.5849893 | 1.26E-05 | 7.06E-05 | 0.079038274 |

| | | | | | | |
|---|---|---|---|---|---|---|
| T2DM | endothelial | *TARS* | -0.513057649 | 0.002236103 | 0.005231999 | 1 |
| T2DM | endothelial | *LNPEP* | -0.257717524 | 0.01444008 | 0.025472722 | 1 |
| T2DM | endothelial | *GNPDA1* | -0.589247559 | 0.000295547 | 0.000947528 | 1 |
| T2DM | endothelial | *C9* | -0.450418293 | 0.000228297 | 0.000767952 | 1 |
| T2DM | endothelial | *ATP6V0E1* | -0.411155826 | 0.001994789 | 0.004741811 | 1 |
| T2DM | endothelial | *CNTN3* | -0.414519368 | 3.05E-07 | 6.16E-06 | 0.001916649 |
| T2DM | endothelial | *ACTR8* | -0.66024039 | 0.002994295 | 0.006673211 | 1 |
| T2DM | endothelial | *HGD* | -1.274189787 | 4.30E-07 | 7.46E-06 | 0.002703208 |
| T2DM | endothelial | *NIT2* | -0.5607392 | 1.92E-06 | 1.82E-05 | 0.012051234 |
| T2DM | endothelial | *FAM162A* | -0.529179205 | 0.001693724 | 0.004135318 | 1 |
| T2DM | endothelial | *PCCB* | -0.55150101 | 2.61E-05 | 0.000128256 | 0.164125125 |
| T2DM | endothelial | *WNT5A* | -0.668041908 | 1.41E-07 | 4.46E-06 | 0.000883574 |
| T2DM | endothelial | *PRKAR2A* | -0.296997973 | 0.007477917 | 0.014537311 | 1 |
| T2DM | endothelial | *HYAL1* | 0.25159696 | 4.40E-05 | 0.000195616 | 0.27658172 |
| T2DM | endothelial | *CYB561D2* | -0.539980198 | 3.98E-06 | 2.98E-05 | 0.025004868 |
| T2DM | endothelial | *IQCG* | -0.449504169 | 0.002544307 | 0.005820983 | 1 |
| T2DM | endothelial | *GBE1* | -0.307907037 | 0.001200715 | 0.00309381 | 1 |
| T2DM | endothelial | *UMPS* | -0.603407379 | 0.000351638 | 0.001097689 | 1 |
| T2DM | endothelial | *SLC41A3* | 0.366698622 | 0.002016478 | 0.004787613 | 1 |
| T2DM | endothelial | *PLXNA1* | -0.392978698 | 3.42E-05 | 0.000159064 | 0.21505435 |
| T2DM | endothelial | *ABTB1* | -0.531882 | 0.003827056 | 0.008245615 | 1 |
| T2DM | endothelial | *EFCC1* | 0.228334219 | 0.000516719 | 0.00151028 | 1 |
| T2DM | endothelial | *NEK11* | -0.472153287 | 7.04E-05 | 0.000285852 | 0.44249833 |
| T2DM | endothelial | *PLSCR4* | 0.194089604 | 0.018671623 | 0.03170948 | 1 |
| T2DM | endothelial | *HEMK1* | -0.319526773 | 0.013725288 | 0.024424253 | 1 |
| T2DM | endothelial | *COMMD2* | -0.349301458 | 0.024291447 | 0.039833158 | 1 |
| T2DM | endothelial | *ABCC5* | -0.501563248 | 6.47E-06 | 4.23E-05 | 0.040669753 |
| T2DM | endothelial | *ARHGEF26* | -0.419563278 | 8.49E-06 | 5.23E-05 | 0.053376031 |
| T2DM | endothelial | *KLHL24* | -0.284714964 | 0.008446551 | 0.01615069 | 1 |
| T2DM | endothelial | *PLCH1* | -0.723664337 | 5.10E-05 | 0.000219669 | 0.320935832 |
| T2DM | endothelial | *ZBTB47* | -0.508143839 | 0.002638057 | 0.00600705 | 1 |
| T2DM | endothelial | *CLCN2* | -0.979124197 | 0.002709266 | 0.006118231 | 1 |
| T2DM | endothelial | *NEK4* | -0.378688341 | 0.029297453 | 0.046880399 | 1 |
| T2DM | endothelial | *MOB1A* | -0.356467608 | 0.028699266 | 0.046087428 | 1 |
| T2DM | endothelial | *LMAN2L* | -0.313014647 | 0.014095634 | 0.024977241 | 1 |
| T2DM | endothelial | *KCNIP3* | -0.804585032 | 5.19E-06 | 3.62E-05 | 0.032655814 |
| T2DM | endothelial | *FAHD2A* | -0.426617965 | 0.028397182 | 0.045695695 | 1 |
| T2DM | endothelial | *TFCP2L1* | -0.585249787 | 0.000191667 | 0.000663188 | 1 |
| T2DM | endothelial | *TP53I3* | -0.483378679 | 0.002783038 | 0.006266819 | 1 |
| T2DM | endothelial | *DNAJC27* | -0.699403718 | 1.50E-06 | 1.58E-05 | 0.009455298 |
| T2DM | endothelial | *STAM2* | -0.332456526 | 0.013048282 | 0.023405006 | 1 |
| T2DM | endothelial | *CENPA* | -0.649862422 | 0.004547768 | 0.009495787 | 1 |
| T2DM | endothelial | *TANC1* | 0.189008442 | 0.000247294 | 0.000819144 | 1 |
| T2DM | endothelial | *NRBP1* | -0.576982038 | 8.57E-05 | 0.000337511 | 0.539004576 |
| T2DM | endothelial | *ITGB6* | -0.543208524 | 1.28E-06 | 1.41E-05 | 0.008059883 |
| T2DM | endothelial | *ITGA4* | -0.474012756 | 3.88E-05 | 0.000176958 | 0.244102338 |
| T2DM | endothelial | *PPM1G* | -0.53376467 | 0.001147822 | 0.002977306 | 1 |
| T2DM | endothelial | *PCSK4* | -0.498727048 | 7.45E-05 | 0.000299282 | 0.468076879 |
| T2DM | endothelial | *INO80B* | -0.447430714 | 0.006030611 | 0.012105508 | 1 |
| T2DM | endothelial | *MOGS* | -0.367806776 | 2.89E-06 | 2.37E-05 | 0.018152772 |

| | | | | | | |
|---|---|---|---|---|---|---|
| T2DM | endothelial | *GRB14* | 0.230700219 | 7.03E-06 | 4.50E-05 | 0.044214021 |
| T2DM | endothelial | *CLIP4* | -0.360384221 | 4.76E-07 | 7.87E-06 | 0.002990834 |
| T2DM | endothelial | *RTN4* | -0.367457333 | 3.08E-05 | 0.000145933 | 0.193506897 |
| T2DM | endothelial | *HTRA2* | -0.723339171 | 0.003040234 | 0.006766 | 1 |
| T2DM | endothelial | *GALNT3* | -0.327606935 | 0.021810694 | 0.036266551 | 1 |
| T2DM | endothelial | *POLE4* | 0.383459511 | 9.79E-05 | 0.00037882 | 0.615203029 |
| T2DM | endothelial | *MRPL19* | -0.355800226 | 0.008292998 | 0.015929752 | 1 |
| T2DM | endothelial | *LANCL1* | -0.360289512 | 0.018605311 | 0.031631041 | 1 |
| T2DM | endothelial | *EFEMP1* | -0.351099262 | 1.89E-06 | 1.81E-05 | 0.011899561 |
| T2DM | endothelial | *GGCX* | -0.333921418 | 0.002575115 | 0.005879085 | 1 |
| T2DM | endothelial | *SF3B1* | -0.286110402 | 0.01128445 | 0.020689803 | 1 |
| T2DM | endothelial | *ST3GAL5* | -0.461655988 | 2.94E-06 | 2.39E-05 | 0.018483251 |
| T2DM | endothelial | *PLCD4* | -0.754447892 | 0.002111229 | 0.004975041 | 1 |
| T2DM | endothelial | *CHMP3* | -0.486832488 | 0.00110678 | 0.002886074 | 1 |
| T2DM | endothelial | *WNT6* | -1.36911083 | 0.000493462 | 0.001455158 | 1 |
| T2DM | endothelial | *MLPH* | 0.148221374 | 1.01E-06 | 1.19E-05 | 0.00632545 |
| T2DM | endothelial | *HDLBP* | -0.325129848 | 0.002993867 | 0.006673211 | 1 |
| T2DM | endothelial | *PASK* | -0.614259963 | 0.004129146 | 0.008789661 | 1 |
| T2DM | endothelial | *STK25* | -0.462213771 | 0.005743199 | 0.01158824 | 1 |
| T2DM | endothelial | *TPO* | 0.084614509 | 1.04E-06 | 1.22E-05 | 0.006556886 |
| T2DM | endothelial | *TAF1B* | -0.471656169 | 0.000188314 | 0.000654829 | 1 |
| T2DM | endothelial | *SLC25A12* | -0.330471468 | 0.007684841 | 0.014884895 | 1 |
| T2DM | endothelial | *RMDN2* | -0.537479288 | 8.83E-05 | 0.000345792 | 0.554996769 |
| T2DM | endothelial | *SDC1* | 1.249252229 | 1.82E-07 | 4.62E-06 | 0.001145296 |
| T2DM | endothelial | *SOS1* | -0.245814389 | 0.00853131 | 0.016268227 | 1 |
| T2DM | endothelial | *WIPF1* | -0.342705118 | 0.002650053 | 0.006025635 | 1 |
| T2DM | endothelial | *ARID3A* | -0.495014049 | 1.02E-06 | 1.20E-05 | 0.006441755 |
| T2DM | endothelial | *CD207* | 1.140221039 | 3.40E-07 | 6.50E-06 | 0.002139438 |
| T2DM | endothelial | *VAX2* | 0.592534351 | 2.65E-05 | 0.000129396 | 0.166662482 |
| T2DM | endothelial | *BCL9* | -0.39689544 | 0.008082018 | 0.015557761 | 1 |
| T2DM | endothelial | *DHCR24* | -0.684030017 | 1.92E-06 | 1.82E-05 | 0.01206736 |
| T2DM | endothelial | *DNAJC16* | -0.354934413 | 0.009824415 | 0.018376196 | 1 |
| T2DM | endothelial | *MORN1* | -0.367939171 | 0.0143197 | 0.025302967 | 1 |
| T2DM | endothelial | *GPX7* | -0.318947661 | 8.51E-05 | 0.000335593 | 0.534935025 |
| T2DM | endothelial | *SCP2* | -0.363954813 | 0.001261457 | 0.003221276 | 1 |
| T2DM | endothelial | *TPSG1* | -0.510731788 | 0.02107055 | 0.035175398 | 1 |
| T2DM | endothelial | *RALGPS2* | -0.449700873 | 8.87E-06 | 5.42E-05 | 0.055791165 |
| T2DM | endothelial | *WRAP73* | -0.559350074 | 0.01363079 | 0.024269832 | 1 |
| T2DM | endothelial | *ICMT* | -0.653377151 | 0.003871782 | 0.008324862 | 1 |
| T2DM | endothelial | *PARK7* | -0.46237518 | 0.002041317 | 0.004841101 | 1 |
| T2DM | endothelial | *KCNC4* | 0.255284121 | 0.000560627 | 0.00162427 | 1 |
| T2DM | endothelial | *EDEM3* | -0.390129214 | 0.002510439 | 0.005760267 | 1 |
| T2DM | endothelial | *CAPZA1* | -0.371808755 | 0.00209835 | 0.004950215 | 1 |
| T2DM | endothelial | *DOCK7* | -0.402564787 | 4.37E-06 | 3.21E-05 | 0.02746694 |
| T2DM | endothelial | *FBXO6* | -0.525215281 | 0.00744214 | 0.014482599 | 1 |
| T2DM | endothelial | *DNAJC6* | 0.359695694 | 0.000216748 | 0.000736426 | 1 |
| T2DM | endothelial | *LEPR* | 0.216562208 | 0.030069453 | 0.048030146 | 1 |
| T2DM | endothelial | *MIIP* | -0.730285002 | 3.37E-07 | 6.50E-06 | 0.002117594 |
| T2DM | endothelial | *NCF2* | -0.613494922 | 3.19E-06 | 2.53E-05 | 0.020061755 |
| T2DM | endothelial | *RGS2* | -0.587475018 | 2.41E-07 | 5.33E-06 | 0.001515262 |

| | | | | | | |
|---|---|---|---|---|---|---|
| T2DM | endothelial | *OLFML3* | -0.549815091 | 5.24E-07 | 8.32E-06 | 0.003293416 |
| T2DM | endothelial | *TNNI3K* | -0.477512392 | 0.002992192 | 0.006673211 | 1 |
| T2DM | endothelial | *CRYZ* | -0.351100279 | 0.002485283 | 0.005715059 | 1 |
| T2DM | endothelial | *TTF2* | -0.573581196 | 0.000123772 | 0.000461813 | 0.778155214 |
| T2DM | endothelial | *TMEM9* | -0.430798611 | 9.55E-05 | 0.000371309 | 0.600406795 |
| T2DM | endothelial | *WARS2* | -0.363178215 | 0.000712728 | 0.001990636 | 1 |
| T2DM | endothelial | *OSCP1* | -0.468111699 | 2.49E-06 | 2.15E-05 | 0.015638971 |
| T2DM | endothelial | *C1orf109* | -0.760387238 | 2.56E-05 | 0.00012624 | 0.160703998 |
| T2DM | endothelial | *TBCE* | -0.550079558 | 0.00014112 | 0.000514762 | 0.887222616 |
| T2DM | endothelial | *MTR* | -0.31239234 | 0.009584641 | 0.017993025 | 1 |
| T2DM | endothelial | *KCNQ4* | -0.458371284 | 8.40E-06 | 5.20E-05 | 0.052796579 |
| T2DM | endothelial | *PADI2* | -0.833392064 | 8.87E-08 | 4.46E-06 | 0.000557677 |
| T2DM | endothelial | *PLA2G2D* | -4.195561966 | 4.23E-08 | 3.59E-06 | 0.000265648 |
| T2DM | endothelial | *KIF17* | 0.388600426 | 8.21E-06 | 5.11E-05 | 0.051604307 |
| T2DM | endothelial | *NSUN4* | -0.550007876 | 0.000462926 | 0.001381308 | 1 |
| T2DM | endothelial | *TMED5* | -0.470085162 | 2.76E-05 | 0.000132992 | 0.173422056 |
| T2DM | endothelial | *TNFSF4* | -0.759621607 | 5.81E-10 | 2.15E-07 | 3.66E-06 |
| T2DM | endothelial | *IRF6* | 0.173508705 | 1.57E-07 | 4.49E-06 | 0.000988482 |
| T2DM | endothelial | *RCOR3* | -0.296773378 | 0.016086253 | 0.027945365 | 1 |
| T2DM | endothelial | *MTFR1L* | -0.591926704 | 0.000229448 | 0.000769353 | 1 |
| T2DM | endothelial | *RPA2* | -0.562497029 | 0.000376423 | 0.001160084 | 1 |
| T2DM | endothelial | *SLC5A9* | -0.877645923 | 0.00011581 | 0.000435035 | 0.728099217 |
| T2DM | endothelial | *OSBPL9* | -0.316539239 | 0.005461758 | 0.011098278 | 1 |
| T2DM | endothelial | *CD3EAP* | -0.703622288 | 8.95E-06 | 5.45E-05 | 0.056266882 |
| T2DM | endothelial | *COLEC11* | 0.378641682 | 0.024225931 | 0.039736089 | 1 |
| T2DM | endothelial | *A4GNT* | -0.351052161 | 4.31E-05 | 0.000192828 | 0.270730325 |
| T2DM | endothelial | *IFT46* | -0.629392699 | 0.000243554 | 0.000810601 | 1 |
| T2DM | endothelial | *CASC1* | -0.572099004 | 0.000486118 | 0.001440256 | 1 |
| T2DM | endothelial | *LRMP* | 0.25232158 | 0.006316102 | 0.01258616 | 1 |
| T2DM | endothelial | *ATP10B* | -1.169543807 | 0.001954941 | 0.004671278 | 1 |
| T2DM | endothelial | *SPCS2* | -0.362567232 | 0.017165205 | 0.029517954 | 1 |
| T2DM | endothelial | *FILIP1* | 0.20055298 | 6.99E-05 | 0.000284369 | 0.439633847 |
| T2DM | endothelial | *SGIP1* | 0.293187288 | 0.010706535 | 0.019774379 | 1 |
| T2DM | endothelial | *AKAP7* | -0.30326251 | 0.001038539 | 0.002729638 | 1 |
| T2DM | endothelial | *TCF21* | -1.784062766 | 0.007685123 | 0.014884895 | 1 |
| T2DM | endothelial | *MED28* | -0.400179156 | 0.006118444 | 0.012258335 | 1 |
| T2DM | endothelial | *SLC16A7* | -0.379673754 | 4.98E-05 | 0.000215517 | 0.313361764 |
| T2DM | endothelial | *VAMP8* | -0.514025105 | 2.63E-05 | 0.000128665 | 0.165205649 |
| T2DM | endothelial | *FOXO3* | -0.344330339 | 0.000595968 | 0.00171402 | 1 |
| T2DM | endothelial | *TGIF2* | -0.611704386 | 0.017931247 | 0.030634171 | 1 |
| T2DM | endothelial | *ABCG2* | 0.304093115 | 5.99E-06 | 4.01E-05 | 0.037659816 |
| T2DM | endothelial | *SPP1* | -0.472974369 | 0.000211121 | 0.000720193 | 1 |
| T2DM | endothelial | *FAM47E-STBD* | -0.347433966 | 0.000106006 | 0.000403914 | 0.666457946 |
| T2DM | endothelial | *MFSD1* | -0.460798129 | 0.000167335 | 0.000590699 | 1 |
| T2DM | endothelial | *UCHL3* | -0.835536476 | 9.14E-07 | 1.11E-05 | 0.005744447 |
| T2DM | endothelial | *WDR35* | -0.564429633 | 1.66E-05 | 8.88E-05 | 0.104316708 |
| T2DM | endothelial | *DNAH7* | -0.412553559 | 1.98E-08 | 2.76E-06 | 0.000124404 |
| T2DM | endothelial | *TRPM6* | 0.140425037 | 1.04E-06 | 1.21E-05 | 0.006534026 |
| T2DM | endothelial | *GDA* | -0.498306883 | 2.32E-06 | 2.06E-05 | 0.014599546 |
| T2DM | endothelial | *C2orf40* | 0.294945681 | 1.73E-05 | 9.17E-05 | 0.108747084 |

| | | | | | | |
|---|---|---|---|---|---|---|
| T2DM | endothelial | *CPSF3* | -0.486729634 | 0.002630605 | 0.005994423 | 1 |
| T2DM | endothelial | *CCDC92* | -0.527429161 | 2.47E-06 | 2.14E-05 | 0.015515855 |
| T2DM | endothelial | *FKBP15* | -0.422204648 | 0.00043768 | 0.001314712 | 1 |
| T2DM | endothelial | *CTNNAL1* | 0.195361218 | 0.004092253 | 0.008718398 | 1 |
| T2DM | endothelial | *WDR34* | -0.631396584 | 6.76E-05 | 0.000277549 | 0.425205767 |
| T2DM | endothelial | *TRIM32* | -0.675659494 | 1.26E-06 | 1.39E-05 | 0.007893217 |
| T2DM | endothelial | *PHF19* | -0.647415074 | 1.69E-07 | 4.55E-06 | 0.001064593 |
| T2DM | endothelial | *BSPRY* | -2.371710553 | 1.02E-07 | 4.46E-06 | 0.000642758 |
| T2DM | endothelial | *PPP6C* | -0.462489078 | 0.000332535 | 0.001046893 | 1 |
| T2DM | endothelial | *HDHD3* | -0.159536771 | 0.000127645 | 0.000474293 | 0.802503343 |
| T2DM | endothelial | *HSDL2* | -0.263374375 | 0.000374076 | 0.001154548 | 1 |
| T2DM | endothelial | *KDSR* | -0.464407354 | 0.001071637 | 0.002803215 | 1 |
| T2DM | endothelial | *VSX2* | 0.249781766 | 4.96E-05 | 0.00021471 | 0.311681271 |
| T2DM | endothelial | *IRF2BPL* | -0.673067604 | 0.000563825 | 0.001632029 | 1 |
| T2DM | endothelial | *FLVCR2* | -0.640602327 | 3.48E-06 | 2.69E-05 | 0.021863653 |
| T2DM | endothelial | *PPP4R4* | -0.593981174 | 5.36E-07 | 8.34E-06 | 0.003370274 |
| T2DM | endothelial | *ZC2HC1C* | 0.438814512 | 0.023126681 | 0.038152045 | 1 |
| T2DM | endothelial | *ALDH6A1* | -0.326847519 | 0.002203927 | 0.005170183 | 1 |
| T2DM | endothelial | *ESRRB* | -0.314735752 | 0.000139633 | 0.000511281 | 0.877869914 |
| T2DM | endothelial | *GPR75* | -2.075863408 | 5.82E-05 | 0.000245417 | 0.366161455 |
| T2DM | endothelial | *KLHL29* | -0.418247354 | 8.09E-05 | 0.000321166 | 0.508405321 |
| T2DM | endothelial | *CRIPT* | -0.535459752 | 0.001206062 | 0.003104654 | 1 |
| T2DM | endothelial | *EPCAM* | -0.716470366 | 0.002435399 | 0.005620908 | 1 |
| T2DM | endothelial | *SLC17A5* | -0.400354657 | 0.000158813 | 0.000566341 | 0.998459218 |
| T2DM | endothelial | *OGFRL1* | -0.329415159 | 0.006996676 | 0.013733407 | 1 |
| T2DM | endothelial | *CUTC* | -0.422723098 | 0.000585171 | 0.001688377 | 1 |
| T2DM | endothelial | *C10orf88* | -0.495114671 | 0.006945234 | 0.013650048 | 1 |
| T2DM | endothelial | *HELLS* | -0.429258509 | 0.006519127 | 0.012929258 | 1 |
| T2DM | endothelial | *TCTN3* | -0.412876589 | 0.007634566 | 0.014809786 | 1 |
| T2DM | endothelial | *AVPI1* | -0.455261062 | 1.02E-06 | 1.20E-05 | 0.006442096 |
| T2DM | endothelial | *WDR11* | -0.366409238 | 0.002095985 | 0.004948583 | 1 |
| T2DM | endothelial | *C10orf76* | -0.337293828 | 0.003656788 | 0.007919015 | 1 |
| T2DM | endothelial | *KCNIP2* | -0.182171734 | 0.002812248 | 0.006323536 | 1 |
| T2DM | endothelial | *HOXB5* | -0.745223762 | 0.000139104 | 0.000510238 | 0.87454764 |
| T2DM | endothelial | *CD274* | -0.657626345 | 0.001271297 | 0.003244549 | 1 |
| T2DM | endothelial | *LRP11* | -0.439418572 | 0.001962602 | 0.004680107 | 1 |
| T2DM | endothelial | *CCDC170* | -0.258638874 | 5.77E-06 | 3.91E-05 | 0.036283449 |
| T2DM | endothelial | *PCDHB14* | 0.521210398 | 2.44E-05 | 0.000121273 | 0.15316723 |
| T2DM | endothelial | *SEC16B* | 0.739563679 | 0.00020475 | 0.000701124 | 1 |
| T2DM | endothelial | *ACAT2* | -0.604899416 | 3.29E-05 | 0.000154205 | 0.206943063 |
| T2DM | endothelial | *MASTL* | -0.360813419 | 1.42E-06 | 1.52E-05 | 0.00894295 |
| T2DM | endothelial | *MRC1* | -0.373491318 | 1.02E-05 | 5.99E-05 | 0.063888379 |
| T2DM | endothelial | *CCDC77* | -0.710437231 | 4.41E-07 | 7.50E-06 | 0.002769661 |
| T2DM | endothelial | *ENOX1* | -0.462464616 | 1.16E-06 | 1.31E-05 | 0.00728991 |
| T2DM | endothelial | *WBP4* | -0.381195313 | 0.004139799 | 0.008798991 | 1 |
| T2DM | endothelial | *SMAD9* | 0.213698504 | 3.59E-06 | 2.75E-05 | 0.022580806 |
| T2DM | endothelial | *SIL1* | -0.360893434 | 0.001203034 | 0.003098514 | 1 |
| T2DM | endothelial | *ARL1* | -0.422924275 | 0.002224912 | 0.005211633 | 1 |
| T2DM | endothelial | *EPHX2* | -0.260706288 | 0.031017694 | 0.049394184 | 1 |
| T2DM | endothelial | *ZNF706* | -0.353295708 | 0.005344852 | 0.010913636 | 1 |

| | | | | | | |
|---|---|---|---|---|---|---|
| T2DM | endothelial | *CRISPLD1* | 0.576019931 | 0.000995648 | 0.002632313 | 1 |
| T2DM | endothelial | *RDH10* | -0.222016756 | 0.007489746 | 0.014555806 | 1 |
| T2DM | endothelial | *SCPEP1* | -0.197972127 | 0.000998956 | 0.002639322 | 1 |
| T2DM | endothelial | *TRIM6* | -0.549084227 | 1.39E-06 | 1.50E-05 | 0.008718759 |
| T2DM | endothelial | *ECHDC2* | -0.313999602 | 1.68E-07 | 4.54E-06 | 0.001058366 |
| T2DM | endothelial | *PYROXD1* | -0.379962498 | 0.003649051 | 0.007905438 | 1 |
| T2DM | endothelial | *A1BG* | -0.977878201 | 7.15E-05 | 0.000289741 | 0.449654506 |
| T2DM | endothelial | *PDZRN3* | -0.339617029 | 0.000115834 | 0.000435035 | 0.728247846 |
| T2DM | endothelial | *LHX4* | -0.642920738 | 0.003055663 | 0.006795527 | 1 |
| T2DM | endothelial | *CSTA* | -0.351642366 | 1.55E-06 | 1.61E-05 | 0.009765208 |
| T2DM | endothelial | *POPDC2* | 0.600571085 | 0.000109114 | 0.000414501 | 0.685999631 |
| T2DM | endothelial | *B4GALT4* | -0.656925172 | 0.000196425 | 0.000675932 | 1 |
| T2DM | endothelial | *CD80* | -1.119504239 | 6.35E-07 | 9.11E-06 | 0.003995078 |
| T2DM | endothelial | *CAT* | -0.211818053 | 0.000898862 | 0.002421228 | 1 |
| T2DM | endothelial | *POLR3GL* | -0.845871465 | 4.39E-05 | 0.000195571 | 0.276145767 |
| T2DM | endothelial | *TNFSF10* | 0.245056044 | 4.63E-05 | 0.000204066 | 0.29120279 |
| T2DM | endothelial | *ZNF639* | -0.449517858 | 0.016983928 | 0.029230209 | 1 |
| T2DM | endothelial | *LIAS* | -0.484002988 | 0.002482081 | 0.005709786 | 1 |
| T2DM | endothelial | *CLCC1* | -0.562720353 | 0.000749023 | 0.002074496 | 1 |
| T2DM | endothelial | *CXCR4* | -0.856238534 | 0.00026917 | 0.000875338 | 1 |
| T2DM | endothelial | *ACVR2A* | -0.31259701 | 0.012135595 | 0.022019187 | 1 |
| T2DM | endothelial | *FLT3* | -0.723447371 | 5.40E-07 | 8.38E-06 | 0.003394461 |
| T2DM | endothelial | *RASL11A* | -1.561525939 | 4.67E-08 | 3.81E-06 | 0.000293342 |
| T2DM | endothelial | *FMOD* | -0.606383897 | 5.27E-07 | 8.34E-06 | 0.003315824 |
| T2DM | endothelial | *SERAC1* | -0.666885293 | 0.000248206 | 0.000821733 | 1 |
| T2DM | endothelial | *LDB3* | -0.283750299 | 0.00036876 | 0.001141504 | 1 |
| T2DM | endothelial | *OPN4* | -0.881508224 | 2.32E-05 | 0.000116701 | 0.145759168 |
| T2DM | endothelial | *FAM35A* | -0.269818973 | 0.017968996 | 0.030684548 | 1 |
| T2DM | endothelial | *NAA60* | -0.684336459 | 0.00608284 | 0.012190887 | 1 |
| T2DM | endothelial | *RWDD3* | -0.862442969 | 0.00121918 | 0.00312729 | 1 |
| T2DM | endothelial | *CCDC18* | -0.427970669 | 5.57E-06 | 3.81E-05 | 0.03501857 |
| T2DM | endothelial | *BBS9* | -0.245513081 | 0.031387394 | 0.049944962 | 1 |
| T2DM | endothelial | *KIAA0087* | 0.592295654 | 6.58E-05 | 0.000270357 | 0.413375375 |
| T2DM | endothelial | *KLHL7* | -0.321495408 | 0.028396145 | 0.045695695 | 1 |
| T2DM | endothelial | *CBX3* | -0.375132208 | 0.013988503 | 0.024815383 | 1 |
| T2DM | endothelial | *FAM126A* | -0.362485586 | 0.000714318 | 0.001994192 | 1 |
| T2DM | endothelial | *POLM* | -0.181972601 | 0.001410325 | 0.003550945 | 1 |
| T2DM | endothelial | *GLIPR2* | -0.485827658 | 4.12E-07 | 7.27E-06 | 0.002588408 |
| T2DM | endothelial | *DCAF10* | -0.272143268 | 0.024529507 | 0.040202558 | 1 |
| T2DM | endothelial | *BICC1* | -0.258903844 | 9.64E-06 | 5.76E-05 | 0.060612346 |
| T2DM | endothelial | *CISD1* | -0.260096413 | 0.020247906 | 0.03393724 | 1 |
| T2DM | endothelial | *SLC25A16* | -0.211508719 | 0.007934534 | 0.015297276 | 1 |
| T2DM | endothelial | *ZWINT* | -0.321719133 | 0.001223959 | 0.00313571 | 1 |
| T2DM | endothelial | *RBM19* | -0.409374005 | 0.010496797 | 0.019450417 | 1 |
| T2DM | endothelial | *ACADS* | -0.576142765 | 8.04E-07 | 1.04E-05 | 0.005057122 |
| T2DM | endothelial | *SSPN* | -0.231440818 | 0.013750417 | 0.024462046 | 1 |
| T2DM | endothelial | *CCDC91* | -0.315855583 | 0.001411148 | 0.003551596 | 1 |
| T2DM | endothelial | *EBPL* | 0.373173968 | 4.52E-05 | 0.000199822 | 0.283946551 |
| T2DM | endothelial | *OPTN* | -0.468466524 | 4.28E-05 | 0.000192073 | 0.269094837 |
| T2DM | endothelial | *ARHGAP9* | -0.699917981 | 1.40E-06 | 1.50E-05 | 0.008774995 |

| | | | | | | |
|---|---|---|---|---|---|---|
| T2DM | endothelial | *ORMDL2* | 0.393344587 | 0.023060003 | 0.038062021 | 1 |
| T2DM | endothelial | *PDE1B* | -0.473524078 | 9.07E-08 | 4.46E-06 | 0.000570488 |
| T2DM | endothelial | *IKZF4* | -0.750698223 | 0.00010371 | 0.00039709 | 0.652021969 |
| T2DM | endothelial | *SARDH* | -1.031910734 | 1.16E-07 | 4.46E-06 | 0.000731207 |
| T2DM | endothelial | *DBH* | -1.380555316 | 0.000374905 | 0.001156539 | 1 |
| T2DM | endothelial | *STIL* | -0.563100746 | 0.015948429 | 0.027752641 | 1 |
| T2DM | endothelial | *ACVR1C* | -0.317134783 | 3.53E-06 | 2.71E-05 | 0.022198447 |
| T2DM | endothelial | *SLC36A1* | -0.215566643 | 0.0030287 | 0.006742718 | 1 |
| T2DM | endothelial | *C4BPB* | -0.429327019 | 3.10E-06 | 2.48E-05 | 0.019474132 |
| T2DM | endothelial | *RAB38* | -1.024621251 | 3.56E-07 | 6.67E-06 | 0.002239481 |
| T2DM | endothelial | *AGO2* | -0.321583763 | 0.021406923 | 0.03567064 | 1 |
| T2DM | endothelial | *CHPF* | -0.382694905 | 7.68E-06 | 4.83E-05 | 0.048286697 |
| T2DM | endothelial | *MOGAT1* | -0.3965476 | 5.77E-07 | 8.63E-06 | 0.003625467 |
| T2DM | endothelial | *FAM124B* | 0.168956061 | 4.90E-07 | 8.02E-06 | 0.003081512 |
| T2DM | endothelial | *FAM210B* | -0.405554866 | 0.001435367 | 0.003596712 | 1 |
| T2DM | endothelial | *SNX21* | -0.439130692 | 5.65E-07 | 8.58E-06 | 0.003553502 |
| T2DM | endothelial | *WFDC3* | -0.168071825 | 0.000100833 | 0.000388442 | 0.633936816 |
| T2DM | endothelial | *SDC4* | -0.785878234 | 3.87E-08 | 3.45E-06 | 0.000243617 |
| T2DM | endothelial | *VAPB* | -0.279361811 | 0.00196301 | 0.004680107 | 1 |
| T2DM | endothelial | *TOX2* | 0.108694754 | 0.000389162 | 0.001191749 | 1 |
| T2DM | endothelial | *CDH26* | -0.804073272 | 2.61E-05 | 0.000128256 | 0.163893281 |
| T2DM | endothelial | *STX16* | -0.46418951 | 0.000644467 | 0.001823475 | 1 |
| T2DM | endothelial | *RNF114* | -0.491433428 | 0.000437198 | 0.001314521 | 1 |
| T2DM | endothelial | *KCNK15* | -1.139449038 | 2.79E-06 | 2.32E-05 | 0.017538769 |
| T2DM | endothelial | *PEPD* | -0.336583151 | 0.000999139 | 0.002639322 | 1 |
| T2DM | endothelial | *CHST8* | -0.521710484 | 1.49E-07 | 4.49E-06 | 0.000938581 |
| T2DM | endothelial | *STAMBP* | -0.410839439 | 0.005738642 | 0.011586012 | 1 |
| T2DM | endothelial | *ATP8A1* | 0.191905464 | 2.62E-06 | 2.23E-05 | 0.016455614 |
| T2DM | endothelial | *HIF3A* | 0.167800916 | 1.15E-06 | 1.30E-05 | 0.007234351 |
| T2DM | endothelial | *MRS2* | -0.329083449 | 0.010867588 | 0.020018906 | 1 |
| T2DM | endothelial | *PEX6* | -0.208663001 | 0.00338606 | 0.007414722 | 1 |
| T2DM | endothelial | *NQO2* | -0.510132507 | 2.01E-05 | 0.000103513 | 0.126078963 |
| T2DM | endothelial | *RPS10* | -1.540372578 | 0.000799958 | 0.002192388 | 1 |
| T2DM | endothelial | *HIST1H2BJ* | 0.492440265 | 0.003819285 | 0.008231692 | 1 |
| T2DM | endothelial | *MED20* | -0.511412436 | 0.001332924 | 0.003374986 | 1 |
| T2DM | endothelial | *TBCC* | -0.644048506 | 0.00011424 | 0.000430593 | 0.718229221 |
| T2DM | endothelial | *TCP11* | -0.43519737 | 0.004309073 | 0.009103205 | 1 |
| T2DM | endothelial | *KLHDC3* | -0.546091888 | 0.001878759 | 0.004516923 | 1 |
| T2DM | endothelial | *KLHL31* | -0.210220374 | 0.000190171 | 0.000659461 | 1 |
| T2DM | endothelial | *COL21A1* | 0.176752904 | 1.41E-05 | 7.78E-05 | 0.088925455 |
| T2DM | endothelial | *CPNE5* | 0.195045366 | 1.71E-05 | 9.08E-05 | 0.107214989 |
| T2DM | endothelial | *NUP153* | -0.327619294 | 0.009966143 | 0.018578823 | 1 |
| T2DM | endothelial | *EEF1E1* | -0.838471186 | 0.008420448 | 0.016110577 | 1 |
| T2DM | endothelial | *EREG* | -1.700055713 | 9.04E-08 | 4.46E-06 | 0.000568535 |
| T2DM | endothelial | *MYRF* | -0.340594452 | 0.016937961 | 0.029159081 | 1 |
| T2DM | endothelial | *MT1G* | -4.379442071 | 5.90E-07 | 8.79E-06 | 0.003710966 |
| T2DM | endothelial | *KIF25* | 0.364414475 | 3.04E-07 | 6.16E-06 | 0.001913027 |
| T2DM | endothelial | *GRK4* | 0.145257306 | 0.004298308 | 0.009083517 | 1 |
| T2DM | endothelial | *TEKT3* | -0.249971175 | 0.004951184 | 0.010195904 | 1 |
| T2DM | endothelial | *MRPS7* | -0.642493264 | 0.000399986 | 0.001217779 | 1 |

| | | | | | | |
|---|---|---|---|---|---|---|
| T2DM | endothelial | *GGA3* | -0.522886903 | 0.000632225 | 0.001796582 | 1 |
| T2DM | endothelial | *MIF4GD* | -0.8268976 | 0.003340868 | 0.007335813 | 1 |
| T2DM | endothelial | *NT5C* | -0.503490096 | 6.37E-07 | 9.11E-06 | 0.004007012 |
| T2DM | endothelial | *DDX31* | -0.348278901 | 0.012158095 | 0.022048263 | 1 |
| T2DM | endothelial | *PPP1R12C* | -0.584074004 | 0.000119295 | 0.000446168 | 0.750009039 |
| T2DM | endothelial | *OPRL1* | -0.895232855 | 2.79E-10 | 1.25E-07 | 1.75E-06 |
| T2DM | endothelial | *PLGLB2* | -2.911612083 | 0.001797404 | 0.004349607 | 1 |
| T2DM | endothelial | *PAX8* | -0.32670626 | 0.001466455 | 0.003661478 | 1 |
| T2DM | endothelial | *INSIG2* | -0.476828371 | 0.007004103 | 0.013739406 | 1 |
| T2DM | endothelial | *PSD4* | -0.717582265 | 1.28E-06 | 1.41E-05 | 0.008041034 |
| T2DM | endothelial | *SLC25A23* | -0.264615708 | 0.007537859 | 0.014640259 | 1 |
| T2DM | endothelial | *SH2D3A* | -1.027683188 | 4.74E-05 | 0.000207519 | 0.297997602 |
| T2DM | endothelial | *TRIP10* | -0.451860322 | 0.000479348 | 0.0014249 | 1 |
| T2DM | endothelial | *SNRPD2* | -0.564933293 | 0.006881979 | 0.013567577 | 1 |
| T2DM | endothelial | *RTN2* | -0.915940715 | 0.000891788 | 0.002405264 | 1 |
| T2DM | endothelial | *SYMPK* | -0.485709776 | 0.000437036 | 0.001314521 | 1 |
| T2DM | endothelial | *GPCPD1* | -0.303836485 | 0.012090175 | 0.021963763 | 1 |
| T2DM | endothelial | *SDCBP2* | 0.550508815 | 3.05E-06 | 2.46E-05 | 0.019172134 |
| T2DM | endothelial | *PANK2* | -0.365658871 | 0.010931646 | 0.020107448 | 1 |
| T2DM | endothelial | *BMP2* | -1.122615824 | 2.35E-06 | 2.08E-05 | 0.014767141 |
| T2DM | endothelial | *PCSK2* | -1.259112417 | 0.001014941 | 0.002676567 | 1 |
| T2DM | endothelial | *MKKS* | -0.36125823 | 0.023235272 | 0.038311081 | 1 |
| T2DM | endothelial | *MGME1* | -0.470511642 | 0.001580434 | 0.003895018 | 1 |
| T2DM | endothelial | *ITPA* | -0.575369836 | 6.38E-06 | 4.19E-05 | 0.04012774 |
| T2DM | endothelial | *TMEM74B* | 0.730898153 | 7.94E-05 | 0.000315556 | 0.499208801 |
| T2DM | endothelial | *FAM110A* | -0.854517348 | 6.21E-07 | 9.01E-06 | 0.003903233 |
| T2DM | endothelial | *NCLN* | -0.444534615 | 0.012083278 | 0.021962293 | 1 |
| T2DM | endothelial | *UROD* | -0.511255893 | 0.003906448 | 0.008385059 | 1 |
| T2DM | endothelial | *XRCC3* | 0.501043427 | 0.000264617 | 0.000863782 | 1 |
| T2DM | endothelial | *MCF2L* | 0.143387278 | 4.16E-07 | 7.31E-06 | 0.0026165 |
| T2DM | endothelial | *F10* | -0.486721572 | 3.25E-07 | 6.36E-06 | 0.002040519 |
| T2DM | endothelial | *PCID2* | -0.502113108 | 8.71E-06 | 5.34E-05 | 0.054776687 |
| T2DM | endothelial | *PROZ* | -2.286388608 | 2.82E-07 | 5.91E-06 | 0.001771847 |
| T2DM | endothelial | *IGFLR1* | -1.539586862 | 8.67E-08 | 4.46E-06 | 0.000544953 |
| T2DM | endothelial | *COX6B1* | -0.519362622 | 0.003505693 | 0.007629038 | 1 |
| T2DM | endothelial | *PRDX5* | -0.50466494 | 0.0176576 | 0.030199492 | 1 |
| T2DM | endothelial | *SCAF1* | -0.771531466 | 0.00020928 | 0.000715076 | 1 |
| T2DM | endothelial | *TSKS* | -0.543798286 | 2.68E-07 | 5.73E-06 | 0.001685422 |
| T2DM | endothelial | *SBDS* | -0.555328612 | 0.000987609 | 0.002616222 | 1 |
| T2DM | endothelial | *DNAJC8* | -0.428634288 | 0.005517364 | 0.011200408 | 1 |
| T2DM | endothelial | *ATG14* | -0.415553695 | 0.000724729 | 0.002018774 | 1 |
| T2DM | endothelial | *RHOJ* | 0.215021552 | 1.05E-05 | 6.16E-05 | 0.066261534 |
| T2DM | endothelial | *HSPA2* | 0.504817583 | 0.001734731 | 0.004217422 | 1 |
| T2DM | endothelial | *ZBTB1* | -0.39458419 | 0.000379091 | 0.001166021 | 1 |
| T2DM | endothelial | *RHOT1* | -0.249748321 | 0.025571318 | 0.041703469 | 1 |
| T2DM | endothelial | *AIF1L* | 0.221600181 | 2.70E-06 | 2.27E-05 | 0.016955462 |
| T2DM | endothelial | *MAP2K2* | -0.485400729 | 0.002369014 | 0.005495938 | 1 |
| T2DM | endothelial | *IPPK* | -0.628493119 | 0.000307666 | 0.00097939 | 1 |
| T2DM | endothelial | *FGD3* | -0.635958941 | 1.64E-05 | 8.78E-05 | 0.102805276 |
| T2DM | endothelial | *ABHD8* | -0.392774592 | 0.005124132 | 0.010503886 | 1 |

| | | | | | | |
|---|---|---|---|---|---|---|
| T2DM | endothelial | *MASP1* | -0.453071805 | 1.24E-07 | 4.46E-06 | 0.000778273 |
| T2DM | endothelial | *ATP13A4* | -0.414614541 | 0.00043329 | 0.001305893 | 1 |
| T2DM | endothelial | *HRASLS* | -0.491134405 | 1.09E-06 | 1.26E-05 | 0.00686945 |
| T2DM | endothelial | *HELB* | -0.40662841 | 0.012325763 | 0.022280641 | 1 |
| T2DM | endothelial | *RAB3IP* | -0.775527199 | 4.43E-06 | 3.24E-05 | 0.02787888 |
| T2DM | endothelial | *LRRC61* | -0.228424508 | 0.000302574 | 0.000965627 | 1 |
| T2DM | endothelial | *IDUA* | -0.791263841 | 0.000142653 | 0.000519616 | 0.896857179 |
| T2DM | endothelial | *TMEM175* | -0.384333347 | 0.006237744 | 0.012457655 | 1 |
| T2DM | endothelial | *UBR4* | -0.290242907 | 0.003892512 | 0.008360855 | 1 |
| T2DM | endothelial | *WFIKKN1* | -2.423623739 | 4.04E-07 | 7.20E-06 | 0.002541337 |
| T2DM | endothelial | *WDR24* | -1.455817627 | 4.89E-06 | 3.49E-05 | 0.030754579 |
| T2DM | endothelial | *TICAM1* | -0.574528917 | 0.008885391 | 0.016876874 | 1 |
| T2DM | endothelial | *AAMP* | -0.707026975 | 8.72E-05 | 0.000341824 | 0.547944064 |
| T2DM | endothelial | *PNKD* | -0.492381096 | 0.001145492 | 0.002973454 | 1 |
| T2DM | endothelial | *ZNF835* | -0.502201818 | 0.000153714 | 0.000551283 | 0.966398623 |
| T2DM | endothelial | *POR* | -0.474309957 | 0.000112603 | 0.000425188 | 0.707937809 |
| T2DM | endothelial | *FGL2* | -0.297470523 | 0.012929717 | 0.023218832 | 1 |
| T2DM | endothelial | *STYXL1* | -0.230127284 | 0.013256132 | 0.023710185 | 1 |
| T2DM | endothelial | *STEAP4* | -0.492525517 | 0.000233148 | 0.000779681 | 1 |
| T2DM | endothelial | *SGCE* | -0.425733438 | 0.001232234 | 0.003150491 | 1 |
| T2DM | endothelial | *RBM48* | -0.610734107 | 0.000107556 | 0.000409229 | 0.676204281 |
| T2DM | endothelial | *SRD5A3* | -0.42255987 | 0.016040846 | 0.02788189 | 1 |
| T2DM | endothelial | *VPREB3* | -2.947465508 | 8.82E-06 | 5.39E-05 | 0.055444809 |
| T2DM | endothelial | *ATF4* | -0.554189229 | 0.000245011 | 0.000814157 | 1 |
| T2DM | endothelial | *A4GALT* | 0.212117143 | 0.011180932 | 0.020505987 | 1 |
| T2DM | endothelial | *BAIAP2L2* | -0.405833291 | 7.15E-05 | 0.000289741 | 0.44967845 |
| T2DM | endothelial | *TST* | -0.353068532 | 0.0004577 | 0.00136766 | 1 |
| T2DM | endothelial | *APOL5* | -1.269842757 | 1.38E-05 | 7.62E-05 | 0.086659171 |
| T2DM | endothelial | *CDHR3* | -0.344611975 | 0.000148322 | 0.000534689 | 0.932498046 |
| T2DM | endothelial | *PRKRIP1* | -0.355562249 | 0.002909602 | 0.006507531 | 1 |
| T2DM | endothelial | *FOXP2* | -0.387306406 | 1.61E-07 | 4.49E-06 | 0.001010853 |
| T2DM | endothelial | *STRIP2* | -0.304554177 | 0.020398333 | 0.034151837 | 1 |
| T2DM | endothelial | *IRF5* | -0.184604191 | 0.014160519 | 0.025049854 | 1 |
| T2DM | endothelial | *KLHDC10* | -0.262752523 | 0.02803566 | 0.04526456 | 1 |
| T2DM | endothelial | *OPN1SW* | -0.397446898 | 4.70E-05 | 0.00020632 | 0.295450083 |
| T2DM | endothelial | *HOXD1* | 0.41086133 | 3.16E-07 | 6.29E-06 | 0.00198752 |
| T2DM | endothelial | *HOXD3* | 0.269253955 | 1.17E-05 | 6.68E-05 | 0.07386525 |
| T2DM | endothelial | *PDE11A* | -0.354249327 | 0.000193262 | 0.000666869 | 1 |
| T2DM | endothelial | *CHN1* | 0.152630867 | 3.69E-06 | 2.80E-05 | 0.023207696 |
| T2DM | endothelial | *GAD1* | -0.959628699 | 1.22E-07 | 4.46E-06 | 0.000767738 |
| T2DM | endothelial | *HOXD9* | 0.494883536 | 0.001440147 | 0.003605816 | 1 |
| T2DM | endothelial | *TWSG1* | -0.550730766 | 0.000101018 | 0.000388916 | 0.635099745 |
| T2DM | endothelial | *WDFY4* | -0.407185688 | 2.89E-06 | 2.37E-05 | 0.018157824 |
| T2DM | endothelial | *EIF2AK4* | -0.411616466 | 0.000163046 | 0.000579135 | 1 |
| T2DM | endothelial | *ANAPC13* | -0.211750546 | 0.006999066 | 0.013733811 | 1 |
| T2DM | endothelial | *SUMF2* | -0.483950854 | 0.000252386 | 0.000832065 | 1 |
| T2DM | endothelial | *DCTD* | -0.433085393 | 0.000299221 | 0.000956869 | 1 |
| T2DM | endothelial | *RPAIN* | -0.502265361 | 0.000191993 | 0.000663782 | 1 |
| T2DM | endothelial | *PLD2* | 0.482105901 | 0.000162096 | 0.000576086 | 1 |
| T2DM | endothelial | *ATP1B2* | -0.721332334 | 0.000993092 | 0.002626659 | 1 |

| | | | | | | |
|---|---|---|---|---|---|---|
| T2DM | endothelial | *CCL4* | -0.415598752 | 5.30E-05 | 0.000227458 | 0.333448722 |
| T2DM | endothelial | *PHF20L1* | -0.290349685 | 0.019753566 | 0.033259418 | 1 |
| T2DM | endothelial | *SLC44A2* | 0.316058321 | 0.002263374 | 0.005280086 | 1 |
| T2DM | endothelial | *CDKN2D* | -0.628877184 | 0.006517564 | 0.012929258 | 1 |
| T2DM | endothelial | *MTUS1* | 0.238220707 | 0.000608721 | 0.001741933 | 1 |
| T2DM | endothelial | *RIPK3* | 0.470960021 | 0.00019376 | 0.000667857 | 1 |
| T2DM | endothelial | *ADCY4* | 0.117131034 | 6.57E-07 | 9.28E-06 | 0.004129396 |
| T2DM | endothelial | *BCL2L2* | -0.613310502 | 1.78E-05 | 9.34E-05 | 0.111701419 |
| T2DM | endothelial | *DTD2* | -0.264278589 | 0.002669954 | 0.006055556 | 1 |
| T2DM | endothelial | *PARP2* | -0.473769997 | 0.000485687 | 0.001439659 | 1 |
| T2DM | endothelial | *TEP1* | -0.526025752 | 0.000787156 | 0.00216296 | 1 |
| T2DM | endothelial | *EPB41L4A* | 0.21278983 | 7.25E-05 | 0.000292755 | 0.455819497 |
| T2DM | endothelial | *CDO1* | -0.474354801 | 4.31E-07 | 7.46E-06 | 0.002709679 |
| T2DM | endothelial | *REEP5* | -0.365098911 | 0.00396027 | 0.008494785 | 1 |
| T2DM | endothelial | *SEC14L1* | 0.218807922 | 0.000329386 | 0.00103802 | 1 |
| T2DM | endothelial | *TMEM8A* | -0.482697126 | 0.01476413 | 0.025986027 | 1 |
| T2DM | endothelial | *SHC2* | 0.422985133 | 0.000133847 | 0.000492969 | 0.841497708 |
| T2DM | endothelial | *ERMARD* | -0.534053429 | 0.000481694 | 0.001429844 | 1 |
| T2DM | endothelial | *PHF10* | -0.372029085 | 0.024851213 | 0.040666209 | 1 |
| T2DM | endothelial | *GALNT8* | -0.402275889 | 0.007444979 | 0.014482599 | 1 |
| T2DM | endothelial | *DOCK6* | 0.27499812 | 0.000236463 | 0.000789507 | 1 |
| T2DM | endothelial | *ECSIT* | -0.438154753 | 0.016207502 | 0.028109398 | 1 |
| T2DM | endothelial | *ELOF1* | -1.052475964 | 2.26E-05 | 0.000114097 | 0.14182232 |
| T2DM | endothelial | *THEM6* | 0.242689859 | 0.023680238 | 0.038952814 | 1 |
| T2DM | endothelial | *TOMM40* | -0.633483753 | 0.000180521 | 0.000632137 | 1 |
| T2DM | endothelial | *DPP6* | -0.672088346 | 5.73E-09 | 1.22E-06 | 3.60E-05 |
| T2DM | endothelial | *FAM98C* | -0.573898612 | 0.000375441 | 0.001157625 | 1 |
| T2DM | endothelial | *ATP8B3* | -0.255426497 | 0.00287737 | 0.006444612 | 1 |
| T2DM | endothelial | *GTPBP3* | -0.767492128 | 4.02E-05 | 0.000182145 | 0.25299889 |
| T2DM | endothelial | *PLVAP* | 0.095711391 | 2.30E-05 | 0.000116007 | 0.144776397 |
| T2DM | endothelial | *BST2* | 0.229954534 | 6.31E-06 | 4.16E-05 | 0.039652516 |
| T2DM | endothelial | *DDA1* | -0.662758742 | 0.000295394 | 0.000947523 | 1 |
| T2DM | endothelial | *MRPL34* | -0.274121141 | 0.000214241 | 0.000729651 | 1 |
| T2DM | endothelial | *LSM7* | -0.362501114 | 0.008323134 | 0.015977876 | 1 |
| T2DM | endothelial | *RTN4IP1* | -0.450114834 | 0.002864388 | 0.00642011 | 1 |
| T2DM | endothelial | *ACTN4* | 0.235045602 | 0.017989231 | 0.030697063 | 1 |
| T2DM | endothelial | *STK33* | 0.160005717 | 2.96E-05 | 0.000141156 | 0.185903005 |
| T2DM | endothelial | *CACNG6* | 0.918425006 | 5.30E-05 | 0.000227458 | 0.333479477 |
| T2DM | endothelial | *PXDN* | -0.41758779 | 0.000189727 | 0.000658649 | 1 |
| T2DM | endothelial | *PGPEP1* | -0.335013858 | 0.007090177 | 0.013886586 | 1 |
| T2DM | endothelial | *HRC* | -0.27321453 | 0.024799826 | 0.040593416 | 1 |
| T2DM | endothelial | *TRPM4* | 0.341708448 | 0.00141188 | 0.003552018 | 1 |
| T2DM | endothelial | *CAMSAP1* | -0.42437854 | 0.010717796 | 0.019783554 | 1 |
| T2DM | endothelial | *UBAC1* | -0.47912831 | 0.000911967 | 0.00245023 | 1 |
| T2DM | endothelial | *TNNT3* | -0.564481426 | 3.06E-05 | 0.000144957 | 0.192068014 |
| T2DM | endothelial | *ATXN10* | -0.29172598 | 0.015213481 | 0.026671732 | 1 |
| T2DM | endothelial | *TUBGCP2* | -0.402287261 | 0.002586419 | 0.005900152 | 1 |
| T2DM | endothelial | *PAK4* | 0.331199916 | 0.02829629 | 0.045580009 | 1 |
| T2DM | endothelial | *CEP85* | -0.38809671 | 0.0011572 | 0.002998894 | 1 |
| T2DM | endothelial | *POMT1* | -0.456224126 | 0.00532413 | 0.010877721 | 1 |

| | | | | | | |
|---|---|---|---|---|---|---|
| T2DM | endothelial | *UCK1* | -0.419394887 | 0.00362285 | 0.007864937 | 1 |
| T2DM | endothelial | *YIPF2* | -0.654711229 | 6.83E-05 | 0.000279414 | 0.429179393 |
| T2DM | endothelial | *MAP3K10* | -0.469822275 | 0.007907312 | 0.015254149 | 1 |
| T2DM | endothelial | *SMPDL3B* | 1.562025803 | 1.79E-06 | 1.74E-05 | 0.011268305 |
| T2DM | endothelial | *ATPIF1* | -0.468221135 | 0.001160371 | 0.003005433 | 1 |
| T2DM | endothelial | *THEMIS2* | -0.325989216 | 0.001534593 | 0.003802912 | 1 |
| T2DM | endothelial | *CLIP1* | -0.34950369 | 0.001618022 | 0.003975188 | 1 |
| T2DM | endothelial | *PPAN* | -1.268933382 | 2.70E-05 | 0.000131067 | 0.169981204 |
| T2DM | endothelial | *ZNF236* | -0.44810119 | 0.000518932 | 0.001516044 | 1 |
| T2DM | endothelial | *PKDREJ* | -0.464421366 | 0.002501544 | 0.005748248 | 1 |
| T2DM | endothelial | *EPS8L1* | 2.125998575 | 0.00987335 | 0.018443647 | 1 |
| T2DM | endothelial | *LILRB2* | -0.661093749 | 2.08E-07 | 4.93E-06 | 0.001306701 |
| T2DM | endothelial | *COX4I2* | -1.738960729 | 2.30E-06 | 2.05E-05 | 0.014455463 |
| T2DM | endothelial | *ATP6V1E1* | -0.416590758 | 0.007421875 | 0.014459661 | 1 |
| T2DM | endothelial | *CHMP1A* | -0.430448268 | 0.004987266 | 0.010260125 | 1 |
| T2DM | endothelial | *PPT1* | -0.512490694 | 0.001787399 | 0.004327062 | 1 |
| T2DM | endothelial | *HAUS8* | -0.721867505 | 7.06E-08 | 4.46E-06 | 0.000443964 |
| T2DM | endothelial | *RFTN1* | -0.326616089 | 0.000704402 | 0.001970007 | 1 |
| T2DM | endothelial | *KCNC3* | -0.370524523 | 0.004736298 | 0.009824184 | 1 |
| T2DM | endothelial | *KIF3A* | -0.391705745 | 0.003844976 | 0.008275715 | 1 |
| T2DM | endothelial | *AOC3* | -0.374719735 | 0.006671771 | 0.013198687 | 1 |
| T2DM | endothelial | *ACLY* | -0.345614688 | 0.025733977 | 0.041892676 | 1 |
| T2DM | endothelial | *AOC2* | -0.883755413 | 2.32E-07 | 5.24E-06 | 0.001456933 |
| T2DM | endothelial | *ACAP3* | -0.492017488 | 2.95E-05 | 0.0001411 | 0.18568761 |
| T2DM | endothelial | *ANO1* | -0.626522189 | 0.000142875 | 0.000520123 | 0.898252221 |
| T2DM | endothelial | *PPFIA1* | -0.341446371 | 0.002222236 | 0.005207664 | 1 |
| T2DM | endothelial | *NINJ1* | -0.766384542 | 1.63E-06 | 1.65E-05 | 0.010264033 |
| T2DM | endothelial | *CKMT2* | -0.367927706 | 0.009342548 | 0.01762803 | 1 |
| T2DM | endothelial | *ZCCHC9* | -0.479787027 | 0.001888426 | 0.004534964 | 1 |
| T2DM | endothelial | *KHDRBS3* | -0.44751377 | 2.35E-06 | 2.08E-05 | 0.014795318 |
| T2DM | endothelial | *CHD1L* | -0.473911504 | 0.000197568 | 0.000679491 | 1 |
| T2DM | endothelial | *RBM8A* | -0.517303028 | 6.96E-05 | 0.000283943 | 0.437556885 |
| T2DM | endothelial | *ZNF304* | -0.409295749 | 0.027777646 | 0.044882565 | 1 |
| T2DM | endothelial | *ZSCAN5A* | -0.456102162 | 0.001783922 | 0.004320308 | 1 |
| T2DM | endothelial | *RHPN2* | -0.857140344 | 5.00E-08 | 3.98E-06 | 0.000314275 |
| T2DM | endothelial | *C19orf12* | -0.273702973 | 0.000254381 | 0.00083645 | 1 |
| T2DM | endothelial | *GCH1* | 0.286754338 | 3.91E-06 | 2.95E-05 | 0.024595715 |
| T2DM | endothelial | *DNAJB1* | -0.510737455 | 3.49E-05 | 0.000161259 | 0.219312798 |
| T2DM | endothelial | *FBXW9* | -0.482584512 | 0.000443331 | 0.001328451 | 1 |
| T2DM | endothelial | *MATN3* | -1.041868781 | 1.11E-07 | 4.46E-06 | 0.000698084 |
| T2DM | endothelial | *CCT6B* | -0.528464493 | 5.32E-05 | 0.000227776 | 0.334375869 |
| T2DM | endothelial | *ENOSF1* | -0.332700626 | 0.001071883 | 0.002803215 | 1 |
| T2DM | endothelial | *EMILIN2* | -0.32954515 | 5.38E-05 | 0.000229966 | 0.338477146 |
| T2DM | endothelial | *TRIM5* | 0.253942822 | 0.011974741 | 0.021771311 | 1 |
| T2DM | endothelial | *MRPL35* | 0.308624805 | 0.004423588 | 0.009273458 | 1 |
| T2DM | endothelial | *RPA1* | -0.419095332 | 0.001983852 | 0.004724423 | 1 |
| T2DM | endothelial | *SERPINF1* | -0.37088677 | 6.05E-06 | 4.03E-05 | 0.038009248 |
| T2DM | endothelial | *TMEM128* | -0.515868502 | 0.010962097 | 0.020157562 | 1 |
| T2DM | endothelial | *FIGNL1* | -0.680021831 | 0.000203733 | 0.000698021 | 1 |
| T2DM | endothelial | *KDM6B* | -0.583133055 | 0.001835004 | 0.004426964 | 1 |

| | | | | | | |
|---|---|---|---|---|---|---|
| T2DM | endothelial | *XAF1* | 0.205213218 | 5.46E-06 | 3.75E-05 | 0.034345997 |
| T2DM | endothelial | *PRMT7* | -0.35769142 | 0.000334687 | 0.001050512 | 1 |
| T2DM | endothelial | *TERF2* | -0.349636859 | 0.031191137 | 0.049657806 | 1 |
| T2DM | endothelial | *PCNA* | -0.676478616 | 9.26E-05 | 0.000361095 | 0.582445579 |
| T2DM | endothelial | *IGHMBP2* | -0.357023669 | 0.000348539 | 0.00108943 | 1 |
| T2DM | endothelial | *MMACHC* | -0.535238003 | 0.000743414 | 0.002061426 | 1 |
| T2DM | endothelial | *LPIN3* | -0.569067926 | 7.20E-07 | 9.76E-06 | 0.004525893 |
| T2DM | endothelial | *DMGDH* | -0.464888427 | 0.001115787 | 0.002905316 | 1 |
| T2DM | endothelial | *BHMT2* | -0.132869289 | 0.0001464 | 0.000529281 | 0.920419251 |
| T2DM | endothelial | *KANK4* | -0.235681367 | 8.26E-05 | 0.00032704 | 0.519339363 |
| T2DM | endothelial | *USPL1* | -0.375576658 | 0.007101729 | 0.013900551 | 1 |
| T2DM | endothelial | *WASF3* | -0.47726202 | 7.37E-06 | 4.67E-05 | 0.046348891 |
| T2DM | endothelial | *PEMT* | -0.470497582 | 0.000351519 | 0.001097689 | 1 |
| T2DM | endothelial | *SCO1* | -0.482507306 | 0.004630182 | 0.009635867 | 1 |
| T2DM | endothelial | *DSTYK* | -0.526050389 | 0.000161623 | 0.000575055 | 1 |
| T2DM | endothelial | *TMCC2* | -0.841030716 | 6.89E-07 | 9.54E-06 | 0.004330753 |
| T2DM | endothelial | *TRPC4* | -0.639293659 | 1.50E-07 | 4.49E-06 | 0.000946002 |
| T2DM | endothelial | *RFC3* | 0.248154405 | 0.015201361 | 0.026658565 | 1 |
| T2DM | endothelial | *BTBD2* | -0.473528379 | 0.001719501 | 0.004185252 | 1 |
| T2DM | endothelial | *PRAM1* | -0.654458808 | 2.91E-08 | 3.21E-06 | 0.000182833 |
| T2DM | endothelial | *ZNF414* | -0.861249701 | 0.000217323 | 0.00073735 | 1 |
| T2DM | endothelial | *RTN3* | -0.384162087 | 2.23E-05 | 0.000113056 | 0.140302963 |
| T2DM | endothelial | *GSTT2B* | 0.21481035 | 0.017596515 | 0.030103208 | 1 |
| T2DM | endothelial | *SLC2A11* | -0.728707718 | 3.72E-07 | 6.87E-06 | 0.002340525 |
| T2DM | endothelial | *C1QTNF6* | -0.794095369 | 8.72E-06 | 5.34E-05 | 0.054795373 |
| T2DM | endothelial | *FAM83F* | 1.002923537 | 0.004624224 | 0.009626654 | 1 |
| T2DM | endothelial | *GIMAP4* | 0.185668188 | 0.000348645 | 0.00108943 | 1 |
| T2DM | endothelial | *ADCK2* | -0.270163907 | 0.002099999 | 0.004952247 | 1 |
| T2DM | endothelial | *AGAP3* | -0.524564935 | 8.98E-05 | 0.00035142 | 0.564731793 |
| T2DM | endothelial | *ACTR3B* | -0.392913715 | 0.022511631 | 0.037274328 | 1 |
| T2DM | endothelial | *SFTPD* | 0.155791666 | 0.002758522 | 0.006217053 | 1 |
| T2DM | endothelial | *DYDC2* | -1.667348756 | 4.63E-07 | 7.72E-06 | 0.002910385 |
| T2DM | endothelial | *TMTC1* | 0.203510418 | 0.000245279 | 0.000814616 | 1 |
| T2DM | endothelial | *SPINK5* | 0.72588864 | 0.00014796 | 0.000533691 | 0.930224056 |
| T2DM | endothelial | *SWAP70* | 0.258729273 | 0.001958027 | 0.004673545 | 1 |
| T2DM | endothelial | *AMPD3* | -0.401642468 | 1.43E-07 | 4.46E-06 | 0.000899338 |
| T2DM | endothelial | *RRAS2* | -0.161807841 | 0.02503066 | 0.040895987 | 1 |
| T2DM | endothelial | *HSD17B4* | -0.38990432 | 0.000792509 | 0.002175212 | 1 |
| T2DM | endothelial | *DPF2* | -0.482007947 | 0.000515438 | 0.001508607 | 1 |
| T2DM | endothelial | *GSC* | -0.807930556 | 1.17E-07 | 4.46E-06 | 0.000735889 |
| T2DM | endothelial | *CATSPERB* | -0.336608822 | 3.33E-05 | 0.00015534 | 0.209553411 |
| T2DM | endothelial | *ELP3* | -0.395170238 | 0.004658109 | 0.009685085 | 1 |
| T2DM | endothelial | *MRO* | -0.46757979 | 1.27E-06 | 1.40E-05 | 0.007987962 |
| T2DM | endothelial | *THUMPD3* | -0.522874508 | 0.000250285 | 0.000826441 | 1 |
| T2DM | endothelial | *EDEM1* | -0.596290983 | 4.27E-05 | 0.000191908 | 0.268478957 |
| T2DM | endothelial | *CNTN6* | -0.367332317 | 1.93E-07 | 4.73E-06 | 0.00121488 |
| T2DM | endothelial | *GSTM5* | -0.282114939 | 0.00058864 | 0.001696828 | 1 |
| T2DM | endothelial | *SYT6* | 0.228517374 | 5.76E-07 | 8.63E-06 | 0.003620137 |
| T2DM | endothelial | *VAV3* | -0.371461275 | 0.002575314 | 0.005879085 | 1 |
| T2DM | endothelial | *PSRC1* | -0.478186286 | 0.000210696 | 0.000719526 | 1 |

| | | | | | | |
|---|---|---|---|---|---|---|
| T2DM | endothelial | *WNT2B* | 0.261381611 | 0.001419965 | 0.003565224 | 1 |
| T2DM | endothelial | *CEPT1* | -0.408200934 | 7.02E-05 | 0.000285488 | 0.44164933 |
| T2DM | endothelial | *NAPG* | -0.374916966 | 0.009083623 | 0.017206609 | 1 |
| T2DM | endothelial | *SPIRE1* | -0.435113716 | 3.78E-05 | 0.000173199 | 0.237802494 |
| T2DM | endothelial | *FKBP11* | -0.539874265 | 0.000841134 | 0.002291251 | 1 |
| T2DM | endothelial | *LPIN1* | -0.405428828 | 2.79E-06 | 2.32E-05 | 0.01751766 |
| T2DM | endothelial | *LDHA* | -0.467494908 | 0.001424586 | 0.003573971 | 1 |
| T2DM | endothelial | *ANO3* | -0.269273354 | 5.27E-06 | 3.65E-05 | 0.033116501 |
| T2DM | endothelial | *FST* | -0.540140075 | 9.12E-08 | 4.46E-06 | 0.000573069 |
| T2DM | endothelial | *TIMM17A* | -0.490399968 | 0.00072416 | 0.002018082 | 1 |
| T2DM | endothelial | *NARS* | -0.414088152 | 0.009845177 | 0.018399711 | 1 |
| T2DM | endothelial | *ECHDC3* | -0.418458088 | 7.25E-07 | 9.76E-06 | 0.004557845 |
| T2DM | endothelial | *TMEM241* | -0.369810962 | 0.002173936 | 0.005105541 | 1 |
| T2DM | endothelial | *EMP1* | -0.209543663 | 0.021755865 | 0.036194529 | 1 |
| T2DM | endothelial | *PRH2* | -0.860051791 | 0.003627622 | 0.007869861 | 1 |
| T2DM | endothelial | *DDB2* | -0.389951308 | 0.001477725 | 0.003683766 | 1 |
| T2DM | endothelial | *ACP2* | -0.385332968 | 0.011814351 | 0.021529514 | 1 |
| T2DM | endothelial | *HOOK1* | -1.188513308 | 5.41E-06 | 3.72E-05 | 0.03403463 |
| T2DM | endothelial | *FHOD3* | -0.303881061 | 3.39E-07 | 6.50E-06 | 0.002133349 |
| T2DM | endothelial | *TPGS2* | -0.327098676 | 0.023912023 | 0.03925193 | 1 |
| T2DM | endothelial | *SLC43A3* | -0.669453572 | 2.78E-06 | 2.32E-05 | 0.017482703 |
| T2DM | endothelial | *TIMM10* | -0.733081244 | 5.01E-05 | 0.00021625 | 0.315248669 |
| T2DM | endothelial | *DHX34* | -0.568974479 | 8.49E-06 | 5.23E-05 | 0.053351365 |
| T2DM | endothelial | *APLNR* | 0.14759111 | 5.97E-07 | 8.81E-06 | 0.003750803 |
| T2DM | endothelial | *FADS2* | -0.587036041 | 1.34E-05 | 7.43E-05 | 0.083953046 |
| T2DM | endothelial | *CLDN10* | 0.314207132 | 2.20E-06 | 1.99E-05 | 0.013814377 |
| T2DM | endothelial | *DZIP1* | -0.307393182 | 0.004060595 | 0.00865682 | 1 |
| T2DM | endothelial | *BIVM* | -0.719948643 | 0.000159931 | 0.000570004 | 1 |
| T2DM | endothelial | *ERCC5* | -1.124310653 | 5.81E-06 | 3.92E-05 | 0.036514881 |
| T2DM | endothelial | *CARS2* | -0.426571084 | 0.000404092 | 0.001227902 | 1 |
| T2DM | endothelial | *KLB* | -0.225388788 | 0.000255488 | 0.000839212 | 1 |
| T2DM | endothelial | *WDR36* | -0.401216577 | 0.005569585 | 0.011295477 | 1 |
| T2DM | endothelial | *UBQLN1* | -0.363911596 | 0.005583785 | 0.011313328 | 1 |
| T2DM | endothelial | *CTSL* | -0.468370315 | 3.30E-05 | 0.000154501 | 0.207494561 |
| T2DM | endothelial | *USP30* | -0.309723764 | 0.002717314 | 0.006134202 | 1 |
| T2DM | endothelial | *HRK* | 0.59205 | 3.35E-06 | 2.62E-05 | 0.021049951 |
| T2DM | endothelial | *DTX1* | -0.336574039 | 1.11E-07 | 4.46E-06 | 0.000699171 |
| T2DM | endothelial | *CCDC146* | -0.39426195 | 0.00013635 | 0.000501012 | 0.85723069 |
| T2DM | endothelial | *POM121C* | -0.601281591 | 4.83E-06 | 3.47E-05 | 0.030389053 |
| T2DM | endothelial | *SRPK2* | -0.25335942 | 0.017428462 | 0.029913387 | 1 |
| T2DM | endothelial | *MDFIC* | -0.318958549 | 0.000618338 | 0.00176624 | 1 |
| T2DM | endothelial | *MTO1* | -0.416195466 | 0.00085612 | 0.002325023 | 1 |
| T2DM | endothelial | *KHDC1* | -0.248461609 | 9.35E-05 | 0.000364214 | 0.587840761 |
| T2DM | endothelial | *EPHA7* | 0.104003054 | 9.54E-06 | 5.72E-05 | 0.05997685 |
| T2DM | endothelial | *PRR5L* | -0.458686527 | 4.42E-05 | 0.000196337 | 0.278013704 |
| T2DM | endothelial | *ITGA7* | -0.246545994 | 2.44E-06 | 2.13E-05 | 0.015350642 |
| T2DM | endothelial | *RDH5* | -0.337553193 | 2.15E-06 | 1.96E-05 | 0.013497429 |
| T2DM | endothelial | *BLOC1S1* | -0.595927872 | 1.96E-05 | 0.000101607 | 0.123248832 |
| T2DM | endothelial | *PPP1R1A* | -0.630858627 | 9.60E-05 | 0.000372405 | 0.603295634 |
| T2DM | endothelial | *B4GALNT1* | -1.08772032 | 2.30E-07 | 5.23E-06 | 0.00144439 |

| | | | | | | |
|---|---|---|---|---|---|---|
| T2DM | endothelial | *FAIM2* | -0.769941701 | 5.99E-08 | 4.43E-06 | 0.000376586 |
| T2DM | endothelial | *KRT7* | 0.76481198 | 0.00040448 | 0.001228485 | 1 |
| T2DM | endothelial | *HNRNPA1* | -0.53719607 | 0.001739315 | 0.004226933 | 1 |
| T2DM | endothelial | *SLC26A10* | -1.26389523 | 3.78E-07 | 6.87E-06 | 0.002375263 |
| T2DM | endothelial | *MAP7* | -0.43163886 | 5.52E-05 | 0.000234867 | 0.346898106 |
| T2DM | endothelial | *PKIB* | -0.644504885 | 7.90E-06 | 4.94E-05 | 0.049638794 |
| T2DM | endothelial | *SMPD2* | -0.419316692 | 0.000594822 | 0.001712293 | 1 |
| T2DM | endothelial | *MICAL1* | -0.540395722 | 0.000163994 | 0.000582173 | 1 |
| T2DM | endothelial | *PRADC1* | -0.855181078 | 0.002422232 | 0.005594626 | 1 |
| T2DM | endothelial | *SMYD5* | -0.994339014 | 6.44E-07 | 9.14E-06 | 0.004048041 |
| T2DM | endothelial | *DYSF* | 0.165182216 | 1.11E-05 | 6.39E-05 | 0.069527083 |
| T2DM | endothelial | *KLHL36* | -0.360561123 | 0.019923956 | 0.033465646 | 1 |
| T2DM | endothelial | *MPHOSPH6* | -0.382775978 | 0.026766689 | 0.043382876 | 1 |
| T2DM | endothelial | *FHOD1* | -0.652344663 | 2.88E-05 | 0.000138128 | 0.18122334 |
| T2DM | endothelial | *RNASEL* | -0.343490633 | 0.012109929 | 0.021985309 | 1 |
| T2DM | endothelial | *SLC19A3* | -0.163689114 | 0.000904847 | 0.002434222 | 1 |
| T2DM | endothelial | *TMBIM1* | -0.489741358 | 0.000160915 | 0.000572861 | 1 |
| T2DM | endothelial | *ARMC9* | -0.399951517 | 0.01550834 | 0.027076072 | 1 |
| T2DM | endothelial | *GCC2* | -0.283143083 | 0.013062617 | 0.023423611 | 1 |
| T2DM | endothelial | *USP44* | -0.420784591 | 0.000107596 | 0.000409229 | 0.676455273 |
| T2DM | endothelial | *APPL2* | -0.361035718 | 0.002742072 | 0.006185649 | 1 |
| T2DM | endothelial | *SLC41A2* | -0.475487452 | 3.44E-05 | 0.000159739 | 0.216446318 |
| T2DM | endothelial | *NEK3* | 0.200689043 | 0.02377723 | 0.039079574 | 1 |
| T2DM | endothelial | *RNASEH2B* | -0.337573058 | 0.008148429 | 0.015666414 | 1 |
| T2DM | endothelial | *TBC1D4* | -0.378012646 | 6.46E-05 | 0.000267136 | 0.406313546 |
| T2DM | endothelial | *LRCH1* | 0.240638673 | 0.00777527 | 0.01503171 | 1 |
| T2DM | endothelial | *SUCLA2* | -0.226655017 | 0.030243679 | 0.048292315 | 1 |
| T2DM | endothelial | *GPNMB* | -0.410026057 | 4.75E-06 | 3.42E-05 | 0.029862182 |
| T2DM | endothelial | *KDELR2* | -0.530988678 | 0.001516924 | 0.003763905 | 1 |
| T2DM | endothelial | *NUPL2* | -0.451564403 | 0.000943796 | 0.002519087 | 1 |
| T2DM | endothelial | *ZDHHC4* | -0.538370823 | 8.18E-05 | 0.000324298 | 0.51457288 |
| T2DM | endothelial | *AOAH* | -0.489374238 | 1.38E-07 | 4.46E-06 | 0.000865444 |
| T2DM | endothelial | *TBRG4* | -0.763863138 | 1.98E-05 | 0.000102395 | 0.124307573 |
| T2DM | endothelial | *DDX56* | -0.71184212 | 2.93E-05 | 0.000140139 | 0.184282386 |
| T2DM | endothelial | *HUS1* | 0.305880768 | 0.004021177 | 0.008599026 | 1 |
| T2DM | endothelial | *NACAD* | -1.679119133 | 4.32E-10 | 1.70E-07 | 2.71E-06 |
| T2DM | endothelial | *ALPK3* | -0.600334663 | 3.66E-06 | 2.79E-05 | 0.022992253 |
| T2DM | endothelial | *TM6SF1* | 0.234268751 | 2.31E-06 | 2.05E-05 | 0.014522231 |
| T2DM | endothelial | *CIB2* | -0.454600349 | 0.000293318 | 0.000942787 | 1 |
| T2DM | endothelial | *RSAD1* | -0.651643583 | 0.000487731 | 0.001444355 | 1 |
| T2DM | endothelial | *BRIP1* | -0.4776584 | 4.99E-07 | 8.08E-06 | 0.003134337 |
| T2DM | endothelial | *MRPL47* | -0.644567029 | 0.000608224 | 0.001741304 | 1 |
| T2DM | endothelial | *BLK* | -0.997481529 | 6.89E-07 | 9.54E-06 | 0.004331415 |
| T2DM | endothelial | *RPS6KC1* | -0.314002453 | 0.006328081 | 0.012602041 | 1 |
| T2DM | endothelial | *CBWD2* | -0.548922446 | 0.000760534 | 0.002099901 | 1 |
| T2DM | endothelial | *IL1RN* | -0.6925816 | 1.50E-06 | 1.58E-05 | 0.009448745 |
| T2DM | endothelial | *UGGT1* | -0.384815331 | 0.006802733 | 0.013432407 | 1 |
| T2DM | endothelial | *GYPC* | -0.440915129 | 4.46E-05 | 0.000197889 | 0.280408697 |
| T2DM | endothelial | *LRRC8A* | 0.319333202 | 0.002609793 | 0.005949155 | 1 |
| T2DM | endothelial | *TXN* | -0.534923012 | 0.000871438 | 0.002358471 | 1 |

| | | | | | | |
|---|---|---|---|---|---|---|
| T2DM | endothelial | *TOR1A* | -0.58917544 | 3.25E-06 | 2.56E-05 | 0.020439292 |
| T2DM | endothelial | *ZNF189* | -0.530582632 | 0.000109282 | 0.000414888 | 0.687054788 |
| T2DM | endothelial | *ALDOB* | -0.61817505 | 0.000391709 | 0.001196883 | 1 |
| T2DM | endothelial | *FPGS* | -0.429532877 | 0.008873585 | 0.016869739 | 1 |
| T2DM | endothelial | *TEX10* | -0.385223458 | 0.013891784 | 0.024675606 | 1 |
| T2DM | endothelial | *GARNL3* | -0.499511091 | 2.11E-05 | 0.000107898 | 0.132823002 |
| T2DM | endothelial | *GABBR2* | -1.57639012 | 0.000843048 | 0.002294478 | 1 |
| T2DM | endothelial | *PSMB7* | -0.417062365 | 0.002087314 | 0.004931582 | 1 |
| T2DM | endothelial | *RABEPK* | -0.550789508 | 0.000257677 | 0.000844197 | 1 |
| T2DM | endothelial | *GOLGA1* | -0.405965074 | 0.003228069 | 0.007118508 | 1 |
| T2DM | endothelial | *LMX1B* | -0.911654433 | 8.73E-08 | 4.46E-06 | 0.000548586 |
| T2DM | endothelial | *DERL1* | -0.4695164 | 0.004791778 | 0.009909838 | 1 |
| T2DM | endothelial | *APTX* | -0.351035399 | 0.007818812 | 0.015111242 | 1 |
| T2DM | endothelial | *TLN1* | -0.337629818 | 0.009141105 | 0.017289448 | 1 |
| T2DM | endothelial | *SPAG8* | -0.546511093 | 2.25E-05 | 0.000113839 | 0.141388504 |
| T2DM | endothelial | *CD72* | -0.559560229 | 5.06E-06 | 3.56E-05 | 0.031839 |
| T2DM | endothelial | *TMEM8B* | -0.767712539 | 3.07E-06 | 2.47E-05 | 0.019307151 |
| T2DM | endothelial | *GRHPR* | -0.454257902 | 0.000442082 | 0.001326036 | 1 |
| T2DM | endothelial | *ARHGEF39* | -0.41186395 | 1.13E-05 | 6.47E-05 | 0.070759518 |
| T2DM | endothelial | *DENND4C* | -0.273842228 | 0.009399834 | 0.017720167 | 1 |
| T2DM | endothelial | *PPIL1* | -1.287061278 | 5.07E-06 | 3.56E-05 | 0.031898355 |
| T2DM | endothelial | *TMEM63B* | -0.417381967 | 0.000764616 | 0.002108396 | 1 |
| T2DM | endothelial | *FRS3* | 0.516897292 | 0.00176115 | 0.004271741 | 1 |
| T2DM | endothelial | *SLC22A23* | -0.321269116 | 0.013232065 | 0.023687356 | 1 |
| T2DM | endothelial | *LRRC1* | 0.184767014 | 9.91E-06 | 5.89E-05 | 0.06233299 |
| T2DM | endothelial | *BPHL* | -0.212250221 | 0.000628413 | 0.001787058 | 1 |
| T2DM | endothelial | *TUBB2B* | -0.303351867 | 0.001648617 | 0.00403616 | 1 |
| T2DM | endothelial | *UQCC2* | -0.675503875 | 8.15E-06 | 5.08E-05 | 0.051249937 |
| T2DM | endothelial | *TCF19* | 0.416432204 | 0.009075929 | 0.017197217 | 1 |
| T2DM | endothelial | *FLOT1* | 0.337831462 | 0.00065503 | 0.001846714 | 1 |
| T2DM | endothelial | *NRM* | 0.368480659 | 0.002096084 | 0.004948583 | 1 |
| T2DM | endothelial | *MTCH1* | -0.457041571 | 0.002708617 | 0.006118231 | 1 |
| T2DM | endothelial | *FAM8A1* | -0.676999651 | 0.000223003 | 0.000753773 | 1 |
| T2DM | endothelial | *FGFBP2* | -0.397627114 | 0.02603702 | 0.042309317 | 1 |
| T2DM | endothelial | *CCDC90B* | -0.401547435 | 0.012988487 | 0.023311053 | 1 |
| T2DM | endothelial | *SYTL2* | -0.611885319 | 1.55E-07 | 4.49E-06 | 0.000971719 |
| T2DM | endothelial | *PRCP* | 0.327150319 | 0.00305541 | 0.006795527 | 1 |
| T2DM | endothelial | *NARS2* | -0.435299614 | 0.000923105 | 0.002472796 | 1 |
| T2DM | endothelial | *PI15* | -2.559749169 | 0.000496103 | 0.001460207 | 1 |
| T2DM | endothelial | *TTPA* | -0.176855313 | 1.63E-06 | 1.65E-05 | 0.010222918 |
| T2DM | endothelial | *GGH* | -0.164600041 | 0.013485285 | 0.024051628 | 1 |
| T2DM | endothelial | *SULF1* | 0.166240583 | 8.27E-06 | 5.14E-05 | 0.05201922 |
| T2DM | endothelial | *TGS1* | -0.302700453 | 0.011572052 | 0.021136981 | 1 |
| T2DM | endothelial | *DDX60* | 0.287560323 | 5.79E-05 | 0.000244298 | 0.363759515 |
| T2DM | endothelial | *DCUN1D5* | -0.491734685 | 2.74E-08 | 3.19E-06 | 0.000172116 |
| T2DM | endothelial | *TRIM29* | -0.442954442 | 4.46E-06 | 3.25E-05 | 0.028022225 |
| T2DM | endothelial | *POU2F3* | -0.303854606 | 0.008376945 | 0.016046878 | 1 |
| T2DM | endothelial | *PPP2R1B* | -0.384133419 | 7.23E-06 | 4.60E-05 | 0.045480353 |
| T2DM | endothelial | *FDX1* | -0.525119037 | 6.30E-05 | 0.000261112 | 0.395845305 |
| T2DM | endothelial | *FXYD6* | 0.495563313 | 0.009570098 | 0.017971088 | 1 |

| | | | | | | |
|---|---|---|---|---|---|---|
| T2DM | endothelial | *TMPRSS13* | -0.881833087 | 0.001408051 | 0.003546643 | 1 |
| T2DM | endothelial | *NDUFAF1* | -0.631250787 | 4.93E-06 | 3.50E-05 | 0.030968376 |
| T2DM | endothelial | *ITGA11* | -0.368079495 | 7.48E-08 | 4.46E-06 | 0.000470454 |
| T2DM | endothelial | *PAQR5* | -0.550639401 | 2.07E-07 | 4.93E-06 | 0.001303418 |
| T2DM | endothelial | *ITPKA* | -1.817724615 | 1.35E-05 | 7.48E-05 | 0.084631805 |
| T2DM | endothelial | *PLCB2* | -0.139554107 | 0.030248948 | 0.048292315 | 1 |
| T2DM | endothelial | *RSL24D1* | -0.376514562 | 0.009669425 | 0.018135941 | 1 |
| T2DM | endothelial | *SPTBN5* | 0.201391142 | 0.013942543 | 0.024747817 | 1 |
| T2DM | endothelial | *RABGGTB* | -0.50424451 | 0.0043769 | 0.009201699 | 1 |
| T2DM | endothelial | *SLC44A5* | 0.246075273 | 1.19E-05 | 6.78E-05 | 0.075053075 |
| T2DM | endothelial | *DNASE2B* | -2.088290135 | 4.07E-07 | 7.23E-06 | 0.002559984 |
| T2DM | endothelial | *DBT* | -0.367887261 | 0.007445166 | 0.014482599 | 1 |
| T2DM | endothelial | *KHK* | -0.126636199 | 0.018993484 | 0.032186532 | 1 |
| T2DM | endothelial | *ADCY3* | -0.550818282 | 5.94E-07 | 8.81E-06 | 0.003734621 |
| T2DM | endothelial | *DYNC2LI1* | -0.372875412 | 0.000152631 | 0.000548024 | 0.959589619 |
| T2DM | endothelial | *LHCGR* | -0.448361822 | 6.79E-08 | 4.46E-06 | 0.000426695 |
| T2DM | endothelial | *PREB* | -0.461567622 | 5.01E-05 | 0.00021625 | 0.315292846 |
| T2DM | endothelial | *CENPO* | -0.931877328 | 8.64E-06 | 5.30E-05 | 0.054307163 |
| T2DM | endothelial | *MYOF* | 0.245584472 | 0.003182524 | 0.007033619 | 1 |
| T2DM | endothelial | *STAMBPL1* | -0.388237161 | 4.53E-06 | 3.29E-05 | 0.028468233 |
| T2DM | endothelial | *ATAD1* | -0.410901614 | 0.003168016 | 0.007008204 | 1 |
| T2DM | endothelial | *BTBD16* | -0.740821699 | 1.53E-08 | 2.38E-06 | 9.62E-05 |
| T2DM | endothelial | *TACC2* | -0.338318009 | 0.001067536 | 0.002797616 | 1 |
| T2DM | endothelial | *PLCE1* | -0.4440184 | 0.000431007 | 0.001299637 | 1 |
| T2DM | endothelial | *RBP4* | -0.473756752 | 0.000133201 | 0.000491453 | 0.837436028 |
| T2DM | endothelial | *DNAJC13* | -0.311327912 | 0.016262672 | 0.028173992 | 1 |
| T2DM | endothelial | *FAM149B1* | -0.351426021 | 0.004194831 | 0.008900743 | 1 |
| T2DM | endothelial | *ASCC1* | -0.391158962 | 0.001744386 | 0.00423598 | 1 |
| T2DM | endothelial | *DNA2* | -0.119319962 | 0.021643194 | 0.036026148 | 1 |
| T2DM | endothelial | *ATIC* | -0.464645612 | 1.54E-06 | 1.60E-05 | 0.009679538 |
| T2DM | endothelial | *SMARCAL1* | -0.414368817 | 0.006839774 | 0.013492834 | 1 |
| T2DM | endothelial | *STAT4* | -0.484775108 | 6.61E-06 | 4.30E-05 | 0.041529186 |
| T2DM | endothelial | *ASNSD1* | -0.464280965 | 0.00170616 | 0.004160834 | 1 |
| T2DM | endothelial | *METTL5* | -0.44064927 | 0.013377215 | 0.023913151 | 1 |
| T2DM | endothelial | *NAB1* | -0.322684251 | 0.030704317 | 0.048966022 | 1 |
| T2DM | endothelial | *MDH1B* | 0.886576708 | 0.000254071 | 0.000836306 | 1 |
| T2DM | endothelial | *COX17* | 0.348192745 | 0.01746516 | 0.029960016 | 1 |
| T2DM | endothelial | *SPPL2A* | -0.360808183 | 0.003709872 | 0.00802338 | 1 |
| T2DM | endothelial | *UBL7* | -0.584618284 | 0.009415956 | 0.01773992 | 1 |
| T2DM | endothelial | *ARHGAP24* | -0.325504046 | 2.70E-05 | 0.000131067 | 0.169870218 |
| T2DM | endothelial | *FAM13A* | -0.266668704 | 0.002820266 | 0.006334767 | 1 |
| T2DM | endothelial | *HERC3* | -0.295333521 | 0.020258323 | 0.033945649 | 1 |
| T2DM | endothelial | *PRKG2* | -0.471352928 | 1.13E-07 | 4.46E-06 | 0.000707544 |
| T2DM | endothelial | *PRDM5* | 0.308936363 | 0.000638777 | 0.001809005 | 1 |
| T2DM | endothelial | *NAAA* | -0.378800031 | 0.002284045 | 0.005326332 | 1 |
| T2DM | endothelial | *BMP2K* | -0.304342137 | 0.001581773 | 0.003896789 | 1 |
| T2DM | endothelial | *SCARB2* | -0.332372676 | 0.009039709 | 0.017149261 | 1 |
| T2DM | endothelial | *ANXA3* | 0.122947563 | 8.65E-07 | 1.08E-05 | 0.005436132 |
| T2DM | endothelial | *ENPEP* | -0.526038712 | 5.16E-07 | 8.26E-06 | 0.003245664 |
| T2DM | endothelial | *PAPSS1* | -0.276080209 | 0.012091074 | 0.021963763 | 1 |

| | | | | | | |
|---|---|---|---|---|---|---|
| T2DM | endothelial | *RNF185* | -0.465895254 | 0.000589556 | 0.001698689 | 1 |
| T2DM | endothelial | *PARVG* | -0.639714385 | 4.68E-05 | 0.000206039 | 0.294223516 |
| T2DM | endothelial | *B4GALNT3* | -0.969738607 | 3.04E-06 | 2.45E-05 | 0.019101163 |
| T2DM | endothelial | *ERP27* | -1.160412997 | 1.80E-05 | 9.45E-05 | 0.113177352 |
| T2DM | endothelial | *KIF21A* | -0.26400765 | 0.000597271 | 0.0017151 | 1 |
| T2DM | endothelial | *CPNE8* | 0.188246924 | 0.00215278 | 0.005065317 | 1 |
| T2DM | endothelial | *ALG10* | -0.96192824 | 0.00020239 | 0.000693798 | 1 |
| T2DM | endothelial | *SLCO1C1* | -0.50035315 | 6.85E-06 | 4.41E-05 | 0.043081091 |
| T2DM | endothelial | *PRICKLE1* | -0.375506962 | 8.19E-05 | 0.000324298 | 0.514661146 |
| T2DM | endothelial | *CD27* | -0.835157656 | 1.81E-05 | 9.48E-05 | 0.113970053 |
| T2DM | endothelial | *PEX5* | -0.523121881 | 0.000908818 | 0.002443857 | 1 |
| T2DM | endothelial | *PPFIA2* | -0.289239216 | 9.29E-06 | 5.61E-05 | 0.058406606 |
| T2DM | endothelial | *GLIPR1* | -0.488046861 | 7.82E-05 | 0.000312041 | 0.491776442 |
| T2DM | endothelial | *LGR5* | -0.570988703 | 1.55E-05 | 8.43E-05 | 0.09757865 |
| T2DM | endothelial | *AMDHD1* | 0.412490814 | 0.000811499 | 0.002223047 | 1 |
| T2DM | endothelial | *GAS2L3* | -0.63360511 | 7.64E-06 | 4.81E-05 | 0.048041515 |
| T2DM | endothelial | *TMEM132B* | -0.536645489 | 0.004616737 | 0.009614252 | 1 |
| T2DM | endothelial | *TDG* | -0.38291825 | 0.008774127 | 0.016710978 | 1 |
| T2DM | endothelial | *MMAB* | -0.542625276 | 0.000403381 | 0.001226927 | 1 |
| T2DM | endothelial | *GLTP* | -0.424253316 | 0.029009525 | 0.046526246 | 1 |
| T2DM | endothelial | *SLC46A3* | -0.260268185 | 0.014394721 | 0.025399834 | 1 |
| T2DM | endothelial | *CCDC65* | -0.485124079 | 0.000155724 | 0.000556587 | 0.979035845 |
| T2DM | endothelial | *ACVRL1* | 0.295434335 | 2.50E-05 | 0.000123889 | 0.156967169 |
| T2DM | endothelial | *GPR84* | -3.290256344 | 2.09E-08 | 2.80E-06 | 0.000131621 |
| T2DM | endothelial | *BRCA2* | -0.734923191 | 2.23E-05 | 0.000112967 | 0.140078521 |
| T2DM | endothelial | *KANSL2* | -0.293150008 | 0.013036886 | 0.02339124 | 1 |
| T2DM | endothelial | *CERS5* | -0.303035079 | 0.030653244 | 0.048900519 | 1 |
| T2DM | endothelial | *ZNF740* | -0.500200081 | 0.00098547 | 0.002612651 | 1 |
| T2DM | endothelial | *WDFY2* | -0.399680951 | 0.000155175 | 0.000555572 | 0.975583868 |
| T2DM | endothelial | *HNRNPA1L2* | 0.755010557 | 0.000139422 | 0.000511106 | 0.876545933 |
| T2DM | endothelial | *VPS33A* | -0.593539825 | 0.029253057 | 0.046833199 | 1 |
| T2DM | endothelial | *RHOF* | -0.588609983 | 3.58E-08 | 3.41E-06 | 0.000225208 |
| T2DM | endothelial | *METTL21C* | -1.251093555 | 0.000715872 | 0.001997642 | 1 |
| T2DM | endothelial | *GRTP1* | -0.253165912 | 0.00026892 | 0.000875102 | 1 |
| T2DM | endothelial | *TSSK4* | -1.617021152 | 0.000443732 | 0.001328451 | 1 |
| T2DM | endothelial | *TMX1* | -0.455765033 | 0.006470775 | 0.012849577 | 1 |
| T2DM | endothelial | *RTN1* | -0.375611615 | 2.79E-06 | 2.32E-05 | 0.017566797 |
| T2DM | endothelial | *RAB15* | -0.448155447 | 0.000112422 | 0.000425015 | 0.706800006 |
| T2DM | endothelial | *STON2* | -0.47089777 | 6.04E-05 | 0.000252299 | 0.379709428 |
| T2DM | endothelial | *AK7* | 0.310423528 | 1.36E-06 | 1.47E-05 | 0.008520073 |
| T2DM | endothelial | *SLC24A4* | -0.366582805 | 4.30E-05 | 0.000192477 | 0.27004555 |
| T2DM | endothelial | *DUOXA1* | -0.410173618 | 3.48E-05 | 0.000161198 | 0.219067841 |
| T2DM | endothelial | *SORD* | -0.803098056 | 0.00011595 | 0.000435212 | 0.728980363 |
| T2DM | endothelial | *ZSCAN29* | -0.672650945 | 1.32E-06 | 1.44E-05 | 0.008319149 |
| T2DM | endothelial | *SRP14* | 0.360230611 | 8.73E-05 | 0.00034222 | 0.548920664 |
| T2DM | endothelial | *ANP32A* | -0.44129631 | 0.001663478 | 0.004065679 | 1 |
| T2DM | endothelial | *COMMD4* | -0.59836854 | 0.004290002 | 0.009069013 | 1 |
| T2DM | endothelial | *PSTPIP1* | -0.504114049 | 4.25E-05 | 0.000191137 | 0.26690574 |
| T2DM | endothelial | *MAN2C1* | -0.246128866 | 7.97E-08 | 4.46E-06 | 0.000501164 |
| T2DM | endothelial | *DNAJA4* | -0.68896228 | 7.23E-07 | 9.76E-06 | 0.004543289 |

| | | | | | | |
|---|---|---|---|---|---|---|
| T2DM | endothelial | *IGF1R* | 0.230211492 | 0.00338952 | 0.007414722 | 1 |
| T2DM | endothelial | *CYP11A1* | -0.272517108 | 0.004850652 | 0.010015122 | 1 |
| T2DM | endothelial | *BBS4* | -0.402017561 | 0.005464834 | 0.011100941 | 1 |
| T2DM | endothelial | *ADAMTS17* | -0.557203493 | 3.52E-06 | 2.71E-05 | 0.022134569 |
| T2DM | endothelial | *ULK3* | -0.661187969 | 0.001190634 | 0.003075396 | 1 |
| T2DM | endothelial | *SCAMP2* | -0.607015862 | 2.21E-05 | 0.000112264 | 0.139094781 |
| T2DM | endothelial | *HAPLN3* | 0.317327347 | 1.30E-05 | 7.27E-05 | 0.081701836 |
| T2DM | endothelial | *POLG* | -0.386880626 | 0.021482924 | 0.035787796 | 1 |
| T2DM | endothelial | *WDR93* | -0.241270069 | 1.44E-06 | 1.53E-05 | 0.009022661 |
| T2DM | endothelial | *TICRR* | -1.063405181 | 6.01E-07 | 8.83E-06 | 0.003778572 |
| T2DM | endothelial | *MFGE8* | -0.524263451 | 5.21E-06 | 3.63E-05 | 0.032738409 |
| T2DM | endothelial | *UNC45A* | -0.283372344 | 0.018493643 | 0.031462298 | 1 |
| T2DM | endothelial | *ST8SIA2* | -2.644462202 | 8.83E-07 | 1.09E-05 | 0.005551485 |
| T2DM | endothelial | *CRTC3* | -0.370672853 | 0.001500607 | 0.003735795 | 1 |
| T2DM | endothelial | *SH3GL3* | -0.459190729 | 6.50E-06 | 4.25E-05 | 0.040862377 |
| T2DM | endothelial | *SEC11A* | -0.385191595 | 0.003143925 | 0.006962259 | 1 |
| T2DM | endothelial | *PARN* | -0.269163192 | 0.023598083 | 0.038849715 | 1 |
| T2DM | endothelial | *TXNL4B* | -0.546171892 | 0.00318398 | 0.007033619 | 1 |
| T2DM | endothelial | *MARVELD3* | -0.822922652 | 3.80E-05 | 0.000173828 | 0.239013465 |
| T2DM | endothelial | *ZFHX3* | -0.373816035 | 3.32E-05 | 0.000155155 | 0.208684107 |
| T2DM | endothelial | *ADAMTS18* | 0.123623261 | 1.24E-06 | 1.38E-05 | 0.007792062 |
| T2DM | endothelial | *GCSH* | -0.359197231 | 0.000282329 | 0.000913537 | 1 |
| T2DM | endothelial | *CMTM3* | -0.545514587 | 6.12E-07 | 8.97E-06 | 0.00384968 |
| T2DM | endothelial | *TLDC1* | -0.540049827 | 0.002940041 | 0.006566266 | 1 |
| T2DM | endothelial | *NDUFB10* | -0.634479357 | 0.001438173 | 0.003602307 | 1 |
| T2DM | endothelial | *DEF8* | -0.573113617 | 1.10E-05 | 6.38E-05 | 0.069315326 |
| T2DM | endothelial | *GALNS* | -0.620167806 | 1.89E-06 | 1.81E-05 | 0.011883859 |
| T2DM | endothelial | *COPS3* | -0.622106453 | 0.000130871 | 0.000485418 | 0.822783609 |
| T2DM | endothelial | *RASL10B* | -0.877259529 | 1.28E-05 | 7.15E-05 | 0.080268243 |
| T2DM | endothelial | *UNC45B* | -1.030898283 | 0.01448943 | 0.025545443 | 1 |
| T2DM | endothelial | *VPS53* | -0.403287919 | 0.000587233 | 0.001693548 | 1 |
| T2DM | endothelial | *SPATA22* | -0.208389259 | 0.001933922 | 0.004627714 | 1 |
| T2DM | endothelial | *SGSM2* | -0.445955942 | 0.005819204 | 0.011718557 | 1 |
| T2DM | endothelial | *NPEPPS* | -0.29434032 | 0.016290752 | 0.028191566 | 1 |
| T2DM | endothelial | *LRRC46* | -0.964042221 | 9.57E-07 | 1.15E-05 | 0.006016505 |
| T2DM | endothelial | *RHBDL3* | 0.362067271 | 0.000595338 | 0.001712994 | 1 |
| T2DM | endothelial | *ARSG* | -0.557669259 | 0.000101288 | 0.000389716 | 0.636795369 |
| T2DM | endothelial | *ABCA8* | -0.282045416 | 5.06E-06 | 3.56E-05 | 0.03182776 |
| T2DM | endothelial | *IMPA2* | -0.542052934 | 9.18E-07 | 1.11E-05 | 0.005771494 |
| T2DM | endothelial | *C18orf21* | -0.625180246 | 0.000235841 | 0.000787848 | 1 |
| T2DM | endothelial | *ASXL3* | 0.2704766 | 5.24E-06 | 3.64E-05 | 0.032939785 |
| T2DM | endothelial | *ADCYAP1* | -0.636814266 | 0.023782162 | 0.039079574 | 1 |
| T2DM | endothelial | *ESCO1* | -0.360571188 | 0.00693405 | 0.013640292 | 1 |
| T2DM | endothelial | *GREB1L* | -0.344996503 | 3.21E-07 | 6.33E-06 | 0.002017997 |
| T2DM | endothelial | *C18orf8* | -0.659909048 | 2.43E-06 | 2.13E-05 | 0.015304926 |
| T2DM | endothelial | *NPC1* | -0.534055239 | 2.52E-05 | 0.000125029 | 0.158661505 |
| T2DM | endothelial | *SLC14A1* | 0.483101901 | 0.000636052 | 0.001802947 | 1 |
| T2DM | endothelial | *SLC13A5* | 1.06098794 | 0.000792653 | 0.002175212 | 1 |
| T2DM | endothelial | *CCDC40* | -0.2757926 | 0.022949498 | 0.037907329 | 1 |
| T2DM | endothelial | *TMC6* | 0.240663711 | 0.016122289 | 0.028000229 | 1 |

| | | | | | | |
|---|---|---|---|---|---|---|
| T2DM | endothelial | *CARD14* | -0.585350023 | 0.014463419 | 0.025506737 | 1 |
| T2DM | endothelial | *EIF4A3* | -0.516620523 | 0.000193367 | 0.000666869 | 1 |
| T2DM | endothelial | *TBCD* | 0.275180663 | 0.000701806 | 0.001965531 | 1 |
| T2DM | endothelial | *FN3KRP* | -0.306348621 | 0.000104 | 0.000397716 | 0.653845492 |
| T2DM | endothelial | *NARF* | -0.411885895 | 0.004502648 | 0.009423484 | 1 |
| T2DM | endothelial | *TRIM65* | -0.3747437 | 0.01063031 | 0.019656694 | 1 |
| T2DM | endothelial | *CBX8* | -0.972072968 | 7.46E-05 | 0.000299884 | 0.469318437 |
| T2DM | endothelial | *SECTM1* | 0.704994734 | 0.013584828 | 0.024201703 | 1 |
| T2DM | endothelial | *RNF157* | -0.19225693 | 0.000989071 | 0.002618227 | 1 |
| T2DM | endothelial | *RNF165* | 0.259703342 | 5.89E-06 | 3.96E-05 | 0.037030651 |
| T2DM | endothelial | *SMAD4* | -0.338506132 | 0.020069656 | 0.033683376 | 1 |
| T2DM | endothelial | *GRB7* | 0.520821184 | 0.000729844 | 0.002030324 | 1 |
| T2DM | endothelial | *NFIC* | -0.369209348 | 6.42E-05 | 0.000265598 | 0.403709067 |
| T2DM | endothelial | *TPGS1* | -0.545430493 | 0.003425429 | 0.007482861 | 1 |
| T2DM | endothelial | *PFKL* | -0.330306092 | 0.020140293 | 0.03377488 | 1 |
| T2DM | endothelial | *TMEM91* | -0.380804507 | 0.023559719 | 0.038815501 | 1 |
| T2DM | endothelial | *IFITM3* | 0.297256748 | 0.013873436 | 0.024659964 | 1 |
| T2DM | endothelial | *COL6A1* | -0.357627666 | 1.11E-06 | 1.27E-05 | 0.007004946 |
| T2DM | endothelial | *TRPM2* | -0.15852888 | 0.013459185 | 0.024025525 | 1 |
| T2DM | endothelial | *DOPEY2* | -0.425760268 | 0.000943841 | 0.002519087 | 1 |
| T2DM | endothelial | *AKT1* | -0.661264217 | 0.001332021 | 0.003374058 | 1 |
| T2DM | endothelial | *SAE1* | -0.369132255 | 0.001638831 | 0.004016892 | 1 |
| T2DM | endothelial | *GEMIN7* | -1.035203667 | 3.71E-09 | 8.63E-07 | 2.33E-05 |
| T2DM | endothelial | *WTIP* | -0.492660056 | 0.001337594 | 0.003385449 | 1 |
| T2DM | endothelial | *MYO1F* | -0.358290366 | 0.003437958 | 0.007507622 | 1 |
| T2DM | endothelial | *CACNG8* | 0.238426034 | 6.17E-07 | 8.98E-06 | 0.003879813 |
| T2DM | endothelial | *CTU1* | -0.897621066 | 4.55E-05 | 0.000200893 | 0.286071851 |
| T2DM | endothelial | *RCN3* | -0.571556204 | 1.99E-06 | 1.87E-05 | 0.012511934 |
| T2DM | endothelial | *PEX14* | -0.44172576 | 0.000141159 | 0.000514762 | 0.887464967 |
| T2DM | endothelial | *SH3BGRL3* | -0.46252801 | 0.003964515 | 0.008500992 | 1 |
| T2DM | endothelial | *CNKSR1* | -0.15583227 | 0.005986805 | 0.012029097 | 1 |
| T2DM | endothelial | *ZNF593* | -0.58763098 | 3.04E-07 | 6.16E-06 | 0.001908692 |
| T2DM | endothelial | *C1orf216* | -0.659363811 | 0.000337301 | 0.001057663 | 1 |
| T2DM | endothelial | *MAP3K6* | 0.379053459 | 0.028263325 | 0.04555025 | 1 |
| T2DM | endothelial | *GPN2* | -0.598049361 | 0.002660694 | 0.006041524 | 1 |
| T2DM | endothelial | *NBPF3* | -0.240010184 | 0.000615461 | 0.00175882 | 1 |
| T2DM | endothelial | *MOB3C* | -0.841374682 | 3.09E-05 | 0.000146425 | 0.194306588 |
| T2DM | endothelial | *SYPL2* | -0.235527428 | 0.000200554 | 0.000687881 | 1 |
| T2DM | endothelial | *ZNF697* | -0.578393561 | 7.27E-07 | 9.76E-06 | 0.004571101 |
| T2DM | endothelial | *CELSR2* | -0.847438388 | 0.000155482 | 0.000556355 | 0.977515127 |
| T2DM | endothelial | *GPR161* | 0.422425986 | 0.004635222 | 0.009643163 | 1 |
| T2DM | endothelial | *ATP1B1* | -0.658833928 | 3.19E-06 | 2.53E-05 | 0.020041766 |
| T2DM | endothelial | *CREG1* | -0.514685815 | 8.89E-06 | 5.42E-05 | 0.055891791 |
| T2DM | endothelial | *GPA33* | -1.103707491 | 1.16E-06 | 1.31E-05 | 0.007312602 |
| T2DM | endothelial | *RXRG* | -1.443355922 | 0.017475642 | 0.029968537 | 1 |
| T2DM | endothelial | *TMCO1* | -0.343450551 | 0.019074403 | 0.032297541 | 1 |
| T2DM | endothelial | *XCL2* | -0.564414971 | 1.77E-05 | 9.34E-05 | 0.111135831 |
| T2DM | endothelial | *ADCY10* | -0.762379486 | 0.001911347 | 0.004577766 | 1 |
| T2DM | endothelial | *RRNAD1* | -0.5775015 | 9.35E-06 | 5.64E-05 | 0.058798099 |
| T2DM | endothelial | *RGL1* | -0.378224088 | 4.98E-06 | 3.52E-05 | 0.0313187 |

| | | | | | | |
|---|---|---|---|---|---|---|
| T2DM | endothelial | *TUFT1* | -0.448107992 | 8.49E-07 | 1.08E-05 | 0.005337179 |
| T2DM | endothelial | *SNX27* | -0.463760248 | 0.000821014 | 0.0022413 | 1 |
| T2DM | endothelial | *SELENBP1* | -0.405291026 | 4.05E-06 | 3.02E-05 | 0.025441999 |
| T2DM | endothelial | *SEMA6C* | 0.35028517 | 1.24E-05 | 7.00E-05 | 0.078102451 |
| T2DM | endothelial | *ARNT* | -0.390865516 | 0.000850067 | 0.002309582 | 1 |
| T2DM | endothelial | *GABPB2* | -0.25653124 | 0.003021324 | 0.006728679 | 1 |
| T2DM | endothelial | *IKBKE* | -0.646874079 | 4.45E-07 | 7.52E-06 | 0.002799227 |
| T2DM | endothelial | *DYRK3* | 0.419406955 | 3.91E-05 | 0.000177807 | 0.246084788 |
| T2DM | endothelial | *EIF2D* | -0.338075279 | 0.017876775 | 0.030557717 | 1 |
| T2DM | endothelial | *HHIPL2* | -0.709959578 | 1.60E-05 | 8.66E-05 | 0.100784164 |
| T2DM | endothelial | *RAB13* | 0.40544836 | 0.004788168 | 0.009908891 | 1 |
| T2DM | endothelial | *S100A8* | -0.951733999 | 1.86E-07 | 4.62E-06 | 0.00116914 |
| T2DM | endothelial | *KCNN3* | 0.151740528 | 9.14E-07 | 1.11E-05 | 0.005745346 |
| T2DM | endothelial | *FLG* | -0.199637923 | 0.001213985 | 0.003117778 | 1 |
| T2DM | endothelial | *GALNT2* | -0.323440897 | 0.014258209 | 0.025208481 | 1 |
| T2DM | endothelial | *SCCPDH* | -0.542998091 | 4.29E-05 | 0.000192466 | 0.269836796 |
| T2DM | endothelial | *CEP170* | -0.314882485 | 0.007014311 | 0.013755137 | 1 |
| T2DM | endothelial | *SDE2* | -0.552870891 | 0.012271881 | 0.022208784 | 1 |
| T2DM | endothelial | *FBXO28* | -0.330088802 | 0.017504276 | 0.029999884 | 1 |
| T2DM | endothelial | *CNIH4* | -0.535796645 | 0.000884446 | 0.002389563 | 1 |
| T2DM | endothelial | *GUK1* | 0.335818972 | 0.008487262 | 0.016213739 | 1 |
| T2DM | endothelial | *CDC42BPA* | 0.20918589 | 0.004771715 | 0.009881347 | 1 |
| T2DM | endothelial | *CNIH3* | -0.356415739 | 0.005811668 | 0.011707131 | 1 |
| T2DM | endothelial | *PSEN2* | -0.707685977 | 3.11E-06 | 2.48E-05 | 0.019537399 |
| T2DM | endothelial | *PYCR2* | -0.531181889 | 0.002137631 | 0.005031557 | 1 |
| T2DM | endothelial | *PLEKHA6* | -0.42300185 | 0.002407492 | 0.005570815 | 1 |
| T2DM | endothelial | *EML4* | -0.275811747 | 0.018428599 | 0.031373031 | 1 |
| T2DM | endothelial | *WDPCP* | -0.187600369 | 0.002303369 | 0.005365426 | 1 |
| T2DM | endothelial | *ASXL2* | -0.291539824 | 0.018918085 | 0.032067404 | 1 |
| T2DM | endothelial | *CIAO1* | -0.481880651 | 0.00079812 | 0.002188302 | 1 |
| T2DM | endothelial | *ZNF514* | -0.222403181 | 0.000249894 | 0.000826016 | 1 |
| T2DM | endothelial | *TEX261* | -0.873263671 | 0.000134865 | 0.000496425 | 0.847893392 |
| T2DM | endothelial | *THNSL2* | -0.121174604 | 0.000701864 | 0.001965531 | 1 |
| T2DM | endothelial | *RALB* | 0.290402954 | 0.00401556 | 0.008592861 | 1 |
| T2DM | endothelial | *TMEM177* | -0.857230073 | 0.003476463 | 0.007586436 | 1 |
| T2DM | endothelial | *RABL2A* | -0.415096719 | 0.019952476 | 0.033504599 | 1 |
| T2DM | endothelial | *FBLN7* | -0.28978941 | 0.003061921 | 0.006807035 | 1 |
| T2DM | endothelial | *ZC3H8* | -0.328611133 | 0.012609424 | 0.022734572 | 1 |
| T2DM | endothelial | *FAHD2B* | -0.586876521 | 0.000403826 | 0.001227685 | 1 |
| T2DM | endothelial | *LYG1* | -0.399408626 | 0.000374022 | 0.001154548 | 1 |
| T2DM | endothelial | *AFF3* | -0.292882406 | 2.36E-06 | 2.08E-05 | 0.014860117 |
| T2DM | endothelial | *THSD7B* | -0.244998223 | 0.000661328 | 0.001861966 | 1 |
| T2DM | endothelial | *GPR17* | 0.160385198 | 2.06E-05 | 0.000105763 | 0.129592826 |
| T2DM | endothelial | *POLR2D* | -0.712101716 | 0.000206542 | 0.000706489 | 1 |
| T2DM | endothelial | *ZNF385B* | -0.394498899 | 0.000237626 | 0.000792549 | 1 |
| T2DM | endothelial | *TMEFF2* | 0.354854377 | 4.88E-05 | 0.000212657 | 0.307076087 |
| T2DM | endothelial | *GULP1* | 0.254287708 | 0.000683682 | 0.001918889 | 1 |
| T2DM | endothelial | *HSPD1* | -0.369863282 | 0.012536288 | 0.02261674 | 1 |
| T2DM | endothelial | *PTH2R* | -0.301235363 | 3.03E-05 | 0.000144041 | 0.190422771 |
| T2DM | endothelial | *SUMF1* | -0.309084998 | 0.001635097 | 0.004009304 | 1 |

| | | | | | | |
|---|---|---|---|---|---|---|
| T2DM | endothelial | *ESPNL* | -2.892233165 | 0.006121563 | 0.012260677 | 1 |
| T2DM | endothelial | *FANCD2* | -0.639230558 | 0.009436757 | 0.017767994 | 1 |
| T2DM | endothelial | *OSBPL10* | 0.273042927 | 6.36E-05 | 0.0002636 | 0.399881163 |
| T2DM | endothelial | *FAM198A* | -0.360036451 | 5.78E-05 | 0.000244298 | 0.363538022 |
| T2DM | endothelial | *SLC25A38* | -0.933219508 | 2.07E-05 | 0.000106004 | 0.130067355 |
| T2DM | endothelial | *CTDSPL* | -0.421939004 | 0.001033635 | 0.002720591 | 1 |
| T2DM | endothelial | *STAC* | -0.478784593 | 2.04E-07 | 4.92E-06 | 0.001283249 |
| T2DM | endothelial | *ARL6IP5* | -0.40747362 | 0.001276712 | 0.003256263 | 1 |
| T2DM | endothelial | *TMF1* | -0.44268825 | 0.000737013 | 0.002046645 | 1 |
| T2DM | endothelial | *LIMD1* | -0.458580884 | 8.41E-05 | 0.000332238 | 0.528922411 |
| T2DM | endothelial | *NXPE3* | 0.318733009 | 0.000129214 | 0.00047984 | 0.812368902 |
| T2DM | endothelial | *PLA1A* | 0.160844778 | 0.000286898 | 0.000924989 | 1 |
| T2DM | endothelial | *ADPRH* | 0.219136715 | 0.030798861 | 0.049095447 | 1 |
| T2DM | endothelial | *BOC* | -0.45769884 | 0.000145547 | 0.000527407 | 0.915050864 |
| T2DM | endothelial | *AGTR1* | -0.434287849 | 1.40E-06 | 1.51E-05 | 0.008824655 |
| T2DM | endothelial | *EIF2A* | -0.250693064 | 0.001458778 | 0.003643759 | 1 |
| T2DM | endothelial | *TCTA* | -0.414410411 | 0.005059286 | 0.010384502 | 1 |
| T2DM | endothelial | *EAF2* | -0.526473098 | 1.32E-05 | 7.36E-05 | 0.0829543 |
| T2DM | endothelial | *MUC4* | -0.212910402 | 0.002210443 | 0.005183533 | 1 |
| T2DM | endothelial | *DGKQ* | -0.380447372 | 0.000779443 | 0.002144577 | 1 |
| T2DM | endothelial | *ATP10D* | 0.274478916 | 0.004533957 | 0.009473243 | 1 |
| T2DM | endothelial | *PYURF* | -0.779978331 | 6.63E-05 | 0.000272347 | 0.416690414 |
| T2DM | endothelial | *CISD2* | -0.517288312 | 0.000147514 | 0.000532389 | 0.927421338 |
| T2DM | endothelial | *TIFA* | 0.630992598 | 0.006144493 | 0.012294853 | 1 |
| T2DM | endothelial | *USP53* | -0.312284182 | 0.003813425 | 0.008221881 | 1 |
| T2DM | endothelial | *MARCH1* | -0.222481287 | 0.024665967 | 0.040405141 | 1 |
| T2DM | endothelial | *PDGFC* | -0.407938646 | 9.86E-07 | 1.17E-05 | 0.006197907 |
| T2DM | endothelial | *NDUFS6* | -0.546184452 | 0.002896499 | 0.006485146 | 1 |
| T2DM | endothelial | *NKD2* | -1.64600946 | 0.003145167 | 0.006962558 | 1 |
| T2DM | endothelial | *FAM105A* | -0.442063735 | 0.003186227 | 0.007036112 | 1 |
| T2DM | endothelial | *BHMT* | -0.290001688 | 2.86E-06 | 2.36E-05 | 0.017985982 |
| T2DM | endothelial | *CRHBP* | 0.259198533 | 0.000181393 | 0.000633567 | 1 |
| T2DM | endothelial | *GTF2H2* | -0.497572825 | 0.001422968 | 0.003571336 | 1 |
| T2DM | endothelial | *SPATA9* | -0.326955803 | 5.20E-06 | 3.62E-05 | 0.032665799 |
| T2DM | endothelial | *YIPF5* | -0.400500715 | 0.031438131 | 0.049975103 | 1 |
| T2DM | endothelial | *SLC25A48* | -0.333196259 | 0.000243879 | 0.000810824 | 1 |
| T2DM | endothelial | *GABRB2* | -0.378290715 | 7.76E-07 | 1.03E-05 | 0.004876949 |
| T2DM | endothelial | *PCYOX1L* | -0.473164362 | 0.002051257 | 0.004859177 | 1 |
| T2DM | endothelial | *MYLK4* | 0.455195942 | 0.023721459 | 0.039010413 | 1 |
| T2DM | endothelial | *GFOD1* | 0.182957744 | 1.38E-05 | 7.61E-05 | 0.086528199 |
| T2DM | endothelial | *PSD2* | 2.19258331 | 5.33E-07 | 8.34E-06 | 0.003350222 |
| T2DM | endothelial | *ZMAT2* | -0.42861557 | 0.006350064 | 0.012633813 | 1 |
| T2DM | endothelial | *KLHL3* | 0.22957846 | 1.06E-05 | 6.17E-05 | 0.066412674 |
| T2DM | endothelial | *TRIM7* | 0.470981799 | 0.00835632 | 0.01601713 | 1 |
| T2DM | endothelial | *HIGD2A* | -0.705027385 | 2.56E-06 | 2.19E-05 | 0.01610015 |
| T2DM | endothelial | *MUT* | -0.425375355 | 0.016417996 | 0.028372716 | 1 |
| T2DM | endothelial | *FGD2* | -0.513979277 | 3.91E-07 | 7.01E-06 | 0.002459105 |
| T2DM | endothelial | *TPBG* | 0.370173964 | 0.019552968 | 0.033010072 | 1 |
| T2DM | endothelial | *PM20D2* | -0.577184169 | 0.00390516 | 0.008385059 | 1 |
| T2DM | endothelial | *RNF217* | -0.337391112 | 0.006283614 | 0.012529362 | 1 |

| | | | | | | |
|---|---|---|---|---|---|---|
| T2DM | endothelial | *AIG1* | -0.342899778 | 0.00118794 | 0.003069699 | 1 |
| T2DM | endothelial | *DYNLT1* | -0.471149735 | 0.001542926 | 0.003820551 | 1 |
| T2DM | endothelial | *VIP* | 0.193114007 | 2.57E-07 | 5.51E-06 | 0.001614644 |
| T2DM | endothelial | *SLC22A3* | -0.405460085 | 2.73E-07 | 5.76E-06 | 0.001714429 |
| T2DM | endothelial | *GNA12* | -0.406792688 | 0.000763223 | 0.002105478 | 1 |
| T2DM | endothelial | *SDK1* | -0.195854646 | 0.002031904 | 0.004822416 | 1 |
| T2DM | endothelial | *CCZ1B* | -0.341501222 | 0.00663593 | 0.013148469 | 1 |
| T2DM | endothelial | *RBAK* | -0.829407828 | 0.000164146 | 0.000582385 | 1 |
| T2DM | endothelial | *EGFR* | -0.406032687 | 2.03E-06 | 1.89E-05 | 0.012748303 |
| T2DM | endothelial | *PSPH* | -0.24559445 | 0.015314731 | 0.026805043 | 1 |
| T2DM | endothelial | *GIGYF1* | -0.426454099 | 0.009055728 | 0.017164112 | 1 |
| T2DM | endothelial | *TRIM4* | -0.208480498 | 0.001181539 | 0.003054414 | 1 |
| T2DM | endothelial | *CSGALNACT1* | 0.214832978 | 0.002129077 | 0.005013298 | 1 |
| T2DM | endothelial | *CHMP7* | -0.492119568 | 6.99E-05 | 0.000284292 | 0.439230945 |
| T2DM | endothelial | *DOCK5* | -0.362426084 | 0.008516362 | 0.016250371 | 1 |
| T2DM | endothelial | *ST18* | -0.51093075 | 3.76E-07 | 6.87E-06 | 0.00236647 |
| T2DM | endothelial | *RGS20* | -0.295941902 | 0.016937752 | 0.029159081 | 1 |
| T2DM | endothelial | *TACC1* | 0.191905389 | 0.007134832 | 0.013956655 | 1 |
| T2DM | endothelial | *ADHFE1* | -0.239554575 | 2.42E-07 | 5.33E-06 | 0.001518461 |
| T2DM | endothelial | *SLC26A7* | -0.257598187 | 0.005441049 | 0.011063349 | 1 |
| T2DM | endothelial | *ATP6V0D2* | -0.507158723 | 4.90E-06 | 3.49E-05 | 0.030817621 |
| T2DM | endothelial | *MTDH* | -0.270211084 | 0.023463065 | 0.038676531 | 1 |
| T2DM | endothelial | *MAL2* | -0.421585862 | 9.95E-08 | 4.46E-06 | 0.000625642 |
| T2DM | endothelial | *UTP23* | -0.518623131 | 0.001038377 | 0.002729638 | 1 |
| T2DM | endothelial | *SLC39A4* | -1.061981348 | 1.03E-09 | 3.61E-07 | 6.50E-06 |
| T2DM | endothelial | *CER1* | -0.642823835 | 4.31E-05 | 0.000193017 | 0.271188587 |
| T2DM | endothelial | *PLIN2* | -0.478919717 | 5.59E-06 | 3.81E-05 | 0.035128181 |
| T2DM | endothelial | *HAUS6* | -0.485999229 | 0.004761355 | 0.009863143 | 1 |
| T2DM | endothelial | *C9orf72* | -0.326161736 | 0.006711267 | 0.013272643 | 1 |
| T2DM | endothelial | *SIGMAR1* | -0.498898457 | 0.001068062 | 0.002797616 | 1 |
| T2DM | endothelial | *ZNF462* | 0.261760636 | 0.009234315 | 0.017444753 | 1 |
| T2DM | endothelial | *ALAD* | -0.180388921 | 0.009967611 | 0.018578823 | 1 |
| T2DM | endothelial | *ASTN2* | -0.479261034 | 3.30E-05 | 0.000154522 | 0.207677556 |
| T2DM | endothelial | *WDR31* | -0.633775809 | 4.74E-05 | 0.00020754 | 0.298234812 |
| T2DM | endothelial | *SURF1* | -0.38203091 | 0.000739431 | 0.002052451 | 1 |
| T2DM | endothelial | *SURF6* | -0.687282046 | 0.000144399 | 0.000524459 | 0.907839084 |
| T2DM | endothelial | *CIZ1* | -0.49982303 | 6.00E-05 | 0.000251584 | 0.377376115 |
| T2DM | endothelial | *SH3GLB2* | -0.594142985 | 0.000190124 | 0.000659461 | 1 |
| T2DM | endothelial | *LRSAM1* | -0.403168975 | 0.010722002 | 0.019785509 | 1 |
| T2DM | endothelial | *INPP5E* | -0.641778335 | 0.001846491 | 0.004451261 | 1 |
| T2DM | endothelial | *DPH7* | -0.57833708 | 0.000987897 | 0.002616222 | 1 |
| T2DM | endothelial | *SLC39A12* | -0.684981264 | 2.10E-05 | 0.00010748 | 0.132092496 |
| T2DM | endothelial | *PARD3* | -0.298078994 | 0.001008715 | 0.002662382 | 1 |
| T2DM | endothelial | *NRBF2* | -0.43307807 | 4.61E-06 | 3.35E-05 | 0.028980798 |
| T2DM | endothelial | *GLUD1* | -0.364218426 | 0.002452027 | 0.005649702 | 1 |
| T2DM | endothelial | *HTR7* | -0.523485592 | 1.65E-07 | 4.50E-06 | 0.001038838 |
| T2DM | endothelial | *RPP30* | -0.362061143 | 0.022195009 | 0.036817948 | 1 |
| T2DM | endothelial | *PLEKHS1* | -1.272318076 | 6.78E-05 | 0.000278021 | 0.426206369 |
| T2DM | endothelial | *FUOM* | -0.800448707 | 1.27E-06 | 1.40E-05 | 0.007954241 |
| T2DM | endothelial | *PAOX* | -0.952696383 | 2.96E-05 | 0.000141214 | 0.186120167 |

| | | | | | | |
|---|---|---|---|---|---|---|
| T2DM | endothelial | *GSTO1* | -0.42355326 | 0.014393038 | 0.025399834 | 1 |
| T2DM | endothelial | *PDCD11* | -0.374538244 | 0.023624183 | 0.038880952 | 1 |
| T2DM | endothelial | *RGS10* | -0.664312082 | 3.06E-07 | 6.16E-06 | 0.001920849 |
| T2DM | endothelial | *GAS2* | 0.401308414 | 0.001544394 | 0.003822678 | 1 |
| T2DM | endothelial | *LIN7C* | -0.626736266 | 0.000418004 | 0.001264066 | 1 |
| T2DM | endothelial | *HSD17B12* | -0.184362471 | 0.007552966 | 0.014665069 | 1 |
| T2DM | endothelial | *APIP* | -0.19472408 | 2.17E-06 | 1.98E-05 | 0.013631564 |
| T2DM | endothelial | *EIF3M* | -0.460069783 | 0.001964471 | 0.004681815 | 1 |
| T2DM | endothelial | *GLYAT* | -0.23474335 | 0.000211469 | 0.000720991 | 1 |
| T2DM | endothelial | *PTPRJ* | -0.30339662 | 0.004400694 | 0.009228539 | 1 |
| T2DM | endothelial | *ENDOD1* | 0.353099112 | 3.89E-05 | 0.000176982 | 0.24441173 |
| T2DM | endothelial | *CCDC82* | -0.243303896 | 0.00974161 | 0.018260436 | 1 |
| T2DM | endothelial | *DRD2* | -1.895938899 | 3.08E-06 | 2.47E-05 | 0.019377151 |
| T2DM | endothelial | *C11orf52* | -1.66412255 | 1.44E-07 | 4.46E-06 | 0.000904041 |
| T2DM | endothelial | *GLB1L2* | 0.727121588 | 0.008334292 | 0.015984654 | 1 |
| T2DM | endothelial | *GRIK4* | 0.209656734 | 1.70E-06 | 1.68E-05 | 0.010678476 |
| T2DM | endothelial | *ADAM33* | -0.363786935 | 9.19E-08 | 4.46E-06 | 0.000577861 |
| T2DM | endothelial | *MTA2* | 0.400792348 | 0.005664197 | 0.011461477 | 1 |
| T2DM | endothelial | *TMC2* | -0.863564928 | 1.58E-07 | 4.49E-06 | 0.000991633 |
| T2DM | endothelial | *ROM1* | 0.175474356 | 0.006229172 | 0.012444488 | 1 |
| T2DM | endothelial | *MS4A3* | -4.463478751 | 1.20E-07 | 4.46E-06 | 0.000754137 |
| T2DM | endothelial | *PLCH2* | -1.263969206 | 1.51E-07 | 4.49E-06 | 0.000947559 |
| T2DM | endothelial | *CPSF7* | -0.450812802 | 0.001139799 | 0.002961123 | 1 |
| T2DM | endothelial | *CCDC15* | -0.483405297 | 1.14E-05 | 6.52E-05 | 0.071640094 |
| T2DM | endothelial | *CHEK1* | -0.276771774 | 0.005325533 | 0.010877721 | 1 |
| T2DM | endothelial | *FEZ1* | -0.747355316 | 5.27E-06 | 3.65E-05 | 0.033101455 |
| T2DM | endothelial | *CNBD2* | -0.782225695 | 5.51E-07 | 8.50E-06 | 0.003466877 |
| T2DM | endothelial | *CABLES2* | -0.377547119 | 0.008535652 | 0.016271573 | 1 |
| T2DM | endothelial | *TRPT1* | -0.579842423 | 0.005054975 | 0.010379042 | 1 |
| T2DM | endothelial | *DOC2A* | -0.6307187 | 0.008305838 | 0.015949543 | 1 |
| T2DM | endothelial | *TAOK2* | 0.318530292 | 5.10E-05 | 0.000219632 | 0.320662064 |
| T2DM | endothelial | *MPP7* | -0.30613185 | 0.000369878 | 0.001144401 | 1 |
| T2DM | endothelial | *ARID5B* | -0.307796537 | 0.001886575 | 0.004532249 | 1 |
| T2DM | endothelial | *LYPD6B* | 0.651053986 | 0.002414658 | 0.005583286 | 1 |
| T2DM | endothelial | *WDR17* | -0.639996491 | 1.39E-06 | 1.50E-05 | 0.008737247 |
| T2DM | endothelial | *CCDC102B* | -0.457961986 | 2.01E-05 | 0.000103522 | 0.126193662 |
| T2DM | endothelial | *RGS18* | -0.62243269 | 2.73E-07 | 5.76E-06 | 0.001715647 |
| T2DM | endothelial | *PPP1R1C* | -0.267585585 | 0.021883407 | 0.036377836 | 1 |
| T2DM | endothelial | *C11orf53* | 2.655827227 | 0.000143243 | 0.000520862 | 0.900570379 |
| T2DM | endothelial | *CCT5* | -0.469941679 | 0.001375877 | 0.00347395 | 1 |
| T2DM | endothelial | *FAM173B* | -0.595978935 | 1.92E-05 | 0.000100043 | 0.120826272 |
| T2DM | endothelial | *DOCK1* | 0.204412007 | 0.006165014 | 0.012328068 | 1 |
| T2DM | endothelial | *TEX12* | -2.468586473 | 1.07E-07 | 4.46E-06 | 0.000672048 |
| T2DM | endothelial | *ABCB9* | 0.210411546 | 5.42E-05 | 0.000231027 | 0.340534525 |
| T2DM | endothelial | *PRSS53* | -0.660836001 | 0.003189649 | 0.007041195 | 1 |
| T2DM | endothelial | *GPR158* | -0.404552796 | 4.58E-05 | 0.000202243 | 0.288196578 |
| T2DM | endothelial | *THRB* | -0.342147281 | 6.90E-05 | 0.000281527 | 0.433552015 |
| T2DM | endothelial | *TMEM86A* | -0.352490225 | 0.000257312 | 0.00084344 | 1 |
| T2DM | endothelial | *BTBD11* | -0.648408015 | 8.00E-07 | 1.04E-05 | 0.005029403 |
| T2DM | endothelial | *UBE3B* | -0.333046693 | 0.003986971 | 0.008537495 | 1 |

| | | | | | | |
|---|---|---|---|---|---|---|
| T2DM | endothelial | *ANK3* | -0.309928368 | 0.001802071 | 0.004359224 | 1 |
| T2DM | endothelial | *IPMK* | -0.500522207 | 9.46E-05 | 0.000368267 | 0.594751652 |
| T2DM | endothelial | *PLBD2* | -0.415853234 | 0.000154674 | 0.00055441 | 0.972435785 |
| T2DM | endothelial | *DLG5* | -0.41666767 | 1.49E-05 | 8.12E-05 | 0.093553422 |
| T2DM | endothelial | *TMEM18* | -0.648789313 | 0.002232845 | 0.005228012 | 1 |
| T2DM | endothelial | *KCTD14* | -1.686283618 | 0.00398569 | 0.008537495 | 1 |
| T2DM | endothelial | *THRSP* | -0.341285198 | 4.54E-05 | 0.000200691 | 0.285583282 |
| T2DM | endothelial | *ADAMTS12* | -0.313282905 | 1.01E-06 | 1.20E-05 | 0.006350667 |
| T2DM | endothelial | *VIPAS39* | -0.629015334 | 4.33E-05 | 0.000193487 | 0.272042054 |
| T2DM | endothelial | *PTPRO* | -0.522914611 | 5.59E-07 | 8.53E-06 | 0.003514946 |
| T2DM | endothelial | *VPS26B* | -0.514744919 | 0.002361006 | 0.005481406 | 1 |
| T2DM | endothelial | *NCAPD3* | -0.572571907 | 7.40E-05 | 0.000297784 | 0.465436475 |
| T2DM | endothelial | *QDPR* | -0.156990392 | 0.004283443 | 0.009058192 | 1 |
| T2DM | endothelial | *FAM160B1* | -0.314255076 | 0.027625562 | 0.044659785 | 1 |
| T2DM | endothelial | *ANO4* | -0.381949214 | 3.90E-07 | 7.01E-06 | 0.002455029 |
| T2DM | endothelial | *AKR1C2* | -0.48974508 | 5.46E-07 | 8.44E-06 | 0.003434898 |
| T2DM | endothelial | *ADAM8* | -0.636900401 | 6.39E-07 | 9.11E-06 | 0.004016954 |
| T2DM | endothelial | *PIGF* | -0.408261752 | 0.004580374 | 0.009553485 | 1 |
| T2DM | endothelial | *INPP1* | 0.311865405 | 1.78E-05 | 9.34E-05 | 0.111619661 |
| T2DM | endothelial | *MFSD6* | 0.264865618 | 0.019749438 | 0.033259418 | 1 |
| T2DM | endothelial | *RNF144A* | 0.234205613 | 0.022952556 | 0.037907329 | 1 |
| T2DM | endothelial | *ADAM17* | -0.349343302 | 0.004091256 | 0.008718398 | 1 |
| T2DM | endothelial | *TMEM45B* | -1.454765846 | 2.70E-05 | 0.000131067 | 0.169846867 |
| T2DM | endothelial | *ACSL1* | -0.21986176 | 0.0009183 | 0.002461985 | 1 |
| T2DM | endothelial | *GUF1* | -0.409869177 | 0.010094411 | 0.018792882 | 1 |
| T2DM | endothelial | *GABRA2* | 0.156309652 | 3.03E-06 | 2.45E-05 | 0.019058346 |
| T2DM | endothelial | *CENPJ* | -0.426373509 | 0.002727315 | 0.006154568 | 1 |
| T2DM | endothelial | *PARP8* | -0.286781695 | 0.001445905 | 0.003615912 | 1 |
| T2DM | endothelial | *CACUL1* | -0.374531525 | 0.002222389 | 0.005207664 | 1 |
| T2DM | endothelial | *BEND6* | -0.290453785 | 2.61E-05 | 0.000128256 | 0.1640626 |
| T2DM | endothelial | *PTPN14* | 0.221143332 | 0.000384562 | 0.001180538 | 1 |
| T2DM | endothelial | *ARL14EP* | -0.716728773 | 3.35E-07 | 6.48E-06 | 0.002105363 |
| T2DM | endothelial | *EPG5* | -0.388384842 | 0.0012082 | 0.003105459 | 1 |
| T2DM | endothelial | *SPC25* | 0.09375816 | 5.61E-06 | 3.82E-05 | 0.035275077 |
| T2DM | endothelial | *G6PC2* | 6.775295889 | 8.30E-07 | 1.06E-05 | 0.005220899 |
| T2DM | endothelial | *POC5* | -0.558809361 | 1.92E-05 | 0.000100047 | 0.120975567 |
| T2DM | endothelial | *GUCY1A2* | -0.382223336 | 1.90E-06 | 1.81E-05 | 0.011921051 |
| T2DM | endothelial | *ZNF547* | -0.781789069 | 0.001935146 | 0.004627714 | 1 |
| T2DM | endothelial | *ZNF773* | -0.611053499 | 1.17E-06 | 1.31E-05 | 0.007334327 |
| T2DM | endothelial | *NMT2* | -0.309243196 | 0.002939121 | 0.006566266 | 1 |
| T2DM | endothelial | *IGSF10* | -0.436177926 | 8.76E-07 | 1.08E-05 | 0.005505647 |
| T2DM | endothelial | *SPEF2* | -0.371661981 | 0.004581479 | 0.009553485 | 1 |
| T2DM | endothelial | *SPARCL1* | 0.141847626 | 1.01E-05 | 5.95E-05 | 0.063416652 |
| T2DM | endothelial | *CCNO* | 1.593364066 | 0.000179491 | 0.000629369 | 1 |
| T2DM | endothelial | *CLEC4F* | 0.590688012 | 1.69E-06 | 1.68E-05 | 0.010603758 |
| T2DM | endothelial | *PELO* | 0.509017913 | 0.010833102 | 0.019962264 | 1 |
| T2DM | endothelial | *RASGRP3* | 0.215032954 | 0.003114318 | 0.006910575 | 1 |
| T2DM | endothelial | *GPR180* | -0.384686725 | 0.000113439 | 0.000427829 | 0.713191329 |
| T2DM | endothelial | *TCTEX1D1* | 0.208373266 | 1.76E-06 | 1.72E-05 | 0.011068095 |
| T2DM | endothelial | *IFIT5* | 0.391199547 | 0.002649412 | 0.006025635 | 1 |

| | | | | | | |
|---|---|---|---|---|---|---|
| T2DM | endothelial | *PRDM8* | -0.3883746 | 2.67E-06 | 2.26E-05 | 0.016764767 |
| T2DM | endothelial | *HHEX* | 0.249347732 | 6.02E-06 | 4.02E-05 | 0.037853597 |
| T2DM | endothelial | *RAB3C* | 0.097162065 | 8.89E-07 | 1.09E-05 | 0.005589262 |
| T2DM | endothelial | *RAD17* | -0.286103301 | 0.009925376 | 0.018516821 | 1 |
| T2DM | endothelial | *SRP19* | -0.495081434 | 0.017553024 | 0.030053339 | 1 |
| T2DM | endothelial | *BANK1* | -0.449122753 | 1.11E-06 | 1.27E-05 | 0.00695868 |
| T2DM | endothelial | *CLGN* | 1.070459655 | 7.84E-07 | 1.03E-05 | 0.004926378 |
| T2DM | endothelial | *TMEM87B* | 0.292713353 | 0.000858901 | 0.002328551 | 1 |
| T2DM | endothelial | *PTPRR* | 0.134594372 | 6.43E-07 | 9.14E-06 | 0.004041288 |
| T2DM | endothelial | *FRMD1* | -0.902743641 | 9.71E-07 | 1.16E-05 | 0.006103011 |
| T2DM | endothelial | *INO80C* | -0.765515685 | 0.001898197 | 0.004554949 | 1 |
| T2DM | endothelial | *C16orf89* | -0.380934942 | 0.000160138 | 0.000570418 | 1 |
| T2DM | endothelial | *TEX29* | -0.557848858 | 4.82E-05 | 0.00021046 | 0.30285215 |
| T2DM | endothelial | *TUBGCP5* | -0.453485486 | 0.002314762 | 0.005385977 | 1 |
| T2DM | endothelial | *LURAP1L* | -0.187726671 | 0.007650454 | 0.014831455 | 1 |
| T2DM | endothelial | *ZDHHC7* | -0.451217808 | 1.04E-05 | 6.06E-05 | 0.065116158 |
| T2DM | endothelial | *FAM92B* | -4.643037592 | 1.02E-07 | 4.46E-06 | 0.000639811 |
| T2DM | endothelial | *C7orf31* | 0.381780124 | 0.030043779 | 0.048001332 | 1 |
| T2DM | endothelial | *JAZF1* | -0.533006642 | 2.42E-07 | 5.33E-06 | 0.001524557 |
| T2DM | endothelial | *MCOLN2* | -0.351239124 | 0.009168569 | 0.017336178 | 1 |
| T2DM | endothelial | *CACNA2D1* | -0.347918048 | 2.64E-05 | 0.000129231 | 0.16619071 |
| T2DM | endothelial | *ZUFSP* | -0.539759442 | 0.000437571 | 0.001314712 | 1 |
| T2DM | endothelial | *ANKRD29* | 0.134159389 | 6.99E-07 | 9.61E-06 | 0.00439627 |
| T2DM | endothelial | *THY1* | -0.527335328 | 2.67E-06 | 2.27E-05 | 0.016813182 |
| T2DM | endothelial | *UBASH3B* | -0.564195357 | 4.32E-06 | 3.17E-05 | 0.027144752 |
| T2DM | endothelial | *ROBO4* | 0.169050876 | 1.42E-06 | 1.52E-05 | 0.008955105 |
| T2DM | endothelial | *PRKCA* | -0.414277648 | 0.00028874 | 0.000929975 | 1 |
| T2DM | endothelial | *LRRK1* | -0.374515208 | 0.006490411 | 0.012884502 | 1 |
| T2DM | endothelial | *CEP112* | 0.204163289 | 0.003663732 | 0.007929047 | 1 |
| T2DM | endothelial | *ENPP3* | -0.586147357 | 0.001129051 | 0.002936841 | 1 |
| T2DM | endothelial | *UCHL1* | -0.504813339 | 1.26E-07 | 4.46E-06 | 0.000792831 |
| T2DM | endothelial | *MIA3* | -0.402507336 | 0.002320119 | 0.005395439 | 1 |
| T2DM | endothelial | *LONRF1* | -0.593831437 | 1.85E-06 | 1.78E-05 | 0.011626895 |
| T2DM | endothelial | *CCSAP* | -0.68206268 | 1.23E-05 | 6.94E-05 | 0.077343403 |
| T2DM | endothelial | *GBP5* | -0.512026368 | 0.015908252 | 0.027697363 | 1 |
| T2DM | endothelial | *FAM69A* | -0.407325591 | 3.48E-06 | 2.69E-05 | 0.021902381 |
| T2DM | endothelial | *PDLIM3* | 0.374430264 | 7.16E-05 | 0.000289778 | 0.450025036 |
| T2DM | endothelial | *PDE1C* | -0.448980268 | 6.72E-07 | 9.43E-06 | 0.004226079 |
| T2DM | endothelial | *RABGEF1* | -0.24542594 | 0.00017742 | 0.0006235 | 1 |
| T2DM | endothelial | *FLCN* | -0.42935395 | 8.37E-06 | 5.19E-05 | 0.052643737 |
| T2DM | endothelial | *DPH3* | -0.804534241 | 0.000141238 | 0.000514762 | 0.887964944 |
| T2DM | endothelial | *CXXC1* | -0.628657846 | 0.002494117 | 0.005733279 | 1 |
| T2DM | endothelial | *APCDD1* | -0.224248887 | 0.000236653 | 0.000789723 | 1 |
| T2DM | endothelial | *ANKRD40* | -0.57946557 | 0.000260115 | 0.000850414 | 1 |
| T2DM | endothelial | *CYP2U1* | -0.633276783 | 1.69E-06 | 1.68E-05 | 0.010654745 |
| T2DM | endothelial | *FBXL18* | -0.598781076 | 7.86E-05 | 0.000313335 | 0.494442326 |
| T2DM | endothelial | *PROM2* | -2.102557069 | 2.40E-05 | 0.000119809 | 0.150719308 |
| T2DM | endothelial | *GTF3C6* | -0.45068027 | 0.010670529 | 0.01971367 | 1 |
| T2DM | endothelial | *AGPAT5* | 0.304442526 | 0.008073573 | 0.015546265 | 1 |
| T2DM | endothelial | *GOLGA7B* | -1.043358944 | 8.08E-08 | 4.46E-06 | 0.000508218 |

| | | | | | | |
|---|---|---|---|---|---|---|
| T2DM | endothelial | *TRMT44* | -0.456424747 | 0.008331074 | 0.015983357 | 1 |
| T2DM | endothelial | *C16orf87* | -0.511371785 | 0.000158344 | 0.000564987 | 0.995507916 |
| T2DM | endothelial | *HEATR3* | -0.220779749 | 0.005745167 | 0.01158824 | 1 |
| T2DM | endothelial | *CNOT8* | -0.384752217 | 0.006349631 | 0.012633813 | 1 |
| T2DM | endothelial | *LRGUK* | -0.255894261 | 0.004855613 | 0.010019252 | 1 |
| T2DM | endothelial | *SETD9* | -0.324690146 | 6.15E-06 | 4.08E-05 | 0.038672485 |
| T2DM | endothelial | *KDM8* | -0.724965858 | 0.010208946 | 0.018989244 | 1 |
| T2DM | endothelial | *FAM126B* | -0.333364588 | 0.006302019 | 0.012562078 | 1 |
| T2DM | endothelial | *ALS2CR12* | -0.594916868 | 2.40E-05 | 0.000119852 | 0.151013792 |
| T2DM | endothelial | *SPAG17* | -0.353080417 | 5.35E-07 | 8.34E-06 | 0.00336656 |
| T2DM | endothelial | *DEPTOR* | -0.376537166 | 0.013076464 | 0.023435498 | 1 |
| T2DM | endothelial | *RRAGA* | -0.804648415 | 0.000433993 | 0.001306761 | 1 |
| T2DM | endothelial | *WIF1* | -0.81368243 | 0.025794281 | 0.041979975 | 1 |
| T2DM | endothelial | *UGT2B4* | -0.689652147 | 1.16E-07 | 4.46E-06 | 0.000731795 |
| T2DM | endothelial | *MMP16* | 0.167219788 | 0.005745275 | 0.01158824 | 1 |
| T2DM | endothelial | *KCNMA1* | -0.385068223 | 7.13E-06 | 4.54E-05 | 0.044810124 |
| T2DM | endothelial | *BATF* | -0.978338771 | 8.96E-07 | 1.09E-05 | 0.005633189 |
| T2DM | endothelial | *DCK* | -0.578431106 | 0.002529479 | 0.005795493 | 1 |
| T2DM | endothelial | *NDUFAF6* | -0.321551207 | 0.008530859 | 0.016268227 | 1 |
| T2DM | endothelial | *DRAM2* | -0.389552202 | 0.000179996 | 0.000630787 | 1 |
| T2DM | endothelial | *C8orf37* | -0.375338233 | 0.0149196 | 0.02621563 | 1 |
| T2DM | endothelial | *ADAMTSL3* | -0.229580287 | 0.001842155 | 0.004442512 | 1 |
| T2DM | endothelial | *WHAMM* | -0.435616096 | 8.03E-06 | 5.02E-05 | 0.05049144 |
| T2DM | endothelial | *CXCL13* | 0.74055053 | 0.014898313 | 0.026185544 | 1 |
| T2DM | endothelial | *N6AMT1* | -0.199382577 | 0.022015395 | 0.036587573 | 1 |
| T2DM | endothelial | *ANKRD9* | -0.60601129 | 1.88E-08 | 2.69E-06 | 0.000118264 |
| T2DM | endothelial | *SFXN2* | -0.396172682 | 0.009095563 | 0.017218851 | 1 |
| T2DM | endothelial | *C14orf2* | -0.472760255 | 6.48E-05 | 0.000267794 | 0.407581845 |
| T2DM | endothelial | *TDRD9* | -0.39901001 | 1.07E-05 | 6.21E-05 | 0.067014278 |
| T2DM | endothelial | *SH3RF2* | 0.175228078 | 2.01E-06 | 1.88E-05 | 0.01266376 |
| T2DM | endothelial | *GDF6* | -0.70095143 | 2.25E-06 | 2.01E-05 | 0.014120922 |
| T2DM | endothelial | *ATAD2* | -0.551136665 | 2.89E-06 | 2.37E-05 | 0.018164153 |
| T2DM | endothelial | *SASS6* | -0.630829969 | 0.000225396 | 0.000759821 | 1 |
| T2DM | endothelial | *ITGAD* | -0.338745714 | 0.000126774 | 0.000471615 | 0.797028727 |
| T2DM | endothelial | *MPV17L* | -0.235728917 | 0.008434661 | 0.016132861 | 1 |
| T2DM | endothelial | *RPUSD3* | -0.71268745 | 2.54E-05 | 0.000125672 | 0.159603618 |
| T2DM | endothelial | *GHRL* | -0.666030072 | 0.001284883 | 0.003274445 | 1 |
| T2DM | endothelial | *TIMP4* | 0.45249431 | 0.012907681 | 0.023185884 | 1 |
| T2DM | endothelial | *CPT2* | -0.685308045 | 3.06E-06 | 2.46E-05 | 0.019219468 |
| T2DM | endothelial | *TMED6* | -3.79805604 | 5.87E-06 | 3.95E-05 | 0.03689721 |
| T2DM | endothelial | *CLEC18A* | -0.593929575 | 0.000561407 | 0.001625779 | 1 |
| T2DM | endothelial | *IL34* | -0.62823694 | 8.96E-07 | 1.09E-05 | 0.005630208 |
| T2DM | endothelial | *RNF111* | -0.310088168 | 0.002676564 | 0.006066171 | 1 |
| T2DM | endothelial | *KCNJ15* | -0.47570438 | 0.002474095 | 0.005693498 | 1 |
| T2DM | endothelial | *MX1* | 0.123613514 | 0.001074856 | 0.00280982 | 1 |
| T2DM | endothelial | *CREB3L1* | -0.406154315 | 0.011156012 | 0.020478203 | 1 |
| T2DM | endothelial | *SLC38A10* | -0.480198542 | 0.000477048 | 0.001418733 | 1 |
| T2DM | endothelial | *C9orf43* | -0.649970125 | 4.42E-06 | 3.23E-05 | 0.027818817 |
| T2DM | endothelial | *WDR19* | -0.491747082 | 5.47E-05 | 0.000233124 | 0.343857595 |
| T2DM | endothelial | *SLC37A3* | -0.493120374 | 0.000140135 | 0.000512822 | 0.881028371 |

| | | | | | | |
|---|---|---|---|---|---|---|
| T2DM | endothelial | *AP3S2* | 0.484891354 | 1.41E-07 | 4.46E-06 | 0.000886366 |
| T2DM | endothelial | *FAM213B* | 0.388893091 | 3.95E-06 | 2.96E-05 | 0.024815966 |
| T2DM | endothelial | *PEX10* | -0.731478965 | 0.000300281 | 0.000958793 | 1 |
| T2DM | endothelial | *RADIL* | -0.527043157 | 1.63E-07 | 4.50E-06 | 0.001027175 |
| T2DM | endothelial | *WDR66* | -0.359123075 | 1.59E-05 | 8.58E-05 | 0.099724205 |
| T2DM | endothelial | *UBXN11* | -0.419814472 | 0.000197811 | 0.000679583 | 1 |
| T2DM | endothelial | *RHPN1* | 0.410322045 | 0.000166816 | 0.00058989 | 1 |
| T2DM | endothelial | *TPRG1L* | -0.650399222 | 0.013542631 | 0.02414021 | 1 |
| T2DM | endothelial | *LRRC43* | -0.683825982 | 1.30E-07 | 4.46E-06 | 0.000818888 |
| T2DM | endothelial | *XKR8* | -0.734483076 | 0.000817846 | 0.002236538 | 1 |
| T2DM | endothelial | *MRAS* | -0.355519323 | 1.78E-05 | 9.34E-05 | 0.111755632 |
| T2DM | endothelial | *ABHD3* | -0.274490056 | 0.006542211 | 0.01296686 | 1 |
| T2DM | endothelial | *CLSTN2* | -0.319041707 | 2.39E-07 | 5.33E-06 | 0.001502365 |
| T2DM | endothelial | *COLEC12* | -0.381870119 | 1.94E-06 | 1.83E-05 | 0.012190764 |
| T2DM | endothelial | *RNF207* | -0.944814005 | 0.000127057 | 0.000472389 | 0.798809346 |
| T2DM | endothelial | *AUTS2* | -0.316721347 | 6.03E-05 | 0.000252274 | 0.379236903 |
| T2DM | endothelial | *TSPAN33* | -0.358112389 | 2.92E-06 | 2.38E-05 | 0.018338671 |
| T2DM | endothelial | *AHCYL2* | -0.26125284 | 0.025861923 | 0.042068283 | 1 |
| T2DM | endothelial | *CD1D* | -0.868693915 | 6.96E-08 | 4.46E-06 | 0.00043774 |
| T2DM | endothelial | *FAM86C1* | -1.848351906 | 3.63E-05 | 0.000166897 | 0.227980955 |
| T2DM | endothelial | *NCF1* | -0.75488304 | 1.42E-07 | 4.46E-06 | 0.000891131 |
| T2DM | endothelial | *ZFAND2B* | -0.468081005 | 0.00696333 | 0.013676493 | 1 |
| T2DM | endothelial | *GDPD5* | -0.536251043 | 2.33E-06 | 2.06E-05 | 0.014622626 |
| T2DM | endothelial | *DYNC1I1* | -0.54243314 | 1.03E-05 | 6.05E-05 | 0.064922449 |
| T2DM | endothelial | *TMED4* | -0.50457475 | 0.002040032 | 0.00483988 | 1 |
| T2DM | endothelial | *COPG2* | -0.183335145 | 6.66E-06 | 4.33E-05 | 0.041864106 |
| T2DM | endothelial | *ELK4* | 0.315001607 | 0.01089833 | 0.020057904 | 1 |
| T2DM | endothelial | *DUSP23* | 0.54369312 | 7.52E-05 | 0.000301823 | 0.472655455 |
| T2DM | endothelial | *RNF166* | -0.601814121 | 6.33E-06 | 4.17E-05 | 0.039767785 |
| T2DM | endothelial | *F11R* | 0.284948324 | 0.01673456 | 0.028848417 | 1 |
| T2DM | endothelial | *SPATA2L* | -0.983086679 | 2.29E-06 | 2.04E-05 | 0.014406605 |
| T2DM | endothelial | *FGF17* | 0.911742459 | 0.018311562 | 0.03119913 | 1 |
| T2DM | endothelial | *CDA* | 0.309510264 | 0.029583697 | 0.047302315 | 1 |
| T2DM | endothelial | *FAM160B2* | -0.382525413 | 0.018332476 | 0.031226301 | 1 |
| T2DM | endothelial | *RAPGEF6* | -0.948766435 | 4.00E-05 | 0.000181119 | 0.251393699 |
| T2DM | endothelial | *ALG8* | -0.466187421 | 0.002291452 | 0.005341622 | 1 |
| T2DM | endothelial | *IFNAR2* | 0.254648708 | 0.00103577 | 0.002724639 | 1 |
| T2DM | endothelial | *MRPL10* | -0.493829513 | 0.000318033 | 0.001007292 | 1 |
| T2DM | endothelial | *SV2A* | -1.678089792 | 0.006820863 | 0.013463571 | 1 |
| T2DM | endothelial | *CLIC6* | -0.299928771 | 2.06E-06 | 1.91E-05 | 0.012963538 |
| T2DM | endothelial | *SIM2* | -0.506365371 | 1.47E-05 | 8.06E-05 | 0.092634914 |
| T2DM | endothelial | *HLCS* | -0.18889848 | 0.01346916 | 0.024036506 | 1 |
| T2DM | endothelial | *ARHGAP27* | 0.335599401 | 0.01190175 | 0.02166994 | 1 |
| T2DM | endothelial | *PADI4* | -1.254998773 | 1.30E-06 | 1.42E-05 | 0.008164628 |
| T2DM | endothelial | *PSMD4* | -0.552044263 | 0.000258582 | 0.000846281 | 1 |
| T2DM | endothelial | *STARD9* | 0.242259101 | 0.00063239 | 0.001796582 | 1 |
| T2DM | endothelial | *THEM4* | -0.407259984 | 0.00072269 | 0.002015772 | 1 |
| T2DM | endothelial | *TCHH* | -1.352775258 | 1.54E-08 | 2.38E-06 | 9.69E-05 |
| T2DM | endothelial | *MED8* | -0.356899511 | 0.00097244 | 0.002580886 | 1 |
| T2DM | endothelial | *ACE* | 0.326007063 | 4.78E-05 | 0.00020899 | 0.300527122 |

| | | | | | | |
|---|---|---|---|---|---|---|
| T2DM | endothelial | *TEPP* | -3.357020633 | 1.83E-05 | 9.57E-05 | 0.115240844 |
| T2DM | endothelial | *ATP6V0D1* | -0.498788052 | 0.000151977 | 0.000545989 | 0.955480131 |
| T2DM | endothelial | *C16orf86* | -0.486809112 | 0.000749537 | 0.002075006 | 1 |
| T2DM | endothelial | *CCDC117* | -0.711411286 | 2.87E-05 | 0.000137723 | 0.180416804 |
| T2DM | endothelial | *CCDC107* | -0.533480664 | 4.84E-07 | 7.96E-06 | 0.003041431 |
| T2DM | endothelial | *NPR2* | -0.678412546 | 0.000667388 | 0.001877345 | 1 |
| T2DM | endothelial | *CALM3* | -0.62706929 | 0.000392311 | 0.001197891 | 1 |
| T2DM | endothelial | *DFFA* | -0.318583189 | 0.028080988 | 0.045314603 | 1 |
| T2DM | endothelial | *ATAD3B* | -0.476484688 | 0.007094012 | 0.013889771 | 1 |
| T2DM | endothelial | *SSU72* | -0.499852103 | 0.003492492 | 0.007605575 | 1 |
| T2DM | endothelial | *NR2F6* | -0.507999108 | 0.000634393 | 0.001800646 | 1 |
| T2DM | endothelial | *ANKLE1* | -0.607860366 | 0.00013303 | 0.000491399 | 0.836361543 |
| T2DM | endothelial | *CILP2* | -1.780441856 | 1.80E-07 | 4.62E-06 | 0.001129624 |
| T2DM | endothelial | *ABCG1* | 0.254480289 | 5.25E-05 | 0.000225449 | 0.329831819 |
| T2DM | endothelial | *SLC37A1* | 0.376947363 | 0.000120541 | 0.000450561 | 0.757843095 |
| T2DM | endothelial | *CSTB* | -0.466540042 | 0.000140442 | 0.000513647 | 0.882959798 |
| T2DM | endothelial | *C21orf33* | -1.10291748 | 9.86E-07 | 1.17E-05 | 0.006201121 |
| T2DM | endothelial | *LRRC3* | -0.492450483 | 0.002950835 | 0.006588032 | 1 |
| T2DM | endothelial | *ITGB2* | -0.306715527 | 0.000231568 | 0.000774811 | 1 |
| T2DM | endothelial | *FAM207A* | -0.499870889 | 0.00044551 | 0.001333136 | 1 |
| T2DM | endothelial | *LSS* | -0.413854635 | 6.29E-06 | 4.15E-05 | 0.039531035 |
| T2DM | endothelial | *PCNT* | -0.246691731 | 0.00334178 | 0.007335813 | 1 |
| T2DM | endothelial | *ADAMTS13* | -0.456236287 | 0.001104583 | 0.00288154 | 1 |
| T2DM | endothelial | *ZNF714* | -0.390526261 | 0.006252785 | 0.012479765 | 1 |
| T2DM | endothelial | *GPSM1* | -0.795652455 | 1.14E-05 | 6.51E-05 | 0.071428809 |
| T2DM | endothelial | *C19orf47* | -0.689059527 | 0.005143621 | 0.0105404 | 1 |
| T2DM | endothelial | *RDH13* | -0.483729305 | 0.025711699 | 0.041868178 | 1 |
| T2DM | endothelial | *MPZL3* | 0.311213627 | 2.77E-06 | 2.32E-05 | 0.017434772 |
| T2DM | endothelial | *PCSK7* | -0.385367675 | 0.007700056 | 0.014904635 | 1 |
| T2DM | endothelial | *S100A1* | -0.680061463 | 0.015378821 | 0.02690224 | 1 |
| T2DM | endothelial | *ZBTB7B* | -0.678221235 | 4.84E-05 | 0.000211205 | 0.304345959 |
| T2DM | endothelial | *VPS11* | -0.680376885 | 1.57E-05 | 8.53E-05 | 0.098960643 |
| T2DM | endothelial | *FDPS* | 0.293563735 | 0.025427439 | 0.041490348 | 1 |
| T2DM | endothelial | *PAQR6* | -0.898931799 | 2.43E-08 | 3.06E-06 | 0.000153038 |
| T2DM | endothelial | *PMF1* | -0.519450375 | 0.000371007 | 0.00114733 | 1 |
| T2DM | endothelial | *FCRL3* | -0.28441118 | 0.004138822 | 0.008798991 | 1 |
| T2DM | endothelial | *NACC1* | -0.84669879 | 0.000200163 | 0.000686914 | 1 |
| T2DM | endothelial | *LY6K* | 1.207125766 | 0.000146275 | 0.000529131 | 0.919630502 |
| T2DM | endothelial | *LY6E* | 0.396644719 | 0.000322745 | 0.001021183 | 1 |
| T2DM | endothelial | *MUM1* | -0.303523737 | 0.013401366 | 0.023942707 | 1 |
| T2DM | endothelial | *ZNF333* | -0.445044393 | 0.004393327 | 0.009216166 | 1 |
| T2DM | endothelial | *PPP1R16A* | -0.206232668 | 0.028420275 | 0.045721154 | 1 |
| T2DM | endothelial | *ORAI2* | 0.428521703 | 0.000953869 | 0.002540015 | 1 |
| T2DM | endothelial | *FBXL13* | -0.406812891 | 0.005025242 | 0.010328112 | 1 |
| T2DM | endothelial | *MFSD12* | -0.531336449 | 0.003610282 | 0.007843069 | 1 |
| T2DM | endothelial | *YDJC* | -1.823952784 | 0.000607308 | 0.001739474 | 1 |
| T2DM | endothelial | *CCDC116* | -0.596201051 | 0.02617476 | 0.042500184 | 1 |
| T2DM | endothelial | *DVL3* | -0.449232341 | 0.003843852 | 0.008275715 | 1 |
| T2DM | endothelial | *AP2M1* | -0.437685756 | 0.005208645 | 0.010656281 | 1 |
| T2DM | endothelial | *FBXO27* | -0.233425708 | 0.000114402 | 0.000430865 | 0.719247511 |

| | | | | | | |
|---|---|---|---|---|---|---|
| T2DM | endothelial | *DMKN* | 0.276383202 | 3.37E-06 | 2.63E-05 | 0.021190581 |
| T2DM | endothelial | *BDH1* | -0.846464935 | 8.70E-07 | 1.08E-05 | 0.005466831 |
| T2DM | endothelial | *COX7A1* | -0.447587891 | 0.000346908 | 0.00108508 | 1 |
| T2DM | endothelial | *DUSP14* | -0.425342136 | 5.17E-06 | 3.61E-05 | 0.032495758 |
| T2DM | endothelial | *IKZF3* | -0.643839993 | 1.35E-05 | 7.49E-05 | 0.084750036 |
| T2DM | endothelial | *ACOX1* | -0.508131112 | 9.42E-06 | 5.66E-05 | 0.059237686 |
| T2DM | endothelial | *TMEM143* | -0.366723035 | 0.000651582 | 0.001838641 | 1 |
| T2DM | endothelial | *CCL16* | 0.164250586 | 5.01E-05 | 0.00021625 | 0.315119071 |
| T2DM | endothelial | *SIGLEC11* | -0.709408913 | 6.17E-07 | 8.98E-06 | 0.003880176 |
| T2DM | endothelial | *RACGAP1* | -0.72812795 | 9.59E-05 | 0.000372253 | 0.602677605 |
| T2DM | endothelial | *LARP4* | -0.252187365 | 0.023994049 | 0.039376294 | 1 |
| T2DM | endothelial | *SPC24* | -1.671933509 | 7.07E-06 | 4.52E-05 | 0.044455768 |
| T2DM | endothelial | *IP6K3* | -1.092291957 | 0.000215305 | 0.000732774 | 1 |
| T2DM | endothelial | *SCIMP* | -0.579420084 | 1.39E-07 | 4.46E-06 | 0.000873091 |
| T2DM | endothelial | *ASGR2* | -0.989042491 | 2.84E-07 | 5.92E-06 | 0.001782791 |
| T2DM | endothelial | *SENP3* | -1.074954535 | 1.26E-05 | 7.07E-05 | 0.079138354 |
| T2DM | endothelial | *SNRNP25* | -0.625170507 | 0.022170227 | 0.036786544 | 1 |
| T2DM | endothelial | *WDR90* | -0.36747236 | 0.00010366 | 0.00039709 | 0.651710339 |
| T2DM | endothelial | *ZG16B* | -3.217138959 | 1.29E-07 | 4.46E-06 | 0.000811692 |
| T2DM | endothelial | *TTC9C* | -0.374125076 | 0.019252097 | 0.032545829 | 1 |
| T2DM | endothelial | *NXF1* | -0.472728644 | 0.000887883 | 0.002395761 | 1 |
| T2DM | endothelial | *TPCN2* | -0.45265242 | 0.000180582 | 0.000632137 | 1 |
| T2DM | endothelial | *CYP4A22* | -1.20661272 | 4.73E-05 | 0.000207323 | 0.297454357 |
| T2DM | endothelial | *PDZK1IP1* | 4.7177844 | 0.020876817 | 0.034870496 | 1 |
| T2DM | endothelial | *COA7* | -0.909979238 | 0.000899841 | 0.002422827 | 1 |
| T2DM | endothelial | *SLC1A7* | -0.136199214 | 0.013862218 | 0.024646993 | 1 |
| T2DM | endothelial | *C1orf123* | -0.629556441 | 0.000440563 | 0.001322108 | 1 |
| T2DM | endothelial | *ACOT11* | -0.595918105 | 1.82E-05 | 9.52E-05 | 0.114531675 |
| T2DM | endothelial | *USP24* | -0.288830655 | 0.002701976 | 0.00610835 | 1 |
| T2DM | endothelial | *RBP7* | 0.211818376 | 3.22E-06 | 2.54E-05 | 0.02022953 |
| T2DM | endothelial | *FBLIM1* | 0.3221256 | 9.14E-06 | 5.53E-05 | 0.05744862 |
| T2DM | endothelial | *SLC25A34* | -1.185969085 | 8.52E-07 | 1.08E-05 | 0.005358272 |
| T2DM | endothelial | *PDPN* | -0.349706801 | 8.90E-07 | 1.09E-05 | 0.00559258 |
| T2DM | endothelial | *LRRC38* | -1.523587462 | 1.08E-06 | 1.25E-05 | 0.006776779 |
| T2DM | endothelial | *LAPTM5* | -0.329967198 | 0.019537866 | 0.032993436 | 1 |
| T2DM | endothelial | *TSSK3* | -0.326841696 | 0.010023748 | 0.018672386 | 1 |
| T2DM | endothelial | *CAMK2N1* | -0.606285902 | 1.62E-07 | 4.50E-06 | 0.00101607 |
| T2DM | endothelial | *TTLL10* | 0.285784547 | 5.80E-07 | 8.66E-06 | 0.00364745 |
| T2DM | endothelial | *OMA1* | -0.293890647 | 0.005582486 | 0.011313328 | 1 |
| T2DM | endothelial | *TM2D1* | -0.287064264 | 0.029051944 | 0.046582396 | 1 |
| T2DM | endothelial | *NEXN* | -0.484441528 | 5.41E-05 | 0.000230964 | 0.340210696 |
| T2DM | endothelial | *B3GALT2* | 0.605434259 | 0.011760523 | 0.021450075 | 1 |
| T2DM | endothelial | *HFM1* | -0.58351222 | 0.000556423 | 0.001613575 | 1 |
| T2DM | endothelial | *KCNT2* | 0.267362262 | 0.010833664 | 0.019962264 | 1 |
| T2DM | endothelial | *VCAM1* | 0.139670381 | 1.17E-06 | 1.32E-05 | 0.007381726 |
| T2DM | endothelial | *EXTL2* | -0.505416818 | 4.11E-08 | 3.59E-06 | 0.000258643 |
| T2DM | endothelial | *ARPC5* | -0.296642771 | 0.015504978 | 0.027076072 | 1 |
| T2DM | endothelial | *ZNF496* | 0.387239341 | 0.009442161 | 0.017767994 | 1 |
| T2DM | endothelial | *TRIM58* | 0.396830922 | 0.003340245 | 0.007335813 | 1 |
| T2DM | endothelial | *IGSF8* | -0.758140549 | 1.68E-06 | 1.68E-05 | 0.010546847 |

| | | | | | | |
|---|---|---|---|---|---|---|
| T2DM | endothelial | *VANGL2* | 0.755588246 | 2.62E-05 | 0.000128614 | 0.165012115 |
| T2DM | endothelial | *FCRLB* | -0.711613701 | 1.00E-05 | 5.94E-05 | 0.063032171 |
| T2DM | endothelial | *LMX1A* | -0.176102577 | 0.006192041 | 0.012378175 | 1 |
| T2DM | endothelial | *TDRD5* | -0.384241248 | 0.002413929 | 0.005583286 | 1 |
| T2DM | endothelial | *SNED1* | -0.238405729 | 0.028917234 | 0.046401901 | 1 |
| T2DM | endothelial | *C1orf115* | 0.338353061 | 1.59E-06 | 1.62E-05 | 0.010011147 |
| T2DM | endothelial | *ACP6* | -0.443181227 | 0.001325701 | 0.003360507 | 1 |
| T2DM | endothelial | *HAAO* | -0.557404958 | 3.79E-05 | 0.000173603 | 0.238531138 |
| T2DM | endothelial | *B3GALNT2* | -0.410350684 | 0.000324912 | 0.001027008 | 1 |
| T2DM | endothelial | *FAM84A* | -1.110078845 | 1.60E-07 | 4.49E-06 | 0.001006688 |
| T2DM | endothelial | *FRZB* | -0.680286502 | 2.28E-05 | 0.000115049 | 0.143351523 |
| T2DM | endothelial | *FBXO41* | -0.765241984 | 3.90E-08 | 3.45E-06 | 0.000244883 |
| T2DM | endothelial | *CCDC74A* | -1.296053565 | 1.05E-07 | 4.46E-06 | 0.000658655 |
| T2DM | endothelial | *NOSTRIN* | 0.122851264 | 1.85E-06 | 1.78E-05 | 0.011650134 |
| T2DM | endothelial | *SGPP2* | -0.815627141 | 6.84E-06 | 4.41E-05 | 0.043009485 |
| T2DM | endothelial | *BBS5* | -0.538101818 | 0.000250244 | 0.000826441 | 1 |
| T2DM | endothelial | *HPGDS* | -0.440195167 | 2.13E-05 | 0.000108622 | 0.134147998 |
| T2DM | endothelial | *C1QTNF7* | -0.398456577 | 4.85E-06 | 3.47E-05 | 0.030494588 |
| T2DM | endothelial | *LYSMD1* | -0.785174843 | 0.000413947 | 0.001253607 | 1 |
| T2DM | endothelial | *RNF149* | -0.390881508 | 0.000443733 | 0.001328451 | 1 |
| T2DM | endothelial | *DHX57* | -0.376782718 | 0.009044868 | 0.017153872 | 1 |
| T2DM | endothelial | *S100A9* | -0.695901418 | 0.001306154 | 0.003320578 | 1 |
| T2DM | endothelial | *S100A12* | -2.385202199 | 0.018479129 | 0.031450537 | 1 |
| T2DM | endothelial | *TDRD10* | 0.334413004 | 0.000258278 | 0.000845728 | 1 |
| T2DM | endothelial | *DCAF16* | -0.369666634 | 0.027466133 | 0.044413471 | 1 |
| T2DM | endothelial | *NIPAL1* | -0.716508902 | 5.10E-07 | 8.20E-06 | 0.003208137 |
| T2DM | endothelial | *ANTXR2* | -0.329221473 | 1.68E-05 | 8.97E-05 | 0.105523381 |
| T2DM | endothelial | *PBXIP1* | -0.64208029 | 0.000147363 | 0.00053215 | 0.926473184 |
| T2DM | endothelial | *DCST2* | 0.66860205 | 1.81E-05 | 9.48E-05 | 0.113978725 |
| T2DM | endothelial | *LMOD3* | -0.806181905 | 2.71E-07 | 5.76E-06 | 0.001702011 |
| T2DM | endothelial | *NBPF10* | -0.504140701 | 2.33E-07 | 5.25E-06 | 0.001466627 |
| T2DM | endothelial | *SLC22A15* | -0.45099395 | 5.50E-06 | 3.77E-05 | 0.03460197 |
| T2DM | endothelial | *LRRC58* | -0.548727781 | 0.000360459 | 0.001119115 | 1 |
| T2DM | endothelial | *FSTL1* | -0.295956766 | 0.000677577 | 0.001903451 | 1 |
| T2DM | endothelial | *LMOD1* | -0.452281416 | 5.68E-09 | 1.22E-06 | 3.57E-05 |
| T2DM | endothelial | *IGFBP7* | 0.200097206 | 0.005285901 | 0.010807304 | 1 |
| T2DM | endothelial | *PYHIN1* | -0.605261551 | 2.28E-08 | 2.92E-06 | 0.000143206 |
| T2DM | endothelial | *AIM2* | 0.165176627 | 9.11E-07 | 1.11E-05 | 0.005728014 |
| T2DM | endothelial | *EFHB* | -0.076986698 | 0.028875612 | 0.04634989 | 1 |
| T2DM | endothelial | *CD200R1* | -0.481449167 | 4.91E-06 | 3.49E-05 | 0.030859661 |
| T2DM | endothelial | *CADPS* | -0.688080748 | 2.96E-07 | 6.09E-06 | 0.001858532 |
| T2DM | endothelial | *PPM1K* | -0.501189403 | 0.001279251 | 0.003261417 | 1 |
| T2DM | endothelial | *TIPARP* | -0.373327585 | 0.006377007 | 0.012679393 | 1 |
| T2DM | endothelial | *RPP14* | -0.700180625 | 0.000290067 | 0.00093377 | 1 |
| T2DM | endothelial | *DNASE1L3* | 0.22771172 | 6.87E-07 | 9.54E-06 | 0.004321269 |
| T2DM | endothelial | *CRELD1* | -0.340906077 | 0.001366919 | 0.00345272 | 1 |
| T2DM | endothelial | *PCOLCE2* | -0.325560688 | 1.96E-06 | 1.84E-05 | 0.012306215 |
| T2DM | endothelial | *TTC14* | -0.230863673 | 0.026728819 | 0.043332667 | 1 |
| T2DM | endothelial | *RCHY1* | -0.445332925 | 0.002012973 | 0.004782902 | 1 |
| T2DM | endothelial | *CCDC158* | -0.486578608 | 8.63E-06 | 5.30E-05 | 0.054255128 |

| | | | | | | |
|---|---|---|---|---|---|---|
| T2DM | endothelial | *GYG1* | -0.430983482 | 0.022026083 | 0.036595662 | 1 |
| T2DM | endothelial | *HPS3* | -0.331827516 | 0.02956858 | 0.04729017 | 1 |
| T2DM | endothelial | *PLB1* | -0.740167852 | 8.05E-07 | 1.04E-05 | 0.005060678 |
| T2DM | endothelial | *CDCP1* | -0.557358577 | 1.83E-07 | 4.62E-06 | 0.001152729 |
| T2DM | endothelial | *LZTFL1* | -0.669407098 | 0.000216978 | 0.000736577 | 1 |
| T2DM | endothelial | *FYCO1* | -0.508292837 | 0.000948113 | 0.002526829 | 1 |
| T2DM | endothelial | *LRRC2* | -0.335650651 | 0.000294089 | 0.00094478 | 1 |
| T2DM | endothelial | *DTX3L* | 0.408543804 | 0.000830885 | 0.002265433 | 1 |
| T2DM | endothelial | *TPRA1* | -0.674993341 | 0.000109616 | 0.000415905 | 0.689155121 |
| T2DM | endothelial | *MEAF6* | -0.401209308 | 0.021220023 | 0.035396732 | 1 |
| T2DM | endothelial | *DNALI1* | -0.728324779 | 7.79E-05 | 0.000310951 | 0.489748383 |
| T2DM | endothelial | *RFT1* | -0.839379587 | 4.96E-07 | 8.08E-06 | 0.003120496 |
| T2DM | endothelial | *SFMBT1* | -0.587902877 | 3.18E-05 | 0.000149831 | 0.199874003 |
| T2DM | endothelial | *GNL3* | -0.468664477 | 0.001206882 | 0.003104654 | 1 |
| T2DM | endothelial | *FAM208A* | -0.272829785 | 0.025040302 | 0.040901112 | 1 |
| T2DM | endothelial | *SLC51A* | -0.462936193 | 0.005679419 | 0.01147844 | 1 |
| T2DM | endothelial | *PIGX* | -0.474566336 | 0.001695452 | 0.004136324 | 1 |
| T2DM | endothelial | *ABLIM2* | 0.293528313 | 9.81E-06 | 5.84E-05 | 0.061661686 |
| T2DM | endothelial | *EXO5* | -0.690323647 | 5.16E-06 | 3.61E-05 | 0.032450454 |
| T2DM | endothelial | *C1orf50* | -0.795401476 | 5.93E-05 | 0.000248585 | 0.372629068 |
| T2DM | endothelial | *ERMAP* | -0.404515419 | 0.000132185 | 0.000489715 | 0.831046969 |
| T2DM | endothelial | *SLC9B2* | 0.292516609 | 0.006498282 | 0.012896054 | 1 |
| T2DM | endothelial | *BDH2* | -0.456447934 | 3.62E-06 | 2.77E-05 | 0.022767668 |
| T2DM | endothelial | *CDC25A* | -0.474469561 | 4.22E-06 | 3.11E-05 | 0.026502537 |
| T2DM | endothelial | *ZNF589* | -0.343691747 | 6.20E-05 | 0.000257618 | 0.389518658 |
| T2DM | endothelial | *ATRIP* | -1.539174442 | 0.022539982 | 0.037291807 | 1 |
| T2DM | endothelial | *SHISA5* | 0.343491481 | 0.001064157 | 0.002792302 | 1 |
| T2DM | endothelial | *APEH* | -0.503774065 | 1.72E-05 | 9.12E-05 | 0.108057943 |
| T2DM | endothelial | *RNF123* | 0.228069487 | 0.020386001 | 0.034141394 | 1 |
| T2DM | endothelial | *RAD54L2* | -0.29222955 | 0.027452969 | 0.044403606 | 1 |
| T2DM | endothelial | *GRM2* | 8.772951981 | 6.77E-07 | 9.48E-06 | 0.004257501 |
| T2DM | endothelial | *PPM1M* | -0.450196483 | 1.09E-05 | 6.32E-05 | 0.068442433 |
| T2DM | endothelial | *ANXA5* | -0.372993303 | 0.000333109 | 0.001047651 | 1 |
| T2DM | endothelial | *MAP9* | -0.613611557 | 4.23E-07 | 7.37E-06 | 0.002661674 |
| T2DM | endothelial | *CEP44* | -0.442301978 | 0.003234758 | 0.007125762 | 1 |
| T2DM | endothelial | *TMEM144* | -0.510274346 | 1.16E-05 | 6.60E-05 | 0.07278353 |
| T2DM | endothelial | *NPY1R* | -0.366525726 | 4.08E-07 | 7.23E-06 | 0.002568044 |
| T2DM | endothelial | *IL15* | 0.267141187 | 6.03E-05 | 0.000252274 | 0.379106463 |
| T2DM | endothelial | *ITGA2* | 0.220214279 | 1.81E-06 | 1.75E-05 | 0.011389007 |
| T2DM | endothelial | *NDUFAF2* | -0.375736913 | 0.001803869 | 0.004361895 | 1 |
| T2DM | endothelial | *LMBRD2* | -0.475158226 | 0.003003545 | 0.006691456 | 1 |
| T2DM | endothelial | *PGGT1B* | -0.347553195 | 0.022391325 | 0.037124013 | 1 |
| T2DM | endothelial | *CMBL* | -0.568426136 | 2.55E-07 | 5.50E-06 | 0.001604332 |
| T2DM | endothelial | *C5orf63* | -0.769293105 | 5.70E-05 | 0.000241382 | 0.358210578 |
| T2DM | endothelial | *PRRC1* | -0.442719412 | 0.003321475 | 0.007303991 | 1 |
| T2DM | endothelial | *F2RL1* | -0.595820066 | 0.000516285 | 0.001509713 | 1 |
| T2DM | endothelial | *WDR41* | -0.456892902 | 1.76E-05 | 9.32E-05 | 0.110774227 |
| T2DM | endothelial | *NDUFS4* | -0.299751427 | 0.022038257 | 0.036597636 | 1 |
| T2DM | endothelial | *RHOBTB3* | -0.24393949 | 0.000294828 | 0.000946671 | 1 |
| T2DM | endothelial | *TIGD6* | -0.983498808 | 5.20E-07 | 8.28E-06 | 0.00326959 |

| | | | | | | |
|---|---|---|---|---|---|---|
| T2DM | endothelial | *SERINC5* | -0.279763567 | 0.02152206 | 0.035834001 | 1 |
| T2DM | endothelial | *ENPP6* | 0.920074546 | 0.006153892 | 0.012309743 | 1 |
| T2DM | endothelial | *PRIMPOL* | -0.422303532 | 0.000819836 | 0.002240031 | 1 |
| T2DM | endothelial | *ANKRA2* | -0.499732589 | 0.001861422 | 0.004480382 | 1 |
| T2DM | endothelial | *UBLCP1* | -0.316577428 | 0.012695262 | 0.02285656 | 1 |
| T2DM | endothelial | *UTP15* | -0.733880453 | 0.000573037 | 0.001655645 | 1 |
| T2DM | endothelial | *TLR3* | 0.469245713 | 0.001825838 | 0.004406544 | 1 |
| T2DM | endothelial | *CCDC127* | -0.739156003 | 0.000334518 | 0.001050508 | 1 |
| T2DM | endothelial | *GDF9* | -0.559217819 | 0.010384762 | 0.01927066 | 1 |
| T2DM | endothelial | *GRIK2* | -0.315112207 | 0.008500324 | 0.01622883 | 1 |
| T2DM | endothelial | *TXLNB* | -0.731188251 | 1.77E-06 | 1.72E-05 | 0.011107584 |
| T2DM | endothelial | *TMEM200A* | -0.811760533 | 0.000107696 | 0.000409362 | 0.677083993 |
| T2DM | endothelial | *STXBP5* | -0.256439691 | 0.028135672 | 0.045391063 | 1 |
| T2DM | endothelial | *PI16* | -0.44276569 | 4.90E-06 | 3.49E-05 | 0.030796901 |
| T2DM | endothelial | *DAGLB* | -0.407698201 | 0.001043316 | 0.002741048 | 1 |
| T2DM | endothelial | *SAP30L* | -0.569134985 | 0.001207393 | 0.003104654 | 1 |
| T2DM | endothelial | *HCN1* | -0.641760406 | 6.91E-06 | 4.44E-05 | 0.043471504 |
| T2DM | endothelial | *SLU7* | -0.454049016 | 0.004331999 | 0.009139356 | 1 |
| T2DM | endothelial | *RP9* | -0.650492475 | 4.70E-05 | 0.00020632 | 0.295357032 |
| T2DM | endothelial | *KCNK5* | 0.761523079 | 0.000170996 | 0.000602318 | 1 |
| T2DM | endothelial | *USP49* | -0.502254376 | 7.70E-05 | 0.000308153 | 0.484109125 |
| T2DM | endothelial | *ZNF704* | -0.273839756 | 0.011345326 | 0.020789386 | 1 |
| T2DM | endothelial | *FNDC1* | -0.362439004 | 3.53E-05 | 0.000162895 | 0.221862633 |
| T2DM | endothelial | *CHMP4C* | 0.203900428 | 0.001090004 | 0.00284469 | 1 |
| T2DM | endothelial | *CTSB* | -0.233829464 | 0.001442571 | 0.003610447 | 1 |
| T2DM | endothelial | *PEX2* | -0.524434728 | 0.000875077 | 0.002367301 | 1 |
| T2DM | endothelial | *DEFB1* | -0.451342331 | 6.51E-06 | 4.25E-05 | 0.040940796 |
| T2DM | endothelial | *GPER1* | 0.273242349 | 0.001659386 | 0.004059361 | 1 |
| T2DM | endothelial | *INTS1* | -0.640947299 | 0.000529922 | 0.001543131 | 1 |
| T2DM | endothelial | *SLC4A2* | -0.737140848 | 7.18E-05 | 0.000290472 | 0.451648449 |
| T2DM | endothelial | *FASTK* | -0.672066552 | 4.96E-05 | 0.00021471 | 0.311974259 |
| T2DM | endothelial | *COX6C* | -0.462998198 | 0.002548044 | 0.005827412 | 1 |
| T2DM | endothelial | *YWHAZ* | -0.213696226 | 0.019313298 | 0.032640512 | 1 |
| T2DM | endothelial | *FREM1* | -0.352052086 | 1.68E-07 | 4.54E-06 | 0.001053193 |
| T2DM | endothelial | *RPP25L* | -0.836318382 | 0.00024662 | 0.000818124 | 1 |
| T2DM | endothelial | *SYK* | -0.4842832 | 2.84E-06 | 2.34E-05 | 0.017826404 |
| T2DM | endothelial | *TMC1* | 0.43153927 | 0.000397614 | 0.001211143 | 1 |
| T2DM | endothelial | *C9orf64* | -0.40509057 | 0.0013174 | 0.00334511 | 1 |
| T2DM | endothelial | *SVEP1* | -0.307621027 | 7.57E-07 | 1.01E-05 | 0.004758267 |
| T2DM | endothelial | *ZNF367* | -0.349178378 | 0.012627085 | 0.022759887 | 1 |
| T2DM | endothelial | *NOL6* | -0.563512921 | 0.022092386 | 0.036667062 | 1 |
| T2DM | endothelial | *TRMT10B* | -0.359034795 | 0.011483913 | 0.021012619 | 1 |
| T2DM | endothelial | *VCP* | -0.378577619 | 0.001572323 | 0.003878069 | 1 |
| T2DM | endothelial | *LRFN5* | -0.239084762 | 0.000252911 | 0.000833334 | 1 |
| T2DM | endothelial | *CFL2* | -0.559462287 | 0.000155643 | 0.000556587 | 0.978528492 |
| T2DM | endothelial | *ZCCHC24* | -0.446557798 | 0.000469455 | 0.001400124 | 1 |
| T2DM | endothelial | *PGM2L1* | 0.434779522 | 0.000533069 | 0.001551576 | 1 |
| T2DM | endothelial | *GJB2* | 0.38704467 | 4.18E-06 | 3.09E-05 | 0.02624906 |
| T2DM | endothelial | *REEP3* | -0.361491539 | 0.003478975 | 0.007586651 | 1 |
| T2DM | endothelial | *LRR1* | -0.642452952 | 0.000396371 | 0.001209058 | 1 |

| | | | | | | |
|---|---|---|---|---|---|---|
| T2DM | endothelial | *OXGR1* | 1.223250551 | 0.016442302 | 0.028406912 | 1 |
| T2DM | endothelial | *ATP5C1* | -0.392897019 | 0.004379113 | 0.009201699 | 1 |
| T2DM | endothelial | *VDAC2* | -0.500839625 | 0.003065839 | 0.006813337 | 1 |
| T2DM | endothelial | *SLC18A2* | -0.503131586 | 0.004867052 | 0.010035801 | 1 |
| T2DM | endothelial | *SNAPC4* | -0.260443092 | 0.015238878 | 0.026702014 | 1 |
| T2DM | endothelial | *SDCCAG3* | -0.613986175 | 9.15E-05 | 0.000357294 | 0.575243433 |
| T2DM | endothelial | *AK8* | -0.805230099 | 1.98E-05 | 0.000102645 | 0.124714243 |
| T2DM | endothelial | *ZMYND19* | -0.845193946 | 1.65E-07 | 4.50E-06 | 0.00103916 |
| T2DM | endothelial | *STOX1* | -0.148625221 | 0.000507108 | 0.001488415 | 1 |
| T2DM | endothelial | *RET* | -0.478972445 | 8.77E-08 | 4.46E-06 | 0.00055165 |
| T2DM | endothelial | *STK32C* | -0.608554031 | 0.000413902 | 0.001253607 | 1 |
| T2DM | endothelial | *NSMF* | -0.83346707 | 6.85E-05 | 0.000280112 | 0.43053278 |
| T2DM | endothelial | *VWA2* | 0.2886092 | 1.29E-06 | 1.42E-05 | 0.008137418 |
| T2DM | endothelial | *TRUB1* | -0.432184408 | 0.000169289 | 0.00059726 | 1 |
| T2DM | endothelial | *HSPA12A* | -0.211757016 | 0.000451664 | 0.001350266 | 1 |
| T2DM | endothelial | *UBTD1* | -0.579270123 | 1.43E-05 | 7.86E-05 | 0.090041214 |
| T2DM | endothelial | *ARHGAP42* | -0.371067028 | 6.84E-07 | 9.54E-06 | 0.004301178 |
| T2DM | endothelial | *OTOGL* | -0.903119965 | 0.000180947 | 0.000632495 | 1 |
| T2DM | endothelial | *PACSIN3* | -0.182406285 | 0.02108914 | 0.035197086 | 1 |
| T2DM | endothelial | *TTC7B* | -0.332702602 | 0.003625939 | 0.007868925 | 1 |
| T2DM | endothelial | *SLC39A13* | -0.757932089 | 0.000601122 | 0.001723327 | 1 |
| T2DM | endothelial | *CLMN* | -0.388722744 | 0.000412036 | 0.001249625 | 1 |
| T2DM | endothelial | *PTER* | -0.508305452 | 0.000315327 | 0.001000322 | 1 |
| T2DM | endothelial | *ABTB2* | -0.528674398 | 5.97E-07 | 8.81E-06 | 0.003754161 |
| T2DM | endothelial | *LIPC* | -0.475575433 | 5.99E-06 | 4.01E-05 | 0.037676117 |
| T2DM | endothelial | *JAM3* | 0.331644302 | 0.00107916 | 0.002818728 | 1 |
| T2DM | endothelial | *GPT2* | -0.332517651 | 2.56E-05 | 0.000126416 | 0.16105441 |
| T2DM | endothelial | *HIF1AN* | -0.608662228 | 0.0002305 | 0.000772469 | 1 |
| T2DM | endothelial | *NDUFB8* | -0.726450079 | 0.000857476 | 0.002325726 | 1 |
| T2DM | endothelial | *SPINT1* | -0.786476062 | 2.19E-06 | 1.99E-05 | 0.013787458 |
| T2DM | endothelial | *AVPR1A* | -0.253293352 | 0.004594355 | 0.009577159 | 1 |
| T2DM | endothelial | *TRMT61A* | -0.85037535 | 1.30E-12 | 4.10E-09 | 8.20E-09 |
| T2DM | endothelial | *BTRC* | -0.281709939 | 0.017337255 | 0.029775706 | 1 |
| T2DM | endothelial | *ASPG* | -1.312970732 | 6.34E-07 | 9.11E-06 | 0.003986936 |
| T2DM | endothelial | *GABRB3* | 0.327165628 | 7.79E-06 | 4.89E-05 | 0.048996309 |
| T2DM | endothelial | *TBATA* | -1.411063903 | 0.000533969 | 0.001553477 | 1 |
| T2DM | endothelial | *PCBD1* | -0.447757776 | 5.80E-09 | 1.22E-06 | 3.65E-05 |
| T2DM | endothelial | *FAM227B* | -0.281838411 | 0.009799707 | 0.018351804 | 1 |
| T2DM | endothelial | *STXBP4* | -0.4002665 | 0.002665941 | 0.00605082 | 1 |
| T2DM | endothelial | *C2* | -0.273152925 | 0.019897346 | 0.033447758 | 1 |
| T2DM | endothelial | *TAF10* | -0.497895177 | 0.016847873 | 0.029019884 | 1 |
| T2DM | endothelial | *DCHS1* | 0.401034217 | 1.53E-05 | 8.34E-05 | 0.096292403 |
| T2DM | endothelial | *NETO1* | 0.514356816 | 5.56E-06 | 3.80E-05 | 0.034938504 |
| T2DM | endothelial | *USP54* | 0.302925193 | 0.000893941 | 0.002410037 | 1 |
| T2DM | endothelial | *C11orf74* | -0.578338771 | 0.000838677 | 0.002285549 | 1 |
| T2DM | endothelial | *WDR88* | -0.643563427 | 2.00E-07 | 4.87E-06 | 0.001260278 |
| T2DM | endothelial | *CYB5R2* | -0.454860519 | 2.68E-05 | 0.00013071 | 0.168633352 |
| T2DM | endothelial | *TUB* | -0.438521635 | 8.18E-09 | 1.61E-06 | 5.14E-05 |
| T2DM | endothelial | *XRRA1* | -0.182617318 | 0.02863803 | 0.046000842 | 1 |
| T2DM | endothelial | *AKIP1* | -0.747606195 | 0.000343227 | 0.001074636 | 1 |

| | | | | | | |
|---|---|---|---|---|---|---|
| T2DM | endothelial | *C16orf46* | -3.792310066 | 7.19E-08 | 4.46E-06 | 0.000451826 |
| T2DM | endothelial | *TMEM41B* | -0.431047735 | 0.002371346 | 0.005499319 | 1 |
| T2DM | endothelial | *ZNF143* | -0.451197755 | 0.004602237 | 0.009587232 | 1 |
| T2DM | endothelial | *CCDC68* | 0.11179269 | 7.00E-07 | 9.61E-06 | 0.004403 |
| T2DM | endothelial | *CLEC4E* | -0.252674844 | 0.005042323 | 0.010356447 | 1 |
| T2DM | endothelial | *CLEC4D* | -0.907827083 | 2.12E-05 | 0.00010833 | 0.133570986 |
| T2DM | endothelial | *BEAN1* | -0.579960273 | 1.38E-05 | 7.61E-05 | 0.086552181 |
| T2DM | endothelial | *TK2* | -0.47665136 | 1.44E-06 | 1.53E-05 | 0.009029916 |
| T2DM | endothelial | *SEC11C* | 0.429555174 | 6.78E-05 | 0.000278032 | 0.426500652 |
| T2DM | endothelial | *GALR1* | 0.202434959 | 7.27E-06 | 4.62E-05 | 0.045726068 |
| T2DM | endothelial | *CENPV* | -0.635002202 | 0.000145364 | 0.000527048 | 0.913901261 |
| T2DM | endothelial | *SPDYE6* | -0.644129879 | 0.000197801 | 0.000679583 | 1 |
| T2DM | endothelial | *ZNF592* | -0.417802055 | 0.001555927 | 0.003842339 | 1 |
| T2DM | endothelial | *ACSM1* | 0.464993511 | 0.012176578 | 0.022074436 | 1 |
| T2DM | endothelial | *AP1G1* | -0.359506868 | 0.005394838 | 0.011000716 | 1 |
| T2DM | endothelial | *SAAL1* | -0.690480592 | 0.001163362 | 0.003009901 | 1 |
| T2DM | endothelial | *YPEL4* | -0.59783781 | 4.47E-07 | 7.53E-06 | 0.002808334 |
| T2DM | endothelial | *ANKDD1A* | -0.397330223 | 0.002457765 | 0.005660061 | 1 |
| T2DM | endothelial | *TSC22D4* | -0.446087476 | 0.014113141 | 0.024994174 | 1 |
| T2DM | endothelial | *MS4A6E* | -0.325458685 | 2.42E-06 | 2.12E-05 | 0.015206826 |
| T2DM | endothelial | *MS4A7* | -0.42249372 | 7.23E-07 | 9.76E-06 | 0.00454785 |
| T2DM | endothelial | *MS4A14* | -0.328217966 | 2.39E-06 | 2.10E-05 | 0.015035186 |
| T2DM | endothelial | *DIS3L* | -0.469258509 | 0.000220874 | 0.000747785 | 1 |
| T2DM | endothelial | *CCNDBP1* | -0.563666682 | 0.000381868 | 0.001172839 | 1 |
| T2DM | endothelial | *SMAD3* | -0.29600768 | 0.008393619 | 0.016064136 | 1 |
| T2DM | endothelial | *RCCD1* | -0.533872476 | 6.37E-07 | 9.11E-06 | 0.004003283 |
| T2DM | endothelial | *EVA1C* | 0.22203568 | 3.25E-06 | 2.56E-05 | 0.02041903 |
| T2DM | endothelial | *DUSP18* | 0.315728503 | 0.013524644 | 0.024114985 | 1 |
| T2DM | endothelial | *PBX3* | -0.369306739 | 0.000124472 | 0.000463599 | 0.782554916 |
| T2DM | endothelial | *SNRPD1* | -0.604269157 | 0.001319638 | 0.003349441 | 1 |
| T2DM | endothelial | *PIP5KL1* | 0.194740822 | 0.009441568 | 0.017767994 | 1 |
| T2DM | endothelial | *ENDOG* | -0.582466222 | 7.00E-06 | 4.49E-05 | 0.044038486 |
| T2DM | endothelial | *ISLR2* | 0.260367826 | 2.96E-06 | 2.41E-05 | 0.018599312 |
| T2DM | endothelial | *SP2* | -0.638338842 | 0.000228586 | 0.000768467 | 1 |
| T2DM | endothelial | *GPRC5B* | 0.277002542 | 8.36E-05 | 0.000330665 | 0.525756816 |
| T2DM | endothelial | *RNF214* | -0.356607743 | 0.015297709 | 0.02678672 | 1 |
| T2DM | endothelial | *ENGASE* | -0.373393843 | 0.001556004 | 0.003842339 | 1 |
| T2DM | endothelial | *MYO5B* | -0.16570241 | 0.018228683 | 0.03106634 | 1 |
| T2DM | endothelial | *RRM1* | -0.313764665 | 0.000521615 | 0.001521762 | 1 |
| T2DM | endothelial | *ZNF23* | -0.729194181 | 0.000515667 | 0.001508607 | 1 |
| T2DM | endothelial | *VKORC1* | -0.582839813 | 2.77E-05 | 0.000133545 | 0.174409874 |
| T2DM | endothelial | *PROCA1* | -0.616835344 | 1.28E-06 | 1.41E-05 | 0.008040792 |
| T2DM | endothelial | *TP53I13* | -0.354778282 | 0.000298318 | 0.000954465 | 1 |
| T2DM | endothelial | *ZNF701* | -0.404652507 | 0.010370143 | 0.019249215 | 1 |
| T2DM | endothelial | *SERTAD3* | -1.047742651 | 0.000145965 | 0.000528314 | 0.917681173 |
| T2DM | endothelial | *TMC4* | -0.913384375 | 1.63E-05 | 8.74E-05 | 0.102272524 |
| T2DM | endothelial | *TTYH1* | -0.685361888 | 1.27E-05 | 7.10E-05 | 0.079642398 |
| T2DM | endothelial | *LENG8* | -0.387393394 | 0.000842198 | 0.002293157 | 1 |
| T2DM | endothelial | *CDC42EP5* | 0.284690335 | 9.13E-06 | 5.53E-05 | 0.057410732 |
| T2DM | endothelial | *PPP1R14A* | 0.505824638 | 0.002424975 | 0.005598905 | 1 |

| | | | | | | |
|---|---|---|---|---|---|---|
| T2DM | endothelial | *C19orf33* | 0.406394934 | 2.04E-05 | 0.000104749 | 0.128212249 |
| T2DM | endothelial | *DNAAF3* | 0.671114557 | 0.011870475 | 0.021619258 | 1 |
| T2DM | endothelial | *PSCA* | -0.339781188 | 0.000325287 | 0.001027679 | 1 |
| T2DM | endothelial | *UBXN6* | -0.669340675 | 1.78E-06 | 1.74E-05 | 0.011219795 |
| T2DM | endothelial | *ZNF444* | -0.474053657 | 0.029505051 | 0.047200574 | 1 |
| T2DM | endothelial | *FAM57A* | -0.28489558 | 0.029165071 | 0.046711383 | 1 |
| T2DM | endothelial | *KIFC2* | -0.42334398 | 1.56E-06 | 1.61E-05 | 0.009806988 |
| T2DM | endothelial | *SERPINF2* | -0.444542672 | 0.001220038 | 0.003128214 | 1 |
| T2DM | endothelial | *CD320* | 0.201710026 | 1.94E-06 | 1.83E-05 | 0.012185089 |
| T2DM | endothelial | *SPRYD3* | -0.79101164 | 4.93E-05 | 0.000214 | 0.309712418 |
| T2DM | endothelial | *SOAT2* | -0.380554144 | 0.000297398 | 0.000952492 | 1 |
| T2DM | endothelial | *ZNF558* | -0.284541571 | 0.00595055 | 0.011967726 | 1 |
| T2DM | endothelial | *NUDT8* | -0.349326421 | 0.000924526 | 0.002475509 | 1 |
| T2DM | endothelial | *MIS12* | -0.644084251 | 2.63E-05 | 0.000128691 | 0.16536759 |
| T2DM | endothelial | *CD300C* | -0.359132478 | 0.030850348 | 0.049152594 | 1 |
| T2DM | endothelial | *HID1* | 0.142750585 | 6.32E-07 | 9.11E-06 | 0.003971846 |
| T2DM | endothelial | *ATP5H* | -0.518940405 | 0.005867009 | 0.011811042 | 1 |
| T2DM | endothelial | *SRP68* | -0.601114605 | 0.000252318 | 0.000832065 | 1 |
| T2DM | endothelial | *MGAT5B* | 0.926722426 | 0.000239649 | 0.000798872 | 1 |
| T2DM | endothelial | *TMC8* | -0.378054653 | 0.004382657 | 0.009206069 | 1 |
| T2DM | endothelial | *GSDMA* | -0.751150357 | 8.16E-07 | 1.05E-05 | 0.005127998 |
| T2DM | endothelial | *KRT24* | -0.654536986 | 1.44E-06 | 1.53E-05 | 0.009070951 |
| T2DM | endothelial | *MLST8* | -0.867552778 | 0.007981755 | 0.015378883 | 1 |
| T2DM | endothelial | *ECI1* | -0.432701935 | 0.002083937 | 0.004927307 | 1 |
| T2DM | endothelial | *KCTD5* | -0.79071687 | 4.25E-05 | 0.000191137 | 0.267017956 |
| T2DM | endothelial | *SDHAF2* | -0.889096702 | 0.001044708 | 0.002743558 | 1 |
| T2DM | endothelial | *VWCE* | 0.474841237 | 0.00141676 | 0.00356002 | 1 |
| T2DM | endothelial | *BSCL2* | 1.050239706 | 0.02501124 | 0.040874881 | 1 |
| T2DM | endothelial | *TRANK1* | -0.308494113 | 0.025712269 | 0.041868178 | 1 |
| T2DM | endothelial | *NAALADL1* | 0.588619361 | 0.018866831 | 0.032015052 | 1 |
| T2DM | endothelial | *SCARA5* | -0.428958443 | 9.53E-08 | 4.46E-06 | 0.000599428 |
| T2DM | endothelial | *COPS6* | -0.342054624 | 0.028214846 | 0.045495444 | 1 |
| T2DM | endothelial | *NUDT16L1* | -0.927512852 | 5.40E-06 | 3.72E-05 | 0.033929987 |
| T2DM | endothelial | *KIAA1586* | -0.556103993 | 0.000144553 | 0.000524638 | 0.908804314 |
| T2DM | endothelial | *ZCCHC4* | -0.405607394 | 0.012525688 | 0.022609531 | 1 |
| T2DM | endothelial | *PTGDR* | -0.36297994 | 0.012287439 | 0.02223054 | 1 |
| T2DM | endothelial | *NT5DC2* | -0.466496332 | 0.011693768 | 0.021340703 | 1 |
| T2DM | endothelial | *COA6* | -0.905678722 | 8.41E-05 | 0.000332188 | 0.528510599 |
| T2DM | endothelial | *KIF5C* | 0.424575759 | 0.009210963 | 0.017411102 | 1 |
| T2DM | endothelial | *ACOX2* | -0.267668302 | 0.000610071 | 0.001745003 | 1 |
| T2DM | endothelial | *FAM107A* | 0.164008205 | 4.19E-06 | 3.09E-05 | 0.026314159 |
| T2DM | endothelial | *CX3CR1* | -0.358061124 | 0.004187689 | 0.008891592 | 1 |
| T2DM | endothelial | *SCN11A* | 0.120770669 | 0.000152804 | 0.000548333 | 0.960680119 |
| T2DM | endothelial | *DTYMK* | -0.687560759 | 0.000473329 | 0.001409673 | 1 |
| T2DM | endothelial | *ING5* | -0.520161585 | 2.28E-05 | 0.000114896 | 0.143045455 |
| T2DM | endothelial | *MLKL* | 0.187149177 | 0.012434761 | 0.022466401 | 1 |
| T2DM | endothelial | *RHOH* | -0.510808497 | 0.000228952 | 0.000768511 | 1 |
| T2DM | endothelial | *KLHL30* | -0.490077205 | 2.56E-06 | 2.19E-05 | 0.016083727 |
| T2DM | endothelial | *BMP1* | -0.477501183 | 5.82E-05 | 0.000245357 | 0.365827112 |
| T2DM | endothelial | *ATXN2L* | -0.36793924 | 0.013882164 | 0.024668503 | 1 |

| | | | | | | |
|---|---|---|---|---|---|---|
| T2DM | endothelial | *TRAPPC11* | -0.315975402 | 0.025940598 | 0.042185345 | 1 |
| T2DM | endothelial | *SLC20A2* | -0.376969526 | 0.001338989 | 0.003387614 | 1 |
| T2DM | endothelial | *ZSWIM1* | -0.578511383 | 0.028468585 | 0.045787157 | 1 |
| T2DM | endothelial | *NBPF9* | -0.665475329 | 4.44E-07 | 7.52E-06 | 0.002790819 |
| T2DM | endothelial | *WFDC13* | -1.688214157 | 1.29E-05 | 7.22E-05 | 0.081108738 |
| T2DM | endothelial | *NDUFS5* | -0.567840899 | 0.000273471 | 0.000887615 | 1 |
| T2DM | endothelial | *ZNF30* | 0.346635926 | 0.0002846 | 0.000918994 | 1 |
| T2DM | endothelial | *FAM84B* | 0.179514444 | 5.55E-07 | 8.50E-06 | 0.003491575 |
| T2DM | endothelial | *TMEM208* | -0.571930348 | 0.000904584 | 0.002434222 | 1 |
| T2DM | endothelial | *LRP1B* | -0.175033303 | 0.015815192 | 0.027558235 | 1 |
| T2DM | endothelial | *NPNT* | 0.245266255 | 3.48E-06 | 2.69E-05 | 0.021874407 |
| T2DM | endothelial | *CNNM3* | -0.796086621 | 5.31E-05 | 0.000227458 | 0.333681053 |
| T2DM | endothelial | *ABHD15* | -0.297092079 | 0.014881439 | 0.026170519 | 1 |
| T2DM | endothelial | *SNTB2* | -0.33740974 | 0.00793856 | 0.015300345 | 1 |
| T2DM | endothelial | *ZNF507* | -0.388412296 | 0.004881601 | 0.010059202 | 1 |
| T2DM | endothelial | *STX18* | -0.29763616 | 0.031415842 | 0.049975103 | 1 |
| T2DM | endothelial | *TNIP2* | -0.350096259 | 0.01594883 | 0.027752641 | 1 |
| T2DM | endothelial | *BTNL3* | 0.127883325 | 4.96E-06 | 3.52E-05 | 0.031188818 |
| T2DM | endothelial | *ZNF608* | 0.288836976 | 0.002574783 | 0.005879085 | 1 |
| T2DM | endothelial | *INPP5D* | 0.243724666 | 0.030925288 | 0.04925951 | 1 |
| T2DM | endothelial | *LETM1* | -0.601274135 | 0.000944407 | 0.002519087 | 1 |
| T2DM | endothelial | *TMEM129* | -0.45637375 | 4.90E-05 | 0.000213142 | 0.30820351 |
| T2DM | endothelial | *CPLX1* | 0.137405252 | 1.71E-06 | 1.69E-05 | 0.010765586 |
| T2DM | endothelial | *SIGLEC7* | -0.516252062 | 1.68E-06 | 1.68E-05 | 0.010590964 |
| T2DM | endothelial | *MFSD7* | -0.797002493 | 3.45E-06 | 2.68E-05 | 0.02168938 |
| T2DM | endothelial | *COL4A3* | -0.628456996 | 1.53E-05 | 8.34E-05 | 0.096370801 |
| T2DM | endothelial | *IRS1* | -0.53507951 | 3.36E-31 | 2.11E-27 | 2.11E-27 |
| T2DM | endothelial | *PARM1* | 0.194442355 | 6.50E-06 | 4.25E-05 | 0.040889021 |
| T2DM | endothelial | *FAM110B* | -0.460712789 | 1.93E-06 | 1.83E-05 | 0.012123682 |
| T2DM | endothelial | *ATF5* | -0.816870741 | 1.17E-06 | 1.32E-05 | 0.007361421 |
| T2DM | endothelial | *CPT1C* | 0.543836252 | 2.35E-05 | 0.000117835 | 0.147528993 |
| T2DM | endothelial | *PCSK9* | -2.519623723 | 1.03E-05 | 6.05E-05 | 0.064965615 |
| T2DM | endothelial | *XPO6* | -0.38820796 | 0.004342815 | 0.009149009 | 1 |
| T2DM | endothelial | *SH3TC2* | 0.253445398 | 1.48E-05 | 8.06E-05 | 0.092791432 |
| T2DM | endothelial | *GPRIN1* | -1.431443562 | 0.009091982 | 0.017217256 | 1 |
| T2DM | endothelial | *PGM2* | 0.414529171 | 0.022450361 | 0.037192469 | 1 |
| T2DM | endothelial | *STK32A* | -0.275119317 | 9.14E-08 | 4.46E-06 | 0.000574536 |
| T2DM | endothelial | *P2RY12* | -0.362293733 | 8.92E-07 | 1.09E-05 | 0.005606202 |
| T2DM | endothelial | *SLC33A1* | -0.420542894 | 0.006393127 | 0.01270341 | 1 |
| T2DM | endothelial | *SNUPN* | -0.666442268 | 1.23E-06 | 1.37E-05 | 0.00775636 |
| T2DM | endothelial | *RNASE6* | -0.790044879 | 1.72E-07 | 4.57E-06 | 0.001083796 |
| T2DM | endothelial | *NPR1* | 0.351950148 | 0.000636856 | 0.001804377 | 1 |
| T2DM | endothelial | *CD52* | -0.464112871 | 0.000123177 | 0.000459864 | 0.774410708 |
| T2DM | endothelial | *CLIC4* | 0.245874274 | 0.001765035 | 0.004277862 | 1 |
| T2DM | endothelial | *SLC38A11* | -0.695335774 | 9.92E-08 | 4.46E-06 | 0.000623452 |
| T2DM | endothelial | *CCDC8* | -0.83312614 | 1.14E-07 | 4.46E-06 | 0.000715806 |
| T2DM | endothelial | *METTL15* | -0.262893746 | 0.010788524 | 0.019902421 | 1 |
| T2DM | endothelial | *HINT1* | -0.432622512 | 0.007058321 | 0.013828503 | 1 |
| T2DM | endothelial | *DTWD2* | -0.398168981 | 0.004714345 | 0.009781877 | 1 |
| T2DM | endothelial | *INO80E* | -0.41677256 | 0.005781996 | 0.011651093 | 1 |

| | | | | | | |
|---|---|---|---|---|---|---|
| T2DM | endothelial | *DFFB* | -0.510037543 | 0.015670551 | 0.02732892 | 1 |
| T2DM | endothelial | *C15orf40* | -0.294723123 | 0.011756187 | 0.021448389 | 1 |
| T2DM | endothelial | *LRRC45* | -0.490959078 | 3.23E-08 | 3.24E-06 | 0.000202888 |
| T2DM | endothelial | *CHRNA5* | -0.803332396 | 8.67E-05 | 0.000340357 | 0.544898949 |
| T2DM | endothelial | *FASN* | -0.247327845 | 0.001115498 | 0.002905316 | 1 |
| T2DM | endothelial | *DCXR* | -0.729598513 | 8.69E-07 | 1.08E-05 | 0.005462661 |
| T2DM | endothelial | *LIMS1* | -0.313371148 | 0.000912415 | 0.002450386 | 1 |
| T2DM | endothelial | *UGP2* | -0.303266809 | 0.00035777 | 0.001114618 | 1 |
| T2DM | endothelial | *CSGALNACT2* | -0.446674146 | 0.000736121 | 0.002045914 | 1 |
| T2DM | endothelial | *CTNND2* | 0.207163869 | 0.008338289 | 0.015987443 | 1 |
| T2DM | endothelial | *CALML6* | -0.692068428 | 9.87E-05 | 0.00038115 | 0.620314852 |
| T2DM | endothelial | *OTUD3* | -0.396017223 | 8.59E-05 | 0.000337886 | 0.53994152 |
| T2DM | endothelial | *GUSB* | -0.483305532 | 0.001858652 | 0.004477143 | 1 |
| T2DM | endothelial | *BRD3* | -0.590810911 | 0.000628469 | 0.001787058 | 1 |
| T2DM | endothelial | *ZNF747* | -1.165152059 | 0.000157631 | 0.000562918 | 0.991023907 |
| T2DM | endothelial | *MAP3K2* | -0.259011686 | 0.029669033 | 0.0474267 | 1 |
| T2DM | endothelial | *PUSL1* | -1.240370923 | 7.22E-07 | 9.76E-06 | 0.004540559 |
| T2DM | endothelial | *MYO7B* | 0.320188071 | 4.96E-05 | 0.00021471 | 0.311954583 |
| T2DM | endothelial | *CNTROB* | -0.662562466 | 0.002908928 | 0.006507531 | 1 |
| T2DM | endothelial | *RNF150* | -0.270635876 | 2.34E-05 | 0.000117419 | 0.146890794 |
| T2DM | endothelial | *ANKK1* | -0.292205199 | 0.005892371 | 0.011858302 | 1 |
| T2DM | endothelial | *ZNF212* | -0.599993787 | 0.000597707 | 0.0017151 | 1 |
| T2DM | endothelial | *GLB1* | -0.479481541 | 9.58E-05 | 0.000372253 | 0.602418756 |
| T2DM | endothelial | *FAXDC2* | -0.35045401 | 0.003480783 | 0.007587963 | 1 |
| T2DM | endothelial | *CRTAP* | -0.520267294 | 0.000359491 | 0.001117763 | 1 |
| T2DM | endothelial | *ELP5* | -0.477551828 | 0.01864983 | 0.031681027 | 1 |
| T2DM | endothelial | *NFRKB* | -0.456178475 | 3.16E-05 | 0.000149136 | 0.198500613 |
| T2DM | endothelial | *SEMA3E* | 0.233927872 | 5.03E-06 | 3.54E-05 | 0.031611564 |
| T2DM | endothelial | *ADORA2B* | -0.211165995 | 0.020403102 | 0.034151837 | 1 |
| T2DM | endothelial | *METTL7B* | -0.956311621 | 1.28E-07 | 4.46E-06 | 0.000802918 |
| T2DM | endothelial | *NFXL1* | -0.369754805 | 0.019693898 | 0.033221234 | 1 |
| T2DM | endothelial | *CD14* | -0.42098602 | 0.000157674 | 0.000562918 | 0.991298136 |
| T2DM | endothelial | *KRT72* | -1.185003936 | 0.00089842 | 0.002421074 | 1 |
| T2DM | endothelial | *HSD17B13* | 0.169925417 | 7.88E-05 | 0.000313826 | 0.495531558 |
| T2DM | endothelial | *TMC7* | 0.125140961 | 5.07E-06 | 3.56E-05 | 0.031868962 |
| T2DM | endothelial | *ARL6IP1* | 0.260717789 | 0.012660597 | 0.022807212 | 1 |
| T2DM | endothelial | *SMAGP* | 0.253177957 | 4.68E-06 | 3.39E-05 | 0.029394796 |
| T2DM | endothelial | *IRX2* | -0.303182554 | 1.77E-05 | 9.34E-05 | 0.111589581 |
| T2DM | endothelial | *EMB* | -0.455862774 | 2.86E-08 | 3.21E-06 | 0.000179917 |
| T2DM | endothelial | *DLGAP1* | 0.226077482 | 6.85E-06 | 4.41E-05 | 0.043041418 |
| T2DM | endothelial | *NUDCD2* | -0.577767298 | 0.002541377 | 0.005816395 | 1 |
| T2DM | endothelial | *HSPA4* | -0.375815949 | 0.01974803 | 0.033259418 | 1 |
| T2DM | endothelial | *COMMD5* | -0.810574481 | 1.63E-06 | 1.65E-05 | 0.010266554 |
| T2DM | endothelial | *GTSF1* | 1.25145978 | 9.20E-06 | 5.56E-05 | 0.057834641 |
| T2DM | endothelial | *RNF34* | -0.378566553 | 0.006945523 | 0.013650048 | 1 |
| T2DM | endothelial | *ACYP2* | -0.340661328 | 0.000255466 | 0.000839212 | 1 |
| T2DM | endothelial | *TRABD* | -0.377168513 | 0.003205621 | 0.007071488 | 1 |
| T2DM | endothelial | *TMEM133* | -0.686365965 | 1.40E-07 | 4.46E-06 | 0.000880205 |
| T2DM | endothelial | *TTLL6* | 1.130725555 | 4.73E-05 | 0.000207323 | 0.297508424 |
| T2DM | endothelial | *GPR37* | -0.353596396 | 0.005433267 | 0.011054675 | 1 |

| | | | | | | |
|---|---|---|---|---|---|---|
| T2DM | endothelial | *HTRA3* | -0.293886114 | 0.000886755 | 0.002394678 | 1 |
| T2DM | endothelial | *FOXN2* | -0.410436243 | 0.00024372 | 0.000810722 | 1 |
| T2DM | endothelial | *MTSS1* | -0.344821656 | 0.000597563 | 0.0017151 | 1 |
| T2DM | endothelial | *TSEN34* | -0.986714992 | 2.75E-06 | 2.31E-05 | 0.017303577 |
| T2DM | endothelial | *NDUFA3* | -0.689054791 | 0.000388871 | 0.00119144 | 1 |
| T2DM | endothelial | *OSCAR* | -0.428411497 | 0.00013954 | 0.000511241 | 0.877288945 |
| T2DM | endothelial | *FPR1* | -0.356810827 | 6.85E-06 | 4.41E-05 | 0.043086551 |
| T2DM | endothelial | *SOX7* | 0.168690611 | 6.72E-07 | 9.43E-06 | 0.004226769 |
| T2DM | endothelial | *C11orf24* | -0.650891324 | 2.94E-07 | 6.09E-06 | 0.001850527 |
| T2DM | endothelial | *ALK* | -0.424636406 | 1.00E-07 | 4.46E-06 | 0.000630134 |
| T2DM | endothelial | *GIMAP8* | 0.16627816 | 2.65E-05 | 0.000129396 | 0.166643796 |
| T2DM | endothelial | *KCNMB3* | 0.365920329 | 0.000635195 | 0.001802107 | 1 |
| T2DM | endothelial | *ATP6V0E2* | -0.613540737 | 2.24E-07 | 5.17E-06 | 0.00141033 |
| T2DM | endothelial | *GRIK1* | -0.489287857 | 0.001069015 | 0.002798043 | 1 |
| T2DM | endothelial | *GAA* | -0.226496331 | 0.011367119 | 0.020817092 | 1 |
| T2DM | endothelial | *KCND3* | -0.40057722 | 0.012312186 | 0.022268904 | 1 |
| T2DM | endothelial | *ZNF581* | -1.147375357 | 0.009566792 | 0.017970249 | 1 |
| T2DM | endothelial | *DLK2* | -1.148771133 | 0.001079949 | 0.002819619 | 1 |
| T2DM | endothelial | *ZNF561* | -0.503587519 | 2.06E-05 | 0.000105763 | 0.129665123 |
| T2DM | endothelial | *HOPX* | -0.901784225 | 2.14E-06 | 1.96E-05 | 0.013473789 |
| T2DM | endothelial | *LRRC8C* | 0.393655489 | 0.000363233 | 0.001127168 | 1 |
| T2DM | endothelial | *RSL1D1* | -0.549445749 | 0.001624656 | 0.003988369 | 1 |
| T2DM | endothelial | *COL24A1* | -0.140456346 | 0.000483417 | 0.001434282 | 1 |
| T2DM | endothelial | *TBCA* | -0.317932881 | 0.007676524 | 0.014877407 | 1 |
| T2DM | endothelial | *MAP6* | -0.720494439 | 9.13E-06 | 5.53E-05 | 0.057392046 |
| T2DM | endothelial | *PLRG1* | -0.545398502 | 0.001725677 | 0.004198657 | 1 |
| T2DM | endothelial | *NMUR1* | -0.6452079 | 3.77E-07 | 6.87E-06 | 0.002368015 |
| T2DM | endothelial | *P2RY6* | -0.599623445 | 1.12E-07 | 4.46E-06 | 0.000702884 |
| T2DM | endothelial | *ZIK1* | -1.03484608 | 4.86E-08 | 3.92E-06 | 0.000305657 |
| T2DM | endothelial | *RGS19* | -0.695369408 | 3.19E-12 | 6.68E-09 | 2.00E-08 |
| T2DM | endothelial | *TCEA2* | 0.37028774 | 2.14E-05 | 0.000108798 | 0.134474277 |
| T2DM | endothelial | *ANO5* | -0.626829149 | 4.32E-07 | 7.46E-06 | 0.002715157 |
| T2DM | endothelial | *GPHN* | -0.200922025 | 0.029816571 | 0.047650428 | 1 |
| T2DM | endothelial | *VAT1L* | 0.382412931 | 9.03E-06 | 5.49E-05 | 0.056741317 |
| T2DM | endothelial | *CAMTA1* | 0.236315936 | 0.011501005 | 0.021035132 | 1 |
| T2DM | endothelial | *SPATA5L1* | -0.382908624 | 0.005540629 | 0.011244008 | 1 |
| T2DM | endothelial | *SYCE1* | -0.430980737 | 3.98E-05 | 0.000180535 | 0.250401668 |
| T2DM | endothelial | *NXNL1* | -0.521761344 | 0.000385636 | 0.001183256 | 1 |
| T2DM | endothelial | *BCL2* | -0.181805467 | 0.000193228 | 0.000666869 | 1 |
| T2DM | endothelial | *RHNO1* | -0.548364629 | 0.010346727 | 0.019217097 | 1 |
| T2DM | endothelial | *KNDC1* | -0.488760516 | 5.33E-06 | 3.68E-05 | 0.033507973 |
| T2DM | endothelial | *METTL18* | -0.922142851 | 0.000438069 | 0.001315254 | 1 |
| T2DM | endothelial | *PWWP2B* | -0.732877393 | 0.001217001 | 0.003122974 | 1 |
| T2DM | endothelial | *ZNF570* | -0.501252336 | 0.028163459 | 0.045424234 | 1 |
| T2DM | endothelial | *FAM90A1* | -2.454798836 | 0.000728316 | 0.002027867 | 1 |
| T2DM | endothelial | *TRAPPC12* | -0.365532883 | 0.006449436 | 0.01281125 | 1 |
| T2DM | endothelial | *C3AR1* | -0.565202334 | 1.30E-07 | 4.46E-06 | 0.000820134 |
| T2DM | endothelial | *AQP4* | -0.526734569 | 1.16E-07 | 4.46E-06 | 0.000727532 |
| T2DM | endothelial | *TLN2* | -0.177196682 | 0.010614149 | 0.019638362 | 1 |
| T2DM | endothelial | *TVP23B* | -0.562680554 | 0.000663295 | 0.001866668 | 1 |

| | | | | | | |
|---|---|---|---|---|---|---|
| T2DM | endothelial | *SRGAP2C* | -0.500904966 | 0.00045987 | 0.001373492 | 1 |
| T2DM | endothelial | *ATPAF2* | -0.560200905 | 0.00466432 | 0.00968917 | 1 |
| T2DM | endothelial | *CYP4F22* | -0.772556905 | 0.003488853 | 0.007600283 | 1 |
| T2DM | endothelial | *ZNF554* | -0.627652292 | 3.85E-08 | 3.45E-06 | 0.000242202 |
| T2DM | endothelial | *ANKRD20A4* | -1.437198427 | 0.015190701 | 0.026647304 | 1 |
| T2DM | endothelial | *LAMB2* | -0.598531743 | 4.17E-07 | 7.31E-06 | 0.002620809 |
| T2DM | endothelial | *ORMDL3* | -0.262716357 | 0.000143818 | 0.00052265 | 0.904185262 |
| T2DM | endothelial | *SERF1A* | 2.363056026 | 0.00019205 | 0.000663782 | 1 |
| T2DM | endothelial | *LRRC15* | -0.844962934 | 2.52E-05 | 0.000124997 | 0.15849617 |
| T2DM | endothelial | *MOB3A* | -0.573756517 | 8.67E-07 | 1.08E-05 | 0.005453092 |
| T2DM | endothelial | *FRMD3* | 0.276519233 | 0.00026806 | 0.00087321 | 1 |
| T2DM | endothelial | *MTBP* | -0.524216049 | 0.001419948 | 0.003565224 | 1 |
| T2DM | endothelial | *ISG20* | 0.232704426 | 0.031331579 | 0.049868769 | 1 |
| T2DM | endothelial | *MBOAT1* | -0.513831047 | 2.74E-05 | 0.000132643 | 0.172568668 |
| T2DM | endothelial | *C5orf34* | -0.434588742 | 0.014089641 | 0.02497366 | 1 |
| T2DM | endothelial | *C1QTNF4* | -0.176872802 | 1.58E-05 | 8.55E-05 | 0.099273176 |
| T2DM | endothelial | *CSDC2* | -0.56296338 | 1.54E-06 | 1.60E-05 | 0.00969147 |
| T2DM | endothelial | *RCAN2* | -0.484653552 | 1.37E-05 | 7.59E-05 | 0.086133144 |
| T2DM | endothelial | *IL16* | -0.368244656 | 1.40E-06 | 1.50E-05 | 0.008777212 |
| T2DM | endothelial | *GNB2* | -0.44689227 | 0.017548522 | 0.030053339 | 1 |
| T2DM | endothelial | *PRSS27* | 1.82023348 | 0.000757418 | 0.002094057 | 1 |
| T2DM | endothelial | *ZNF24* | -0.441766049 | 0.003761388 | 0.008121292 | 1 |
| T2DM | endothelial | *MANEA* | -0.464535785 | 0.011007091 | 0.02023438 | 1 |
| T2DM | endothelial | *CARNS1* | 0.900642924 | 2.13E-05 | 0.000108622 | 0.134146933 |
| T2DM | endothelial | *CTSW* | -0.162574858 | 3.01E-07 | 6.15E-06 | 0.001895293 |
| T2DM | endothelial | *SNTG2* | 0.173154772 | 3.72E-05 | 0.000170795 | 0.233988828 |
| T2DM | endothelial | *PDE3A* | -0.308187075 | 0.000335185 | 0.001051551 | 1 |
| T2DM | endothelial | *SMPDL3A* | -0.61612619 | 4.47E-06 | 3.25E-05 | 0.028106176 |
| T2DM | endothelial | *EFEMP2* | -0.415519406 | 0.000364098 | 0.001129295 | 1 |
| T2DM | endothelial | *AGAP5* | -25.71654629 | 4.98E-06 | 3.52E-05 | 0.031336099 |
| T2DM | endothelial | *ZMAT3* | -0.606868799 | 2.56E-06 | 2.19E-05 | 0.016089569 |
| T2DM | endothelial | *SLFN11* | 0.251159425 | 0.028548305 | 0.045894985 | 1 |
| T2DM | endothelial | *CCL19* | -2.453062091 | 5.34E-08 | 4.14E-06 | 0.000335514 |
| T2DM | endothelial | *FUT10* | -0.220344927 | 0.01729365 | 0.029714451 | 1 |
| T2DM | endothelial | *EFCAB12* | -0.30838112 | 0.000626658 | 0.001784324 | 1 |
| T2DM | endothelial | *FAM192A* | -0.341124084 | 0.000282783 | 0.000914537 | 1 |
| T2DM | endothelial | *RAB43* | 1.340511801 | 1.19E-07 | 4.46E-06 | 0.00074527 |
| T2DM | endothelial | *CBWD1* | -0.574832865 | 0.000378634 | 0.001165185 | 1 |
| T2DM | endothelial | *HOXC5* | -0.513912098 | 4.36E-05 | 0.000194825 | 0.274313811 |
| T2DM | endothelial | *RAB37* | -0.742762692 | 0.000620708 | 0.001770595 | 1 |
| T2DM | endothelial | *SNX32* | 0.574418187 | 1.42E-05 | 7.81E-05 | 0.089453765 |
| T2DM | endothelial | *CYP7B1* | -0.403047881 | 1.54E-06 | 1.60E-05 | 0.009656331 |
| T2DM | endothelial | *METAP1D* | -0.526316799 | 3.37E-05 | 0.000157042 | 0.212007096 |
| T2DM | endothelial | *NADSYN1* | -0.410919794 | 0.00667097 | 0.013198687 | 1 |
| T2DM | endothelial | *RNASEH2C* | -0.603282139 | 5.52E-05 | 0.000234886 | 0.347161242 |
| T2DM | endothelial | *MYEOV* | -0.21944948 | 1.58E-06 | 1.61E-05 | 0.009905091 |
| T2DM | endothelial | *ANKRD13D* | -0.413580885 | 0.004898672 | 0.010091072 | 1 |
| T2DM | endothelial | *MRGPRF* | -0.590580976 | 2.80E-06 | 2.32E-05 | 0.017597166 |
| T2DM | endothelial | *ADH6* | -0.224798996 | 0.00032593 | 0.001029194 | 1 |
| T2DM | endothelial | *GXYLT2* | -0.421610335 | 9.20E-08 | 4.46E-06 | 0.000578671 |

| | | | | | | |
|---|---|---|---|---|---|---|
| T2DM | endothelial | *HPSE2* | -0.231772407 | 0.000512889 | 0.001502579 | 1 |
| T2DM | endothelial | *DCAKD* | -0.21157271 | 0.003387532 | 0.007414722 | 1 |
| T2DM | endothelial | *BNC2* | -0.28906889 | 0.001112809 | 0.002900592 | 1 |
| T2DM | endothelial | *NOC3L* | -0.398453547 | 0.014139656 | 0.025027031 | 1 |
| T2DM | endothelial | *PARP15* | -0.395903752 | 1.71E-05 | 9.11E-05 | 0.107765365 |
| T2DM | endothelial | *AHSA2* | -0.37128081 | 0.001360354 | 0.003440285 | 1 |
| T2DM | endothelial | *ABLIM3* | 0.139564979 | 2.72E-06 | 2.29E-05 | 0.017103218 |
| T2DM | endothelial | *GLRX* | -0.3809847 | 7.61E-07 | 1.02E-05 | 0.004781617 |
| T2DM | endothelial | *IQCB1* | -0.227189537 | 0.027280735 | 0.044147743 | 1 |
| T2DM | endothelial | *DMRT2* | -0.344368252 | 2.48E-06 | 2.15E-05 | 0.015590613 |
| T2DM | endothelial | *SLC2A14* | -0.37845335 | 0.001824063 | 0.004403948 | 1 |
| T2DM | endothelial | *MZT2A* | -0.329322902 | 0.015425661 | 0.026969168 | 1 |
| T2DM | endothelial | *MAP3K11* | 0.398968488 | 0.000421916 | 0.00127467 | 1 |
| T2DM | endothelial | *C1QA* | -0.550759839 | 3.98E-05 | 0.000180453 | 0.250108145 |
| T2DM | endothelial | *NDNF* | 0.276881117 | 6.46E-06 | 4.23E-05 | 0.040627797 |
| T2DM | endothelial | *OLR1* | -0.846128997 | 5.34E-07 | 8.34E-06 | 0.003358493 |
| T2DM | endothelial | *ARV1* | -0.478138775 | 0.007928405 | 0.015290147 | 1 |
| T2DM | endothelial | *CCDC36* | -0.417876696 | 0.000682789 | 0.001917237 | 1 |
| T2DM | endothelial | *THAP2* | -0.599707265 | 0.00026927 | 0.000875338 | 1 |
| T2DM | endothelial | *PPP1R14B* | -0.887967025 | 0.000433911 | 0.001306761 | 1 |
| T2DM | endothelial | *ZNF417* | -0.89907233 | 5.83E-06 | 3.93E-05 | 0.036662553 |
| T2DM | endothelial | *VEGFB* | -0.606807301 | 3.35E-10 | 1.40E-07 | 2.11E-06 |
| T2DM | endothelial | *TNFRSF10D* | 0.170338613 | 0.000120649 | 0.000450697 | 0.758523128 |
| T2DM | endothelial | *GMPPB* | -0.522208025 | 0.001955592 | 0.004671278 | 1 |
| T2DM | endothelial | *CSPG4* | -0.4403841 | 8.79E-06 | 5.38E-05 | 0.055263233 |
| T2DM | endothelial | *SNX33* | -0.599024581 | 3.36E-06 | 2.63E-05 | 0.021152095 |
| T2DM | endothelial | *NUDT18* | -1.270108239 | 7.71E-07 | 1.03E-05 | 0.004844638 |
| T2DM | endothelial | *SULT1B1* | 0.532874626 | 0.000104677 | 0.000399577 | 0.658103993 |
| T2DM | endothelial | *NUDT4* | -0.326012515 | 0.020178981 | 0.033830734 | 1 |
| T2DM | endothelial | *SCAI* | -0.403864116 | 0.000789281 | 0.002167851 | 1 |
| T2DM | endothelial | *C1orf100* | 0.819784098 | 0.000391361 | 0.001196735 | 1 |
| T2DM | endothelial | *AGFG1* | -0.324372077 | 0.000416436 | 0.00125993 | 1 |
| T2DM | endothelial | *TUBB8* | 0.668925519 | 6.24E-05 | 0.000259153 | 0.392097754 |
| T2DM | endothelial | *GPR160* | 0.178713932 | 1.08E-06 | 1.25E-05 | 0.006818296 |
| T2DM | endothelial | *HOXB2* | -0.628677569 | 2.37E-05 | 0.000118867 | 0.149295511 |
| T2DM | endothelial | *XXYLT1* | -0.458362855 | 0.001016406 | 0.002679306 | 1 |
| T2DM | endothelial | *UBXN2A* | -0.427528861 | 0.000132859 | 0.000491305 | 0.835286696 |
| T2DM | endothelial | *CCS* | -0.484640134 | 0.00010595 | 0.000403914 | 0.66610568 |
| T2DM | endothelial | *CEP19* | -0.306147613 | 0.025443076 | 0.041505091 | 1 |
| T2DM | endothelial | *CTSF* | -0.310148802 | 0.001606783 | 0.00395221 | 1 |
| T2DM | endothelial | *LEMD3* | -0.308617958 | 0.027082304 | 0.043837911 | 1 |
| T2DM | endothelial | *C16orf91* | -0.777772862 | 0.011089411 | 0.020373796 | 1 |
| T2DM | endothelial | *SOCS7* | -0.564080983 | 2.33E-05 | 0.000117361 | 0.146701794 |
| T2DM | endothelial | *TLR6* | -0.513508594 | 0.000102317 | 0.000392956 | 0.643268177 |
| T2DM | endothelial | *FAM53A* | -0.940655154 | 1.45E-07 | 4.46E-06 | 0.000910527 |
| T2DM | endothelial | *CTU2* | -0.684756927 | 4.04E-05 | 0.00018293 | 0.254272541 |
| T2DM | endothelial | *MGA* | -0.344289053 | 0.001231256 | 0.003149396 | 1 |
| T2DM | endothelial | *SNX31* | -0.372997454 | 0.007377267 | 0.014386129 | 1 |
| T2DM | endothelial | *GLIS1* | -0.25028797 | 9.78E-05 | 0.00037882 | 0.61508919 |
| T2DM | endothelial | *LIG4* | -0.389034934 | 0.000172843 | 0.000608095 | 1 |

| | | | | | | |
|---|---|---|---|---|---|---|
| T2DM | endothelial | *LINGO2* | -0.627641591 | 0.00024226 | 0.000806722 | 1 |
| T2DM | endothelial | *ANKRD36C* | -0.272473016 | 0.02827208 | 0.045552683 | 1 |
| T2DM | endothelial | *TMEM81* | -1.624036155 | 0.002170436 | 0.005099227 | 1 |
| T2DM | endothelial | *ZNF497* | -1.42402361 | 1.68E-05 | 8.99E-05 | 0.105815653 |
| T2DM | endothelial | *KLF14* | -2.663784845 | 4.00E-11 | 3.59E-08 | 2.51E-07 |
| T2DM | endothelial | *ZNF266* | -0.318656522 | 0.000387192 | 0.001186873 | 1 |
| T2DM | endothelial | *BRSK2* | 0.561078204 | 2.14E-06 | 1.96E-05 | 0.013480809 |
| T2DM | endothelial | *SH3PXD2B* | 0.226526616 | 0.010327484 | 0.019187024 | 1 |
| T2DM | endothelial | *LARP7* | -0.37550718 | 0.00899021 | 0.017065655 | 1 |
| T2DM | endothelial | *FGFBP3* | -0.920420937 | 5.07E-07 | 8.20E-06 | 0.003189456 |
| T2DM | endothelial | *C4orf32* | 0.189324375 | 1.00E-05 | 5.94E-05 | 0.063138548 |
| T2DM | endothelial | *HRAS* | -0.540361156 | 0.015359861 | 0.02687655 | 1 |
| T2DM | endothelial | *C4orf26* | -1.350575069 | 0.000960051 | 0.002553232 | 1 |
| T2DM | endothelial | *BTC* | 0.527609118 | 0.000393701 | 0.001201554 | 1 |
| T2DM | endothelial | *NLRP6* | -2.877053852 | 2.38E-10 | 1.15E-07 | 1.49E-06 |
| T2DM | endothelial | *NDUFA11* | -0.651407265 | 0.000229234 | 0.000769048 | 1 |
| T2DM | endothelial | *RSRC1* | -0.254722635 | 0.012193466 | 0.022092312 | 1 |
| T2DM | endothelial | *PTDSS2* | -0.391561301 | 0.0145751 | 0.025689277 | 1 |
| T2DM | endothelial | *FUT1* | 0.343805457 | 3.51E-07 | 6.64E-06 | 0.002205512 |
| T2DM | endothelial | *DHX36* | -0.322471862 | 0.011519844 | 0.021059977 | 1 |
| T2DM | endothelial | *SLC22A1* | -0.681559291 | 2.12E-06 | 1.95E-05 | 0.013308155 |
| T2DM | endothelial | *ATR* | -0.270661728 | 0.019922733 | 0.033465646 | 1 |
| T2DM | endothelial | *PACS1* | -0.308187842 | 0.005673734 | 0.01147708 | 1 |
| T2DM | endothelial | *CADM2* | 0.103906446 | 2.70E-06 | 2.27E-05 | 0.016970161 |
| T2DM | endothelial | *ARHGAP1* | -0.504780064 | 1.07E-05 | 6.22E-05 | 0.067324313 |
| T2DM | endothelial | *CHST1* | 0.453884245 | 1.75E-06 | 1.72E-05 | 0.011001058 |
| T2DM | endothelial | *TP53I11* | 0.343400713 | 6.25E-06 | 4.14E-05 | 0.039298973 |
| T2DM | endothelial | *PHYHD1* | -0.468382232 | 0.000429721 | 0.00129638 | 1 |
| T2DM | endothelial | *ZNF519* | -0.490382122 | 0.000252145 | 0.000832065 | 1 |
| T2DM | endothelial | *CHRNA7* | 0.560614292 | 1.19E-06 | 1.34E-05 | 0.007511041 |
| T2DM | endothelial | *NRIP3* | -0.580638362 | 6.80E-05 | 0.000278439 | 0.427404523 |
| T2DM | endothelial | *SCUBE2* | 0.286169057 | 0.000360092 | 0.001118527 | 1 |
| T2DM | endothelial | *ZNF25* | -0.358514955 | 0.000404905 | 0.001229183 | 1 |
| T2DM | endothelial | *CLTB* | -0.392370304 | 0.002689762 | 0.006089497 | 1 |
| T2DM | endothelial | *PCSK1* | -0.580118652 | 8.78E-07 | 1.08E-05 | 0.005521853 |
| T2DM | endothelial | *LPL* | -0.252921808 | 3.66E-06 | 2.79E-05 | 0.023041324 |
| T2DM | endothelial | *TBC1D10C* | -0.417612794 | 4.99E-05 | 0.000215534 | 0.313601505 |
| T2DM | endothelial | *MCTP1* | 0.145061556 | 1.70E-05 | 9.06E-05 | 0.106755668 |
| T2DM | endothelial | *LIPT2* | -1.264300756 | 0.000190612 | 0.000659898 | 1 |
| T2DM | endothelial | *UCP3* | -0.757816213 | 0.002404085 | 0.00556498 | 1 |
| T2DM | endothelial | *UCP2* | -0.572100762 | 1.17E-07 | 4.46E-06 | 0.000735309 |
| T2DM | endothelial | *MRPL48* | -0.319105853 | 0.003762442 | 0.008121292 | 1 |
| T2DM | endothelial | *RPS6KB2* | -0.637298718 | 0.001684474 | 0.004114331 | 1 |
| T2DM | endothelial | *TEX26* | -0.570037965 | 0.000101689 | 0.000390782 | 0.639318766 |
| T2DM | endothelial | *GPR156* | -0.701963897 | 6.98E-06 | 4.48E-05 | 0.043880106 |
| T2DM | endothelial | *B3GNTL1* | -0.559628568 | 1.63E-05 | 8.74E-05 | 0.102262564 |
| T2DM | endothelial | *CTDNEP1* | -0.693670353 | 4.18E-05 | 0.00018836 | 0.262574056 |
| T2DM | endothelial | *ETV4* | -0.364532169 | 0.004041216 | 0.008630137 | 1 |
| T2DM | endothelial | *A2M* | 0.20311551 | 5.09E-05 | 0.000219375 | 0.320068628 |
| T2DM | endothelial | *ARL4D* | -0.597916223 | 7.34E-06 | 4.65E-05 | 0.046142477 |

| | | | | | | |
|---|---|---|---|---|---|---|
| T2DM | endothelial | *LRRN1* | -0.755532329 | 2.06E-06 | 1.91E-05 | 0.012937397 |
| T2DM | endothelial | *SSNA1* | -0.679395805 | 0.00338614 | 0.007414722 | 1 |
| T2DM | endothelial | *CHMP6* | -0.652567834 | 0.001067438 | 0.002797616 | 1 |
| T2DM | endothelial | *CCDC57* | -0.382103007 | 0.001994925 | 0.004741811 | 1 |
| T2DM | endothelial | *CIDEA* | -0.103830624 | 0.003632097 | 0.007874135 | 1 |
| T2DM | endothelial | *SLC35G1* | -0.42004885 | 9.09E-06 | 5.52E-05 | 0.057120607 |
| T2DM | endothelial | *B3GNT4* | -2.05364941 | 1.24E-07 | 4.46E-06 | 0.000780104 |
| T2DM | endothelial | *CDC26* | -0.526412263 | 6.02E-06 | 4.02E-05 | 0.037878783 |
| T2DM | endothelial | *CRLF3* | -0.274124772 | 0.023206339 | 0.038273414 | 1 |
| T2DM | endothelial | *SPRYD4* | -0.353417654 | 1.14E-05 | 6.52E-05 | 0.071744673 |
| T2DM | endothelial | *PHLDB3* | -0.415941677 | 0.010514494 | 0.019470426 | 1 |
| T2DM | endothelial | *PRR15* | 0.672999978 | 1.93E-05 | 0.000100162 | 0.121295737 |
| T2DM | endothelial | *RNF152* | 0.329476878 | 0.002857124 | 0.006406113 | 1 |
| T2DM | endothelial | *LRRC37A* | -0.470279237 | 7.89E-05 | 0.000314134 | 0.496331367 |
| T2DM | endothelial | *ACSF3* | -0.474856677 | 0.000298172 | 0.000954465 | 1 |
| T2DM | endothelial | *BOK* | -0.496295771 | 7.30E-05 | 0.000294411 | 0.458987101 |
| T2DM | endothelial | *ZNF843* | -1.593018491 | 2.36E-06 | 2.08E-05 | 0.014831709 |
| T2DM | endothelial | *C8orf59* | -0.336552532 | 0.020651719 | 0.03454959 | 1 |
| T2DM | endothelial | *RUFY1* | -0.332390033 | 0.002233568 | 0.005228012 | 1 |
| T2DM | endothelial | *BASP1* | -0.379353719 | 5.22E-05 | 0.000224488 | 0.32820215 |
| T2DM | endothelial | *LRRC37A3* | -0.502012781 | 0.00455233 | 0.009502158 | 1 |
| T2DM | endothelial | *IRX5* | -0.44123831 | 6.10E-06 | 4.06E-05 | 0.038336088 |
| T2DM | endothelial | *PNMA1* | -0.89254353 | 4.80E-07 | 7.93E-06 | 0.003019891 |
| T2DM | endothelial | *GCNT4* | 1.095686448 | 4.17E-06 | 3.09E-05 | 0.026218119 |
| T2DM | endothelial | *MUC20* | -0.897981797 | 3.46E-06 | 2.68E-05 | 0.021779218 |
| T2DM | endothelial | *THAP4* | -0.454387898 | 0.003999626 | 0.008561678 | 1 |
| T2DM | endothelial | *DPP7* | -0.536996432 | 0.000118313 | 0.000443021 | 0.743831765 |
| T2DM | endothelial | *SEC24C* | -0.595365645 | 0.000332712 | 0.001046927 | 1 |
| T2DM | endothelial | *TMEM80* | -0.299534811 | 0.004152342 | 0.008819519 | 1 |
| T2DM | endothelial | *POLE* | -0.550868611 | 0.000985718 | 0.002612651 | 1 |
| T2DM | endothelial | *DSCAML1* | -0.430964985 | 9.70E-06 | 5.79E-05 | 0.061010476 |
| T2DM | endothelial | *EPS8L2* | -0.524025195 | 1.35E-08 | 2.24E-06 | 8.49E-05 |
| T2DM | endothelial | *TALDO1* | -0.340237004 | 0.004802941 | 0.009923132 | 1 |
| T2DM | endothelial | *RIMKLA* | 0.337583304 | 1.45E-05 | 7.93E-05 | 0.09101514 |
| T2DM | endothelial | *B3GNT8* | -2.334235729 | 2.12E-05 | 0.000108163 | 0.133257394 |
| T2DM | endothelial | *AP006621.1* | -0.288877241 | 0.012109874 | 0.021985309 | 1 |
| T2DM | endothelial | *MAN1B1* | -0.619390223 | 0.000303 | 0.000966004 | 1 |
| T2DM | endothelial | *FBXO39* | 0.226444662 | 0.000118084 | 0.000442429 | 0.742395946 |
| T2DM | endothelial | *TOP3A* | -0.448978162 | 0.002758959 | 0.006217053 | 1 |
| T2DM | endothelial | *C8orf31* | -0.595548734 | 1.31E-07 | 4.46E-06 | 0.000823622 |
| T2DM | endothelial | *AC024940.1* | -0.488737891 | 0.028003565 | 0.045224354 | 1 |
| T2DM | endothelial | *PPFIA3* | 0.299967119 | 3.04E-06 | 2.45E-05 | 0.019101523 |
| T2DM | endothelial | *MAGEF1* | -0.664298164 | 1.91E-07 | 4.69E-06 | 0.001199808 |
| T2DM | endothelial | *SAMD9L* | 0.210612095 | 0.020337162 | 0.034068676 | 1 |
| T2DM | endothelial | *PAWR* | 0.278010653 | 0.017723622 | 0.030304164 | 1 |
| T2DM | endothelial | *NIM1K* | -0.669391678 | 0.002660884 | 0.006041524 | 1 |
| T2DM | endothelial | *ACOT4* | -0.426876681 | 1.42E-05 | 7.81E-05 | 0.089353648 |
| T2DM | endothelial | *ARIH2* | -0.283577578 | 0.00451065 | 0.009437089 | 1 |
| T2DM | endothelial | *RBM44* | 0.298327155 | 1.73E-06 | 1.70E-05 | 0.010851095 |
| T2DM | endothelial | *IRX3* | -0.333577959 | 0.000413822 | 0.001253607 | 1 |

| | | | | | | |
|---|---|---|---|---|---|---|
| T2DM | endothelial | *TBL1XR1* | -0.300902856 | 0.002787409 | 0.006274415 | 1 |
| T2DM | endothelial | *SAMD12* | 0.174419437 | 5.61E-06 | 3.82E-05 | 0.03527682 |
| T2DM | endothelial | *C18orf32* | -0.521345171 | 0.011915693 | 0.021689045 | 1 |
| T2DM | endothelial | *C12orf54* | -1.969755736 | 0.000205111 | 0.000701976 | 1 |
| T2DM | endothelial | *GBA* | -0.793402057 | 6.69E-05 | 0.000274789 | 0.420701843 |
| T2DM | endothelial | *IL17RA* | -0.254857903 | 0.019920883 | 0.033465646 | 1 |
| T2DM | endothelial | *CD163L1* | -0.289152365 | 0.000234858 | 0.000784982 | 1 |
| T2DM | endothelial | *FAM87B* | -0.948770929 | 7.66E-08 | 4.46E-06 | 0.00048132 |
| T2DM | endothelial | *ZCCHC3* | -0.817020924 | 0.000535552 | 0.001557361 | 1 |
| T2DM | endothelial | *ZNF619* | 0.595448995 | 0.011103409 | 0.020393553 | 1 |
| T2DM | endothelial | *ZBTB41* | -0.472196483 | 0.000636064 | 0.001802947 | 1 |
| T2DM | endothelial | *CENPBD1* | -0.697996663 | 7.22E-06 | 4.60E-05 | 0.045400313 |
| T2DM | endothelial | *BET1L* | -0.441293435 | 0.005019001 | 0.010318659 | 1 |
| T2DM | endothelial | *RIC8A* | -0.821479476 | 0.000316063 | 0.001001557 | 1 |
| T2DM | endothelial | *IMP3* | -0.555689432 | 0.004350513 | 0.009153841 | 1 |
| T2DM | endothelial | *C2orf73* | -1.006011764 | 7.92E-07 | 1.03E-05 | 0.004982278 |
| T2DM | endothelial | *FAM26E* | 0.431745957 | 1.10E-05 | 6.36E-05 | 0.068953897 |
| T2DM | endothelial | *ALS2CL* | 0.253912157 | 7.84E-07 | 1.03E-05 | 0.004931143 |
| T2DM | endothelial | *MLF1* | -0.558973671 | 0.000132927 | 0.000491305 | 0.835709639 |
| T2DM | endothelial | *GRAMD1C* | 0.163514236 | 1.06E-06 | 1.22E-05 | 0.006636383 |
| T2DM | endothelial | *STAP2* | -0.879206444 | 8.70E-05 | 0.000341294 | 0.546753135 |
| T2DM | endothelial | *TSSK6* | -1.088396575 | 7.58E-05 | 0.000304063 | 0.476771237 |
| T2DM | endothelial | *ZNF114* | 0.457345931 | 1.04E-05 | 6.06E-05 | 0.065174015 |
| T2DM | endothelial | *ZNF518B* | -0.704332113 | 0.000519596 | 0.001517278 | 1 |
| T2DM | endothelial | *ZNF454* | 0.307479498 | 4.92E-06 | 3.49E-05 | 0.030905356 |
| T2DM | endothelial | *SH2B1* | -0.374640949 | 8.39E-06 | 5.20E-05 | 0.052730483 |
| T2DM | endothelial | *ZC3H12D* | -0.629520874 | 2.12E-07 | 4.94E-06 | 0.00133029 |
| T2DM | endothelial | *KDELC2* | -0.585435695 | 5.38E-05 | 0.000229966 | 0.33851062 |
| T2DM | endothelial | *PLEC* | 0.301312175 | 0.011014248 | 0.020241618 | 1 |
| T2DM | endothelial | *SH2D4B* | 1.181380236 | 0.000492137 | 0.001452614 | 1 |
| T2DM | endothelial | *PRSS36* | -0.716077789 | 1.39E-07 | 4.46E-06 | 0.000872355 |
| T2DM | endothelial | *TMPRSS9* | -0.470671543 | 0.000341546 | 0.00107044 | 1 |
| T2DM | endothelial | *AQP11* | -0.405167879 | 0.028621285 | 0.045985694 | 1 |
| T2DM | endothelial | *TMEM11* | -0.653069512 | 0.002835008 | 0.006361061 | 1 |
| T2DM | endothelial | *FAM220A* | -0.327181457 | 0.02149059 | 0.035791083 | 1 |
| T2DM | endothelial | *AC006111.1* | -2.368203321 | 0.004387374 | 0.009209824 | 1 |
| T2DM | endothelial | *LINC00843* | -0.921347777 | 1.15E-05 | 6.55E-05 | 0.072168583 |
| T2DM | endothelial | *COX14* | -0.425319786 | 3.98E-06 | 2.98E-05 | 0.025034427 |
| T2DM | endothelial | *P4HTM* | -0.519350817 | 0.000224533 | 0.000757725 | 1 |
| T2DM | endothelial | *CTXN1* | -1.442538825 | 8.54E-05 | 0.000336333 | 0.536787369 |
| T2DM | endothelial | *CA8* | 0.149043254 | 3.42E-06 | 2.66E-05 | 0.021513276 |
| T2DM | endothelial | *ERBB4* | -0.356711155 | 4.49E-05 | 0.000199031 | 0.282424872 |
| T2DM | endothelial | *GPR35* | -0.165748205 | 0.024889299 | 0.040717935 | 1 |
| T2DM | endothelial | *PARP10* | -0.346137341 | 0.015300002 | 0.02678672 | 1 |
| T2DM | endothelial | *KCTD12* | 0.191711144 | 0.000119077 | 0.000445616 | 0.74863563 |
| T2DM | endothelial | *STX19* | 0.560412405 | 0.00035897 | 0.001117249 | 1 |
| T2DM | endothelial | *FAM219B* | -0.518367665 | 1.60E-05 | 8.64E-05 | 0.10049512 |
| T2DM | endothelial | *MPI* | -0.497424614 | 0.007368526 | 0.014376175 | 1 |
| T2DM | endothelial | *TRIM73* | -0.906925667 | 0.000501365 | 0.001474312 | 1 |
| T2DM | endothelial | *HYI* | -0.161140197 | 0.030831985 | 0.049135789 | 1 |

| | | | | | | |
|---|---|---|---|---|---|---|
| T2DM | endothelial | *C17orf62* | -0.494789016 | 0.005425591 | 0.01104388 | 1 |
| T2DM | endothelial | *C14orf39* | -0.233232842 | 2.43E-06 | 2.12E-05 | 0.015255318 |
| T2DM | endothelial | *TRIML2* | -4.979113525 | 5.60E-08 | 4.24E-06 | 0.000352126 |
| T2DM | endothelial | *FARSA* | -0.956538227 | 4.34E-07 | 7.47E-06 | 0.002727443 |
| T2DM | endothelial | *SPTY2D1* | -0.563382266 | 0.00060518 | 0.00173417 | 1 |
| T2DM | endothelial | *TCAIM* | -0.252347195 | 0.025415932 | 0.041482338 | 1 |
| T2DM | endothelial | *FUCA1* | -0.407003398 | 0.002158422 | 0.005072897 | 1 |
| T2DM | endothelial | *ZNF664* | -0.699730339 | 4.08E-06 | 3.03E-05 | 0.025631975 |
| T2DM | endothelial | *CDH4* | -0.493790549 | 0.000328504 | 0.001035758 | 1 |
| T2DM | endothelial | *HLA-DQB1* | -0.235998641 | 3.43E-05 | 0.000159409 | 0.215839964 |
| T2DM | endothelial | *ARID3B* | -0.587655876 | 0.000343148 | 0.001074636 | 1 |
| T2DM | endothelial | *LINC00174* | -0.21319723 | 0.002065369 | 0.004888921 | 1 |
| T2DM | endothelial | *GEMIN4* | -0.405038807 | 1.10E-05 | 6.38E-05 | 0.069322856 |
| T2DM | endothelial | *FJX1* | -0.551523484 | 0.001525994 | 0.003783094 | 1 |
| T2DM | endothelial | *CIITA* | 0.332523497 | 0.00157941 | 0.00389402 | 1 |
| T2DM | endothelial | *PLD6* | -0.286816399 | 0.022878408 | 0.037811922 | 1 |
| T2DM | endothelial | *MAF1* | -0.835008934 | 6.14E-05 | 0.00025589 | 0.38588201 |
| T2DM | endothelial | *FCER1A* | -0.595809549 | 2.57E-05 | 0.000126696 | 0.16153764 |
| T2DM | endothelial | *RPRML* | 1.145992898 | 1.58E-07 | 4.49E-06 | 0.000996152 |
| T2DM | endothelial | *PCED1B* | -0.293136257 | 0.004758376 | 0.009860221 | 1 |
| T2DM | endothelial | *PIPOX* | -0.769978478 | 1.03E-07 | 4.46E-06 | 0.000646357 |
| T2DM | endothelial | *CDH5* | 0.112974607 | 5.69E-07 | 8.58E-06 | 0.003578203 |
| T2DM | endothelial | *GIPC3* | 0.370561708 | 7.74E-05 | 0.000309204 | 0.486377797 |
| T2DM | endothelial | *CITED4* | -0.864442678 | 1.01E-05 | 5.96E-05 | 0.06354306 |
| T2DM | endothelial | *TIGD5* | -0.576656355 | 8.06E-06 | 5.03E-05 | 0.050653767 |
| T2DM | endothelial | *ZNF154* | 0.403966868 | 0.004875239 | 0.010049387 | 1 |
| T2DM | endothelial | *R3HDM2* | -0.298654034 | 0.006130837 | 0.012275341 | 1 |
| T2DM | endothelial | *B3GNT3* | 1.662959829 | 9.31E-09 | 1.77E-06 | 5.85E-05 |
| T2DM | endothelial | *SEPT1* | -0.652433748 | 1.73E-07 | 4.57E-06 | 0.001089791 |
| T2DM | endothelial | *LYNX1* | -0.36010881 | 0.018365325 | 0.03127378 | 1 |
| T2DM | endothelial | *TDRP* | 0.261191854 | 3.95E-06 | 2.96E-05 | 0.024836825 |
| T2DM | endothelial | *FGD6* | -0.342810788 | 0.022404728 | 0.037126654 | 1 |
| T2DM | endothelial | *HCLS1* | -0.540210732 | 0.000131316 | 0.000486782 | 0.825581428 |
| T2DM | endothelial | *ZNF609* | -0.299115282 | 0.00318298 | 0.007033619 | 1 |
| T2DM | endothelial | *DEFB124* | -1.316302599 | 4.63E-07 | 7.72E-06 | 0.002912204 |
| T2DM | endothelial | *CYP8B1* | -0.499279275 | 6.15E-06 | 4.08E-05 | 0.038641792 |
| T2DM | endothelial | *RNF182* | -0.483875209 | 1.05E-08 | 1.89E-06 | 6.63E-05 |
| T2DM | endothelial | *MB21D2* | -0.431230773 | 0.002154526 | 0.005067529 | 1 |
| T2DM | endothelial | *SSTR2* | -0.866555563 | 1.29E-07 | 4.46E-06 | 0.000813433 |
| T2DM | endothelial | *ZNF594* | -0.612545845 | 1.62E-05 | 8.74E-05 | 0.102135658 |
| T2DM | endothelial | *SHISA2* | 2.600382722 | 5.74E-07 | 8.63E-06 | 0.003608021 |
| T2DM | endothelial | *OR51E1* | 0.25891653 | 6.25E-05 | 0.000259423 | 0.393025169 |
| T2DM | endothelial | *CAPS2* | -0.424843165 | 0.000285691 | 0.000922044 | 1 |
| T2DM | endothelial | *ZNF792* | 0.288499524 | 4.20E-05 | 0.000189206 | 0.263941778 |
| T2DM | endothelial | *D2HGDH* | -0.406823143 | 0.000315355 | 0.001000322 | 1 |
| T2DM | endothelial | *CMTR2* | -0.562734822 | 0.000277533 | 0.000899407 | 1 |
| T2DM | endothelial | *MRPL14* | -0.55801022 | 0.002043889 | 0.004845373 | 1 |
| T2DM | endothelial | *BBS12* | -0.807695117 | 0.001467071 | 0.003661563 | 1 |
| T2DM | endothelial | *LSMEM1* | -0.411747094 | 1.20E-05 | 6.78E-05 | 0.075152558 |
| T2DM | endothelial | *AEN* | -0.939502096 | 0.005961618 | 0.011986151 | 1 |

| | | | | | | |
|---|---|---|---|---|---|---|
| T2DM | endothelial | *FKRP* | -0.208046286 | 0.017593896 | 0.030103208 | 1 |
| T2DM | endothelial | *ANKRD34A* | -0.938387832 | 6.63E-08 | 4.46E-06 | 0.000416569 |
| T2DM | endothelial | *SLC26A11* | -0.412036381 | 0.021702922 | 0.036116006 | 1 |
| T2DM | endothelial | *HIGD1A* | -0.539851087 | 0.000359994 | 0.001118527 | 1 |
| T2DM | endothelial | *ADIPOQ* | -0.156894599 | 0.000633308 | 0.001798378 | 1 |
| T2DM | endothelial | *HIST3H2A* | -0.497569573 | 0.002830569 | 0.006355638 | 1 |
| T2DM | endothelial | *TMEM132C* | -0.272794685 | 3.14E-05 | 0.00014865 | 0.197704951 |
| T2DM | endothelial | *SLC25A41* | -1.48644223 | 9.16E-08 | 4.46E-06 | 0.000575626 |
| T2DM | endothelial | *TMEM132E* | 0.906974219 | 1.24E-05 | 6.96E-05 | 0.077649407 |
| T2DM | endothelial | *CCL13* | -0.586292061 | 1.85E-06 | 1.78E-05 | 0.011612143 |
| T2DM | endothelial | *ACBD4* | -0.625234711 | 2.03E-05 | 0.000104461 | 0.127755993 |
| T2DM | endothelial | *SGSH* | -0.835007232 | 9.98E-05 | 0.000384962 | 0.627488193 |
| T2DM | endothelial | *MRPS23* | -0.661024428 | 0.001516284 | 0.003763905 | 1 |
| T2DM | endothelial | *GPR135* | -0.332711789 | 2.37E-05 | 0.000118867 | 0.14929752 |
| T2DM | endothelial | *ZFP41* | -0.301819592 | 0.005444179 | 0.011066134 | 1 |
| T2DM | endothelial | *ATG9B* | 0.177164691 | 6.00E-07 | 8.83E-06 | 0.003769683 |
| T2DM | endothelial | *AMIGO1* | -0.243345382 | 0.005998432 | 0.012048608 | 1 |
| T2DM | endothelial | *ODF3L2* | 0.483148923 | 0.026001439 | 0.042273351 | 1 |
| T2DM | endothelial | *CLDN7* | -0.447004861 | 9.61E-06 | 5.75E-05 | 0.060416458 |
| T2DM | endothelial | *SNRPE* | -0.760338004 | 0.000266359 | 0.000868569 | 1 |
| T2DM | endothelial | *IDH2* | -0.599445441 | 5.53E-06 | 3.79E-05 | 0.034751839 |
| T2DM | endothelial | *PTCHD3* | 0.205753542 | 0.00039179 | 0.001196883 | 1 |
| T2DM | endothelial | *FAM181B* | 0.946782787 | 0.000210956 | 0.00072002 | 1 |
| T2DM | endothelial | *NOP10* | -0.485179209 | 0.006337111 | 0.012616028 | 1 |
| T2DM | endothelial | *ZNF708* | -0.399932055 | 0.000828991 | 0.002262095 | 1 |
| T2DM | endothelial | *IST1* | -0.386675326 | 0.004499912 | 0.009420895 | 1 |
| T2DM | endothelial | *ERCC6L2* | -0.266795923 | 0.019817459 | 0.033358062 | 1 |
| T2DM | endothelial | *MRPL41* | -0.590881844 | 4.40E-05 | 0.000195616 | 0.276601382 |
| T2DM | endothelial | *HHIPL1* | -0.982705372 | 8.28E-07 | 1.06E-05 | 0.005207358 |
| T2DM | endothelial | *BACE2* | 0.215357326 | 0.000400523 | 0.001218822 | 1 |
| T2DM | endothelial | *SYNM* | -0.68183005 | 2.24E-06 | 2.01E-05 | 0.014099011 |
| T2DM | endothelial | *IZUMO1* | 0.314029597 | 9.81E-07 | 1.17E-05 | 0.006168202 |
| T2DM | endothelial | *FBXL6* | -0.82698958 | 0.000846143 | 0.002301906 | 1 |
| T2DM | endothelial | *CACNB4* | -0.266488334 | 2.86E-05 | 0.000137374 | 0.179822026 |
| T2DM | endothelial | *TRAPPC6B* | -0.455967288 | 0.000281522 | 0.000911395 | 1 |
| T2DM | endothelial | *NPLOC4* | -0.323521093 | 0.001652869 | 0.004044992 | 1 |
| T2DM | endothelial | *CAPN12* | 0.409069563 | 1.10E-06 | 1.26E-05 | 0.006921798 |
| T2DM | endothelial | *FES* | 0.390966425 | 0.003487476 | 0.007599918 | 1 |
| T2DM | endothelial | *GLRX5* | -0.644694544 | 0.001326136 | 0.003360507 | 1 |
| T2DM | endothelial | *CAV3* | -0.224301756 | 0.001298478 | 0.003302856 | 1 |
| T2DM | endothelial | *MFSD5* | -0.558388209 | 0.002549852 | 0.005829426 | 1 |
| T2DM | endothelial | *ADI1* | -0.594668916 | 2.82E-06 | 2.33E-05 | 0.017727448 |
| T2DM | endothelial | *SPNS3* | 0.398533448 | 2.52E-06 | 2.17E-05 | 0.015853261 |
| T2DM | endothelial | *CLEC4G* | -0.732441304 | 1.80E-07 | 4.62E-06 | 0.001128753 |
| T2DM | endothelial | *SATB1* | -0.622719281 | 0.000283647 | 0.000916859 | 1 |
| T2DM | endothelial | *ACTL10* | -0.788670733 | 9.31E-06 | 5.62E-05 | 0.058520153 |
| T2DM | endothelial | *SKA2* | -0.56889129 | 0.000137185 | 0.000503491 | 0.862479662 |
| T2DM | endothelial | *KCNB2* | -0.240315871 | 8.68E-07 | 1.08E-05 | 0.005454734 |
| T2DM | endothelial | *BRICD5* | -0.313372561 | 0.010825444 | 0.019958817 | 1 |
| T2DM | endothelial | *TSKU* | -0.536773268 | 1.14E-06 | 1.30E-05 | 0.007178884 |

| | | | | | | |
|---|---|---|---|---|---|---|
| T2DM | endothelial | *GRID1* | -0.590721488 | 1.33E-08 | 2.24E-06 | 8.35E-05 |
| T2DM | endothelial | *PLCXD3* | 0.125165306 | 9.73E-07 | 1.17E-05 | 0.00611632 |
| T2DM | endothelial | *GPIHBP1* | 0.121713713 | 1.64E-05 | 8.78E-05 | 0.102900056 |
| T2DM | endothelial | *SLC25A18* | -0.359870683 | 1.00E-05 | 5.94E-05 | 0.063141981 |
| T2DM | endothelial | *CEP63* | -0.262364463 | 0.000880564 | 0.00238112 | 1 |
| T2DM | endothelial | *HMGN4* | -0.650662118 | 1.93E-05 | 0.000100047 | 0.121056653 |
| T2DM | endothelial | *ZNF320* | 0.185601834 | 0.001221152 | 0.003129794 | 1 |
| T2DM | endothelial | *WBP2NL* | -0.574183613 | 0.000475109 | 0.001413634 | 1 |
| T2DM | endothelial | *BEGAIN* | -1.813525434 | 2.14E-07 | 4.97E-06 | 0.001346834 |
| T2DM | endothelial | *ARHGEF37* | 0.388053166 | 0.001639643 | 0.004017316 | 1 |
| T2DM | endothelial | *FAM43B* | -0.990417285 | 3.68E-06 | 2.80E-05 | 0.023119933 |
| T2DM | endothelial | *PTGDR2* | -4.797757297 | 0.001136128 | 0.002954027 | 1 |
| T2DM | endothelial | *GPR19* | 1.382354538 | 0.005398607 | 0.011001958 | 1 |
| T2DM | endothelial | *C2CD4C* | 0.220214068 | 3.10E-07 | 6.22E-06 | 0.001945986 |
| T2DM | endothelial | *CTNNA3* | -0.401109504 | 4.86E-05 | 0.000211649 | 0.305410159 |
| T2DM | endothelial | *CCBE1* | -0.235495181 | 1.07E-05 | 6.22E-05 | 0.067261006 |
| T2DM | endothelial | *ZNF623* | -0.626026329 | 6.53E-05 | 0.000268847 | 0.41026095 |
| T2DM | endothelial | *CCDC125* | -0.47411763 | 4.85E-05 | 0.000211484 | 0.304959388 |
| T2DM | endothelial | *FHL3* | -1.03562927 | 2.71E-05 | 0.000131368 | 0.17051621 |
| T2DM | endothelial | *LRIT3* | -0.810445645 | 0.003368046 | 0.007381952 | 1 |
| T2DM | endothelial | *GRIN2A* | -0.472860286 | 0.00030299 | 0.000966004 | 1 |
| T2DM | endothelial | *URAD* | -1.408689476 | 3.16E-08 | 3.24E-06 | 0.000198703 |
| T2DM | endothelial | *GPR132* | -0.702652826 | 6.64E-07 | 9.37E-06 | 0.004177661 |
| T2DM | endothelial | *ACSM5* | -0.264558761 | 0.000290281 | 0.00093398 | 1 |
| T2DM | endothelial | *AC131971.1* | -1.472054331 | 2.88E-05 | 0.000137985 | 0.180897794 |
| T2DM | endothelial | *PCBP3* | -0.107568058 | 2.01E-05 | 0.000103663 | 0.126469453 |
| T2DM | endothelial | *TANGO2* | -0.486304438 | 0.000103956 | 0.000397716 | 0.653569032 |
| T2DM | endothelial | *SFXN4* | -0.633630781 | 8.52E-05 | 0.000336001 | 0.535921764 |
| T2DM | endothelial | *FAM167B* | 0.517322561 | 1.95E-06 | 1.83E-05 | 0.012250021 |
| T2DM | endothelial | *MRPL54* | -0.364354536 | 0.000645341 | 0.001825128 | 1 |
| T2DM | endothelial | *DGCR6* | -0.517563493 | 0.001147923 | 0.002977306 | 1 |
| T2DM | endothelial | *RP1L1* | 0.537842261 | 2.78E-09 | 7.82E-07 | 1.75E-05 |
| T2DM | endothelial | *FAM19A1* | -0.49082124 | 7.85E-07 | 1.03E-05 | 0.004935344 |
| T2DM | endothelial | *TRMT12* | -0.683842177 | 0.001049133 | 0.002754028 | 1 |
| T2DM | endothelial | *FIGLA* | 0.887266596 | 1.38E-05 | 7.63E-05 | 0.086864958 |
| T2DM | endothelial | *KREMEN1* | -0.143269822 | 0.00922148 | 0.017425742 | 1 |
| T2DM | endothelial | *AIFM3* | -0.601128293 | 1.92E-05 | 0.000100043 | 0.120770396 |
| T2DM | endothelial | *KCTD16* | -0.452853058 | 0.001900856 | 0.004556631 | 1 |
| T2DM | endothelial | *B3GALT5* | -0.756064119 | 0.000155097 | 0.000555572 | 0.975096636 |
| T2DM | endothelial | *ZNF703* | -0.79120229 | 4.96E-11 | 3.69E-08 | 3.12E-07 |
| T2DM | endothelial | *KCTD8* | -0.264252015 | 2.27E-05 | 0.000114753 | 0.142752649 |
| T2DM | endothelial | *C9orf66* | -0.705851847 | 2.12E-07 | 4.94E-06 | 0.001334358 |
| T2DM | endothelial | *TUBA8* | 0.398084865 | 0.018045826 | 0.030779736 | 1 |
| T2DM | endothelial | *NPIPA5* | -0.132144078 | 0.007242772 | 0.014145792 | 1 |
| T2DM | endothelial | *FAM162B* | -0.317383011 | 0.007823653 | 0.015115952 | 1 |
| T2DM | endothelial | *BTBD9* | -0.248812156 | 0.016143736 | 0.028029734 | 1 |
| T2DM | endothelial | *NUDT14* | 0.357802584 | 6.11E-05 | 0.000255251 | 0.384407709 |
| T2DM | endothelial | *ANKRD45* | 1.355662899 | 0.001775383 | 0.004301284 | 1 |
| T2DM | endothelial | *MAATS1* | 0.356935446 | 0.000105545 | 0.000402646 | 0.663560097 |
| T2DM | endothelial | *GPR39* | -0.442306217 | 1.31E-07 | 4.46E-06 | 0.000823318 |

| | | | | | | |
|---|---|---|---|---|---|---|
| T2DM | endothelial | *DENND5A* | -0.297660154 | 0.019340998 | 0.032678541 | 1 |
| T2DM | endothelial | *TMEM236* | -0.764021666 | 1.56E-07 | 4.49E-06 | 0.000980209 |
| T2DM | endothelial | *TBX1* | 0.394064963 | 1.77E-05 | 9.34E-05 | 0.111296155 |
| T2DM | endothelial | *UQCR10* | -0.757209822 | 3.11E-05 | 0.000147246 | 0.195542321 |
| T2DM | endothelial | *EIF3C* | 0.429066914 | 9.89E-05 | 0.000381831 | 0.622002245 |
| T2DM | endothelial | *CNTN2* | -1.134509941 | 6.95E-07 | 9.58E-06 | 0.004366591 |
| T2DM | endothelial | *CRELD2* | -0.716084105 | 0.0001664 | 0.000589054 | 1 |
| T2DM | endothelial | *UBE2F* | -0.774471943 | 1.56E-06 | 1.61E-05 | 0.009816764 |
| T2DM | endothelial | *GOLGA6L4* | -0.89248231 | 1.22E-05 | 6.91E-05 | 0.076919333 |
| T2DM | endothelial | *PCDH9* | -0.287623076 | 3.11E-06 | 2.48E-05 | 0.019578504 |
| T2DM | endothelial | *OAF* | -0.334335413 | 0.000813247 | 0.002226867 | 1 |
| T2DM | endothelial | *TACSTD2* | 0.2188539 | 0.002815247 | 0.006325754 | 1 |
| T2DM | endothelial | *GDF3* | 0.554410806 | 0.000288194 | 0.000928691 | 1 |
| T2DM | endothelial | *PKP3* | -0.393719344 | 0.026562829 | 0.043085786 | 1 |
| T2DM | endothelial | *PLA2G6* | -0.186135711 | 0.030516515 | 0.048694754 | 1 |
| T2DM | endothelial | *A3GALT2* | -0.304393571 | 0.001519456 | 0.003768371 | 1 |
| T2DM | endothelial | *THAP7* | -0.905739914 | 6.36E-06 | 4.19E-05 | 0.040014182 |
| T2DM | endothelial | *CCR10* | -0.865537931 | 1.77E-06 | 1.73E-05 | 0.011130927 |
| T2DM | endothelial | *TXNRD2* | -0.559596527 | 0.000626054 | 0.001783418 | 1 |
| T2DM | endothelial | *ZFP1* | -0.620266504 | 0.006988428 | 0.013721501 | 1 |
| T2DM | endothelial | *LPAR5* | -0.517520727 | 6.28E-06 | 4.15E-05 | 0.039489981 |
| T2DM | endothelial | *TMEM173* | 0.381813709 | 0.000178833 | 0.000627411 | 1 |
| T2DM | endothelial | *NELL2* | -0.244600183 | 0.026721904 | 0.043332631 | 1 |
| T2DM | endothelial | *ZNF93* | -0.828994101 | 1.92E-05 | 0.000100043 | 0.120852319 |
| T2DM | endothelial | *SEPT5* | -2.256109166 | 6.94E-07 | 9.58E-06 | 0.004364411 |
| T2DM | endothelial | *UBE2G2* | -0.404570521 | 0.000759095 | 0.002097773 | 1 |
| T2DM | endothelial | *TUSC5* | -0.227325208 | 0.004345609 | 0.009149009 | 1 |
| T2DM | endothelial | *BTBD6* | -0.468105303 | 0.000109728 | 0.00041608 | 0.689860252 |
| T2DM | endothelial | *H1FX* | -0.560606821 | 0.002177749 | 0.005110678 | 1 |
| T2DM | endothelial | *NUTM2A* | -0.558660168 | 0.010516259 | 0.019470426 | 1 |
| T2DM | endothelial | *FAM227A* | -0.440083916 | 3.21E-05 | 0.000150625 | 0.201687305 |
| T2DM | endothelial | *NOC4L* | -0.321320516 | 0.008388927 | 0.016060043 | 1 |
| T2DM | endothelial | *SORCS2* | -0.505490772 | 3.11E-06 | 2.48E-05 | 0.019528789 |
| T2DM | endothelial | *TMEM106A* | -0.490474312 | 0.001750228 | 0.004246886 | 1 |
| T2DM | endothelial | *UBOX5* | -0.586129612 | 6.37E-05 | 0.000263978 | 0.400718707 |
| T2DM | endothelial | *CIB1* | -0.415685502 | 0.001325175 | 0.003360507 | 1 |
| T2DM | endothelial | *FLRT2* | -0.30094572 | 0.000220564 | 0.000747539 | 1 |
| T2DM | endothelial | *ADSSL1* | -0.773954245 | 8.63E-07 | 1.08E-05 | 0.005423455 |
| T2DM | endothelial | *C6orf120* | -0.819723518 | 7.72E-06 | 4.85E-05 | 0.048509617 |
| T2DM | endothelial | *SIGIRR* | 0.302708531 | 0.01141491 | 0.020898527 | 1 |
| T2DM | endothelial | *KIAA0825* | -0.37123265 | 0.001934581 | 0.004627714 | 1 |
| T2DM | endothelial | *KLHL33* | -0.265874527 | 0.008018719 | 0.015445369 | 1 |
| T2DM | endothelial | *RBM11* | 0.886611703 | 4.54E-05 | 0.000200579 | 0.285223552 |
| T2DM | endothelial | *TMEM105* | -3.821213756 | 2.30E-05 | 0.000115795 | 0.144396056 |
| T2DM | endothelial | *TCN2* | 0.188171436 | 0.002674165 | 0.00606292 | 1 |
| T2DM | endothelial | *ATP6V0A2* | -0.386034157 | 0.008812907 | 0.016769596 | 1 |
| T2DM | endothelial | *TNFAIP8L1* | 0.448105639 | 3.27E-06 | 2.57E-05 | 0.020569045 |
| T2DM | endothelial | *MAPK11* | 0.24221706 | 3.93E-06 | 2.96E-05 | 0.024714134 |
| T2DM | endothelial | *SH3BGR* | -0.100638508 | 0.014371582 | 0.025373248 | 1 |
| T2DM | endothelial | *FAM174B* | 0.293357555 | 0.00628068 | 0.012527486 | 1 |

| | | | | | | |
|---|---|---|---|---|---|---|
| T2DM | endothelial | *SV2B* | 0.208174777 | 3.21E-06 | 2.54E-05 | 0.020161676 |
| T2DM | endothelial | *PDE6G* | 0.562939865 | 0.027431416 | 0.044380163 | 1 |
| T2DM | endothelial | *TLCD2* | -0.551217227 | 5.37E-05 | 0.000229856 | 0.337830149 |
| T2DM | endothelial | *LSAMP* | -0.355246044 | 2.06E-06 | 1.91E-05 | 0.01297765 |
| T2DM | endothelial | *SP1* | -0.375694124 | 0.001206962 | 0.003104654 | 1 |
| T2DM | endothelial | *PDIA2* | -0.414371798 | 0.00159006 | 0.00391567 | 1 |
| T2DM | endothelial | *PBX1* | -0.249964682 | 0.007387644 | 0.014397432 | 1 |
| T2DM | endothelial | *UBE2L3* | -0.446543187 | 0.001952069 | 0.00466641 | 1 |
| T2DM | endothelial | *PMEL* | 0.265387504 | 0.003368669 | 0.007381952 | 1 |
| T2DM | endothelial | *SNAI3* | -0.569312249 | 7.69E-05 | 0.000307771 | 0.483201094 |
| T2DM | endothelial | *MYBL1* | -0.340404896 | 4.96E-05 | 0.00021471 | 0.311913494 |
| T2DM | endothelial | *ADARB2* | 0.133496905 | 9.92E-07 | 1.18E-05 | 0.006238933 |
| T2DM | endothelial | *SRL* | 0.211195091 | 0.000315036 | 0.00100032 | 1 |
| T2DM | endothelial | *KCNIP4* | -0.289594032 | 1.11E-05 | 6.42E-05 | 0.069993288 |
| T2DM | endothelial | *PIGP* | -0.125670127 | 0.005436161 | 0.011056987 | 1 |
| T2DM | endothelial | *ARL17A* | -0.376744004 | 0.006365891 | 0.012661297 | 1 |
| T2DM | endothelial | *IFITM1* | 0.595158828 | 3.40E-05 | 0.000158406 | 0.214006891 |
| T2DM | endothelial | *LINC00839* | 0.904117167 | 3.06E-09 | 7.82E-07 | 1.92E-05 |
| T2DM | endothelial | *RTN4RL1* | -0.64600728 | 5.29E-06 | 3.65E-05 | 0.033244714 |
| T2DM | endothelial | *PLK5* | -0.634703009 | 0.000326686 | 0.001031062 | 1 |
| T2DM | endothelial | *RASA3* | -0.490902557 | 3.19E-08 | 3.24E-06 | 0.000200412 |
| T2DM | endothelial | *ZNF566* | -0.558787816 | 3.91E-05 | 0.000177807 | 0.245970388 |
| T2DM | endothelial | *ZNF284* | -0.924340211 | 1.32E-07 | 4.46E-06 | 0.000830287 |
| T2DM | endothelial | *C2orf76* | -0.444650914 | 0.001717136 | 0.004182734 | 1 |
| T2DM | endothelial | *WWOX* | -0.208486642 | 0.012839588 | 0.023083354 | 1 |
| T2DM | endothelial | *CYP4Z1* | -0.517177448 | 1.88E-06 | 1.81E-05 | 0.011840924 |
| T2DM | endothelial | *CCDC84* | -0.351354836 | 0.009275953 | 0.017518148 | 1 |
| T2DM | endothelial | *FFAR4* | -0.258706799 | 2.58E-05 | 0.000127236 | 0.162353352 |
| T2DM | endothelial | *BPIFB4* | 5.975343207 | 1.76E-07 | 4.59E-06 | 0.001105092 |
| T2DM | endothelial | *CYP4F12* | 0.239702607 | 0.002399342 | 0.005556046 | 1 |
| T2DM | endothelial | *KLHL32* | -0.372966348 | 0.008570838 | 0.016333695 | 1 |
| T2DM | endothelial | *NBPF12* | -0.492855051 | 0.000816914 | 0.002235346 | 1 |
| T2DM | endothelial | *CYP4X1* | -0.416884972 | 0.000420108 | 0.001269817 | 1 |
| T2DM | endothelial | *CCDC30* | -0.346645629 | 0.015984005 | 0.027806154 | 1 |
| T2DM | endothelial | *FCAR* | -0.389080302 | 0.011798207 | 0.021506328 | 1 |
| T2DM | endothelial | *PCLO* | -0.286217278 | 3.88E-05 | 0.000176958 | 0.244040611 |
| T2DM | endothelial | *RGS7BP* | -0.581801982 | 0.00313733 | 0.006950104 | 1 |
| T2DM | endothelial | *ZNF396* | -0.735399343 | 2.53E-07 | 5.49E-06 | 0.00159355 |
| T2DM | endothelial | *CLCNKA* | -0.736546354 | 7.62E-05 | 0.000305495 | 0.479321994 |
| T2DM | endothelial | *SMYD4* | -0.422947443 | 0.00027456 | 0.000890692 | 1 |
| T2DM | endothelial | *NF2* | -0.385291088 | 0.00508305 | 0.010429875 | 1 |
| T2DM | endothelial | *MIR22HG* | -0.436845728 | 1.20E-06 | 1.34E-05 | 0.007532557 |
| T2DM | endothelial | *PDE2A* | 0.163294721 | 2.12E-06 | 1.95E-05 | 0.013312307 |
| T2DM | endothelial | *C17orf58* | -0.237829444 | 0.011585732 | 0.021155823 | 1 |
| T2DM | endothelial | *CYP27C1* | -0.202387063 | 0.000223508 | 0.00075467 | 1 |
| T2DM | endothelial | *BCR* | 0.246121227 | 0.007639557 | 0.014814897 | 1 |
| T2DM | endothelial | *LILRB4* | -0.279490131 | 0.00065657 | 0.001849396 | 1 |
| T2DM | endothelial | *KRT16* | -0.367947183 | 0.000280616 | 0.00090893 | 1 |
| T2DM | endothelial | *RTN4RL2* | -1.432513413 | 8.11E-06 | 5.06E-05 | 0.051013065 |
| T2DM | endothelial | *TMEM232* | -0.397456061 | 0.000506722 | 0.001488415 | 1 |

| | | | | | | |
|---|---|---|---|---|---|---|
| T2DM | endothelial | *EFCAB6* | -0.437721356 | 0.000166987 | 0.000590113 | 1 |
| T2DM | endothelial | *KANK3* | 0.178002435 | 2.79E-06 | 2.32E-05 | 0.017559536 |
| T2DM | endothelial | *EMID1* | -0.979038404 | 2.00E-06 | 1.87E-05 | 0.012546479 |
| T2DM | endothelial | *CYP4A11* | -0.386160596 | 0.004216153 | 0.008933924 | 1 |
| T2DM | endothelial | *PLCD1* | -0.337753531 | 0.02105159 | 0.035153079 | 1 |
| T2DM | endothelial | *ENTPD5* | -0.492066636 | 0.001298658 | 0.003302856 | 1 |
| T2DM | endothelial | *MRPS21* | -0.433982695 | 0.001366164 | 0.003452199 | 1 |
| T2DM | endothelial | *ANGPTL5* | -0.477755313 | 1.51E-07 | 4.49E-06 | 0.000948616 |
| T2DM | endothelial | *MT1X* | -0.494188172 | 0.000136487 | 0.000501224 | 0.858095484 |
| T2DM | endothelial | *WDR86* | -0.486896402 | 3.49E-07 | 6.64E-06 | 0.002193369 |
| T2DM | endothelial | *EPOR* | -0.557042695 | 1.81E-05 | 9.48E-05 | 0.113626899 |
| T2DM | endothelial | *FPR3* | -0.398597144 | 3.08E-06 | 2.47E-05 | 0.019337887 |
| T2DM | endothelial | *GJA4* | 0.209282081 | 4.62E-07 | 7.72E-06 | 0.002906575 |
| T2DM | endothelial | *IFT140* | 0.344335226 | 0.005396247 | 0.011000716 | 1 |
| T2DM | endothelial | *TLR5* | -0.539379248 | 5.19E-07 | 8.28E-06 | 0.003261633 |
| T2DM | endothelial | *PLEKHN1* | -0.481715392 | 0.003128634 | 0.006933282 | 1 |
| T2DM | endothelial | *ZNF385C* | -1.345078069 | 3.03E-05 | 0.000144052 | 0.190580961 |
| T2DM | endothelial | *TET3* | -0.374627438 | 0.010028412 | 0.018675542 | 1 |
| T2DM | endothelial | *DHRS4L2* | -0.38017942 | 0.000133334 | 0.000491656 | 0.838273867 |
| T2DM | endothelial | *TRPV2* | -0.689469081 | 1.71E-06 | 1.69E-05 | 0.010757849 |
| T2DM | endothelial | *C2orf88* | -0.348048499 | 0.004745822 | 0.009840694 | 1 |
| T2DM | endothelial | *FANCA* | -0.356543542 | 0.004384946 | 0.009207802 | 1 |
| T2DM | endothelial | *ADH1A* | -0.585878511 | 9.70E-08 | 4.46E-06 | 0.000609841 |
| T2DM | endothelial | *FAM69C* | 0.479716746 | 2.14E-06 | 1.96E-05 | 0.01346223 |
| T2DM | endothelial | *ZFP69* | -0.50038883 | 0.003317184 | 0.007299662 | 1 |
| T2DM | endothelial | *TMEM220* | -0.432679105 | 0.001141354 | 0.002963937 | 1 |
| T2DM | endothelial | *GFRAL* | 0.562922179 | 5.64E-05 | 0.000239236 | 0.354547404 |
| T2DM | endothelial | *C2orf66* | 1.244024753 | 0.016471445 | 0.028435152 | 1 |
| T2DM | endothelial | *CPSF4L* | 0.5290166 | 0.004133097 | 0.008793496 | 1 |
| T2DM | endothelial | *KLHL17* | -0.686171432 | 6.30E-07 | 9.11E-06 | 0.003962078 |
| T2DM | endothelial | *OR7D2* | -0.622967854 | 0.013565781 | 0.024174622 | 1 |
| T2DM | endothelial | *MORN2* | -0.290177857 | 0.006822799 | 0.013463571 | 1 |
| T2DM | endothelial | *S100A3* | -0.458034714 | 0.000640765 | 0.001813818 | 1 |
| T2DM | endothelial | *C6orf132* | -0.4486914 | 4.38E-07 | 7.49E-06 | 0.002750807 |
| T2DM | endothelial | *MAPK12* | 0.288578463 | 1.94E-06 | 1.83E-05 | 0.012180802 |
| T2DM | endothelial | *AGRN* | 0.237976393 | 4.98E-07 | 8.08E-06 | 0.00312967 |
| T2DM | endothelial | *FAM166A* | -1.422105868 | 1.17E-07 | 4.46E-06 | 0.000732529 |
| T2DM | endothelial | *ZC3H6* | -0.353241303 | 0.003398904 | 0.007429148 | 1 |
| T2DM | endothelial | *LINC00265* | -0.57931637 | 1.13E-05 | 6.50E-05 | 0.071307014 |
| T2DM | endothelial | *LAMTOR4* | -0.362021608 | 0.018909317 | 0.032061186 | 1 |
| T2DM | endothelial | *PRKAR1B* | 0.27975376 | 7.23E-05 | 0.000292012 | 0.454371209 |
| T2DM | endothelial | *NUTM2B* | -8.54122547 | 2.99E-09 | 7.82E-07 | 1.88E-05 |
| T2DM | endothelial | *NCR3LG1* | -1.24292842 | 0.018168888 | 0.03097283 | 1 |
| T2DM | endothelial | *ZNF793* | 0.247384049 | 5.28E-06 | 3.65E-05 | 0.033197632 |
| T2DM | endothelial | *AGAP4* | -0.757333107 | 0.002014728 | 0.004785265 | 1 |
| T2DM | endothelial | *RUFY4* | -0.873657734 | 1.03E-07 | 4.46E-06 | 0.000649061 |
| T2DM | endothelial | *CENPP* | -0.199013017 | 0.004139878 | 0.008798991 | 1 |
| T2DM | endothelial | *PLSCR1* | 0.255572962 | 5.75E-05 | 0.0002433 | 0.361343652 |
| T2DM | endothelial | *ZP3* | -0.471871429 | 1.63E-05 | 8.74E-05 | 0.102205828 |
| T2DM | endothelial | *JAKMIP3* | -0.24683327 | 0.001259284 | 0.003217033 | 1 |

| | | | | | | |
|---|---|---|---|---|---|---|
| T2DM | endothelial | *PDCD1* | -0.81401835 | 0.012228144 | 0.022141357 | 1 |
| T2DM | endothelial | *CERKL* | -0.85318269 | 0.000116044 | 0.000435303 | 0.729567697 |
| T2DM | endothelial | *C19orf54* | -0.375433383 | 0.011831826 | 0.021555111 | 1 |
| T2DM | endothelial | *C22orf34* | 0.210270262 | 3.65E-05 | 0.000167961 | 0.229602987 |
| T2DM | endothelial | *COL25A1* | -0.412463778 | 0.0002631 | 0.000859282 | 1 |
| T2DM | endothelial | *NBR1* | -0.335130573 | 0.026135977 | 0.042448176 | 1 |
| T2DM | endothelial | *NDOR1* | -0.79268367 | 3.17E-05 | 0.000149596 | 0.199412036 |
| T2DM | endothelial | *FAM72B* | 1.6972052 | 0.000315925 | 0.001001557 | 1 |
| T2DM | endothelial | *SUMO2* | -0.447589693 | 0.007169159 | 0.014019441 | 1 |
| T2DM | endothelial | *GOLGA8M* | 0.477566444 | 0.000208085 | 0.000711382 | 1 |
| T2DM | endothelial | *DPYD* | -0.214793336 | 0.026112951 | 0.042421738 | 1 |
| T2DM | endothelial | *UROS* | -0.444017985 | 0.003393286 | 0.007420379 | 1 |
| T2DM | endothelial | *NOXA1* | -0.69719541 | 0.000778566 | 0.002143523 | 1 |
| T2DM | endothelial | *TMEM201* | 0.705616312 | 0.007429128 | 0.014469309 | 1 |
| T2DM | endothelial | *LINC00910* | -0.837976013 | 1.75E-06 | 1.72E-05 | 0.011031328 |
| T2DM | endothelial | *BEND4* | 0.387390863 | 3.76E-07 | 6.87E-06 | 0.002362957 |
| T2DM | endothelial | *ZNF563* | -0.413138971 | 0.001598826 | 0.003935715 | 1 |
| T2DM | endothelial | *FBF1* | -0.178593776 | 0.005354354 | 0.010928299 | 1 |
| T2DM | endothelial | *FAM196A* | 0.262105782 | 0.000111397 | 0.000421644 | 0.700350036 |
| T2DM | endothelial | *UTS2B* | -0.310571684 | 0.000124038 | 0.00046253 | 0.779825181 |
| T2DM | endothelial | *C9orf152* | -1.502617663 | 0.014725483 | 0.025932524 | 1 |
| T2DM | endothelial | *NELFB* | -0.549371173 | 0.003840083 | 0.008270847 | 1 |
| T2DM | endothelial | *SLC15A5* | -0.184384364 | 0.010581782 | 0.01958424 | 1 |
| T2DM | endothelial | *ADAT2* | -0.363439657 | 0.004666583 | 0.00968917 | 1 |
| T2DM | endothelial | *ANKDD1B* | -0.303727749 | 0.000777045 | 0.002140789 | 1 |
| T2DM | endothelial | *SF3B3* | -0.386973648 | 0.01045372 | 0.019387179 | 1 |
| T2DM | endothelial | *SP6* | 3.496312325 | 5.65E-06 | 3.84E-05 | 0.035503911 |
| T2DM | endothelial | *PLAC9* | -0.41391231 | 5.27E-06 | 3.65E-05 | 0.033162978 |
| T2DM | endothelial | *ZNF527* | -0.845986892 | 5.37E-05 | 0.000229856 | 0.337888879 |
| T2DM | endothelial | *LINC00994* | -0.787351402 | 4.62E-06 | 3.35E-05 | 0.029015637 |
| T2DM | endothelial | *C15orf61* | -0.783056847 | 1.72E-05 | 9.12E-05 | 0.108108794 |
| T2DM | endothelial | *PNRC2* | -0.636004912 | 0.000140696 | 0.000514277 | 0.884555844 |
| T2DM | endothelial | *ZKSCAN3* | -0.574729944 | 0.002969498 | 0.006627346 | 1 |
| T2DM | endothelial | *SLC35E2B* | -0.21386193 | 0.019537839 | 0.032993436 | 1 |
| T2DM | endothelial | *BLOC1S2* | -0.330302894 | 0.000102621 | 0.00039364 | 0.645176438 |
| T2DM | endothelial | *AKR1C3* | -0.455138991 | 0.000144616 | 0.000524638 | 0.90919812 |
| T2DM | endothelial | *ZNF79* | -0.400900904 | 0.002378673 | 0.005514276 | 1 |
| T2DM | endothelial | *S100A4* | -0.269664332 | 0.000174306 | 0.000612899 | 1 |
| T2DM | endothelial | *COLCA1* | -0.689603643 | 4.37E-08 | 3.67E-06 | 0.000275055 |
| T2DM | endothelial | *ACADSB* | -0.525571469 | 5.81E-05 | 0.000245129 | 0.365241971 |
| T2DM | endothelial | *SEMA4A* | -0.419422188 | 0.008211272 | 0.015777589 | 1 |
| T2DM | endothelial | *HRCT1* | 0.306447611 | 0.024915121 | 0.040738977 | 1 |
| T2DM | endothelial | *SIRPB2* | -0.649967011 | 4.87E-07 | 7.99E-06 | 0.003062071 |
| T2DM | endothelial | *SRGAP3* | -0.202945401 | 0.007469776 | 0.014525976 | 1 |
| T2DM | endothelial | *SUPT5H* | -0.429739532 | 0.005041061 | 0.010356447 | 1 |
| T2DM | endothelial | *XPNPEP3* | -0.304131883 | 0.000166824 | 0.00058989 | 1 |
| T2DM | endothelial | *GRM7* | -0.214088312 | 3.17E-05 | 0.000149496 | 0.19912867 |
| T2DM | endothelial | *POM121* | -0.351741291 | 0.009925511 | 0.018516821 | 1 |
| T2DM | endothelial | *GIMAP5* | 0.263769831 | 1.56E-06 | 1.61E-05 | 0.009781245 |
| T2DM | endothelial | *STK31* | 0.588504974 | 0.000183367 | 0.000638914 | 1 |

| | | | | | | |
|---|---|---|---|---|---|---|
| T2DM | endothelial | *ZKSCAN7* | -0.615819522 | 2.16E-07 | 4.99E-06 | 0.001356399 |
| T2DM | endothelial | *ZNF565* | -0.283335088 | 0.021398223 | 0.035665597 | 1 |
| T2DM | endothelial | *NTNG2* | -1.015033361 | 2.78E-06 | 2.32E-05 | 0.017485508 |
| T2DM | endothelial | *WDR5* | -0.575137058 | 0.000215392 | 0.000732774 | 1 |
| T2DM | endothelial | *ASB13* | -0.71960679 | 0.000110261 | 0.000417848 | 0.693209789 |
| T2DM | endothelial | *SLC35F1* | -0.609455322 | 0.000736425 | 0.002045914 | 1 |
| T2DM | endothelial | *INCA1* | -0.821829816 | 0.004516744 | 0.009443555 | 1 |
| T2DM | endothelial | *ZNF124* | -0.375257443 | 0.016356436 | 0.028274104 | 1 |
| T2DM | endothelial | *PIK3R4* | -0.280038384 | 0.021313364 | 0.035543002 | 1 |
| T2DM | endothelial | *ZNF775* | -0.623059377 | 7.22E-07 | 9.76E-06 | 0.004537999 |
| T2DM | endothelial | *ZNF605* | -0.640047174 | 3.43E-05 | 0.000159409 | 0.215779615 |
| T2DM | endothelial | *RFX8* | -0.487384049 | 0.000649982 | 0.001835777 | 1 |
| T2DM | endothelial | *MYL6B* | -0.657566458 | 0.004056134 | 0.008651492 | 1 |
| T2DM | endothelial | *SULT1A1* | -0.389268045 | 2.02E-08 | 2.76E-06 | 0.000127107 |
| T2DM | endothelial | *ANAPC7* | -0.448073974 | 0.004596401 | 0.009578247 | 1 |
| T2DM | endothelial | *C1orf186* | -0.853837543 | 1.17E-05 | 6.68E-05 | 0.073794819 |
| T2DM | endothelial | *MYO18A* | 0.278846503 | 0.00264896 | 0.006025635 | 1 |
| T2DM | endothelial | *SPTSSB* | 0.78062037 | 7.43E-06 | 4.70E-05 | 0.046710515 |
| T2DM | endothelial | *SULF2* | 0.199379564 | 5.79E-06 | 3.92E-05 | 0.03639079 |
| T2DM | endothelial | *XRCC2* | -0.915874202 | 1.81E-06 | 1.75E-05 | 0.01139237 |
| T2DM | endothelial | *HDAC2* | -0.345570492 | 0.03145058 | 0.049982255 | 1 |
| T2DM | endothelial | *ADH1B* | -0.319625542 | 1.22E-06 | 1.36E-05 | 0.007639838 |
| T2DM | endothelial | *FAM180B* | -0.614979744 | 8.19E-11 | 4.68E-08 | 5.15E-07 |
| T2DM | endothelial | *ERI2* | -0.441016875 | 0.000471824 | 0.001405857 | 1 |
| T2DM | endothelial | *TRPV1* | -0.363209628 | 0.000389702 | 0.001192569 | 1 |
| T2DM | endothelial | *AMZ2* | -0.512426285 | 0.002900369 | 0.006491498 | 1 |
| T2DM | endothelial | *ZNF431* | -0.395174737 | 0.001443837 | 0.003612179 | 1 |
| T2DM | endothelial | *DAPK1* | -0.23808907 | 0.019880559 | 0.03343742 | 1 |
| T2DM | endothelial | *HLA-DQA1* | -0.267924382 | 0.001431614 | 0.003590169 | 1 |
| T2DM | endothelial | *GM2A* | -0.303583189 | 0.022780669 | 0.037680101 | 1 |
| T2DM | endothelial | *STRN3* | -0.264217785 | 0.018618017 | 0.031644086 | 1 |
| T2DM | endothelial | *ZNF239* | -0.280102918 | 0.006079224 | 0.012187525 | 1 |
| T2DM | endothelial | *PPTC7* | -0.503866713 | 1.53E-05 | 8.35E-05 | 0.096498462 |
| T2DM | endothelial | *NHLRC2* | -0.332633021 | 0.010273137 | 0.019091697 | 1 |
| T2DM | endothelial | *ZFP28* | 0.454937084 | 0.004217813 | 0.00893443 | 1 |
| T2DM | endothelial | *KIAA1211L* | 0.16097101 | 1.54E-06 | 1.60E-05 | 0.009686116 |
| T2DM | endothelial | *LAMB3* | -0.522547986 | 7.90E-07 | 1.03E-05 | 0.004967105 |
| T2DM | endothelial | *KPNA5* | -0.32804798 | 0.016472216 | 0.028435152 | 1 |
| T2DM | endothelial | *HCAR1* | -0.366100481 | 0.000909443 | 0.002444492 | 1 |
| T2DM | endothelial | *SRGAP1* | -0.330419418 | 0.002422063 | 0.005594626 | 1 |
| T2DM | endothelial | *FAM3C* | -0.389289378 | 0.012522519 | 0.022609531 | 1 |
| T2DM | endothelial | *HIST1H3E* | 0.310000274 | 0.011673872 | 0.021310579 | 1 |
| T2DM | endothelial | *ANXA4* | -0.273881137 | 0.020704431 | 0.03461935 | 1 |
| T2DM | endothelial | *ZNF138* | -0.321543988 | 0.014016067 | 0.024857268 | 1 |
| T2DM | endothelial | *ZNF429* | -0.450435114 | 0.000304506 | 0.000970315 | 1 |
| T2DM | endothelial | *ZNF100* | -0.282818843 | 0.007847548 | 0.015152805 | 1 |
| T2DM | endothelial | *GMFB* | -0.528261265 | 1.91E-06 | 1.82E-05 | 0.012019072 |
| T2DM | endothelial | *ZMYM1* | -0.431424751 | 0.003633954 | 0.007875446 | 1 |
| T2DM | endothelial | *IGF2R* | -0.381323864 | 0.001872771 | 0.004504251 | 1 |
| T2DM | endothelial | *GAL3ST4* | -0.586870077 | 1.01E-05 | 5.95E-05 | 0.063380643 |

| | | | | | | |
|---|---|---|---|---|---|---|
| T2DM | endothelial | *SLC25A29* | 0.343463508 | 2.02E-05 | 0.000104068 | 0.127067581 |
| T2DM | endothelial | *ZNF682* | -0.54530668 | 0.00132027 | 0.003349692 | 1 |
| T2DM | endothelial | *ZNF772* | -0.272874447 | 0.001116007 | 0.002905316 | 1 |
| T2DM | endothelial | *ADAM32* | -0.367919017 | 0.017548989 | 0.030053339 | 1 |
| T2DM | endothelial | *LRRC8B* | -0.410776501 | 0.00112406 | 0.002925068 | 1 |
| T2DM | endothelial | *ABCB8* | -0.408051334 | 2.90E-05 | 0.000138611 | 0.182134801 |
| T2DM | endothelial | *SND1* | -0.345731458 | 0.001469805 | 0.003666931 | 1 |
| T2DM | endothelial | *ENTPD4* | -0.349727759 | 0.016317261 | 0.028218023 | 1 |
| T2DM | endothelial | *C1D* | -0.419380959 | 0.005688963 | 0.011493094 | 1 |
| T2DM | endothelial | *TBC1D9B* | -0.380402901 | 0.001865548 | 0.004488596 | 1 |
| T2DM | endothelial | *FAM110D* | 0.150507353 | 5.54E-07 | 8.50E-06 | 0.003480053 |
| T2DM | endothelial | *KANK2* | -0.490985005 | 6.62E-06 | 4.31E-05 | 0.041612573 |
| T2DM | endothelial | *C6orf141* | 0.119092115 | 3.34E-06 | 2.62E-05 | 0.021001045 |
| T2DM | endothelial | *CCL4L2* | -0.341950544 | 0.000198787 | 0.000682564 | 1 |
| T2DM | endothelial | *GTF2E2* | -0.376160597 | 0.002442735 | 0.005633704 | 1 |
| T2DM | endothelial | *ZNF720* | -0.365973275 | 0.007764099 | 0.01501473 | 1 |
| T2DM | endothelial | *DDI2* | -0.461557521 | 1.00E-05 | 5.94E-05 | 0.063012679 |
| T2DM | endothelial | *C5AR1* | -0.445131663 | 9.65E-06 | 5.76E-05 | 0.060697286 |
| T2DM | endothelial | *DCHS2* | -0.25418932 | 2.48E-06 | 2.15E-05 | 0.015608896 |
| T2DM | endothelial | *VEPH1* | -0.515917928 | 0.000181602 | 0.000633944 | 1 |
| T2DM | endothelial | *SHPK* | -0.260044901 | 0.001415596 | 0.003558518 | 1 |
| T2DM | endothelial | *PCDHB11* | -1.221205226 | 1.59E-07 | 4.49E-06 | 0.000999337 |
| T2DM | endothelial | *SLC2A10* | -0.39922194 | 0.00031459 | 0.000999407 | 1 |
| T2DM | endothelial | *ZNF665* | -0.264885175 | 0.026036281 | 0.042309317 | 1 |
| T2DM | endothelial | *MYO5A* | -0.268455547 | 0.005298401 | 0.01082934 | 1 |
| T2DM | endothelial | *ATG7* | -0.28159615 | 0.003759301 | 0.008121292 | 1 |
| T2DM | endothelial | *TTC30A* | -0.81201067 | 1.51E-07 | 4.49E-06 | 0.000951751 |
| T2DM | endothelial | *RAB40C* | -0.617336791 | 2.69E-05 | 0.00013075 | 0.168928435 |
| T2DM | endothelial | *ZNF624* | -0.722988737 | 0.000165145 | 0.000585599 | 1 |
| T2DM | endothelial | *HHLA3* | -0.810653707 | 4.37E-07 | 7.49E-06 | 0.002747064 |
| T2DM | endothelial | *HOXA4* | -0.896456456 | 1.25E-07 | 4.46E-06 | 0.000784408 |
| T2DM | endothelial | *BCO2* | -0.662711995 | 2.70E-06 | 2.27E-05 | 0.016952635 |
| T2DM | endothelial | *ENTPD6* | -0.641468623 | 1.06E-05 | 6.18E-05 | 0.066570406 |
| T2DM | endothelial | *CCDC154* | -0.644985793 | 4.93E-05 | 0.000214 | 0.309872452 |
| T2DM | endothelial | *C5orf42* | -0.307791225 | 0.002119009 | 0.004991461 | 1 |
| T2DM | endothelial | *MFAP5* | -0.38028385 | 7.28E-06 | 4.62E-05 | 0.045742138 |
| T2DM | endothelial | *MPEG1* | -0.202016964 | 0.002533136 | 0.005801759 | 1 |
| T2DM | endothelial | *PDCD1LG2* | -0.269967549 | 0.006757157 | 0.013354997 | 1 |
| T2DM | endothelial | *NMB* | -0.788371342 | 0.001033799 | 0.002720591 | 1 |
| T2DM | endothelial | *KLHL14* | 0.351274171 | 2.50E-06 | 2.16E-05 | 0.015747581 |
| T2DM | endothelial | *FAM114A1* | -0.401114432 | 0.000765708 | 0.00211048 | 1 |
| T2DM | endothelial | *RPS26* | -0.143797717 | 0.016665998 | 0.028745989 | 1 |
| T2DM | endothelial | *C14orf178* | -0.267706688 | 0.002474072 | 0.005693498 | 1 |
| T2DM | endothelial | *PSAP* | -0.328122609 | 0.003657837 | 0.007919015 | 1 |
| T2DM | endothelial | *S100A10* | -0.298021655 | 0.014886916 | 0.026172831 | 1 |
| T2DM | endothelial | *MAP1LC3C* | -0.415267322 | 4.05E-06 | 3.02E-05 | 0.025441709 |
| T2DM | endothelial | *ZNF780A* | -0.211847207 | 0.002436321 | 0.005620973 | 1 |
| T2DM | endothelial | *AC122129.1* | -1.73400544 | 2.48E-07 | 5.42E-06 | 0.001556746 |
| T2DM | endothelial | *OCLN* | 0.223499622 | 2.90E-05 | 0.000138611 | 0.182035891 |
| T2DM | endothelial | *ADH5* | -0.467754841 | 9.85E-06 | 5.86E-05 | 0.061914067 |

| | | | | | | |
|---|---|---|---|---|---|---|
| T2DM | endothelial | *ZNF311* | -0.283565027 | 0.015744599 | 0.027450443 | 1 |
| T2DM | endothelial | *LEKR1* | -0.529984392 | 7.88E-07 | 1.03E-05 | 0.004954377 |
| T2DM | endothelial | *NOL8* | -0.456594788 | 0.001661838 | 0.004063778 | 1 |
| T2DM | endothelial | *ZNF335* | -0.335361588 | 0.021373953 | 0.035634591 | 1 |
| T2DM | endothelial | *AGAP9* | -0.859415011 | 0.000178509 | 0.000626627 | 1 |
| T2DM | endothelial | *ZNF273* | -0.407458321 | 0.010270518 | 0.019091697 | 1 |
| T2DM | endothelial | *ZNF84* | -0.371140105 | 0.009747388 | 0.018265821 | 1 |
| T2DM | endothelial | *ZNF667* | -0.521829376 | 8.15E-05 | 0.000323298 | 0.512427215 |
| T2DM | endothelial | *PRIM1* | -0.661830317 | 0.000497846 | 0.00146465 | 1 |
| T2DM | endothelial | *SULT1C4* | 0.281241973 | 8.35E-07 | 1.07E-05 | 0.005252002 |
| T2DM | endothelial | *ADH4* | -0.177470933 | 0.00269882 | 0.006105606 | 1 |
| T2DM | endothelial | *ZNF248* | -0.398350364 | 0.000149452 | 0.000537839 | 0.93960476 |
| T2DM | endothelial | *HIBCH* | -0.237797812 | 0.024910099 | 0.040738977 | 1 |
| T2DM | endothelial | *SOWAHC* | -1.151838181 | 1.13E-07 | 4.46E-06 | 0.00070922 |
| T2DM | endothelial | *AC135178.1* | 0.452668928 | 3.89E-07 | 7.01E-06 | 0.002448688 |
| T2DM | endothelial | *SVIP* | 0.287349157 | 1.71E-05 | 9.10E-05 | 0.107528875 |
| T2DM | endothelial | *DDRGK1* | -0.507048294 | 0.008516776 | 0.016250371 | 1 |
| T2DM | endothelial | *ZNF334* | -0.319907305 | 0.000979737 | 0.002598991 | 1 |
| T2DM | endothelial | *HSD17B11* | 0.294684132 | 0.009814804 | 0.018368684 | 1 |
| T2DM | endothelial | *SULT1C2* | 0.2807133 | 4.41E-05 | 0.000196158 | 0.277563542 |
| T2DM | endothelial | *QRICH1* | -0.229659472 | 0.02506766 | 0.040935163 | 1 |
| T2DM | endothelial | *SLC29A3* | -0.728474156 | 3.33E-06 | 2.61E-05 | 0.020922459 |
| T2DM | endothelial | *ANTXRL* | -13.20216435 | 9.85E-08 | 4.46E-06 | 0.00061912 |
| T2DM | endothelial | *STYX* | -0.386615612 | 0.026926857 | 0.043619982 | 1 |
| T2DM | endothelial | *TMEM116* | -0.357927531 | 2.44E-06 | 2.13E-05 | 0.015353184 |
| T2DM | endothelial | *MYL4* | 0.485826679 | 7.15E-06 | 4.56E-05 | 0.044975521 |
| T2DM | endothelial | *GFPT1* | -0.399017242 | 6.97E-05 | 0.000284107 | 0.438295033 |
| T2DM | endothelial | *UVRAG* | -0.22044959 | 0.005661366 | 0.011459436 | 1 |
| T2DM | endothelial | *MGEA5* | -0.313282016 | 0.001559893 | 0.00385043 | 1 |
| T2DM | endothelial | *MT1F* | -1.490842804 | 1.07E-05 | 6.21E-05 | 0.067001812 |
| T2DM | endothelial | *ZNF587* | -0.499608311 | 0.001743851 | 0.00423598 | 1 |
| T2DM | endothelial | *SH3BGRL2* | 0.174744392 | 2.10E-05 | 0.00010748 | 0.132075935 |
| T2DM | endothelial | *HLA-DRB5* | -0.175659877 | 2.54E-06 | 2.18E-05 | 0.01594405 |
| T2DM | endothelial | *CNGA1* | -0.289326493 | 0.001226272 | 0.003140354 | 1 |
| T2DM | endothelial | *MAFK* | -0.691270267 | 0.000116242 | 0.000435785 | 0.730811489 |
| T2DM | endothelial | *HIST1H4E* | 0.682161723 | 0.004233554 | 0.008958719 | 1 |
| T2DM | endothelial | *C20orf203* | 0.383116364 | 1.99E-05 | 0.000102943 | 0.125178421 |
| T2DM | endothelial | *ZNF627* | -0.575414933 | 2.84E-05 | 0.000136353 | 0.178350333 |
| T2DM | endothelial | *ARC* | -2.928420532 | 2.30E-07 | 5.23E-06 | 0.001448403 |
| T2DM | endothelial | *MMP17* | -0.470131927 | 1.22E-07 | 4.46E-06 | 0.000767283 |
| T2DM | endothelial | *MDM4* | -0.276585164 | 0.020729115 | 0.034651408 | 1 |
| T2DM | endothelial | *ZNF534* | -0.714915562 | 0.008459434 | 0.016170405 | 1 |
| T2DM | endothelial | *FAM19A2* | -0.405147423 | 0.002050678 | 0.004859177 | 1 |
| T2DM | endothelial | *TUSC1* | -1.187545863 | 5.88E-05 | 0.000246743 | 0.369373858 |
| T2DM | endothelial | *PAPSS2* | -0.350256818 | 0.000870059 | 0.002355753 | 1 |
| T2DM | endothelial | *FAN1* | -0.361280917 | 0.028330832 | 0.04562396 | 1 |
| T2DM | endothelial | *ANKRD13B* | -0.307981766 | 0.003113357 | 0.006910575 | 1 |
| T2DM | endothelial | *ZNF652* | -0.328081828 | 0.011128108 | 0.020432949 | 1 |
| T2DM | endothelial | *RASSF9* | 0.248693349 | 7.11E-06 | 4.54E-05 | 0.044729342 |
| T2DM | endothelial | *FAM169A* | 0.417064484 | 0.002986032 | 0.006661881 | 1 |

| | | | | | | |
|---|---|---|---|---|---|---|
| T2DM | endothelial | *TMEM184B* | 0.27585832 | 0.022954223 | 0.037907329 | 1 |
| T2DM | endothelial | *ALPK2* | -0.33392492 | 3.64E-06 | 2.78E-05 | 0.022865485 |
| T2DM | endothelial | *PNP* | 0.123990965 | 0.015778121 | 0.02750126 | 1 |
| T2DM | endothelial | *SFT2D1* | -0.519715964 | 9.49E-05 | 0.000369082 | 0.596436165 |
| T2DM | endothelial | *ARHGAP11A* | -0.680604644 | 0.000135252 | 0.000497558 | 0.850326993 |
| T2DM | endothelial | *HMGN2* | -0.333930069 | 0.010517189 | 0.019470426 | 1 |
| T2DM | endothelial | *DENND4B* | -0.495783098 | 0.001068407 | 0.002797616 | 1 |
| T2DM | endothelial | *CD3E* | -0.37051134 | 0.00451235 | 0.009437506 | 1 |
| T2DM | endothelial | *TSEN15* | -0.461857804 | 0.004449652 | 0.009318774 | 1 |
| T2DM | endothelial | *PRMT6* | -2.141799704 | 7.79E-08 | 4.46E-06 | 0.000489973 |
| T2DM | endothelial | *C1orf174* | -0.544853618 | 0.000887101 | 0.002394678 | 1 |
| T2DM | endothelial | *RASGEF1A* | -1.306238331 | 1.87E-07 | 4.63E-06 | 0.001176495 |
| T2DM | endothelial | *DCLRE1A* | -0.587336192 | 0.000167075 | 0.000590113 | 1 |
| T2DM | endothelial | *APRT* | -0.668364431 | 0.001614732 | 0.003968655 | 1 |
| T2DM | endothelial | *TBKBP1* | 0.204175403 | 0.024929489 | 0.040751872 | 1 |
| T2DM | endothelial | *ZFP2* | 0.407819495 | 0.001315361 | 0.003341281 | 1 |
| T2DM | endothelial | *L3MBTL3* | -0.404195641 | 0.015090751 | 0.026479361 | 1 |
| T2DM | endothelial | *NAGA* | -0.700020529 | 1.12E-05 | 6.45E-05 | 0.070486779 |
| T2DM | endothelial | *SMG5* | -0.397436873 | 0.012231101 | 0.022141357 | 1 |
| T2DM | endothelial | *TGM2* | 0.133357149 | 9.41E-06 | 5.66E-05 | 0.059155658 |
| T2DM | endothelial | *AL590822.1* | -0.29824453 | 1.24E-05 | 7.00E-05 | 0.078149571 |
| T2DM | endothelial | *AP001007.1* | -2.571243633 | 1.84E-07 | 4.62E-06 | 0.001154929 |
| T2DM | endothelial | *AC105020.1* | -1.313585226 | 0.01395356 | 0.024760381 | 1 |
| T2DM | endothelial | *EFCAB2* | -0.274996202 | 0.025695508 | 0.041868178 | 1 |
| T2DM | endothelial | *TATDN3* | -0.228256127 | 0.009884185 | 0.018456154 | 1 |
| T2DM | endothelial | *CENPW* | -1.558406232 | 5.28E-11 | 3.69E-08 | 3.32E-07 |
| T2DM | endothelial | *FAM229B* | -0.218679183 | 0.018496048 | 0.031462298 | 1 |
| T2DM | endothelial | *DDO* | -0.858569439 | 1.31E-05 | 7.29E-05 | 0.082055984 |
| T2DM | endothelial | *HIST2H2BF* | -1.353138422 | 1.22E-07 | 4.46E-06 | 0.000766299 |
| T2DM | endothelial | *C6orf163* | -0.943209626 | 0.000295026 | 0.000946826 | 1 |
| T2DM | endothelial | *OOEP* | -0.480094929 | 1.39E-05 | 7.66E-05 | 0.087346384 |
| T2DM | endothelial | *FOXO6* | -0.623954959 | 5.87E-05 | 0.000246613 | 0.368933459 |
| T2DM | endothelial | *INPP5B* | -0.154365502 | 0.001363962 | 0.00344802 | 1 |
| T2DM | endothelial | *NEU4* | -1.082040258 | 0.018881492 | 0.032031285 | 1 |
| T2DM | endothelial | *TRAF3IP1* | -0.5120873 | 0.000270925 | 0.000879805 | 1 |
| T2DM | endothelial | *ASAH2B* | 0.394486761 | 0.00115664 | 0.002998677 | 1 |
| T2DM | endothelial | *C10orf128* | 0.257516577 | 8.25E-06 | 5.13E-05 | 0.051891146 |
| T2DM | endothelial | *SYT15* | 0.274900369 | 1.34E-07 | 4.46E-06 | 0.000845335 |
| T2DM | endothelial | *RING1* | -0.458179371 | 0.000569686 | 0.001647476 | 1 |
| T2DM | endothelial | *HLA-DRA* | 0.481219263 | 2.27E-08 | 2.92E-06 | 0.00014256 |
| T2DM | endothelial | *AGER* | 0.476650616 | 0.000469829 | 0.001400577 | 1 |
| T2DM | endothelial | *PRRT1* | 0.548095628 | 2.66E-08 | 3.16E-06 | 0.000167267 |
| T2DM | endothelial | *DXO* | -0.314419116 | 0.013246353 | 0.023706182 | 1 |
| T2DM | endothelial | *SKIV2L* | -0.12656134 | 0.009379525 | 0.017692491 | 1 |
| T2DM | endothelial | *LAYN* | 0.360009177 | 8.10E-05 | 0.000321357 | 0.509029192 |
| T2DM | endothelial | *C6orf48* | -0.380545951 | 0.018534017 | 0.031518356 | 1 |
| T2DM | endothelial | *GPANK1* | -0.398558259 | 0.001449858 | 0.003624356 | 1 |
| T2DM | endothelial | *NFKBIL1* | -0.337106558 | 0.012536876 | 0.02261674 | 1 |
| T2DM | endothelial | *MICA* | -0.149570843 | 0.017450793 | 0.029943542 | 1 |
| T2DM | endothelial | *HLA-C* | 0.128083934 | 0.023678855 | 0.038952814 | 1 |

| | | | | | | |
|---|---|---|---|---|---|---|
| T2DM | endothelial | *PSORS1C1* | 0.069303117 | 0.003478402 | 0.007586651 | 1 |
| T2DM | endothelial | *ABCF1* | -0.337454656 | 0.01153355 | 0.021078903 | 1 |
| T2DM | endothelial | *LILRB3* | -0.350476019 | 0.002262264 | 0.005280086 | 1 |
| T2DM | endothelial | *LINC01123* | 2.127825299 | 1.36E-05 | 7.52E-05 | 0.085234161 |
| T2DM | endothelial | *HLA-E* | 0.203015123 | 0.000147159 | 0.000531719 | 0.925190483 |
| T2DM | endothelial | *ZNF468* | -0.856264339 | 0.003461847 | 0.007557163 | 1 |
| T2DM | endothelial | *HLA-G* | 0.421357678 | 4.27E-05 | 0.000191956 | 0.268738568 |
| T2DM | endothelial | *HLA-F* | 0.26188721 | 0.000573588 | 0.001656477 | 1 |
| T2DM | endothelial | *FAM153C* | -1.049655189 | 0.011948494 | 0.021742455 | 1 |
| T2DM | endothelial | *GABBR1* | -0.408439024 | 0.009110115 | 0.017241208 | 1 |
| T2DM | endothelial | *TRIM27* | -0.285475902 | 0.026839635 | 0.043489893 | 1 |
| T2DM | endothelial | *MALRD1* | -0.220730284 | 0.000333503 | 0.001048367 | 1 |
| T2DM | endothelial | *FAM216A* | -0.445075491 | 0.01178538 | 0.021489177 | 1 |
| T2DM | endothelial | *ZNF783* | -0.327247407 | 0.031425188 | 0.049975103 | 1 |
| T2DM | endothelial | *ZNF425* | -0.374461375 | 0.001500973 | 0.003735795 | 1 |
| T2DM | endothelial | *LRRC10B* | -2.106554941 | 5.75E-05 | 0.0002433 | 0.361543784 |
| T2DM | endothelial | *C12orf73* | -0.523941846 | 0.000539339 | 0.001566925 | 1 |
| T2DM | endothelial | *PCDHGA1* | -0.38968166 | 0.013179288 | 0.02360632 | 1 |
| T2DM | endothelial | *PCDHA5* | -2.597659806 | 0.000226393 | 0.000762362 | 1 |
| T2DM | endothelial | *PCDHA4* | 1.439252827 | 0.00070068 | 0.001963964 | 1 |
| T2DM | endothelial | *CLLU1OS* | -0.528303167 | 0.004098665 | 0.008729102 | 1 |
| T2DM | endothelial | *TMEM231* | 0.298801916 | 0.000450323 | 0.001346899 | 1 |
| T2DM | endothelial | *FAM71F2* | -0.602611944 | 2.04E-07 | 4.92E-06 | 0.001285479 |
| T2DM | endothelial | *AC016586.1* | -2.22653049 | 0.000372537 | 0.001151496 | 1 |
| T2DM | endothelial | *LINC00654* | -0.644717184 | 5.41E-06 | 3.72E-05 | 0.034008577 |
| T2DM | endothelial | *TMEM170B* | -0.143046755 | 0.010713739 | 0.019781873 | 1 |
| T2DM | endothelial | *SAP25* | -0.37400036 | 0.001821863 | 0.004401131 | 1 |
| T2DM | endothelial | *PRR13* | -0.609899485 | 0.003630191 | 0.007872719 | 1 |
| T2DM | endothelial | *MT1A* | -0.875525085 | 1.29E-07 | 4.46E-06 | 0.000810522 |
| T2DM | endothelial | *MT1M* | -1.055536953 | 0.000378304 | 0.001164738 | 1 |
| T2DM | endothelial | *CFI* | 0.141004901 | 2.42E-06 | 2.12E-05 | 0.015218484 |
| T2DM | endothelial | *EXOC3L4* | -0.652721632 | 3.83E-08 | 3.45E-06 | 0.000240514 |
| T2DM | endothelial | *TP53TG3D* | -1.001356434 | 0.006664664 | 0.013197085 | 1 |
| T2DM | endothelial | *NAP1L4* | -0.305828127 | 0.009429973 | 0.017761007 | 1 |
| T2DM | endothelial | *SMN2* | -0.27053834 | 0.001404178 | 0.003538304 | 1 |
| T2DM | endothelial | *SERF1B* | -0.902438381 | 0.00051455 | 0.001506741 | 1 |
| T2DM | endothelial | *DENND6B* | 0.497509988 | 0.012813562 | 0.023049747 | 1 |
| T2DM | endothelial | *CDPF1* | -0.38283309 | 0.003673027 | 0.007946428 | 1 |
| T2DM | endothelial | *LINC00487* | 0.555561676 | 5.02E-06 | 3.54E-05 | 0.031567979 |
| T2DM | endothelial | *RFPL3S* | -0.415778335 | 0.000113122 | 0.00042689 | 0.711199335 |
| T2DM | endothelial | *C1QTNF9B* | 4.154094431 | 5.02E-06 | 3.54E-05 | 0.031591395 |
| T2DM | endothelial | *NYNRIN* | 0.383864453 | 4.39E-06 | 3.21E-05 | 0.027570949 |
| T2DM | endothelial | *GOLGA8O* | -1.254399295 | 1.55E-06 | 1.60E-05 | 0.009742225 |
| T2DM | endothelial | *HCG27* | 0.167789445 | 0.011502908 | 0.021035132 | 1 |
| T2DM | endothelial | *HLA-A* | 0.187035524 | 0.010922985 | 0.020097398 | 1 |
| T2DM | endothelial | *LNP1* | -0.585311767 | 1.74E-07 | 4.57E-06 | 0.001095097 |
| T2DM | endothelial | *VGLL3* | -0.253316041 | 3.65E-07 | 6.76E-06 | 0.002292132 |
| T2DM | endothelial | *COLQ* | -0.611324161 | 2.99E-07 | 6.14E-06 | 0.0018791 |
| T2DM | endothelial | *METTL6* | -0.376696805 | 0.003153855 | 0.006979334 | 1 |
| T2DM | endothelial | *SLC48A1* | 0.206401576 | 3.87E-06 | 2.92E-05 | 0.024310064 |

| | | | | | | |
|---|---|---|---|---|---|---|
| T2DM | endothelial | *TAS2R14* | 0.343296617 | 0.001549559 | 0.003832447 | 1 |
| T2DM | endothelial | *ZRSR1* | -2.259327792 | 0.016637639 | 0.02870495 | 1 |
| T2DM | endothelial | *AC084018.1* | -0.229745885 | 0.0056145 | 0.011371895 | 1 |
| T2DM | endothelial | *C17orf51* | -0.50059109 | 1.65E-07 | 4.50E-06 | 0.001036891 |
| T2DM | endothelial | *OR2A42* | -0.274378191 | 0.024634799 | 0.040364603 | 1 |
| T2DM | endothelial | *RNF208* | -1.079077124 | 1.13E-07 | 4.46E-06 | 0.000707482 |
| T2DM | endothelial | *MAP10* | 0.509296083 | 0.004344606 | 0.009149009 | 1 |
| T2DM | endothelial | *ZNF580* | -0.406417083 | 0.000796245 | 0.002184116 | 1 |
| T2DM | endothelial | *NUP62* | -0.703620072 | 0.000240096 | 0.000799938 | 1 |
| T2DM | endothelial | *FAM106A* | -0.254314607 | 0.008492544 | 0.016218902 | 1 |
| T2DM | endothelial | *ZNF254* | -0.339740405 | 0.00959019 | 0.017998067 | 1 |
| T2DM | endothelial | *LINGO4* | -2.887232788 | 0.000490712 | 0.001449087 | 1 |
| T2DM | endothelial | *GIMAP1* | 0.253074609 | 4.21E-06 | 3.11E-05 | 0.026484098 |
| T2DM | endothelial | *CCDC183* | -1.402390098 | 7.35E-08 | 4.46E-06 | 0.000462285 |
| T2DM | endothelial | *ARHGEF35* | -0.805684832 | 0.000115007 | 0.000432561 | 0.723049171 |
| T2DM | endothelial | *DNLZ* | 0.273610492 | 0.000489013 | 0.001446107 | 1 |
| T2DM | endothelial | *GSTM2* | -0.307234892 | 0.000305395 | 0.000972652 | 1 |
| T2DM | endothelial | *ARHGAP19* | 0.714221041 | 0.006273841 | 0.012517816 | 1 |
| T2DM | endothelial | *CCL14* | 0.103114042 | 3.70E-05 | 0.000169903 | 0.232426701 |
| T2DM | endothelial | *RBMXL1* | -0.707437922 | 0.003351903 | 0.007350336 | 1 |
| T2DM | endothelial | *SRA1* | -0.344489404 | 0.003980957 | 0.008530428 | 1 |
| T2DM | endothelial | *TMEM110* | -0.843749427 | 0.002511375 | 0.005760312 | 1 |
| T2DM | endothelial | *LBH* | -0.73545387 | 5.76E-06 | 3.91E-05 | 0.036215316 |
| T2DM | endothelial | *PPP1CB* | -0.309887499 | 0.015993578 | 0.027808631 | 1 |
| T2DM | endothelial | *ATF6B* | 0.279238257 | 0.013252734 | 0.023710185 | 1 |
| T2DM | endothelial | *SLC35F6* | -0.535807377 | 0.014151733 | 0.025041358 | 1 |
| T2DM | endothelial | *CLIC1* | -0.393602792 | 0.015457437 | 0.027009702 | 1 |
| T2DM | endothelial | *EMP2* | 0.24948139 | 0.000480084 | 0.001426411 | 1 |
| T2DM | endothelial | *PPM1N* | -0.747435952 | 9.18E-05 | 0.000358355 | 0.577309892 |
| T2DM | endothelial | *LTB4R* | -0.262544053 | 0.019003794 | 0.032195325 | 1 |
| T2DM | endothelial | *GALT* | -0.390227999 | 0.0291255 | 0.046664632 | 1 |
| T2DM | endothelial | *ITGA1* | 0.238553925 | 0.001901072 | 0.004556631 | 1 |
| T2DM | endothelial | *NUDT19* | -0.541777252 | 2.40E-05 | 0.000119852 | 0.150938156 |
| T2DM | endothelial | *ZNF726* | -1.01662101 | 1.69E-06 | 1.68E-05 | 0.010655278 |
| T2DM | endothelial | *AP1G2* | 0.46672657 | 0.014753914 | 0.025975318 | 1 |
| T2DM | endothelial | *ZNF90* | -0.56205429 | 0.003399659 | 0.007429148 | 1 |
| T2DM | endothelial | *TM6SF2* | -10.88848935 | 1.32E-05 | 7.34E-05 | 0.082692941 |
| T2DM | endothelial | *REPIN1* | -0.563410416 | 7.32E-05 | 0.000295175 | 0.46047241 |
| T2DM | endothelial | *SMIM7* | -0.379602139 | 0.009710589 | 0.018207717 | 1 |
| T2DM | endothelial | *TSPAN4* | -0.432498078 | 0.003123878 | 0.006925932 | 1 |
| T2DM | endothelial | *CPNE1* | -0.151977529 | 0.026972592 | 0.043682814 | 1 |
| T2DM | endothelial | *AC079776.2* | -0.722583731 | 8.42E-07 | 1.07E-05 | 0.00529115 |
| T2DM | endothelial | *LYRM4* | -0.353703605 | 0.013894011 | 0.024675606 | 1 |
| T2DM | endothelial | *PRCD* | 0.501522124 | 3.29E-05 | 0.000154205 | 0.206860315 |
| T2DM | endothelial | *C17orf67* | 0.23969576 | 0.00023128 | 0.000774671 | 1 |
| T2DM | endothelial | *COLCA2* | -0.476272274 | 1.74E-07 | 4.57E-06 | 0.00109408 |
| T2DM | endothelial | *NEURL1B* | 0.182843488 | 1.45E-06 | 1.54E-05 | 0.009128986 |
| T2DM | endothelial | *VSTM5* | 0.246674363 | 0.0104971 | 0.019450417 | 1 |
| T2DM | endothelial | *PPME1* | -0.352224839 | 0.000869255 | 0.002354592 | 1 |
| T2DM | endothelial | *CPEB1* | -0.37876335 | 8.17E-08 | 4.46E-06 | 0.000513793 |

| | | | | | | |
|---|---|---|---|---|---|---|
| T2DM | endothelial | *C10orf105* | 0.498193512 | 3.47E-05 | 0.00016082 | 0.218321528 |
| T2DM | endothelial | *ARHGEF33* | -0.250166551 | 0.014221493 | 0.02515064 | 1 |
| T2DM | endothelial | *AC005082.1* | -1.491191922 | 2.90E-07 | 6.03E-06 | 0.001826315 |
| T2DM | endothelial | *ACSM4* | -0.179142051 | 0.011345377 | 0.020789386 | 1 |
| T2DM | endothelial | *COL28A1* | -0.679320047 | 0.004440135 | 0.009301942 | 1 |
| T2DM | endothelial | *PHB2* | -0.724776524 | 0.000443015 | 0.001328201 | 1 |
| T2DM | endothelial | *GRID2IP* | 0.431646126 | 6.17E-05 | 0.000256549 | 0.387645847 |
| T2DM | endothelial | *UBXN2B* | -0.299687413 | 0.018099568 | 0.030863027 | 1 |
| T2DM | endothelial | *UBE2QL1* | -0.556415461 | 1.13E-07 | 4.46E-06 | 0.000713222 |
| T2DM | endothelial | *KCNU1* | -0.75065867 | 1.51E-11 | 1.90E-08 | 9.49E-08 |
| T2DM | endothelial | *MYL5* | -0.981477237 | 2.73E-07 | 5.76E-06 | 0.001714667 |
| T2DM | endothelial | *NPEPL1* | -0.622793513 | 3.77E-06 | 2.86E-05 | 0.023708255 |
| T2DM | endothelial | *LINC01139* | -0.496204961 | 5.77E-05 | 0.000243838 | 0.362587318 |
| T2DM | endothelial | *TSTD1* | -0.651802532 | 6.17E-07 | 8.98E-06 | 0.003877801 |
| T2DM | endothelial | *LINC01138* | -0.462784303 | 7.87E-07 | 1.03E-05 | 0.004949409 |
| T2DM | endothelial | *CYB5RL* | -0.683422963 | 5.58E-05 | 0.000237345 | 0.351033637 |
| T2DM | endothelial | *ATAD3C* | 0.782557999 | 2.59E-06 | 2.22E-05 | 0.016311177 |
| T2DM | endothelial | *AP000350.4* | -0.111009056 | 0.017970495 | 0.030684548 | 1 |
| T2DM | endothelial | *RNASEK* | -0.452462586 | 0.019202595 | 0.032470876 | 1 |
| T2DM | endothelial | *NBPF1* | -0.463035015 | 0.0004279 | 0.001291507 | 1 |
| T2DM | endothelial | *RNU6ATAC35I* | -3.04855221 | 1.86E-07 | 4.62E-06 | 0.001168884 |
| T2DM | endothelial | *C6orf226* | -0.672638119 | 4.38E-05 | 0.000195348 | 0.275527522 |
| T2DM | endothelial | *PPP3R1* | -0.48698218 | 0.003298219 | 0.007260469 | 1 |
| T2DM | endothelial | *FANCG* | -0.399321016 | 0.000267318 | 0.000871243 | 1 |
| T2DM | endothelial | *C19orf73* | -1.613476204 | 8.10E-07 | 1.04E-05 | 0.005091584 |
| T2DM | endothelial | *OR2A25* | -8.725061586 | 2.25E-06 | 2.01E-05 | 0.014120101 |
| T2DM | endothelial | *TIGD1* | -0.31413453 | 0.002515691 | 0.005766004 | 1 |
| T2DM | endothelial | *XKR9* | -0.309171254 | 1.12E-06 | 1.27E-05 | 0.007019535 |
| T2DM | endothelial | *C1orf229* | -0.576884658 | 2.91E-06 | 2.38E-05 | 0.018293695 |
| T2DM | endothelial | *SLC12A8* | -0.572349426 | 3.13E-08 | 3.24E-06 | 0.000196898 |
| T2DM | endothelial | *APOL6* | -0.448901561 | 4.81E-06 | 3.46E-05 | 0.030233659 |
| T2DM | endothelial | *TIAF1* | 0.536034737 | 3.24E-06 | 2.55E-05 | 0.020351359 |
| T2DM | endothelial | *FAM185A* | -0.47040107 | 0.000732293 | 0.002036235 | 1 |
| T2DM | endothelial | *AC062017.1* | -0.566215368 | 1.90E-09 | 6.14E-07 | 1.20E-05 |
| T2DM | endothelial | *ADRA2B* | -1.249952741 | 1.27E-07 | 4.46E-06 | 0.000800229 |
| T2DM | endothelial | *EXOSC6* | -0.395027043 | 0.024778728 | 0.040579282 | 1 |
| T2DM | endothelial | *VPS52* | -0.406755674 | 0.021804963 | 0.036266551 | 1 |
| T2DM | endothelial | *LINC00205* | -0.771031654 | 2.26E-06 | 2.02E-05 | 0.014193303 |
| T2DM | endothelial | *OSER1-AS1* | -0.701886928 | 0.000187798 | 0.000653757 | 1 |
| T2DM | endothelial | *LINC01133* | -0.460876787 | 1.90E-07 | 4.69E-06 | 0.001197577 |
| T2DM | endothelial | *LINC01117* | 0.280312644 | 8.53E-06 | 5.25E-05 | 0.053612772 |
| T2DM | endothelial | *RASAL2-AS1* | -1.857180923 | 5.55E-07 | 8.50E-06 | 0.003488498 |
| T2DM | endothelial | *AC069213.1* | -1.26307288 | 0.000312715 | 0.000993952 | 1 |
| T2DM | endothelial | *LINC00240* | -0.24472341 | 0.0114839 | 0.021012619 | 1 |
| T2DM | endothelial | *LINC00863* | -0.409143705 | 0.001891976 | 0.004541754 | 1 |
| T2DM | endothelial | *LINC00184* | -0.819236262 | 1.69E-08 | 2.53E-06 | 0.000106059 |
| T2DM | endothelial | *TMEM233* | 0.269455613 | 0.000103347 | 0.000396184 | 0.649741214 |
| T2DM | endothelial | *AL049840.1* | -0.48446664 | 2.46E-06 | 2.14E-05 | 0.015477465 |
| T2DM | endothelial | *LINC00237* | -1.810134254 | 1.08E-07 | 4.46E-06 | 0.000681077 |
| T2DM | endothelial | *PLEKHM1* | -0.384088596 | 0.006141656 | 0.012293088 | 1 |

| | | | | | | |
|---|---|---|---|---|---|---|
| T2DM | endothelial | *AP000350.6* | -0.326722593 | 0.000415727 | 0.001258389 | 1 |
| T2DM | endothelial | *SFTA1P* | -1.864494002 | 8.43E-07 | 1.07E-05 | 0.005299185 |
| T2DM | endothelial | *FAM225B* | -0.393161002 | 0.001289715 | 0.003285429 | 1 |
| T2DM | endothelial | *LINC00626* | -5.546877621 | 6.83E-06 | 4.41E-05 | 0.042971269 |
| T2DM | endothelial | *ERCC6* | -0.433840005 | 0.003192282 | 0.007044533 | 1 |
| T2DM | endothelial | *ELFN1* | 0.248531881 | 5.60E-07 | 8.53E-06 | 0.003521257 |
| T2DM | endothelial | *ZNF503-AS1* | -0.851042679 | 6.74E-06 | 4.37E-05 | 0.042348971 |
| T2DM | endothelial | *LINC00937* | -0.77322963 | 1.23E-07 | 4.46E-06 | 0.000772097 |
| T2DM | endothelial | *AC010536.1* | -0.557084316 | 0.004339583 | 0.009149009 | 1 |
| T2DM | endothelial | *AC064836.3* | 0.816545773 | 1.13E-05 | 6.48E-05 | 0.070847054 |
| T2DM | endothelial | *TAS2R46* | -0.676959099 | 0.000525512 | 0.001532418 | 1 |
| T2DM | endothelial | *AC093901.1* | -0.73097739 | 0.001709743 | 0.004167954 | 1 |
| T2DM | endothelial | *AC009299.3* | -0.58400144 | 0.005176574 | 0.010597565 | 1 |
| T2DM | endothelial | *SLC2A1-AS1* | -0.658652841 | 1.35E-06 | 1.47E-05 | 0.008512318 |
| T2DM | endothelial | *LINC01068* | -0.510625747 | 0.000124918 | 0.000464986 | 0.785361108 |
| T2DM | endothelial | *AC108463.2* | -0.62642718 | 0.000150436 | 0.000540761 | 0.945791308 |
| T2DM | endothelial | *C19orf24* | -0.718221557 | 5.48E-05 | 0.000233609 | 0.344806667 |
| T2DM | endothelial | *LINC01013* | 0.543605414 | 2.91E-06 | 2.38E-05 | 0.018277193 |
| T2DM | endothelial | *AC073133.2* | 2.182658309 | 2.75E-05 | 0.00013285 | 0.172970349 |
| T2DM | endothelial | *ARL17B* | -0.487417627 | 2.77E-06 | 2.32E-05 | 0.017396402 |
| T2DM | endothelial | *DHFR* | -0.234058974 | 0.000165631 | 0.000586991 | 1 |
| T2DM | endothelial | *LINC00954* | -0.801452709 | 1.81E-07 | 4.62E-06 | 0.001140403 |
| T2DM | endothelial | *AC067945.3* | -0.558989993 | 0.00049449 | 0.001457504 | 1 |
| T2DM | endothelial | *PATL2* | -0.646679495 | 2.56E-05 | 0.00012624 | 0.16069938 |
| T2DM | endothelial | *U91324.1* | -0.757980143 | 1.54E-07 | 4.49E-06 | 0.00096811 |
| T2DM | endothelial | *ZNF688* | -0.641422195 | 0.01309306 | 0.023458555 | 1 |
| T2DM | endothelial | *CISD3* | 0.601755784 | 0.000367593 | 0.001139012 | 1 |
| T2DM | endothelial | *PSMG3-AS1* | -0.572918118 | 3.51E-06 | 2.70E-05 | 0.022044569 |
| T2DM | endothelial | *PPP5D1* | -0.55480552 | 3.55E-05 | 0.000163789 | 0.223407826 |
| T2DM | endothelial | *AC112229.4* | 0.232270388 | 0.004047612 | 0.008637929 | 1 |
| T2DM | endothelial | *KLHL7-AS1* | -1.624244747 | 2.08E-07 | 4.93E-06 | 0.001310211 |
| T2DM | endothelial | *AC092171.4* | -0.653075294 | 5.93E-06 | 3.98E-05 | 0.037252833 |
| T2DM | endothelial | *HLA-DPA1* | 0.235505164 | 0.019741684 | 0.033259418 | 1 |
| T2DM | endothelial | *DIRC3* | -0.328076497 | 1.02E-06 | 1.20E-05 | 0.00639108 |
| T2DM | endothelial | *TAPBP* | 0.27091426 | 0.020815726 | 0.034777696 | 1 |
| T2DM | endothelial | *LINC00865* | -0.602227675 | 4.45E-06 | 3.24E-05 | 0.027954858 |
| T2DM | endothelial | *AC009495.3* | -1.000800252 | 0.002504024 | 0.005749743 | 1 |
| T2DM | endothelial | *HLA-DQB2* | 0.194766269 | 3.13E-07 | 6.27E-06 | 0.001969381 |
| T2DM | endothelial | *LYRM9* | -0.833001714 | 3.03E-05 | 0.000144041 | 0.190342765 |
| T2DM | endothelial | *LINC00342* | -0.73856261 | 5.67E-05 | 0.000240304 | 0.356370136 |
| T2DM | endothelial | *SNHG15* | -0.648014075 | 0.002262521 | 0.005280086 | 1 |
| T2DM | endothelial | *LINC00702* | -0.458862451 | 8.43E-08 | 4.46E-06 | 0.000529775 |
| T2DM | endothelial | *AC123768.3* | -0.881636041 | 1.77E-05 | 9.34E-05 | 0.111467869 |
| T2DM | endothelial | *PINLYP* | -0.25197121 | 0.001403028 | 0.003536822 | 1 |
| T2DM | endothelial | *HLA-B* | 0.291020708 | 6.41E-06 | 4.20E-05 | 0.040271388 |
| T2DM | endothelial | *C19orf81* | -0.810835295 | 8.63E-05 | 0.000339291 | 0.542525667 |
| T2DM | endothelial | *ZSCAN31* | 0.246004284 | 0.00025983 | 0.000849922 | 1 |
| T2DM | endothelial | *LINC00957* | 0.89573162 | 0.0001328 | 0.000491305 | 0.834910459 |
| T2DM | endothelial | *KIAA0040* | -0.450098795 | 0.005677772 | 0.01147844 | 1 |
| T2DM | endothelial | *LINC00941* | -2.447627556 | 1.06E-06 | 1.23E-05 | 0.006672937 |

| | | | | | | |
|---|---|---|---|---|---|---|
| T2DM | endothelial | *LINC00545* | -2.230675659 | 0.004801532 | 0.009923132 | 1 |
| T2DM | endothelial | *ZBTB22* | -0.299769994 | 0.021144056 | 0.035279374 | 1 |
| T2DM | endothelial | *B3GNT9* | -0.790507831 | 1.09E-06 | 1.26E-05 | 0.006862733 |
| T2DM | endothelial | *CDKN2AIPNL* | -0.285327617 | 0.023906378 | 0.03925193 | 1 |
| T2DM | endothelial | *GS1-124K5.4* | 0.101961853 | 0.020421903 | 0.03417421 | 1 |
| T2DM | endothelial | *ZNF737* | -0.377986483 | 0.027661572 | 0.044706505 | 1 |
| T2DM | endothelial | *HLA-DQA2* | 0.416143208 | 2.96E-11 | 3.11E-08 | 1.86E-07 |
| T2DM | endothelial | *AC073257.2* | -2.233009378 | 0.004666573 | 0.00968917 | 1 |
| T2DM | endothelial | *IRGM* | -0.266306308 | 0.013262919 | 0.023715578 | 1 |
| T2DM | endothelial | *LRRC37A2* | -0.430079864 | 0.003230983 | 0.007120524 | 1 |
| T2DM | endothelial | *TXNDC5* | 1.438781212 | 1.02E-05 | 6.01E-05 | 0.06434223 |
| T2DM | endothelial | *ASB14* | 0.32861567 | 0.023584548 | 0.038846229 | 1 |
| T2DM | endothelial | *NME1* | -0.172889953 | 0.019012399 | 0.032201227 | 1 |
| T2DM | endothelial | *CDRT4* | -0.95471148 | 2.47E-05 | 0.000122827 | 0.155498601 |
| T2DM | endothelial | *ADSL* | -0.608981651 | 0.000225938 | 0.00076124 | 1 |
| T2DM | endothelial | *LILRA4* | -5.242074889 | 0.006956125 | 0.013666612 | 1 |
| T2DM | endothelial | *LILRA2* | -0.271955499 | 0.000189482 | 0.000658161 | 1 |
| T2DM | endothelial | *LY6G5B* | -0.154390997 | 0.000991953 | 0.002624752 | 1 |
| T2DM | endothelial | *PSMB9* | 0.224225566 | 0.000703818 | 0.001969249 | 1 |
| T2DM | endothelial | *COX19* | -0.565189113 | 0.000124116 | 0.000462546 | 0.780314396 |
| T2DM | endothelial | *AQP1* | 0.192668264 | 6.73E-06 | 4.37E-05 | 0.042312417 |
| T2DM | endothelial | *C1QTNF9* | 0.344231631 | 3.23E-07 | 6.35E-06 | 0.002032215 |
| T2DM | endothelial | *ARHGEF25* | -0.586221049 | 4.34E-05 | 0.000193879 | 0.272787715 |
| T2DM | endothelial | *LINC00886* | -0.346691241 | 0.014337014 | 0.025326441 | 1 |
| T2DM | endothelial | *MIF* | -0.12068735 | 0.017246904 | 0.029642232 | 1 |
| T2DM | endothelial | *CD302* | -0.585939632 | 0.000434786 | 0.001308521 | 1 |
| T2DM | endothelial | *ATP5J2* | -0.568614049 | 0.001602806 | 0.00394397 | 1 |
| T2DM | endothelial | *ARHGAP8* | -1.134745499 | 7.97E-08 | 4.46E-06 | 0.000501308 |
| T2DM | endothelial | *PLEKHO2* | -0.642590523 | 0.001194323 | 0.003082392 | 1 |
| T2DM | endothelial | *PWP2* | -1.712520414 | 0.022960696 | 0.037908061 | 1 |
| T2DM | endothelial | *AKAP2* | 0.711603135 | 1.38E-06 | 1.50E-05 | 0.008670057 |
| T2DM | endothelial | *LINC00969* | -0.298110649 | 0.001019524 | 0.0026864 | 1 |
| T2DM | endothelial | *ARHGDIG* | -0.536435491 | 7.53E-05 | 0.000302081 | 0.473361539 |
| T2DM | endothelial | *C22orf39* | -0.571874896 | 0.027016 | 0.043741847 | 1 |
| T2DM | endothelial | *EIF6* | -0.602859022 | 0.000327089 | 0.001031816 | 1 |
| T2DM | endothelial | *DECR2* | -0.773623807 | 3.64E-07 | 6.76E-06 | 0.002288357 |
| T2DM | endothelial | *AP5Z1* | -0.542164305 | 0.000753221 | 0.002083752 | 1 |
| T2DM | endothelial | *ZNF709* | 0.431843092 | 0.005414172 | 0.011026531 | 1 |
| T2DM | endothelial | *VSIG8* | -0.876506443 | 1.76E-06 | 1.72E-05 | 0.011075103 |
| T2DM | endothelial | *C7orf73* | -0.487421411 | 0.000196264 | 0.000675746 | 1 |
| T2DM | endothelial | *FGF14-IT1* | -0.684191982 | 1.35E-07 | 4.46E-06 | 0.000846591 |
| T2DM | endothelial | *TICAM2* | -0.975903666 | 2.24E-06 | 2.01E-05 | 0.014064424 |
| T2DM | endothelial | *PALM2* | -0.225387476 | 7.27E-05 | 0.000293541 | 0.457336339 |
| T2DM | endothelial | *NAT6* | -0.43433521 | 0.000583462 | 0.001684219 | 1 |
| T2DM | endothelial | *UPK3B* | -0.433924812 | 0.001196535 | 0.003084303 | 1 |
| T2DM | endothelial | *PLA2G4B* | 0.354603789 | 0.001271603 | 0.003244549 | 1 |
| T2DM | endothelial | *JMJD7* | 0.491797463 | 5.86E-05 | 0.000246551 | 0.368593548 |
| T2DM | endothelial | *RN7SL832P* | -0.379500605 | 0.019153337 | 0.032413736 | 1 |
| T2DM | endothelial | *TMEM199* | -0.685268509 | 0.000268519 | 0.000874253 | 1 |
| T2DM | endothelial | *LILRA6* | -0.217953779 | 0.00311509 | 0.006910575 | 1 |

| | | | | | | |
|---|---|---|---|---|---|---|
| T2DM | endothelial | *APOBEC3C* | -0.753597181 | 1.74E-07 | 4.57E-06 | 0.001096036 |
| T2DM | endothelial | *CCDC13* | -0.378577242 | 5.62E-05 | 0.000238732 | 0.353323498 |
| T2DM | endothelial | *ASPRV1* | 0.93160587 | 0.01680937 | 0.028969437 | 1 |
| T2DM | endothelial | *OR2AE1* | 1.763803773 | 4.71E-06 | 3.40E-05 | 0.029591144 |
| T2DM | endothelial | *CRYBB2* | -0.793987981 | 0.000300028 | 0.000958472 | 1 |
| T2DM | endothelial | *LINC00847* | -0.841549099 | 3.16E-06 | 2.51E-05 | 0.019893583 |
| T2DM | endothelial | *NEAT1* | -0.227742175 | 0.001749438 | 0.004246609 | 1 |
| T2DM | endothelial | *RAB30-AS1* | -0.394287473 | 0.000933722 | 0.002495881 | 1 |
| T2DM | endothelial | *LINC00920* | -0.690577145 | 0.003522268 | 0.007659806 | 1 |
| T2DM | endothelial | *UBAP1L* | -0.617976467 | 2.42E-05 | 0.000120476 | 0.152041247 |
| T2DM | endothelial | *DNAJC3-AS1* | -0.607503119 | 0.002534664 | 0.005803145 | 1 |
| T2DM | endothelial | *LINC00926* | -0.712011579 | 0.002443719 | 0.005633905 | 1 |
| T2DM | endothelial | *BCKDHA* | -0.867525747 | 0.00515336 | 0.010554479 | 1 |
| T2DM | endothelial | *CDK11B* | -0.39721765 | 0.018725845 | 0.031792975 | 1 |
| T2DM | endothelial | *PCDHAC1* | -1.068549201 | 0.000652877 | 0.001841472 | 1 |
| T2DM | endothelial | *ABHD14A* | -0.608378978 | 4.14E-05 | 0.000186946 | 0.260415835 |
| T2DM | endothelial | *SRP14-AS1* | -0.221700926 | 7.62E-06 | 4.81E-05 | 0.047906746 |
| T2DM | endothelial | *C5orf17* | -0.576084014 | 0.000916928 | 0.002460404 | 1 |
| T2DM | endothelial | *FMN1* | -0.243327532 | 0.00154022 | 0.003815353 | 1 |
| T2DM | endothelial | *NAIP* | -0.872624363 | 8.57E-07 | 1.08E-05 | 0.00538674 |
| T2DM | endothelial | *ZNF286B* | -1.587070858 | 2.34E-07 | 5.25E-06 | 0.001470148 |
| T2DM | endothelial | *LINC00942* | -0.673431878 | 0.005240287 | 0.010717529 | 1 |
| T2DM | endothelial | *LINC00603* | -0.523451436 | 4.20E-07 | 7.34E-06 | 0.002642694 |
| T2DM | endothelial | *YJEFN3* | -0.528525714 | 2.63E-06 | 2.23E-05 | 0.016518083 |
| T2DM | endothelial | *DNAH10OS* | -2.28645068 | 3.11E-09 | 7.82E-07 | 1.95E-05 |
| T2DM | endothelial | *LINC00989* | -0.608288269 | 1.83E-07 | 4.62E-06 | 0.001151358 |
| T2DM | endothelial | *LINC01021* | -1.19231754 | 1.36E-07 | 4.46E-06 | 0.000854068 |
| T2DM | endothelial | *FAM218A* | -0.639122003 | 1.18E-07 | 4.46E-06 | 0.000740059 |
| T2DM | endothelial | *GLI4* | -0.550373901 | 3.45E-06 | 2.68E-05 | 0.021712223 |
| T2DM | endothelial | *SNHG18* | -0.514486957 | 7.18E-09 | 1.46E-06 | 4.51E-05 |
| T2DM | endothelial | *SHANK3* | 0.112548663 | 7.80E-07 | 1.03E-05 | 0.004903243 |
| T2DM | endothelial | *ZNF550* | -0.478593434 | 0.010362417 | 0.019240554 | 1 |
| T2DM | endothelial | *CCDC71L* | -0.603502675 | 0.000397101 | 0.001210168 | 1 |
| T2DM | endothelial | *TMEM200B* | -0.250157012 | 0.004141762 | 0.008800019 | 1 |
| T2DM | endothelial | *TRNP1* | -1.614642534 | 1.09E-07 | 4.46E-06 | 0.000687704 |
| T2DM | endothelial | *ALG11* | -0.342578195 | 0.002701842 | 0.00610835 | 1 |
| T2DM | endothelial | *ATXN7L3B* | -0.414313313 | 0.0239066 | 0.03925193 | 1 |
| T2DM | endothelial | *ARMS2* | -0.750419702 | 0.000923119 | 0.002472796 | 1 |
| T2DM | endothelial | *FPGT* | -0.77161808 | 2.36E-05 | 0.000118517 | 0.148610706 |
| T2DM | endothelial | *EID3* | -0.228788599 | 0.009559477 | 0.017961875 | 1 |
| T2DM | endothelial | *SNHG9* | -0.20847781 | 0.006865024 | 0.013538396 | 1 |
| T2DM | endothelial | *TAS2R43* | 0.156130174 | 0.022874341 | 0.037811922 | 1 |
| T2DM | endothelial | *EBLN2* | -0.628015222 | 0.001230881 | 0.003149396 | 1 |
| T2DM | endothelial | *LINC00944* | -0.427452502 | 0.004850647 | 0.010015122 | 1 |
| T2DM | endothelial | *PGA5* | 3.225625471 | 5.30E-05 | 0.000227458 | 0.33338811 |
| T2DM | endothelial | *ZNF253* | -0.489170608 | 0.001136689 | 0.002954263 | 1 |
| T2DM | endothelial | *ZBED6* | -1.411496232 | 1.64E-07 | 4.50E-06 | 0.001033274 |
| T2DM | endothelial | *FNTB* | -1.101855133 | 2.61E-06 | 2.23E-05 | 0.016414228 |
| T2DM | endothelial | *CUX1* | -0.257758359 | 0.016830597 | 0.028998072 | 1 |
| T2DM | endothelial | *TEN1* | -0.626040418 | 0.020095656 | 0.033718012 | 1 |

| | | | | | | |
|---|---|---|---|---|---|---|
| T2DM | endothelial | *C17orf49* | -0.961800237 | 7.78E-05 | 0.000310905 | 0.489364549 |
| T2DM | endothelial | *ZNF578* | 0.337716958 | 0.000248356 | 0.000821797 | 1 |
| T2DM | endothelial | *LINC00640* | -0.225689892 | 0.00084885 | 0.002307271 | 1 |
| T2DM | endothelial | *LINC00638* | -0.983619681 | 5.13E-08 | 4.03E-06 | 0.000322567 |
| T2DM | endothelial | *CEP95* | -0.382791073 | 0.001917485 | 0.004590719 | 1 |
| T2DM | endothelial | *LINC00639* | 0.099463597 | 1.59E-07 | 4.49E-06 | 0.001002316 |
| T2DM | endothelial | *LINC00648* | -0.398366132 | 6.62E-08 | 4.46E-06 | 0.000416431 |
| T2DM | endothelial | *LINC00924* | -0.514975227 | 7.28E-07 | 9.76E-06 | 0.004579064 |
| T2DM | endothelial | *ITGB3* | 0.361206202 | 1.13E-05 | 6.50E-05 | 0.071335266 |
| T2DM | endothelial | *UBE2Q2L* | -1.357754777 | 2.85E-07 | 5.93E-06 | 0.00179014 |
| T2DM | endothelial | *AC083843.1* | -6.989801476 | 1.69E-10 | 8.84E-08 | 1.06E-06 |
| T2DM | endothelial | *PWRN1* | -0.415205013 | 0.022508611 | 0.037274328 | 1 |
| T2DM | endothelial | *LINC00562* | -0.189897103 | 0.009905005 | 0.018489538 | 1 |
| T2DM | endothelial | *AC009120.5* | -0.769415188 | 1.42E-07 | 4.46E-06 | 0.000891777 |
| T2DM | endothelial | *LINC00565* | -2.317788814 | 0.000220734 | 0.000747713 | 1 |
| T2DM | endothelial | *TMEM178B* | 0.30699227 | 5.83E-05 | 0.000245696 | 0.366823699 |
| T2DM | endothelial | *HPR* | -1.178477307 | 6.24E-05 | 0.000259174 | 0.392390163 |
| T2DM | endothelial | *GOLGA8H* | -0.282353644 | 0.002686671 | 0.006084691 | 1 |
| T2DM | endothelial | *LINC00662* | -0.408491484 | 0.00359858 | 0.00782035 | 1 |
| T2DM | endothelial | *AC123768.4* | -1.40621829 | 0.00557319 | 0.011299144 | 1 |
| T2DM | endothelial | *LINC00667* | -0.243073723 | 0.004215592 | 0.008933924 | 1 |
| T2DM | endothelial | *LINC00672* | -0.51888594 | 4.90E-05 | 0.000213088 | 0.307912378 |
| T2DM | endothelial | *LINC00909* | -0.362884473 | 0.007341153 | 0.014329038 | 1 |
| T2DM | endothelial | *DYNLL2* | -0.888634152 | 1.77E-05 | 9.34E-05 | 0.111114658 |
| T2DM | endothelial | *LINC00526* | -0.843072783 | 1.37E-05 | 7.59E-05 | 0.086162977 |
| T2DM | endothelial | *LINC00668* | 7.423687808 | 0.000270017 | 0.00087731 | 1 |
| T2DM | endothelial | *MYO15B* | 0.266306666 | 0.018859018 | 0.032010434 | 1 |
| T2DM | endothelial | *ILF3-AS1* | -0.322108488 | 0.009454249 | 0.017785417 | 1 |
| T2DM | endothelial | *AC139100.2* | -0.973972982 | 1.56E-07 | 4.49E-06 | 0.000978585 |
| T2DM | endothelial | *LINC01140* | -0.443871124 | 1.64E-05 | 8.80E-05 | 0.103215289 |
| T2DM | endothelial | *LINC00906* | -1.816802656 | 1.33E-07 | 4.46E-06 | 0.000835415 |
| T2DM | endothelial | *ZNF582-AS1* | 0.569243568 | 0.01426981 | 0.025221899 | 1 |
| T2DM | endothelial | *ZNF285* | 0.177829163 | 0.00691016 | 0.013601733 | 1 |
| T2DM | endothelial | *AL132989.1* | -1.774096833 | 1.05E-06 | 1.22E-05 | 0.006578026 |
| T2DM | endothelial | *LINC00664* | -0.87136025 | 0.020751958 | 0.034680373 | 1 |
| T2DM | endothelial | *FBXO17* | -0.402122035 | 3.75E-07 | 6.87E-06 | 0.002357121 |
| T2DM | endothelial | *AC009060.1* | -0.814628654 | 0.005648293 | 0.011436656 | 1 |
| T2DM | endothelial | *HCG17* | -0.452008958 | 0.00052962 | 0.001542967 | 1 |
| T2DM | endothelial | *YTHDF3-AS1* | -0.78056126 | 5.81E-06 | 3.92E-05 | 0.036515689 |
| T2DM | endothelial | *LINC01050* | -1.980346865 | 1.70E-06 | 1.69E-05 | 0.010702328 |
| T2DM | endothelial | *MILR1* | -0.174089572 | 0.003862727 | 0.008308232 | 1 |
| T2DM | endothelial | *GTF2H5* | -0.392102479 | 0.001957776 | 0.004673545 | 1 |
| T2DM | endothelial | *FGF14-AS2* | -0.624033366 | 0.005496337 | 0.011161328 | 1 |
| T2DM | endothelial | *LINC01023* | -0.663627639 | 7.87E-06 | 4.94E-05 | 0.049506475 |
| T2DM | endothelial | *AP000347.2* | -0.30158094 | 0.000917621 | 0.002461213 | 1 |
| T2DM | endothelial | *DOC2B* | 0.092724505 | 2.11E-05 | 0.000107757 | 0.13254101 |
| T2DM | endothelial | *AP000350.5* | -0.133774921 | 0.00749233 | 0.014556328 | 1 |
| T2DM | endothelial | *PAXIP1-AS1* | -0.553866614 | 6.12E-06 | 4.07E-05 | 0.038465306 |
| T2DM | LEC | *C1orf112* | 0.902141284 | 0.000323172 | 0.001159063 | 1 |
| T2DM | LEC | *CFH* | 0.779241453 | 0.000338227 | 0.001185962 | 1 |

| | | | | | | |
|---|---|---|---|---|---|---|
| T2DM | LEC | *NIPAL3* | -0.779887858 | 0.00306586 | 0.005619552 | 1 |
| T2DM | LEC | *ENPP4* | 0.813611221 | 0.00109957 | 0.00252115 | 1 |
| T2DM | LEC | *SEMA3F* | -0.814287738 | 8.93E-05 | 0.000908725 | 0.561579853 |
| T2DM | LEC | *RAD52* | 0.814875443 | 0.000304252 | 0.001126076 | 1 |
| T2DM | LEC | *BAD* | 0.544017597 | 0.000111504 | 0.000908725 | 0.701025852 |
| T2DM | LEC | *HS3ST1* | -0.145833031 | 0.000143618 | 0.000908725 | 0.902928832 |
| T2DM | LEC | *WNT16* | -1.450760165 | 6.64E-05 | 0.000908725 | 0.417522952 |
| T2DM | LEC | *HECW1* | 0.445748081 | 0.000239858 | 0.00100947 | 1 |
| T2DM | LEC | *MAD1L1* | 0.896643859 | 0.002629132 | 0.004969739 | 1 |
| T2DM | LEC | *LASP1* | 0.987902522 | 0.002947438 | 0.005451763 | 1 |
| T2DM | LEC | *SNX11* | 0.700972371 | 6.40E-06 | 0.000600399 | 0.040226716 |
| T2DM | LEC | *TMEM176A* | 0.515822881 | 8.58E-06 | 0.000673907 | 0.053942467 |
| T2DM | LEC | *CYP26B1* | 0.629103196 | 9.14E-05 | 0.000908725 | 0.574436695 |
| T2DM | LEC | *ICA1* | -0.614589983 | 0.002603335 | 0.004923938 | 1 |
| T2DM | LEC | *ALS2* | 0.978535327 | 0.002159283 | 0.004264975 | 1 |
| T2DM | LEC | *POLDIP2* | -1.01291032 | 0.011665838 | 0.017685827 | 1 |
| T2DM | LEC | *CD38* | 0.940651374 | 0.008429265 | 0.013342092 | 1 |
| T2DM | LEC | *RBM6* | -0.425766145 | 0.001719906 | 0.003568662 | 1 |
| T2DM | LEC | *RECQL* | 1.006322814 | 0.000411561 | 0.001311277 | 1 |
| T2DM | LEC | *ARHGAP33* | 0.602244104 | 0.001475917 | 0.003159376 | 1 |
| T2DM | LEC | *PDK4* | 0.651748983 | 0.00036947 | 0.001240843 | 1 |
| T2DM | LEC | *SLC22A16* | 0.723282686 | 0.003871656 | 0.006852787 | 1 |
| T2DM | LEC | *CDC27* | 0.650611792 | 0.02231637 | 0.031451024 | 1 |
| T2DM | LEC | *MEOX1* | 0.533455114 | 0.000345172 | 0.001196042 | 1 |
| T2DM | LEC | *FAM214B* | 1.268704941 | 0.000341108 | 0.001190094 | 1 |
| T2DM | LEC | *COPZ2* | 0.735774095 | 0.000109914 | 0.000908725 | 0.691027857 |
| T2DM | LEC | *GCFC2* | 0.85496495 | 0.008782668 | 0.013821436 | 1 |
| T2DM | LEC | *ABCB4* | 0.775984723 | 0.00013028 | 0.000908725 | 0.819069023 |
| T2DM | LEC | *ZNF195* | -1.041161658 | 0.00270636 | 0.005091229 | 1 |
| T2DM | LEC | *PDK2* | 1.125657532 | 0.000737184 | 0.001903545 | 1 |
| T2DM | LEC | *ASB4* | 1.336627877 | 0.000411716 | 0.001311277 | 1 |
| T2DM | LEC | *ABCC8* | -1.490304271 | 0.00049667 | 0.001454972 | 1 |
| T2DM | LEC | *CCL18* | 0.51980879 | 5.36E-05 | 0.000908725 | 0.336935615 |
| T2DM | LEC | *SYNRG* | 0.82135265 | 0.008739199 | 0.013759916 | 1 |
| T2DM | LEC | *TMEM132A* | 1.076243997 | 7.83E-05 | 0.000908725 | 0.49201563 |
| T2DM | LEC | *TNFRSF12A* | 0.565750627 | 0.000203434 | 0.000964732 | 1 |
| T2DM | LEC | *BAIAP2L1* | 0.562023277 | 0.00014711 | 0.000908725 | 0.924879598 |
| T2DM | LEC | *FARP2* | 0.831121848 | 0.001992845 | 0.004001984 | 1 |
| T2DM | LEC | *IFRD1* | 0.637985328 | 0.000904756 | 0.002195681 | 1 |
| T2DM | LEC | *GTF2IRD1* | 0.595139175 | 0.001126936 | 0.002568874 | 1 |
| T2DM | LEC | *VPS41* | 0.644581302 | 0.030373339 | 0.041440361 | 1 |
| T2DM | LEC | *ARHGAP44* | -0.811001525 | 0.000646297 | 0.001740903 | 1 |
| T2DM | LEC | *ELAC2* | 1.07005469 | 0.004993866 | 0.008545574 | 1 |
| T2DM | LEC | *CCDC124* | 0.913353705 | 0.005233143 | 0.008882498 | 1 |
| T2DM | LEC | *CEACAM21* | 0.438662012 | 0.000407316 | 0.00130653 | 1 |
| T2DM | LEC | *NOS2* | -1.505989159 | 0.000234235 | 0.001001608 | 1 |
| T2DM | LEC | *CD79B* | -1.000870437 | 0.000200116 | 0.000962607 | 1 |
| T2DM | LEC | *PAX6* | -0.45346747 | 0.000226116 | 0.000992757 | 1 |
| T2DM | LEC | *RPUSD1* | 1.533232267 | 0.000124703 | 0.000908725 | 0.784005267 |
| T2DM | LEC | *BAIAP3* | -1.127856294 | 0.000595245 | 0.001651502 | 1 |

| | | | | | | |
|---|---|---|---|---|---|---|
| T2DM | LEC | *TSR3* | 0.950449241 | 4.82E-05 | 0.000908725 | 0.302998212 |
| T2DM | LEC | *PIGQ* | 0.710358688 | 0.000463309 | 0.001396368 | 1 |
| T2DM | LEC | *PSMB1* | 0.958288072 | 0.000905325 | 0.002195681 | 1 |
| T2DM | LEC | *CDK11A* | 1.321834291 | 0.005328821 | 0.009028291 | 1 |
| T2DM | LEC | *DLEC1* | 0.573477258 | 0.000126087 | 0.000908725 | 0.792708189 |
| T2DM | LEC | *SS18L2* | -0.913038278 | 0.000521723 | 0.001506001 | 1 |
| T2DM | LEC | *MPND* | 1.647333002 | 0.00032862 | 0.001168572 | 1 |
| T2DM | LEC | *MGST1* | 0.405526364 | 0.000133642 | 0.000908725 | 0.840204288 |
| T2DM | LEC | *MMP25* | 0.5281488 | 5.65E-05 | 0.000908725 | 0.355273712 |
| T2DM | LEC | *IL32* | 0.894613674 | 0.00028053 | 0.00108013 | 1 |
| T2DM | LEC | *PKD1* | 1.011338672 | 0.000323181 | 0.001159063 | 1 |
| T2DM | LEC | *MAPK8IP2* | -1.521465357 | 5.65E-05 | 0.000908725 | 0.355177328 |
| T2DM | LEC | *SEC62* | 0.839004939 | 0.0069911 | 0.011366187 | 1 |
| T2DM | LEC | *MLXIPL* | 0.328010568 | 7.91E-06 | 0.000664383 | 0.049727317 |
| T2DM | LEC | *METTL13* | 1.244333007 | 0.000217121 | 0.000982034 | 1 |
| T2DM | LEC | *DYRK4* | 0.574290693 | 2.91E-05 | 0.000908725 | 0.182939827 |
| T2DM | LEC | *NCAPD2* | 0.46388157 | 0.007747366 | 0.012428602 | 1 |
| T2DM | LEC | *ZNF200* | -0.705520424 | 0.003946138 | 0.006945512 | 1 |
| T2DM | LEC | *CD4* | 0.381992507 | 0.00011549 | 0.000908725 | 0.726084245 |
| T2DM | LEC | *LRRC23* | 0.584779035 | 0.000123324 | 0.000908725 | 0.775340172 |
| T2DM | LEC | *SCMH1* | 0.652717578 | 0.033659429 | 0.04545035 | 1 |
| T2DM | LEC | *CLCN6* | 0.987056165 | 0.007506142 | 0.012103389 | 1 |
| T2DM | LEC | *MRC2* | 0.413687256 | 0.000197538 | 0.000956093 | 1 |
| T2DM | LEC | *TSPAN9* | -0.544169456 | 8.04E-05 | 0.000908725 | 0.50556397 |
| T2DM | LEC | *MKS1* | 1.288720739 | 0.000172561 | 0.000910705 | 1 |
| T2DM | LEC | *TTC19* | 0.789698945 | 0.008299868 | 0.013180417 | 1 |
| T2DM | LEC | *DPF1* | -0.65893485 | 6.66E-05 | 0.000908725 | 0.418429542 |
| T2DM | LEC | *LARS2* | 1.026405655 | 0.013146738 | 0.019623347 | 1 |
| T2DM | LEC | *ZBTB32* | 1.078012374 | 0.000168838 | 0.000910705 | 1 |
| T2DM | LEC | *TMEM159* | 0.513382317 | 0.000262277 | 0.00104809 | 1 |
| T2DM | LEC | *ERCC1* | 0.441783947 | 0.000156815 | 0.000910705 | 0.985896177 |
| T2DM | LEC | *LTF* | -0.476224603 | 0.00037195 | 0.001243195 | 1 |
| T2DM | LEC | *ALOX5* | 0.862560126 | 0.000237009 | 0.001005449 | 1 |
| T2DM | LEC | *MAP4K5* | -0.626866502 | 0.022826228 | 0.032097628 | 1 |
| T2DM | LEC | *PSMC4* | 1.271825046 | 0.003659445 | 0.006523089 | 1 |
| T2DM | LEC | *MAN2B2* | 0.833425799 | 0.000358375 | 0.001223183 | 1 |
| T2DM | LEC | *CLDN11* | -0.185143484 | 0.000184185 | 0.000929091 | 1 |
| T2DM | LEC | *SLC25A39* | 0.854388016 | 0.000148776 | 0.000910197 | 0.935354153 |
| T2DM | LEC | *ANGEL1* | 0.867004823 | 6.83E-05 | 0.000908725 | 0.429500749 |
| T2DM | LEC | *DDX11* | 0.541292982 | 0.002952535 | 0.005459584 | 1 |
| T2DM | LEC | *HEBP1* | -1.162556035 | 0.001952025 | 0.003941034 | 1 |
| T2DM | LEC | *TACC3* | 0.818032744 | 0.001105634 | 0.002530657 | 1 |
| T2DM | LEC | *ZC3H3* | 0.846324642 | 0.025863412 | 0.035910616 | 1 |
| T2DM | LEC | *MDH1* | 0.748651916 | 0.001497593 | 0.003194871 | 1 |
| T2DM | LEC | *SLC30A9* | 0.825978015 | 0.003450417 | 0.006205026 | 1 |
| T2DM | LEC | *MTMR11* | 0.420789738 | 0.0214204 | 0.030324263 | 1 |
| T2DM | LEC | *COX15* | 0.863408101 | 0.001710333 | 0.003554665 | 1 |
| T2DM | LEC | *CCDC88C* | 0.610705661 | 0.000284583 | 0.001090187 | 1 |
| T2DM | LEC | *NUDCD3* | 0.535726145 | 0.014592011 | 0.02152005 | 1 |
| T2DM | LEC | *CHDH* | -0.69737608 | 0.000233949 | 0.001001608 | 1 |

| | | | | | | |
|---|---|---|---|---|---|---|
| T2DM | LEC | *IL20RA* | -0.375050118 | 0.000142889 | 0.000908725 | 0.898340364 |
| T2DM | LEC | *WWTR1* | -0.414833942 | 0.000739216 | 0.001906254 | 1 |
| T2DM | LEC | *TTC27* | 0.751899781 | 0.007659801 | 0.012319563 | 1 |
| T2DM | LEC | *VSIG2* | 1.103641385 | 0.000137619 | 0.000908725 | 0.865207702 |
| T2DM | LEC | *PHLDB1* | 0.302451405 | 0.01053282 | 0.016187948 | 1 |
| T2DM | LEC | *MARCO* | 0.520073183 | 3.26E-05 | 0.000908725 | 0.204784707 |
| T2DM | LEC | *MNAT1* | 0.819884379 | 0.001742277 | 0.003605114 | 1 |
| T2DM | LEC | *SLC7A9* | -0.493870552 | 0.0142507 | 0.021099406 | 1 |
| T2DM | LEC | *OSBPL5* | -0.783641116 | 0.016907087 | 0.024537132 | 1 |
| T2DM | LEC | *AQR* | 0.924166142 | 0.003191933 | 0.00581672 | 1 |
| T2DM | LEC | *NLRP2* | 0.620870499 | 0.000539355 | 0.001540629 | 1 |
| T2DM | LEC | *RNH1* | 0.735083795 | 0.000830343 | 0.002064201 | 1 |
| T2DM | LEC | *NDUFS1* | 0.834988775 | 0.003172901 | 0.005790429 | 1 |
| T2DM | LEC | *BIRC3* | 0.960257464 | 0.002860274 | 0.005328161 | 1 |
| T2DM | LEC | *AKAP11* | 0.931310137 | 0.004384382 | 0.007616637 | 1 |
| T2DM | LEC | *GLRX2* | 0.719916737 | 0.001508172 | 0.003208758 | 1 |
| T2DM | LEC | *SNAPC1* | 0.499886222 | 0.00018439 | 0.000929091 | 1 |
| T2DM | LEC | *GCLM* | 0.75917298 | 0.001043676 | 0.002427703 | 1 |
| T2DM | LEC | *UBR2* | 0.734050572 | 0.0113313 | 0.017232676 | 1 |
| T2DM | LEC | *HSF2* | 1.282188276 | 0.000485166 | 0.001436081 | 1 |
| T2DM | LEC | *NR1H3* | 1.34211048 | 0.000293569 | 0.001109196 | 1 |
| T2DM | LEC | *TYMP* | 0.729159607 | 0.000192733 | 0.000946191 | 1 |
| T2DM | LEC | *KPNA6* | 0.983760683 | 0.000331381 | 0.001173744 | 1 |
| T2DM | LEC | *VIM* | 0.599584558 | 0.031266555 | 0.042575879 | 1 |
| T2DM | LEC | *FAS* | 0.66449386 | 0.000120338 | 0.000908725 | 0.756565389 |
| T2DM | LEC | *RNASET2* | 0.675250733 | 0.000408744 | 0.001307728 | 1 |
| T2DM | LEC | *CD44* | 0.512655385 | 0.000270175 | 0.001066753 | 1 |
| T2DM | LEC | *MIPEP* | 0.8573301 | 0.000360567 | 0.001224756 | 1 |
| T2DM | LEC | *INSRR* | 0.96359083 | 0.017668814 | 0.025501339 | 1 |
| T2DM | LEC | *B4GALT7* | -0.789270368 | 0.002076305 | 0.004127009 | 1 |
| T2DM | LEC | *VRK2* | 0.75243211 | 0.022250988 | 0.031365911 | 1 |
| T2DM | LEC | *SNX1* | 0.678709499 | 0.003297033 | 0.005982236 | 1 |
| T2DM | LEC | *ARNTL2* | -0.818451722 | 0.001526685 | 0.003240468 | 1 |
| T2DM | LEC | *SLC39A9* | 0.747334431 | 0.001176848 | 0.002649067 | 1 |
| T2DM | LEC | *ANK1* | 0.451720343 | 8.42E-05 | 0.000908725 | 0.529446516 |
| T2DM | LEC | *RABEP1* | 0.783321194 | 0.001032115 | 0.002411901 | 1 |
| T2DM | LEC | *BAK1* | 0.846079901 | 2.66E-06 | 0.000388325 | 0.01669798 |
| T2DM | LEC | *GRN* | 0.751559785 | 0.000264632 | 0.001053693 | 1 |
| T2DM | LEC | *ARID4A* | 0.853828889 | 0.004640834 | 0.008000254 | 1 |
| T2DM | LEC | *ABCF2* | 0.978454381 | 8.49E-05 | 0.000908725 | 0.533924 |
| T2DM | LEC | *FUT8* | 0.717248938 | 0.000596482 | 0.001654204 | 1 |
| T2DM | LEC | *PIAS1* | 0.722684166 | 0.004335523 | 0.007548445 | 1 |
| T2DM | LEC | *SLC4A7* | 0.539007212 | 0.000240135 | 0.00100947 | 1 |
| T2DM | LEC | *ASTE1* | 0.74107334 | 0.016380125 | 0.023855932 | 1 |
| T2DM | LEC | *MYOC* | 0.459712521 | 4.87E-05 | 0.000908725 | 0.30628813 |
| T2DM | LEC | *VCL* | 0.614587156 | 0.009101904 | 0.01425247 | 1 |
| T2DM | LEC | *ADSS* | 1.04138685 | 0.001718002 | 0.003568245 | 1 |
| T2DM | LEC | *STAP1* | 1.144007455 | 0.001916675 | 0.003894678 | 1 |
| T2DM | LEC | *TBC1D23* | 0.873231118 | 0.00472477 | 0.008124899 | 1 |
| T2DM | LEC | *CUL3* | 0.840797456 | 0.001812151 | 0.003717127 | 1 |

| | | | | | | |
|---|---|---|---|---|---|---|
| T2DM | LEC | *MYOM2* | 0.656341845 | 0.000160591 | 0.000910705 | 1 |
| T2DM | LEC | *NSUN2* | 0.880733425 | 0.003645677 | 0.006505818 | 1 |
| T2DM | LEC | *AGA* | 1.306344427 | 0.000397134 | 0.001287664 | 1 |
| T2DM | LEC | *BOD1L1* | 0.703440228 | 0.018832625 | 0.027044475 | 1 |
| T2DM | LEC | *MAT2B* | 1.284628829 | 0.000427979 | 0.001336216 | 1 |
| T2DM | LEC | *CLEC16A* | 0.778509716 | 0.00064195 | 0.001733394 | 1 |
| T2DM | LEC | *SKIV2L2* | 0.840328821 | 0.002283522 | 0.004447492 | 1 |
| T2DM | LEC | *DNAH5* | 1.34287896 | 0.003711634 | 0.00660675 | 1 |
| T2DM | LEC | *C6* | 0.340704663 | 0.00022629 | 0.000992757 | 1 |
| T2DM | LEC | *RAI14* | -0.225570269 | 0.000204112 | 0.000964732 | 1 |
| T2DM | LEC | *PHLPP2* | 0.997167278 | 0.001336148 | 0.002925936 | 1 |
| T2DM | LEC | *SPDL1* | -0.673598225 | 0.002863803 | 0.005331237 | 1 |
| T2DM | LEC | *PQLC2* | 1.518414336 | 0.000106025 | 0.000908725 | 0.666577177 |
| T2DM | LEC | *INPP4A* | 0.643761609 | 0.013079294 | 0.019541236 | 1 |
| T2DM | LEC | *RAB27B* | 0.854962532 | 3.16E-06 | 0.000432123 | 0.019877643 |
| T2DM | LEC | *PSMA4* | 1.020083795 | 0.00122299 | 0.00272472 | 1 |
| T2DM | LEC | *THAP3* | 0.740902042 | 0.000283612 | 0.001087982 | 1 |
| T2DM | LEC | *TDP1* | 1.231535539 | 0.004148467 | 0.007244836 | 1 |
| T2DM | LEC | *SPATA7* | 0.833928491 | 0.000860824 | 0.002114889 | 1 |
| T2DM | LEC | *MED17* | 1.503974346 | 4.59E-05 | 0.000908725 | 0.288544889 |
| T2DM | LEC | *RETSAT* | 0.432430621 | 0.000752577 | 0.001926061 | 1 |
| T2DM | LEC | *CAPG* | 0.495236009 | 0.000533749 | 0.001530525 | 1 |
| T2DM | LEC | *AP2S1* | 1.247077371 | 0.002047607 | 0.00408717 | 1 |
| T2DM | LEC | *LCP2* | 0.48842253 | 0.000143105 | 0.000908725 | 0.899700061 |
| T2DM | LEC | *CUL7* | 0.612613996 | 0.000231628 | 0.001001608 | 1 |
| T2DM | LEC | *EPHA3* | 0.768581556 | 0.000253659 | 0.001032203 | 1 |
| T2DM | LEC | *DSG2* | 0.515577663 | 8.28E-05 | 0.000908725 | 0.520761212 |
| T2DM | LEC | *WDR37* | 0.788459929 | 0.000741152 | 0.001906556 | 1 |
| T2DM | LEC | *YTHDC2* | 0.778037403 | 0.024368543 | 0.034075852 | 1 |
| T2DM | LEC | *ATP6V1H* | 0.894509551 | 0.002303632 | 0.004474185 | 1 |
| T2DM | LEC | *CP* | 0.53734303 | 0.000111967 | 0.000908725 | 0.703939117 |
| T2DM | LEC | *ANO2* | -0.237618504 | 0.000332243 | 0.001173862 | 1 |
| T2DM | LEC | *TSPAN17* | 0.57500935 | 1.30E-05 | 0.000893669 | 0.081958023 |
| T2DM | LEC | *CC2D2A* | 0.823737653 | 0.001438532 | 0.003095348 | 1 |
| T2DM | LEC | *RRM2B* | 1.089094866 | 0.003506997 | 0.006294173 | 1 |
| T2DM | LEC | *SNX29* | 0.587615502 | 0.000264958 | 0.001053693 | 1 |
| T2DM | LEC | *LMO3* | 0.427034793 | 0.000215923 | 0.000979981 | 1 |
| T2DM | LEC | *CELF2* | 0.522443039 | 0.000385214 | 0.001268529 | 1 |
| T2DM | LEC | *ADAMTS6* | -0.573508808 | 0.031554421 | 0.042912102 | 1 |
| T2DM | LEC | *UTS2* | 0.529538672 | 0.000553342 | 0.001565645 | 1 |
| T2DM | LEC | *EPN3* | 5.000453406 | 5.51E-06 | 0.000549402 | 0.034612299 |
| T2DM | LEC | *RFC2* | 0.69487587 | 6.48E-05 | 0.000908725 | 0.407530662 |
| T2DM | LEC | *JKAMP* | 0.736862371 | 0.001448755 | 0.003113956 | 1 |
| T2DM | LEC | *DKK3* | -0.104147969 | 0.00015849 | 0.000910705 | 0.996427432 |
| T2DM | LEC | *ARHGEF5* | 2.195847289 | 0.005670134 | 0.009511241 | 1 |
| T2DM | LEC | *NFE2L3* | 1.485730735 | 0.000401568 | 0.001294696 | 1 |
| T2DM | LEC | *MCUR1* | 0.891761914 | 0.000968273 | 0.002298049 | 1 |
| T2DM | LEC | *LIMA1* | 0.309836209 | 0.008748302 | 0.0137708 | 1 |
| T2DM | LEC | *PTGER3* | 0.496019973 | 0.00018788 | 0.000936299 | 1 |
| T2DM | LEC | *MAPK9* | 1.044748886 | 0.000144448 | 0.000908725 | 0.908141601 |

| | | | | | | |
|---|---|---|---|---|---|---|
| T2DM | LEC | *FAM160A2* | 0.63870696 | 0.035705793 | 0.047986814 | 1 |
| T2DM | LEC | *HERPUD1* | 0.894987291 | 0.001016664 | 0.002384605 | 1 |
| T2DM | LEC | *PIK3CB* | 0.642539906 | 0.030845109 | 0.042047529 | 1 |
| T2DM | LEC | *CYBA* | 0.786556654 | 0.000246617 | 0.001019088 | 1 |
| T2DM | LEC | *THOC3* | 0.764587066 | 0.00011832 | 0.000908725 | 0.743879317 |
| T2DM | LEC | *RRP12* | 0.600801474 | 8.01E-05 | 0.000908725 | 0.50372535 |
| T2DM | LEC | *ALX4* | 2.669690396 | 0.000500161 | 0.001463103 | 1 |
| T2DM | LEC | *FOXN3* | 0.639256507 | 0.000842397 | 0.0020815 | 1 |
| T2DM | LEC | *AKR7A2* | 1.289243245 | 0.001498252 | 0.003194871 | 1 |
| T2DM | LEC | *MRTO4* | 1.181784758 | 0.014029985 | 0.020803423 | 1 |
| T2DM | LEC | *MCF2L2* | 1.057799281 | 0.006169517 | 0.010201933 | 1 |
| T2DM | LEC | *ANAPC4* | 1.103720494 | 0.002141277 | 0.004234731 | 1 |
| T2DM | LEC | *LY75* | 1.055274847 | 0.000784407 | 0.001980697 | 1 |
| T2DM | LEC | *OPN3* | 0.968101593 | 0.000766115 | 0.001948143 | 1 |
| T2DM | LEC | *SDCCAG8* | 0.638952055 | 0.001246101 | 0.00276535 | 1 |
| T2DM | LEC | *TBC1D22A* | 0.735241218 | 0.006940208 | 0.01129513 | 1 |
| T2DM | LEC | *PLEKHH1* | 0.709627168 | 0.006447541 | 0.010598452 | 1 |
| T2DM | LEC | *CHRDL2* | -0.413114582 | 0.000741012 | 0.001906556 | 1 |
| T2DM | LEC | *GALC* | 0.801275228 | 0.001693849 | 0.003532083 | 1 |
| T2DM | LEC | *GINM1* | 0.996998866 | 0.005022905 | 0.008590589 | 1 |
| T2DM | LEC | *MRPL43* | 0.841661952 | 0.007204253 | 0.011670482 | 1 |
| T2DM | LEC | *ITIH4* | 0.807686588 | 5.44E-08 | 1.90E-05 | 0.000342138 |
| T2DM | LEC | *TRAF1* | 0.867776776 | 0.000754551 | 0.001929184 | 1 |
| T2DM | LEC | *IL17RB* | -1.625605506 | 0.000133138 | 0.000908725 | 0.837036789 |
| T2DM | LEC | *TRAF3IP2* | 1.080172624 | 0.007374211 | 0.011905923 | 1 |
| T2DM | LEC | *DCBLD2* | 0.881158264 | 0.00467983 | 0.008058639 | 1 |
| T2DM | LEC | *GDI2* | 0.919657971 | 0.004359079 | 0.007578963 | 1 |
| T2DM | LEC | *TMCC3* | 1.008206113 | 0.008331105 | 0.01320667 | 1 |
| T2DM | LEC | *PITHD1* | 0.733586753 | 0.000108103 | 0.000908725 | 0.679642562 |
| T2DM | LEC | *ATP11B* | 0.648901913 | 0.010317395 | 0.015910096 | 1 |
| T2DM | LEC | *CDK14* | 0.648416329 | 0.000418369 | 0.001322531 | 1 |
| T2DM | LEC | *ZC3H11A* | 1.916603833 | 0.00017714 | 0.000918726 | 1 |
| T2DM | LEC | *RIOK2* | 1.16231775 | 0.012080414 | 0.018208957 | 1 |
| T2DM | LEC | *DNAJC25* | -0.775506702 | 0.000925219 | 0.002222312 | 1 |
| T2DM | LEC | *PIGV* | 0.583094768 | 0.033409898 | 0.045132796 | 1 |
| T2DM | LEC | *QSER1* | 0.800028558 | 0.000380502 | 0.00125962 | 1 |
| T2DM | LEC | *SFSWAP* | 0.87640367 | 0.003212555 | 0.00584752 | 1 |
| T2DM | LEC | *TNK2* | 0.733549039 | 0.028976229 | 0.039793143 | 1 |
| T2DM | LEC | *VMP1* | 0.712576007 | 0.00091693 | 0.00221423 | 1 |
| T2DM | LEC | *EIF4B* | 0.644144596 | 0.013032174 | 0.019489363 | 1 |
| T2DM | LEC | *SPHK2* | 0.783222987 | 7.28E-05 | 0.000908725 | 0.457547709 |
| T2DM | LEC | *ISOC2* | 1.431487484 | 0.000250413 | 0.001023586 | 1 |
| T2DM | LEC | *EPN1* | 1.16324075 | 0.000316196 | 0.001151139 | 1 |
| T2DM | LEC | *ADCK1* | 0.776883965 | 0.00016219 | 0.000910705 | 1 |
| T2DM | LEC | *CASP8* | 0.866027069 | 0.00013551 | 0.000908725 | 0.851949873 |
| T2DM | LEC | *DMRT3* | 0.625320939 | 8.09E-05 | 0.000908725 | 0.508621737 |
| T2DM | LEC | *ATP2C2* | -0.923082351 | 0.001177415 | 0.002649395 | 1 |
| T2DM | LEC | *TAF2* | 0.886898682 | 0.007328948 | 0.011845012 | 1 |
| T2DM | LEC | *TMEM161A* | 0.899302132 | 0.000126537 | 0.000908725 | 0.795536164 |
| T2DM | LEC | *LPAR2* | 0.492377927 | 4.49E-05 | 0.000908725 | 0.282161736 |

| | | | | | | |
|---|---|---|---|---|---|---|
| T2DM | LEC | *SUGP2* | 1.102340072 | 0.000319227 | 0.00115467 | 1 |
| T2DM | LEC | *ABCA7* | 0.658432582 | 0.000277631 | 0.00108013 | 1 |
| T2DM | LEC | *BTBD1* | 1.130110517 | 0.001187214 | 0.002664263 | 1 |
| T2DM | LEC | *CHI3L2* | 0.675998231 | 8.15E-05 | 0.000908725 | 0.512640745 |
| T2DM | LEC | *PMS1* | 0.877869704 | 0.000865872 | 0.002122726 | 1 |
| T2DM | LEC | *NTHL1* | -0.691383267 | 0.0020671 | 0.004116521 | 1 |
| T2DM | LEC | *IPO5* | 1.139281931 | 0.001055224 | 0.002445543 | 1 |
| T2DM | LEC | *MCM10* | 0.527344564 | 0.005027788 | 0.008596601 | 1 |
| T2DM | LEC | *ANKRD44* | 0.640655684 | 0.000297443 | 0.00111609 | 1 |
| T2DM | LEC | *ZC3H15* | 0.801485687 | 0.001123633 | 0.002563238 | 1 |
| T2DM | LEC | *MAP2K4* | 0.864427864 | 0.003469808 | 0.006235577 | 1 |
| T2DM | LEC | *TMEM206* | 1.054575895 | 0.000928433 | 0.002227034 | 1 |
| T2DM | LEC | *SEC61A2* | 1.108637963 | 0.000606771 | 0.001670213 | 1 |
| T2DM | LEC | *ASB1* | 0.703317676 | 0.000424704 | 0.0013333 | 1 |
| T2DM | LEC | *TBC1D1* | 0.898227217 | 0.012760803 | 0.01911536 | 1 |
| T2DM | LEC | *TIE1* | -0.202409679 | 0.000183971 | 0.000929091 | 1 |
| T2DM | LEC | *NFYC* | 0.925137949 | 0.006129042 | 0.010151024 | 1 |
| T2DM | LEC | *ZMYND12* | 0.576359142 | 0.000113289 | 0.000908725 | 0.712250781 |
| T2DM | LEC | *NGEF* | 1.187688191 | 0.000112545 | 0.000908725 | 0.707572321 |
| T2DM | LEC | *CD84* | 0.598353757 | 4.86E-05 | 0.000908725 | 0.30536809 |
| T2DM | LEC | *ZNRD1* | 0.868034443 | 0.003741428 | 0.006652251 | 1 |
| T2DM | LEC | *EML1* | 0.554994941 | 0.001375899 | 0.002994506 | 1 |
| T2DM | LEC | *ZFAT* | 0.473709687 | 0.000130653 | 0.000908725 | 0.821417176 |
| T2DM | LEC | *MTFR1* | 0.74614439 | 0.006146452 | 0.010169143 | 1 |
| T2DM | LEC | *STAG3* | 1.167159848 | 0.00106892 | 0.002469789 | 1 |
| T2DM | LEC | *MYO9A* | 0.657343277 | 0.013617249 | 0.020243946 | 1 |
| T2DM | LEC | *SP100* | 0.691100524 | 0.028722846 | 0.039479784 | 1 |
| T2DM | LEC | *STOML1* | 1.653852374 | 0.000278781 | 0.00108013 | 1 |
| T2DM | LEC | *TP53BP1* | 0.99989394 | 0.002909343 | 0.005393159 | 1 |
| T2DM | LEC | *RRP15* | 0.514960246 | 0.004808622 | 0.008251038 | 1 |
| T2DM | LEC | *SYT1* | -0.255893254 | 7.16E-05 | 0.000908725 | 0.450188738 |
| T2DM | LEC | *ROGDI* | 0.593057484 | 0.000106031 | 0.000908725 | 0.666618215 |
| T2DM | LEC | *RASSF1* | 0.90225992 | 0.024427748 | 0.034143453 | 1 |
| T2DM | LEC | *FGFR3* | -0.176718741 | 0.008697713 | 0.013704892 | 1 |
| T2DM | LEC | *IFI35* | 0.769316358 | 3.50E-05 | 0.000908725 | 0.219914065 |
| T2DM | LEC | *HEATR6* | 0.912232384 | 0.000151765 | 0.000910197 | 0.954146608 |
| T2DM | LEC | *SRBD1* | 0.821992359 | 0.001345927 | 0.002942227 | 1 |
| T2DM | LEC | *CYFIP1* | 0.791250624 | 0.0220014 | 0.031055861 | 1 |
| T2DM | LEC | *PSME4* | 0.781658498 | 0.006664621 | 0.010914423 | 1 |
| T2DM | LEC | *PITX1* | 2.417035977 | 0.000149581 | 0.000910197 | 0.940416514 |
| T2DM | LEC | *SDK2* | -0.824810291 | 0.000437658 | 0.001345504 | 1 |
| T2DM | LEC | *NUCKS1* | 0.775802656 | 0.027960318 | 0.03858352 | 1 |
| T2DM | LEC | *DNAJA2* | 0.999514054 | 0.000324651 | 0.001162346 | 1 |
| T2DM | LEC | *KCNAB2* | 0.533207958 | 0.000697778 | 0.00183247 | 1 |
| T2DM | LEC | *GAL* | -0.594914022 | 0.000182549 | 0.000929091 | 1 |
| T2DM | LEC | *CLEC2D* | 0.703707313 | 0.002330381 | 0.004514979 | 1 |
| T2DM | LEC | *NEDD4* | -0.528792423 | 0.021299365 | 0.03017895 | 1 |
| T2DM | LEC | *PIGB* | 0.824450155 | 0.008235493 | 0.013094725 | 1 |
| T2DM | LEC | *GNB5* | -0.572514034 | 0.014128209 | 0.020934256 | 1 |
| T2DM | LEC | *PFN2* | 0.988324392 | 0.001465764 | 0.003144067 | 1 |

| | | | | | | |
|---|---|---|---|---|---|---|
| T2DM | LEC | *DAPP1* | 0.618848271 | 0.000339058 | 0.001186892 | 1 |
| T2DM | LEC | *FGF10* | 0.687424733 | 0.000558494 | 0.001575965 | 1 |
| T2DM | LEC | *TMEM260* | 0.782077892 | 0.000411501 | 0.001311277 | 1 |
| T2DM | LEC | *CLTCL1* | 0.927255135 | 0.008156151 | 0.012991568 | 1 |
| T2DM | LEC | *MNT* | 0.399379763 | 0.000120725 | 0.000908725 | 0.758995387 |
| T2DM | LEC | *ZXDC* | 0.785815083 | 0.022482728 | 0.031671277 | 1 |
| T2DM | LEC | *POLB* | -0.801983191 | 0.002699894 | 0.005080585 | 1 |
| T2DM | LEC | *WIPI1* | 0.865968041 | 0.000302705 | 0.001124766 | 1 |
| T2DM | LEC | *NDST1* | 1.056132752 | 0.00475766 | 0.008173126 | 1 |
| T2DM | LEC | *ASNS* | 1.136650439 | 1.42E-05 | 0.000902248 | 0.08938897 |
| T2DM | LEC | *ST6GALNAC2* | 0.456397449 | 0.000109022 | 0.000908725 | 0.685419572 |
| T2DM | LEC | *EIF2B3* | 0.643113937 | 0.0002852 | 0.001090506 | 1 |
| T2DM | LEC | *CAMK2A* | 1.954621939 | 0.000220957 | 0.00098409 | 1 |
| T2DM | LEC | *OSBPL3* | 0.825479404 | 0.002464668 | 0.004718443 | 1 |
| T2DM | LEC | *RAD18* | 1.080650917 | 0.006782005 | 0.011080682 | 1 |
| T2DM | LEC | *SNX13* | 0.819041951 | 0.005389495 | 0.009103642 | 1 |
| T2DM | LEC | *VASH1* | 0.83168282 | 0.000408353 | 0.001307186 | 1 |
| T2DM | LEC | *TRIP13* | 1.019745771 | 0.000113419 | 0.000908725 | 0.713063346 |
| T2DM | LEC | *TCF3* | -1.027658738 | 0.003240982 | 0.005890736 | 1 |
| T2DM | LEC | *HLTF* | 0.581656798 | 0.01078985 | 0.016545314 | 1 |
| T2DM | LEC | *CYBRD1* | 0.745274118 | 0.000169015 | 0.000910705 | 1 |
| T2DM | LEC | *AFF4* | 0.694053798 | 0.012017295 | 0.018131206 | 1 |
| T2DM | LEC | *TRHDE* | 0.331024873 | 0.000127143 | 0.000908725 | 0.799345842 |
| T2DM | LEC | *NFATC3* | 0.820769188 | 0.008195831 | 0.013048162 | 1 |
| T2DM | LEC | *TRNT1* | 1.006593897 | 0.001963875 | 0.003957334 | 1 |
| T2DM | LEC | *ACADVL* | 0.622178299 | 0.000219268 | 0.00098409 | 1 |
| T2DM | LEC | *NDE1* | 0.821047037 | 0.003100748 | 0.005674 | 1 |
| T2DM | LEC | *AP1M1* | 0.787907615 | 0.006938459 | 0.01129513 | 1 |
| T2DM | LEC | *XRCC1* | 0.671762122 | 0.000112045 | 0.000908725 | 0.704426794 |
| T2DM | LEC | *MOV10L1* | -0.179890641 | 9.78E-05 | 0.000908725 | 0.614692456 |
| T2DM | LEC | *TP63* | -0.755941484 | 0.005632467 | 0.009468268 | 1 |
| T2DM | LEC | *ALPK1* | 0.705153545 | 0.000252271 | 0.001027887 | 1 |
| T2DM | LEC | *LLGL2* | 0.562987137 | 8.12E-05 | 0.000908725 | 0.510404111 |
| T2DM | LEC | *PDE8A* | 0.741475481 | 0.001090399 | 0.00250469 | 1 |
| T2DM | LEC | *NLE1* | 0.654407156 | 7.38E-05 | 0.000908725 | 0.464135427 |
| T2DM | LEC | *SDHA* | 1.267510538 | 4.69E-05 | 0.000908725 | 0.294787617 |
| T2DM | LEC | *GSDMB* | 0.452793211 | 0.000327073 | 0.00116703 | 1 |
| T2DM | LEC | *ADAM11* | 0.501478525 | 0.019838586 | 0.028365974 | 1 |
| T2DM | LEC | *PICALM* | 0.647402134 | 0.001217759 | 0.002719735 | 1 |
| T2DM | LEC | *GLI2* | 0.356711669 | 0.000476303 | 0.001418531 | 1 |
| T2DM | LEC | *CLNS1A* | 0.875844928 | 0.000374576 | 0.00124799 | 1 |
| T2DM | LEC | *ATP2A3* | 0.4782379 | 0.000106372 | 0.000908725 | 0.668761984 |
| T2DM | LEC | *CA12* | 0.339563467 | 0.000264917 | 0.001053693 | 1 |
| T2DM | LEC | *NTN4* | -0.659248554 | 0.00286142 | 0.005328716 | 1 |
| T2DM | LEC | *LMAN1* | 0.914082621 | 0.020750328 | 0.02949521 | 1 |
| T2DM | LEC | *TIPIN* | 0.391954779 | 0.019388539 | 0.027754041 | 1 |
| T2DM | LEC | *SRI* | 0.95746453 | 0.000343544 | 0.001193551 | 1 |
| T2DM | LEC | *TTC38* | 0.883509004 | 0.000977534 | 0.002312173 | 1 |
| T2DM | LEC | *GRAMD4* | 0.928403207 | 0.000326229 | 0.001165342 | 1 |
| T2DM | LEC | *CELSR1* | -0.123533557 | 0.000140515 | 0.000908725 | 0.883416196 |

| | | | | | | |
|---|---|---|---|---|---|---|
| T2DM | LEC | *ADD2* | -1.038236058 | 0.000423452 | 0.00133112 | 1 |
| T2DM | LEC | *MARK3* | 0.538701273 | 0.018135895 | 0.026127491 | 1 |
| T2DM | LEC | *SLC25A3* | 1.165776735 | 0.000487032 | 0.00143852 | 1 |
| T2DM | LEC | *FSCN1* | -0.353113411 | 0.00019363 | 0.000948095 | 1 |
| T2DM | LEC | *MOCOS* | 0.568628416 | 0.000181419 | 0.000928057 | 1 |
| T2DM | LEC | *PLD1* | -0.617791955 | 0.010055548 | 0.015525352 | 1 |
| T2DM | LEC | *SEC31B* | 0.430252982 | 0.001243716 | 0.002762982 | 1 |
| T2DM | LEC | *TUBA3D* | 1.430388852 | 0.00017086 | 0.000910705 | 1 |
| T2DM | LEC | *MKRN2* | 1.040391215 | 0.0159614 | 0.023309947 | 1 |
| T2DM | LEC | *MCM6* | 1.108191106 | 6.82E-05 | 0.000908725 | 0.429076917 |
| T2DM | LEC | *FMO4* | 1.077052351 | 0.000918266 | 0.002214476 | 1 |
| T2DM | LEC | *RGS11* | 0.403681626 | 0.005863642 | 0.009783631 | 1 |
| T2DM | LEC | *TPD52* | 0.90666067 | 0.002670087 | 0.005038066 | 1 |
| T2DM | LEC | *GPATCH1* | 1.059150683 | 0.000283757 | 0.001087982 | 1 |
| T2DM | LEC | *ICAM3* | 1.005898127 | 0.000176322 | 0.000918425 | 1 |
| T2DM | LEC | *NT5C2* | 0.826562916 | 0.005692389 | 0.009543481 | 1 |
| T2DM | LEC | *CAMSAP3* | -1.168461594 | 0.000146575 | 0.000908725 | 0.921519843 |
| T2DM | LEC | *XAB2* | -1.943933448 | 9.46E-05 | 0.000908725 | 0.594800194 |
| T2DM | LEC | *STXBP2* | 1.793791185 | 2.03E-06 | 0.000326655 | 0.01273953 |
| T2DM | LEC | *MAP2K7* | 1.030441425 | 0.000354999 | 0.00121562 | 1 |
| T2DM | LEC | *NMRK2* | 2.089041383 | 0.002724437 | 0.005117578 | 1 |
| T2DM | LEC | *CTTNBP2* | -1.020653926 | 0.000129724 | 0.000908725 | 0.815577239 |
| T2DM | LEC | *TM9SF3* | 0.819404748 | 0.011251586 | 0.017136319 | 1 |
| T2DM | LEC | *DNAJC10* | 0.901796251 | 0.024415212 | 0.03413352 | 1 |
| T2DM | LEC | *APBB1IP* | 0.495542732 | 0.00014169 | 0.000908725 | 0.890805001 |
| T2DM | LEC | *LRCH4* | 0.402477789 | 0.015799076 | 0.023105092 | 1 |
| T2DM | LEC | *SIRT6* | 1.876831202 | 0.002886363 | 0.005360875 | 1 |
| T2DM | LEC | *CAPZB* | 0.762729243 | 0.009555256 | 0.014858742 | 1 |
| T2DM | LEC | *GPR137B* | 0.577607392 | 0.00440012 | 0.007631337 | 1 |
| T2DM | LEC | *NAALAD2* | 0.91884709 | 0.000160127 | 0.000910705 | 1 |
| T2DM | LEC | *FBLN1* | 0.465741679 | 7.79E-05 | 0.000908725 | 0.489547379 |
| T2DM | LEC | *PIAS2* | 0.785470396 | 0.004251356 | 0.007408058 | 1 |
| T2DM | LEC | *AMPH* | 0.479033963 | 0.000560174 | 0.001579288 | 1 |
| T2DM | LEC | *MCCC1* | 0.577357688 | 0.000137755 | 0.000908725 | 0.86606797 |
| T2DM | LEC | *FAP* | 0.294983863 | 0.000118922 | 0.000908725 | 0.747661644 |
| T2DM | LEC | *ACER3* | 0.55205095 | 0.006097675 | 0.010104398 | 1 |
| T2DM | LEC | *N4BP2* | 0.875164532 | 0.000914836 | 0.002211224 | 1 |
| T2DM | LEC | *SYNJ2* | 0.913579399 | 0.000479539 | 0.001424968 | 1 |
| T2DM | LEC | *ZCWPW1* | 0.649831307 | 6.45E-05 | 0.000908725 | 0.405232042 |
| T2DM | LEC | *FGF20* | -4.407692687 | 0.00022097 | 0.00098409 | 1 |
| T2DM | LEC | *PCM1* | 0.70375008 | 0.00434877 | 0.007567317 | 1 |
| T2DM | LEC | *MYH7B* | 1.642904563 | 0.000616275 | 0.001691929 | 1 |
| T2DM | LEC | *FKBP7* | 0.560511629 | 0.007231123 | 0.011704961 | 1 |
| T2DM | LEC | *OSBPL6* | -0.73371959 | 0.001501934 | 0.003199816 | 1 |
| T2DM | LEC | *XRCC5* | 0.805687048 | 0.009253571 | 0.014453963 | 1 |
| T2DM | LEC | *REXO1* | 1.15862981 | 0.000507848 | 0.0014768 | 1 |
| T2DM | LEC | *SAR1A* | 0.695689533 | 0.000246166 | 0.00101886 | 1 |
| T2DM | LEC | *RAPGEF3* | 0.29085646 | 0.03642492 | 0.048848864 | 1 |
| T2DM | LEC | *FDFT1* | 0.93640053 | 0.000421981 | 0.001328489 | 1 |
| T2DM | LEC | *SCGN* | 3.298604119 | 9.20E-05 | 0.000908725 | 0.578429645 |

| | | | | | | |
|---|---|---|---|---|---|---|
| T2DM | LEC | *DDX1* | 0.96281673 | 0.002594837 | 0.004911782 | 1 |
| T2DM | LEC | *DNM2* | 0.883241172 | 0.001353556 | 0.002954794 | 1 |
| T2DM | LEC | *RIMS1* | 0.877497644 | 0.000185324 | 0.000930618 | 1 |
| T2DM | LEC | *DDX43* | 0.664453847 | 0.002176104 | 0.004286532 | 1 |
| T2DM | LEC | *DCT* | 0.62326719 | 0.000109509 | 0.000908725 | 0.688480924 |
| T2DM | LEC | *SLC35C2* | 0.877706019 | 0.000285216 | 0.001090506 | 1 |
| T2DM | LEC | *SCTR* | 0.406994637 | 0.000104256 | 0.000908725 | 0.655457087 |
| T2DM | LEC | *RIF1* | 0.681103559 | 0.009268332 | 0.014473424 | 1 |
| T2DM | LEC | *KCNN2* | -2.370734396 | 0.000109704 | 0.000908725 | 0.689710838 |
| T2DM | LEC | *PSEN1* | 0.766717501 | 0.002557684 | 0.00485512 | 1 |
| T2DM | LEC | *CPOX* | 1.042012902 | 0.003972011 | 0.006975994 | 1 |
| T2DM | LEC | *MOK* | 1.063732192 | 0.000801444 | 0.002011447 | 1 |
| T2DM | LEC | *RSBN1* | 0.870071096 | 0.004571363 | 0.007895649 | 1 |
| T2DM | LEC | *COL4A4* | 0.355364826 | 0.000215216 | 0.000978911 | 1 |
| T2DM | LEC | *STK17B* | 0.600579207 | 0.00016613 | 0.000910705 | 1 |
| T2DM | LEC | *ZNF506* | -0.927124656 | 0.002068246 | 0.004117499 | 1 |
| T2DM | LEC | *KIAA0141* | -0.876950831 | 0.01154673 | 0.017522156 | 1 |
| T2DM | LEC | *PCDHGA2* | 1.158047265 | 0.008318206 | 0.013194242 | 1 |
| T2DM | LEC | *WDR70* | 0.71976257 | 0.002301604 | 0.004471627 | 1 |
| T2DM | LEC | *C5orf22* | 1.228217179 | 0.011865077 | 0.017944609 | 1 |
| T2DM | LEC | *GEMIN5* | 0.759714381 | 0.011149403 | 0.017013664 | 1 |
| T2DM | LEC | *ERC1* | 0.674338944 | 0.001373349 | 0.002991769 | 1 |
| T2DM | LEC | *DOPEY1* | 0.643177115 | 0.007801159 | 0.012502138 | 1 |
| T2DM | LEC | *ULK2* | 0.685082595 | 0.037053278 | 0.049649181 | 1 |
| T2DM | LEC | *PLOD1* | 1.003247809 | 0.000168582 | 0.000910705 | 1 |
| T2DM | LEC | *TDRD3* | 0.686506635 | 0.030542201 | 0.041661709 | 1 |
| T2DM | LEC | *PDS5B* | 0.720009873 | 0.021885913 | 0.030913668 | 1 |
| T2DM | LEC | *CYLD* | 0.866726913 | 0.00097054 | 0.002300824 | 1 |
| T2DM | LEC | *ZNF324* | 0.887458855 | 0.000152712 | 0.000910197 | 0.960098514 |
| T2DM | LEC | *ZNF586* | 0.941941736 | 0.001158089 | 0.00261903 | 1 |
| T2DM | LEC | *RPS5* | 1.04916405 | 0.007716102 | 0.012384654 | 1 |
| T2DM | LEC | *CHMP2B* | 0.819341124 | 0.001389756 | 0.003013935 | 1 |
| T2DM | LEC | *PPIE* | 0.945225601 | 0.001701679 | 0.003544883 | 1 |
| T2DM | LEC | *STARD7* | -0.769983134 | 0.029825647 | 0.040817119 | 1 |
| T2DM | LEC | *NOA1* | -0.833964012 | 0.000477499 | 0.001420746 | 1 |
| T2DM | LEC | *GSTP1* | 1.211047452 | 0.004711721 | 0.008104675 | 1 |
| T2DM | LEC | *WBP11* | 0.841798295 | 0.000211483 | 0.0009784 | 1 |
| T2DM | LEC | *COL16A1* | 0.428445784 | 0.001618728 | 0.003397977 | 1 |
| T2DM | LEC | *APOB* | 0.653837615 | 0.000270997 | 0.001067753 | 1 |
| T2DM | LEC | *NCOA1* | 0.689945358 | 0.00439736 | 0.007630748 | 1 |
| T2DM | LEC | *CD59* | -0.559629908 | 0.002148413 | 0.004246171 | 1 |
| T2DM | LEC | *CD82* | 1.168979159 | 8.37E-06 | 0.000673907 | 0.052597869 |
| T2DM | LEC | *TAF9* | -1.07641201 | 0.00154661 | 0.003275412 | 1 |
| T2DM | LEC | *FCN1* | 0.504702642 | 0.000353728 | 0.001215214 | 1 |
| T2DM | LEC | *SCAMP1* | 1.021270739 | 0.000534917 | 0.001531431 | 1 |
| T2DM | LEC | *HACE1* | 0.80090373 | 0.001150324 | 0.002606816 | 1 |
| T2DM | LEC | *SEH1L* | 1.011731753 | 0.000473515 | 0.001412904 | 1 |
| T2DM | LEC | *OVGP1* | -1.351652321 | 1.73E-05 | 0.000908725 | 0.108624686 |
| T2DM | LEC | *SLC25A24* | 0.858247091 | 0.010826636 | 0.016585541 | 1 |
| T2DM | LEC | *PILRA* | 0.323746202 | 1.99E-05 | 0.000908725 | 0.125066597 |

| | | | | | | |
|---|---|---|---|---|---|---|
| T2DM | LEC | *ABCB1* | 0.439993996 | 9.58E-05 | 0.000908725 | 0.602021326 |
| T2DM | LEC | *ZNF213* | 0.909922886 | 0.002486688 | 0.00474903 | 1 |
| T2DM | LEC | *RRN3* | 1.055850332 | 0.010931572 | 0.016729989 | 1 |
| T2DM | LEC | *EPS15* | 0.697983378 | 0.004199516 | 0.007327882 | 1 |
| T2DM | LEC | *ATG16L1* | 1.024258141 | 0.000108076 | 0.000908725 | 0.67947103 |
| T2DM | LEC | *USP40* | 0.778289532 | 0.003790727 | 0.006728487 | 1 |
| T2DM | LEC | *MAST2* | 0.803276878 | 0.002334158 | 0.004519511 | 1 |
| T2DM | LEC | *B4GALT1* | 0.815931226 | 0.000413825 | 0.001314663 | 1 |
| T2DM | LEC | *CHMP5* | 1.108492377 | 0.000195878 | 0.000953164 | 1 |
| T2DM | LEC | *FOLH1* | 0.428397038 | 0.003938508 | 0.006937909 | 1 |
| T2DM | LEC | *EIF2AK1* | 0.952959183 | 0.000105838 | 0.000908725 | 0.665402191 |
| T2DM | LEC | *EPDR1* | 0.806002257 | 0.000174195 | 0.000911117 | 1 |
| T2DM | LEC | *SEPHS1* | -0.782993122 | 0.013340614 | 0.01988441 | 1 |
| T2DM | LEC | *ITPKC* | 0.566497801 | 0.000135747 | 0.000908725 | 0.853442708 |
| T2DM | LEC | *CEACAM6* | -4.487622684 | 0.000912949 | 0.002208614 | 1 |
| T2DM | LEC | *TMED2* | 0.996140502 | 0.000418616 | 0.001322531 | 1 |
| T2DM | LEC | *ACOX3* | 0.884401987 | 0.026635016 | 0.036916743 | 1 |
| T2DM | LEC | *HSD17B14* | 0.442646642 | 0.000387781 | 0.00126979 | 1 |
| T2DM | LEC | *TRIP6* | 0.479810076 | 0.000290665 | 0.001103508 | 1 |
| T2DM | LEC | *ACHE* | -0.754746002 | 0.000173289 | 0.000910705 | 1 |
| T2DM | LEC | *PIGS* | 0.717527945 | 0.000875777 | 0.002140751 | 1 |
| T2DM | LEC | *ATXN7L3* | 0.695844303 | 8.20E-05 | 0.000908725 | 0.515586186 |
| T2DM | LEC | *PGS1* | 0.747381619 | 0.000354684 | 0.001215214 | 1 |
| T2DM | LEC | *LPCAT2* | 0.547906775 | 0.000365122 | 0.001235026 | 1 |
| T2DM | LEC | *TXNDC16* | 0.880064674 | 0.001761011 | 0.003637139 | 1 |
| T2DM | LEC | *GMCL1* | 1.009907463 | 0.010528431 | 0.016187881 | 1 |
| T2DM | LEC | *PTHLH* | 1.300055653 | 0.000354106 | 0.001215214 | 1 |
| T2DM | LEC | *PHACTR3* | 0.387162108 | 5.76E-05 | 0.000908725 | 0.362038832 |
| T2DM | LEC | *METTL2A* | 0.5246467 | 0.000712263 | 0.001861013 | 1 |
| T2DM | LEC | *GP6* | 0.798451271 | 0.000975531 | 0.002310484 | 1 |
| T2DM | LEC | *DDX18* | 0.995557812 | 0.002909306 | 0.005393159 | 1 |
| T2DM | LEC | *DOCK3* | 0.513732956 | 0.003709186 | 0.006604263 | 1 |
| T2DM | LEC | *C3orf18* | 0.699372289 | 1.53E-06 | 0.000282266 | 0.009597054 |
| T2DM | LEC | *NSFL1C* | 0.682532745 | 0.000179563 | 0.000923826 | 1 |
| T2DM | LEC | *CPXM1* | 0.350893269 | 0.00193893 | 0.003923416 | 1 |
| T2DM | LEC | *MAVS* | 1.007554838 | 0.000999458 | 0.002351644 | 1 |
| T2DM | LEC | *LZTS3* | 1.349431622 | 0.001196773 | 0.002682393 | 1 |
| T2DM | LEC | *TESC* | 1.244078558 | 0.000160817 | 0.000910705 | 1 |
| T2DM | LEC | *RBBP9* | 1.684670742 | 0.001656429 | 0.003465548 | 1 |
| T2DM | LEC | *ANAPC5* | 0.733602044 | 0.032030549 | 0.043474971 | 1 |
| T2DM | LEC | *SLC23A2* | 0.855503057 | 0.000747984 | 0.00191907 | 1 |
| T2DM | LEC | *TMEM230* | 0.907798705 | 0.000197545 | 0.000956093 | 1 |
| T2DM | LEC | *DZANK1* | -0.990414984 | 0.031016975 | 0.042263485 | 1 |
| T2DM | LEC | *TASP1* | 0.723323428 | 0.008684171 | 0.013686985 | 1 |
| T2DM | LEC | *OAS1* | 0.775175491 | 0.000146787 | 0.000908725 | 0.922851472 |
| T2DM | LEC | *KIF16B* | 0.844713383 | 0.00161786 | 0.003397289 | 1 |
| T2DM | LEC | *FUS* | 0.76168566 | 0.012144188 | 0.018291929 | 1 |
| T2DM | LEC | *FXYD3* | 1.850544743 | 0.000234437 | 0.001001608 | 1 |
| T2DM | LEC | *GANAB* | 1.060193229 | 0.003715725 | 0.006612161 | 1 |
| T2DM | LEC | *GMIP* | 0.592560241 | 0.000116049 | 0.000908725 | 0.72960205 |

| | | | | | | |
|---|---|---|---|---|---|---|
| T2DM | LEC | *NECAP1* | 0.812552894 | 0.000171638 | 0.000910705 | 1 |
| T2DM | LEC | *ANKRD24* | 0.512189725 | 8.25E-05 | 0.000908725 | 0.518641874 |
| T2DM | LEC | *GPATCH2L* | 0.81868111 | 0.003875054 | 0.006856872 | 1 |
| T2DM | LEC | *SLC9A1* | 0.810837104 | 0.000112594 | 0.000908725 | 0.707879089 |
| T2DM | LEC | *SPTLC1* | 0.77824972 | 0.001473347 | 0.003156024 | 1 |
| T2DM | LEC | *RGS1* | 0.510970289 | 0.000106304 | 0.000908725 | 0.668333997 |
| T2DM | LEC | *YPEL3* | 0.814961676 | 0.000278756 | 0.00108013 | 1 |
| T2DM | LEC | *MRPS33* | 0.624690802 | 0.000468842 | 0.001404291 | 1 |
| T2DM | LEC | *STRN4* | 1.24929553 | 0.000572733 | 0.001603699 | 1 |
| T2DM | LEC | *LYZ* | 0.403727967 | 5.60E-05 | 0.000908725 | 0.352016459 |
| T2DM | LEC | *MUL1* | 0.852752573 | 0.001996218 | 0.004005817 | 1 |
| T2DM | LEC | *GNPTG* | 0.685268597 | 0.000144051 | 0.000908725 | 0.905647555 |
| T2DM | LEC | *GOLGA3* | 0.854484816 | 0.000105929 | 0.000908725 | 0.665976439 |
| T2DM | LEC | *CERS4* | 1.050492512 | 0.007918531 | 0.012667635 | 1 |
| T2DM | LEC | *PDPR* | 0.92012417 | 0.000112927 | 0.000908725 | 0.709971938 |
| T2DM | LEC | *EXOC1* | 0.802443464 | 0.009026664 | 0.01415934 | 1 |
| T2DM | LEC | *NLRC4* | 0.546450815 | 0.000117601 | 0.000908725 | 0.739358478 |
| T2DM | LEC | *SLC26A4* | -0.100076646 | 0.000127878 | 0.000908725 | 0.803967229 |
| T2DM | LEC | *DLD* | 0.915077158 | 0.000297998 | 0.00111718 | 1 |
| T2DM | LEC | *TXNL1* | 0.984086082 | 0.000305855 | 0.001128721 | 1 |
| T2DM | LEC | *IL5RA* | -1.104068089 | 0.000213871 | 0.0009784 | 1 |
| T2DM | LEC | *ITGA6* | -0.300861305 | 0.000185983 | 0.000932435 | 1 |
| T2DM | LEC | *RAPGEF4* | -0.201320669 | 0.000208067 | 0.000972609 | 1 |
| T2DM | LEC | *SEL1L3* | 1.290296735 | 0.002511589 | 0.004790764 | 1 |
| T2DM | LEC | *TF* | 1.292834463 | 0.00063309 | 0.001718583 | 1 |
| T2DM | LEC | *MYO15A* | 1.887574574 | 4.20E-05 | 0.000908725 | 0.263978961 |
| T2DM | LEC | *ALKBH5* | 0.4939577 | 0.009371746 | 0.014613137 | 1 |
| T2DM | LEC | *NLRP1* | 0.638918656 | 0.00037071 | 0.001241526 | 1 |
| T2DM | LEC | *SPAG7* | 0.652779005 | 0.000996342 | 0.002345876 | 1 |
| T2DM | LEC | *TMEM101* | 1.491158265 | 0.008855689 | 0.013925461 | 1 |
| T2DM | LEC | *CMA1* | -0.779462575 | 0.000255377 | 0.001034507 | 1 |
| T2DM | LEC | *PPP2R3C* | 0.941834934 | 0.000624427 | 0.001705358 | 1 |
| T2DM | LEC | *OSGEP* | 1.083801491 | 0.001148313 | 0.002603477 | 1 |
| T2DM | LEC | *SLC22A17* | 0.8693075 | 0.009125388 | 0.014285686 | 1 |
| T2DM | LEC | *SCFD1* | 0.743655591 | 0.000345266 | 0.001196042 | 1 |
| T2DM | LEC | *RPGRIP1* | -0.660133237 | 0.004094154 | 0.007161922 | 1 |
| T2DM | LEC | *TOX4* | 0.87206936 | 0.000618923 | 0.001697248 | 1 |
| T2DM | LEC | *TGM1* | 0.749391045 | 0.010118111 | 0.01561811 | 1 |
| T2DM | LEC | *AGO1* | 0.61141739 | 0.000242991 | 0.001015577 | 1 |
| T2DM | LEC | *RFFL* | 0.602305456 | 0.000112414 | 0.000908725 | 0.706748413 |
| T2DM | LEC | *MFSD11* | 0.735768972 | 0.000668117 | 0.001781361 | 1 |
| T2DM | LEC | *NUP50* | 1.069155691 | 0.000188419 | 0.000937634 | 1 |
| T2DM | LEC | *COMT* | 0.897191829 | 0.000493416 | 0.001449584 | 1 |
| T2DM | LEC | *VNN3* | 1.9343839 | 3.15E-05 | 0.000908725 | 0.198337159 |
| T2DM | LEC | *SEC22C* | 1.115059934 | 0.008541377 | 0.013488982 | 1 |
| T2DM | LEC | *FMO2* | 0.708726468 | 0.000147547 | 0.000908725 | 0.927626398 |
| T2DM | LEC | *SUCO* | 0.876519266 | 0.001211362 | 0.002708334 | 1 |
| T2DM | LEC | *MSH2* | 1.155110604 | 0.002424031 | 0.004659089 | 1 |
| T2DM | LEC | *MAP3K1* | -0.444737908 | 0.000170341 | 0.000910705 | 1 |
| T2DM | LEC | *TMEM38B* | 1.459218632 | 0.003463735 | 0.006227195 | 1 |

| | | | | | | |
|---|---|---|---|---|---|---|
| T2DM | LEC | *CWF19L1* | -0.528732658 | 0.03612081 | 0.048503104 | 1 |
| T2DM | LEC | *IKZF5* | 0.852745514 | 0.000225752 | 0.000992757 | 1 |
| T2DM | LEC | *BLNK* | 0.417130291 | 0.000170381 | 0.000910705 | 1 |
| T2DM | LEC | *CREM* | 0.919953333 | 0.000737257 | 0.001903545 | 1 |
| T2DM | LEC | *TPSD1* | 3.236815465 | 3.86E-05 | 0.000908725 | 0.242825526 |
| T2DM | LEC | *ITPR3* | 1.137111081 | 0.01062045 | 0.016305438 | 1 |
| T2DM | LEC | *DSP* | -0.066149857 | 0.000192492 | 0.000946191 | 1 |
| T2DM | LEC | *IL12RB1* | 0.61018421 | 6.96E-05 | 0.000908725 | 0.437767255 |
| T2DM | LEC | *CDC7* | 0.511332332 | 0.011527217 | 0.017500993 | 1 |
| T2DM | LEC | *PCSK5* | 0.580703967 | 0.000297529 | 0.00111609 | 1 |
| T2DM | LEC | *NRP1* | 0.506112769 | 0.000255161 | 0.001034507 | 1 |
| T2DM | LEC | *PRTFDC1* | 0.523105093 | 0.00013763 | 0.000908725 | 0.865279514 |
| T2DM | LEC | *PALMD* | 0.727528082 | 0.000449534 | 0.001369955 | 1 |
| T2DM | LEC | *TSPAN15* | 0.844050712 | 0.000487134 | 0.00143852 | 1 |
| T2DM | LEC | *OCEL1* | 0.706597322 | 7.23E-05 | 0.000908725 | 0.454388724 |
| T2DM | LEC | *CATSPERG* | 0.651222268 | 0.000979872 | 0.00231596 | 1 |
| T2DM | LEC | *STX1B* | 0.552813621 | 0.001633218 | 0.003424963 | 1 |
| T2DM | LEC | *HSD3B7* | 0.621042163 | 0.000113594 | 0.000908725 | 0.714168296 |
| T2DM | LEC | *MARCH2* | 1.263076325 | 8.55E-05 | 0.000908725 | 0.537699879 |
| T2DM | LEC | *MTAP* | 0.861395641 | 0.000218694 | 0.00098409 | 1 |
| T2DM | LEC | *CEP170B* | 0.44845931 | 0.000276682 | 0.00108013 | 1 |
| T2DM | LEC | *POLR2E* | 0.910166538 | 0.00050817 | 0.001477052 | 1 |
| T2DM | LEC | *MADCAM1* | 0.91252568 | 0.000133396 | 0.000908725 | 0.838657559 |
| T2DM | LEC | *TRMT2A* | 0.636937724 | 0.00024687 | 0.001019088 | 1 |
| T2DM | LEC | *MED15* | 0.834513426 | 0.000803659 | 0.002014594 | 1 |
| T2DM | LEC | *SNAP29* | 0.637639077 | 0.016380795 | 0.023855932 | 1 |
| T2DM | LEC | *LZTR1* | 1.109887632 | 0.000492807 | 0.001449584 | 1 |
| T2DM | LEC | *CECR2* | 0.581906848 | 0.002299215 | 0.004468366 | 1 |
| T2DM | LEC | *P2RX6* | 0.49300943 | 0.00016753 | 0.000910705 | 1 |
| T2DM | LEC | *SLC7A4* | 1.494262936 | 0.000700303 | 0.001836115 | 1 |
| T2DM | LEC | *DDTL* | 0.546311925 | 0.000519582 | 0.001501892 | 1 |
| T2DM | LEC | *GGT5* | -0.602783059 | 0.000657195 | 0.001758954 | 1 |
| T2DM | LEC | *PPIL2* | 0.581761082 | 0.000301501 | 0.001123614 | 1 |
| T2DM | LEC | *UPB1* | 0.654496579 | 0.000773837 | 0.001960158 | 1 |
| T2DM | LEC | *YPEL1* | -0.842086231 | 0.001922558 | 0.00390159 | 1 |
| T2DM | LEC | *PPM1F* | -0.566042658 | 0.000555818 | 0.001569824 | 1 |
| T2DM | LEC | *TOP3B* | 0.652845358 | 0.000212128 | 0.0009784 | 1 |
| T2DM | LEC | *CRYBB3* | -0.26377023 | 0.000129337 | 0.000908725 | 0.813141161 |
| T2DM | LEC | *CYTH4* | 0.296822633 | 0.000108541 | 0.000908725 | 0.682395411 |
| T2DM | LEC | *CARD10* | -0.156828121 | 0.000164542 | 0.000910705 | 1 |
| T2DM | LEC | *LRP5L* | 1.052679834 | 0.005370295 | 0.00908097 | 1 |
| T2DM | LEC | *SLC25A1* | 0.809875393 | 8.53E-05 | 0.000908725 | 0.536253868 |
| T2DM | LEC | *LGALS2* | 0.637150004 | 0.000162665 | 0.000910705 | 1 |
| T2DM | LEC | *SH3BP1* | 0.797063764 | 0.000123964 | 0.000908725 | 0.779361434 |
| T2DM | LEC | *HPS4* | 0.909495573 | 0.000314332 | 0.001146958 | 1 |
| T2DM | LEC | *PIK3IP1* | 0.746557708 | 0.004205542 | 0.007334326 | 1 |
| T2DM | LEC | *TFIP11* | 0.99466139 | 0.000134857 | 0.000908725 | 0.847843261 |
| T2DM | LEC | *GCAT* | 0.582668265 | 0.000277748 | 0.00108013 | 1 |
| T2DM | LEC | *ANKRD54* | 0.471677635 | 0.030788311 | 0.041979204 | 1 |
| T2DM | LEC | *CCDC134* | -0.96160629 | 0.00238793 | 0.004608016 | 1 |

| | | | | | | |
|---|---|---|---|---|---|---|
| T2DM | LEC | *CBY1* | 0.48171059 | 0.003951429 | 0.006948989 | 1 |
| T2DM | LEC | *FBXO7* | 0.914903081 | 0.000903732 | 0.002194577 | 1 |
| T2DM | LEC | *POLDIP3* | 1.10860121 | 0.030084927 | 0.04110931 | 1 |
| T2DM | LEC | *PPP6R2* | 0.61829803 | 0.00885763 | 0.013925461 | 1 |
| T2DM | LEC | *CYB5R3* | -0.730016789 | 0.033610924 | 0.045394604 | 1 |
| T2DM | LEC | *RHBDD3* | 1.052719649 | 0.000257993 | 0.001039746 | 1 |
| T2DM | LEC | *TTLL1* | -0.694198063 | 0.000339548 | 0.001187188 | 1 |
| T2DM | LEC | *AP1B1* | 0.71549853 | 0.000787021 | 0.00198635 | 1 |
| T2DM | LEC | *HMGXB4* | 0.690360085 | 0.000172832 | 0.000910705 | 1 |
| T2DM | LEC | *TOM1* | 0.635049978 | 0.000232233 | 0.001001608 | 1 |
| T2DM | LEC | *NEFH* | 1.525715002 | 0.026901554 | 0.03725332 | 1 |
| T2DM | LEC | *CHKB* | -0.631125724 | 0.011921724 | 0.018015145 | 1 |
| T2DM | LEC | *BIK* | -0.241279612 | 0.000143773 | 0.000908725 | 0.903900077 |
| T2DM | LEC | *MCAT* | -0.97654453 | 0.000310826 | 0.001138389 | 1 |
| T2DM | LEC | *THOC5* | 0.838561741 | 0.002447501 | 0.004695587 | 1 |
| T2DM | LEC | *RASD2* | -2.103408568 | 0.000309727 | 0.001136749 | 1 |
| T2DM | LEC | *TTLL12* | -1.568318049 | 0.011276484 | 0.017165921 | 1 |
| T2DM | LEC | *MTMR3* | 0.561665817 | 0.014762349 | 0.021750853 | 1 |
| T2DM | LEC | *APOL4* | 0.483244713 | 0.000426027 | 0.001334543 | 1 |
| T2DM | LEC | *SAMM50* | 0.729128455 | 0.000120344 | 0.000908725 | 0.756602458 |
| T2DM | LEC | *TXN2* | 1.301862844 | 8.14E-05 | 0.000908725 | 0.511789546 |
| T2DM | LEC | *FOXRED2* | 0.461558017 | 2.75E-05 | 0.000908725 | 0.173013832 |
| T2DM | LEC | *IFT27* | 1.080127738 | 0.000279472 | 0.00108013 | 1 |
| T2DM | LEC | *KIAA0930* | 0.616865467 | 9.94E-05 | 0.000908725 | 0.624954336 |
| T2DM | LEC | *NCF4* | 0.375706425 | 9.21E-05 | 0.000908725 | 0.578813282 |
| T2DM | LEC | *SLC25A17* | 0.952386319 | 0.010134228 | 0.015639149 | 1 |
| T2DM | LEC | *KCTD17* | -0.70764847 | 0.000181632 | 0.00092839 | 1 |
| T2DM | LEC | *EP300* | 0.812338782 | 0.004383001 | 0.007616342 | 1 |
| T2DM | LEC | *ACO2* | 0.916696218 | 0.000371739 | 0.00124315 | 1 |
| T2DM | LEC | *DESI1* | 1.075556984 | 0.000436848 | 0.001344329 | 1 |
| T2DM | LEC | *KHNYN* | -0.520660506 | 0.019094667 | 0.027364526 | 1 |
| T2DM | LEC | *VCPKMT* | 0.920156488 | 0.002452355 | 0.004702029 | 1 |
| T2DM | LEC | *SOS2* | 0.774820194 | 0.003346241 | 0.006061024 | 1 |
| T2DM | LEC | *PYGL* | 0.386699839 | 0.000427971 | 0.001336216 | 1 |
| T2DM | LEC | *DDHD1* | 0.497206741 | 0.000190869 | 0.000942652 | 1 |
| T2DM | LEC | *CNIH1* | 0.994310023 | 0.000392076 | 0.00127792 | 1 |
| T2DM | LEC | *PIGH* | 0.695457235 | 3.89E-05 | 0.000908725 | 0.244738073 |
| T2DM | LEC | *VTI1B* | 1.16966179 | 0.000678539 | 0.001799318 | 1 |
| T2DM | LEC | *ISM2* | 2.744345823 | 1.43E-06 | 0.000272781 | 0.009001771 |
| T2DM | LEC | *PPM1A* | 0.982913217 | 0.002747838 | 0.005153836 | 1 |
| T2DM | LEC | *SLC10A1* | 0.85204148 | 0.000345667 | 0.001196042 | 1 |
| T2DM | LEC | *DICER1* | 0.762204877 | 0.001934566 | 0.003918369 | 1 |
| T2DM | LEC | *MTHFD1* | 0.595364221 | 0.000162915 | 0.000910705 | 1 |
| T2DM | LEC | *TELO2* | 1.098293605 | 0.000224793 | 0.000992757 | 1 |
| T2DM | LEC | *GSKIP* | -1.079580269 | 0.004582763 | 0.007910992 | 1 |
| T2DM | LEC | *VRK1* | 1.060918764 | 0.014719951 | 0.021698553 | 1 |
| T2DM | LEC | *PSMB5* | 1.241822703 | 0.000859086 | 0.002111443 | 1 |
| T2DM | LEC | *PCK2* | 0.768632898 | 0.000206779 | 0.000972342 | 1 |
| T2DM | LEC | *KIAA0391* | -1.075830179 | 0.000435726 | 0.00134367 | 1 |
| T2DM | LEC | *PLTP* | 0.50421944 | 6.57E-06 | 0.000607202 | 0.041289719 |

| | | | | | | |
|---|---|---|---|---|---|---|
| T2DM | LEC | *GSS* | 1.371235617 | 0.000776826 | 0.001966144 | 1 |
| T2DM | LEC | *PYGB* | 0.610070181 | 0.00101688 | 0.002384605 | 1 |
| T2DM | LEC | *ABHD12* | 0.890845109 | 0.000752717 | 0.001926061 | 1 |
| T2DM | LEC | *IFT52* | 0.996318122 | 8.88E-05 | 0.000908725 | 0.558067766 |
| T2DM | LEC | *DOK5* | 0.599039554 | 0.000131631 | 0.000908725 | 0.827564739 |
| T2DM | LEC | *CSTF1* | 1.025903725 | 0.000370111 | 0.001241526 | 1 |
| T2DM | LEC | *BMP7* | 0.959443467 | 0.000135746 | 0.000908725 | 0.8534339 |
| T2DM | LEC | *TPD52L2* | 1.02882846 | 0.001868937 | 0.003810873 | 1 |
| T2DM | LEC | *NELFCD* | 1.021967717 | 0.000635073 | 0.001720377 | 1 |
| T2DM | LEC | *CTSZ* | 0.797046206 | 0.004388552 | 0.007621774 | 1 |
| T2DM | LEC | *MTG2* | 0.626517553 | 0.00035898 | 0.001223256 | 1 |
| T2DM | LEC | *TCFL5* | -0.583938256 | 0.000302023 | 0.001123941 | 1 |
| T2DM | LEC | *GID8* | 1.422008714 | 0.001858721 | 0.003795316 | 1 |
| T2DM | LEC | *SLC17A9* | 0.57059565 | 6.92E-05 | 0.000908725 | 0.435369031 |
| T2DM | LEC | *NKAIN4* | 0.960667881 | 0.001132246 | 0.002576341 | 1 |
| T2DM | LEC | *PTK6* | -0.504651803 | 0.000150348 | 0.000910197 | 0.945240576 |
| T2DM | LEC | *GMEB2* | 0.702061664 | 0.000199689 | 0.000961874 | 1 |
| T2DM | LEC | *CDC25B* | -0.829132432 | 0.001058799 | 0.002451812 | 1 |
| T2DM | LEC | *RNF24* | 0.635667462 | 0.000425661 | 0.001334064 | 1 |
| T2DM | LEC | *NDUFAF5* | 0.262535761 | 0.017446103 | 0.025226231 | 1 |
| T2DM | LEC | *TRIB3* | 1.415683149 | 0.001430169 | 0.003082438 | 1 |
| T2DM | LEC | *SLC52A3* | 1.226059961 | 0.002310899 | 0.004485526 | 1 |
| T2DM | LEC | *ANGPT4* | 1.472671775 | 0.002215345 | 0.00433485 | 1 |
| T2DM | LEC | *RSPO4* | 0.629914995 | 0.000204547 | 0.000964732 | 1 |
| T2DM | LEC | *SIRPB1* | 0.466924935 | 0.031387442 | 0.042712738 | 1 |
| T2DM | LEC | *SEC23B* | 0.732432548 | 0.000633761 | 0.001718869 | 1 |
| T2DM | LEC | *CCM2L* | -0.264892347 | 0.000169268 | 0.000910705 | 1 |
| T2DM | LEC | *MYL9* | 0.77611536 | 0.00013953 | 0.000908725 | 0.877223654 |
| T2DM | LEC | *MROH8* | 0.574180667 | 0.003918139 | 0.006911618 | 1 |
| T2DM | LEC | *MANBAL* | 0.443769498 | 3.32E-05 | 0.000908725 | 0.208572635 |
| T2DM | LEC | *IDH3B* | 1.138551875 | 0.000190091 | 0.000942374 | 1 |
| T2DM | LEC | *MAPRE1* | -0.932173684 | 0.032474414 | 0.044048897 | 1 |
| T2DM | LEC | *TTI1* | 1.039155195 | 0.000678572 | 0.001799318 | 1 |
| T2DM | LEC | *RPRD1B* | 0.872612273 | 0.009652896 | 0.014986869 | 1 |
| T2DM | LEC | *PXMP4* | 0.859541126 | 0.003623699 | 0.006470376 | 1 |
| T2DM | LEC | *BPI* | 1.469782076 | 0.037162395 | 0.049752977 | 1 |
| T2DM | LEC | *MAP1LC3A* | 1.170780017 | 2.17E-05 | 0.000908725 | 0.136186937 |
| T2DM | LEC | *PIGU* | 0.824021891 | 0.000120905 | 0.000908725 | 0.760130804 |
| T2DM | LEC | *APMAP* | 0.639810115 | 0.000134429 | 0.000908725 | 0.845152414 |
| T2DM | LEC | *RBFA* | 1.006263191 | 0.000154561 | 0.000910197 | 0.971723911 |
| T2DM | LEC | *LPIN2* | 0.991307437 | 0.003801187 | 0.006743245 | 1 |
| T2DM | LEC | *SMCHD1* | 0.71280216 | 0.012584267 | 0.018891425 | 1 |
| T2DM | LEC | *CEP192* | 0.515795107 | 0.028545379 | 0.03928739 | 1 |
| T2DM | LEC | *LIPG* | 1.339137669 | 0.000259167 | 0.001042471 | 1 |
| T2DM | LEC | *POLI* | 0.254823224 | 0.027454234 | 0.037960142 | 1 |
| T2DM | LEC | *NALCN* | -0.055084962 | 0.000138652 | 0.000908725 | 0.871704692 |
| T2DM | LEC | *HTR2A* | 2.410843653 | 0.000165688 | 0.000910705 | 1 |
| T2DM | LEC | *KLF5* | 1.035120444 | 0.005574956 | 0.009376604 | 1 |
| T2DM | LEC | *DNAJC3* | 0.846981923 | 0.005197971 | 0.00883108 | 1 |
| T2DM | LEC | *SGCG* | 0.350338905 | 0.000136926 | 0.000908725 | 0.860850868 |

| | | | | | | |
|---|---|---|---|---|---|---|
| T2DM | LEC | *PARP4* | 0.851128139 | 0.004116384 | 0.007196803 | 1 |
| T2DM | LEC | *RGCC* | 0.319211951 | 0.021157076 | 0.030012305 | 1 |
| T2DM | LEC | *VWA8* | 0.79140448 | 0.000351258 | 0.001209397 | 1 |
| T2DM | LEC | *KATNAL1* | 0.675561627 | 0.003982945 | 0.006990725 | 1 |
| T2DM | LEC | *TSC22D1* | 0.832168352 | 0.012709572 | 0.019052236 | 1 |
| T2DM | LEC | *CLN5* | 0.737046399 | 0.011924496 | 0.018015145 | 1 |
| T2DM | LEC | *MGRN1* | 0.706708098 | 0.000403481 | 0.001298865 | 1 |
| T2DM | LEC | *CORO1A* | 0.586674786 | 0.001369339 | 0.002984067 | 1 |
| T2DM | LEC | *GDPD3* | 1.275880002 | 0.000376695 | 0.001251655 | 1 |
| T2DM | LEC | *ELMO3* | -0.630748216 | 0.001466469 | 0.003144506 | 1 |
| T2DM | LEC | *NFAT5* | -0.523813947 | 0.007192101 | 0.011653799 | 1 |
| T2DM | LEC | *PLLP* | 0.529033278 | 0.000137708 | 0.000908725 | 0.865770783 |
| T2DM | LEC | *DHODH* | 0.803430504 | 6.89E-05 | 0.000908725 | 0.432964095 |
| T2DM | LEC | *CTCF* | 0.949684618 | 0.013098141 | 0.019564746 | 1 |
| T2DM | LEC | *ACD* | -0.901486724 | 0.028216018 | 0.038893687 | 1 |
| T2DM | LEC | *USB1* | 1.525960031 | 0.002839924 | 0.005293389 | 1 |
| T2DM | LEC | *SLC7A6OS* | 0.390645096 | 0.002368131 | 0.004572616 | 1 |
| T2DM | LEC | *MON1B* | 1.121623251 | 0.000801959 | 0.002011482 | 1 |
| T2DM | LEC | *HCFC1R1* | 1.362481661 | 0.000365857 | 0.001235973 | 1 |
| T2DM | LEC | *NPRL3* | 0.782227005 | 0.000322952 | 0.001159063 | 1 |
| T2DM | LEC | *NAGPA* | 1.148276383 | 0.000715195 | 0.00186342 | 1 |
| T2DM | LEC | *COTL1* | 0.705327461 | 0.000402219 | 0.001296129 | 1 |
| T2DM | LEC | *CRISPLD2* | 0.666721553 | 0.00022095 | 0.00098409 | 1 |
| T2DM | LEC | *ZNF500* | 0.65219717 | 0.012814272 | 0.019186313 | 1 |
| T2DM | LEC | *NME4* | 1.492828974 | 0.011846397 | 0.017923492 | 1 |
| T2DM | LEC | *NOMO3* | 0.649058636 | 0.000925404 | 0.002222312 | 1 |
| T2DM | LEC | *LMF1* | 0.974779207 | 0.000551146 | 0.001560836 | 1 |
| T2DM | LEC | *NARFL* | 1.247508783 | 0.000322815 | 0.001159063 | 1 |
| T2DM | LEC | *MTHFSD* | 0.410346347 | 0.001959495 | 0.003951134 | 1 |
| T2DM | LEC | *CLCN7* | 1.013635765 | 0.007340666 | 0.011857854 | 1 |
| T2DM | LEC | *HAGHL* | 0.620818515 | 0.000186492 | 0.000932902 | 1 |
| T2DM | LEC | *SLC7A5* | -0.653915354 | 0.000167821 | 0.000910705 | 1 |
| T2DM | LEC | *FBXO31* | 0.819380679 | 0.002040411 | 0.004077579 | 1 |
| T2DM | LEC | *RHBDL1* | 0.574393709 | 0.023175112 | 0.032566368 | 1 |
| T2DM | LEC | *NUBP1* | 1.216635678 | 0.029445461 | 0.040358319 | 1 |
| T2DM | LEC | *CLUAP1* | 0.830761676 | 1.68E-05 | 0.000908725 | 0.105558985 |
| T2DM | LEC | *EARS2* | 0.429877562 | 0.00360315 | 0.006441002 | 1 |
| T2DM | LEC | *CPPED1* | 0.896579853 | 0.000959088 | 0.002281418 | 1 |
| T2DM | LEC | *BFAR* | 1.078380346 | 0.005746611 | 0.009621556 | 1 |
| T2DM | LEC | *RRN3P2* | 0.761642607 | 0.001920301 | 0.003898267 | 1 |
| T2DM | LEC | *RBL2* | 1.07194761 | 2.30E-06 | 0.000361341 | 0.014453652 |
| T2DM | LEC | *STX4* | 0.972954281 | 0.000460921 | 0.001393179 | 1 |
| T2DM | LEC | *KAT8* | 1.141519356 | 6.05E-05 | 0.000908725 | 0.380249094 |
| T2DM | LEC | *NOMO1* | 1.006220609 | 0.000562401 | 0.001582728 | 1 |
| T2DM | LEC | *SYT17* | -0.97995943 | 0.0006128 | 0.001683862 | 1 |
| T2DM | LEC | *TMC5* | 0.472316181 | 0.000229687 | 0.000997266 | 1 |
| T2DM | LEC | *RNF40* | -0.960258564 | 0.035048328 | 0.047163707 | 1 |
| T2DM | LEC | *IQCH* | 0.65245516 | 0.000157324 | 0.000910705 | 0.989093956 |
| T2DM | LEC | *LACTB* | 0.831185936 | 0.000537505 | 0.00153674 | 1 |
| T2DM | LEC | *CSK* | 1.400569837 | 0.003224761 | 0.005864643 | 1 |

| | | | | | | |
|---|---|---|---|---|---|---|
| T2DM | LEC | *HERC1* | 0.748921308 | 0.001421054 | 0.003070161 | 1 |
| T2DM | LEC | *RASL12* | -0.850969716 | 0.000381806 | 0.001262046 | 1 |
| T2DM | LEC | *AP3B2* | 0.27796813 | 0.01975698 | 0.028262146 | 1 |
| T2DM | LEC | *IGDCC4* | 0.299253972 | 0.0024505 | 0.004699907 | 1 |
| T2DM | LEC | *TTC23* | 0.920771037 | 0.000926971 | 0.002225226 | 1 |
| T2DM | LEC | *FAH* | 0.590059763 | 0.000137668 | 0.000908725 | 0.865519295 |
| T2DM | LEC | *HOMER2* | 0.628944064 | 0.000199529 | 0.000961874 | 1 |
| T2DM | LEC | *EHD4* | 0.994977928 | 0.000649298 | 0.001745996 | 1 |
| T2DM | LEC | *ATP8B4* | 0.462786612 | 0.000140886 | 0.000908725 | 0.885752484 |
| T2DM | LEC | *DTWD1* | 0.997771826 | 0.000304972 | 0.001127196 | 1 |
| T2DM | LEC | *DNAJC17* | 0.970461283 | 0.000504713 | 0.001471086 | 1 |
| T2DM | LEC | *PDGFRL* | 0.458409778 | 5.15E-05 | 0.000908725 | 0.323782545 |
| T2DM | LEC | *RIPK2* | 0.562542758 | 0.000168609 | 0.000910705 | 1 |
| T2DM | LEC | *EYA1* | -0.099944462 | 0.000764562 | 0.001947414 | 1 |
| T2DM | LEC | *NBN* | 0.843763191 | 0.001055023 | 0.002445543 | 1 |
| T2DM | LEC | *CPQ* | 0.57837693 | 0.000404597 | 0.001300425 | 1 |
| T2DM | LEC | *IKBKB* | 0.979450636 | 0.004546471 | 0.007856971 | 1 |
| T2DM | LEC | *PLAT* | 1.006787727 | 0.000208688 | 0.000972609 | 1 |
| T2DM | LEC | *GDAP1* | -0.527151927 | 0.000399537 | 0.001290795 | 1 |
| T2DM | LEC | *RAB2A* | 0.760034768 | 0.00031863 | 0.001153268 | 1 |
| T2DM | LEC | *SPAG1* | 0.707886482 | 0.005102136 | 0.008697703 | 1 |
| T2DM | LEC | *CHRAC1* | 1.331471839 | 0.000100313 | 0.000908725 | 0.630667026 |
| T2DM | LEC | *SNX16* | -0.786155767 | 0.010806054 | 0.016558046 | 1 |
| T2DM | LEC | *GSDMD* | 1.012940497 | 0.000789381 | 0.001989911 | 1 |
| T2DM | LEC | *TSTA3* | -0.967121396 | 0.000298673 | 0.001118132 | 1 |
| T2DM | LEC | *ERI1* | 0.813147335 | 0.000225406 | 0.000992757 | 1 |
| T2DM | LEC | *MTMR9* | 0.957824884 | 0.000719758 | 0.001872204 | 1 |
| T2DM | LEC | *LEPROTL1* | 1.03147301 | 0.000435779 | 0.00134367 | 1 |
| T2DM | LEC | *TNFRSF10A* | 0.769706075 | 0.001174538 | 0.002644815 | 1 |
| T2DM | LEC | *TUSC3* | 0.699537703 | 0.000152627 | 0.000910197 | 0.959563367 |
| T2DM | LEC | *KLHDC4* | 0.774167084 | 0.004575394 | 0.00790044 | 1 |
| T2DM | LEC | *MCM4* | 0.897678593 | 0.000428254 | 0.001336216 | 1 |
| T2DM | LEC | *MAN2B1* | 0.82930974 | 0.000128715 | 0.000908725 | 0.809234009 |
| T2DM | LEC | *TULP2* | 0.962077322 | 6.00E-05 | 0.000908725 | 0.377330271 |
| T2DM | LEC | *NUCB1* | 0.875894612 | 0.000486187 | 0.001437751 | 1 |
| T2DM | LEC | *NFKBIB* | 0.583075403 | 0.00016524 | 0.000910705 | 1 |
| T2DM | LEC | *TUBB4A* | 3.567860725 | 0.010802701 | 0.016556943 | 1 |
| T2DM | LEC | *SARS2* | -0.654971547 | 6.55E-05 | 0.000908725 | 0.411625092 |
| T2DM | LEC | *SNRNP70* | 0.662900082 | 0.012788632 | 0.019152485 | 1 |
| T2DM | LEC | *FCGRT* | 0.563294906 | 0.004432503 | 0.007674785 | 1 |
| T2DM | LEC | *PEX11G* | 0.526621585 | 0.000127993 | 0.000908725 | 0.804690339 |
| T2DM | LEC | *CD37* | 1.12351303 | 0.002508192 | 0.004785736 | 1 |
| T2DM | LEC | *AMH* | -1.12296593 | 0.001381098 | 0.003000964 | 1 |
| T2DM | LEC | *DKKL1* | 1.16111974 | 0.000422913 | 0.001330091 | 1 |
| T2DM | LEC | *STX10* | 0.96501557 | 9.76E-05 | 0.000908725 | 0.613573136 |
| T2DM | LEC | *FCER2* | 0.596648651 | 0.000184622 | 0.000929091 | 1 |
| T2DM | LEC | *IL4I1* | 0.777803849 | 5.18E-05 | 0.000908725 | 0.325575066 |
| T2DM | LEC | *TLE6* | 1.680004913 | 0.000421276 | 0.001326934 | 1 |
| T2DM | LEC | *AES* | 1.003347413 | 0.000370659 | 0.001241526 | 1 |
| T2DM | LEC | *SGTA* | 1.360483356 | 0.001135677 | 0.002582278 | 1 |

| | | | | | | |
|---|---|---|---|---|---|---|
| T2DM | LEC | *C19orf53* | -1.108641825 | 0.008377001 | 0.013272733 | 1 |
| T2DM | LEC | *TIMM44* | 0.818001743 | 0.000109558 | 0.000908725 | 0.688789994 |
| T2DM | LEC | *CCDC61* | -1.648212037 | 9.15E-05 | 0.000908725 | 0.575297441 |
| T2DM | LEC | *IL27RA* | 1.318725127 | 8.44E-05 | 0.000908725 | 0.530379 |
| T2DM | LEC | *C19orf44* | 0.687261441 | 0.008310533 | 0.01318736 | 1 |
| T2DM | LEC | *OLFM2* | 0.314482572 | 0.002420145 | 0.004657315 | 1 |
| T2DM | LEC | *RASAL3* | 0.59139454 | 0.000153363 | 0.000910197 | 0.964194222 |
| T2DM | LEC | *SYDE1* | -1.206249395 | 0.00069683 | 0.001830745 | 1 |
| T2DM | LEC | *AURKC* | 0.806892348 | 0.000917805 | 0.002214476 | 1 |
| T2DM | LEC | *CCNE1* | 0.822102243 | 0.000142703 | 0.000908725 | 0.897175017 |
| T2DM | LEC | *PDCD5* | 1.128928925 | 0.00042724 | 0.001336216 | 1 |
| T2DM | LEC | *ANKRD27* | 0.84240104 | 0.001344002 | 0.002939041 | 1 |
| T2DM | LEC | *PRX* | 0.606171867 | 0.00022547 | 0.000992757 | 1 |
| T2DM | LEC | *CCDC94* | 0.667930568 | 0.000124638 | 0.000908725 | 0.783601804 |
| T2DM | LEC | *TBCB* | 0.804962703 | 0.000256201 | 0.001035475 | 1 |
| T2DM | LEC | *POLR2I* | 1.747319482 | 0.000516537 | 0.001495765 | 1 |
| T2DM | LEC | *SLC1A5* | 0.643136604 | 0.000115367 | 0.000908725 | 0.725315163 |
| T2DM | LEC | *CACTIN* | -0.733750845 | 0.000292888 | 0.001109196 | 1 |
| T2DM | LEC | *CCDC9* | 0.970029034 | 4.75E-05 | 0.000908725 | 0.298321214 |
| T2DM | LEC | *FZR1* | 0.904530148 | 0.000116421 | 0.000908725 | 0.731939541 |
| T2DM | LEC | *TGFB1* | 0.848701008 | 0.03141298 | 0.042738239 | 1 |
| T2DM | LEC | *CEACAM4* | -1.503956691 | 0.000186659 | 0.000932902 | 1 |
| T2DM | LEC | *PLIN3* | 0.72322263 | 0.000200639 | 0.000962913 | 1 |
| T2DM | LEC | *MRPL4* | 1.385550874 | 0.011737848 | 0.017782132 | 1 |
| T2DM | LEC | *ICAM5* | 2.18346341 | 0.005138658 | 0.008752842 | 1 |
| T2DM | LEC | *CD33* | 0.460917832 | 9.72E-05 | 0.000908725 | 0.61106904 |
| T2DM | LEC | *BABAM1* | 1.101053261 | 0.000120462 | 0.000908725 | 0.757346199 |
| T2DM | LEC | *TYK2* | 0.773667674 | 5.92E-05 | 0.000908725 | 0.372268841 |
| T2DM | LEC | *PTPRS* | 0.604264778 | 0.00013192 | 0.000908725 | 0.829380921 |
| T2DM | LEC | *GRWD1* | 0.473658311 | 0.001946784 | 0.00393298 | 1 |
| T2DM | LEC | *GRIN2D* | 1.246743713 | 0.000189689 | 0.000942003 | 1 |
| T2DM | LEC | *CARD8* | 0.744727364 | 0.013060231 | 0.019522033 | 1 |
| T2DM | LEC | *LIG1* | 1.33098156 | 0.000759644 | 0.001936692 | 1 |
| T2DM | LEC | *PLA2G4C* | -0.56136138 | 0.0023271 | 0.004510011 | 1 |
| T2DM | LEC | *CAPS* | 0.918963359 | 3.58E-05 | 0.000908725 | 0.225382375 |
| T2DM | LEC | *BCAT2* | 1.312744905 | 0.001252116 | 0.002774782 | 1 |
| T2DM | LEC | *MIER2* | 1.161096215 | 0.009819347 | 0.015197989 | 1 |
| T2DM | LEC | *TNPO2* | 1.060731254 | 0.000214607 | 0.000978911 | 1 |
| T2DM | LEC | *LILRB5* | 0.357137784 | 0.000112209 | 0.000908725 | 0.705457164 |
| T2DM | LEC | *MAST1* | 0.991365524 | 0.00010894 | 0.000908725 | 0.684905778 |
| T2DM | LEC | *TFPT* | 1.467981881 | 0.006030113 | 0.010018848 | 1 |
| T2DM | LEC | *RPL18A* | 0.929754814 | 0.018682951 | 0.026860214 | 1 |
| T2DM | LEC | *ELL* | 1.181765448 | 0.000343345 | 0.001193551 | 1 |
| T2DM | LEC | *UPK1A* | 1.899908577 | 9.82E-05 | 0.000908725 | 0.617608775 |
| T2DM | LEC | *COPE* | 1.311286441 | 0.002876452 | 0.005348345 | 1 |
| T2DM | LEC | *LSR* | 0.66994122 | 0.012379512 | 0.018601814 | 1 |
| T2DM | LEC | *HPN* | 0.449205457 | 0.000431351 | 0.00134099 | 1 |
| T2DM | LEC | *ZNF14* | 1.168772311 | 0.000598453 | 0.001657479 | 1 |
| T2DM | LEC | *ATP13A1* | 2.04340878 | 1.78E-08 | 8.62E-06 | 0.000112 |
| T2DM | LEC | *ZNF85* | 1.428773566 | 0.004277384 | 0.007449283 | 1 |

| | | | | | | |
|---|---|---|---|---|---|---|
| T2DM | LEC | *SMG9* | 0.632366878 | 0.00011157 | 0.000908725 | 0.701442354 |
| T2DM | LEC | *RASA4* | 1.002430656 | 0.000472926 | 0.001412844 | 1 |
| T2DM | LEC | *PON3* | 0.618820882 | 0.000107914 | 0.000908725 | 0.678456517 |
| T2DM | LEC | *WDR91* | 0.846980332 | 0.000214922 | 0.000978911 | 1 |
| T2DM | LEC | *STEAP1B* | 1.18083138 | 0.001521474 | 0.00323159 | 1 |
| T2DM | LEC | *PTN* | 0.462362393 | 8.28E-05 | 0.000908725 | 0.520281915 |
| T2DM | LEC | *MPP6* | 0.502282972 | 0.000131988 | 0.000908725 | 0.829811206 |
| T2DM | LEC | *DFNA5* | 0.562834893 | 0.000168064 | 0.000910705 | 1 |
| T2DM | LEC | *OGDH* | 0.866884786 | 0.003518229 | 0.006311569 | 1 |
| T2DM | LEC | *ADAP1* | 1.052227245 | 0.000110102 | 0.000908725 | 0.692208976 |
| T2DM | LEC | *H2AFV* | 1.068845658 | 0.002462283 | 0.004715313 | 1 |
| T2DM | LEC | *MET* | -0.512605013 | 0.000375833 | 0.001250548 | 1 |
| T2DM | LEC | *RNF32* | -0.742379093 | 0.000540829 | 0.00154361 | 1 |
| T2DM | LEC | *LMBR1* | -0.676671192 | 0.009195593 | 0.014370543 | 1 |
| T2DM | LEC | *HOXA2* | 3.054289051 | 0.000630465 | 0.00171385 | 1 |
| T2DM | LEC | *HOXA5* | 0.43582999 | 0.000417574 | 0.001322531 | 1 |
| T2DM | LEC | *CPED1* | 0.535053276 | 0.000255558 | 0.001034574 | 1 |
| T2DM | LEC | *HIBADH* | 0.731597747 | 0.00081858 | 0.002039798 | 1 |
| T2DM | LEC | *CPVL* | 0.605427837 | 1.63E-05 | 0.000908725 | 0.102655214 |
| T2DM | LEC | *ABHD11* | -0.698517422 | 0.000439927 | 0.00134918 | 1 |
| T2DM | LEC | *PLEKHA8* | 0.859571067 | 0.000561063 | 0.001579965 | 1 |
| T2DM | LEC | *EPHB6* | 0.625381958 | 6.89E-05 | 0.000908725 | 0.432959239 |
| T2DM | LEC | *CHCHD2* | 0.853002721 | 0.009231997 | 0.014423848 | 1 |
| T2DM | LEC | *CCL24* | -0.947290321 | 0.000566382 | 0.001590372 | 1 |
| T2DM | LEC | *NPTX2* | 9.743704374 | 0.036276182 | 0.048659774 | 1 |
| T2DM | LEC | *CYP3A5* | 1.042052927 | 0.000483226 | 0.001433038 | 1 |
| T2DM | LEC | *ZKSCAN1* | 0.929578461 | 0.005085438 | 0.008681007 | 1 |
| T2DM | LEC | *SNX8* | 0.996502521 | 0.000723819 | 0.001881173 | 1 |
| T2DM | LEC | *AIMP2* | 1.188636376 | 0.01232742 | 0.01853683 | 1 |
| T2DM | LEC | *TFR2* | 1.864249644 | 5.39E-05 | 0.000908725 | 0.338730887 |
| T2DM | LEC | *RBM28* | 0.56352648 | 0.024544144 | 0.034290896 | 1 |
| T2DM | LEC | *AP1S1* | 0.567892215 | 0.000107176 | 0.000908725 | 0.673816628 |
| T2DM | LEC | *C1GALT1* | 0.912829075 | 0.00121479 | 0.002714068 | 1 |
| T2DM | LEC | *PLOD3* | 1.003085217 | 0.033226094 | 0.044923108 | 1 |
| T2DM | LEC | *RPA3* | 0.760248489 | 0.00029085 | 0.001103544 | 1 |
| T2DM | LEC | *CEP41* | 0.894314887 | 0.029705419 | 0.04067664 | 1 |
| T2DM | LEC | *ZNF862* | 0.946270776 | 0.001724286 | 0.003574213 | 1 |
| T2DM | LEC | *MEST* | 0.284456592 | 0.002527148 | 0.004814599 | 1 |
| T2DM | LEC | *ACTR3C* | 0.897464002 | 0.00019083 | 0.000942652 | 1 |
| T2DM | LEC | *POU6F2* | 0.855596789 | 0.000188511 | 0.000937634 | 1 |
| T2DM | LEC | *TSPAN13* | 0.929668594 | 0.00017756 | 0.000918726 | 1 |
| T2DM | LEC | *RARRES2* | 0.825519014 | 0.000257845 | 0.001039746 | 1 |
| T2DM | LEC | *TMEM176B* | 0.443300612 | 6.76E-06 | 0.000610811 | 0.042494639 |
| T2DM | LEC | *MRPL32* | 1.06050767 | 0.000322907 | 0.001159063 | 1 |
| T2DM | LEC | *COA1* | 0.579682315 | 0.026836689 | 0.037179873 | 1 |
| T2DM | LEC | *BLVRA* | 0.606829539 | 0.002577652 | 0.004887123 | 1 |
| T2DM | LEC | *URGCP* | 0.845022089 | 0.000349317 | 0.001204034 | 1 |
| T2DM | LEC | *POLD2* | 0.677450958 | 0.00545664 | 0.009199758 | 1 |
| T2DM | LEC | *GCK* | -8.21886867 | 0.000321563 | 0.001158549 | 1 |
| T2DM | LEC | *BCL7B* | 0.807147765 | 7.03E-05 | 0.000908725 | 0.442263699 |

| | | | | | | |
|---|---|---|---|---|---|---|
| T2DM | LEC | *TBL2* | 0.479999771 | 0.004085053 | 0.007151972 | 1 |
| T2DM | LEC | *CLIP2* | -0.825974428 | 0.002877064 | 0.005348345 | 1 |
| T2DM | LEC | *LIMK1* | 0.927946585 | 0.000165717 | 0.000910705 | 1 |
| T2DM | LEC | *SPATA6L* | -0.304011221 | 0.000162696 | 0.000910705 | 1 |
| T2DM | LEC | *TMEM245* | 0.722031075 | 0.00866641 | 0.013665844 | 1 |
| T2DM | LEC | *MEGF9* | 0.536158383 | 0.000946963 | 0.002256401 | 1 |
| T2DM | LEC | *TRIM14* | 0.531257177 | 0.00070787 | 0.00185124 | 1 |
| T2DM | LEC | *CORO2A* | 1.110037109 | 0.002367876 | 0.004572616 | 1 |
| T2DM | LEC | *C5* | 0.477480324 | 0.000209002 | 0.000972609 | 1 |
| T2DM | LEC | *OGN* | 0.838154355 | 0.001315182 | 0.002885048 | 1 |
| T2DM | LEC | *ECM2* | 0.454584384 | 0.000200955 | 0.00096305 | 1 |
| T2DM | LEC | *PTGR1* | 0.52509308 | 0.000753395 | 0.001927011 | 1 |
| T2DM | LEC | *SUSD1* | 0.688450482 | 0.00086767 | 0.002125064 | 1 |
| T2DM | LEC | *TNFSF8* | 0.413662007 | 0.000133274 | 0.000908725 | 0.837892696 |
| T2DM | LEC | *AK1* | 1.139483904 | 0.000909176 | 0.002203542 | 1 |
| T2DM | LEC | *KDM4C* | 0.76765515 | 0.002444599 | 0.004692884 | 1 |
| T2DM | LEC | *DOCK8* | 0.663960758 | 0.000169815 | 0.000910705 | 1 |
| T2DM | LEC | *ELAVL2* | 0.749534151 | 0.032982926 | 0.044642336 | 1 |
| T2DM | LEC | *FUBP3* | 0.906690227 | 0.018869161 | 0.027090755 | 1 |
| T2DM | LEC | *RGP1* | 1.245432751 | 8.73E-05 | 0.000908725 | 0.548758348 |
| T2DM | LEC | *MPDZ* | -0.465505071 | 0.000766701 | 0.001948143 | 1 |
| T2DM | LEC | *PTGDS* | 0.859690045 | 0.000113583 | 0.000908725 | 0.714095797 |
| T2DM | LEC | *SHB* | 1.135417304 | 8.02E-05 | 0.000908725 | 0.504215408 |
| T2DM | LEC | *DVL1* | 1.418559321 | 0.008267799 | 0.013142769 | 1 |
| T2DM | LEC | *PDLIM1* | -0.68070626 | 0.015536438 | 0.022763362 | 1 |
| T2DM | LEC | *CCNJ* | -0.685446374 | 0.002054698 | 0.004094417 | 1 |
| T2DM | LEC | *ATRNL1* | 0.636009924 | 0.000432439 | 0.001341345 | 1 |
| T2DM | LEC | *HPS1* | 0.867267588 | 0.000317968 | 0.001152864 | 1 |
| T2DM | LEC | *DNMBP* | -0.680017019 | 0.028496297 | 0.039228426 | 1 |
| T2DM | LEC | *TRDMT1* | 1.101402412 | 0.0031902 | 0.005816714 | 1 |
| T2DM | LEC | *DDX50* | 0.820299849 | 0.000386763 | 0.001268529 | 1 |
| T2DM | LEC | *MAPK8* | 0.709972265 | 0.024043176 | 0.033635836 | 1 |
| T2DM | LEC | *ATE1* | 0.771940254 | 0.000931409 | 0.00222992 | 1 |
| T2DM | LEC | *PLEKHA1* | 0.921341444 | 0.000565611 | 0.001588918 | 1 |
| T2DM | LEC | *SPOCK2* | 0.413917341 | 0.000322875 | 0.001159063 | 1 |
| T2DM | LEC | *MICU1* | 0.889051645 | 0.002290467 | 0.004459637 | 1 |
| T2DM | LEC | *CCSER2* | 0.794054518 | 0.011260749 | 0.017146121 | 1 |
| T2DM | LEC | *BMPR1A* | 0.488571396 | 0.000321296 | 0.001158249 | 1 |
| T2DM | LEC | *LIPA* | 0.555979546 | 0.000172423 | 0.000910705 | 1 |
| T2DM | LEC | *SFXN3* | -0.907114671 | 0.035516167 | 0.047772816 | 1 |
| T2DM | LEC | *KAZALD1* | -2.287202489 | 0.000111207 | 0.000908725 | 0.699159835 |
| T2DM | LEC | *ANKRD26* | 0.794334852 | 0.002592734 | 0.004911273 | 1 |
| T2DM | LEC | *ACBD5* | 0.697175771 | 6.15E-05 | 0.000908725 | 0.386435071 |
| T2DM | LEC | *MTPAP* | 1.112573625 | 0.000236367 | 0.001005438 | 1 |
| T2DM | LEC | *MAP3K8* | 0.737943014 | 0.002243885 | 0.004381152 | 1 |
| T2DM | LEC | *SHOC2* | 0.86103602 | 0.003794385 | 0.006733079 | 1 |
| T2DM | LEC | *TFAM* | 1.023834546 | 0.02761203 | 0.038144766 | 1 |
| T2DM | LEC | *TADA2A* | 0.460874137 | 0.014035877 | 0.020807252 | 1 |
| T2DM | LEC | *MLLT6* | 0.978183289 | 0.020732019 | 0.029475848 | 1 |
| T2DM | LEC | *PSMB3* | 1.57529935 | 0.000434165 | 0.001343306 | 1 |

| | | | | | | |
|---|---|---|---|---|---|---|
| T2DM | LEC | *CASC3* | 0.904748774 | 0.002926224 | 0.005420498 | 1 |
| T2DM | LEC | *RAPGEFL1* | 0.565119427 | 0.000738054 | 0.001904039 | 1 |
| T2DM | LEC | *RGS9* | -1.396964147 | 0.005237916 | 0.0088882 | 1 |
| T2DM | LEC | *WNT3* | -2.139678225 | 0.001052209 | 0.002443904 | 1 |
| T2DM | LEC | *ASPA* | 0.89741704 | 0.000693258 | 0.001825172 | 1 |
| T2DM | LEC | *TRIM37* | 1.066013922 | 0.003306821 | 0.005994804 | 1 |
| T2DM | LEC | *CBX1* | 0.983270195 | 0.017512785 | 0.025311006 | 1 |
| T2DM | LEC | *PIGL* | 0.830320002 | 0.000894298 | 0.002177557 | 1 |
| T2DM | LEC | *GALK1* | -1.126257064 | 0.000740433 | 0.001906556 | 1 |
| T2DM | LEC | *HOXB6* | 0.659061136 | 0.00313727 | 0.005732059 | 1 |
| T2DM | LEC | *RNF167* | 1.572291412 | 0.001125953 | 0.002567596 | 1 |
| T2DM | LEC | *CHRNE* | -0.357336282 | 0.033874004 | 0.045710638 | 1 |
| T2DM | LEC | *C1QBP* | 0.722940244 | 7.57E-05 | 0.000908725 | 0.476007948 |
| T2DM | LEC | *GOSR1* | 0.814275861 | 0.025763101 | 0.03580073 | 1 |
| T2DM | LEC | *AKAP10* | 0.77098255 | 0.001575416 | 0.003323704 | 1 |
| T2DM | LEC | *SMARCD2* | 0.950358114 | 0.00106285 | 0.002458476 | 1 |
| T2DM | LEC | *ICAM2* | 0.666901379 | 1.74E-05 | 0.000908725 | 0.109298237 |
| T2DM | LEC | *UTP6* | 1.011771758 | 0.001937856 | 0.003922505 | 1 |
| T2DM | LEC | *DDX5* | 0.569021101 | 0.007605174 | 0.012237964 | 1 |
| T2DM | LEC | *C17orf75* | 1.077550482 | 0.00037594 | 0.001250548 | 1 |
| T2DM | LEC | *CYTH1* | 0.845653273 | 5.92E-05 | 0.000908725 | 0.371999526 |
| T2DM | LEC | *PEX12* | -0.982435254 | 0.030244931 | 0.041283083 | 1 |
| T2DM | LEC | *DHX58* | 0.67248963 | 0.019901846 | 0.028437024 | 1 |
| T2DM | LEC | *RAB5C* | 0.882801281 | 9.47E-05 | 0.000908725 | 0.595670922 |
| T2DM | LEC | *NAGLU* | 0.767955775 | 0.000261313 | 0.001045752 | 1 |
| T2DM | LEC | *HSD17B1* | -0.489311194 | 0.001381389 | 0.003000964 | 1 |
| T2DM | LEC | *EZH1* | 0.939253606 | 0.000771669 | 0.001957823 | 1 |
| T2DM | LEC | *DLX4* | 4.280899753 | 0.000120219 | 0.000908725 | 0.755814786 |
| T2DM | LEC | *VAT1* | 1.733350776 | 4.91E-05 | 0.000908725 | 0.308714142 |
| T2DM | LEC | *LRRC59* | -1.201660638 | 0.036208533 | 0.048589765 | 1 |
| T2DM | LEC | *ABCC3* | 0.360613672 | 3.08E-05 | 0.000908725 | 0.193947479 |
| T2DM | LEC | *MPP2* | 1.136566259 | 0.000109866 | 0.000908725 | 0.690730319 |
| T2DM | LEC | *EFTUD2* | 0.994079282 | 0.000232609 | 0.001001608 | 1 |
| T2DM | LEC | *EFNB3* | 5.848419839 | 0.000133327 | 0.000908725 | 0.838225896 |
| T2DM | LEC | *FAM20A* | 0.350364335 | 0.000114038 | 0.000908725 | 0.716958925 |
| T2DM | LEC | *DPH1* | 1.140006381 | 0.000141388 | 0.000908725 | 0.888906086 |
| T2DM | LEC | *DHRS7B* | 0.843955827 | 0.0007009 | 0.001836832 | 1 |
| T2DM | LEC | *RCVRN* | -1.033303927 | 0.000326998 | 0.00116703 | 1 |
| T2DM | LEC | *SLC9A3R1* | 1.334715263 | 7.81E-05 | 0.000908725 | 0.491136354 |
| T2DM | LEC | *TMEM104* | 1.289739798 | 0.000439502 | 0.001348591 | 1 |
| T2DM | LEC | *TMEM97* | 0.989496937 | 0.000147813 | 0.000908725 | 0.929297831 |
| T2DM | LEC | *RAB34* | 0.890931715 | 0.001001772 | 0.002355325 | 1 |
| T2DM | LEC | *CWH43* | 4.464436645 | 2.69E-05 | 0.000908725 | 0.168820038 |
| T2DM | LEC | *KIAA1211* | 0.536825299 | 0.000459677 | 0.001391616 | 1 |
| T2DM | LEC | *AREG* | 0.567958068 | 0.016039622 | 0.023413305 | 1 |
| T2DM | LEC | *UBE2D3* | 0.790665925 | 0.001321695 | 0.002897314 | 1 |
| T2DM | LEC | *NDUFC1* | 1.071634985 | 0.000356466 | 0.00121733 | 1 |
| T2DM | LEC | *TBC1D9* | 0.723530276 | 0.000603233 | 0.00166412 | 1 |
| T2DM | LEC | *CPE* | 0.896112388 | 0.017194084 | 0.024907651 | 1 |
| T2DM | LEC | *WFS1* | -0.345534644 | 0.000963798 | 0.002290907 | 1 |

| | | | | | | |
|---|---|---|---|---|---|---|
| T2DM | LEC | *GRPEL1* | 1.732123458 | 0.007971115 | 0.012732317 | 1 |
| T2DM | LEC | *AADAT* | 1.551659139 | 0.005182354 | 0.008812946 | 1 |
| T2DM | LEC | *CPZ* | 8.536604336 | 0.000116359 | 0.000908725 | 0.731548982 |
| T2DM | LEC | *SLC2A9* | 0.465706532 | 3.80E-05 | 0.000908725 | 0.238640894 |
| T2DM | LEC | *RAPGEF2* | 0.756853091 | 0.016552826 | 0.024067441 | 1 |
| T2DM | LEC | *UFSP2* | 1.011230768 | 0.001220206 | 0.002722298 | 1 |
| T2DM | LEC | *FAM149A* | 0.731993908 | 3.29E-06 | 0.000436409 | 0.020714781 |
| T2DM | LEC | *UGDH* | 0.572518616 | 0.000124005 | 0.000908725 | 0.779617585 |
| T2DM | LEC | *DDX25* | 1.578160203 | 0.000280958 | 0.001081019 | 1 |
| T2DM | LEC | *CTSC* | 0.611923933 | 0.000683449 | 0.001808437 | 1 |
| T2DM | LEC | *ELP4* | 0.799918882 | 0.005996738 | 0.009969517 | 1 |
| T2DM | LEC | *SC5D* | 1.325522613 | 0.014996887 | 0.022050189 | 1 |
| T2DM | LEC | *C11orf63* | 0.579244742 | 0.000138142 | 0.000908725 | 0.868498708 |
| T2DM | LEC | *DTX4* | -0.326557241 | 0.000221676 | 0.000985627 | 1 |
| T2DM | LEC | *PUS3* | -0.47299141 | 0.000601092 | 0.001659669 | 1 |
| T2DM | LEC | *DCPS* | 1.16075249 | 0.000136287 | 0.000908725 | 0.856834314 |
| T2DM | LEC | *FOXRED1* | 0.671708 | 0.000110493 | 0.000908725 | 0.69466769 |
| T2DM | LEC | *MS4A4A* | 0.311719415 | 0.000100821 | 0.000908725 | 0.633860722 |
| T2DM | LEC | *CCDC86* | -0.781372715 | 0.000169527 | 0.000910705 | 1 |
| T2DM | LEC | *HPX* | -0.568210516 | 0.000996245 | 0.002345876 | 1 |
| T2DM | LEC | *CHORDC1* | 0.894573622 | 0.000393984 | 0.001282395 | 1 |
| T2DM | LEC | *FOLR3* | 0.439299465 | 0.021533819 | 0.030471105 | 1 |
| T2DM | LEC | *EIF4G2* | 0.81897386 | 0.030306914 | 0.041358708 | 1 |
| T2DM | LEC | *GALNT18* | -0.205924694 | 0.000170859 | 0.000910705 | 1 |
| T2DM | LEC | *CBL* | 0.923784906 | 0.000800533 | 0.002010768 | 1 |
| T2DM | LEC | *KIAA1549L* | 0.649280498 | 0.000155691 | 0.000910705 | 0.978828753 |
| T2DM | LEC | *CD5* | 0.597205835 | 0.000147164 | 0.000908725 | 0.92522052 |
| T2DM | LEC | *ACCS* | 0.542669043 | 0.00029823 | 0.001117384 | 1 |
| T2DM | LEC | *NAA40* | 1.261153692 | 0.00112733 | 0.002568874 | 1 |
| T2DM | LEC | *CARS* | 0.930987202 | 3.13E-05 | 0.000908725 | 0.196685726 |
| T2DM | LEC | *SLC22A18* | 0.365555554 | 0.000416008 | 0.001318933 | 1 |
| T2DM | LEC | *SLC35F2* | 0.424325851 | 0.00158979 | 0.003352906 | 1 |
| T2DM | LEC | *ELMOD1* | -1.475076766 | 0.000170458 | 0.000910705 | 1 |
| T2DM | LEC | *AIP* | 1.146331474 | 0.000809108 | 0.002022608 | 1 |
| T2DM | LEC | *NDUFS8* | 1.314982952 | 0.006437254 | 0.010586193 | 1 |
| T2DM | LEC | *TCIRG1* | 0.9050023 | 7.11E-05 | 0.000908725 | 0.446754405 |
| T2DM | LEC | *GTF2H1* | 0.7568119 | 0.001887827 | 0.00384227 | 1 |
| T2DM | LEC | *PTPN5* | 0.634115798 | 0.000318405 | 0.001153119 | 1 |
| T2DM | LEC | *PPFIBP1* | -0.120166938 | 0.000183367 | 0.000929091 | 1 |
| T2DM | LEC | *COQ5* | 0.440402422 | 0.000149016 | 0.000910197 | 0.936863008 |
| T2DM | LEC | *ASIC1* | 0.562393213 | 0.00196474 | 0.003957808 | 1 |
| T2DM | LEC | *CAPRIN2* | 0.902353029 | 0.000980943 | 0.00231762 | 1 |
| T2DM | LEC | *CYP27B1* | 3.525835752 | 0.000103291 | 0.000908725 | 0.649389292 |
| T2DM | LEC | *ACSS3* | 0.39067859 | 0.002095495 | 0.004157936 | 1 |
| T2DM | LEC | *GLI1* | -2.38719554 | 0.00753212 | 0.012132831 | 1 |
| T2DM | LEC | *METAP2* | 0.785365686 | 0.001420747 | 0.003070161 | 1 |
| T2DM | LEC | *LTA4H* | 0.507295088 | 0.00012129 | 0.000908725 | 0.762547358 |
| T2DM | LEC | *WNT5B* | -0.749358437 | 0.002991188 | 0.00550999 | 1 |
| T2DM | LEC | *TRPV4* | 1.070942417 | 0.003432374 | 0.006183194 | 1 |
| T2DM | LEC | *ITFG2* | 0.766914058 | 0.00037565 | 0.001250548 | 1 |

| | | | | | | |
|---|---|---|---|---|---|---|
| T2DM | LEC | *PRR4* | 0.995737799 | 4.87E-06 | 0.000529126 | 0.030606233 |
| T2DM | LEC | *GPN3* | 1.339961406 | 0.000547192 | 0.00155496 | 1 |
| T2DM | LEC | *VPS29* | 0.891609229 | 0.010331957 | 0.01592474 | 1 |
| T2DM | LEC | *AKAP3* | 0.502028164 | 0.001486543 | 0.003179957 | 1 |
| T2DM | LEC | *KCNA1* | 1.043292601 | 0.000227133 | 0.000992757 | 1 |
| T2DM | LEC | *CREBL2* | 1.262207209 | 0.000984568 | 0.002321822 | 1 |
| T2DM | LEC | *SCNN1A* | 0.627580014 | 0.000168277 | 0.000910705 | 1 |
| T2DM | LEC | *LTBR* | 0.827376473 | 0.001303114 | 0.002864572 | 1 |
| T2DM | LEC | *CDK2AP1* | 0.966788633 | 3.09E-06 | 0.000431731 | 0.019427908 |
| T2DM | LEC | *MGP* | 0.642436177 | 3.87E-05 | 0.000908725 | 0.243339768 |
| T2DM | LEC | *RASAL1* | 1.854558625 | 0.004138363 | 0.007231208 | 1 |
| T2DM | LEC | *ARHGDIB* | 0.811421834 | 0.000497224 | 0.001455329 | 1 |
| T2DM | LEC | *EIF2B1* | 0.838972604 | 0.000233151 | 0.001001608 | 1 |
| T2DM | LEC | *ENDOU* | -1.374592407 | 2.48E-06 | 0.000371603 | 0.015607335 |
| T2DM | LEC | *STX2* | 0.693794771 | 0.032992415 | 0.044645569 | 1 |
| T2DM | LEC | *NUP107* | 0.893180038 | 0.005475301 | 0.00922627 | 1 |
| T2DM | LEC | *CNOT2* | 0.743762116 | 0.005423178 | 0.009148249 | 1 |
| T2DM | LEC | *KRR1* | 0.777263987 | 0.02939939 | 0.040303961 | 1 |
| T2DM | LEC | *COPS7A* | 0.771474094 | 0.001795698 | 0.003688191 | 1 |
| T2DM | LEC | *GNB3* | -0.352247561 | 0.007820856 | 0.01253051 | 1 |
| T2DM | LEC | *CDCA3* | -0.909825809 | 0.00356715 | 0.006382093 | 1 |
| T2DM | LEC | *CHPT1* | 0.317554701 | 0.005291103 | 0.008971188 | 1 |
| T2DM | LEC | *GNPTAB* | 0.854130252 | 0.00039112 | 0.001277842 | 1 |
| T2DM | LEC | *ATN1* | 0.82640465 | 0.00029511 | 0.00111174 | 1 |
| T2DM | LEC | *NT5DC3* | 0.757799281 | 0.014081698 | 0.020870259 | 1 |
| T2DM | LEC | *ST8SIA1* | 0.481643875 | 0.000149879 | 0.000910197 | 0.942291402 |
| T2DM | LEC | *C2CD5* | 0.823613794 | 0.001762766 | 0.003638382 | 1 |
| T2DM | LEC | *PHC1* | 1.595790439 | 0.000228178 | 0.000994837 | 1 |
| T2DM | LEC | *COL12A1* | 0.535947295 | 0.000223954 | 0.000992757 | 1 |
| T2DM | LEC | *BTN3A3* | 0.420635221 | 0.031721329 | 0.043092479 | 1 |
| T2DM | LEC | *RSPH4A* | -0.430766871 | 0.001280321 | 0.002822591 | 1 |
| T2DM | LEC | *SMIM8* | 0.695740254 | 0.000223446 | 0.000991392 | 1 |
| T2DM | LEC | *MCM9* | 0.907177752 | 0.00485902 | 0.008330695 | 1 |
| T2DM | LEC | *FAM184A* | 0.969525037 | 0.003942932 | 0.006943757 | 1 |
| T2DM | LEC | *RNGTT* | 0.846824326 | 0.002539029 | 0.00483138 | 1 |
| T2DM | LEC | *GABRR2* | 0.527816209 | 0.000203518 | 0.000964732 | 1 |
| T2DM | LEC | *HDDC2* | 0.656534552 | 0.000253058 | 0.001030426 | 1 |
| T2DM | LEC | *TPD52L1* | 0.813683985 | 0.005170018 | 0.008798793 | 1 |
| T2DM | LEC | *UST* | 0.384106741 | 0.000120332 | 0.000908725 | 0.756527152 |
| T2DM | LEC | *MTRF1L* | 1.171662035 | 0.000831212 | 0.002064728 | 1 |
| T2DM | LEC | *TULP1* | 5.489558748 | 0.000111718 | 0.000908725 | 0.702368465 |
| T2DM | LEC | *SLC26A8* | 0.536663151 | 0.000144742 | 0.000908725 | 0.909992296 |
| T2DM | LEC | *MRPL18* | -0.694251544 | 0.033335823 | 0.045042407 | 1 |
| T2DM | LEC | *RNF8* | 0.818201714 | 0.006591374 | 0.010811366 | 1 |
| T2DM | LEC | *PHACTR1* | 0.676916173 | 0.00017688 | 0.000918726 | 1 |
| T2DM | LEC | *CD83* | 0.379754729 | 0.001119313 | 0.002556164 | 1 |
| T2DM | LEC | *SAYSD1* | 1.046689303 | 9.51E-05 | 0.000908725 | 0.597852849 |
| T2DM | LEC | *CAP2* | 0.443047173 | 0.001086228 | 0.00249742 | 1 |
| T2DM | LEC | *GPR63* | -0.377434187 | 0.000366301 | 0.001236807 | 1 |
| T2DM | LEC | *ASCC3* | 0.738692595 | 0.017841456 | 0.025738695 | 1 |

| | | | | | | |
|---|---|---|---|---|---|---|
| T2DM | LEC | *GPLD1* | 0.607122389 | 0.00013406 | 0.000908725 | 0.84283587 |
| T2DM | LEC | *ALDH5A1* | 0.516280046 | 2.99E-05 | 0.000908725 | 0.187740464 |
| T2DM | LEC | *VNN1* | 0.390163465 | 0.000387784 | 0.00126979 | 1 |
| T2DM | LEC | *VNN2* | 1.973152435 | 0.000682219 | 0.001805941 | 1 |
| T2DM | LEC | *ACOT13* | 1.005257921 | 0.00074959 | 0.001920977 | 1 |
| T2DM | LEC | *C6orf62* | 0.842092251 | 0.000731883 | 0.00189512 | 1 |
| T2DM | LEC | *B3GAT2* | 1.028028942 | 0.00155219 | 0.003283501 | 1 |
| T2DM | LEC | *PERP* | -0.610075233 | 0.002217688 | 0.004338085 | 1 |
| T2DM | LEC | *SLC16A10* | 0.348259085 | 5.90E-05 | 0.000908725 | 0.370987999 |
| T2DM | LEC | *PHACTR2* | 0.701505245 | 0.000667106 | 0.00177942 | 1 |
| T2DM | LEC | *TFEB* | -0.712425844 | 0.002605284 | 0.004926141 | 1 |
| T2DM | LEC | *FAM120B* | 0.879095343 | 0.000911311 | 0.002207017 | 1 |
| T2DM | LEC | *TBP* | 1.151487579 | 0.007993794 | 0.012758818 | 1 |
| T2DM | LEC | *PRPH2* | -0.946430905 | 0.024961089 | 0.03479609 | 1 |
| T2DM | LEC | *DNPH1* | 0.739100282 | 0.000479407 | 0.001424968 | 1 |
| T2DM | LEC | *DUSP22* | 1.017908871 | 0.004249585 | 0.007407026 | 1 |
| T2DM | LEC | *EXOC2* | 0.771230074 | 0.03011439 | 0.041122756 | 1 |
| T2DM | LEC | *SENP6* | 0.729114013 | 0.006820668 | 0.011140956 | 1 |
| T2DM | LEC | *TTK* | 0.870795734 | 0.005198392 | 0.00883108 | 1 |
| T2DM | LEC | *CLIC5* | 0.350898435 | 0.000129848 | 0.000908725 | 0.816355355 |
| T2DM | LEC | *FBRSL1* | 0.907486857 | 0.000125727 | 0.000908725 | 0.790447264 |
| T2DM | LEC | *LY86* | 0.451108725 | 0.000109975 | 0.000908725 | 0.691411155 |
| T2DM | LEC | *TBX18* | 0.838613744 | 0.000964171 | 0.002290907 | 1 |
| T2DM | LEC | *PCDHB2* | -0.481388744 | 0.020882663 | 0.029676605 | 1 |
| T2DM | LEC | *NUDT12* | 0.798616618 | 0.000386617 | 0.001268529 | 1 |
| T2DM | LEC | *CEP72* | -1.066546241 | 0.009601806 | 0.014923747 | 1 |
| T2DM | LEC | *C7* | 0.916836399 | 0.000685564 | 0.001813268 | 1 |
| T2DM | LEC | *GHR* | 0.508832205 | 3.33E-05 | 0.000908725 | 0.209422412 |
| T2DM | LEC | *DAP* | 1.052828499 | 0.002566717 | 0.004869328 | 1 |
| T2DM | LEC | *PFDN1* | 0.710993771 | 0.031987584 | 0.043435409 | 1 |
| T2DM | LEC | *LOX* | -0.850552481 | 0.032012218 | 0.043459472 | 1 |
| T2DM | LEC | *APBB3* | 0.713189134 | 0.001961958 | 0.00395474 | 1 |
| T2DM | LEC | *TMCO6* | -1.422564221 | 0.00259814 | 0.00491559 | 1 |
| T2DM | LEC | *SPARC* | 0.617843939 | 0.000518592 | 0.001499719 | 1 |
| T2DM | LEC | *PCDHB6* | 2.09014653 | 4.50E-05 | 0.000908725 | 0.282978877 |
| T2DM | LEC | *PDE8B* | 0.830052714 | 0.000581993 | 0.001623223 | 1 |
| T2DM | LEC | *CLK4* | 0.868425659 | 0.001658229 | 0.00346816 | 1 |
| T2DM | LEC | *PCDHB15* | -0.479792457 | 0.000260995 | 0.001045146 | 1 |
| T2DM | LEC | *HAVCR1* | 0.578158857 | 0.000198628 | 0.000959857 | 1 |
| T2DM | LEC | *RNF130* | 0.587312453 | 0.000273584 | 0.00107497 | 1 |
| T2DM | LEC | *ARSB* | 0.741270777 | 0.000514212 | 0.001491856 | 1 |
| T2DM | LEC | *CLINT1* | 0.759467148 | 0.000187778 | 0.000936299 | 1 |
| T2DM | LEC | *BTNL8* | 0.365357662 | 0.001883608 | 0.003834923 | 1 |
| T2DM | LEC | *RASGRF2* | 0.516399752 | 0.000190363 | 0.000942374 | 1 |
| T2DM | LEC | *POLR3G* | 0.482201603 | 0.000277047 | 0.00108013 | 1 |
| T2DM | LEC | *DROSHA* | 0.886670802 | 0.000441399 | 0.001351717 | 1 |
| T2DM | LEC | *SLC27A6* | 0.7130693 | 0.000134917 | 0.000908725 | 0.84822092 |
| T2DM | LEC | *TARS* | 1.248724479 | 0.002721122 | 0.005112879 | 1 |
| T2DM | LEC | *LNPEP* | 0.868207553 | 0.000942255 | 0.00224989 | 1 |
| T2DM | LEC | *GNPDA1* | 1.167847294 | 0.013635944 | 0.020266946 | 1 |

| | | | | | | |
|---|---|---|---|---|---|---|
| T2DM | LEC | *SKP1* | 0.99340905 | 0.001304652 | 0.002865949 | 1 |
| T2DM | LEC | *TRAPPC13* | 0.924985568 | 0.001374816 | 0.002993927 | 1 |
| T2DM | LEC | *C9* | 0.702451098 | 0.001133448 | 0.002578143 | 1 |
| T2DM | LEC | *ATP6V0E1* | 0.785272898 | 0.023350765 | 0.032776571 | 1 |
| T2DM | LEC | *CNTN3* | 0.501442737 | 0.000132068 | 0.000908725 | 0.830314125 |
| T2DM | LEC | *ACTR8* | 0.886770933 | 0.00019279 | 0.000946191 | 1 |
| T2DM | LEC | *HGD* | -0.946303639 | 0.000245598 | 0.001017515 | 1 |
| T2DM | LEC | *NPHP3* | 0.555067715 | 0.001055053 | 0.002445543 | 1 |
| T2DM | LEC | *NIT2* | 0.483017321 | 0.000139127 | 0.000908725 | 0.874693792 |
| T2DM | LEC | *PCCB* | 1.0504711 | 0.002171226 | 0.004280495 | 1 |
| T2DM | LEC | *RBP1* | 1.292157087 | 0.001155033 | 0.002614 | 1 |
| T2DM | LEC | *RNF7* | 0.905499495 | 0.002775035 | 0.005193646 | 1 |
| T2DM | LEC | *BCHE* | -0.213124321 | 0.000118917 | 0.000908725 | 0.747630859 |
| T2DM | LEC | *PDCD10* | 0.947877281 | 0.005341492 | 0.009044428 | 1 |
| T2DM | LEC | *WNT5A* | -0.829697857 | 0.000131173 | 0.000908725 | 0.824684703 |
| T2DM | LEC | *PRKAR2A* | 0.56962019 | 0.008734595 | 0.013756112 | 1 |
| T2DM | LEC | *HYAL1* | -0.457580624 | 0.001539163 | 0.003262549 | 1 |
| T2DM | LEC | *CYB561D2* | 0.603743627 | 0.00273474 | 0.005133864 | 1 |
| T2DM | LEC | *C3orf14* | 0.485186981 | 0.000417834 | 0.001322531 | 1 |
| T2DM | LEC | *IFT57* | 1.19357324 | 0.003956539 | 0.006955376 | 1 |
| T2DM | LEC | *IQCG* | 0.795218606 | 0.000278409 | 0.00108013 | 1 |
| T2DM | LEC | *GBE1* | 0.558319609 | 0.000360589 | 0.001224756 | 1 |
| T2DM | LEC | *UMPS* | 0.740115961 | 0.025562161 | 0.035555157 | 1 |
| T2DM | LEC | *NCBP2* | 1.270807926 | 0.000473334 | 0.001412904 | 1 |
| T2DM | LEC | *SLC41A3* | -0.457601845 | 0.00022532 | 0.000992757 | 1 |
| T2DM | LEC | *PLXNA1* | 0.520346828 | 0.000106389 | 0.000908725 | 0.668868448 |
| T2DM | LEC | *ABTB1* | 1.326145502 | 0.003122831 | 0.005707336 | 1 |
| T2DM | LEC | *EFCC1* | -0.127468742 | 7.59E-05 | 0.000908725 | 0.477255778 |
| T2DM | LEC | *NEK11* | 0.880816564 | 0.010702535 | 0.016423441 | 1 |
| T2DM | LEC | *PLSCR4* | -0.345262927 | 0.00058119 | 0.001621811 | 1 |
| T2DM | LEC | *HEMK1* | 0.606243723 | 2.96E-05 | 0.000908725 | 0.185923522 |
| T2DM | LEC | *WDR48* | 0.801640825 | 0.028662486 | 0.039422676 | 1 |
| T2DM | LEC | *COMMD2* | 0.888813389 | 0.000757555 | 0.001932935 | 1 |
| T2DM | LEC | *ABCC5* | 0.704953323 | 0.002168677 | 0.004276812 | 1 |
| T2DM | LEC | *AADAC* | 0.1353018 | 0.031470331 | 0.042807003 | 1 |
| T2DM | LEC | *ARHGEF26* | 0.444475044 | 9.77E-05 | 0.000908725 | 0.614329987 |
| T2DM | LEC | *KLHL24* | 0.74600822 | 0.000337237 | 0.001184473 | 1 |
| T2DM | LEC | *PLCH1* | 0.589419387 | 4.61E-05 | 0.000908725 | 0.289621291 |
| T2DM | LEC | *CLCN2* | 2.360865511 | 0.000269569 | 0.001065229 | 1 |
| T2DM | LEC | *NEK4* | 1.106227191 | 0.001761586 | 0.003637139 | 1 |
| T2DM | LEC | *LMAN2L* | 1.079177185 | 0.001152887 | 0.002610082 | 1 |
| T2DM | LEC | *PIKFYVE* | 0.709732661 | 0.000784466 | 0.001980697 | 1 |
| T2DM | LEC | *KCNIP3* | 2.005242208 | 7.34E-05 | 0.000908725 | 0.461188011 |
| T2DM | LEC | *FAHD2A* | 1.364817471 | 0.000590666 | 0.001641698 | 1 |
| T2DM | LEC | *NCL* | 0.865034574 | 0.006366149 | 0.010488464 | 1 |
| T2DM | LEC | *ACTR1B* | 1.253979759 | 0.000584916 | 0.001630039 | 1 |
| T2DM | LEC | *SLC35F5* | 0.852347237 | 0.012246136 | 0.018432238 | 1 |
| T2DM | LEC | *ACTR3* | 0.807394381 | 0.015913025 | 0.023250102 | 1 |
| T2DM | LEC | *STEAP3* | 0.371325948 | 0.001042666 | 0.002427203 | 1 |
| T2DM | LEC | *TFCP2L1* | 0.729746167 | 6.79E-05 | 0.000908725 | 0.427075749 |

| | | | | | | |
|---|---|---|---|---|---|---|
| T2DM | LEC | *DNAJC27* | -1.312268878 | 0.000752367 | 0.001926061 | 1 |
| T2DM | LEC | *STAM2* | 0.897144389 | 0.003425061 | 0.006175325 | 1 |
| T2DM | LEC | *GPD2* | 0.712351456 | 0.000656301 | 0.00175806 | 1 |
| T2DM | LEC | *CENPA* | -0.561389148 | 0.00025009 | 0.001023586 | 1 |
| T2DM | LEC | *TANC1* | -0.47424029 | 0.00026673 | 0.001057634 | 1 |
| T2DM | LEC | *ITGB6* | 0.420959959 | 0.013959955 | 0.02071424 | 1 |
| T2DM | LEC | *ITGA4* | 0.740373851 | 0.000169345 | 0.000910705 | 1 |
| T2DM | LEC | *PPM1G* | 0.659545305 | 0.013503379 | 0.020098425 | 1 |
| T2DM | LEC | *PCSK4* | 0.680880514 | 0.000447236 | 0.001364277 | 1 |
| T2DM | LEC | *GRB14* | 0.388895807 | 0.000137688 | 0.000908725 | 0.865642716 |
| T2DM | LEC | *CLIP4* | 0.427106471 | 0.00015175 | 0.000910197 | 0.954054526 |
| T2DM | LEC | *HTRA2* | -1.483037768 | 0.001357534 | 0.00296245 | 1 |
| T2DM | LEC | *GALNT3* | 0.686899926 | 0.00053483 | 0.001531431 | 1 |
| T2DM | LEC | *POLE4* | 1.269209737 | 0.000516304 | 0.001495765 | 1 |
| T2DM | LEC | *MRPL19* | 0.748479844 | 0.017012728 | 0.024667671 | 1 |
| T2DM | LEC | *LANCL1* | 0.962033864 | 0.001529407 | 0.00324515 | 1 |
| T2DM | LEC | *WDR75* | 1.01632042 | 0.002095833 | 0.004157936 | 1 |
| T2DM | LEC | *EFEMP1* | -0.406839587 | 0.000547698 | 0.001555274 | 1 |
| T2DM | LEC | *DNAH6* | 0.194909613 | 0.011482503 | 0.017437317 | 1 |
| T2DM | LEC | *GGCX* | 0.47596223 | 0.000463081 | 0.001396351 | 1 |
| T2DM | LEC | *GNLY* | 0.352994094 | 0.000117618 | 0.000908725 | 0.739465881 |
| T2DM | LEC | *SF3B1* | 0.775657782 | 0.004062028 | 0.007117606 | 1 |
| T2DM | LEC | *ST3GAL5* | 0.615246614 | 0.00047512 | 0.00141635 | 1 |
| T2DM | LEC | *PLCD4* | 1.556619014 | 0.01364792 | 0.020279951 | 1 |
| T2DM | LEC | *CHMP3* | -1.315456687 | 0.002121886 | 0.004206969 | 1 |
| T2DM | LEC | *ZNF142* | 0.853229104 | 0.000342394 | 0.001192594 | 1 |
| T2DM | LEC | *WNT6* | 3.744288158 | 0.000329485 | 0.001170986 | 1 |
| T2DM | LEC | *HDLBP* | 0.71017213 | 0.016775043 | 0.024362368 | 1 |
| T2DM | LEC | *STK25* | -0.920810617 | 0.012005448 | 0.018117679 | 1 |
| T2DM | LEC | *TPO* | 0.30992481 | 0.000444471 | 0.001357774 | 1 |
| T2DM | LEC | *TAF1B* | 0.840193939 | 0.001504512 | 0.003202916 | 1 |
| T2DM | LEC | *PRKD3* | 0.847086773 | 0.004980619 | 0.008525224 | 1 |
| T2DM | LEC | *SLC25A12* | 0.627295004 | 0.001032114 | 0.002411901 | 1 |
| T2DM | LEC | *SDC1* | 3.228077055 | 3.57E-06 | 0.000439835 | 0.022431571 |
| T2DM | LEC | *SOS1* | 0.5411014 | 0.001279948 | 0.002822591 | 1 |
| T2DM | LEC | *WIPF1* | 0.725122855 | 0.003036939 | 0.005580596 | 1 |
| T2DM | LEC | *COX7A2L* | -0.712595852 | 0.001023924 | 0.002398438 | 1 |
| T2DM | LEC | *PNO1* | -1.104063688 | 0.009324011 | 0.014549531 | 1 |
| T2DM | LEC | *THADA* | 0.774728179 | 0.003195206 | 0.005820998 | 1 |
| T2DM | LEC | *ARID3A* | 0.661465301 | 0.000170171 | 0.000910705 | 1 |
| T2DM | LEC | *CD207* | 8.97098646 | 4.09E-05 | 0.000908725 | 0.257340511 |
| T2DM | LEC | *GRIN3B* | 1.32791289 | 0.001739928 | 0.003603072 | 1 |
| T2DM | LEC | *VAX2* | 0.593661451 | 0.000109666 | 0.000908725 | 0.689472604 |
| T2DM | LEC | *MSH6* | 0.998448093 | 0.001555544 | 0.003288402 | 1 |
| T2DM | LEC | *PLEKHA3* | 1.121284604 | 0.000956356 | 0.00227578 | 1 |
| T2DM | LEC | *ALMS1* | 0.800736944 | 0.002134668 | 0.004225649 | 1 |
| T2DM | LEC | *BCL9* | 0.808660784 | 5.81E-05 | 0.000908725 | 0.365327511 |
| T2DM | LEC | *DHCR24* | 0.93184364 | 0.000144353 | 0.000908725 | 0.907545444 |
| T2DM | LEC | *SCP2* | 0.841400067 | 0.000864501 | 0.002121437 | 1 |
| T2DM | LEC | *CEP104* | 0.859398319 | 0.018490137 | 0.026589088 | 1 |

| | | | | | | |
|---|---|---|---|---|---|---|
| T2DM | LEC | *FAM20B* | 1.050240056 | 0.00944965 | 0.01471636 | 1 |
| T2DM | LEC | *TMEM59* | 0.908309585 | 0.005345106 | 0.009048111 | 1 |
| T2DM | LEC | *WRAP73* | 1.312209128 | 0.025013997 | 0.034854388 | 1 |
| T2DM | LEC | *MRPL37* | -0.685255869 | 0.020936588 | 0.029733077 | 1 |
| T2DM | LEC | *ICMT* | 0.825298141 | 0.000436343 | 0.00134367 | 1 |
| T2DM | LEC | *KCNC4* | -0.994572259 | 0.001877693 | 0.003825359 | 1 |
| T2DM | LEC | *EDEM3* | 0.848800986 | 0.002184799 | 0.004295588 | 1 |
| T2DM | LEC | *CAPZA1* | 0.708768435 | 0.006667672 | 0.010916577 | 1 |
| T2DM | LEC | *DOCK7* | 0.692056227 | 0.00046816 | 0.001403402 | 1 |
| T2DM | LEC | *FBXO6* | 0.625629447 | 0.000135528 | 0.000908725 | 0.852065578 |
| T2DM | LEC | *C1orf21* | -0.621095345 | 0.001613924 | 0.00339129 | 1 |
| T2DM | LEC | *DNAJC6* | 0.573274931 | 0.000224766 | 0.000992757 | 1 |
| T2DM | LEC | *LEPR* | 0.820778089 | 0.007639064 | 0.012289353 | 1 |
| T2DM | LEC | *MIIP* | 1.020144427 | 0.002036459 | 0.004070976 | 1 |
| T2DM | LEC | *NCF2* | 0.557435485 | 0.000125118 | 0.000908725 | 0.786616085 |
| T2DM | LEC | *RGS2* | 0.788754308 | 0.000119293 | 0.000908725 | 0.749994344 |
| T2DM | LEC | *OLFML3* | 0.593550729 | 0.00015114 | 0.000910197 | 0.950216693 |
| T2DM | LEC | *CFHR3* | -0.457559939 | 0.000437648 | 0.001345504 | 1 |
| T2DM | LEC | *TTF2* | 0.58567204 | 0.000114315 | 0.000908725 | 0.718699611 |
| T2DM | LEC | *TMEM9* | -0.721305873 | 0.015795498 | 0.023105092 | 1 |
| T2DM | LEC | *WARS2* | 0.571786871 | 0.024872052 | 0.034689274 | 1 |
| T2DM | LEC | *OSCP1* | 1.174362474 | 0.000292592 | 0.001108817 | 1 |
| T2DM | LEC | *C1orf109* | 0.510084196 | 0.000270321 | 0.001066753 | 1 |
| T2DM | LEC | *TBCE* | 0.648005809 | 0.00090231 | 0.002192819 | 1 |
| T2DM | LEC | *MTR* | 0.772678774 | 0.000734529 | 0.001898843 | 1 |
| T2DM | LEC | *KCNQ4* | -0.51991317 | 3.36E-05 | 0.000908725 | 0.211427948 |
| T2DM | LEC | *ACADM* | 0.934300248 | 0.007213899 | 0.011683096 | 1 |
| T2DM | LEC | *PADI2* | 0.781284151 | 0.000139159 | 0.000908725 | 0.87489086 |
| T2DM | LEC | *CTBS* | 0.781798357 | 0.030870436 | 0.042072931 | 1 |
| T2DM | LEC | *KLHL12* | 0.710701188 | 0.00019655 | 0.000955691 | 1 |
| T2DM | LEC | *PLA2G2D* | -2.629070738 | 1.46E-05 | 0.000902248 | 0.091611666 |
| T2DM | LEC | *RBBP5* | 0.96699006 | 0.010923789 | 0.016722147 | 1 |
| T2DM | LEC | *KIF17* | 0.78680052 | 0.000492883 | 0.001449584 | 1 |
| T2DM | LEC | *CD46* | 0.719844902 | 0.000591248 | 0.001642588 | 1 |
| T2DM | LEC | *FAAH* | 0.451997632 | 3.26E-05 | 0.000908725 | 0.205187077 |
| T2DM | LEC | *TMED5* | 0.921711048 | 0.003380864 | 0.00611317 | 1 |
| T2DM | LEC | *CNN3* | -0.757517456 | 0.011717654 | 0.017755818 | 1 |
| T2DM | LEC | *PRRC2C* | 0.779244463 | 0.004600272 | 0.007933414 | 1 |
| T2DM | LEC | *ABCD3* | 0.938531197 | 0.003100981 | 0.005674 | 1 |
| T2DM | LEC | *VAMP4* | 1.143609723 | 0.001130266 | 0.002573698 | 1 |
| T2DM | LEC | *PTBP2* | 0.671630755 | 0.012290691 | 0.018490446 | 1 |
| T2DM | LEC | *TNFSF4* | 0.457410966 | 0.001708294 | 0.003552777 | 1 |
| T2DM | LEC | *DARS2* | -1.14881806 | 0.00532909 | 0.009028291 | 1 |
| T2DM | LEC | *IRF6* | 0.701579219 | 0.000367202 | 0.001237855 | 1 |
| T2DM | LEC | *DIEXF* | -1.178441001 | 0.001498596 | 0.003194871 | 1 |
| T2DM | LEC | *SERPINC1* | 1.170667381 | 8.83E-06 | 0.000676784 | 0.055496258 |
| T2DM | LEC | *RCAN3* | 0.667581133 | 0.000152216 | 0.000910197 | 0.956982222 |
| T2DM | LEC | *RCOR3* | 0.788827512 | 0.000939314 | 0.002244571 | 1 |
| T2DM | LEC | *MTFR1L* | 1.026782032 | 6.55E-05 | 0.000908725 | 0.411754837 |
| T2DM | LEC | *NSL1* | 0.832232605 | 0.020260422 | 0.028870642 | 1 |

| | | | | | | |
|---|---|---|---|---|---|---|
| T2DM | LEC | *RPA2* | 1.158725972 | 0.001662625 | 0.003475042 | 1 |
| T2DM | LEC | *TXNDC12* | 1.005869289 | 0.000532132 | 0.001527633 | 1 |
| T2DM | LEC | *ESYT2* | 0.682824151 | 0.002964445 | 0.005473558 | 1 |
| T2DM | LEC | *KMT2A* | 0.76811223 | 0.015607153 | 0.022861643 | 1 |
| T2DM | LEC | *IFT46* | -0.987797728 | 0.0145423 | 0.021456804 | 1 |
| T2DM | LEC | *DDX59* | -0.866574915 | 0.002824742 | 0.005268215 | 1 |
| T2DM | LEC | *ATF6* | 0.798878467 | 0.010958561 | 0.016759054 | 1 |
| T2DM | LEC | *CASC1* | 0.420399848 | 0.000650074 | 0.001746587 | 1 |
| T2DM | LEC | *LRMP* | 0.664089428 | 0.02170006 | 0.030671825 | 1 |
| T2DM | LEC | *ATP10B* | -1.166074801 | 0.00037269 | 0.001243243 | 1 |
| T2DM | LEC | *SPCS2* | 0.981635149 | 0.002352164 | 0.004545974 | 1 |
| T2DM | LEC | *ELOVL4* | 0.863628004 | 1.08E-06 | 0.000241487 | 0.006761646 |
| T2DM | LEC | *FILIP1* | -0.653314335 | 0.002414869 | 0.004652293 | 1 |
| T2DM | LEC | *SGIP1* | -0.137767 | 0.000162948 | 0.000910705 | 1 |
| T2DM | LEC | *AKAP7* | 0.432316221 | 0.008209538 | 0.013065728 | 1 |
| T2DM | LEC | *SLC16A7* | 0.635804688 | 0.000428589 | 0.001336576 | 1 |
| T2DM | LEC | *VAMP8* | 0.664405953 | 0.00025399 | 0.001032458 | 1 |
| T2DM | LEC | *FOXO3* | 0.645313211 | 0.000515464 | 0.001494799 | 1 |
| T2DM | LEC | *TGIF2* | 1.209010046 | 0.000163637 | 0.000910705 | 1 |
| T2DM | LEC | *ABCG2* | -0.858782457 | 0.000465684 | 0.001398163 | 1 |
| T2DM | LEC | *MFSD1* | 0.594929121 | 0.00015407 | 0.000910197 | 0.968635596 |
| T2DM | LEC | *RAB3GAP2* | 0.838696183 | 0.002956759 | 0.005464546 | 1 |
| T2DM | LEC | *UCHL3* | 2.040506813 | 0.005634526 | 0.009469197 | 1 |
| T2DM | LEC | *WDR35* | 1.002083812 | 0.009452413 | 0.014717018 | 1 |
| T2DM | LEC | *NDUFB3* | 1.103370704 | 0.00111741 | 0.002553675 | 1 |
| T2DM | LEC | *TRPM6* | 0.860272006 | 0.006028966 | 0.010018848 | 1 |
| T2DM | LEC | *GDA* | 0.847684658 | 0.000162931 | 0.000910705 | 1 |
| T2DM | LEC | *C2orf40* | 0.749659394 | 0.000332188 | 0.001173862 | 1 |
| T2DM | LEC | *CPSF3* | 1.095469248 | 0.000389953 | 0.001274903 | 1 |
| T2DM | LEC | *PIGZ* | 0.38401432 | 0.00477872 | 0.00820645 | 1 |
| T2DM | LEC | *SENP5* | 0.923095931 | 0.001934191 | 0.003918369 | 1 |
| T2DM | LEC | *FKBP15* | 0.828109461 | 0.0001525 | 0.000910197 | 0.95876489 |
| T2DM | LEC | *CNTRL* | 0.837112101 | 0.001276185 | 0.002817196 | 1 |
| T2DM | LEC | *TRIM32* | 1.774550131 | 0.00848142 | 0.013411139 | 1 |
| T2DM | LEC | *PHF19* | 0.498866111 | 0.000137836 | 0.000908725 | 0.866577732 |
| T2DM | LEC | *BSPRY* | 1.833375375 | 0.000125559 | 0.000908725 | 0.789386746 |
| T2DM | LEC | *PPP6C* | 0.881021167 | 0.001927589 | 0.003910536 | 1 |
| T2DM | LEC | *HSDL2* | 0.517326246 | 0.000611159 | 0.001680086 | 1 |
| T2DM | LEC | *INVS* | 0.724129216 | 0.013197093 | 0.019689161 | 1 |
| T2DM | LEC | *KDSR* | 0.721236948 | 0.000216317 | 0.000979981 | 1 |
| T2DM | LEC | *DCAF4* | 1.234588407 | 2.08E-05 | 0.000908725 | 0.130914475 |
| T2DM | LEC | *IFI27L2* | 0.872808618 | 0.00390583 | 0.006899678 | 1 |
| T2DM | LEC | *IRF2BPL* | -0.888631394 | 0.003558731 | 0.006368842 | 1 |
| T2DM | LEC | *PPP4R4* | 0.577073629 | 0.000794381 | 0.00199931 | 1 |
| T2DM | LEC | *ZC2HC1C* | 1.233369455 | 0.000236275 | 0.001005438 | 1 |
| T2DM | LEC | *ALDH6A1* | 0.458052764 | 0.000839051 | 0.002076816 | 1 |
| T2DM | LEC | *EIF2B2* | 0.773594727 | 0.008505021 | 0.013441696 | 1 |
| T2DM | LEC | *COQ6* | 1.129166723 | 0.000226822 | 0.000992757 | 1 |
| T2DM | LEC | *GPR75* | -0.813887601 | 0.00022876 | 0.000995303 | 1 |
| T2DM | LEC | *KLHL29* | 0.83064373 | 0.015897708 | 0.023233121 | 1 |

| | | | | | | |
|---|---|---|---|---|---|---|
| T2DM | LEC | *ATAD2B* | 0.784394163 | 0.005093722 | 0.008690428 | 1 |
| T2DM | LEC | *ATL2* | 0.848288102 | 0.001071995 | 0.002473259 | 1 |
| T2DM | LEC | *CRIPT* | 0.868274911 | 0.002392304 | 0.00461504 | 1 |
| T2DM | LEC | *EPCAM* | -0.708884181 | 2.73E-05 | 0.000908725 | 0.171768658 |
| T2DM | LEC | *SLC17A5* | 0.724999289 | 0.002014841 | 0.004034174 | 1 |
| T2DM | LEC | *OGFRL1* | 0.93262758 | 0.000270463 | 0.001066753 | 1 |
| T2DM | LEC | *CUTC* | 0.930009298 | 0.002210472 | 0.004329357 | 1 |
| T2DM | LEC | *PYROXD2* | 0.37363365 | 0.019036382 | 0.027312126 | 1 |
| T2DM | LEC | *MXI1* | 0.964869986 | 0.00599725 | 0.009969517 | 1 |
| T2DM | LEC | *HELLS* | 0.879814205 | 0.000199272 | 0.000961874 | 1 |
| T2DM | LEC | *TCTN3* | -0.459580958 | 0.000423777 | 0.001331479 | 1 |
| T2DM | LEC | *AVPI1* | 0.726740521 | 0.000316577 | 0.001151139 | 1 |
| T2DM | LEC | *WDR11* | 0.811703566 | 0.00651405 | 0.0106957 | 1 |
| T2DM | LEC | *C10orf76* | 0.819982522 | 0.002017559 | 0.004037045 | 1 |
| T2DM | LEC | *KCNIP2* | 0.326801905 | 5.65E-05 | 0.000908725 | 0.355415028 |
| T2DM | LEC | *C10orf95* | 0.573823994 | 0.00281602 | 0.005255066 | 1 |
| T2DM | LEC | *KANSL1* | 0.68910754 | 0.003885882 | 0.00687023 | 1 |
| T2DM | LEC | *HOXB5* | -1.440314747 | 0.000334345 | 0.001176051 | 1 |
| T2DM | LEC | *CD274* | -1.100515943 | 0.014990781 | 0.022050189 | 1 |
| T2DM | LEC | *LRP11* | 0.59661049 | 0.002394891 | 0.004618613 | 1 |
| T2DM | LEC | *CCDC170* | 0.308966934 | 0.000142659 | 0.000908725 | 0.896899687 |
| T2DM | LEC | *CYSTM1* | 0.737795851 | 0.000599197 | 0.001658076 | 1 |
| T2DM | LEC | *WDR55* | 1.107277934 | 0.018461064 | 0.026553354 | 1 |
| T2DM | LEC | *PCDHB14* | -0.841526314 | 0.000334465 | 0.001176051 | 1 |
| T2DM | LEC | *PCDHB12* | -0.615910175 | 0.00103369 | 0.00241412 | 1 |
| T2DM | LEC | *SEC16B* | 0.94860267 | 9.94E-05 | 0.000908725 | 0.624957615 |
| T2DM | LEC | *GORAB* | 1.188959288 | 0.001731978 | 0.003588894 | 1 |
| T2DM | LEC | *ACAT2* | 1.388395575 | 0.002679159 | 0.005050637 | 1 |
| T2DM | LEC | *SNX19* | 0.770126418 | 0.000688681 | 0.001819172 | 1 |
| T2DM | LEC | *MRC1* | 0.444361321 | 0.000133344 | 0.000908725 | 0.838335367 |
| T2DM | LEC | *CCDC77* | 0.630898755 | 0.000901619 | 0.002191988 | 1 |
| T2DM | LEC | *ENOX1* | 0.808658703 | 0.000809857 | 0.002022874 | 1 |
| T2DM | LEC | *DNAJC15* | 1.007169367 | 0.002195352 | 0.004306451 | 1 |
| T2DM | LEC | *WBP4* | 0.891245235 | 0.018945433 | 0.02719405 | 1 |
| T2DM | LEC | *SMAD9* | -0.161811892 | 0.000156945 | 0.000910705 | 0.986710788 |
| T2DM | LEC | *SIL1* | 0.757546572 | 0.000541939 | 0.001545201 | 1 |
| T2DM | LEC | *ZFP30* | 0.783042208 | 0.005170372 | 0.008798793 | 1 |
| T2DM | LEC | *APAF1* | 0.981004736 | 0.003020053 | 0.0055534 | 1 |
| T2DM | LEC | *ADRA1A* | 0.362163914 | 9.11E-06 | 0.000681925 | 0.057281683 |
| T2DM | LEC | *EPHX2* | 0.500780025 | 0.033775017 | 0.045596636 | 1 |
| T2DM | LEC | *CRISPLD1* | 0.461465992 | 0.00010577 | 0.000908725 | 0.664978311 |
| T2DM | LEC | *RDH10* | 0.777472198 | 0.000167039 | 0.000910705 | 1 |
| T2DM | LEC | *SCPEP1* | 0.339741771 | 0.000631008 | 0.001714412 | 1 |
| T2DM | LEC | *TEX14* | 0.831955073 | 3.56E-06 | 0.000439835 | 0.022373491 |
| T2DM | LEC | *TRIM6* | 0.698260033 | 0.028359413 | 0.039057093 | 1 |
| T2DM | LEC | *PRB2* | 1.576032029 | 3.89E-05 | 0.000908725 | 0.244805901 |
| T2DM | LEC | *PYROXD1* | 0.615901542 | 0.009943554 | 0.015359982 | 1 |
| T2DM | LEC | *A1BG* | 1.225385209 | 3.36E-05 | 0.000908725 | 0.211342421 |
| T2DM | LEC | *ZNF211* | 0.949519085 | 0.000127789 | 0.000908725 | 0.803412399 |
| T2DM | LEC | *PDZRN3* | 0.629055417 | 0.000756808 | 0.001932596 | 1 |

| | | | | | | |
|---|---|---|---|---|---|---|
| T2DM | LEC | *LHX4* | 1.1400721 | 0.021603425 | 0.030542103 | 1 |
| T2DM | LEC | *SEC22A* | 0.826265775 | 1.54E-05 | 0.000908725 | 0.096521676 |
| T2DM | LEC | *CSTA* | 0.409909864 | 0.00033005 | 0.001172329 | 1 |
| T2DM | LEC | *POPDC2* | 2.04092016 | 0.00813067 | 0.012957547 | 1 |
| T2DM | LEC | *CD80* | -1.969402213 | 0.005741043 | 0.009614795 | 1 |
| T2DM | LEC | *CAT* | 0.346222317 | 0.003043944 | 0.005589157 | 1 |
| T2DM | LEC | *PILRB* | 0.308254451 | 0.002047811 | 0.00408717 | 1 |
| T2DM | LEC | *POLR3GL* | -1.250453748 | 0.00905691 | 0.014199698 | 1 |
| T2DM | LEC | *TNFSF10* | 0.889920686 | 0.000727895 | 0.001887164 | 1 |
| T2DM | LEC | *ZNF639* | 1.383597411 | 0.000621 | 0.001699706 | 1 |
| T2DM | LEC | *LIAS* | 0.931150619 | 0.000823021 | 0.002047095 | 1 |
| T2DM | LEC | *CLCC1* | 1.096054097 | 0.002441733 | 0.004689632 | 1 |
| T2DM | LEC | *GPSM2* | 1.039736217 | 0.001778027 | 0.003661947 | 1 |
| T2DM | LEC | *CXCR4* | 0.566557086 | 0.000310898 | 0.001138389 | 1 |
| T2DM | LEC | *ACVR2A* | 0.827824904 | 0.000921324 | 0.002218447 | 1 |
| T2DM | LEC | *POLK* | 0.808405968 | 0.004977148 | 0.008521605 | 1 |
| T2DM | LEC | *FLT3* | -0.798125074 | 0.000508693 | 0.001477891 | 1 |
| T2DM | LEC | *MTIF3* | 1.013234748 | 0.00445052 | 0.00769962 | 1 |
| T2DM | LEC | *RASL11A* | 0.77444411 | 0.000715685 | 0.001863923 | 1 |
| T2DM | LEC | *FMOD* | -0.995569957 | 0.023613032 | 0.03311513 | 1 |
| T2DM | LEC | *SERAC1* | 0.686461397 | 0.00027707 | 0.00108013 | 1 |
| T2DM | LEC | *LDB3* | 0.480530827 | 5.49E-05 | 0.000908725 | 0.345211294 |
| T2DM | LEC | *OPN4* | 1.742208461 | 0.000792296 | 0.001996459 | 1 |
| T2DM | LEC | *FAM35A* | 0.761638037 | 0.001245088 | 0.002764077 | 1 |
| T2DM | LEC | *NAA60* | 1.499334411 | 0.000395861 | 0.001286191 | 1 |
| T2DM | LEC | *SPATA1* | 0.962994314 | 0.009665484 | 0.015000468 | 1 |
| T2DM | LEC | *LRRC39* | 0.505572814 | 0.001226476 | 0.002729718 | 1 |
| T2DM | LEC | *CCDC18* | 0.665709677 | 0.00037063 | 0.001241526 | 1 |
| T2DM | LEC | *RPAP2* | 0.789522425 | 0.014411025 | 0.021308116 | 1 |
| T2DM | LEC | *PMS2* | 1.011113591 | 1.75E-05 | 0.000908725 | 0.110166484 |
| T2DM | LEC | *KIAA0087* | 1.067204777 | 0.000843567 | 0.002083073 | 1 |
| T2DM | LEC | *HERPUD2* | 0.891591623 | 0.00084059 | 0.002079807 | 1 |
| T2DM | LEC | *FKBP9* | 0.900053061 | 0.000887655 | 0.002164702 | 1 |
| T2DM | LEC | *CCZ1* | 1.062559482 | 0.00015327 | 0.000910197 | 0.963605804 |
| T2DM | LEC | *SMU1* | 0.679811321 | 0.000135431 | 0.000908725 | 0.851452635 |
| T2DM | LEC | *GLIPR2* | 0.821972388 | 0.002969986 | 0.005482179 | 1 |
| T2DM | LEC | *SLC25A51* | 0.441602796 | 1.34E-05 | 0.000893669 | 0.084004927 |
| T2DM | LEC | *DCAF10* | 0.73199843 | 0.024766749 | 0.03457117 | 1 |
| T2DM | LEC | *TRIM24* | -0.680426466 | 0.001742634 | 0.003605114 | 1 |
| T2DM | LEC | *BICC1* | 0.331284862 | 0.000168408 | 0.000910705 | 1 |
| T2DM | LEC | *SLC25A16* | 0.626814233 | 9.81E-05 | 0.000908725 | 0.617016399 |
| T2DM | LEC | *RBM19* | 1.124753843 | 0.002684236 | 0.005057419 | 1 |
| T2DM | LEC | *ACADS* | 0.567791784 | 0.000192408 | 0.000946191 | 1 |
| T2DM | LEC | *SSPN* | 0.705374848 | 1.09E-05 | 0.000799063 | 0.068719385 |
| T2DM | LEC | *CCDC91* | 0.574899748 | 0.000236752 | 0.001005449 | 1 |
| T2DM | LEC | *PKN1* | 1.017938177 | 0.013232929 | 0.019737941 | 1 |
| T2DM | LEC | *EBPL* | -0.33546954 | 0.005646575 | 0.009481843 | 1 |
| T2DM | LEC | *NLN* | 1.042672339 | 0.000302664 | 0.001124766 | 1 |
| T2DM | LEC | *CENPK* | 0.329116227 | 0.00048909 | 0.001442942 | 1 |
| T2DM | LEC | *OPTN* | 0.579354043 | 0.032570632 | 0.044150834 | 1 |

| | | | | | | |
|---|---|---|---|---|---|---|
| T2DM | LEC | *ATF1* | 1.056067791 | 0.000302125 | 0.001123941 | 1 |
| T2DM | LEC | *TSFM* | 1.045527138 | 8.11E-05 | 0.000908725 | 0.510009054 |
| T2DM | LEC | *ARHGAP9* | 0.437304499 | 0.000128645 | 0.000908725 | 0.808792138 |
| T2DM | LEC | *ORMDL2* | 0.99528203 | 0.000208464 | 0.000972609 | 1 |
| T2DM | LEC | *PDE1B* | 0.468302699 | 0.000248149 | 0.001023026 | 1 |
| T2DM | LEC | *IKZF4* | 0.763010819 | 0.001032239 | 0.002411901 | 1 |
| T2DM | LEC | *SARDH* | 0.96423699 | 0.000110955 | 0.000908725 | 0.697576416 |
| T2DM | LEC | *STIL* | 0.530232058 | 0.000106411 | 0.000908725 | 0.669006985 |
| T2DM | LEC | *ACVR1C* | 0.574560147 | 8.01E-05 | 0.000908725 | 0.50354879 |
| T2DM | LEC | *SLC36A1* | 0.496098324 | 0.001385407 | 0.003006836 | 1 |
| T2DM | LEC | *PLA2G12A* | 1.025071315 | 0.00098692 | 0.002326496 | 1 |
| T2DM | LEC | *C4BPB* | 0.518237351 | 0.002189824 | 0.004298291 | 1 |
| T2DM | LEC | *RAB38* | 0.666373938 | 0.000156402 | 0.000910705 | 0.983296374 |
| T2DM | LEC | *CKS2* | 1.319682558 | 0.000700335 | 0.001836115 | 1 |
| T2DM | LEC | *CHPF* | -0.699422548 | 0.007185562 | 0.011646205 | 1 |
| T2DM | LEC | *MOGAT1* | 0.898274402 | 0.000337745 | 0.001185596 | 1 |
| T2DM | LEC | *FAM210B* | 1.215109704 | 0.000303482 | 0.001125658 | 1 |
| T2DM | LEC | *SNX21* | 0.452364753 | 0.000124502 | 0.000908725 | 0.782745473 |
| T2DM | LEC | *SDC4* | -1.211737073 | 0.000240269 | 0.00100947 | 1 |
| T2DM | LEC | *TOX2* | -0.327994963 | 0.004917039 | 0.008425573 | 1 |
| T2DM | LEC | *ARFGEF2* | 0.879620102 | 0.004694312 | 0.008081363 | 1 |
| T2DM | LEC | *CSE1L* | 0.870064164 | 0.003979058 | 0.006985852 | 1 |
| T2DM | LEC | *CDH26* | 1.396137236 | 0.000699728 | 0.001836115 | 1 |
| T2DM | LEC | *STX16* | 0.93118079 | 0.000878546 | 0.002146684 | 1 |
| T2DM | LEC | *RNF114* | 0.898346335 | 0.002031285 | 0.004061925 | 1 |
| T2DM | LEC | *KCNK15* | 1.218317044 | 0.000213788 | 0.0009784 | 1 |
| T2DM | LEC | *NEURL2* | 0.761022072 | 0.006608626 | 0.01083401 | 1 |
| T2DM | LEC | *FASTKD3* | 1.360737891 | 0.000129941 | 0.000908725 | 0.816938629 |
| T2DM | LEC | *CHST8* | -0.465650362 | 0.000184554 | 0.000929091 | 1 |
| T2DM | LEC | *STAMBP* | 1.032871456 | 0.003914119 | 0.006910437 | 1 |
| T2DM | LEC | *ATP8A1* | 0.753724435 | 8.74E-05 | 0.000908725 | 0.549528221 |
| T2DM | LEC | *HIF3A* | -0.229787492 | 0.000429762 | 0.001338907 | 1 |
| T2DM | LEC | *ZNF45* | 1.204253882 | 0.000431497 | 0.00134099 | 1 |
| T2DM | LEC | *SERPINB6* | 0.975190894 | 0.002535299 | 0.004827202 | 1 |
| T2DM | LEC | *NQO2* | 0.813385275 | 0.000307953 | 0.001132221 | 1 |
| T2DM | LEC | *ZNF391* | 0.335647465 | 0.002075744 | 0.004127009 | 1 |
| T2DM | LEC | *RPS10* | -1.765382705 | 0.000712495 | 0.001861013 | 1 |
| T2DM | LEC | *HIST1H2BJ* | 1.122669308 | 0.000237894 | 0.001007844 | 1 |
| T2DM | LEC | *MED20* | 0.784063508 | 0.018319467 | 0.026367786 | 1 |
| T2DM | LEC | *TBCC* | 0.54733569 | 0.001179296 | 0.00265078 | 1 |
| T2DM | LEC | *TCP11* | -1.100548216 | 0.00020843 | 0.000972609 | 1 |
| T2DM | LEC | *MEA1* | -0.815168082 | 0.001061369 | 0.002455954 | 1 |
| T2DM | LEC | *KLHL31* | 0.389116707 | 0.032689983 | 0.044303066 | 1 |
| T2DM | LEC | *COL21A1* | 0.51912009 | 0.000441736 | 0.001351816 | 1 |
| T2DM | LEC | *GLO1* | 1.371658618 | 0.000382931 | 0.001263772 | 1 |
| T2DM | LEC | *CPNE5* | -0.315315323 | 6.56E-05 | 0.000908725 | 0.412363242 |
| T2DM | LEC | *RIOK1* | 1.094291049 | 0.000240365 | 0.00100947 | 1 |
| T2DM | LEC | *RPP40* | 1.150894915 | 0.0217919 | 0.030794712 | 1 |
| T2DM | LEC | *NUP153* | 0.913904724 | 0.002752035 | 0.005160169 | 1 |
| T2DM | LEC | *EEF1E1* | -1.565314994 | 0.02989611 | 0.040895744 | 1 |

| | | | | | | |
|---|---|---|---|---|---|---|
| T2DM | LEC | *EREG* | -0.645138216 | 0.000138819 | 0.000908725 | 0.872756938 |
| T2DM | LEC | *CNOT1* | 0.902655757 | 0.003761447 | 0.006682715 | 1 |
| T2DM | LEC | *BBS2* | 0.911312886 | 0.000409721 | 0.001308237 | 1 |
| T2DM | LEC | *MT1G* | 2.656649849 | 0.00033395 | 0.001176051 | 1 |
| T2DM | LEC | *GOT2* | 1.399963413 | 0.000327519 | 0.001167883 | 1 |
| T2DM | LEC | *TMTC4* | -0.920772942 | 0.005966629 | 0.009934374 | 1 |
| T2DM | LEC | *KIF25* | -0.591411576 | 0.000458138 | 0.001388103 | 1 |
| T2DM | LEC | *GRK4* | 0.460043442 | 0.000844976 | 0.002084721 | 1 |
| T2DM | LEC | *TEKT3* | 0.514089928 | 0.001224139 | 0.002726235 | 1 |
| T2DM | LEC | *MRPS7* | 1.349627099 | 0.005016285 | 0.0085816 | 1 |
| T2DM | LEC | *NUP85* | 0.963024583 | 0.014779775 | 0.021771425 | 1 |
| T2DM | LEC | *MIF4GD* | -1.308719126 | 0.001223025 | 0.00272472 | 1 |
| T2DM | LEC | *NT5C* | 0.606787725 | 0.000101608 | 0.000908725 | 0.638808089 |
| T2DM | LEC | *DDX31* | 0.722913409 | 0.020241943 | 0.028850849 | 1 |
| T2DM | LEC | *OPRL1* | -0.550233797 | 1.17E-06 | 0.000254094 | 0.007368738 |
| T2DM | LEC | *PPDPF* | 1.583125108 | 0.000935613 | 0.002237428 | 1 |
| T2DM | LEC | *CHCHD5* | -1.025446055 | 0.004073657 | 0.007134006 | 1 |
| T2DM | LEC | *PAX8* | 0.510403247 | 0.000260673 | 0.001044521 | 1 |
| T2DM | LEC | *INSIG2* | 1.08482501 | 0.027081728 | 0.037469812 | 1 |
| T2DM | LEC | *PSD4* | 0.697207308 | 0.000141696 | 0.000908725 | 0.890845867 |
| T2DM | LEC | *CLPP* | 1.386328674 | 0.005564486 | 0.009364005 | 1 |
| T2DM | LEC | *SH2D3A* | 0.903587607 | 0.0001803 | 0.00092505 | 1 |
| T2DM | LEC | *TRIP10* | 1.04134237 | 0.001427961 | 0.003079791 | 1 |
| T2DM | LEC | *GPR108* | 0.833263486 | 0.02382037 | 0.033361254 | 1 |
| T2DM | LEC | *RTN2* | 1.039348341 | 0.000288923 | 0.001098223 | 1 |
| T2DM | LEC | *SYMPK* | 0.857338657 | 0.005648993 | 0.009482412 | 1 |
| T2DM | LEC | *GPCPD1* | 0.768335433 | 0.000806744 | 0.002020322 | 1 |
| T2DM | LEC | *SDCBP2* | 1.183239592 | 7.64E-05 | 0.000908725 | 0.480256628 |
| T2DM | LEC | *PANK2* | 0.905133756 | 0.000576454 | 0.001612173 | 1 |
| T2DM | LEC | *PSMF1* | 0.719681728 | 0.014906422 | 0.021937423 | 1 |
| T2DM | LEC | *RBCK1* | 0.884737264 | 0.009766595 | 0.015131242 | 1 |
| T2DM | LEC | *RRBP1* | 0.70695871 | 0.035661192 | 0.047937121 | 1 |
| T2DM | LEC | *BMP2* | -1.734386394 | 0.006845652 | 0.011173056 | 1 |
| T2DM | LEC | *PCSK2* | -2.068872728 | 0.002593671 | 0.004911569 | 1 |
| T2DM | LEC | *MKKS* | 0.997710019 | 0.001307437 | 0.002871064 | 1 |
| T2DM | LEC | *DSTN* | 0.837023518 | 0.011226343 | 0.017106161 | 1 |
| T2DM | LEC | *MGME1* | 0.870131345 | 0.000788402 | 0.001989039 | 1 |
| T2DM | LEC | *ITPA* | 0.988316967 | 0.000287944 | 0.001097202 | 1 |
| T2DM | LEC | *TMEM74B* | -0.515683249 | 0.000511061 | 0.001484083 | 1 |
| T2DM | LEC | *FAM110A* | 0.473775846 | 0.000108028 | 0.000908725 | 0.679172019 |
| T2DM | LEC | *ZNF436* | 0.468250613 | 7.93E-06 | 0.000664383 | 0.049828702 |
| T2DM | LEC | *GDF5* | 0.782049317 | 0.00039141 | 0.001277842 | 1 |
| T2DM | LEC | *MMP24* | 0.920713353 | 0.000293574 | 0.001109196 | 1 |
| T2DM | LEC | *ST3GAL3* | 0.781817948 | 0.005689031 | 0.009540394 | 1 |
| T2DM | LEC | *KLC1* | 1.015058073 | 0.002151459 | 0.004250856 | 1 |
| T2DM | LEC | *MCF2L* | 1.158303514 | 0.000851224 | 0.002095399 | 1 |
| T2DM | LEC | *F10* | 0.552198949 | 0.000124766 | 0.000908725 | 0.784403984 |
| T2DM | LEC | *PROZ* | -0.497758253 | 0.000137911 | 0.000908725 | 0.86704569 |
| T2DM | LEC | *IGFLR1* | 1.386272818 | 0.000124569 | 0.000908725 | 0.783167836 |
| T2DM | LEC | *CAPNS1* | -0.663628774 | 0.002980736 | 0.005495568 | 1 |

| | | | | | | |
|---|---|---|---|---|---|---|
| T2DM | LEC | *PRDX5* | 1.113741 | 0.008211022 | 0.013065728 | 1 |
| T2DM | LEC | *SCAF1* | 1.07790251 | 0.000202183 | 0.00096305 | 1 |
| T2DM | LEC | *PRR12* | 1.36572578 | 1.27E-05 | 0.000887194 | 0.079847441 |
| T2DM | LEC | *TSKS* | 0.610831924 | 8.51E-05 | 0.000908725 | 0.535021444 |
| T2DM | LEC | *FLRT1* | 0.575735789 | 0.000686787 | 0.00181574 | 1 |
| T2DM | LEC | *SBDS* | 1.188816341 | 0.011999535 | 0.018113104 | 1 |
| T2DM | LEC | *TRAP1* | 0.324778877 | 0.003103968 | 0.005677814 | 1 |
| T2DM | LEC | *DNAJC8* | 0.7196006 | 0.001262603 | 0.002792115 | 1 |
| T2DM | LEC | *ZNF384* | 1.296735718 | 0.009551192 | 0.014856098 | 1 |
| T2DM | LEC | *ATG14* | 0.857569918 | 0.016298574 | 0.023758205 | 1 |
| T2DM | LEC | *RHOJ* | -0.098056269 | 0.000144655 | 0.000908725 | 0.909443386 |
| T2DM | LEC | *L3HYPDH* | 0.520701078 | 0.007518119 | 0.012113403 | 1 |
| T2DM | LEC | *ZBTB1* | 0.78111137 | 0.014458488 | 0.021351573 | 1 |
| T2DM | LEC | *PZP* | 0.473705659 | 8.28E-05 | 0.000908725 | 0.520864336 |
| T2DM | LEC | *RHOT1* | 0.633759176 | 0.000150288 | 0.000910197 | 0.94486287 |
| T2DM | LEC | *WDR60* | 0.667245621 | 0.00282577 | 0.005268569 | 1 |
| T2DM | LEC | *AIF1L* | 0.426973183 | 0.000105679 | 0.000908725 | 0.664405171 |
| T2DM | LEC | *MAP2K2* | 0.874834154 | 0.021288883 | 0.030172048 | 1 |
| T2DM | LEC | *CANX* | 0.957779775 | 0.003041612 | 0.005586507 | 1 |
| T2DM | LEC | *FGD3* | 0.5899023 | 0.000123504 | 0.000908725 | 0.776470893 |
| T2DM | LEC | *ABHD8* | 0.902783212 | 0.007944213 | 0.012692571 | 1 |
| T2DM | LEC | *MASP1* | 0.423464213 | 0.000307573 | 0.00113215 | 1 |
| T2DM | LEC | *ATP13A4* | 0.559304601 | 0.000174123 | 0.000911117 | 1 |
| T2DM | LEC | *HELB* | 0.837073524 | 0.003154463 | 0.005760124 | 1 |
| T2DM | LEC | *RAP1B* | 0.690394167 | 0.015234805 | 0.022347461 | 1 |
| T2DM | LEC | *RAB3IP* | 0.788101963 | 0.00141412 | 0.003057281 | 1 |
| T2DM | LEC | *TAS2R4* | -0.627676014 | 0.000158072 | 0.000910705 | 0.993798414 |
| T2DM | LEC | *TMEM175* | 0.843344382 | 5.31E-05 | 0.000908725 | 0.333776463 |
| T2DM | LEC | *EMC1* | 0.902459742 | 0.018086748 | 0.026074612 | 1 |
| T2DM | LEC | *SIN3B* | 0.947015751 | 0.018233481 | 0.026250034 | 1 |
| T2DM | LEC | *WFIKKN1* | -5.386417964 | 0.000582212 | 0.001623223 | 1 |
| T2DM | LEC | *WDR24* | 0.621503984 | 0.000217953 | 0.00098409 | 1 |
| T2DM | LEC | *CHTF18* | 0.496557045 | 7.08E-05 | 0.000908725 | 0.445262248 |
| T2DM | LEC | *SMARCA4* | 0.867402152 | 0.006973128 | 0.011342835 | 1 |
| T2DM | LEC | *TICAM1* | 1.046750485 | 0.000194553 | 0.000949652 | 1 |
| T2DM | LEC | *PNKD* | 0.882002424 | 0.002912843 | 0.005397302 | 1 |
| T2DM | LEC | *ZNF835* | 0.562120405 | 0.001138784 | 0.002587136 | 1 |
| T2DM | LEC | *POR* | 0.889244977 | 0.016671727 | 0.024223515 | 1 |
| T2DM | LEC | *STEAP4* | 0.930065782 | 0.000220939 | 0.00098409 | 1 |
| T2DM | LEC | *GNAI1* | 0.693214672 | 0.004400127 | 0.007631337 | 1 |
| T2DM | LEC | *SGCE* | -0.686009068 | 0.023725553 | 0.03325068 | 1 |
| T2DM | LEC | *RBM48* | 0.78482805 | 0.000297149 | 0.001115996 | 1 |
| T2DM | LEC | *SRD5A3* | -0.822831072 | 0.003259948 | 0.005923354 | 1 |
| T2DM | LEC | *VPREB3* | 5.888782081 | 0.000249211 | 0.001023586 | 1 |
| T2DM | LEC | *MGAT3* | 0.602055405 | 0.000125084 | 0.000908725 | 0.78640002 |
| T2DM | LEC | *ATF4* | 1.097762447 | 0.001640312 | 0.003438694 | 1 |
| T2DM | LEC | *A4GALT* | -0.491796529 | 0.009693918 | 0.015040884 | 1 |
| T2DM | LEC | *BAIAP2L2* | 0.703455573 | 1.83E-05 | 0.000908725 | 0.114790559 |
| T2DM | LEC | *TST* | 0.462432467 | 0.000123285 | 0.000908725 | 0.775089687 |
| T2DM | LEC | *RIBC2* | 1.260282104 | 8.44E-06 | 0.000673907 | 0.053057734 |

| | | | | | | |
|---|---|---|---|---|---|---|
| T2DM | LEC | *EMC4* | 1.486153109 | 2.70E-05 | 0.000908725 | 0.169829152 |
| T2DM | LEC | *SPECC1* | 0.741274073 | 0.000372721 | 0.001243243 | 1 |
| T2DM | LEC | *CDHR3* | 0.676954948 | 0.00015425 | 0.000910197 | 0.969767009 |
| T2DM | LEC | *PRKRIP1* | 0.642575079 | 0.010356193 | 0.015954272 | 1 |
| T2DM | LEC | *FOXP2* | -0.22460103 | 8.86E-05 | 0.000908725 | 0.556730201 |
| T2DM | LEC | *FLNC* | -0.324776685 | 0.000294683 | 0.00111174 | 1 |
| T2DM | LEC | *IRF5* | 0.446954755 | 9.69E-05 | 0.000908725 | 0.609455842 |
| T2DM | LEC | *KLHDC10* | 0.824768185 | 0.002823165 | 0.005266837 | 1 |
| T2DM | LEC | *OPN1SW* | -0.945679079 | 6.49E-05 | 0.000908725 | 0.408292154 |
| T2DM | LEC | *HOXD1* | -2.778094226 | 0.001989472 | 0.003998661 | 1 |
| T2DM | LEC | *HOXD3* | -0.372146015 | 0.000334084 | 0.001176051 | 1 |
| T2DM | LEC | *CHN1* | -0.649710157 | 0.003521348 | 0.006312722 | 1 |
| T2DM | LEC | *GAD1* | -0.188798783 | 0.000133259 | 0.000908725 | 0.837800682 |
| T2DM | LEC | *ORMDL1* | 0.7246966 | 0.000812373 | 0.002026742 | 1 |
| T2DM | LEC | *HOXD9* | -0.092677587 | 0.000141652 | 0.000908725 | 0.890568776 |
| T2DM | LEC | *PSMG2* | 0.786508236 | 0.003720562 | 0.006618895 | 1 |
| T2DM | LEC | *WDFY4* | 0.656353042 | 0.00134345 | 0.002938854 | 1 |
| T2DM | LEC | *EIF2AK4* | 0.452000162 | 0.023986985 | 0.033564695 | 1 |
| T2DM | LEC | *TMOD2* | -0.831900937 | 0.007490475 | 0.012081225 | 1 |
| T2DM | LEC | *KNSTRN* | 0.321198091 | 0.000793521 | 0.001997943 | 1 |
| T2DM | LEC | *ARPP19* | 0.987930473 | 0.005133898 | 0.008747104 | 1 |
| T2DM | LEC | *VPS13C* | 0.649985004 | 0.018103576 | 0.026086908 | 1 |
| T2DM | LEC | *ANAPC13* | 0.402496069 | 0.003662232 | 0.006524355 | 1 |
| T2DM | LEC | *SERGEF* | 1.011548845 | 0.008906347 | 0.013995052 | 1 |
| T2DM | LEC | *RPAIN* | 0.668228724 | 0.009721384 | 0.015076059 | 1 |
| T2DM | LEC | *PLD2* | -0.57114912 | 0.002418993 | 0.004656525 | 1 |
| T2DM | LEC | *ATP1B2* | 1.21486585 | 0.000332348 | 0.001173862 | 1 |
| T2DM | LEC | *CCL4* | 0.344974901 | 0.000145455 | 0.000908725 | 0.914477881 |
| T2DM | LEC | *PUS7L* | 0.943428271 | 0.004652881 | 0.008018823 | 1 |
| T2DM | LEC | *KRI1* | 0.628022709 | 0.003806358 | 0.006750514 | 1 |
| T2DM | LEC | *CDKN2D* | 1.278165701 | 0.012023597 | 0.018136361 | 1 |
| T2DM | LEC | *MTUS1* | -0.69943201 | 0.002306062 | 0.00447752 | 1 |
| T2DM | LEC | *RIPK3* | -0.677170944 | 0.00083334 | 0.002067564 | 1 |
| T2DM | LEC | *ADCY4* | -0.338658342 | 0.000281448 | 0.00108224 | 1 |
| T2DM | LEC | *BCL2L2* | 0.856478844 | 0.000219207 | 0.00098409 | 1 |
| T2DM | LEC | *DTD2* | 0.567369695 | 0.003412555 | 0.006159843 | 1 |
| T2DM | LEC | *PARP2* | 0.696183513 | 0.000288862 | 0.001098223 | 1 |
| T2DM | LEC | *MIS18BP1* | 0.683686448 | 0.007778451 | 0.012472105 | 1 |
| T2DM | LEC | *RNASE1* | 0.540926543 | 0.000680475 | 0.001802884 | 1 |
| T2DM | LEC | *DAD1* | 0.979183123 | 0.001672505 | 0.003492209 | 1 |
| T2DM | LEC | *CDO1* | 0.783212135 | 0.000201715 | 0.00096305 | 1 |
| T2DM | LEC | *REEP5* | 0.875416563 | 0.002799089 | 0.005234273 | 1 |
| T2DM | LEC | *QRICH2* | -0.656441727 | 0.001591881 | 0.003356188 | 1 |
| T2DM | LEC | *RHBDF2* | 0.683735973 | 0.000109368 | 0.000908725 | 0.687598095 |
| T2DM | LEC | *AANAT* | -0.501377239 | 0.001700183 | 0.003544115 | 1 |
| T2DM | LEC | *TMEM8A* | 1.266830428 | 0.003537988 | 0.006337131 | 1 |
| T2DM | LEC | *SHC2* | -0.233785994 | 0.000525301 | 0.001513551 | 1 |
| T2DM | LEC | *ERMARD* | 0.632694637 | 0.000411642 | 0.001311277 | 1 |
| T2DM | LEC | *PHF10* | 1.0950057 | 0.005397829 | 0.00911527 | 1 |
| T2DM | LEC | *GALNT8* | -0.985294372 | 0.000114511 | 0.000908725 | 0.719929182 |

| | | | | | | |
|---|---|---|---|---|---|---|
| T2DM | LEC | *DOCK6* | -0.55605566 | 0.004181917 | 0.007299199 | 1 |
| T2DM | LEC | *ECSIT* | 0.857566962 | 0.000152684 | 0.000910197 | 0.959923079 |
| T2DM | LEC | *ELOF1* | 1.797379201 | 0.001409127 | 0.003047535 | 1 |
| T2DM | LEC | *THEM6* | -0.565022305 | 0.000109628 | 0.000908725 | 0.689228488 |
| T2DM | LEC | *TOMM40* | 1.105109607 | 0.001044777 | 0.002429183 | 1 |
| T2DM | LEC | *DPP6* | 0.359054349 | 0.000424781 | 0.0013333 | 1 |
| T2DM | LEC | *FAM98C* | 0.82632042 | 0.000474197 | 0.001414267 | 1 |
| T2DM | LEC | *ATP8B3* | 0.402295512 | 0.009300577 | 0.014516566 | 1 |
| T2DM | LEC | *BST2* | 0.77930304 | 0.000184504 | 0.000929091 | 1 |
| T2DM | LEC | *DDA1* | 0.960464182 | 0.000436107 | 0.00134367 | 1 |
| T2DM | LEC | *LSM7* | 0.992532244 | 0.002385331 | 0.004604414 | 1 |
| T2DM | LEC | *TULP4* | 0.800243317 | 0.007906482 | 0.012651578 | 1 |
| T2DM | LEC | *RTN4IP1* | 1.155557709 | 0.001997029 | 0.004006165 | 1 |
| T2DM | LEC | *RSPH3* | 0.538050555 | 0.005637546 | 0.00947174 | 1 |
| T2DM | LEC | *STK33* | -0.35022307 | 5.89E-05 | 0.000908725 | 0.369995011 |
| T2DM | LEC | *CACNG6* | 1.843170102 | 0.000387691 | 0.00126979 | 1 |
| T2DM | LEC | *PGPEP1* | 1.144011603 | 0.002277053 | 0.004437642 | 1 |
| T2DM | LEC | *HRC* | -0.594875116 | 0.017651744 | 0.02548381 | 1 |
| T2DM | LEC | *TRPM4* | 0.975210628 | 0.003919185 | 0.006911618 | 1 |
| T2DM | LEC | *CAMSAP1* | 0.948088931 | 0.000385798 | 0.001268529 | 1 |
| T2DM | LEC | *UBAC1* | 0.988256479 | 0.00293492 | 0.005431805 | 1 |
| T2DM | LEC | *TNNT3* | 0.620457172 | 0.015154666 | 0.022235096 | 1 |
| T2DM | LEC | *ATXN10* | 0.801438289 | 0.005795214 | 0.009690029 | 1 |
| T2DM | LEC | *TUBGCP2* | 0.888615354 | 0.000266974 | 0.001057634 | 1 |
| T2DM | LEC | *CYP2E1* | 0.647210811 | 0.000118716 | 0.000908725 | 0.746367477 |
| T2DM | LEC | *HBZ* | 4.477216027 | 0.002472785 | 0.004730177 | 1 |
| T2DM | LEC | *PAK4* | -0.80662441 | 0.000945594 | 0.002255291 | 1 |
| T2DM | LEC | *POMT1* | 0.679578712 | 0.000169147 | 0.000910705 | 1 |
| T2DM | LEC | *MAP3K10* | 0.53930631 | 0.000220906 | 0.00098409 | 1 |
| T2DM | LEC | *SMPDL3B* | 2.796277091 | 9.95E-05 | 0.000908725 | 0.62559102 |
| T2DM | LEC | *THEMIS2* | 0.415121224 | 4.84E-05 | 0.000908725 | 0.304242489 |
| T2DM | LEC | *CLIP1* | 0.75197399 | 0.000541889 | 0.001545201 | 1 |
| T2DM | LEC | *PPAN* | 2.344535138 | 0.000688953 | 0.001819172 | 1 |
| T2DM | LEC | *ZNF236* | 0.656769795 | 0.001559098 | 0.003294807 | 1 |
| T2DM | LEC | *C12orf65* | 1.682232041 | 0.000670389 | 0.001785148 | 1 |
| T2DM | LEC | *PKDREJ* | -1.432932508 | 0.000133263 | 0.000908725 | 0.837824772 |
| T2DM | LEC | *HSD17B3* | 0.729144465 | 0.007538133 | 0.012139406 | 1 |
| T2DM | LEC | *HABP4* | 1.003080794 | 0.008416708 | 0.013325571 | 1 |
| T2DM | LEC | *EPS8L1* | -1.019646581 | 0.000120128 | 0.000908725 | 0.755245177 |
| T2DM | LEC | *LILRB2* | 0.432531679 | 0.000110776 | 0.000908725 | 0.696447007 |
| T2DM | LEC | *COX4I2* | 1.040090188 | 0.000147247 | 0.000908725 | 0.925739953 |
| T2DM | LEC | *ATP6V1E1* | 0.79135317 | 0.007339949 | 0.011857854 | 1 |
| T2DM | LEC | *HAUS8* | 0.981502007 | 0.005721533 | 0.009584672 | 1 |
| T2DM | LEC | *RFTN1* | 0.68789255 | 0.000293532 | 0.001109196 | 1 |
| T2DM | LEC | *PDLIM4* | -0.387483288 | 0.000850499 | 0.002094432 | 1 |
| T2DM | LEC | *KIF3A* | 0.901232421 | 0.001849218 | 0.003780825 | 1 |
| T2DM | LEC | *AOC3* | 0.677328262 | 0.000887167 | 0.002164385 | 1 |
| T2DM | LEC | *AOC2* | 1.108267746 | 0.000116537 | 0.000908725 | 0.732669945 |
| T2DM | LEC | *EXOC4* | 0.591299091 | 0.035589943 | 0.047861813 | 1 |
| T2DM | LEC | *ACAP3* | 0.851443795 | 7.34E-05 | 0.000908725 | 0.46159023 |

| | | | | | | |
|---|---|---|---|---|---|---|
| T2DM | LEC | *C1orf159* | 0.769300185 | 0.000191504 | 0.000945045 | 1 |
| T2DM | LEC | *ANO1* | -0.909989473 | 0.034643758 | 0.046659235 | 1 |
| T2DM | LEC | *PPFIA1* | 0.616850166 | 0.000891771 | 0.002172246 | 1 |
| T2DM | LEC | *NINJ1* | 1.204309098 | 0.000585529 | 0.001631023 | 1 |
| T2DM | LEC | *NPHP4* | 1.197750659 | 0.000642051 | 0.001733394 | 1 |
| T2DM | LEC | *ZCCHC9* | 0.849764009 | 0.000166901 | 0.000910705 | 1 |
| T2DM | LEC | *KHDRBS3* | -0.577342461 | 0.001057346 | 0.002449348 | 1 |
| T2DM | LEC | *RBM8A* | 0.661753929 | 0.008534978 | 0.013482263 | 1 |
| T2DM | LEC | *ZNF304* | 0.674880963 | 0.000841944 | 0.0020815 | 1 |
| T2DM | LEC | *ZSCAN5A* | 0.958802848 | 0.003834223 | 0.006796098 | 1 |
| T2DM | LEC | *RHPN2* | -0.263578719 | 7.32E-05 | 0.000908725 | 0.46023561 |
| T2DM | LEC | *C19orf12* | 0.501204676 | 0.00942581 | 0.01469015 | 1 |
| T2DM | LEC | *ACTR10* | 0.866729655 | 0.009347491 | 0.014582549 | 1 |
| T2DM | LEC | *GCH1* | 0.850303972 | 4.47E-05 | 0.000908725 | 0.280730224 |
| T2DM | LEC | *DNAJB1* | 0.727317176 | 0.000280434 | 0.00108013 | 1 |
| T2DM | LEC | *FBXW9* | 0.617747571 | 3.45E-05 | 0.000908725 | 0.216775168 |
| T2DM | LEC | *C19orf57* | 0.71408468 | 8.95E-05 | 0.000908725 | 0.562743991 |
| T2DM | LEC | *MATN3* | -3.246828214 | 0.000901295 | 0.002191988 | 1 |
| T2DM | LEC | *CCT6B* | 0.779103793 | 0.012341747 | 0.018553937 | 1 |
| T2DM | LEC | *RAF1* | 0.81022189 | 0.00122226 | 0.00272472 | 1 |
| T2DM | LEC | *EMILIN2* | 0.522532564 | 0.000131432 | 0.000908725 | 0.826313168 |
| T2DM | LEC | *TRIM5* | 0.91319047 | 0.004413817 | 0.007650859 | 1 |
| T2DM | LEC | *CNGA4* | 0.71538695 | 0.000157137 | 0.000910705 | 0.987922515 |
| T2DM | LEC | *TRIM22* | 0.866628427 | 0.001367859 | 0.002981875 | 1 |
| T2DM | LEC | *IMMT* | 0.998969554 | 0.000500586 | 0.001463127 | 1 |
| T2DM | LEC | *CARD6* | -0.967818276 | 0.014395901 | 0.021295772 | 1 |
| T2DM | LEC | *INPP5K* | 0.756475435 | 0.006033439 | 0.010019078 | 1 |
| T2DM | LEC | *MYBBP1A* | 1.149084036 | 0.016406971 | 0.02387746 | 1 |
| T2DM | LEC | *RPA1* | 0.833179731 | 0.002986355 | 0.0055027 | 1 |
| T2DM | LEC | *SERPINF1* | 0.522336982 | 0.000360447 | 0.001224756 | 1 |
| T2DM | LEC | *TMEM128* | -0.781418528 | 0.000369904 | 0.001241526 | 1 |
| T2DM | LEC | *SEC61G* | 0.985084667 | 0.007364344 | 0.011893046 | 1 |
| T2DM | LEC | *FIGNL1* | 1.447776949 | 0.001928246 | 0.003910608 | 1 |
| T2DM | LEC | *WBP2* | 1.052913559 | 0.036149564 | 0.048531349 | 1 |
| T2DM | LEC | *KDM6B* | 1.024704888 | 0.00052267 | 0.00150666 | 1 |
| T2DM | LEC | *XAF1* | 0.669996708 | 8.67E-05 | 0.000908725 | 0.544783971 |
| T2DM | LEC | *REEP2* | -0.65805974 | 0.002296901 | 0.00446598 | 1 |
| T2DM | LEC | *PRMT7* | 0.483784421 | 0.013446962 | 0.020028678 | 1 |
| T2DM | LEC | *TERF2* | 1.166575682 | 0.001781398 | 0.00366649 | 1 |
| T2DM | LEC | *BTBD3* | -0.756138873 | 0.00146491 | 0.003143306 | 1 |
| T2DM | LEC | *PCNA* | 0.922640884 | 0.000116118 | 0.000908725 | 0.730031895 |
| T2DM | LEC | *PTPRA* | 0.715987705 | 0.016196307 | 0.023620084 | 1 |
| T2DM | LEC | *IGHMBP2* | 0.41369011 | 0.026333646 | 0.036531252 | 1 |
| T2DM | LEC | *MMACHC* | 0.469612367 | 0.000664022 | 0.001773452 | 1 |
| T2DM | LEC | *CTNNBL1* | 0.887188508 | 0.000467423 | 0.001402582 | 1 |
| T2DM | LEC | *LPIN3* | 0.556751174 | 0.000124687 | 0.000908725 | 0.783904191 |
| T2DM | LEC | *SERINC3* | 0.50440159 | 0.025243541 | 0.035158649 | 1 |
| T2DM | LEC | *KANK4* | 0.44701239 | 0.003305413 | 0.005994804 | 1 |
| T2DM | LEC | *RSG1* | -0.489144466 | 0.00025405 | 0.001032458 | 1 |
| T2DM | LEC | *CASP9* | 0.611071918 | 5.07E-05 | 0.000908725 | 0.318768577 |

| | | | | | | |
|---|---|---|---|---|---|---|
| T2DM | LEC | *ATP8A2* | 1.913249597 | 0.001565454 | 0.003307126 | 1 |
| T2DM | LEC | *USPL1* | 0.9253724 | 0.002176219 | 0.004286532 | 1 |
| T2DM | LEC | *WASF3* | -0.704786016 | 0.023466237 | 0.032931302 | 1 |
| T2DM | LEC | *RNF17* | 1.072460128 | 0.006160901 | 0.010190366 | 1 |
| T2DM | LEC | *PEMT* | 0.587175801 | 0.000293372 | 0.001109196 | 1 |
| T2DM | LEC | *SCO1* | 0.933789478 | 0.002231743 | 0.004360152 | 1 |
| T2DM | LEC | *TMCC2* | -1.673445079 | 7.00E-06 | 0.000610811 | 0.04397841 |
| T2DM | LEC | *COG6* | 0.78583217 | 0.001986438 | 0.003993838 | 1 |
| T2DM | LEC | *TRPC4* | -1.090823783 | 0.000624692 | 0.001705358 | 1 |
| T2DM | LEC | *RFC3* | 0.858911636 | 0.005860407 | 0.009780828 | 1 |
| T2DM | LEC | *BTBD2* | 0.8705442 | 0.00304552 | 0.005590419 | 1 |
| T2DM | LEC | *PRAM1* | 0.474173887 | 0.000976531 | 0.002311539 | 1 |
| T2DM | LEC | *ZNF414* | 0.863982701 | 0.000130222 | 0.000908725 | 0.818703823 |
| T2DM | LEC | *CNDP2* | 1.251652272 | 0.000605237 | 0.001668064 | 1 |
| T2DM | LEC | *RTN3* | 0.697409586 | 0.000580708 | 0.001621185 | 1 |
| T2DM | LEC | *SLC2A11* | 1.047645666 | 0.000214975 | 0.000978911 | 1 |
| T2DM | LEC | *C1QTNF6* | 0.936758976 | 0.000112781 | 0.000908725 | 0.709052095 |
| T2DM | LEC | *FAM83F* | -0.63994149 | 0.000156181 | 0.000910705 | 0.981907442 |
| T2DM | LEC | *GIMAP4* | -0.690523746 | 0.001977585 | 0.003979858 | 1 |
| T2DM | LEC | *AGAP3* | 0.899111901 | 0.001079751 | 0.002487502 | 1 |
| T2DM | LEC | *ACTR3B* | 0.868925352 | 0.000630529 | 0.00171385 | 1 |
| T2DM | LEC | *SFTPD* | 0.432665759 | 0.002275793 | 0.004436562 | 1 |
| T2DM | LEC | *DYDC2* | 1.980643188 | 0.000280092 | 0.00108013 | 1 |
| T2DM | LEC | *TMTC1* | -0.530959743 | 0.020373342 | 0.029018396 | 1 |
| T2DM | LEC | *IPO8* | 0.988777655 | 0.007047886 | 0.011452053 | 1 |
| T2DM | LEC | *SPINK5* | -0.544706357 | 0.000418429 | 0.001322531 | 1 |
| T2DM | LEC | *LRRCC1* | 0.87961256 | 8.36E-05 | 0.000908725 | 0.525856737 |
| T2DM | LEC | *SWAP70* | -0.651807501 | 0.033149407 | 0.044829065 | 1 |
| T2DM | LEC | *AMPD3* | 0.36766778 | 0.000325879 | 0.001164755 | 1 |
| T2DM | LEC | *MICAL2* | -0.371984588 | 0.000969868 | 0.002300824 | 1 |
| T2DM | LEC | *RRAS2* | 0.528701316 | 0.011028506 | 0.016857821 | 1 |
| T2DM | LEC | *HSD17B4* | 0.661036771 | 0.000811029 | 0.002024995 | 1 |
| T2DM | LEC | *GSC* | 1.083053918 | 0.000111197 | 0.000908725 | 0.699094608 |
| T2DM | LEC | *CATSPERB* | -0.294166346 | 0.000127213 | 0.000908725 | 0.799789055 |
| T2DM | LEC | *MED6* | 0.841199681 | 0.000493885 | 0.001449978 | 1 |
| T2DM | LEC | *ELP3* | -0.731228031 | 0.010943394 | 0.016744006 | 1 |
| T2DM | LEC | *MRO* | 0.62008069 | 0.00029588 | 0.001112688 | 1 |
| T2DM | LEC | *THUMPD3* | 0.750120358 | 0.024578547 | 0.034323707 | 1 |
| T2DM | LEC | *EDEM1* | 1.227754476 | 0.007393359 | 0.011930711 | 1 |
| T2DM | LEC | *CNTN6* | 0.538770079 | 0.000245318 | 0.001017515 | 1 |
| T2DM | LEC | *KATNBL1* | 0.820964228 | 0.000677975 | 0.001799252 | 1 |
| T2DM | LEC | *GSTM5* | 0.444821301 | 0.000213349 | 0.0009784 | 1 |
| T2DM | LEC | *GSTM3* | 0.450366648 | 0.001288915 | 0.002837328 | 1 |
| T2DM | LEC | *SYT6* | 1.199440253 | 0.000114594 | 0.000908725 | 0.720452888 |
| T2DM | LEC | *VAV3* | -0.20321511 | 0.000214293 | 0.0009784 | 1 |
| T2DM | LEC | *PSRC1* | 0.868134386 | 0.002932808 | 0.005429494 | 1 |
| T2DM | LEC | *WNT2B* | 0.824478794 | 0.002485784 | 0.004748747 | 1 |
| T2DM | LEC | *CEPT1* | 0.546860445 | 0.000230199 | 0.0009988 | 1 |
| T2DM | LEC | *AP4B1* | 1.604414636 | 0.001106126 | 0.002530657 | 1 |
| T2DM | LEC | *NAPG* | 0.985454792 | 0.002526311 | 0.004814465 | 1 |

| | | | | | | |
|---|---|---|---|---|---|---|
| T2DM | LEC | *SPIRE1* | 0.474819106 | 0.001073732 | 0.002476359 | 1 |
| T2DM | LEC | *PPHLN1* | 0.861112981 | 0.000438271 | 0.001346072 | 1 |
| T2DM | LEC | *YWHAQ* | 0.891103113 | 0.019077248 | 0.027358271 | 1 |
| T2DM | LEC | *LPIN1* | 0.539637607 | 0.011848263 | 0.017923492 | 1 |
| T2DM | LEC | *LDHA* | 0.866599299 | 0.000319398 | 0.00115467 | 1 |
| T2DM | LEC | *ANO3* | 0.388002989 | 0.000431325 | 0.00134099 | 1 |
| T2DM | LEC | *FST* | 1.091557356 | 0.001919214 | 0.00389732 | 1 |
| T2DM | LEC | *NARS* | 0.967369501 | 0.000442134 | 0.001351994 | 1 |
| T2DM | LEC | *KIAA1468* | 0.910586745 | 0.002054634 | 0.004094417 | 1 |
| T2DM | LEC | *FBXO18* | 0.953673976 | 0.011196875 | 0.017073672 | 1 |
| T2DM | LEC | *ECHDC3* | 0.800133694 | 0.007229935 | 0.011704961 | 1 |
| T2DM | LEC | *IL15RA* | 1.151302759 | 0.001998142 | 0.004007118 | 1 |
| T2DM | LEC | *TMEM241* | 0.845221048 | 0.003867529 | 0.00684934 | 1 |
| T2DM | LEC | *EMP1* | 0.386917386 | 0.000520636 | 0.001503555 | 1 |
| T2DM | LEC | *KLRD1* | 1.010786855 | 0.001509765 | 0.003211059 | 1 |
| T2DM | LEC | *PRH2* | -1.63288244 | 1.43E-05 | 0.000902248 | 0.090167401 |
| T2DM | LEC | *DDB2* | 1.112899892 | 0.005406308 | 0.009124687 | 1 |
| T2DM | LEC | *ACP2* | 1.148239007 | 4.62E-05 | 0.000908725 | 0.29071909 |
| T2DM | LEC | *ZCCHC11* | 0.70752345 | 0.010547007 | 0.016200594 | 1 |
| T2DM | LEC | *PRPF38A* | -0.903299018 | 0.002638318 | 0.004984107 | 1 |
| T2DM | LEC | *FHOD3* | 0.41225647 | 0.005386467 | 0.009100973 | 1 |
| T2DM | LEC | *TIMM10* | -1.304951206 | 9.23E-05 | 0.000908725 | 0.580122339 |
| T2DM | LEC | *DHX34* | 0.5327471 | 0.001446688 | 0.003110578 | 1 |
| T2DM | LEC | *APLNR* | 0.747675606 | 7.45E-05 | 0.000908725 | 0.468602438 |
| T2DM | LEC | *TMEM165* | 0.823295399 | 0.001455473 | 0.003127328 | 1 |
| T2DM | LEC | *CLOCK* | 0.843957386 | 0.003420883 | 0.006173103 | 1 |
| T2DM | LEC | *CLDN10* | -0.280322175 | 0.000154457 | 0.000910197 | 0.971070043 |
| T2DM | LEC | *DZIP1* | 0.471735877 | 0.00046515 | 0.001398163 | 1 |
| T2DM | LEC | *BIVM* | 1.088029595 | 0.000272486 | 0.001071675 | 1 |
| T2DM | LEC | *ERCC5* | 1.208126609 | 0.000131585 | 0.000908725 | 0.827272543 |
| T2DM | LEC | *KLB* | 0.419136565 | 0.000201782 | 0.00096305 | 1 |
| T2DM | LEC | *OSTF1* | 0.962674429 | 0.000909854 | 0.002204335 | 1 |
| T2DM | LEC | *UBQLN1* | 0.869422542 | 0.013744526 | 0.020413852 | 1 |
| T2DM | LEC | *CTSL* | 0.823421347 | 0.000459934 | 0.001391616 | 1 |
| T2DM | LEC | *TMEM2* | 0.619830318 | 0.00028358 | 0.001087982 | 1 |
| T2DM | LEC | *AGTPBP1* | 0.798703936 | 0.0023195 | 0.004499444 | 1 |
| T2DM | LEC | *ADAM19* | -0.175534275 | 0.003424153 | 0.006175325 | 1 |
| T2DM | LEC | *USP30* | 0.679942448 | 0.000107484 | 0.000908725 | 0.675751371 |
| T2DM | LEC | *HRK* | -1.39888895 | 0.00027988 | 0.00108013 | 1 |
| T2DM | LEC | *P2RX4* | 1.275119346 | 8.68E-06 | 0.000673907 | 0.054586434 |
| T2DM | LEC | *DTX1* | -0.326390895 | 8.15E-05 | 0.000908725 | 0.512398806 |
| T2DM | LEC | *DMTF1* | 0.702688291 | 0.011886508 | 0.017968375 | 1 |
| T2DM | LEC | *CCDC146* | 0.859066654 | 0.000241311 | 0.00101202 | 1 |
| T2DM | LEC | *TMEM60* | 1.094493268 | 0.020591529 | 0.029302613 | 1 |
| T2DM | LEC | *POM121C* | 0.610930425 | 0.007174622 | 0.011631472 | 1 |
| T2DM | LEC | *SRPK2* | 0.716886488 | 0.006175851 | 0.010209722 | 1 |
| T2DM | LEC | *ANKRD6* | -0.711472863 | 0.002513372 | 0.004792711 | 1 |
| T2DM | LEC | *KHDC1* | 0.465866912 | 0.001312812 | 0.002880856 | 1 |
| T2DM | LEC | *PRR5L* | -0.652993583 | 0.023802681 | 0.033343907 | 1 |
| T2DM | LEC | *NAT10* | 1.064525045 | 0.00644808 | 0.010598452 | 1 |

| | | | | | | |
|---|---|---|---|---|---|---|
| T2DM | LEC | *RDH5* | 0.41426686 | 9.99E-05 | 0.000908725 | 0.627836864 |
| T2DM | LEC | *BLOC1S1* | 1.004936953 | 0.027662606 | 0.03820624 | 1 |
| T2DM | LEC | *PPP1R1A* | 0.673302371 | 0.000108568 | 0.000908725 | 0.682568344 |
| T2DM | LEC | *B4GALNT1* | 1.548977032 | 0.002322005 | 0.004502914 | 1 |
| T2DM | LEC | *FAIM2* | 0.823952266 | 0.000149204 | 0.000910197 | 0.938048138 |
| T2DM | LEC | *KRT7* | -0.670647855 | 0.00220661 | 0.004324487 | 1 |
| T2DM | LEC | *SLC26A10* | 1.178353588 | 0.000669039 | 0.001782308 | 1 |
| T2DM | LEC | *MAP7* | 0.408058518 | 9.56E-05 | 0.000908725 | 0.601007705 |
| T2DM | LEC | *PKIB* | 1.025527845 | 0.001719053 | 0.003568493 | 1 |
| T2DM | LEC | *SMPD2* | 0.745834848 | 0.016975627 | 0.024625235 | 1 |
| T2DM | LEC | *MICAL1* | 0.875958353 | 2.51E-05 | 0.000908725 | 0.158102001 |
| T2DM | LEC | *PRADC1* | 1.626510654 | 0.011280322 | 0.017167607 | 1 |
| T2DM | LEC | *SMYD5* | 0.841896595 | 0.003857296 | 0.006833142 | 1 |
| T2DM | LEC | *DYSF* | -0.147673721 | 0.000125323 | 0.000908725 | 0.787905 |
| T2DM | LEC | *KCNMB4* | 0.820657338 | 0.000234698 | 0.001001608 | 1 |
| T2DM | LEC | *USP15* | 0.717486063 | 0.007663804 | 0.012322848 | 1 |
| T2DM | LEC | *KLHL36* | 1.155280422 | 0.003913947 | 0.006910437 | 1 |
| T2DM | LEC | *MPHOSPH6* | 1.217883769 | 7.41E-05 | 0.000908725 | 0.465642725 |
| T2DM | LEC | *FHOD1* | 0.979646847 | 0.001160604 | 0.00262189 | 1 |
| T2DM | LEC | *CCDC102A* | -0.426159713 | 0.000119659 | 0.000908725 | 0.752299095 |
| T2DM | LEC | *AGT* | 0.459580796 | 0.00218299 | 0.004294261 | 1 |
| T2DM | LEC | *ZNF670* | -0.593016804 | 0.022598923 | 0.031806453 | 1 |
| T2DM | LEC | *NTPCR* | 1.074680187 | 0.007859952 | 0.012586734 | 1 |
| T2DM | LEC | *RNASEL* | 1.086219153 | 7.57E-05 | 0.000908725 | 0.475959983 |
| T2DM | LEC | *PIGC* | 0.89345305 | 0.003924024 | 0.00691821 | 1 |
| T2DM | LEC | *SP110* | 0.888545702 | 0.013520274 | 0.020114047 | 1 |
| T2DM | LEC | *SLC19A3* | 0.312548479 | 0.000714059 | 0.001861576 | 1 |
| T2DM | LEC | *ARMC9* | 0.673547076 | 0.00016852 | 0.000910705 | 1 |
| T2DM | LEC | *TMEM127* | 0.674408807 | 0.000169391 | 0.000910705 | 1 |
| T2DM | LEC | *MRPS9* | 0.913731327 | 0.000546693 | 0.001554691 | 1 |
| T2DM | LEC | *USP44* | -0.687265454 | 0.000147761 | 0.000908725 | 0.928976336 |
| T2DM | LEC | *APPL2* | 0.706917999 | 0.000460879 | 0.001393179 | 1 |
| T2DM | LEC | *DRAM1* | 0.767102494 | 0.007259854 | 0.011745421 | 1 |
| T2DM | LEC | *SLC41A2* | 0.630777445 | 0.000219384 | 0.00098409 | 1 |
| T2DM | LEC | *NEK3* | 0.646179873 | 0.007846929 | 0.012569081 | 1 |
| T2DM | LEC | *RNASEH2B* | 0.783341337 | 0.000377136 | 0.001251877 | 1 |
| T2DM | LEC | *CKAP2* | 0.389702824 | 0.014460792 | 0.021351573 | 1 |
| T2DM | LEC | *TBC1D4* | 0.705476888 | 0.002769824 | 0.005187336 | 1 |
| T2DM | LEC | *LRCH1* | 0.864610333 | 0.00305891 | 0.005610083 | 1 |
| T2DM | LEC | *RCBTB1* | -0.987589151 | 0.001679739 | 0.003506149 | 1 |
| T2DM | LEC | *RCBTB2* | 0.866602064 | 0.032900832 | 0.044540813 | 1 |
| T2DM | LEC | *SETDB2* | 0.730181999 | 0.000399052 | 0.001290555 | 1 |
| T2DM | LEC | *GPNMB* | 0.344368588 | 0.000170445 | 0.000910705 | 1 |
| T2DM | LEC | *KDELR2* | 1.058125171 | 0.028029464 | 0.038661966 | 1 |
| T2DM | LEC | *ZDHHC4* | 0.986145169 | 0.000554931 | 0.001568727 | 1 |
| T2DM | LEC | *AOAH* | 0.336014498 | 0.003686347 | 0.006565458 | 1 |
| T2DM | LEC | *DDX56* | 0.662005913 | 0.000130141 | 0.000908725 | 0.818193703 |
| T2DM | LEC | *HUS1* | 0.755645417 | 0.00016067 | 0.000910705 | 1 |
| T2DM | LEC | *NACAD* | -4.411259767 | 1.36E-06 | 0.000269656 | 0.008538808 |
| T2DM | LEC | *MTHFS* | 0.369558901 | 0.002979709 | 0.005495568 | 1 |

| | | | | | | |
|---|---|---|---|---|---|---|
| T2DM | LEC | *ALPK3* | -0.826129601 | 0.000493168 | 0.001449584 | 1 |
| T2DM | LEC | *CIB2* | -0.964348619 | 0.000184872 | 0.000929091 | 1 |
| T2DM | LEC | *RSAD1* | -1.029146041 | 0.022082304 | 0.031147789 | 1 |
| T2DM | LEC | *NMT1* | 0.719202941 | 0.000890811 | 0.002170748 | 1 |
| T2DM | LEC | *MYCBPAP* | 0.493854538 | 0.026425016 | 0.036641834 | 1 |
| T2DM | LEC | *BRIP1* | 0.446846627 | 8.20E-05 | 0.000908725 | 0.515683061 |
| T2DM | LEC | *RTP4* | 0.734462596 | 0.028627288 | 0.039382879 | 1 |
| T2DM | LEC | *MRPL47* | 1.173720157 | 0.003277949 | 0.00595225 | 1 |
| T2DM | LEC | *BLK* | -0.621317129 | 0.000104428 | 0.000908725 | 0.656537792 |
| T2DM | LEC | *RPS6KC1* | 0.768526646 | 0.006926903 | 0.011279316 | 1 |
| T2DM | LEC | *CBWD2* | 0.894728179 | 0.000338114 | 0.001185962 | 1 |
| T2DM | LEC | *IL1RN* | 1.283237513 | 0.000505771 | 0.001472919 | 1 |
| T2DM | LEC | *SAP130* | 0.84545943 | 0.014850614 | 0.02186553 | 1 |
| T2DM | LEC | *UGGT1* | 0.955191606 | 0.00306831 | 0.005622402 | 1 |
| T2DM | LEC | *NIPSNAP3A* | 0.75058332 | 0.008823498 | 0.013879005 | 1 |
| T2DM | LEC | *TXN* | -0.882319984 | 0.01446456 | 0.021352123 | 1 |
| T2DM | LEC | *TOR1B* | 0.923488411 | 0.003397591 | 0.00613812 | 1 |
| T2DM | LEC | *TOR1A* | 0.609248712 | 0.011321139 | 0.017221388 | 1 |
| T2DM | LEC | *SLC2A8* | 1.010674422 | 0.002653694 | 0.005011647 | 1 |
| T2DM | LEC | *ZFP37* | 1.037333385 | 0.000143316 | 0.000908725 | 0.90102728 |
| T2DM | LEC | *ZNF189* | 0.61014794 | 0.000171923 | 0.000910705 | 1 |
| T2DM | LEC | *ALDOB* | -0.693042677 | 0.0004034 | 0.001298865 | 1 |
| T2DM | LEC | *STX17* | 0.744861305 | 0.000352294 | 0.001211637 | 1 |
| T2DM | LEC | *PRPF4* | 1.139255503 | 5.70E-06 | 0.000559501 | 0.035866817 |
| T2DM | LEC | *FPGS* | 0.948247774 | 0.006109247 | 0.010120906 | 1 |
| T2DM | LEC | *TEX10* | 1.00019045 | 0.01534548 | 0.022499308 | 1 |
| T2DM | LEC | *GARNL3* | 0.635535234 | 0.000183655 | 0.000929091 | 1 |
| T2DM | LEC | *GABBR2* | 1.092271529 | 0.00010532 | 0.000908725 | 0.662145391 |
| T2DM | LEC | *PSMB7* | 0.919715957 | 0.00076265 | 0.001943568 | 1 |
| T2DM | LEC | *RABEPK* | 0.680592436 | 0.000171637 | 0.000910705 | 1 |
| T2DM | LEC | *GOLGA1* | 0.908636522 | 0.001068808 | 0.002469789 | 1 |
| T2DM | LEC | *NOV* | 0.690028153 | 9.49E-05 | 0.000908725 | 0.596468733 |
| T2DM | LEC | *IL11RA* | 0.707014814 | 0.00012531 | 0.000908725 | 0.787824792 |
| T2DM | LEC | *APTX* | 0.769352097 | 0.000462145 | 0.001395537 | 1 |
| T2DM | LEC | *TLN1* | 1.065561921 | 0.002439101 | 0.004686622 | 1 |
| T2DM | LEC | *SPAG8* | 1.205943435 | 6.47E-05 | 0.000908725 | 0.406638426 |
| T2DM | LEC | *CD72* | 0.673090192 | 0.000215339 | 0.000978911 | 1 |
| T2DM | LEC | *TMEM8B* | 0.508161773 | 0.000137971 | 0.000908725 | 0.867424488 |
| T2DM | LEC | *GRHPR* | -0.878857652 | 0.002548545 | 0.004846552 | 1 |
| T2DM | LEC | *ARHGEF39* | 0.548849155 | 0.002562548 | 0.004862885 | 1 |
| T2DM | LEC | *DENND4C* | 0.724138078 | 0.000809679 | 0.002022874 | 1 |
| T2DM | LEC | *PPIL1* | 1.88133612 | 0.000182703 | 0.000929091 | 1 |
| T2DM | LEC | *KIF13A* | 0.709476539 | 0.00222913 | 0.0043564 | 1 |
| T2DM | LEC | *ZSCAN9* | 0.74008023 | 0.000342682 | 0.001192937 | 1 |
| T2DM | LEC | *TMEM63B* | 1.014209905 | 0.020357652 | 0.029002619 | 1 |
| T2DM | LEC | *SLC22A23* | -0.296013974 | 0.000279477 | 0.00108013 | 1 |
| T2DM | LEC | *LRRC1* | -0.180296036 | 0.000134125 | 0.000908725 | 0.843245609 |
| T2DM | LEC | *FOXF2* | 1.344611582 | 0.004355405 | 0.00757467 | 1 |
| T2DM | LEC | *BPHL* | 0.470270722 | 0.000245016 | 0.001017515 | 1 |
| T2DM | LEC | *RIPK1* | 0.891344097 | 0.008512885 | 0.013450744 | 1 |

| | | | | | | |
|---|---|---|---|---|---|---|
| T2DM | LEC | *TUBB2B* | 0.75952236 | 0.000306981 | 0.001131296 | 1 |
| T2DM | LEC | *UQCC2* | 1.205069249 | 0.000765089 | 0.001947414 | 1 |
| T2DM | LEC | *TCF19* | 1.200201268 | 0.008099927 | 0.012911826 | 1 |
| T2DM | LEC | *FLOT1* | 1.18513446 | 0.000641964 | 0.001733394 | 1 |
| T2DM | LEC | *TPMT* | 1.212063773 | 0.000362085 | 0.001227184 | 1 |
| T2DM | LEC | *NRM* | 0.656478158 | 0.000905583 | 0.002195681 | 1 |
| T2DM | LEC | *MTCH1* | 1.078716762 | 0.001803837 | 0.003702489 | 1 |
| T2DM | LEC | *VARS2* | 0.379780378 | 0.000465449 | 0.001398163 | 1 |
| T2DM | LEC | *FAM8A1* | 1.013681657 | 0.000236972 | 0.001005449 | 1 |
| T2DM | LEC | *FGFBP2* | 0.530098785 | 0.000232039 | 0.001001608 | 1 |
| T2DM | LEC | *FHDC1* | -0.741938677 | 0.000347453 | 0.001200239 | 1 |
| T2DM | LEC | *MYO7A* | 0.596944776 | 0.014430539 | 0.021323957 | 1 |
| T2DM | LEC | *CCDC90B* | 1.058076096 | 0.00902308 | 0.014157251 | 1 |
| T2DM | LEC | *SYTL2* | 0.615946598 | 0.001515948 | 0.00322203 | 1 |
| T2DM | LEC | *RAB30* | 0.845042903 | 0.000928873 | 0.002227231 | 1 |
| T2DM | LEC | *NARS2* | 0.663781935 | 0.000212044 | 0.0009784 | 1 |
| T2DM | LEC | *PI15* | 2.243966463 | 0.000587095 | 0.001632494 | 1 |
| T2DM | LEC | *SULF1* | 0.744742932 | 0.000507392 | 0.0014768 | 1 |
| T2DM | LEC | *NEK1* | 0.79731754 | 0.000386794 | 0.001268529 | 1 |
| T2DM | LEC | *DDX60* | -0.809381306 | 0.004736461 | 0.008142776 | 1 |
| T2DM | LEC | *MMP27* | 0.316357698 | 0.006757816 | 0.011046233 | 1 |
| T2DM | LEC | *TRIM29* | -0.28070278 | 0.000100825 | 0.000908725 | 0.633887749 |
| T2DM | LEC | *SLC37A4* | 1.005808658 | 0.000153839 | 0.000910197 | 0.967183808 |
| T2DM | LEC | *POU2F3* | -0.645063601 | 0.002673627 | 0.005043233 | 1 |
| T2DM | LEC | *PPP2R1B* | 0.504277528 | 0.000206069 | 0.000970455 | 1 |
| T2DM | LEC | *FXYD6* | -0.296700075 | 0.000158329 | 0.000910705 | 0.995415821 |
| T2DM | LEC | *TMPRSS13* | 1.711301045 | 0.000159535 | 0.000910705 | 1 |
| T2DM | LEC | *CASP1* | 1.149935366 | 0.00024559 | 0.001017515 | 1 |
| T2DM | LEC | *ITGA11* | 0.303502043 | 0.000180455 | 0.00092505 | 1 |
| T2DM | LEC | *PAQR5* | -1.036442405 | 0.001081854 | 0.002491435 | 1 |
| T2DM | LEC | *RMDN3* | 1.009286567 | 2.93E-05 | 0.000908725 | 0.184029683 |
| T2DM | LEC | *ITPKA* | -1.581839097 | 9.26E-05 | 0.000908725 | 0.582080259 |
| T2DM | LEC | *PLCB2* | 0.412888509 | 0.00047671 | 0.001419069 | 1 |
| T2DM | LEC | *SPTBN5* | 1.016444918 | 3.01E-08 | 1.35E-05 | 0.00018934 |
| T2DM | LEC | *RABGGTB* | 1.130906635 | 0.001351339 | 0.002952003 | 1 |
| T2DM | LEC | *SLC44A5* | -0.154694207 | 0.000144821 | 0.000908725 | 0.910492574 |
| T2DM | LEC | *DNASE2B* | 2.37969851 | 8.29E-05 | 0.000908725 | 0.520907942 |
| T2DM | LEC | *DBT* | 0.728841432 | 0.000444672 | 0.001357774 | 1 |
| T2DM | LEC | *RTCA* | 0.889551718 | 0.017786573 | 0.025665409 | 1 |
| T2DM | LEC | *IFT172* | 1.198707271 | 0.000127471 | 0.000908725 | 0.801408704 |
| T2DM | LEC | *ADCY3* | 0.749212537 | 0.00030399 | 0.001126076 | 1 |
| T2DM | LEC | *LHCGR* | 0.480270854 | 0.000189987 | 0.000942374 | 1 |
| T2DM | LEC | *PREB* | 0.635510253 | 0.003650146 | 0.006510203 | 1 |
| T2DM | LEC | *ABCG5* | 1.079997559 | 5.31E-05 | 0.000908725 | 0.334023716 |
| T2DM | LEC | *CENPO* | 1.04719357 | 0.000137784 | 0.000908725 | 0.866250923 |
| T2DM | LEC | *LRPPRC* | 0.813846734 | 0.001106131 | 0.002530657 | 1 |
| T2DM | LEC | *DTNB* | 0.732604138 | 0.015045487 | 0.022100696 | 1 |
| T2DM | LEC | *CYP2C9* | 0.449900373 | 0.013114397 | 0.019584374 | 1 |
| T2DM | LEC | *CYP2C8* | -0.52408478 | 0.000268696 | 0.001062766 | 1 |
| T2DM | LEC | *MYOF* | 0.920218177 | 3.00E-05 | 0.000908725 | 0.188362703 |

| | | | | | | |
|---|---|---|---|---|---|---|
| T2DM | LEC | *STAMBPL1* | 0.910121933 | 0.003761751 | 0.006682715 | 1 |
| T2DM | LEC | *ATAD1* | 0.892724182 | 0.019952503 | 0.028477046 | 1 |
| T2DM | LEC | *BTBD16* | 0.51103002 | 0.001711214 | 0.003555322 | 1 |
| T2DM | LEC | *TACC2* | 0.873044428 | 0.000516749 | 0.001495765 | 1 |
| T2DM | LEC | *CALHM2* | 0.898484481 | 0.000905557 | 0.002195681 | 1 |
| T2DM | LEC | *ARL3* | 1.021971807 | 0.000306543 | 0.001130344 | 1 |
| T2DM | LEC | *PLCE1* | -0.392258002 | 0.000201329 | 0.00096305 | 1 |
| T2DM | LEC | *RBP4* | 0.954231117 | 0.000619822 | 0.001697248 | 1 |
| T2DM | LEC | *DNAJC13* | 0.762068631 | 0.033056943 | 0.044713641 | 1 |
| T2DM | LEC | *FAM149B1* | 0.61032943 | 0.001150718 | 0.002606816 | 1 |
| T2DM | LEC | *ASCC1* | 0.682906011 | 0.015656612 | 0.02292341 | 1 |
| T2DM | LEC | *DNA2* | 0.232243368 | 0.034776767 | 0.046818316 | 1 |
| T2DM | LEC | *ATIC* | 0.47168686 | 0.014473039 | 0.021359623 | 1 |
| T2DM | LEC | *SMARCAL1* | 0.960841927 | 0.000766924 | 0.001948143 | 1 |
| T2DM | LEC | *BARD1* | 0.904133193 | 0.000320403 | 0.001156359 | 1 |
| T2DM | LEC | *STAT4* | 0.365788048 | 0.000640812 | 0.001733394 | 1 |
| T2DM | LEC | *CARF* | 1.099486253 | 0.000430602 | 0.001340859 | 1 |
| T2DM | LEC | *ASNSD1* | 0.65881235 | 0.019655332 | 0.028123138 | 1 |
| T2DM | LEC | *METTL5* | -0.896486313 | 0.021054006 | 0.02987958 | 1 |
| T2DM | LEC | *FASTKD1* | 0.862386086 | 0.005995541 | 0.009969517 | 1 |
| T2DM | LEC | *MDH1B* | 0.994287529 | 0.00022638 | 0.000992757 | 1 |
| T2DM | LEC | *ITGAV* | 0.824428611 | 0.001076036 | 0.002480222 | 1 |
| T2DM | LEC | *SLC35A5* | 0.712113165 | 0.003278613 | 0.00595225 | 1 |
| T2DM | LEC | *COX17* | 0.973623903 | 0.000197541 | 0.000956093 | 1 |
| T2DM | LEC | *USP8* | 0.759758965 | 0.013873044 | 0.020595 | 1 |
| T2DM | LEC | *SPPL2A* | 0.71767603 | 0.026998487 | 0.037371089 | 1 |
| T2DM | LEC | *UBL7* | 1.457839313 | 0.000399983 | 0.001291573 | 1 |
| T2DM | LEC | *ARHGAP24* | 0.448590484 | 0.000212395 | 0.0009784 | 1 |
| T2DM | LEC | *FAM13A* | 0.516114734 | 6.11E-05 | 0.000908725 | 0.383949012 |
| T2DM | LEC | *HERC3* | 0.740239066 | 0.000451631 | 0.001373685 | 1 |
| T2DM | LEC | *PRKG2* | 0.306430634 | 0.000138093 | 0.000908725 | 0.868188561 |
| T2DM | LEC | *MMRN1* | -0.025389013 | 6.06E-05 | 0.000908725 | 0.38088994 |
| T2DM | LEC | *PRDM5* | -0.62339662 | 0.010248321 | 0.015807458 | 1 |
| T2DM | LEC | *TRPC3* | 0.706479237 | 0.000133947 | 0.000908725 | 0.84212473 |
| T2DM | LEC | *NAAA* | 1.027726352 | 0.007921377 | 0.012668964 | 1 |
| T2DM | LEC | *BMP2K* | -0.554225667 | 0.021162709 | 0.030013523 | 1 |
| T2DM | LEC | *G3BP2* | 0.763989431 | 0.017608381 | 0.025431632 | 1 |
| T2DM | LEC | *SCARB2* | 0.798446727 | 0.018741594 | 0.026926051 | 1 |
| T2DM | LEC | *CNOT6L* | 0.546620815 | 0.032206098 | 0.043694377 | 1 |
| T2DM | LEC | *ANXA3* | -1.083334266 | 0.022591069 | 0.031802519 | 1 |
| T2DM | LEC | *PPA2* | 0.91610732 | 0.003283905 | 0.005960137 | 1 |
| T2DM | LEC | *GSTCD* | 0.916353638 | 0.000464156 | 0.001396913 | 1 |
| T2DM | LEC | *ENPEP* | -1.050521906 | 0.007133453 | 0.01157667 | 1 |
| T2DM | LEC | *PAPSS1* | 0.774467797 | 0.000201905 | 0.00096305 | 1 |
| T2DM | LEC | *RNF185* | 0.774769816 | 0.003363842 | 0.006087645 | 1 |
| T2DM | LEC | *PARVG* | 0.576876251 | 0.000295182 | 0.00111174 | 1 |
| T2DM | LEC | *ERP27* | 1.007793567 | 0.000529576 | 0.001522379 | 1 |
| T2DM | LEC | *ETV6* | 0.766390984 | 0.00361047 | 0.006450418 | 1 |
| T2DM | LEC | *KIF21A* | 0.619297762 | 6.23E-05 | 0.000908725 | 0.391466799 |
| T2DM | LEC | *ALG10* | -1.197591519 | 0.001151029 | 0.002606816 | 1 |

| | | | | | | |
|---|---|---|---|---|---|---|
| T2DM | LEC | *SLCO1C1* | 0.52464259 | 4.13E-05 | 0.000908725 | 0.259926686 |
| T2DM | LEC | *PRICKLE1* | 0.70600834 | 9.63E-05 | 0.000908725 | 0.605568016 |
| T2DM | LEC | *C1RL* | 1.151444433 | 0.000113 | 0.000908725 | 0.710431137 |
| T2DM | LEC | *CLSTN3* | 0.756624078 | 7.50E-05 | 0.000908725 | 0.471476428 |
| T2DM | LEC | *KLRG1* | 0.45330686 | 0.022377676 | 0.031530357 | 1 |
| T2DM | LEC | *VAMP1* | 0.549823061 | 4.00E-05 | 0.000908725 | 0.251543945 |
| T2DM | LEC | *CD27* | 1.023538196 | 0.000229536 | 0.000997266 | 1 |
| T2DM | LEC | *PEX5* | 0.527365551 | 0.000287957 | 0.001097202 | 1 |
| T2DM | LEC | *PIANP* | -1.641088153 | 0.002550106 | 0.004848054 | 1 |
| T2DM | LEC | *PPFIA2* | 0.557330376 | 0.005835323 | 0.009744138 | 1 |
| T2DM | LEC | *GLIPR1* | 0.541736307 | 0.000183242 | 0.000929091 | 1 |
| T2DM | LEC | *LGR5* | 0.526991745 | 0.000354687 | 0.001215214 | 1 |
| T2DM | LEC | *GAS2L3* | 0.600491695 | 0.000128444 | 0.000908725 | 0.80752817 |
| T2DM | LEC | *TMEM132B* | 0.954484804 | 0.000238876 | 0.001009285 | 1 |
| T2DM | LEC | *TDG* | 0.941491782 | 0.016320491 | 0.023779125 | 1 |
| T2DM | LEC | *RITA1* | -0.617542 | 0.013178607 | 0.019666248 | 1 |
| T2DM | LEC | *MMAB* | 0.731410717 | 0.017334125 | 0.025089275 | 1 |
| T2DM | LEC | *SLC46A3* | 0.734650443 | 6.99E-05 | 0.000908725 | 0.43960368 |
| T2DM | LEC | *SUOX* | 0.57761058 | 0.000355969 | 0.001216909 | 1 |
| T2DM | LEC | *CCDC65* | 0.65667795 | 0.01425309 | 0.021099406 | 1 |
| T2DM | LEC | *BRCA2* | 1.078817218 | 0.000486961 | 0.00143852 | 1 |
| T2DM | LEC | *CERS5* | 0.782458278 | 0.006399972 | 0.010538664 | 1 |
| T2DM | LEC | *LMBR1L* | 0.925869188 | 0.012257105 | 0.018444332 | 1 |
| T2DM | LEC | *ZNF740* | 1.174444983 | 0.007934683 | 0.012683791 | 1 |
| T2DM | LEC | *WDFY2* | 0.702309914 | 0.000707163 | 0.00185016 | 1 |
| T2DM | LEC | *HNRNPA1L2* | -1.69882193 | 0.000334185 | 0.001176051 | 1 |
| T2DM | LEC | *LPAR6* | 0.882312296 | 0.002461302 | 0.004714872 | 1 |
| T2DM | LEC | *VPS33A* | -1.322825618 | 0.008948079 | 0.014053603 | 1 |
| T2DM | LEC | *RHOF* | 0.811949606 | 0.011782529 | 0.017841224 | 1 |
| T2DM | LEC | *METTL21C* | -1.807427313 | 0.000333087 | 0.00117581 | 1 |
| T2DM | LEC | *GRTP1* | 0.493833317 | 0.002213993 | 0.004333817 | 1 |
| T2DM | LEC | *CBLN3* | 0.250533859 | 0.004397132 | 0.007630748 | 1 |
| T2DM | LEC | *TSSK4* | 1.991771484 | 0.000178801 | 0.000922168 | 1 |
| T2DM | LEC | *TMX1* | 1.108315318 | 0.002871467 | 0.005341098 | 1 |
| T2DM | LEC | *RTN1* | 0.364211404 | 0.000107685 | 0.000908725 | 0.677016024 |
| T2DM | LEC | *C14orf37* | 0.680865175 | 0.010608192 | 0.016290597 | 1 |
| T2DM | LEC | *RAB15* | 0.585545698 | 0.009697527 | 0.015042772 | 1 |
| T2DM | LEC | *STON2* | -0.161931491 | 0.000165313 | 0.000910705 | 1 |
| T2DM | LEC | *EFCAB11* | 0.940290297 | 0.000398226 | 0.001289522 | 1 |
| T2DM | LEC | *AK7* | 2.815889311 | 0.002928961 | 0.005423969 | 1 |
| T2DM | LEC | *FBLN5* | 0.594499311 | 0.000132407 | 0.000908725 | 0.832440383 |
| T2DM | LEC | *C14orf79* | 0.622178234 | 8.85E-05 | 0.000908725 | 0.556518364 |
| T2DM | LEC | *NIPA2* | 0.873904709 | 0.000635119 | 0.001720377 | 1 |
| T2DM | LEC | *DUOXA1* | 0.819833188 | 3.34E-05 | 0.000908725 | 0.209855142 |
| T2DM | LEC | *SORD* | 0.519859036 | 0.000103174 | 0.000908725 | 0.648654414 |
| T2DM | LEC | *ZSCAN29* | 0.918974192 | 0.000299126 | 0.001118744 | 1 |
| T2DM | LEC | *SRP14* | 1.271251078 | 0.000924767 | 0.002222312 | 1 |
| T2DM | LEC | *BAHD1* | 1.000818731 | 0.000285672 | 0.001091143 | 1 |
| T2DM | LEC | *ANP32A* | 0.948616791 | 0.002168535 | 0.004276812 | 1 |
| T2DM | LEC | *COMMD4* | 0.571863931 | 0.000225344 | 0.000992757 | 1 |

| | | | | | | |
|---|---|---|---|---|---|---|
| T2DM | LEC | *UBE2Q2* | 0.775209848 | 0.000255349 | 0.001034507 | 1 |
| T2DM | LEC | *PSTPIP1* | 0.735275522 | 0.000107245 | 0.000908725 | 0.674247973 |
| T2DM | LEC | *HMG20A* | 1.215395967 | 3.80E-05 | 0.000908725 | 0.23896901 |
| T2DM | LEC | *SCAPER* | 0.589055474 | 0.03379895 | 0.045619149 | 1 |
| T2DM | LEC | *DNAJA4* | 1.112714182 | 0.000141717 | 0.000908725 | 0.890972885 |
| T2DM | LEC | *TPM1* | 0.833553803 | 0.001530928 | 0.003247282 | 1 |
| T2DM | LEC | *CYP11A1* | 0.885318218 | 0.000295309 | 0.00111174 | 1 |
| T2DM | LEC | *BBS4* | 0.810247838 | 0.000116962 | 0.000908725 | 0.73533777 |
| T2DM | LEC | *ADAMTS17* | 0.32930587 | 0.000155375 | 0.000910705 | 0.976843361 |
| T2DM | LEC | *ULK3* | 1.351683716 | 0.001187413 | 0.002664263 | 1 |
| T2DM | LEC | *HAPLN3* | 1.201059825 | 0.001790594 | 0.00368252 | 1 |
| T2DM | LEC | *POLG* | 0.950216544 | 0.001121578 | 0.002559477 | 1 |
| T2DM | LEC | *WDR93* | 0.233428073 | 0.001218779 | 0.002720662 | 1 |
| T2DM | LEC | *TICRR* | 0.739612715 | 0.000173085 | 0.000910705 | 1 |
| T2DM | LEC | *MFGE8* | 0.604325848 | 0.000208698 | 0.000972609 | 1 |
| T2DM | LEC | *UNC45A* | 0.857303927 | 0.003869329 | 0.006850598 | 1 |
| T2DM | LEC | *ST8SIA2* | 2.917631774 | 0.000538232 | 0.001538119 | 1 |
| T2DM | LEC | *IQGAP1* | 0.657506665 | 0.002538382 | 0.00483138 | 1 |
| T2DM | LEC | *CRTC3* | 0.82185171 | 0.002053709 | 0.004094417 | 1 |
| T2DM | LEC | *SEC11A* | 0.828644461 | 0.018142126 | 0.026130481 | 1 |
| T2DM | LEC | *TGFB1I1* | -0.61219206 | 0.000548538 | 0.001555553 | 1 |
| T2DM | LEC | *PARN* | 0.791225759 | 0.002279052 | 0.00444016 | 1 |
| T2DM | LEC | *FTO* | 0.695351757 | 0.005772826 | 0.009655162 | 1 |
| T2DM | LEC | *MARVELD3* | -0.568731387 | 0.000259715 | 0.001042963 | 1 |
| T2DM | LEC | *KIFC3* | 0.843230486 | 0.000262753 | 0.001048732 | 1 |
| T2DM | LEC | *ADAMTS18* | -0.701264147 | 0.026538956 | 0.036791712 | 1 |
| T2DM | LEC | *GCSH* | 1.183696906 | 0.005751375 | 0.009626968 | 1 |
| T2DM | LEC | *CMTM3* | 0.736688928 | 0.000144924 | 0.000908725 | 0.911136721 |
| T2DM | LEC | *MAP1LC3B* | 1.00150598 | 0.001024619 | 0.002399174 | 1 |
| T2DM | LEC | *TLDC1* | 0.848593099 | 0.000218706 | 0.00098409 | 1 |
| T2DM | LEC | *NDUFB10* | 1.31308351 | 0.009756642 | 0.015119549 | 1 |
| T2DM | LEC | *TIGD7* | 0.879446557 | 1.42E-05 | 0.000902248 | 0.089526644 |
| T2DM | LEC | *DEF8* | 0.685584872 | 0.011315424 | 0.017216861 | 1 |
| T2DM | LEC | *GALNS* | 0.893500748 | 0.008437652 | 0.0133511 | 1 |
| T2DM | LEC | *COPS3* | 1.010074486 | 0.000315944 | 0.001151139 | 1 |
| T2DM | LEC | *MYO19* | 1.365745833 | 0.000468322 | 0.001403402 | 1 |
| T2DM | LEC | *DDX52* | 0.879150305 | 0.028339496 | 0.039038215 | 1 |
| T2DM | LEC | *RASL10B* | 0.829787693 | 0.022695661 | 0.031921168 | 1 |
| T2DM | LEC | *UNC45B* | 1.850753279 | 0.001945334 | 0.003931313 | 1 |
| T2DM | LEC | *TOM1L1* | 0.791378711 | 0.000182699 | 0.000929091 | 1 |
| T2DM | LEC | *C17orf80* | 0.623091483 | 0.002459003 | 0.004711903 | 1 |
| T2DM | LEC | *VPS53* | 0.526873999 | 0.013910267 | 0.020645384 | 1 |
| T2DM | LEC | *SPATA22* | 0.86993216 | 2.48E-05 | 0.000908725 | 0.155779617 |
| T2DM | LEC | *SGSM2* | 0.853017367 | 0.002135915 | 0.004226786 | 1 |
| T2DM | LEC | *RHBDL3* | 0.606405886 | 0.000725516 | 0.001883287 | 1 |
| T2DM | LEC | *ARSG* | 0.764318038 | 0.0295945 | 0.040544916 | 1 |
| T2DM | LEC | *ABCA8* | 0.436812355 | 0.000136758 | 0.000908725 | 0.859800632 |
| T2DM | LEC | *G6PC3* | -1.265050476 | 0.0021872 | 0.004297165 | 1 |
| T2DM | LEC | *IMPA2* | 0.525928268 | 0.000103601 | 0.000908725 | 0.651339878 |
| T2DM | LEC | *GNAL* | 0.812805296 | 0.000272564 | 0.001071675 | 1 |

| | | | | | | |
|---|---|---|---|---|---|---|
| T2DM | LEC | *C18orf21* | 1.902566589 | 0.000655553 | 0.001756803 | 1 |
| T2DM | LEC | *GALNT1* | -0.665026971 | 0.004547783 | 0.007857079 | 1 |
| T2DM | LEC | *ASXL3* | 1.31703098 | 0.000427254 | 0.001336216 | 1 |
| T2DM | LEC | *ADCYAP1* | 2.249954974 | 0.002291714 | 0.004460684 | 1 |
| T2DM | LEC | *ESCO1* | 0.806966596 | 0.002188855 | 0.00429773 | 1 |
| T2DM | LEC | *GREB1L* | 0.404136018 | 0.001719257 | 0.003568493 | 1 |
| T2DM | LEC | *C18orf8* | 0.507867639 | 0.004665162 | 0.008037166 | 1 |
| T2DM | LEC | *NPC1* | 0.578720241 | 0.021407951 | 0.030313466 | 1 |
| T2DM | LEC | *SLC13A5* | 2.268840604 | 0.015136557 | 0.02221716 | 1 |
| T2DM | LEC | *CCDC40* | 0.94375824 | 0.000646023 | 0.001740903 | 1 |
| T2DM | LEC | *TMC6* | 0.831252986 | 3.56E-05 | 0.000908725 | 0.223659146 |
| T2DM | LEC | *CARD14* | 1.527221122 | 0.012684439 | 0.019019095 | 1 |
| T2DM | LEC | *EIF4A3* | 1.227770606 | 0.000258853 | 0.001042471 | 1 |
| T2DM | LEC | *FN3KRP* | -0.819358785 | 9.09E-06 | 0.000681925 | 0.057117603 |
| T2DM | LEC | *CBX8* | -1.622871441 | 0.000494243 | 0.001449978 | 1 |
| T2DM | LEC | *SECTM1* | 2.067390277 | 0.000408301 | 0.001307186 | 1 |
| T2DM | LEC | *RNF157* | 0.320384022 | 0.018772739 | 0.026964636 | 1 |
| T2DM | LEC | *WDR45B* | 0.973881566 | 0.00012888 | 0.000908725 | 0.810269601 |
| T2DM | LEC | *RNF165* | -0.824605455 | 0.000830957 | 0.002064728 | 1 |
| T2DM | LEC | *MAPK4* | 0.661392986 | 3.79E-05 | 0.000908725 | 0.238331131 |
| T2DM | LEC | *SMAD4* | 0.899946215 | 0.00557471 | 0.009376604 | 1 |
| T2DM | LEC | *PIP4K2B* | 0.994476289 | 0.002267507 | 0.00442178 | 1 |
| T2DM | LEC | *GRB7* | 0.970926066 | 0.000653931 | 0.001753205 | 1 |
| T2DM | LEC | *NFIC* | 0.587543232 | 0.020494236 | 0.029177369 | 1 |
| T2DM | LEC | *TPGS1* | 1.074035167 | 0.000134012 | 0.000908725 | 0.84253518 |
| T2DM | LEC | *PRDM15* | 0.715257628 | 0.008087121 | 0.012894682 | 1 |
| T2DM | LEC | *PFKL* | 0.613860779 | 0.002042269 | 0.0040787 | 1 |
| T2DM | LEC | *MVB12A* | 0.783051757 | 0.000313746 | 0.001145484 | 1 |
| T2DM | LEC | *DUS3L* | 0.671530032 | 3.40E-06 | 0.000436409 | 0.021384055 |
| T2DM | LEC | *TMEM91* | -1.082338547 | 4.57E-05 | 0.000908725 | 0.28726372 |
| T2DM | LEC | *SIRT3* | 0.86968194 | 4.92E-06 | 0.000529126 | 0.030930672 |
| T2DM | LEC | *IFITM3* | 0.820492799 | 0.000243508 | 0.001016559 | 1 |
| T2DM | LEC | *COL6A1* | 0.45612752 | 0.006731032 | 0.011006907 | 1 |
| T2DM | LEC | *IFNAR1* | 0.497734453 | 0.01991099 | 0.028443625 | 1 |
| T2DM | LEC | *TRPM2* | 0.307243816 | 1.96E-05 | 0.000908725 | 0.123136208 |
| T2DM | LEC | *TMEM50B* | 0.510639315 | 0.01032067 | 0.015911244 | 1 |
| T2DM | LEC | *AKT1* | 1.321779238 | 0.000579307 | 0.001617994 | 1 |
| T2DM | LEC | *SAE1* | 0.641599914 | 0.000106864 | 0.000908725 | 0.671854123 |
| T2DM | LEC | *NTN5* | 0.819492317 | 0.000399422 | 0.001290795 | 1 |
| T2DM | LEC | *GEMIN7* | -1.792241112 | 4.79E-05 | 0.000908725 | 0.301461546 |
| T2DM | LEC | *CAPN10* | 0.913060201 | 0.000234186 | 0.001001608 | 1 |
| T2DM | LEC | *MYO1F* | 0.496812056 | 7.96E-05 | 0.000908725 | 0.500399307 |
| T2DM | LEC | *CACNG8* | 0.993548749 | 0.000102053 | 0.000908725 | 0.641606733 |
| T2DM | LEC | *EVI5L* | 1.427253272 | 0.006541158 | 0.010734601 | 1 |
| T2DM | LEC | *CTU1* | -1.423280781 | 0.007475886 | 0.01206079 | 1 |
| T2DM | LEC | *RCN3* | 0.553829811 | 0.000125793 | 0.000908725 | 0.790863241 |
| T2DM | LEC | *ARHGEF19* | -0.369411823 | 0.000177504 | 0.000918726 | 1 |
| T2DM | LEC | *PEX14* | 0.788604691 | 0.003114414 | 0.005693609 | 1 |
| T2DM | LEC | *IL22RA1* | 0.74644201 | 0.01275645 | 0.019113394 | 1 |
| T2DM | LEC | *ZNF593* | 0.668809988 | 0.020964201 | 0.029758847 | 1 |

| | | | | | | |
|---|---|---|---|---|---|---|
| T2DM | LEC | *C1orf216* | 1.519216294 | 0.001781633 | 0.00366649 | 1 |
| T2DM | LEC | *MAP3K6* | 1.151834398 | 0.005843496 | 0.009755193 | 1 |
| T2DM | LEC | *ITGB3BP* | 0.693197942 | 0.002297277 | 0.00446598 | 1 |
| T2DM | LEC | *PIGK* | 0.889369769 | 0.00041848 | 0.001322531 | 1 |
| T2DM | LEC | *CYP4B1* | 0.642516229 | 3.10E-05 | 0.000908725 | 0.194627736 |
| T2DM | LEC | *SYPL2* | 0.429080004 | 0.000180217 | 0.00092505 | 1 |
| T2DM | LEC | *ZNF697* | 0.683326721 | 0.000147865 | 0.000908725 | 0.92962556 |
| T2DM | LEC | *CELSR2* | 1.167966719 | 0.000505811 | 0.001472919 | 1 |
| T2DM | LEC | *ATP1B1* | 0.657921378 | 0.000271057 | 0.001067753 | 1 |
| T2DM | LEC | *TIPRL* | 0.952945348 | 0.000853992 | 0.002101389 | 1 |
| T2DM | LEC | *CREG1* | 0.587388678 | 0.00030554 | 0.001128632 | 1 |
| T2DM | LEC | *GPA33* | -0.823789194 | 0.000137626 | 0.000908725 | 0.865254497 |
| T2DM | LEC | *RXRG* | 3.705760287 | 0.000236895 | 0.001005449 | 1 |
| T2DM | LEC | *TMCO1* | 0.900915414 | 0.015000487 | 0.022050189 | 1 |
| T2DM | LEC | *MGST3* | 0.805999017 | 0.002714672 | 0.00510381 | 1 |
| T2DM | LEC | *ADCY10* | 0.704832214 | 0.000242425 | 0.001014056 | 1 |
| T2DM | LEC | *PPOX* | 1.26060892 | 0.001643086 | 0.003443361 | 1 |
| T2DM | LEC | *RGL1* | 0.562117982 | 0.000200556 | 0.000962913 | 1 |
| T2DM | LEC | *TUFT1* | 0.488349821 | 0.006989329 | 0.011366187 | 1 |
| T2DM | LEC | *SNX27* | 0.966149474 | 0.001850191 | 0.003781584 | 1 |
| T2DM | LEC | *SELENBP1* | 0.700756365 | 0.000409595 | 0.001308237 | 1 |
| T2DM | LEC | *CERS2* | 0.954863158 | 0.000319568 | 0.00115467 | 1 |
| T2DM | LEC | *ENSA* | 0.887907206 | 0.00376109 | 0.006682715 | 1 |
| T2DM | LEC | *SEMA6C* | 1.360808888 | 0.024869041 | 0.034689274 | 1 |
| T2DM | LEC | *ARNT* | 0.691815019 | 0.00133267 | 0.002919337 | 1 |
| T2DM | LEC | *POGZ* | 0.692897487 | 0.006710832 | 0.010981519 | 1 |
| T2DM | LEC | *GABPB2* | 0.706368013 | 0.000163736 | 0.000910705 | 1 |
| T2DM | LEC | *IKBKE* | 1.437164521 | 0.000302026 | 0.001123941 | 1 |
| T2DM | LEC | *DYRK3* | 0.814966904 | 0.0001246 | 0.000908725 | 0.783363009 |
| T2DM | LEC | *EIF2D* | 0.718995184 | 0.002716047 | 0.00510487 | 1 |
| T2DM | LEC | *HHIPL2* | 1.40338126 | 0.006707161 | 0.01097837 | 1 |
| T2DM | LEC | *TP53BP2* | 1.094280816 | 0.001703691 | 0.003546723 | 1 |
| T2DM | LEC | *RAB13* | 0.636191843 | 0.000265747 | 0.00105572 | 1 |
| T2DM | LEC | *S100A8* | -1.703800803 | 0.001611585 | 0.00338864 | 1 |
| T2DM | LEC | *HAX1* | 0.88776256 | 0.000127713 | 0.000908725 | 0.802931776 |
| T2DM | LEC | *CREB3L4* | -0.785541671 | 0.028005391 | 0.038637237 | 1 |
| T2DM | LEC | *KCNN3* | -0.124955346 | 0.000159283 | 0.000910705 | 1 |
| T2DM | LEC | *GATAD2B* | 0.651339398 | 0.031186528 | 0.042476105 | 1 |
| T2DM | LEC | *RIT1* | 1.121581651 | 0.000204524 | 0.000964732 | 1 |
| T2DM | LEC | *GALNT2* | 0.837658249 | 0.000844801 | 0.002084721 | 1 |
| T2DM | LEC | *CEP170* | 0.777822351 | 0.01267749 | 0.019013211 | 1 |
| T2DM | LEC | *SNAP47* | 1.427029974 | 0.00067556 | 0.001793599 | 1 |
| T2DM | LEC | *PSEN2* | 1.016932723 | 0.015015961 | 0.022067637 | 1 |
| T2DM | LEC | *PYCR2* | -0.775074914 | 0.002865317 | 0.005331237 | 1 |
| T2DM | LEC | *WNT9A* | -0.487190784 | 0.01243009 | 0.01866889 | 1 |
| T2DM | LEC | *PLEKHA6* | 0.992663642 | 3.03E-05 | 0.000908725 | 0.190704836 |
| T2DM | LEC | *ABCG8* | 2.050507314 | 1.48E-05 | 0.000902894 | 0.093252907 |
| T2DM | LEC | *EML4* | 0.74951129 | 0.000821329 | 0.002045028 | 1 |
| T2DM | LEC | *CHAC2* | -0.930112611 | 0.001542086 | 0.003267643 | 1 |
| T2DM | LEC | *WDPCP* | 0.37065353 | 0.004596541 | 0.007930588 | 1 |

| | | | | | | |
|---|---|---|---|---|---|---|
| T2DM | LEC | *ASXL2* | 0.824261008 | 0.003446147 | 0.006199573 | 1 |
| T2DM | LEC | *CIAO1* | 0.798011548 | 0.007934663 | 0.012683791 | 1 |
| T2DM | LEC | *EXOC6B* | 0.649795979 | 0.00143862 | 0.003095348 | 1 |
| T2DM | LEC | *TEX261* | 0.825660701 | 0.000123025 | 0.000908725 | 0.773457176 |
| T2DM | LEC | *THNSL2* | 0.250603995 | 0.001660411 | 0.003471567 | 1 |
| T2DM | LEC | *RABL2A* | 0.936131132 | 0.027700076 | 0.038241189 | 1 |
| T2DM | LEC | *SLC20A1* | 0.761983495 | 0.000130235 | 0.000908725 | 0.818785895 |
| T2DM | LEC | *FBLN7* | -0.440400382 | 0.019889821 | 0.028426303 | 1 |
| T2DM | LEC | *FAHD2B* | -0.724396174 | 0.000169028 | 0.000910705 | 1 |
| T2DM | LEC | *AFF3* | 0.391327183 | 0.000773051 | 0.001959414 | 1 |
| T2DM | LEC | *UBXN4* | 0.807680378 | 0.001968474 | 0.003964061 | 1 |
| T2DM | LEC | *THSD7B* | 0.556045849 | 8.66E-06 | 0.000673907 | 0.054462672 |
| T2DM | LEC | *GPR17* | 0.395286818 | 0.000833221 | 0.002067564 | 1 |
| T2DM | LEC | *POLR2D* | 0.454861164 | 0.000121565 | 0.000908725 | 0.764276922 |
| T2DM | LEC | *ZNF385B* | 0.249468642 | 8.17E-05 | 0.000908725 | 0.513575709 |
| T2DM | LEC | *TMEFF2* | 1.2800927 | 0.003945129 | 0.006945512 | 1 |
| T2DM | LEC | *FAM171B* | -0.206657714 | 0.00017734 | 0.000918726 | 1 |
| T2DM | LEC | *HSPD1* | 0.973135943 | 0.000249056 | 0.001023586 | 1 |
| T2DM | LEC | *METTL21A* | 0.787359179 | 0.000236309 | 0.001005438 | 1 |
| T2DM | LEC | *PTH2R* | 0.339054959 | 9.85E-05 | 0.000908725 | 0.619213487 |
| T2DM | LEC | *NBEAL1* | 0.827374932 | 0.000202959 | 0.000964479 | 1 |
| T2DM | LEC | *SUMF1* | 0.688049669 | 0.000934457 | 0.002235514 | 1 |
| T2DM | LEC | *ESPNL* | -0.74001827 | 0.000193119 | 0.000946328 | 1 |
| T2DM | LEC | *TAMM41* | 1.104422188 | 0.000854614 | 0.002102096 | 1 |
| T2DM | LEC | *FAM198A* | 1.040116534 | 0.002757379 | 0.005167107 | 1 |
| T2DM | LEC | *CSRNP1* | 0.854044302 | 0.000118785 | 0.000908725 | 0.746800136 |
| T2DM | LEC | *SLC25A38* | 0.615991819 | 0.000221149 | 0.00098409 | 1 |
| T2DM | LEC | *CTDSPL* | -0.744557075 | 0.001236006 | 0.002747797 | 1 |
| T2DM | LEC | *STAC* | 1.325798091 | 0.003260813 | 0.005923354 | 1 |
| T2DM | LEC | *ARL6IP5* | 0.88999489 | 0.001005646 | 0.002361784 | 1 |
| T2DM | LEC | *TMF1* | 0.882548664 | 0.004703329 | 0.00809467 | 1 |
| T2DM | LEC | *LIMD1* | 0.895192527 | 0.000471155 | 0.001409207 | 1 |
| T2DM | LEC | *COL8A1* | 0.447415108 | 0.000630274 | 0.00171385 | 1 |
| T2DM | LEC | *NXPE3* | -1.05265014 | 0.004529485 | 0.00782977 | 1 |
| T2DM | LEC | *PLA1A* | -0.269997206 | 1.66E-05 | 0.000908725 | 0.104569223 |
| T2DM | LEC | *RABL3* | 1.195090624 | 0.00235214 | 0.004545974 | 1 |
| T2DM | LEC | *IGSF11* | 0.485609553 | 8.01E-05 | 0.000908725 | 0.503438645 |
| T2DM | LEC | *BOC* | 0.42498582 | 0.000165457 | 0.000910705 | 1 |
| T2DM | LEC | *AGTR1* | 0.490698433 | 0.00023381 | 0.001001608 | 1 |
| T2DM | LEC | *AMT* | 0.440416739 | 0.002325371 | 0.004508051 | 1 |
| T2DM | LEC | *TCTA* | 1.204516552 | 0.001242845 | 0.002762023 | 1 |
| T2DM | LEC | *EAF2* | 0.864887364 | 0.035021707 | 0.047137973 | 1 |
| T2DM | LEC | *LYAR* | -1.099381384 | 0.022927103 | 0.032232267 | 1 |
| T2DM | LEC | *ATP10D* | 0.965506145 | 0.001723897 | 0.003574213 | 1 |
| T2DM | LEC | *SLC10A4* | 3.636744869 | 0.000126211 | 0.000908725 | 0.79349157 |
| T2DM | LEC | *PYURF* | 1.221626244 | 0.007881448 | 0.012617943 | 1 |
| T2DM | LEC | *CISD2* | 1.062933172 | 7.52E-05 | 0.000908725 | 0.472616903 |
| T2DM | LEC | *TIFA* | 0.58254148 | 0.00014933 | 0.000910197 | 0.93884056 |
| T2DM | LEC | *CCNA2* | 0.703279256 | 0.000382688 | 0.001263636 | 1 |
| T2DM | LEC | *USP53* | 0.786475024 | 0.000661844 | 0.001769892 | 1 |

| | | | | | | |
|---|---|---|---|---|---|---|
| T2DM | LEC | *MARCH1* | 0.481450442 | 0.000415959 | 0.001318933 | 1 |
| T2DM | LEC | *PDGFC* | 0.478531191 | 0.000617508 | 0.001694577 | 1 |
| T2DM | LEC | *CBR4* | 0.78092489 | 0.007254403 | 0.011739623 | 1 |
| T2DM | LEC | *FAM105A* | 0.477089208 | 0.000550407 | 0.001559445 | 1 |
| T2DM | LEC | *LHFPL2* | 0.853681879 | 0.002398782 | 0.004624699 | 1 |
| T2DM | LEC | *SSBP2* | 0.654845804 | 0.011085235 | 0.016923961 | 1 |
| T2DM | LEC | *BHMT* | -0.720578787 | 1.43E-05 | 0.000902248 | 0.090166054 |
| T2DM | LEC | *CRHBP* | 0.367384935 | 0.000408938 | 0.001307728 | 1 |
| T2DM | LEC | *RASA1* | 0.640915474 | 0.005384305 | 0.009099765 | 1 |
| T2DM | LEC | *GTF2H2* | 1.042556204 | 0.000548465 | 0.001555553 | 1 |
| T2DM | LEC | *SPATA9* | 0.264388112 | 5.99E-05 | 0.000908725 | 0.376858236 |
| T2DM | LEC | *TSLP* | 0.570029203 | 0.011416377 | 0.017349472 | 1 |
| T2DM | LEC | *FEM1C* | 0.986848445 | 0.030067458 | 0.041094371 | 1 |
| T2DM | LEC | *YIPF5* | -0.920827454 | 0.017493458 | 0.025288887 | 1 |
| T2DM | LEC | *GABRB2* | 0.299187494 | 0.000592534 | 0.001645434 | 1 |
| T2DM | LEC | *MYLK4* | 1.422563569 | 0.001474334 | 0.003157062 | 1 |
| T2DM | LEC | *TBC1D7* | -0.895828966 | 0.031770851 | 0.043150431 | 1 |
| T2DM | LEC | *GFOD1* | 0.586861346 | 0.000924876 | 0.002222312 | 1 |
| T2DM | LEC | *PSD2* | -1.761899923 | 0.007942001 | 0.012692263 | 1 |
| T2DM | LEC | *ZMAT2* | 1.057321165 | 0.002405022 | 0.004635307 | 1 |
| T2DM | LEC | *KLHL3* | -0.209173355 | 0.00022658 | 0.000992757 | 1 |
| T2DM | LEC | *TRIM7* | 0.913479905 | 0.000417238 | 0.001322166 | 1 |
| T2DM | LEC | *HIGD2A* | 0.765199256 | 0.00084631 | 0.002086542 | 1 |
| T2DM | LEC | *FAM193B* | 0.932318378 | 0.011202364 | 0.0170779 | 1 |
| T2DM | LEC | *MUT* | 1.133457293 | 0.014211966 | 0.021048441 | 1 |
| T2DM | LEC | *DOK3* | 0.83392761 | 0.003541276 | 0.006341214 | 1 |
| T2DM | LEC | *FGD2* | 0.37087989 | 0.000122551 | 0.000908725 | 0.770476693 |
| T2DM | LEC | *RPL7L1* | 1.048030588 | 0.000691151 | 0.001821616 | 1 |
| T2DM | LEC | *MMS22L* | 0.805837155 | 0.001395119 | 0.00302348 | 1 |
| T2DM | LEC | *RNF217* | 0.723777711 | 0.000428261 | 0.001336216 | 1 |
| T2DM | LEC | *ABRACL* | 0.942675196 | 0.01741207 | 0.025194403 | 1 |
| T2DM | LEC | *AIG1* | 0.646067881 | 0.000425443 | 0.001334043 | 1 |
| T2DM | LEC | *DYNLT1* | 0.783430373 | 0.000737142 | 0.001903545 | 1 |
| T2DM | LEC | *WTAP* | 0.696807659 | 0.011457548 | 0.017403624 | 1 |
| T2DM | LEC | *VIP* | -0.267845586 | 0.000732807 | 0.00189673 | 1 |
| T2DM | LEC | *SLC22A3* | 0.90540332 | 0.000435682 | 0.00134367 | 1 |
| T2DM | LEC | *GNA12* | 0.827535705 | 0.001055314 | 0.002445543 | 1 |
| T2DM | LEC | *C7orf50* | 1.141928772 | 0.000365776 | 0.001235973 | 1 |
| T2DM | LEC | *EGFR* | 0.48232667 | 0.016830743 | 0.024437617 | 1 |
| T2DM | LEC | *POMZP3* | 0.659719461 | 0.004879552 | 0.008363615 | 1 |
| T2DM | LEC | *PSPH* | 1.075960962 | 0.000593739 | 0.001648051 | 1 |
| T2DM | LEC | *GIGYF1* | 0.783792636 | 0.021208569 | 0.03007178 | 1 |
| T2DM | LEC | *TMEM209* | 1.319746329 | 0.000276335 | 0.00108013 | 1 |
| T2DM | LEC | *EPHA1* | -0.914441921 | 0.001901141 | 0.003865613 | 1 |
| T2DM | LEC | *NCAPG2* | 1.181307685 | 0.0064303 | 0.010580291 | 1 |
| T2DM | LEC | *FBXO25* | -0.790724047 | 0.005113778 | 0.008715186 | 1 |
| T2DM | LEC | *CSGALNACT1* | -0.27338166 | 0.000164098 | 0.000910705 | 1 |
| T2DM | LEC | *CCDC25* | 1.557957703 | 0.000372755 | 0.001243243 | 1 |
| T2DM | LEC | *BIN3* | 0.835583311 | 0.000199729 | 0.000961874 | 1 |
| T2DM | LEC | *SLC25A37* | 0.894685168 | 0.013361041 | 0.019905418 | 1 |

| | | | | | | |
|---|---|---|---|---|---|---|
| T2DM | LEC | *CHMP7* | 0.737862925 | 6.33E-05 | 0.000908725 | 0.398033704 |
| T2DM | LEC | *DOCK5* | -0.203522973 | 0.000186818 | 0.000932902 | 1 |
| T2DM | LEC | *ST18* | 0.335842578 | 0.000167645 | 0.000910705 | 1 |
| T2DM | LEC | *RGS20* | -0.546773279 | 0.000841286 | 0.00208071 | 1 |
| T2DM | LEC | *MTDH* | 0.732261548 | 0.02159372 | 0.030535249 | 1 |
| T2DM | LEC | *UTP23* | 1.193791048 | 0.002025886 | 0.004052417 | 1 |
| T2DM | LEC | *FAM135B* | -1.634795426 | 0.001515048 | 0.003221207 | 1 |
| T2DM | LEC | *ZNF7* | 1.316467374 | 0.000120475 | 0.000908725 | 0.757428145 |
| T2DM | LEC | *SLC39A4* | -1.68744524 | 6.94E-06 | 0.000610811 | 0.043615076 |
| T2DM | LEC | *CER1* | 1.269807929 | 0.002176334 | 0.004286532 | 1 |
| T2DM | LEC | *PLIN2* | 0.6292696 | 0.000197144 | 0.000956093 | 1 |
| T2DM | LEC | *HAUS6* | 0.957253786 | 0.000317905 | 0.001152864 | 1 |
| T2DM | LEC | *C9orf72* | 0.638511631 | 0.000133282 | 0.000908725 | 0.837946373 |
| T2DM | LEC | *SIGMAR1* | 0.673082937 | 0.001066557 | 0.002466144 | 1 |
| T2DM | LEC | *CBWD5* | 0.8753129 | 0.000180062 | 0.00092505 | 1 |
| T2DM | LEC | *ASTN2* | 0.920902276 | 0.000930936 | 0.002229636 | 1 |
| T2DM | LEC | *WDR31* | 1.448285733 | 0.000279517 | 0.00108013 | 1 |
| T2DM | LEC | *POLE3* | 1.125729387 | 0.00173251 | 0.003588894 | 1 |
| T2DM | LEC | *SURF6* | 1.336639714 | 0.00517512 | 0.008803025 | 1 |
| T2DM | LEC | *ASB6* | 0.999438125 | 0.00032463 | 0.001162346 | 1 |
| T2DM | LEC | *NTMT1* | 0.855172704 | 0.000248786 | 0.001023586 | 1 |
| T2DM | LEC | *CIZ1* | 1.010164149 | 0.00071363 | 0.001861576 | 1 |
| T2DM | LEC | *LRSAM1* | 0.702360671 | 0.000420746 | 0.001326594 | 1 |
| T2DM | LEC | *INPP5E* | 1.032031645 | 0.001143004 | 0.002594247 | 1 |
| T2DM | LEC | *SEC16A* | 0.755678972 | 1.68E-05 | 0.000908725 | 0.105352142 |
| T2DM | LEC | *SLC39A12* | 0.812168801 | 9.44E-05 | 0.000908725 | 0.59331247 |
| T2DM | LEC | *GLUD1* | 0.702125763 | 0.000164185 | 0.000910705 | 1 |
| T2DM | LEC | *HTR7* | 0.466794841 | 0.000100667 | 0.000908725 | 0.632891649 |
| T2DM | LEC | *FRA10AC1* | 0.88170164 | 0.000866545 | 0.002123137 | 1 |
| T2DM | LEC | *DNAJB12* | 1.035089671 | 0.000454605 | 0.001380727 | 1 |
| T2DM | LEC | *PAOX* | 0.701175697 | 0.00012295 | 0.000908725 | 0.772988981 |
| T2DM | LEC | *GSTO1* | 0.890442865 | 0.010468037 | 0.016102899 | 1 |
| T2DM | LEC | *PDCD11* | 1.089524072 | 0.001019448 | 0.002389734 | 1 |
| T2DM | LEC | *RGS10* | 0.629605366 | 0.000275403 | 0.001079463 | 1 |
| T2DM | LEC | *GAS2* | -1.024054946 | 0.005466412 | 0.009213763 | 1 |
| T2DM | LEC | *LIN7C* | 1.255040256 | 0.0010032 | 0.002356919 | 1 |
| T2DM | LEC | *ZNF215* | 0.419083107 | 0.000343618 | 0.001193551 | 1 |
| T2DM | LEC | *HSD17B12* | -0.993590812 | 3.08E-06 | 0.000431731 | 0.019378771 |
| T2DM | LEC | *APIP* | -0.285328022 | 0.009896162 | 0.015305578 | 1 |
| T2DM | LEC | *EIF3M* | 0.634026215 | 0.030678559 | 0.041838633 | 1 |
| T2DM | LEC | *GLYAT* | 0.319616635 | 2.19E-05 | 0.000908725 | 0.137422556 |
| T2DM | LEC | *PTPRJ* | 0.715845593 | 0.000119114 | 0.000908725 | 0.748866811 |
| T2DM | LEC | *C11orf49* | 0.914319992 | 0.001595142 | 0.003359685 | 1 |
| T2DM | LEC | *ENDOD1* | -0.835052726 | 0.0009894 | 0.002331469 | 1 |
| T2DM | LEC | *SERPINH1* | 0.930435151 | 0.014415893 | 0.021310303 | 1 |
| T2DM | LEC | *CAPN5* | 0.449780392 | 6.75E-05 | 0.000908725 | 0.424105132 |
| T2DM | LEC | *INTS4* | 0.900554708 | 0.003931564 | 0.006927619 | 1 |
| T2DM | LEC | *ZC3H12C* | 0.936077471 | 0.000847434 | 0.002087704 | 1 |
| T2DM | LEC | *TTC12* | 0.534167578 | 0.006036605 | 0.010020662 | 1 |
| T2DM | LEC | *DRD2* | 1.959360548 | 0.000105889 | 0.000908725 | 0.665726634 |

| | | | | | | |
|---|---|---|---|---|---|---|
| T2DM | LEC | *C11orf52* | 3.174478635 | 0.00010983 | 0.000908725 | 0.690502537 |
| T2DM | LEC | *GRIK4* | 0.698433107 | 0.003198745 | 0.005824709 | 1 |
| T2DM | LEC | *ADAM33* | 0.375547084 | 0.000163413 | 0.000910705 | 1 |
| T2DM | LEC | *TMC2* | 2.314352446 | 0.033015709 | 0.044667476 | 1 |
| T2DM | LEC | *INCENP* | 0.800963706 | 0.000117314 | 0.000908725 | 0.737555005 |
| T2DM | LEC | *MS4A3* | 8.113543248 | 0.000125307 | 0.000908725 | 0.787806381 |
| T2DM | LEC | *PLCH2* | 1.437104833 | 0.000101546 | 0.000908725 | 0.638421158 |
| T2DM | LEC | *CPSF7* | 0.988743795 | 0.004087652 | 0.007154403 | 1 |
| T2DM | LEC | *CHEK1* | 0.635150148 | 0.011224929 | 0.017106161 | 1 |
| T2DM | LEC | *FEZ1* | 0.772841746 | 6.96E-05 | 0.000908725 | 0.437766838 |
| T2DM | LEC | *KIAA1755* | 0.644317835 | 0.000400625 | 0.001292438 | 1 |
| T2DM | LEC | *CNBD2* | 0.919452197 | 0.000108301 | 0.000908725 | 0.68088839 |
| T2DM | LEC | *CABLES2* | 0.879923977 | 0.023276538 | 0.03269428 | 1 |
| T2DM | LEC | *TBX6* | 0.730345848 | 0.000173066 | 0.000910705 | 1 |
| T2DM | LEC | *DOC2A* | 0.594355544 | 0.032713868 | 0.044325881 | 1 |
| T2DM | LEC | *TAOK2* | -1.033826948 | 0.001611041 | 0.00338863 | 1 |
| T2DM | LEC | *MPP7* | -0.110648238 | 6.19E-05 | 0.000908725 | 0.388904152 |
| T2DM | LEC | *CWC15* | 0.85162801 | 0.009190257 | 0.014365775 | 1 |
| T2DM | LEC | *ARID5B* | 0.741752334 | 0.00054734 | 0.00155496 | 1 |
| T2DM | LEC | *LYPD6B* | -0.369765804 | 0.000127981 | 0.000908725 | 0.804613708 |
| T2DM | LEC | *PDCD4* | 0.74174649 | 0.000425402 | 0.001334043 | 1 |
| T2DM | LEC | *WDR17* | 0.605869518 | 0.000120789 | 0.000908725 | 0.759402234 |
| T2DM | LEC | *CCDC102B* | -0.498137656 | 0.000690813 | 0.001821616 | 1 |
| T2DM | LEC | *CNDP1* | 0.872805472 | 0.023938769 | 0.033512145 | 1 |
| T2DM | LEC | *FSIP1* | 1.207549916 | 0.000405243 | 0.001301208 | 1 |
| T2DM | LEC | *RGS18* | 0.774955359 | 0.000103579 | 0.000908725 | 0.651198377 |
| T2DM | LEC | *MTMR12* | 0.873002844 | 0.019566745 | 0.02800276 | 1 |
| T2DM | LEC | *PPP1R1C* | -0.3770658 | 0.002733664 | 0.005133378 | 1 |
| T2DM | LEC | *C11orf53* | -0.958576392 | 0.000120897 | 0.000908725 | 0.76007984 |
| T2DM | LEC | *CCT5* | 1.125285559 | 0.002741539 | 0.00514509 | 1 |
| T2DM | LEC | *TEX12* | 2.79297275 | 0.000114744 | 0.000908725 | 0.721397528 |
| T2DM | LEC | *PIP4K2A* | 0.742224987 | 0.003048931 | 0.005595047 | 1 |
| T2DM | LEC | *ABCB9* | 0.620781146 | 0.018103521 | 0.026086908 | 1 |
| T2DM | LEC | *RILPL2* | 0.537388379 | 0.000361546 | 0.00122668 | 1 |
| T2DM | LEC | *DHX37* | 1.134926884 | 0.000244108 | 0.001017397 | 1 |
| T2DM | LEC | *PRSS53* | -1.363456843 | 4.63E-06 | 0.000522362 | 0.029099666 |
| T2DM | LEC | *GPR158* | -0.479822019 | 0.000599173 | 0.001658076 | 1 |
| T2DM | LEC | *THRB* | 0.584321777 | 0.00045137 | 0.001373553 | 1 |
| T2DM | LEC | *TMEM86A* | 0.472468168 | 0.010387075 | 0.01599401 | 1 |
| T2DM | LEC | *BTBD11* | 0.645975203 | 0.000121757 | 0.000908725 | 0.765487018 |
| T2DM | LEC | *ANK3* | 0.478454416 | 3.52E-05 | 0.000908725 | 0.221395768 |
| T2DM | LEC | *IPMK* | 0.688350216 | 0.000202454 | 0.00096305 | 1 |
| T2DM | LEC | *PLBD2* | 1.027420387 | 0.000368702 | 0.001239589 | 1 |
| T2DM | LEC | *DLG5* | 0.542578896 | 0.010533624 | 0.016187948 | 1 |
| T2DM | LEC | *GXYLT1* | 1.04961498 | 0.001500865 | 0.003198622 | 1 |
| T2DM | LEC | *FAM177A1* | 1.090346704 | 0.002695979 | 0.005074737 | 1 |
| T2DM | LEC | *EXT2* | 0.546713992 | 0.012079695 | 0.018208957 | 1 |
| T2DM | LEC | *NDUFC2* | 1.391878619 | 0.001815436 | 0.003721436 | 1 |
| T2DM | LEC | *ME3* | 0.810900339 | 0.003653821 | 0.00651491 | 1 |
| T2DM | LEC | *VIPAS39* | 0.788255825 | 0.001340328 | 0.002933047 | 1 |

| | | | | | | |
|---|---|---|---|---|---|---|
| T2DM | LEC | *C4orf33* | 0.795882771 | 0.000249034 | 0.001023586 | 1 |
| T2DM | LEC | *PTPRO* | 0.404610746 | 0.000115605 | 0.000908725 | 0.726806189 |
| T2DM | LEC | *VPS26B* | 1.024528704 | 0.000872043 | 0.002134942 | 1 |
| T2DM | LEC | *NCAPD3* | 0.682486763 | 0.000624182 | 0.001705358 | 1 |
| T2DM | LEC | *VTI1A* | 0.752821982 | 0.002078698 | 0.004130459 | 1 |
| T2DM | LEC | *ANO4* | 0.696747644 | 0.000160911 | 0.000910705 | 1 |
| T2DM | LEC | *AKR1C2* | 0.723522281 | 0.001010184 | 0.002371557 | 1 |
| T2DM | LEC | *ADAM8* | 0.656629307 | 0.000143831 | 0.000908725 | 0.904264201 |
| T2DM | LEC | *ITIH2* | -1.508845612 | 0.001173791 | 0.002644078 | 1 |
| T2DM | LEC | *KIN* | 1.029004956 | 0.002314858 | 0.004491824 | 1 |
| T2DM | LEC | *PIGF* | 0.919170089 | 0.003220379 | 0.005858369 | 1 |
| T2DM | LEC | *INPP1* | 1.084024303 | 0.028707525 | 0.039475986 | 1 |
| T2DM | LEC | *RNF144A* | 0.886100272 | 0.006308581 | 0.010401797 | 1 |
| T2DM | LEC | *ASAP2* | 0.824309177 | 0.001472803 | 0.003156024 | 1 |
| T2DM | LEC | *ADAM17* | 0.754068135 | 0.000555577 | 0.001569824 | 1 |
| T2DM | LEC | *TMEM45B* | 2.067961714 | 0.000239906 | 0.00100947 | 1 |
| T2DM | LEC | *CENPU* | 0.705390255 | 0.000419571 | 0.001324883 | 1 |
| T2DM | LEC | *ACSL1* | 0.439459842 | 0.000174612 | 0.000911424 | 1 |
| T2DM | LEC | *GABRA2* | 0.671631468 | 0.009397858 | 0.014650219 | 1 |
| T2DM | LEC | *CENPJ* | 1.060166378 | 0.000946945 | 0.002256401 | 1 |
| T2DM | LEC | *PARP8* | 0.657996936 | 0.000419825 | 0.001325019 | 1 |
| T2DM | LEC | *CACUL1* | 0.75558688 | 0.000917108 | 0.00221423 | 1 |
| T2DM | LEC | *BEND6* | 0.326741224 | 0.000808714 | 0.002022429 | 1 |
| T2DM | LEC | *PTPN14* | -0.331995967 | 0.000278022 | 0.00108013 | 1 |
| T2DM | LEC | *RNF219* | 0.878360262 | 0.00051819 | 0.001499244 | 1 |
| T2DM | LEC | *ARL14EP* | 0.51115153 | 0.020054479 | 0.028603111 | 1 |
| T2DM | LEC | *HAUS1* | -0.65885637 | 0.027345032 | 0.03781747 | 1 |
| T2DM | LEC | *SPC25* | 0.287169028 | 0.030043843 | 0.041079957 | 1 |
| T2DM | LEC | *G6PC2* | -20.07375246 | 0.000214188 | 0.0009784 | 1 |
| T2DM | LEC | *TGOLN2* | 0.890398847 | 0.001742228 | 0.003605114 | 1 |
| T2DM | LEC | *POC5* | -1.244861955 | 0.000213341 | 0.0009784 | 1 |
| T2DM | LEC | *GUCY1A2* | 0.657200952 | 0.037135863 | 0.049732774 | 1 |
| T2DM | LEC | *CWF19L2* | 0.79202325 | 0.013978816 | 0.020737333 | 1 |
| T2DM | LEC | *ZNF547* | 1.189971127 | 0.000690905 | 0.001821616 | 1 |
| T2DM | LEC | *ZNF773* | -0.932019488 | 0.019084982 | 0.02736312 | 1 |
| T2DM | LEC | *ZNF837* | -0.627009753 | 0.003966911 | 0.006970366 | 1 |
| T2DM | LEC | *CCDC50* | 0.569725833 | 0.012373385 | 0.018597053 | 1 |
| T2DM | LEC | *ZFP36L2* | 0.892820314 | 0.000345664 | 0.001196042 | 1 |
| T2DM | LEC | *PLEKHH2* | 0.339948334 | 0.00149123 | 0.003184566 | 1 |
| T2DM | LEC | *TMEM123* | -0.775481779 | 0.01742031 | 0.025200526 | 1 |
| T2DM | LEC | *IGSF10* | 0.320822286 | 0.000117438 | 0.000908725 | 0.738332402 |
| T2DM | LEC | *SPEF2* | 0.821568504 | 0.001778067 | 0.003661947 | 1 |
| T2DM | LEC | *SPARCL1* | 0.490337504 | 0.000391674 | 0.00127787 | 1 |
| T2DM | LEC | *MBNL1* | 0.561036908 | 0.00807766 | 0.012882863 | 1 |
| T2DM | LEC | *CLEC4F* | 1.146468723 | 0.000107073 | 0.000908725 | 0.673168854 |
| T2DM | LEC | *PELO* | 0.589613871 | 0.000109272 | 0.000908725 | 0.686991048 |
| T2DM | LEC | *RASGRP3* | -0.09921825 | 0.000185754 | 0.000932031 | 1 |
| T2DM | LEC | *SAR1B* | 1.238308105 | 2.23E-05 | 0.000908725 | 0.139980633 |
| T2DM | LEC | *GPR180* | 0.682802059 | 0.006725949 | 0.011003394 | 1 |
| T2DM | LEC | *TCTEX1D1* | -0.418683388 | 0.003432376 | 0.006183194 | 1 |

| | | | | | | |
|---|---|---|---|---|---|---|
| T2DM | LEC | *WDR78* | 0.657695717 | 0.000700191 | 0.001836115 | 1 |
| T2DM | LEC | *IFIT5* | 0.477339862 | 0.000103786 | 0.000908725 | 0.652500583 |
| T2DM | LEC | *PRDM8* | -0.71449879 | 0.002264686 | 0.004417648 | 1 |
| T2DM | LEC | *HHEX* | -0.289451891 | 0.000141667 | 0.000908725 | 0.890659588 |
| T2DM | LEC | *ZNF117* | 0.307120548 | 0.00258393 | 0.004896073 | 1 |
| T2DM | LEC | *RAB3C* | 0.915916947 | 0.002961283 | 0.005470934 | 1 |
| T2DM | LEC | *MED21* | 0.932771094 | 0.000280099 | 0.00108013 | 1 |
| T2DM | LEC | *SRP19* | 0.617521999 | 9.29E-05 | 0.000908725 | 0.583813937 |
| T2DM | LEC | *CARHSP1* | 0.933368291 | 0.008391234 | 0.013291934 | 1 |
| T2DM | LEC | *TEKT5* | 1.07957662 | 5.19E-05 | 0.000908725 | 0.326051785 |
| T2DM | LEC | *BANK1* | 0.617905268 | 0.000166213 | 0.000910705 | 1 |
| T2DM | LEC | *CAST* | 0.576282078 | 0.014949454 | 0.021995604 | 1 |
| T2DM | LEC | *SCOC* | 1.177368673 | 0.000351637 | 0.001210038 | 1 |
| T2DM | LEC | *CLGN* | 2.427810087 | 7.15E-05 | 0.000908725 | 0.449740567 |
| T2DM | LEC | *AHCTF1* | 0.858649206 | 0.000755712 | 0.001930582 | 1 |
| T2DM | LEC | *PTPRR* | 0.903286251 | 0.000117315 | 0.000908725 | 0.73755818 |
| T2DM | LEC | *INO80C* | -0.205291989 | 0.000146458 | 0.000908725 | 0.920782508 |
| T2DM | LEC | *LPCAT1* | 0.759700383 | 0.000154324 | 0.000910197 | 0.970233892 |
| T2DM | LEC | *NMRAL1* | 1.081544958 | 0.000140165 | 0.000908725 | 0.88121675 |
| T2DM | LEC | *C16orf89* | 0.438458697 | 0.000126229 | 0.000908725 | 0.793600385 |
| T2DM | LEC | *ADPRHL1* | 0.55482078 | 0.001254147 | 0.002778302 | 1 |
| T2DM | LEC | *TUBGCP5* | 0.970792107 | 0.01837924 | 0.026447764 | 1 |
| T2DM | LEC | *LURAP1L* | 0.496101819 | 0.000251265 | 0.001025781 | 1 |
| T2DM | LEC | *ZDHHC7* | 0.783431412 | 0.000179402 | 0.00092375 | 1 |
| T2DM | LEC | *FAM92B* | 5.224633275 | 0.000117076 | 0.000908725 | 0.736056868 |
| T2DM | LEC | *C7orf31* | -0.594625296 | 0.000491192 | 0.001447869 | 1 |
| T2DM | LEC | *FBXO36* | 1.307782397 | 0.000649785 | 0.001746558 | 1 |
| T2DM | LEC | *CACNA2D1* | 0.62199026 | 0.004430489 | 0.007673411 | 1 |
| T2DM | LEC | *ZUFSP* | 1.097023098 | 0.000902716 | 0.002192958 | 1 |
| T2DM | LEC | *HS3ST3A1* | 0.625094948 | 0.000143496 | 0.000908725 | 0.90216216 |
| T2DM | LEC | *PPP2R5E* | 0.838062718 | 0.00778727 | 0.012483062 | 1 |
| T2DM | LEC | *ANKRD29* | 0.745968036 | 4.98E-05 | 0.000908725 | 0.313296927 |
| T2DM | LEC | *THY1* | 0.453454174 | 0.000102832 | 0.000908725 | 0.646504444 |
| T2DM | LEC | *UBASH3B* | 0.550387608 | 0.000173392 | 0.000910705 | 1 |
| T2DM | LEC | *ROBO4* | -0.461985806 | 0.00058612 | 0.001631948 | 1 |
| T2DM | LEC | *PRKCA* | 0.81559956 | 0.001039872 | 0.00242502 | 1 |
| T2DM | LEC | *LRRK1* | 0.902165195 | 0.000963911 | 0.002290907 | 1 |
| T2DM | LEC | *GAL3ST2* | 3.91492467 | 0.005982768 | 0.009953338 | 1 |
| T2DM | LEC | *ENPP3* | -0.436977448 | 0.000245024 | 0.001017515 | 1 |
| T2DM | LEC | *UCHL1* | 0.462600241 | 0.000122009 | 0.000908725 | 0.76706906 |
| T2DM | LEC | *MIA3* | 0.768987343 | 0.00130084 | 0.002860574 | 1 |
| T2DM | LEC | *DISP1* | 0.860570859 | 0.005561129 | 0.009360862 | 1 |
| T2DM | LEC | *FAM167A* | 2.088090424 | 0.001652357 | 0.003458323 | 1 |
| T2DM | LEC | *NEIL2* | 0.680875183 | 4.62E-05 | 0.000908725 | 0.290425781 |
| T2DM | LEC | *CCSAP* | 0.506747521 | 0.00014596 | 0.000908725 | 0.917651748 |
| T2DM | LEC | *FAM69A* | 0.513575319 | 0.00053716 | 0.001536454 | 1 |
| T2DM | LEC | *ATP5G3* | 1.154897521 | 0.000158203 | 0.000910705 | 0.99462051 |
| T2DM | LEC | *PDLIM3* | -0.406942285 | 0.000929223 | 0.002227231 | 1 |
| T2DM | LEC | *NCAM2* | 0.619252522 | 0.001259995 | 0.002787506 | 1 |
| T2DM | LEC | *L3MBTL4* | -0.572828267 | 0.008984071 | 0.014103085 | 1 |

| | | | | | | |
|---|---|---|---|---|---|---|
| T2DM | LEC | *PDE1C* | -0.712823044 | 0.001593532 | 0.003357751 | 1 |
| T2DM | LEC | *MRPL39* | 0.639718838 | 0.006249967 | 0.010315973 | 1 |
| T2DM | LEC | *TSEN2* | 1.283621869 | 0.000207023 | 0.000972609 | 1 |
| T2DM | LEC | *FLCN* | 0.864581685 | 0.001973584 | 0.003973078 | 1 |
| T2DM | LEC | *DPH3* | 0.705642582 | 0.000117821 | 0.000908725 | 0.74074146 |
| T2DM | LEC | *APCDD1* | 0.47062318 | 0.016547769 | 0.024065655 | 1 |
| T2DM | LEC | *EPHB1* | -0.795993454 | 0.001160077 | 0.002621641 | 1 |
| T2DM | LEC | *ANKRD40* | 0.902464139 | 0.003095101 | 0.005666541 | 1 |
| T2DM | LEC | *CYP2U1* | 1.000832695 | 0.000215655 | 0.00097964 | 1 |
| T2DM | LEC | *FBXL18* | 0.733709484 | 0.000125993 | 0.000908725 | 0.792117325 |
| T2DM | LEC | *PROM2* | 1.784405083 | 0.00025504 | 0.001034507 | 1 |
| T2DM | LEC | *CDK19* | 0.811304564 | 0.004367023 | 0.007590675 | 1 |
| T2DM | LEC | *GTF3C6* | 1.021035484 | 0.000983038 | 0.002320899 | 1 |
| T2DM | LEC | *GOLGA7B* | 0.665063699 | 0.000856473 | 0.002105844 | 1 |
| T2DM | LEC | *TRMT44* | 1.177741627 | 0.007284888 | 0.011779858 | 1 |
| T2DM | LEC | *HEATR3* | 0.491373953 | 0.00360201 | 0.006440795 | 1 |
| T2DM | LEC | *CNOT8* | 0.776786623 | 0.000355984 | 0.001216909 | 1 |
| T2DM | LEC | *PDIA4* | 1.323454813 | 0.002814854 | 0.005254449 | 1 |
| T2DM | LEC | *KDM8* | -1.492862843 | 0.018716762 | 0.026897848 | 1 |
| T2DM | LEC | *FAM126B* | 0.807921466 | 0.003786255 | 0.006722448 | 1 |
| T2DM | LEC | *ALS2CR12* | 0.61764759 | 0.001459553 | 0.003133308 | 1 |
| T2DM | LEC | *SPAG17* | 0.503387794 | 0.000129351 | 0.000908725 | 0.813232027 |
| T2DM | LEC | *DEPTOR* | 0.875245924 | 0.000456158 | 0.001383438 | 1 |
| T2DM | LEC | *WIF1* | -1.75368626 | 0.028166248 | 0.038833597 | 1 |
| T2DM | LEC | *UGT2B4* | 0.684409787 | 0.000113478 | 0.000908725 | 0.713435065 |
| T2DM | LEC | *MMP16* | -0.624768327 | 0.002522423 | 0.004808512 | 1 |
| T2DM | LEC | *KCNMA1* | 0.572998275 | 0.000415502 | 0.001318656 | 1 |
| T2DM | LEC | *BATF* | 0.50060651 | 0.000114588 | 0.000908725 | 0.720412946 |
| T2DM | LEC | *DRAM2* | 0.748431405 | 0.000217275 | 0.000982034 | 1 |
| T2DM | LEC | *C8orf37* | -0.825992481 | 0.000341573 | 0.001191055 | 1 |
| T2DM | LEC | *ADAMTSL3* | 0.545783218 | 0.005101322 | 0.008697703 | 1 |
| T2DM | LEC | *CXCL13* | -1.353028625 | 0.000483487 | 0.001433135 | 1 |
| T2DM | LEC | *N6AMT1* | 0.414112954 | 0.003616511 | 0.006459376 | 1 |
| T2DM | LEC | *CCT8* | 0.545841583 | 0.012402822 | 0.018632388 | 1 |
| T2DM | LEC | *ANKRD9* | 0.448583559 | 0.000295943 | 0.001112688 | 1 |
| T2DM | LEC | *SFXN2* | 1.098775864 | 0.000124508 | 0.000908725 | 0.782781943 |
| T2DM | LEC | *C14orf2* | 0.541277767 | 0.025532087 | 0.035521184 | 1 |
| T2DM | LEC | *SH3RF2* | 0.751358039 | 0.00233546 | 0.004520639 | 1 |
| T2DM | LEC | *NODAL* | 1.66914184 | 0.000259787 | 0.001042963 | 1 |
| T2DM | LEC | *MED19* | 0.439174265 | 0.000404675 | 0.001300425 | 1 |
| T2DM | LEC | *WDYHV1* | -0.957274914 | 0.003238631 | 0.005888164 | 1 |
| T2DM | LEC | *ATAD2* | 0.691469712 | 0.000166594 | 0.000910705 | 1 |
| T2DM | LEC | *SASS6* | 0.875728285 | 0.001172319 | 0.00264171 | 1 |
| T2DM | LEC | *ITGAD* | 0.489113725 | 0.000241924 | 0.001012636 | 1 |
| T2DM | LEC | *VPS8* | 0.812754442 | 9.48E-05 | 0.000908725 | 0.596273189 |
| T2DM | LEC | *MPV17L* | 0.696484563 | 0.000642131 | 0.001733394 | 1 |
| T2DM | LEC | *RPUSD3* | 1.404150472 | 0.011649068 | 0.017664663 | 1 |
| T2DM | LEC | *GHRL* | 0.550744845 | 0.000112534 | 0.000908725 | 0.707503925 |
| T2DM | LEC | *EXOG* | 0.944337969 | 0.000970947 | 0.002300922 | 1 |
| T2DM | LEC | *SLC6A1* | 0.747787459 | 0.001282413 | 0.002825984 | 1 |

| | | | | | | |
|---|---|---|---|---|---|---|
| T2DM | LEC | *FCHO2* | 0.713578919 | 0.000619323 | 0.001697248 | 1 |
| T2DM | LEC | *TIMP4* | 2.138147452 | 1.33E-05 | 0.000893669 | 0.083881551 |
| T2DM | LEC | *CPT2* | 0.821526513 | 0.000106013 | 0.000908725 | 0.666502628 |
| T2DM | LEC | *LRP8* | 1.29400999 | 0.000804548 | 0.002016019 | 1 |
| T2DM | LEC | *STEAP2* | 0.428646248 | 2.49E-05 | 0.000908725 | 0.156684123 |
| T2DM | LEC | *GATAD1* | 0.821634729 | 0.004389774 | 0.007621792 | 1 |
| T2DM | LEC | *SUSD3* | 0.660478493 | 0.001488562 | 0.00318103 | 1 |
| T2DM | LEC | *TMED6* | -5.307361843 | 0.002072033 | 0.004122483 | 1 |
| T2DM | LEC | *CLEC18A* | -2.538999006 | 1.35E-06 | 0.000269656 | 0.008488715 |
| T2DM | LEC | *IL34* | -0.851455763 | 0.0011028 | 0.002527636 | 1 |
| T2DM | LEC | *DHRS1* | 0.552958959 | 0.009538401 | 0.014839873 | 1 |
| T2DM | LEC | *AASDH* | 0.839634957 | 0.012129145 | 0.018278029 | 1 |
| T2DM | LEC | *RNF111* | 0.709343176 | 0.001645964 | 0.003448243 | 1 |
| T2DM | LEC | *MX1* | 0.255643063 | 0.023286028 | 0.032700303 | 1 |
| T2DM | LEC | *CREB3L1* | 0.580595016 | 5.92E-05 | 0.000908725 | 0.371955836 |
| T2DM | LEC | *C2CD2* | 0.4060133 | 0.000808318 | 0.002022243 | 1 |
| T2DM | LEC | *SLC38A10* | 0.914950174 | 0.000873445 | 0.002137544 | 1 |
| T2DM | LEC | *C9orf43* | 0.593927952 | 0.000983913 | 0.002321149 | 1 |
| T2DM | LEC | *WDR19* | 0.844983319 | 0.000515901 | 0.001495377 | 1 |
| T2DM | LEC | *SLC37A3* | 0.908740872 | 0.021243939 | 0.03011514 | 1 |
| T2DM | LEC | *AP3S2* | 1.558318632 | 5.32E-10 | 3.71E-07 | 3.34E-06 |
| T2DM | LEC | *SPPL3* | 0.629608899 | 0.027890389 | 0.038495472 | 1 |
| T2DM | LEC | *FAM213B* | 1.069995315 | 0.003325941 | 0.006025991 | 1 |
| T2DM | LEC | *RADIL* | -0.150519336 | 0.000131474 | 0.000908725 | 0.826574571 |
| T2DM | LEC | *LDLRAP1* | 1.248651625 | 0.000527285 | 0.00151718 | 1 |
| T2DM | LEC | *TRIM63* | 1.143447054 | 3.26E-07 | 9.30E-05 | 0.002046803 |
| T2DM | LEC | *UBXN11* | 0.569026277 | 0.000833768 | 0.002067811 | 1 |
| T2DM | LEC | *GALNT14* | -1.988304747 | 0.009432377 | 0.014693101 | 1 |
| T2DM | LEC | *RHPN1* | -0.733488459 | 0.001688751 | 0.003522619 | 1 |
| T2DM | LEC | *TPRG1L* | -0.993903642 | 0.000276525 | 0.00108013 | 1 |
| T2DM | LEC | *LRRC43* | 3.956131615 | 0.034329476 | 0.046275604 | 1 |
| T2DM | LEC | *XKR8* | 0.62499204 | 0.000279399 | 0.00108013 | 1 |
| T2DM | LEC | *EYA3* | 0.791899321 | 0.004475754 | 0.007741147 | 1 |
| T2DM | LEC | *ABHD3* | 0.720848813 | 8.76E-05 | 0.000908725 | 0.550775936 |
| T2DM | LEC | *CLSTN2* | 0.543066857 | 0.000327856 | 0.001167883 | 1 |
| T2DM | LEC | *COLEC12* | -0.309626005 | 0.000317416 | 0.001152864 | 1 |
| T2DM | LEC | *RNF207* | 1.881792193 | 0.002475477 | 0.004733372 | 1 |
| T2DM | LEC | *MITD1* | 0.828753353 | 0.010536328 | 0.016188147 | 1 |
| T2DM | LEC | *AHCYL2* | 0.816340094 | 0.002194317 | 0.004305765 | 1 |
| T2DM | LEC | *CD1D* | 0.71427634 | 0.001495617 | 0.003191766 | 1 |
| T2DM | LEC | *CD1E* | 2.889200382 | 4.37E-07 | 0.000119417 | 0.00274659 |
| T2DM | LEC | *NCF1* | 0.577192563 | 0.000111245 | 0.000908725 | 0.69939701 |
| T2DM | LEC | *PPP1R9A* | -0.373339832 | 0.000328055 | 0.001167883 | 1 |
| T2DM | LEC | *DYNC1I1* | -0.341489117 | 0.000251587 | 0.001026403 | 1 |
| T2DM | LEC | *TMED4* | 1.062972389 | 0.000712286 | 0.001861013 | 1 |
| T2DM | LEC | *ELK4* | -0.797520035 | 0.031597386 | 0.042942664 | 1 |
| T2DM | LEC | *DUSP23* | 1.294562175 | 0.000135397 | 0.000908725 | 0.851239261 |
| T2DM | LEC | *RNF166* | 1.042447431 | 0.000147813 | 0.000908725 | 0.929297929 |
| T2DM | LEC | *SPATA2L* | 0.914147559 | 0.000218167 | 0.00098409 | 1 |
| T2DM | LEC | *NIT1* | 0.74247286 | 0.000128533 | 0.000908725 | 0.808088534 |

| | | | | | | |
|---|---|---|---|---|---|---|
| T2DM | LEC | *ZNF276* | 0.69391977 | 0.00229707 | 0.00446598 | 1 |
| T2DM | LEC | *FGF17* | -1.327963595 | 0.001118432 | 0.002555081 | 1 |
| T2DM | LEC | *CDA* | 0.496586612 | 0.000119561 | 0.000908725 | 0.751682067 |
| T2DM | LEC | *B4GALT3* | 1.420562672 | 0.001434194 | 0.003088996 | 1 |
| T2DM | LEC | *DMTN* | -0.227638238 | 0.000951307 | 0.002265481 | 1 |
| T2DM | LEC | *FAM160B2* | 0.9667566 | 0.007518133 | 0.012113403 | 1 |
| T2DM | LEC | *NDUFS2* | 0.665339012 | 0.000638279 | 0.001727446 | 1 |
| T2DM | LEC | *FCER1G* | 0.80062399 | 0.000110668 | 0.000908725 | 0.6957685 |
| T2DM | LEC | *CCAR2* | 1.238169305 | 0.001791729 | 0.00368365 | 1 |
| T2DM | LEC | *CDC42SE2* | 0.796777825 | 0.00889881 | 0.013986705 | 1 |
| T2DM | LEC | *EPB41* | 0.637771294 | 0.000173298 | 0.000910705 | 1 |
| T2DM | LEC | *ALG8* | 0.835799376 | 0.000630107 | 0.00171385 | 1 |
| T2DM | LEC | *IFNAR2* | 0.754477723 | 0.000184179 | 0.000929091 | 1 |
| T2DM | LEC | *MRPL10* | 0.787659577 | 0.002146937 | 0.004244589 | 1 |
| T2DM | LEC | *SV2A* | -0.411187024 | 0.00016312 | 0.000910705 | 1 |
| T2DM | LEC | *UBE2Z* | 0.902083959 | 0.011871807 | 0.01795047 | 1 |
| T2DM | LEC | *CLIC6* | 0.521000708 | 0.000238685 | 0.001009156 | 1 |
| T2DM | LEC | *CBR1* | 0.524653303 | 0.000224491 | 0.000992757 | 1 |
| T2DM | LEC | *CBR3* | 0.820853133 | 0.00301706 | 0.00554952 | 1 |
| T2DM | LEC | *SIM2* | 1.182424904 | 0.000325471 | 0.001163956 | 1 |
| T2DM | LEC | *HLCS* | 0.428040894 | 0.014404871 | 0.021304029 | 1 |
| T2DM | LEC | *ARHGAP27* | -0.646482296 | 0.0006337 | 0.001718869 | 1 |
| T2DM | LEC | *PADI4* | 3.434010045 | 0.000177695 | 0.000918726 | 1 |
| T2DM | LEC | *PSMD4* | 1.027779816 | 0.000586511 | 0.001632314 | 1 |
| T2DM | LEC | *STARD9* | -0.56011781 | 0.005406244 | 0.009124687 | 1 |
| T2DM | LEC | *TCHH* | 0.981933371 | 0.001980622 | 0.003984694 | 1 |
| T2DM | LEC | *MED8* | 0.813797174 | 0.00012808 | 0.000908725 | 0.805240115 |
| T2DM | LEC | *CCDC17* | -1.289998501 | 0.00835899 | 0.013247535 | 1 |
| T2DM | LEC | *ACE* | 0.327517489 | 0.000118561 | 0.000908725 | 0.745394388 |
| T2DM | LEC | *TEPP* | -1.412699411 | 7.00E-05 | 0.000908725 | 0.440309791 |
| T2DM | LEC | *ATP6V0D1* | 0.724019535 | 0.000206032 | 0.000970455 | 1 |
| T2DM | LEC | *ZFYVE28* | 0.724201858 | 0.002350017 | 0.004544618 | 1 |
| T2DM | LEC | *C16orf86* | 1.24325163 | 0.000368576 | 0.001239589 | 1 |
| T2DM | LEC | *LYPD5* | -0.376180137 | 4.56E-05 | 0.000908725 | 0.286888617 |
| T2DM | LEC | *CCDC117* | 1.281372239 | 0.01112998 | 0.016988148 | 1 |
| T2DM | LEC | *CCDC107* | 0.487936856 | 0.00082079 | 0.002044494 | 1 |
| T2DM | LEC | *NPR2* | 1.252047943 | 7.37E-05 | 0.000908725 | 0.463305374 |
| T2DM | LEC | *ZNF235* | 0.742110362 | 0.007110939 | 0.011546094 | 1 |
| T2DM | LEC | *CALM3* | 1.253614604 | 0.001032357 | 0.002411901 | 1 |
| T2DM | LEC | *DFFA* | 1.220840348 | 0.000917988 | 0.002214476 | 1 |
| T2DM | LEC | *ATAD3B* | 1.197848315 | 0.00318376 | 0.005806876 | 1 |
| T2DM | LEC | *SSU72* | 0.9246953 | 0.000234554 | 0.001001608 | 1 |
| T2DM | LEC | *NR2F6* | 0.60546199 | 4.14E-05 | 0.000908725 | 0.260532366 |
| T2DM | LEC | *ANKLE1* | 1.175594161 | 0.005197262 | 0.00883108 | 1 |
| T2DM | LEC | *CILP2* | 3.406436989 | 0.000114406 | 0.000908725 | 0.719269344 |
| T2DM | LEC | *ABCG1* | -0.375323399 | 0.000728211 | 0.001887164 | 1 |
| T2DM | LEC | *RSPH1* | -0.604749523 | 2.57E-05 | 0.000908725 | 0.161444236 |
| T2DM | LEC | *NDUFV3* | 0.251463821 | 0.026419852 | 0.036641834 | 1 |
| T2DM | LEC | *U2AF1* | 0.529307101 | 0.000945039 | 0.002254824 | 1 |
| T2DM | LEC | *HSF2BP* | 0.937491498 | 3.57E-05 | 0.000908725 | 0.224674103 |

| | | | | | | |
|---|---|---|---|---|---|---|
| T2DM | LEC | *C21orf33* | 1.057509777 | 0.009002032 | 0.014127752 | 1 |
| T2DM | LEC | *ZNF66* | 1.288698633 | 0.00045667 | 0.001384323 | 1 |
| T2DM | LEC | *LRRC3* | 0.811037334 | 0.000317509 | 0.001152864 | 1 |
| T2DM | LEC | *ITGB2* | 0.447376799 | 0.001918703 | 0.00389732 | 1 |
| T2DM | LEC | *FAM207A* | 1.221986042 | 0.00049238 | 0.001449584 | 1 |
| T2DM | LEC | *SPATC1L* | -0.185444534 | 0.033328023 | 0.045041548 | 1 |
| T2DM | LEC | *LSS* | 0.416577966 | 0.000445612 | 0.001359981 | 1 |
| T2DM | LEC | *PCNT* | 0.525304974 | 0.000673832 | 0.001791283 | 1 |
| T2DM | LEC | *S100B* | 0.216155448 | 0.026891367 | 0.037247416 | 1 |
| T2DM | LEC | *ZNF714* | 0.973176801 | 0.009836487 | 0.015220772 | 1 |
| T2DM | LEC | *GPSM1* | -0.451524983 | 4.10E-05 | 0.000908725 | 0.258049124 |
| T2DM | LEC | *C19orf47* | 0.799143112 | 0.00013859 | 0.000908725 | 0.871315209 |
| T2DM | LEC | *HIPK4* | 0.776182689 | 0.00043642 | 0.00134367 | 1 |
| T2DM | LEC | *RDH13* | 0.803465079 | 0.000277415 | 0.00108013 | 1 |
| T2DM | LEC | *MPZL3* | 0.548904653 | 0.000106581 | 0.000908725 | 0.670076369 |
| T2DM | LEC | *PCSK7* | 0.789101677 | 0.012930526 | 0.019351158 | 1 |
| T2DM | LEC | *S100A1* | -0.675292873 | 0.000535386 | 0.001532077 | 1 |
| T2DM | LEC | *ZBTB7B* | 0.718656521 | 6.54E-05 | 0.000908725 | 0.411441689 |
| T2DM | LEC | *VPS11* | 0.949946078 | 0.009735566 | 0.015090607 | 1 |
| T2DM | LEC | *PMF1* | 0.89128426 | 0.000130537 | 0.000908725 | 0.82068577 |
| T2DM | LEC | *SLC25A44* | -1.184916427 | 0.001388515 | 0.003012283 | 1 |
| T2DM | LEC | *NBEAL2* | 0.981568041 | 0.000287012 | 0.001094929 | 1 |
| T2DM | LEC | *FCRL3* | 0.420314794 | 0.008489395 | 0.013420374 | 1 |
| T2DM | LEC | *LY6K* | 2.896652968 | 0.000241203 | 0.00101202 | 1 |
| T2DM | LEC | *VPS28* | 0.964989395 | 0.009518609 | 0.014812746 | 1 |
| T2DM | LEC | *TONSL* | -0.653366036 | 0.000244699 | 0.001017515 | 1 |
| T2DM | LEC | *MUM1* | 0.943771931 | 0.000394081 | 0.001282395 | 1 |
| T2DM | LEC | *ZNF333* | 1.038775503 | 0.002139277 | 0.004232107 | 1 |
| T2DM | LEC | *ORAI2* | 1.008514523 | 0.00033382 | 0.001176051 | 1 |
| T2DM | LEC | *LRWD1* | -0.503082789 | 0.003361972 | 0.006086011 | 1 |
| T2DM | LEC | *FBXL13* | 0.85166097 | 0.003069338 | 0.005622647 | 1 |
| T2DM | LEC | *SCGB3A1* | -0.599683507 | 0.000136027 | 0.000908725 | 0.855198991 |
| T2DM | LEC | *MFSD12* | 0.751540198 | 0.000206707 | 0.000972342 | 1 |
| T2DM | LEC | *YDJC* | 1.255930131 | 0.0001066 | 0.000908725 | 0.670193899 |
| T2DM | LEC | *DVL3* | 1.163804395 | 0.000209443 | 0.00097394 | 1 |
| T2DM | LEC | *AP2M1* | 1.032525606 | 0.000101288 | 0.000908725 | 0.636800109 |
| T2DM | LEC | *FBXO27* | 0.42799719 | 0.001646672 | 0.003448576 | 1 |
| T2DM | LEC | *DMKN* | -0.368278532 | 0.000599618 | 0.001658512 | 1 |
| T2DM | LEC | *BDH1* | 0.512366605 | 0.000109223 | 0.000908725 | 0.686683805 |
| T2DM | LEC | *COX7A1* | -0.768831101 | 0.022214046 | 0.031320858 | 1 |
| T2DM | LEC | *DUSP14* | 0.62066812 | 6.48E-05 | 0.000908725 | 0.407389371 |
| T2DM | LEC | *IKZF3* | 0.761847681 | 0.000143021 | 0.000908725 | 0.899172838 |
| T2DM | LEC | *ACOX1* | 0.582442021 | 0.000453693 | 0.001379287 | 1 |
| T2DM | LEC | *PRPSAP1* | -0.878931566 | 0.002423926 | 0.004659089 | 1 |
| T2DM | LEC | *CYGB* | -1.566873521 | 0.00633552 | 0.010440738 | 1 |
| T2DM | LEC | *ZNF577* | 0.644703176 | 0.03653627 | 0.048987743 | 1 |
| T2DM | LEC | *TMEM143* | 0.878182387 | 5.91E-05 | 0.000908725 | 0.371628162 |
| T2DM | LEC | *ALDH16A1* | 0.985640218 | 0.000114552 | 0.000908725 | 0.720186814 |
| T2DM | LEC | *SIGLEC11* | 0.772063653 | 0.00076648 | 0.001948143 | 1 |
| T2DM | LEC | *MPP3* | 0.650779108 | 0.000202505 | 0.00096305 | 1 |

| | | | | | | |
|---|---|---|---|---|---|---|
| T2DM | LEC | *NAGS* | -2.227723295 | 0.001705373 | 0.00354905 | 1 |
| T2DM | LEC | *ASB16* | -0.868596441 | 0.00342383 | 0.006175325 | 1 |
| T2DM | LEC | *PLCD3* | -0.58103336 | 0.016444114 | 0.023925976 | 1 |
| T2DM | LEC | *RACGAP1* | 0.660001598 | 7.47E-05 | 0.000908725 | 0.469505802 |
| T2DM | LEC | *LARP4* | 0.657967243 | 0.00031089 | 0.001138389 | 1 |
| T2DM | LEC | *SPC24* | 1.148335469 | 0.000300526 | 0.001121312 | 1 |
| T2DM | LEC | *IP6K3* | -0.350061638 | 5.59E-05 | 0.000908725 | 0.351286803 |
| T2DM | LEC | *LEMD2* | 1.061345739 | 0.011766776 | 0.017821662 | 1 |
| T2DM | LEC | *SCIMP* | 0.69466825 | 0.004921412 | 0.008430767 | 1 |
| T2DM | LEC | *ASGR2* | 1.513935668 | 7.03E-05 | 0.000908725 | 0.44189334 |
| T2DM | LEC | *TNFSF13* | 1.696476864 | 0.002161532 | 0.004267386 | 1 |
| T2DM | LEC | *SENP3* | 1.535015585 | 0.009588399 | 0.014906594 | 1 |
| T2DM | LEC | *SNRNP25* | -1.556835372 | 0.00867925 | 0.013682659 | 1 |
| T2DM | LEC | *JMJD8* | 1.128260072 | 2.01E-05 | 0.000908725 | 0.126310777 |
| T2DM | LEC | *MEIOB* | 1.051933853 | 9.92E-05 | 0.000908725 | 0.62339974 |
| T2DM | LEC | *FLYWCH2* | 1.042800545 | 0.000184396 | 0.000929091 | 1 |
| T2DM | LEC | *ZG16B* | 6.088624961 | 0.000120813 | 0.000908725 | 0.759550943 |
| T2DM | LEC | *RPL29* | 0.955575342 | 0.002812075 | 0.005252381 | 1 |
| T2DM | LEC | *TPCN2* | 0.938111628 | 0.00069139 | 0.001821616 | 1 |
| T2DM | LEC | *CYP4A22* | 3.117789548 | 2.09E-05 | 0.000908725 | 0.131140389 |
| T2DM | LEC | *PDZK1IP1* | 9.931743656 | 0.002127118 | 0.004213356 | 1 |
| T2DM | LEC | *COA7* | 2.217862721 | 0.001360162 | 0.002967155 | 1 |
| T2DM | LEC | *SLC1A7* | 0.421120368 | 0.000846631 | 0.002086542 | 1 |
| T2DM | LEC | *C1orf123* | -1.392232623 | 0.002675639 | 0.005045513 | 1 |
| T2DM | LEC | *USP24* | 0.5694568 | 0.021303353 | 0.03017895 | 1 |
| T2DM | LEC | *ZSWIM5* | -0.517637575 | 0.00115656 | 0.002616514 | 1 |
| T2DM | LEC | *RBP7* | 0.519065119 | 0.000108988 | 0.000908725 | 0.685210533 |
| T2DM | LEC | *FBLIM1* | 0.996535654 | 0.000207209 | 0.000972609 | 1 |
| T2DM | LEC | *SLC25A34* | 2.000634473 | 0.025216697 | 0.035129044 | 1 |
| T2DM | LEC | *PDPN* | -0.139505569 | 0.000138347 | 0.000908725 | 0.869790119 |
| T2DM | LEC | *LRRC38* | 3.384674218 | 0.000142324 | 0.000908725 | 0.894791188 |
| T2DM | LEC | *LAPTM5* | -0.585790631 | 0.037139395 | 0.049732774 | 1 |
| T2DM | LEC | *SYNC* | 0.863673553 | 0.005212797 | 0.008850352 | 1 |
| T2DM | LEC | *RBBP4* | 0.88319017 | 0.006236875 | 0.010297067 | 1 |
| T2DM | LEC | *CAMK2N1* | 0.534376812 | 9.79E-05 | 0.000908725 | 0.615218191 |
| T2DM | LEC | *TTLL10* | 1.164858999 | 7.88E-05 | 0.000908725 | 0.495301728 |
| T2DM | LEC | *OMA1* | 0.614842359 | 0.008328091 | 0.013205223 | 1 |
| T2DM | LEC | *TM2D1* | 0.863509507 | 0.001569261 | 0.003312943 | 1 |
| T2DM | LEC | *FUBP1* | 0.747140744 | 0.025613734 | 0.035611133 | 1 |
| T2DM | LEC | *NEXN* | 0.95641053 | 0.003402658 | 0.006145508 | 1 |
| T2DM | LEC | *B3GALT2* | -0.872812714 | 0.000238508 | 0.001009087 | 1 |
| T2DM | LEC | *NTNG1* | 0.356040577 | 9.36E-05 | 0.000908725 | 0.58857763 |
| T2DM | LEC | *HFM1* | -0.527634604 | 0.000360182 | 0.001224756 | 1 |
| T2DM | LEC | *KCNT2* | -0.572742879 | 0.002332135 | 0.004516984 | 1 |
| T2DM | LEC | *VCAM1* | 0.503780451 | 0.000121958 | 0.000908725 | 0.766750253 |
| T2DM | LEC | *EXTL2* | 0.411188522 | 0.006435654 | 0.010586193 | 1 |
| T2DM | LEC | *ARPC5* | 0.755883986 | 0.02095124 | 0.029747164 | 1 |
| T2DM | LEC | *IGSF8* | 1.451607231 | 0.00065054 | 0.001747094 | 1 |
| T2DM | LEC | *VANGL2* | 1.018283188 | 0.000103871 | 0.000908725 | 0.65303957 |
| T2DM | LEC | *FCRLB* | -1.404247818 | 0.006052599 | 0.010037639 | 1 |

| | | | | | | |
|---|---|---|---|---|---|---|
| T2DM | LEC | *KLHDC9* | -0.919560872 | 0.008293696 | 0.013173944 | 1 |
| T2DM | LEC | *LMX1A* | 0.530019448 | 2.44E-06 | 0.000371603 | 0.015331271 |
| T2DM | LEC | *TDRD5* | -0.739971457 | 0.025493184 | 0.03547491 | 1 |
| T2DM | LEC | *BROX* | 0.609879464 | 0.015138908 | 0.02221716 | 1 |
| T2DM | LEC | *ACP6* | 0.913235809 | 0.000656626 | 0.00175818 | 1 |
| T2DM | LEC | *PM20D1* | -0.941031393 | 0.000241818 | 0.001012636 | 1 |
| T2DM | LEC | *HAAO* | 0.691312707 | 7.71E-05 | 0.000908725 | 0.484536709 |
| T2DM | LEC | *B3GALNT2* | 0.702608597 | 0.004825578 | 0.008277875 | 1 |
| T2DM | LEC | *PUS10* | 1.01886802 | 0.015828699 | 0.023143031 | 1 |
| T2DM | LEC | *RFTN2* | -0.70607704 | 0.002473054 | 0.004730177 | 1 |
| T2DM | LEC | *DISC1* | 0.716309423 | 0.001257685 | 0.002784177 | 1 |
| T2DM | LEC | *FAM84A* | -0.351904398 | 0.00014652 | 0.000908725 | 0.921169651 |
| T2DM | LEC | *CLHC1* | 0.379093739 | 0.0046008 | 0.007933414 | 1 |
| T2DM | LEC | *FRZB* | 1.433460363 | 0.000644913 | 0.001740158 | 1 |
| T2DM | LEC | *NUP35* | 1.048091231 | 0.001394695 | 0.00302348 | 1 |
| T2DM | LEC | *FBXO41* | -0.896030528 | 0.000117065 | 0.000908725 | 0.73598996 |
| T2DM | LEC | *SMC6* | 0.786607753 | 0.000668907 | 0.001782308 | 1 |
| T2DM | LEC | *CCDC74A* | 1.803057353 | 0.000120556 | 0.000908725 | 0.75793367 |
| T2DM | LEC | *NOSTRIN* | 0.472279657 | 0.000223167 | 0.000990853 | 1 |
| T2DM | LEC | *SGPP2* | -1.066754366 | 0.000746626 | 0.001918282 | 1 |
| T2DM | LEC | *BBS5* | 0.555806451 | 8.25E-05 | 0.000908725 | 0.518496567 |
| T2DM | LEC | *HPGDS* | 0.699254587 | 0.000201112 | 0.00096305 | 1 |
| T2DM | LEC | *PDLIM5* | 0.724785085 | 0.001853268 | 0.003786642 | 1 |
| T2DM | LEC | *CTSS* | 0.424028168 | 0.000120274 | 0.000908725 | 0.756162141 |
| T2DM | LEC | *PACRGL* | 0.900223447 | 0.000550171 | 0.001559445 | 1 |
| T2DM | LEC | *C1QTNF7* | 0.650827947 | 0.000320024 | 0.001155653 | 1 |
| T2DM | LEC | *LYSMD1* | 1.0742546 | 0.000713496 | 0.001861576 | 1 |
| T2DM | LEC | *ERCC3* | 0.667820319 | 3.00E-05 | 0.000908725 | 0.188826344 |
| T2DM | LEC | *RNF149* | 0.752131566 | 0.000663496 | 0.001773452 | 1 |
| T2DM | LEC | *DHX57* | 0.965806554 | 0.002049599 | 0.004089441 | 1 |
| T2DM | LEC | *S100A12* | 3.77732257 | 0.000330799 | 0.001172764 | 1 |
| T2DM | LEC | *TDRD10* | -0.776248059 | 0.001504917 | 0.003202916 | 1 |
| T2DM | LEC | *GNPDA2* | 0.543189522 | 0.023489989 | 0.032952261 | 1 |
| T2DM | LEC | *NIPAL1* | 0.781948933 | 0.000113444 | 0.000908725 | 0.713223343 |
| T2DM | LEC | *ANTXR2* | 0.733948131 | 0.001901061 | 0.003865613 | 1 |
| T2DM | LEC | *CGGBP1* | 0.403607177 | 0.005694248 | 0.009544052 | 1 |
| T2DM | LEC | *PBXIP1* | 1.110779369 | 0.000361771 | 0.001226783 | 1 |
| T2DM | LEC | *DCST2* | 1.443618648 | 8.61E-05 | 0.000908725 | 0.541278798 |
| T2DM | LEC | *LMOD3* | 1.301557629 | 0.000174688 | 0.000911424 | 1 |
| T2DM | LEC | *NBPF10* | 0.618851024 | 0.000392096 | 0.00127792 | 1 |
| T2DM | LEC | *SLC22A15* | 0.416926091 | 6.69E-05 | 0.000908725 | 0.420333839 |
| T2DM | LEC | *LRRC58* | 1.113498811 | 0.00142362 | 0.003073591 | 1 |
| T2DM | LEC | *FSTL1* | 0.597086087 | 0.001052274 | 0.002443904 | 1 |
| T2DM | LEC | *LMOD1* | 0.339469393 | 0.02956812 | 0.040517604 | 1 |
| T2DM | LEC | *TMEM183A* | -1.183086988 | 0.00847743 | 0.013408202 | 1 |
| T2DM | LEC | *IGFBP7* | -0.340280377 | 0.000339908 | 0.001187224 | 1 |
| T2DM | LEC | *ARPC2* | 0.821184211 | 0.005871284 | 0.009793781 | 1 |
| T2DM | LEC | *CCT3* | 0.849549295 | 0.000484652 | 0.001435914 | 1 |
| T2DM | LEC | *GLB1L* | 0.804522791 | 0.030113588 | 0.041122756 | 1 |
| T2DM | LEC | *SERPINI1* | -1.008353951 | 0.01645927 | 0.023942487 | 1 |

| | | | | | | |
|---|---|---|---|---|---|---|
| T2DM | LEC | *NUAK2* | 0.780371015 | 0.003199097 | 0.005824709 | 1 |
| T2DM | LEC | *PYHIN1* | 0.31283439 | 0.000432549 | 0.001341345 | 1 |
| T2DM | LEC | *AIM2* | 1.891163765 | 0.008026488 | 0.012807748 | 1 |
| T2DM | LEC | *CD200R1* | 0.439545443 | 0.000368653 | 0.001239589 | 1 |
| T2DM | LEC | *GTPBP8* | 1.250013428 | 0.008274428 | 0.013149982 | 1 |
| T2DM | LEC | *SPICE1* | 0.967162876 | 0.001473117 | 0.003156024 | 1 |
| T2DM | LEC | *CADPS* | 0.711428252 | 0.000123235 | 0.000908725 | 0.774775676 |
| T2DM | LEC | *PPM1K* | 0.761656616 | 0.000491915 | 0.001449236 | 1 |
| T2DM | LEC | *TIPARP* | 0.755529882 | 6.12E-05 | 0.000908725 | 0.384723946 |
| T2DM | LEC | *DCLK3* | -1.345388897 | 0.016107126 | 0.023500929 | 1 |
| T2DM | LEC | *RPL9* | -0.434868582 | 0.000513241 | 0.001489726 | 1 |
| T2DM | LEC | *RPP14* | 0.923632288 | 0.000168609 | 0.000910705 | 1 |
| T2DM | LEC | *DNASE1L3* | 0.775240191 | 5.66E-05 | 0.000908725 | 0.356036401 |
| T2DM | LEC | *CRELD1* | 0.856439762 | 0.004440502 | 0.00768652 | 1 |
| T2DM | LEC | *PCOLCE2* | 0.49933706 | 0.000371348 | 0.001242505 | 1 |
| T2DM | LEC | *TTC14* | 0.639049724 | 0.012820374 | 0.019190879 | 1 |
| T2DM | LEC | *RCHY1* | 0.788053316 | 0.003426568 | 0.00617627 | 1 |
| T2DM | LEC | *CCDC158* | 0.828190696 | 5.15E-05 | 0.000908725 | 0.323701184 |
| T2DM | LEC | *GYG1* | 0.754067952 | 9.23E-05 | 0.000908725 | 0.580279806 |
| T2DM | LEC | *HPS3* | 0.937394925 | 0.001249718 | 0.002772398 | 1 |
| T2DM | LEC | *RYK* | 0.829368771 | 0.006409416 | 0.010551452 | 1 |
| T2DM | LEC | *PLB1* | 0.395730319 | 0.000107935 | 0.000908725 | 0.678589809 |
| T2DM | LEC | *KIAA1143* | 0.529755216 | 0.008032331 | 0.01281382 | 1 |
| T2DM | LEC | *CDCP1* | 0.916136972 | 0.000136024 | 0.000908725 | 0.855181331 |
| T2DM | LEC | *FYCO1* | 0.803232493 | 0.000278577 | 0.00108013 | 1 |
| T2DM | LEC | *LRRC2* | 0.374237875 | 0.000103647 | 0.000908725 | 0.65162958 |
| T2DM | LEC | *DTX3L* | 0.930742648 | 4.02E-05 | 0.000908725 | 0.252777982 |
| T2DM | LEC | *NMNAT3* | 0.975964431 | 8.54E-05 | 0.000908725 | 0.537164165 |
| T2DM | LEC | *TPRA1* | 1.282749305 | 0.019103634 | 0.027371136 | 1 |
| T2DM | LEC | *MEAF6* | 1.16595389 | 0.008396943 | 0.013297628 | 1 |
| T2DM | LEC | *DNALI1* | 0.999681948 | 3.23E-05 | 0.000908725 | 0.203285143 |
| T2DM | LEC | *RPN1* | 1.036887259 | 0.001001253 | 0.002354986 | 1 |
| T2DM | LEC | *RFC4* | 1.230015397 | 0.000104822 | 0.000908725 | 0.65901347 |
| T2DM | LEC | *SFMBT1* | 1.083647519 | 0.000757169 | 0.001932733 | 1 |
| T2DM | LEC | *GNL3* | 0.855466624 | 0.02833388 | 0.039038215 | 1 |
| T2DM | LEC | *UVSSA* | 0.928442916 | 0.000455438 | 0.001382587 | 1 |
| T2DM | LEC | *FAM208A* | 0.666318217 | 0.025580725 | 0.035573107 | 1 |
| T2DM | LEC | *SLC51A* | 0.817221002 | 0.000358723 | 0.001223256 | 1 |
| T2DM | LEC | *PIGX* | 0.703084784 | 0.000749814 | 0.001920977 | 1 |
| T2DM | LEC | *ABLIM2* | -0.854308067 | 0.001255656 | 0.002780665 | 1 |
| T2DM | LEC | *EXO5* | 0.660502632 | 0.00712792 | 0.011570677 | 1 |
| T2DM | LEC | *C1orf50* | -1.132377677 | 0.00098361 | 0.002321149 | 1 |
| T2DM | LEC | *ERMAP* | 0.469467199 | 0.000531835 | 0.001527476 | 1 |
| T2DM | LEC | *DNAJB14* | 0.937444869 | 0.003004458 | 0.005527957 | 1 |
| T2DM | LEC | *SLC9B2* | 0.869223154 | 0.01669988 | 0.024258814 | 1 |
| T2DM | LEC | *ATRIP* | 1.050857423 | 0.00019442 | 0.000949652 | 1 |
| T2DM | LEC | *SHISA5* | 1.056848553 | 0.022662461 | 0.031881605 | 1 |
| T2DM | LEC | *RNF123* | 1.521010762 | 7.47E-10 | 4.70E-07 | 4.70E-06 |
| T2DM | LEC | *MST1R* | 0.976752119 | 1.92E-05 | 0.000908725 | 0.120597881 |
| T2DM | LEC | *POC1A* | 0.626505406 | 8.38E-05 | 0.000908725 | 0.52704363 |

| | | | | | | |
|---|---|---|---|---|---|---|
| T2DM | LEC | *PPM1M* | 0.467390065 | 0.001105483 | 0.002530657 | 1 |
| T2DM | LEC | *C4orf3* | 1.205675547 | 0.003214384 | 0.005849155 | 1 |
| T2DM | LEC | *ANXA5* | 0.796443754 | 0.000648173 | 0.001744462 | 1 |
| T2DM | LEC | *MAP9* | 1.234778206 | 0.000577075 | 0.001613193 | 1 |
| T2DM | LEC | *CEP44* | 0.975478482 | 0.000777817 | 0.001967609 | 1 |
| T2DM | LEC | *C4orf45* | 0.716750935 | 0.000564459 | 0.001587631 | 1 |
| T2DM | LEC | *TMEM144* | 0.597813489 | 0.0001581 | 0.000910705 | 0.993975198 |
| T2DM | LEC | *NPY1R* | 0.857396539 | 0.009145835 | 0.014310569 | 1 |
| T2DM | LEC | *IL15* | 1.043541003 | 0.002979987 | 0.005495568 | 1 |
| T2DM | LEC | *ARFIP1* | 0.69156053 | 0.028901718 | 0.039699607 | 1 |
| T2DM | LEC | *ITGA2* | -0.293965045 | 0.000184581 | 0.000929091 | 1 |
| T2DM | LEC | *NDUFAF2* | 0.665618597 | 0.006887103 | 0.01122905 | 1 |
| T2DM | LEC | *LMBRD2* | -0.849407555 | 0.010793775 | 0.016547297 | 1 |
| T2DM | LEC | *PGGT1B* | 0.976334712 | 0.008319067 | 0.013194242 | 1 |
| T2DM | LEC | *CMBL* | 1.351732388 | 0.001030113 | 0.002410242 | 1 |
| T2DM | LEC | *C5orf63* | -0.98859161 | 0.000882841 | 0.002154666 | 1 |
| T2DM | LEC | *PRRC1* | 0.926330494 | 0.000110931 | 0.000908725 | 0.69742289 |
| T2DM | LEC | *WDR41* | 0.522135265 | 0.002764424 | 0.005178765 | 1 |
| T2DM | LEC | *RHOBTB3* | 0.460666789 | 3.79E-05 | 0.000908725 | 0.238452189 |
| T2DM | LEC | *TIGD6* | -1.420123789 | 0.000925289 | 0.002222312 | 1 |
| T2DM | LEC | *SERINC5* | 0.82648405 | 0.003571531 | 0.006388112 | 1 |
| T2DM | LEC | *ENPP6* | -1.112040091 | 0.000330918 | 0.001172764 | 1 |
| T2DM | LEC | *ERAP1* | 0.646955006 | 0.004045906 | 0.007091332 | 1 |
| T2DM | LEC | *ANKRA2* | 1.11973396 | 0.008439203 | 0.0133511 | 1 |
| T2DM | LEC | *UTP15* | 0.780752922 | 0.000210908 | 0.0009784 | 1 |
| T2DM | LEC | *TLR3* | 1.602021567 | 0.003174321 | 0.00579134 | 1 |
| T2DM | LEC | *CCDC127* | 0.857335635 | 0.000138874 | 0.000908725 | 0.873102642 |
| T2DM | LEC | *GDF9* | 1.46780107 | 0.011532492 | 0.017504775 | 1 |
| T2DM | LEC | *MB21D1* | 0.742643756 | 0.000152792 | 0.000910197 | 0.960604399 |
| T2DM | LEC | *TXLNB* | -0.531345759 | 0.000212887 | 0.0009784 | 1 |
| T2DM | LEC | *PDSS2* | 0.724706304 | 0.02185311 | 0.03087427 | 1 |
| T2DM | LEC | *STXBP5* | 0.760801972 | 0.001136105 | 0.002582317 | 1 |
| T2DM | LEC | *PI16* | 0.373580225 | 0.000107291 | 0.000908725 | 0.67453656 |
| T2DM | LEC | *DAGLB* | 0.647099639 | 0.009611367 | 0.014934914 | 1 |
| T2DM | LEC | *STK17A* | 0.76653874 | 0.001956805 | 0.003948148 | 1 |
| T2DM | LEC | *SAP30L* | 1.157150948 | 0.005661285 | 0.009498931 | 1 |
| T2DM | LEC | *HCN1* | 1.509807954 | 0.000122582 | 0.000908725 | 0.770674121 |
| T2DM | LEC | *SLU7* | 1.030208621 | 0.005381205 | 0.009096971 | 1 |
| T2DM | LEC | *CAMLG* | 1.497583841 | 0.000183106 | 0.000929091 | 1 |
| T2DM | LEC | *KCNK5* | -1.588314021 | 0.000861977 | 0.002116068 | 1 |
| T2DM | LEC | *KIAA1324L* | -0.527454791 | 0.000296091 | 0.001112688 | 1 |
| T2DM | LEC | *USP49* | 0.983349448 | 0.000672252 | 0.001787838 | 1 |
| T2DM | LEC | *ZNF704* | 0.748367734 | 0.001295641 | 0.002851136 | 1 |
| T2DM | LEC | *FNDC1* | 0.402972267 | 0.000121085 | 0.000908725 | 0.76126425 |
| T2DM | LEC | *CHMP4C* | -0.348136531 | 0.000256275 | 0.001035475 | 1 |
| T2DM | LEC | *BRI3* | 0.907754182 | 0.000811599 | 0.002025615 | 1 |
| T2DM | LEC | *LMTK2* | 1.053344386 | 0.001985343 | 0.003992915 | 1 |
| T2DM | LEC | *CTSB* | 0.40142078 | 0.002504722 | 0.004780567 | 1 |
| T2DM | LEC | *TNFRSF11B* | 1.682704864 | 0.003763616 | 0.00668414 | 1 |
| T2DM | LEC | *OXR1* | 0.7757568 | 0.005938419 | 0.009892644 | 1 |

| | | | | | | |
|---|---|---|---|---|---|---|
| T2DM | LEC | *GPER1* | 0.685402912 | 0.000691302 | 0.001821616 | 1 |
| T2DM | LEC | *TMEM184A* | 0.539627217 | 0.000316538 | 0.001151139 | 1 |
| T2DM | LEC | *INTS1* | 1.089700102 | 0.000367149 | 0.001237855 | 1 |
| T2DM | LEC | *SLC4A2* | 0.736098993 | 0.000129964 | 0.000908725 | 0.817083283 |
| T2DM | LEC | *FASTK* | 1.056528927 | 0.000355823 | 0.001216909 | 1 |
| T2DM | LEC | *INTS8* | 0.928794563 | 0.020013498 | 0.028551137 | 1 |
| T2DM | LEC | *FREM1* | 0.484335934 | 0.000302968 | 0.001125081 | 1 |
| T2DM | LEC | *RPP25L* | 1.22178039 | 0.000542359 | 0.001545699 | 1 |
| T2DM | LEC | *FAM219A* | 0.90980248 | 0.000674366 | 0.001791945 | 1 |
| T2DM | LEC | *CCDC171* | 0.770365993 | 0.003534883 | 0.006333373 | 1 |
| T2DM | LEC | *SYK* | 0.302881539 | 0.000127754 | 0.000908725 | 0.803186338 |
| T2DM | LEC | *NIPSNAP3B* | 0.429673524 | 0.000915159 | 0.002211224 | 1 |
| T2DM | LEC | *METTL2B* | 0.418007649 | 0.000835142 | 0.002070401 | 1 |
| T2DM | LEC | *TMC1* | 0.725321557 | 9.76E-05 | 0.000908725 | 0.613505836 |
| T2DM | LEC | *HGSNAT* | 0.710027174 | 0.024759657 | 0.034568946 | 1 |
| T2DM | LEC | *C9orf64* | 1.027278446 | 0.000154228 | 0.000910197 | 0.969632298 |
| T2DM | LEC | *HNRNPK* | 0.947239608 | 0.001186615 | 0.002664263 | 1 |
| T2DM | LEC | *SVEP1* | 0.585519875 | 0.000212562 | 0.0009784 | 1 |
| T2DM | LEC | *ZNF367* | 0.82037633 | 4.72E-05 | 0.000908725 | 0.296475385 |
| T2DM | LEC | *NOL6* | 0.579673341 | 9.79E-05 | 0.000908725 | 0.615289813 |
| T2DM | LEC | *TRMT10B* | 0.958784507 | 0.002551169 | 0.00484861 | 1 |
| T2DM | LEC | *VCP* | 0.845899101 | 0.002684366 | 0.005057419 | 1 |
| T2DM | LEC | *PIGO* | 0.771895165 | 0.000184104 | 0.000929091 | 1 |
| T2DM | LEC | *LRFN5* | 0.438595233 | 6.17E-05 | 0.000908725 | 0.388101111 |
| T2DM | LEC | *WRN* | 0.726854117 | 0.03412822 | 0.046024049 | 1 |
| T2DM | LEC | *MARCH8* | 1.018290299 | 0.004803908 | 0.008245201 | 1 |
| T2DM | LEC | *CFL2* | 1.223106404 | 0.001438282 | 0.003095348 | 1 |
| T2DM | LEC | *ZCCHC24* | 0.891781043 | 0.000802097 | 0.002011482 | 1 |
| T2DM | LEC | *PGM2L1* | -0.356567652 | 0.000462808 | 0.001396351 | 1 |
| T2DM | LEC | *CRYL1* | 0.658526173 | 0.000724403 | 0.001881173 | 1 |
| T2DM | LEC | *REEP3* | -0.85171291 | 0.004443588 | 0.007689743 | 1 |
| T2DM | LEC | *HEPACAM* | 0.273479111 | 0.000130152 | 0.000908725 | 0.818263778 |
| T2DM | LEC | *LRR1* | -0.904734539 | 0.00343413 | 0.006183509 | 1 |
| T2DM | LEC | *C10orf25* | 0.704169949 | 0.000213121 | 0.0009784 | 1 |
| T2DM | LEC | *KLHDC2* | 0.9603201 | 0.00288286 | 0.005355951 | 1 |
| T2DM | LEC | *NOXRED1* | -0.973943218 | 0.002963637 | 0.005473558 | 1 |
| T2DM | LEC | *AKR1E2* | 0.613569093 | 0.014563516 | 0.021483066 | 1 |
| T2DM | LEC | *OXGR1* | -0.554164869 | 0.000104658 | 0.000908725 | 0.657987769 |
| T2DM | LEC | *TAF3* | 0.809014231 | 0.000288167 | 0.00109734 | 1 |
| T2DM | LEC | *SLC18A2* | 0.940124443 | 5.96E-05 | 0.000908725 | 0.374994411 |
| T2DM | LEC | *PDZD8* | 0.910116151 | 4.93E-05 | 0.000908725 | 0.310229422 |
| T2DM | LEC | *SNAPC4* | 0.713693391 | 0.006144106 | 0.010167937 | 1 |
| T2DM | LEC | *SDCCAG3* | 1.238008468 | 0.000671588 | 0.001786827 | 1 |
| T2DM | LEC | *AK8* | 0.43423089 | 0.00013731 | 0.000908725 | 0.863268196 |
| T2DM | LEC | *TSC1* | 0.998406814 | 0.007988505 | 0.012753614 | 1 |
| T2DM | LEC | *ZMYND19* | 0.607753533 | 0.000109641 | 0.000908725 | 0.689312833 |
| T2DM | LEC | *STOX1* | 0.22131303 | 0.025680656 | 0.035696282 | 1 |
| T2DM | LEC | *RET* | -1.484302334 | 0.000393978 | 0.001282395 | 1 |
| T2DM | LEC | *STK32C* | 1.117241216 | 0.000932022 | 0.002230538 | 1 |
| T2DM | LEC | *NSMF* | -1.090193078 | 0.001613266 | 0.003391041 | 1 |

| | | | | | | |
|---|---|---|---|---|---|---|
| T2DM | LEC | *VWA2* | 0.81804662 | 0.000240305 | 0.00100947 | 1 |
| T2DM | LEC | *TRUB1* | 0.605797089 | 0.016633129 | 0.024173201 | 1 |
| T2DM | LEC | *HSPA12A* | 0.500148319 | 0.0007243 | 0.001881173 | 1 |
| T2DM | LEC | *UBTD1* | -0.718181402 | 0.001200315 | 0.002687458 | 1 |
| T2DM | LEC | *ARHGAP42* | 0.832244731 | 0.002890747 | 0.005367432 | 1 |
| T2DM | LEC | *OTOGL* | -1.488413039 | 0.000571774 | 0.001603363 | 1 |
| T2DM | LEC | *PACSIN3* | 0.417888403 | 0.022654512 | 0.031877555 | 1 |
| T2DM | LEC | *TTC7B* | 0.677029287 | 0.007162028 | 0.01161746 | 1 |
| T2DM | LEC | *SLC39A13* | 0.958243197 | 9.71E-05 | 0.000908725 | 0.610194747 |
| T2DM | LEC | *PSMC3* | 0.707338252 | 8.64E-05 | 0.000908725 | 0.542935765 |
| T2DM | LEC | *CPSF2* | 0.500976499 | 0.017000603 | 0.024655776 | 1 |
| T2DM | LEC | *IFI27L1* | 1.245330439 | 0.011054118 | 0.016887183 | 1 |
| T2DM | LEC | *CLMN* | 0.668038725 | 0.00062251 | 0.001703099 | 1 |
| T2DM | LEC | *PTER* | 0.829273712 | 0.000783086 | 0.001978803 | 1 |
| T2DM | LEC | *C1QL3* | 0.426627986 | 4.24E-05 | 0.000908725 | 0.266752882 |
| T2DM | LEC | *ABTB2* | 0.597985469 | 0.000121951 | 0.000908725 | 0.766704025 |
| T2DM | LEC | *LIPC* | -0.438459477 | 0.000432678 | 0.001341345 | 1 |
| T2DM | LEC | *JAM3* | 0.470718408 | 8.02E-05 | 0.000908725 | 0.504135506 |
| T2DM | LEC | *GPT2* | 0.440449522 | 0.000308529 | 0.001133676 | 1 |
| T2DM | LEC | *IKBIP* | 0.696118194 | 0.000211876 | 0.0009784 | 1 |
| T2DM | LEC | *HIF1AN* | 1.216281437 | 0.003916823 | 0.006911618 | 1 |
| T2DM | LEC | *NDUFB8* | 1.337433323 | 0.000627322 | 0.001711418 | 1 |
| T2DM | LEC | *SPINT1* | 0.958707569 | 0.000845229 | 0.002084721 | 1 |
| T2DM | LEC | *AVPR1A* | -0.386749771 | 0.000842598 | 0.0020815 | 1 |
| T2DM | LEC | *BTRC* | 0.788072753 | 0.006084412 | 0.010085077 | 1 |
| T2DM | LEC | *LARP6* | 0.98255729 | 0.000482332 | 0.001432414 | 1 |
| T2DM | LEC | *ASPG* | 1.537917519 | 6.62E-05 | 0.000908725 | 0.41613143 |
| T2DM | LEC | *TBATA* | 2.324371668 | 0.029710057 | 0.04067664 | 1 |
| T2DM | LEC | *PCBD1* | -0.999553356 | 1.01E-06 | 0.000241487 | 0.006325489 |
| T2DM | LEC | *COX11* | 0.797399176 | 0.027516931 | 0.038030104 | 1 |
| T2DM | LEC | *ZNF202* | 1.541664887 | 0.024873429 | 0.034689274 | 1 |
| T2DM | LEC | *FAM227B* | 0.865341331 | 0.000407147 | 0.00130653 | 1 |
| T2DM | LEC | *C2* | 1.118927173 | 1.99E-07 | 5.95E-05 | 0.001249437 |
| T2DM | LEC | *APBB1* | 0.581173918 | 0.000115433 | 0.000908725 | 0.725724692 |
| T2DM | LEC | *ILK* | 1.039264623 | 0.002064532 | 0.00411271 | 1 |
| T2DM | LEC | *TAF10* | 0.894931366 | 0.000437877 | 0.001345521 | 1 |
| T2DM | LEC | *DCHS1* | 1.659372809 | 0.001546795 | 0.003275412 | 1 |
| T2DM | LEC | *NETO1* | 0.708028955 | 5.56E-05 | 0.000908725 | 0.349347631 |
| T2DM | LEC | *USP54* | 1.006888952 | 0.001479744 | 0.003166491 | 1 |
| T2DM | LEC | *C11orf74* | -1.001410506 | 0.014458923 | 0.021351573 | 1 |
| T2DM | LEC | *WDR88* | 1.331163543 | 0.001397175 | 0.003025848 | 1 |
| T2DM | LEC | *CYB5R2* | 0.803542037 | 0.000244195 | 0.001017397 | 1 |
| T2DM | LEC | *RIC3* | 0.867150994 | 0.000354328 | 0.001215214 | 1 |
| T2DM | LEC | *LMO1* | 1.426753966 | 0.000111981 | 0.000908725 | 0.7040251 |
| T2DM | LEC | *TRIM66* | 0.274737343 | 0.004124658 | 0.007209265 | 1 |
| T2DM | LEC | *CDYL2* | 0.577826488 | 0.028768876 | 0.039534409 | 1 |
| T2DM | LEC | *PRTG* | -0.911586239 | 0.001336827 | 0.002926403 | 1 |
| T2DM | LEC | *C16orf46* | 5.104250155 | 0.000201528 | 0.00096305 | 1 |
| T2DM | LEC | *TMEM41B* | 0.992041309 | 0.002897805 | 0.005378931 | 1 |
| T2DM | LEC | *ZNF143* | 1.140209906 | 0.000806908 | 0.002020322 | 1 |

| | | | | | | |
|---|---|---|---|---|---|---|
| T2DM | LEC | *TMX3* | 0.876379493 | 0.011366858 | 0.017282572 | 1 |
| T2DM | LEC | *CLEC4E* | 0.628491057 | 1.77E-05 | 0.000908725 | 0.111546801 |
| T2DM | LEC | *ZNF3* | 1.042172735 | 0.026185355 | 0.036333553 | 1 |
| T2DM | LEC | *CLEC4D* | 1.841262942 | 2.78E-05 | 0.000908725 | 0.174678976 |
| T2DM | LEC | *BEAN1* | 1.04404523 | 0.000359491 | 0.001224333 | 1 |
| T2DM | LEC | *TK2* | 0.474245789 | 0.011016621 | 0.016843749 | 1 |
| T2DM | LEC | *SEC11C* | 1.323767549 | 0.000298785 | 0.001118132 | 1 |
| T2DM | LEC | *GALR1* | 0.497958653 | 0.001422063 | 0.003071284 | 1 |
| T2DM | LEC | *CENPV* | 0.866877119 | 0.000280555 | 0.00108013 | 1 |
| T2DM | LEC | *CHRFAM7A* | -0.662735731 | 0.014431913 | 0.021323957 | 1 |
| T2DM | LEC | *SPDYE6* | 0.95566427 | 9.63E-05 | 0.000908725 | 0.605384477 |
| T2DM | LEC | *ATF7IP2* | 0.731932623 | 0.005049396 | 0.008628854 | 1 |
| T2DM | LEC | *ZNF592* | 0.774195853 | 0.00177185 | 0.003654731 | 1 |
| T2DM | LEC | *ACSM1* | -0.283503825 | 0.000228721 | 0.000995303 | 1 |
| T2DM | LEC | *AP1G1* | 0.818981695 | 0.001766014 | 0.003643889 | 1 |
| T2DM | LEC | *SLFN5* | 0.741415216 | 0.002185031 | 0.004295588 | 1 |
| T2DM | LEC | *CATSPER2* | 1.155385269 | 0.001832514 | 0.003751678 | 1 |
| T2DM | LEC | *C16orf45* | 0.867404418 | 0.001425689 | 0.003075946 | 1 |
| T2DM | LEC | *YPEL4* | 0.764213096 | 0.000358827 | 0.001223256 | 1 |
| T2DM | LEC | *FAM111A* | 0.889896309 | 0.000913025 | 0.002208614 | 1 |
| T2DM | LEC | *SCNN1G* | -0.998137914 | 0.000144832 | 0.000908725 | 0.910556678 |
| T2DM | LEC | *ANKDD1A* | -0.719139964 | 0.002294856 | 0.004465417 | 1 |
| T2DM | LEC | *DCTN5* | 0.63591161 | 0.00266852 | 0.005036622 | 1 |
| T2DM | LEC | *STX3* | -0.723417771 | 0.001042767 | 0.002427203 | 1 |
| T2DM | LEC | *MTMR10* | 0.965266103 | 0.002468232 | 0.004723827 | 1 |
| T2DM | LEC | *YWHAB* | 0.945131268 | 0.002554646 | 0.00485375 | 1 |
| T2DM | LEC | *NYAP1* | -0.300091839 | 2.96E-05 | 0.000908725 | 0.186264656 |
| T2DM | LEC | *MS4A6E* | 0.435405294 | 0.003883443 | 0.006867849 | 1 |
| T2DM | LEC | *MS4A7* | 0.231315141 | 0.000324477 | 0.001162346 | 1 |
| T2DM | LEC | *MS4A14* | 0.299170183 | 0.00170783 | 0.003552777 | 1 |
| T2DM | LEC | *DIS3L* | 0.960453161 | 0.000766495 | 0.001948143 | 1 |
| T2DM | LEC | *CCNDBP1* | 1.156678223 | 0.008652386 | 0.013647153 | 1 |
| T2DM | LEC | *SMAD3* | 0.670384671 | 0.000354527 | 0.001215214 | 1 |
| T2DM | LEC | *AKTIP* | 0.602006504 | 6.22E-05 | 0.000908725 | 0.391146712 |
| T2DM | LEC | *CNPY4* | 1.041574087 | 6.10E-05 | 0.000908725 | 0.38352678 |
| T2DM | LEC | *NKX3-1* | 2.81114247 | 0.002163534 | 0.004269347 | 1 |
| T2DM | LEC | *DUSP18* | -0.959020251 | 0.004151134 | 0.007247481 | 1 |
| T2DM | LEC | *TEF* | 0.815777172 | 8.39E-06 | 0.000673907 | 0.052769115 |
| T2DM | LEC | *MEI1* | 0.735630768 | 0.002595346 | 0.004911782 | 1 |
| T2DM | LEC | *PBX3* | 0.548055984 | 0.000661357 | 0.001769341 | 1 |
| T2DM | LEC | *SNRPD1* | 1.38989537 | 0.002814329 | 0.005254449 | 1 |
| T2DM | LEC | *SAMD14* | -1.302336081 | 0.006900271 | 0.011246648 | 1 |
| T2DM | LEC | *URM1* | 0.893949101 | 0.008217396 | 0.013072563 | 1 |
| T2DM | LEC | *CERCAM* | 0.658581914 | 0.006313786 | 0.010407649 | 1 |
| T2DM | LEC | *ENDOG* | 0.559625401 | 0.002686797 | 0.005058968 | 1 |
| T2DM | LEC | *ISLR2* | -0.19652367 | 0.000154619 | 0.000910197 | 0.972090039 |
| T2DM | LEC | *SP2* | 0.935584048 | 0.001859906 | 0.003796504 | 1 |
| T2DM | LEC | *COQ7* | 0.838617575 | 0.000167391 | 0.000910705 | 1 |
| T2DM | LEC | *GPRC5B* | -0.633294507 | 0.00050101 | 0.001463685 | 1 |
| T2DM | LEC | *KATNAL2* | 0.515590961 | 0.00021991 | 0.00098409 | 1 |

| | | | | | | |
|---|---|---|---|---|---|---|
| T2DM | LEC | *CCL23* | -0.34489608 | 0.015735254 | 0.023027827 | 1 |
| T2DM | LEC | *ENGASE* | 1.014040737 | 0.006274243 | 0.010350608 | 1 |
| T2DM | LEC | *RBFOX3* | -1.634792828 | 0.008140841 | 0.012970469 | 1 |
| T2DM | LEC | *OR51E2* | 0.713506573 | 0.012720648 | 0.019064293 | 1 |
| T2DM | LEC | *TRIM68* | -0.8504133 | 0.000189189 | 0.000940261 | 1 |
| T2DM | LEC | *FN3K* | 0.72175035 | 0.000106497 | 0.000908725 | 0.669546946 |
| T2DM | LEC | *ZNF23* | -1.226692678 | 0.00398586 | 0.006993889 | 1 |
| T2DM | LEC | *ZNF229* | 2.002920248 | 1.80E-05 | 0.000908725 | 0.112929052 |
| T2DM | LEC | *ZNF646* | -0.65928865 | 0.001708887 | 0.003552836 | 1 |
| T2DM | LEC | *VKORC1* | -1.042278428 | 0.007974354 | 0.012734255 | 1 |
| T2DM | LEC | *SMG8* | 1.141578857 | 0.00010144 | 0.000908725 | 0.637755806 |
| T2DM | LEC | *RAB8A* | 0.779883666 | 9.23E-05 | 0.000908725 | 0.580099255 |
| T2DM | LEC | *GATAD2A* | 0.997497032 | 0.000124118 | 0.000908725 | 0.780331601 |
| T2DM | LEC | *PROCA1* | 0.536740446 | 0.000305923 | 0.001128721 | 1 |
| T2DM | LEC | *ZNF641* | 0.703065934 | 1.74E-06 | 0.000289824 | 0.010929065 |
| T2DM | LEC | *CACNB3* | -1.250141496 | 0.001442958 | 0.003103619 | 1 |
| T2DM | LEC | *TP53I13* | 0.577931294 | 5.88E-05 | 0.000908725 | 0.369389056 |
| T2DM | LEC | *ZNF528* | 1.104836739 | 0.003888232 | 0.006872452 | 1 |
| T2DM | LEC | *SERTAD3* | 0.635280667 | 0.000109663 | 0.000908725 | 0.689451588 |
| T2DM | LEC | *TMC4* | 1.625449459 | 0.001201226 | 0.002688539 | 1 |
| T2DM | LEC | *TTYH1* | 1.531380731 | 9.26E-05 | 0.000908725 | 0.582131073 |
| T2DM | LEC | *LENG8* | 0.755293382 | 0.029039533 | 0.039854081 | 1 |
| T2DM | LEC | *CDC42EP5* | -0.635689907 | 0.000194354 | 0.000949652 | 1 |
| T2DM | LEC | *PPP1R14A* | 0.878508074 | 0.000122555 | 0.000908725 | 0.770504191 |
| T2DM | LEC | *C19orf33* | 0.735113611 | 0.036029216 | 0.048390446 | 1 |
| T2DM | LEC | *DNAAF3* | -1.812095712 | 0.020625549 | 0.029342179 | 1 |
| T2DM | LEC | *CHAF1A* | 0.369904073 | 0.000748599 | 0.001919429 | 1 |
| T2DM | LEC | *KIFC2* | 0.36151955 | 0.011630953 | 0.017641449 | 1 |
| T2DM | LEC | *SERPINF2* | 0.75845673 | 0.000100642 | 0.000908725 | 0.632735319 |
| T2DM | LEC | *SRR* | 0.413552292 | 0.028721653 | 0.039479784 | 1 |
| T2DM | LEC | *CYB5D2* | 0.754422147 | 7.78E-05 | 0.000908725 | 0.488934815 |
| T2DM | LEC | *C19orf48* | 0.608035309 | 0.000174637 | 0.000911424 | 1 |
| T2DM | LEC | *ZNF83* | 0.661783425 | 0.036841938 | 0.049376523 | 1 |
| T2DM | LEC | *CD320* | 1.317500548 | 0.000391462 | 0.001277842 | 1 |
| T2DM | LEC | *SPRYD3* | 1.500217892 | 0.011034301 | 0.01686258 | 1 |
| T2DM | LEC | *SOAT2* | 0.921393947 | 0.002662129 | 0.005026068 | 1 |
| T2DM | LEC | *ZNF558* | 0.822227695 | 0.002227617 | 0.004354797 | 1 |
| T2DM | LEC | *NDUFV1* | 1.102188183 | 0.001854026 | 0.00378696 | 1 |
| T2DM | LEC | *NUDT8* | -0.487266277 | 0.000572497 | 0.001603699 | 1 |
| T2DM | LEC | *PRDX2* | 1.130195414 | 1.89E-05 | 0.000908725 | 0.118975832 |
| T2DM | LEC | *MIS12* | 0.707369497 | 0.000212041 | 0.0009784 | 1 |
| T2DM | LEC | *CD300C* | 0.826628763 | 0.025260534 | 0.035174525 | 1 |
| T2DM | LEC | *HID1* | -0.164492632 | 0.000155111 | 0.000910705 | 0.975181383 |
| T2DM | LEC | *ATP5H* | 1.01348082 | 0.000331846 | 0.001173862 | 1 |
| T2DM | LEC | *SRP68* | 0.824140435 | 0.000476119 | 0.001418531 | 1 |
| T2DM | LEC | *MGAT5B* | 0.991751626 | 0.000105081 | 0.000908725 | 0.660642807 |
| T2DM | LEC | *TMC8* | 0.747165789 | 1.31E-05 | 0.000893669 | 0.08262725 |
| T2DM | LEC | *TMEM68* | 0.845953401 | 0.001169165 | 0.002636492 | 1 |
| T2DM | LEC | *CYP7A1* | 1.71209098 | 0.01073664 | 0.016471756 | 1 |
| T2DM | LEC | *GSDMA* | -1.372643336 | 0.001183433 | 0.002659129 | 1 |

| | | | | | | |
|---|---|---|---|---|---|---|
| T2DM | LEC | *ZNF598* | 0.748634512 | 0.000273744 | 0.00107497 | 1 |
| T2DM | LEC | *MLST8* | -1.696109586 | 0.002583819 | 0.004896073 | 1 |
| T2DM | LEC | *ECI1* | 0.701141343 | 0.001178947 | 0.00265078 | 1 |
| T2DM | LEC | *ABCA3* | -0.702579058 | 0.003160844 | 0.005770101 | 1 |
| T2DM | LEC | *KCTD5* | -0.917102234 | 0.0015377 | 0.003260546 | 1 |
| T2DM | LEC | *SDHAF2* | -1.457203429 | 0.006829261 | 0.011152094 | 1 |
| T2DM | LEC | *VWCE* | -1.245839088 | 0.013003908 | 0.019456347 | 1 |
| T2DM | LEC | *HRASLS5* | 0.317700152 | 7.19E-05 | 0.000908725 | 0.45200188 |
| T2DM | LEC | *TRANK1* | 0.81974208 | 0.00193703 | 0.003922096 | 1 |
| T2DM | LEC | *TTC21A* | 1.271872849 | 0.000440524 | 0.001350355 | 1 |
| T2DM | LEC | *RPSA* | 0.916188835 | 0.000193067 | 0.000946328 | 1 |
| T2DM | LEC | *LTBP3* | 1.439287887 | 3.36E-06 | 0.000436409 | 0.021107084 |
| T2DM | LEC | *NAALADL1* | -0.78369986 | 0.000200426 | 0.000962913 | 1 |
| T2DM | LEC | *SCARA5* | 0.363620709 | 0.000247163 | 0.00101963 | 1 |
| T2DM | LEC | *RNF187* | 1.013993845 | 0.000135868 | 0.000908725 | 0.85420237 |
| T2DM | LEC | *PTGDR* | 0.365766558 | 0.000153961 | 0.000910197 | 0.967954925 |
| T2DM | LEC | *GLYCTK* | 1.510387745 | 2.23E-05 | 0.000908725 | 0.140250291 |
| T2DM | LEC | *POLR2J3* | 1.059286521 | 2.30E-05 | 0.000908725 | 0.144654869 |
| T2DM | LEC | *COA6* | 0.556826516 | 0.00010949 | 0.000908725 | 0.688364929 |
| T2DM | LEC | *PXK* | 1.135954704 | 0.002257379 | 0.004404761 | 1 |
| T2DM | LEC | *ACOX2* | 0.370620815 | 0.002346258 | 0.004540142 | 1 |
| T2DM | LEC | *FAM107A* | 0.432418297 | 0.000428194 | 0.001336216 | 1 |
| T2DM | LEC | *CX3CR1* | 0.368530807 | 0.000400662 | 0.001292438 | 1 |
| T2DM | LEC | *DTYMK* | 0.712949894 | 9.85E-05 | 0.000908725 | 0.619526177 |
| T2DM | LEC | *TAP1* | 0.534019932 | 0.026996538 | 0.037371089 | 1 |
| T2DM | LEC | *ING5* | 0.946943195 | 0.000794986 | 0.002000018 | 1 |
| T2DM | LEC | *MLKL* | 0.455930153 | 0.035853396 | 0.048174888 | 1 |
| T2DM | LEC | *RFWD3* | 1.111083264 | 3.59E-05 | 0.000908725 | 0.225695559 |
| T2DM | LEC | *RHOH* | 0.387540039 | 7.69E-05 | 0.000908725 | 0.483663818 |
| T2DM | LEC | *KLHL30* | 0.604683488 | 0.000970522 | 0.002300824 | 1 |
| T2DM | LEC | *REEP4* | 1.188570451 | 0.000234782 | 0.001001608 | 1 |
| T2DM | LEC | *BMP1* | 0.801308833 | 0.021984696 | 0.031039251 | 1 |
| T2DM | LEC | *ATXN2L* | 0.907742818 | 0.003080875 | 0.005642139 | 1 |
| T2DM | LEC | *FNTA* | 1.044006842 | 0.029868675 | 0.040867108 | 1 |
| T2DM | LEC | *TRAPPC11* | 0.927944005 | 0.005584121 | 0.009389508 | 1 |
| T2DM | LEC | *TMEM223* | -1.15843063 | 0.005824673 | 0.009731523 | 1 |
| T2DM | LEC | *SLC20A2* | 0.862581435 | 0.001190266 | 0.002668759 | 1 |
| T2DM | LEC | *ZSWIM1* | 1.002181131 | 2.15E-05 | 0.000908725 | 0.135363851 |
| T2DM | LEC | *NBPF9* | 0.499637539 | 0.000114559 | 0.000908725 | 0.720231543 |
| T2DM | LEC | *WFDC13* | 8.337859515 | 0.0008163 | 0.002034923 | 1 |
| T2DM | LEC | *ZNF30* | 0.868114327 | 0.019957512 | 0.02847773 | 1 |
| T2DM | LEC | *SLC16A4* | 0.880501683 | 0.002982662 | 0.005497508 | 1 |
| T2DM | LEC | *LRP1B* | 0.357537258 | 0.002204492 | 0.004321685 | 1 |
| T2DM | LEC | *NPNT* | 0.467873344 | 0.000124569 | 0.000908725 | 0.783165095 |
| T2DM | LEC | *CNNM3* | 1.119310303 | 0.000288789 | 0.001098223 | 1 |
| T2DM | LEC | *GSTM4* | 0.723129615 | 4.01E-05 | 0.000908725 | 0.251805287 |
| T2DM | LEC | *TCTN2* | 0.452911133 | 0.006197167 | 0.010239576 | 1 |
| T2DM | LEC | *ABHD15* | 0.555353028 | 2.19E-05 | 0.000908725 | 0.137797495 |
| T2DM | LEC | *ADAL* | 0.290190768 | 0.02050423 | 0.029184989 | 1 |
| T2DM | LEC | *ZNF507* | 0.578987882 | 0.008823665 | 0.013879005 | 1 |

| | | | | | | |
|---|---|---|---|---|---|---|
| T2DM | LEC | *STX18* | 0.887097983 | 0.011192347 | 0.017070908 | 1 |
| T2DM | LEC | *ZBTB49* | 1.390450678 | 0.000996634 | 0.002345876 | 1 |
| T2DM | LEC | *TNIP2* | -0.899400387 | 0.000619832 | 0.001697248 | 1 |
| T2DM | LEC | *BTNL3* | 0.302920715 | 0.005523387 | 0.009299821 | 1 |
| T2DM | LEC | *INPP5D* | 0.893620006 | 0.000663821 | 0.001773452 | 1 |
| T2DM | LEC | *LETM1* | 1.242711726 | 0.000979452 | 0.00231584 | 1 |
| T2DM | LEC | *TMEM129* | 0.499241944 | 0.007162283 | 0.01161746 | 1 |
| T2DM | LEC | *CEP120* | 0.920084808 | 0.007906087 | 0.012651578 | 1 |
| T2DM | LEC | *STXBP6* | -0.170363213 | 0.000182076 | 0.000929091 | 1 |
| T2DM | LEC | *MFF* | 0.785711729 | 0.005097464 | 0.008694454 | 1 |
| T2DM | LEC | *SIGLEC7* | 0.649849728 | 0.000482591 | 0.001432508 | 1 |
| T2DM | LEC | *FEM1B* | 1.209779976 | 8.30E-05 | 0.000908725 | 0.521818704 |
| T2DM | LEC | *COL4A3* | 0.454059398 | 0.000262398 | 0.00104809 | 1 |
| T2DM | LEC | *IRS1* | -0.849043282 | 1.85E-12 | 3.87E-09 | 1.16E-08 |
| T2DM | LEC | *C8orf46* | 0.276513165 | 1.37E-05 | 0.000902248 | 0.086104583 |
| T2DM | LEC | *HSPBAP1* | 0.789928324 | 0.009075745 | 0.014218591 | 1 |
| T2DM | LEC | *ATF5* | 0.411474265 | 0.000121352 | 0.000908725 | 0.762937533 |
| T2DM | LEC | *CPT1C* | -0.422277257 | 0.000190277 | 0.000942374 | 1 |
| T2DM | LEC | *PCSK9* | 3.172567107 | 0.006949513 | 0.011307348 | 1 |
| T2DM | LEC | *XPO6* | 0.987455281 | 0.001251363 | 0.002774089 | 1 |
| T2DM | LEC | *RGS14* | -0.466363196 | 0.017335424 | 0.025089275 | 1 |
| T2DM | LEC | *RAB24* | 0.722324231 | 0.003828051 | 0.006787072 | 1 |
| T2DM | LEC | *CXCL10* | 0.523875675 | 0.002415315 | 0.004652293 | 1 |
| T2DM | LEC | *SH3TC2* | -0.826102278 | 0.016171721 | 0.0235897 | 1 |
| T2DM | LEC | *GPRIN1* | -1.177823709 | 0.00030746 | 0.00113215 | 1 |
| T2DM | LEC | *PGM2* | -1.084524652 | 0.016100713 | 0.023497025 | 1 |
| T2DM | LEC | *STK32A* | 0.279917026 | 0.001165032 | 0.002628115 | 1 |
| T2DM | LEC | *P2RY12* | 0.294084396 | 0.000219438 | 0.00098409 | 1 |
| T2DM | LEC | *SLC33A1* | 1.03100255 | 0.009045572 | 0.014185461 | 1 |
| T2DM | LEC | *SNUPN* | 0.771081931 | 0.002576261 | 0.004885959 | 1 |
| T2DM | LEC | *PTAFR* | 0.423621058 | 0.000117869 | 0.000908725 | 0.741042382 |
| T2DM | LEC | *RNASE6* | 0.620052401 | 0.000101497 | 0.000908725 | 0.638109602 |
| T2DM | LEC | *NPR1* | 0.711678722 | 0.000152497 | 0.000910197 | 0.958751604 |
| T2DM | LEC | *SLC38A11* | 0.464166429 | 0.000168536 | 0.000910705 | 1 |
| T2DM | LEC | *CCDC8* | 0.928537184 | 0.000150716 | 0.000910197 | 0.947550599 |
| T2DM | LEC | *CLIC3* | 0.849832995 | 0.000157597 | 0.000910705 | 0.990814843 |
| T2DM | LEC | *DFFB* | 0.807090129 | 0.000264974 | 0.001053693 | 1 |
| T2DM | LEC | *C15orf40* | 0.612569882 | 0.000396746 | 0.00128761 | 1 |
| T2DM | LEC | *RGPD8* | 0.361509903 | 0.001376034 | 0.002994506 | 1 |
| T2DM | LEC | *HEXDC* | 1.107812602 | 0.001093381 | 0.002510624 | 1 |
| T2DM | LEC | *LRRC45* | -1.053145908 | 0.00083576 | 0.002071116 | 1 |
| T2DM | LEC | *FASN* | 0.715394066 | 0.000262259 | 0.00104809 | 1 |
| T2DM | LEC | *MT1E* | 0.550935104 | 0.000250137 | 0.001023586 | 1 |
| T2DM | LEC | *DCXR* | 1.405871662 | 0.008305701 | 0.013186349 | 1 |
| T2DM | LEC | *LIMS1* | 0.541944455 | 0.037114428 | 0.049720521 | 1 |
| T2DM | LEC | *CSGALNACT2* | 0.819058214 | 0.000338953 | 0.001186892 | 1 |
| T2DM | LEC | *CTNND2* | 0.46133852 | 0.00086604 | 0.002122726 | 1 |
| T2DM | LEC | *BRD3* | 1.069393588 | 0.001250323 | 0.002772761 | 1 |
| T2DM | LEC | *ZFPM2* | -0.433139148 | 0.001801002 | 0.003697877 | 1 |
| T2DM | LEC | *ZNF747* | -0.879346106 | 0.000447582 | 0.001364669 | 1 |

| | | | | | | |
|---|---|---|---|---|---|---|
| T2DM | LEC | *MAP3K2* | 0.759760128 | 0.011172753 | 0.017045158 | 1 |
| T2DM | LEC | *PUSL1* | 0.816166814 | 0.000172764 | 0.000910705 | 1 |
| T2DM | LEC | *MYO7B* | 0.600380648 | 0.000268777 | 0.001062766 | 1 |
| T2DM | LEC | *CNTROB* | 1.432005583 | 0.003397179 | 0.00613812 | 1 |
| T2DM | LEC | *KCNAB3* | -0.983620286 | 0.000533272 | 0.001530205 | 1 |
| T2DM | LEC | *NIPA1* | 0.763841178 | 0.000207445 | 0.000972609 | 1 |
| T2DM | LEC | *RNF150* | 0.509419487 | 0.00011929 | 0.000908725 | 0.749976824 |
| T2DM | LEC | *CHRNB1* | 0.518560254 | 0.000152882 | 0.000910197 | 0.961167973 |
| T2DM | LEC | *PDCD6IP* | 0.645553087 | 0.03280479 | 0.044429925 | 1 |
| T2DM | LEC | *ZNF212* | 1.168410339 | 0.009812432 | 0.015191026 | 1 |
| T2DM | LEC | *FAM161A* | 1.060712848 | 0.000836788 | 0.002072846 | 1 |
| T2DM | LEC | *GLB1* | 0.699728005 | 0.001110214 | 0.002539075 | 1 |
| T2DM | LEC | *FAXDC2* | 0.785240681 | 0.016633254 | 0.024173201 | 1 |
| T2DM | LEC | *CRTAP* | 0.982168244 | 0.016320016 | 0.023779125 | 1 |
| T2DM | LEC | *ELP5* | 0.956076981 | 0.001723997 | 0.003574213 | 1 |
| T2DM | LEC | *STX8* | 0.736231897 | 0.000936225 | 0.002238041 | 1 |
| T2DM | LEC | *NFRKB* | 0.847468088 | 0.000172696 | 0.000910705 | 1 |
| T2DM | LEC | *SP7* | 1.862770337 | 0.028601243 | 0.039355661 | 1 |
| T2DM | LEC | *SEMA3E* | 0.882135638 | 0.000121072 | 0.000908725 | 0.761181929 |
| T2DM | LEC | *MGMT* | 0.694867904 | 0.001993035 | 0.004001984 | 1 |
| T2DM | LEC | *METTL7B* | 1.751953694 | 0.000115105 | 0.000908725 | 0.723664194 |
| T2DM | LEC | *NFXL1* | 0.933805814 | 0.031618073 | 0.042961493 | 1 |
| T2DM | LEC | *CD14* | 0.516492377 | 0.0001848 | 0.000929091 | 1 |
| T2DM | LEC | *HSD17B13* | 0.369558153 | 0.032549034 | 0.044131071 | 1 |
| T2DM | LEC | *ARL6IP1* | -0.755366588 | 0.000230522 | 0.00099951 | 1 |
| T2DM | LEC | *SERPINB9* | -0.602096341 | 0.000450235 | 0.001370763 | 1 |
| T2DM | LEC | *SMAGP* | -0.259484966 | 0.000161345 | 0.000910705 | 1 |
| T2DM | LEC | *IRX2* | 0.457912666 | 9.68E-05 | 0.000908725 | 0.608735067 |
| T2DM | LEC | *STAT2* | 0.780380172 | 0.000121026 | 0.000908725 | 0.760889888 |
| T2DM | LEC | *NUDCD2* | 0.52684684 | 0.000105865 | 0.000908725 | 0.66557421 |
| T2DM | LEC | *HSPA4* | 1.084480006 | 0.003521096 | 0.006312722 | 1 |
| T2DM | LEC | *SLC26A5* | -0.7288493 | 0.029966836 | 0.040983576 | 1 |
| T2DM | LEC | *COMMD5* | 0.742723503 | 0.000652068 | 0.001749701 | 1 |
| T2DM | LEC | *GTSF1* | 1.483720988 | 9.94E-05 | 0.000908725 | 0.624729944 |
| T2DM | LEC | *ARMC10* | 0.937187477 | 0.010443239 | 0.016068684 | 1 |
| T2DM | LEC | *ACYP2* | 0.641734058 | 0.000561168 | 0.001579965 | 1 |
| T2DM | LEC | *TRABD* | 0.940399601 | 0.028458769 | 0.039185344 | 1 |
| T2DM | LEC | *TMEM133* | 0.994396127 | 0.000105143 | 0.000908725 | 0.661032696 |
| T2DM | LEC | *TTLL6* | -0.405420204 | 0.0001217 | 0.000908725 | 0.765125115 |
| T2DM | LEC | *KCNS3* | 0.75679907 | 0.000147155 | 0.000908725 | 0.925162223 |
| T2DM | LEC | *GPR37* | -0.445608346 | 0.000147144 | 0.000908725 | 0.925096795 |
| T2DM | LEC | *AKAP13* | 0.661326764 | 0.000411121 | 0.001311277 | 1 |
| T2DM | LEC | *HTRA3* | 0.516459585 | 6.26E-05 | 0.000908725 | 0.393666354 |
| T2DM | LEC | *FOXN2* | 0.763396986 | 0.000627668 | 0.001711418 | 1 |
| T2DM | LEC | *MTSS1* | 0.490384836 | 0.000166176 | 0.000910705 | 1 |
| T2DM | LEC | *RPS9* | 0.844468541 | 0.007049379 | 0.011452053 | 1 |
| T2DM | LEC | *NDUFA3* | 1.070478069 | 0.00033086 | 0.001172764 | 1 |
| T2DM | LEC | *OSCAR* | 0.496186452 | 8.50E-05 | 0.000908725 | 0.534602043 |
| T2DM | LEC | *PAQR8* | 1.052786 | 0.000652714 | 0.001750688 | 1 |
| T2DM | LEC | *NUDT6* | 0.839420537 | 8.58E-05 | 0.000908725 | 0.539223359 |

| | | | | | | |
|---|---|---|---|---|---|---|
| T2DM | LEC | *ZNF160* | 0.780600903 | 0.000610381 | 0.001678682 | 1 |
| T2DM | LEC | *ZNF415* | -0.325532745 | 0.000464115 | 0.001396913 | 1 |
| T2DM | LEC | *TSNARE1* | 0.797317343 | 0.000327965 | 0.001167883 | 1 |
| T2DM | LEC | *FPR1* | 0.315867483 | 0.000208013 | 0.000972609 | 1 |
| T2DM | LEC | *C11orf24* | -1.228572494 | 0.005477716 | 0.009227867 | 1 |
| T2DM | LEC | *ALK* | 0.443439868 | 0.000250564 | 0.001023586 | 1 |
| T2DM | LEC | *MFN1* | 1.022800213 | 0.000528859 | 0.001521015 | 1 |
| T2DM | LEC | *GIMAP8* | -0.492656359 | 0.000128692 | 0.000908725 | 0.80908805 |
| T2DM | LEC | *ATP6V0E2* | 0.549940665 | 0.000107532 | 0.000908725 | 0.67605079 |
| T2DM | LEC | *GRIK1* | -0.679724187 | 0.003552176 | 0.006358921 | 1 |
| T2DM | LEC | *CANT1* | -0.961227951 | 0.017327206 | 0.025088932 | 1 |
| T2DM | LEC | *CHST11* | 0.507581908 | 0.01791883 | 0.025844387 | 1 |
| T2DM | LEC | *LURAP1* | -0.574270861 | 0.000921316 | 0.002218447 | 1 |
| T2DM | LEC | *KCND3* | -0.693304788 | 0.030066279 | 0.041094371 | 1 |
| T2DM | LEC | *MRPL36* | 1.10041083 | 1.46E-05 | 0.000902248 | 0.092029282 |
| T2DM | LEC | *ZNF581* | 1.897103849 | 0.000430872 | 0.00134099 | 1 |
| T2DM | LEC | *MCC* | 0.556945769 | 0.000145935 | 0.000908725 | 0.917493689 |
| T2DM | LEC | *DLK2* | 1.264250588 | 0.001566667 | 0.003308577 | 1 |
| T2DM | LEC | *HOPX* | 0.70165626 | 7.63E-05 | 0.000908725 | 0.479871808 |
| T2DM | LEC | *RSL1D1* | 1.250978055 | 0.000221174 | 0.00098409 | 1 |
| T2DM | LEC | *PPID* | 0.94254552 | 0.000344906 | 0.001196042 | 1 |
| T2DM | LEC | *ETFDH* | 0.299742616 | 0.034549492 | 0.046542245 | 1 |
| T2DM | LEC | *TBCA* | 0.684882663 | 0.000730077 | 0.001891221 | 1 |
| T2DM | LEC | *MAP6* | -0.311756471 | 0.000124193 | 0.000908725 | 0.780800944 |
| T2DM | LEC | *ZNF584* | 0.505630098 | 1.49E-05 | 0.000902894 | 0.093900957 |
| T2DM | LEC | *NMUR1* | 0.497293131 | 9.29E-05 | 0.000908725 | 0.584019653 |
| T2DM | LEC | *P2RY6* | 0.411688999 | 0.000128673 | 0.000908725 | 0.808966702 |
| T2DM | LEC | *ZIK1* | -1.83608447 | 0.000142662 | 0.000908725 | 0.89691877 |
| T2DM | LEC | *TCEA2* | -0.842495723 | 0.00067545 | 0.001793599 | 1 |
| T2DM | LEC | *VAT1L* | 0.505600379 | 0.000129115 | 0.000908725 | 0.811746884 |
| T2DM | LEC | *LGALS4* | -0.759707398 | 0.003733053 | 0.006639238 | 1 |
| T2DM | LEC | *LRRC34* | 0.557176651 | 0.000747195 | 0.001918961 | 1 |
| T2DM | LEC | *SPATA5L1* | 0.575638789 | 0.031134541 | 0.042414487 | 1 |
| T2DM | LEC | *SYCE1* | 0.745573385 | 0.000463906 | 0.001396913 | 1 |
| T2DM | LEC | *NXNL1* | 0.86776274 | 0.006226116 | 0.010282004 | 1 |
| T2DM | LEC | *BCL2* | 0.31183229 | 0.000967895 | 0.00229802 | 1 |
| T2DM | LEC | *RHNO1* | 0.87134669 | 0.000727954 | 0.001887164 | 1 |
| T2DM | LEC | *PWWP2B* | 1.101028013 | 0.000436158 | 0.00134367 | 1 |
| T2DM | LEC | *NINJ2* | 0.59464575 | 0.000168504 | 0.000910705 | 1 |
| T2DM | LEC | *FAM90A1* | -1.840817306 | 0.000285332 | 0.001090506 | 1 |
| T2DM | LEC | *TRAPPC12* | 0.772999104 | 0.020709617 | 0.029450659 | 1 |
| T2DM | LEC | *C3AR1* | 0.508290351 | 0.000465294 | 0.001398163 | 1 |
| T2DM | LEC | *AQP4* | 0.522531089 | 0.000112956 | 0.000908725 | 0.710157448 |
| T2DM | LEC | *TLN2* | 0.406493862 | 0.000365184 | 0.001235026 | 1 |
| T2DM | LEC | *SRGAP2C* | 0.999072119 | 0.000771307 | 0.001957692 | 1 |
| T2DM | LEC | *ATPAF2* | 1.360511941 | 0.005498652 | 0.009260654 | 1 |
| T2DM | LEC | *CYP4F22* | -1.83428074 | 0.006558518 | 0.010760282 | 1 |
| T2DM | LEC | *C20orf196* | 0.908956875 | 0.001036853 | 0.00241971 | 1 |
| T2DM | LEC | *ANKRD20A4* | -0.626749061 | 0.000166208 | 0.000910705 | 1 |
| T2DM | LEC | *ORMDL3* | 0.41481431 | 0.006615016 | 0.010841659 | 1 |

| | | | | | | |
|---|---|---|---|---|---|---|
| T2DM | LEC | *SERF1A* | -1.328368909 | 0.000154825 | 0.000910555 | 0.973382826 |
| T2DM | LEC | *LRRC15* | -0.134784641 | 0.000120526 | 0.000908725 | 0.757750046 |
| T2DM | LEC | *NME6* | 0.831291365 | 0.000123798 | 0.000908725 | 0.77831664 |
| T2DM | LEC | *FRMD3* | 0.883582917 | 0.000880936 | 0.002150853 | 1 |
| T2DM | LEC | *MTBP* | 0.872635203 | 0.000303663 | 0.001125666 | 1 |
| T2DM | LEC | *MALT1* | 0.899425515 | 0.006511392 | 0.010694128 | 1 |
| T2DM | LEC | *MBOAT1* | -0.697291975 | 0.001352914 | 0.002954418 | 1 |
| T2DM | LEC | *C5orf34* | 0.906615907 | 0.000600615 | 0.00165908 | 1 |
| T2DM | LEC | *ZNF131* | 0.800503115 | 0.005895766 | 0.009829404 | 1 |
| T2DM | LEC | *HINFP* | 1.326496082 | 0.009177448 | 0.014352889 | 1 |
| T2DM | LEC | *SPTLC3* | 0.432711148 | 0.000118932 | 0.000908725 | 0.747724716 |
| T2DM | LEC | *COPRS* | 1.590724219 | 0.012300227 | 0.018500366 | 1 |
| T2DM | LEC | *CLEC12A* | 0.176141388 | 0.009937767 | 0.015355583 | 1 |
| T2DM | LEC | *SUCLG2* | 0.821028287 | 0.000556666 | 0.001571513 | 1 |
| T2DM | LEC | *CSDC2* | -1.233883709 | 0.004022641 | 0.007056457 | 1 |
| T2DM | LEC | *RCAN2* | 0.895104042 | 0.001070189 | 0.002470212 | 1 |
| T2DM | LEC | *IL16* | 0.36755349 | 0.000227132 | 0.000992757 | 1 |
| T2DM | LEC | *GNB2* | 0.893630016 | 0.001088734 | 0.00250178 | 1 |
| T2DM | LEC | *PRSS27* | 5.733982638 | 4.01E-05 | 0.000908725 | 0.252183237 |
| T2DM | LEC | *FGGY* | 1.006179056 | 0.00029974 | 0.001119043 | 1 |
| T2DM | LEC | *ZNF24* | 0.955810341 | 0.029110593 | 0.039942885 | 1 |
| T2DM | LEC | *MANEA* | 1.07482926 | 0.000216603 | 0.000980405 | 1 |
| T2DM | LEC | *FIBP* | 0.693029614 | 3.30E-05 | 0.000908725 | 0.207611129 |
| T2DM | LEC | *CARNS1* | 3.044383107 | 0.00391769 | 0.006911618 | 1 |
| T2DM | LEC | *NIPAL4* | -0.544753336 | 0.001287967 | 0.002836234 | 1 |
| T2DM | LEC | *SNTG2* | -0.148171791 | 0.000142025 | 0.000908725 | 0.892908643 |
| T2DM | LEC | *PDE3A* | 0.962765711 | 0.000543623 | 0.001548598 | 1 |
| T2DM | LEC | *MRPL52* | 1.30242222 | 0.000208492 | 0.000972609 | 1 |
| T2DM | LEC | *SMPDL3A* | 0.922673704 | 0.000874943 | 0.002140375 | 1 |
| T2DM | LEC | *AGAP5* | -4.367058197 | 0.000141004 | 0.000908725 | 0.88649233 |
| T2DM | LEC | *ZMAT3* | 1.237724919 | 0.006759163 | 0.011046233 | 1 |
| T2DM | LEC | *ZFAND4* | 0.968154967 | 0.015767485 | 0.023069626 | 1 |
| T2DM | LEC | *ZNF738* | 0.862754256 | 7.20E-05 | 0.000908725 | 0.452813445 |
| T2DM | LEC | *SLFN11* | 0.795778707 | 0.000694457 | 0.001826037 | 1 |
| T2DM | LEC | *CCL19* | 2.837324774 | 0.000186383 | 0.000932902 | 1 |
| T2DM | LEC | *CFL1* | 0.969951498 | 0.029302614 | 0.040180051 | 1 |
| T2DM | LEC | *EFCAB12* | 0.629458434 | 0.024212594 | 0.033865312 | 1 |
| T2DM | LEC | *FAM192A* | 0.624193006 | 0.00186937 | 0.003810873 | 1 |
| T2DM | LEC | *CBWD1* | 1.175253147 | 0.026771535 | 0.037097783 | 1 |
| T2DM | LEC | *RAB37* | 0.945409484 | 8.52E-05 | 0.000908725 | 0.535714616 |
| T2DM | LEC | *DCP2* | 0.867335168 | 0.012629202 | 0.018949831 | 1 |
| T2DM | LEC | *SNX32* | 1.490565283 | 1.05E-06 | 0.000241487 | 0.006593492 |
| T2DM | LEC | *METAP1D* | 0.969564291 | 0.000745818 | 0.00191699 | 1 |
| T2DM | LEC | *ANKRD13D* | 0.879177804 | 0.000522113 | 0.001506436 | 1 |
| T2DM | LEC | *LCLAT1* | -0.762499013 | 0.01796426 | 0.025903968 | 1 |
| T2DM | LEC | *ADH6* | 0.26920604 | 0.021342827 | 0.03022806 | 1 |
| T2DM | LEC | *GXYLT2* | 0.45580746 | 0.000257562 | 0.00103934 | 1 |
| T2DM | LEC | *HPSE2* | 0.972645546 | 0.000203473 | 0.000964732 | 1 |
| T2DM | LEC | *HECTD4* | 0.810875836 | 0.002498034 | 0.00476925 | 1 |
| T2DM | LEC | *BNC2* | 0.562667767 | 0.000361447 | 0.00122668 | 1 |

| | | | | | | |
|---|---|---|---|---|---|---|
| T2DM | LEC | *NOC3L* | 1.023734956 | 0.010408381 | 0.016022893 | 1 |
| T2DM | LEC | *PARP15* | 0.651600977 | 0.006635929 | 0.010870267 | 1 |
| T2DM | LEC | *GLRX* | 0.533931371 | 0.000165408 | 0.000910705 | 1 |
| T2DM | LEC | *GOLGB1* | 0.747746109 | 0.000748153 | 0.00191907 | 1 |
| T2DM | LEC | *DMRT2* | 0.35206975 | 0.000197284 | 0.000956093 | 1 |
| T2DM | LEC | *SLC2A14* | -0.811824941 | 0.0054121 | 0.00913201 | 1 |
| T2DM | LEC | *C1QA* | 0.592772745 | 0.000140676 | 0.000908725 | 0.884427002 |
| T2DM | LEC | *NDNF* | -0.864889889 | 0.000215144 | 0.000978911 | 1 |
| T2DM | LEC | *GLIPR1L1* | 0.91218138 | 0.000180537 | 0.00092505 | 1 |
| T2DM | LEC | *ARV1* | 0.910620276 | 0.00079788 | 0.002004905 | 1 |
| T2DM | LEC | *CCDC36* | 0.840859843 | 0.000166872 | 0.000910705 | 1 |
| T2DM | LEC | *THAP2* | 0.809676744 | 0.000367693 | 0.001238845 | 1 |
| T2DM | LEC | *PPP1R14B* | 1.033201369 | 0.000232729 | 0.001001608 | 1 |
| T2DM | LEC | *ZNF417* | 0.881465992 | 0.000525595 | 0.001513704 | 1 |
| T2DM | LEC | *PEAK1* | 0.664003792 | 0.001260075 | 0.002787506 | 1 |
| T2DM | LEC | *MST1* | 0.546017156 | 0.009914042 | 0.01532946 | 1 |
| T2DM | LEC | *TNFRSF10C* | 0.67523297 | 0.000801266 | 0.002011447 | 1 |
| T2DM | LEC | *CSPG4* | 1.044490769 | 0.001615229 | 0.003392899 | 1 |
| T2DM | LEC | *NUDT18* | -1.504881519 | 0.000340622 | 0.001189058 | 1 |
| T2DM | LEC | *SULT1B1* | 1.035791428 | 0.000130926 | 0.000908725 | 0.823134844 |
| T2DM | LEC | *NUDT4* | -0.686349685 | 0.001076196 | 0.002480222 | 1 |
| T2DM | LEC | *SCAI* | 0.737374532 | 0.000463066 | 0.001396351 | 1 |
| T2DM | LEC | *C1orf100* | -1.428727894 | 0.000605459 | 0.001668064 | 1 |
| T2DM | LEC | *AGFG1* | 0.685259228 | 0.000396912 | 0.00128761 | 1 |
| T2DM | LEC | *RNF213* | 0.741711076 | 0.006923239 | 0.01127627 | 1 |
| T2DM | LEC | *PLK3* | 0.891668808 | 0.000106548 | 0.000908725 | 0.669866131 |
| T2DM | LEC | *TUBB8* | 1.255763193 | 0.001097453 | 0.002517215 | 1 |
| T2DM | LEC | *GPR160* | 1.391847799 | 0.000299384 | 0.001118805 | 1 |
| T2DM | LEC | *GOLIM4* | 0.801383614 | 0.01684927 | 0.024458869 | 1 |
| T2DM | LEC | *HOXB2* | 0.368182164 | 0.009728897 | 0.01508399 | 1 |
| T2DM | LEC | *UBXN2A* | 0.691025096 | 0.006037581 | 0.010020662 | 1 |
| T2DM | LEC | *CCS* | 0.89412957 | 0.000176469 | 0.000918426 | 1 |
| T2DM | LEC | *NRROS* | 0.576380905 | 0.000234098 | 0.001001608 | 1 |
| T2DM | LEC | *CEP19* | 0.793770795 | 0.028286155 | 0.038981819 | 1 |
| T2DM | LEC | *SLC25A30* | 1.316117888 | 0.000378069 | 0.001253649 | 1 |
| T2DM | LEC | *CTSF* | 0.550647982 | 0.002214128 | 0.004333817 | 1 |
| T2DM | LEC | *C16orf91* | 1.09686787 | 3.92E-05 | 0.000908725 | 0.246189306 |
| T2DM | LEC | *SOCS7* | 0.992700972 | 0.000561006 | 0.001579965 | 1 |
| T2DM | LEC | *TLR6* | 0.863177604 | 0.000113354 | 0.000908725 | 0.712659002 |
| T2DM | LEC | *FAM53A* | 0.481816789 | 0.000225709 | 0.000992757 | 1 |
| T2DM | LEC | *SNX31* | 1.249912849 | 0.005148143 | 0.008766623 | 1 |
| T2DM | LEC | *RALGAPA1* | 0.632082592 | 0.020090994 | 0.028648691 | 1 |
| T2DM | LEC | *LIG4* | 0.892065982 | 8.02E-05 | 0.000908725 | 0.504255788 |
| T2DM | LEC | *LINGO2* | 0.440342629 | 6.74E-05 | 0.000908725 | 0.423647459 |
| T2DM | LEC | *BBS1* | 0.948864997 | 0.000141218 | 0.000908725 | 0.887835821 |
| T2DM | LEC | *ANKRD36C* | 0.878909443 | 0.001867009 | 0.003808528 | 1 |
| T2DM | LEC | *TMEM81* | 1.955733831 | 1.76E-05 | 0.000908725 | 0.110607711 |
| T2DM | LEC | *IL20RB* | 0.9410112 | 4.00E-06 | 0.000465666 | 0.02514597 |
| T2DM | LEC | *ZNF497* | 2.532605216 | 0.00032284 | 0.001159063 | 1 |
| T2DM | LEC | *KLF14* | -0.758643971 | 4.45E-05 | 0.000908725 | 0.2795986 |

| | | | | | | |
|---|---|---|---|---|---|---|
| T2DM | LEC | *BRSK2* | -0.678493379 | 0.000755128 | 0.001929875 | 1 |
| T2DM | LEC | *SH3PXD2B* | 0.624262166 | 1.46E-05 | 0.000902248 | 0.091747643 |
| T2DM | LEC | *LARP7* | 0.960049679 | 0.001828996 | 0.00374679 | 1 |
| T2DM | LEC | *FGFBP3* | -1.02944571 | 0.000315929 | 0.001151139 | 1 |
| T2DM | LEC | *HRAS* | 0.771512927 | 0.00029222 | 0.001108073 | 1 |
| T2DM | LEC | *SRP72* | 0.748452053 | 0.005649931 | 0.009482412 | 1 |
| T2DM | LEC | *C4orf26* | 1.81992327 | 0.005210196 | 0.008848326 | 1 |
| T2DM | LEC | *BTC* | 0.768679714 | 6.21E-05 | 0.000908725 | 0.390592898 |
| T2DM | LEC | *NLRP6* | -3.811792053 | 1.01E-06 | 0.000241487 | 0.006332576 |
| T2DM | LEC | *NDUFA11* | 1.419860843 | 0.007764081 | 0.012452239 | 1 |
| T2DM | LEC | *PTDSS2* | 0.84259654 | 0.015524489 | 0.022751157 | 1 |
| T2DM | LEC | *FUT1* | -1.064277651 | 0.002804516 | 0.005241376 | 1 |
| T2DM | LEC | *FBXW8* | 0.905615618 | 0.000807975 | 0.002022189 | 1 |
| T2DM | LEC | *CHST2* | -1.635558266 | 0.00109589 | 0.002514547 | 1 |
| T2DM | LEC | *ATR* | 0.823713818 | 0.002859148 | 0.005327642 | 1 |
| T2DM | LEC | *TVP23C* | 0.571666992 | 0.000130961 | 0.000908725 | 0.823351241 |
| T2DM | LEC | *PACS1* | 0.662043069 | 0.009064686 | 0.014208347 | 1 |
| T2DM | LEC | *SH3BP5L* | 0.686162811 | 4.45E-05 | 0.000908725 | 0.279957102 |
| T2DM | LEC | *CADM2* | 0.562377643 | 0.008584508 | 0.01354618 | 1 |
| T2DM | LEC | *ARHGAP1* | 0.601616681 | 0.000307925 | 0.001132221 | 1 |
| T2DM | LEC | *CHST1* | -0.3243631 | 0.000172438 | 0.000910705 | 1 |
| T2DM | LEC | *GOLGA8A* | 0.729737243 | 0.000578436 | 0.00161628 | 1 |
| T2DM | LEC | *TP53I11* | 1.289637485 | 0.022517056 | 0.031712529 | 1 |
| T2DM | LEC | *PHYHD1* | 0.875902424 | 8.91E-05 | 0.000908725 | 0.560150265 |
| T2DM | LEC | *ZNF519* | 0.85820219 | 0.000671309 | 0.001786827 | 1 |
| T2DM | LEC | *CHRNA7* | -0.686366236 | 0.023908163 | 0.033476753 | 1 |
| T2DM | LEC | *TMEM9B* | 1.15204479 | 5.74E-05 | 0.000908725 | 0.360683006 |
| T2DM | LEC | *NRIP3* | 0.504816835 | 0.034231116 | 0.046152911 | 1 |
| T2DM | LEC | *SMAD2* | 0.68061374 | 0.008231203 | 0.013091216 | 1 |
| T2DM | LEC | *EIF3F* | 0.93310723 | 0.00100238 | 0.002355874 | 1 |
| T2DM | LEC | *CLTB* | 1.059774977 | 0.001620479 | 0.003400518 | 1 |
| T2DM | LEC | *PCSK1* | 0.489155315 | 8.44E-05 | 0.000908725 | 0.530444159 |
| T2DM | LEC | *PPP2R2D* | 0.81753386 | 0.003957274 | 0.006955376 | 1 |
| T2DM | LEC | *MCTP1* | 0.538582252 | 0.003000204 | 0.005521746 | 1 |
| T2DM | LEC | *LRRC25* | 0.568639646 | 7.63E-05 | 0.000908725 | 0.479922963 |
| T2DM | LEC | *UBQLNL* | -1.700708035 | 0.007672187 | 0.012333173 | 1 |
| T2DM | LEC | *LIPT2* | 1.412798895 | 0.000339595 | 0.001187188 | 1 |
| T2DM | LEC | *KCNE3* | 0.654388114 | 9.87E-05 | 0.000908725 | 0.620818354 |
| T2DM | LEC | *UCP3* | 0.891526489 | 0.002975914 | 0.00549151 | 1 |
| T2DM | LEC | *UCP2* | 0.937367435 | 0.001607095 | 0.003382592 | 1 |
| T2DM | LEC | *PAAF1* | 0.304066451 | 0.023236731 | 0.03264566 | 1 |
| T2DM | LEC | *RAB6A* | 0.705608072 | 0.025336724 | 0.035272804 | 1 |
| T2DM | LEC | *TOM1L2* | 0.880073975 | 0.002017209 | 0.004037045 | 1 |
| T2DM | LEC | *TEX26* | 1.181199839 | 8.43E-05 | 0.000908725 | 0.530196879 |
| T2DM | LEC | *GPR156* | 0.764449615 | 5.85E-05 | 0.000908725 | 0.36757444 |
| T2DM | LEC | *B3GNTL1* | 0.698252879 | 0.000144008 | 0.000908725 | 0.905380134 |
| T2DM | LEC | *LINC01106* | -2.6196408 | 5.53E-05 | 0.000908725 | 0.347920331 |
| T2DM | LEC | *MSRA* | 0.748922773 | 0.002556963 | 0.00485512 | 1 |
| T2DM | LEC | *CTDNEP1* | 0.879260696 | 9.39E-05 | 0.000908725 | 0.590230522 |
| T2DM | LEC | *ETV4* | 0.841165566 | 1.90E-05 | 0.000908725 | 0.119261278 |

| | | | | | | |
|---|---|---|---|---|---|---|
| T2DM | LEC | *SWI5* | 0.868465372 | 0.003927725 | 0.006922794 | 1 |
| T2DM | LEC | *A2M* | 0.658374746 | 0.000347681 | 0.001200369 | 1 |
| T2DM | LEC | *ARL4D* | 0.657773182 | 0.000162458 | 0.000910705 | 1 |
| T2DM | LEC | *LRRN1* | 1.462176923 | 0.000783006 | 0.001978803 | 1 |
| T2DM | LEC | *B3GALT6* | -0.783065121 | 0.000349155 | 0.001204034 | 1 |
| T2DM | LEC | *ZNF613* | 0.729826245 | 3.13E-05 | 0.000908725 | 0.196552985 |
| T2DM | LEC | *IP6K1* | 0.915943741 | 0.014895638 | 0.021926687 | 1 |
| T2DM | LEC | *SSNA1* | 1.389086084 | 0.031585124 | 0.042942664 | 1 |
| T2DM | LEC | *CHMP6* | 1.657982477 | 0.002787807 | 0.0052148 | 1 |
| T2DM | LEC | *UFSP1* | 0.636319703 | 0.021574593 | 0.030515065 | 1 |
| T2DM | LEC | *TCP11L1* | 0.995016133 | 0.000126828 | 0.000908725 | 0.797368016 |
| T2DM | LEC | *CIDEA* | 0.213265785 | 0.005949456 | 0.009908406 | 1 |
| T2DM | LEC | *SMIM19* | 0.795704522 | 0.000120227 | 0.000908725 | 0.755870153 |
| T2DM | LEC | *SLC35G1* | 0.752403199 | 0.001991414 | 0.004001284 | 1 |
| T2DM | LEC | *B3GNT4* | 3.295358332 | 0.000111608 | 0.000908725 | 0.70168084 |
| T2DM | LEC | *CDC26* | -0.875550985 | 2.15E-05 | 0.000908725 | 0.135478133 |
| T2DM | LEC | *CRLF3* | 0.935759038 | 0.000666792 | 0.001779338 | 1 |
| T2DM | LEC | *RNPEP* | 0.873983554 | 0.000364731 | 0.001234821 | 1 |
| T2DM | LEC | *SPRYD4* | 0.637607641 | 0.005808454 | 0.009709585 | 1 |
| T2DM | LEC | *SLCO3A1* | 0.837434175 | 0.004976487 | 0.008521605 | 1 |
| T2DM | LEC | *WDR25* | 0.970614536 | 0.000449838 | 0.001370217 | 1 |
| T2DM | LEC | *PLA2G16* | 0.401311928 | 0.001212236 | 0.002709324 | 1 |
| T2DM | LEC | *PHLDB3* | -0.421578068 | 0.000412587 | 0.001312173 | 1 |
| T2DM | LEC | *RMDN1* | 0.924052746 | 0.001230557 | 0.00273665 | 1 |
| T2DM | LEC | *HORMAD2* | 1.154181315 | 5.78E-06 | 0.000559501 | 0.036367562 |
| T2DM | LEC | *RNF152* | -0.161979654 | 0.000142437 | 0.000908725 | 0.895499587 |
| T2DM | LEC | *LRRC37A* | 0.615861066 | 0.001999923 | 0.004009413 | 1 |
| T2DM | LEC | *BOK* | 0.774279836 | 0.000105536 | 0.000908725 | 0.66350189 |
| T2DM | LEC | *ZNF843* | 0.654376028 | 0.000192715 | 0.000946191 | 1 |
| T2DM | LEC | *C8orf59* | 1.109684136 | 0.00086569 | 0.002122726 | 1 |
| T2DM | LEC | *PFN4* | 1.024908921 | 0.000484957 | 0.001436081 | 1 |
| T2DM | LEC | *RUFY1* | 0.721477404 | 0.006499033 | 0.010679409 | 1 |
| T2DM | LEC | *BASP1* | 0.687037258 | 0.000470809 | 0.001408842 | 1 |
| T2DM | LEC | *LRRC37A3* | 0.808255479 | 0.000153726 | 0.000910197 | 0.966473978 |
| T2DM | LEC | *VSIG10* | 0.537450022 | 0.000239304 | 0.00100947 | 1 |
| T2DM | LEC | *IRX5* | 0.606547124 | 0.000114087 | 0.000908725 | 0.717266899 |
| T2DM | LEC | *PNMA1* | 1.507686296 | 0.001668069 | 0.00348526 | 1 |
| T2DM | LEC | *ANKLE2* | 1.08092217 | 0.001955478 | 0.003946739 | 1 |
| T2DM | LEC | *GCNT4* | 6.250406817 | 0.009937808 | 0.015355583 | 1 |
| T2DM | LEC | *MUC20* | 1.863166572 | 0.001162173 | 0.00262449 | 1 |
| T2DM | LEC | *THAP4* | 0.924820533 | 0.000120672 | 0.000908725 | 0.758665884 |
| T2DM | LEC | *SHMT1* | 0.584415103 | 7.81E-05 | 0.000908725 | 0.49089891 |
| T2DM | LEC | *DPP7* | 1.075932184 | 0.003037506 | 0.005580596 | 1 |
| T2DM | LEC | *SEC24C* | 0.984944122 | 0.013044154 | 0.019502639 | 1 |
| T2DM | LEC | *MTHFR* | 1.001462955 | 0.004229389 | 0.007373868 | 1 |
| T2DM | LEC | *C19orf18* | -0.909733973 | 0.011824387 | 0.017895984 | 1 |
| T2DM | LEC | *WDR73* | 1.008694087 | 0.000147272 | 0.000908725 | 0.925900437 |
| T2DM | LEC | *POLE* | 1.200496652 | 0.000461381 | 0.0013939 | 1 |
| T2DM | LEC | *DSCAML1* | 1.090170001 | 0.001227889 | 0.002731684 | 1 |
| T2DM | LEC | *TALDO1* | 0.727259957 | 7.31E-05 | 0.000908725 | 0.459806531 |

| | | | | | | |
|---|---|---|---|---|---|---|
| T2DM | LEC | *RIMKLA* | -0.590475564 | 0.005309272 | 0.008999566 | 1 |
| T2DM | LEC | *B3GNT8* | -2.629586842 | 0.000887986 | 0.002164702 | 1 |
| T2DM | LEC | *AP006621.1* | 0.595203761 | 3.54E-05 | 0.000908725 | 0.222803514 |
| T2DM | LEC | *MAN1B1* | 1.240234909 | 0.001300315 | 0.00286042 | 1 |
| T2DM | LEC | *FBXO39* | -0.3237857 | 0.00081332 | 0.0020283 | 1 |
| T2DM | LEC | *C8orf31* | 0.709749196 | 0.000112565 | 0.000908725 | 0.707693056 |
| T2DM | LEC | *AC024940.1* | -0.498313715 | 0.000212653 | 0.0009784 | 1 |
| T2DM | LEC | *TIMM22* | 1.213889351 | 7.30E-05 | 0.000908725 | 0.459226206 |
| T2DM | LEC | *PPFIA3* | 0.947145639 | 0.011926043 | 0.018015145 | 1 |
| T2DM | LEC | *MAGEF1* | -1.419794393 | 0.027023562 | 0.037397565 | 1 |
| T2DM | LEC | *NAP1L5* | 0.708335729 | 0.000140446 | 0.000908725 | 0.882985944 |
| T2DM | LEC | *NIM1K* | -0.493213613 | 0.000408315 | 0.001307186 | 1 |
| T2DM | LEC | *ACOT4* | -0.731623863 | 0.000467602 | 0.001402582 | 1 |
| T2DM | LEC | *ARIH2* | 0.678706871 | 0.016401972 | 0.02387746 | 1 |
| T2DM | LEC | *IRX3* | -0.398587884 | 0.001632 | 0.003423552 | 1 |
| T2DM | LEC | *ATOX1* | 1.289300419 | 0.000168743 | 0.000910705 | 1 |
| T2DM | LEC | *TBL1XR1* | 0.632183573 | 0.011055773 | 0.016887183 | 1 |
| T2DM | LEC | *SAMD12* | 0.707635272 | 0.007383564 | 0.011917963 | 1 |
| T2DM | LEC | *C18orf32* | 0.832484042 | 5.72E-05 | 0.000908725 | 0.359724004 |
| T2DM | LEC | *RPLP2* | 0.847636777 | 0.007597447 | 0.012228661 | 1 |
| T2DM | LEC | *C12orf54* | -0.921818278 | 2.60E-05 | 0.000908725 | 0.163222011 |
| T2DM | LEC | *ACAD9* | 0.401330879 | 0.009938264 | 0.015355583 | 1 |
| T2DM | LEC | *IL17RA* | 0.860727059 | 0.001086439 | 0.00249742 | 1 |
| T2DM | LEC | *CD163L1* | 0.274509592 | 0.000117209 | 0.000908725 | 0.736893079 |
| T2DM | LEC | *THAP5* | 0.920091841 | 0.000186743 | 0.000932902 | 1 |
| T2DM | LEC | *CD151* | 0.915374552 | 0.009287693 | 0.014500056 | 1 |
| T2DM | LEC | *POLR2L* | 1.10449141 | 0.000930597 | 0.002229636 | 1 |
| T2DM | LEC | *FAM87B* | 1.187486171 | 0.001163295 | 0.002626081 | 1 |
| T2DM | LEC | *PCDHB9* | 1.027528152 | 0.00020396 | 0.000964732 | 1 |
| T2DM | LEC | *ZNF619* | 2.000872578 | 0.009777543 | 0.015144472 | 1 |
| T2DM | LEC | *ZBTB41* | 0.678743825 | 0.0001467 | 0.000908725 | 0.922304593 |
| T2DM | LEC | *ZNF354C* | 1.226971074 | 0.017273118 | 0.025016376 | 1 |
| T2DM | LEC | *CENPBD1* | -0.383852817 | 8.79E-05 | 0.000908725 | 0.552546811 |
| T2DM | LEC | *BET1L* | 1.217696079 | 2.11E-05 | 0.000908725 | 0.132657693 |
| T2DM | LEC | *RIC8A* | 0.722410125 | 0.002115762 | 0.004196149 | 1 |
| T2DM | LEC | *IMP3* | 1.671368348 | 3.15E-05 | 0.000908725 | 0.197822355 |
| T2DM | LEC | *ODF3B* | 1.263212491 | 0.003065279 | 0.005619552 | 1 |
| T2DM | LEC | *DPY19L2* | -0.462155582 | 0.00024879 | 0.001023586 | 1 |
| T2DM | LEC | *FAM26E* | 0.584768176 | 7.93E-05 | 0.000908725 | 0.498430657 |
| T2DM | LEC | *MLF1* | 0.683959379 | 0.000147469 | 0.000908725 | 0.927136538 |
| T2DM | LEC | *PRSS42* | 1.706467852 | 4.21E-05 | 0.000908725 | 0.264824971 |
| T2DM | LEC | *GRAMD1C* | 0.828730817 | 0.000111866 | 0.000908725 | 0.703304679 |
| T2DM | LEC | *STAP2* | -0.669324238 | 0.000142412 | 0.000908725 | 0.895341945 |
| T2DM | LEC | *GOLGA8Q* | 19.90109601 | 0.000742934 | 0.001910358 | 1 |
| T2DM | LEC | *DALRD3* | 1.177238011 | 0.002185751 | 0.00429566 | 1 |
| T2DM | LEC | *ZNF114* | 0.780328616 | 0.00028239 | 0.001085201 | 1 |
| T2DM | LEC | *ZNF518B* | -0.633761041 | 0.000177314 | 0.000918726 | 1 |
| T2DM | LEC | *LCORL* | 0.739109428 | 0.025783175 | 0.035815029 | 1 |
| T2DM | LEC | *ZNF454* | -0.632311993 | 0.005815104 | 0.009718117 | 1 |
| T2DM | LEC | *ZC3H12D* | 0.486769298 | 0.000107755 | 0.000908725 | 0.677454963 |

| | | | | | | |
|---|---|---|---|---|---|---|
| T2DM | LEC | *KDELC2* | 0.80728714 | 0.000609978 | 0.001678308 | 1 |
| T2DM | LEC | *PLEC* | -0.804363111 | 0.010923231 | 0.016722147 | 1 |
| T2DM | LEC | *SH2D4B* | -1.155970091 | 0.000218298 | 0.00098409 | 1 |
| T2DM | LEC | *RNF212* | 0.581726857 | 0.000380871 | 0.00125962 | 1 |
| T2DM | LEC | *PRSS36* | 0.594881181 | 0.000109246 | 0.000908725 | 0.686832131 |
| T2DM | LEC | *AQP11* | 0.928405988 | 0.000244127 | 0.001017397 | 1 |
| T2DM | LEC | *KCNG2* | 0.53488791 | 0.00152009 | 0.003229743 | 1 |
| T2DM | LEC | *ZFAND2A* | -0.858220251 | 0.000548105 | 0.001555553 | 1 |
| T2DM | LEC | *ZNF223* | -0.440224657 | 0.007698906 | 0.012369799 | 1 |
| T2DM | LEC | *FAM220A* | 0.780638742 | 0.005354858 | 0.00905974 | 1 |
| T2DM | LEC | *BEND3* | 1.130039369 | 9.86E-05 | 0.000908725 | 0.620005637 |
| T2DM | LEC | *NT5DC1* | 0.758586026 | 0.000914552 | 0.002211224 | 1 |
| T2DM | LEC | *AC006111.1* | 2.114479534 | 5.53E-05 | 0.000908725 | 0.347806827 |
| T2DM | LEC | *LINC00843* | 1.76697577 | 0.00136447 | 0.00297552 | 1 |
| T2DM | LEC | *COX14* | 0.995466584 | 0.004409046 | 0.007644697 | 1 |
| T2DM | LEC | *P4HTM* | 0.426240511 | 0.000103028 | 0.000908725 | 0.647734335 |
| T2DM | LEC | *CA8* | -0.512234091 | 0.002356907 | 0.004553742 | 1 |
| T2DM | LEC | *ERBB4* | 0.605012189 | 0.000459962 | 0.001391616 | 1 |
| T2DM | LEC | *CSRNP3* | -0.349062683 | 0.000277496 | 0.00108013 | 1 |
| T2DM | LEC | *ZNF713* | -0.590598367 | 0.000919953 | 0.00221763 | 1 |
| T2DM | LEC | *PARP10* | 0.72433311 | 0.000104386 | 0.000908725 | 0.656277472 |
| T2DM | LEC | *RPP25* | -0.635972374 | 0.000385123 | 0.001268529 | 1 |
| T2DM | LEC | *STX19* | 0.690408653 | 5.06E-06 | 0.000529126 | 0.031781421 |
| T2DM | LEC | *FAM219B* | 0.909159894 | 0.0002672 | 0.001057861 | 1 |
| T2DM | LEC | *CPNE7* | -1.648704222 | 0.029174606 | 0.040021983 | 1 |
| T2DM | LEC | *GDPD4* | 3.261522459 | 0.007708087 | 0.012378224 | 1 |
| T2DM | LEC | *MPI* | 0.932327143 | 0.00013278 | 0.000908725 | 0.834786605 |
| T2DM | LEC | *TRIM73* | -1.774944749 | 1.60E-06 | 0.000288231 | 0.010088091 |
| T2DM | LEC | *EFCAB13* | 0.751735956 | 0.001139459 | 0.002587136 | 1 |
| T2DM | LEC | *HYI* | 0.338570226 | 0.0192587 | 0.02758074 | 1 |
| T2DM | LEC | *C17orf62* | 0.905940969 | 0.000250094 | 0.001023586 | 1 |
| T2DM | LEC | *TUFM* | 0.973336601 | 0.000706993 | 0.00185016 | 1 |
| T2DM | LEC | *RMI1* | 1.246706036 | 0.000129545 | 0.000908725 | 0.814446917 |
| T2DM | LEC | *SNX18* | 0.815612462 | 0.000120391 | 0.000908725 | 0.75689923 |
| T2DM | LEC | *C14orf39* | 0.343170454 | 0.000493382 | 0.001449584 | 1 |
| T2DM | LEC | *TRIML2* | 8.343224419 | 0.00043952 | 0.001348591 | 1 |
| T2DM | LEC | *SPTY2D1* | 1.066759237 | 0.034679016 | 0.046696718 | 1 |
| T2DM | LEC | *TCAIM* | 0.584969061 | 0.031368946 | 0.042696809 | 1 |
| T2DM | LEC | *FUCA1* | 0.919005299 | 0.000103592 | 0.000908725 | 0.651285344 |
| T2DM | LEC | *ZNF664* | 0.53049736 | 0.000156475 | 0.000910705 | 0.983761153 |
| T2DM | LEC | *CALR* | 1.37472395 | 0.000173032 | 0.000910705 | 1 |
| T2DM | LEC | *CDH4* | 0.58146718 | 0.000260419 | 0.001044168 | 1 |
| T2DM | LEC | *HLA-DQB1* | -0.831550569 | 1.72E-07 | 5.41E-05 | 0.001081209 |
| T2DM | LEC | *ARID3B* | 0.775713099 | 0.01181937 | 0.017892699 | 1 |
| T2DM | LEC | *EGR3* | -1.493231868 | 0.00241891 | 0.004656525 | 1 |
| T2DM | LEC | *GEMIN4* | 0.611448986 | 0.00603342 | 0.010019078 | 1 |
| T2DM | LEC | *FJX1* | 0.861556086 | 0.011248485 | 0.017135747 | 1 |
| T2DM | LEC | *CIITA* | 0.653489236 | 9.60E-05 | 0.000908725 | 0.603587595 |
| T2DM | LEC | *PLD6* | -0.524950145 | 0.007324063 | 0.01184016 | 1 |
| T2DM | LEC | *ZBTB42* | 1.515592979 | 0.000151593 | 0.000910197 | 0.953064899 |

| | | | | | | |
|---|---|---|---|---|---|---|
| T2DM | LEC | *LACC1* | 1.419301378 | 0.013141521 | 0.019620219 | 1 |
| T2DM | LEC | *FCER1A* | 0.687309619 | 0.00013172 | 0.000908725 | 0.828125421 |
| T2DM | LEC | *PCED1B* | 0.568428024 | 0.001202673 | 0.00269082 | 1 |
| T2DM | LEC | *PIPOX* | 0.656990281 | 0.001814245 | 0.003720209 | 1 |
| T2DM | LEC | *FOXS1* | 1.263693016 | 0.000727789 | 0.001887164 | 1 |
| T2DM | LEC | *CDH5* | -0.308282137 | 0.001487365 | 0.003180633 | 1 |
| T2DM | LEC | *SERTAD2* | -0.702811168 | 0.009454981 | 0.014717373 | 1 |
| T2DM | LEC | *GIPC3* | -0.822933501 | 0.000619269 | 0.001697248 | 1 |
| T2DM | LEC | *CITED4* | 0.615571891 | 0.000255984 | 0.001035475 | 1 |
| T2DM | LEC | *TIGD5* | -0.482543532 | 0.000245635 | 0.001017515 | 1 |
| T2DM | LEC | *PDXDC1* | 0.720908665 | 0.027687978 | 0.038232883 | 1 |
| T2DM | LEC | *R3HDM2* | 0.757174215 | 0.004852251 | 0.008321358 | 1 |
| T2DM | LEC | *B3GNT3* | 4.080626276 | 1.48E-07 | 4.90E-05 | 0.000931463 |
| T2DM | LEC | *SSC5D* | 0.33046762 | 0.03660515 | 0.049069633 | 1 |
| T2DM | LEC | *ZADH2* | 1.019069113 | 0.002956956 | 0.005464546 | 1 |
| T2DM | LEC | *TMEM86B* | 0.609692358 | 0.000370858 | 0.001241526 | 1 |
| T2DM | LEC | *SEPT1* | 0.756346736 | 0.000150613 | 0.000910197 | 0.946905438 |
| T2DM | LEC | *LYNX1* | -0.857202469 | 5.73E-05 | 0.000908725 | 0.360393336 |
| T2DM | LEC | *TDRP* | -0.301739417 | 0.000246846 | 0.001019088 | 1 |
| T2DM | LEC | *ZNF816* | 0.964681546 | 0.002074365 | 0.004125762 | 1 |
| T2DM | LEC | *FGD6* | -0.522699152 | 0.000606722 | 0.001670213 | 1 |
| T2DM | LEC | *HCLS1* | 0.693761352 | 0.000145736 | 0.000908725 | 0.916245284 |
| T2DM | LEC | *ZNF609* | 0.68473655 | 0.002994927 | 0.005513648 | 1 |
| T2DM | LEC | *CCDC66* | 0.846545326 | 0.000441861 | 0.001351816 | 1 |
| T2DM | LEC | *DEFB124* | 1.519365015 | 0.001940262 | 0.003924678 | 1 |
| T2DM | LEC | *CYP8B1* | -0.139510671 | 0.000139932 | 0.000908725 | 0.879752992 |
| T2DM | LEC | *GLIPR1L2* | 0.371778103 | 0.01043359 | 0.016057768 | 1 |
| T2DM | LEC | *RNF182* | 0.408966393 | 0.02453357 | 0.034283742 | 1 |
| T2DM | LEC | *MB21D2* | 0.746055067 | 0.000486087 | 0.001437751 | 1 |
| T2DM | LEC | *SSTR2* | 0.726642919 | 0.000109629 | 0.000908725 | 0.689235092 |
| T2DM | LEC | *ZNF594* | 1.092380141 | 0.01351386 | 0.020109264 | 1 |
| T2DM | LEC | *PCGF5* | 0.830002618 | 0.001836885 | 0.003759277 | 1 |
| T2DM | LEC | *SHISA2* | -1.580245787 | 0.000137812 | 0.000908725 | 0.866426504 |
| T2DM | LEC | *CHST13* | 1.638356058 | 0.00122252 | 0.00272472 | 1 |
| T2DM | LEC | *SRSF8* | -1.022354736 | 0.001069401 | 0.002469994 | 1 |
| T2DM | LEC | *SLC36A4* | 0.746540642 | 0.015031785 | 0.022085728 | 1 |
| T2DM | LEC | *OR51E1* | 0.581913489 | 0.00174954 | 0.003614642 | 1 |
| T2DM | LEC | *PSMG4* | -0.701395389 | 0.002198072 | 0.004310443 | 1 |
| T2DM | LEC | *ZNF443* | 0.732888672 | 0.024973473 | 0.034805636 | 1 |
| T2DM | LEC | *CAPS2* | 0.618909374 | 0.000171876 | 0.000910705 | 1 |
| T2DM | LEC | *ZNF792* | 0.770705691 | 0.009099282 | 0.014251915 | 1 |
| T2DM | LEC | *D2HGDH* | 0.744191236 | 0.00011286 | 0.000908725 | 0.709550827 |
| T2DM | LEC | *CMTR2* | 0.991491739 | 0.000412203 | 0.001312162 | 1 |
| T2DM | LEC | *LRRC57* | 1.233851673 | 0.027165717 | 0.037577747 | 1 |
| T2DM | LEC | *MRPL14* | -0.879617104 | 0.000470572 | 0.001408802 | 1 |
| T2DM | LEC | *BBS12* | 0.823892576 | 0.000123176 | 0.000908725 | 0.774407188 |
| T2DM | LEC | *LSMEM1* | 0.906363085 | 0.002534163 | 0.004826502 | 1 |
| T2DM | LEC | *AEN* | 1.97831217 | 0.010379497 | 0.015986257 | 1 |
| T2DM | LEC | *TRAPPC5* | 0.958551948 | 0.001523344 | 0.003234469 | 1 |
| T2DM | LEC | *ANKRD34A* | -1.753544958 | 9.69E-05 | 0.000908725 | 0.609059686 |

| | | | | | | |
|---|---|---|---|---|---|---|
| T2DM | LEC | *SLC26A11* | 0.852517835 | 0.000130778 | 0.000908725 | 0.822201306 |
| T2DM | LEC | *HIGD1A* | 0.532668696 | 0.000274416 | 0.001076939 | 1 |
| T2DM | LEC | *HIST3H2A* | 0.969023824 | 0.000488584 | 0.001442126 | 1 |
| T2DM | LEC | *POLR2A* | 0.838986076 | 0.003636208 | 0.006490868 | 1 |
| T2DM | LEC | *TMEM132C* | 0.439179717 | 6.53E-05 | 0.000908725 | 0.410435369 |
| T2DM | LEC | *SLC25A41* | -1.03251702 | 0.000131641 | 0.000908725 | 0.827626733 |
| T2DM | LEC | *TMEM136* | 0.895058474 | 0.024573772 | 0.034323707 | 1 |
| T2DM | LEC | *TMEM132E* | -2.048194489 | 0.000195858 | 0.000953164 | 1 |
| T2DM | LEC | *ZNF322* | -0.751776057 | 0.003489054 | 0.006263758 | 1 |
| T2DM | LEC | *CCL13* | 0.669513587 | 0.001272437 | 0.002809909 | 1 |
| T2DM | LEC | *OGFOD3* | 0.428138689 | 0.012473987 | 0.018730345 | 1 |
| T2DM | LEC | *ACBD4* | 0.625564945 | 0.000108481 | 0.000908725 | 0.682020528 |
| T2DM | LEC | *SGSH* | 0.991348338 | 0.000124979 | 0.000908725 | 0.785741805 |
| T2DM | LEC | *SETD2* | 0.783491231 | 0.000764974 | 0.001947414 | 1 |
| T2DM | LEC | *MRPS23* | 1.553365792 | 0.004758015 | 0.008173126 | 1 |
| T2DM | LEC | *GPR135* | 0.531922398 | 0.00060045 | 0.00165908 | 1 |
| T2DM | LEC | *ZFP41* | -0.621709833 | 0.001376917 | 0.00299539 | 1 |
| T2DM | LEC | *ATG9B* | -1.135230055 | 0.000227227 | 0.000992757 | 1 |
| T2DM | LEC | *C3orf58* | 0.942475922 | 0.000531263 | 0.001526532 | 1 |
| T2DM | LEC | *ODF3L2* | 1.413295084 | 0.000861206 | 0.002115 | 1 |
| T2DM | LEC | *RELL1* | 0.723835507 | 0.000257482 | 0.00103934 | 1 |
| T2DM | LEC | *CLDN7* | 0.622083913 | 0.004666082 | 0.008037166 | 1 |
| T2DM | LEC | *ADO* | 1.121836444 | 0.000107559 | 0.000908725 | 0.676222363 |
| T2DM | LEC | *SNRPE* | 0.755134698 | 0.000202282 | 0.00096305 | 1 |
| T2DM | LEC | *CHST15* | -0.413293408 | 0.000286834 | 0.001094915 | 1 |
| T2DM | LEC | *IDH2* | 0.794627483 | 0.000412624 | 0.001312173 | 1 |
| T2DM | LEC | *WRB* | -0.461852084 | 0.010339396 | 0.015932299 | 1 |
| T2DM | LEC | *TNRC18* | 0.846731757 | 0.003029915 | 0.005569906 | 1 |
| T2DM | LEC | *FAM181B* | 2.067639739 | 0.000385019 | 0.001268529 | 1 |
| T2DM | LEC | *NOP10* | 0.542021098 | 0.001093941 | 0.002510992 | 1 |
| T2DM | LEC | *TDRKH* | 0.586456997 | 0.000875482 | 0.002140751 | 1 |
| T2DM | LEC | *ZNF708* | 0.739722551 | 0.005268814 | 0.008938218 | 1 |
| T2DM | LEC | *IST1* | 0.796598207 | 0.002179155 | 0.004290745 | 1 |
| T2DM | LEC | *ERCC6L2* | 0.774684205 | 0.002248255 | 0.004388321 | 1 |
| T2DM | LEC | *FAM153B* | 0.738813471 | 0.000350343 | 0.001206906 | 1 |
| T2DM | LEC | *BACE2* | -0.307660764 | 0.000432262 | 0.001341345 | 1 |
| T2DM | LEC | *SYNM* | -0.39255299 | 0.000222524 | 0.0009887 | 1 |
| T2DM | LEC | *IZUMO1* | -0.361872487 | 0.007715191 | 0.012384654 | 1 |
| T2DM | LEC | *C8orf33* | 0.992063991 | 0.02972818 | 0.040692591 | 1 |
| T2DM | LEC | *FBXL6* | 1.981237656 | 0.002181387 | 0.004292451 | 1 |
| T2DM | LEC | *KBTBD3* | 0.580011456 | 0.000110813 | 0.000908725 | 0.696682475 |
| T2DM | LEC | *CLN8* | 0.742306974 | 0.000404793 | 0.001300425 | 1 |
| T2DM | LEC | *CACNB4* | 0.363045963 | 0.00037687 | 0.001251655 | 1 |
| T2DM | LEC | *TRAPPC6B* | 0.843586377 | 0.010510529 | 0.016164309 | 1 |
| T2DM | LEC | *NPLOC4* | 0.534952044 | 0.015113358 | 0.022190024 | 1 |
| T2DM | LEC | *TSHZ2* | -0.485225563 | 0.009068728 | 0.014211139 | 1 |
| T2DM | LEC | *CAPN12* | 1.047336747 | 1.94E-05 | 0.000908725 | 0.122175141 |
| T2DM | LEC | *FES* | 0.920765845 | 0.000316499 | 0.001151139 | 1 |
| T2DM | LEC | *CAV3* | 0.43369697 | 0.001381273 | 0.003000964 | 1 |
| T2DM | LEC | *ADI1* | 0.87048969 | 0.000636835 | 0.001724281 | 1 |

| | | | | | | |
|---|---|---|---|---|---|---|
| T2DM | LEC | *CLEC4G* | 1.760089142 | 0.000142882 | 0.000908725 | 0.898298676 |
| T2DM | LEC | *SATB1* | 0.91464576 | 0.000624237 | 0.001705358 | 1 |
| T2DM | LEC | *ACTL10* | -0.405424103 | 0.000113069 | 0.000908725 | 0.710862703 |
| T2DM | LEC | *TRAK1* | 0.788057424 | 0.005054608 | 0.008635413 | 1 |
| T2DM | LEC | *SKA2* | 0.80603277 | 0.000213237 | 0.0009784 | 1 |
| T2DM | LEC | *TTC3* | 0.562133767 | 7.11E-06 | 0.000612246 | 0.044693994 |
| T2DM | LEC | *KCNB2* | 0.22968236 | 0.000455875 | 0.001383246 | 1 |
| T2DM | LEC | *BRICD5* | -0.790403578 | 0.014756295 | 0.021747029 | 1 |
| T2DM | LEC | *TSKU* | 1.167323604 | 0.000627729 | 0.001711418 | 1 |
| T2DM | LEC | *ACBD3* | 0.877371806 | 0.001054506 | 0.002445543 | 1 |
| T2DM | LEC | *PLCXD3* | -0.450290594 | 0.000795516 | 0.002000018 | 1 |
| T2DM | LEC | *GPIHBP1* | -0.674829129 | 0.002710299 | 0.005097114 | 1 |
| T2DM | LEC | *VMO1* | 0.604859675 | 8.35E-05 | 0.000908725 | 0.524973981 |
| T2DM | LEC | *COL18A1* | -0.589061971 | 0.000630261 | 0.00171385 | 1 |
| T2DM | LEC | *SLC25A18* | 0.344357734 | 0.000603553 | 0.00166427 | 1 |
| T2DM | LEC | *LENG9* | 0.794768469 | 0.000164685 | 0.000910705 | 1 |
| T2DM | LEC | *CEP63* | 0.44317332 | 0.006387906 | 0.010521552 | 1 |
| T2DM | LEC | *HMGN4* | 0.964682209 | 0.01347691 | 0.02006542 | 1 |
| T2DM | LEC | *ZNF320* | 0.600644368 | 0.000101414 | 0.000908725 | 0.637588859 |
| T2DM | LEC | *LYSMD4* | 0.569119687 | 0.002745252 | 0.005150523 | 1 |
| T2DM | LEC | *WBP2NL* | 1.14779319 | 0.000977258 | 0.002312173 | 1 |
| T2DM | LEC | *BEGAIN* | 4.194562996 | 0.000534 | 0.001530525 | 1 |
| T2DM | LEC | *FAM43B* | 1.420630193 | 0.000174133 | 0.000911117 | 1 |
| T2DM | LEC | *CSMD1* | 0.652124629 | 0.000737608 | 0.00190367 | 1 |
| T2DM | LEC | *PTGDR2* | -0.978261582 | 0.000159249 | 0.000910705 | 1 |
| T2DM | LEC | *GPR19* | 1.227547257 | 3.63E-05 | 0.000908725 | 0.228109323 |
| T2DM | LEC | *FANCF* | -1.391855925 | 0.001040808 | 0.002426236 | 1 |
| T2DM | LEC | *C2CD4C* | 1.329631474 | 0.000144792 | 0.000908725 | 0.910310052 |
| T2DM | LEC | *RUVBL2* | 1.328736476 | 0.000441056 | 0.001351326 | 1 |
| T2DM | LEC | *GDPGP1* | -0.670508059 | 0.021429995 | 0.030331017 | 1 |
| T2DM | LEC | *CTNNA3* | 0.688940272 | 0.01909205 | 0.027364526 | 1 |
| T2DM | LEC | *PLGLB1* | 1.430307081 | 0.000234829 | 0.001001608 | 1 |
| T2DM | LEC | *CCBE1* | 0.58578546 | 0.000157455 | 0.000910705 | 0.98991695 |
| T2DM | LEC | *ZNF623* | 0.795324625 | 0.000259532 | 0.001042963 | 1 |
| T2DM | LEC | *CCDC125* | 0.714762825 | 0.000228757 | 0.000995303 | 1 |
| T2DM | LEC | *FHL3* | 0.648327366 | 0.000473045 | 0.001412844 | 1 |
| T2DM | LEC | *C19orf71* | 1.057249394 | 0.000422211 | 0.001328547 | 1 |
| T2DM | LEC | *LRIT3* | 1.202509281 | 0.00021359 | 0.0009784 | 1 |
| T2DM | LEC | *NPIPA1* | 0.492875426 | 0.001178069 | 0.002649919 | 1 |
| T2DM | LEC | *SF3A3* | 1.152466536 | 0.003110709 | 0.00568849 | 1 |
| T2DM | LEC | *GRIN2A* | 0.873392871 | 0.000693835 | 0.001825927 | 1 |
| T2DM | LEC | *URAD* | 1.749866282 | 0.001104616 | 0.002530657 | 1 |
| T2DM | LEC | *GPR132* | 0.869985116 | 0.000246331 | 0.00101887 | 1 |
| T2DM | LEC | *MX2* | 0.609793097 | 0.000496378 | 0.001454972 | 1 |
| T2DM | LEC | *PSMG1* | 0.431596841 | 0.014319185 | 0.021187271 | 1 |
| T2DM | LEC | *ACSM5* | 0.705412609 | 4.44E-05 | 0.000908725 | 0.279270029 |
| T2DM | LEC | *AC131971.1* | -1.100100737 | 0.000231132 | 0.001001467 | 1 |
| T2DM | LEC | *SERHL2* | 0.87574843 | 0.000101546 | 0.000908725 | 0.638417578 |
| T2DM | LEC | *PCBP3* | 0.112759888 | 0.03276384 | 0.044384026 | 1 |
| T2DM | LEC | *TANGO2* | 0.752963964 | 0.000149873 | 0.000910197 | 0.942252088 |

| | | | | | | |
|---|---|---|---|---|---|---|
| T2DM | LEC | *SFXN4* | 0.873968391 | 0.002126991 | 0.004213356 | 1 |
| T2DM | LEC | *FAM167B* | 1.911179972 | 0.001494096 | 0.003189604 | 1 |
| T2DM | LEC | *MRPL54* | -0.641889181 | 0.00132232 | 0.002897675 | 1 |
| T2DM | LEC | *NDUFB1* | 1.023243815 | 0.000436287 | 0.00134367 | 1 |
| T2DM | LEC | *FAM19A1* | 0.389924431 | 0.000111652 | 0.000908725 | 0.701956529 |
| T2DM | LEC | *TRMT12* | 0.6698743 | 0.000502903 | 0.001467078 | 1 |
| T2DM | LEC | *BMP8A* | 1.689736435 | 3.91E-08 | 1.47E-05 | 0.000245929 |
| T2DM | LEC | *CMTM4* | 0.963459118 | 0.001943015 | 0.00392789 | 1 |
| T2DM | LEC | *CBX6* | 1.403168204 | 0.001277078 | 0.002818179 | 1 |
| T2DM | LEC | *AIFM3* | 1.550717625 | 0.023718019 | 0.033247533 | 1 |
| T2DM | LEC | *KCTD16* | 1.380165616 | 0.00255689 | 0.00485512 | 1 |
| T2DM | LEC | *B3GALT5* | 1.505608328 | 0.001283941 | 0.00282836 | 1 |
| T2DM | LEC | *ZNF703* | 0.451499097 | 0.015698134 | 0.022978852 | 1 |
| T2DM | LEC | *KCTD8* | 0.305035794 | 5.15E-05 | 0.000908725 | 0.32396617 |
| T2DM | LEC | *C9orf66* | 1.453322992 | 7.72E-05 | 0.000908725 | 0.485380819 |
| T2DM | LEC | *TUBA8* | 0.798257992 | 2.06E-05 | 0.000908725 | 0.129413733 |
| T2DM | LEC | *FAM162B* | -0.313174867 | 0.000300705 | 0.001121313 | 1 |
| T2DM | LEC | *BTBD9* | 0.668471114 | 0.000407734 | 0.001307186 | 1 |
| T2DM | LEC | *NUDT14* | 0.872768282 | 0.001128217 | 0.002569964 | 1 |
| T2DM | LEC | *ANKRD45* | 1.157374432 | 0.00011443 | 0.000908725 | 0.719422977 |
| T2DM | LEC | *MAATS1* | 0.50904049 | 5.85E-05 | 0.000908725 | 0.367669137 |
| T2DM | LEC | *GPR39* | 0.523338686 | 0.000146147 | 0.000908725 | 0.918823311 |
| T2DM | LEC | *NPIPA7* | -1.957864666 | 9.08E-05 | 0.000908725 | 0.571155975 |
| T2DM | LEC | *PP2D1* | 0.940377402 | 2.59E-05 | 0.000908725 | 0.162837105 |
| T2DM | LEC | *DENND5A* | 0.78923866 | 0.008952341 | 0.014056786 | 1 |
| T2DM | LEC | *TMEM236* | 0.593652252 | 0.000114948 | 0.000908725 | 0.722677556 |
| T2DM | LEC | *VPS33B* | 1.31338886 | 0.00102246 | 0.002395903 | 1 |
| T2DM | LEC | *TBX1* | -0.061747625 | 0.000133703 | 0.000908725 | 0.840589772 |
| T2DM | LEC | *CNTN2* | 0.770021986 | 9.25E-05 | 0.000908725 | 0.581298388 |
| T2DM | LEC | *CRELD2* | 0.705819811 | 0.000119922 | 0.000908725 | 0.753947037 |
| T2DM | LEC | *UBE2F* | 0.91136323 | 0.015870997 | 0.023199479 | 1 |
| T2DM | LEC | *GOLGA6L4* | -0.651667561 | 0.000204374 | 0.000964732 | 1 |
| T2DM | LEC | *PCDH9* | 0.318552806 | 0.006135555 | 0.01015646 | 1 |
| T2DM | LEC | *OAF* | -0.505571868 | 0.000576301 | 0.001612173 | 1 |
| T2DM | LEC | *CLECL1* | 0.767835312 | 0.000119809 | 0.000908725 | 0.75324032 |
| T2DM | LEC | *MROH7* | 1.963423022 | 3.99E-06 | 0.000465666 | 0.025064728 |
| T2DM | LEC | *GDF3* | -0.455838884 | 0.000166663 | 0.000910705 | 1 |
| T2DM | LEC | *CSF1* | 0.719822665 | 2.76E-05 | 0.000908725 | 0.173658711 |
| T2DM | LEC | *PLA2G6* | 0.793701231 | 3.96E-05 | 0.000908725 | 0.248784981 |
| T2DM | LEC | *SS18L1* | 0.669859439 | 0.000116387 | 0.000908725 | 0.731722758 |
| T2DM | LEC | *THAP7* | 1.208580511 | 0.000280452 | 0.00108013 | 1 |
| T2DM | LEC | *CCR10* | 0.765028164 | 7.30E-05 | 0.000908725 | 0.45909651 |
| T2DM | LEC | *WDR27* | 0.880737048 | 0.000572315 | 0.001603699 | 1 |
| T2DM | LEC | *TXNRD2* | -0.842580358 | 0.005769316 | 0.009651859 | 1 |
| T2DM | LEC | *PROS1* | 0.841602685 | 0.01506596 | 0.0221256 | 1 |
| T2DM | LEC | *LPAR5* | 0.395920588 | 0.007025409 | 0.011419015 | 1 |
| T2DM | LEC | *TMEM173* | -0.852103893 | 0.008310375 | 0.01318736 | 1 |
| T2DM | LEC | *SNN* | 1.222241326 | 0.000113863 | 0.000908725 | 0.715858438 |
| T2DM | LEC | *NELL2* | 0.310332487 | 0.000121826 | 0.000908725 | 0.765921294 |
| T2DM | LEC | *KRBA2* | -1.184478636 | 0.000339709 | 0.001187188 | 1 |

| | | | | | | |
|---|---|---|---|---|---|---|
| T2DM | LEC | *SEPT5* | 1.753516145 | 9.91E-05 | 0.000908725 | 0.623287284 |
| T2DM | LEC | *ATL3* | 0.666196966 | 0.001208411 | 0.002702697 | 1 |
| T2DM | LEC | *NDUFA12* | 1.112901301 | 0.000502148 | 0.001466241 | 1 |
| T2DM | LEC | *UBE2G2* | 0.725934084 | 0.002237079 | 0.004369218 | 1 |
| T2DM | LEC | *OSBP2* | -0.185344828 | 0.000177679 | 0.000918726 | 1 |
| T2DM | LEC | *TUSC5* | 0.540236323 | 2.80E-05 | 0.000908725 | 0.176158335 |
| T2DM | LEC | *TMED9* | 1.275215099 | 0.000271967 | 0.001070668 | 1 |
| T2DM | LEC | *H1FX* | 0.754908532 | 0.000284728 | 0.001090187 | 1 |
| T2DM | LEC | *RBM43* | 1.000798345 | 0.000444286 | 0.001357774 | 1 |
| T2DM | LEC | *NUTM2A* | 1.32465174 | 0.011979222 | 0.018086784 | 1 |
| T2DM | LEC | *NDUFA6* | 0.462838507 | 0.003950275 | 0.006948903 | 1 |
| T2DM | LEC | *SORCS2* | 0.989431438 | 0.000174108 | 0.000911117 | 1 |
| T2DM | LEC | *TMEM106A* | 0.847698837 | 4.91E-05 | 0.000908725 | 0.308909782 |
| T2DM | LEC | *BRF1* | 0.884913073 | 0.00033341 | 0.001176051 | 1 |
| T2DM | LEC | *CIB1* | 0.705904608 | 0.019149881 | 0.027431146 | 1 |
| T2DM | LEC | *FLRT2* | -0.254364022 | 0.000147289 | 0.000908725 | 0.926005802 |
| T2DM | LEC | *ADSSL1* | 2.344483587 | 0.000233653 | 0.001001608 | 1 |
| T2DM | LEC | *C6orf120* | 0.62958123 | 0.000317663 | 0.001152864 | 1 |
| T2DM | LEC | *SIGIRR* | 0.814208784 | 0.025978963 | 0.036055131 | 1 |
| T2DM | LEC | *IFITM2* | 0.990367576 | 0.003372734 | 0.006101979 | 1 |
| T2DM | LEC | *GP1BA* | 1.426828979 | 3.11E-05 | 0.000908725 | 0.195652586 |
| T2DM | LEC | *RBM11* | 2.054640321 | 0.001408176 | 0.003046525 | 1 |
| T2DM | LEC | *CCDC137* | 0.963579699 | 8.88E-05 | 0.000908725 | 0.558016323 |
| T2DM | LEC | *TMEM105* | 8.757232887 | 0.000201697 | 0.00096305 | 1 |
| T2DM | LEC | *GAS2L1* | -1.00887179 | 0.000243112 | 0.001015577 | 1 |
| T2DM | LEC | *ATP6V0A2* | 0.879157005 | 0.02486907 | 0.034689274 | 1 |
| T2DM | LEC | *HS6ST3* | -0.596671989 | 0.000245167 | 0.001017515 | 1 |
| T2DM | LEC | *TNFAIP8L1* | -0.350906563 | 0.00038256 | 0.001263636 | 1 |
| T2DM | LEC | *MAPK11* | 0.678331865 | 0.000265987 | 0.00105572 | 1 |
| T2DM | LEC | *SP140L* | 0.828536435 | 0.035441084 | 0.047682024 | 1 |
| T2DM | LEC | *SH3BGR* | -0.320239621 | 0.001794195 | 0.003687515 | 1 |
| T2DM | LEC | *FAM174B* | -0.717383147 | 0.02396806 | 0.03354568 | 1 |
| T2DM | LEC | *MUC1* | 0.851215053 | 3.04E-05 | 0.000908725 | 0.191417547 |
| T2DM | LEC | *SV2B* | -0.197927437 | 0.000141429 | 0.000908725 | 0.889166669 |
| T2DM | LEC | *PDE6G* | 1.657471437 | 8.74E-05 | 0.000908725 | 0.549557658 |
| T2DM | LEC | *TLCD2* | 0.977440381 | 0.001046712 | 0.002432782 | 1 |
| T2DM | LEC | *LSAMP* | 0.685562887 | 0.000235536 | 0.001003942 | 1 |
| T2DM | LEC | *AHNAK2* | 1.007969672 | 0.000429619 | 0.001338907 | 1 |
| T2DM | LEC | *SP1* | 0.708647264 | 0.000343128 | 0.001193551 | 1 |
| T2DM | LEC | *PDIA2* | 1.091846882 | 3.74E-05 | 0.000908725 | 0.234997793 |
| T2DM | LEC | *PSMD13* | 1.000035675 | 0.008281493 | 0.013157884 | 1 |
| T2DM | LEC | *PBX1* | 0.583586465 | 0.018167809 | 0.026161478 | 1 |
| T2DM | LEC | *UBE2L3* | 1.096690322 | 0.011600054 | 0.017598827 | 1 |
| T2DM | LEC | *PMEL* | 0.50645087 | 0.000696377 | 0.001830319 | 1 |
| T2DM | LEC | *SNAI3* | 1.338014386 | 0.000770979 | 0.00195765 | 1 |
| T2DM | LEC | *MYBL1* | 0.253289391 | 0.004029974 | 0.00706735 | 1 |
| T2DM | LEC | *C16orf52* | 0.837140138 | 0.017428995 | 0.02520729 | 1 |
| T2DM | LEC | *ADARB2* | 0.433121937 | 0.000115976 | 0.000908725 | 0.729140877 |
| T2DM | LEC | *SRL* | 0.396261484 | 0.000520015 | 0.00150245 | 1 |
| T2DM | LEC | *KCNIP4* | 0.38354836 | 0.01904861 | 0.027323435 | 1 |

| | | | | | | |
|---|---|---|---|---|---|---|
| T2DM | LEC | *SLC52A2* | -1.130262322 | 0.001549103 | 0.003278092 | 1 |
| T2DM | LEC | *PIGP* | -0.336434101 | 0.005829384 | 0.009736806 | 1 |
| T2DM | LEC | *ARL17A* | 0.827194865 | 0.000694138 | 0.00182596 | 1 |
| T2DM | LEC | *NPIPB4* | 0.531760433 | 0.000135834 | 0.000908725 | 0.853985874 |
| T2DM | LEC | *ZNF829* | 0.624286302 | 0.000131826 | 0.000908725 | 0.828792777 |
| T2DM | LEC | *TRIM69* | 0.663754145 | 0.019937951 | 0.028463984 | 1 |
| T2DM | LEC | *IFITM1* | 1.17099138 | 0.000280428 | 0.00108013 | 1 |
| T2DM | LEC | *POMK* | 0.679004722 | 6.90E-05 | 0.000908725 | 0.433615996 |
| T2DM | LEC | *LINC00839* | -0.14536866 | 0.000344859 | 0.001196042 | 1 |
| T2DM | LEC | *PTCH1* | -0.854127699 | 0.001931009 | 0.003914949 | 1 |
| T2DM | LEC | *RTN4RL1* | 1.216940338 | 0.000975717 | 0.002310484 | 1 |
| T2DM | LEC | *PLK5* | 0.920034393 | 0.009796633 | 0.015170304 | 1 |
| T2DM | LEC | *RASA3* | 0.449537617 | 0.002126765 | 0.004213356 | 1 |
| T2DM | LEC | *ZNF566* | 0.611770153 | 0.000758173 | 0.001933725 | 1 |
| T2DM | LEC | *ZNF529* | 1.037634879 | 0.001861491 | 0.003798505 | 1 |
| T2DM | LEC | *ZNF284* | 0.764181714 | 0.000109519 | 0.000908725 | 0.688547866 |
| T2DM | LEC | *GSAP* | 0.880345 | 0.000131378 | 0.000908725 | 0.825973118 |
| T2DM | LEC | *C2orf76* | 0.718633045 | 0.006731598 | 0.011006907 | 1 |
| T2DM | LEC | *WWOX* | 0.593465582 | 0.005447114 | 0.009186161 | 1 |
| T2DM | LEC | *CYP4Z1* | 0.896128381 | 5.39E-05 | 0.000908725 | 0.338694237 |
| T2DM | LEC | *CCDC84* | 0.904526711 | 0.001185648 | 0.002663155 | 1 |
| T2DM | LEC | *FFAR4* | 0.348972579 | 0.007510162 | 0.012106766 | 1 |
| T2DM | LEC | *BPIFB4* | 28.91776807 | 3.01E-05 | 0.000908725 | 0.189326144 |
| T2DM | LEC | *CYP4F12* | 0.629030773 | 0.002775672 | 0.005193646 | 1 |
| T2DM | LEC | *KLHL32* | 0.448741403 | 0.000143927 | 0.000908725 | 0.904866836 |
| T2DM | LEC | *NBPF12* | 0.829097919 | 0.000793165 | 0.001997848 | 1 |
| T2DM | LEC | *GPAT2* | -0.307918513 | 0.000144335 | 0.000908725 | 0.907435249 |
| T2DM | LEC | *TOR3A* | 1.021764851 | 0.000149562 | 0.000910197 | 0.940294296 |
| T2DM | LEC | *CYP4X1* | -0.402996685 | 0.001197749 | 0.002683623 | 1 |
| T2DM | LEC | *GOLGA8R* | -1.233589314 | 0.001189301 | 0.002667547 | 1 |
| T2DM | LEC | *CCDC30* | 0.808700013 | 0.001144083 | 0.00259576 | 1 |
| T2DM | LEC | *GLDN* | 0.609467143 | 0.000634016 | 0.001718869 | 1 |
| T2DM | LEC | *FCAR* | 0.748042266 | 0.000154341 | 0.000910197 | 0.970339739 |
| T2DM | LEC | *KPNA4* | 0.883703749 | 0.006060521 | 0.010048127 | 1 |
| T2DM | LEC | *ZNF501* | 1.028536292 | 0.010687612 | 0.016404546 | 1 |
| T2DM | LEC | *PCLO* | -0.257062973 | 0.000507723 | 0.0014768 | 1 |
| T2DM | LEC | *RGS7BP* | 0.544188146 | 0.018415655 | 0.026494102 | 1 |
| T2DM | LEC | *CLCNKA* | 1.41914935 | 0.016255394 | 0.023700756 | 1 |
| T2DM | LEC | *SMYD4* | 0.806503402 | 0.005171233 | 0.008798793 | 1 |
| T2DM | LEC | *CEACAM19* | -0.798943825 | 0.010214519 | 0.015759185 | 1 |
| T2DM | LEC | *NF2* | 0.867246019 | 0.034484311 | 0.046464394 | 1 |
| T2DM | LEC | *MIR22HG* | 0.691109828 | 0.000688506 | 0.001819172 | 1 |
| T2DM | LEC | *PDE2A* | -0.368974916 | 0.000347981 | 0.001200743 | 1 |
| T2DM | LEC | *C17orf58* | 0.767037162 | 0.000233042 | 0.001001608 | 1 |
| T2DM | LEC | *CYP27C1* | 0.437113702 | 0.000740495 | 0.001906556 | 1 |
| T2DM | LEC | *BCR* | -0.513674452 | 0.000435653 | 0.00134367 | 1 |
| T2DM | LEC | *HYAL3* | -0.707073773 | 0.004787735 | 0.008219686 | 1 |
| T2DM | LEC | *LILRB4* | 0.392036123 | 6.04E-05 | 0.000908725 | 0.379500089 |
| T2DM | LEC | *KRT16* | 0.7594107 | 0.001652426 | 0.003458323 | 1 |
| T2DM | LEC | *TRABD2A* | 0.29415052 | 0.000299499 | 0.001118805 | 1 |

| | | | | | | |
|---|---|---|---|---|---|---|
| T2DM | LEC | *QRFPR* | 1.540774294 | 0.035637721 | 0.047915815 | 1 |
| T2DM | LEC | *TMEM17* | -0.778152752 | 0.002226417 | 0.004353805 | 1 |
| T2DM | LEC | *TNFRSF18* | 1.166512705 | 0.00010431 | 0.000908725 | 0.655794717 |
| T2DM | LEC | *RTN4RL2* | -0.698344893 | 0.000207737 | 0.000972609 | 1 |
| T2DM | LEC | *TMEM232* | 0.862260038 | 0.003147893 | 0.005749797 | 1 |
| T2DM | LEC | *EFCAB6* | 0.843093922 | 5.01E-05 | 0.000908725 | 0.314995027 |
| T2DM | LEC | *KANK3* | -0.199364144 | 0.000153059 | 0.000910197 | 0.962281129 |
| T2DM | LEC | *EMID1* | -0.268196832 | 0.000132478 | 0.000908725 | 0.83289073 |
| T2DM | LEC | *CYP4A11* | 0.987458156 | 0.0002783 | 0.00108013 | 1 |
| T2DM | LEC | *TMEM216* | 1.51168552 | 0.000103661 | 0.000908725 | 0.651719661 |
| T2DM | LEC | *RPS19BP1* | 0.902327044 | 0.000197304 | 0.000956093 | 1 |
| T2DM | LEC | *C3orf70* | -0.33939364 | 0.002754004 | 0.005162321 | 1 |
| T2DM | LEC | *PLCD1* | 0.943384924 | 0.006901474 | 0.011246648 | 1 |
| T2DM | LEC | *ENTPD5* | 1.166459765 | 0.000838899 | 0.002076816 | 1 |
| T2DM | LEC | *HEATR4* | 0.603993768 | 0.000172228 | 0.000910705 | 1 |
| T2DM | LEC | *MRPS21* | 0.918679427 | 0.000433749 | 0.001342678 | 1 |
| T2DM | LEC | *ANGPTL5* | 0.373959315 | 0.000108093 | 0.000908725 | 0.679582976 |
| T2DM | LEC | *TSPYL4* | -0.783935435 | 0.009431834 | 0.014693101 | 1 |
| T2DM | LEC | *MT1X* | 0.366771495 | 8.57E-05 | 0.000908725 | 0.538757884 |
| T2DM | LEC | *FNBP1* | 0.571298461 | 0.037266855 | 0.049871588 | 1 |
| T2DM | LEC | *WDR86* | 0.512309979 | 0.000308814 | 0.001134063 | 1 |
| T2DM | LEC | *EPOR* | 0.60689611 | 5.54E-05 | 0.000908725 | 0.34845068 |
| T2DM | LEC | *FPR3* | 0.262893341 | 0.000138144 | 0.000908725 | 0.868512862 |
| T2DM | LEC | *GJA4* | -0.403320093 | 0.00035615 | 0.001216909 | 1 |
| T2DM | LEC | *TLR5* | 0.493258219 | 0.000105062 | 0.000908725 | 0.660527596 |
| T2DM | LEC | *PLEKHN1* | -0.902490047 | 0.002898651 | 0.005378931 | 1 |
| T2DM | LEC | *ZNF385C* | 0.694358397 | 0.000128918 | 0.000908725 | 0.810505257 |
| T2DM | LEC | *TET3* | 0.853216284 | 0.000432013 | 0.001341345 | 1 |
| T2DM | LEC | *C17orf97* | 0.256209662 | 0.022086344 | 0.031147789 | 1 |
| T2DM | LEC | *TRPV2* | 0.752201371 | 0.000132317 | 0.000908725 | 0.831878684 |
| T2DM | LEC | *C2orf88* | 0.884827102 | 0.001552704 | 0.003283501 | 1 |
| T2DM | LEC | *FANCA* | 0.652243997 | 0.002188343 | 0.00429773 | 1 |
| T2DM | LEC | *ADH1A* | 0.475186543 | 0.023326449 | 0.032749752 | 1 |
| T2DM | LEC | *FAM69C* | 1.068157628 | 0.000113553 | 0.000908725 | 0.713905163 |
| T2DM | LEC | *DNAH17* | -1.371626685 | 0.001457567 | 0.003130757 | 1 |
| T2DM | LEC | *ZFP69* | 1.035613051 | 0.00819123 | 0.013044139 | 1 |
| T2DM | LEC | *TMEM220* | 0.92780419 | 0.001082741 | 0.002491652 | 1 |
| T2DM | LEC | *GFRAL* | 0.79428023 | 3.39E-05 | 0.000908725 | 0.212975696 |
| T2DM | LEC | *C2orf66* | 3.552106627 | 0.009866209 | 0.015263006 | 1 |
| T2DM | LEC | *CPSF4L* | -0.498012846 | 9.70E-05 | 0.000908725 | 0.609905136 |
| T2DM | LEC | *KLHL17* | -0.929007296 | 0.000496809 | 0.001454972 | 1 |
| T2DM | LEC | *ZSCAN23* | 0.303802592 | 0.001795599 | 0.003688191 | 1 |
| T2DM | LEC | *OR7D2* | -1.218712905 | 0.025767187 | 0.03580073 | 1 |
| T2DM | LEC | *MORN2* | 0.475632513 | 0.003661343 | 0.006524355 | 1 |
| T2DM | LEC | *C6orf132* | 0.520982522 | 0.00015768 | 0.000910705 | 0.991332288 |
| T2DM | LEC | *MAPK12* | -0.523495246 | 0.000156806 | 0.000910705 | 0.985842366 |
| T2DM | LEC | *AGRN* | -0.298410586 | 0.000220376 | 0.00098409 | 1 |
| T2DM | LEC | *FAM166A* | 1.935779613 | 0.000114212 | 0.000908725 | 0.718053764 |
| T2DM | LEC | *ZNF626* | -1.018418209 | 0.006873897 | 0.011212884 | 1 |
| T2DM | LEC | *SMTNL2* | 2.782196292 | 0.000110506 | 0.000908725 | 0.694750987 |

| | | | | | | |
|---|---|---|---|---|---|---|
| T2DM | LEC | *ZC3H6* | 0.817117975 | 0.005198636 | 0.00883108 | 1 |
| T2DM | LEC | *LINC00265* | 0.963355969 | 8.37E-05 | 0.000908725 | 0.526307668 |
| T2DM | LEC | *LAMTOR4* | 0.993133841 | 0.014155997 | 0.020970488 | 1 |
| T2DM | LEC | *PRKAR1B* | -0.214282578 | 0.000144631 | 0.000908725 | 0.909295478 |
| T2DM | LEC | *NUTM2B* | -6.789984133 | 1.01E-05 | 0.000749023 | 0.06366698 |
| T2DM | LEC | *NCR3LG1* | 2.020007398 | 0.000126919 | 0.000908725 | 0.797938021 |
| T2DM | LEC | *HYKK* | 0.843185001 | 6.57E-05 | 0.000908725 | 0.413000824 |
| T2DM | LEC | *RUFY4* | 0.994830266 | 0.000116801 | 0.000908725 | 0.73432628 |
| T2DM | LEC | *CENPP* | 0.422715111 | 0.001011445 | 0.002373631 | 1 |
| T2DM | LEC | *PLSCR1* | 0.981365742 | 0.004750163 | 0.008164099 | 1 |
| T2DM | LEC | *ZNF559* | 0.397383691 | 0.018717685 | 0.026897848 | 1 |
| T2DM | LEC | *ZP3* | -0.84565218 | 0.001775544 | 0.003661149 | 1 |
| T2DM | LEC | *JAKMIP3* | 0.566256735 | 5.85E-05 | 0.000908725 | 0.367888771 |
| T2DM | LEC | *PDCD1* | 2.846006981 | 0.000940511 | 0.002246579 | 1 |
| T2DM | LEC | *CERKL* | -1.371244317 | 0.029018702 | 0.039834187 | 1 |
| T2DM | LEC | *C19orf54* | 0.549227076 | 0.000124767 | 0.000908725 | 0.784408139 |
| T2DM | LEC | *COL25A1* | 0.689840919 | 0.019924195 | 0.028456023 | 1 |
| T2DM | LEC | *SRSF10* | 0.87009884 | 0.005352058 | 0.00905744 | 1 |
| T2DM | LEC | *NBR1* | 0.996783182 | 0.002940677 | 0.005440859 | 1 |
| T2DM | LEC | *NDOR1* | 0.893684588 | 0.000148668 | 0.000910197 | 0.934678588 |
| T2DM | LEC | *FAM72B* | 3.093378951 | 0.00013042 | 0.000908725 | 0.819949883 |
| T2DM | LEC | *SUMO2* | 1.281183699 | 0.001832576 | 0.003751678 | 1 |
| T2DM | LEC | *GOLGA8M* | 1.480582284 | 0.011425162 | 0.017358626 | 1 |
| T2DM | LEC | *DPYD* | 0.582894517 | 0.00052598 | 0.001514119 | 1 |
| T2DM | LEC | *S100A16* | -0.950132816 | 0.004418165 | 0.007656285 | 1 |
| T2DM | LEC | *RHCE* | 0.552490625 | 0.000265877 | 0.00105572 | 1 |
| T2DM | LEC | *PARVB* | 0.562560182 | 0.012047554 | 0.01816814 | 1 |
| T2DM | LEC | *SLC4A5* | -0.588218811 | 0.000170103 | 0.000910705 | 1 |
| T2DM | LEC | *UROS* | 1.291338825 | 0.000642002 | 0.001733394 | 1 |
| T2DM | LEC | *FAM221A* | 0.987634839 | 0.001113681 | 0.002546078 | 1 |
| T2DM | LEC | *NOXA1* | 1.166702851 | 0.000283806 | 0.001087982 | 1 |
| T2DM | LEC | *SKOR1* | 2.037961924 | 6.90E-05 | 0.000908725 | 0.433998799 |
| T2DM | LEC | *TMEM201* | 1.65323199 | 6.28E-06 | 0.000598155 | 0.039478243 |
| T2DM | LEC | *ZDHHC11* | 0.958564222 | 0.001684024 | 0.003513925 | 1 |
| T2DM | LEC | *FAM26F* | 0.480660164 | 0.000479598 | 0.001424968 | 1 |
| T2DM | LEC | *LINC00910* | -1.362797433 | 0.009654338 | 0.014986869 | 1 |
| T2DM | LEC | *BEND4* | 2.313995873 | 4.48E-05 | 0.000908725 | 0.281347816 |
| T2DM | LEC | *ZNF563* | 0.600259302 | 8.69E-05 | 0.000908725 | 0.546198853 |
| T2DM | LEC | *FBF1* | 0.367079744 | 0.002799883 | 0.005234273 | 1 |
| T2DM | LEC | *LRRK2* | 0.970193829 | 0.00068052 | 0.001802884 | 1 |
| T2DM | LEC | *FAM196A* | 0.916932255 | 0.005933397 | 0.009886897 | 1 |
| T2DM | LEC | *UTS2B* | 0.475405563 | 0.000714191 | 0.001861576 | 1 |
| T2DM | LEC | *C9orf152* | 1.100611494 | 0.000111152 | 0.000908725 | 0.698810611 |
| T2DM | LEC | *NELFB* | 0.940824613 | 0.000210755 | 0.0009784 | 1 |
| T2DM | LEC | *SLC15A5* | 0.404271741 | 4.01E-05 | 0.000908725 | 0.252068623 |
| T2DM | LEC | *LIPI* | -0.484988481 | 0.004422696 | 0.007662025 | 1 |
| T2DM | LEC | *ADAT2* | 1.068474676 | 7.07E-05 | 0.000908725 | 0.444190405 |
| T2DM | LEC | *ZNF567* | 1.237534334 | 0.002072061 | 0.004122483 | 1 |
| T2DM | LEC | *ANKDD1B* | -0.318517514 | 7.44E-05 | 0.000908725 | 0.467580144 |
| T2DM | LEC | *LITAF* | 0.67914914 | 0.000250437 | 0.001023586 | 1 |

| | | | | | | |
|---|---|---|---|---|---|---|
| T2DM | LEC | *VSTM1* | 1.363896923 | 0.001903527 | 0.003869212 | 1 |
| T2DM | LEC | *SF3B3* | 0.938692232 | 0.00071301 | 0.001861576 | 1 |
| T2DM | LEC | *SP6* | -1.951562101 | 9.17E-05 | 0.000908725 | 0.57645883 |
| T2DM | LEC | *PLAC9* | 0.469563893 | 0.021937856 | 0.030980076 | 1 |
| T2DM | LEC | *CLDN4* | 0.757053576 | 0.016955497 | 0.024601709 | 1 |
| T2DM | LEC | *ZNF573* | 1.065263881 | 0.00277098 | 0.005187955 | 1 |
| T2DM | LEC | *S100A13* | 0.973764565 | 0.000166641 | 0.000910705 | 1 |
| T2DM | LEC | *C15orf61* | 1.594103107 | 0.000414641 | 0.001316589 | 1 |
| T2DM | LEC | *PNRC2* | 0.975341774 | 0.000421181 | 0.001326934 | 1 |
| T2DM | LEC | *ZKSCAN3* | 1.123264801 | 0.033254761 | 0.0449522 | 1 |
| T2DM | LEC | *SLC35E2B* | 0.635623358 | 0.001131953 | 0.002576341 | 1 |
| T2DM | LEC | *BLOC1S2* | 0.275507906 | 0.019853862 | 0.028381362 | 1 |
| T2DM | LEC | *WDSUB1* | 1.034541214 | 0.000880747 | 0.002150853 | 1 |
| T2DM | LEC | *ZNF79* | 0.606054096 | 0.012152652 | 0.018300292 | 1 |
| T2DM | LEC | *COLCA1* | 0.723861811 | 0.001385526 | 0.003006836 | 1 |
| T2DM | LEC | *ACADSB* | 0.563870459 | 0.00290975 | 0.005393159 | 1 |
| T2DM | LEC | *SEMA4A* | 0.433494907 | 0.000101703 | 0.000908725 | 0.639408166 |
| T2DM | LEC | *MPHOSPH8* | 0.785053027 | 0.003379263 | 0.006112033 | 1 |
| T2DM | LEC | *SIRPB2* | 0.386373224 | 0.000110695 | 0.000908725 | 0.695939832 |
| T2DM | LEC | *SRGAP3* | -0.23455545 | 0.001402803 | 0.003036991 | 1 |
| T2DM | LEC | *SUPT5H* | 0.950774332 | 0.014624 | 0.021562169 | 1 |
| T2DM | LEC | *ZNF107* | 0.746730257 | 0.004140374 | 0.007232712 | 1 |
| T2DM | LEC | *PPIA* | 0.969559372 | 0.000522622 | 0.00150666 | 1 |
| T2DM | LEC | *GTF2IRD2* | 1.271737899 | 6.02E-05 | 0.000908725 | 0.378294983 |
| T2DM | LEC | *GRM7* | 0.437424088 | 0.004354102 | 0.007574499 | 1 |
| T2DM | LEC | *POM121* | 0.698820021 | 0.004200981 | 0.007328405 | 1 |
| T2DM | LEC | *GIMAP5* | -0.290253275 | 0.000276108 | 0.00108013 | 1 |
| T2DM | LEC | *STK31* | 0.689919684 | 0.000111618 | 0.000908725 | 0.70174196 |
| T2DM | LEC | *ZKSCAN7* | 0.43281797 | 0.000112958 | 0.000908725 | 0.710168421 |
| T2DM | LEC | *ZNF565* | 0.765412344 | 0.00914405 | 0.014310569 | 1 |
| T2DM | LEC | *NTNG2* | -1.605794864 | 0.007718001 | 0.012384654 | 1 |
| T2DM | LEC | *WDR5* | 1.368910395 | 0.000795617 | 0.002000018 | 1 |
| T2DM | LEC | *LONP1* | 0.847888843 | 0.011931041 | 0.018018365 | 1 |
| T2DM | LEC | *ASB13* | -1.074371979 | 0.006049565 | 0.010035255 | 1 |
| T2DM | LEC | *SLC35F1* | 1.07677585 | 0.000388485 | 0.001271424 | 1 |
| T2DM | LEC | *INCA1* | 0.884444902 | 3.87E-05 | 0.000908725 | 0.24308068 |
| T2DM | LEC | *ZNF774* | -1.244251375 | 0.00177826 | 0.003661947 | 1 |
| T2DM | LEC | *ZNF124* | 0.803250216 | 0.000262892 | 0.001048732 | 1 |
| T2DM | LEC | *ZNF569* | 0.868433666 | 0.017566813 | 0.025383257 | 1 |
| T2DM | LEC | *PIK3R4* | -0.625855007 | 0.017117213 | 0.024807727 | 1 |
| T2DM | LEC | *ZNF775* | -0.966212904 | 0.012135673 | 0.018283484 | 1 |
| T2DM | LEC | *RFX8* | 0.38446386 | 7.55E-05 | 0.000908725 | 0.474388873 |
| T2DM | LEC | *MYL6B* | -0.608317928 | 0.000311989 | 0.001141055 | 1 |
| T2DM | LEC | *ZNF799* | 0.665382317 | 0.00019263 | 0.000946191 | 1 |
| T2DM | LEC | *C20orf96* | 0.969387225 | 0.000143046 | 0.000908725 | 0.899332692 |
| T2DM | LEC | *SULT1A1* | 0.273053825 | 0.003409818 | 0.006156671 | 1 |
| T2DM | LEC | *ANAPC7* | 1.118835644 | 0.002485457 | 0.004748747 | 1 |
| T2DM | LEC | *TPK1* | 0.961227825 | 0.000194953 | 0.000950868 | 1 |
| T2DM | LEC | *C1orf186* | -1.231469432 | 0.003878993 | 0.00686191 | 1 |
| T2DM | LEC | *SPTSSB* | 1.420726089 | 0.000226807 | 0.000992757 | 1 |

| | | | | | | |
|---|---|---|---|---|---|---|
| T2DM | LEC | *SULF2* | -0.311731144 | 0.000404095 | 0.001300178 | 1 |
| T2DM | LEC | *XRCC2* | -0.740046607 | 0.000360424 | 0.001224756 | 1 |
| T2DM | LEC | *ADH1B* | 0.570317504 | 0.000233749 | 0.001001608 | 1 |
| T2DM | LEC | *HRH1* | 0.368411578 | 0.000118196 | 0.000908725 | 0.743095439 |
| T2DM | LEC | *ZNF502* | 0.919078306 | 0.009185481 | 0.01436188 | 1 |
| T2DM | LEC | *TRAPPC4* | 0.751642711 | 0.000740796 | 0.001906556 | 1 |
| T2DM | LEC | *TECPR2* | 0.980593576 | 0.00187612 | 0.003823393 | 1 |
| T2DM | LEC | *ZFP62* | -1.085026678 | 0.005901647 | 0.0098366 | 1 |
| T2DM | LEC | *ERI2* | 0.907987108 | 0.00024568 | 0.001017515 | 1 |
| T2DM | LEC | *TRPV1* | 0.564928559 | 1.51E-05 | 0.00090697 | 0.095231883 |
| T2DM | LEC | *ZNF33B* | -0.824767867 | 1.20E-05 | 0.000851536 | 0.075295201 |
| T2DM | LEC | *AMZ2* | 0.827849933 | 0.001840385 | 0.003765213 | 1 |
| T2DM | LEC | *ZNF431* | 0.811296165 | 0.003838581 | 0.006801904 | 1 |
| T2DM | LEC | *NF1* | 0.731319531 | 0.002002795 | 0.004013889 | 1 |
| T2DM | LEC | *DAPK1* | -0.500568368 | 0.034399051 | 0.04635945 | 1 |
| T2DM | LEC | *GM2A* | 0.772045779 | 0.002907604 | 0.005393159 | 1 |
| T2DM | LEC | *TLE1* | -0.644214045 | 0.00126982 | 0.002805116 | 1 |
| T2DM | LEC | *STRN3* | 0.663364528 | 0.027545895 | 0.038061768 | 1 |
| T2DM | LEC | *MVB12B* | -0.692002233 | 0.013858768 | 0.020578667 | 1 |
| T2DM | LEC | *C6orf106* | 0.841089045 | 8.49E-05 | 0.000908725 | 0.533969372 |
| T2DM | LEC | *PPTC7* | 0.51289026 | 0.000135528 | 0.000908725 | 0.852064828 |
| T2DM | LEC | *NHLRC2* | 0.87125522 | 0.002053761 | 0.004094417 | 1 |
| T2DM | LEC | *LAMB3* | -0.808460787 | 0.000775919 | 0.001964639 | 1 |
| T2DM | LEC | *KPNA5* | 0.909693262 | 0.004480847 | 0.007747824 | 1 |
| T2DM | LEC | *HCAR1* | 0.569929436 | 9.05E-05 | 0.000908725 | 0.569070015 |
| T2DM | LEC | *SRGAP1* | 0.660413751 | 0.000328241 | 0.001167885 | 1 |
| T2DM | LEC | *SLC39A10* | 0.766489712 | 0.001948126 | 0.003934427 | 1 |
| T2DM | LEC | *HIST1H3E* | 0.897851224 | 0.004254677 | 0.007411792 | 1 |
| T2DM | LEC | *ZNF585A* | 0.687660871 | 1.76E-05 | 0.000908725 | 0.110929204 |
| T2DM | LEC | *FUT11* | 1.397623977 | 0.00661939 | 0.010846 | 1 |
| T2DM | LEC | *ZNF138* | 0.8734022 | 0.000249734 | 0.001023586 | 1 |
| T2DM | LEC | *ZNF429* | 0.768660082 | 0.011687881 | 0.017714973 | 1 |
| T2DM | LEC | *ZNF470* | 1.090425162 | 0.025446405 | 0.035417655 | 1 |
| T2DM | LEC | *ZNF763* | 0.360386233 | 0.017652616 | 0.02548381 | 1 |
| T2DM | LEC | *ZMYM1* | 0.95646737 | 0.000772681 | 0.001959414 | 1 |
| T2DM | LEC | *IGF2R* | 0.735230231 | 0.009649078 | 0.014986104 | 1 |
| T2DM | LEC | *GAL3ST4* | 0.797972502 | 2.23E-05 | 0.000908725 | 0.139910498 |
| T2DM | LEC | *SLC25A29* | 1.091843512 | 7.89E-05 | 0.000908725 | 0.495914339 |
| T2DM | LEC | *ZNF772* | -0.664876401 | 0.001503504 | 0.003202077 | 1 |
| T2DM | LEC | *ZNF257* | -0.927216556 | 0.003191005 | 0.005816714 | 1 |
| T2DM | LEC | *ADAM32* | -0.804236247 | 0.00159369 | 0.003357751 | 1 |
| T2DM | LEC | *LRRC8B* | 0.648129528 | 0.000347176 | 0.001199942 | 1 |
| T2DM | LEC | *SND1* | 0.590788181 | 0.000920279 | 0.00221763 | 1 |
| T2DM | LEC | *SULT1A2* | 0.330385814 | 3.65E-05 | 0.000908725 | 0.22975602 |
| T2DM | LEC | *NEK5* | 0.738390053 | 0.002207803 | 0.004325478 | 1 |
| T2DM | LEC | *PIWIL2* | 1.09964541 | 0.000380159 | 0.001259252 | 1 |
| T2DM | LEC | *C1D* | 0.613877793 | 0.00200891 | 0.004024861 | 1 |
| T2DM | LEC | *FAM110D* | 0.695312922 | 8.33E-05 | 0.000908725 | 0.523570902 |
| T2DM | LEC | *C6orf141* | -0.063289817 | 0.000108063 | 0.000908725 | 0.679393934 |
| T2DM | LEC | *CCL4L2* | 0.323226602 | 0.001747461 | 0.003611535 | 1 |

| | | | | | | |
|---|---|---|---|---|---|---|
| T2DM | LEC | *GTF2E2* | 0.912178183 | 0.000187945 | 0.000936299 | 1 |
| T2DM | LEC | *ZNF165* | -1.167845156 | 0.001845604 | 0.003774663 | 1 |
| T2DM | LEC | *ZNF720* | -0.843094375 | 0.01370591 | 0.020361308 | 1 |
| T2DM | LEC | *DDI2* | 0.673197799 | 0.000347021 | 0.001199942 | 1 |
| T2DM | LEC | *ZNF786* | 0.820243989 | 0.000615932 | 0.001691727 | 1 |
| T2DM | LEC | *SLC22A5* | 0.724232268 | 0.000398313 | 0.001289522 | 1 |
| T2DM | LEC | *C5AR1* | 0.665148185 | 5.45E-05 | 0.000908725 | 0.342657392 |
| T2DM | LEC | *VEPH1* | -0.701944245 | 0.001778135 | 0.003661947 | 1 |
| T2DM | LEC | *SHPK* | 0.470799321 | 0.000100872 | 0.000908725 | 0.634184607 |
| T2DM | LEC | *MAP3K5* | 0.719384058 | 0.000716509 | 0.001865296 | 1 |
| T2DM | LEC | *STMN3* | 1.185521292 | 0.00054089 | 0.00154361 | 1 |
| T2DM | LEC | *GYPE* | 1.059720849 | 3.50E-05 | 0.000908725 | 0.220158778 |
| T2DM | LEC | *PCDHB11* | -2.214618737 | 0.003947821 | 0.006946529 | 1 |
| T2DM | LEC | *SLC2A10* | 1.355208474 | 0.002347637 | 0.004541414 | 1 |
| T2DM | LEC | *ZNF665* | 0.652292271 | 0.002089196 | 0.004148697 | 1 |
| T2DM | LEC | *RPF2* | 0.832152118 | 0.000220916 | 0.00098409 | 1 |
| T2DM | LEC | *MYO5A* | 0.547241106 | 0.000171268 | 0.000910705 | 1 |
| T2DM | LEC | *ATG7* | 0.51969822 | 0.000409665 | 0.001308237 | 1 |
| T2DM | LEC | *TTC30A* | -1.486971695 | 5.26E-05 | 0.000908725 | 0.330653401 |
| T2DM | LEC | *PIGN* | 0.596902701 | 0.00613315 | 0.010155153 | 1 |
| T2DM | LEC | *ZNF624* | 1.2658534 | 0.02812345 | 0.038783094 | 1 |
| T2DM | LEC | *HHLA3* | 0.837835568 | 0.013069751 | 0.01953162 | 1 |
| T2DM | LEC | *HOXA4* | 0.699399106 | 0.000172255 | 0.000910705 | 1 |
| T2DM | LEC | *BCO2* | -0.476366818 | 0.000211992 | 0.0009784 | 1 |
| T2DM | LEC | *ENTPD6* | 0.768780723 | 0.002423437 | 0.004659089 | 1 |
| T2DM | LEC | *CCDC154* | 1.216054915 | 1.87E-05 | 0.000908725 | 0.117293808 |
| T2DM | LEC | *ZNF841* | -1.088831394 | 0.036226817 | 0.048603926 | 1 |
| T2DM | LEC | *MFAP5* | 0.542598737 | 0.000227758 | 0.000994385 | 1 |
| T2DM | LEC | *CDC42SE1* | 0.997242547 | 0.009628317 | 0.014957556 | 1 |
| T2DM | LEC | *MPEG1* | 0.376715394 | 0.001139299 | 0.002587136 | 1 |
| T2DM | LEC | *PDCD1LG2* | 0.61604747 | 0.000433452 | 0.001342678 | 1 |
| T2DM | LEC | *NMB* | 1.995805509 | 0.005089365 | 0.008685351 | 1 |
| T2DM | LEC | *KLHL14* | -0.076454372 | 0.000137713 | 0.000908725 | 0.865801739 |
| T2DM | LEC | *FAM114A1* | 0.888766214 | 0.007168524 | 0.011624584 | 1 |
| T2DM | LEC | *PHF2* | -0.959075055 | 0.003325689 | 0.006025991 | 1 |
| T2DM | LEC | *RPS26* | -0.391339405 | 0.015000582 | 0.022050189 | 1 |
| T2DM | LEC | *C14orf178* | 0.72174631 | 0.006505961 | 0.010688 | 1 |
| T2DM | LEC | *PSAP* | 0.623101865 | 0.000386364 | 0.001268529 | 1 |
| T2DM | LEC | *MAP1LC3C* | 0.644619399 | 0.000192365 | 0.000946191 | 1 |
| T2DM | LEC | *TAF13* | 1.114166268 | 0.000372747 | 0.001243243 | 1 |
| T2DM | LEC | *AC122129.1* | 1.061543407 | 0.000111546 | 0.000908725 | 0.701288024 |
| T2DM | LEC | *CCDC180* | 0.879076481 | 0.000313617 | 0.001145484 | 1 |
| T2DM | LEC | *OCLN* | -0.492220029 | 0.03391335 | 0.045744096 | 1 |
| T2DM | LEC | *ZNF181* | 1.028586175 | 9.16E-05 | 0.000908725 | 0.576077184 |
| T2DM | LEC | *SGTB* | 1.07093384 | 0.00019981 | 0.000961874 | 1 |
| T2DM | LEC | *ADH5* | 0.626043317 | 0.000773232 | 0.001959414 | 1 |
| T2DM | LEC | *HRNR* | 0.22534334 | 0.019773839 | 0.028279828 | 1 |
| T2DM | LEC | *ZNF677* | 1.359251824 | 0.015479475 | 0.022690478 | 1 |
| T2DM | LEC | *ZNF311* | 0.492880021 | 0.004343198 | 0.007559713 | 1 |
| T2DM | LEC | *FCHSD1* | 1.158797161 | 0.000897559 | 0.00218465 | 1 |

| | | | | | | |
|---|---|---|---|---|---|---|
| T2DM | LEC | *ZNF71* | -0.988108937 | 0.003481144 | 0.006253129 | 1 |
| T2DM | LEC | *LEKR1* | 0.607654572 | 0.001025952 | 0.0024014 | 1 |
| T2DM | LEC | *NOL8* | 0.974852621 | 0.001809709 | 0.003713328 | 1 |
| T2DM | LEC | *ZNF335* | 0.62837444 | 0.000232185 | 0.001001608 | 1 |
| T2DM | LEC | *AGAP9* | 1.472667796 | 0.000159089 | 0.000910705 | 1 |
| T2DM | LEC | *MAK16* | 0.973891137 | 0.000295117 | 0.00111174 | 1 |
| T2DM | LEC | *SULT1C4* | -0.368855463 | 0.000367923 | 0.001238957 | 1 |
| T2DM | LEC | *ZNF649* | 0.841562482 | 0.003446398 | 0.006199573 | 1 |
| T2DM | LEC | *HIBCH* | 0.57339749 | 0.020892096 | 0.029683302 | 1 |
| T2DM | LEC | *ZNF544* | 0.847662622 | 0.020628667 | 0.029342179 | 1 |
| T2DM | LEC | *SOWAHC* | -0.91265747 | 0.000178275 | 0.000920211 | 1 |
| T2DM | LEC | *AC135178.1* | 3.098773216 | 0.00038876 | 0.001271661 | 1 |
| T2DM | LEC | *MIER1* | 0.918182266 | 0.001041884 | 0.002427203 | 1 |
| T2DM | LEC | *SVIP* | -0.603592513 | 0.000491221 | 0.001447869 | 1 |
| T2DM | LEC | *ZNF251* | 0.958744301 | 0.000947136 | 0.002256401 | 1 |
| T2DM | LEC | *DDRGK1* | 0.681344254 | 0.000144484 | 0.000908725 | 0.908371573 |
| T2DM | LEC | *ZNF334* | 0.653065878 | 0.000107294 | 0.000908725 | 0.674559433 |
| T2DM | LEC | *HSD17B11* | 1.069178275 | 0.001548902 | 0.003278092 | 1 |
| T2DM | LEC | *SULT1C2* | 0.545460053 | 4.71E-05 | 0.000908725 | 0.29625174 |
| T2DM | LEC | *RPS6KL1* | -0.305697372 | 0.001147042 | 0.002601535 | 1 |
| T2DM | LEC | *DDX42* | 0.812404353 | 0.037195284 | 0.049786407 | 1 |
| T2DM | LEC | *SLC29A3* | 0.525065666 | 0.000204274 | 0.000964732 | 1 |
| T2DM | LEC | *ANTXRL* | -8.69574327 | 0.000145283 | 0.000908725 | 0.913395646 |
| T2DM | LEC | *STYX* | 0.671533715 | 0.000128715 | 0.000908725 | 0.809229603 |
| T2DM | LEC | *ZKSCAN8* | 1.124291485 | 0.009169337 | 0.014343773 | 1 |
| T2DM | LEC | *GFPT1* | 0.740901608 | 0.000216103 | 0.000979981 | 1 |
| T2DM | LEC | *UVRAG* | 0.548381812 | 0.000718166 | 0.001868836 | 1 |
| T2DM | LEC | *MGEA5* | 0.602088333 | 0.003434518 | 0.006183509 | 1 |
| T2DM | LEC | *MT1F* | -0.807446302 | 9.46E-05 | 0.000908725 | 0.59482008 |
| T2DM | LEC | *ZNF587* | 1.059216378 | 0.006283183 | 0.010362636 | 1 |
| T2DM | LEC | *SH3BGRL2* | -0.61216307 | 0.000330257 | 0.001172403 | 1 |
| T2DM | LEC | *ZNF808* | 1.051515769 | 0.014003597 | 0.020769195 | 1 |
| T2DM | LEC | *TMA16* | 1.13166163 | 0.001996081 | 0.004005817 | 1 |
| T2DM | LEC | *HLA-DRB5* | -0.701955206 | 1.66E-12 | 3.87E-09 | 1.04E-08 |
| T2DM | LEC | *C20orf203* | -1.321283121 | 0.023619063 | 0.033116202 | 1 |
| T2DM | LEC | *ZNF627* | 1.039182815 | 0.000396363 | 0.001287157 | 1 |
| T2DM | LEC | *ARC* | 8.384840914 | 0.00188254 | 0.00383399 | 1 |
| T2DM | LEC | *LRBA* | 0.708749947 | 0.00305221 | 0.005599429 | 1 |
| T2DM | LEC | *MMP17* | 0.897349676 | 0.000153315 | 0.000910197 | 0.963890392 |
| T2DM | LEC | *CCDC69* | 0.868660471 | 0.000648826 | 0.001745472 | 1 |
| T2DM | LEC | *MDM4* | 0.771914757 | 0.001082674 | 0.002491652 | 1 |
| T2DM | LEC | *ZNF534* | -1.295924751 | 8.59E-05 | 0.000908725 | 0.539815828 |
| T2DM | LEC | *FAM19A2* | 0.715167355 | 0.000600198 | 0.00165908 | 1 |
| T2DM | LEC | *TTC37* | 0.797744427 | 0.002808347 | 0.005246977 | 1 |
| T2DM | LEC | *PAPSS2* | 0.366373352 | 0.000129858 | 0.000908725 | 0.816414763 |
| T2DM | LEC | *FAN1* | 1.046053588 | 0.000822616 | 0.002047095 | 1 |
| T2DM | LEC | *CEP290* | 0.807281949 | 0.020385409 | 0.029029007 | 1 |
| T2DM | LEC | *ECI2* | 1.13329729 | 0.001701416 | 0.003544883 | 1 |
| T2DM | LEC | *ZNF652* | 0.814679103 | 0.00074815 | 0.00191907 | 1 |
| T2DM | LEC | *RASSF9* | -0.171230145 | 0.000159725 | 0.000910705 | 1 |

| | | | | | | |
|---|---|---|---|---|---|---|
| T2DM | LEC | *FAM169A* | -0.317594598 | 0.000322373 | 0.001159063 | 1 |
| T2DM | LEC | *SCAMP5* | 0.664923761 | 0.000623111 | 0.001704001 | 1 |
| T2DM | LEC | *ALPK2* | 0.441184467 | 0.000153555 | 0.000910197 | 0.965399936 |
| T2DM | LEC | *SFT2D1* | 1.0723496 | 0.001429151 | 0.003081302 | 1 |
| T2DM | LEC | *ARHGAP11A* | 0.675567987 | 7.11E-05 | 0.000908725 | 0.447318433 |
| T2DM | LEC | *HMGN2* | 1.148464567 | 0.002041594 | 0.004078648 | 1 |
| T2DM | LEC | *DENND4B* | 0.762761738 | 0.017040837 | 0.024702731 | 1 |
| T2DM | LEC | *CD3E* | 0.721265197 | 0.000559543 | 0.001578217 | 1 |
| T2DM | LEC | *TSEN15* | 0.843148183 | 0.000249971 | 0.001023586 | 1 |
| T2DM | LEC | *TYW1* | 0.6886374 | 0.001381262 | 0.003000964 | 1 |
| T2DM | LEC | *ITPRIPL1* | -0.625542247 | 0.000680777 | 0.001802884 | 1 |
| T2DM | LEC | *PRMT6* | 1.477754985 | 0.000554687 | 0.001568727 | 1 |
| T2DM | LEC | *SHISA4* | 1.645482027 | 0.000332032 | 0.001173862 | 1 |
| T2DM | LEC | *CAPZA2* | 0.728431772 | 0.000336364 | 0.001182068 | 1 |
| T2DM | LEC | *C1orf174* | 1.096814645 | 0.019938824 | 0.028463984 | 1 |
| T2DM | LEC | *RASGEF1A* | 2.095087784 | 0.000116267 | 0.000908725 | 0.730971354 |
| T2DM | LEC | *DCLRE1A* | 1.397659595 | 0.027505674 | 0.038022905 | 1 |
| T2DM | LEC | *APRT* | 1.503096056 | 0.002540535 | 0.004832781 | 1 |
| T2DM | LEC | *TBKBP1* | -0.837891042 | 0.000190677 | 0.000942652 | 1 |
| T2DM | LEC | *L3MBTL3* | -0.884496779 | 0.002133339 | 0.004224347 | 1 |
| T2DM | LEC | *MFAP3L* | -0.906626485 | 0.000646025 | 0.001740903 | 1 |
| T2DM | LEC | *NAGA* | 1.195208871 | 0.002447208 | 0.004695587 | 1 |
| T2DM | LEC | *PJA2* | 0.738836006 | 0.006426921 | 0.0105775 | 1 |
| T2DM | LEC | *AL590822.1* | 0.569194874 | 0.014827522 | 0.021836643 | 1 |
| T2DM | LEC | *AP001007.1* | -3.434971326 | 0.000363395 | 0.001230962 | 1 |
| T2DM | LEC | *ZNF525* | 0.736013298 | 0.000165274 | 0.000910705 | 1 |
| T2DM | LEC | *EFCAB2* | 0.801397594 | 0.00389099 | 0.006875394 | 1 |
| T2DM | LEC | *CHML* | 0.87411899 | 4.65E-06 | 0.000522362 | 0.029252287 |
| T2DM | LEC | *CAPN8* | 1.334841268 | 0.000572914 | 0.001603699 | 1 |
| T2DM | LEC | *TATDN3* | 0.494940736 | 0.006517512 | 0.010698589 | 1 |
| T2DM | LEC | *ECT2L* | -0.998089716 | 0.006044424 | 0.010029373 | 1 |
| T2DM | LEC | *CENPW* | -3.369632726 | 3.36E-08 | 1.41E-05 | 0.000211353 |
| T2DM | LEC | *DDO* | -0.91758135 | 0.000965225 | 0.002292547 | 1 |
| T2DM | LEC | *HIST2H2BF* | 1.40880064 | 0.000109695 | 0.000908725 | 0.689651509 |
| T2DM | LEC | *RBM20* | -1.315442515 | 0.013478013 | 0.02006542 | 1 |
| T2DM | LEC | *C6orf163* | -1.505032981 | 0.022548926 | 0.031750301 | 1 |
| T2DM | LEC | *PCMTD2* | 1.105679303 | 0.003518697 | 0.006311569 | 1 |
| T2DM | LEC | *OOEP* | -0.528446318 | 9.72E-05 | 0.000908725 | 0.61087941 |
| T2DM | LEC | *SYS1* | 1.756308097 | 0.008586214 | 0.01354618 | 1 |
| T2DM | LEC | *NEU4* | 3.826027059 | 0.001790394 | 0.00368252 | 1 |
| T2DM | LEC | *TRAF3IP1* | 0.872745721 | 0.015941918 | 0.023286904 | 1 |
| T2DM | LEC | *GIGYF2* | 0.753572263 | 0.01085909 | 0.016631206 | 1 |
| T2DM | LEC | *C2orf72* | -0.559481776 | 0.000105465 | 0.000908725 | 0.663058042 |
| T2DM | LEC | *AGAP6* | 0.678267594 | 2.00E-05 | 0.000908725 | 0.125551981 |
| T2DM | LEC | *C10orf128* | 0.863848624 | 0.00683125 | 0.011152445 | 1 |
| T2DM | LEC | *SYT15* | -0.811116405 | 0.003485798 | 0.0062597 | 1 |
| T2DM | LEC | *RING1* | 0.786483734 | 0.023769365 | 0.033304658 | 1 |
| T2DM | LEC | *OXLD1* | 0.723473675 | 5.13E-06 | 0.000529126 | 0.032276668 |
| T2DM | LEC | *HLA-DOA* | 0.522791931 | 0.028982472 | 0.039793143 | 1 |
| T2DM | LEC | *HLA-DMA* | 0.725881702 | 0.001382931 | 0.003003276 | 1 |

| | | | | | | |
|---|---|---|---|---|---|---|
| T2DM | LEC | *TAP2* | -0.523465213 | 0.01352749 | 0.020120021 | 1 |
| T2DM | LEC | *HLA-DRA* | 1.090890917 | 3.67E-11 | 3.84E-08 | 2.31E-07 |
| T2DM | LEC | *COL15A1* | 0.549039718 | 0.001245013 | 0.002764077 | 1 |
| T2DM | LEC | *NOTCH4* | -0.247071914 | 3.98E-08 | 1.47E-05 | 0.000249952 |
| T2DM | LEC | *STK19* | 0.943656314 | 0.009352026 | 0.014586005 | 1 |
| T2DM | LEC | *NELFE* | 1.192444137 | 1.37E-06 | 0.000269656 | 0.008628978 |
| T2DM | LEC | *LAYN* | -0.106632635 | 0.000123151 | 0.000908725 | 0.774247311 |
| T2DM | LEC | *C6orf48* | 0.877481174 | 0.025891044 | 0.035941045 | 1 |
| T2DM | LEC | *CARD16* | -0.711380929 | 0.000259126 | 0.001042471 | 1 |
| T2DM | LEC | *MSH5* | 0.948705549 | 3.35E-05 | 0.000908725 | 0.210389618 |
| T2DM | LEC | *BAG6* | 1.218045574 | 5.35E-06 | 0.000542914 | 0.033660663 |
| T2DM | LEC | *NFKBIL1* | 1.195121227 | 2.62E-05 | 0.000908725 | 0.164526837 |
| T2DM | LEC | *MICB* | 0.267970699 | 0.005644463 | 0.009480828 | 1 |
| T2DM | LEC | *HLA-C* | 0.356272056 | 0.031300902 | 0.042613419 | 1 |
| T2DM | LEC | *CCHCR1* | 0.388329429 | 0.010746542 | 0.016482925 | 1 |
| T2DM | LEC | *ABCF1* | 0.790399599 | 0.005038236 | 0.008612124 | 1 |
| T2DM | LEC | *LILRB3* | 0.465718462 | 0.000132491 | 0.000908725 | 0.832970563 |
| T2DM | LEC | *ZNF468* | 0.720724407 | 0.000107534 | 0.000908725 | 0.676068698 |
| T2DM | LEC | *HCG9* | -0.482129598 | 0.0058817 | 0.009808554 | 1 |
| T2DM | LEC | *HLA-G* | 0.831549721 | 3.64E-05 | 0.000908725 | 0.229029863 |
| T2DM | LEC | *HLA-F* | 0.717277714 | 0.005974648 | 0.00994246 | 1 |
| T2DM | LEC | *FAM153C* | -1.239470524 | 0.000232521 | 0.001001608 | 1 |
| T2DM | LEC | *TRIM27* | 0.663877744 | 0.032181836 | 0.043670883 | 1 |
| T2DM | LEC | *MALRD1* | 0.423391935 | 0.001702312 | 0.003545027 | 1 |
| T2DM | LEC | *ATXN2* | 0.695446419 | 0.006875404 | 0.011212884 | 1 |
| T2DM | LEC | *FAM216A* | 1.090656601 | 0.000318226 | 0.001153119 | 1 |
| T2DM | LEC | *ZNF783* | 1.009213723 | 0.000733328 | 0.001897299 | 1 |
| T2DM | LEC | *ZNF425* | 0.564625176 | 0.024892014 | 0.034707494 | 1 |
| T2DM | LEC | *LRRC10B* | 4.213003885 | 0.002455057 | 0.004705775 | 1 |
| T2DM | LEC | *PCDHGA1* | -0.988601327 | 0.013587083 | 0.020203877 | 1 |
| T2DM | LEC | *PCDHA5* | -2.882784647 | 0.000546751 | 0.001554691 | 1 |
| T2DM | LEC | *PCDHA4* | 2.836557244 | 0.000398915 | 0.001290555 | 1 |
| T2DM | LEC | *TRIM13* | 0.772865654 | 0.000740732 | 0.001906556 | 1 |
| T2DM | LEC | *SPIRE2* | 0.844998257 | 0.000274773 | 0.001077667 | 1 |
| T2DM | LEC | *SLFN12L* | 0.306640552 | 0.000105401 | 0.000908725 | 0.662653879 |
| T2DM | LEC | *CLLU1OS* | 1.248995108 | 0.002180729 | 0.004292451 | 1 |
| T2DM | LEC | *SYCE1L* | 1.016253977 | 3.14E-05 | 0.000908725 | 0.197497532 |
| T2DM | LEC | *FAM71F2* | 0.506415372 | 0.000127959 | 0.000908725 | 0.804477222 |
| T2DM | LEC | *AC016586.1* | 6.998803493 | 6.95E-06 | 0.000610811 | 0.043697436 |
| T2DM | LEC | *TMEM170B* | 0.341391029 | 0.022965936 | 0.032279643 | 1 |
| T2DM | LEC | *PRR13* | 0.789190588 | 0.000630068 | 0.00171385 | 1 |
| T2DM | LEC | *MT1A* | 4.529366501 | 0.00121904 | 0.002720662 | 1 |
| T2DM | LEC | *MT1M* | 0.940763531 | 0.000117642 | 0.000908725 | 0.739612367 |
| T2DM | LEC | *CFI* | -0.310042875 | 0.000168204 | 0.000910705 | 1 |
| T2DM | LEC | *SAMD9* | 1.100798643 | 0.029634651 | 0.040591078 | 1 |
| T2DM | LEC | *EXOC3L4* | 0.886992549 | 0.000228119 | 0.000994837 | 1 |
| T2DM | LEC | *ATP6AP1L* | 0.753563741 | 4.95E-05 | 0.000908725 | 0.311241427 |
| T2DM | LEC | *NAP1L4* | 0.629044456 | 0.036163223 | 0.048539321 | 1 |
| T2DM | LEC | *SMN2* | 0.329059805 | 0.007564202 | 0.012178268 | 1 |
| T2DM | LEC | *SERF1B* | -0.672648506 | 1.87E-05 | 0.000908725 | 0.117416646 |

| | | | | | | |
|---|---|---|---|---|---|---|
| T2DM | LEC | *DENND6B* | 1.292679567 | 0.00017282 | 0.000910705 | 1 |
| T2DM | LEC | *EIF3CL* | 2.667413422 | 5.06E-06 | 0.000529126 | 0.031812924 |
| T2DM | LEC | *LCMT1* | 0.907474113 | 0.000545268 | 0.001551879 | 1 |
| T2DM | LEC | *CDPF1* | 0.533489952 | 0.001430813 | 0.00308277 | 1 |
| T2DM | LEC | *LIN52* | 1.024418471 | 3.91E-06 | 0.000465666 | 0.024569553 |
| T2DM | LEC | *C17orf107* | 0.802905189 | 0.010954926 | 0.016757571 | 1 |
| T2DM | LEC | *RFPL3S* | -0.864520716 | 0.000178173 | 0.000920211 | 1 |
| T2DM | LEC | *C1QTNF9B* | 5.522295096 | 0.00010842 | 0.000908725 | 0.681637423 |
| T2DM | LEC | *RNPS1* | 1.023451757 | 0.006222521 | 0.010278767 | 1 |
| T2DM | LEC | *NYNRIN* | 0.781481109 | 6.63E-05 | 0.000908725 | 0.416896538 |
| T2DM | LEC | *GOLGA8O* | 2.173065039 | 0.000823136 | 0.002047095 | 1 |
| T2DM | LEC | *HCG27* | 0.923636862 | 1.84E-11 | 2.32E-08 | 1.16E-07 |
| T2DM | LEC | *LNP1* | -1.001868773 | 0.000569079 | 0.001596519 | 1 |
| T2DM | LEC | *VGLL3* | 0.283324516 | 0.000564648 | 0.001587631 | 1 |
| T2DM | LEC | *METTL6* | 1.069251738 | 0.0009435 | 0.002252007 | 1 |
| T2DM | LEC | *SACM1L* | 0.922246209 | 0.007279259 | 0.011773784 | 1 |
| T2DM | LEC | *SLC48A1* | 0.884323949 | 0.000289428 | 0.001099475 | 1 |
| T2DM | LEC | *TAS2R14* | 0.965409448 | 0.001226571 | 0.002729718 | 1 |
| T2DM | LEC | *ZRSR1* | 2.392394498 | 4.91E-05 | 0.000908725 | 0.308455247 |
| T2DM | LEC | *C17orf51* | 0.999351605 | 0.001959541 | 0.003951134 | 1 |
| T2DM | LEC | *OR2A42* | 0.461828066 | 0.020168855 | 0.028753195 | 1 |
| T2DM | LEC | *RNF208* | -3.80135406 | 0.007087406 | 0.011510856 | 1 |
| T2DM | LEC | *MAP10* | 0.735344464 | 7.34E-05 | 0.000908725 | 0.461484425 |
| T2DM | LEC | *ZNF580* | 1.09977325 | 0.000534114 | 0.001530525 | 1 |
| T2DM | LEC | *NUP62* | 0.918957503 | 0.000212342 | 0.0009784 | 1 |
| T2DM | LEC | *TCTEX1D2* | -0.384282236 | 0.00116428 | 0.002627361 | 1 |
| T2DM | LEC | *AC022498.1* | -0.689700266 | 0.00039487 | 0.001284297 | 1 |
| T2DM | LEC | *LINGO4* | 5.220100875 | 0.01759551 | 0.025418881 | 1 |
| T2DM | LEC | *ASIC3* | 1.037725637 | 0.000384188 | 0.001267255 | 1 |
| T2DM | LEC | *GIMAP1* | -0.597625786 | 0.000722591 | 0.001878796 | 1 |
| T2DM | LEC | *CCDC183* | 1.400356803 | 0.000213606 | 0.0009784 | 1 |
| T2DM | LEC | *ARHGEF35* | -0.997914371 | 0.000107501 | 0.000908725 | 0.675855663 |
| T2DM | LEC | *DNLZ* | 0.643222635 | 0.001896773 | 0.003859227 | 1 |
| T2DM | LEC | *MXD3* | 0.666876568 | 9.90E-05 | 0.000908725 | 0.622212753 |
| T2DM | LEC | *GSTM2* | 0.680355908 | 0.001397116 | 0.003025848 | 1 |
| T2DM | LEC | *SIPA1* | 1.77726261 | 4.68E-05 | 0.000908725 | 0.294084056 |
| T2DM | LEC | *ERV3-1* | 0.315182084 | 0.000169827 | 0.000910705 | 1 |
| T2DM | LEC | *RBMXL1* | 1.360931144 | 0.000927818 | 0.002226408 | 1 |
| T2DM | LEC | *TMEM110* | 1.31563938 | 5.98E-05 | 0.000908725 | 0.37584679 |
| T2DM | LEC | *HEXA* | 1.121830406 | 0.001459752 | 0.003133308 | 1 |
| T2DM | LEC | *LEPROT* | 0.87639016 | 0.004066695 | 0.007123797 | 1 |
| T2DM | LEC | *LBH* | 0.764772195 | 0.000113924 | 0.000908725 | 0.716239885 |
| T2DM | LEC | *PPP1CB* | 0.807007166 | 0.003532726 | 0.006331313 | 1 |
| T2DM | LEC | *NCKIPSD* | 0.526992543 | 8.48E-05 | 0.000908725 | 0.533195314 |
| T2DM | LEC | *ATF6B* | 1.199091655 | 0.000142436 | 0.000908725 | 0.895496154 |
| T2DM | LEC | *SLC35F6* | 1.348087458 | 0.02090076 | 0.029688901 | 1 |
| T2DM | LEC | *ATP6V1G2* | 0.519730671 | 0.002479034 | 0.004738731 | 1 |
| T2DM | LEC | *KLRK1* | 1.623507819 | 0.000922396 | 0.002219328 | 1 |
| T2DM | LEC | *PPM1N* | 0.660806215 | 3.60E-05 | 0.000908725 | 0.226583321 |
| T2DM | LEC | *SLC23A3* | 0.575668176 | 0.00105958 | 0.002452718 | 1 |

| | | | | | | |
|---|---|---|---|---|---|---|
| T2DM | LEC | *LTB4R* | 0.536916536 | 0.012676593 | 0.019013211 | 1 |
| T2DM | LEC | *LTB4R2* | 0.818281637 | 0.004040129 | 0.007083182 | 1 |
| T2DM | LEC | *GALT* | 1.140878055 | 0.000180859 | 0.000925947 | 1 |
| T2DM | LEC | *NUDT19* | -0.989733934 | 0.004104102 | 0.007177326 | 1 |
| T2DM | LEC | *ZNF726* | 1.623108983 | 0.001171501 | 0.002640812 | 1 |
| T2DM | LEC | *TAX1BP3* | -0.509271267 | 0.000310575 | 0.001138389 | 1 |
| T2DM | LEC | *AP1G2* | -0.618434357 | 0.000597773 | 0.001656323 | 1 |
| T2DM | LEC | *ZNF90* | 1.1635451 | 0.008548298 | 0.013493134 | 1 |
| T2DM | LEC | *TM6SF2* | 6.485234649 | 9.66E-05 | 0.000908725 | 0.607165387 |
| T2DM | LEC | *SMIM7* | 0.698563574 | 7.19E-05 | 0.000908725 | 0.451769626 |
| T2DM | LEC | *AC079776.2* | -1.522928507 | 0.001310415 | 0.002876599 | 1 |
| T2DM | LEC | *PAXIP1-AS2* | 0.840057519 | 0.000214232 | 0.0009784 | 1 |
| T2DM | LEC | *PRCD* | 0.48267218 | 6.09E-05 | 0.000908725 | 0.383108417 |
| T2DM | LEC | *C17orf67* | -0.68004205 | 0.002686278 | 0.005058968 | 1 |
| T2DM | LEC | *COLCA2* | 0.507161067 | 0.001070281 | 0.002470212 | 1 |
| T2DM | LEC | *MBLAC1* | 0.604032423 | 0.000691617 | 0.001821616 | 1 |
| T2DM | LEC | *NEURL1B* | 0.782142953 | 0.000105729 | 0.000908725 | 0.664717927 |
| T2DM | LEC | *VSTM5* | 0.573290885 | 0.000494216 | 0.001449978 | 1 |
| T2DM | LEC | *STARD10* | 1.012600226 | 2.80E-10 | 2.20E-07 | 1.76E-06 |
| T2DM | LEC | *CPEB1* | 0.460622485 | 0.000203743 | 0.000964732 | 1 |
| T2DM | LEC | *C10orf105* | -1.185922671 | 0.000433739 | 0.001342678 | 1 |
| T2DM | LEC | *ARHGEF33* | 0.786591159 | 0.000373074 | 0.001243646 | 1 |
| T2DM | LEC | *AC005082.1* | 1.856064439 | 0.000131047 | 0.000908725 | 0.82389153 |
| T2DM | LEC | *ACSM4* | 0.4355727 | 0.017151751 | 0.024852053 | 1 |
| T2DM | LEC | *COL28A1* | 1.465909092 | 0.000552577 | 0.001564182 | 1 |
| T2DM | LEC | *PHB2* | 0.712034902 | 0.000647932 | 0.001744462 | 1 |
| T2DM | LEC | *UBXN2B* | 0.853040305 | 0.015253324 | 0.022369407 | 1 |
| T2DM | LEC | *UBE2QL1* | -1.052998873 | 0.000303422 | 0.001125658 | 1 |
| T2DM | LEC | *MYL5* | 0.594089794 | 0.00011194 | 0.000908725 | 0.703765723 |
| T2DM | LEC | *NPEPL1* | 0.527195102 | 0.000107793 | 0.000908725 | 0.677695527 |
| T2DM | LEC | *LINC01139* | -0.427941958 | 0.00015713 | 0.000910705 | 0.987873208 |
| T2DM | LEC | *TSTD1* | 0.452871405 | 9.90E-05 | 0.000908725 | 0.622314035 |
| T2DM | LEC | *LINC01138* | 0.764233514 | 0.000233093 | 0.001001608 | 1 |
| T2DM | LEC | *CYB5RL* | -0.374711481 | 0.000212501 | 0.0009784 | 1 |
| T2DM | LEC | *ATAD3C* | -0.651312642 | 0.001269232 | 0.002804801 | 1 |
| T2DM | LEC | *RNASEK* | 1.210294538 | 0.0015958 | 0.003359946 | 1 |
| T2DM | LEC | *NBPF1* | 0.581358022 | 0.000229181 | 0.000996446 | 1 |
| T2DM | LEC | *FAM228B* | 0.485863708 | 0.003391476 | 0.006130594 | 1 |
| T2DM | LEC | *RNU6ATAC35l* | 6.15583807 | 0.000172528 | 0.000910705 | 1 |
| T2DM | LEC | *C6orf226* | -1.001689166 | 0.000380701 | 0.00125962 | 1 |
| T2DM | LEC | *PPP3R1* | 0.888555442 | 0.000219692 | 0.00098409 | 1 |
| T2DM | LEC | *FANCG* | -0.933221005 | 0.000241455 | 0.00101202 | 1 |
| T2DM | LEC | *AP4M1* | 0.638257877 | 0.001940802 | 0.003924678 | 1 |
| T2DM | LEC | *C19orf73* | 3.329709463 | 9.18E-05 | 0.000908725 | 0.577388558 |
| T2DM | LEC | *OR2A25* | 17.35722777 | 0.000345395 | 0.001196042 | 1 |
| T2DM | LEC | *C1orf229* | 1.229166368 | 0.006922179 | 0.01127627 | 1 |
| T2DM | LEC | *SLC12A8* | 0.548918821 | 0.013358923 | 0.019905418 | 1 |
| T2DM | LEC | *APOL6* | 0.605473615 | 0.000194514 | 0.000949652 | 1 |
| T2DM | LEC | *TIAF1* | 1.005732247 | 3.53E-05 | 0.000908725 | 0.221730403 |
| T2DM | LEC | *FAM185A* | 0.999052453 | 0.000412834 | 0.001312177 | 1 |

| | | | | | | |
|---|---|---|---|---|---|---|
| T2DM | LEC | *AC062017.1* | 0.369699834 | 0.001320567 | 0.002895852 | 1 |
| T2DM | LEC | *ADRA2B* | -4.534443591 | 0.001347745 | 0.002945177 | 1 |
| T2DM | LEC | *VPS52* | 1.139732077 | 0.005081016 | 0.008675814 | 1 |
| T2DM | LEC | *LINC00205* | 0.714415563 | 0.005358168 | 0.0090629 | 1 |
| T2DM | LEC | *AC147651.3* | 1.057852548 | 0.000565136 | 0.001588293 | 1 |
| T2DM | LEC | *HLA-DPB1* | 0.611252415 | 0.001035465 | 0.002417367 | 1 |
| T2DM | LEC | *OSER1-AS1* | 0.981108544 | 0.000166739 | 0.000910705 | 1 |
| T2DM | LEC | *LINC01133* | 0.33369051 | 0.000110764 | 0.000908725 | 0.696373511 |
| T2DM | LEC | *LINC01117* | -0.384781007 | 0.000278976 | 0.00108013 | 1 |
| T2DM | LEC | *RASAL2-AS1* | -1.011961268 | 0.000105454 | 0.000908725 | 0.662986168 |
| T2DM | LEC | *AC069213.1* | -0.503812702 | 9.64E-05 | 0.000908725 | 0.606084445 |
| T2DM | LEC | *LINC00240* | 0.589381883 | 3.98E-05 | 0.000908725 | 0.249925243 |
| T2DM | LEC | *LINC00863* | 0.690373145 | 5.12E-05 | 0.000908725 | 0.321847631 |
| T2DM | LEC | *LINC00184* | 0.949315532 | 0.000702922 | 0.001841362 | 1 |
| T2DM | LEC | *TMEM233* | 0.324201438 | 0.00014617 | 0.000908725 | 0.918970195 |
| T2DM | LEC | *LINC00237* | -2.711585273 | 0.000218673 | 0.00098409 | 1 |
| T2DM | LEC | *PLEKHM1* | 0.977405547 | 6.11E-05 | 0.000908725 | 0.384257967 |
| T2DM | LEC | *AP000350.6* | -0.572001013 | 0.000239454 | 0.00100947 | 1 |
| T2DM | LEC | *SFTA1P* | 1.523425668 | 0.000239101 | 0.00100947 | 1 |
| T2DM | LEC | *FAM225B* | 0.645735885 | 0.012229481 | 0.018411577 | 1 |
| T2DM | LEC | *SLC26A6* | -0.743634908 | 0.005969254 | 0.009936114 | 1 |
| T2DM | LEC | *MIAT* | 0.584826618 | 0.000251744 | 0.001026403 | 1 |
| T2DM | LEC | *LINC00626* | -3.152313861 | 0.000264576 | 0.001053693 | 1 |
| T2DM | LEC | *ERCC6* | 1.054777673 | 0.003306017 | 0.005994804 | 1 |
| T2DM | LEC | *NOL7* | 1.206995879 | 0.002442159 | 0.004689632 | 1 |
| T2DM | LEC | *ELFN1* | 1.168977491 | 9.79E-05 | 0.000908725 | 0.615765512 |
| T2DM | LEC | *ZNF503-AS1* | 1.664510899 | 0.001488069 | 0.00318103 | 1 |
| T2DM | LEC | *LINC00937* | 0.524188123 | 0.000161936 | 0.000910705 | 1 |
| T2DM | LEC | *AC064836.3* | 0.917543353 | 0.000107631 | 0.000908725 | 0.676678852 |
| T2DM | LEC | *TMEM185B* | -1.022759305 | 0.000156599 | 0.000910705 | 0.984535379 |
| T2DM | LEC | *HSBP1L1* | 1.301015798 | 0.004708339 | 0.008101075 | 1 |
| T2DM | LEC | *TAS2R46* | -1.072263635 | 3.87E-05 | 0.000908725 | 0.243198879 |
| T2DM | LEC | *AC093901.1* | 0.836134923 | 9.46E-05 | 0.000908725 | 0.594937254 |
| T2DM | LEC | *AC017074.2* | 5.295551676 | 0.00201343 | 0.004032633 | 1 |
| T2DM | LEC | *AC009299.3* | 0.837646658 | 0.000254417 | 0.00103328 | 1 |
| T2DM | LEC | *SLC2A1-AS1* | -1.115805608 | 0.001159312 | 0.002620854 | 1 |
| T2DM | LEC | *LINC01068* | 1.184383725 | 9.75E-05 | 0.000908725 | 0.613077013 |
| T2DM | LEC | *AC108463.2* | -0.329163308 | 0.000312331 | 0.001141643 | 1 |
| T2DM | LEC | *C19orf24* | 0.702206766 | 0.002634852 | 0.004979054 | 1 |
| T2DM | LEC | *LINC01013* | -1.80751295 | 0.00343764 | 0.006187359 | 1 |
| T2DM | LEC | *AC073133.2* | 4.938994309 | 0.001816557 | 0.003722521 | 1 |
| T2DM | LEC | *PROB1* | 2.315697557 | 0.005076649 | 0.008670713 | 1 |
| T2DM | LEC | *ARL17B* | 0.573719518 | 0.032820161 | 0.04444117 | 1 |
| T2DM | LEC | *AC067945.3* | 0.931025298 | 0.000502351 | 0.001466241 | 1 |
| T2DM | LEC | *RPL41* | 1.237897272 | 0.000225339 | 0.000992757 | 1 |
| T2DM | LEC | *LINC00200* | 5.329370255 | 0.000545146 | 0.001551879 | 1 |
| T2DM | LEC | *PATL2* | 0.938578636 | 0.000170895 | 0.000910705 | 1 |
| T2DM | LEC | *ZNF492* | 0.540772355 | 0.008707878 | 0.013717471 | 1 |
| T2DM | LEC | *U91324.1* | 2.71497095 | 0.000121893 | 0.000908725 | 0.766343835 |
| T2DM | LEC | *ZNF688* | 1.240147631 | 8.96E-05 | 0.000908725 | 0.563118701 |

| | | | | | | |
|---|---|---|---|---|---|---|
| T2DM | LEC | *CISD3* | -0.688036233 | 0.000386501 | 0.001268529 | 1 |
| T2DM | LEC | *KLHL7-AS1* | 1.087119915 | 0.000107763 | 0.000908725 | 0.677503708 |
| T2DM | LEC | *AC092171.4* | 1.408023942 | 0.000369048 | 0.001240089 | 1 |
| T2DM | LEC | *HLA-DPA1* | 0.724483866 | 0.00108614 | 0.00249742 | 1 |
| T2DM | LEC | *DIRC3* | 0.519698725 | 0.000788836 | 0.001989336 | 1 |
| T2DM | LEC | *LINC00865* | 0.607450503 | 0.0004206 | 0.001326594 | 1 |
| T2DM | LEC | *HLA-DQB2* | 0.576945894 | 1.58E-08 | 8.30E-06 | 9.96E-05 |
| T2DM | LEC | *GOLGA8N* | 0.727481725 | 0.000983069 | 0.002320899 | 1 |
| T2DM | LEC | *TNF* | 1.587155793 | 7.18E-07 | 0.000188181 | 0.004516346 |
| T2DM | LEC | *LYRM9* | 0.564089686 | 0.000198356 | 0.000959281 | 1 |
| T2DM | LEC | *LINC00342* | 1.369068451 | 0.006177683 | 0.010210067 | 1 |
| T2DM | LEC | *SNHG15* | 1.187614381 | 0.002421913 | 0.004659089 | 1 |
| T2DM | LEC | *LINC00702* | 0.588745999 | 0.000207759 | 0.000972609 | 1 |
| T2DM | LEC | *LINC00472* | 0.768219374 | 0.000135883 | 0.000908725 | 0.854299077 |
| T2DM | LEC | *AC073133.1* | -1.044067027 | 0.000143994 | 0.000908725 | 0.905290302 |
| T2DM | LEC | *TRIM26* | 0.971481429 | 0.015615194 | 0.022868093 | 1 |
| T2DM | LEC | *LINC01114* | -3.888123663 | 0.006264134 | 0.010336644 | 1 |
| T2DM | LEC | *HLA-B* | 0.837553173 | 1.75E-06 | 0.000289824 | 0.011013327 |
| T2DM | LEC | *C19orf81* | -1.395624819 | 0.021571934 | 0.030515065 | 1 |
| T2DM | LEC | *ANKRD65* | -0.578988479 | 0.000734517 | 0.001898843 | 1 |
| T2DM | LEC | *LINC00957* | 1.168854682 | 0.000100891 | 0.000908725 | 0.634301603 |
| T2DM | LEC | *KIAA0040* | 1.050281864 | 0.001304165 | 0.00286588 | 1 |
| T2DM | LEC | *LINC00941* | -0.585183048 | 0.000145214 | 0.000908725 | 0.912962587 |
| T2DM | LEC | *LINC00545* | 4.933928716 | 0.019023544 | 0.027299936 | 1 |
| T2DM | LEC | *ZBTB22* | 0.854626992 | 0.00287849 | 0.005349414 | 1 |
| T2DM | LEC | *B3GNT9* | 1.10801201 | 0.000216353 | 0.000979981 | 1 |
| T2DM | LEC | *GS1-124K5.4* | 0.405572621 | 0.000496871 | 0.001454972 | 1 |
| T2DM | LEC | *ZNF737* | 1.113428639 | 0.005698977 | 0.009549432 | 1 |
| T2DM | LEC | *HLA-DQA2* | 1.482894961 | 2.28E-22 | 1.43E-18 | 1.43E-18 |
| T2DM | LEC | *AC073257.2* | 5.221540688 | 2.46E-05 | 0.000908725 | 0.154636275 |
| T2DM | LEC | *LRRC37A2* | 0.754826541 | 7.99E-05 | 0.000908725 | 0.502192225 |
| T2DM | LEC | *TXNDC5* | -0.744992016 | 0.000266916 | 0.001057634 | 1 |
| T2DM | LEC | *ASB14* | 1.368053596 | 0.00038578 | 0.001268529 | 1 |
| T2DM | LEC | *ADSL* | 0.512201543 | 0.000379525 | 0.001257815 | 1 |
| T2DM | LEC | *LILRA4* | 15.01670109 | 0.003609217 | 0.006450014 | 1 |
| T2DM | LEC | *PSMB9* | 0.597244979 | 0.003470381 | 0.006235577 | 1 |
| T2DM | LEC | *COX19* | 0.691386337 | 0.000161135 | 0.000910705 | 1 |
| T2DM | LEC | *L1TD1* | 0.401474563 | 0.011372089 | 0.017286345 | 1 |
| T2DM | LEC | *AQP1* | 0.970251114 | 0.013252679 | 0.019762712 | 1 |
| T2DM | LEC | *C1QTNF9* | 2.154007684 | 0.000195377 | 0.000952199 | 1 |
| T2DM | LEC | *ARHGEF25* | 1.383091369 | 0.000167989 | 0.000910705 | 1 |
| T2DM | LEC | *LINC00886* | 0.773066272 | 0.023491655 | 0.032952261 | 1 |
| T2DM | LEC | *NSUN6* | 0.790448512 | 0.000597459 | 0.001656184 | 1 |
| T2DM | LEC | *CD302* | 1.145422156 | 0.000376204 | 0.001250763 | 1 |
| T2DM | LEC | *ARHGAP8* | 1.810131245 | 0.001042177 | 0.002427203 | 1 |
| T2DM | LEC | *ATP5O* | -0.920996658 | 0.028876966 | 0.039674275 | 1 |
| T2DM | LEC | *PLEKHO2* | 1.361591013 | 0.001574184 | 0.003322221 | 1 |
| T2DM | LEC | *PWP2* | 1.7871297 | 0.000901568 | 0.002191988 | 1 |
| T2DM | LEC | *AKAP2* | -0.400889844 | 0.000141351 | 0.000908725 | 0.888675667 |
| T2DM | LEC | *AMACR* | 0.286920523 | 0.006867248 | 0.011205396 | 1 |

| | | | | | | |
|---|---|---|---|---|---|---|
| T2DM | LEC | *ARHGDIG* | 0.693643104 | 0.000398322 | 0.001289522 | 1 |
| T2DM | LEC | *TCP10L* | 3.903344502 | 0.000320883 | 0.001157423 | 1 |
| T2DM | LEC | *C22orf39* | 1.001165424 | 5.56E-05 | 0.000908725 | 0.349821004 |
| T2DM | LEC | *MRPL20* | 1.223179747 | 0.000367137 | 0.001237855 | 1 |
| T2DM | LEC | *DECR2* | 0.541022427 | 9.57E-05 | 0.000908725 | 0.601718476 |
| T2DM | LEC | *CCDC169* | 0.75251322 | 0.01303144 | 0.019489363 | 1 |
| T2DM | LEC | *AP5Z1* | 0.899264132 | 0.000152698 | 0.000910197 | 0.960012187 |
| T2DM | LEC | *STRC* | -3.127907191 | 0.000435034 | 0.00134367 | 1 |
| T2DM | LEC | *PSG4* | 0.756747017 | 0.000168385 | 0.000910705 | 1 |
| T2DM | LEC | *VSIG8* | 0.678519741 | 0.000325104 | 0.001163305 | 1 |
| T2DM | LEC | *C7orf73* | 0.807036436 | 0.000341785 | 0.001191132 | 1 |
| T2DM | LEC | *FGF14-IT1* | 4.625113923 | 0.000568405 | 0.001595341 | 1 |
| T2DM | LEC | *TICAM2* | 1.418167089 | 0.000385692 | 0.001268529 | 1 |
| T2DM | LEC | *PALM2* | 1.055312456 | 1.69E-05 | 0.000908725 | 0.106415461 |
| T2DM | LEC | *NAT6* | 1.186568909 | 1.72E-06 | 0.000289824 | 0.010831763 |
| T2DM | LEC | *IL10RB* | 1.104411767 | 0.000156363 | 0.000910705 | 0.983051904 |
| T2DM | LEC | *WDR92* | 1.010564523 | 3.60E-05 | 0.000908725 | 0.226409661 |
| T2DM | LEC | *PLA2G4B* | 1.920339218 | 7.95E-11 | 7.14E-08 | 5.00E-07 |
| T2DM | LEC | *LEFTY1* | -0.729818254 | 9.57E-05 | 0.000908725 | 0.601577526 |
| T2DM | LEC | *TTC4* | 0.478320377 | 0.000921722 | 0.002218555 | 1 |
| T2DM | LEC | *JMJD7* | 2.044462183 | 3.64E-12 | 5.72E-09 | 2.29E-08 |
| T2DM | LEC | *APOBEC3D* | 2.230776413 | 0.000311335 | 0.001139327 | 1 |
| T2DM | LEC | *RN7SL832P* | 0.700928913 | 0.00029457 | 0.00111174 | 1 |
| T2DM | LEC | *P2RY11* | 0.974450672 | 0.001406351 | 0.003043624 | 1 |
| T2DM | LEC | *DBNDD2* | -1.093589643 | 0.005757877 | 0.009635287 | 1 |
| T2DM | LEC | *LILRA6* | 0.443032682 | 0.000160786 | 0.000910705 | 1 |
| T2DM | LEC | *APOBEC3C* | 1.307899534 | 0.000182766 | 0.000929091 | 1 |
| T2DM | LEC | *ASPRV1* | -1.662462326 | 0.000238239 | 0.001008626 | 1 |
| T2DM | LEC | *UBE2V1* | -0.708103638 | 0.000338738 | 0.001186892 | 1 |
| T2DM | LEC | *N4BP2L2* | 0.612769914 | 0.012593533 | 0.018900822 | 1 |
| T2DM | LEC | *LINC00847* | 0.500128218 | 0.000102833 | 0.000908725 | 0.646510381 |
| T2DM | LEC | *NEAT1* | 0.477828695 | 0.000956165 | 0.00227578 | 1 |
| T2DM | LEC | *ZNF585B* | 0.817093383 | 0.008926434 | 0.014023111 | 1 |
| T2DM | LEC | *RAB30-AS1* | 0.665604678 | 0.019372954 | 0.027738046 | 1 |
| T2DM | LEC | *LINC00920* | -0.441844357 | 0.000219612 | 0.00098409 | 1 |
| T2DM | LEC | *UBAP1L* | -1.030701904 | 0.007703523 | 0.012374054 | 1 |
| T2DM | LEC | *PGAM5* | -0.746039377 | 0.000260415 | 0.001044168 | 1 |
| T2DM | LEC | *DNAJC3-AS1* | -0.994190507 | 0.007682311 | 0.012346291 | 1 |
| T2DM | LEC | *BCKDHA* | 2.307105329 | 0.000250009 | 0.001023586 | 1 |
| T2DM | LEC | *CDK11B* | 1.139431398 | 0.002994409 | 0.005513648 | 1 |
| T2DM | LEC | *PCDHAC1* | 1.775328794 | 0.032491817 | 0.044062997 | 1 |
| T2DM | LEC | *ABHD14A* | 1.063045545 | 0.00011438 | 0.000908725 | 0.719104125 |
| T2DM | LEC | *CCDC153* | 0.92385519 | 0.000154557 | 0.000910197 | 0.971702521 |
| T2DM | LEC | *FMN1* | 0.369477293 | 3.26E-05 | 0.000908725 | 0.205199863 |
| T2DM | LEC | *LINC01060* | -0.661597081 | 0.00999012 | 0.015428122 | 1 |
| T2DM | LEC | *NAIP* | 0.59558412 | 0.000586822 | 0.001632455 | 1 |
| T2DM | LEC | *ZNF286B* | 0.580507996 | 0.000645791 | 0.001740903 | 1 |
| T2DM | LEC | *LINC00942* | 1.97428446 | 0.000482844 | 0.00143258 | 1 |
| T2DM | LEC | *PDCD6* | 1.100682324 | 0.001280425 | 0.002822591 | 1 |
| T2DM | LEC | *LINC00603* | 0.70482982 | 0.00030449 | 0.001126076 | 1 |

| | | | | | | |
|---|---|---|---|---|---|---|
| T2DM | LEC | *YJEFN3* | 0.696578021 | 7.71E-05 | 0.000908725 | 0.484982181 |
| T2DM | LEC | *DNAH10OS* | 2.570141016 | 0.00166981 | 0.003487739 | 1 |
| T2DM | LEC | *PCDHA10* | 1.010149917 | 9.15E-05 | 0.000908725 | 0.575278641 |
| T2DM | LEC | *LINC00989* | -1.055880189 | 0.001198479 | 0.002684303 | 1 |
| T2DM | LEC | *LINC01021* | 1.909207746 | 0.000135559 | 0.000908725 | 0.852256852 |
| T2DM | LEC | *FAM218A* | 0.867687671 | 0.000112544 | 0.000908725 | 0.707564496 |
| T2DM | LEC | *ATP6V1E2* | 0.570591535 | 0.001120114 | 0.002557065 | 1 |
| T2DM | LEC | *GLI4* | 1.183914778 | 0.001745997 | 0.003609696 | 1 |
| T2DM | LEC | *SHANK3* | -0.205845958 | 0.000354093 | 0.001215214 | 1 |
| T2DM | LEC | *ZNF550* | 1.231904034 | 0.00208673 | 0.00414511 | 1 |
| T2DM | LEC | *CCDC71L* | -0.950245911 | 0.016013429 | 0.023380499 | 1 |
| T2DM | LEC | *TMEM200B* | 1.14151389 | 0.00105479 | 0.002445543 | 1 |
| T2DM | LEC | *PCDHGB6* | 0.899748013 | 6.46E-05 | 0.000908725 | 0.405912681 |
| T2DM | LEC | *TRNP1* | 2.029318798 | 0.000112473 | 0.000908725 | 0.70711705 |
| T2DM | LEC | *PCDHGA5* | 0.439193073 | 0.000454255 | 0.001380328 | 1 |
| T2DM | LEC | *ATXN7L3B* | 1.227353901 | 0.005336001 | 0.009037565 | 1 |
| T2DM | LEC | *PRKDC* | 0.794938944 | 0.014315527 | 0.021186846 | 1 |
| T2DM | LEC | *PCDHGA6* | -0.434936937 | 0.025794635 | 0.035823033 | 1 |
| T2DM | LEC | *PCDHGA11* | 0.646165611 | 0.003972333 | 0.006975994 | 1 |
| T2DM | LEC | *CLDN23* | 0.975165863 | 6.73E-05 | 0.000908725 | 0.423016136 |
| T2DM | LEC | *ZNF260* | 1.105285065 | 0.033897777 | 0.045732902 | 1 |
| T2DM | LEC | *ARMS2* | 1.540690655 | 0.00077803 | 0.001967609 | 1 |
| T2DM | LEC | *FPGT* | 0.898350267 | 0.00015792 | 0.000910705 | 0.992844928 |
| T2DM | LEC | *MPV17L2* | -0.457090524 | 0.000601381 | 0.001659739 | 1 |
| T2DM | LEC | *DPP3* | 0.620769005 | 0.000500345 | 0.001463103 | 1 |
| T2DM | LEC | *KRTAP5-9* | 3.658929974 | 0.008544856 | 0.013491088 | 1 |
| T2DM | LEC | *EID3* | 0.551863799 | 0.0010399 | 0.00242502 | 1 |
| T2DM | LEC | *SNHG9* | 0.692016757 | 0.001743261 | 0.003605225 | 1 |
| T2DM | LEC | *EBLN2* | 0.936257433 | 3.90E-05 | 0.000908725 | 0.245497663 |
| T2DM | LEC | *POLR2M* | 1.084953697 | 0.000208949 | 0.000972609 | 1 |
| T2DM | LEC | *TAS2R20* | -0.229969595 | 0.004596638 | 0.007930588 | 1 |
| T2DM | LEC | *LINC00346* | 1.069645253 | 0.00193371 | 0.003918369 | 1 |
| T2DM | LEC | *CTSO* | 1.010283466 | 0.000395375 | 0.001285276 | 1 |
| T2DM | LEC | *ZNF432* | 0.924936428 | 0.002865287 | 0.005331237 | 1 |
| T2DM | LEC | *LINC00944* | 0.640818395 | 0.000378056 | 0.001253649 | 1 |
| T2DM | LEC | *ZNF10* | 0.585050483 | 0.029260726 | 0.040131367 | 1 |
| T2DM | LEC | *HMBS* | 1.211017679 | 9.93E-09 | 5.68E-06 | 6.24E-05 |
| T2DM | LEC | *TAS2R31* | -0.587740255 | 0.000465662 | 0.001398163 | 1 |
| T2DM | LEC | *PGA5* | 6.172177327 | 0.000912975 | 0.002208614 | 1 |
| T2DM | LEC | *HP* | 1.08326875 | 0.013272156 | 0.019787064 | 1 |
| T2DM | LEC | *KIAA1147* | 0.874815913 | 0.030204362 | 0.041236661 | 1 |
| T2DM | LEC | *GATC* | 0.963975522 | 0.003360065 | 0.006084311 | 1 |
| T2DM | LEC | *ZBED6* | -2.927842583 | 0.002090577 | 0.004150129 | 1 |
| T2DM | LEC | *CUX1* | 0.636520473 | 0.036016378 | 0.048383541 | 1 |
| T2DM | LEC | *RTEL1* | 1.290003179 | 1.21E-05 | 0.000851536 | 0.075786694 |
| T2DM | LEC | *ZNF578* | -0.662782926 | 0.011073134 | 0.016909593 | 1 |
| T2DM | LEC | *PDF* | -2.617507536 | 0.000472345 | 0.001412093 | 1 |
| T2DM | LEC | *LINC00640* | 0.400943812 | 0.001424572 | 0.00307459 | 1 |
| T2DM | LEC | *LINC00638* | 1.580748655 | 0.000665276 | 0.001776048 | 1 |
| T2DM | LEC | *CEP95* | 0.821201849 | 0.001043754 | 0.002427703 | 1 |

| | | | | | | |
|---|---|---|---|---|---|---|
| T2DM | LEC | *LINC00639* | 0.3323042 | 1.15E-05 | 0.000830158 | 0.072223786 |
| T2DM | LEC | *LINC00648* | 0.548235768 | 0.000174893 | 0.000911734 | 1 |
| T2DM | LEC | *LINC00924* | -0.122288142 | 0.000148274 | 0.000910197 | 0.93219596 |
| T2DM | LEC | *ITGB3* | 0.649413505 | 4.81E-05 | 0.000908725 | 0.302264854 |
| T2DM | LEC | *UBE2Q2L* | -1.188176522 | 9.92E-05 | 0.000908725 | 0.623792369 |
| T2DM | LEC | *IQCH-AS1* | 0.750070189 | 0.00126767 | 0.002802335 | 1 |
| T2DM | LEC | *AC083843.1* | -3.899691776 | 1.61E-05 | 0.000908725 | 0.101369185 |
| T2DM | LEC | *SH3RF3-AS1* | -1.203342265 | 0.000503105 | 0.001467078 | 1 |
| T2DM | LEC | *HOXB7* | 1.115303643 | 0.001609006 | 0.003385483 | 1 |
| T2DM | LEC | *FRRS1L* | 1.113174561 | 6.77E-05 | 0.000908725 | 0.425381023 |
| T2DM | LEC | *LINC00562* | 0.56126425 | 0.00070609 | 0.001848892 | 1 |
| T2DM | LEC | *AC009120.5* | -0.960128319 | 0.000304327 | 0.001126076 | 1 |
| T2DM | LEC | *TMEM178B* | 0.66732231 | 0.000226248 | 0.000992757 | 1 |
| T2DM | LEC | *LINC01003* | 0.624806488 | 0.000174055 | 0.000911117 | 1 |
| T2DM | LEC | *HPR* | -0.69201399 | 0.000226595 | 0.000992757 | 1 |
| T2DM | LEC | *GOLGA8H* | 0.528675622 | 0.016405136 | 0.02387746 | 1 |
| T2DM | LEC | *LINC00662* | 0.881619376 | 0.022018128 | 0.031072496 | 1 |
| T2DM | LEC | *AC123768.4* | -2.26295598 | 0.000831961 | 0.002065773 | 1 |
| T2DM | LEC | *LINC00667* | 0.614434031 | 0.004088718 | 0.007154403 | 1 |
| T2DM | LEC | *LINC00672* | 0.723445035 | 0.005274667 | 0.008945733 | 1 |
| T2DM | LEC | *LINC00909* | 0.873565926 | 0.0315927 | 0.042942664 | 1 |
| T2DM | LEC | *DYNLL2* | 1.053859599 | 0.001490156 | 0.003183353 | 1 |
| T2DM | LEC | *LINC00526* | 0.663955958 | 0.000231534 | 0.001001608 | 1 |
| T2DM | LEC | *STRADA* | 0.647489619 | 0.022054018 | 0.031116161 | 1 |
| T2DM | LEC | *LINC00663* | 0.229881363 | 0.021118661 | 0.029964573 | 1 |
| T2DM | LEC | *ILF3-AS1* | 0.777991316 | 3.29E-05 | 0.000908725 | 0.206535891 |
| T2DM | LEC | *AC139100.2* | -0.865801374 | 0.000153393 | 0.000910197 | 0.96438136 |
| T2DM | LEC | *LINC01140* | 0.732294687 | 2.47E-05 | 0.000908725 | 0.154993382 |
| T2DM | LEC | *LINC00906* | 3.434251356 | 0.000115469 | 0.000908725 | 0.725954649 |
| T2DM | LEC | *ZNF582-AS1* | -0.931524713 | 0.000426838 | 0.001336216 | 1 |
| T2DM | LEC | *AC137932.1* | 1.788185151 | 0.000631597 | 0.001715271 | 1 |
| T2DM | LEC | *AC003005.2* | 1.161643399 | 0.000237557 | 0.001007093 | 1 |
| T2DM | LEC | *AL132989.1* | -0.554064125 | 0.000166393 | 0.000910705 | 1 |
| T2DM | LEC | *LINC00664* | -1.931995882 | 0.003646652 | 0.006505818 | 1 |
| T2DM | LEC | *AC009060.1* | 0.809234973 | 0.006594517 | 0.010813701 | 1 |
| T2DM | LEC | *SNHG8* | 0.982929991 | 0.002161862 | 0.004267386 | 1 |
| T2DM | LEC | *YTHDF3-AS1* | 0.903562985 | 7.78E-05 | 0.000908725 | 0.489180709 |
| T2DM | LEC | *LINC01050* | 4.207205083 | 3.28E-05 | 0.000908725 | 0.205905334 |
| T2DM | LEC | *MILR1* | 0.348118886 | 9.53E-05 | 0.000908725 | 0.598935508 |
| T2DM | LEC | *FGF14-AS2* | 1.473643474 | 0.000628762 | 0.001713493 | 1 |
| T2DM | LEC | *CASC15* | -0.208420476 | 0.000179037 | 0.000922628 | 1 |
| T2DM | LEC | *LINC01023* | 0.806438808 | 4.70E-05 | 0.000908725 | 0.295608142 |
| T2DM | LEC | *DOC2B* | -0.257165379 | 2.61E-05 | 0.000908725 | 0.164077718 |
| T2DM | LEC | *POLR2J4* | 0.664660674 | 0.000115366 | 0.000908725 | 0.725306538 |
| T2DM | LEC | *AC144652.1* | 0.879115992 | 0.000651672 | 0.001749386 | 1 |
| T2DM | LEC | *PAXIP1-AS1* | -0.825783334 | 0.030093624 | 0.041112258 | 1 |
| T2DM | macrophage | *CFH* | 0.816244653 | 3.79E-06 | 0.000108369 | 0.023841125 |
| T2DM | macrophage | *SEMA3F* | -0.217301599 | 4.13E-05 | 0.000586182 | 0.259678597 |
| T2DM | macrophage | *BAD* | 1.298032146 | 0.007869322 | 0.033470855 | 1 |
| T2DM | macrophage | *HS3ST1* | -0.601527042 | 0.000927067 | 0.006440298 | 1 |

| | | | | | | |
|---|---|---|---|---|---|---|
| T2DM | macrophage | *CYP26B1* | 0.931397295 | 7.27E-05 | 0.0009036 | 0.457221583 |
| T2DM | macrophage | *ALS2* | 0.738498771 | 0.005038376 | 0.023568655 | 1 |
| T2DM | macrophage | *POLDIP2* | 1.094512223 | 0.000463952 | 0.003789843 | 1 |
| T2DM | macrophage | *RBM6* | -0.204259149 | 1.84E-07 | 2.90E-05 | 0.001158485 |
| T2DM | macrophage | *SLC22A16* | -0.265225244 | 0.001473364 | 0.009281601 | 1 |
| T2DM | macrophage | *CIAPIN1* | 1.218519556 | 0.000300723 | 0.002644258 | 1 |
| T2DM | macrophage | *COPZ2* | 0.215304591 | 0.005174074 | 0.02408848 | 1 |
| T2DM | macrophage | *GCFC2* | 0.744086711 | 0.001668889 | 0.010256408 | 1 |
| T2DM | macrophage | *WDR54* | 0.862486517 | 0.00162187 | 0.010016404 | 1 |
| T2DM | macrophage | *ABCB4* | 1.151490157 | 0.000208636 | 0.002027346 | 1 |
| T2DM | macrophage | *PDK2* | 0.580484452 | 0.00349752 | 0.018025187 | 1 |
| T2DM | macrophage | *GGNBP2* | -0.399901664 | 0.000989056 | 0.006722381 | 1 |
| T2DM | macrophage | *ASB4* | 0.255657775 | 0.011648588 | 0.044984444 | 1 |
| T2DM | macrophage | *ABCC8* | 1.35731029 | 0.001676225 | 0.010289005 | 1 |
| T2DM | macrophage | *TMEM132A* | 1.947673426 | 3.79E-05 | 0.000552152 | 0.238529822 |
| T2DM | macrophage | *ARHGAP44* | 0.587774711 | 0.002719746 | 0.014907624 | 1 |
| T2DM | macrophage | *NOS2* | 5.726700641 | 8.30E-06 | 0.000186258 | 0.052152131 |
| T2DM | macrophage | *BAIAP3* | 0.711582917 | 0.011164817 | 0.043550002 | 1 |
| T2DM | macrophage | *TSR3* | 1.695299083 | 2.23E-05 | 0.000379264 | 0.139948304 |
| T2DM | macrophage | *MGST1* | 0.498870656 | 0.000953072 | 0.006584578 | 1 |
| T2DM | macrophage | *MMP25* | 0.538400161 | 0.001609902 | 0.009962062 | 1 |
| T2DM | macrophage | *PKD1* | 0.430183779 | 0.001663822 | 0.010235276 | 1 |
| T2DM | macrophage | *MLXIPL* | 0.118157289 | 0.001339757 | 0.008612526 | 1 |
| T2DM | macrophage | *CD4* | -0.479665766 | 1.29E-05 | 0.000257003 | 0.080927118 |
| T2DM | macrophage | *SCMH1* | 0.41126307 | 0.007109775 | 0.030827004 | 1 |
| T2DM | macrophage | *MKS1* | 1.323787653 | 0.000484235 | 0.003918131 | 1 |
| T2DM | macrophage | *TTC19* | 0.41505248 | 0.008308833 | 0.034732469 | 1 |
| T2DM | macrophage | *SYT7* | 0.775947441 | 9.52E-05 | 0.001102229 | 0.598510387 |
| T2DM | macrophage | *LARS2* | 0.538357924 | 0.00854226 | 0.035500042 | 1 |
| T2DM | macrophage | *PPP5C* | 0.688904103 | 0.000800205 | 0.005762761 | 1 |
| T2DM | macrophage | *CEP68* | 0.68201273 | 1.19E-10 | 6.78E-08 | 7.45E-07 |
| T2DM | macrophage | *ZBTB32* | 1.61410439 | 3.17E-05 | 0.000487893 | 0.199060167 |
| T2DM | macrophage | *ALOX5* | -0.492930238 | 0.000100252 | 0.001148054 | 0.630281439 |
| T2DM | macrophage | *PSMC4* | 0.389905111 | 0.008863234 | 0.036515828 | 1 |
| T2DM | macrophage | *CLDN11* | 0.42332429 | 0.002204348 | 0.012796617 | 1 |
| T2DM | macrophage | *TACC3* | -0.408411026 | 0.008284765 | 0.034654901 | 1 |
| T2DM | macrophage | *CHDH* | 1.450647692 | 1.58E-05 | 0.000294107 | 0.099408204 |
| T2DM | macrophage | *IL20RA* | 1.864453933 | 4.87E-07 | 3.87E-05 | 0.003059619 |
| T2DM | macrophage | *WWTR1* | 0.72607395 | 4.25E-05 | 0.000599701 | 0.267466663 |
| T2DM | macrophage | *MNAT1* | 0.711717282 | 0.001712337 | 0.010462064 | 1 |
| T2DM | macrophage | *OSBPL5* | 0.975879 | 0.00032404 | 0.002817761 | 1 |
| T2DM | macrophage | *HSF2* | 0.881348826 | 0.009811011 | 0.039489006 | 1 |
| T2DM | macrophage | *RNASET2* | -0.322685194 | 0.003959227 | 0.019849811 | 1 |
| T2DM | macrophage | *ARNTL2* | 1.183099246 | 2.43E-05 | 0.000407339 | 0.152752167 |
| T2DM | macrophage | *ANK1* | 1.504449976 | 5.30E-06 | 0.000135329 | 0.033290843 |
| T2DM | macrophage | *GRN* | -0.555704137 | 0.004074158 | 0.020248403 | 1 |
| T2DM | macrophage | *FUT8* | 0.572188525 | 0.012779687 | 0.048178716 | 1 |
| T2DM | macrophage | *TUBG2* | -0.260640314 | 4.60E-07 | 3.80E-05 | 0.002889783 |
| T2DM | macrophage | *MAT2B* | 0.971142158 | 0.001280951 | 0.008310981 | 1 |
| T2DM | macrophage | *DNAH5* | 0.612205144 | 0.006773465 | 0.029675802 | 1 |

| | | | | | | |
|---|---|---|---|---|---|---|
| T2DM | macrophage | *C6* | 0.48741095 | 0.007109316 | 0.030827004 | 1 |
| T2DM | macrophage | *RAB27B* | 0.322791151 | 0.000294121 | 0.002608095 | 1 |
| T2DM | macrophage | *MED17* | 0.809471429 | 0.000481108 | 0.003897846 | 1 |
| T2DM | macrophage | *LCP2* | -0.417419971 | 0.000347152 | 0.002981622 | 1 |
| T2DM | macrophage | *EPHA3* | 0.947363272 | 2.05E-05 | 0.000356591 | 0.128729389 |
| T2DM | macrophage | *DSG2* | 0.921989352 | 1.34E-05 | 0.000265687 | 0.084488557 |
| T2DM | macrophage | *TSPAN17* | 0.340422749 | 0.002419556 | 0.013679628 | 1 |
| T2DM | macrophage | *CC2D2A* | 0.757044971 | 0.000748965 | 0.005437349 | 1 |
| T2DM | macrophage | *EPN3* | 1.291207618 | 0.000457094 | 0.003756536 | 1 |
| T2DM | macrophage | *DKK3* | 0.915537818 | 0.000161351 | 0.00162827 | 1 |
| T2DM | macrophage | *ARHGEF5* | 1.658611659 | 7.96E-06 | 0.000180097 | 0.050066969 |
| T2DM | macrophage | *PTGER3* | 0.602299536 | 0.000278969 | 0.002493278 | 1 |
| T2DM | macrophage | *ALX4* | 2.074014392 | 1.43E-06 | 6.22E-05 | 0.008967502 |
| T2DM | macrophage | *MCF2L2* | -0.575400009 | 0.002475574 | 0.013933694 | 1 |
| T2DM | macrophage | *ATP9A* | 0.65757324 | 0.000914806 | 0.006380743 | 1 |
| T2DM | macrophage | *GINM1* | 0.625717331 | 0.010913795 | 0.042830856 | 1 |
| T2DM | macrophage | *ITIH4* | 0.18113777 | 2.92E-06 | 9.41E-05 | 0.018356701 |
| T2DM | macrophage | *IL17RB* | 1.110539236 | 0.004900802 | 0.023114286 | 1 |
| T2DM | macrophage | *TRAF3IP2* | 0.941417745 | 0.000258027 | 0.00238211 | 1 |
| T2DM | macrophage | *DCBLD2* | 0.64230616 | 0.010210548 | 0.040680426 | 1 |
| T2DM | macrophage | *NCKAP1* | 0.65416262 | 6.02E-05 | 0.000776918 | 0.378359225 |
| T2DM | macrophage | *SPHK2* | 0.659760748 | 0.008044948 | 0.033878241 | 1 |
| T2DM | macrophage | *ISOC2* | 1.792592528 | 1.89E-05 | 0.00033478 | 0.118846974 |
| T2DM | macrophage | *DMRT3* | 1.394847439 | 8.06E-07 | 4.61E-05 | 0.00506552 |
| T2DM | macrophage | *ATP2C2* | 1.062137597 | 0.001145599 | 0.00758944 | 1 |
| T2DM | macrophage | *TMEM161A* | 0.822170311 | 0.002039728 | 0.012086494 | 1 |
| T2DM | macrophage | *SUGP2* | 0.494825565 | 0.001359355 | 0.008729588 | 1 |
| T2DM | macrophage | *NTHL1* | 0.431654778 | 0.007989092 | 0.033754988 | 1 |
| T2DM | macrophage | *IPO5* | 0.925473116 | 0.000931956 | 0.006463856 | 1 |
| T2DM | macrophage | *NDUFB4* | 0.823562512 | 0.010780864 | 0.042441634 | 1 |
| T2DM | macrophage | *SEC61A2* | 0.999023941 | 0.003776051 | 0.019097377 | 1 |
| T2DM | macrophage | *PPP2R5A* | 0.622016616 | 0.006052867 | 0.027142922 | 1 |
| T2DM | macrophage | *NGEF* | 2.645733981 | 1.88E-06 | 7.17E-05 | 0.0118277 |
| T2DM | macrophage | *CD84* | -0.21764647 | 0.002395709 | 0.013637966 | 1 |
| T2DM | macrophage | *STOML1* | 1.955508725 | 3.33E-05 | 0.000502535 | 0.209054719 |
| T2DM | macrophage | *TP53BP1* | 0.553236255 | 0.004428009 | 0.021510192 | 1 |
| T2DM | macrophage | *SYT1* | -0.27285376 | 0.007124555 | 0.030869798 | 1 |
| T2DM | macrophage | *PITX1* | 2.847526731 | 9.26E-07 | 4.80E-05 | 0.005819685 |
| T2DM | macrophage | *SDK2* | 0.98144843 | 0.002405029 | 0.013657666 | 1 |
| T2DM | macrophage | *DNAJA2* | 0.897872833 | 0.012564578 | 0.047672602 | 1 |
| T2DM | macrophage | *KCNAB2* | -0.468682675 | 8.76E-06 | 0.000192632 | 0.055092838 |
| T2DM | macrophage | *GAL* | -1.442005378 | 8.20E-05 | 0.000985306 | 0.51531483 |
| T2DM | macrophage | *NEDD4* | 0.438416458 | 0.002527646 | 0.014150766 | 1 |
| T2DM | macrophage | *GNB5* | 0.362116407 | 0.006592039 | 0.029042853 | 1 |
| T2DM | macrophage | *DAPP1* | -0.285423135 | 0.00222719 | 0.012893503 | 1 |
| T2DM | macrophage | *FGF10* | 0.207971503 | 0.003811886 | 0.019233809 | 1 |
| T2DM | macrophage | *CLTCL1* | 0.393411136 | 0.002938927 | 0.015846767 | 1 |
| T2DM | macrophage | *WIPI1* | 0.958106701 | 0.000878608 | 0.00617185 | 1 |
| T2DM | macrophage | *NDST1* | 0.791898957 | 0.000215805 | 0.002074564 | 1 |
| T2DM | macrophage | *ASNS* | 1.034395469 | 7.72E-05 | 0.000948133 | 0.485444195 |

| | | | | | | |
|---|---|---|---|---|---|---|
| T2DM | macrophage | *TESK2* | -0.58131433 | 0.005118807 | 0.02387384 | 1 |
| T2DM | macrophage | *CAMK2A* | 2.796992788 | 0.000100014 | 0.001147421 | 0.628786573 |
| T2DM | macrophage | *OSBPL3* | -0.484339442 | 5.15E-05 | 0.000696188 | 0.323727286 |
| T2DM | macrophage | *TRIP13* | 3.552843158 | 0.002691175 | 0.014802639 | 1 |
| T2DM | macrophage | *CYBRD1* | 0.371503844 | 0.005942123 | 0.026818468 | 1 |
| T2DM | macrophage | *LIMS2* | 0.481093332 | 0.000832021 | 0.005910641 | 1 |
| T2DM | macrophage | *TRHDE* | 0.696909751 | 5.98E-05 | 0.000776326 | 0.375935287 |
| T2DM | macrophage | *TP63* | 0.831684169 | 0.000439824 | 0.003635299 | 1 |
| T2DM | macrophage | *NLE1* | 0.615960454 | 0.010131418 | 0.040488293 | 1 |
| T2DM | macrophage | *SDHA* | 0.551258589 | 4.97E-05 | 0.000674748 | 0.312408236 |
| T2DM | macrophage | *SMARCE1* | 0.666043608 | 0.003114485 | 0.016551789 | 1 |
| T2DM | macrophage | *ATP2A3* | 0.476663708 | 0.00215189 | 0.01254608 | 1 |
| T2DM | macrophage | *NTN4* | 0.744415832 | 0.000170561 | 0.00171023 | 1 |
| T2DM | macrophage | *LMAN1* | 0.788606295 | 0.003119326 | 0.016563516 | 1 |
| T2DM | macrophage | *SRI* | 0.912743616 | 0.004010777 | 0.020028402 | 1 |
| T2DM | macrophage | *CELSR1* | -0.913088235 | 0.012808726 | 0.048249526 | 1 |
| T2DM | macrophage | *SLC25A3* | 0.561478785 | 0.013364198 | 0.049834349 | 1 |
| T2DM | macrophage | *BCAP29* | 0.553590108 | 0.012425215 | 0.047229337 | 1 |
| T2DM | macrophage | *RBMS2* | 0.691266512 | 0.001733734 | 0.010562001 | 1 |
| T2DM | macrophage | *CAMSAP3* | 10.19628654 | 0.00030334 | 0.002663548 | 1 |
| T2DM | macrophage | *XAB2* | -0.555426496 | 0.00864436 | 0.035801773 | 1 |
| T2DM | macrophage | *CTTNBP2* | -0.694517121 | 5.79E-07 | 4.13E-05 | 0.00363703 |
| T2DM | macrophage | *LRCH4* | -0.472594884 | 1.47E-05 | 0.000282513 | 0.092664212 |
| T2DM | macrophage | *GPR137B* | -0.190257905 | 0.011169487 | 0.043550002 | 1 |
| T2DM | macrophage | *NAALAD2* | 0.356663154 | 0.000956652 | 0.00659701 | 1 |
| T2DM | macrophage | *TULP3* | 0.900987705 | 0.000357638 | 0.003046705 | 1 |
| T2DM | macrophage | *SYNJ2* | 0.534989927 | 0.003777375 | 0.019097377 | 1 |
| T2DM | macrophage | *FGF20* | -1.926829926 | 1.52E-06 | 6.42E-05 | 0.009539485 |
| T2DM | macrophage | *MYH7B* | 0.581100967 | 0.00191695 | 0.011423566 | 1 |
| T2DM | macrophage | *FDFT1* | 0.770546792 | 0.008549675 | 0.035503174 | 1 |
| T2DM | macrophage | *RIMS1* | 0.74395631 | 0.001493719 | 0.009381629 | 1 |
| T2DM | macrophage | *DCT* | 1.877564242 | 5.22E-06 | 0.000134402 | 0.032836079 |
| T2DM | macrophage | *SCTR* | -0.609644881 | 0.000141796 | 0.001479525 | 0.891472168 |
| T2DM | macrophage | *MOK* | 1.074957828 | 4.01E-05 | 0.000576252 | 0.252223175 |
| T2DM | macrophage | *MAGI3* | 0.709525716 | 0.001247587 | 0.008132722 | 1 |
| T2DM | macrophage | *STK17B* | -0.456041041 | 0.010606057 | 0.0419901 | 1 |
| T2DM | macrophage | *C5orf22* | 0.882693159 | 0.0064858 | 0.028655111 | 1 |
| T2DM | macrophage | *COBLL1* | -0.269952421 | 0.001043425 | 0.007046204 | 1 |
| T2DM | macrophage | *ZNF324* | 1.229010608 | 0.005520824 | 0.025261586 | 1 |
| T2DM | macrophage | *ZNF446* | 1.803164405 | 1.33E-05 | 0.000263397 | 0.083496734 |
| T2DM | macrophage | *GSTP1* | 0.844430123 | 0.005236932 | 0.02429859 | 1 |
| T2DM | macrophage | *APOB* | 0.543054856 | 0.007452862 | 0.032102302 | 1 |
| T2DM | macrophage | *CD82* | 0.430847922 | 0.002067101 | 0.01221416 | 1 |
| T2DM | macrophage | *ZNF213* | 0.345571268 | 0.005271557 | 0.024423196 | 1 |
| T2DM | macrophage | *CPNE3* | 0.49722448 | 6.71E-05 | 0.000849692 | 0.422053093 |
| T2DM | macrophage | *CTTN* | 0.842123622 | 1.16E-05 | 0.000236032 | 0.072769727 |
| T2DM | macrophage | *ATG16L1* | 0.978133148 | 0.001372544 | 0.008796314 | 1 |
| T2DM | macrophage | *CHMP5* | 0.704068852 | 0.008045206 | 0.033878241 | 1 |
| T2DM | macrophage | *EIF2AK1* | 0.709084465 | 0.001441585 | 0.009155971 | 1 |
| T2DM | macrophage | *EPDR1* | 0.186053537 | 0.003414062 | 0.017783272 | 1 |

| | | | | | | |
|---|---|---|---|---|---|---|
| T2DM | macrophage | *SNX10* | -0.084017459 | 0.001435618 | 0.009135358 | 1 |
| T2DM | macrophage | *SEPHS1* | 0.582266143 | 0.007410061 | 0.031952713 | 1 |
| T2DM | macrophage | *ITPKC* | 0.762238421 | 0.000664762 | 0.004969511 | 1 |
| T2DM | macrophage | *HSD17B14* | -0.627591177 | 1.97E-06 | 7.42E-05 | 0.012393854 |
| T2DM | macrophage | *GMCL1* | 0.699592227 | 0.002947433 | 0.015878757 | 1 |
| T2DM | macrophage | *KLHL42* | 0.625652374 | 0.002044875 | 0.012105581 | 1 |
| T2DM | macrophage | *PTHLH* | 0.549359199 | 0.002640778 | 0.014630053 | 1 |
| T2DM | macrophage | *PHACTR3* | 0.244299272 | 0.007341731 | 0.031701553 | 1 |
| T2DM | macrophage | *ASAP3* | 0.682917673 | 0.000109057 | 0.001215675 | 0.685640729 |
| T2DM | macrophage | *DOCK3* | -0.221717103 | 0.000456293 | 0.00375486 | 1 |
| T2DM | macrophage | *C3orf18* | 1.033349835 | 6.69E-11 | 4.67E-08 | 4.20E-07 |
| T2DM | macrophage | *ZNF343* | 0.795434582 | 0.000185287 | 0.001846118 | 1 |
| T2DM | macrophage | *LZTS3* | 0.743868862 | 0.001745614 | 0.010603553 | 1 |
| T2DM | macrophage | *TESC* | 1.753355623 | 0.000874221 | 0.006154791 | 1 |
| T2DM | macrophage | *TMEM230* | 0.473408146 | 0.004985308 | 0.023424988 | 1 |
| T2DM | macrophage | *OAS1* | -0.751479302 | 0.000193881 | 0.001904576 | 1 |
| T2DM | macrophage | *GMIP* | -0.752361096 | 2.59E-05 | 0.000417424 | 0.162640819 |
| T2DM | macrophage | *MLF2* | 1.012728 | 0.002092989 | 0.012307926 | 1 |
| T2DM | macrophage | *SLC9A1* | 0.939263131 | 0.006122016 | 0.027355446 | 1 |
| T2DM | macrophage | *RGS1* | -0.452751473 | 3.00E-06 | 9.57E-05 | 0.01884921 |
| T2DM | macrophage | *YPEL3* | 0.192410188 | 0.004474084 | 0.021654015 | 1 |
| T2DM | macrophage | *GOLGA3* | 0.419059085 | 0.010088494 | 0.040424706 | 1 |
| T2DM | macrophage | *PABPC4* | -0.751690828 | 2.83E-10 | 1.48E-07 | 1.78E-06 |
| T2DM | macrophage | *CERS4* | 0.748219884 | 0.006324399 | 0.028020789 | 1 |
| T2DM | macrophage | *NLRC4* | -0.762361238 | 0.000134042 | 0.001418721 | 0.842720008 |
| T2DM | macrophage | *SLC26A4* | 1.75279244 | 9.74E-05 | 0.001124126 | 0.612648638 |
| T2DM | macrophage | *IL5RA* | 2.999729666 | 1.24E-05 | 0.000250222 | 0.078069178 |
| T2DM | macrophage | *RAPGEF4* | 0.790537622 | 9.29E-06 | 0.00020003 | 0.058408683 |
| T2DM | macrophage | *TF* | 0.763128983 | 0.000994437 | 0.00675165 | 1 |
| T2DM | macrophage | *MYO15A* | 0.781998086 | 0.001501655 | 0.00940579 | 1 |
| T2DM | macrophage | *ZFHX4* | 0.623052693 | 0.000245775 | 0.002289166 | 1 |
| T2DM | macrophage | *VNN3* | 2.391344426 | 2.37E-05 | 0.000399358 | 0.148960527 |
| T2DM | macrophage | *SEC22C* | 0.762342461 | 0.003074442 | 0.016422275 | 1 |
| T2DM | macrophage | *FMO2* | 0.289737565 | 0.001760644 | 0.010655495 | 1 |
| T2DM | macrophage | *MAP3K1* | -0.565903232 | 0.003029337 | 0.016236522 | 1 |
| T2DM | macrophage | *CWF19L1* | -0.240447944 | 0.001135341 | 0.007553318 | 1 |
| T2DM | macrophage | *BLNK* | -0.285901999 | 2.06E-05 | 0.000357193 | 0.12930382 |
| T2DM | macrophage | *CREM* | -0.483614418 | 0.009513706 | 0.038638674 | 1 |
| T2DM | macrophage | *TPSD1* | 4.564179195 | 2.56E-08 | 8.47E-06 | 0.000160966 |
| T2DM | macrophage | *ITPR3* | 1.094633002 | 1.66E-05 | 0.000303568 | 0.104427347 |
| T2DM | macrophage | *PCSK5* | 0.69133084 | 6.72E-05 | 0.000849692 | 0.422296877 |
| T2DM | macrophage | *PRTFDC1* | 0.81448037 | 0.001205265 | 0.007934558 | 1 |
| T2DM | macrophage | *PALMD* | 0.65514946 | 1.25E-05 | 0.000251018 | 0.078568776 |
| T2DM | macrophage | *CDC34* | 0.941691722 | 0.009707782 | 0.03919899 | 1 |
| T2DM | macrophage | *MADCAM1* | 1.070038331 | 0.000892252 | 0.006253722 | 1 |
| T2DM | macrophage | *LZTR1* | 0.954392672 | 0.006160794 | 0.027485775 | 1 |
| T2DM | macrophage | *P2RX6* | 0.775579949 | 0.00144177 | 0.009155971 | 1 |
| T2DM | macrophage | *GGT5* | 1.320784517 | 1.56E-06 | 6.42E-05 | 0.009809675 |
| T2DM | macrophage | *CYTH4* | -0.379260937 | 7.09E-06 | 0.000165711 | 0.044565272 |
| T2DM | macrophage | *CARD10* | 0.583327262 | 0.000469485 | 0.003828343 | 1 |

| | | | | | | |
|---|---|---|---|---|---|---|
| T2DM | macrophage | *LRP5L* | -0.63349361 | 0.004458721 | 0.021629615 | 1 |
| T2DM | macrophage | *SLC25A1* | 0.803265047 | 0.003020579 | 0.016203395 | 1 |
| T2DM | macrophage | *GGA1* | -0.378371258 | 0.004522103 | 0.021769113 | 1 |
| T2DM | macrophage | *SH3BP1* | -0.72080975 | 1.10E-05 | 0.000227343 | 0.069339684 |
| T2DM | macrophage | *TIMP3* | 0.621490911 | 0.000812744 | 0.005819727 | 1 |
| T2DM | macrophage | *CYB5R3* | 0.649949266 | 0.004495751 | 0.021692086 | 1 |
| T2DM | macrophage | *RASL10A* | -0.344698705 | 0.000716364 | 0.005279931 | 1 |
| T2DM | macrophage | *AP1B1* | -0.514288128 | 7.46E-05 | 0.000920979 | 0.468778278 |
| T2DM | macrophage | *BIK* | 4.087313876 | 3.76E-05 | 0.000548284 | 0.236097317 |
| T2DM | macrophage | *RBFOX2* | 0.710583337 | 0.00011635 | 0.001274378 | 0.731493153 |
| T2DM | macrophage | *SAMM50* | 1.006218191 | 0.002523316 | 0.014139118 | 1 |
| T2DM | macrophage | *IFT27* | 1.290531469 | 0.00029825 | 0.002629867 | 1 |
| T2DM | macrophage | *NCF4* | -0.219984509 | 0.000296333 | 0.002620315 | 1 |
| T2DM | macrophage | *KCTD17* | -2.039671937 | 0.001248302 | 0.008132722 | 1 |
| T2DM | macrophage | *DESI1* | 0.247411693 | 0.00426701 | 0.021007591 | 1 |
| T2DM | macrophage | *ISM2* | 0.650696804 | 0.001050736 | 0.007080362 | 1 |
| T2DM | macrophage | *ZFYVE21* | 0.665445005 | 0.004293066 | 0.021069873 | 1 |
| T2DM | macrophage | *MTHFD1* | 0.664793286 | 0.013178026 | 0.049286287 | 1 |
| T2DM | macrophage | *PSMB5* | 0.425423254 | 0.000698358 | 0.005171465 | 1 |
| T2DM | macrophage | *NINL* | 0.556267021 | 0.001429579 | 0.009115379 | 1 |
| T2DM | macrophage | *IFT52* | 0.523061739 | 0.003416923 | 0.017783272 | 1 |
| T2DM | macrophage | *DOK5* | 1.089526586 | 8.43E-06 | 0.000188037 | 0.053026354 |
| T2DM | macrophage | *BMP7* | 1.752940915 | 1.88E-05 | 0.000334233 | 0.117984406 |
| T2DM | macrophage | *CTSZ* | -0.421001956 | 0.000260197 | 0.002384638 | 1 |
| T2DM | macrophage | *DIDO1* | 0.522024113 | 0.003428572 | 0.017807627 | 1 |
| T2DM | macrophage | *SLC17A9* | -0.444967361 | 0.000119727 | 0.001304542 | 0.752720999 |
| T2DM | macrophage | *PTK6* | -2.387263881 | 5.91E-05 | 0.000774578 | 0.371797283 |
| T2DM | macrophage | *CCM2L* | 1.396867059 | 9.40E-07 | 4.80E-05 | 0.005912901 |
| T2DM | macrophage | *MYL9* | 0.474200648 | 0.001038505 | 0.00702052 | 1 |
| T2DM | macrophage | *BPI* | 0.477576629 | 0.01190344 | 0.045799834 | 1 |
| T2DM | macrophage | *MAP1LC3A* | 0.392514385 | 6.09E-05 | 0.000783447 | 0.382579802 |
| T2DM | macrophage | *CELF4* | -0.700994021 | 1.45E-05 | 0.000280658 | 0.091449138 |
| T2DM | macrophage | *RBFA* | 0.569961405 | 0.003345023 | 0.017495973 | 1 |
| T2DM | macrophage | *METTL4* | 0.844509246 | 0.013146704 | 0.049229431 | 1 |
| T2DM | macrophage | *LIPG* | 0.878105956 | 0.002413189 | 0.013668215 | 1 |
| T2DM | macrophage | *NALCN* | 1.074872247 | 1.05E-05 | 0.000217794 | 0.065773684 |
| T2DM | macrophage | *HTR2A* | 2.454021923 | 6.94E-07 | 4.47E-05 | 0.004364672 |
| T2DM | macrophage | *UGGT2* | 0.502361355 | 0.002569769 | 0.014343922 | 1 |
| T2DM | macrophage | *DHRS12* | 0.882615047 | 0.001737096 | 0.010562017 | 1 |
| T2DM | macrophage | *MEDAG* | 1.064205313 | 3.59E-06 | 0.000105956 | 0.022568534 |
| T2DM | macrophage | *PLLP* | 0.611739948 | 0.006475567 | 0.028630023 | 1 |
| T2DM | macrophage | *DHODH* | 0.170552925 | 0.00580132 | 0.026277306 | 1 |
| T2DM | macrophage | *SETD6* | 0.791941975 | 0.002406982 | 0.013657666 | 1 |
| T2DM | macrophage | *COG4* | -0.424300498 | 0.004468913 | 0.021645654 | 1 |
| T2DM | macrophage | *HCFC1R1* | 1.299397374 | 0.005330027 | 0.024675907 | 1 |
| T2DM | macrophage | *NME4* | 0.888882123 | 0.003322996 | 0.017409731 | 1 |
| T2DM | macrophage | *LMF1* | 0.730595503 | 0.004404526 | 0.021432861 | 1 |
| T2DM | macrophage | *CLCN7* | -0.659803946 | 0.007863431 | 0.033470855 | 1 |
| T2DM | macrophage | *HAGHL* | -0.800392892 | 0.00067773 | 0.005053357 | 1 |
| T2DM | macrophage | *SLC7A5* | 3.579971751 | 5.68E-05 | 0.000746677 | 0.356911762 |

| | | | | | | |
|---|---|---|---|---|---|---|
| T2DM | macrophage | *CLUAP1* | 0.250421653 | 0.001770846 | 0.010704694 | 1 |
| T2DM | macrophage | *RBL2* | 0.499458734 | 4.48E-06 | 0.000122018 | 0.028186185 |
| T2DM | macrophage | *SYT17* | -0.437940881 | 0.000279069 | 0.002493278 | 1 |
| T2DM | macrophage | *IQCH* | 0.773290732 | 0.004632555 | 0.022198837 | 1 |
| T2DM | macrophage | *LACTB* | -0.600239735 | 0.009165724 | 0.037540657 | 1 |
| T2DM | macrophage | *RASL12* | 1.593614131 | 1.05E-06 | 5.15E-05 | 0.006591135 |
| T2DM | macrophage | *FAH* | 0.896333197 | 0.008067044 | 0.033924751 | 1 |
| T2DM | macrophage | *ATP8B4* | -0.40387698 | 6.49E-05 | 0.000826369 | 0.408226504 |
| T2DM | macrophage | *DTWD1* | 0.654095976 | 0.000119428 | 0.001303547 | 0.750843263 |
| T2DM | macrophage | *PDGFRL* | 0.67333576 | 4.40E-05 | 0.000612685 | 0.276539698 |
| T2DM | macrophage | *EYA1* | 0.335247221 | 3.11E-07 | 3.61E-05 | 0.001957895 |
| T2DM | macrophage | *NIPAL2* | 0.490200772 | 0.004909257 | 0.023125646 | 1 |
| T2DM | macrophage | *PLAT* | 1.112458315 | 7.69E-07 | 4.48E-05 | 0.004835091 |
| T2DM | macrophage | *RAB2A* | 0.280700348 | 0.007854166 | 0.033470855 | 1 |
| T2DM | macrophage | *CHRAC1* | 0.855848146 | 0.006866374 | 0.02998038 | 1 |
| T2DM | macrophage | *TUSC3* | 0.96087992 | 2.31E-06 | 8.26E-05 | 0.014529914 |
| T2DM | macrophage | *ARHGEF10* | 0.32258156 | 0.001677466 | 0.010289005 | 1 |
| T2DM | macrophage | *FCGRT* | -0.336405227 | 0.000723142 | 0.005311211 | 1 |
| T2DM | macrophage | *FCER2* | -0.475973793 | 5.26E-07 | 3.92E-05 | 0.003307123 |
| T2DM | macrophage | *TLE6* | 0.377249816 | 0.002173378 | 0.012640175 | 1 |
| T2DM | macrophage | *C19orf53* | 0.856071363 | 0.007911729 | 0.033563453 | 1 |
| T2DM | macrophage | *CCDC61* | -0.434554486 | 0.008451681 | 0.035235889 | 1 |
| T2DM | macrophage | *ZNF419* | 0.146852037 | 0.011700351 | 0.045156602 | 1 |
| T2DM | macrophage | *SYDE1* | 0.614427437 | 0.006647235 | 0.029245041 | 1 |
| T2DM | macrophage | *PDCD5* | 0.452104552 | 0.009420834 | 0.038360612 | 1 |
| T2DM | macrophage | *POLR2I* | 0.487524583 | 0.012177767 | 0.046485503 | 1 |
| T2DM | macrophage | *CCDC9* | 1.030070506 | 0.000850915 | 0.006024442 | 1 |
| T2DM | macrophage | *CEACAM4* | -1.322582004 | 1.99E-06 | 7.44E-05 | 0.012492331 |
| T2DM | macrophage | *MRPL4* | 0.710876409 | 0.005744291 | 0.026037748 | 1 |
| T2DM | macrophage | *ICAM5* | -1.657428029 | 0.006744038 | 0.029588115 | 1 |
| T2DM | macrophage | *CD33* | -0.323018912 | 0.000142755 | 0.00148347 | 0.897499082 |
| T2DM | macrophage | *BABAM1* | 1.629673837 | 0.000294583 | 0.002608515 | 1 |
| T2DM | macrophage | *PTPRS* | 0.360264625 | 0.000975214 | 0.006649858 | 1 |
| T2DM | macrophage | *BCAT2* | 1.076636297 | 0.005163157 | 0.024062837 | 1 |
| T2DM | macrophage | *PPP2R1A* | 0.862535479 | 0.009676819 | 0.039164328 | 1 |
| T2DM | macrophage | *TNPO2* | 0.705513275 | 0.001549436 | 0.009666361 | 1 |
| T2DM | macrophage | *GCDH* | -0.469273446 | 0.012879385 | 0.048428645 | 1 |
| T2DM | macrophage | *LILRB5* | -0.295875515 | 4.24E-06 | 0.000116858 | 0.02664357 |
| T2DM | macrophage | *MAST1* | 3.211327472 | 1.62E-06 | 6.59E-05 | 0.010207451 |
| T2DM | macrophage | *HPN* | -0.57930749 | 2.36E-07 | 3.14E-05 | 0.001481474 |
| T2DM | macrophage | *ZNF14* | 1.01464118 | 0.006190289 | 0.027543061 | 1 |
| T2DM | macrophage | *ATP13A1* | 0.655396691 | 2.97E-07 | 3.52E-05 | 0.001865777 |
| T2DM | macrophage | *PON3* | 1.438644701 | 9.58E-06 | 0.000204901 | 0.060240918 |
| T2DM | macrophage | *WDR91* | -0.644446152 | 0.000476532 | 0.003865751 | 1 |
| T2DM | macrophage | *STEAP1B* | 0.309913395 | 0.006410214 | 0.028380998 | 1 |
| T2DM | macrophage | *PTN* | 1.169133959 | 6.34E-07 | 4.29E-05 | 0.003987079 |
| T2DM | macrophage | *MPP6* | 0.94957867 | 2.50E-05 | 0.000409886 | 0.156986258 |
| T2DM | macrophage | *ADAP1* | -1.593485874 | 3.73E-05 | 0.000546549 | 0.234469439 |
| T2DM | macrophage | *CAV1* | 0.357967459 | 0.003166645 | 0.016758161 | 1 |
| T2DM | macrophage | *MET* | 0.683242643 | 0.000261 | 0.002388507 | 1 |

| | | | | | | |
|---|---|---|---|---|---|---|
| T2DM | macrophage | *HOXA2* | 1.263451626 | 0.000242312 | 0.002263623 | 1 |
| T2DM | macrophage | *NOD1* | 0.703443451 | 0.005966388 | 0.026908667 | 1 |
| T2DM | macrophage | *CRHR2* | 1.617901804 | 0.000113006 | 0.001250082 | 0.710465762 |
| T2DM | macrophage | *NPTX2* | -4.776814155 | 4.99E-06 | 0.000129744 | 0.031397963 |
| T2DM | macrophage | *CYP3A5* | 1.120187233 | 3.09E-05 | 0.000478466 | 0.193976403 |
| T2DM | macrophage | *SNX8* | -0.621776154 | 0.000731443 | 0.005342734 | 1 |
| T2DM | macrophage | *TFR2* | 0.598188623 | 0.009418445 | 0.038360612 | 1 |
| T2DM | macrophage | *LSM5* | 1.220815883 | 0.000334132 | 0.002889528 | 1 |
| T2DM | macrophage | *AP1S1* | 2.296331176 | 0.000100894 | 0.001150456 | 0.634321739 |
| T2DM | macrophage | *POU6F2* | 1.158128746 | 6.82E-06 | 0.000160518 | 0.042858291 |
| T2DM | macrophage | *TSPAN13* | 1.32066403 | 4.10E-05 | 0.000584458 | 0.257707259 |
| T2DM | macrophage | *RARRES2* | 0.564398834 | 0.00288258 | 0.015582788 | 1 |
| T2DM | macrophage | *MRPL32* | 0.343599504 | 0.0009703 | 0.006626591 | 1 |
| T2DM | macrophage | *URGCP* | 0.728705839 | 0.003363995 | 0.01758058 | 1 |
| T2DM | macrophage | *LIMK1* | -0.747720693 | 0.000101298 | 0.001151643 | 0.636858708 |
| T2DM | macrophage | *SLC1A1* | 0.14596114 | 0.010881577 | 0.042731089 | 1 |
| T2DM | macrophage | *TRIM14* | -0.524140235 | 2.30E-06 | 8.26E-05 | 0.014487595 |
| T2DM | macrophage | *CORO2A* | -0.764476348 | 0.000494039 | 0.003971896 | 1 |
| T2DM | macrophage | *TNFSF8* | -0.576627659 | 5.64E-06 | 0.000141335 | 0.035467453 |
| T2DM | macrophage | *MPDZ* | 0.773721107 | 1.64E-05 | 0.000300683 | 0.103134353 |
| T2DM | macrophage | *PTGDS* | 1.897543214 | 5.19E-06 | 0.000134193 | 0.0326089 |
| T2DM | macrophage | *DVL1* | 0.722241939 | 0.011360452 | 0.04408837 | 1 |
| T2DM | macrophage | *NSMCE4A* | 0.735038891 | 0.011629129 | 0.044936897 | 1 |
| T2DM | macrophage | *PITRM1* | 0.81665347 | 0.011142492 | 0.043538127 | 1 |
| T2DM | macrophage | *PSMB3* | 0.866007242 | 0.000443657 | 0.003660465 | 1 |
| T2DM | macrophage | *RGS9* | 1.629456019 | 1.77E-06 | 6.90E-05 | 0.01113226 |
| T2DM | macrophage | *ASPA* | 0.880963649 | 9.23E-06 | 0.000199597 | 0.058058682 |
| T2DM | macrophage | *PIGL* | 0.633701446 | 0.013044811 | 0.048933609 | 1 |
| T2DM | macrophage | *GALK1* | 1.897127424 | 0.000212817 | 0.002056243 | 1 |
| T2DM | macrophage | *RNF167* | 0.801593108 | 0.00214472 | 0.012523855 | 1 |
| T2DM | macrophage | *C1QBP* | 1.220008457 | 0.000824414 | 0.005883187 | 1 |
| T2DM | macrophage | *DRG2* | 1.130143052 | 0.007214508 | 0.031216525 | 1 |
| T2DM | macrophage | *ICAM2* | 0.597474153 | 0.000133847 | 0.001418721 | 0.841498112 |
| T2DM | macrophage | *SYNGR2* | -0.363383615 | 0.005622015 | 0.025647368 | 1 |
| T2DM | macrophage | *C17orf75* | 0.723350481 | 0.004033762 | 0.020111229 | 1 |
| T2DM | macrophage | *HSD17B1* | -0.190460129 | 2.06E-06 | 7.62E-05 | 0.012974209 |
| T2DM | macrophage | *EZH1* | 0.646960261 | 0.000661393 | 0.004956109 | 1 |
| T2DM | macrophage | *DLX4* | -9.70147829 | 4.52E-05 | 0.000625016 | 0.284382457 |
| T2DM | macrophage | *VAT1* | 0.768112677 | 4.23E-05 | 0.000597667 | 0.265961664 |
| T2DM | macrophage | *LRRC59* | 1.262148124 | 0.008640094 | 0.035801773 | 1 |
| T2DM | macrophage | *MPP2* | 1.604234489 | 3.91E-07 | 3.80E-05 | 0.002458308 |
| T2DM | macrophage | *EFNB3* | 2.817553928 | 0.001843721 | 0.01103056 | 1 |
| T2DM | macrophage | *FAM20A* | -0.468761417 | 0.000112126 | 0.001243274 | 0.704936421 |
| T2DM | macrophage | *DPH1* | 1.550864875 | 0.001756419 | 0.010648606 | 1 |
| T2DM | macrophage | *MYH3* | 0.603454042 | 0.001240336 | 0.00812291 | 1 |
| T2DM | macrophage | *CPE* | 0.785679406 | 0.000153375 | 0.001560306 | 0.964269095 |
| T2DM | macrophage | *WFS1* | 2.431724047 | 3.23E-16 | 5.07E-13 | 2.03E-12 |
| T2DM | macrophage | *GRPEL1* | 1.515835065 | 0.000504218 | 0.004029778 | 1 |
| T2DM | macrophage | *AADAT* | 0.780597633 | 0.000187734 | 0.00186459 | 1 |
| T2DM | macrophage | *CPZ* | 12.15880123 | 6.27E-07 | 4.28E-05 | 0.003941631 |

| | | | | | | |
|---|---|---|---|---|---|---|
| T2DM | macrophage | *SLC2A9* | -0.291490205 | 0.000657657 | 0.004933998 | 1 |
| T2DM | macrophage | *SH3D19* | 0.339481519 | 0.000628671 | 0.004779269 | 1 |
| T2DM | macrophage | *FAM149A* | 0.231259379 | 0.000103617 | 0.001171038 | 0.651440304 |
| T2DM | macrophage | *UGDH* | 0.829941189 | 0.002612425 | 0.01452194 | 1 |
| T2DM | macrophage | *CTSC* | -0.338917954 | 8.59E-05 | 0.001026506 | 0.539942129 |
| T2DM | macrophage | *SC5D* | 1.255828511 | 0.000612146 | 0.004676258 | 1 |
| T2DM | macrophage | *C11orf63* | 0.91355695 | 0.001830601 | 0.010971393 | 1 |
| T2DM | macrophage | *NRXN2* | -0.855767368 | 6.37E-06 | 0.000154182 | 0.040075435 |
| T2DM | macrophage | *MS4A4A* | -0.255633634 | 0.00010054 | 0.00114926 | 0.632092967 |
| T2DM | macrophage | *MADD* | -0.497721444 | 0.004814883 | 0.022896096 | 1 |
| T2DM | macrophage | *CARS* | 0.716571399 | 0.000105581 | 0.00118958 | 0.663785588 |
| T2DM | macrophage | *C11orf21* | -0.314420495 | 0.005466408 | 0.025122301 | 1 |
| T2DM | macrophage | *NDUFS8* | 1.084773032 | 0.003689841 | 0.018799054 | 1 |
| T2DM | macrophage | *PPFIBP1* | 0.485955924 | 0.002627252 | 0.014591459 | 1 |
| T2DM | macrophage | *COQ5* | 0.096153481 | 0.001549816 | 0.009666361 | 1 |
| T2DM | macrophage | *CYP27B1* | -2.079390476 | 9.41E-06 | 0.000201911 | 0.059159782 |
| T2DM | macrophage | *GLI1* | 2.558951905 | 2.47E-06 | 8.38E-05 | 0.015504856 |
| T2DM | macrophage | *TRPV4* | -0.501551799 | 2.11E-05 | 0.000364056 | 0.132516481 |
| T2DM | macrophage | *PRR4* | 0.359129138 | 0.000274824 | 0.002482495 | 1 |
| T2DM | macrophage | *KCNA1* | 0.838746745 | 0.004999535 | 0.023474292 | 1 |
| T2DM | macrophage | *CDK2AP1* | 0.195608338 | 0.000193826 | 0.001904576 | 1 |
| T2DM | macrophage | *MGP* | 0.31604025 | 0.001628566 | 0.010047884 | 1 |
| T2DM | macrophage | *ENDOU* | -0.342075352 | 0.002237695 | 0.012942397 | 1 |
| T2DM | macrophage | *MDM1* | 0.365554931 | 0.011204719 | 0.043645641 | 1 |
| T2DM | macrophage | *GNB3* | -0.082824395 | 0.011064374 | 0.043286697 | 1 |
| T2DM | macrophage | *CDCA3* | -0.200590076 | 0.009364575 | 0.038205765 | 1 |
| T2DM | macrophage | *PHC1* | 1.726110226 | 1.04E-05 | 0.000217536 | 0.065478352 |
| T2DM | macrophage | *COL12A1* | 0.716418333 | 0.000115202 | 0.001265032 | 0.724275652 |
| T2DM | macrophage | *FAM184A* | 0.456872682 | 0.000256906 | 0.002375244 | 1 |
| T2DM | macrophage | *MTRF1L* | 0.303161116 | 0.007454964 | 0.032102302 | 1 |
| T2DM | macrophage | *TULP1* | 7.865325395 | 4.65E-07 | 3.80E-05 | 0.002921758 |
| T2DM | macrophage | *CD83* | -0.354418835 | 2.40E-07 | 3.14E-05 | 0.001506329 |
| T2DM | macrophage | *BVES* | -0.244648037 | 0.000618311 | 0.004711901 | 1 |
| T2DM | macrophage | *GPLD1* | 0.147028124 | 0.011832662 | 0.045574405 | 1 |
| T2DM | macrophage | *ALDH5A1* | 0.556382603 | 5.95E-05 | 0.00077563 | 0.373991399 |
| T2DM | macrophage | *VNN1* | -0.153653872 | 0.0034301 | 0.017807627 | 1 |
| T2DM | macrophage | *SLC16A10* | -0.202018189 | 0.003747647 | 0.019031873 | 1 |
| T2DM | macrophage | *FAM120B* | 0.363483179 | 0.00806344 | 0.033924751 | 1 |
| T2DM | macrophage | *TTK* | -0.414710482 | 0.003614719 | 0.018506303 | 1 |
| T2DM | macrophage | *LY86* | -0.557974439 | 2.47E-05 | 0.000408348 | 0.15526028 |
| T2DM | macrophage | *TBX18* | 0.321471476 | 0.010626751 | 0.042030045 | 1 |
| T2DM | macrophage | *C7* | 0.737809193 | 0.000750433 | 0.005441719 | 1 |
| T2DM | macrophage | *GHR* | 0.156862977 | 0.002664673 | 0.014708339 | 1 |
| T2DM | macrophage | *NME5* | 0.293320109 | 0.002749481 | 0.015018234 | 1 |
| T2DM | macrophage | *LOX* | 1.095922495 | 5.25E-05 | 0.000702298 | 0.330080264 |
| T2DM | macrophage | *SPARC* | 0.567029571 | 0.000829136 | 0.005896808 | 1 |
| T2DM | macrophage | *PCDHB6* | 0.475970596 | 0.004427879 | 0.021510192 | 1 |
| T2DM | macrophage | *RNF130* | -0.422169048 | 0.010685042 | 0.04219652 | 1 |
| T2DM | macrophage | *CLINT1* | 0.5892914 | 0.007820902 | 0.033380861 | 1 |
| T2DM | macrophage | *RASGRF2* | 0.721391923 | 0.000121702 | 0.001323776 | 0.76514273 |

| | | | | | | |
|---|---|---|---|---|---|---|
| T2DM | macrophage | *SLC27A6* | 1.810808095 | 6.82E-05 | 0.000859407 | 0.428844271 |
| T2DM | macrophage | *TARS* | 1.049121439 | 0.001804183 | 0.010864844 | 1 |
| T2DM | macrophage | *LNPEP* | 0.483383127 | 0.003250427 | 0.017143821 | 1 |
| T2DM | macrophage | *SLC12A7* | -0.76501688 | 5.03E-05 | 0.000682019 | 0.316456643 |
| T2DM | macrophage | *GNPDA1* | -0.66062119 | 0.011776391 | 0.045422192 | 1 |
| T2DM | macrophage | *C9* | -0.284822568 | 0.000728502 | 0.005331889 | 1 |
| T2DM | macrophage | *SMAD5* | 0.691266287 | 0.004103484 | 0.020345901 | 1 |
| T2DM | macrophage | *CNTN3* | 0.906223797 | 5.71E-06 | 0.000142573 | 0.03592839 |
| T2DM | macrophage | *NPHP3* | 0.133076884 | 0.002683875 | 0.014788362 | 1 |
| T2DM | macrophage | *IFT57* | 1.0852422 | 0.000452307 | 0.003726935 | 1 |
| T2DM | macrophage | *NCBP2* | 0.733039256 | 0.010766766 | 0.042412692 | 1 |
| T2DM | macrophage | *PLSCR4* | 0.405099782 | 0.001796356 | 0.010828083 | 1 |
| T2DM | macrophage | *ABCC5* | -0.544227108 | 6.88E-05 | 0.000864931 | 0.432465475 |
| T2DM | macrophage | *ARHGEF26* | 0.511059655 | 0.000166529 | 0.001675148 | 1 |
| T2DM | macrophage | *PLCH1* | 1.089049777 | 0.000195077 | 0.001913338 | 1 |
| T2DM | macrophage | *ZBTB47* | 0.547070902 | 0.003568758 | 0.01831574 | 1 |
| T2DM | macrophage | *NKTR* | 0.539142125 | 0.00428132 | 0.021028641 | 1 |
| T2DM | macrophage | *CLCN2* | 1.796083364 | 2.42E-06 | 8.38E-05 | 0.015240438 |
| T2DM | macrophage | *LMAN2L* | 0.420737933 | 0.00270529 | 0.014851852 | 1 |
| T2DM | macrophage | *KCNIP3* | 1.08770016 | 8.86E-05 | 0.001048551 | 0.556780641 |
| T2DM | macrophage | *FAHD2A* | 0.544043757 | 0.00609906 | 0.027272253 | 1 |
| T2DM | macrophage | *TFCP2L1* | -0.343980853 | 0.00026459 | 0.002410836 | 1 |
| T2DM | macrophage | *NRBP1* | -0.522131978 | 0.002280663 | 0.013130981 | 1 |
| T2DM | macrophage | *PPM1G* | -0.797391668 | 0.000632965 | 0.004801556 | 1 |
| T2DM | macrophage | *GRB14* | 0.624016876 | 0.000811855 | 0.005819727 | 1 |
| T2DM | macrophage | *POLE4* | 1.300453428 | 0.000494999 | 0.003974529 | 1 |
| T2DM | macrophage | *LANCL1* | 0.760661978 | 0.00534295 | 0.024705859 | 1 |
| T2DM | macrophage | *PLCD4* | 1.229796354 | 2.49E-05 | 0.000409886 | 0.156856746 |
| T2DM | macrophage | *CHMP3* | -0.596156769 | 2.80E-05 | 0.000446916 | 0.176084708 |
| T2DM | macrophage | *MLPH* | 1.495365216 | 0.000220895 | 0.002116401 | 1 |
| T2DM | macrophage | *SDC1* | 0.871457888 | 0.000291299 | 0.002586722 | 1 |
| T2DM | macrophage | *ARID3A* | -0.632433703 | 0.00280501 | 0.015268482 | 1 |
| T2DM | macrophage | *CD207* | 11.16523001 | 5.55E-08 | 1.34E-05 | 0.00034875 |
| T2DM | macrophage | *VAX2* | 1.694137228 | 1.06E-06 | 5.18E-05 | 0.006687245 |
| T2DM | macrophage | *PLEKHA3* | 0.864826748 | 0.010201727 | 0.040671056 | 1 |
| T2DM | macrophage | *BCL9* | 0.803964637 | 7.74E-06 | 0.00017637 | 0.04863335 |
| T2DM | macrophage | *GPX7* | -0.148020098 | 0.006800039 | 0.029750764 | 1 |
| T2DM | macrophage | *RALGPS2* | 0.532959165 | 0.01004607 | 0.040306089 | 1 |
| T2DM | macrophage | *MRPL37* | 0.737924615 | 0.00010764 | 0.001206299 | 0.676733515 |
| T2DM | macrophage | *PARK7* | 0.671864069 | 0.005404157 | 0.024889909 | 1 |
| T2DM | macrophage | *LEPR* | 0.463790968 | 0.009268486 | 0.037862879 | 1 |
| T2DM | macrophage | *NCF2* | -0.443417758 | 8.88E-06 | 0.000193762 | 0.05580335 |
| T2DM | macrophage | *RGS2* | -0.824206103 | 0.000278106 | 0.002493278 | 1 |
| T2DM | macrophage | *OLFML3* | 0.712594557 | 0.002737742 | 0.014980142 | 1 |
| T2DM | macrophage | *TNNI3K* | 0.46616659 | 0.003760718 | 0.019067447 | 1 |
| T2DM | macrophage | *ACADM* | 0.543052075 | 0.008829485 | 0.036424521 | 1 |
| T2DM | macrophage | *RBBP5* | 0.365306677 | 0.00787393 | 0.033470855 | 1 |
| T2DM | macrophage | *CD46* | 0.570204216 | 0.005846103 | 0.026461085 | 1 |
| T2DM | macrophage | *FAAH* | 0.091056813 | 0.008283872 | 0.034654901 | 1 |
| T2DM | macrophage | *CNN3* | 0.924263213 | 1.42E-05 | 0.000277162 | 0.089523436 |

| | | | | | | |
|---|---|---|---|---|---|---|
| T2DM | macrophage | *VAMP4* | 0.505316069 | 0.006047421 | 0.027137855 | 1 |
| T2DM | macrophage | *PTBP2* | 0.310829641 | 0.005729249 | 0.026024802 | 1 |
| T2DM | macrophage | *SERPINC1* | 0.256358644 | 0.001083184 | 0.00724466 | 1 |
| T2DM | macrophage | *SLC5A9* | 1.543469023 | 4.06E-06 | 0.000114426 | 0.025524279 |
| T2DM | macrophage | *CD3EAP* | -0.396272571 | 0.003465287 | 0.017931076 | 1 |
| T2DM | macrophage | *ATP10B* | -0.905260472 | 4.35E-05 | 0.000608022 | 0.273609791 |
| T2DM | macrophage | *ELOVL4* | 0.248213745 | 0.000227878 | 0.002164148 | 1 |
| T2DM | macrophage | *SGIP1* | 0.626995624 | 0.000731683 | 0.005342734 | 1 |
| T2DM | macrophage | *SLC16A7* | 0.562336973 | 0.007134725 | 0.030892573 | 1 |
| T2DM | macrophage | *FOXO3* | -0.418125143 | 0.005407924 | 0.024889909 | 1 |
| T2DM | macrophage | *ABCG2* | -0.962484231 | 0.000161769 | 0.001629878 | 1 |
| T2DM | macrophage | *SPP1* | -0.611736562 | 0.000265074 | 0.002411751 | 1 |
| T2DM | macrophage | *MFSD1* | -0.48906315 | 0.000475939 | 0.003865751 | 1 |
| T2DM | macrophage | *WDR35* | 0.700255007 | 0.011490877 | 0.044484695 | 1 |
| T2DM | macrophage | *DNAH7* | -0.184342573 | 0.000545287 | 0.004285274 | 1 |
| T2DM | macrophage | *CTNNAL1* | 0.143033637 | 0.007083921 | 0.030757327 | 1 |
| T2DM | macrophage | *BSPRY* | 3.665647014 | 2.44E-06 | 8.38E-05 | 0.01531258 |
| T2DM | macrophage | *KDSR* | 0.798262939 | 0.005701482 | 0.025918451 | 1 |
| T2DM | macrophage | *DCAF4* | 0.415989851 | 0.001813099 | 0.010887255 | 1 |
| T2DM | macrophage | *IRF2BPL* | 1.153978066 | 0.000123724 | 0.001341128 | 0.777854167 |
| T2DM | macrophage | *FLVCR2* | -0.658117904 | 0.002312627 | 0.013290204 | 1 |
| T2DM | macrophage | *ZC2HC1C* | 1.16932267 | 0.001244423 | 0.008131648 | 1 |
| T2DM | macrophage | *COQ6* | 1.068972065 | 0.002091763 | 0.012307926 | 1 |
| T2DM | macrophage | *KLHL29* | 0.711842664 | 1.59E-05 | 0.000295695 | 0.100240517 |
| T2DM | macrophage | *ATL2* | 0.609345024 | 0.008004745 | 0.033798411 | 1 |
| T2DM | macrophage | *CNRIP1* | -0.514716421 | 0.005627281 | 0.025647368 | 1 |
| T2DM | macrophage | *CUTC* | 0.559696912 | 0.006186864 | 0.027543061 | 1 |
| T2DM | macrophage | *C10orf88* | 0.903581053 | 0.013147163 | 0.049229431 | 1 |
| T2DM | macrophage | *PCDHB14* | 1.113099917 | 4.02E-07 | 3.80E-05 | 0.002529501 |
| T2DM | macrophage | *MRC1* | -0.251373196 | 2.19E-05 | 0.000375669 | 0.137870448 |
| T2DM | macrophage | *ENOX1* | 0.703539738 | 0.000424729 | 0.003536786 | 1 |
| T2DM | macrophage | *WBP4* | 0.536591632 | 0.009180325 | 0.037575977 | 1 |
| T2DM | macrophage | *SMAD9* | 1.13869822 | 4.79E-06 | 0.000127086 | 0.030119458 |
| T2DM | macrophage | *ADRA1A* | 0.0846473 | 4.61E-05 | 0.000629611 | 0.289866824 |
| T2DM | macrophage | *CRISPLD1* | 1.212007899 | 4.71E-07 | 3.80E-05 | 0.002961018 |
| T2DM | macrophage | *TEX14* | 0.447972322 | 0.007773256 | 0.03324521 | 1 |
| T2DM | macrophage | *PRB2* | 1.364864098 | 3.01E-06 | 9.57E-05 | 0.018946247 |
| T2DM | macrophage | *ZNF211* | 0.207084744 | 0.0069874 | 0.030422289 | 1 |
| T2DM | macrophage | *PDZRN3* | 0.640772563 | 5.94E-05 | 0.00077563 | 0.37367507 |
| T2DM | macrophage | *SEC22A* | 0.444677634 | 0.000425592 | 0.0035383 | 1 |
| T2DM | macrophage | *B4GALT4* | 1.020700432 | 0.004921307 | 0.023158874 | 1 |
| T2DM | macrophage | *POLR3GL* | 1.03885157 | 0.002462876 | 0.013887084 | 1 |
| T2DM | macrophage | *TNFSF10* | 0.678450645 | 0.001692028 | 0.010358113 | 1 |
| T2DM | macrophage | *ZNF639* | 0.842663418 | 0.002017948 | 0.011968717 | 1 |
| T2DM | macrophage | *GPSM2* | 0.618800775 | 0.000517527 | 0.004108194 | 1 |
| T2DM | macrophage | *FLT3* | 2.269220935 | 5.62E-05 | 0.000741335 | 0.353616755 |
| T2DM | macrophage | *MTIF3* | 0.561618479 | 0.003478058 | 0.017982362 | 1 |
| T2DM | macrophage | *FMOD* | 0.586634994 | 0.010040946 | 0.040306089 | 1 |
| T2DM | macrophage | *ODF2L* | 0.461934696 | 0.001078576 | 0.007229217 | 1 |
| T2DM | macrophage | *RPAP2* | 0.656187247 | 0.005593258 | 0.025537265 | 1 |

| | | | | | | |
|---|---|---|---|---|---|---|
| T2DM | macrophage | *PMS2* | 1.23983231 | 7.11E-07 | 4.47E-05 | 0.004468412 |
| T2DM | macrophage | *KLHL7* | 0.774066994 | 0.005344356 | 0.024705859 | 1 |
| T2DM | macrophage | *FKBP9* | 0.507554407 | 0.002063521 | 0.012204474 | 1 |
| T2DM | macrophage | *SLC25A51* | 0.113100359 | 0.010629517 | 0.042030045 | 1 |
| T2DM | macrophage | *SSPN* | 0.385964181 | 0.000137027 | 0.001443028 | 0.861487736 |
| T2DM | macrophage | *EBPL* | 0.270784384 | 0.003129121 | 0.016601507 | 1 |
| T2DM | macrophage | *ATF1* | 0.578555583 | 0.011168744 | 0.043550002 | 1 |
| T2DM | macrophage | *TSFM* | 0.519542088 | 0.008591889 | 0.035640853 | 1 |
| T2DM | macrophage | *ARHGAP9* | -0.514213292 | 1.39E-05 | 0.000273082 | 0.087659183 |
| T2DM | macrophage | *SARDH* | 3.350270007 | 3.14E-06 | 9.77E-05 | 0.019740244 |
| T2DM | macrophage | *TTC21B* | 0.714357831 | 0.004301361 | 0.021080579 | 1 |
| T2DM | macrophage | *ACVR1C* | 0.544180403 | 7.36E-05 | 0.000910738 | 0.46265485 |
| T2DM | macrophage | *C4BPB* | -0.264772944 | 1.06E-05 | 0.000219177 | 0.066410693 |
| T2DM | macrophage | *RAB38* | -0.571142181 | 1.54E-06 | 6.42E-05 | 0.009677816 |
| T2DM | macrophage | *DNPEP* | 0.935478811 | 0.00660345 | 0.029072754 | 1 |
| T2DM | macrophage | *FAM210B* | 0.535723785 | 0.00018993 | 0.001880457 | 1 |
| T2DM | macrophage | *SNX21* | 0.923187684 | 1.29E-05 | 0.000257003 | 0.080956082 |
| T2DM | macrophage | *HIF3A* | 0.352652976 | 0.000287714 | 0.00256212 | 1 |
| T2DM | macrophage | *RPS10* | 1.878894635 | 0.001244985 | 0.008131648 | 1 |
| T2DM | macrophage | *HIST1H2BJ* | 0.305043004 | 0.00788932 | 0.033513617 | 1 |
| T2DM | macrophage | *GLO1* | 1.069189702 | 1.46E-05 | 0.000281609 | 0.092086135 |
| T2DM | macrophage | *RPP40* | 0.377409911 | 0.003274692 | 0.017228444 | 1 |
| T2DM | macrophage | *EREG* | -4.425883837 | 0.001049011 | 0.007076324 | 1 |
| T2DM | macrophage | *MT1G* | -2.560091393 | 1.67E-06 | 6.69E-05 | 0.010511398 |
| T2DM | macrophage | *TMTC4* | 0.898299189 | 0.012896618 | 0.048464459 | 1 |
| T2DM | macrophage | *KIF25* | 1.125236964 | 0.000103749 | 0.001171038 | 0.652268256 |
| T2DM | macrophage | *GRK4* | 0.136337433 | 8.07E-05 | 0.000979583 | 0.507424123 |
| T2DM | macrophage | *OPRL1* | -0.511125135 | 1.66E-06 | 6.69E-05 | 0.010464013 |
| T2DM | macrophage | *PPDPF* | 1.260348597 | 8.73E-05 | 0.001039579 | 0.54889768 |
| T2DM | macrophage | *PSD4* | -0.629973504 | 0.000389958 | 0.003273248 | 1 |
| T2DM | macrophage | *CLPP* | 0.460471273 | 0.007031002 | 0.030569784 | 1 |
| T2DM | macrophage | *SH2D3A* | 1.016792897 | 0.000489838 | 0.003943161 | 1 |
| T2DM | macrophage | *TRIP10* | 0.878544501 | 4.13E-05 | 0.000586182 | 0.259486667 |
| T2DM | macrophage | *RTN2* | -0.953476802 | 0.010189365 | 0.040647548 | 1 |
| T2DM | macrophage | *PCSK2* | 0.920991488 | 0.002401746 | 0.0136526 | 1 |
| T2DM | macrophage | *FAM110A* | -0.809816671 | 2.46E-05 | 0.000408348 | 0.154437664 |
| T2DM | macrophage | *ZNF436* | 0.677819491 | 1.18E-07 | 2.05E-05 | 0.000741475 |
| T2DM | macrophage | *UROD* | 0.907901775 | 9.04E-05 | 0.001064332 | 0.568353148 |
| T2DM | macrophage | *KLC1* | 0.595554353 | 0.008499926 | 0.035390088 | 1 |
| T2DM | macrophage | *XRCC3* | 0.749365878 | 0.004663195 | 0.022311647 | 1 |
| T2DM | macrophage | *MCF2L* | 0.433109653 | 9.82E-05 | 0.001130317 | 0.617153276 |
| T2DM | macrophage | *F10* | 1.142998493 | 7.92E-05 | 0.000967461 | 0.498242186 |
| T2DM | macrophage | *PROZ* | 3.610753957 | 0.000115178 | 0.001265032 | 0.724125514 |
| T2DM | macrophage | *IGFLR1* | -2.251847006 | 6.58E-06 | 0.000156638 | 0.041352333 |
| T2DM | macrophage | *PRR12* | 0.455807189 | 0.000154419 | 0.001568385 | 0.970830425 |
| T2DM | macrophage | *TSKS* | 1.629482352 | 5.23E-05 | 0.000700784 | 0.328667543 |
| T2DM | macrophage | *SBDS* | 0.598899477 | 0.012730461 | 0.048127727 | 1 |
| T2DM | macrophage | *ATG14* | 0.632395474 | 0.013064754 | 0.048979196 | 1 |
| T2DM | macrophage | *RHOJ* | 0.913269934 | 1.79E-05 | 0.000320978 | 0.112663109 |
| T2DM | macrophage | *PZP* | 0.906873996 | 6.11E-05 | 0.000783447 | 0.383889009 |

| | | | | | | |
|---|---|---|---|---|---|---|
| T2DM | macrophage | *AIF1L* | 1.202945405 | 1.53E-06 | 6.42E-05 | 0.009592193 |
| T2DM | macrophage | *FGD3* | -0.623231816 | 6.92E-05 | 0.00086873 | 0.435233871 |
| T2DM | macrophage | *HRASLS* | 0.657504171 | 0.003562871 | 0.018300468 | 1 |
| T2DM | macrophage | *WFIKKN1* | -1.276514448 | 0.002094716 | 0.012307926 | 1 |
| T2DM | macrophage | *SGCE* | 0.799496961 | 3.56E-05 | 0.000528152 | 0.223649848 |
| T2DM | macrophage | *MGAT3* | 1.20150592 | 2.50E-06 | 8.38E-05 | 0.015727841 |
| T2DM | macrophage | *BAIAP2L2* | 0.859728093 | 9.01E-07 | 4.80E-05 | 0.00566263 |
| T2DM | macrophage | *EMC4* | 0.721869206 | 0.000253598 | 0.002355054 | 1 |
| T2DM | macrophage | *CDHR3* | 0.41977282 | 0.000404647 | 0.003392018 | 1 |
| T2DM | macrophage | *HOXD1* | 1.728614645 | 0.000485525 | 0.003922479 | 1 |
| T2DM | macrophage | *HOXD3* | 0.594382544 | 0.000213381 | 0.002057553 | 1 |
| T2DM | macrophage | *WDFY4* | -0.301034817 | 0.000352808 | 0.003015336 | 1 |
| T2DM | macrophage | *EIF2AK4* | -0.330680155 | 0.004484619 | 0.021671638 | 1 |
| T2DM | macrophage | *CALML4* | -0.963938102 | 3.47E-05 | 0.00052152 | 0.217995238 |
| T2DM | macrophage | *ACKR4* | 0.540044909 | 0.001563961 | 0.009725639 | 1 |
| T2DM | macrophage | *CCL4* | -0.24231268 | 0.00865098 | 0.035805604 | 1 |
| T2DM | macrophage | *SLC44A2* | 0.947731504 | 8.85E-05 | 0.001048551 | 0.556415409 |
| T2DM | macrophage | *NGDN* | 0.945171039 | 0.00722374 | 0.031234977 | 1 |
| T2DM | macrophage | *RNASE1* | -0.383461264 | 0.000210214 | 0.002039533 | 1 |
| T2DM | macrophage | *TEP1* | -0.588908593 | 0.005553587 | 0.025374566 | 1 |
| T2DM | macrophage | *EPB41L4A* | 0.75904144 | 2.22E-05 | 0.000378833 | 0.13941044 |
| T2DM | macrophage | *CDO1* | 0.850896839 | 9.70E-05 | 0.001120682 | 0.609650853 |
| T2DM | macrophage | *SHC2* | 0.424501191 | 9.34E-05 | 0.001087022 | 0.586991882 |
| T2DM | macrophage | *PHF10* | 0.867044248 | 0.005398754 | 0.024889909 | 1 |
| T2DM | macrophage | *DPP6* | -0.491625083 | 3.49E-08 | 1.04E-05 | 0.000219326 |
| T2DM | macrophage | *GTPBP3* | -0.531242246 | 0.002252185 | 0.013014236 | 1 |
| T2DM | macrophage | *PXDN* | 0.778275829 | 1.54E-05 | 0.000291188 | 0.09677067 |
| T2DM | macrophage | *CYP2E1* | 0.497718677 | 0.005666219 | 0.025795452 | 1 |
| T2DM | macrophage | *HBZ* | 0.931807669 | 0.011272367 | 0.043881962 | 1 |
| T2DM | macrophage | *SMPDL3B* | 3.984270915 | 3.16E-07 | 3.61E-05 | 0.0019847 |
| T2DM | macrophage | *THEMIS2* | -0.19453336 | 0.004691961 | 0.022415166 | 1 |
| T2DM | macrophage | *PPAN* | 2.411134599 | 3.64E-05 | 0.000536366 | 0.22902849 |
| T2DM | macrophage | *C12orf65* | 0.933921856 | 3.12E-06 | 9.77E-05 | 0.019631202 |
| T2DM | macrophage | *PKDREJ* | -0.26697647 | 0.008662201 | 0.03582846 | 1 |
| T2DM | macrophage | *HABP4* | 0.75031792 | 0.001059227 | 0.007114704 | 1 |
| T2DM | macrophage | *LILRB2* | -0.53690545 | 4.97E-06 | 0.000129744 | 0.031268956 |
| T2DM | macrophage | *COX4I2* | 1.502273749 | 1.54E-05 | 0.000291188 | 0.096965665 |
| T2DM | macrophage | *HAUS8* | -0.593163519 | 0.007936048 | 0.033621251 | 1 |
| T2DM | macrophage | *RFTN1* | 0.575370477 | 0.000488108 | 0.003934273 | 1 |
| T2DM | macrophage | *PDLIM4* | 0.499825766 | 0.000946361 | 0.006545402 | 1 |
| T2DM | macrophage | *KIF3A* | 0.3848837 | 0.011939944 | 0.045884124 | 1 |
| T2DM | macrophage | *AOC2* | 1.51135495 | 8.62E-06 | 0.000190089 | 0.054175495 |
| T2DM | macrophage | *NINJ1* | -0.569350915 | 1.06E-05 | 0.000220204 | 0.066941961 |
| T2DM | macrophage | *ZCCHC9* | 0.598733245 | 0.009342436 | 0.038140191 | 1 |
| T2DM | macrophage | *KHDRBS3* | 0.67422633 | 0.000279323 | 0.002493278 | 1 |
| T2DM | macrophage | *GCH1* | 0.537137925 | 0.002505874 | 0.014053906 | 1 |
| T2DM | macrophage | *FBXW9* | 0.453391087 | 0.005474225 | 0.025139847 | 1 |
| T2DM | macrophage | *MATN3* | 3.149876613 | 1.04E-05 | 0.000217536 | 0.065310645 |
| T2DM | macrophage | *CNGA4* | 0.693855267 | 0.001432395 | 0.009124078 | 1 |
| T2DM | macrophage | *MUTYH* | 1.125569296 | 0.001171445 | 0.007744349 | 1 |

| | | | | | | |
|---|---|---|---|---|---|---|
| T2DM | macrophage | *LPIN3* | 1.179663231 | 3.09E-05 | 0.000478466 | 0.194257171 |
| T2DM | macrophage | *ATP8A2* | 1.698243792 | 0.001461545 | 0.009244199 | 1 |
| T2DM | macrophage | *WASF3* | 0.829541469 | 1.52E-05 | 0.000287932 | 0.095305621 |
| T2DM | macrophage | *TMCC2* | -0.544577323 | 6.50E-06 | 0.000155672 | 0.040874004 |
| T2DM | macrophage | *TRPC4* | 1.386126159 | 1.30E-06 | 5.95E-05 | 0.008151892 |
| T2DM | macrophage | *PRAM1* | -0.561386538 | 6.46E-06 | 0.000155636 | 0.040620897 |
| T2DM | macrophage | *ZNF414* | 1.651757762 | 4.22E-05 | 0.000597415 | 0.265252445 |
| T2DM | macrophage | *SLC2A11* | 1.244816263 | 0.001752254 | 0.010633613 | 1 |
| T2DM | macrophage | *C1QTNF6* | 1.571162424 | 1.49E-06 | 6.36E-05 | 0.00934934 |
| T2DM | macrophage | *FAM83F* | 1.895292196 | 2.50E-06 | 8.38E-05 | 0.015748104 |
| T2DM | macrophage | *SFTPD* | 0.105271529 | 0.003088686 | 0.016478053 | 1 |
| T2DM | macrophage | *TMTC1* | 0.679679542 | 6.60E-05 | 0.000837871 | 0.414746264 |
| T2DM | macrophage | *SPINK5* | 0.472929016 | 0.009395449 | 0.038306867 | 1 |
| T2DM | macrophage | *GSC* | 2.256853151 | 5.07E-07 | 3.89E-05 | 0.003187908 |
| T2DM | macrophage | *ADAMDEC1* | -0.529461909 | 0.003259716 | 0.017164014 | 1 |
| T2DM | macrophage | *SYT6* | 1.963897447 | 0.004279111 | 0.021028641 | 1 |
| T2DM | macrophage | *VAV3* | -0.380972458 | 0.001885814 | 0.011248685 | 1 |
| T2DM | macrophage | *AP4B1* | 0.733060322 | 0.002159331 | 0.012570105 | 1 |
| T2DM | macrophage | *SPIRE1* | -0.387011289 | 0.005888918 | 0.026607186 | 1 |
| T2DM | macrophage | *LDHA* | 0.988895903 | 0.000187712 | 0.00186459 | 1 |
| T2DM | macrophage | *IL15RA* | 1.086621381 | 0.000212917 | 0.002056243 | 1 |
| T2DM | macrophage | *KLRD1* | 0.935231247 | 4.31E-05 | 0.000604534 | 0.270831276 |
| T2DM | macrophage | *TPGS2* | 0.688066005 | 0.000624059 | 0.00474995 | 1 |
| T2DM | macrophage | *SLC43A3* | -0.526788928 | 0.001140347 | 0.007570605 | 1 |
| T2DM | macrophage | *DHX34* | -0.413034163 | 0.002309957 | 0.013287007 | 1 |
| T2DM | macrophage | *APLNR* | 0.965820399 | 5.19E-05 | 0.000698131 | 0.326253737 |
| T2DM | macrophage | *BIVM* | 1.324466857 | 2.85E-05 | 0.000450977 | 0.179038033 |
| T2DM | macrophage | *CCDC146* | 0.742948861 | 0.001160511 | 0.007680139 | 1 |
| T2DM | macrophage | *NT5E* | 0.581422095 | 1.37E-05 | 0.000270852 | 0.086401923 |
| T2DM | macrophage | *RDH5* | 0.476832521 | 0.00089996 | 0.006300722 | 1 |
| T2DM | macrophage | *PPP1R1A* | 0.866835059 | 0.001376491 | 0.008812624 | 1 |
| T2DM | macrophage | *FAIM2* | 1.618633169 | 3.56E-05 | 0.000528152 | 0.223936473 |
| T2DM | macrophage | *KRT7* | 0.393533188 | 0.012377607 | 0.04707684 | 1 |
| T2DM | macrophage | *ARMC9* | -0.62485032 | 0.003544754 | 0.01823721 | 1 |
| T2DM | macrophage | *RCBTB1* | -0.318390605 | 0.007969992 | 0.033696932 | 1 |
| T2DM | macrophage | *GPNMB* | -0.389566659 | 0.002075465 | 0.012252064 | 1 |
| T2DM | macrophage | *KDELR2* | 1.012743909 | 0.002011416 | 0.011951008 | 1 |
| T2DM | macrophage | *AOAH* | -0.480417339 | 1.34E-06 | 6.01E-05 | 0.008453406 |
| T2DM | macrophage | *IL1RN* | -0.578738237 | 1.45E-05 | 0.000280658 | 0.091036466 |
| T2DM | macrophage | *ZFP37* | 0.393527169 | 0.002411922 | 0.013668215 | 1 |
| T2DM | macrophage | *PRPF4* | 0.289432855 | 0.007615488 | 0.032703945 | 1 |
| T2DM | macrophage | *GABBR2* | 3.234249299 | 0.000307477 | 0.002693507 | 1 |
| T2DM | macrophage | *RABEPK* | 0.819620556 | 0.008671311 | 0.035842561 | 1 |
| T2DM | macrophage | *LMX1B* | 2.471443019 | 0.000231751 | 0.002194305 | 1 |
| T2DM | macrophage | *NOV* | 0.573199652 | 0.001088841 | 0.007274751 | 1 |
| T2DM | macrophage | *SPAG8* | 0.449334939 | 0.012770435 | 0.048178716 | 1 |
| T2DM | macrophage | *TMEM8B* | 0.844790707 | 0.000307609 | 0.002693507 | 1 |
| T2DM | macrophage | *KIF13A* | 0.543647814 | 0.008019909 | 0.033823988 | 1 |
| T2DM | macrophage | *FRS3* | 1.448240136 | 0.00014247 | 0.001482962 | 0.895708974 |
| T2DM | macrophage | *FLOT1* | 0.615276008 | 0.000869219 | 0.006133313 | 1 |

| | | | | | | |
|---|---|---|---|---|---|---|
| T2DM | macrophage | *FAM8A1* | 1.299399973 | 0.000132339 | 0.00140781 | 0.832015847 |
| T2DM | macrophage | *FHDC1* | 1.605628408 | 4.72E-06 | 0.00012579 | 0.029686485 |
| T2DM | macrophage | *MYO7A* | -0.19608418 | 0.002465447 | 0.013889124 | 1 |
| T2DM | macrophage | *RAB30* | 0.596855021 | 0.000965139 | 0.006619669 | 1 |
| T2DM | macrophage | *SULF1* | 0.464694931 | 0.000256383 | 0.002373898 | 1 |
| T2DM | macrophage | *NEK1* | 0.575045854 | 0.00480457 | 0.022866261 | 1 |
| T2DM | macrophage | *PPP2R1B* | 0.624534349 | 0.000611215 | 0.004674827 | 1 |
| T2DM | macrophage | *FXYD6* | -0.469692568 | 6.23E-05 | 0.000796134 | 0.391697775 |
| T2DM | macrophage | *CASP1* | -0.81931879 | 0.00091731 | 0.006380743 | 1 |
| T2DM | macrophage | *PAQR5* | -0.65548981 | 0.000205344 | 0.001998447 | 1 |
| T2DM | macrophage | *RMDN3* | 0.707500608 | 0.000513192 | 0.004089275 | 1 |
| T2DM | macrophage | *SPTBN5* | 0.17969699 | 0.00128086 | 0.008310981 | 1 |
| T2DM | macrophage | *DNASE2B* | -1.072044402 | 4.23E-06 | 0.000116858 | 0.026588815 |
| T2DM | macrophage | *IFT172* | 0.965077641 | 1.98E-05 | 0.000349157 | 0.124649076 |
| T2DM | macrophage | *LHCGR* | 0.540259518 | 0.004835758 | 0.022945213 | 1 |
| T2DM | macrophage | *ABCG5* | 0.401554524 | 0.004880774 | 0.023037106 | 1 |
| T2DM | macrophage | *CENPO* | 1.372944377 | 0.010602147 | 0.0419901 | 1 |
| T2DM | macrophage | *MYOF* | 0.315038522 | 0.000643761 | 0.004858734 | 1 |
| T2DM | macrophage | *STAMBPL1* | 0.474541613 | 0.005255777 | 0.024368046 | 1 |
| T2DM | macrophage | *TACC2* | 0.472402417 | 0.000145702 | 0.001506599 | 0.916026906 |
| T2DM | macrophage | *RBP4* | 0.669496697 | 0.00693256 | 0.030225384 | 1 |
| T2DM | macrophage | *CARF* | 0.437833797 | 3.98E-05 | 0.000574019 | 0.250272266 |
| T2DM | macrophage | *METTL5* | 1.056872139 | 0.001245551 | 0.008131648 | 1 |
| T2DM | macrophage | *MDH1B* | 0.955945965 | 0.000214622 | 0.002066354 | 1 |
| T2DM | macrophage | *PRKG2* | 0.66621549 | 0.001257426 | 0.008183685 | 1 |
| T2DM | macrophage | *BBS7* | 0.90716559 | 0.004467048 | 0.021645654 | 1 |
| T2DM | macrophage | *PRDM5* | 0.794886689 | 1.78E-06 | 6.90E-05 | 0.011171715 |
| T2DM | macrophage | *TRPC3* | 0.577740853 | 0.006027573 | 0.027087457 | 1 |
| T2DM | macrophage | *NUP54* | 0.444665686 | 0.0060937 | 0.027267679 | 1 |
| T2DM | macrophage | *BMP2K* | -0.274373257 | 0.003929814 | 0.019718068 | 1 |
| T2DM | macrophage | *ANXA3* | 1.158074072 | 9.47E-07 | 4.80E-05 | 0.005954661 |
| T2DM | macrophage | *PARVG* | -0.486793967 | 3.12E-05 | 0.000482056 | 0.196196914 |
| T2DM | macrophage | *ERP27* | -0.947426487 | 0.001500778 | 0.00940579 | 1 |
| T2DM | macrophage | *KIF21A* | 0.273317423 | 0.003053421 | 0.016323858 | 1 |
| T2DM | macrophage | *SLCO1C1* | 0.829346092 | 7.09E-06 | 0.000165711 | 0.044576389 |
| T2DM | macrophage | *ZCRB1* | 0.346237687 | 0.008878675 | 0.036555489 | 1 |
| T2DM | macrophage | *PRICKLE1* | 0.739128812 | 2.39E-06 | 8.38E-05 | 0.015049369 |
| T2DM | macrophage | *C1RL* | 0.564229083 | 6.16E-06 | 0.000150633 | 0.03871277 |
| T2DM | macrophage | *CLSTN3* | 0.525103138 | 0.000341045 | 0.002945261 | 1 |
| T2DM | macrophage | *GLIPR1* | -0.395305305 | 0.001311378 | 0.008482131 | 1 |
| T2DM | macrophage | *GAS2L3* | -0.378371923 | 5.64E-06 | 0.000141335 | 0.035475168 |
| T2DM | macrophage | *GPR84* | -2.055341499 | 6.27E-08 | 1.41E-05 | 0.000394384 |
| T2DM | macrophage | *BRCA2* | -0.709924293 | 7.07E-05 | 0.000882182 | 0.444619798 |
| T2DM | macrophage | *VPS33A* | 1.226102191 | 0.004864132 | 0.023010382 | 1 |
| T2DM | macrophage | *RTN1* | -0.390722615 | 0.003007714 | 0.016148163 | 1 |
| T2DM | macrophage | *STON2* | -0.520210408 | 0.00095697 | 0.00659701 | 1 |
| T2DM | macrophage | *AK7* | 2.055352985 | 9.43E-07 | 4.80E-05 | 0.005929834 |
| T2DM | macrophage | *FBLN5* | 0.873714673 | 2.92E-05 | 0.000457252 | 0.183398122 |
| T2DM | macrophage | *C14orf79* | 1.463358262 | 0.000136008 | 0.001434697 | 0.85507961 |
| T2DM | macrophage | *DUOXA1* | 0.431344344 | 0.002954393 | 0.015902626 | 1 |

| | | | | | | |
|---|---|---|---|---|---|---|
| T2DM | macrophage | *SRP14* | 0.602164138 | 0.001662797 | 0.010235276 | 1 |
| T2DM | macrophage | *PSTPIP1* | -0.332117314 | 0.002186023 | 0.012701965 | 1 |
| T2DM | macrophage | *HMG20A* | 0.96890077 | 1.14E-05 | 0.000234117 | 0.071873946 |
| T2DM | macrophage | *MAN2C1* | -0.125927922 | 0.000106663 | 0.001199625 | 0.670590567 |
| T2DM | macrophage | *DNAJA4* | 1.172606607 | 0.009090518 | 0.037256899 | 1 |
| T2DM | macrophage | *ADAMTS17* | 0.513958922 | 0.006913324 | 0.030162436 | 1 |
| T2DM | macrophage | *SCAMP2* | -0.611758562 | 8.90E-05 | 0.001051356 | 0.559321548 |
| T2DM | macrophage | *MFGE8* | 0.674411492 | 0.00095916 | 0.006597169 | 1 |
| T2DM | macrophage | *CRTC3* | 0.596170807 | 0.013326979 | 0.049725055 | 1 |
| T2DM | macrophage | *ADAMTS18* | 0.334906346 | 0.009957659 | 0.040053615 | 1 |
| T2DM | macrophage | *GCSH* | 0.378148259 | 0.001611821 | 0.009964127 | 1 |
| T2DM | macrophage | *TLDC1* | 0.855270456 | 0.004182819 | 0.020674043 | 1 |
| T2DM | macrophage | *NDUFB10* | 0.835818496 | 0.010950782 | 0.042922424 | 1 |
| T2DM | macrophage | *TIGD7* | 0.372995342 | 0.001446204 | 0.009174859 | 1 |
| T2DM | macrophage | *DEF8* | -0.348660887 | 0.006771559 | 0.029675802 | 1 |
| T2DM | macrophage | *MYO19* | 0.975027105 | 3.21E-05 | 0.000489553 | 0.201695845 |
| T2DM | macrophage | *TOM1L1* | 0.875055147 | 0.00026173 | 0.002391709 | 1 |
| T2DM | macrophage | *VPS53* | -0.390098343 | 0.011469089 | 0.044427704 | 1 |
| T2DM | macrophage | *SPATA22* | 0.311398771 | 9.28E-05 | 0.001082672 | 0.583560284 |
| T2DM | macrophage | *LRRC46* | -0.36383373 | 0.006201524 | 0.027554049 | 1 |
| T2DM | macrophage | *GNAL* | 0.814554309 | 0.000108474 | 0.001213482 | 0.681976634 |
| T2DM | macrophage | *C18orf21* | 0.676192134 | 0.011098716 | 0.043394045 | 1 |
| T2DM | macrophage | *ASXL3* | 0.968769029 | 2.07E-06 | 7.62E-05 | 0.012989113 |
| T2DM | macrophage | *C18orf8* | -0.485517765 | 0.005075125 | 0.02371491 | 1 |
| T2DM | macrophage | *NPC1* | -0.621709331 | 0.001561238 | 0.009718321 | 1 |
| T2DM | macrophage | *CARD14* | -0.884651941 | 0.000243776 | 0.002273913 | 1 |
| T2DM | macrophage | *FN3KRP* | -0.158545391 | 0.005186389 | 0.024117476 | 1 |
| T2DM | macrophage | *SECTM1* | -0.579951898 | 0.009250017 | 0.037812 | 1 |
| T2DM | macrophage | *RNF165* | 0.760120342 | 0.000275656 | 0.002486443 | 1 |
| T2DM | macrophage | *DUS3L* | 0.285737348 | 0.003443297 | 0.017861391 | 1 |
| T2DM | macrophage | *TMEM91* | -0.403871023 | 0.002332838 | 0.013394109 | 1 |
| T2DM | macrophage | *SIRT3* | 0.678930168 | 3.29E-06 | 0.000100599 | 0.020668345 |
| T2DM | macrophage | *IFITM3* | 0.960018524 | 0.001195922 | 0.007889571 | 1 |
| T2DM | macrophage | *DOPEY2* | -0.440059533 | 0.002796625 | 0.015236032 | 1 |
| T2DM | macrophage | *GEMIN7* | -1.041349966 | 9.72E-06 | 0.000207124 | 0.061101527 |
| T2DM | macrophage | *WTIP* | 0.63262692 | 0.000706393 | 0.005212549 | 1 |
| T2DM | macrophage | *MYO1F* | -0.313780516 | 0.011022856 | 0.043151117 | 1 |
| T2DM | macrophage | *CACNG8* | 2.424289328 | 3.55E-07 | 3.80E-05 | 0.002229376 |
| T2DM | macrophage | *EVI5L* | 0.834348032 | 0.007501345 | 0.032279914 | 1 |
| T2DM | macrophage | *RCN3* | -0.5345427 | 3.19E-05 | 0.000489383 | 0.200608762 |
| T2DM | macrophage | *CYP4B1* | 0.199778244 | 0.00345538 | 0.017894541 | 1 |
| T2DM | macrophage | *ATP1B1* | -0.416301839 | 3.32E-06 | 0.00010084 | 0.020873927 |
| T2DM | macrophage | *CREG1* | -0.439425761 | 0.007039444 | 0.030585338 | 1 |
| T2DM | macrophage | *RXRG* | 1.81696117 | 1.18E-06 | 5.60E-05 | 0.007446358 |
| T2DM | macrophage | *RGL1* | -0.393611994 | 0.00022947 | 0.002175988 | 1 |
| T2DM | macrophage | *TARS2* | 0.829934499 | 5.44E-05 | 0.000724055 | 0.342046938 |
| T2DM | macrophage | *DYRK3* | 0.869743908 | 3.27E-05 | 0.000495853 | 0.205497296 |
| T2DM | macrophage | *KCNN3* | 0.938350131 | 6.95E-05 | 0.00087095 | 0.437217015 |
| T2DM | macrophage | *SCCPDH* | 0.763198814 | 0.00284426 | 0.015409039 | 1 |
| T2DM | macrophage | *SNAP47* | 0.485155005 | 0.001862279 | 0.011129416 | 1 |

| | | | | | | |
|---|---|---|---|---|---|---|
| T2DM | macrophage | *GUK1* | 0.825909151 | 0.008355254 | 0.034856987 | 1 |
| T2DM | macrophage | *CDC42BPA* | 0.54913679 | 0.00072739 | 0.005329953 | 1 |
| T2DM | macrophage | *WNT9A* | 0.734116777 | 9.10E-08 | 1.68E-05 | 0.000571916 |
| T2DM | macrophage | *PLEKHA6* | 0.752242573 | 7.66E-07 | 4.48E-05 | 0.004814351 |
| T2DM | macrophage | *CHAC2* | 0.885520284 | 0.001809404 | 0.010885858 | 1 |
| T2DM | macrophage | *FBLN7* | 0.287939429 | 0.002690549 | 0.014802639 | 1 |
| T2DM | macrophage | *THSD7B* | 0.451648644 | 4.94E-06 | 0.000129527 | 0.031086565 |
| T2DM | macrophage | *ZNF385B* | 0.529739859 | 6.74E-05 | 0.000850616 | 0.423606537 |
| T2DM | macrophage | *TMEFF2* | 0.970985659 | 8.92E-07 | 4.80E-05 | 0.005609493 |
| T2DM | macrophage | *GULP1* | 0.583031493 | 0.000191571 | 0.001890748 | 1 |
| T2DM | macrophage | *NBEAL1* | 0.325977718 | 1.01E-05 | 0.000212236 | 0.063246231 |
| T2DM | macrophage | *ESPNL* | 4.40072067 | 8.37E-07 | 4.67E-05 | 0.005263199 |
| T2DM | macrophage | *TAMM41* | 0.691206624 | 0.003651241 | 0.018659934 | 1 |
| T2DM | macrophage | *OSBPL10* | 0.641485935 | 0.000124953 | 0.001347477 | 0.785579273 |
| T2DM | macrophage | *STAC* | 1.111962606 | 3.58E-06 | 0.000105956 | 0.022487462 |
| T2DM | macrophage | *ARL6IP5* | 0.673324548 | 0.010136563 | 0.040488293 | 1 |
| T2DM | macrophage | *PLA1A* | -0.243629949 | 0.004020694 | 0.020061989 | 1 |
| T2DM | macrophage | *RABL3* | 0.865354713 | 0.006150722 | 0.027464197 | 1 |
| T2DM | macrophage | *BOC* | 0.375529204 | 0.006679936 | 0.029368362 | 1 |
| T2DM | macrophage | *TCTA* | 0.611557707 | 0.000486021 | 0.003922479 | 1 |
| T2DM | macrophage | *EAF2* | -0.388785559 | 0.005414646 | 0.02490262 | 1 |
| T2DM | macrophage | *USP53* | 0.778598629 | 1.57E-05 | 0.000293149 | 0.098498065 |
| T2DM | macrophage | *MARCH1* | -0.201221761 | 0.0089997 | 0.036932842 | 1 |
| T2DM | macrophage | *PDGFC* | -0.508308103 | 1.16E-05 | 0.000236032 | 0.072933752 |
| T2DM | macrophage | *CBR4* | 0.584166764 | 0.010130469 | 0.040488293 | 1 |
| T2DM | macrophage | *FAM105A* | -0.38723418 | 7.52E-06 | 0.000173209 | 0.047286126 |
| T2DM | macrophage | *BHMT* | -0.122870348 | 0.003297506 | 0.017333965 | 1 |
| T2DM | macrophage | *CRHBP* | -0.406683919 | 1.57E-05 | 0.00029382 | 0.099017311 |
| T2DM | macrophage | *SPATA9* | 0.548473966 | 0.000586846 | 0.004518593 | 1 |
| T2DM | macrophage | *YIPF5* | 0.957647796 | 0.005356059 | 0.024741767 | 1 |
| T2DM | macrophage | *GABRB2* | -0.239283983 | 8.80E-06 | 0.000192708 | 0.055307178 |
| T2DM | macrophage | *PSD2* | 1.674625311 | 0.000158643 | 0.001606097 | 0.997385965 |
| T2DM | macrophage | *KLHL3* | 0.345789653 | 0.011956114 | 0.045918196 | 1 |
| T2DM | macrophage | *MUT* | 0.993996717 | 0.002843967 | 0.015409039 | 1 |
| T2DM | macrophage | *DOK3* | -0.207293698 | 0.001603088 | 0.009939464 | 1 |
| T2DM | macrophage | *MLIP* | 0.594408607 | 0.000300645 | 0.002644258 | 1 |
| T2DM | macrophage | *FGD2* | -0.366212495 | 6.51E-06 | 0.000155672 | 0.040941788 |
| T2DM | macrophage | *TPBG* | 1.288090465 | 0.000259916 | 0.002384638 | 1 |
| T2DM | macrophage | *IRAK1BP1* | 1.03966845 | 4.01E-05 | 0.000576252 | 0.252398441 |
| T2DM | macrophage | *PM20D2* | 0.908350272 | 0.008174517 | 0.034328914 | 1 |
| T2DM | macrophage | *SLC22A3* | 0.911535219 | 4.65E-06 | 0.00012484 | 0.029212545 |
| T2DM | macrophage | *C7orf50* | 0.724437974 | 0.003534459 | 0.018199137 | 1 |
| T2DM | macrophage | *PSPH* | 0.30636071 | 0.003106915 | 0.016525526 | 1 |
| T2DM | macrophage | *TMEM209* | 0.678890364 | 0.005030799 | 0.02356828 | 1 |
| T2DM | macrophage | *FBXO25* | 0.728307807 | 0.010936431 | 0.042892916 | 1 |
| T2DM | macrophage | *CSGALNACT1* | -0.386717894 | 0.002349854 | 0.013467212 | 1 |
| T2DM | macrophage | *CCDC25* | 1.020928223 | 2.83E-06 | 9.17E-05 | 0.017795716 |
| T2DM | macrophage | *SLC25A37* | 0.806283772 | 0.000368787 | 0.003119001 | 1 |
| T2DM | macrophage | *DOCK5* | -0.559748467 | 0.00272463 | 0.014921384 | 1 |
| T2DM | macrophage | *ST18* | -0.300121912 | 4.09E-06 | 0.000114426 | 0.025716362 |

| | | | | | | |
|---|---|---|---|---|---|---|
| T2DM | macrophage | *ATP6V0D2* | -0.309438084 | 3.86E-05 | 0.000559827 | 0.242965069 |
| T2DM | macrophage | *ZNF7* | 0.613879548 | 0.003416765 | 0.017783272 | 1 |
| T2DM | macrophage | *SLC39A4* | -0.655620281 | 0.001126351 | 0.007501447 | 1 |
| T2DM | macrophage | *CBWD5* | 1.052749444 | 0.006081642 | 0.027233107 | 1 |
| T2DM | macrophage | *ZNF462* | 0.859095262 | 0.000459007 | 0.003767332 | 1 |
| T2DM | macrophage | *WDR31* | 1.423521315 | 3.79E-06 | 0.000108369 | 0.023802641 |
| T2DM | macrophage | *SH3GLB2* | 0.727537155 | 0.003674306 | 0.018750294 | 1 |
| T2DM | macrophage | *SEC16A* | 0.457154214 | 0.00208649 | 0.012294058 | 1 |
| T2DM | macrophage | *SLC39A12* | -0.401304912 | 0.000379299 | 0.003196589 | 1 |
| T2DM | macrophage | *PARD3* | 0.316225882 | 0.008092132 | 0.034007508 | 1 |
| T2DM | macrophage | *ADIRF* | 0.734545537 | 0.002845538 | 0.015409039 | 1 |
| T2DM | macrophage | *HTR7* | -0.462249197 | 4.58E-06 | 0.000123484 | 0.028771704 |
| T2DM | macrophage | *FRA10AC1* | 0.328164358 | 0.012782265 | 0.048178716 | 1 |
| T2DM | macrophage | *GSTO1* | -0.750642612 | 0.002571274 | 0.014343922 | 1 |
| T2DM | macrophage | *RGS10* | -0.585207455 | 2.63E-06 | 8.69E-05 | 0.016512934 |
| T2DM | macrophage | *GAS2* | 1.22188101 | 0.000694911 | 0.005152008 | 1 |
| T2DM | macrophage | *LIN7C* | 0.968476923 | 0.010752766 | 0.0423841 | 1 |
| T2DM | macrophage | *HSD17B12* | -0.290576904 | 2.80E-07 | 3.38E-05 | 0.001758046 |
| T2DM | macrophage | *APIP* | -0.099601912 | 0.000461475 | 0.003782653 | 1 |
| T2DM | macrophage | *GLYAT* | 0.291313987 | 0.000507717 | 0.004050782 | 1 |
| T2DM | macrophage | *C11orf49* | 0.502189584 | 0.013219303 | 0.049352588 | 1 |
| T2DM | macrophage | *ENDOD1* | 1.251648536 | 0.001015339 | 0.006871299 | 1 |
| T2DM | macrophage | *SERPINH1* | 0.453723936 | 0.002790585 | 0.015216311 | 1 |
| T2DM | macrophage | *CAPN5* | 0.814290752 | 8.13E-05 | 0.000982587 | 0.510945029 |
| T2DM | macrophage | *DRD2* | 1.985351105 | 0.000273813 | 0.00247692 | 1 |
| T2DM | macrophage | *C11orf52* | 5.788963695 | 4.01E-07 | 3.80E-05 | 0.002522919 |
| T2DM | macrophage | *P4HA3* | 0.425153449 | 0.007807098 | 0.03334458 | 1 |
| T2DM | macrophage | *TMC2* | 2.301135279 | 5.58E-06 | 0.000140968 | 0.035101015 |
| T2DM | macrophage | *EML3* | 0.644559407 | 0.003422047 | 0.017795213 | 1 |
| T2DM | macrophage | *MS4A3* | -11.1714103 | 3.14E-06 | 9.77E-05 | 0.01973892 |
| T2DM | macrophage | *PLCH2* | 2.982978772 | 2.76E-07 | 3.38E-05 | 0.001737907 |
| T2DM | macrophage | *FEZ1* | 1.465362693 | 0.000152422 | 0.00155565 | 0.958280209 |
| T2DM | macrophage | *DUSP15* | -0.447502931 | 0.004622629 | 0.02216817 | 1 |
| T2DM | macrophage | *DOC2A* | -0.790592314 | 4.50E-08 | 1.18E-05 | 0.000283022 |
| T2DM | macrophage | *ARID5B* | 0.400534666 | 0.012333923 | 0.046939087 | 1 |
| T2DM | macrophage | *LYPD6B* | 1.038875119 | 0.000678036 | 0.005053357 | 1 |
| T2DM | macrophage | *WDR17* | -0.569704674 | 0.003497809 | 0.018025187 | 1 |
| T2DM | macrophage | *CCDC102B* | 0.691823892 | 0.00574183 | 0.026037748 | 1 |
| T2DM | macrophage | *RGS18* | -0.620073407 | 1.91E-05 | 0.000336892 | 0.119933692 |
| T2DM | macrophage | *PPP1R1C* | 0.386813883 | 0.000735732 | 0.00535364 | 1 |
| T2DM | macrophage | *C11orf53* | 2.920824623 | 0.000760868 | 0.005498363 | 1 |
| T2DM | macrophage | *TEX12* | 10.50831949 | 2.53E-05 | 0.000412577 | 0.158842205 |
| T2DM | macrophage | *BTBD11* | 1.34470981 | 2.03E-05 | 0.000354745 | 0.127708034 |
| T2DM | macrophage | *ANK3* | 0.352761284 | 0.006076018 | 0.027227319 | 1 |
| T2DM | macrophage | *IPMK* | -0.58105023 | 0.001718698 | 0.010480555 | 1 |
| T2DM | macrophage | *FAM177A1* | 0.814132846 | 0.00214541 | 0.012523855 | 1 |
| T2DM | macrophage | *MIPOL1* | 0.494308657 | 0.000238512 | 0.002241446 | 1 |
| T2DM | macrophage | *ADAMTS12* | 0.626444304 | 8.30E-05 | 0.000996094 | 0.521953 |
| T2DM | macrophage | *PTPRO* | -0.518282798 | 0.000114074 | 0.001258217 | 0.717183744 |
| T2DM | macrophage | *FAM160B1* | 0.88100367 | 0.000345231 | 0.002969178 | 1 |

| | | | | | | |
|---|---|---|---|---|---|---|
| T2DM | macrophage | *ANO4* | 0.463417826 | 0.006293012 | 0.027901389 | 1 |
| T2DM | macrophage | *AKR1C2* | 0.57349322 | 0.00264428 | 0.014630053 | 1 |
| T2DM | macrophage | *RNF144A* | 0.751558054 | 1.76E-05 | 0.000317813 | 0.110916744 |
| T2DM | macrophage | *ASAP2* | 0.643040436 | 0.002851079 | 0.015425761 | 1 |
| T2DM | macrophage | *PTPN14* | 0.32458015 | 0.011347744 | 0.044066256 | 1 |
| T2DM | macrophage | *ARL14EP* | -0.792843056 | 1.38E-05 | 0.000272055 | 0.087057646 |
| T2DM | macrophage | *G6PC2* | 28.62242163 | 4.16E-07 | 3.80E-05 | 0.002613293 |
| T2DM | macrophage | *POC5* | -0.377066665 | 0.003212404 | 0.016971749 | 1 |
| T2DM | macrophage | *TADA1* | 1.185262501 | 0.001816388 | 0.010896597 | 1 |
| T2DM | macrophage | *CLEC4F* | 4.26240966 | 2.89E-05 | 0.000456382 | 0.181640202 |
| T2DM | macrophage | *PELO* | 1.513020819 | 2.15E-06 | 7.84E-05 | 0.013488789 |
| T2DM | macrophage | *SAR1B* | 0.830989439 | 2.81E-06 | 9.17E-05 | 0.017688718 |
| T2DM | macrophage | *RAB3C* | 0.324368346 | 0.01016884 | 0.040591429 | 1 |
| T2DM | macrophage | *MED21* | 0.905655451 | 0.00434669 | 0.021250108 | 1 |
| T2DM | macrophage | *BANK1* | 0.893991546 | 0.000497153 | 0.003981657 | 1 |
| T2DM | macrophage | *CLGN* | 5.272132399 | 8.12E-08 | 1.55E-05 | 0.000510581 |
| T2DM | macrophage | *AHCTF1* | 0.657529485 | 0.010541776 | 0.041814604 | 1 |
| T2DM | macrophage | *PTPRR* | 0.98593896 | 0.000264264 | 0.002410836 | 1 |
| T2DM | macrophage | *PLA2R1* | 0.76366403 | 7.99E-08 | 1.55E-05 | 0.00050251 |
| T2DM | macrophage | *FRMD1* | 1.389671925 | 1.02E-06 | 5.04E-05 | 0.00640155 |
| T2DM | macrophage | *TUBGCP5* | 0.778722803 | 0.009749672 | 0.039342869 | 1 |
| T2DM | macrophage | *FAM92B* | 11.95896611 | 1.10E-06 | 5.29E-05 | 0.006924827 |
| T2DM | macrophage | *JAZF1* | -0.754155599 | 2.91E-05 | 0.000457209 | 0.182727159 |
| T2DM | macrophage | *FBXO36* | 0.860754146 | 0.003162887 | 0.016752374 | 1 |
| T2DM | macrophage | *CACNA2D1* | 0.508256718 | 0.000702371 | 0.005188959 | 1 |
| T2DM | macrophage | *ANKRD29* | 0.670401528 | 0.000260163 | 0.002384638 | 1 |
| T2DM | macrophage | *THY1* | 0.956450856 | 0.000296977 | 0.00262232 | 1 |
| T2DM | macrophage | *UBASH3B* | -0.603761492 | 0.00035979 | 0.003060894 | 1 |
| T2DM | macrophage | *LRRK1* | -0.541144733 | 0.003801878 | 0.019198718 | 1 |
| T2DM | macrophage | *CEP112* | 0.337702955 | 0.00348106 | 0.017983095 | 1 |
| T2DM | macrophage | *ENPP3* | 0.676665064 | 0.002153208 | 0.01254608 | 1 |
| T2DM | macrophage | *UCHL1* | 1.149638009 | 0.000237868 | 0.00223874 | 1 |
| T2DM | macrophage | *CCSAP* | -0.560927232 | 0.000258479 | 0.002382785 | 1 |
| T2DM | macrophage | *ATP5G3* | 0.321209254 | 0.008334644 | 0.034794095 | 1 |
| T2DM | macrophage | *PDLIM3* | 0.419827141 | 0.000803602 | 0.005775736 | 1 |
| T2DM | macrophage | *PDE1C* | 0.755084125 | 0.000148468 | 0.001526639 | 0.93342047 |
| T2DM | macrophage | *TSEN2* | 0.700367185 | 8.73E-09 | 3.23E-06 | 5.49E-05 |
| T2DM | macrophage | *CYP2U1* | 0.991882689 | 0.010503547 | 0.041715603 | 1 |
| T2DM | macrophage | *FBXL18* | 0.889072369 | 0.010871724 | 0.042719079 | 1 |
| T2DM | macrophage | *GOLGA7B* | -0.644520742 | 8.39E-07 | 4.67E-05 | 0.005274479 |
| T2DM | macrophage | *SPAG17* | 0.675940189 | 4.50E-05 | 0.000623084 | 0.282880185 |
| T2DM | macrophage | *DEPTOR* | -0.534502414 | 0.011837602 | 0.045574405 | 1 |
| T2DM | macrophage | *FAM161B* | 0.786394415 | 0.008237901 | 0.034550825 | 1 |
| T2DM | macrophage | *UGT2B4* | 1.538560989 | 7.11E-07 | 4.47E-05 | 0.004472438 |
| T2DM | macrophage | *KCNMA1* | -0.313846553 | 9.27E-05 | 0.001082672 | 0.582493033 |
| T2DM | macrophage | *BATF* | -1.199808137 | 0.000149876 | 0.001534644 | 0.942271401 |
| T2DM | macrophage | *SH3RF2* | 0.205397113 | 0.00681799 | 0.029808557 | 1 |
| T2DM | macrophage | *GDF6* | 1.566180232 | 2.11E-07 | 3.06E-05 | 0.001328509 |
| T2DM | macrophage | *NODAL* | 1.511638405 | 9.61E-07 | 4.83E-05 | 0.006041675 |
| T2DM | macrophage | *GHRL* | 1.133032716 | 0.007553021 | 0.032480057 | 1 |

| | | | | | | |
|---|---|---|---|---|---|---|
| T2DM | macrophage | *TIMP4* | 1.153623053 | 4.58E-09 | 1.92E-06 | 2.88E-05 |
| T2DM | macrophage | *CPT2* | 1.059552029 | 0.00245403 | 0.013849627 | 1 |
| T2DM | macrophage | *LRP8* | -1.132408944 | 0.00069472 | 0.005152008 | 1 |
| T2DM | macrophage | *STEAP2* | 0.125740239 | 0.003374605 | 0.01762138 | 1 |
| T2DM | macrophage | *TMED6* | 5.022868601 | 1.55E-06 | 6.42E-05 | 0.009728563 |
| T2DM | macrophage | *CLEC18A* | -0.666993357 | 2.42E-08 | 8.46E-06 | 0.000152256 |
| T2DM | macrophage | *IL34* | 1.305246872 | 5.59E-07 | 4.04E-05 | 0.003511454 |
| T2DM | macrophage | *KCNJ15* | -0.295791427 | 0.007964311 | 0.033695573 | 1 |
| T2DM | macrophage | *CREB3L1* | 0.398303036 | 0.003983671 | 0.019940556 | 1 |
| T2DM | macrophage | *WDR19* | 0.819361481 | 0.001288779 | 0.008353149 | 1 |
| T2DM | macrophage | *SLC37A3* | 0.904388088 | 0.000141905 | 0.001479525 | 0.892153717 |
| T2DM | macrophage | *AP3S2* | 0.433102655 | 7.55E-11 | 4.75E-08 | 4.75E-07 |
| T2DM | macrophage | *C12orf43* | 0.529265807 | 0.000124186 | 0.001341512 | 0.780760188 |
| T2DM | macrophage | *RADIL* | 0.671054764 | 0.001144959 | 0.00758944 | 1 |
| T2DM | macrophage | *LDLRAP1* | 1.278066482 | 0.000271974 | 0.002463833 | 1 |
| T2DM | macrophage | *TRIM63* | 0.321457977 | 0.000222902 | 0.002129406 | 1 |
| T2DM | macrophage | *LRRC43* | 4.185705761 | 1.42E-05 | 0.000277162 | 0.089376788 |
| T2DM | macrophage | *CLSTN2* | 0.488063799 | 0.000757281 | 0.005478743 | 1 |
| T2DM | macrophage | *COLEC12* | -0.356085331 | 2.65E-05 | 0.000426599 | 0.166800168 |
| T2DM | macrophage | *RNF207* | 1.601206786 | 3.64E-06 | 0.000106444 | 0.022885451 |
| T2DM | macrophage | *TSPAN33* | -0.207332111 | 0.000212918 | 0.002056243 | 1 |
| T2DM | macrophage | *CD1D* | -0.564412311 | 0.00063313 | 0.004801556 | 1 |
| T2DM | macrophage | *FAM86C1* | 1.661842599 | 0.002504499 | 0.014053906 | 1 |
| T2DM | macrophage | *CD1E* | 0.972738736 | 2.93E-05 | 0.000457252 | 0.184272379 |
| T2DM | macrophage | *NCF1* | -0.492045697 | 2.48E-06 | 8.38E-05 | 0.015607042 |
| T2DM | macrophage | *PPP1R9A* | 0.428600406 | 0.005492268 | 0.025149227 | 1 |
| T2DM | macrophage | *GDPD5* | 0.752751023 | 0.000202755 | 0.00197631 | 1 |
| T2DM | macrophage | *DYNC1I1* | 1.000806367 | 0.000200102 | 0.001959569 | 1 |
| T2DM | macrophage | *TMED4* | 0.836910731 | 0.012173258 | 0.046485503 | 1 |
| T2DM | macrophage | *SPATA2L* | -0.805276692 | 0.005037291 | 0.023568655 | 1 |
| T2DM | macrophage | *DMTN* | 0.234255814 | 0.003003215 | 0.016137789 | 1 |
| T2DM | macrophage | *EPB41* | 0.282633499 | 0.01129224 | 0.043904954 | 1 |
| T2DM | macrophage | *SV2A* | 1.763947403 | 0.000315774 | 0.002757318 | 1 |
| T2DM | macrophage | *PADI4* | 1.673865545 | 0.00058058 | 0.004490856 | 1 |
| T2DM | macrophage | *STARD9* | 0.500283012 | 0.003095368 | 0.016478053 | 1 |
| T2DM | macrophage | *THEM4* | 0.422495942 | 0.00884641 | 0.036470412 | 1 |
| T2DM | macrophage | *TCHH* | -0.845877108 | 4.62E-07 | 3.80E-05 | 0.00290216 |
| T2DM | macrophage | *NPR2* | 1.083876589 | 6.03E-07 | 4.22E-05 | 0.003792102 |
| T2DM | macrophage | *DFFA* | 0.53494898 | 0.001082165 | 0.00724466 | 1 |
| T2DM | macrophage | *NR2F6* | 0.642993965 | 0.00036497 | 0.003092411 | 1 |
| T2DM | macrophage | *VAV2* | -0.691263824 | 0.003379279 | 0.017631141 | 1 |
| T2DM | macrophage | *DIP2A* | -0.478557818 | 0.000717685 | 0.005283475 | 1 |
| T2DM | macrophage | *ZNF208* | 0.34020159 | 0.006166926 | 0.027485775 | 1 |
| T2DM | macrophage | *MPZL3* | 1.787506876 | 0.0011803 | 0.007794689 | 1 |
| T2DM | macrophage | *S100A1* | 0.687299527 | 0.004662074 | 0.022311647 | 1 |
| T2DM | macrophage | *FDPS* | 0.656052122 | 0.000221167 | 0.002116401 | 1 |
| T2DM | macrophage | *LY6K* | 2.35320439 | 0.000352996 | 0.003015336 | 1 |
| T2DM | macrophage | *VPS28* | 0.418047624 | 0.000564959 | 0.004395913 | 1 |
| T2DM | macrophage | *MUM1* | 0.376162208 | 0.01003599 | 0.040306089 | 1 |
| T2DM | macrophage | *MFSD12* | -0.722646521 | 0.000112084 | 0.001243274 | 0.704672824 |

| | | | | | | |
|---|---|---|---|---|---|---|
| T2DM | macrophage | *YDJC* | -2.043868128 | 0.000220139 | 0.002112997 | 1 |
| T2DM | macrophage | *DVL3* | 0.824026531 | 0.000582741 | 0.00449533 | 1 |
| T2DM | macrophage | *AP2M1* | 0.672184222 | 0.007666513 | 0.032881373 | 1 |
| T2DM | macrophage | *BDH1* | 1.780528161 | 4.40E-05 | 0.000612685 | 0.276933631 |
| T2DM | macrophage | *DUSP14* | 0.55155138 | 0.000330042 | 0.002862031 | 1 |
| T2DM | macrophage | *PGAP3* | 1.314692435 | 0.010533722 | 0.041809034 | 1 |
| T2DM | macrophage | *IKZF3* | 0.702634776 | 0.002013061 | 0.011951008 | 1 |
| T2DM | macrophage | *TMEM143* | 0.354007092 | 0.005483607 | 0.025146197 | 1 |
| T2DM | macrophage | *SIGLEC11* | -0.424153532 | 4.67E-06 | 0.000124967 | 0.029367138 |
| T2DM | macrophage | *MPP3* | 0.615238011 | 0.004005778 | 0.020019339 | 1 |
| T2DM | macrophage | *ASGR2* | -1.368083915 | 2.91E-05 | 0.000457209 | 0.18288348 |
| T2DM | macrophage | *JMJD8* | 0.32209299 | 0.003978638 | 0.019931233 | 1 |
| T2DM | macrophage | *FLYWCH2* | 1.039816318 | 0.000529586 | 0.004182799 | 1 |
| T2DM | macrophage | *ZG16B* | -3.188066444 | 1.35E-06 | 6.01E-05 | 0.008477985 |
| T2DM | macrophage | *CYP4A22* | 1.542866423 | 0.000232815 | 0.0021989 | 1 |
| T2DM | macrophage | *ACOT11* | -0.433878428 | 0.000580733 | 0.004490856 | 1 |
| T2DM | macrophage | *ZSWIM5* | 0.439799907 | 0.002675544 | 0.014755392 | 1 |
| T2DM | macrophage | *RBP7* | 1.037911035 | 0.005981282 | 0.0269565 | 1 |
| T2DM | macrophage | *SLC25A34* | -1.176171409 | 1.63E-05 | 0.000300039 | 0.102514154 |
| T2DM | macrophage | *LRRC38* | 1.914788695 | 0.000122701 | 0.001332332 | 0.771420236 |
| T2DM | macrophage | *PEF1* | 1.545016683 | 0.000440029 | 0.003635299 | 1 |
| T2DM | macrophage | *TTLL10* | 1.149180146 | 0.000965827 | 0.006619669 | 1 |
| T2DM | macrophage | *NFIA* | 0.477545482 | 0.007782456 | 0.03326193 | 1 |
| T2DM | macrophage | *NEXN* | 0.903139955 | 5.20E-05 | 0.000698131 | 0.326725537 |
| T2DM | macrophage | *B3GALT2* | 1.524554222 | 3.66E-05 | 0.000537915 | 0.230227608 |
| T2DM | macrophage | *HFM1* | 0.954868015 | 8.15E-06 | 0.000183759 | 0.051268819 |
| T2DM | macrophage | *KCNT2* | 0.793966505 | 3.13E-06 | 9.77E-05 | 0.019653418 |
| T2DM | macrophage | *IGSF8* | 1.505354142 | 0.000102675 | 0.001163091 | 0.645515335 |
| T2DM | macrophage | *VANGL2* | 0.609991662 | 0.004040546 | 0.02012909 | 1 |
| T2DM | macrophage | *LMX1A* | 0.305428729 | 2.85E-05 | 0.000450977 | 0.178968047 |
| T2DM | macrophage | *SNED1* | 0.680786148 | 0.000177635 | 0.001778329 | 1 |
| T2DM | macrophage | *C1orf115* | 0.288420553 | 0.006694414 | 0.029390907 | 1 |
| T2DM | macrophage | *RFTN2* | 0.876753587 | 4.45E-06 | 0.000121677 | 0.027985816 |
| T2DM | macrophage | *DISC1* | -0.573861713 | 3.76E-05 | 0.000548284 | 0.236310611 |
| T2DM | macrophage | *FAM84A* | 1.866707307 | 6.08E-07 | 4.22E-05 | 0.00382444 |
| T2DM | macrophage | *FRZB* | 0.548312444 | 0.003210539 | 0.016971749 | 1 |
| T2DM | macrophage | *FBXO41* | -0.741374229 | 7.97E-07 | 4.60E-05 | 0.005012816 |
| T2DM | macrophage | *CCDC74A* | 3.86672804 | 1.30E-06 | 5.95E-05 | 0.008150773 |
| T2DM | macrophage | *NOSTRIN* | 0.336491628 | 0.012841412 | 0.048343687 | 1 |
| T2DM | macrophage | *HPGDS* | -0.30103702 | 4.59E-05 | 0.00062858 | 0.288364029 |
| T2DM | macrophage | *C1QTNF7* | 0.558252476 | 0.000414278 | 0.003458917 | 1 |
| T2DM | macrophage | *NIPAL1* | -0.961367753 | 0.000959387 | 0.006597169 | 1 |
| T2DM | macrophage | *PBXIP1* | 1.108601009 | 0.001384059 | 0.008852066 | 1 |
| T2DM | macrophage | *DCST2* | 2.222425395 | 0.000590037 | 0.004525383 | 1 |
| T2DM | macrophage | *LMOD3* | 1.916467932 | 1.73E-05 | 0.000314983 | 0.108984162 |
| T2DM | macrophage | *SERPINI1* | 0.809452272 | 0.002362944 | 0.013524503 | 1 |
| T2DM | macrophage | *SPTA1* | -0.09582269 | 0.004803843 | 0.022866261 | 1 |
| T2DM | macrophage | *AIM2* | 1.703552155 | 2.78E-06 | 9.11E-05 | 0.017490406 |
| T2DM | macrophage | *CD200R1* | -0.301071763 | 1.51E-05 | 0.000287022 | 0.094717303 |
| T2DM | macrophage | *CADPS* | 1.447017785 | 5.55E-06 | 0.000140968 | 0.034878673 |

| | | | | | | |
|---|---|---|---|---|---|---|
| T2DM | macrophage | *DNASE1L3* | 0.946833481 | 9.07E-06 | 0.000197296 | 0.057018457 |
| T2DM | macrophage | *CCDC158* | 0.69562161 | 8.85E-05 | 0.001048551 | 0.556670783 |
| T2DM | macrophage | *PLB1* | -0.487896242 | 5.57E-06 | 0.000140968 | 0.035022383 |
| T2DM | macrophage | *CDCP1* | -0.319299016 | 2.44E-06 | 8.38E-05 | 0.015367755 |
| T2DM | macrophage | *LZTFL1* | 1.211704145 | 0.001005149 | 0.006817015 | 1 |
| T2DM | macrophage | *FYCO1* | 0.718893794 | 0.00654322 | 0.028884355 | 1 |
| T2DM | macrophage | *NMNAT3* | 0.623298789 | 0.001706655 | 0.010437489 | 1 |
| T2DM | macrophage | *DNALI1* | 1.0447236 | 2.56E-05 | 0.00041543 | 0.160771453 |
| T2DM | macrophage | *RFT1* | -0.639554132 | 0.000141257 | 0.001477671 | 0.888080085 |
| T2DM | macrophage | *PIGX* | -0.595515597 | 2.59E-05 | 0.000417424 | 0.162795522 |
| T2DM | macrophage | *ABLIM2* | 1.240559618 | 0.00054625 | 0.004287486 | 1 |
| T2DM | macrophage | *CDC25A* | -0.20894717 | 0.008538397 | 0.035500042 | 1 |
| T2DM | macrophage | *ATRIP* | 2.035745754 | 0.000907869 | 0.006349027 | 1 |
| T2DM | macrophage | *SHISA5* | 0.789953782 | 0.00616867 | 0.027485775 | 1 |
| T2DM | macrophage | *RNF123* | 0.376363389 | 3.34E-07 | 3.75E-05 | 0.002099816 |
| T2DM | macrophage | *MST1R* | 0.282394968 | 0.001327303 | 0.008558721 | 1 |
| T2DM | macrophage | *GRM2* | -4.521205443 | 1.62E-06 | 6.59E-05 | 0.010169646 |
| T2DM | macrophage | *MAP9* | 0.995521758 | 2.09E-05 | 0.000361931 | 0.131381069 |
| T2DM | macrophage | *CEP44* | 0.764986366 | 0.011547019 | 0.044674527 | 1 |
| T2DM | macrophage | *NPY1R* | 0.825145613 | 3.93E-05 | 0.000568132 | 0.247137213 |
| T2DM | macrophage | *ITGA2* | 1.094907169 | 1.76E-05 | 0.000317813 | 0.110853166 |
| T2DM | macrophage | *CMBL* | 1.295268982 | 7.81E-05 | 0.000957443 | 0.491168212 |
| T2DM | macrophage | *C5orf63* | 1.018812494 | 0.0002027 | 0.00197631 | 1 |
| T2DM | macrophage | *PRRC1* | 0.82560283 | 0.0032287 | 0.017043526 | 1 |
| T2DM | macrophage | *RHOBTB3* | 0.279099222 | 0.004573892 | 0.021967958 | 1 |
| T2DM | macrophage | *TIGD6* | 1.625307151 | 0.000966575 | 0.006619669 | 1 |
| T2DM | macrophage | *ANKRA2* | 0.861139673 | 0.004061224 | 0.020200091 | 1 |
| T2DM | macrophage | *TLR3* | 1.12517773 | 0.000259991 | 0.002384638 | 1 |
| T2DM | macrophage | *CCDC127* | 0.685891975 | 0.004853357 | 0.0229767 | 1 |
| T2DM | macrophage | *MB21D1* | -0.711404538 | 0.007333722 | 0.031688734 | 1 |
| T2DM | macrophage | *TXLNB* | -0.476715345 | 2.15E-05 | 0.000370143 | 0.135472158 |
| T2DM | macrophage | *TMEM200A* | 1.119903997 | 8.48E-06 | 0.000188394 | 0.05331543 |
| T2DM | macrophage | *PI16* | 0.82936866 | 0.000521113 | 0.004131449 | 1 |
| T2DM | macrophage | *HCN1* | 0.858268948 | 0.000278193 | 0.002493278 | 1 |
| T2DM | macrophage | *CAMLG* | 0.609259219 | 5.59E-05 | 0.000737916 | 0.351248212 |
| T2DM | macrophage | *KCNK5* | 1.871260905 | 1.20E-06 | 5.64E-05 | 0.007551238 |
| T2DM | macrophage | *ZNF704* | 0.573048323 | 0.001866553 | 0.011144366 | 1 |
| T2DM | macrophage | *LMTK2* | 0.713673947 | 0.001237859 | 0.008121141 | 1 |
| T2DM | macrophage | *TNFRSF11B* | 0.931492386 | 1.60E-05 | 0.000296065 | 0.100662143 |
| T2DM | macrophage | *SUN1* | 0.632263089 | 2.13E-05 | 0.000367723 | 0.134218982 |
| T2DM | macrophage | *DCAF13* | 0.902179091 | 0.012746549 | 0.0481385 | 1 |
| T2DM | macrophage | *INTS8* | 0.456614234 | 0.012043588 | 0.046197645 | 1 |
| T2DM | macrophage | *FREM1* | 0.523183206 | 0.002115323 | 0.012405815 | 1 |
| T2DM | macrophage | *SYK* | -0.342656457 | 6.05E-06 | 0.000149044 | 0.038006156 |
| T2DM | macrophage | *TMC1* | 1.173664285 | 1.32E-06 | 5.98E-05 | 0.008309138 |
| T2DM | macrophage | *SVEP1* | 0.699478483 | 2.31E-05 | 0.000391051 | 0.145471052 |
| T2DM | macrophage | *LRFN5* | 0.203383405 | 0.003094305 | 0.016478053 | 1 |
| T2DM | macrophage | *CFL2* | 0.552916063 | 0.012748624 | 0.0481385 | 1 |
| T2DM | macrophage | *ZCCHC24* | 0.74752455 | 0.004479536 | 0.021663727 | 1 |
| T2DM | macrophage | *CRYL1* | -0.291675843 | 0.008899561 | 0.036617502 | 1 |

| | | | | | | |
|---|---|---|---|---|---|---|
| T2DM | macrophage | *KLHDC2* | 0.48755756 | 0.007727846 | 0.033118585 | 1 |
| T2DM | macrophage | *SLC18A2* | 0.803122497 | 0.000932514 | 0.006463856 | 1 |
| T2DM | macrophage | *PDZD8* | 0.378053903 | 0.005198497 | 0.024155914 | 1 |
| T2DM | macrophage | *RET* | 1.134000846 | 0.000588514 | 0.004523214 | 1 |
| T2DM | macrophage | *NSMF* | 1.39619038 | 0.002451346 | 0.013846912 | 1 |
| T2DM | macrophage | *ARHGAP42* | 0.7863018 | 7.67E-05 | 0.000945536 | 0.482223197 |
| T2DM | macrophage | *SLC39A13* | 1.330470436 | 0.002886384 | 0.015589945 | 1 |
| T2DM | macrophage | *IFI27L1* | 0.838035552 | 0.001333874 | 0.008592279 | 1 |
| T2DM | macrophage | *ABTB2* | 1.184410037 | 2.77E-05 | 0.000444329 | 0.174176785 |
| T2DM | macrophage | *LIPC* | 0.495226414 | 0.001760945 | 0.010655495 | 1 |
| T2DM | macrophage | *TCP11L2* | 0.852509561 | 0.002555811 | 0.014283006 | 1 |
| T2DM | macrophage | *JAM3* | 0.737958368 | 4.75E-05 | 0.000646766 | 0.298805664 |
| T2DM | macrophage | *SPINT1* | -0.494169509 | 0.002813194 | 0.01528656 | 1 |
| T2DM | macrophage | *TRMT61A* | -0.437876093 | 0.000853253 | 0.006034197 | 1 |
| T2DM | macrophage | *LARP6* | 0.526326515 | 0.000341533 | 0.00294543 | 1 |
| T2DM | macrophage | *ASPG* | 2.735317282 | 9.01E-07 | 4.80E-05 | 0.005664751 |
| T2DM | macrophage | *PCBD1* | -0.193190987 | 0.000826207 | 0.005889302 | 1 |
| T2DM | macrophage | *ZNF202* | 1.262352041 | 0.000833306 | 0.005913085 | 1 |
| T2DM | macrophage | *FAM227B* | 0.384458204 | 0.004186832 | 0.020677623 | 1 |
| T2DM | macrophage | *C2* | 1.031847423 | 2.19E-07 | 3.06E-05 | 0.001377874 |
| T2DM | macrophage | *APBB1* | 0.740256554 | 0.000967875 | 0.006621362 | 1 |
| T2DM | macrophage | *ILK* | 0.752972262 | 0.003311926 | 0.017380698 | 1 |
| T2DM | macrophage | *NETO1* | 0.56682415 | 0.002823065 | 0.015326953 | 1 |
| T2DM | macrophage | *WDR88* | 0.893378688 | 0.003703868 | 0.018855239 | 1 |
| T2DM | macrophage | *LMO1* | -0.502106479 | 1.63E-05 | 0.000300039 | 0.102613437 |
| T2DM | macrophage | *PRTG* | 1.036735202 | 8.21E-07 | 4.65E-05 | 0.005162097 |
| T2DM | macrophage | *C16orf46* | 4.861771473 | 0.000200667 | 0.001962041 | 1 |
| T2DM | macrophage | *C16orf45* | 0.649095675 | 0.003039817 | 0.016264961 | 1 |
| T2DM | macrophage | *MS4A6E* | -0.21153503 | 0.000411515 | 0.003445001 | 1 |
| T2DM | macrophage | *MS4A7* | -0.375831619 | 5.67E-08 | 1.34E-05 | 0.000356554 |
| T2DM | macrophage | *MS4A14* | -0.20979075 | 4.59E-05 | 0.00062858 | 0.288518024 |
| T2DM | macrophage | *SMAD3* | 0.50784003 | 0.001238774 | 0.008121141 | 1 |
| T2DM | macrophage | *AKTIP* | 1.60412066 | 2.44E-07 | 3.14E-05 | 0.001536614 |
| T2DM | macrophage | *EVA1C* | 0.319909104 | 0.001232699 | 0.008098201 | 1 |
| T2DM | macrophage | *CNPY4* | 0.235543706 | 0.008264349 | 0.034615566 | 1 |
| T2DM | macrophage | *TEF* | 0.31288745 | 7.69E-05 | 0.00094639 | 0.483605283 |
| T2DM | macrophage | *COQ7* | 1.16574059 | 0.001587637 | 0.009853378 | 1 |
| T2DM | macrophage | *RBFOX3* | 2.615175349 | 8.37E-06 | 0.000187252 | 0.052617795 |
| T2DM | macrophage | *VKORC1* | -0.286199559 | 0.012595228 | 0.047677251 | 1 |
| T2DM | macrophage | *SMG8* | 4.167465189 | 0.001464936 | 0.009247041 | 1 |
| T2DM | macrophage | *ZNF641* | 0.239306497 | 0.004107449 | 0.020349515 | 1 |
| T2DM | macrophage | *CDC42EP5* | 0.483532735 | 0.012706361 | 0.048065517 | 1 |
| T2DM | macrophage | *PPP1R14A* | 1.524616468 | 8.05E-05 | 0.000979583 | 0.506386886 |
| T2DM | macrophage | *ZNF444* | 1.33596171 | 0.000319325 | 0.002780608 | 1 |
| T2DM | macrophage | *CYB5D2* | 1.164128548 | 4.33E-05 | 0.000606009 | 0.272097888 |
| T2DM | macrophage | *CD320* | 1.361239059 | 3.66E-06 | 0.000106585 | 0.023037026 |
| T2DM | macrophage | *SPRYD3* | 1.460840701 | 0.000129297 | 0.001381946 | 0.812889779 |
| T2DM | macrophage | *PRDX2* | 1.454528771 | 3.81E-08 | 1.04E-05 | 0.000239723 |
| T2DM | macrophage | *GSDMA* | -0.554163981 | 3.27E-05 | 0.000495853 | 0.20577897 |
| T2DM | macrophage | *BSCL2* | -1.10064061 | 0.009509466 | 0.038638674 | 1 |

| | | | | | | |
|---|---|---|---|---|---|---|
| T2DM | macrophage | *LTBP3* | 0.676928839 | 8.42E-09 | 3.23E-06 | 5.30E-05 |
| T2DM | macrophage | *NAALADL1* | 0.681437328 | 0.00405137 | 0.020167031 | 1 |
| T2DM | macrophage | *UBTD2* | 0.573792867 | 0.003305863 | 0.017363378 | 1 |
| T2DM | macrophage | *PXK* | 0.716418596 | 3.85E-06 | 0.000109505 | 0.024200516 |
| T2DM | macrophage | *TAP1* | -0.513992118 | 0.008964879 | 0.036838037 | 1 |
| T2DM | macrophage | *RHOH* | 0.541293048 | 0.002645836 | 0.014630053 | 1 |
| T2DM | macrophage | *SLC20A2* | 0.641797327 | 0.005891077 | 0.026607186 | 1 |
| T2DM | macrophage | *ZSWIM1* | 0.709495125 | 0.003039013 | 0.016264961 | 1 |
| T2DM | macrophage | *WFDC13* | 2.538194766 | 4.07E-05 | 0.000582519 | 0.255725975 |
| T2DM | macrophage | *SLC16A4* | 0.261775732 | 0.008261705 | 0.034615566 | 1 |
| T2DM | macrophage | *NPNT* | 0.697975734 | 0.005395251 | 0.024889909 | 1 |
| T2DM | macrophage | *SNTB2* | 0.667739928 | 0.007370878 | 0.031805566 | 1 |
| T2DM | macrophage | *ZNF608* | 0.710972567 | 3.63E-05 | 0.000536296 | 0.228462201 |
| T2DM | macrophage | *INPP5D* | -0.553266529 | 0.001054231 | 0.007088718 | 1 |
| T2DM | macrophage | *LETM1* | 1.065983444 | 0.003740958 | 0.0190223 | 1 |
| T2DM | macrophage | *STXBP6* | 0.722343503 | 0.000224068 | 0.002134422 | 1 |
| T2DM | macrophage | *LGALS9* | -0.331468976 | 0.000363616 | 0.003085096 | 1 |
| T2DM | macrophage | *SIGLEC7* | -0.370853804 | 0.000423837 | 0.003534038 | 1 |
| T2DM | macrophage | *IRS1* | -0.354428013 | 1.72E-24 | 1.08E-20 | 1.08E-20 |
| T2DM | macrophage | *C8orf46* | 0.046032659 | 0.008021563 | 0.033823988 | 1 |
| T2DM | macrophage | *FAM110B* | 0.848145957 | 0.001053017 | 0.007088131 | 1 |
| T2DM | macrophage | *ATF5* | -0.833898749 | 0.005176322 | 0.02408848 | 1 |
| T2DM | macrophage | *PTAFR* | -0.346012134 | 0.000515969 | 0.0041062 | 1 |
| T2DM | macrophage | *RNASE6* | -0.754364493 | 6.77E-06 | 0.000160089 | 0.04258361 |
| T2DM | macrophage | *NPR1* | 1.001118721 | 2.48E-06 | 8.38E-05 | 0.015565964 |
| T2DM | macrophage | *SLC38A11* | 0.888084465 | 0.001316289 | 0.008505145 | 1 |
| T2DM | macrophage | *CCDC8* | 1.158823261 | 0.001812929 | 0.010887255 | 1 |
| T2DM | macrophage | *INO80E* | -0.378513575 | 0.009809831 | 0.039489006 | 1 |
| T2DM | macrophage | *UGP2* | 0.34227222 | 0.007930329 | 0.033619677 | 1 |
| T2DM | macrophage | *AVEN* | 0.757862489 | 0.008746808 | 0.036130869 | 1 |
| T2DM | macrophage | *ZFPM2* | 0.587834166 | 0.000143222 | 0.001485869 | 0.900436677 |
| T2DM | macrophage | *ZNF747* | 1.87967269 | 6.01E-05 | 0.000776918 | 0.37781981 |
| T2DM | macrophage | *SEMA3E* | 1.019383874 | 1.27E-07 | 2.09E-05 | 0.000795923 |
| T2DM | macrophage | *METTL7B* | 3.535153108 | 3.44E-06 | 0.000103127 | 0.021656731 |
| T2DM | macrophage | *CD14* | -0.289545031 | 0.000637246 | 0.004821136 | 1 |
| T2DM | macrophage | *EMB* | -0.327586878 | 3.64E-08 | 1.04E-05 | 0.000228676 |
| T2DM | macrophage | *DLGAP1* | 0.877625757 | 9.45E-05 | 0.001097803 | 0.593911283 |
| T2DM | macrophage | *NUDCD2* | 0.985065314 | 0.001485434 | 0.009347503 | 1 |
| T2DM | macrophage | *SLC26A5* | 1.249159913 | 8.55E-06 | 0.000189238 | 0.05374358 |
| T2DM | macrophage | *COMMD5* | -0.718680272 | 0.006786213 | 0.029710947 | 1 |
| T2DM | macrophage | *GTSF1* | -2.159374331 | 0.002604986 | 0.014493403 | 1 |
| T2DM | macrophage | *TMEM133* | 3.029056398 | 6.11E-07 | 4.22E-05 | 0.003844262 |
| T2DM | macrophage | *KCNS3* | 0.198527438 | 0.000538093 | 0.004234034 | 1 |
| T2DM | macrophage | *MTSS1* | -0.375299686 | 0.000587194 | 0.004518593 | 1 |
| T2DM | macrophage | *TSEN34* | -0.950218386 | 0.007859473 | 0.033470855 | 1 |
| T2DM | macrophage | *OSCAR* | -0.255544232 | 0.006546875 | 0.028884355 | 1 |
| T2DM | macrophage | *PAQR8* | -0.726646481 | 0.000504443 | 0.004029778 | 1 |
| T2DM | macrophage | *NUDT6* | 0.818839565 | 1.89E-05 | 0.00033478 | 0.118728513 |
| T2DM | macrophage | *FPR1* | -0.227785179 | 0.000290128 | 0.002579963 | 1 |
| T2DM | macrophage | *SOX7* | 1.020101248 | 7.31E-05 | 0.000906005 | 0.459344673 |

| | | | | | | |
|---|---|---|---|---|---|---|
| T2DM | macrophage | *MFN1* | 0.588629122 | 0.001137146 | 0.007557334 | 1 |
| T2DM | macrophage | *GIMAP8* | -0.259455861 | 0.010716658 | 0.042268273 | 1 |
| T2DM | macrophage | *KCNMB3* | 0.390630366 | 0.000496851 | 0.003981657 | 1 |
| T2DM | macrophage | *ATP6V0E2* | 1.076478829 | 0.0035727 | 0.018321018 | 1 |
| T2DM | macrophage | *CHST11* | -0.311316163 | 0.000552987 | 0.004318792 | 1 |
| T2DM | macrophage | *KCND3* | 0.794810647 | 1.31E-06 | 5.97E-05 | 0.008242332 |
| T2DM | macrophage | *MRPL36* | 0.572172126 | 0.000616216 | 0.00470164 | 1 |
| T2DM | macrophage | *MCC* | 0.663979855 | 0.000641787 | 0.004849659 | 1 |
| T2DM | macrophage | *ZNF561* | -0.298588226 | 0.003683111 | 0.018779984 | 1 |
| T2DM | macrophage | *HOPX* | 1.303542814 | 0.000141239 | 0.001477671 | 0.887970335 |
| T2DM | macrophage | *ZNF584* | 0.187704669 | 0.009195644 | 0.037614194 | 1 |
| T2DM | macrophage | *NMUR1* | 1.352576838 | 7.66E-07 | 4.48E-05 | 0.004813968 |
| T2DM | macrophage | *P2RY6* | -0.306666789 | 1.44E-06 | 6.22E-05 | 0.009081296 |
| T2DM | macrophage | *RGS19* | -0.47009291 | 1.59E-09 | 7.15E-07 | 1.00E-05 |
| T2DM | macrophage | *TCEA2* | 1.490650506 | 1.75E-05 | 0.000316778 | 0.109921889 |
| T2DM | macrophage | *ANO5* | -0.78100247 | 8.54E-05 | 0.001022897 | 0.537020708 |
| T2DM | macrophage | *VAT1L* | 0.458938399 | 0.004818122 | 0.022896096 | 1 |
| T2DM | macrophage | *NINJ2* | 0.747940291 | 0.000655835 | 0.004926208 | 1 |
| T2DM | macrophage | *FAM90A1* | 2.59744508 | 4.41E-07 | 3.80E-05 | 0.002774211 |
| T2DM | macrophage | *C3AR1* | -0.393787735 | 3.61E-06 | 0.000106131 | 0.022711951 |
| T2DM | macrophage | *AQP4* | 1.59338554 | 2.40E-06 | 8.38E-05 | 0.015092727 |
| T2DM | macrophage | *SRGAP2C* | -0.467630616 | 0.003778772 | 0.019097377 | 1 |
| T2DM | macrophage | *ANKRD20A4* | 3.079561284 | 1.84E-06 | 7.11E-05 | 0.011587785 |
| T2DM | macrophage | *SERF1A* | 4.350802308 | 1.44E-06 | 6.22E-05 | 0.009045023 |
| T2DM | macrophage | *MOB3A* | -0.612508434 | 0.001394613 | 0.0089105 | 1 |
| T2DM | macrophage | *C1QTNF4* | -0.103034755 | 0.000166896 | 0.001676162 | 1 |
| T2DM | macrophage | *HINFP* | 0.99514181 | 0.011600746 | 0.04485479 | 1 |
| T2DM | macrophage | *COPRS* | 1.199261257 | 0.000118365 | 0.001294193 | 0.744160802 |
| T2DM | macrophage | *RCAN2* | 0.824665392 | 0.000191087 | 0.001888934 | 1 |
| T2DM | macrophage | *FGGY* | 0.544459098 | 0.004910567 | 0.023125646 | 1 |
| T2DM | macrophage | *FIBP* | 1.027005204 | 0.004490313 | 0.021682489 | 1 |
| T2DM | macrophage | *CARNS1* | 1.043998121 | 0.003919829 | 0.019683676 | 1 |
| T2DM | macrophage | *CTSW* | -0.069559887 | 0.001714119 | 0.010462784 | 1 |
| T2DM | macrophage | *SNTG2* | 0.360185854 | 0.011829354 | 0.045574405 | 1 |
| T2DM | macrophage | *PDE3A* | 0.680752499 | 1.70E-06 | 6.69E-05 | 0.010687126 |
| T2DM | macrophage | *SMPDL3A* | -0.444968297 | 0.004365546 | 0.02130915 | 1 |
| T2DM | macrophage | *AGAP5* | -15.35690277 | 4.10E-06 | 0.000114426 | 0.025745818 |
| T2DM | macrophage | *CCL19* | 4.696900114 | 0.000256273 | 0.002373898 | 1 |
| T2DM | macrophage | *SNX32* | 0.426594815 | 0.000107073 | 0.001202087 | 0.673168827 |
| T2DM | macrophage | *METAP1D* | 0.916488134 | 0.000605472 | 0.004636546 | 1 |
| T2DM | macrophage | *MRGPRF* | 0.697416623 | 0.001688325 | 0.010345516 | 1 |
| T2DM | macrophage | *HPSE2* | 0.411095572 | 1.68E-05 | 0.000305604 | 0.105433452 |
| T2DM | macrophage | *BNC2* | 0.355902051 | 0.012437554 | 0.047247674 | 1 |
| T2DM | macrophage | *VANGL1* | 0.904689087 | 9.26E-05 | 0.001082672 | 0.582261465 |
| T2DM | macrophage | *GLRX* | 0.750648291 | 0.001069329 | 0.007174891 | 1 |
| T2DM | macrophage | *C1QA* | -0.466504399 | 2.47E-05 | 0.000408348 | 0.155464163 |
| T2DM | macrophage | *NDNF* | 0.779197774 | 0.000820816 | 0.005864173 | 1 |
| T2DM | macrophage | *OLR1* | -0.921230587 | 5.46E-05 | 0.000724055 | 0.343202174 |
| T2DM | macrophage | *GLIPR1L1* | 0.476145205 | 0.001364752 | 0.0087553 | 1 |
| T2DM | macrophage | *CCDC36* | 0.68458272 | 1.13E-05 | 0.000233163 | 0.07134777 |

| | | | | | | |
|---|---|---|---|---|---|---|
| T2DM | macrophage | *VEGFB* | -0.326891139 | 0.000184354 | 0.001839734 | 1 |
| T2DM | macrophage | *C1orf100* | 2.340671328 | 6.98E-05 | 0.000872883 | 0.439060215 |
| T2DM | macrophage | *GPR160* | 0.870170086 | 0.009630843 | 0.039013603 | 1 |
| T2DM | macrophage | *USMG5* | 0.876209014 | 0.000549408 | 0.004301529 | 1 |
| T2DM | macrophage | *CCS* | 0.688062633 | 0.004001791 | 0.02001532 | 1 |
| T2DM | macrophage | *C16orf91* | 1.861198191 | 5.76E-05 | 0.000755578 | 0.361921948 |
| T2DM | macrophage | *SOCS7* | 0.83497747 | 0.009070713 | 0.037199982 | 1 |
| T2DM | macrophage | *TLR6* | -0.301425004 | 0.001094744 | 0.007306428 | 1 |
| T2DM | macrophage | *LINGO2* | 1.116794515 | 1.70E-06 | 6.69E-05 | 0.010710287 |
| T2DM | macrophage | *BBS1* | 0.586954215 | 0.010556932 | 0.041848316 | 1 |
| T2DM | macrophage | *IL20RB* | 0.310899455 | 0.002144672 | 0.012523855 | 1 |
| T2DM | macrophage | *KLF14* | -2.464370183 | 0.004343589 | 0.021250108 | 1 |
| T2DM | macrophage | *ANGEL2* | 0.708641042 | 0.012303971 | 0.046853461 | 1 |
| T2DM | macrophage | *SH3PXD2B* | 0.305604026 | 0.000138261 | 0.001453593 | 0.869248897 |
| T2DM | macrophage | *FGFBP3* | 1.108744151 | 0.001582646 | 0.009832112 | 1 |
| T2DM | macrophage | *C4orf32* | 0.624097655 | 0.004936939 | 0.02321506 | 1 |
| T2DM | macrophage | *BTC* | 0.863688855 | 1.46E-05 | 0.000280658 | 0.091494462 |
| T2DM | macrophage | *NLRP6* | -2.503843494 | 7.82E-08 | 1.55E-05 | 0.000491653 |
| T2DM | macrophage | *NDUFA11* | 1.053276111 | 0.001653677 | 0.01019281 | 1 |
| T2DM | macrophage | *FBXW8* | 0.754258469 | 0.006228848 | 0.02765591 | 1 |
| T2DM | macrophage | *CHST2* | 3.007297683 | 2.36E-06 | 8.37E-05 | 0.014820042 |
| T2DM | macrophage | *PHYHD1* | 0.524011906 | 0.000557005 | 0.004344776 | 1 |
| T2DM | macrophage | *ZNF519* | 0.583804265 | 0.01253047 | 0.0475719 | 1 |
| T2DM | macrophage | *BANF1* | 0.223070476 | 0.004871951 | 0.023014001 | 1 |
| T2DM | macrophage | *CHRNA7* | 0.479697933 | 0.001967741 | 0.01171514 | 1 |
| T2DM | macrophage | *TMEM9B* | 0.909118265 | 0.002269422 | 0.013101794 | 1 |
| T2DM | macrophage | *NRIP3* | -0.389618935 | 1.19E-06 | 5.60E-05 | 0.007453045 |
| T2DM | macrophage | *UCP2* | -0.622298458 | 3.55E-05 | 0.000528152 | 0.223033171 |
| T2DM | macrophage | *TEX26* | 0.566390416 | 0.002397002 | 0.013637966 | 1 |
| T2DM | macrophage | *ETV4* | 0.425410586 | 0.00146094 | 0.009244199 | 1 |
| T2DM | macrophage | *LRRN1* | 1.450413262 | 0.000362722 | 0.003081669 | 1 |
| T2DM | macrophage | *GPX2* | 1.329576882 | 0.00010101 | 0.001150456 | 0.635051949 |
| T2DM | macrophage | *COX8A* | 1.140757586 | 0.00954787 | 0.038752395 | 1 |
| T2DM | macrophage | *B3GNT4* | 7.832561929 | 7.30E-07 | 4.48E-05 | 0.004587787 |
| T2DM | macrophage | *CDC26* | -0.245829031 | 0.008470584 | 0.035291294 | 1 |
| T2DM | macrophage | *HORMAD2* | 0.28308829 | 0.000193172 | 0.001903564 | 1 |
| T2DM | macrophage | *RNF152* | 0.819674097 | 0.00088298 | 0.006195643 | 1 |
| T2DM | macrophage | *BOK* | 0.655878246 | 0.000100011 | 0.001147421 | 0.628770783 |
| T2DM | macrophage | *C8orf59* | 0.834373751 | 0.000316222 | 0.002757407 | 1 |
| T2DM | macrophage | *PFN4* | 0.2301137 | 0.005491861 | 0.025149227 | 1 |
| T2DM | macrophage | *IRX5* | 0.422092631 | 0.002393489 | 0.013637966 | 1 |
| T2DM | macrophage | *PNMA1* | 0.904303782 | 0.00502119 | 0.023558376 | 1 |
| T2DM | macrophage | *GCNT4* | 2.889812268 | 4.35E-07 | 3.80E-05 | 0.002731923 |
| T2DM | macrophage | *WDR73* | 0.436606095 | 0.002705366 | 0.014851852 | 1 |
| T2DM | macrophage | *DSCAML1* | -0.406605073 | 0.004094965 | 0.020335739 | 1 |
| T2DM | macrophage | *AP006621.1* | 0.32332106 | 0.001208309 | 0.007946273 | 1 |
| T2DM | macrophage | *MAN1B1* | 0.952310715 | 0.009686739 | 0.039164328 | 1 |
| T2DM | macrophage | *C8orf31* | 1.596833018 | 1.42E-06 | 6.22E-05 | 0.008946717 |
| T2DM | macrophage | *TIMM22* | 1.090004186 | 0.002995987 | 0.016112721 | 1 |
| T2DM | macrophage | *MAGEF1* | 1.647386317 | 4.86E-06 | 0.000128317 | 0.030539397 |

| | | | | | | |
|---|---|---|---|---|---|---|
| T2DM | macrophage | *PAWR* | 0.98245302 | 1.77E-05 | 0.00031823 | 0.111380415 |
| T2DM | macrophage | *IRX3* | 0.433553492 | 0.000145939 | 0.001506599 | 0.917518619 |
| T2DM | macrophage | *C18orf32* | 0.646469677 | 0.004143894 | 0.020513909 | 1 |
| T2DM | macrophage | *CD163L1* | -0.228565325 | 3.00E-05 | 0.000466318 | 0.188392668 |
| T2DM | macrophage | *ZNF619* | 0.956029598 | 0.003653631 | 0.018659934 | 1 |
| T2DM | macrophage | *ZNF354C* | 1.2364285 | 0.000130982 | 0.001395739 | 0.823486047 |
| T2DM | macrophage | *BET1L* | 0.608564753 | 0.002078041 | 0.012255766 | 1 |
| T2DM | macrophage | *IMP3* | 1.755998542 | 2.17E-07 | 3.06E-05 | 0.00136408 |
| T2DM | macrophage | *C2orf73* | 1.499709799 | 1.00E-06 | 5.00E-05 | 0.006295501 |
| T2DM | macrophage | *FAM26E* | 0.463249721 | 0.002367444 | 0.01353102 | 1 |
| T2DM | macrophage | *PRSS42* | 0.399536359 | 0.00668738 | 0.029380545 | 1 |
| T2DM | macrophage | *C2orf69* | 0.978239501 | 0.006039516 | 0.027121741 | 1 |
| T2DM | macrophage | *GRAMD1C* | 1.396310225 | 0.000133611 | 0.001418721 | 0.840010021 |
| T2DM | macrophage | *GOLGA8Q* | 3.862310742 | 0.01205092 | 0.046197645 | 1 |
| T2DM | macrophage | *DALRD3* | 0.386435662 | 0.010853585 | 0.042674477 | 1 |
| T2DM | macrophage | *ZC3H12D* | -0.656021987 | 1.18E-05 | 0.00023889 | 0.074294761 |
| T2DM | macrophage | *SH2D4B* | -1.927534213 | 2.93E-05 | 0.000457252 | 0.183953912 |
| T2DM | macrophage | *PRSS36* | -0.40967511 | 2.07E-06 | 7.62E-05 | 0.013031958 |
| T2DM | macrophage | *AQP11* | 0.7487923 | 0.004402695 | 0.021432861 | 1 |
| T2DM | macrophage | *TMEM11* | 1.200659505 | 0.005481888 | 0.025146197 | 1 |
| T2DM | macrophage | *BEND3* | 1.794300477 | 0.000989057 | 0.006722381 | 1 |
| T2DM | macrophage | *LINC00843* | 1.430818133 | 0.005700234 | 0.025918451 | 1 |
| T2DM | macrophage | *COX14* | 0.467416205 | 0.004721091 | 0.022537205 | 1 |
| T2DM | macrophage | *P4HTM* | 0.489796007 | 0.006439425 | 0.028490266 | 1 |
| T2DM | macrophage | *ERBB4* | 0.65758224 | 0.000159436 | 0.001611535 | 1 |
| T2DM | macrophage | *CSRNP3* | 0.768650566 | 0.000735325 | 0.00535364 | 1 |
| T2DM | macrophage | *STX19* | 0.929620162 | 7.41E-07 | 4.48E-05 | 0.004660236 |
| T2DM | macrophage | *FAM219B* | 0.68775853 | 0.008594168 | 0.035640853 | 1 |
| T2DM | macrophage | *GDPD4* | -0.862572561 | 4.55E-06 | 0.000123286 | 0.028602415 |
| T2DM | macrophage | *TRIM73* | -0.503697427 | 0.003777644 | 0.019097377 | 1 |
| T2DM | macrophage | *TRIML2* | -7.143472871 | 2.40E-06 | 8.38E-05 | 0.015060188 |
| T2DM | macrophage | *FARSA* | -0.678980702 | 0.000917479 | 0.006380743 | 1 |
| T2DM | macrophage | *CALR* | 0.584027506 | 0.007024449 | 0.030562429 | 1 |
| T2DM | macrophage | *NSUN7* | -0.113799641 | 0.004872218 | 0.023014001 | 1 |
| T2DM | macrophage | *HLA-DQB1* | -0.313056098 | 5.89E-16 | 7.40E-13 | 3.70E-12 |
| T2DM | macrophage | *EGR3* | -1.01952699 | 4.28E-05 | 0.000601316 | 0.268788061 |
| T2DM | macrophage | *ZBTB42* | 1.1158916 | 0.001463366 | 0.009246414 | 1 |
| T2DM | macrophage | *LACC1* | -0.547426694 | 0.000155877 | 0.001580643 | 0.979998844 |
| T2DM | macrophage | *MAF1* | 1.403521035 | 0.003136836 | 0.016628404 | 1 |
| T2DM | macrophage | *CDH5* | 0.198251586 | 0.012148075 | 0.046456781 | 1 |
| T2DM | macrophage | *B3GNT3* | 0.738626375 | 0.006010933 | 0.027051348 | 1 |
| T2DM | macrophage | *SEPT1* | 1.320102793 | 0.000725306 | 0.005320888 | 1 |
| T2DM | macrophage | *TDRP* | 0.904269345 | 2.52E-05 | 0.000412233 | 0.158297571 |
| T2DM | macrophage | *HCLS1* | -0.512775033 | 8.16E-05 | 0.000984184 | 0.512760062 |
| T2DM | macrophage | *CCDC66* | 0.318971819 | 0.012857324 | 0.048374623 | 1 |
| T2DM | macrophage | *SSTR2* | 2.365778734 | 1.56E-06 | 6.42E-05 | 0.009819212 |
| T2DM | macrophage | *SHISA2* | 14.92545459 | 2.07E-07 | 3.06E-05 | 0.001304377 |
| T2DM | macrophage | *CHST13* | -1.513970644 | 5.52E-07 | 4.03E-05 | 0.003469873 |
| T2DM | macrophage | *SRSF8* | 1.477942665 | 0.000253144 | 0.002354311 | 1 |
| T2DM | macrophage | *CAPS2* | 0.765264096 | 0.000960141 | 0.006597169 | 1 |

| | | | | | | |
|---|---|---|---|---|---|---|
| T2DM | macrophage | *KCTD2* | 1.090626344 | 0.003752265 | 0.019039942 | 1 |
| T2DM | macrophage | *LRRC57* | 0.767421931 | 0.002638109 | 0.014630053 | 1 |
| T2DM | macrophage | *ZFP82* | 0.819659503 | 0.002600508 | 0.014481303 | 1 |
| T2DM | macrophage | *RPH3AL* | -0.292014376 | 0.009772792 | 0.039401807 | 1 |
| T2DM | macrophage | *SLC26A11* | -0.548578241 | 0.00147287 | 0.009281601 | 1 |
| T2DM | macrophage | *TMEM132C* | 0.349994271 | 0.000328412 | 0.002851836 | 1 |
| T2DM | macrophage | *SLC25A41* | 3.035033181 | 2.01E-05 | 0.000352846 | 0.126490284 |
| T2DM | macrophage | *TMEM136* | 0.294452708 | 0.007948096 | 0.033649616 | 1 |
| T2DM | macrophage | *CCL13* | -0.391247856 | 0.000124134 | 0.001341512 | 0.780432037 |
| T2DM | macrophage | *ACBD4* | 0.83324601 | 0.001013947 | 0.006869274 | 1 |
| T2DM | macrophage | *ATG9B* | 2.550476306 | 3.25E-05 | 0.000494113 | 0.204068741 |
| T2DM | macrophage | *CLDN7* | -0.271221172 | 0.000234304 | 0.002208501 | 1 |
| T2DM | macrophage | *CHST15* | -0.522016046 | 0.000351044 | 0.003006827 | 1 |
| T2DM | macrophage | *HHIPL1* | -0.836083356 | 5.29E-05 | 0.000706099 | 0.332572676 |
| T2DM | macrophage | *SYNM* | 1.222774901 | 4.05E-06 | 0.000114426 | 0.025478395 |
| T2DM | macrophage | *KBTBD3* | 1.172757479 | 0.002588606 | 0.014427807 | 1 |
| T2DM | macrophage | *CLN8* | -0.382774051 | 0.007583198 | 0.032587537 | 1 |
| T2DM | macrophage | *TSHZ2* | 0.400196547 | 0.002272993 | 0.013110371 | 1 |
| T2DM | macrophage | *CAPN12* | 1.237513005 | 1.12E-07 | 2.00E-05 | 0.000701108 |
| T2DM | macrophage | *FES* | -0.894766142 | 0.002211407 | 0.012814501 | 1 |
| T2DM | macrophage | *SPNS3* | 0.901784991 | 0.004572239 | 0.021967958 | 1 |
| T2DM | macrophage | *CLEC4G* | -0.489336696 | 1.87E-06 | 7.15E-05 | 0.011726518 |
| T2DM | macrophage | *SKA2* | 0.942332218 | 0.003256983 | 0.017163998 | 1 |
| T2DM | macrophage | *TTC3* | 0.134023643 | 0.001099293 | 0.007329009 | 1 |
| T2DM | macrophage | *TSKU* | 1.092717517 | 2.68E-06 | 8.82E-05 | 0.01684829 |
| T2DM | macrophage | *ZNF320* | 0.134858928 | 0.000528756 | 0.004181499 | 1 |
| T2DM | macrophage | *C12orf60* | 0.631071929 | 0.002754748 | 0.015033939 | 1 |
| T2DM | macrophage | *GAS6* | -0.621026108 | 8.09E-05 | 0.000980389 | 0.508821738 |
| T2DM | macrophage | *BEGAIN* | 2.54152711 | 7.57E-07 | 4.48E-05 | 0.004758215 |
| T2DM | macrophage | *FANCF* | 1.796432834 | 0.005629612 | 0.025647368 | 1 |
| T2DM | macrophage | *C2CD4C* | 0.409884869 | 0.012109536 | 0.046365807 | 1 |
| T2DM | macrophage | *RUVBL2* | 0.401239031 | 0.012918951 | 0.048519382 | 1 |
| T2DM | macrophage | *CCBE1* | 0.188308496 | 0.013177383 | 0.049286287 | 1 |
| T2DM | macrophage | *FHL3* | -0.584203288 | 0.009472917 | 0.038547723 | 1 |
| T2DM | macrophage | *C19orf71* | -1.061200955 | 0.000464161 | 0.003789843 | 1 |
| T2DM | macrophage | *GPR132* | -0.556134762 | 9.92E-06 | 0.000210661 | 0.062355791 |
| T2DM | macrophage | *SERHL2* | 1.20328793 | 3.19E-05 | 0.000489383 | 0.20064704 |
| T2DM | macrophage | *RP1L1* | 0.699065641 | 6.75E-07 | 4.42E-05 | 0.004244714 |
| T2DM | macrophage | *FAM19A1* | 1.001699607 | 9.24E-06 | 0.000199597 | 0.058082652 |
| T2DM | macrophage | *BMP8A* | 2.08991586 | 2.95E-15 | 3.09E-12 | 1.86E-11 |
| T2DM | macrophage | *KCTD16* | 0.645620451 | 5.49E-05 | 0.000726902 | 0.345278659 |
| T2DM | macrophage | *ZNF703* | -0.429267116 | 0.002418107 | 0.013679628 | 1 |
| T2DM | macrophage | *KCTD8* | 0.316736139 | 0.000376222 | 0.003174909 | 1 |
| T2DM | macrophage | *TUBA8* | 0.522792884 | 0.00043914 | 0.003635299 | 1 |
| T2DM | macrophage | *ANKRD45* | 2.487569185 | 2.47E-05 | 0.000408348 | 0.155580452 |
| T2DM | macrophage | *GPR39* | 0.75179729 | 0.000552715 | 0.004318792 | 1 |
| T2DM | macrophage | *PP2D1* | 0.578945439 | 0.000516645 | 0.004106384 | 1 |
| T2DM | macrophage | *TMEM236* | -0.646560097 | 4.32E-06 | 0.00011848 | 0.027132 |
| T2DM | macrophage | *TBX1* | -0.961750031 | 0.004518577 | 0.021768808 | 1 |
| T2DM | macrophage | *EIF3C* | 0.253845404 | 0.000570053 | 0.004424593 | 1 |

| | | | | | | |
|---|---|---|---|---|---|---|
| T2DM | macrophage | *CNTN2* | 2.274117662 | 0.000240283 | 0.002254711 | 1 |
| T2DM | macrophage | *CSF1* | 0.311649083 | 0.001532643 | 0.009587788 | 1 |
| T2DM | macrophage | *PLA2G6* | 0.228383882 | 0.002938974 | 0.015846767 | 1 |
| T2DM | macrophage | *CCR10* | 1.297981684 | 1.17E-05 | 0.00023767 | 0.073677564 |
| T2DM | macrophage | *WDR27* | 0.690271792 | 0.001320368 | 0.008522744 | 1 |
| T2DM | macrophage | *LPAR5* | -0.318511612 | 8.18E-05 | 0.000984838 | 0.514085531 |
| T2DM | macrophage | *ZNF93* | -0.893172403 | 0.006563941 | 0.028939337 | 1 |
| T2DM | macrophage | *SEPT5* | 2.332662743 | 4.08E-07 | 3.80E-05 | 0.002567709 |
| T2DM | macrophage | *NDUFA12* | 0.931589492 | 0.010453964 | 0.041544925 | 1 |
| T2DM | macrophage | *TUSC5* | 0.219932121 | 0.005402749 | 0.024889909 | 1 |
| T2DM | macrophage | *ZFP90* | 0.844530773 | 0.004404263 | 0.021432861 | 1 |
| T2DM | macrophage | *NDUFA6* | 0.114113864 | 0.004842543 | 0.022954579 | 1 |
| T2DM | macrophage | *TMEM106A* | -0.381592051 | 0.000909827 | 0.00635565 | 1 |
| T2DM | macrophage | *BRF1* | 0.893191746 | 0.003495158 | 0.018025187 | 1 |
| T2DM | macrophage | *ADSSL1* | 1.517731119 | 7.74E-06 | 0.00017637 | 0.048678124 |
| T2DM | macrophage | *HS6ST3* | 0.746171895 | 0.000472434 | 0.0038474 | 1 |
| T2DM | macrophage | *MAPK11* | 0.565693317 | 0.009493233 | 0.038605405 | 1 |
| T2DM | macrophage | *MUC1* | 0.250575494 | 0.009776812 | 0.039401807 | 1 |
| T2DM | macrophage | *AHNAK2* | 0.951591348 | 0.000223203 | 0.002129406 | 1 |
| T2DM | macrophage | *PDIA2* | 0.369167477 | 0.011922552 | 0.045845313 | 1 |
| T2DM | macrophage | *PBX1* | 0.488079643 | 0.000582166 | 0.00449533 | 1 |
| T2DM | macrophage | *ADARB2* | 0.886907266 | 0.000662658 | 0.004959678 | 1 |
| T2DM | macrophage | *SLC52A2* | 1.287059947 | 0.010706168 | 0.042253408 | 1 |
| T2DM | macrophage | *PIGP* | -0.08108226 | 0.007760341 | 0.033212569 | 1 |
| T2DM | macrophage | *LINC00839* | 0.573141103 | 4.46E-05 | 0.000618718 | 0.280279308 |
| T2DM | macrophage | *RTN4RL1* | 1.16354089 | 0.000633927 | 0.004801806 | 1 |
| T2DM | macrophage | *CYP4Z1* | 0.916687336 | 4.07E-07 | 3.80E-05 | 0.002561251 |
| T2DM | macrophage | *BPIFB4* | 16.22151584 | 6.10E-05 | 0.000783447 | 0.383519854 |
| T2DM | macrophage | *CYP4X1* | 0.69655249 | 3.44E-06 | 0.000103127 | 0.021611954 |
| T2DM | macrophage | *PCLO* | 0.263808388 | 0.006234107 | 0.027659723 | 1 |
| T2DM | macrophage | *LILRB4* | -0.157254688 | 0.00620079 | 0.027554049 | 1 |
| T2DM | macrophage | *TMEM17* | 0.43354977 | 0.007667265 | 0.032881373 | 1 |
| T2DM | macrophage | *TNFRSF18* | 4.842272227 | 3.39E-05 | 0.000511013 | 0.213092426 |
| T2DM | macrophage | *RTN4RL2* | 1.276784154 | 0.000568175 | 0.004415472 | 1 |
| T2DM | macrophage | *TMEM232* | 0.375854891 | 0.010132211 | 0.040488293 | 1 |
| T2DM | macrophage | *EFCAB6* | 0.773509984 | 0.000113138 | 0.001250082 | 0.71129662 |
| T2DM | macrophage | *KANK3* | 0.962809999 | 0.000111836 | 0.001243274 | 0.703115805 |
| T2DM | macrophage | *EMID1* | -0.748923706 | 3.20E-05 | 0.000489553 | 0.201443707 |
| T2DM | macrophage | *CYP4A11* | 0.279631636 | 0.01321586 | 0.049352588 | 1 |
| T2DM | macrophage | *TMEM216* | 0.42399668 | 0.001736447 | 0.010562017 | 1 |
| T2DM | macrophage | *ENTPD5* | 1.080911295 | 0.000385888 | 0.003246985 | 1 |
| T2DM | macrophage | *ANGPTL5* | 1.469171765 | 2.41E-05 | 0.000405572 | 0.151683868 |
| T2DM | macrophage | *MT1X* | 0.40847684 | 0.005028448 | 0.02356828 | 1 |
| T2DM | macrophage | *FPR3* | -0.27072773 | 3.82E-05 | 0.000554106 | 0.239927849 |
| T2DM | macrophage | *GJA4* | 0.606349695 | 0.002530363 | 0.014153372 | 1 |
| T2DM | macrophage | *IFT140* | 1.127810389 | 2.57E-05 | 0.000416737 | 0.161694003 |
| T2DM | macrophage | *TLR5* | -0.600295098 | 0.000241478 | 0.002259181 | 1 |
| T2DM | macrophage | *TET3* | -0.477017338 | 5.45E-05 | 0.000724055 | 0.34257676 |
| T2DM | macrophage | *TRPV2* | -0.841126731 | 0.000872991 | 0.006153018 | 1 |
| T2DM | macrophage | *C2orf88* | 0.409115148 | 0.00970002 | 0.03919282 | 1 |

| | | | | | | |
|---|---|---|---|---|---|---|
| T2DM | macrophage | *FAM69C* | 0.486809923 | 0.01213277 | 0.046426492 | 1 |
| T2DM | macrophage | *MCRS1* | 0.6759591 | 0.000277594 | 0.002493278 | 1 |
| T2DM | macrophage | *GFRAL* | 0.415917923 | 0.000369101 | 0.003119001 | 1 |
| T2DM | macrophage | *C2orf66* | 1.050200641 | 0.00213266 | 0.012484202 | 1 |
| T2DM | macrophage | *C6orf132* | 0.418561003 | 0.002812234 | 0.01528656 | 1 |
| T2DM | macrophage | *FAM166A* | 4.585882406 | 7.48E-07 | 4.48E-05 | 0.004703116 |
| T2DM | macrophage | *SMTNL2* | 5.882030365 | 6.67E-07 | 4.41E-05 | 0.004193739 |
| T2DM | macrophage | *LINC00265* | 0.760143616 | 0.004362596 | 0.02130915 | 1 |
| T2DM | macrophage | *NUTM2B* | -5.920892567 | 6.18E-05 | 0.000791569 | 0.388660471 |
| T2DM | macrophage | *NCR3LG1* | -1.974285322 | 0.001203592 | 0.007931851 | 1 |
| T2DM | macrophage | *ZNF793* | 0.144196129 | 0.008179531 | 0.034328914 | 1 |
| T2DM | macrophage | *HYKK* | 0.560594265 | 0.000464066 | 0.003789843 | 1 |
| T2DM | macrophage | *JAKMIP3* | 0.192205396 | 0.010082021 | 0.040424531 | 1 |
| T2DM | macrophage | *FAM72B* | -1.284114972 | 3.19E-06 | 9.87E-05 | 0.020040374 |
| T2DM | macrophage | *GOLGA8M* | 0.888310536 | 2.57E-06 | 8.54E-05 | 0.016138155 |
| T2DM | macrophage | *S100A16* | 0.719574299 | 0.000741215 | 0.005387303 | 1 |
| T2DM | macrophage | *UROS* | 0.615954993 | 0.003898361 | 0.019622897 | 1 |
| T2DM | macrophage | *NOXA1* | -0.868563621 | 0.000866694 | 0.006122367 | 1 |
| T2DM | macrophage | *SKOR1* | 2.042435793 | 6.25E-05 | 0.00079716 | 0.392999738 |
| T2DM | macrophage | *TMEM201* | 0.931094603 | 0.00026599 | 0.002416586 | 1 |
| T2DM | macrophage | *BEND4* | 2.243830504 | 7.16E-06 | 0.00016667 | 0.045000872 |
| T2DM | macrophage | *FAM196A* | 0.544279106 | 0.006972999 | 0.030380625 | 1 |
| T2DM | macrophage | *C9orf152* | 2.255713181 | 7.50E-06 | 0.000173209 | 0.04714361 |
| T2DM | macrophage | *ADAT2* | 0.436258071 | 0.007905186 | 0.033558343 | 1 |
| T2DM | macrophage | *ALKBH2* | 0.641787113 | 1.56E-05 | 0.000292939 | 0.097841562 |
| T2DM | macrophage | *S100A13* | 0.856676031 | 0.000701004 | 0.005184958 | 1 |
| T2DM | macrophage | *C15orf61* | 1.356217567 | 2.46E-05 | 0.000408348 | 0.154670518 |
| T2DM | macrophage | *BLOC1S2* | -0.156689093 | 0.012593943 | 0.047677251 | 1 |
| T2DM | macrophage | *HLA-DRB1* | -0.562241988 | 8.94E-19 | 2.81E-15 | 5.62E-15 |
| T2DM | macrophage | *AKR1C3* | 0.531014339 | 0.000934407 | 0.006469846 | 1 |
| T2DM | macrophage | *SEMA4A* | -0.285970509 | 0.008939813 | 0.036759061 | 1 |
| T2DM | macrophage | *SIRPB2* | -0.425088191 | 5.24E-06 | 0.000134402 | 0.032928413 |
| T2DM | macrophage | *GTF2IRD2* | 0.385138265 | 0.000153277 | 0.001560306 | 0.963651695 |
| T2DM | macrophage | *NTNG2* | 1.441563655 | 0.001338356 | 0.008612326 | 1 |
| T2DM | macrophage | *WDR5* | 1.047755155 | 0.00145694 | 0.00923365 | 1 |
| T2DM | macrophage | *ASB13* | 1.148459495 | 0.00475652 | 0.022689103 | 1 |
| T2DM | macrophage | *SLC35F1* | 0.856005375 | 0.000682997 | 0.00507565 | 1 |
| T2DM | macrophage | *ZNF34* | 1.249210906 | 0.000778962 | 0.00561621 | 1 |
| T2DM | macrophage | *INCA1* | 1.311422476 | 5.83E-06 | 0.000144835 | 0.036643129 |
| T2DM | macrophage | *ZNF605* | 1.202451827 | 5.99E-05 | 0.000776326 | 0.376517933 |
| T2DM | macrophage | *RFX8* | 0.488048499 | 0.000827383 | 0.005891008 | 1 |
| T2DM | macrophage | *MYL6B* | 0.927048365 | 0.003742737 | 0.0190223 | 1 |
| T2DM | macrophage | *SULT1A1* | -0.182638872 | 0.012005101 | 0.046078189 | 1 |
| T2DM | macrophage | *C1orf186* | 0.7532967 | 0.004584385 | 0.022001548 | 1 |
| T2DM | macrophage | *ADH1B* | 0.70726233 | 4.10E-05 | 0.000584458 | 0.257746134 |
| T2DM | macrophage | *HRH1* | -0.398841535 | 3.56E-05 | 0.000528152 | 0.223873543 |
| T2DM | macrophage | *FAM180B* | -0.338554737 | 0.000803844 | 0.005775736 | 1 |
| T2DM | macrophage | *ERI2* | 0.526573377 | 0.012162837 | 0.046484958 | 1 |
| T2DM | macrophage | *TRPV1* | 0.601312632 | 6.73E-06 | 0.000159629 | 0.042301774 |
| T2DM | macrophage | *ZNF33B* | -0.36751096 | 0.000435617 | 0.003613092 | 1 |

| | | | | | | |
|---|---|---|---|---|---|---|
| T2DM | macrophage | *DAPK1* | -0.245886993 | 0.003909532 | 0.019657985 | 1 |
| T2DM | macrophage | *HLA-DQA1* | -0.379062063 | 4.03E-10 | 1.95E-07 | 2.53E-06 |
| T2DM | macrophage | *MVB12B* | -0.506320788 | 0.010388523 | 0.041337117 | 1 |
| T2DM | macrophage | *C6orf106* | 0.212149914 | 0.00600601 | 0.027048558 | 1 |
| T2DM | macrophage | *ZFP28* | 0.946128025 | 0.003587469 | 0.018381756 | 1 |
| T2DM | macrophage | *LAMB3* | 0.980293596 | 0.000109051 | 0.001215675 | 0.685602585 |
| T2DM | macrophage | *ZNF585A* | 0.264836297 | 0.011173239 | 0.043550002 | 1 |
| T2DM | macrophage | *FUT11* | 0.799180859 | 0.00047422 | 0.003856951 | 1 |
| T2DM | macrophage | *ZNF138* | 0.545565851 | 0.004152294 | 0.020539318 | 1 |
| T2DM | macrophage | *ARRDC1* | -0.746459674 | 0.007833508 | 0.033411982 | 1 |
| T2DM | macrophage | *GAL3ST4* | 1.212549927 | 3.63E-07 | 3.80E-05 | 0.002282608 |
| T2DM | macrophage | *SLC25A29* | 0.520261464 | 0.002742508 | 0.014993175 | 1 |
| T2DM | macrophage | *ZNF682* | 0.746564938 | 0.004507322 | 0.021731238 | 1 |
| T2DM | macrophage | *PIWIL2* | 0.371545913 | 0.004829863 | 0.022934553 | 1 |
| T2DM | macrophage | *TBC1D9B* | -0.353936585 | 0.001305855 | 0.008455111 | 1 |
| T2DM | macrophage | *FAM110D* | 1.101487076 | 2.56E-05 | 0.00041543 | 0.160658691 |
| T2DM | macrophage | *CCL4L2* | -0.210997521 | 0.000765869 | 0.005528152 | 1 |
| T2DM | macrophage | *GTF2E2* | 0.661148448 | 0.002707209 | 0.014851852 | 1 |
| T2DM | macrophage | *RAMP2-AS1* | 0.716791742 | 1.20E-07 | 2.05E-05 | 0.000757127 |
| T2DM | macrophage | *ZNF517* | 1.876471663 | 1.56E-05 | 0.000293149 | 0.098259545 |
| T2DM | macrophage | *SLC22A5* | 0.169212668 | 0.004277474 | 0.021028641 | 1 |
| T2DM | macrophage | *C5AR1* | -0.366210864 | 0.000820784 | 0.005864173 | 1 |
| T2DM | macrophage | *STMN3* | 0.465113821 | 0.012216864 | 0.046578184 | 1 |
| T2DM | macrophage | *GYPE* | 0.302480746 | 0.001609372 | 0.009962062 | 1 |
| T2DM | macrophage | *SLC2A10* | 0.636164965 | 3.48E-05 | 0.000522015 | 0.218724148 |
| T2DM | macrophage | *ATG7* | -0.322756964 | 0.002645093 | 0.014630053 | 1 |
| T2DM | macrophage | *RAB40C* | 1.040058463 | 0.001786847 | 0.010781101 | 1 |
| T2DM | macrophage | *HOXA4* | 0.907446622 | 0.000534071 | 0.004207653 | 1 |
| T2DM | macrophage | *MFAP5* | 0.639641516 | 0.002118882 | 0.012415106 | 1 |
| T2DM | macrophage | *NMB* | 1.383314872 | 0.000413994 | 0.003458917 | 1 |
| T2DM | macrophage | *FAM114A1* | 0.70928711 | 0.000755642 | 0.005473185 | 1 |
| T2DM | macrophage | *AC122129.1* | 2.063042999 | 5.14E-07 | 3.90E-05 | 0.003232915 |
| T2DM | macrophage | *ZNF181* | 0.637211395 | 0.004304656 | 0.021080579 | 1 |
| T2DM | macrophage | *ZNF677* | 1.283518153 | 0.000151496 | 0.001548703 | 0.952452296 |
| T2DM | macrophage | *FCHSD1* | -0.827286224 | 0.005872305 | 0.026560563 | 1 |
| T2DM | macrophage | *ZNF273* | 0.627230187 | 0.001411584 | 0.009009774 | 1 |
| T2DM | macrophage | *PRIM1* | -0.680199882 | 0.000681769 | 0.005072526 | 1 |
| T2DM | macrophage | *SVIP* | 1.176649049 | 0.000344477 | 0.002966752 | 1 |
| T2DM | macrophage | *ZNF251* | 0.584031337 | 0.010111158 | 0.040488293 | 1 |
| T2DM | macrophage | *DDRGK1* | 0.925417689 | 0.010009236 | 0.040235336 | 1 |
| T2DM | macrophage | *DDX42* | 0.477292686 | 0.010839936 | 0.042647484 | 1 |
| T2DM | macrophage | *SLC29A3* | -0.493625354 | 1.51E-05 | 0.000287022 | 0.094654062 |
| T2DM | macrophage | *ANTXRL* | 21.81056316 | 9.13E-07 | 4.80E-05 | 0.005738871 |
| T2DM | macrophage | *ZKSCAN8* | 0.508032803 | 0.011302047 | 0.043915929 | 1 |
| T2DM | macrophage | *GFPT1* | 0.565642915 | 0.005077186 | 0.02371491 | 1 |
| T2DM | macrophage | *ZNF568* | 0.911534844 | 4.93E-06 | 0.000129527 | 0.030965506 |
| T2DM | macrophage | *TMA16* | 0.803041177 | 0.005226382 | 0.02426755 | 1 |
| T2DM | macrophage | *HLA-DRB5* | -0.164992717 | 1.80E-13 | 1.61E-10 | 1.13E-09 |
| T2DM | macrophage | *ZNF627* | -0.582036342 | 0.002211505 | 0.012814501 | 1 |
| T2DM | macrophage | *ARC* | -2.386832824 | 5.94E-06 | 0.000146917 | 0.03731703 |

| | | | | | | |
|---|---|---|---|---|---|---|
| T2DM | macrophage | *MMP17* | 1.418182133 | 7.43E-06 | 0.000172484 | 0.046743095 |
| T2DM | macrophage | *CCDC69* | 0.458515732 | 0.004672259 | 0.022338018 | 1 |
| T2DM | macrophage | *TUSC1* | 1.926499362 | 3.30E-06 | 0.000100599 | 0.020723364 |
| T2DM | macrophage | *PAPSS2* | -0.363125645 | 0.002659629 | 0.014693397 | 1 |
| T2DM | macrophage | *FAN1* | 0.946538124 | 0.001843983 | 0.01103056 | 1 |
| T2DM | macrophage | *ECI2* | 1.03889882 | 0.000267817 | 0.002429673 | 1 |
| T2DM | macrophage | *APCDD1L* | 1.493670025 | 7.70E-06 | 0.00017637 | 0.048418395 |
| T2DM | macrophage | *RASSF9* | 1.0942461 | 3.68E-06 | 0.000106585 | 0.023128908 |
| T2DM | macrophage | *TMEM184B* | 0.917566591 | 0.000241236 | 0.002259181 | 1 |
| T2DM | macrophage | *HMGN2* | 0.410528542 | 0.012214244 | 0.046578184 | 1 |
| T2DM | macrophage | *DENND4B* | -0.605781593 | 5.96E-05 | 0.00077563 | 0.374629134 |
| T2DM | macrophage | *TSEN15* | 0.868339122 | 0.006866828 | 0.02998038 | 1 |
| T2DM | macrophage | *ITPRIPL1* | 0.431412265 | 0.012101695 | 0.046364019 | 1 |
| T2DM | macrophage | *RASGEF1A* | 3.741277017 | 1.70E-06 | 6.69E-05 | 0.010699098 |
| T2DM | macrophage | *APRT* | 1.270772842 | 0.002348591 | 0.013467212 | 1 |
| T2DM | macrophage | *ZFP2* | 0.747991676 | 0.000648052 | 0.004885253 | 1 |
| T2DM | macrophage | *MFAP3L* | 1.556109714 | 1.42E-07 | 2.29E-05 | 0.000893406 |
| T2DM | macrophage | *NAGA* | -0.472265052 | 0.006025501 | 0.027087457 | 1 |
| T2DM | macrophage | *AP001007.1* | 3.297913903 | 2.36E-07 | 3.14E-05 | 0.001484148 |
| T2DM | macrophage | *HIST2H2BF* | -2.871033771 | 4.56E-05 | 0.000627754 | 0.286883716 |
| T2DM | macrophage | *RBM20* | 0.64838681 | 0.004103109 | 0.020345901 | 1 |
| T2DM | macrophage | *PCMTD2* | 0.856992012 | 0.000590236 | 0.004525383 | 1 |
| T2DM | macrophage | *SYS1* | 1.676208185 | 0.000129041 | 0.001381946 | 0.811280632 |
| T2DM | macrophage | *ASAH2B* | 0.320466895 | 0.000311403 | 0.00272294 | 1 |
| T2DM | macrophage | *AGAP6* | 0.161945209 | 0.012595774 | 0.047677251 | 1 |
| T2DM | macrophage | *SYT15* | 0.722143731 | 8.92E-05 | 0.001051669 | 0.560539317 |
| T2DM | macrophage | *ZDBF2* | 0.95583144 | 1.08E-06 | 5.25E-05 | 0.006820502 |
| T2DM | macrophage | *OXLD1* | 0.580701637 | 0.000148335 | 0.001526639 | 0.932580502 |
| T2DM | macrophage | *HLA-DMA* | -0.692183781 | 6.23E-06 | 0.000151884 | 0.039186112 |
| T2DM | macrophage | *HLA-DRA* | 0.350066747 | 3.77E-06 | 0.000108369 | 0.023698575 |
| T2DM | macrophage | *NOTCH4* | -0.056161959 | 6.76E-08 | 1.42E-05 | 0.000424963 |
| T2DM | macrophage | *AGER* | 0.352709424 | 0.003561306 | 0.018300468 | 1 |
| T2DM | macrophage | *PRRT1* | 0.434045294 | 7.41E-07 | 4.48E-05 | 0.004658305 |
| T2DM | macrophage | *NELFE* | 0.406876048 | 0.000101732 | 0.001154492 | 0.639588507 |
| T2DM | macrophage | *BAG6* | 0.387083031 | 0.000734915 | 0.00535364 | 1 |
| T2DM | macrophage | *NFKBIL1* | 0.486753469 | 0.000279587 | 0.002493278 | 1 |
| T2DM | macrophage | *PSORS1C1* | 0.043350879 | 0.005540442 | 0.025332915 | 1 |
| T2DM | macrophage | *LILRB3* | -0.217025251 | 0.004791401 | 0.022838163 | 1 |
| T2DM | macrophage | *FAM153C* | -0.762521644 | 0.003322033 | 0.017409731 | 1 |
| T2DM | macrophage | *PCDHA4* | 1.224231317 | 0.004571988 | 0.021967958 | 1 |
| T2DM | macrophage | *SPIRE2* | 0.17066778 | 0.008981556 | 0.036882456 | 1 |
| T2DM | macrophage | *FAM71F2* | 1.227538343 | 0.000720773 | 0.005300003 | 1 |
| T2DM | macrophage | *AC016586.1* | 2.730420934 | 0.00065578 | 0.004926208 | 1 |
| T2DM | macrophage | *MT1A* | 1.773097043 | 4.61E-07 | 3.80E-05 | 0.002900673 |
| T2DM | macrophage | *MT1M* | 1.859639121 | 3.49E-06 | 0.000103861 | 0.021914594 |
| T2DM | macrophage | *ATP6AP1L* | 0.319204872 | 0.004305307 | 0.021080579 | 1 |
| T2DM | macrophage | *EIF3CL* | 1.489819213 | 4.62E-05 | 0.000629611 | 0.290250674 |
| T2DM | macrophage | *LIN52* | 0.565687654 | 1.02E-05 | 0.000214771 | 0.064216622 |
| T2DM | macrophage | *C1QTNF9B* | 7.974609262 | 4.29E-07 | 3.80E-05 | 0.002698894 |
| T2DM | macrophage | *HCG27* | 0.344389207 | 3.65E-11 | 2.87E-08 | 2.30E-07 |

| | | | | | | |
|---|---|---|---|---|---|---|
| T2DM | macrophage | *METTL6* | 0.505939587 | 0.003337191 | 0.017469543 | 1 |
| T2DM | macrophage | *TAS2R14* | 0.22101967 | 0.004254797 | 0.020963876 | 1 |
| T2DM | macrophage | *ZRSR1* | 3.749973375 | 7.09E-05 | 0.000882435 | 0.445629665 |
| T2DM | macrophage | *RNF208* | 4.57005452 | 0.000225456 | 0.00214439 | 1 |
| T2DM | macrophage | *FGFR1OP* | 1.028389453 | 4.26E-07 | 3.80E-05 | 0.002680781 |
| T2DM | macrophage | *CCDC183* | 1.420702012 | 0.000386312 | 0.003246985 | 1 |
| T2DM | macrophage | *DNLZ* | 0.197042356 | 0.000144993 | 0.001501769 | 0.911573584 |
| T2DM | macrophage | *LBH* | 1.315761547 | 0.000188167 | 0.001865939 | 1 |
| T2DM | macrophage | *ATF6B* | 0.424925138 | 2.01E-05 | 0.000352846 | 0.126671715 |
| T2DM | macrophage | *PPM1N* | -0.460434106 | 0.003911586 | 0.019657985 | 1 |
| T2DM | macrophage | *GALT* | 0.558045054 | 0.008814308 | 0.036385788 | 1 |
| T2DM | macrophage | *ITGA1* | 0.468698787 | 0.001975902 | 0.011752598 | 1 |
| T2DM | macrophage | *TAX1BP3* | 1.060421762 | 0.000349785 | 0.003000135 | 1 |
| T2DM | macrophage | *TM6SF2* | 8.289933391 | 9.45E-07 | 4.80E-05 | 0.005943913 |
| T2DM | macrophage | *AC079776.2* | 1.608091972 | 3.59E-05 | 0.000531261 | 0.225785903 |
| T2DM | macrophage | *PRCD* | 0.596055177 | 0.000561003 | 0.004370542 | 1 |
| T2DM | macrophage | *NEURL1B* | 1.245522889 | 7.95E-06 | 0.000180097 | 0.049951288 |
| T2DM | macrophage | *STARD10* | 0.244522727 | 2.08E-07 | 3.06E-05 | 0.001310285 |
| T2DM | macrophage | *COL28A1* | 0.975738356 | 4.56E-05 | 0.000627754 | 0.286563846 |
| T2DM | macrophage | *UBE2QL1* | 1.498411427 | 6.66E-07 | 4.41E-05 | 0.004186282 |
| T2DM | macrophage | *GOLGA8B* | 1.038166544 | 3.51E-05 | 0.000525775 | 0.220825293 |
| T2DM | macrophage | *LINC01138* | 0.949701354 | 0.00054776 | 0.004293973 | 1 |
| T2DM | macrophage | *ATAD3C* | 0.694951506 | 0.000354909 | 0.003027565 | 1 |
| T2DM | macrophage | *RNU6ATAC35I* | 3.217888139 | 3.87E-07 | 3.80E-05 | 0.002430938 |
| T2DM | macrophage | *C19orf73* | 2.338621575 | 0.000184045 | 0.001839577 | 1 |
| T2DM | macrophage | *APOL6* | 0.775819279 | 0.001772481 | 0.010704694 | 1 |
| T2DM | macrophage | *FAM185A* | 0.747565889 | 0.001561072 | 0.009718321 | 1 |
| T2DM | macrophage | *ADRA2B* | -1.945707214 | 4.20E-06 | 0.000116858 | 0.026419443 |
| T2DM | macrophage | *AC147651.3* | -0.770479361 | 2.79E-05 | 0.000445745 | 0.175177938 |
| T2DM | macrophage | *OSER1-AS1* | 0.796153599 | 0.002114995 | 0.012405815 | 1 |
| T2DM | macrophage | *LINC01133* | 1.189675568 | 2.31E-05 | 0.000391048 | 0.145078842 |
| T2DM | macrophage | *LINC01117* | 0.720082582 | 1.33E-05 | 0.000263397 | 0.083494519 |
| T2DM | macrophage | *LINC00240* | 0.294447036 | 9.47E-05 | 0.001099029 | 0.595673539 |
| T2DM | macrophage | *LINC00863* | 0.339120708 | 0.009591045 | 0.038902516 | 1 |
| T2DM | macrophage | *LINC00237* | -2.442583403 | 3.30E-06 | 0.000100599 | 0.020716784 |
| T2DM | macrophage | *PLEKHM1* | 0.527501242 | 0.012596137 | 0.047677251 | 1 |
| T2DM | macrophage | *SFTA1P* | 1.067559729 | 0.008543274 | 0.035500042 | 1 |
| T2DM | macrophage | *SLC26A6* | 0.873818968 | 8.70E-05 | 0.00103773 | 0.546883508 |
| T2DM | macrophage | *ERCC6* | 0.773620373 | 0.004845034 | 0.022954579 | 1 |
| T2DM | macrophage | *NOL7* | 0.895569329 | 0.003852562 | 0.0194079 | 1 |
| T2DM | macrophage | *ELFN1* | 3.709052999 | 4.13E-07 | 3.80E-05 | 0.002599245 |
| T2DM | macrophage | *ZNF503-AS1* | 1.372314538 | 1.94E-06 | 7.34E-05 | 0.012179397 |
| T2DM | macrophage | *AC064836.3* | 2.613288004 | 7.08E-07 | 4.47E-05 | 0.004451013 |
| T2DM | macrophage | *C14orf132* | 0.661583114 | 0.000115296 | 0.001265032 | 0.724863542 |
| T2DM | macrophage | *SLC2A1-AS1* | 0.530553187 | 0.002379474 | 0.013575091 | 1 |
| T2DM | macrophage | *AC108463.2* | 0.868213142 | 0.000125958 | 0.001353669 | 0.791896167 |
| T2DM | macrophage | *HCG20* | 1.275846662 | 0.000678389 | 0.005053357 | 1 |
| T2DM | macrophage | *LINC00954* | 2.327224307 | 8.07E-05 | 0.000979583 | 0.507254545 |
| T2DM | macrophage | *RPL41* | 1.20195867 | 0.002372447 | 0.013547298 | 1 |
| T2DM | macrophage | *PATL2* | -0.594131186 | 0.01227393 | 0.046767391 | 1 |

| | | | | | | |
|---|---|---|---|---|---|---|
| T2DM | macrophage | *U91324.1* | 3.013200708 | 2.72E-07 | 3.38E-05 | 0.001710263 |
| T2DM | macrophage | *CISD3* | 1.866245246 | 0.00014941 | 0.001532369 | 0.939342447 |
| T2DM | macrophage | *KLHL7-AS1* | 2.066289664 | 3.88E-07 | 3.80E-05 | 0.00243787 |
| T2DM | macrophage | *LINC00865* | -0.361939765 | 2.27E-05 | 0.000385077 | 0.142478465 |
| T2DM | macrophage | *AC009495.3* | 1.025040431 | 0.000876252 | 0.006162188 | 1 |
| T2DM | macrophage | *HLA-DQB2* | 0.138114047 | 5.77E-08 | 1.34E-05 | 0.000362977 |
| T2DM | macrophage | *LINC00665* | 0.225390091 | 0.009685558 | 0.039164328 | 1 |
| T2DM | macrophage | *LINC01114* | -1.146450761 | 0.002490207 | 0.013991 | 1 |
| T2DM | macrophage | *HLA-B* | 0.336403679 | 2.46E-05 | 0.000408348 | 0.154651798 |
| T2DM | macrophage | *C19orf81* | 0.594898014 | 0.01128654 | 0.043904954 | 1 |
| T2DM | macrophage | *ANKRD65* | 0.484914325 | 0.003446908 | 0.017865385 | 1 |
| T2DM | macrophage | *ZSCAN31* | 0.167808483 | 0.008328805 | 0.034792824 | 1 |
| T2DM | macrophage | *LINC00941* | 2.242262294 | 5.19E-05 | 0.000698131 | 0.326103737 |
| T2DM | macrophage | *GS1-124K5.4* | 0.086522847 | 0.003090734 | 0.016478053 | 1 |
| T2DM | macrophage | *ZNF737* | 0.920232263 | 0.001539104 | 0.009618634 | 1 |
| T2DM | macrophage | *HLA-DQA2* | 0.365568318 | 2.34E-16 | 4.90E-13 | 1.47E-12 |
| T2DM | macrophage | *TXNDC5* | 2.139075396 | 2.95E-06 | 9.46E-05 | 0.018537062 |
| T2DM | macrophage | *ASB14* | 0.508846725 | 0.001267023 | 0.008237616 | 1 |
| T2DM | macrophage | *LILRA2* | -0.145566094 | 0.004233922 | 0.020889324 | 1 |
| T2DM | macrophage | *LY6G5B* | -0.107557243 | 0.000651228 | 0.00490332 | 1 |
| T2DM | macrophage | *AQP1* | 0.64700012 | 0.002364153 | 0.013524503 | 1 |
| T2DM | macrophage | *ARHGEF25* | 0.51722856 | 0.005733156 | 0.026024802 | 1 |
| T2DM | macrophage | *ARHGAP8* | -0.614073334 | 8.88E-07 | 4.80E-05 | 0.005581946 |
| T2DM | macrophage | *PLEKHO2* | -0.488545365 | 0.003823546 | 0.019277171 | 1 |
| T2DM | macrophage | *PWP2* | -2.036139619 | 7.84E-05 | 0.000959518 | 0.49319207 |
| T2DM | macrophage | *AKAP2* | 2.835018639 | 3.68E-07 | 3.80E-05 | 0.002315808 |
| T2DM | macrophage | *C22orf39* | 1.639830501 | 0.000148609 | 0.001526639 | 0.934303245 |
| T2DM | macrophage | *MRPL20* | 0.715222487 | 0.007735149 | 0.033127302 | 1 |
| T2DM | macrophage | *DECR2* | 2.202710807 | 2.81E-05 | 0.000447025 | 0.176575043 |
| T2DM | macrophage | *PSG4* | 0.331259153 | 0.010961206 | 0.042936511 | 1 |
| T2DM | macrophage | *C7orf73* | 0.713258323 | 0.013018936 | 0.048865702 | 1 |
| T2DM | macrophage | *FGF14-IT1* | 2.468193075 | 5.01E-07 | 3.89E-05 | 0.003149451 |
| T2DM | macrophage | *PALM2* | 0.390229551 | 6.38E-06 | 0.000154182 | 0.040087261 |
| T2DM | macrophage | *NAT6* | 0.859577909 | 6.11E-06 | 0.00015016 | 0.038440836 |
| T2DM | macrophage | *WDR92* | 0.654965474 | 0.000806744 | 0.005789951 | 1 |
| T2DM | macrophage | *NME1-NME2* | 0.504093794 | 0.001502054 | 0.00940579 | 1 |
| T2DM | macrophage | *PLA2G4B* | 0.424699541 | 3.13E-08 | 9.84E-06 | 0.000196774 |
| T2DM | macrophage | *JMJD7* | 0.448635612 | 6.77E-08 | 1.42E-05 | 0.000425891 |
| T2DM | macrophage | *UBE2V1* | 1.6737783 | 0.000232936 | 0.0021989 | 1 |
| T2DM | macrophage | *LINC00847* | -0.873693887 | 0.010660808 | 0.042127278 | 1 |
| T2DM | macrophage | *ZNF585B* | 0.249467838 | 0.002280743 | 0.013130981 | 1 |
| T2DM | macrophage | *LINC00920* | -0.436831823 | 0.000426037 | 0.0035383 | 1 |
| T2DM | macrophage | *PGAM5* | 2.107450694 | 0.000533045 | 0.004204835 | 1 |
| T2DM | macrophage | *ABHD14A* | 0.954692997 | 0.000332457 | 0.002879008 | 1 |
| T2DM | macrophage | *SRP14-AS1* | -0.087992387 | 0.011453082 | 0.044393048 | 1 |
| T2DM | macrophage | *NAIP* | -0.565944031 | 2.19E-06 | 7.90E-05 | 0.013747249 |
| T2DM | macrophage | *ZNF286B* | -1.109321354 | 0.000834595 | 0.00591556 | 1 |
| T2DM | macrophage | *PCDHA10* | 0.990842907 | 0.000279083 | 0.002493278 | 1 |
| T2DM | macrophage | *LINC00989* | 1.275462937 | 0.000523242 | 0.004143101 | 1 |
| T2DM | macrophage | *LINC01021* | 1.599917405 | 4.68E-07 | 3.80E-05 | 0.002940885 |

| | | | | | | |
|---|---|---|---|---|---|---|
| T2DM | macrophage | *FAM218A* | 2.341000292 | 8.86E-07 | 4.80E-05 | 0.005573199 |
| T2DM | macrophage | *CCDC71L* | 0.597867099 | 0.010410242 | 0.041397339 | 1 |
| T2DM | macrophage | *TMEM200B* | 0.356917615 | 9.20E-05 | 0.001081586 | 0.578648356 |
| T2DM | macrophage | *TRNP1* | 7.128101687 | 1.82E-05 | 0.000324545 | 0.114239939 |
| T2DM | macrophage | *PCDHGA6* | 0.257758871 | 0.005110962 | 0.02385495 | 1 |
| T2DM | macrophage | *ZNF260* | 1.031767886 | 0.000128884 | 0.001381946 | 0.810293602 |
| T2DM | macrophage | *EBLN2* | 0.816222561 | 0.003619649 | 0.018516465 | 1 |
| T2DM | macrophage | *POLR2M* | 1.364183149 | 0.000970748 | 0.006626591 | 1 |
| T2DM | macrophage | *TAS2R20* | 0.201140926 | 0.004430682 | 0.021510192 | 1 |
| T2DM | macrophage | *CTSO* | 0.872526594 | 0.011390032 | 0.0441759 | 1 |
| T2DM | macrophage | *HMBS* | 0.341359275 | 1.25E-06 | 5.80E-05 | 0.007831367 |
| T2DM | macrophage | *AC093668.2* | 0.319807329 | 0.013212568 | 0.049352588 | 1 |
| T2DM | macrophage | *ZBED6* | 3.839310766 | 0.000125708 | 0.001353293 | 0.790323186 |
| T2DM | macrophage | *FNTB* | 1.816147017 | 1.00E-05 | 0.000212236 | 0.063159957 |
| T2DM | macrophage | *OVCH1-AS1* | 0.806089045 | 0.010385498 | 0.041337117 | 1 |
| T2DM | macrophage | *RTEL1* | 0.405898675 | 0.004236343 | 0.020889324 | 1 |
| T2DM | macrophage | *LINC00639* | 0.323995568 | 0.000983945 | 0.006702124 | 1 |
| T2DM | macrophage | *LINC00648* | 0.922498653 | 9.24E-05 | 0.001082672 | 0.580916993 |
| T2DM | macrophage | *LINC00924* | 0.773211013 | 0.000572272 | 0.004436344 | 1 |
| T2DM | macrophage | *FRRS1L* | 2.937112137 | 4.93E-07 | 3.88E-05 | 0.003100197 |
| T2DM | macrophage | *AC009120.5* | 1.964195276 | 3.34E-06 | 0.000100897 | 0.020986564 |
| T2DM | macrophage | *TMEM178B* | 0.80161359 | 0.000129468 | 0.001381946 | 0.813966297 |
| T2DM | macrophage | *HPR* | 0.866903996 | 0.000917065 | 0.006380743 | 1 |
| T2DM | macrophage | *ILF3-AS1* | 0.458196914 | 0.001486799 | 0.009347503 | 1 |
| T2DM | macrophage | *AC139100.2* | 7.496422546 | 0.000139329 | 0.001462378 | 0.875964206 |
| T2DM | macrophage | *LINC01140* | 0.908406442 | 2.16E-06 | 7.85E-05 | 0.013577445 |
| T2DM | macrophage | *LINC00906* | 4.928817636 | 5.30E-07 | 3.92E-05 | 0.003334528 |
| T2DM | macrophage | *ZNF582-AS1* | 0.701213138 | 0.002481468 | 0.013954371 | 1 |
| T2DM | macrophage | *FBXO17* | 0.957469834 | 1.42E-06 | 6.22E-05 | 0.008921023 |
| T2DM | macrophage | *AC009060.1* | -0.87973978 | 0.004383847 | 0.021381884 | 1 |
| T2DM | macrophage | *LINC01023* | 0.764146388 | 0.009600433 | 0.038915487 | 1 |
| T2DM | macrophage | *POLR2J4* | 1.552691778 | 0.000134607 | 0.001422306 | 0.846272177 |
| T2DM | mast_cell | *SCYL3* | 2.079438344 | 0.000158567 | 0.000291625 | 0.996910677 |
| T2DM | mast_cell | *C1orf112* | 3.516087065 | 2.69E-09 | 7.78E-08 | 1.69E-05 |
| T2DM | mast_cell | *CFH* | 1.87011747 | 1.27E-09 | 6.77E-08 | 8.00E-06 |
| T2DM | mast_cell | *NFYA* | 3.850806678 | 0.000104699 | 0.000198865 | 0.658241619 |
| T2DM | mast_cell | *STPG1* | 1.888122188 | 7.45E-05 | 0.000144995 | 0.468187496 |
| T2DM | mast_cell | *NIPAL3* | -1.814411072 | 0.014316887 | 0.01929067 | 1 |
| T2DM | mast_cell | *ENPP4* | 2.222420171 | 3.80E-06 | 9.90E-06 | 0.023870607 |
| T2DM | mast_cell | *KRIT1* | 3.556660071 | 5.13E-08 | 2.77E-07 | 0.000322279 |
| T2DM | mast_cell | *RAD52* | 4.591320247 | 9.19E-08 | 4.28E-07 | 0.000577633 |
| T2DM | mast_cell | *HS3ST1* | 5.050004559 | 5.44E-07 | 1.83E-06 | 0.003418434 |
| T2DM | mast_cell | *WNT16* | 1.936630946 | 0.004128588 | 0.006032171 | 1 |
| T2DM | mast_cell | *HECW1* | 1.466601858 | 1.10E-05 | 2.54E-05 | 0.069406139 |
| T2DM | mast_cell | *MAD1L1* | 2.785880141 | 0.00741836 | 0.010454882 | 1 |
| T2DM | mast_cell | *LASP1* | 2.792822398 | 0.00658784 | 0.009359943 | 1 |
| T2DM | mast_cell | *SNX11* | 1.576860237 | 0.002363242 | 0.003599249 | 1 |
| T2DM | mast_cell | *TMEM176A* | 2.039306673 | 4.75E-05 | 9.64E-05 | 0.29855312 |
| T2DM | mast_cell | *M6PR* | 3.52129095 | 0.000130443 | 0.000243566 | 0.820092259 |
| T2DM | mast_cell | *CYP26B1* | 3.409651803 | 5.00E-09 | 7.84E-08 | 3.14E-05 |

| | | | | | | |
|---|---|---|---|---|---|---|
| T2DM | mast_cell | *ALS2* | -1.368341392 | 1.43E-05 | 3.20E-05 | 0.090101352 |
| T2DM | mast_cell | *POLDIP2* | 6.037359743 | 1.58E-08 | 1.24E-07 | 9.95E-05 |
| T2DM | mast_cell | *CD38* | 1.748745018 | 6.19E-08 | 3.21E-07 | 0.000388918 |
| T2DM | mast_cell | *RBM6* | -1.025605244 | 3.32E-05 | 6.96E-05 | 0.208575317 |
| T2DM | mast_cell | *RECQL* | 5.090870101 | 1.83E-07 | 7.40E-07 | 0.001148721 |
| T2DM | mast_cell | *ARHGAP33* | -2.464450467 | 0.000130405 | 0.000243566 | 0.819853128 |
| T2DM | mast_cell | *PDK4* | 2.790739203 | 0.001037461 | 0.001653782 | 1 |
| T2DM | mast_cell | *SLC22A16* | -1.749123839 | 1.33E-05 | 3.01E-05 | 0.083860667 |
| T2DM | mast_cell | *CDC27* | 1.899418669 | 0.00295653 | 0.004438324 | 1 |
| T2DM | mast_cell | *MEOX1* | 1.691786404 | 5.79E-06 | 1.43E-05 | 0.036383128 |
| T2DM | mast_cell | *LIG3* | 1.28986539 | 0.009004415 | 0.012527276 | 1 |
| T2DM | mast_cell | *CIAPIN1* | 3.95450641 | 1.40E-09 | 6.85E-08 | 8.80E-06 |
| T2DM | mast_cell | *FAM214B* | 4.41299387 | 6.09E-09 | 8.15E-08 | 3.83E-05 |
| T2DM | mast_cell | *GCFC2* | 3.925764258 | 7.88E-08 | 3.85E-07 | 0.000495513 |
| T2DM | mast_cell | *WDR54* | 6.310238405 | 5.61E-05 | 0.000112448 | 0.352637369 |
| T2DM | mast_cell | *ABCB4* | 2.58762932 | 8.99E-09 | 9.66E-08 | 5.65E-05 |
| T2DM | mast_cell | *ASB4* | 2.278380663 | 0.000392505 | 0.000672759 | 1 |
| T2DM | mast_cell | *TMEM98* | 1.796923282 | 1.40E-05 | 3.13E-05 | 0.087910265 |
| T2DM | mast_cell | *ABCC8* | 6.626151089 | 1.85E-08 | 1.36E-07 | 0.000116557 |
| T2DM | mast_cell | *SYNRG* | 2.997386577 | 0.000988864 | 0.001583139 | 1 |
| T2DM | mast_cell | *ZNF263* | 4.967217912 | 3.79E-08 | 2.20E-07 | 0.00023797 |
| T2DM | mast_cell | *SPATA20* | 3.129443432 | 0.002839844 | 0.004271315 | 1 |
| T2DM | mast_cell | *DLX6* | 7.711455465 | 0.008755845 | 0.012211179 | 1 |
| T2DM | mast_cell | *MAP3K9* | 3.781273755 | 4.35E-07 | 1.51E-06 | 0.002736908 |
| T2DM | mast_cell | *RALA* | 4.16462003 | 4.07E-07 | 1.43E-06 | 0.002560527 |
| T2DM | mast_cell | *BAIAP2L1* | 3.053846366 | 7.60E-07 | 2.43E-06 | 0.004775483 |
| T2DM | mast_cell | *FARP2* | 2.291482211 | 0.00421662 | 0.006155072 | 1 |
| T2DM | mast_cell | *IFRD1* | 2.147022475 | 0.000285851 | 0.000501296 | 1 |
| T2DM | mast_cell | *GTF2IRD1* | 1.544483624 | 0.002120411 | 0.003250676 | 1 |
| T2DM | mast_cell | *ARHGAP44* | 3.108710858 | 7.37E-08 | 3.66E-07 | 0.000463056 |
| T2DM | mast_cell | *ELAC2* | 2.357653458 | 0.012520024 | 0.016993391 | 1 |
| T2DM | mast_cell | *CCDC124* | 1.819634091 | 1.37E-07 | 5.83E-07 | 0.000859916 |
| T2DM | mast_cell | *CEACAM21* | 2.19415493 | 0.003612998 | 0.005335898 | 1 |
| T2DM | mast_cell | *NOS2* | 10.31992261 | 4.83E-09 | 7.84E-08 | 3.04E-05 |
| T2DM | mast_cell | *TRAPPC6A* | 5.240835289 | 1.23E-07 | 5.36E-07 | 0.000772954 |
| T2DM | mast_cell | *RPUSD1* | 2.933411001 | 1.44E-08 | 1.19E-07 | 9.06E-05 |
| T2DM | mast_cell | *BAIAP3* | 5.214109354 | 0.005079066 | 0.007322193 | 1 |
| T2DM | mast_cell | *PIGQ* | 2.966146666 | 2.99E-06 | 8.00E-06 | 0.018804063 |
| T2DM | mast_cell | *PSMB1* | 3.363828226 | 0.002489424 | 0.003780437 | 1 |
| T2DM | mast_cell | *DLEC1* | 3.514564929 | 9.28E-08 | 4.31E-07 | 0.000583125 |
| T2DM | mast_cell | *MGST1* | 2.869184302 | 6.47E-07 | 2.12E-06 | 0.004067237 |
| T2DM | mast_cell | *NFIX* | 2.42035633 | 5.20E-06 | 1.31E-05 | 0.032717886 |
| T2DM | mast_cell | *MMP25* | 2.99917261 | 6.10E-09 | 8.15E-08 | 3.84E-05 |
| T2DM | mast_cell | *IL32* | 2.631057945 | 3.12E-08 | 1.90E-07 | 0.000195971 |
| T2DM | mast_cell | *PKD1* | 3.357419965 | 1.19E-07 | 5.24E-07 | 0.000746299 |
| T2DM | mast_cell | *MAPK8IP2* | 4.179241442 | 0.006902019 | 0.009760008 | 1 |
| T2DM | mast_cell | *SEC62* | 3.596136039 | 5.55E-05 | 0.000111362 | 0.348896817 |
| T2DM | mast_cell | *RPS20* | 2.581323945 | 0.01354491 | 0.018317239 | 1 |
| T2DM | mast_cell | *MLXIPL* | 1.053197025 | 8.00E-09 | 9.17E-08 | 5.03E-05 |
| T2DM | mast_cell | *SPRTN* | 4.18417916 | 3.86E-09 | 7.78E-08 | 2.43E-05 |

| | | | | | | |
|---|---|---|---|---|---|---|
| T2DM | mast_cell | *METTL13* | 5.688854953 | 8.60E-07 | 2.71E-06 | 0.005407998 |
| T2DM | mast_cell | *DYRK4* | 2.831971196 | 4.74E-08 | 2.61E-07 | 0.000298182 |
| T2DM | mast_cell | *STARD3NL* | 4.90768017 | 1.17E-06 | 3.49E-06 | 0.007346455 |
| T2DM | mast_cell | *ZNF200* | 4.334690342 | 8.00E-09 | 9.17E-08 | 5.03E-05 |
| T2DM | mast_cell | *CD4* | 5.432218391 | 7.14E-07 | 2.30E-06 | 0.004491308 |
| T2DM | mast_cell | *LRRC23* | 1.769632134 | 0.020030717 | 0.026517818 | 1 |
| T2DM | mast_cell | *SCMH1* | 2.677042218 | 0.000167308 | 0.000305954 | 1 |
| T2DM | mast_cell | *CLCN6* | 3.131292953 | 0.004941333 | 0.007136724 | 1 |
| T2DM | mast_cell | *MRC2* | 1.929773042 | 1.69E-07 | 6.94E-07 | 0.00106125 |
| T2DM | mast_cell | *TSPAN9* | 1.210001441 | 0.024649805 | 0.032185529 | 1 |
| T2DM | mast_cell | *MKS1* | 4.371800082 | 1.94E-09 | 7.78E-08 | 1.22E-05 |
| T2DM | mast_cell | *UTP18* | 3.91405426 | 4.35E-09 | 7.78E-08 | 2.73E-05 |
| T2DM | mast_cell | *TTC19* | 2.556160184 | 0.000563646 | 0.000941456 | 1 |
| T2DM | mast_cell | *SYT7* | 5.477251988 | 7.46E-08 | 3.69E-07 | 0.000469168 |
| T2DM | mast_cell | *LARS2* | 4.586291626 | 1.67E-07 | 6.91E-07 | 0.001052446 |
| T2DM | mast_cell | *PPP5C* | 3.401246022 | 0.001131058 | 0.001797059 | 1 |
| T2DM | mast_cell | *CEP68* | 3.627415487 | 2.73E-09 | 7.78E-08 | 1.71E-05 |
| T2DM | mast_cell | *ZBTB32* | -3.634161266 | 6.29E-05 | 0.000124571 | 0.395638133 |
| T2DM | mast_cell | *TMEM159* | 2.31966103 | 1.97E-07 | 7.87E-07 | 0.00123748 |
| T2DM | mast_cell | *LTF* | 3.674810638 | 5.33E-06 | 1.33E-05 | 0.03349425 |
| T2DM | mast_cell | *ALOX5* | -0.647066047 | 2.89E-07 | 1.07E-06 | 0.001819699 |
| T2DM | mast_cell | *MAP4K5* | 3.194916966 | 3.11E-05 | 6.55E-05 | 0.195542326 |
| T2DM | mast_cell | *PSMC4* | 2.94778914 | 1.77E-10 | 2.59E-08 | 1.11E-06 |
| T2DM | mast_cell | *MAN2B2* | -2.267788412 | 0.00019392 | 0.000350639 | 1 |
| T2DM | mast_cell | *CLDN11* | 2.50736173 | 4.47E-08 | 2.49E-07 | 0.000280784 |
| T2DM | mast_cell | *ANGEL1* | 6.007534867 | 1.37E-06 | 4.01E-06 | 0.008614705 |
| T2DM | mast_cell | *RNF14* | 2.828926883 | 0.009093135 | 0.012647907 | 1 |
| T2DM | mast_cell | *DDX11* | 1.30316591 | 0.006461636 | 0.009193099 | 1 |
| T2DM | mast_cell | *HEBP1* | 2.326887376 | 2.79E-07 | 1.04E-06 | 0.001756535 |
| T2DM | mast_cell | *TACC3* | 2.987053827 | 0.000473727 | 0.00079998 | 1 |
| T2DM | mast_cell | *ZC3H3* | 1.915826507 | 0.016488314 | 0.022055751 | 1 |
| T2DM | mast_cell | *MDH1* | 2.591745685 | 2.28E-07 | 8.85E-07 | 0.001430681 |
| T2DM | mast_cell | *SLC30A9* | 3.396509211 | 1.62E-05 | 3.58E-05 | 0.102059028 |
| T2DM | mast_cell | *MTMR11* | 1.011031162 | 0.000398466 | 0.000682046 | 1 |
| T2DM | mast_cell | *COX15* | 2.409742394 | 3.56E-08 | 2.10E-07 | 0.000223962 |
| T2DM | mast_cell | *NPC1L1* | 3.435593414 | 1.02E-05 | 2.37E-05 | 0.064230226 |
| T2DM | mast_cell | *NUDCD3* | 1.583696549 | 0.001374504 | 0.002163622 | 1 |
| T2DM | mast_cell | *CHDH* | 4.633575166 | 9.93E-09 | 1.04E-07 | 6.24E-05 |
| T2DM | mast_cell | *IL20RA* | 4.043557764 | 1.46E-08 | 1.19E-07 | 9.20E-05 |
| T2DM | mast_cell | *GLT8D1* | 2.622873911 | 0.000219968 | 0.000393663 | 1 |
| T2DM | mast_cell | *RUFY3* | 2.311841688 | 0.000476047 | 0.000802819 | 1 |
| T2DM | mast_cell | *WWTR1* | 3.355762496 | 1.06E-08 | 1.06E-07 | 6.66E-05 |
| T2DM | mast_cell | *TTC27* | 2.625062 | 0.000268915 | 0.000473443 | 1 |
| T2DM | mast_cell | *VSIG2* | 3.777158169 | 8.52E-08 | 4.06E-07 | 0.000535606 |
| T2DM | mast_cell | *MARCO* | 1.498881553 | 2.13E-07 | 8.38E-07 | 0.001337093 |
| T2DM | mast_cell | *MNAT1* | 3.797013651 | 4.13E-07 | 1.45E-06 | 0.002597869 |
| T2DM | mast_cell | *OSBPL5* | 4.766901904 | 4.00E-06 | 1.04E-05 | 0.025151955 |
| T2DM | mast_cell | *AQR* | 3.920862778 | 4.40E-08 | 2.47E-07 | 0.000276527 |
| T2DM | mast_cell | *CPS1* | 1.213772201 | 0.005070698 | 0.007312401 | 1 |
| T2DM | mast_cell | *NLRP2* | 2.032778512 | 3.80E-05 | 7.88E-05 | 0.238815066 |

| | | | | | | |
|---|---|---|---|---|---|---|
| T2DM | mast_cell | *ZDHHC6* | 3.774196193 | 3.75E-05 | 7.79E-05 | 0.235796676 |
| T2DM | mast_cell | *RNH1* | 1.720033135 | 0.015675848 | 0.021022623 | 1 |
| T2DM | mast_cell | *NDUFS1* | 2.244534506 | 0.010577073 | 0.014528744 | 1 |
| T2DM | mast_cell | *BIRC3* | -1.149744067 | 8.51E-07 | 2.68E-06 | 0.005348703 |
| T2DM | mast_cell | *AKAP11* | 4.284547341 | 4.24E-07 | 1.48E-06 | 0.002663229 |
| T2DM | mast_cell | *SNAPC1* | 3.927402046 | 3.03E-07 | 1.12E-06 | 0.001904447 |
| T2DM | mast_cell | *GCLM* | 2.26077295 | 3.05E-07 | 1.12E-06 | 0.001915661 |
| T2DM | mast_cell | *UBR2* | 2.70888058 | 0.000990195 | 0.001584866 | 1 |
| T2DM | mast_cell | *HSF2* | 4.620837536 | 1.43E-09 | 6.91E-08 | 8.98E-06 |
| T2DM | mast_cell | *NR1H3* | 4.202847298 | 3.89E-11 | 9.78E-09 | 2.44E-07 |
| T2DM | mast_cell | *TYMP* | 2.223739249 | 0.006790191 | 0.009617254 | 1 |
| T2DM | mast_cell | *TOMM34* | 5.311804729 | 0.00321545 | 0.00480065 | 1 |
| T2DM | mast_cell | *KPNA6* | 2.198418572 | 0.015725658 | 0.021084925 | 1 |
| T2DM | mast_cell | *FAS* | 2.042926594 | 0.000291816 | 0.000511044 | 1 |
| T2DM | mast_cell | *RNASET2* | 1.666540617 | 0.038010291 | 0.04844328 | 1 |
| T2DM | mast_cell | *CD44* | -1.44931654 | 0.000234035 | 0.000416584 | 1 |
| T2DM | mast_cell | *MIPEP* | 4.265497596 | 1.69E-08 | 1.29E-07 | 0.000106073 |
| T2DM | mast_cell | *INSRR* | 2.175609343 | 0.006653981 | 0.009447511 | 1 |
| T2DM | mast_cell | *B4GALT7* | 2.235056681 | 8.06E-09 | 9.17E-08 | 5.07E-05 |
| T2DM | mast_cell | *VRK2* | 2.416964287 | 0.005065115 | 0.007307108 | 1 |
| T2DM | mast_cell | *VEZT* | 3.075467982 | 6.09E-05 | 0.000120976 | 0.382648513 |
| T2DM | mast_cell | *SNX1* | 1.630479587 | 0.024313699 | 0.031773067 | 1 |
| T2DM | mast_cell | *ARNTL2* | 5.537368813 | 4.29E-08 | 2.42E-07 | 0.000269875 |
| T2DM | mast_cell | *SLC39A9* | 2.950027714 | 1.40E-05 | 3.14E-05 | 0.088196972 |
| T2DM | mast_cell | *ANK1* | -2.657324058 | 0.03235393 | 0.041537505 | 1 |
| T2DM | mast_cell | *RABEP1* | 2.560405389 | 0.000178876 | 0.000325687 | 1 |
| T2DM | mast_cell | *GRN* | 4.368322805 | 6.55E-06 | 1.60E-05 | 0.041155178 |
| T2DM | mast_cell | *ARID4A* | 2.772427281 | 0.00307217 | 0.004596557 | 1 |
| T2DM | mast_cell | *IFT88* | 2.443602302 | 0.002911316 | 0.004372537 | 1 |
| T2DM | mast_cell | *ZCCHC8* | 4.311213243 | 2.95E-06 | 7.92E-06 | 0.018566318 |
| T2DM | mast_cell | *ABCF2* | 3.447496347 | 1.77E-08 | 1.33E-07 | 0.000111243 |
| T2DM | mast_cell | *FUT8* | 3.335734807 | 5.24E-05 | 0.000105612 | 0.329540197 |
| T2DM | mast_cell | *PIAS1* | 1.847125584 | 0.021105222 | 0.027793994 | 1 |
| T2DM | mast_cell | *SLC4A7* | 3.523497806 | 3.72E-07 | 1.32E-06 | 0.002336905 |
| T2DM | mast_cell | *ASTE1* | 2.131042513 | 8.83E-06 | 2.08E-05 | 0.055490114 |
| T2DM | mast_cell | *MYOC* | 1.779628926 | 1.44E-06 | 4.19E-06 | 0.009075154 |
| T2DM | mast_cell | *SH3YL1* | 1.109523582 | 0.037549456 | 0.047865658 | 1 |
| T2DM | mast_cell | *VCL* | 2.296638023 | 0.000680296 | 0.001117591 | 1 |
| T2DM | mast_cell | *ADSS* | 4.742131589 | 2.11E-07 | 8.32E-07 | 0.001325319 |
| T2DM | mast_cell | *STAP1* | -0.357477952 | 7.88E-08 | 3.85E-07 | 0.00049566 |
| T2DM | mast_cell | *TBC1D23* | 2.987321755 | 0.001675411 | 0.002607901 | 1 |
| T2DM | mast_cell | *CUL3* | 3.249968193 | 0.000119742 | 0.000225193 | 0.752818999 |
| T2DM | mast_cell | *MYOM2* | 2.279893975 | 6.72E-07 | 2.19E-06 | 0.004227114 |
| T2DM | mast_cell | *TUBG2* | -1.268894055 | 0.000108208 | 0.000205097 | 0.680306037 |
| T2DM | mast_cell | *NSUN2* | 3.573006978 | 0.00024695 | 0.000437961 | 1 |
| T2DM | mast_cell | *MFAP3* | 2.151670468 | 1.11E-05 | 2.55E-05 | 0.069556599 |
| T2DM | mast_cell | *MRI1* | 1.335885178 | 0.027856742 | 0.036006443 | 1 |
| T2DM | mast_cell | *AGA* | 4.705943192 | 3.64E-09 | 7.78E-08 | 2.29E-05 |
| T2DM | mast_cell | *BOD1L1* | 2.258235776 | 0.004315136 | 0.006288211 | 1 |
| T2DM | mast_cell | *CLEC16A* | 3.954089223 | 8.85E-07 | 2.77E-06 | 0.00556222 |

| | | | | | | |
|---|---|---|---|---|---|---|
| T2DM | mast_cell | *SKIV2L2* | 3.00022099 | 0.000456275 | 0.000772791 | 1 |
| T2DM | mast_cell | *DNAH5* | 3.075142443 | 6.76E-08 | 3.42E-07 | 0.000425176 |
| T2DM | mast_cell | *C6* | 3.385462867 | 0.000110753 | 0.000209478 | 0.69630425 |
| T2DM | mast_cell | *RAI14* | 2.207140977 | 0.000993042 | 0.001587865 | 1 |
| T2DM | mast_cell | *PHLPP2* | 3.385824206 | 0.000102334 | 0.000194785 | 0.64337334 |
| T2DM | mast_cell | *SPDL1* | 1.187090121 | 0.026105307 | 0.033881929 | 1 |
| T2DM | mast_cell | *CTNS* | 0.966208138 | 0.028970928 | 0.037377432 | 1 |
| T2DM | mast_cell | *INPP4A* | 2.035891728 | 0.000530756 | 0.000888409 | 1 |
| T2DM | mast_cell | *RAB27B* | -0.13948926 | 1.33E-07 | 5.71E-07 | 0.000839081 |
| T2DM | mast_cell | *PSMA4* | 3.69416469 | 1.83E-08 | 1.35E-07 | 0.000114855 |
| T2DM | mast_cell | *THAP3* | 2.90498371 | 3.40E-08 | 2.03E-07 | 0.000213642 |
| T2DM | mast_cell | *TDP1* | 4.282720256 | 0.001572515 | 0.00245442 | 1 |
| T2DM | mast_cell | *SPATA7* | 3.614213732 | 0.000280315 | 0.000492137 | 1 |
| T2DM | mast_cell | *MED17* | 3.649150921 | 1.01E-10 | 1.77E-08 | 6.37E-07 |
| T2DM | mast_cell | *RETSAT* | 1.694995357 | 1.27E-05 | 2.89E-05 | 0.080100161 |
| T2DM | mast_cell | *CAPG* | -2.001368615 | 4.62E-06 | 1.17E-05 | 0.029030369 |
| T2DM | mast_cell | *AP2S1* | 5.144929668 | 0.000158068 | 0.000291088 | 0.993774565 |
| T2DM | mast_cell | *LCP2* | -1.52475288 | 2.67E-05 | 5.69E-05 | 0.1680042 |
| T2DM | mast_cell | *TRIT1* | 2.071208366 | 0.00103364 | 0.00164811 | 1 |
| T2DM | mast_cell | *CUL7* | 1.89715312 | 0.004506111 | 0.0065533 | 1 |
| T2DM | mast_cell | *CTNNA1* | 3.164482095 | 9.89E-06 | 2.31E-05 | 0.062202212 |
| T2DM | mast_cell | *CNTLN* | 2.860388735 | 0.001010957 | 0.001614809 | 1 |
| T2DM | mast_cell | *EPHA3* | 2.605869202 | 3.57E-09 | 7.78E-08 | 2.24E-05 |
| T2DM | mast_cell | *WDR37* | 2.402653548 | 0.009800674 | 0.013563029 | 1 |
| T2DM | mast_cell | *YTHDC2* | 3.386715543 | 0.000141082 | 0.000261956 | 0.886982298 |
| T2DM | mast_cell | *ATP6V1H* | 3.759178138 | 6.63E-05 | 0.000130481 | 0.416626745 |
| T2DM | mast_cell | *POLR2B* | 3.827448631 | 6.96E-05 | 0.000136173 | 0.437324756 |
| T2DM | mast_cell | *CP* | 4.483927496 | 3.88E-09 | 7.78E-08 | 2.44E-05 |
| T2DM | mast_cell | *DTNBP1* | 2.192543374 | 0.033843584 | 0.043370284 | 1 |
| T2DM | mast_cell | *ANO2* | 1.167679219 | 0.013949529 | 0.018827971 | 1 |
| T2DM | mast_cell | *GOPC* | 2.355373304 | 0.013931231 | 0.018807311 | 1 |
| T2DM | mast_cell | *TSPAN17* | 2.903724017 | 1.08E-09 | 6.34E-08 | 6.81E-06 |
| T2DM | mast_cell | *LMO3* | 1.510010384 | 0.00100038 | 0.001599133 | 1 |
| T2DM | mast_cell | *CELF2* | 1.693133106 | 0.013501578 | 0.018262569 | 1 |
| T2DM | mast_cell | *ERCC8* | 3.87216557 | 1.16E-08 | 1.09E-07 | 7.29E-05 |
| T2DM | mast_cell | *ADAMTS6* | 2.049145112 | 6.16E-06 | 1.52E-05 | 0.038714315 |
| T2DM | mast_cell | *H6PD* | 1.842992446 | 0.000240072 | 0.000426605 | 1 |
| T2DM | mast_cell | *UTS2* | 1.736286421 | 2.05E-06 | 5.73E-06 | 0.012877431 |
| T2DM | mast_cell | *EPN3* | 10.86865706 | 2.60E-07 | 9.81E-07 | 0.001633351 |
| T2DM | mast_cell | *RCN1* | 2.411763712 | 0.012657738 | 0.017164161 | 1 |
| T2DM | mast_cell | *RFC2* | 6.165827995 | 2.35E-06 | 6.46E-06 | 0.014773396 |
| T2DM | mast_cell | *JKAMP* | 2.257209621 | 0.000763479 | 0.001244489 | 1 |
| T2DM | mast_cell | *DKK3* | 1.798812991 | 4.07E-09 | 7.78E-08 | 2.56E-05 |
| T2DM | mast_cell | *ARHGEF5* | 2.399348908 | 8.58E-09 | 9.42E-08 | 5.40E-05 |
| T2DM | mast_cell | *NFE2L3* | -2.223983606 | 0.000257935 | 0.000455389 | 1 |
| T2DM | mast_cell | *MCUR1* | 2.74913631 | 0.004760265 | 0.006892627 | 1 |
| T2DM | mast_cell | *LIMA1* | 0.821367802 | 0.002283924 | 0.003483511 | 1 |
| T2DM | mast_cell | *MAPK9* | 4.045421347 | 2.02E-09 | 7.78E-08 | 1.27E-05 |
| T2DM | mast_cell | *FAM160A2* | 1.713216617 | 0.003484801 | 0.005167204 | 1 |
| T2DM | mast_cell | *HERPUD1* | 3.502530304 | 0.000791578 | 0.001287953 | 1 |

| | | | | | | |
|---|---|---|---|---|---|---|
| T2DM | mast_cell | *PIK3CB* | 1.745231213 | 0.024744417 | 0.03229565 | 1 |
| T2DM | mast_cell | *CYBA* | 4.42896205 | 2.68E-07 | 1.01E-06 | 0.001683842 |
| T2DM | mast_cell | *THOC3* | 5.655854885 | 2.38E-08 | 1.57E-07 | 0.000149348 |
| T2DM | mast_cell | *RRP12* | 3.58801301 | 7.53E-09 | 9.02E-08 | 4.74E-05 |
| T2DM | mast_cell | *ALX4* | 2.734741742 | 1.44E-08 | 1.19E-07 | 9.03E-05 |
| T2DM | mast_cell | *FOXN3* | 2.991273298 | 1.44E-05 | 3.21E-05 | 0.090428726 |
| T2DM | mast_cell | *AKR7A2* | 3.547737344 | 1.64E-08 | 1.26E-07 | 0.000102904 |
| T2DM | mast_cell | *MRTO4* | 3.179823856 | 5.52E-08 | 2.93E-07 | 0.000347209 |
| T2DM | mast_cell | *MCF2L2* | -1.45284293 | 1.85E-06 | 5.26E-06 | 0.011655491 |
| T2DM | mast_cell | *LY75* | 2.754852749 | 0.00274282 | 0.004135278 | 1 |
| T2DM | mast_cell | *OPN3* | 1.37241234 | 0.008247007 | 0.011537368 | 1 |
| T2DM | mast_cell | *SDCCAG8* | 1.765200118 | 0.00169352 | 0.002634784 | 1 |
| T2DM | mast_cell | *TBC1D22A* | 2.453645044 | 0.001979956 | 0.003051724 | 1 |
| T2DM | mast_cell | *PLEKHH1* | 1.757592232 | 2.81E-06 | 7.57E-06 | 0.017672619 |
| T2DM | mast_cell | *ATP9A* | 3.211433063 | 5.25E-08 | 2.82E-07 | 0.000329948 |
| T2DM | mast_cell | *CHRDL2* | -1.698739044 | 0.000594705 | 0.000990703 | 1 |
| T2DM | mast_cell | *FAM114A2* | 2.610021266 | 0.009104021 | 0.012654649 | 1 |
| T2DM | mast_cell | *GINM1* | 4.263492723 | 6.78E-06 | 1.65E-05 | 0.042644832 |
| T2DM | mast_cell | *MRPL43* | 2.572521359 | 6.22E-05 | 0.000123342 | 0.391117688 |
| T2DM | mast_cell | *ITIH4* | 0.604775907 | 0.011566684 | 0.015808639 | 1 |
| T2DM | mast_cell | *TRAF1* | 2.195298443 | 4.18E-08 | 2.37E-07 | 0.000262993 |
| T2DM | mast_cell | *PCGF2* | 0.942768105 | 0.001462132 | 0.002292946 | 1 |
| T2DM | mast_cell | *IL17RB* | -5.313611056 | 1.63E-06 | 4.69E-06 | 0.01024282 |
| T2DM | mast_cell | *GDI2* | 3.109182737 | 0.000214443 | 0.000384213 | 1 |
| T2DM | mast_cell | *PITHD1* | 3.643600635 | 0.002540476 | 0.003849596 | 1 |
| T2DM | mast_cell | *ATP11B* | 2.147939751 | 1.47E-05 | 3.28E-05 | 0.092711432 |
| T2DM | mast_cell | *CDK14* | 3.156333462 | 1.89E-08 | 1.38E-07 | 0.00011892 |
| T2DM | mast_cell | *ZC3H11A* | 8.083153898 | 7.40E-06 | 1.78E-05 | 0.046536282 |
| T2DM | mast_cell | *DNAJC25* | 4.168380448 | 2.12E-05 | 4.60E-05 | 0.133284497 |
| T2DM | mast_cell | *YBX3* | 4.015195242 | 3.00E-06 | 8.02E-06 | 0.018858851 |
| T2DM | mast_cell | *OGFR* | 3.536860285 | 4.88E-06 | 1.23E-05 | 0.030696194 |
| T2DM | mast_cell | *QSER1* | 4.22616357 | 6.87E-08 | 3.46E-07 | 0.000431794 |
| T2DM | mast_cell | *NCKAP1* | 3.49338817 | 8.51E-08 | 4.06E-07 | 0.000535048 |
| T2DM | mast_cell | *SFSWAP* | 2.239170677 | 0.016466287 | 0.022035664 | 1 |
| T2DM | mast_cell | *TNK2* | 2.364464331 | 0.00010681 | 0.00020269 | 0.671513402 |
| T2DM | mast_cell | *SLC6A16* | 2.181591856 | 0.002558359 | 0.003875761 | 1 |
| T2DM | mast_cell | *SPHK2* | 4.036851757 | 1.49E-09 | 6.92E-08 | 9.38E-06 |
| T2DM | mast_cell | *ISOC2* | 6.90965219 | 7.16E-09 | 8.82E-08 | 4.50E-05 |
| T2DM | mast_cell | *EPN1* | 5.958418749 | 4.60E-07 | 1.58E-06 | 0.002890422 |
| T2DM | mast_cell | *ADCK1* | 3.377128155 | 0.003052749 | 0.004569675 | 1 |
| T2DM | mast_cell | *HAGH* | 1.107426251 | 0.032693681 | 0.041956557 | 1 |
| T2DM | mast_cell | *CASP8* | 4.244060312 | 1.10E-06 | 3.32E-06 | 0.006899078 |
| T2DM | mast_cell | *TM7SF3* | 3.705656334 | 1.32E-06 | 3.90E-06 | 0.00832084 |
| T2DM | mast_cell | *DMRT3* | 6.604535565 | 0.003044197 | 0.004561218 | 1 |
| T2DM | mast_cell | *ATP2C2* | -3.139522357 | 0.000908466 | 0.001466374 | 1 |
| T2DM | mast_cell | *CDON* | 1.637399792 | 0.000350213 | 0.000604721 | 1 |
| T2DM | mast_cell | *TAF2* | 3.625545399 | 2.65E-07 | 9.95E-07 | 0.001663334 |
| T2DM | mast_cell | *TMEM161A* | 5.550111108 | 1.96E-06 | 5.54E-06 | 0.012350301 |
| T2DM | mast_cell | *SUGP2* | 3.642352202 | 1.76E-06 | 5.03E-06 | 0.011080815 |
| T2DM | mast_cell | *DDX20* | 7.457121372 | 2.24E-07 | 8.75E-07 | 0.001410062 |

| | | | | | | |
|---|---|---|---|---|---|---|
| T2DM | mast_cell | *BTBD1* | 4.990647967 | 1.05E-07 | 4.75E-07 | 0.000658227 |
| T2DM | mast_cell | *CHI3L2* | 1.487126683 | 0.023296077 | 0.030519366 | 1 |
| T2DM | mast_cell | *PMS1* | 3.680030009 | 0.000157204 | 0.000289667 | 0.988342114 |
| T2DM | mast_cell | *ANKS1A* | 3.428223381 | 6.49E-07 | 2.12E-06 | 0.004080934 |
| T2DM | mast_cell | *NTHL1* | 3.132154366 | 1.99E-08 | 1.41E-07 | 0.000125335 |
| T2DM | mast_cell | *UHRF1BP1* | 0.782896658 | 0.026234769 | 0.034042929 | 1 |
| T2DM | mast_cell | *IPO5* | 5.137468334 | 7.48E-07 | 2.40E-06 | 0.004702757 |
| T2DM | mast_cell | *ADAT1* | 1.828202289 | 3.85E-06 | 1.00E-05 | 0.024212614 |
| T2DM | mast_cell | *NDUFB4* | 5.325611443 | 5.46E-05 | 0.000109607 | 0.343070063 |
| T2DM | mast_cell | *ZC3H15* | 3.665284565 | 4.56E-05 | 9.30E-05 | 0.286607926 |
| T2DM | mast_cell | *MAP2K4* | 3.375640576 | 0.000181592 | 0.000329772 | 1 |
| T2DM | mast_cell | *SEC61A2* | 5.617345874 | 1.08E-06 | 3.27E-06 | 0.006780325 |
| T2DM | mast_cell | *ASB1* | -2.981053136 | 0.003359635 | 0.004993387 | 1 |
| T2DM | mast_cell | *PPP2R5A* | 3.61058993 | 3.28E-06 | 8.68E-06 | 0.020603727 |
| T2DM | mast_cell | *TIE1* | 1.806826073 | 3.28E-05 | 6.88E-05 | 0.206204541 |
| T2DM | mast_cell | *NFYC* | 3.400293502 | 0.000465437 | 0.000786403 | 1 |
| T2DM | mast_cell | *ZMYND12* | 1.703809953 | 1.50E-05 | 3.34E-05 | 0.094496322 |
| T2DM | mast_cell | *NGEF* | -6.31249984 | 0.000713162 | 0.001168529 | 1 |
| T2DM | mast_cell | *CD84* | -0.949229529 | 1.09E-05 | 2.51E-05 | 0.068435612 |
| T2DM | mast_cell | *ZNRD1* | 2.455101808 | 6.40E-05 | 0.000126562 | 0.402594518 |
| T2DM | mast_cell | *ATXN3* | 2.600892848 | 4.48E-07 | 1.55E-06 | 0.002815107 |
| T2DM | mast_cell | *EML1* | 2.034962034 | 0.000121545 | 0.00022831 | 0.764154716 |
| T2DM | mast_cell | *ZFAT* | 4.031414014 | 3.56E-06 | 9.37E-06 | 0.022402538 |
| T2DM | mast_cell | *MTFR1* | 2.779327497 | 0.001026591 | 0.001638116 | 1 |
| T2DM | mast_cell | *STAG3* | 2.961929122 | 5.48E-08 | 2.92E-07 | 0.000344632 |
| T2DM | mast_cell | *MYO9A* | 2.705793212 | 5.77E-05 | 0.000115246 | 0.363025928 |
| T2DM | mast_cell | *SP100* | 2.925349187 | 0.000437564 | 0.000744509 | 1 |
| T2DM | mast_cell | *STOML1* | 1.935340134 | 1.48E-08 | 1.20E-07 | 9.30E-05 |
| T2DM | mast_cell | *METTL22* | 3.23618427 | 0.020439585 | 0.026996569 | 1 |
| T2DM | mast_cell | *RRP15* | 1.189818102 | 0.001181317 | 0.001873125 | 1 |
| T2DM | mast_cell | *DHX8* | 3.389890013 | 7.76E-05 | 0.000150533 | 0.48757602 |
| T2DM | mast_cell | *SYT1* | 1.571326493 | 4.34E-05 | 8.89E-05 | 0.272624514 |
| T2DM | mast_cell | *ROGDI* | 2.22350413 | 1.38E-08 | 1.18E-07 | 8.68E-05 |
| T2DM | mast_cell | *HDAC4* | 3.842469463 | 1.46E-06 | 4.24E-06 | 0.009186769 |
| T2DM | mast_cell | *FGFR3* | 0.615168957 | 0.007850397 | 0.011019301 | 1 |
| T2DM | mast_cell | *IFI35* | 1.794729943 | 2.96E-09 | 7.78E-08 | 1.86E-05 |
| T2DM | mast_cell | *HEATR6* | 5.941300435 | 3.50E-07 | 1.26E-06 | 0.002201164 |
| T2DM | mast_cell | *SRBD1* | 3.196915804 | 0.000451469 | 0.000765476 | 1 |
| T2DM | mast_cell | *CYFIP1* | 3.236648076 | 0.000518815 | 0.000870275 | 1 |
| T2DM | mast_cell | *PSME4* | 3.189853595 | 3.56E-05 | 7.44E-05 | 0.224037374 |
| T2DM | mast_cell | *PITX1* | 16.20514938 | 0.001417731 | 0.002226649 | 1 |
| T2DM | mast_cell | *SDK2* | -0.509484779 | 1.46E-07 | 6.16E-07 | 0.000917496 |
| T2DM | mast_cell | *NUCKS1* | 2.45938183 | 0.007129586 | 0.010063698 | 1 |
| T2DM | mast_cell | *DNAJA2* | 5.551378597 | 9.05E-08 | 4.24E-07 | 0.000568766 |
| T2DM | mast_cell | *GAL* | 2.629542422 | 1.49E-08 | 1.20E-07 | 9.34E-05 |
| T2DM | mast_cell | *NEDD4* | 3.372101526 | 3.62E-07 | 1.29E-06 | 0.002276522 |
| T2DM | mast_cell | *PIGB* | 2.439102114 | 0.010779516 | 0.014790663 | 1 |
| T2DM | mast_cell | *GNB5* | 2.873499298 | 0.000181459 | 0.000329721 | 1 |
| T2DM | mast_cell | *PFN2* | 3.264194263 | 2.54E-07 | 9.64E-07 | 0.001597783 |
| T2DM | mast_cell | *DAPP1* | 2.357497508 | 0.000251647 | 0.000445538 | 1 |

| | | | | | | |
|---|---|---|---|---|---|---|
| T2DM | mast_cell | *FGF10* | 1.590607308 | 6.37E-08 | 3.27E-07 | 0.000400513 |
| T2DM | mast_cell | *TMEM260* | 4.016803637 | 1.74E-05 | 3.81E-05 | 0.109183723 |
| T2DM | mast_cell | *CLTCL1* | 3.013563152 | 8.51E-08 | 4.06E-07 | 0.0005351 |
| T2DM | mast_cell | *DGCR2* | 4.162204541 | 4.55E-06 | 1.16E-05 | 0.02863146 |
| T2DM | mast_cell | *MNT* | 2.333765427 | 0.004100399 | 0.005995165 | 1 |
| T2DM | mast_cell | *ZXDC* | 2.36604978 | 0.010625437 | 0.014591988 | 1 |
| T2DM | mast_cell | *POLB* | 3.167125647 | 3.73E-05 | 7.76E-05 | 0.234593378 |
| T2DM | mast_cell | *NDST1* | 3.890779338 | 1.20E-08 | 1.11E-07 | 7.56E-05 |
| T2DM | mast_cell | *AP3M2* | 4.682689407 | 1.79E-07 | 7.27E-07 | 0.001122965 |
| T2DM | mast_cell | *ST6GALNAC2* | -1.983317578 | 5.75E-05 | 0.000114859 | 0.361521246 |
| T2DM | mast_cell | *TESK2* | -1.429453577 | 5.35E-06 | 1.34E-05 | 0.033645722 |
| T2DM | mast_cell | *EIF2B3* | 3.129348564 | 8.21E-09 | 9.23E-08 | 5.16E-05 |
| T2DM | mast_cell | *CAMK2A* | 7.30025632 | 7.21E-08 | 3.59E-07 | 0.000453481 |
| T2DM | mast_cell | *CDC42* | 4.351430627 | 1.19E-07 | 5.24E-07 | 0.000747894 |
| T2DM | mast_cell | *OSBPL3* | 3.367518014 | 3.59E-07 | 1.28E-06 | 0.002255596 |
| T2DM | mast_cell | *SNX13* | 3.236874203 | 0.000229996 | 0.000410092 | 1 |
| T2DM | mast_cell | *VASH1* | 3.93255293 | 4.44E-08 | 2.48E-07 | 0.000279377 |
| T2DM | mast_cell | *TRIP13* | -2.756086134 | 3.82E-07 | 1.35E-06 | 0.002404364 |
| T2DM | mast_cell | *TCF3* | 4.040753492 | 9.07E-08 | 4.25E-07 | 0.000570266 |
| T2DM | mast_cell | *MBD3* | 2.784273942 | 0.00072014 | 0.001179348 | 1 |
| T2DM | mast_cell | *CPSF1* | -2.551897285 | 0.006807599 | 0.009635158 | 1 |
| T2DM | mast_cell | *CYBRD1* | 3.062832933 | 1.16E-06 | 3.47E-06 | 0.007293416 |
| T2DM | mast_cell | *LIMS2* | 2.805618096 | 2.86E-08 | 1.79E-07 | 0.000179923 |
| T2DM | mast_cell | *TRHDE* | 2.123697143 | 4.50E-09 | 7.78E-08 | 2.83E-05 |
| T2DM | mast_cell | *NFATC3* | 2.175001173 | 0.003838416 | 0.005639664 | 1 |
| T2DM | mast_cell | *TRNT1* | 3.43739722 | 0.001783333 | 0.002766982 | 1 |
| T2DM | mast_cell | *ACADVL* | 4.101904436 | 6.41E-06 | 1.57E-05 | 0.04027052 |
| T2DM | mast_cell | *CRMP1* | 2.721774424 | 2.73E-06 | 7.39E-06 | 0.017175198 |
| T2DM | mast_cell | *DERL2* | 5.099984067 | 2.47E-07 | 9.40E-07 | 0.001550182 |
| T2DM | mast_cell | *NDE1* | 3.334923856 | 2.48E-06 | 6.76E-06 | 0.015610187 |
| T2DM | mast_cell | *AP1M1* | 2.42247128 | 2.31E-06 | 6.36E-06 | 0.014491847 |
| T2DM | mast_cell | *PVR* | 2.920100099 | 0.000634844 | 0.001049229 | 1 |
| T2DM | mast_cell | *XRCC1* | 4.820180559 | 0.000974554 | 0.001562617 | 1 |
| T2DM | mast_cell | *MOV10L1* | 1.112224036 | 9.36E-05 | 0.00017937 | 0.588691191 |
| T2DM | mast_cell | *TP63* | 1.987244222 | 3.14E-09 | 7.78E-08 | 1.97E-05 |
| T2DM | mast_cell | *ALPK1* | 3.692178903 | 2.00E-05 | 4.34E-05 | 0.125468045 |
| T2DM | mast_cell | *LLGL2* | 2.529973467 | 1.37E-07 | 5.86E-07 | 0.000864412 |
| T2DM | mast_cell | *PDE8A* | 2.495470974 | 0.000139476 | 0.000259051 | 0.876888181 |
| T2DM | mast_cell | *SDHA* | 4.0053593 | 4.66E-09 | 7.80E-08 | 2.93E-05 |
| T2DM | mast_cell | *SMARCE1* | 4.742940081 | 7.78E-07 | 2.48E-06 | 0.004893528 |
| T2DM | mast_cell | *GSDMB* | 2.061061861 | 7.36E-07 | 2.37E-06 | 0.004629967 |
| T2DM | mast_cell | *ADAM11* | 0.973472485 | 0.014230567 | 0.019178472 | 1 |
| T2DM | mast_cell | *PICALM* | 2.998936485 | 6.75E-06 | 1.65E-05 | 0.042447929 |
| T2DM | mast_cell | *GLI2* | 1.174271422 | 2.93E-05 | 6.20E-05 | 0.184208434 |
| T2DM | mast_cell | *CLNS1A* | 2.261754188 | 3.14E-05 | 6.61E-05 | 0.197296545 |
| T2DM | mast_cell | *ATP2A3* | -1.75174304 | 0.000513539 | 0.000862114 | 1 |
| T2DM | mast_cell | *CA12* | 1.682761866 | 5.72E-06 | 1.42E-05 | 0.035989092 |
| T2DM | mast_cell | *NTN4* | 2.314564499 | 5.23E-09 | 7.96E-08 | 3.29E-05 |
| T2DM | mast_cell | *LMAN1* | 4.018427266 | 9.65E-05 | 0.000184496 | 0.606622578 |
| T2DM | mast_cell | *ENO1* | 4.412692204 | 1.07E-07 | 4.85E-07 | 0.000673465 |

| | | | | | | |
|---|---|---|---|---|---|---|
| T2DM | mast_cell | *TIPIN* | 0.973675928 | 0.000295867 | 0.00051785 | 1 |
| T2DM | mast_cell | *SRI* | 5.278476277 | 9.17E-07 | 2.86E-06 | 0.005762465 |
| T2DM | mast_cell | *EIF4G3* | 2.375363028 | 0.001614661 | 0.002517078 | 1 |
| T2DM | mast_cell | *TTC38* | 3.421417008 | 1.38E-07 | 5.89E-07 | 0.000869906 |
| T2DM | mast_cell | *CELSR1* | 7.313705465 | 2.17E-07 | 8.53E-07 | 0.001365515 |
| T2DM | mast_cell | *ADD2* | 2.68761366 | 1.10E-08 | 1.07E-07 | 6.94E-05 |
| T2DM | mast_cell | *VPS9D1* | 4.08405575 | 4.94E-09 | 7.84E-08 | 3.11E-05 |
| T2DM | mast_cell | *MARK3* | 1.913430501 | 0.008609144 | 0.012019918 | 1 |
| T2DM | mast_cell | *SLC25A3* | 4.753572065 | 1.96E-08 | 1.40E-07 | 0.000123115 |
| T2DM | mast_cell | *MOCOS* | 1.338723284 | 1.06E-07 | 4.79E-07 | 0.000663973 |
| T2DM | mast_cell | *BCAP29* | 4.047090686 | 6.87E-07 | 2.23E-06 | 0.004319083 |
| T2DM | mast_cell | *SEC31B* | 0.753321488 | 0.00135887 | 0.002141157 | 1 |
| T2DM | mast_cell | *MKRN2* | 3.579757312 | 1.06E-07 | 4.81E-07 | 0.00066703 |
| T2DM | mast_cell | *MCM6* | 2.846081033 | 1.25E-09 | 6.74E-08 | 7.89E-06 |
| T2DM | mast_cell | *RBMS2* | 3.89724835 | 1.44E-07 | 6.08E-07 | 0.000904178 |
| T2DM | mast_cell | *MLH1* | 2.149069012 | 0.006488254 | 0.009224706 | 1 |
| T2DM | mast_cell | *RGS11* | 1.065575266 | 0.000608571 | 0.001010321 | 1 |
| T2DM | mast_cell | *GPATCH1* | 3.377156671 | 3.21E-09 | 7.78E-08 | 2.02E-05 |
| T2DM | mast_cell | *ICAM3* | 2.633033083 | 1.15E-07 | 5.13E-07 | 0.000723904 |
| T2DM | mast_cell | *NT5C2* | 2.609161752 | 0.001343157 | 0.002117459 | 1 |
| T2DM | mast_cell | *CAMSAP3* | 10.25944153 | 1.37E-08 | 1.17E-07 | 8.62E-05 |
| T2DM | mast_cell | *XAB2* | 2.30116265 | 1.97E-05 | 4.30E-05 | 0.12405147 |
| T2DM | mast_cell | *MAP2K7* | 5.624230056 | 1.23E-07 | 5.38E-07 | 0.000776294 |
| T2DM | mast_cell | *NMRK2* | -2.748215902 | 9.35E-07 | 2.91E-06 | 0.005878591 |
| T2DM | mast_cell | *CTTNBP2* | -2.370004179 | 2.72E-06 | 7.37E-06 | 0.01709684 |
| T2DM | mast_cell | *TM9SF3* | 3.604156425 | 7.06E-06 | 1.71E-05 | 0.044365402 |
| T2DM | mast_cell | *DNAJC10* | 4.596850799 | 5.30E-07 | 1.78E-06 | 0.003332386 |
| T2DM | mast_cell | *DYNC1I2* | 3.935693339 | 1.92E-07 | 7.71E-07 | 0.00120612 |
| T2DM | mast_cell | *APBB1IP* | 2.753275387 | 5.04E-09 | 7.84E-08 | 3.17E-05 |
| T2DM | mast_cell | *LRCH4* | -2.826115172 | 5.81E-05 | 0.00011595 | 0.365474344 |
| T2DM | mast_cell | *SIRT6* | 8.643095196 | 3.27E-07 | 1.19E-06 | 0.002053159 |
| T2DM | mast_cell | *NAALAD2* | 1.77301858 | 0.007104264 | 0.01003246 | 1 |
| T2DM | mast_cell | *FBLN1* | 1.964317124 | 8.04E-09 | 9.17E-08 | 5.05E-05 |
| T2DM | mast_cell | *PIAS2* | 3.373178791 | 5.63E-06 | 1.40E-05 | 0.035393089 |
| T2DM | mast_cell | *AMPH* | 1.344352833 | 1.07E-05 | 2.47E-05 | 0.067342732 |
| T2DM | mast_cell | *MCCC1* | 2.557621055 | 4.31E-07 | 1.50E-06 | 0.002708954 |
| T2DM | mast_cell | *FAP* | 1.688990034 | 5.95E-07 | 1.98E-06 | 0.00374313 |
| T2DM | mast_cell | *ACER3* | -1.584597529 | 0.001601891 | 0.00249779 | 1 |
| T2DM | mast_cell | *PIK3C3* | 3.481926985 | 8.23E-05 | 0.000158843 | 0.517137446 |
| T2DM | mast_cell | *N4BP2* | 3.681351012 | 3.54E-07 | 1.27E-06 | 0.00222814 |
| T2DM | mast_cell | *TULP3* | 4.454183927 | 6.05E-08 | 3.15E-07 | 0.000380108 |
| T2DM | mast_cell | *SYNJ2* | 3.914466392 | 2.13E-06 | 5.93E-06 | 0.013380702 |
| T2DM | mast_cell | *ZCWPW1* | 3.469691195 | 1.44E-06 | 4.18E-06 | 0.009039803 |
| T2DM | mast_cell | *FGF20* | -15.29248814 | 1.23E-06 | 3.64E-06 | 0.007723276 |
| T2DM | mast_cell | *PCM1* | 2.631505154 | 0.000419501 | 0.000715714 | 1 |
| T2DM | mast_cell | *MYH7B* | 4.778251867 | 2.42E-06 | 6.61E-06 | 0.015216144 |
| T2DM | mast_cell | *OSBPL6* | 4.745642172 | 5.29E-09 | 7.96E-08 | 3.33E-05 |
| T2DM | mast_cell | *XRCC5* | 2.839982914 | 0.001470324 | 0.002305218 | 1 |
| T2DM | mast_cell | *LXN* | -1.738490405 | 0.024799874 | 0.032354598 | 1 |
| T2DM | mast_cell | *REXO1* | 3.74192462 | 3.81E-09 | 7.78E-08 | 2.39E-05 |

| | | | | | | |
|---|---|---|---|---|---|---|
| T2DM | mast_cell | *SAR1A* | 3.623730583 | 1.78E-08 | 1.33E-07 | 0.000111645 |
| T2DM | mast_cell | *RAPGEF3* | 0.579953935 | 0.011594437 | 0.015843127 | 1 |
| T2DM | mast_cell | *FDFT1* | 4.749693695 | 1.09E-06 | 3.30E-06 | 0.006853637 |
| T2DM | mast_cell | *SCGN* | 7.580800197 | 5.20E-08 | 2.80E-07 | 0.000326951 |
| T2DM | mast_cell | *DDX1* | 4.385101388 | 1.68E-07 | 6.92E-07 | 0.00105808 |
| T2DM | mast_cell | *DNM2* | 2.474354652 | 0.011953103 | 0.016287204 | 1 |
| T2DM | mast_cell | *STX7* | 2.54174387 | 0.000357633 | 0.000616687 | 1 |
| T2DM | mast_cell | *DDX43* | -2.235027395 | 0.021891435 | 0.028751087 | 1 |
| T2DM | mast_cell | *DCT* | 2.335638964 | 1.46E-08 | 1.19E-07 | 9.20E-05 |
| T2DM | mast_cell | *SLC35C2* | 2.638965361 | 0.033017038 | 0.042345597 | 1 |
| T2DM | mast_cell | *SCTR* | 5.09530431 | 1.06E-08 | 1.06E-07 | 6.69E-05 |
| T2DM | mast_cell | *RIF1* | 1.622261762 | 0.022913023 | 0.030055117 | 1 |
| T2DM | mast_cell | *KCNN2* | 3.395636606 | 1.39E-07 | 5.91E-07 | 0.000876196 |
| T2DM | mast_cell | *PSEN1* | 2.711864409 | 0.000485692 | 0.000817768 | 1 |
| T2DM | mast_cell | *CPOX* | 4.105716686 | 1.61E-06 | 4.64E-06 | 0.010122621 |
| T2DM | mast_cell | *MOK* | 4.282311328 | 7.85E-09 | 9.16E-08 | 4.94E-05 |
| T2DM | mast_cell | *RSBN1* | 3.810897496 | 3.64E-06 | 9.53E-06 | 0.022859234 |
| T2DM | mast_cell | *MAGI3* | 3.506042812 | 2.40E-05 | 5.15E-05 | 0.150875253 |
| T2DM | mast_cell | *COL4A4* | 1.591288703 | 2.23E-07 | 8.70E-07 | 0.001399993 |
| T2DM | mast_cell | *PKP1* | -1.438051761 | 0.009604738 | 0.013303966 | 1 |
| T2DM | mast_cell | *STK17B* | 4.090810025 | 1.04E-06 | 3.18E-06 | 0.006531979 |
| T2DM | mast_cell | *ZNF506* | 3.670448425 | 2.42E-06 | 6.62E-06 | 0.015242654 |
| T2DM | mast_cell | *KIAA0141* | 3.873443942 | 1.04E-07 | 4.74E-07 | 0.000654566 |
| T2DM | mast_cell | *PCDHA6* | -5.484294715 | 1.07E-08 | 1.06E-07 | 6.73E-05 |
| T2DM | mast_cell | *PCDHGA2* | 3.243340531 | 0.000196095 | 0.000354267 | 1 |
| T2DM | mast_cell | *WDR70* | 2.724323517 | 0.000316643 | 0.000550992 | 1 |
| T2DM | mast_cell | *PGR* | 1.746043448 | 6.79E-06 | 1.65E-05 | 0.04268168 |
| T2DM | mast_cell | *C5orf22* | 4.929115992 | 5.85E-09 | 8.14E-08 | 3.68E-05 |
| T2DM | mast_cell | *CCNT2* | 2.645781588 | 2.34E-07 | 9.01E-07 | 0.001468946 |
| T2DM | mast_cell | *GEMIN5* | 1.887059132 | 0.011218234 | 0.015355766 | 1 |
| T2DM | mast_cell | *GSK3B* | 3.47972227 | 5.21E-07 | 1.76E-06 | 0.003274607 |
| T2DM | mast_cell | *ERC1* | 2.748434743 | 0.000181588 | 0.000329772 | 1 |
| T2DM | mast_cell | *DOPEY1* | 1.619350855 | 0.000658382 | 0.001085612 | 1 |
| T2DM | mast_cell | *LYRM2* | 2.252169105 | 2.53E-07 | 9.62E-07 | 0.001593168 |
| T2DM | mast_cell | *ULK2* | 2.89888889 | 1.12E-05 | 2.58E-05 | 0.070684928 |
| T2DM | mast_cell | *PLOD1* | 6.404774225 | 4.70E-07 | 1.61E-06 | 0.002955294 |
| T2DM | mast_cell | *PIBF1* | 2.776871637 | 0.000129705 | 0.000242335 | 0.81545785 |
| T2DM | mast_cell | *PDS5B* | 2.551104464 | 0.003406804 | 0.005059905 | 1 |
| T2DM | mast_cell | *OXCT1* | 1.748569533 | 5.89E-07 | 1.96E-06 | 0.003700174 |
| T2DM | mast_cell | *CYLD* | 2.630544434 | 0.011281728 | 0.015439318 | 1 |
| T2DM | mast_cell | *ZNF324* | -2.95233078 | 0.000338891 | 0.000586448 | 1 |
| T2DM | mast_cell | *ZNF586* | 3.477013214 | 3.54E-07 | 1.27E-06 | 0.00222865 |
| T2DM | mast_cell | *ZNF446* | 6.394606184 | 2.88E-09 | 7.78E-08 | 1.81E-05 |
| T2DM | mast_cell | *RPS5* | 4.500110006 | 4.52E-05 | 9.23E-05 | 0.284198436 |
| T2DM | mast_cell | *CHMP2B* | 2.235910223 | 0.011141944 | 0.015257984 | 1 |
| T2DM | mast_cell | *PPIE* | 3.057872543 | 3.27E-05 | 6.87E-05 | 0.205801888 |
| T2DM | mast_cell | *ZMPSTE24* | 4.645890221 | 4.45E-05 | 9.09E-05 | 0.279526144 |
| T2DM | mast_cell | *STARD7* | 5.294370217 | 7.49E-07 | 2.40E-06 | 0.004711649 |
| T2DM | mast_cell | *NOA1* | 2.606123414 | 3.77E-05 | 7.84E-05 | 0.237220247 |
| T2DM | mast_cell | *GSTP1* | 5.176493214 | 6.09E-06 | 1.50E-05 | 0.038260638 |

| | | | | | | |
|---|---|---|---|---|---|---|
| T2DM | mast_cell | *WBP11* | 4.523914352 | 0.000373747 | 0.00064271 | 1 |
| T2DM | mast_cell | *COL16A1* | 1.565253372 | 0.005000869 | 0.007217959 | 1 |
| T2DM | mast_cell | *APOB* | 1.703628131 | 6.70E-08 | 3.39E-07 | 0.000421154 |
| T2DM | mast_cell | *NCOA1* | 2.062906571 | 0.006614093 | 0.009392999 | 1 |
| T2DM | mast_cell | *CD59* | 3.818996123 | 1.68E-08 | 1.29E-07 | 0.000105696 |
| T2DM | mast_cell | *CD82* | -0.522783262 | 9.30E-07 | 2.90E-06 | 0.00584867 |
| T2DM | mast_cell | *TAF9* | 4.255568045 | 3.57E-09 | 7.78E-08 | 2.24E-05 |
| T2DM | mast_cell | *FCN1* | 1.628797871 | 3.39E-07 | 1.23E-06 | 0.002130191 |
| T2DM | mast_cell | *SCAMP1* | 4.255365113 | 2.75E-06 | 7.44E-06 | 0.017308407 |
| T2DM | mast_cell | *HACE1* | 3.45155352 | 2.21E-07 | 8.66E-07 | 0.001391381 |
| T2DM | mast_cell | *SEH1L* | 3.883458436 | 4.30E-08 | 2.43E-07 | 0.00027039 |
| T2DM | mast_cell | *OVGP1* | 2.162397225 | 0.002676956 | 0.004045678 | 1 |
| T2DM | mast_cell | *SLC25A24* | 3.613303853 | 1.39E-07 | 5.89E-07 | 0.000871216 |
| T2DM | mast_cell | *PILRA* | 0.673286064 | 9.72E-05 | 0.000185645 | 0.610957315 |
| T2DM | mast_cell | *ZNF213* | 2.888511065 | 6.72E-06 | 1.64E-05 | 0.042260112 |
| T2DM | mast_cell | *CPNE3* | 3.328084792 | 8.18E-07 | 2.59E-06 | 0.005143368 |
| T2DM | mast_cell | *RRN3* | 3.875534783 | 5.26E-05 | 0.000105829 | 0.330505175 |
| T2DM | mast_cell | *CTTN* | 3.856980757 | 2.26E-08 | 1.52E-07 | 0.000141959 |
| T2DM | mast_cell | *MTIF2* | 3.10092398 | 1.61E-05 | 3.56E-05 | 0.10143833 |
| T2DM | mast_cell | *EPS15* | 2.518126844 | 4.53E-06 | 1.16E-05 | 0.028489033 |
| T2DM | mast_cell | *ATG16L1* | 5.627733612 | 1.60E-06 | 4.62E-06 | 0.010073564 |
| T2DM | mast_cell | *USP40* | 3.284226225 | 1.55E-06 | 4.47E-06 | 0.00971748 |
| T2DM | mast_cell | *MAST2* | 3.082121169 | 0.000318712 | 0.000554133 | 1 |
| T2DM | mast_cell | *B4GALT1* | 3.610502191 | 0.000145335 | 0.000269296 | 0.913721507 |
| T2DM | mast_cell | *CHMP5* | 5.28703715 | 2.17E-07 | 8.53E-07 | 0.001366357 |
| T2DM | mast_cell | *FOLH1* | 0.638790101 | 0.020629259 | 0.027224213 | 1 |
| T2DM | mast_cell | *EIF2AK1* | 4.773941339 | 2.33E-08 | 1.55E-07 | 0.00014668 |
| T2DM | mast_cell | *EPDR1* | 1.073397193 | 0.007464375 | 0.010508142 | 1 |
| T2DM | mast_cell | *ITPKC* | 4.325269387 | 4.43E-05 | 9.07E-05 | 0.278662031 |
| T2DM | mast_cell | *TMED2* | 4.44583393 | 1.00E-08 | 1.04E-07 | 6.29E-05 |
| T2DM | mast_cell | *ACOX3* | 3.159422401 | 0.000261792 | 0.00046155 | 1 |
| T2DM | mast_cell | *HSD17B14* | 2.9443274 | 0.000202383 | 0.000364371 | 1 |
| T2DM | mast_cell | *TRIP6* | 2.932080535 | 0.00038522 | 0.000660813 | 1 |
| T2DM | mast_cell | *PIGS* | 1.814998572 | 1.43E-05 | 3.20E-05 | 0.090158776 |
| T2DM | mast_cell | *ATXN7L3* | 3.303005351 | 2.47E-09 | 7.78E-08 | 1.55E-05 |
| T2DM | mast_cell | *PGS1* | 4.362088641 | 6.46E-06 | 1.58E-05 | 0.040595087 |
| T2DM | mast_cell | *PSMC5* | 1.824976948 | 0.009470909 | 0.013135584 | 1 |
| T2DM | mast_cell | *LPCAT2* | 3.019860412 | 0.000171281 | 0.000312855 | 1 |
| T2DM | mast_cell | *TXNDC16* | 3.158281394 | 5.03E-07 | 1.71E-06 | 0.003163101 |
| T2DM | mast_cell | *GMCL1* | 4.255576153 | 1.87E-08 | 1.37E-07 | 0.000117597 |
| T2DM | mast_cell | *PTHLH* | 2.818675068 | 0.020896539 | 0.027548027 | 1 |
| T2DM | mast_cell | *PHACTR3* | 1.458464672 | 1.15E-08 | 1.09E-07 | 7.24E-05 |
| T2DM | mast_cell | *ERGIC2* | 3.170837837 | 0.002407679 | 0.003662492 | 1 |
| T2DM | mast_cell | *METTL2A* | 2.064527282 | 0.010703208 | 0.014689166 | 1 |
| T2DM | mast_cell | *ALG6* | 3.491842247 | 5.75E-06 | 1.43E-05 | 0.036176596 |
| T2DM | mast_cell | *DDX18* | 3.933515218 | 7.97E-06 | 1.91E-05 | 0.050117767 |
| T2DM | mast_cell | *ASAP3* | 3.036587244 | 2.07E-08 | 1.44E-07 | 0.000129889 |
| T2DM | mast_cell | *EDEM2* | -2.226960256 | 3.49E-05 | 7.29E-05 | 0.219698003 |
| T2DM | mast_cell | *DOCK3* | 1.518294908 | 0.000252378 | 0.000446707 | 1 |
| T2DM | mast_cell | *FKBP1A* | 4.631500724 | 0.001039778 | 0.001657055 | 1 |

| | | | | | | |
|---|---|---|---|---|---|---|
| T2DM | mast_cell | *NSFL1C* | 4.51034683 | 1.03E-06 | 3.16E-06 | 0.006482124 |
| T2DM | mast_cell | *ZNF343* | 3.89665168 | 1.85E-09 | 7.58E-08 | 1.17E-05 |
| T2DM | mast_cell | *MAVS* | 3.261597885 | 0.000447848 | 0.000760157 | 1 |
| T2DM | mast_cell | *LZTS3* | -2.886806929 | 5.63E-07 | 1.88E-06 | 0.003540174 |
| T2DM | mast_cell | *XRN2* | 2.797230849 | 0.003938482 | 0.00577185 | 1 |
| T2DM | mast_cell | *TESC* | -0.975398862 | 2.79E-07 | 1.04E-06 | 0.001755667 |
| T2DM | mast_cell | *ANAPC5* | 2.74832547 | 0.002740762 | 0.004133166 | 1 |
| T2DM | mast_cell | *SLC23A2* | 4.010761851 | 2.24E-05 | 4.84E-05 | 0.140715462 |
| T2DM | mast_cell | *TMEM230* | 3.498582917 | 9.77E-09 | 1.03E-07 | 6.14E-05 |
| T2DM | mast_cell | *DZANK1* | 3.844819517 | 0.00130622 | 0.002060779 | 1 |
| T2DM | mast_cell | *TASP1* | 2.942369161 | 6.53E-06 | 1.60E-05 | 0.04103732 |
| T2DM | mast_cell | *OAS1* | 5.4818456 | 0.000200141 | 0.000360644 | 1 |
| T2DM | mast_cell | *KIF16B* | 3.876685221 | 8.67E-06 | 2.04E-05 | 0.054533518 |
| T2DM | mast_cell | *FUS* | 3.383584867 | 3.46E-05 | 7.23E-05 | 0.217423027 |
| T2DM | mast_cell | *FXYD5* | 5.104555489 | 0.000113417 | 0.000214002 | 0.713055356 |
| T2DM | mast_cell | *FXYD3* | 5.799074495 | 7.89E-07 | 2.51E-06 | 0.004957519 |
| T2DM | mast_cell | *KCNH4* | 2.118913334 | 5.08E-07 | 1.72E-06 | 0.003191465 |
| T2DM | mast_cell | *GANAB* | 4.302587549 | 2.99E-08 | 1.84E-07 | 0.000187838 |
| T2DM | mast_cell | *GMIP* | 7.502382571 | 1.18E-07 | 5.22E-07 | 0.000742758 |
| T2DM | mast_cell | *NECAP1* | 4.064391815 | 1.30E-06 | 3.83E-06 | 0.008160868 |
| T2DM | mast_cell | *ANKRD24* | 3.078060816 | 1.19E-07 | 5.24E-07 | 0.000747339 |
| T2DM | mast_cell | *GPATCH2L* | 2.877240836 | 7.93E-05 | 0.000153623 | 0.498831717 |
| T2DM | mast_cell | *SLC9A1* | -1.963354283 | 3.17E-05 | 6.67E-05 | 0.199498592 |
| T2DM | mast_cell | *SPTLC1* | 2.652491493 | 0.000681969 | 0.001120047 | 1 |
| T2DM | mast_cell | *PAPOLA* | 3.759745971 | 3.78E-07 | 1.34E-06 | 0.002376399 |
| T2DM | mast_cell | *RGS1* | -3.524644436 | 4.13E-05 | 8.50E-05 | 0.259719285 |
| T2DM | mast_cell | *YPEL3* | 1.970448482 | 1.32E-07 | 5.68E-07 | 0.000829694 |
| T2DM | mast_cell | *MRPS33* | 3.372604697 | 2.23E-06 | 6.18E-06 | 0.014021999 |
| T2DM | mast_cell | *STRN4* | 5.558551632 | 2.02E-05 | 4.40E-05 | 0.127087127 |
| T2DM | mast_cell | *MUL1* | 2.052372309 | 3.49E-07 | 1.26E-06 | 0.002191182 |
| T2DM | mast_cell | *SPG21* | 2.554642767 | 1.02E-06 | 3.14E-06 | 0.006427497 |
| T2DM | mast_cell | *GNPTG* | 4.550270086 | 7.57E-09 | 9.02E-08 | 4.76E-05 |
| T2DM | mast_cell | *ZNF268* | 4.969351546 | 6.89E-05 | 0.000135086 | 0.433355438 |
| T2DM | mast_cell | *GOLGA3* | 2.824506781 | 0.000752593 | 0.001228337 | 1 |
| T2DM | mast_cell | *CERS4* | -1.917636561 | 4.10E-05 | 8.43E-05 | 0.257455756 |
| T2DM | mast_cell | *PDPR* | 4.811323144 | 1.21E-06 | 3.59E-06 | 0.00758591 |
| T2DM | mast_cell | *EXOC1* | 3.225757214 | 0.00023566 | 0.000419359 | 1 |
| T2DM | mast_cell | *NLRC4* | 4.907368929 | 5.46E-09 | 8.00E-08 | 3.43E-05 |
| T2DM | mast_cell | *LAMB1* | 1.43530389 | 0.004325647 | 0.006299593 | 1 |
| T2DM | mast_cell | *SLC26A4* | 8.514302233 | 3.07E-08 | 1.88E-07 | 0.000192798 |
| T2DM | mast_cell | *DLD* | 3.74053897 | 0.000480625 | 0.000810104 | 1 |
| T2DM | mast_cell | *TXNL1* | 4.79356262 | 1.99E-06 | 5.58E-06 | 0.012480016 |
| T2DM | mast_cell | *IL5RA* | -0.321044049 | 9.11E-08 | 4.26E-07 | 0.000572445 |
| T2DM | mast_cell | *RAPGEF4* | 3.516660911 | 7.93E-09 | 9.16E-08 | 4.98E-05 |
| T2DM | mast_cell | *SEL1L3* | -1.880133224 | 0.023139594 | 0.030333325 | 1 |
| T2DM | mast_cell | *TF* | 3.063207469 | 1.42E-08 | 1.19E-07 | 8.93E-05 |
| T2DM | mast_cell | *MYO15A* | -2.062032573 | 0.008038222 | 0.011267849 | 1 |
| T2DM | mast_cell | *NLRP1* | 3.481588181 | 2.00E-07 | 7.98E-07 | 0.001257482 |
| T2DM | mast_cell | *ZFHX4* | 3.409708094 | 9.41E-06 | 2.20E-05 | 0.059178338 |
| T2DM | mast_cell | *RGS17* | -1.224006446 | 3.11E-07 | 1.14E-06 | 0.001956397 |

| | | | | | | |
|---|---|---|---|---|---|---|
| T2DM | mast_cell | *TMEM101* | 2.46657041 | 3.58E-08 | 2.10E-07 | 0.000225152 |
| T2DM | mast_cell | *CMA1* | -0.197081938 | 8.31E-08 | 4.00E-07 | 0.000522566 |
| T2DM | mast_cell | *PPP2R3C* | 4.192505536 | 4.19E-08 | 2.37E-07 | 0.000263145 |
| T2DM | mast_cell | *HAUS4* | 0.76446372 | 0.02972848 | 0.038299785 | 1 |
| T2DM | mast_cell | *SLC22A17* | 1.837012462 | 1.64E-06 | 4.71E-06 | 0.010288263 |
| T2DM | mast_cell | *SCFD1* | 2.720512905 | 9.77E-05 | 0.000186497 | 0.614319803 |
| T2DM | mast_cell | *RPGRIP1* | 4.290307027 | 4.66E-08 | 2.58E-07 | 0.000293132 |
| T2DM | mast_cell | *TOX4* | 4.104569587 | 1.32E-07 | 5.68E-07 | 0.000828524 |
| T2DM | mast_cell | *TGM1* | 1.734814087 | 0.000147656 | 0.000273436 | 0.928315536 |
| T2DM | mast_cell | *AGO1* | 3.560801072 | 0.001114669 | 0.001772364 | 1 |
| T2DM | mast_cell | *RFFL* | 5.755906684 | 5.16E-09 | 7.89E-08 | 3.24E-05 |
| T2DM | mast_cell | *MFSD11* | 2.791095319 | 4.00E-06 | 1.03E-05 | 0.025127705 |
| T2DM | mast_cell | *DPYSL2* | 3.850234514 | 8.61E-07 | 2.71E-06 | 0.005410797 |
| T2DM | mast_cell | *COMT* | 4.070841454 | 1.21E-08 | 1.11E-07 | 7.63E-05 |
| T2DM | mast_cell | *FMO2* | 2.19875999 | 3.99E-08 | 2.30E-07 | 0.000251053 |
| T2DM | mast_cell | *MSH2* | 3.675534254 | 0.003599115 | 0.005319143 | 1 |
| T2DM | mast_cell | *TMEM38B* | 4.91662081 | 0.001852354 | 0.002865588 | 1 |
| T2DM | mast_cell | *PDE6C* | 3.77675642 | 2.30E-06 | 6.35E-06 | 0.014457984 |
| T2DM | mast_cell | *IKZF5* | 4.546861984 | 4.04E-05 | 8.33E-05 | 0.25405548 |
| T2DM | mast_cell | *BLNK* | 2.449327672 | 6.60E-07 | 2.15E-06 | 0.004146575 |
| T2DM | mast_cell | *WAC* | 2.721508162 | 0.000227408 | 0.000406005 | 1 |
| T2DM | mast_cell | *CREM* | 2.927327696 | 0.004318506 | 0.00629065 | 1 |
| T2DM | mast_cell | *TPSD1* | -0.370517353 | 8.84E-08 | 4.17E-07 | 0.000555842 |
| T2DM | mast_cell | *ITPR3* | 3.879091706 | 4.81E-09 | 7.84E-08 | 3.02E-05 |
| T2DM | mast_cell | *CDC7* | 1.095086958 | 0.013775975 | 0.018613702 | 1 |
| T2DM | mast_cell | *PCSK5* | 3.339075129 | 8.55E-07 | 2.70E-06 | 0.005377533 |
| T2DM | mast_cell | *RAB18* | 2.334535594 | 0.000668737 | 0.001099751 | 1 |
| T2DM | mast_cell | *NRP1* | 3.001310415 | 7.01E-06 | 1.70E-05 | 0.044068308 |
| T2DM | mast_cell | *PRTFDC1* | 3.38094181 | 1.16E-08 | 1.09E-07 | 7.27E-05 |
| T2DM | mast_cell | *PALMD* | 3.23402873 | 1.79E-08 | 1.34E-07 | 0.00011265 |
| T2DM | mast_cell | *TSPAN15* | 3.389856479 | 1.04E-08 | 1.05E-07 | 6.54E-05 |
| T2DM | mast_cell | *STX1B* | 1.851222202 | 1.44E-05 | 3.21E-05 | 0.090639904 |
| T2DM | mast_cell | *HSD3B7* | 2.986917818 | 1.22E-07 | 5.34E-07 | 0.000767773 |
| T2DM | mast_cell | *ATP5D* | 5.071850042 | 6.25E-05 | 0.000123825 | 0.39314289 |
| T2DM | mast_cell | *MARCH2* | 5.228735658 | 5.55E-09 | 8.00E-08 | 3.49E-05 |
| T2DM | mast_cell | *CDC34* | 5.664300334 | 0.000275073 | 0.000483744 | 1 |
| T2DM | mast_cell | *MTAP* | 5.062933125 | 1.96E-08 | 1.40E-07 | 0.000123315 |
| T2DM | mast_cell | *CEP170B* | 1.520743517 | 2.88E-06 | 7.74E-06 | 0.018114294 |
| T2DM | mast_cell | *POLR2E* | -2.607341423 | 0.001646508 | 0.002564816 | 1 |
| T2DM | mast_cell | *MADCAM1* | 2.232110426 | 6.31E-09 | 8.24E-08 | 3.97E-05 |
| T2DM | mast_cell | *TRMT2A* | 3.414453095 | 1.09E-07 | 4.92E-07 | 0.000686223 |
| T2DM | mast_cell | *ZDHHC8* | 3.468225578 | 2.45E-06 | 6.68E-06 | 0.01540847 |
| T2DM | mast_cell | *MED15* | 3.786487046 | 2.31E-07 | 8.94E-07 | 0.001451891 |
| T2DM | mast_cell | *SNAP29* | 1.865743541 | 1.90E-05 | 4.15E-05 | 0.119626517 |
| T2DM | mast_cell | *LZTR1* | 4.803381568 | 9.97E-09 | 1.04E-07 | 6.27E-05 |
| T2DM | mast_cell | *CECR2* | 1.620834568 | 5.12E-06 | 1.29E-05 | 0.032162015 |
| T2DM | mast_cell | *SMARCB1* | 1.475168739 | 0.026536002 | 0.034405412 | 1 |
| T2DM | mast_cell | *SLC7A4* | -2.83484487 | 1.99E-05 | 4.34E-05 | 0.12523998 |
| T2DM | mast_cell | *DDTL* | 1.124359553 | 0.001437215 | 0.002254997 | 1 |
| T2DM | mast_cell | *DDT* | 3.530236323 | 3.27E-08 | 1.98E-07 | 0.000205802 |

| | | | | | | |
|---|---|---|---|---|---|---|
| T2DM | mast_cell | *GGT5* | 4.183678446 | 6.69E-10 | 5.76E-08 | 4.20E-06 |
| T2DM | mast_cell | *SPECC1L* | 1.912081037 | 4.44E-06 | 1.14E-05 | 0.027938287 |
| T2DM | mast_cell | *UPB1* | 1.450188183 | 0.000270205 | 0.000475582 | 1 |
| T2DM | mast_cell | *YPEL1* | 2.731902588 | 4.77E-09 | 7.84E-08 | 3.00E-05 |
| T2DM | mast_cell | *PRODH* | -9.317205425 | 5.21E-07 | 1.76E-06 | 0.003276866 |
| T2DM | mast_cell | *PPM1F* | 4.791020572 | 9.44E-07 | 2.93E-06 | 0.005933557 |
| T2DM | mast_cell | *CRYBB3* | 3.071004856 | 1.27E-05 | 2.88E-05 | 0.079724522 |
| T2DM | mast_cell | *CYTH4* | 4.077546095 | 4.61E-09 | 7.79E-08 | 2.90E-05 |
| T2DM | mast_cell | *LRP5L* | 3.484172591 | 0.002562816 | 0.003881577 | 1 |
| T2DM | mast_cell | *SLC25A1* | 4.791358634 | 5.03E-09 | 7.84E-08 | 3.16E-05 |
| T2DM | mast_cell | *SH3BP1* | 3.363195702 | 6.96E-09 | 8.77E-08 | 4.38E-05 |
| T2DM | mast_cell | *HPS4* | 4.870991984 | 1.79E-07 | 7.27E-07 | 0.001125042 |
| T2DM | mast_cell | *TRIOBP* | 2.55689872 | 2.22E-05 | 4.81E-05 | 0.13975075 |
| T2DM | mast_cell | *EIF3L* | 2.853615809 | 0.000166721 | 0.000304968 | 1 |
| T2DM | mast_cell | *CCDC134* | -1.988737819 | 9.95E-06 | 2.32E-05 | 0.062534129 |
| T2DM | mast_cell | *KDELR3* | -1.505572322 | 0.003709247 | 0.005467769 | 1 |
| T2DM | mast_cell | *CYP2D6* | 1.123295488 | 0.036262935 | 0.046315096 | 1 |
| T2DM | mast_cell | *DDX17* | 1.937092612 | 0.002526336 | 0.003830939 | 1 |
| T2DM | mast_cell | *CBY1* | 0.905391739 | 0.006106675 | 0.008723623 | 1 |
| T2DM | mast_cell | *FBXO7* | 3.902677936 | 1.37E-06 | 4.01E-06 | 0.008613243 |
| T2DM | mast_cell | *POLDIP3* | 5.566346962 | 1.12E-05 | 2.58E-05 | 0.070431451 |
| T2DM | mast_cell | *PPP6R2* | 2.338047242 | 0.00256351 | 0.003881692 | 1 |
| T2DM | mast_cell | *CYB5R3* | 3.027483771 | 0.003406675 | 0.005059905 | 1 |
| T2DM | mast_cell | *RHBDD3* | 5.021617059 | 5.41E-08 | 2.89E-07 | 0.000340148 |
| T2DM | mast_cell | *TTLL1* | 6.06856574 | 2.63E-05 | 5.60E-05 | 0.165119777 |
| T2DM | mast_cell | *RASL10A* | 2.614950415 | 3.43E-07 | 1.24E-06 | 0.002156696 |
| T2DM | mast_cell | *AP1B1* | 2.762158816 | 0.00370539 | 0.005463365 | 1 |
| T2DM | mast_cell | *HMGXB4* | 4.420817184 | 3.59E-05 | 7.48E-05 | 0.225641875 |
| T2DM | mast_cell | *TOM1* | 1.720305111 | 0.00157785 | 0.002462136 | 1 |
| T2DM | mast_cell | *NEFH* | 3.017078531 | 0.019039956 | 0.025296747 | 1 |
| T2DM | mast_cell | *CHKB* | -1.052372007 | 0.030535455 | 0.039275042 | 1 |
| T2DM | mast_cell | *BIK* | -5.187077341 | 2.04E-06 | 5.71E-06 | 0.012844822 |
| T2DM | mast_cell | *MCAT* | 3.530104853 | 1.17E-05 | 2.68E-05 | 0.07377165 |
| T2DM | mast_cell | *THOC5* | 2.310415308 | 0.007292489 | 0.010284405 | 1 |
| T2DM | mast_cell | *TTLL12* | 2.667684795 | 1.22E-08 | 1.12E-07 | 7.70E-05 |
| T2DM | mast_cell | *RBFOX2* | 3.384188405 | 1.04E-07 | 4.74E-07 | 0.000655024 |
| T2DM | mast_cell | *MTMR3* | 1.473380146 | 0.009885105 | 0.013662823 | 1 |
| T2DM | mast_cell | *SAMM50* | 5.631698194 | 4.25E-06 | 1.09E-05 | 0.026739147 |
| T2DM | mast_cell | *TXN2* | 3.300200609 | 2.44E-09 | 7.78E-08 | 1.53E-05 |
| T2DM | mast_cell | *IFT27* | 6.336624038 | 2.75E-07 | 1.03E-06 | 0.0017284 |
| T2DM | mast_cell | *KIAA0930* | 3.631841877 | 2.41E-06 | 6.58E-06 | 0.015126682 |
| T2DM | mast_cell | *NCF4* | 2.301255872 | 1.05E-05 | 2.42E-05 | 0.065705636 |
| T2DM | mast_cell | *SLC25A17* | 4.192631753 | 1.76E-05 | 3.85E-05 | 0.11038492 |
| T2DM | mast_cell | *FAM118A* | 1.423426739 | 0.000729936 | 0.001193216 | 1 |
| T2DM | mast_cell | *KCTD17* | 14.83168131 | 6.55E-08 | 3.34E-07 | 0.000412052 |
| T2DM | mast_cell | *EP300* | 3.454653884 | 8.56E-06 | 2.02E-05 | 0.053832116 |
| T2DM | mast_cell | *ACO2* | 2.999223755 | 3.86E-09 | 7.78E-08 | 2.43E-05 |
| T2DM | mast_cell | *POLR3H* | 1.08590074 | 0.013745726 | 0.018576823 | 1 |
| T2DM | mast_cell | *DESI1* | 1.607790389 | 0.002826769 | 0.004254703 | 1 |
| T2DM | mast_cell | *HDAC10* | 3.104427525 | 0.000345012 | 0.000596232 | 1 |

| | | | | | | |
|---|---|---|---|---|---|---|
| T2DM | mast_cell | *RBM23* | 4.182164933 | 1.81E-05 | 3.95E-05 | 0.113598577 |
| T2DM | mast_cell | *VCPKMT* | 2.747262085 | 7.71E-05 | 0.000149768 | 0.484499991 |
| T2DM | mast_cell | *SOS2* | 2.138258723 | 0.012161006 | 0.01654539 | 1 |
| T2DM | mast_cell | *PYGL* | 1.009310863 | 0.000161457 | 0.000296373 | 1 |
| T2DM | mast_cell | *TRIM9* | 1.325151666 | 0.009250252 | 0.012843713 | 1 |
| T2DM | mast_cell | *DDHD1* | 3.446532818 | 9.15E-07 | 2.85E-06 | 0.005751296 |
| T2DM | mast_cell | *CNIH1* | 3.975380268 | 6.40E-08 | 3.28E-07 | 0.000402179 |
| T2DM | mast_cell | *PIGH* | 3.835377596 | 1.77E-07 | 7.21E-07 | 0.001111937 |
| T2DM | mast_cell | *VTI1B* | 4.767694868 | 3.61E-08 | 2.11E-07 | 0.000226662 |
| T2DM | mast_cell | *TMED8* | -2.445922842 | 3.63E-05 | 7.56E-05 | 0.228448548 |
| T2DM | mast_cell | *DAAM1* | 2.588600393 | 1.10E-06 | 3.33E-06 | 0.006930002 |
| T2DM | mast_cell | *PPM1A* | 3.702218195 | 1.42E-06 | 4.14E-06 | 0.008926022 |
| T2DM | mast_cell | *DICER1* | 1.870344154 | 0.01528957 | 0.020539642 | 1 |
| T2DM | mast_cell | *ZFYVE21* | 4.846075574 | 1.38E-06 | 4.04E-06 | 0.008679477 |
| T2DM | mast_cell | *MTHFD1* | 3.939395215 | 2.09E-08 | 1.45E-07 | 0.00013167 |
| T2DM | mast_cell | *TELO2* | 3.368557507 | 5.39E-09 | 8.00E-08 | 3.39E-05 |
| T2DM | mast_cell | *VRK1* | 5.084190108 | 2.81E-06 | 7.57E-06 | 0.017666434 |
| T2DM | mast_cell | *PAPLN* | 4.124213268 | 8.95E-09 | 9.66E-08 | 5.63E-05 |
| T2DM | mast_cell | *PSMB5* | 3.498921175 | 6.87E-07 | 2.23E-06 | 0.004316714 |
| T2DM | mast_cell | *CCNB1IP1* | 3.486732383 | 9.58E-07 | 2.96E-06 | 0.006024064 |
| T2DM | mast_cell | *TRIP11* | 2.137219452 | 0.023518196 | 0.030784696 | 1 |
| T2DM | mast_cell | *PCK2* | 2.445454505 | 2.29E-08 | 1.53E-07 | 0.000143962 |
| T2DM | mast_cell | *KIAA0391* | 8.149444285 | 1.71E-08 | 1.30E-07 | 0.0001077 |
| T2DM | mast_cell | *PLTP* | 2.239470549 | 5.86E-08 | 3.08E-07 | 0.000368701 |
| T2DM | mast_cell | *GSS* | 3.993479624 | 6.12E-09 | 8.15E-08 | 3.85E-05 |
| T2DM | mast_cell | *PYGB* | 2.109279904 | 3.75E-06 | 9.79E-06 | 0.023563422 |
| T2DM | mast_cell | *ABHD12* | 2.992102767 | 3.57E-05 | 7.44E-05 | 0.224305635 |
| T2DM | mast_cell | *NINL* | 3.609423251 | 2.02E-09 | 7.78E-08 | 1.27E-05 |
| T2DM | mast_cell | *CD40* | 3.497616048 | 4.86E-09 | 7.84E-08 | 3.06E-05 |
| T2DM | mast_cell | *UQCC1* | 1.695618963 | 0.007702657 | 0.010828847 | 1 |
| T2DM | mast_cell | *IFT52* | 4.215445407 | 2.05E-07 | 8.13E-07 | 0.001286565 |
| T2DM | mast_cell | *NDRG3* | 4.083862765 | 3.94E-09 | 7.78E-08 | 2.48E-05 |
| T2DM | mast_cell | *DOK5* | 3.328673191 | 4.01E-09 | 7.78E-08 | 2.52E-05 |
| T2DM | mast_cell | *CSTF1* | 4.209369415 | 3.28E-05 | 6.89E-05 | 0.20639471 |
| T2DM | mast_cell | *BMP7* | 1.727200161 | 1.49E-08 | 1.20E-07 | 9.34E-05 |
| T2DM | mast_cell | *TPD52L2* | 3.995313605 | 1.22E-05 | 2.78E-05 | 0.076514056 |
| T2DM | mast_cell | *NELFCD* | 5.050890871 | 3.69E-06 | 9.65E-06 | 0.02320088 |
| T2DM | mast_cell | *CTSZ* | 2.644956015 | 0.000899549 | 0.001453846 | 1 |
| T2DM | mast_cell | *PRPF6* | 2.608305124 | 0.012619617 | 0.017117482 | 1 |
| T2DM | mast_cell | *MTG2* | 1.929370431 | 0.014139529 | 0.019063955 | 1 |
| T2DM | mast_cell | *TCFL5* | 2.153153307 | 8.07E-05 | 0.000156169 | 0.507550558 |
| T2DM | mast_cell | *DIDO1* | 3.73420807 | 3.56E-06 | 9.35E-06 | 0.022359829 |
| T2DM | mast_cell | *GID8* | 4.572396118 | 0.000113008 | 0.000213358 | 0.710482374 |
| T2DM | mast_cell | *SLC17A9* | 3.266502757 | 3.34E-09 | 7.78E-08 | 2.10E-05 |
| T2DM | mast_cell | *BIRC7* | -1.202112292 | 1.28E-08 | 1.13E-07 | 8.07E-05 |
| T2DM | mast_cell | *NKAIN4* | 3.099104378 | 1.18E-05 | 2.69E-05 | 0.073951091 |
| T2DM | mast_cell | *PTK6* | -4.176160962 | 1.13E-06 | 3.39E-06 | 0.007110481 |
| T2DM | mast_cell | *GMEB2* | 4.220663995 | 4.60E-08 | 2.55E-07 | 0.000289459 |
| T2DM | mast_cell | *CDC25B* | 2.05109136 | 4.60E-08 | 2.55E-07 | 0.000289471 |
| T2DM | mast_cell | *RNF24* | 3.182813835 | 6.25E-07 | 2.06E-06 | 0.003928434 |

| | | | | | | |
|---|---|---|---|---|---|---|
| T2DM | mast_cell | *NDUFAF5* | 0.539409277 | 0.00659574 | 0.00936905 | 1 |
| T2DM | mast_cell | *TRIB3* | -3.628392619 | 0.008876086 | 0.012362418 | 1 |
| T2DM | mast_cell | *SLC52A3* | 2.943742271 | 0.000355838 | 0.000613759 | 1 |
| T2DM | mast_cell | *ANGPT4* | 3.372042752 | 5.98E-08 | 3.13E-07 | 0.000376124 |
| T2DM | mast_cell | *RSPO4* | 2.696208302 | 3.95E-07 | 1.39E-06 | 0.002481733 |
| T2DM | mast_cell | *SIRPB1* | 0.768993545 | 0.039201622 | 0.049860529 | 1 |
| T2DM | mast_cell | *SEC23B* | 1.747495047 | 0.007720459 | 0.010849022 | 1 |
| T2DM | mast_cell | *CCM2L* | 1.784915654 | 1.10E-08 | 1.06E-07 | 6.90E-05 |
| T2DM | mast_cell | *MYL9* | 3.33679621 | 5.54E-08 | 2.94E-07 | 0.00034849 |
| T2DM | mast_cell | *MROH8* | 1.672277097 | 6.24E-06 | 1.53E-05 | 0.03922218 |
| T2DM | mast_cell | *MANBAL* | 1.475547143 | 0.001570544 | 0.002451952 | 1 |
| T2DM | mast_cell | *IDH3B* | 5.360189011 | 0.00062428 | 0.001034215 | 1 |
| T2DM | mast_cell | *MAPRE1* | 3.824707013 | 0.00376215 | 0.005540556 | 1 |
| T2DM | mast_cell | *RPRD1B* | 3.896127405 | 1.18E-05 | 2.69E-05 | 0.074032415 |
| T2DM | mast_cell | *PXMP4* | 2.762886718 | 0.000930025 | 0.001495796 | 1 |
| T2DM | mast_cell | *BPI* | -2.640293715 | 3.06E-05 | 6.46E-05 | 0.192653152 |
| T2DM | mast_cell | *ACTR5* | 4.795033787 | 4.23E-08 | 2.39E-07 | 0.000265949 |
| T2DM | mast_cell | *DHX35* | 4.839805347 | 1.11E-07 | 4.99E-07 | 0.000699445 |
| T2DM | mast_cell | *MAP1LC3A* | 1.904792112 | 3.09E-10 | 3.81E-08 | 1.94E-06 |
| T2DM | mast_cell | *PIGU* | 4.138176015 | 9.28E-08 | 4.31E-07 | 0.000583214 |
| T2DM | mast_cell | *APMAP* | 2.646705517 | 3.08E-09 | 7.78E-08 | 1.94E-05 |
| T2DM | mast_cell | *CELF4* | -3.190743974 | 0.000345239 | 0.00059646 | 1 |
| T2DM | mast_cell | *RBFA* | 4.228710547 | 6.53E-08 | 3.33E-07 | 0.000410773 |
| T2DM | mast_cell | *LPIN2* | 4.173522309 | 1.17E-08 | 1.09E-07 | 7.34E-05 |
| T2DM | mast_cell | *SMCHD1* | 3.222562106 | 2.19E-05 | 4.73E-05 | 0.137378991 |
| T2DM | mast_cell | *MYL12A* | 4.485596638 | 6.84E-08 | 3.44E-07 | 0.000429814 |
| T2DM | mast_cell | *LIPG* | 4.630347171 | 2.72E-07 | 1.02E-06 | 0.001710437 |
| T2DM | mast_cell | *POLI* | 0.433274344 | 0.026794283 | 0.034697354 | 1 |
| T2DM | mast_cell | *NALCN* | 2.356740044 | 3.32E-09 | 7.78E-08 | 2.09E-05 |
| T2DM | mast_cell | *HTR2A* | 3.599934795 | 1.59E-08 | 1.24E-07 | 1.00E-04 |
| T2DM | mast_cell | *KLF5* | 2.44085292 | 3.51E-06 | 9.25E-06 | 0.022073602 |
| T2DM | mast_cell | *STK24* | 3.554146494 | 1.31E-05 | 2.95E-05 | 0.082108265 |
| T2DM | mast_cell | *DNAJC3* | 3.585626657 | 8.63E-05 | 0.000166145 | 0.542428528 |
| T2DM | mast_cell | *UGGT2* | 2.745676937 | 0.000197318 | 0.000355862 | 1 |
| T2DM | mast_cell | *ARHGEF7* | 3.358355966 | 4.77E-07 | 1.63E-06 | 0.002996613 |
| T2DM | mast_cell | *SGCG* | 1.450468134 | 1.43E-08 | 1.19E-07 | 9.02E-05 |
| T2DM | mast_cell | *RGCC* | 0.563404094 | 0.020240055 | 0.026761141 | 1 |
| T2DM | mast_cell | *VWA8* | 3.296299991 | 3.25E-06 | 8.60E-06 | 0.020417793 |
| T2DM | mast_cell | *KATNAL1* | 2.812946196 | 3.89E-05 | 8.06E-05 | 0.244783854 |
| T2DM | mast_cell | *DHRS12* | 5.092976297 | 1.05E-07 | 4.76E-07 | 0.000659398 |
| T2DM | mast_cell | *MEDAG* | 2.481308622 | 2.55E-09 | 7.78E-08 | 1.61E-05 |
| T2DM | mast_cell | *TSC22D1* | 3.914922794 | 7.19E-08 | 3.58E-07 | 0.00045187 |
| T2DM | mast_cell | *CLN5* | 1.897088633 | 0.004722574 | 0.006841203 | 1 |
| T2DM | mast_cell | *MGRN1* | 1.737872285 | 0.006566146 | 0.00933123 | 1 |
| T2DM | mast_cell | *GDPD3* | 4.344383123 | 1.67E-05 | 3.67E-05 | 0.104912003 |
| T2DM | mast_cell | *NFAT5* | 2.988452972 | 7.24E-06 | 1.75E-05 | 0.045542778 |
| T2DM | mast_cell | *LONP2* | 1.973065867 | 0.017956878 | 0.023908279 | 1 |
| T2DM | mast_cell | *PLLP* | -2.579216436 | 0.014833118 | 0.019956305 | 1 |
| T2DM | mast_cell | *DHODH* | 1.565777996 | 4.71E-05 | 9.56E-05 | 0.295851024 |
| T2DM | mast_cell | *CTCF* | 4.259491499 | 6.88E-08 | 3.46E-07 | 0.00043247 |

| | | | | | | |
|---|---|---|---|---|---|---|
| T2DM | mast_cell | *ACD* | 1.956196757 | 0.000818911 | 0.001329673 | 1 |
| T2DM | mast_cell | *USB1* | 4.69563209 | 2.15E-08 | 1.48E-07 | 0.000135339 |
| T2DM | mast_cell | *SETD6* | 5.536468514 | 5.59E-08 | 2.96E-07 | 0.000351667 |
| T2DM | mast_cell | *SLC7A6OS* | 1.154265727 | 0.000394191 | 0.000675465 | 1 |
| T2DM | mast_cell | *HCFC1R1* | 6.899538155 | 2.40E-08 | 1.58E-07 | 0.000151115 |
| T2DM | mast_cell | *NPRL3* | 2.063104797 | 0.000924346 | 0.001487568 | 1 |
| T2DM | mast_cell | *NAGPA* | 4.43546492 | 1.79E-06 | 5.10E-06 | 0.011266358 |
| T2DM | mast_cell | *COTL1* | 3.660519309 | 2.23E-07 | 8.70E-07 | 0.001399697 |
| T2DM | mast_cell | *CRISPLD2* | 2.115191696 | 4.74E-07 | 1.62E-06 | 0.002983091 |
| T2DM | mast_cell | *NME4* | 5.767940699 | 2.44E-07 | 9.32E-07 | 0.001532803 |
| T2DM | mast_cell | *LMF1* | 4.577185897 | 9.15E-08 | 4.27E-07 | 0.000575282 |
| T2DM | mast_cell | *NARFL* | 3.888794506 | 1.72E-08 | 1.30E-07 | 0.000107849 |
| T2DM | mast_cell | *CLCN7* | -2.312710768 | 0.012191493 | 0.016576107 | 1 |
| T2DM | mast_cell | *HAGHL* | -3.586572857 | 8.38E-06 | 1.99E-05 | 0.05268426 |
| T2DM | mast_cell | *SLC7A5* | 8.294961864 | 3.68E-09 | 7.78E-08 | 2.31E-05 |
| T2DM | mast_cell | *STUB1* | 2.833846656 | 5.91E-07 | 1.97E-06 | 0.003717124 |
| T2DM | mast_cell | *RHBDL1* | 1.300021803 | 0.011911901 | 0.016238101 | 1 |
| T2DM | mast_cell | *NUBP1* | 4.424968231 | 2.01E-09 | 7.78E-08 | 1.27E-05 |
| T2DM | mast_cell | *UBE2I* | 3.136808989 | 0.003045211 | 0.00456165 | 1 |
| T2DM | mast_cell | *CLUAP1* | 2.009789551 | 2.15E-09 | 7.78E-08 | 1.35E-05 |
| T2DM | mast_cell | *CPPED1* | -1.813686843 | 0.00018676 | 0.000338473 | 1 |
| T2DM | mast_cell | *HMOX2* | 3.991059543 | 8.09E-08 | 3.92E-07 | 0.000508351 |
| T2DM | mast_cell | *BFAR* | -1.998785238 | 0.015531307 | 0.020842119 | 1 |
| T2DM | mast_cell | *RRN3P2* | 2.757305639 | 0.000124387 | 0.000233091 | 0.782020912 |
| T2DM | mast_cell | *RBL2* | 3.514288772 | 3.86E-10 | 4.05E-08 | 2.43E-06 |
| T2DM | mast_cell | *STX4* | 4.025178405 | 7.49E-08 | 3.69E-07 | 0.000470713 |
| T2DM | mast_cell | *KAT8* | 5.384196012 | 6.98E-10 | 5.79E-08 | 4.39E-06 |
| T2DM | mast_cell | *TMC5* | 2.895621233 | 2.57E-07 | 9.72E-07 | 0.001615963 |
| T2DM | mast_cell | *RNF40* | 3.918279203 | 6.38E-08 | 3.27E-07 | 0.000401042 |
| T2DM | mast_cell | *KNOP1* | 3.217908317 | 1.25E-05 | 2.83E-05 | 0.078330616 |
| T2DM | mast_cell | *LACTB* | 4.380824631 | 1.98E-06 | 5.58E-06 | 0.012468388 |
| T2DM | mast_cell | *HERC1* | 1.931682157 | 0.014562954 | 0.019613816 | 1 |
| T2DM | mast_cell | *RASL12* | 2.03915107 | 1.09E-08 | 1.06E-07 | 6.86E-05 |
| T2DM | mast_cell | *AP3B2* | 0.584017882 | 0.0245155 | 0.032023468 | 1 |
| T2DM | mast_cell | *IGDCC4* | 0.347697945 | 0.028217776 | 0.036458109 | 1 |
| T2DM | mast_cell | *CTSH* | 1.277314206 | 1.84E-06 | 5.22E-06 | 0.011557447 |
| T2DM | mast_cell | *TTC23* | 3.832196384 | 5.49E-07 | 1.84E-06 | 0.003453977 |
| T2DM | mast_cell | *FAH* | 1.982520888 | 6.10E-09 | 8.15E-08 | 3.83E-05 |
| T2DM | mast_cell | *RPAP1* | 3.962514748 | 0.000516798 | 0.000867354 | 1 |
| T2DM | mast_cell | *EHD4* | 4.004148862 | 1.84E-08 | 1.36E-07 | 0.000115925 |
| T2DM | mast_cell | *ATP8B4* | 2.797899715 | 0.002588545 | 0.003918656 | 1 |
| T2DM | mast_cell | *DTWD1* | 3.458614799 | 1.39E-05 | 3.12E-05 | 0.087358467 |
| T2DM | mast_cell | *DNAJC17* | 4.702586721 | 3.41E-05 | 7.13E-05 | 0.214091394 |
| T2DM | mast_cell | *PDGFRL* | 2.201287641 | 1.11E-09 | 6.34E-08 | 6.97E-06 |
| T2DM | mast_cell | *ZFAND1* | 2.083513104 | 0.003932582 | 0.005764548 | 1 |
| T2DM | mast_cell | *RIPK2* | 4.033757147 | 7.70E-07 | 2.46E-06 | 0.004840628 |
| T2DM | mast_cell | *EYA1* | 1.958507162 | 3.63E-12 | 1.52E-09 | 2.28E-08 |
| T2DM | mast_cell | *CPQ* | 2.405800959 | 1.48E-06 | 4.29E-06 | 0.009312716 |
| T2DM | mast_cell | *NIPAL2* | 3.684630507 | 9.39E-10 | 6.13E-08 | 5.90E-06 |
| T2DM | mast_cell | *IKBKB* | 4.198701563 | 1.08E-05 | 2.50E-05 | 0.068086519 |

| | | | | | | |
|---|---|---|---|---|---|---|
| T2DM | mast_cell | *PLAT* | 4.774637187 | 3.25E-09 | 7.78E-08 | 2.04E-05 |
| T2DM | mast_cell | *GDAP1* | -1.695433101 | 9.94E-06 | 2.32E-05 | 0.062472728 |
| T2DM | mast_cell | *RAB2A* | 2.569648665 | 7.88E-07 | 2.51E-06 | 0.004957232 |
| T2DM | mast_cell | *ZC2HC1A* | 3.47244487 | 5.10E-05 | 0.000102969 | 0.320645443 |
| T2DM | mast_cell | *SPAG1* | 2.129623356 | 6.08E-05 | 0.000120807 | 0.381990305 |
| T2DM | mast_cell | *CHRAC1* | 3.364156292 | 2.63E-09 | 7.78E-08 | 1.65E-05 |
| T2DM | mast_cell | *SNX16* | 4.138063866 | 8.54E-06 | 2.02E-05 | 0.053681719 |
| T2DM | mast_cell | *GSDMD* | 2.874024766 | 1.05E-05 | 2.44E-05 | 0.066175343 |
| T2DM | mast_cell | *TSTA3* | 2.251589251 | 4.81E-06 | 1.22E-05 | 0.030267276 |
| T2DM | mast_cell | *ERI1* | 4.898891083 | 9.37E-06 | 2.19E-05 | 0.058887214 |
| T2DM | mast_cell | *MTMR9* | 4.569120263 | 8.50E-06 | 2.01E-05 | 0.053439637 |
| T2DM | mast_cell | *LEPROTL1* | -1.930459384 | 0.013438265 | 0.018180842 | 1 |
| T2DM | mast_cell | *TNFRSF10A* | 0.99356405 | 0.038252783 | 0.048722699 | 1 |
| T2DM | mast_cell | *ERICH1* | 2.114467864 | 0.021590698 | 0.0283917 | 1 |
| T2DM | mast_cell | *TUSC3* | 2.270857017 | 3.94E-09 | 7.78E-08 | 2.48E-05 |
| T2DM | mast_cell | *ARHGEF10* | 2.614633136 | 3.09E-07 | 1.13E-06 | 0.001943095 |
| T2DM | mast_cell | *KLHDC4* | 2.058692282 | 0.001954034 | 0.003013988 | 1 |
| T2DM | mast_cell | *MCM4* | -2.025971354 | 8.99E-05 | 0.000172654 | 0.565097151 |
| T2DM | mast_cell | *MAN2B1* | 2.81214722 | 2.81E-06 | 7.57E-06 | 0.017684257 |
| T2DM | mast_cell | *TULP2* | -0.660229426 | 3.44E-07 | 1.24E-06 | 0.002160858 |
| T2DM | mast_cell | *NUCB1* | 2.047386192 | 0.025223682 | 0.032839364 | 1 |
| T2DM | mast_cell | *NFKBIB* | 5.649863694 | 0.00052376 | 0.000877632 | 1 |
| T2DM | mast_cell | *TUBB4A* | 6.57511063 | 0.010949205 | 0.015003848 | 1 |
| T2DM | mast_cell | *SARS2* | 1.091057284 | 0.018951673 | 0.025195426 | 1 |
| T2DM | mast_cell | *SNRNP70* | 2.072653542 | 0.00679036 | 0.009617254 | 1 |
| T2DM | mast_cell | *PPP1R37* | -2.354162947 | 0.018934942 | 0.025183834 | 1 |
| T2DM | mast_cell | *FCGRT* | 1.768766925 | 0.002086438 | 0.003204846 | 1 |
| T2DM | mast_cell | *ERCC2* | 6.547218655 | 2.20E-08 | 1.50E-07 | 0.000138191 |
| T2DM | mast_cell | *PLEKHJ1* | 5.496119601 | 7.41E-08 | 3.67E-07 | 0.000465704 |
| T2DM | mast_cell | *CD37* | -2.076911135 | 0.000196683 | 0.000355125 | 1 |
| T2DM | mast_cell | *AMH* | 2.260636399 | 0.023281646 | 0.030506817 | 1 |
| T2DM | mast_cell | *DKKL1* | -1.86373673 | 0.000334076 | 0.000579084 | 1 |
| T2DM | mast_cell | *STX10* | 4.282068815 | 0.002004898 | 0.00308714 | 1 |
| T2DM | mast_cell | *FCER2* | -5.209803479 | 0.010115942 | 0.013941018 | 1 |
| T2DM | mast_cell | *IL4I1* | 2.486337705 | 1.24E-07 | 5.40E-07 | 0.000780384 |
| T2DM | mast_cell | *AES* | 2.715430934 | 0.021111544 | 0.027796497 | 1 |
| T2DM | mast_cell | *SGTA* | 4.641590256 | 1.57E-08 | 1.24E-07 | 9.90E-05 |
| T2DM | mast_cell | *C19orf53* | -2.701494101 | 0.003553105 | 0.005261039 | 1 |
| T2DM | mast_cell | *IL27RA* | 4.843728405 | 1.05E-08 | 1.05E-07 | 6.57E-05 |
| T2DM | mast_cell | *C19orf44* | 1.465792672 | 0.001942549 | 0.002998071 | 1 |
| T2DM | mast_cell | *RASAL3* | -2.098824638 | 0.000848844 | 0.001376852 | 1 |
| T2DM | mast_cell | *ZNF419* | 1.620502536 | 4.50E-06 | 1.15E-05 | 0.028313615 |
| T2DM | mast_cell | *SYDE1* | 2.362797182 | 4.42E-08 | 2.47E-07 | 0.000277748 |
| T2DM | mast_cell | *AURKC* | 1.696365496 | 0.000240252 | 0.000426806 | 1 |
| T2DM | mast_cell | *CCNE1* | -5.190509389 | 0.029033854 | 0.037450931 | 1 |
| T2DM | mast_cell | *PDCD5* | 2.959669233 | 5.79E-09 | 8.14E-08 | 3.64E-05 |
| T2DM | mast_cell | *ANKRD27* | -1.396967529 | 4.74E-06 | 1.20E-05 | 0.029823267 |
| T2DM | mast_cell | *PRX* | 3.125176629 | 0.005957756 | 0.008526386 | 1 |
| T2DM | mast_cell | *TBCB* | 3.220876952 | 0.001901413 | 0.002937146 | 1 |
| T2DM | mast_cell | *POLR2I* | 4.906441508 | 3.07E-06 | 8.20E-06 | 0.019327635 |

| | | | | | | |
|---|---|---|---|---|---|---|
| T2DM | mast_cell | *CLIP3* | -1.365801806 | 0.001412232 | 0.002219675 | 1 |
| T2DM | mast_cell | *SLC1A5* | 3.267145147 | 3.00E-08 | 1.84E-07 | 0.000188633 |
| T2DM | mast_cell | *PRKD2* | 2.056683468 | 4.01E-05 | 8.28E-05 | 0.252108855 |
| T2DM | mast_cell | *CACTIN* | 7.828831476 | 0.000501069 | 0.00084253 | 1 |
| T2DM | mast_cell | *CCDC9* | 2.991246013 | 7.60E-10 | 5.88E-08 | 4.78E-06 |
| T2DM | mast_cell | *FZR1* | 5.134547869 | 2.42E-09 | 7.78E-08 | 1.52E-05 |
| T2DM | mast_cell | *CEACAM4* | 6.156996205 | 1.26E-08 | 1.13E-07 | 7.92E-05 |
| T2DM | mast_cell | *PLIN3* | 3.646107543 | 2.24E-08 | 1.52E-07 | 0.000141026 |
| T2DM | mast_cell | *MRPL4* | 5.503480029 | 3.79E-06 | 9.89E-06 | 0.023829188 |
| T2DM | mast_cell | *ICAM5* | -15.17222938 | 1.13E-07 | 5.04E-07 | 0.000709669 |
| T2DM | mast_cell | *CD33* | -1.529665938 | 6.23E-06 | 1.53E-05 | 0.039166669 |
| T2DM | mast_cell | *BABAM1* | 5.802343367 | 1.61E-09 | 7.17E-08 | 1.01E-05 |
| T2DM | mast_cell | *TYK2* | 3.890841028 | 6.91E-10 | 5.79E-08 | 4.35E-06 |
| T2DM | mast_cell | *RABAC1* | 3.2776333 | 1.01E-07 | 4.61E-07 | 0.000633605 |
| T2DM | mast_cell | *PTPRS* | 2.18202877 | 0.000338753 | 0.000586382 | 1 |
| T2DM | mast_cell | *CYTH2* | 5.406068536 | 2.15E-08 | 1.48E-07 | 0.000135268 |
| T2DM | mast_cell | *GRIN2D* | -2.594987492 | 3.90E-06 | 1.01E-05 | 0.024526418 |
| T2DM | mast_cell | *CCDC114* | 1.245132364 | 0.02644663 | 0.03429661 | 1 |
| T2DM | mast_cell | *CARD8* | 2.076044966 | 0.0183909 | 0.024470601 | 1 |
| T2DM | mast_cell | *LIG1* | 5.861931321 | 2.36E-07 | 9.08E-07 | 0.001481361 |
| T2DM | mast_cell | *ZNF175* | -2.206748645 | 1.66E-05 | 3.65E-05 | 0.104338987 |
| T2DM | mast_cell | *PLA2G4C* | 2.381138005 | 5.12E-06 | 1.29E-05 | 0.03218445 |
| T2DM | mast_cell | *CAPS* | 3.812906703 | 6.38E-06 | 1.56E-05 | 0.040082167 |
| T2DM | mast_cell | *MIER2* | 2.590704455 | 2.92E-08 | 1.81E-07 | 0.000183455 |
| T2DM | mast_cell | *PPP2R1A* | 5.567241047 | 7.04E-07 | 2.27E-06 | 0.004428682 |
| T2DM | mast_cell | *TNPO2* | 4.683385844 | 6.09E-07 | 2.02E-06 | 0.003828751 |
| T2DM | mast_cell | *LILRB5* | 2.946634911 | 4.01E-09 | 7.78E-08 | 2.52E-05 |
| T2DM | mast_cell | *PRPF31* | 2.541824587 | 0.000974934 | 0.001562827 | 1 |
| T2DM | mast_cell | *RPL18A* | 1.565016832 | 0.026768691 | 0.0346785 | 1 |
| T2DM | mast_cell | *PDE4C* | 2.661078424 | 3.14E-07 | 1.15E-06 | 0.0019737 |
| T2DM | mast_cell | *ELL* | 4.257169565 | 1.34E-05 | 3.01E-05 | 0.083942395 |
| T2DM | mast_cell | *UPK1A* | 6.278206524 | 7.16E-08 | 3.57E-07 | 0.000450125 |
| T2DM | mast_cell | *COPE* | 4.814021961 | 1.71E-08 | 1.30E-07 | 0.000107798 |
| T2DM | mast_cell | *LSR* | -1.570497287 | 5.08E-05 | 0.000102659 | 0.31947332 |
| T2DM | mast_cell | *KXD1* | 5.920537955 | 3.97E-05 | 8.20E-05 | 0.249548361 |
| T2DM | mast_cell | *HPN* | 1.853623446 | 6.14E-07 | 2.03E-06 | 0.003862027 |
| T2DM | mast_cell | *ZNF14* | 5.986962747 | 9.93E-07 | 3.05E-06 | 0.006240714 |
| T2DM | mast_cell | *ATP13A1* | 4.462265242 | 9.61E-15 | 8.63E-12 | 6.04E-11 |
| T2DM | mast_cell | *SIPA1L3* | 3.4908978 | 7.89E-05 | 0.000152788 | 0.495795646 |
| T2DM | mast_cell | *ZNF85* | 4.156077878 | 9.27E-10 | 6.13E-08 | 5.83E-06 |
| T2DM | mast_cell | *ETHE1* | 2.453040978 | 1.47E-08 | 1.19E-07 | 9.22E-05 |
| T2DM | mast_cell | *SMG9* | 3.614681362 | 1.27E-08 | 1.13E-07 | 7.99E-05 |
| T2DM | mast_cell | *GTPBP10* | 1.948473798 | 9.71E-05 | 0.000185543 | 0.610250247 |
| T2DM | mast_cell | *RASA4* | 1.769028051 | 0.000849064 | 0.001376855 | 1 |
| T2DM | mast_cell | *PON3* | 1.787770965 | 1.45E-08 | 1.19E-07 | 9.12E-05 |
| T2DM | mast_cell | *PON2* | 4.262732777 | 3.45E-08 | 2.05E-07 | 0.000217087 |
| T2DM | mast_cell | *STEAP1B* | 2.819243635 | 1.82E-07 | 7.39E-07 | 0.001146396 |
| T2DM | mast_cell | *PTN* | 4.086307949 | 1.77E-09 | 7.51E-08 | 1.11E-05 |
| T2DM | mast_cell | *MPP6* | 2.132994758 | 4.43E-09 | 7.78E-08 | 2.79E-05 |
| T2DM | mast_cell | *DFNA5* | 2.543185719 | 1.55E-05 | 3.43E-05 | 0.097336106 |

| | | | | | | |
|---|---|---|---|---|---|---|
| T2DM | mast_cell | *OGDH* | 3.578195298 | 8.99E-05 | 0.000172699 | 0.565416816 |
| T2DM | mast_cell | *ADAP1* | 8.571499199 | 0.023891315 | 0.031234081 | 1 |
| T2DM | mast_cell | *CAV1* | 2.468332398 | 7.96E-07 | 2.53E-06 | 0.005001609 |
| T2DM | mast_cell | *MET* | 2.331588446 | 1.18E-08 | 1.10E-07 | 7.45E-05 |
| T2DM | mast_cell | *LMBR1* | 3.00368616 | 0.000300912 | 0.000525801 | 1 |
| T2DM | mast_cell | *HOXA2* | 4.216613111 | 2.08E-08 | 1.44E-07 | 0.000130573 |
| T2DM | mast_cell | *HOXA5* | 2.321923979 | 0.000102771 | 0.000195558 | 0.646124053 |
| T2DM | mast_cell | *HOXA6* | 2.775694672 | 2.47E-05 | 5.29E-05 | 0.155409643 |
| T2DM | mast_cell | *BRAT1* | 2.331156159 | 2.72E-05 | 5.78E-05 | 0.170794257 |
| T2DM | mast_cell | *CPED1* | 3.037206324 | 6.23E-05 | 0.000123498 | 0.391734185 |
| T2DM | mast_cell | *HIBADH* | 3.554650911 | 1.11E-06 | 3.35E-06 | 0.00700083 |
| T2DM | mast_cell | *CPVL* | 3.545145631 | 7.36E-12 | 2.89E-09 | 4.63E-08 |
| T2DM | mast_cell | *PLEKHA8* | 3.487344029 | 2.64E-08 | 1.69E-07 | 0.00016588 |
| T2DM | mast_cell | *NOD1* | 3.972591073 | 3.48E-05 | 7.26E-05 | 0.21855986 |
| T2DM | mast_cell | *CRHR2* | -2.959447325 | 4.25E-06 | 1.09E-05 | 0.026716132 |
| T2DM | mast_cell | *EPHB6* | 3.167499644 | 4.31E-08 | 2.43E-07 | 0.000271167 |
| T2DM | mast_cell | *NPTX2* | 21.18568449 | 0.02154681 | 0.028339915 | 1 |
| T2DM | mast_cell | *CYP3A5* | 5.259353464 | 1.27E-05 | 2.89E-05 | 0.080158726 |
| T2DM | mast_cell | *ZKSCAN1* | 4.215499317 | 1.05E-07 | 4.75E-07 | 0.000658556 |
| T2DM | mast_cell | *EIF3B* | 3.594592369 | 0.000260922 | 0.000460274 | 1 |
| T2DM | mast_cell | *SNX8* | 3.523144306 | 0.002862169 | 0.004302835 | 1 |
| T2DM | mast_cell | *TAF6* | 3.527250692 | 6.02E-08 | 3.15E-07 | 0.000378673 |
| T2DM | mast_cell | *AIMP2* | 5.600580273 | 1.95E-06 | 5.49E-06 | 0.012239623 |
| T2DM | mast_cell | *TFR2* | 5.714383164 | 1.12E-07 | 5.03E-07 | 0.000706226 |
| T2DM | mast_cell | *USP42* | 4.866866544 | 4.12E-09 | 7.78E-08 | 2.59E-05 |
| T2DM | mast_cell | *LSM5* | 5.235442004 | 0.000205314 | 0.000369014 | 1 |
| T2DM | mast_cell | *AP1S1* | 2.129250599 | 1.40E-08 | 1.19E-07 | 8.82E-05 |
| T2DM | mast_cell | *C1GALT1* | 3.887017245 | 1.98E-08 | 1.41E-07 | 0.000124628 |
| T2DM | mast_cell | *PLOD3* | 3.380520651 | 2.19E-08 | 1.50E-07 | 0.000137996 |
| T2DM | mast_cell | *PHF14* | 3.382923863 | 3.12E-06 | 8.30E-06 | 0.019605449 |
| T2DM | mast_cell | *CEP41* | 2.885181974 | 4.50E-05 | 9.20E-05 | 0.282868196 |
| T2DM | mast_cell | *ZNF862* | 3.390806589 | 6.31E-05 | 0.000124868 | 0.396704104 |
| T2DM | mast_cell | *MEST* | 0.973317164 | 0.00039954 | 0.000683698 | 1 |
| T2DM | mast_cell | *ACTR3C* | 2.053213713 | 3.21E-08 | 1.94E-07 | 0.000202 |
| T2DM | mast_cell | *POU6F2* | 2.751254688 | 4.59E-09 | 7.79E-08 | 2.88E-05 |
| T2DM | mast_cell | *TSPAN13* | 6.350040622 | 3.69E-08 | 2.16E-07 | 0.000232034 |
| T2DM | mast_cell | *RARRES2* | 3.940121337 | 4.40E-07 | 1.52E-06 | 0.002764596 |
| T2DM | mast_cell | *TMEM176B* | 1.770329437 | 4.46E-07 | 1.54E-06 | 0.002804553 |
| T2DM | mast_cell | *MRPL32* | 2.699620112 | 1.42E-08 | 1.19E-07 | 8.93E-05 |
| T2DM | mast_cell | *URGCP* | 2.674131124 | 4.98E-09 | 7.84E-08 | 3.13E-05 |
| T2DM | mast_cell | *GCK* | 73.3765265 | 1.14E-08 | 1.09E-07 | 7.18E-05 |
| T2DM | mast_cell | *BCL7B* | 2.403699149 | 1.81E-05 | 3.96E-05 | 0.113693097 |
| T2DM | mast_cell | *TBL2* | 1.080493948 | 0.000627866 | 0.001039335 | 1 |
| T2DM | mast_cell | *CLIP2* | 5.33534724 | 1.81E-07 | 7.33E-07 | 0.001134999 |
| T2DM | mast_cell | *LIMK1* | 4.332374846 | 3.38E-09 | 7.78E-08 | 2.13E-05 |
| T2DM | mast_cell | *SPATA6L* | 1.971899842 | 0.030546927 | 0.039281761 | 1 |
| T2DM | mast_cell | *SLC1A1* | 1.530232762 | 5.80E-06 | 1.44E-05 | 0.036453402 |
| T2DM | mast_cell | *FKTN* | 3.936635221 | 4.07E-05 | 8.39E-05 | 0.255981866 |
| T2DM | mast_cell | *NMRK1* | 3.366121575 | 2.96E-08 | 1.83E-07 | 0.000185806 |
| T2DM | mast_cell | *TMEM245* | 1.694070815 | 0.035359973 | 0.045258174 | 1 |

| | | | | | | |
|---|---|---|---|---|---|---|
| T2DM | mast_cell | *MEGF9* | 2.029438115 | 8.96E-07 | 2.80E-06 | 0.00563398 |
| T2DM | mast_cell | *TRIM14* | 2.885767709 | 8.79E-05 | 0.000169047 | 0.552836395 |
| T2DM | mast_cell | *OGN* | 2.854596253 | 0.000484395 | 0.000815802 | 1 |
| T2DM | mast_cell | *ECM2* | 2.451117329 | 9.73E-05 | 0.000185882 | 0.611921999 |
| T2DM | mast_cell | *PTGR1* | 1.305241235 | 0.003349583 | 0.00498198 | 1 |
| T2DM | mast_cell | *TNFSF8* | 3.961647625 | 2.02E-08 | 1.42E-07 | 0.000127083 |
| T2DM | mast_cell | *PLGRKT* | -1.320789709 | 0.001970298 | 0.003038328 | 1 |
| T2DM | mast_cell | *KDM4C* | 2.952498504 | 0.000340637 | 0.000589157 | 1 |
| T2DM | mast_cell | *DOCK8* | 2.40869389 | 1.31E-06 | 3.86E-06 | 0.008224079 |
| T2DM | mast_cell | *ELAVL2* | 1.958791494 | 4.25E-05 | 8.75E-05 | 0.26742256 |
| T2DM | mast_cell | *RGP1* | 5.675657079 | 0.000224368 | 0.000400967 | 1 |
| T2DM | mast_cell | *MPDZ* | 3.59183418 | 3.11E-08 | 1.90E-07 | 0.000195621 |
| T2DM | mast_cell | *PTGDS* | 3.013176218 | 6.62E-09 | 8.54E-08 | 4.16E-05 |
| T2DM | mast_cell | *SHB* | -3.06943665 | 1.17E-06 | 3.48E-06 | 0.00733117 |
| T2DM | mast_cell | *DVL1* | 2.316768377 | 1.05E-08 | 1.05E-07 | 6.59E-05 |
| T2DM | mast_cell | *PDLIM1* | 2.595259498 | 6.78E-05 | 0.000133163 | 0.426253908 |
| T2DM | mast_cell | *CCNJ* | 3.105561588 | 0.007149889 | 0.010087825 | 1 |
| T2DM | mast_cell | *ATRNL1* | 2.203263433 | 2.71E-05 | 5.77E-05 | 0.170440067 |
| T2DM | mast_cell | *HPS1* | 4.429144063 | 8.64E-08 | 4.10E-07 | 0.000543168 |
| T2DM | mast_cell | *DNMBP* | 4.001885097 | 3.40E-08 | 2.03E-07 | 0.000214005 |
| T2DM | mast_cell | *TRDMT1* | 2.615426684 | 0.027182684 | 0.035171338 | 1 |
| T2DM | mast_cell | *DDX50* | 4.290371565 | 8.36E-06 | 1.98E-05 | 0.052561807 |
| T2DM | mast_cell | *MAPK8* | 3.297015632 | 3.78E-05 | 7.84E-05 | 0.237498205 |
| T2DM | mast_cell | *ATE1* | 2.353193058 | 0.001133511 | 0.001800501 | 1 |
| T2DM | mast_cell | *NSMCE4A* | 4.133797884 | 4.22E-08 | 2.39E-07 | 0.000265564 |
| T2DM | mast_cell | *PLEKHA1* | 3.852566838 | 3.48E-09 | 7.78E-08 | 2.19E-05 |
| T2DM | mast_cell | *SPOCK2* | 1.332066039 | 0.020190901 | 0.026707383 | 1 |
| T2DM | mast_cell | *MICU1* | 3.984535669 | 2.42E-07 | 9.26E-07 | 0.001520933 |
| T2DM | mast_cell | *CCSER2* | 3.683673535 | 5.63E-08 | 2.97E-07 | 0.000353834 |
| T2DM | mast_cell | *LIPA* | 2.45523983 | 0.000361161 | 0.000622599 | 1 |
| T2DM | mast_cell | *SFXN3* | 4.64194878 | 5.27E-08 | 2.83E-07 | 0.000331226 |
| T2DM | mast_cell | *KAZALD1* | 3.07446291 | 1.25E-06 | 3.69E-06 | 0.007845852 |
| T2DM | mast_cell | *TNKS2* | 3.766429382 | 9.42E-06 | 2.20E-05 | 0.059194541 |
| T2DM | mast_cell | *ANKRD26* | 2.281807727 | 0.000272289 | 0.000478982 | 1 |
| T2DM | mast_cell | *ACBD5* | 4.841004745 | 9.11E-07 | 2.84E-06 | 0.005728548 |
| T2DM | mast_cell | *MTPAP* | 3.559620547 | 1.50E-09 | 6.92E-08 | 9.42E-06 |
| T2DM | mast_cell | *MAP3K8* | 2.745382895 | 0.002447117 | 0.003718884 | 1 |
| T2DM | mast_cell | *SHOC2* | 3.895188639 | 3.09E-07 | 1.13E-06 | 0.001941723 |
| T2DM | mast_cell | *TFAM* | 4.208381369 | 0.000627598 | 0.001039335 | 1 |
| T2DM | mast_cell | *MLLT6* | 4.664120234 | 1.00E-05 | 2.33E-05 | 0.062976795 |
| T2DM | mast_cell | *PSMB3* | 4.263627193 | 8.83E-11 | 1.63E-08 | 5.55E-07 |
| T2DM | mast_cell | *CASC3* | 3.118210243 | 0.001281038 | 0.002024098 | 1 |
| T2DM | mast_cell | *RAPGEFL1* | 2.009860889 | 1.71E-05 | 3.75E-05 | 0.107366058 |
| T2DM | mast_cell | *RGS9* | 7.716792817 | 0.000180139 | 0.000327635 | 1 |
| T2DM | mast_cell | *WNT3* | 2.554959068 | 3.91E-07 | 1.38E-06 | 0.002455512 |
| T2DM | mast_cell | *ASPA* | 4.473700133 | 4.58E-07 | 1.58E-06 | 0.002878132 |
| T2DM | mast_cell | *SEPT4* | 1.904964051 | 0.001075879 | 0.001714154 | 1 |
| T2DM | mast_cell | *TRIM37* | 4.300286513 | 2.33E-05 | 5.03E-05 | 0.146699467 |
| T2DM | mast_cell | *PNPO* | 2.533034055 | 2.32E-05 | 5.01E-05 | 0.145963132 |
| T2DM | mast_cell | *RPS6KB1* | 4.33632726 | 8.08E-06 | 1.93E-05 | 0.050815287 |

| | | | | | | |
|---|---|---|---|---|---|---|
| T2DM | mast_cell | *TRIM16L* | 1.244421505 | 7.08E-07 | 2.28E-06 | 0.004451572 |
| T2DM | mast_cell | *CDK5RAP3* | -2.152243987 | 7.39E-05 | 0.000144071 | 0.464773632 |
| T2DM | mast_cell | *CBX1* | 2.862931871 | 0.001295599 | 0.002045563 | 1 |
| T2DM | mast_cell | *RECQL5* | 2.286807483 | 0.000260869 | 0.000460274 | 1 |
| T2DM | mast_cell | *PIGL* | 3.677386517 | 2.08E-08 | 1.44E-07 | 0.000130473 |
| T2DM | mast_cell | *GALK1* | -3.255369395 | 2.66E-05 | 5.68E-05 | 0.167455853 |
| T2DM | mast_cell | *MED13* | 2.356976275 | 0.000921271 | 0.001483615 | 1 |
| T2DM | mast_cell | *HOXB6* | 2.376075087 | 0.000428953 | 0.000731442 | 1 |
| T2DM | mast_cell | *RNF167* | 5.755004984 | 1.84E-06 | 5.23E-06 | 0.011573698 |
| T2DM | mast_cell | *GOSR1* | 4.148126182 | 1.19E-06 | 3.54E-06 | 0.007478313 |
| T2DM | mast_cell | *DRG2* | 6.603696304 | 9.33E-08 | 4.33E-07 | 0.000586809 |
| T2DM | mast_cell | *AKAP10* | 3.446175346 | 3.53E-06 | 9.30E-06 | 0.022220393 |
| T2DM | mast_cell | *SMARCD2* | 4.167497831 | 0.000474613 | 0.00080126 | 1 |
| T2DM | mast_cell | *ICAM2* | 4.157226926 | 8.51E-10 | 6.08E-08 | 5.35E-06 |
| T2DM | mast_cell | *SYNGR2* | -1.872049119 | 0.014796813 | 0.019911722 | 1 |
| T2DM | mast_cell | *UTP6* | 4.626556144 | 2.56E-06 | 6.97E-06 | 0.016118813 |
| T2DM | mast_cell | *DDX5* | 1.568905342 | 0.001543849 | 0.002411473 | 1 |
| T2DM | mast_cell | *C17orf75* | 3.910513261 | 1.69E-08 | 1.29E-07 | 0.000106158 |
| T2DM | mast_cell | *CYTH1* | 3.791048288 | 8.53E-08 | 4.06E-07 | 0.00053606 |
| T2DM | mast_cell | *NAGLU* | 4.537187772 | 9.51E-08 | 4.40E-07 | 0.000597595 |
| T2DM | mast_cell | *HSD17B1* | -1.447635012 | 1.87E-08 | 1.37E-07 | 0.000117256 |
| T2DM | mast_cell | *EZH1* | 3.620059031 | 1.82E-09 | 7.58E-08 | 1.14E-05 |
| T2DM | mast_cell | *DLX4* | 16.10444353 | 1.71E-08 | 1.30E-07 | 0.000107779 |
| T2DM | mast_cell | *VAT1* | 5.318007516 | 9.81E-08 | 4.52E-07 | 0.000616534 |
| T2DM | mast_cell | *ALOX12* | 3.08655099 | 8.42E-08 | 4.03E-07 | 0.000529177 |
| T2DM | mast_cell | *ABCC3* | 1.340846728 | 0.000517573 | 0.000868422 | 1 |
| T2DM | mast_cell | *MPP2* | 3.045601892 | 1.00E-08 | 1.04E-07 | 6.30E-05 |
| T2DM | mast_cell | *EFTUD2* | 4.230273421 | 1.73E-05 | 3.80E-05 | 0.108894323 |
| T2DM | mast_cell | *EFNB3* | 18.79458476 | 1.87E-07 | 7.55E-07 | 0.001175028 |
| T2DM | mast_cell | *FAM20A* | 2.796567398 | 3.92E-09 | 7.78E-08 | 2.47E-05 |
| T2DM | mast_cell | *YWHAE* | 3.787763458 | 8.17E-06 | 1.94E-05 | 0.051341407 |
| T2DM | mast_cell | *DPH1* | 4.214409866 | 3.25E-09 | 7.78E-08 | 2.04E-05 |
| T2DM | mast_cell | *DHRS7B* | 2.892506505 | 0.004313538 | 0.006287784 | 1 |
| T2DM | mast_cell | *RCVRN* | -2.376007229 | 5.28E-07 | 1.78E-06 | 0.003321046 |
| T2DM | mast_cell | *SLC9A3R1* | 2.661669716 | 0.023082748 | 0.030271431 | 1 |
| T2DM | mast_cell | *MYH3* | 2.759826906 | 0.02056471 | 0.027144727 | 1 |
| T2DM | mast_cell | *TMEM97* | 2.311948478 | 2.31E-07 | 8.94E-07 | 0.001452116 |
| T2DM | mast_cell | *RAB34* | 2.35050431 | 2.48E-07 | 9.45E-07 | 0.001561153 |
| T2DM | mast_cell | *SLAIN2* | 3.057808015 | 0.000302062 | 0.000527664 | 1 |
| T2DM | mast_cell | *OCIAD1* | 3.314434728 | 2.33E-06 | 6.41E-06 | 0.014618225 |
| T2DM | mast_cell | *KIAA1211* | 1.114180966 | 0.000298415 | 0.000521873 | 1 |
| T2DM | mast_cell | *AREG* | -0.94668196 | 0.005618074 | 0.008064117 | 1 |
| T2DM | mast_cell | *UBE2D3* | 3.21959852 | 7.87E-05 | 0.000152633 | 0.49498755 |
| T2DM | mast_cell | *NDUFC1* | 4.797477483 | 0.000576137 | 0.000961299 | 1 |
| T2DM | mast_cell | *TBC1D9* | 3.605829395 | 1.02E-07 | 4.66E-07 | 0.000641127 |
| T2DM | mast_cell | *CPE* | 4.239184403 | 1.12E-07 | 5.03E-07 | 0.000706194 |
| T2DM | mast_cell | *WFS1* | 3.885478952 | 1.03E-12 | 5.00E-10 | 6.49E-09 |
| T2DM | mast_cell | *GRPEL1* | 7.09158981 | 7.56E-05 | 0.000147089 | 0.475391037 |
| T2DM | mast_cell | *AADAT* | 2.441172586 | 1.39E-08 | 1.18E-07 | 8.74E-05 |
| T2DM | mast_cell | *CPZ* | 32.16567057 | 1.56E-08 | 1.23E-07 | 9.80E-05 |

| | | | | | | |
|---|---|---|---|---|---|---|
| T2DM | mast_cell | *TRIM2* | 2.059196246 | 0.000111203 | 0.000210202 | 0.699131116 |
| T2DM | mast_cell | *SLC2A9* | 2.529030189 | 2.17E-10 | 2.96E-08 | 1.36E-06 |
| T2DM | mast_cell | *SH3D19* | 2.152728412 | 6.29E-08 | 3.25E-07 | 0.000395425 |
| T2DM | mast_cell | *STIM2* | 3.804629334 | 3.15E-06 | 8.36E-06 | 0.019779311 |
| T2DM | mast_cell | *RAPGEF2* | -1.161412849 | 0.014989136 | 0.020153271 | 1 |
| T2DM | mast_cell | *LRP2BP* | 2.632824208 | 8.75E-06 | 2.06E-05 | 0.054997373 |
| T2DM | mast_cell | *UFSP2* | 4.384585663 | 5.61E-05 | 0.000112526 | 0.35299251 |
| T2DM | mast_cell | *KLF3* | 3.951689033 | 5.03E-08 | 2.73E-07 | 0.000315952 |
| T2DM | mast_cell | *KLHL5* | 1.867425809 | 0.002891796 | 0.004344259 | 1 |
| T2DM | mast_cell | *FAM149A* | 1.860081687 | 6.87E-08 | 3.46E-07 | 0.000432099 |
| T2DM | mast_cell | *UGDH* | 3.127752807 | 4.03E-09 | 7.78E-08 | 2.54E-05 |
| T2DM | mast_cell | *DDX25* | 3.637464075 | 2.74E-05 | 5.83E-05 | 0.17229479 |
| T2DM | mast_cell | *HTATIP2* | 1.422242888 | 0.012055305 | 0.01641579 | 1 |
| T2DM | mast_cell | *CTSC* | 2.50565241 | 0.000222144 | 0.000397445 | 1 |
| T2DM | mast_cell | *SC5D* | 5.828897665 | 1.49E-08 | 1.20E-07 | 9.38E-05 |
| T2DM | mast_cell | *C11orf63* | 3.209031499 | 4.04E-09 | 7.78E-08 | 2.54E-05 |
| T2DM | mast_cell | *DTX4* | 5.144820509 | 1.95E-08 | 1.40E-07 | 0.000122523 |
| T2DM | mast_cell | *PUS3* | -2.032399014 | 0.004560237 | 0.006625886 | 1 |
| T2DM | mast_cell | *DCPS* | 4.888643554 | 8.27E-09 | 9.23E-08 | 5.20E-05 |
| T2DM | mast_cell | *FOXRED1* | 2.587814412 | 2.09E-06 | 5.83E-06 | 0.013134641 |
| T2DM | mast_cell | *NRXN2* | 11.35255017 | 5.95E-05 | 0.000118494 | 0.374202933 |
| T2DM | mast_cell | *MS4A4A* | 1.670625043 | 1.36E-06 | 4.00E-06 | 0.008575409 |
| T2DM | mast_cell | *CCDC86* | 18.67537543 | 4.41E-08 | 2.47E-07 | 0.000277509 |
| T2DM | mast_cell | *PRPF19* | 2.821039018 | 2.44E-09 | 7.78E-08 | 1.54E-05 |
| T2DM | mast_cell | *HPX* | 2.134110421 | 1.61E-05 | 3.55E-05 | 0.101035836 |
| T2DM | mast_cell | *FOLR3* | -0.651007025 | 0.037519193 | 0.047836781 | 1 |
| T2DM | mast_cell | *PANX1* | 3.305803712 | 3.69E-07 | 1.31E-06 | 0.002322137 |
| T2DM | mast_cell | *EIF4G2* | 4.181077589 | 5.37E-06 | 1.34E-05 | 0.033786224 |
| T2DM | mast_cell | *CBL* | 3.449831666 | 6.22E-07 | 2.05E-06 | 0.003908645 |
| T2DM | mast_cell | *MADD* | 3.850329908 | 0.000632078 | 0.001044931 | 1 |
| T2DM | mast_cell | *NAA40* | 4.154705952 | 1.00E-07 | 4.60E-07 | 0.000630712 |
| T2DM | mast_cell | *CARS* | 4.524555557 | 8.85E-10 | 6.08E-08 | 5.57E-06 |
| T2DM | mast_cell | *SLC35F2* | 1.468745504 | 3.16E-05 | 6.64E-05 | 0.198537908 |
| T2DM | mast_cell | *C11orf21* | -2.970055637 | 5.55E-07 | 1.86E-06 | 0.00348833 |
| T2DM | mast_cell | *ELMOD1* | 3.74086989 | 0.009835945 | 0.013608844 | 1 |
| T2DM | mast_cell | *RPS13* | 2.558031573 | 0.004315831 | 0.006288211 | 1 |
| T2DM | mast_cell | *AIP* | 3.798434985 | 4.44E-05 | 9.09E-05 | 0.279268158 |
| T2DM | mast_cell | *NDUFS8* | 5.683052869 | 2.63E-08 | 1.69E-07 | 0.000165254 |
| T2DM | mast_cell | *TCIRG1* | 3.317088343 | 0.003885923 | 0.005704133 | 1 |
| T2DM | mast_cell | *GTF2H1* | 2.335012493 | 0.006465211 | 0.009196105 | 1 |
| T2DM | mast_cell | *PTPN5* | 1.397238234 | 3.86E-05 | 8.00E-05 | 0.242615117 |
| T2DM | mast_cell | *PPFIBP1* | 3.357532234 | 1.88E-06 | 5.32E-06 | 0.011830104 |
| T2DM | mast_cell | *PRDM4* | 2.752624407 | 0.014180304 | 0.019114831 | 1 |
| T2DM | mast_cell | *COQ5* | 0.905086596 | 4.87E-07 | 1.66E-06 | 0.00306193 |
| T2DM | mast_cell | *ASIC1* | 1.812322506 | 0.000603489 | 0.001003208 | 1 |
| T2DM | mast_cell | *CAPRIN2* | 3.864376525 | 0.000153456 | 0.000283175 | 0.964775885 |
| T2DM | mast_cell | *CYP27B1* | 12.91152089 | 1.85E-08 | 1.36E-07 | 0.000116414 |
| T2DM | mast_cell | *ACSS3* | 0.702327121 | 0.009864291 | 0.013644619 | 1 |
| T2DM | mast_cell | *GLI1* | 7.783376646 | 4.15E-09 | 7.78E-08 | 2.61E-05 |
| T2DM | mast_cell | *LTA4H* | 4.522560014 | 1.67E-08 | 1.28E-07 | 0.000105095 |

| | | | | | | |
|---|---|---|---|---|---|---|
| T2DM | mast_cell | *WNT5B* | 2.295524259 | 2.19E-09 | 7.78E-08 | 1.38E-05 |
| T2DM | mast_cell | *TRPV4* | 2.295945582 | 3.63E-07 | 1.29E-06 | 0.00228293 |
| T2DM | mast_cell | *ITFG2* | 3.748893431 | 4.31E-05 | 8.84E-05 | 0.270802133 |
| T2DM | mast_cell | *PRR4* | 2.821788457 | 1.11E-06 | 3.34E-06 | 0.006967492 |
| T2DM | mast_cell | *PARP11* | 4.75423959 | 1.09E-07 | 4.92E-07 | 0.00068795 |
| T2DM | mast_cell | *ARPC3* | 4.725318634 | 1.16E-07 | 5.15E-07 | 0.000729545 |
| T2DM | mast_cell | *GPN3* | 6.092300141 | 2.15E-06 | 5.98E-06 | 0.013498909 |
| T2DM | mast_cell | *VPS29* | 3.838895371 | 6.36E-05 | 0.000125841 | 0.400047266 |
| T2DM | mast_cell | *AKAP3* | 1.497854946 | 3.25E-05 | 6.83E-05 | 0.204391742 |
| T2DM | mast_cell | *KCNA1* | 3.732954303 | 1.27E-07 | 5.48E-07 | 0.00079595 |
| T2DM | mast_cell | *CREBL2* | 4.058411622 | 1.81E-10 | 2.59E-08 | 1.14E-06 |
| T2DM | mast_cell | *NAA25* | 3.634060186 | 4.76E-05 | 9.65E-05 | 0.299085141 |
| T2DM | mast_cell | *SCNN1A* | 3.227590951 | 1.05E-06 | 3.21E-06 | 0.006609429 |
| T2DM | mast_cell | *LTBR* | 2.448601449 | 0.001899951 | 0.002935609 | 1 |
| T2DM | mast_cell | *CDK2AP1* | 1.684925511 | 4.50E-07 | 1.55E-06 | 0.002828005 |
| T2DM | mast_cell | *MGP* | 2.596648226 | 6.33E-08 | 3.26E-07 | 0.000397835 |
| T2DM | mast_cell | *RASAL1* | -4.420683587 | 4.64E-06 | 1.18E-05 | 0.029173849 |
| T2DM | mast_cell | *ARHGDIB* | -1.769876904 | 0.000277786 | 0.000487969 | 1 |
| T2DM | mast_cell | *EIF2B1* | 5.248601785 | 1.14E-06 | 3.41E-06 | 0.007163816 |
| T2DM | mast_cell | *ENDOU* | -3.093274008 | 3.97E-08 | 2.29E-07 | 0.000249797 |
| T2DM | mast_cell | *STX2* | 2.553114577 | 4.04E-05 | 8.33E-05 | 0.254058767 |
| T2DM | mast_cell | *MDM1* | 3.124723006 | 6.07E-08 | 3.16E-07 | 0.000381689 |
| T2DM | mast_cell | *NUP107* | 2.476162626 | 0.007006354 | 0.009900865 | 1 |
| T2DM | mast_cell | *CNOT2* | 3.394354953 | 4.24E-06 | 1.09E-05 | 0.026663842 |
| T2DM | mast_cell | *COPS7A* | 2.053165485 | 0.026837728 | 0.034739303 | 1 |
| T2DM | mast_cell | *GNPTAB* | -1.388789555 | 7.94E-05 | 0.000153623 | 0.498968965 |
| T2DM | mast_cell | *SPSB2* | 3.644827976 | 3.29E-08 | 1.99E-07 | 0.000207086 |
| T2DM | mast_cell | *ATN1* | 3.484051158 | 7.73E-08 | 3.79E-07 | 0.000486187 |
| T2DM | mast_cell | *C12orf57* | 3.698730743 | 1.32E-05 | 2.98E-05 | 0.08295306 |
| T2DM | mast_cell | *NT5DC3* | 2.258030858 | 0.000160121 | 0.000294351 | 1 |
| T2DM | mast_cell | *ST8SIA1* | -0.320523889 | 1.49E-07 | 6.26E-07 | 0.000937725 |
| T2DM | mast_cell | *C2CD5* | 3.438577928 | 8.96E-05 | 0.000172087 | 0.563069773 |
| T2DM | mast_cell | *PHC1* | 4.328515418 | 1.01E-09 | 6.18E-08 | 6.36E-06 |
| T2DM | mast_cell | *COL12A1* | 3.515779507 | 4.64E-06 | 1.18E-05 | 0.029181205 |
| T2DM | mast_cell | *TDP2* | 3.661379023 | 0.001829945 | 0.002834409 | 1 |
| T2DM | mast_cell | *RWDD1* | 2.662362294 | 3.52E-07 | 1.26E-06 | 0.002214765 |
| T2DM | mast_cell | *RSPH4A* | 1.192609186 | 0.0006623 | 0.001091163 | 1 |
| T2DM | mast_cell | *SMIM8* | 3.626816533 | 6.36E-06 | 1.56E-05 | 0.039983728 |
| T2DM | mast_cell | *MCM9* | 3.591115179 | 0.000150787 | 0.000278577 | 0.947998076 |
| T2DM | mast_cell | *FAM184A* | 2.821772873 | 9.76E-09 | 1.03E-07 | 6.13E-05 |
| T2DM | mast_cell | *RNGTT* | 3.836109724 | 1.50E-07 | 6.29E-07 | 0.000942481 |
| T2DM | mast_cell | *GABRR2* | 1.869428518 | 1.13E-07 | 5.04E-07 | 0.000708875 |
| T2DM | mast_cell | *TPD52L1* | 2.118619399 | 0.000353606 | 0.000610244 | 1 |
| T2DM | mast_cell | *UST* | 2.367981685 | 1.26E-08 | 1.13E-07 | 7.92E-05 |
| T2DM | mast_cell | *MTRF1L* | 2.495000231 | 0.00025377 | 0.000449044 | 1 |
| T2DM | mast_cell | *TULP1* | 9.564930482 | 1.17E-08 | 1.10E-07 | 7.37E-05 |
| T2DM | mast_cell | *SLC26A8* | 1.554136819 | 3.88E-05 | 8.04E-05 | 0.244108162 |
| T2DM | mast_cell | *RNF8* | 2.17461222 | 0.005437539 | 0.007812113 | 1 |
| T2DM | mast_cell | *PHACTR1* | 3.694777906 | 9.43E-09 | 1.00E-07 | 5.93E-05 |
| T2DM | mast_cell | *MDGA1* | 4.027770997 | 0.000973847 | 0.001561882 | 1 |

| | | | | | | |
|---|---|---|---|---|---|---|
| T2DM | mast_cell | *CD83* | -1.671410543 | 6.09E-07 | 2.02E-06 | 0.00383161 |
| T2DM | mast_cell | *MDN1* | 2.38053871 | 0.002839411 | 0.004271315 | 1 |
| T2DM | mast_cell | *SAYSD1* | 4.689534053 | 2.74E-09 | 7.78E-08 | 1.72E-05 |
| T2DM | mast_cell | *CAP2* | 1.848279402 | 6.89E-06 | 1.67E-05 | 0.043294929 |
| T2DM | mast_cell | *GPR63* | -1.584471433 | 0.000616517 | 0.001022163 | 1 |
| T2DM | mast_cell | *ASCC3* | 3.427409584 | 1.63E-05 | 3.60E-05 | 0.102699059 |
| T2DM | mast_cell | *GPLD1* | 1.300690317 | 2.77E-08 | 1.75E-07 | 0.000174096 |
| T2DM | mast_cell | *ALDH5A1* | 1.894080792 | 0.022077759 | 0.028983686 | 1 |
| T2DM | mast_cell | *VNN1* | 0.891625349 | 0.030840182 | 0.039650762 | 1 |
| T2DM | mast_cell | *VNN2* | 3.791935673 | 1.60E-09 | 7.17E-08 | 1.00E-05 |
| T2DM | mast_cell | *ACOT13* | 4.76758204 | 1.37E-05 | 3.08E-05 | 0.086170146 |
| T2DM | mast_cell | *C6orf62* | 3.399735123 | 5.05E-08 | 2.74E-07 | 0.000317638 |
| T2DM | mast_cell | *B3GAT2* | -2.606753015 | 0.003891712 | 0.005711296 | 1 |
| T2DM | mast_cell | *PEX7* | 1.24008727 | 0.005364772 | 0.007716385 | 1 |
| T2DM | mast_cell | *SLC16A10* | -1.044557918 | 0.035630791 | 0.045595519 | 1 |
| T2DM | mast_cell | *PACRG* | 3.110062115 | 1.56E-08 | 1.23E-07 | 9.82E-05 |
| T2DM | mast_cell | *TFEB* | 5.414695092 | 5.44E-09 | 8.00E-08 | 3.42E-05 |
| T2DM | mast_cell | *FAM120B* | 1.813405986 | 0.008212579 | 0.01149432 | 1 |
| T2DM | mast_cell | *TBP* | 3.700223737 | 4.06E-09 | 7.78E-08 | 2.55E-05 |
| T2DM | mast_cell | *GUCA1B* | 3.297652502 | 0.021094201 | 0.027785301 | 1 |
| T2DM | mast_cell | *DNPH1* | 1.697704831 | 0.000333089 | 0.000578011 | 1 |
| T2DM | mast_cell | *DUSP22* | 4.590464241 | 6.04E-06 | 1.49E-05 | 0.037984529 |
| T2DM | mast_cell | *EXOC2* | 3.97180336 | 1.44E-06 | 4.18E-06 | 0.009031838 |
| T2DM | mast_cell | *TMEM30A* | 4.23123078 | 1.68E-07 | 6.92E-07 | 0.001057593 |
| T2DM | mast_cell | *SENP6* | 2.974681627 | 0.000133919 | 0.000249319 | 0.84195071 |
| T2DM | mast_cell | *TTK* | 2.058288217 | 0.01080342 | 0.014816995 | 1 |
| T2DM | mast_cell | *BTN2A1* | 2.500583698 | 6.54E-05 | 0.000128991 | 0.41096418 |
| T2DM | mast_cell | *LAMA4* | 1.753773274 | 3.30E-06 | 8.73E-06 | 0.020764597 |
| T2DM | mast_cell | *CLIC5* | 2.005340935 | 2.43E-07 | 9.30E-07 | 0.001528568 |
| T2DM | mast_cell | *FBRSL1* | 2.80137283 | 0.015640604 | 0.020979833 | 1 |
| T2DM | mast_cell | *LY86* | 4.392338496 | 5.55E-09 | 8.00E-08 | 3.49E-05 |
| T2DM | mast_cell | *TBX18* | 2.739029773 | 4.78E-05 | 9.69E-05 | 0.300436257 |
| T2DM | mast_cell | *NUDT12* | 3.08855354 | 0.020136335 | 0.02665203 | 1 |
| T2DM | mast_cell | *CEP72* | -2.326008019 | 0.000158484 | 0.000291621 | 0.996389412 |
| T2DM | mast_cell | *C7* | 4.46028432 | 1.01E-06 | 3.09E-06 | 0.006330448 |
| T2DM | mast_cell | *PAPD7* | 4.054244657 | 5.42E-06 | 1.35E-05 | 0.034087445 |
| T2DM | mast_cell | *GHR* | 1.418231983 | 6.87E-07 | 2.23E-06 | 0.004317972 |
| T2DM | mast_cell | *NME5* | 2.342610167 | 3.93E-05 | 8.13E-05 | 0.247116218 |
| T2DM | mast_cell | *NNT* | 1.025006248 | 0.033710634 | 0.043208717 | 1 |
| T2DM | mast_cell | *PFDN1* | 2.007768171 | 0.005669763 | 0.008136453 | 1 |
| T2DM | mast_cell | *LOX* | 5.312257178 | 8.48E-07 | 2.68E-06 | 0.005332942 |
| T2DM | mast_cell | *APBB3* | 1.9146046 | 1.22E-06 | 3.61E-06 | 0.00764115 |
| T2DM | mast_cell | *TMCO6* | 2.725895328 | 1.75E-07 | 7.15E-07 | 0.001101631 |
| T2DM | mast_cell | *SPARC* | 2.62678107 | 0.000240572 | 0.000427252 | 1 |
| T2DM | mast_cell | *PCDHB6* | 4.47541362 | 3.58E-06 | 9.39E-06 | 0.022499263 |
| T2DM | mast_cell | *PDE8B* | 2.46238957 | 1.37E-08 | 1.17E-07 | 8.60E-05 |
| T2DM | mast_cell | *CLK4* | 3.89407949 | 2.04E-05 | 4.44E-05 | 0.128515366 |
| T2DM | mast_cell | *HAVCR1* | 1.687480309 | 1.52E-06 | 4.39E-06 | 0.009529794 |
| T2DM | mast_cell | *ARSB* | 2.458025149 | 7.55E-05 | 0.00014689 | 0.474601813 |
| T2DM | mast_cell | *CLINT1* | 3.878438322 | 9.92E-09 | 1.04E-07 | 6.23E-05 |

| | | | | | | |
|---|---|---|---|---|---|---|
| T2DM | mast_cell | *TTC1* | 4.271914216 | 5.27E-06 | 1.32E-05 | 0.033129084 |
| T2DM | mast_cell | *MSH3* | 1.005614951 | 0.036051451 | 0.046058823 | 1 |
| T2DM | mast_cell | *RASGRF2* | 3.205788264 | 2.23E-08 | 1.51E-07 | 0.000139901 |
| T2DM | mast_cell | *POLR3G* | 2.882089864 | 0.000272272 | 0.000478982 | 1 |
| T2DM | mast_cell | *DROSHA* | 3.317475249 | 6.01E-06 | 1.48E-05 | 0.037777121 |
| T2DM | mast_cell | *SLC27A6* | 4.560550225 | 1.55E-08 | 1.23E-07 | 9.76E-05 |
| T2DM | mast_cell | *TARS* | 4.142511074 | 2.04E-09 | 7.78E-08 | 1.28E-05 |
| T2DM | mast_cell | *LNPEP* | 3.348667251 | 4.55E-09 | 7.78E-08 | 2.86E-05 |
| T2DM | mast_cell | *SLC12A7* | 4.247005563 | 1.22E-05 | 2.78E-05 | 0.076806048 |
| T2DM | mast_cell | *IL4* | 2.530330004 | 0.016701119 | 0.022326161 | 1 |
| T2DM | mast_cell | *GNPDA1* | 5.513071165 | 8.33E-08 | 4.00E-07 | 0.000523841 |
| T2DM | mast_cell | *SKP1* | 2.631426422 | 0.014686306 | 0.019767245 | 1 |
| T2DM | mast_cell | *PPWD1* | 1.343950128 | 0.00478051 | 0.006920347 | 1 |
| T2DM | mast_cell | *TRIM23* | 3.672026006 | 0.000254204 | 0.000449559 | 1 |
| T2DM | mast_cell | *TRAPPC13* | 3.238819784 | 1.40E-05 | 3.14E-05 | 0.088111516 |
| T2DM | mast_cell | *C9* | 2.013347849 | 2.23E-06 | 6.18E-06 | 0.014025316 |
| T2DM | mast_cell | *SMAD5* | 3.085930807 | 0.001436286 | 0.002254101 | 1 |
| T2DM | mast_cell | *HMGXB3* | 4.171649663 | 1.62E-08 | 1.26E-07 | 0.00010213 |
| T2DM | mast_cell | *ATP6V0E1* | 3.442892538 | 0.000130977 | 0.000244419 | 0.823453016 |
| T2DM | mast_cell | *BNIP1* | 0.849272905 | 0.015527634 | 0.020841639 | 1 |
| T2DM | mast_cell | *CNTN3* | 3.300264743 | 4.96E-09 | 7.84E-08 | 3.12E-05 |
| T2DM | mast_cell | *ACTR8* | 5.21493963 | 1.58E-08 | 1.24E-07 | 9.93E-05 |
| T2DM | mast_cell | *HGD* | 6.683914955 | 3.65E-09 | 7.78E-08 | 2.29E-05 |
| T2DM | mast_cell | *NPHP3* | 1.069903158 | 0.000163749 | 0.00030023 | 1 |
| T2DM | mast_cell | *NIT2* | 3.414583931 | 2.53E-08 | 1.64E-07 | 0.000159307 |
| T2DM | mast_cell | *RBP1* | 2.857834465 | 4.17E-07 | 1.46E-06 | 0.0026241 |
| T2DM | mast_cell | *WNT5A* | 2.814410812 | 0.017772967 | 0.023678459 | 1 |
| T2DM | mast_cell | *HYAL1* | 1.457190892 | 0.000261238 | 0.000460702 | 1 |
| T2DM | mast_cell | *CYB561D2* | 1.41613764 | 0.034333435 | 0.043971305 | 1 |
| T2DM | mast_cell | *C3orf14* | 1.692442037 | 3.60E-06 | 9.44E-06 | 0.022625601 |
| T2DM | mast_cell | *IFT57* | 5.190574442 | 4.75E-08 | 2.61E-07 | 0.000298392 |
| T2DM | mast_cell | *IQCG* | 4.747268255 | 4.82E-07 | 1.65E-06 | 0.003030435 |
| T2DM | mast_cell | *NCBP2* | 5.469753367 | 2.42E-08 | 1.59E-07 | 0.000152446 |
| T2DM | mast_cell | *PLXNA1* | 2.281209841 | 3.96E-06 | 1.03E-05 | 0.024915472 |
| T2DM | mast_cell | *ABTB1* | 4.571271666 | 2.06E-08 | 1.44E-07 | 0.000129664 |
| T2DM | mast_cell | *PLSCR4* | 2.779996208 | 7.57E-07 | 2.42E-06 | 0.004756609 |
| T2DM | mast_cell | *HEMK1* | 3.42228495 | 5.34E-06 | 1.34E-05 | 0.033584454 |
| T2DM | mast_cell | *WDR48* | 3.438885343 | 4.17E-07 | 1.46E-06 | 0.002620742 |
| T2DM | mast_cell | *COMMD2* | 3.96740944 | 2.84E-06 | 7.63E-06 | 0.017842816 |
| T2DM | mast_cell | *GORASP1* | 6.039635691 | 6.92E-08 | 3.47E-07 | 0.00043491 |
| T2DM | mast_cell | *ABCC5* | 3.021356789 | 4.71E-06 | 1.19E-05 | 0.029621796 |
| T2DM | mast_cell | *ARHGEF26* | 3.145867292 | 8.77E-05 | 0.000168777 | 0.55156321 |
| T2DM | mast_cell | *KLHL24* | 1.847231923 | 0.010024643 | 0.013833391 | 1 |
| T2DM | mast_cell | *PLCH1* | 7.141163379 | 1.08E-09 | 6.34E-08 | 6.78E-06 |
| T2DM | mast_cell | *ZBTB47* | 3.418710251 | 9.82E-10 | 6.17E-08 | 6.17E-06 |
| T2DM | mast_cell | *NKTR* | 2.386596865 | 0.001672075 | 0.002603351 | 1 |
| T2DM | mast_cell | *CLCN2* | 2.26054571 | 8.32E-09 | 9.26E-08 | 5.23E-05 |
| T2DM | mast_cell | *NEK4* | 4.477759908 | 3.40E-08 | 2.03E-07 | 0.000213581 |
| T2DM | mast_cell | *MOB1A* | 5.003756147 | 1.57E-06 | 4.53E-06 | 0.009869732 |
| T2DM | mast_cell | *LMAN2L* | 2.545987005 | 0.001502434 | 0.002349702 | 1 |

| | | | | | | |
|---|---|---|---|---|---|---|
| T2DM | mast_cell | *PIKFYVE* | 2.3027685 | 0.007520454 | 0.01058216 | 1 |
| T2DM | mast_cell | *KCNIP3* | 2.350465566 | 6.17E-09 | 8.15E-08 | 3.88E-05 |
| T2DM | mast_cell | *FAHD2A* | 3.885855806 | 1.74E-08 | 1.31E-07 | 0.000109693 |
| T2DM | mast_cell | *NCL* | 3.651795545 | 2.81E-07 | 1.05E-06 | 0.001769782 |
| T2DM | mast_cell | *ACTR1B* | 4.740109196 | 1.65E-08 | 1.27E-07 | 0.000103864 |
| T2DM | mast_cell | *SLC35F5* | 3.060437675 | 0.000156697 | 0.000288816 | 0.985150931 |
| T2DM | mast_cell | *ACTR3* | 3.915303119 | 2.13E-06 | 5.94E-06 | 0.013404919 |
| T2DM | mast_cell | *STEAP3* | 0.839761207 | 0.004517545 | 0.006568411 | 1 |
| T2DM | mast_cell | *TFCP2L1* | 3.031555808 | 9.06E-09 | 9.71E-08 | 5.69E-05 |
| T2DM | mast_cell | *TP53I3* | 4.346175458 | 2.44E-05 | 5.24E-05 | 0.15356504 |
| T2DM | mast_cell | *STAM2* | 4.117803506 | 4.28E-07 | 1.49E-06 | 0.002693279 |
| T2DM | mast_cell | *GPD2* | 2.705480842 | 0.00279423 | 0.004209759 | 1 |
| T2DM | mast_cell | *CENPA* | 5.725802025 | 2.35E-05 | 5.07E-05 | 0.148021357 |
| T2DM | mast_cell | *TANC1* | 2.080077164 | 0.000436861 | 0.000743516 | 1 |
| T2DM | mast_cell | *ITGB6* | -2.091949331 | 3.43E-06 | 9.03E-06 | 0.021538394 |
| T2DM | mast_cell | *PPM1G* | 2.253952335 | 0.014128682 | 0.019053416 | 1 |
| T2DM | mast_cell | *PCSK4* | 2.203234394 | 0.001666583 | 0.002595445 | 1 |
| T2DM | mast_cell | *GRB14* | 2.693383466 | 0.024943188 | 0.032521324 | 1 |
| T2DM | mast_cell | *CLIP4* | 3.468380752 | 5.28E-05 | 0.000106181 | 0.331817087 |
| T2DM | mast_cell | *SPTBN1* | 2.104726176 | 0.000117012 | 0.000220388 | 0.735654384 |
| T2DM | mast_cell | *HTRA2* | 4.768130758 | 6.02E-06 | 1.49E-05 | 0.037838839 |
| T2DM | mast_cell | *GALNT3* | -1.204822825 | 2.00E-06 | 5.63E-06 | 0.012594627 |
| T2DM | mast_cell | *POLE4* | 4.841137396 | 4.47E-09 | 7.78E-08 | 2.81E-05 |
| T2DM | mast_cell | *MRPL19* | 2.826597143 | 0.000451717 | 0.000765691 | 1 |
| T2DM | mast_cell | *LANCL1* | 4.481788832 | 6.19E-07 | 2.04E-06 | 0.003890619 |
| T2DM | mast_cell | *WDR75* | 4.508417429 | 8.04E-08 | 3.90E-07 | 0.000505188 |
| T2DM | mast_cell | *EFEMP1* | 2.408682282 | 2.41E-06 | 6.58E-06 | 0.015139586 |
| T2DM | mast_cell | *GLS* | 3.079571017 | 3.63E-05 | 7.55E-05 | 0.228056488 |
| T2DM | mast_cell | *DNAH6* | 0.421992314 | 0.000622362 | 0.00103131 | 1 |
| T2DM | mast_cell | *GGCX* | 1.780314526 | 0.00024162 | 0.000428993 | 1 |
| T2DM | mast_cell | *GNLY* | 1.802603107 | 8.24E-09 | 9.23E-08 | 5.18E-05 |
| T2DM | mast_cell | *SF3B1* | 3.289535115 | 2.70E-05 | 5.76E-05 | 0.170040488 |
| T2DM | mast_cell | *ST3GAL5* | 3.504769519 | 2.02E-06 | 5.67E-06 | 0.012726204 |
| T2DM | mast_cell | *PLCD4* | 5.255330382 | 1.33E-08 | 1.15E-07 | 8.39E-05 |
| T2DM | mast_cell | *CHMP3* | -3.329696126 | 3.15E-06 | 8.37E-06 | 0.019807818 |
| T2DM | mast_cell | *ZNF142* | 2.604375179 | 0.004903982 | 0.007089293 | 1 |
| T2DM | mast_cell | *WNT6* | -5.307009733 | 0.006795725 | 0.009622685 | 1 |
| T2DM | mast_cell | *MLPH* | -0.31470157 | 1.21E-07 | 5.31E-07 | 0.000760769 |
| T2DM | mast_cell | *HDLBP* | 2.412285061 | 0.00063114 | 0.001043656 | 1 |
| T2DM | mast_cell | *PASK* | 2.657108066 | 0.000726673 | 0.001189118 | 1 |
| T2DM | mast_cell | *STK25* | 4.074553183 | 2.65E-07 | 9.97E-07 | 0.001667989 |
| T2DM | mast_cell | *TPO* | 1.143090678 | 0.000334626 | 0.000579717 | 1 |
| T2DM | mast_cell | *TAF1B* | 3.084371177 | 2.82E-08 | 1.78E-07 | 0.000177556 |
| T2DM | mast_cell | *STRN* | 2.82736644 | 0.003361252 | 0.004994609 | 1 |
| T2DM | mast_cell | *PRKD3* | 3.311766329 | 0.000105698 | 0.000200641 | 0.664524577 |
| T2DM | mast_cell | *RMDN2* | 4.675219092 | 4.53E-08 | 2.52E-07 | 0.000284878 |
| T2DM | mast_cell | *SDC1* | -2.16280217 | 2.55E-05 | 5.44E-05 | 0.16016311 |
| T2DM | mast_cell | *SOS1* | 2.287841563 | 3.03E-05 | 6.40E-05 | 0.190744185 |
| T2DM | mast_cell | *COX7A2L* | 4.186078199 | 8.98E-09 | 9.66E-08 | 5.65E-05 |
| T2DM | mast_cell | *ORC4* | 2.38058067 | 0.006716457 | 0.009523313 | 1 |

| | | | | | | |
|---|---|---|---|---|---|---|
| T2DM | mast_cell | *THADA* | 3.548744573 | 1.53E-06 | 4.41E-06 | 0.009590829 |
| T2DM | mast_cell | *ARID3A* | 3.613671616 | 0.002149818 | 0.003290142 | 1 |
| T2DM | mast_cell | *CD207* | -7.419470435 | 2.23E-07 | 8.72E-07 | 0.001405125 |
| T2DM | mast_cell | *GRIN3B* | 2.799266196 | 1.42E-05 | 3.17E-05 | 0.089124764 |
| T2DM | mast_cell | *VAX2* | -4.76423196 | 0.002276288 | 0.003472707 | 1 |
| T2DM | mast_cell | *MSH6* | 3.836727054 | 0.000797344 | 0.001296663 | 1 |
| T2DM | mast_cell | *PLEKHA3* | 4.34505622 | 5.77E-09 | 8.14E-08 | 3.63E-05 |
| T2DM | mast_cell | *BCL9* | 4.802288468 | 6.49E-08 | 3.32E-07 | 0.000408009 |
| T2DM | mast_cell | *DHCR24* | 2.728354656 | 8.50E-09 | 9.37E-08 | 5.34E-05 |
| T2DM | mast_cell | *MORN1* | 2.844631186 | 0.000521843 | 0.00087512 | 1 |
| T2DM | mast_cell | *GPX7* | -1.205170825 | 0.000733596 | 0.001198576 | 1 |
| T2DM | mast_cell | *SCP2* | 3.439748256 | 1.48E-05 | 3.29E-05 | 0.092923576 |
| T2DM | mast_cell | *TPSG1* | -0.544690985 | 3.89E-08 | 2.25E-07 | 0.000244833 |
| T2DM | mast_cell | *RALGPS2* | 3.554392475 | 0.000125187 | 0.00023445 | 0.787049021 |
| T2DM | mast_cell | *FAM20B* | 4.0809539 | 7.85E-09 | 9.16E-08 | 4.94E-05 |
| T2DM | mast_cell | *TMEM59* | 3.936045451 | 1.04E-05 | 2.40E-05 | 0.065165004 |
| T2DM | mast_cell | *WRAP73* | 4.430147438 | 0.006004744 | 0.008589721 | 1 |
| T2DM | mast_cell | *MRPL37* | 3.690362928 | 3.59E-10 | 4.05E-08 | 2.26E-06 |
| T2DM | mast_cell | *ICMT* | 2.745990515 | 1.01E-07 | 4.61E-07 | 0.0006321 |
| T2DM | mast_cell | *QSOX1* | 4.848441636 | 4.17E-08 | 2.37E-07 | 0.000262013 |
| T2DM | mast_cell | *PARK7* | 4.179459799 | 1.08E-08 | 1.06E-07 | 6.79E-05 |
| T2DM | mast_cell | *KCNC4* | -1.719905591 | 0.005979718 | 0.008555869 | 1 |
| T2DM | mast_cell | *EDEM3* | 3.654603539 | 5.83E-08 | 3.06E-07 | 0.00036652 |
| T2DM | mast_cell | *RAP1A* | 2.230193476 | 0.009003539 | 0.012527276 | 1 |
| T2DM | mast_cell | *CAPZA1* | 2.694162389 | 0.000667255 | 0.001098459 | 1 |
| T2DM | mast_cell | *DOCK7* | 3.29954342 | 0.000101452 | 0.000193164 | 0.637826351 |
| T2DM | mast_cell | *FBXO6* | 2.090936926 | 4.45E-08 | 2.48E-07 | 0.000279511 |
| T2DM | mast_cell | *DNAJC6* | 2.898490799 | 1.33E-07 | 5.70E-07 | 0.00083603 |
| T2DM | mast_cell | *LEPR* | 1.983213363 | 0.018928028 | 0.025179964 | 1 |
| T2DM | mast_cell | *MIIP* | 3.194878614 | 5.58E-07 | 1.87E-06 | 0.003508592 |
| T2DM | mast_cell | *SMG7* | 3.890113367 | 2.37E-06 | 6.50E-06 | 0.014887826 |
| T2DM | mast_cell | *NCF2* | 2.751358771 | 4.08E-09 | 7.78E-08 | 2.57E-05 |
| T2DM | mast_cell | *OLFML3* | 2.751561458 | 1.18E-08 | 1.10E-07 | 7.45E-05 |
| T2DM | mast_cell | *TNNI3K* | 3.434300742 | 1.39E-08 | 1.18E-07 | 8.73E-05 |
| T2DM | mast_cell | *CFHR3* | 1.499166171 | 0.001211637 | 0.001919749 | 1 |
| T2DM | mast_cell | *TTF2* | 4.512688391 | 0.000213492 | 0.000382618 | 1 |
| T2DM | mast_cell | *WARS2* | 1.85909123 | 0.000475093 | 0.000801424 | 1 |
| T2DM | mast_cell | *OSCP1* | -1.667951944 | 0.035774735 | 0.045742476 | 1 |
| T2DM | mast_cell | *GNPAT* | 2.292988283 | 0.008466505 | 0.011831278 | 1 |
| T2DM | mast_cell | *C1orf109* | 3.931820041 | 0.000208142 | 0.000373668 | 1 |
| T2DM | mast_cell | *TBCE* | 2.464754372 | 0.011925969 | 0.016253754 | 1 |
| T2DM | mast_cell | *MTR* | 3.642117323 | 8.33E-07 | 2.63E-06 | 0.00523966 |
| T2DM | mast_cell | *ACADM* | 3.893854255 | 2.30E-08 | 1.54E-07 | 0.000144859 |
| T2DM | mast_cell | *PADI2* | -0.506762483 | 1.01E-07 | 4.63E-07 | 0.000636575 |
| T2DM | mast_cell | *CTBS* | 3.167072252 | 1.08E-06 | 3.27E-06 | 0.006782572 |
| T2DM | mast_cell | *KLHL12* | 3.776666875 | 1.08E-07 | 4.87E-07 | 0.000677757 |
| T2DM | mast_cell | *ZNHIT6* | 2.097716704 | 0.001276036 | 0.002016701 | 1 |
| T2DM | mast_cell | *RBBP5* | 3.248353897 | 5.49E-09 | 8.00E-08 | 3.45E-05 |
| T2DM | mast_cell | *KIF17* | -2.826972542 | 0.000229274 | 0.000409037 | 1 |
| T2DM | mast_cell | *CD46* | 3.3177094 | 1.36E-05 | 3.05E-05 | 0.085205288 |

| | | | | | | |
|---|---|---|---|---|---|---|
| T2DM | mast_cell | *PRDX1* | 3.036708643 | 0.00324586 | 0.004840304 | 1 |
| T2DM | mast_cell | *FAAH* | 0.881197899 | 2.98E-05 | 6.30E-05 | 0.187522633 |
| T2DM | mast_cell | *NSUN4* | 2.570741761 | 0.001343019 | 0.002117459 | 1 |
| T2DM | mast_cell | *TMED5* | 4.239464548 | 6.28E-07 | 2.06E-06 | 0.003947363 |
| T2DM | mast_cell | *DR1* | 5.578774022 | 1.83E-07 | 7.40E-07 | 0.001149641 |
| T2DM | mast_cell | *CNN3* | 4.394100421 | 4.80E-08 | 2.63E-07 | 0.000301884 |
| T2DM | mast_cell | *PRRC2C* | 2.456153151 | 0.002189286 | 0.003346472 | 1 |
| T2DM | mast_cell | *ABCD3* | 3.814304152 | 1.43E-08 | 1.19E-07 | 8.96E-05 |
| T2DM | mast_cell | *VAMP4* | 4.173821205 | 2.61E-08 | 1.68E-07 | 0.00016424 |
| T2DM | mast_cell | *PTBP2* | 2.586560969 | 2.61E-06 | 7.08E-06 | 0.01638071 |
| T2DM | mast_cell | *TNFSF4* | 1.576986236 | 1.30E-05 | 2.94E-05 | 0.081734446 |
| T2DM | mast_cell | *DARS2* | 3.449746659 | 6.08E-07 | 2.01E-06 | 0.003823487 |
| T2DM | mast_cell | *DIEXF* | 3.326337409 | 1.22E-06 | 3.61E-06 | 0.007640245 |
| T2DM | mast_cell | *SERPINC1* | 2.304982659 | 5.14E-07 | 1.74E-06 | 0.003233435 |
| T2DM | mast_cell | *RCAN3* | -2.015712423 | 0.000149257 | 0.000276157 | 0.938380578 |
| T2DM | mast_cell | *RCOR3* | 3.800466831 | 1.84E-06 | 5.23E-06 | 0.011586341 |
| T2DM | mast_cell | *MTFR1L* | 3.272462544 | 0.012358336 | 0.016795689 | 1 |
| T2DM | mast_cell | *NSL1* | 2.253522141 | 0.014984174 | 0.020150909 | 1 |
| T2DM | mast_cell | *RPA2* | 5.272589624 | 2.92E-07 | 1.08E-06 | 0.001834494 |
| T2DM | mast_cell | *SLC5A9* | 7.21242824 | 0.000143481 | 0.000266096 | 0.902066791 |
| T2DM | mast_cell | *OSBPL9* | 2.381511272 | 0.005405057 | 0.007770774 | 1 |
| T2DM | mast_cell | *TXNDC12* | 4.989592685 | 2.87E-07 | 1.07E-06 | 0.001806811 |
| T2DM | mast_cell | *ESYT2* | 2.038347193 | 0.006371374 | 0.009070839 | 1 |
| T2DM | mast_cell | *CD3EAP* | 1.376371447 | 0.005481003 | 0.007872759 | 1 |
| T2DM | mast_cell | *COLEC11* | -2.389107228 | 9.26E-05 | 0.000177525 | 0.582283274 |
| T2DM | mast_cell | *KMT2A* | 3.350718985 | 9.32E-05 | 0.000178582 | 0.585927648 |
| T2DM | mast_cell | *IFT46* | 3.649390931 | 1.88E-07 | 7.59E-07 | 0.001183689 |
| T2DM | mast_cell | *CAMSAP2* | 3.578756156 | 2.30E-07 | 8.92E-07 | 0.001445709 |
| T2DM | mast_cell | *ATF6* | 3.688008274 | 3.69E-06 | 9.65E-06 | 0.023182853 |
| T2DM | mast_cell | *LRMP* | -1.065316314 | 0.025851347 | 0.033586985 | 1 |
| T2DM | mast_cell | *ATP10B* | 3.305277472 | 1.83E-08 | 1.35E-07 | 0.000114839 |
| T2DM | mast_cell | *SPCS2* | 3.044583444 | 0.004148017 | 0.00605915 | 1 |
| T2DM | mast_cell | *ELOVL4* | 2.00894946 | 1.81E-06 | 5.14E-06 | 0.011358845 |
| T2DM | mast_cell | *FILIP1* | 1.703414491 | 3.21E-07 | 1.17E-06 | 0.002019528 |
| T2DM | mast_cell | *TCF21* | 8.205125046 | 0.010084356 | 0.013906635 | 1 |
| T2DM | mast_cell | *MED28* | 3.133642577 | 2.64E-05 | 5.63E-05 | 0.165973801 |
| T2DM | mast_cell | *SLC16A7* | 3.021921929 | 1.58E-08 | 1.24E-07 | 9.94E-05 |
| T2DM | mast_cell | *FOXO3* | 3.23974559 | 1.08E-05 | 2.50E-05 | 0.068024103 |
| T2DM | mast_cell | *TGIF2* | -1.758429313 | 1.81E-06 | 5.15E-06 | 0.011381184 |
| T2DM | mast_cell | *ABCG2* | -3.250084816 | 0.00533233 | 0.007671478 | 1 |
| T2DM | mast_cell | *SPP1* | 3.602055032 | 3.93E-07 | 1.39E-06 | 0.002468905 |
| T2DM | mast_cell | *FAM47E-STBL* | 1.26533926 | 7.36E-07 | 2.37E-06 | 0.004627267 |
| T2DM | mast_cell | *MFSD1* | 4.316410421 | 4.40E-07 | 1.52E-06 | 0.002764799 |
| T2DM | mast_cell | *RAB3GAP2* | 3.84174289 | 3.52E-07 | 1.26E-06 | 0.002210307 |
| T2DM | mast_cell | *UBN1* | 3.617153228 | 2.12E-06 | 5.92E-06 | 0.013353313 |
| T2DM | mast_cell | *UCHL3* | -3.580391238 | 0.00061353 | 0.001017747 | 1 |
| T2DM | mast_cell | *WDR35* | 4.394284647 | 2.20E-07 | 8.62E-07 | 0.001384308 |
| T2DM | mast_cell | *DNAH7* | -1.231510612 | 0.000365475 | 0.00062969 | 1 |
| T2DM | mast_cell | *NDUFB3* | 3.752753204 | 2.73E-05 | 5.81E-05 | 0.171714655 |
| T2DM | mast_cell | *C2orf40* | 2.371354645 | 3.10E-06 | 8.26E-06 | 0.019486752 |

| | | | | | | |
|---|---|---|---|---|---|---|
| T2DM | mast_cell | *CPSF3* | 5.090213271 | 7.76E-09 | 9.13E-08 | 4.88E-05 |
| T2DM | mast_cell | *PIGZ* | 0.796926644 | 0.010033136 | 0.013842074 | 1 |
| T2DM | mast_cell | *SENP5* | 3.862955979 | 1.01E-05 | 2.35E-05 | 0.063495759 |
| T2DM | mast_cell | *PTBP3* | 2.429660384 | 0.011451641 | 0.015654809 | 1 |
| T2DM | mast_cell | *FKBP15* | 4.325888377 | 2.05E-08 | 1.43E-07 | 0.000128907 |
| T2DM | mast_cell | *CTNNAL1* | 1.153534741 | 0.000540194 | 0.000903725 | 1 |
| T2DM | mast_cell | *WDR34* | 1.466221544 | 1.87E-06 | 5.30E-06 | 0.011757828 |
| T2DM | mast_cell | *TRIM32* | 4.546451571 | 2.22E-09 | 7.78E-08 | 1.39E-05 |
| T2DM | mast_cell | *PHF19* | 4.467248346 | 2.51E-08 | 1.64E-07 | 0.000157858 |
| T2DM | mast_cell | *BSPRY* | -0.338191473 | 8.27E-08 | 3.99E-07 | 0.000520067 |
| T2DM | mast_cell | *PPP6C* | 3.669083211 | 7.42E-08 | 3.67E-07 | 0.000466488 |
| T2DM | mast_cell | *HDHD3* | -0.411052711 | 0.030268535 | 0.038963611 | 1 |
| T2DM | mast_cell | *RBM18* | 3.763130764 | 7.56E-09 | 9.02E-08 | 4.75E-05 |
| T2DM | mast_cell | *HSDL2* | 1.654975304 | 9.60E-05 | 0.000183647 | 0.603464891 |
| T2DM | mast_cell | *INVS* | 3.185988471 | 1.16E-05 | 2.66E-05 | 0.073038867 |
| T2DM | mast_cell | *KDSR* | 3.907597239 | 0.000322291 | 0.000560045 | 1 |
| T2DM | mast_cell | *DCAF4* | 1.979720059 | 9.92E-10 | 6.17E-08 | 6.24E-06 |
| T2DM | mast_cell | *IFI27L2* | 2.260799169 | 2.64E-06 | 7.17E-06 | 0.016612025 |
| T2DM | mast_cell | *DNAL1* | 2.039981946 | 7.50E-09 | 9.02E-08 | 4.71E-05 |
| T2DM | mast_cell | *IRF2BPL* | 6.78364006 | 3.19E-06 | 8.47E-06 | 0.020065104 |
| T2DM | mast_cell | *TTLL5* | 3.594185536 | 0.000109102 | 0.000206674 | 0.685925764 |
| T2DM | mast_cell | *FLVCR2* | 3.828929932 | 0.004333745 | 0.006308464 | 1 |
| T2DM | mast_cell | *PPP4R4* | 1.855375699 | 6.17E-06 | 1.52E-05 | 0.038815519 |
| T2DM | mast_cell | *ALDH6A1* | 1.610262514 | 2.06E-05 | 4.48E-05 | 0.12977014 |
| T2DM | mast_cell | *EIF2B2* | -1.072950122 | 0.026408083 | 0.034253686 | 1 |
| T2DM | mast_cell | *COQ6* | 5.979767714 | 1.78E-08 | 1.33E-07 | 0.000111794 |
| T2DM | mast_cell | *GPR75* | 8.366124564 | 7.75E-09 | 9.13E-08 | 4.87E-05 |
| T2DM | mast_cell | *KLHL29* | 3.292776616 | 7.79E-09 | 9.15E-08 | 4.90E-05 |
| T2DM | mast_cell | *ATAD2B* | 2.955796601 | 0.0004949 | 0.000832381 | 1 |
| T2DM | mast_cell | *ATL2* | 3.477887415 | 1.62E-08 | 1.26E-07 | 0.000102137 |
| T2DM | mast_cell | *CNRIP1* | -2.095715768 | 0.006087672 | 0.008698453 | 1 |
| T2DM | mast_cell | *EPCAM* | -3.396207008 | 2.55E-07 | 9.66E-07 | 0.00160205 |
| T2DM | mast_cell | *OGFRL1* | 3.472813328 | 1.90E-06 | 5.36E-06 | 0.01192459 |
| T2DM | mast_cell | *CUTC* | 3.785789148 | 7.99E-08 | 3.88E-07 | 0.000502312 |
| T2DM | mast_cell | *PYROXD2* | 0.830061438 | 0.002085987 | 0.003204846 | 1 |
| T2DM | mast_cell | *MXI1* | 3.935835028 | 5.08E-08 | 2.75E-07 | 0.000319128 |
| T2DM | mast_cell | *HELLS* | 2.113802438 | 0.029454349 | 0.037962176 | 1 |
| T2DM | mast_cell | *TCTN3* | 4.497358781 | 9.34E-07 | 2.90E-06 | 0.005870578 |
| T2DM | mast_cell | *WDR11* | 3.190007214 | 0.000175936 | 0.000320891 | 1 |
| T2DM | mast_cell | *C10orf76* | 3.329649349 | 6.79E-05 | 0.0001334 | 0.427148284 |
| T2DM | mast_cell | *KCNIP2* | 1.444351152 | 7.01E-07 | 2.27E-06 | 0.004409084 |
| T2DM | mast_cell | *C10orf95* | 1.332968728 | 0.006472423 | 0.00920428 | 1 |
| T2DM | mast_cell | *KANSL1* | 2.589884448 | 5.94E-05 | 0.000118257 | 0.373336223 |
| T2DM | mast_cell | *HOXB5* | 2.153562269 | 0.014569983 | 0.01961908 | 1 |
| T2DM | mast_cell | *CAAP1* | 2.187180643 | 0.021815686 | 0.028657588 | 1 |
| T2DM | mast_cell | *CD274* | -2.342613167 | 0.010388807 | 0.014285746 | 1 |
| T2DM | mast_cell | *NUP43* | 2.455043784 | 0.000417053 | 0.000711924 | 1 |
| T2DM | mast_cell | *LRP11* | 2.403538839 | 5.26E-05 | 0.000105829 | 0.330445347 |
| T2DM | mast_cell | *CCDC170* | 1.523210631 | 3.11E-06 | 8.28E-06 | 0.019541406 |
| T2DM | mast_cell | *CYSTM1* | 3.55774013 | 1.53E-05 | 3.40E-05 | 0.096482992 |

| | | | | | | |
|---|---|---|---|---|---|---|
| T2DM | mast_cell | *PCDHB10* | 1.622706267 | 0.000701831 | 0.001150563 | 1 |
| T2DM | mast_cell | *PCDHB14* | 3.163102778 | 6.80E-09 | 8.67E-08 | 4.28E-05 |
| T2DM | mast_cell | *SEC16B* | 4.582022765 | 0.000663203 | 0.001092078 | 1 |
| T2DM | mast_cell | *GORAB* | 4.803966092 | 7.49E-08 | 3.69E-07 | 0.000470673 |
| T2DM | mast_cell | *ACAT2* | 4.391798779 | 0.000197665 | 0.000356386 | 1 |
| T2DM | mast_cell | *SNX19* | 2.871766475 | 1.11E-06 | 3.34E-06 | 0.006979392 |
| T2DM | mast_cell | *MRC1* | 1.975697061 | 6.48E-09 | 8.40E-08 | 4.07E-05 |
| T2DM | mast_cell | *IQSEC3* | 7.690307973 | 1.14E-06 | 3.42E-06 | 0.007180651 |
| T2DM | mast_cell | *CCDC77* | 2.989094093 | 7.02E-06 | 1.70E-05 | 0.044135747 |
| T2DM | mast_cell | *ENOX1* | 3.680353088 | 8.91E-08 | 4.20E-07 | 0.000560332 |
| T2DM | mast_cell | *DNAJC15* | 4.543926427 | 3.69E-06 | 9.65E-06 | 0.023173363 |
| T2DM | mast_cell | *WBP4* | 3.967410592 | 9.84E-08 | 4.53E-07 | 0.000618584 |
| T2DM | mast_cell | *SMAD9* | 2.725650823 | 3.76E-09 | 7.78E-08 | 2.36E-05 |
| T2DM | mast_cell | *SIL1* | 3.598203437 | 2.31E-06 | 6.38E-06 | 0.014544461 |
| T2DM | mast_cell | *ZFP30* | 1.341920496 | 0.030505608 | 0.039244681 | 1 |
| T2DM | mast_cell | *ARL1* | 3.18945769 | 4.22E-07 | 1.47E-06 | 0.002652212 |
| T2DM | mast_cell | *APAF1* | 3.606003221 | 8.00E-08 | 3.88E-07 | 0.000502698 |
| T2DM | mast_cell | *ADRA1A* | 0.712151379 | 1.93E-08 | 1.39E-07 | 0.000121038 |
| T2DM | mast_cell | *EPHX2* | 1.546207097 | 5.67E-05 | 0.000113546 | 0.35664778 |
| T2DM | mast_cell | *ZNF706* | 1.944733546 | 0.027095752 | 0.035066075 | 1 |
| T2DM | mast_cell | *CRISPLD1* | 3.162973864 | 5.88E-09 | 8.14E-08 | 3.70E-05 |
| T2DM | mast_cell | *RDH10* | 1.458694972 | 0.002861143 | 0.004302322 | 1 |
| T2DM | mast_cell | *SCPEP1* | 0.779831026 | 0.010680127 | 0.01466069 | 1 |
| T2DM | mast_cell | *TBX4* | 2.564892028 | 0.005414824 | 0.007783036 | 1 |
| T2DM | mast_cell | *TEX14* | 2.875449838 | 0.003307217 | 0.004923627 | 1 |
| T2DM | mast_cell | *TRIM6* | -1.677114944 | 0.015245383 | 0.020489038 | 1 |
| T2DM | mast_cell | *CEP89* | 0.979229831 | 0.016386577 | 0.021938332 | 1 |
| T2DM | mast_cell | *PRB2* | 6.381141711 | 8.26E-05 | 0.000159512 | 0.519529273 |
| T2DM | mast_cell | *A1BG* | -1.958332871 | 1.55E-06 | 4.49E-06 | 0.009768405 |
| T2DM | mast_cell | *ZNF211* | 2.109492347 | 1.66E-06 | 4.77E-06 | 0.010450216 |
| T2DM | mast_cell | *PDZRN3* | 2.889076151 | 2.99E-08 | 1.84E-07 | 0.000187684 |
| T2DM | mast_cell | *SEC22A* | 3.429087569 | 2.39E-08 | 1.58E-07 | 0.000150494 |
| T2DM | mast_cell | *CSTA* | 1.862231353 | 5.17E-05 | 0.000104229 | 0.324778508 |
| T2DM | mast_cell | *B4GALT4* | 5.679884556 | 2.97E-05 | 6.28E-05 | 0.186732153 |
| T2DM | mast_cell | *CD80* | 2.838127177 | 1.28E-08 | 1.13E-07 | 8.04E-05 |
| T2DM | mast_cell | *PILRB* | 0.610356981 | 0.000178124 | 0.000324504 | 1 |
| T2DM | mast_cell | *TMEM39B* | 3.507057941 | 7.39E-08 | 3.67E-07 | 0.000464862 |
| T2DM | mast_cell | *RNF115* | 3.329312818 | 0.000161436 | 0.000296373 | 1 |
| T2DM | mast_cell | *POLR3GL* | 5.05596861 | 7.10E-09 | 8.80E-08 | 4.46E-05 |
| T2DM | mast_cell | *TNFSF10* | 3.054832591 | 0.002083565 | 0.003201998 | 1 |
| T2DM | mast_cell | *ZNF639* | 4.291558556 | 0.000920469 | 0.001483243 | 1 |
| T2DM | mast_cell | *LIAS* | 4.123690252 | 5.09E-05 | 0.000102891 | 0.320298198 |
| T2DM | mast_cell | *CLCC1* | -2.037827872 | 0.020161436 | 0.026679636 | 1 |
| T2DM | mast_cell | *GPSM2* | 4.054772152 | 1.17E-07 | 5.19E-07 | 0.00073676 |
| T2DM | mast_cell | *ACVR2A* | 3.42622362 | 0.000135795 | 0.000252661 | 0.853740881 |
| T2DM | mast_cell | *POLK* | 3.522264014 | 1.30E-05 | 2.95E-05 | 0.082039997 |
| T2DM | mast_cell | *FLT3* | 2.005165555 | 6.17E-09 | 8.15E-08 | 3.88E-05 |
| T2DM | mast_cell | *MTIF3* | 4.070690875 | 1.10E-06 | 3.33E-06 | 0.006942802 |
| T2DM | mast_cell | *RASL11A* | 4.577523505 | 0.000596106 | 0.00099251 | 1 |
| T2DM | mast_cell | *FMOD* | 2.088269982 | 2.17E-08 | 1.48E-07 | 0.000136686 |

| | | | | | | |
|---|---|---|---|---|---|---|
| T2DM | mast_cell | *SERAC1* | 3.814917589 | 0.003700712 | 0.005457747 | 1 |
| T2DM | mast_cell | *LDB3* | 1.895542942 | 6.81E-05 | 0.000133574 | 0.427836573 |
| T2DM | mast_cell | *OPN4* | -2.64536413 | 1.75E-06 | 5.01E-06 | 0.011027076 |
| T2DM | mast_cell | *FAM35A* | 1.594944701 | 0.031032603 | 0.039889996 | 1 |
| T2DM | mast_cell | *FAM213A* | 1.909043307 | 0.000163852 | 0.000300332 | 1 |
| T2DM | mast_cell | *NAA60* | 4.295161567 | 0.019189636 | 0.025468703 | 1 |
| T2DM | mast_cell | *ODF2L* | 3.324392763 | 6.98E-08 | 3.49E-07 | 0.000438625 |
| T2DM | mast_cell | *SPATA1* | 4.479997494 | 7.88E-07 | 2.51E-06 | 0.004951828 |
| T2DM | mast_cell | *LRRC39* | 1.267452274 | 0.000334215 | 0.000579166 | 1 |
| T2DM | mast_cell | *CCDC18* | 2.734072652 | 0.00199186 | 0.003069319 | 1 |
| T2DM | mast_cell | *RPAP2* | 3.476203955 | 6.60E-05 | 0.000130132 | 0.41524803 |
| T2DM | mast_cell | *BBS9* | 2.218880119 | 0.003735136 | 0.005502062 | 1 |
| T2DM | mast_cell | *PMS2* | 6.903674056 | 6.61E-10 | 5.76E-08 | 4.15E-06 |
| T2DM | mast_cell | *KIAA0087* | 3.645472313 | 3.78E-06 | 9.86E-06 | 0.023750054 |
| T2DM | mast_cell | *KLHL7* | 3.957554669 | 1.29E-08 | 1.13E-07 | 8.13E-05 |
| T2DM | mast_cell | *CBX3* | 2.076408783 | 0.03323615 | 0.042617923 | 1 |
| T2DM | mast_cell | *FAM126A* | 3.319188145 | 2.29E-06 | 6.34E-06 | 0.014422 |
| T2DM | mast_cell | *FKBP9* | 3.482421809 | 1.60E-07 | 6.64E-07 | 0.001005835 |
| T2DM | mast_cell | *CCZ1* | 3.388384379 | 1.97E-06 | 5.55E-06 | 0.012393199 |
| T2DM | mast_cell | *SMU1* | 4.851703086 | 1.22E-07 | 5.35E-07 | 0.000769769 |
| T2DM | mast_cell | *GLIPR2* | 3.378749979 | 7.42E-07 | 2.38E-06 | 0.004661862 |
| T2DM | mast_cell | *DCAF10* | 3.506211468 | 3.64E-08 | 2.13E-07 | 0.000228643 |
| T2DM | mast_cell | *TRIM24* | -1.841784589 | 0.000312062 | 0.000543622 | 1 |
| T2DM | mast_cell | *BICC1* | 1.470219454 | 1.76E-06 | 5.03E-06 | 0.011079331 |
| T2DM | mast_cell | *SLC25A16* | 1.870127992 | 1.75E-08 | 1.32E-07 | 0.000110286 |
| T2DM | mast_cell | *ZWINT* | 1.512547793 | 0.00171494 | 0.002665471 | 1 |
| T2DM | mast_cell | *RBM19* | 4.353657475 | 1.25E-08 | 1.13E-07 | 7.87E-05 |
| T2DM | mast_cell | *ACADS* | 4.397902456 | 1.57E-05 | 3.47E-05 | 0.098565074 |
| T2DM | mast_cell | *CCDC91* | 3.187029488 | 1.42E-06 | 4.14E-06 | 0.008923984 |
| T2DM | mast_cell | *EBPL* | 2.455020504 | 7.52E-06 | 1.80E-05 | 0.047269886 |
| T2DM | mast_cell | *NLN* | 2.898738383 | 2.13E-07 | 8.39E-07 | 0.001339033 |
| T2DM | mast_cell | *OPTN* | 2.402560318 | 0.000225757 | 0.000403219 | 1 |
| T2DM | mast_cell | *ATF1* | 4.55021644 | 1.83E-08 | 1.35E-07 | 0.000114915 |
| T2DM | mast_cell | *TSFM* | 4.632050672 | 4.56E-08 | 2.54E-07 | 0.000286675 |
| T2DM | mast_cell | *ARHGAP9* | 4.470296368 | 5.97E-08 | 3.12E-07 | 0.000375281 |
| T2DM | mast_cell | *SPATS2* | 3.845178905 | 1.76E-06 | 5.02E-06 | 0.011059688 |
| T2DM | mast_cell | *ORMDL2* | 3.577384619 | 1.72E-08 | 1.30E-07 | 0.000107891 |
| T2DM | mast_cell | *PDE1B* | 3.395269095 | 1.38E-05 | 3.11E-05 | 0.087018241 |
| T2DM | mast_cell | *IKZF4* | 2.771137245 | 5.18E-07 | 1.75E-06 | 0.003258499 |
| T2DM | mast_cell | *SARDH* | -4.450832866 | 1.88E-06 | 5.31E-06 | 0.011792361 |
| T2DM | mast_cell | *STIL* | 5.0432669 | 6.51E-07 | 2.13E-06 | 0.004094435 |
| T2DM | mast_cell | *TTC21B* | 3.815613792 | 2.78E-05 | 5.91E-05 | 0.174937083 |
| T2DM | mast_cell | *ACVR1C* | 2.42721794 | 1.49E-09 | 6.92E-08 | 9.34E-06 |
| T2DM | mast_cell | *SLC36A1* | -0.739437141 | 0.019088419 | 0.025355777 | 1 |
| T2DM | mast_cell | *PLA2G12A* | 2.802489469 | 0.01693754 | 0.02261336 | 1 |
| T2DM | mast_cell | *C4BPB* | 1.559522753 | 5.71E-05 | 0.000114254 | 0.35921367 |
| T2DM | mast_cell | *RAB38* | -0.963838143 | 1.72E-07 | 7.03E-07 | 0.001078912 |
| T2DM | mast_cell | *AGO2* | 4.321231842 | 7.43E-07 | 2.38E-06 | 0.004668187 |
| T2DM | mast_cell | *CKS2* | 4.019959324 | 7.31E-07 | 2.35E-06 | 0.00459589 |
| T2DM | mast_cell | *CHPF* | 1.329688109 | 0.006114473 | 0.008731006 | 1 |

| | | | | | | |
|---|---|---|---|---|---|---|
| T2DM | mast_cell | *DNPEP* | 5.53590028 | 2.03E-08 | 1.43E-07 | 0.000127559 |
| T2DM | mast_cell | *MOGAT1* | 3.728113188 | 5.36E-06 | 1.34E-05 | 0.03371197 |
| T2DM | mast_cell | *FAM210B* | 3.895841756 | 8.65E-07 | 2.72E-06 | 0.005435632 |
| T2DM | mast_cell | *SNX21* | 3.312412663 | 4.95E-09 | 7.84E-08 | 3.11E-05 |
| T2DM | mast_cell | *SDC4* | 2.005457289 | 7.62E-08 | 3.74E-07 | 0.000479352 |
| T2DM | mast_cell | *TOX2* | -0.610152557 | 0.003342059 | 0.004971965 | 1 |
| T2DM | mast_cell | *ARFGEF2* | 3.92851425 | 4.00E-06 | 1.03E-05 | 0.025134197 |
| T2DM | mast_cell | *CDH26* | 3.426096227 | 7.70E-09 | 9.13E-08 | 4.84E-05 |
| T2DM | mast_cell | *STX16* | 4.020920648 | 1.48E-08 | 1.20E-07 | 9.29E-05 |
| T2DM | mast_cell | *RNF114* | 3.486632747 | 2.85E-07 | 1.06E-06 | 0.001794355 |
| T2DM | mast_cell | *NEURL2* | 1.081051039 | 0.020275573 | 0.026802466 | 1 |
| T2DM | mast_cell | *FASTKD3* | 5.196657822 | 6.82E-05 | 0.000133808 | 0.428853681 |
| T2DM | mast_cell | *PEPD* | 3.438556402 | 2.28E-05 | 4.93E-05 | 0.143505053 |
| T2DM | mast_cell | *CHST8* | -1.214582857 | 2.65E-07 | 9.96E-07 | 0.001665433 |
| T2DM | mast_cell | *STAMBP* | 4.483496418 | 7.33E-06 | 1.76E-05 | 0.046104274 |
| T2DM | mast_cell | *ATP8A1* | 1.994838565 | 0.001373451 | 0.002162506 | 1 |
| T2DM | mast_cell | *HIF3A* | 2.575812199 | 4.06E-06 | 1.05E-05 | 0.025549451 |
| T2DM | mast_cell | *ZNF45* | 4.011871861 | 3.32E-09 | 7.78E-08 | 2.09E-05 |
| T2DM | mast_cell | *BTN2A2* | -0.784659808 | 0.000141272 | 0.000262232 | 0.888179875 |
| T2DM | mast_cell | *MRS2* | 2.955357214 | 2.32E-05 | 5.01E-05 | 0.146017675 |
| T2DM | mast_cell | *SNRPC* | 1.413541034 | 0.014434742 | 0.019445302 | 1 |
| T2DM | mast_cell | *HIST1H1D* | -1.168940862 | 4.36E-07 | 1.51E-06 | 0.002743231 |
| T2DM | mast_cell | *NQO2* | 4.375334785 | 1.06E-05 | 2.45E-05 | 0.066547542 |
| T2DM | mast_cell | *ZNF391* | 0.586181072 | 0.005944716 | 0.008509661 | 1 |
| T2DM | mast_cell | *RPS10* | 3.317112539 | 1.37E-08 | 1.17E-07 | 8.64E-05 |
| T2DM | mast_cell | *HIST1H2BJ* | 2.510754081 | 2.33E-05 | 5.03E-05 | 0.146727192 |
| T2DM | mast_cell | *MED20* | 2.060330189 | 0.002529833 | 0.003835316 | 1 |
| T2DM | mast_cell | *TBCC* | -2.285814975 | 0.00319413 | 0.004771084 | 1 |
| T2DM | mast_cell | *TCP11* | -1.979649841 | 2.64E-07 | 9.95E-07 | 0.001661415 |
| T2DM | mast_cell | *KLHDC3* | 3.029038908 | 3.84E-08 | 2.23E-07 | 0.000241588 |
| T2DM | mast_cell | *KLHL31* | 0.979652785 | 7.44E-07 | 2.39E-06 | 0.004679562 |
| T2DM | mast_cell | *COL21A1* | 0.935168601 | 5.53E-08 | 2.93E-07 | 0.000347534 |
| T2DM | mast_cell | *GLO1* | 2.94529262 | 0.007605187 | 0.010696602 | 1 |
| T2DM | mast_cell | *CPNE5* | -0.34865647 | 5.77E-08 | 3.03E-07 | 0.000362973 |
| T2DM | mast_cell | *RIOK1* | 3.580371756 | 2.31E-07 | 8.94E-07 | 0.001451359 |
| T2DM | mast_cell | *RPP40* | 3.354797124 | 1.13E-05 | 2.60E-05 | 0.071037394 |
| T2DM | mast_cell | *NUP153* | 3.860175298 | 2.39E-08 | 1.58E-07 | 0.000150377 |
| T2DM | mast_cell | *EEF1E1* | 7.462002179 | 0.000599919 | 0.00099833 | 1 |
| T2DM | mast_cell | *EREG* | 6.062320791 | 1.53E-08 | 1.21E-07 | 9.61E-05 |
| T2DM | mast_cell | *CNOT1* | 3.833648219 | 4.84E-08 | 2.65E-07 | 0.000304369 |
| T2DM | mast_cell | *BBS2* | 2.033659992 | 0.000363408 | 0.000626301 | 1 |
| T2DM | mast_cell | *MT1G* | 17.89911649 | 0.000627823 | 0.001039335 | 1 |
| T2DM | mast_cell | *GOT2* | 6.314549861 | 5.41E-07 | 1.82E-06 | 0.003399051 |
| T2DM | mast_cell | *CLYBL* | 4.314231737 | 3.19E-06 | 8.46E-06 | 0.020033621 |
| T2DM | mast_cell | *KIF25* | 6.666146826 | 4.04E-09 | 7.78E-08 | 2.54E-05 |
| T2DM | mast_cell | *GRK4* | 0.983066572 | 1.14E-05 | 2.62E-05 | 0.071862389 |
| T2DM | mast_cell | *MRPS7* | 3.897370711 | 0.003047758 | 0.00456329 | 1 |
| T2DM | mast_cell | *GGA3* | 4.70980388 | 4.35E-05 | 8.91E-05 | 0.273292278 |
| T2DM | mast_cell | *NUP85* | 2.905227725 | 0.003443145 | 0.005107847 | 1 |
| T2DM | mast_cell | *MIF4GD* | 3.219248383 | 8.51E-09 | 9.37E-08 | 5.35E-05 |

| | | | | | | |
|---|---|---|---|---|---|---|
| T2DM | mast_cell | *NT5C* | 5.74750812 | 1.90E-08 | 1.38E-07 | 0.000119463 |
| T2DM | mast_cell | *DDX31* | 2.406905611 | 0.000185435 | 0.000336169 | 1 |
| T2DM | mast_cell | *PPP1R12C* | 3.813772736 | 3.35E-07 | 1.21E-06 | 0.002103807 |
| T2DM | mast_cell | *OPRL1* | -2.830622625 | 4.84E-05 | 9.81E-05 | 0.304404349 |
| T2DM | mast_cell | *CHCHD5* | 3.822254278 | 7.77E-05 | 0.000150643 | 0.48823247 |
| T2DM | mast_cell | *PAX8* | 1.918904749 | 6.80E-06 | 1.65E-05 | 0.042730248 |
| T2DM | mast_cell | *INSIG2* | 3.014178349 | 3.70E-09 | 7.78E-08 | 2.32E-05 |
| T2DM | mast_cell | *PSD4* | 4.921203539 | 0.000444173 | 0.000754531 | 1 |
| T2DM | mast_cell | *SLC25A23* | 1.911548518 | 1.29E-06 | 3.81E-06 | 0.008114888 |
| T2DM | mast_cell | *CLPP* | 3.470554349 | 2.75E-09 | 7.78E-08 | 1.73E-05 |
| T2DM | mast_cell | *SH2D3A* | 6.615308525 | 6.20E-07 | 2.04E-06 | 0.003895363 |
| T2DM | mast_cell | *TRIP10* | 3.835685889 | 3.77E-09 | 7.78E-08 | 2.37E-05 |
| T2DM | mast_cell | *SNRPD2* | 5.712435485 | 4.74E-08 | 2.61E-07 | 0.00029797 |
| T2DM | mast_cell | *RTN2* | 2.157442537 | 2.70E-08 | 1.73E-07 | 0.000169961 |
| T2DM | mast_cell | *GPCPD1* | 3.682211626 | 6.52E-06 | 1.59E-05 | 0.040972885 |
| T2DM | mast_cell | *SDCBP2* | -2.667389912 | 0.025961536 | 0.033716211 | 1 |
| T2DM | mast_cell | *PANK2* | 4.529790236 | 1.79E-07 | 7.27E-07 | 0.001123337 |
| T2DM | mast_cell | *DTD1* | 4.11795997 | 1.89E-05 | 4.12E-05 | 0.118636374 |
| T2DM | mast_cell | *RBCK1* | 2.880247585 | 2.50E-07 | 9.52E-07 | 0.001574864 |
| T2DM | mast_cell | *RRBP1* | 2.440146751 | 0.003351514 | 0.004983673 | 1 |
| T2DM | mast_cell | *BMP2* | 4.566703336 | 4.24E-09 | 7.78E-08 | 2.67E-05 |
| T2DM | mast_cell | *PCSK2* | 3.582765011 | 4.77E-08 | 2.62E-07 | 0.000299864 |
| T2DM | mast_cell | *MKKS* | 3.584235904 | 0.001592305 | 0.002483458 | 1 |
| T2DM | mast_cell | *DSTN* | 3.29602565 | 0.000426915 | 0.000728165 | 1 |
| T2DM | mast_cell | *ITPA* | 5.547713818 | 2.95E-07 | 1.09E-06 | 0.001853153 |
| T2DM | mast_cell | *TMEM74B* | 3.932221809 | 6.32E-05 | 0.000125045 | 0.397393845 |
| T2DM | mast_cell | *FAM110A* | -2.722341975 | 8.22E-06 | 1.95E-05 | 0.051649965 |
| T2DM | mast_cell | *MRPS26* | 0.90741697 | 0.000668333 | 0.001099374 | 1 |
| T2DM | mast_cell | *ZNF436* | 2.136764152 | 2.42E-11 | 7.21E-09 | 1.52E-07 |
| T2DM | mast_cell | *GDF5* | 1.941758971 | 2.26E-05 | 4.89E-05 | 0.142165234 |
| T2DM | mast_cell | *MMP24* | 3.198578763 | 9.99E-05 | 0.000190385 | 0.627890033 |
| T2DM | mast_cell | *UROD* | 4.80476645 | 0.000179774 | 0.000327131 | 1 |
| T2DM | mast_cell | *KLC1* | 4.098787461 | 6.22E-06 | 1.53E-05 | 0.03908965 |
| T2DM | mast_cell | *XRCC3* | 4.129535824 | 5.28E-10 | 4.99E-08 | 3.32E-06 |
| T2DM | mast_cell | *MCF2L* | 3.06343534 | 1.02E-08 | 1.04E-07 | 6.42E-05 |
| T2DM | mast_cell | *F10* | 4.957691065 | 2.13E-08 | 1.47E-07 | 0.000134089 |
| T2DM | mast_cell | *PCID2* | 2.974288923 | 6.68E-07 | 2.18E-06 | 0.004201642 |
| T2DM | mast_cell | *PROZ* | 9.703831517 | 1.74E-08 | 1.31E-07 | 0.00010951 |
| T2DM | mast_cell | *IGFLR1* | 5.212185291 | 1.88E-08 | 1.37E-07 | 0.00011812 |
| T2DM | mast_cell | *CAPNS1* | 4.796255227 | 3.00E-07 | 1.11E-06 | 0.001888619 |
| T2DM | mast_cell | *COX6B1* | 3.233704687 | 0.017182577 | 0.022925904 | 1 |
| T2DM | mast_cell | *SCAF1* | 2.113533392 | 7.91E-09 | 9.16E-08 | 4.97E-05 |
| T2DM | mast_cell | *PRR12* | 3.748552055 | 3.00E-08 | 1.84E-07 | 0.000188346 |
| T2DM | mast_cell | *TSKS* | 5.935839553 | 1.74E-09 | 7.51E-08 | 1.10E-05 |
| T2DM | mast_cell | *SBDS* | 4.510012619 | 7.17E-05 | 0.000140094 | 0.450540704 |
| T2DM | mast_cell | *BECN1* | 5.704254468 | 5.89E-09 | 8.14E-08 | 3.71E-05 |
| T2DM | mast_cell | *TRAP1* | 0.68010539 | 0.004746734 | 0.006874619 | 1 |
| T2DM | mast_cell | *DNAJC8* | 3.151084198 | 2.40E-05 | 5.16E-05 | 0.151151212 |
| T2DM | mast_cell | *ZNF384* | 4.510939098 | 0.000412328 | 0.000704431 | 1 |
| T2DM | mast_cell | *EMG1* | 4.416650376 | 9.56E-07 | 2.96E-06 | 0.006012701 |

| | | | | | | |
|---|---|---|---|---|---|---|
| T2DM | mast_cell | *ATG14* | 4.156947001 | 1.82E-07 | 7.38E-07 | 0.001144025 |
| T2DM | mast_cell | *RHOJ* | 4.739813847 | 6.70E-07 | 2.18E-06 | 0.004213593 |
| T2DM | mast_cell | *HSPA2* | 3.549581476 | 9.63E-07 | 2.97E-06 | 0.00605305 |
| T2DM | mast_cell | *ZBTB1* | 3.191496325 | 0.000105618 | 0.00020055 | 0.664022128 |
| T2DM | mast_cell | *PZP* | -0.434163207 | 2.41E-07 | 9.23E-07 | 0.001512526 |
| T2DM | mast_cell | *RHOT1* | 2.794233416 | 2.91E-05 | 6.16E-05 | 0.183047993 |
| T2DM | mast_cell | *WDR60* | 2.040352632 | 0.000914075 | 0.00147467 | 1 |
| T2DM | mast_cell | *AIF1L* | 1.81839267 | 8.85E-09 | 9.64E-08 | 5.56E-05 |
| T2DM | mast_cell | *MAP2K2* | 3.364317861 | 1.51E-05 | 3.36E-05 | 0.095157411 |
| T2DM | mast_cell | *MASP1* | 1.597840001 | 6.23E-07 | 2.05E-06 | 0.003916868 |
| T2DM | mast_cell | *ATP13A4* | -1.533364025 | 3.49E-05 | 7.29E-05 | 0.2194036 |
| T2DM | mast_cell | *HRASLS* | 2.846253946 | 1.88E-08 | 1.37E-07 | 0.000118293 |
| T2DM | mast_cell | *HELB* | 3.27549457 | 8.15E-07 | 2.58E-06 | 0.005123954 |
| T2DM | mast_cell | *RAP1B* | 3.343277661 | 9.39E-07 | 2.92E-06 | 0.005904612 |
| T2DM | mast_cell | *RAB3IP* | 3.272297934 | 2.21E-05 | 4.78E-05 | 0.138876264 |
| T2DM | mast_cell | *TAS2R4* | 1.661651037 | 0.000115927 | 0.000218475 | 0.728831664 |
| T2DM | mast_cell | *LRRC61* | 1.293371873 | 8.09E-07 | 2.57E-06 | 0.005086117 |
| T2DM | mast_cell | *IDUA* | 4.176053148 | 3.71E-09 | 7.78E-08 | 2.33E-05 |
| T2DM | mast_cell | *TMEM175* | 4.021725894 | 1.01E-08 | 1.04E-07 | 6.36E-05 |
| T2DM | mast_cell | *EMC1* | 3.255304404 | 0.000110114 | 0.000208332 | 0.692286904 |
| T2DM | mast_cell | *PLA2G5* | 1.936959648 | 1.77E-05 | 3.88E-05 | 0.111291935 |
| T2DM | mast_cell | *SIN3B* | 4.26529729 | 2.07E-08 | 1.44E-07 | 0.000130372 |
| T2DM | mast_cell | *WDR24* | 6.212550317 | 3.23E-06 | 8.57E-06 | 0.020307832 |
| T2DM | mast_cell | *CHTF18* | 3.535583861 | 1.78E-09 | 7.51E-08 | 1.12E-05 |
| T2DM | mast_cell | *SMARCA4* | 3.159190562 | 0.000897491 | 0.001450894 | 1 |
| T2DM | mast_cell | *TICAM1* | 4.067736682 | 3.59E-09 | 7.78E-08 | 2.26E-05 |
| T2DM | mast_cell | *AAMP* | 3.730374494 | 0.002219353 | 0.003390783 | 1 |
| T2DM | mast_cell | *ZNF835* | 1.791032156 | 0.000505594 | 0.000849911 | 1 |
| T2DM | mast_cell | *HIP1* | 3.211618298 | 2.37E-08 | 1.57E-07 | 0.000149064 |
| T2DM | mast_cell | *PTPN12* | 3.438724594 | 9.10E-07 | 2.84E-06 | 0.005722053 |
| T2DM | mast_cell | *POR* | 4.257119891 | 6.28E-06 | 1.54E-05 | 0.039452844 |
| T2DM | mast_cell | *FGL2* | 3.410202181 | 7.54E-07 | 2.41E-06 | 0.004737995 |
| T2DM | mast_cell | *STYXL1* | 1.047942009 | 0.013970414 | 0.018852113 | 1 |
| T2DM | mast_cell | *STEAP4* | 2.684009492 | 6.41E-09 | 8.33E-08 | 4.03E-05 |
| T2DM | mast_cell | *SGCE* | 4.237283968 | 8.12E-07 | 2.57E-06 | 0.005102876 |
| T2DM | mast_cell | *RBM48* | 2.858255141 | 1.24E-08 | 1.12E-07 | 7.78E-05 |
| T2DM | mast_cell | *CASD1* | 4.073451609 | 5.17E-06 | 1.30E-05 | 0.032482235 |
| T2DM | mast_cell | *SRD5A3* | -2.094972499 | 0.002042074 | 0.003142075 | 1 |
| T2DM | mast_cell | *DGCR6L* | 2.839465484 | 0.000193472 | 0.000349931 | 1 |
| T2DM | mast_cell | *VPREB3* | 17.42011615 | 1.77E-06 | 5.05E-06 | 0.011130823 |
| T2DM | mast_cell | *MGAT3* | 2.760497103 | 7.72E-09 | 9.13E-08 | 4.85E-05 |
| T2DM | mast_cell | *ATF4* | 3.883022978 | 0.000554532 | 0.000926726 | 1 |
| T2DM | mast_cell | *CDC42EP1* | 3.915455903 | 5.85E-06 | 1.45E-05 | 0.036775165 |
| T2DM | mast_cell | *TPST2* | 3.173153508 | 0.002657042 | 0.004019447 | 1 |
| T2DM | mast_cell | *BAIAP2L2* | 4.500529486 | 9.56E-11 | 1.72E-08 | 6.01E-07 |
| T2DM | mast_cell | *TST* | 1.618021147 | 9.60E-08 | 4.43E-07 | 0.000603272 |
| T2DM | mast_cell | *RIBC2* | 2.49536592 | 1.09E-06 | 3.30E-06 | 0.006851695 |
| T2DM | mast_cell | *EMC4* | 5.264317971 | 4.04E-07 | 1.42E-06 | 0.002538589 |
| T2DM | mast_cell | *SPECC1* | -1.577953457 | 0.00022751 | 0.000406005 | 1 |
| T2DM | mast_cell | *POT1* | 3.39316384 | 7.93E-05 | 0.000153599 | 0.498581396 |

| | | | | | | |
|---|---|---|---|---|---|---|
| T2DM | mast_cell | *CDHR3* | 2.615458677 | 1.52E-08 | 1.21E-07 | 9.56E-05 |
| T2DM | mast_cell | *PRKRIP1* | 1.899514441 | 0.008667282 | 0.012095716 | 1 |
| T2DM | mast_cell | *FOXP2* | 1.777007543 | 0.000205415 | 0.000369089 | 1 |
| T2DM | mast_cell | *STRIP2* | 2.155105163 | 6.83E-08 | 3.44E-07 | 0.000429217 |
| T2DM | mast_cell | *FLNC* | 2.356222711 | 8.31E-07 | 2.63E-06 | 0.005225297 |
| T2DM | mast_cell | *IRF5* | 1.322398652 | 0.000314066 | 0.000546961 | 1 |
| T2DM | mast_cell | *KLHDC10* | 3.556975991 | 9.82E-07 | 3.03E-06 | 0.006173086 |
| T2DM | mast_cell | *HOXD1* | 10.07409646 | 8.34E-08 | 4.01E-07 | 0.000524564 |
| T2DM | mast_cell | *HOXD3* | 3.962313309 | 2.06E-07 | 8.18E-07 | 0.001296989 |
| T2DM | mast_cell | *CHN1* | 2.655274708 | 0.000203858 | 0.000366711 | 1 |
| T2DM | mast_cell | *GAD1* | 6.368500839 | 7.14E-09 | 8.82E-08 | 4.49E-05 |
| T2DM | mast_cell | *ORMDL1* | 1.548483805 | 0.001009638 | 0.001613112 | 1 |
| T2DM | mast_cell | *HOXD9* | 2.557324422 | 0.000693737 | 0.001138482 | 1 |
| T2DM | mast_cell | *PSMG2* | 2.2303704 | 0.000529843 | 0.000887117 | 1 |
| T2DM | mast_cell | *WDFY4* | 1.709258905 | 0.010204899 | 0.01405745 | 1 |
| T2DM | mast_cell | *TMOD2* | 2.707696097 | 0.000770557 | 0.001255376 | 1 |
| T2DM | mast_cell | *IVD* | -1.349181208 | 0.017491332 | 0.023318068 | 1 |
| T2DM | mast_cell | *ARPP19* | 4.288141827 | 5.73E-07 | 1.91E-06 | 0.003604511 |
| T2DM | mast_cell | *VPS13C* | 2.11293491 | 0.005498612 | 0.007896249 | 1 |
| T2DM | mast_cell | *CALML4* | -2.892930141 | 7.16E-07 | 2.30E-06 | 0.004498385 |
| T2DM | mast_cell | *ACKR4* | 3.217607347 | 1.42E-08 | 1.19E-07 | 8.91E-05 |
| T2DM | mast_cell | *SUMF2* | 3.643797673 | 2.01E-08 | 1.42E-07 | 0.000126182 |
| T2DM | mast_cell | *SPCS3* | 4.324274607 | 1.42E-05 | 3.17E-05 | 0.089014434 |
| T2DM | mast_cell | *SERGEF* | 3.031400349 | 0.004479319 | 0.006515844 | 1 |
| T2DM | mast_cell | *RPAIN* | 2.382630692 | 0.000474903 | 0.000801319 | 1 |
| T2DM | mast_cell | *PLD2* | 3.099120771 | 8.13E-10 | 6.02E-08 | 5.11E-06 |
| T2DM | mast_cell | *ATP1B2* | 3.644471156 | 2.09E-06 | 5.83E-06 | 0.013129525 |
| T2DM | mast_cell | *PHF20L1* | 3.19137503 | 0.000246204 | 0.000436762 | 1 |
| T2DM | mast_cell | *PUS7L* | 3.408769647 | 2.75E-05 | 5.85E-05 | 0.173109518 |
| T2DM | mast_cell | *KRI1* | 1.560492352 | 0.000103022 | 0.000195917 | 0.64770009 |
| T2DM | mast_cell | *SLC44A2* | 4.588249078 | 2.73E-08 | 1.74E-07 | 0.000171784 |
| T2DM | mast_cell | *CDKN2D* | 3.354260638 | 1.41E-06 | 4.13E-06 | 0.008882104 |
| T2DM | mast_cell | *MTUS1* | 3.020777486 | 1.12E-06 | 3.37E-06 | 0.007057583 |
| T2DM | mast_cell | *NGDN* | 5.275454151 | 4.12E-05 | 8.49E-05 | 0.259217673 |
| T2DM | mast_cell | *RIPK3* | 3.172673446 | 7.03E-05 | 0.000137646 | 0.442256535 |
| T2DM | mast_cell | *ADCY4* | 2.670791303 | 2.18E-06 | 6.07E-06 | 0.013718425 |
| T2DM | mast_cell | *BCL2L2* | 5.38457833 | 8.81E-08 | 4.16E-07 | 0.000554128 |
| T2DM | mast_cell | *DTD2* | 1.293844665 | 4.13E-06 | 1.07E-05 | 0.025976882 |
| T2DM | mast_cell | *PARP2* | 2.762038453 | 5.86E-07 | 1.95E-06 | 0.003685953 |
| T2DM | mast_cell | *MIS18BP1* | 1.929072094 | 0.0002552 | 0.000451066 | 1 |
| T2DM | mast_cell | *RNASE1* | 2.410918081 | 0.000152155 | 0.000280939 | 0.956598634 |
| T2DM | mast_cell | *EPB41L4A* | 2.278965954 | 0.009968204 | 0.013764572 | 1 |
| T2DM | mast_cell | *CDO1* | 1.984570914 | 7.90E-09 | 9.16E-08 | 4.97E-05 |
| T2DM | mast_cell | *REEP5* | 3.273185738 | 0.000542202 | 0.000906843 | 1 |
| T2DM | mast_cell | *QRICH2* | 3.374182736 | 7.06E-09 | 8.80E-08 | 4.44E-05 |
| T2DM | mast_cell | *SEC14L1* | 1.721493687 | 0.009951944 | 0.013745139 | 1 |
| T2DM | mast_cell | *RHBDF2* | 1.712058208 | 0.002245648 | 0.00342929 | 1 |
| T2DM | mast_cell | *AANAT* | 1.680443306 | 0.010043529 | 0.013853371 | 1 |
| T2DM | mast_cell | *TMEM8A* | 4.632249348 | 4.70E-07 | 1.61E-06 | 0.002957354 |
| T2DM | mast_cell | *SHC2* | 2.08916485 | 1.41E-09 | 6.85E-08 | 8.84E-06 |

| | | | | | | |
|---|---|---|---|---|---|---|
| T2DM | mast_cell | *ERMARD* | 3.713258653 | 2.81E-05 | 5.95E-05 | 0.176582271 |
| T2DM | mast_cell | *PHF10* | 4.301673675 | 1.19E-08 | 1.10E-07 | 7.50E-05 |
| T2DM | mast_cell | *GALNT8* | 1.274135286 | 0.029902101 | 0.038515573 | 1 |
| T2DM | mast_cell | *DOCK6* | 2.708409264 | 1.46E-07 | 6.16E-07 | 0.000919712 |
| T2DM | mast_cell | *ECSIT* | 4.878142063 | 3.86E-07 | 1.36E-06 | 0.002425805 |
| T2DM | mast_cell | *ELOF1* | 3.60818726 | 9.99E-09 | 1.04E-07 | 6.28E-05 |
| T2DM | mast_cell | *CDC16* | 1.931342603 | 0.003164848 | 0.00472847 | 1 |
| T2DM | mast_cell | *TOMM40* | 3.129693554 | 0.006275657 | 0.008950784 | 1 |
| T2DM | mast_cell | *FAM98C* | 2.387361261 | 0.006068708 | 0.008673328 | 1 |
| T2DM | mast_cell | *RPL36* | 4.339215831 | 1.69E-06 | 4.86E-06 | 0.010648604 |
| T2DM | mast_cell | *ATP8B3* | 0.904848171 | 0.004078307 | 0.005967028 | 1 |
| T2DM | mast_cell | *GTPBP3* | -3.298048017 | 4.92E-07 | 1.67E-06 | 0.003091333 |
| T2DM | mast_cell | *PLVAP* | 0.481104367 | 0.007967291 | 0.011173402 | 1 |
| T2DM | mast_cell | *BST2* | 5.48127636 | 6.82E-06 | 1.66E-05 | 0.042863711 |
| T2DM | mast_cell | *DDA1* | 3.346160007 | 1.13E-07 | 5.04E-07 | 0.000709306 |
| T2DM | mast_cell | *LSM7* | 2.911284821 | 5.24E-05 | 0.000105612 | 0.329614591 |
| T2DM | mast_cell | *TULP4* | 2.848374247 | 0.000158497 | 0.000291621 | 0.996470503 |
| T2DM | mast_cell | *RTN4IP1* | 3.629309334 | 8.28E-05 | 0.000159722 | 0.520375261 |
| T2DM | mast_cell | *QRSL1* | 0.953469996 | 0.015624553 | 0.020962775 | 1 |
| T2DM | mast_cell | *RSPH3* | 1.231470476 | 0.002821777 | 0.004248206 | 1 |
| T2DM | mast_cell | *ACTN4* | 3.455026897 | 0.000145201 | 0.000269126 | 0.912876897 |
| T2DM | mast_cell | *STK33* | 1.350367866 | 7.24E-05 | 0.000141342 | 0.45512185 |
| T2DM | mast_cell | *CACNG6* | 4.736668408 | 1.45E-05 | 3.22E-05 | 0.090880412 |
| T2DM | mast_cell | *PXDN* | 2.925887851 | 5.54E-09 | 8.00E-08 | 3.48E-05 |
| T2DM | mast_cell | *KIAA1683* | -0.915050866 | 0.007920848 | 0.011110748 | 1 |
| T2DM | mast_cell | *LSM4* | -1.089565042 | 0.028964028 | 0.037376199 | 1 |
| T2DM | mast_cell | *HRC* | 2.009975091 | 4.85E-05 | 9.83E-05 | 0.30500683 |
| T2DM | mast_cell | *TRPM4* | 2.984222897 | 4.06E-07 | 1.43E-06 | 0.002552705 |
| T2DM | mast_cell | *UBAC1* | 3.620884084 | 1.64E-07 | 6.77E-07 | 0.001029352 |
| T2DM | mast_cell | *TNNT3* | 1.661613118 | 3.95E-05 | 8.16E-05 | 0.248350544 |
| T2DM | mast_cell | *ATXN10* | 3.656047443 | 7.90E-07 | 2.51E-06 | 0.004966017 |
| T2DM | mast_cell | *CYP2E1* | 2.251648865 | 3.30E-08 | 1.99E-07 | 0.000207208 |
| T2DM | mast_cell | *HBZ* | 7.072676715 | 0.003506206 | 0.005197718 | 1 |
| T2DM | mast_cell | *ZNF337* | 2.367550786 | 2.36E-06 | 6.49E-06 | 0.014847452 |
| T2DM | mast_cell | *CEP85* | 1.391762739 | 0.017891517 | 0.023826301 | 1 |
| T2DM | mast_cell | *POMT1* | 4.089356011 | 2.58E-06 | 7.01E-06 | 0.016201174 |
| T2DM | mast_cell | *UCK1* | 1.548652763 | 7.29E-06 | 1.76E-05 | 0.045852882 |
| T2DM | mast_cell | *YIPF2* | 2.424674459 | 0.009709801 | 0.013443189 | 1 |
| T2DM | mast_cell | *TMEM160* | 5.652036333 | 0.005001023 | 0.007217959 | 1 |
| T2DM | mast_cell | *MAP3K10* | 3.594637261 | 1.44E-05 | 3.20E-05 | 0.090235376 |
| T2DM | mast_cell | *SMPDL3B* | -9.686466318 | 1.07E-06 | 3.25E-06 | 0.006722355 |
| T2DM | mast_cell | *ATPIF1* | -1.946129456 | 0.004084253 | 0.005974336 | 1 |
| T2DM | mast_cell | *THEMIS2* | 2.52701829 | 1.26E-07 | 5.47E-07 | 0.000794826 |
| T2DM | mast_cell | *CLIP1* | 3.517254935 | 6.98E-06 | 1.69E-05 | 0.043879303 |
| T2DM | mast_cell | *PPAN* | 2.816927239 | 9.58E-09 | 1.01E-07 | 6.03E-05 |
| T2DM | mast_cell | *ZNF236* | 2.662326521 | 0.000650479 | 0.001073375 | 1 |
| T2DM | mast_cell | *C12orf65* | 4.645197442 | 8.31E-12 | 3.07E-09 | 5.23E-08 |
| T2DM | mast_cell | *PKDREJ* | -1.794736697 | 0.007053659 | 0.009965473 | 1 |
| T2DM | mast_cell | *HSD17B3* | 1.950421427 | 0.002157811 | 0.003300768 | 1 |
| T2DM | mast_cell | *HABP4* | 4.356744562 | 4.88E-07 | 1.66E-06 | 0.003067361 |

| | | | | | | |
|---|---|---|---|---|---|---|
| T2DM | mast_cell | *EPS8L1* | -13.64862727 | 4.66E-06 | 1.18E-05 | 0.029315612 |
| T2DM | mast_cell | *LILRB2* | 2.679588733 | 6.90E-09 | 8.77E-08 | 4.34E-05 |
| T2DM | mast_cell | *COX4I2* | 4.545726369 | 1.93E-08 | 1.39E-07 | 0.000121066 |
| T2DM | mast_cell | *HAUS8* | 3.19945787 | 2.81E-06 | 7.57E-06 | 0.017647645 |
| T2DM | mast_cell | *TBC1D5* | 2.788786459 | 3.06E-05 | 6.46E-05 | 0.192633376 |
| T2DM | mast_cell | *RFTN1* | 3.32725603 | 2.74E-07 | 1.03E-06 | 0.001722984 |
| T2DM | mast_cell | *KCNC3* | 1.761564046 | 4.04E-05 | 8.33E-05 | 0.254161403 |
| T2DM | mast_cell | *PDLIM4* | 3.533521417 | 2.53E-09 | 7.78E-08 | 1.59E-05 |
| T2DM | mast_cell | *KIF3A* | 3.149840451 | 8.82E-07 | 2.76E-06 | 0.005543114 |
| T2DM | mast_cell | *AOC3* | 2.691947106 | 0.00011462 | 0.000216142 | 0.72061618 |
| T2DM | mast_cell | *ACLY* | 3.815282746 | 9.44E-06 | 2.21E-05 | 0.059378389 |
| T2DM | mast_cell | *AOC2* | 7.241319412 | 1.22E-08 | 1.11E-07 | 7.65E-05 |
| T2DM | mast_cell | *EXOC4* | 2.509491599 | 0.000244203 | 0.000433458 | 1 |
| T2DM | mast_cell | *ACAP3* | 4.393539231 | 1.13E-05 | 2.60E-05 | 0.071017001 |
| T2DM | mast_cell | *C1orf159* | 3.817242016 | 0.002119507 | 0.003250084 | 1 |
| T2DM | mast_cell | *ANO1* | 3.637760384 | 5.50E-09 | 8.00E-08 | 3.46E-05 |
| T2DM | mast_cell | *PPFIA1* | 2.152059889 | 0.003335511 | 0.004964573 | 1 |
| T2DM | mast_cell | *NINJ1* | 3.850661845 | 2.52E-08 | 1.64E-07 | 0.000158637 |
| T2DM | mast_cell | *NPHP4* | 5.774271847 | 1.28E-05 | 2.90E-05 | 0.080498297 |
| T2DM | mast_cell | *MAP1B* | 0.998953737 | 0.001032552 | 0.001646792 | 1 |
| T2DM | mast_cell | *CKMT2* | 2.216617727 | 0.002062114 | 0.003171358 | 1 |
| T2DM | mast_cell | *ZCCHC9* | 4.399535844 | 3.93E-08 | 2.27E-07 | 0.000247131 |
| T2DM | mast_cell | *KHDRBS3* | 3.921203519 | 1.10E-07 | 4.93E-07 | 0.000689131 |
| T2DM | mast_cell | *CHD1L* | 2.870489288 | 1.33E-07 | 5.70E-07 | 0.000836689 |
| T2DM | mast_cell | *RBM8A* | 2.058398783 | 8.42E-05 | 0.000162385 | 0.529536303 |
| T2DM | mast_cell | *ZNF304* | 1.77660491 | 6.76E-07 | 2.20E-06 | 0.004247501 |
| T2DM | mast_cell | *ZSCAN5A* | 4.192705677 | 1.31E-05 | 2.95E-05 | 0.082051944 |
| T2DM | mast_cell | *RHPN2* | -3.312477337 | 0.000291713 | 0.000511006 | 1 |
| T2DM | mast_cell | *C19orf12* | 1.476561405 | 2.38E-06 | 6.53E-06 | 0.014976207 |
| T2DM | mast_cell | *ACTR10* | 3.642834911 | 5.41E-05 | 0.000108629 | 0.339898768 |
| T2DM | mast_cell | *GCH1* | 4.219441651 | 1.01E-08 | 1.04E-07 | 6.35E-05 |
| T2DM | mast_cell | *LGALS3* | -1.602845378 | 0.00390727 | 0.005731452 | 1 |
| T2DM | mast_cell | *MATN3* | 7.563882295 | 1.09E-08 | 1.06E-07 | 6.87E-05 |
| T2DM | mast_cell | *RAF1* | 2.968727644 | 0.00152724 | 0.002386123 | 1 |
| T2DM | mast_cell | *ENOSF1* | 2.562913459 | 2.33E-05 | 5.02E-05 | 0.146363332 |
| T2DM | mast_cell | *TRIM5* | 3.961862797 | 8.38E-06 | 1.99E-05 | 0.052688796 |
| T2DM | mast_cell | *CNGA4* | 4.35277775 | 3.33E-07 | 1.21E-06 | 0.002094264 |
| T2DM | mast_cell | *TRIM22* | 3.409229876 | 0.000375844 | 0.000645963 | 1 |
| T2DM | mast_cell | *RRP8* | 3.442631559 | 0.001792692 | 0.002780817 | 1 |
| T2DM | mast_cell | *IMMT* | 2.183529486 | 0.006498013 | 0.009236493 | 1 |
| T2DM | mast_cell | *MRPL35* | 2.164396378 | 1.23E-05 | 2.81E-05 | 0.077541507 |
| T2DM | mast_cell | *CARD6* | 4.250307266 | 2.95E-08 | 1.82E-07 | 0.000185165 |
| T2DM | mast_cell | *INPP5K* | 2.26542631 | 1.12E-06 | 3.37E-06 | 0.007063685 |
| T2DM | mast_cell | *MYBBP1A* | 2.323623488 | 0.026263808 | 0.034073578 | 1 |
| T2DM | mast_cell | *RPA1* | 2.481260336 | 0.003528867 | 0.005226727 | 1 |
| T2DM | mast_cell | *SERPINF1* | 1.872918909 | 0.003528408 | 0.005226727 | 1 |
| T2DM | mast_cell | *SEC61G* | 1.984827704 | 0.012339417 | 0.016773603 | 1 |
| T2DM | mast_cell | *FIGNL1* | 4.147111282 | 0.003883441 | 0.005701819 | 1 |
| T2DM | mast_cell | *WBP2* | 5.324132399 | 5.87E-08 | 3.08E-07 | 0.000368899 |
| T2DM | mast_cell | *KDM6B* | 3.444982639 | 0.009385102 | 0.01302232 | 1 |

| | | | | | | |
|---|---|---|---|---|---|---|
| T2DM | mast_cell | *XAF1* | 3.012558096 | 6.44E-08 | 3.30E-07 | 0.000404994 |
| T2DM | mast_cell | *TERF2* | 4.518101668 | 6.19E-09 | 8.16E-08 | 3.89E-05 |
| T2DM | mast_cell | *BTBD3* | 3.913717663 | 1.26E-08 | 1.13E-07 | 7.92E-05 |
| T2DM | mast_cell | *PCNA* | 2.808058003 | 1.35E-08 | 1.16E-07 | 8.46E-05 |
| T2DM | mast_cell | *PTPRA* | 3.042470516 | 0.000160954 | 0.000295623 | 1 |
| T2DM | mast_cell | *MMACHC* | -2.220288488 | 5.26E-06 | 1.32E-05 | 0.033095099 |
| T2DM | mast_cell | *CTNNBL1* | -1.305813285 | 6.91E-06 | 1.68E-05 | 0.043436723 |
| T2DM | mast_cell | *LPIN3* | 4.200188224 | 5.50E-09 | 8.00E-08 | 3.46E-05 |
| T2DM | mast_cell | *DMGDH* | 2.909072644 | 6.47E-07 | 2.12E-06 | 0.004068334 |
| T2DM | mast_cell | *BHMT2* | 0.439418377 | 0.00459092 | 0.006665846 | 1 |
| T2DM | mast_cell | *KANK4* | 1.034284441 | 1.64E-06 | 4.71E-06 | 0.010309969 |
| T2DM | mast_cell | *RSG1* | -2.231232268 | 0.016927669 | 0.022604981 | 1 |
| T2DM | mast_cell | *CASP9* | 1.574384518 | 0.001397415 | 0.002198035 | 1 |
| T2DM | mast_cell | *ATP8A2* | -1.938235556 | 1.32E-06 | 3.88E-06 | 0.008279383 |
| T2DM | mast_cell | *USPL1* | 4.250849392 | 3.76E-07 | 1.34E-06 | 0.002366454 |
| T2DM | mast_cell | *POMP* | 3.601690887 | 0.00240425 | 0.003658161 | 1 |
| T2DM | mast_cell | *WASF3* | 4.023827673 | 5.72E-08 | 3.01E-07 | 0.000359321 |
| T2DM | mast_cell | *PEMT* | 2.56609722 | 3.82E-07 | 1.35E-06 | 0.002400083 |
| T2DM | mast_cell | *SCO1* | 3.471079337 | 2.07E-07 | 8.20E-07 | 0.001301468 |
| T2DM | mast_cell | *CHI3L1* | 0.880343503 | 6.97E-06 | 1.69E-05 | 0.043806407 |
| T2DM | mast_cell | *DSTYK* | 2.483766082 | 0.003417783 | 0.005074508 | 1 |
| T2DM | mast_cell | *TMCC2* | -3.176116487 | 4.41E-08 | 2.47E-07 | 0.000277163 |
| T2DM | mast_cell | *COG6* | 2.493775128 | 0.000907388 | 0.001465763 | 1 |
| T2DM | mast_cell | *TRPC4* | 8.072750589 | 1.16E-08 | 1.09E-07 | 7.32E-05 |
| T2DM | mast_cell | *RFC3* | 3.382532691 | 6.08E-06 | 1.50E-05 | 0.038239873 |
| T2DM | mast_cell | *BTBD2* | 3.11147507 | 0.000637505 | 0.001053072 | 1 |
| T2DM | mast_cell | *PRAM1* | 2.221908257 | 0.00013485 | 0.000250977 | 0.847799543 |
| T2DM | mast_cell | *CNDP2* | 5.246496796 | 5.84E-09 | 8.14E-08 | 3.67E-05 |
| T2DM | mast_cell | *RTN3* | 3.433486746 | 2.73E-06 | 7.39E-06 | 0.017164652 |
| T2DM | mast_cell | *SLC2A11* | 3.041698853 | 8.38E-09 | 9.31E-08 | 5.27E-05 |
| T2DM | mast_cell | *C1QTNF6* | 3.391381943 | 2.53E-09 | 7.78E-08 | 1.59E-05 |
| T2DM | mast_cell | *FAM83F* | -1.077915968 | 1.42E-07 | 6.00E-07 | 0.000890502 |
| T2DM | mast_cell | *AGAP3* | 2.83479398 | 0.006895717 | 0.00975329 | 1 |
| T2DM | mast_cell | *ACTR3B* | 3.562344237 | 8.38E-08 | 4.02E-07 | 0.000526742 |
| T2DM | mast_cell | *SFTPD* | 0.891976383 | 7.27E-05 | 0.00014185 | 0.45689972 |
| T2DM | mast_cell | *DYDC2* | 9.404483424 | 0.000102939 | 0.000195818 | 0.647177973 |
| T2DM | mast_cell | *TMTC1* | 3.28817171 | 2.92E-07 | 1.08E-06 | 0.001835211 |
| T2DM | mast_cell | *IPO8* | 4.320477641 | 2.29E-05 | 4.94E-05 | 0.143708165 |
| T2DM | mast_cell | *LARS* | 1.043851386 | 0.003144055 | 0.004699637 | 1 |
| T2DM | mast_cell | *SPINK5* | 3.207821832 | 5.14E-08 | 2.78E-07 | 0.000322897 |
| T2DM | mast_cell | *LRRCC1* | 3.159075909 | 2.30E-07 | 8.93E-07 | 0.001448823 |
| T2DM | mast_cell | *SWAP70* | 3.935648629 | 1.42E-07 | 6.02E-07 | 0.000894777 |
| T2DM | mast_cell | *AMPD3* | 1.449088159 | 1.67E-07 | 6.90E-07 | 0.001051126 |
| T2DM | mast_cell | *SBF2* | 2.973625873 | 4.64E-06 | 1.18E-05 | 0.029166622 |
| T2DM | mast_cell | *MICAL2* | 2.659965169 | 5.63E-05 | 0.000112851 | 0.35412733 |
| T2DM | mast_cell | *RRAS2* | 1.469816943 | 5.98E-05 | 0.00011892 | 0.375667224 |
| T2DM | mast_cell | *DPF2* | 3.375123773 | 1.53E-07 | 6.41E-07 | 0.000963974 |
| T2DM | mast_cell | *GSC* | 4.574788186 | 1.28E-08 | 1.13E-07 | 8.02E-05 |
| T2DM | mast_cell | *CATSPERB* | 1.005986134 | 0.012826973 | 0.017379995 | 1 |
| T2DM | mast_cell | *COX16* | -1.041185942 | 6.54E-07 | 2.14E-06 | 0.00411126 |

| | | | | | | |
|---|---|---|---|---|---|---|
| T2DM | mast_cell | *EIF2S1* | 4.7066535 | 7.08E-09 | 8.80E-08 | 4.45E-05 |
| T2DM | mast_cell | *ELP3* | 4.23801895 | 9.05E-08 | 4.24E-07 | 0.000569113 |
| T2DM | mast_cell | *ADAMDEC1* | -3.366217354 | 7.86E-07 | 2.50E-06 | 0.004939575 |
| T2DM | mast_cell | *MRO* | 2.097943317 | 4.34E-07 | 1.51E-06 | 0.002731051 |
| T2DM | mast_cell | *THUMPD3* | 2.208183549 | 0.006304085 | 0.008985215 | 1 |
| T2DM | mast_cell | *CNTN6* | 1.870023602 | 7.48E-08 | 3.69E-07 | 0.000470406 |
| T2DM | mast_cell | *KATNBL1* | 3.866420835 | 1.36E-06 | 3.98E-06 | 0.008525138 |
| T2DM | mast_cell | *GSTM5* | 1.358626314 | 0.003015856 | 0.004524144 | 1 |
| T2DM | mast_cell | *SYT6* | 4.420642896 | 2.07E-08 | 1.44E-07 | 0.000130363 |
| T2DM | mast_cell | *PSRC1* | 2.025566472 | 0.000614886 | 0.001019727 | 1 |
| T2DM | mast_cell | *WNT2B* | 2.298066745 | 0.004999809 | 0.007217959 | 1 |
| T2DM | mast_cell | *CEPT1* | 3.271076891 | 2.53E-05 | 5.40E-05 | 0.158832297 |
| T2DM | mast_cell | *AP4B1* | 5.174713016 | 1.50E-05 | 3.34E-05 | 0.09456282 |
| T2DM | mast_cell | *NAPG* | 3.866756846 | 4.20E-08 | 2.38E-07 | 0.000264028 |
| T2DM | mast_cell | *PPHLN1* | 3.122892361 | 4.50E-05 | 9.21E-05 | 0.283210904 |
| T2DM | mast_cell | *FKBP11* | 3.438970228 | 1.32E-07 | 5.69E-07 | 0.000830605 |
| T2DM | mast_cell | *YWHAQ* | 4.248774188 | 5.33E-08 | 2.85E-07 | 0.000334994 |
| T2DM | mast_cell | *LPIN1* | 1.187268601 | 0.037028673 | 0.047240111 | 1 |
| T2DM | mast_cell | *ANO3* | 1.265607892 | 7.85E-07 | 2.50E-06 | 0.004937481 |
| T2DM | mast_cell | *FST* | 1.56310812 | 6.92E-08 | 3.47E-07 | 0.00043503 |
| T2DM | mast_cell | *TIMM17A* | 3.961065929 | 7.48E-10 | 5.88E-08 | 4.70E-06 |
| T2DM | mast_cell | *FBXO18* | 3.869356799 | 8.14E-09 | 9.22E-08 | 5.12E-05 |
| T2DM | mast_cell | *ECHDC3* | 2.455917639 | 1.44E-06 | 4.18E-06 | 0.009041896 |
| T2DM | mast_cell | *IL15RA* | 2.631449599 | 3.89E-09 | 7.78E-08 | 2.44E-05 |
| T2DM | mast_cell | *TMEM241* | 3.752795514 | 1.07E-05 | 2.47E-05 | 0.067326445 |
| T2DM | mast_cell | *EMP1* | 1.586425841 | 0.002508513 | 0.003807585 | 1 |
| T2DM | mast_cell | *KLRD1* | 3.403670579 | 2.14E-09 | 7.78E-08 | 1.35E-05 |
| T2DM | mast_cell | *PRH2* | 2.525480863 | 0.000176861 | 0.000322484 | 1 |
| T2DM | mast_cell | *DDB2* | 4.527096775 | 1.12E-09 | 6.34E-08 | 7.03E-06 |
| T2DM | mast_cell | *ACP2* | 4.007018676 | 2.83E-08 | 1.78E-07 | 0.000178092 |
| T2DM | mast_cell | *HOOK1* | 5.42288431 | 2.17E-07 | 8.53E-07 | 0.001366948 |
| T2DM | mast_cell | *ZCCHC11* | 2.617840112 | 0.000671062 | 0.001102998 | 1 |
| T2DM | mast_cell | *FHOD3* | 1.28330644 | 8.76E-05 | 0.000168485 | 0.550439556 |
| T2DM | mast_cell | *TPGS2* | 4.439883857 | 6.93E-07 | 2.24E-06 | 0.004353835 |
| T2DM | mast_cell | *SLC43A3* | 2.417428795 | 0.008511611 | 0.011889024 | 1 |
| T2DM | mast_cell | *TIMM10* | 2.205173136 | 0.00098192 | 0.001573624 | 1 |
| T2DM | mast_cell | *DHX34* | 1.984264649 | 0.004895865 | 0.007079186 | 1 |
| T2DM | mast_cell | *APLNR* | 3.298278167 | 5.16E-09 | 7.89E-08 | 3.24E-05 |
| T2DM | mast_cell | *FADS2* | 5.366968652 | 6.11E-09 | 8.15E-08 | 3.84E-05 |
| T2DM | mast_cell | *TMEM165* | 3.107342463 | 1.06E-05 | 2.45E-05 | 0.06672103 |
| T2DM | mast_cell | *CLOCK* | 3.785715571 | 9.31E-08 | 4.33E-07 | 0.000585435 |
| T2DM | mast_cell | *DZIP1* | 1.659051251 | 2.52E-05 | 5.38E-05 | 0.158201897 |
| T2DM | mast_cell | *BIVM* | 4.216121167 | 4.48E-09 | 7.78E-08 | 2.81E-05 |
| T2DM | mast_cell | *ERCC5* | 4.644633547 | 4.53E-09 | 7.78E-08 | 2.85E-05 |
| T2DM | mast_cell | *CARS2* | 1.609092796 | 0.035727683 | 0.045691608 | 1 |
| T2DM | mast_cell | *KLB* | 1.513175768 | 6.18E-06 | 1.52E-05 | 0.038856391 |
| T2DM | mast_cell | *WDR36* | 4.506779479 | 5.69E-06 | 1.41E-05 | 0.03577718 |
| T2DM | mast_cell | *OSTF1* | 4.4475851 | 6.17E-06 | 1.52E-05 | 0.038816303 |
| T2DM | mast_cell | *UBQLN1* | 4.135926543 | 2.17E-06 | 6.05E-06 | 0.013656671 |
| T2DM | mast_cell | *NAA35* | 3.912551765 | 4.07E-08 | 2.34E-07 | 0.000255993 |

| | | | | | | |
|---|---|---|---|---|---|---|
| T2DM | mast_cell | *CTSL* | 2.645130844 | 0.008013107 | 0.011235149 | 1 |
| T2DM | mast_cell | *TMEM2* | 3.702695951 | 4.26E-07 | 1.48E-06 | 0.002679661 |
| T2DM | mast_cell | *GOLM1* | 3.06831398 | 0.000109399 | 0.000207165 | 0.687788665 |
| T2DM | mast_cell | *ADAM19* | 0.994932394 | 9.44E-05 | 0.000180723 | 0.593494668 |
| T2DM | mast_cell | *USP30* | 2.549438796 | 1.25E-06 | 3.70E-06 | 0.007874282 |
| T2DM | mast_cell | *HRK* | 4.929843388 | 3.06E-06 | 8.16E-06 | 0.019215755 |
| T2DM | mast_cell | *P2RX4* | 3.391574231 | 1.14E-06 | 3.41E-06 | 0.007163848 |
| T2DM | mast_cell | *DMTF1* | 2.712114652 | 4.65E-06 | 1.18E-05 | 0.029212846 |
| T2DM | mast_cell | *CCDC146* | 4.235182462 | 2.02E-08 | 1.42E-07 | 0.000126851 |
| T2DM | mast_cell | *POM121C* | 2.286797562 | 0.001154517 | 0.001832016 | 1 |
| T2DM | mast_cell | *SRPK2* | 3.166941767 | 1.57E-05 | 3.48E-05 | 0.098843575 |
| T2DM | mast_cell | *TES* | 2.486484326 | 1.23E-06 | 3.65E-06 | 0.007756059 |
| T2DM | mast_cell | *MDFIC* | 2.452779319 | 1.13E-07 | 5.05E-07 | 0.000711184 |
| T2DM | mast_cell | *MTO1* | 2.757362453 | 2.35E-06 | 6.45E-06 | 0.014745927 |
| T2DM | mast_cell | *ANKRD6* | 4.116477819 | 2.22E-07 | 8.67E-07 | 0.001394915 |
| T2DM | mast_cell | *SNX14* | 3.650332067 | 1.17E-06 | 3.49E-06 | 0.007343904 |
| T2DM | mast_cell | *NT5E* | 3.857747765 | 1.06E-09 | 6.34E-08 | 6.63E-06 |
| T2DM | mast_cell | *PRR5L* | 2.996275957 | 6.62E-08 | 3.36E-07 | 0.000416433 |
| T2DM | mast_cell | *NAT10* | 3.6482711 | 4.42E-08 | 2.47E-07 | 0.000278011 |
| T2DM | mast_cell | *ATP5G2* | 4.633370483 | 2.90E-08 | 1.81E-07 | 0.000182632 |
| T2DM | mast_cell | *ITGA7* | 0.589469191 | 0.011620715 | 0.015872134 | 1 |
| T2DM | mast_cell | *RDH5* | 1.895721268 | 4.89E-09 | 7.84E-08 | 3.07E-05 |
| T2DM | mast_cell | *PPP1R1A* | 2.105975302 | 5.69E-09 | 8.09E-08 | 3.57E-05 |
| T2DM | mast_cell | *B4GALNT1* | 4.238347027 | 0.000181232 | 0.000329499 | 1 |
| T2DM | mast_cell | *FAIM2* | -1.857461749 | 1.92E-07 | 7.71E-07 | 0.001205066 |
| T2DM | mast_cell | *KRT7* | 3.713587157 | 6.88E-05 | 0.00013488 | 0.43242569 |
| T2DM | mast_cell | *HNRNPA1* | 5.006419088 | 2.37E-07 | 9.12E-07 | 0.001490584 |
| T2DM | mast_cell | *SLC26A10* | -2.892631772 | 1.34E-07 | 5.72E-07 | 0.000841382 |
| T2DM | mast_cell | *MAP7* | 2.396868518 | 0.004603136 | 0.006678956 | 1 |
| T2DM | mast_cell | *PKIB* | -2.314205619 | 6.17E-05 | 0.000122421 | 0.388074837 |
| T2DM | mast_cell | *PRADC1* | -3.58109809 | 2.46E-05 | 5.28E-05 | 0.154837237 |
| T2DM | mast_cell | *SMYD5* | 2.314779426 | 1.87E-06 | 5.30E-06 | 0.011752933 |
| T2DM | mast_cell | *KCNMB4* | 2.887603956 | 2.05E-08 | 1.43E-07 | 0.000128784 |
| T2DM | mast_cell | *USP15* | 2.607708519 | 0.000868977 | 0.001406968 | 1 |
| T2DM | mast_cell | *KLHL36* | 2.299620878 | 0.025958249 | 0.033716211 | 1 |
| T2DM | mast_cell | *MPHOSPH6* | 6.278923652 | 8.68E-09 | 9.51E-08 | 5.46E-05 |
| T2DM | mast_cell | *CCDC102A* | 2.697388555 | 7.30E-06 | 1.76E-05 | 0.045911964 |
| T2DM | mast_cell | *AGT* | 0.759192722 | 0.002483276 | 0.003772012 | 1 |
| T2DM | mast_cell | *RNASEL* | 3.535565619 | 8.88E-08 | 4.19E-07 | 0.000558166 |
| T2DM | mast_cell | *PIGC* | 3.044173282 | 0.000147798 | 0.000273618 | 0.929207582 |
| T2DM | mast_cell | *SP110* | 3.238978918 | 0.00074991 | 0.001224593 | 1 |
| T2DM | mast_cell | *SLC19A3* | 0.907748001 | 1.12E-05 | 2.58E-05 | 0.070601604 |
| T2DM | mast_cell | *ARMC9* | 3.424797612 | 5.35E-09 | 8.00E-08 | 3.36E-05 |
| T2DM | mast_cell | *TMEM127* | 4.359322129 | 4.68E-08 | 2.59E-07 | 0.000294067 |
| T2DM | mast_cell | *GCC2* | 2.618261973 | 7.23E-05 | 0.000141128 | 0.454290269 |
| T2DM | mast_cell | *MRPS9* | 1.466092747 | 0.020200563 | 0.026714543 | 1 |
| T2DM | mast_cell | *ANKRD36* | 3.604799325 | 2.35E-05 | 5.06E-05 | 0.147653261 |
| T2DM | mast_cell | *USP44* | 1.411028532 | 0.000460651 | 0.000779363 | 1 |
| T2DM | mast_cell | *DRAM1* | 2.304682131 | 0.000740031 | 0.001208774 | 1 |
| T2DM | mast_cell | *SLC41A2* | 3.957341433 | 6.24E-08 | 3.23E-07 | 0.000392602 |

| | | | | | | |
|---|---|---|---|---|---|---|
| T2DM | mast_cell | *NEK3* | 1.689622617 | 0.000882946 | 0.001427748 | 1 |
| T2DM | mast_cell | *TBC1D4* | 3.175571799 | 2.79E-06 | 7.52E-06 | 0.017526461 |
| T2DM | mast_cell | *RCBTB1* | 1.540186826 | 0.009547864 | 0.013236477 | 1 |
| T2DM | mast_cell | *RCBTB2* | 3.999068111 | 1.94E-08 | 1.39E-07 | 0.000121874 |
| T2DM | mast_cell | *SETDB2* | 3.254547681 | 1.04E-05 | 2.40E-05 | 0.065133651 |
| T2DM | mast_cell | *SPDYE1* | 2.543416745 | 0.014880799 | 0.020016171 | 1 |
| T2DM | mast_cell | *GPNMB* | 2.943222261 | 1.80E-05 | 3.93E-05 | 0.11293887 |
| T2DM | mast_cell | *KDELR2* | 3.893454073 | 0.002724236 | 0.004110216 | 1 |
| T2DM | mast_cell | *NUPL2* | 3.017408272 | 0.001618743 | 0.002522816 | 1 |
| T2DM | mast_cell | *TBRG4* | 2.163273318 | 2.80E-08 | 1.76E-07 | 0.000175795 |
| T2DM | mast_cell | *DDX56* | 3.042109467 | 4.37E-09 | 7.78E-08 | 2.74E-05 |
| T2DM | mast_cell | *HUS1* | 3.2673149 | 0.000609916 | 0.001012286 | 1 |
| T2DM | mast_cell | *NACAD* | 4.076765826 | 0.000136728 | 0.000254172 | 0.859609253 |
| T2DM | mast_cell | *TTC5* | 2.306641844 | 0.015424687 | 0.020712304 | 1 |
| T2DM | mast_cell | *ALPK3* | 3.16771655 | 9.78E-05 | 0.000186568 | 0.614741379 |
| T2DM | mast_cell | *RSAD1* | -2.61847078 | 0.001086056 | 0.001728178 | 1 |
| T2DM | mast_cell | *NMT1* | 1.683710534 | 7.71E-05 | 0.000149772 | 0.48466175 |
| T2DM | mast_cell | *MYCBPAP* | -0.791934919 | 0.024737035 | 0.032292719 | 1 |
| T2DM | mast_cell | *BRIP1* | 3.908660761 | 0.009166167 | 0.012729776 | 1 |
| T2DM | mast_cell | *RTP4* | 1.918267587 | 0.00058833 | 0.000981122 | 1 |
| T2DM | mast_cell | *MRPL47* | 4.793083442 | 1.21E-05 | 2.75E-05 | 0.075891156 |
| T2DM | mast_cell | *EPRS* | 4.734373845 | 3.52E-07 | 1.26E-06 | 0.002212009 |
| T2DM | mast_cell | *RPS6KC1* | 3.199077805 | 9.05E-05 | 0.000173753 | 0.569214484 |
| T2DM | mast_cell | *CBWD2* | 4.458204128 | 2.14E-07 | 8.44E-07 | 0.001347684 |
| T2DM | mast_cell | *IL1RN* | 2.443299909 | 5.93E-09 | 8.14E-08 | 3.73E-05 |
| T2DM | mast_cell | *UGGT1* | 3.583437405 | 0.000601448 | 0.001000345 | 1 |
| T2DM | mast_cell | *GYPC* | 3.525527818 | 1.70E-06 | 4.87E-06 | 0.010694871 |
| T2DM | mast_cell | *NIPSNAP3A* | 2.233371847 | 1.05E-06 | 3.21E-06 | 0.006621479 |
| T2DM | mast_cell | *TXN* | -2.185330416 | 0.000690341 | 0.001133204 | 1 |
| T2DM | mast_cell | *ODF2* | 5.109583662 | 9.94E-08 | 4.57E-07 | 0.000624839 |
| T2DM | mast_cell | *ZFP37* | 3.193148095 | 1.87E-07 | 7.55E-07 | 0.001174795 |
| T2DM | mast_cell | *ZNF189* | 3.83367482 | 1.69E-06 | 4.86E-06 | 0.010655003 |
| T2DM | mast_cell | *PRPF4* | 3.261414941 | 5.30E-08 | 2.84E-07 | 0.000333419 |
| T2DM | mast_cell | *FPGS* | 3.468596203 | 0.000434161 | 0.000739119 | 1 |
| T2DM | mast_cell | *TEX10* | 4.773225928 | 4.74E-06 | 1.20E-05 | 0.029801808 |
| T2DM | mast_cell | *GARNL3* | 3.047204248 | 4.42E-09 | 7.78E-08 | 2.78E-05 |
| T2DM | mast_cell | *GABBR2* | 6.25499095 | 4.26E-09 | 7.78E-08 | 2.68E-05 |
| T2DM | mast_cell | *PSMB7* | 4.439705581 | 6.33E-07 | 2.08E-06 | 0.003979487 |
| T2DM | mast_cell | *RABEPK* | 4.578094704 | 2.73E-08 | 1.74E-07 | 0.000171497 |
| T2DM | mast_cell | *GOLGA1* | 3.777656875 | 0.000165696 | 0.000303535 | 1 |
| T2DM | mast_cell | *LMX1B* | 7.199816292 | 1.39E-08 | 1.18E-07 | 8.76E-05 |
| T2DM | mast_cell | *DERL1* | 4.243133332 | 4.73E-07 | 1.62E-06 | 0.002972277 |
| T2DM | mast_cell | *APTX* | 3.517008214 | 4.98E-06 | 1.26E-05 | 0.031340521 |
| T2DM | mast_cell | *TLN1* | 4.181508147 | 3.35E-06 | 8.84E-06 | 0.021067735 |
| T2DM | mast_cell | *SPAG8* | -2.028187065 | 7.82E-05 | 0.000151581 | 0.491427028 |
| T2DM | mast_cell | *CD72* | 3.734969897 | 0.000122854 | 0.000230356 | 0.772385231 |
| T2DM | mast_cell | *TMEM8B* | 2.312535474 | 1.57E-08 | 1.24E-07 | 9.90E-05 |
| T2DM | mast_cell | *GRHPR* | 1.713538948 | 0.003908568 | 0.00573202 | 1 |
| T2DM | mast_cell | *ARHGEF39* | 1.487043676 | 0.000181891 | 0.000330128 | 1 |
| T2DM | mast_cell | *DENND4C* | 2.239183849 | 0.004566904 | 0.00663404 | 1 |

| | | | | | | |
|---|---|---|---|---|---|---|
| T2DM | mast_cell | *PPIL1* | 6.876626496 | 2.44E-09 | 7.78E-08 | 1.53E-05 |
| T2DM | mast_cell | *KIF13A* | 3.19079979 | 7.76E-06 | 1.86E-05 | 0.048790538 |
| T2DM | mast_cell | *FRS3* | 4.713423085 | 1.63E-09 | 7.17E-08 | 1.03E-05 |
| T2DM | mast_cell | *SLC22A23* | 3.878446837 | 4.16E-08 | 2.37E-07 | 0.0002618 |
| T2DM | mast_cell | *BPHL* | 0.931539201 | 0.009885825 | 0.013662823 | 1 |
| T2DM | mast_cell | *RIPK1* | 3.578861026 | 0.000239532 | 0.000426008 | 1 |
| T2DM | mast_cell | *TUBB2B* | 1.329547481 | 0.012445128 | 0.016902684 | 1 |
| T2DM | mast_cell | *UQCC2* | 2.971385245 | 6.76E-09 | 8.65E-08 | 4.25E-05 |
| T2DM | mast_cell | *TCF19* | 2.811208683 | 3.53E-06 | 9.28E-06 | 0.022178291 |
| T2DM | mast_cell | *FLOT1* | 2.3583528 | 0.010563985 | 0.014513936 | 1 |
| T2DM | mast_cell | *TPMT* | 3.799666559 | 0.003769338 | 0.005548543 | 1 |
| T2DM | mast_cell | *NRM* | -1.641599643 | 0.015060028 | 0.020244258 | 1 |
| T2DM | mast_cell | *MTCH1* | 4.959484231 | 6.07E-07 | 2.01E-06 | 0.003816982 |
| T2DM | mast_cell | *FAM8A1* | 5.713633643 | 1.91E-08 | 1.38E-07 | 0.000120197 |
| T2DM | mast_cell | *FGFBP2* | 1.82458978 | 2.51E-07 | 9.53E-07 | 0.001576791 |
| T2DM | mast_cell | *FHDC1* | 6.493535353 | 5.90E-09 | 8.14E-08 | 3.71E-05 |
| T2DM | mast_cell | *MYO7A* | 1.550056164 | 5.92E-05 | 0.000117974 | 0.372091452 |
| T2DM | mast_cell | *ARRB1* | 2.577762519 | 0.004690688 | 0.006801281 | 1 |
| T2DM | mast_cell | *CCDC90B* | 4.716799266 | 1.05E-06 | 3.21E-06 | 0.006609096 |
| T2DM | mast_cell | *SYTL2* | -0.605851986 | 1.07E-07 | 4.85E-07 | 0.000673278 |
| T2DM | mast_cell | *RAB30* | 3.731167692 | 1.29E-05 | 2.93E-05 | 0.081265889 |
| T2DM | mast_cell | *PRCP* | 4.205559987 | 5.59E-06 | 1.39E-05 | 0.035117679 |
| T2DM | mast_cell | *NARS2* | 4.144477008 | 1.45E-05 | 3.23E-05 | 0.091317568 |
| T2DM | mast_cell | *PI15* | 7.480331248 | 2.28E-06 | 6.30E-06 | 0.014316936 |
| T2DM | mast_cell | *SULF1* | 2.06264248 | 4.64E-09 | 7.80E-08 | 2.92E-05 |
| T2DM | mast_cell | *TGS1* | 2.171352671 | 0.001850495 | 0.002863416 | 1 |
| T2DM | mast_cell | *NEK1* | 3.646555495 | 3.28E-06 | 8.68E-06 | 0.020635423 |
| T2DM | mast_cell | *DDX60* | 5.073142799 | 1.79E-06 | 5.10E-06 | 0.011261249 |
| T2DM | mast_cell | *MMP27* | 0.671130393 | 0.002432974 | 0.003699179 | 1 |
| T2DM | mast_cell | *DCUN1D5* | 1.626420934 | 5.06E-05 | 0.000102192 | 0.317817843 |
| T2DM | mast_cell | *TRIM29* | 1.661647048 | 2.34E-05 | 5.04E-05 | 0.14723778 |
| T2DM | mast_cell | *SLC37A4* | 6.380073084 | 0.001872759 | 0.00289573 | 1 |
| T2DM | mast_cell | *POU2F3* | 1.353384818 | 0.01137985 | 0.015563437 | 1 |
| T2DM | mast_cell | *PPP2R1B* | 2.513628374 | 7.52E-09 | 9.02E-08 | 4.73E-05 |
| T2DM | mast_cell | *FDX1* | 4.027202727 | 6.81E-05 | 0.000133621 | 0.428121888 |
| T2DM | mast_cell | *FXYD6* | -2.686275325 | 0.003421337 | 0.005077891 | 1 |
| T2DM | mast_cell | *TMPRSS13* | 4.972113262 | 1.07E-06 | 3.26E-06 | 0.00675485 |
| T2DM | mast_cell | *MAP2K5* | 3.64256228 | 1.07E-06 | 3.25E-06 | 0.006733615 |
| T2DM | mast_cell | *ITGA11* | 2.348454752 | 1.20E-07 | 5.26E-07 | 0.00075282 |
| T2DM | mast_cell | *PAQR5* | -0.406332917 | 1.25E-07 | 5.42E-07 | 0.000783703 |
| T2DM | mast_cell | *RMDN3* | 4.452965662 | 2.92E-10 | 3.68E-08 | 1.83E-06 |
| T2DM | mast_cell | *ITPKA* | 5.864075559 | 0.007457291 | 0.010505038 | 1 |
| T2DM | mast_cell | *UACA* | 3.414366519 | 2.00E-06 | 5.62E-06 | 0.012587196 |
| T2DM | mast_cell | *RSL24D1* | 2.264337714 | 0.001930423 | 0.002980493 | 1 |
| T2DM | mast_cell | *SPTBN5* | 1.884130824 | 1.36E-11 | 4.76E-09 | 8.57E-08 |
| T2DM | mast_cell | *RABGGTB* | 4.976188936 | 6.25E-07 | 2.06E-06 | 0.003926654 |
| T2DM | mast_cell | *DNASE2B* | 8.721400317 | 1.27E-08 | 1.13E-07 | 7.99E-05 |
| T2DM | mast_cell | *DBT* | 3.073989711 | 4.04E-07 | 1.42E-06 | 0.00254017 |
| T2DM | mast_cell | *RTCA* | 2.424362024 | 0.001118479 | 0.001777971 | 1 |
| T2DM | mast_cell | *IFT172* | 5.229497275 | 3.71E-09 | 7.78E-08 | 2.33E-05 |

| | | | | | | |
|---|---|---|---|---|---|---|
| T2DM | mast_cell | *ADCY3* | 3.633538287 | 2.31E-08 | 1.54E-07 | 0.000144962 |
| T2DM | mast_cell | *LHCGR* | 1.680047003 | 4.79E-08 | 2.63E-07 | 0.00030132 |
| T2DM | mast_cell | *THUMPD2* | 3.612372206 | 0.000156121 | 0.000287839 | 0.981529958 |
| T2DM | mast_cell | *PREB* | 1.463500068 | 0.015850351 | 0.021238525 | 1 |
| T2DM | mast_cell | *ABCG5* | 3.071777728 | 2.90E-08 | 1.81E-07 | 0.000182616 |
| T2DM | mast_cell | *ATRAID* | 4.367114695 | 5.03E-07 | 1.71E-06 | 0.003159758 |
| T2DM | mast_cell | *CENPO* | 8.465366272 | 8.40E-08 | 4.03E-07 | 0.000528407 |
| T2DM | mast_cell | *LRPPRC* | 3.858155103 | 3.59E-06 | 9.41E-06 | 0.022552533 |
| T2DM | mast_cell | *DTNB* | 1.677723987 | 0.020526179 | 0.027105248 | 1 |
| T2DM | mast_cell | *CYP2C9* | 0.881911896 | 0.008449852 | 0.011813257 | 1 |
| T2DM | mast_cell | *CYP2C8* | 2.389944975 | 2.35E-05 | 5.07E-05 | 0.148057038 |
| T2DM | mast_cell | *MYOF* | 2.710579139 | 1.69E-08 | 1.29E-07 | 0.000106376 |
| T2DM | mast_cell | *STAMBPL1* | 3.399620125 | 6.30E-08 | 3.25E-07 | 0.000395866 |
| T2DM | mast_cell | *CH25H* | -0.846826162 | 0.029293725 | 0.037762897 | 1 |
| T2DM | mast_cell | *ATAD1* | 4.374371897 | 2.03E-06 | 5.70E-06 | 0.012791908 |
| T2DM | mast_cell | *BTBD16* | 1.890552057 | 2.35E-05 | 5.06E-05 | 0.147656993 |
| T2DM | mast_cell | *TACC2* | 3.170894932 | 1.02E-06 | 3.14E-06 | 0.00642944 |
| T2DM | mast_cell | *CALHM2* | 2.780301319 | 1.68E-06 | 4.81E-06 | 0.010545849 |
| T2DM | mast_cell | *PLCE1* | 2.988071302 | 2.81E-05 | 5.95E-05 | 0.176607934 |
| T2DM | mast_cell | *RBP4* | 2.97670647 | 2.40E-09 | 7.78E-08 | 1.51E-05 |
| T2DM | mast_cell | *ASCC1* | 1.688692505 | 0.02882534 | 0.037212508 | 1 |
| T2DM | mast_cell | *RPS24* | 2.407797349 | 0.016764487 | 0.022401345 | 1 |
| T2DM | mast_cell | *BARD1* | 5.205300175 | 5.56E-06 | 1.38E-05 | 0.034978638 |
| T2DM | mast_cell | *CARF* | 3.398119357 | 1.48E-09 | 6.92E-08 | 9.33E-06 |
| T2DM | mast_cell | *METTL5* | 4.129024532 | 4.60E-09 | 7.79E-08 | 2.89E-05 |
| T2DM | mast_cell | *NAB1* | 3.925618535 | 1.01E-08 | 1.04E-07 | 6.36E-05 |
| T2DM | mast_cell | *FASTKD1* | 2.833901473 | 0.001343137 | 0.002117459 | 1 |
| T2DM | mast_cell | *MDH1B* | 3.60421905 | 1.13E-07 | 5.05E-07 | 0.000712261 |
| T2DM | mast_cell | *FAM117B* | 3.778831492 | 4.37E-09 | 7.78E-08 | 2.75E-05 |
| T2DM | mast_cell | *ITGAV* | 3.236307403 | 5.19E-08 | 2.80E-07 | 0.000326595 |
| T2DM | mast_cell | *SLC35A5* | 1.903374296 | 1.84E-06 | 5.23E-06 | 0.011570127 |
| T2DM | mast_cell | *SENP7* | 2.875897906 | 0.000877892 | 0.00141994 | 1 |
| T2DM | mast_cell | *USP8* | 3.005507566 | 9.71E-05 | 0.000185608 | 0.610651931 |
| T2DM | mast_cell | *SPPL2A* | 2.845938648 | 0.001178891 | 0.00186975 | 1 |
| T2DM | mast_cell | *PPCDC* | 1.99050422 | 0.000307338 | 0.000536433 | 1 |
| T2DM | mast_cell | *ARHGAP24* | 2.121343586 | 0.002443591 | 0.003714423 | 1 |
| T2DM | mast_cell | *FAM13A* | 3.176252874 | 3.18E-06 | 8.44E-06 | 0.019987753 |
| T2DM | mast_cell | *AP1AR* | 4.02226388 | 1.98E-08 | 1.41E-07 | 0.000124674 |
| T2DM | mast_cell | *PRKG2* | -0.39418966 | 1.35E-07 | 5.75E-07 | 0.000845683 |
| T2DM | mast_cell | *BBS7* | 4.240020435 | 7.85E-09 | 9.16E-08 | 4.93E-05 |
| T2DM | mast_cell | *KIAA1109* | 2.977207867 | 0.000136213 | 0.000253364 | 0.856369968 |
| T2DM | mast_cell | *MMRN1* | -0.56286732 | 0.014016567 | 0.018910335 | 1 |
| T2DM | mast_cell | *PRDM5* | 4.38002105 | 4.78E-08 | 2.63E-07 | 0.000300573 |
| T2DM | mast_cell | *TRPC3* | 2.535265135 | 4.20E-08 | 2.38E-07 | 0.000264129 |
| T2DM | mast_cell | *NAAA* | 2.284467565 | 0.008260338 | 0.011553448 | 1 |
| T2DM | mast_cell | *NUP54* | 3.498113183 | 8.58E-06 | 2.03E-05 | 0.053917636 |
| T2DM | mast_cell | *BMP2K* | -0.87231408 | 9.45E-07 | 2.93E-06 | 0.005938103 |
| T2DM | mast_cell | *SCARB2* | 3.926359977 | 1.08E-06 | 3.28E-06 | 0.006797465 |
| T2DM | mast_cell | *ANXA3* | 6.648003372 | 6.15E-08 | 3.20E-07 | 0.000386476 |
| T2DM | mast_cell | *PPA2* | 1.661029498 | 0.037359274 | 0.047642547 | 1 |

| | | | | | | |
|---|---|---|---|---|---|---|
| T2DM | mast_cell | *GSTCD* | 3.891706798 | 6.04E-08 | 3.15E-07 | 0.000379455 |
| T2DM | mast_cell | *PAPSS1* | 3.057644235 | 2.25E-06 | 6.23E-06 | 0.014155343 |
| T2DM | mast_cell | *SLC39A8* | 2.295623602 | 8.88E-06 | 2.09E-05 | 0.055804354 |
| T2DM | mast_cell | *FBN2* | 0.535412117 | 0.001418342 | 0.002227052 | 1 |
| T2DM | mast_cell | *RNF185* | 2.974022825 | 0.000349354 | 0.000603404 | 1 |
| T2DM | mast_cell | *B4GALNT3* | 3.62988226 | 0.000177114 | 0.000322851 | 1 |
| T2DM | mast_cell | *ERP27* | -4.196606728 | 1.90E-05 | 4.15E-05 | 0.119580813 |
| T2DM | mast_cell | *KIF21A* | 2.216646836 | 2.26E-08 | 1.52E-07 | 0.000141879 |
| T2DM | mast_cell | *CPNE8* | 1.732345044 | 0.001881028 | 0.002907086 | 1 |
| T2DM | mast_cell | *ALG10* | 6.764459066 | 3.94E-09 | 7.78E-08 | 2.48E-05 |
| T2DM | mast_cell | *SLCO1C1* | 2.280748758 | 2.13E-09 | 7.78E-08 | 1.34E-05 |
| T2DM | mast_cell | *ZCRB1* | 3.291678434 | 1.68E-07 | 6.91E-07 | 0.001053655 |
| T2DM | mast_cell | *PRICKLE1* | 2.823475979 | 6.60E-10 | 5.76E-08 | 4.15E-06 |
| T2DM | mast_cell | *C1RL* | 3.434743177 | 3.67E-10 | 4.05E-08 | 2.31E-06 |
| T2DM | mast_cell | *CLSTN3* | 3.874020934 | 5.87E-07 | 1.96E-06 | 0.003690698 |
| T2DM | mast_cell | *KLRG1* | -1.205305491 | 1.38E-06 | 4.03E-06 | 0.008657098 |
| T2DM | mast_cell | *VAMP1* | 1.538436267 | 0.012683164 | 0.017192551 | 1 |
| T2DM | mast_cell | *CD27* | 3.161450719 | 5.91E-08 | 3.10E-07 | 0.000371782 |
| T2DM | mast_cell | *PEX5* | 2.55623484 | 0.000960724 | 0.001542802 | 1 |
| T2DM | mast_cell | *PIANP* | 3.816555436 | 0.027711781 | 0.035826438 | 1 |
| T2DM | mast_cell | *PPFIA2* | 1.471017282 | 0.001086709 | 0.00172878 | 1 |
| T2DM | mast_cell | *GLIPR1* | 3.207512365 | 9.41E-07 | 2.92E-06 | 0.005914706 |
| T2DM | mast_cell | *LGR5* | 1.731800256 | 5.51E-07 | 1.85E-06 | 0.003462985 |
| T2DM | mast_cell | *POC1B* | 4.7923842 | 2.06E-07 | 8.17E-07 | 0.001295446 |
| T2DM | mast_cell | *AMDHD1* | 1.986333517 | 8.03E-08 | 3.90E-07 | 0.000504841 |
| T2DM | mast_cell | *GAS2L3* | 5.730749534 | 2.41E-07 | 9.24E-07 | 0.001515122 |
| T2DM | mast_cell | *TMEM132B* | 2.652373049 | 1.70E-07 | 6.97E-07 | 0.001067092 |
| T2DM | mast_cell | *TDG* | 4.031767763 | 6.86E-06 | 1.67E-05 | 0.043153262 |
| T2DM | mast_cell | *GLTP* | 3.952334701 | 8.98E-09 | 9.66E-08 | 5.64E-05 |
| T2DM | mast_cell | *MTMR6* | 3.134836107 | 0.00021585 | 0.000386514 | 1 |
| T2DM | mast_cell | *SLC46A3* | 2.275423322 | 0.000372279 | 0.000640536 | 1 |
| T2DM | mast_cell | *ACVRL1* | 2.927616164 | 1.95E-07 | 7.83E-07 | 0.001228565 |
| T2DM | mast_cell | *GPR84* | -13.21470961 | 1.59E-08 | 1.24E-07 | 9.99E-05 |
| T2DM | mast_cell | *BRCA2* | -2.700240621 | 0.000256418 | 0.000452964 | 1 |
| T2DM | mast_cell | *KANSL2* | 1.886370911 | 0.002702106 | 0.004078785 | 1 |
| T2DM | mast_cell | *CERS5* | 3.181800671 | 0.000111625 | 0.00021081 | 0.701785792 |
| T2DM | mast_cell | *LMBR1L* | 3.868375631 | 6.52E-06 | 1.60E-05 | 0.041014729 |
| T2DM | mast_cell | *ZNF740* | 3.129413349 | 0.002521558 | 0.003824616 | 1 |
| T2DM | mast_cell | *HNRNPA1L2* | 3.321138571 | 1.26E-07 | 5.45E-07 | 0.000789681 |
| T2DM | mast_cell | *LPAR6* | 3.923640327 | 5.03E-06 | 1.27E-05 | 0.031598795 |
| T2DM | mast_cell | *VPS33A* | 7.415940635 | 4.93E-05 | 9.98E-05 | 0.309743072 |
| T2DM | mast_cell | *METTL21C* | 4.030490911 | 0.025082213 | 0.032682254 | 1 |
| T2DM | mast_cell | *CBLN3* | 0.413678602 | 0.006680961 | 0.009479395 | 1 |
| T2DM | mast_cell | *TSSK4* | 6.622591181 | 2.11E-07 | 8.32E-07 | 0.001323824 |
| T2DM | mast_cell | *TMX1* | 5.087161585 | 1.34E-06 | 3.95E-06 | 0.008455323 |
| T2DM | mast_cell | *RTN1* | 3.190390646 | 1.16E-08 | 1.09E-07 | 7.28E-05 |
| T2DM | mast_cell | *C14orf37* | 2.381574024 | 0.000714533 | 0.001170471 | 1 |
| T2DM | mast_cell | *RAB15* | 1.204075802 | 7.34E-07 | 2.36E-06 | 0.00461311 |
| T2DM | mast_cell | *STON2* | 2.559251521 | 5.92E-09 | 8.14E-08 | 3.72E-05 |
| T2DM | mast_cell | *EFCAB11* | 2.863660671 | 1.19E-07 | 5.24E-07 | 0.000748383 |

| | | | | | | |
|---|---|---|---|---|---|---|
| T2DM | mast_cell | *PTGR2* | -1.050671872 | 0.00020739 | 0.000372426 | 1 |
| T2DM | mast_cell | *AK7* | 4.976139993 | 5.06E-09 | 7.84E-08 | 3.18E-05 |
| T2DM | mast_cell | *SLC24A4* | 1.393449151 | 3.74E-07 | 1.33E-06 | 0.002350016 |
| T2DM | mast_cell | *FBLN5* | 3.678849409 | 6.12E-09 | 8.15E-08 | 3.85E-05 |
| T2DM | mast_cell | *C14orf79* | 7.978996078 | 6.57E-08 | 3.34E-07 | 0.000413032 |
| T2DM | mast_cell | *NIPA2* | 4.323214654 | 5.13E-07 | 1.74E-06 | 0.003223648 |
| T2DM | mast_cell | *DUOXA1* | 2.765540815 | 0.003720911 | 0.005483677 | 1 |
| T2DM | mast_cell | *MFAP1* | 3.349003645 | 2.74E-06 | 7.40E-06 | 0.01721546 |
| T2DM | mast_cell | *TCF12* | 2.770143282 | 6.12E-05 | 0.000121569 | 0.38476585 |
| T2DM | mast_cell | *SORD* | 5.092335626 | 3.15E-07 | 1.15E-06 | 0.001981409 |
| T2DM | mast_cell | *ZSCAN29* | 2.38284963 | 1.18E-08 | 1.10E-07 | 7.40E-05 |
| T2DM | mast_cell | *LYSMD2* | 2.811234547 | 2.47E-06 | 6.75E-06 | 0.015558442 |
| T2DM | mast_cell | *SRP14* | 3.082994253 | 0.001252853 | 0.001982055 | 1 |
| T2DM | mast_cell | *BAHD1* | 4.51450966 | 9.99E-08 | 4.59E-07 | 0.000628018 |
| T2DM | mast_cell | *ANP32A* | 2.404259476 | 0.024607638 | 0.032137146 | 1 |
| T2DM | mast_cell | *UBE2Q2* | 2.22978206 | 0.031352808 | 0.040285123 | 1 |
| T2DM | mast_cell | *PSTPIP1* | 3.79259706 | 1.44E-06 | 4.19E-06 | 0.0090628 |
| T2DM | mast_cell | *HMG20A* | 5.311447933 | 3.25E-09 | 7.78E-08 | 2.04E-05 |
| T2DM | mast_cell | *SCAPER* | 2.376445734 | 0.000262791 | 0.000463051 | 1 |
| T2DM | mast_cell | *MAN2C1* | -0.640553934 | 0.002188261 | 0.003345719 | 1 |
| T2DM | mast_cell | *DNAJA4* | -2.890067879 | 0.000125108 | 0.000234372 | 0.786552134 |
| T2DM | mast_cell | *TPM1* | 3.254063256 | 6.95E-09 | 8.77E-08 | 4.37E-05 |
| T2DM | mast_cell | *IGF1R* | 3.310952756 | 1.30E-08 | 1.13E-07 | 8.15E-05 |
| T2DM | mast_cell | *CYP11A1* | 2.089639894 | 8.47E-06 | 2.00E-05 | 0.053234801 |
| T2DM | mast_cell | *BBS4* | -1.656139887 | 0.010269245 | 0.014133701 | 1 |
| T2DM | mast_cell | *ADAMTS17* | 2.983925531 | 9.24E-08 | 4.30E-07 | 0.000580844 |
| T2DM | mast_cell | *HAPLN3* | 2.468790262 | 8.12E-08 | 3.93E-07 | 0.000510262 |
| T2DM | mast_cell | *POLG* | 4.191031317 | 0.000109765 | 0.000207734 | 0.690093098 |
| T2DM | mast_cell | *WDR93* | 0.770495293 | 0.000155566 | 0.000286901 | 0.978043882 |
| T2DM | mast_cell | *TICRR* | 3.594605969 | 1.22E-07 | 5.33E-07 | 0.000764806 |
| T2DM | mast_cell | *MFGE8* | 3.430940383 | 3.54E-08 | 2.09E-07 | 0.000222458 |
| T2DM | mast_cell | *UNC45A* | 2.057687112 | 0.000964954 | 0.001548409 | 1 |
| T2DM | mast_cell | *ST8SIA2* | 9.662029718 | 7.87E-06 | 1.88E-05 | 0.04945393 |
| T2DM | mast_cell | *IQGAP1* | 2.09346935 | 0.001305296 | 0.002059838 | 1 |
| T2DM | mast_cell | *CRTC3* | 3.559996228 | 3.17E-05 | 6.67E-05 | 0.199411728 |
| T2DM | mast_cell | *SH3GL3* | 1.253163685 | 0.004306197 | 0.006278958 | 1 |
| T2DM | mast_cell | *SEC11A* | 3.919363267 | 1.33E-07 | 5.70E-07 | 0.000835739 |
| T2DM | mast_cell | *TGFB1I1* | 2.202651071 | 8.06E-08 | 3.91E-07 | 0.000506828 |
| T2DM | mast_cell | *PARN* | 2.900026635 | 0.00013662 | 0.000254045 | 0.858926859 |
| T2DM | mast_cell | *FTO* | 2.122481613 | 0.007126222 | 0.010061207 | 1 |
| T2DM | mast_cell | *TXNL4B* | 2.890908995 | 1.57E-08 | 1.24E-07 | 9.89E-05 |
| T2DM | mast_cell | *MARVELD3* | 4.661305933 | 1.48E-07 | 6.24E-07 | 0.000932583 |
| T2DM | mast_cell | *ZFHX3* | 2.256095648 | 0.005784442 | 0.008293451 | 1 |
| T2DM | mast_cell | *KIFC3* | 4.720000653 | 1.16E-07 | 5.14E-07 | 0.00072721 |
| T2DM | mast_cell | *ADAMTS18* | 1.822755547 | 3.58E-08 | 2.10E-07 | 0.000225128 |
| T2DM | mast_cell | *GCSH* | 3.189947982 | 1.53E-07 | 6.41E-07 | 0.000963615 |
| T2DM | mast_cell | *CMTM3* | 5.0989054 | 0.000169889 | 0.000310491 | 1 |
| T2DM | mast_cell | *MAP1LC3B* | 4.051535035 | 3.65E-07 | 1.30E-06 | 0.002291618 |
| T2DM | mast_cell | *TLDC1* | 3.561734649 | 7.05E-09 | 8.80E-08 | 4.43E-05 |
| T2DM | mast_cell | *TIGD7* | 2.823412154 | 1.11E-09 | 6.34E-08 | 7.00E-06 |

| | | | | | | |
|---|---|---|---|---|---|---|
| T2DM | mast_cell | *DEF8* | 1.970209623 | 4.17E-06 | 1.08E-05 | 0.026219612 |
| T2DM | mast_cell | *COPS3* | 5.723763819 | 2.26E-07 | 8.80E-07 | 0.001419713 |
| T2DM | mast_cell | *RANBP10* | 3.876932587 | 1.51E-08 | 1.21E-07 | 9.48E-05 |
| T2DM | mast_cell | *MYO19* | 5.521925344 | 6.65E-11 | 1.31E-08 | 4.18E-07 |
| T2DM | mast_cell | *DDX52* | 3.76842443 | 0.000137426 | 0.000255394 | 0.863997598 |
| T2DM | mast_cell | *RASL10B* | 2.093009452 | 0.008790432 | 0.012251263 | 1 |
| T2DM | mast_cell | *UNC45B* | -5.061562794 | 4.15E-06 | 1.07E-05 | 0.026078254 |
| T2DM | mast_cell | *TOM1L1* | 3.72947023 | 2.69E-09 | 7.78E-08 | 1.69E-05 |
| T2DM | mast_cell | *C17orf80* | 1.581181876 | 0.004601227 | 0.006677727 | 1 |
| T2DM | mast_cell | *SPATA22* | 2.356904952 | 7.66E-08 | 3.76E-07 | 0.000481878 |
| T2DM | mast_cell | *SGSM2* | 3.146676631 | 0.000628607 | 0.001040287 | 1 |
| T2DM | mast_cell | *NPEPPS* | 3.474663406 | 2.84E-05 | 6.01E-05 | 0.178283034 |
| T2DM | mast_cell | *SCRN2* | 0.44347602 | 0.02018129 | 0.026700289 | 1 |
| T2DM | mast_cell | *RHBDL3* | -1.893471518 | 1.04E-06 | 3.19E-06 | 0.006559566 |
| T2DM | mast_cell | *ABCA8* | 2.036668632 | 6.25E-05 | 0.000123744 | 0.392763825 |
| T2DM | mast_cell | *IMPA2* | 4.49378173 | 3.13E-09 | 7.78E-08 | 1.97E-05 |
| T2DM | mast_cell | *GNAL* | 4.397124685 | 1.26E-07 | 5.46E-07 | 0.000790605 |
| T2DM | mast_cell | *RPRD1A* | 2.3054602 | 0.003633462 | 0.00536486 | 1 |
| T2DM | mast_cell | *C18orf21* | 4.70331408 | 7.65E-10 | 5.88E-08 | 4.81E-06 |
| T2DM | mast_cell | *GALNT1* | 3.350986343 | 2.25E-05 | 4.86E-05 | 0.141156169 |
| T2DM | mast_cell | *ASXL3* | 1.892385185 | 7.07E-09 | 8.80E-08 | 4.44E-05 |
| T2DM | mast_cell | *ADCYAP1* | -0.797316647 | 1.57E-07 | 6.53E-07 | 0.000983961 |
| T2DM | mast_cell | *ESCO1* | 3.350329949 | 0.000277443 | 0.000487503 | 1 |
| T2DM | mast_cell | *GREB1L* | 1.483932072 | 0.00090818 | 0.001466288 | 1 |
| T2DM | mast_cell | *C18orf8* | 2.028027871 | 0.002275902 | 0.003472707 | 1 |
| T2DM | mast_cell | *NPC1* | 2.26936673 | 1.07E-05 | 2.48E-05 | 0.06753756 |
| T2DM | mast_cell | *SLC14A1* | 2.682808968 | 0.002657787 | 0.004019606 | 1 |
| T2DM | mast_cell | *SLC13A5* | 5.157614381 | 0.00673074 | 0.009539527 | 1 |
| T2DM | mast_cell | *CCDC40* | 2.491528595 | 7.88E-08 | 3.85E-07 | 0.000495496 |
| T2DM | mast_cell | *CARD14* | -2.5592389 | 2.02E-05 | 4.39E-05 | 0.126889779 |
| T2DM | mast_cell | *EIF4A3* | 3.334652414 | 8.92E-09 | 9.66E-08 | 5.61E-05 |
| T2DM | mast_cell | *TBCD* | 2.628169776 | 0.003605996 | 0.005327074 | 1 |
| T2DM | mast_cell | *NARF* | 2.698673676 | 0.002664647 | 0.004029013 | 1 |
| T2DM | mast_cell | *TRIM65* | 2.487135013 | 9.07E-07 | 2.83E-06 | 0.005703097 |
| T2DM | mast_cell | *CBX8* | 3.155865707 | 2.72E-06 | 7.37E-06 | 0.017115791 |
| T2DM | mast_cell | *SECTM1* | 2.789151135 | 1.42E-08 | 1.19E-07 | 8.91E-05 |
| T2DM | mast_cell | *RNF157* | 0.791786127 | 3.46E-05 | 7.23E-05 | 0.217467192 |
| T2DM | mast_cell | *WDR45B* | 3.309345952 | 0.000467698 | 0.00079001 | 1 |
| T2DM | mast_cell | *RNF165* | 4.873147388 | 2.49E-05 | 5.34E-05 | 0.15683411 |
| T2DM | mast_cell | *SMAD4* | 2.954664844 | 0.003595549 | 0.005315122 | 1 |
| T2DM | mast_cell | *PIP4K2B* | 3.720456544 | 1.04E-07 | 4.74E-07 | 0.000654105 |
| T2DM | mast_cell | *GRB7* | 2.839333977 | 1.07E-05 | 2.47E-05 | 0.067099072 |
| T2DM | mast_cell | *NFIC* | 2.021690364 | 0.000918796 | 0.001481147 | 1 |
| T2DM | mast_cell | *TPGS1* | 3.91505099 | 0.000698917 | 0.001146085 | 1 |
| T2DM | mast_cell | *PRDM15* | 1.739401725 | 0.000189324 | 0.000342724 | 1 |
| T2DM | mast_cell | *PFKL* | 2.176778567 | 0.000115556 | 0.000217841 | 0.72649926 |
| T2DM | mast_cell | *MVB12A* | 2.967771893 | 8.72E-07 | 2.74E-06 | 0.00547938 |
| T2DM | mast_cell | *DUS3L* | 2.442099592 | 7.28E-05 | 0.00014214 | 0.457975178 |
| T2DM | mast_cell | *DPP9* | 3.080702576 | 1.08E-07 | 4.87E-07 | 0.000679059 |
| T2DM | mast_cell | *CCDC97* | 5.416094097 | 5.93E-05 | 0.000118117 | 0.372659794 |

| | | | | | | |
|---|---|---|---|---|---|---|
| T2DM | mast_cell | *SIRT3* | 4.28632681 | 1.66E-07 | 6.85E-07 | 0.001042294 |
| T2DM | mast_cell | *IFITM3* | 4.875183412 | 8.14E-06 | 1.94E-05 | 0.051174971 |
| T2DM | mast_cell | *COL6A1* | 1.13109454 | 0.03026196 | 0.038963126 | 1 |
| T2DM | mast_cell | *IFNAR1* | 1.329596932 | 0.026042643 | 0.033807577 | 1 |
| T2DM | mast_cell | *TRPM2* | 1.046767091 | 0.010973435 | 0.015033774 | 1 |
| T2DM | mast_cell | *TMEM50B* | 1.078665027 | 0.002134565 | 0.003269186 | 1 |
| T2DM | mast_cell | *DOPEY2* | 3.300813978 | 4.98E-05 | 0.000100851 | 0.313344723 |
| T2DM | mast_cell | *URB1* | 1.953078462 | 0.000120802 | 0.000226983 | 0.759484454 |
| T2DM | mast_cell | *AKT1* | 4.281304894 | 3.53E-09 | 7.78E-08 | 2.22E-05 |
| T2DM | mast_cell | *SAE1* | 3.428481764 | 2.87E-07 | 1.07E-06 | 0.001806977 |
| T2DM | mast_cell | *NTN5* | -2.522919214 | 0.000439394 | 0.000747017 | 1 |
| T2DM | mast_cell | *GEMIN7* | 3.41091851 | 4.30E-05 | 8.84E-05 | 0.270520282 |
| T2DM | mast_cell | *WTIP* | 2.768685878 | 1.44E-08 | 1.19E-07 | 9.03E-05 |
| T2DM | mast_cell | *MYO1F* | 3.255732972 | 0.000229594 | 0.000409492 | 1 |
| T2DM | mast_cell | *CACNG8* | -1.210670033 | 1.57E-07 | 6.53E-07 | 0.0009847 |
| T2DM | mast_cell | *FBN3* | -1.768742846 | 0.00141529 | 0.002223371 | 1 |
| T2DM | mast_cell | *EVI5L* | 3.904873077 | 5.28E-09 | 7.96E-08 | 3.32E-05 |
| T2DM | mast_cell | *CTU1* | 5.419393598 | 2.97E-05 | 6.27E-05 | 0.186414199 |
| T2DM | mast_cell | *RCN3* | 3.918591762 | 4.87E-09 | 7.84E-08 | 3.06E-05 |
| T2DM | mast_cell | *RERE* | 1.816371707 | 0.006267646 | 0.008943415 | 1 |
| T2DM | mast_cell | *PEX14* | 3.477317585 | 2.41E-06 | 6.58E-06 | 0.015138918 |
| T2DM | mast_cell | *SH3BGRL3* | 2.504327943 | 0.036913143 | 0.047121407 | 1 |
| T2DM | mast_cell | *IL22RA1* | 1.616571448 | 0.006422265 | 0.009139154 | 1 |
| T2DM | mast_cell | *ZNF593* | 1.39998587 | 0.037152902 | 0.047388983 | 1 |
| T2DM | mast_cell | *C1orf216* | 3.96858316 | 8.68E-08 | 4.11E-07 | 0.000545492 |
| T2DM | mast_cell | *GPN2* | 5.035643098 | 0.000118723 | 0.00022341 | 0.74641361 |
| T2DM | mast_cell | *ITGB3BP* | 1.982116689 | 7.88E-05 | 0.000152666 | 0.495249649 |
| T2DM | mast_cell | *SERBP1* | 4.558946223 | 2.26E-07 | 8.81E-07 | 0.001421274 |
| T2DM | mast_cell | *PIGK* | 4.048349154 | 0.000276468 | 0.000486061 | 1 |
| T2DM | mast_cell | *MOB3C* | 4.538820032 | 2.81E-09 | 7.78E-08 | 1.76E-05 |
| T2DM | mast_cell | *SYPL2* | 1.483079864 | 2.37E-05 | 5.09E-05 | 0.148754044 |
| T2DM | mast_cell | *ZNF697* | 4.327454727 | 1.93E-08 | 1.39E-07 | 0.000121119 |
| T2DM | mast_cell | *GPR161* | 4.23663534 | 2.40E-08 | 1.58E-07 | 0.000150927 |
| T2DM | mast_cell | *TIPRL* | 4.558062804 | 4.07E-07 | 1.43E-06 | 0.002557077 |
| T2DM | mast_cell | *NME7* | 3.213897042 | 1.75E-05 | 3.83E-05 | 0.109877055 |
| T2DM | mast_cell | *GPA33* | 3.329599361 | 2.55E-07 | 9.67E-07 | 0.001604829 |
| T2DM | mast_cell | *RXRG* | 4.31406454 | 1.12E-08 | 1.07E-07 | 7.05E-05 |
| T2DM | mast_cell | *TMCO1* | 3.081237847 | 0.00386418 | 0.005676192 | 1 |
| T2DM | mast_cell | *XCL2* | 1.786830253 | 0.008647788 | 0.01207119 | 1 |
| T2DM | mast_cell | *MGST3* | 3.655486441 | 4.20E-07 | 1.47E-06 | 0.002642925 |
| T2DM | mast_cell | *ADCY10* | 4.285962881 | 1.57E-07 | 6.54E-07 | 0.000988205 |
| T2DM | mast_cell | *RFWD2* | 1.729981569 | 0.025849408 | 0.033586985 | 1 |
| T2DM | mast_cell | *PPOX* | 2.39107127 | 0.009098988 | 0.01265045 | 1 |
| T2DM | mast_cell | *RGS5* | 3.286404609 | 6.45E-07 | 2.11E-06 | 0.004055296 |
| T2DM | mast_cell | *RGL1* | 3.291297127 | 1.88E-08 | 1.37E-07 | 0.000118035 |
| T2DM | mast_cell | *TARS2* | 5.410623082 | 6.86E-07 | 2.23E-06 | 0.004314952 |
| T2DM | mast_cell | *SNX27* | 4.357551449 | 8.09E-06 | 1.93E-05 | 0.05087094 |
| T2DM | mast_cell | *CERS2* | 2.666784458 | 0.019710034 | 0.026131797 | 1 |
| T2DM | mast_cell | *ENSA* | 3.000793268 | 0.002236464 | 0.003416095 | 1 |
| T2DM | mast_cell | *SEMA6C* | 2.229354303 | 3.34E-08 | 2.00E-07 | 0.000209811 |

| | | | | | | |
|---|---|---|---|---|---|---|
| T2DM | mast_cell | *ARNT* | 2.512489419 | 0.001711946 | 0.002661475 | 1 |
| T2DM | mast_cell | *POGZ* | 2.117521579 | 0.002534408 | 0.003841326 | 1 |
| T2DM | mast_cell | *HORMAD1* | 0.426249772 | 0.00238789 | 0.003634147 | 1 |
| T2DM | mast_cell | *GABPB2* | 2.172802302 | 1.69E-05 | 3.71E-05 | 0.106142914 |
| T2DM | mast_cell | *IKBKE* | 3.601179745 | 5.58E-09 | 8.01E-08 | 3.51E-05 |
| T2DM | mast_cell | *DYRK3* | 4.93863566 | 3.07E-08 | 1.88E-07 | 0.000192728 |
| T2DM | mast_cell | *HHIPL2* | -2.944538696 | 3.84E-05 | 7.96E-05 | 0.241216576 |
| T2DM | mast_cell | *TP53BP2* | 3.028695044 | 0.014065333 | 0.018972055 | 1 |
| T2DM | mast_cell | *RAB13* | 4.911248198 | 2.81E-06 | 7.57E-06 | 0.01768099 |
| T2DM | mast_cell | *HAX1* | 2.757982211 | 0.009144262 | 0.01270216 | 1 |
| T2DM | mast_cell | *KCNN3* | -2.159752588 | 0.000139452 | 0.000259051 | 0.876732382 |
| T2DM | mast_cell | *GATAD2B* | 3.00805782 | 2.83E-06 | 7.62E-06 | 0.017813347 |
| T2DM | mast_cell | *RIT1* | 4.669082653 | 4.30E-09 | 7.78E-08 | 2.70E-05 |
| T2DM | mast_cell | *GALNT2* | 3.306502621 | 0.000592819 | 0.000988084 | 1 |
| T2DM | mast_cell | *SCCPDH* | 4.218137773 | 3.59E-08 | 2.11E-07 | 0.000225768 |
| T2DM | mast_cell | *CEP170* | 3.287558947 | 1.41E-05 | 3.14E-05 | 0.088338179 |
| T2DM | mast_cell | *SNAP47* | 3.940469781 | 2.62E-09 | 7.78E-08 | 1.64E-05 |
| T2DM | mast_cell | *NVL* | 4.172291359 | 7.56E-08 | 3.72E-07 | 0.000475044 |
| T2DM | mast_cell | *SDE2* | 4.606526446 | 9.33E-07 | 2.90E-06 | 0.005865034 |
| T2DM | mast_cell | *FBXO28* | 3.810491159 | 0.000117864 | 0.000221926 | 0.74101026 |
| T2DM | mast_cell | *CNIH4* | 4.445773587 | 0.000131667 | 0.000245344 | 0.827791063 |
| T2DM | mast_cell | *GUK1* | 5.174930534 | 7.94E-08 | 3.87E-07 | 0.000498965 |
| T2DM | mast_cell | *CDC42BPA* | 3.149445271 | 5.38E-07 | 1.81E-06 | 0.003379343 |
| T2DM | mast_cell | *CNIH3* | 2.143083068 | 0.020726579 | 0.027341167 | 1 |
| T2DM | mast_cell | *PSEN2* | 2.440640522 | 3.83E-07 | 1.36E-06 | 0.002410156 |
| T2DM | mast_cell | *PYCR2* | 2.867820153 | 1.18E-08 | 1.10E-07 | 7.39E-05 |
| T2DM | mast_cell | *WNT9A* | 3.476061073 | 2.85E-11 | 7.54E-09 | 1.79E-07 |
| T2DM | mast_cell | *ABCG8* | 6.22120639 | 1.03E-08 | 1.04E-07 | 6.49E-05 |
| T2DM | mast_cell | *EML4* | 3.61531731 | 1.93E-07 | 7.75E-07 | 0.001214604 |
| T2DM | mast_cell | *CHAC2* | 5.904861508 | 2.28E-08 | 1.53E-07 | 0.000143473 |
| T2DM | mast_cell | *WDPCP* | 1.104583725 | 0.000158352 | 0.000291525 | 0.995559161 |
| T2DM | mast_cell | *ASXL2* | 3.628623685 | 5.35E-06 | 1.34E-05 | 0.033625253 |
| T2DM | mast_cell | *CIAO1* | 2.592901538 | 0.002375698 | 0.003616468 | 1 |
| T2DM | mast_cell | *TEX261* | 5.247507471 | 3.60E-09 | 7.78E-08 | 2.27E-05 |
| T2DM | mast_cell | *RALB* | -2.038009739 | 0.004810114 | 0.006961598 | 1 |
| T2DM | mast_cell | *TMEM177* | 6.550198485 | 5.28E-06 | 1.32E-05 | 0.033195901 |
| T2DM | mast_cell | *RABL2A* | 3.448571163 | 4.34E-06 | 1.11E-05 | 0.027299791 |
| T2DM | mast_cell | *SLC20A1* | 3.458588893 | 2.16E-09 | 7.78E-08 | 1.36E-05 |
| T2DM | mast_cell | *FBLN7* | 2.157588358 | 0.000372852 | 0.000641346 | 1 |
| T2DM | mast_cell | *LIPT1* | 1.972727637 | 2.98E-06 | 7.99E-06 | 0.018761441 |
| T2DM | mast_cell | *FAHD2B* | 1.831319501 | 6.25E-07 | 2.06E-06 | 0.00392776 |
| T2DM | mast_cell | *AFF3* | 1.566823954 | 7.42E-05 | 0.000144648 | 0.466780607 |
| T2DM | mast_cell | *UBXN4* | 3.544718062 | 9.20E-06 | 2.16E-05 | 0.057854301 |
| T2DM | mast_cell | *THSD7B* | 1.969282366 | 5.56E-11 | 1.16E-08 | 3.49E-07 |
| T2DM | mast_cell | *GPR17* | 0.773716253 | 0.000783666 | 0.001275409 | 1 |
| T2DM | mast_cell | *POLR2D* | 5.430492839 | 1.16E-07 | 5.15E-07 | 0.000730639 |
| T2DM | mast_cell | *ZNF385B* | 1.076329326 | 7.10E-09 | 8.80E-08 | 4.46E-05 |
| T2DM | mast_cell | *TMEFF2* | -0.466566138 | 3.33E-07 | 1.21E-06 | 0.002093731 |
| T2DM | mast_cell | *GULP1* | 2.56013188 | 2.85E-09 | 7.78E-08 | 1.79E-05 |
| T2DM | mast_cell | *FAM171B* | 2.339630549 | 0.000667427 | 0.001098459 | 1 |

| | | | | | | |
|---|---|---|---|---|---|---|
| T2DM | mast_cell | *HSPD1* | 4.934475054 | 4.92E-07 | 1.68E-06 | 0.003095595 |
| T2DM | mast_cell | *PTH2R* | -1.096403659 | 8.39E-05 | 0.000161722 | 0.527214037 |
| T2DM | mast_cell | *NBEAL1* | 2.11758356 | 1.02E-06 | 3.13E-06 | 0.006407819 |
| T2DM | mast_cell | *SUMF1* | 3.11045647 | 3.71E-06 | 9.70E-06 | 0.023323936 |
| T2DM | mast_cell | *ESPNL* | 18.06925447 | 9.56E-10 | 6.13E-08 | 6.01E-06 |
| T2DM | mast_cell | *TAMM41* | 4.357717438 | 2.23E-09 | 7.78E-08 | 1.40E-05 |
| T2DM | mast_cell | *OSBPL10* | 3.341296291 | 1.08E-09 | 6.34E-08 | 6.79E-06 |
| T2DM | mast_cell | *FAM198A* | 1.827736108 | 3.40E-08 | 2.03E-07 | 0.000213881 |
| T2DM | mast_cell | *CSRNP1* | 4.659897069 | 5.03E-06 | 1.27E-05 | 0.031640368 |
| T2DM | mast_cell | *SLC25A38* | 2.208272225 | 4.86E-08 | 2.66E-07 | 0.000305423 |
| T2DM | mast_cell | *CTDSPL* | 2.925756763 | 0.000177179 | 0.000322876 | 1 |
| T2DM | mast_cell | *STAC* | 3.091309342 | 4.08E-09 | 7.78E-08 | 2.56E-05 |
| T2DM | mast_cell | *ARL6IP5* | 2.799017327 | 0.004838284 | 0.006999146 | 1 |
| T2DM | mast_cell | *TMF1* | 3.968141151 | 4.34E-06 | 1.11E-05 | 0.027272175 |
| T2DM | mast_cell | *COL8A1* | 1.875964026 | 2.70E-06 | 7.30E-06 | 0.016946686 |
| T2DM | mast_cell | *NXPE3* | -2.597443875 | 0.000658412 | 0.001085612 | 1 |
| T2DM | mast_cell | *RABL3* | 4.895521985 | 1.11E-08 | 1.07E-07 | 6.97E-05 |
| T2DM | mast_cell | *ADPRH* | -1.115167296 | 0.011149604 | 0.015265149 | 1 |
| T2DM | mast_cell | *BOC* | 2.676387329 | 2.76E-07 | 1.03E-06 | 0.001733673 |
| T2DM | mast_cell | *AGTR1* | 2.033718543 | 5.74E-08 | 3.02E-07 | 0.000360987 |
| T2DM | mast_cell | *EIF2A* | 1.220178708 | 0.001005084 | 0.001606244 | 1 |
| T2DM | mast_cell | *AMT* | 0.983742066 | 0.000196869 | 0.000355258 | 1 |
| T2DM | mast_cell | *TCTA* | 3.264381889 | 0.001029705 | 0.001642669 | 1 |
| T2DM | mast_cell | *EAF2* | 3.238781177 | 3.44E-07 | 1.24E-06 | 0.002162681 |
| T2DM | mast_cell | *SLC26A1* | -1.286122345 | 0.000483167 | 0.000813995 | 1 |
| T2DM | mast_cell | *LYAR* | -2.588055119 | 0.017437986 | 0.023261749 | 1 |
| T2DM | mast_cell | *CENPC* | 1.497700013 | 0.022756534 | 0.029856079 | 1 |
| T2DM | mast_cell | *ATP10D* | 3.279302354 | 5.01E-07 | 1.70E-06 | 0.003152803 |
| T2DM | mast_cell | *SLC10A4* | 6.439030178 | 9.42E-05 | 0.000180363 | 0.592132592 |
| T2DM | mast_cell | *PYURF* | 3.047845746 | 0.028635791 | 0.036990594 | 1 |
| T2DM | mast_cell | *CISD2* | 5.356752392 | 8.43E-08 | 4.03E-07 | 0.000529775 |
| T2DM | mast_cell | *TIFA* | 4.081439306 | 3.30E-08 | 1.99E-07 | 0.000207296 |
| T2DM | mast_cell | *CCNA2* | -3.04210158 | 0.003832993 | 0.005634329 | 1 |
| T2DM | mast_cell | *USP53* | 3.302049304 | 4.98E-09 | 7.84E-08 | 3.13E-05 |
| T2DM | mast_cell | *MARCH1* | 1.824440349 | 8.13E-05 | 0.000157214 | 0.511260045 |
| T2DM | mast_cell | *PDGFC* | 2.630706195 | 0.000232082 | 0.000413343 | 1 |
| T2DM | mast_cell | *CBR4* | 3.30478049 | 5.27E-05 | 0.000106154 | 0.331625736 |
| T2DM | mast_cell | *NDUFS6* | 4.560809247 | 6.65E-05 | 0.000130968 | 0.418313161 |
| T2DM | mast_cell | *FAM105A* | 2.303632597 | 2.76E-05 | 5.86E-05 | 0.173404386 |
| T2DM | mast_cell | *LHFPL2* | 2.965755696 | 2.74E-08 | 1.74E-07 | 0.000172326 |
| T2DM | mast_cell | *SSBP2* | 2.575381345 | 0.000403194 | 0.000689763 | 1 |
| T2DM | mast_cell | *CRHBP* | 1.904756162 | 0.004813849 | 0.006965401 | 1 |
| T2DM | mast_cell | *RASA1* | 1.850064558 | 0.002252276 | 0.003438577 | 1 |
| T2DM | mast_cell | *GTF2H2* | 2.28505658 | 5.04E-08 | 2.74E-07 | 0.000316846 |
| T2DM | mast_cell | *SPATA9* | 2.585870442 | 5.92E-10 | 5.40E-08 | 3.72E-06 |
| T2DM | mast_cell | *TSLP* | -1.388390735 | 0.000479044 | 0.000807657 | 1 |
| T2DM | mast_cell | *FEM1C* | 4.381140847 | 3.82E-08 | 2.22E-07 | 0.000240305 |
| T2DM | mast_cell | *YIPF5* | 4.143849435 | 3.70E-09 | 7.78E-08 | 2.32E-05 |
| T2DM | mast_cell | *GABRB2* | 1.090291479 | 2.37E-06 | 6.51E-06 | 0.014922502 |
| T2DM | mast_cell | *PCYOX1L* | 1.890929452 | 0.000630764 | 0.001043309 | 1 |

| | | | | | | |
|---|---|---|---|---|---|---|
| T2DM | mast_cell | *MYLK4* | 3.472763143 | 6.41E-08 | 3.29E-07 | 0.000403095 |
| T2DM | mast_cell | *GFOD1* | 1.518403878 | 0.000396479 | 0.0006792 | 1 |
| T2DM | mast_cell | *ZMAT2* | 3.184127623 | 6.44E-05 | 0.000127275 | 0.404988855 |
| T2DM | mast_cell | *KLHL3* | 1.666654186 | 0.038457708 | 0.048963874 | 1 |
| T2DM | mast_cell | *TRIM7* | 2.710241302 | 4.56E-07 | 1.57E-06 | 0.002866279 |
| T2DM | mast_cell | *HIGD2A* | 3.324820076 | 1.00E-05 | 2.33E-05 | 0.062884481 |
| T2DM | mast_cell | *FAM193B* | 4.2387318 | 1.27E-05 | 2.89E-05 | 0.080094936 |
| T2DM | mast_cell | *MUT* | 3.34742594 | 3.57E-09 | 7.78E-08 | 2.24E-05 |
| T2DM | mast_cell | *DOK3* | -1.3364918 | 0.000991032 | 0.001585802 | 1 |
| T2DM | mast_cell | *MLIP* | 3.304557189 | 9.68E-09 | 1.02E-07 | 6.08E-05 |
| T2DM | mast_cell | *FGD2* | 2.397879061 | 1.05E-08 | 1.05E-07 | 6.58E-05 |
| T2DM | mast_cell | *TPBG* | 5.95795119 | 3.88E-08 | 2.25E-07 | 0.000243691 |
| T2DM | mast_cell | *IRAK1BP1* | 4.781866233 | 3.94E-08 | 2.27E-07 | 0.000247406 |
| T2DM | mast_cell | *PM20D2* | 5.769356054 | 3.12E-06 | 8.30E-06 | 0.01958774 |
| T2DM | mast_cell | *RNF217* | 3.544600349 | 2.25E-08 | 1.52E-07 | 0.000141444 |
| T2DM | mast_cell | *ABRACL* | 2.531090769 | 0.002371426 | 0.00361084 | 1 |
| T2DM | mast_cell | *SLC18B1* | 0.677584029 | 0.006114621 | 0.008731006 | 1 |
| T2DM | mast_cell | *AIG1* | 3.044359588 | 2.23E-06 | 6.18E-06 | 0.013991192 |
| T2DM | mast_cell | *PNLDC1* | 2.734683944 | 6.92E-05 | 0.000135591 | 0.435111516 |
| T2DM | mast_cell | *WTAP* | 2.423710071 | 0.0001432 | 0.000265654 | 0.900299918 |
| T2DM | mast_cell | *VIP* | 1.24397597 | 0.021039611 | 0.027725013 | 1 |
| T2DM | mast_cell | *SLC22A3* | 2.923522163 | 3.62E-09 | 7.78E-08 | 2.28E-05 |
| T2DM | mast_cell | *GNA12* | 3.204024495 | 8.89E-08 | 4.19E-07 | 0.000558935 |
| T2DM | mast_cell | *C7orf50* | 4.864489075 | 8.06E-09 | 9.17E-08 | 5.07E-05 |
| T2DM | mast_cell | *CCZ1B* | 1.866871388 | 0.003022342 | 0.004532792 | 1 |
| T2DM | mast_cell | *EGFR* | 1.814241768 | 0.000166428 | 0.00030461 | 1 |
| T2DM | mast_cell | *CCT6A* | 4.812493701 | 2.32E-07 | 8.96E-07 | 0.001457946 |
| T2DM | mast_cell | *PSPH* | 2.16573279 | 1.62E-09 | 7.17E-08 | 1.02E-05 |
| T2DM | mast_cell | *GIGYF1* | 2.931238297 | 0.001233333 | 0.00195314 | 1 |
| T2DM | mast_cell | *TMEM209* | 4.24445278 | 4.55E-10 | 4.47E-08 | 2.86E-06 |
| T2DM | mast_cell | *EPHA1* | -1.934685888 | 6.43E-06 | 1.57E-05 | 0.040420466 |
| T2DM | mast_cell | *NOM1* | 4.907067788 | 4.16E-08 | 2.37E-07 | 0.00026141 |
| T2DM | mast_cell | *NCAPG2* | 2.796253849 | 0.002925746 | 0.004393161 | 1 |
| T2DM | mast_cell | *FBXO25* | 3.035908987 | 0.008953791 | 0.01246512 | 1 |
| T2DM | mast_cell | *CCDC25* | 3.546119748 | 2.48E-11 | 7.21E-09 | 1.56E-07 |
| T2DM | mast_cell | *BIN3* | 4.361315679 | 3.46E-07 | 1.25E-06 | 0.002177705 |
| T2DM | mast_cell | *SLC25A37* | 4.073194487 | 1.28E-05 | 2.90E-05 | 0.080401334 |
| T2DM | mast_cell | *CHMP7* | 3.730395875 | 4.96E-06 | 1.25E-05 | 0.031208933 |
| T2DM | mast_cell | *DOCK5* | -1.79746855 | 0.001880384 | 0.002906804 | 1 |
| T2DM | mast_cell | *ST18* | 1.726985115 | 7.11E-08 | 3.55E-07 | 0.000447176 |
| T2DM | mast_cell | *TACC1* | 2.748379269 | 7.69E-05 | 0.000149538 | 0.483606318 |
| T2DM | mast_cell | *SLC26A7* | 1.374995049 | 0.000608252 | 0.00101024 | 1 |
| T2DM | mast_cell | *ATP6V0D2* | 1.975876112 | 4.31E-05 | 8.84E-05 | 0.270795856 |
| T2DM | mast_cell | *MAL2* | 1.627861688 | 2.37E-06 | 6.50E-06 | 0.014884217 |
| T2DM | mast_cell | *UTP23* | 4.904422569 | 1.97E-08 | 1.40E-07 | 0.000123644 |
| T2DM | mast_cell | *FAM135B* | -4.497779595 | 0.000235739 | 0.000419381 | 1 |
| T2DM | mast_cell | *ZNF7* | 4.984969812 | 1.62E-07 | 6.70E-07 | 0.001016444 |
| T2DM | mast_cell | *SLC39A4* | 2.372957877 | 0.001192904 | 0.001891021 | 1 |
| T2DM | mast_cell | *CER1* | 3.167894701 | 4.37E-05 | 8.95E-05 | 0.274620977 |
| T2DM | mast_cell | *PLIN2* | -1.828214261 | 0.002688507 | 0.004060208 | 1 |

| | | | | | | |
|---|---|---|---|---|---|---|
| T2DM | mast_cell | *HAUS6* | 4.955474562 | 6.70E-05 | 0.000131649 | 0.421012318 |
| T2DM | mast_cell | *C9orf72* | 3.122879789 | 2.54E-07 | 9.62E-07 | 0.001594567 |
| T2DM | mast_cell | *CBWD5* | 3.401778106 | 3.57E-09 | 7.78E-08 | 2.24E-05 |
| T2DM | mast_cell | *ZNF462* | 3.503463283 | 6.73E-09 | 8.65E-08 | 4.23E-05 |
| T2DM | mast_cell | *ALAD* | 1.104697618 | 0.000450103 | 0.000763466 | 1 |
| T2DM | mast_cell | *ASTN2* | 4.183873378 | 6.65E-06 | 1.62E-05 | 0.04179363 |
| T2DM | mast_cell | *WDR31* | 6.704809572 | 1.11E-08 | 1.07E-07 | 6.95E-05 |
| T2DM | mast_cell | *POLE3* | -2.544395787 | 7.20E-05 | 0.000140679 | 0.452703792 |
| T2DM | mast_cell | *SURF6* | 5.981222025 | 4.02E-06 | 1.04E-05 | 0.025298522 |
| T2DM | mast_cell | *ASB6* | 3.808268154 | 9.60E-06 | 2.24E-05 | 0.060343453 |
| T2DM | mast_cell | *NTMT1* | 2.886792564 | 0.000851304 | 0.001380131 | 1 |
| T2DM | mast_cell | *CIZ1* | 3.847964645 | 1.23E-05 | 2.80E-05 | 0.077452843 |
| T2DM | mast_cell | *SH3GLB2* | 5.144758092 | 5.36E-08 | 2.86E-07 | 0.000336815 |
| T2DM | mast_cell | *INPP5E* | 2.578880993 | 7.56E-08 | 3.72E-07 | 0.000475347 |
| T2DM | mast_cell | *DPH7* | 4.705583728 | 8.80E-05 | 0.000169047 | 0.552951396 |
| T2DM | mast_cell | *SLC39A12* | 3.803198158 | 0.000670874 | 0.001102977 | 1 |
| T2DM | mast_cell | *RSU1* | 2.939638392 | 0.000986101 | 0.001579452 | 1 |
| T2DM | mast_cell | *PARD3* | 2.353378573 | 1.49E-05 | 3.32E-05 | 0.093832865 |
| T2DM | mast_cell | *GLUD1* | 3.898593417 | 8.30E-09 | 9.26E-08 | 5.22E-05 |
| T2DM | mast_cell | *HTR7* | 3.408860186 | 3.55E-09 | 7.78E-08 | 2.23E-05 |
| T2DM | mast_cell | *RPP30* | 4.019819784 | 0.000166312 | 0.000304486 | 1 |
| T2DM | mast_cell | *FRA10AC1* | 2.632936361 | 4.41E-09 | 7.78E-08 | 2.77E-05 |
| T2DM | mast_cell | *DNAJB12* | 3.279915451 | 0.006798379 | 0.009624276 | 1 |
| T2DM | mast_cell | *FUOM* | -3.055615839 | 2.21E-05 | 4.78E-05 | 0.138679055 |
| T2DM | mast_cell | *LRRC27* | 1.331474516 | 4.42E-06 | 1.13E-05 | 0.027816682 |
| T2DM | mast_cell | *PAOX* | 4.874044952 | 0.015927304 | 0.021337089 | 1 |
| T2DM | mast_cell | *GSTO1* | 3.749360328 | 8.21E-05 | 0.000158672 | 0.516318586 |
| T2DM | mast_cell | *PDCD11* | 4.160082445 | 8.25E-07 | 2.61E-06 | 0.005189757 |
| T2DM | mast_cell | *LIN7C* | 2.933061953 | 3.77E-09 | 7.78E-08 | 2.37E-05 |
| T2DM | mast_cell | *ZNF215* | 0.971528289 | 0.000378798 | 0.000650506 | 1 |
| T2DM | mast_cell | *HSD17B12* | -1.142676595 | 0.001129921 | 0.001795706 | 1 |
| T2DM | mast_cell | *EIF3M* | 2.540397639 | 5.21E-05 | 0.000105112 | 0.32773874 |
| T2DM | mast_cell | *GLYAT* | 1.369778684 | 8.80E-10 | 6.08E-08 | 5.53E-06 |
| T2DM | mast_cell | *PTPRJ* | 3.50075292 | 2.10E-09 | 7.78E-08 | 1.32E-05 |
| T2DM | mast_cell | *ENDOD1* | 4.808515705 | 0.001821012 | 0.002821962 | 1 |
| T2DM | mast_cell | *SERPINH1* | 3.148806584 | 0.00012239 | 0.000229622 | 0.769464656 |
| T2DM | mast_cell | *CAPN5* | 5.080502837 | 4.33E-05 | 8.87E-05 | 0.271995764 |
| T2DM | mast_cell | *INTS4* | 2.919563925 | 6.72E-05 | 0.000131997 | 0.422258294 |
| T2DM | mast_cell | *ZC3H12C* | 3.099430971 | 2.79E-09 | 7.78E-08 | 1.75E-05 |
| T2DM | mast_cell | *TTC12* | 1.296908079 | 3.59E-05 | 7.48E-05 | 0.225466313 |
| T2DM | mast_cell | *DRD2* | -0.597832636 | 9.56E-08 | 4.42E-07 | 0.000601184 |
| T2DM | mast_cell | *C11orf52* | 11.94317716 | 1.40E-08 | 1.19E-07 | 8.81E-05 |
| T2DM | mast_cell | *NPAT* | 2.229583634 | 0.019374613 | 0.025703354 | 1 |
| T2DM | mast_cell | *GLB1L2* | -3.379471445 | 0.00021145 | 0.000379175 | 1 |
| T2DM | mast_cell | *P4HA3* | 4.144836006 | 2.17E-08 | 1.48E-07 | 0.000136633 |
| T2DM | mast_cell | *GRIK4* | 1.632530566 | 0.000180155 | 0.000327635 | 1 |
| T2DM | mast_cell | *ADAM33* | 2.536570152 | 5.91E-08 | 3.09E-07 | 0.000371274 |
| T2DM | mast_cell | *MTA2* | 2.350870365 | 0.012164548 | 0.016546628 | 1 |
| T2DM | mast_cell | *TMC2* | 11.15839592 | 2.77E-08 | 1.75E-07 | 0.000173927 |
| T2DM | mast_cell | *ROM1* | -0.980834851 | 0.00194704 | 0.003003936 | 1 |

| | | | | | | |
|---|---|---|---|---|---|---|
| T2DM | mast_cell | *EML3* | 4.225028597 | 2.33E-08 | 1.55E-07 | 0.000146683 |
| T2DM | mast_cell | *INCENP* | 5.16597476 | 3.36E-09 | 7.78E-08 | 2.11E-05 |
| T2DM | mast_cell | *MS4A3* | -4.558971623 | 9.87E-08 | 4.54E-07 | 0.000620756 |
| T2DM | mast_cell | *PLCH2* | 5.421287717 | 1.12E-08 | 1.07E-07 | 7.05E-05 |
| T2DM | mast_cell | *CPSF7* | 3.10333442 | 0.004306484 | 0.006278958 | 1 |
| T2DM | mast_cell | *FEZ1* | 2.216918917 | 4.04E-09 | 7.78E-08 | 2.54E-05 |
| T2DM | mast_cell | *DUSP15* | -3.525912269 | 8.02E-09 | 9.17E-08 | 5.04E-05 |
| T2DM | mast_cell | *COMMD7* | 4.631596565 | 0.000266889 | 0.000470008 | 1 |
| T2DM | mast_cell | *KIAA1755* | 1.36037805 | 1.36E-07 | 5.80E-07 | 0.000854988 |
| T2DM | mast_cell | *CNBD2* | 4.703518795 | 2.48E-09 | 7.78E-08 | 1.56E-05 |
| T2DM | mast_cell | *CABLES2* | 1.910926969 | 4.82E-07 | 1.65E-06 | 0.003031506 |
| T2DM | mast_cell | *TRPT1* | 4.09092463 | 9.82E-06 | 2.29E-05 | 0.061720487 |
| T2DM | mast_cell | *TBX6* | 1.508605127 | 0.000311587 | 0.000543095 | 1 |
| T2DM | mast_cell | *TAOK2* | 4.066091395 | 1.33E-07 | 5.70E-07 | 0.000835648 |
| T2DM | mast_cell | *MPP7* | 0.829142334 | 0.021195631 | 0.027889689 | 1 |
| T2DM | mast_cell | *CWC15* | 2.14806594 | 0.009502651 | 0.013176702 | 1 |
| T2DM | mast_cell | *ARID5B* | 3.128904637 | 2.04E-06 | 5.71E-06 | 0.012825276 |
| T2DM | mast_cell | *TMCO3* | 4.723256314 | 1.90E-07 | 7.65E-07 | 0.001192631 |
| T2DM | mast_cell | *LYPD6B* | 2.494649616 | 4.12E-08 | 2.35E-07 | 0.000258827 |
| T2DM | mast_cell | *WDR17* | -2.370312325 | 0.00507111 | 0.007312401 | 1 |
| T2DM | mast_cell | *CCDC102B* | 3.60007695 | 1.82E-08 | 1.35E-07 | 0.000114722 |
| T2DM | mast_cell | *CNDP1* | 2.163434641 | 7.08E-05 | 0.000138488 | 0.445239268 |
| T2DM | mast_cell | *FSIP1* | -0.603951455 | 1.24E-07 | 5.40E-07 | 0.000780846 |
| T2DM | mast_cell | *RGS18* | 2.759140866 | 5.06E-09 | 7.84E-08 | 3.18E-05 |
| T2DM | mast_cell | *PRSS23* | 2.26922654 | 8.78E-06 | 2.07E-05 | 0.055228575 |
| T2DM | mast_cell | *MTMR12* | 3.609002176 | 6.91E-09 | 8.77E-08 | 4.34E-05 |
| T2DM | mast_cell | *PPP1R1C* | 2.848999664 | 1.28E-09 | 6.77E-08 | 8.05E-06 |
| T2DM | mast_cell | *C11orf53* | 11.21951829 | 1.08E-07 | 4.88E-07 | 0.000680232 |
| T2DM | mast_cell | *CCT5* | 3.624465451 | 0.00066201 | 0.001090972 | 1 |
| T2DM | mast_cell | *DOCK1* | 2.995916786 | 2.04E-07 | 8.10E-07 | 0.001281953 |
| T2DM | mast_cell | *TEX12* | -3.831717152 | 1.59E-07 | 6.61E-07 | 0.001000496 |
| T2DM | mast_cell | *CRIM1* | 2.156263695 | 0.004013989 | 0.005878394 | 1 |
| T2DM | mast_cell | *ABCB9* | 2.114402847 | 0.000667741 | 0.001098688 | 1 |
| T2DM | mast_cell | *DHX37* | 6.501748867 | 2.87E-08 | 1.79E-07 | 0.000180139 |
| T2DM | mast_cell | *GPR158* | 2.067870255 | 3.52E-07 | 1.26E-06 | 0.002213087 |
| T2DM | mast_cell | *THRB* | 2.654045606 | 0.000245606 | 0.000435824 | 1 |
| T2DM | mast_cell | *BTBD11* | 3.931321861 | 3.18E-09 | 7.78E-08 | 2.00E-05 |
| T2DM | mast_cell | *UBE3B* | 1.941183353 | 0.000509838 | 0.00085636 | 1 |
| T2DM | mast_cell | *ANK3* | 2.109652918 | 7.86E-10 | 5.88E-08 | 4.94E-06 |
| T2DM | mast_cell | *IPMK* | 4.520336584 | 1.57E-07 | 6.55E-07 | 0.00098988 |
| T2DM | mast_cell | *FAM177A1* | 3.058668641 | 0.00330303 | 0.004918558 | 1 |
| T2DM | mast_cell | *MIPOL1* | 3.010313254 | 3.73E-08 | 2.17E-07 | 0.000234612 |
| T2DM | mast_cell | *TMEM18* | 3.369635003 | 3.12E-09 | 7.78E-08 | 1.96E-05 |
| T2DM | mast_cell | *NDUFC2* | 5.565055086 | 8.69E-07 | 2.73E-06 | 0.005461623 |
| T2DM | mast_cell | *ME3* | 3.041365175 | 1.92E-07 | 7.72E-07 | 0.001208956 |
| T2DM | mast_cell | *ADAMTS12* | 2.899823273 | 7.30E-09 | 8.89E-08 | 4.59E-05 |
| T2DM | mast_cell | *VIPAS39* | 3.566609681 | 4.67E-06 | 1.18E-05 | 0.029360124 |
| T2DM | mast_cell | *ANKRD50* | 4.78590786 | 6.48E-06 | 1.59E-05 | 0.040739721 |
| T2DM | mast_cell | *C4orf33* | -2.270365602 | 0.002698277 | 0.004073984 | 1 |
| T2DM | mast_cell | *PTPRO* | 3.98925025 | 2.61E-08 | 1.68E-07 | 0.000164141 |

| | | | | | | |
|---|---|---|---|---|---|---|
| T2DM | mast_cell | *VPS26B* | 4.501846491 | 5.73E-05 | 0.000114561 | 0.360410127 |
| T2DM | mast_cell | *NCAPD3* | 3.403026288 | 7.26E-06 | 1.75E-05 | 0.045674092 |
| T2DM | mast_cell | *VTI1A* | 2.182922615 | 0.00296335 | 0.004447501 | 1 |
| T2DM | mast_cell | *FAM160B1* | 3.347612373 | 5.35E-09 | 8.00E-08 | 3.37E-05 |
| T2DM | mast_cell | *ANO4* | 1.629484299 | 1.41E-08 | 1.19E-07 | 8.88E-05 |
| T2DM | mast_cell | *TEX9* | 4.247991314 | 1.33E-05 | 3.00E-05 | 0.083515299 |
| T2DM | mast_cell | *AKR1C2* | 2.469347489 | 4.34E-08 | 2.44E-07 | 0.000272767 |
| T2DM | mast_cell | *ADAM8* | 2.12581994 | 2.05E-08 | 1.43E-07 | 0.000128938 |
| T2DM | mast_cell | *ITIH2* | 2.814294847 | 1.29E-08 | 1.13E-07 | 8.10E-05 |
| T2DM | mast_cell | *KIN* | 3.863436976 | 5.41E-08 | 2.89E-07 | 0.000340313 |
| T2DM | mast_cell | *PIGF* | 4.106767424 | 1.70E-06 | 4.87E-06 | 0.010699612 |
| T2DM | mast_cell | *INPP1* | 3.85412897 | 2.99E-09 | 7.78E-08 | 1.88E-05 |
| T2DM | mast_cell | *RNF144A* | 3.797981359 | 4.71E-08 | 2.60E-07 | 0.000296219 |
| T2DM | mast_cell | *ASAP2* | 3.705292349 | 8.94E-08 | 4.21E-07 | 0.000561898 |
| T2DM | mast_cell | *ADAM17* | 3.707628158 | 1.44E-05 | 3.21E-05 | 0.090651774 |
| T2DM | mast_cell | *TMEM45B* | 7.016566716 | 0.000855604 | 0.001386744 | 1 |
| T2DM | mast_cell | *ACSL1* | 1.46696336 | 0.000725533 | 0.001187565 | 1 |
| T2DM | mast_cell | *BICD1* | 3.793499228 | 2.24E-06 | 6.21E-06 | 0.014095019 |
| T2DM | mast_cell | *GUF1* | 2.918604006 | 0.005838347 | 0.008365015 | 1 |
| T2DM | mast_cell | *GABRA2* | -1.572319797 | 1.92E-06 | 5.41E-06 | 0.012051197 |
| T2DM | mast_cell | *CENPJ* | 2.866886081 | 0.002000911 | 0.003081755 | 1 |
| T2DM | mast_cell | *PARP8* | 2.520927435 | 3.28E-07 | 1.19E-06 | 0.002063387 |
| T2DM | mast_cell | *CACUL1* | 3.025861035 | 0.000905292 | 0.001462752 | 1 |
| T2DM | mast_cell | *BEND6* | 0.983333949 | 0.002087185 | 0.00320521 | 1 |
| T2DM | mast_cell | *RABGAP1L* | 2.066053701 | 0.003700545 | 0.005457747 | 1 |
| T2DM | mast_cell | *PTPN14* | 2.546052602 | 1.04E-06 | 3.18E-06 | 0.006545528 |
| T2DM | mast_cell | *RNF219* | 4.719741173 | 1.34E-06 | 3.94E-06 | 0.008422306 |
| T2DM | mast_cell | *ARL14EP* | 1.676411379 | 0.021614364 | 0.028410937 | 1 |
| T2DM | mast_cell | *G6PC2* | -51.73854295 | 4.65E-07 | 1.60E-06 | 0.002922981 |
| T2DM | mast_cell | *TADA1* | 3.606518742 | 3.34E-09 | 7.78E-08 | 2.10E-05 |
| T2DM | mast_cell | *GUCY1A2* | 2.333420555 | 1.92E-06 | 5.42E-06 | 0.012090209 |
| T2DM | mast_cell | *CWF19L2* | 3.351843 | 5.75E-05 | 0.000114859 | 0.36157567 |
| T2DM | mast_cell | *ZNF837* | 1.883392837 | 3.60E-05 | 7.50E-05 | 0.226448495 |
| T2DM | mast_cell | *CCDC50* | 1.203545802 | 0.021595432 | 0.028391986 | 1 |
| T2DM | mast_cell | *ZFP36L2* | 3.518460226 | 5.71E-05 | 0.000114189 | 0.358896388 |
| T2DM | mast_cell | *PLEKHH2* | 0.598898002 | 0.003216414 | 0.004800949 | 1 |
| T2DM | mast_cell | *TMEM123* | 4.331674535 | 6.07E-07 | 2.01E-06 | 0.003819299 |
| T2DM | mast_cell | *IGSF10* | 1.772934788 | 0.035089868 | 0.044921604 | 1 |
| T2DM | mast_cell | *SPEF2* | 2.710584029 | 6.18E-07 | 2.04E-06 | 0.003887578 |
| T2DM | mast_cell | *SPARCL1* | 1.674374809 | 7.97E-08 | 3.88E-07 | 0.000500847 |
| T2DM | mast_cell | *MBNL1* | 2.417569029 | 5.06E-05 | 0.000102287 | 0.318213751 |
| T2DM | mast_cell | *CCNO* | 7.36037674 | 0.001773864 | 0.00275365 | 1 |
| T2DM | mast_cell | *CLEC4F* | -4.399968434 | 1.22E-06 | 3.61E-06 | 0.007649271 |
| T2DM | mast_cell | *PELO* | 3.785339517 | 4.17E-09 | 7.78E-08 | 2.62E-05 |
| T2DM | mast_cell | *RASGRP3* | 1.595385824 | 0.000209548 | 0.000376086 | 1 |
| T2DM | mast_cell | *SAR1B* | 4.669362001 | 5.73E-11 | 1.16E-08 | 3.61E-07 |
| T2DM | mast_cell | *GPR180* | 2.038822874 | 0.000172963 | 0.000315652 | 1 |
| T2DM | mast_cell | *TCTEX1D1* | -0.855839971 | 6.24E-06 | 1.53E-05 | 0.039229825 |
| T2DM | mast_cell | *WDR78* | 1.939508492 | 0.025074331 | 0.032678755 | 1 |
| T2DM | mast_cell | *PRDM8* | 1.432099543 | 2.36E-06 | 6.48E-06 | 0.01482687 |

| | | | | | | |
|---|---|---|---|---|---|---|
| T2DM | mast_cell | *HHEX* | 2.783978561 | 0.000804792 | 0.001307762 | 1 |
| T2DM | mast_cell | *ZNF117* | 0.562089379 | 0.003903112 | 0.00572669 | 1 |
| T2DM | mast_cell | *MED21* | 5.255939882 | 7.43E-07 | 2.38E-06 | 0.0046696 |
| T2DM | mast_cell | *SRP19* | 5.910972206 | 1.52E-07 | 6.35E-07 | 0.000953753 |
| T2DM | mast_cell | *TEKT5* | 4.01604853 | 3.21E-09 | 7.78E-08 | 2.02E-05 |
| T2DM | mast_cell | *BANK1* | 2.03040803 | 3.79E-09 | 7.78E-08 | 2.38E-05 |
| T2DM | mast_cell | *SCOC* | 3.596572229 | 8.01E-09 | 9.17E-08 | 5.04E-05 |
| T2DM | mast_cell | *CLGN* | -1.651378661 | 1.39E-07 | 5.91E-07 | 0.000875152 |
| T2DM | mast_cell | *SYCP2L* | -0.666333115 | 2.56E-07 | 9.69E-07 | 0.001609651 |
| T2DM | mast_cell | *AHCTF1* | 3.77906693 | 1.08E-08 | 1.06E-07 | 6.78E-05 |
| T2DM | mast_cell | *TMEM87B* | 2.747555477 | 0.002075644 | 0.003191385 | 1 |
| T2DM | mast_cell | *PTPRR* | 3.737746368 | 1.09E-08 | 1.06E-07 | 6.84E-05 |
| T2DM | mast_cell | *PLA2R1* | 3.40000411 | 1.63E-10 | 2.44E-08 | 1.02E-06 |
| T2DM | mast_cell | *SLC25A27* | 0.856084903 | 0.012905333 | 0.017482402 | 1 |
| T2DM | mast_cell | *FRMD1* | 1.760164091 | 1.45E-08 | 1.19E-07 | 9.14E-05 |
| T2DM | mast_cell | *ASAP1* | 2.912447023 | 7.23E-06 | 1.74E-05 | 0.045457064 |
| T2DM | mast_cell | *INO80C* | -3.099175402 | 0.000205012 | 0.000368682 | 1 |
| T2DM | mast_cell | *LPCAT1* | 3.767563894 | 4.35E-09 | 7.78E-08 | 2.74E-05 |
| T2DM | mast_cell | *TEX29* | 3.050035145 | 1.62E-05 | 3.57E-05 | 0.101636281 |
| T2DM | mast_cell | *ADPRHL1* | 1.582489364 | 4.68E-05 | 9.53E-05 | 0.294478192 |
| T2DM | mast_cell | *TUBGCP5* | 4.235037848 | 1.36E-08 | 1.17E-07 | 8.57E-05 |
| T2DM | mast_cell | *LURAP1L* | 1.359096636 | 0.000372031 | 0.000640459 | 1 |
| T2DM | mast_cell | *CFDP1* | 3.791463389 | 1.93E-07 | 7.75E-07 | 0.001213693 |
| T2DM | mast_cell | *ZDHHC7* | 4.335055377 | 2.91E-08 | 1.81E-07 | 0.000182674 |
| T2DM | mast_cell | *FAM92B* | -2.527434044 | 9.69E-08 | 4.47E-07 | 0.000608929 |
| T2DM | mast_cell | *C7orf31* | 3.700605706 | 8.19E-09 | 9.23E-08 | 5.15E-05 |
| T2DM | mast_cell | *FBXO36* | 4.662646533 | 4.28E-09 | 7.78E-08 | 2.69E-05 |
| T2DM | mast_cell | *CACNA2D1* | 2.79050718 | 2.94E-06 | 7.89E-06 | 0.018479613 |
| T2DM | mast_cell | *ZUFSP* | 4.546041303 | 8.71E-06 | 2.05E-05 | 0.054784833 |
| T2DM | mast_cell | *HS3ST3A1* | 1.934386281 | 4.60E-07 | 1.58E-06 | 0.002892822 |
| T2DM | mast_cell | *PPP2R5E* | 3.383499368 | 9.61E-07 | 2.97E-06 | 0.006042652 |
| T2DM | mast_cell | *ANKRD29* | 1.849462055 | 1.51E-09 | 6.93E-08 | 9.50E-06 |
| T2DM | mast_cell | *THY1* | 2.793325559 | 2.90E-09 | 7.78E-08 | 1.82E-05 |
| T2DM | mast_cell | *UBASH3B* | 4.492248447 | 9.40E-06 | 2.20E-05 | 0.05911401 |
| T2DM | mast_cell | *ROBO4* | 3.456666324 | 2.57E-07 | 9.73E-07 | 0.001618376 |
| T2DM | mast_cell | *ROBO3* | 2.377893412 | 2.42E-06 | 6.61E-06 | 0.015210338 |
| T2DM | mast_cell | *CC2D1B* | 1.49217248 | 0.001724041 | 0.002678954 | 1 |
| T2DM | mast_cell | *PRKCA* | -1.088221692 | 4.53E-06 | 1.16E-05 | 0.02848982 |
| T2DM | mast_cell | *LRRK1* | 4.31421613 | 1.94E-07 | 7.76E-07 | 0.001217003 |
| T2DM | mast_cell | *CEP112* | 2.435094211 | 2.29E-07 | 8.88E-07 | 0.001438416 |
| T2DM | mast_cell | *GAL3ST2* | 6.835207343 | 0.005142349 | 0.007408329 | 1 |
| T2DM | mast_cell | *ENPP3* | -0.283504501 | 1.16E-07 | 5.14E-07 | 0.000726773 |
| T2DM | mast_cell | *UCHL1* | 6.640838296 | 5.43E-07 | 1.82E-06 | 0.003411782 |
| T2DM | mast_cell | *MIA3* | 3.313521526 | 0.0002332 | 0.000415217 | 1 |
| T2DM | mast_cell | *DISP1* | 2.194871928 | 7.37E-05 | 0.000143756 | 0.463612509 |
| T2DM | mast_cell | *FAM167A* | 4.83013611 | 0.000163365 | 0.000299613 | 1 |
| T2DM | mast_cell | *NEIL2* | 1.824593635 | 7.37E-08 | 3.66E-07 | 0.000463284 |
| T2DM | mast_cell | *CCSAP* | 3.706185563 | 8.51E-08 | 4.06E-07 | 0.000534725 |
| T2DM | mast_cell | *GBP5* | -2.370692487 | 0.004926096 | 0.007116352 | 1 |
| T2DM | mast_cell | *FAM69A* | 2.338112814 | 5.48E-07 | 1.84E-06 | 0.003446401 |

| | | | | | | |
|---|---|---|---|---|---|---|
| T2DM | mast_cell | *ATP5G3* | 2.871780937 | 4.55E-05 | 9.28E-05 | 0.285864939 |
| T2DM | mast_cell | *PDLIM3* | 1.906476817 | 5.39E-09 | 8.00E-08 | 3.39E-05 |
| T2DM | mast_cell | *NCAM2* | 1.887865998 | 3.47E-05 | 7.26E-05 | 0.218397163 |
| T2DM | mast_cell | *L3MBTL4* | 3.613540095 | 1.24E-08 | 1.12E-07 | 7.77E-05 |
| T2DM | mast_cell | *PDE1C* | 2.049944517 | 6.14E-09 | 8.15E-08 | 3.86E-05 |
| T2DM | mast_cell | *MRPL39* | 1.16614213 | 0.011604102 | 0.015852887 | 1 |
| T2DM | mast_cell | *TSEN2* | 3.932751987 | 1.02E-08 | 1.04E-07 | 6.43E-05 |
| T2DM | mast_cell | *FLCN* | 2.615245678 | 0.000565399 | 0.000944133 | 1 |
| T2DM | mast_cell | *DPH3* | 4.132267705 | 3.39E-09 | 7.78E-08 | 2.13E-05 |
| T2DM | mast_cell | *CXXC1* | 6.530618418 | 7.12E-08 | 3.55E-07 | 0.000447832 |
| T2DM | mast_cell | *PPP4R1* | 4.031712115 | 1.37E-05 | 3.09E-05 | 0.086430848 |
| T2DM | mast_cell | *APCDD1* | 1.326902117 | 3.99E-06 | 1.03E-05 | 0.025085598 |
| T2DM | mast_cell | *MPPE1* | 0.986123578 | 0.028813489 | 0.037204848 | 1 |
| T2DM | mast_cell | *EPHB1* | 4.239031668 | 0.000126557 | 0.000236734 | 0.795661469 |
| T2DM | mast_cell | *ANKRD40* | 3.507205406 | 7.18E-06 | 1.73E-05 | 0.045120953 |
| T2DM | mast_cell | *CYP2U1* | 5.237850336 | 7.19E-09 | 8.83E-08 | 4.52E-05 |
| T2DM | mast_cell | *FBXL18* | 5.014233761 | 9.24E-05 | 0.00017713 | 0.58080966 |
| T2DM | mast_cell | *PROM2* | 6.203156845 | 2.41E-07 | 9.24E-07 | 0.001514026 |
| T2DM | mast_cell | *CDK19* | 3.055084975 | 0.00045961 | 0.00077802 | 1 |
| T2DM | mast_cell | *GTF3C6* | 3.34837946 | 4.63E-07 | 1.59E-06 | 0.002907822 |
| T2DM | mast_cell | *TTC39B* | -1.475569848 | 0.000196822 | 0.000355258 | 1 |
| T2DM | mast_cell | *AGPAT5* | -1.923887238 | 0.000107939 | 0.000204648 | 0.678611641 |
| T2DM | mast_cell | *GOLGA7B* | 2.710897329 | 1.20E-05 | 2.74E-05 | 0.075401038 |
| T2DM | mast_cell | *TRMT44* | 5.009885942 | 1.04E-07 | 4.72E-07 | 0.000651211 |
| T2DM | mast_cell | *C16orf87* | 3.239701576 | 3.20E-09 | 7.78E-08 | 2.01E-05 |
| T2DM | mast_cell | *HEATR3* | 1.216360604 | 0.001493983 | 0.002339272 | 1 |
| T2DM | mast_cell | *CNOT8* | 3.866026631 | 2.00E-06 | 5.62E-06 | 0.012560958 |
| T2DM | mast_cell | *ZKSCAN2* | 1.226006941 | 0.012431506 | 0.016891481 | 1 |
| T2DM | mast_cell | *PDIA4* | 5.482685429 | 1.70E-08 | 1.30E-07 | 0.00010716 |
| T2DM | mast_cell | *FAM126B* | 3.711334307 | 1.12E-06 | 3.37E-06 | 0.007050197 |
| T2DM | mast_cell | *ALS2CR12* | 2.352936907 | 0.00061798 | 0.001024319 | 1 |
| T2DM | mast_cell | *SPAG17* | 3.46062403 | 3.46E-08 | 2.05E-07 | 0.000217619 |
| T2DM | mast_cell | *DEPTOR* | 4.491687744 | 1.98E-07 | 7.89E-07 | 0.001242326 |
| T2DM | mast_cell | *RRAGA* | 5.803128195 | 3.52E-07 | 1.26E-06 | 0.002214851 |
| T2DM | mast_cell | *FAM161B* | 2.634367844 | 7.85E-10 | 5.88E-08 | 4.94E-06 |
| T2DM | mast_cell | *WIF1* | 6.533553043 | 5.10E-06 | 1.28E-05 | 0.032040666 |
| T2DM | mast_cell | *UGT2B4* | 2.35872133 | 1.50E-08 | 1.21E-07 | 9.46E-05 |
| T2DM | mast_cell | *MMP16* | 1.314366782 | 0.000813742 | 0.001321622 | 1 |
| T2DM | mast_cell | *KCNMA1* | -1.43728368 | 0.000368146 | 0.000634119 | 1 |
| T2DM | mast_cell | *BATF* | -0.648853761 | 1.54E-07 | 6.45E-07 | 0.000970789 |
| T2DM | mast_cell | *DCK* | 3.654496462 | 6.89E-05 | 0.000135026 | 0.433029652 |
| T2DM | mast_cell | *DPY19L4* | 3.60267919 | 0.000109437 | 0.000207176 | 0.688031256 |
| T2DM | mast_cell | *DRAM2* | 3.496146405 | 1.03E-06 | 3.16E-06 | 0.006479819 |
| T2DM | mast_cell | *C8orf37* | 2.036361606 | 0.000438508 | 0.000745915 | 1 |
| T2DM | mast_cell | *CXCL13* | -4.272544139 | 2.47E-05 | 5.29E-05 | 0.155401511 |
| T2DM | mast_cell | *N6AMT1* | 0.941484962 | 0.008112674 | 0.011367145 | 1 |
| T2DM | mast_cell | *CCT8* | 1.230079203 | 0.001485624 | 0.002328626 | 1 |
| T2DM | mast_cell | *ANKRD9* | 3.644149085 | 6.89E-06 | 1.67E-05 | 0.043321705 |
| T2DM | mast_cell | *SFXN2* | 3.523987741 | 5.53E-06 | 1.38E-05 | 0.034762601 |
| T2DM | mast_cell | *TDRD9* | 1.495506244 | 0.003030673 | 0.004544203 | 1 |

| | | | | | | |
|---|---|---|---|---|---|---|
| T2DM | mast_cell | *SH3RF2* | 1.508984176 | 0.001738521 | 0.002699452 | 1 |
| T2DM | mast_cell | *GDF6* | 4.148349232 | 1.84E-09 | 7.58E-08 | 1.16E-05 |
| T2DM | mast_cell | *NODAL* | 6.350890511 | 4.41E-09 | 7.78E-08 | 2.77E-05 |
| T2DM | mast_cell | *MED19* | 1.236731725 | 0.006057505 | 0.008659284 | 1 |
| T2DM | mast_cell | *WDYHV1* | 2.181942558 | 9.17E-05 | 0.000176002 | 0.576759499 |
| T2DM | mast_cell | *ATAD2* | 4.379144508 | 9.94E-09 | 1.04E-07 | 6.25E-05 |
| T2DM | mast_cell | *ITGAD* | -1.281082531 | 0.000276909 | 0.0004867 | 1 |
| T2DM | mast_cell | *VPS8* | 3.198830704 | 9.24E-06 | 2.17E-05 | 0.058104487 |
| T2DM | mast_cell | *RPUSD3* | 6.337055919 | 1.65E-05 | 3.64E-05 | 0.103860645 |
| T2DM | mast_cell | *GHRL* | 6.178014761 | 2.00E-08 | 1.42E-07 | 0.000125707 |
| T2DM | mast_cell | *EXOG* | 4.245910988 | 8.69E-08 | 4.12E-07 | 0.000546219 |
| T2DM | mast_cell | *NTAN1* | 3.278354568 | 1.56E-08 | 1.23E-07 | 9.79E-05 |
| T2DM | mast_cell | *SLC6A1* | -2.094398023 | 0.038634767 | 0.049169389 | 1 |
| T2DM | mast_cell | *FCHO2* | 3.58940022 | 1.04E-06 | 3.19E-06 | 0.006562181 |
| T2DM | mast_cell | *TIMP4* | 2.115595175 | 0.035671415 | 0.045628929 | 1 |
| T2DM | mast_cell | *CPT2* | 5.193120048 | 0.000909928 | 0.001468357 | 1 |
| T2DM | mast_cell | *NECAP2* | 4.601972701 | 5.74E-08 | 3.02E-07 | 0.000361096 |
| T2DM | mast_cell | *LRP8* | 4.772436059 | 1.02E-07 | 4.67E-07 | 0.000642628 |
| T2DM | mast_cell | *STEAP2* | 1.181478283 | 4.77E-07 | 1.63E-06 | 0.00299856 |
| T2DM | mast_cell | *GATAD1* | 2.655634912 | 1.33E-05 | 3.01E-05 | 0.083768134 |
| T2DM | mast_cell | *SUSD3* | -0.862553278 | 0.02672717 | 0.034631847 | 1 |
| T2DM | mast_cell | *TMED6* | 22.41837301 | 5.13E-09 | 7.89E-08 | 3.23E-05 |
| T2DM | mast_cell | *CLEC18A* | -3.76605225 | 5.50E-14 | 4.32E-11 | 3.46E-10 |
| T2DM | mast_cell | *IL34* | 2.460355645 | 3.56E-09 | 7.78E-08 | 2.24E-05 |
| T2DM | mast_cell | *DHRS1* | 0.961234607 | 0.03216718 | 0.041314619 | 1 |
| T2DM | mast_cell | *RNF111* | 3.286061302 | 1.96E-07 | 7.85E-07 | 0.001233708 |
| T2DM | mast_cell | *DYRK1A* | 2.868577608 | 0.000250109 | 0.000443064 | 1 |
| T2DM | mast_cell | *KCNJ15* | -1.650432536 | 0.021459018 | 0.028230351 | 1 |
| T2DM | mast_cell | *C2CD2* | 1.152758282 | 0.000446478 | 0.000758241 | 1 |
| T2DM | mast_cell | *SLC38A10* | 3.808819482 | 2.16E-08 | 1.48E-07 | 0.000135856 |
| T2DM | mast_cell | *C9orf43* | 2.173157783 | 0.000205143 | 0.000368812 | 1 |
| T2DM | mast_cell | *SLC37A3* | 4.190612925 | 3.11E-08 | 1.90E-07 | 0.00019533 |
| T2DM | mast_cell | *AP3S2* | 2.735524628 | 1.13E-10 | 1.92E-08 | 7.10E-07 |
| T2DM | mast_cell | *SPPL3* | 2.205598296 | 0.004247846 | 0.006199212 | 1 |
| T2DM | mast_cell | *FAM213B* | -1.262625682 | 0.006844699 | 0.009685487 | 1 |
| T2DM | mast_cell | *C12orf43* | 4.413182638 | 1.44E-08 | 1.19E-07 | 9.08E-05 |
| T2DM | mast_cell | *PEX10* | -2.952135327 | 1.75E-05 | 3.83E-05 | 0.109725708 |
| T2DM | mast_cell | *TRIM63* | -0.221316253 | 3.58E-07 | 1.28E-06 | 0.002251648 |
| T2DM | mast_cell | *WDR66* | -1.040739151 | 0.019393104 | 0.025722457 | 1 |
| T2DM | mast_cell | *GALNT14* | -1.024712023 | 2.03E-07 | 8.07E-07 | 0.001275362 |
| T2DM | mast_cell | *HPD* | 2.109717422 | 0.00052332 | 0.00087713 | 1 |
| T2DM | mast_cell | *RHPN1* | 2.501128715 | 1.89E-05 | 4.12E-05 | 0.118626803 |
| T2DM | mast_cell | *TPRG1L* | 4.998859795 | 0.001283804 | 0.00202745 | 1 |
| T2DM | mast_cell | *LRRC43* | 4.075443505 | 1.44E-08 | 1.19E-07 | 9.03E-05 |
| T2DM | mast_cell | *XKR8* | -2.568441733 | 0.011877881 | 0.016202264 | 1 |
| T2DM | mast_cell | *EYA3* | 2.704970024 | 0.000865109 | 0.001401066 | 1 |
| T2DM | mast_cell | *MRAS* | 1.042597041 | 0.002141702 | 0.003279318 | 1 |
| T2DM | mast_cell | *ABHD3* | 2.16092233 | 0.001486798 | 0.002329886 | 1 |
| T2DM | mast_cell | *CLSTN2* | 2.064472012 | 1.51E-08 | 1.21E-07 | 9.52E-05 |
| T2DM | mast_cell | *COLEC12* | 2.206650872 | 3.80E-09 | 7.78E-08 | 2.39E-05 |

| | | | | | | |
|---|---|---|---|---|---|---|
| T2DM | mast_cell | *RNF207* | 8.648026849 | 1.32E-06 | 3.88E-06 | 0.008284336 |
| T2DM | mast_cell | *AUTS2* | 2.786954717 | 5.47E-06 | 1.36E-05 | 0.034379487 |
| T2DM | mast_cell | *MITD1* | 1.917904685 | 0.024793053 | 0.032352413 | 1 |
| T2DM | mast_cell | *TSPAN33* | -1.213889763 | 0.001161062 | 0.001841937 | 1 |
| T2DM | mast_cell | *AHCYL2* | 3.547769152 | 3.98E-07 | 1.40E-06 | 0.002502841 |
| T2DM | mast_cell | *CD1D* | 2.301787967 | 1.99E-05 | 4.33E-05 | 0.125087816 |
| T2DM | mast_cell | *FAM86C1* | 3.234056791 | 2.89E-08 | 1.80E-07 | 0.000181719 |
| T2DM | mast_cell | *CD1E* | 7.505774139 | 2.56E-10 | 3.36E-08 | 1.61E-06 |
| T2DM | mast_cell | *NCF1* | 3.168551055 | 8.53E-09 | 9.38E-08 | 5.36E-05 |
| T2DM | mast_cell | *PPP1R9A* | 2.181758856 | 2.83E-08 | 1.78E-07 | 0.000177749 |
| T2DM | mast_cell | *ZFAND2B* | 2.934674616 | 9.88E-08 | 4.54E-07 | 0.000620949 |
| T2DM | mast_cell | *GDPD5* | 3.524779866 | 2.82E-08 | 1.78E-07 | 0.000177477 |
| T2DM | mast_cell | *DYNC1I1* | 4.867148676 | 1.00E-07 | 4.60E-07 | 0.000630229 |
| T2DM | mast_cell | *TMED4* | 3.691007201 | 0.003427606 | 0.005085995 | 1 |
| T2DM | mast_cell | *ELK4* | 3.761600154 | 0.001488734 | 0.002332337 | 1 |
| T2DM | mast_cell | *RNF166* | 3.052884791 | 0.020696588 | 0.027307335 | 1 |
| T2DM | mast_cell | *SPATA2L* | -3.163257227 | 8.09E-06 | 1.93E-05 | 0.050840878 |
| T2DM | mast_cell | *NIT1* | 1.706937981 | 0.003800344 | 0.005590258 | 1 |
| T2DM | mast_cell | *ZNF276* | 2.58906552 | 0.011655566 | 0.015916278 | 1 |
| T2DM | mast_cell | *FGF17* | 5.98686024 | 1.60E-05 | 3.54E-05 | 0.100734029 |
| T2DM | mast_cell | *CDA* | 2.566023374 | 2.13E-05 | 4.62E-05 | 0.133799527 |
| T2DM | mast_cell | *B4GALT3* | 3.015443381 | 0.001401353 | 0.002203127 | 1 |
| T2DM | mast_cell | *DMTN* | 1.495519169 | 4.27E-09 | 7.78E-08 | 2.69E-05 |
| T2DM | mast_cell | *FAM160B2* | 2.791664294 | 3.66E-08 | 2.14E-07 | 0.000230374 |
| T2DM | mast_cell | *NDUFS2* | 2.092237969 | 0.002129986 | 0.003264559 | 1 |
| T2DM | mast_cell | *SLC5A11* | 0.583556671 | 0.008125136 | 0.01138207 | 1 |
| T2DM | mast_cell | *FCER1G* | 1.708184173 | 0.000756999 | 0.001235207 | 1 |
| T2DM | mast_cell | *CCAR2* | 5.272747588 | 1.02E-05 | 2.37E-05 | 0.064267869 |
| T2DM | mast_cell | *RAPGEF6* | -3.570428712 | 0.001972738 | 0.003041345 | 1 |
| T2DM | mast_cell | *EPB41* | 2.42541527 | 1.78E-08 | 1.33E-07 | 0.000112082 |
| T2DM | mast_cell | *ALG8* | 3.206756444 | 1.62E-08 | 1.26E-07 | 0.000101911 |
| T2DM | mast_cell | *IFNAR2* | 2.059061223 | 9.89E-05 | 0.000188557 | 0.621670842 |
| T2DM | mast_cell | *MRPL10* | 2.03202849 | 0.0015704 | 0.002451952 | 1 |
| T2DM | mast_cell | *SV2A* | 7.110552037 | 1.23E-08 | 1.12E-07 | 7.76E-05 |
| T2DM | mast_cell | *RCAN1* | 3.359669002 | 0.000288587 | 0.000505929 | 1 |
| T2DM | mast_cell | *UBE2Z* | 3.977319279 | 3.85E-06 | 1.00E-05 | 0.024195221 |
| T2DM | mast_cell | *CLIC6* | -1.263879827 | 0.000874903 | 0.001415834 | 1 |
| T2DM | mast_cell | *CBR1* | 1.279838353 | 0.000455774 | 0.000772359 | 1 |
| T2DM | mast_cell | *CBR3* | -1.306036043 | 0.026614545 | 0.034493021 | 1 |
| T2DM | mast_cell | *SIM2* | 2.868457317 | 0.001399571 | 0.002200876 | 1 |
| T2DM | mast_cell | *ARHGAP27* | 4.228471649 | 4.69E-09 | 7.82E-08 | 2.95E-05 |
| T2DM | mast_cell | *PADI4* | 4.299107114 | 7.51E-09 | 9.02E-08 | 4.72E-05 |
| T2DM | mast_cell | *PSMD4* | 5.080809506 | 7.29E-08 | 3.62E-07 | 0.000458234 |
| T2DM | mast_cell | *STARD9* | 2.987313699 | 2.21E-08 | 1.50E-07 | 0.000139005 |
| T2DM | mast_cell | *THEM4* | 3.35919327 | 1.42E-08 | 1.19E-07 | 8.93E-05 |
| T2DM | mast_cell | *TCHH* | 3.142259001 | 3.78E-05 | 7.85E-05 | 0.237894615 |
| T2DM | mast_cell | *MED8* | 2.687586544 | 8.61E-06 | 2.03E-05 | 0.054118208 |
| T2DM | mast_cell | *RSPRY1* | 3.223065705 | 0.00046331 | 0.000783441 | 1 |
| T2DM | mast_cell | *CCDC17* | 2.179840798 | 7.75E-05 | 0.000150533 | 0.487491369 |
| T2DM | mast_cell | *ACE* | 2.569699774 | 1.03E-08 | 1.04E-07 | 6.46E-05 |

| | | | | | | |
|---|---|---|---|---|---|---|
| T2DM | mast_cell | *TEPP* | 9.515042393 | 0.000157466 | 0.000290064 | 0.989988191 |
| T2DM | mast_cell | *EFCAB14* | 3.449136871 | 0.001795461 | 0.002784426 | 1 |
| T2DM | mast_cell | *ATP6V0D1* | 3.342794821 | 0.000104398 | 0.000198414 | 0.656353289 |
| T2DM | mast_cell | *C16orf86* | -1.862755296 | 0.000210778 | 0.000378078 | 1 |
| T2DM | mast_cell | *LYPD5* | -1.9118004 | 2.33E-07 | 9.00E-07 | 0.001467343 |
| T2DM | mast_cell | *CCDC117* | 5.871835884 | 8.72E-07 | 2.74E-06 | 0.005484785 |
| T2DM | mast_cell | *CCDC107* | 2.338940103 | 0.000282377 | 0.000495618 | 1 |
| T2DM | mast_cell | *NPR2* | 4.488095871 | 7.85E-10 | 5.88E-08 | 4.94E-06 |
| T2DM | mast_cell | *CALM3* | 5.260899123 | 3.92E-06 | 1.02E-05 | 0.024657238 |
| T2DM | mast_cell | *DFFA* | 2.720270181 | 0.003131255 | 0.004682731 | 1 |
| T2DM | mast_cell | *ATAD3B* | 3.105748337 | 2.71E-08 | 1.73E-07 | 0.000170282 |
| T2DM | mast_cell | *SSU72* | 5.228578744 | 3.18E-08 | 1.93E-07 | 0.000200045 |
| T2DM | mast_cell | *NR2F6* | 4.587051894 | 2.02E-08 | 1.42E-07 | 0.000127167 |
| T2DM | mast_cell | *CILP2* | 12.47856779 | 2.46E-08 | 1.61E-07 | 0.000154524 |
| T2DM | mast_cell | *ABCG1* | 2.127237853 | 4.23E-07 | 1.48E-06 | 0.002660619 |
| T2DM | mast_cell | *SLC37A1* | 2.591105308 | 4.79E-09 | 7.84E-08 | 3.01E-05 |
| T2DM | mast_cell | *U2AF1* | 1.938056694 | 2.40E-05 | 5.15E-05 | 0.150854888 |
| T2DM | mast_cell | *HSF2BP* | 2.543880405 | 1.16E-08 | 1.09E-07 | 7.27E-05 |
| T2DM | mast_cell | *ICOSLG* | -2.440466271 | 0.002682937 | 0.004053743 | 1 |
| T2DM | mast_cell | *C21orf2* | 1.032307252 | 0.001513265 | 0.002365464 | 1 |
| T2DM | mast_cell | *ZNF66* | 4.400122877 | 9.15E-06 | 2.15E-05 | 0.057494892 |
| T2DM | mast_cell | *LRRC3* | 2.441859328 | 2.41E-06 | 6.58E-06 | 0.015134627 |
| T2DM | mast_cell | *ITGB2* | 1.098466371 | 0.017456992 | 0.023282162 | 1 |
| T2DM | mast_cell | *FAM207A* | 3.575863729 | 7.28E-06 | 1.75E-05 | 0.045793892 |
| T2DM | mast_cell | *SPATC1L* | -0.380470516 | 0.005901236 | 0.008451269 | 1 |
| T2DM | mast_cell | *LSS* | 2.05566875 | 0.000457694 | 0.000774986 | 1 |
| T2DM | mast_cell | *VAV2* | 4.001304264 | 1.26E-07 | 5.47E-07 | 0.000794567 |
| T2DM | mast_cell | *PCNT* | 1.318567832 | 0.003694808 | 0.005451597 | 1 |
| T2DM | mast_cell | *ZNF208* | 2.729537991 | 4.54E-06 | 1.16E-05 | 0.028549494 |
| T2DM | mast_cell | *ADAMTS13* | 2.319944931 | 0.000255338 | 0.000451184 | 1 |
| T2DM | mast_cell | *FCN2* | 1.332881803 | 0.002043459 | 0.003143437 | 1 |
| T2DM | mast_cell | *ZNF714* | 3.73548572 | 7.51E-06 | 1.80E-05 | 0.04720883 |
| T2DM | mast_cell | *GPSM1* | -3.106963466 | 4.21E-06 | 1.09E-05 | 0.02649241 |
| T2DM | mast_cell | *C19orf47* | 2.968185791 | 2.27E-08 | 1.52E-07 | 0.000142658 |
| T2DM | mast_cell | *HIPK4* | 2.693954716 | 9.04E-07 | 2.82E-06 | 0.005681003 |
| T2DM | mast_cell | *RDH13* | 3.226208782 | 2.98E-08 | 1.84E-07 | 0.000187077 |
| T2DM | mast_cell | *MPZL3* | -3.211579018 | 2.32E-05 | 5.01E-05 | 0.145928 |
| T2DM | mast_cell | *VPS11* | 3.292637678 | 4.26E-06 | 1.10E-05 | 0.026812649 |
| T2DM | mast_cell | *FDPS* | 4.619404614 | 2.60E-08 | 1.68E-07 | 0.000163513 |
| T2DM | mast_cell | *PAQR6* | -3.219256084 | 0.005914285 | 0.008468027 | 1 |
| T2DM | mast_cell | *SLC25A44* | 4.321350973 | 4.56E-07 | 1.57E-06 | 0.002869263 |
| T2DM | mast_cell | *NBEAL2* | -2.096639578 | 0.003784847 | 0.005568765 | 1 |
| T2DM | mast_cell | *FCRL3* | 1.04509772 | 0.003354535 | 0.004986986 | 1 |
| T2DM | mast_cell | *NACC1* | 5.199387617 | 6.31E-09 | 8.24E-08 | 3.97E-05 |
| T2DM | mast_cell | *LY6K* | -1.999438505 | 2.27E-07 | 8.82E-07 | 0.00142432 |
| T2DM | mast_cell | *VPS28* | 2.793519304 | 7.75E-05 | 0.000150491 | 0.487138316 |
| T2DM | mast_cell | *TONSL* | -1.623824698 | 5.19E-07 | 1.75E-06 | 0.003262121 |
| T2DM | mast_cell | *MUM1* | 1.950521584 | 0.013180204 | 0.017839385 | 1 |
| T2DM | mast_cell | *ZNF333* | 4.293546803 | 5.18E-06 | 1.30E-05 | 0.032581544 |
| T2DM | mast_cell | *LRWD1* | -1.171074902 | 0.0017755 | 0.002755509 | 1 |

| | | | | | | |
|---|---|---|---|---|---|---|
| T2DM | mast_cell | *MFSD12* | 4.230552167 | 1.21E-08 | 1.11E-07 | 7.63E-05 |
| T2DM | mast_cell | *YDJC* | 12.23633029 | 3.17E-09 | 7.78E-08 | 1.99E-05 |
| T2DM | mast_cell | *CCDC116* | -3.922273674 | 1.30E-05 | 2.94E-05 | 0.081803282 |
| T2DM | mast_cell | *DVL3* | 3.722337649 | 7.47E-10 | 5.88E-08 | 4.70E-06 |
| T2DM | mast_cell | *AP2M1* | 5.180020711 | 1.45E-06 | 4.21E-06 | 0.009115293 |
| T2DM | mast_cell | *ABCF3* | 1.52007648 | 0.004839727 | 0.006999624 | 1 |
| T2DM | mast_cell | *FBXO27* | 1.100867425 | 5.51E-05 | 0.000110698 | 0.346596157 |
| T2DM | mast_cell | *DMKN* | 1.692865581 | 7.65E-05 | 0.000148768 | 0.480967964 |
| T2DM | mast_cell | *U2AF1L4* | 0.781403667 | 0.000638663 | 0.001054497 | 1 |
| T2DM | mast_cell | *BDH1* | 6.339668573 | 4.88E-09 | 7.84E-08 | 3.07E-05 |
| T2DM | mast_cell | *COX7A1* | 1.446897411 | 0.010487963 | 0.014412639 | 1 |
| T2DM | mast_cell | *DUSP14* | 2.733482969 | 0.004542861 | 0.006602166 | 1 |
| T2DM | mast_cell | *IKZF3* | 2.950052362 | 8.19E-09 | 9.23E-08 | 5.15E-05 |
| T2DM | mast_cell | *ACOX1* | 3.343087106 | 1.29E-06 | 3.80E-06 | 0.008087834 |
| T2DM | mast_cell | *PRPSAP1* | 3.623049306 | 2.79E-05 | 5.92E-05 | 0.175353772 |
| T2DM | mast_cell | *ZNF577* | 2.098032365 | 0.000239854 | 0.00042646 | 1 |
| T2DM | mast_cell | *TMEM143* | 2.354405875 | 2.88E-09 | 7.78E-08 | 1.81E-05 |
| T2DM | mast_cell | *CCL16* | -0.849666418 | 0.003340767 | 0.004971219 | 1 |
| T2DM | mast_cell | *SIGLEC11* | 2.482819564 | 5.55E-06 | 1.38E-05 | 0.034893541 |
| T2DM | mast_cell | *MPP3* | 4.271429749 | 6.29E-07 | 2.07E-06 | 0.003952376 |
| T2DM | mast_cell | *PLCD3* | 2.202448364 | 1.67E-06 | 4.79E-06 | 0.010485636 |
| T2DM | mast_cell | *RACGAP1* | 5.098574796 | 1.83E-09 | 7.58E-08 | 1.15E-05 |
| T2DM | mast_cell | *LARP4* | 2.631391021 | 3.27E-07 | 1.19E-06 | 0.002054067 |
| T2DM | mast_cell | *SPC24* | 2.826082055 | 6.73E-08 | 3.41E-07 | 0.000423282 |
| T2DM | mast_cell | *IP6K3* | -5.164512076 | 1.36E-06 | 3.98E-06 | 0.008531948 |
| T2DM | mast_cell | *LEMD2* | 2.647166614 | 0.010790049 | 0.014801885 | 1 |
| T2DM | mast_cell | *ASGR2* | 5.7040734 | 5.41E-09 | 8.00E-08 | 3.40E-05 |
| T2DM | mast_cell | *TNFSF13* | -2.219874135 | 0.000248977 | 0.000441183 | 1 |
| T2DM | mast_cell | *SENP3* | 5.235839956 | 0.000200115 | 0.000360644 | 1 |
| T2DM | mast_cell | *SNRNP25* | -3.435002784 | 8.55E-05 | 0.000164809 | 0.537608467 |
| T2DM | mast_cell | *WDR90* | 1.442722926 | 9.00E-05 | 0.000172767 | 0.565812485 |
| T2DM | mast_cell | *JMJD8* | 3.056541179 | 2.99E-06 | 8.00E-06 | 0.018805155 |
| T2DM | mast_cell | *FLYWCH2* | 5.955349591 | 8.22E-08 | 3.97E-07 | 0.000516678 |
| T2DM | mast_cell | *ZG16B* | 43.142991 | 8.50E-09 | 9.37E-08 | 5.34E-05 |
| T2DM | mast_cell | *NXF1* | 3.257889729 | 1.40E-09 | 6.85E-08 | 8.78E-06 |
| T2DM | mast_cell | *RPL29* | 2.693110867 | 0.004671406 | 0.006774885 | 1 |
| T2DM | mast_cell | *TPCN2* | 3.33061289 | 6.08E-05 | 0.000120807 | 0.38198909 |
| T2DM | mast_cell | *PDZK1IP1* | 28.09566438 | 8.79E-05 | 0.000169047 | 0.552739157 |
| T2DM | mast_cell | *COA7* | 3.509021224 | 4.34E-09 | 7.78E-08 | 2.73E-05 |
| T2DM | mast_cell | *SLC1A7* | 0.612392279 | 0.036266775 | 0.046315096 | 1 |
| T2DM | mast_cell | *ACOT11* | -0.8939984 | 1.17E-07 | 5.17E-07 | 0.000733067 |
| T2DM | mast_cell | *USP24* | 2.066023601 | 0.000284142 | 0.00049845 | 1 |
| T2DM | mast_cell | *NOL9* | 4.507616163 | 1.44E-07 | 6.08E-07 | 0.000904026 |
| T2DM | mast_cell | *ZSWIM5* | 2.763112869 | 2.99E-08 | 1.84E-07 | 0.00018792 |
| T2DM | mast_cell | *RBP7* | 2.606918875 | 4.01E-09 | 7.78E-08 | 2.52E-05 |
| T2DM | mast_cell | *FBLIM1* | 2.146668484 | 1.28E-08 | 1.13E-07 | 8.05E-05 |
| T2DM | mast_cell | *SLC25A34* | 3.839526191 | 1.49E-08 | 1.20E-07 | 9.38E-05 |
| T2DM | mast_cell | *PDPN* | 1.772325279 | 3.80E-07 | 1.35E-06 | 0.002386343 |
| T2DM | mast_cell | *LRRC38* | 6.277308376 | 1.71E-08 | 1.30E-07 | 0.000107674 |
| T2DM | mast_cell | *PEF1* | 5.779715916 | 0.000869717 | 0.001407804 | 1 |

| | | | | | | |
|---|---|---|---|---|---|---|
| T2DM | mast_cell | *SYNC* | 2.621214354 | 0.000727899 | 0.001190815 | 1 |
| T2DM | mast_cell | *RBBP4* | 3.792395569 | 1.18E-06 | 3.53E-06 | 0.007440974 |
| T2DM | mast_cell | *TSSK3* | 2.360022302 | 0.001239696 | 0.001962722 | 1 |
| T2DM | mast_cell | *CAMK2N1* | 4.010389216 | 3.86E-09 | 7.78E-08 | 2.43E-05 |
| T2DM | mast_cell | *TTLL10* | 4.143894341 | 1.84E-08 | 1.36E-07 | 0.000115846 |
| T2DM | mast_cell | *NFIA* | 2.738893306 | 3.00E-05 | 6.34E-05 | 0.188815245 |
| T2DM | mast_cell | *OMA1* | 1.166445921 | 0.021184593 | 0.027881 | 1 |
| T2DM | mast_cell | *TM2D1* | 3.998983567 | 2.88E-06 | 7.73E-06 | 0.018092674 |
| T2DM | mast_cell | *FUBP1* | 3.345654333 | 6.66E-05 | 0.000131141 | 0.41899549 |
| T2DM | mast_cell | *NEXN* | 4.353981448 | 1.67E-07 | 6.89E-07 | 0.001048641 |
| T2DM | mast_cell | *B3GALT2* | 3.565418724 | 1.19E-08 | 1.10E-07 | 7.48E-05 |
| T2DM | mast_cell | *NTNG1* | 1.084420055 | 0.003240857 | 0.004833989 | 1 |
| T2DM | mast_cell | *KCNT2* | 3.968649511 | 1.52E-08 | 1.21E-07 | 9.54E-05 |
| T2DM | mast_cell | *VCAM1* | 1.883883453 | 1.99E-08 | 1.41E-07 | 0.000125159 |
| T2DM | mast_cell | *ARPC5* | 2.832642322 | 2.70E-05 | 5.76E-05 | 0.169901501 |
| T2DM | mast_cell | *ZNF496* | 5.833136931 | 3.34E-06 | 8.81E-06 | 0.020971857 |
| T2DM | mast_cell | *TRIM58* | 2.005802469 | 0.003529101 | 0.005226727 | 1 |
| T2DM | mast_cell | *IGSF8* | 7.191946534 | 4.59E-06 | 1.17E-05 | 0.028858823 |
| T2DM | mast_cell | *VANGL2* | 3.732865875 | 7.85E-08 | 3.84E-07 | 0.000493702 |
| T2DM | mast_cell | *LMX1A* | 1.743641599 | 7.58E-11 | 1.44E-08 | 4.77E-07 |
| T2DM | mast_cell | *TDRD5* | 1.769434266 | 3.44E-05 | 7.19E-05 | 0.2160798 |
| T2DM | mast_cell | *SNED1* | 3.332843031 | 7.03E-07 | 2.27E-06 | 0.004417874 |
| T2DM | mast_cell | *C1orf115* | 2.227016124 | 2.25E-08 | 1.52E-07 | 0.000141559 |
| T2DM | mast_cell | *ACP6* | 2.584505795 | 1.26E-07 | 5.47E-07 | 0.000794447 |
| T2DM | mast_cell | *PM20D1* | 2.177773844 | 2.05E-08 | 1.43E-07 | 0.000128677 |
| T2DM | mast_cell | *HAAO* | 4.112283583 | 2.01E-09 | 7.78E-08 | 1.26E-05 |
| T2DM | mast_cell | *B3GALNT2* | 2.432056761 | 2.79E-05 | 5.93E-05 | 0.175672416 |
| T2DM | mast_cell | *PUS10* | 4.859708626 | 4.73E-06 | 1.20E-05 | 0.029733244 |
| T2DM | mast_cell | *RFTN2* | 3.60122788 | 6.07E-09 | 8.15E-08 | 3.81E-05 |
| T2DM | mast_cell | *DISC1* | -1.751288368 | 0.009722833 | 0.013458267 | 1 |
| T2DM | mast_cell | *FAM84A* | 3.123060422 | 1.46E-08 | 1.19E-07 | 9.17E-05 |
| T2DM | mast_cell | *CLHC1* | 0.82654789 | 0.00011988 | 0.000225385 | 0.753687516 |
| T2DM | mast_cell | *FRZB* | 4.098576843 | 1.57E-07 | 6.54E-07 | 0.000987097 |
| T2DM | mast_cell | *NUP35* | 3.851042386 | 3.14E-08 | 1.91E-07 | 0.0001973 |
| T2DM | mast_cell | *FBXO41* | -2.679034367 | 3.98E-07 | 1.40E-06 | 0.00249994 |
| T2DM | mast_cell | *CCDC74A* | 6.735911762 | 1.91E-08 | 1.38E-07 | 0.000120283 |
| T2DM | mast_cell | *H3F3A* | 1.84873962 | 0.021936085 | 0.028803711 | 1 |
| T2DM | mast_cell | *NOSTRIN* | 2.421981023 | 4.23E-07 | 1.48E-06 | 0.002658704 |
| T2DM | mast_cell | *BBS5* | 2.213838965 | 2.29E-08 | 1.53E-07 | 0.000143979 |
| T2DM | mast_cell | *HPGDS* | -0.251325759 | 1.22E-07 | 5.33E-07 | 0.000765311 |
| T2DM | mast_cell | *PDLIM5* | 3.274980363 | 7.09E-06 | 1.71E-05 | 0.044573381 |
| T2DM | mast_cell | *RPRD2* | 2.781742205 | 9.22E-05 | 0.000176893 | 0.579856859 |
| T2DM | mast_cell | *CTSS* | 0.946142655 | 0.004925754 | 0.007116352 | 1 |
| T2DM | mast_cell | *PACRGL* | 3.317174637 | 5.87E-07 | 1.96E-06 | 0.003692454 |
| T2DM | mast_cell | *C1QTNF7* | 3.347138871 | 7.11E-06 | 1.72E-05 | 0.044712475 |
| T2DM | mast_cell | *LYSMD1* | 3.208087069 | 0.015522794 | 0.020839592 | 1 |
| T2DM | mast_cell | *ERCC3* | 2.703443891 | 2.40E-07 | 9.23E-07 | 0.001510146 |
| T2DM | mast_cell | *RNF149* | 3.431812668 | 6.13E-05 | 0.000121764 | 0.385562172 |
| T2DM | mast_cell | *DHX57* | 3.05085919 | 7.94E-09 | 9.16E-08 | 4.99E-05 |
| T2DM | mast_cell | *S100A9* | 4.536746835 | 9.86E-07 | 3.04E-06 | 0.006197445 |

| | | | | | | |
|---|---|---|---|---|---|---|
| T2DM | mast_cell | *S100A12* | 12.06385213 | 1.27E-06 | 3.75E-06 | 0.00798356 |
| T2DM | mast_cell | *TDRD10* | 2.343977091 | 4.97E-09 | 7.84E-08 | 3.13E-05 |
| T2DM | mast_cell | *DCAF16* | 4.131229156 | 1.93E-08 | 1.39E-07 | 0.000121303 |
| T2DM | mast_cell | *NIPAL1* | 7.573097352 | 1.27E-08 | 1.13E-07 | 7.98E-05 |
| T2DM | mast_cell | *ANTXR2* | 1.790494 | 0.007459123 | 0.010505266 | 1 |
| T2DM | mast_cell | *CGGBP1* | 0.760862399 | 0.009266951 | 0.012864059 | 1 |
| T2DM | mast_cell | *DCST2* | 4.071907864 | 3.48E-09 | 7.78E-08 | 2.19E-05 |
| T2DM | mast_cell | *LMOD3* | 4.301613103 | 4.55E-09 | 7.78E-08 | 2.86E-05 |
| T2DM | mast_cell | *NBPF10* | 2.071472893 | 1.14E-06 | 3.40E-06 | 0.007137444 |
| T2DM | mast_cell | *SLC22A15* | 2.686086217 | 1.76E-09 | 7.51E-08 | 1.11E-05 |
| T2DM | mast_cell | *LRRC58* | 5.129369063 | 5.54E-06 | 1.38E-05 | 0.034854961 |
| T2DM | mast_cell | *FSTL1* | 2.264367538 | 2.53E-05 | 5.41E-05 | 0.159088776 |
| T2DM | mast_cell | *IGFBP7* | 2.358747894 | 8.59E-06 | 2.03E-05 | 0.053978413 |
| T2DM | mast_cell | *ARPC2* | 3.741382283 | 4.29E-06 | 1.10E-05 | 0.026969841 |
| T2DM | mast_cell | *CCT3* | 1.944840914 | 0.00960501 | 0.013303966 | 1 |
| T2DM | mast_cell | *GLB1L* | -1.617393363 | 0.007902175 | 0.011089503 | 1 |
| T2DM | mast_cell | *NUAK2* | 1.7875931 | 3.64E-05 | 7.58E-05 | 0.229075309 |
| T2DM | mast_cell | *PYHIN1* | 1.823259647 | 3.80E-06 | 9.90E-06 | 0.023882213 |
| T2DM | mast_cell | *AIM2* | -3.420386646 | 0.000396992 | 0.000679893 | 1 |
| T2DM | mast_cell | *CD200R1* | -1.015472192 | 5.10E-07 | 1.73E-06 | 0.003208604 |
| T2DM | mast_cell | *SPICE1* | 2.241749998 | 0.012769484 | 0.01730583 | 1 |
| T2DM | mast_cell | *CADPS* | -0.488041635 | 1.47E-07 | 6.20E-07 | 0.000925776 |
| T2DM | mast_cell | *PPM1K* | 3.995538512 | 6.88E-07 | 2.23E-06 | 0.004323811 |
| T2DM | mast_cell | *TIPARP* | 4.592027438 | 3.47E-07 | 1.25E-06 | 0.002184142 |
| T2DM | mast_cell | *DCLK3* | -3.111427303 | 0.005786837 | 0.008294994 | 1 |
| T2DM | mast_cell | *RPP14* | 5.523845404 | 2.49E-07 | 9.48E-07 | 0.001567795 |
| T2DM | mast_cell | *DNASE1L3* | 2.276318388 | 2.91E-09 | 7.78E-08 | 1.83E-05 |
| T2DM | mast_cell | *CRELD1* | 2.512554286 | 8.07E-07 | 2.56E-06 | 0.005074826 |
| T2DM | mast_cell | *PCOLCE2* | 1.5217324 | 0.025089527 | 0.03268501 | 1 |
| T2DM | mast_cell | *TTC14* | 1.824842791 | 0.004594862 | 0.006670029 | 1 |
| T2DM | mast_cell | *RCHY1* | 2.595077623 | 0.003809616 | 0.005602585 | 1 |
| T2DM | mast_cell | *CCDC158* | 1.920189382 | 4.51E-09 | 7.78E-08 | 2.84E-05 |
| T2DM | mast_cell | *GYG1* | 2.677334282 | 2.56E-09 | 7.78E-08 | 1.61E-05 |
| T2DM | mast_cell | *HPS3* | 3.980930518 | 1.44E-05 | 3.21E-05 | 0.09046615 |
| T2DM | mast_cell | *RYK* | 3.369193577 | 9.56E-05 | 0.000182982 | 0.601094494 |
| T2DM | mast_cell | *PLB1* | 2.875016132 | 5.02E-09 | 7.84E-08 | 3.16E-05 |
| T2DM | mast_cell | *CDCP1* | 7.885993513 | 6.59E-07 | 2.15E-06 | 0.004143576 |
| T2DM | mast_cell | *LZTFL1* | 5.83893807 | 3.69E-05 | 7.68E-05 | 0.232175711 |
| T2DM | mast_cell | *FYCO1* | 4.590460676 | 3.26E-07 | 1.19E-06 | 0.00204709 |
| T2DM | mast_cell | *LRRC2* | 2.127656422 | 1.55E-05 | 3.44E-05 | 0.097561541 |
| T2DM | mast_cell | *DTX3L* | 4.431840927 | 7.45E-06 | 1.79E-05 | 0.04682431 |
| T2DM | mast_cell | *NMNAT3* | 3.53329766 | 3.63E-10 | 4.05E-08 | 2.28E-06 |
| T2DM | mast_cell | *SMIM12* | 5.198730642 | 2.92E-07 | 1.08E-06 | 0.001834537 |
| T2DM | mast_cell | *TPRA1* | 4.11335375 | 4.88E-09 | 7.84E-08 | 3.07E-05 |
| T2DM | mast_cell | *YEATS2* | 4.289295359 | 6.61E-08 | 3.36E-07 | 0.000415884 |
| T2DM | mast_cell | *MEAF6* | 4.610929735 | 0.00022522 | 0.000402376 | 1 |
| T2DM | mast_cell | *DNALI1* | 2.788835681 | 2.30E-09 | 7.78E-08 | 1.44E-05 |
| T2DM | mast_cell | *RPN1* | 3.698407029 | 0.002110114 | 0.00323726 | 1 |
| T2DM | mast_cell | *SFMBT1* | 4.764685366 | 2.52E-08 | 1.64E-07 | 0.000158712 |
| T2DM | mast_cell | *GNL3* | 3.924206845 | 6.68E-05 | 0.000131372 | 0.419996991 |

| | | | | | | |
|---|---|---|---|---|---|---|
| T2DM | mast_cell | *UVSSA* | 2.581290719 | 3.88E-05 | 8.03E-05 | 0.243793679 |
| T2DM | mast_cell | *FAM208A* | 1.887778577 | 0.015260552 | 0.020505042 | 1 |
| T2DM | mast_cell | *LRPAP1* | 3.810545371 | 3.47E-08 | 2.06E-07 | 0.000218107 |
| T2DM | mast_cell | *SLC51A* | 3.630817163 | 0.000331268 | 0.000575168 | 1 |
| T2DM | mast_cell | *PIGX* | 3.632686199 | 6.95E-06 | 1.69E-05 | 0.043704592 |
| T2DM | mast_cell | *ABLIM2* | 4.379590987 | 3.36E-09 | 7.78E-08 | 2.11E-05 |
| T2DM | mast_cell | *EXO5* | -2.280557595 | 1.06E-06 | 3.22E-06 | 0.00665199 |
| T2DM | mast_cell | *ERMAP* | 1.88198055 | 0.000317562 | 0.000552285 | 1 |
| T2DM | mast_cell | *METAP1* | 3.149576962 | 5.34E-08 | 2.86E-07 | 0.000335865 |
| T2DM | mast_cell | *DNAJB14* | 3.89123695 | 6.96E-08 | 3.48E-07 | 0.000437599 |
| T2DM | mast_cell | *SLC9B2* | 2.799793625 | 7.02E-07 | 2.27E-06 | 0.004416234 |
| T2DM | mast_cell | *CDC25A* | -1.155900751 | 0.026782225 | 0.034688885 | 1 |
| T2DM | mast_cell | *ATRIP* | 9.530864082 | 1.46E-07 | 6.16E-07 | 0.000918731 |
| T2DM | mast_cell | *SHISA5* | 4.851299982 | 1.62E-07 | 6.71E-07 | 0.001017205 |
| T2DM | mast_cell | *APEH* | -1.974648566 | 0.000404431 | 0.000691692 | 1 |
| T2DM | mast_cell | *RNF123* | 3.102573979 | 1.44E-13 | 9.62E-11 | 9.03E-10 |
| T2DM | mast_cell | *MST1R* | 2.245563886 | 4.18E-05 | 8.59E-05 | 0.262515355 |
| T2DM | mast_cell | *RAD54L2* | 2.806188622 | 0.000316589 | 0.000550992 | 1 |
| T2DM | mast_cell | *TEX264* | 2.169000907 | 4.55E-05 | 9.28E-05 | 0.285929019 |
| T2DM | mast_cell | *POC1A* | 2.918662696 | 2.40E-08 | 1.58E-07 | 0.000150782 |
| T2DM | mast_cell | *C4orf3* | 4.071022919 | 0.000543502 | 0.000908775 | 1 |
| T2DM | mast_cell | *ANXA5* | 3.161864106 | 3.36E-05 | 7.04E-05 | 0.21106774 |
| T2DM | mast_cell | *MAP9* | -2.722168882 | 0.000460179 | 0.000778775 | 1 |
| T2DM | mast_cell | *CEP44* | 4.506669432 | 3.34E-08 | 2.01E-07 | 0.00021029 |
| T2DM | mast_cell | *C4orf45* | -2.487012572 | 0.000712716 | 0.001168104 | 1 |
| T2DM | mast_cell | *TMEM144* | 4.308575185 | 3.21E-08 | 1.94E-07 | 0.000201523 |
| T2DM | mast_cell | *NPY1R* | 2.04927618 | 4.08E-09 | 7.78E-08 | 2.57E-05 |
| T2DM | mast_cell | *IL15* | 2.364530603 | 4.67E-09 | 7.80E-08 | 2.93E-05 |
| T2DM | mast_cell | *ARFIP1* | 2.891795375 | 0.000194306 | 0.000351237 | 1 |
| T2DM | mast_cell | *OTUD4* | 4.51193451 | 5.59E-06 | 1.39E-05 | 0.035130429 |
| T2DM | mast_cell | *ITGA2* | 4.991071537 | 6.63E-09 | 8.54E-08 | 4.17E-05 |
| T2DM | mast_cell | *NDUFAF2* | 2.307579074 | 0.000927953 | 0.001492846 | 1 |
| T2DM | mast_cell | *LMBRD2* | 4.096234354 | 0.000510036 | 0.000856462 | 1 |
| T2DM | mast_cell | *PGGT1B* | 2.365287007 | 0.034333567 | 0.043971305 | 1 |
| T2DM | mast_cell | *C5orf63* | 5.95414337 | 5.57E-06 | 1.39E-05 | 0.035022434 |
| T2DM | mast_cell | *PRRC1* | 4.518376135 | 1.09E-09 | 6.34E-08 | 6.88E-06 |
| T2DM | mast_cell | *NDUFS4* | 3.492713341 | 0.000165191 | 0.000302697 | 1 |
| T2DM | mast_cell | *SERINC5* | 3.770331532 | 1.10E-06 | 3.32E-06 | 0.006912296 |
| T2DM | mast_cell | *ENPP6* | 6.311863639 | 5.56E-05 | 0.000111559 | 0.349624528 |
| T2DM | mast_cell | *CASP3* | 5.158009801 | 6.60E-06 | 1.61E-05 | 0.041472944 |
| T2DM | mast_cell | *PRIMPOL* | -1.91715202 | 0.002687332 | 0.004059408 | 1 |
| T2DM | mast_cell | *ERAP2* | 1.111420733 | 0.002594691 | 0.003927015 | 1 |
| T2DM | mast_cell | *RICTOR* | 2.852061387 | 0.000801613 | 0.001302932 | 1 |
| T2DM | mast_cell | *ANKRA2* | 4.740653708 | 2.32E-08 | 1.54E-07 | 0.000145641 |
| T2DM | mast_cell | *UBLCP1* | 2.235150623 | 0.003507221 | 0.005197995 | 1 |
| T2DM | mast_cell | *UTP15* | -2.661076662 | 0.010827042 | 0.014846154 | 1 |
| T2DM | mast_cell | *TLR3* | 3.243177652 | 3.84E-09 | 7.78E-08 | 2.42E-05 |
| T2DM | mast_cell | *KLKB1* | 1.803591117 | 5.69E-09 | 8.09E-08 | 3.57E-05 |
| T2DM | mast_cell | *NSA2* | 0.810985942 | 0.013815515 | 0.018663116 | 1 |
| T2DM | mast_cell | *CCDC127* | 2.282910536 | 1.36E-08 | 1.17E-07 | 8.53E-05 |

| | | | | | | |
|---|---|---|---|---|---|---|
| T2DM | mast_cell | *GDF9* | 3.489926853 | 6.96E-05 | 0.000136173 | 0.437389014 |
| T2DM | mast_cell | *GRIK2* | 1.683511403 | 2.42E-05 | 5.19E-05 | 0.152164494 |
| T2DM | mast_cell | *MB21D1* | 5.294082272 | 6.13E-05 | 0.000121764 | 0.385625633 |
| T2DM | mast_cell | *TXLNB* | -1.990476918 | 1.49E-06 | 4.30E-06 | 0.009337239 |
| T2DM | mast_cell | *DCBLD1* | 3.885751031 | 5.58E-05 | 0.000111973 | 0.351036358 |
| T2DM | mast_cell | *PDSS2* | 3.305458795 | 6.76E-05 | 0.00013291 | 0.425311374 |
| T2DM | mast_cell | *STXBP5* | -1.101956802 | 9.52E-06 | 2.23E-05 | 0.059841187 |
| T2DM | mast_cell | *PI16* | 2.813184209 | 3.08E-09 | 7.78E-08 | 1.94E-05 |
| T2DM | mast_cell | *DAGLB* | 1.336540706 | 0.03879904 | 0.049368461 | 1 |
| T2DM | mast_cell | *STK17A* | 3.330546565 | 4.52E-05 | 9.24E-05 | 0.284472375 |
| T2DM | mast_cell | *GALNT10* | 3.51531361 | 4.24E-06 | 1.09E-05 | 0.02665866 |
| T2DM | mast_cell | *SAP30L* | 4.402853977 | 2.06E-07 | 8.17E-07 | 0.001294924 |
| T2DM | mast_cell | *SLU7* | 3.785806469 | 2.44E-07 | 9.33E-07 | 0.001534429 |
| T2DM | mast_cell | *RP9* | 3.257867278 | 0.001433822 | 0.002250797 | 1 |
| T2DM | mast_cell | *CAMLG* | 4.02409499 | 2.22E-06 | 6.16E-06 | 0.013939552 |
| T2DM | mast_cell | *KCNK5* | 11.90703777 | 8.09E-08 | 3.92E-07 | 0.000508882 |
| T2DM | mast_cell | *KIAA1324L* | 1.331137439 | 0.005771343 | 0.008276559 | 1 |
| T2DM | mast_cell | *USP49* | 3.465207129 | 0.001397145 | 0.002198035 | 1 |
| T2DM | mast_cell | *ZNF704* | 1.653804873 | 0.036050892 | 0.046058823 | 1 |
| T2DM | mast_cell | *FNDC1* | 1.770721813 | 7.78E-08 | 3.81E-07 | 0.000489042 |
| T2DM | mast_cell | *BRI3* | 3.545259974 | 6.20E-06 | 1.52E-05 | 0.038983299 |
| T2DM | mast_cell | *LMTK2* | 4.239826122 | 2.94E-08 | 1.82E-07 | 0.000184676 |
| T2DM | mast_cell | *CTSB* | 0.897697452 | 0.017581534 | 0.023428381 | 1 |
| T2DM | mast_cell | *PEX2* | 4.20118998 | 6.20E-08 | 3.22E-07 | 0.000389777 |
| T2DM | mast_cell | *DEFB1* | 1.379166148 | 0.000447528 | 0.000759819 | 1 |
| T2DM | mast_cell | *SUN1* | 3.737454607 | 1.42E-08 | 1.19E-07 | 8.91E-05 |
| T2DM | mast_cell | *OXR1* | 3.407146128 | 3.97E-07 | 1.40E-06 | 0.002495032 |
| T2DM | mast_cell | *TMEM184A* | 3.237619669 | 8.23E-05 | 0.000158843 | 0.517193171 |
| T2DM | mast_cell | *INTS1* | 4.965165582 | 9.25E-09 | 9.89E-08 | 5.82E-05 |
| T2DM | mast_cell | *SLC4A2* | 5.791613791 | 1.07E-08 | 1.06E-07 | 6.72E-05 |
| T2DM | mast_cell | *FASTK* | 3.395677253 | 0.012135785 | 0.016518225 | 1 |
| T2DM | mast_cell | *ALDH7A1* | 1.332820892 | 0.033290553 | 0.042678978 | 1 |
| T2DM | mast_cell | *COX6C* | 4.944069344 | 2.17E-07 | 8.53E-07 | 0.001364546 |
| T2DM | mast_cell | *SLC25A32* | 3.181319673 | 1.38E-07 | 5.87E-07 | 0.000867575 |
| T2DM | mast_cell | *DCAF13* | 6.102544953 | 2.52E-08 | 1.64E-07 | 0.000158541 |
| T2DM | mast_cell | *INTS8* | 3.325960604 | 0.000174732 | 0.000318787 | 1 |
| T2DM | mast_cell | *FREM1* | -1.597215419 | 0.006959556 | 0.009836944 | 1 |
| T2DM | mast_cell | *RPP25L* | 4.40711034 | 6.56E-05 | 0.000129405 | 0.412544535 |
| T2DM | mast_cell | *FAM219A* | 3.838057116 | 8.93E-09 | 9.66E-08 | 5.61E-05 |
| T2DM | mast_cell | *NUDT2* | 0.666650961 | 0.020026931 | 0.026517818 | 1 |
| T2DM | mast_cell | *TMEM65* | 3.578135223 | 1.33E-07 | 5.70E-07 | 0.00083362 |
| T2DM | mast_cell | *CCDC171* | 3.486853449 | 1.40E-05 | 3.14E-05 | 0.088011095 |
| T2DM | mast_cell | *SYK* | 3.898545945 | 3.72E-06 | 9.73E-06 | 0.023409406 |
| T2DM | mast_cell | *METTL2B* | 0.808547193 | 0.000196661 | 0.000355125 | 1 |
| T2DM | mast_cell | *HGSNAT* | 2.846063799 | 0.000405473 | 0.000693285 | 1 |
| T2DM | mast_cell | *HNRNPK* | 3.568897677 | 0.000439724 | 0.000747377 | 1 |
| T2DM | mast_cell | *SVEP1* | 3.272598721 | 1.09E-05 | 2.51E-05 | 0.068359811 |
| T2DM | mast_cell | *ZNF367* | 3.206969513 | 1.74E-07 | 7.12E-07 | 0.001095163 |
| T2DM | mast_cell | *NOL6* | 5.756414468 | 3.87E-05 | 8.03E-05 | 0.243576089 |
| T2DM | mast_cell | *AQP3* | -2.420055012 | 0.000951414 | 0.001529414 | 1 |

| | | | | | | |
|---|---|---|---|---|---|---|
| T2DM | mast_cell | *TRMT10B* | 3.681274043 | 0.000369805 | 0.000636802 | 1 |
| T2DM | mast_cell | *VCP* | 3.187167425 | 9.88E-07 | 3.04E-06 | 0.006214231 |
| T2DM | mast_cell | *PIGO* | 4.323746338 | 5.46E-08 | 2.91E-07 | 0.000343214 |
| T2DM | mast_cell | *LRFN5* | 0.978012003 | 1.03E-08 | 1.04E-07 | 6.48E-05 |
| T2DM | mast_cell | *WRN* | 3.616053669 | 7.54E-06 | 1.81E-05 | 0.047417696 |
| T2DM | mast_cell | *MARCH8* | -1.85167499 | 0.002289256 | 0.003490796 | 1 |
| T2DM | mast_cell | *CFL2* | 2.831009878 | 5.57E-09 | 8.01E-08 | 3.50E-05 |
| T2DM | mast_cell | *ZCCHC24* | 4.195606434 | 8.22E-06 | 1.95E-05 | 0.051696727 |
| T2DM | mast_cell | *PGM2L1* | 3.706890538 | 6.67E-10 | 5.76E-08 | 4.19E-06 |
| T2DM | mast_cell | *CRYL1* | 2.367150374 | 0.00031473 | 0.000547966 | 1 |
| T2DM | mast_cell | *REEP3* | 2.264546924 | 0.00035296 | 0.000609297 | 1 |
| T2DM | mast_cell | *HEPACAM* | 0.434843214 | 0.006307197 | 0.008987613 | 1 |
| T2DM | mast_cell | *LRR1* | 5.026398603 | 1.18E-05 | 2.70E-05 | 0.074240436 |
| T2DM | mast_cell | *RPL36AL* | 2.937383214 | 4.63E-07 | 1.59E-06 | 0.002911588 |
| T2DM | mast_cell | *NOXRED1* | -2.326870519 | 0.000316965 | 0.0005514 | 1 |
| T2DM | mast_cell | *AKR1E2* | 1.506139411 | 0.023735246 | 0.031055877 | 1 |
| T2DM | mast_cell | *ATP5C1* | 3.23760162 | 1.85E-06 | 5.25E-06 | 0.011641404 |
| T2DM | mast_cell | *TAF3* | 3.021634664 | 0.000800498 | 0.001301456 | 1 |
| T2DM | mast_cell | *VDAC2* | 4.062399377 | 1.00E-09 | 6.17E-08 | 6.30E-06 |
| T2DM | mast_cell | *SLC18A2* | -0.169428623 | 1.10E-07 | 4.94E-07 | 0.000690619 |
| T2DM | mast_cell | *PDZD8* | 2.877976083 | 4.26E-05 | 8.76E-05 | 0.267808234 |
| T2DM | mast_cell | *CLEC1B* | -0.594576978 | 0.000638703 | 0.001054497 | 1 |
| T2DM | mast_cell | *SNAPC4* | 1.275228053 | 0.019987563 | 0.026477415 | 1 |
| T2DM | mast_cell | *SDCCAG3* | 5.623305598 | 1.23E-05 | 2.80E-05 | 0.077303296 |
| T2DM | mast_cell | *AK8* | -2.437613541 | 0.02819558 | 0.03643692 | 1 |
| T2DM | mast_cell | *TSC1* | 4.148516843 | 5.32E-06 | 1.33E-05 | 0.033426147 |
| T2DM | mast_cell | *ZMYND19* | 2.280421882 | 1.46E-08 | 1.19E-07 | 9.17E-05 |
| T2DM | mast_cell | *STOX1* | 0.435573085 | 0.029059655 | 0.037468838 | 1 |
| T2DM | mast_cell | *RET* | -1.906031198 | 3.23E-07 | 1.18E-06 | 0.00202981 |
| T2DM | mast_cell | *STK32C* | -2.401871137 | 0.001683801 | 0.002620311 | 1 |
| T2DM | mast_cell | *NSMF* | 5.217117772 | 3.74E-09 | 7.78E-08 | 2.35E-05 |
| T2DM | mast_cell | *CASP7* | 2.027518959 | 0.000131015 | 0.000244419 | 0.823693074 |
| T2DM | mast_cell | *VWA2* | 2.926218675 | 4.64E-05 | 9.46E-05 | 0.291978004 |
| T2DM | mast_cell | *METTL3* | 4.340013355 | 5.13E-09 | 7.89E-08 | 3.22E-05 |
| T2DM | mast_cell | *HSPA12A* | 1.286912064 | 0.000149378 | 0.000276217 | 0.939138834 |
| T2DM | mast_cell | *UBTD1* | 2.861079438 | 3.65E-09 | 7.78E-08 | 2.29E-05 |
| T2DM | mast_cell | *ARHGAP42* | 3.824805387 | 3.80E-07 | 1.35E-06 | 0.002391 |
| T2DM | mast_cell | *OTOGL* | 3.414864888 | 3.01E-05 | 6.36E-05 | 0.189266595 |
| T2DM | mast_cell | *PACSIN3* | 0.759336506 | 0.023807265 | 0.031130673 | 1 |
| T2DM | mast_cell | *TTC7B* | -1.534752228 | 0.003164134 | 0.00472847 | 1 |
| T2DM | mast_cell | *SLC39A13* | 2.487384233 | 3.80E-09 | 7.78E-08 | 2.39E-05 |
| T2DM | mast_cell | *PSMC3* | 3.134698399 | 7.01E-07 | 2.27E-06 | 0.00440833 |
| T2DM | mast_cell | *TC2N* | 0.875436766 | 0.000203612 | 0.000366373 | 1 |
| T2DM | mast_cell | *CPSF2* | 1.288666781 | 0.000298783 | 0.000522371 | 1 |
| T2DM | mast_cell | *CLMN* | 2.81179523 | 1.47E-07 | 6.20E-07 | 0.000926247 |
| T2DM | mast_cell | *PTER* | 2.845582218 | 0.007642475 | 0.010746642 | 1 |
| T2DM | mast_cell | *ABTB2* | 2.983289965 | 0.029057476 | 0.037468838 | 1 |
| T2DM | mast_cell | *LIPC* | 2.660930766 | 1.21E-08 | 1.11E-07 | 7.60E-05 |
| T2DM | mast_cell | *CEP57* | 1.817957741 | 0.01265949 | 0.017164161 | 1 |
| T2DM | mast_cell | *TCP11L2* | 3.608185118 | 5.46E-09 | 8.00E-08 | 3.43E-05 |

| | | | | | | |
|---|---|---|---|---|---|---|
| T2DM | mast_cell | *JAM3* | 3.622988379 | 3.80E-09 | 7.78E-08 | 2.39E-05 |
| T2DM | mast_cell | *GPT2* | 1.29159753 | 1.56E-07 | 6.50E-07 | 0.000978197 |
| T2DM | mast_cell | *IKBIP* | 3.412318843 | 3.94E-07 | 1.39E-06 | 0.002477263 |
| T2DM | mast_cell | *HIF1AN* | 4.62810237 | 1.31E-08 | 1.14E-07 | 8.23E-05 |
| T2DM | mast_cell | *ZFYVE19* | 1.791017378 | 4.65E-05 | 9.47E-05 | 0.292222349 |
| T2DM | mast_cell | *SPINT1* | -2.859590799 | 2.34E-06 | 6.45E-06 | 0.014722456 |
| T2DM | mast_cell | *AVPR1A* | 1.010817369 | 0.03296383 | 0.042285982 | 1 |
| T2DM | mast_cell | *BTRC* | 3.064459491 | 0.000181299 | 0.000329525 | 1 |
| T2DM | mast_cell | *LARP6* | 2.873311829 | 2.48E-08 | 1.62E-07 | 0.000156016 |
| T2DM | mast_cell | *ASPG* | 5.707935186 | 6.00E-09 | 8.15E-08 | 3.77E-05 |
| T2DM | mast_cell | *GABRB3* | 2.816078511 | 5.07E-07 | 1.72E-06 | 0.003184474 |
| T2DM | mast_cell | *TBATA* | 4.395199121 | 0.025518389 | 0.033195554 | 1 |
| T2DM | mast_cell | *PCBD1* | -1.487904749 | 6.09E-05 | 0.000121016 | 0.382894176 |
| T2DM | mast_cell | *COX11* | 3.012123755 | 2.18E-05 | 4.72E-05 | 0.136934599 |
| T2DM | mast_cell | *ZNF202* | 3.573220569 | 4.65E-10 | 4.49E-08 | 2.92E-06 |
| T2DM | mast_cell | *FAM227B* | 2.218654615 | 0.002151899 | 0.003292526 | 1 |
| T2DM | mast_cell | *STXBP4* | 3.005192482 | 3.37E-05 | 7.06E-05 | 0.211772992 |
| T2DM | mast_cell | *C2* | 7.974933753 | 6.82E-16 | 1.07E-12 | 4.29E-12 |
| T2DM | mast_cell | *APBB1* | 1.984904877 | 1.14E-08 | 1.09E-07 | 7.19E-05 |
| T2DM | mast_cell | *ILK* | 3.796217079 | 2.33E-09 | 7.78E-08 | 1.46E-05 |
| T2DM | mast_cell | *TAF10* | 4.818696235 | 1.69E-05 | 3.71E-05 | 0.106127891 |
| T2DM | mast_cell | *DCHS1* | 3.351734899 | 5.10E-09 | 7.88E-08 | 3.21E-05 |
| T2DM | mast_cell | *NETO1* | 3.922321392 | 3.55E-08 | 2.09E-07 | 0.000223105 |
| T2DM | mast_cell | *USP54* | 3.028033563 | 0.000442656 | 0.000752157 | 1 |
| T2DM | mast_cell | *C11orf74* | 3.857910252 | 2.47E-07 | 9.41E-07 | 0.001554163 |
| T2DM | mast_cell | *WDR88* | 3.062732041 | 1.39E-08 | 1.18E-07 | 8.72E-05 |
| T2DM | mast_cell | *CYB5R2* | -1.560394022 | 0.020026627 | 0.026517818 | 1 |
| T2DM | mast_cell | *TUB* | -4.035634796 | 7.22E-06 | 1.74E-05 | 0.045382362 |
| T2DM | mast_cell | *RIC3* | -2.359305626 | 0.00010464 | 0.000198813 | 0.657873427 |
| T2DM | mast_cell | *LMO1* | 3.428464188 | 2.26E-08 | 1.52E-07 | 0.000141852 |
| T2DM | mast_cell | *TRIM66* | 0.504160766 | 0.002105888 | 0.003231565 | 1 |
| T2DM | mast_cell | *RPL27A* | 3.658604163 | 0.000485898 | 0.000817895 | 1 |
| T2DM | mast_cell | *CDYL2* | 1.777292146 | 8.81E-07 | 2.76E-06 | 0.005540466 |
| T2DM | mast_cell | *PRTG* | 2.730582893 | 4.32E-09 | 7.78E-08 | 2.71E-05 |
| T2DM | mast_cell | *AKIP1* | 6.369475111 | 1.61E-08 | 1.25E-07 | 0.000100976 |
| T2DM | mast_cell | *C16orf46* | 18.54749194 | 9.17E-08 | 4.28E-07 | 0.000576409 |
| T2DM | mast_cell | *TMEM41B* | 4.516506627 | 1.73E-07 | 7.08E-07 | 0.001087914 |
| T2DM | mast_cell | *ZNF143* | 4.548285453 | 7.04E-09 | 8.80E-08 | 4.43E-05 |
| T2DM | mast_cell | *TMX3* | 4.06300077 | 4.17E-07 | 1.46E-06 | 0.002619494 |
| T2DM | mast_cell | *CCDC68* | -0.84152447 | 9.49E-07 | 2.94E-06 | 0.005964942 |
| T2DM | mast_cell | *CLEC4E* | 1.912870252 | 1.32E-08 | 1.14E-07 | 8.27E-05 |
| T2DM | mast_cell | *CLEC4D* | 6.219911392 | 4.74E-09 | 7.84E-08 | 2.98E-05 |
| T2DM | mast_cell | *BEAN1* | 5.325104377 | 9.02E-08 | 4.23E-07 | 0.000567151 |
| T2DM | mast_cell | *SEC11C* | 6.415328135 | 0.000224291 | 0.000400944 | 1 |
| T2DM | mast_cell | *GALR1* | 1.617676174 | 8.11E-05 | 0.000156809 | 0.509784794 |
| T2DM | mast_cell | *CENPV* | 3.522144194 | 1.75E-07 | 7.16E-07 | 0.00110332 |
| T2DM | mast_cell | *SPDYE6* | 2.639295833 | 3.39E-08 | 2.03E-07 | 0.00021295 |
| T2DM | mast_cell | *ATF7IP2* | 2.812825023 | 1.28E-06 | 3.77E-06 | 0.008022049 |
| T2DM | mast_cell | *AP1G1* | 3.53387169 | 5.50E-08 | 2.92E-07 | 0.000345503 |
| T2DM | mast_cell | *SLFN5* | 2.439632672 | 0.005306264 | 0.007637473 | 1 |

| | | | | | | |
|---|---|---|---|---|---|---|
| T2DM | mast_cell | *CATSPER2* | 2.654766722 | 0.006694962 | 0.009497117 | 1 |
| T2DM | mast_cell | *C16orf45* | 3.443426981 | 0.000154598 | 0.000285199 | 0.971959249 |
| T2DM | mast_cell | *LDHC* | 1.358738653 | 3.09E-06 | 8.25E-06 | 0.019439492 |
| T2DM | mast_cell | *FAM111A* | 3.177782519 | 5.68E-07 | 1.90E-06 | 0.003568727 |
| T2DM | mast_cell | *SCNN1G* | 6.790484563 | 1.03E-05 | 2.40E-05 | 0.065065125 |
| T2DM | mast_cell | *ANKDD1A* | 2.77356282 | 1.71E-06 | 4.88E-06 | 0.010721307 |
| T2DM | mast_cell | *DCTN5* | 1.414477582 | 0.003043035 | 0.004560562 | 1 |
| T2DM | mast_cell | *STX3* | -0.283562263 | 1.24E-07 | 5.39E-07 | 0.000778437 |
| T2DM | mast_cell | *MTMR10* | 3.26578827 | 2.11E-05 | 4.59E-05 | 0.132913969 |
| T2DM | mast_cell | *YWHAB* | 3.937830295 | 1.04E-05 | 2.41E-05 | 0.065302617 |
| T2DM | mast_cell | *NYAP1* | -1.109619643 | 0.010646562 | 0.014617806 | 1 |
| T2DM | mast_cell | *TSC22D4* | -2.065277683 | 0.012463238 | 0.016923624 | 1 |
| T2DM | mast_cell | *MS4A6E* | 1.140943908 | 0.000126169 | 0.000236078 | 0.793222238 |
| T2DM | mast_cell | *MS4A7* | 1.37034144 | 1.03E-06 | 3.16E-06 | 0.006475932 |
| T2DM | mast_cell | *DIS3L* | 3.791888022 | 6.75E-06 | 1.64E-05 | 0.042414262 |
| T2DM | mast_cell | *CCNDBP1* | 4.240951866 | 5.25E-09 | 7.96E-08 | 3.30E-05 |
| T2DM | mast_cell | *SMAD3* | 3.188948092 | 1.40E-07 | 5.92E-07 | 0.000879265 |
| T2DM | mast_cell | *RCCD1* | 3.767101336 | 0.003579072 | 0.005294777 | 1 |
| T2DM | mast_cell | *AKTIP* | 4.369744768 | 3.21E-10 | 3.88E-08 | 2.02E-06 |
| T2DM | mast_cell | *NKX3-1* | 8.760985655 | 4.67E-05 | 9.50E-05 | 0.293366182 |
| T2DM | mast_cell | *DUSP18* | 3.889143839 | 2.37E-06 | 6.51E-06 | 0.014925888 |
| T2DM | mast_cell | *TEF* | 2.347543854 | 8.61E-07 | 2.71E-06 | 0.005410617 |
| T2DM | mast_cell | *SNRPD1* | 5.743989971 | 3.89E-05 | 8.04E-05 | 0.244280486 |
| T2DM | mast_cell | *SAMD14* | -1.765089508 | 8.73E-07 | 2.74E-06 | 0.00548851 |
| T2DM | mast_cell | *PIP5KL1* | 0.851525014 | 0.010112212 | 0.013938934 | 1 |
| T2DM | mast_cell | *URM1* | 2.624254551 | 6.47E-07 | 2.12E-06 | 0.004068771 |
| T2DM | mast_cell | *ISLR2* | 2.566095712 | 6.96E-05 | 0.000136173 | 0.437365325 |
| T2DM | mast_cell | *SP2* | 3.085804583 | 1.04E-07 | 4.74E-07 | 0.000654642 |
| T2DM | mast_cell | *COQ7* | 3.337633926 | 3.74E-09 | 7.78E-08 | 2.35E-05 |
| T2DM | mast_cell | *GPRC5B* | 2.968586747 | 1.14E-06 | 3.42E-06 | 0.0071816 |
| T2DM | mast_cell | *KATNAL2* | 0.962231788 | 0.017803632 | 0.023714287 | 1 |
| T2DM | mast_cell | *HDHD2* | 1.611545143 | 3.91E-05 | 8.08E-05 | 0.245624342 |
| T2DM | mast_cell | *RNF214* | 3.929284293 | 0.000189705 | 0.000343315 | 1 |
| T2DM | mast_cell | *ENGASE* | 3.020041156 | 4.28E-07 | 1.49E-06 | 0.002692869 |
| T2DM | mast_cell | *RBFOX3* | 7.827880195 | 0.003876724 | 0.005693288 | 1 |
| T2DM | mast_cell | *MYO5B* | 0.855037933 | 0.02562893 | 0.03332556 | 1 |
| T2DM | mast_cell | *RRM1* | 1.058570669 | 0.001414877 | 0.002223277 | 1 |
| T2DM | mast_cell | *TRIM68* | 1.903144026 | 2.48E-06 | 6.75E-06 | 0.015579597 |
| T2DM | mast_cell | *FN3K* | 4.034373131 | 1.29E-08 | 1.13E-07 | 8.14E-05 |
| T2DM | mast_cell | *ZNF23* | 2.161202874 | 1.18E-07 | 5.22E-07 | 0.000742737 |
| T2DM | mast_cell | *ZNF229* | -3.237631728 | 0.005589267 | 0.0080246 | 1 |
| T2DM | mast_cell | *ZNF646* | 4.240954756 | 0.000217925 | 0.000390119 | 1 |
| T2DM | mast_cell | *VKORC1* | 1.138805743 | 0.024869685 | 0.032438944 | 1 |
| T2DM | mast_cell | *SMG8* | 4.281013166 | 1.24E-08 | 1.13E-07 | 7.82E-05 |
| T2DM | mast_cell | *RAB8A* | 5.120160005 | 7.58E-08 | 3.73E-07 | 0.000476675 |
| T2DM | mast_cell | *GATAD2A* | 4.193217721 | 1.17E-05 | 2.68E-05 | 0.073603086 |
| T2DM | mast_cell | *RPL13* | 2.878226724 | 0.001825407 | 0.002828076 | 1 |
| T2DM | mast_cell | *ZNF641* | 2.44431356 | 1.51E-06 | 4.39E-06 | 0.009520754 |
| T2DM | mast_cell | *CACNB3* | -2.59122026 | 0.019461418 | 0.025807622 | 1 |
| T2DM | mast_cell | *TP53I13* | 2.967401297 | 2.33E-07 | 9.00E-07 | 0.001467301 |

| | | | | | | |
|---|---|---|---|---|---|---|
| T2DM | mast_cell | *ZNF528* | 3.057998676 | 0.010852794 | 0.01487822 | 1 |
| T2DM | mast_cell | *ZNF701* | 3.497673514 | 2.49E-05 | 5.34E-05 | 0.156805224 |
| T2DM | mast_cell | *SERTAD3* | 2.411563562 | 1.00E-08 | 1.04E-07 | 6.29E-05 |
| T2DM | mast_cell | *TMC4* | 4.439387346 | 0.012010299 | 0.016358048 | 1 |
| T2DM | mast_cell | *TTYH1* | 1.723538808 | 6.22E-09 | 8.17E-08 | 3.91E-05 |
| T2DM | mast_cell | *LENG8* | 2.144109145 | 0.011752455 | 0.016034654 | 1 |
| T2DM | mast_cell | *CDC42EP5* | -2.904660256 | 0.001707332 | 0.002654958 | 1 |
| T2DM | mast_cell | *ZNF283* | 3.425510904 | 5.19E-07 | 1.75E-06 | 0.003260322 |
| T2DM | mast_cell | *SPINT2* | 1.224504361 | 0.008228652 | 0.011514253 | 1 |
| T2DM | mast_cell | *PSCA* | 3.552096705 | 0.000227497 | 0.000406005 | 1 |
| T2DM | mast_cell | *UBXN6* | 2.887191619 | 1.50E-05 | 3.33E-05 | 0.09410672 |
| T2DM | mast_cell | *FAM57A* | 1.486765577 | 5.82E-05 | 0.000116053 | 0.365913766 |
| T2DM | mast_cell | *GLOD4* | 3.810853522 | 7.92E-09 | 9.16E-08 | 4.98E-05 |
| T2DM | mast_cell | *KIFC2* | 0.927797952 | 0.006920984 | 0.009784625 | 1 |
| T2DM | mast_cell | *SRR* | 0.784616699 | 0.015819323 | 0.021205988 | 1 |
| T2DM | mast_cell | *CYB5D2* | 4.032839862 | 4.53E-10 | 4.47E-08 | 2.85E-06 |
| T2DM | mast_cell | *ZNF83* | 3.085777167 | 6.67E-06 | 1.63E-05 | 0.041953346 |
| T2DM | mast_cell | *CD320* | 4.840416938 | 3.36E-09 | 7.78E-08 | 2.12E-05 |
| T2DM | mast_cell | *SPRYD3* | 6.501011204 | 2.44E-08 | 1.60E-07 | 0.000153691 |
| T2DM | mast_cell | *SOAT2* | 1.680577136 | 0.008183225 | 0.011458338 | 1 |
| T2DM | mast_cell | *ZNF558* | 1.362331714 | 0.016601127 | 0.022197211 | 1 |
| T2DM | mast_cell | *NDUFV1* | 4.040626994 | 5.52E-05 | 0.000110835 | 0.347136306 |
| T2DM | mast_cell | *CDK2AP2* | 5.64942951 | 2.14E-07 | 8.44E-07 | 0.001348391 |
| T2DM | mast_cell | *PRDX2* | 6.54583224 | 4.31E-11 | 1.04E-08 | 2.71E-07 |
| T2DM | mast_cell | *ZNF232* | -0.944140564 | 0.004172333 | 0.006091839 | 1 |
| T2DM | mast_cell | *MIS12* | 2.582303818 | 9.47E-09 | 1.01E-07 | 5.95E-05 |
| T2DM | mast_cell | *HID1* | 2.400338424 | 2.66E-06 | 7.22E-06 | 0.016737485 |
| T2DM | mast_cell | *ATP5H* | 2.924970291 | 0.023722739 | 0.031045974 | 1 |
| T2DM | mast_cell | *SRP68* | 2.695060905 | 4.14E-08 | 2.36E-07 | 0.000260575 |
| T2DM | mast_cell | *MGAT5B* | -3.561619604 | 0.011285193 | 0.015440698 | 1 |
| T2DM | mast_cell | *TMC8* | -0.958616788 | 1.17E-05 | 2.69E-05 | 0.073820317 |
| T2DM | mast_cell | *TMEM68* | 3.460421542 | 4.45E-07 | 1.54E-06 | 0.002799565 |
| T2DM | mast_cell | *CYP7A1* | 3.876462737 | 0.000161547 | 0.000296452 | 1 |
| T2DM | mast_cell | *GSDMA* | 4.551083269 | 4.13E-08 | 2.36E-07 | 0.000259361 |
| T2DM | mast_cell | *ZNF598* | 2.044851483 | 4.71E-08 | 2.60E-07 | 0.000296279 |
| T2DM | mast_cell | *MLST8* | 6.189261107 | 0.005255166 | 0.007569124 | 1 |
| T2DM | mast_cell | *ECI1* | 2.40226847 | 0.000338977 | 0.000586448 | 1 |
| T2DM | mast_cell | *ABCA3* | 2.971512856 | 1.21E-08 | 1.11E-07 | 7.62E-05 |
| T2DM | mast_cell | *KCTD5* | -2.712360415 | 0.01252999 | 0.017003248 | 1 |
| T2DM | mast_cell | *SDHAF2* | 4.294021416 | 4.04E-09 | 7.78E-08 | 2.54E-05 |
| T2DM | mast_cell | *DDB1* | 2.550093389 | 0.025445277 | 0.033107297 | 1 |
| T2DM | mast_cell | *VPS37C* | 3.588077222 | 1.63E-08 | 1.26E-07 | 0.000102438 |
| T2DM | mast_cell | *VWCE* | -2.835817826 | 0.000150323 | 0.000277802 | 0.945082304 |
| T2DM | mast_cell | *BSCL2* | -2.809153153 | 2.20E-08 | 1.50E-07 | 0.000138065 |
| T2DM | mast_cell | *HRASLS5* | 0.703017429 | 1.61E-05 | 3.55E-05 | 0.101172132 |
| T2DM | mast_cell | *TRANK1* | -1.610585889 | 0.023097585 | 0.030284571 | 1 |
| T2DM | mast_cell | *TTC21A* | 4.850531121 | 6.09E-09 | 8.15E-08 | 3.83E-05 |
| T2DM | mast_cell | *RPSA* | 1.390333123 | 0.023802894 | 0.031130673 | 1 |
| T2DM | mast_cell | *LTBP3* | 3.692423739 | 7.15E-13 | 3.75E-10 | 4.50E-09 |
| T2DM | mast_cell | *NAALADL1* | 2.962968298 | 6.68E-08 | 3.39E-07 | 0.000420092 |

| | | | | | | |
|---|---|---|---|---|---|---|
| T2DM | mast_cell | *SCARA5* | 2.409184451 | 1.63E-07 | 6.74E-07 | 0.001023265 |
| T2DM | mast_cell | *COPS6* | 2.581957494 | 0.008070297 | 0.01131029 | 1 |
| T2DM | mast_cell | *NUDT16L1* | 6.155425201 | 4.66E-05 | 9.48E-05 | 0.292725461 |
| T2DM | mast_cell | *KIAA1586* | 2.92036581 | 0.000455901 | 0.000772365 | 1 |
| T2DM | mast_cell | *ZCCHC4* | 4.650793767 | 1.49E-05 | 3.30E-05 | 0.093427926 |
| T2DM | mast_cell | *GLYCTK* | 2.623804025 | 5.32E-10 | 4.99E-08 | 3.34E-06 |
| T2DM | mast_cell | *UBTD2* | 4.146723984 | 4.89E-08 | 2.67E-07 | 0.000307567 |
| T2DM | mast_cell | *POLR2J3* | 2.492668598 | 1.20E-07 | 5.28E-07 | 0.000755 |
| T2DM | mast_cell | *NT5DC2* | -2.194244056 | 4.60E-05 | 9.38E-05 | 0.289340168 |
| T2DM | mast_cell | *KIF5C* | 3.772066917 | 1.01E-05 | 2.35E-05 | 0.063574362 |
| T2DM | mast_cell | *PXK* | 4.329431442 | 2.13E-08 | 1.47E-07 | 0.00013364 |
| T2DM | mast_cell | *ACOX2* | 0.914400387 | 0.009095457 | 0.012648338 | 1 |
| T2DM | mast_cell | *CX3CR1* | 1.695932913 | 7.18E-06 | 1.73E-05 | 0.045154169 |
| T2DM | mast_cell | *SCN11A* | 0.678561899 | 6.37E-05 | 0.00012593 | 0.400458315 |
| T2DM | mast_cell | *DTYMK* | 4.82502384 | 1.46E-08 | 1.19E-07 | 9.17E-05 |
| T2DM | mast_cell | *ING5* | 4.594210569 | 4.62E-06 | 1.17E-05 | 0.029026406 |
| T2DM | mast_cell | *MLKL* | 1.357060193 | 6.17E-05 | 0.000122396 | 0.387873054 |
| T2DM | mast_cell | *RFWD3* | 3.629348038 | 2.77E-09 | 7.78E-08 | 1.74E-05 |
| T2DM | mast_cell | *RHOH* | -0.520206426 | 2.94E-07 | 1.09E-06 | 0.001851248 |
| T2DM | mast_cell | *KLHL30* | 1.869604449 | 1.12E-05 | 2.58E-05 | 0.070509913 |
| T2DM | mast_cell | *REEP4* | 2.36912416 | 1.71E-08 | 1.30E-07 | 0.000107416 |
| T2DM | mast_cell | *BMP1* | 2.989276749 | 3.87E-06 | 1.00E-05 | 0.024307892 |
| T2DM | mast_cell | *ATXN2L* | 3.796757428 | 9.88E-05 | 0.000188471 | 0.621200779 |
| T2DM | mast_cell | *FNTA* | 3.764005701 | 2.38E-09 | 7.78E-08 | 1.50E-05 |
| T2DM | mast_cell | *SERINC2* | 2.797656244 | 1.21E-06 | 3.59E-06 | 0.007584716 |
| T2DM | mast_cell | *TRAPPC11* | 3.474040016 | 1.02E-08 | 1.04E-07 | 6.39E-05 |
| T2DM | mast_cell | *TMEM223* | -2.366151654 | 0.01373014 | 0.018559748 | 1 |
| T2DM | mast_cell | *ZSWIM1* | 6.559749976 | 1.22E-06 | 3.61E-06 | 0.007647267 |
| T2DM | mast_cell | *NBPF9* | 4.603625982 | 0.007092132 | 0.010017577 | 1 |
| T2DM | mast_cell | *WFDC13* | 13.67560276 | 8.82E-09 | 9.63E-08 | 5.55E-05 |
| T2DM | mast_cell | *ZNF30* | 2.5084129 | 0.000260077 | 0.000459041 | 1 |
| T2DM | mast_cell | *SLC16A4* | 2.078684616 | 0.000374066 | 0.000643083 | 1 |
| T2DM | mast_cell | *LRP1B* | -0.978127982 | 0.000414096 | 0.000707259 | 1 |
| T2DM | mast_cell | *DNAJC21* | 4.602312646 | 6.21E-08 | 3.22E-07 | 0.000390421 |
| T2DM | mast_cell | *NPNT* | 1.438571222 | 1.86E-08 | 1.37E-07 | 0.000117019 |
| T2DM | mast_cell | *ABHD15* | 3.053331962 | 4.48E-09 | 7.78E-08 | 2.82E-05 |
| T2DM | mast_cell | *ADAL* | 0.676496578 | 0.00061227 | 0.001015926 | 1 |
| T2DM | mast_cell | *SNTB2* | 3.635475191 | 1.80E-08 | 1.34E-07 | 0.00011299 |
| T2DM | mast_cell | *ZNF507* | 2.118626346 | 0.000732729 | 0.00119747 | 1 |
| T2DM | mast_cell | *STX18* | 3.129766238 | 0.001817961 | 0.002817928 | 1 |
| T2DM | mast_cell | *ZBTB49* | 4.913372236 | 3.72E-09 | 7.78E-08 | 2.34E-05 |
| T2DM | mast_cell | *TNIP2* | 1.749780976 | 0.004332058 | 0.006307469 | 1 |
| T2DM | mast_cell | *BTNL3* | 0.590172404 | 0.002819849 | 0.004246321 | 1 |
| T2DM | mast_cell | *ZNF608* | 2.215906767 | 5.95E-09 | 8.15E-08 | 3.74E-05 |
| T2DM | mast_cell | *INPP5D* | -1.76588439 | 0.000254887 | 0.000450639 | 1 |
| T2DM | mast_cell | *LETM1* | 3.089051475 | 3.72E-09 | 7.78E-08 | 2.34E-05 |
| T2DM | mast_cell | *CEP120* | 3.835253558 | 1.23E-05 | 2.80E-05 | 0.077187798 |
| T2DM | mast_cell | *STXBP6* | -0.887877564 | 1.21E-06 | 3.60E-06 | 0.007625672 |
| T2DM | mast_cell | *MFF* | 3.306046019 | 4.70E-05 | 9.55E-05 | 0.295498252 |
| T2DM | mast_cell | *CPLX1* | 1.618889853 | 3.05E-05 | 6.43E-05 | 0.191693732 |

| | | | | | | |
|---|---|---|---|---|---|---|
| T2DM | mast_cell | *SIGLEC7* | -2.330799331 | 0.000250754 | 0.000444081 | 1 |
| T2DM | mast_cell | *FEM1B* | 4.357434253 | 9.98E-10 | 6.17E-08 | 6.27E-06 |
| T2DM | mast_cell | *MFSD7* | 4.498683131 | 1.15E-05 | 2.64E-05 | 0.072328841 |
| T2DM | mast_cell | *COL4A3* | 2.170497125 | 3.04E-07 | 1.12E-06 | 0.001908555 |
| T2DM | mast_cell | *IRS1* | -2.334333399 | 2.08E-25 | 1.31E-21 | 1.31E-21 |
| T2DM | mast_cell | *UPF3A* | 1.58279209 | 0.005324665 | 0.007662204 | 1 |
| T2DM | mast_cell | *C8orf46* | 0.389469392 | 0.000570825 | 0.000952942 | 1 |
| T2DM | mast_cell | *HSPBAP1* | 2.204500525 | 0.000207195 | 0.000372181 | 1 |
| T2DM | mast_cell | *PARM1* | 1.341580446 | 0.022642511 | 0.029712683 | 1 |
| T2DM | mast_cell | *FAM110B* | 3.77130972 | 1.09E-08 | 1.06E-07 | 6.83E-05 |
| T2DM | mast_cell | *ATF5* | 6.09253933 | 1.50E-07 | 6.28E-07 | 0.000941489 |
| T2DM | mast_cell | *PCSK9* | 8.535970166 | 0.000757211 | 0.001235233 | 1 |
| T2DM | mast_cell | *XPO6* | 4.548987174 | 9.65E-07 | 2.98E-06 | 0.006068553 |
| T2DM | mast_cell | *RGS14* | 1.210007795 | 0.000384052 | 0.000658989 | 1 |
| T2DM | mast_cell | *RAB24* | 2.110979933 | 1.57E-05 | 3.47E-05 | 0.098435063 |
| T2DM | mast_cell | *SH3TC2* | -1.778812202 | 5.35E-05 | 0.000107619 | 0.336417151 |
| T2DM | mast_cell | *GPRIN1* | 9.668451228 | 8.92E-06 | 2.10E-05 | 0.0560981 |
| T2DM | mast_cell | *STK32A* | 0.845425697 | 0.02118034 | 0.027881 | 1 |
| T2DM | mast_cell | *P2RY12* | 1.508192491 | 9.73E-07 | 3.00E-06 | 0.006114304 |
| T2DM | mast_cell | *PTK2* | 2.991296489 | 8.66E-06 | 2.04E-05 | 0.054470355 |
| T2DM | mast_cell | *RNASE6* | 2.576027432 | 1.02E-08 | 1.04E-07 | 6.40E-05 |
| T2DM | mast_cell | *NPR1* | 3.673030563 | 3.95E-09 | 7.78E-08 | 2.48E-05 |
| T2DM | mast_cell | *CLIC4* | 3.498202005 | 4.50E-07 | 1.56E-06 | 0.002832042 |
| T2DM | mast_cell | *SLC38A11* | -0.474053236 | 1.18E-07 | 5.22E-07 | 0.000743344 |
| T2DM | mast_cell | *CCDC8* | 3.402025629 | 4.18E-08 | 2.37E-07 | 0.000262583 |
| T2DM | mast_cell | *METTL15* | 2.080052032 | 0.000122125 | 0.000229193 | 0.767797843 |
| T2DM | mast_cell | *DTWD2* | 3.797733607 | 4.71E-05 | 9.57E-05 | 0.296186757 |
| T2DM | mast_cell | *INO80E* | -2.091282053 | 0.010101133 | 0.013926716 | 1 |
| T2DM | mast_cell | *DFFB* | 4.79152191 | 1.24E-07 | 5.40E-07 | 0.00078028 |
| T2DM | mast_cell | *LUZP1* | 3.357778463 | 7.83E-06 | 1.87E-05 | 0.049222908 |
| T2DM | mast_cell | *HEXDC* | 3.686701589 | 0.000522955 | 0.000876751 | 1 |
| T2DM | mast_cell | *CHRNA5* | 2.044283094 | 1.42E-06 | 4.15E-06 | 0.008950725 |
| T2DM | mast_cell | *FASN* | 1.867184951 | 9.56E-08 | 4.42E-07 | 0.000601349 |
| T2DM | mast_cell | *MT1E* | -3.239938271 | 0.008780829 | 0.012240592 | 1 |
| T2DM | mast_cell | *DCXR* | 4.118696305 | 2.15E-08 | 1.48E-07 | 0.000135239 |
| T2DM | mast_cell | *LIMS1* | 2.324813119 | 5.09E-06 | 1.28E-05 | 0.031984764 |
| T2DM | mast_cell | *UGP2* | 2.645458193 | 8.05E-06 | 1.92E-05 | 0.050632611 |
| T2DM | mast_cell | *CSGALNACT2* | 3.880281879 | 0.000126659 | 0.000236785 | 0.79630764 |
| T2DM | mast_cell | *AVEN* | 4.182646482 | 1.11E-08 | 1.07E-07 | 6.96E-05 |
| T2DM | mast_cell | *CTNND2* | 1.259955077 | 2.03E-06 | 5.68E-06 | 0.012743109 |
| T2DM | mast_cell | *TPST1* | 1.179075839 | 0.004263869 | 0.006221151 | 1 |
| T2DM | mast_cell | *GUSB* | 4.98040144 | 4.73E-07 | 1.62E-06 | 0.002974896 |
| T2DM | mast_cell | *ZFPM2* | 2.820791176 | 1.02E-07 | 4.68E-07 | 0.000644334 |
| T2DM | mast_cell | *ZNF747* | -4.100008825 | 0.00081014 | 0.001316111 | 1 |
| T2DM | mast_cell | *MAP3K2* | 3.570561685 | 3.24E-07 | 1.18E-06 | 0.002037704 |
| T2DM | mast_cell | *PUSL1* | 3.545467261 | 7.61E-08 | 3.74E-07 | 0.000478466 |
| T2DM | mast_cell | *MYO7B* | 2.628839187 | 0.001807533 | 0.002802456 | 1 |
| T2DM | mast_cell | *CNTROB* | 4.903873887 | 0.002671212 | 0.004037968 | 1 |
| T2DM | mast_cell | *KCNAB3* | 1.277528187 | 0.021674957 | 0.028484626 | 1 |
| T2DM | mast_cell | *NIPA1* | 5.428312766 | 1.04E-06 | 3.19E-06 | 0.006552327 |

| | | | | | | |
|---|---|---|---|---|---|---|
| T2DM | mast_cell | *RNF150* | 2.292792481 | 3.45E-07 | 1.25E-06 | 0.002171697 |
| T2DM | mast_cell | *HOXD4* | -2.450775993 | 0.001522908 | 0.002379946 | 1 |
| T2DM | mast_cell | *CHRNB1* | -1.124869606 | 0.01091571 | 0.01496121 | 1 |
| T2DM | mast_cell | *ANKK1* | -1.038866705 | 3.49E-08 | 2.07E-07 | 0.000219396 |
| T2DM | mast_cell | *PDCD6IP* | 2.553179012 | 0.000376311 | 0.000646412 | 1 |
| T2DM | mast_cell | *ZNF212* | 3.990580301 | 6.94E-08 | 3.48E-07 | 0.000436311 |
| T2DM | mast_cell | *FAM161A* | 3.370866291 | 1.14E-06 | 3.42E-06 | 0.007173883 |
| T2DM | mast_cell | *GLB1* | 3.36931033 | 8.05E-06 | 1.92E-05 | 0.050612883 |
| T2DM | mast_cell | *FAXDC2* | 3.257373872 | 9.12E-08 | 4.26E-07 | 0.000573538 |
| T2DM | mast_cell | *ELP5* | 3.813140594 | 2.75E-05 | 5.85E-05 | 0.173145438 |
| T2DM | mast_cell | *STX8* | 3.196638558 | 6.05E-05 | 0.000120347 | 0.380295136 |
| T2DM | mast_cell | *NFRKB* | 3.818884414 | 1.50E-07 | 6.30E-07 | 0.000944585 |
| T2DM | mast_cell | *SP7* | 4.549988486 | 0.010487921 | 0.014412639 | 1 |
| T2DM | mast_cell | *SEMA3E* | 5.575655021 | 1.09E-07 | 4.91E-07 | 0.000684772 |
| T2DM | mast_cell | *DCLK2* | 2.271263156 | 0.00058482 | 0.000975527 | 1 |
| T2DM | mast_cell | *ADORA2B* | 0.725010779 | 0.032286464 | 0.041459355 | 1 |
| T2DM | mast_cell | *MGMT* | 2.523060979 | 4.51E-05 | 9.22E-05 | 0.283557854 |
| T2DM | mast_cell | *METTL7B* | 4.856793188 | 1.50E-08 | 1.21E-07 | 9.43E-05 |
| T2DM | mast_cell | *NFXL1* | 4.570915279 | 1.35E-05 | 3.04E-05 | 0.084987756 |
| T2DM | mast_cell | *CD14* | 2.558797739 | 1.65E-05 | 3.64E-05 | 0.103714113 |
| T2DM | mast_cell | *KRT72* | -5.639853794 | 6.24E-05 | 0.000123716 | 0.392550641 |
| T2DM | mast_cell | *HSD17B13* | 0.924936857 | 0.003055734 | 0.004573054 | 1 |
| T2DM | mast_cell | *TMC7* | 1.375493791 | 1.73E-05 | 3.80E-05 | 0.108932394 |
| T2DM | mast_cell | *ARL6IP1* | -1.940762671 | 0.016477998 | 0.022046643 | 1 |
| T2DM | mast_cell | *SERPINB9* | 4.002956655 | 1.45E-06 | 4.20E-06 | 0.00909162 |
| T2DM | mast_cell | *SMAGP* | -1.757481833 | 4.88E-05 | 9.89E-05 | 0.30706877 |
| T2DM | mast_cell | *IRX2* | 2.07570236 | 8.28E-07 | 2.62E-06 | 0.005205929 |
| T2DM | mast_cell | *STAT2* | 3.900307294 | 1.44E-07 | 6.09E-07 | 0.000906308 |
| T2DM | mast_cell | *NUDCD2* | 5.017942824 | 9.75E-09 | 1.03E-07 | 6.13E-05 |
| T2DM | mast_cell | *HSPA4* | 3.062208795 | 0.010266031 | 0.014133701 | 1 |
| T2DM | mast_cell | *SLC26A5* | 5.408408195 | 1.44E-09 | 6.91E-08 | 9.05E-06 |
| T2DM | mast_cell | *COMMD5* | 3.731223279 | 2.34E-06 | 6.45E-06 | 0.014710576 |
| T2DM | mast_cell | *GTSF1* | 20.07667285 | 4.02E-05 | 8.30E-05 | 0.252684719 |
| T2DM | mast_cell | *RNF34* | 2.833978661 | 0.000490082 | 0.000824496 | 1 |
| T2DM | mast_cell | *ACYP2* | 3.155274844 | 5.06E-06 | 1.28E-05 | 0.031824694 |
| T2DM | mast_cell | *TRABD* | 1.87573367 | 0.030362667 | 0.039068786 | 1 |
| T2DM | mast_cell | *TMEM133* | 3.738523403 | 1.28E-08 | 1.13E-07 | 8.07E-05 |
| T2DM | mast_cell | *TTLL6* | -6.052857777 | 5.68E-07 | 1.90E-06 | 0.003570746 |
| T2DM | mast_cell | *KCNS3* | 1.646223981 | 7.57E-07 | 2.42E-06 | 0.004756555 |
| T2DM | mast_cell | *HTRA3* | 2.309170679 | 2.38E-06 | 6.53E-06 | 0.014993405 |
| T2DM | mast_cell | *FOXN2* | 2.596304207 | 0.004912744 | 0.007100327 | 1 |
| T2DM | mast_cell | *KBTBD2* | 3.337574205 | 4.66E-07 | 1.60E-06 | 0.002931899 |
| T2DM | mast_cell | *MTSS1* | 3.267996886 | 2.22E-06 | 6.16E-06 | 0.013939083 |
| T2DM | mast_cell | *RPS9* | 2.615784088 | 0.000532351 | 0.000890842 | 1 |
| T2DM | mast_cell | *TSEN34* | 3.436436807 | 4.69E-08 | 2.60E-07 | 0.000295105 |
| T2DM | mast_cell | *NDUFA3* | 4.057038953 | 1.52E-08 | 1.21E-07 | 9.55E-05 |
| T2DM | mast_cell | *OSCAR* | 2.977262141 | 8.05E-07 | 2.56E-06 | 0.005060704 |
| T2DM | mast_cell | *PAQR8* | 3.04609855 | 0.036854158 | 0.047055664 | 1 |
| T2DM | mast_cell | *NUDT6* | 2.317959272 | 1.23E-09 | 6.68E-08 | 7.74E-06 |
| T2DM | mast_cell | *ZNF160* | 3.493141999 | 3.24E-06 | 8.58E-06 | 0.020349742 |

| | | | | | | |
|---|---|---|---|---|---|---|
| T2DM | mast_cell | *ZNF415* | -0.816869369 | 0.005293558 | 0.007620929 | 1 |
| T2DM | mast_cell | *TSNARE1* | 2.178443069 | 3.23E-05 | 6.80E-05 | 0.203314184 |
| T2DM | mast_cell | *FPR1* | 1.628290301 | 0.000648294 | 0.001070051 | 1 |
| T2DM | mast_cell | *FEZ2* | 2.997710895 | 0.000449255 | 0.000762339 | 1 |
| T2DM | mast_cell | *SOX7* | 7.030840711 | 1.61E-05 | 3.56E-05 | 0.101438329 |
| T2DM | mast_cell | *TRMT61B* | 1.147442013 | 0.001101897 | 0.001752498 | 1 |
| T2DM | mast_cell | *MFN1* | 3.935688602 | 5.37E-09 | 8.00E-08 | 3.38E-05 |
| T2DM | mast_cell | *KCNMB3* | 1.421164733 | 0.032523669 | 0.041746897 | 1 |
| T2DM | mast_cell | *ATP6V0E2* | 2.050044689 | 1.55E-08 | 1.23E-07 | 9.76E-05 |
| T2DM | mast_cell | *CLDN20* | -1.509122433 | 2.05E-05 | 4.45E-05 | 0.128856414 |
| T2DM | mast_cell | *CANT1* | 3.690148908 | 8.68E-06 | 2.05E-05 | 0.054584777 |
| T2DM | mast_cell | *CHST11* | -1.472819493 | 0.000160188 | 0.000294387 | 1 |
| T2DM | mast_cell | *KCND3* | 2.509246779 | 1.14E-09 | 6.34E-08 | 7.17E-06 |
| T2DM | mast_cell | *ZNF581* | 3.150456193 | 3.32E-08 | 2.00E-07 | 0.000208912 |
| T2DM | mast_cell | *MCC* | 3.453011237 | 1.95E-08 | 1.39E-07 | 0.000122295 |
| T2DM | mast_cell | *DLK2* | -2.595546683 | 1.07E-07 | 4.86E-07 | 0.000675681 |
| T2DM | mast_cell | *ZNF561* | 1.115960819 | 0.020331827 | 0.026871179 | 1 |
| T2DM | mast_cell | *LRRC8C* | 5.411674738 | 1.03E-08 | 1.04E-07 | 6.44E-05 |
| T2DM | mast_cell | *PPID* | 4.213255625 | 3.45E-08 | 2.05E-07 | 0.00021688 |
| T2DM | mast_cell | *ETFDH* | 0.588542164 | 0.020367935 | 0.026913242 | 1 |
| T2DM | mast_cell | *TBCA* | 3.168934837 | 1.27E-05 | 2.89E-05 | 0.080105951 |
| T2DM | mast_cell | *MAP6* | 1.859038557 | 1.70E-07 | 6.99E-07 | 0.001071126 |
| T2DM | mast_cell | *BCL2L1* | 2.986009909 | 0.001449817 | 0.002274201 | 1 |
| T2DM | mast_cell | *PLRG1* | 4.636823199 | 2.56E-08 | 1.66E-07 | 0.000161062 |
| T2DM | mast_cell | *NMUR1* | 1.866895005 | 9.97E-09 | 1.04E-07 | 6.27E-05 |
| T2DM | mast_cell | *P2RY6* | 1.541655935 | 2.13E-08 | 1.47E-07 | 0.00013399 |
| T2DM | mast_cell | *ZIK1* | -5.446543 | 0.000181893 | 0.000330128 | 1 |
| T2DM | mast_cell | *RGS19* | -1.663120807 | 0.000957023 | 0.001537644 | 1 |
| T2DM | mast_cell | *TCEA2* | 2.983668183 | 4.54E-09 | 7.78E-08 | 2.86E-05 |
| T2DM | mast_cell | *ANO5* | 2.377156333 | 6.15E-09 | 8.15E-08 | 3.87E-05 |
| T2DM | mast_cell | *VAT1L* | 3.418405161 | 0.00099158 | 0.001586276 | 1 |
| T2DM | mast_cell | *CAMTA1* | 3.056890136 | 0.001011551 | 0.001615347 | 1 |
| T2DM | mast_cell | *LRRC34* | 0.96514445 | 0.007711346 | 0.01083864 | 1 |
| T2DM | mast_cell | *SYCE1* | 2.035126123 | 1.38E-05 | 3.09E-05 | 0.086671541 |
| T2DM | mast_cell | *NXNL1* | 2.022997163 | 0.001493256 | 0.00233884 | 1 |
| T2DM | mast_cell | *BCL2* | 0.88484766 | 0.000508869 | 0.00085496 | 1 |
| T2DM | mast_cell | *RHNO1* | 2.745709725 | 5.05E-05 | 0.000102021 | 0.317184028 |
| T2DM | mast_cell | *METTL18* | 2.376240159 | 3.21E-07 | 1.17E-06 | 0.002017453 |
| T2DM | mast_cell | *ZNF570* | 4.051563382 | 5.02E-05 | 0.000101508 | 0.315486429 |
| T2DM | mast_cell | *NINJ2* | 2.957033724 | 7.92E-09 | 9.16E-08 | 4.98E-05 |
| T2DM | mast_cell | *FAM90A1* | 5.4131322 | 1.16E-08 | 1.09E-07 | 7.27E-05 |
| T2DM | mast_cell | *AQP4* | 1.904346241 | 1.42E-08 | 1.19E-07 | 8.92E-05 |
| T2DM | mast_cell | *TLN2* | 1.433741436 | 4.58E-05 | 9.34E-05 | 0.28804449 |
| T2DM | mast_cell | *TVP23B* | 3.531889209 | 1.21E-08 | 1.11E-07 | 7.62E-05 |
| T2DM | mast_cell | *SRGAP2C* | 2.909026181 | 6.17E-09 | 8.15E-08 | 3.88E-05 |
| T2DM | mast_cell | *ATPAF2* | 6.095561937 | 4.34E-06 | 1.11E-05 | 0.027278199 |
| T2DM | mast_cell | *CYP4F22* | -1.13313887 | 4.18E-08 | 2.37E-07 | 0.000263033 |
| T2DM | mast_cell | *C20orf196* | 4.303605719 | 1.34E-06 | 3.93E-06 | 0.008402796 |
| T2DM | mast_cell | *ZNF554* | 1.441982431 | 0.030342028 | 0.039050221 | 1 |
| T2DM | mast_cell | *ANKRD20A4* | -1.89679388 | 2.61E-07 | 9.86E-07 | 0.001643154 |

| | | | | | | |
|---|---|---|---|---|---|---|
| T2DM | mast_cell | *SERF1A* | 7.75886126 | 1.80E-08 | 1.34E-07 | 0.000113049 |
| T2DM | mast_cell | *LRRC15* | -3.601107071 | 0.000372263 | 0.000640536 | 1 |
| T2DM | mast_cell | *NME6* | 1.324181784 | 0.007824137 | 0.010988923 | 1 |
| T2DM | mast_cell | *FRMD3* | 4.190210352 | 1.44E-06 | 4.19E-06 | 0.009066208 |
| T2DM | mast_cell | *MTBP* | 4.94445197 | 2.63E-06 | 7.13E-06 | 0.016512454 |
| T2DM | mast_cell | *MALT1* | 3.953030579 | 1.28E-05 | 2.90E-05 | 0.080411594 |
| T2DM | mast_cell | *ISG20* | 1.33868402 | 0.011883886 | 0.016206939 | 1 |
| T2DM | mast_cell | *CXCR6* | 2.361574859 | 1.57E-07 | 6.53E-07 | 0.000985127 |
| T2DM | mast_cell | *C5orf34* | 3.273583212 | 1.78E-07 | 7.25E-07 | 0.00111842 |
| T2DM | mast_cell | *C1QTNF4* | -0.601114731 | 0.00077083 | 0.001255494 | 1 |
| T2DM | mast_cell | *ZNF131* | 3.185438409 | 0.000194617 | 0.000351697 | 1 |
| T2DM | mast_cell | *HINFP* | 6.223405255 | 2.99E-07 | 1.10E-06 | 0.001877649 |
| T2DM | mast_cell | *SPTLC3* | 4.681521282 | 1.91E-06 | 5.38E-06 | 0.011981267 |
| T2DM | mast_cell | *COPRS* | -2.198531247 | 0.028657073 | 0.03701048 | 1 |
| T2DM | mast_cell | *SUCLG2* | 3.18130321 | 3.49E-05 | 7.29E-05 | 0.219694871 |
| T2DM | mast_cell | *RCAN2* | 1.714294708 | 5.80E-09 | 8.14E-08 | 3.65E-05 |
| T2DM | mast_cell | *IL16* | 2.209219914 | 6.90E-06 | 1.68E-05 | 0.043388769 |
| T2DM | mast_cell | *GNB2* | 4.161197852 | 4.27E-06 | 1.10E-05 | 0.02687515 |
| T2DM | mast_cell | *PRSS27* | 10.38443025 | 9.00E-10 | 6.08E-08 | 5.66E-06 |
| T2DM | mast_cell | *ZNF24* | 4.864650611 | 5.15E-06 | 1.29E-05 | 0.03238735 |
| T2DM | mast_cell | *FIBP* | 2.63217487 | 8.47E-10 | 6.08E-08 | 5.33E-06 |
| T2DM | mast_cell | *CARNS1* | 4.254118187 | 3.57E-08 | 2.10E-07 | 0.00022468 |
| T2DM | mast_cell | *NIPAL4* | 2.169590003 | 0.035666199 | 0.045628929 | 1 |
| T2DM | mast_cell | *SNTG2* | 2.658942435 | 2.11E-06 | 5.89E-06 | 0.013267437 |
| T2DM | mast_cell | *PDE3A* | -1.071191487 | 0.000729189 | 0.001192503 | 1 |
| T2DM | mast_cell | *MRPL52* | 1.819457017 | 0.026828095 | 0.034733985 | 1 |
| T2DM | mast_cell | *SMPDL3A* | 2.67910822 | 1.74E-07 | 7.09E-07 | 0.001091126 |
| T2DM | mast_cell | *RAD9A* | 0.907538232 | 0.006330739 | 0.009015571 | 1 |
| T2DM | mast_cell | *EFEMP2* | 2.710689872 | 5.66E-05 | 0.000113264 | 0.355648297 |
| T2DM | mast_cell | *AGAP5* | 46.26988581 | 1.53E-08 | 1.21E-07 | 9.60E-05 |
| T2DM | mast_cell | *ZFAND4* | 2.697708846 | 0.006672038 | 0.009468872 | 1 |
| T2DM | mast_cell | *SLFN11* | 2.600998032 | 2.59E-07 | 9.80E-07 | 0.001631278 |
| T2DM | mast_cell | *CCL19* | 7.344020194 | 3.02E-08 | 1.85E-07 | 0.000189708 |
| T2DM | mast_cell | *EFCAB12* | 1.205376274 | 0.01674656 | 0.022382147 | 1 |
| T2DM | mast_cell | *FAM192A* | 2.057876492 | 4.54E-06 | 1.16E-05 | 0.028546491 |
| T2DM | mast_cell | *RAB43* | -9.048961951 | 9.03E-07 | 2.82E-06 | 0.005678218 |
| T2DM | mast_cell | *CBWD1* | 3.870991343 | 1.36E-09 | 6.85E-08 | 8.56E-06 |
| T2DM | mast_cell | *HOXC5* | 2.300661035 | 0.000608361 | 0.00101024 | 1 |
| T2DM | mast_cell | *RAB37* | -1.428254515 | 1.37E-06 | 4.02E-06 | 0.008626288 |
| T2DM | mast_cell | *DCP2* | 4.072399462 | 3.53E-06 | 9.28E-06 | 0.022164592 |
| T2DM | mast_cell | *SNX32* | 1.721944426 | 0.011714147 | 0.015989328 | 1 |
| T2DM | mast_cell | *METAP1D* | 4.72877005 | 1.57E-06 | 4.54E-06 | 0.009896369 |
| T2DM | mast_cell | *NADSYN1* | 3.544652148 | 6.93E-07 | 2.24E-06 | 0.0043543 |
| T2DM | mast_cell | *RNASEH2C* | 1.488952581 | 0.008211267 | 0.01149432 | 1 |
| T2DM | mast_cell | *MYEOV* | 0.377646959 | 0.020762716 | 0.027377348 | 1 |
| T2DM | mast_cell | *MRGPRF* | 3.646696627 | 8.45E-09 | 9.37E-08 | 5.31E-05 |
| T2DM | mast_cell | *ADH6* | 0.622633251 | 0.006732445 | 0.009539527 | 1 |
| T2DM | mast_cell | *GXYLT2* | 2.882555592 | 5.32E-07 | 1.79E-06 | 0.003342802 |
| T2DM | mast_cell | *HECTD4* | 3.45141769 | 1.16E-05 | 2.65E-05 | 0.072699807 |
| T2DM | mast_cell | *TRMT112* | 3.0737199 | 6.48E-05 | 0.000128077 | 0.407668411 |

| | | | | | | |
|---|---|---|---|---|---|---|
| T2DM | mast_cell | *NOC3L* | 3.724641733 | 3.36E-08 | 2.01E-07 | 0.000210982 |
| T2DM | mast_cell | *ABLIM3* | 4.202287509 | 2.25E-06 | 6.23E-06 | 0.01414361 |
| T2DM | mast_cell | *VANGL1* | 4.617782277 | 0.000416554 | 0.000711265 | 1 |
| T2DM | mast_cell | *IQCB1* | 1.468313369 | 0.009994925 | 0.013795411 | 1 |
| T2DM | mast_cell | *GOLGB1* | 3.583735053 | 3.00E-06 | 8.03E-06 | 0.018884645 |
| T2DM | mast_cell | *DMRT2* | 1.5192225 | 0.008927461 | 0.012431217 | 1 |
| T2DM | mast_cell | *ZNF483* | 1.284144238 | 0.000222748 | 0.000398299 | 1 |
| T2DM | mast_cell | *SLC2A14* | -1.528040687 | 0.00261217 | 0.003952517 | 1 |
| T2DM | mast_cell | *MZT2A* | 2.850193072 | 6.94E-08 | 3.48E-07 | 0.000436391 |
| T2DM | mast_cell | *MAP3K11* | 6.032325677 | 2.33E-07 | 9.00E-07 | 0.001465942 |
| T2DM | mast_cell | *C1QA* | 2.91872647 | 3.57E-09 | 7.78E-08 | 2.25E-05 |
| T2DM | mast_cell | *NDNF* | 2.282460492 | 4.36E-08 | 2.45E-07 | 0.000274186 |
| T2DM | mast_cell | *OLR1* | 4.74122044 | 3.84E-09 | 7.78E-08 | 2.42E-05 |
| T2DM | mast_cell | *GLIPR1L1* | 2.601129669 | 3.81E-09 | 7.78E-08 | 2.40E-05 |
| T2DM | mast_cell | *ARV1* | 3.592005429 | 3.53E-07 | 1.27E-06 | 0.00222018 |
| T2DM | mast_cell | *CCDC36* | 3.229073694 | 2.85E-09 | 7.78E-08 | 1.79E-05 |
| T2DM | mast_cell | *THAP2* | 2.502061238 | 1.32E-07 | 5.70E-07 | 0.000832981 |
| T2DM | mast_cell | *PPP1R14B* | 1.721402536 | 2.01E-08 | 1.42E-07 | 0.000126199 |
| T2DM | mast_cell | *SMARCC1* | 1.885704392 | 0.038339461 | 0.04882321 | 1 |
| T2DM | mast_cell | *PEAK1* | 3.087297793 | 7.78E-06 | 1.86E-05 | 0.048904993 |
| T2DM | mast_cell | *MST1* | 1.29858944 | 7.42E-06 | 1.78E-05 | 0.04662872 |
| T2DM | mast_cell | *TNFRSF10C* | 0.993727709 | 0.018315074 | 0.024380027 | 1 |
| T2DM | mast_cell | *GMPPB* | -1.79163905 | 0.000162025 | 0.000297242 | 1 |
| T2DM | mast_cell | *CSPG4* | 2.703428425 | 5.01E-08 | 2.73E-07 | 0.00031515 |
| T2DM | mast_cell | *SNX33* | -2.26383078 | 0.000183213 | 0.000332427 | 1 |
| T2DM | mast_cell | *NUDT18* | -4.947258937 | 0.003587053 | 0.005305058 | 1 |
| T2DM | mast_cell | *SULT1B1* | 3.161066093 | 0.000100604 | 0.000191607 | 0.632494516 |
| T2DM | mast_cell | *NUDT4* | 2.854877542 | 4.71E-05 | 9.57E-05 | 0.296088619 |
| T2DM | mast_cell | *SCAI* | 3.833557935 | 8.16E-06 | 1.94E-05 | 0.051316782 |
| T2DM | mast_cell | *C1orf100* | 9.19240875 | 5.27E-09 | 7.96E-08 | 3.32E-05 |
| T2DM | mast_cell | *AGFG1* | 2.818464001 | 1.89E-06 | 5.36E-06 | 0.011913129 |
| T2DM | mast_cell | *RNF213* | 3.433134406 | 9.41E-07 | 2.92E-06 | 0.005916511 |
| T2DM | mast_cell | *TUBB8* | 5.094814978 | 0.003281804 | 0.004888107 | 1 |
| T2DM | mast_cell | *GPR160* | 5.16170916 | 6.23E-09 | 8.17E-08 | 3.91E-05 |
| T2DM | mast_cell | *GOLIM4* | 2.868375429 | 0.000429538 | 0.000731845 | 1 |
| T2DM | mast_cell | *USMG5* | 5.379433012 | 3.31E-09 | 7.78E-08 | 2.08E-05 |
| T2DM | mast_cell | *HOXB2* | 1.203423296 | 0.013887734 | 0.018752617 | 1 |
| T2DM | mast_cell | *XXYLT1* | 4.098440851 | 2.82E-06 | 7.57E-06 | 0.017702125 |
| T2DM | mast_cell | *UBXN2A* | 2.580283333 | 0.00029931 | 0.000523148 | 1 |
| T2DM | mast_cell | *CCS* | 4.665133124 | 1.39E-07 | 5.91E-07 | 0.00087528 |
| T2DM | mast_cell | *NRROS* | 1.624983143 | 3.68E-05 | 7.65E-05 | 0.2310516 |
| T2DM | mast_cell | *LEMD3* | 4.365016499 | 6.00E-07 | 1.99E-06 | 0.003774538 |
| T2DM | mast_cell | *C16orf91* | 3.106473799 | 1.40E-09 | 6.85E-08 | 8.81E-06 |
| T2DM | mast_cell | *SOCS7* | 4.291374635 | 9.29E-09 | 9.92E-08 | 5.84E-05 |
| T2DM | mast_cell | *TLR6* | 1.719693767 | 1.64E-08 | 1.26E-07 | 0.000103207 |
| T2DM | mast_cell | *CTU2* | 3.178589328 | 0.00030952 | 0.000539653 | 1 |
| T2DM | mast_cell | *SNX31* | 2.103855062 | 0.009591392 | 0.013293889 | 1 |
| T2DM | mast_cell | *DDX23* | 3.906053338 | 0.002132653 | 0.003267851 | 1 |
| T2DM | mast_cell | *GLIS1* | 1.021572807 | 3.85E-06 | 1.00E-05 | 0.024193511 |
| T2DM | mast_cell | *RALGAPA1* | 1.779351434 | 0.011909577 | 0.016238101 | 1 |

| | | | | | | |
|---|---|---|---|---|---|---|
| T2DM | mast_cell | *LIG4* | 3.360542943 | 1.17E-06 | 3.49E-06 | 0.007351484 |
| T2DM | mast_cell | *SNAPC5* | 3.950072231 | 0.000151704 | 0.000280189 | 0.953763225 |
| T2DM | mast_cell | *LINGO2* | -2.219019433 | 0.002458581 | 0.003735404 | 1 |
| T2DM | mast_cell | *BBS1* | 2.988229913 | 1.83E-08 | 1.35E-07 | 0.000114973 |
| T2DM | mast_cell | *ANKRD36C* | 3.311649171 | 9.64E-05 | 0.000184348 | 0.605952081 |
| T2DM | mast_cell | *TMEM81* | -3.663642656 | 5.93E-06 | 1.47E-05 | 0.037286224 |
| T2DM | mast_cell | *IL20RB* | 3.019067056 | 2.18E-07 | 8.54E-07 | 0.00137147 |
| T2DM | mast_cell | *ZNF497* | 7.277924784 | 4.50E-06 | 1.15E-05 | 0.028322365 |
| T2DM | mast_cell | *KLF14* | 7.423409702 | 0.001077635 | 0.001716516 | 1 |
| T2DM | mast_cell | *ANGEL2* | 4.489708642 | 7.74E-07 | 2.47E-06 | 0.004868811 |
| T2DM | mast_cell | *BRSK2* | 3.316646735 | 2.79E-08 | 1.76E-07 | 0.000175303 |
| T2DM | mast_cell | *SH3PXD2B* | 1.831781857 | 4.00E-10 | 4.13E-08 | 2.52E-06 |
| T2DM | mast_cell | *FGFBP3* | 2.730664643 | 1.97E-08 | 1.41E-07 | 0.000124094 |
| T2DM | mast_cell | *C4orf32* | -1.69799072 | 0.007737018 | 0.010869862 | 1 |
| T2DM | mast_cell | *HRAS* | 2.454916382 | 8.22E-08 | 3.97E-07 | 0.000517028 |
| T2DM | mast_cell | *SRP72* | 1.774702588 | 0.003956715 | 0.005797219 | 1 |
| T2DM | mast_cell | *C4orf26* | -5.679685786 | 4.59E-06 | 1.17E-05 | 0.028868132 |
| T2DM | mast_cell | *BTC* | 1.728443716 | 5.91E-09 | 8.14E-08 | 3.72E-05 |
| T2DM | mast_cell | *NDUFA11* | 3.7367693 | 0.010139629 | 0.013970599 | 1 |
| T2DM | mast_cell | *PTDSS2* | 2.516865568 | 9.57E-07 | 2.96E-06 | 0.006015664 |
| T2DM | mast_cell | *FUT1* | -2.217443172 | 8.41E-06 | 1.99E-05 | 0.0528435 |
| T2DM | mast_cell | *DHX36* | 2.029816141 | 0.026570631 | 0.034443208 | 1 |
| T2DM | mast_cell | *CHST2* | -2.451606987 | 1.13E-06 | 3.40E-06 | 0.007134107 |
| T2DM | mast_cell | *ATR* | 3.49523661 | 1.94E-08 | 1.39E-07 | 0.000121935 |
| T2DM | mast_cell | *TVP23C* | -1.953378253 | 1.36E-06 | 4.00E-06 | 0.008566538 |
| T2DM | mast_cell | *PACS1* | 2.673225136 | 0.00047489 | 0.000801319 | 1 |
| T2DM | mast_cell | *CADM2* | -1.062276488 | 0.010362791 | 0.014253089 | 1 |
| T2DM | mast_cell | *PSMD2* | 2.512902751 | 1.73E-07 | 7.07E-07 | 0.001085688 |
| T2DM | mast_cell | *ARHGAP1* | 3.154243114 | 3.65E-06 | 9.56E-06 | 0.022940697 |
| T2DM | mast_cell | *CHST1* | 3.432833362 | 0.000908137 | 0.001466288 | 1 |
| T2DM | mast_cell | *GOLGA8A* | 2.732273733 | 7.83E-07 | 2.50E-06 | 0.0049237 |
| T2DM | mast_cell | *TP53I11* | 4.972527405 | 1.17E-09 | 6.44E-08 | 7.37E-06 |
| T2DM | mast_cell | *PHYHD1* | 2.784022031 | 0.003229555 | 0.004818275 | 1 |
| T2DM | mast_cell | *ZNF519* | 2.618556474 | 0.01198853 | 0.016331936 | 1 |
| T2DM | mast_cell | *BANF1* | 2.0671211 | 1.05E-08 | 1.06E-07 | 6.62E-05 |
| T2DM | mast_cell | *TMEM9B* | 5.718800081 | 2.33E-10 | 3.12E-08 | 1.47E-06 |
| T2DM | mast_cell | *NRIP3* | 1.43679295 | 0.002079415 | 0.003196402 | 1 |
| T2DM | mast_cell | *SCUBE2* | 2.887027527 | 2.40E-06 | 6.56E-06 | 0.01507087 |
| T2DM | mast_cell | *SMAD2* | 1.853802959 | 0.005267451 | 0.007585081 | 1 |
| T2DM | mast_cell | *EIF3F* | 4.106655573 | 1.02E-05 | 2.36E-05 | 0.063828449 |
| T2DM | mast_cell | *CLTB* | 2.269089918 | 0.007464508 | 0.010508142 | 1 |
| T2DM | mast_cell | *PCSK1* | -1.134999019 | 5.68E-07 | 1.90E-06 | 0.003568447 |
| T2DM | mast_cell | *TBC1D10C* | 1.748897529 | 0.00458091 | 0.006652848 | 1 |
| T2DM | mast_cell | *PPP2R2D* | 2.208337098 | 0.002173451 | 0.003323883 | 1 |
| T2DM | mast_cell | *LRRC25* | 1.269053983 | 6.86E-06 | 1.67E-05 | 0.043130779 |
| T2DM | mast_cell | *UBQLNL* | 4.365406369 | 5.59E-06 | 1.39E-05 | 0.03516097 |
| T2DM | mast_cell | *LIPT2* | 4.601254948 | 1.03E-06 | 3.16E-06 | 0.006478183 |
| T2DM | mast_cell | *KCNE3* | -2.468111965 | 0.01343491 | 0.018180215 | 1 |
| T2DM | mast_cell | *UCP3* | 2.395780139 | 6.17E-06 | 1.52E-05 | 0.038806374 |
| T2DM | mast_cell | *UCP2* | 2.554210712 | 0.030118928 | 0.038786912 | 1 |

| | | | | | | |
|---|---|---|---|---|---|---|
| T2DM | mast_cell | *PAAF1* | 0.549786523 | 0.015936914 | 0.021345415 | 1 |
| T2DM | mast_cell | *MRPL48* | 1.203271805 | 0.01918414 | 0.025466784 | 1 |
| T2DM | mast_cell | *RAB6A* | 2.747083691 | 0.000860102 | 0.001393676 | 1 |
| T2DM | mast_cell | *RPS6KB2* | 6.373810574 | 4.71E-07 | 1.61E-06 | 0.002961377 |
| T2DM | mast_cell | *RMI2* | 1.592181229 | 0.001499011 | 0.0023461 | 1 |
| T2DM | mast_cell | *TOM1L2* | 3.889622195 | 1.05E-05 | 2.42E-05 | 0.065768444 |
| T2DM | mast_cell | *TEX26* | 3.785024021 | 6.70E-08 | 3.39E-07 | 0.000421376 |
| T2DM | mast_cell | *B3GNTL1* | 4.54107998 | 1.41E-06 | 4.13E-06 | 0.008889069 |
| T2DM | mast_cell | *TTLL11* | 1.982833203 | 0.000222487 | 0.000397944 | 1 |
| T2DM | mast_cell | *MSRA* | -1.416633619 | 1.18E-05 | 2.70E-05 | 0.074243198 |
| T2DM | mast_cell | *CTDNEP1* | 3.465548301 | 1.62E-09 | 7.17E-08 | 1.02E-05 |
| T2DM | mast_cell | *ETV4* | 3.533786952 | 1.09E-08 | 1.06E-07 | 6.85E-05 |
| T2DM | mast_cell | *SWI5* | 2.522941164 | 0.000308618 | 0.000538368 | 1 |
| T2DM | mast_cell | *ZDHHC21* | 3.214444087 | 2.76E-06 | 7.46E-06 | 0.017365435 |
| T2DM | mast_cell | *A2M* | -1.2437617 | 2.31E-05 | 4.98E-05 | 0.144992269 |
| T2DM | mast_cell | *ARL4D* | 2.136458666 | 4.09E-08 | 2.34E-07 | 0.000257343 |
| T2DM | mast_cell | *LRRN1* | 3.37437828 | 4.21E-09 | 7.78E-08 | 2.64E-05 |
| T2DM | mast_cell | *B3GALT6* | -2.641309597 | 3.74E-06 | 9.78E-06 | 0.023535358 |
| T2DM | mast_cell | *CHMP6* | 3.608769681 | 2.12E-09 | 7.78E-08 | 1.33E-05 |
| T2DM | mast_cell | *UFSP1* | -0.981305315 | 0.019277837 | 0.025580363 | 1 |
| T2DM | mast_cell | *TCP11L1* | 3.502781377 | 9.96E-07 | 3.06E-06 | 0.006264273 |
| T2DM | mast_cell | *GPX2* | -3.193994325 | 0.012441165 | 0.016900952 | 1 |
| T2DM | mast_cell | *CCDC57* | 2.622323859 | 3.37E-05 | 7.06E-05 | 0.211697995 |
| T2DM | mast_cell | *CIDEA* | 0.46228682 | 0.002343646 | 0.003571135 | 1 |
| T2DM | mast_cell | *SMIM19* | 2.028682677 | 5.71E-05 | 0.000114185 | 0.358770553 |
| T2DM | mast_cell | *SLC35G1* | 1.69887393 | 1.27E-07 | 5.50E-07 | 0.000800906 |
| T2DM | mast_cell | *COX8A* | -2.896230994 | 0.001927149 | 0.00297617 | 1 |
| T2DM | mast_cell | *PRR18* | -4.350595368 | 2.32E-06 | 6.40E-06 | 0.014583399 |
| T2DM | mast_cell | *B3GNT4* | -7.221450802 | 2.86E-07 | 1.06E-06 | 0.001798758 |
| T2DM | mast_cell | *CRLF3* | 3.7831682 | 3.11E-08 | 1.90E-07 | 0.000195814 |
| T2DM | mast_cell | *RNPEP* | 2.982958479 | 1.53E-08 | 1.21E-07 | 9.59E-05 |
| T2DM | mast_cell | *WDR25* | 4.625240719 | 1.07E-06 | 3.25E-06 | 0.006721605 |
| T2DM | mast_cell | *PLA2G16* | 0.911243273 | 0.000172938 | 0.000315652 | 1 |
| T2DM | mast_cell | *PHLDB3* | 3.471824486 | 3.60E-06 | 9.43E-06 | 0.022614947 |
| T2DM | mast_cell | *RMDN1* | 3.598796899 | 4.53E-08 | 2.52E-07 | 0.000284713 |
| T2DM | mast_cell | *RNF152* | 3.000771126 | 1.29E-08 | 1.13E-07 | 8.09E-05 |
| T2DM | mast_cell | *BOK* | 4.019576218 | 3.51E-08 | 2.07E-07 | 0.000220692 |
| T2DM | mast_cell | *ZNF843* | 8.363071061 | 8.32E-05 | 0.000160548 | 0.523226131 |
| T2DM | mast_cell | *C8orf59* | 4.718900048 | 3.66E-07 | 1.30E-06 | 0.002299065 |
| T2DM | mast_cell | *PFN4* | 2.140882072 | 1.41E-08 | 1.19E-07 | 8.88E-05 |
| T2DM | mast_cell | *RUFY1* | 2.099980444 | 0.001942761 | 0.002998071 | 1 |
| T2DM | mast_cell | *LRRC37A3* | 3.768375984 | 2.86E-09 | 7.78E-08 | 1.80E-05 |
| T2DM | mast_cell | *VSIG10* | 0.901767745 | 0.027481201 | 0.035535646 | 1 |
| T2DM | mast_cell | *IRX5* | 3.256133923 | 8.57E-05 | 0.00016518 | 0.538983806 |
| T2DM | mast_cell | *PNMA1* | 4.721224103 | 6.36E-08 | 3.27E-07 | 0.00039973 |
| T2DM | mast_cell | *ANKLE2* | 4.315573542 | 5.73E-05 | 0.000114538 | 0.360223013 |
| T2DM | mast_cell | *GCNT4* | -1.006362804 | 1.12E-07 | 5.01E-07 | 0.000702534 |
| T2DM | mast_cell | *THAP4* | 3.150309235 | 0.002509761 | 0.003808225 | 1 |
| T2DM | mast_cell | *DPP7* | 4.94747773 | 8.65E-07 | 2.72E-06 | 0.00543755 |
| T2DM | mast_cell | *SEC24C* | 4.267495288 | 4.98E-05 | 0.000100825 | 0.313163025 |

| | | | | | | |
|---|---|---|---|---|---|---|
| T2DM | mast_cell | *MTHFR* | 4.166355878 | 1.59E-05 | 3.51E-05 | 0.099720952 |
| T2DM | mast_cell | *WDR73* | 3.344817823 | 2.62E-09 | 7.78E-08 | 1.65E-05 |
| T2DM | mast_cell | *DSCAML1* | 2.001155968 | 1.36E-09 | 6.85E-08 | 8.57E-06 |
| T2DM | mast_cell | *EPS8L2* | 1.910119973 | 0.020737525 | 0.027349868 | 1 |
| T2DM | mast_cell | *TALDO1* | 3.191627219 | 5.91E-07 | 1.97E-06 | 0.003718544 |
| T2DM | mast_cell | *RIMKLA* | 1.990634548 | 1.17E-05 | 2.68E-05 | 0.073563001 |
| T2DM | mast_cell | *B3GNT8* | 7.771780175 | 4.90E-09 | 7.84E-08 | 3.08E-05 |
| T2DM | mast_cell | *AP006621.1* | 1.355927593 | 5.28E-09 | 7.96E-08 | 3.32E-05 |
| T2DM | mast_cell | *FBXO39* | 0.832491224 | 0.019731194 | 0.026154336 | 1 |
| T2DM | mast_cell | *TOP3A* | 4.166701888 | 2.97E-06 | 7.96E-06 | 0.018685199 |
| T2DM | mast_cell | *ZBTB38* | 2.852222731 | 0.000185398 | 0.000336169 | 1 |
| T2DM | mast_cell | *C8orf31* | 3.598197337 | 5.84E-09 | 8.14E-08 | 3.67E-05 |
| T2DM | mast_cell | *AC024940.1* | 3.21444342 | 4.43E-05 | 9.07E-05 | 0.278502441 |
| T2DM | mast_cell | *TIMM22* | -2.60281665 | 0.00200699 | 0.003089605 | 1 |
| T2DM | mast_cell | *PPFIA3* | 1.932787222 | 2.47E-07 | 9.41E-07 | 0.001554204 |
| T2DM | mast_cell | *MAGEF1* | -3.194572483 | 3.93E-05 | 8.13E-05 | 0.247382166 |
| T2DM | mast_cell | *SAMD9L* | 1.380769167 | 0.025027659 | 0.032624693 | 1 |
| T2DM | mast_cell | *PAWR* | 2.806975341 | 2.96E-09 | 7.78E-08 | 1.86E-05 |
| T2DM | mast_cell | *NAP1L5* | -2.397312294 | 2.77E-06 | 7.47E-06 | 0.017398888 |
| T2DM | mast_cell | *NIM1K* | 2.190852547 | 6.12E-09 | 8.15E-08 | 3.84E-05 |
| T2DM | mast_cell | *ACOT4* | 1.725169305 | 1.35E-05 | 3.04E-05 | 0.085023892 |
| T2DM | mast_cell | *ARIH2* | 2.492194177 | 1.41E-05 | 3.16E-05 | 0.088856455 |
| T2DM | mast_cell | *ATOX1* | 4.783954249 | 5.63E-05 | 0.000112854 | 0.354248155 |
| T2DM | mast_cell | *TBL1XR1* | 2.785077281 | 7.01E-06 | 1.70E-05 | 0.044043518 |
| T2DM | mast_cell | *SAMD12* | 2.027931199 | 2.29E-06 | 6.34E-06 | 0.014423677 |
| T2DM | mast_cell | *C18orf32* | 5.500212686 | 4.39E-07 | 1.52E-06 | 0.002757313 |
| T2DM | mast_cell | *RPLP2* | 2.232046957 | 0.018364446 | 0.024440573 | 1 |
| T2DM | mast_cell | *GBA* | 3.098690792 | 0.00066159 | 0.001090565 | 1 |
| T2DM | mast_cell | *ACAD9* | 0.604927663 | 0.033994184 | 0.043554399 | 1 |
| T2DM | mast_cell | *IL17RA* | 2.154697255 | 0.000766249 | 0.001248679 | 1 |
| T2DM | mast_cell | *CD163L1* | 1.130704817 | 3.60E-08 | 2.11E-07 | 0.000226518 |
| T2DM | mast_cell | *THAP5* | 5.75997864 | 8.60E-08 | 4.09E-07 | 0.000540893 |
| T2DM | mast_cell | *POLR2L* | 3.28048881 | 0.001282542 | 0.002025964 | 1 |
| T2DM | mast_cell | *FAM87B* | 3.763320171 | 9.26E-06 | 2.17E-05 | 0.058203446 |
| T2DM | mast_cell | *ZCCHC3* | -2.807800705 | 0.031781586 | 0.040827714 | 1 |
| T2DM | mast_cell | *PCDHB9* | 2.093040369 | 2.00E-07 | 7.99E-07 | 0.001259546 |
| T2DM | mast_cell | *ZNF619* | 2.099551278 | 1.09E-08 | 1.06E-07 | 6.83E-05 |
| T2DM | mast_cell | *ZNF354C* | 3.483380422 | 2.61E-09 | 7.78E-08 | 1.64E-05 |
| T2DM | mast_cell | *CENPBD1* | 1.800483074 | 0.038034148 | 0.048463861 | 1 |
| T2DM | mast_cell | *BET1L* | 3.655718455 | 2.11E-10 | 2.95E-08 | 1.33E-06 |
| T2DM | mast_cell | *RIC8A* | 3.035922612 | 2.89E-05 | 6.11E-05 | 0.181441193 |
| T2DM | mast_cell | *IMP3* | 3.964631901 | 0.008852525 | 0.012332334 | 1 |
| T2DM | mast_cell | *ODF3B* | 3.862509047 | 5.23E-05 | 0.000105475 | 0.328977472 |
| T2DM | mast_cell | *DPY19L2* | -1.081101022 | 0.008839347 | 0.012316705 | 1 |
| T2DM | mast_cell | *C2orf73* | 5.280868713 | 4.37E-09 | 7.78E-08 | 2.75E-05 |
| T2DM | mast_cell | *FAM26E* | 3.597001266 | 4.36E-07 | 1.51E-06 | 0.002740597 |
| T2DM | mast_cell | *ALS2CL* | 2.291350318 | 2.07E-08 | 1.44E-07 | 0.000129943 |
| T2DM | mast_cell | *MLF1* | 4.403084094 | 7.51E-08 | 3.70E-07 | 0.000472254 |
| T2DM | mast_cell | *PRSS42* | 3.811189938 | 2.95E-06 | 7.92E-06 | 0.018567072 |
| T2DM | mast_cell | *C2orf69* | 5.670848802 | 5.48E-06 | 1.37E-05 | 0.034473238 |

| | | | | | | |
|---|---|---|---|---|---|---|
| T2DM | mast_cell | *GRAMD1C* | 4.849022293 | 3.61E-09 | 7.78E-08 | 2.27E-05 |
| T2DM | mast_cell | *STAP2* | 3.135762969 | 0.006033824 | 0.008629356 | 1 |
| T2DM | mast_cell | *TSSK6* | 2.935446046 | 0.000636645 | 0.001051929 | 1 |
| T2DM | mast_cell | *GOLGA8Q* | 32.71608414 | 0.000914793 | 0.00147545 | 1 |
| T2DM | mast_cell | *DALRD3* | 3.380794845 | 1.29E-08 | 1.13E-07 | 8.12E-05 |
| T2DM | mast_cell | *ZNF114* | -5.478257244 | 5.53E-06 | 1.38E-05 | 0.034786357 |
| T2DM | mast_cell | *ZNF518B* | 3.275937798 | 2.61E-07 | 9.84E-07 | 0.001640092 |
| T2DM | mast_cell | *ZNF454* | 1.87268788 | 1.29E-07 | 5.58E-07 | 0.000812428 |
| T2DM | mast_cell | *ZC3H12D* | 4.060492465 | 3.62E-09 | 7.78E-08 | 2.28E-05 |
| T2DM | mast_cell | *KDELC2* | 3.403753408 | 1.38E-06 | 4.04E-06 | 0.008680578 |
| T2DM | mast_cell | *PLEC* | 4.418433444 | 1.72E-07 | 7.03E-07 | 0.001078695 |
| T2DM | mast_cell | *SH2D4B* | -7.906906337 | 0.001143216 | 0.001814541 | 1 |
| T2DM | mast_cell | *RNF212* | 1.316030399 | 8.60E-06 | 2.03E-05 | 0.054085172 |
| T2DM | mast_cell | *PRSS36* | 2.424648886 | 1.31E-08 | 1.14E-07 | 8.23E-05 |
| T2DM | mast_cell | *GEN1* | 0.960608467 | 0.010267745 | 0.014133701 | 1 |
| T2DM | mast_cell | *TMPRSS9* | 2.582667649 | 0.001366048 | 0.002151928 | 1 |
| T2DM | mast_cell | *AQP11* | 2.750723803 | 0.029680336 | 0.038245598 | 1 |
| T2DM | mast_cell | *TMEM11* | 6.788632609 | 1.10E-07 | 4.96E-07 | 0.000694483 |
| T2DM | mast_cell | *KCNG2* | -0.736658224 | 0.023304089 | 0.030523502 | 1 |
| T2DM | mast_cell | *ZFAND2A* | 2.635443322 | 7.22E-08 | 3.59E-07 | 0.000453976 |
| T2DM | mast_cell | *ZNF223* | -0.782169545 | 0.003137041 | 0.004690269 | 1 |
| T2DM | mast_cell | *FAM220A* | 1.904015319 | 5.49E-06 | 1.37E-05 | 0.034499408 |
| T2DM | mast_cell | *BEND3* | -4.806018841 | 0.001508805 | 0.002359079 | 1 |
| T2DM | mast_cell | *NT5DC1* | 2.84427232 | 0.000960305 | 0.001542524 | 1 |
| T2DM | mast_cell | *AC006111.1* | 9.488791029 | 4.20E-09 | 7.78E-08 | 2.64E-05 |
| T2DM | mast_cell | *COX14* | 2.415650487 | 7.71E-09 | 9.13E-08 | 4.84E-05 |
| T2DM | mast_cell | *P4HTM* | 2.928562008 | 0.004096802 | 0.005991299 | 1 |
| T2DM | mast_cell | *CTXN1* | -6.266150284 | 2.18E-05 | 4.72E-05 | 0.136965747 |
| T2DM | mast_cell | *CA8* | 1.159190997 | 0.00740654 | 0.010440564 | 1 |
| T2DM | mast_cell | *ERBB4* | 2.532380096 | 6.76E-09 | 8.65E-08 | 4.25E-05 |
| T2DM | mast_cell | *CSRNP3* | 4.348288363 | 5.52E-09 | 8.00E-08 | 3.47E-05 |
| T2DM | mast_cell | *PARP10* | 3.297480926 | 9.64E-07 | 2.98E-06 | 0.006060644 |
| T2DM | mast_cell | *RPP25* | 1.197237273 | 0.012498114 | 0.016967316 | 1 |
| T2DM | mast_cell | *STX19* | 4.265758938 | 4.31E-05 | 8.84E-05 | 0.270714642 |
| T2DM | mast_cell | *FAM219B* | 3.082575784 | 2.92E-09 | 7.78E-08 | 1.83E-05 |
| T2DM | mast_cell | *CPNE7* | 2.410529576 | 0.032957891 | 0.042285982 | 1 |
| T2DM | mast_cell | *GDPD4* | -5.721552531 | 4.61E-05 | 9.40E-05 | 0.289873376 |
| T2DM | mast_cell | *MPI* | 2.871387538 | 0.010207998 | 0.014058638 | 1 |
| T2DM | mast_cell | *EFCAB13* | 2.322080404 | 0.000608237 | 0.00101024 | 1 |
| T2DM | mast_cell | *C17orf62* | 4.244803942 | 0.000335346 | 0.000580805 | 1 |
| T2DM | mast_cell | *GAK* | 3.77530439 | 1.52E-06 | 4.40E-06 | 0.009558104 |
| T2DM | mast_cell | *TUFM* | 2.364863975 | 5.78E-05 | 0.00011526 | 0.363185778 |
| T2DM | mast_cell | *RMI1* | 2.42117547 | 1.32E-09 | 6.84E-08 | 8.28E-06 |
| T2DM | mast_cell | *CTC1* | 2.224203118 | 0.00304773 | 0.00456329 | 1 |
| T2DM | mast_cell | *SNX18* | 3.988893656 | 4.43E-09 | 7.78E-08 | 2.79E-05 |
| T2DM | mast_cell | *C14orf39* | 1.093905157 | 0.00096259 | 0.001545404 | 1 |
| T2DM | mast_cell | *TRIML2* | 29.31733381 | 6.80E-07 | 2.21E-06 | 0.004277904 |
| T2DM | mast_cell | *FARSA* | 1.965588106 | 0.010322572 | 0.014200878 | 1 |
| T2DM | mast_cell | *SPTY2D1* | 5.265466664 | 1.88E-05 | 4.10E-05 | 0.117992725 |
| T2DM | mast_cell | *TCAIM* | 1.827856591 | 0.003003239 | 0.004506293 | 1 |

| | | | | | | |
|---|---|---|---|---|---|---|
| T2DM | mast_cell | *FUCA1* | 3.314868325 | 2.32E-05 | 5.00E-05 | 0.145723017 |
| T2DM | mast_cell | *ZNF664* | 5.114622655 | 9.39E-08 | 4.36E-07 | 0.000590615 |
| T2DM | mast_cell | *CALR* | 5.027767581 | 2.52E-11 | 7.21E-09 | 1.59E-07 |
| T2DM | mast_cell | *CDH4* | -1.819474565 | 0.000280181 | 0.000492039 | 1 |
| T2DM | mast_cell | *HLA-DQB1* | -1.986378359 | 1.53E-15 | 1.93E-12 | 9.64E-12 |
| T2DM | mast_cell | *EGR3* | -0.759446453 | 1.83E-07 | 7.40E-07 | 0.001149553 |
| T2DM | mast_cell | *GEMIN4* | 1.427692678 | 3.86E-06 | 1.00E-05 | 0.024253138 |
| T2DM | mast_cell | *FJX1* | 2.002050252 | 0.003455416 | 0.005124841 | 1 |
| T2DM | mast_cell | *CIITA* | 2.817663817 | 3.36E-09 | 7.78E-08 | 2.12E-05 |
| T2DM | mast_cell | *PLD6* | -1.311642745 | 0.003965883 | 0.005809297 | 1 |
| T2DM | mast_cell | *ZBTB42* | 8.188888741 | 0.000411642 | 0.00070345 | 1 |
| T2DM | mast_cell | *LACC1* | 4.591954191 | 5.26E-08 | 2.83E-07 | 0.000330723 |
| T2DM | mast_cell | *MAF1* | 5.882893272 | 3.02E-09 | 7.78E-08 | 1.90E-05 |
| T2DM | mast_cell | *FCER1A* | -0.6960155 | 2.11E-07 | 8.32E-07 | 0.001325886 |
| T2DM | mast_cell | *PCED1B* | 2.043920463 | 0.003765138 | 0.005543659 | 1 |
| T2DM | mast_cell | *PIPOX* | 2.519025178 | 6.55E-05 | 0.000129285 | 0.412029879 |
| T2DM | mast_cell | *FOXS1* | 3.008013716 | 7.76E-05 | 0.000150643 | 0.488112233 |
| T2DM | mast_cell | *CDH5* | 1.951263312 | 2.49E-06 | 6.79E-06 | 0.015671624 |
| T2DM | mast_cell | *SERTAD2* | 4.178264605 | 1.19E-07 | 5.24E-07 | 0.000746659 |
| T2DM | mast_cell | *CITED4* | 2.129894853 | 1.49E-07 | 6.26E-07 | 0.000937409 |
| T2DM | mast_cell | *TIGD5* | -2.130736959 | 2.63E-05 | 5.61E-05 | 0.165345115 |
| T2DM | mast_cell | *PDXDC1* | 2.774093779 | 0.000985651 | 0.001579202 | 1 |
| T2DM | mast_cell | *ZNF154* | 2.121864323 | 8.51E-09 | 9.37E-08 | 5.35E-05 |
| T2DM | mast_cell | *R3HDM2* | 2.693812847 | 0.000955814 | 0.001536095 | 1 |
| T2DM | mast_cell | *B3GNT3* | -2.320294468 | 0.023802897 | 0.031130673 | 1 |
| T2DM | mast_cell | *SSC5D* | 0.794102925 | 0.001992915 | 0.003070192 | 1 |
| T2DM | mast_cell | *ZADH2* | 3.964050885 | 0.000376248 | 0.000646412 | 1 |
| T2DM | mast_cell | *TMEM86B* | 1.65928945 | 2.01E-06 | 5.63E-06 | 0.012613893 |
| T2DM | mast_cell | *SEPT1* | 1.986448951 | 1.33E-08 | 1.15E-07 | 8.35E-05 |
| T2DM | mast_cell | *EXOC3* | 2.226849614 | 0.003557136 | 0.005265768 | 1 |
| T2DM | mast_cell | *TDRD6* | 1.14814455 | 0.000675423 | 0.001109876 | 1 |
| T2DM | mast_cell | *TDRP* | 5.32860704 | 2.60E-08 | 1.68E-07 | 0.000163511 |
| T2DM | mast_cell | *ZNF816* | 2.24487633 | 9.87E-07 | 3.04E-06 | 0.006205601 |
| T2DM | mast_cell | *FGD6* | 4.265336854 | 1.90E-08 | 1.38E-07 | 0.000119613 |
| T2DM | mast_cell | *HCLS1* | 5.186691765 | 6.32E-07 | 2.08E-06 | 0.003975096 |
| T2DM | mast_cell | *ZNF609* | 2.626467823 | 0.000230919 | 0.000411621 | 1 |
| T2DM | mast_cell | *CCDC66* | 2.567437656 | 0.00010008 | 0.000190725 | 0.629200783 |
| T2DM | mast_cell | *DEFB124* | -1.701377049 | 6.28E-08 | 3.25E-07 | 0.000394707 |
| T2DM | mast_cell | *MCFD2* | 4.728930844 | 1.91E-07 | 7.67E-07 | 0.001198626 |
| T2DM | mast_cell | *CYP8B1* | 2.627051615 | 3.62E-07 | 1.29E-06 | 0.002275227 |
| T2DM | mast_cell | *RNF182* | 1.129885402 | 0.005432881 | 0.007807206 | 1 |
| T2DM | mast_cell | *MB21D2* | 3.681796645 | 2.75E-06 | 7.43E-06 | 0.017276849 |
| T2DM | mast_cell | *SSTR2* | 3.216256386 | 1.14E-08 | 1.09E-07 | 7.19E-05 |
| T2DM | mast_cell | *PCGF5* | 2.896810711 | 0.001725158 | 0.002680027 | 1 |
| T2DM | mast_cell | *SHISA2* | 39.17431641 | 8.26E-09 | 9.23E-08 | 5.19E-05 |
| T2DM | mast_cell | *CHST13* | -2.729191778 | 4.04E-08 | 2.32E-07 | 0.000254112 |
| T2DM | mast_cell | *SRSF8* | 6.742457438 | 0.000144264 | 0.000267468 | 0.90698509 |
| T2DM | mast_cell | *SLC36A4* | 2.934899442 | 0.000465064 | 0.000785983 | 1 |
| T2DM | mast_cell | *ZDHHC20* | 3.904931585 | 8.37E-06 | 1.99E-05 | 0.052615097 |
| T2DM | mast_cell | *OR51E1* | 1.341559259 | 0.000307567 | 0.000536684 | 1 |

| | | | | | | |
|---|---|---|---|---|---|---|
| T2DM | mast_cell | *PSMG4* | 2.947196093 | 0.000795856 | 0.001294579 | 1 |
| T2DM | mast_cell | *CAPS2* | 3.644648206 | 1.11E-08 | 1.07E-07 | 6.99E-05 |
| T2DM | mast_cell | *ZNF792* | -2.17753857 | 5.11E-07 | 1.73E-06 | 0.00321093 |
| T2DM | mast_cell | *KCTD2* | 5.404752537 | 1.20E-08 | 1.11E-07 | 7.54E-05 |
| T2DM | mast_cell | *D2HGDH* | 3.803516141 | 1.22E-07 | 5.34E-07 | 0.000768121 |
| T2DM | mast_cell | *LRRC57* | 5.467388663 | 4.30E-06 | 1.10E-05 | 0.027047821 |
| T2DM | mast_cell | *MRPL14* | 3.056623133 | 0.000294535 | 0.000515661 | 1 |
| T2DM | mast_cell | *BBS12* | 2.229544616 | 2.80E-08 | 1.76E-07 | 0.000175736 |
| T2DM | mast_cell | *ZFP82* | 3.205420826 | 1.40E-09 | 6.85E-08 | 8.78E-06 |
| T2DM | mast_cell | *LSMEM1* | 2.018561769 | 1.22E-07 | 5.34E-07 | 0.000768495 |
| T2DM | mast_cell | *AEN* | 2.375044 | 3.99E-08 | 2.30E-07 | 0.000250715 |
| T2DM | mast_cell | *TRAPPC5* | 2.667395481 | 8.18E-05 | 0.000158012 | 0.514012774 |
| T2DM | mast_cell | *RPH3AL* | 3.126498853 | 2.86E-05 | 6.05E-05 | 0.17965095 |
| T2DM | mast_cell | *ANKRD34A* | -3.667504512 | 0.004032661 | 0.005904364 | 1 |
| T2DM | mast_cell | *HIGD1A* | 2.208333328 | 2.83E-05 | 5.99E-05 | 0.177705458 |
| T2DM | mast_cell | *F2R* | 1.399435161 | 4.24E-06 | 1.09E-05 | 0.026638194 |
| T2DM | mast_cell | *HIST3H2A* | 2.618601855 | 1.35E-05 | 3.04E-05 | 0.08508365 |
| T2DM | mast_cell | *TMEM132C* | 2.259338096 | 2.12E-08 | 1.47E-07 | 0.000133497 |
| T2DM | mast_cell | *TMEM136* | -1.366287017 | 0.002087828 | 0.003205415 | 1 |
| T2DM | mast_cell | *TMEM132E* | 5.942649868 | 0.002257806 | 0.003446182 | 1 |
| T2DM | mast_cell | *DDX60L* | 3.649741251 | 3.53E-07 | 1.27E-06 | 0.002222234 |
| T2DM | mast_cell | *OGFOD3* | 1.098081571 | 0.000630393 | 0.001042968 | 1 |
| T2DM | mast_cell | *ACBD4* | 3.264849552 | 4.32E-09 | 7.78E-08 | 2.71E-05 |
| T2DM | mast_cell | *SGSH* | 6.908284266 | 1.09E-08 | 1.06E-07 | 6.87E-05 |
| T2DM | mast_cell | *SETD2* | 3.099744272 | 2.59E-05 | 5.54E-05 | 0.163124821 |
| T2DM | mast_cell | *MRPS23* | 4.390593986 | 1.94E-09 | 7.78E-08 | 1.22E-05 |
| T2DM | mast_cell | *GPR135* | 1.821341042 | 3.68E-05 | 7.66E-05 | 0.231585345 |
| T2DM | mast_cell | *ZFP41* | 1.320984127 | 0.003418251 | 0.005074508 | 1 |
| T2DM | mast_cell | *ATG9B* | -6.866683481 | 0.007604057 | 0.010696602 | 1 |
| T2DM | mast_cell | *HKR1* | 3.896749467 | 3.05E-07 | 1.12E-06 | 0.001919229 |
| T2DM | mast_cell | *C3orf58* | 3.767132268 | 2.69E-06 | 7.30E-06 | 0.016919341 |
| T2DM | mast_cell | *ODF3L2* | 3.16954874 | 0.000150221 | 0.000277694 | 0.944436562 |
| T2DM | mast_cell | *RELL1* | 3.42100103 | 0.000483192 | 0.000813995 | 1 |
| T2DM | mast_cell | *RFX7* | 2.661081889 | 0.003121279 | 0.004668923 | 1 |
| T2DM | mast_cell | *CLDN7* | 1.504948953 | 5.95E-06 | 1.47E-05 | 0.037433572 |
| T2DM | mast_cell | *SNRPE* | 5.290842283 | 2.18E-07 | 8.53E-07 | 0.001368136 |
| T2DM | mast_cell | *CHST15* | 3.690852107 | 1.11E-07 | 4.98E-07 | 0.000698233 |
| T2DM | mast_cell | *TNRC18* | 3.81635134 | 3.28E-06 | 8.68E-06 | 0.020632031 |
| T2DM | mast_cell | *FAM181B* | 5.356274148 | 1.82E-05 | 3.97E-05 | 0.114259605 |
| T2DM | mast_cell | *NOP10* | 2.505719234 | 0.0001313 | 0.000244879 | 0.825486096 |
| T2DM | mast_cell | *FAM89A* | 1.302988853 | 0.002789942 | 0.004204307 | 1 |
| T2DM | mast_cell | *ZNF708* | 2.882432327 | 9.97E-06 | 2.32E-05 | 0.062711188 |
| T2DM | mast_cell | *IST1* | 3.507339785 | 6.78E-06 | 1.65E-05 | 0.042625822 |
| T2DM | mast_cell | *ERCC6L2* | 2.906725612 | 1.64E-06 | 4.71E-06 | 0.010305313 |
| T2DM | mast_cell | *HHIPL1* | 1.904903743 | 3.94E-08 | 2.27E-07 | 0.000247806 |
| T2DM | mast_cell | *FAM153B* | 2.889911498 | 1.75E-06 | 5.00E-06 | 0.011004555 |
| T2DM | mast_cell | *BACE2* | -0.64641071 | 9.03E-07 | 2.82E-06 | 0.005677543 |
| T2DM | mast_cell | *SYNM* | 1.868568102 | 7.98E-09 | 9.17E-08 | 5.02E-05 |
| T2DM | mast_cell | *IZUMO1* | -0.908623408 | 0.00721466 | 0.010176927 | 1 |
| T2DM | mast_cell | *C8orf33* | 4.243391908 | 4.38E-06 | 1.12E-05 | 0.027529778 |

| | | | | | | |
|---|---|---|---|---|---|---|
| T2DM | mast_cell | *FBXL6* | 2.470412876 | 9.34E-09 | 9.95E-08 | 5.87E-05 |
| T2DM | mast_cell | *KBTBD3* | 2.688499877 | 4.79E-09 | 7.84E-08 | 3.01E-05 |
| T2DM | mast_cell | *CLN8* | 3.141162301 | 0.000123201 | 0.000230938 | 0.774567672 |
| T2DM | mast_cell | *CACNB4* | 1.195128976 | 1.52E-06 | 4.40E-06 | 0.009565884 |
| T2DM | mast_cell | *TRAPPC6B* | 3.204038122 | 7.99E-07 | 2.54E-06 | 0.005025225 |
| T2DM | mast_cell | *NPLOC4* | 1.262467361 | 0.027479054 | 0.035535646 | 1 |
| T2DM | mast_cell | *TSHZ2* | 2.487635727 | 6.08E-06 | 1.50E-05 | 0.038237468 |
| T2DM | mast_cell | *CAPN12* | 5.798706534 | 0.000213278 | 0.000382344 | 1 |
| T2DM | mast_cell | *GLRX5* | 3.178632093 | 2.07E-08 | 1.44E-07 | 0.000129956 |
| T2DM | mast_cell | *CAV3* | 1.052601102 | 0.000333954 | 0.000579032 | 1 |
| T2DM | mast_cell | *MXRA7* | 1.176169581 | 0.000210331 | 0.000377383 | 1 |
| T2DM | mast_cell | *MFSD5* | -2.110901312 | 0.009138695 | 0.012697818 | 1 |
| T2DM | mast_cell | *ADI1* | 4.603474144 | 7.05E-06 | 1.71E-05 | 0.04434434 |
| T2DM | mast_cell | *SPNS3* | 3.225012498 | 3.51E-08 | 2.07E-07 | 0.000220742 |
| T2DM | mast_cell | *CLEC4G* | 4.343775911 | 2.60E-08 | 1.68E-07 | 0.00016375 |
| T2DM | mast_cell | *SATB1* | 3.879420803 | 4.42E-08 | 2.47E-07 | 0.000277671 |
| T2DM | mast_cell | *ACTL10* | -3.36710586 | 2.40E-05 | 5.15E-05 | 0.150743022 |
| T2DM | mast_cell | *TRAK1* | 2.880225808 | 0.000187313 | 0.000339376 | 1 |
| T2DM | mast_cell | *TTC3* | 1.224991122 | 6.82E-07 | 2.22E-06 | 0.004288174 |
| T2DM | mast_cell | *KCNB2* | 0.94104494 | 0.000107844 | 0.00020453 | 0.678016057 |
| T2DM | mast_cell | *TSKU* | 3.389856461 | 0.011436949 | 0.015638125 | 1 |
| T2DM | mast_cell | *PLCXD3* | 1.666778504 | 0.002739371 | 0.00413206 | 1 |
| T2DM | mast_cell | *GPIHBP1* | -1.387317395 | 1.22E-05 | 2.78E-05 | 0.076561789 |
| T2DM | mast_cell | *COL18A1* | 2.035683326 | 0.000336216 | 0.000582151 | 1 |
| T2DM | mast_cell | *SLC25A18* | 1.33313867 | 0.000506467 | 0.000851151 | 1 |
| T2DM | mast_cell | *ZNF721* | 2.05003869 | 0.003274824 | 0.004880023 | 1 |
| T2DM | mast_cell | *LENG9* | 2.946632597 | 1.14E-06 | 3.42E-06 | 0.007190435 |
| T2DM | mast_cell | *HMGN4* | 3.625393114 | 2.37E-05 | 5.10E-05 | 0.149238115 |
| T2DM | mast_cell | *ZNF320* | 1.255654262 | 1.68E-07 | 6.92E-07 | 0.001056208 |
| T2DM | mast_cell | *C12orf60* | 3.226300006 | 3.43E-08 | 2.04E-07 | 0.000215861 |
| T2DM | mast_cell | *LYSMD4* | 1.455868342 | 4.44E-06 | 1.14E-05 | 0.02790793 |
| T2DM | mast_cell | *WBP2NL* | 5.550191654 | 9.37E-07 | 2.91E-06 | 0.005892615 |
| T2DM | mast_cell | *GAS6* | 2.642385867 | 0.000603224 | 0.001003034 | 1 |
| T2DM | mast_cell | *BEGAIN* | -0.910006543 | 1.16E-07 | 5.15E-07 | 0.000730809 |
| T2DM | mast_cell | *ARHGEF37* | 6.096759925 | 2.94E-07 | 1.09E-06 | 0.001848867 |
| T2DM | mast_cell | *FAM43B* | 6.62181723 | 4.94E-06 | 1.25E-05 | 0.031077691 |
| T2DM | mast_cell | *CSMD1* | 2.958425528 | 6.17E-06 | 1.52E-05 | 0.038762655 |
| T2DM | mast_cell | *PTGDR2* | 38.24504764 | 3.99E-06 | 1.03E-05 | 0.025082855 |
| T2DM | mast_cell | *GPR19* | 5.662481027 | 8.97E-10 | 6.08E-08 | 5.64E-06 |
| T2DM | mast_cell | *FANCF* | 8.730140331 | 0.000590947 | 0.000985225 | 1 |
| T2DM | mast_cell | *SMDT1* | 1.024758039 | 0.000776991 | 0.001264873 | 1 |
| T2DM | mast_cell | *C2CD4C* | 3.400588236 | 3.53E-07 | 1.27E-06 | 0.002221498 |
| T2DM | mast_cell | *RUVBL2* | -1.709232279 | 0.001501372 | 0.002349209 | 1 |
| T2DM | mast_cell | *GDPGP1* | 2.719342692 | 2.08E-08 | 1.44E-07 | 0.0001308 |
| T2DM | mast_cell | *CTNNA3* | 1.820523429 | 0.006699842 | 0.009501896 | 1 |
| T2DM | mast_cell | *CCBE1* | 1.323791622 | 1.07E-08 | 1.06E-07 | 6.72E-05 |
| T2DM | mast_cell | *ZNF623* | 3.579996998 | 0.000964351 | 0.001547836 | 1 |
| T2DM | mast_cell | *CCDC125* | 3.307566041 | 7.93E-09 | 9.16E-08 | 4.99E-05 |
| T2DM | mast_cell | *FHL3* | -3.414689802 | 2.11E-06 | 5.88E-06 | 0.013242931 |
| T2DM | mast_cell | *C19orf71* | 3.460626163 | 8.31E-08 | 4.00E-07 | 0.000522182 |

| | | | | | | |
|---|---|---|---|---|---|---|
| T2DM | mast_cell | *LRIT3* | 3.475837518 | 2.73E-07 | 1.02E-06 | 0.001717333 |
| T2DM | mast_cell | *SF3A3* | 3.81726275 | 2.36E-08 | 1.56E-07 | 0.000148076 |
| T2DM | mast_cell | *GRIN2A* | 1.943420099 | 1.27E-07 | 5.50E-07 | 0.000801425 |
| T2DM | mast_cell | *URAD* | 5.783227006 | 8.00E-06 | 1.91E-05 | 0.050279958 |
| T2DM | mast_cell | *GPR132* | 4.008473431 | 3.73E-08 | 2.17E-07 | 0.000234553 |
| T2DM | mast_cell | *MX2* | 2.728513134 | 3.30E-06 | 8.73E-06 | 0.02077212 |
| T2DM | mast_cell | *ACSM5* | 2.262046202 | 1.28E-05 | 2.90E-05 | 0.080473965 |
| T2DM | mast_cell | *AC131971.1* | -4.216757402 | 2.01E-06 | 5.65E-06 | 0.012665564 |
| T2DM | mast_cell | *SERHL2* | 1.922815861 | 6.36E-09 | 8.30E-08 | 4.00E-05 |
| T2DM | mast_cell | *TANGO2* | 4.167641378 | 3.14E-06 | 8.36E-06 | 0.019755949 |
| T2DM | mast_cell | *FAM167B* | 5.443359444 | 4.68E-05 | 9.53E-05 | 0.294513508 |
| T2DM | mast_cell | *RP1L1* | 4.718376269 | 2.62E-09 | 7.78E-08 | 1.64E-05 |
| T2DM | mast_cell | *NDUFB1* | 2.540299746 | 7.29E-09 | 8.89E-08 | 4.58E-05 |
| T2DM | mast_cell | *FAM19A1* | 1.843167663 | 5.69E-09 | 8.09E-08 | 3.58E-05 |
| T2DM | mast_cell | *TRMT12* | 2.378756868 | 3.51E-07 | 1.26E-06 | 0.002203669 |
| T2DM | mast_cell | *CMTM4* | 2.025052143 | 3.26E-08 | 1.97E-07 | 0.000204707 |
| T2DM | mast_cell | *FIGLA* | -7.067049849 | 0.000355576 | 0.000613475 | 1 |
| T2DM | mast_cell | *CBX6* | 5.509492902 | 4.79E-09 | 7.84E-08 | 3.01E-05 |
| T2DM | mast_cell | *KREMEN1* | -0.691307591 | 1.33E-06 | 3.93E-06 | 0.00838554 |
| T2DM | mast_cell | *KCTD16* | 2.898319838 | 1.18E-09 | 6.44E-08 | 7.40E-06 |
| T2DM | mast_cell | *B3GALT5* | 3.74038084 | 0.000119211 | 0.00022426 | 0.74947724 |
| T2DM | mast_cell | *ZNF703* | -2.397721462 | 8.58E-10 | 6.08E-08 | 5.39E-06 |
| T2DM | mast_cell | *KCTD8* | 1.553466795 | 4.29E-09 | 7.78E-08 | 2.70E-05 |
| T2DM | mast_cell | *C9orf66* | 5.978154635 | 4.40E-09 | 7.78E-08 | 2.77E-05 |
| T2DM | mast_cell | *TUBA8* | 3.269855301 | 0.000450162 | 0.000763466 | 1 |
| T2DM | mast_cell | *BTBD9* | 2.800813327 | 0.000121735 | 0.000228531 | 0.76534975 |
| T2DM | mast_cell | *ANKRD45* | 4.303429512 | 1.83E-08 | 1.36E-07 | 0.000115355 |
| T2DM | mast_cell | *GPR39* | 1.987349149 | 1.53E-08 | 1.21E-07 | 9.60E-05 |
| T2DM | mast_cell | *FAM3B* | 0.44310479 | 0.018940351 | 0.0251857 | 1 |
| T2DM | mast_cell | *NPIPA7* | 2.798906434 | 0.019801825 | 0.026242426 | 1 |
| T2DM | mast_cell | *PP2D1* | 2.69264962 | 0.006316998 | 0.008999539 | 1 |
| T2DM | mast_cell | *DENND5A* | 3.558614127 | 1.69E-05 | 3.71E-05 | 0.106203029 |
| T2DM | mast_cell | *TMEM236* | 2.0979727 | 1.61E-08 | 1.25E-07 | 0.000101307 |
| T2DM | mast_cell | *VPS33B* | 4.69521669 | 1.78E-07 | 7.26E-07 | 0.001121143 |
| T2DM | mast_cell | *TBX1* | -2.62013673 | 2.43E-05 | 5.22E-05 | 0.153044629 |
| T2DM | mast_cell | *UQCR10* | 3.871194489 | 3.72E-08 | 2.17E-07 | 0.000234187 |
| T2DM | mast_cell | *EIF3C* | 1.131079782 | 0.01639652 | 0.021946971 | 1 |
| T2DM | mast_cell | *CNTN2* | 3.543934957 | 7.73E-09 | 9.13E-08 | 4.86E-05 |
| T2DM | mast_cell | *CRELD2* | 3.574990876 | 6.56E-09 | 8.49E-08 | 4.13E-05 |
| T2DM | mast_cell | *UBE2F* | 2.725759641 | 0.003606176 | 0.005327074 | 1 |
| T2DM | mast_cell | *GOLGA6L4* | 4.088826245 | 3.20E-07 | 1.17E-06 | 0.002013838 |
| T2DM | mast_cell | *PCDH9* | 1.107938951 | 0.000683152 | 0.001121696 | 1 |
| T2DM | mast_cell | *ACOT1* | 1.068300244 | 0.002886717 | 0.004337664 | 1 |
| T2DM | mast_cell | *OAF* | -1.10204098 | 0.02542221 | 0.033084131 | 1 |
| T2DM | mast_cell | *TM2D3* | 3.515276398 | 0.000920805 | 0.001483243 | 1 |
| T2DM | mast_cell | *TACSTD2* | -1.225781684 | 0.002510145 | 0.003808225 | 1 |
| T2DM | mast_cell | *CLECL1* | 1.549850287 | 8.43E-06 | 2.00E-05 | 0.053021408 |
| T2DM | mast_cell | *MROH7* | -0.713805554 | 3.58E-07 | 1.28E-06 | 0.002250595 |
| T2DM | mast_cell | *CSF1* | -0.581377526 | 2.21E-06 | 6.14E-06 | 0.013892834 |
| T2DM | mast_cell | *PLA2G6* | 2.103901187 | 2.38E-07 | 9.17E-07 | 0.001498533 |

| | | | | | | |
|---|---|---|---|---|---|---|
| T2DM | mast_cell | *SS18L1* | -1.688464306 | 0.019142506 | 0.025416882 | 1 |
| T2DM | mast_cell | *THAP7* | 4.847153914 | 8.94E-09 | 9.66E-08 | 5.62E-05 |
| T2DM | mast_cell | *WDR27* | 1.926129111 | 0.038555446 | 0.049078373 | 1 |
| T2DM | mast_cell | *TXNRD2* | 5.361980236 | 2.79E-08 | 1.76E-07 | 0.00017511 |
| T2DM | mast_cell | *HDDC3* | 2.806026046 | 0.000598799 | 0.00099673 | 1 |
| T2DM | mast_cell | *LPAR5* | -1.001680115 | 1.98E-08 | 1.41E-07 | 0.000124472 |
| T2DM | mast_cell | *TMEM173* | -1.912981292 | 0.015325907 | 0.020584059 | 1 |
| T2DM | mast_cell | *SNN* | 4.7014442 | 0.01697914 | 0.022659277 | 1 |
| T2DM | mast_cell | *NELL2* | 1.490741873 | 1.90E-07 | 7.67E-07 | 0.001196615 |
| T2DM | mast_cell | *KRBA2* | 2.444981542 | 0.000178613 | 0.000325302 | 1 |
| T2DM | mast_cell | *ZNF93* | 6.705488433 | 8.18E-09 | 9.23E-08 | 5.15E-05 |
| T2DM | mast_cell | *SEPT5* | 13.47661451 | 4.27E-09 | 7.78E-08 | 2.68E-05 |
| T2DM | mast_cell | *ATL3* | 2.790853308 | 4.07E-05 | 8.39E-05 | 0.256081276 |
| T2DM | mast_cell | *NDUFA12* | 5.289321585 | 2.12E-08 | 1.46E-07 | 0.000133081 |
| T2DM | mast_cell | *UBE2G2* | 3.08157205 | 2.14E-05 | 4.65E-05 | 0.13475049 |
| T2DM | mast_cell | *OSBP2* | 4.532944454 | 1.25E-08 | 1.13E-07 | 7.84E-05 |
| T2DM | mast_cell | *TUSC5* | 1.589242241 | 3.83E-09 | 7.78E-08 | 2.41E-05 |
| T2DM | mast_cell | *RBM43* | 3.619518383 | 3.18E-08 | 1.93E-07 | 0.000199786 |
| T2DM | mast_cell | *NUTM2A* | 2.658734303 | 0.006671539 | 0.009468872 | 1 |
| T2DM | mast_cell | *ZFP90* | 4.793008333 | 1.82E-05 | 3.97E-05 | 0.114140158 |
| T2DM | mast_cell | *NOC4L* | 1.583877306 | 0.001245073 | 0.001970739 | 1 |
| T2DM | mast_cell | *NDUFA6* | 1.137675215 | 4.22E-06 | 1.09E-05 | 0.026531804 |
| T2DM | mast_cell | *SORCS2* | 2.230389579 | 7.50E-09 | 9.02E-08 | 4.72E-05 |
| T2DM | mast_cell | *TMEM106A* | 5.18070039 | 7.57E-06 | 1.81E-05 | 0.047563854 |
| T2DM | mast_cell | *SIVA1* | -2.212284682 | 0.00013048 | 0.000243566 | 0.820330572 |
| T2DM | mast_cell | *BRF1* | 4.141687106 | 8.23E-09 | 9.23E-08 | 5.18E-05 |
| T2DM | mast_cell | *CIB1* | 2.094111058 | 7.19E-05 | 0.000140487 | 0.451947506 |
| T2DM | mast_cell | *FLRT2* | 1.447711383 | 0.000296028 | 0.000517986 | 1 |
| T2DM | mast_cell | *ADSSL1* | -2.684520029 | 5.75E-06 | 1.43E-05 | 0.036155898 |
| T2DM | mast_cell | *C6orf120* | 2.932076008 | 0.014595699 | 0.019649499 | 1 |
| T2DM | mast_cell | *IFITM2* | 3.381398635 | 0.000158592 | 0.000291625 | 0.997066676 |
| T2DM | mast_cell | *GP1BA* | 3.212305087 | 1.05E-06 | 3.21E-06 | 0.006614127 |
| T2DM | mast_cell | *KIAA0825* | 2.854581388 | 2.26E-08 | 1.52E-07 | 0.000141992 |
| T2DM | mast_cell | *KLHL33* | -1.12315089 | 0.008539129 | 0.011924812 | 1 |
| T2DM | mast_cell | *RBM11* | 3.994641334 | 0.000750114 | 0.001224608 | 1 |
| T2DM | mast_cell | *CCDC137* | 3.007008039 | 8.50E-06 | 2.01E-05 | 0.05342358 |
| T2DM | mast_cell | *CDK10* | 2.129880791 | 4.54E-06 | 1.16E-05 | 0.028551378 |
| T2DM | mast_cell | *TMEM105* | -15.17126644 | 2.49E-05 | 5.32E-05 | 0.156249473 |
| T2DM | mast_cell | *TCN2* | 0.996537729 | 0.016951824 | 0.022627626 | 1 |
| T2DM | mast_cell | *GAS2L1* | -2.707042791 | 0.031155386 | 0.040039638 | 1 |
| T2DM | mast_cell | *ATP6V0A2* | -0.833628297 | 6.44E-07 | 2.11E-06 | 0.004051576 |
| T2DM | mast_cell | *TNFAIP8L1* | 6.99710873 | 6.92E-09 | 8.77E-08 | 4.35E-05 |
| T2DM | mast_cell | *MAPK11* | 4.533804756 | 8.23E-06 | 1.96E-05 | 0.051749649 |
| T2DM | mast_cell | *SP140L* | 3.263472593 | 0.001017497 | 0.001624017 | 1 |
| T2DM | mast_cell | *PDE6G* | 3.603101483 | 0.000846853 | 0.001373977 | 1 |
| T2DM | mast_cell | *TLCD2* | 1.277110356 | 2.69E-08 | 1.72E-07 | 0.000168922 |
| T2DM | mast_cell | *LSAMP* | 3.074362744 | 2.56E-06 | 6.97E-06 | 0.01611308 |
| T2DM | mast_cell | *AHNAK2* | 5.328373544 | 7.63E-07 | 2.44E-06 | 0.004794362 |
| T2DM | mast_cell | *SP1* | 3.034305755 | 2.34E-06 | 6.45E-06 | 0.014726924 |
| T2DM | mast_cell | *PDIA2* | 3.278193461 | 4.06E-08 | 2.33E-07 | 0.00025522 |

| | | | | | | |
|---|---|---|---|---|---|---|
| T2DM | mast_cell | *PCGF3* | 2.795971686 | 0.003276363 | 0.00488116 | 1 |
| T2DM | mast_cell | *PSMD13* | 2.667191268 | 5.58E-08 | 2.96E-07 | 0.000351071 |
| T2DM | mast_cell | *UBE2L3* | 4.086476784 | 9.47E-10 | 6.13E-08 | 5.95E-06 |
| T2DM | mast_cell | *PMEL* | 1.270375568 | 0.019927912 | 0.026403958 | 1 |
| T2DM | mast_cell | *SNAI3* | 3.28170547 | 0.001232592 | 0.001952459 | 1 |
| T2DM | mast_cell | *MYBL1* | 0.858528519 | 0.001012051 | 0.001615735 | 1 |
| T2DM | mast_cell | *C16orf52* | 3.218817328 | 0.000876571 | 0.001418168 | 1 |
| T2DM | mast_cell | *ADARB2* | 4.972526852 | 6.54E-08 | 3.34E-07 | 0.00041121 |
| T2DM | mast_cell | *SRL* | 1.146551584 | 8.15E-06 | 1.94E-05 | 0.051232023 |
| T2DM | mast_cell | *SLC52A2* | 4.970524783 | 3.42E-09 | 7.78E-08 | 2.15E-05 |
| T2DM | mast_cell | *ARL17A* | 2.499829849 | 2.18E-07 | 8.53E-07 | 0.001368662 |
| T2DM | mast_cell | *NPIPB4* | 2.739451409 | 2.35E-08 | 1.56E-07 | 0.00014792 |
| T2DM | mast_cell | *POMK* | 5.604699086 | 0.000120089 | 0.000225709 | 0.754996959 |
| T2DM | mast_cell | *PTCH1* | 2.592048541 | 0.000152973 | 0.000282367 | 0.961743035 |
| T2DM | mast_cell | *RTN4RL1* | -2.284927809 | 0.016571894 | 0.022162837 | 1 |
| T2DM | mast_cell | *RASA3* | 2.082535458 | 0.000555958 | 0.000928861 | 1 |
| T2DM | mast_cell | *ZNF566* | 3.034818075 | 3.05E-06 | 8.15E-06 | 0.019174677 |
| T2DM | mast_cell | *ZNF529* | 2.663667853 | 0.00276385 | 0.004165985 | 1 |
| T2DM | mast_cell | *ZNF284* | 8.022159087 | 3.97E-09 | 7.78E-08 | 2.49E-05 |
| T2DM | mast_cell | *GSAP* | 3.948616669 | 5.56E-08 | 2.95E-07 | 0.000349756 |
| T2DM | mast_cell | *C2orf76* | 2.530106973 | 0.000288654 | 0.000505929 | 1 |
| T2DM | mast_cell | *WWOX* | 1.900982881 | 0.000835341 | 0.001355651 | 1 |
| T2DM | mast_cell | *CYP4Z1* | 1.989181291 | 3.59E-09 | 7.78E-08 | 2.26E-05 |
| T2DM | mast_cell | *CCDC84* | 4.266298526 | 9.26E-07 | 2.88E-06 | 0.005820996 |
| T2DM | mast_cell | *POLR1D* | 3.652019975 | 2.61E-09 | 7.78E-08 | 1.64E-05 |
| T2DM | mast_cell | *FFAR4* | 0.870983317 | 7.54E-06 | 1.81E-05 | 0.047392152 |
| T2DM | mast_cell | *BPIFB4* | 112.5359455 | 4.68E-05 | 9.53E-05 | 0.29440612 |
| T2DM | mast_cell | *ZNF749* | 1.106664942 | 0.002207781 | 0.003373922 | 1 |
| T2DM | mast_cell | *NBPF12* | 3.909475948 | 9.53E-06 | 2.23E-05 | 0.059908246 |
| T2DM | mast_cell | *GPAT2* | 3.409898499 | 1.50E-05 | 3.33E-05 | 0.094066175 |
| T2DM | mast_cell | *TOR3A* | 6.675352182 | 1.89E-08 | 1.38E-07 | 0.000118842 |
| T2DM | mast_cell | *CYP4X1* | 4.469173572 | 7.41E-09 | 8.98E-08 | 4.66E-05 |
| T2DM | mast_cell | *CCDC30* | 3.472812627 | 5.36E-05 | 0.000107735 | 0.33688861 |
| T2DM | mast_cell | *GLDN* | 1.991408943 | 3.99E-07 | 1.40E-06 | 0.00251102 |
| T2DM | mast_cell | *FCAR* | 2.272722913 | 6.07E-07 | 2.01E-06 | 0.003818638 |
| T2DM | mast_cell | *KPNA4* | 2.200258114 | 0.024492655 | 0.032000275 | 1 |
| T2DM | mast_cell | *ZNF501* | 3.006272946 | 4.33E-06 | 1.11E-05 | 0.027233658 |
| T2DM | mast_cell | *RPS23* | 2.39773412 | 0.001136623 | 0.001804988 | 1 |
| T2DM | mast_cell | *PCLO* | 1.756845732 | 1.51E-08 | 1.21E-07 | 9.46E-05 |
| T2DM | mast_cell | *RGS7BP* | 1.663726636 | 0.000113082 | 0.000213433 | 0.71094595 |
| T2DM | mast_cell | *ZNF396* | 1.665441491 | 2.57E-05 | 5.49E-05 | 0.161733874 |
| T2DM | mast_cell | *CLCNKA* | 3.538557461 | 0.000332383 | 0.000576944 | 1 |
| T2DM | mast_cell | *SEPT10* | 2.217369387 | 0.005370486 | 0.007722837 | 1 |
| T2DM | mast_cell | *SMYD4* | 2.595466261 | 0.000114212 | 0.000215436 | 0.718049249 |
| T2DM | mast_cell | *NF2* | 2.344950106 | 0.011063011 | 0.015153192 | 1 |
| T2DM | mast_cell | *MIR22HG* | 2.823137156 | 1.16E-06 | 3.47E-06 | 0.007313971 |
| T2DM | mast_cell | *PDE2A* | 1.746853651 | 0.008966002 | 0.012479357 | 1 |
| T2DM | mast_cell | *C17orf58* | 2.041964422 | 8.31E-07 | 2.63E-06 | 0.00522368 |
| T2DM | mast_cell | *CYP27C1* | 1.11422102 | 5.22E-06 | 1.31E-05 | 0.032799219 |
| T2DM | mast_cell | *BCR* | 2.508673859 | 0.001272799 | 0.002012091 | 1 |

| | | | | | | |
|---|---|---|---|---|---|---|
| T2DM | mast_cell | *FOXI2* | -2.037266022 | 4.73E-06 | 1.20E-05 | 0.02974441 |
| T2DM | mast_cell | *HYAL3* | -1.476433807 | 2.21E-08 | 1.50E-07 | 0.000138869 |
| T2DM | mast_cell | *ZSCAN30* | 3.611473771 | 4.37E-09 | 7.78E-08 | 2.74E-05 |
| T2DM | mast_cell | *LILRB4* | 1.74397918 | 1.23E-07 | 5.36E-07 | 0.000771148 |
| T2DM | mast_cell | *TRABD2A* | 0.96814717 | 0.001081828 | 0.001722323 | 1 |
| T2DM | mast_cell | *TMEM17* | 3.294037272 | 4.92E-08 | 2.68E-07 | 0.000309007 |
| T2DM | mast_cell | *TNFRSF18* | -3.265350098 | 4.92E-07 | 1.67E-06 | 0.003091482 |
| T2DM | mast_cell | *RTN4RL2* | -4.465408652 | 1.01E-05 | 2.36E-05 | 0.063779315 |
| T2DM | mast_cell | *TMEM232* | 2.532679639 | 2.87E-08 | 1.79E-07 | 0.000180623 |
| T2DM | mast_cell | *EFCAB6* | 4.123801186 | 3.57E-05 | 7.45E-05 | 0.224666789 |
| T2DM | mast_cell | *KANK3* | 3.219465616 | 9.84E-09 | 1.03E-07 | 6.19E-05 |
| T2DM | mast_cell | *EMID1* | -0.97822276 | 1.73E-07 | 7.07E-07 | 0.001085178 |
| T2DM | mast_cell | *RHD* | 2.199755287 | 9.74E-05 | 0.00018596 | 0.612365218 |
| T2DM | mast_cell | *CYP4A11* | 2.19004108 | 0.002413147 | 0.003669922 | 1 |
| T2DM | mast_cell | *RPS19BP1* | 2.632941325 | 0.025377316 | 0.033032544 | 1 |
| T2DM | mast_cell | *C3orf70* | 2.615709263 | 1.68E-06 | 4.83E-06 | 0.010592385 |
| T2DM | mast_cell | *HEATR4* | 1.307905387 | 0.000320655 | 0.000557357 | 1 |
| T2DM | mast_cell | *MRPS21* | 3.815516449 | 2.64E-07 | 9.95E-07 | 0.001660044 |
| T2DM | mast_cell | *ANGPTL5* | 2.593143674 | 6.06E-09 | 8.15E-08 | 3.81E-05 |
| T2DM | mast_cell | *FNBP1* | 1.849086075 | 0.000993079 | 0.001587865 | 1 |
| T2DM | mast_cell | *WDR86* | 2.345680463 | 0.004520405 | 0.006571049 | 1 |
| T2DM | mast_cell | *EPOR* | -1.336472527 | 3.34E-06 | 8.82E-06 | 0.021019649 |
| T2DM | mast_cell | *FPR3* | 2.303677721 | 1.09E-06 | 3.30E-06 | 0.006870281 |
| T2DM | mast_cell | *GJA4* | 2.152334866 | 6.77E-09 | 8.65E-08 | 4.26E-05 |
| T2DM | mast_cell | *IFT140* | 3.620366944 | 8.77E-10 | 6.08E-08 | 5.51E-06 |
| T2DM | mast_cell | *TLR5* | 5.648403148 | 1.06E-06 | 3.23E-06 | 0.0066706 |
| T2DM | mast_cell | *PLEKHN1* | 2.01119487 | 0.009139116 | 0.012697818 | 1 |
| T2DM | mast_cell | *ZNF385C* | 2.57665751 | 2.50E-08 | 1.63E-07 | 0.000157437 |
| T2DM | mast_cell | *TET3* | 4.122711247 | 1.74E-08 | 1.31E-07 | 0.000109277 |
| T2DM | mast_cell | *C17orf97* | 0.597930864 | 0.00059568 | 0.000992064 | 1 |
| T2DM | mast_cell | *TRPV2* | -2.572548407 | 0.000100449 | 0.000191371 | 0.63152355 |
| T2DM | mast_cell | *C2orf88* | 2.738850934 | 0.000725535 | 0.001187565 | 1 |
| T2DM | mast_cell | *FANCA* | 1.847539276 | 1.83E-05 | 3.99E-05 | 0.114909965 |
| T2DM | mast_cell | *ADH1A* | 1.330877873 | 0.002100249 | 0.003223698 | 1 |
| T2DM | mast_cell | *SEMA4D* | -1.82594105 | 8.63E-06 | 2.04E-05 | 0.054259661 |
| T2DM | mast_cell | *FAM69C* | 4.314190265 | 3.10E-06 | 8.26E-06 | 0.019481946 |
| T2DM | mast_cell | *DNAH17* | -3.191383261 | 2.15E-05 | 4.66E-05 | 0.134950846 |
| T2DM | mast_cell | *ZFP69* | 2.959205349 | 1.81E-06 | 5.15E-06 | 0.011387343 |
| T2DM | mast_cell | *TMEM220* | 3.882140535 | 5.93E-05 | 0.000118144 | 0.372863068 |
| T2DM | mast_cell | *CCDC157* | 6.257268949 | 4.60E-08 | 2.55E-07 | 0.000289355 |
| T2DM | mast_cell | *GFRAL* | 2.501114918 | 2.23E-08 | 1.51E-07 | 0.000140416 |
| T2DM | mast_cell | *LDLRAD2* | -1.836041567 | 1.35E-05 | 3.04E-05 | 0.084932254 |
| T2DM | mast_cell | *C2orf66* | 8.631543096 | 1.19E-06 | 3.54E-06 | 0.007476366 |
| T2DM | mast_cell | *CPSF4L* | -3.3768148 | 0.000262447 | 0.000462575 | 1 |
| T2DM | mast_cell | *KLHL17* | 2.099510284 | 3.33E-08 | 2.00E-07 | 0.000209511 |
| T2DM | mast_cell | *ZSCAN23* | 0.567687154 | 0.004363624 | 0.006350487 | 1 |
| T2DM | mast_cell | *OR7D2* | 4.085744975 | 0.000214996 | 0.000385093 | 1 |
| T2DM | mast_cell | *S100A3* | -2.095328631 | 0.000142777 | 0.000264946 | 0.897637803 |
| T2DM | mast_cell | *MAPK12* | -1.080540155 | 1.59E-07 | 6.60E-07 | 0.00099916 |
| T2DM | mast_cell | *AGRN* | 4.647665405 | 3.50E-08 | 2.07E-07 | 0.000220081 |

| | | | | | | |
|---|---|---|---|---|---|---|
| T2DM | mast_cell | *FAM166A* | -3.349038022 | 2.03E-07 | 8.07E-07 | 0.001276611 |
| T2DM | mast_cell | *ZNF626* | 3.905791854 | 0.000148507 | 0.00027485 | 0.933664195 |
| T2DM | mast_cell | *SMTNL2* | 10.54825279 | 1.25E-08 | 1.13E-07 | 7.88E-05 |
| T2DM | mast_cell | *ZC3H6* | 3.627459384 | 2.47E-07 | 9.40E-07 | 0.001550798 |
| T2DM | mast_cell | *LINC00265* | 2.472596072 | 2.25E-09 | 7.78E-08 | 1.41E-05 |
| T2DM | mast_cell | *LAMTOR4* | 3.567538561 | 3.51E-07 | 1.26E-06 | 0.002207649 |
| T2DM | mast_cell | *PRKAR1B* | -1.700939055 | 0.019008224 | 0.025259925 | 1 |
| T2DM | mast_cell | *NCR3LG1* | 12.81010154 | 2.93E-09 | 7.78E-08 | 1.84E-05 |
| T2DM | mast_cell | *HYKK* | 1.883803048 | 5.70E-09 | 8.09E-08 | 3.58E-05 |
| T2DM | mast_cell | *RUFY4* | 18.6208656 | 1.27E-07 | 5.49E-07 | 0.000798355 |
| T2DM | mast_cell | *ZNF669* | -3.04082314 | 0.017486584 | 0.023316682 | 1 |
| T2DM | mast_cell | *PLSCR1* | 3.511146661 | 0.000697767 | 0.001144498 | 1 |
| T2DM | mast_cell | *JAKMIP3* | 1.540464828 | 6.99E-08 | 3.50E-07 | 0.000439682 |
| T2DM | mast_cell | *PDCD1* | 5.725895733 | 0.000344808 | 0.000596042 | 1 |
| T2DM | mast_cell | *CERKL* | 4.338635262 | 3.36E-09 | 7.78E-08 | 2.11E-05 |
| T2DM | mast_cell | *C19orf54* | 2.335072032 | 0.001080845 | 0.001721194 | 1 |
| T2DM | mast_cell | *C22orf34* | -1.312616447 | 0.000118376 | 0.000222822 | 0.744226928 |
| T2DM | mast_cell | *COL25A1* | 2.604638408 | 1.86E-05 | 4.06E-05 | 0.117005812 |
| T2DM | mast_cell | *NBR1* | 4.373013707 | 5.67E-08 | 2.99E-07 | 0.000356556 |
| T2DM | mast_cell | *NDOR1* | 5.141675564 | 4.16E-09 | 7.78E-08 | 2.62E-05 |
| T2DM | mast_cell | *FAM72B* | -3.120235079 | 3.16E-07 | 1.16E-06 | 0.00198923 |
| T2DM | mast_cell | *GOLGA8M* | 1.682922458 | 5.83E-09 | 8.14E-08 | 3.67E-05 |
| T2DM | mast_cell | *DPYD* | 1.778999876 | 0.012170786 | 0.016551532 | 1 |
| T2DM | mast_cell | *S100A16* | 4.656104759 | 6.67E-08 | 3.39E-07 | 0.000419567 |
| T2DM | mast_cell | *SLC4A5* | 2.815730596 | 4.91E-08 | 2.68E-07 | 0.000308707 |
| T2DM | mast_cell | *UROS* | 3.492236928 | 1.31E-09 | 6.84E-08 | 8.24E-06 |
| T2DM | mast_cell | *FAM221A* | 2.959826333 | 6.57E-07 | 2.15E-06 | 0.00413307 |
| T2DM | mast_cell | *FSIP2* | 1.262860145 | 0.001259624 | 0.001992266 | 1 |
| T2DM | mast_cell | *NOXA1* | 6.159170827 | 1.65E-08 | 1.27E-07 | 0.000103711 |
| T2DM | mast_cell | *SKOR1* | 7.318987274 | 1.06E-08 | 1.06E-07 | 6.66E-05 |
| T2DM | mast_cell | *TMEM201* | -2.061189977 | 0.000833889 | 0.001353643 | 1 |
| T2DM | mast_cell | *ZDHHC11* | -2.48842943 | 0.000488518 | 0.000822086 | 1 |
| T2DM | mast_cell | *LINC00910* | -3.195326966 | 0.000328878 | 0.000571176 | 1 |
| T2DM | mast_cell | *BEND4* | 6.83401282 | 5.62E-09 | 8.05E-08 | 3.53E-05 |
| T2DM | mast_cell | *ZNF563* | 3.125114037 | 9.86E-06 | 2.30E-05 | 0.061970886 |
| T2DM | mast_cell | *MSL1* | 1.46042011 | 0.000922098 | 0.001484565 | 1 |
| T2DM | mast_cell | *LRRK2* | 3.479743563 | 7.72E-08 | 3.79E-07 | 0.000485538 |
| T2DM | mast_cell | *FAM196A* | 3.546967216 | 2.85E-07 | 1.06E-06 | 0.001793044 |
| T2DM | mast_cell | *UTS2B* | 1.216865945 | 0.007915176 | 0.011105269 | 1 |
| T2DM | mast_cell | *C9orf152* | -2.740596338 | 4.12E-07 | 1.44E-06 | 0.002588119 |
| T2DM | mast_cell | *NOC2L* | -0.838308527 | 0.023316547 | 0.030533457 | 1 |
| T2DM | mast_cell | *NELFB* | 2.888668526 | 3.75E-09 | 7.78E-08 | 2.36E-05 |
| T2DM | mast_cell | *SLC15A5* | 1.46947733 | 6.08E-07 | 2.01E-06 | 0.003819597 |
| T2DM | mast_cell | *LIPI* | 1.194103913 | 0.023248742 | 0.030470052 | 1 |
| T2DM | mast_cell | *ADAT2* | 4.11661833 | 3.89E-08 | 2.25E-07 | 0.00024466 |
| T2DM | mast_cell | *ZNF567* | 4.002025195 | 2.72E-09 | 7.78E-08 | 1.71E-05 |
| T2DM | mast_cell | *ANKDD1B* | -1.331640197 | 1.58E-05 | 3.49E-05 | 0.099080754 |
| T2DM | mast_cell | *ALKBH2* | 3.560922099 | 0.000179162 | 0.000326113 | 1 |
| T2DM | mast_cell | *LITAF* | -1.628245321 | 0.003040143 | 0.004557315 | 1 |
| T2DM | mast_cell | *VSTM1* | 3.019973723 | 4.56E-06 | 1.16E-05 | 0.028657912 |

| | | | | | | |
|---|---|---|---|---|---|---|
| T2DM | mast_cell | *SF3B3* | 4.478609256 | 5.62E-08 | 2.97E-07 | 0.000353468 |
| T2DM | mast_cell | *PLAC9* | 1.474347728 | 0.005766881 | 0.008272047 | 1 |
| T2DM | mast_cell | *ZNF573* | 4.701529168 | 8.10E-07 | 2.57E-06 | 0.005094462 |
| T2DM | mast_cell | *FAM47E* | 1.522792417 | 1.16E-05 | 2.66E-05 | 0.072873787 |
| T2DM | mast_cell | *ZNF527* | 4.518425077 | 0.000131519 | 0.000245141 | 0.826860146 |
| T2DM | mast_cell | *ZNF600* | 2.217895006 | 1.20E-06 | 3.57E-06 | 0.007553112 |
| T2DM | mast_cell | *PNRC2* | 3.497478192 | 0.009381678 | 0.013020444 | 1 |
| T2DM | mast_cell | *ZKSCAN3* | 3.655642018 | 3.39E-08 | 2.03E-07 | 0.000213075 |
| T2DM | mast_cell | *RRP7A* | 2.766499496 | 1.79E-06 | 5.09E-06 | 0.011223232 |
| T2DM | mast_cell | *SLC35E2B* | 1.391834772 | 0.002490343 | 0.003780919 | 1 |
| T2DM | mast_cell | *HLA-DRB1* | -3.399045722 | 6.61E-17 | 1.39E-13 | 4.16E-13 |
| T2DM | mast_cell | *AKR1C3* | 3.483295425 | 3.69E-06 | 9.65E-06 | 0.023207813 |
| T2DM | mast_cell | *COLCA1* | 2.464976033 | 1.85E-05 | 4.05E-05 | 0.116547432 |
| T2DM | mast_cell | *ACADSB* | 1.872494987 | 0.004043685 | 0.005917748 | 1 |
| T2DM | mast_cell | *SEMA4A* | -1.809095428 | 0.0003335 | 0.000578481 | 1 |
| T2DM | mast_cell | *HRCT1* | 2.372858869 | 2.61E-06 | 7.10E-06 | 0.016430261 |
| T2DM | mast_cell | *MPHOSPH8* | 3.50741263 | 2.01E-06 | 5.64E-06 | 0.012643232 |
| T2DM | mast_cell | *SIRPB2* | 3.460785266 | 5.00E-09 | 7.84E-08 | 3.14E-05 |
| T2DM | mast_cell | *SRGAP3* | 1.239947241 | 0.007833382 | 0.010997873 | 1 |
| T2DM | mast_cell | *SUPT5H* | 4.241275325 | 3.41E-07 | 1.24E-06 | 0.002144541 |
| T2DM | mast_cell | *PPIA* | 3.326620696 | 0.007511015 | 0.010571245 | 1 |
| T2DM | mast_cell | *ZNF471* | 2.751307109 | 9.01E-06 | 2.12E-05 | 0.056655457 |
| T2DM | mast_cell | *GTF2IRD2* | 3.130558582 | 9.13E-10 | 6.11E-08 | 5.74E-06 |
| T2DM | mast_cell | *GRM7* | 1.005736316 | 0.001208204 | 0.001914792 | 1 |
| T2DM | mast_cell | *SUPT3H* | 2.709063144 | 0.002316082 | 0.003530846 | 1 |
| T2DM | mast_cell | *STK31* | 6.896645636 | 4.86E-09 | 7.84E-08 | 3.06E-05 |
| T2DM | mast_cell | *ZKSCAN7* | 3.439984821 | 1.02E-08 | 1.04E-07 | 6.41E-05 |
| T2DM | mast_cell | *NTNG2* | -1.058218405 | 2.87E-07 | 1.07E-06 | 0.001805745 |
| T2DM | mast_cell | *WDR5* | 6.04992914 | 1.07E-06 | 3.24E-06 | 0.00670168 |
| T2DM | mast_cell | *ASB13* | 2.23326626 | 6.40E-09 | 8.33E-08 | 4.02E-05 |
| T2DM | mast_cell | *SLC35F1* | 4.665378311 | 0.000303961 | 0.000530835 | 1 |
| T2DM | mast_cell | *ZNF34* | -2.46789937 | 7.44E-05 | 0.000144964 | 0.467944916 |
| T2DM | mast_cell | *ZNF781* | 1.038246964 | 0.018983859 | 0.02523288 | 1 |
| T2DM | mast_cell | *INCA1* | -3.062915565 | 0.001582655 | 0.00246902 | 1 |
| T2DM | mast_cell | *ZNF774* | 2.597752261 | 7.24E-09 | 8.86E-08 | 4.55E-05 |
| T2DM | mast_cell | *EVL* | 1.619875237 | 0.016862416 | 0.022522628 | 1 |
| T2DM | mast_cell | *ZNF124* | 4.410275146 | 6.16E-05 | 0.000122274 | 0.387364088 |
| T2DM | mast_cell | *ZNF569* | 3.239526045 | 1.77E-06 | 5.05E-06 | 0.011122039 |
| T2DM | mast_cell | *PIK3R4* | 2.312881069 | 2.23E-06 | 6.18E-06 | 0.014017122 |
| T2DM | mast_cell | *ZNF605* | 5.130095833 | 1.29E-08 | 1.13E-07 | 8.12E-05 |
| T2DM | mast_cell | *RFX8* | 3.103444361 | 2.63E-08 | 1.69E-07 | 0.000165157 |
| T2DM | mast_cell | *MYL6B* | 5.643538325 | 0.000231768 | 0.000412901 | 1 |
| T2DM | mast_cell | *C20orf96* | 3.160142603 | 5.19E-09 | 7.92E-08 | 3.26E-05 |
| T2DM | mast_cell | *ANAPC7* | 4.43466762 | 2.08E-07 | 8.24E-07 | 0.001310427 |
| T2DM | mast_cell | *TPK1* | 1.980216188 | 0.009870748 | 0.013647986 | 1 |
| T2DM | mast_cell | *C1orf186* | -0.156246499 | 8.96E-08 | 4.21E-07 | 0.000563159 |
| T2DM | mast_cell | *MYO18A* | 3.041239435 | 4.35E-05 | 8.92E-05 | 0.273606617 |
| T2DM | mast_cell | *SPTSSB* | 4.360389427 | 8.75E-08 | 4.13E-07 | 0.000549879 |
| T2DM | mast_cell | *SULF2* | 3.180786475 | 7.18E-06 | 1.73E-05 | 0.045157481 |
| T2DM | mast_cell | *XRCC2* | 10.82859265 | 3.15E-06 | 8.38E-06 | 0.01983279 |

| | | | | | | |
|---|---|---|---|---|---|---|
| T2DM | mast_cell | *HDAC2* | 4.434286636 | 1.16E-07 | 5.14E-07 | 0.000727274 |
| T2DM | mast_cell | *ADH1B* | 3.072633521 | 1.34E-08 | 1.16E-07 | 8.45E-05 |
| T2DM | mast_cell | *HRH1* | 2.866026852 | 4.54E-09 | 7.78E-08 | 2.85E-05 |
| T2DM | mast_cell | *ZNF502* | 2.456209837 | 6.74E-06 | 1.64E-05 | 0.042379596 |
| T2DM | mast_cell | *TRAPPC4* | 1.44558405 | 0.005090739 | 0.00733734 | 1 |
| T2DM | mast_cell | *TECPR2* | 3.957545328 | 1.12E-08 | 1.07E-07 | 7.01E-05 |
| T2DM | mast_cell | *FAM180B* | 0.936218162 | 0.022491093 | 0.029520146 | 1 |
| T2DM | mast_cell | *ZFP62* | 6.198322188 | 1.35E-06 | 3.98E-06 | 0.008506882 |
| T2DM | mast_cell | *TOMM7* | 5.088170396 | 4.04E-06 | 1.04E-05 | 0.025402346 |
| T2DM | mast_cell | *TRPV1* | 2.250721665 | 0.00745243 | 0.010500545 | 1 |
| T2DM | mast_cell | *ZNF33B* | -2.298572499 | 1.05E-05 | 2.44E-05 | 0.06629241 |
| T2DM | mast_cell | *AMZ2* | 2.916247163 | 0.000187752 | 0.000340073 | 1 |
| T2DM | mast_cell | *ZNF431* | 3.025949109 | 0.000553167 | 0.00092469 | 1 |
| T2DM | mast_cell | *NF1* | 2.885217076 | 4.38E-05 | 8.98E-05 | 0.275626676 |
| T2DM | mast_cell | *DAPK1* | 1.552473722 | 0.009430937 | 0.013083032 | 1 |
| T2DM | mast_cell | *HLA-DQA1* | -2.309115315 | 4.72E-11 | 1.10E-08 | 2.97E-07 |
| T2DM | mast_cell | *GM2A* | -1.491474872 | 0.006331119 | 0.009015571 | 1 |
| T2DM | mast_cell | *TLE1* | 2.045897549 | 2.74E-05 | 5.83E-05 | 0.172382817 |
| T2DM | mast_cell | *STRN3* | 3.263356671 | 1.19E-05 | 2.73E-05 | 0.075075101 |
| T2DM | mast_cell | *ZNF239* | 1.925367001 | 8.32E-06 | 1.98E-05 | 0.05228549 |
| T2DM | mast_cell | *MVB12B* | 4.074652129 | 5.58E-06 | 1.39E-05 | 0.035084971 |
| T2DM | mast_cell | *C6orf106* | 1.663641659 | 0.000309526 | 0.000539653 | 1 |
| T2DM | mast_cell | *PPTC7* | 3.696747019 | 0.000131364 | 0.000244924 | 0.825882368 |
| T2DM | mast_cell | *NHLRC2* | 3.801813379 | 4.75E-06 | 1.20E-05 | 0.029889863 |
| T2DM | mast_cell | *ZFP28* | 3.318663333 | 1.02E-08 | 1.04E-07 | 6.41E-05 |
| T2DM | mast_cell | *KIAA1211L* | 0.929326372 | 0.008300906 | 0.011607606 | 1 |
| T2DM | mast_cell | *LAMB3* | 2.834522911 | 5.04E-09 | 7.84E-08 | 3.17E-05 |
| T2DM | mast_cell | *KPNA5* | 4.144030238 | 2.45E-07 | 9.35E-07 | 0.001539006 |
| T2DM | mast_cell | *ANKRD36B* | 1.874343694 | 0.016848335 | 0.022508602 | 1 |
| T2DM | mast_cell | *HCAR1* | 1.855650134 | 8.54E-08 | 4.06E-07 | 0.000536597 |
| T2DM | mast_cell | *SRGAP1* | 3.289642163 | 1.69E-08 | 1.29E-07 | 0.000106358 |
| T2DM | mast_cell | *FAM3C* | 3.523066708 | 6.93E-06 | 1.68E-05 | 0.043565147 |
| T2DM | mast_cell | *SLC39A10* | 2.230104365 | 0.0251942 | 0.032814571 | 1 |
| T2DM | mast_cell | *AP2A1* | 1.818538672 | 0.035793401 | 0.045747736 | 1 |
| T2DM | mast_cell | *PCDHB16* | 1.448369135 | 2.80E-06 | 7.54E-06 | 0.01757912 |
| T2DM | mast_cell | *ZNF585A* | 2.515900027 | 7.31E-05 | 0.000142683 | 0.459866037 |
| T2DM | mast_cell | *FUT11* | 2.846350752 | 0.011689052 | 0.015958539 | 1 |
| T2DM | mast_cell | *ANXA4* | 2.607868732 | 0.002721064 | 0.004106417 | 1 |
| T2DM | mast_cell | *ZNF138* | 2.484446157 | 0.006290502 | 0.008967887 | 1 |
| T2DM | mast_cell | *ZNF429* | 3.124846142 | 8.66E-05 | 0.000166658 | 0.544306508 |
| T2DM | mast_cell | *ZNF470* | 2.883363333 | 5.30E-07 | 1.78E-06 | 0.003330589 |
| T2DM | mast_cell | *GMFB* | 4.495442563 | 0.000202548 | 0.000364563 | 1 |
| T2DM | mast_cell | *ZNF763* | 0.740805364 | 0.0120701 | 0.016432378 | 1 |
| T2DM | mast_cell | *ZMYM1* | 4.530044824 | 1.33E-05 | 3.00E-05 | 0.083579951 |
| T2DM | mast_cell | *ARRDC1* | 4.798050135 | 1.08E-05 | 2.48E-05 | 0.067639016 |
| T2DM | mast_cell | *IGF2R* | 2.885876331 | 0.000391646 | 0.000671469 | 1 |
| T2DM | mast_cell | *SLC25A29* | 2.095470376 | 0.025726755 | 0.033445845 | 1 |
| T2DM | mast_cell | *ZNF682* | 3.334596569 | 1.24E-08 | 1.12E-07 | 7.79E-05 |
| T2DM | mast_cell | *ZNF772* | -1.342656554 | 1.13E-05 | 2.60E-05 | 0.071318742 |
| T2DM | mast_cell | *ZNF257* | -1.975397487 | 0.020961488 | 0.027627856 | 1 |

| | | | | | | |
|---|---|---|---|---|---|---|
| T2DM | mast_cell | *ADAM32* | 3.357041385 | 1.52E-05 | 3.36E-05 | 0.09529399 |
| T2DM | mast_cell | *LRRC8B* | 3.120936479 | 2.37E-06 | 6.50E-06 | 0.014884361 |
| T2DM | mast_cell | *SND1* | 2.424990915 | 0.000864272 | 0.001400072 | 1 |
| T2DM | mast_cell | *PIWIL2* | 3.294813467 | 4.38E-07 | 1.52E-06 | 0.00275621 |
| T2DM | mast_cell | *ENTPD4* | 3.567002533 | 2.00E-05 | 4.35E-05 | 0.125658004 |
| T2DM | mast_cell | *C1D* | 2.651991038 | 0.000122755 | 0.000230238 | 0.771757686 |
| T2DM | mast_cell | *FAM110D* | 2.550442775 | 1.17E-08 | 1.10E-07 | 7.37E-05 |
| T2DM | mast_cell | *KANK2* | 4.387352245 | 2.26E-06 | 6.25E-06 | 0.014217429 |
| T2DM | mast_cell | *CCL4L2* | -1.381529141 | 8.18E-06 | 1.95E-05 | 0.051445551 |
| T2DM | mast_cell | *GTF2E2* | 4.321697608 | 5.94E-07 | 1.98E-06 | 0.003737009 |
| T2DM | mast_cell | *ZNF165* | -2.220769299 | 0.000924436 | 0.001487568 | 1 |
| T2DM | mast_cell | *RAMP2-AS1* | 3.152364241 | 1.56E-10 | 2.39E-08 | 9.81E-07 |
| T2DM | mast_cell | *ZNF720* | 1.798827877 | 0.024125296 | 0.031533417 | 1 |
| T2DM | mast_cell | *DDI2* | 3.490024287 | 1.33E-07 | 5.70E-07 | 0.000835798 |
| T2DM | mast_cell | *MRPL21* | 1.200556343 | 4.00E-06 | 1.04E-05 | 0.025176122 |
| T2DM | mast_cell | *ZNF786* | 3.846402703 | 3.00E-06 | 8.02E-06 | 0.018871789 |
| T2DM | mast_cell | *ZNF517* | 5.352035487 | 0.01911921 | 0.025391313 | 1 |
| T2DM | mast_cell | *SLC22A5* | 1.606081607 | 3.85E-06 | 1.00E-05 | 0.024192242 |
| T2DM | mast_cell | *C5AR1* | 5.276161532 | 2.95E-08 | 1.83E-07 | 0.000185428 |
| T2DM | mast_cell | *DCHS2* | 0.760946708 | 0.000183783 | 0.000333366 | 1 |
| T2DM | mast_cell | *VEPH1* | 3.021132175 | 8.62E-08 | 4.10E-07 | 0.000542142 |
| T2DM | mast_cell | *SHPK* | 1.6206183 | 6.68E-05 | 0.000131357 | 0.419816126 |
| T2DM | mast_cell | *MAP3K5* | 2.636189484 | 0.000166227 | 0.000304419 | 1 |
| T2DM | mast_cell | *STMN3* | 3.245970143 | 0.003590629 | 0.005309098 | 1 |
| T2DM | mast_cell | *GYPE* | 1.363847364 | 0.028909725 | 0.037313784 | 1 |
| T2DM | mast_cell | *SLC2A10* | 3.358286455 | 0.000333545 | 0.000578481 | 1 |
| T2DM | mast_cell | *ZNF665* | 1.76622934 | 0.000655278 | 0.00108101 | 1 |
| T2DM | mast_cell | *RPF2* | 2.977981283 | 1.10E-08 | 1.06E-07 | 6.91E-05 |
| T2DM | mast_cell | *MYO5A* | 1.720093077 | 0.00782526 | 0.010988923 | 1 |
| T2DM | mast_cell | *ATG7* | 2.554887015 | 3.86E-05 | 8.01E-05 | 0.242793683 |
| T2DM | mast_cell | *TTC30A* | -2.729579195 | 0.000942855 | 0.001516043 | 1 |
| T2DM | mast_cell | *RAB40C* | 3.897863619 | 4.24E-09 | 7.78E-08 | 2.67E-05 |
| T2DM | mast_cell | *PIGN* | 1.371929018 | 0.003686903 | 0.005441211 | 1 |
| T2DM | mast_cell | *ZNF624* | 6.075615883 | 1.21E-07 | 5.31E-07 | 0.000760003 |
| T2DM | mast_cell | *HHLA3* | 2.486541457 | 1.14E-05 | 2.63E-05 | 0.071974369 |
| T2DM | mast_cell | *HOXA4* | 6.460223885 | 2.44E-06 | 6.67E-06 | 0.015358447 |
| T2DM | mast_cell | *BCO2* | 3.988968463 | 6.32E-08 | 3.26E-07 | 0.000397338 |
| T2DM | mast_cell | *ENTPD6* | 2.695200711 | 2.98E-06 | 7.98E-06 | 0.018733423 |
| T2DM | mast_cell | *CCDC154* | 6.909634993 | 1.78E-09 | 7.51E-08 | 1.12E-05 |
| T2DM | mast_cell | *C5orf42* | 2.08319863 | 0.000397806 | 0.000681102 | 1 |
| T2DM | mast_cell | *ZNF841* | 3.974329783 | 3.86E-06 | 1.00E-05 | 0.024298155 |
| T2DM | mast_cell | *MFAP5* | 3.375574455 | 5.81E-06 | 1.44E-05 | 0.036523932 |
| T2DM | mast_cell | *CDC42SE1* | 3.968112028 | 6.53E-08 | 3.33E-07 | 0.000410523 |
| T2DM | mast_cell | *MPEG1* | 0.906369081 | 0.004719204 | 0.006837897 | 1 |
| T2DM | mast_cell | *NMB* | 2.073265773 | 5.50E-09 | 8.00E-08 | 3.46E-05 |
| T2DM | mast_cell | *KLHL14* | 2.265923045 | 8.45E-08 | 4.04E-07 | 0.000531297 |
| T2DM | mast_cell | *FAM114A1* | 3.79705112 | 3.07E-08 | 1.88E-07 | 0.000193323 |
| T2DM | mast_cell | *PSAP* | 2.695693574 | 0.001353769 | 0.002133655 | 1 |
| T2DM | mast_cell | *MAP1LC3C* | 2.047516523 | 4.93E-07 | 1.68E-06 | 0.003099093 |
| T2DM | mast_cell | *TAF13* | 3.174799198 | 2.09E-07 | 8.25E-07 | 0.001313131 |

| | | | | | | |
|---|---|---|---|---|---|---|
| T2DM | mast_cell | *AC122129.1* | -2.835481669 | 3.87E-07 | 1.37E-06 | 0.002431607 |
| T2DM | mast_cell | *CCDC180* | 1.889817527 | 0.008778668 | 0.012240294 | 1 |
| T2DM | mast_cell | *OCLN* | 1.12096676 | 0.008454958 | 0.011817768 | 1 |
| T2DM | mast_cell | *ZNF181* | 2.404776532 | 3.42E-09 | 7.78E-08 | 2.15E-05 |
| T2DM | mast_cell | *SGTB* | 4.02550658 | 0.001494276 | 0.002339272 | 1 |
| T2DM | mast_cell | *ADH5* | 2.764541026 | 1.24E-05 | 2.82E-05 | 0.078024775 |
| T2DM | mast_cell | *ZNF677* | 5.997212861 | 3.16E-08 | 1.92E-07 | 0.000198749 |
| T2DM | mast_cell | *ZNF311* | 1.295254162 | 4.79E-05 | 9.72E-05 | 0.301335429 |
| T2DM | mast_cell | *FCHSD1* | 5.327421059 | 8.63E-05 | 0.000166145 | 0.54246224 |
| T2DM | mast_cell | *VPS13A* | 2.420644196 | 1.42E-05 | 3.17E-05 | 0.089061287 |
| T2DM | mast_cell | *LEKR1* | -1.878337401 | 0.004149795 | 0.00606034 | 1 |
| T2DM | mast_cell | *NOL8* | 4.293592364 | 1.36E-07 | 5.79E-07 | 0.00085245 |
| T2DM | mast_cell | *MRPL42* | 2.050099861 | 5.04E-06 | 1.27E-05 | 0.031670844 |
| T2DM | mast_cell | *ZNF335* | 3.16506055 | 4.01E-05 | 8.27E-05 | 0.25184449 |
| T2DM | mast_cell | *ZNF273* | 4.396023622 | 1.46E-06 | 4.24E-06 | 0.009189719 |
| T2DM | mast_cell | *ZNF667* | 1.577795638 | 3.41E-05 | 7.14E-05 | 0.214325736 |
| T2DM | mast_cell | *SULT1C4* | 4.243010861 | 1.46E-08 | 1.19E-07 | 9.17E-05 |
| T2DM | mast_cell | *SFI1* | 5.007065813 | 4.09E-08 | 2.34E-07 | 0.000257278 |
| T2DM | mast_cell | *ZNF649* | 2.672614742 | 3.22E-05 | 6.76E-05 | 0.202251791 |
| T2DM | mast_cell | *ADH4* | 0.54689372 | 0.006283329 | 0.008959694 | 1 |
| T2DM | mast_cell | *ZNF248* | 1.583904975 | 0.00503038 | 0.007258664 | 1 |
| T2DM | mast_cell | *HIBCH* | 1.832609966 | 0.006269786 | 0.00894444 | 1 |
| T2DM | mast_cell | *ZNF544* | 2.34807587 | 0.000306762 | 0.000535576 | 1 |
| T2DM | mast_cell | *SOWAHC* | 5.469034343 | 2.65E-08 | 1.70E-07 | 0.000166781 |
| T2DM | mast_cell | *AC135178.1* | 7.998286462 | 1.35E-05 | 3.04E-05 | 0.085066647 |
| T2DM | mast_cell | *MIER1* | 3.484162918 | 0.000131914 | 0.000245659 | 0.829344908 |
| T2DM | mast_cell | *SVIP* | -2.341098946 | 0.006385342 | 0.009088668 | 1 |
| T2DM | mast_cell | *ZNF251* | 4.122180439 | 2.93E-08 | 1.82E-07 | 0.000184123 |
| T2DM | mast_cell | *DDRGK1* | 5.648713061 | 9.03E-08 | 4.23E-07 | 0.00056741 |
| T2DM | mast_cell | *ZNF334* | 1.993333804 | 0.000247318 | 0.000438491 | 1 |
| T2DM | mast_cell | *HSD17B11* | 4.517970538 | 2.62E-05 | 5.59E-05 | 0.164772816 |
| T2DM | mast_cell | *SULT1C2* | 1.491493168 | 4.98E-08 | 2.71E-07 | 0.00031295 |
| T2DM | mast_cell | *RPS6KL1* | 1.312690714 | 0.013699955 | 0.018522928 | 1 |
| T2DM | mast_cell | *DDX42* | 4.197618528 | 8.71E-08 | 4.12E-07 | 0.000547555 |
| T2DM | mast_cell | *ANTXRL* | -66.65133575 | 1.10E-06 | 3.33E-06 | 0.006924503 |
| T2DM | mast_cell | *STYX* | 3.356111877 | 2.45E-09 | 7.78E-08 | 1.54E-05 |
| T2DM | mast_cell | *ZKSCAN8* | 4.131103161 | 7.58E-09 | 9.02E-08 | 4.76E-05 |
| T2DM | mast_cell | *ZNF813* | -3.211041972 | 0.000160812 | 0.000295448 | 1 |
| T2DM | mast_cell | *GFPT1* | 3.8096976 | 2.39E-07 | 9.18E-07 | 0.001501037 |
| T2DM | mast_cell | *UVRAG* | 2.066794946 | 5.54E-07 | 1.85E-06 | 0.003480403 |
| T2DM | mast_cell | *MGEA5* | 2.514629094 | 4.68E-07 | 1.61E-06 | 0.002943461 |
| T2DM | mast_cell | *MT1F* | -4.013175068 | 2.90E-07 | 1.07E-06 | 0.001820432 |
| T2DM | mast_cell | *ZNF568* | 4.093320914 | 2.88E-11 | 7.54E-09 | 1.81E-07 |
| T2DM | mast_cell | *ZNF587* | 3.876073607 | 7.15E-07 | 2.30E-06 | 0.004493966 |
| T2DM | mast_cell | *ZNF808* | 4.021393151 | 4.32E-05 | 8.87E-05 | 0.271722194 |
| T2DM | mast_cell | *TMA16* | 3.773233251 | 0.000965206 | 0.001548418 | 1 |
| T2DM | mast_cell | *HLA-DRB5* | -1.103636079 | 1.53E-13 | 9.62E-11 | 9.62E-10 |
| T2DM | mast_cell | *CNGA1* | 1.821107137 | 0.000574922 | 0.000959526 | 1 |
| T2DM | mast_cell | *MAFK* | 3.807441385 | 3.62E-09 | 7.78E-08 | 2.27E-05 |
| T2DM | mast_cell | *C20orf203* | -2.849339408 | 1.63E-05 | 3.59E-05 | 0.102221303 |

| | | | | | | |
|---|---|---|---|---|---|---|
| T2DM | mast_cell | *ZNF627* | 5.079718244 | 1.36E-05 | 3.06E-05 | 0.085476259 |
| T2DM | mast_cell | *ARC* | 8.764774473 | 1.87E-08 | 1.37E-07 | 0.000117784 |
| T2DM | mast_cell | *TLK1* | 3.639775467 | 1.63E-06 | 4.69E-06 | 0.01024975 |
| T2DM | mast_cell | *LRBA* | 2.649397625 | 0.000526848 | 0.000882338 | 1 |
| T2DM | mast_cell | *CCDC69* | 3.44091488 | 9.55E-06 | 2.23E-05 | 0.060037333 |
| T2DM | mast_cell | *MDM4* | 3.679196152 | 2.95E-07 | 1.09E-06 | 0.001851585 |
| T2DM | mast_cell | *ZNF534* | 2.70909711 | 3.03E-05 | 6.40E-05 | 0.190678299 |
| T2DM | mast_cell | *NCOA6* | 2.621390628 | 0.000291448 | 0.000510684 | 1 |
| T2DM | mast_cell | *FAM19A2* | 3.504054965 | 4.76E-07 | 1.63E-06 | 0.002991393 |
| T2DM | mast_cell | *TTC37* | 3.52447683 | 2.63E-06 | 7.15E-06 | 0.016563396 |
| T2DM | mast_cell | *TUSC1* | -4.362298118 | 0.000284149 | 0.00049845 | 1 |
| T2DM | mast_cell | *PAPSS2* | 2.736451877 | 0.00025749 | 0.000454731 | 1 |
| T2DM | mast_cell | *FAN1* | 3.555015326 | 3.66E-09 | 7.78E-08 | 2.30E-05 |
| T2DM | mast_cell | *CEP290* | 3.775812356 | 2.05E-06 | 5.74E-06 | 0.012908592 |
| T2DM | mast_cell | *ANKRD13B* | 1.511302375 | 0.000253996 | 0.000449318 | 1 |
| T2DM | mast_cell | *ZNF652* | 4.008863643 | 2.45E-07 | 9.37E-07 | 0.001542623 |
| T2DM | mast_cell | *APCDD1L* | 4.320039208 | 3.55E-09 | 7.78E-08 | 2.23E-05 |
| T2DM | mast_cell | *RCSD1* | -0.932927039 | 0.027299702 | 0.035315478 | 1 |
| T2DM | mast_cell | *RASSF9* | 4.995106456 | 7.54E-09 | 9.02E-08 | 4.74E-05 |
| T2DM | mast_cell | *FAM169A* | 2.818894222 | 5.01E-07 | 1.70E-06 | 0.003148699 |
| T2DM | mast_cell | *TMEM184B* | 4.303568933 | 1.29E-08 | 1.13E-07 | 8.08E-05 |
| T2DM | mast_cell | *SCAMP5* | 1.680728042 | 7.02E-07 | 2.27E-06 | 0.004410969 |
| T2DM | mast_cell | *ALPK2* | 1.455609447 | 0.024894297 | 0.032464311 | 1 |
| T2DM | mast_cell | *SFT2D1* | 3.200323789 | 0.005093525 | 0.007339673 | 1 |
| T2DM | mast_cell | *ARHGAP11A* | -1.646432501 | 2.92E-06 | 7.83E-06 | 0.018333859 |
| T2DM | mast_cell | *HMGN2* | 2.379706104 | 0.006732192 | 0.009539527 | 1 |
| T2DM | mast_cell | *DENND4B* | 2.946741631 | 1.43E-05 | 3.19E-05 | 0.089800581 |
| T2DM | mast_cell | *CD3E* | 1.649869462 | 1.87E-07 | 7.56E-07 | 0.001178069 |
| T2DM | mast_cell | *TSEN15* | 5.142485124 | 1.63E-06 | 4.69E-06 | 0.010238005 |
| T2DM | mast_cell | *TYW1* | 1.593254138 | 0.001140056 | 0.001809983 | 1 |
| T2DM | mast_cell | *ITPRIPL1* | 2.567729977 | 2.05E-08 | 1.43E-07 | 0.000128842 |
| T2DM | mast_cell | *PRMT6* | 3.85549999 | 1.97E-07 | 7.88E-07 | 0.001239316 |
| T2DM | mast_cell | *SHISA4* | 3.464100011 | 6.43E-08 | 3.29E-07 | 0.000404131 |
| T2DM | mast_cell | *CAPZA2* | 3.936127657 | 4.19E-06 | 1.08E-05 | 0.026314326 |
| T2DM | mast_cell | *C1orf174* | 3.636928875 | 6.88E-07 | 2.23E-06 | 0.004323028 |
| T2DM | mast_cell | *RASGEF1A* | 3.796022218 | 1.20E-08 | 1.11E-07 | 7.55E-05 |
| T2DM | mast_cell | *KIAA0753* | 3.438655188 | 1.14E-05 | 2.61E-05 | 0.07141346 |
| T2DM | mast_cell | *DCLRE1A* | 4.682797772 | 1.05E-06 | 3.21E-06 | 0.006622354 |
| T2DM | mast_cell | *APRT* | 4.795674603 | 0.003579259 | 0.005294777 | 1 |
| T2DM | mast_cell | *TBKBP1* | 1.373992804 | 0.003199576 | 0.004778084 | 1 |
| T2DM | mast_cell | *ZFP2* | 3.044650937 | 4.00E-09 | 7.78E-08 | 2.52E-05 |
| T2DM | mast_cell | *L3MBTL3* | -2.071529471 | 0.008505449 | 0.011883058 | 1 |
| T2DM | mast_cell | *MFAP3L* | 10.03668669 | 2.37E-07 | 9.11E-07 | 0.001487948 |
| T2DM | mast_cell | *NAGA* | 3.847528948 | 7.50E-05 | 0.00014596 | 0.471450824 |
| T2DM | mast_cell | *SMG5* | 3.778010748 | 5.39E-05 | 0.000108316 | 0.338812798 |
| T2DM | mast_cell | *PJA2* | 2.97255572 | 0.000111306 | 0.000210271 | 0.699782607 |
| T2DM | mast_cell | *AP001007.1* | 8.569660497 | 1.06E-08 | 1.06E-07 | 6.68E-05 |
| T2DM | mast_cell | *ZNF525* | -2.587610324 | 0.000131783 | 0.000245487 | 0.828517914 |
| T2DM | mast_cell | *AC105020.1* | 3.9576067 | 2.69E-07 | 1.01E-06 | 0.001691414 |
| T2DM | mast_cell | *EFCAB2* | 2.63005911 | 0.002361803 | 0.003597929 | 1 |

| | | | | | | |
|---|---|---|---|---|---|---|
| T2DM | mast_cell | *CHML* | 2.844726254 | 1.39E-07 | 5.91E-07 | 0.00087644 |
| T2DM | mast_cell | *CAPN8* | -4.955806198 | 0.001262417 | 0.001996181 | 1 |
| T2DM | mast_cell | *TATDN3* | 1.040657233 | 0.013860039 | 0.018719241 | 1 |
| T2DM | mast_cell | *ECT2L* | 2.924707326 | 0.000757857 | 0.001235645 | 1 |
| T2DM | mast_cell | *CENPW* | -5.385176064 | 1.28E-05 | 2.91E-05 | 0.080742317 |
| T2DM | mast_cell | *DDO* | 7.133400738 | 3.18E-07 | 1.16E-06 | 0.002000245 |
| T2DM | mast_cell | *HIST2H2BF* | 5.321065678 | 1.31E-08 | 1.14E-07 | 8.25E-05 |
| T2DM | mast_cell | *RBM20* | 3.372301176 | 6.34E-08 | 3.27E-07 | 0.000398858 |
| T2DM | mast_cell | *PCMTD2* | 4.855218458 | 1.15E-07 | 5.13E-07 | 0.000723361 |
| T2DM | mast_cell | *OOEP* | 1.449069588 | 0.000391204 | 0.000670894 | 1 |
| T2DM | mast_cell | *FOXO6* | -1.636799367 | 0.036981732 | 0.0471898 | 1 |
| T2DM | mast_cell | *SYS1* | 4.757751263 | 3.44E-09 | 7.78E-08 | 2.17E-05 |
| T2DM | mast_cell | *NEU4* | 7.039928724 | 0.001502189 | 0.002349702 | 1 |
| T2DM | mast_cell | *GIGYF2* | 2.717612236 | 0.001370719 | 0.002158745 | 1 |
| T2DM | mast_cell | *RUFY2* | 2.776253871 | 0.001845092 | 0.002856463 | 1 |
| T2DM | mast_cell | *ASAH2B* | 1.907183045 | 1.46E-09 | 6.92E-08 | 9.17E-06 |
| T2DM | mast_cell | *AGAP6* | 1.550852034 | 0.000125456 | 0.000234884 | 0.788739092 |
| T2DM | mast_cell | *SYT15* | 5.25163323 | 1.18E-07 | 5.22E-07 | 0.000743542 |
| T2DM | mast_cell | *RING1* | 2.494361992 | 3.98E-06 | 1.03E-05 | 0.025022466 |
| T2DM | mast_cell | *HSD17B8* | -1.822543504 | 1.70E-05 | 3.73E-05 | 0.106783444 |
| T2DM | mast_cell | *OXLD1* | 3.953297595 | 2.02E-11 | 6.69E-09 | 1.27E-07 |
| T2DM | mast_cell | *HLA-DMA* | -2.501694647 | 0.005760615 | 0.008264945 | 1 |
| T2DM | mast_cell | *HLA-DRA* | 2.334665782 | 2.73E-07 | 1.02E-06 | 0.001716002 |
| T2DM | mast_cell | *COL15A1* | 2.204692964 | 4.60E-06 | 1.17E-05 | 0.028904069 |
| T2DM | mast_cell | *NOTCH4* | -0.34512479 | 8.49E-08 | 4.06E-07 | 0.000533587 |
| T2DM | mast_cell | *AGER* | 1.970389912 | 2.16E-08 | 1.48E-07 | 0.000135516 |
| T2DM | mast_cell | *PRRT1* | 2.782652369 | 3.50E-06 | 9.23E-06 | 0.022022798 |
| T2DM | mast_cell | *MRPL38* | 2.144804008 | 6.58E-05 | 0.000129743 | 0.41375066 |
| T2DM | mast_cell | *STK19* | 2.788159197 | 2.02E-07 | 8.04E-07 | 0.001269499 |
| T2DM | mast_cell | *NELFE* | 3.37298441 | 3.88E-08 | 2.25E-07 | 0.000244165 |
| T2DM | mast_cell | *LAYN* | 2.478899497 | 0.000126731 | 0.000236848 | 0.796756142 |
| T2DM | mast_cell | *CARD16* | 3.014380751 | 4.83E-05 | 9.80E-05 | 0.303799531 |
| T2DM | mast_cell | *MSH5* | 1.908145199 | 0.000191285 | 0.000346074 | 1 |
| T2DM | mast_cell | *GPANK1* | 2.460218855 | 0.000430912 | 0.000733787 | 1 |
| T2DM | mast_cell | *BAG6* | 3.557595386 | 3.29E-10 | 3.90E-08 | 2.07E-06 |
| T2DM | mast_cell | *NFKBIL1* | 3.529005954 | 2.48E-08 | 1.62E-07 | 0.000155851 |
| T2DM | mast_cell | *ZNF814* | 4.06137725 | 8.70E-07 | 2.73E-06 | 0.005469857 |
| T2DM | mast_cell | *PSORS1C1* | 0.428249256 | 1.31E-05 | 2.96E-05 | 0.082329939 |
| T2DM | mast_cell | *ABCF1* | 2.513599397 | 0.000298148 | 0.000521552 | 1 |
| T2DM | mast_cell | *LILRB3* | 2.139183184 | 1.39E-06 | 4.05E-06 | 0.008717807 |
| T2DM | mast_cell | *HLA-E* | 1.630490971 | 0.003250517 | 0.004846099 | 1 |
| T2DM | mast_cell | *ZNF468* | 4.798919114 | 2.87E-08 | 1.79E-07 | 0.000180261 |
| T2DM | mast_cell | *TBC1D8* | 3.291611817 | 2.25E-08 | 1.52E-07 | 0.00014152 |
| T2DM | mast_cell | *FAM153C* | -4.399093078 | 7.51E-06 | 1.80E-05 | 0.047238409 |
| T2DM | mast_cell | *GABBR1* | 4.939196405 | 2.86E-08 | 1.79E-07 | 0.000179523 |
| T2DM | mast_cell | *MALRD1* | -0.815139624 | 0.000524443 | 0.000878543 | 1 |
| T2DM | mast_cell | *MROH6* | 1.555278593 | 0.000166691 | 0.000304968 | 1 |
| T2DM | mast_cell | *ATXN2* | 2.766360471 | 0.000109106 | 0.000206674 | 0.685950017 |
| T2DM | mast_cell | *FAM216A* | 3.869040604 | 5.41E-09 | 8.00E-08 | 3.40E-05 |
| T2DM | mast_cell | *ZNF155* | 2.867714382 | 7.33E-06 | 1.76E-05 | 0.04605683 |

| | | | | | | |
|---|---|---|---|---|---|---|
| T2DM | mast_cell | *ZNF783* | 3.247398563 | 4.79E-08 | 2.63E-07 | 0.000301185 |
| T2DM | mast_cell | *LRRC10B* | -4.221856191 | 2.89E-07 | 1.07E-06 | 0.001817141 |
| T2DM | mast_cell | *PCDHGA1* | 1.413931112 | 0.010310021 | 0.014186715 | 1 |
| T2DM | mast_cell | *PCDHA5* | 13.04856353 | 7.96E-06 | 1.90E-05 | 0.050045422 |
| T2DM | mast_cell | *PCDHA4* | 7.171935693 | 3.32E-07 | 1.20E-06 | 0.002084477 |
| T2DM | mast_cell | *TRIM13* | 2.900601302 | 0.000757505 | 0.001235391 | 1 |
| T2DM | mast_cell | *SPIRE2* | 1.778090959 | 7.75E-08 | 3.79E-07 | 0.000486979 |
| T2DM | mast_cell | *SLFN12L* | 2.744162029 | 1.58E-07 | 6.56E-07 | 0.000991497 |
| T2DM | mast_cell | *SYCE1L* | 2.59623059 | 0.00012589 | 0.000235627 | 0.791471853 |
| T2DM | mast_cell | *TMEM231* | 1.72518416 | 3.08E-05 | 6.48E-05 | 0.193554235 |
| T2DM | mast_cell | *FAM71F2* | 2.460935393 | 8.96E-09 | 9.66E-08 | 5.63E-05 |
| T2DM | mast_cell | *AC016586.1* | 20.05303932 | 7.16E-09 | 8.82E-08 | 4.50E-05 |
| T2DM | mast_cell | *LINC00654* | 4.203710629 | 8.17E-08 | 3.95E-07 | 0.000513772 |
| T2DM | mast_cell | *TMEM170B* | 0.745419122 | 0.00466295 | 0.006764183 | 1 |
| T2DM | mast_cell | *NT5M* | -1.27683739 | 6.49E-05 | 0.000128165 | 0.408123361 |
| T2DM | mast_cell | *PRR13* | 2.685887783 | 3.58E-07 | 1.28E-06 | 0.002248137 |
| T2DM | mast_cell | *MT1A* | -1.965330151 | 2.30E-07 | 8.92E-07 | 0.001445087 |
| T2DM | mast_cell | *MT1M* | -2.427777951 | 2.21E-06 | 6.15E-06 | 0.01390348 |
| T2DM | mast_cell | *CFI* | 2.335728594 | 6.62E-05 | 0.000130387 | 0.416195064 |
| T2DM | mast_cell | *EXOC3L4* | 2.926167582 | 8.74E-08 | 4.13E-07 | 0.000549396 |
| T2DM | mast_cell | *TP53TG3D* | -4.30042094 | 2.79E-07 | 1.04E-06 | 0.001756183 |
| T2DM | mast_cell | *ATP6AP1L* | 2.466564189 | 4.59E-09 | 7.79E-08 | 2.89E-05 |
| T2DM | mast_cell | *NAP1L4* | 1.788288116 | 0.02056467 | 0.027144727 | 1 |
| T2DM | mast_cell | *CPT1B* | -1.408260314 | 0.00072931 | 0.001192503 | 1 |
| T2DM | mast_cell | *DENND6B* | 2.335117235 | 7.06E-09 | 8.80E-08 | 4.44E-05 |
| T2DM | mast_cell | *EIF3CL* | 9.578255235 | 2.93E-10 | 3.68E-08 | 1.84E-06 |
| T2DM | mast_cell | *LCMT1* | 4.585703893 | 1.12E-06 | 3.37E-06 | 0.00706135 |
| T2DM | mast_cell | *CDPF1* | 1.572665711 | 8.04E-05 | 0.000155559 | 0.505411622 |
| T2DM | mast_cell | *LIN52* | 3.989482054 | 1.31E-10 | 2.13E-08 | 8.25E-07 |
| T2DM | mast_cell | *C17orf107* | 1.309631487 | 0.011328187 | 0.015496151 | 1 |
| T2DM | mast_cell | *LINC00487* | -3.345383488 | 0.002828191 | 0.004255825 | 1 |
| T2DM | mast_cell | *RFPL3S* | 0.991642856 | 0.020424392 | 0.026982171 | 1 |
| T2DM | mast_cell | *C1QTNF9B* | -3.109976659 | 1.03E-07 | 4.71E-07 | 0.000649714 |
| T2DM | mast_cell | *RNPS1* | 3.754628091 | 2.36E-08 | 1.56E-07 | 0.00014835 |
| T2DM | mast_cell | *NYNRIN* | 3.502998353 | 1.86E-09 | 7.58E-08 | 1.17E-05 |
| T2DM | mast_cell | *ZDHHC11B* | -1.625985721 | 1.69E-05 | 3.72E-05 | 0.106354332 |
| T2DM | mast_cell | *GOLGA8O* | -2.820069492 | 1.80E-07 | 7.30E-07 | 0.001130263 |
| T2DM | mast_cell | *HCG27* | 2.513875522 | 2.73E-15 | 2.86E-12 | 1.72E-11 |
| T2DM | mast_cell | *LNP1* | 2.206723238 | 2.74E-08 | 1.74E-07 | 0.000172345 |
| T2DM | mast_cell | *VGLL3* | 1.08233317 | 0.000608246 | 0.00101024 | 1 |
| T2DM | mast_cell | *COLQ* | 3.071752783 | 8.29E-08 | 3.99E-07 | 0.000521122 |
| T2DM | mast_cell | *METTL6* | 3.820077644 | 2.61E-08 | 1.68E-07 | 0.00016393 |
| T2DM | mast_cell | *SACM1L* | 3.963261114 | 5.81E-06 | 1.44E-05 | 0.03653141 |
| T2DM | mast_cell | *SLC48A1* | 3.090713264 | 1.22E-08 | 1.11E-07 | 7.65E-05 |
| T2DM | mast_cell | *TAS2R14* | 2.219089273 | 2.71E-09 | 7.78E-08 | 1.70E-05 |
| T2DM | mast_cell | *ZRSR1* | 8.81671198 | 3.14E-09 | 7.78E-08 | 1.97E-05 |
| T2DM | mast_cell | *AC084018.1* | -0.753528968 | 0.0380976 | 0.048534875 | 1 |
| T2DM | mast_cell | *C17orf51* | 2.023606752 | 2.89E-07 | 1.07E-06 | 0.001816296 |
| T2DM | mast_cell | *OR2A42* | 1.128917738 | 0.00615744 | 0.008788156 | 1 |
| T2DM | mast_cell | *RNF208* | 4.425106196 | 1.52E-08 | 1.21E-07 | 9.57E-05 |

| | | | | | | |
|---|---|---|---|---|---|---|
| T2DM | mast_cell | *MAP10* | 3.471687771 | 0.000106977 | 0.000202946 | 0.67256304 |
| T2DM | mast_cell | *ZNF580* | 3.305806989 | 8.42E-06 | 1.99E-05 | 0.052935595 |
| T2DM | mast_cell | *FGFR1OP* | 6.459597986 | 5.48E-10 | 5.06E-08 | 3.44E-06 |
| T2DM | mast_cell | *ZNF254* | 3.896759199 | 1.36E-05 | 3.06E-05 | 0.085678397 |
| T2DM | mast_cell | *TCTEX1D2* | 2.512356606 | 1.92E-08 | 1.38E-07 | 0.000120495 |
| T2DM | mast_cell | *LINGO4* | 10.71518318 | 0.009609676 | 0.013307496 | 1 |
| T2DM | mast_cell | *ASIC3* | 2.006837309 | 0.000146081 | 0.000270598 | 0.918408155 |
| T2DM | mast_cell | *GIMAP1* | 3.131324787 | 0.000342532 | 0.000592271 | 1 |
| T2DM | mast_cell | *CCDC183* | -2.495739574 | 1.35E-07 | 5.77E-07 | 0.000850032 |
| T2DM | mast_cell | *ARHGEF35* | 2.649560237 | 0.00325572 | 0.004852705 | 1 |
| T2DM | mast_cell | *DNLZ* | 1.409425988 | 3.46E-05 | 7.23E-05 | 0.217221606 |
| T2DM | mast_cell | *SUPT4H1* | 5.410979373 | 5.37E-08 | 2.87E-07 | 0.000337567 |
| T2DM | mast_cell | *MXD3* | 2.231320526 | 3.32E-05 | 6.96E-05 | 0.208486602 |
| T2DM | mast_cell | *GSTM2* | -1.021199673 | 0.013147956 | 0.017799569 | 1 |
| T2DM | mast_cell | *ARHGAP19* | 6.533052965 | 0.000417686 | 0.000712811 | 1 |
| T2DM | mast_cell | *LCAT* | 1.924170076 | 0.000126644 | 0.000236785 | 0.796209965 |
| T2DM | mast_cell | *SIPA1* | 4.721528983 | 7.00E-10 | 5.79E-08 | 4.40E-06 |
| T2DM | mast_cell | *ERV3-1* | 0.51980326 | 0.010465826 | 0.014388508 | 1 |
| T2DM | mast_cell | *SRA1* | 1.35120493 | 0.035059182 | 0.044891462 | 1 |
| T2DM | mast_cell | *LEPROT* | 3.688150748 | 2.92E-05 | 6.17E-05 | 0.183295393 |
| T2DM | mast_cell | *LBH* | 6.932706521 | 1.10E-06 | 3.33E-06 | 0.006940288 |
| T2DM | mast_cell | *PPP1CB* | 2.306758022 | 0.009866143 | 0.013644619 | 1 |
| T2DM | mast_cell | *NCKIPSD* | 4.685311314 | 2.42E-07 | 9.26E-07 | 0.001520765 |
| T2DM | mast_cell | *ATF6B* | 3.370189811 | 5.18E-11 | 1.16E-08 | 3.26E-07 |
| T2DM | mast_cell | *SLC35F6* | 5.423983837 | 1.26E-08 | 1.13E-07 | 7.91E-05 |
| T2DM | mast_cell | *EMP2* | 2.665591246 | 4.76E-06 | 1.20E-05 | 0.029909148 |
| T2DM | mast_cell | *PPM1N* | 6.275355411 | 1.35E-10 | 2.13E-08 | 8.51E-07 |
| T2DM | mast_cell | *LTB4R2* | -1.87129357 | 0.038888446 | 0.049472209 | 1 |
| T2DM | mast_cell | *ITGA1* | 2.762094829 | 2.77E-08 | 1.75E-07 | 0.000174233 |
| T2DM | mast_cell | *ZNF726* | -3.783201653 | 0.000103661 | 0.000197073 | 0.651719384 |
| T2DM | mast_cell | *TAX1BP3* | 5.123638925 | 0.002868356 | 0.004311105 | 1 |
| T2DM | mast_cell | *AP1G2* | -2.4282568 | 0.000149318 | 0.000276188 | 0.938763364 |
| T2DM | mast_cell | *ZNF90* | 3.526691885 | 2.00E-07 | 7.99E-07 | 0.001258462 |
| T2DM | mast_cell | *TM6SF2* | 23.99721461 | 1.33E-08 | 1.15E-07 | 8.37E-05 |
| T2DM | mast_cell | *REPIN1* | 3.398237356 | 1.70E-05 | 3.74E-05 | 0.107059402 |
| T2DM | mast_cell | *SMIM7* | 3.296519524 | 1.40E-09 | 6.85E-08 | 8.78E-06 |
| T2DM | mast_cell | *AC079776.2* | 4.057908828 | 6.10E-09 | 8.15E-08 | 3.83E-05 |
| T2DM | mast_cell | *WEE2* | 1.665258126 | 0.000600639 | 0.000999263 | 1 |
| T2DM | mast_cell | *PAXIP1-AS2* | 4.909775784 | 2.29E-08 | 1.53E-07 | 0.000143689 |
| T2DM | mast_cell | *LYRM4* | 4.074036921 | 4.94E-07 | 1.68E-06 | 0.00310344 |
| T2DM | mast_cell | *MBLAC1* | 1.172602435 | 0.000311749 | 0.000543227 | 1 |
| T2DM | mast_cell | *NEURL1B* | 1.489788775 | 9.55E-09 | 1.01E-07 | 6.00E-05 |
| T2DM | mast_cell | *VSTM5* | 1.394668832 | 0.001619306 | 0.002523067 | 1 |
| T2DM | mast_cell | *PPME1* | 1.441005549 | 0.00372452 | 0.00548771 | 1 |
| T2DM | mast_cell | *STARD10* | 0.812190041 | 0.004476786 | 0.006513666 | 1 |
| T2DM | mast_cell | *CPEB1* | -2.399193063 | 0.000429455 | 0.000731845 | 1 |
| T2DM | mast_cell | *C10orf105* | -3.075765971 | 5.16E-05 | 0.000104123 | 0.324344062 |
| T2DM | mast_cell | *ARHGEF33* | 1.950204675 | 1.02E-05 | 2.37E-05 | 0.06403477 |
| T2DM | mast_cell | *AC005082.1* | -4.369471004 | 5.58E-07 | 1.87E-06 | 0.003508004 |
| T2DM | mast_cell | *ACSM4* | 1.017069722 | 0.000201832 | 0.000363483 | 1 |

| | | | | | | |
|---|---|---|---|---|---|---|
| T2DM | mast_cell | *COL28A1* | 5.531914475 | 0.000382257 | 0.000656087 | 1 |
| T2DM | mast_cell | *PHB2* | 3.418963043 | 1.88E-06 | 5.32E-06 | 0.011815828 |
| T2DM | mast_cell | *GRID2IP* | 3.725420712 | 8.02E-06 | 1.92E-05 | 0.05043743 |
| T2DM | mast_cell | *UBXN2B* | 2.23392043 | 0.002318843 | 0.003534197 | 1 |
| T2DM | mast_cell | *UBE2QL1* | 2.418200307 | 1.54E-08 | 1.22E-07 | 9.65E-05 |
| T2DM | mast_cell | *GOLGA8B* | 3.27948599 | 2.26E-09 | 7.78E-08 | 1.42E-05 |
| T2DM | mast_cell | *KCNU1* | 1.393244139 | 0.000464845 | 0.000785824 | 1 |
| T2DM | mast_cell | *MYL5* | 11.38494615 | 4.19E-07 | 1.46E-06 | 0.002634911 |
| T2DM | mast_cell | *NPEPL1* | 5.458011442 | 6.81E-08 | 3.43E-07 | 0.000427945 |
| T2DM | mast_cell | *LINC01139* | 1.620573986 | 0.02580809 | 0.033544648 | 1 |
| T2DM | mast_cell | *TSTD1* | 7.46179119 | 8.98E-08 | 4.22E-07 | 0.000564459 |
| T2DM | mast_cell | *LINC01138* | 2.827993581 | 3.38E-09 | 7.78E-08 | 2.13E-05 |
| T2DM | mast_cell | *CYB5RL* | 3.183059272 | 9.58E-08 | 4.43E-07 | 0.000602501 |
| T2DM | mast_cell | *ATAD3C* | -2.176657803 | 4.31E-05 | 8.84E-05 | 0.27084289 |
| T2DM | mast_cell | *CCDC7* | 2.211168075 | 0.006151057 | 0.008781039 | 1 |
| T2DM | mast_cell | *AC016773.1* | -1.122409481 | 0.000662536 | 0.001091267 | 1 |
| T2DM | mast_cell | *RNASEK* | 3.762216388 | 7.36E-09 | 8.93E-08 | 4.63E-05 |
| T2DM | mast_cell | *FAM228B* | 0.864326408 | 0.003577869 | 0.005294777 | 1 |
| T2DM | mast_cell | *RNU6ATAC35P* | 8.411191623 | 1.26E-08 | 1.13E-07 | 7.90E-05 |
| T2DM | mast_cell | *PPP3R1* | 3.429492169 | 2.71E-07 | 1.02E-06 | 0.001705471 |
| T2DM | mast_cell | *FANCG* | -1.662711015 | 1.09E-06 | 3.30E-06 | 0.006841501 |
| T2DM | mast_cell | *C19orf73* | 11.33087369 | 5.17E-08 | 2.79E-07 | 0.00032513 |
| T2DM | mast_cell | *OR2A25* | -22.09849359 | 3.20E-07 | 1.17E-06 | 0.002010155 |
| T2DM | mast_cell | *XKR9* | 1.038586173 | 2.36E-05 | 5.08E-05 | 0.14865551 |
| T2DM | mast_cell | *C1orf229* | 3.13760681 | 0.000197071 | 0.00035552 | 1 |
| T2DM | mast_cell | *SLC12A8* | 1.67100656 | 0.000172147 | 0.000314345 | 1 |
| T2DM | mast_cell | *APOL6* | 2.908099287 | 4.42E-09 | 7.78E-08 | 2.78E-05 |
| T2DM | mast_cell | *UBA52* | 2.301585306 | 0.008681658 | 0.01211309 | 1 |
| T2DM | mast_cell | *TIAF1* | 2.696023235 | 2.07E-07 | 8.21E-07 | 0.001304365 |
| T2DM | mast_cell | *BTBD19* | 3.555939706 | 3.84E-08 | 2.23E-07 | 0.000241455 |
| T2DM | mast_cell | *FAM185A* | 4.239199495 | 1.09E-08 | 1.06E-07 | 6.87E-05 |
| T2DM | mast_cell | *AC062017.1* | 1.691198598 | 0.000771874 | 0.001256869 | 1 |
| T2DM | mast_cell | *ADRA2B* | 6.38817114 | 1.30E-08 | 1.13E-07 | 8.15E-05 |
| T2DM | mast_cell | *EXOSC6* | 2.522538202 | 0.000189218 | 0.00034263 | 1 |
| T2DM | mast_cell | *VPS52* | 3.224485846 | 1.32E-08 | 1.14E-07 | 8.30E-05 |
| T2DM | mast_cell | *LINC00205* | 1.894941587 | 1.47E-05 | 3.27E-05 | 0.092355862 |
| T2DM | mast_cell | *AC147651.3* | -1.59715182 | 9.40E-08 | 4.36E-07 | 0.000590684 |
| T2DM | mast_cell | *OSER1-AS1* | 5.537330504 | 1.98E-06 | 5.58E-06 | 0.012458636 |
| T2DM | mast_cell | *LINC01133* | 1.703163979 | 9.07E-09 | 9.71E-08 | 5.70E-05 |
| T2DM | mast_cell | *LINC01117* | 2.266438974 | 7.14E-09 | 8.82E-08 | 4.49E-05 |
| T2DM | mast_cell | *RASAL2-AS1* | 5.802939764 | 1.13E-06 | 3.39E-06 | 0.007101828 |
| T2DM | mast_cell | *AC069213.1* | -4.608419982 | 0.0046921 | 0.00680176 | 1 |
| T2DM | mast_cell | *LINC00240* | 1.907466907 | 6.58E-08 | 3.35E-07 | 0.000413941 |
| T2DM | mast_cell | *LINC00863* | 3.190587597 | 1.21E-06 | 3.58E-06 | 0.007577989 |
| T2DM | mast_cell | *LINC00184* | 3.323108725 | 1.87E-06 | 5.30E-06 | 0.011761186 |
| T2DM | mast_cell | *TMEM233* | 1.571370408 | 2.81E-07 | 1.05E-06 | 0.001766182 |
| T2DM | mast_cell | *LINC00237* | 8.044854648 | 1.63E-08 | 1.26E-07 | 0.000102627 |
| T2DM | mast_cell | *PLEKHM1* | 4.756368043 | 1.85E-07 | 7.49E-07 | 0.001165007 |
| T2DM | mast_cell | *AP000350.6* | 1.13520603 | 0.000121695 | 0.000228523 | 0.765094478 |
| T2DM | mast_cell | *SFTA1P* | 5.394391645 | 1.94E-07 | 7.77E-07 | 0.001218829 |

| | | | | | | |
|---|---|---|---|---|---|---|
| T2DM | mast_cell | *FAM225B* | 1.390849783 | 0.006057309 | 0.008659284 | 1 |
| T2DM | mast_cell | *LINC00626* | 35.16556713 | 8.73E-09 | 9.55E-08 | 5.49E-05 |
| T2DM | mast_cell | *ERCC6* | 4.731036159 | 3.48E-07 | 1.26E-06 | 0.002187497 |
| T2DM | mast_cell | *LINC00115* | 1.695979771 | 0.021085177 | 0.027779235 | 1 |
| T2DM | mast_cell | *ZNF503-AS1* | 1.510034559 | 1.42E-08 | 1.19E-07 | 8.96E-05 |
| T2DM | mast_cell | *AC064836.3* | 9.419400174 | 3.84E-09 | 7.78E-08 | 2.42E-05 |
| T2DM | mast_cell | *TMEM185B* | -2.26471038 | 3.13E-06 | 8.32E-06 | 0.01965942 |
| T2DM | mast_cell | *TAS2R46* | -2.606900765 | 0.002134179 | 0.003269186 | 1 |
| T2DM | mast_cell | *AC093901.1* | 2.970581531 | 4.09E-08 | 2.34E-07 | 0.000257391 |
| T2DM | mast_cell | *C14orf132* | 2.046368879 | 7.31E-09 | 8.89E-08 | 4.60E-05 |
| T2DM | mast_cell | *AC017074.2* | 9.182983078 | 0.001834612 | 0.002840937 | 1 |
| T2DM | mast_cell | *AC009299.3* | 5.032448723 | 6.33E-06 | 1.55E-05 | 0.039784134 |
| T2DM | mast_cell | *SLC2A1-AS1* | 2.046298413 | 4.58E-08 | 2.54E-07 | 0.000287803 |
| T2DM | mast_cell | *LINC01068* | -1.646427764 | 5.43E-06 | 1.36E-05 | 0.03414022 |
| T2DM | mast_cell | *AC108463.2* | 4.725124858 | 1.72E-09 | 7.51E-08 | 1.08E-05 |
| T2DM | mast_cell | *HCG20* | -5.870440528 | 0.000168377 | 0.000307818 | 1 |
| T2DM | mast_cell | *C19orf24* | 2.302040495 | 0.006857842 | 0.009701902 | 1 |
| T2DM | mast_cell | *LINC01013* | 4.582108115 | 2.75E-09 | 7.78E-08 | 1.73E-05 |
| T2DM | mast_cell | *AC073133.2* | -6.453842503 | 9.16E-08 | 4.28E-07 | 0.000575867 |
| T2DM | mast_cell | *PROB1* | 5.624012115 | 0.001698097 | 0.002641251 | 1 |
| T2DM | mast_cell | *LINC00954* | -2.161404982 | 9.76E-07 | 3.01E-06 | 0.006138767 |
| T2DM | mast_cell | *AC067945.3* | 2.758623981 | 5.77E-05 | 0.000115214 | 0.362807482 |
| T2DM | mast_cell | *RPL41* | 3.308283561 | 5.76E-09 | 8.14E-08 | 3.62E-05 |
| T2DM | mast_cell | *PATL2* | 4.136747762 | 3.53E-09 | 7.78E-08 | 2.22E-05 |
| T2DM | mast_cell | *ZNF492* | 1.351590621 | 0.001845853 | 0.002856936 | 1 |
| T2DM | mast_cell | *U91324.1* | 4.076352847 | 1.07E-08 | 1.06E-07 | 6.75E-05 |
| T2DM | mast_cell | *CISD3* | 5.406144561 | 1.14E-09 | 6.34E-08 | 7.16E-06 |
| T2DM | mast_cell | *PSMG3-AS1* | 4.681938652 | 1.07E-06 | 3.24E-06 | 0.006703152 |
| T2DM | mast_cell | *PPP5D1* | 4.297483929 | 8.64E-08 | 4.10E-07 | 0.000543288 |
| T2DM | mast_cell | *AC112229.4* | 0.866534877 | 0.021781183 | 0.028618245 | 1 |
| T2DM | mast_cell | *AC092171.4* | 4.138323764 | 0.000438706 | 0.00074605 | 1 |
| T2DM | mast_cell | *DIRC3* | 2.471216091 | 1.24E-06 | 3.67E-06 | 0.007790958 |
| T2DM | mast_cell | *TAPBP* | 3.616952911 | 1.63E-07 | 6.75E-07 | 0.001025339 |
| T2DM | mast_cell | *AC107021.1* | 1.724582851 | 1.92E-06 | 5.42E-06 | 0.012072171 |
| T2DM | mast_cell | *LINC00865* | 2.04736712 | 8.87E-07 | 2.78E-06 | 0.005578998 |
| T2DM | mast_cell | *AC009495.3* | -4.052894344 | 0.000326253 | 0.000566773 | 1 |
| T2DM | mast_cell | *HLA-DQB2* | 0.916033465 | 5.41E-11 | 1.16E-08 | 3.40E-07 |
| T2DM | mast_cell | *GOLGA8N* | 1.412208705 | 0.001083474 | 0.001724507 | 1 |
| T2DM | mast_cell | *LINC00665* | 2.121764575 | 1.59E-05 | 3.51E-05 | 0.099820463 |
| T2DM | mast_cell | *TNF* | -1.130529944 | 0.007148151 | 0.010087638 | 1 |
| T2DM | mast_cell | *LYRM9* | -2.821637623 | 0.009600487 | 0.013303562 | 1 |
| T2DM | mast_cell | *LINC00342* | 5.613739226 | 1.10E-06 | 3.33E-06 | 0.006929095 |
| T2DM | mast_cell | *SNHG15* | 2.819034895 | 6.08E-08 | 3.17E-07 | 0.000382336 |
| T2DM | mast_cell | *LINC00702* | 1.562923859 | 5.29E-08 | 2.84E-07 | 0.000332876 |
| T2DM | mast_cell | *LINC00472* | 2.702420983 | 7.96E-08 | 3.88E-07 | 0.000500696 |
| T2DM | mast_cell | *RPS28* | 4.319862843 | 0.009942306 | 0.013734845 | 1 |
| T2DM | mast_cell | *AC123768.3* | 5.330170073 | 5.24E-08 | 2.82E-07 | 0.000329475 |
| T2DM | mast_cell | *TRIM26* | 3.81145806 | 2.32E-07 | 8.96E-07 | 0.001456451 |
| T2DM | mast_cell | *ZNF736* | 1.88294093 | 0.001861997 | 0.002879797 | 1 |
| T2DM | mast_cell | *HLA-B* | 2.520220421 | 4.93E-08 | 2.69E-07 | 0.000309916 |

| | | | | | | |
|---|---|---|---|---|---|---|
| T2DM | mast_cell | *ANKRD65* | 3.81069827 | 3.22E-07 | 1.17E-06 | 0.00202306 |
| T2DM | mast_cell | *ZSCAN31* | 1.364904158 | 0.000461906 | 0.000781275 | 1 |
| T2DM | mast_cell | *LINC00957* | 5.633315107 | 5.16E-06 | 1.30E-05 | 0.032409843 |
| T2DM | mast_cell | *KIAA0040* | 2.819330427 | 0.012152636 | 0.016537581 | 1 |
| T2DM | mast_cell | *B3GALT4* | -2.436191556 | 0.000264216 | 0.000465432 | 1 |
| T2DM | mast_cell | *LINC00941* | 6.270594146 | 2.65E-08 | 1.70E-07 | 0.000166624 |
| T2DM | mast_cell | *LINC00545* | -9.951645913 | 0.001738514 | 0.002699452 | 1 |
| T2DM | mast_cell | *CDKN2AIPNL* | 1.103499448 | 0.035785463 | 0.04574689 | 1 |
| T2DM | mast_cell | *ZNF737* | 4.087575479 | 1.15E-08 | 1.09E-07 | 7.25E-05 |
| T2DM | mast_cell | *LINC00959* | 3.209690638 | 1.26E-05 | 2.87E-05 | 0.079299709 |
| T2DM | mast_cell | *HLA-DQA2* | 2.667035688 | 6.67E-20 | 2.10E-16 | 4.20E-16 |
| T2DM | mast_cell | *AC073257.2* | 16.14379957 | 1.60E-08 | 1.25E-07 | 0.000100686 |
| T2DM | mast_cell | *LRRC37A2* | 4.261790532 | 5.43E-08 | 2.89E-07 | 0.000341604 |
| T2DM | mast_cell | *TXNDC5* | 7.929523838 | 2.10E-09 | 7.78E-08 | 1.32E-05 |
| T2DM | mast_cell | *ASB14* | 2.28461931 | 3.77E-10 | 4.05E-08 | 2.37E-06 |
| T2DM | mast_cell | *CDRT4* | 2.889520683 | 3.90E-06 | 1.01E-05 | 0.024503302 |
| T2DM | mast_cell | *APOBEC3G* | -0.787079835 | 0.004122335 | 0.006024436 | 1 |
| T2DM | mast_cell | *ADSL* | 2.17639303 | 2.00E-06 | 5.62E-06 | 0.012562361 |
| T2DM | mast_cell | *LILRA4* | -18.62632448 | 3.34E-06 | 8.82E-06 | 0.020995757 |
| T2DM | mast_cell | *LY6G5B* | -0.549999678 | 0.007311942 | 0.010309527 | 1 |
| T2DM | mast_cell | *COX19* | 5.207754451 | 6.63E-07 | 2.16E-06 | 0.004165775 |
| T2DM | mast_cell | *L1TD1* | 0.861613461 | 0.004696394 | 0.006806416 | 1 |
| T2DM | mast_cell | *AQP1* | 4.11363526 | 9.87E-06 | 2.30E-05 | 0.062044959 |
| T2DM | mast_cell | *C1QTNF9* | -3.181647124 | 0.023468682 | 0.030726282 | 1 |
| T2DM | mast_cell | *ARHGEF25* | 2.478614926 | 1.43E-08 | 1.19E-07 | 8.96E-05 |
| T2DM | mast_cell | *LINC00886* | 2.394094426 | 0.009973899 | 0.013769412 | 1 |
| T2DM | mast_cell | *TDGF1* | -1.987952732 | 0.000379652 | 0.000651795 | 1 |
| T2DM | mast_cell | *ARHGAP8* | 2.861143704 | 6.34E-08 | 3.27E-07 | 0.000398405 |
| T2DM | mast_cell | *ATP5O* | 4.17990874 | 0.000116624 | 0.000219724 | 0.733217983 |
| T2DM | mast_cell | *PWP2* | 6.170634185 | 3.48E-06 | 9.16E-06 | 0.021854823 |
| T2DM | mast_cell | *PI4KA* | 2.71050725 | 0.002033312 | 0.003129359 | 1 |
| T2DM | mast_cell | *AKAP2* | 18.57610553 | 2.23E-09 | 7.78E-08 | 1.40E-05 |
| T2DM | mast_cell | *AMACR* | 0.529035216 | 0.005882609 | 0.008426512 | 1 |
| T2DM | mast_cell | *ARHGDIG* | 2.055316102 | 0.008151518 | 0.011416484 | 1 |
| T2DM | mast_cell | *TCP10L* | 10.77838283 | 5.40E-06 | 1.35E-05 | 0.033960423 |
| T2DM | mast_cell | *ARFGAP3* | 4.228247803 | 2.43E-08 | 1.59E-07 | 0.000152466 |
| T2DM | mast_cell | *C22orf39* | 8.653672259 | 1.62E-09 | 7.17E-08 | 1.02E-05 |
| T2DM | mast_cell | *EIF6* | -2.002170714 | 0.015841515 | 0.021231209 | 1 |
| T2DM | mast_cell | *MRPL20* | 5.162252046 | 4.86E-09 | 7.84E-08 | 3.06E-05 |
| T2DM | mast_cell | *DECR2* | 1.944221773 | 1.08E-08 | 1.06E-07 | 6.81E-05 |
| T2DM | mast_cell | *AP5Z1* | 4.830062883 | 7.23E-09 | 8.86E-08 | 4.55E-05 |
| T2DM | mast_cell | *STRC* | -5.767645014 | 0.002145184 | 0.00328385 | 1 |
| T2DM | mast_cell | *PSG4* | 2.406534383 | 3.82E-07 | 1.35E-06 | 0.002399343 |
| T2DM | mast_cell | *STON1* | 1.670511102 | 0.000594068 | 0.000989903 | 1 |
| T2DM | mast_cell | *VSIG8* | -2.239769837 | 5.47E-07 | 1.83E-06 | 0.003436175 |
| T2DM | mast_cell | *C7orf73* | 4.318886886 | 1.80E-05 | 3.93E-05 | 0.112941619 |
| T2DM | mast_cell | *FGF14-IT1* | 4.677782463 | 1.61E-08 | 1.25E-07 | 0.000100947 |
| T2DM | mast_cell | *TICAM2* | -3.346765043 | 1.55E-06 | 4.47E-06 | 0.009724293 |
| T2DM | mast_cell | *PALM2* | 1.470719729 | 3.69E-10 | 4.05E-08 | 2.32E-06 |
| T2DM | mast_cell | *NAT6* | -1.562492653 | 0.003777484 | 0.005559233 | 1 |

| | | | | | | |
|---|---|---|---|---|---|---|
| T2DM | mast_cell | *UPK3B* | 1.468221435 | 0.003837605 | 0.005639664 | 1 |
| T2DM | mast_cell | *IL10RB* | 3.916867853 | 1.04E-05 | 2.41E-05 | 0.065315123 |
| T2DM | mast_cell | *WDR92* | 3.604721069 | 7.37E-10 | 5.88E-08 | 4.63E-06 |
| T2DM | mast_cell | *NME1-NME2* | 4.655615506 | 1.69E-07 | 6.94E-07 | 0.001061158 |
| T2DM | mast_cell | *PLA2G4B* | 3.104059388 | 3.80E-10 | 4.05E-08 | 2.39E-06 |
| T2DM | mast_cell | *TTC4* | 1.020313779 | 8.99E-06 | 2.11E-05 | 0.056490093 |
| T2DM | mast_cell | *JMJD7* | 3.709154386 | 5.14E-13 | 2.94E-10 | 3.23E-09 |
| T2DM | mast_cell | *APOBEC3D* | 5.850994585 | 9.14E-06 | 2.14E-05 | 0.057434482 |
| T2DM | mast_cell | *LINC01011* | -2.198585361 | 0.000920622 | 0.001483243 | 1 |
| T2DM | mast_cell | *TMEM199* | -2.542109847 | 0.003818845 | 0.005614845 | 1 |
| T2DM | mast_cell | *P2RY11* | -3.03186236 | 0.000915207 | 0.001475739 | 1 |
| T2DM | mast_cell | *LILRA6* | 1.41328107 | 1.07E-05 | 2.48E-05 | 0.067501847 |
| T2DM | mast_cell | *APOBEC3C* | 2.809283616 | 9.54E-09 | 1.01E-07 | 6.00E-05 |
| T2DM | mast_cell | *ASPRV1* | 4.896139528 | 6.16E-07 | 2.04E-06 | 0.003874633 |
| T2DM | mast_cell | *OR2AE1* | -3.378089285 | 4.29E-06 | 1.10E-05 | 0.02696088 |
| T2DM | mast_cell | *UBE2V1* | 6.568130848 | 5.89E-09 | 8.14E-08 | 3.70E-05 |
| T2DM | mast_cell | *CRYBB2* | -2.637143329 | 2.69E-08 | 1.72E-07 | 0.000169392 |
| T2DM | mast_cell | *N4BP2L2* | 2.656898694 | 5.21E-05 | 0.000105112 | 0.327721853 |
| T2DM | mast_cell | *LINC00847* | 4.628980352 | 2.95E-09 | 7.78E-08 | 1.86E-05 |
| T2DM | mast_cell | *NEAT1* | 1.749521723 | 0.001384396 | 0.002178648 | 1 |
| T2DM | mast_cell | *ZNF585B* | 2.052421014 | 6.49E-05 | 0.000128165 | 0.408204692 |
| T2DM | mast_cell | *RAB30-AS1* | 2.399128558 | 0.001301234 | 0.002053943 | 1 |
| T2DM | mast_cell | *LINC00920* | 3.440063505 | 9.56E-07 | 2.96E-06 | 0.006009214 |
| T2DM | mast_cell | *UBAP1L* | 3.180861544 | 7.36E-05 | 0.000143514 | 0.462690391 |
| T2DM | mast_cell | *PGAM5* | 2.065657222 | 1.37E-08 | 1.17E-07 | 8.62E-05 |
| T2DM | mast_cell | *DNAJC3-AS1* | 5.418811138 | 1.12E-08 | 1.07E-07 | 7.03E-05 |
| T2DM | mast_cell | *LINC00926* | 1.850812019 | 1.44E-08 | 1.19E-07 | 9.07E-05 |
| T2DM | mast_cell | *BCKDHA* | 4.36316074 | 5.17E-08 | 2.79E-07 | 0.000325242 |
| T2DM | mast_cell | *PCDHAC1* | 3.374044651 | 0.025211896 | 0.032830818 | 1 |
| T2DM | mast_cell | *ABHD14A* | 4.857899641 | 0.000231009 | 0.000411665 | 1 |
| T2DM | mast_cell | *C5orf17* | 5.625902543 | 1.31E-07 | 5.66E-07 | 0.000825829 |
| T2DM | mast_cell | *FMN1* | -0.837516608 | 0.000171097 | 0.000312609 | 1 |
| T2DM | mast_cell | *ZNF286B* | -3.898456858 | 0.036923574 | 0.047125154 | 1 |
| T2DM | mast_cell | *LINC00942* | 4.448600956 | 6.60E-05 | 0.000130132 | 0.415252281 |
| T2DM | mast_cell | *PDCD6* | 4.734597844 | 6.80E-08 | 3.43E-07 | 0.000427763 |
| T2DM | mast_cell | *LINC00603* | 2.305396681 | 3.03E-07 | 1.11E-06 | 0.001902204 |
| T2DM | mast_cell | *YJEFN3* | 2.521462848 | 6.94E-09 | 8.77E-08 | 4.36E-05 |
| T2DM | mast_cell | *DNAH10OS* | 9.076964767 | 9.57E-06 | 2.23E-05 | 0.060142019 |
| T2DM | mast_cell | *PCDHA10* | 4.677590438 | 2.82E-08 | 1.78E-07 | 0.000177199 |
| T2DM | mast_cell | *ZNF718* | 4.361772702 | 2.62E-07 | 9.86E-07 | 0.001645254 |
| T2DM | mast_cell | *LINC00989* | 6.826006513 | 2.28E-07 | 8.87E-07 | 0.001434548 |
| T2DM | mast_cell | *LINC01021* | 10.76906652 | 0.013029969 | 0.01764744 | 1 |
| T2DM | mast_cell | *FAM218A* | -4.581865949 | 3.58E-06 | 9.39E-06 | 0.02249728 |
| T2DM | mast_cell | *GLI4* | 2.570487062 | 1.76E-08 | 1.32E-07 | 0.000110513 |
| T2DM | mast_cell | *SNHG18* | -2.814158472 | 0.000200348 | 0.000360913 | 1 |
| T2DM | mast_cell | *ZNF345* | 2.933005244 | 0.001252213 | 0.001981541 | 1 |
| T2DM | mast_cell | *SHANK3* | 1.220056881 | 0.011732549 | 0.016010969 | 1 |
| T2DM | mast_cell | *ZNF550* | 4.216563598 | 0.000239963 | 0.000426532 | 1 |
| T2DM | mast_cell | *CCDC71L* | 4.444121931 | 9.67E-08 | 4.46E-07 | 0.000607959 |
| T2DM | mast_cell | *TMEM200B* | 2.4780219 | 2.28E-09 | 7.78E-08 | 1.43E-05 |

| | | | | | | |
|---|---|---|---|---|---|---|
| T2DM | mast_cell | *TRNP1* | 16.09640347 | 4.02E-09 | 7.78E-08 | 2.53E-05 |
| T2DM | mast_cell | *ALG11* | 1.198802444 | 0.004300205 | 0.006272712 | 1 |
| T2DM | mast_cell | *ATXN7L3B* | 5.651567889 | 3.17E-07 | 1.16E-06 | 0.001991417 |
| T2DM | mast_cell | *PRKDC* | 2.81919965 | 0.002117033 | 0.003247082 | 1 |
| T2DM | mast_cell | *PCDHGA6* | 2.408883277 | 3.57E-06 | 9.37E-06 | 0.022420834 |
| T2DM | mast_cell | *PCDHGA11* | 1.687497423 | 0.000430629 | 0.000733504 | 1 |
| T2DM | mast_cell | *CLDN23* | -2.346697944 | 2.24E-06 | 6.21E-06 | 0.014082049 |
| T2DM | mast_cell | *ZNF260* | 4.730050198 | 4.36E-10 | 4.42E-08 | 2.74E-06 |
| T2DM | mast_cell | *ARMS2* | -2.861984498 | 1.34E-05 | 3.01E-05 | 0.084096845 |
| T2DM | mast_cell | *FPGT* | 6.928422462 | 1.44E-06 | 4.18E-06 | 0.009027171 |
| T2DM | mast_cell | *MPV17L2* | -1.879629327 | 0.003911475 | 0.005734944 | 1 |
| T2DM | mast_cell | *DPP3* | 1.586009081 | 1.39E-05 | 3.11E-05 | 0.087209519 |
| T2DM | mast_cell | *KRTAP5-9* | 6.360934532 | 0.008743744 | 0.012197009 | 1 |
| T2DM | mast_cell | *EBLN2* | 6.307678756 | 8.82E-06 | 2.08E-05 | 0.055477284 |
| T2DM | mast_cell | *POLR2M* | 6.607500758 | 3.14E-08 | 1.91E-07 | 0.000197327 |
| T2DM | mast_cell | *TAS2R20* | 1.815793472 | 7.05E-05 | 0.000137967 | 0.443425305 |
| T2DM | mast_cell | *LINC00346* | -2.30415934 | 3.95E-07 | 1.39E-06 | 0.002482359 |
| T2DM | mast_cell | *CTSO* | 3.842690657 | 0.003222002 | 0.004808147 | 1 |
| T2DM | mast_cell | *ZNF432* | 2.997053864 | 0.002801564 | 0.004219797 | 1 |
| T2DM | mast_cell | *LINC00944* | 2.069991047 | 9.93E-07 | 3.05E-06 | 0.006245496 |
| T2DM | mast_cell | *ZNF10* | 1.451395291 | 0.009917007 | 0.013702907 | 1 |
| T2DM | mast_cell | *HMBS* | 2.476047825 | 1.99E-12 | 8.94E-10 | 1.25E-08 |
| T2DM | mast_cell | *AC093668.2* | 3.22136732 | 4.02E-05 | 8.30E-05 | 0.25275351 |
| T2DM | mast_cell | *PGA5* | 17.17219933 | 3.07E-05 | 6.47E-05 | 0.193212817 |
| T2DM | mast_cell | *ZNF253* | 4.827591857 | 2.28E-07 | 8.87E-07 | 0.001433679 |
| T2DM | mast_cell | *HP* | 2.484756628 | 0.005807603 | 0.008322863 | 1 |
| T2DM | mast_cell | *KIAA1147* | 3.33822982 | 0.000429346 | 0.000731845 | 1 |
| T2DM | mast_cell | *GATC* | 3.172675975 | 7.86E-06 | 1.88E-05 | 0.049387919 |
| T2DM | mast_cell | *ZBED6* | -4.26964964 | 1.38E-06 | 4.04E-06 | 0.008681608 |
| T2DM | mast_cell | *OVCH1-AS1* | 4.123956916 | 8.08E-09 | 9.17E-08 | 5.08E-05 |
| T2DM | mast_cell | *CUX1* | 2.161470206 | 0.00368215 | 0.005435473 | 1 |
| T2DM | mast_cell | *CHURC1* | 1.708060231 | 0.000696451 | 0.001142637 | 1 |
| T2DM | mast_cell | *CEP95* | 2.631371649 | 0.004112224 | 0.006011056 | 1 |
| T2DM | mast_cell | *LINC00639* | 2.896080991 | 1.32E-10 | 2.13E-08 | 8.29E-07 |
| T2DM | mast_cell | *LINC00648* | 1.613087331 | 2.26E-08 | 1.52E-07 | 0.000141894 |
| T2DM | mast_cell | *LINC00924* | 1.98440017 | 1.08E-08 | 1.06E-07 | 6.77E-05 |
| T2DM | mast_cell | *ITGB3* | 1.839281519 | 2.07E-07 | 8.18E-07 | 0.00129832 |
| T2DM | mast_cell | *IQCH-AS1* | 1.376618548 | 0.017567464 | 0.023414595 | 1 |
| T2DM | mast_cell | *AC083843.1* | 13.41453549 | 0.013147235 | 0.017799569 | 1 |
| T2DM | mast_cell | *PWRN1* | 2.419926284 | 1.72E-06 | 4.91E-06 | 0.010787003 |
| T2DM | mast_cell | *HOXB7* | 4.12053003 | 8.32E-06 | 1.98E-05 | 0.052315834 |
| T2DM | mast_cell | *FRRS1L* | 5.067401516 | 3.55E-09 | 7.78E-08 | 2.23E-05 |
| T2DM | mast_cell | *LINC00562* | 1.227587269 | 6.45E-07 | 2.11E-06 | 0.00405457 |
| T2DM | mast_cell | *AC009120.5* | 2.205692812 | 1.47E-08 | 1.20E-07 | 9.26E-05 |
| T2DM | mast_cell | *LINC01003* | 1.436928768 | 4.75E-08 | 2.62E-07 | 0.000298927 |
| T2DM | mast_cell | *HPR* | 3.884465102 | 5.07E-08 | 2.75E-07 | 0.000318669 |
| T2DM | mast_cell | *LINC00662* | 3.634366899 | 0.000407696 | 0.000696896 | 1 |
| T2DM | mast_cell | *AC123768.4* | 6.035525465 | 0.012585257 | 0.017074559 | 1 |
| T2DM | mast_cell | *LINC00667* | 1.585099591 | 0.000110855 | 0.000209608 | 0.696946775 |
| T2DM | mast_cell | *LINC00672* | 1.941793546 | 0.000111288 | 0.000210271 | 0.699668668 |

| | | | | | | |
|---|---|---|---|---|---|---|
| T2DM | mast_cell | *LINC00909* | 3.529466957 | 6.24E-08 | 3.23E-07 | 0.000392067 |
| T2DM | mast_cell | *DYNLL2* | -3.554722621 | 0.00098631 | 0.001579452 | 1 |
| T2DM | mast_cell | *LINC00526* | 2.325222912 | 2.02E-07 | 8.03E-07 | 0.001267538 |
| T2DM | mast_cell | *LINC00663* | 0.50482172 | 0.004041093 | 0.005915333 | 1 |
| T2DM | mast_cell | *ILF3-AS1* | 3.489384404 | 3.59E-09 | 7.78E-08 | 2.26E-05 |
| T2DM | mast_cell | *AC139100.2* | 20.88916631 | 1.96E-09 | 7.78E-08 | 1.23E-05 |
| T2DM | mast_cell | *LINC01140* | -1.459307739 | 8.01E-06 | 1.91E-05 | 0.050342529 |
| T2DM | mast_cell | *LINC00906* | -19.78728602 | 5.13E-06 | 1.29E-05 | 0.032253023 |
| T2DM | mast_cell | *ZNF285* | 0.646697429 | 0.025538272 | 0.033214546 | 1 |
| T2DM | mast_cell | *AC137932.1* | -5.606307832 | 0.000247813 | 0.000439244 | 1 |
| T2DM | mast_cell | *AC003005.2* | 3.14872364 | 3.89E-06 | 1.01E-05 | 0.02447267 |
| T2DM | mast_cell | *AL132989.1* | 3.796613839 | 1.09E-08 | 1.06E-07 | 6.84E-05 |
| T2DM | mast_cell | *LINC00664* | 5.153199032 | 2.03E-06 | 5.68E-06 | 0.012760561 |
| T2DM | mast_cell | *AC026740.1* | -6.375843287 | 5.04E-06 | 1.27E-05 | 0.031655157 |
| T2DM | mast_cell | *FBXO17* | 5.115940715 | 6.35E-08 | 3.27E-07 | 0.000399441 |
| T2DM | mast_cell | *AC009060.1* | -1.811962038 | 1.33E-08 | 1.15E-07 | 8.33E-05 |
| T2DM | mast_cell | *SNHG8* | 2.001659853 | 0.026017242 | 0.033781578 | 1 |
| T2DM | mast_cell | *YTHDF3-AS1* | 3.198964057 | 1.91E-08 | 1.38E-07 | 0.000120122 |
| T2DM | mast_cell | *LINC01050* | 15.09011342 | 2.39E-09 | 7.78E-08 | 1.50E-05 |
| T2DM | mast_cell | *MILR1* | -0.61444703 | 0.016122868 | 0.021589877 | 1 |
| T2DM | mast_cell | *GTF2H5* | 2.503980887 | 2.76E-08 | 1.75E-07 | 0.000173353 |
| T2DM | mast_cell | *CASC15* | 2.839552787 | 3.30E-06 | 8.72E-06 | 0.020717332 |
| T2DM | mast_cell | *LINC01023* | 2.217962314 | 4.64E-09 | 7.80E-08 | 2.92E-05 |
| T2DM | mast_cell | *POLR2J4* | 3.566056117 | 3.97E-09 | 7.78E-08 | 2.50E-05 |
| T2DM | mast_cell | *AC144652.1* | 1.790208958 | 3.07E-08 | 1.88E-07 | 0.000192724 |
| T2DM | monocyte | *SCYL3* | 1.158091171 | 0.002777054 | 0.004736663 | 1 |
| T2DM | monocyte | *C1orf112* | 2.150055105 | 7.48E-05 | 0.000433477 | 0.470322336 |
| T2DM | monocyte | *CFH* | 0.584654945 | 0.000661516 | 0.001633259 | 1 |
| T2DM | monocyte | *NFYA* | 2.215419121 | 0.000390303 | 0.001132887 | 1 |
| T2DM | monocyte | *STPG1* | 0.894036593 | 0.004219878 | 0.006816642 | 1 |
| T2DM | monocyte | *NIPAL3* | 1.749092083 | 0.000390047 | 0.001132668 | 1 |
| T2DM | monocyte | *ENPP4* | 1.491865933 | 0.000604463 | 0.001529222 | 1 |
| T2DM | monocyte | *KRIT1* | 2.104731664 | 6.38E-05 | 0.000411623 | 0.400920561 |
| T2DM | monocyte | *RAD52* | 1.619080231 | 0.00019132 | 0.000714856 | 1 |
| T2DM | monocyte | *BAD* | 1.408445693 | 0.000771376 | 0.001822488 | 1 |
| T2DM | monocyte | *LASP1* | 3.142867164 | 1.80E-05 | 0.000362859 | 0.112925381 |
| T2DM | monocyte | *SNX11* | 1.708541408 | 0.004402849 | 0.00706862 | 1 |
| T2DM | monocyte | *TMEM176A* | 1.632843173 | 0.006465911 | 0.009922183 | 1 |
| T2DM | monocyte | *M6PR* | 3.977528419 | 3.62E-05 | 0.000365043 | 0.227845014 |
| T2DM | monocyte | *CYP26B1* | 3.399030465 | 7.66E-05 | 0.000437295 | 0.481713487 |
| T2DM | monocyte | *ICA1* | 1.05528505 | 0.000450142 | 0.001247813 | 1 |
| T2DM | monocyte | *ALS2* | 1.851419374 | 8.70E-05 | 0.000461034 | 0.546741005 |
| T2DM | monocyte | *POLDIP2* | 1.316010854 | 0.000552349 | 0.001445117 | 1 |
| T2DM | monocyte | *RBM6* | -0.814758013 | 0.003047756 | 0.005137063 | 1 |
| T2DM | monocyte | *RECQL* | 2.869029228 | 2.26E-05 | 0.000362859 | 0.14228812 |
| T2DM | monocyte | *PDK4* | 1.928110384 | 5.48E-05 | 0.000388233 | 0.344362934 |
| T2DM | monocyte | *SLC22A16* | -0.419591273 | 0.007725836 | 0.011600748 | 1 |
| T2DM | monocyte | *CDC27* | 2.432634087 | 0.00030975 | 0.000971846 | 1 |
| T2DM | monocyte | *MEOX1* | 0.683945577 | 0.002843244 | 0.00483513 | 1 |
| T2DM | monocyte | *CIAPIN1* | 2.553452798 | 8.07E-05 | 0.000443644 | 0.507299305 |

| | | | | | | |
|---|---|---|---|---|---|---|
| T2DM | monocyte | *COPZ2* | 1.288389542 | 0.000221898 | 0.000777632 | 1 |
| T2DM | monocyte | *GCFC2* | 1.757398821 | 8.10E-05 | 0.000444359 | 0.509235203 |
| T2DM | monocyte | *WDR54* | 6.259186786 | 0.000321262 | 0.000995944 | 1 |
| T2DM | monocyte | *ABCB4* | 5.563378813 | 0.003408466 | 0.005667554 | 1 |
| T2DM | monocyte | *ZNF195* | 0.687293892 | 0.013457748 | 0.018953597 | 1 |
| T2DM | monocyte | *PDK2* | 1.039683596 | 0.000998898 | 0.002204307 | 1 |
| T2DM | monocyte | *ASB4* | 2.193834361 | 0.003255919 | 0.005446627 | 1 |
| T2DM | monocyte | *TMEM98* | 0.718883764 | 0.002914749 | 0.004942025 | 1 |
| T2DM | monocyte | *ABCC8* | 1.863814464 | 0.001824395 | 0.003390473 | 1 |
| T2DM | monocyte | *CCL18* | 0.980374773 | 0.000791588 | 0.001858369 | 1 |
| T2DM | monocyte | *SYNRG* | 2.400282827 | 3.56E-05 | 0.000365043 | 0.223551823 |
| T2DM | monocyte | *ZNF263* | 4.715373241 | 3.46E-05 | 0.000365043 | 0.217652257 |
| T2DM | monocyte | *SPATA20* | 2.069671945 | 0.000700849 | 0.001698293 | 1 |
| T2DM | monocyte | *TNFRSF12A* | 1.518971257 | 0.003687565 | 0.006051786 | 1 |
| T2DM | monocyte | *MAP3K9* | 1.836527228 | 0.000262303 | 0.000866124 | 1 |
| T2DM | monocyte | *RALA* | 2.259563634 | 5.33E-05 | 0.000385235 | 0.33525252 |
| T2DM | monocyte | *BAIAP2L1* | 1.661340279 | 0.000566564 | 0.00146765 | 1 |
| T2DM | monocyte | *FARP2* | 2.438341393 | 6.28E-05 | 0.000408289 | 0.395096403 |
| T2DM | monocyte | *IFRD1* | 1.178292443 | 0.00061343 | 0.001543731 | 1 |
| T2DM | monocyte | *GTF2IRD1* | 0.893208959 | 0.001848009 | 0.00341618 | 1 |
| T2DM | monocyte | *VPS41* | 2.429883654 | 0.000457829 | 0.001261891 | 1 |
| T2DM | monocyte | *ARHGAP44* | 0.892622097 | 0.001593814 | 0.003095555 | 1 |
| T2DM | monocyte | *ELAC2* | 2.633143433 | 8.67E-05 | 0.00046101 | 0.544913702 |
| T2DM | monocyte | *CCDC124* | 1.696382486 | 0.002925984 | 0.004958399 | 1 |
| T2DM | monocyte | *NOS2* | 7.417707113 | 0.000559921 | 0.001458252 | 1 |
| T2DM | monocyte | *TRAPPC6A* | 2.563637884 | 0.000203008 | 0.000735552 | 1 |
| T2DM | monocyte | *CD79B* | -1.11228349 | 0.005169869 | 0.008156328 | 1 |
| T2DM | monocyte | *RPUSD1* | 2.063049871 | 0.001200676 | 0.002510852 | 1 |
| T2DM | monocyte | *TSR3* | 2.108411313 | 0.000158681 | 0.000639029 | 0.997625343 |
| T2DM | monocyte | *PIGQ* | 2.482164695 | 0.004524963 | 0.007239269 | 1 |
| T2DM | monocyte | *PSMB1* | 2.459563508 | 0.000122669 | 0.000553734 | 0.771218843 |
| T2DM | monocyte | *DLEC1* | 1.780256721 | 0.000557217 | 0.001451812 | 1 |
| T2DM | monocyte | *SS18L2* | 3.155395551 | 0.002627549 | 0.004529587 | 1 |
| T2DM | monocyte | *MPND* | 2.345148752 | 0.000196629 | 0.000724198 | 1 |
| T2DM | monocyte | *MGST1* | 0.789871839 | 0.000296689 | 0.000946439 | 1 |
| T2DM | monocyte | *NFIX* | 1.156910802 | 0.000316895 | 0.000986784 | 1 |
| T2DM | monocyte | *MMP25* | 1.320432493 | 0.000650297 | 0.001611517 | 1 |
| T2DM | monocyte | *IL32* | 1.011844581 | 0.001359191 | 0.00275475 | 1 |
| T2DM | monocyte | *PKD1* | 1.928102946 | 5.04E-05 | 0.000383643 | 0.316634352 |
| T2DM | monocyte | *MED24* | 1.513373984 | 0.001923832 | 0.003508886 | 1 |
| T2DM | monocyte | *SEC62* | 2.791548547 | 3.42E-05 | 0.000365043 | 0.215327847 |
| T2DM | monocyte | *RPS20* | 3.288076822 | 4.89E-05 | 0.000381666 | 0.307623048 |
| T2DM | monocyte | *POMT2* | 1.131266482 | 0.002062231 | 0.003713907 | 1 |
| T2DM | monocyte | *MLXIPL* | 0.536700819 | 6.74E-05 | 0.000421481 | 0.423673581 |
| T2DM | monocyte | *SPRTN* | 5.209491072 | 2.97E-05 | 0.000362859 | 0.186517384 |
| T2DM | monocyte | *METTL13* | 2.365787739 | 0.000188059 | 0.000707135 | 1 |
| T2DM | monocyte | *DYRK4* | 2.720336208 | 0.00023713 | 0.000809797 | 1 |
| T2DM | monocyte | *STARD3NL* | 2.346759859 | 6.12E-05 | 0.000403714 | 0.384739098 |
| T2DM | monocyte | *ZNF200* | 4.735429626 | 2.48E-05 | 0.000362859 | 0.155715246 |
| T2DM | monocyte | *CD4* | -0.445257414 | 0.023645935 | 0.031436243 | 1 |

| | | | | | | |
|---|---|---|---|---|---|---|
| T2DM | monocyte | *LRRC23* | 2.182217301 | 0.011820658 | 0.016917022 | 1 |
| T2DM | monocyte | *HFE* | 1.131843489 | 0.017475858 | 0.023916134 | 1 |
| T2DM | monocyte | *SCMH1* | 1.501352436 | 7.78E-05 | 0.000438718 | 0.489222746 |
| T2DM | monocyte | *CLCN6* | 2.675749831 | 0.000157313 | 0.000635622 | 0.989028538 |
| T2DM | monocyte | *MRC2* | 0.54777733 | 0.001468211 | 0.002916135 | 1 |
| T2DM | monocyte | *TSPAN9* | 0.663300069 | 0.00625885 | 0.009656293 | 1 |
| T2DM | monocyte | *MKS1* | 1.237414037 | 0.000739201 | 0.001771097 | 1 |
| T2DM | monocyte | *UTP18* | 4.967760326 | 0.000194754 | 0.000722248 | 1 |
| T2DM | monocyte | *TTC19* | 2.041872858 | 2.28E-05 | 0.000362859 | 0.143635447 |
| T2DM | monocyte | *DPF1* | -1.660037517 | 0.004954285 | 0.007861582 | 1 |
| T2DM | monocyte | *SYT7* | 2.055434556 | 0.000449273 | 0.001246505 | 1 |
| T2DM | monocyte | *LARS2* | 3.563866454 | 1.69E-06 | 0.000169734 | 0.010650569 |
| T2DM | monocyte | *PPP5C* | 2.581909547 | 1.75E-05 | 0.000362859 | 0.10984451 |
| T2DM | monocyte | *CEP68* | 2.51753886 | 6.85E-10 | 7.18E-07 | 4.31E-06 |
| T2DM | monocyte | *ZBTB32* | 0.927870199 | 0.001971941 | 0.003582174 | 1 |
| T2DM | monocyte | *TMEM159* | 1.380139382 | 0.001276473 | 0.002626773 | 1 |
| T2DM | monocyte | *ERCC1* | 1.57831288 | 0.00019438 | 0.000721836 | 1 |
| T2DM | monocyte | *LTF* | 2.977706901 | 0.012723937 | 0.018024817 | 1 |
| T2DM | monocyte | *ALOX5* | -0.244703212 | 0.012203568 | 0.017362262 | 1 |
| T2DM | monocyte | *MAP4K5* | 2.052506625 | 0.000124967 | 0.000556889 | 0.785669145 |
| T2DM | monocyte | *PSMC4* | 4.232500803 | 1.48E-06 | 0.000157658 | 0.009320594 |
| T2DM | monocyte | *MAN2B2* | 4.056012441 | 0.000277665 | 0.000902161 | 1 |
| T2DM | monocyte | *CLDN11* | 1.562674849 | 0.000374805 | 0.001099195 | 1 |
| T2DM | monocyte | *SLC25A39* | 2.391545961 | 0.000341183 | 0.001031258 | 1 |
| T2DM | monocyte | *ANGEL1* | 4.944162288 | 0.000262826 | 0.000866941 | 1 |
| T2DM | monocyte | *RNF14* | 2.660915033 | 4.28E-05 | 0.000377787 | 0.269361935 |
| T2DM | monocyte | *HEBP1* | 2.400490761 | 0.002908642 | 0.004933 | 1 |
| T2DM | monocyte | *ZC3H3* | 2.30887481 | 0.0099931 | 0.014566802 | 1 |
| T2DM | monocyte | *MDH1* | 1.060123573 | 0.000727536 | 0.001749147 | 1 |
| T2DM | monocyte | *SLC30A9* | 2.925632175 | 4.47E-05 | 0.000379941 | 0.280944217 |
| T2DM | monocyte | *COX15* | 1.689999995 | 0.000505519 | 0.001355309 | 1 |
| T2DM | monocyte | *NPC1L1* | 1.967958289 | 0.019117071 | 0.025897226 | 1 |
| T2DM | monocyte | *NUDCD3* | 1.341845457 | 0.004333871 | 0.006973905 | 1 |
| T2DM | monocyte | *CHDH* | 2.131383529 | 0.001342762 | 0.002727608 | 1 |
| T2DM | monocyte | *IL20RA* | 4.560488285 | 0.000575185 | 0.00148083 | 1 |
| T2DM | monocyte | *GLT8D1* | 1.473785389 | 0.000896352 | 0.00202711 | 1 |
| T2DM | monocyte | *RUFY3* | 2.053727897 | 0.000231995 | 0.00079821 | 1 |
| T2DM | monocyte | *WWTR1* | 1.677388083 | 7.20E-05 | 0.000429362 | 0.452976749 |
| T2DM | monocyte | *TTC27* | 2.058392704 | 0.000324262 | 0.001000803 | 1 |
| T2DM | monocyte | *VSIG2* | 2.169389414 | 0.00168107 | 0.003212428 | 1 |
| T2DM | monocyte | *PHLDB1* | 0.441447073 | 0.013146299 | 0.01855653 | 1 |
| T2DM | monocyte | *MARCO* | 0.885931571 | 0.000291437 | 0.0009334 | 1 |
| T2DM | monocyte | *MNAT1* | 1.344358815 | 0.000146362 | 0.000609994 | 0.920176455 |
| T2DM | monocyte | *OSBPL5* | 4.356029552 | 0.000180423 | 0.000688301 | 1 |
| T2DM | monocyte | *AQR* | 3.512521916 | 1.43E-05 | 0.000362859 | 0.089752849 |
| T2DM | monocyte | *CPS1* | 0.645318775 | 0.011566712 | 0.016583791 | 1 |
| T2DM | monocyte | *ZDHHC6* | 3.359963152 | 0.001814121 | 0.003381376 | 1 |
| T2DM | monocyte | *RNH1* | 2.233899252 | 0.000735923 | 0.001764586 | 1 |
| T2DM | monocyte | *NDUFS1* | 3.000735169 | 7.32E-05 | 0.000431915 | 0.460337457 |
| T2DM | monocyte | *AKAP11* | 3.026548653 | 2.53E-05 | 0.000362859 | 0.159014177 |

| | | | | | | |
|---|---|---|---|---|---|---|
| T2DM | monocyte | *GLRX2* | 0.84035343 | 0.003198911 | 0.005365943 | 1 |
| T2DM | monocyte | *SNAPC1* | 3.651415281 | 0.002466871 | 0.004289054 | 1 |
| T2DM | monocyte | *GCLM* | 1.262765169 | 0.000444738 | 0.001238293 | 1 |
| T2DM | monocyte | *UBR2* | 3.085730687 | 2.66E-05 | 0.000362859 | 0.167160509 |
| T2DM | monocyte | *HSF2* | 2.288147494 | 7.21E-05 | 0.000429365 | 0.453409246 |
| T2DM | monocyte | *NR1H3* | 1.651363392 | 2.06E-05 | 0.000362859 | 0.129223208 |
| T2DM | monocyte | *TOMM34* | 1.356550886 | 0.000894797 | 0.00202711 | 1 |
| T2DM | monocyte | *KPNA6* | 2.22338497 | 2.12E-05 | 0.000362859 | 0.133442214 |
| T2DM | monocyte | *VIM* | 2.164284513 | 2.36E-05 | 0.000362859 | 0.148080508 |
| T2DM | monocyte | *FAS* | 1.611037316 | 0.020101336 | 0.027142847 | 1 |
| T2DM | monocyte | *RNASET2* | -0.390314445 | 0.020174178 | 0.027217579 | 1 |
| T2DM | monocyte | *MIPEP* | 2.34042477 | 6.91E-05 | 0.000425443 | 0.434573465 |
| T2DM | monocyte | *B4GALT7* | 4.079579348 | 0.001907919 | 0.003493037 | 1 |
| T2DM | monocyte | *VRK2* | 1.869958679 | 0.028924174 | 0.037688348 | 1 |
| T2DM | monocyte | *VEZT* | 2.162896017 | 9.54E-05 | 0.000480415 | 0.599558143 |
| T2DM | monocyte | *SNX1* | 2.01513303 | 0.000414771 | 0.001179826 | 1 |
| T2DM | monocyte | *ARNTL2* | 0.995753987 | 0.000829316 | 0.001921116 | 1 |
| T2DM | monocyte | *ANK1* | 1.890199984 | 0.000297964 | 0.000948026 | 1 |
| T2DM | monocyte | *RABEP1* | 2.759718635 | 8.98E-05 | 0.000465291 | 0.564831195 |
| T2DM | monocyte | *GRN* | 3.003663389 | 0.006429167 | 0.009880268 | 1 |
| T2DM | monocyte | *ARID4A* | 3.571241518 | 0.000148206 | 0.000613409 | 0.93176881 |
| T2DM | monocyte | *IFT88* | 1.900724815 | 0.000146507 | 0.000609994 | 0.921091462 |
| T2DM | monocyte | *ZCCHC8* | 2.793422885 | 3.55E-05 | 0.000365043 | 0.22339507 |
| T2DM | monocyte | *FUT8* | 1.840313736 | 6.16E-05 | 0.000405482 | 0.387235308 |
| T2DM | monocyte | *UBA6* | 1.786813586 | 0.004709882 | 0.007507868 | 1 |
| T2DM | monocyte | *PIAS1* | 2.002147594 | 0.012722831 | 0.018024817 | 1 |
| T2DM | monocyte | *SLC4A7* | 2.848370566 | 4.41E-05 | 0.000378997 | 0.277046584 |
| T2DM | monocyte | *ASTE1* | 1.726229646 | 0.003894076 | 0.006350728 | 1 |
| T2DM | monocyte | *MYOC* | 0.876402235 | 0.000402614 | 0.001158461 | 1 |
| T2DM | monocyte | *VCL* | 1.874852915 | 0.019984118 | 0.026996164 | 1 |
| T2DM | monocyte | *ADSS* | 3.517286357 | 0.002456931 | 0.004274135 | 1 |
| T2DM | monocyte | *STAP1* | 1.411815553 | 0.007864385 | 0.011777843 | 1 |
| T2DM | monocyte | *TBC1D23* | 3.209569504 | 3.27E-05 | 0.000365043 | 0.205623904 |
| T2DM | monocyte | *CUL3* | 3.426413676 | 1.47E-05 | 0.000362859 | 0.092320775 |
| T2DM | monocyte | *MYOM2* | 0.882195222 | 0.002310419 | 0.004054035 | 1 |
| T2DM | monocyte | *NSUN2* | 2.704720975 | 0.000427596 | 0.001205513 | 1 |
| T2DM | monocyte | *MFAP3* | 1.286964687 | 0.000885992 | 0.002012922 | 1 |
| T2DM | monocyte | *MRI1* | 1.151655235 | 0.007441177 | 0.011226945 | 1 |
| T2DM | monocyte | *AGA* | 3.863418472 | 4.98E-05 | 0.000383513 | 0.313002173 |
| T2DM | monocyte | *BOD1L1* | 2.694686081 | 0.000984167 | 0.002175619 | 1 |
| T2DM | monocyte | *MAT2B* | 3.630477991 | 1.75E-05 | 0.000362859 | 0.110190281 |
| T2DM | monocyte | *CLEC16A* | 2.525106718 | 0.009835288 | 0.014369504 | 1 |
| T2DM | monocyte | *SKIV2L2* | 1.904577693 | 5.11E-05 | 0.000384204 | 0.321312831 |
| T2DM | monocyte | *DNAH5* | 0.817874993 | 0.002627315 | 0.004529587 | 1 |
| T2DM | monocyte | *C6* | 0.584586697 | 0.0025125 | 0.004357542 | 1 |
| T2DM | monocyte | *RAI14* | 1.274526941 | 0.000712052 | 0.001720472 | 1 |
| T2DM | monocyte | *PHLPP2* | 3.470675524 | 0.000242066 | 0.000820147 | 1 |
| T2DM | monocyte | *PQLC2* | 2.927120593 | 0.000135686 | 0.000582688 | 0.853055551 |
| T2DM | monocyte | *CTNS* | 1.248931697 | 0.003205277 | 0.005375188 | 1 |
| T2DM | monocyte | *RAB27B* | 0.887184879 | 0.000101062 | 0.00049639 | 0.635379519 |

| | | | | | | |
|---|---|---|---|---|---|---|
| T2DM | monocyte | *PSMA4* | 3.642232998 | 7.06E-05 | 0.000427286 | 0.443863151 |
| T2DM | monocyte | *LSG1* | 1.088497819 | 0.00505109 | 0.007993004 | 1 |
| T2DM | monocyte | *THAP3* | 4.053338571 | 0.000421249 | 0.001193507 | 1 |
| T2DM | monocyte | *TDP1* | 4.767129307 | 3.82E-05 | 0.00036941 | 0.240142621 |
| T2DM | monocyte | *SPATA7* | 1.913957823 | 0.000141052 | 0.000595965 | 0.886795908 |
| T2DM | monocyte | *MED17* | 5.423275765 | 6.32E-07 | 8.64E-05 | 0.003974527 |
| T2DM | monocyte | *RETSAT* | 0.662890677 | 0.002266706 | 0.003990697 | 1 |
| T2DM | monocyte | *CAPG* | 1.467099246 | 0.008351541 | 0.012436319 | 1 |
| T2DM | monocyte | *AP2S1* | 3.918026503 | 6.52E-05 | 0.000417158 | 0.41006659 |
| T2DM | monocyte | *LCP2* | -0.444024173 | 0.034844567 | 0.04469859 | 1 |
| T2DM | monocyte | *CUL7* | 1.701338167 | 0.001383903 | 0.00279313 | 1 |
| T2DM | monocyte | *CTNNA1* | 1.907221076 | 5.26E-05 | 0.000385235 | 0.330404185 |
| T2DM | monocyte | *CNTLN* | 2.766000268 | 2.33E-05 | 0.000362859 | 0.14653102 |
| T2DM | monocyte | *EPHA3* | 0.482397818 | 0.001683053 | 0.003212508 | 1 |
| T2DM | monocyte | *DSG2* | 0.768364531 | 0.001327606 | 0.002703173 | 1 |
| T2DM | monocyte | *WDR37* | 3.072479838 | 5.05E-05 | 0.000383728 | 0.317625539 |
| T2DM | monocyte | *YTHDC2* | 3.419863058 | 8.52E-06 | 0.000362859 | 0.053550292 |
| T2DM | monocyte | *ATP6V1H* | 1.991179581 | 7.99E-05 | 0.000442726 | 0.502050955 |
| T2DM | monocyte | *POLR2B* | 3.14408483 | 2.42E-05 | 0.000362859 | 0.15187894 |
| T2DM | monocyte | *CP* | 3.571705358 | 3.81E-05 | 0.00036941 | 0.23937759 |
| T2DM | monocyte | *ANO2* | 0.478602554 | 0.001916238 | 0.003500607 | 1 |
| T2DM | monocyte | *GOPC* | 3.325664241 | 0.000223263 | 0.00077951 | 1 |
| T2DM | monocyte | *TSPAN17* | 2.074884994 | 1.10E-05 | 0.000362859 | 0.069074586 |
| T2DM | monocyte | *CC2D2A* | 2.376078409 | 3.80E-05 | 0.00036941 | 0.239015286 |
| T2DM | monocyte | *RRM2B* | 3.860983782 | 0.000259571 | 0.000859358 | 1 |
| T2DM | monocyte | *SNX29* | 2.997190205 | 2.24E-05 | 0.000362859 | 0.140646885 |
| T2DM | monocyte | *LMO3* | 0.682860687 | 0.001148421 | 0.002434297 | 1 |
| T2DM | monocyte | *CELF2* | 2.21615922 | 2.49E-05 | 0.000362859 | 0.156415268 |
| T2DM | monocyte | *ERCC8* | 3.127074237 | 9.00E-06 | 0.000362859 | 0.056594724 |
| T2DM | monocyte | *ADAMTS6* | 0.780928794 | 0.00158993 | 0.003092788 | 1 |
| T2DM | monocyte | *H6PD* | 1.556381064 | 0.012677789 | 0.017971873 | 1 |
| T2DM | monocyte | *UTS2* | 1.122927409 | 0.01212582 | 0.017271189 | 1 |
| T2DM | monocyte | *EPN3* | 9.161359143 | 7.78E-05 | 0.000438718 | 0.488995043 |
| T2DM | monocyte | *RCN1* | 1.559975502 | 0.002026772 | 0.003661797 | 1 |
| T2DM | monocyte | *JKAMP* | 1.684937957 | 0.000387831 | 0.001127275 | 1 |
| T2DM | monocyte | *DKK3* | 1.047115714 | 0.000686959 | 0.001675945 | 1 |
| T2DM | monocyte | *ARHGEF5* | 1.042306176 | 0.00153833 | 0.003011046 | 1 |
| T2DM | monocyte | *NFE2L3* | 4.003875282 | 0.006730443 | 0.010282939 | 1 |
| T2DM | monocyte | *MCUR1* | 1.713841391 | 0.000600023 | 0.001523326 | 1 |
| T2DM | monocyte | *LIMA1* | 0.532292595 | 0.00305125 | 0.005141573 | 1 |
| T2DM | monocyte | *PTGER3* | 0.728923698 | 0.000576365 | 0.001482303 | 1 |
| T2DM | monocyte | *MAPK9* | 1.977533433 | 3.65E-05 | 0.000365396 | 0.22942068 |
| T2DM | monocyte | *FAM160A2* | 1.768471996 | 0.008948555 | 0.013218883 | 1 |
| T2DM | monocyte | *HERPUD1* | 1.999261528 | 0.000124697 | 0.000556889 | 0.783968934 |
| T2DM | monocyte | *PIK3CB* | 2.392124367 | 0.000529933 | 0.00140183 | 1 |
| T2DM | monocyte | *THOC3* | 1.07121942 | 0.001642633 | 0.00315721 | 1 |
| T2DM | monocyte | *ALX4* | 12.50273916 | 5.88E-05 | 0.000398946 | 0.36943694 |
| T2DM | monocyte | *FOXN3* | 2.401207801 | 4.68E-05 | 0.000379941 | 0.293993301 |
| T2DM | monocyte | *AKR7A2* | 2.679635735 | 0.000183496 | 0.000696642 | 1 |
| T2DM | monocyte | *MRTO4* | 2.557015966 | 0.000536793 | 0.001412959 | 1 |

| | | | | | | |
|---|---|---|---|---|---|---|
| T2DM | monocyte | *MCF2L2* | 1.351783772 | 0.000750474 | 0.00178992 | 1 |
| T2DM | monocyte | *ANAPC4* | 3.468043461 | 3.85E-06 | 0.000269285 | 0.024235681 |
| T2DM | monocyte | *OPN3* | 1.302297669 | 0.000102648 | 0.000500426 | 0.64534495 |
| T2DM | monocyte | *SDCCAG8* | 1.938681174 | 8.99E-05 | 0.000465291 | 0.565320374 |
| T2DM | monocyte | *PLEKHH1* | 0.711403509 | 0.005750221 | 0.008959514 | 1 |
| T2DM | monocyte | *ATP9A* | 0.991269039 | 0.000601999 | 0.001524272 | 1 |
| T2DM | monocyte | *CHRDL2* | 2.419951222 | 0.001098381 | 0.002357638 | 1 |
| T2DM | monocyte | *GALC* | 2.242725503 | 0.037784141 | 0.048135541 | 1 |
| T2DM | monocyte | *FAM114A2* | 3.311943359 | 0.002071637 | 0.003726575 | 1 |
| T2DM | monocyte | *GINM1* | 2.807472093 | 4.36E-05 | 0.000378997 | 0.274192283 |
| T2DM | monocyte | *MRPL43* | 1.953224417 | 0.0308649 | 0.039927495 | 1 |
| T2DM | monocyte | *ITIH4* | 1.279271659 | 1.85E-05 | 0.000362859 | 0.116150238 |
| T2DM | monocyte | *TRAF1* | 1.07285722 | 0.001459577 | 0.002905834 | 1 |
| T2DM | monocyte | *PCGF2* | 0.777808305 | 0.004025649 | 0.006538169 | 1 |
| T2DM | monocyte | *IL17RB* | 3.808126547 | 0.002578629 | 0.004462385 | 1 |
| T2DM | monocyte | *TRAF3IP2* | 1.904916695 | 0.000110768 | 0.000523558 | 0.696397078 |
| T2DM | monocyte | *DCBLD2* | 1.062529149 | 0.000426985 | 0.001204331 | 1 |
| T2DM | monocyte | *MSH4* | 1.307423573 | 0.016574146 | 0.022816216 | 1 |
| T2DM | monocyte | *TMCC3* | 3.558453574 | 0.000168086 | 0.000658686 | 1 |
| T2DM | monocyte | *PITHD1* | 2.547070035 | 9.99E-05 | 0.000492855 | 0.627896831 |
| T2DM | monocyte | *ATP11B* | 1.523361602 | 7.33E-05 | 0.000431915 | 0.460885568 |
| T2DM | monocyte | *LAMC2* | 0.533450989 | 0.029967965 | 0.038879199 | 1 |
| T2DM | monocyte | *CDK14* | 1.272348557 | 0.000158124 | 0.000638077 | 0.994124132 |
| T2DM | monocyte | *ZC3H11A* | -0.665469924 | 0.014739729 | 0.02055565 | 1 |
| T2DM | monocyte | *RIOK2* | 2.00936068 | 0.000158625 | 0.000639029 | 0.997277012 |
| T2DM | monocyte | *DNAJC25* | 2.664060785 | 0.000487362 | 0.001325599 | 1 |
| T2DM | monocyte | *YBX3* | 3.773862707 | 8.74E-05 | 0.000461034 | 0.549330044 |
| T2DM | monocyte | *QSER1* | 3.735528835 | 0.000504473 | 0.00135308 | 1 |
| T2DM | monocyte | *NCKAP1* | 1.201089662 | 0.000166584 | 0.000656212 | 1 |
| T2DM | monocyte | *SFSWAP* | 3.048012522 | 2.50E-05 | 0.000362859 | 0.157288597 |
| T2DM | monocyte | *SLC6A16* | 1.257395752 | 0.002715897 | 0.004661437 | 1 |
| T2DM | monocyte | *SPHK2* | 5.160581175 | 1.79E-05 | 0.000362859 | 0.112628914 |
| T2DM | monocyte | *ISOC2* | 2.423369966 | 0.000358477 | 0.001066609 | 1 |
| T2DM | monocyte | *ADCK1* | 2.548344283 | 0.000147098 | 0.000610432 | 0.924805235 |
| T2DM | monocyte | *TM7SF3* | 2.649924214 | 0.005816046 | 0.009046383 | 1 |
| T2DM | monocyte | *DMRT3* | 1.508125969 | 0.001010817 | 0.002220478 | 1 |
| T2DM | monocyte | *ST3GAL6* | 0.772957517 | 0.010101776 | 0.014704761 | 1 |
| T2DM | monocyte | *ATP2C2* | 1.759421702 | 0.000613802 | 0.001543731 | 1 |
| T2DM | monocyte | *CDON* | 0.527470101 | 0.001663217 | 0.003184119 | 1 |
| T2DM | monocyte | *TAF2* | 3.199646447 | 1.52E-05 | 0.000362859 | 0.095865833 |
| T2DM | monocyte | *TMEM161A* | 3.677719456 | 4.69E-05 | 0.000379941 | 0.294599711 |
| T2DM | monocyte | *SUGP2* | 3.388343007 | 1.34E-06 | 0.000147316 | 0.008397021 |
| T2DM | monocyte | *DDX20* | 3.071425555 | 0.000126451 | 0.00055907 | 0.794997739 |
| T2DM | monocyte | *BTBD1* | 4.603851196 | 3.11E-06 | 0.000235686 | 0.019561957 |
| T2DM | monocyte | *CHI3L2* | 1.446987027 | 0.001516126 | 0.002978363 | 1 |
| T2DM | monocyte | *PMS1* | 3.785391845 | 2.80E-05 | 0.000362859 | 0.176316475 |
| T2DM | monocyte | *TAF11* | 1.387723104 | 0.011910815 | 0.017028633 | 1 |
| T2DM | monocyte | *ANKS1A* | 2.813660694 | 4.87E-05 | 0.000381649 | 0.306464447 |
| T2DM | monocyte | *NTHL1* | 2.425939426 | 0.000248449 | 0.000834848 | 1 |
| T2DM | monocyte | *IPO5* | 4.757061045 | 5.33E-05 | 0.000385235 | 0.33485905 |

| | | | | | | |
|---|---|---|---|---|---|---|
| T2DM | monocyte | *ADAT1* | 1.551931989 | 0.001482866 | 0.002935383 | 1 |
| T2DM | monocyte | *NDUFB4* | 3.721167922 | 1.13E-05 | 0.000362859 | 0.071348911 |
| T2DM | monocyte | *ZC3H15* | 3.282006059 | 0.00016879 | 0.000659529 | 1 |
| T2DM | monocyte | *MAP2K4* | 1.953435292 | 5.88E-05 | 0.000398946 | 0.36975461 |
| T2DM | monocyte | *TMEM206* | 4.284748591 | 0.00016722 | 0.000657069 | 1 |
| T2DM | monocyte | *SEC61A2* | 2.508877242 | 9.30E-05 | 0.000474395 | 0.584728292 |
| T2DM | monocyte | *PPP2R5A* | 1.920602746 | 5.78E-05 | 0.000396862 | 0.36330729 |
| T2DM | monocyte | *TIE1* | 0.582841629 | 0.002476415 | 0.004300889 | 1 |
| T2DM | monocyte | *NFYC* | 3.415679437 | 0.000969194 | 0.002152556 | 1 |
| T2DM | monocyte | *ZMYND12* | 1.352069327 | 0.014631544 | 0.020429667 | 1 |
| T2DM | monocyte | *NGEF* | 8.581481199 | 0.016766443 | 0.023045612 | 1 |
| T2DM | monocyte | *CD84* | 1.871643188 | 0.021294995 | 0.028619417 | 1 |
| T2DM | monocyte | *ZNRD1* | 1.147669832 | 0.005573332 | 0.008714136 | 1 |
| T2DM | monocyte | *ATXN3* | 2.251134541 | 2.94E-05 | 0.000362859 | 0.184539628 |
| T2DM | monocyte | *GOLGA5* | 1.309607477 | 0.000456262 | 0.001258123 | 1 |
| T2DM | monocyte | *EML1* | 0.580788443 | 0.002957912 | 0.005003065 | 1 |
| T2DM | monocyte | *ZFAT* | 3.559103352 | 0.000136154 | 0.000583903 | 0.856002481 |
| T2DM | monocyte | *MTFR1* | 1.278477683 | 0.000729532 | 0.001752605 | 1 |
| T2DM | monocyte | *STAG3* | 2.445232899 | 0.00013848 | 0.000589065 | 0.870624371 |
| T2DM | monocyte | *MYO9A* | 1.465730142 | 0.000118787 | 0.000544237 | 0.746816366 |
| T2DM | monocyte | *SP100* | 3.279702797 | 0.000111847 | 0.000524745 | 0.703181221 |
| T2DM | monocyte | *METTL22* | 3.386770467 | 0.016306707 | 0.022472658 | 1 |
| T2DM | monocyte | *TP53BP1* | 2.219857832 | 1.84E-05 | 0.000362859 | 0.115437139 |
| T2DM | monocyte | *RRP15* | 1.048828489 | 0.002304867 | 0.00404655 | 1 |
| T2DM | monocyte | *DHX8* | 3.193726589 | 0.001196114 | 0.002504666 | 1 |
| T2DM | monocyte | *SYT1* | 0.463800707 | 0.005576165 | 0.008716397 | 1 |
| T2DM | monocyte | *ROGDI* | -0.756719453 | 0.019757172 | 0.026729791 | 1 |
| T2DM | monocyte | *HDAC4* | 3.534020541 | 8.70E-05 | 0.000461034 | 0.547243903 |
| T2DM | monocyte | *RASSF1* | 3.531803539 | 0.000462167 | 0.001271543 | 1 |
| T2DM | monocyte | *FGFR3* | 0.707412024 | 0.00442121 | 0.007092662 | 1 |
| T2DM | monocyte | *IFI35* | 1.65870403 | 0.000120664 | 0.00054853 | 0.758616653 |
| T2DM | monocyte | *HEATR6* | 5.540516445 | 4.22E-05 | 0.000374514 | 0.265155742 |
| T2DM | monocyte | *SRBD1* | 2.245695788 | 0.015688453 | 0.021691952 | 1 |
| T2DM | monocyte | *CYFIP1* | 1.982853245 | 7.08E-05 | 0.000427286 | 0.445258405 |
| T2DM | monocyte | *PSME4* | 2.081246107 | 7.25E-05 | 0.000431378 | 0.455966764 |
| T2DM | monocyte | *PITX1* | 18.96455412 | 0.000193166 | 0.000718174 | 1 |
| T2DM | monocyte | *SDK2* | 1.000254514 | 0.001648376 | 0.003165345 | 1 |
| T2DM | monocyte | *NUCKS1* | 1.726543721 | 6.86E-05 | 0.000423977 | 0.431081525 |
| T2DM | monocyte | *DNAJA2* | 5.079819465 | 3.82E-05 | 0.00036941 | 0.240172882 |
| T2DM | monocyte | *KCNAB2* | -0.715901073 | 0.03128986 | 0.040443945 | 1 |
| T2DM | monocyte | *CLEC2D* | 2.345030059 | 0.001449722 | 0.002892752 | 1 |
| T2DM | monocyte | *NEDD4* | 1.74963812 | 4.71E-05 | 0.000379941 | 0.296219751 |
| T2DM | monocyte | *PIGB* | 3.018086777 | 0.000882883 | 0.00201039 | 1 |
| T2DM | monocyte | *GNB5* | 2.950603728 | 8.25E-05 | 0.000448526 | 0.518944057 |
| T2DM | monocyte | *DAPP1* | -0.198850499 | 0.010190907 | 0.01482078 | 1 |
| T2DM | monocyte | *FGF10* | 0.909985375 | 0.000366638 | 0.00108397 | 1 |
| T2DM | monocyte | *TMEM260* | 3.456471308 | 2.33E-05 | 0.000362859 | 0.146642023 |
| T2DM | monocyte | *CLTCL1* | 1.703565887 | 0.000125072 | 0.000556889 | 0.786327296 |
| T2DM | monocyte | *DGCR2* | 3.0568512 | 7.19E-05 | 0.000428995 | 0.452160859 |
| T2DM | monocyte | *ZXDC* | 2.813369155 | 0.003559056 | 0.005880626 | 1 |

| | | | | | | |
|---|---|---|---|---|---|---|
| T2DM | monocyte | *POLB* | 2.343163156 | 0.000511514 | 0.001369046 | 1 |
| T2DM | monocyte | *WIPI1* | 1.569512413 | 0.000218221 | 0.000769035 | 1 |
| T2DM | monocyte | *NDST1* | 2.398315154 | 5.96E-05 | 0.000399203 | 0.374835384 |
| T2DM | monocyte | *AP3M2* | 2.612059514 | 9.76E-05 | 0.000485446 | 0.613603714 |
| T2DM | monocyte | *TESK2* | 3.690837757 | 7.85E-05 | 0.000440255 | 0.493802416 |
| T2DM | monocyte | *EIF2B3* | 2.597657712 | 3.03E-05 | 0.000362859 | 0.190620771 |
| T2DM | monocyte | *CAMK2A* | 3.716981575 | 0.003624433 | 0.005973525 | 1 |
| T2DM | monocyte | *CDC42* | 2.776674342 | 0.007965869 | 0.011921308 | 1 |
| T2DM | monocyte | *OSBPL3* | 2.955280839 | 0.002956027 | 0.005002531 | 1 |
| T2DM | monocyte | *RAD18* | 1.364353882 | 0.000346559 | 0.001039015 | 1 |
| T2DM | monocyte | *SNX13* | 3.058180096 | 2.39E-05 | 0.000362859 | 0.150000363 |
| T2DM | monocyte | *TRIP13* | 14.64967642 | 0.002709926 | 0.004652459 | 1 |
| T2DM | monocyte | *TCF3* | 3.718402441 | 0.006816436 | 0.010401682 | 1 |
| T2DM | monocyte | *HLTF* | 1.324043217 | 0.004402155 | 0.00706862 | 1 |
| T2DM | monocyte | *CPSF1* | 2.272061541 | 0.000202512 | 0.000735524 | 1 |
| T2DM | monocyte | *CYBRD1* | 1.990053533 | 1.34E-05 | 0.000362859 | 0.084421456 |
| T2DM | monocyte | *EPN2* | 1.514713144 | 0.000177819 | 0.000681674 | 1 |
| T2DM | monocyte | *LIMS2* | 0.672854679 | 0.001148105 | 0.002434297 | 1 |
| T2DM | monocyte | *AFF4* | 2.114018113 | 0.000110932 | 0.000523558 | 0.697431137 |
| T2DM | monocyte | *TRHDE* | 0.528546122 | 0.001032286 | 0.002255032 | 1 |
| T2DM | monocyte | *NFATC3* | 3.40500917 | 6.15E-06 | 0.000322038 | 0.038659246 |
| T2DM | monocyte | *TRNT1* | 3.303464804 | 2.39E-05 | 0.000362859 | 0.150178252 |
| T2DM | monocyte | *ACADVL* | 2.298348715 | 9.01E-05 | 0.000465303 | 0.566414334 |
| T2DM | monocyte | *CRMP1* | 0.776005269 | 0.002344111 | 0.004107421 | 1 |
| T2DM | monocyte | *DERL2* | 1.904641855 | 0.000279566 | 0.000906931 | 1 |
| T2DM | monocyte | *AP1M1* | 2.136750416 | 0.003672817 | 0.006033707 | 1 |
| T2DM | monocyte | *TP63* | 0.822086311 | 0.000804291 | 0.001880468 | 1 |
| T2DM | monocyte | *ALPK1* | 3.629444172 | 1.72E-05 | 0.000362859 | 0.107857392 |
| T2DM | monocyte | *LLGL2* | 1.57197348 | 0.000387806 | 0.001127275 | 1 |
| T2DM | monocyte | *PDE8A* | 2.195347116 | 2.08E-05 | 0.000362859 | 0.130558055 |
| T2DM | monocyte | *NLE1* | 1.304001364 | 0.000997994 | 0.002203086 | 1 |
| T2DM | monocyte | *SDHA* | 3.867023253 | 2.25E-07 | 4.94E-05 | 0.001416774 |
| T2DM | monocyte | *SMARCE1* | 4.507064406 | 2.60E-05 | 0.000362859 | 0.163327143 |
| T2DM | monocyte | *GSDMB* | 0.990817695 | 0.001339399 | 0.002721813 | 1 |
| T2DM | monocyte | *PICALM* | 2.422091159 | 0.001385333 | 0.002795118 | 1 |
| T2DM | monocyte | *GLI2* | 0.504360152 | 0.005688643 | 0.008872363 | 1 |
| T2DM | monocyte | *CLNS1A* | 2.105528917 | 1.36E-05 | 0.000362859 | 0.085468792 |
| T2DM | monocyte | *ATP2A3* | 2.326012638 | 0.000142356 | 0.000598657 | 0.894992758 |
| T2DM | monocyte | *CA12* | 0.636197703 | 0.004176416 | 0.006755115 | 1 |
| T2DM | monocyte | *NTN4* | 0.53935392 | 0.001395389 | 0.002809098 | 1 |
| T2DM | monocyte | *LMAN1* | 1.600904606 | 0.000162361 | 0.000646053 | 1 |
| T2DM | monocyte | *TIPIN* | 0.980790755 | 0.000315613 | 0.000983768 | 1 |
| T2DM | monocyte | *SRI* | 2.903246134 | 4.55E-05 | 0.000379941 | 0.285785444 |
| T2DM | monocyte | *EIF4G3* | 2.426590085 | 4.66E-05 | 0.000379941 | 0.293172794 |
| T2DM | monocyte | *TTC38* | 0.746987023 | 0.001301147 | 0.002670687 | 1 |
| T2DM | monocyte | *GRAMD4* | 4.611868121 | 7.95E-05 | 0.000442619 | 0.499820733 |
| T2DM | monocyte | *CELSR1* | 2.832388189 | 0.000176921 | 0.000679891 | 1 |
| T2DM | monocyte | *ADD2* | -0.560801102 | 0.01474433 | 0.02055565 | 1 |
| T2DM | monocyte | *VPS9D1* | 5.38664028 | 9.50E-05 | 0.000480034 | 0.597161909 |
| T2DM | monocyte | *MARK3* | 1.897445446 | 0.003029626 | 0.005110614 | 1 |

| | | | | | | |
|---|---|---|---|---|---|---|
| T2DM | monocyte | *SLC25A3* | 4.484018844 | 2.53E-05 | 0.000362859 | 0.159096855 |
| T2DM | monocyte | *FSCN1* | 1.183258729 | 0.001309703 | 0.002683867 | 1 |
| T2DM | monocyte | *MOCOS* | 1.422159995 | 0.006490009 | 0.009951875 | 1 |
| T2DM | monocyte | *PLD1* | 1.795989418 | 7.13E-05 | 0.000427313 | 0.44825184 |
| T2DM | monocyte | *BCAP29* | 2.709286319 | 2.60E-05 | 0.000362859 | 0.163713421 |
| T2DM | monocyte | *SEC31B* | 0.881403284 | 0.000322081 | 0.000997498 | 1 |
| T2DM | monocyte | *TUBA3D* | -1.323594884 | 0.015523767 | 0.021502076 | 1 |
| T2DM | monocyte | *MKRN2* | 1.383591443 | 0.001061444 | 0.002301931 | 1 |
| T2DM | monocyte | *MCM6* | 1.927305429 | 0.000115311 | 0.000534632 | 0.72496051 |
| T2DM | monocyte | *RBMS2* | 1.699414202 | 0.000105412 | 0.000507835 | 0.662724185 |
| T2DM | monocyte | *MLH1* | 1.712421449 | 0.000495695 | 0.001337525 | 1 |
| T2DM | monocyte | *FMO4* | 1.847950569 | 0.000181193 | 0.000690399 | 1 |
| T2DM | monocyte | *RGS11* | 0.692714382 | 0.0231421 | 0.030838147 | 1 |
| T2DM | monocyte | *TPD52* | 1.867609334 | 0.000283926 | 0.000915489 | 1 |
| T2DM | monocyte | *GPATCH1* | 2.680483336 | 4.09E-05 | 0.000371379 | 0.256994285 |
| T2DM | monocyte | *NT5C2* | 2.735302721 | 0.001134596 | 0.002413127 | 1 |
| T2DM | monocyte | *CAMSAP3* | 5.237839354 | 0.001947681 | 0.003544611 | 1 |
| T2DM | monocyte | *XAB2* | 1.244927174 | 0.020698543 | 0.027883382 | 1 |
| T2DM | monocyte | *STXBP2* | -0.29387854 | 0.024603298 | 0.032543853 | 1 |
| T2DM | monocyte | *MAP2K7* | 2.243544136 | 0.00013093 | 0.000572307 | 0.823159052 |
| T2DM | monocyte | *CTTNBP2* | 1.267557381 | 0.027106571 | 0.035474399 | 1 |
| T2DM | monocyte | *TM9SF3* | 3.301349016 | 3.86E-05 | 0.00036941 | 0.242772058 |
| T2DM | monocyte | *DNAJC10* | 4.048196763 | 0.000277543 | 0.000902161 | 1 |
| T2DM | monocyte | *DYNC1I2* | 3.654418573 | 8.78E-05 | 0.00046125 | 0.552182647 |
| T2DM | monocyte | *APBB1IP* | 1.872016755 | 0.015654852 | 0.021655017 | 1 |
| T2DM | monocyte | *CAPZB* | 3.140303988 | 0.000198238 | 0.000726217 | 1 |
| T2DM | monocyte | *NAALAD2* | 1.924787647 | 2.70E-05 | 0.000362859 | 0.169748123 |
| T2DM | monocyte | *FGFR1* | 1.037313316 | 0.0005718 | 0.001475743 | 1 |
| T2DM | monocyte | *FBLN1* | 0.539887451 | 0.00058685 | 0.001496766 | 1 |
| T2DM | monocyte | *PIAS2* | 1.877674911 | 0.000152061 | 0.0006224 | 0.956006834 |
| T2DM | monocyte | *AMPH* | 0.517753581 | 0.002116132 | 0.003791428 | 1 |
| T2DM | monocyte | *MCCC1* | 1.37987827 | 0.000139368 | 0.00059075 | 0.876206237 |
| T2DM | monocyte | *FAP* | 0.49747858 | 0.001895497 | 0.003476368 | 1 |
| T2DM | monocyte | *ACER3* | 1.45351873 | 0.001840899 | 0.003406162 | 1 |
| T2DM | monocyte | *PIK3C3* | 3.055847786 | 4.44E-05 | 0.000379941 | 0.279113859 |
| T2DM | monocyte | *N4BP2* | 2.982623034 | 0.000101668 | 0.000497422 | 0.639187125 |
| T2DM | monocyte | *TULP3* | 2.24908642 | 4.68E-05 | 0.000379941 | 0.293944311 |
| T2DM | monocyte | *SYNJ2* | 2.671660945 | 0.000121716 | 0.000552114 | 0.765229407 |
| T2DM | monocyte | *FGF20* | 7.168258614 | 0.00187837 | 0.003452005 | 1 |
| T2DM | monocyte | *PCM1* | 2.395082759 | 5.43E-05 | 0.000387662 | 0.341405008 |
| T2DM | monocyte | *MYH7B* | 2.663136588 | 9.00E-05 | 0.000465291 | 0.565794214 |
| T2DM | monocyte | *FKBP7* | 1.056878302 | 0.008869708 | 0.01311782 | 1 |
| T2DM | monocyte | *OSBPL6* | 2.162998761 | 0.000109834 | 0.000521153 | 0.690527402 |
| T2DM | monocyte | *XRCC5* | 3.4874276 | 4.13E-05 | 0.000373192 | 0.259371515 |
| T2DM | monocyte | *LXN* | 2.682127277 | 5.58E-05 | 0.000390942 | 0.350674673 |
| T2DM | monocyte | *SAR1A* | 1.600782627 | 0.000168155 | 0.000658686 | 1 |
| T2DM | monocyte | *CDC14A* | 2.25254673 | 0.00031829 | 0.000989488 | 1 |
| T2DM | monocyte | *RAPGEF3* | 0.519488667 | 0.002113769 | 0.003788275 | 1 |
| T2DM | monocyte | *FDFT1* | 2.717915144 | 3.12E-05 | 0.000362859 | 0.195975602 |
| T2DM | monocyte | *SCGN* | 4.472760057 | 0.001092511 | 0.00234747 | 1 |

| | | | | | | |
|---|---|---|---|---|---|---|
| T2DM | monocyte | *DDX1* | 4.054188765 | 5.06E-05 | 0.000383728 | 0.318381047 |
| T2DM | monocyte | *DNM2* | 3.415304932 | 0.000759276 | 0.001805434 | 1 |
| T2DM | monocyte | *STX7* | 2.911510524 | 1.37E-05 | 0.000362859 | 0.086242246 |
| T2DM | monocyte | *RABL2B* | 1.871921119 | 0.000242965 | 0.000821174 | 1 |
| T2DM | monocyte | *DDX43* | 0.754725261 | 0.00856026 | 0.012707995 | 1 |
| T2DM | monocyte | *DCT* | 5.260194264 | 0.000102093 | 0.000498724 | 0.641857227 |
| T2DM | monocyte | *SLC35C2* | 4.716601319 | 0.000124969 | 0.000556889 | 0.785678513 |
| T2DM | monocyte | *SCTR* | 0.588431631 | 0.001713432 | 0.003250557 | 1 |
| T2DM | monocyte | *RIF1* | 2.212012487 | 0.000604684 | 0.001529222 | 1 |
| T2DM | monocyte | *KCNN2* | 1.467120879 | 0.004252041 | 0.006861545 | 1 |
| T2DM | monocyte | *CPOX* | 3.301649224 | 0.000406695 | 0.001166997 | 1 |
| T2DM | monocyte | *MOK* | 1.038475475 | 0.000636954 | 0.00158721 | 1 |
| T2DM | monocyte | *RSBN1* | 2.155946847 | 0.00022263 | 0.000778207 | 1 |
| T2DM | monocyte | *MAGI3* | 1.391159929 | 0.000147748 | 0.000612266 | 0.928891718 |
| T2DM | monocyte | *COL4A4* | 1.48147949 | 0.003013624 | 0.00508635 | 1 |
| T2DM | monocyte | *PKP1* | 1.052446465 | 0.032896346 | 0.0423463 | 1 |
| T2DM | monocyte | *ZNF506* | 1.331948754 | 0.00106582 | 0.002308237 | 1 |
| T2DM | monocyte | *PCDHGA2* | -1.319799151 | 0.002944045 | 0.004984974 | 1 |
| T2DM | monocyte | *HSPB11* | 2.923714211 | 1.97E-05 | 0.000362859 | 0.12399134 |
| T2DM | monocyte | *WDR70* | 2.369340161 | 4.00E-05 | 0.000371009 | 0.251627104 |
| T2DM | monocyte | *PGR* | 0.653906281 | 0.00374704 | 0.006131609 | 1 |
| T2DM | monocyte | *C5orf22* | 5.000319003 | 1.74E-05 | 0.000362859 | 0.109159199 |
| T2DM | monocyte | *CCNT2* | 2.555790677 | 4.95E-05 | 0.000383513 | 0.311049771 |
| T2DM | monocyte | *GEMIN5* | 2.001113457 | 0.001035003 | 0.002259396 | 1 |
| T2DM | monocyte | *GSK3B* | 3.047958247 | 1.93E-05 | 0.000362859 | 0.121412407 |
| T2DM | monocyte | *ERC1* | 1.833909534 | 4.23E-05 | 0.00037505 | 0.266210193 |
| T2DM | monocyte | *DOPEY1* | 1.986632414 | 0.000123123 | 0.000553734 | 0.774076045 |
| T2DM | monocyte | *LYRM2* | 1.948844495 | 0.005489854 | 0.008598583 | 1 |
| T2DM | monocyte | *PLOD1* | 3.350222627 | 4.38E-05 | 0.000378997 | 0.275143896 |
| T2DM | monocyte | *PIBF1* | 2.398822773 | 7.12E-05 | 0.000427286 | 0.447778825 |
| T2DM | monocyte | *TDRD3* | 1.947815951 | 5.21E-05 | 0.000385178 | 0.327786166 |
| T2DM | monocyte | *PDS5B* | 3.093869991 | 4.12E-05 | 0.000373192 | 0.259071976 |
| T2DM | monocyte | *OXCT1* | 1.041662148 | 0.00116603 | 0.00246001 | 1 |
| T2DM | monocyte | *ZNF324* | 1.984893384 | 0.000336366 | 0.00102161 | 1 |
| T2DM | monocyte | *ZNF586* | 2.346742938 | 2.20E-05 | 0.000362859 | 0.138512979 |
| T2DM | monocyte | *ZNF446* | 7.764772464 | 2.55E-05 | 0.000362859 | 0.16050208 |
| T2DM | monocyte | *ZNF264* | 2.075155086 | 0.000137938 | 0.000587547 | 0.867218922 |
| T2DM | monocyte | *RPS5* | 3.957809004 | 4.56E-05 | 0.000379941 | 0.286926568 |
| T2DM | monocyte | *CHMP2B* | 2.606787271 | 0.003464481 | 0.005750051 | 1 |
| T2DM | monocyte | *PPIE* | 1.151538772 | 0.001148927 | 0.002434548 | 1 |
| T2DM | monocyte | *ZMPSTE24* | 4.520508234 | 5.94E-05 | 0.000399203 | 0.373541222 |
| T2DM | monocyte | *STARD7* | 3.809015393 | 0.002187255 | 0.003883679 | 1 |
| T2DM | monocyte | *NOA1* | 1.933694308 | 0.034981424 | 0.044846699 | 1 |
| T2DM | monocyte | *GSTP1* | 2.625277549 | 4.17E-05 | 0.00037367 | 0.261944925 |
| T2DM | monocyte | *WBP11* | 4.685556189 | 2.44E-05 | 0.000362859 | 0.153518792 |
| T2DM | monocyte | *COL16A1* | 0.476276125 | 0.006356235 | 0.009784929 | 1 |
| T2DM | monocyte | *APOB* | 1.045955319 | 0.003097804 | 0.005213034 | 1 |
| T2DM | monocyte | *NCOA1* | 2.819477604 | 3.18E-05 | 0.000363519 | 0.200143777 |
| T2DM | monocyte | *CD59* | 1.900634312 | 6.00E-05 | 0.000399998 | 0.377456538 |
| T2DM | monocyte | *CD82* | 3.119486627 | 4.79E-06 | 0.000292389 | 0.030116088 |

| | | | | | | |
|---|---|---|---|---|---|---|
| T2DM | monocyte | *TAF9* | 3.782621117 | 6.76E-05 | 0.0004219 | 0.425275023 |
| T2DM | monocyte | *FCN1* | -0.056045923 | 0.006544774 | 0.010028514 | 1 |
| T2DM | monocyte | *SCAMP1* | 2.778985943 | 0.005190047 | 0.008177901 | 1 |
| T2DM | monocyte | *HACE1* | 2.483024815 | 0.00013787 | 0.000587547 | 0.8667892 |
| T2DM | monocyte | *SEH1L* | 2.860843038 | 0.000122836 | 0.000553734 | 0.77227147 |
| T2DM | monocyte | *SLC25A24* | 2.099173431 | 4.73E-05 | 0.000379941 | 0.297284596 |
| T2DM | monocyte | *PILRA* | -0.151278491 | 0.028240147 | 0.036881139 | 1 |
| T2DM | monocyte | *ABCB1* | 0.997946678 | 0.000948627 | 0.002117152 | 1 |
| T2DM | monocyte | *ZNF213* | 2.26566504 | 6.25E-05 | 0.000407346 | 0.393089241 |
| T2DM | monocyte | *CPNE3* | 1.690612315 | 9.96E-06 | 0.000362859 | 0.062598084 |
| T2DM | monocyte | *RRN3* | 2.211005244 | 0.000130645 | 0.000572307 | 0.821364002 |
| T2DM | monocyte | *CTTN* | 0.812931078 | 0.000672415 | 0.001649073 | 1 |
| T2DM | monocyte | *MTIF2* | 2.169265386 | 0.001967221 | 0.003575576 | 1 |
| T2DM | monocyte | *EPS15* | 2.118449563 | 1.63E-05 | 0.000362859 | 0.102475297 |
| T2DM | monocyte | *ATG16L1* | 5.346171303 | 2.58E-06 | 0.000207301 | 0.016240955 |
| T2DM | monocyte | *USP40* | 1.599779149 | 0.000154431 | 0.000629232 | 0.970905413 |
| T2DM | monocyte | *MAST2* | 3.100192506 | 0.000251054 | 0.000841352 | 1 |
| T2DM | monocyte | *B4GALT1* | 3.863553351 | 3.51E-05 | 0.000365043 | 0.220636164 |
| T2DM | monocyte | *CHMP5* | 3.672971347 | 1.53E-05 | 0.000362859 | 0.096169901 |
| T2DM | monocyte | *FOLH1* | 0.68032274 | 0.024266509 | 0.032172826 | 1 |
| T2DM | monocyte | *EIF2AK1* | 2.20949308 | 2.12E-05 | 0.000362859 | 0.133190515 |
| T2DM | monocyte | *EPDR1* | 1.82889198 | 3.55E-05 | 0.000365043 | 0.22297281 |
| T2DM | monocyte | *SNX10* | -0.584935813 | 0.00069807 | 0.00169385 | 1 |
| T2DM | monocyte | *SEPHS1* | 4.04366391 | 2.43E-05 | 0.000362859 | 0.153067808 |
| T2DM | monocyte | *ITPKC* | 1.395325944 | 0.000313828 | 0.000980148 | 1 |
| T2DM | monocyte | *TMED2* | 1.706867478 | 0.000240286 | 0.000817468 | 1 |
| T2DM | monocyte | *ACOX3* | 1.543597486 | 0.000193164 | 0.000718174 | 1 |
| T2DM | monocyte | *HSD17B14* | 1.103428369 | 0.007006331 | 0.010647524 | 1 |
| T2DM | monocyte | *TRIP6* | 2.248321621 | 0.003163973 | 0.005313008 | 1 |
| T2DM | monocyte | *SRRT* | 1.848562957 | 0.001479236 | 0.002929121 | 1 |
| T2DM | monocyte | *PIGS* | 1.518615565 | 0.003308889 | 0.005522427 | 1 |
| T2DM | monocyte | *PGS1* | 2.880457891 | 0.015065357 | 0.020945577 | 1 |
| T2DM | monocyte | *PSMC5* | 0.817601676 | 0.004556775 | 0.007282268 | 1 |
| T2DM | monocyte | *OGFOD1* | 1.28601241 | 0.002890648 | 0.004907779 | 1 |
| T2DM | monocyte | *TXNDC16* | 2.660002917 | 0.000195385 | 0.000722579 | 1 |
| T2DM | monocyte | *GMCL1* | 4.241865773 | 3.67E-05 | 0.000365515 | 0.2308404 |
| T2DM | monocyte | *KLHL42* | 3.373053728 | 8.85E-06 | 0.000362859 | 0.055633747 |
| T2DM | monocyte | *PTHLH* | 0.959429394 | 0.001672719 | 0.003198414 | 1 |
| T2DM | monocyte | *PHACTR3* | 0.652113217 | 0.000938355 | 0.002099444 | 1 |
| T2DM | monocyte | *ERGIC2* | 2.059260541 | 0.000133618 | 0.000578424 | 0.840058373 |
| T2DM | monocyte | *METTL2A* | 0.944467308 | 0.00375035 | 0.006135429 | 1 |
| T2DM | monocyte | *ALG6* | 2.8973632 | 0.009235094 | 0.013606993 | 1 |
| T2DM | monocyte | *GP6* | -0.810998004 | 0.01146727 | 0.016463742 | 1 |
| T2DM | monocyte | *DDX18* | 3.829661731 | 1.16E-05 | 0.000362859 | 0.072874634 |
| T2DM | monocyte | *ASAP3* | 0.690567047 | 0.001369484 | 0.002771144 | 1 |
| T2DM | monocyte | *TPX2* | 0.722820127 | 0.025889755 | 0.034044947 | 1 |
| T2DM | monocyte | *DOCK3* | 0.781669524 | 0.002685734 | 0.004619757 | 1 |
| T2DM | monocyte | *C3orf18* | 7.3693133 | 8.49E-10 | 7.62E-07 | 5.34E-06 |
| T2DM | monocyte | *FKBP1A* | 4.137659311 | 4.15E-05 | 0.00037367 | 0.261201345 |
| T2DM | monocyte | *NSFL1C* | 3.57679718 | 5.45E-05 | 0.000387662 | 0.34234422 |

| | | | | | | |
|---|---|---|---|---|---|---|
| T2DM | monocyte | *ZNF343* | 2.86385599 | 4.17E-05 | 0.00037367 | 0.262313556 |
| T2DM | monocyte | *MAVS* | 2.891971269 | 2.10E-05 | 0.000362859 | 0.131808978 |
| T2DM | monocyte | *LZTS3* | 1.413592848 | 0.003365491 | 0.005603508 | 1 |
| T2DM | monocyte | *XRN2* | 3.737833655 | 7.71E-05 | 0.00043763 | 0.484476188 |
| T2DM | monocyte | *TESC* | -0.629162374 | 0.021427135 | 0.028772404 | 1 |
| T2DM | monocyte | *RBBP9* | 4.270970997 | 0.000206696 | 0.000743876 | 1 |
| T2DM | monocyte | *ANAPC5* | 2.286656428 | 7.74E-05 | 0.000437657 | 0.486674051 |
| T2DM | monocyte | *SLC23A2* | 3.875330084 | 9.16E-05 | 0.000469636 | 0.575589241 |
| T2DM | monocyte | *TMEM230* | 2.580861816 | 2.12E-05 | 0.000362859 | 0.133585049 |
| T2DM | monocyte | *DZANK1* | 1.444950354 | 0.000845431 | 0.001950526 | 1 |
| T2DM | monocyte | *TASP1* | 1.741792484 | 0.000222086 | 0.000777859 | 1 |
| T2DM | monocyte | *OAS1* | -0.503113667 | 0.023621484 | 0.031410379 | 1 |
| T2DM | monocyte | *KIF16B* | 2.118342576 | 7.58E-05 | 0.000435317 | 0.476671759 |
| T2DM | monocyte | *FUS* | 3.083041176 | 2.05E-05 | 0.000362859 | 0.129162435 |
| T2DM | monocyte | *ZNF302* | 0.950031036 | 0.014406237 | 0.020140541 | 1 |
| T2DM | monocyte | *FXYD3* | 3.269845871 | 0.005081875 | 0.008031611 | 1 |
| T2DM | monocyte | *KCNH4* | 1.413813908 | 0.001236207 | 0.002563289 | 1 |
| T2DM | monocyte | *GANAB* | 2.625672873 | 5.88E-05 | 0.000398946 | 0.369915372 |
| T2DM | monocyte | *GMIP* | -0.453849545 | 0.016026208 | 0.022129754 | 1 |
| T2DM | monocyte | *MLF2* | 3.250375862 | 6.99E-05 | 0.000427286 | 0.439562906 |
| T2DM | monocyte | *NECAP1* | 3.668133931 | 0.001151944 | 0.002436759 | 1 |
| T2DM | monocyte | *ANKRD24* | 2.084023886 | 0.000601886 | 0.001524272 | 1 |
| T2DM | monocyte | *GPATCH2L* | 2.494849274 | 4.87E-05 | 0.000381649 | 0.306115699 |
| T2DM | monocyte | *SLC9A1* | 4.368075398 | 0.002526279 | 0.004380231 | 1 |
| T2DM | monocyte | *SPTLC1* | 2.757239867 | 0.000567213 | 0.00146776 | 1 |
| T2DM | monocyte | *PAPOLA* | 3.260654234 | 2.90E-05 | 0.000362859 | 0.182407201 |
| T2DM | monocyte | *RGS1* | 1.366330459 | 0.001185221 | 0.002488805 | 1 |
| T2DM | monocyte | *YPEL3* | 1.94717148 | 2.26E-05 | 0.000362859 | 0.142361529 |
| T2DM | monocyte | *STRN4* | 4.880404103 | 0.000900839 | 0.002036525 | 1 |
| T2DM | monocyte | *LYZ* | -0.212615703 | 0.016684103 | 0.022952507 | 1 |
| T2DM | monocyte | *SPG21* | 2.221520899 | 0.021912355 | 0.029348738 | 1 |
| T2DM | monocyte | *GNPTG* | 4.586829509 | 0.000377424 | 0.001105714 | 1 |
| T2DM | monocyte | *ZNF268* | 2.268829072 | 0.001446016 | 0.002888039 | 1 |
| T2DM | monocyte | *GOLGA3* | 2.954348549 | 9.62E-06 | 0.000362859 | 0.060510513 |
| T2DM | monocyte | *PABPC4* | -2.746335509 | 0.000464816 | 0.001277145 | 1 |
| T2DM | monocyte | *CERS4* | 1.179715616 | 0.000579413 | 0.001487717 | 1 |
| T2DM | monocyte | *USP48* | 1.974395973 | 0.000452825 | 0.001253041 | 1 |
| T2DM | monocyte | *PDPR* | 4.528093296 | 4.39E-05 | 0.000378997 | 0.276210747 |
| T2DM | monocyte | *EXOC1* | 2.958403108 | 2.34E-05 | 0.000362859 | 0.146843091 |
| T2DM | monocyte | *NLRC4* | -0.327404523 | 0.013994696 | 0.019630668 | 1 |
| T2DM | monocyte | *LAMB1* | 0.877683727 | 0.00567099 | 0.008851419 | 1 |
| T2DM | monocyte | *SLC26A4* | 6.719295729 | 0.000436581 | 0.001224257 | 1 |
| T2DM | monocyte | *DLD* | 1.686799579 | 0.000130988 | 0.000572307 | 0.823520586 |
| T2DM | monocyte | *TXNL1* | 2.432022154 | 2.43E-05 | 0.000362859 | 0.152685628 |
| T2DM | monocyte | *IL5RA* | 1.719610221 | 0.002066225 | 0.003718969 | 1 |
| T2DM | monocyte | *ITGA6* | 0.873785051 | 0.000938884 | 0.002099819 | 1 |
| T2DM | monocyte | *RAPGEF4* | 0.702570829 | 0.000771125 | 0.001822488 | 1 |
| T2DM | monocyte | *SEL1L3* | 2.131162336 | 7.68E-05 | 0.000437295 | 0.482617731 |
| T2DM | monocyte | *TF* | 0.762914677 | 0.001855129 | 0.003422505 | 1 |
| T2DM | monocyte | *MYO15A* | 2.140195383 | 9.76E-05 | 0.000485446 | 0.613520702 |

| | | | | | | |
|---|---|---|---|---|---|---|
| T2DM | monocyte | *ZFHX4* | 0.509547237 | 0.001396928 | 0.002810396 | 1 |
| T2DM | monocyte | *RGS17* | 0.988614665 | 0.003872644 | 0.006319053 | 1 |
| T2DM | monocyte | *CMA1* | 2.419216335 | 0.005468909 | 0.008574322 | 1 |
| T2DM | monocyte | *PPP2R3C* | 3.990421095 | 2.65E-05 | 0.000362859 | 0.166430903 |
| T2DM | monocyte | *OSGEP* | 2.216823459 | 0.000114113 | 0.000530249 | 0.717427022 |
| T2DM | monocyte | *SLC22A17* | 0.951233497 | 0.009839172 | 0.014369504 | 1 |
| T2DM | monocyte | *SCFD1* | 2.733552224 | 1.35E-05 | 0.000362859 | 0.084771272 |
| T2DM | monocyte | *RPGRIP1* | 3.849876978 | 8.43E-05 | 0.000454809 | 0.530307786 |
| T2DM | monocyte | *TOX4* | 3.20260736 | 4.67E-05 | 0.000379941 | 0.293463827 |
| T2DM | monocyte | *RFFL* | 6.670309739 | 3.63E-05 | 0.000365043 | 0.228316662 |
| T2DM | monocyte | *MFSD11* | 2.338388191 | 9.68E-05 | 0.000485026 | 0.608707438 |
| T2DM | monocyte | *DPYSL2* | 2.658719035 | 3.02E-05 | 0.000362859 | 0.190092736 |
| T2DM | monocyte | *COMT* | 3.560171953 | 2.45E-05 | 0.000362859 | 0.154021606 |
| T2DM | monocyte | *VNN3* | -0.266390582 | 0.007980335 | 0.011940116 | 1 |
| T2DM | monocyte | *SEC22C* | 2.964682301 | 1.97E-05 | 0.000362859 | 0.123801142 |
| T2DM | monocyte | *FMO2* | 0.970089167 | 0.000207594 | 0.000745798 | 1 |
| T2DM | monocyte | *MSH2* | 2.077914986 | 0.000123218 | 0.000553734 | 0.774674111 |
| T2DM | monocyte | *MAP3K1* | -0.506853573 | 0.032363924 | 0.041729285 | 1 |
| T2DM | monocyte | *TMEM38B* | 2.900519925 | 9.74E-05 | 0.000485446 | 0.612547185 |
| T2DM | monocyte | *PDE6C* | 2.651151587 | 0.001152296 | 0.002436759 | 1 |
| T2DM | monocyte | *IKZF5* | 2.746329923 | 6.61E-05 | 0.00041942 | 0.415645614 |
| T2DM | monocyte | *BLNK* | 1.647204112 | 0.001038581 | 0.002264062 | 1 |
| T2DM | monocyte | *WAC* | 2.64074971 | 0.000944839 | 0.002110809 | 1 |
| T2DM | monocyte | *CREM* | 2.84914752 | 2.04E-05 | 0.000362859 | 0.127997126 |
| T2DM | monocyte | *TPSD1* | 6.399583359 | 0.000664974 | 0.00163686 | 1 |
| T2DM | monocyte | *ITPR3* | 1.430582366 | 0.000331675 | 0.001015705 | 1 |
| T2DM | monocyte | *DSP* | 1.267721599 | 0.022343504 | 0.029881645 | 1 |
| T2DM | monocyte | *IFT74* | 1.592542337 | 0.000866575 | 0.001984033 | 1 |
| T2DM | monocyte | *IL12RB1* | -0.510914158 | 0.025187925 | 0.033219317 | 1 |
| T2DM | monocyte | *PCSK5* | 1.130591169 | 0.000180349 | 0.000688301 | 1 |
| T2DM | monocyte | *RAB18* | 2.745740304 | 4.97E-05 | 0.000383513 | 0.312561531 |
| T2DM | monocyte | *NRP1* | 1.147315524 | 0.000149266 | 0.000617391 | 0.9384347 |
| T2DM | monocyte | *PRTFDC1* | 0.604688998 | 0.001894727 | 0.003476368 | 1 |
| T2DM | monocyte | *PALMD* | 0.615889949 | 0.000516939 | 0.001380375 | 1 |
| T2DM | monocyte | *TSPAN15* | 0.788349154 | 0.0008879 | 0.002015244 | 1 |
| T2DM | monocyte | *OCEL1* | 2.341478983 | 0.000904996 | 0.002042944 | 1 |
| T2DM | monocyte | *CATSPERG* | 1.367493086 | 0.00388793 | 0.006342351 | 1 |
| T2DM | monocyte | *STX1B* | 0.95081964 | 0.021999359 | 0.029452719 | 1 |
| T2DM | monocyte | *HSD3B7* | 1.434090561 | 0.000619912 | 0.001553363 | 1 |
| T2DM | monocyte | *ATP5D* | 1.971267753 | 0.000500289 | 0.001345878 | 1 |
| T2DM | monocyte | *MARCH2* | 5.408080573 | 4.87E-07 | 7.60E-05 | 0.003060303 |
| T2DM | monocyte | *CDC34* | 3.645625593 | 0.032059951 | 0.041379781 | 1 |
| T2DM | monocyte | *MTAP* | 2.166820895 | 0.000165041 | 0.000652176 | 1 |
| T2DM | monocyte | *CEP170B* | 1.208742984 | 0.005119584 | 0.00808308 | 1 |
| T2DM | monocyte | *POLR2E* | 2.614633342 | 0.00021786 | 0.000768284 | 1 |
| T2DM | monocyte | *MADCAM1* | 0.725890228 | 0.001623939 | 0.003136622 | 1 |
| T2DM | monocyte | *ZDHHC8* | 2.583581344 | 0.000147832 | 0.000612266 | 0.929420053 |
| T2DM | monocyte | *MED15* | 3.194516394 | 3.11E-05 | 0.000362859 | 0.195332191 |
| T2DM | monocyte | *LZTR1* | 2.713736272 | 8.82E-05 | 0.000461502 | 0.55426425 |
| T2DM | monocyte | *CECR2* | 0.548643138 | 0.003535771 | 0.005849839 | 1 |

| | | | | | | |
|---|---|---|---|---|---|---|
| T2DM | monocyte | *SMARCB1* | 1.996705571 | 0.003296707 | 0.005503558 | 1 |
| T2DM | monocyte | *P2RX6* | 1.363382949 | 0.002359735 | 0.004131344 | 1 |
| T2DM | monocyte | *SLC7A4* | 2.417939582 | 0.013211313 | 0.01863993 | 1 |
| T2DM | monocyte | *DDT* | 3.822900508 | 0.000309095 | 0.000971153 | 1 |
| T2DM | monocyte | *GGT5* | 0.807145591 | 0.001114017 | 0.002381442 | 1 |
| T2DM | monocyte | *SPECC1L* | 1.188441562 | 0.005187936 | 0.008176624 | 1 |
| T2DM | monocyte | *UPB1* | 1.112004109 | 0.015058015 | 0.020939999 | 1 |
| T2DM | monocyte | *YPEL1* | 1.924524047 | 0.000228845 | 0.000790087 | 1 |
| T2DM | monocyte | *TOP3B* | 2.873584594 | 0.002798348 | 0.004769102 | 1 |
| T2DM | monocyte | *CRYBB3* | 1.834307592 | 0.009822492 | 0.01435472 | 1 |
| T2DM | monocyte | *CYTH4* | -0.470747752 | 0.029217195 | 0.038054383 | 1 |
| T2DM | monocyte | *CARD10* | 0.986496214 | 0.001496717 | 0.002953704 | 1 |
| T2DM | monocyte | *SLC25A1* | 2.665734105 | 0.000107551 | 0.000515377 | 0.676174163 |
| T2DM | monocyte | *LGALS2* | -0.487699157 | 0.018840908 | 0.02556719 | 1 |
| T2DM | monocyte | *GGA1* | 1.654582626 | 0.014988575 | 0.020857275 | 1 |
| T2DM | monocyte | *SH3BP1* | -0.681367277 | 0.021416833 | 0.028764715 | 1 |
| T2DM | monocyte | *HPS4* | 2.811088195 | 3.27E-05 | 0.000365043 | 0.205563326 |
| T2DM | monocyte | *PIK3IP1* | 1.946885923 | 0.00260223 | 0.004497037 | 1 |
| T2DM | monocyte | *TRIOBP* | 2.264354937 | 0.000147053 | 0.000610432 | 0.924525022 |
| T2DM | monocyte | *TFIP11* | 2.588907022 | 3.86E-05 | 0.00036941 | 0.242937934 |
| T2DM | monocyte | *GCAT* | 1.068636898 | 0.00453354 | 0.007248822 | 1 |
| T2DM | monocyte | *ANKRD54* | 0.696826183 | 0.025723062 | 0.03386825 | 1 |
| T2DM | monocyte | *EIF3L* | 2.245770731 | 0.005779438 | 0.008998347 | 1 |
| T2DM | monocyte | *SLC16A8* | 1.492772814 | 0.006004721 | 0.009307614 | 1 |
| T2DM | monocyte | *KDELR3* | 1.670523565 | 0.000836392 | 0.001933944 | 1 |
| T2DM | monocyte | *DDX17* | 1.969067437 | 6.66E-05 | 0.000420641 | 0.418958843 |
| T2DM | monocyte | *FBXO7* | 2.784246611 | 5.85E-05 | 0.000397957 | 0.367712488 |
| T2DM | monocyte | *POLDIP3* | 2.862694513 | 7.00E-05 | 0.000427286 | 0.439872453 |
| T2DM | monocyte | *TIMP3* | 1.394467837 | 8.13E-05 | 0.000445492 | 0.511298741 |
| T2DM | monocyte | *PPP6R2* | 2.231228198 | 0.001498684 | 0.002955529 | 1 |
| T2DM | monocyte | *CYB5R3* | 2.598818415 | 7.70E-05 | 0.00043763 | 0.483883496 |
| T2DM | monocyte | *RHBDD3* | 0.854954152 | 0.001103135 | 0.002364792 | 1 |
| T2DM | monocyte | *TTLL1* | 3.127758788 | 0.000169974 | 0.000662097 | 1 |
| T2DM | monocyte | *RASL10A* | 2.282401073 | 0.030633684 | 0.039677374 | 1 |
| T2DM | monocyte | *AP1B1* | 2.744382878 | 0.000521336 | 0.001387654 | 1 |
| T2DM | monocyte | *HMGXB4* | 3.07690447 | 2.83E-05 | 0.000362859 | 0.177670293 |
| T2DM | monocyte | *TOM1* | 1.619772075 | 0.000242795 | 0.000821174 | 1 |
| T2DM | monocyte | *CHKB* | 0.749271763 | 0.005890181 | 0.009143597 | 1 |
| T2DM | monocyte | *BIK* | 3.780903398 | 0.0013225 | 0.002699532 | 1 |
| T2DM | monocyte | *MCAT* | 1.34719734 | 0.003737389 | 0.006119001 | 1 |
| T2DM | monocyte | *THOC5* | 2.207408164 | 0.020001934 | 0.027014428 | 1 |
| T2DM | monocyte | *ARSA* | 0.927803866 | 0.005102092 | 0.00805951 | 1 |
| T2DM | monocyte | *TTLL12* | 2.553847691 | 0.000453381 | 0.001253852 | 1 |
| T2DM | monocyte | *RBFOX2* | 1.324429553 | 0.000136909 | 0.000584748 | 0.860748805 |
| T2DM | monocyte | *MTMR3* | 1.785447035 | 0.002016798 | 0.00365091 | 1 |
| T2DM | monocyte | *APOL4* | 0.941346301 | 0.011934661 | 0.017056879 | 1 |
| T2DM | monocyte | *SAMM50* | 2.819609821 | 5.03E-05 | 0.000383643 | 0.316450694 |
| T2DM | monocyte | *TXN2* | 3.490926167 | 2.07E-05 | 0.000362859 | 0.130137195 |
| T2DM | monocyte | *FOXRED2* | 2.059983588 | 0.000310046 | 0.000972198 | 1 |
| T2DM | monocyte | *IFT27* | 3.44613951 | 5.27E-05 | 0.000385235 | 0.33101984 |

| | | | | | | |
|---|---|---|---|---|---|---|
| T2DM | monocyte | *NCF4* | 0.929552294 | 0.001316175 | 0.002691604 | 1 |
| T2DM | monocyte | *SLC25A17* | 2.312456813 | 6.80E-05 | 0.000422315 | 0.427382549 |
| T2DM | monocyte | *KCTD17* | -0.649112211 | 0.01189665 | 0.017014158 | 1 |
| T2DM | monocyte | *EP300* | 3.008367537 | 2.58E-05 | 0.000362859 | 0.162062711 |
| T2DM | monocyte | *ACO2* | 2.490852841 | 3.56E-05 | 0.000365043 | 0.223774629 |
| T2DM | monocyte | *POLR3H* | 0.919540598 | 0.02151975 | 0.028865852 | 1 |
| T2DM | monocyte | *DESI1* | 2.57374442 | 4.14E-05 | 0.000373192 | 0.260075403 |
| T2DM | monocyte | *ZBED4* | 3.989437271 | 2.62E-05 | 0.000362859 | 0.164410198 |
| T2DM | monocyte | *HDAC10* | 2.48482451 | 0.004435137 | 0.007111376 | 1 |
| T2DM | monocyte | *SDR39U1* | 0.591737809 | 0.032097423 | 0.041419643 | 1 |
| T2DM | monocyte | *RBM23* | 3.827585555 | 3.97E-05 | 0.000371009 | 0.249560652 |
| T2DM | monocyte | *VCPKMT* | 2.394804337 | 0.000356598 | 0.001062714 | 1 |
| T2DM | monocyte | *SOS2* | 2.876996498 | 2.40E-05 | 0.000362859 | 0.150820657 |
| T2DM | monocyte | *PYGL* | 0.745522114 | 0.001198718 | 0.002507933 | 1 |
| T2DM | monocyte | *TRIM9* | 0.978098423 | 0.002878681 | 0.004888781 | 1 |
| T2DM | monocyte | *DDHD1* | 3.112214245 | 0.000183057 | 0.000695815 | 1 |
| T2DM | monocyte | *CNIH1* | 3.365171346 | 8.92E-05 | 0.000464259 | 0.560824669 |
| T2DM | monocyte | *PIGH* | 2.687731188 | 3.60E-05 | 0.000365043 | 0.226265041 |
| T2DM | monocyte | *VTI1B* | 2.664198013 | 0.000105786 | 0.000509246 | 0.665075748 |
| T2DM | monocyte | *GSTZ1* | 1.757815411 | 0.008286226 | 0.01235076 | 1 |
| T2DM | monocyte | *TMED8* | 1.947523667 | 0.006618004 | 0.010133315 | 1 |
| T2DM | monocyte | *DAAM1* | 0.982283233 | 0.000396837 | 0.001146558 | 1 |
| T2DM | monocyte | *PPM1A* | 2.745035904 | 5.71E-06 | 0.000322038 | 0.035917341 |
| T2DM | monocyte | *SLC10A1* | 1.643844323 | 0.000571653 | 0.001475743 | 1 |
| T2DM | monocyte | *DICER1* | 2.819011383 | 0.000407537 | 0.001168881 | 1 |
| T2DM | monocyte | *ZFYVE21* | 2.682260862 | 4.52E-05 | 0.000379941 | 0.284040136 |
| T2DM | monocyte | *MTHFD1* | 0.858886213 | 0.000791133 | 0.001857996 | 1 |
| T2DM | monocyte | *VRK1* | 4.615251983 | 3.75E-05 | 0.000367486 | 0.235558388 |
| T2DM | monocyte | *PAPLN* | 2.185667283 | 7.54E-05 | 0.000435317 | 0.474242993 |
| T2DM | monocyte | *PSMB5* | 2.180185136 | 1.04E-05 | 0.000362859 | 0.065684162 |
| T2DM | monocyte | *CCNB1IP1* | 2.994966654 | 0.000492581 | 0.001333243 | 1 |
| T2DM | monocyte | *TRIP11* | 2.481287478 | 3.56E-05 | 0.000365043 | 0.223516992 |
| T2DM | monocyte | *KIAA0391* | 1.158617043 | 0.001795407 | 0.003358443 | 1 |
| T2DM | monocyte | *EMC9* | 0.847313739 | 0.006872857 | 0.010472529 | 1 |
| T2DM | monocyte | *PLTP* | 1.043910348 | 3.30E-05 | 0.000365043 | 0.207377282 |
| T2DM | monocyte | *GSS* | 1.634464002 | 0.000123616 | 0.000554584 | 0.777172097 |
| T2DM | monocyte | *PYGB* | 1.323772943 | 0.001510904 | 0.002973099 | 1 |
| T2DM | monocyte | *ABHD12* | 2.873507214 | 6.55E-05 | 0.000417788 | 0.411778347 |
| T2DM | monocyte | *PROCR* | 0.757240835 | 0.014614209 | 0.020413137 | 1 |
| T2DM | monocyte | *NINL* | 1.317113648 | 0.000129466 | 0.000569596 | 0.813952287 |
| T2DM | monocyte | *CD40* | 3.053247205 | 6.66E-05 | 0.000420641 | 0.418763954 |
| T2DM | monocyte | *UQCC1* | 1.189235747 | 0.004401525 | 0.00706862 | 1 |
| T2DM | monocyte | *IFT52* | 3.417751817 | 6.27E-06 | 0.000322862 | 0.039389162 |
| T2DM | monocyte | *NDRG3* | 2.529300952 | 0.032415695 | 0.041787466 | 1 |
| T2DM | monocyte | *DOK5* | 1.368754965 | 0.0004107 | 0.00117346 | 1 |
| T2DM | monocyte | *BMP7* | 0.878036543 | 0.001748503 | 0.003296203 | 1 |
| T2DM | monocyte | *NELFCD* | 3.334904367 | 3.55E-05 | 0.000365043 | 0.222924315 |
| T2DM | monocyte | *PRPF6* | 2.892535452 | 0.000125166 | 0.000556913 | 0.786917829 |
| T2DM | monocyte | *MTG2* | 1.601528434 | 0.000512762 | 0.001370635 | 1 |
| T2DM | monocyte | *DIDO1* | 2.655856337 | 4.32E-06 | 0.000277801 | 0.027188739 |

| | | | | | | |
|---|---|---|---|---|---|---|
| T2DM | monocyte | *GID8* | 4.546497461 | 0.000411568 | 0.001175081 | 1 |
| T2DM | monocyte | *SLC17A9* | -0.681359091 | 0.033286112 | 0.042804735 | 1 |
| T2DM | monocyte | *BIRC7* | -2.255126724 | 0.020736986 | 0.027917223 | 1 |
| T2DM | monocyte | *NKAIN4* | 1.653555837 | 0.017054375 | 0.023386887 | 1 |
| T2DM | monocyte | *PTK6* | -1.064554908 | 0.013287201 | 0.018742794 | 1 |
| T2DM | monocyte | *GMEB2* | 4.498000212 | 0.000235745 | 0.000805942 | 1 |
| T2DM | monocyte | *CDC25B* | 3.000873009 | 0.005996978 | 0.009297904 | 1 |
| T2DM | monocyte | *SLC52A3* | 2.054115788 | 0.020150138 | 0.027197063 | 1 |
| T2DM | monocyte | *ANGPT4* | 0.978731448 | 0.00271736 | 0.004662675 | 1 |
| T2DM | monocyte | *RSPO4* | 2.220607506 | 0.004068326 | 0.006600662 | 1 |
| T2DM | monocyte | *SIRPB1* | -0.227141888 | 0.009927917 | 0.014481859 | 1 |
| T2DM | monocyte | *CCM2L* | 0.865464209 | 0.001473225 | 0.002921819 | 1 |
| T2DM | monocyte | *MYL9* | 1.62375213 | 0.000116485 | 0.000538484 | 0.732338357 |
| T2DM | monocyte | *MROH8* | 1.17328108 | 0.011979537 | 0.017101671 | 1 |
| T2DM | monocyte | *IDH3B* | 2.130890809 | 0.000198466 | 0.000726217 | 1 |
| T2DM | monocyte | *MAPRE1* | 4.538639911 | 0.00117567 | 0.002474535 | 1 |
| T2DM | monocyte | *TTI1* | 2.496311733 | 9.98E-05 | 0.000492855 | 0.627479085 |
| T2DM | monocyte | *RPRD1B* | 2.146267824 | 0.00017727 | 0.000680399 | 1 |
| T2DM | monocyte | *BPI* | 3.458178467 | 0.002775457 | 0.004735223 | 1 |
| T2DM | monocyte | *ACTR5* | 3.808384845 | 0.000177169 | 0.000680399 | 1 |
| T2DM | monocyte | *DHX35* | 4.263827585 | 5.31E-05 | 0.000385235 | 0.333730208 |
| T2DM | monocyte | *DNTTIP1* | 1.217949349 | 0.02686758 | 0.035227628 | 1 |
| T2DM | monocyte | *MAP1LC3A* | 1.548853703 | 1.99E-05 | 0.000362859 | 0.124837324 |
| T2DM | monocyte | *PIGU* | 3.80538418 | 2.86E-05 | 0.000362859 | 0.179522405 |
| T2DM | monocyte | *APMAP* | 1.122277472 | 0.000426415 | 0.001203701 | 1 |
| T2DM | monocyte | *CELF4* | 1.686542879 | 0.00080916 | 0.00188834 | 1 |
| T2DM | monocyte | *RBFA* | 4.280782241 | 0.000314564 | 0.000981471 | 1 |
| T2DM | monocyte | *METTL4* | 2.327248275 | 9.08E-05 | 0.000467343 | 0.570625727 |
| T2DM | monocyte | *LPIN2* | 3.033342656 | 1.78E-05 | 0.000362859 | 0.111619493 |
| T2DM | monocyte | *SMCHD1* | 2.879492096 | 0.000598619 | 0.001521227 | 1 |
| T2DM | monocyte | *MYL12A* | 3.6862096 | 0.00177394 | 0.003333161 | 1 |
| T2DM | monocyte | *CEP76* | 1.98161836 | 0.001190623 | 0.002495148 | 1 |
| T2DM | monocyte | *POLI* | 0.50454595 | 0.018180377 | 0.024767071 | 1 |
| T2DM | monocyte | *NALCN* | 0.754738365 | 0.001587081 | 0.003089932 | 1 |
| T2DM | monocyte | *HTR2A* | 1.881325901 | 0.001783625 | 0.003344363 | 1 |
| T2DM | monocyte | *KLF5* | 1.575430365 | 0.005330039 | 0.008371211 | 1 |
| T2DM | monocyte | *STK24* | 2.529369498 | 0.000125801 | 0.000558159 | 0.790910789 |
| T2DM | monocyte | *DNAJC3* | 3.13496643 | 7.65E-05 | 0.000437295 | 0.480979865 |
| T2DM | monocyte | *UGGT2* | 1.762536821 | 2.33E-05 | 0.000362859 | 0.14631218 |
| T2DM | monocyte | *ARHGEF7* | 1.909510904 | 6.74E-05 | 0.000421481 | 0.423599929 |
| T2DM | monocyte | *SGCG* | 0.919290799 | 0.000793752 | 0.001861364 | 1 |
| T2DM | monocyte | *PARP4* | 3.548578454 | 0.000156049 | 0.000632549 | 0.98108295 |
| T2DM | monocyte | *RGCC* | 0.502058923 | 0.001797167 | 0.003359735 | 1 |
| T2DM | monocyte | *VWA8* | 2.225861851 | 1.09E-05 | 0.000362859 | 0.068502581 |
| T2DM | monocyte | *KATNAL1* | 1.314041356 | 0.0007068 | 0.001710412 | 1 |
| T2DM | monocyte | *DHRS12* | 1.775927002 | 0.000169971 | 0.000662097 | 1 |
| T2DM | monocyte | *MEDAG* | 0.529342505 | 0.001579556 | 0.003079277 | 1 |
| T2DM | monocyte | *TSC22D1* | 2.551273328 | 1.15E-05 | 0.000362859 | 0.072035656 |
| T2DM | monocyte | *CLN5* | 1.929292131 | 0.002367054 | 0.004139546 | 1 |
| T2DM | monocyte | *MGRN1* | 2.195856406 | 0.001149438 | 0.002434811 | 1 |

| | | | | | | |
|---|---|---|---|---|---|---|
| T2DM | monocyte | *CORO1A* | -0.560310106 | 0.007435721 | 0.011224534 | 1 |
| T2DM | monocyte | *GDPD3* | 3.593717481 | 0.000161853 | 0.000644849 | 1 |
| T2DM | monocyte | *NFAT5* | 2.011352947 | 4.60E-05 | 0.000379941 | 0.289502529 |
| T2DM | monocyte | *LONP2* | 1.749413962 | 0.000194794 | 0.000722248 | 1 |
| T2DM | monocyte | *PLLP* | 1.478872395 | 0.001030606 | 0.002252927 | 1 |
| T2DM | monocyte | *DHODH* | 1.392685693 | 0.002956801 | 0.005002531 | 1 |
| T2DM | monocyte | *CTCF* | 3.652530991 | 0.000361516 | 0.001073112 | 1 |
| T2DM | monocyte | *ACD* | -0.844737498 | 0.016344872 | 0.022520317 | 1 |
| T2DM | monocyte | *SETD6* | 1.344571073 | 0.000562197 | 0.00146183 | 1 |
| T2DM | monocyte | *SLC7A6OS* | 0.697637043 | 0.006141967 | 0.009489936 | 1 |
| T2DM | monocyte | *NPRL3* | 2.319625756 | 7.66E-05 | 0.000437295 | 0.481627258 |
| T2DM | monocyte | *NAGPA* | 3.514818184 | 0.000177623 | 0.00068134 | 1 |
| T2DM | monocyte | *COTL1* | -0.194499033 | 0.01026769 | 0.014922091 | 1 |
| T2DM | monocyte | *CRISPLD2* | 1.598238623 | 0.000647364 | 0.001606948 | 1 |
| T2DM | monocyte | *ZNF500* | 1.291320715 | 0.002487768 | 0.004319414 | 1 |
| T2DM | monocyte | *NME4* | 1.359671348 | 0.001142368 | 0.002424735 | 1 |
| T2DM | monocyte | *NOMO3* | 1.266332016 | 0.009498899 | 0.013943399 | 1 |
| T2DM | monocyte | *LMF1* | 2.435446543 | 2.33E-05 | 0.000362859 | 0.146587582 |
| T2DM | monocyte | *NARFL* | 3.162798776 | 0.000128971 | 0.000567814 | 0.810838145 |
| T2DM | monocyte | *MTHFSD* | 0.744303772 | 0.022966696 | 0.030636881 | 1 |
| T2DM | monocyte | *CLCN7* | 2.807130263 | 0.030775358 | 0.039836251 | 1 |
| T2DM | monocyte | *HAGHL* | 3.808853402 | 0.005882526 | 0.009133969 | 1 |
| T2DM | monocyte | *FBXO31* | 1.649342692 | 0.000599973 | 0.001523326 | 1 |
| T2DM | monocyte | *STUB1* | 2.546746367 | 0.002995842 | 0.005060413 | 1 |
| T2DM | monocyte | *NUBP1* | 3.799750284 | 0.008965845 | 0.013241312 | 1 |
| T2DM | monocyte | *UBE2I* | 2.700384768 | 2.66E-05 | 0.000362859 | 0.167048249 |
| T2DM | monocyte | *CLUAP1* | 1.879141166 | 2.56E-05 | 0.000362859 | 0.160893002 |
| T2DM | monocyte | *EARS2* | 0.664057881 | 0.012726604 | 0.018024817 | 1 |
| T2DM | monocyte | *CPPED1* | -0.268194319 | 0.015752689 | 0.021775979 | 1 |
| T2DM | monocyte | *HMOX2* | 3.676048365 | 1.12E-05 | 0.000362859 | 0.070716645 |
| T2DM | monocyte | *BFAR* | 2.860680266 | 7.36E-05 | 0.000431915 | 0.46258063 |
| T2DM | monocyte | *RBL2* | 2.967225353 | 1.89E-08 | 9.91E-06 | 0.00011893 |
| T2DM | monocyte | *STX4* | 1.284515133 | 0.000485213 | 0.001322296 | 1 |
| T2DM | monocyte | *KAT8* | 5.342803422 | 2.55E-05 | 0.000362859 | 0.160156623 |
| T2DM | monocyte | *SYT17* | -1.009484326 | 0.038405997 | 0.048828817 | 1 |
| T2DM | monocyte | *TMC5* | -1.098289737 | 0.019582735 | 0.02652233 | 1 |
| T2DM | monocyte | *RNF40* | 2.25968113 | 0.000325681 | 0.001003705 | 1 |
| T2DM | monocyte | *KNOP1* | 1.657773961 | 0.000535169 | 0.001410737 | 1 |
| T2DM | monocyte | *IQCH* | 2.624443102 | 6.83E-05 | 0.000423957 | 0.429524808 |
| T2DM | monocyte | *CSK* | -0.496709116 | 0.023928894 | 0.031772113 | 1 |
| T2DM | monocyte | *HERC1* | 2.989635808 | 1.15E-05 | 0.000362859 | 0.072309032 |
| T2DM | monocyte | *TRIP4* | 1.520687882 | 0.006445655 | 0.009900765 | 1 |
| T2DM | monocyte | *RASL12* | 1.058401502 | 0.001607662 | 0.003112833 | 1 |
| T2DM | monocyte | *IGDCC4* | 0.544605409 | 0.00429439 | 0.006919229 | 1 |
| T2DM | monocyte | *TTC23* | 1.447106685 | 0.000207471 | 0.00074578 | 1 |
| T2DM | monocyte | *FAH* | 1.372024654 | 0.000417624 | 0.00118535 | 1 |
| T2DM | monocyte | *RPAP1* | 2.812538507 | 0.000802572 | 0.001877146 | 1 |
| T2DM | monocyte | *HOMER2* | 1.172147262 | 0.000869297 | 0.001988093 | 1 |
| T2DM | monocyte | *EHD4* | 1.891828792 | 9.74E-05 | 0.000485446 | 0.612305947 |
| T2DM | monocyte | *ATP8B4* | 3.337160753 | 0.00049015 | 0.001329984 | 1 |

| | | | | | | |
|---|---|---|---|---|---|---|
| T2DM | monocyte | *DTWD1* | 1.604456702 | 2.01E-05 | 0.000362859 | 0.126657275 |
| T2DM | monocyte | *DNAJC17* | 3.403506737 | 2.84E-05 | 0.000362859 | 0.178304592 |
| T2DM | monocyte | *PDGFRL* | 0.639262037 | 0.000764967 | 0.001813827 | 1 |
| T2DM | monocyte | *ZFAND1* | 0.971038288 | 0.000999773 | 0.002205463 | 1 |
| T2DM | monocyte | *EYA1* | 1.178543183 | 2.27E-06 | 0.000190588 | 0.014244181 |
| T2DM | monocyte | *NBN* | 3.679101389 | 0.000225171 | 0.000782857 | 1 |
| T2DM | monocyte | *CPQ* | 2.011781227 | 5.71E-05 | 0.000395506 | 0.358818136 |
| T2DM | monocyte | *NIPAL2* | 3.373848645 | 1.78E-06 | 0.000169734 | 0.011202428 |
| T2DM | monocyte | *IKBKB* | 3.527854813 | 6.16E-06 | 0.000322038 | 0.03871179 |
| T2DM | monocyte | *PLAT* | 0.941346565 | 0.000640194 | 0.001593662 | 1 |
| T2DM | monocyte | *GDAP1* | 3.319120708 | 0.000109533 | 0.000520508 | 0.688631974 |
| T2DM | monocyte | *RAB2A* | 1.75701818 | 9.36E-06 | 0.000362859 | 0.058847804 |
| T2DM | monocyte | *ZC2HC1A* | 1.925448342 | 9.30E-05 | 0.000474395 | 0.584928897 |
| T2DM | monocyte | *SPAG1* | 1.154445088 | 0.000680284 | 0.001664181 | 1 |
| T2DM | monocyte | *SNX16* | 2.04519405 | 0.0001876 | 0.000706541 | 1 |
| T2DM | monocyte | *ERI1* | 2.309155954 | 8.00E-05 | 0.000443003 | 0.503251804 |
| T2DM | monocyte | *MTMR9* | 3.468684139 | 2.18E-05 | 0.000362859 | 0.137315896 |
| T2DM | monocyte | *LEPROTL1* | 3.934805195 | 0.001822781 | 0.003388476 | 1 |
| T2DM | monocyte | *ERICH1* | 3.058548877 | 0.001273643 | 0.002623654 | 1 |
| T2DM | monocyte | *TUSC3* | 0.666599411 | 0.001141352 | 0.002424215 | 1 |
| T2DM | monocyte | *ARHGEF10* | 1.218342503 | 0.00022389 | 0.000779936 | 1 |
| T2DM | monocyte | *KLHDC4* | 2.045756888 | 0.002118185 | 0.003794025 | 1 |
| T2DM | monocyte | *MCM4* | 1.863306366 | 0.001302924 | 0.002672589 | 1 |
| T2DM | monocyte | *NUCB1* | 1.849829955 | 0.000173453 | 0.000671081 | 1 |
| T2DM | monocyte | *NFKBIB* | 1.936905078 | 0.000895921 | 0.00202711 | 1 |
| T2DM | monocyte | *SNRNP70* | 2.093123867 | 0.000601931 | 0.001524272 | 1 |
| T2DM | monocyte | *FCGRT* | 1.588756371 | 0.011979602 | 0.017101671 | 1 |
| T2DM | monocyte | *ERCC2* | 3.373316135 | 0.000195294 | 0.000722579 | 1 |
| T2DM | monocyte | *PLEKHJ1* | 2.094115862 | 0.000332572 | 0.001017459 | 1 |
| T2DM | monocyte | *CD37* | -0.378081845 | 0.016789333 | 0.023072031 | 1 |
| T2DM | monocyte | *DKKL1* | 2.003236586 | 0.00518689 | 0.008176624 | 1 |
| T2DM | monocyte | *FCER2* | 2.860464621 | 0.001517988 | 0.002978363 | 1 |
| T2DM | monocyte | *DMPK* | 0.944905682 | 0.006436534 | 0.009889171 | 1 |
| T2DM | monocyte | *TLE6* | 2.295440423 | 0.013630857 | 0.019163059 | 1 |
| T2DM | monocyte | *AES* | 2.823188417 | 2.92E-05 | 0.000362859 | 0.183438358 |
| T2DM | monocyte | *C19orf53* | 6.058228767 | 0.000204349 | 0.000739207 | 1 |
| T2DM | monocyte | *TIMM44* | 3.568611122 | 1.67E-05 | 0.000362859 | 0.105118864 |
| T2DM | monocyte | *IL27RA* | -0.44554553 | 0.023273349 | 0.030993338 | 1 |
| T2DM | monocyte | *C19orf44* | 1.444062311 | 0.00156753 | 0.003058679 | 1 |
| T2DM | monocyte | *OLFM2* | 0.511328988 | 0.01733896 | 0.02375464 | 1 |
| T2DM | monocyte | *RASAL3* | -0.51598194 | 0.020353187 | 0.027441666 | 1 |
| T2DM | monocyte | *ZNF419* | 1.405104184 | 5.75E-05 | 0.000396678 | 0.361373473 |
| T2DM | monocyte | *SYDE1* | 2.152845433 | 0.001582062 | 0.00308225 | 1 |
| T2DM | monocyte | *AURKC* | 1.354980748 | 0.008027965 | 0.011999956 | 1 |
| T2DM | monocyte | *PDCD5* | 2.244218075 | 0.000103493 | 0.000502828 | 0.650659032 |
| T2DM | monocyte | *ANKRD27* | 2.390043893 | 0.00015666 | 0.000634614 | 0.984921116 |
| T2DM | monocyte | *RPS16* | 2.822120366 | 0.000797269 | 0.001867522 | 1 |
| T2DM | monocyte | *PRX* | 2.828772096 | 0.010667875 | 0.01540752 | 1 |
| T2DM | monocyte | *TBCB* | 3.067053417 | 0.000356056 | 0.001061919 | 1 |
| T2DM | monocyte | *POLR2I* | 4.878858567 | 0.000105215 | 0.000507835 | 0.661487853 |

| | | | | | | |
|---|---|---|---|---|---|---|
| T2DM | monocyte | *SLC1A5* | 3.192671193 | 0.000139442 | 0.00059075 | 0.876673034 |
| T2DM | monocyte | *CACTIN* | 1.952397923 | 0.000617985 | 0.001550389 | 1 |
| T2DM | monocyte | *CEACAM4* | -0.270670861 | 0.007096919 | 0.01077217 | 1 |
| T2DM | monocyte | *PLIN3* | 3.627680221 | 0.00015031 | 0.000618751 | 0.944998917 |
| T2DM | monocyte | *MRPL4* | 3.88668081 | 1.63E-05 | 0.000362859 | 0.102511768 |
| T2DM | monocyte | *ICAM5* | -6.514683333 | 0.001430223 | 0.002866707 | 1 |
| T2DM | monocyte | *CD33* | -0.35955594 | 0.015576663 | 0.021556346 | 1 |
| T2DM | monocyte | *BABAM1* | 1.631854501 | 0.00045838 | 0.001262855 | 1 |
| T2DM | monocyte | *RABAC1* | 3.533651868 | 0.000670926 | 0.001646414 | 1 |
| T2DM | monocyte | *PTPRS* | 0.818568373 | 0.000287043 | 0.000922809 | 1 |
| T2DM | monocyte | *CYTH2* | 2.273436677 | 0.000179082 | 0.000684948 | 1 |
| T2DM | monocyte | *CARD8* | 2.291708599 | 0.014632542 | 0.020429667 | 1 |
| T2DM | monocyte | *ZNF175* | 1.013286808 | 0.02279102 | 0.03043292 | 1 |
| T2DM | monocyte | *PLA2G4C* | 1.909785827 | 0.002188231 | 0.00388408 | 1 |
| T2DM | monocyte | *CAPS* | 2.03110615 | 7.47E-05 | 0.000433477 | 0.469915006 |
| T2DM | monocyte | *BCAT2* | 3.058481754 | 7.05E-05 | 0.000427286 | 0.443350488 |
| T2DM | monocyte | *MIER2* | 1.756439636 | 0.000824246 | 0.001915724 | 1 |
| T2DM | monocyte | *PPP2R1A* | 4.49989524 | 0.000585216 | 0.001495631 | 1 |
| T2DM | monocyte | *TNPO2* | 1.91472097 | 5.20E-05 | 0.000384775 | 0.32705844 |
| T2DM | monocyte | *GCDH* | 0.99320974 | 0.034574855 | 0.044370711 | 1 |
| T2DM | monocyte | *LILRB5* | 0.673255114 | 0.001276828 | 0.002626773 | 1 |
| T2DM | monocyte | *MAST1* | 3.806881512 | 0.000636828 | 0.00158721 | 1 |
| T2DM | monocyte | *PRPF31* | 1.653769232 | 0.003013208 | 0.00508635 | 1 |
| T2DM | monocyte | *TFPT* | 2.133604851 | 0.000461756 | 0.001271042 | 1 |
| T2DM | monocyte | *RPL18A* | 1.06330315 | 0.000877422 | 0.002002136 | 1 |
| T2DM | monocyte | *PDE4C* | 1.054831278 | 0.002159956 | 0.003852382 | 1 |
| T2DM | monocyte | *ELL* | 3.200255392 | 5.09E-06 | 0.000298974 | 0.031990213 |
| T2DM | monocyte | *COPE* | 3.02076682 | 0.000103824 | 0.000504047 | 0.65274052 |
| T2DM | monocyte | *LSR* | 1.348343228 | 0.008661035 | 0.012830331 | 1 |
| T2DM | monocyte | *KXD1* | 4.694249813 | 2.01E-05 | 0.000362859 | 0.126056533 |
| T2DM | monocyte | *HPN* | 0.822487682 | 0.007339409 | 0.011094702 | 1 |
| T2DM | monocyte | *ZNF14* | 2.334385157 | 0.000149916 | 0.000618049 | 0.942524619 |
| T2DM | monocyte | *ATP13A1* | 5.416787213 | 1.44E-09 | 1.13E-06 | 9.06E-06 |
| T2DM | monocyte | *SIPA1L3* | 2.886774089 | 0.002625581 | 0.004528678 | 1 |
| T2DM | monocyte | *ZNF85* | 4.737224598 | 6.98E-06 | 0.000338043 | 0.043914078 |
| T2DM | monocyte | *ETHE1* | 3.106418349 | 0.000247423 | 0.000833181 | 1 |
| T2DM | monocyte | *GTPBP10* | 1.680278291 | 0.005943856 | 0.009224503 | 1 |
| T2DM | monocyte | *RASA4* | 1.707387258 | 0.00116987 | 0.00246563 | 1 |
| T2DM | monocyte | *PON3* | 0.914917081 | 0.001700676 | 0.003233153 | 1 |
| T2DM | monocyte | *PON2* | 1.997779396 | 9.23E-05 | 0.00047287 | 0.580240662 |
| T2DM | monocyte | *HBP1* | 2.337388819 | 7.12E-05 | 0.000427286 | 0.447399356 |
| T2DM | monocyte | *WDR91* | 1.182877169 | 0.000629554 | 0.001574064 | 1 |
| T2DM | monocyte | *STEAP1B* | 1.008215275 | 0.000556276 | 0.001451164 | 1 |
| T2DM | monocyte | *PTN* | 0.910745023 | 0.001321557 | 0.002698483 | 1 |
| T2DM | monocyte | *MPP6* | 1.079056728 | 0.000476906 | 0.00130418 | 1 |
| T2DM | monocyte | *DFNA5* | 1.549701857 | 0.000757845 | 0.001803396 | 1 |
| T2DM | monocyte | *ADAP1* | -0.456216781 | 0.009714435 | 0.014216633 | 1 |
| T2DM | monocyte | *H2AFV* | 2.403863078 | 5.29E-05 | 0.000385235 | 0.332821799 |
| T2DM | monocyte | *CAV1* | 0.767188211 | 0.000581645 | 0.00149014 | 1 |
| T2DM | monocyte | *MET* | 0.74602617 | 0.001224031 | 0.002549862 | 1 |

| | | | | | | |
|---|---|---|---|---|---|---|
| T2DM | monocyte | *RNF32* | 1.161009412 | 0.014112298 | 0.019777641 | 1 |
| T2DM | monocyte | *LMBR1* | 2.899498406 | 0.000186719 | 0.000704201 | 1 |
| T2DM | monocyte | *HOXA2* | 7.983278789 | 0.00672998 | 0.010282939 | 1 |
| T2DM | monocyte | *HOXA5* | 1.1384695 | 0.003158863 | 0.005305844 | 1 |
| T2DM | monocyte | *HOXA6* | 1.2915988 | 0.002825202 | 0.004805748 | 1 |
| T2DM | monocyte | *BRAT1* | 1.392254201 | 0.001662876 | 0.003184119 | 1 |
| T2DM | monocyte | *CPED1* | 1.590174334 | 8.81E-05 | 0.000461329 | 0.553595241 |
| T2DM | monocyte | *HIBADH* | 1.705158442 | 9.75E-05 | 0.000485446 | 0.61293613 |
| T2DM | monocyte | *CPVL* | -0.237890858 | 0.032487647 | 0.041863053 | 1 |
| T2DM | monocyte | *PLEKHA8* | 2.409338787 | 0.000164615 | 0.000651311 | 1 |
| T2DM | monocyte | *NOD1* | 2.796837526 | 2.72E-05 | 0.000362859 | 0.171266228 |
| T2DM | monocyte | *EPHB6* | 3.313863542 | 0.000482993 | 0.001317386 | 1 |
| T2DM | monocyte | *CHCHD2* | 1.678837238 | 0.000610731 | 0.001538945 | 1 |
| T2DM | monocyte | *CYP3A5* | 1.53504261 | 0.000314307 | 0.000981157 | 1 |
| T2DM | monocyte | *ZKSCAN1* | 2.9877666 | 1.47E-05 | 0.000362859 | 0.092547139 |
| T2DM | monocyte | *EIF3B* | 2.693498681 | 0.000190105 | 0.000713121 | 1 |
| T2DM | monocyte | *SNX8* | 2.736394994 | 5.16E-05 | 0.000384775 | 0.324707835 |
| T2DM | monocyte | *TAF6* | 3.584848867 | 0.0026938 | 0.004632363 | 1 |
| T2DM | monocyte | *AIMP2* | 1.583195654 | 0.000346047 | 0.001038471 | 1 |
| T2DM | monocyte | *RBM28* | 1.048214625 | 0.002806783 | 0.00477959 | 1 |
| T2DM | monocyte | *USP42* | 4.466419853 | 4.33E-06 | 0.000277801 | 0.027224538 |
| T2DM | monocyte | *LSM5* | 2.004643526 | 0.000213127 | 0.000760026 | 1 |
| T2DM | monocyte | *AP1S1* | 4.196059396 | 0.000127934 | 0.00056466 | 0.804318916 |
| T2DM | monocyte | *C1GALT1* | 3.787683475 | 3.01E-05 | 0.000362859 | 0.189014277 |
| T2DM | monocyte | *PLOD3* | 2.587433348 | 0.000186045 | 0.000703043 | 1 |
| T2DM | monocyte | *RPA3* | 1.939415828 | 0.00010977 | 0.000521153 | 0.690125926 |
| T2DM | monocyte | *ZNHIT1* | 1.478702989 | 0.007153855 | 0.01084863 | 1 |
| T2DM | monocyte | *PHF14* | 2.655395012 | 2.34E-05 | 0.000362859 | 0.146895021 |
| T2DM | monocyte | *CEP41* | 1.347458732 | 0.000751889 | 0.001791936 | 1 |
| T2DM | monocyte | *ZNF862* | 2.173340458 | 6.94E-05 | 0.00042643 | 0.436321727 |
| T2DM | monocyte | *MEST* | 0.437447991 | 0.013328349 | 0.018788191 | 1 |
| T2DM | monocyte | *ACTR3C* | 1.250993136 | 0.000825888 | 0.001917414 | 1 |
| T2DM | monocyte | *POU6F2* | 1.173799661 | 0.000713303 | 0.001722834 | 1 |
| T2DM | monocyte | *TSPAN13* | 2.441748429 | 0.000327149 | 0.001006043 | 1 |
| T2DM | monocyte | *RARRES2* | 1.971484156 | 8.07E-05 | 0.000443644 | 0.507479864 |
| T2DM | monocyte | *MRPL32* | 1.486355978 | 6.50E-05 | 0.000415842 | 0.408356517 |
| T2DM | monocyte | *COA1* | 1.935522856 | 0.000272404 | 0.00089059 | 1 |
| T2DM | monocyte | *URGCP* | 3.930230477 | 4.58E-05 | 0.000379941 | 0.288058887 |
| T2DM | monocyte | *POLD2* | 0.960045129 | 0.009791452 | 0.014312686 | 1 |
| T2DM | monocyte | *GCK* | -4.985471959 | 0.01442358 | 0.020160304 | 1 |
| T2DM | monocyte | *YKT6* | 6.009672202 | 0.000160802 | 0.000643059 | 1 |
| T2DM | monocyte | *CLIP2* | 4.694450671 | 3.05E-05 | 0.000362859 | 0.191603529 |
| T2DM | monocyte | *LIMK1* | 4.988082671 | 2.49E-05 | 0.000362859 | 0.156238383 |
| T2DM | monocyte | *SPATA6L* | 0.838334486 | 0.00182624 | 0.003390895 | 1 |
| T2DM | monocyte | *SLC1A1* | 0.934062831 | 8.16E-05 | 0.000445905 | 0.513236334 |
| T2DM | monocyte | *FKTN* | 2.093343036 | 0.000144762 | 0.000606338 | 0.910120161 |
| T2DM | monocyte | *NMRK1* | 1.624248694 | 5.15E-05 | 0.000384775 | 0.32357953 |
| T2DM | monocyte | *TMEM245* | 1.532741715 | 0.000252496 | 0.000843844 | 1 |
| T2DM | monocyte | *CORO2A* | 3.25685649 | 0.029924318 | 0.038830586 | 1 |
| T2DM | monocyte | *C5* | 1.939602282 | 0.001278582 | 0.002628661 | 1 |

| | | | | | | |
|---|---|---|---|---|---|---|
| T2DM | monocyte | *OGN* | 1.712337123 | 0.000340618 | 0.001030047 | 1 |
| T2DM | monocyte | *ECM2* | 1.389231512 | 0.000233908 | 0.000801844 | 1 |
| T2DM | monocyte | *PTGR1* | 0.804880997 | 0.000953923 | 0.002125952 | 1 |
| T2DM | monocyte | *TNFSF8* | -0.643135465 | 0.02640976 | 0.034685223 | 1 |
| T2DM | monocyte | *AK1* | 1.026916911 | 0.002157157 | 0.00385176 | 1 |
| T2DM | monocyte | *PLGRKT* | 1.032935481 | 0.007068839 | 0.010734732 | 1 |
| T2DM | monocyte | *KDM4C* | 3.090502628 | 0.00033367 | 0.001020323 | 1 |
| T2DM | monocyte | *ELAVL2* | -1.7604051 | 0.004530747 | 0.007246199 | 1 |
| T2DM | monocyte | *FUBP3* | 1.96150864 | 9.81E-05 | 0.000486996 | 0.617023821 |
| T2DM | monocyte | *RGP1* | 2.96269771 | 4.46E-05 | 0.000379941 | 0.280222131 |
| T2DM | monocyte | *MPDZ* | 0.837583447 | 0.000525272 | 0.001392823 | 1 |
| T2DM | monocyte | *PTGDS* | 0.968773364 | 0.001742838 | 0.00328999 | 1 |
| T2DM | monocyte | *SHB* | 7.715085824 | 0.000109036 | 0.000518963 | 0.685509873 |
| T2DM | monocyte | *DVL1* | 2.892601528 | 0.000160415 | 0.000642374 | 1 |
| T2DM | monocyte | *PDLIM1* | 1.396836453 | 0.000623498 | 0.001561104 | 1 |
| T2DM | monocyte | *CCNJ* | 3.105510144 | 0.011778791 | 0.016860942 | 1 |
| T2DM | monocyte | *ATRNL1* | 1.165839671 | 0.001607644 | 0.003112833 | 1 |
| T2DM | monocyte | *HPS1* | 3.550771273 | 0.004053544 | 0.006580075 | 1 |
| T2DM | monocyte | *DNMBP* | 3.241232177 | 2.51E-05 | 0.000362859 | 0.157612563 |
| T2DM | monocyte | *TRDMT1* | 2.440340641 | 0.000122 | 0.000552602 | 0.767011269 |
| T2DM | monocyte | *DDX50* | 3.206494105 | 2.37E-05 | 0.000362859 | 0.148836537 |
| T2DM | monocyte | *MAPK8* | 2.319712984 | 2.63E-05 | 0.000362859 | 0.165261891 |
| T2DM | monocyte | *ATE1* | 2.212649097 | 3.06E-05 | 0.000362859 | 0.192108215 |
| T2DM | monocyte | *NSMCE4A* | 1.739618791 | 0.000239836 | 0.000816378 | 1 |
| T2DM | monocyte | *PLEKHA1* | 2.439657416 | 2.20E-05 | 0.000362859 | 0.138523078 |
| T2DM | monocyte | *SPOCK2* | 0.99379115 | 0.006230488 | 0.009619617 | 1 |
| T2DM | monocyte | *MICU1* | 3.440074051 | 0.000134289 | 0.00057865 | 0.844274253 |
| T2DM | monocyte | *CCSER2* | 2.612093895 | 1.89E-05 | 0.000362859 | 0.11877856 |
| T2DM | monocyte | *BMPR1A* | 0.977515472 | 0.00057463 | 0.001480084 | 1 |
| T2DM | monocyte | *LIPA* | 1.583174747 | 0.000202075 | 0.000734787 | 1 |
| T2DM | monocyte | *SFXN3* | 3.192482014 | 8.52E-05 | 0.000457699 | 0.535507731 |
| T2DM | monocyte | *TNKS2* | 3.285965997 | 7.05E-05 | 0.000427286 | 0.442947875 |
| T2DM | monocyte | *ANKRD26* | 1.571130816 | 9.29E-05 | 0.00047433 | 0.583837339 |
| T2DM | monocyte | *ACBD5* | 4.656127247 | 4.27E-06 | 0.000277801 | 0.026860698 |
| T2DM | monocyte | *MTPAP* | 4.118089143 | 1.06E-05 | 0.000362859 | 0.066479104 |
| T2DM | monocyte | *PITRM1* | 2.089966078 | 0.000145655 | 0.000608331 | 0.915731561 |
| T2DM | monocyte | *MAP3K8* | 2.855430678 | 0.002875773 | 0.004885163 | 1 |
| T2DM | monocyte | *SHOC2* | 3.599749481 | 4.79E-05 | 0.000379941 | 0.301293371 |
| T2DM | monocyte | *TFAM* | 2.575799094 | 0.000620602 | 0.001554472 | 1 |
| T2DM | monocyte | *RPL28* | 1.608360328 | 0.020156666 | 0.027200034 | 1 |
| T2DM | monocyte | *TADA2A* | 0.992792404 | 0.009567228 | 0.014027323 | 1 |
| T2DM | monocyte | *MLLT6* | 4.128069213 | 1.50E-05 | 0.000362859 | 0.094115355 |
| T2DM | monocyte | *PSMB3* | 5.68917866 | 5.42E-07 | 8.11E-05 | 0.003407724 |
| T2DM | monocyte | *CASC3* | 2.679744824 | 9.62E-05 | 0.000482554 | 0.604640545 |
| T2DM | monocyte | *RAPGEFL1* | 0.944559296 | 0.004988139 | 0.007903334 | 1 |
| T2DM | monocyte | *RGS9* | 2.122047805 | 0.000436885 | 0.001224565 | 1 |
| T2DM | monocyte | *WNT3* | 1.308584224 | 0.006346212 | 0.00977668 | 1 |
| T2DM | monocyte | *ASPA* | 0.802559811 | 0.000808589 | 0.001887709 | 1 |
| T2DM | monocyte | *TRIM37* | 2.639378629 | 1.98E-05 | 0.000362859 | 0.124321327 |
| T2DM | monocyte | *PNPO* | 2.195046911 | 0.002364381 | 0.004136023 | 1 |

| | | | | | | |
|---|---|---|---|---|---|---|
| T2DM | monocyte | *RPS6KB1* | 3.530782593 | 2.46E-05 | 0.000362859 | 0.154644887 |
| T2DM | monocyte | *TRIM16L* | 1.066170294 | 0.016517161 | 0.022743413 | 1 |
| T2DM | monocyte | *CDK5RAP3* | 2.291947697 | 0.00140243 | 0.002820562 | 1 |
| T2DM | monocyte | *CBX1* | 3.104996594 | 7.46E-05 | 0.000433444 | 0.46898685 |
| T2DM | monocyte | *RECQL5* | 1.044880207 | 0.003170462 | 0.005322481 | 1 |
| T2DM | monocyte | *PIGL* | 2.410850564 | 4.01E-05 | 0.000371009 | 0.251870905 |
| T2DM | monocyte | *GALK1* | 6.993001157 | 2.37E-05 | 0.000362859 | 0.149286067 |
| T2DM | monocyte | *CAMTA2* | 2.911018225 | 0.0050811 | 0.008031611 | 1 |
| T2DM | monocyte | *MED13* | 2.235184599 | 0.000149743 | 0.00061774 | 0.941436158 |
| T2DM | monocyte | *HOXB6* | 1.577396104 | 0.018208583 | 0.024794751 | 1 |
| T2DM | monocyte | *RNF167* | 4.002414252 | 0.003550012 | 0.005867226 | 1 |
| T2DM | monocyte | *NUP88* | 2.438472861 | 0.000114282 | 0.000530644 | 0.718492124 |
| T2DM | monocyte | *C1QBP* | 6.705645896 | 7.54E-05 | 0.000435317 | 0.474155776 |
| T2DM | monocyte | *GOSR1* | 3.12338912 | 2.79E-05 | 0.000362859 | 0.175367335 |
| T2DM | monocyte | *DRG2* | 2.628225435 | 0.000141949 | 0.000598067 | 0.892435224 |
| T2DM | monocyte | *AKAP10* | 2.611965267 | 0.003631606 | 0.005981637 | 1 |
| T2DM | monocyte | *UTP6* | 4.190456257 | 3.73E-05 | 0.000366571 | 0.23423872 |
| T2DM | monocyte | *DDX5* | 1.491161375 | 7.56E-05 | 0.000435317 | 0.475204958 |
| T2DM | monocyte | *C17orf75* | 3.283309247 | 0.000133259 | 0.000578193 | 0.837801238 |
| T2DM | monocyte | *CYTH1* | 2.675449811 | 0.000610499 | 0.001538945 | 1 |
| T2DM | monocyte | *NAGLU* | 2.492263832 | 0.000186284 | 0.000703402 | 1 |
| T2DM | monocyte | *EZH1* | 2.416830217 | 9.13E-06 | 0.000362859 | 0.057408157 |
| T2DM | monocyte | *DLX4* | -1.722777739 | 0.008394741 | 0.012485861 | 1 |
| T2DM | monocyte | *VAT1* | 1.44810592 | 6.78E-05 | 0.000422229 | 0.426451426 |
| T2DM | monocyte | *MPP2* | 1.280095342 | 0.001616053 | 0.00312427 | 1 |
| T2DM | monocyte | *EFTUD2* | 4.451640258 | 2.27E-05 | 0.000362859 | 0.142641865 |
| T2DM | monocyte | *EFNB3* | 10.88344922 | 0.002244232 | 0.003961621 | 1 |
| T2DM | monocyte | *FAM20A* | 1.207468673 | 0.000281418 | 0.000911031 | 1 |
| T2DM | monocyte | *YWHAE* | 3.222236309 | 2.48E-05 | 0.000362859 | 0.155783751 |
| T2DM | monocyte | *DPH1* | 6.351627563 | 1.97E-05 | 0.000362859 | 0.124027032 |
| T2DM | monocyte | *DHRS7B* | 2.881625282 | 0.005466815 | 0.008573178 | 1 |
| T2DM | monocyte | *RCVRN* | 3.20071589 | 0.010353459 | 0.0150259 | 1 |
| T2DM | monocyte | *MYH3* | 2.111114614 | 0.000234542 | 0.000803155 | 1 |
| T2DM | monocyte | *SLAIN2* | 3.282817279 | 0.000314861 | 0.000981911 | 1 |
| T2DM | monocyte | *OCIAD1* | 1.782736117 | 0.000336274 | 0.00102161 | 1 |
| T2DM | monocyte | *CWH43* | 8.717482704 | 0.000530007 | 0.00140183 | 1 |
| T2DM | monocyte | *UBE2D3* | 3.305133822 | 1.38E-05 | 0.000362859 | 0.086569696 |
| T2DM | monocyte | *NDUFC1* | 5.156326966 | 0.000409349 | 0.001172717 | 1 |
| T2DM | monocyte | *TBC1D9* | 3.403237142 | 0.00010188 | 0.000498069 | 0.640516786 |
| T2DM | monocyte | *CPE* | 1.184070022 | 0.000230217 | 0.000794388 | 1 |
| T2DM | monocyte | *WFS1* | 3.398870039 | 3.98E-07 | 6.81E-05 | 0.002500485 |
| T2DM | monocyte | *GRPEL1* | 6.486017295 | 1.56E-05 | 0.000362859 | 0.09829969 |
| T2DM | monocyte | *AADAT* | 3.51618962 | 0.000274444 | 0.000895398 | 1 |
| T2DM | monocyte | *CPZ* | -2.176547532 | 0.007613493 | 0.011451203 | 1 |
| T2DM | monocyte | *TRIM2* | 0.938512238 | 0.001058196 | 0.002296761 | 1 |
| T2DM | monocyte | *SH3D19* | 0.69393111 | 0.00037168 | 0.00109424 | 1 |
| T2DM | monocyte | *STIM2* | 3.195588753 | 0.000714164 | 0.001724252 | 1 |
| T2DM | monocyte | *RAPGEF2* | 2.089180886 | 1.03E-05 | 0.000362859 | 0.064532923 |
| T2DM | monocyte | *LRP2BP* | 0.887034484 | 0.00102029 | 0.002237378 | 1 |
| T2DM | monocyte | *UFSP2* | 1.841858618 | 0.000174573 | 0.000674165 | 1 |

| | | | | | | |
|---|---|---|---|---|---|---|
| T2DM | monocyte | *KLHL5* | 1.302990216 | 0.000576081 | 0.001482303 | 1 |
| T2DM | monocyte | *FAM149A* | 0.968447406 | 4.20E-05 | 0.000374514 | 0.264349317 |
| T2DM | monocyte | *UGDH* | 1.080745542 | 0.000565894 | 0.001466519 | 1 |
| T2DM | monocyte | *HTATIP2* | 1.571627456 | 0.016037813 | 0.022136055 | 1 |
| T2DM | monocyte | *CTSC* | 2.253193974 | 0.000770167 | 0.001822488 | 1 |
| T2DM | monocyte | *ELP4* | 3.245179981 | 6.65E-05 | 0.000420396 | 0.417873636 |
| T2DM | monocyte | *SC5D* | 3.136944227 | 6.24E-05 | 0.000407194 | 0.392346729 |
| T2DM | monocyte | *C11orf63* | 0.847328179 | 0.000864937 | 0.00198108 | 1 |
| T2DM | monocyte | *DTX4* | 1.398945806 | 0.000492835 | 0.001333243 | 1 |
| T2DM | monocyte | *PUS3* | 5.121979244 | 0.000110945 | 0.000523558 | 0.697509346 |
| T2DM | monocyte | *DCPS* | 4.2041741 | 1.81E-05 | 0.000362859 | 0.113977708 |
| T2DM | monocyte | *FOXRED1* | 1.194581807 | 0.000239288 | 0.000815394 | 1 |
| T2DM | monocyte | *NRXN2* | 8.087996757 | 3.91E-05 | 0.000371009 | 0.245665507 |
| T2DM | monocyte | *MS4A4A* | 1.164297255 | 0.001408445 | 0.002830149 | 1 |
| T2DM | monocyte | *PRPF19* | 3.797624096 | 3.61E-05 | 0.000365043 | 0.22694389 |
| T2DM | monocyte | *HPX* | 1.438745823 | 0.005333203 | 0.008374087 | 1 |
| T2DM | monocyte | *CHORDC1* | 3.51267798 | 2.92E-05 | 0.000362859 | 0.183844003 |
| T2DM | monocyte | *EIF4G2* | 3.439926765 | 0.001084178 | 0.002333526 | 1 |
| T2DM | monocyte | *GALNT18* | 0.457260435 | 0.003176806 | 0.005331709 | 1 |
| T2DM | monocyte | *CD5* | 1.128967516 | 0.002159735 | 0.003852382 | 1 |
| T2DM | monocyte | *ACCS* | 1.239028103 | 0.000971439 | 0.002154039 | 1 |
| T2DM | monocyte | *MADD* | 2.704035726 | 3.61E-05 | 0.000365043 | 0.227166104 |
| T2DM | monocyte | *NAA40* | 3.556620723 | 0.000211381 | 0.000755944 | 1 |
| T2DM | monocyte | *CARS* | 3.702982606 | 3.97E-07 | 6.81E-05 | 0.002495038 |
| T2DM | monocyte | *SLC35F2* | 0.969913171 | 0.037421209 | 0.047728165 | 1 |
| T2DM | monocyte | *C11orf21* | -0.405046246 | 0.002761377 | 0.00471376 | 1 |
| T2DM | monocyte | *ELMOD1* | 1.684204271 | 0.009759589 | 0.014273164 | 1 |
| T2DM | monocyte | *RPS13* | 3.057454169 | 0.001593445 | 0.003095555 | 1 |
| T2DM | monocyte | *AIP* | 2.581467711 | 0.00115694 | 0.002444761 | 1 |
| T2DM | monocyte | *NDUFS8* | 4.624607609 | 2.41E-05 | 0.000362859 | 0.151727703 |
| T2DM | monocyte | *GTF2H1* | 2.416315701 | 0.000232137 | 0.00079821 | 1 |
| T2DM | monocyte | *PPFIBP1* | 1.544832397 | 7.55E-05 | 0.000435317 | 0.474527107 |
| T2DM | monocyte | *PRDM4* | 3.479809593 | 5.71E-05 | 0.000395506 | 0.359119334 |
| T2DM | monocyte | *COQ5* | 0.839398844 | 0.00018747 | 0.000706541 | 1 |
| T2DM | monocyte | *ASIC1* | -1.648216013 | 0.012505465 | 0.017751605 | 1 |
| T2DM | monocyte | *CAPRIN2* | 3.80913992 | 0.000254809 | 0.000848508 | 1 |
| T2DM | monocyte | *CYP27B1* | -3.14179815 | 0.014924941 | 0.02078713 | 1 |
| T2DM | monocyte | *ACSS3* | 0.529989156 | 0.001106878 | 0.002369404 | 1 |
| T2DM | monocyte | *GLI1* | -1.098353018 | 0.022794436 | 0.03043292 | 1 |
| T2DM | monocyte | *METAP2* | 2.542127047 | 0.000186077 | 0.000703043 | 1 |
| T2DM | monocyte | *LTA4H* | -0.428578638 | 0.030362455 | 0.039350392 | 1 |
| T2DM | monocyte | *WNT5B* | 3.439973113 | 1.61E-05 | 0.000362859 | 0.101238066 |
| T2DM | monocyte | *TRPV4* | 1.150227886 | 0.004339298 | 0.006979065 | 1 |
| T2DM | monocyte | *ITFG2* | 3.504706814 | 5.17E-05 | 0.000384775 | 0.324977174 |
| T2DM | monocyte | *PRR4* | 3.010978729 | 1.31E-06 | 0.000146927 | 0.008227896 |
| T2DM | monocyte | *PARP11* | 3.209032409 | 1.32E-05 | 0.000362859 | 0.083100591 |
| T2DM | monocyte | *GPN3* | 5.647068487 | 9.49E-05 | 0.000479779 | 0.596365387 |
| T2DM | monocyte | *VPS29* | 3.59516858 | 0.000555086 | 0.001449263 | 1 |
| T2DM | monocyte | *KCNA1* | 2.41177912 | 0.003330289 | 0.00555372 | 1 |
| T2DM | monocyte | *CREBL2* | 3.766006121 | 1.06E-06 | 0.000125709 | 0.006636634 |

| | | | | | | |
|---|---|---|---|---|---|---|
| T2DM | monocyte | *NAA25* | 3.036944556 | 3.98E-05 | 0.000371009 | 0.249966529 |
| T2DM | monocyte | *SCNN1A* | 1.165432904 | 0.003361569 | 0.005599944 | 1 |
| T2DM | monocyte | *LTBR* | 2.587825807 | 0.003492263 | 0.005786994 | 1 |
| T2DM | monocyte | *CDK2AP1* | 1.678163883 | 2.79E-06 | 0.000219074 | 0.017525953 |
| T2DM | monocyte | *MGP* | 1.215089186 | 2.95E-05 | 0.000362859 | 0.185552481 |
| T2DM | monocyte | *RASAL1* | 2.745641079 | 0.027325502 | 0.035753472 | 1 |
| T2DM | monocyte | *EIF2B1* | 2.818024699 | 5.38E-05 | 0.000385603 | 0.338174214 |
| T2DM | monocyte | *STX2* | 2.418855903 | 0.001150241 | 0.00243487 | 1 |
| T2DM | monocyte | *CAND1* | 1.427454047 | 0.001071757 | 0.002317103 | 1 |
| T2DM | monocyte | *MDM1* | 3.107125003 | 0.000344523 | 0.001037263 | 1 |
| T2DM | monocyte | *NUP107* | 2.562715945 | 2.94E-05 | 0.000362859 | 0.184553056 |
| T2DM | monocyte | *CNOT2* | 3.046272798 | 0.000103328 | 0.000502417 | 0.649624696 |
| T2DM | monocyte | *KRR1* | 3.051260371 | 0.001489056 | 0.002943636 | 1 |
| T2DM | monocyte | *GNB3* | -0.563427411 | 0.025508593 | 0.033606983 | 1 |
| T2DM | monocyte | *CDCA3* | -1.379594282 | 0.000562641 | 0.00146183 | 1 |
| T2DM | monocyte | *GNPTAB* | 2.456672083 | 1.94E-05 | 0.000362859 | 0.121695949 |
| T2DM | monocyte | *ATN1* | 1.727610205 | 0.000213568 | 0.000760736 | 1 |
| T2DM | monocyte | *NT5DC3* | 1.255107238 | 0.001133704 | 0.002412863 | 1 |
| T2DM | monocyte | *PRKAB1* | 1.813581312 | 0.017462949 | 0.023903671 | 1 |
| T2DM | monocyte | *ST8SIA1* | 0.82479093 | 0.000950812 | 0.002120522 | 1 |
| T2DM | monocyte | *C2CD5* | 3.478516178 | 4.06E-05 | 0.00037107 | 0.255382734 |
| T2DM | monocyte | *PHC1* | 1.994327953 | 0.000243753 | 0.000822585 | 1 |
| T2DM | monocyte | *COL12A1* | 0.867333409 | 0.000496525 | 0.001338052 | 1 |
| T2DM | monocyte | *TDP2* | 2.463405723 | 0.000119297 | 0.000545328 | 0.750019199 |
| T2DM | monocyte | *RWDD1* | 2.083354155 | 0.000765057 | 0.001813827 | 1 |
| T2DM | monocyte | *RSPH4A* | 0.858138313 | 0.025232923 | 0.033271684 | 1 |
| T2DM | monocyte | *SMIM8* | 1.276578291 | 0.000259176 | 0.000858504 | 1 |
| T2DM | monocyte | *MCM9* | 3.186162998 | 1.71E-05 | 0.000362859 | 0.107706835 |
| T2DM | monocyte | *FAM184A* | 1.232093783 | 0.000160999 | 0.000643075 | 1 |
| T2DM | monocyte | *RNGTT* | 1.918032145 | 0.035064018 | 0.044943422 | 1 |
| T2DM | monocyte | *HDDC2* | 2.726120366 | 0.001526557 | 0.002992894 | 1 |
| T2DM | monocyte | *TPD52L1* | 0.724225491 | 0.001105194 | 0.002367413 | 1 |
| T2DM | monocyte | *UST* | 0.901929427 | 0.000422239 | 0.001195374 | 1 |
| T2DM | monocyte | *MTRF1L* | 2.714834201 | 1.23E-05 | 0.000362859 | 0.077309164 |
| T2DM | monocyte | *TULP1* | 4.989117497 | 0.001603228 | 0.003109994 | 1 |
| T2DM | monocyte | *MAPK14* | 2.390315055 | 0.005232763 | 0.008239014 | 1 |
| T2DM | monocyte | *MRPL18* | 2.235195013 | 0.014155878 | 0.019821381 | 1 |
| T2DM | monocyte | *RNF8* | 1.849789164 | 0.001041241 | 0.00226774 | 1 |
| T2DM | monocyte | *PHACTR1* | 1.877267665 | 5.09E-05 | 0.000383728 | 0.320087466 |
| T2DM | monocyte | *MDGA1* | 2.253153689 | 0.001996513 | 0.003616865 | 1 |
| T2DM | monocyte | *MDN1* | 2.690510333 | 0.000186048 | 0.000703043 | 1 |
| T2DM | monocyte | *SAYSD1* | 2.043828909 | 0.00028013 | 0.00090829 | 1 |
| T2DM | monocyte | *CAP2* | 0.566671414 | 0.003490394 | 0.005786948 | 1 |
| T2DM | monocyte | *GPR63* | 1.177866734 | 0.004246971 | 0.006855123 | 1 |
| T2DM | monocyte | *ASCC3* | 2.998641023 | 0.000746655 | 0.001782842 | 1 |
| T2DM | monocyte | *WASF1* | 0.552555656 | 0.010466092 | 0.015171852 | 1 |
| T2DM | monocyte | *GPLD1* | 0.755646265 | 0.000334506 | 0.00102039 | 1 |
| T2DM | monocyte | *ALDH5A1* | 1.949815527 | 2.66E-05 | 0.000362859 | 0.16698287 |
| T2DM | monocyte | *VNN1* | -0.470416791 | 0.029922893 | 0.038830586 | 1 |
| T2DM | monocyte | *VNN2* | -0.277203553 | 0.015024693 | 0.020898285 | 1 |

| | | | | | | |
|---|---|---|---|---|---|---|
| T2DM | monocyte | *ACOT13* | 1.633179419 | 0.000210639 | 0.00075415 | 1 |
| T2DM | monocyte | *C6orf62* | 2.11267737 | 3.21E-05 | 0.000363519 | 0.202045023 |
| T2DM | monocyte | *B3GAT2* | 1.811598318 | 0.001692667 | 0.003223856 | 1 |
| T2DM | monocyte | *HBS1L* | 1.539092118 | 0.000902797 | 0.002039484 | 1 |
| T2DM | monocyte | *PEX7* | 1.229519722 | 0.020782157 | 0.027966057 | 1 |
| T2DM | monocyte | *PERP* | 1.097867042 | 0.004161817 | 0.006739231 | 1 |
| T2DM | monocyte | *SLC16A10* | 0.896105486 | 0.000617262 | 0.001549812 | 1 |
| T2DM | monocyte | *PHACTR2* | 2.331346717 | 0.007346108 | 0.01109949 | 1 |
| T2DM | monocyte | *PACRG* | 1.032559149 | 0.001469102 | 0.002916135 | 1 |
| T2DM | monocyte | *FAM120B* | 2.318812065 | 5.64E-06 | 0.000322038 | 0.035487672 |
| T2DM | monocyte | *TBP* | 3.561033792 | 2.32E-05 | 0.000362859 | 0.145750168 |
| T2DM | monocyte | *GUCA1B* | 1.313736066 | 0.001207397 | 0.002521895 | 1 |
| T2DM | monocyte | *PRPH2* | 0.894492638 | 0.036170854 | 0.046258372 | 1 |
| T2DM | monocyte | *DNPH1* | 1.591059509 | 0.003491984 | 0.005786994 | 1 |
| T2DM | monocyte | *DUSP22* | 2.600204768 | 5.92E-05 | 0.000399203 | 0.371941059 |
| T2DM | monocyte | *EXOC2* | 3.084532278 | 3.46E-05 | 0.000365043 | 0.21780196 |
| T2DM | monocyte | *TMEM30A* | 2.823885312 | 3.71E-05 | 0.000366571 | 0.233108731 |
| T2DM | monocyte | *SENP6* | 3.125440106 | 7.42E-05 | 0.000432877 | 0.466641523 |
| T2DM | monocyte | *TTK* | -1.545596739 | 0.006067469 | 0.009390984 | 1 |
| T2DM | monocyte | *LAMA4* | 0.717709367 | 0.000719713 | 0.001732325 | 1 |
| T2DM | monocyte | *CLIC5* | 0.593941567 | 0.001832267 | 0.003395067 | 1 |
| T2DM | monocyte | *LY86* | -0.214742715 | 0.010394272 | 0.015075037 | 1 |
| T2DM | monocyte | *TBX18* | 0.626527632 | 0.001348183 | 0.002736638 | 1 |
| T2DM | monocyte | *NUDT12* | 3.05801345 | 0.000199372 | 0.000726217 | 1 |
| T2DM | monocyte | *CEP72* | 1.881160251 | 0.018503553 | 0.02515283 | 1 |
| T2DM | monocyte | *C7* | 0.974808371 | 0.001189005 | 0.002493421 | 1 |
| T2DM | monocyte | *PAPD7* | 2.744922327 | 3.47E-05 | 0.000365043 | 0.218437141 |
| T2DM | monocyte | *GHR* | 0.677201511 | 9.16E-05 | 0.000469636 | 0.575774343 |
| T2DM | monocyte | *DAP* | 3.101145878 | 0.007705335 | 0.011572728 | 1 |
| T2DM | monocyte | *NME5* | 0.710961425 | 0.001156273 | 0.002444346 | 1 |
| T2DM | monocyte | *NNT* | 1.074450709 | 0.010555602 | 0.015269919 | 1 |
| T2DM | monocyte | *PFDN1* | 2.011993663 | 6.84E-05 | 0.000423957 | 0.429892339 |
| T2DM | monocyte | *LOX* | 0.933696416 | 0.00082569 | 0.001917414 | 1 |
| T2DM | monocyte | *SPARC* | 1.048387886 | 0.000195069 | 0.000722579 | 1 |
| T2DM | monocyte | *IK* | 2.306971727 | 0.001029447 | 0.002251178 | 1 |
| T2DM | monocyte | *PDE8B* | 0.960830997 | 0.000425096 | 0.001201159 | 1 |
| T2DM | monocyte | *CLK4* | 1.758682766 | 0.000126329 | 0.00055907 | 0.794231772 |
| T2DM | monocyte | *HAVCR1* | 1.017611719 | 0.004134642 | 0.006697886 | 1 |
| T2DM | monocyte | *RNF130* | 1.864174997 | 0.02388679 | 0.031722909 | 1 |
| T2DM | monocyte | *ARSB* | 1.960413416 | 0.011091766 | 0.015972041 | 1 |
| T2DM | monocyte | *CLINT1* | 2.730603397 | 8.08E-06 | 0.000355191 | 0.050804083 |
| T2DM | monocyte | *BTNL8* | 0.581554639 | 0.037426412 | 0.047728165 | 1 |
| T2DM | monocyte | *TTC1* | 3.308517323 | 1.86E-05 | 0.000362859 | 0.116630281 |
| T2DM | monocyte | *MSH3* | 0.974980937 | 0.019881499 | 0.02687566 | 1 |
| T2DM | monocyte | *RASGRF2* | 0.818652044 | 0.000583688 | 0.001493548 | 1 |
| T2DM | monocyte | *POLR3G* | 1.147231634 | 0.001608391 | 0.003112938 | 1 |
| T2DM | monocyte | *DROSHA* | 2.142921044 | 3.07E-05 | 0.000362859 | 0.19272403 |
| T2DM | monocyte | *TARS* | 3.89188178 | 1.45E-05 | 0.000362859 | 0.091196157 |
| T2DM | monocyte | *LNPEP* | 2.626958242 | 3.77E-06 | 0.000266654 | 0.023732219 |
| T2DM | monocyte | *GNPDA1* | 2.809212992 | 6.26E-05 | 0.00040772 | 0.393857378 |

| | | | | | | |
|---|---|---|---|---|---|---|
| T2DM | monocyte | *SKP1* | 2.9908304 | 3.19E-05 | 0.000363519 | 0.200709634 |
| T2DM | monocyte | *PPWD1* | 1.136603761 | 0.039335922 | 0.049930334 | 1 |
| T2DM | monocyte | *TRIM23* | 2.241836272 | 0.00016692 | 0.000656301 | 1 |
| T2DM | monocyte | *TRAPPC13* | 2.047134746 | 8.74E-05 | 0.000461034 | 0.549330615 |
| T2DM | monocyte | *C9* | 0.745592054 | 0.00322334 | 0.005402597 | 1 |
| T2DM | monocyte | *TXNDC15* | 2.947475456 | 0.00011353 | 0.000529891 | 0.713762891 |
| T2DM | monocyte | *SMAD5* | 1.970805582 | 4.65E-05 | 0.000379941 | 0.29253936 |
| T2DM | monocyte | *HMGXB3* | 1.689310424 | 0.000131519 | 0.00057341 | 0.826857105 |
| T2DM | monocyte | *ATP6V0E1* | 2.877119954 | 0.004727526 | 0.007532174 | 1 |
| T2DM | monocyte | *BNIP1* | 0.829162053 | 0.004121374 | 0.006680744 | 1 |
| T2DM | monocyte | *CNTN3* | 0.781845176 | 0.000923983 | 0.002074672 | 1 |
| T2DM | monocyte | *ACTR8* | 1.312532793 | 0.000852193 | 0.001959669 | 1 |
| T2DM | monocyte | *HGD* | 2.400653819 | 0.000911973 | 0.002054308 | 1 |
| T2DM | monocyte | *NPHP3* | 1.295437271 | 1.29E-05 | 0.000362859 | 0.08126578 |
| T2DM | monocyte | *NIT2* | 2.068830227 | 0.000252058 | 0.000842919 | 1 |
| T2DM | monocyte | *FAM162A* | 2.233941327 | 0.00501247 | 0.007937884 | 1 |
| T2DM | monocyte | *PCCB* | 2.448045266 | 0.000143738 | 0.000603259 | 0.903681746 |
| T2DM | monocyte | *RBP1* | 2.190633568 | 0.004484081 | 0.007180698 | 1 |
| T2DM | monocyte | *RNF7* | 1.819791448 | 0.000743101 | 0.001777289 | 1 |
| T2DM | monocyte | *BCHE* | 0.791634869 | 0.007742961 | 0.011616834 | 1 |
| T2DM | monocyte | *PDCD10* | 2.751259561 | 5.50E-05 | 0.000388972 | 0.345796328 |
| T2DM | monocyte | *WNT5A* | 1.082258347 | 0.004812927 | 0.007660474 | 1 |
| T2DM | monocyte | *PRKAR2A* | 1.560290796 | 0.006021463 | 0.009328964 | 1 |
| T2DM | monocyte | *HYAL1* | 1.053528992 | 0.004594717 | 0.007337309 | 1 |
| T2DM | monocyte | *C3orf14* | 1.287562914 | 0.015773577 | 0.021800061 | 1 |
| T2DM | monocyte | *IFT57* | 1.495273565 | 0.000347443 | 0.001041169 | 1 |
| T2DM | monocyte | *IQCG* | 2.861302745 | 4.31E-05 | 0.000378034 | 0.270676194 |
| T2DM | monocyte | *GBE1* | 0.874913644 | 0.00031368 | 0.000980148 | 1 |
| T2DM | monocyte | *UMPS* | 1.586999324 | 0.00518205 | 0.008173494 | 1 |
| T2DM | monocyte | *NCBP2* | 5.317074637 | 1.44E-05 | 0.000362859 | 0.09039003 |
| T2DM | monocyte | *SLC41A3* | 3.044318892 | 0.000664017 | 0.001636486 | 1 |
| T2DM | monocyte | *PLXNA1* | 1.993759795 | 0.001832921 | 0.003395279 | 1 |
| T2DM | monocyte | *ABTB1* | 4.86747213 | 0.000152702 | 0.000624618 | 0.960038496 |
| T2DM | monocyte | *EFCC1* | 0.650181377 | 0.024621102 | 0.032560553 | 1 |
| T2DM | monocyte | *NEK11* | 1.79304715 | 0.000132216 | 0.000575651 | 0.831239411 |
| T2DM | monocyte | *PLSCR4* | 1.333246109 | 0.000168027 | 0.000658686 | 1 |
| T2DM | monocyte | *HEMK1* | 2.266881328 | 3.20E-05 | 0.000363519 | 0.201113705 |
| T2DM | monocyte | *WDR48* | 3.214316893 | 4.96E-05 | 0.000383513 | 0.31194332 |
| T2DM | monocyte | *COMMD2* | 1.820389617 | 0.000252052 | 0.000842919 | 1 |
| T2DM | monocyte | *GORASP1* | 5.285462667 | 1.86E-05 | 0.000362859 | 0.117138492 |
| T2DM | monocyte | *ABCC5* | 1.063082401 | 0.001158007 | 0.002445545 | 1 |
| T2DM | monocyte | *ARHGEF26* | 1.171555602 | 0.000312588 | 0.000978124 | 1 |
| T2DM | monocyte | *KLHL24* | 1.716720586 | 6.06E-05 | 0.000402465 | 0.381134506 |
| T2DM | monocyte | *PLCH1* | 1.371725363 | 0.000523421 | 0.001390262 | 1 |
| T2DM | monocyte | *ZBTB47* | 4.180799667 | 2.00E-05 | 0.000362859 | 0.125782422 |
| T2DM | monocyte | *NKTR* | 3.081648133 | 4.07E-06 | 0.000271729 | 0.02558528 |
| T2DM | monocyte | *CLCN2* | 1.209417409 | 0.001302762 | 0.002672589 | 1 |
| T2DM | monocyte | *NEK4* | 3.074107103 | 0.015385663 | 0.021343703 | 1 |
| T2DM | monocyte | *LMAN2L* | 1.869172327 | 7.00E-05 | 0.000427286 | 0.440221908 |
| T2DM | monocyte | *PIKFYVE* | 2.937612896 | 0.001009137 | 0.002219887 | 1 |

| | | | | | | |
|---|---|---|---|---|---|---|
| T2DM | monocyte | *KCNIP3* | 1.258327162 | 0.001063423 | 0.002304102 | 1 |
| T2DM | monocyte | *FAHD2A* | 1.823266322 | 0.000339017 | 0.001028171 | 1 |
| T2DM | monocyte | *NCL* | 2.542000421 | 0.00015502 | 0.000629999 | 0.974608065 |
| T2DM | monocyte | *ACTR1B* | 2.955519755 | 5.57E-05 | 0.000390932 | 0.349884383 |
| T2DM | monocyte | *SLC35F5* | 2.148537717 | 1.53E-05 | 0.000362859 | 0.096417389 |
| T2DM | monocyte | *STEAP3* | 0.65822366 | 0.016863859 | 0.02316938 | 1 |
| T2DM | monocyte | *TFCP2L1* | 1.253216316 | 0.000835318 | 0.001932173 | 1 |
| T2DM | monocyte | *TP53I3* | 1.248663166 | 0.000523118 | 0.001390262 | 1 |
| T2DM | monocyte | *DNAJC27* | 1.296069143 | 0.00908989 | 0.013411907 | 1 |
| T2DM | monocyte | *STAM2* | 3.774930813 | 0.000113847 | 0.000529958 | 0.715753983 |
| T2DM | monocyte | *GPD2* | 3.012531258 | 5.04E-05 | 0.000383643 | 0.316888778 |
| T2DM | monocyte | *CENPA* | 5.309392882 | 0.000299225 | 0.000951556 | 1 |
| T2DM | monocyte | *TANC1* | 0.88389247 | 0.00197267 | 0.003582373 | 1 |
| T2DM | monocyte | *ITGA4* | -0.191363431 | 0.012388388 | 0.017597334 | 1 |
| T2DM | monocyte | *PPM1G* | 1.91217377 | 0.032714564 | 0.042129551 | 1 |
| T2DM | monocyte | *INO80B* | 1.429138182 | 0.001196359 | 0.002504666 | 1 |
| T2DM | monocyte | *GRB14* | 0.720838451 | 0.001849478 | 0.003416441 | 1 |
| T2DM | monocyte | *CLIP4* | 2.416984306 | 5.30E-05 | 0.000385235 | 0.333107123 |
| T2DM | monocyte | *SPTBN1* | 0.920154201 | 0.000495018 | 0.00133742 | 1 |
| T2DM | monocyte | *RTN4* | 1.218512471 | 0.007286862 | 0.011025872 | 1 |
| T2DM | monocyte | *HTRA2* | 3.432595824 | 0.001722121 | 0.00326212 | 1 |
| T2DM | monocyte | *POLE4* | 1.755524232 | 0.000317112 | 0.000986971 | 1 |
| T2DM | monocyte | *MRPL19* | 1.149922799 | 0.000799299 | 0.001870187 | 1 |
| T2DM | monocyte | *LANCL1* | 1.777365393 | 0.000190663 | 0.000713511 | 1 |
| T2DM | monocyte | *WDR75* | 3.035666409 | 4.16E-05 | 0.00037367 | 0.261663965 |
| T2DM | monocyte | *EFEMP1* | 0.971163377 | 0.000702509 | 0.001701339 | 1 |
| T2DM | monocyte | *GLS* | 2.175087978 | 3.05E-05 | 0.000362859 | 0.191943655 |
| T2DM | monocyte | *DNAH6* | 0.395295205 | 0.006688547 | 0.01022909 | 1 |
| T2DM | monocyte | *GNLY* | 0.575059048 | 0.001728165 | 0.003269629 | 1 |
| T2DM | monocyte | *SF3B1* | 2.592005307 | 2.13E-05 | 0.000362859 | 0.134171681 |
| T2DM | monocyte | *ST3GAL5* | 2.970198668 | 0.000172549 | 0.000669641 | 1 |
| T2DM | monocyte | *PLCD4* | 0.906673435 | 0.001698024 | 0.003231077 | 1 |
| T2DM | monocyte | *CHMP3* | -1.931642186 | 0.002245045 | 0.003961621 | 1 |
| T2DM | monocyte | *ZNF142* | 3.179982596 | 0.001092524 | 0.00234747 | 1 |
| T2DM | monocyte | *WNT6* | -1.727450877 | 0.017247905 | 0.023635043 | 1 |
| T2DM | monocyte | *MLPH* | 0.751246715 | 0.001643203 | 0.003157341 | 1 |
| T2DM | monocyte | *HDLBP* | 2.602349679 | 0.000413657 | 0.00117837 | 1 |
| T2DM | monocyte | *PASK* | -0.966533586 | 0.010246294 | 0.01489444 | 1 |
| T2DM | monocyte | *STK25* | 2.291899672 | 0.000232214 | 0.00079821 | 1 |
| T2DM | monocyte | *TPO* | 0.459427653 | 0.005009036 | 0.007934444 | 1 |
| T2DM | monocyte | *TAF1B* | 3.338309589 | 0.000152812 | 0.000624661 | 0.960728854 |
| T2DM | monocyte | *STRN* | 2.696054321 | 3.04E-05 | 0.000362859 | 0.191247241 |
| T2DM | monocyte | *CEBPZ* | 2.345749893 | 0.000267692 | 0.000879761 | 1 |
| T2DM | monocyte | *PRKD3* | 2.753667114 | 3.04E-05 | 0.000362859 | 0.190969096 |
| T2DM | monocyte | *SLC25A12* | 2.327007317 | 0.000518182 | 0.00138236 | 1 |
| T2DM | monocyte | *RMDN2* | 3.297397208 | 3.16E-05 | 0.000363519 | 0.19863248 |
| T2DM | monocyte | *SDC1* | 5.905434312 | 6.99E-05 | 0.000427286 | 0.43946825 |
| T2DM | monocyte | *SOS1* | 1.413423971 | 0.000220273 | 0.000773036 | 1 |
| T2DM | monocyte | *COX7A2L* | 2.772607346 | 6.85E-05 | 0.000423977 | 0.430582416 |
| T2DM | monocyte | *PNO1* | 1.372661596 | 0.000613858 | 0.001543731 | 1 |

| | | | | | | |
|---|---|---|---|---|---|---|
| T2DM | monocyte | *ORC4* | 2.476744161 | 6.73E-05 | 0.000421481 | 0.423417252 |
| T2DM | monocyte | *THADA* | 2.642456113 | 2.37E-05 | 0.000362859 | 0.149210656 |
| T2DM | monocyte | *CD207* | 18.45443916 | 0.000520245 | 0.001385338 | 1 |
| T2DM | monocyte | *VAX2* | 1.164597076 | 0.001719682 | 0.003261056 | 1 |
| T2DM | monocyte | *MSH6* | 4.039274259 | 8.53E-05 | 0.000458095 | 0.536565227 |
| T2DM | monocyte | *PLEKHA3* | 3.67781448 | 2.43E-05 | 0.000362859 | 0.152798837 |
| T2DM | monocyte | *ALMS1* | 2.774699095 | 0.000118437 | 0.000544237 | 0.74461169 |
| T2DM | monocyte | *BCL9* | 1.728658857 | 8.18E-05 | 0.000446179 | 0.514444715 |
| T2DM | monocyte | *DHCR24* | 3.445242484 | 0.000110077 | 0.000521673 | 0.692052261 |
| T2DM | monocyte | *DNAJC16* | 2.63519017 | 0.00108297 | 0.002332521 | 1 |
| T2DM | monocyte | *MORN1* | 2.134378557 | 0.029610932 | 0.038495436 | 1 |
| T2DM | monocyte | *SCP2* | 2.018347813 | 4.22E-05 | 0.000374514 | 0.265057389 |
| T2DM | monocyte | *RALGPS2* | 2.484543806 | 3.94E-05 | 0.000371009 | 0.24751162 |
| T2DM | monocyte | *CEP104* | 2.660893962 | 0.002708568 | 0.004651398 | 1 |
| T2DM | monocyte | *FAM20B* | 4.227061337 | 2.55E-05 | 0.000362859 | 0.160632051 |
| T2DM | monocyte | *TMEM59* | 3.434428763 | 1.98E-05 | 0.000362859 | 0.124641753 |
| T2DM | monocyte | *MRPL37* | 4.565233735 | 0.000179408 | 0.000685677 | 1 |
| T2DM | monocyte | *ICMT* | 2.039729123 | 0.001351861 | 0.002741661 | 1 |
| T2DM | monocyte | *QSOX1* | 4.88301255 | 0.000143272 | 0.000601705 | 0.900751873 |
| T2DM | monocyte | *PARK7* | 3.480783454 | 2.53E-05 | 0.000362859 | 0.159352053 |
| T2DM | monocyte | *KCNC4* | 0.634357264 | 0.008215326 | 0.012253798 | 1 |
| T2DM | monocyte | *EDEM3* | 3.574568622 | 0.000115945 | 0.000536386 | 0.728948214 |
| T2DM | monocyte | *RAP1A* | 2.578445047 | 0.003268011 | 0.005461096 | 1 |
| T2DM | monocyte | *DOCK7* | 1.687148471 | 9.52E-05 | 0.000480165 | 0.598765666 |
| T2DM | monocyte | *FBXO6* | 2.925478406 | 0.005820798 | 0.009051535 | 1 |
| T2DM | monocyte | *C1orf21* | 0.735940687 | 0.004962897 | 0.007869706 | 1 |
| T2DM | monocyte | *DNAJC6* | 0.934973739 | 0.002469672 | 0.004292736 | 1 |
| T2DM | monocyte | *LEPR* | 2.678209244 | 4.00E-05 | 0.000371009 | 0.251530137 |
| T2DM | monocyte | *MFN2* | 3.70775343 | 5.17E-05 | 0.000384775 | 0.325249805 |
| T2DM | monocyte | *SMG7* | 3.112783555 | 0.001651653 | 0.003169364 | 1 |
| T2DM | monocyte | *NCF2* | -0.250534295 | 0.012443222 | 0.017668908 | 1 |
| T2DM | monocyte | *RGS2* | -0.178303985 | 0.008889185 | 0.013140443 | 1 |
| T2DM | monocyte | *OLFML3* | 1.822617182 | 0.000363679 | 0.001076982 | 1 |
| T2DM | monocyte | *CFHR3* | 1.057100835 | 0.035979765 | 0.046023353 | 1 |
| T2DM | monocyte | *CRYZ* | 1.038997012 | 0.006963331 | 0.010584734 | 1 |
| T2DM | monocyte | *WARS2* | 1.51017904 | 0.014882865 | 0.02073312 | 1 |
| T2DM | monocyte | *OSCP1* | 1.95161183 | 0.000130751 | 0.000572307 | 0.822031886 |
| T2DM | monocyte | *GNPAT* | 1.824698578 | 0.000910756 | 0.002052303 | 1 |
| T2DM | monocyte | *C1orf109* | 1.730567714 | 0.001504269 | 0.002964682 | 1 |
| T2DM | monocyte | *TBCE* | 2.693090902 | 0.001610699 | 0.003114877 | 1 |
| T2DM | monocyte | *LGALS8* | 1.11208309 | 0.005806257 | 0.009035628 | 1 |
| T2DM | monocyte | *MTR* | 2.444199852 | 3.72E-05 | 0.000366571 | 0.233705294 |
| T2DM | monocyte | *ACADM* | 1.90493375 | 4.48E-05 | 0.000379941 | 0.281658799 |
| T2DM | monocyte | *PADI2* | -0.481321119 | 0.009328242 | 0.013724937 | 1 |
| T2DM | monocyte | *KLHL12* | 3.764770169 | 0.001082062 | 0.002331366 | 1 |
| T2DM | monocyte | *ZNHIT6* | 1.564803973 | 0.001455794 | 0.002900975 | 1 |
| T2DM | monocyte | *RBBP5* | 3.463614561 | 2.72E-05 | 0.000362859 | 0.17076275 |
| T2DM | monocyte | *KIF17* | 1.789175822 | 0.010509216 | 0.015220327 | 1 |
| T2DM | monocyte | *CD46* | 1.721536584 | 4.72E-05 | 0.000379941 | 0.297026686 |
| T2DM | monocyte | *PRDX1* | 2.402315071 | 8.78E-05 | 0.00046125 | 0.551976015 |

| | | | | | | |
|---|---|---|---|---|---|---|
| T2DM | monocyte | *FAAH* | 0.814066153 | 0.001765532 | 0.003319348 | 1 |
| T2DM | monocyte | *NSUN4* | 2.064421993 | 0.010588489 | 0.015304675 | 1 |
| T2DM | monocyte | *TMED5* | 2.108377383 | 7.07E-05 | 0.000427286 | 0.444255031 |
| T2DM | monocyte | *DR1* | 4.633166785 | 0.000406643 | 0.001166997 | 1 |
| T2DM | monocyte | *CNN3* | 1.537468282 | 0.000202752 | 0.000735545 | 1 |
| T2DM | monocyte | *PRRC2C* | 2.952558189 | 1.26E-05 | 0.000362859 | 0.079514128 |
| T2DM | monocyte | *ABCD3* | 2.270618295 | 5.65E-05 | 0.000394257 | 0.355003309 |
| T2DM | monocyte | *VAMP4* | 3.894041054 | 1.04E-05 | 0.000362859 | 0.065683387 |
| T2DM | monocyte | *PTBP2* | 2.148214826 | 8.74E-06 | 0.000362859 | 0.054974884 |
| T2DM | monocyte | *TNFSF4* | 0.770571841 | 0.026949482 | 0.03531292 | 1 |
| T2DM | monocyte | *DARS2* | 1.192943591 | 0.001712456 | 0.003249687 | 1 |
| T2DM | monocyte | *IRF6* | 1.133655658 | 0.004702936 | 0.007500443 | 1 |
| T2DM | monocyte | *SERPINC1* | 1.999167935 | 9.75E-05 | 0.000485446 | 0.613193718 |
| T2DM | monocyte | *RCAN3* | 2.843668414 | 0.016120835 | 0.02224088 | 1 |
| T2DM | monocyte | *RCOR3* | 2.464661623 | 3.31E-05 | 0.000365043 | 0.208238858 |
| T2DM | monocyte | *MTFR1L* | 5.033221475 | 0.000381558 | 0.001113675 | 1 |
| T2DM | monocyte | *NSL1* | 3.145876412 | 7.67E-05 | 0.000437295 | 0.482441346 |
| T2DM | monocyte | *SLC5A9* | 6.46016122 | 1.08E-05 | 0.000362859 | 0.068148669 |
| T2DM | monocyte | *OSBPL9* | 2.161382644 | 4.78E-05 | 0.000379941 | 0.300622035 |
| T2DM | monocyte | *TXNDC12* | 2.538523946 | 5.09E-05 | 0.000383728 | 0.320198877 |
| T2DM | monocyte | *ESYT2* | 1.658654142 | 0.000195257 | 0.000722579 | 1 |
| T2DM | monocyte | *COLEC11* | 1.248909257 | 0.007514674 | 0.011324246 | 1 |
| T2DM | monocyte | *KMT2A* | 3.134216174 | 2.44E-05 | 0.000362859 | 0.153311704 |
| T2DM | monocyte | *CAMSAP2* | 2.293479866 | 3.56E-05 | 0.000365043 | 0.22370146 |
| T2DM | monocyte | *ATF6* | 3.445099973 | 9.98E-05 | 0.000492855 | 0.627430097 |
| T2DM | monocyte | *LRMP* | -0.434417562 | 0.033806097 | 0.043437345 | 1 |
| T2DM | monocyte | *ATP10B* | 1.431179928 | 0.002025326 | 0.003661077 | 1 |
| T2DM | monocyte | *SPCS2* | 3.046953248 | 7.44E-05 | 0.000433444 | 0.467985382 |
| T2DM | monocyte | *ELOVL4* | 1.69764107 | 9.50E-06 | 0.000362859 | 0.059743737 |
| T2DM | monocyte | *FILIP1* | 0.704904471 | 0.002110849 | 0.00378412 | 1 |
| T2DM | monocyte | *SGIP1* | 0.950999141 | 0.000688757 | 0.001678379 | 1 |
| T2DM | monocyte | *MED28* | 2.681704737 | 0.00585351 | 0.009095654 | 1 |
| T2DM | monocyte | *SLC16A7* | 1.164145744 | 0.00020629 | 0.000743876 | 1 |
| T2DM | monocyte | *VAMP8* | 2.560119365 | 0.001874388 | 0.003446704 | 1 |
| T2DM | monocyte | *FOXO3* | 2.130340233 | 3.56E-05 | 0.000365043 | 0.224113582 |
| T2DM | monocyte | *TGIF2* | 3.657918397 | 0.000200102 | 0.000728454 | 1 |
| T2DM | monocyte | *ABCG2* | 3.201981036 | 0.000157546 | 0.000636155 | 0.990493449 |
| T2DM | monocyte | *SPP1* | 2.900859791 | 0.000135969 | 0.000583505 | 0.854835273 |
| T2DM | monocyte | *FAM47E-STBL* | 0.71287053 | 0.004525263 | 0.007239269 | 1 |
| T2DM | monocyte | *MFSD1* | 3.504017884 | 2.19E-05 | 0.000362859 | 0.137606175 |
| T2DM | monocyte | *RAB3GAP2* | 3.374203191 | 2.28E-05 | 0.000362859 | 0.14336859 |
| T2DM | monocyte | *UBN1* | 3.013777816 | 5.44E-05 | 0.000387662 | 0.342004931 |
| T2DM | monocyte | *WDR35* | 1.685287661 | 0.000287699 | 0.000923782 | 1 |
| T2DM | monocyte | *NDUFB3* | 2.997234904 | 0.000633155 | 0.001579622 | 1 |
| T2DM | monocyte | *TRPM6* | 0.879210535 | 0.001994505 | 0.00361576 | 1 |
| T2DM | monocyte | *GDA* | 3.382854957 | 0.001724088 | 0.003264862 | 1 |
| T2DM | monocyte | *C2orf40* | 1.025698544 | 0.002401977 | 0.004195951 | 1 |
| T2DM | monocyte | *CPSF3* | 1.902494371 | 0.00011944 | 0.000545328 | 0.750916425 |
| T2DM | monocyte | *PIGZ* | 0.720156192 | 0.015451111 | 0.021420316 | 1 |
| T2DM | monocyte | *SENP5* | 3.600115146 | 6.48E-06 | 0.000331164 | 0.040733195 |

| | | | | | | |
|---|---|---|---|---|---|---|
| T2DM | monocyte | *CCDC92* | 0.856534588 | 0.015292488 | 0.021233187 | 1 |
| T2DM | monocyte | *PTBP3* | 3.222148745 | 0.001131537 | 0.002409882 | 1 |
| T2DM | monocyte | *FKBP15* | 2.961221546 | 0.003128689 | 0.005259377 | 1 |
| T2DM | monocyte | *CTNNAL1* | 1.039645002 | 0.000171425 | 0.000666827 | 1 |
| T2DM | monocyte | *TRIM32* | 6.573480635 | 2.10E-05 | 0.000362859 | 0.132102537 |
| T2DM | monocyte | *PHF19* | -0.73824965 | 0.028804617 | 0.03754813 | 1 |
| T2DM | monocyte | *BSPRY* | 3.502773309 | 0.00227712 | 0.004004546 | 1 |
| T2DM | monocyte | *PPP6C* | 1.709152098 | 0.000206704 | 0.000743876 | 1 |
| T2DM | monocyte | *RBM18* | 4.155318527 | 5.37E-05 | 0.000385603 | 0.337576281 |
| T2DM | monocyte | *HSDL2* | 0.881214214 | 0.000519292 | 0.001383972 | 1 |
| T2DM | monocyte | *INVS* | 1.62262658 | 8.48E-05 | 0.000456188 | 0.532827274 |
| T2DM | monocyte | *KDSR* | 1.830187845 | 0.000129756 | 0.00057004 | 0.815774031 |
| T2DM | monocyte | *DCAF4* | 3.164690237 | 1.06E-05 | 0.000362859 | 0.066441295 |
| T2DM | monocyte | *IFI27L2* | 1.932637513 | 0.014295228 | 0.020007591 | 1 |
| T2DM | monocyte | *DNAL1* | 1.835353832 | 0.000183418 | 0.000696642 | 1 |
| T2DM | monocyte | *IRF2BPL* | 2.879978452 | 5.29E-05 | 0.000385235 | 0.332682548 |
| T2DM | monocyte | *TTLL5* | 2.146047265 | 5.47E-05 | 0.000388233 | 0.344050764 |
| T2DM | monocyte | *FLVCR2* | -0.447943448 | 0.021190592 | 0.028488824 | 1 |
| T2DM | monocyte | *PPP4R4* | 1.010849218 | 0.012349612 | 0.017550183 | 1 |
| T2DM | monocyte | *ZC2HC1C* | 2.216121582 | 0.000267152 | 0.000878904 | 1 |
| T2DM | monocyte | *ALDH6A1* | 0.889582649 | 0.004300092 | 0.006926641 | 1 |
| T2DM | monocyte | *EIF2B2* | 1.569137526 | 0.001210535 | 0.002527609 | 1 |
| T2DM | monocyte | *COQ6* | 2.506998052 | 6.95E-05 | 0.00042643 | 0.436663933 |
| T2DM | monocyte | *GPR75* | 9.571319966 | 0.000268281 | 0.000880774 | 1 |
| T2DM | monocyte | *KLHL29* | 0.898261834 | 0.000437911 | 0.001226618 | 1 |
| T2DM | monocyte | *ATAD2B* | 3.208256765 | 3.12E-05 | 0.000362859 | 0.196081931 |
| T2DM | monocyte | *ATL2* | 2.456086829 | 3.79E-05 | 0.00036941 | 0.238108512 |
| T2DM | monocyte | *CNRIP1* | 1.372778755 | 0.001841505 | 0.003406162 | 1 |
| T2DM | monocyte | *CRIPT* | 1.246434394 | 0.001508647 | 0.002969588 | 1 |
| T2DM | monocyte | *SLC17A5* | 2.059426515 | 0.001592435 | 0.003095555 | 1 |
| T2DM | monocyte | *OGFRL1* | 2.21568738 | 0.009932126 | 0.014484639 | 1 |
| T2DM | monocyte | *CUTC* | 1.980370289 | 7.58E-05 | 0.000435317 | 0.476292544 |
| T2DM | monocyte | *PYROXD2* | 0.828367069 | 0.000770857 | 0.001822488 | 1 |
| T2DM | monocyte | *MXI1* | 3.807278586 | 0.000769516 | 0.001822488 | 1 |
| T2DM | monocyte | *C10orf88* | 3.591880892 | 3.75E-05 | 0.000367486 | 0.235527885 |
| T2DM | monocyte | *HELLS* | 2.756393227 | 0.000175999 | 0.000677754 | 1 |
| T2DM | monocyte | *TCTN3* | 2.671241504 | 0.018074591 | 0.024638976 | 1 |
| T2DM | monocyte | *AVPI1* | 1.868912141 | 0.001247148 | 0.002578368 | 1 |
| T2DM | monocyte | *WDR11* | 2.865631989 | 0.002036052 | 0.00367202 | 1 |
| T2DM | monocyte | *C10orf76* | 2.906093481 | 2.32E-05 | 0.000362859 | 0.146054781 |
| T2DM | monocyte | *KCNIP2* | 0.645349214 | 0.000565089 | 0.001465478 | 1 |
| T2DM | monocyte | *KANSL1* | 2.291413059 | 1.65E-05 | 0.000362859 | 0.103966461 |
| T2DM | monocyte | *HOXB5* | 1.123866023 | 0.012588522 | 0.017861462 | 1 |
| T2DM | monocyte | *HOXB3* | 0.691037571 | 0.038076491 | 0.048442494 | 1 |
| T2DM | monocyte | *CAAP1* | 3.609963378 | 7.96E-05 | 0.000442619 | 0.500159059 |
| T2DM | monocyte | *CD274* | 0.846742998 | 0.00210376 | 0.003774641 | 1 |
| T2DM | monocyte | *NUP43* | 2.417281563 | 0.00077123 | 0.001822488 | 1 |
| T2DM | monocyte | *LRP11* | 1.342809237 | 0.00365322 | 0.006008899 | 1 |
| T2DM | monocyte | *CCDC170* | 1.191274437 | 0.002438491 | 0.004246756 | 1 |
| T2DM | monocyte | *CYSTM1* | 2.380720312 | 3.35E-05 | 0.000365043 | 0.210793082 |

| | | | | | | |
|---|---|---|---|---|---|---|
| T2DM | monocyte | *PCDHB10* | 1.175509662 | 0.028910111 | 0.037677833 | 1 |
| T2DM | monocyte | *PCDHB14* | 1.271595359 | 0.001266844 | 0.00261136 | 1 |
| T2DM | monocyte | *SEC16B* | 1.8219895 | 0.001657547 | 0.003176913 | 1 |
| T2DM | monocyte | *GORAB* | 4.731651803 | 0.000108143 | 0.000517029 | 0.679892934 |
| T2DM | monocyte | *ACAT2* | 3.676550511 | 6.76E-05 | 0.0004219 | 0.425242528 |
| T2DM | monocyte | *SNX19* | 2.377944006 | 0.00019891 | 0.000726217 | 1 |
| T2DM | monocyte | *MRC1* | 0.841770824 | 0.000489482 | 0.001329318 | 1 |
| T2DM | monocyte | *IQSEC3* | 0.915215635 | 0.001971991 | 0.003582174 | 1 |
| T2DM | monocyte | *ENOX1* | 0.926685977 | 0.000746377 | 0.001782842 | 1 |
| T2DM | monocyte | *DNAJC15* | 3.748706825 | 2.42E-05 | 0.000362859 | 0.152102037 |
| T2DM | monocyte | *WBP4* | 2.191547472 | 5.16E-05 | 0.000384775 | 0.324357493 |
| T2DM | monocyte | *SMAD9* | 0.728096103 | 0.001348514 | 0.002736638 | 1 |
| T2DM | monocyte | *SIL1* | 2.435060904 | 2.63E-05 | 0.000362859 | 0.165337463 |
| T2DM | monocyte | *ZFP30* | 1.497618762 | 0.00028386 | 0.000915489 | 1 |
| T2DM | monocyte | *ARL1* | 2.629426144 | 0.006468662 | 0.009923982 | 1 |
| T2DM | monocyte | *ADRA1A* | 0.679887778 | 3.63E-07 | 6.70E-05 | 0.002279098 |
| T2DM | monocyte | *EPHX2* | 0.539864439 | 0.007623704 | 0.011463819 | 1 |
| T2DM | monocyte | *ZNF706* | 1.947474254 | 0.0001501 | 0.000618401 | 0.943680323 |
| T2DM | monocyte | *CRISPLD1* | 0.908714876 | 0.001628131 | 0.003142787 | 1 |
| T2DM | monocyte | *RDH10* | 0.978420787 | 0.000273732 | 0.000893537 | 1 |
| T2DM | monocyte | *TEX14* | 5.980407267 | 1.91E-07 | 4.50E-05 | 0.001199348 |
| T2DM | monocyte | *TRIM6* | 1.165118163 | 0.0167106 | 0.022978902 | 1 |
| T2DM | monocyte | *PRB2* | 0.930053647 | 0.00087535 | 0.001998302 | 1 |
| T2DM | monocyte | *PYROXD1* | 1.671547709 | 0.020648912 | 0.027822483 | 1 |
| T2DM | monocyte | *ZSCAN18* | 1.862279029 | 0.000119673 | 0.000545998 | 0.752384854 |
| T2DM | monocyte | *ZNF211* | 1.887473882 | 5.54E-05 | 0.000390932 | 0.348391085 |
| T2DM | monocyte | *PDZRN3* | 0.687229012 | 0.000531527 | 0.001405262 | 1 |
| T2DM | monocyte | *LHX4* | 0.935492921 | 0.002502392 | 0.004342407 | 1 |
| T2DM | monocyte | *SEC22A* | 2.643275362 | 6.14E-07 | 8.58E-05 | 0.003858999 |
| T2DM | monocyte | *CSTA* | -0.61269162 | 0.026104618 | 0.034305964 | 1 |
| T2DM | monocyte | *POPDC2* | 6.144648271 | 3.66E-05 | 0.000365495 | 0.229896601 |
| T2DM | monocyte | *B4GALT4* | 3.91474941 | 3.67E-05 | 0.000365515 | 0.231005169 |
| T2DM | monocyte | *CD80* | -0.919554099 | 0.021799864 | 0.029216744 | 1 |
| T2DM | monocyte | *CAT* | 0.715925092 | 0.012161213 | 0.017313756 | 1 |
| T2DM | monocyte | *PILRB* | 0.424422863 | 0.013488411 | 0.018988849 | 1 |
| T2DM | monocyte | *GJB6* | -30.44695366 | 0.00089603 | 0.00202711 | 1 |
| T2DM | monocyte | *TMEM39B* | 4.077678472 | 0.001323392 | 0.002699599 | 1 |
| T2DM | monocyte | *RNF115* | 2.270210461 | 4.21E-05 | 0.000374514 | 0.264901995 |
| T2DM | monocyte | *POLR3GL* | 3.910965131 | 7.95E-05 | 0.000442619 | 0.500085061 |
| T2DM | monocyte | *TNFSF10* | 2.304455389 | 0.022708306 | 0.030337254 | 1 |
| T2DM | monocyte | *ZNF639* | 1.917336573 | 0.000155814 | 0.000632001 | 0.979602063 |
| T2DM | monocyte | *LIAS* | 1.284070432 | 0.000601503 | 0.001524272 | 1 |
| T2DM | monocyte | *CLCC1* | 2.015669798 | 0.000361508 | 0.001073112 | 1 |
| T2DM | monocyte | *GPSM2* | 1.602647724 | 4.72E-05 | 0.000379941 | 0.296874876 |
| T2DM | monocyte | *CXCR4* | -0.721214909 | 0.01272999 | 0.018025551 | 1 |
| T2DM | monocyte | *ACVR2A* | 3.643578107 | 7.10E-05 | 0.000427286 | 0.446295775 |
| T2DM | monocyte | *POLK* | 3.106998571 | 2.61E-05 | 0.000362859 | 0.16377912 |
| T2DM | monocyte | *FLT3* | -0.580922282 | 0.018981553 | 0.025730276 | 1 |
| T2DM | monocyte | *MTIF3* | 1.453356423 | 7.08E-05 | 0.000427286 | 0.445422452 |
| T2DM | monocyte | *RASL11A* | 1.145362045 | 0.008288872 | 0.012351775 | 1 |

| | | | | | | |
|---|---|---|---|---|---|---|
| T2DM | monocyte | *FMOD* | 2.056162047 | 0.000465254 | 0.001277315 | 1 |
| T2DM | monocyte | *SERAC1* | 2.727880405 | 0.00033536 | 0.00102039 | 1 |
| T2DM | monocyte | *LDB3* | 0.737446649 | 0.00102826 | 0.002249366 | 1 |
| T2DM | monocyte | *FAM35A* | 2.528479743 | 0.00011109 | 0.000523558 | 0.698425762 |
| T2DM | monocyte | *FAM213A* | 0.585351167 | 0.002734455 | 0.004680512 | 1 |
| T2DM | monocyte | *NAA60* | 4.907494125 | 3.95E-05 | 0.000371009 | 0.248124185 |
| T2DM | monocyte | *ODF2L* | 1.995365985 | 1.65E-05 | 0.000362859 | 0.103719942 |
| T2DM | monocyte | *SPATA1* | 2.002898872 | 6.73E-05 | 0.000421481 | 0.4231795 |
| T2DM | monocyte | *LRRC39* | 0.944066602 | 0.005186458 | 0.008176624 | 1 |
| T2DM | monocyte | *RWDD3* | 1.130576968 | 0.001559409 | 0.003043777 | 1 |
| T2DM | monocyte | *CCDC18* | 1.849871667 | 0.000108662 | 0.000518843 | 0.683154971 |
| T2DM | monocyte | *RPAP2* | 1.742356641 | 8.71E-05 | 0.000461034 | 0.547415391 |
| T2DM | monocyte | *BBS9* | 1.735599303 | 0.000309623 | 0.000971846 | 1 |
| T2DM | monocyte | *PMS2* | 1.588443871 | 8.45E-05 | 0.000455349 | 0.531391977 |
| T2DM | monocyte | *KIAA0087* | 1.872713948 | 0.014173302 | 0.01984136 | 1 |
| T2DM | monocyte | *KLHL7* | 2.49011573 | 4.94E-05 | 0.000383513 | 0.310740581 |
| T2DM | monocyte | *HERPUD2* | 4.013153148 | 0.000133681 | 0.000578424 | 0.840449736 |
| T2DM | monocyte | *FAM126A* | 2.759328498 | 4.00E-05 | 0.000371009 | 0.251377693 |
| T2DM | monocyte | *FKBP9* | 0.894050137 | 0.000692097 | 0.00168521 | 1 |
| T2DM | monocyte | *CCZ1* | 1.977568177 | 8.86E-05 | 0.000462594 | 0.556880864 |
| T2DM | monocyte | *SMU1* | 4.074187958 | 0.000496528 | 0.001338052 | 1 |
| T2DM | monocyte | *GLIPR2* | 3.070181547 | 0.000605454 | 0.001529335 | 1 |
| T2DM | monocyte | *SLC25A51* | 1.04692016 | 0.000523067 | 0.001390262 | 1 |
| T2DM | monocyte | *DCAF10* | 2.37057097 | 3.40E-05 | 0.000365043 | 0.213917597 |
| T2DM | monocyte | *TRIM24* | 3.220481693 | 0.001914204 | 0.003499449 | 1 |
| T2DM | monocyte | *BICC1* | 0.372923132 | 0.002020976 | 0.003655316 | 1 |
| T2DM | monocyte | *SLC25A16* | 1.170803934 | 0.000101579 | 0.000497371 | 0.638624467 |
| T2DM | monocyte | *ZWINT* | 1.091776524 | 0.012893164 | 0.018236068 | 1 |
| T2DM | monocyte | *ACADS* | 1.351738217 | 0.000770844 | 0.001822488 | 1 |
| T2DM | monocyte | *SSPN* | 1.159206402 | 4.68E-05 | 0.000379941 | 0.294218063 |
| T2DM | monocyte | *CCDC91* | 2.48316905 | 1.98E-05 | 0.000362859 | 0.12453779 |
| T2DM | monocyte | *EBPL* | 1.582449908 | 2.81E-05 | 0.000362859 | 0.176931298 |
| T2DM | monocyte | *OPTN* | 1.135441787 | 0.001665449 | 0.003186451 | 1 |
| T2DM | monocyte | *ATF1* | 2.898538634 | 9.50E-06 | 0.000362859 | 0.059706772 |
| T2DM | monocyte | *TSFM* | 3.532660935 | 2.25E-05 | 0.000362859 | 0.141476335 |
| T2DM | monocyte | *SPATS2* | 2.212864482 | 7.66E-05 | 0.000437295 | 0.481845609 |
| T2DM | monocyte | *ORMDL2* | 1.938388297 | 0.000402271 | 0.001158461 | 1 |
| T2DM | monocyte | *PDE1B* | 2.842520079 | 0.004224414 | 0.006822216 | 1 |
| T2DM | monocyte | *IKZF4* | 0.778431801 | 0.003624781 | 0.005973525 | 1 |
| T2DM | monocyte | *SARDH* | 7.331995783 | 0.000139625 | 0.000591126 | 0.877821468 |
| T2DM | monocyte | *NDUFAF4* | 1.125355741 | 0.006061568 | 0.009384162 | 1 |
| T2DM | monocyte | *TTC21B* | 3.552624461 | 1.46E-05 | 0.000362859 | 0.091994015 |
| T2DM | monocyte | *NMI* | 4.459337867 | 0.000301203 | 0.000955067 | 1 |
| T2DM | monocyte | *ACVR1C* | 0.509462929 | 0.000847582 | 0.001953354 | 1 |
| T2DM | monocyte | *SLC36A1* | 0.912817659 | 0.030798685 | 0.03985635 | 1 |
| T2DM | monocyte | *PLA2G12A* | 2.0034286 | 0.000134252 | 0.00057865 | 0.844044463 |
| T2DM | monocyte | *C4BPB* | 0.842736897 | 0.029678507 | 0.038559366 | 1 |
| T2DM | monocyte | *RAB38* | 2.15892135 | 0.000920379 | 0.002069536 | 1 |
| T2DM | monocyte | *AGO2* | 3.876599197 | 0.000197121 | 0.000724426 | 1 |
| T2DM | monocyte | *CKS2* | -1.206946534 | 0.021185267 | 0.028488824 | 1 |

| | | | | | | |
|---|---|---|---|---|---|---|
| T2DM | monocyte | *CHPF* | 0.762518371 | 0.01016242 | 0.014786192 | 1 |
| T2DM | monocyte | *DNPEP* | 3.431251796 | 3.04E-05 | 0.000362859 | 0.191231508 |
| T2DM | monocyte | *MOGAT1* | 0.613655105 | 0.001898462 | 0.003477941 | 1 |
| T2DM | monocyte | *FAM124B* | 1.697873854 | 0.004216971 | 0.006815047 | 1 |
| T2DM | monocyte | *FAM210B* | 1.370194329 | 0.000175846 | 0.000677754 | 1 |
| T2DM | monocyte | *SNX21* | 0.957937759 | 0.000606063 | 0.001530249 | 1 |
| T2DM | monocyte | *SLC12A5* | -2.214989408 | 0.000164199 | 0.000650483 | 1 |
| T2DM | monocyte | *SDC4* | 2.990595919 | 0.00254347 | 0.004407607 | 1 |
| T2DM | monocyte | *ARFGEF2* | 2.813966643 | 1.72E-05 | 0.000362859 | 0.108343511 |
| T2DM | monocyte | *STX16* | 3.85721096 | 2.65E-05 | 0.000362859 | 0.166512655 |
| T2DM | monocyte | *RNF114* | 2.640757562 | 0.000227437 | 0.000787811 | 1 |
| T2DM | monocyte | *KCNK15* | 2.284638501 | 0.003507375 | 0.005808974 | 1 |
| T2DM | monocyte | *NEURL2* | 1.040424118 | 0.000163894 | 0.000650094 | 1 |
| T2DM | monocyte | *FASTKD3* | 3.877978446 | 8.73E-05 | 0.000461034 | 0.548914706 |
| T2DM | monocyte | *PEPD* | 1.457442214 | 3.37E-05 | 0.000365043 | 0.211693843 |
| T2DM | monocyte | *STAMBP* | 2.734626724 | 4.17E-05 | 0.00037367 | 0.262387194 |
| T2DM | monocyte | *ATP8A1* | 2.317913883 | 9.73E-06 | 0.000362859 | 0.061192942 |
| T2DM | monocyte | *USP22* | 2.475393831 | 3.59E-05 | 0.000365043 | 0.225782462 |
| T2DM | monocyte | *HIF3A* | 0.856106839 | 0.000293094 | 0.000937676 | 1 |
| T2DM | monocyte | *ZNF45* | 3.250227958 | 4.82E-05 | 0.00038013 | 0.30334382 |
| T2DM | monocyte | *MRS2* | 2.05773233 | 0.000172124 | 0.000668817 | 1 |
| T2DM | monocyte | *SERPINB6* | 2.667685668 | 4.01E-05 | 0.000371009 | 0.251915409 |
| T2DM | monocyte | *HIST1H1D* | -0.84196693 | 0.012751452 | 0.018051876 | 1 |
| T2DM | monocyte | *NQO2* | 2.594283995 | 5.96E-05 | 0.000399203 | 0.374851699 |
| T2DM | monocyte | *ZNF391* | 0.614291536 | 0.002621757 | 0.004523936 | 1 |
| T2DM | monocyte | *RPS10* | 5.468990062 | 0.000230567 | 0.000794721 | 1 |
| T2DM | monocyte | *MED20* | 1.215205613 | 0.007642859 | 0.011489874 | 1 |
| T2DM | monocyte | *TBCC* | 1.213791485 | 0.032287554 | 0.041656444 | 1 |
| T2DM | monocyte | *KLHDC3* | 1.895602275 | 0.000472896 | 0.001295467 | 1 |
| T2DM | monocyte | *GNMT* | -0.728081776 | 0.011151041 | 0.01605372 | 1 |
| T2DM | monocyte | *KLHL31* | 0.47030989 | 0.002646899 | 0.004561692 | 1 |
| T2DM | monocyte | *COL21A1* | 0.473384903 | 0.002672074 | 0.004601295 | 1 |
| T2DM | monocyte | *GLO1* | 4.830267449 | 4.31E-07 | 6.94E-05 | 0.002707351 |
| T2DM | monocyte | *CPNE5* | 1.052998878 | 0.038028427 | 0.048426667 | 1 |
| T2DM | monocyte | *RIOK1* | 3.484005786 | 0.00019869 | 0.000726217 | 1 |
| T2DM | monocyte | *NUP153* | 3.121676427 | 1.41E-05 | 0.000362859 | 0.08895603 |
| T2DM | monocyte | *EEF1E1* | 1.915056817 | 0.000783495 | 0.001843127 | 1 |
| T2DM | monocyte | *EREG* | -0.137028078 | 0.006688689 | 0.01022909 | 1 |
| T2DM | monocyte | *CNOT1* | 3.280577888 | 1.18E-05 | 0.000362859 | 0.074124078 |
| T2DM | monocyte | *BBS2* | 1.826389937 | 2.62E-05 | 0.000362859 | 0.164973564 |
| T2DM | monocyte | *MT1G* | 4.971718177 | 0.005088968 | 0.0080408 | 1 |
| T2DM | monocyte | *CLYBL* | 3.476159539 | 3.01E-05 | 0.000362859 | 0.189072848 |
| T2DM | monocyte | *TMTC4* | 2.907274335 | 6.19E-05 | 0.000406647 | 0.389416358 |
| T2DM | monocyte | *GRK4* | 0.92585709 | 1.28E-06 | 0.000146309 | 0.008046968 |
| T2DM | monocyte | *TEKT3* | 0.993203827 | 0.030648493 | 0.039688378 | 1 |
| T2DM | monocyte | *MRPS7* | 2.237252395 | 0.000215648 | 0.000765509 | 1 |
| T2DM | monocyte | *NUP85* | 3.031588971 | 7.18E-05 | 0.000428995 | 0.451530159 |
| T2DM | monocyte | *MIF4GD* | 1.531180017 | 0.001001992 | 0.002209584 | 1 |
| T2DM | monocyte | *NT5C* | 4.187068926 | 1.53E-05 | 0.000362859 | 0.095895875 |
| T2DM | monocyte | *DDX31* | 1.718649641 | 0.002334444 | 0.004093782 | 1 |

| | | | | | | |
|---|---|---|---|---|---|---|
| T2DM | monocyte | *PPP1R12C* | 2.947923737 | 0.015404493 | 0.021365111 | 1 |
| T2DM | monocyte | *OPRL1* | -1.519143585 | 2.14E-05 | 0.000362859 | 0.134308639 |
| T2DM | monocyte | *PPDPF* | 6.000923468 | 4.63E-06 | 0.000287938 | 0.029081785 |
| T2DM | monocyte | *PLGLB2* | -7.280164549 | 0.001919921 | 0.00350277 | 1 |
| T2DM | monocyte | *CHCHD5* | 1.024013745 | 0.002055887 | 0.003704603 | 1 |
| T2DM | monocyte | *PAX8* | 1.152479514 | 0.001460633 | 0.002906013 | 1 |
| T2DM | monocyte | *INSIG2* | 4.193970391 | 0.002103748 | 0.003774641 | 1 |
| T2DM | monocyte | *POLR1B* | 0.968734895 | 0.015368128 | 0.02132879 | 1 |
| T2DM | monocyte | *SLC25A23* | 1.156329735 | 0.001031468 | 0.002254028 | 1 |
| T2DM | monocyte | *GTF2F1* | 3.94052703 | 0.000155568 | 0.000631819 | 0.978055805 |
| T2DM | monocyte | *CLPP* | 2.305666837 | 6.34E-05 | 0.000409778 | 0.398304569 |
| T2DM | monocyte | *SH2D3A* | 1.656326056 | 0.0012267 | 0.00255373 | 1 |
| T2DM | monocyte | *TRIP10* | 0.905286601 | 0.000502289 | 0.001349618 | 1 |
| T2DM | monocyte | *GPR108* | 2.296931641 | 0.008211107 | 0.012250411 | 1 |
| T2DM | monocyte | *SNRPD2* | 4.863403787 | 4.73E-05 | 0.000379941 | 0.297606611 |
| T2DM | monocyte | *RTN2* | 6.036558307 | 0.001204578 | 0.00251758 | 1 |
| T2DM | monocyte | *SDCBP2* | 1.990390602 | 0.000663064 | 0.001634778 | 1 |
| T2DM | monocyte | *PSMF1* | 2.115060086 | 0.025940861 | 0.034105017 | 1 |
| T2DM | monocyte | *DTD1* | 3.042369761 | 5.78E-05 | 0.000396862 | 0.363399612 |
| T2DM | monocyte | *RBCK1* | 2.798970322 | 0.000438927 | 0.00122857 | 1 |
| T2DM | monocyte | *RRBP1* | 1.681287074 | 0.000196399 | 0.000723774 | 1 |
| T2DM | monocyte | *BMP2* | 1.947191774 | 0.000795898 | 0.001865701 | 1 |
| T2DM | monocyte | *PCSK2* | 1.889243127 | 0.002547824 | 0.004413936 | 1 |
| T2DM | monocyte | *MKKS* | 3.434468658 | 4.54E-05 | 0.000379941 | 0.285702206 |
| T2DM | monocyte | *DSTN* | 1.511871017 | 0.000153529 | 0.000625966 | 0.96523919 |
| T2DM | monocyte | *MGME1* | 2.442183388 | 0.00023538 | 0.000805134 | 1 |
| T2DM | monocyte | *FAM110A* | -0.349081921 | 0.011474183 | 0.016466147 | 1 |
| T2DM | monocyte | *MRPS26* | 0.669166714 | 0.036925267 | 0.047165614 | 1 |
| T2DM | monocyte | *NCLN* | 2.767250971 | 0.006936948 | 0.010552284 | 1 |
| T2DM | monocyte | *ZNF436* | 4.351894615 | 3.24E-08 | 1.36E-05 | 0.000203902 |
| T2DM | monocyte | *MMP24* | 2.402614262 | 3.36E-05 | 0.000365043 | 0.211112593 |
| T2DM | monocyte | *UROD* | 5.714538429 | 1.65E-06 | 0.000169672 | 0.010349977 |
| T2DM | monocyte | *ST3GAL3* | 1.839029019 | 9.45E-05 | 0.000478875 | 0.593992028 |
| T2DM | monocyte | *KLC1* | 2.575228436 | 1.77E-05 | 0.000362859 | 0.111463497 |
| T2DM | monocyte | *XRCC3* | 2.975834051 | 2.68E-05 | 0.000362859 | 0.168542806 |
| T2DM | monocyte | *MCF2L* | 0.64692742 | 0.000525066 | 0.001392823 | 1 |
| T2DM | monocyte | *F10* | 0.851123876 | 0.001005696 | 0.002214803 | 1 |
| T2DM | monocyte | *PCID2* | 2.125485552 | 0.000466205 | 0.001278809 | 1 |
| T2DM | monocyte | *PROZ* | 4.930357954 | 0.002196692 | 0.003897996 | 1 |
| T2DM | monocyte | *IGFLR1* | -1.719544141 | 0.015891444 | 0.021958134 | 1 |
| T2DM | monocyte | *COX6B1* | 2.660570207 | 8.72E-05 | 0.000461034 | 0.548528388 |
| T2DM | monocyte | *PRDX5* | 1.746194873 | 0.001391524 | 0.002804151 | 1 |
| T2DM | monocyte | *PRR12* | 2.581416282 | 4.12E-07 | 6.81E-05 | 0.002588137 |
| T2DM | monocyte | *FLRT1* | 0.813906445 | 0.004905848 | 0.007796529 | 1 |
| T2DM | monocyte | *SBDS* | 4.552612807 | 0.000176041 | 0.000677754 | 1 |
| T2DM | monocyte | *BECN1* | 5.59511019 | 1.24E-05 | 0.000362859 | 0.078121545 |
| T2DM | monocyte | *TRAP1* | 0.632430463 | 0.023368884 | 0.031107384 | 1 |
| T2DM | monocyte | *DNAJC8* | 2.219166402 | 0.001576429 | 0.003074135 | 1 |
| T2DM | monocyte | *ZNF384* | 4.33552544 | 0.001670336 | 0.003194829 | 1 |
| T2DM | monocyte | *EMG1* | 4.043432243 | 0.000473801 | 0.001297381 | 1 |

| | | | | | | |
|---|---|---|---|---|---|---|
| T2DM | monocyte | *ATG14* | 1.631990328 | 0.00011169 | 0.000524745 | 0.702198033 |
| T2DM | monocyte | *RHOJ* | 1.052954519 | 0.000576464 | 0.001482303 | 1 |
| T2DM | monocyte | *L3HYPDH* | 0.830114249 | 0.002163522 | 0.003856553 | 1 |
| T2DM | monocyte | *HSPA2* | -0.797087606 | 0.017237676 | 0.023626176 | 1 |
| T2DM | monocyte | *ZBTB1* | 2.140939617 | 6.24E-05 | 0.000407194 | 0.392398406 |
| T2DM | monocyte | *PZP* | 1.472918155 | 0.000230948 | 0.000795163 | 1 |
| T2DM | monocyte | *RHOT1* | 2.49968563 | 4.14E-05 | 0.000373192 | 0.260114844 |
| T2DM | monocyte | *WDR60* | 1.377097056 | 0.000450709 | 0.001248286 | 1 |
| T2DM | monocyte | *AIF1L* | 1.708919948 | 0.000323668 | 0.000999459 | 1 |
| T2DM | monocyte | *MAP2K2* | 2.919920961 | 0.001246138 | 0.002577977 | 1 |
| T2DM | monocyte | *CANX* | 3.899299298 | 0.000151108 | 0.0006205 | 0.950014228 |
| T2DM | monocyte | *IPPK* | 3.44887711 | 0.001255209 | 0.002590775 | 1 |
| T2DM | monocyte | *FGD3* | -0.420603331 | 0.021521747 | 0.028865852 | 1 |
| T2DM | monocyte | *MASP1* | 0.777538497 | 0.005530009 | 0.00865932 | 1 |
| T2DM | monocyte | *ATP13A4* | 1.037824641 | 0.002917314 | 0.00494504 | 1 |
| T2DM | monocyte | *HRASLS* | 1.557284302 | 0.001312539 | 0.00268705 | 1 |
| T2DM | monocyte | *HELB* | 1.672309836 | 0.000866918 | 0.001984096 | 1 |
| T2DM | monocyte | *RAP1B* | 2.38123963 | 0.003620667 | 0.00597281 | 1 |
| T2DM | monocyte | *RAB3IP* | 1.119811168 | 0.00358235 | 0.005914453 | 1 |
| T2DM | monocyte | *YEATS4* | 2.009700927 | 0.001829287 | 0.003393752 | 1 |
| T2DM | monocyte | *TAS2R4* | 0.832333328 | 0.013180167 | 0.018600159 | 1 |
| T2DM | monocyte | *LRRC61* | 0.811688838 | 0.002766073 | 0.004720495 | 1 |
| T2DM | monocyte | *IDUA* | 2.620595791 | 0.000171395 | 0.000666827 | 1 |
| T2DM | monocyte | *TMEM175* | 4.121573236 | 3.02E-05 | 0.000362859 | 0.189805647 |
| T2DM | monocyte | *EMC1* | 3.039781954 | 0.001419374 | 0.002847353 | 1 |
| T2DM | monocyte | *PLA2G5* | 0.968493045 | 0.00102268 | 0.002239495 | 1 |
| T2DM | monocyte | *SIN3B* | 3.678653664 | 2.37E-05 | 0.000362859 | 0.148870186 |
| T2DM | monocyte | *CHTF18* | 2.448404367 | 0.000134835 | 0.000580224 | 0.847707992 |
| T2DM | monocyte | *TICAM1* | 6.204675562 | 5.36E-05 | 0.000385603 | 0.337145826 |
| T2DM | monocyte | *PNKD* | 1.53568878 | 0.000603732 | 0.001528045 | 1 |
| T2DM | monocyte | *HIP1* | 1.90805789 | 4.82E-05 | 0.000379967 | 0.302833674 |
| T2DM | monocyte | *PTPN12* | 2.974625572 | 3.96E-05 | 0.000371009 | 0.249039869 |
| T2DM | monocyte | *POR* | 1.847399791 | 0.00011011 | 0.000521673 | 0.692259723 |
| T2DM | monocyte | *FGL2* | -0.293973163 | 0.030769298 | 0.039836251 | 1 |
| T2DM | monocyte | *STEAP4* | 0.939373557 | 0.000779899 | 0.001837789 | 1 |
| T2DM | monocyte | *GNAI1* | 0.855364977 | 0.00104135 | 0.00226774 | 1 |
| T2DM | monocyte | *SGCE* | 1.368648565 | 0.00021576 | 0.000765509 | 1 |
| T2DM | monocyte | *RBM48* | 1.784174743 | 0.000340038 | 0.001029894 | 1 |
| T2DM | monocyte | *CASD1* | 2.287040747 | 5.80E-05 | 0.000397161 | 0.364594111 |
| T2DM | monocyte | *SRD5A3* | 1.599657498 | 0.000373937 | 0.001098059 | 1 |
| T2DM | monocyte | *VPREB3* | 10.58962357 | 0.003904621 | 0.006366274 | 1 |
| T2DM | monocyte | *MGAT3* | 0.788445002 | 0.001870097 | 0.003444321 | 1 |
| T2DM | monocyte | *ATF4* | 1.297200749 | 0.00049546 | 0.001337467 | 1 |
| T2DM | monocyte | *A4GALT* | 0.887010725 | 0.002589069 | 0.004479218 | 1 |
| T2DM | monocyte | *CDC42EP1* | 3.265778601 | 0.00054917 | 0.001440035 | 1 |
| T2DM | monocyte | *TPST2* | 2.068605794 | 8.95E-05 | 0.000464456 | 0.562455626 |
| T2DM | monocyte | *BAIAP2L2* | 1.449379152 | 6.72E-05 | 0.000421481 | 0.422182583 |
| T2DM | monocyte | *TST* | 1.642931756 | 0.003529268 | 0.005840618 | 1 |
| T2DM | monocyte | *APOL5* | -3.710492587 | 0.018242883 | 0.024826708 | 1 |
| T2DM | monocyte | *EMC4* | 3.277750072 | 2.99E-06 | 0.000230511 | 0.01877521 |

| | | | | | | |
|---|---|---|---|---|---|---|
| T2DM | monocyte | *SPECC1* | 3.004881468 | 0.002245681 | 0.003961621 | 1 |
| T2DM | monocyte | *POT1* | 3.128955957 | 0.000105412 | 0.000507835 | 0.662725243 |
| T2DM | monocyte | *CDHR3* | 0.930256788 | 0.00027826 | 0.000903626 | 1 |
| T2DM | monocyte | *PRKRIP1* | 1.953811019 | 0.003157653 | 0.005305228 | 1 |
| T2DM | monocyte | *FOXP2* | 0.608565202 | 0.003664553 | 0.006021706 | 1 |
| T2DM | monocyte | *STRIP2* | 2.029006181 | 0.001460171 | 0.002906012 | 1 |
| T2DM | monocyte | *FLNC* | 1.190357084 | 0.001610642 | 0.003114877 | 1 |
| T2DM | monocyte | *IRF5* | -0.270358471 | 0.026561668 | 0.034870162 | 1 |
| T2DM | monocyte | *KLHDC10* | 2.498388524 | 2.87E-05 | 0.000362859 | 0.180503684 |
| T2DM | monocyte | *HOXD1* | 6.176392722 | 0.001056611 | 0.002294615 | 1 |
| T2DM | monocyte | *HOXD3* | 0.70096762 | 0.001246037 | 0.002577977 | 1 |
| T2DM | monocyte | *CHN1* | 1.030036697 | 0.000697628 | 0.001693432 | 1 |
| T2DM | monocyte | *GAD1* | 2.130121236 | 0.002124916 | 0.003804997 | 1 |
| T2DM | monocyte | *ORMDL1* | 1.581818581 | 0.000125385 | 0.000557491 | 0.788292601 |
| T2DM | monocyte | *HOXD9* | 1.629913346 | 0.00745744 | 0.011248782 | 1 |
| T2DM | monocyte | *PSMG2* | 2.032091643 | 0.000417993 | 0.00118535 | 1 |
| T2DM | monocyte | *TWSG1* | 4.581681353 | 0.00016672 | 0.000656301 | 1 |
| T2DM | monocyte | *ARPP19* | 3.519198586 | 9.26E-05 | 0.000473501 | 0.581933026 |
| T2DM | monocyte | *VPS13C* | 2.776815692 | 0.000304588 | 0.000963252 | 1 |
| T2DM | monocyte | *CALML4* | -0.689203686 | 0.011276483 | 0.016219457 | 1 |
| T2DM | monocyte | *ACKR4* | 2.74094104 | 9.61E-05 | 0.000482554 | 0.604223881 |
| T2DM | monocyte | *SUMF2* | 3.795043407 | 4.67E-05 | 0.000379941 | 0.293445032 |
| T2DM | monocyte | *SPCS3* | 3.979029358 | 0.000199345 | 0.000726217 | 1 |
| T2DM | monocyte | *SERGEF* | 4.43550464 | 2.62E-05 | 0.000362859 | 0.164409031 |
| T2DM | monocyte | *RPAIN* | 1.768853553 | 0.004075695 | 0.006610912 | 1 |
| T2DM | monocyte | *USP6* | 0.675911902 | 0.029756371 | 0.038652542 | 1 |
| T2DM | monocyte | *ATP1B2* | 2.141047239 | 0.005641709 | 0.008810091 | 1 |
| T2DM | monocyte | *CCL4* | 1.838461452 | 0.00511214 | 0.008073355 | 1 |
| T2DM | monocyte | *PHF20L1* | 2.782086413 | 4.67E-05 | 0.000379941 | 0.293550153 |
| T2DM | monocyte | *PUS7L* | 2.845349268 | 0.000115896 | 0.000536386 | 0.728635324 |
| T2DM | monocyte | *KRI1* | 1.218524879 | 0.016726479 | 0.022995708 | 1 |
| T2DM | monocyte | *CDKN2D* | -0.832671746 | 0.019882089 | 0.02687566 | 1 |
| T2DM | monocyte | *MTUS1* | 1.213445243 | 0.000595557 | 0.001515557 | 1 |
| T2DM | monocyte | *NGDN* | 1.941669431 | 0.000159539 | 0.000640496 | 1 |
| T2DM | monocyte | *RIPK3* | 2.542132276 | 0.003949735 | 0.006434824 | 1 |
| T2DM | monocyte | *ADCY4* | 0.758434534 | 0.001825249 | 0.003390895 | 1 |
| T2DM | monocyte | *RAB2B* | 1.553814001 | 0.000239165 | 0.000815394 | 1 |
| T2DM | monocyte | *BCL2L2* | 3.340982705 | 4.06E-05 | 0.00037107 | 0.255139886 |
| T2DM | monocyte | *PARP2* | 2.394542749 | 0.000827822 | 0.001920485 | 1 |
| T2DM | monocyte | *RNASE1* | 1.002622604 | 0.001275224 | 0.00262605 | 1 |
| T2DM | monocyte | *DAD1* | 1.647609515 | 0.000214536 | 0.00076289 | 1 |
| T2DM | monocyte | *EPB41L4A* | 1.095636981 | 0.000253625 | 0.00084636 | 1 |
| T2DM | monocyte | *CDO1* | 1.438260682 | 0.000434241 | 0.001219871 | 1 |
| T2DM | monocyte | *REEP5* | 2.992372088 | 2.52E-05 | 0.000362859 | 0.158415907 |
| T2DM | monocyte | *QRICH2* | 1.521670781 | 0.000447425 | 0.001242475 | 1 |
| T2DM | monocyte | *SEC14L1* | 1.786931278 | 0.018010629 | 0.024562434 | 1 |
| T2DM | monocyte | *TMEM8A* | 3.966584493 | 0.003690851 | 0.006055423 | 1 |
| T2DM | monocyte | *SHC2* | 1.623422891 | 7.37E-05 | 0.000432034 | 0.463217288 |
| T2DM | monocyte | *ERMARD* | 2.785879884 | 0.000440198 | 0.001228968 | 1 |
| T2DM | monocyte | *PHF10* | 2.816793399 | 5.36E-05 | 0.000385603 | 0.336947417 |

| | | | | | | |
|---|---|---|---|---|---|---|
| T2DM | monocyte | *DOCK6* | 1.606606693 | 0.000166919 | 0.000656301 | 1 |
| T2DM | monocyte | *ECSIT* | 4.60366252 | 9.59E-05 | 0.000482439 | 0.603048518 |
| T2DM | monocyte | *ELOF1* | 1.609476674 | 0.001003003 | 0.002211039 | 1 |
| T2DM | monocyte | *CDC16* | 1.95824834 | 0.000755249 | 0.001797898 | 1 |
| T2DM | monocyte | *THEM6* | 1.064762539 | 0.018644044 | 0.025332852 | 1 |
| T2DM | monocyte | *TOMM40* | 3.71690571 | 0.001074326 | 0.00232186 | 1 |
| T2DM | monocyte | *DPP6* | 0.856673247 | 0.005561278 | 0.008701781 | 1 |
| T2DM | monocyte | *RPL36* | 3.050354709 | 0.004271808 | 0.006889907 | 1 |
| T2DM | monocyte | *GTPBP3* | 1.046198026 | 0.032993387 | 0.042445145 | 1 |
| T2DM | monocyte | *PLVAP* | 0.326475427 | 0.034368965 | 0.044124501 | 1 |
| T2DM | monocyte | *BST2* | 4.990866477 | 3.05E-05 | 0.000362859 | 0.191916823 |
| T2DM | monocyte | *DDA1* | 2.353703097 | 0.000640876 | 0.001593879 | 1 |
| T2DM | monocyte | *LSM7* | 1.391590032 | 0.000433452 | 0.001218264 | 1 |
| T2DM | monocyte | *TULP4* | 1.675338511 | 0.000169836 | 0.000662097 | 1 |
| T2DM | monocyte | *RTN4IP1* | 2.379390282 | 0.000136248 | 0.000583905 | 0.85658858 |
| T2DM | monocyte | *RSPH3* | 1.132570917 | 0.016036982 | 0.022136055 | 1 |
| T2DM | monocyte | *ACTN4* | 1.668719482 | 0.000151802 | 0.000622149 | 0.954376835 |
| T2DM | monocyte | *STK33* | 0.541319401 | 0.010793714 | 0.01557853 | 1 |
| T2DM | monocyte | *NDUFA10* | 1.004787229 | 0.013704119 | 0.019253139 | 1 |
| T2DM | monocyte | *PXDN* | 0.783013343 | 0.00063536 | 0.001584495 | 1 |
| T2DM | monocyte | *PGPEP1* | 1.793616125 | 0.033526549 | 0.04309577 | 1 |
| T2DM | monocyte | *KIAA1683* | 0.991812119 | 0.000429463 | 0.001210234 | 1 |
| T2DM | monocyte | *HRC* | 1.443559988 | 0.006356121 | 0.009784929 | 1 |
| T2DM | monocyte | *TRPM4* | 2.32457108 | 0.000496174 | 0.001338052 | 1 |
| T2DM | monocyte | *CAMSAP1* | 2.8105483 | 5.32E-05 | 0.000385235 | 0.3346381 |
| T2DM | monocyte | *UBAC1* | 3.471691854 | 0.000302191 | 0.000957114 | 1 |
| T2DM | monocyte | *TNNT3* | 0.750722315 | 0.012085109 | 0.017232724 | 1 |
| T2DM | monocyte | *ATXN10* | 2.614921508 | 2.90E-05 | 0.000362859 | 0.18213618 |
| T2DM | monocyte | *TUBGCP2* | 3.345494838 | 3.10E-05 | 0.000362859 | 0.194640601 |
| T2DM | monocyte | *CYP2E1* | 2.041732402 | 0.001303744 | 0.002673399 | 1 |
| T2DM | monocyte | *HBZ* | 7.468747374 | 0.00880285 | 0.013022004 | 1 |
| T2DM | monocyte | *PAK4* | 1.399061181 | 0.000524988 | 0.001392823 | 1 |
| T2DM | monocyte | *ZNF337* | 1.496234765 | 0.000718255 | 0.001730618 | 1 |
| T2DM | monocyte | *POMT1* | 1.953946426 | 0.000263734 | 0.000869479 | 1 |
| T2DM | monocyte | *YIPF2* | 1.668247226 | 0.003646203 | 0.006000031 | 1 |
| T2DM | monocyte | *TMEM160* | 5.546379441 | 2.98E-05 | 0.000362859 | 0.187197768 |
| T2DM | monocyte | *MAP3K10* | 1.100197999 | 0.00153194 | 0.003000408 | 1 |
| T2DM | monocyte | *ARHGEF16* | 1.356368193 | 0.003273966 | 0.005468498 | 1 |
| T2DM | monocyte | *SMPDL3B* | 5.471328642 | 0.001606355 | 0.003112582 | 1 |
| T2DM | monocyte | *ATPIF1* | 1.868839048 | 0.024586281 | 0.032528188 | 1 |
| T2DM | monocyte | *THEMIS2* | -0.276468275 | 0.021575264 | 0.028928063 | 1 |
| T2DM | monocyte | *CLIP1* | 3.349207925 | 2.59E-05 | 0.000362859 | 0.162815057 |
| T2DM | monocyte | *ZNF317* | 2.984079133 | 0.018198016 | 0.024785729 | 1 |
| T2DM | monocyte | *PPAN* | 1.308133111 | 0.001518044 | 0.002978363 | 1 |
| T2DM | monocyte | *ZNF236* | 2.430471719 | 0.003139364 | 0.005275911 | 1 |
| T2DM | monocyte | *C12orf65* | 2.638609953 | 3.36E-06 | 0.000245788 | 0.021137732 |
| T2DM | monocyte | *HSD17B3* | 0.872101473 | 0.003709323 | 0.006079383 | 1 |
| T2DM | monocyte | *HABP4* | 1.612287408 | 0.000304998 | 0.000964063 | 1 |
| T2DM | monocyte | *EPS8L1* | 10.99984572 | 0.004428578 | 0.007102671 | 1 |
| T2DM | monocyte | *LILRB2* | -0.113013673 | 0.007700545 | 0.011568297 | 1 |

| | | | | | | |
|---|---|---|---|---|---|---|
| T2DM | monocyte | *COX4I2* | 1.688306067 | 0.002214438 | 0.003920634 | 1 |
| T2DM | monocyte | *ATP6V1E1* | 2.587706799 | 0.003089904 | 0.005201132 | 1 |
| T2DM | monocyte | *CHMP1A* | 3.613932262 | 0.000182129 | 0.000693125 | 1 |
| T2DM | monocyte | *TBC1D5* | 2.431452845 | 3.31E-05 | 0.000365043 | 0.208308051 |
| T2DM | monocyte | *RFTN1* | 1.36497472 | 8.33E-05 | 0.000450242 | 0.523631641 |
| T2DM | monocyte | *KCNC3* | -0.404549885 | 0.010072097 | 0.014664955 | 1 |
| T2DM | monocyte | *PDLIM4* | 1.763392096 | 0.000179767 | 0.000686631 | 1 |
| T2DM | monocyte | *KIF3A* | 1.488354649 | 0.000396225 | 0.001146371 | 1 |
| T2DM | monocyte | *AOC3* | 0.98272369 | 0.001238739 | 0.002566892 | 1 |
| T2DM | monocyte | *ACLY* | 2.388661462 | 0.000138748 | 0.0005894 | 0.872311329 |
| T2DM | monocyte | *AOC2* | 1.468337808 | 0.00184978 | 0.003416441 | 1 |
| T2DM | monocyte | *EXOC4* | 2.402388008 | 2.13E-06 | 0.000188222 | 0.013389774 |
| T2DM | monocyte | *ACAP3* | 4.321522102 | 1.10E-05 | 0.000362859 | 0.068954929 |
| T2DM | monocyte | *ANO1* | 3.845317314 | 4.57E-05 | 0.000379941 | 0.287275089 |
| T2DM | monocyte | *PPFIA1* | 2.057836084 | 0.000302049 | 0.000957114 | 1 |
| T2DM | monocyte | *NPHP4* | 3.974905809 | 3.00E-05 | 0.000362859 | 0.188785635 |
| T2DM | monocyte | *MAP1B* | 0.675061784 | 0.000401535 | 0.001156944 | 1 |
| T2DM | monocyte | *CKMT2* | 1.532023609 | 0.001943063 | 0.003537804 | 1 |
| T2DM | monocyte | *ZCCHC9* | 2.436338036 | 7.29E-05 | 0.000431726 | 0.458078178 |
| T2DM | monocyte | *KHDRBS3* | 0.700236986 | 0.001005221 | 0.002214803 | 1 |
| T2DM | monocyte | *CHD1L* | 2.919430471 | 0.000895866 | 0.00202711 | 1 |
| T2DM | monocyte | *RBM8A* | 1.439754217 | 0.011538146 | 0.016546607 | 1 |
| T2DM | monocyte | *ZNF304* | 1.508178684 | 0.01143458 | 0.016424309 | 1 |
| T2DM | monocyte | *ZSCAN5A* | 3.507147152 | 9.45E-05 | 0.000478875 | 0.594283808 |
| T2DM | monocyte | *ZNF132* | 2.852326394 | 0.00077771 | 0.001834694 | 1 |
| T2DM | monocyte | *RHPN2* | 1.201719328 | 0.023062658 | 0.030745321 | 1 |
| T2DM | monocyte | *C19orf12* | 0.728721319 | 0.002187388 | 0.003883679 | 1 |
| T2DM | monocyte | *ACTR10* | 2.000799082 | 0.00011209 | 0.000525118 | 0.704708881 |
| T2DM | monocyte | *GCH1* | -0.231473039 | 0.015342116 | 0.021297391 | 1 |
| T2DM | monocyte | *LGALS3* | 2.108340606 | 7.10E-05 | 0.000427286 | 0.446124042 |
| T2DM | monocyte | *DNAJB1* | 1.735281203 | 0.001103218 | 0.002364792 | 1 |
| T2DM | monocyte | *FBXW9* | 1.729744291 | 0.000238641 | 0.000814072 | 1 |
| T2DM | monocyte | *C19orf57* | 5.722873278 | 8.49E-05 | 0.000456772 | 0.533966959 |
| T2DM | monocyte | *MATN3* | 2.574922102 | 0.001975891 | 0.003586151 | 1 |
| T2DM | monocyte | *CCT6B* | 1.481889823 | 0.004175979 | 0.006755115 | 1 |
| T2DM | monocyte | *RAF1* | 3.538795525 | 9.52E-05 | 0.000480165 | 0.59843548 |
| T2DM | monocyte | *ENOSF1* | 1.172749844 | 0.000409574 | 0.001172717 | 1 |
| T2DM | monocyte | *TRIM5* | 3.338613917 | 5.84E-05 | 0.000397957 | 0.366913457 |
| T2DM | monocyte | *TRIM22* | 3.376480227 | 0.001251725 | 0.002585282 | 1 |
| T2DM | monocyte | *RRP8* | 2.668919603 | 0.000349163 | 0.001045826 | 1 |
| T2DM | monocyte | *IMMT* | 2.964856085 | 8.38E-06 | 0.000361025 | 0.052709674 |
| T2DM | monocyte | *MRPL35* | 2.245966091 | 3.10E-05 | 0.000362859 | 0.195207014 |
| T2DM | monocyte | *CARD6* | 2.69311958 | 0.00011676 | 0.000539362 | 0.734072251 |
| T2DM | monocyte | *INPP5K* | 1.284206138 | 0.00220648 | 0.003909848 | 1 |
| T2DM | monocyte | *MYBBP1A* | 1.541961359 | 0.000678112 | 0.001661104 | 1 |
| T2DM | monocyte | *RPA1* | 2.707829218 | 0.003646591 | 0.006000031 | 1 |
| T2DM | monocyte | *SERPINF1* | 0.924309867 | 0.000845735 | 0.001950526 | 1 |
| T2DM | monocyte | *TMEM128* | 2.930590756 | 0.00035477 | 0.001060094 | 1 |
| T2DM | monocyte | *FIGNL1* | 3.040303605 | 8.76E-05 | 0.00046125 | 0.551005007 |
| T2DM | monocyte | *WBP2* | 3.709549372 | 3.59E-05 | 0.000365043 | 0.225881741 |

| | | | | | | |
|---|---|---|---|---|---|---|
| T2DM | monocyte | *XAF1* | 1.884202575 | 3.34E-05 | 0.000365043 | 0.210164596 |
| T2DM | monocyte | *TERF2* | 4.16875171 | 1.15E-05 | 0.000362859 | 0.072316189 |
| T2DM | monocyte | *BTBD3* | 4.399551822 | 9.38E-05 | 0.000477414 | 0.589606531 |
| T2DM | monocyte | *PTPRA* | 2.428873198 | 3.22E-05 | 0.000363519 | 0.202314888 |
| T2DM | monocyte | *MMACHC* | 1.365418493 | 0.033509526 | 0.043082697 | 1 |
| T2DM | monocyte | *MUTYH* | 6.007232171 | 4.01E-06 | 0.000271729 | 0.025203397 |
| T2DM | monocyte | *CTNNBL1* | 4.100055902 | 4.88E-05 | 0.000381666 | 0.307016689 |
| T2DM | monocyte | *LPIN3* | 1.580240604 | 0.000342918 | 0.001035674 | 1 |
| T2DM | monocyte | *SERINC3* | 1.193457138 | 0.037714615 | 0.048056706 | 1 |
| T2DM | monocyte | *DMGDH* | 1.032083191 | 0.001432651 | 0.002870323 | 1 |
| T2DM | monocyte | *BHMT2* | 0.28544444 | 0.034979967 | 0.044846699 | 1 |
| T2DM | monocyte | *KANK4* | 0.473427575 | 0.003853735 | 0.006291465 | 1 |
| T2DM | monocyte | *RSG1* | -0.981155353 | 0.021524275 | 0.028865852 | 1 |
| T2DM | monocyte | *CASP9* | 1.474327594 | 0.000211865 | 0.000756388 | 1 |
| T2DM | monocyte | *USPL1* | 3.387128423 | 3.39E-05 | 0.000365043 | 0.213067098 |
| T2DM | monocyte | *POMP* | 3.866356873 | 3.54E-05 | 0.000365043 | 0.222584768 |
| T2DM | monocyte | *WASF3* | 1.072079092 | 0.000442661 | 0.001234698 | 1 |
| T2DM | monocyte | *PEMT* | 0.846845408 | 0.00165781 | 0.003176913 | 1 |
| T2DM | monocyte | *SCO1* | 2.621163214 | 0.000407778 | 0.001169038 | 1 |
| T2DM | monocyte | *CHI3L1* | 0.726009315 | 0.001633058 | 0.003150365 | 1 |
| T2DM | monocyte | *DSTYK* | 2.208131823 | 0.00783282 | 0.011733367 | 1 |
| T2DM | monocyte | *COG6* | 2.268888089 | 8.30E-05 | 0.000449548 | 0.522016652 |
| T2DM | monocyte | *TRPC4* | 1.346060643 | 0.001178837 | 0.002479541 | 1 |
| T2DM | monocyte | *RFC3* | 3.151338424 | 0.000243234 | 0.000821275 | 1 |
| T2DM | monocyte | *BTBD2* | 2.686561431 | 0.000576819 | 0.00148261 | 1 |
| T2DM | monocyte | *PRAM1* | -0.102049161 | 0.00581327 | 0.009044303 | 1 |
| T2DM | monocyte | *ZNF414* | 7.090590744 | 3.04E-05 | 0.000362859 | 0.190899554 |
| T2DM | monocyte | *RTN3* | 1.825038299 | 5.95E-05 | 0.000399203 | 0.373792842 |
| T2DM | monocyte | *SLC2A11* | 4.336748436 | 9.15E-05 | 0.000469636 | 0.575444679 |
| T2DM | monocyte | *C1QTNF6* | 0.97409965 | 0.001144329 | 0.002428079 | 1 |
| T2DM | monocyte | *FAM83F* | 1.649030582 | 0.002105299 | 0.003776324 | 1 |
| T2DM | monocyte | *GIMAP4* | -0.763552197 | 0.025559125 | 0.033659452 | 1 |
| T2DM | monocyte | *AGAP3* | 1.800631275 | 0.000257325 | 0.000854171 | 1 |
| T2DM | monocyte | *SFTPD* | 0.641863529 | 0.019898792 | 0.026892456 | 1 |
| T2DM | monocyte | *TMTC1* | 0.95944644 | 0.000270312 | 0.000886054 | 1 |
| T2DM | monocyte | *IPO8* | 3.867518653 | 2.70E-05 | 0.000362859 | 0.169769095 |
| T2DM | monocyte | *LARS* | 0.863155661 | 0.038629202 | 0.049102667 | 1 |
| T2DM | monocyte | *SPINK5* | 2.461437432 | 0.000884561 | 0.00201134 | 1 |
| T2DM | monocyte | *IMPA1* | 1.155001013 | 0.010637678 | 0.015370968 | 1 |
| T2DM | monocyte | *LRRCC1* | 1.615234678 | 0.000161774 | 0.000644849 | 1 |
| T2DM | monocyte | *SWAP70* | 3.046693061 | 2.97E-05 | 0.000362859 | 0.186613263 |
| T2DM | monocyte | *AMPD3* | 1.868900965 | 0.006091116 | 0.009420626 | 1 |
| T2DM | monocyte | *SBF2* | 2.777971272 | 8.80E-05 | 0.000461329 | 0.553497436 |
| T2DM | monocyte | *RRAS2* | 0.892778594 | 0.000334852 | 0.00102039 | 1 |
| T2DM | monocyte | *HSD17B4* | 1.834972217 | 0.000219739 | 0.000772651 | 1 |
| T2DM | monocyte | *DPF2* | 1.484061811 | 0.0003873 | 0.001126773 | 1 |
| T2DM | monocyte | *GSC* | 15.66123302 | 2.95E-05 | 0.000362859 | 0.185257035 |
| T2DM | monocyte | *CATSPERB* | 0.70313921 | 0.006663176 | 0.010199997 | 1 |
| T2DM | monocyte | *MED6* | 1.451674911 | 0.000158237 | 0.000638126 | 0.994838067 |
| T2DM | monocyte | *EIF2S1* | 3.514550577 | 1.38E-05 | 0.000362859 | 0.086476332 |

| | | | | | | |
|---|---|---|---|---|---|---|
| T2DM | monocyte | *ELP3* | 2.253748944 | 7.19E-05 | 0.000428995 | 0.452158216 |
| T2DM | monocyte | *MRO* | 1.1518591 | 0.004410228 | 0.007078658 | 1 |
| T2DM | monocyte | *THUMPD3* | 1.610352444 | 0.002451412 | 0.004266895 | 1 |
| T2DM | monocyte | *CNTN6* | 0.641356167 | 0.002471815 | 0.004294086 | 1 |
| T2DM | monocyte | *KATNBL1* | 2.525382383 | 1.82E-05 | 0.000362859 | 0.114127204 |
| T2DM | monocyte | *GSTM5* | 0.818593553 | 0.00333726 | 0.00556387 | 1 |
| T2DM | monocyte | *GSTM3* | 0.681984059 | 0.001506236 | 0.002966699 | 1 |
| T2DM | monocyte | *SYT6* | 2.384606624 | 0.00162546 | 0.003138596 | 1 |
| T2DM | monocyte | *PSRC1* | -0.926549112 | 0.023765946 | 0.031580784 | 1 |
| T2DM | monocyte | *WNT2B* | 1.507838792 | 0.00053326 | 0.001408658 | 1 |
| T2DM | monocyte | *CEPT1* | 2.028927811 | 8.15E-05 | 0.000445803 | 0.512227252 |
| T2DM | monocyte | *AP4B1* | 5.530390147 | 6.99E-06 | 0.000338043 | 0.043945623 |
| T2DM | monocyte | *NAPG* | 3.234107837 | 3.89E-05 | 0.000370888 | 0.24478594 |
| T2DM | monocyte | *SPIRE1* | 1.304457465 | 0.001697745 | 0.003231077 | 1 |
| T2DM | monocyte | *PPHLN1* | 2.706475201 | 7.35E-06 | 0.000349439 | 0.046187286 |
| T2DM | monocyte | *YWHAQ* | 2.906786478 | 1.66E-05 | 0.000362859 | 0.104308084 |
| T2DM | monocyte | *LPIN1* | 1.278799457 | 0.006682637 | 0.010224809 | 1 |
| T2DM | monocyte | *LDHA* | 1.702789899 | 0.000157224 | 0.000635622 | 0.98846615 |
| T2DM | monocyte | *ANO3* | 0.441846888 | 0.002431446 | 0.004238009 | 1 |
| T2DM | monocyte | *FST* | 3.102311722 | 0.006478313 | 0.009936363 | 1 |
| T2DM | monocyte | *TIMM17A* | 3.382292784 | 9.92E-06 | 0.000362859 | 0.062384788 |
| T2DM | monocyte | *NARS* | 4.588147011 | 4.39E-05 | 0.000378997 | 0.275869876 |
| T2DM | monocyte | *KIAA1468* | 3.266814494 | 0.000524742 | 0.001392823 | 1 |
| T2DM | monocyte | *FBXO18* | 3.881387534 | 1.85E-05 | 0.000362859 | 0.11650921 |
| T2DM | monocyte | *ECHDC3* | 0.798702785 | 0.002239001 | 0.003956323 | 1 |
| T2DM | monocyte | *IL15RA* | 3.100547188 | 3.93E-05 | 0.000371009 | 0.247367025 |
| T2DM | monocyte | *TMEM241* | 1.35480795 | 0.0001838 | 0.000697374 | 1 |
| T2DM | monocyte | *EMP1* | 0.941728191 | 0.000977497 | 0.002164679 | 1 |
| T2DM | monocyte | *KLRD1* | 0.766386134 | 0.00071873 | 0.001730623 | 1 |
| T2DM | monocyte | *DDB2* | 4.665113238 | 2.08E-06 | 0.000188095 | 0.013069599 |
| T2DM | monocyte | *ACP2* | 3.900368056 | 0.000272364 | 0.00089059 | 1 |
| T2DM | monocyte | *HOOK1* | -1.630554034 | 0.0346871 | 0.044505673 | 1 |
| T2DM | monocyte | *ZCCHC11* | 1.508161364 | 0.000149721 | 0.00061774 | 0.94129509 |
| T2DM | monocyte | *PRPF38A* | 2.385021122 | 0.000236598 | 0.000808419 | 1 |
| T2DM | monocyte | *FHOD3* | 0.4886462 | 0.00607647 | 0.009400288 | 1 |
| T2DM | monocyte | *TPGS2* | 2.167851327 | 3.48E-05 | 0.000365043 | 0.218695469 |
| T2DM | monocyte | *SLC43A3* | 2.229413069 | 0.024542765 | 0.03249797 | 1 |
| T2DM | monocyte | *APLNR* | 1.116472234 | 0.001032682 | 0.002255114 | 1 |
| T2DM | monocyte | *FADS2* | 3.038556035 | 3.91E-05 | 0.000371009 | 0.246124052 |
| T2DM | monocyte | *TMEM165* | 3.032830296 | 9.26E-06 | 0.000362859 | 0.058225103 |
| T2DM | monocyte | *CLOCK* | 3.402666227 | 2.82E-05 | 0.000362859 | 0.177385285 |
| T2DM | monocyte | *CLDN10* | 1.469837994 | 0.002150106 | 0.003840261 | 1 |
| T2DM | monocyte | *DZIP1* | 0.678191532 | 0.003473022 | 0.005762704 | 1 |
| T2DM | monocyte | *BIVM* | 2.153351038 | 0.000224087 | 0.000780086 | 1 |
| T2DM | monocyte | *ERCC5* | 1.532030251 | 0.001100488 | 0.002360549 | 1 |
| T2DM | monocyte | *CARS2* | 2.041799321 | 0.003703799 | 0.006073497 | 1 |
| T2DM | monocyte | *STT3A* | 1.803573666 | 0.000210532 | 0.00075415 | 1 |
| T2DM | monocyte | *KLB* | 0.435301107 | 0.001469431 | 0.002916135 | 1 |
| T2DM | monocyte | *WDR36* | 1.415246448 | 0.000225257 | 0.000782857 | 1 |
| T2DM | monocyte | *UBQLN1* | 2.155289735 | 5.67E-05 | 0.000394331 | 0.356474993 |

| | | | | | | |
|---|---|---|---|---|---|---|
| T2DM | monocyte | *NAA35* | 2.995822881 | 2.56E-05 | 0.000362859 | 0.160902814 |
| T2DM | monocyte | *CTSL* | 1.674644684 | 0.000131333 | 0.000572999 | 0.825691251 |
| T2DM | monocyte | *TMEM2* | 2.810660126 | 3.19E-05 | 0.000363519 | 0.200576106 |
| T2DM | monocyte | *GOLM1* | 1.720852721 | 0.000931906 | 0.002088305 | 1 |
| T2DM | monocyte | *ADAM19* | 0.898428894 | 0.001077205 | 0.002326482 | 1 |
| T2DM | monocyte | *USP30* | 1.210341108 | 0.000323574 | 0.000999459 | 1 |
| T2DM | monocyte | *HRK* | 3.16019135 | 0.009148449 | 0.013491977 | 1 |
| T2DM | monocyte | *DTX1* | 0.526363287 | 0.009951466 | 0.014509477 | 1 |
| T2DM | monocyte | *DMTF1* | 1.799590972 | 7.12E-05 | 0.000427286 | 0.447770158 |
| T2DM | monocyte | *CCDC146* | 1.503214346 | 6.00E-05 | 0.000399998 | 0.377031611 |
| T2DM | monocyte | *TMEM60* | 2.648018648 | 0.000308373 | 0.000969854 | 1 |
| T2DM | monocyte | *POM121C* | 1.431582886 | 0.007755107 | 0.011628037 | 1 |
| T2DM | monocyte | *SRPK2* | 2.912088106 | 2.55E-05 | 0.000362859 | 0.160334794 |
| T2DM | monocyte | *MDFIC* | 1.436453207 | 0.000175686 | 0.000677718 | 1 |
| T2DM | monocyte | *MTO1* | 1.641810293 | 0.000573572 | 0.001479101 | 1 |
| T2DM | monocyte | *ANKRD6* | 3.473807029 | 5.00E-05 | 0.000383513 | 0.314097203 |
| T2DM | monocyte | *KHDC1* | 0.891656496 | 0.022839898 | 0.030480671 | 1 |
| T2DM | monocyte | *SNX14* | 2.719829104 | 4.87E-05 | 0.000381649 | 0.306034648 |
| T2DM | monocyte | *NT5E* | 1.939790979 | 1.52E-05 | 0.000362859 | 0.095549117 |
| T2DM | monocyte | *PRR5L* | 3.059877084 | 0.000489708 | 0.001329359 | 1 |
| T2DM | monocyte | *NAT10* | 2.873476943 | 6.91E-05 | 0.000425443 | 0.434580601 |
| T2DM | monocyte | *ATP5G2* | 3.869204279 | 2.29E-05 | 0.000362859 | 0.143782413 |
| T2DM | monocyte | *ITGA7* | 0.380305851 | 0.032324453 | 0.041695494 | 1 |
| T2DM | monocyte | *RDH5* | 0.607830292 | 0.000864263 | 0.001980904 | 1 |
| T2DM | monocyte | *BLOC1S1* | 2.917649018 | 0.010208297 | 0.014842637 | 1 |
| T2DM | monocyte | *PPP1R1A* | 2.541153674 | 0.000123672 | 0.000554584 | 0.777527237 |
| T2DM | monocyte | *B4GALNT1* | 2.402859087 | 0.037411602 | 0.047728165 | 1 |
| T2DM | monocyte | *FAIM2* | 1.593604578 | 0.002411071 | 0.004207162 | 1 |
| T2DM | monocyte | *KRT7* | 3.161192321 | 0.002504095 | 0.004344162 | 1 |
| T2DM | monocyte | *SLC26A10* | 2.092721583 | 0.010533818 | 0.015241913 | 1 |
| T2DM | monocyte | *MAP7* | 1.371910502 | 0.000241809 | 0.000819987 | 1 |
| T2DM | monocyte | *AHI1* | 0.846547724 | 0.003456362 | 0.005739605 | 1 |
| T2DM | monocyte | *PKIB* | 1.093409005 | 0.001606542 | 0.003112582 | 1 |
| T2DM | monocyte | *PRADC1* | 1.53392316 | 0.001450821 | 0.002893817 | 1 |
| T2DM | monocyte | *KCNMB4* | 3.911889948 | 0.000322901 | 0.00099909 | 1 |
| T2DM | monocyte | *KLHL36* | 2.799852248 | 3.33E-05 | 0.000365043 | 0.209669265 |
| T2DM | monocyte | *FHOD1* | 2.095247955 | 0.000554594 | 0.00144858 | 1 |
| T2DM | monocyte | *CCDC102A* | 0.817207845 | 0.004504498 | 0.007211556 | 1 |
| T2DM | monocyte | *ZNF670* | 0.887358706 | 0.030213304 | 0.039181321 | 1 |
| T2DM | monocyte | *NTPCR* | 3.71441005 | 3.87E-05 | 0.00036941 | 0.243441002 |
| T2DM | monocyte | *PIGC* | 2.019675908 | 0.000115937 | 0.000536386 | 0.728896912 |
| T2DM | monocyte | *USP37* | 3.520535467 | 1.92E-05 | 0.000362859 | 0.120503416 |
| T2DM | monocyte | *SLC19A3* | 0.514015011 | 0.002865221 | 0.004869868 | 1 |
| T2DM | monocyte | *ARMC9* | 1.053822764 | 0.000615465 | 0.001547152 | 1 |
| T2DM | monocyte | *GCC2* | 2.265097457 | 0.000486334 | 0.001323627 | 1 |
| T2DM | monocyte | *MRPS9* | 2.444096485 | 0.000405556 | 0.001164794 | 1 |
| T2DM | monocyte | *ANKRD36* | 2.067296953 | 6.09E-05 | 0.000403612 | 0.382624163 |
| T2DM | monocyte | *USP44* | 0.708457168 | 0.016916284 | 0.023221108 | 1 |
| T2DM | monocyte | *APPL2* | 3.233503814 | 3.94E-05 | 0.000371009 | 0.247497209 |
| T2DM | monocyte | *DRAM1* | 2.275237802 | 0.001636679 | 0.003153478 | 1 |

| | | | | | | |
|---|---|---|---|---|---|---|
| T2DM | monocyte | *SLC41A2* | 0.742394197 | 0.00087022 | 0.001988233 | 1 |
| T2DM | monocyte | *NEK3* | 1.34049031 | 0.000696129 | 0.0016911 | 1 |
| T2DM | monocyte | *VPS36* | 2.272592448 | 0.00012643 | 0.00055907 | 0.79486842 |
| T2DM | monocyte | *RNASEH2B* | 2.960171691 | 3.86E-05 | 0.00036941 | 0.24251462 |
| T2DM | monocyte | *TBC1D4* | 1.067221068 | 0.00030124 | 0.000955067 | 1 |
| T2DM | monocyte | *LRCH1* | 2.084345158 | 3.34E-05 | 0.000365043 | 0.209937287 |
| T2DM | monocyte | *SUCLA2* | 1.15851452 | 0.004414495 | 0.007083699 | 1 |
| T2DM | monocyte | *MED4* | 2.456401567 | 0.00073515 | 0.001763407 | 1 |
| T2DM | monocyte | *SETDB2* | 2.801699538 | 0.000176407 | 0.000678746 | 1 |
| T2DM | monocyte | *C7orf25* | -0.947312886 | 0.008244662 | 0.012294636 | 1 |
| T2DM | monocyte | *SPDYE1* | 1.931500856 | 0.000196299 | 0.000723774 | 1 |
| T2DM | monocyte | *CHST12* | 3.115203568 | 0.000184875 | 0.00070061 | 1 |
| T2DM | monocyte | *GPNMB* | 0.967942589 | 0.000668378 | 0.001643367 | 1 |
| T2DM | monocyte | *KDELR2* | 4.280205575 | 2.32E-05 | 0.000362859 | 0.145949628 |
| T2DM | monocyte | *NUPL2* | 1.991080458 | 0.000220802 | 0.000774224 | 1 |
| T2DM | monocyte | *ZDHHC4* | 2.423365061 | 0.018365431 | 0.024986683 | 1 |
| T2DM | monocyte | *AOAH* | -0.191344254 | 0.006154289 | 0.009506637 | 1 |
| T2DM | monocyte | *TBRG4* | 3.297453 | 0.02514914 | 0.033186829 | 1 |
| T2DM | monocyte | *DDX56* | 5.689168801 | 4.07E-05 | 0.00037107 | 0.256038196 |
| T2DM | monocyte | *HUS1* | 3.378845676 | 2.71E-05 | 0.000362859 | 0.170388774 |
| T2DM | monocyte | *DBNL* | 3.273478645 | 0.000422288 | 0.001195374 | 1 |
| T2DM | monocyte | *TTC5* | 2.478177976 | 0.016999008 | 0.02331939 | 1 |
| T2DM | monocyte | *ALPK3* | 0.695192177 | 0.002466051 | 0.004288815 | 1 |
| T2DM | monocyte | *TM6SF1* | 1.930686581 | 3.85E-05 | 0.00036941 | 0.242271948 |
| T2DM | monocyte | *RSAD1* | 1.153217672 | 0.000868438 | 0.001986853 | 1 |
| T2DM | monocyte | *NMT1* | 1.668580891 | 0.000121495 | 0.000551508 | 0.763838618 |
| T2DM | monocyte | *MYCBPAP* | 0.826307008 | 0.009289099 | 0.013676947 | 1 |
| T2DM | monocyte | *BRIP1* | 3.761354087 | 2.28E-05 | 0.000362859 | 0.143161214 |
| T2DM | monocyte | *BLK* | 1.277060234 | 0.010891656 | 0.015705468 | 1 |
| T2DM | monocyte | *EPRS* | 2.369897227 | 5.78E-05 | 0.000396862 | 0.363525304 |
| T2DM | monocyte | *RPS6KC1* | 2.965652363 | 3.45E-05 | 0.000365043 | 0.216916064 |
| T2DM | monocyte | *IL1RN* | 5.575820123 | 2.72E-05 | 0.000362859 | 0.170946655 |
| T2DM | monocyte | *UGGT1* | 3.686465662 | 3.27E-05 | 0.000365043 | 0.205800553 |
| T2DM | monocyte | *GYPC* | 1.345164365 | 0.000284373 | 0.000916377 | 1 |
| T2DM | monocyte | *LRRC8A* | 1.303360568 | 0.000840238 | 0.001942125 | 1 |
| T2DM | monocyte | *TXN* | 1.750984133 | 0.000296863 | 0.000946439 | 1 |
| T2DM | monocyte | *ODF2* | 3.308609434 | 4.75E-05 | 0.000379941 | 0.298904147 |
| T2DM | monocyte | *TOR1B* | 2.105571507 | 0.012216567 | 0.017376823 | 1 |
| T2DM | monocyte | *SLC2A8* | 2.059192048 | 0.036829065 | 0.047052292 | 1 |
| T2DM | monocyte | *ZFP37* | 1.268087318 | 0.000574151 | 0.001479988 | 1 |
| T2DM | monocyte | *ZNF189* | 1.9285541 | 0.000260785 | 0.00086247 | 1 |
| T2DM | monocyte | *ALDOB* | 2.240963299 | 0.003495701 | 0.005791165 | 1 |
| T2DM | monocyte | *STX17* | 1.766410938 | 0.000161233 | 0.000643434 | 1 |
| T2DM | monocyte | *PRPF4* | 1.663756841 | 0.018687462 | 0.02538636 | 1 |
| T2DM | monocyte | *FPGS* | 2.574478757 | 0.000486165 | 0.001323627 | 1 |
| T2DM | monocyte | *TEX10* | 2.455371697 | 6.49E-05 | 0.000415842 | 0.40833928 |
| T2DM | monocyte | *GARNL3* | 2.180321389 | 7.57E-05 | 0.000435317 | 0.475911433 |
| T2DM | monocyte | *GABBR2* | 12.57205939 | 1.82E-05 | 0.000362859 | 0.114654936 |
| T2DM | monocyte | *PSMB7* | 2.736356509 | 3.07E-05 | 0.000362859 | 0.192879236 |
| T2DM | monocyte | *RABEPK* | 1.303307125 | 0.000413193 | 0.001178344 | 1 |

| | | | | | | |
|---|---|---|---|---|---|---|
| T2DM | monocyte | *GOLGA1* | 2.184429754 | 7.93E-05 | 0.000442263 | 0.498430872 |
| T2DM | monocyte | *LMX1B* | 1.947783173 | 0.002182707 | 0.003878654 | 1 |
| T2DM | monocyte | *DERL1* | 2.663269711 | 0.000218615 | 0.00076956 | 1 |
| T2DM | monocyte | *NOV* | 3.720487656 | 0.004289219 | 0.00691444 | 1 |
| T2DM | monocyte | *POLR1E* | 1.479193371 | 0.002208823 | 0.003912896 | 1 |
| T2DM | monocyte | *IL11RA* | 2.17573305 | 0.000241441 | 0.0008199 | 1 |
| T2DM | monocyte | *APTX* | 2.147768214 | 9.06E-05 | 0.000467143 | 0.56991479 |
| T2DM | monocyte | *TLN1* | 3.984170442 | 3.19E-06 | 0.000238747 | 0.020054711 |
| T2DM | monocyte | *CD72* | 3.52810284 | 0.001443951 | 0.002885606 | 1 |
| T2DM | monocyte | *TMEM8B* | 2.989940682 | 0.000267678 | 0.000879761 | 1 |
| T2DM | monocyte | *GRHPR* | 1.301328114 | 0.018520261 | 0.025170099 | 1 |
| T2DM | monocyte | *DENND4C* | 1.669248375 | 4.81E-05 | 0.000379967 | 0.302429732 |
| T2DM | monocyte | *PPIL1* | 7.167268085 | 0.013390847 | 0.018863601 | 1 |
| T2DM | monocyte | *KIF13A* | 2.100188958 | 3.39E-05 | 0.000365043 | 0.212908348 |
| T2DM | monocyte | *TMEM63B* | 3.584994754 | 1.85E-05 | 0.000362859 | 0.116380757 |
| T2DM | monocyte | *FRS3* | 3.261827561 | 7.97E-05 | 0.000442726 | 0.501246873 |
| T2DM | monocyte | *SLC22A23* | 1.271222523 | 0.000228305 | 0.000788692 | 1 |
| T2DM | monocyte | *LRRC1* | 0.767629437 | 0.002379013 | 0.004156991 | 1 |
| T2DM | monocyte | *FOXF2* | 2.139456531 | 0.034211766 | 0.043940628 | 1 |
| T2DM | monocyte | *BPHL* | 0.707418859 | 0.001106529 | 0.002369404 | 1 |
| T2DM | monocyte | *RIPK1* | 2.200356606 | 6.13E-05 | 0.000404 | 0.385415761 |
| T2DM | monocyte | *TUBB2B* | 1.245176049 | 0.006816051 | 0.010401682 | 1 |
| T2DM | monocyte | *UQCC2* | 4.418296025 | 8.06E-05 | 0.000443644 | 0.506433973 |
| T2DM | monocyte | *TCF19* | 3.164096629 | 0.000145787 | 0.000608331 | 0.916565918 |
| T2DM | monocyte | *FLOT1* | 3.130640602 | 2.16E-06 | 0.000188222 | 0.013572021 |
| T2DM | monocyte | *VARS2* | 0.58105699 | 0.001223567 | 0.00254974 | 1 |
| T2DM | monocyte | *FAM8A1* | 2.654257448 | 0.00010877 | 0.000518843 | 0.683835661 |
| T2DM | monocyte | *FGFBP2* | 0.979987107 | 0.003994972 | 0.006500101 | 1 |
| T2DM | monocyte | *FHDC1* | 0.855064963 | 0.001749501 | 0.003297097 | 1 |
| T2DM | monocyte | *MYO7A* | 1.102533573 | 0.012658715 | 0.017948882 | 1 |
| T2DM | monocyte | *CCDC90B* | 2.548405431 | 7.76E-05 | 0.000438618 | 0.488181617 |
| T2DM | monocyte | *SYTL2* | 0.513471637 | 0.003810042 | 0.006228219 | 1 |
| T2DM | monocyte | *RAB30* | 0.904347712 | 0.000755182 | 0.001797898 | 1 |
| T2DM | monocyte | *PRCP* | 4.337823057 | 1.50E-06 | 0.000157658 | 0.009459457 |
| T2DM | monocyte | *NARS2* | 2.291497626 | 5.62E-05 | 0.000392825 | 0.353149744 |
| T2DM | monocyte | *PI15* | 4.025866358 | 0.009279833 | 0.013666505 | 1 |
| T2DM | monocyte | *SULF1* | 0.778428238 | 0.000441711 | 0.001232596 | 1 |
| T2DM | monocyte | *TGS1* | 2.023278985 | 0.000344654 | 0.001037263 | 1 |
| T2DM | monocyte | *NEK1* | 2.499334991 | 1.55E-05 | 0.000362859 | 0.09763529 |
| T2DM | monocyte | *DCUN1D5* | 0.742294493 | 0.007489 | 0.01129097 | 1 |
| T2DM | monocyte | *TRIM29* | 0.945467884 | 0.016192833 | 0.02233041 | 1 |
| T2DM | monocyte | *SLC37A4* | 5.357698844 | 8.61E-05 | 0.000459567 | 0.54156379 |
| T2DM | monocyte | *POU2F3* | 1.147585621 | 0.026933149 | 0.035298876 | 1 |
| T2DM | monocyte | *PPP2R1B* | 0.632198539 | 0.000812734 | 0.001894571 | 1 |
| T2DM | monocyte | *FDX1* | 1.620888784 | 0.000241623 | 0.0008199 | 1 |
| T2DM | monocyte | *FXYD6* | 1.055085329 | 0.002175322 | 0.003869907 | 1 |
| T2DM | monocyte | *CASP1* | -0.420988965 | 0.015667268 | 0.021667425 | 1 |
| T2DM | monocyte | *MAP2K5* | 2.877618169 | 9.07E-06 | 0.000362859 | 0.057001178 |
| T2DM | monocyte | *ITGA11* | 0.476441009 | 0.002404276 | 0.004197179 | 1 |
| T2DM | monocyte | *PAQR5* | 1.428035746 | 0.00105967 | 0.002298877 | 1 |

| | | | | | | |
|---|---|---|---|---|---|---|
| T2DM | monocyte | *RMDN3* | 1.910532114 | 3.95E-05 | 0.000371009 | 0.248166689 |
| T2DM | monocyte | *ITPKA* | -4.225230915 | 0.007926912 | 0.011865832 | 1 |
| T2DM | monocyte | *UACA* | 1.485157579 | 0.000101437 | 0.000497371 | 0.637736707 |
| T2DM | monocyte | *PLCB2* | -0.237677105 | 0.024832802 | 0.032799123 | 1 |
| T2DM | monocyte | *SPTBN5* | 1.458664965 | 7.47E-07 | 1.00E-04 | 0.004697971 |
| T2DM | monocyte | *RABGGTB* | 3.963400928 | 6.58E-05 | 0.000418344 | 0.413436655 |
| T2DM | monocyte | *SLC44A5* | 1.311555467 | 0.002252851 | 0.003972981 | 1 |
| T2DM | monocyte | *DNASE2B* | -1.733099147 | 0.013342128 | 0.018803397 | 1 |
| T2DM | monocyte | *DBT* | 2.497407285 | 0.00018192 | 0.000692752 | 1 |
| T2DM | monocyte | *RTCA* | 3.027806569 | 6.65E-06 | 0.000332508 | 0.041798186 |
| T2DM | monocyte | *IFT172* | 1.815252254 | 3.08E-05 | 0.000362859 | 0.193440974 |
| T2DM | monocyte | *ADCY3* | 1.589961074 | 0.000432541 | 0.001217273 | 1 |
| T2DM | monocyte | *DYNC2LI1* | 1.097539579 | 0.004773561 | 0.007599741 | 1 |
| T2DM | monocyte | *LHCGR* | 1.086472915 | 0.002031677 | 0.003666233 | 1 |
| T2DM | monocyte | *THUMPD2* | 2.330117931 | 8.13E-05 | 0.000445492 | 0.51142473 |
| T2DM | monocyte | *PREB* | 1.323951897 | 0.012118076 | 0.017264071 | 1 |
| T2DM | monocyte | *ABCG5* | 1.214419644 | 0.000678762 | 0.001661104 | 1 |
| T2DM | monocyte | *ATRAID* | 1.422645945 | 0.000371767 | 0.00109424 | 1 |
| T2DM | monocyte | *CENPO* | 6.535901916 | 2.08E-05 | 0.000362859 | 0.130551864 |
| T2DM | monocyte | *LRPPRC* | 2.632476983 | 3.63E-05 | 0.000365043 | 0.228516747 |
| T2DM | monocyte | *DTNB* | 2.435316488 | 0.000307524 | 0.000968153 | 1 |
| T2DM | monocyte | *CYP2C8* | 1.537217416 | 0.008121026 | 0.012121768 | 1 |
| T2DM | monocyte | *MYOF* | 2.673232547 | 9.88E-07 | 0.000122893 | 0.006214657 |
| T2DM | monocyte | *STAMBPL1* | 0.964214795 | 0.000465724 | 0.001278048 | 1 |
| T2DM | monocyte | *CH25H* | 1.082456093 | 0.021705443 | 0.029096401 | 1 |
| T2DM | monocyte | *ATAD1* | 2.585391861 | 4.72E-05 | 0.000379941 | 0.296936926 |
| T2DM | monocyte | *BTBD16* | 0.884066034 | 0.017797827 | 0.024314415 | 1 |
| T2DM | monocyte | *TACC2* | 1.361908243 | 0.000121354 | 0.000551268 | 0.762954624 |
| T2DM | monocyte | *PLCE1* | 1.449097646 | 0.000714787 | 0.001725092 | 1 |
| T2DM | monocyte | *RBP4* | 1.102678233 | 0.000372765 | 0.001095639 | 1 |
| T2DM | monocyte | *DNAJC13* | 3.342974332 | 2.55E-05 | 0.000362859 | 0.160159402 |
| T2DM | monocyte | *FAM149B1* | 1.455709353 | 0.000808071 | 0.001887709 | 1 |
| T2DM | monocyte | *ASCC1* | 1.852884973 | 0.001540072 | 0.00301258 | 1 |
| T2DM | monocyte | *RPS24* | 3.461665357 | 4.07E-05 | 0.00037107 | 0.255596039 |
| T2DM | monocyte | *SMARCAL1* | 3.163568979 | 3.61E-05 | 0.000365043 | 0.227149766 |
| T2DM | monocyte | *STAT4* | 0.632578779 | 0.004816669 | 0.00766449 | 1 |
| T2DM | monocyte | *CARF* | 3.210931431 | 7.29E-08 | 2.08E-05 | 0.000458579 |
| T2DM | monocyte | *ASNSD1* | 1.341931406 | 0.005988669 | 0.009287312 | 1 |
| T2DM | monocyte | *METTL5* | 1.475383381 | 0.000352373 | 0.001053935 | 1 |
| T2DM | monocyte | *NAB1* | 3.558096942 | 2.51E-05 | 0.000362859 | 0.157713828 |
| T2DM | monocyte | *FASTKD1* | 3.019301413 | 0.001829935 | 0.003393752 | 1 |
| T2DM | monocyte | *MDH1B* | -1.949517539 | 0.022745606 | 0.03038063 | 1 |
| T2DM | monocyte | *ITGAV* | 1.746903194 | 6.19E-05 | 0.000406647 | 0.388861503 |
| T2DM | monocyte | *SLC35A5* | 1.661447514 | 0.00112316 | 0.00239631 | 1 |
| T2DM | monocyte | *SENP7* | 2.488430714 | 2.30E-05 | 0.000362859 | 0.144716384 |
| T2DM | monocyte | *COX17* | 5.123283119 | 2.07E-05 | 0.000362859 | 0.130014056 |
| T2DM | monocyte | *USP8* | 2.158979026 | 5.30E-05 | 0.000385235 | 0.333055765 |
| T2DM | monocyte | *SPPL2A* | 3.04147346 | 0.000847939 | 0.001953461 | 1 |
| T2DM | monocyte | *PPCDC* | 1.976455828 | 0.00049523 | 0.00133742 | 1 |
| T2DM | monocyte | *UBL7* | 2.118750945 | 0.000539611 | 0.001418877 | 1 |

| | | | | | | |
|---|---|---|---|---|---|---|
| T2DM | monocyte | *ARHGAP24* | 1.386304042 | 8.07E-05 | 0.000443644 | 0.50752911 |
| T2DM | monocyte | *FAM13A* | 1.277279122 | 3.19E-05 | 0.000363519 | 0.200799318 |
| T2DM | monocyte | *HERC3* | 1.896145909 | 0.037086599 | 0.047347847 | 1 |
| T2DM | monocyte | *AP1AR* | 2.4065339 | 0.000120114 | 0.000547214 | 0.755155241 |
| T2DM | monocyte | *PRKG2* | 0.586808858 | 0.002245779 | 0.003961621 | 1 |
| T2DM | monocyte | *BBS7* | 4.54309397 | 0.000345427 | 0.001037992 | 1 |
| T2DM | monocyte | *KIAA1109* | 3.004661876 | 3.86E-05 | 0.00036941 | 0.242826748 |
| T2DM | monocyte | *PRDM5* | 0.907738749 | 0.000394677 | 0.001142946 | 1 |
| T2DM | monocyte | *TRPC3* | 1.350101364 | 0.002304695 | 0.00404655 | 1 |
| T2DM | monocyte | *NAAA* | -0.341194467 | 0.031380252 | 0.04053578 | 1 |
| T2DM | monocyte | *NUP54* | 2.882707295 | 4.11E-06 | 0.000271729 | 0.025814288 |
| T2DM | monocyte | *BMP2K* | 1.441145407 | 0.022846278 | 0.030482715 | 1 |
| T2DM | monocyte | *G3BP2* | 2.018281551 | 0.001006597 | 0.002215852 | 1 |
| T2DM | monocyte | *SCARB2* | 2.479162408 | 4.07E-05 | 0.00037107 | 0.25583394 |
| T2DM | monocyte | *CNOT6L* | 1.75815257 | 0.011319473 | 0.016277568 | 1 |
| T2DM | monocyte | *ANXA3* | 1.792426089 | 0.000282483 | 0.000912628 | 1 |
| T2DM | monocyte | *PPA2* | 3.039353053 | 3.33E-06 | 0.000245788 | 0.020953738 |
| T2DM | monocyte | *ENPEP* | 0.646992494 | 0.002622113 | 0.004523936 | 1 |
| T2DM | monocyte | *PAPSS1* | 2.968502776 | 2.24E-05 | 0.000362859 | 0.141084264 |
| T2DM | monocyte | *FBN2* | -0.245227923 | 0.024688651 | 0.032636154 | 1 |
| T2DM | monocyte | *RNF185* | 1.643583347 | 0.000682868 | 0.001668508 | 1 |
| T2DM | monocyte | *PARVG* | -0.371319524 | 0.011977856 | 0.017101671 | 1 |
| T2DM | monocyte | *B4GALNT3* | 1.135317984 | 0.006340612 | 0.009772843 | 1 |
| T2DM | monocyte | *ERP27* | -0.365825753 | 0.008366043 | 0.012454963 | 1 |
| T2DM | monocyte | *KIF21A* | 0.874364736 | 0.000271449 | 0.000888419 | 1 |
| T2DM | monocyte | *ALG10* | 4.745044246 | 0.025115442 | 0.033158501 | 1 |
| T2DM | monocyte | *SLCO1C1* | 3.121090049 | 5.08E-05 | 0.000383728 | 0.319082409 |
| T2DM | monocyte | *ZCRB1* | 1.617559961 | 0.000107733 | 0.000515853 | 0.677315233 |
| T2DM | monocyte | *PRICKLE1* | 0.998316106 | 0.000192265 | 0.000716946 | 1 |
| T2DM | monocyte | *C1RL* | 2.021143538 | 5.39E-06 | 0.000310813 | 0.033878565 |
| T2DM | monocyte | *CLSTN3* | 3.596180945 | 0.000790729 | 0.001857739 | 1 |
| T2DM | monocyte | *KLRG1* | 0.836220115 | 0.019756606 | 0.026729791 | 1 |
| T2DM | monocyte | *VAMP1* | 2.076383054 | 0.000659204 | 0.001631017 | 1 |
| T2DM | monocyte | *CD27* | 4.02808035 | 0.001179665 | 0.002479769 | 1 |
| T2DM | monocyte | *PPFIA2* | 0.600466112 | 0.001735372 | 0.003279315 | 1 |
| T2DM | monocyte | *LGR5* | 0.950423368 | 0.004587673 | 0.007328604 | 1 |
| T2DM | monocyte | *POC1B* | 4.359761071 | 4.71E-05 | 0.000379941 | 0.295817528 |
| T2DM | monocyte | *SNRPF* | 1.687953852 | 0.006451252 | 0.009906942 | 1 |
| T2DM | monocyte | *AMDHD1* | 1.20208328 | 0.001052031 | 0.002286248 | 1 |
| T2DM | monocyte | *GAS2L3* | 0.804926295 | 0.001186551 | 0.002489935 | 1 |
| T2DM | monocyte | *TMEM132B* | 1.074989724 | 0.002230823 | 0.00394409 | 1 |
| T2DM | monocyte | *TDG* | 2.537820595 | 6.92E-05 | 0.000425443 | 0.434803085 |
| T2DM | monocyte | *MMAB* | 0.823250342 | 0.004601006 | 0.007343622 | 1 |
| T2DM | monocyte | *SLC46A3* | 1.712244201 | 4.19E-05 | 0.000374152 | 0.263402937 |
| T2DM | monocyte | *SUOX* | 1.620993696 | 0.023256823 | 0.030977891 | 1 |
| T2DM | monocyte | *CCDC65* | 1.729750988 | 0.004755034 | 0.007572163 | 1 |
| T2DM | monocyte | *ACVRL1* | 2.549278079 | 0.000258559 | 0.000857815 | 1 |
| T2DM | monocyte | *BRCA2* | 3.207753908 | 0.000163773 | 0.000650094 | 1 |
| T2DM | monocyte | *KANSL2* | 1.940462194 | 0.006138658 | 0.009487154 | 1 |
| T2DM | monocyte | *CERS5* | 2.551700489 | 0.000556228 | 0.001451164 | 1 |

| | | | | | | |
|---|---|---|---|---|---|---|
| T2DM | monocyte | *LMBR1L* | 2.928861123 | 0.013491838 | 0.018988849 | 1 |
| T2DM | monocyte | *ZNF740* | 2.529198156 | 8.86E-05 | 0.000462594 | 0.556963667 |
| T2DM | monocyte | *HNRNPA1L2* | 1.75085379 | 0.003622447 | 0.00597281 | 1 |
| T2DM | monocyte | *LPAR6* | 3.251990502 | 3.35E-05 | 0.000365043 | 0.210421762 |
| T2DM | monocyte | *VPS33A* | 1.111291173 | 0.001185062 | 0.002488805 | 1 |
| T2DM | monocyte | *RHOF* | 1.270606893 | 0.00607458 | 0.009399676 | 1 |
| T2DM | monocyte | *GRTP1* | 0.596706005 | 0.005441862 | 0.008536175 | 1 |
| T2DM | monocyte | *CBLN3* | 0.422072082 | 0.008620556 | 0.012782414 | 1 |
| T2DM | monocyte | *TMX1* | 4.011319302 | 0.001898011 | 0.003477941 | 1 |
| T2DM | monocyte | *C14orf37* | 2.063900402 | 0.000363848 | 0.001076982 | 1 |
| T2DM | monocyte | *STON2* | 1.767477182 | 0.000161293 | 0.000643434 | 1 |
| T2DM | monocyte | *EFCAB11* | 1.252486659 | 0.001698675 | 0.003231337 | 1 |
| T2DM | monocyte | *SLC24A4* | -0.123295215 | 0.007040893 | 0.010694877 | 1 |
| T2DM | monocyte | *FBLN5* | 0.695380365 | 0.000834134 | 0.001930854 | 1 |
| T2DM | monocyte | *C14orf79* | 3.051426378 | 0.000108124 | 0.000517029 | 0.67977383 |
| T2DM | monocyte | *NIPA2* | 3.141140792 | 0.010584438 | 0.015304591 | 1 |
| T2DM | monocyte | *DUOXA1* | 1.609748351 | 0.000523375 | 0.001390262 | 1 |
| T2DM | monocyte | *TCF12* | 2.150419539 | 2.70E-05 | 0.000362859 | 0.16963428 |
| T2DM | monocyte | *SORD* | 3.081670213 | 0.000413238 | 0.001178344 | 1 |
| T2DM | monocyte | *ZSCAN29* | 1.342937109 | 0.000797045 | 0.001867522 | 1 |
| T2DM | monocyte | *SRP14* | 2.173627407 | 1.32E-05 | 0.000362859 | 0.083157288 |
| T2DM | monocyte | *ANP32A* | 2.196695749 | 0.037662168 | 0.047999604 | 1 |
| T2DM | monocyte | *COMMD4* | 2.622055164 | 0.000494938 | 0.00133742 | 1 |
| T2DM | monocyte | *UBE2Q2* | 3.8622632 | 2.05E-05 | 0.000362859 | 0.12864571 |
| T2DM | monocyte | *HMG20A* | 2.543257829 | 1.06E-06 | 0.000125709 | 0.006662579 |
| T2DM | monocyte | *SCAPER* | 2.096481793 | 0.000112584 | 0.000527042 | 0.707817839 |
| T2DM | monocyte | *NEIL1* | 1.153510214 | 0.004885201 | 0.007765679 | 1 |
| T2DM | monocyte | *MAN2C1* | -0.501978038 | 0.031302614 | 0.040452114 | 1 |
| T2DM | monocyte | *DNAJA4* | 2.398813543 | 0.00020624 | 0.000743876 | 1 |
| T2DM | monocyte | *TPM1* | 1.205633127 | 0.000216603 | 0.000766771 | 1 |
| T2DM | monocyte | *IGF1R* | 1.480166981 | 9.64E-05 | 0.000483327 | 0.606092463 |
| T2DM | monocyte | *CYP11A1* | 1.236420174 | 0.000976257 | 0.002162694 | 1 |
| T2DM | monocyte | *BBS4* | 2.728653379 | 1.98E-05 | 0.000362859 | 0.124401259 |
| T2DM | monocyte | *ADAMTS17* | 1.105019981 | 0.000920892 | 0.002069949 | 1 |
| T2DM | monocyte | *ULK3* | 2.992150561 | 0.000111566 | 0.000524745 | 0.701417725 |
| T2DM | monocyte | *HAPLN3* | 0.903207775 | 0.002371383 | 0.004145963 | 1 |
| T2DM | monocyte | *POLG* | 3.569413839 | 0.003864674 | 0.006307686 | 1 |
| T2DM | monocyte | *WDR93* | 0.394001678 | 0.019075548 | 0.025846545 | 1 |
| T2DM | monocyte | *TICRR* | -1.278159778 | 0.023946605 | 0.031788916 | 1 |
| T2DM | monocyte | *MFGE8* | 1.072949799 | 0.000680599 | 0.001664305 | 1 |
| T2DM | monocyte | *UNC45A* | 1.541207724 | 0.000615993 | 0.00154786 | 1 |
| T2DM | monocyte | *ST8SIA2* | 4.447662649 | 0.00860634 | 0.012767356 | 1 |
| T2DM | monocyte | *IQGAP1* | 2.30827951 | 0.000811815 | 0.001893131 | 1 |
| T2DM | monocyte | *CRTC3* | 1.74335735 | 8.61E-05 | 0.000459567 | 0.54134323 |
| T2DM | monocyte | *SEC11A* | 3.527249609 | 0.000462343 | 0.001271543 | 1 |
| T2DM | monocyte | *TGFB1I1* | 2.638687813 | 0.00308414 | 0.005193932 | 1 |
| T2DM | monocyte | *PARN* | 2.030383532 | 9.01E-05 | 0.000465303 | 0.566738757 |
| T2DM | monocyte | *FTO* | 1.627544192 | 9.45E-05 | 0.000478875 | 0.594145686 |
| T2DM | monocyte | *TXNL4B* | 3.365785623 | 0.00015072 | 0.000619333 | 0.947578816 |
| T2DM | monocyte | *MARVELD3* | 4.654626056 | 0.001817804 | 0.00338523 | 1 |

| | | | | | | |
|---|---|---|---|---|---|---|
| T2DM | monocyte | *ZFHX3* | 2.725139395 | 1.90E-05 | 0.000362859 | 0.119457748 |
| T2DM | monocyte | *KIFC3* | 1.647430406 | 0.000277421 | 0.000902161 | 1 |
| T2DM | monocyte | *ADAMTS18* | 2.579302336 | 0.001045405 | 0.002274995 | 1 |
| T2DM | monocyte | *NUDT7* | 0.611594178 | 0.009292896 | 0.013679335 | 1 |
| T2DM | monocyte | *GCSH* | 2.03023082 | 0.000133576 | 0.000578424 | 0.839792793 |
| T2DM | monocyte | *CMTM3* | 2.310064108 | 0.000136701 | 0.000584748 | 0.85944086 |
| T2DM | monocyte | *MAP1LC3B* | 2.597235233 | 5.96E-05 | 0.000399203 | 0.374644427 |
| T2DM | monocyte | *TLDC1* | 2.760706089 | 0.000106609 | 0.000511748 | 0.670252653 |
| T2DM | monocyte | *RPS2* | 2.373490206 | 0.029652323 | 0.03853331 | 1 |
| T2DM | monocyte | *NDUFB10* | 2.420483473 | 0.00019769 | 0.000725134 | 1 |
| T2DM | monocyte | *TIGD7* | 2.022994208 | 1.48E-05 | 0.000362859 | 0.092896229 |
| T2DM | monocyte | *GALNS* | 2.760970496 | 0.002491144 | 0.004324081 | 1 |
| T2DM | monocyte | *COPS3* | 2.295101828 | 0.000125614 | 0.000558116 | 0.789733764 |
| T2DM | monocyte | *RANBP10* | 3.511362661 | 2.15E-05 | 0.000362859 | 0.135140803 |
| T2DM | monocyte | *PRPSAP2* | 2.597030164 | 7.06E-05 | 0.000427286 | 0.443923657 |
| T2DM | monocyte | *MYO19* | 4.089017244 | 9.16E-08 | 2.50E-05 | 0.000575854 |
| T2DM | monocyte | *DDX52* | 3.786977797 | 6.48E-05 | 0.000415426 | 0.40711751 |
| T2DM | monocyte | *UNC45B* | 3.116582171 | 0.021193293 | 0.028488824 | 1 |
| T2DM | monocyte | *TOM1L1* | 1.260999935 | 0.000216209 | 0.000766669 | 1 |
| T2DM | monocyte | *C17orf80* | 1.424148001 | 0.009708068 | 0.014216616 | 1 |
| T2DM | monocyte | *SPATA22* | 0.86868286 | 0.000103289 | 0.000502417 | 0.649377765 |
| T2DM | monocyte | *SGSM2* | 2.188724209 | 0.000870429 | 0.001988233 | 1 |
| T2DM | monocyte | *NPEPPS* | 3.034192726 | 4.36E-05 | 0.000378997 | 0.274210656 |
| T2DM | monocyte | *ABCA8* | 0.539245736 | 0.000828817 | 0.001920669 | 1 |
| T2DM | monocyte | *G6PC3* | 3.616233806 | 0.034307414 | 0.044054475 | 1 |
| T2DM | monocyte | *GNAL* | 1.686720787 | 0.000198469 | 0.000726217 | 1 |
| T2DM | monocyte | *RPRD1A* | 2.415953821 | 0.00350975 | 0.005811377 | 1 |
| T2DM | monocyte | *C18orf21* | 6.268331538 | 1.06E-05 | 0.000362859 | 0.06654143 |
| T2DM | monocyte | *GALNT1* | 2.787281166 | 0.000132541 | 0.000576268 | 0.833283475 |
| T2DM | monocyte | *ASXL3* | 1.118884745 | 0.000760225 | 0.001806324 | 1 |
| T2DM | monocyte | *ADCYAP1* | 3.774269329 | 0.012994037 | 0.018362218 | 1 |
| T2DM | monocyte | *ESCO1* | 2.640399406 | 0.000104514 | 0.000506226 | 0.657081251 |
| T2DM | monocyte | *GREB1L* | 0.611881177 | 0.003853389 | 0.006291465 | 1 |
| T2DM | monocyte | *NPC1* | 1.855973396 | 0.018243899 | 0.024826708 | 1 |
| T2DM | monocyte | *CCDC40* | 1.407260386 | 0.000280511 | 0.000909057 | 1 |
| T2DM | monocyte | *EIF4A3* | 3.650491886 | 0.000763705 | 0.001813651 | 1 |
| T2DM | monocyte | *TBCD* | 3.552590323 | 9.59E-05 | 0.000482439 | 0.602920261 |
| T2DM | monocyte | *NARF* | 2.80823346 | 0.000978988 | 0.002167218 | 1 |
| T2DM | monocyte | *TRIM65* | 1.0249717 | 0.001571998 | 0.003066446 | 1 |
| T2DM | monocyte | *CBX8* | -1.799036362 | 0.036333301 | 0.046456674 | 1 |
| T2DM | monocyte | *SECTM1* | -0.422104227 | 0.011521368 | 0.016526315 | 1 |
| T2DM | monocyte | *RNF157* | 0.515841783 | 0.020983208 | 0.028224525 | 1 |
| T2DM | monocyte | *WDR45B* | 4.024012934 | 7.28E-05 | 0.000431726 | 0.457484462 |
| T2DM | monocyte | *RNF165* | 2.07594176 | 0.000316473 | 0.000985959 | 1 |
| T2DM | monocyte | *MAPK4* | 1.273054501 | 0.000607596 | 0.001532888 | 1 |
| T2DM | monocyte | *SMAD4* | 3.503375979 | 0.000147232 | 0.000610586 | 0.925648855 |
| T2DM | monocyte | *NT5C3B* | 0.749100285 | 0.009712758 | 0.014216633 | 1 |
| T2DM | monocyte | *PIP4K2B* | 3.802095362 | 0.000776396 | 0.001832661 | 1 |
| T2DM | monocyte | *GRB7* | 1.681341863 | 0.010337694 | 0.015006483 | 1 |
| T2DM | monocyte | *FKBP10* | 0.542591041 | 0.031978412 | 0.041283013 | 1 |

| | | | | | | |
|---|---|---|---|---|---|---|
| T2DM | monocyte | *NFIC* | 1.030595973 | 0.000793366 | 0.001861364 | 1 |
| T2DM | monocyte | *TPGS1* | 2.089126647 | 0.000250238 | 0.000839513 | 1 |
| T2DM | monocyte | *PRDM15* | 1.626396267 | 0.000431022 | 0.001214084 | 1 |
| T2DM | monocyte | *MVB12A* | 1.615662431 | 0.000535096 | 0.001410737 | 1 |
| T2DM | monocyte | *DUS3L* | 2.118565874 | 0.002426762 | 0.004232192 | 1 |
| T2DM | monocyte | *DPP9* | 2.972211543 | 0.000378859 | 0.001107855 | 1 |
| T2DM | monocyte | *SIRT3* | 4.446378701 | 1.86E-06 | 0.000174546 | 0.01169461 |
| T2DM | monocyte | *IFITM3* | 2.044320121 | 0.000111081 | 0.000523558 | 0.698367213 |
| T2DM | monocyte | *COL6A1* | 0.77494234 | 0.003116623 | 0.005241896 | 1 |
| T2DM | monocyte | *TRPM2* | 1.140032246 | 0.014973333 | 0.020840679 | 1 |
| T2DM | monocyte | *TMEM50B* | 1.230099754 | 0.002822269 | 0.004802058 | 1 |
| T2DM | monocyte | *DOPEY2* | 3.011621751 | 0.000581632 | 0.00149014 | 1 |
| T2DM | monocyte | *URB1* | 1.588942403 | 0.000584278 | 0.001494448 | 1 |
| T2DM | monocyte | *AKT1* | 5.213945171 | 2.47E-05 | 0.000362859 | 0.155382756 |
| T2DM | monocyte | *SAE1* | 2.903583284 | 2.70E-05 | 0.000362859 | 0.16950848 |
| T2DM | monocyte | *NTN5* | 1.601950423 | 0.007335718 | 0.011091789 | 1 |
| T2DM | monocyte | *WTIP* | 0.698814226 | 0.001468788 | 0.002916135 | 1 |
| T2DM | monocyte | *CAPN10* | 2.014921883 | 0.000591881 | 0.001507761 | 1 |
| T2DM | monocyte | *MYO1F* | -0.251657126 | 0.019926349 | 0.026923911 | 1 |
| T2DM | monocyte | *CACNG8* | 1.980200566 | 0.001459619 | 0.002905834 | 1 |
| T2DM | monocyte | *EVI5L* | 4.083424038 | 4.52E-05 | 0.000379941 | 0.284405049 |
| T2DM | monocyte | *RPS11* | 2.924958959 | 0.000119016 | 0.000544578 | 0.748250779 |
| T2DM | monocyte | *RCN3* | 1.458864832 | 0.000357117 | 0.00106357 | 1 |
| T2DM | monocyte | *RERE* | 1.966956479 | 2.59E-05 | 0.000362859 | 0.162743749 |
| T2DM | monocyte | *PEX14* | 2.261810389 | 0.000133131 | 0.000578036 | 0.836995564 |
| T2DM | monocyte | *SH3BGRL3* | 3.987883235 | 0.001196806 | 0.002504767 | 1 |
| T2DM | monocyte | *C1orf216* | 0.970169529 | 0.00153372 | 0.003002958 | 1 |
| T2DM | monocyte | *GPN2* | 1.274082197 | 0.000992057 | 0.002191518 | 1 |
| T2DM | monocyte | *HSPG2* | 0.632327216 | 0.012370431 | 0.017575797 | 1 |
| T2DM | monocyte | *ITGB3BP* | 1.683902504 | 0.000282747 | 0.000913009 | 1 |
| T2DM | monocyte | *SERBP1* | 1.650555232 | 0.000165315 | 0.000652847 | 1 |
| T2DM | monocyte | *PIGK* | 4.243041611 | 4.75E-05 | 0.000379941 | 0.298615813 |
| T2DM | monocyte | *RPS8* | 2.279896641 | 0.000142958 | 0.000600787 | 0.898777391 |
| T2DM | monocyte | *MOB3C* | 5.898028002 | 2.13E-05 | 0.000362859 | 0.133832657 |
| T2DM | monocyte | *CYP4B1* | 0.681802496 | 0.000410146 | 0.00117346 | 1 |
| T2DM | monocyte | *SYPL2* | 1.194030543 | 0.004728929 | 0.0075325 | 1 |
| T2DM | monocyte | *ZNF697* | 1.257219706 | 0.00090651 | 0.0020442 | 1 |
| T2DM | monocyte | *CELSR2* | 2.068441273 | 0.00752881 | 0.01134283 | 1 |
| T2DM | monocyte | *GPR161* | 0.714591948 | 0.001871493 | 0.003444403 | 1 |
| T2DM | monocyte | *ATP1B1* | 1.665863255 | 0.000765114 | 0.001813827 | 1 |
| T2DM | monocyte | *TIPRL* | 2.163032545 | 0.00016855 | 0.000659148 | 1 |
| T2DM | monocyte | *NME7* | 2.733335236 | 0.000886836 | 0.002013557 | 1 |
| T2DM | monocyte | *CREG1* | 2.296261703 | 0.000394369 | 0.001142581 | 1 |
| T2DM | monocyte | *GPA33* | -1.458515972 | 0.016687929 | 0.022952748 | 1 |
| T2DM | monocyte | *RXRG* | 3.026198346 | 0.000910223 | 0.002051836 | 1 |
| T2DM | monocyte | *TMCO1* | 2.058571348 | 0.000118409 | 0.000544237 | 0.744435303 |
| T2DM | monocyte | *MGST3* | 1.753461718 | 0.000111896 | 0.000524745 | 0.703488154 |
| T2DM | monocyte | *ADCY10* | 1.5507736 | 0.001435723 | 0.002874671 | 1 |
| T2DM | monocyte | *PPOX* | 3.064007228 | 3.09E-05 | 0.000362859 | 0.194004495 |
| T2DM | monocyte | *RGS5* | 0.824068672 | 0.001272677 | 0.002622523 | 1 |

| | | | | | | |
|---|---|---|---|---|---|---|
| T2DM | monocyte | *PIGM* | 5.141435929 | 3.38E-05 | 0.000365043 | 0.21228529 |
| T2DM | monocyte | *RGL1* | 1.244107762 | 0.000174107 | 0.000673193 | 1 |
| T2DM | monocyte | *TUFT1* | 0.681572494 | 0.022519616 | 0.030104365 | 1 |
| T2DM | monocyte | *TARS2* | 1.415876311 | 7.86E-05 | 0.000440257 | 0.494408446 |
| T2DM | monocyte | *SELENBP1* | 0.697542774 | 0.001476136 | 0.002925747 | 1 |
| T2DM | monocyte | *CERS2* | 4.039846451 | 0.000264784 | 0.000872484 | 1 |
| T2DM | monocyte | *ENSA* | 2.813513811 | 3.61E-05 | 0.000365043 | 0.22713398 |
| T2DM | monocyte | *SEMA6C* | 0.977798865 | 0.002019471 | 0.003654695 | 1 |
| T2DM | monocyte | *ARNT* | 2.081805922 | 0.000293146 | 0.000937676 | 1 |
| T2DM | monocyte | *POGZ* | 2.393458973 | 0.000133397 | 0.000578389 | 0.838664677 |
| T2DM | monocyte | *GABPB2* | 1.583749399 | 7.47E-05 | 0.000433477 | 0.469773366 |
| T2DM | monocyte | *DYRK3* | 1.49099742 | 0.000610183 | 0.001538797 | 1 |
| T2DM | monocyte | *EIF2D* | 1.851506597 | 0.020241772 | 0.027297302 | 1 |
| T2DM | monocyte | *HHIPL2* | 2.09857941 | 0.018464443 | 0.025105094 | 1 |
| T2DM | monocyte | *TP53BP2* | 4.193090194 | 3.14E-05 | 0.000362859 | 0.19715724 |
| T2DM | monocyte | *RAB13* | 0.943482539 | 0.002242756 | 0.003960732 | 1 |
| T2DM | monocyte | *S100A8* | -0.239812389 | 0.007118271 | 0.010799366 | 1 |
| T2DM | monocyte | *HAX1* | 4.061649654 | 0.000197152 | 0.000724426 | 1 |
| T2DM | monocyte | *KCNN3* | 0.952880834 | 0.000844531 | 0.001949181 | 1 |
| T2DM | monocyte | *GATAD2B* | 2.350221471 | 8.07E-05 | 0.000443644 | 0.507333371 |
| T2DM | monocyte | *GALNT2* | 2.943089306 | 2.66E-05 | 0.000362859 | 0.167229585 |
| T2DM | monocyte | *SCCPDH* | 1.953924567 | 0.000164006 | 0.000650129 | 1 |
| T2DM | monocyte | *CEP170* | 2.540808755 | 3.80E-05 | 0.00036941 | 0.238846168 |
| T2DM | monocyte | *SNAP47* | 2.114262437 | 0.025847733 | 0.034003913 | 1 |
| T2DM | monocyte | *NVL* | 2.74381033 | 3.44E-05 | 0.000365043 | 0.216045173 |
| T2DM | monocyte | *SDE2* | 3.062356155 | 0.000662451 | 0.001633907 | 1 |
| T2DM | monocyte | *FBXO28* | 3.706070094 | 2.77E-05 | 0.000362859 | 0.174298796 |
| T2DM | monocyte | *CNIH4* | 4.175092743 | 0.000947428 | 0.002115227 | 1 |
| T2DM | monocyte | *GUK1* | 4.52032626 | 1.90E-05 | 0.000362859 | 0.119725874 |
| T2DM | monocyte | *CDC42BPA* | 1.331249331 | 7.74E-05 | 0.000437657 | 0.486454677 |
| T2DM | monocyte | *CNIH3* | 3.253515338 | 1.38E-05 | 0.000362859 | 0.086640143 |
| T2DM | monocyte | *PSEN2* | 1.62523538 | 0.002732683 | 0.004680512 | 1 |
| T2DM | monocyte | *WNT9A* | 1.218084197 | 0.000100658 | 0.000495511 | 0.632839227 |
| T2DM | monocyte | *PLEKHA6* | 0.788327276 | 0.000528298 | 0.001398488 | 1 |
| T2DM | monocyte | *ABCG8* | 4.092004065 | 0.000261469 | 0.000864279 | 1 |
| T2DM | monocyte | *EML4* | 2.814168126 | 0.001588933 | 0.003091805 | 1 |
| T2DM | monocyte | *CHAC2* | 3.635605541 | 8.99E-05 | 0.000465291 | 0.565424932 |
| T2DM | monocyte | *WDPCP* | 0.625048895 | 0.00152859 | 0.00299478 | 1 |
| T2DM | monocyte | *ASXL2* | 3.245207725 | 4.99E-05 | 0.000383513 | 0.313537302 |
| T2DM | monocyte | *CIAO1* | 1.374520921 | 0.001235873 | 0.002563289 | 1 |
| T2DM | monocyte | *EXOC6B* | 1.458478151 | 7.35E-05 | 0.000431915 | 0.462286892 |
| T2DM | monocyte | *THNSL2* | 0.41380255 | 0.033780841 | 0.043413767 | 1 |
| T2DM | monocyte | *TMEM177* | 1.20558921 | 0.001914144 | 0.003499449 | 1 |
| T2DM | monocyte | *RABL2A* | 1.642017783 | 0.001217197 | 0.002539833 | 1 |
| T2DM | monocyte | *FBLN7* | 1.43966871 | 0.000158766 | 0.000639029 | 0.998163666 |
| T2DM | monocyte | *ZC3H8* | 1.836860199 | 0.00018571 | 0.000702925 | 1 |
| T2DM | monocyte | *LIPT1* | 1.066735095 | 0.00159342 | 0.003095555 | 1 |
| T2DM | monocyte | *FAHD2B* | 1.305791009 | 0.00739498 | 0.011170649 | 1 |
| T2DM | monocyte | *AFF3* | 1.075511616 | 0.001266231 | 0.002610954 | 1 |
| T2DM | monocyte | *UBXN4* | 2.794361904 | 4.36E-05 | 0.000378997 | 0.274235702 |

| | | | | | | |
|---|---|---|---|---|---|---|
| T2DM | monocyte | *THSD7B* | 1.002955015 | 4.83E-05 | 0.000380416 | 0.303952566 |
| T2DM | monocyte | *GPR17* | 0.610285925 | 0.003355426 | 0.005592673 | 1 |
| T2DM | monocyte | *ZNF385B* | 0.497423867 | 0.00115915 | 0.002447137 | 1 |
| T2DM | monocyte | *TMEFF2* | 0.838281865 | 0.000562922 | 0.00146183 | 1 |
| T2DM | monocyte | *GULP1* | 0.799089113 | 0.000413845 | 0.00117837 | 1 |
| T2DM | monocyte | *FAM171B* | 1.901522306 | 0.002470375 | 0.004292771 | 1 |
| T2DM | monocyte | *HSPD1* | 3.065422285 | 1.05E-05 | 0.000362859 | 0.066235517 |
| T2DM | monocyte | *METTL21A* | 2.271181278 | 6.10E-05 | 0.000403714 | 0.383714384 |
| T2DM | monocyte | *PTH2R* | 1.156203917 | 0.000678595 | 0.001661104 | 1 |
| T2DM | monocyte | *NBEAL1* | 1.818300164 | 1.76E-07 | 4.42E-05 | 0.001103797 |
| T2DM | monocyte | *SUMF1* | 2.759308034 | 2.35E-05 | 0.000362859 | 0.147859768 |
| T2DM | monocyte | *ESPNL* | 10.20027024 | 0.000326997 | 0.001006043 | 1 |
| T2DM | monocyte | *FANCD2* | 7.052683077 | 0.000163674 | 0.000650094 | 1 |
| T2DM | monocyte | *TAMM41* | 2.966845856 | 6.72E-06 | 0.000332508 | 0.042228567 |
| T2DM | monocyte | *OSBPL10* | 1.146681579 | 0.000120483 | 0.000548498 | 0.757475965 |
| T2DM | monocyte | *FAM198A* | 0.907215058 | 0.001222191 | 0.002548562 | 1 |
| T2DM | monocyte | *CSRNP1* | 2.321194742 | 0.000278512 | 0.000903978 | 1 |
| T2DM | monocyte | *SLC25A38* | 4.404686374 | 0.00130002 | 0.002669245 | 1 |
| T2DM | monocyte | *CTDSPL* | 2.414855192 | 0.000834474 | 0.00193093 | 1 |
| T2DM | monocyte | *STAC* | 0.981904917 | 0.000958258 | 0.002133345 | 1 |
| T2DM | monocyte | *ARL6IP5* | 3.439329465 | 1.81E-05 | 0.000362859 | 0.113657992 |
| T2DM | monocyte | *TMF1* | 3.574326489 | 0.000132057 | 0.00057536 | 0.830244903 |
| T2DM | monocyte | *COL8A1* | 0.710383057 | 0.001223352 | 0.00254974 | 1 |
| T2DM | monocyte | *NXPE3* | 2.953450568 | 0.028411621 | 0.037081972 | 1 |
| T2DM | monocyte | *RABL3* | 1.477877364 | 0.000265669 | 0.00087494 | 1 |
| T2DM | monocyte | *IGSF11* | 2.490767329 | 0.000419415 | 0.001188848 | 1 |
| T2DM | monocyte | *BOC* | 0.461061217 | 0.002200777 | 0.003901942 | 1 |
| T2DM | monocyte | *AGTR1* | 0.488400227 | 0.002259281 | 0.00398097 | 1 |
| T2DM | monocyte | *EIF2A* | 1.08242856 | 0.010589365 | 0.015304675 | 1 |
| T2DM | monocyte | *AMT* | 0.778247272 | 0.004451444 | 0.007133884 | 1 |
| T2DM | monocyte | *TCTA* | 2.239001112 | 7.01E-05 | 0.000427286 | 0.440850755 |
| T2DM | monocyte | *EAF2* | -0.3473095 | 0.016898619 | 0.023206993 | 1 |
| T2DM | monocyte | *LYAR* | 2.746193865 | 0.000124661 | 0.000556889 | 0.783744761 |
| T2DM | monocyte | *CENPC* | 1.645656094 | 0.00032772 | 0.001006043 | 1 |
| T2DM | monocyte | *ATP10D* | 2.770164339 | 0.000770814 | 0.001822488 | 1 |
| T2DM | monocyte | *SLC10A4* | 5.956819133 | 0.001693832 | 0.003225051 | 1 |
| T2DM | monocyte | *PYURF* | 1.029480831 | 0.001880721 | 0.003455316 | 1 |
| T2DM | monocyte | *CISD2* | 5.092292088 | 4.74E-06 | 0.000291975 | 0.029781399 |
| T2DM | monocyte | *TIFA* | 3.65776998 | 0.000640304 | 0.001593662 | 1 |
| T2DM | monocyte | *CCNA2* | -1.110063754 | 0.023804639 | 0.031620487 | 1 |
| T2DM | monocyte | *USP53* | 0.820780718 | 0.000344986 | 0.001037595 | 1 |
| T2DM | monocyte | *MARCH1* | -0.317419915 | 0.024815132 | 0.032782672 | 1 |
| T2DM | monocyte | *PDGFC* | 1.104723146 | 0.000718453 | 0.001730618 | 1 |
| T2DM | monocyte | *CBR4* | 1.595006637 | 7.66E-05 | 0.000437295 | 0.481414006 |
| T2DM | monocyte | *CYP4V2* | 1.021706986 | 0.017855643 | 0.024382803 | 1 |
| T2DM | monocyte | *FAM105A* | -0.561519814 | 0.014745662 | 0.02055565 | 1 |
| T2DM | monocyte | *LHFPL2* | 2.074729002 | 0.000199246 | 0.000726217 | 1 |
| T2DM | monocyte | *SSBP2* | 2.002426861 | 3.22E-05 | 0.000363519 | 0.202465411 |
| T2DM | monocyte | *CRHBP* | 0.815916683 | 0.007210535 | 0.010924689 | 1 |
| T2DM | monocyte | *RASA1* | 1.96108075 | 5.83E-05 | 0.000397957 | 0.366480631 |

| | | | | | | |
|---|---|---|---|---|---|---|
| T2DM | monocyte | *GTF2H2* | 2.764220427 | 0.002975338 | 0.005028481 | 1 |
| T2DM | monocyte | *SPATA9* | 0.623261367 | 0.000698562 | 0.00169439 | 1 |
| T2DM | monocyte | *FEM1C* | 1.833466018 | 0.000211817 | 0.000756388 | 1 |
| T2DM | monocyte | *COMMD10* | 2.346440333 | 0.000242737 | 0.000821174 | 1 |
| T2DM | monocyte | *YIPF5* | 2.304455784 | 0.00011403 | 0.000530249 | 0.71690742 |
| T2DM | monocyte | *GABRB2* | 0.533867125 | 0.009839465 | 0.014369504 | 1 |
| T2DM | monocyte | *PCYOX1L* | -0.996584288 | 0.014738371 | 0.02055565 | 1 |
| T2DM | monocyte | *TBC1D7* | 2.973953946 | 4.70E-05 | 0.000379941 | 0.29533797 |
| T2DM | monocyte | *GFOD1* | 0.888217866 | 0.000496835 | 0.001338304 | 1 |
| T2DM | monocyte | *PSD2* | 9.094933127 | 0.008875983 | 0.013124013 | 1 |
| T2DM | monocyte | *ZMAT2* | 2.544946144 | 0.011473771 | 0.016466147 | 1 |
| T2DM | monocyte | *KLHL3* | 1.418229647 | 0.000255772 | 0.000851264 | 1 |
| T2DM | monocyte | *FAM193B* | 2.328697837 | 5.38E-05 | 0.000385603 | 0.338172698 |
| T2DM | monocyte | *MUT* | 4.854268678 | 3.14E-05 | 0.000362859 | 0.197343273 |
| T2DM | monocyte | *DOK3* | -0.358968411 | 0.018143007 | 0.024726877 | 1 |
| T2DM | monocyte | *ABT1* | 1.925036352 | 0.000971664 | 0.002154039 | 1 |
| T2DM | monocyte | *MLIP* | 1.089429488 | 0.000675137 | 0.00165481 | 1 |
| T2DM | monocyte | *FGD2* | -0.173462505 | 0.009645346 | 0.014131972 | 1 |
| T2DM | monocyte | *RPL7L1* | 2.512440001 | 8.06E-05 | 0.000443644 | 0.506491178 |
| T2DM | monocyte | *TPBG* | 1.144026887 | 0.000771201 | 0.001822488 | 1 |
| T2DM | monocyte | *IRAK1BP1* | 1.231303457 | 0.000239567 | 0.000815904 | 1 |
| T2DM | monocyte | *PHIP* | 3.160775712 | 0.000159941 | 0.000641295 | 1 |
| T2DM | monocyte | *MMS22L* | 3.23783862 | 0.00053605 | 0.001412368 | 1 |
| T2DM | monocyte | *PM20D2* | 3.658878875 | 9.40E-05 | 0.000478278 | 0.591152174 |
| T2DM | monocyte | *RNF217* | 1.72281747 | 9.44E-05 | 0.000478875 | 0.593486388 |
| T2DM | monocyte | *ABRACL* | -0.712898881 | 0.023437399 | 0.031185382 | 1 |
| T2DM | monocyte | *SLC18B1* | 0.582203208 | 0.024162845 | 0.032055668 | 1 |
| T2DM | monocyte | *AIG1* | 1.489772454 | 0.00013541 | 0.000582302 | 0.851325687 |
| T2DM | monocyte | *DYNLT1* | 3.285466492 | 0.00165367 | 0.003171637 | 1 |
| T2DM | monocyte | *PNLDC1* | 1.867261573 | 0.017887512 | 0.024415716 | 1 |
| T2DM | monocyte | *WTAP* | 2.373416958 | 0.000362451 | 0.001073858 | 1 |
| T2DM | monocyte | *SLC22A3* | 0.518122368 | 0.001687184 | 0.003219219 | 1 |
| T2DM | monocyte | *GNA12* | 2.415743191 | 7.38E-05 | 0.000432034 | 0.464190623 |
| T2DM | monocyte | *C7orf50* | 2.216670422 | 3.12E-05 | 0.000362859 | 0.196308462 |
| T2DM | monocyte | *SDK1* | 0.512558598 | 0.00493028 | 0.007831397 | 1 |
| T2DM | monocyte | *CCZ1B* | 1.772213911 | 0.009634005 | 0.014118646 | 1 |
| T2DM | monocyte | *EGFR* | 0.581654186 | 0.003258097 | 0.00544778 | 1 |
| T2DM | monocyte | *POMZP3* | 1.179891916 | 0.004121935 | 0.006680744 | 1 |
| T2DM | monocyte | *CCT6A* | 1.945025756 | 0.000127985 | 0.00056466 | 0.804640851 |
| T2DM | monocyte | *PSPH* | 1.047205248 | 0.000189049 | 0.000709582 | 1 |
| T2DM | monocyte | *GIGYF1* | 1.894761643 | 0.001892649 | 0.003474186 | 1 |
| T2DM | monocyte | *TMEM209* | 2.473005632 | 2.39E-05 | 0.000362859 | 0.150164835 |
| T2DM | monocyte | *AGBL3* | 1.70484811 | 0.000173454 | 0.000671081 | 1 |
| T2DM | monocyte | *EPHA1* | 2.090914648 | 0.002266434 | 0.003990697 | 1 |
| T2DM | monocyte | *NCAPG2* | 2.538330901 | 3.71E-05 | 0.000366571 | 0.233504493 |
| T2DM | monocyte | *FBXO25* | 4.381626113 | 0.000176895 | 0.000679891 | 1 |
| T2DM | monocyte | *CSGALNACT1* | 0.938190361 | 0.001082013 | 0.002331366 | 1 |
| T2DM | monocyte | *CCDC25* | 5.81643627 | 2.28E-07 | 4.94E-05 | 0.00143199 |
| T2DM | monocyte | *BIN3* | 3.871527379 | 2.81E-05 | 0.000362859 | 0.176702236 |
| T2DM | monocyte | *SLC25A37* | 2.819070406 | 3.53E-05 | 0.000365043 | 0.222243709 |

| | | | | | | |
|---|---|---|---|---|---|---|
| T2DM | monocyte | *CHMP7* | 2.161940642 | 0.00011827 | 0.000544237 | 0.743563444 |
| T2DM | monocyte | *DOCK5* | -0.325974266 | 0.021991179 | 0.029448039 | 1 |
| T2DM | monocyte | *RGS20* | 1.174271114 | 0.003541374 | 0.005856028 | 1 |
| T2DM | monocyte | *TACC1* | 1.279322597 | 0.000215472 | 0.000765353 | 1 |
| T2DM | monocyte | *SLC26A7* | 0.781359298 | 0.003399512 | 0.005655658 | 1 |
| T2DM | monocyte | *ATP6V0D2* | 1.302726639 | 0.016578936 | 0.022817813 | 1 |
| T2DM | monocyte | *MTDH* | 3.201441286 | 2.90E-05 | 0.000362859 | 0.182465205 |
| T2DM | monocyte | *MAL2* | 0.618304246 | 0.006012204 | 0.009316916 | 1 |
| T2DM | monocyte | *UTP23* | 4.839340612 | 4.39E-05 | 0.000378997 | 0.276064779 |
| T2DM | monocyte | *ZNF7* | 2.554887508 | 3.93E-06 | 0.000271625 | 0.024717904 |
| T2DM | monocyte | *SLC39A4* | -0.846917569 | 0.002362795 | 0.004135085 | 1 |
| T2DM | monocyte | *AK3* | 1.898590701 | 0.000217295 | 0.000767755 | 1 |
| T2DM | monocyte | *CER1* | 1.958060265 | 0.015230462 | 0.02115641 | 1 |
| T2DM | monocyte | *PLIN2* | 1.734829794 | 0.000137893 | 0.000587547 | 0.866931454 |
| T2DM | monocyte | *HAUS6* | 4.83144121 | 3.08E-05 | 0.000362859 | 0.193387535 |
| T2DM | monocyte | *SIGMAR1* | 1.235187409 | 0.007087749 | 0.010760849 | 1 |
| T2DM | monocyte | *CBWD5* | 5.532080705 | 3.38E-05 | 0.000365043 | 0.212363987 |
| T2DM | monocyte | *ZNF462* | 0.886405341 | 0.000580192 | 0.001488843 | 1 |
| T2DM | monocyte | *ALAD* | 0.919874911 | 0.017526733 | 0.023980537 | 1 |
| T2DM | monocyte | *ASTN2* | 2.146810569 | 6.86E-05 | 0.000423977 | 0.431184589 |
| T2DM | monocyte | *WDR31* | 3.053232788 | 2.75E-05 | 0.000362859 | 0.173165441 |
| T2DM | monocyte | *POLE3* | 2.621898262 | 0.001659202 | 0.003178368 | 1 |
| T2DM | monocyte | *SURF6* | 1.218434626 | 0.000753361 | 0.001794764 | 1 |
| T2DM | monocyte | *ASB6* | 1.918017428 | 0.001617767 | 0.003126622 | 1 |
| T2DM | monocyte | *NTMT1* | 2.623236796 | 0.000492038 | 0.001333243 | 1 |
| T2DM | monocyte | *CIZ1* | 2.188043308 | 0.000122832 | 0.000553734 | 0.772241651 |
| T2DM | monocyte | *SH3GLB2* | 2.476324964 | 0.000111279 | 0.000524053 | 0.699610241 |
| T2DM | monocyte | *SEC16A* | 4.187942955 | 2.09E-06 | 0.000188095 | 0.013166635 |
| T2DM | monocyte | *DPH7* | 4.573619079 | 0.000494587 | 0.001337406 | 1 |
| T2DM | monocyte | *SLC39A12* | 1.551767631 | 0.00163632 | 0.003153478 | 1 |
| T2DM | monocyte | *RSU1* | 3.440698065 | 0.000165551 | 0.000653372 | 1 |
| T2DM | monocyte | *PARD3* | 0.801928702 | 0.00066233 | 0.001633907 | 1 |
| T2DM | monocyte | *NRBF2* | 0.797471283 | 0.037147283 | 0.047410671 | 1 |
| T2DM | monocyte | *ADIRF* | 2.017301795 | 5.44E-05 | 0.000387662 | 0.342044106 |
| T2DM | monocyte | *GLUD1* | 2.838770397 | 1.00E-05 | 0.000362859 | 0.063125664 |
| T2DM | monocyte | *HTR7* | -0.235044509 | 0.009563698 | 0.014025419 | 1 |
| T2DM | monocyte | *RPP30* | 3.164075463 | 3.37E-05 | 0.000365043 | 0.211829588 |
| T2DM | monocyte | *FRA10AC1* | 1.737180585 | 4.54E-05 | 0.000379941 | 0.285496646 |
| T2DM | monocyte | *DNAJB12* | 4.0967703 | 3.12E-05 | 0.000362859 | 0.196148971 |
| T2DM | monocyte | *PLEKHS1* | -2.984631419 | 0.003455207 | 0.005739605 | 1 |
| T2DM | monocyte | *FUOM* | -1.09287484 | 0.036815515 | 0.047044542 | 1 |
| T2DM | monocyte | *LRRC27* | 1.054112371 | 0.016928593 | 0.023232932 | 1 |
| T2DM | monocyte | *PDCD11* | 3.554290975 | 8.23E-05 | 0.000447645 | 0.517477066 |
| T2DM | monocyte | *GAS2* | 1.237221654 | 0.000661669 | 0.001633259 | 1 |
| T2DM | monocyte | *LIN7C* | 1.600408371 | 0.000345095 | 0.001037595 | 1 |
| T2DM | monocyte | *ZNF215* | 0.745170268 | 0.005471267 | 0.00857588 | 1 |
| T2DM | monocyte | *HSD17B12* | -1.049254472 | 0.006373409 | 0.009808965 | 1 |
| T2DM | monocyte | *APIP* | -0.445180225 | 0.032959329 | 0.042410009 | 1 |
| T2DM | monocyte | *EIF3M* | 1.99424861 | 0.017168481 | 0.023536467 | 1 |
| T2DM | monocyte | *GLYAT* | 1.111298426 | 4.99E-05 | 0.000383513 | 0.313963328 |

| | | | | | | |
|---|---|---|---|---|---|---|
| T2DM | monocyte | *C11orf49* | 2.539211758 | 1.90E-05 | 0.000362859 | 0.119459371 |
| T2DM | monocyte | *ENDOD1* | 1.890466078 | 0.000266441 | 0.000877024 | 1 |
| T2DM | monocyte | *SERPINH1* | 1.31454888 | 0.000215132 | 0.000764578 | 1 |
| T2DM | monocyte | *CAPN5* | 0.903506788 | 0.000904508 | 0.002042616 | 1 |
| T2DM | monocyte | *INTS4* | 2.790035848 | 4.40E-05 | 0.000378997 | 0.276391548 |
| T2DM | monocyte | *RPS3* | 2.27268206 | 0.000619597 | 0.001553363 | 1 |
| T2DM | monocyte | *ZC3H12C* | 1.270001097 | 0.000250171 | 0.000839513 | 1 |
| T2DM | monocyte | *TTC12* | 1.1511904 | 0.001497147 | 0.002953704 | 1 |
| T2DM | monocyte | *DRD2* | -1.980351577 | 0.018895763 | 0.025636094 | 1 |
| T2DM | monocyte | *C11orf52* | 7.335105671 | 0.001517322 | 0.002978363 | 1 |
| T2DM | monocyte | *NPAT* | 3.354317461 | 5.74E-05 | 0.000396663 | 0.3605671 |
| T2DM | monocyte | *GLB1L2* | -1.505384716 | 0.025736243 | 0.03387851 | 1 |
| T2DM | monocyte | *P4HA3* | 2.145222715 | 1.39E-05 | 0.000362859 | 0.087586183 |
| T2DM | monocyte | *GRIK4* | 1.357403759 | 0.00534069 | 0.00838375 | 1 |
| T2DM | monocyte | *ADAM33* | 0.537737649 | 0.002310374 | 0.004054035 | 1 |
| T2DM | monocyte | *MTA2* | 1.396070473 | 0.002708437 | 0.004651398 | 1 |
| T2DM | monocyte | *TMC2* | 1.467416371 | 0.001477452 | 0.002926509 | 1 |
| T2DM | monocyte | *EML3* | 1.414226409 | 0.00038265 | 0.001115309 | 1 |
| T2DM | monocyte | *MS4A3* | -3.787481069 | 0.00865191 | 0.012822857 | 1 |
| T2DM | monocyte | *PLCH2* | 2.776919799 | 0.001761961 | 0.003315608 | 1 |
| T2DM | monocyte | *CPSF7* | 2.015308543 | 9.52E-05 | 0.000480165 | 0.598215795 |
| T2DM | monocyte | *CHEK1* | 1.068652299 | 0.007038541 | 0.010693887 | 1 |
| T2DM | monocyte | *FEZ1* | 1.131252312 | 0.000828174 | 0.001920594 | 1 |
| T2DM | monocyte | *COMMD7* | 3.616032555 | 3.10E-05 | 0.000362859 | 0.195048364 |
| T2DM | monocyte | *KIAA1755* | 0.789113472 | 0.001911839 | 0.003497157 | 1 |
| T2DM | monocyte | *TRPT1* | 1.425268264 | 0.00137419 | 0.0027762 | 1 |
| T2DM | monocyte | *TBX6* | 1.272350804 | 0.001513584 | 0.00297567 | 1 |
| T2DM | monocyte | *TAOK2* | 3.035216387 | 0.000269371 | 0.000883892 | 1 |
| T2DM | monocyte | *MPP7* | -0.840626572 | 0.035367129 | 0.045304226 | 1 |
| T2DM | monocyte | *ARID5B* | 1.99958359 | 2.48E-05 | 0.000362859 | 0.155651195 |
| T2DM | monocyte | *TMCO3* | 3.164827161 | 2.96E-05 | 0.000362859 | 0.185875865 |
| T2DM | monocyte | *SAP18* | 1.899639128 | 0.000190792 | 0.000713569 | 1 |
| T2DM | monocyte | *LYPD6B* | 1.163538917 | 0.002749711 | 0.004698948 | 1 |
| T2DM | monocyte | *PDCD4* | 3.009923358 | 1.52E-05 | 0.000362859 | 0.09551843 |
| T2DM | monocyte | *WDR17* | 4.435786556 | 0.000307203 | 0.000967849 | 1 |
| T2DM | monocyte | *CCDC102B* | 1.602051352 | 0.00014964 | 0.00061774 | 0.940786788 |
| T2DM | monocyte | *RGS18* | -0.104663043 | 0.007650874 | 0.011499173 | 1 |
| T2DM | monocyte | *PRSS23* | 0.846681691 | 0.00057466 | 0.001480084 | 1 |
| T2DM | monocyte | *MTMR12* | 2.279387586 | 4.36E-05 | 0.000378997 | 0.274041627 |
| T2DM | monocyte | *PPP1R1C* | 1.843623234 | 0.027576902 | 0.036044903 | 1 |
| T2DM | monocyte | *C11orf53* | 5.839926209 | 0.003601411 | 0.005942801 | 1 |
| T2DM | monocyte | *CCT5* | 3.708033343 | 2.37E-05 | 0.000362859 | 0.148874765 |
| T2DM | monocyte | *FAM173B* | 1.545552289 | 0.000776556 | 0.001832661 | 1 |
| T2DM | monocyte | *DOCK1* | 1.537347728 | 0.000113775 | 0.000529958 | 0.715301051 |
| T2DM | monocyte | *TEX12* | 5.439276244 | 0.001871164 | 0.003444403 | 1 |
| T2DM | monocyte | *CRIM1* | 0.886309384 | 0.000630595 | 0.001575722 | 1 |
| T2DM | monocyte | *ABCB9* | 0.606707947 | 0.010330505 | 0.014999511 | 1 |
| T2DM | monocyte | *DHX37* | 4.47132532 | 3.21E-05 | 0.000363519 | 0.202111636 |
| T2DM | monocyte | *GPR158* | 0.889454464 | 0.002270122 | 0.003993357 | 1 |
| T2DM | monocyte | *THRB* | 1.049650277 | 0.000201201 | 0.000732033 | 1 |

| | | | | | | |
|---|---|---|---|---|---|---|
| T2DM | monocyte | *TMEM86A* | -0.544558936 | 0.007282141 | 0.011021381 | 1 |
| T2DM | monocyte | *BTBD11* | -0.302039617 | 0.012793021 | 0.018106647 | 1 |
| T2DM | monocyte | *UBE3B* | 1.380837314 | 0.00218268 | 0.003878654 | 1 |
| T2DM | monocyte | *ANK3* | 1.15462298 | 0.000113881 | 0.000529958 | 0.715972969 |
| T2DM | monocyte | *PLBD2* | 2.261579195 | 6.74E-05 | 0.000421481 | 0.42400976 |
| T2DM | monocyte | *DLG5* | 1.012793863 | 0.0052356 | 0.008241416 | 1 |
| T2DM | monocyte | *GXYLT1* | 3.752298765 | 0.001734431 | 0.003278522 | 1 |
| T2DM | monocyte | *FAM177A1* | 3.318871916 | 4.54E-06 | 0.000285742 | 0.028574237 |
| T2DM | monocyte | *MIPOL1* | 1.193198254 | 0.000174566 | 0.000674165 | 1 |
| T2DM | monocyte | *THRSP* | 0.716866934 | 0.010524351 | 0.015233247 | 1 |
| T2DM | monocyte | *NDUFC2* | 5.090555573 | 0.000346239 | 0.001038551 | 1 |
| T2DM | monocyte | *ME3* | 1.43344111 | 0.000129839 | 0.00057004 | 0.816297799 |
| T2DM | monocyte | *ADAMTS12* | 0.72598056 | 0.00051193 | 0.001369576 | 1 |
| T2DM | monocyte | *VIPAS39* | 1.638040466 | 0.00114573 | 0.00243023 | 1 |
| T2DM | monocyte | *ANKRD50* | 3.543924604 | 0.003256541 | 0.005446627 | 1 |
| T2DM | monocyte | *VPS26B* | 1.111290128 | 0.000758699 | 0.001804745 | 1 |
| T2DM | monocyte | *NCAPD3* | 2.916606446 | 0.005588015 | 0.00873275 | 1 |
| T2DM | monocyte | *VTI1A* | 3.068549393 | 3.59E-05 | 0.000365043 | 0.225524217 |
| T2DM | monocyte | *FAM160B1* | 1.971132769 | 8.99E-05 | 0.000465291 | 0.565105664 |
| T2DM | monocyte | *ANO4* | 0.771671637 | 0.001323329 | 0.002699599 | 1 |
| T2DM | monocyte | *TEX9* | 1.748339745 | 0.000283952 | 0.000915489 | 1 |
| T2DM | monocyte | *AKR1C2* | 0.798802499 | 0.001326965 | 0.002703173 | 1 |
| T2DM | monocyte | *ITIH2* | 5.342909314 | 0.03328652 | 0.042804735 | 1 |
| T2DM | monocyte | *KIN* | 3.603093557 | 6.91E-06 | 0.000338043 | 0.043453166 |
| T2DM | monocyte | *PIGF* | 2.27614086 | 0.000136875 | 0.000584748 | 0.860531689 |
| T2DM | monocyte | *INPP1* | 2.199479747 | 8.70E-05 | 0.000461034 | 0.547086717 |
| T2DM | monocyte | *MFSD6* | 4.147201795 | 4.34E-05 | 0.000378997 | 0.272597144 |
| T2DM | monocyte | *RNF144A* | 1.054388723 | 0.000345557 | 0.001037992 | 1 |
| T2DM | monocyte | *ASAP2* | 1.322661146 | 0.000141194 | 0.000596094 | 0.887687963 |
| T2DM | monocyte | *ADAM17* | 3.062147959 | 2.41E-05 | 0.000362859 | 0.1515624 |
| T2DM | monocyte | *TMEM45B* | 3.842842834 | 0.001846237 | 0.003413909 | 1 |
| T2DM | monocyte | *ACSL1* | 1.02204154 | 0.000248103 | 0.000834576 | 1 |
| T2DM | monocyte | *BICD1* | 2.143248463 | 6.74E-05 | 0.000421481 | 0.423682652 |
| T2DM | monocyte | *GUF1* | 3.582952419 | 0.000306122 | 0.000965629 | 1 |
| T2DM | monocyte | *GABRA2* | 0.902236599 | 0.009448097 | 0.013881792 | 1 |
| T2DM | monocyte | *CENPJ* | 2.965505545 | 6.12E-05 | 0.000403714 | 0.384719501 |
| T2DM | monocyte | *PARP8* | 2.227149489 | 0.001136295 | 0.002415108 | 1 |
| T2DM | monocyte | *CACUL1* | 2.763856023 | 5.04E-05 | 0.000383643 | 0.316755141 |
| T2DM | monocyte | *BEND6* | 0.540211897 | 0.015207213 | 0.021128785 | 1 |
| T2DM | monocyte | *RABGAP1L* | 2.960580547 | 2.10E-05 | 0.000362859 | 0.13230489 |
| T2DM | monocyte | *PTPN14* | 0.846005131 | 0.000861588 | 0.001977101 | 1 |
| T2DM | monocyte | *RNF219* | 2.729423979 | 0.000122148 | 0.000552874 | 0.767941554 |
| T2DM | monocyte | *ARL14EP* | 1.051826015 | 0.020705796 | 0.027887176 | 1 |
| T2DM | monocyte | *EPG5* | 1.786581739 | 0.002326856 | 0.004081736 | 1 |
| T2DM | monocyte | *SPC25* | 0.504725568 | 0.011761233 | 0.016839644 | 1 |
| T2DM | monocyte | *G6PC2* | -5.294944559 | 0.00779373 | 0.011680377 | 1 |
| T2DM | monocyte | *TGOLN2* | 2.439735888 | 0.021485452 | 0.028832238 | 1 |
| T2DM | monocyte | *ATG10* | 0.592210968 | 0.009189875 | 0.013546716 | 1 |
| T2DM | monocyte | *TADA1* | 4.17035169 | 0.006822534 | 0.010407854 | 1 |
| T2DM | monocyte | *GUCY1A2* | 0.591495403 | 0.002070414 | 0.003725441 | 1 |

| | | | | | | |
|---|---|---|---|---|---|---|
| T2DM | monocyte | *CWF19L2* | 2.390220951 | 1.74E-05 | 0.000362859 | 0.109357512 |
| T2DM | monocyte | *ZNF547* | 1.371123765 | 0.003268666 | 0.005461096 | 1 |
| T2DM | monocyte | *NMT2* | 0.931558935 | 0.002453426 | 0.004269219 | 1 |
| T2DM | monocyte | *CCDC50* | 1.47575887 | 0.000700982 | 0.001698293 | 1 |
| T2DM | monocyte | *ZFP36L2* | 3.975631508 | 8.13E-06 | 0.000355191 | 0.051082733 |
| T2DM | monocyte | *PLEKHH2* | 0.601075489 | 0.001245321 | 0.002577977 | 1 |
| T2DM | monocyte | *TMEM123* | 3.681653297 | 5.70E-05 | 0.000395506 | 0.358490108 |
| T2DM | monocyte | *IGSF10* | 1.08136043 | 0.001320889 | 0.002697995 | 1 |
| T2DM | monocyte | *SPEF2* | 2.496946447 | 0.000687873 | 0.001677082 | 1 |
| T2DM | monocyte | *SPARCL1* | 0.484409458 | 0.001526676 | 0.002992894 | 1 |
| T2DM | monocyte | *MBNL1* | 2.328863544 | 0.000287523 | 0.000923687 | 1 |
| T2DM | monocyte | *CLEC4F* | -0.123423386 | 0.006886688 | 0.010488519 | 1 |
| T2DM | monocyte | *PELO* | 7.152781743 | 0.000591117 | 0.001506427 | 1 |
| T2DM | monocyte | *RASGRP3* | 0.937144009 | 0.000772948 | 0.001825517 | 1 |
| T2DM | monocyte | *SAR1B* | 4.772326796 | 4.30E-08 | 1.50E-05 | 0.000270494 |
| T2DM | monocyte | *GPR180* | 1.269201696 | 0.001508093 | 0.002969427 | 1 |
| T2DM | monocyte | *WDR78* | 1.849760287 | 0.000842999 | 0.001946359 | 1 |
| T2DM | monocyte | *PRDM8* | 0.76552675 | 0.007186085 | 0.010894361 | 1 |
| T2DM | monocyte | *ZNF117* | 0.527306535 | 0.001025077 | 0.002243961 | 1 |
| T2DM | monocyte | *RAB3C* | 0.656488425 | 0.001760263 | 0.003313404 | 1 |
| T2DM | monocyte | *RAD17* | 1.163549105 | 0.039149169 | 0.049733446 | 1 |
| T2DM | monocyte | *MED21* | 1.755668361 | 0.000168022 | 0.000658686 | 1 |
| T2DM | monocyte | *SRP19* | 2.112688413 | 0.000193942 | 0.000720633 | 1 |
| T2DM | monocyte | *CENPH* | 1.652530378 | 0.000323071 | 0.00099909 | 1 |
| T2DM | monocyte | *CARHSP1* | 1.370275352 | 0.000410648 | 0.00117346 | 1 |
| T2DM | monocyte | *TEKT5* | 2.18043149 | 0.000708494 | 0.001713194 | 1 |
| T2DM | monocyte | *BANK1* | 0.970873787 | 0.000527205 | 0.001396582 | 1 |
| T2DM | monocyte | *CAST* | 1.908769643 | 0.000553988 | 0.0014482 | 1 |
| T2DM | monocyte | *SCOC* | 4.069724661 | 6.29E-05 | 0.000408289 | 0.39563196 |
| T2DM | monocyte | *CLGN* | 4.793832507 | 0.0011403 | 0.0024228 | 1 |
| T2DM | monocyte | *SYCP2L* | 1.478589802 | 0.000716169 | 0.001727103 | 1 |
| T2DM | monocyte | *AHCTF1* | 3.147128849 | 1.50E-05 | 0.000362859 | 0.094495586 |
| T2DM | monocyte | *PTPRR* | 1.025881926 | 0.001505979 | 0.002966699 | 1 |
| T2DM | monocyte | *PLA2R1* | 1.140951291 | 7.58E-05 | 0.000435317 | 0.476365466 |
| T2DM | monocyte | *SLC25A27* | 0.663878208 | 0.008516452 | 0.012651922 | 1 |
| T2DM | monocyte | *FRMD1* | 0.913293032 | 0.001800965 | 0.003361444 | 1 |
| T2DM | monocyte | *ASAP1* | 2.616225508 | 0.000101399 | 0.000497371 | 0.637495666 |
| T2DM | monocyte | *INO80C* | 1.697738548 | 0.001096893 | 0.002355248 | 1 |
| T2DM | monocyte | *NMRAL1* | 2.732174897 | 0.000104823 | 0.000507331 | 0.659023241 |
| T2DM | monocyte | *C16orf89* | 0.980430819 | 0.00214685 | 0.003836625 | 1 |
| T2DM | monocyte | *TEX29* | 1.679173244 | 0.001752046 | 0.003299914 | 1 |
| T2DM | monocyte | *ADPRHL1* | -0.956374022 | 0.029906792 | 0.038830586 | 1 |
| T2DM | monocyte | *TUBGCP5* | 2.148477661 | 9.11E-05 | 0.00046854 | 0.572555595 |
| T2DM | monocyte | *LURAP1L* | 0.836607224 | 0.000492788 | 0.001333243 | 1 |
| T2DM | monocyte | *CFDP1* | 2.377422551 | 3.52E-05 | 0.000365043 | 0.221586467 |
| T2DM | monocyte | *FAM92B* | 10.13724527 | 0.001947929 | 0.003544611 | 1 |
| T2DM | monocyte | *C7orf31* | 1.767367324 | 0.000414425 | 0.001179488 | 1 |
| T2DM | monocyte | *FBXO36* | 1.315465286 | 0.00042657 | 0.001203701 | 1 |
| T2DM | monocyte | *MCOLN2* | 1.321811488 | 0.000343138 | 0.001035674 | 1 |
| T2DM | monocyte | *CACNA2D1* | 0.794538788 | 0.000497161 | 0.001338608 | 1 |

| | | | | | | |
|---|---|---|---|---|---|---|
| T2DM | monocyte | *ZUFSP* | 3.599018946 | 6.64E-05 | 0.000420249 | 0.417307708 |
| T2DM | monocyte | *HS3ST3A1* | 1.125177886 | 0.003320876 | 0.005540963 | 1 |
| T2DM | monocyte | *PPP2R5E* | 2.579246689 | 3.50E-06 | 0.000249851 | 0.021986897 |
| T2DM | monocyte | *ANKRD29* | 0.58150482 | 0.000896096 | 0.00202711 | 1 |
| T2DM | monocyte | *THY1* | 1.140961816 | 0.000484255 | 0.001320257 | 1 |
| T2DM | monocyte | *TBCEL* | 2.222862829 | 8.00E-05 | 0.000443003 | 0.50313006 |
| T2DM | monocyte | *UBASH3B* | -0.232445405 | 0.011963004 | 0.017089617 | 1 |
| T2DM | monocyte | *ROBO4* | 0.80887272 | 0.000863739 | 0.001980904 | 1 |
| T2DM | monocyte | *ROBO3* | 1.483820915 | 0.002429328 | 0.004235492 | 1 |
| T2DM | monocyte | *CC2D1B* | 1.107585838 | 0.029255367 | 0.038096207 | 1 |
| T2DM | monocyte | *CEP112* | 0.961469044 | 0.000320854 | 0.00099517 | 1 |
| T2DM | monocyte | *GAL3ST2* | 5.851957715 | 0.037638057 | 0.047978602 | 1 |
| T2DM | monocyte | *ENPP3* | 0.762957297 | 0.00164011 | 0.00315622 | 1 |
| T2DM | monocyte | *UCHL1* | 1.03078551 | 0.001435735 | 0.002874671 | 1 |
| T2DM | monocyte | *MIA3* | 2.14236425 | 0.000180594 | 0.000688537 | 1 |
| T2DM | monocyte | *DISP1* | 1.925574739 | 0.005770566 | 0.008986934 | 1 |
| T2DM | monocyte | *NEIL2* | 0.887451078 | 0.000611479 | 0.001540211 | 1 |
| T2DM | monocyte | *LONRF1* | 1.857935022 | 0.012647102 | 0.01793646 | 1 |
| T2DM | monocyte | *CCSAP* | -0.593677019 | 0.026974 | 0.035337682 | 1 |
| T2DM | monocyte | *GBP5* | -0.669138106 | 0.032485389 | 0.041863053 | 1 |
| T2DM | monocyte | *FAM69A* | 1.149413504 | 0.00068338 | 0.001668508 | 1 |
| T2DM | monocyte | *ATP5G3* | 1.935997389 | 1.90E-05 | 0.000362859 | 0.11930045 |
| T2DM | monocyte | *PDLIM3* | 0.705103846 | 0.000846739 | 0.001952127 | 1 |
| T2DM | monocyte | *NCAM2* | 1.398534535 | 0.001890498 | 0.003471251 | 1 |
| T2DM | monocyte | *L3MBTL4* | 1.9635217 | 3.51E-05 | 0.000365043 | 0.220733373 |
| T2DM | monocyte | *PDE1C* | 0.693961729 | 0.001293655 | 0.002657912 | 1 |
| T2DM | monocyte | *MRPL39* | 1.142812065 | 0.001157246 | 0.002444761 | 1 |
| T2DM | monocyte | *TSEN2* | 1.653162643 | 8.04E-07 | 0.000105311 | 0.005054947 |
| T2DM | monocyte | *FLCN* | 2.471390195 | 0.000308764 | 0.0009706 | 1 |
| T2DM | monocyte | *DPH3* | 2.725708686 | 0.000151981 | 0.0006224 | 0.955505492 |
| T2DM | monocyte | *CXXC1* | 2.528880057 | 0.000123208 | 0.000553734 | 0.77460905 |
| T2DM | monocyte | *PPP4R1* | 2.147836082 | 6.24E-05 | 0.000407194 | 0.392535366 |
| T2DM | monocyte | *APCDD1* | 0.777121501 | 0.001125755 | 0.002400821 | 1 |
| T2DM | monocyte | *EPHB1* | 1.217196175 | 0.000700677 | 0.001698293 | 1 |
| T2DM | monocyte | *ANKRD40* | 2.632266472 | 0.000797808 | 0.00186809 | 1 |
| T2DM | monocyte | *CYP2U1* | 3.218639824 | 5.42E-05 | 0.00038765 | 0.34074408 |
| T2DM | monocyte | *FBXL18* | 3.94079779 | 1.84E-05 | 0.000362859 | 0.115416599 |
| T2DM | monocyte | *PROM2* | 4.249595688 | 0.003549763 | 0.005867226 | 1 |
| T2DM | monocyte | *AK9* | 2.105734824 | 0.00012505 | 0.000556889 | 0.786188104 |
| T2DM | monocyte | *CDK19* | 2.816888359 | 0.002302615 | 0.004044857 | 1 |
| T2DM | monocyte | *GTF3C6* | 2.352534081 | 0.000579516 | 0.001487717 | 1 |
| T2DM | monocyte | *TTC39B* | 2.60705125 | 2.89E-05 | 0.000362859 | 0.181498815 |
| T2DM | monocyte | *MMS19* | 2.855485577 | 0.000197309 | 0.000724581 | 1 |
| T2DM | monocyte | *GOLGA7B* | 1.206499819 | 0.012847017 | 0.018174887 | 1 |
| T2DM | monocyte | *TRMT44* | 2.405977557 | 9.29E-05 | 0.00047433 | 0.583899837 |
| T2DM | monocyte | *CNOT8* | 1.41108821 | 0.0002968 | 0.000946439 | 1 |
| T2DM | monocyte | *LRGUK* | 0.822300427 | 0.003118082 | 0.005242947 | 1 |
| T2DM | monocyte | *ZKSCAN2* | 1.188128605 | 0.000515533 | 0.001377457 | 1 |
| T2DM | monocyte | *PDIA4* | 5.074327678 | 2.39E-05 | 0.000362859 | 0.150170596 |
| T2DM | monocyte | *KDM8* | 2.366715816 | 0.000551586 | 0.001444324 | 1 |

| | | | | | | |
|---|---|---|---|---|---|---|
| T2DM | monocyte | *FAM126B* | 2.899009628 | 3.97E-05 | 0.000371009 | 0.249558965 |
| T2DM | monocyte | *ALS2CR12* | 1.776226607 | 0.014369177 | 0.020093197 | 1 |
| T2DM | monocyte | *SPAG17* | 0.41137673 | 0.001657937 | 0.003176913 | 1 |
| T2DM | monocyte | *DEPTOR* | 1.379045793 | 0.000293219 | 0.000937676 | 1 |
| T2DM | monocyte | *RRAGA* | 4.673872442 | 0.000206434 | 0.000743876 | 1 |
| T2DM | monocyte | *FAM161B* | 1.343235089 | 0.000343419 | 0.001035977 | 1 |
| T2DM | monocyte | *UGT2B4* | 1.953219722 | 0.000938129 | 0.002099444 | 1 |
| T2DM | monocyte | *MMP16* | 0.92872663 | 0.014805453 | 0.020634424 | 1 |
| T2DM | monocyte | *KCNMA1* | 0.553011643 | 0.001447775 | 0.002890492 | 1 |
| T2DM | monocyte | *BATF* | 2.560290752 | 0.000423495 | 0.001197173 | 1 |
| T2DM | monocyte | *DPY19L4* | 1.898876594 | 0.00024115 | 0.0008199 | 1 |
| T2DM | monocyte | *NDUFAF6* | 1.114294699 | 0.022331399 | 0.029872151 | 1 |
| T2DM | monocyte | *DRAM2* | 2.754084084 | 2.54E-05 | 0.000362859 | 0.15957032 |
| T2DM | monocyte | *ADAMTSL3* | 0.490476848 | 0.0011359 | 0.002415085 | 1 |
| T2DM | monocyte | *N6AMT1* | 0.613056827 | 0.014511331 | 0.020278448 | 1 |
| T2DM | monocyte | *RWDD2B* | 1.584053424 | 0.001449827 | 0.002892752 | 1 |
| T2DM | monocyte | *SFXN2* | 1.810408932 | 0.000234546 | 0.000803155 | 1 |
| T2DM | monocyte | *SH3RF2* | 0.567862005 | 0.000764319 | 0.001813827 | 1 |
| T2DM | monocyte | *GDF6* | 2.295517874 | 0.00049239 | 0.001333243 | 1 |
| T2DM | monocyte | *ATAD2* | 3.876909186 | 2.75E-05 | 0.000362859 | 0.17311437 |
| T2DM | monocyte | *SASS6* | 2.867328202 | 0.000884574 | 0.00201134 | 1 |
| T2DM | monocyte | *VPS8* | 3.166635536 | 5.39E-05 | 0.000385932 | 0.338848188 |
| T2DM | monocyte | *MPV17L* | 1.001845317 | 0.004078248 | 0.006613346 | 1 |
| T2DM | monocyte | *RPUSD3* | 3.315451225 | 0.029210727 | 0.038053842 | 1 |
| T2DM | monocyte | *GHRL* | 1.818551577 | 0.000334228 | 0.00102039 | 1 |
| T2DM | monocyte | *EXOG* | 4.141181551 | 0.00032941 | 0.001010243 | 1 |
| T2DM | monocyte | *NTAN1* | 1.454279479 | 0.000396588 | 0.001146558 | 1 |
| T2DM | monocyte | *SLC6A1* | 1.218935171 | 0.024654882 | 0.032598369 | 1 |
| T2DM | monocyte | *FCHO2* | 2.191379645 | 3.50E-05 | 0.000365043 | 0.219949808 |
| T2DM | monocyte | *TIMP4* | 0.855535527 | 0.000222681 | 0.000778207 | 1 |
| T2DM | monocyte | *NECAP2* | 1.666499099 | 0.000328132 | 0.001006817 | 1 |
| T2DM | monocyte | *STEAP2* | 0.793034474 | 0.000118854 | 0.000544237 | 0.747237435 |
| T2DM | monocyte | *GATAD1* | 1.650712951 | 5.58E-05 | 0.000390942 | 0.350539944 |
| T2DM | monocyte | *TMED6* | -1.047588453 | 0.009887031 | 0.014432264 | 1 |
| T2DM | monocyte | *CLEC18A* | -4.824937359 | 3.31E-09 | 1.89E-06 | 2.08E-05 |
| T2DM | monocyte | *IL34* | 1.366295689 | 0.000563866 | 0.001463676 | 1 |
| T2DM | monocyte | *DHRS1* | 1.069238518 | 0.01397777 | 0.019611301 | 1 |
| T2DM | monocyte | *AASDH* | 2.826774376 | 0.001477028 | 0.002926509 | 1 |
| T2DM | monocyte | *RNF111* | 2.370863994 | 2.74E-05 | 0.000362859 | 0.172282658 |
| T2DM | monocyte | *DYRK1A* | 3.093516177 | 6.72E-05 | 0.000421481 | 0.422330344 |
| T2DM | monocyte | *KCNJ15* | -1.958731592 | 0.000222591 | 0.000778207 | 1 |
| T2DM | monocyte | *LCA5L* | 0.59258813 | 0.015001326 | 0.0208704 | 1 |
| T2DM | monocyte | *CREB3L1* | 0.933381164 | 0.001075667 | 0.002323959 | 1 |
| T2DM | monocyte | *C2CD2* | 0.876167118 | 0.001636203 | 0.003153478 | 1 |
| T2DM | monocyte | *SLC38A10* | 3.90274872 | 0.000372454 | 0.001095237 | 1 |
| T2DM | monocyte | *C9orf43* | 1.019484373 | 0.015900996 | 0.021966504 | 1 |
| T2DM | monocyte | *WDR19* | 1.30089309 | 0.000300812 | 0.00095467 | 1 |
| T2DM | monocyte | *SLC37A3* | 2.165471644 | 7.50E-05 | 0.000434165 | 0.471503622 |
| T2DM | monocyte | *AP3S2* | 3.168850807 | 1.80E-13 | 1.09E-09 | 1.13E-09 |
| T2DM | monocyte | *SPPL3* | 2.586299966 | 0.00048806 | 0.00132603 | 1 |

| | | | | | | |
|---|---|---|---|---|---|---|
| T2DM | monocyte | *FAM213B* | 1.68212376 | 0.030265554 | 0.039240985 | 1 |
| T2DM | monocyte | *C12orf43* | 4.161130318 | 9.97E-07 | 0.000122893 | 0.006267546 |
| T2DM | monocyte | *PEX10* | 0.87970272 | 0.00348695 | 0.005782763 | 1 |
| T2DM | monocyte | *RADIL* | 0.723556152 | 0.002055385 | 0.003704603 | 1 |
| T2DM | monocyte | *LDLRAP1* | 4.49281244 | 0.006823791 | 0.010407854 | 1 |
| T2DM | monocyte | *PAFAH2* | 2.059684688 | 0.004511199 | 0.007220444 | 1 |
| T2DM | monocyte | *TRIM63* | 2.18256723 | 6.70E-06 | 0.000332508 | 0.042107591 |
| T2DM | monocyte | *WDR66* | 0.56443885 | 0.016145409 | 0.022269896 | 1 |
| T2DM | monocyte | *UBXN11* | -0.524667854 | 0.018227015 | 0.024814475 | 1 |
| T2DM | monocyte | *GALNT14* | 1.219903321 | 0.00183109 | 0.003394888 | 1 |
| T2DM | monocyte | *HPD* | 1.340405205 | 0.000254608 | 0.000848508 | 1 |
| T2DM | monocyte | *RHPN1* | 1.845441596 | 0.010574651 | 0.015293957 | 1 |
| T2DM | monocyte | *TPRG1L* | 2.72307427 | 0.00097424 | 0.002158986 | 1 |
| T2DM | monocyte | *LRRC43* | 2.287245487 | 0.001721997 | 0.00326212 | 1 |
| T2DM | monocyte | *XKR8* | 1.108259776 | 0.004325354 | 0.006961981 | 1 |
| T2DM | monocyte | *EYA3* | 2.934857555 | 0.000227129 | 0.000787188 | 1 |
| T2DM | monocyte | *MRAS* | 0.620985841 | 0.02933683 | 0.038186469 | 1 |
| T2DM | monocyte | *ABHD3* | 1.747894457 | 0.015074168 | 0.020950812 | 1 |
| T2DM | monocyte | *CLSTN2* | 0.712808648 | 0.000879772 | 0.002006213 | 1 |
| T2DM | monocyte | *COLEC12* | 0.729415091 | 0.000678295 | 0.001661104 | 1 |
| T2DM | monocyte | *AUTS2* | 1.180716507 | 0.000124826 | 0.000556889 | 0.784783828 |
| T2DM | monocyte | *MITD1* | 2.553105451 | 0.000450074 | 0.001247813 | 1 |
| T2DM | monocyte | *AHCYL2* | 2.374347004 | 0.005686029 | 0.008870487 | 1 |
| T2DM | monocyte | *B4GALT5* | 1.721893424 | 0.00015964 | 0.000640496 | 1 |
| T2DM | monocyte | *CD1D* | -0.198817781 | 0.005623788 | 0.008786252 | 1 |
| T2DM | monocyte | *FAM86C1* | 10.35612327 | 0.000969648 | 0.002152556 | 1 |
| T2DM | monocyte | *NCF1* | -0.303321602 | 0.009488433 | 0.013931289 | 1 |
| T2DM | monocyte | *PPP1R9A* | 0.997549188 | 0.000554338 | 0.001448513 | 1 |
| T2DM | monocyte | *ZFAND2B* | 2.219097578 | 0.000500837 | 0.001346776 | 1 |
| T2DM | monocyte | *GDPD5* | 1.593950918 | 0.000334558 | 0.00102039 | 1 |
| T2DM | monocyte | *DYNC1I1* | 1.58314809 | 0.000243074 | 0.000821174 | 1 |
| T2DM | monocyte | *TMED4* | 2.495874101 | 7.85E-05 | 0.000440255 | 0.493466833 |
| T2DM | monocyte | *ELK4* | 4.419080276 | 0.000335255 | 0.00102039 | 1 |
| T2DM | monocyte | *DUSP23* | 1.984518709 | 0.00047282 | 0.001295467 | 1 |
| T2DM | monocyte | *RNF166* | -0.461663484 | 0.023497629 | 0.031258907 | 1 |
| T2DM | monocyte | *F11R* | 1.183967846 | 0.001021381 | 0.002238696 | 1 |
| T2DM | monocyte | *NIT1* | 1.835960815 | 7.72E-05 | 0.00043763 | 0.485226059 |
| T2DM | monocyte | *ZNF276* | 2.421880506 | 0.007737714 | 0.011613036 | 1 |
| T2DM | monocyte | *CDA* | -0.365673767 | 0.014943188 | 0.020807934 | 1 |
| T2DM | monocyte | *DMTN* | 1.153107549 | 0.000209374 | 0.000750879 | 1 |
| T2DM | monocyte | *FAM160B2* | 1.677200082 | 0.000467709 | 0.001282377 | 1 |
| T2DM | monocyte | *NDUFS2* | 1.880423843 | 0.001126354 | 0.002401253 | 1 |
| T2DM | monocyte | *FCER1G* | -0.244501223 | 0.028318664 | 0.036968322 | 1 |
| T2DM | monocyte | *CCAR2* | 2.504445687 | 8.55E-05 | 0.000458095 | 0.537345745 |
| T2DM | monocyte | *CDC42SE2* | 2.210019342 | 0.013612234 | 0.01914544 | 1 |
| T2DM | monocyte | *RAPGEF6* | 2.030318698 | 0.000660508 | 0.001632317 | 1 |
| T2DM | monocyte | *EPB41* | 1.439448136 | 5.02E-05 | 0.000383643 | 0.315851927 |
| T2DM | monocyte | *ALG8* | 3.336883367 | 8.28E-05 | 0.000448982 | 0.520819368 |
| T2DM | monocyte | *MRPL10* | 0.804673477 | 0.003682666 | 0.006046728 | 1 |
| T2DM | monocyte | *RCAN1* | 3.748763757 | 7.35E-05 | 0.000431915 | 0.462027335 |

| | | | | | | |
|---|---|---|---|---|---|---|
| T2DM | monocyte | *CLIC6* | 0.62765324 | 0.00253847 | 0.004400154 | 1 |
| T2DM | monocyte | *CBR3* | 1.714751098 | 0.000384344 | 0.001119209 | 1 |
| T2DM | monocyte | *SIM2* | 2.495806759 | 0.002937944 | 0.004977326 | 1 |
| T2DM | monocyte | *HLCS* | 0.926237916 | 0.005478437 | 0.008584978 | 1 |
| T2DM | monocyte | *ARHGAP27* | -0.53957398 | 0.037505556 | 0.047819393 | 1 |
| T2DM | monocyte | *PADI4* | -0.463906719 | 0.009516715 | 0.01396629 | 1 |
| T2DM | monocyte | *PSMD4* | 3.737775002 | 2.55E-05 | 0.000362859 | 0.16062524 |
| T2DM | monocyte | *STARD9* | 1.213490724 | 0.00017857 | 0.000684139 | 1 |
| T2DM | monocyte | *THEM4* | 1.171267058 | 0.000273218 | 0.000892323 | 1 |
| T2DM | monocyte | *MED8* | 1.140940077 | 0.000380705 | 0.001112734 | 1 |
| T2DM | monocyte | *RSPRY1* | 2.177935877 | 0.00028091 | 0.000909882 | 1 |
| T2DM | monocyte | *ACE* | 1.07917154 | 0.000960716 | 0.002136547 | 1 |
| T2DM | monocyte | *TEPP* | -3.329714257 | 0.005016328 | 0.007941993 | 1 |
| T2DM | monocyte | *EFCAB14* | 3.647990398 | 2.30E-05 | 0.000362859 | 0.14431683 |
| T2DM | monocyte | *ZFYVE28* | 1.653992995 | 0.001100484 | 0.002360549 | 1 |
| T2DM | monocyte | *CCDC117* | 3.602529301 | 8.04E-05 | 0.000443644 | 0.50525938 |
| T2DM | monocyte | *CCDC107* | 1.866867028 | 0.013302521 | 0.018758897 | 1 |
| T2DM | monocyte | *NPR2* | 1.22274553 | 0.000584676 | 0.001494858 | 1 |
| T2DM | monocyte | *ZNF235* | 1.453138736 | 0.000586497 | 0.001496766 | 1 |
| T2DM | monocyte | *CALM3* | 4.194731698 | 6.21E-05 | 0.000406647 | 0.390471843 |
| T2DM | monocyte | *DFFA* | 4.00196483 | 2.87E-05 | 0.000362859 | 0.180431279 |
| T2DM | monocyte | *ATAD3B* | 2.784543175 | 0.000196828 | 0.000724272 | 1 |
| T2DM | monocyte | *SSU72* | 4.957436386 | 2.56E-05 | 0.000362859 | 0.160855945 |
| T2DM | monocyte | *NR2F6* | 1.540427495 | 0.00019798 | 0.000725771 | 1 |
| T2DM | monocyte | *ANKLE1* | 1.847906968 | 0.038733253 | 0.049224977 | 1 |
| T2DM | monocyte | *CILP2* | 6.530825828 | 0.002063036 | 0.003714292 | 1 |
| T2DM | monocyte | *ABCG1* | 2.029575383 | 0.002169382 | 0.003863712 | 1 |
| T2DM | monocyte | *U2AF1* | -1.104592269 | 0.011573428 | 0.016589635 | 1 |
| T2DM | monocyte | *HSF2BP* | 2.628197904 | 0.000340076 | 0.001029894 | 1 |
| T2DM | monocyte | *CSTB* | 1.495475959 | 0.031322685 | 0.040469732 | 1 |
| T2DM | monocyte | *ZNF66* | 3.11448612 | 1.47E-05 | 0.000362859 | 0.092715762 |
| T2DM | monocyte | *LRRC3* | 1.667556996 | 0.007754603 | 0.011628037 | 1 |
| T2DM | monocyte | *ITGB2* | -0.444400916 | 0.029622177 | 0.038502093 | 1 |
| T2DM | monocyte | *FTCD* | -0.747488313 | 0.018991078 | 0.025737639 | 1 |
| T2DM | monocyte | *LSS* | 1.027086407 | 0.001701795 | 0.003233153 | 1 |
| T2DM | monocyte | *PCNT* | 1.220704708 | 0.001895472 | 0.003476368 | 1 |
| T2DM | monocyte | *S100B* | 0.339859237 | 0.030883743 | 0.039943652 | 1 |
| T2DM | monocyte | *ZNF208* | 1.729547991 | 0.000518849 | 0.001383377 | 1 |
| T2DM | monocyte | *ADAMTS13* | 0.99362854 | 0.002976545 | 0.005029168 | 1 |
| T2DM | monocyte | *FCN2* | 1.168457991 | 0.017538337 | 0.023991193 | 1 |
| T2DM | monocyte | *ZNF714* | 3.222495424 | 8.22E-05 | 0.000447645 | 0.5170148 |
| T2DM | monocyte | *GPSM1* | 0.727523904 | 0.016400267 | 0.022591691 | 1 |
| T2DM | monocyte | *C19orf47* | 4.73962172 | 0.001190568 | 0.002495148 | 1 |
| T2DM | monocyte | *HIPK4* | 2.073352744 | 0.008545169 | 0.012690659 | 1 |
| T2DM | monocyte | *RDH13* | 2.341183976 | 0.000439996 | 0.001228968 | 1 |
| T2DM | monocyte | *PCSK7* | 3.003391382 | 0.001538827 | 0.003011081 | 1 |
| T2DM | monocyte | *S100A1* | -0.92636722 | 0.029580411 | 0.038463711 | 1 |
| T2DM | monocyte | *VPS11* | 1.687899698 | 0.001690481 | 0.003223856 | 1 |
| T2DM | monocyte | *FDPS* | 2.756494223 | 1.22E-06 | 0.00014194 | 0.007664784 |
| T2DM | monocyte | *PAQR6* | -0.8620145 | 0.002373353 | 0.004148254 | 1 |

| | | | | | | |
|---|---|---|---|---|---|---|
| T2DM | monocyte | *SLC25A44* | 2.283444279 | 0.000825387 | 0.001917414 | 1 |
| T2DM | monocyte | *NBEAL2* | -0.489141332 | 0.02670847 | 0.035048169 | 1 |
| T2DM | monocyte | *LY6K* | 2.140854439 | 0.001986264 | 0.003602898 | 1 |
| T2DM | monocyte | *LY6E* | 2.37122889 | 0.000453517 | 0.001253852 | 1 |
| T2DM | monocyte | *VPS28* | 3.726581585 | 1.40E-06 | 0.000152128 | 0.008823448 |
| T2DM | monocyte | *TONSL* | 1.294474569 | 0.001503595 | 0.002964284 | 1 |
| T2DM | monocyte | *MUM1* | 1.896257127 | 5.20E-05 | 0.000384775 | 0.32699806 |
| T2DM | monocyte | *ZNF333* | 3.979971087 | 0.000175709 | 0.000677718 | 1 |
| T2DM | monocyte | *PPP1R16A* | 0.834752935 | 0.003275189 | 0.005469087 | 1 |
| T2DM | monocyte | *RPL8* | 3.734646984 | 0.000860258 | 0.001975366 | 1 |
| T2DM | monocyte | *FBXL13* | 1.866215205 | 0.00029708 | 0.000946652 | 1 |
| T2DM | monocyte | *SCGB3A1* | -1.925567851 | 0.015377308 | 0.021336821 | 1 |
| T2DM | monocyte | *MFSD12* | 4.071711376 | 5.46E-05 | 0.000387662 | 0.343042715 |
| T2DM | monocyte | *DVL3* | 4.036662665 | 6.03E-06 | 0.000322038 | 0.037902701 |
| T2DM | monocyte | *AP2M1* | 3.862901971 | 5.03E-06 | 0.000298974 | 0.031594825 |
| T2DM | monocyte | *ABCF3* | 1.487365675 | 0.017953039 | 0.024489208 | 1 |
| T2DM | monocyte | *FBXO27* | 0.57078693 | 0.004956792 | 0.007863576 | 1 |
| T2DM | monocyte | *DMKN* | 1.160683312 | 0.008163617 | 0.01218245 | 1 |
| T2DM | monocyte | *BDH1* | 2.797718328 | 0.000217244 | 0.000767755 | 1 |
| T2DM | monocyte | *DUSP14* | 0.659859213 | 0.001127747 | 0.002402624 | 1 |
| T2DM | monocyte | *LRRC56* | -0.610513387 | 0.006831297 | 0.010416775 | 1 |
| T2DM | monocyte | *PGAP3* | 7.461639129 | 0.000109064 | 0.000518963 | 0.685683787 |
| T2DM | monocyte | *IKZF3* | 0.819348253 | 0.001391933 | 0.002804151 | 1 |
| T2DM | monocyte | *ACOX1* | 1.585641031 | 0.000474597 | 0.001298995 | 1 |
| T2DM | monocyte | *CYGB* | -1.20082218 | 0.038216972 | 0.048598322 | 1 |
| T2DM | monocyte | *ZNF577* | 1.404800157 | 0.000337695 | 0.001024656 | 1 |
| T2DM | monocyte | *TMEM143* | 1.256904575 | 0.00019625 | 0.000723774 | 1 |
| T2DM | monocyte | *SIGLEC11* | -0.847599939 | 0.008614301 | 0.012776153 | 1 |
| T2DM | monocyte | *MPP3* | 1.17289756 | 0.001245516 | 0.002577977 | 1 |
| T2DM | monocyte | *NAGS* | -1.747353706 | 0.011883271 | 0.016998891 | 1 |
| T2DM | monocyte | *ASB16* | 2.00431933 | 0.001017807 | 0.002233491 | 1 |
| T2DM | monocyte | *DBF4B* | 0.656709988 | 0.02828673 | 0.036934303 | 1 |
| T2DM | monocyte | *PLCD3* | 1.586680496 | 0.00120493 | 0.00251758 | 1 |
| T2DM | monocyte | *LARP4* | 2.093066158 | 7.55E-05 | 0.000435317 | 0.474913575 |
| T2DM | monocyte | *SPC24* | 4.387630624 | 0.00202947 | 0.003664354 | 1 |
| T2DM | monocyte | *LEMD2* | 2.435350696 | 0.000188799 | 0.000709469 | 1 |
| T2DM | monocyte | *SCIMP* | -0.302093529 | 0.008657342 | 0.012827884 | 1 |
| T2DM | monocyte | *ASGR2* | -0.219423944 | 0.007326333 | 0.01108293 | 1 |
| T2DM | monocyte | *SNRNP25* | 3.225120167 | 0.004013903 | 0.006522463 | 1 |
| T2DM | monocyte | *JMJD8* | 1.435719909 | 9.70E-05 | 0.000485446 | 0.6099777 |
| T2DM | monocyte | *MEIOB* | 2.931582897 | 0.0005345 | 0.001410156 | 1 |
| T2DM | monocyte | *ZG16B* | -8.921681928 | 0.020117063 | 0.027158251 | 1 |
| T2DM | monocyte | *TTC9C* | 3.043242285 | 0.000233867 | 0.000801844 | 1 |
| T2DM | monocyte | *NXF1* | 3.548957725 | 1.08E-05 | 0.000362859 | 0.06786961 |
| T2DM | monocyte | *RPL29* | 3.143501936 | 6.87E-05 | 0.000424475 | 0.432115676 |
| T2DM | monocyte | *TPCN2* | 2.911103903 | 7.89E-05 | 0.000441133 | 0.495833966 |
| T2DM | monocyte | *CYP4A22* | 5.938835934 | 0.000448068 | 0.001243711 | 1 |
| T2DM | monocyte | *PDZK1IP1* | -14.31097399 | 0.017551334 | 0.024003749 | 1 |
| T2DM | monocyte | *COA7* | -0.716836625 | 0.025770096 | 0.033915971 | 1 |
| T2DM | monocyte | *SLC1A7* | 0.681687067 | 0.005832183 | 0.009066997 | 1 |

| | | | | | | |
|---|---|---|---|---|---|---|
| T2DM | monocyte | *C1orf123* | 1.544825036 | 0.035503295 | 0.045450869 | 1 |
| T2DM | monocyte | *ACOT11* | 1.01226312 | 0.006343767 | 0.009775309 | 1 |
| T2DM | monocyte | *USP24* | 1.587063346 | 0.000444714 | 0.001238293 | 1 |
| T2DM | monocyte | *NOL9* | 2.457320244 | 5.66E-05 | 0.000394257 | 0.355620019 |
| T2DM | monocyte | *ZSWIM5* | 1.261456669 | 0.000320059 | 0.000993684 | 1 |
| T2DM | monocyte | *RBP7* | 1.862443475 | 0.000206235 | 0.000743876 | 1 |
| T2DM | monocyte | *FBLIM1* | 0.948295912 | 0.001058329 | 0.002296761 | 1 |
| T2DM | monocyte | *SLC25A34* | 6.964455534 | 0.007436008 | 0.011224534 | 1 |
| T2DM | monocyte | *PDPN* | 0.965745232 | 0.002759187 | 0.00471258 | 1 |
| T2DM | monocyte | *PEF1* | 1.505278603 | 0.000422489 | 0.001195403 | 1 |
| T2DM | monocyte | *SYNC* | 1.458230795 | 0.000413463 | 0.001178352 | 1 |
| T2DM | monocyte | *RBBP4* | 3.197415942 | 0.001819313 | 0.003386941 | 1 |
| T2DM | monocyte | *TSSK3* | 1.068204979 | 0.000579445 | 0.001487717 | 1 |
| T2DM | monocyte | *CAMK2N1* | 3.421956489 | 0.000160378 | 0.000642374 | 1 |
| T2DM | monocyte | *TTLL10* | 2.311249883 | 0.001176418 | 0.002475281 | 1 |
| T2DM | monocyte | *NFIA* | 1.498303115 | 4.18E-05 | 0.00037367 | 0.262689845 |
| T2DM | monocyte | *OMA1* | 1.240931952 | 0.009475287 | 0.013918488 | 1 |
| T2DM | monocyte | *TM2D1* | 3.078818337 | 2.39E-05 | 0.000362859 | 0.150442441 |
| T2DM | monocyte | *FUBP1* | 2.335565948 | 4.41E-05 | 0.000379147 | 0.277535504 |
| T2DM | monocyte | *NEXN* | 1.021269895 | 0.000479674 | 0.001309918 | 1 |
| T2DM | monocyte | *B3GALT2* | 7.460774439 | 0.000118052 | 0.000544132 | 0.742195957 |
| T2DM | monocyte | *NTNG1* | 0.663202048 | 0.00178059 | 0.003341663 | 1 |
| T2DM | monocyte | *HFM1* | 1.952650444 | 0.000220012 | 0.000772859 | 1 |
| T2DM | monocyte | *KCNT2* | 0.676891941 | 0.000731319 | 0.001755557 | 1 |
| T2DM | monocyte | *VCAM1* | 1.144496189 | 0.000982783 | 0.002174088 | 1 |
| T2DM | monocyte | *ZNF496* | 5.335438236 | 2.52E-05 | 0.000362859 | 0.158177277 |
| T2DM | monocyte | *TRIM58* | 1.239222394 | 0.006377866 | 0.009812078 | 1 |
| T2DM | monocyte | *IGSF8* | 1.415437173 | 0.000692472 | 0.001685365 | 1 |
| T2DM | monocyte | *VANGL2* | 1.976312342 | 0.001483695 | 0.0029361 | 1 |
| T2DM | monocyte | *FCRLB* | -0.88046761 | 0.01877918 | 0.0254897 | 1 |
| T2DM | monocyte | *LMX1A* | 0.872064336 | 4.98E-05 | 0.000383513 | 0.313396742 |
| T2DM | monocyte | *FLVCR1* | 1.8212872 | 0.00048557 | 0.001322695 | 1 |
| T2DM | monocyte | *TDRD5* | 1.050947099 | 0.016078161 | 0.022186874 | 1 |
| T2DM | monocyte | *SNED1* | 2.515659606 | 2.51E-05 | 0.000362859 | 0.157593344 |
| T2DM | monocyte | *C1orf115* | 0.959708539 | 0.000616844 | 0.001549381 | 1 |
| T2DM | monocyte | *BROX* | 1.39130005 | 0.015586959 | 0.021565848 | 1 |
| T2DM | monocyte | *ACP6* | 1.174652743 | 0.00086497 | 0.00198108 | 1 |
| T2DM | monocyte | *CNST* | 1.651927264 | 0.000144677 | 0.000606338 | 0.909586506 |
| T2DM | monocyte | *PM20D1* | -0.929495673 | 0.021850499 | 0.029278364 | 1 |
| T2DM | monocyte | *HAAO* | 1.317482044 | 0.000349343 | 0.001045868 | 1 |
| T2DM | monocyte | *B3GALNT2* | 1.71433712 | 0.000993059 | 0.002192962 | 1 |
| T2DM | monocyte | *PUS10* | 2.171560549 | 7.17E-05 | 0.000428945 | 0.450820916 |
| T2DM | monocyte | *RFTN2* | 1.032110211 | 0.000512493 | 0.0013705 | 1 |
| T2DM | monocyte | *FAM84A* | 1.605520875 | 0.001909982 | 0.003494778 | 1 |
| T2DM | monocyte | *CLHC1* | 0.864901007 | 0.000192459 | 0.000717244 | 1 |
| T2DM | monocyte | *FRZB* | 1.043388468 | 0.001494377 | 0.002950737 | 1 |
| T2DM | monocyte | *NUP35* | 3.07491438 | 0.000144858 | 0.000606338 | 0.91071987 |
| T2DM | monocyte | *FBXO41* | -0.432086958 | 0.008465083 | 0.01258092 | 1 |
| T2DM | monocyte | *SMC6* | 2.271307577 | 8.42E-05 | 0.000454787 | 0.529372532 |
| T2DM | monocyte | *CCDC74A* | 4.745360544 | 0.001316476 | 0.002691604 | 1 |

| | | | | | | |
|---|---|---|---|---|---|---|
| T2DM | monocyte | *H3F3A* | 3.545987491 | 5.93E-05 | 0.000399203 | 0.373038534 |
| T2DM | monocyte | *NOSTRIN* | 0.65819945 | 0.001948628 | 0.003544856 | 1 |
| T2DM | monocyte | *SGPP2* | 1.628010234 | 0.001869867 | 0.003444321 | 1 |
| T2DM | monocyte | *BBS5* | 0.968072999 | 0.001492204 | 0.002947372 | 1 |
| T2DM | monocyte | *HPGDS* | 0.557063465 | 0.0016392 | 0.003155435 | 1 |
| T2DM | monocyte | *PDLIM5* | 2.962276853 | 2.65E-05 | 0.000362859 | 0.166372389 |
| T2DM | monocyte | *RPRD2* | 2.077115883 | 0.000149562 | 0.00061774 | 0.940298699 |
| T2DM | monocyte | *CTSS* | -0.182155466 | 0.021466293 | 0.028818831 | 1 |
| T2DM | monocyte | *PACRGL* | 2.602592768 | 4.93E-05 | 0.000383513 | 0.310130889 |
| T2DM | monocyte | *C1QTNF7* | 1.779823199 | 0.000118609 | 0.000544237 | 0.745696009 |
| T2DM | monocyte | *LYSMD1* | 3.597024533 | 0.015417918 | 0.021379014 | 1 |
| T2DM | monocyte | *ERCC3* | 2.082098958 | 0.002081748 | 0.003742623 | 1 |
| T2DM | monocyte | *DHX57* | 3.75314535 | 5.97E-05 | 0.00039944 | 0.375473471 |
| T2DM | monocyte | *S100A9* | -0.27586586 | 0.010051303 | 0.014638068 | 1 |
| T2DM | monocyte | *S100A12* | 6.693622709 | 0.006349816 | 0.009779837 | 1 |
| T2DM | monocyte | *TDRD10* | 1.036971858 | 0.000692697 | 0.001685365 | 1 |
| T2DM | monocyte | *DCAF16* | 1.743517654 | 0.000254701 | 0.000848508 | 1 |
| T2DM | monocyte | *GNPDA2* | 1.104994579 | 0.001331245 | 0.002707712 | 1 |
| T2DM | monocyte | *NIPAL1* | 1.391497697 | 0.000869943 | 0.001988233 | 1 |
| T2DM | monocyte | *ANTXR2* | 1.966367524 | 5.56E-05 | 0.000390932 | 0.349655629 |
| T2DM | monocyte | *CGGBP1* | 0.85786451 | 0.001463791 | 0.002910453 | 1 |
| T2DM | monocyte | *PBXIP1* | 2.30661097 | 0.000130785 | 0.000572307 | 0.822243405 |
| T2DM | monocyte | *DCST2* | 3.570633089 | 0.000173438 | 0.000671081 | 1 |
| T2DM | monocyte | *NBPF10* | -0.796785356 | 0.012270528 | 0.017445683 | 1 |
| T2DM | monocyte | *SLC22A15* | -0.372697628 | 0.027973698 | 0.036540752 | 1 |
| T2DM | monocyte | *SLC15A2* | -0.460363734 | 0.037909223 | 0.048285106 | 1 |
| T2DM | monocyte | *LRRC58* | 2.588678629 | 6.91E-05 | 0.000425443 | 0.434628758 |
| T2DM | monocyte | *FSTL1* | 0.913002655 | 0.000397203 | 0.001147063 | 1 |
| T2DM | monocyte | *LMOD1* | 0.371074224 | 0.017652598 | 0.024136991 | 1 |
| T2DM | monocyte | *TMEM183A* | 4.078490337 | 4.81E-05 | 0.000379967 | 0.302389448 |
| T2DM | monocyte | *IGFBP7* | 0.840271645 | 0.000741087 | 0.001774262 | 1 |
| T2DM | monocyte | *CCT3* | 2.661803725 | 0.000744645 | 0.001779393 | 1 |
| T2DM | monocyte | *GLB1L* | 1.637633263 | 0.003626744 | 0.005975193 | 1 |
| T2DM | monocyte | *SERPINI1* | 1.148384162 | 0.000905854 | 0.002043453 | 1 |
| T2DM | monocyte | *NUAK2* | -0.499289298 | 0.024555148 | 0.032500677 | 1 |
| T2DM | monocyte | *PYHIN1* | 0.506341613 | 0.006239688 | 0.009631456 | 1 |
| T2DM | monocyte | *AIM2* | 1.489688736 | 0.000683334 | 0.001668508 | 1 |
| T2DM | monocyte | *ICA1L* | 1.052098929 | 0.01402987 | 0.01967123 | 1 |
| T2DM | monocyte | *CD200R1* | 0.551414041 | 0.003663526 | 0.006021591 | 1 |
| T2DM | monocyte | *GTPBP8* | 2.331385273 | 0.000100642 | 0.000495511 | 0.632734472 |
| T2DM | monocyte | *SPICE1* | 1.976835569 | 8.91E-05 | 0.000464119 | 0.560191418 |
| T2DM | monocyte | *CADPS* | 1.576167261 | 0.00066521 | 0.00163686 | 1 |
| T2DM | monocyte | *PPM1K* | 1.672647607 | 0.000446746 | 0.001241137 | 1 |
| T2DM | monocyte | *GMPS* | 4.431978697 | 3.26E-05 | 0.000365043 | 0.205116272 |
| T2DM | monocyte | *TIPARP* | 1.174278724 | 0.000182624 | 0.000694589 | 1 |
| T2DM | monocyte | *DCLK3* | 3.872639775 | 0.001231932 | 0.002559536 | 1 |
| T2DM | monocyte | *RPP14* | 4.180408216 | 0.006395904 | 0.009836362 | 1 |
| T2DM | monocyte | *DNASE1L3* | 1.106666624 | 0.000666706 | 0.001639898 | 1 |
| T2DM | monocyte | *CRELD1* | 1.749468419 | 0.000297468 | 0.00094709 | 1 |
| T2DM | monocyte | *PCOLCE2* | 0.626153518 | 0.000859465 | 0.001974948 | 1 |

| | | | | | | |
|---|---|---|---|---|---|---|
| T2DM | monocyte | *TTC14* | 1.645882531 | 0.000172838 | 0.000670346 | 1 |
| T2DM | monocyte | *RCHY1* | 2.362211269 | 0.019740131 | 0.026718235 | 1 |
| T2DM | monocyte | *CCDC158* | 0.902629703 | 0.000931838 | 0.002088305 | 1 |
| T2DM | monocyte | *GYG1* | 5.323547451 | 4.54E-05 | 0.000379941 | 0.285665496 |
| T2DM | monocyte | *HPS3* | 2.940004158 | 0.007495667 | 0.011298312 | 1 |
| T2DM | monocyte | *RYK* | 2.754158566 | 2.38E-05 | 0.000362859 | 0.149570273 |
| T2DM | monocyte | *PLB1* | -0.4227387 | 0.015466699 | 0.021431204 | 1 |
| T2DM | monocyte | *CDCP1* | 6.919924404 | 4.03E-05 | 0.00037107 | 0.253299843 |
| T2DM | monocyte | *LZTFL1* | 1.647762808 | 0.000368316 | 0.001087648 | 1 |
| T2DM | monocyte | *FYCO1* | 3.210308891 | 6.01E-05 | 0.000399998 | 0.377917104 |
| T2DM | monocyte | *LRRC2* | 0.851032705 | 0.000969969 | 0.002152556 | 1 |
| T2DM | monocyte | *DTX3L* | 4.689122592 | 1.01E-05 | 0.000362859 | 0.063512762 |
| T2DM | monocyte | *NMNAT3* | 2.660274403 | 8.33E-06 | 0.000361025 | 0.052384059 |
| T2DM | monocyte | *SMIM12* | 1.436871564 | 0.000325955 | 0.001004055 | 1 |
| T2DM | monocyte | *TPRA1* | 2.072769718 | 0.000213341 | 0.00076036 | 1 |
| T2DM | monocyte | *YEATS2* | 3.842683216 | 0.000335378 | 0.00102039 | 1 |
| T2DM | monocyte | *MEAF6* | 2.756095088 | 5.26E-05 | 0.000385235 | 0.330978721 |
| T2DM | monocyte | *DNALI1* | 1.375688413 | 0.000682373 | 0.001667993 | 1 |
| T2DM | monocyte | *RPN1* | 2.447086196 | 8.84E-05 | 0.000462503 | 0.555928313 |
| T2DM | monocyte | *RFC4* | 3.51081483 | 8.28E-05 | 0.000448982 | 0.520377776 |
| T2DM | monocyte | *SFMBT1* | 2.150402161 | 0.000125762 | 0.000558159 | 0.790665548 |
| T2DM | monocyte | *GNL3* | 1.805700526 | 0.000258974 | 0.000858355 | 1 |
| T2DM | monocyte | *UVSSA* | 2.761651155 | 5.12E-05 | 0.000384204 | 0.321963182 |
| T2DM | monocyte | *FAM208A* | 2.322992428 | 0.000606685 | 0.001531204 | 1 |
| T2DM | monocyte | *LRPAP1* | 3.842543848 | 9.81E-05 | 0.000486996 | 0.616601131 |
| T2DM | monocyte | *SLC51A* | 1.254010906 | 0.00079372 | 0.001861364 | 1 |
| T2DM | monocyte | *PIGX* | -0.536130308 | 0.028581865 | 0.037288688 | 1 |
| T2DM | monocyte | *ABLIM2* | 0.728088665 | 0.001466264 | 0.002913527 | 1 |
| T2DM | monocyte | *C1orf50* | 1.481222492 | 0.002905054 | 0.004929575 | 1 |
| T2DM | monocyte | *ERMAP* | 1.539419933 | 0.012948538 | 0.018306152 | 1 |
| T2DM | monocyte | *METAP1* | 1.524519333 | 0.000560482 | 0.001459111 | 1 |
| T2DM | monocyte | *DNAJB14* | 3.643252494 | 8.06E-06 | 0.000355191 | 0.050653525 |
| T2DM | monocyte | *SLC9B2* | 1.11462926 | 0.001683156 | 0.003212508 | 1 |
| T2DM | monocyte | *ATRIP* | -2.171141295 | 0.01877975 | 0.0254897 | 1 |
| T2DM | monocyte | *SHISA5* | 4.115148591 | 0.001987884 | 0.003604795 | 1 |
| T2DM | monocyte | *RNF123* | 2.777411546 | 3.60E-10 | 4.53E-07 | 2.26E-06 |
| T2DM | monocyte | *RAD54L2* | 2.506100466 | 0.000289298 | 0.000927969 | 1 |
| T2DM | monocyte | *C4orf3* | 3.336027996 | 5.46E-05 | 0.000387662 | 0.343080809 |
| T2DM | monocyte | *ANXA5* | 2.726847431 | 2.82E-05 | 0.000362859 | 0.177567139 |
| T2DM | monocyte | *MAP9* | 0.794542686 | 0.001600692 | 0.003106528 | 1 |
| T2DM | monocyte | *CEP44* | 1.744423302 | 0.000160892 | 0.000643059 | 1 |
| T2DM | monocyte | *C4orf45* | 1.541117113 | 0.00114207 | 0.002424735 | 1 |
| T2DM | monocyte | *TMEM144* | 1.184375737 | 0.000390735 | 0.00113362 | 1 |
| T2DM | monocyte | *NPY1R* | 1.075116723 | 0.000372314 | 0.001095237 | 1 |
| T2DM | monocyte | *IL15* | -0.42204191 | 0.030272863 | 0.039242369 | 1 |
| T2DM | monocyte | *ARFIP1* | 2.139328708 | 6.78E-05 | 0.000422229 | 0.42622653 |
| T2DM | monocyte | *ITGA2* | 1.490400969 | 0.000619792 | 0.001553363 | 1 |
| T2DM | monocyte | *NDUFAF2* | 1.975501179 | 0.001581675 | 0.00308225 | 1 |
| T2DM | monocyte | *LMBRD2* | 2.152903755 | 0.000134149 | 0.00057865 | 0.843394619 |
| T2DM | monocyte | *CMBL* | 1.294639849 | 0.000743199 | 0.001777289 | 1 |

| | | | | | | |
|---|---|---|---|---|---|---|
| T2DM | monocyte | *C5orf63* | 1.024888601 | 0.00134777 | 0.002736638 | 1 |
| T2DM | monocyte | *PRRC1* | 1.490277592 | 0.000139364 | 0.00059075 | 0.876179172 |
| T2DM | monocyte | *WDR41* | 1.595528495 | 0.003708425 | 0.006079383 | 1 |
| T2DM | monocyte | *NDUFS4* | 2.662832003 | 3.12E-05 | 0.000362859 | 0.195842867 |
| T2DM | monocyte | *RHOBTB3* | 0.971138826 | 7.91E-05 | 0.000441994 | 0.497242964 |
| T2DM | monocyte | *TIGD6* | 5.556611619 | 0.02855624 | 0.037262989 | 1 |
| T2DM | monocyte | *SERINC5* | 2.660246044 | 2.45E-05 | 0.000362859 | 0.153853662 |
| T2DM | monocyte | *ENPP6* | 2.413499674 | 0.003084801 | 0.005193932 | 1 |
| T2DM | monocyte | *CASP3* | 4.432460651 | 5.78E-05 | 0.000396862 | 0.36348795 |
| T2DM | monocyte | *PRIMPOL* | 1.648047809 | 0.002704191 | 0.004646419 | 1 |
| T2DM | monocyte | *ERAP1* | 1.534064337 | 0.000396672 | 0.001146558 | 1 |
| T2DM | monocyte | *RICTOR* | 2.921868571 | 2.50E-05 | 0.000362859 | 0.156959315 |
| T2DM | monocyte | *ANKRA2* | 2.729600852 | 5.70E-05 | 0.000395506 | 0.358541917 |
| T2DM | monocyte | *UBLCP1* | 2.997468784 | 0.000257212 | 0.000854171 | 1 |
| T2DM | monocyte | *TLR3* | 3.306358684 | 7.06E-05 | 0.000427286 | 0.443968392 |
| T2DM | monocyte | *KLKB1* | 1.018786617 | 8.89E-05 | 0.000463596 | 0.559096437 |
| T2DM | monocyte | *NSA2* | 0.893556823 | 0.016517643 | 0.022743413 | 1 |
| T2DM | monocyte | *CCDC127* | 1.380458569 | 0.000912428 | 0.002054597 | 1 |
| T2DM | monocyte | *GDF9* | 2.424963992 | 0.009305575 | 0.013694792 | 1 |
| T2DM | monocyte | *GRIK2* | 0.741133307 | 0.005405208 | 0.008481024 | 1 |
| T2DM | monocyte | *TXLNB* | 1.808866438 | 0.001317397 | 0.002691736 | 1 |
| T2DM | monocyte | *DCBLD1* | 3.789565718 | 7.56E-06 | 0.000349439 | 0.047523766 |
| T2DM | monocyte | *TMEM200A* | 1.274995751 | 0.000808417 | 0.001887709 | 1 |
| T2DM | monocyte | *PDSS2* | 3.293301163 | 0.000162106 | 0.000645445 | 1 |
| T2DM | monocyte | *STXBP5* | 3.269746686 | 4.30E-05 | 0.000378034 | 0.270240035 |
| T2DM | monocyte | *PI16* | 0.573060016 | 0.001108974 | 0.002373084 | 1 |
| T2DM | monocyte | *STK17A* | 1.710553157 | 0.000343126 | 0.001035674 | 1 |
| T2DM | monocyte | *GALNT10* | 3.129494662 | 0.000361243 | 0.001073112 | 1 |
| T2DM | monocyte | *SAP30L* | 3.022120135 | 0.000179109 | 0.000684948 | 1 |
| T2DM | monocyte | *RPS14* | 2.720758119 | 4.53E-05 | 0.000379941 | 0.285042046 |
| T2DM | monocyte | *HCN1* | 1.095708098 | 0.001081864 | 0.002331366 | 1 |
| T2DM | monocyte | *SLU7* | 2.4906815 | 0.000190614 | 0.000713511 | 1 |
| T2DM | monocyte | *RP9* | 1.07081238 | 0.001874037 | 0.003446704 | 1 |
| T2DM | monocyte | *CAMLG* | 4.058895089 | 3.62E-07 | 6.70E-05 | 0.002274359 |
| T2DM | monocyte | *KCNK5* | 1.276122737 | 0.001743879 | 0.003290446 | 1 |
| T2DM | monocyte | *KIAA1324L* | 0.845874615 | 0.005963611 | 0.009250734 | 1 |
| T2DM | monocyte | *USP49* | 3.136176077 | 2.70E-05 | 0.000362859 | 0.169571788 |
| T2DM | monocyte | *ZNF704* | 1.249822434 | 0.000125038 | 0.000556889 | 0.786112228 |
| T2DM | monocyte | *FNDC1* | 0.655981954 | 0.001265021 | 0.002609313 | 1 |
| T2DM | monocyte | *BRI3* | 3.455354931 | 4.74E-05 | 0.000379941 | 0.297817935 |
| T2DM | monocyte | *LMTK2* | 3.734899492 | 1.23E-05 | 0.000362859 | 0.077435436 |
| T2DM | monocyte | *PEX2* | 3.580378289 | 7.39E-05 | 0.000432034 | 0.464690135 |
| T2DM | monocyte | *TNFRSF11B* | 1.504488433 | 0.000651167 | 0.001613036 | 1 |
| T2DM | monocyte | *SUN1* | 1.473240982 | 2.32E-05 | 0.000362859 | 0.145957123 |
| T2DM | monocyte | *OXR1* | 2.956440899 | 0.000281735 | 0.000911031 | 1 |
| T2DM | monocyte | *GPER1* | 0.806946828 | 0.002741563 | 0.004688849 | 1 |
| T2DM | monocyte | *SLC4A2* | 3.038297243 | 8.63E-05 | 0.000459567 | 0.542748494 |
| T2DM | monocyte | *FASTK* | 2.141378702 | 0.000258995 | 0.000858355 | 1 |
| T2DM | monocyte | *ALDH7A1* | 1.808945751 | 0.000371471 | 0.00109424 | 1 |
| T2DM | monocyte | *COX6C* | 3.02071239 | 3.72E-05 | 0.000366571 | 0.233922226 |

| | | | | | | |
|---|---|---|---|---|---|---|
| T2DM | monocyte | *YWHAZ* | 1.371455222 | 0.001825888 | 0.003390895 | 1 |
| T2DM | monocyte | *SLC25A32* | 1.902605397 | 0.000440216 | 0.001228968 | 1 |
| T2DM | monocyte | *DCAF13* | 1.253546939 | 0.000453947 | 0.001253938 | 1 |
| T2DM | monocyte | *INTS8* | 3.790971048 | 2.60E-06 | 0.000207301 | 0.01633558 |
| T2DM | monocyte | *FREM1* | 0.93859699 | 0.000694839 | 0.001689923 | 1 |
| T2DM | monocyte | *RPP25L* | 2.187709016 | 0.001641311 | 0.003156599 | 1 |
| T2DM | monocyte | *FAM219A* | 1.672133087 | 0.000151213 | 0.0006205 | 0.950673935 |
| T2DM | monocyte | *NUDT2* | 0.686346481 | 0.018755678 | 0.025468024 | 1 |
| T2DM | monocyte | *TMEM65* | 2.859490116 | 0.005074181 | 0.008023486 | 1 |
| T2DM | monocyte | *CCDC171* | 1.353515891 | 0.000502348 | 0.001349618 | 1 |
| T2DM | monocyte | *SYK* | -0.304208779 | 0.01803725 | 0.024593405 | 1 |
| T2DM | monocyte | *NIPSNAP3B* | 0.516468406 | 0.005304354 | 0.008335034 | 1 |
| T2DM | monocyte | *METTL2B* | 0.799154875 | 0.000600208 | 0.001523326 | 1 |
| T2DM | monocyte | *TMC1* | 0.88641215 | 0.001009265 | 0.002219887 | 1 |
| T2DM | monocyte | *HGSNAT* | 2.282324202 | 0.00010268 | 0.000500426 | 0.645549603 |
| T2DM | monocyte | *C9orf64* | 2.893179588 | 0.001455703 | 0.002900975 | 1 |
| T2DM | monocyte | *HNRNPK* | 2.277189087 | 5.59E-05 | 0.000391656 | 0.351706859 |
| T2DM | monocyte | *SVEP1* | 0.498522186 | 0.000960554 | 0.002136547 | 1 |
| T2DM | monocyte | *ZNF367* | 2.682576274 | 0.000134294 | 0.00057865 | 0.844308917 |
| T2DM | monocyte | *NOL6* | 1.052544562 | 0.001331134 | 0.002707712 | 1 |
| T2DM | monocyte | *TRMT10B* | 2.754466432 | 3.65E-05 | 0.000365396 | 0.229468497 |
| T2DM | monocyte | *VCP* | 1.49841957 | 0.000228253 | 0.000788692 | 1 |
| T2DM | monocyte | *PIGO* | 5.081815928 | 0.000561657 | 0.001461564 | 1 |
| T2DM | monocyte | *LRFN5* | 0.550050808 | 0.000870623 | 0.001988233 | 1 |
| T2DM | monocyte | *WRN* | 1.898061403 | 6.12E-05 | 0.000403714 | 0.384597398 |
| T2DM | monocyte | *MARCH8* | 1.837518373 | 0.000108718 | 0.000518843 | 0.683510005 |
| T2DM | monocyte | *CFL2* | 1.646173733 | 0.000271175 | 0.000888419 | 1 |
| T2DM | monocyte | *ZCCHC24* | 1.987985865 | 7.98E-05 | 0.000442726 | 0.501625687 |
| T2DM | monocyte | *PGM2L1* | 2.523489095 | 2.98E-05 | 0.000362859 | 0.187106435 |
| T2DM | monocyte | *CRYL1* | 1.898059216 | 0.001812749 | 0.00337982 | 1 |
| T2DM | monocyte | *REEP3* | 1.636245409 | 0.005846771 | 0.009087429 | 1 |
| T2DM | monocyte | *HEPACAM* | 0.50974746 | 0.001721011 | 0.003261983 | 1 |
| T2DM | monocyte | *RPL36AL* | 2.154769492 | 0.011513246 | 0.016518435 | 1 |
| T2DM | monocyte | *C10orf25* | 1.289803422 | 0.001225022 | 0.002551081 | 1 |
| T2DM | monocyte | *KLHDC2* | 1.971804094 | 5.30E-05 | 0.000385235 | 0.33289859 |
| T2DM | monocyte | *NOXRED1* | 3.475759636 | 0.000333989 | 0.00102039 | 1 |
| T2DM | monocyte | *AKR1E2* | 0.944456479 | 0.010155698 | 0.014779832 | 1 |
| T2DM | monocyte | *ATP5C1* | 2.491284037 | 0.000323419 | 0.000999459 | 1 |
| T2DM | monocyte | *TAF3* | 2.384471861 | 0.007986093 | 0.011943046 | 1 |
| T2DM | monocyte | *VDAC2* | 1.685556246 | 0.000163876 | 0.000650094 | 1 |
| T2DM | monocyte | *SLC18A2* | 0.736934083 | 0.001062031 | 0.002302409 | 1 |
| T2DM | monocyte | *PDZD8* | 3.231895944 | 4.95E-05 | 0.000383513 | 0.311457933 |
| T2DM | monocyte | *QSOX2* | 4.084123365 | 7.26E-05 | 0.000431404 | 0.456425458 |
| T2DM | monocyte | *CLEC1B* | -0.46505194 | 0.000723969 | 0.001741904 | 1 |
| T2DM | monocyte | *SNAPC4* | 1.516557168 | 0.009927408 | 0.014481859 | 1 |
| T2DM | monocyte | *SDCCAG3* | 3.166269364 | 3.42E-05 | 0.000365043 | 0.21517865 |
| T2DM | monocyte | *AK8* | 1.539045765 | 0.001150098 | 0.00243487 | 1 |
| T2DM | monocyte | *TSC1* | 2.054711089 | 6.46E-05 | 0.000415426 | 0.406342239 |
| T2DM | monocyte | *STOX1* | 0.334324244 | 0.037317044 | 0.047617669 | 1 |
| T2DM | monocyte | *RET* | 6.860832605 | 0.001167624 | 0.002461722 | 1 |

| | | | | | | |
|---|---|---|---|---|---|---|
| T2DM | monocyte | *NSMF* | 3.510964059 | 8.78E-05 | 0.00046125 | 0.552086691 |
| T2DM | monocyte | *CASP7* | 1.979950401 | 0.001229216 | 0.002555581 | 1 |
| T2DM | monocyte | *VWA2* | 1.500930938 | 0.003953007 | 0.006438486 | 1 |
| T2DM | monocyte | *METTL3* | 3.694371482 | 2.21E-05 | 0.000362859 | 0.139150709 |
| T2DM | monocyte | *TRUB1* | 1.235796159 | 0.005277019 | 0.008298304 | 1 |
| T2DM | monocyte | *HSPA12A* | 0.769921771 | 0.000760058 | 0.001806324 | 1 |
| T2DM | monocyte | *UBTD1* | 2.602030243 | 7.33E-05 | 0.000431915 | 0.460926036 |
| T2DM | monocyte | *ARHGAP42* | 0.830043927 | 0.000567306 | 0.00146776 | 1 |
| T2DM | monocyte | *OTOGL* | 1.606527413 | 0.007994543 | 0.01195284 | 1 |
| T2DM | monocyte | *TTC7B* | 1.008489064 | 0.00088458 | 0.00201134 | 1 |
| T2DM | monocyte | *SLC39A13* | 2.069519658 | 0.000228131 | 0.000788692 | 1 |
| T2DM | monocyte | *PSMC3* | 2.856495402 | 0.000817938 | 0.001903879 | 1 |
| T2DM | monocyte | *TC2N* | 0.685628029 | 0.016223692 | 0.022363155 | 1 |
| T2DM | monocyte | *IFI27L1* | 0.824404855 | 0.001104828 | 0.002367413 | 1 |
| T2DM | monocyte | *CLMN* | 1.242480863 | 0.000345742 | 0.001038052 | 1 |
| T2DM | monocyte | *PTER* | 3.624929778 | 0.000779863 | 0.001837789 | 1 |
| T2DM | monocyte | *C1QL3* | 1.230457344 | 0.000640133 | 0.001593662 | 1 |
| T2DM | monocyte | *ABTB2* | 1.332232321 | 0.00045055 | 0.001248286 | 1 |
| T2DM | monocyte | *LIPC* | 1.208169939 | 0.000744591 | 0.001779393 | 1 |
| T2DM | monocyte | *CEP57* | 1.765634035 | 0.006912036 | 0.010522027 | 1 |
| T2DM | monocyte | *TCP11L2* | 1.766832405 | 0.000141662 | 0.000597338 | 0.89063024 |
| T2DM | monocyte | *JAM3* | 1.204221803 | 0.000276487 | 0.000900194 | 1 |
| T2DM | monocyte | *GPT2* | 0.590897358 | 0.00213929 | 0.003825289 | 1 |
| T2DM | monocyte | *HIF1AN* | 3.563742117 | 4.78E-05 | 0.000379941 | 0.300315281 |
| T2DM | monocyte | *NDUFB8* | 2.963831863 | 0.000191413 | 0.000714856 | 1 |
| T2DM | monocyte | *ZFYVE19* | 1.162028737 | 0.004555543 | 0.00728215 | 1 |
| T2DM | monocyte | *AVPR1A* | 0.881529099 | 0.003540609 | 0.005856028 | 1 |
| T2DM | monocyte | *BTRC* | 1.833848436 | 7.45E-05 | 0.000433444 | 0.468528666 |
| T2DM | monocyte | *LARP6* | 2.264626637 | 0.000146812 | 0.000610432 | 0.923007331 |
| T2DM | monocyte | *ASPG* | 3.028437533 | 0.00108968 | 0.002343763 | 1 |
| T2DM | monocyte | *GABRB3* | 2.6596384 | 0.002740914 | 0.004688849 | 1 |
| T2DM | monocyte | *PCBD1* | -0.895656247 | 0.032924951 | 0.042374446 | 1 |
| T2DM | monocyte | *COX11* | 1.290198591 | 0.000687958 | 0.001677082 | 1 |
| T2DM | monocyte | *ZNF202* | 2.725461343 | 4.26E-05 | 0.000376656 | 0.26780243 |
| T2DM | monocyte | *FAM227B* | 1.515423293 | 6.36E-05 | 0.000411242 | 0.400138871 |
| T2DM | monocyte | *STXBP4* | 1.646421167 | 0.000695742 | 0.001690811 | 1 |
| T2DM | monocyte | *C2* | 2.441999739 | 1.93E-07 | 4.50E-05 | 0.001215847 |
| T2DM | monocyte | *APBB1* | 1.436151416 | 0.000613196 | 0.001543731 | 1 |
| T2DM | monocyte | *NUDT13* | 0.522359219 | 0.026559433 | 0.034870162 | 1 |
| T2DM | monocyte | *ILK* | 2.081274332 | 4.62E-05 | 0.000379941 | 0.290552511 |
| T2DM | monocyte | *DCHS1* | 0.83482061 | 0.001489376 | 0.002943636 | 1 |
| T2DM | monocyte | *NETO1* | 3.139337184 | 0.000217798 | 0.000768284 | 1 |
| T2DM | monocyte | *USP54* | 1.815237775 | 7.85E-05 | 0.000440255 | 0.493730944 |
| T2DM | monocyte | *C11orf74* | 2.894180429 | 0.000365086 | 0.00108014 | 1 |
| T2DM | monocyte | *WDR88* | 1.853565503 | 0.000595905 | 0.001515557 | 1 |
| T2DM | monocyte | *CYB5R2* | 1.397739428 | 0.005280406 | 0.008301553 | 1 |
| T2DM | monocyte | *TUB* | 0.679514047 | 0.005258422 | 0.008273198 | 1 |
| T2DM | monocyte | *RIC3* | 0.748475149 | 0.001936925 | 0.00352765 | 1 |
| T2DM | monocyte | *LMO1* | 1.851591714 | 0.001608706 | 0.003112938 | 1 |
| T2DM | monocyte | *TRIM66* | 0.576938236 | 0.002174552 | 0.003869632 | 1 |

| | | | | | | |
|---|---|---|---|---|---|---|
| T2DM | monocyte | *RPL27A* | 3.984205216 | 0.000226022 | 0.000784218 | 1 |
| T2DM | monocyte | *PRTG* | 1.322574966 | 0.000617569 | 0.001549963 | 1 |
| T2DM | monocyte | *AKIP1* | 2.395076636 | 0.00020318 | 0.000735552 | 1 |
| T2DM | monocyte | *C16orf46* | -1.081687491 | 0.006904276 | 0.010512759 | 1 |
| T2DM | monocyte | *TMEM41B* | 3.091425781 | 3.01E-05 | 0.000362859 | 0.189059121 |
| T2DM | monocyte | *ZNF143* | 3.122080663 | 0.008936928 | 0.01320481 | 1 |
| T2DM | monocyte | *TMX3* | 3.205958253 | 4.81E-05 | 0.000379967 | 0.302515879 |
| T2DM | monocyte | *CCDC68* | 0.840600493 | 0.001977792 | 0.003588566 | 1 |
| T2DM | monocyte | *CLEC4E* | -0.115159159 | 0.010289199 | 0.014949894 | 1 |
| T2DM | monocyte | *ZNF3* | 1.825175057 | 0.000225619 | 0.000783249 | 1 |
| T2DM | monocyte | *CLEC4D* | -0.210594256 | 0.00790914 | 0.01184205 | 1 |
| T2DM | monocyte | *ZSCAN21* | 1.246263787 | 0.01039451 | 0.015075037 | 1 |
| T2DM | monocyte | *RIMKLB* | 0.756362351 | 0.012822273 | 0.018143964 | 1 |
| T2DM | monocyte | *BEAN1* | 3.988034398 | 2.23E-05 | 0.000362859 | 0.140266673 |
| T2DM | monocyte | *TK2* | 0.931010563 | 0.013130631 | 0.01854274 | 1 |
| T2DM | monocyte | *SEC11C* | 2.696514528 | 0.000146507 | 0.000609994 | 0.921088394 |
| T2DM | monocyte | *GALR1* | 0.970993822 | 0.024229587 | 0.03213065 | 1 |
| T2DM | monocyte | *CENPV* | 2.499607703 | 0.000397403 | 0.001147063 | 1 |
| T2DM | monocyte | *SPDYE6* | 1.931658144 | 0.001028261 | 0.002249366 | 1 |
| T2DM | monocyte | *ATF7IP2* | 1.976516607 | 0.000382218 | 0.001114602 | 1 |
| T2DM | monocyte | *ZNF592* | 1.961206146 | 0.000479838 | 0.001309918 | 1 |
| T2DM | monocyte | *ACSM1* | 1.650426508 | 0.001516167 | 0.002978363 | 1 |
| T2DM | monocyte | *AP1G1* | 3.316639411 | 6.22E-05 | 0.000406647 | 0.390787475 |
| T2DM | monocyte | *SLFN5* | 1.48524726 | 0.000312521 | 0.000978124 | 1 |
| T2DM | monocyte | *CATSPER2* | 3.345855149 | 7.09E-05 | 0.000427286 | 0.446000606 |
| T2DM | monocyte | *C16orf45* | 1.537429216 | 0.000164336 | 0.000650619 | 1 |
| T2DM | monocyte | *SAAL1* | 1.003382623 | 0.004167017 | 0.006743381 | 1 |
| T2DM | monocyte | *YPEL4* | 1.98581613 | 0.024808091 | 0.032780259 | 1 |
| T2DM | monocyte | *LDHC* | 0.937149628 | 0.002219969 | 0.003929319 | 1 |
| T2DM | monocyte | *FAM111A* | 2.870179004 | 0.000827097 | 0.001919512 | 1 |
| T2DM | monocyte | *ANKDD1A* | 1.815063645 | 0.000586731 | 0.001496766 | 1 |
| T2DM | monocyte | *DCTN5* | 1.394012724 | 0.012686297 | 0.01797988 | 1 |
| T2DM | monocyte | *STX3* | 2.01650712 | 0.000340575 | 0.001030047 | 1 |
| T2DM | monocyte | *MTMR10* | 2.871425018 | 1.56E-05 | 0.000362859 | 0.097958859 |
| T2DM | monocyte | *YWHAB* | 2.713190809 | 0.008387548 | 0.012481069 | 1 |
| T2DM | monocyte | *TSC22D4* | 5.36575069 | 1.60E-05 | 0.000362859 | 0.100832194 |
| T2DM | monocyte | *MS4A7* | -0.315025888 | 0.009998607 | 0.014568074 | 1 |
| T2DM | monocyte | *MS4A14* | -0.269091953 | 0.005557306 | 0.00869773 | 1 |
| T2DM | monocyte | *DIS3L* | 2.229600651 | 0.000121999 | 0.000552602 | 0.767005036 |
| T2DM | monocyte | *CCNDBP1* | 3.814599994 | 2.52E-05 | 0.000362859 | 0.158375597 |
| T2DM | monocyte | *SMAD3* | 1.532403487 | 0.000106631 | 0.000511748 | 0.670390437 |
| T2DM | monocyte | *RCCD1* | 1.830006346 | 0.000580777 | 0.001489736 | 1 |
| T2DM | monocyte | *AKTIP* | 6.764032171 | 1.75E-06 | 0.000169734 | 0.010972729 |
| T2DM | monocyte | *EVA1C* | 1.743438371 | 1.50E-05 | 0.000362859 | 0.094050399 |
| T2DM | monocyte | *CNPY4* | 2.035172485 | 0.002289558 | 0.004025295 | 1 |
| T2DM | monocyte | *NUDT21* | 3.713536876 | 4.58E-05 | 0.000379941 | 0.287784062 |
| T2DM | monocyte | *NKX3-1* | -3.621844475 | 0.004944431 | 0.007851891 | 1 |
| T2DM | monocyte | *DUSP18* | 3.46312011 | 0.002608359 | 0.00450639 | 1 |
| T2DM | monocyte | *TEF* | 1.760623284 | 7.55E-06 | 0.000349439 | 0.047479798 |
| T2DM | monocyte | *PBX3* | 2.012139034 | 0.000144545 | 0.000606241 | 0.908755717 |

| | | | | | | |
|---|---|---|---|---|---|---|
| T2DM | monocyte | *SNRPD1* | 3.342725038 | 4.54E-05 | 0.000379941 | 0.28543387 |
| T2DM | monocyte | *SAMD14* | 2.926562868 | 0.005725484 | 0.008923182 | 1 |
| T2DM | monocyte | *CERCAM* | 1.164537847 | 0.002174522 | 0.003869632 | 1 |
| T2DM | monocyte | *ENDOG* | -1.143936306 | 0.012984182 | 0.018352417 | 1 |
| T2DM | monocyte | *ISLR2* | 1.133346382 | 0.003808351 | 0.006227074 | 1 |
| T2DM | monocyte | *COQ7* | 1.732200863 | 0.000306221 | 0.000965629 | 1 |
| T2DM | monocyte | *GPRC5B* | 1.342558393 | 0.001005768 | 0.002214803 | 1 |
| T2DM | monocyte | *KATNAL2* | 1.118629959 | 0.000643549 | 0.00159984 | 1 |
| T2DM | monocyte | *HDHD2* | 1.443630594 | 0.000935313 | 0.002094129 | 1 |
| T2DM | monocyte | *RNF214* | 3.090142702 | 3.25E-05 | 0.000365043 | 0.204216701 |
| T2DM | monocyte | *RBFOX3* | 1.744018877 | 0.000728481 | 0.00175075 | 1 |
| T2DM | monocyte | *MYO5B* | 0.652261431 | 0.001038076 | 0.002263746 | 1 |
| T2DM | monocyte | *TRIM68* | 1.42599255 | 0.025397665 | 0.033474868 | 1 |
| T2DM | monocyte | *FN3K* | 1.545845563 | 0.000399225 | 0.001150816 | 1 |
| T2DM | monocyte | *ZNF23* | 1.174326588 | 0.002723224 | 0.004668914 | 1 |
| T2DM | monocyte | *ZNF229* | 3.903228618 | 0.000300646 | 0.000954626 | 1 |
| T2DM | monocyte | *GATAD2A* | 1.872941318 | 0.029443872 | 0.038309939 | 1 |
| T2DM | monocyte | *RPL13* | 3.191519343 | 0.0008953 | 0.00202711 | 1 |
| T2DM | monocyte | *ZNF641* | 2.443051394 | 4.65E-05 | 0.000379941 | 0.292305668 |
| T2DM | monocyte | *TP53I13* | 1.67336877 | 0.000101508 | 0.000497371 | 0.638177692 |
| T2DM | monocyte | *ZNF528* | 1.136493426 | 0.000647402 | 0.001606948 | 1 |
| T2DM | monocyte | *ZNF701* | 2.788482458 | 0.000928888 | 0.002083453 | 1 |
| T2DM | monocyte | *SERTAD3* | 3.60325907 | 0.000145355 | 0.000607712 | 0.913847106 |
| T2DM | monocyte | *TTYH1* | 3.723128323 | 4.31E-05 | 0.000378034 | 0.270975474 |
| T2DM | monocyte | *LENG8* | 2.119969897 | 8.56E-05 | 0.0004583 | 0.538044606 |
| T2DM | monocyte | *CDC42EP5* | 1.139387572 | 0.002295666 | 0.004034905 | 1 |
| T2DM | monocyte | *ZNF283* | 3.035117597 | 0.003008867 | 0.005081049 | 1 |
| T2DM | monocyte | *PPP1R14A* | 0.797113008 | 0.001691691 | 0.003223856 | 1 |
| T2DM | monocyte | *PSCA* | 1.777773996 | 5.80E-05 | 0.000397161 | 0.364511867 |
| T2DM | monocyte | *UBXN6* | 1.963718378 | 0.004236378 | 0.006839781 | 1 |
| T2DM | monocyte | *ZNF444* | 5.273320367 | 2.70E-05 | 0.000362859 | 0.169585234 |
| T2DM | monocyte | *FAM57A* | 0.795963677 | 0.01891104 | 0.025645753 | 1 |
| T2DM | monocyte | *GLOD4* | 3.444585904 | 4.74E-05 | 0.000379941 | 0.298273086 |
| T2DM | monocyte | *SRR* | 0.892256595 | 0.009757679 | 0.014273164 | 1 |
| T2DM | monocyte | *CYB5D2* | 2.68622799 | 1.62E-05 | 0.000362859 | 0.10192259 |
| T2DM | monocyte | *ZNF83* | 1.832927616 | 0.000142154 | 0.000598208 | 0.893722217 |
| T2DM | monocyte | *CD320* | 1.266162962 | 0.000649961 | 0.001611319 | 1 |
| T2DM | monocyte | *SPRYD3* | 1.36684036 | 0.000697589 | 0.001693432 | 1 |
| T2DM | monocyte | *SOAT2* | 1.559363511 | 0.006302783 | 0.009721687 | 1 |
| T2DM | monocyte | *ZNF558* | 1.799779123 | 0.001797156 | 0.003359735 | 1 |
| T2DM | monocyte | *NDUFV1* | 3.622960125 | 1.72E-05 | 0.000362859 | 0.108062029 |
| T2DM | monocyte | *NUDT8* | 0.925309719 | 0.006044747 | 0.009360424 | 1 |
| T2DM | monocyte | *PRDX2* | 1.427349378 | 0.000126825 | 0.000560331 | 0.797350365 |
| T2DM | monocyte | *ZNF232* | 0.795043208 | 0.0254401 | 0.033523771 | 1 |
| T2DM | monocyte | *MIS12* | 3.113614873 | 0.021401696 | 0.028750526 | 1 |
| T2DM | monocyte | *CD300C* | -0.276689749 | 0.006573909 | 0.010070703 | 1 |
| T2DM | monocyte | *HID1* | 1.093343591 | 0.002134426 | 0.003817678 | 1 |
| T2DM | monocyte | *ATP5H* | 1.447178292 | 0.000534395 | 0.001410156 | 1 |
| T2DM | monocyte | *MGAT5B* | 1.938850392 | 0.001546052 | 0.003021457 | 1 |
| T2DM | monocyte | *TMEM68* | 2.886280124 | 0.000398967 | 0.001150599 | 1 |

| | | | | | | |
|---|---|---|---|---|---|---|
| T2DM | monocyte | *CYP7A1* | 3.329678694 | 0.004012368 | 0.006521654 | 1 |
| T2DM | monocyte | *GSDMA* | 1.754025543 | 0.001917099 | 0.003500668 | 1 |
| T2DM | monocyte | *TMEM99* | 1.117899194 | 0.01467272 | 0.020476668 | 1 |
| T2DM | monocyte | *ZNF598* | 1.691483771 | 0.001420887 | 0.00284948 | 1 |
| T2DM | monocyte | *ECI1* | 1.182555099 | 0.002093879 | 0.00376134 | 1 |
| T2DM | monocyte | *ABCA3* | 2.017977131 | 0.000198646 | 0.000726217 | 1 |
| T2DM | monocyte | *KCTD5* | 4.164692415 | 2.10E-05 | 0.000362859 | 0.13219403 |
| T2DM | monocyte | *SDHAF2* | 2.218107373 | 0.000413273 | 0.001178344 | 1 |
| T2DM | monocyte | *DDB1* | 1.85976877 | 0.000134172 | 0.00057865 | 0.843539091 |
| T2DM | monocyte | *VWCE* | 2.147940702 | 0.001379376 | 0.002785781 | 1 |
| T2DM | monocyte | *BSCL2* | -0.937928531 | 0.003729411 | 0.006107531 | 1 |
| T2DM | monocyte | *HRASLS5* | 0.596463904 | 0.000882379 | 0.00200997 | 1 |
| T2DM | monocyte | *TRANK1* | 3.280023357 | 0.000242947 | 0.000821174 | 1 |
| T2DM | monocyte | *TTC21A* | 4.772855584 | 0.00185524 | 0.003422505 | 1 |
| T2DM | monocyte | *RPSA* | 2.406688703 | 4.78E-05 | 0.000379941 | 0.300385517 |
| T2DM | monocyte | *LTBP3* | 1.657137091 | 2.04E-06 | 0.000188095 | 0.012849736 |
| T2DM | monocyte | *SCARA5* | 0.510811701 | 0.002423505 | 0.004227685 | 1 |
| T2DM | monocyte | *COPS6* | 2.309849677 | 0.00027644 | 0.000900194 | 1 |
| T2DM | monocyte | *KIAA1586* | 2.425121215 | 0.004595957 | 0.007337426 | 1 |
| T2DM | monocyte | *ZCCHC4* | 3.119943875 | 2.00E-05 | 0.000362859 | 0.125457353 |
| T2DM | monocyte | *PTGDR* | 0.729947349 | 0.001919389 | 0.00350277 | 1 |
| T2DM | monocyte | *GLYCTK* | 5.595331344 | 4.06E-06 | 0.000271729 | 0.025541548 |
| T2DM | monocyte | *UBTD2* | 3.82525613 | 1.19E-05 | 0.000362859 | 0.074900853 |
| T2DM | monocyte | *NT5DC2* | 1.680783207 | 0.005488373 | 0.008598405 | 1 |
| T2DM | monocyte | *KIF5C* | 2.893302089 | 0.022331654 | 0.029872151 | 1 |
| T2DM | monocyte | *PDHB* | 1.073784199 | 0.003528086 | 0.005840199 | 1 |
| T2DM | monocyte | *PXK* | 3.98075045 | 3.61E-08 | 1.42E-05 | 0.000226848 |
| T2DM | monocyte | *ACOX2* | 0.583972212 | 0.012343645 | 0.017545669 | 1 |
| T2DM | monocyte | *FAM107A* | 0.791240558 | 0.006733851 | 0.010285647 | 1 |
| T2DM | monocyte | *CX3CR1* | -0.246749354 | 0.007274545 | 0.011012537 | 1 |
| T2DM | monocyte | *SCN11A* | 0.445999909 | 0.011404969 | 0.01638552 | 1 |
| T2DM | monocyte | *SEPT2* | 0.869479319 | 0.015469191 | 0.021431204 | 1 |
| T2DM | monocyte | *DTYMK* | 2.547362178 | 0.000150601 | 0.000619248 | 0.94683083 |
| T2DM | monocyte | *ING5* | 1.699644067 | 0.00016921 | 0.000660586 | 1 |
| T2DM | monocyte | *RFWD3* | 3.042512707 | 3.47E-06 | 0.000249851 | 0.021840378 |
| T2DM | monocyte | *RHOH* | 3.305690563 | 5.99E-05 | 0.000399998 | 0.37669615 |
| T2DM | monocyte | *KLHL30* | 1.026754278 | 0.015551139 | 0.021530502 | 1 |
| T2DM | monocyte | *BMP1* | 0.711975244 | 0.001632263 | 0.003149797 | 1 |
| T2DM | monocyte | *ATXN2L* | 1.901426753 | 0.000251874 | 0.000842919 | 1 |
| T2DM | monocyte | *FNTA* | 4.873212252 | 2.95E-05 | 0.000362859 | 0.185387638 |
| T2DM | monocyte | *SERINC2* | 1.250008001 | 0.002733864 | 0.004680512 | 1 |
| T2DM | monocyte | *TRAPPC11* | 2.363937806 | 0.022169447 | 0.029667798 | 1 |
| T2DM | monocyte | *TMEM223* | 1.531080011 | 0.001744997 | 0.003291122 | 1 |
| T2DM | monocyte | *SLC20A2* | 2.005172733 | 4.50E-05 | 0.000379941 | 0.283215349 |
| T2DM | monocyte | *ZSWIM1* | 2.011618147 | 0.000340333 | 0.001030047 | 1 |
| T2DM | monocyte | *NBPF9* | 1.550221119 | 0.000568727 | 0.001470224 | 1 |
| T2DM | monocyte | *WFDC13* | 8.300984246 | 0.000403923 | 0.001161162 | 1 |
| T2DM | monocyte | *NDUFS5* | 2.229509549 | 0.001475897 | 0.002925747 | 1 |
| T2DM | monocyte | *ZNF30* | 2.35337734 | 0.000362077 | 0.001073764 | 1 |
| T2DM | monocyte | *FAM84B* | 1.443096423 | 0.005770678 | 0.008986934 | 1 |

| | | | | | | |
|---|---|---|---|---|---|---|
| T2DM | monocyte | *SLC16A4* | 1.185966643 | 8.78E-05 | 0.00046125 | 0.552279248 |
| T2DM | monocyte | *TMEM208* | 1.545290344 | 0.001727036 | 0.003268475 | 1 |
| T2DM | monocyte | *LRP1B* | 0.556927417 | 0.002720825 | 0.004666073 | 1 |
| T2DM | monocyte | *DNAJC21* | 4.454864904 | 0.000123217 | 0.000553734 | 0.774668175 |
| T2DM | monocyte | *NPNT* | 0.75662268 | 0.00180644 | 0.003369055 | 1 |
| T2DM | monocyte | *CNNM3* | 3.238624963 | 0.000222281 | 0.000778106 | 1 |
| T2DM | monocyte | *GSTM4* | 1.243233297 | 0.000206547 | 0.000743876 | 1 |
| T2DM | monocyte | *TCTN2* | 0.782629042 | 0.031520039 | 0.040707988 | 1 |
| T2DM | monocyte | *ABHD15* | 2.820856407 | 1.93E-05 | 0.000362859 | 0.121653137 |
| T2DM | monocyte | *ADAL* | 0.59497524 | 0.011952651 | 0.017078709 | 1 |
| T2DM | monocyte | *SNTB2* | 1.883334104 | 7.68E-05 | 0.000437295 | 0.482714221 |
| T2DM | monocyte | *ZNF507* | 0.880252808 | 0.004964411 | 0.007869706 | 1 |
| T2DM | monocyte | *STX18* | 2.163525645 | 6.55E-05 | 0.000417788 | 0.411913439 |
| T2DM | monocyte | *ZBTB49* | 2.530983966 | 0.000128086 | 0.000564711 | 0.805278191 |
| T2DM | monocyte | *GFM1* | 2.076179496 | 0.000860276 | 0.001975366 | 1 |
| T2DM | monocyte | *TNIP2* | 1.144522185 | 0.013347626 | 0.01880693 | 1 |
| T2DM | monocyte | *BTNL3* | 0.488093085 | 0.029519697 | 0.038400649 | 1 |
| T2DM | monocyte | *ZNF608* | 0.954958723 | 0.000605058 | 0.001529335 | 1 |
| T2DM | monocyte | *LETM1* | 2.073270856 | 0.000187789 | 0.000706541 | 1 |
| T2DM | monocyte | *CEP120* | 2.485816584 | 0.000111678 | 0.000524745 | 0.702120032 |
| T2DM | monocyte | *STXBP6* | 1.476708158 | 0.00022392 | 0.000779936 | 1 |
| T2DM | monocyte | *MFF* | 1.482331426 | 0.000439098 | 0.00122857 | 1 |
| T2DM | monocyte | *CPLX1* | 0.851184437 | 0.006927093 | 0.010539843 | 1 |
| T2DM | monocyte | *SIGLEC7* | -0.319358437 | 0.00774394 | 0.011616834 | 1 |
| T2DM | monocyte | *FEM1B* | 2.742248858 | 8.09E-06 | 0.000355191 | 0.050857306 |
| T2DM | monocyte | *COL4A3* | 0.841052178 | 0.002570039 | 0.004448743 | 1 |
| T2DM | monocyte | *IRS1* | -0.713930808 | 2.74E-05 | 0.000362859 | 0.172497035 |
| T2DM | monocyte | *UPF3A* | 1.038883831 | 0.000573419 | 0.001479101 | 1 |
| T2DM | monocyte | *C8orf46* | 0.451588398 | 0.000557158 | 0.001451812 | 1 |
| T2DM | monocyte | *HSPBAP1* | 2.417212727 | 0.000281844 | 0.000911031 | 1 |
| T2DM | monocyte | *PARM1* | 0.555350751 | 0.00186291 | 0.003434638 | 1 |
| T2DM | monocyte | *FAM110B* | 1.147430291 | 0.000381119 | 0.001112943 | 1 |
| T2DM | monocyte | *ATF5* | 4.199519353 | 0.000129683 | 0.00057004 | 0.815316924 |
| T2DM | monocyte | *PCSK9* | -5.657547245 | 0.009868014 | 0.014407851 | 1 |
| T2DM | monocyte | *RGS14* | -0.363807144 | 0.014874804 | 0.020726483 | 1 |
| T2DM | monocyte | *RAB24* | -1.254249316 | 0.006401127 | 0.009839581 | 1 |
| T2DM | monocyte | *CXCL10* | 0.832534762 | 0.026734912 | 0.035068307 | 1 |
| T2DM | monocyte | *SH3TC2* | 1.373795658 | 0.022020427 | 0.029474649 | 1 |
| T2DM | monocyte | *GPRIN1* | 7.532297117 | 0.015075787 | 0.020950812 | 1 |
| T2DM | monocyte | *STK32A* | 0.577457311 | 0.023055849 | 0.030742762 | 1 |
| T2DM | monocyte | *SLC33A1* | 4.427190139 | 0.000118476 | 0.000544237 | 0.744861702 |
| T2DM | monocyte | *SNUPN* | 0.977884152 | 0.002614577 | 0.004514652 | 1 |
| T2DM | monocyte | *PTK2* | 1.309709426 | 0.00015571 | 0.000631986 | 0.978946528 |
| T2DM | monocyte | *RNASE6* | -0.912319006 | 0.024577125 | 0.032522919 | 1 |
| T2DM | monocyte | *NPR1* | 1.211546798 | 0.000502538 | 0.001349618 | 1 |
| T2DM | monocyte | *CLIC4* | 1.909732454 | 0.000120635 | 0.00054853 | 0.758432432 |
| T2DM | monocyte | *SLC38A11* | 0.745351045 | 0.002600076 | 0.004494551 | 1 |
| T2DM | monocyte | *CCDC8* | 1.774639302 | 0.002654364 | 0.004573303 | 1 |
| T2DM | monocyte | *METTL15* | 1.615674847 | 0.000849566 | 0.001955271 | 1 |
| T2DM | monocyte | *HINT1* | 2.549251683 | 0.00135157 | 0.002741661 | 1 |

| | | | | | | |
|---|---|---|---|---|---|---|
| T2DM | monocyte | *DTWD2* | 2.921514933 | 3.62E-05 | 0.000365043 | 0.227491475 |
| T2DM | monocyte | *CLIC3* | 4.38307054 | 0.002164853 | 0.003857404 | 1 |
| T2DM | monocyte | *DFFB* | 2.70044914 | 0.000275542 | 0.000898513 | 1 |
| T2DM | monocyte | *C15orf40* | 1.727161652 | 0.001404943 | 0.002824713 | 1 |
| T2DM | monocyte | *RGPD8* | 0.617025662 | 0.013849369 | 0.019439826 | 1 |
| T2DM | monocyte | *LUZP1* | 1.990502558 | 0.000244083 | 0.000823255 | 1 |
| T2DM | monocyte | *HEXDC* | 3.153472877 | 2.98E-05 | 0.000362859 | 0.187429279 |
| T2DM | monocyte | *FASN* | 1.156598194 | 0.000171506 | 0.000666827 | 1 |
| T2DM | monocyte | *MT1E* | 0.751486352 | 0.00311002 | 0.005232191 | 1 |
| T2DM | monocyte | *DCXR* | 5.096488157 | 0.000378762 | 0.001107855 | 1 |
| T2DM | monocyte | *LIMS1* | 1.53565244 | 0.000302816 | 0.000958611 | 1 |
| T2DM | monocyte | *UGP2* | 1.062322417 | 0.000156814 | 0.000634753 | 0.985889758 |
| T2DM | monocyte | *AVEN* | 2.633978781 | 3.85E-05 | 0.00036941 | 0.242047556 |
| T2DM | monocyte | *TPST1* | 0.613624208 | 0.003678691 | 0.00604178 | 1 |
| T2DM | monocyte | *GUSB* | 3.222670187 | 3.79E-05 | 0.00036941 | 0.238432405 |
| T2DM | monocyte | *BRD3* | 3.954777057 | 9.88E-05 | 0.000489328 | 0.620957249 |
| T2DM | monocyte | *ZFPM2* | 0.976806131 | 0.000216363 | 0.000766771 | 1 |
| T2DM | monocyte | *MAP3K2* | 1.998263719 | 0.021353318 | 0.028691667 | 1 |
| T2DM | monocyte | *PUSL1* | -1.021817933 | 0.014154847 | 0.019821381 | 1 |
| T2DM | monocyte | *CNTROB* | 5.71913389 | 0.000140296 | 0.000593167 | 0.882039145 |
| T2DM | monocyte | *KCNAB3* | -0.941039242 | 0.004952228 | 0.007860302 | 1 |
| T2DM | monocyte | *NIPA1* | 1.395430728 | 0.000498949 | 0.001342849 | 1 |
| T2DM | monocyte | *RNF150* | 0.718892293 | 0.000350698 | 0.001049423 | 1 |
| T2DM | monocyte | *PDCD6IP* | 2.535858991 | 5.89E-05 | 0.000398946 | 0.370222121 |
| T2DM | monocyte | *FAM161A* | 0.872521435 | 0.001778272 | 0.00333926 | 1 |
| T2DM | monocyte | *GLB1* | 2.710856466 | 0.000336559 | 0.001021702 | 1 |
| T2DM | monocyte | *FAXDC2* | 1.550918516 | 0.000151301 | 0.0006205 | 0.95122675 |
| T2DM | monocyte | *CRTAP* | 3.070872179 | 5.31E-05 | 0.000385235 | 0.334035128 |
| T2DM | monocyte | *ELP5* | 1.922397125 | 0.001798418 | 0.00336093 | 1 |
| T2DM | monocyte | *STX8* | 2.471847418 | 3.41E-05 | 0.000365043 | 0.214218884 |
| T2DM | monocyte | *NFRKB* | 1.334384866 | 0.000281742 | 0.000911031 | 1 |
| T2DM | monocyte | *SEMA3E* | 0.541641419 | 0.001317132 | 0.002691736 | 1 |
| T2DM | monocyte | *DCLK2* | 0.914056793 | 0.002081644 | 0.003742623 | 1 |
| T2DM | monocyte | *GPRC5C* | 1.23309729 | 0.00035956 | 0.001068819 | 1 |
| T2DM | monocyte | *ADORA2B* | -0.794418955 | 0.011447759 | 0.016439484 | 1 |
| T2DM | monocyte | *MGMT* | 1.847790657 | 0.000154551 | 0.000629314 | 0.971661391 |
| T2DM | monocyte | *METTL7B* | 2.506161812 | 0.001870353 | 0.003444321 | 1 |
| T2DM | monocyte | *TMC7* | 0.604508952 | 0.014589274 | 0.020382837 | 1 |
| T2DM | monocyte | *SMAGP* | 0.908829613 | 0.002803854 | 0.004775895 | 1 |
| T2DM | monocyte | *IRX2* | 0.854711793 | 0.001439452 | 0.002879962 | 1 |
| T2DM | monocyte | *DLGAP1* | 1.099535029 | 0.000284621 | 0.000916707 | 1 |
| T2DM | monocyte | *STAT2* | 2.982513298 | 0.001109783 | 0.002374008 | 1 |
| T2DM | monocyte | *NUDCD2* | 4.935047428 | 0.000941601 | 0.002104459 | 1 |
| T2DM | monocyte | *HSPA4* | 2.515029152 | 6.47E-05 | 0.000415426 | 0.406999649 |
| T2DM | monocyte | *SLC26A5* | 1.369239231 | 0.000190219 | 0.000713122 | 1 |
| T2DM | monocyte | *COMMD5* | 2.717238618 | 0.002270001 | 0.003993357 | 1 |
| T2DM | monocyte | *ARMC10* | 2.455889491 | 0.000130993 | 0.000572307 | 0.823549924 |
| T2DM | monocyte | *RNF34* | 2.608765785 | 0.003697141 | 0.00606416 | 1 |
| T2DM | monocyte | *ACYP2* | 1.710527052 | 5.01E-05 | 0.000383643 | 0.314699001 |
| T2DM | monocyte | *TRABD* | 3.34809245 | 9.39E-06 | 0.000362859 | 0.059050548 |

| | | | | | | |
|---|---|---|---|---|---|---|
| T2DM | monocyte | *TMEM133* | 3.834413111 | 0.000605458 | 0.001529335 | 1 |
| T2DM | monocyte | *TTLL6* | 2.374229396 | 0.003045934 | 0.005135368 | 1 |
| T2DM | monocyte | *KCNS3* | 0.843354738 | 0.000118823 | 0.000544237 | 0.747039163 |
| T2DM | monocyte | *AKAP13* | 1.882590146 | 0.008266428 | 0.012324172 | 1 |
| T2DM | monocyte | *HTRA3* | 0.548642673 | 0.000952488 | 0.002123508 | 1 |
| T2DM | monocyte | *FOXN2* | 2.808769464 | 2.73E-05 | 0.000362859 | 0.171572817 |
| T2DM | monocyte | *KBTBD2* | 2.754089597 | 0.00039569 | 0.001145351 | 1 |
| T2DM | monocyte | *TSEN34* | 4.706262729 | 0.013136742 | 0.018547203 | 1 |
| T2DM | monocyte | *NDUFA3* | 4.294559592 | 0.000142026 | 0.000598067 | 0.892914605 |
| T2DM | monocyte | *OSCAR* | -0.138082632 | 0.008898268 | 0.013150779 | 1 |
| T2DM | monocyte | *PAQR8* | -1.479900124 | 0.027566237 | 0.036038456 | 1 |
| T2DM | monocyte | *NUDT6* | 2.668817313 | 1.55E-05 | 0.000362859 | 0.097415954 |
| T2DM | monocyte | *ZNF160* | 2.873664591 | 0.000102828 | 0.000500758 | 0.646478173 |
| T2DM | monocyte | *ZNF415* | 0.691030619 | 0.029924035 | 0.038830586 | 1 |
| T2DM | monocyte | *TSNARE1* | 2.095960299 | 0.000309779 | 0.000971846 | 1 |
| T2DM | monocyte | *FPR1* | -0.480533934 | 0.027546928 | 0.036020703 | 1 |
| T2DM | monocyte | *FEZ2* | 2.129667398 | 4.27E-05 | 0.000376681 | 0.268197053 |
| T2DM | monocyte | *SOX7* | 1.22488656 | 0.001619753 | 0.003129498 | 1 |
| T2DM | monocyte | *ALK* | 1.268490339 | 0.001834435 | 0.003397083 | 1 |
| T2DM | monocyte | *TRMT61B* | 1.150290002 | 0.001488417 | 0.002943636 | 1 |
| T2DM | monocyte | *MFN1* | 2.21942979 | 1.27E-05 | 0.000362859 | 0.079775641 |
| T2DM | monocyte | *KCNMB3* | 1.271994056 | 6.01E-05 | 0.000399998 | 0.377998091 |
| T2DM | monocyte | *ATP6V0E2* | 4.334996268 | 0.000162738 | 0.000647145 | 1 |
| T2DM | monocyte | *GRIK1* | 3.833142745 | 0.00086166 | 0.001977101 | 1 |
| T2DM | monocyte | *CLDN20* | 2.143599772 | 0.00035567 | 0.001061525 | 1 |
| T2DM | monocyte | *CANT1* | 2.497157283 | 0.000730266 | 0.001753699 | 1 |
| T2DM | monocyte | *KCND3* | 0.624122827 | 0.001037721 | 0.002263746 | 1 |
| T2DM | monocyte | *MRPL36* | 1.961211854 | 9.76E-05 | 0.000485446 | 0.613333459 |
| T2DM | monocyte | *MCC* | 1.447998128 | 8.60E-05 | 0.000459567 | 0.540864959 |
| T2DM | monocyte | *DLK2* | 2.088575298 | 0.024373302 | 0.032287178 | 1 |
| T2DM | monocyte | *HOPX* | 1.372427624 | 0.001276092 | 0.002626773 | 1 |
| T2DM | monocyte | *RSL1D1* | 4.480543545 | 7.20E-06 | 0.000345566 | 0.045269207 |
| T2DM | monocyte | *PPID* | 4.380835005 | 0.000126126 | 0.00055907 | 0.792953075 |
| T2DM | monocyte | *ETFDH* | 0.661908648 | 0.01173123 | 0.016800511 | 1 |
| T2DM | monocyte | *TBCA* | 2.771421465 | 0.000134377 | 0.00057865 | 0.844828877 |
| T2DM | monocyte | *MAP6* | 0.975230816 | 0.004010756 | 0.006520719 | 1 |
| T2DM | monocyte | *BCL2L1* | 3.031697306 | 1.15E-05 | 0.000362859 | 0.072470189 |
| T2DM | monocyte | *PLRG1* | 1.584955195 | 0.000371562 | 0.00109424 | 1 |
| T2DM | monocyte | *ZNF584* | 1.818414212 | 0.000311743 | 0.000976546 | 1 |
| T2DM | monocyte | *NMUR1* | 0.972438629 | 0.00151363 | 0.00297567 | 1 |
| T2DM | monocyte | *P2RY6* | 0.845801306 | 0.002026889 | 0.003661797 | 1 |
| T2DM | monocyte | *RGS19* | -0.386902891 | 0.000526835 | 0.001396379 | 1 |
| T2DM | monocyte | *TCEA2* | 4.537469654 | 0.009520792 | 0.013969012 | 1 |
| T2DM | monocyte | *ANO5* | 1.314414253 | 0.000747822 | 0.001784951 | 1 |
| T2DM | monocyte | *GPHN* | 0.923545669 | 0.009161021 | 0.01350735 | 1 |
| T2DM | monocyte | *VAT1L* | 0.988504739 | 0.001817234 | 0.003385171 | 1 |
| T2DM | monocyte | *CAMTA1* | 2.541240744 | 4.58E-05 | 0.000379941 | 0.287736088 |
| T2DM | monocyte | *LGALS4* | 0.968128951 | 0.004881677 | 0.007762039 | 1 |
| T2DM | monocyte | *LRRC34* | 0.67178784 | 0.001018621 | 0.002234498 | 1 |
| T2DM | monocyte | *SYCE1* | 1.241759177 | 0.009995443 | 0.014566841 | 1 |

| | | | | | | |
|---|---|---|---|---|---|---|
| T2DM | monocyte | *BCL2* | 0.602506218 | 0.002345537 | 0.004108774 | 1 |
| T2DM | monocyte | *KNDC1* | -0.148732679 | 0.004322242 | 0.006958754 | 1 |
| T2DM | monocyte | *METTL18* | 1.242571295 | 0.005209062 | 0.008205806 | 1 |
| T2DM | monocyte | *ZNF570* | 1.430664508 | 0.002619088 | 0.004521199 | 1 |
| T2DM | monocyte | *NINJ2* | 2.454026862 | 7.81E-05 | 0.000439826 | 0.491285435 |
| T2DM | monocyte | *FAM90A1* | 2.434461619 | 0.001762704 | 0.003316013 | 1 |
| T2DM | monocyte | *TRAPPC12* | 3.250973806 | 0.000464571 | 0.001277112 | 1 |
| T2DM | monocyte | *C3AR1* | -0.655671663 | 0.017361787 | 0.023770374 | 1 |
| T2DM | monocyte | *AQP4* | 0.935424897 | 0.001836469 | 0.003399846 | 1 |
| T2DM | monocyte | *TLN2* | 0.674428558 | 0.000549996 | 0.00144136 | 1 |
| T2DM | monocyte | *TVP23B* | 4.145929183 | 7.36E-05 | 0.000431915 | 0.462421732 |
| T2DM | monocyte | *ATPAF2* | 4.604159043 | 2.82E-05 | 0.000362859 | 0.177304293 |
| T2DM | monocyte | *CYP4F22* | -0.355851477 | 0.005222327 | 0.008224642 | 1 |
| T2DM | monocyte | *C20orf196* | 2.975775275 | 0.00955772 | 0.014019922 | 1 |
| T2DM | monocyte | *LAMB2* | 0.723500903 | 0.022498284 | 0.030082243 | 1 |
| T2DM | monocyte | *ORMDL3* | 0.51547247 | 0.009260134 | 0.01364069 | 1 |
| T2DM | monocyte | *SERF1A* | 4.033886891 | 0.00193177 | 0.003520301 | 1 |
| T2DM | monocyte | *NME6* | 1.786808612 | 0.000849969 | 0.001955271 | 1 |
| T2DM | monocyte | *FRMD3* | 1.907505473 | 8.79E-05 | 0.00046125 | 0.552577815 |
| T2DM | monocyte | *MTBP* | 1.759600148 | 0.000256181 | 0.000851723 | 1 |
| T2DM | monocyte | *MALT1* | 3.193075805 | 0.002030904 | 0.003665889 | 1 |
| T2DM | monocyte | *CXCR6* | 1.609404649 | 0.000503892 | 0.0013521 | 1 |
| T2DM | monocyte | *C1QTNF4* | -0.875306475 | 2.81E-05 | 0.000362859 | 0.17697733 |
| T2DM | monocyte | *ZNF131* | 3.217120026 | 0.000323036 | 0.00099909 | 1 |
| T2DM | monocyte | *HINFP* | 2.249816041 | 0.000166348 | 0.000655961 | 1 |
| T2DM | monocyte | *SPTLC3* | 0.818186719 | 0.00081753 | 0.001903633 | 1 |
| T2DM | monocyte | *COPRS* | 2.08644165 | 0.000159333 | 0.000640079 | 1 |
| T2DM | monocyte | *CLEC12A* | -0.16148452 | 0.002020674 | 0.003655316 | 1 |
| T2DM | monocyte | *SUCLG2* | 3.119400643 | 0.000198979 | 0.000726217 | 1 |
| T2DM | monocyte | *CSDC2* | 1.867631339 | 0.002719863 | 0.004665696 | 1 |
| T2DM | monocyte | *RCAN2* | 0.440188522 | 0.001820265 | 0.003386941 | 1 |
| T2DM | monocyte | *IL16* | 1.153686185 | 0.001016614 | 0.002231652 | 1 |
| T2DM | monocyte | *GNB2* | 2.822740651 | 0.035750404 | 0.045748584 | 1 |
| T2DM | monocyte | *PRSS27* | 3.6898653 | 0.000356661 | 0.001062714 | 1 |
| T2DM | monocyte | *FGGY* | 2.665469364 | 4.93E-06 | 0.000297779 | 0.030969024 |
| T2DM | monocyte | *ZNF24* | 4.608440482 | 5.20E-05 | 0.000384775 | 0.327028575 |
| T2DM | monocyte | *MANEA* | 2.600489074 | 0.03930432 | 0.049900294 | 1 |
| T2DM | monocyte | *FIBP* | 6.777939921 | 8.35E-07 | 0.000107191 | 0.005252364 |
| T2DM | monocyte | *CARNS1* | 2.275416469 | 0.002061059 | 0.003712859 | 1 |
| T2DM | monocyte | *SNTG2* | 0.524700204 | 0.001858678 | 0.003427841 | 1 |
| T2DM | monocyte | *PDE3A* | 0.757599326 | 0.000223426 | 0.00077951 | 1 |
| T2DM | monocyte | *MRPL52* | 2.866273096 | 0.00234242 | 0.004105602 | 1 |
| T2DM | monocyte | *RAD9A* | 0.985239765 | 0.006858183 | 0.010452702 | 1 |
| T2DM | monocyte | *EFEMP2* | 0.761933266 | 0.00158678 | 0.003089932 | 1 |
| T2DM | monocyte | *AGAP5* | -10.69495367 | 0.012115575 | 0.017264071 | 1 |
| T2DM | monocyte | *ZMAT3* | 2.579970401 | 9.47E-05 | 0.000479275 | 0.595259795 |
| T2DM | monocyte | *ZFAND4* | 2.566707448 | 6.69E-05 | 0.000421481 | 0.420773182 |
| T2DM | monocyte | *CCL19* | -3.19508114 | 0.018452425 | 0.02509418 | 1 |
| T2DM | monocyte | *CFL1* | 3.824133661 | 0.002363187 | 0.004135085 | 1 |
| T2DM | monocyte | *EFCAB12* | 0.994797603 | 0.008120832 | 0.012121768 | 1 |

| | | | | | | |
|---|---|---|---|---|---|---|
| T2DM | monocyte | *RAB37* | -0.195783307 | 0.009482461 | 0.013925773 | 1 |
| T2DM | monocyte | *DCP2* | 2.551439454 | 0.01213011 | 0.017273387 | 1 |
| T2DM | monocyte | *SNX32* | 1.70768079 | 2.73E-05 | 0.000362859 | 0.171926203 |
| T2DM | monocyte | *CYP7B1* | 0.842295932 | 0.001233432 | 0.002561806 | 1 |
| T2DM | monocyte | *METAP1D* | 1.768871675 | 0.000154659 | 0.000629348 | 0.972342542 |
| T2DM | monocyte | *NADSYN1* | 3.31717866 | 0.001304311 | 0.002673689 | 1 |
| T2DM | monocyte | *MRGPRF* | 1.356793514 | 0.0006651 | 0.00163686 | 1 |
| T2DM | monocyte | *LCLAT1* | 2.976797414 | 6.63E-05 | 0.000420249 | 0.417028057 |
| T2DM | monocyte | *GXYLT2* | 0.883707042 | 0.001455296 | 0.002900975 | 1 |
| T2DM | monocyte | *HPSE2* | 0.732134625 | 0.000138482 | 0.000589065 | 0.870638211 |
| T2DM | monocyte | *HECTD4* | 2.455434917 | 2.43E-05 | 0.000362859 | 0.153037133 |
| T2DM | monocyte | *BNC2* | 0.887727612 | 0.000271457 | 0.000888419 | 1 |
| T2DM | monocyte | *TRMT112* | 2.408789137 | 0.000438008 | 0.001226618 | 1 |
| T2DM | monocyte | *NOC3L* | 4.019997217 | 0.000223314 | 0.00077951 | 1 |
| T2DM | monocyte | *PARP15* | 1.25710523 | 0.006725825 | 0.01028088 | 1 |
| T2DM | monocyte | *AHSA2* | 1.471754383 | 0.001725589 | 0.00326672 | 1 |
| T2DM | monocyte | *ABLIM3* | 1.335413931 | 0.000230512 | 0.000794721 | 1 |
| T2DM | monocyte | *VANGL1* | 1.011548537 | 0.000732105 | 0.001756772 | 1 |
| T2DM | monocyte | *GLRX* | 1.318613544 | 0.00020597 | 0.000743876 | 1 |
| T2DM | monocyte | *IQCB1* | 1.720262449 | 0.002734197 | 0.004680512 | 1 |
| T2DM | monocyte | *GOLGB1* | 1.994128233 | 3.95E-05 | 0.000371009 | 0.248213875 |
| T2DM | monocyte | *DMRT2* | 0.752933126 | 0.003247402 | 0.005435681 | 1 |
| T2DM | monocyte | *ZNF483* | 1.009625116 | 0.010378107 | 0.015058195 | 1 |
| T2DM | monocyte | *MZT2A* | 3.00460164 | 0.000196216 | 0.000723774 | 1 |
| T2DM | monocyte | *MAP3K11* | 4.537102302 | 1.59E-05 | 0.000362859 | 0.099810354 |
| T2DM | monocyte | *C1QA* | 2.981886203 | 4.04E-05 | 0.00037107 | 0.254302714 |
| T2DM | monocyte | *NDNF* | 1.302863321 | 0.001702051 | 0.003233153 | 1 |
| T2DM | monocyte | *OLR1* | -0.304546288 | 0.008380067 | 0.012472888 | 1 |
| T2DM | monocyte | *GLIPR1L1* | 1.37480639 | 0.000166416 | 0.000655961 | 1 |
| T2DM | monocyte | *ARV1* | 2.101851635 | 0.00063084 | 0.001575722 | 1 |
| T2DM | monocyte | *CCDC36* | 0.801827946 | 0.000567208 | 0.00146776 | 1 |
| T2DM | monocyte | *THAP2* | 2.808470985 | 0.00137161 | 0.002774286 | 1 |
| T2DM | monocyte | *PPP1R14B* | 5.126861869 | 0.000105273 | 0.000507835 | 0.661851805 |
| T2DM | monocyte | *SMARCC1* | 3.007698152 | 1.67E-05 | 0.000362859 | 0.104919202 |
| T2DM | monocyte | *PEAK1* | 1.916229389 | 4.62E-05 | 0.000379941 | 0.290560559 |
| T2DM | monocyte | *MST1* | 1.034776129 | 0.00658347 | 0.010082893 | 1 |
| T2DM | monocyte | *GMPPB* | 1.635947552 | 0.000669223 | 0.00164416 | 1 |
| T2DM | monocyte | *CSPG4* | 1.545770055 | 0.000433399 | 0.001218264 | 1 |
| T2DM | monocyte | *SNX33* | 1.07948772 | 0.001799202 | 0.00336093 | 1 |
| T2DM | monocyte | *NUDT18* | -0.717506641 | 0.010953095 | 0.01578903 | 1 |
| T2DM | monocyte | *SULT1B1* | -0.283942793 | 0.012615955 | 0.017896324 | 1 |
| T2DM | monocyte | *NUDT4* | 1.469311873 | 0.000967477 | 0.002150822 | 1 |
| T2DM | monocyte | *SCAI* | 1.910116004 | 8.18E-05 | 0.000446179 | 0.514328539 |
| T2DM | monocyte | *AGFG1* | 2.095493541 | 4.79E-05 | 0.000379941 | 0.301291798 |
| T2DM | monocyte | *HAP1* | -1.259502992 | 0.025172039 | 0.033205331 | 1 |
| T2DM | monocyte | *RNF213* | 2.817855369 | 0.001071285 | 0.00231688 | 1 |
| T2DM | monocyte | *PLK3* | 1.867578488 | 0.000291127 | 0.000933359 | 1 |
| T2DM | monocyte | *DPY19L1* | 3.545880178 | 5.20E-05 | 0.000384775 | 0.326799562 |
| T2DM | monocyte | *TUBB8* | -1.477178145 | 0.006463416 | 0.009920775 | 1 |
| T2DM | monocyte | *GPR160* | 2.773339094 | 0.00013937 | 0.00059075 | 0.876216483 |

| | | | | | | |
|---|---|---|---|---|---|---|
| T2DM | monocyte | *GOLIM4* | 1.974045518 | 0.000232392 | 0.000798387 | 1 |
| T2DM | monocyte | *USMG5* | 3.988790205 | 2.19E-06 | 0.000188222 | 0.013740221 |
| T2DM | monocyte | *XXYLT1* | 2.07873105 | 0.000136719 | 0.000584748 | 0.859549793 |
| T2DM | monocyte | *UBXN2A* | 1.596884087 | 0.000533656 | 0.001409112 | 1 |
| T2DM | monocyte | *CCS* | 1.993969642 | 6.32E-05 | 0.00040959 | 0.397302267 |
| T2DM | monocyte | *CTSF* | 1.189722025 | 0.010471337 | 0.015175956 | 1 |
| T2DM | monocyte | *LEMD3* | 2.807761802 | 2.21E-05 | 0.000362859 | 0.139028354 |
| T2DM | monocyte | *C16orf91* | 2.690190398 | 0.000132676 | 0.000576456 | 0.834132453 |
| T2DM | monocyte | *SOCS7* | 2.926287362 | 4.62E-05 | 0.000379941 | 0.290671959 |
| T2DM | monocyte | *RGMB* | 1.958254308 | 0.000446461 | 0.001240893 | 1 |
| T2DM | monocyte | *FAM53A* | 1.172091753 | 0.002811516 | 0.004786353 | 1 |
| T2DM | monocyte | *CTU2* | 2.312661644 | 0.005536249 | 0.008666931 | 1 |
| T2DM | monocyte | *MGA* | 2.033376806 | 8.23E-05 | 0.000447645 | 0.517187624 |
| T2DM | monocyte | *SNX31* | 1.559027076 | 0.000925774 | 0.002077951 | 1 |
| T2DM | monocyte | *DDX23* | 4.251714367 | 9.06E-05 | 0.000467143 | 0.569658426 |
| T2DM | monocyte | *GLIS1* | 0.594501547 | 0.006378553 | 0.009812078 | 1 |
| T2DM | monocyte | *RALGAPA1* | 1.96078717 | 9.97E-05 | 0.000492855 | 0.626527255 |
| T2DM | monocyte | *LIG4* | 2.286690258 | 7.15E-05 | 0.000428217 | 0.44962806 |
| T2DM | monocyte | *LINGO2* | 1.4417539 | 0.000308273 | 0.000969854 | 1 |
| T2DM | monocyte | *BBS1* | 3.134331906 | 0.000327556 | 0.001006043 | 1 |
| T2DM | monocyte | *ANKRD36C* | 2.121892309 | 3.31E-05 | 0.000365043 | 0.208399133 |
| T2DM | monocyte | *TMEM81* | -1.223924681 | 0.028702161 | 0.037422332 | 1 |
| T2DM | monocyte | *IL20RB* | 2.699476024 | 2.43E-06 | 0.000200808 | 0.015261381 |
| T2DM | monocyte | *ZNF497* | -2.082893045 | 0.022813255 | 0.030451579 | 1 |
| T2DM | monocyte | *KLF14* | -6.255349341 | 0.000276383 | 0.000900194 | 1 |
| T2DM | monocyte | *ANGEL2* | 2.487376875 | 4.91E-05 | 0.000382804 | 0.308922435 |
| T2DM | monocyte | *BRSK2* | 1.943111457 | 0.000905303 | 0.002042944 | 1 |
| T2DM | monocyte | *SH3PXD2B* | 0.769378847 | 0.000113681 | 0.000529958 | 0.714712409 |
| T2DM | monocyte | *LARP7* | 2.264229161 | 6.79E-05 | 0.000422315 | 0.427041731 |
| T2DM | monocyte | *FGFBP3* | 1.410820311 | 0.001804101 | 0.003365693 | 1 |
| T2DM | monocyte | *C4orf32* | 1.214892907 | 0.000332528 | 0.001017459 | 1 |
| T2DM | monocyte | *HRAS* | 2.333653004 | 0.001414407 | 0.002840771 | 1 |
| T2DM | monocyte | *SRP72* | 2.039127176 | 0.002696235 | 0.004635283 | 1 |
| T2DM | monocyte | *BTC* | 1.520586054 | 0.000410463 | 0.00117346 | 1 |
| T2DM | monocyte | *RSRC1* | 1.787754527 | 0.003071774 | 0.005174771 | 1 |
| T2DM | monocyte | *PTDSS2* | 1.559363265 | 0.00217791 | 0.003873415 | 1 |
| T2DM | monocyte | *FUT1* | 2.670459525 | 0.001559263 | 0.003043777 | 1 |
| T2DM | monocyte | *DHX36* | 2.726724216 | 0.000296556 | 0.000946439 | 1 |
| T2DM | monocyte | *FBXW8* | 3.551798294 | 2.35E-05 | 0.000362859 | 0.147523823 |
| T2DM | monocyte | *CHST2* | -0.368220743 | 0.009709901 | 0.014216616 | 1 |
| T2DM | monocyte | *ATR* | 2.569143178 | 1.53E-05 | 0.000362859 | 0.09631564 |
| T2DM | monocyte | *TVP23C* | 1.419365942 | 0.001682506 | 0.003212508 | 1 |
| T2DM | monocyte | *PACS1* | 2.189118521 | 0.000160532 | 0.000642436 | 1 |
| T2DM | monocyte | *SH3BP5L* | 2.270941078 | 8.93E-05 | 0.000464416 | 0.561478938 |
| T2DM | monocyte | *CADM2* | 0.557356277 | 0.002299635 | 0.00404075 | 1 |
| T2DM | monocyte | *PSMD2* | 1.536316813 | 0.000187683 | 0.000706541 | 1 |
| T2DM | monocyte | *ARHGAP1* | 2.780367722 | 0.002404682 | 0.004197179 | 1 |
| T2DM | monocyte | *GOLGA8A* | 1.152022749 | 0.000595826 | 0.001515557 | 1 |
| T2DM | monocyte | *TP53I11* | 1.505023869 | 0.000227559 | 0.000787811 | 1 |
| T2DM | monocyte | *PHYHD1* | 0.777143437 | 0.000923429 | 0.002074169 | 1 |

| | | | | | | |
|---|---|---|---|---|---|---|
| T2DM | monocyte | *ZNF519* | 1.890410744 | 0.00017243 | 0.000669592 | 1 |
| T2DM | monocyte | *BANF1* | 2.153460011 | 0.000122966 | 0.000553734 | 0.773085449 |
| T2DM | monocyte | *CHRNA7* | 1.638721299 | 0.000270292 | 0.000886054 | 1 |
| T2DM | monocyte | *TMEM9B* | 5.334102723 | 5.62E-07 | 8.21E-05 | 0.003530663 |
| T2DM | monocyte | *SCUBE2* | 0.781618692 | 0.002744769 | 0.004693055 | 1 |
| T2DM | monocyte | *SMAD2* | 2.400028737 | 0.000233379 | 0.000800901 | 1 |
| T2DM | monocyte | *EIF3F* | 2.398981003 | 0.000374849 | 0.001099195 | 1 |
| T2DM | monocyte | *PCSK1* | 0.963282069 | 0.001315573 | 0.00269151 | 1 |
| T2DM | monocyte | *LPL* | 0.326254869 | 0.038079021 | 0.048442494 | 1 |
| T2DM | monocyte | *PPP2R2D* | 2.756837526 | 8.73E-05 | 0.000461034 | 0.548862378 |
| T2DM | monocyte | *MCTP1* | 1.732925005 | 0.00272965 | 0.004677381 | 1 |
| T2DM | monocyte | *LRRC25* | -0.136033984 | 0.014967407 | 0.020837044 | 1 |
| T2DM | monocyte | *UBQLNL* | 2.132382235 | 0.005313482 | 0.008347292 | 1 |
| T2DM | monocyte | *LIPT2* | 2.634657589 | 0.004397558 | 0.007065537 | 1 |
| T2DM | monocyte | *UCP3* | 1.448122227 | 0.0310491 | 0.040149258 | 1 |
| T2DM | monocyte | *PAAF1* | 0.657505821 | 0.008996562 | 0.013280439 | 1 |
| T2DM | monocyte | *RAB6A* | 3.004033204 | 8.31E-05 | 0.000449548 | 0.52237468 |
| T2DM | monocyte | *RPS6KB2* | 5.671194829 | 2.44E-05 | 0.000362859 | 0.153154299 |
| T2DM | monocyte | *RMI2* | 0.916010511 | 0.009761241 | 0.014273164 | 1 |
| T2DM | monocyte | *TOM1L2* | 2.094464644 | 5.28E-05 | 0.000385235 | 0.332042731 |
| T2DM | monocyte | *TEX26* | 4.083019429 | 0.003716047 | 0.006088816 | 1 |
| T2DM | monocyte | *GPR156* | 1.058255113 | 0.001078974 | 0.002327903 | 1 |
| T2DM | monocyte | *TTLL11* | 1.320536024 | 0.000452167 | 0.001251773 | 1 |
| T2DM | monocyte | *LINC01106* | -3.348228295 | 0.00439737 | 0.007065537 | 1 |
| T2DM | monocyte | *MSRA* | 2.932777274 | 0.000294138 | 0.000940136 | 1 |
| T2DM | monocyte | *ETV4* | 2.319497051 | 4.06E-05 | 0.00037107 | 0.255026979 |
| T2DM | monocyte | *SWI5* | 2.006874754 | 0.025152734 | 0.033186829 | 1 |
| T2DM | monocyte | *ZDHHC21* | 1.383987728 | 0.000335267 | 0.00102039 | 1 |
| T2DM | monocyte | *A2M* | 1.070034469 | 0.000261976 | 0.000865499 | 1 |
| T2DM | monocyte | *ARL4D* | 2.409053637 | 0.001221656 | 0.002548292 | 1 |
| T2DM | monocyte | *LRRN1* | 2.806531823 | 0.000140176 | 0.00059306 | 0.88128645 |
| T2DM | monocyte | *B3GALT6* | 1.560288087 | 0.001383147 | 0.0027925 | 1 |
| T2DM | monocyte | *ZNF613* | 1.706146492 | 0.000212479 | 0.000758148 | 1 |
| T2DM | monocyte | *IP6K1* | 3.710948027 | 0.000168588 | 0.000659148 | 1 |
| T2DM | monocyte | *SSNA1* | 3.038554883 | 0.000138725 | 0.0005894 | 0.872164132 |
| T2DM | monocyte | *CHMP6* | 1.467158924 | 0.000414919 | 0.001179826 | 1 |
| T2DM | monocyte | *TMEM39A* | 2.525725335 | 3.50E-05 | 0.000365043 | 0.220100945 |
| T2DM | monocyte | *TCP11L1* | 3.39543119 | 7.56E-06 | 0.000349439 | 0.047510551 |
| T2DM | monocyte | *GPX2* | 1.529390322 | 0.000798134 | 0.001868157 | 1 |
| T2DM | monocyte | *CCDC57* | 1.717716094 | 0.000585883 | 0.001496493 | 1 |
| T2DM | monocyte | *CIDEA* | 0.313787151 | 0.011577364 | 0.016591494 | 1 |
| T2DM | monocyte | *SMIM19* | 1.535001169 | 5.84E-05 | 0.000397957 | 0.367186571 |
| T2DM | monocyte | *SLC35G1* | 1.510638217 | 0.00176409 | 0.003317629 | 1 |
| T2DM | monocyte | *ZNF135* | 0.996798851 | 0.004264976 | 0.006880653 | 1 |
| T2DM | monocyte | *COX8A* | 6.338189612 | 3.59E-05 | 0.000365043 | 0.225640445 |
| T2DM | monocyte | *B3GNT4* | 6.417819289 | 0.001827193 | 0.003391662 | 1 |
| T2DM | monocyte | *CRLF3* | 3.516374663 | 7.71E-05 | 0.00043763 | 0.484437519 |
| T2DM | monocyte | *RNPEP* | 2.660815482 | 0.015575209 | 0.021556346 | 1 |
| T2DM | monocyte | *SPRYD4* | 1.196646958 | 0.011912188 | 0.017028633 | 1 |
| T2DM | monocyte | *WDR25* | 4.058141926 | 4.69E-05 | 0.000379941 | 0.294584815 |

| | | | | | | |
|---|---|---|---|---|---|---|
| T2DM | monocyte | *PLA2G16* | 0.634441561 | 0.00051127 | 0.001368977 | 1 |
| T2DM | monocyte | *RMDN1* | 1.773398402 | 4.37E-05 | 0.000378997 | 0.274720382 |
| T2DM | monocyte | *HORMAD2* | 1.495493501 | 2.69E-05 | 0.000362859 | 0.168966492 |
| T2DM | monocyte | *RNF152* | 0.767692565 | 0.001497289 | 0.002953704 | 1 |
| T2DM | monocyte | *ACSF3* | 2.000327454 | 0.014115202 | 0.019777641 | 1 |
| T2DM | monocyte | *BOK* | 0.644764837 | 0.001173033 | 0.002469812 | 1 |
| T2DM | monocyte | *ZNF843* | 2.916600961 | 0.001821574 | 0.003387234 | 1 |
| T2DM | monocyte | *C8orf59* | 2.559938331 | 1.84E-05 | 0.000362859 | 0.115615072 |
| T2DM | monocyte | *PFN4* | 1.104413161 | 0.000205216 | 0.000741918 | 1 |
| T2DM | monocyte | *RUFY1* | 2.310367265 | 0.000553344 | 0.00144712 | 1 |
| T2DM | monocyte | *LRRC37A3* | 4.851977878 | 1.84E-05 | 0.000362859 | 0.115874208 |
| T2DM | monocyte | *VSIG10* | 0.887431617 | 0.00049184 | 0.001333243 | 1 |
| T2DM | monocyte | *IRX5* | 0.944725809 | 0.001641019 | 0.003156599 | 1 |
| T2DM | monocyte | *PNMA1* | 1.125436235 | 0.002132766 | 0.003815795 | 1 |
| T2DM | monocyte | *ANKLE2* | 1.70630924 | 0.000104356 | 0.000505851 | 0.656088882 |
| T2DM | monocyte | *GCNT4* | 2.964166453 | 0.001682205 | 0.003212508 | 1 |
| T2DM | monocyte | *MUC20* | 1.991081062 | 0.000971274 | 0.002154039 | 1 |
| T2DM | monocyte | *THAP4* | 3.244335809 | 4.62E-05 | 0.000379941 | 0.2906872 |
| T2DM | monocyte | *SHMT1* | 1.127712845 | 0.000251005 | 0.000841352 | 1 |
| T2DM | monocyte | *DPP7* | 2.142477196 | 0.000100805 | 0.000495511 | 0.633758544 |
| T2DM | monocyte | *SEC24C* | 3.780282963 | 0.001324985 | 0.002701972 | 1 |
| T2DM | monocyte | *MTHFR* | 2.52446708 | 0.035441121 | 0.045389759 | 1 |
| T2DM | monocyte | *C19orf18* | 0.94250147 | 0.002052616 | 0.003700831 | 1 |
| T2DM | monocyte | *WDR73* | 2.695883419 | 5.78E-06 | 0.000322038 | 0.036315437 |
| T2DM | monocyte | *POLE* | 4.206970691 | 9.68E-06 | 0.000362859 | 0.060856882 |
| T2DM | monocyte | *DSCAML1* | 0.749461186 | 0.000601598 | 0.001524272 | 1 |
| T2DM | monocyte | *EPS8L2* | 0.789208395 | 0.010684105 | 0.015427416 | 1 |
| T2DM | monocyte | *RIMKLA* | 1.349295321 | 0.004608169 | 0.007353187 | 1 |
| T2DM | monocyte | *B3GNT8* | 5.944407658 | 0.000287103 | 0.000922809 | 1 |
| T2DM | monocyte | *AP006621.1* | 1.285877747 | 0.000260673 | 0.00086247 | 1 |
| T2DM | monocyte | *MAN1B1* | 4.766099907 | 2.42E-05 | 0.000362859 | 0.152238346 |
| T2DM | monocyte | *TOP3A* | 3.931653922 | 0.000214266 | 0.000762361 | 1 |
| T2DM | monocyte | *ZBTB38* | 1.446316825 | 0.000565259 | 0.001465478 | 1 |
| T2DM | monocyte | *C8orf31* | 6.210079313 | 3.22E-05 | 0.000363519 | 0.202480314 |
| T2DM | monocyte | *TIMM22* | 1.38549656 | 0.000459758 | 0.001266097 | 1 |
| T2DM | monocyte | *PPFIA3* | 1.276260272 | 0.002108024 | 0.003780134 | 1 |
| T2DM | monocyte | *MAGEF1* | 1.200586183 | 0.001250242 | 0.002583916 | 1 |
| T2DM | monocyte | *PAWR* | 0.936241055 | 0.000632522 | 0.001578678 | 1 |
| T2DM | monocyte | *NAP1L5* | 3.978801691 | 0.001045909 | 0.002275253 | 1 |
| T2DM | monocyte | *ACOT4* | 0.948035345 | 0.00967851 | 0.014177258 | 1 |
| T2DM | monocyte | *ARIH2* | 1.754612928 | 0.000112977 | 0.000528063 | 0.710289237 |
| T2DM | monocyte | *IRX3* | 1.403730457 | 0.000213727 | 0.000760874 | 1 |
| T2DM | monocyte | *ATOX1* | 5.558110292 | 1.71E-06 | 0.000169734 | 0.010735165 |
| T2DM | monocyte | *TBL1XR1* | 1.932486248 | 0.000111927 | 0.000524745 | 0.703683621 |
| T2DM | monocyte | *SAMD12* | 1.468627496 | 0.000586029 | 0.001496493 | 1 |
| T2DM | monocyte | *C18orf32* | 4.657164259 | 0.001295339 | 0.002660502 | 1 |
| T2DM | monocyte | *RPLP2* | 2.302271521 | 4.24E-05 | 0.00037505 | 0.266285592 |
| T2DM | monocyte | *C12orf54* | -1.7969454 | 0.002204381 | 0.00390723 | 1 |
| T2DM | monocyte | *GBA* | 0.909812505 | 0.004823954 | 0.00767414 | 1 |
| T2DM | monocyte | *ACAD9* | 0.778272822 | 0.009383072 | 0.013792699 | 1 |

| | | | | | | |
|---|---|---|---|---|---|---|
| T2DM | monocyte | *CD163L1* | 0.525099194 | 0.001651127 | 0.003169364 | 1 |
| T2DM | monocyte | *THAP5* | 3.480286408 | 7.10E-05 | 0.000427286 | 0.446180578 |
| T2DM | monocyte | *CD151* | 2.095503196 | 0.000135536 | 0.000582442 | 0.85211231 |
| T2DM | monocyte | *POLR2L* | 3.445100057 | 5.09E-05 | 0.000383728 | 0.320043864 |
| T2DM | monocyte | *ZCCHC3* | 1.118403112 | 0.001914772 | 0.003499468 | 1 |
| T2DM | monocyte | *PCDHB9* | 1.417401053 | 0.000669185 | 0.00164416 | 1 |
| T2DM | monocyte | *ZNF619* | 2.208230485 | 0.000417791 | 0.00118535 | 1 |
| T2DM | monocyte | *ZBTB41* | 2.862079988 | 4.89E-05 | 0.000381666 | 0.307456928 |
| T2DM | monocyte | *ZNF354C* | 4.553000731 | 1.64E-05 | 0.000362859 | 0.102956908 |
| T2DM | monocyte | *CENPBD1* | -0.856396236 | 0.008338338 | 0.0124196 | 1 |
| T2DM | monocyte | *BET1L* | 1.748482249 | 7.73E-05 | 0.00043763 | 0.485748953 |
| T2DM | monocyte | *RIC8A* | 2.173918981 | 0.024018081 | 0.031877069 | 1 |
| T2DM | monocyte | *IMP3* | 3.218906526 | 8.14E-06 | 0.000355191 | 0.051147517 |
| T2DM | monocyte | *DPY19L2* | 0.552204271 | 0.011220077 | 0.016145713 | 1 |
| T2DM | monocyte | *C2orf73* | 1.799237793 | 0.000830938 | 0.001924166 | 1 |
| T2DM | monocyte | *FAM26E* | 1.458365077 | 0.000922624 | 0.002073102 | 1 |
| T2DM | monocyte | *ALS2CL* | 1.305963908 | 0.000150382 | 0.000618751 | 0.945452097 |
| T2DM | monocyte | *MLF1* | 1.205830238 | 0.000536237 | 0.001412368 | 1 |
| T2DM | monocyte | *PRSS42* | 3.273578493 | 0.002803337 | 0.004775895 | 1 |
| T2DM | monocyte | *C2orf69* | 1.532378431 | 0.000318392 | 0.000989488 | 1 |
| T2DM | monocyte | *GRAMD1C* | 0.801861558 | 0.001591921 | 0.003095555 | 1 |
| T2DM | monocyte | *STAP2* | 1.154635907 | 0.004290916 | 0.006915403 | 1 |
| T2DM | monocyte | *GOLGA8Q* | 34.32806497 | 0.002895031 | 0.004913893 | 1 |
| T2DM | monocyte | *NDUFV2* | 6.08211959 | 0.000717851 | 0.001730495 | 1 |
| T2DM | monocyte | *DALRD3* | 1.574066909 | 0.000137118 | 0.000585241 | 0.862059656 |
| T2DM | monocyte | *ZNF114* | 4.458268114 | 0.007613328 | 0.011451203 | 1 |
| T2DM | monocyte | *ZNF518B* | 2.066521327 | 0.001368796 | 0.002771144 | 1 |
| T2DM | monocyte | *LCORL* | 3.401606347 | 0.000164916 | 0.000652094 | 1 |
| T2DM | monocyte | *ZNF454* | 0.947527611 | 0.001392049 | 0.002804151 | 1 |
| T2DM | monocyte | *ZC3H12D* | 5.04658766 | 0.01126123 | 0.016201225 | 1 |
| T2DM | monocyte | *KDELC2* | 2.908295122 | 0.010320249 | 0.014988081 | 1 |
| T2DM | monocyte | *SH2D4B* | -0.935213773 | 0.013668687 | 0.019211946 | 1 |
| T2DM | monocyte | *RNF212* | 1.003984487 | 0.000849858 | 0.001955271 | 1 |
| T2DM | monocyte | *PRSS36* | 1.159025148 | 0.00179021 | 0.00335271 | 1 |
| T2DM | monocyte | *TMPRSS9* | 0.934283793 | 0.00123572 | 0.002563289 | 1 |
| T2DM | monocyte | *AQP11* | 2.28496334 | 0.000159083 | 0.000639485 | 1 |
| T2DM | monocyte | *TMEM11* | 1.368468539 | 0.000669911 | 0.001644565 | 1 |
| T2DM | monocyte | *ZFAND2A* | 3.459846649 | 0.002223561 | 0.00393432 | 1 |
| T2DM | monocyte | *ZNF223* | -0.743631293 | 0.018149599 | 0.0247305 | 1 |
| T2DM | monocyte | *BEND3* | 10.08519183 | 5.07E-05 | 0.000383728 | 0.318559747 |
| T2DM | monocyte | *NT5DC1* | 2.206785934 | 6.43E-05 | 0.000414227 | 0.404466302 |
| T2DM | monocyte | *AC006111.1* | 4.164447967 | 0.00095568 | 0.002129115 | 1 |
| T2DM | monocyte | *LINC00843* | 2.11744981 | 0.000397559 | 0.001147063 | 1 |
| T2DM | monocyte | *COX14* | 0.770221635 | 0.000884911 | 0.002011365 | 1 |
| T2DM | monocyte | *P4HTM* | 1.777467528 | 0.000285353 | 0.000918593 | 1 |
| T2DM | monocyte | *CA8* | 0.704190655 | 0.005692681 | 0.008876459 | 1 |
| T2DM | monocyte | *ERBB4* | 0.748854587 | 0.000480514 | 0.001311194 | 1 |
| T2DM | monocyte | *CSRNP3* | 0.70935385 | 0.001415851 | 0.002842099 | 1 |
| T2DM | monocyte | *ZNF713* | 1.251492081 | 0.003641865 | 0.005995393 | 1 |
| T2DM | monocyte | *PARP10* | 3.10521803 | 0.000439291 | 0.00122857 | 1 |

| | | | | | | |
|---|---|---|---|---|---|---|
| T2DM | monocyte | *KCTD12* | 1.219206473 | 0.035167078 | 0.04506633 | 1 |
| T2DM | monocyte | *RPP25* | -0.863953505 | 0.014094649 | 0.019757649 | 1 |
| T2DM | monocyte | *STX19* | 1.508717323 | 2.89E-05 | 0.000362859 | 0.181718388 |
| T2DM | monocyte | *FAM219B* | 0.991418833 | 0.000562706 | 0.00146183 | 1 |
| T2DM | monocyte | *GDPD4* | 1.633392162 | 0.001816953 | 0.003385171 | 1 |
| T2DM | monocyte | *MPI* | 0.978838282 | 0.001068043 | 0.002310922 | 1 |
| T2DM | monocyte | *EFCAB13* | 1.177999513 | 0.000248365 | 0.000834848 | 1 |
| T2DM | monocyte | *GAK* | 3.15761909 | 0.003268242 | 0.005461096 | 1 |
| T2DM | monocyte | *TUFM* | 2.094277963 | 0.003635811 | 0.005986993 | 1 |
| T2DM | monocyte | *RMI1* | 2.844746336 | 0.002747437 | 0.00469634 | 1 |
| T2DM | monocyte | *CTC1* | 2.635529207 | 0.002235956 | 0.003952054 | 1 |
| T2DM | monocyte | *C14orf39* | 0.611726169 | 0.007208096 | 0.010924689 | 1 |
| T2DM | monocyte | *TRIML2* | 15.16384461 | 0.007439172 | 0.011226613 | 1 |
| T2DM | monocyte | *SPTY2D1* | 2.659495827 | 6.20E-05 | 0.000406647 | 0.38985477 |
| T2DM | monocyte | *TCAIM* | 1.816366405 | 0.001781945 | 0.003343207 | 1 |
| T2DM | monocyte | *FUCA1* | 1.437687206 | 0.000247616 | 0.000833385 | 1 |
| T2DM | monocyte | *ZNF664* | 2.0495938 | 0.000339972 | 0.001029894 | 1 |
| T2DM | monocyte | *CALR* | 5.330195761 | 6.07E-08 | 1.82E-05 | 0.000381399 |
| T2DM | monocyte | *HLA-DQB1* | -1.038103447 | 2.40E-09 | 1.51E-06 | 1.51E-05 |
| T2DM | monocyte | *ARID3B* | 2.141837325 | 0.004911174 | 0.00780302 | 1 |
| T2DM | monocyte | *EGR3* | 0.996224442 | 0.001792009 | 0.00335478 | 1 |
| T2DM | monocyte | *GEMIN4* | 0.730283892 | 0.007557489 | 0.011383309 | 1 |
| T2DM | monocyte | *FJX1* | -0.763852396 | 0.006400023 | 0.009839581 | 1 |
| T2DM | monocyte | *CIITA* | -0.36076189 | 0.030579674 | 0.03961558 | 1 |
| T2DM | monocyte | *PLD6* | 1.716326647 | 0.000358107 | 0.001066013 | 1 |
| T2DM | monocyte | *ZBTB42* | -1.737249859 | 0.025353377 | 0.033423502 | 1 |
| T2DM | monocyte | *MAF1* | 3.753410554 | 5.77E-05 | 0.000396862 | 0.362548681 |
| T2DM | monocyte | *PCED1B* | 1.239221534 | 0.000391769 | 0.001135569 | 1 |
| T2DM | monocyte | *FOXS1* | 2.056241576 | 0.011659149 | 0.016701087 | 1 |
| T2DM | monocyte | *CDH5* | 0.589736398 | 0.000818473 | 0.001904418 | 1 |
| T2DM | monocyte | *SERTAD2* | 3.46988241 | 2.48E-05 | 0.000362859 | 0.155984932 |
| T2DM | monocyte | *TIGD5* | 1.893096549 | 0.002083831 | 0.003745296 | 1 |
| T2DM | monocyte | *PDXDC1* | 1.889515823 | 5.66E-05 | 0.000394257 | 0.355600857 |
| T2DM | monocyte | *ZNF154* | 1.189345215 | 0.000527356 | 0.001396582 | 1 |
| T2DM | monocyte | *R3HDM2* | 3.176766115 | 0.000117414 | 0.000541588 | 0.73818454 |
| T2DM | monocyte | *SSC5D* | 0.534939773 | 0.00743221 | 0.011224191 | 1 |
| T2DM | monocyte | *ZADH2* | 2.438033548 | 0.000123858 | 0.000554793 | 0.778693265 |
| T2DM | monocyte | *SEPT1* | 1.322074899 | 0.00104344 | 0.002271506 | 1 |
| T2DM | monocyte | *EXOC3* | 2.423693009 | 0.001418725 | 0.002846959 | 1 |
| T2DM | monocyte | *TDRD6* | 0.80780841 | 0.001898567 | 0.003477941 | 1 |
| T2DM | monocyte | *TDRP* | 1.409615682 | 0.000370577 | 0.001093812 | 1 |
| T2DM | monocyte | *FGD6* | 3.560757457 | 0.001841299 | 0.003406162 | 1 |
| T2DM | monocyte | *HCLS1* | -0.389932216 | 0.024884473 | 0.032860466 | 1 |
| T2DM | monocyte | *ZNF609* | 2.635395287 | 5.96E-05 | 0.000399203 | 0.374780563 |
| T2DM | monocyte | *CCDC66* | 1.721185738 | 2.46E-05 | 0.000362859 | 0.154456791 |
| T2DM | monocyte | *MCFD2* | 2.935032864 | 3.72E-05 | 0.000366571 | 0.233913279 |
| T2DM | monocyte | *CYP8B1* | 2.472464663 | 0.005253153 | 0.008266977 | 1 |
| T2DM | monocyte | *MB21D2* | 1.428256556 | 0.000417883 | 0.00118535 | 1 |
| T2DM | monocyte | *SSTR2* | 1.417688397 | 0.001778779 | 0.00333926 | 1 |
| T2DM | monocyte | *ZNF594* | 1.401334696 | 0.000540624 | 0.001420353 | 1 |

| | | | | | | |
|---|---|---|---|---|---|---|
| T2DM | monocyte | *PCGF5* | 3.218894807 | 3.87E-05 | 0.00036941 | 0.243099633 |
| T2DM | monocyte | *SHISA2* | 20.12374769 | 0.001541481 | 0.003014399 | 1 |
| T2DM | monocyte | *CHST13* | 3.458230169 | 0.031721284 | 0.04095948 | 1 |
| T2DM | monocyte | *SRSF8* | 1.169717698 | 0.001021995 | 0.002238775 | 1 |
| T2DM | monocyte | *ZDHHC20* | 3.003687786 | 0.002437034 | 0.004245396 | 1 |
| T2DM | monocyte | *OR51E1* | 0.922401778 | 0.021909088 | 0.029348738 | 1 |
| T2DM | monocyte | *PSMG4* | 1.813521223 | 0.000317822 | 0.000988691 | 1 |
| T2DM | monocyte | *CAPS2* | 1.452496867 | 0.000145818 | 0.000608331 | 0.916755149 |
| T2DM | monocyte | *KCTD2* | 3.052053346 | 6.55E-05 | 0.000417788 | 0.411938807 |
| T2DM | monocyte | *D2HGDH* | 2.099424555 | 4.40E-05 | 0.000378997 | 0.276897809 |
| T2DM | monocyte | *CMTR2* | 1.967258239 | 0.000153354 | 0.000625657 | 0.964136869 |
| T2DM | monocyte | *LRRC57* | 5.054551639 | 0.000202722 | 0.000735545 | 1 |
| T2DM | monocyte | *MRPL14* | 2.301879033 | 0.006026611 | 0.00933464 | 1 |
| T2DM | monocyte | *BBS12* | 1.235842104 | 0.001875355 | 0.003447473 | 1 |
| T2DM | monocyte | *ZFP82* | 2.439052915 | 5.51E-05 | 0.000389085 | 0.346285854 |
| T2DM | monocyte | *LSMEM1* | 1.153740281 | 0.001167472 | 0.002461722 | 1 |
| T2DM | monocyte | *AEN* | 2.062664979 | 0.001663811 | 0.003184287 | 1 |
| T2DM | monocyte | *TRAPPC5* | -0.962959581 | 0.008723541 | 0.01291684 | 1 |
| T2DM | monocyte | *RPH3AL* | 1.039410628 | 0.00045382 | 0.001253938 | 1 |
| T2DM | monocyte | *ANKRD34A* | 1.247062732 | 0.014352755 | 0.020074699 | 1 |
| T2DM | monocyte | *SLC26A11* | 0.996822023 | 0.000661398 | 0.001633259 | 1 |
| T2DM | monocyte | *HIGD1A* | 1.027263494 | 0.005945234 | 0.009224503 | 1 |
| T2DM | monocyte | *F2R* | 0.936686569 | 0.007253644 | 0.010986074 | 1 |
| T2DM | monocyte | *HIST3H2A* | 1.666672659 | 0.007735781 | 0.011612907 | 1 |
| T2DM | monocyte | *POLR2A* | 3.199757619 | 0.000518248 | 0.00138236 | 1 |
| T2DM | monocyte | *TMEM132C* | 0.701448516 | 0.000355755 | 0.001061525 | 1 |
| T2DM | monocyte | *SLC25A41* | -2.522579484 | 0.012444397 | 0.017668908 | 1 |
| T2DM | monocyte | *TMEM136* | 0.863187192 | 0.000939191 | 0.002099819 | 1 |
| T2DM | monocyte | *ZNF322* | 1.756831209 | 0.018921742 | 0.025654732 | 1 |
| T2DM | monocyte | *CCL13* | 1.189673851 | 0.012111922 | 0.017263127 | 1 |
| T2DM | monocyte | *DDX60L* | 2.327255394 | 0.015455163 | 0.02142121 | 1 |
| T2DM | monocyte | *ACBD4* | 1.41229801 | 0.000417092 | 0.001184932 | 1 |
| T2DM | monocyte | *SETD2* | 2.734637879 | 6.56E-06 | 0.000332508 | 0.041246466 |
| T2DM | monocyte | *MRPS23* | 1.423943165 | 0.000519987 | 0.001385238 | 1 |
| T2DM | monocyte | *GPR135* | 1.011334088 | 0.00125167 | 0.002585282 | 1 |
| T2DM | monocyte | *ZFP41* | 0.868308965 | 0.026876077 | 0.035231422 | 1 |
| T2DM | monocyte | *ATG9B* | 1.752268252 | 0.001691201 | 0.003223856 | 1 |
| T2DM | monocyte | *HKR1* | 1.850838832 | 0.000185275 | 0.0007017 | 1 |
| T2DM | monocyte | *ZBTB20* | 1.599456077 | 3.36E-05 | 0.000365043 | 0.211297139 |
| T2DM | monocyte | *ODF3L2* | 2.320142703 | 0.010845314 | 0.015643823 | 1 |
| T2DM | monocyte | *RELL1* | 2.516594769 | 3.52E-05 | 0.000365043 | 0.221266897 |
| T2DM | monocyte | *RFX7* | 3.413096944 | 3.04E-05 | 0.000362859 | 0.191120301 |
| T2DM | monocyte | *ADO* | 8.431772617 | 7.02E-05 | 0.000427286 | 0.441245112 |
| T2DM | monocyte | *SNRPE* | 3.557239959 | 0.00019909 | 0.000726217 | 1 |
| T2DM | monocyte | *CHST15* | -0.257566656 | 0.013489742 | 0.018988849 | 1 |
| T2DM | monocyte | *IDH2* | 2.065509657 | 0.000325277 | 0.001002952 | 1 |
| T2DM | monocyte | *TNRC18* | 2.063471991 | 0.022702707 | 0.030337254 | 1 |
| T2DM | monocyte | *FAM181B* | 3.582296795 | 0.005882085 | 0.009133969 | 1 |
| T2DM | monocyte | *NOP10* | 1.195038781 | 0.006747739 | 0.010304357 | 1 |
| T2DM | monocyte | *FAM89A* | 0.684131372 | 0.009414067 | 0.013835026 | 1 |

| | | | | | | |
|---|---|---|---|---|---|---|
| T2DM | monocyte | *TDRKH* | 1.161582148 | 0.00906792 | 0.013382633 | 1 |
| T2DM | monocyte | *ZNF708* | 2.568656217 | 0.001700571 | 0.003233153 | 1 |
| T2DM | monocyte | *IST1* | 2.704520718 | 0.000217886 | 0.000768284 | 1 |
| T2DM | monocyte | *ERCC6L2* | 2.0049115 | 0.000110652 | 0.000523558 | 0.695670528 |
| T2DM | monocyte | *MRPL41* | 2.69195506 | 0.004287811 | 0.006913944 | 1 |
| T2DM | monocyte | *HHIPL1* | 1.01375346 | 0.002435124 | 0.004243244 | 1 |
| T2DM | monocyte | *FAM153B* | -1.111111863 | 0.029886959 | 0.038814152 | 1 |
| T2DM | monocyte | *BACE2* | 1.088879653 | 0.000763885 | 0.001813651 | 1 |
| T2DM | monocyte | *SYNM* | 1.127781794 | 0.000863955 | 0.001980904 | 1 |
| T2DM | monocyte | *IZUMO1* | 1.169795015 | 0.032779299 | 0.042204271 | 1 |
| T2DM | monocyte | *C8orf33* | 2.144953823 | 0.000252603 | 0.000843844 | 1 |
| T2DM | monocyte | *FBXL6* | 1.936559557 | 0.000543059 | 0.001426153 | 1 |
| T2DM | monocyte | *KBTBD3* | 5.580014546 | 1.88E-05 | 0.000362859 | 0.118449136 |
| T2DM | monocyte | *CLN8* | 2.210594849 | 0.000969933 | 0.002152556 | 1 |
| T2DM | monocyte | *CACNB4* | 0.695469259 | 0.002969393 | 0.005019783 | 1 |
| T2DM | monocyte | *TRAPPC6B* | 2.626690439 | 0.00020721 | 0.000745268 | 1 |
| T2DM | monocyte | *NPLOC4* | 1.190062151 | 0.003256088 | 0.005446627 | 1 |
| T2DM | monocyte | *TSHZ2* | 0.724208717 | 0.000543845 | 0.001427623 | 1 |
| T2DM | monocyte | *CAPN12* | 2.340782419 | 5.56E-05 | 0.000390932 | 0.349376168 |
| T2DM | monocyte | *GLRX5* | 4.136317121 | 0.000306261 | 0.000965629 | 1 |
| T2DM | monocyte | *CAV3* | 0.591067459 | 0.008064163 | 0.012051199 | 1 |
| T2DM | monocyte | *MXRA7* | 0.909792008 | 0.01985123 | 0.026845489 | 1 |
| T2DM | monocyte | *MFSD5* | 3.398145321 | 0.004311496 | 0.006943232 | 1 |
| T2DM | monocyte | *ADI1* | 2.149280515 | 0.000242117 | 0.000820147 | 1 |
| T2DM | monocyte | *SPNS3* | -1.271803523 | 0.027066637 | 0.03544469 | 1 |
| T2DM | monocyte | *CLEC4G* | 8.909602097 | 0.001236592 | 0.002563289 | 1 |
| T2DM | monocyte | *SATB1* | 1.692273928 | 0.00051109 | 0.001368977 | 1 |
| T2DM | monocyte | *TRAK1* | 2.799193322 | 2.53E-05 | 0.000362859 | 0.159306231 |
| T2DM | monocyte | *SKA2* | 4.805181108 | 5.25E-05 | 0.000385235 | 0.330172727 |
| T2DM | monocyte | *TTC3* | 1.158070992 | 5.04E-06 | 0.000298974 | 0.031712392 |
| T2DM | monocyte | *KCNB2* | 0.406276701 | 0.008726106 | 0.012917596 | 1 |
| T2DM | monocyte | *TSKU* | 0.574805484 | 0.001651981 | 0.003169364 | 1 |
| T2DM | monocyte | *GRID1* | 0.547838193 | 0.007341791 | 0.011095635 | 1 |
| T2DM | monocyte | *ACBD3* | 2.027151529 | 8.02E-05 | 0.000443644 | 0.504511379 |
| T2DM | monocyte | *PLCXD3* | 1.304999959 | 0.001729967 | 0.003271069 | 1 |
| T2DM | monocyte | *GPIHBP1* | 0.535923044 | 0.032630345 | 0.042029703 | 1 |
| T2DM | monocyte | *COL18A1* | 0.765549926 | 0.00192889 | 0.003516205 | 1 |
| T2DM | monocyte | *SLC25A18* | 0.759804974 | 0.01784467 | 0.024373113 | 1 |
| T2DM | monocyte | *ZNF721* | 1.940243952 | 3.67E-05 | 0.000365515 | 0.230449635 |
| T2DM | monocyte | *LENG9* | 2.496458042 | 0.000409621 | 0.001172717 | 1 |
| T2DM | monocyte | *CEP63* | 0.964341489 | 0.010649932 | 0.015385138 | 1 |
| T2DM | monocyte | *HMGN4* | 2.935398284 | 0.006455712 | 0.00991137 | 1 |
| T2DM | monocyte | *ZNF320* | 1.148048737 | 0.000122717 | 0.000553734 | 0.771521556 |
| T2DM | monocyte | *C12orf60* | 1.462132366 | 0.000690739 | 0.001682556 | 1 |
| T2DM | monocyte | *LYSMD4* | 1.12571446 | 0.027066957 | 0.03544469 | 1 |
| T2DM | monocyte | *GAS6* | 1.376772503 | 0.002814413 | 0.004789988 | 1 |
| T2DM | monocyte | *BEGAIN* | -1.199873928 | 0.014313944 | 0.020029327 | 1 |
| T2DM | monocyte | *ARHGEF37* | 2.527593913 | 5.49E-05 | 0.000388828 | 0.34527963 |
| T2DM | monocyte | *FAM43B* | 2.696858458 | 0.002568259 | 0.004446886 | 1 |
| T2DM | monocyte | *CSMD1* | 0.930125431 | 0.001126721 | 0.002401253 | 1 |

| | | | | | | |
|---|---|---|---|---|---|---|
| T2DM | monocyte | *PTGDR2* | 26.49245691 | 0.002239919 | 0.003956834 | 1 |
| T2DM | monocyte | *FANCF* | 1.432205463 | 0.00118199 | 0.00248368 | 1 |
| T2DM | monocyte | *C2CD4C* | 2.213862016 | 0.001186448 | 0.002489935 | 1 |
| T2DM | monocyte | *RUVBL2* | 3.831111646 | 0.000126198 | 0.00055907 | 0.793407727 |
| T2DM | monocyte | *CTNNA3* | 0.46448546 | 0.00221026 | 0.003914339 | 1 |
| T2DM | monocyte | *PTTG1IP* | 2.70959437 | 0.000739531 | 0.001771212 | 1 |
| T2DM | monocyte | *PLGLB1* | 2.006266987 | 0.024312864 | 0.032220695 | 1 |
| T2DM | monocyte | *CCBE1* | 0.519238483 | 0.000916787 | 0.002062936 | 1 |
| T2DM | monocyte | *CCDC125* | 1.693072705 | 0.000145379 | 0.000607712 | 0.913998727 |
| T2DM | monocyte | *FHL3* | 1.220116019 | 0.006942229 | 0.01055521 | 1 |
| T2DM | monocyte | *C19orf71* | 0.996865941 | 0.003042324 | 0.005130657 | 1 |
| T2DM | monocyte | *LRIT3* | 1.981216931 | 0.002738258 | 0.004685746 | 1 |
| T2DM | monocyte | *NPIPA1* | 0.867399337 | 0.002264632 | 0.00398928 | 1 |
| T2DM | monocyte | *SF3A3* | 3.751134724 | 0.000389245 | 0.00113086 | 1 |
| T2DM | monocyte | *GRIN2A* | 0.861496677 | 0.002352744 | 0.004120252 | 1 |
| T2DM | monocyte | *URAD* | 2.989591379 | 0.017941615 | 0.024479397 | 1 |
| T2DM | monocyte | *GPR132* | -0.701959051 | 0.026713985 | 0.035048169 | 1 |
| T2DM | monocyte | *MX2* | -0.463140555 | 0.023324581 | 0.031054986 | 1 |
| T2DM | monocyte | *KLRC4* | -0.659612752 | 0.0045881 | 0.007328604 | 1 |
| T2DM | monocyte | *ACSM5* | 0.61346493 | 0.000664703 | 0.00163686 | 1 |
| T2DM | monocyte | *AC131971.1* | 3.638367666 | 0.003727646 | 0.00610623 | 1 |
| T2DM | monocyte | *SERHL2* | 0.928324101 | 0.001122764 | 0.00239631 | 1 |
| T2DM | monocyte | *TANGO2* | 3.823516477 | 7.29E-05 | 0.000431726 | 0.458630734 |
| T2DM | monocyte | *SFXN4* | 1.864066681 | 0.00096064 | 0.002136547 | 1 |
| T2DM | monocyte | *FAM167B* | 3.259507647 | 0.017941954 | 0.024479397 | 1 |
| T2DM | monocyte | *RP1L1* | 3.426985101 | 2.27E-06 | 0.000190588 | 0.014294128 |
| T2DM | monocyte | *NDUFB1* | 2.979809463 | 0.000570677 | 0.001474055 | 1 |
| T2DM | monocyte | *FAM19A1* | 1.104585912 | 0.000532054 | 0.001406063 | 1 |
| T2DM | monocyte | *BMP8A* | 6.93760601 | 2.70E-05 | 0.000362859 | 0.169805065 |
| T2DM | monocyte | *AIFM3* | -1.000849798 | 0.027105462 | 0.035474399 | 1 |
| T2DM | monocyte | *KCTD16* | 0.935456854 | 0.000487706 | 0.00132564 | 1 |
| T2DM | monocyte | *B3GALT5* | 2.455426293 | 0.016905864 | 0.023211873 | 1 |
| T2DM | monocyte | *ZNF703* | -0.503637564 | 0.002093954 | 0.00376134 | 1 |
| T2DM | monocyte | *KCTD8* | 0.512388736 | 0.000842186 | 0.001945197 | 1 |
| T2DM | monocyte | *C9orf66* | -1.646396836 | 0.026338756 | 0.034599197 | 1 |
| T2DM | monocyte | *TUBA8* | 2.814331017 | 3.78E-05 | 0.00036941 | 0.237903901 |
| T2DM | monocyte | *FAM162B* | 0.97652797 | 0.002161687 | 0.003854376 | 1 |
| T2DM | monocyte | *BTBD9* | 2.941467368 | 1.94E-05 | 0.000362859 | 0.122089751 |
| T2DM | monocyte | *NUDT14* | 1.507380946 | 0.004016805 | 0.006525492 | 1 |
| T2DM | monocyte | *ANKRD45* | 9.397108268 | 0.000173277 | 0.000671081 | 1 |
| T2DM | monocyte | *MAATS1* | 0.861902065 | 0.001009842 | 0.002219887 | 1 |
| T2DM | monocyte | *GPR39* | 1.221592293 | 0.001079558 | 0.002328364 | 1 |
| T2DM | monocyte | *NPIPA7* | -3.098473967 | 0.009599229 | 0.014070961 | 1 |
| T2DM | monocyte | *TTC32* | 1.270724575 | 0.000849904 | 0.001955271 | 1 |
| T2DM | monocyte | *PP2D1* | 1.153599081 | 0.000268029 | 0.000880406 | 1 |
| T2DM | monocyte | *TMEM236* | 1.090522705 | 0.001848656 | 0.003416373 | 1 |
| T2DM | monocyte | *TBX1* | 1.600342591 | 0.000695368 | 0.001690555 | 1 |
| T2DM | monocyte | *UQCR10* | 3.518366154 | 0.000225578 | 0.000783249 | 1 |
| T2DM | monocyte | *EIF3C* | 2.270665612 | 6.20E-06 | 0.000322038 | 0.038966617 |
| T2DM | monocyte | *NIPSNAP1* | 2.271797755 | 0.00044618 | 0.00124066 | 1 |

| | | | | | | |
|---|---|---|---|---|---|---|
| T2DM | monocyte | *CNTN2* | 1.512677317 | 0.001471421 | 0.002919162 | 1 |
| T2DM | monocyte | *CRELD2* | 4.600191362 | 0.003362718 | 0.005600374 | 1 |
| T2DM | monocyte | *UBE2F* | 2.221529891 | 0.024807311 | 0.032780259 | 1 |
| T2DM | monocyte | *GOLGA6L4* | 1.257470526 | 0.002402981 | 0.004196539 | 1 |
| T2DM | monocyte | *PCDH9* | 0.465718906 | 0.005145338 | 0.008119664 | 1 |
| T2DM | monocyte | *ACOT1* | 0.883289305 | 0.006836056 | 0.010421505 | 1 |
| T2DM | monocyte | *OAF* | 1.288471791 | 0.003687672 | 0.006051786 | 1 |
| T2DM | monocyte | *TM2D3* | 3.327930786 | 0.003959845 | 0.006447953 | 1 |
| T2DM | monocyte | *CLECL1* | 1.240727228 | 0.007211303 | 0.010924689 | 1 |
| T2DM | monocyte | *MROH7* | 3.383580593 | 9.74E-05 | 0.000485446 | 0.612568067 |
| T2DM | monocyte | *CSF1* | 1.342053542 | 4.37E-05 | 0.000378997 | 0.274673438 |
| T2DM | monocyte | *PLA2G6* | 1.132439079 | 8.16E-05 | 0.000445905 | 0.512929462 |
| T2DM | monocyte | *SS18L1* | 4.091418285 | 0.000272806 | 0.000891441 | 1 |
| T2DM | monocyte | *THAP7* | 6.775525102 | 0.00014707 | 0.000610432 | 0.9246321 |
| T2DM | monocyte | *WDR27* | 1.504733294 | 0.000109125 | 0.000518963 | 0.686068967 |
| T2DM | monocyte | *TXNRD2* | 4.201418024 | 0.001820339 | 0.003386941 | 1 |
| T2DM | monocyte | *PROS1* | 0.98282572 | 0.000582349 | 0.001490729 | 1 |
| T2DM | monocyte | *HDDC3* | 2.182169571 | 0.002784963 | 0.004748864 | 1 |
| T2DM | monocyte | *ZFP1* | 1.461212049 | 0.005405354 | 0.008481024 | 1 |
| T2DM | monocyte | *TMEM173* | 2.38406673 | 0.000105106 | 0.000507835 | 0.660799406 |
| T2DM | monocyte | *NELL2* | 1.035161358 | 0.001518271 | 0.002978363 | 1 |
| T2DM | monocyte | *KRBA2* | 1.267246024 | 0.019843656 | 0.026841021 | 1 |
| T2DM | monocyte | *ZNF93* | -0.546774478 | 0.021480302 | 0.028831482 | 1 |
| T2DM | monocyte | *SEPT5* | 2.743875307 | 0.001511544 | 0.002973428 | 1 |
| T2DM | monocyte | *ATL3* | 2.19284285 | 0.000569011 | 0.001470353 | 1 |
| T2DM | monocyte | *NDUFA12* | 4.806162422 | 0.000738865 | 0.001770966 | 1 |
| T2DM | monocyte | *UBE2G2* | 1.566220831 | 0.000327357 | 0.001006043 | 1 |
| T2DM | monocyte | *OSBP2* | 1.370951101 | 0.000358788 | 0.001067029 | 1 |
| T2DM | monocyte | *TUSC5* | 1.055580753 | 0.000158919 | 0.000639235 | 0.999123909 |
| T2DM | monocyte | *TMED9* | 2.775784212 | 2.45E-05 | 0.000362859 | 0.154209246 |
| T2DM | monocyte | *H1FX* | 2.827109558 | 0.000335477 | 0.00102039 | 1 |
| T2DM | monocyte | *RBM43* | 1.09717465 | 0.001260108 | 0.002600033 | 1 |
| T2DM | monocyte | *ZFP90* | 2.133146968 | 6.09E-05 | 0.000403663 | 0.383075935 |
| T2DM | monocyte | *FAM227A* | 0.796420655 | 0.011878915 | 0.016996527 | 1 |
| T2DM | monocyte | *NOC4L* | 1.063265161 | 0.023769726 | 0.031580784 | 1 |
| T2DM | monocyte | *NDUFA6* | 1.051161001 | 5.12E-05 | 0.000384204 | 0.321781978 |
| T2DM | monocyte | *SORCS2* | 0.784121831 | 0.001372673 | 0.002774917 | 1 |
| T2DM | monocyte | *TMEM106A* | 3.988207287 | 0.00146549 | 0.00291291 | 1 |
| T2DM | monocyte | *SIVA1* | 1.439371601 | 0.001227716 | 0.002554388 | 1 |
| T2DM | monocyte | *UBOX5* | 1.685593352 | 0.001200912 | 0.002510852 | 1 |
| T2DM | monocyte | *MAFF* | 0.779625532 | 0.002130005 | 0.003813025 | 1 |
| T2DM | monocyte | *BRF1* | 4.080431137 | 2.38E-05 | 0.000362859 | 0.149781337 |
| T2DM | monocyte | *FLRT2* | 0.739113498 | 0.002199635 | 0.003901017 | 1 |
| T2DM | monocyte | *ADSSL1* | 1.292515365 | 0.001357572 | 0.002752355 | 1 |
| T2DM | monocyte | *DDX51* | 1.318045145 | 0.017861167 | 0.024385051 | 1 |
| T2DM | monocyte | *SIGIRR* | 2.021395743 | 0.001336495 | 0.002717512 | 1 |
| T2DM | monocyte | *IFITM2* | 2.357817377 | 0.00022034 | 0.000773036 | 1 |
| T2DM | monocyte | *GP1BA* | 1.746337028 | 0.000217029 | 0.000767755 | 1 |
| T2DM | monocyte | *KIAA0825* | 2.282420783 | 8.63E-05 | 0.000459567 | 0.542367004 |
| T2DM | monocyte | *KLHL33* | 1.374213056 | 0.002907306 | 0.004932065 | 1 |

| | | | | | | |
|---|---|---|---|---|---|---|
| T2DM | monocyte | *CCDC137* | 1.980956569 | 9.15E-05 | 0.000469636 | 0.575318085 |
| T2DM | monocyte | *CDK10* | 1.815577008 | 0.001151665 | 0.002436759 | 1 |
| T2DM | monocyte | *TMEM105* | 15.27202154 | 0.003997317 | 0.006502234 | 1 |
| T2DM | monocyte | *TCN2* | 0.768786831 | 0.032628395 | 0.042029703 | 1 |
| T2DM | monocyte | *GAS2L1* | 2.249394598 | 0.001600946 | 0.003106528 | 1 |
| T2DM | monocyte | *ATP6V0A2* | 1.969963391 | 0.000230799 | 0.000795088 | 1 |
| T2DM | monocyte | *HS6ST3* | 0.774482137 | 0.002102026 | 0.003773683 | 1 |
| T2DM | monocyte | *TNFAIP8L1* | 5.817197287 | 0.000307273 | 0.000967849 | 1 |
| T2DM | monocyte | *MAPK11* | 1.347046799 | 0.002702227 | 0.004644314 | 1 |
| T2DM | monocyte | *FAM174B* | 1.59670116 | 0.000209487 | 0.000750879 | 1 |
| T2DM | monocyte | *MUC1* | 2.06847832 | 0.000640898 | 0.001593879 | 1 |
| T2DM | monocyte | *SV2B* | 1.260470633 | 0.001365941 | 0.002767539 | 1 |
| T2DM | monocyte | *PDE6G* | 3.477954158 | 0.001442723 | 0.00288407 | 1 |
| T2DM | monocyte | *TLCD2* | 1.307968842 | 0.000884036 | 0.00201134 | 1 |
| T2DM | monocyte | *LSAMP* | 0.571618787 | 0.001236063 | 0.002563289 | 1 |
| T2DM | monocyte | *AHNAK2* | 0.868432155 | 0.001279806 | 0.002630317 | 1 |
| T2DM | monocyte | *SP1* | 2.521031672 | 0.000153327 | 0.000625657 | 0.96396414 |
| T2DM | monocyte | *PDIA2* | 2.131429162 | 0.000431633 | 0.001215259 | 1 |
| T2DM | monocyte | *PCGF3* | 2.796123594 | 4.06E-05 | 0.00037107 | 0.255213369 |
| T2DM | monocyte | *PSMD13* | 3.539944369 | 0.000683997 | 0.001669367 | 1 |
| T2DM | monocyte | *PBX1* | 0.809880852 | 0.000366725 | 0.00108397 | 1 |
| T2DM | monocyte | *UBE2L3* | 3.330854572 | 7.87E-06 | 0.000355191 | 0.049455077 |
| T2DM | monocyte | *SNAI3* | 3.152012535 | 0.00044593 | 0.001240515 | 1 |
| T2DM | monocyte | *C6orf201* | 0.629163295 | 0.015276108 | 0.02121513 | 1 |
| T2DM | monocyte | *MYBL1* | 0.40746263 | 0.034409529 | 0.04416756 | 1 |
| T2DM | monocyte | *C16orf52* | 2.663449522 | 3.04E-05 | 0.000362859 | 0.191334461 |
| T2DM | monocyte | *ADARB2* | 0.761093086 | 0.001800592 | 0.003361444 | 1 |
| T2DM | monocyte | *SRL* | 0.795508595 | 0.011646859 | 0.016687283 | 1 |
| T2DM | monocyte | *KCNIP4* | 0.442950252 | 0.003979894 | 0.006477244 | 1 |
| T2DM | monocyte | *SLC52A2* | 1.694461006 | 0.000562792 | 0.00146183 | 1 |
| T2DM | monocyte | *ARL17A* | 2.467489203 | 0.003243568 | 0.005432155 | 1 |
| T2DM | monocyte | *ZNF829* | 1.392853018 | 0.000926432 | 0.002078686 | 1 |
| T2DM | monocyte | *IFITM1* | 2.548100521 | 0.004068197 | 0.006600662 | 1 |
| T2DM | monocyte | *FFAR3* | -3.708455746 | 0.007581035 | 0.011414856 | 1 |
| T2DM | monocyte | *POMK* | 3.329933135 | 2.92E-05 | 0.000362859 | 0.183561646 |
| T2DM | monocyte | *LINC00839* | 3.664717743 | 2.60E-06 | 0.000207301 | 0.016376806 |
| T2DM | monocyte | *PTCH1* | 0.91059704 | 0.002184447 | 0.00388065 | 1 |
| T2DM | monocyte | *RTN4RL1* | 0.596936937 | 0.00179941 | 0.00336093 | 1 |
| T2DM | monocyte | *RASA3* | 1.543053907 | 0.017728542 | 0.024225026 | 1 |
| T2DM | monocyte | *ZNF566* | 1.953175406 | 0.001040016 | 0.002266405 | 1 |
| T2DM | monocyte | *ZNF529* | 2.374619377 | 3.29E-05 | 0.000365043 | 0.206741516 |
| T2DM | monocyte | *ZNF284* | 4.805294578 | 0.000218853 | 0.000769966 | 1 |
| T2DM | monocyte | *GSAP* | 3.589127902 | 5.28E-05 | 0.000385235 | 0.331856774 |
| T2DM | monocyte | *WWOX* | 1.643893799 | 3.02E-05 | 0.000362859 | 0.189791173 |
| T2DM | monocyte | *CYP4Z1* | 1.811912051 | 0.000218398 | 0.000769226 | 1 |
| T2DM | monocyte | *CCDC84* | 2.322973363 | 4.56E-05 | 0.000379941 | 0.286667446 |
| T2DM | monocyte | *POLR1D* | 3.845239952 | 1.42E-05 | 0.000362859 | 0.089306854 |
| T2DM | monocyte | *FFAR4* | 0.465298567 | 0.011356684 | 0.016323611 | 1 |
| T2DM | monocyte | *BPIFB4* | 57.66031056 | 0.00037167 | 0.00109424 | 1 |
| T2DM | monocyte | *CYP4F12* | 0.962362591 | 0.002595687 | 0.004489432 | 1 |

| | | | | | | |
|---|---|---|---|---|---|---|
| T2DM | monocyte | *KLHL32* | 1.06925162 | 0.001998756 | 0.003619292 | 1 |
| T2DM | monocyte | *NBPF12* | 2.487700303 | 0.000203046 | 0.000735552 | 1 |
| T2DM | monocyte | *GPAT2* | 2.081799387 | 0.009352373 | 0.013754004 | 1 |
| T2DM | monocyte | *TOR3A* | 3.574546371 | 4.51E-05 | 0.000379941 | 0.283856281 |
| T2DM | monocyte | *CYP4X1* | 1.246928942 | 0.000168479 | 0.000659148 | 1 |
| T2DM | monocyte | *CCDC30* | 2.126965431 | 0.000245057 | 0.000826098 | 1 |
| T2DM | monocyte | *GLDN* | 0.625145809 | 0.002171193 | 0.003865842 | 1 |
| T2DM | monocyte | *FCAR* | -0.174274126 | 0.008411306 | 0.012507541 | 1 |
| T2DM | monocyte | *KPNA4* | 3.437659722 | 0.000629619 | 0.001574064 | 1 |
| T2DM | monocyte | *ZNF197* | 1.891134239 | 0.000816877 | 0.001902817 | 1 |
| T2DM | monocyte | *RPS23* | 2.131788122 | 0.001277611 | 0.002627525 | 1 |
| T2DM | monocyte | *PCLO* | 0.596424393 | 0.000741454 | 0.001774465 | 1 |
| T2DM | monocyte | *RGS7BP* | 0.677299007 | 0.027105018 | 0.035474399 | 1 |
| T2DM | monocyte | *ZNF396* | -0.740012239 | 0.004162309 | 0.006739231 | 1 |
| T2DM | monocyte | *TMEM222* | 1.937520947 | 0.00107795 | 0.002327292 | 1 |
| T2DM | monocyte | *SEPT10* | 1.681965969 | 0.000123557 | 0.000554584 | 0.776801526 |
| T2DM | monocyte | *SMYD4* | 1.769175625 | 0.000582143 | 0.001490729 | 1 |
| T2DM | monocyte | *NF2* | 3.487729091 | 5.20E-05 | 0.000384775 | 0.326979376 |
| T2DM | monocyte | *MIR22HG* | 1.047744353 | 0.000983322 | 0.002174515 | 1 |
| T2DM | monocyte | *PDE2A* | 0.744326425 | 0.001775791 | 0.003335643 | 1 |
| T2DM | monocyte | *C17orf58* | 1.811123123 | 0.000440178 | 0.001228968 | 1 |
| T2DM | monocyte | *CYP27C1* | 0.678656364 | 0.003380866 | 0.005626126 | 1 |
| T2DM | monocyte | *BCR* | 1.208128659 | 0.00074863 | 0.0017862 | 1 |
| T2DM | monocyte | *ZSCAN30* | 2.094373737 | 0.000104044 | 0.000504723 | 0.654121584 |
| T2DM | monocyte | *KRT16* | 1.188132903 | 0.023050162 | 0.030741699 | 1 |
| T2DM | monocyte | *TMEM17* | 1.213629177 | 0.001089365 | 0.002343763 | 1 |
| T2DM | monocyte | *TNFRSF18* | 3.645890478 | 0.000902346 | 0.002039198 | 1 |
| T2DM | monocyte | *RTN4RL2* | 2.286134797 | 0.001853364 | 0.00342105 | 1 |
| T2DM | monocyte | *TMEM232* | 1.210746538 | 0.00036175 | 0.001073298 | 1 |
| T2DM | monocyte | *EFCAB6* | 1.485537579 | 9.38E-05 | 0.000477414 | 0.589504052 |
| T2DM | monocyte | *KANK3* | 0.944452497 | 0.001123259 | 0.00239631 | 1 |
| T2DM | monocyte | *RHD* | 1.622777811 | 0.000152926 | 0.00062472 | 0.961443439 |
| T2DM | monocyte | *CYP4A11* | 1.49712746 | 0.001793192 | 0.003355298 | 1 |
| T2DM | monocyte | *TMEM216* | 2.730869066 | 2.01E-05 | 0.000362859 | 0.12658467 |
| T2DM | monocyte | *RPS19BP1* | 2.833964194 | 0.029347252 | 0.038192129 | 1 |
| T2DM | monocyte | *C3orf70* | 0.959576415 | 0.000305741 | 0.000965441 | 1 |
| T2DM | monocyte | *PLCD1* | 1.646899574 | 0.000646544 | 0.001606649 | 1 |
| T2DM | monocyte | *ENTPD5* | 2.579804781 | 3.69E-05 | 0.000366137 | 0.231764463 |
| T2DM | monocyte | *HEATR4* | 1.028947285 | 0.000377778 | 0.001106237 | 1 |
| T2DM | monocyte | *MRPS21* | 3.022788472 | 5.67E-05 | 0.000394331 | 0.356462011 |
| T2DM | monocyte | *ANGPTL5* | 0.984242008 | 0.001162637 | 0.002453676 | 1 |
| T2DM | monocyte | *TSPYL4* | 1.888105554 | 0.000436007 | 0.001223193 | 1 |
| T2DM | monocyte | *MT1X* | 2.171681563 | 0.000241653 | 0.0008199 | 1 |
| T2DM | monocyte | *WDR86* | 0.834403072 | 0.0026745 | 0.004602951 | 1 |
| T2DM | monocyte | *EPOR* | 4.566070818 | 1.79E-05 | 0.000362859 | 0.112789753 |
| T2DM | monocyte | *GJA4* | 1.1652769 | 0.000886236 | 0.002012922 | 1 |
| T2DM | monocyte | *IFT140* | 2.210463207 | 5.02E-05 | 0.000383643 | 0.315553456 |
| T2DM | monocyte | *ZNF385C* | -1.151349758 | 0.025547812 | 0.033651602 | 1 |
| T2DM | monocyte | *DHRS4L2* | -0.81086293 | 0.020756148 | 0.027937038 | 1 |
| T2DM | monocyte | *C2orf88* | 1.53292232 | 0.000192654 | 0.000717543 | 1 |

| | | | | | | |
|---|---|---|---|---|---|---|
| T2DM | monocyte | *KBTBD12* | 0.692645117 | 0.003015254 | 0.005087735 | 1 |
| T2DM | monocyte | *FANCA* | 1.441997526 | 0.028685549 | 0.037408431 | 1 |
| T2DM | monocyte | *SEMA4D* | -0.455205822 | 0.022705155 | 0.030337254 | 1 |
| T2DM | monocyte | *DNAH17* | 2.068817165 | 0.000932048 | 0.002088305 | 1 |
| T2DM | monocyte | *MCRS1* | 3.576766167 | 6.03E-07 | 8.58E-05 | 0.003793663 |
| T2DM | monocyte | *ZFP69* | 2.088175681 | 0.001490144 | 0.002944228 | 1 |
| T2DM | monocyte | *TMEM220* | 1.941042239 | 9.60E-05 | 0.000482554 | 0.603817495 |
| T2DM | monocyte | *CCDC157* | 2.704872685 | 0.000659789 | 0.001631182 | 1 |
| T2DM | monocyte | *GFRAL* | 2.951023259 | 0.000751439 | 0.001791541 | 1 |
| T2DM | monocyte | *LDLRAD2* | 1.185524784 | 0.024300468 | 0.032211057 | 1 |
| T2DM | monocyte | *C2orf66* | 6.39464869 | 0.000709754 | 0.001715579 | 1 |
| T2DM | monocyte | *KLHL17* | 2.797449494 | 0.000811801 | 0.001893131 | 1 |
| T2DM | monocyte | *ZSCAN23* | 0.587686153 | 0.005795675 | 0.009021393 | 1 |
| T2DM | monocyte | *OR7D2* | 3.004448309 | 0.013065045 | 0.018458413 | 1 |
| T2DM | monocyte | *TREML4* | -0.284406881 | 0.00653381 | 0.010014155 | 1 |
| T2DM | monocyte | *C6orf132* | 0.733803377 | 0.002025075 | 0.003661077 | 1 |
| T2DM | monocyte | *MAPK12* | 1.492909301 | 0.004123502 | 0.006681561 | 1 |
| T2DM | monocyte | *FAM166A* | 3.748529241 | 0.001928963 | 0.003516205 | 1 |
| T2DM | monocyte | *ZNF626* | 1.552920991 | 0.000828766 | 0.001920669 | 1 |
| T2DM | monocyte | *SMTNL2* | 5.401975227 | 0.001851351 | 0.003418339 | 1 |
| T2DM | monocyte | *ZC3H6* | 3.163294043 | 4.76E-05 | 0.000379941 | 0.299320457 |
| T2DM | monocyte | *LINC00265* | 3.566506037 | 0.009352257 | 0.013754004 | 1 |
| T2DM | monocyte | *LAMTOR4* | 3.366472588 | 0.000821339 | 0.001910379 | 1 |
| T2DM | monocyte | *PRKAR1B* | 1.123292724 | 0.001396672 | 0.002810396 | 1 |
| T2DM | monocyte | *ZNF793* | 1.012750608 | 0.000193116 | 0.000718174 | 1 |
| T2DM | monocyte | *AGAP4* | 4.283506885 | 0.000305309 | 0.000964562 | 1 |
| T2DM | monocyte | *HYKK* | 1.792458635 | 0.000197472 | 0.000724754 | 1 |
| T2DM | monocyte | *RUFY4* | -0.191441986 | 0.007255317 | 0.010986074 | 1 |
| T2DM | monocyte | *ZNF669* | 2.48254055 | 0.001787052 | 0.003347794 | 1 |
| T2DM | monocyte | *CENPP* | 1.050787278 | 0.004704031 | 0.007500443 | 1 |
| T2DM | monocyte | *ZNF559* | 0.696129631 | 0.007818062 | 0.011714051 | 1 |
| T2DM | monocyte | *JAKMIP3* | 1.234220197 | 0.000715322 | 0.001725722 | 1 |
| T2DM | monocyte | *PDCD1* | 4.633159383 | 0.009362927 | 0.013766306 | 1 |
| T2DM | monocyte | *CERKL* | -0.411342326 | 0.012060974 | 0.017206114 | 1 |
| T2DM | monocyte | *C19orf54* | 1.931255795 | 0.020178326 | 0.027217579 | 1 |
| T2DM | monocyte | *C22orf34* | 1.840518136 | 0.012097872 | 0.017247012 | 1 |
| T2DM | monocyte | *COL25A1* | 0.696164077 | 0.001745284 | 0.003291122 | 1 |
| T2DM | monocyte | *SRSF10* | 2.514907052 | 2.91E-05 | 0.000362859 | 0.182765893 |
| T2DM | monocyte | *NBR1* | 3.969262805 | 0.000454996 | 0.001255734 | 1 |
| T2DM | monocyte | *FAM72B* | 3.564943288 | 0.001227829 | 0.002554388 | 1 |
| T2DM | monocyte | *SUMO2* | 4.945502627 | 5.23E-06 | 0.000304341 | 0.032868794 |
| T2DM | monocyte | *GOLGA8M* | 1.659947304 | 0.000296556 | 0.000946439 | 1 |
| T2DM | monocyte | *S100A16* | 1.676693614 | 0.00038113 | 0.001112943 | 1 |
| T2DM | monocyte | *PARVB* | 1.151143997 | 0.00392063 | 0.006390718 | 1 |
| T2DM | monocyte | *SLC4A5* | 1.481637137 | 0.00255149 | 0.004419069 | 1 |
| T2DM | monocyte | *UROS* | 2.047770978 | 6.20E-05 | 0.000406647 | 0.390003693 |
| T2DM | monocyte | *FAM221A* | 2.104936735 | 0.000416733 | 0.001184448 | 1 |
| T2DM | monocyte | *FSIP2* | 0.903412715 | 0.000491781 | 0.001333243 | 1 |
| T2DM | monocyte | *NOXA1* | 1.736213747 | 0.00040254 | 0.001158461 | 1 |
| T2DM | monocyte | *SKOR1* | -1.699003559 | 0.023528702 | 0.031293622 | 1 |

| | | | | | | |
|---|---|---|---|---|---|---|
| T2DM | monocyte | *TMEM201* | 7.196114407 | 3.00E-07 | 6.09E-05 | 0.001888488 |
| T2DM | monocyte | *ZDHHC11* | 0.914412645 | 0.00362213 | 0.00597281 | 1 |
| T2DM | monocyte | *LINC00910* | 3.140274098 | 0.005065868 | 0.008012355 | 1 |
| T2DM | monocyte | *BEND4* | -0.90036584 | 0.013510717 | 0.01900691 | 1 |
| T2DM | monocyte | *ZNF563* | 2.224153229 | 0.000184287 | 0.0006988 | 1 |
| T2DM | monocyte | *MSL1* | 1.208754897 | 0.023136566 | 0.030837309 | 1 |
| T2DM | monocyte | *FAM196A* | 1.189853498 | 0.000588422 | 0.001500165 | 1 |
| T2DM | monocyte | *C9orf152* | 2.062609287 | 0.001766383 | 0.003319955 | 1 |
| T2DM | monocyte | *NELFB* | 2.736927437 | 8.43E-05 | 0.000454809 | 0.530282196 |
| T2DM | monocyte | *SLC15A5* | 0.764451715 | 0.000785855 | 0.001846979 | 1 |
| T2DM | monocyte | *LIPI* | 1.368486313 | 0.003428374 | 0.005699151 | 1 |
| T2DM | monocyte | *ADAT2* | 1.98103256 | 3.19E-05 | 0.000363519 | 0.200605456 |
| T2DM | monocyte | *ZNF567* | 3.507675944 | 2.66E-05 | 0.000362859 | 0.167434302 |
| T2DM | monocyte | *ALKBH2* | 1.626924216 | 6.29E-05 | 0.000408289 | 0.395398107 |
| T2DM | monocyte | *LITAF* | 2.814787633 | 6.02E-05 | 0.000400332 | 0.378714111 |
| T2DM | monocyte | *VSTM1* | -0.131230368 | 0.00841466 | 0.012509569 | 1 |
| T2DM | monocyte | *SF3B3* | 3.389055854 | 0.003403696 | 0.005661121 | 1 |
| T2DM | monocyte | *PLAC9* | 0.825572157 | 0.004458789 | 0.007143834 | 1 |
| T2DM | monocyte | *CLDN4* | 1.346734977 | 0.016884264 | 0.023192346 | 1 |
| T2DM | monocyte | *ZNF573* | 1.752889301 | 0.000224225 | 0.000780135 | 1 |
| T2DM | monocyte | *FAM47E* | 1.032365399 | 0.002256392 | 0.003976993 | 1 |
| T2DM | monocyte | *ZNF527* | 0.943659797 | 0.002860946 | 0.004863917 | 1 |
| T2DM | monocyte | *S100A13* | 1.411943691 | 0.000208878 | 0.000749656 | 1 |
| T2DM | monocyte | *ZNF33A* | 3.10998378 | 5.27E-05 | 0.000385235 | 0.331477872 |
| T2DM | monocyte | *ZNF600* | 0.966834347 | 0.001115115 | 0.00238217 | 1 |
| T2DM | monocyte | *C15orf61* | 6.861845916 | 1.50E-05 | 0.000362859 | 0.094557744 |
| T2DM | monocyte | *ZKSCAN3* | 3.166109411 | 0.001046247 | 0.002275253 | 1 |
| T2DM | monocyte | *RRP7A* | 2.227837952 | 0.003325831 | 0.005547757 | 1 |
| T2DM | monocyte | *SLC35E2B* | 1.430523967 | 0.003227146 | 0.005406093 | 1 |
| T2DM | monocyte | *HLA-DRB1* | -1.564251845 | 2.95E-08 | 1.33E-05 | 0.000185669 |
| T2DM | monocyte | *AKR1C3* | 0.86858522 | 0.000783629 | 0.001843127 | 1 |
| T2DM | monocyte | *WDSUB1* | 1.354239287 | 7.41E-05 | 0.000432843 | 0.466171701 |
| T2DM | monocyte | *COLCA1* | 1.203949332 | 0.018909608 | 0.025645753 | 1 |
| T2DM | monocyte | *ACADSB* | 1.445096201 | 0.017055651 | 0.023386887 | 1 |
| T2DM | monocyte | *SEMA4A* | -0.564998919 | 0.039167077 | 0.049746144 | 1 |
| T2DM | monocyte | *HRCT1* | 1.316388863 | 0.006328101 | 0.009755952 | 1 |
| T2DM | monocyte | *MPHOSPH8* | 2.710731077 | 2.65E-05 | 0.000362859 | 0.166360235 |
| T2DM | monocyte | *SIRPB2* | -0.162132631 | 0.008695262 | 0.012878001 | 1 |
| T2DM | monocyte | *SRGAP3* | 1.351137248 | 8.74E-05 | 0.000461034 | 0.549552037 |
| T2DM | monocyte | *SUPT5H* | 2.297536239 | 0.000136803 | 0.000584748 | 0.860080043 |
| T2DM | monocyte | *ZNF107* | 1.760191953 | 0.000889811 | 0.002018853 | 1 |
| T2DM | monocyte | *PPIA* | 3.838421249 | 7.30E-05 | 0.000431726 | 0.458647777 |
| T2DM | monocyte | *ZNF471* | 1.537204154 | 0.000537136 | 0.001412959 | 1 |
| T2DM | monocyte | *GTF2IRD2* | 1.841652295 | 5.99E-06 | 0.000322038 | 0.037641048 |
| T2DM | monocyte | *GRM7* | 0.871020746 | 0.010524213 | 0.015233247 | 1 |
| T2DM | monocyte | *SUPT3H* | 2.077408372 | 5.56E-05 | 0.000390932 | 0.34981584 |
| T2DM | monocyte | *GIMAP5* | 2.864563893 | 0.000106798 | 0.00051216 | 0.671441193 |
| T2DM | monocyte | *STK31* | 2.665116125 | 0.000310626 | 0.000973531 | 1 |
| T2DM | monocyte | *ZKSCAN7* | 1.898520251 | 0.00053693 | 0.001412959 | 1 |
| T2DM | monocyte | *ZNF565* | 1.479175545 | 0.000326721 | 0.001005923 | 1 |

| | | | | | | |
|---|---|---|---|---|---|---|
| T2DM | monocyte | *NTNG2* | 4.4046593 | 0.000122615 | 0.000553734 | 0.770883067 |
| T2DM | monocyte | *WDR5* | 2.691443749 | 3.56E-05 | 0.000365043 | 0.223975299 |
| T2DM | monocyte | *LONP1* | 2.413263825 | 0.00118788 | 0.002491892 | 1 |
| T2DM | monocyte | *ASB13* | 4.026605852 | 0.011378357 | 0.016351024 | 1 |
| T2DM | monocyte | *SLC35F1* | 0.943763149 | 0.001122856 | 0.00239631 | 1 |
| T2DM | monocyte | *ZNF34* | 4.38693902 | 1.97E-05 | 0.000362859 | 0.124147563 |
| T2DM | monocyte | *INCA1* | 1.999439087 | 0.000378827 | 0.001107855 | 1 |
| T2DM | monocyte | *ZNF774* | 1.363681773 | 0.001310765 | 0.002685167 | 1 |
| T2DM | monocyte | *EVL* | 2.23313769 | 0.001708613 | 0.003243372 | 1 |
| T2DM | monocyte | *ZNF124* | 2.983185884 | 0.010507012 | 0.015220327 | 1 |
| T2DM | monocyte | *ZNF569* | 1.018035121 | 0.00123487 | 0.002563289 | 1 |
| T2DM | monocyte | *PIK3R4* | 1.9506676 | 0.002148436 | 0.003838368 | 1 |
| T2DM | monocyte | *ZNF775* | -1.647913241 | 0.000917495 | 0.00206379 | 1 |
| T2DM | monocyte | *ZNF605* | 1.165519472 | 0.000629676 | 0.001574064 | 1 |
| T2DM | monocyte | *RFX8* | 0.750109646 | 0.001339477 | 0.002721813 | 1 |
| T2DM | monocyte | *MYL6B* | 1.068020874 | 0.001247105 | 0.002578368 | 1 |
| T2DM | monocyte | *ZNF799* | -1.01872818 | 0.036688346 | 0.04689157 | 1 |
| T2DM | monocyte | *C20orf96* | 2.250146656 | 0.000288746 | 0.000926668 | 1 |
| T2DM | monocyte | *ANAPC7* | 4.229175462 | 0.000208907 | 0.000749656 | 1 |
| T2DM | monocyte | *C1orf186* | 1.111864542 | 0.001638339 | 0.003155126 | 1 |
| T2DM | monocyte | *SULF2* | -0.37635802 | 0.035489043 | 0.045441877 | 1 |
| T2DM | monocyte | *XRCC2* | 7.335842712 | 9.40E-06 | 0.000362859 | 0.059112258 |
| T2DM | monocyte | *MYO6* | 0.867674531 | 0.00439039 | 0.007057627 | 1 |
| T2DM | monocyte | *HDAC2* | 3.986825041 | 0.000725876 | 0.001745823 | 1 |
| T2DM | monocyte | *ADH1B* | 0.64611611 | 0.000596944 | 0.001517586 | 1 |
| T2DM | monocyte | *HRH1* | 0.782872368 | 0.000659528 | 0.001631176 | 1 |
| T2DM | monocyte | *TRAPPC4* | 1.651111291 | 0.001311899 | 0.002686616 | 1 |
| T2DM | monocyte | *TECPR2* | 3.158817304 | 2.73E-05 | 0.000362859 | 0.171821584 |
| T2DM | monocyte | *ZFP62* | 1.965455137 | 0.000321789 | 0.000997087 | 1 |
| T2DM | monocyte | *ERI2* | 1.218449184 | 0.000262796 | 0.000866941 | 1 |
| T2DM | monocyte | *TOMM7* | 3.121927082 | 4.04E-05 | 0.00037107 | 0.254081622 |
| T2DM | monocyte | *TRPV1* | 3.452051943 | 1.76E-06 | 0.000169734 | 0.011038739 |
| T2DM | monocyte | *ZNF431* | 2.442258853 | 0.000105316 | 0.000507835 | 0.662120702 |
| T2DM | monocyte | *NF1* | 2.142919515 | 2.69E-05 | 0.000362859 | 0.169115931 |
| T2DM | monocyte | *HLA-DQA1* | -0.791774003 | 5.10E-05 | 0.000383728 | 0.320413207 |
| T2DM | monocyte | *GM2A* | 1.288518604 | 0.001408547 | 0.002830149 | 1 |
| T2DM | monocyte | *TLE1* | 0.830771896 | 0.002165226 | 0.003857404 | 1 |
| T2DM | monocyte | *STRN3* | 2.350526632 | 4.00E-05 | 0.000371009 | 0.251303269 |
| T2DM | monocyte | *ZNF239* | 0.9114871 | 0.001446087 | 0.002888039 | 1 |
| T2DM | monocyte | *MVB12B* | 1.385276648 | 0.000220044 | 0.000772859 | 1 |
| T2DM | monocyte | *C6orf106* | 2.138159549 | 5.79E-06 | 0.000322038 | 0.036402321 |
| T2DM | monocyte | *PPTC7* | 3.337380974 | 4.43E-05 | 0.000379884 | 0.27845492 |
| T2DM | monocyte | *NHLRC2* | 2.722307759 | 4.31E-05 | 0.000378034 | 0.271050603 |
| T2DM | monocyte | *KIAA1211L* | 0.774517678 | 0.00202808 | 0.003662895 | 1 |
| T2DM | monocyte | *LAMB3* | 1.163552331 | 0.001083536 | 0.002332942 | 1 |
| T2DM | monocyte | *KPNA5* | 1.810493407 | 0.000112973 | 0.000528063 | 0.710258867 |
| T2DM | monocyte | *ANKRD36B* | 1.945822029 | 0.000251504 | 0.000842412 | 1 |
| T2DM | monocyte | *HCAR1* | 1.073924418 | 0.001605357 | 0.003112582 | 1 |
| T2DM | monocyte | *SRGAP1* | 1.942956113 | 4.08E-05 | 0.000371379 | 0.256625329 |
| T2DM | monocyte | *FAM3C* | 2.777871644 | 0.000672534 | 0.001649073 | 1 |

| | | | | | | |
|---|---|---|---|---|---|---|
| T2DM | monocyte | *SLC39A10* | 2.220497011 | 0.00021655 | 0.000766771 | 1 |
| T2DM | monocyte | *PCDHB16* | 1.129468884 | 0.020479732 | 0.027606362 | 1 |
| T2DM | monocyte | *HIST1H3E* | 1.473351358 | 0.00276011 | 0.004712876 | 1 |
| T2DM | monocyte | *ZNF585A* | 1.279880751 | 0.000178757 | 0.000684437 | 1 |
| T2DM | monocyte | *FUT11* | 2.331040165 | 1.75E-05 | 0.000362859 | 0.10985513 |
| T2DM | monocyte | *ANXA4* | 1.926619902 | 5.74E-05 | 0.000396678 | 0.361089262 |
| T2DM | monocyte | *WDR5B* | 1.069374879 | 0.000517064 | 0.001380375 | 1 |
| T2DM | monocyte | *ZNF138* | 1.731911948 | 7.12E-05 | 0.000427286 | 0.447795905 |
| T2DM | monocyte | *ZNF429* | 1.387407258 | 0.000545557 | 0.001431518 | 1 |
| T2DM | monocyte | *ZNF470* | 2.204654372 | 0.002962132 | 0.005008856 | 1 |
| T2DM | monocyte | *GMFB* | 4.083828723 | 2.96E-05 | 0.000362859 | 0.185904193 |
| T2DM | monocyte | *ZMYM1* | 3.828467111 | 2.42E-05 | 0.000362859 | 0.152039004 |
| T2DM | monocyte | *IGF2R* | 2.01185538 | 0.000254421 | 0.000848508 | 1 |
| T2DM | monocyte | *SLC25A29* | 3.082967317 | 8.01E-06 | 0.000355191 | 0.050353236 |
| T2DM | monocyte | *ZNF682* | 2.286470519 | 0.000211187 | 0.000755683 | 1 |
| T2DM | monocyte | *ZNF257* | 3.413139248 | 0.003923427 | 0.00639362 | 1 |
| T2DM | monocyte | *ADAM32* | 1.144653878 | 0.001063544 | 0.002304102 | 1 |
| T2DM | monocyte | *LRRC8B* | 2.109750293 | 0.000211759 | 0.000756388 | 1 |
| T2DM | monocyte | *SND1* | 2.40056392 | 0.000300205 | 0.000953709 | 1 |
| T2DM | monocyte | *SULT1A2* | 0.581406002 | 0.000841554 | 0.00194445 | 1 |
| T2DM | monocyte | *NEK5* | 1.431867749 | 0.002797269 | 0.004768554 | 1 |
| T2DM | monocyte | *PIWIL2* | 0.897662144 | 0.000949697 | 0.002118789 | 1 |
| T2DM | monocyte | *ENTPD4* | 2.504118217 | 0.01415262 | 0.019821381 | 1 |
| T2DM | monocyte | *C1D* | 2.035116282 | 0.007984551 | 0.011943046 | 1 |
| T2DM | monocyte | *FAM110D* | 1.377920806 | 0.00131356 | 0.002688266 | 1 |
| T2DM | monocyte | *KANK2* | 1.589361009 | 0.000188906 | 0.000709469 | 1 |
| T2DM | monocyte | *C6orf141* | 0.676157968 | 0.031181106 | 0.040311662 | 1 |
| T2DM | monocyte | *GTF2E2* | 3.484920421 | 4.49E-06 | 0.000285323 | 0.028247006 |
| T2DM | monocyte | *RAMP2-AS1* | 1.123971558 | 0.000190493 | 0.000713511 | 1 |
| T2DM | monocyte | *ZNF720* | 1.169378233 | 0.002440185 | 0.00424853 | 1 |
| T2DM | monocyte | *DDI2* | 2.060347324 | 0.000115063 | 0.000533875 | 0.723401048 |
| T2DM | monocyte | *ZNF786* | 3.367225459 | 0.00215953 | 0.003852382 | 1 |
| T2DM | monocyte | *SLC22A5* | 1.079908834 | 0.00010044 | 0.000495267 | 0.631464943 |
| T2DM | monocyte | *VEPH1* | 1.116208016 | 0.000982084 | 0.002173305 | 1 |
| T2DM | monocyte | *SHPK* | 0.944905028 | 0.001648173 | 0.003165345 | 1 |
| T2DM | monocyte | *STMN3* | 3.715121339 | 0.002143831 | 0.00383232 | 1 |
| T2DM | monocyte | *GYPE* | 1.373893666 | 0.000130179 | 0.000571134 | 0.818434812 |
| T2DM | monocyte | *PCDHB11* | -1.336587428 | 0.004947231 | 0.007854354 | 1 |
| T2DM | monocyte | *SLC2A10* | 0.874070961 | 0.000403726 | 0.001161128 | 1 |
| T2DM | monocyte | *ZNF665* | 1.399604007 | 0.01766825 | 0.024153139 | 1 |
| T2DM | monocyte | *RPF2* | 1.444338621 | 0.000297519 | 0.00094709 | 1 |
| T2DM | monocyte | *MYO5A* | 1.667860495 | 5.30E-05 | 0.000385235 | 0.333311187 |
| T2DM | monocyte | *ATG7* | 1.804618861 | 0.012588538 | 0.017861462 | 1 |
| T2DM | monocyte | *RAB40C* | 1.92308923 | 0.000194836 | 0.000722248 | 1 |
| T2DM | monocyte | *PIGN* | 1.525756805 | 0.004337924 | 0.00697864 | 1 |
| T2DM | monocyte | *ZNF624* | 5.637438429 | 6.44E-05 | 0.000414227 | 0.40470024 |
| T2DM | monocyte | *HOXA4* | 1.332565295 | 0.002943481 | 0.004984974 | 1 |
| T2DM | monocyte | *BCO2* | 0.820248624 | 0.001784445 | 0.003344903 | 1 |
| T2DM | monocyte | *ENTPD6* | 2.324912256 | 0.017359291 | 0.023770374 | 1 |
| T2DM | monocyte | *CCDC154* | 4.724629881 | 1.31E-05 | 0.000362859 | 0.082139393 |

| | | | | | | |
|---|---|---|---|---|---|---|
| T2DM | monocyte | *C5orf42* | 1.433758629 | 0.000454867 | 0.001255734 | 1 |
| T2DM | monocyte | *ZNF841* | 1.71903988 | 0.001430389 | 0.002866707 | 1 |
| T2DM | monocyte | *MFAP5* | 0.839043048 | 0.000434753 | 0.001220221 | 1 |
| T2DM | monocyte | *MPEG1* | -0.367216506 | 0.025887187 | 0.034044947 | 1 |
| T2DM | monocyte | *PDCD1LG2* | 0.983159262 | 0.001067962 | 0.002310922 | 1 |
| T2DM | monocyte | *NMB* | 4.205597834 | 3.18E-05 | 0.000363519 | 0.19979311 |
| T2DM | monocyte | *KLHL14* | 1.265279367 | 0.001918312 | 0.003501866 | 1 |
| T2DM | monocyte | *FAM114A1* | 1.390193072 | 0.000155005 | 0.000629999 | 0.97451865 |
| T2DM | monocyte | *PHF2* | 3.745743049 | 0.000443647 | 0.001236351 | 1 |
| T2DM | monocyte | *S100A10* | 1.954518989 | 0.002158063 | 0.003852284 | 1 |
| T2DM | monocyte | *MAP1LC3C* | 1.155980889 | 0.004166786 | 0.006743381 | 1 |
| T2DM | monocyte | *TAF13* | 1.960549325 | 0.027649055 | 0.036124192 | 1 |
| T2DM | monocyte | *AC122129.1* | 2.073680017 | 0.001792382 | 0.00335478 | 1 |
| T2DM | monocyte | *CCDC180* | 1.48527962 | 0.001996841 | 0.003616865 | 1 |
| T2DM | monocyte | *OCLN* | 0.768493221 | 0.008094299 | 0.012087615 | 1 |
| T2DM | monocyte | *ZNF181* | 5.026399716 | 5.35E-05 | 0.000385603 | 0.336319083 |
| T2DM | monocyte | *SGTB* | 4.170253513 | 0.002132705 | 0.003815795 | 1 |
| T2DM | monocyte | *ADH5* | 1.473766615 | 0.000783189 | 0.001843127 | 1 |
| T2DM | monocyte | *ZNF677* | 1.4150374 | 0.000581573 | 0.00149014 | 1 |
| T2DM | monocyte | *FCHSD1* | 3.103758138 | 0.001009545 | 0.002219887 | 1 |
| T2DM | monocyte | *RPL12* | 1.414913675 | 0.038975101 | 0.049522324 | 1 |
| T2DM | monocyte | *VPS13A* | 2.099248229 | 2.64E-05 | 0.000362859 | 0.166048016 |
| T2DM | monocyte | *LEKR1* | 0.560273979 | 0.003803395 | 0.00622059 | 1 |
| T2DM | monocyte | *NOL8* | 3.978290813 | 7.79E-05 | 0.000438718 | 0.489609214 |
| T2DM | monocyte | *MRPL42* | 1.702329381 | 0.008466657 | 0.01258092 | 1 |
| T2DM | monocyte | *ZNF335* | 1.775728111 | 0.000362426 | 0.001073858 | 1 |
| T2DM | monocyte | *AGAP9* | 3.901404402 | 0.033876961 | 0.043519504 | 1 |
| T2DM | monocyte | *ZNF273* | 1.434089594 | 0.00027755 | 0.000902161 | 1 |
| T2DM | monocyte | *ZNF84* | 1.558046371 | 0.00035262 | 0.001054172 | 1 |
| T2DM | monocyte | *MAK16* | 2.912514552 | 0.000203222 | 0.000735552 | 1 |
| T2DM | monocyte | *ZNF667* | 0.654856794 | 0.010525406 | 0.015233247 | 1 |
| T2DM | monocyte | *PRIM1* | 1.021391947 | 0.012085099 | 0.017232724 | 1 |
| T2DM | monocyte | *SULT1C4* | 1.023978436 | 0.000804857 | 0.001881092 | 1 |
| T2DM | monocyte | *ZNF248* | 0.909412029 | 0.004034643 | 0.006551084 | 1 |
| T2DM | monocyte | *HIBCH* | 1.251202531 | 0.000556747 | 0.001451792 | 1 |
| T2DM | monocyte | *ZNF544* | 2.084311868 | 0.000232855 | 0.000799542 | 1 |
| T2DM | monocyte | *SOWAHC* | 2.30287474 | 0.001214078 | 0.002534167 | 1 |
| T2DM | monocyte | *AC135178.1* | 5.632188421 | 0.003965019 | 0.006454705 | 1 |
| T2DM | monocyte | *MIER1* | 3.810095521 | 7.52E-06 | 0.000349439 | 0.04728725 |
| T2DM | monocyte | *SVIP* | 1.23056894 | 0.000552077 | 0.001445007 | 1 |
| T2DM | monocyte | *ZNF251* | 3.09971211 | 1.13E-05 | 0.000362859 | 0.071272765 |
| T2DM | monocyte | *DDRGK1* | 1.167057079 | 0.000703918 | 0.001704095 | 1 |
| T2DM | monocyte | *ZNF334* | 1.451799774 | 0.000782583 | 0.001842733 | 1 |
| T2DM | monocyte | *HSD17B11* | 2.74711013 | 0.013917421 | 0.019530988 | 1 |
| T2DM | monocyte | *SULT1C2* | 0.832489589 | 0.000319421 | 0.000992193 | 1 |
| T2DM | monocyte | *RPS6KL1* | 0.864478257 | 0.000957288 | 0.002131941 | 1 |
| T2DM | monocyte | *DDX42* | 2.721711401 | 5.87E-06 | 0.000322038 | 0.036908064 |
| T2DM | monocyte | *SLC29A3* | 2.293993029 | 0.001371919 | 0.002774286 | 1 |
| T2DM | monocyte | *ANTXRL* | -7.231783109 | 0.008596538 | 0.012755826 | 1 |
| T2DM | monocyte | *STYX* | 5.090638451 | 5.26E-05 | 0.000385235 | 0.330609821 |

| | | | | | | |
|---|---|---|---|---|---|---|
| T2DM | monocyte | *ZKSCAN8* | 4.294895968 | 3.83E-05 | 0.00036941 | 0.240546108 |
| T2DM | monocyte | *GFPT1* | 2.146222484 | 2.80E-05 | 0.000362859 | 0.17596389 |
| T2DM | monocyte | *UVRAG* | 1.228884757 | 0.000110898 | 0.000523558 | 0.697216675 |
| T2DM | monocyte | *MGEA5* | 1.184182059 | 0.000355268 | 0.001061078 | 1 |
| T2DM | monocyte | *ZNF568* | 3.802837884 | 2.61E-07 | 5.47E-05 | 0.001641266 |
| T2DM | monocyte | *ZNF587* | 1.85993305 | 0.000313241 | 0.000979286 | 1 |
| T2DM | monocyte | *FLVCR1-AS1* | 0.690535216 | 0.010011816 | 0.01458394 | 1 |
| T2DM | monocyte | *SH3BGRL2* | 0.926743881 | 0.004963437 | 0.007869706 | 1 |
| T2DM | monocyte | *ZNF808* | 4.112710211 | 3.95E-05 | 0.000371009 | 0.248083216 |
| T2DM | monocyte | *TMA16* | 4.161128739 | 8.86E-06 | 0.000362859 | 0.055705726 |
| T2DM | monocyte | *HLA-DRB5* | -0.814234412 | 2.76E-10 | 4.33E-07 | 1.73E-06 |
| T2DM | monocyte | *CNGA1* | 1.281692924 | 0.000913886 | 0.002057143 | 1 |
| T2DM | monocyte | *MAFK* | 4.085682515 | 3.48E-05 | 0.000365043 | 0.218563968 |
| T2DM | monocyte | *HIST1H4E* | 2.342633664 | 0.003564441 | 0.005886431 | 1 |
| T2DM | monocyte | *C20orf203* | 3.186215847 | 0.006668537 | 0.010205719 | 1 |
| T2DM | monocyte | *ZNF627* | 1.189057195 | 0.000506102 | 0.001356293 | 1 |
| T2DM | monocyte | *ARC* | 4.548053982 | 0.001975829 | 0.003586151 | 1 |
| T2DM | monocyte | *TLK1* | 2.610268593 | 2.66E-05 | 0.000362859 | 0.167111988 |
| T2DM | monocyte | *LRBA* | 2.103649267 | 3.42E-05 | 0.000365043 | 0.215295736 |
| T2DM | monocyte | *CCDC69* | 2.895047894 | 0.0001343 | 0.00057865 | 0.844343159 |
| T2DM | monocyte | *MDM4* | 3.047086202 | 2.41E-05 | 0.000362859 | 0.151584555 |
| T2DM | monocyte | *NCOA6* | 2.466586002 | 0.000241563 | 0.0008199 | 1 |
| T2DM | monocyte | *FAM19A2* | 2.978811628 | 0.002665496 | 0.004591226 | 1 |
| T2DM | monocyte | *TTC37* | 2.204911714 | 3.43E-05 | 0.000365043 | 0.215947536 |
| T2DM | monocyte | *TUSC1* | 9.348322132 | 3.14E-05 | 0.000362859 | 0.197395047 |
| T2DM | monocyte | *PAPSS2* | 1.538642904 | 0.000131128 | 0.0005725 | 0.824399886 |
| T2DM | monocyte | *FAN1* | 2.308774646 | 8.09E-05 | 0.000444291 | 0.508713744 |
| T2DM | monocyte | *CEP290* | 3.264935685 | 0.000117208 | 0.000541031 | 0.73688454 |
| T2DM | monocyte | *ANKRD13B* | 0.882660511 | 0.010293534 | 0.014952738 | 1 |
| T2DM | monocyte | *ECI2* | 2.522851572 | 6.57E-05 | 0.000418224 | 0.412787509 |
| T2DM | monocyte | *ZNF652* | 3.685894701 | 0.000108971 | 0.000518963 | 0.685103755 |
| T2DM | monocyte | *APCDD1L* | 5.563043585 | 2.40E-05 | 0.000362859 | 0.150645593 |
| T2DM | monocyte | *RCSD1* | -0.365561723 | 0.037085474 | 0.047347847 | 1 |
| T2DM | monocyte | *RASSF9* | 0.799558422 | 0.001179734 | 0.002479769 | 1 |
| T2DM | monocyte | *FAM169A* | 1.66232571 | 0.000877666 | 0.002002136 | 1 |
| T2DM | monocyte | *TMEM184B* | 1.968732669 | 0.000105988 | 0.000509831 | 0.666348879 |
| T2DM | monocyte | *SCAMP5* | 0.981573507 | 0.001518316 | 0.002978363 | 1 |
| T2DM | monocyte | *ALPK2* | 0.820610313 | 0.002945579 | 0.00498623 | 1 |
| T2DM | monocyte | *SFT2D1* | 3.117561332 | 2.75E-05 | 0.000362859 | 0.173085938 |
| T2DM | monocyte | *ARHGAP11A* | 1.592479924 | 0.000785396 | 0.001846592 | 1 |
| T2DM | monocyte | *HMGN2* | 1.586909234 | 7.73E-05 | 0.00043763 | 0.485768907 |
| T2DM | monocyte | *CD3E* | 0.969924384 | 0.003431417 | 0.005702701 | 1 |
| T2DM | monocyte | *TSEN15* | 2.09803143 | 0.000119425 | 0.000545328 | 0.750827842 |
| T2DM | monocyte | *TYW1* | 1.613970787 | 8.62E-05 | 0.000459567 | 0.542174609 |
| T2DM | monocyte | *ITPRIPL1* | 1.453725984 | 0.001010359 | 0.002220247 | 1 |
| T2DM | monocyte | *PRMT6* | -0.803406005 | 0.007582064 | 0.011414856 | 1 |
| T2DM | monocyte | *CAPZA2* | 2.054438094 | 0.025961906 | 0.034125549 | 1 |
| T2DM | monocyte | *C1orf174* | 3.330398534 | 0.002335025 | 0.004093782 | 1 |
| T2DM | monocyte | *RASGEF1A* | -0.919912469 | 0.013507524 | 0.019006671 | 1 |
| T2DM | monocyte | *KIAA0753* | 1.627772365 | 0.000669645 | 0.001644554 | 1 |

| | | | | | | |
|---|---|---|---|---|---|---|
| T2DM | monocyte | *DCLRE1A* | 2.401162625 | 0.000336291 | 0.00102161 | 1 |
| T2DM | monocyte | *APRT* | 1.731528088 | 0.00039163 | 0.001135569 | 1 |
| T2DM | monocyte | *TBKBP1* | 0.76623995 | 0.030079231 | 0.0390155 | 1 |
| T2DM | monocyte | *ZFP2* | 2.808502084 | 7.68E-05 | 0.000437295 | 0.4827732 |
| T2DM | monocyte | *MFAP3L* | 0.966604511 | 0.001462226 | 0.002908262 | 1 |
| T2DM | monocyte | *NAGA* | -0.610971188 | 0.020847882 | 0.028048499 | 1 |
| T2DM | monocyte | *SMG5* | 3.442617193 | 0.000405527 | 0.001164794 | 1 |
| T2DM | monocyte | *TGM2* | 0.597129502 | 0.011046483 | 0.015910479 | 1 |
| T2DM | monocyte | *PJA2* | 2.754913084 | 0.000238225 | 0.000813095 | 1 |
| T2DM | monocyte | *AL590822.1* | 0.76641285 | 0.006243621 | 0.009635162 | 1 |
| T2DM | monocyte | *AP001007.1* | 4.460847368 | 0.001559047 | 0.003043777 | 1 |
| T2DM | monocyte | *ZNF525* | 1.157099352 | 0.00111081 | 0.002375395 | 1 |
| T2DM | monocyte | *AC105020.1* | 2.035841871 | 0.0055694 | 0.008711412 | 1 |
| T2DM | monocyte | *EFCAB2* | 1.763136168 | 0.000113054 | 0.000528063 | 0.710772923 |
| T2DM | monocyte | *CAPN8* | 2.309710945 | 0.010988857 | 0.015834735 | 1 |
| T2DM | monocyte | *ECT2L* | 2.200628696 | 0.001995314 | 0.003616182 | 1 |
| T2DM | monocyte | *DDO* | 1.940389444 | 0.000568262 | 0.001469626 | 1 |
| T2DM | monocyte | *HIST2H2BF* | -2.356331895 | 0.027584068 | 0.036046776 | 1 |
| T2DM | monocyte | *RBM20* | 0.84244433 | 0.002598281 | 0.004492682 | 1 |
| T2DM | monocyte | *C6orf163* | 1.203381852 | 0.001702201 | 0.003233153 | 1 |
| T2DM | monocyte | *PCMTD2* | 2.306793855 | 1.82E-05 | 0.000362859 | 0.114571374 |
| T2DM | monocyte | *NEU4* | 5.688879687 | 0.023167331 | 0.030865228 | 1 |
| T2DM | monocyte | *TRAF3IP1* | 1.613460149 | 0.000478475 | 0.001307333 | 1 |
| T2DM | monocyte | *GIGYF2* | 3.286470185 | 5.90E-05 | 0.000399203 | 0.370967735 |
| T2DM | monocyte | *RUFY2* | 2.117038775 | 7.97E-05 | 0.000442726 | 0.500948532 |
| T2DM | monocyte | *PHACTR4* | 1.729162876 | 0.004449325 | 0.007132307 | 1 |
| T2DM | monocyte | *ASAH2B* | 1.586064952 | 7.06E-05 | 0.000427286 | 0.443703261 |
| T2DM | monocyte | *AGAP6* | 1.804501297 | 9.92E-05 | 0.000490923 | 0.623471989 |
| T2DM | monocyte | *C10orf128* | 1.103264608 | 0.000334757 | 0.00102039 | 1 |
| T2DM | monocyte | *ZDBF2* | 1.373522541 | 0.000253272 | 0.000845631 | 1 |
| T2DM | monocyte | *RING1* | 1.872915422 | 0.001752975 | 0.003300676 | 1 |
| T2DM | monocyte | *HSD17B8* | 0.899492563 | 0.038204528 | 0.048592326 | 1 |
| T2DM | monocyte | *OXLD1* | 1.692589468 | 2.11E-05 | 0.000362859 | 0.132546127 |
| T2DM | monocyte | *HLA-DOA* | -0.778580258 | 0.019693377 | 0.026660694 | 1 |
| T2DM | monocyte | *HLA-DMA* | 1.825673469 | 0.00268059 | 0.004612171 | 1 |
| T2DM | monocyte | *COL15A1* | 0.937083363 | 0.001134347 | 0.002413127 | 1 |
| T2DM | monocyte | *NOTCH4* | -0.366627731 | 6.14E-06 | 0.000322038 | 0.038576169 |
| T2DM | monocyte | *PRRT1* | 1.360641453 | 1.67E-05 | 0.000362859 | 0.104924422 |
| T2DM | monocyte | *MRPL38* | 1.46549409 | 0.009912836 | 0.014466574 | 1 |
| T2DM | monocyte | *STK19* | 1.517700584 | 0.000102398 | 0.000499824 | 0.643773131 |
| T2DM | monocyte | *NELFE* | 3.18392764 | 5.15E-08 | 1.70E-05 | 0.000323527 |
| T2DM | monocyte | *LAYN* | 1.05560204 | 0.007111525 | 0.010791734 | 1 |
| T2DM | monocyte | *CARD16* | -0.758053371 | 0.016220864 | 0.022363155 | 1 |
| T2DM | monocyte | *BAG6* | 3.282486817 | 4.10E-07 | 6.81E-05 | 0.002580552 |
| T2DM | monocyte | *NFKBIL1* | 1.813677835 | 0.027528222 | 0.036003731 | 1 |
| T2DM | monocyte | *ZNF814* | 1.294416237 | 0.00032767 | 0.001006043 | 1 |
| T2DM | monocyte | *CCHCR1* | 0.915921272 | 0.006878062 | 0.01047792 | 1 |
| T2DM | monocyte | *PSORS1C1* | 0.373451911 | 0.001366453 | 0.002767683 | 1 |
| T2DM | monocyte | *ABCF1* | 1.746855552 | 0.000256112 | 0.000851723 | 1 |
| T2DM | monocyte | *LILRB3* | -0.249187694 | 0.012221861 | 0.017380421 | 1 |

| | | | | | | |
|---|---|---|---|---|---|---|
| T2DM | monocyte | *HLA-E* | 2.256816904 | 4.59E-05 | 0.000379941 | 0.288732659 |
| T2DM | monocyte | *ZFP57* | 0.603818083 | 0.032345548 | 0.041714146 | 1 |
| T2DM | monocyte | *FAM153C* | 2.806811328 | 0.003455912 | 0.005739605 | 1 |
| T2DM | monocyte | *TRIM27* | 1.949919969 | 0.004711127 | 0.007507948 | 1 |
| T2DM | monocyte | *MALRD1* | 0.603072053 | 0.004364517 | 0.007017831 | 1 |
| T2DM | monocyte | *MROH6* | -0.47611409 | 0.010846417 | 0.015643823 | 1 |
| T2DM | monocyte | *ATXN2* | 2.153736662 | 5.34E-05 | 0.000385235 | 0.335539911 |
| T2DM | monocyte | *FAM216A* | 3.516949705 | 2.72E-05 | 0.000362859 | 0.171294683 |
| T2DM | monocyte | *ZNF155* | 2.158907521 | 0.000821911 | 0.001911003 | 1 |
| T2DM | monocyte | *ZNF783* | 3.160729554 | 0.000120102 | 0.000547214 | 0.755083104 |
| T2DM | monocyte | *ZNF425* | 1.01386868 | 0.013820121 | 0.019403105 | 1 |
| T2DM | monocyte | *LRRC10B* | -0.958229434 | 0.00733504 | 0.011091789 | 1 |
| T2DM | monocyte | *C12orf73* | 1.033642693 | 0.007700041 | 0.011568297 | 1 |
| T2DM | monocyte | *PCDHA4* | 3.925138496 | 0.004217806 | 0.006815047 | 1 |
| T2DM | monocyte | *TRIM13* | 3.236572787 | 0.000299392 | 0.000951607 | 1 |
| T2DM | monocyte | *SPIRE2* | 1.673781201 | 4.46E-05 | 0.000379941 | 0.280525729 |
| T2DM | monocyte | *SLFN12L* | 2.03404204 | 0.025779138 | 0.03392077 | 1 |
| T2DM | monocyte | *SYCE1L* | 1.651747212 | 0.000228316 | 0.000788692 | 1 |
| T2DM | monocyte | *TMEM231* | 0.997176156 | 0.009762145 | 0.014273164 | 1 |
| T2DM | monocyte | *FAM71F2* | 0.697386686 | 0.001955376 | 0.003555075 | 1 |
| T2DM | monocyte | *AC016586.1* | 13.51586368 | 0.000132371 | 0.000575926 | 0.832213621 |
| T2DM | monocyte | *LINC00654* | 4.306365818 | 0.007605602 | 0.011444811 | 1 |
| T2DM | monocyte | *NT5M* | 1.268500071 | 0.024198377 | 0.032096032 | 1 |
| T2DM | monocyte | *MT1A* | 2.294599261 | 0.001691271 | 0.003223856 | 1 |
| T2DM | monocyte | *MT1M* | 1.016385338 | 0.001742277 | 0.00328999 | 1 |
| T2DM | monocyte | *CFI* | 1.313472245 | 0.002179483 | 0.003875115 | 1 |
| T2DM | monocyte | *EXOC3L4* | 1.506352034 | 0.003742401 | 0.006125611 | 1 |
| T2DM | monocyte | *ATP6AP1L* | 1.544930665 | 5.93E-05 | 0.000399203 | 0.372559612 |
| T2DM | monocyte | *NAP1L4* | 1.962539892 | 0.00034447 | 0.001037263 | 1 |
| T2DM | monocyte | *SMN2* | -0.860831821 | 0.024551682 | 0.032500677 | 1 |
| T2DM | monocyte | *DENND6B* | 7.697540496 | 9.45E-05 | 0.000478875 | 0.594027827 |
| T2DM | monocyte | *EIF3CL* | 3.122479456 | 0.00018668 | 0.000704201 | 1 |
| T2DM | monocyte | *LCMT1* | 3.158165949 | 3.81E-05 | 0.00036941 | 0.239411936 |
| T2DM | monocyte | *LIN52* | 2.974459298 | 5.40E-08 | 1.70E-05 | 0.000339569 |
| T2DM | monocyte | *C17orf107* | 1.348311874 | 0.026702289 | 0.035047452 | 1 |
| T2DM | monocyte | *RFPL3S* | -0.698733728 | 0.003476021 | 0.005766159 | 1 |
| T2DM | monocyte | *C1QTNF9B* | 10.81792782 | 0.001720004 | 0.003261056 | 1 |
| T2DM | monocyte | *RNPS1* | 3.69756977 | 1.99E-05 | 0.000362859 | 0.124844832 |
| T2DM | monocyte | *NYNRIN* | 1.326405576 | 0.000595239 | 0.001515557 | 1 |
| T2DM | monocyte | *ZDHHC11B* | 0.89544828 | 0.00136923 | 0.002771144 | 1 |
| T2DM | monocyte | *GOLGA8O* | 3.824243444 | 0.010494374 | 0.015205838 | 1 |
| T2DM | monocyte | *HCG27* | 2.480834831 | 3.48E-13 | 1.09E-09 | 2.19E-09 |
| T2DM | monocyte | *HLA-A* | 1.300236694 | 0.020577831 | 0.027732652 | 1 |
| T2DM | monocyte | *LNP1* | 3.599824815 | 0.034921711 | 0.044788413 | 1 |
| T2DM | monocyte | *VGLL3* | 0.49707829 | 0.005065458 | 0.008012355 | 1 |
| T2DM | monocyte | *COLQ* | 0.790134374 | 0.001896628 | 0.003477428 | 1 |
| T2DM | monocyte | *METTL6* | 3.03814194 | 1.72E-05 | 0.000362859 | 0.107852483 |
| T2DM | monocyte | *SACM1L* | 3.673926929 | 0.000246782 | 0.000831467 | 1 |
| T2DM | monocyte | *SLC48A1* | 0.967841223 | 0.001228402 | 0.002554734 | 1 |
| T2DM | monocyte | *TAS2R14* | 2.186826079 | 7.86E-05 | 0.000440255 | 0.49396564 |

| | | | | | | |
|---|---|---|---|---|---|---|
| T2DM | monocyte | *ZRSR1* | 4.828054729 | 0.000654941 | 0.001621107 | 1 |
| T2DM | monocyte | *C17orf51* | 1.706520943 | 0.005721622 | 0.008919374 | 1 |
| T2DM | monocyte | *RNF208* | 3.431502916 | 0.001114832 | 0.00238217 | 1 |
| T2DM | monocyte | *MAP10* | 3.022499111 | 0.002098849 | 0.003769056 | 1 |
| T2DM | monocyte | *ZNF580* | 1.510572281 | 0.000234977 | 0.000804193 | 1 |
| T2DM | monocyte | *FGFR1OP* | 1.87988325 | 2.57E-05 | 0.000362859 | 0.161699358 |
| T2DM | monocyte | *FAM106A* | -0.896006379 | 0.035591352 | 0.045554322 | 1 |
| T2DM | monocyte | *ZNF254* | 2.786298201 | 2.97E-05 | 0.000362859 | 0.186811189 |
| T2DM | monocyte | *TCTEX1D2* | 1.584882068 | 0.000433475 | 0.001218264 | 1 |
| T2DM | monocyte | *LINGO4* | -4.166349432 | 0.010822095 | 0.015615908 | 1 |
| T2DM | monocyte | *CCDC183* | 2.986356676 | 0.00319563 | 0.00536187 | 1 |
| T2DM | monocyte | *DNLZ* | 1.488421154 | 0.00021737 | 0.000767755 | 1 |
| T2DM | monocyte | *SUPT4H1* | 3.909595028 | 1.45E-05 | 0.000362859 | 0.09130804 |
| T2DM | monocyte | *GSTM2* | 1.425940825 | 0.001600067 | 0.003106528 | 1 |
| T2DM | monocyte | *LCAT* | 1.182746355 | 0.001244152 | 0.00257726 | 1 |
| T2DM | monocyte | *SIPA1* | 2.536277688 | 0.006786631 | 0.010361231 | 1 |
| T2DM | monocyte | *ERV3-1* | 0.577134293 | 0.000632526 | 0.001578678 | 1 |
| T2DM | monocyte | *RBMXL1* | 2.401718597 | 0.000426358 | 0.001203701 | 1 |
| T2DM | monocyte | *SRA1* | 1.53508506 | 0.006712888 | 0.010263601 | 1 |
| T2DM | monocyte | *HEXA* | 3.609769919 | 2.45E-05 | 0.000362859 | 0.153985817 |
| T2DM | monocyte | *LEPROT* | 2.352607939 | 0.000123895 | 0.000554793 | 0.778929038 |
| T2DM | monocyte | *LBH* | 0.715681911 | 0.001678522 | 0.003208534 | 1 |
| T2DM | monocyte | *PPP1CB* | 2.744056716 | 0.003657143 | 0.006012672 | 1 |
| T2DM | monocyte | *NCKIPSD* | 2.862614053 | 8.54E-05 | 0.000458095 | 0.537116794 |
| T2DM | monocyte | *ATF6B* | 3.51444961 | 3.97E-08 | 1.47E-05 | 0.000249496 |
| T2DM | monocyte | *SLC35F6* | 5.919788181 | 7.92E-05 | 0.000442227 | 0.49794726 |
| T2DM | monocyte | *KLRK1* | 2.755355584 | 0.005625046 | 0.008786252 | 1 |
| T2DM | monocyte | *EMP2* | 1.10194875 | 0.001078462 | 0.002327598 | 1 |
| T2DM | monocyte | *PPM1N* | -0.371842654 | 0.01597213 | 0.022059926 | 1 |
| T2DM | monocyte | *SLC23A3* | 0.986766306 | 0.039294977 | 0.04989851 | 1 |
| T2DM | monocyte | *LTB4R2* | -1.235772751 | 0.00898096 | 0.01326052 | 1 |
| T2DM | monocyte | *GALT* | 2.385914189 | 9.24E-05 | 0.00047287 | 0.580683897 |
| T2DM | monocyte | *ITGA1* | 0.976696595 | 0.000373099 | 0.001096108 | 1 |
| T2DM | monocyte | *ZNF726* | 1.156812209 | 0.001783322 | 0.003344363 | 1 |
| T2DM | monocyte | *TAX1BP3* | 3.670968426 | 7.36E-05 | 0.000431915 | 0.462473822 |
| T2DM | monocyte | *AP1G2* | 3.548489613 | 2.97E-05 | 0.000362859 | 0.186704453 |
| T2DM | monocyte | *ZNF90* | 1.792035532 | 0.000813687 | 0.00189609 | 1 |
| T2DM | monocyte | *TM6SF2* | 12.81886942 | 0.001437695 | 0.002877679 | 1 |
| T2DM | monocyte | *REPIN1* | 0.944803882 | 0.001587479 | 0.003089932 | 1 |
| T2DM | monocyte | *SMIM7* | 2.56044578 | 0.01368335 | 0.019228257 | 1 |
| T2DM | monocyte | *TSPAN4* | 1.44122496 | 0.000376959 | 0.001104869 | 1 |
| T2DM | monocyte | *AC079776.2* | 2.164010976 | 0.001048761 | 0.00227993 | 1 |
| T2DM | monocyte | *WEE2* | 0.905910591 | 0.027411565 | 0.035858616 | 1 |
| T2DM | monocyte | *PAXIP1-AS2* | 5.11184591 | 7.46E-05 | 0.000433444 | 0.468801127 |
| T2DM | monocyte | *LYRM4* | 2.835961078 | 6.58E-05 | 0.000418344 | 0.413742541 |
| T2DM | monocyte | *PRCD* | 3.63559028 | 5.81E-05 | 0.000397753 | 0.365535072 |
| T2DM | monocyte | *C17orf67* | 1.327860152 | 0.006939822 | 0.010554103 | 1 |
| T2DM | monocyte | *COLCA2* | 1.014831326 | 0.02073098 | 0.027915115 | 1 |
| T2DM | monocyte | *NEURL1B* | 0.703620885 | 0.001489087 | 0.002943636 | 1 |
| T2DM | monocyte | *PPME1* | 1.098394548 | 0.012021602 | 0.017153838 | 1 |

| | | | | | | |
|---|---|---|---|---|---|---|
| T2DM | monocyte | *STARD10* | 1.456022812 | 2.54E-08 | 1.23E-05 | 0.000159507 |
| T2DM | monocyte | *CPEB1* | 0.856190072 | 0.001869005 | 0.003444321 | 1 |
| T2DM | monocyte | *C10orf105* | -0.484781152 | 0.008625513 | 0.012786749 | 1 |
| T2DM | monocyte | *ARHGEF33* | 0.898463885 | 0.001090373 | 0.002344452 | 1 |
| T2DM | monocyte | *AC005082.1* | -0.595548081 | 0.008079115 | 0.012070674 | 1 |
| T2DM | monocyte | *TBC1D26* | -14.67123671 | 0.013304584 | 0.018758897 | 1 |
| T2DM | monocyte | *ACSM4* | 0.742890601 | 0.024318066 | 0.032220797 | 1 |
| T2DM | monocyte | *COL28A1* | 0.890906895 | 0.001606349 | 0.003112582 | 1 |
| T2DM | monocyte | *PHB2* | 2.985952099 | 0.008589354 | 0.012748174 | 1 |
| T2DM | monocyte | *UBXN2B* | 2.048386784 | 0.01470237 | 0.020513493 | 1 |
| T2DM | monocyte | *UBE2QL1* | 1.249725588 | 0.001885546 | 0.00346317 | 1 |
| T2DM | monocyte | *GOLGA8B* | 1.134329014 | 0.000434538 | 0.001220161 | 1 |
| T2DM | monocyte | *NPEPL1* | 2.975552911 | 6.73E-05 | 0.000421481 | 0.422928508 |
| T2DM | monocyte | *LINC01139* | 1.301135313 | 0.015537494 | 0.021516349 | 1 |
| T2DM | monocyte | *TSTD1* | 1.364974965 | 0.000648271 | 0.001608397 | 1 |
| T2DM | monocyte | *LINC01138* | 2.456627375 | 6.33E-05 | 0.000409778 | 0.398083886 |
| T2DM | monocyte | *CYB5RL* | 1.385354183 | 0.001327097 | 0.002703173 | 1 |
| T2DM | monocyte | *ATAD3C* | 2.623652614 | 0.000374544 | 0.001099195 | 1 |
| T2DM | monocyte | *CCDC7* | 1.333765738 | 0.000312714 | 0.000978124 | 1 |
| T2DM | monocyte | *PAM16* | 1.020998915 | 0.005685376 | 0.008870487 | 1 |
| T2DM | monocyte | *RNASEK* | 3.815881539 | 0.00809121 | 0.012085873 | 1 |
| T2DM | monocyte | *NBPF1* | 2.428168097 | 0.0005651 | 0.001465478 | 1 |
| T2DM | monocyte | *FAM228B* | 0.972108981 | 0.004460478 | 0.00714472 | 1 |
| T2DM | monocyte | *RNU6ATAC35I* | 4.317058653 | 0.001821226 | 0.003387234 | 1 |
| T2DM | monocyte | *C6orf226* | 1.772984095 | 0.010705991 | 0.015455468 | 1 |
| T2DM | monocyte | *PPP3R1* | 3.148155434 | 0.000550691 | 0.001442582 | 1 |
| T2DM | monocyte | *AP4M1* | 1.348324508 | 0.000600414 | 0.001523326 | 1 |
| T2DM | monocyte | *C19orf73* | 6.270817178 | 0.001829938 | 0.003393752 | 1 |
| T2DM | monocyte | *TIGD1* | 0.897687928 | 0.026803601 | 0.035151072 | 1 |
| T2DM | monocyte | *C1orf229* | 2.108817871 | 0.006532693 | 0.010014155 | 1 |
| T2DM | monocyte | *SLC12A8* | 0.736174913 | 0.018411268 | 0.025043626 | 1 |
| T2DM | monocyte | *APOL6* | 1.50260594 | 0.000169271 | 0.000660586 | 1 |
| T2DM | monocyte | *UBA52* | 2.815760132 | 7.91E-06 | 0.000355191 | 0.049707549 |
| T2DM | monocyte | *BTBD19* | 1.855364224 | 0.000343733 | 0.001035977 | 1 |
| T2DM | monocyte | *FAM185A* | 2.326550264 | 5.09E-05 | 0.000383728 | 0.319799353 |
| T2DM | monocyte | *AC062017.1* | 0.83459983 | 0.030524925 | 0.039552804 | 1 |
| T2DM | monocyte | *ADRA2B* | 3.271932176 | 0.001873767 | 0.003446704 | 1 |
| T2DM | monocyte | *EXOSC6* | 1.767817718 | 0.016995378 | 0.02331939 | 1 |
| T2DM | monocyte | *VPS52* | 2.551769749 | 0.000167856 | 0.000658686 | 1 |
| T2DM | monocyte | *AC147651.3* | -1.762315923 | 0.01220016 | 0.017361342 | 1 |
| T2DM | monocyte | *OSER1-AS1* | 5.146780628 | 0.000257137 | 0.000854171 | 1 |
| T2DM | monocyte | *LINC01133* | 0.59774112 | 0.001741686 | 0.00328999 | 1 |
| T2DM | monocyte | *LINC01117* | 0.688601977 | 0.001542876 | 0.003016188 | 1 |
| T2DM | monocyte | *RASAL2-AS1* | -2.849998024 | 0.014003061 | 0.01963802 | 1 |
| T2DM | monocyte | *LINC00240* | 0.747059036 | 0.000286309 | 0.0009212 | 1 |
| T2DM | monocyte | *LINC00863* | 2.956493241 | 0.000778314 | 0.001835432 | 1 |
| T2DM | monocyte | *LINC00184* | 1.673976331 | 0.01219536 | 0.01735844 | 1 |
| T2DM | monocyte | *TMEM233* | 1.06252781 | 0.001704041 | 0.003235672 | 1 |
| T2DM | monocyte | *LINC00237* | 4.150076268 | 0.001954845 | 0.003555075 | 1 |
| T2DM | monocyte | *PLEKHM1* | 4.272474525 | 7.10E-05 | 0.000427286 | 0.446251853 |

| | | | | | | |
|---|---|---|---|---|---|---|
| T2DM | monocyte | *SLC26A6* | 3.086686053 | 2.04E-05 | 0.000362859 | 0.128545673 |
| T2DM | monocyte | *MIAT* | 2.122204997 | 0.029579533 | 0.038463711 | 1 |
| T2DM | monocyte | *ERCC6* | 2.526555068 | 4.77E-05 | 0.000379941 | 0.299578573 |
| T2DM | monocyte | *LINC00115* | 1.06068618 | 0.001385833 | 0.00279523 | 1 |
| T2DM | monocyte | *NOL7* | 2.042459425 | 6.44E-05 | 0.000414227 | 0.404601662 |
| T2DM | monocyte | *ELFN1* | 2.338698966 | 0.001373432 | 0.00277556 | 1 |
| T2DM | monocyte | *ZNF503-AS1* | 0.866524989 | 0.001558491 | 0.003043777 | 1 |
| T2DM | monocyte | *LINC00937* | -0.243894001 | 0.008391716 | 0.012484316 | 1 |
| T2DM | monocyte | *AC064836.3* | 1.809030183 | 0.001638538 | 0.003155126 | 1 |
| T2DM | monocyte | *HSBP1L1* | 1.156615582 | 0.001751471 | 0.003299819 | 1 |
| T2DM | monocyte | *AC093901.1* | -0.998136109 | 0.03806471 | 0.048442494 | 1 |
| T2DM | monocyte | *C14orf132* | 0.755780363 | 0.001230797 | 0.002558022 | 1 |
| T2DM | monocyte | *AC009299.3* | 2.687225958 | 0.000329806 | 0.001010965 | 1 |
| T2DM | monocyte | *SLC2A1-AS1* | 1.23860334 | 0.002226679 | 0.003937872 | 1 |
| T2DM | monocyte | *LINC01068* | 2.124199362 | 0.001936238 | 0.00352742 | 1 |
| T2DM | monocyte | *AC108463.2* | 5.354859254 | 0.003245044 | 0.00543318 | 1 |
| T2DM | monocyte | *HCG20* | 2.678594596 | 0.005124539 | 0.008088872 | 1 |
| T2DM | monocyte | *C19orf24* | 1.016835091 | 0.006125283 | 0.009468812 | 1 |
| T2DM | monocyte | *LINC01013* | 1.780017871 | 0.000890722 | 0.002020191 | 1 |
| T2DM | monocyte | *AC073133.2* | 8.19647716 | 0.02413583 | 0.032026585 | 1 |
| T2DM | monocyte | *PROB1* | -2.095700267 | 0.008325904 | 0.012404018 | 1 |
| T2DM | monocyte | *ARL17B* | 1.445543159 | 0.012903684 | 0.018246842 | 1 |
| T2DM | monocyte | *LINC00954* | 2.633593877 | 0.000477982 | 0.001306553 | 1 |
| T2DM | monocyte | *AC067945.3* | -1.045159653 | 0.035248146 | 0.045161013 | 1 |
| T2DM | monocyte | *LINC00200* | 8.499402426 | 0.005570204 | 0.008711412 | 1 |
| T2DM | monocyte | *U91324.1* | 2.097231146 | 0.001692691 | 0.003223856 | 1 |
| T2DM | monocyte | *ZNF688* | 7.554226534 | 3.44E-05 | 0.000365043 | 0.216060189 |
| T2DM | monocyte | *CISD3* | 1.254290834 | 0.000945114 | 0.002110809 | 1 |
| T2DM | monocyte | *PSMG3-AS1* | 3.011044916 | 8.95E-05 | 0.000464456 | 0.562418457 |
| T2DM | monocyte | *PPP5D1* | 1.279370682 | 0.000540031 | 0.001419388 | 1 |
| T2DM | monocyte | *KLHL7-AS1* | 2.122847759 | 0.001743114 | 0.00328999 | 1 |
| T2DM | monocyte | *AC092171.4* | 1.403800777 | 0.00098579 | 0.002178439 | 1 |
| T2DM | monocyte | *RPS18* | 1.941873145 | 0.005286735 | 0.008309426 | 1 |
| T2DM | monocyte | *DIRC3* | 0.664530106 | 0.000854514 | 0.001964289 | 1 |
| T2DM | monocyte | *TAPBP* | 2.463106997 | 0.003562537 | 0.005884832 | 1 |
| T2DM | monocyte | *AC107021.1* | 0.523465794 | 0.002199489 | 0.003901017 | 1 |
| T2DM | monocyte | *LINC00865* | 1.110174424 | 0.006325471 | 0.00975429 | 1 |
| T2DM | monocyte | *HLA-DQB2* | 0.991433934 | 4.96E-07 | 7.60E-05 | 0.003118045 |
| T2DM | monocyte | *LINC00665* | 1.545993487 | 0.000156896 | 0.000634753 | 0.98640675 |
| T2DM | monocyte | *LYRM9* | 1.965475124 | 0.001389978 | 0.002802692 | 1 |
| T2DM | monocyte | *LINC00342* | 2.209822822 | 0.000320698 | 0.00099517 | 1 |
| T2DM | monocyte | *LINC00702* | 0.816399527 | 0.0027533 | 0.004703803 | 1 |
| T2DM | monocyte | *LINC00472* | 1.767590566 | 0.000226598 | 0.000785783 | 1 |
| T2DM | monocyte | *AC073133.1* | -1.262234996 | 0.036532854 | 0.046702329 | 1 |
| T2DM | monocyte | *RPS28* | 1.185088485 | 0.000881676 | 0.002009097 | 1 |
| T2DM | monocyte | *AC123768.3* | 1.378543727 | 0.001193117 | 0.002499543 | 1 |
| T2DM | monocyte | *TRIM26* | 3.801372746 | 0.000118805 | 0.000544237 | 0.74692487 |
| T2DM | monocyte | *LINC01114* | -4.588953427 | 0.018751091 | 0.025467295 | 1 |
| T2DM | monocyte | *ZNF736* | 2.168658015 | 0.001036684 | 0.002262281 | 1 |
| T2DM | monocyte | *HLA-B* | 2.690146691 | 3.42E-07 | 6.70E-05 | 0.002149514 |

| | | | | | | |
|---|---|---|---|---|---|---|
| T2DM | monocyte | *C19orf81* | 4.227014325 | 0.009123425 | 0.013458229 | 1 |
| T2DM | monocyte | *ANKRD65* | 2.021095236 | 0.000892323 | 0.002023092 | 1 |
| T2DM | monocyte | *ZSCAN31* | 1.229123665 | 0.0001955 | 0.000722579 | 1 |
| T2DM | monocyte | *KIAA0040* | 2.006920198 | 0.000196879 | 0.000724272 | 1 |
| T2DM | monocyte | *B3GALT4* | 2.157482875 | 0.000304576 | 0.000963252 | 1 |
| T2DM | monocyte | *LINC00941* | 3.255192797 | 0.002224045 | 0.00393432 | 1 |
| T2DM | monocyte | *ZBTB22* | 1.080097388 | 0.002253938 | 0.003973782 | 1 |
| T2DM | monocyte | *B3GNT9* | 1.752792681 | 0.00267442 | 0.004602951 | 1 |
| T2DM | monocyte | *GS1-124K5.4* | 0.770048037 | 8.87E-05 | 0.000463045 | 0.557969769 |
| T2DM | monocyte | *ZNF737* | 1.379600327 | 0.000410813 | 0.00117346 | 1 |
| T2DM | monocyte | *LINC00959* | 2.129792567 | 0.000549261 | 0.001440035 | 1 |
| T2DM | monocyte | *HLA-DQA2* | 0.512655726 | 0.007591658 | 0.011426564 | 1 |
| T2DM | monocyte | *AC073257.2* | 10.19343619 | 0.000371276 | 0.00109424 | 1 |
| T2DM | monocyte | *LRRC37A2* | 4.17839483 | 9.79E-05 | 0.000486788 | 0.615786548 |
| T2DM | monocyte | *ASB14* | 2.336117737 | 1.00E-05 | 0.000362859 | 0.063000434 |
| T2DM | monocyte | *NME1* | 0.747526583 | 0.023701567 | 0.031503542 | 1 |
| T2DM | monocyte | *TNFSF12* | 0.778529255 | 0.004631048 | 0.00738782 | 1 |
| T2DM | monocyte | *CDRT4* | -1.372739168 | 0.013129614 | 0.01854274 | 1 |
| T2DM | monocyte | *ADSL* | 1.445464048 | 0.007766608 | 0.011642505 | 1 |
| T2DM | monocyte | *LILRA4* | 24.0268081 | 0.024414766 | 0.032335292 | 1 |
| T2DM | monocyte | *LILRA2* | -0.206191096 | 0.003653892 | 0.006008899 | 1 |
| T2DM | monocyte | *LY6G5B* | -0.843587832 | 0.000383142 | 0.001116226 | 1 |
| T2DM | monocyte | *COX19* | 2.718587783 | 5.84E-05 | 0.000397957 | 0.367408443 |
| T2DM | monocyte | *L1TD1* | -0.275058221 | 0.010433837 | 0.015128583 | 1 |
| T2DM | monocyte | *AQP1* | 0.991642371 | 0.000647431 | 0.001606948 | 1 |
| T2DM | monocyte | *C1QTNF9* | 4.672007217 | 0.005874127 | 0.009125435 | 1 |
| T2DM | monocyte | *ARHGEF25* | 1.363853812 | 0.001170277 | 0.00246566 | 1 |
| T2DM | monocyte | *LINC00886* | 1.697697242 | 0.000503294 | 0.001351072 | 1 |
| T2DM | monocyte | *NSUN6* | 2.751890279 | 3.33E-05 | 0.000365043 | 0.209143229 |
| T2DM | monocyte | *HLA-DOB* | 0.567879215 | 0.025143921 | 0.033186829 | 1 |
| T2DM | monocyte | *ATP5J2* | 1.907349027 | 0.00038223 | 0.001114602 | 1 |
| T2DM | monocyte | *ARHGAP8* | 1.464315131 | 0.003366969 | 0.005604483 | 1 |
| T2DM | monocyte | *ATP5O* | 1.936283796 | 0.000464989 | 0.001277145 | 1 |
| T2DM | monocyte | *PWP2* | 3.045199862 | 0.017344719 | 0.023757353 | 1 |
| T2DM | monocyte | *PI4KA* | 2.047386502 | 0.029276652 | 0.03811603 | 1 |
| T2DM | monocyte | *AKAP2* | 2.845096685 | 0.001729246 | 0.003270688 | 1 |
| T2DM | monocyte | *LINC00969* | 1.115873086 | 0.002609223 | 0.004506644 | 1 |
| T2DM | monocyte | *ARHGDIG* | 1.250638908 | 0.006096912 | 0.009427271 | 1 |
| T2DM | monocyte | *ARFGAP3* | 3.07424869 | 3.02E-05 | 0.000362859 | 0.18980772 |
| T2DM | monocyte | *C22orf39* | 8.031085104 | 4.50E-05 | 0.000379941 | 0.283208353 |
| T2DM | monocyte | *EIF6* | 1.209762267 | 0.001527905 | 0.00299437 | 1 |
| T2DM | monocyte | *MRPL20* | 3.495423725 | 5.99E-06 | 0.000322038 | 0.037637493 |
| T2DM | monocyte | *DECR2* | 3.742643677 | 0.000100796 | 0.000495511 | 0.633704873 |
| T2DM | monocyte | *AP5Z1* | 4.823388661 | 1.37E-05 | 0.000362859 | 0.086344309 |
| T2DM | monocyte | *PSG4* | 1.410140916 | 0.002726015 | 0.004672425 | 1 |
| T2DM | monocyte | *STON1* | 0.720550844 | 0.006921776 | 0.010534303 | 1 |
| T2DM | monocyte | *VSIG8* | 1.254199004 | 0.005266223 | 0.008283399 | 1 |
| T2DM | monocyte | *C7orf73* | 2.725661685 | 4.02E-05 | 0.00037107 | 0.252825447 |
| T2DM | monocyte | *FGF14-IT1* | 2.442308674 | 0.001801289 | 0.003361444 | 1 |
| T2DM | monocyte | *PALM2* | 0.66773006 | 0.000250203 | 0.000839513 | 1 |

| | | | | | | |
|---|---|---|---|---|---|---|
| T2DM | monocyte | *NAT6* | 1.484338561 | 9.83E-05 | 0.000487362 | 0.617974463 |
| T2DM | monocyte | *WDR92* | 1.96431373 | 7.30E-05 | 0.000431726 | 0.458925006 |
| T2DM | monocyte | *NME1-NME2* | 3.859297666 | 7.84E-05 | 0.000440255 | 0.492976366 |
| T2DM | monocyte | *PLA2G4B* | 3.460432239 | 1.78E-09 | 1.24E-06 | 1.12E-05 |
| T2DM | monocyte | *TTC4* | 0.853784176 | 0.003998892 | 0.006503112 | 1 |
| T2DM | monocyte | *JMJD7* | 3.922298979 | 1.45E-10 | 3.04E-07 | 9.11E-07 |
| T2DM | monocyte | *APOBEC3D* | -0.937881404 | 0.013628046 | 0.019163059 | 1 |
| T2DM | monocyte | *RN7SL832P* | 1.26355446 | 0.004862075 | 0.007732828 | 1 |
| T2DM | monocyte | *TMEM199* | 2.349139929 | 0.000445488 | 0.001239833 | 1 |
| T2DM | monocyte | *P2RY11* | 1.969651349 | 0.007154195 | 0.01084863 | 1 |
| T2DM | monocyte | *LILRA6* | -0.215804916 | 0.014642774 | 0.020439414 | 1 |
| T2DM | monocyte | *APOBEC3C* | 8.001698799 | 4.48E-05 | 0.000379941 | 0.281485129 |
| T2DM | monocyte | *ASPRV1* | 2.679635473 | 0.005030703 | 0.007962747 | 1 |
| T2DM | monocyte | *UBE2V1* | 1.912615959 | 0.000475622 | 0.001301235 | 1 |
| T2DM | monocyte | *N4BP2L2* | 1.765321584 | 5.13E-05 | 0.000384272 | 0.322404142 |
| T2DM | monocyte | *NEAT1* | 1.315292952 | 6.60E-05 | 0.00041942 | 0.415253685 |
| T2DM | monocyte | *ZNF585B* | 1.429426163 | 4.59E-05 | 0.000379941 | 0.288472809 |
| T2DM | monocyte | *RAB30-AS1* | 1.518830413 | 0.000881194 | 0.002008725 | 1 |
| T2DM | monocyte | *LINC00920* | 1.99668992 | 0.004214213 | 0.006812743 | 1 |
| T2DM | monocyte | *UBAP1L* | 2.558224949 | 0.003846897 | 0.006286831 | 1 |
| T2DM | monocyte | *PGAM5* | 4.02952246 | 0.000147012 | 0.000610432 | 0.924262733 |
| T2DM | monocyte | *DNAJC3-AS1* | 4.212129623 | 3.30E-05 | 0.000365043 | 0.207654679 |
| T2DM | monocyte | *LINC00926* | 1.739584469 | 0.000654355 | 0.001620296 | 1 |
| T2DM | monocyte | *BCKDHA* | 2.519398986 | 0.00132789 | 0.002703173 | 1 |
| T2DM | monocyte | *CDK11B* | 2.197804949 | 0.000106465 | 0.00051173 | 0.669342702 |
| T2DM | monocyte | *ABHD14A* | 2.050055266 | 0.000128592 | 0.000566542 | 0.808455412 |
| T2DM | monocyte | *CCDC153* | 1.401957609 | 0.001171041 | 0.002466443 | 1 |
| T2DM | monocyte | *C5orf17* | 1.370040493 | 0.000455812 | 0.001257432 | 1 |
| T2DM | monocyte | *FMN1* | 2.055776492 | 2.40E-05 | 0.000362859 | 0.151118958 |
| T2DM | monocyte | *LINC01060* | 1.067430912 | 0.00418259 | 0.006763359 | 1 |
| T2DM | monocyte | *NAIP* | -0.616528694 | 0.008546564 | 0.012690659 | 1 |
| T2DM | monocyte | *ZNF286B* | 1.185726357 | 0.011355549 | 0.016323611 | 1 |
| T2DM | monocyte | *PDCD6* | 3.870168512 | 9.90E-06 | 0.000362859 | 0.062214073 |
| T2DM | monocyte | *LINC00603* | 2.017545841 | 0.003358079 | 0.005595612 | 1 |
| T2DM | monocyte | *DNAH10OS* | -8.749934558 | 0.001786391 | 0.003347554 | 1 |
| T2DM | monocyte | *PCDHA10* | 1.977924569 | 0.001392486 | 0.002804151 | 1 |
| T2DM | monocyte | *ZNF718* | 2.489001237 | 5.93E-05 | 0.000399203 | 0.372532138 |
| T2DM | monocyte | *LINC00989* | 0.71582121 | 0.002033066 | 0.003667685 | 1 |
| T2DM | monocyte | *LINC01021* | 1.81367926 | 0.001831945 | 0.003395067 | 1 |
| T2DM | monocyte | *FAM218A* | 6.075884711 | 0.000195628 | 0.00072263 | 1 |
| T2DM | monocyte | *ATP6V1E2* | 0.905979614 | 0.001327722 | 0.002703173 | 1 |
| T2DM | monocyte | *GLI4* | 1.772582322 | 0.00048748 | 0.001325599 | 1 |
| T2DM | monocyte | *SNHG18* | 1.057299032 | 0.037090527 | 0.047347847 | 1 |
| T2DM | monocyte | *ZNF345* | 1.34984667 | 0.001015222 | 0.002229375 | 1 |
| T2DM | monocyte | *SHANK3* | 0.530979909 | 0.00096986 | 0.002152556 | 1 |
| T2DM | monocyte | *ZNF550* | 1.883139912 | 0.000343674 | 0.001035977 | 1 |
| T2DM | monocyte | *CCDC71L* | 1.082873583 | 0.000934341 | 0.002092697 | 1 |
| T2DM | monocyte | *TMEM200B* | 1.017771977 | 0.000191591 | 0.000714856 | 1 |
| T2DM | monocyte | *PCDHGB6* | 1.680814269 | 0.000324548 | 0.001001195 | 1 |
| T2DM | monocyte | *TRNP1* | -1.020218908 | 0.009201744 | 0.013561033 | 1 |

| | | | | | | |
|---|---|---|---|---|---|---|
| T2DM | monocyte | *PCDHGA5* | 0.796199234 | 0.007482254 | 0.011283505 | 1 |
| T2DM | monocyte | *ATXN7L3B* | 1.531007177 | 0.000330941 | 0.001013952 | 1 |
| T2DM | monocyte | *PRKDC* | 3.415930906 | 4.14E-05 | 0.000373192 | 0.260031615 |
| T2DM | monocyte | *PCDHGA6* | 1.416771065 | 0.000191559 | 0.000714856 | 1 |
| T2DM | monocyte | *CLDN23* | 1.911320198 | 0.001094908 | 0.00235179 | 1 |
| T2DM | monocyte | *ZNF260* | 2.094307228 | 4.87E-05 | 0.000381649 | 0.30632822 |
| T2DM | monocyte | *SIGLEC14* | -0.397277503 | 0.010992958 | 0.015837013 | 1 |
| T2DM | monocyte | *ARMS2* | 2.672129013 | 0.008800777 | 0.013022001 | 1 |
| T2DM | monocyte | *FPGT* | 4.08187836 | 0.014336904 | 0.020056991 | 1 |
| T2DM | monocyte | *MPV17L2* | 2.593931658 | 0.000367139 | 0.001084682 | 1 |
| T2DM | monocyte | *DPP3* | 1.171067436 | 0.026125224 | 0.03432587 | 1 |
| T2DM | monocyte | *EID3* | 0.746669824 | 0.003851434 | 0.006290976 | 1 |
| T2DM | monocyte | *SNHG9* | 1.140337306 | 0.000628222 | 0.001572307 | 1 |
| T2DM | monocyte | *EBLN2* | 5.695425021 | 3.01E-06 | 0.000230511 | 0.018901867 |
| T2DM | monocyte | *POLR2M* | 6.142489024 | 2.59E-05 | 0.000362859 | 0.162642787 |
| T2DM | monocyte | *TAS2R20* | 1.827590325 | 0.000648556 | 0.001608469 | 1 |
| T2DM | monocyte | *LINC00346* | 1.870836889 | 0.017681953 | 0.024166618 | 1 |
| T2DM | monocyte | *CTSO* | 4.112543925 | 2.29E-05 | 0.000362859 | 0.143844611 |
| T2DM | monocyte | *ZNF432* | 2.409946306 | 0.00070717 | 0.001710649 | 1 |
| T2DM | monocyte | *LINC00944* | 1.686433759 | 0.035878234 | 0.04590282 | 1 |
| T2DM | monocyte | *ZNF10* | 0.982755681 | 0.013734376 | 0.019291336 | 1 |
| T2DM | monocyte | *HMBS* | 2.755217554 | 1.18E-07 | 3.10E-05 | 0.000744957 |
| T2DM | monocyte | *TAS2R31* | 1.040413855 | 0.019666017 | 0.026629388 | 1 |
| T2DM | monocyte | *AC093668.2* | 2.797551403 | 0.00346085 | 0.005745542 | 1 |
| T2DM | monocyte | *CLEC12B* | -0.391457845 | 0.000780342 | 0.001838145 | 1 |
| T2DM | monocyte | *PGA5* | 10.66391296 | 0.011984389 | 0.01710462 | 1 |
| T2DM | monocyte | *ZNF253* | 2.383336243 | 7.39E-05 | 0.000432034 | 0.464673486 |
| T2DM | monocyte | *HP* | -1.155641643 | 0.004969511 | 0.007875804 | 1 |
| T2DM | monocyte | *KIAA1147* | 1.711130968 | 8.58E-05 | 0.000459295 | 0.539671084 |
| T2DM | monocyte | *GATC* | 2.061511745 | 0.001916509 | 0.003500607 | 1 |
| T2DM | monocyte | *FNTB* | 1.158810622 | 0.001642188 | 0.00315721 | 1 |
| T2DM | monocyte | *OVCH1-AS1* | 4.904507304 | 8.28E-05 | 0.000448982 | 0.520436 |
| T2DM | monocyte | *CHURC1* | 1.517729488 | 0.001414738 | 0.002840771 | 1 |
| T2DM | monocyte | *RTEL1* | 3.380933431 | 0.001442108 | 0.002883756 | 1 |
| T2DM | monocyte | *ZNF578* | 1.592157809 | 0.030803564 | 0.03985635 | 1 |
| T2DM | monocyte | *LINC00640* | 0.691885458 | 0.01375097 | 0.01931033 | 1 |
| T2DM | monocyte | *LINC00638* | 2.790650405 | 0.010954628 | 0.01578903 | 1 |
| T2DM | monocyte | *CEP95* | 2.054907801 | 7.39E-05 | 0.000432034 | 0.464868559 |
| T2DM | monocyte | *LINC00639* | 0.566166848 | 0.000223819 | 0.000779936 | 1 |
| T2DM | monocyte | *LINC00648* | 0.651433845 | 0.002269436 | 0.003993357 | 1 |
| T2DM | monocyte | *LINC00924* | 1.037471136 | 0.001457152 | 0.00290276 | 1 |
| T2DM | monocyte | *ITGB3* | 1.751962333 | 0.002873572 | 0.004882743 | 1 |
| T2DM | monocyte | *UBE2Q2L* | 2.767993514 | 0.005651509 | 0.008823203 | 1 |
| T2DM | monocyte | *IQCH-AS1* | 1.947890875 | 0.001633617 | 0.003150476 | 1 |
| T2DM | monocyte | *PWRN1* | 0.822274347 | 0.003589316 | 0.005924397 | 1 |
| T2DM | monocyte | *HOXB7* | 3.363204753 | 0.016675076 | 0.022945108 | 1 |
| T2DM | monocyte | *FRRS1L* | 2.218502762 | 0.001021603 | 0.002238696 | 1 |
| T2DM | monocyte | *LINC00562* | 1.059181168 | 0.010179449 | 0.014807542 | 1 |
| T2DM | monocyte | *AC009120.5* | 9.580743917 | 5.17E-05 | 0.000384775 | 0.324993804 |
| T2DM | monocyte | *LINC00565* | -5.651897926 | 0.001439752 | 0.002879962 | 1 |

| | | | | | | |
|---|---|---|---|---|---|---|
| T2DM | monocyte | *TMEM178B* | 1.005991903 | 0.001132204 | 0.002410486 | 1 |
| T2DM | monocyte | *LINC01003* | 1.917379266 | 0.001253615 | 0.002588334 | 1 |
| T2DM | monocyte | *HPR* | 1.813580227 | 0.001909691 | 0.003494778 | 1 |
| T2DM | monocyte | *GOLGA8H* | 0.793306588 | 0.01121836 | 0.016145713 | 1 |
| T2DM | monocyte | *LINC00662* | 2.614914435 | 4.95E-05 | 0.000383513 | 0.311011571 |
| T2DM | monocyte | *LINC00667* | 0.888496782 | 0.001068163 | 0.002310922 | 1 |
| T2DM | monocyte | *LINC00909* | 1.004047065 | 0.000422763 | 0.001195642 | 1 |
| T2DM | monocyte | *DYNLL2* | 2.528554855 | 0.001328154 | 0.002703173 | 1 |
| T2DM | monocyte | *ILF3-AS1* | 2.305989464 | 2.25E-05 | 0.000362859 | 0.141162369 |
| T2DM | monocyte | *AC139100.2* | 3.621287884 | 0.001900326 | 0.003480148 | 1 |
| T2DM | monocyte | *LINC01140* | 1.376666878 | 0.000385507 | 0.001122076 | 1 |
| T2DM | monocyte | *LINC00906* | -5.319010953 | 0.023412404 | 0.031158717 | 1 |
| T2DM | monocyte | *ZNF582-AS1* | 2.633697731 | 0.001054447 | 0.002290709 | 1 |
| T2DM | monocyte | *AC137932.1* | 3.171683251 | 0.008778078 | 0.012991472 | 1 |
| T2DM | monocyte | *AC003005.2* | 2.088305311 | 0.003226848 | 0.005406093 | 1 |
| T2DM | monocyte | *AL132989.1* | -1.581516527 | 0.028656775 | 0.037378661 | 1 |
| T2DM | monocyte | *LINC00664* | 3.147091836 | 0.003850751 | 0.006290976 | 1 |
| T2DM | monocyte | *FBXO17* | 1.539129987 | 0.000202465 | 0.000735524 | 1 |
| T2DM | monocyte | *SNHG8* | 2.636719014 | 0.000521974 | 0.001388764 | 1 |
| T2DM | monocyte | *HCG17* | 1.889723672 | 0.003213036 | 0.005386762 | 1 |
| T2DM | monocyte | *GTF2H5* | 1.943957027 | 0.000291309 | 0.0009334 | 1 |
| T2DM | monocyte | *FGF14-AS2* | 2.918000983 | 0.038035769 | 0.048426667 | 1 |
| T2DM | monocyte | *CASC15* | 1.734139003 | 0.000442886 | 0.001234778 | 1 |
| T2DM | monocyte | *LINC01023* | 5.248984166 | 1.87E-05 | 0.000362859 | 0.117315909 |
| T2DM | monocyte | *AP000347.2* | -0.981422429 | 0.038073027 | 0.048442494 | 1 |
| T2DM | monocyte | *POLR2J4* | 6.669480426 | 2.47E-05 | 0.000362859 | 0.155230318 |
| T2DM | monocyte | *AC144652.1* | 1.250376225 | 0.000141272 | 0.000596094 | 0.888179355 |
| T2DM | monocyte | *PAXIP1-AS1* | 1.216721352 | 0.006188282 | 0.009556799 | 1 |
| T2DM | neutrophil | *SEMA3F* | -1.526017666 | 6.60E-05 | 0.037702573 | 0.414728308 |
| T2DM | neutrophil | *CDK2AP1* | 3.783658184 | 4.08E-05 | 0.028508489 | 0.256576405 |
| T2DM | neutrophil | *CLEC18A* | -11.49141072 | 1.15E-06 | 0.00120087 | 0.007205223 |
| T2DM | neutrophil | *AP3S2* | 3.902726008 | 2.21E-07 | 0.000462412 | 0.001387237 |
| T2DM | neutrophil | *HLA-DQB1* | -3.000258431 | 6.77E-07 | 0.000866682 | 0.004255975 |
| T2DM | neutrophil | *EIF3C* | 4.301414927 | 6.43E-05 | 0.037702573 | 0.404450901 |
| T2DM | neutrophil | *NOTCH4* | -1.079865019 | 1.69E-07 | 0.000462412 | 0.001062305 |
| T2DM | neutrophil | *HCG27* | 2.184671034 | 3.20E-06 | 0.002516808 | 0.02013446 |
| T2DM | neutrophil | *STARD10* | 4.027659494 | 1.42E-06 | 0.001272416 | 0.008906912 |
| T2DM | neutrophil | *HLA-DQB2* | 1.613967408 | 6.89E-07 | 0.000866682 | 0.00433341 |
| T2DM | neutrophil | *HLA-DQA2* | 2.823419916 | 1.91E-10 | 1.20E-06 | 1.20E-06 |
| T2DM | nk_cell | *SEMA3F* | -0.978944788 | 3.28E-07 | 0.000121136 | 0.002059315 |
| T2DM | nk_cell | *RBM6* | -0.730618357 | 1.09E-07 | 5.72E-05 | 0.000686017 |
| T2DM | nk_cell | *GGNBP2* | -1.157925924 | 0.000250119 | 0.03520144 | 1 |
| T2DM | nk_cell | *TUBG2* | -1.022738802 | 1.72E-08 | 1.20E-05 | 0.000107996 |
| T2DM | nk_cell | *ITIH4* | 0.606399953 | 2.16E-05 | 0.005033482 | 0.135904022 |
| T2DM | nk_cell | *COBLL1* | -0.672720651 | 8.14E-05 | 0.014224023 | 0.512064822 |
| T2DM | nk_cell | *PABPC4* | -1.943550676 | 7.86E-07 | 0.000235223 | 0.004939692 |
| T2DM | nk_cell | *CWF19L1* | -0.912398579 | 0.00015996 | 0.024528488 | 1 |
| T2DM | nk_cell | *GCDH* | -1.942790847 | 0.000383978 | 0.048948718 | 1 |
| T2DM | nk_cell | *ATP13A1* | 1.427292092 | 0.000389285 | 0.048948718 | 1 |
| T2DM | nk_cell | *HSD17B1* | -0.71879808 | 1.02E-06 | 0.000290532 | 0.006391712 |

| | | | | | | |
|---|---|---|---|---|---|---|
| T2DM | nk_cell | *CHMP3* | -1.162537439 | 0.000253556 | 0.03520144 | 1 |
| T2DM | nk_cell | *CD3EAP* | -1.492909728 | 0.000129687 | 0.021456394 | 0.815342967 |
| T2DM | nk_cell | *DNAH7* | -0.719292061 | 1.34E-05 | 0.003365326 | 0.084133155 |
| T2DM | nk_cell | *OPRL1* | -1.101960466 | 2.25E-05 | 0.005045653 | 0.141278294 |
| T2DM | nk_cell | *MAN2C1* | -0.489988514 | 1.71E-05 | 0.004134993 | 0.107509805 |
| T2DM | nk_cell | *LRRC46* | -1.748800473 | 2.87E-05 | 0.006125879 | 0.180319691 |
| T2DM | nk_cell | *BHMT* | -0.55670964 | 3.29E-05 | 0.006676435 | 0.206969481 |
| T2DM | nk_cell | *HSD17B12* | -1.06984988 | 2.08E-08 | 1.31E-05 | 0.0001306 |
| T2DM | nk_cell | *APIP* | -0.401285462 | 7.41E-05 | 0.013707859 | 0.466067199 |
| T2DM | nk_cell | *CLEC18A* | -2.506705813 | 1.45E-07 | 7.01E-05 | 0.000910901 |
| T2DM | nk_cell | *KCNJ15* | -1.328629738 | 0.00018822 | 0.028174748 | 1 |
| T2DM | nk_cell | *AP3S2* | 1.174940962 | 1.89E-07 | 8.51E-05 | 0.001190884 |
| T2DM | nk_cell | *CD1E* | 2.981945002 | 0.000257558 | 0.03520144 | 1 |
| T2DM | nk_cell | *RFT1* | -1.81344871 | 0.000102114 | 0.017351062 | 0.641989278 |
| T2DM | nk_cell | *RNF123* | 1.248384672 | 5.89E-07 | 0.000194779 | 0.003700804 |
| T2DM | nk_cell | *PCBD1* | -0.698009629 | 5.63E-05 | 0.010780216 | 0.35422902 |
| T2DM | nk_cell | *IRS1* | -1.099043512 | 3.30E-23 | 2.08E-19 | 2.08E-19 |
| T2DM | nk_cell | *RGS19* | -1.038109131 | 8.12E-05 | 0.014224023 | 0.510750853 |
| T2DM | nk_cell | *VEGFB* | -1.175465868 | 1.22E-05 | 0.003200038 | 0.0768009 |
| T2DM | nk_cell | *HOXB2* | -1.558577709 | 0.000143332 | 0.023105835 | 0.901127561 |
| T2DM | nk_cell | *HLA-DQB1* | -1.221608571 | 5.83E-16 | 1.83E-12 | 3.66E-12 |
| T2DM | nk_cell | *BMP8A* | 5.726380034 | 2.99E-09 | 3.13E-06 | 1.88E-05 |
| T2DM | nk_cell | *ZNF703* | -1.752679539 | 5.66E-05 | 0.010780216 | 0.355747115 |
| T2DM | nk_cell | *EIF3C* | 0.9856892 | 0.000264381 | 0.035365135 | 1 |
| T2DM | nk_cell | *HLA-DRB1* | -1.67549758 | 1.85E-12 | 2.91E-09 | 1.16E-08 |
| T2DM | nk_cell | *FAM180B* | -1.071184057 | 0.000149408 | 0.023483198 | 0.93932792 |
| T2DM | nk_cell | *HLA-DQA1* | -1.294572743 | 1.62E-08 | 1.20E-05 | 0.000101863 |
| T2DM | nk_cell | *HLA-DRB5* | -0.590640432 | 2.25E-11 | 2.83E-08 | 1.42E-07 |
| T2DM | nk_cell | *HLA-DRA* | 1.337056087 | 3.80E-07 | 0.000132702 | 0.002388644 |
| T2DM | nk_cell | *NOTCH4* | -0.203177979 | 9.92E-08 | 5.67E-05 | 0.000623532 |
| T2DM | nk_cell | *AGER* | 1.110280392 | 0.000363603 | 0.047624427 | 1 |
| T2DM | nk_cell | *HCG27* | 0.760391739 | 4.48E-06 | 0.001223259 | 0.028134957 |
| T2DM | nk_cell | *DNLZ* | 0.702357367 | 0.000245291 | 0.03520144 | 1 |
| T2DM | nk_cell | *STARD10* | 0.858305156 | 6.80E-07 | 0.000213816 | 0.004276321 |
| T2DM | nk_cell | *HLA-DQB2* | 0.469818458 | 2.16E-07 | 9.05E-05 | 0.001357788 |
| T2DM | nk_cell | *HLA-DQA2* | 1.205108076 | 1.22E-14 | 2.56E-11 | 7.68E-11 |
| T2DM | nk_cell | *PLA2G4B* | 1.560089662 | 1.64E-08 | 1.20E-05 | 0.000102989 |
| T2DM | nk_cell | *JMJD7* | 1.545905907 | 2.39E-07 | 9.41E-05 | 0.001504884 |
| T2DM | nk_cell | *HMBS* | 1.069368286 | 2.92E-05 | 0.006125879 | 0.183776366 |
| T2DM | pericyte | *C1orf112* | 4.387517007 | 4.07E-05 | 0.000607238 | 0.255832035 |
| T2DM | pericyte | *CFH* | 3.159159459 | 2.88E-06 | 0.000234693 | 0.018123381 |
| T2DM | pericyte | *NIPAL3* | 1.540370368 | 0.001639892 | 0.004686364 | 1 |
| T2DM | pericyte | *ENPP4* | 0.943138525 | 0.005658915 | 0.012035724 | 1 |
| T2DM | pericyte | *SEMA3F* | -1.099161479 | 4.54E-07 | 6.21E-05 | 0.002851237 |
| T2DM | pericyte | *KRIT1* | 2.81910969 | 0.001138313 | 0.003599887 | 1 |
| T2DM | pericyte | *RAD52* | 3.892363987 | 4.71E-05 | 0.000607238 | 0.296028947 |
| T2DM | pericyte | *BAD* | 5.089304619 | 5.94E-05 | 0.00062437 | 0.373736818 |
| T2DM | pericyte | *HS3ST1* | 2.680500977 | 0.006715209 | 0.013842137 | 1 |
| T2DM | pericyte | *HECW1* | 0.554599613 | 0.012050828 | 0.022772334 | 1 |
| T2DM | pericyte | *MAD1L1* | 3.065064269 | 0.002071577 | 0.005535064 | 1 |

| | | | | | | |
|---|---|---|---|---|---|---|
| T2DM | pericyte | *LASP1* | 3.046102522 | 3.22E-05 | 0.000599418 | 0.20234708 |
| T2DM | pericyte | *SNX11* | 1.266656383 | 0.000427151 | 0.001772607 | 1 |
| T2DM | pericyte | *TMEM176A* | 1.096075735 | 0.002736156 | 0.006900205 | 1 |
| T2DM | pericyte | *M6PR* | 2.600339206 | 0.000156432 | 0.000945663 | 0.983489139 |
| T2DM | pericyte | *ICA1* | 2.277897578 | 0.000236992 | 0.001209392 | 1 |
| T2DM | pericyte | *ALS2* | 3.678083958 | 2.81E-05 | 0.000573841 | 0.17643949 |
| T2DM | pericyte | *POLDIP2* | 1.476172718 | 0.000899732 | 0.003037923 | 1 |
| T2DM | pericyte | *CD38* | 0.892210196 | 0.00912201 | 0.01789394 | 1 |
| T2DM | pericyte | *RBM6* | -0.779143663 | 1.11E-05 | 0.000426258 | 0.06948002 |
| T2DM | pericyte | *RECQL* | 3.598279909 | 1.16E-05 | 0.000446471 | 0.073221226 |
| T2DM | pericyte | *PDK4* | 2.437482747 | 5.32E-05 | 0.000619691 | 0.334266041 |
| T2DM | pericyte | *CDC27* | 1.578075326 | 0.00069972 | 0.002522443 | 1 |
| T2DM | pericyte | *CIAPIN1* | 1.752330568 | 0.00047436 | 0.001905624 | 1 |
| T2DM | pericyte | *FAM214B* | 1.935772727 | 0.000495941 | 0.001972158 | 1 |
| T2DM | pericyte | *COPZ2* | 0.935889201 | 0.001529579 | 0.004462395 | 1 |
| T2DM | pericyte | *GCFC2* | 3.483068986 | 0.000103496 | 0.000753104 | 0.650681955 |
| T2DM | pericyte | *WDR54* | 1.896972436 | 0.002646955 | 0.006720028 | 1 |
| T2DM | pericyte | *ABCB4* | 2.181148087 | 0.001183512 | 0.003700641 | 1 |
| T2DM | pericyte | *PDK2* | 1.744303474 | 0.001317752 | 0.003986867 | 1 |
| T2DM | pericyte | *SYNRG* | 3.037083475 | 0.000689352 | 0.00249078 | 1 |
| T2DM | pericyte | *ZNF263* | 2.564518881 | 0.000152227 | 0.000937005 | 0.957050085 |
| T2DM | pericyte | *SPATA20* | 1.603208054 | 0.006440705 | 0.013377177 | 1 |
| T2DM | pericyte | *TNFRSF12A* | 0.916082212 | 0.019503496 | 0.034375799 | 1 |
| T2DM | pericyte | *MAP3K9* | 1.342737741 | 0.000981137 | 0.003246531 | 1 |
| T2DM | pericyte | *RALA* | 2.343569137 | 9.23E-05 | 0.00071031 | 0.580323491 |
| T2DM | pericyte | *FARP2* | 2.061768547 | 0.002210092 | 0.005824542 | 1 |
| T2DM | pericyte | *IFRD1* | 1.099271636 | 0.00561929 | 0.011979816 | 1 |
| T2DM | pericyte | *VPS41* | 1.551812034 | 0.012358991 | 0.023284679 | 1 |
| T2DM | pericyte | *ELAC2* | 2.149605265 | 0.002560324 | 0.006541705 | 1 |
| T2DM | pericyte | *NOS2* | 5.683233991 | 0.001904076 | 0.005211549 | 1 |
| T2DM | pericyte | *TRAPPC6A* | 2.734006131 | 0.023238508 | 0.039841969 | 1 |
| T2DM | pericyte | *TSR3* | 3.921342174 | 1.42E-05 | 0.000473503 | 0.089018601 |
| T2DM | pericyte | *PIGQ* | 1.18210012 | 0.020231863 | 0.03545087 | 1 |
| T2DM | pericyte | *CDK11A* | 1.05218312 | 0.011221217 | 0.021486231 | 1 |
| T2DM | pericyte | *DLEC1* | 1.356953033 | 0.003037929 | 0.007484839 | 1 |
| T2DM | pericyte | *MPND* | 2.518073032 | 0.000846144 | 0.002911416 | 1 |
| T2DM | pericyte | *MGST1* | 1.849340349 | 5.15E-05 | 0.000616617 | 0.323847073 |
| T2DM | pericyte | *NFIX* | 1.288727519 | 0.007960311 | 0.015984183 | 1 |
| T2DM | pericyte | *MMP25* | 1.426221664 | 0.001351137 | 0.004043123 | 1 |
| T2DM | pericyte | *PKD1* | 1.881176096 | 0.000203602 | 0.001095342 | 1 |
| T2DM | pericyte | *MAPK8IP2* | -2.325469012 | 0.012956559 | 0.024265084 | 1 |
| T2DM | pericyte | *RHOBTB2* | 1.516503273 | 0.022284474 | 0.038500272 | 1 |
| T2DM | pericyte | *SEC62* | 3.082914857 | 0.000795726 | 0.002774668 | 1 |
| T2DM | pericyte | *RPS20* | 3.022407479 | 0.000187038 | 0.001043394 | 1 |
| T2DM | pericyte | *MLXIPL* | 0.600369858 | 0.000237724 | 0.001211238 | 1 |
| T2DM | pericyte | *SPRTN* | 2.262831107 | 0.000274995 | 0.001323244 | 1 |
| T2DM | pericyte | *METTL13* | 3.630499825 | 0.001360244 | 0.004060709 | 1 |
| T2DM | pericyte | *STARD3NL* | 2.932419961 | 6.45E-05 | 0.000632025 | 0.405416338 |
| T2DM | pericyte | *ZNF200* | 2.915206537 | 0.000316695 | 0.001431389 | 1 |
| T2DM | pericyte | *CD4* | 1.52057257 | 0.00099829 | 0.003284638 | 1 |

| | | | | | | |
|---|---|---|---|---|---|---|
| T2DM | pericyte | *SCMH1* | 2.200858501 | 0.000299031 | 0.001384395 | 1 |
| T2DM | pericyte | *CLCN6* | 2.873858896 | 0.000913159 | 0.003067079 | 1 |
| T2DM | pericyte | *MRC2* | 1.088107365 | 0.015329263 | 0.028016011 | 1 |
| T2DM | pericyte | *MKS1* | 2.02869467 | 0.000541928 | 0.002104259 | 1 |
| T2DM | pericyte | *UTP18* | 4.031589447 | 4.72E-05 | 0.000607238 | 0.297039713 |
| T2DM | pericyte | *TTC19* | 2.087785423 | 0.001201998 | 0.00373589 | 1 |
| T2DM | pericyte | *SYT7* | 1.868599251 | 0.001195824 | 0.00372924 | 1 |
| T2DM | pericyte | *LARS2* | 2.508539726 | 6.86E-06 | 0.000347886 | 0.043137812 |
| T2DM | pericyte | *PPP5C* | 3.073744736 | 1.49E-05 | 0.000478946 | 0.093873354 |
| T2DM | pericyte | *CEP68* | 3.287821023 | 4.42E-13 | 5.56E-10 | 2.78E-09 |
| T2DM | pericyte | *ZBTB32* | 4.521463425 | 0.026705776 | 0.044880838 | 1 |
| T2DM | pericyte | *ALOX5* | 1.059117044 | 0.002288074 | 0.005984165 | 1 |
| T2DM | pericyte | *MAP4K5* | 2.485858736 | 0.00091352 | 0.003067079 | 1 |
| T2DM | pericyte | *PSMC4* | 2.277192239 | 1.36E-05 | 0.000465252 | 0.085606312 |
| T2DM | pericyte | *MAN2B2* | 2.740441056 | 0.001299523 | 0.003943098 | 1 |
| T2DM | pericyte | *CLDN11* | 1.52968793 | 0.02427571 | 0.041316023 | 1 |
| T2DM | pericyte | *ANGEL1* | 3.28208966 | 6.69E-06 | 0.000344883 | 0.042064233 |
| T2DM | pericyte | *RNF14* | 2.706262926 | 9.48E-05 | 0.000717456 | 0.595911873 |
| T2DM | pericyte | *ZC3H3* | 1.391807457 | 0.001068176 | 0.003435102 | 1 |
| T2DM | pericyte | *MDH1* | 1.454945599 | 0.023896791 | 0.040814759 | 1 |
| T2DM | pericyte | *SLC30A9* | 2.461140265 | 0.000151578 | 0.000935202 | 0.9529712 |
| T2DM | pericyte | *MTMR11* | 0.453399995 | 0.021214338 | 0.0369254 | 1 |
| T2DM | pericyte | *COX15* | 2.052167706 | 0.00459067 | 0.010300337 | 1 |
| T2DM | pericyte | *CCDC88C* | 1.261758959 | 0.004575319 | 0.010269558 | 1 |
| T2DM | pericyte | *IL20RA* | 8.927687308 | 0.000281015 | 0.001336306 | 1 |
| T2DM | pericyte | *GLT8D1* | 1.083917249 | 0.009375084 | 0.018316083 | 1 |
| T2DM | pericyte | *RUFY3* | 1.529927122 | 0.007990997 | 0.016025326 | 1 |
| T2DM | pericyte | *WWTR1* | 3.072064535 | 3.49E-05 | 0.000599418 | 0.219387136 |
| T2DM | pericyte | *TTC27* | 1.651096006 | 0.004532067 | 0.010190668 | 1 |
| T2DM | pericyte | *VSIG2* | 2.246990035 | 0.022190076 | 0.03841107 | 1 |
| T2DM | pericyte | *MARCO* | 0.628128332 | 0.023230576 | 0.039839234 | 1 |
| T2DM | pericyte | *MNAT1* | 2.792619874 | 2.64E-05 | 0.000562456 | 0.166021081 |
| T2DM | pericyte | *OSBPL5* | 4.044896845 | 0.000289594 | 0.001357702 | 1 |
| T2DM | pericyte | *AQR* | 3.32840275 | 4.07E-05 | 0.000607238 | 0.256004115 |
| T2DM | pericyte | *ZDHHC6* | 2.416300464 | 0.002176061 | 0.005755127 | 1 |
| T2DM | pericyte | *RNH1* | 1.21904901 | 0.02652144 | 0.044630699 | 1 |
| T2DM | pericyte | *NDUFS1* | 2.304548957 | 0.00057515 | 0.002189103 | 1 |
| T2DM | pericyte | *BIRC3* | 0.763251723 | 0.005466964 | 0.011746685 | 1 |
| T2DM | pericyte | *AKAP11* | 2.886713705 | 4.75E-05 | 0.000607238 | 0.29832869 |
| T2DM | pericyte | *SNAPC1* | 1.377973121 | 0.002458528 | 0.00633474 | 1 |
| T2DM | pericyte | *GCLM* | 1.148704315 | 0.006150466 | 0.012876451 | 1 |
| T2DM | pericyte | *UBR2* | 3.056937961 | 6.01E-05 | 0.00062437 | 0.377913246 |
| T2DM | pericyte | *HSF2* | 2.907373513 | 4.76E-05 | 0.000607238 | 0.298971601 |
| T2DM | pericyte | *NR1H3* | 1.409246794 | 0.021358265 | 0.037145066 | 1 |
| T2DM | pericyte | *TYMP* | 1.802703726 | 0.006815875 | 0.014003728 | 1 |
| T2DM | pericyte | *TOMM34* | 4.44211549 | 0.000178277 | 0.001021719 | 1 |
| T2DM | pericyte | *KPNA6* | 2.323044885 | 2.31E-05 | 0.000548455 | 0.145055395 |
| T2DM | pericyte | *VIM* | 2.657022295 | 6.14E-05 | 0.000627415 | 0.385860401 |
| T2DM | pericyte | *RNASET2* | 1.148060489 | 0.008737919 | 0.017264392 | 1 |
| T2DM | pericyte | *CD44* | 2.38492336 | 0.001011103 | 0.003317749 | 1 |

| | | | | | | |
|---|---|---|---|---|---|---|
| T2DM | pericyte | *MIPEP* | 4.099510128 | 6.24E-05 | 0.000630549 | 0.392474616 |
| T2DM | pericyte | *B4GALT7* | 2.395473467 | 0.003344212 | 0.00805558 | 1 |
| T2DM | pericyte | *VRK2* | 2.864493467 | 0.000160882 | 0.000958177 | 1 |
| T2DM | pericyte | *VEZT* | 2.235361504 | 0.000760796 | 0.002694718 | 1 |
| T2DM | pericyte | *SNX1* | 1.349800807 | 0.018645755 | 0.033077274 | 1 |
| T2DM | pericyte | *ARNTL2* | 2.70238579 | 0.000162484 | 0.000963715 | 1 |
| T2DM | pericyte | *SLC39A9* | 1.355185085 | 0.004121083 | 0.009459276 | 1 |
| T2DM | pericyte | *ANK1* | 0.920724234 | 0.002847791 | 0.007107606 | 1 |
| T2DM | pericyte | *RABEP1* | 1.943316626 | 0.000302242 | 0.00139207 | 1 |
| T2DM | pericyte | *BAK1* | 1.136197027 | 0.000171771 | 0.000999928 | 1 |
| T2DM | pericyte | *GRN* | 3.045386405 | 6.69E-05 | 0.000632837 | 0.4204818 |
| T2DM | pericyte | *ARID4A* | 3.349987773 | 0.000204811 | 0.001098675 | 1 |
| T2DM | pericyte | *IFT88* | 2.033775346 | 0.003326274 | 0.008037003 | 1 |
| T2DM | pericyte | *ZCCHC8* | 3.413375657 | 6.11E-05 | 0.000627415 | 0.38390072 |
| T2DM | pericyte | *FUT8* | 3.187709904 | 8.70E-05 | 0.000695771 | 0.546876037 |
| T2DM | pericyte | *PIAS1* | 2.815802478 | 6.19E-05 | 0.000628694 | 0.389385131 |
| T2DM | pericyte | *SLC4A7* | 2.873851187 | 0.000160679 | 0.000958177 | 1 |
| T2DM | pericyte | *MYOC* | 0.843991425 | 0.006883198 | 0.014114353 | 1 |
| T2DM | pericyte | *ADSS* | 4.1260021 | 0.000104814 | 0.000759174 | 0.658963041 |
| T2DM | pericyte | *TBC1D23* | 2.99341601 | 6.77E-05 | 0.000632837 | 0.42589204 |
| T2DM | pericyte | *CUL3* | 2.94978779 | 0.000347529 | 0.001521528 | 1 |
| T2DM | pericyte | *TUBG2* | -0.798431998 | 0.000221674 | 0.001158399 | 1 |
| T2DM | pericyte | *NSUN2* | 1.432429437 | 0.003552621 | 0.008447551 | 1 |
| T2DM | pericyte | *MFAP3* | 0.953417178 | 0.026272982 | 0.044259979 | 1 |
| T2DM | pericyte | *AGA* | 6.059949623 | 5.68E-05 | 0.00062437 | 0.357349869 |
| T2DM | pericyte | *BOD1L1* | 1.75022173 | 0.010516998 | 0.020294772 | 1 |
| T2DM | pericyte | *MAT2B* | 3.993982906 | 1.87E-05 | 0.000512131 | 0.117351399 |
| T2DM | pericyte | *CLEC16A* | 2.919753558 | 7.36E-05 | 0.000648165 | 0.462916809 |
| T2DM | pericyte | *SKIV2L2* | 3.023631906 | 0.000161721 | 0.000961913 | 1 |
| T2DM | pericyte | *DNAH5* | 1.049042577 | 0.009194629 | 0.018013908 | 1 |
| T2DM | pericyte | *C6* | 0.665618425 | 0.010014431 | 0.019438322 | 1 |
| T2DM | pericyte | *RAI14* | 1.210405151 | 0.018338568 | 0.032633619 | 1 |
| T2DM | pericyte | *PHLPP2* | 2.300269684 | 0.000327623 | 0.001467071 | 1 |
| T2DM | pericyte | *RAB27B* | 1.389718077 | 2.81E-05 | 0.000573841 | 0.176743117 |
| T2DM | pericyte | *PSMA4* | 3.111905234 | 0.003448479 | 0.008259273 | 1 |
| T2DM | pericyte | *THAP3* | 0.997439914 | 0.007353766 | 0.014923539 | 1 |
| T2DM | pericyte | *TDP1* | 4.745073368 | 0.000211034 | 0.00112039 | 1 |
| T2DM | pericyte | *SPATA7* | 2.530605679 | 0.000207345 | 0.001105665 | 1 |
| T2DM | pericyte | *MED17* | 3.152923669 | 3.33E-06 | 0.000258649 | 0.020950529 |
| T2DM | pericyte | *AP2S1* | 4.408542124 | 0.000239151 | 0.001217445 | 1 |
| T2DM | pericyte | *LCP2* | 1.221989637 | 0.001439143 | 0.004243851 | 1 |
| T2DM | pericyte | *TRIT1* | 1.180127025 | 0.015009545 | 0.027503646 | 1 |
| T2DM | pericyte | *CTNNA1* | 2.290723383 | 8.92E-05 | 0.000702125 | 0.561071961 |
| T2DM | pericyte | *CNTLN* | 3.222189816 | 7.22E-05 | 0.00064687 | 0.453902891 |
| T2DM | pericyte | *WDR37* | 2.215299295 | 0.000176349 | 0.001018057 | 1 |
| T2DM | pericyte | *YTHDC2* | 3.26137756 | 8.26E-06 | 0.000379198 | 0.051950149 |
| T2DM | pericyte | *ATP6V1H* | 2.83680632 | 6.20E-05 | 0.000628694 | 0.389790121 |
| T2DM | pericyte | *POLR2B* | 3.137394191 | 0.001280019 | 0.003902185 | 1 |
| T2DM | pericyte | *CP* | 4.025174958 | 0.004343927 | 0.009877133 | 1 |
| T2DM | pericyte | *DTNBP1* | 3.430877743 | 3.86E-05 | 0.000607238 | 0.242911537 |

| | | | | | | |
|---|---|---|---|---|---|---|
| T2DM | pericyte | *GOPC* | 2.049003307 | 0.021170019 | 0.036868672 | 1 |
| T2DM | pericyte | *TSPAN17* | 1.696891857 | 0.000824739 | 0.002855045 | 1 |
| T2DM | pericyte | *CC2D2A* | 2.839950236 | 4.52E-05 | 0.000607238 | 0.284464374 |
| T2DM | pericyte | *RRM2B* | 2.31293796 | 0.000810758 | 0.002819267 | 1 |
| T2DM | pericyte | *SNX29* | 2.80040628 | 4.18E-05 | 0.000607238 | 0.262868784 |
| T2DM | pericyte | *CELF2* | 2.140182561 | 4.47E-05 | 0.000607238 | 0.281118188 |
| T2DM | pericyte | *ERCC8* | 2.97785536 | 2.58E-05 | 0.000562456 | 0.162062113 |
| T2DM | pericyte | *EPN3* | 5.528834278 | 0.0005553 | 0.002141821 | 1 |
| T2DM | pericyte | *RFC2* | 4.563281466 | 3.46E-05 | 0.000599418 | 0.217426887 |
| T2DM | pericyte | *JKAMP* | 1.324729436 | 0.009121778 | 0.01789394 | 1 |
| T2DM | pericyte | *DKK3* | 1.926752905 | 0.000345091 | 0.001515074 | 1 |
| T2DM | pericyte | *ARHGEF5* | 1.056955263 | 0.00388039 | 0.009045612 | 1 |
| T2DM | pericyte | *NFE2L3* | 5.799513523 | 3.56E-05 | 0.00060096 | 0.223897326 |
| T2DM | pericyte | *MCUR1* | 1.768093448 | 0.005815178 | 0.012318067 | 1 |
| T2DM | pericyte | *PTGER3* | 2.303578725 | 0.000107132 | 0.000770639 | 0.673538623 |
| T2DM | pericyte | *MAPK9* | 3.04908354 | 1.26E-05 | 0.000458996 | 0.079406286 |
| T2DM | pericyte | *FAM160A2* | 0.977433958 | 0.012162244 | 0.02296217 | 1 |
| T2DM | pericyte | *HERPUD1* | 2.844824246 | 0.000136182 | 0.000878628 | 0.856176487 |
| T2DM | pericyte | *PIK3CB* | 1.5203817 | 0.00426294 | 0.009731948 | 1 |
| T2DM | pericyte | *CYBA* | 3.287397087 | 2.77E-05 | 0.000570694 | 0.174061657 |
| T2DM | pericyte | *THOC3* | 0.993688174 | 0.005233183 | 0.011356929 | 1 |
| T2DM | pericyte | *RRP12* | 1.759752405 | 0.000267041 | 0.0012953 | 1 |
| T2DM | pericyte | *FOXN3* | 2.473897875 | 0.000311178 | 0.001420754 | 1 |
| T2DM | pericyte | *MCF2L2* | 2.410464098 | 0.002325541 | 0.006059128 | 1 |
| T2DM | pericyte | *ANAPC4* | 3.754704391 | 5.02E-05 | 0.000610424 | 0.315382173 |
| T2DM | pericyte | *OPN3* | 1.169811575 | 0.00099206 | 0.003275777 | 1 |
| T2DM | pericyte | *SDCCAG8* | 1.371577509 | 0.001838147 | 0.005086852 | 1 |
| T2DM | pericyte | *TBC1D22A* | 2.916926352 | 3.09E-05 | 0.000599418 | 0.194254895 |
| T2DM | pericyte | *ATP9A* | 1.42365341 | 0.000975927 | 0.003232694 | 1 |
| T2DM | pericyte | *CHRDL2* | 1.514015853 | 0.00636097 | 0.013246579 | 1 |
| T2DM | pericyte | *GALC* | 2.034436731 | 0.009862528 | 0.019173072 | 1 |
| T2DM | pericyte | *FAM114A2* | 2.161279778 | 0.003968347 | 0.009199483 | 1 |
| T2DM | pericyte | *GINM1* | 2.224358351 | 0.018660381 | 0.033093883 | 1 |
| T2DM | pericyte | *ITIH4* | 0.820292449 | 3.86E-06 | 0.000279804 | 0.024288032 |
| T2DM | pericyte | *TRAF1* | 1.933557409 | 0.022811902 | 0.039260451 | 1 |
| T2DM | pericyte | *IL17RB* | 1.377576663 | 0.017342192 | 0.031115971 | 1 |
| T2DM | pericyte | *TRAF3IP2* | 2.575194173 | 7.71E-05 | 0.000661364 | 0.484829976 |
| T2DM | pericyte | *GDI2* | 2.126657906 | 0.003821272 | 0.008937625 | 1 |
| T2DM | pericyte | *TMCC3* | 2.725879793 | 0.003604958 | 0.008533272 | 1 |
| T2DM | pericyte | *PITHD1* | 2.262633085 | 0.000321047 | 0.001444826 | 1 |
| T2DM | pericyte | *ATP11B* | 1.280520693 | 0.001772008 | 0.004962413 | 1 |
| T2DM | pericyte | *ZC3H11A* | 1.464843199 | 0.004677675 | 0.010447086 | 1 |
| T2DM | pericyte | *RIOK2* | 4.117938728 | 0.000144945 | 0.00091218 | 0.911268207 |
| T2DM | pericyte | *YBX3* | 3.463322039 | 0.000114032 | 0.000791304 | 0.716921129 |
| T2DM | pericyte | *OGFR* | 2.031925199 | 0.001405195 | 0.004173103 | 1 |
| T2DM | pericyte | *QSER1* | 3.822896134 | 8.44E-05 | 0.000682391 | 0.5303453 |
| T2DM | pericyte | *NCKAP1* | 2.389619275 | 0.00151228 | 0.004420133 | 1 |
| T2DM | pericyte | *SFSWAP* | 2.700082695 | 6.68E-05 | 0.000632837 | 0.41967447 |
| T2DM | pericyte | *VMP1* | 2.797861836 | 0.00059558 | 0.002240819 | 1 |
| T2DM | pericyte | *SPHK2* | 2.574764881 | 0.00010239 | 0.000750264 | 0.643726099 |

| | | | | | | |
|---|---|---|---|---|---|---|
| T2DM | pericyte | *ISOC2* | 5.311087101 | 9.52E-05 | 0.000718199 | 0.59825974 |
| T2DM | pericyte | *EPN1* | 4.937896214 | 2.33E-05 | 0.000548455 | 0.146178613 |
| T2DM | pericyte | *ADCK1* | 2.579505137 | 0.008117256 | 0.016216457 | 1 |
| T2DM | pericyte | *CASP8* | 2.110283839 | 6.71E-05 | 0.000632837 | 0.421587523 |
| T2DM | pericyte | *TM7SF3* | 2.568083681 | 0.000243324 | 0.001224801 | 1 |
| T2DM | pericyte | *TAF2* | 2.582524483 | 0.000326816 | 0.001464497 | 1 |
| T2DM | pericyte | *TMEM161A* | 1.737972543 | 0.000711038 | 0.002550083 | 1 |
| T2DM | pericyte | *LPAR2* | 2.932393395 | 0.000982773 | 0.003250233 | 1 |
| T2DM | pericyte | *SUGP2* | 2.771792055 | 2.52E-06 | 0.000216894 | 0.015833269 |
| T2DM | pericyte | *ABCA7* | 1.87862323 | 0.028497052 | 0.047281329 | 1 |
| T2DM | pericyte | *DDX20* | 3.435576046 | 0.000197913 | 0.001074506 | 1 |
| T2DM | pericyte | *BTBD1* | 3.832640836 | 1.58E-06 | 0.000159839 | 0.009910042 |
| T2DM | pericyte | *PMS1* | 3.235299635 | 3.33E-05 | 0.000599418 | 0.209091773 |
| T2DM | pericyte | *ANKS1A* | 2.70995591 | 0.000159117 | 0.000954278 | 1 |
| T2DM | pericyte | *NTHL1* | 1.828685447 | 0.002006767 | 0.005412502 | 1 |
| T2DM | pericyte | *IPO5* | 4.262915426 | 8.79E-05 | 0.000700657 | 0.552818119 |
| T2DM | pericyte | *ANKRD44* | 2.606788142 | 0.000102588 | 0.000750837 | 0.64496913 |
| T2DM | pericyte | *ADAT1* | 0.786193762 | 0.024793726 | 0.042061024 | 1 |
| T2DM | pericyte | *NDUFB4* | 4.186787251 | 8.90E-06 | 0.000385718 | 0.055929097 |
| T2DM | pericyte | *ZC3H15* | 2.653681875 | 0.001050077 | 0.003394258 | 1 |
| T2DM | pericyte | *MAP2K4* | 3.40870149 | 6.38E-05 | 0.000632025 | 0.401401029 |
| T2DM | pericyte | *TMEM206* | 1.354024424 | 0.001750759 | 0.004918241 | 1 |
| T2DM | pericyte | *SEC61A2* | 2.643138992 | 0.000165753 | 0.000976653 | 1 |
| T2DM | pericyte | *PPP2R5A* | 1.804186801 | 0.000138988 | 0.000891653 | 0.873820315 |
| T2DM | pericyte | *NFYC* | 3.068623834 | 7.73E-05 | 0.000661364 | 0.48589667 |
| T2DM | pericyte | *ZMYND12* | 0.705141974 | 0.01178051 | 0.022375925 | 1 |
| T2DM | pericyte | *NGEF* | 1.547400785 | 0.004569613 | 0.010263196 | 1 |
| T2DM | pericyte | *CD84* | 1.261010508 | 0.000464592 | 0.00188404 | 1 |
| T2DM | pericyte | *ATXN3* | 1.602056773 | 0.000677364 | 0.00245877 | 1 |
| T2DM | pericyte | *GOLGA5* | 3.513976408 | 0.001327108 | 0.003999773 | 1 |
| T2DM | pericyte | *MTFR1* | 1.889257142 | 0.006185968 | 0.012942157 | 1 |
| T2DM | pericyte | *MYO9A* | 2.114644447 | 0.000433391 | 0.001790229 | 1 |
| T2DM | pericyte | *SP100* | 2.9066827 | 0.000395583 | 0.001672514 | 1 |
| T2DM | pericyte | *METTL22* | 1.643034589 | 0.000868772 | 0.002970075 | 1 |
| T2DM | pericyte | *TP53BP1* | 2.854893582 | 0.000267159 | 0.0012953 | 1 |
| T2DM | pericyte | *DHX8* | 1.934740512 | 0.010889347 | 0.020916994 | 1 |
| T2DM | pericyte | *ROGDI* | 2.580875836 | 0.001122967 | 0.003562106 | 1 |
| T2DM | pericyte | *HDAC4* | 3.044944781 | 4.51E-05 | 0.000607238 | 0.283466454 |
| T2DM | pericyte | *RASSF1* | 2.451507408 | 0.001266905 | 0.003877815 | 1 |
| T2DM | pericyte | *FGFR3* | 0.3558911 | 0.026899128 | 0.045097284 | 1 |
| T2DM | pericyte | *HEATR6* | 4.593586293 | 3.01E-05 | 0.000596445 | 0.189073117 |
| T2DM | pericyte | *SRBD1* | 2.525098277 | 6.27E-05 | 0.000630549 | 0.394093319 |
| T2DM | pericyte | *CYFIP1* | 3.370800792 | 0.000190401 | 0.001051889 | 1 |
| T2DM | pericyte | *PSME4* | 2.347118648 | 0.000349497 | 0.001525896 | 1 |
| T2DM | pericyte | *SDK2* | 3.578845384 | 0.000707392 | 0.002541358 | 1 |
| T2DM | pericyte | *NUCKS1* | 3.248106118 | 1.75E-05 | 0.000512131 | 0.110187957 |
| T2DM | pericyte | *KCNAB2* | 0.92328833 | 0.025753345 | 0.043551715 | 1 |
| T2DM | pericyte | *GAL* | 1.366075345 | 0.005196397 | 0.011296594 | 1 |
| T2DM | pericyte | *CLEC2D* | 1.430851467 | 0.007944411 | 0.015962452 | 1 |
| T2DM | pericyte | *PIGB* | 1.840427172 | 0.00427081 | 0.009742592 | 1 |

| | | | | | | |
|---|---|---|---|---|---|---|
| T2DM | pericyte | *PFN2* | 1.941467992 | 0.016234434 | 0.029439251 | 1 |
| T2DM | pericyte | *DAPP1* | 0.592149964 | 0.010467525 | 0.020211711 | 1 |
| T2DM | pericyte | *TMEM260* | 3.626162441 | 5.36E-05 | 0.000619691 | 0.336987428 |
| T2DM | pericyte | *DGCR2* | 3.233782215 | 0.000792911 | 0.002767925 | 1 |
| T2DM | pericyte | *MNT* | 1.339115139 | 0.014063399 | 0.025966694 | 1 |
| T2DM | pericyte | *ZXDC* | 2.619657398 | 0.00014598 | 0.00091686 | 0.917776378 |
| T2DM | pericyte | *POLB* | 1.685453922 | 0.008097946 | 0.016183022 | 1 |
| T2DM | pericyte | *WIPI1* | 4.197604644 | 4.40E-05 | 0.000607238 | 0.276870736 |
| T2DM | pericyte | *NDST1* | 2.603866454 | 0.000126579 | 0.00084164 | 0.795803026 |
| T2DM | pericyte | *ASNS* | 1.889209357 | 0.000297631 | 0.001381518 | 1 |
| T2DM | pericyte | *AP3M2* | 3.797093603 | 0.000473637 | 0.001905155 | 1 |
| T2DM | pericyte | *ST6GALNAC2* | 1.80198603 | 0.004486135 | 0.010112704 | 1 |
| T2DM | pericyte | *EIF2B3* | 2.75428719 | 4.88E-05 | 0.000607238 | 0.306644774 |
| T2DM | pericyte | *CAMK2A* | -0.446057266 | 0.021160415 | 0.036868672 | 1 |
| T2DM | pericyte | *CDC42* | 3.07819056 | 3.19E-05 | 0.000599418 | 0.200431638 |
| T2DM | pericyte | *OSBPL3* | 1.853082322 | 0.001499163 | 0.004392003 | 1 |
| T2DM | pericyte | *RAD18* | 2.879070817 | 0.000115726 | 0.000796114 | 0.727569737 |
| T2DM | pericyte | *SNX13* | 2.664106231 | 4.72E-05 | 0.000607238 | 0.296700919 |
| T2DM | pericyte | *TRIP13* | 1.991367851 | 0.005242314 | 0.011372819 | 1 |
| T2DM | pericyte | *TCF3* | 2.757523749 | 0.002196078 | 0.005798716 | 1 |
| T2DM | pericyte | *CPSF1* | 3.039535634 | 0.000256946 | 0.001267988 | 1 |
| T2DM | pericyte | *CYBRD1* | 2.083552822 | 2.67E-05 | 0.000562456 | 0.167779657 |
| T2DM | pericyte | *EPN2* | 2.902826981 | 0.000276201 | 0.001324545 | 1 |
| T2DM | pericyte | *AFF4* | 2.012318039 | 0.001575782 | 0.004551324 | 1 |
| T2DM | pericyte | *TRHDE* | 1.420031931 | 0.000283072 | 0.001339905 | 1 |
| T2DM | pericyte | *NFATC3* | 2.674675191 | 2.37E-05 | 0.000548455 | 0.149157726 |
| T2DM | pericyte | *TRNT1* | 4.011542984 | 5.25E-05 | 0.000619691 | 0.330198935 |
| T2DM | pericyte | *ACADVL* | 3.273906586 | 0.000314012 | 0.001424384 | 1 |
| T2DM | pericyte | *DERL2* | 2.545675517 | 0.000399817 | 0.001684752 | 1 |
| T2DM | pericyte | *NDE1* | 1.862039692 | 0.01836077 | 0.032654643 | 1 |
| T2DM | pericyte | *AP1M1* | 1.1527768 | 0.01314375 | 0.024542547 | 1 |
| T2DM | pericyte | *PVR* | 1.630498271 | 0.003702936 | 0.008711864 | 1 |
| T2DM | pericyte | *XRCC1* | 3.643841176 | 0.000232456 | 0.00119595 | 1 |
| T2DM | pericyte | *TP63* | 1.004497244 | 0.001253681 | 0.003850462 | 1 |
| T2DM | pericyte | *ALPK1* | 2.317092148 | 4.28E-05 | 0.000607238 | 0.268818294 |
| T2DM | pericyte | *LLGL2* | 1.311243522 | 0.004610865 | 0.010341958 | 1 |
| T2DM | pericyte | *PDE8A* | 1.943600246 | 0.000624635 | 0.002316644 | 1 |
| T2DM | pericyte | *NLE1* | 1.279576347 | 0.002428084 | 0.006276878 | 1 |
| T2DM | pericyte | *SDHA* | 2.776191324 | 1.81E-06 | 0.000177356 | 0.011350803 |
| T2DM | pericyte | *SMARCE1* | 2.139144389 | 0.000116546 | 0.000798466 | 0.732722112 |
| T2DM | pericyte | *GSDMB* | 0.97311851 | 0.015975181 | 0.029069408 | 1 |
| T2DM | pericyte | *PICALM* | 2.655858977 | 6.41E-05 | 0.000632025 | 0.402985673 |
| T2DM | pericyte | *CLNS1A* | 1.475776267 | 0.000184053 | 0.001035729 | 1 |
| T2DM | pericyte | *ATP2A3* | 0.781748348 | 0.002977925 | 0.007365152 | 1 |
| T2DM | pericyte | *NTN4* | 1.48409342 | 0.000517999 | 0.002034139 | 1 |
| T2DM | pericyte | *LMAN1* | 3.524444373 | 0.000608034 | 0.002274991 | 1 |
| T2DM | pericyte | *ENO1* | 3.423241642 | 0.000285987 | 0.001346398 | 1 |
| T2DM | pericyte | *TIPIN* | 0.513750937 | 0.008014522 | 0.016067378 | 1 |
| T2DM | pericyte | *SRI* | 2.803917025 | 0.013995638 | 0.025866618 | 1 |
| T2DM | pericyte | *EIF4G3* | 2.385787057 | 0.000931915 | 0.003118121 | 1 |

| | | | | | | |
|---|---|---|---|---|---|---|
| T2DM | pericyte | *TTC38* | 1.197225806 | 0.002097529 | 0.005578327 | 1 |
| T2DM | pericyte | *GRAMD4* | 3.564854186 | 9.83E-05 | 0.000733473 | 0.618318055 |
| T2DM | pericyte | *CELSR1* | 3.502389114 | 0.000234524 | 0.001198741 | 1 |
| T2DM | pericyte | *ADD2* | 7.633258405 | 0.000125866 | 0.00084164 | 0.7913166 |
| T2DM | pericyte | *VPS9D1* | 4.63051572 | 0.000483264 | 0.001933981 | 1 |
| T2DM | pericyte | *SLC25A3* | 2.863119278 | 0.001037803 | 0.003365197 | 1 |
| T2DM | pericyte | *FSCN1* | 2.418617506 | 0.004903462 | 0.01080928 | 1 |
| T2DM | pericyte | *PLD1* | 2.773447465 | 0.001410157 | 0.004182358 | 1 |
| T2DM | pericyte | *BCAP29* | 3.170720416 | 6.23E-05 | 0.000630549 | 0.391721316 |
| T2DM | pericyte | *SEC31B* | 0.443803694 | 0.007955208 | 0.01597904 | 1 |
| T2DM | pericyte | *MKRN2* | 1.143294427 | 0.005372319 | 0.01159484 | 1 |
| T2DM | pericyte | *MCM6* | 2.663574542 | 7.23E-05 | 0.00064687 | 0.454742897 |
| T2DM | pericyte | *RBMS2* | 2.671728939 | 0.002729955 | 0.006895615 | 1 |
| T2DM | pericyte | *MLH1* | 1.339179906 | 0.006102636 | 0.012810441 | 1 |
| T2DM | pericyte | *FMO4* | 2.385413637 | 0.000192561 | 0.001061026 | 1 |
| T2DM | pericyte | *TPD52* | 0.792348406 | 0.00278941 | 0.006989646 | 1 |
| T2DM | pericyte | *GPATCH1* | 2.781894467 | 0.000580568 | 0.002202711 | 1 |
| T2DM | pericyte | *ICAM3* | 1.018418947 | 0.00186737 | 0.005138542 | 1 |
| T2DM | pericyte | *NT5C2* | 3.074949798 | 5.33E-06 | 0.000328654 | 0.033522717 |
| T2DM | pericyte | *CAMSAP3* | 5.274181231 | 0.005540567 | 0.011852175 | 1 |
| T2DM | pericyte | *STXBP2* | 2.667406761 | 1.26E-05 | 0.000458996 | 0.079202509 |
| T2DM | pericyte | *MAP2K7* | 2.175055947 | 0.000312059 | 0.001422709 | 1 |
| T2DM | pericyte | *TM9SF3* | 2.697732402 | 0.000116895 | 0.000799696 | 0.73492053 |
| T2DM | pericyte | *DNAJC10* | 3.893479734 | 3.59E-05 | 0.00060096 | 0.225998944 |
| T2DM | pericyte | *DYNC1I2* | 3.116902713 | 0.000221288 | 0.001158398 | 1 |
| T2DM | pericyte | *APBB1IP* | 1.251762976 | 0.000237739 | 0.001211238 | 1 |
| T2DM | pericyte | *SIRT6* | 5.507456925 | 0.005311296 | 0.011494705 | 1 |
| T2DM | pericyte | *CAPZB* | 2.603894683 | 0.000264493 | 0.001293055 | 1 |
| T2DM | pericyte | *NAALAD2* | 1.499916771 | 0.000221796 | 0.001158399 | 1 |
| T2DM | pericyte | *FGFR1* | 1.61394848 | 0.00197638 | 0.005351206 | 1 |
| T2DM | pericyte | *FBLN1* | 1.039885127 | 0.000370696 | 0.001595186 | 1 |
| T2DM | pericyte | *PIAS2* | 2.243012402 | 0.000623748 | 0.002315491 | 1 |
| T2DM | pericyte | *AMPH* | 0.553154873 | 0.026841142 | 0.045012072 | 1 |
| T2DM | pericyte | *MCCC1* | 1.435245799 | 0.000764708 | 0.002704005 | 1 |
| T2DM | pericyte | *ACER3* | 0.967873585 | 0.026215422 | 0.044186691 | 1 |
| T2DM | pericyte | *PIK3C3* | 3.016684359 | 0.000630556 | 0.002333317 | 1 |
| T2DM | pericyte | *N4BP2* | 2.167075161 | 0.000541066 | 0.0021024 | 1 |
| T2DM | pericyte | *TULP3* | 3.856194776 | 2.07E-05 | 0.000527586 | 0.130013529 |
| T2DM | pericyte | *ZCWPW1* | 1.40482578 | 0.000922504 | 0.003093217 | 1 |
| T2DM | pericyte | *FGF20* | 17.62885518 | 0.001226306 | 0.003789498 | 1 |
| T2DM | pericyte | *PCM1* | 2.272130981 | 0.001637461 | 0.004681545 | 1 |
| T2DM | pericyte | *MYH7B* | 2.373100621 | 0.000377691 | 0.001619456 | 1 |
| T2DM | pericyte | *OSBPL6* | 4.667398774 | 0.000180932 | 0.001033172 | 1 |
| T2DM | pericyte | *XRCC5* | 3.321258373 | 7.58E-05 | 0.000657143 | 0.47642893 |
| T2DM | pericyte | *LXN* | 2.433873236 | 0.000170792 | 0.000995155 | 1 |
| T2DM | pericyte | *REXO1* | 3.423844511 | 0.011492901 | 0.021935602 | 1 |
| T2DM | pericyte | *SAR1A* | 2.462135576 | 0.000139775 | 0.000894176 | 0.87876583 |
| T2DM | pericyte | *CDC14A* | 1.231185415 | 0.001734942 | 0.004885952 | 1 |
| T2DM | pericyte | *FDFT1* | 3.076270974 | 3.27E-05 | 0.000599418 | 0.205476714 |
| T2DM | pericyte | *SCGN* | -0.387116852 | 0.028191525 | 0.046851736 | 1 |

| | | | | | | |
|---|---|---|---|---|---|---|
| T2DM | pericyte | *DDX1* | 3.760514811 | 0.00022888 | 0.001183363 | 1 |
| T2DM | pericyte | *DNM2* | 3.481729241 | 5.75E-05 | 0.00062437 | 0.361618875 |
| T2DM | pericyte | *STX7* | 2.253359918 | 8.61E-05 | 0.000692254 | 0.541342964 |
| T2DM | pericyte | *RABL2B* | 1.640777562 | 0.000782055 | 0.002742726 | 1 |
| T2DM | pericyte | *DCT* | 1.22336836 | 0.004739431 | 0.010547082 | 1 |
| T2DM | pericyte | *SLC35C2* | 3.412802994 | 5.73E-05 | 0.00062437 | 0.360547905 |
| T2DM | pericyte | *SCTR* | 1.333097338 | 0.001072872 | 0.003444917 | 1 |
| T2DM | pericyte | *RIF1* | 1.36794219 | 0.008645811 | 0.017130859 | 1 |
| T2DM | pericyte | *KCNN2* | 1.806147816 | 0.023878798 | 0.04079511 | 1 |
| T2DM | pericyte | *PSEN1* | 2.072758673 | 0.002949935 | 0.007310303 | 1 |
| T2DM | pericyte | *CPOX* | 1.941325953 | 0.003382769 | 0.00812666 | 1 |
| T2DM | pericyte | *MOK* | 1.46800227 | 0.000724095 | 0.002582631 | 1 |
| T2DM | pericyte | *RSBN1* | 2.30641954 | 0.001079526 | 0.003457454 | 1 |
| T2DM | pericyte | *MAGI3* | 2.829799695 | 4.03E-05 | 0.000607238 | 0.253119915 |
| T2DM | pericyte | *STK17B* | 2.094262383 | 8.09E-05 | 0.000671315 | 0.508731182 |
| T2DM | pericyte | *KIAA0141* | 2.28197028 | 0.000792057 | 0.002766478 | 1 |
| T2DM | pericyte | *HSPB11* | 3.070685525 | 2.41E-05 | 0.000553241 | 0.151588082 |
| T2DM | pericyte | *WDR70* | 2.50472165 | 0.000201186 | 0.001088517 | 1 |
| T2DM | pericyte | *C5orf22* | 1.400485953 | 0.000783529 | 0.002745844 | 1 |
| T2DM | pericyte | *CCNT2* | 1.498973216 | 0.000684674 | 0.002476722 | 1 |
| T2DM | pericyte | *GEMIN5* | 1.122432922 | 0.010934283 | 0.020984078 | 1 |
| T2DM | pericyte | *GSK3B* | 2.958052946 | 0.000121249 | 0.000821438 | 0.762294657 |
| T2DM | pericyte | *ERC1* | 2.764816945 | 0.000116158 | 0.000798129 | 0.730288114 |
| T2DM | pericyte | *DOPEY1* | 1.076372047 | 0.002851162 | 0.007113198 | 1 |
| T2DM | pericyte | *ULK2* | 1.416628548 | 0.02931993 | 0.048470787 | 1 |
| T2DM | pericyte | *PIBF1* | 2.197806805 | 0.000617262 | 0.002296876 | 1 |
| T2DM | pericyte | *TDRD3* | 2.347806462 | 5.69E-05 | 0.00062437 | 0.357490473 |
| T2DM | pericyte | *PDS5B* | 2.229474283 | 0.007526783 | 0.015205939 | 1 |
| T2DM | pericyte | *CYLD* | 2.985622205 | 0.000181886 | 0.001034031 | 1 |
| T2DM | pericyte | *ZNF586* | 2.121381507 | 6.59E-05 | 0.000632701 | 0.414029001 |
| T2DM | pericyte | *ZNF446* | 1.108069815 | 0.003292815 | 0.007971477 | 1 |
| T2DM | pericyte | *ZNF264* | 2.951125534 | 0.000173287 | 0.001004107 | 1 |
| T2DM | pericyte | *RPS5* | 3.729357436 | 0.000127424 | 0.000844709 | 0.801117795 |
| T2DM | pericyte | *CHMP2B* | 1.4234289 | 0.028042189 | 0.04666523 | 1 |
| T2DM | pericyte | *ZMPSTE24* | 2.11529025 | 0.000315122 | 0.001426579 | 1 |
| T2DM | pericyte | *STARD7* | 3.564483707 | 1.86E-05 | 0.000512131 | 0.117215417 |
| T2DM | pericyte | *GSTP1* | 4.292709956 | 0.00019368 | 0.001063462 | 1 |
| T2DM | pericyte | *WBP11* | 4.677828064 | 0.000177771 | 0.001020443 | 1 |
| T2DM | pericyte | *APOB* | 0.794561363 | 0.014922021 | 0.02737518 | 1 |
| T2DM | pericyte | *NCOA1* | 2.740882562 | 0.000147802 | 0.000921857 | 0.929232037 |
| T2DM | pericyte | *CD59* | 2.334010707 | 0.004385658 | 0.009950428 | 1 |
| T2DM | pericyte | *CD82* | 2.06389687 | 2.23E-05 | 0.000547486 | 0.140318571 |
| T2DM | pericyte | *TAF9* | 4.996603577 | 0.003873054 | 0.009031857 | 1 |
| T2DM | pericyte | *SCAMP1* | 3.417756467 | 8.37E-06 | 0.000381367 | 0.052628649 |
| T2DM | pericyte | *HACE1* | 2.312461582 | 0.0010238 | 0.003343213 | 1 |
| T2DM | pericyte | *SEH1L* | 2.658867018 | 0.001910545 | 0.005226978 | 1 |
| T2DM | pericyte | *OVGP1* | -1.803350602 | 0.007379889 | 0.014962064 | 1 |
| T2DM | pericyte | *SLC25A24* | 2.620312638 | 0.000348576 | 0.001522932 | 1 |
| T2DM | pericyte | *PILRA* | 0.283316794 | 0.020100905 | 0.035270553 | 1 |
| T2DM | pericyte | *ABCB1* | 1.031590521 | 0.011601926 | 0.022111163 | 1 |

| | | | | | | |
|---|---|---|---|---|---|---|
| T2DM | pericyte | *CPNE3* | 2.406833162 | 5.01E-06 | 0.000320172 | 0.031482417 |
| T2DM | pericyte | *RRN3* | 2.544215155 | 0.0022846 | 0.005982209 | 1 |
| T2DM | pericyte | *CTTN* | 3.598329051 | 4.80E-05 | 0.000607238 | 0.302034183 |
| T2DM | pericyte | *EPS15* | 1.613341557 | 0.000995448 | 0.003283509 | 1 |
| T2DM | pericyte | *ATG16L1* | 3.344798048 | 6.69E-06 | 0.000344883 | 0.042075741 |
| T2DM | pericyte | *USP40* | 2.324005082 | 0.000726028 | 0.002587607 | 1 |
| T2DM | pericyte | *MAST2* | 1.777344633 | 0.000896063 | 0.003035317 | 1 |
| T2DM | pericyte | *B4GALT1* | 2.921455366 | 3.03E-05 | 0.000598882 | 0.19044436 |
| T2DM | pericyte | *CHMP5* | 2.390453608 | 0.016647104 | 0.030040281 | 1 |
| T2DM | pericyte | *EIF2AK1* | 2.50118512 | 1.32E-05 | 0.000462556 | 0.082969889 |
| T2DM | pericyte | *EPDR1* | 1.019820854 | 0.000251004 | 0.00125243 | 1 |
| T2DM | pericyte | *SEPHS1* | 1.872749754 | 0.000313012 | 0.001424252 | 1 |
| T2DM | pericyte | *ITPKC* | 1.172088897 | 0.001245819 | 0.003833353 | 1 |
| T2DM | pericyte | *TMED2* | 4.016446224 | 5.76E-05 | 0.00062437 | 0.362415654 |
| T2DM | pericyte | *ACOX3* | 1.487741974 | 0.000547575 | 0.002121028 | 1 |
| T2DM | pericyte | *ACHE* | -0.64860156 | 0.005785607 | 0.012267829 | 1 |
| T2DM | pericyte | *ATXN7L3* | 1.797759326 | 0.000652512 | 0.002392036 | 1 |
| T2DM | pericyte | *PGS1* | 2.384917983 | 0.000576867 | 0.002192723 | 1 |
| T2DM | pericyte | *LPCAT2* | 1.846234855 | 0.010048913 | 0.019487204 | 1 |
| T2DM | pericyte | *TXNDC16* | 1.649301615 | 0.00183829 | 0.005086852 | 1 |
| T2DM | pericyte | *GMCL1* | 3.099426065 | 0.001098072 | 0.003511486 | 1 |
| T2DM | pericyte | *PHACTR3* | 1.047537652 | 0.003527108 | 0.008418727 | 1 |
| T2DM | pericyte | *ERGIC2* | 2.687984616 | 0.006728196 | 0.013859818 | 1 |
| T2DM | pericyte | *ALG6* | 1.836525637 | 0.025976508 | 0.04385454 | 1 |
| T2DM | pericyte | *DDX18* | 3.206901492 | 2.35E-05 | 0.000548455 | 0.147577887 |
| T2DM | pericyte | *ASAP3* | 2.704516089 | 0.005859281 | 0.01238645 | 1 |
| T2DM | pericyte | *C3orf18* | 1.697207586 | 2.22E-06 | 0.000202161 | 0.01393874 |
| T2DM | pericyte | *FKBP1A* | 3.916589557 | 0.00011134 | 0.00078265 | 0.699996859 |
| T2DM | pericyte | *NSFL1C* | 3.12613433 | 0.000304929 | 0.001399049 | 1 |
| T2DM | pericyte | *ZNF343* | 2.011465777 | 0.000210242 | 0.001117319 | 1 |
| T2DM | pericyte | *MAVS* | 2.522525538 | 9.10E-05 | 0.000704357 | 0.572342973 |
| T2DM | pericyte | *LZTS3* | 1.170816097 | 0.019197364 | 0.033912286 | 1 |
| T2DM | pericyte | *XRN2* | 3.350788304 | 4.76E-05 | 0.000607238 | 0.299477004 |
| T2DM | pericyte | *RBBP9* | 2.980347232 | 0.004384892 | 0.009950428 | 1 |
| T2DM | pericyte | *ANAPC5* | 1.977343891 | 0.02164721 | 0.037584749 | 1 |
| T2DM | pericyte | *SLC23A2* | 2.854552939 | 0.003483696 | 0.008334776 | 1 |
| T2DM | pericyte | *TMEM230* | 2.326643844 | 7.98E-05 | 0.000667567 | 0.502010175 |
| T2DM | pericyte | *DZANK1* | 2.206236113 | 0.003044725 | 0.007494982 | 1 |
| T2DM | pericyte | *TASP1* | 1.680098121 | 0.001761958 | 0.004938667 | 1 |
| T2DM | pericyte | *OAS1* | 5.246270961 | 0.000184199 | 0.001035729 | 1 |
| T2DM | pericyte | *KIF16B* | 3.318780286 | 0.00021776 | 0.001144699 | 1 |
| T2DM | pericyte | *FUS* | 2.996350981 | 0.000351531 | 0.001530521 | 1 |
| T2DM | pericyte | *FXYD5* | 3.945955243 | 0.001269241 | 0.003881185 | 1 |
| T2DM | pericyte | *GANAB* | 3.043891574 | 9.55E-05 | 0.000719108 | 0.600455163 |
| T2DM | pericyte | *GMIP* | 1.843764717 | 0.001260476 | 0.003865666 | 1 |
| T2DM | pericyte | *MLF2* | 2.907313518 | 0.000229706 | 0.001186657 | 1 |
| T2DM | pericyte | *NECAP1* | 2.173100825 | 0.001094432 | 0.003501625 | 1 |
| T2DM | pericyte | *ANKRD24* | 1.290869259 | 0.003546438 | 0.008443811 | 1 |
| T2DM | pericyte | *GPATCH2L* | 2.053894761 | 0.000650895 | 0.0023875 | 1 |
| T2DM | pericyte | *SLC9A1* | 2.965277835 | 7.30E-05 | 0.000646906 | 0.459065034 |

| | | | | | | |
|---|---|---|---|---|---|---|
| T2DM | pericyte | *SPTLC1* | 1.657843033 | 0.011628309 | 0.022140272 | 1 |
| T2DM | pericyte | *PAPOLA* | 3.070153832 | 0.000338737 | 0.001503983 | 1 |
| T2DM | pericyte | *RGS1* | 1.004198194 | 0.004802938 | 0.010636164 | 1 |
| T2DM | pericyte | *YPEL3* | 1.039388072 | 0.000655731 | 0.002401036 | 1 |
| T2DM | pericyte | *MRPS33* | 1.497039286 | 0.030117465 | 0.049528774 | 1 |
| T2DM | pericyte | *STRN4* | 3.666787826 | 0.004002165 | 0.009253972 | 1 |
| T2DM | pericyte | *LYZ* | 2.227717611 | 0.001086383 | 0.003477641 | 1 |
| T2DM | pericyte | *GNPTG* | 3.984539046 | 2.83E-05 | 0.000575064 | 0.177694867 |
| T2DM | pericyte | *ZNF268* | 1.132618405 | 0.01028591 | 0.019879347 | 1 |
| T2DM | pericyte | *GOLGA3* | 2.075205065 | 6.31E-05 | 0.000632025 | 0.396618424 |
| T2DM | pericyte | *CERS4* | 3.539333 | 0.000315177 | 0.001426579 | 1 |
| T2DM | pericyte | *USP48* | 1.448322559 | 0.011442767 | 0.021866468 | 1 |
| T2DM | pericyte | *PDPR* | 3.120560224 | 6.21E-06 | 0.000343901 | 0.039017989 |
| T2DM | pericyte | *EXOC1* | 3.194201952 | 4.62E-05 | 0.000607238 | 0.290651347 |
| T2DM | pericyte | *NLRC4* | 1.53840759 | 0.001183711 | 0.003700641 | 1 |
| T2DM | pericyte | *SLC26A4* | 1.351847343 | 0.008971688 | 0.017637587 | 1 |
| T2DM | pericyte | *DLD* | 2.971279908 | 3.27E-05 | 0.000599418 | 0.205440044 |
| T2DM | pericyte | *TXNL1* | 4.422419605 | 5.86E-06 | 0.000331752 | 0.036824482 |
| T2DM | pericyte | *IL5RA* | 1.723673935 | 0.006107027 | 0.012815381 | 1 |
| T2DM | pericyte | *RAPGEF4* | 1.14726846 | 0.000613632 | 0.00229011 | 1 |
| T2DM | pericyte | *SEL1L3* | 1.22642687 | 0.000732215 | 0.002605225 | 1 |
| T2DM | pericyte | *TF* | 1.275755638 | 0.002974 | 0.007358338 | 1 |
| T2DM | pericyte | *MYO15A* | 1.05214053 | 0.000997357 | 0.003284638 | 1 |
| T2DM | pericyte | *NLRP1* | 2.8210923 | 0.000183692 | 0.001035729 | 1 |
| T2DM | pericyte | *ZFHX4* | 2.288853795 | 0.005227895 | 0.01134937 | 1 |
| T2DM | pericyte | *RGS17* | 0.659789749 | 0.029052608 | 0.048041491 | 1 |
| T2DM | pericyte | *PPP2R3C* | 3.477712091 | 6.61E-05 | 0.000632701 | 0.415684491 |
| T2DM | pericyte | *OSGEP* | 2.415186936 | 0.000193656 | 0.001063462 | 1 |
| T2DM | pericyte | *SLC22A17* | -1.22688441 | 0.022534884 | 0.038877497 | 1 |
| T2DM | pericyte | *SCFD1* | 2.275279383 | 0.000216553 | 0.001141214 | 1 |
| T2DM | pericyte | *RPGRIP1* | 2.162685757 | 0.000476776 | 0.001911666 | 1 |
| T2DM | pericyte | *TOX4* | 1.925219692 | 0.000359564 | 0.00155902 | 1 |
| T2DM | pericyte | *AGO1* | 2.079181987 | 0.000972739 | 0.003223833 | 1 |
| T2DM | pericyte | *RFFL* | 1.326627297 | 0.001914357 | 0.005235129 | 1 |
| T2DM | pericyte | *MFSD11* | 1.548322189 | 0.001247126 | 0.003834074 | 1 |
| T2DM | pericyte | *NUP50* | 2.091101496 | 0.000159223 | 0.000954278 | 1 |
| T2DM | pericyte | *COMT* | 2.926531 | 6.64E-05 | 0.000632837 | 0.417469389 |
| T2DM | pericyte | *VNN3* | 4.002472432 | 0.000754537 | 0.002675563 | 1 |
| T2DM | pericyte | *SEC22C* | 4.061502624 | 3.38E-05 | 0.000599418 | 0.212736505 |
| T2DM | pericyte | *FMO2* | 1.301130695 | 0.001267597 | 0.003878045 | 1 |
| T2DM | pericyte | *SUCO* | 3.502670291 | 0.000177528 | 0.001020443 | 1 |
| T2DM | pericyte | *MSH2* | 2.047979552 | 0.000268506 | 0.001300536 | 1 |
| T2DM | pericyte | *TMEM38B* | 2.216807138 | 0.000456109 | 0.001859636 | 1 |
| T2DM | pericyte | *PDE6C* | 1.614312089 | 0.010616301 | 0.020436216 | 1 |
| T2DM | pericyte | *IKZF5* | 3.192660458 | 0.000160808 | 0.000958177 | 1 |
| T2DM | pericyte | *BLNK* | 0.972501305 | 0.008839901 | 0.017438487 | 1 |
| T2DM | pericyte | *WAC* | 2.224006111 | 0.001334716 | 0.004015006 | 1 |
| T2DM | pericyte | *CREM* | 2.420587795 | 5.24E-05 | 0.000619691 | 0.329630485 |
| T2DM | pericyte | *ITPR3* | 2.754627821 | 0.000108073 | 0.000774167 | 0.679453032 |
| T2DM | pericyte | *DSP* | 0.755519368 | 0.017147282 | 0.030810221 | 1 |

| | | | | | | |
|---|---|---|---|---|---|---|
| T2DM | pericyte | *IFT74* | 0.89474928 | 0.027350112 | 0.045694965 | 1 |
| T2DM | pericyte | *IL12RB1* | 1.799317773 | 0.006133931 | 0.012850392 | 1 |
| T2DM | pericyte | *PCSK5* | 2.005552037 | 0.007200368 | 0.014654813 | 1 |
| T2DM | pericyte | *RAB18* | 1.761915136 | 0.000620074 | 0.002305385 | 1 |
| T2DM | pericyte | *NRP1* | 2.03809311 | 0.003787618 | 0.008875421 | 1 |
| T2DM | pericyte | *PALMD* | 1.446636168 | 7.34E-05 | 0.000646906 | 0.461244219 |
| T2DM | pericyte | *HSD3B7* | 1.568357654 | 0.003608339 | 0.008537124 | 1 |
| T2DM | pericyte | *ATP5D* | 3.606286377 | 0.002981573 | 0.007371274 | 1 |
| T2DM | pericyte | *MARCH2* | 3.150003767 | 1.97E-06 | 0.000187928 | 0.012403218 |
| T2DM | pericyte | *MTAP* | 4.278001861 | 0.001454843 | 0.00428613 | 1 |
| T2DM | pericyte | *POLR2E* | 1.574672735 | 0.001858396 | 0.005124384 | 1 |
| T2DM | pericyte | *MADCAM1* | 2.054682348 | 0.000608281 | 0.002274991 | 1 |
| T2DM | pericyte | *ZDHHC8* | 1.428801069 | 0.001545861 | 0.004496702 | 1 |
| T2DM | pericyte | *MED15* | 2.33350051 | 0.000126274 | 0.00084164 | 0.793881739 |
| T2DM | pericyte | *SERPIND1* | 1.058044507 | 0.005024692 | 0.011007052 | 1 |
| T2DM | pericyte | *SNAP29* | 0.717316901 | 0.003616178 | 0.008550174 | 1 |
| T2DM | pericyte | *SMARCB1* | 1.072058576 | 0.021517343 | 0.037390695 | 1 |
| T2DM | pericyte | *P2RX6* | 2.461257207 | 0.012412659 | 0.023365917 | 1 |
| T2DM | pericyte | *DDT* | 3.193573512 | 0.008894698 | 0.017510884 | 1 |
| T2DM | pericyte | *SUSD2* | -0.335659395 | 0.027303876 | 0.045641975 | 1 |
| T2DM | pericyte | *PPIL2* | 0.807387847 | 0.016050904 | 0.029165327 | 1 |
| T2DM | pericyte | *YPEL1* | 1.874138425 | 0.000610042 | 0.002280221 | 1 |
| T2DM | pericyte | *PRODH* | -3.473247215 | 0.003912418 | 0.009100027 | 1 |
| T2DM | pericyte | *PPM1F* | 3.393710985 | 0.000183682 | 0.001035729 | 1 |
| T2DM | pericyte | *CYTH4* | 1.055536188 | 0.00172972 | 0.004874385 | 1 |
| T2DM | pericyte | *CARD10* | 0.967265576 | 0.00350644 | 0.008372574 | 1 |
| T2DM | pericyte | *SLC25A1* | 1.776830601 | 0.000685781 | 0.002479301 | 1 |
| T2DM | pericyte | *SH3BP1* | 6.173114737 | 0.000887233 | 0.003018117 | 1 |
| T2DM | pericyte | *HPS4* | 3.789503774 | 2.75E-05 | 0.000569342 | 0.172660577 |
| T2DM | pericyte | *TRIOBP* | 1.432513626 | 0.001786078 | 0.004988689 | 1 |
| T2DM | pericyte | *TFIP11* | 1.251336395 | 0.000472811 | 0.001905155 | 1 |
| T2DM | pericyte | *CCDC134* | 3.011828483 | 9.37E-05 | 0.000716959 | 0.589115279 |
| T2DM | pericyte | *DDX17* | 1.723013939 | 0.002860704 | 0.007128517 | 1 |
| T2DM | pericyte | *FBXO7* | 2.23044276 | 0.000281417 | 0.001336306 | 1 |
| T2DM | pericyte | *TIMP3* | 2.814147299 | 7.24E-05 | 0.00064687 | 0.454910283 |
| T2DM | pericyte | *PPP6R2* | 1.4433981 | 0.010676527 | 0.02054586 | 1 |
| T2DM | pericyte | *CYB5R3* | 2.622773818 | 0.000242786 | 0.001224801 | 1 |
| T2DM | pericyte | *RHBDD3* | 3.526425005 | 0.000182399 | 0.001034031 | 1 |
| T2DM | pericyte | *HMGXB4* | 4.001342871 | 0.000111829 | 0.000783039 | 0.703069737 |
| T2DM | pericyte | *TOM1* | 1.108035373 | 0.005093689 | 0.011134918 | 1 |
| T2DM | pericyte | *BIK* | 3.943101787 | 0.002765923 | 0.006950183 | 1 |
| T2DM | pericyte | *TSPO* | 2.205841483 | 0.004918387 | 0.010830788 | 1 |
| T2DM | pericyte | *RASD2* | -1.01867525 | 0.01326851 | 0.024724102 | 1 |
| T2DM | pericyte | *TTLL12* | 1.337781332 | 0.003983152 | 0.009226999 | 1 |
| T2DM | pericyte | *RBFOX2* | 2.755262033 | 0.000615441 | 0.002292225 | 1 |
| T2DM | pericyte | *MTMR3* | 0.961230424 | 0.02300024 | 0.039476525 | 1 |
| T2DM | pericyte | *SAMM50* | 2.571754573 | 0.000115186 | 0.000794697 | 0.724175123 |
| T2DM | pericyte | *TXN2* | 1.82040299 | 0.00316928 | 0.007750006 | 1 |
| T2DM | pericyte | *FOXRED2* | 0.785186304 | 0.001512 | 0.004420133 | 1 |
| T2DM | pericyte | *IFT27* | 1.330500985 | 0.001461455 | 0.004301576 | 1 |

| | | | | | | |
|---|---|---|---|---|---|---|
| T2DM | pericyte | *KIAA0930* | 1.903502207 | 0.000357081 | 0.001551465 | 1 |
| T2DM | pericyte | *SLC25A17* | 3.542517345 | 0.000128454 | 0.000850095 | 0.807589777 |
| T2DM | pericyte | *EP300* | 2.803393953 | 0.000281367 | 0.001336306 | 1 |
| T2DM | pericyte | *ACO2* | 2.199384488 | 0.002900148 | 0.007206811 | 1 |
| T2DM | pericyte | *DESI1* | 1.53067602 | 1.47E-05 | 0.000476496 | 0.092117803 |
| T2DM | pericyte | *ZBED4* | 4.479869227 | 5.17E-05 | 0.000616661 | 0.325159956 |
| T2DM | pericyte | *HDAC10* | 1.142329085 | 0.017975612 | 0.032096754 | 1 |
| T2DM | pericyte | *RBM23* | 3.346949683 | 0.000296749 | 0.00138095 | 1 |
| T2DM | pericyte | *SOS2* | 3.090998417 | 0.000115738 | 0.000796114 | 0.727647788 |
| T2DM | pericyte | *PYGL* | 0.436445715 | 0.028565628 | 0.04737328 | 1 |
| T2DM | pericyte | *TRIM9* | 0.669052293 | 0.017932147 | 0.032046735 | 1 |
| T2DM | pericyte | *DDHD1* | 2.489901838 | 0.000727555 | 0.00259158 | 1 |
| T2DM | pericyte | *PIGH* | 1.768078659 | 0.008193656 | 0.016337938 | 1 |
| T2DM | pericyte | *VTI1B* | 1.435956711 | 0.001202116 | 0.00373589 | 1 |
| T2DM | pericyte | *DAAM1* | 1.616551147 | 0.001233556 | 0.003805096 | 1 |
| T2DM | pericyte | *ISM2* | 3.104398192 | 0.000212118 | 0.001122547 | 1 |
| T2DM | pericyte | *PPM1A* | 2.805396382 | 1.92E-05 | 0.000512152 | 0.120952533 |
| T2DM | pericyte | *DICER1* | 2.084041291 | 0.001984934 | 0.005369741 | 1 |
| T2DM | pericyte | *ZFYVE21* | 2.674293017 | 8.44E-05 | 0.000682391 | 0.530900387 |
| T2DM | pericyte | *MTHFD1* | 1.552411833 | 0.000594231 | 0.002237251 | 1 |
| T2DM | pericyte | *TELO2* | 4.984514655 | 0.000234432 | 0.001198741 | 1 |
| T2DM | pericyte | *GSKIP* | 2.141118744 | 0.003531452 | 0.008419041 | 1 |
| T2DM | pericyte | *VRK1* | 3.426624462 | 5.29E-05 | 0.000619691 | 0.332682737 |
| T2DM | pericyte | *PAPLN* | 1.806598426 | 0.000340764 | 0.00150554 | 1 |
| T2DM | pericyte | *PSMB5* | 1.937006289 | 2.04E-05 | 0.000524717 | 0.128066948 |
| T2DM | pericyte | *CCNB1IP1* | 1.935829938 | 0.006865994 | 0.014088284 | 1 |
| T2DM | pericyte | *TRIP11* | 3.349944063 | 5.31E-05 | 0.000619691 | 0.33383904 |
| T2DM | pericyte | *PLTP* | 1.238455872 | 3.28E-05 | 0.000599418 | 0.206342293 |
| T2DM | pericyte | *GSS* | 2.046342415 | 0.000129114 | 0.000852666 | 0.811737899 |
| T2DM | pericyte | *PYGB* | 0.867725002 | 0.019829796 | 0.034833732 | 1 |
| T2DM | pericyte | *ABHD12* | 1.857876797 | 0.000331252 | 0.001477009 | 1 |
| T2DM | pericyte | *NINL* | 2.061129839 | 4.15E-05 | 0.000607238 | 0.260957486 |
| T2DM | pericyte | *CD40* | 2.744728471 | 0.012955893 | 0.024265084 | 1 |
| T2DM | pericyte | *IFT52* | 1.80599944 | 0.011686412 | 0.022237431 | 1 |
| T2DM | pericyte | *NDRG3* | 4.16385362 | 8.76E-05 | 0.000699239 | 0.551000185 |
| T2DM | pericyte | *DOK5* | 2.858920292 | 0.000119671 | 0.000813238 | 0.75237468 |
| T2DM | pericyte | *CSTF1* | 1.432613672 | 0.003721192 | 0.008739516 | 1 |
| T2DM | pericyte | *TPD52L2* | 1.878124937 | 0.000878253 | 0.002995813 | 1 |
| T2DM | pericyte | *NELFCD* | 4.284597544 | 6.46E-05 | 0.000632025 | 0.406078249 |
| T2DM | pericyte | *CTSZ* | 1.551407841 | 0.010088119 | 0.019557201 | 1 |
| T2DM | pericyte | *PRPF6* | 2.602794552 | 0.001013422 | 0.003321888 | 1 |
| T2DM | pericyte | *MTG2* | 1.174517656 | 0.003397066 | 0.008148553 | 1 |
| T2DM | pericyte | *DIDO1* | 2.513809171 | 5.65E-06 | 0.000331374 | 0.035502129 |
| T2DM | pericyte | *GID8* | 1.693015212 | 0.000225782 | 0.001172167 | 1 |
| T2DM | pericyte | *SLC17A9* | 4.135658249 | 0.000350236 | 0.001528062 | 1 |
| T2DM | pericyte | *BIRC7* | -1.400009343 | 0.002573922 | 0.006564807 | 1 |
| T2DM | pericyte | *NKAIN4* | -0.884913279 | 0.015458136 | 0.028234666 | 1 |
| T2DM | pericyte | *PTK6* | 6.491084474 | 0.000777646 | 0.002735903 | 1 |
| T2DM | pericyte | *GMEB2* | 2.718187414 | 0.001151818 | 0.003624363 | 1 |
| T2DM | pericyte | *TRIB3* | 1.026487898 | 0.01714106 | 0.030807846 | 1 |

| | | | | | | |
|---|---|---|---|---|---|---|
| T2DM | pericyte | *MYL9* | 1.451537024 | 0.025402728 | 0.043047696 | 1 |
| T2DM | pericyte | *MANBAL* | 0.945139069 | 0.001190322 | 0.003713923 | 1 |
| T2DM | pericyte | *IDH3B* | 1.549292889 | 0.001019159 | 0.003333742 | 1 |
| T2DM | pericyte | *MAPRE1* | 2.75816104 | 0.000348058 | 0.001522785 | 1 |
| T2DM | pericyte | *TTI1* | 2.490724986 | 0.000247407 | 0.001238415 | 1 |
| T2DM | pericyte | *RPRD1B* | 2.846126477 | 0.002045812 | 0.005482185 | 1 |
| T2DM | pericyte | *ACTR5* | 1.958180297 | 0.001945329 | 0.00529329 | 1 |
| T2DM | pericyte | *DHX35* | 3.219432331 | 0.000149051 | 0.000925057 | 0.937082331 |
| T2DM | pericyte | *MAP1LC3A* | 1.679454883 | 3.16E-05 | 0.000599418 | 0.19856361 |
| T2DM | pericyte | *PIGU* | 1.923273755 | 0.000163263 | 0.000967422 | 1 |
| T2DM | pericyte | *APMAP* | 2.945013624 | 9.09E-05 | 0.000704357 | 0.571296315 |
| T2DM | pericyte | *CELF4* | 0.846831353 | 0.00636066 | 0.013246579 | 1 |
| T2DM | pericyte | *METTL4* | 3.990064031 | 3.23E-05 | 0.000599418 | 0.202782378 |
| T2DM | pericyte | *LPIN2* | 3.309645281 | 1.89E-05 | 0.000512131 | 0.118789691 |
| T2DM | pericyte | *SMCHD1* | 2.911668951 | 6.74E-05 | 0.000632837 | 0.423566133 |
| T2DM | pericyte | *MYL12A* | 3.510485273 | 0.001185282 | 0.003703712 | 1 |
| T2DM | pericyte | *CEP76* | 1.371398117 | 0.024611844 | 0.041789672 | 1 |
| T2DM | pericyte | *NALCN* | 0.885180401 | 0.003172854 | 0.007752714 | 1 |
| T2DM | pericyte | *HTR2A* | 1.960822799 | 0.004800183 | 0.010636164 | 1 |
| T2DM | pericyte | *STK24* | 1.963087584 | 0.026171736 | 0.044124888 | 1 |
| T2DM | pericyte | *DNAJC3* | 2.976223319 | 0.000339498 | 0.00150511 | 1 |
| T2DM | pericyte | *UGGT2* | 2.6648249 | 5.16E-05 | 0.000616617 | 0.32434046 |
| T2DM | pericyte | *ARHGEF7* | 2.667847027 | 0.000487039 | 0.001946607 | 1 |
| T2DM | pericyte | *SGCG* | 1.510538638 | 0.001280453 | 0.003902185 | 1 |
| T2DM | pericyte | *PARP4* | 3.287876182 | 4.31E-05 | 0.000607238 | 0.270898525 |
| T2DM | pericyte | *VWA8* | 2.68821643 | 2.00E-05 | 0.000518274 | 0.125940561 |
| T2DM | pericyte | *DHRS12* | 4.137032264 | 4.08E-05 | 0.000607238 | 0.256640119 |
| T2DM | pericyte | *MEDAG* | 1.957073462 | 0.000195585 | 0.001065597 | 1 |
| T2DM | pericyte | *TSC22D1* | 3.412405719 | 2.54E-05 | 0.000562456 | 0.15979953 |
| T2DM | pericyte | *GDPD3* | 1.181059192 | 0.002249401 | 0.005917149 | 1 |
| T2DM | pericyte | *NFAT5* | 2.535800531 | 0.000187323 | 0.001044058 | 1 |
| T2DM | pericyte | *LONP2* | 2.317543697 | 0.00213311 | 0.005663372 | 1 |
| T2DM | pericyte | *PLLP* | 1.515753095 | 0.003802331 | 0.008906578 | 1 |
| T2DM | pericyte | *DHODH* | 0.909106645 | 0.000549283 | 0.002123828 | 1 |
| T2DM | pericyte | *CTCF* | 3.366929114 | 1.91E-05 | 0.000512152 | 0.120023515 |
| T2DM | pericyte | *USB1* | 3.583809539 | 0.001214085 | 0.00376564 | 1 |
| T2DM | pericyte | *SETD6* | 3.810683211 | 0.000519801 | 0.002038672 | 1 |
| T2DM | pericyte | *MON1B* | 2.250376646 | 0.021210764 | 0.0369254 | 1 |
| T2DM | pericyte | *HCFC1R1* | 1.092136134 | 0.002927204 | 0.007259697 | 1 |
| T2DM | pericyte | *NPRL3* | 1.413168521 | 0.002519248 | 0.006459425 | 1 |
| T2DM | pericyte | *NAGPA* | 2.027868148 | 0.001635093 | 0.004681162 | 1 |
| T2DM | pericyte | *COTL1* | 1.700993707 | 0.002082233 | 0.005553279 | 1 |
| T2DM | pericyte | *CRISPLD2* | 0.883006717 | 0.002076348 | 0.005545454 | 1 |
| T2DM | pericyte | *LMF1* | 4.025009443 | 9.82E-06 | 0.000406319 | 0.061760424 |
| T2DM | pericyte | *NARFL* | 1.800262903 | 0.001032327 | 0.00335812 | 1 |
| T2DM | pericyte | *CLCN7* | 1.368797176 | 0.003992586 | 0.009242042 | 1 |
| T2DM | pericyte | *SLC7A5* | 10.16462397 | 0.011713534 | 0.022272851 | 1 |
| T2DM | pericyte | *FBXO31* | 1.708726789 | 0.005657844 | 0.012035724 | 1 |
| T2DM | pericyte | *NUBP1* | 2.314297167 | 0.00020869 | 0.001111258 | 1 |
| T2DM | pericyte | *UBE2I* | 3.433376733 | 3.21E-05 | 0.000599418 | 0.201644148 |

| | | | | | | |
|---|---|---|---|---|---|---|
| T2DM | pericyte | *CLUAP1* | 1.245481452 | 0.000218299 | 0.001146081 | 1 |
| T2DM | pericyte | *CPPED1* | 1.14397071 | 0.001325599 | 0.003998058 | 1 |
| T2DM | pericyte | *HMOX2* | 2.353198369 | 0.000740455 | 0.00263008 | 1 |
| T2DM | pericyte | *BFAR* | 2.766511493 | 0.000215204 | 0.00113601 | 1 |
| T2DM | pericyte | *RBL2* | 2.501540209 | 3.71E-08 | 8.64E-06 | 0.000233184 |
| T2DM | pericyte | *STX4* | 2.767842545 | 0.002895828 | 0.007198921 | 1 |
| T2DM | pericyte | *KAT8* | 4.745604116 | 2.23E-06 | 0.000202161 | 0.013997287 |
| T2DM | pericyte | *NOMO1* | 2.600144423 | 0.005821812 | 0.012320515 | 1 |
| T2DM | pericyte | *TMC5* | 0.789372562 | 0.019939588 | 0.035007035 | 1 |
| T2DM | pericyte | *IQCH* | 3.424038554 | 9.45E-05 | 0.000717456 | 0.594012148 |
| T2DM | pericyte | *LACTB* | 2.239035576 | 0.000757228 | 0.002683591 | 1 |
| T2DM | pericyte | *CSK* | 5.553485448 | 0.000139769 | 0.000894176 | 0.8787265 |
| T2DM | pericyte | *HERC1* | 2.570464518 | 0.000707053 | 0.002541358 | 1 |
| T2DM | pericyte | *AP3B2* | -0.377265919 | 0.020551063 | 0.035900121 | 1 |
| T2DM | pericyte | *CTSH* | 0.553320608 | 0.013923544 | 0.025765202 | 1 |
| T2DM | pericyte | *TTC23* | 2.439013515 | 0.000224137 | 0.001166517 | 1 |
| T2DM | pericyte | *FAH* | 1.233495732 | 0.001336844 | 0.004019481 | 1 |
| T2DM | pericyte | *RPAP1* | 1.844820526 | 0.007765561 | 0.015653121 | 1 |
| T2DM | pericyte | *EHD4* | 3.295407672 | 7.21E-05 | 0.00064687 | 0.453050934 |
| T2DM | pericyte | *ATP8B4* | 2.680059334 | 0.000142395 | 0.000905195 | 0.89523751 |
| T2DM | pericyte | *DTWD1* | 3.217290399 | 2.25E-06 | 0.000202161 | 0.014151276 |
| T2DM | pericyte | *PDGFRL* | 1.096333645 | 0.000394101 | 0.001670746 | 1 |
| T2DM | pericyte | *ZFAND1* | 1.304567176 | 0.007773393 | 0.015663886 | 1 |
| T2DM | pericyte | *RIPK2* | 2.71117989 | 0.001860464 | 0.005125652 | 1 |
| T2DM | pericyte | *EYA1* | 0.870745452 | 3.79E-06 | 0.000279804 | 0.023815743 |
| T2DM | pericyte | *NBN* | 2.278069509 | 0.000245863 | 0.001234577 | 1 |
| T2DM | pericyte | *CPQ* | 1.551261054 | 0.000343876 | 0.001511465 | 1 |
| T2DM | pericyte | *NIPAL2* | 3.042921269 | 4.25E-06 | 0.000287028 | 0.026693594 |
| T2DM | pericyte | *IKBKB* | 4.126046479 | 1.06E-05 | 0.000413298 | 0.066540919 |
| T2DM | pericyte | *PLAT* | 5.17170769 | 4.19E-06 | 0.000287028 | 0.026357614 |
| T2DM | pericyte | *GDAP1* | 2.464534155 | 0.01296135 | 0.024266827 | 1 |
| T2DM | pericyte | *RAB2A* | 1.707980656 | 2.81E-05 | 0.000573841 | 0.176499735 |
| T2DM | pericyte | *ZC2HC1A* | 2.518968579 | 0.000193053 | 0.001062757 | 1 |
| T2DM | pericyte | *SPAG1* | 1.002203048 | 0.022902109 | 0.03932957 | 1 |
| T2DM | pericyte | *SNX16* | 1.915178884 | 0.000623897 | 0.002315491 | 1 |
| T2DM | pericyte | *GSDMD* | 1.476962055 | 0.00193744 | 0.005279882 | 1 |
| T2DM | pericyte | *ERI1* | 3.411901391 | 4.59E-05 | 0.000607238 | 0.288724386 |
| T2DM | pericyte | *MTMR9* | 3.477513208 | 3.34E-05 | 0.000599418 | 0.210277533 |
| T2DM | pericyte | *LEPROTL1* | 2.487933775 | 9.45E-05 | 0.000717456 | 0.594414761 |
| T2DM | pericyte | *TNFRSF10A* | 0.89925274 | 0.006118956 | 0.012831848 | 1 |
| T2DM | pericyte | *ERICH1* | 3.393916452 | 8.87E-05 | 0.000702125 | 0.557633833 |
| T2DM | pericyte | *ARHGEF10* | 1.360600615 | 0.001559543 | 0.004518363 | 1 |
| T2DM | pericyte | *KLHDC4* | 1.091020959 | 0.019393463 | 0.034201039 | 1 |
| T2DM | pericyte | *MAN2B1* | 1.427907791 | 0.002741665 | 0.006905789 | 1 |
| T2DM | pericyte | *TULP2* | 1.337563273 | 0.003068508 | 0.007535824 | 1 |
| T2DM | pericyte | *NUCB1* | 1.690384283 | 0.000672464 | 0.002446108 | 1 |
| T2DM | pericyte | *NFKBIB* | 1.068963314 | 0.006574211 | 0.013618473 | 1 |
| T2DM | pericyte | *SARS2* | -0.755514199 | 0.022600538 | 0.038949996 | 1 |
| T2DM | pericyte | *ERCC2* | 2.126160723 | 0.001575987 | 0.004551324 | 1 |
| T2DM | pericyte | *PLEKHJ1* | 1.596409078 | 0.001656731 | 0.004725893 | 1 |

| | | | | | | |
|---|---|---|---|---|---|---|
| T2DM | pericyte | *CD37* | 1.894481622 | 0.002779669 | 0.006973574 | 1 |
| T2DM | pericyte | *STX10* | 1.336058602 | 0.001034245 | 0.003362097 | 1 |
| T2DM | pericyte | *FCER2* | 1.235318117 | 0.012072271 | 0.022806 | 1 |
| T2DM | pericyte | *IL4I1* | 1.24082984 | 0.015268164 | 0.027912459 | 1 |
| T2DM | pericyte | *AES* | 4.458552394 | 6.02E-05 | 0.00062437 | 0.378428807 |
| T2DM | pericyte | *SGTA* | 1.674850521 | 0.001063705 | 0.003424226 | 1 |
| T2DM | pericyte | *TIMM44* | 2.918067464 | 0.000189351 | 0.001048037 | 1 |
| T2DM | pericyte | *IL27RA* | 1.752356975 | 0.00056077 | 0.002157627 | 1 |
| T2DM | pericyte | *RASAL3* | 0.786274142 | 0.008452978 | 0.016781252 | 1 |
| T2DM | pericyte | *ZNF419* | 0.789732319 | 0.000299641 | 0.001384824 | 1 |
| T2DM | pericyte | *CCNE1* | 2.234152109 | 0.005790051 | 0.012273111 | 1 |
| T2DM | pericyte | *ANKRD27* | 2.033219402 | 0.000904363 | 0.003045383 | 1 |
| T2DM | pericyte | *PRX* | 1.154365205 | 0.023996793 | 0.040962226 | 1 |
| T2DM | pericyte | *CCDC94* | 1.314238706 | 0.002780832 | 0.006973709 | 1 |
| T2DM | pericyte | *TBCB* | 1.924392812 | 0.022494359 | 0.038830872 | 1 |
| T2DM | pericyte | *POLR2I* | 2.803760922 | 0.000679172 | 0.002463909 | 1 |
| T2DM | pericyte | *SLC1A5* | 2.181888743 | 0.001489494 | 0.004367745 | 1 |
| T2DM | pericyte | *CACTIN* | 1.40526747 | 0.002860325 | 0.007128517 | 1 |
| T2DM | pericyte | *CCDC9* | 3.474922715 | 1.31E-05 | 0.000462556 | 0.082667079 |
| T2DM | pericyte | *FZR1* | 6.367598192 | 2.48E-05 | 0.000558632 | 0.155833875 |
| T2DM | pericyte | *TGFB1* | 2.025736783 | 0.000195933 | 0.001065597 | 1 |
| T2DM | pericyte | *CEACAM4* | 3.151590466 | 0.005724821 | 0.01214713 | 1 |
| T2DM | pericyte | *PLIN3* | 2.197349243 | 0.000897633 | 0.00303638 | 1 |
| T2DM | pericyte | *MRPL4* | 2.869352766 | 9.94E-05 | 0.000737891 | 0.624993879 |
| T2DM | pericyte | *CD33* | 1.337303495 | 0.005127924 | 0.011198076 | 1 |
| T2DM | pericyte | *BABAM1* | 2.188198536 | 0.00044716 | 0.001832657 | 1 |
| T2DM | pericyte | *TYK2* | 3.769467282 | 3.12E-05 | 0.000599418 | 0.195860438 |
| T2DM | pericyte | *RABAC1* | 2.314169173 | 0.004277238 | 0.009753717 | 1 |
| T2DM | pericyte | *PTPRS* | 1.459444583 | 0.000191634 | 0.001057775 | 1 |
| T2DM | pericyte | *GRWD1* | -0.560090909 | 0.026780404 | 0.044922199 | 1 |
| T2DM | pericyte | *GRIN2D* | 1.736748412 | 0.002061708 | 0.005518076 | 1 |
| T2DM | pericyte | *CARD8* | 2.035505882 | 0.00372128 | 0.008739516 | 1 |
| T2DM | pericyte | *PLA2G4C* | 0.946689196 | 0.007493622 | 0.015153418 | 1 |
| T2DM | pericyte | *CAPS* | 2.029344795 | 0.000251212 | 0.001252473 | 1 |
| T2DM | pericyte | *BCAT2* | 1.930459925 | 0.000574368 | 0.002188517 | 1 |
| T2DM | pericyte | *PPP2R1A* | 4.906341123 | 9.11E-06 | 0.000390594 | 0.057281021 |
| T2DM | pericyte | *TNPO2* | 2.512318107 | 3.29E-05 | 0.000599418 | 0.207132626 |
| T2DM | pericyte | *LILRB5* | 1.20024605 | 0.001188421 | 0.003709834 | 1 |
| T2DM | pericyte | *RPL18A* | 1.116821678 | 0.022014545 | 0.038128222 | 1 |
| T2DM | pericyte | *ELL* | 2.200341269 | 2.59E-05 | 0.000562456 | 0.162790989 |
| T2DM | pericyte | *UPK1A* | 2.960739318 | 0.006199694 | 0.012959391 | 1 |
| T2DM | pericyte | *COPE* | 3.983724328 | 0.000209225 | 0.001112855 | 1 |
| T2DM | pericyte | *KXD1* | 1.843888454 | 0.000580789 | 0.002202711 | 1 |
| T2DM | pericyte | *ATP13A1* | 3.206952123 | 1.80E-08 | 5.64E-06 | 0.000113187 |
| T2DM | pericyte | *SIPA1L3* | 3.078846164 | 8.31E-05 | 0.000681373 | 0.522730617 |
| T2DM | pericyte | *SMG9* | 0.835469464 | 0.003108333 | 0.007618749 | 1 |
| T2DM | pericyte | *GTPBP10* | 0.920490181 | 0.013486412 | 0.025063278 | 1 |
| T2DM | pericyte | *PON2* | 3.898652213 | 5.42E-05 | 0.00062437 | 0.341022286 |
| T2DM | pericyte | *HBP1* | 3.00297446 | 0.000226531 | 0.001175081 | 1 |
| T2DM | pericyte | *DLX5* | -1.637204306 | 0.017942735 | 0.032047151 | 1 |

| | | | | | | |
|---|---|---|---|---|---|---|
| T2DM | pericyte | *STEAP1B* | 1.308802638 | 0.001318758 | 0.00398799 | 1 |
| T2DM | pericyte | *PTN* | 1.068790344 | 0.002499342 | 0.006423036 | 1 |
| T2DM | pericyte | *MPP6* | 1.052989842 | 0.001259305 | 0.003865666 | 1 |
| T2DM | pericyte | *DFNA5* | 0.807613927 | 0.00667032 | 0.013772185 | 1 |
| T2DM | pericyte | *OGDH* | 3.260241607 | 4.62E-05 | 0.000607238 | 0.29038507 |
| T2DM | pericyte | *ADAP1* | 1.700186742 | 0.004740902 | 0.010547082 | 1 |
| T2DM | pericyte | *H2AFV* | 2.87717371 | 6.02E-05 | 0.00062437 | 0.378588126 |
| T2DM | pericyte | *CAV1* | 1.496345605 | 0.001171942 | 0.003672981 | 1 |
| T2DM | pericyte | *LMBR1* | 2.36239163 | 0.002152903 | 0.00570869 | 1 |
| T2DM | pericyte | *HOXA2* | 4.146612362 | 0.005579707 | 0.01191563 | 1 |
| T2DM | pericyte | *HOXA6* | 1.058863154 | 0.030256392 | 0.049718227 | 1 |
| T2DM | pericyte | *CPED1* | 1.848555618 | 0.00012657 | 0.00084164 | 0.795742722 |
| T2DM | pericyte | *HIBADH* | 2.518374931 | 9.25E-05 | 0.000711161 | 0.581729635 |
| T2DM | pericyte | *CPVL* | 3.835651899 | 9.40E-09 | 3.48E-06 | 5.91E-05 |
| T2DM | pericyte | *PLEKHA8* | 3.158535379 | 0.001425198 | 0.004216575 | 1 |
| T2DM | pericyte | *NOD1* | 3.687675644 | 8.44E-05 | 0.000682391 | 0.530358668 |
| T2DM | pericyte | *EPHB6* | 1.787951294 | 0.008781347 | 0.017333855 | 1 |
| T2DM | pericyte | *NPTX2* | -9.443357548 | 0.013114693 | 0.024495566 | 1 |
| T2DM | pericyte | *CYP3A5* | 3.134431625 | 8.96E-05 | 0.000702125 | 0.563052082 |
| T2DM | pericyte | *ZKSCAN1* | 2.507961187 | 3.56E-05 | 0.00060096 | 0.223627487 |
| T2DM | pericyte | *SNX8* | 2.328833916 | 0.000168819 | 0.000990075 | 1 |
| T2DM | pericyte | *TAF6* | 1.356218981 | 0.001867595 | 0.005138542 | 1 |
| T2DM | pericyte | *AIMP2* | 3.613823959 | 8.95E-05 | 0.000702125 | 0.562634523 |
| T2DM | pericyte | *TFR2* | 3.008096633 | 0.002563824 | 0.006541705 | 1 |
| T2DM | pericyte | *USP42* | 4.662414655 | 6.48E-06 | 0.000343901 | 0.040745762 |
| T2DM | pericyte | *LSM5* | 3.240043225 | 0.000113517 | 0.000790347 | 0.7136836 |
| T2DM | pericyte | *AP1S1* | 5.816825725 | 0.014873197 | 0.027293576 | 1 |
| T2DM | pericyte | *C1GALT1* | 3.304127213 | 2.84E-05 | 0.000576866 | 0.178828496 |
| T2DM | pericyte | *PLOD3* | 2.962357847 | 0.009788455 | 0.019052095 | 1 |
| T2DM | pericyte | *RPA3* | 3.619376329 | 0.000340637 | 0.00150554 | 1 |
| T2DM | pericyte | *PHF14* | 2.980326938 | 7.11E-05 | 0.000642058 | 0.44687238 |
| T2DM | pericyte | *ZNF862* | 2.065952894 | 0.006775453 | 0.01394346 | 1 |
| T2DM | pericyte | *ACTR3C* | 1.170656846 | 0.004793642 | 0.010630556 | 1 |
| T2DM | pericyte | *POU6F2* | 0.635998932 | 0.004891875 | 0.01078752 | 1 |
| T2DM | pericyte | *TSPAN13* | 0.824171711 | 0.00555099 | 0.011866397 | 1 |
| T2DM | pericyte | *RARRES2* | 2.373313797 | 0.000133439 | 0.000871003 | 0.838932153 |
| T2DM | pericyte | *TMEM176B* | 0.968510743 | 0.000253786 | 0.001257649 | 1 |
| T2DM | pericyte | *MRPL32* | 1.471104827 | 0.000211966 | 0.001122547 | 1 |
| T2DM | pericyte | *COA1* | 1.057644949 | 0.018583581 | 0.033003476 | 1 |
| T2DM | pericyte | *URGCP* | 2.915832924 | 0.000126532 | 0.00084164 | 0.795504605 |
| T2DM | pericyte | *GCK* | -2.036100131 | 0.028648255 | 0.047485257 | 1 |
| T2DM | pericyte | *BCL7B* | 1.189812006 | 0.005355632 | 0.011570878 | 1 |
| T2DM | pericyte | *YKT6* | 1.018247 | 0.002437925 | 0.006291968 | 1 |
| T2DM | pericyte | *SPATA6L* | 0.971056956 | 0.008044706 | 0.016112477 | 1 |
| T2DM | pericyte | *SLC1A1* | 0.734845339 | 0.004832586 | 0.01067925 | 1 |
| T2DM | pericyte | *FKTN* | 2.002333493 | 0.000462739 | 0.001879352 | 1 |
| T2DM | pericyte | *NMRK1* | 2.085594177 | 0.000102713 | 0.000750883 | 0.645759018 |
| T2DM | pericyte | *TMEM245* | 1.889055183 | 0.001948269 | 0.00529562 | 1 |
| T2DM | pericyte | *MEGF9* | 0.984732627 | 0.019742029 | 0.03471836 | 1 |
| T2DM | pericyte | *CORO2A* | 1.282500795 | 0.009822493 | 0.019107059 | 1 |

| | | | | | | |
|---|---|---|---|---|---|---|
| T2DM | pericyte | *OGN* | 1.443536918 | 0.002563082 | 0.006541705 | 1 |
| T2DM | pericyte | *ECM2* | 1.35427349 | 0.018535548 | 0.032937533 | 1 |
| T2DM | pericyte | *SUSD1* | 0.937920717 | 0.010023808 | 0.019450518 | 1 |
| T2DM | pericyte | *TNFSF8* | 0.872354072 | 0.005668453 | 0.012051932 | 1 |
| T2DM | pericyte | *AK1* | 1.323428917 | 0.013196953 | 0.024619954 | 1 |
| T2DM | pericyte | *KDM4C* | 2.920922033 | 4.19E-05 | 0.000607238 | 0.263690582 |
| T2DM | pericyte | *DOCK8* | 1.501598235 | 0.000227516 | 0.001178248 | 1 |
| T2DM | pericyte | *FUBP3* | 2.510737295 | 9.32E-05 | 0.000715299 | 0.585829928 |
| T2DM | pericyte | *RGP1* | 5.616980301 | 2.50E-05 | 0.000558632 | 0.15690882 |
| T2DM | pericyte | *MPDZ* | 2.742554307 | 4.29E-05 | 0.000607238 | 0.26981624 |
| T2DM | pericyte | *PTGDS* | 0.97651197 | 0.004733172 | 0.01053734 | 1 |
| T2DM | pericyte | *SHB* | 2.667397922 | 0.002280064 | 0.005975306 | 1 |
| T2DM | pericyte | *DVL1* | 2.208318098 | 0.000969388 | 0.003214419 | 1 |
| T2DM | pericyte | *CCNJ* | 3.030971779 | 0.001101926 | 0.003518983 | 1 |
| T2DM | pericyte | *ATRNL1* | 0.7938258 | 0.015145856 | 0.027729177 | 1 |
| T2DM | pericyte | *HPS1* | 3.405686036 | 0.00077394 | 0.002724402 | 1 |
| T2DM | pericyte | *DNMBP* | 3.70131336 | 9.04E-05 | 0.000703483 | 0.568414478 |
| T2DM | pericyte | *TRDMT1* | 2.785767704 | 0.000286112 | 0.001346398 | 1 |
| T2DM | pericyte | *DDX50* | 3.658883686 | 0.000316317 | 0.001430707 | 1 |
| T2DM | pericyte | *MAPK8* | 3.102920876 | 4.55E-05 | 0.000607238 | 0.285827997 |
| T2DM | pericyte | *ATE1* | 2.133628756 | 0.000385391 | 0.001643795 | 1 |
| T2DM | pericyte | *NSMCE4A* | 2.104572853 | 0.000417913 | 0.001744124 | 1 |
| T2DM | pericyte | *PLEKHA1* | 2.682134362 | 1.90E-05 | 0.000512152 | 0.119355542 |
| T2DM | pericyte | *MICU1* | 3.593982636 | 3.46E-06 | 0.000265476 | 0.021769055 |
| T2DM | pericyte | *CCSER2* | 3.29668184 | 6.77E-05 | 0.000632837 | 0.425899168 |
| T2DM | pericyte | *LIPA* | 1.573638997 | 0.003382067 | 0.00812666 | 1 |
| T2DM | pericyte | *SFXN3* | 1.466829027 | 0.001410366 | 0.004182358 | 1 |
| T2DM | pericyte | *TNKS2* | 2.631134764 | 0.000194305 | 0.001065033 | 1 |
| T2DM | pericyte | *ANKRD26* | 1.548755364 | 0.001859189 | 0.005124384 | 1 |
| T2DM | pericyte | *ACBD5* | 2.818016245 | 1.28E-05 | 0.000461315 | 0.080498825 |
| T2DM | pericyte | *MTPAP* | 2.602603131 | 7.48E-05 | 0.000650766 | 0.470438077 |
| T2DM | pericyte | *PITRM1* | 2.665023285 | 0.000169947 | 0.000993915 | 1 |
| T2DM | pericyte | *MAP3K8* | 2.216434795 | 0.002436866 | 0.006291968 | 1 |
| T2DM | pericyte | *SHOC2* | 3.137897951 | 8.52E-06 | 0.000381879 | 0.053596368 |
| T2DM | pericyte | *MLLT6* | 2.895397103 | 0.005965976 | 0.012569735 | 1 |
| T2DM | pericyte | *PSMB3* | 2.333325373 | 1.62E-05 | 0.00049967 | 0.101932618 |
| T2DM | pericyte | *CASC3* | 2.410124358 | 0.000693376 | 0.002503881 | 1 |
| T2DM | pericyte | *RGS9* | 3.365225083 | 0.000298477 | 0.001383867 | 1 |
| T2DM | pericyte | *ASPA* | 1.562393244 | 0.000456075 | 0.001859636 | 1 |
| T2DM | pericyte | *SEPT4* | -0.192392971 | 0.013268082 | 0.024724102 | 1 |
| T2DM | pericyte | *TRIM37* | 3.835475585 | 7.79E-06 | 0.000364517 | 0.048985389 |
| T2DM | pericyte | *PNPO* | 1.127126538 | 0.014045923 | 0.025942044 | 1 |
| T2DM | pericyte | *RPS6KB1* | 3.032200983 | 0.003067846 | 0.007535824 | 1 |
| T2DM | pericyte | *CDK5RAP3* | 1.460897411 | 0.021134648 | 0.036837685 | 1 |
| T2DM | pericyte | *CBX1* | 2.038714539 | 0.000548276 | 0.002121239 | 1 |
| T2DM | pericyte | *PIGL* | 3.360667197 | 4.46E-05 | 0.000607238 | 0.280705806 |
| T2DM | pericyte | *GALK1* | 3.079605632 | 0.000466003 | 0.001887734 | 1 |
| T2DM | pericyte | *CAMTA2* | 2.442794194 | 0.001709327 | 0.00483425 | 1 |
| T2DM | pericyte | *MED13* | 1.838146826 | 0.003116433 | 0.007629679 | 1 |
| T2DM | pericyte | *RNF167* | 3.603119253 | 3.18E-05 | 0.000599418 | 0.199733595 |

| | | | | | | |
|---|---|---|---|---|---|---|
| T2DM | pericyte | *NUP88* | 2.325160278 | 0.004847408 | 0.01070448 | 1 |
| T2DM | pericyte | *C1QBP* | 4.242186449 | 0.002747825 | 0.006912996 | 1 |
| T2DM | pericyte | *GOSR1* | 3.530669459 | 7.27E-05 | 0.000646895 | 0.457309937 |
| T2DM | pericyte | *DRG2* | 5.096604782 | 4.92E-05 | 0.000607238 | 0.30911 |
| T2DM | pericyte | *AKAP10* | 2.667147585 | 0.000240454 | 0.001220212 | 1 |
| T2DM | pericyte | *SMARCD2* | 2.390986576 | 0.001624 | 0.004655763 | 1 |
| T2DM | pericyte | *ICAM2* | 2.946817794 | 1.30E-05 | 0.000461315 | 0.081652829 |
| T2DM | pericyte | *UTP6* | 3.98328096 | 9.02E-05 | 0.000702451 | 0.56687789 |
| T2DM | pericyte | *DDX5* | 1.17686423 | 0.003018405 | 0.007444768 | 1 |
| T2DM | pericyte | *C17orf75* | 3.320820471 | 0.002391703 | 0.00619812 | 1 |
| T2DM | pericyte | *CYTH1* | 2.882385256 | 3.66E-07 | 5.61E-05 | 0.002301251 |
| T2DM | pericyte | *RAB5C* | 2.332737506 | 0.000621561 | 0.002309548 | 1 |
| T2DM | pericyte | *NAGLU* | 3.392832632 | 0.00097717 | 0.003235107 | 1 |
| T2DM | pericyte | *HSD17B1* | -0.535136706 | 0.001181174 | 0.003698229 | 1 |
| T2DM | pericyte | *EZH1* | 3.139849947 | 3.64E-05 | 0.00060096 | 0.228965589 |
| T2DM | pericyte | *DLX4* | 8.255265219 | 0.006062416 | 0.012743228 | 1 |
| T2DM | pericyte | *VAT1* | 1.854673808 | 1.64E-05 | 0.000502137 | 0.10314433 |
| T2DM | pericyte | *LRRC59* | 6.663568795 | 6.99E-05 | 0.000639553 | 0.439627757 |
| T2DM | pericyte | *ABCC3* | 0.752229969 | 0.001074993 | 0.003448205 | 1 |
| T2DM | pericyte | *MPP2* | 2.492326926 | 0.001338105 | 0.004021351 | 1 |
| T2DM | pericyte | *EFTUD2* | 2.893208787 | 2.15E-05 | 0.000540879 | 0.135237196 |
| T2DM | pericyte | *EFNB3* | -0.304308681 | 0.022220356 | 0.038431741 | 1 |
| T2DM | pericyte | *FAM20A* | 3.296326478 | 3.63E-05 | 0.00060096 | 0.227991508 |
| T2DM | pericyte | *YWHAE* | 3.283798211 | 4.85E-05 | 0.000607238 | 0.305154774 |
| T2DM | pericyte | *DPH1* | 3.737379192 | 0.000165019 | 0.00097324 | 1 |
| T2DM | pericyte | *DHRS7B* | 2.876637515 | 6.01E-05 | 0.00062437 | 0.377749009 |
| T2DM | pericyte | *SLC9A3R1* | 1.332330573 | 0.001047728 | 0.003388409 | 1 |
| T2DM | pericyte | *TMEM104* | 1.49582624 | 0.000730655 | 0.002601147 | 1 |
| T2DM | pericyte | *SLAIN2* | 2.830176614 | 4.70E-05 | 0.000607238 | 0.295297182 |
| T2DM | pericyte | *OCIAD1* | 2.345980657 | 0.005112894 | 0.011173016 | 1 |
| T2DM | pericyte | *CWH43* | 8.392985697 | 0.001006614 | 0.003309186 | 1 |
| T2DM | pericyte | *KIAA1211* | 0.510616515 | 0.027899396 | 0.046464504 | 1 |
| T2DM | pericyte | *UBE2D3* | 2.715759688 | 1.33E-05 | 0.000462556 | 0.083722609 |
| T2DM | pericyte | *TBC1D9* | 2.984886495 | 9.41E-05 | 0.000717456 | 0.591768301 |
| T2DM | pericyte | *WFS1* | 9.808308759 | 6.21E-13 | 5.60E-10 | 3.90E-09 |
| T2DM | pericyte | *AADAT* | 2.435740224 | 0.001998969 | 0.005396101 | 1 |
| T2DM | pericyte | *SLC2A9* | 4.469396568 | 6.84E-07 | 8.78E-05 | 0.004303306 |
| T2DM | pericyte | *SH3D19* | 1.512250519 | 0.000780431 | 0.002741102 | 1 |
| T2DM | pericyte | *STIM2* | 3.146930775 | 0.000139951 | 0.000894176 | 0.879869587 |
| T2DM | pericyte | *RAPGEF2* | 1.860048291 | 0.005545138 | 0.011857919 | 1 |
| T2DM | pericyte | *UFSP2* | 2.374360061 | 0.024056551 | 0.040998519 | 1 |
| T2DM | pericyte | *KLF3* | 3.612246158 | 7.60E-05 | 0.000657799 | 0.477952782 |
| T2DM | pericyte | *FAM149A* | 0.958371197 | 0.000563271 | 0.002159249 | 1 |
| T2DM | pericyte | *UGDH* | 2.626985094 | 0.000123678 | 0.000831605 | 0.777565624 |
| T2DM | pericyte | *ELP4* | 2.761807205 | 0.000106698 | 0.000769275 | 0.670808127 |
| T2DM | pericyte | *SC5D* | 4.680062058 | 3.44E-05 | 0.000599418 | 0.216389102 |
| T2DM | pericyte | *C11orf63* | 2.100781057 | 0.000247385 | 0.001238415 | 1 |
| T2DM | pericyte | *PUS3* | 1.514231585 | 0.000663027 | 0.002422108 | 1 |
| T2DM | pericyte | *DCPS* | 1.796075618 | 0.000415397 | 0.001737591 | 1 |
| T2DM | pericyte | *FOXRED1* | 1.360577023 | 0.002383392 | 0.006181678 | 1 |

| | | | | | | |
|---|---|---|---|---|---|---|
| T2DM | pericyte | *NRXN2* | 4.068834664 | 0.000562451 | 0.002159249 | 1 |
| T2DM | pericyte | *MS4A4A* | 0.646781879 | 0.008652692 | 0.017139091 | 1 |
| T2DM | pericyte | *PRPF19* | 3.225638051 | 0.002447073 | 0.006307809 | 1 |
| T2DM | pericyte | *CHORDC1* | 2.45606854 | 0.000150096 | 0.000927922 | 0.943653169 |
| T2DM | pericyte | *EIF4G2* | 3.601490054 | 4.93E-05 | 0.000607238 | 0.309875982 |
| T2DM | pericyte | *CBL* | 2.453105002 | 0.000107492 | 0.000771465 | 0.675803048 |
| T2DM | pericyte | *KIAA1549L* | 1.062773743 | 0.022836477 | 0.0392717 | 1 |
| T2DM | pericyte | *ACCS* | 0.602277272 | 0.024039782 | 0.040981049 | 1 |
| T2DM | pericyte | *MADD* | 3.366464417 | 0.001288187 | 0.003921953 | 1 |
| T2DM | pericyte | *NAA40* | 1.988201618 | 0.00185251 | 0.005114945 | 1 |
| T2DM | pericyte | *CARS* | 3.399887462 | 3.95E-07 | 5.78E-05 | 0.002485599 |
| T2DM | pericyte | *RPS13* | 2.501809567 | 0.000724221 | 0.002582631 | 1 |
| T2DM | pericyte | *NDUFS8* | 5.149081477 | 0.000248895 | 0.001243879 | 1 |
| T2DM | pericyte | *TCIRG1* | 2.968798865 | 1.97E-05 | 0.000515615 | 0.123691231 |
| T2DM | pericyte | *GTF2H1* | 1.871976756 | 0.00333899 | 0.008049169 | 1 |
| T2DM | pericyte | *PTPN5* | 0.590335527 | 0.022759284 | 0.039212831 | 1 |
| T2DM | pericyte | *PPFIBP1* | 2.115736257 | 0.001955823 | 0.005313697 | 1 |
| T2DM | pericyte | *PRDM4* | 3.068449168 | 0.000379418 | 0.001624931 | 1 |
| T2DM | pericyte | *COQ5* | 0.482569137 | 0.000174581 | 0.001009742 | 1 |
| T2DM | pericyte | *CAPRIN2* | 2.789222033 | 0.001871846 | 0.005147484 | 1 |
| T2DM | pericyte | *CYP27B1* | 6.514682768 | 0.005540483 | 0.011852175 | 1 |
| T2DM | pericyte | *GLI1* | 1.471013218 | 0.00499458 | 0.010952538 | 1 |
| T2DM | pericyte | *METAP2* | 2.140563714 | 0.004097128 | 0.009421597 | 1 |
| T2DM | pericyte | *LTA4H* | 4.265517888 | 0.000114431 | 0.000791693 | 0.719425534 |
| T2DM | pericyte | *ITFG2* | 3.079256393 | 0.000312904 | 0.001424252 | 1 |
| T2DM | pericyte | *PRR4* | 1.84127569 | 3.87E-06 | 0.000279804 | 0.024342925 |
| T2DM | pericyte | *PARP11* | 2.991708079 | 1.86E-05 | 0.000512131 | 0.117074197 |
| T2DM | pericyte | *ARPC3* | 3.032387587 | 0.000216874 | 0.001141949 | 1 |
| T2DM | pericyte | *GPN3* | 3.673025393 | 0.000156162 | 0.000945663 | 0.98179276 |
| T2DM | pericyte | *VPS29* | 3.304586286 | 0.000693879 | 0.002504256 | 1 |
| T2DM | pericyte | *KCNA1* | 1.672073786 | 0.018741355 | 0.033192521 | 1 |
| T2DM | pericyte | *CREBL2* | 3.309512627 | 1.13E-06 | 0.000128924 | 0.007090815 |
| T2DM | pericyte | *NAA25* | 2.887762959 | 0.000155924 | 0.000945663 | 0.980294759 |
| T2DM | pericyte | *LTBR* | 1.263579194 | 0.004104536 | 0.00943518 | 1 |
| T2DM | pericyte | *CDK2AP1* | 1.007406999 | 2.18E-05 | 0.000541395 | 0.136972965 |
| T2DM | pericyte | *MGP* | 1.680407711 | 1.69E-05 | 0.000507168 | 0.10599821 |
| T2DM | pericyte | *EIF2B1* | 3.194857305 | 7.04E-05 | 0.000639553 | 0.44279735 |
| T2DM | pericyte | *ENDOU* | -1.42625182 | 0.000344028 | 0.001511465 | 1 |
| T2DM | pericyte | *STX2* | 1.331124086 | 0.010569565 | 0.020364958 | 1 |
| T2DM | pericyte | *MDM1* | 1.904488538 | 0.001778242 | 0.004977652 | 1 |
| T2DM | pericyte | *NUP107* | 2.725822503 | 5.04E-05 | 0.000610492 | 0.316845169 |
| T2DM | pericyte | *CNOT2* | 2.752250961 | 0.000606522 | 0.002271117 | 1 |
| T2DM | pericyte | *COPS7A* | 0.886986643 | 0.018260732 | 0.032522725 | 1 |
| T2DM | pericyte | *GNB3* | -0.40038233 | 0.007471933 | 0.015119421 | 1 |
| T2DM | pericyte | *CDCA3* | -0.957018015 | 0.001564583 | 0.004530877 | 1 |
| T2DM | pericyte | *GNPTAB* | 3.645511897 | 2.69E-05 | 0.000562456 | 0.169086837 |
| T2DM | pericyte | *SPSB2* | 2.529979122 | 0.005212888 | 0.0113207 | 1 |
| T2DM | pericyte | *ATN1* | 2.20968676 | 0.000850962 | 0.002925096 | 1 |
| T2DM | pericyte | *C12orf57* | 1.072837125 | 0.009791207 | 0.019052095 | 1 |
| T2DM | pericyte | *NT5DC3* | 1.021838823 | 0.015220596 | 0.027841689 | 1 |

| | | | | | | |
|---|---|---|---|---|---|---|
| T2DM | pericyte | *ST8SIA1* | 1.270750783 | 0.001464271 | 0.004305833 | 1 |
| T2DM | pericyte | *C2CD5* | 3.002103861 | 4.05E-05 | 0.000607238 | 0.254656537 |
| T2DM | pericyte | *PHC1* | 7.255482019 | 2.08E-05 | 0.000527586 | 0.130841379 |
| T2DM | pericyte | *TDP2* | 3.413597772 | 0.000383944 | 0.001639846 | 1 |
| T2DM | pericyte | *SMIM8* | 2.60368401 | 0.000266083 | 0.001293785 | 1 |
| T2DM | pericyte | *MCM9* | 3.612171377 | 3.25E-05 | 0.000599418 | 0.204497334 |
| T2DM | pericyte | *FAM184A* | 1.811512934 | 0.000199064 | 0.001077963 | 1 |
| T2DM | pericyte | *RNGTT* | 2.870399784 | 5.02E-05 | 0.000610424 | 0.31558912 |
| T2DM | pericyte | *GABRR2* | 0.839377866 | 0.016346478 | 0.029616804 | 1 |
| T2DM | pericyte | *HDDC2* | 1.112215869 | 0.003491766 | 0.008342281 | 1 |
| T2DM | pericyte | *TPD52L1* | 1.070441856 | 0.019348581 | 0.03414104 | 1 |
| T2DM | pericyte | *UST* | 1.159234171 | 0.000581808 | 0.002203511 | 1 |
| T2DM | pericyte | *MTRF1L* | 1.364618973 | 0.000122546 | 0.000826661 | 0.770447999 |
| T2DM | pericyte | *TULP1* | 5.077642663 | 0.004052476 | 0.009339413 | 1 |
| T2DM | pericyte | *SLC26A8* | 0.616789518 | 0.008677181 | 0.017165966 | 1 |
| T2DM | pericyte | *RNF8* | 1.234782533 | 0.014010336 | 0.025883922 | 1 |
| T2DM | pericyte | *PHACTR1* | 2.858724137 | 0.000955478 | 0.003180036 | 1 |
| T2DM | pericyte | *MDGA1* | 0.936746348 | 0.012600365 | 0.023661438 | 1 |
| T2DM | pericyte | *SAYSD1* | 4.411373628 | 8.37E-05 | 0.000681373 | 0.526039192 |
| T2DM | pericyte | *CAP2* | 0.632654523 | 0.028894275 | 0.047829991 | 1 |
| T2DM | pericyte | *ASCC3* | 3.028697585 | 0.000108238 | 0.000774167 | 0.680492909 |
| T2DM | pericyte | *BVES* | -0.956076836 | 0.00017036 | 0.000994477 | 1 |
| T2DM | pericyte | *GPLD1* | 0.696895412 | 0.002015712 | 0.00542956 | 1 |
| T2DM | pericyte | *ALDH5A1* | 1.555000998 | 7.04E-05 | 0.000639553 | 0.442317386 |
| T2DM | pericyte | *VNN2* | 2.693900146 | 0.000464792 | 0.00188404 | 1 |
| T2DM | pericyte | *ACOT13* | 2.064685684 | 0.000239685 | 0.001219172 | 1 |
| T2DM | pericyte | *C6orf62* | 2.5244561 | 6.91E-05 | 0.000637539 | 0.434314192 |
| T2DM | pericyte | *SLC16A10* | 1.064565052 | 0.001443256 | 0.004253984 | 1 |
| T2DM | pericyte | *PHACTR2* | 2.71982093 | 5.89E-05 | 0.00062437 | 0.370495174 |
| T2DM | pericyte | *TFEB* | 4.670000517 | 5.73E-05 | 0.00062437 | 0.360504071 |
| T2DM | pericyte | *FAM120B* | 2.237164358 | 1.82E-05 | 0.000512131 | 0.114714395 |
| T2DM | pericyte | *TBP* | 3.625515286 | 7.44E-05 | 0.000649647 | 0.46774603 |
| T2DM | pericyte | *DNPH1* | 0.804233196 | 0.020146589 | 0.035321139 | 1 |
| T2DM | pericyte | *DUSP22* | 3.168872182 | 7.32E-05 | 0.000646906 | 0.460454175 |
| T2DM | pericyte | *EXOC2* | 3.379570555 | 0.000140264 | 0.00089527 | 0.881840959 |
| T2DM | pericyte | *TMEM30A* | 1.718410139 | 0.000295284 | 0.001375149 | 1 |
| T2DM | pericyte | *SENP6* | 2.561231452 | 0.00106069 | 0.003418019 | 1 |
| T2DM | pericyte | *CLIC5* | 1.065859082 | 0.018640887 | 0.033077274 | 1 |
| T2DM | pericyte | *FBRSL1* | 2.691709747 | 5.98E-05 | 0.00062437 | 0.375727409 |
| T2DM | pericyte | *LY86* | 4.354430593 | 0.000124149 | 0.000832113 | 0.7805219 |
| T2DM | pericyte | *NUDT12* | 1.503746491 | 0.002444182 | 0.006302941 | 1 |
| T2DM | pericyte | *C7* | 1.896671382 | 0.001709008 | 0.00483425 | 1 |
| T2DM | pericyte | *PAPD7* | 2.994984454 | 0.000102904 | 0.00075118 | 0.646956876 |
| T2DM | pericyte | *GHR* | 0.764553029 | 0.000361207 | 0.001563986 | 1 |
| T2DM | pericyte | *DAP* | 2.841550768 | 0.004423472 | 0.010021754 | 1 |
| T2DM | pericyte | *NME5* | 0.661030576 | 0.003409719 | 0.008172666 | 1 |
| T2DM | pericyte | *PFDN1* | 1.68466609 | 0.003859428 | 0.009008206 | 1 |
| T2DM | pericyte | *LOX* | 3.228004153 | 8.17E-05 | 0.000675975 | 0.513741252 |
| T2DM | pericyte | *SPARC* | 2.485447875 | 5.87E-05 | 0.00062437 | 0.36881115 |
| T2DM | pericyte | *PCDHB6* | 1.999552071 | 0.008957576 | 0.017615351 | 1 |

| | | | | | | |
|---|---|---|---|---|---|---|
| T2DM | pericyte | *CLK4* | 2.034747977 | 0.000195855 | 0.001065597 | 1 |
| T2DM | pericyte | *RNF130* | 2.904670844 | 4.76E-05 | 0.000607238 | 0.299142377 |
| T2DM | pericyte | *ARSB* | 1.290714557 | 0.003854626 | 0.00900224 | 1 |
| T2DM | pericyte | *CLINT1* | 3.111667174 | 5.40E-06 | 0.000329604 | 0.033949234 |
| T2DM | pericyte | *BTNL8* | -0.25642329 | 0.028298552 | 0.04699234 | 1 |
| T2DM | pericyte | *TTC1* | 3.685373261 | 2.99E-05 | 0.00059534 | 0.188084304 |
| T2DM | pericyte | *RASGRF2* | 1.532871054 | 0.000326179 | 0.001462687 | 1 |
| T2DM | pericyte | *POLR3G* | 0.944697741 | 0.010543162 | 0.020320312 | 1 |
| T2DM | pericyte | *DROSHA* | 2.166223717 | 0.000928221 | 0.003107414 | 1 |
| T2DM | pericyte | *CDH6* | -0.255000645 | 0.007431077 | 0.015042688 | 1 |
| T2DM | pericyte | *TARS* | 2.094434676 | 0.000187719 | 0.001045339 | 1 |
| T2DM | pericyte | *LNPEP* | 2.737990092 | 6.51E-06 | 0.000343901 | 0.040924242 |
| T2DM | pericyte | *GNPDA1* | 2.161491987 | 0.000282491 | 0.001338374 | 1 |
| T2DM | pericyte | *SKP1* | 3.586956147 | 6.36E-05 | 0.000632025 | 0.400046078 |
| T2DM | pericyte | *TRIM23* | 2.83913873 | 0.001291939 | 0.003925772 | 1 |
| T2DM | pericyte | *TRAPPC13* | 1.980144011 | 0.002298251 | 0.006000462 | 1 |
| T2DM | pericyte | *TXNDC15* | 1.82052745 | 0.00094234 | 0.003142967 | 1 |
| T2DM | pericyte | *SMAD5* | 3.685967595 | 2.60E-05 | 0.000562456 | 0.163437167 |
| T2DM | pericyte | *HMGXB3* | 3.473626411 | 0.000591303 | 0.00222873 | 1 |
| T2DM | pericyte | *ATP6V0E1* | 2.634429841 | 0.001570443 | 0.00454366 | 1 |
| T2DM | pericyte | *CNTN3* | 2.16083074 | 0.000204623 | 0.001098602 | 1 |
| T2DM | pericyte | *ACTR8* | 1.90245466 | 0.001208254 | 0.003749405 | 1 |
| T2DM | pericyte | *HGD* | 2.423330163 | 0.002336383 | 0.006079819 | 1 |
| T2DM | pericyte | *NPHP3* | 0.684907789 | 0.000701085 | 0.002523526 | 1 |
| T2DM | pericyte | *NIT2* | 2.751259511 | 0.00517749 | 0.01126718 | 1 |
| T2DM | pericyte | *PCCB* | 1.918826673 | 0.000772565 | 0.002724025 | 1 |
| T2DM | pericyte | *PDCD10* | 3.328001121 | 0.000108684 | 0.000775065 | 0.683295592 |
| T2DM | pericyte | *WNT5A* | 0.874816817 | 0.030188077 | 0.049618939 | 1 |
| T2DM | pericyte | *IFT57* | 2.514967094 | 0.000195644 | 0.001065597 | 1 |
| T2DM | pericyte | *IQCG* | 4.04341709 | 6.42E-05 | 0.000632025 | 0.403527807 |
| T2DM | pericyte | *GBE1* | 1.588270819 | 0.000130645 | 0.000858273 | 0.821366963 |
| T2DM | pericyte | *NCBP2* | 3.656445392 | 1.65E-05 | 0.000502137 | 0.103942266 |
| T2DM | pericyte | *SLC41A3* | 1.579647556 | 0.005701168 | 0.012109203 | 1 |
| T2DM | pericyte | *ABTB1* | 3.471190919 | 0.00114838 | 0.003618978 | 1 |
| T2DM | pericyte | *NEK11* | 2.782642762 | 0.000114347 | 0.000791693 | 0.718896934 |
| T2DM | pericyte | *PLSCR4* | 1.702286204 | 0.004134763 | 0.009480399 | 1 |
| T2DM | pericyte | *HEMK1* | 1.89667906 | 0.001540434 | 0.004486927 | 1 |
| T2DM | pericyte | *WDR48* | 2.152516949 | 0.001824718 | 0.005067139 | 1 |
| T2DM | pericyte | *COMMD2* | 2.574601145 | 0.001344221 | 0.004034577 | 1 |
| T2DM | pericyte | *GORASP1* | 4.095210061 | 5.96E-05 | 0.00062437 | 0.374847131 |
| T2DM | pericyte | *ARHGEF26* | 0.851691219 | 0.001681923 | 0.004780403 | 1 |
| T2DM | pericyte | *KLHL24* | 1.876970406 | 0.001271762 | 0.003881628 | 1 |
| T2DM | pericyte | *ZBTB47* | 2.49408958 | 0.00014485 | 0.00091218 | 0.910670982 |
| T2DM | pericyte | *NKTR* | 3.093569007 | 9.71E-06 | 0.000404168 | 0.061029427 |
| T2DM | pericyte | *CLCN2* | 1.885412016 | 0.00131243 | 0.003976505 | 1 |
| T2DM | pericyte | *NEK4* | 2.983564519 | 0.000180321 | 0.001031556 | 1 |
| T2DM | pericyte | *MOB1A* | 4.073123056 | 0.000466312 | 0.001887768 | 1 |
| T2DM | pericyte | *LMAN2L* | 1.372211468 | 0.023754556 | 0.040615962 | 1 |
| T2DM | pericyte | *PIKFYVE* | 2.015156867 | 0.010308607 | 0.01991709 | 1 |
| T2DM | pericyte | *FAHD2A* | 1.184413324 | 0.00174516 | 0.004909093 | 1 |

| | | | | | | |
|---|---|---|---|---|---|---|
| T2DM | pericyte | *SLC35F5* | 2.838534317 | 3.76E-05 | 0.000607238 | 0.236281804 |
| T2DM | pericyte | *ACTR3* | 3.366221426 | 3.64E-05 | 0.00060096 | 0.22859744 |
| T2DM | pericyte | *TFCP2L1* | 1.190419272 | 0.002390599 | 0.006197813 | 1 |
| T2DM | pericyte | *STAM2* | 3.014933131 | 0.001762772 | 0.004938747 | 1 |
| T2DM | pericyte | *GPD2* | 1.788379253 | 0.000313767 | 0.001424297 | 1 |
| T2DM | pericyte | *CENPA* | 2.87068202 | 0.001284683 | 0.003913179 | 1 |
| T2DM | pericyte | *ITGA4* | 0.731947714 | 0.002395921 | 0.006203936 | 1 |
| T2DM | pericyte | *GRB14* | 0.653153629 | 0.006679942 | 0.013785158 | 1 |
| T2DM | pericyte | *CLIP4* | 2.051216159 | 0.000183494 | 0.001035729 | 1 |
| T2DM | pericyte | *SPTBN1* | 1.336876995 | 0.009095876 | 0.017853815 | 1 |
| T2DM | pericyte | *HTRA2* | 0.904906091 | 0.013114408 | 0.024495566 | 1 |
| T2DM | pericyte | *POLE4* | 4.700987911 | 3.36E-05 | 0.000599418 | 0.211510138 |
| T2DM | pericyte | *LANCL1* | 2.182554386 | 0.000277861 | 0.001326433 | 1 |
| T2DM | pericyte | *WDR75* | 3.83454494 | 8.33E-05 | 0.000681373 | 0.523504445 |
| T2DM | pericyte | *EFEMP1* | 0.973947809 | 0.003051372 | 0.007505467 | 1 |
| T2DM | pericyte | *GLS* | 2.855417707 | 5.18E-05 | 0.000616661 | 0.325597058 |
| T2DM | pericyte | *DNAH6* | 0.208318299 | 0.018588406 | 0.033003476 | 1 |
| T2DM | pericyte | *GNLY* | 3.099049421 | 0.001222346 | 0.003783797 | 1 |
| T2DM | pericyte | *SF3B1* | 2.980842835 | 0.000104083 | 0.000755623 | 0.654369348 |
| T2DM | pericyte | *ST3GAL5* | 1.266074001 | 0.001558994 | 0.004518363 | 1 |
| T2DM | pericyte | *PLCD4* | 1.863417026 | 0.001718071 | 0.004847304 | 1 |
| T2DM | pericyte | *CHMP3* | -1.389142365 | 0.025616041 | 0.043362426 | 1 |
| T2DM | pericyte | *ZNF142* | 1.013267814 | 0.002869844 | 0.007148458 | 1 |
| T2DM | pericyte | *MLPH* | 0.759994648 | 0.00431736 | 0.00982383 | 1 |
| T2DM | pericyte | *HDLBP* | 1.721707377 | 0.001920095 | 0.005241701 | 1 |
| T2DM | pericyte | *STK25* | 2.773584016 | 0.002332895 | 0.006073254 | 1 |
| T2DM | pericyte | *STRN* | 3.453178412 | 0.000195618 | 0.001065597 | 1 |
| T2DM | pericyte | *CEBPZ* | 1.781779296 | 0.00612583 | 0.012841978 | 1 |
| T2DM | pericyte | *PRKD3* | 2.836057074 | 0.000542791 | 0.0021052 | 1 |
| T2DM | pericyte | *SLC25A12* | 1.666503662 | 0.00451597 | 0.010159665 | 1 |
| T2DM | pericyte | *RMDN2* | 4.049020807 | 0.000290167 | 0.001359225 | 1 |
| T2DM | pericyte | *SDC1* | 3.634286267 | 0.000470148 | 0.001897189 | 1 |
| T2DM | pericyte | *SOS1* | 1.44442645 | 0.001965074 | 0.005332077 | 1 |
| T2DM | pericyte | *WIPF1* | 2.142947979 | 0.000443529 | 0.001821336 | 1 |
| T2DM | pericyte | *COX7A2L* | 4.199372394 | 9.16E-05 | 0.000706851 | 0.576083177 |
| T2DM | pericyte | *PNO1* | 3.091973955 | 0.000172974 | 0.001003219 | 1 |
| T2DM | pericyte | *ORC4* | 2.345623782 | 0.001916663 | 0.005239156 | 1 |
| T2DM | pericyte | *THADA* | 2.97915723 | 4.05E-05 | 0.000607238 | 0.254764784 |
| T2DM | pericyte | *ARID3A* | 1.925187428 | 0.000329085 | 0.001471521 | 1 |
| T2DM | pericyte | *CD207* | 19.33656189 | 0.000713932 | 0.002559001 | 1 |
| T2DM | pericyte | *GRIN3B* | 1.18661972 | 0.017598064 | 0.031521091 | 1 |
| T2DM | pericyte | *VAX2* | 3.399214189 | 0.000605052 | 0.002267078 | 1 |
| T2DM | pericyte | *MSH6* | 2.350098287 | 0.000409309 | 0.00171784 | 1 |
| T2DM | pericyte | *PLEKHA3* | 4.046731471 | 3.05E-05 | 0.000599418 | 0.19204483 |
| T2DM | pericyte | *ALMS1* | 2.231764773 | 0.003171815 | 0.007752714 | 1 |
| T2DM | pericyte | *BCL9* | 2.797894914 | 0.001388343 | 0.004130862 | 1 |
| T2DM | pericyte | *DHCR24* | 2.022395467 | 0.001141835 | 0.003605585 | 1 |
| T2DM | pericyte | *DNAJC16* | 1.482601261 | 0.005031062 | 0.011017166 | 1 |
| T2DM | pericyte | *MORN1* | 1.540977562 | 0.002841652 | 0.00709792 | 1 |
| T2DM | pericyte | *GPX7* | -0.603182645 | 0.014142342 | 0.026097125 | 1 |

| | | | | | | |
|---|---|---|---|---|---|---|
| T2DM | pericyte | *SCP2* | 2.442519747 | 6.60E-05 | 0.000632701 | 0.415151323 |
| T2DM | pericyte | *TPSG1* | -1.163010276 | 0.006610025 | 0.013675943 | 1 |
| T2DM | pericyte | *RALGPS2* | 2.319272667 | 0.005925837 | 0.012496433 | 1 |
| T2DM | pericyte | *CEP104* | 1.688235084 | 0.001459954 | 0.004299171 | 1 |
| T2DM | pericyte | *FAM20B* | 4.23970794 | 0.000149901 | 0.000927922 | 0.94243029 |
| T2DM | pericyte | *TMEM59* | 3.34862442 | 3.38E-05 | 0.000599418 | 0.212523073 |
| T2DM | pericyte | *WRAP73* | 4.200594577 | 0.000402689 | 0.001695718 | 1 |
| T2DM | pericyte | *MRPL37* | 3.237837284 | 1.79E-05 | 0.000512131 | 0.112842724 |
| T2DM | pericyte | *QSOX1* | 3.780199394 | 0.000231717 | 0.001193695 | 1 |
| T2DM | pericyte | *PARK7* | 3.54307185 | 0.000186639 | 0.001043394 | 1 |
| T2DM | pericyte | *EDEM3* | 2.815766715 | 0.000142757 | 0.000906581 | 0.897515412 |
| T2DM | pericyte | *RAP1A* | 3.04978725 | 0.000150726 | 0.00093086 | 0.947615131 |
| T2DM | pericyte | *CAPZA1* | 1.75588352 | 0.02087283 | 0.036421727 | 1 |
| T2DM | pericyte | *DOCK7* | 2.806296368 | 0.001147198 | 0.003618882 | 1 |
| T2DM | pericyte | *FBXO6* | 1.016274981 | 0.008702297 | 0.017204825 | 1 |
| T2DM | pericyte | *DNAJC6* | 1.791729241 | 0.01712392 | 0.030791957 | 1 |
| T2DM | pericyte | *LEPR* | 1.689865806 | 0.000253625 | 0.001257649 | 1 |
| T2DM | pericyte | *MFN2* | 2.991641256 | 0.000231732 | 0.001193695 | 1 |
| T2DM | pericyte | *SMG7* | 3.210610069 | 0.000123678 | 0.000831605 | 0.777566038 |
| T2DM | pericyte | *NCF2* | 2.105862855 | 0.000391201 | 0.001664062 | 1 |
| T2DM | pericyte | *OLFML3* | 1.426294138 | 0.001809639 | 0.005031932 | 1 |
| T2DM | pericyte | *TNNI3K* | 1.711168264 | 0.001519197 | 0.004436225 | 1 |
| T2DM | pericyte | *TTF2* | 2.89145958 | 9.11E-05 | 0.000704357 | 0.572530045 |
| T2DM | pericyte | *OSCP1* | 2.110680213 | 0.000248835 | 0.001243879 | 1 |
| T2DM | pericyte | *GNPAT* | 1.482745615 | 0.022837177 | 0.0392717 | 1 |
| T2DM | pericyte | *TBCE* | 1.636286079 | 0.016635843 | 0.030037204 | 1 |
| T2DM | pericyte | *MTR* | 2.730177088 | 8.54E-05 | 0.000687866 | 0.537223049 |
| T2DM | pericyte | *ACADM* | 2.099962381 | 5.65E-05 | 0.00062437 | 0.355279585 |
| T2DM | pericyte | *CTBS* | 1.808170343 | 0.010149734 | 0.019640314 | 1 |
| T2DM | pericyte | *KLHL12* | 2.61317387 | 0.00116237 | 0.003652084 | 1 |
| T2DM | pericyte | *RBBP5* | 2.142680833 | 6.95E-05 | 0.000637863 | 0.436936397 |
| T2DM | pericyte | *CD46* | 2.625418391 | 1.87E-05 | 0.000512131 | 0.117881045 |
| T2DM | pericyte | *PRDX1* | 2.759538958 | 0.000475051 | 0.001907181 | 1 |
| T2DM | pericyte | *FAAH* | 0.459797621 | 0.001557151 | 0.004517679 | 1 |
| T2DM | pericyte | *TMED5* | 3.646469302 | 0.000149214 | 0.000925159 | 0.938111522 |
| T2DM | pericyte | *DR1* | 4.53687641 | 1.34E-05 | 0.000464073 | 0.084461246 |
| T2DM | pericyte | *CNN3* | 3.620258928 | 4.13E-05 | 0.000607238 | 0.259651507 |
| T2DM | pericyte | *PRRC2C* | 2.917515293 | 8.66E-05 | 0.000695574 | 0.544634341 |
| T2DM | pericyte | *ABCD3* | 3.737232671 | 6.12E-05 | 0.000627415 | 0.384767986 |
| T2DM | pericyte | *VAMP4* | 2.042649854 | 5.60E-05 | 0.00062437 | 0.352281232 |
| T2DM | pericyte | *PTBP2* | 1.625338891 | 0.000739941 | 0.002629741 | 1 |
| T2DM | pericyte | *TNFSF4* | -1.174627169 | 0.002352114 | 0.00611569 | 1 |
| T2DM | pericyte | *IRF6* | 0.598365765 | 0.020494295 | 0.035840788 | 1 |
| T2DM | pericyte | *SERPINC1* | 1.141304204 | 0.000896964 | 0.00303638 | 1 |
| T2DM | pericyte | *RCAN3* | 1.892298514 | 0.002644392 | 0.006717289 | 1 |
| T2DM | pericyte | *RCOR3* | 2.948650715 | 0.000900394 | 0.003038527 | 1 |
| T2DM | pericyte | *MTFR1L* | 4.226312315 | 8.99E-05 | 0.000702125 | 0.565157182 |
| T2DM | pericyte | *NSL1* | 2.144008018 | 0.000369453 | 0.001590927 | 1 |
| T2DM | pericyte | *RPA2* | 2.786038295 | 0.000136067 | 0.000878628 | 0.855452957 |
| T2DM | pericyte | *SLC5A9* | 6.679456421 | 2.27E-05 | 0.000548455 | 0.142433275 |

| | | | | | | |
|---|---|---|---|---|---|---|
| T2DM | pericyte | *OSBPL9* | 2.790185099 | 6.86E-05 | 0.000635523 | 0.431520163 |
| T2DM | pericyte | *TXNDC12* | 2.402255716 | 0.000112834 | 0.00078821 | 0.70938883 |
| T2DM | pericyte | *ESYT2* | 1.711290282 | 0.011221795 | 0.021486231 | 1 |
| T2DM | pericyte | *CD3EAP* | -1.836069828 | 0.002275348 | 0.005967924 | 1 |
| T2DM | pericyte | *KMT2A* | 3.252298581 | 0.000117896 | 0.000803917 | 0.741211917 |
| T2DM | pericyte | *IFT46* | 2.113957714 | 0.020548337 | 0.035900121 | 1 |
| T2DM | pericyte | *CAMSAP2* | 2.772874886 | 9.95E-05 | 0.000737961 | 0.625791123 |
| T2DM | pericyte | *ATF6* | 2.8912111 | 3.66E-05 | 0.000601794 | 0.229885353 |
| T2DM | pericyte | *ATP10B* | 1.710459351 | 0.005265248 | 0.011417805 | 1 |
| T2DM | pericyte | *SPCS2* | 2.361426031 | 0.000352628 | 0.001534236 | 1 |
| T2DM | pericyte | *ELOVL4* | 1.165613586 | 8.35E-05 | 0.000681373 | 0.524893628 |
| T2DM | pericyte | *MED28* | 1.627703169 | 0.018291599 | 0.032564233 | 1 |
| T2DM | pericyte | *SLC16A7* | 1.56452589 | 0.000204111 | 0.00109679 | 1 |
| T2DM | pericyte | *FOXO3* | 2.721190065 | 4.55E-05 | 0.000607238 | 0.285972383 |
| T2DM | pericyte | *TGIF2* | 1.635486839 | 0.002163505 | 0.005731965 | 1 |
| T2DM | pericyte | *ABCG2* | 2.657997218 | 0.000665114 | 0.002428322 | 1 |
| T2DM | pericyte | *SPP1* | 2.733715269 | 0.001621814 | 0.004653739 | 1 |
| T2DM | pericyte | *MFSD1* | 2.002897226 | 0.000152317 | 0.000937005 | 0.957619273 |
| T2DM | pericyte | *RAB3GAP2* | 2.459451041 | 4.63E-05 | 0.000607238 | 0.291100207 |
| T2DM | pericyte | *UBN1* | 2.061463435 | 0.000281847 | 0.001337337 | 1 |
| T2DM | pericyte | *WDR35* | 2.272554023 | 0.000437088 | 0.00180313 | 1 |
| T2DM | pericyte | *DNAH7* | -0.692667294 | 0.003355451 | 0.008073371 | 1 |
| T2DM | pericyte | *TRPM6* | 0.806942794 | 0.017732716 | 0.03172612 | 1 |
| T2DM | pericyte | *GDA* | 0.977555583 | 0.006407525 | 0.013321465 | 1 |
| T2DM | pericyte | *CPSF3* | 3.825242212 | 1.54E-05 | 0.000488489 | 0.097004353 |
| T2DM | pericyte | *SENP5* | 3.693066366 | 1.87E-05 | 0.000512131 | 0.117818099 |
| T2DM | pericyte | *PTBP3* | 3.606583386 | 6.20E-05 | 0.000628694 | 0.389773876 |
| T2DM | pericyte | *FKBP15* | 3.302447661 | 8.68E-06 | 0.000381879 | 0.054562411 |
| T2DM | pericyte | *CTNNAL1* | 0.706684044 | 0.001960983 | 0.005323272 | 1 |
| T2DM | pericyte | *CNTRL* | 3.241645411 | 0.000384676 | 0.001641859 | 1 |
| T2DM | pericyte | *PHF19* | 1.067024798 | 0.004946507 | 0.010877471 | 1 |
| T2DM | pericyte | *BSPRY* | -1.04932776 | 0.030110679 | 0.049528774 | 1 |
| T2DM | pericyte | *PPP6C* | 2.789853649 | 0.000302501 | 0.00139207 | 1 |
| T2DM | pericyte | *RBM18* | 4.011435992 | 0.000550485 | 0.002127167 | 1 |
| T2DM | pericyte | *HSDL2* | 0.812452906 | 0.006588087 | 0.013642722 | 1 |
| T2DM | pericyte | *INVS* | 2.593693863 | 0.000188064 | 0.001046336 | 1 |
| T2DM | pericyte | *KDSR* | 3.565612327 | 4.08E-05 | 0.000607238 | 0.256198425 |
| T2DM | pericyte | *DCAF4* | 1.700725186 | 0.000111399 | 0.00078265 | 0.700367953 |
| T2DM | pericyte | *DNAL1* | 1.795609431 | 0.000858771 | 0.00294389 | 1 |
| T2DM | pericyte | *IRF2BPL* | 1.0108526 | 0.00163676 | 0.004681545 | 1 |
| T2DM | pericyte | *TTLL5* | 2.620173995 | 5.66E-05 | 0.00062437 | 0.355590689 |
| T2DM | pericyte | *FLVCR2* | 1.751027812 | 0.001078779 | 0.00345682 | 1 |
| T2DM | pericyte | *PPP4R4* | -0.399753819 | 0.019164563 | 0.033873379 | 1 |
| T2DM | pericyte | *COQ6* | 1.971353185 | 0.000269771 | 0.001305657 | 1 |
| T2DM | pericyte | *GPR75* | 7.594476517 | 0.002269367 | 0.005957208 | 1 |
| T2DM | pericyte | *KLHL29* | 2.91398731 | 5.03E-05 | 0.000610492 | 0.316530956 |
| T2DM | pericyte | *ATAD2B* | 3.015360219 | 4.34E-05 | 0.000607238 | 0.272656468 |
| T2DM | pericyte | *ATL2* | 2.649318178 | 6.84E-05 | 0.000635523 | 0.430268598 |
| T2DM | pericyte | *SLC17A5* | 1.11219342 | 0.030066376 | 0.049483589 | 1 |
| T2DM | pericyte | *OGFRL1* | 2.242236345 | 0.000634621 | 0.002337355 | 1 |

| | | | | | | |
|---|---|---|---|---|---|---|
| T2DM | pericyte | *CUTC* | 2.949179062 | 0.000218466 | 0.001146081 | 1 |
| T2DM | pericyte | *MXI1* | 3.44648621 | 0.000813297 | 0.002824311 | 1 |
| T2DM | pericyte | *C10orf88* | 3.974274675 | 5.31E-05 | 0.000619691 | 0.333889629 |
| T2DM | pericyte | *HELLS* | 1.902418614 | 0.001548033 | 0.004499531 | 1 |
| T2DM | pericyte | *AVPI1* | 0.729265289 | 0.009150954 | 0.017945117 | 1 |
| T2DM | pericyte | *WDR11* | 2.50697135 | 0.000162242 | 0.000963715 | 1 |
| T2DM | pericyte | *C10orf76* | 2.903601023 | 4.80E-05 | 0.000607238 | 0.301796456 |
| T2DM | pericyte | *KCNIP2* | 0.654548022 | 0.013175148 | 0.02459387 | 1 |
| T2DM | pericyte | *KANSL1* | 2.217699105 | 0.000134779 | 0.000874555 | 0.847352867 |
| T2DM | pericyte | *CAAP1* | 2.581718598 | 0.001887626 | 0.005174823 | 1 |
| T2DM | pericyte | *CD274* | 1.94349878 | 0.006155627 | 0.012882964 | 1 |
| T2DM | pericyte | *CYSTM1* | 3.113468931 | 0.00025327 | 0.001257649 | 1 |
| T2DM | pericyte | *WDR55* | 1.090476675 | 0.006349096 | 0.01323414 | 1 |
| T2DM | pericyte | *SEC16B* | 1.543950421 | 0.007362609 | 0.014931846 | 1 |
| T2DM | pericyte | *GORAB* | 3.396442739 | 0.000233909 | 0.001198741 | 1 |
| T2DM | pericyte | *ACAT2* | 1.300861491 | 0.00150817 | 0.004412221 | 1 |
| T2DM | pericyte | *SNX19* | 1.516426559 | 0.010905507 | 0.020941639 | 1 |
| T2DM | pericyte | *MRC1* | 1.532753884 | 0.000392929 | 0.00166915 | 1 |
| T2DM | pericyte | *DNAJC15* | 3.091731637 | 0.003202623 | 0.007801197 | 1 |
| T2DM | pericyte | *WBP4* | 2.940214374 | 0.000104014 | 0.000755623 | 0.653937016 |
| T2DM | pericyte | *SIL1* | 2.861827188 | 5.62E-05 | 0.00062437 | 0.353357433 |
| T2DM | pericyte | *ZFP30* | 1.171454999 | 0.007116081 | 0.014506745 | 1 |
| T2DM | pericyte | *ARL1* | 1.52724592 | 0.001789171 | 0.004988689 | 1 |
| T2DM | pericyte | *APAF1* | 2.367579351 | 0.000345809 | 0.00151717 | 1 |
| T2DM | pericyte | *ADRA1A* | 0.408361162 | 1.40E-05 | 0.000470281 | 0.087942579 |
| T2DM | pericyte | *ZNF706* | 2.261785796 | 0.00168564 | 0.004786638 | 1 |
| T2DM | pericyte | *CRISPLD1* | 0.915903885 | 0.004320878 | 0.009828279 | 1 |
| T2DM | pericyte | *RDH10* | 0.823831196 | 0.001431584 | 0.004231421 | 1 |
| T2DM | pericyte | *PRB2* | 4.51101126 | 4.61E-06 | 0.000304712 | 0.029011962 |
| T2DM | pericyte | *A1BG* | 5.626375019 | 0.000276872 | 0.001325163 | 1 |
| T2DM | pericyte | *ZSCAN18* | 3.351381551 | 0.000113026 | 0.000788672 | 0.710593783 |
| T2DM | pericyte | *ZNF211* | 0.937765171 | 0.00039482 | 0.001672514 | 1 |
| T2DM | pericyte | *PDZRN3* | 2.456315027 | 4.55E-05 | 0.000607238 | 0.285850257 |
| T2DM | pericyte | *SEC22A* | 2.443589663 | 5.71E-06 | 0.000331374 | 0.035916736 |
| T2DM | pericyte | *POPDC2* | 4.849311799 | 0.00029775 | 0.001381518 | 1 |
| T2DM | pericyte | *B4GALT4* | 2.540697457 | 0.000265824 | 0.001293785 | 1 |
| T2DM | pericyte | *CD80* | 1.208256636 | 0.005564145 | 0.011886436 | 1 |
| T2DM | pericyte | *TMEM39B* | 1.746179577 | 0.003174397 | 0.007753471 | 1 |
| T2DM | pericyte | *RNF115* | 3.265993365 | 6.12E-05 | 0.000627415 | 0.384967659 |
| T2DM | pericyte | *POLR3GL* | 3.223151642 | 0.000413237 | 0.001732013 | 1 |
| T2DM | pericyte | *TNFSF10* | 3.12041304 | 0.00014163 | 0.000901244 | 0.890429384 |
| T2DM | pericyte | *ZNF639* | 3.300335605 | 4.89E-05 | 0.000607238 | 0.307213686 |
| T2DM | pericyte | *LIAS* | 2.762717508 | 0.004093067 | 0.009415701 | 1 |
| T2DM | pericyte | *CLCC1* | 2.310126905 | 0.025474781 | 0.043146538 | 1 |
| T2DM | pericyte | *GPSM2* | 2.911115026 | 6.83E-06 | 0.000347886 | 0.042963392 |
| T2DM | pericyte | *CXCR4* | 0.934149284 | 0.021765643 | 0.037769968 | 1 |
| T2DM | pericyte | *ACVR2A* | 3.450640008 | 4.55E-05 | 0.000607238 | 0.286046893 |
| T2DM | pericyte | *POLK* | 3.077738132 | 0.000117217 | 0.000800153 | 0.736940861 |
| T2DM | pericyte | *MTIF3* | 2.913426035 | 4.17E-06 | 0.000287028 | 0.02620661 |
| T2DM | pericyte | *RASL11A* | -0.437829306 | 0.015207226 | 0.027825329 | 1 |

| | | | | | | |
|---|---|---|---|---|---|---|
| T2DM | pericyte | *FMOD* | 2.037839739 | 0.002609038 | 0.006640899 | 1 |
| T2DM | pericyte | *OPN4* | -0.117539889 | 0.021884633 | 0.037926929 | 1 |
| T2DM | pericyte | *FAM35A* | 1.817360093 | 0.001386533 | 0.004127429 | 1 |
| T2DM | pericyte | *NAA60* | 4.464052918 | 0.000111068 | 0.00078265 | 0.698287344 |
| T2DM | pericyte | *ODF2L* | 1.940851338 | 2.69E-05 | 0.000562456 | 0.168825161 |
| T2DM | pericyte | *SPATA1* | 3.868726916 | 5.68E-05 | 0.00062437 | 0.35725317 |
| T2DM | pericyte | *RWDD3* | 2.415098096 | 0.001787947 | 0.004988689 | 1 |
| T2DM | pericyte | *CCDC18* | 2.721111086 | 0.00086961 | 0.002971326 | 1 |
| T2DM | pericyte | *RPAP2* | 2.736983775 | 4.67E-05 | 0.000607238 | 0.293700384 |
| T2DM | pericyte | *BBS9* | 1.673974811 | 0.018886072 | 0.033409323 | 1 |
| T2DM | pericyte | *PMS2* | 5.267625577 | 1.43E-06 | 0.000155353 | 0.009010467 |
| T2DM | pericyte | *KIAA0087* | -2.170830192 | 0.016448876 | 0.029759448 | 1 |
| T2DM | pericyte | *KLHL7* | 2.730958475 | 0.007060567 | 0.014416949 | 1 |
| T2DM | pericyte | *HERPUD2* | 3.335833165 | 8.40E-05 | 0.000681639 | 0.528270068 |
| T2DM | pericyte | *FAM126A* | 2.41437936 | 0.00087504 | 0.002988255 | 1 |
| T2DM | pericyte | *FKBP9* | 2.218514918 | 0.000555185 | 0.002141821 | 1 |
| T2DM | pericyte | *CCZ1* | 1.590263572 | 0.010135024 | 0.019629974 | 1 |
| T2DM | pericyte | *SMU1* | 2.741016379 | 4.74E-05 | 0.000607238 | 0.298167108 |
| T2DM | pericyte | *GLIPR2* | 2.157169196 | 0.007881239 | 0.015855792 | 1 |
| T2DM | pericyte | *SLC25A51* | 0.581062491 | 0.001348927 | 0.004041043 | 1 |
| T2DM | pericyte | *DCAF10* | 3.237631766 | 7.61E-05 | 0.000657799 | 0.478220048 |
| T2DM | pericyte | *TRIM24* | 2.290119727 | 0.000519725 | 0.002038672 | 1 |
| T2DM | pericyte | *BICC1* | 0.654262065 | 0.011321899 | 0.021651366 | 1 |
| T2DM | pericyte | *SLC25A16* | 0.983724148 | 0.000553388 | 0.002137072 | 1 |
| T2DM | pericyte | *RBM19* | 2.969617719 | 7.94E-05 | 0.000667567 | 0.499183866 |
| T2DM | pericyte | *ACADS* | 1.4736067 | 0.00261235 | 0.006646638 | 1 |
| T2DM | pericyte | *SSPN* | 1.698323214 | 2.50E-05 | 0.000558632 | 0.156975536 |
| T2DM | pericyte | *CCDC91* | 2.357515016 | 5.25E-05 | 0.000619691 | 0.330215314 |
| T2DM | pericyte | *PKN1* | 3.096814183 | 0.002144018 | 0.005689928 | 1 |
| T2DM | pericyte | *EBPL* | 1.286737849 | 0.001971177 | 0.005341719 | 1 |
| T2DM | pericyte | *NLN* | 1.421547578 | 5.87E-05 | 0.00062437 | 0.369011723 |
| T2DM | pericyte | *ATF1* | 3.384841598 | 8.22E-06 | 0.000379198 | 0.051709549 |
| T2DM | pericyte | *TSFM* | 2.072086872 | 0.000202144 | 0.001092758 | 1 |
| T2DM | pericyte | *ARHGAP9* | 0.736992137 | 0.006322503 | 0.013192308 | 1 |
| T2DM | pericyte | *SPATS2* | 2.765340676 | 0.00194657 | 0.00529329 | 1 |
| T2DM | pericyte | *ORMDL2* | 3.016716056 | 0.001826008 | 0.005068483 | 1 |
| T2DM | pericyte | *IKZF4* | 1.270670343 | 0.028241133 | 0.04690938 | 1 |
| T2DM | pericyte | *STIL* | 1.493977448 | 0.00323082 | 0.007857704 | 1 |
| T2DM | pericyte | *TTC21B* | 3.66095584 | 2.61E-05 | 0.000562456 | 0.163959199 |
| T2DM | pericyte | *NMI* | 2.347801224 | 0.000177882 | 0.001020443 | 1 |
| T2DM | pericyte | *ACVR1C* | 1.015149371 | 0.000361477 | 0.00156408 | 1 |
| T2DM | pericyte | *PLA2G12A* | 3.942260398 | 0.000283414 | 0.001339905 | 1 |
| T2DM | pericyte | *RAB38* | 2.109634346 | 0.003638122 | 0.008590671 | 1 |
| T2DM | pericyte | *AGO2* | 3.189727517 | 0.001359559 | 0.004060594 | 1 |
| T2DM | pericyte | *DNPEP* | 2.936214097 | 8.86E-05 | 0.000702125 | 0.556804445 |
| T2DM | pericyte | *MOGAT1* | 0.724836543 | 0.004629498 | 0.010365261 | 1 |
| T2DM | pericyte | *FAM124B* | 0.864717586 | 0.018033016 | 0.032171842 | 1 |
| T2DM | pericyte | *FAM210B* | 2.343791799 | 0.000320818 | 0.001444826 | 1 |
| T2DM | pericyte | *SNX21* | 3.342442824 | 5.12E-05 | 0.000615408 | 0.321662411 |
| T2DM | pericyte | *SLC12A5* | -0.964902726 | 0.022795491 | 0.039254116 | 1 |

| | | | | | | |
|---|---|---|---|---|---|---|
| T2DM | pericyte | *ARFGEF2* | 2.948618792 | 2.10E-05 | 0.000529508 | 0.13184742 |
| T2DM | pericyte | *CSE1L* | 1.929843006 | 0.012943185 | 0.024254487 | 1 |
| T2DM | pericyte | *STX16* | 3.743459306 | 4.55E-05 | 0.000607238 | 0.286215251 |
| T2DM | pericyte | *KCNK15* | 2.29208747 | 0.027719621 | 0.046189572 | 1 |
| T2DM | pericyte | *NEURL2* | 0.839963501 | 0.000513092 | 0.002021185 | 1 |
| T2DM | pericyte | *FASTKD3* | 1.352483258 | 0.001668125 | 0.004749773 | 1 |
| T2DM | pericyte | *PEPD* | 2.220899679 | 2.74E-06 | 0.000229534 | 0.017215069 |
| T2DM | pericyte | *CHST8* | 3.217905566 | 0.002499947 | 0.006423036 | 1 |
| T2DM | pericyte | *STAMBP* | 2.600989448 | 0.000100753 | 0.000742087 | 0.633434128 |
| T2DM | pericyte | *ATP8A1* | 1.555153435 | 0.001201876 | 0.00373589 | 1 |
| T2DM | pericyte | *USP22* | 3.20708802 | 6.52E-05 | 0.00063262 | 0.410016809 |
| T2DM | pericyte | *HIF3A* | 1.290847553 | 0.006350787 | 0.01323414 | 1 |
| T2DM | pericyte | *ZNF45* | 3.601462051 | 7.29E-05 | 0.000646906 | 0.458312076 |
| T2DM | pericyte | *BTN2A2* | 0.569929813 | 0.008060746 | 0.016139168 | 1 |
| T2DM | pericyte | *MRS2* | 1.632750781 | 0.002810422 | 0.007036688 | 1 |
| T2DM | pericyte | *SERPINB6* | 3.898398315 | 6.07E-05 | 0.000626153 | 0.38132737 |
| T2DM | pericyte | *NQO2* | 2.300925708 | 0.000160941 | 0.000958177 | 1 |
| T2DM | pericyte | *HIST1H2BJ* | 1.359382335 | 0.004843098 | 0.01069872 | 1 |
| T2DM | pericyte | *TCP11* | -0.959035132 | 0.024261573 | 0.041303143 | 1 |
| T2DM | pericyte | *KLHDC3* | 2.528489959 | 0.022254206 | 0.038469121 | 1 |
| T2DM | pericyte | *KLHL31* | 0.419139403 | 0.028836195 | 0.047746421 | 1 |
| T2DM | pericyte | *GLO1* | 4.339911351 | 2.17E-07 | 3.90E-05 | 0.001363263 |
| T2DM | pericyte | *RIOK1* | 1.867023658 | 0.000492561 | 0.001961199 | 1 |
| T2DM | pericyte | *NUP153* | 3.13107057 | 0.000395145 | 0.001672514 | 1 |
| T2DM | pericyte | *EREG* | -0.871172781 | 0.029861667 | 0.049185303 | 1 |
| T2DM | pericyte | *CNOT1* | 3.188897473 | 4.65E-05 | 0.000607238 | 0.292229592 |
| T2DM | pericyte | *BBS2* | 1.138300124 | 0.008228644 | 0.016392106 | 1 |
| T2DM | pericyte | *MT1G* | -2.44845456 | 0.023730214 | 0.04058538 | 1 |
| T2DM | pericyte | *GOT2* | 2.598884763 | 0.000282105 | 0.00133755 | 1 |
| T2DM | pericyte | *CLYBL* | 3.414529131 | 5.89E-05 | 0.00062437 | 0.37030858 |
| T2DM | pericyte | *TMTC4* | 1.712561587 | 0.000605083 | 0.002267078 | 1 |
| T2DM | pericyte | *KIF25* | 2.167758105 | 0.000805738 | 0.002804915 | 1 |
| T2DM | pericyte | *GRK4* | 0.653608723 | 1.46E-05 | 0.000476496 | 0.091762957 |
| T2DM | pericyte | *MRPS7* | 2.422303382 | 0.000531054 | 0.002071177 | 1 |
| T2DM | pericyte | *GGA3* | 2.301799561 | 0.000205441 | 0.001099503 | 1 |
| T2DM | pericyte | *NT5C* | 4.528167478 | 1.87E-05 | 0.000512131 | 0.117620646 |
| T2DM | pericyte | *OPRL1* | -1.834225537 | 2.35E-06 | 0.000207979 | 0.014766484 |
| T2DM | pericyte | *PPDPF* | 3.868934879 | 1.24E-05 | 0.000458996 | 0.07769291 |
| T2DM | pericyte | *CHCHD5* | 1.663948931 | 0.025972161 | 0.04385454 | 1 |
| T2DM | pericyte | *PAX8* | 0.694588427 | 0.011537918 | 0.022008158 | 1 |
| T2DM | pericyte | *INSIG2* | 1.164898505 | 0.001466892 | 0.004311523 | 1 |
| T2DM | pericyte | *PSD4* | 1.05248474 | 0.005478785 | 0.011764044 | 1 |
| T2DM | pericyte | *SLC25A23* | 0.701016459 | 0.005403574 | 0.011637364 | 1 |
| T2DM | pericyte | *GTF2F1* | 2.442301439 | 0.00357278 | 0.008479453 | 1 |
| T2DM | pericyte | *CLPP* | 2.519206914 | 0.000586441 | 0.002215719 | 1 |
| T2DM | pericyte | *TRIP10* | 3.052439042 | 2.44E-05 | 0.000556688 | 0.153089177 |
| T2DM | pericyte | *SYMPK* | 2.336739869 | 0.013006588 | 0.024329789 | 1 |
| T2DM | pericyte | *GPCPD1* | 3.268725902 | 6.67E-05 | 0.000632837 | 0.419530931 |
| T2DM | pericyte | *SDCBP2* | 3.209583698 | 0.00334373 | 0.00805558 | 1 |
| T2DM | pericyte | *PANK2* | 3.412870151 | 0.001020348 | 0.003335894 | 1 |

| | | | | | | |
|---|---|---|---|---|---|---|
| T2DM | pericyte | *DTD1* | 3.08308143 | 0.00028118 | 0.001336306 | 1 |
| T2DM | pericyte | *RBCK1* | 1.622995658 | 0.004461595 | 0.010075449 | 1 |
| T2DM | pericyte | *RRBP1* | 2.118379305 | 0.001151096 | 0.003624363 | 1 |
| T2DM | pericyte | *MKKS* | 3.171647439 | 0.000133691 | 0.000871003 | 0.840518096 |
| T2DM | pericyte | *DSTN* | 3.409285488 | 0.00012975 | 0.000855691 | 0.815740728 |
| T2DM | pericyte | *MGME1* | 2.052046501 | 0.002323614 | 0.006059128 | 1 |
| T2DM | pericyte | *ITPA* | 4.76817233 | 2.41E-05 | 0.000553241 | 0.151218242 |
| T2DM | pericyte | *TMEM74B* | -0.14078368 | 0.027819982 | 0.046344522 | 1 |
| T2DM | pericyte | *ZNF436* | 2.062924879 | 0.000300606 | 0.001387598 | 1 |
| T2DM | pericyte | *MMP24* | 2.352086435 | 0.000673206 | 0.002446108 | 1 |
| T2DM | pericyte | *UROD* | 3.925599833 | 0.000144098 | 0.000909584 | 0.905945406 |
| T2DM | pericyte | *ST3GAL3* | 2.199694301 | 0.000488261 | 0.001948566 | 1 |
| T2DM | pericyte | *KLC1* | 3.493517702 | 5.50E-05 | 0.00062437 | 0.345984279 |
| T2DM | pericyte | *XRCC3* | 2.772798454 | 3.96E-05 | 0.000607238 | 0.248792676 |
| T2DM | pericyte | *MCF2L* | 1.893511251 | 0.000101805 | 0.000747722 | 0.640049858 |
| T2DM | pericyte | *F10* | 1.814535479 | 0.000500062 | 0.001982277 | 1 |
| T2DM | pericyte | *PCID2* | 1.743058897 | 0.007059529 | 0.014416949 | 1 |
| T2DM | pericyte | *CAPNS1* | 2.648717691 | 7.19E-05 | 0.00064687 | 0.451739025 |
| T2DM | pericyte | *SCAF1* | 5.706792171 | 4.11E-05 | 0.000607238 | 0.258106638 |
| T2DM | pericyte | *PRR12* | 2.527616042 | 9.98E-07 | 0.000118335 | 0.006271778 |
| T2DM | pericyte | *TSKS* | 1.331814098 | 0.002014586 | 0.005428933 | 1 |
| T2DM | pericyte | *SBDS* | 2.435141219 | 0.017831442 | 0.031884606 | 1 |
| T2DM | pericyte | *ZNF384* | 1.949723261 | 0.000372067 | 0.001599989 | 1 |
| T2DM | pericyte | *ATG14* | 2.347522822 | 6.50E-05 | 0.00063262 | 0.40841029 |
| T2DM | pericyte | *L3HYPDH* | 0.526582943 | 0.029733804 | 0.048987533 | 1 |
| T2DM | pericyte | *HSPA2* | 0.963566093 | 0.016219199 | 0.02942011 | 1 |
| T2DM | pericyte | *ZBTB1* | 2.461530853 | 0.000154553 | 0.000942457 | 0.97167268 |
| T2DM | pericyte | *PZP* | 1.300657831 | 0.000575436 | 0.002189103 | 1 |
| T2DM | pericyte | *RHOT1* | 1.777120795 | 0.000240209 | 0.001220212 | 1 |
| T2DM | pericyte | *WDR60* | 1.203191005 | 0.017373882 | 0.031163935 | 1 |
| T2DM | pericyte | *MAP2K2* | 1.692112926 | 0.014324759 | 0.026379543 | 1 |
| T2DM | pericyte | *CANX* | 3.018636439 | 3.80E-05 | 0.000607238 | 0.239041983 |
| T2DM | pericyte | *IPPK* | 1.52890596 | 0.006975981 | 0.014268373 | 1 |
| T2DM | pericyte | *FGD3* | 1.177512636 | 0.001580955 | 0.004563575 | 1 |
| T2DM | pericyte | *ABHD8* | 1.120932779 | 0.011501751 | 0.021945831 | 1 |
| T2DM | pericyte | *HRASLS* | 1.437672953 | 0.004305875 | 0.009804795 | 1 |
| T2DM | pericyte | *HELB* | 1.622799405 | 0.007219766 | 0.014689536 | 1 |
| T2DM | pericyte | *RAP1B* | 2.860527627 | 0.000156281 | 0.000945663 | 0.982539073 |
| T2DM | pericyte | *YEATS4* | 1.108283087 | 0.001887518 | 0.005174823 | 1 |
| T2DM | pericyte | *LRRC61* | 0.536934331 | 0.018011598 | 0.032142752 | 1 |
| T2DM | pericyte | *IDUA* | 1.373589048 | 0.001855793 | 0.005119514 | 1 |
| T2DM | pericyte | *TMEM175* | 2.780545489 | 5.61E-05 | 0.00062437 | 0.352875462 |
| T2DM | pericyte | *EMC1* | 1.765997936 | 0.005599863 | 0.011945142 | 1 |
| T2DM | pericyte | *PLA2G5* | 0.737571439 | 0.006327162 | 0.013193654 | 1 |
| T2DM | pericyte | *SIN3B* | 3.430885999 | 4.32E-05 | 0.000607238 | 0.271765142 |
| T2DM | pericyte | *WFIKKN1* | -4.08273846 | 0.002832228 | 0.007077193 | 1 |
| T2DM | pericyte | *WDR24* | 1.069012295 | 0.017000188 | 0.030607154 | 1 |
| T2DM | pericyte | *SMARCA4* | 1.912061052 | 0.004001064 | 0.009253972 | 1 |
| T2DM | pericyte | *TICAM1* | 5.412043641 | 0.000302903 | 0.00139207 | 1 |
| T2DM | pericyte | *AAMP* | 1.490043527 | 0.007897791 | 0.015878929 | 1 |

| | | | | | | |
|---|---|---|---|---|---|---|
| T2DM | pericyte | *PNKD* | 2.36235817 | 0.003205699 | 0.007805666 | 1 |
| T2DM | pericyte | *HIP1* | 2.14218857 | 0.005797054 | 0.012283815 | 1 |
| T2DM | pericyte | *PTPN12* | 2.870581724 | 0.000283454 | 0.001339905 | 1 |
| T2DM | pericyte | *POR* | 3.185697016 | 4.93E-05 | 0.000607238 | 0.309638455 |
| T2DM | pericyte | *FGL2* | 1.735737568 | 8.91E-05 | 0.000702125 | 0.560184958 |
| T2DM | pericyte | *STEAP4* | -0.094448129 | 0.028677188 | 0.047520685 | 1 |
| T2DM | pericyte | *SGCE* | 3.406341876 | 0.000132246 | 0.000864273 | 0.831430408 |
| T2DM | pericyte | *RBM48* | 1.147774011 | 0.002487602 | 0.006402905 | 1 |
| T2DM | pericyte | *CASD1* | 3.237875913 | 6.05E-05 | 0.000625492 | 0.380299396 |
| T2DM | pericyte | *SRD5A3* | 2.235509579 | 0.000852851 | 0.002928387 | 1 |
| T2DM | pericyte | *ATF4* | 2.94303647 | 0.000340072 | 0.00150554 | 1 |
| T2DM | pericyte | *TPST2* | 2.227130583 | 0.000153739 | 0.000941145 | 0.966555505 |
| T2DM | pericyte | *BAIAP2L2* | 3.774659754 | 1.02E-06 | 0.000118642 | 0.006406677 |
| T2DM | pericyte | *RIBC2* | 1.329313571 | 0.000419768 | 0.00174765 | 1 |
| T2DM | pericyte | *EMC4* | 1.518396675 | 8.98E-05 | 0.000702125 | 0.564434086 |
| T2DM | pericyte | *KLHDC10* | 2.646332387 | 6.82E-05 | 0.000635523 | 0.429024508 |
| T2DM | pericyte | *HOXD1* | 4.927757157 | 0.004625169 | 0.010362725 | 1 |
| T2DM | pericyte | *ORMDL1* | 1.022203503 | 0.002529863 | 0.006481356 | 1 |
| T2DM | pericyte | *PSMG2* | 1.253011171 | 0.011887953 | 0.022498364 | 1 |
| T2DM | pericyte | *TWSG1* | 3.099615879 | 6.70E-05 | 0.000632837 | 0.421368344 |
| T2DM | pericyte | *TMOD2* | 1.447260228 | 0.01672132 | 0.030156896 | 1 |
| T2DM | pericyte | *ARPP19* | 2.503481824 | 0.000413962 | 0.001733897 | 1 |
| T2DM | pericyte | *VPS13C* | 2.41763509 | 0.000365667 | 0.00157895 | 1 |
| T2DM | pericyte | *CALML4* | 1.795814296 | 0.00644809 | 0.013383672 | 1 |
| T2DM | pericyte | *ACKR4* | 2.236874864 | 0.004452883 | 0.010066505 | 1 |
| T2DM | pericyte | *SUMF2* | 2.635412186 | 0.004244284 | 0.009699678 | 1 |
| T2DM | pericyte | *SPCS3* | 3.118034036 | 0.003600882 | 0.008526835 | 1 |
| T2DM | pericyte | *SERGEF* | 4.270056381 | 1.29E-05 | 0.000461315 | 0.081293447 |
| T2DM | pericyte | *PLD2* | 2.855221927 | 0.000473517 | 0.001905155 | 1 |
| T2DM | pericyte | *PHF20L1* | 2.952587176 | 0.000222284 | 0.001159749 | 1 |
| T2DM | pericyte | *PUS7L* | 1.710589596 | 0.022539625 | 0.038877497 | 1 |
| T2DM | pericyte | *SLC44A2* | 4.015780917 | 0.000172725 | 0.001002699 | 1 |
| T2DM | pericyte | *NGDN* | 3.553384033 | 3.84E-05 | 0.000607238 | 0.241530445 |
| T2DM | pericyte | *RAB2B* | 2.263321433 | 0.000198461 | 0.001076468 | 1 |
| T2DM | pericyte | *BCL2L2* | 3.521860624 | 6.57E-05 | 0.000632701 | 0.413345081 |
| T2DM | pericyte | *PARP2* | 1.428538312 | 0.018156289 | 0.032382578 | 1 |
| T2DM | pericyte | *MIS18BP1* | 1.190099187 | 0.001973799 | 0.005346521 | 1 |
| T2DM | pericyte | *DAD1* | 2.031661665 | 0.000633243 | 0.002337355 | 1 |
| T2DM | pericyte | *TEP1* | 1.087304268 | 0.008085133 | 0.016172839 | 1 |
| T2DM | pericyte | *EPB41L4A* | 2.914238126 | 2.96E-05 | 0.000593226 | 0.186007679 |
| T2DM | pericyte | *CDO1* | 1.096960338 | 0.00195661 | 0.005313697 | 1 |
| T2DM | pericyte | *REEP5* | 3.461903577 | 5.75E-05 | 0.00062437 | 0.361792923 |
| T2DM | pericyte | *QRICH2* | 2.394508502 | 0.002552925 | 0.006527142 | 1 |
| T2DM | pericyte | *RHBDF2* | 0.974691303 | 0.016020261 | 0.029126483 | 1 |
| T2DM | pericyte | *TMEM8A* | 2.787275351 | 0.004304138 | 0.009804795 | 1 |
| T2DM | pericyte | *SHC2* | 1.410359395 | 0.007075227 | 0.014437505 | 1 |
| T2DM | pericyte | *ERMARD* | 1.907240343 | 0.005369105 | 0.01159484 | 1 |
| T2DM | pericyte | *PHF10* | 2.094377628 | 0.000272989 | 0.001319089 | 1 |
| T2DM | pericyte | *DOCK6* | 1.529846382 | 0.007778846 | 0.01566985 | 1 |
| T2DM | pericyte | *ECSIT* | 3.173854971 | 0.001783507 | 0.004987948 | 1 |

| | | | | | | |
|---|---|---|---|---|---|---|
| T2DM | pericyte | *ELOF1* | 4.440820364 | 0.000448849 | 0.001838378 | 1 |
| T2DM | pericyte | *TOMM40* | 2.242564744 | 0.006197052 | 0.012959391 | 1 |
| T2DM | pericyte | *FAM98C* | 1.034559347 | 0.015597305 | 0.028448 | 1 |
| T2DM | pericyte | *RPL36* | 3.266594946 | 0.000529839 | 0.002070139 | 1 |
| T2DM | pericyte | *BST2* | 3.664793862 | 0.00464133 | 0.010388053 | 1 |
| T2DM | pericyte | *DDA1* | 0.739872408 | 0.007519387 | 0.01519588 | 1 |
| T2DM | pericyte | *LSM7* | 1.520639321 | 0.003843047 | 0.008981873 | 1 |
| T2DM | pericyte | *TULP4* | 1.823044449 | 0.001229627 | 0.003796987 | 1 |
| T2DM | pericyte | *RTN4IP1* | 1.94707269 | 0.012546976 | 0.023582314 | 1 |
| T2DM | pericyte | *ACTN4* | 2.262086233 | 0.010842334 | 0.020852172 | 1 |
| T2DM | pericyte | *PXDN* | 1.87471986 | 0.000159514 | 0.000955112 | 1 |
| T2DM | pericyte | *PGPEP1* | 1.640152023 | 0.004164859 | 0.00953897 | 1 |
| T2DM | pericyte | *KIAA1683* | 0.98549023 | 0.005675265 | 0.01205826 | 1 |
| T2DM | pericyte | *TRPM4* | 1.612108983 | 0.012503489 | 0.023525993 | 1 |
| T2DM | pericyte | *CAMSAP1* | 2.674139595 | 0.000121592 | 0.000822872 | 0.764448175 |
| T2DM | pericyte | *UBAC1* | 1.766656793 | 0.00121515 | 0.003767083 | 1 |
| T2DM | pericyte | *ATXN10* | 3.052547435 | 8.84E-05 | 0.000701359 | 0.555476506 |
| T2DM | pericyte | *TUBGCP2* | 2.715292593 | 0.000723946 | 0.002582631 | 1 |
| T2DM | pericyte | *CYP2E1* | 1.358433863 | 0.005904742 | 0.012460459 | 1 |
| T2DM | pericyte | *HBZ* | 4.12886404 | 0.015462375 | 0.028234666 | 1 |
| T2DM | pericyte | *PAK4* | 4.745911286 | 0.000563299 | 0.002159249 | 1 |
| T2DM | pericyte | *ZNF337* | 1.132452323 | 0.025755571 | 0.043551715 | 1 |
| T2DM | pericyte | *POMT1* | 2.338056737 | 0.006567119 | 0.013608265 | 1 |
| T2DM | pericyte | *TMEM160* | 0.986106044 | 0.003917126 | 0.00910509 | 1 |
| T2DM | pericyte | *ARHGEF16* | 0.724604009 | 0.016931928 | 0.030527969 | 1 |
| T2DM | pericyte | *SMPDL3B* | 5.620736289 | 0.003900168 | 0.009074892 | 1 |
| T2DM | pericyte | *THEMIS2* | 0.901834663 | 0.001402182 | 0.004166124 | 1 |
| T2DM | pericyte | *CLIP1* | 2.845343976 | 3.39E-05 | 0.000599418 | 0.213428457 |
| T2DM | pericyte | *ZNF317* | 1.869254955 | 0.001207501 | 0.003749405 | 1 |
| T2DM | pericyte | *PPAN* | 5.110575944 | 0.000181354 | 0.001034031 | 1 |
| T2DM | pericyte | *C12orf65* | 2.526315459 | 1.43E-06 | 0.000155353 | 0.008983928 |
| T2DM | pericyte | *PKDREJ* | -1.089520927 | 0.020478051 | 0.035840788 | 1 |
| T2DM | pericyte | *HABP4* | 3.096456817 | 0.00044046 | 0.001812287 | 1 |
| T2DM | pericyte | *EPS8L1* | -1.062107547 | 0.022598396 | 0.038949996 | 1 |
| T2DM | pericyte | *LILRB2* | 1.018566787 | 0.003997716 | 0.009250512 | 1 |
| T2DM | pericyte | *COX4I2* | -0.114767978 | 0.022126622 | 0.03831178 | 1 |
| T2DM | pericyte | *CHMP1A* | 2.094570777 | 0.000781389 | 0.002742726 | 1 |
| T2DM | pericyte | *PPT1* | 2.368110745 | 0.00055744 | 0.002148758 | 1 |
| T2DM | pericyte | *TBC1D5* | 2.201994223 | 0.001582971 | 0.004567297 | 1 |
| T2DM | pericyte | *RFTN1* | 2.181280891 | 0.001423737 | 0.004214236 | 1 |
| T2DM | pericyte | *PDLIM4* | 1.683459602 | 0.000292794 | 0.00136659 | 1 |
| T2DM | pericyte | *KIF3A* | 1.806045539 | 0.004788317 | 0.010622495 | 1 |
| T2DM | pericyte | *AOC3* | 1.200859796 | 0.010425363 | 0.020136484 | 1 |
| T2DM | pericyte | *ACLY* | 1.724546279 | 0.000881298 | 0.003001473 | 1 |
| T2DM | pericyte | *DIAPH1* | 2.014644908 | 0.003142986 | 0.007689588 | 1 |
| T2DM | pericyte | *EXOC4* | 2.840124918 | 9.54E-07 | 0.000115401 | 0.006000875 |
| T2DM | pericyte | *C1orf159* | 1.688837853 | 0.00057371 | 0.002187336 | 1 |
| T2DM | pericyte | *PPFIA1* | 1.495174926 | 0.018379306 | 0.032678365 | 1 |
| T2DM | pericyte | *NINJ1* | 3.359781734 | 0.009543129 | 0.018609694 | 1 |
| T2DM | pericyte | *NPHP4* | 3.844281478 | 0.004802 | 0.010636164 | 1 |

| | | | | | | |
|---|---|---|---|---|---|---|
| T2DM | pericyte | *CKMT2* | 1.078379123 | 0.02357434 | 0.040373706 | 1 |
| T2DM | pericyte | *ZCCHC9* | 2.599071896 | 0.000168003 | 0.000986702 | 1 |
| T2DM | pericyte | *CHD1L* | 1.822970947 | 0.007245013 | 0.014736136 | 1 |
| T2DM | pericyte | *ZSCAN5A* | 2.707111292 | 0.000339435 | 0.00150511 | 1 |
| T2DM | pericyte | *ZNF132* | 1.504662695 | 0.001787739 | 0.004988689 | 1 |
| T2DM | pericyte | *RHPN2* | -1.168891594 | 0.011610404 | 0.022113189 | 1 |
| T2DM | pericyte | *C19orf12* | 0.616784526 | 0.027712807 | 0.046189572 | 1 |
| T2DM | pericyte | *ACTR10* | 2.862358186 | 0.000265899 | 0.001293785 | 1 |
| T2DM | pericyte | *GCH1* | 2.089785356 | 0.000107087 | 0.000770639 | 0.673256625 |
| T2DM | pericyte | *LGALS3* | 2.301831557 | 0.006066154 | 0.012746628 | 1 |
| T2DM | pericyte | *DNAJB1* | 0.781907624 | 0.008332983 | 0.016578945 | 1 |
| T2DM | pericyte | *FBXW9* | 2.058352792 | 0.000509097 | 0.002007964 | 1 |
| T2DM | pericyte | *C19orf57* | 2.073597189 | 0.002002638 | 0.005403684 | 1 |
| T2DM | pericyte | *MATN3* | 9.409123433 | 0.000921668 | 0.003092064 | 1 |
| T2DM | pericyte | *RAF1* | 3.057380742 | 8.98E-05 | 0.000702125 | 0.564586462 |
| T2DM | pericyte | *ENOSF1* | 1.488811571 | 0.002959131 | 0.007327316 | 1 |
| T2DM | pericyte | *EMILIN2* | 1.247929927 | 0.001661536 | 0.004735303 | 1 |
| T2DM | pericyte | *TRIM5* | 3.086974219 | 0.000285566 | 0.001346398 | 1 |
| T2DM | pericyte | *CNGA4* | 1.822960489 | 0.01479828 | 0.027164025 | 1 |
| T2DM | pericyte | *TRIM22* | 2.811478952 | 0.001118766 | 0.003550571 | 1 |
| T2DM | pericyte | *RRP8* | 2.177099619 | 0.004960707 | 0.010901071 | 1 |
| T2DM | pericyte | *IMMT* | 2.4022634 | 0.000240983 | 0.00122084 | 1 |
| T2DM | pericyte | *MRPL35* | 1.300104926 | 0.000123808 | 0.000831605 | 0.778382569 |
| T2DM | pericyte | *CARD6* | 2.313476381 | 0.000560132 | 0.002156489 | 1 |
| T2DM | pericyte | *RPA1* | 1.554706863 | 0.00481002 | 0.010648097 | 1 |
| T2DM | pericyte | *TMEM128* | 1.694830374 | 0.003335374 | 0.008049169 | 1 |
| T2DM | pericyte | *FIGNL1* | 1.229852039 | 0.001919259 | 0.005241694 | 1 |
| T2DM | pericyte | *WBP2* | 2.365925076 | 0.000256383 | 0.001266207 | 1 |
| T2DM | pericyte | *KDM6B* | 2.683686598 | 0.00089325 | 0.003029051 | 1 |
| T2DM | pericyte | *XAF1* | 1.553788011 | 0.006659736 | 0.013754849 | 1 |
| T2DM | pericyte | *TERF2* | 2.359464591 | 0.016541466 | 0.029909748 | 1 |
| T2DM | pericyte | *BTBD3* | 2.728999697 | 0.000205665 | 0.001099503 | 1 |
| T2DM | pericyte | *PTPRA* | 2.689219058 | 0.000887625 | 0.003018117 | 1 |
| T2DM | pericyte | *CTNNBL1* | 3.625680635 | 0.00027837 | 0.001327854 | 1 |
| T2DM | pericyte | *LPIN3* | 4.867055555 | 5.92E-05 | 0.00062437 | 0.372381789 |
| T2DM | pericyte | *DMGDH* | 1.442674272 | 0.008324755 | 0.016567817 | 1 |
| T2DM | pericyte | *KANK4* | 0.358233016 | 0.023636583 | 0.040447251 | 1 |
| T2DM | pericyte | *CASP9* | 0.928307614 | 0.008761754 | 0.017306047 | 1 |
| T2DM | pericyte | *ATP8A2* | 2.736605479 | 0.000629149 | 0.002329483 | 1 |
| T2DM | pericyte | *USPL1* | 3.562131672 | 0.00011978 | 0.000813238 | 0.753057988 |
| T2DM | pericyte | *POMP* | 2.502238445 | 0.020688453 | 0.036120051 | 1 |
| T2DM | pericyte | *WASF3* | 1.468491898 | 0.000526703 | 0.00206062 | 1 |
| T2DM | pericyte | *PEMT* | 1.177176257 | 0.006610655 | 0.013675943 | 1 |
| T2DM | pericyte | *SCO1* | 2.153170234 | 0.003685005 | 0.008677014 | 1 |
| T2DM | pericyte | *CHI3L1* | 0.381126887 | 0.014175769 | 0.026151133 | 1 |
| T2DM | pericyte | *COG6* | 1.968173012 | 0.001434847 | 0.004237145 | 1 |
| T2DM | pericyte | *RFC3* | 2.066012303 | 0.00101824 | 0.003332471 | 1 |
| T2DM | pericyte | *BTBD2* | 1.800169704 | 0.009458025 | 0.018466648 | 1 |
| T2DM | pericyte | *RTN3* | 2.496276178 | 5.64E-05 | 0.00062437 | 0.354379807 |
| T2DM | pericyte | *SLC2A11* | 1.338034585 | 0.002828375 | 0.007073187 | 1 |

| | | | | | | |
|---|---|---|---|---|---|---|
| T2DM | pericyte | *C1QTNF6* | 6.869596969 | 1.29E-05 | 0.000461315 | 0.08090515 |
| T2DM | pericyte | *FAM83F* | 11.20495671 | 0.002083694 | 0.005553279 | 1 |
| T2DM | pericyte | *AGAP3* | 2.277318578 | 0.000709281 | 0.002545233 | 1 |
| T2DM | pericyte | *ACTR3B* | 1.465879882 | 0.0025314 | 0.006482653 | 1 |
| T2DM | pericyte | *SFTPD* | 0.487969148 | 0.001710165 | 0.004834445 | 1 |
| T2DM | pericyte | *TMTC1* | 2.667236994 | 3.29E-05 | 0.000599418 | 0.206615836 |
| T2DM | pericyte | *IPO8* | 3.785653404 | 5.99E-05 | 0.00062437 | 0.376352667 |
| T2DM | pericyte | *LRRCC1* | 1.577185912 | 0.000939578 | 0.003136594 | 1 |
| T2DM | pericyte | *SWAP70* | 2.836510199 | 0.00268079 | 0.006790543 | 1 |
| T2DM | pericyte | *AMPD3* | 0.858628746 | 0.024669083 | 0.041872172 | 1 |
| T2DM | pericyte | *SBF2* | 2.632117437 | 9.68E-05 | 0.000726663 | 0.608411943 |
| T2DM | pericyte | *MICAL2* | 1.497104469 | 0.004049004 | 0.009334831 | 1 |
| T2DM | pericyte | *RRAS2* | 0.762064384 | 0.005706316 | 0.012116045 | 1 |
| T2DM | pericyte | *HSD17B4* | 1.592295619 | 0.001436703 | 0.004240634 | 1 |
| T2DM | pericyte | *DPF2* | 1.942430503 | 0.016073539 | 0.029189584 | 1 |
| T2DM | pericyte | *MED6* | 1.430895961 | 0.006054259 | 0.012734401 | 1 |
| T2DM | pericyte | *EIF2S1* | 3.58363276 | 1.82E-05 | 0.000512131 | 0.114118965 |
| T2DM | pericyte | *ELP3* | 3.50065847 | 7.04E-05 | 0.000639553 | 0.442661004 |
| T2DM | pericyte | *MRO* | 0.860830907 | 0.029387982 | 0.048557752 | 1 |
| T2DM | pericyte | *EDEM1* | 4.551000785 | 3.62E-05 | 0.00060096 | 0.227519005 |
| T2DM | pericyte | *KATNBL1* | 3.133284273 | 1.65E-05 | 0.000502137 | 0.103830806 |
| T2DM | pericyte | *GSTM5* | 0.575172701 | 0.025069134 | 0.042516764 | 1 |
| T2DM | pericyte | *GSTM3* | 0.426834166 | 0.012552361 | 0.023585383 | 1 |
| T2DM | pericyte | *SYT6* | 2.326070895 | 0.004819046 | 0.010664323 | 1 |
| T2DM | pericyte | *VAV3* | 1.539818983 | 0.000395949 | 0.001672936 | 1 |
| T2DM | pericyte | *CEPT1* | 2.371969578 | 0.000245537 | 0.001233967 | 1 |
| T2DM | pericyte | *AP4B1* | 2.903630645 | 0.002874767 | 0.007157885 | 1 |
| T2DM | pericyte | *NAPG* | 3.078096836 | 0.000277783 | 0.001326433 | 1 |
| T2DM | pericyte | *PPHLN1* | 2.189947543 | 1.98E-05 | 0.000515615 | 0.124263211 |
| T2DM | pericyte | *FKBP11* | 1.851339957 | 0.003688011 | 0.008680841 | 1 |
| T2DM | pericyte | *YWHAQ* | 3.643003042 | 3.83E-05 | 0.000607238 | 0.241038285 |
| T2DM | pericyte | *LDHA* | 2.443372154 | 0.000106005 | 0.000765156 | 0.666450601 |
| T2DM | pericyte | *TIMM17A* | 2.294568839 | 5.47E-05 | 0.00062437 | 0.343635708 |
| T2DM | pericyte | *NARS* | 4.116345035 | 0.000322086 | 0.001447429 | 1 |
| T2DM | pericyte | *KIAA1468* | 2.877045155 | 1.02E-05 | 0.000407741 | 0.063947357 |
| T2DM | pericyte | *FBXO18* | 3.738664174 | 0.000246051 | 0.001234577 | 1 |
| T2DM | pericyte | *IL15RA* | 3.700469713 | 4.25E-05 | 0.000607238 | 0.266907544 |
| T2DM | pericyte | *TMEM241* | 2.268456951 | 8.38E-05 | 0.000681373 | 0.526600142 |
| T2DM | pericyte | *EMP1* | 0.928537888 | 0.014435274 | 0.026559721 | 1 |
| T2DM | pericyte | *KLRD1* | 1.540293588 | 0.000265085 | 0.001293785 | 1 |
| T2DM | pericyte | *PRH2* | -2.036994633 | 0.003408875 | 0.008172666 | 1 |
| T2DM | pericyte | *DDB2* | 2.864514578 | 2.81E-06 | 0.000232551 | 0.017673859 |
| T2DM | pericyte | *ACP2* | 1.413635246 | 0.000418068 | 0.001744124 | 1 |
| T2DM | pericyte | *HOOK1* | 2.313449725 | 0.011114643 | 0.021310692 | 1 |
| T2DM | pericyte | *ZCCHC11* | 2.131566145 | 0.003532596 | 0.008419041 | 1 |
| T2DM | pericyte | *PRPF38A* | 2.98845143 | 0.002402663 | 0.006218833 | 1 |
| T2DM | pericyte | *TPGS2* | 2.799677166 | 2.31E-05 | 0.000548455 | 0.14551321 |
| T2DM | pericyte | *TIMM10* | -1.030789026 | 0.022337745 | 0.038581704 | 1 |
| T2DM | pericyte | *APLNR* | 1.820203161 | 0.000898889 | 0.003037034 | 1 |
| T2DM | pericyte | *TMEM165* | 2.571171705 | 1.38E-05 | 0.00046738 | 0.086932724 |

| | | | | | | |
|---|---|---|---|---|---|---|
| T2DM | pericyte | *CLOCK* | 3.174106174 | 4.06E-05 | 0.000607238 | 0.255077323 |
| T2DM | pericyte | *CLDN10* | 1.465658711 | 0.009710642 | 0.018912889 | 1 |
| T2DM | pericyte | *BIVM* | 1.395861271 | 0.001513308 | 0.00442108 | 1 |
| T2DM | pericyte | *ERCC5* | 1.978479228 | 0.001819716 | 0.00505548 | 1 |
| T2DM | pericyte | *STT3A* | 2.482465678 | 0.000423262 | 0.00175879 | 1 |
| T2DM | pericyte | *KLB* | 0.610399939 | 0.005657723 | 0.012035724 | 1 |
| T2DM | pericyte | *WDR36* | 2.690606026 | 6.13E-05 | 0.000627415 | 0.385404675 |
| T2DM | pericyte | *OSTF1* | 3.549234025 | 1.81E-05 | 0.000512131 | 0.114046022 |
| T2DM | pericyte | *UBQLN1* | 3.212009089 | 3.60E-05 | 0.00060096 | 0.226378099 |
| T2DM | pericyte | *NAA35* | 3.546542376 | 5.56E-05 | 0.00062437 | 0.349413101 |
| T2DM | pericyte | *CTSL* | 2.726660422 | 7.73E-05 | 0.000661364 | 0.486102491 |
| T2DM | pericyte | *TMEM2* | 2.013770031 | 0.000139922 | 0.000894176 | 0.879691337 |
| T2DM | pericyte | *AGTPBP1* | 2.304269323 | 0.001052433 | 0.003396635 | 1 |
| T2DM | pericyte | *ADAM19* | 0.434215366 | 0.022808416 | 0.039260451 | 1 |
| T2DM | pericyte | *USP30* | 1.11106788 | 0.001391801 | 0.004139192 | 1 |
| T2DM | pericyte | *P2RX4* | 1.605506772 | 6.42E-06 | 0.000343901 | 0.040358826 |
| T2DM | pericyte | *DMTF1* | 1.662709675 | 0.001898846 | 0.005201763 | 1 |
| T2DM | pericyte | *CCDC146* | 3.357515238 | 2.23E-06 | 0.000202161 | 0.014006274 |
| T2DM | pericyte | *TMEM60* | 1.737913352 | 0.01019672 | 0.019713032 | 1 |
| T2DM | pericyte | *SRPK2* | 2.871287669 | 4.72E-05 | 0.000607238 | 0.296510383 |
| T2DM | pericyte | *MDFIC* | 1.771448167 | 0.000786024 | 0.002753053 | 1 |
| T2DM | pericyte | *MTO1* | 1.404690667 | 0.008555648 | 0.016962902 | 1 |
| T2DM | pericyte | *ANKRD6* | 3.155492655 | 0.000205305 | 0.001099503 | 1 |
| T2DM | pericyte | *SNX14* | 2.754826923 | 0.000184556 | 0.001035963 | 1 |
| T2DM | pericyte | *NT5E* | 1.538409314 | 3.46E-05 | 0.000599418 | 0.217730152 |
| T2DM | pericyte | *PRR5L* | 1.673028684 | 0.001347228 | 0.004041042 | 1 |
| T2DM | pericyte | *NAT10* | 2.499723369 | 0.000963396 | 0.003197926 | 1 |
| T2DM | pericyte | *ATP5G2* | 2.991947963 | 7.10E-05 | 0.000642058 | 0.446395138 |
| T2DM | pericyte | *RDH5* | 0.803800343 | 0.001262436 | 0.003869788 | 1 |
| T2DM | pericyte | *PPP1R1A* | 1.816738125 | 0.00061378 | 0.00229011 | 1 |
| T2DM | pericyte | *KRT7* | 1.679791675 | 0.015553321 | 0.02837601 | 1 |
| T2DM | pericyte | *HNRNPA1* | 1.611403676 | 0.000957591 | 0.003185385 | 1 |
| T2DM | pericyte | *SLC26A10* | -1.576763929 | 0.028957916 | 0.04791011 | 1 |
| T2DM | pericyte | *MAP7* | 1.990508658 | 0.001923998 | 0.005250077 | 1 |
| T2DM | pericyte | *MICAL1* | 3.801956331 | 4.00E-06 | 0.000285979 | 0.025166126 |
| T2DM | pericyte | *PRADC1* | 1.833610961 | 0.004827482 | 0.010671722 | 1 |
| T2DM | pericyte | *USP15* | 2.723197367 | 0.000160166 | 0.000957189 | 1 |
| T2DM | pericyte | *KLHL36* | 3.05789363 | 0.000183194 | 0.001035729 | 1 |
| T2DM | pericyte | *MPHOSPH6* | 5.139678825 | 2.84E-07 | 4.69E-05 | 0.001783816 |
| T2DM | pericyte | *NTPCR* | 1.744389888 | 0.000443002 | 0.00182036 | 1 |
| T2DM | pericyte | *RNASEL* | 1.834776539 | 0.000419025 | 0.001746957 | 1 |
| T2DM | pericyte | *PIGC* | 2.011048531 | 0.000655663 | 0.002401036 | 1 |
| T2DM | pericyte | *SP110* | 2.879569958 | 0.000716965 | 0.002566945 | 1 |
| T2DM | pericyte | *USP37* | 3.199938263 | 0.000148302 | 0.000922861 | 0.932373896 |
| T2DM | pericyte | *SLC19A3* | 0.382398876 | 0.029039563 | 0.048032553 | 1 |
| T2DM | pericyte | *ARMC9* | 3.377437905 | 7.64E-05 | 0.00065815 | 0.480449442 |
| T2DM | pericyte | *TMEM127* | 3.1615301 | 0.003643763 | 0.008595999 | 1 |
| T2DM | pericyte | *MRPS9* | 1.394881543 | 0.000746169 | 0.002647383 | 1 |
| T2DM | pericyte | *ANKRD36* | 2.999931905 | 8.27E-05 | 0.000681373 | 0.520186075 |
| T2DM | pericyte | *APPL2* | 2.928337488 | 7.88E-05 | 0.000667567 | 0.495379369 |

| | | | | | | |
|---|---|---|---|---|---|---|
| T2DM | pericyte | *DRAM1* | 1.230227097 | 0.018294396 | 0.032564233 | 1 |
| T2DM | pericyte | *SLC41A2* | 2.131369327 | 0.021443838 | 0.037273268 | 1 |
| T2DM | pericyte | *VPS36* | 1.940447225 | 0.004024275 | 0.009288039 | 1 |
| T2DM | pericyte | *RNASEH2B* | 2.357822308 | 0.000374242 | 0.001607143 | 1 |
| T2DM | pericyte | *TBC1D4* | 2.314708808 | 0.00018982 | 0.0010496 | 1 |
| T2DM | pericyte | *LRCH1* | 2.790804919 | 0.001225389 | 0.003789498 | 1 |
| T2DM | pericyte | *MED4* | 1.68834169 | 0.013371102 | 0.024878401 | 1 |
| T2DM | pericyte | *RCBTB2* | 3.405602596 | 6.69E-05 | 0.000632837 | 0.420421153 |
| T2DM | pericyte | *SETDB2* | 1.764218824 | 0.012037922 | 0.022754785 | 1 |
| T2DM | pericyte | *SPDYE1* | 2.121139873 | 0.000839864 | 0.002894862 | 1 |
| T2DM | pericyte | *CHST12* | 2.132022306 | 0.001427237 | 0.004220622 | 1 |
| T2DM | pericyte | *GPNMB* | 1.420813175 | 0.001600545 | 0.004605322 | 1 |
| T2DM | pericyte | *KDELR2* | 1.801428806 | 0.000464579 | 0.00188404 | 1 |
| T2DM | pericyte | *NUPL2* | 1.891676965 | 0.029974275 | 0.049344924 | 1 |
| T2DM | pericyte | *HUS1* | 2.769298918 | 0.000113652 | 0.000790392 | 0.714531378 |
| T2DM | pericyte | *NACAD* | -5.108765347 | 0.000486707 | 0.00194652 | 1 |
| T2DM | pericyte | *DBNL* | 2.121173213 | 0.003371949 | 0.008106861 | 1 |
| T2DM | pericyte | *ALPK3* | 0.557126775 | 0.012291211 | 0.023177818 | 1 |
| T2DM | pericyte | *TM6SF1* | 2.858064061 | 6.57E-06 | 0.000344284 | 0.041314077 |
| T2DM | pericyte | *RSAD1* | 1.065137028 | 0.002732142 | 0.006895615 | 1 |
| T2DM | pericyte | *NMT1* | 0.908219068 | 0.005142806 | 0.011222778 | 1 |
| T2DM | pericyte | *BRIP1* | 1.072460678 | 0.001540948 | 0.004486927 | 1 |
| T2DM | pericyte | *MRPL47* | 2.603465667 | 0.003078222 | 0.007553778 | 1 |
| T2DM | pericyte | *EPRS* | 2.590971306 | 8.00E-05 | 0.000668213 | 0.503164306 |
| T2DM | pericyte | *RPS6KC1* | 2.932351078 | 0.00030142 | 0.001390335 | 1 |
| T2DM | pericyte | *CBWD2* | 3.007068998 | 0.000854307 | 0.002931784 | 1 |
| T2DM | pericyte | *IL1RN* | 4.435793289 | 6.38E-05 | 0.000632025 | 0.400970264 |
| T2DM | pericyte | *SAP130* | 1.889979886 | 0.007298279 | 0.014820503 | 1 |
| T2DM | pericyte | *UGGT1* | 3.096535328 | 0.002665294 | 0.006759461 | 1 |
| T2DM | pericyte | *GYPC* | 2.226829084 | 0.001148324 | 0.003618978 | 1 |
| T2DM | pericyte | *LRRC8A* | 1.096107522 | 0.005279629 | 0.011434043 | 1 |
| T2DM | pericyte | *TXN* | 2.056613652 | 0.000667679 | 0.002436274 | 1 |
| T2DM | pericyte | *ODF2* | 4.138840255 | 0.000454405 | 0.00185713 | 1 |
| T2DM | pericyte | *ZFP37* | 1.597614857 | 0.005042543 | 0.011037216 | 1 |
| T2DM | pericyte | *ZNF189* | 1.901428726 | 0.001332538 | 0.004010488 | 1 |
| T2DM | pericyte | *STX17* | 2.992619929 | 0.003317222 | 0.008024385 | 1 |
| T2DM | pericyte | *PRPF4* | 1.696688575 | 0.000491105 | 0.00195664 | 1 |
| T2DM | pericyte | *TEX10* | 2.327201107 | 0.000154507 | 0.000942457 | 0.971386686 |
| T2DM | pericyte | *GARNL3* | 2.834360453 | 0.005486826 | 0.011769252 | 1 |
| T2DM | pericyte | *GABBR2* | 1.603296265 | 0.004511022 | 0.010154242 | 1 |
| T2DM | pericyte | *PSMB7* | 2.353419954 | 8.33E-05 | 0.000681373 | 0.523532666 |
| T2DM | pericyte | *RABEPK* | 3.351378113 | 8.35E-05 | 0.000681373 | 0.524703686 |
| T2DM | pericyte | *GOLGA1* | 3.418590637 | 7.81E-05 | 0.000663325 | 0.490860762 |
| T2DM | pericyte | *LMX1B* | 8.871897527 | 0.016440608 | 0.029753051 | 1 |
| T2DM | pericyte | *DERL1* | 3.110792468 | 0.002738568 | 0.00690352 | 1 |
| T2DM | pericyte | *NOV* | 2.302019362 | 0.00398971 | 0.009238788 | 1 |
| T2DM | pericyte | *IL11RA* | 1.661519536 | 0.020483398 | 0.035840788 | 1 |
| T2DM | pericyte | *APTX* | 2.083730267 | 0.005906182 | 0.012460459 | 1 |
| T2DM | pericyte | *TLN1* | 1.99591424 | 0.00885238 | 0.017457626 | 1 |
| T2DM | pericyte | *CD72* | 2.135404255 | 0.002552643 | 0.006527142 | 1 |

| | | | | | | |
|---|---|---|---|---|---|---|
| T2DM | pericyte | *DENND4C* | 2.854612979 | 1.60E-05 | 0.000496985 | 0.100390902 |
| T2DM | pericyte | *PPIL1* | 2.996766205 | 0.000890459 | 0.003026116 | 1 |
| T2DM | pericyte | *KIF13A* | 2.657201675 | 0.000495628 | 0.001972158 | 1 |
| T2DM | pericyte | *FRS3* | 1.712705119 | 0.000935687 | 0.003125752 | 1 |
| T2DM | pericyte | *SLC22A23* | 3.245361417 | 5.36E-05 | 0.000619691 | 0.337112036 |
| T2DM | pericyte | *LRRC1* | 1.020648476 | 0.024203032 | 0.041214642 | 1 |
| T2DM | pericyte | *RIPK1* | 3.549608392 | 6.39E-05 | 0.000632025 | 0.401621995 |
| T2DM | pericyte | *UQCC2* | 2.477727209 | 0.000572854 | 0.002185396 | 1 |
| T2DM | pericyte | *TCF19* | 1.543173616 | 0.001432236 | 0.004231421 | 1 |
| T2DM | pericyte | *FLOT1* | 3.299910149 | 3.29E-06 | 0.000258296 | 0.020663644 |
| T2DM | pericyte | *MTCH1* | 4.054934451 | 3.10E-05 | 0.000599418 | 0.194678476 |
| T2DM | pericyte | *VARS2* | 0.329159175 | 0.006789447 | 0.013963119 | 1 |
| T2DM | pericyte | *FHDC1* | 1.588736552 | 0.001804541 | 0.005024423 | 1 |
| T2DM | pericyte | *ARRB1* | 2.981912237 | 0.000469603 | 0.001897115 | 1 |
| T2DM | pericyte | *RAB30* | 2.190577914 | 0.011785437 | 0.022378448 | 1 |
| T2DM | pericyte | *PRCP* | 2.50806519 | 5.31E-06 | 0.000328654 | 0.033399249 |
| T2DM | pericyte | *NARS2* | 3.366725037 | 0.000838963 | 0.002894862 | 1 |
| T2DM | pericyte | *TGS1* | 1.316373569 | 0.017705502 | 0.031686447 | 1 |
| T2DM | pericyte | *NEK1* | 3.036357358 | 2.35E-05 | 0.000548455 | 0.147548081 |
| T2DM | pericyte | *DDX60* | 4.254076218 | 0.000318644 | 0.001438132 | 1 |
| T2DM | pericyte | *SLC37A4* | 1.535523107 | 0.003055798 | 0.007510479 | 1 |
| T2DM | pericyte | *PPP2R1B* | 1.133927289 | 0.000613074 | 0.00229011 | 1 |
| T2DM | pericyte | *FDX1* | 2.960710107 | 0.00101735 | 0.00333129 | 1 |
| T2DM | pericyte | *TMPRSS13* | 1.988742801 | 0.024002679 | 0.040962226 | 1 |
| T2DM | pericyte | *CASP1* | 3.148308738 | 0.000292071 | 0.001364229 | 1 |
| T2DM | pericyte | *MAP2K5* | 2.860395116 | 9.27E-06 | 0.000393808 | 0.058283556 |
| T2DM | pericyte | *ITGA11* | 0.650822631 | 0.007111399 | 0.014501902 | 1 |
| T2DM | pericyte | *PAQR5* | 2.213122702 | 0.003256976 | 0.007906025 | 1 |
| T2DM | pericyte | *RMDN3* | 3.761320075 | 6.98E-06 | 0.000348151 | 0.04386857 |
| T2DM | pericyte | *UACA* | 2.989693283 | 8.06E-05 | 0.000671194 | 0.506751161 |
| T2DM | pericyte | *SPTBN5* | 1.105473556 | 8.53E-07 | 0.000105105 | 0.00536034 |
| T2DM | pericyte | *RABGGTB* | 3.395110463 | 0.003363189 | 0.008088894 | 1 |
| T2DM | pericyte | *SLC44A5* | 0.806693521 | 0.013308779 | 0.024784447 | 1 |
| T2DM | pericyte | *DNASE2B* | 4.544710233 | 0.004047293 | 0.009334311 | 1 |
| T2DM | pericyte | *DBT* | 1.204764348 | 0.002108497 | 0.005605126 | 1 |
| T2DM | pericyte | *RTCA* | 2.183354111 | 7.97E-05 | 0.000667567 | 0.501216725 |
| T2DM | pericyte | *IFT172* | 4.461178641 | 4.64E-07 | 6.21E-05 | 0.002917009 |
| T2DM | pericyte | *LHCGR* | 0.68856998 | 0.011835746 | 0.022446858 | 1 |
| T2DM | pericyte | *THUMPD2* | 2.633100769 | 0.000202505 | 0.001093325 | 1 |
| T2DM | pericyte | *ATRAID* | 2.778132714 | 0.001266291 | 0.003877815 | 1 |
| T2DM | pericyte | *CENPO* | 4.365052021 | 0.000109845 | 0.000778599 | 0.690596595 |
| T2DM | pericyte | *LRPPRC* | 3.209632335 | 6.92E-05 | 0.000637539 | 0.434801875 |
| T2DM | pericyte | *DTNB* | 1.470830678 | 0.006527334 | 0.013539212 | 1 |
| T2DM | pericyte | *MYOF* | 1.734549762 | 1.73E-05 | 0.000512131 | 0.108634366 |
| T2DM | pericyte | *ATAD1* | 3.602015521 | 0.000350739 | 0.001529192 | 1 |
| T2DM | pericyte | *TACC2* | 1.348974306 | 0.000267184 | 0.0012953 | 1 |
| T2DM | pericyte | *CALHM2* | 0.619897949 | 0.008063166 | 0.016139168 | 1 |
| T2DM | pericyte | *ARL3* | 1.971641192 | 0.013925998 | 0.025765202 | 1 |
| T2DM | pericyte | *PLCE1* | 1.451460117 | 0.006146013 | 0.012871413 | 1 |
| T2DM | pericyte | *RBP4* | 3.62807697 | 4.94E-05 | 0.000607238 | 0.310681364 |

| | | | | | | |
|---|---|---|---|---|---|---|
| T2DM | pericyte | *DNAJC13* | 3.386656625 | 0.000250058 | 0.001248702 | 1 |
| T2DM | pericyte | *RPS24* | 2.989266554 | 0.001968492 | 0.005336743 | 1 |
| T2DM | pericyte | *SMARCAL1* | 2.689705559 | 0.000111356 | 0.00078265 | 0.700098266 |
| T2DM | pericyte | *BARD1* | 3.515129373 | 0.000273531 | 0.001319791 | 1 |
| T2DM | pericyte | *CARF* | 2.107157302 | 7.50E-06 | 0.000356314 | 0.047155701 |
| T2DM | pericyte | *METTL5* | 2.803232162 | 0.000129844 | 0.000855691 | 0.816329 |
| T2DM | pericyte | *NAB1* | 3.064264173 | 0.003552444 | 0.008447551 | 1 |
| T2DM | pericyte | *FASTKD1* | 1.76182635 | 0.001623566 | 0.004655763 | 1 |
| T2DM | pericyte | *MDH1B* | 1.670775422 | 0.016244757 | 0.029449477 | 1 |
| T2DM | pericyte | *FAM117B* | 3.409255459 | 1.95E-05 | 0.000515615 | 0.122833191 |
| T2DM | pericyte | *ITGAV* | 2.397034483 | 0.000168086 | 0.000986702 | 1 |
| T2DM | pericyte | *SLC35A5* | 1.008015237 | 0.000411113 | 0.001724262 | 1 |
| T2DM | pericyte | *SENP7* | 2.895603172 | 0.000521339 | 0.002043031 | 1 |
| T2DM | pericyte | *USP8* | 2.225451945 | 0.001887844 | 0.005174823 | 1 |
| T2DM | pericyte | *SPPL2A* | 2.074892489 | 0.000642816 | 0.002359244 | 1 |
| T2DM | pericyte | *PPCDC* | 1.140641463 | 0.003872282 | 0.009031857 | 1 |
| T2DM | pericyte | *ARHGAP24* | 2.253428037 | 0.000987967 | 0.003265694 | 1 |
| T2DM | pericyte | *FAM13A* | 2.776571863 | 4.60E-07 | 6.21E-05 | 0.002891747 |
| T2DM | pericyte | *HERC3* | 3.353760828 | 0.000195672 | 0.001065597 | 1 |
| T2DM | pericyte | *AP1AR* | 3.513807159 | 0.000276963 | 0.001325163 | 1 |
| T2DM | pericyte | *PRKG2* | 0.854639409 | 0.005205483 | 0.011308525 | 1 |
| T2DM | pericyte | *BBS7* | 4.32412109 | 0.000320089 | 0.001443616 | 1 |
| T2DM | pericyte | *KIAA1109* | 2.814440831 | 0.000277426 | 0.001326373 | 1 |
| T2DM | pericyte | *PRDM5* | 2.521555625 | 2.31E-05 | 0.000548455 | 0.145535365 |
| T2DM | pericyte | *NAAA* | 2.385198268 | 0.000137814 | 0.000885929 | 0.866438164 |
| T2DM | pericyte | *NUP54* | 2.719534631 | 3.76E-05 | 0.000607238 | 0.236126675 |
| T2DM | pericyte | *G3BP2* | 1.457742754 | 0.015928848 | 0.028993824 | 1 |
| T2DM | pericyte | *SCARB2* | 3.360890606 | 0.000127506 | 0.000844709 | 0.801628962 |
| T2DM | pericyte | *PPA2* | 2.823193688 | 5.04E-06 | 0.000320172 | 0.031696994 |
| T2DM | pericyte | *GSTCD* | 2.599693326 | 0.000134793 | 0.000874555 | 0.847444042 |
| T2DM | pericyte | *ENPEP* | -0.140061651 | 0.025868934 | 0.043708139 | 1 |
| T2DM | pericyte | *PAPSS1* | 1.872378481 | 0.000130481 | 0.000858087 | 0.82033129 |
| T2DM | pericyte | *SLC39A8* | 0.992456185 | 0.021255379 | 0.036986595 | 1 |
| T2DM | pericyte | *RNF185* | 1.446499898 | 0.006000741 | 0.012638747 | 1 |
| T2DM | pericyte | *PARVG* | 1.311193554 | 0.01060783 | 0.020426165 | 1 |
| T2DM | pericyte | *ERP27* | 1.65939573 | 0.022007905 | 0.038127225 | 1 |
| T2DM | pericyte | *ETV6* | 2.886112498 | 2.69E-05 | 0.000562456 | 0.168919595 |
| T2DM | pericyte | *KIF21A* | 1.202186153 | 0.004207712 | 0.009623094 | 1 |
| T2DM | pericyte | *ZCRB1* | 1.757106951 | 0.002879709 | 0.007164515 | 1 |
| T2DM | pericyte | *PRICKLE1* | 2.389933531 | 1.02E-05 | 0.000407741 | 0.064423057 |
| T2DM | pericyte | *C1RL* | 2.425833176 | 3.87E-06 | 0.000279804 | 0.024321921 |
| T2DM | pericyte | *CLSTN3* | 1.921121951 | 0.009424237 | 0.018406393 | 1 |
| T2DM | pericyte | *VAMP1* | 1.051287533 | 0.002713613 | 0.006857109 | 1 |
| T2DM | pericyte | *CD27* | 1.540880371 | 0.007166976 | 0.014596301 | 1 |
| T2DM | pericyte | *GLIPR1* | 1.346580591 | 0.002906944 | 0.007217992 | 1 |
| T2DM | pericyte | *POC1B* | 3.378586734 | 6.20E-05 | 0.000628694 | 0.389693153 |
| T2DM | pericyte | *GAS2L3* | 4.586808444 | 0.000683069 | 0.002473764 | 1 |
| T2DM | pericyte | *TDG* | 1.9438292 | 0.025830184 | 0.043654399 | 1 |
| T2DM | pericyte | *MTMR6* | 1.709685287 | 0.023364604 | 0.040047238 | 1 |
| T2DM | pericyte | *SLC46A3* | 1.415903095 | 0.000259668 | 0.00127893 | 1 |

| | | | | | | |
|---|---|---|---|---|---|---|
| T2DM | pericyte | *SUOX* | 0.829764566 | 0.02713073 | 0.045388744 | 1 |
| T2DM | pericyte | *BRCA2* | 1.289526484 | 0.002743788 | 0.006906875 | 1 |
| T2DM | pericyte | *LMBR1L* | 2.303285181 | 0.016401203 | 0.029698838 | 1 |
| T2DM | pericyte | *ZNF740* | 2.481063803 | 0.000892825 | 0.003029051 | 1 |
| T2DM | pericyte | *WDFY2* | 2.267292627 | 0.00268033 | 0.006790543 | 1 |
| T2DM | pericyte | *VPS33A* | 1.138324753 | 0.003038223 | 0.007484839 | 1 |
| T2DM | pericyte | *TSSK4* | 2.841782872 | 0.017932307 | 0.032046735 | 1 |
| T2DM | pericyte | *TMX1* | 2.538261696 | 0.026683092 | 0.044866702 | 1 |
| T2DM | pericyte | *RTN1* | 2.644724419 | 8.53E-05 | 0.000687295 | 0.536089845 |
| T2DM | pericyte | *C14orf37* | 1.517659964 | 0.013541505 | 0.025158227 | 1 |
| T2DM | pericyte | *STON2* | 2.763104016 | 0.000227306 | 0.001178129 | 1 |
| T2DM | pericyte | *EFCAB11* | 0.771614773 | 0.009673384 | 0.018846163 | 1 |
| T2DM | pericyte | *FBLN5* | 3.167531228 | 2.54E-05 | 0.000562456 | 0.159491076 |
| T2DM | pericyte | *C14orf79* | 3.89921909 | 0.013717141 | 0.02543943 | 1 |
| T2DM | pericyte | *NIPA2* | 2.977690052 | 0.001160191 | 0.003647061 | 1 |
| T2DM | pericyte | *DUOXA1* | 1.380744712 | 0.001605824 | 0.004616285 | 1 |
| T2DM | pericyte | *MFAP1* | 1.278020141 | 0.006885343 | 0.014114353 | 1 |
| T2DM | pericyte | *TCF12* | 2.552252923 | 0.000276416 | 0.001324564 | 1 |
| T2DM | pericyte | *SRP14* | 3.412982216 | 1.68E-05 | 0.000506931 | 0.105441557 |
| T2DM | pericyte | *BAHD1* | 2.18753841 | 0.001221706 | 0.003783676 | 1 |
| T2DM | pericyte | *ANP32A* | 3.186799498 | 0.000143088 | 0.000906847 | 0.899591813 |
| T2DM | pericyte | *COMMD4* | 2.4219722 | 0.003574973 | 0.008481455 | 1 |
| T2DM | pericyte | *UBE2Q2* | 3.34220825 | 0.000723817 | 0.002582631 | 1 |
| T2DM | pericyte | *PSTPIP1* | 0.993700287 | 0.002599859 | 0.006620215 | 1 |
| T2DM | pericyte | *HMG20A* | 4.501560961 | 2.07E-09 | 9.32E-07 | 1.30E-05 |
| T2DM | pericyte | *SCAPER* | 1.680469767 | 0.001988907 | 0.005378177 | 1 |
| T2DM | pericyte | *MAN2C1* | -0.519039048 | 0.000462173 | 0.00187827 | 1 |
| T2DM | pericyte | *DNAJA4* | 5.42342114 | 3.43E-05 | 0.000599418 | 0.215716238 |
| T2DM | pericyte | *IGF1R* | 2.791471905 | 0.000743689 | 0.002640076 | 1 |
| T2DM | pericyte | *CYP11A1* | 0.860576533 | 0.004682656 | 0.010454495 | 1 |
| T2DM | pericyte | *BBS4* | 2.554886571 | 2.96E-05 | 0.000593226 | 0.186273038 |
| T2DM | pericyte | *ADAMTS17* | 2.224149647 | 0.005836686 | 0.012346987 | 1 |
| T2DM | pericyte | *ULK3* | 1.170021623 | 0.002378199 | 0.006170755 | 1 |
| T2DM | pericyte | *POLG* | 1.766262879 | 0.00121883 | 0.00377663 | 1 |
| T2DM | pericyte | *TICRR* | 1.238399031 | 0.014260649 | 0.026276877 | 1 |
| T2DM | pericyte | *UNC45A* | 1.120222459 | 0.01382388 | 0.025607169 | 1 |
| T2DM | pericyte | *IQGAP1* | 1.632792079 | 0.003544969 | 0.008443811 | 1 |
| T2DM | pericyte | *CRTC3* | 2.937661357 | 0.000907276 | 0.003050291 | 1 |
| T2DM | pericyte | *SEC11A* | 3.202958696 | 0.000109237 | 0.000776889 | 0.686769914 |
| T2DM | pericyte | *PARN* | 1.985696666 | 0.000406666 | 0.00170903 | 1 |
| T2DM | pericyte | *FTO* | 2.494740133 | 0.000116589 | 0.000798466 | 0.73299223 |
| T2DM | pericyte | *TXNL4B* | 1.09604021 | 0.003532064 | 0.008419041 | 1 |
| T2DM | pericyte | *MARVELD3* | 1.647270314 | 0.006046089 | 0.012721474 | 1 |
| T2DM | pericyte | *ZFHX3* | 3.014223124 | 5.44E-05 | 0.00062437 | 0.341870801 |
| T2DM | pericyte | *KIFC3* | 3.111873827 | 0.005642904 | 0.012018075 | 1 |
| T2DM | pericyte | *GCSH* | 1.366741654 | 0.000790883 | 0.00276545 | 1 |
| T2DM | pericyte | *CMTM3* | 4.904271295 | 0.000155856 | 0.000945663 | 0.979865801 |
| T2DM | pericyte | *MAP1LC3B* | 1.944561577 | 0.000330177 | 0.001474519 | 1 |
| T2DM | pericyte | *TLDC1* | 3.806899735 | 0.00018574 | 0.001039848 | 1 |
| T2DM | pericyte | *TIGD7* | 1.744116531 | 0.000998401 | 0.003284638 | 1 |

| | | | | | | |
|---|---|---|---|---|---|---|
| T2DM | pericyte | *COPS3* | 2.144199061 | 0.000332035 | 0.001479451 | 1 |
| T2DM | pericyte | *RANBP10* | 2.831693109 | 9.20E-05 | 0.000708744 | 0.578335504 |
| T2DM | pericyte | *PRPSAP2* | 1.831915051 | 0.000633917 | 0.002337355 | 1 |
| T2DM | pericyte | *MYO19* | 5.046229136 | 2.25E-09 | 9.44E-07 | 1.42E-05 |
| T2DM | pericyte | *DDX52* | 2.558005695 | 0.000182253 | 0.001034031 | 1 |
| T2DM | pericyte | *TOM1L1* | 2.910355555 | 2.22E-05 | 0.000547486 | 0.139588784 |
| T2DM | pericyte | *NPEPPS* | 2.919965761 | 0.000124088 | 0.000832113 | 0.780143747 |
| T2DM | pericyte | *G6PC3* | 1.450013 | 0.005593806 | 0.01193763 | 1 |
| T2DM | pericyte | *IMPA2* | 2.325298872 | 0.000180617 | 0.00103231 | 1 |
| T2DM | pericyte | *GNAL* | 1.682315307 | 0.000503103 | 0.001990567 | 1 |
| T2DM | pericyte | *C18orf21* | 0.920853379 | 0.001612712 | 0.004631851 | 1 |
| T2DM | pericyte | *GALNT1* | 2.340313203 | 0.000790444 | 0.00276545 | 1 |
| T2DM | pericyte | *ESCO1* | 2.758161517 | 0.000631377 | 0.002333607 | 1 |
| T2DM | pericyte | *SLC14A1* | 1.174564145 | 0.027075504 | 0.045308409 | 1 |
| T2DM | pericyte | *CCDC40* | 1.15237552 | 0.001411637 | 0.004182358 | 1 |
| T2DM | pericyte | *TMC6* | 1.215697673 | 0.000421861 | 0.001754127 | 1 |
| T2DM | pericyte | *CARD14* | 5.68249532 | 0.000108733 | 0.000775065 | 0.683607482 |
| T2DM | pericyte | *TBCD* | 2.666480745 | 7.48E-05 | 0.000650766 | 0.470503805 |
| T2DM | pericyte | *FN3KRP* | -0.605849287 | 0.018904893 | 0.033433211 | 1 |
| T2DM | pericyte | *NARF* | 1.733878685 | 0.022886996 | 0.0393251 | 1 |
| T2DM | pericyte | *TRIM65* | 1.015718617 | 0.012977817 | 0.024290425 | 1 |
| T2DM | pericyte | *SECTM1* | 1.445273717 | 0.003948174 | 0.009166237 | 1 |
| T2DM | pericyte | *WDR45B* | 3.170681005 | 1.98E-05 | 0.000515615 | 0.124186891 |
| T2DM | pericyte | *RNF165* | 2.913782374 | 0.001114711 | 0.003544881 | 1 |
| T2DM | pericyte | *MAPK4* | 0.991445364 | 0.002826246 | 0.007070676 | 1 |
| T2DM | pericyte | *SMAD4* | 2.898319129 | 0.000994117 | 0.003280848 | 1 |
| T2DM | pericyte | *PIP4K2B* | 1.464555926 | 0.001939509 | 0.005283229 | 1 |
| T2DM | pericyte | *NFIC* | 1.129615667 | 0.008343241 | 0.016594102 | 1 |
| T2DM | pericyte | *TPGS1* | 1.62183975 | 0.001201033 | 0.00373589 | 1 |
| T2DM | pericyte | *PRDM15* | 0.884101064 | 0.009487431 | 0.01851831 | 1 |
| T2DM | pericyte | *DUS3L* | 1.728554374 | 2.69E-05 | 0.000562456 | 0.169299266 |
| T2DM | pericyte | *DPP9* | 2.154678156 | 0.026736519 | 0.044912373 | 1 |
| T2DM | pericyte | *CCDC97* | 2.782742768 | 0.000177892 | 0.001020443 | 1 |
| T2DM | pericyte | *SIRT3* | 3.048229115 | 2.30E-07 | 4.01E-05 | 0.001443622 |
| T2DM | pericyte | *IFITM3* | 2.940833306 | 0.008476089 | 0.016815769 | 1 |
| T2DM | pericyte | *TRPM2* | 0.872661365 | 0.003046787 | 0.007497123 | 1 |
| T2DM | pericyte | *DOPEY2* | 1.961266334 | 0.001571657 | 0.004544679 | 1 |
| T2DM | pericyte | *URB1* | 0.923102214 | 0.017163406 | 0.03083038 | 1 |
| T2DM | pericyte | *AKT1* | 3.273767829 | 0.00014772 | 0.000921857 | 0.928714198 |
| T2DM | pericyte | *SAE1* | 1.92820639 | 0.000223849 | 0.00116598 | 1 |
| T2DM | pericyte | *CAPN10* | 1.020669173 | 0.003529029 | 0.008419041 | 1 |
| T2DM | pericyte | *MYO1F* | 1.952328784 | 6.41E-05 | 0.000632025 | 0.402733485 |
| T2DM | pericyte | *CACNG8* | 5.058626253 | 0.000488459 | 0.001948566 | 1 |
| T2DM | pericyte | *RPS11* | 1.820994469 | 0.027408427 | 0.045775001 | 1 |
| T2DM | pericyte | *CTU1* | 0.954823686 | 0.007579834 | 0.015303281 | 1 |
| T2DM | pericyte | *RCN3* | 4.635102187 | 6.94E-05 | 0.000637766 | 0.436210568 |
| T2DM | pericyte | *RERE* | 2.11859913 | 0.00021857 | 0.001146081 | 1 |
| T2DM | pericyte | *ARHGEF19* | -0.29386516 | 0.019872574 | 0.034899126 | 1 |
| T2DM | pericyte | *PEX14* | 1.690850245 | 0.000825134 | 0.002855045 | 1 |
| T2DM | pericyte | *SH3BGRL3* | 3.289616734 | 0.000228044 | 0.00118001 | 1 |

| | | | | | | |
|---|---|---|---|---|---|---|
| T2DM | pericyte | *C1orf216* | 1.72901297 | 0.002713503 | 0.006857109 | 1 |
| T2DM | pericyte | *ITGB3BP* | 0.941706163 | 0.02443145 | 0.041536108 | 1 |
| T2DM | pericyte | *SERBP1* | 3.271110899 | 0.001693897 | 0.004805744 | 1 |
| T2DM | pericyte | *PIGK* | 3.876400943 | 0.000100983 | 0.000742551 | 0.634881253 |
| T2DM | pericyte | *CYP4B1* | 0.900240305 | 0.003385736 | 0.008130681 | 1 |
| T2DM | pericyte | *ATP1B1* | 1.324586494 | 0.00435479 | 0.009894676 | 1 |
| T2DM | pericyte | *TIPRL* | 3.340115038 | 0.002735746 | 0.006900205 | 1 |
| T2DM | pericyte | *NME7* | 1.889766136 | 0.019300397 | 0.034072898 | 1 |
| T2DM | pericyte | *CREG1* | 1.650300628 | 0.003221579 | 0.007838262 | 1 |
| T2DM | pericyte | *GPA33* | 1.416324066 | 0.027041346 | 0.045263297 | 1 |
| T2DM | pericyte | *TMCO1* | 3.567181903 | 0.00033589 | 0.001493474 | 1 |
| T2DM | pericyte | *MGST3* | 2.920056541 | 0.000791743 | 0.002766478 | 1 |
| T2DM | pericyte | *ADCY10* | 1.216402756 | 0.007495965 | 0.015153418 | 1 |
| T2DM | pericyte | *RFWD2* | 2.343010737 | 0.001037482 | 0.003365197 | 1 |
| T2DM | pericyte | *PPOX* | 1.669736943 | 0.000359228 | 0.00155864 | 1 |
| T2DM | pericyte | *PIGM* | 2.32489792 | 0.000287735 | 0.001350999 | 1 |
| T2DM | pericyte | *RGL1* | 2.496532507 | 4.73E-05 | 0.000607238 | 0.297138798 |
| T2DM | pericyte | *TARS2* | 2.208547885 | 8.77E-06 | 0.000382983 | 0.055149552 |
| T2DM | pericyte | *SNX27* | 2.887771972 | 9.88E-05 | 0.00073561 | 0.621366211 |
| T2DM | pericyte | *CERS2* | 2.51484005 | 0.008346271 | 0.016594879 | 1 |
| T2DM | pericyte | *ENSA* | 3.119378924 | 7.02E-05 | 0.000639553 | 0.441520188 |
| T2DM | pericyte | *POGZ* | 1.78058688 | 0.002296527 | 0.006000462 | 1 |
| T2DM | pericyte | *GABPB2* | 1.226662133 | 0.001320817 | 0.003992297 | 1 |
| T2DM | pericyte | *IKBKE* | 0.851365786 | 0.004650886 | 0.010402035 | 1 |
| T2DM | pericyte | *TP53BP2* | 2.547555202 | 0.022928364 | 0.039363906 | 1 |
| T2DM | pericyte | *HAX1* | 2.496336498 | 0.000827419 | 0.002861377 | 1 |
| T2DM | pericyte | *KCNN3* | 1.693013441 | 0.000566994 | 0.002169792 | 1 |
| T2DM | pericyte | *GATAD2B* | 2.006615454 | 0.002845567 | 0.007104876 | 1 |
| T2DM | pericyte | *RIT1* | 4.33374735 | 4.25E-05 | 0.000607238 | 0.267496358 |
| T2DM | pericyte | *GALNT2* | 2.246551057 | 9.91E-05 | 0.000736303 | 0.622912753 |
| T2DM | pericyte | *SCCPDH* | 3.25401549 | 0.000158014 | 0.000950652 | 0.993431336 |
| T2DM | pericyte | *CEP170* | 2.470237485 | 0.000580979 | 0.002202711 | 1 |
| T2DM | pericyte | *SNAP47* | 2.511137758 | 0.000257943 | 0.001271911 | 1 |
| T2DM | pericyte | *NVL* | 3.719963467 | 0.000126641 | 0.00084164 | 0.796191346 |
| T2DM | pericyte | *SDE2* | 1.654056943 | 0.006885434 | 0.014114353 | 1 |
| T2DM | pericyte | *FBXO28* | 2.870068487 | 5.49E-05 | 0.00062437 | 0.3451699 |
| T2DM | pericyte | *CNIH4* | 3.315492915 | 0.000265117 | 0.001293785 | 1 |
| T2DM | pericyte | *GUK1* | 2.735518012 | 0.015729542 | 0.028672552 | 1 |
| T2DM | pericyte | *CNIH3* | 1.746894351 | 0.000144726 | 0.00091218 | 0.909891484 |
| T2DM | pericyte | *WNT9A* | 2.699852729 | 1.83E-06 | 0.000177371 | 0.011529101 |
| T2DM | pericyte | *PLEKHA6* | 2.914606898 | 1.26E-05 | 0.000458996 | 0.079205331 |
| T2DM | pericyte | *ABCG8* | 3.112186844 | 0.001170236 | 0.003671295 | 1 |
| T2DM | pericyte | *EML4* | 2.897149006 | 3.62E-05 | 0.00060096 | 0.227496748 |
| T2DM | pericyte | *WDPCP* | 0.47493755 | 0.030318089 | 0.049793581 | 1 |
| T2DM | pericyte | *ASXL2* | 2.968508849 | 0.000192188 | 0.001059899 | 1 |
| T2DM | pericyte | *EXOC6B* | 2.639445938 | 5.66E-05 | 0.00062437 | 0.355885075 |
| T2DM | pericyte | *TEX261* | 2.34939566 | 0.000627303 | 0.002324014 | 1 |
| T2DM | pericyte | *SLC20A1* | 3.171751128 | 2.89E-05 | 0.000585 | 0.181935146 |
| T2DM | pericyte | *FBLN7* | 1.283485306 | 0.00102643 | 0.003347079 | 1 |
| T2DM | pericyte | *ZC3H8* | 1.86450254 | 0.001478038 | 0.004338199 | 1 |

| | | | | | | |
|---|---|---|---|---|---|---|
| T2DM | pericyte | *UBXN4* | 2.805338957 | 0.000136029 | 0.000878628 | 0.855213576 |
| T2DM | pericyte | *GPR17* | 0.347067662 | 0.026059173 | 0.043970484 | 1 |
| T2DM | pericyte | *POLR2D* | 3.986579316 | 0.000839741 | 0.002894862 | 1 |
| T2DM | pericyte | *ZNF385B* | 0.922980509 | 0.000813556 | 0.002824311 | 1 |
| T2DM | pericyte | *TMEFF2* | 2.142346912 | 3.87E-05 | 0.000607238 | 0.243183663 |
| T2DM | pericyte | *GULP1* | 2.515715568 | 5.83E-05 | 0.00062437 | 0.366326064 |
| T2DM | pericyte | *FAM171B* | 0.856629031 | 0.013929684 | 0.025765202 | 1 |
| T2DM | pericyte | *HSPD1* | 2.843538963 | 1.75E-05 | 0.000512131 | 0.11003538 |
| T2DM | pericyte | *METTL21A* | 4.019404831 | 0.000188341 | 0.001046837 | 1 |
| T2DM | pericyte | *PTH2R* | 0.771940414 | 0.005861797 | 0.012386875 | 1 |
| T2DM | pericyte | *NBEAL1* | 1.200988568 | 0.000158608 | 0.000953314 | 0.997166021 |
| T2DM | pericyte | *SUMF1* | 2.409780434 | 5.88E-05 | 0.00062437 | 0.369441386 |
| T2DM | pericyte | *ESPNL* | 10.12380545 | 0.000377541 | 0.001619456 | 1 |
| T2DM | pericyte | *FANCD2* | 4.286941061 | 0.004968768 | 0.010914971 | 1 |
| T2DM | pericyte | *TAMM41* | 3.3283068 | 0.000241215 | 0.001221028 | 1 |
| T2DM | pericyte | *CSRNP1* | 2.290379756 | 0.001172659 | 0.003673396 | 1 |
| T2DM | pericyte | *ARL6IP5* | 3.336464944 | 2.62E-05 | 0.000562456 | 0.164411032 |
| T2DM | pericyte | *TMF1* | 3.070119715 | 0.001050833 | 0.003394957 | 1 |
| T2DM | pericyte | *COL8A1* | 0.901586322 | 0.021849439 | 0.037885658 | 1 |
| T2DM | pericyte | *NXPE3* | 2.572763368 | 0.005385833 | 0.011612049 | 1 |
| T2DM | pericyte | *PLA1A* | -1.146936661 | 0.003040049 | 0.007486403 | 1 |
| T2DM | pericyte | *RABL3* | 3.796112156 | 2.68E-05 | 0.000562456 | 0.168219162 |
| T2DM | pericyte | *AGTR1* | -0.133426499 | 0.028104637 | 0.046743192 | 1 |
| T2DM | pericyte | *TCTA* | 2.525563609 | 0.000205544 | 0.001099503 | 1 |
| T2DM | pericyte | *EAF2* | 1.648849762 | 0.003492426 | 0.008342281 | 1 |
| T2DM | pericyte | *ILDR1* | -0.945519512 | 0.028153501 | 0.046800915 | 1 |
| T2DM | pericyte | *LYAR* | 2.486194977 | 0.000362221 | 0.001566219 | 1 |
| T2DM | pericyte | *CENPC* | 1.172281293 | 0.02414828 | 0.041132549 | 1 |
| T2DM | pericyte | *ATP10D* | 1.766457995 | 0.003026383 | 0.007461516 | 1 |
| T2DM | pericyte | *CISD2* | 3.705557061 | 1.24E-05 | 0.000458996 | 0.07807591 |
| T2DM | pericyte | *TIFA* | 2.273153135 | 0.00491775 | 0.010830788 | 1 |
| T2DM | pericyte | *CCNA2* | 1.01047957 | 0.025748943 | 0.043551715 | 1 |
| T2DM | pericyte | *USP53* | 3.181600446 | 7.47E-06 | 0.000356314 | 0.046960196 |
| T2DM | pericyte | *PDGFC* | 1.491715593 | 0.003643634 | 0.008595999 | 1 |
| T2DM | pericyte | *CBR4* | 3.21934101 | 2.99E-05 | 0.00059534 | 0.188127427 |
| T2DM | pericyte | *NDUFS6* | 2.798553223 | 0.017942119 | 0.032047151 | 1 |
| T2DM | pericyte | *LHFPL2* | 2.524345921 | 0.011481409 | 0.02192698 | 1 |
| T2DM | pericyte | *SSBP2* | 2.168886267 | 0.002437516 | 0.006291968 | 1 |
| T2DM | pericyte | *RASA1* | 1.590614502 | 0.000906428 | 0.00304947 | 1 |
| T2DM | pericyte | *GTF2H2* | 1.469197818 | 0.012384393 | 0.023325548 | 1 |
| T2DM | pericyte | *SPATA9* | 2.293692503 | 2.68E-05 | 0.000562456 | 0.168221487 |
| T2DM | pericyte | *FEM1C* | 3.045738414 | 0.000182166 | 0.001034031 | 1 |
| T2DM | pericyte | *COMMD10* | 1.718352485 | 0.001350147 | 0.004042083 | 1 |
| T2DM | pericyte | *YIPF5* | 1.835109561 | 0.00047354 | 0.001905155 | 1 |
| T2DM | pericyte | *TBC1D7* | 2.230640627 | 0.00023523 | 0.001201373 | 1 |
| T2DM | pericyte | *PSD2* | 5.896879884 | 0.000264471 | 0.001293055 | 1 |
| T2DM | pericyte | *ZMAT2* | 1.20403416 | 0.004241542 | 0.009696936 | 1 |
| T2DM | pericyte | *KLHL3* | 1.578474539 | 0.001714583 | 0.004842579 | 1 |
| T2DM | pericyte | *TRIM7* | 0.984443869 | 0.022448987 | 0.038763192 | 1 |
| T2DM | pericyte | *FAM193B* | 3.808736815 | 8.95E-05 | 0.000702125 | 0.562594851 |

| | | | | | | |
|---|---|---|---|---|---|---|
| T2DM | pericyte | *MUT* | 3.541787278 | 6.01E-05 | 0.00062437 | 0.37772582 |
| T2DM | pericyte | *FGD2* | 0.775185098 | 0.004405044 | 0.009990804 | 1 |
| T2DM | pericyte | *RPL7L1* | 1.676498559 | 0.001111193 | 0.00353725 | 1 |
| T2DM | pericyte | *TPBG* | 4.012279733 | 6.28E-05 | 0.000630768 | 0.394860895 |
| T2DM | pericyte | *IRAK1BP1* | 4.299773099 | 6.99E-06 | 0.000348151 | 0.043974604 |
| T2DM | pericyte | *PHIP* | 3.014814766 | 1.47E-05 | 0.000476496 | 0.092440238 |
| T2DM | pericyte | *MMS22L* | 2.457790344 | 0.003782025 | 0.00886562 | 1 |
| T2DM | pericyte | *PM20D2* | 2.442421468 | 0.00063415 | 0.002337355 | 1 |
| T2DM | pericyte | *RNF217* | 2.663804671 | 5.31E-05 | 0.000619691 | 0.334066613 |
| T2DM | pericyte | *AIG1* | 1.932048707 | 0.00039055 | 0.001662417 | 1 |
| T2DM | pericyte | *DYNLT1* | 2.100627151 | 0.01871693 | 0.033166105 | 1 |
| T2DM | pericyte | *WTAP* | 1.469891681 | 0.002406268 | 0.006223039 | 1 |
| T2DM | pericyte | *SLC22A3* | 1.186086717 | 0.001107555 | 0.003529245 | 1 |
| T2DM | pericyte | *GNA12* | 2.340592344 | 0.001027481 | 0.003348766 | 1 |
| T2DM | pericyte | *C7orf50* | 4.105642103 | 2.32E-05 | 0.000548455 | 0.145748411 |
| T2DM | pericyte | *RBAK* | 2.5726643 | 0.001024713 | 0.003343213 | 1 |
| T2DM | pericyte | *CCT6A* | 2.913813579 | 8.27E-05 | 0.000681373 | 0.520182507 |
| T2DM | pericyte | *PSPH* | 1.564984747 | 0.000583566 | 0.00220618 | 1 |
| T2DM | pericyte | *TMEM209* | 1.842469234 | 8.82E-05 | 0.000701359 | 0.554474942 |
| T2DM | pericyte | *AGBL3* | 2.231535059 | 0.000305089 | 0.001399049 | 1 |
| T2DM | pericyte | *EPHA1* | 1.307025137 | 0.011323316 | 0.021651366 | 1 |
| T2DM | pericyte | *NOM1* | 2.516423865 | 0.000263052 | 0.001288014 | 1 |
| T2DM | pericyte | *NCAPG2* | 1.720572905 | 0.000241853 | 0.001222377 | 1 |
| T2DM | pericyte | *FBXO25* | 3.622743922 | 6.69E-05 | 0.000632837 | 0.420337745 |
| T2DM | pericyte | *CCDC25* | 3.738513102 | 1.45E-07 | 2.76E-05 | 0.000909392 |
| T2DM | pericyte | *BIN3* | 3.185272916 | 0.000261717 | 0.001284286 | 1 |
| T2DM | pericyte | *SLC25A37* | 2.823401321 | 7.63E-05 | 0.00065815 | 0.480006652 |
| T2DM | pericyte | *CHMP7* | 2.04884526 | 0.002638699 | 0.006705539 | 1 |
| T2DM | pericyte | *DOCK5* | 2.782565083 | 8.70E-05 | 0.000695771 | 0.546722402 |
| T2DM | pericyte | *TACC1* | 1.80226756 | 0.01147373 | 0.021918974 | 1 |
| T2DM | pericyte | *MTDH* | 2.99538342 | 4.32E-05 | 0.000607238 | 0.271512493 |
| T2DM | pericyte | *UTP23* | 3.503050381 | 4.34E-05 | 0.000607238 | 0.27274306 |
| T2DM | pericyte | *FAM135B* | 1.184234061 | 0.005053786 | 0.011055376 | 1 |
| T2DM | pericyte | *ZNF7* | 3.863335666 | 1.53E-07 | 2.83E-05 | 0.000962135 |
| T2DM | pericyte | *AK3* | 1.882159035 | 0.019408926 | 0.03421871 | 1 |
| T2DM | pericyte | *PLIN2* | 3.488026245 | 5.86E-05 | 0.00062437 | 0.368161379 |
| T2DM | pericyte | *HAUS6* | 4.327749241 | 4.01E-05 | 0.000607238 | 0.252270129 |
| T2DM | pericyte | *C9orf72* | 1.729702061 | 0.000480994 | 0.00192612 | 1 |
| T2DM | pericyte | *CBWD5* | 2.385997794 | 0.000299785 | 0.001384824 | 1 |
| T2DM | pericyte | *ZNF462* | 3.716578522 | 0.000164787 | 0.000972788 | 1 |
| T2DM | pericyte | *ASTN2* | 3.331070668 | 6.19E-05 | 0.000628694 | 0.389050389 |
| T2DM | pericyte | *WDR31* | 3.01101284 | 0.011780549 | 0.022375925 | 1 |
| T2DM | pericyte | *POLE3* | 0.915168332 | 0.011736643 | 0.022306008 | 1 |
| T2DM | pericyte | *SURF6* | 3.627579152 | 0.000156432 | 0.000945663 | 0.983489887 |
| T2DM | pericyte | *NTMT1* | 1.551979838 | 0.010144401 | 0.019638275 | 1 |
| T2DM | pericyte | *CIZ1* | 1.944627799 | 0.000547885 | 0.002121028 | 1 |
| T2DM | pericyte | *SH3GLB2* | 3.169239092 | 0.00014855 | 0.000922861 | 0.933935756 |
| T2DM | pericyte | *LRSAM1* | 1.267844293 | 0.002783101 | 0.006976618 | 1 |
| T2DM | pericyte | *SEC16A* | 2.106257858 | 7.40E-06 | 0.000356314 | 0.046498824 |
| T2DM | pericyte | *DPH7* | 3.258255505 | 0.006730618 | 0.013860268 | 1 |

| | | | | | | |
|---|---|---|---|---|---|---|
| T2DM | pericyte | *RSU1* | 2.819837099 | 0.001035192 | 0.003363438 | 1 |
| T2DM | pericyte | *ADIRF* | 2.401484245 | 0.00705654 | 0.014416949 | 1 |
| T2DM | pericyte | *GLUD1* | 3.602213802 | 1.85E-05 | 0.000512131 | 0.116314882 |
| T2DM | pericyte | *HTR7* | 1.105992789 | 0.003332407 | 0.008048729 | 1 |
| T2DM | pericyte | *RPP30* | 3.305704357 | 0.000818752 | 0.002839212 | 1 |
| T2DM | pericyte | *FRA10AC1* | 1.91915017 | 0.00016462 | 0.000972714 | 1 |
| T2DM | pericyte | *DNAJB12* | 3.8296087 | 0.000158949 | 0.000954278 | 0.999310332 |
| T2DM | pericyte | *PLEKHS1* | -1.816005002 | 0.008300352 | 0.016524482 | 1 |
| T2DM | pericyte | *LRRC27* | 0.537696479 | 0.026698865 | 0.044880838 | 1 |
| T2DM | pericyte | *GSTO1* | 1.810202301 | 0.011839981 | 0.022448118 | 1 |
| T2DM | pericyte | *PDCD11* | 2.558600596 | 0.001470206 | 0.004319247 | 1 |
| T2DM | pericyte | *RGS10* | 2.389972148 | 0.005404979 | 0.011637364 | 1 |
| T2DM | pericyte | *GAS2* | 3.418388764 | 0.000108436 | 0.000774699 | 0.681735086 |
| T2DM | pericyte | *LIN7C* | 1.767264985 | 0.000635651 | 0.002339775 | 1 |
| T2DM | pericyte | *HSD17B12* | -1.23326821 | 1.67E-06 | 0.000166159 | 0.010468026 |
| T2DM | pericyte | *APIP* | -0.33143293 | 0.008824193 | 0.017412964 | 1 |
| T2DM | pericyte | *GLYAT* | 0.838979514 | 0.000188676 | 0.001046959 | 1 |
| T2DM | pericyte | *PTPRJ* | 2.065827034 | 1.25E-05 | 0.000458996 | 0.078461241 |
| T2DM | pericyte | *C11orf49* | 3.018295514 | 0.00013142 | 0.000860073 | 0.826238088 |
| T2DM | pericyte | *SERPINH1* | 2.126486807 | 0.000451235 | 0.00184695 | 1 |
| T2DM | pericyte | *CAPN5* | 0.917307388 | 0.001966106 | 0.005332575 | 1 |
| T2DM | pericyte | *INTS4* | 1.972886967 | 0.001062851 | 0.003423228 | 1 |
| T2DM | pericyte | *ZC3H12C* | 3.812063781 | 2.68E-05 | 0.000562456 | 0.168788561 |
| T2DM | pericyte | *DRD2* | 3.561238411 | 0.005522744 | 0.011830151 | 1 |
| T2DM | pericyte | *C11orf52* | 8.570606216 | 0.00339377 | 0.008146862 | 1 |
| T2DM | pericyte | *NPAT* | 2.170872203 | 0.000274748 | 0.001323244 | 1 |
| T2DM | pericyte | *P4HA3* | 1.793486099 | 0.005534949 | 0.011848221 | 1 |
| T2DM | pericyte | *GRIK4* | 0.625690967 | 0.011879782 | 0.022489668 | 1 |
| T2DM | pericyte | *ADAM33* | 1.247001263 | 0.004134355 | 0.009480399 | 1 |
| T2DM | pericyte | *TMC2* | 1.261856602 | 0.004984815 | 0.010942574 | 1 |
| T2DM | pericyte | *EML3* | 2.986154628 | 0.001983329 | 0.005367711 | 1 |
| T2DM | pericyte | *MS4A3* | 15.53033729 | 0.006697278 | 0.013809703 | 1 |
| T2DM | pericyte | *CPSF7* | 3.894411397 | 5.86E-05 | 0.00062437 | 0.368539074 |
| T2DM | pericyte | *FEZ1* | 1.340369024 | 0.001265516 | 0.003877339 | 1 |
| T2DM | pericyte | *COMMD7* | 4.544303761 | 0.000321398 | 0.001445372 | 1 |
| T2DM | pericyte | *KIAA1755* | 0.651252008 | 0.012533916 | 0.023564811 | 1 |
| T2DM | pericyte | *CNBD2* | 1.094043978 | 0.003326257 | 0.008037003 | 1 |
| T2DM | pericyte | *ARID5B* | 2.310051283 | 7.45E-05 | 0.00064967 | 0.468411813 |
| T2DM | pericyte | *TMCO3* | 3.322302126 | 4.56E-05 | 0.000607238 | 0.286486134 |
| T2DM | pericyte | *LYPD6B* | 1.094704161 | 0.011007779 | 0.021112235 | 1 |
| T2DM | pericyte | *PDCD4* | 3.199193472 | 1.87E-05 | 0.000512131 | 0.117255961 |
| T2DM | pericyte | *WDR17* | 2.040948244 | 0.001169833 | 0.003671295 | 1 |
| T2DM | pericyte | *FSIP1* | 0.923993207 | 0.008691697 | 0.017189274 | 1 |
| T2DM | pericyte | *RGS18* | 0.877003184 | 0.004410351 | 0.009999235 | 1 |
| T2DM | pericyte | *PRSS23* | 1.350330333 | 0.0046692 | 0.010435571 | 1 |
| T2DM | pericyte | *MTMR12* | 2.403580538 | 7.80E-05 | 0.000663325 | 0.490344676 |
| T2DM | pericyte | *PPP1R1C* | 1.439971645 | 0.000178942 | 0.0010246 | 1 |
| T2DM | pericyte | *CCT5* | 2.694626807 | 0.000118359 | 0.000806199 | 0.744121691 |
| T2DM | pericyte | *FAM173B* | 1.103702339 | 0.00559068 | 0.011935011 | 1 |
| T2DM | pericyte | *DOCK1* | 1.747431071 | 0.014796606 | 0.027164025 | 1 |

| | | | | | | |
|---|---|---|---|---|---|---|
| T2DM | pericyte | *TEX12* | 5.437350097 | 0.005392233 | 0.011617877 | 1 |
| T2DM | pericyte | *PIP4K2A* | 2.479055196 | 0.000516586 | 0.002031129 | 1 |
| T2DM | pericyte | *CRIM1* | 1.888135464 | 0.001207798 | 0.003749405 | 1 |
| T2DM | pericyte | *ABCB9* | -1.506302949 | 0.027677721 | 0.0461687 | 1 |
| T2DM | pericyte | *GPR158* | 0.760047237 | 0.009199346 | 0.018017535 | 1 |
| T2DM | pericyte | *THRB* | 2.687859891 | 8.08E-05 | 0.000671315 | 0.50773717 |
| T2DM | pericyte | *ANK3* | 0.942964629 | 0.000335895 | 0.001493474 | 1 |
| T2DM | pericyte | *IPMK* | 2.004185692 | 0.000208748 | 0.001111258 | 1 |
| T2DM | pericyte | *PLBD2* | 1.86972628 | 0.004551834 | 0.010231456 | 1 |
| T2DM | pericyte | *GXYLT1* | 3.31062745 | 7.24E-05 | 0.00064687 | 0.455097893 |
| T2DM | pericyte | *FAM177A1* | 4.337919914 | 9.68E-06 | 0.000404168 | 0.060827394 |
| T2DM | pericyte | *MIPOL1* | 2.140509554 | 0.000240471 | 0.001220212 | 1 |
| T2DM | pericyte | *TMEM18* | 4.88353219 | 5.93E-05 | 0.00062437 | 0.372660997 |
| T2DM | pericyte | *NDUFC2* | 1.834005103 | 0.001171735 | 0.003672981 | 1 |
| T2DM | pericyte | *ME3* | 1.674548794 | 0.000233883 | 0.001198741 | 1 |
| T2DM | pericyte | *ANKRD50* | 2.49712799 | 0.000122314 | 0.000825981 | 0.768988312 |
| T2DM | pericyte | *C4orf33* | 2.379406472 | 0.008454022 | 0.016781252 | 1 |
| T2DM | pericyte | *PTPRO* | 1.010873788 | 0.003009434 | 0.007425554 | 1 |
| T2DM | pericyte | *VTI1A* | 2.799760276 | 1.59E-05 | 0.000496416 | 0.099779581 |
| T2DM | pericyte | *FAM160B1* | 3.574042725 | 4.29E-05 | 0.000607238 | 0.269652255 |
| T2DM | pericyte | *TEX9* | 1.861157533 | 0.000793822 | 0.002769565 | 1 |
| T2DM | pericyte | *AKR1C2* | 0.777144487 | 0.004886036 | 0.010782208 | 1 |
| T2DM | pericyte | *ADAM8* | 0.894347149 | 0.006403326 | 0.01331714 | 1 |
| T2DM | pericyte | *ITIH2* | 3.496150587 | 0.001032487 | 0.00335812 | 1 |
| T2DM | pericyte | *KIN* | 2.701597344 | 3.62E-05 | 0.00060096 | 0.227328733 |
| T2DM | pericyte | *PIGF* | 2.805849365 | 0.000382768 | 0.001635935 | 1 |
| T2DM | pericyte | *MFSD6* | 3.573543389 | 5.68E-05 | 0.00062437 | 0.357151458 |
| T2DM | pericyte | *RNF144A* | 1.729314641 | 0.000186876 | 0.001043394 | 1 |
| T2DM | pericyte | *ASAP2* | 1.728095675 | 0.022579754 | 0.038935521 | 1 |
| T2DM | pericyte | *ADAM17* | 2.992599334 | 4.14E-05 | 0.000607238 | 0.260425602 |
| T2DM | pericyte | *ACSL1* | 0.897869401 | 0.002227373 | 0.005866476 | 1 |
| T2DM | pericyte | *BICD1* | 3.104631186 | 0.000439118 | 0.001807948 | 1 |
| T2DM | pericyte | *GUF1* | 2.516950514 | 0.001751922 | 0.00491931 | 1 |
| T2DM | pericyte | *CENPJ* | 1.97856156 | 0.000706598 | 0.002541358 | 1 |
| T2DM | pericyte | *PARP8* | 1.503981631 | 0.000768473 | 0.002714263 | 1 |
| T2DM | pericyte | *CACUL1* | 1.959009791 | 0.00025459 | 0.001259327 | 1 |
| T2DM | pericyte | *RABGAP1L* | 2.630777127 | 2.47E-05 | 0.000558632 | 0.155539553 |
| T2DM | pericyte | *RNF219* | 3.418783327 | 0.002886297 | 0.007178065 | 1 |
| T2DM | pericyte | *G6PC2* | 39.96881367 | 0.004470037 | 0.010087265 | 1 |
| T2DM | pericyte | *TGOLN2* | 2.373584305 | 0.013706293 | 0.025426812 | 1 |
| T2DM | pericyte | *TADA1* | 4.803261862 | 4.13E-05 | 0.000607238 | 0.259850858 |
| T2DM | pericyte | *CWF19L2* | 3.251370162 | 3.45E-05 | 0.000599418 | 0.217119756 |
| T2DM | pericyte | *ZNF547* | 1.201101828 | 0.019365762 | 0.034161769 | 1 |
| T2DM | pericyte | *CCDC50* | 0.902068876 | 0.014954935 | 0.027419562 | 1 |
| T2DM | pericyte | *ZFP36L2* | 3.230640768 | 8.15E-07 | 0.000102524 | 0.005126188 |
| T2DM | pericyte | *TMEM123* | 3.49506751 | 0.000414847 | 0.001736448 | 1 |
| T2DM | pericyte | *SPEF2* | 1.486983815 | 0.006902911 | 0.014145567 | 1 |
| T2DM | pericyte | *MBNL1* | 2.102580559 | 0.000497128 | 0.00197275 | 1 |
| T2DM | pericyte | *CLEC4F* | 2.299025896 | 0.004348944 | 0.009884963 | 1 |
| T2DM | pericyte | *RASGRP3* | 0.715125201 | 0.018585938 | 0.033003476 | 1 |

| | | | | | | |
|---|---|---|---|---|---|---|
| T2DM | pericyte | *SAR1B* | 2.546481816 | 3.91E-07 | 5.78E-05 | 0.00245932 |
| T2DM | pericyte | *IFIT5* | 1.180663579 | 0.01059831 | 0.020414086 | 1 |
| T2DM | pericyte | *RAB3C* | 1.278503362 | 0.006930487 | 0.01419745 | 1 |
| T2DM | pericyte | *MED21* | 3.083631808 | 5.01E-05 | 0.000610424 | 0.315012813 |
| T2DM | pericyte | *CENPH* | 1.004158288 | 0.004066901 | 0.009365791 | 1 |
| T2DM | pericyte | *CARHSP1* | 1.782700064 | 0.000815655 | 0.002830036 | 1 |
| T2DM | pericyte | *TEKT5* | 2.288201355 | 0.00118204 | 0.003699095 | 1 |
| T2DM | pericyte | *BANK1* | 2.681772342 | 6.42E-05 | 0.000632025 | 0.403849514 |
| T2DM | pericyte | *CAST* | 1.228414727 | 0.00730496 | 0.014829281 | 1 |
| T2DM | pericyte | *SCOC* | 2.662567034 | 0.001234417 | 0.003805096 | 1 |
| T2DM | pericyte | *CLGN* | 5.05499787 | 0.002181073 | 0.005761515 | 1 |
| T2DM | pericyte | *SYCP2L* | 1.891241338 | 0.003076948 | 0.0075536 | 1 |
| T2DM | pericyte | *AHCTF1* | 3.423749212 | 1.78E-05 | 0.000512131 | 0.111975564 |
| T2DM | pericyte | *TMEM87B* | 2.277978512 | 0.002439614 | 0.006293743 | 1 |
| T2DM | pericyte | *PLA2R1* | 1.373539411 | 3.86E-05 | 0.000607238 | 0.24274359 |
| T2DM | pericyte | *FRMD1* | 0.918103384 | 0.005000993 | 0.010958955 | 1 |
| T2DM | pericyte | *INO80C* | 2.701361864 | 0.004725622 | 0.010527989 | 1 |
| T2DM | pericyte | *LPCAT1* | 3.371031688 | 0.00012504 | 0.000837193 | 0.786123948 |
| T2DM | pericyte | *NMRAL1* | 1.521878881 | 0.000562294 | 0.002159249 | 1 |
| T2DM | pericyte | *TEX29* | 1.025657088 | 0.013444502 | 0.025007568 | 1 |
| T2DM | pericyte | *LURAP1L* | 0.707421441 | 0.004688295 | 0.010459551 | 1 |
| T2DM | pericyte | *CFDP1* | 3.325047986 | 7.38E-05 | 0.000648165 | 0.464085862 |
| T2DM | pericyte | *ZDHHC7* | 2.438259936 | 4.42E-05 | 0.000607238 | 0.277818262 |
| T2DM | pericyte | *FAM92B* | 10.15125174 | 0.005653284 | 0.012035724 | 1 |
| T2DM | pericyte | *FBXO36* | 4.341724414 | 0.000176704 | 0.001018276 | 1 |
| T2DM | pericyte | *MCOLN2* | 1.02501931 | 0.001129638 | 0.003577852 | 1 |
| T2DM | pericyte | *CACNA2D1* | 1.894639265 | 0.000297629 | 0.001381518 | 1 |
| T2DM | pericyte | *ZUFSP* | 2.550150136 | 0.000419351 | 0.001747155 | 1 |
| T2DM | pericyte | *PPP2R5E* | 2.551449086 | 4.34E-05 | 0.000607238 | 0.272702324 |
| T2DM | pericyte | *TBCEL* | 3.208943829 | 0.002298252 | 0.006000462 | 1 |
| T2DM | pericyte | *UBASH3B* | 1.053844476 | 0.002474451 | 0.006373155 | 1 |
| T2DM | pericyte | *ROBO3* | 0.844258363 | 0.018353931 | 0.032651715 | 1 |
| T2DM | pericyte | *PRKCA* | 2.565569143 | 0.002992264 | 0.007391892 | 1 |
| T2DM | pericyte | *LRRK1* | 3.015297443 | 4.57E-05 | 0.000607238 | 0.287323098 |
| T2DM | pericyte | *CEP112* | 1.483026232 | 0.004058139 | 0.009349037 | 1 |
| T2DM | pericyte | *ENPP3* | 0.640347165 | 0.006016395 | 0.012667474 | 1 |
| T2DM | pericyte | *UCHL1* | 4.285542964 | 0.000517188 | 0.002032225 | 1 |
| T2DM | pericyte | *MIA3* | 2.578786464 | 0.001332577 | 0.004010488 | 1 |
| T2DM | pericyte | *NEIL2* | 1.086781067 | 0.004089784 | 0.009411594 | 1 |
| T2DM | pericyte | *CCSAP* | 2.462097617 | 0.012263258 | 0.023132044 | 1 |
| T2DM | pericyte | *GBP5* | 0.838313366 | 0.005043728 | 0.011037216 | 1 |
| T2DM | pericyte | *FAM69A* | 1.25698245 | 0.006814685 | 0.014003728 | 1 |
| T2DM | pericyte | *ATP5G3* | 1.431297473 | 0.006563453 | 0.013605154 | 1 |
| T2DM | pericyte | *PDLIM3* | 1.554113665 | 0.000831548 | 0.002872496 | 1 |
| T2DM | pericyte | *NCAM2* | 0.728035526 | 0.013226142 | 0.024667089 | 1 |
| T2DM | pericyte | *L3MBTL4* | 2.381806381 | 2.37E-05 | 0.000548455 | 0.149179699 |
| T2DM | pericyte | *TSEN2* | 3.29395365 | 5.24E-10 | 2.74E-07 | 3.29E-06 |
| T2DM | pericyte | *SLFN13* | -0.444499171 | 0.013083426 | 0.024451695 | 1 |
| T2DM | pericyte | *FLCN* | 1.058233921 | 0.002632927 | 0.006693575 | 1 |
| T2DM | pericyte | *DPH3* | 1.712540206 | 0.00113172 | 0.003582639 | 1 |

| | | | | | | |
|---|---|---|---|---|---|---|
| T2DM | pericyte | *PPP4R1* | 3.56968161 | 5.26E-05 | 0.000619691 | 0.330899042 |
| T2DM | pericyte | *APCDD1* | 0.489686831 | 0.008409503 | 0.016710034 | 1 |
| T2DM | pericyte | *EPHB1* | 1.54917414 | 0.001588397 | 0.004578197 | 1 |
| T2DM | pericyte | *ANKRD40* | 1.162705058 | 0.008201952 | 0.016349294 | 1 |
| T2DM | pericyte | *CYP2U1* | 4.711989615 | 3.58E-05 | 0.00060096 | 0.225301764 |
| T2DM | pericyte | *FBXL18* | 4.375541595 | 1.84E-05 | 0.000512131 | 0.115652456 |
| T2DM | pericyte | *PROM2* | 2.659564667 | 0.024000209 | 0.040962226 | 1 |
| T2DM | pericyte | *AK9* | 2.124029134 | 0.003278356 | 0.00794259 | 1 |
| T2DM | pericyte | *CDK19* | 3.013440317 | 0.000347382 | 0.001521528 | 1 |
| T2DM | pericyte | *TTC39B* | 1.925587489 | 0.009370324 | 0.018312474 | 1 |
| T2DM | pericyte | *AGPAT5* | 1.233222294 | 0.00644642 | 0.013383672 | 1 |
| T2DM | pericyte | *MMS19* | 2.747013969 | 0.001697165 | 0.004807262 | 1 |
| T2DM | pericyte | *TRMT44* | 3.79465654 | 0.00011528 | 0.000794697 | 0.724764084 |
| T2DM | pericyte | *C16orf87* | 1.979966249 | 0.000265899 | 0.001293785 | 1 |
| T2DM | pericyte | *CNOT8* | 2.57115844 | 0.001944416 | 0.00529329 | 1 |
| T2DM | pericyte | *ZKSCAN2* | 0.739968687 | 0.002599486 | 0.006620215 | 1 |
| T2DM | pericyte | *PDIA4* | 5.296092478 | 6.77E-05 | 0.000632837 | 0.425778775 |
| T2DM | pericyte | *KDM8* | 4.405040416 | 0.000763244 | 0.002700345 | 1 |
| T2DM | pericyte | *FAM126B* | 2.911810265 | 0.000100802 | 0.000742087 | 0.633742127 |
| T2DM | pericyte | *SPAG17* | 2.874540949 | 0.000259773 | 0.00127893 | 1 |
| T2DM | pericyte | *DEPTOR* | 3.292278221 | 0.001749432 | 0.00491671 | 1 |
| T2DM | pericyte | *RRAGA* | 2.867374805 | 0.001541556 | 0.004486927 | 1 |
| T2DM | pericyte | *FAM161B* | 1.692754471 | 0.000251685 | 0.001253836 | 1 |
| T2DM | pericyte | *WIF1* | 2.647631591 | 0.022795734 | 0.039254116 | 1 |
| T2DM | pericyte | *UGT2B4* | 1.190930823 | 0.005172383 | 0.011261976 | 1 |
| T2DM | pericyte | *KCNMA1* | 1.383398895 | 0.010117147 | 0.019601388 | 1 |
| T2DM | pericyte | *BATF* | 0.972749603 | 0.005485191 | 0.011769252 | 1 |
| T2DM | pericyte | *DPY19L4* | 2.506999656 | 0.001549583 | 0.004501954 | 1 |
| T2DM | pericyte | *DRAM2* | 2.342694676 | 0.000601239 | 0.002255362 | 1 |
| T2DM | pericyte | *ANKRD9* | 0.723105802 | 0.026941613 | 0.045132407 | 1 |
| T2DM | pericyte | *SFXN2* | 0.934927588 | 0.00188463 | 0.005174823 | 1 |
| T2DM | pericyte | *SH3RF2* | 0.800174228 | 0.024493179 | 0.041629796 | 1 |
| T2DM | pericyte | *GDF6* | 2.355498715 | 0.000808831 | 0.002814124 | 1 |
| T2DM | pericyte | *NODAL* | 1.900231565 | 0.001928949 | 0.005261302 | 1 |
| T2DM | pericyte | *ATAD2* | 2.96238187 | 9.48E-05 | 0.000717456 | 0.596205613 |
| T2DM | pericyte | *SASS6* | 1.735715922 | 0.011610568 | 0.022113189 | 1 |
| T2DM | pericyte | *VPS8* | 2.841231462 | 1.52E-06 | 0.000159759 | 0.009535615 |
| T2DM | pericyte | *RPUSD3* | 5.74058492 | 0.000134161 | 0.000872252 | 0.843468003 |
| T2DM | pericyte | *GHRL* | 2.793161125 | 0.000231828 | 0.001193695 | 1 |
| T2DM | pericyte | *EXOG* | 2.954563903 | 0.000339709 | 0.00150511 | 1 |
| T2DM | pericyte | *NTAN1* | 2.432193974 | 0.000537174 | 0.002091155 | 1 |
| T2DM | pericyte | *SLC6A1* | -0.508224076 | 0.016196992 | 0.029388308 | 1 |
| T2DM | pericyte | *FCHO2* | 3.134471127 | 0.000111845 | 0.000783039 | 0.703169075 |
| T2DM | pericyte | *CPT2* | 3.38043285 | 0.000111426 | 0.00078265 | 0.700538051 |
| T2DM | pericyte | *NECAP2* | 3.641028242 | 0.001367536 | 0.004078605 | 1 |
| T2DM | pericyte | *LRP8* | 2.439763082 | 0.005525849 | 0.011832769 | 1 |
| T2DM | pericyte | *STEAP2* | 0.595105608 | 0.001735379 | 0.004885952 | 1 |
| T2DM | pericyte | *GATAD1* | 1.609288294 | 0.00021488 | 0.001135251 | 1 |
| T2DM | pericyte | *CLEC18A* | -3.052306242 | 9.05E-09 | 3.48E-06 | 5.69E-05 |
| T2DM | pericyte | *AASDH* | 1.821865324 | 0.004974094 | 0.010922853 | 1 |

| | | | | | | |
|---|---|---|---|---|---|---|
| T2DM | pericyte | *RNF111* | 2.195520396 | 5.76E-05 | 0.00062437 | 0.362073588 |
| T2DM | pericyte | *DYRK1A* | 2.604487416 | 0.000672863 | 0.002446108 | 1 |
| T2DM | pericyte | *CREB3L1* | 1.524403105 | 0.00199408 | 0.005385215 | 1 |
| T2DM | pericyte | *SLC38A10* | 2.145672814 | 0.000267219 | 0.0012953 | 1 |
| T2DM | pericyte | *WDR19* | 1.425027301 | 0.000581247 | 0.002202711 | 1 |
| T2DM | pericyte | *SLC37A3* | 3.781375845 | 6.08E-05 | 0.000626323 | 0.382057199 |
| T2DM | pericyte | *AP3S2* | 1.771568185 | 2.21E-12 | 1.55E-09 | 1.39E-08 |
| T2DM | pericyte | *SPPL3* | 1.925180152 | 0.004475474 | 0.010095912 | 1 |
| T2DM | pericyte | *FAM213B* | 1.005982149 | 0.007184488 | 0.014627227 | 1 |
| T2DM | pericyte | *PEX10* | 1.16437127 | 0.028233698 | 0.04690938 | 1 |
| T2DM | pericyte | *LDLRAP1* | 3.454015207 | 0.000193213 | 0.001062757 | 1 |
| T2DM | pericyte | *PAFAH2* | 1.028316627 | 0.030265343 | 0.049719941 | 1 |
| T2DM | pericyte | *TRIM63* | 1.479616521 | 3.37E-05 | 0.000599418 | 0.211846455 |
| T2DM | pericyte | *GALNT14* | -0.201956497 | 0.026763896 | 0.044912373 | 1 |
| T2DM | pericyte | *HPD* | 1.057010126 | 0.001046239 | 0.003385334 | 1 |
| T2DM | pericyte | *TPRG1L* | 2.232241454 | 0.009078669 | 0.017831175 | 1 |
| T2DM | pericyte | *XKR8* | 0.960316654 | 0.027008006 | 0.045231576 | 1 |
| T2DM | pericyte | *EYA3* | 2.054044852 | 0.001073439 | 0.003444976 | 1 |
| T2DM | pericyte | *ABHD3* | 1.580707706 | 9.74E-05 | 0.000728999 | 0.612359565 |
| T2DM | pericyte | *CLSTN2* | 0.878744951 | 0.00201305 | 0.005427121 | 1 |
| T2DM | pericyte | *COLEC12* | 1.761941543 | 0.000184223 | 0.001035729 | 1 |
| T2DM | pericyte | *RNF207* | 2.14740297 | 0.001007991 | 0.003309263 | 1 |
| T2DM | pericyte | *AUTS2* | 2.366877984 | 0.000302844 | 0.00139207 | 1 |
| T2DM | pericyte | *AHCYL2* | 2.851073566 | 7.66E-05 | 0.000658626 | 0.481455503 |
| T2DM | pericyte | *B4GALT5* | 2.162030686 | 0.00034121 | 0.001506451 | 1 |
| T2DM | pericyte | *CD1D* | -1.356691332 | 0.01078689 | 0.02075189 | 1 |
| T2DM | pericyte | *FAM86C1* | 3.826693458 | 0.004262201 | 0.009731948 | 1 |
| T2DM | pericyte | *CD1E* | 4.127099826 | 1.25E-05 | 0.000458996 | 0.078328312 |
| T2DM | pericyte | *PPP1R9A* | 0.927062935 | 0.002255018 | 0.005929444 | 1 |
| T2DM | pericyte | *ZFAND2B* | 1.769585988 | 0.008870326 | 0.01748565 | 1 |
| T2DM | pericyte | *GDPD5* | 1.379831042 | 0.001383509 | 0.00412038 | 1 |
| T2DM | pericyte | *DYNC1I1* | 3.612655031 | 0.001696828 | 0.004807262 | 1 |
| T2DM | pericyte | *TMED4* | 3.179278512 | 7.41E-05 | 0.000649163 | 0.465883052 |
| T2DM | pericyte | *ELK4* | 2.684803153 | 0.012223956 | 0.023064829 | 1 |
| T2DM | pericyte | *DUSP23* | 5.032325864 | 6.26E-05 | 0.000630549 | 0.393810245 |
| T2DM | pericyte | *F11R* | 1.065400876 | 0.011854207 | 0.022461544 | 1 |
| T2DM | pericyte | *SPATA2L* | 2.290173852 | 0.005376743 | 0.011600405 | 1 |
| T2DM | pericyte | *NIT1* | 1.08618207 | 0.014208574 | 0.026203961 | 1 |
| T2DM | pericyte | *B4GALT3* | 1.917689402 | 0.001532348 | 0.0044684 | 1 |
| T2DM | pericyte | *DMTN* | 1.019897755 | 0.00110911 | 0.00353241 | 1 |
| T2DM | pericyte | *FAM160B2* | 2.090552375 | 0.002515739 | 0.006455694 | 1 |
| T2DM | pericyte | *FCER1G* | 1.045308962 | 0.001704006 | 0.004823541 | 1 |
| T2DM | pericyte | *CCAR2* | 4.293809964 | 0.000330331 | 0.001474519 | 1 |
| T2DM | pericyte | *CDC42SE2* | 2.963442428 | 4.92E-05 | 0.000607238 | 0.309141474 |
| T2DM | pericyte | *RAPGEF6* | 2.686008791 | 0.001406337 | 0.004174524 | 1 |
| T2DM | pericyte | *EPB41* | 0.997105276 | 0.000298997 | 0.001384395 | 1 |
| T2DM | pericyte | *ALG8* | 3.37576515 | 0.001290153 | 0.003924699 | 1 |
| T2DM | pericyte | *IFNAR2* | 1.33068695 | 0.000631274 | 0.002333607 | 1 |
| T2DM | pericyte | *SV2A* | 3.557596639 | 0.003917491 | 0.00910509 | 1 |
| T2DM | pericyte | *RCAN1* | 2.279774822 | 0.003271691 | 0.007929498 | 1 |

| | | | | | | |
|---|---|---|---|---|---|---|
| T2DM | pericyte | *UBE2Z* | 2.325803342 | 0.003190237 | 0.007774038 | 1 |
| T2DM | pericyte | *CBR1* | 0.597394581 | 0.024416963 | 0.041522706 | 1 |
| T2DM | pericyte | *SIM2* | 1.236083923 | 0.013609002 | 0.025268693 | 1 |
| T2DM | pericyte | *ARHGAP27* | 2.229593835 | 0.000280199 | 0.001334556 | 1 |
| T2DM | pericyte | *PADI4* | 5.677167096 | 0.001007593 | 0.003309263 | 1 |
| T2DM | pericyte | *PSMD4* | 2.708111786 | 0.017108974 | 0.030776571 | 1 |
| T2DM | pericyte | *STARD9* | 2.282841163 | 0.00149584 | 0.00438431 | 1 |
| T2DM | pericyte | *THEM4* | 2.129067151 | 0.000203475 | 0.001095342 | 1 |
| T2DM | pericyte | *TCHH* | -2.432078958 | 0.008876413 | 0.01748565 | 1 |
| T2DM | pericyte | *MED8* | 1.31316748 | 0.001807803 | 0.005029053 | 1 |
| T2DM | pericyte | *RSPRY1* | 2.061701095 | 0.018187821 | 0.032411233 | 1 |
| T2DM | pericyte | *ACE* | 3.715090279 | 0.000472118 | 0.001903916 | 1 |
| T2DM | pericyte | *TEPP* | -3.674263757 | 0.012350015 | 0.023274743 | 1 |
| T2DM | pericyte | *EFCAB14* | 3.323770995 | 4.33E-05 | 0.000607238 | 0.272188224 |
| T2DM | pericyte | *ATP6V0D1* | 1.951836611 | 0.007877095 | 0.015852527 | 1 |
| T2DM | pericyte | *ZFYVE28* | 1.368010035 | 0.013367673 | 0.024878401 | 1 |
| T2DM | pericyte | *CCDC117* | 2.674304507 | 0.000469753 | 0.001897115 | 1 |
| T2DM | pericyte | *NPR2* | 4.570576134 | 2.16E-05 | 0.000540879 | 0.135760511 |
| T2DM | pericyte | *ZNF235* | 1.595174504 | 0.005422998 | 0.011664177 | 1 |
| T2DM | pericyte | *CALM3* | 3.44518698 | 0.000539375 | 0.002097605 | 1 |
| T2DM | pericyte | *DFFA* | 1.50122851 | 0.000203667 | 0.001095342 | 1 |
| T2DM | pericyte | *ATAD3B* | 2.256238474 | 0.004114001 | 0.009450028 | 1 |
| T2DM | pericyte | *SSU72* | 4.093829089 | 3.40E-05 | 0.000599418 | 0.213447275 |
| T2DM | pericyte | *NR2F6* | 2.856041048 | 0.000241969 | 0.001222377 | 1 |
| T2DM | pericyte | *CILP2* | 6.213352596 | 0.006976456 | 0.014268373 | 1 |
| T2DM | pericyte | *RSPH1* | -1.040860478 | 0.0203587 | 0.035650336 | 1 |
| T2DM | pericyte | *ZNF66* | 3.187175363 | 6.03E-05 | 0.00062437 | 0.378992636 |
| T2DM | pericyte | *FAM207A* | 1.310912541 | 0.002324953 | 0.006059128 | 1 |
| T2DM | pericyte | *LSS* | 0.829948638 | 0.017998434 | 0.032128381 | 1 |
| T2DM | pericyte | *VAV2* | 2.340033668 | 0.001358345 | 0.004058896 | 1 |
| T2DM | pericyte | *ZNF208* | 1.284385893 | 0.030417046 | 0.049943057 | 1 |
| T2DM | pericyte | *ADAMTS13* | 0.772486265 | 0.019233741 | 0.033967003 | 1 |
| T2DM | pericyte | *ZNF714* | 2.354206444 | 0.00084652 | 0.002911416 | 1 |
| T2DM | pericyte | *C19orf47* | 2.469124757 | 0.005285621 | 0.011443078 | 1 |
| T2DM | pericyte | *RDH13* | 3.078891477 | 0.004270413 | 0.009742592 | 1 |
| T2DM | pericyte | *MPZL3* | 1.528387805 | 0.002097272 | 0.005578327 | 1 |
| T2DM | pericyte | *PCSK7* | 2.07605491 | 0.00921438 | 0.018041201 | 1 |
| T2DM | pericyte | *S100A1* | 2.575037667 | 0.002355153 | 0.006118532 | 1 |
| T2DM | pericyte | *ZBTB7B* | 2.410979384 | 0.000156128 | 0.000945663 | 0.98157474 |
| T2DM | pericyte | *VPS11* | 1.300636062 | 0.019080264 | 0.033733864 | 1 |
| T2DM | pericyte | *FDPS* | 3.610459527 | 3.42E-07 | 5.38E-05 | 0.002151681 |
| T2DM | pericyte | *PMF1* | 1.806820816 | 0.006091403 | 0.012795406 | 1 |
| T2DM | pericyte | *NBEAL2* | 2.682332863 | 0.00186535 | 0.005136861 | 1 |
| T2DM | pericyte | *NACC1* | 1.984394974 | 0.001115488 | 0.003545538 | 1 |
| T2DM | pericyte | *LY6E* | 1.673862701 | 0.004925873 | 0.01083968 | 1 |
| T2DM | pericyte | *VPS28* | 2.3350258 | 4.66E-06 | 0.000304712 | 0.029269631 |
| T2DM | pericyte | *TONSL* | 3.220362046 | 0.000660099 | 0.002412817 | 1 |
| T2DM | pericyte | *MUM1* | 2.021363968 | 0.000188487 | 0.001046837 | 1 |
| T2DM | pericyte | *RPL8* | 3.525350568 | 0.000348363 | 0.001522932 | 1 |
| T2DM | pericyte | *FBXL13* | 2.703119996 | 0.002064619 | 0.005523514 | 1 |

| | | | | | | |
|---|---|---|---|---|---|---|
| T2DM | pericyte | *MFSD12* | 1.938693332 | 0.000709217 | 0.002545233 | 1 |
| T2DM | pericyte | *YDJC* | 1.819930248 | 0.004649098 | 0.010401736 | 1 |
| T2DM | pericyte | *DVL3* | 3.981253714 | 5.81E-05 | 0.00062437 | 0.365148242 |
| T2DM | pericyte | *AP2M1* | 3.708325007 | 6.10E-06 | 0.000342645 | 0.038376281 |
| T2DM | pericyte | *BDH1* | 2.81484178 | 0.00048821 | 0.001948566 | 1 |
| T2DM | pericyte | *DUSP14* | 1.899431212 | 0.000454609 | 0.00185713 | 1 |
| T2DM | pericyte | *PGAP3* | 3.854709184 | 0.027921879 | 0.046489633 | 1 |
| T2DM | pericyte | *IKZF3* | 1.054496251 | 0.002647608 | 0.006720028 | 1 |
| T2DM | pericyte | *ACOX1* | 1.713992053 | 0.002351758 | 0.00611569 | 1 |
| T2DM | pericyte | *PRPSAP1* | 1.737658566 | 0.003581267 | 0.008493183 | 1 |
| T2DM | pericyte | *CYGB* | -0.386922874 | 0.028644541 | 0.047485257 | 1 |
| T2DM | pericyte | *ZNF577* | 1.118042418 | 0.01241326 | 0.023365917 | 1 |
| T2DM | pericyte | *TMEM143* | 1.520996438 | 0.000273889 | 0.001320506 | 1 |
| T2DM | pericyte | *ALDH16A1* | 3.250205713 | 0.006324418 | 0.013192308 | 1 |
| T2DM | pericyte | *MPP3* | 1.491901767 | 0.00373886 | 0.008774249 | 1 |
| T2DM | pericyte | *ASB16* | 1.443572845 | 0.015984244 | 0.029069408 | 1 |
| T2DM | pericyte | *RACGAP1* | 0.824222359 | 0.0030811 | 0.007557891 | 1 |
| T2DM | pericyte | *LARP4* | 1.565261043 | 0.001197281 | 0.00373193 | 1 |
| T2DM | pericyte | *SPC24* | 2.29723641 | 0.016372387 | 0.0296552 | 1 |
| T2DM | pericyte | *LEMD2* | 2.303172031 | 0.001781797 | 0.004985385 | 1 |
| T2DM | pericyte | *ASGR2* | 3.101676837 | 0.002507638 | 0.006440164 | 1 |
| T2DM | pericyte | *SENP3* | 0.940800702 | 0.013996856 | 0.025866618 | 1 |
| T2DM | pericyte | *SNRNP25* | 0.987096981 | 0.015245291 | 0.027878751 | 1 |
| T2DM | pericyte | *TTC9C* | 2.381821661 | 0.004989522 | 0.010945263 | 1 |
| T2DM | pericyte | *NXF1* | 3.058843767 | 4.62E-05 | 0.000607238 | 0.290278853 |
| T2DM | pericyte | *TPCN2* | 2.022850277 | 0.002172609 | 0.005751238 | 1 |
| T2DM | pericyte | *CYP4A22* | 4.754318473 | 0.001674886 | 0.004766868 | 1 |
| T2DM | pericyte | *PDZK1IP1* | -3.843859871 | 0.021886257 | 0.037926929 | 1 |
| T2DM | pericyte | *COA7* | 1.755882304 | 0.001586247 | 0.004574649 | 1 |
| T2DM | pericyte | *SLC1A7* | 0.389420297 | 0.026116533 | 0.044055445 | 1 |
| T2DM | pericyte | *C1orf123* | -1.218291765 | 0.026572209 | 0.044704169 | 1 |
| T2DM | pericyte | *USP24* | 1.168368639 | 0.016520397 | 0.029880246 | 1 |
| T2DM | pericyte | *NOL9* | 3.285208134 | 0.000150103 | 0.000927922 | 0.943696618 |
| T2DM | pericyte | *ZSWIM5* | 1.988219758 | 0.001325903 | 0.003998058 | 1 |
| T2DM | pericyte | *RBP7* | 5.566355616 | 4.82E-05 | 0.000607238 | 0.302887077 |
| T2DM | pericyte | *SLC25A34* | 1.070044996 | 0.007272025 | 0.014771962 | 1 |
| T2DM | pericyte | *PDPN* | 0.61241674 | 0.017245105 | 0.030959444 | 1 |
| T2DM | pericyte | *LAPTM5* | 2.430758624 | 2.66E-05 | 0.000562456 | 0.167072096 |
| T2DM | pericyte | *PEF1* | 1.508503669 | 0.000824291 | 0.002855045 | 1 |
| T2DM | pericyte | *SYNC* | 1.43820046 | 0.026513924 | 0.044629997 | 1 |
| T2DM | pericyte | *RBBP4* | 2.605285475 | 0.000958609 | 0.003185399 | 1 |
| T2DM | pericyte | *TTLL10* | 2.16780012 | 0.003629753 | 0.00857582 | 1 |
| T2DM | pericyte | *NFIA* | 2.644337701 | 3.24E-05 | 0.000599418 | 0.20369155 |
| T2DM | pericyte | *TM2D1* | 3.511851241 | 6.85E-05 | 0.000635523 | 0.430407697 |
| T2DM | pericyte | *FUBP1* | 2.179358786 | 0.007489985 | 0.015151073 | 1 |
| T2DM | pericyte | *NEXN* | 3.796329586 | 7.98E-05 | 0.000667567 | 0.501460447 |
| T2DM | pericyte | *HFM1* | 1.077343102 | 0.002036605 | 0.005464848 | 1 |
| T2DM | pericyte | *KCNT2* | 3.091761085 | 1.47E-05 | 0.000476496 | 0.092175636 |
| T2DM | pericyte | *VCAM1* | 2.194685064 | 0.007976293 | 0.01600605 | 1 |
| T2DM | pericyte | *ARPC5* | 1.586996907 | 0.007431619 | 0.015042688 | 1 |

| | | | | | | |
|---|---|---|---|---|---|---|
| T2DM | pericyte | *ZNF496* | 4.264281329 | 0.000865227 | 0.002961177 | 1 |
| T2DM | pericyte | *IGSF8* | 2.17948602 | 0.000668518 | 0.00243792 | 1 |
| T2DM | pericyte | *VANGL2* | 2.082553826 | 0.006822461 | 0.01401268 | 1 |
| T2DM | pericyte | *LMX1A* | 1.181922293 | 1.46E-05 | 0.000476496 | 0.092024304 |
| T2DM | pericyte | *FLVCR1* | 2.4465592 | 0.004923607 | 0.010838487 | 1 |
| T2DM | pericyte | *SNED1* | 2.493786447 | 4.49E-05 | 0.000607238 | 0.282362623 |
| T2DM | pericyte | *C1orf115* | 1.179150848 | 0.016955467 | 0.030552887 | 1 |
| T2DM | pericyte | *ACP6* | 1.466268408 | 0.016088316 | 0.02919955 | 1 |
| T2DM | pericyte | *CNST* | 2.827304516 | 8.52E-05 | 0.000687243 | 0.535362371 |
| T2DM | pericyte | *PM20D1* | 1.328142814 | 0.005508521 | 0.011807889 | 1 |
| T2DM | pericyte | *HAAO* | 3.717878879 | 0.000183171 | 0.001035729 | 1 |
| T2DM | pericyte | *B3GALNT2* | 1.121963752 | 0.026762167 | 0.044912373 | 1 |
| T2DM | pericyte | *PUS10* | 3.027377726 | 3.79E-05 | 0.000607238 | 0.238102166 |
| T2DM | pericyte | *RFTN2* | 2.778503998 | 7.37E-05 | 0.000648165 | 0.463477473 |
| T2DM | pericyte | *DISC1* | 2.036441931 | 0.001305953 | 0.003958787 | 1 |
| T2DM | pericyte | *FAM84A* | 1.610023107 | 0.005476652 | 0.011763481 | 1 |
| T2DM | pericyte | *CLHC1* | 0.464209353 | 0.002262136 | 0.005943187 | 1 |
| T2DM | pericyte | *NUP35* | 3.476022482 | 0.000892162 | 0.003028629 | 1 |
| T2DM | pericyte | *FBXO41* | 1.045131266 | 0.01697292 | 0.030570668 | 1 |
| T2DM | pericyte | *H3F3A* | 2.128534211 | 0.000575567 | 0.002189103 | 1 |
| T2DM | pericyte | *SGPP2* | 1.285844412 | 0.008896097 | 0.017510884 | 1 |
| T2DM | pericyte | *HPGDS* | 0.766068674 | 0.00370506 | 0.008711864 | 1 |
| T2DM | pericyte | *PDLIM5* | 2.666780332 | 0.000641627 | 0.002356256 | 1 |
| T2DM | pericyte | *RPRD2* | 1.921460797 | 0.002731332 | 0.006895615 | 1 |
| T2DM | pericyte | *CTSS* | 0.591912025 | 0.007356382 | 0.014924032 | 1 |
| T2DM | pericyte | *PACRGL* | 2.071738243 | 0.000842076 | 0.002900896 | 1 |
| T2DM | pericyte | *C1QTNF7* | 1.675361048 | 0.000415698 | 0.001737696 | 1 |
| T2DM | pericyte | *ERCC3* | 1.5373693 | 0.000567331 | 0.002169792 | 1 |
| T2DM | pericyte | *RNF149* | 2.780861603 | 4.39E-05 | 0.000607238 | 0.276198738 |
| T2DM | pericyte | *DHX57* | 2.816139522 | 0.000211354 | 0.00112039 | 1 |
| T2DM | pericyte | *S100A9* | 0.831785098 | 0.007269996 | 0.014771962 | 1 |
| T2DM | pericyte | *S100A12* | -1.174636806 | 0.026008843 | 0.04389734 | 1 |
| T2DM | pericyte | *TDRD10* | 1.939829515 | 0.000387321 | 0.001650908 | 1 |
| T2DM | pericyte | *DCAF16* | 2.446373349 | 0.029423564 | 0.048578243 | 1 |
| T2DM | pericyte | *NIPAL1* | -0.145121944 | 0.027523143 | 0.045935228 | 1 |
| T2DM | pericyte | *ANTXR2* | 1.55918187 | 0.005508595 | 0.011807889 | 1 |
| T2DM | pericyte | *DCST2* | 1.481992756 | 0.002694847 | 0.006817909 | 1 |
| T2DM | pericyte | *LMOD3* | 6.045663746 | 7.78E-05 | 0.000663325 | 0.488909708 |
| T2DM | pericyte | *SLC15A2* | 1.174813109 | 0.012614274 | 0.023680484 | 1 |
| T2DM | pericyte | *LRRC58* | 4.54457817 | 8.15E-05 | 0.000675424 | 0.512647051 |
| T2DM | pericyte | *FSTL1* | 1.382306216 | 0.001759854 | 0.004937173 | 1 |
| T2DM | pericyte | *LMOD1* | -0.616462003 | 0.004802187 | 0.010636164 | 1 |
| T2DM | pericyte | *TMEM183A* | 1.370418557 | 0.001804011 | 0.005024423 | 1 |
| T2DM | pericyte | *ARPC2* | 3.125310646 | 0.000166265 | 0.00097875 | 1 |
| T2DM | pericyte | *CCT3* | 1.505809019 | 0.005847318 | 0.012365316 | 1 |
| T2DM | pericyte | *GLB1L* | 0.680954098 | 0.017380083 | 0.031166168 | 1 |
| T2DM | pericyte | *SERPINI1* | 2.014425416 | 0.001042073 | 0.003375644 | 1 |
| T2DM | pericyte | *AIM2* | 3.51954744 | 0.000130253 | 0.000857489 | 0.818901883 |
| T2DM | pericyte | *CD200R1* | 0.491584314 | 0.01705859 | 0.030694721 | 1 |
| T2DM | pericyte | *GTPBP8* | 3.378831434 | 6.53E-05 | 0.00063262 | 0.410568574 |

| | | | | | | |
|---|---|---|---|---|---|---|
| T2DM | pericyte | *SPICE1* | 2.213582296 | 0.000353851 | 0.001538493 | 1 |
| T2DM | pericyte | *CADPS* | 2.085343804 | 0.000946831 | 0.003156272 | 1 |
| T2DM | pericyte | *PPM1K* | 2.538720287 | 0.010045896 | 0.019487204 | 1 |
| T2DM | pericyte | *GMPS* | 4.277419946 | 4.68E-05 | 0.000607238 | 0.294045652 |
| T2DM | pericyte | *TIPARP* | 3.454670734 | 4.48E-05 | 0.000607238 | 0.281484161 |
| T2DM | pericyte | *RPP14* | 3.66065873 | 0.002203353 | 0.005811429 | 1 |
| T2DM | pericyte | *DNASE1L3* | 3.051393686 | 0.006062509 | 0.012743228 | 1 |
| T2DM | pericyte | *CRELD1* | 1.276393131 | 0.005189565 | 0.011285644 | 1 |
| T2DM | pericyte | *PCOLCE2* | 1.635010642 | 0.002180774 | 0.005761515 | 1 |
| T2DM | pericyte | *TTC14* | 1.459675017 | 0.002689788 | 0.006810591 | 1 |
| T2DM | pericyte | *CCDC158* | 0.909266042 | 0.001990464 | 0.005380071 | 1 |
| T2DM | pericyte | *GYG1* | 3.241593052 | 6.54E-05 | 0.00063262 | 0.410996968 |
| T2DM | pericyte | *HPS3* | 2.763855176 | 0.00093993 | 0.003136594 | 1 |
| T2DM | pericyte | *RYK* | 3.12421038 | 8.09E-05 | 0.000671315 | 0.508856503 |
| T2DM | pericyte | *PLB1* | 1.446417851 | 0.001787098 | 0.004988689 | 1 |
| T2DM | pericyte | *CDCP1* | 0.99774805 | 0.003622765 | 0.008562529 | 1 |
| T2DM | pericyte | *LZTFL1* | 3.093402233 | 0.000147022 | 0.000919731 | 0.924329554 |
| T2DM | pericyte | *FYCO1* | 3.425563245 | 0.000260833 | 0.001281362 | 1 |
| T2DM | pericyte | *LRRC2* | 0.91095793 | 0.005863423 | 0.012386875 | 1 |
| T2DM | pericyte | *DTX3L* | 3.276802636 | 4.89E-05 | 0.000607238 | 0.307734283 |
| T2DM | pericyte | *NMNAT3* | 3.496097824 | 7.83E-06 | 0.000364517 | 0.049209765 |
| T2DM | pericyte | *YEATS2* | 3.124948419 | 3.49E-05 | 0.000599418 | 0.219225918 |
| T2DM | pericyte | *MEAF6* | 4.612449062 | 0.000159804 | 0.000955936 | 1 |
| T2DM | pericyte | *DNALI1* | 3.42527194 | 0.005385324 | 0.011612049 | 1 |
| T2DM | pericyte | *RPN1* | 3.461374997 | 8.35E-05 | 0.000681373 | 0.524923892 |
| T2DM | pericyte | *RFC4* | 1.473744275 | 0.001199123 | 0.003733971 | 1 |
| T2DM | pericyte | *SFMBT1* | 3.001785138 | 0.000109848 | 0.000778599 | 0.690617236 |
| T2DM | pericyte | *GNL3* | 2.121524308 | 0.000696651 | 0.002512821 | 1 |
| T2DM | pericyte | *UVSSA* | 1.676582433 | 0.000438633 | 0.001807948 | 1 |
| T2DM | pericyte | *SLC51A* | 1.875515937 | 0.002024681 | 0.005437849 | 1 |
| T2DM | pericyte | *PIGX* | 1.859389034 | 0.005266683 | 0.011417805 | 1 |
| T2DM | pericyte | *METAP1* | 2.401407888 | 0.002488038 | 0.006402905 | 1 |
| T2DM | pericyte | *DNAJB14* | 3.110024183 | 8.69E-06 | 0.000381879 | 0.054608711 |
| T2DM | pericyte | *BDH2* | -0.873763557 | 0.013480049 | 0.025058861 | 1 |
| T2DM | pericyte | *ATRIP* | 1.880153036 | 0.013182586 | 0.024600451 | 1 |
| T2DM | pericyte | *SHISA5* | 2.789847467 | 0.000303349 | 0.001393101 | 1 |
| T2DM | pericyte | *RNF123* | 1.863496942 | 2.63E-08 | 6.90E-06 | 0.000165648 |
| T2DM | pericyte | *MST1R* | 1.117857995 | 0.001012984 | 0.003321888 | 1 |
| T2DM | pericyte | *RAD54L2* | 1.765604774 | 0.004754811 | 0.010566806 | 1 |
| T2DM | pericyte | *GRM2* | -1.193073451 | 0.020066921 | 0.035220752 | 1 |
| T2DM | pericyte | *POC1A* | 1.134736745 | 0.004280527 | 0.009757678 | 1 |
| T2DM | pericyte | *C4orf3* | 3.347148877 | 0.000333555 | 0.001485171 | 1 |
| T2DM | pericyte | *ANXA5* | 2.434281324 | 0.000368323 | 0.001587148 | 1 |
| T2DM | pericyte | *CEP44* | 4.180453221 | 7.84E-05 | 0.000665088 | 0.492830571 |
| T2DM | pericyte | *TMEM144* | 2.264524952 | 0.000152051 | 0.000937005 | 0.955945733 |
| T2DM | pericyte | *NPY1R* | 2.06385494 | 0.00014795 | 0.000921868 | 0.930164611 |
| T2DM | pericyte | *IL15* | 2.962245753 | 0.000152904 | 0.000938389 | 0.961309567 |
| T2DM | pericyte | *ARFIP1* | 2.296046397 | 0.001271855 | 0.003881628 | 1 |
| T2DM | pericyte | *OTUD4* | 3.529900748 | 4.05E-05 | 0.000607238 | 0.254360059 |
| T2DM | pericyte | *ITGA2* | 1.514988224 | 0.001438641 | 0.004243851 | 1 |

| | | | | | | |
|---|---|---|---|---|---|---|
| T2DM | pericyte | *LMBRD2* | 2.856315255 | 0.00014621 | 0.000917289 | 0.919223161 |
| T2DM | pericyte | *PGGT1B* | 3.443939714 | 9.42E-05 | 0.000717456 | 0.592080982 |
| T2DM | pericyte | *CMBL* | 1.489441816 | 0.001546345 | 0.004496702 | 1 |
| T2DM | pericyte | *C5orf63* | 2.324538543 | 0.001226599 | 0.003789498 | 1 |
| T2DM | pericyte | *PRRC1* | 3.964714183 | 0.00018315 | 0.001035729 | 1 |
| T2DM | pericyte | *NDUFS4* | 3.293283052 | 0.000340495 | 0.00150554 | 1 |
| T2DM | pericyte | *RHOBTB3* | 1.206043444 | 0.00411302 | 0.009450028 | 1 |
| T2DM | pericyte | *SERINC5* | 2.765734972 | 3.61E-05 | 0.00060096 | 0.227131784 |
| T2DM | pericyte | *ENPP6* | 1.948247972 | 0.015860955 | 0.028886971 | 1 |
| T2DM | pericyte | *CASP3* | 3.040848175 | 0.000219809 | 0.001151615 | 1 |
| T2DM | pericyte | *ERAP2* | 0.638863832 | 0.016575575 | 0.029954194 | 1 |
| T2DM | pericyte | *RICTOR* | 3.114511962 | 7.43E-05 | 0.00064946 | 0.466961661 |
| T2DM | pericyte | *ANKRA2* | 2.987832909 | 9.13E-05 | 0.000705517 | 0.574290599 |
| T2DM | pericyte | *UTP15* | 0.797220957 | 0.007833098 | 0.015774084 | 1 |
| T2DM | pericyte | *TLR3* | 4.44095759 | 9.99E-05 | 0.000739101 | 0.628235482 |
| T2DM | pericyte | *CCDC127* | 1.382696226 | 0.0027967 | 0.007005119 | 1 |
| T2DM | pericyte | *MB21D1* | 3.363270655 | 7.79E-05 | 0.000663325 | 0.489680015 |
| T2DM | pericyte | *TXLNB* | 1.126104992 | 0.006411829 | 0.013326007 | 1 |
| T2DM | pericyte | *PDSS2* | 3.043475231 | 4.67E-05 | 0.000607238 | 0.293653366 |
| T2DM | pericyte | *STXBP5* | 2.904892104 | 4.30E-05 | 0.000607238 | 0.270150432 |
| T2DM | pericyte | *PI16* | 2.334368339 | 6.60E-05 | 0.000632701 | 0.414864448 |
| T2DM | pericyte | *STK17A* | 1.961062312 | 0.002228272 | 0.005866476 | 1 |
| T2DM | pericyte | *GALNT10* | 2.407361258 | 0.002957018 | 0.007324969 | 1 |
| T2DM | pericyte | *RPS14* | 2.60369945 | 0.002176824 | 0.005755127 | 1 |
| T2DM | pericyte | *SLU7* | 1.904782183 | 0.001313065 | 0.003976511 | 1 |
| T2DM | pericyte | *CAMLG* | 2.687560949 | 2.39E-06 | 0.000208914 | 0.015041843 |
| T2DM | pericyte | *USP49* | 2.848624026 | 7.32E-05 | 0.000646906 | 0.460419843 |
| T2DM | pericyte | *ZNF704* | 2.016126961 | 5.60E-05 | 0.00062437 | 0.351931938 |
| T2DM | pericyte | *FNDC1* | 0.905595938 | 0.005273424 | 0.01142454 | 1 |
| T2DM | pericyte | *BRI3* | 2.528713592 | 0.000252753 | 0.001256173 | 1 |
| T2DM | pericyte | *LMTK2* | 3.475539706 | 4.53E-05 | 0.000607238 | 0.284901301 |
| T2DM | pericyte | *PEX2* | 3.268933585 | 0.002282051 | 0.005978024 | 1 |
| T2DM | pericyte | *TNFRSF11B* | 1.048398953 | 0.00324698 | 0.007889627 | 1 |
| T2DM | pericyte | *SUN1* | 2.688997014 | 1.57E-06 | 0.000159839 | 0.009864212 |
| T2DM | pericyte | *OXR1* | 2.767008866 | 3.29E-05 | 0.000599418 | 0.207037662 |
| T2DM | pericyte | *INTS1* | 3.475866547 | 0.00011154 | 0.00078265 | 0.701254653 |
| T2DM | pericyte | *SLC4A2* | 5.621201435 | 0.000306034 | 0.001401337 | 1 |
| T2DM | pericyte | *FASTK* | 3.54190838 | 0.000680944 | 0.002468911 | 1 |
| T2DM | pericyte | *ALDH7A1* | 1.12614186 | 0.004447384 | 0.010061426 | 1 |
| T2DM | pericyte | *COX6C* | 3.758790875 | 3.96E-05 | 0.000607238 | 0.249261949 |
| T2DM | pericyte | *INTS8* | 3.056128036 | 1.38E-05 | 0.00046738 | 0.086486518 |
| T2DM | pericyte | *FREM1* | 1.3326196 | 0.001535088 | 0.004474316 | 1 |
| T2DM | pericyte | *FAM219A* | 3.207652226 | 0.002168603 | 0.005743054 | 1 |
| T2DM | pericyte | *TMEM65* | 2.948486294 | 0.000562652 | 0.002159249 | 1 |
| T2DM | pericyte | *CCDC171* | 2.117598121 | 0.021850389 | 0.037885658 | 1 |
| T2DM | pericyte | *SYK* | 1.366125781 | 0.001106725 | 0.003528386 | 1 |
| T2DM | pericyte | *METTL2B* | 0.423448347 | 0.005822213 | 0.012320515 | 1 |
| T2DM | pericyte | *TMC1* | 5.4641812 | 1.71E-05 | 0.00051051 | 0.107207084 |
| T2DM | pericyte | *HGSNAT* | 2.318904426 | 0.001695025 | 0.004806775 | 1 |
| T2DM | pericyte | *C9orf64* | 1.02180421 | 0.001559142 | 0.004518363 | 1 |

| | | | | | | |
|---|---|---|---|---|---|---|
| T2DM | pericyte | *HNRNPK* | 3.222861077 | 0.001032431 | 0.00335812 | 1 |
| T2DM | pericyte | *SVEP1* | 1.632199828 | 0.000100292 | 0.000740845 | 0.630538871 |
| T2DM | pericyte | *ZNF367* | 1.685147045 | 0.002285733 | 0.005982683 | 1 |
| T2DM | pericyte | *AQP3* | 1.256443804 | 0.015501665 | 0.028298191 | 1 |
| T2DM | pericyte | *TRMT10B* | 3.729484717 | 0.00025385 | 0.001257649 | 1 |
| T2DM | pericyte | *VCP* | 1.96772348 | 0.002859325 | 0.007128517 | 1 |
| T2DM | pericyte | *PIGO* | 3.336334986 | 0.006835732 | 0.014035351 | 1 |
| T2DM | pericyte | *WRN* | 2.59485498 | 5.65E-05 | 0.00062437 | 0.354908041 |
| T2DM | pericyte | *MARCH8* | 2.711346593 | 5.43E-05 | 0.00062437 | 0.341506871 |
| T2DM | pericyte | *CFL2* | 2.833809462 | 0.002772854 | 0.006962033 | 1 |
| T2DM | pericyte | *ZCCHC24* | 3.228683658 | 0.001153967 | 0.003629309 | 1 |
| T2DM | pericyte | *CRYL1* | 1.136554221 | 0.026160775 | 0.044118238 | 1 |
| T2DM | pericyte | *HEPACAM* | 0.268505174 | 0.009597027 | 0.018703195 | 1 |
| T2DM | pericyte | *LRR1* | 2.871252663 | 0.00541266 | 0.011646864 | 1 |
| T2DM | pericyte | *RPL36AL* | 1.532358156 | 0.006480962 | 0.013447462 | 1 |
| T2DM | pericyte | *KLHDC2* | 2.627614273 | 0.000475795 | 0.00190895 | 1 |
| T2DM | pericyte | *NOXRED1* | 1.087715665 | 0.003547019 | 0.008443811 | 1 |
| T2DM | pericyte | *TAF3* | 3.082650971 | 0.000275299 | 0.001323244 | 1 |
| T2DM | pericyte | *VDAC2* | 4.411541648 | 1.45E-05 | 0.000476496 | 0.091162941 |
| T2DM | pericyte | *SLC18A2* | 1.087002334 | 0.001051547 | 0.003395518 | 1 |
| T2DM | pericyte | *PDZD8* | 2.067174351 | 1.05E-05 | 0.000413298 | 0.066112985 |
| T2DM | pericyte | *QSOX2* | 5.636901807 | 0.002513355 | 0.00645221 | 1 |
| T2DM | pericyte | *SDCCAG3* | 5.224357758 | 5.56E-05 | 0.00062437 | 0.34972968 |
| T2DM | pericyte | *TSC1* | 2.139452315 | 0.011485408 | 0.021927956 | 1 |
| T2DM | pericyte | *ZMYND19* | 2.251762925 | 0.001789326 | 0.004988689 | 1 |
| T2DM | pericyte | *STK32C* | 1.193193341 | 0.002393704 | 0.006200749 | 1 |
| T2DM | pericyte | *NSMF* | 3.767926579 | 0.000146866 | 0.000919668 | 0.923346243 |
| T2DM | pericyte | *CASP7* | 1.00306405 | 0.014089361 | 0.026006992 | 1 |
| T2DM | pericyte | *METTL3* | 3.993329521 | 5.12E-05 | 0.000615408 | 0.321858321 |
| T2DM | pericyte | *UBTD1* | 2.896401357 | 0.000109148 | 0.000776889 | 0.686212508 |
| T2DM | pericyte | *SLC39A13* | 6.74871397 | 8.67E-06 | 0.000381879 | 0.054516923 |
| T2DM | pericyte | *IFI27L1* | 1.765169366 | 0.00061742 | 0.002296876 | 1 |
| T2DM | pericyte | *PTER* | 2.687921127 | 0.000868306 | 0.002970075 | 1 |
| T2DM | pericyte | *C1QL3* | 0.692763978 | 0.004742861 | 0.010547706 | 1 |
| T2DM | pericyte | *ABTB2* | 2.806133333 | 0.000127321 | 0.000844709 | 0.800465641 |
| T2DM | pericyte | *LIPC* | 1.577590062 | 0.001888189 | 0.005174823 | 1 |
| T2DM | pericyte | *TCP11L2* | 2.186220722 | 0.000157001 | 0.000947957 | 0.987064687 |
| T2DM | pericyte | *GPT2* | 0.552849663 | 0.013776681 | 0.025542316 | 1 |
| T2DM | pericyte | *IKBIP* | 2.009211996 | 0.003321303 | 0.008031166 | 1 |
| T2DM | pericyte | *HIF1AN* | 3.249738323 | 0.000136372 | 0.000878628 | 0.857373134 |
| T2DM | pericyte | *NDUFB8* | 4.314299432 | 0.002200069 | 0.005806816 | 1 |
| T2DM | pericyte | *AVPR1A* | -0.098178301 | 0.028502662 | 0.047281329 | 1 |
| T2DM | pericyte | *BTRC* | 2.762314087 | 0.000279788 | 0.001333604 | 1 |
| T2DM | pericyte | *LARP6* | 1.797272351 | 0.001275037 | 0.003889452 | 1 |
| T2DM | pericyte | *PCBD1* | -0.939465902 | 0.000367673 | 0.001585434 | 1 |
| T2DM | pericyte | *FAM227B* | 1.955945673 | 0.000153099 | 0.000938389 | 0.962532515 |
| T2DM | pericyte | *STXBP4* | 1.612474736 | 0.017294027 | 0.031038409 | 1 |
| T2DM | pericyte | *C2* | 3.515104387 | 5.09E-11 | 2.91E-08 | 3.20E-07 |
| T2DM | pericyte | *APBB1* | 3.146069871 | 0.002273537 | 0.005965663 | 1 |
| T2DM | pericyte | *ILK* | 3.988148985 | 1.61E-05 | 0.00049967 | 0.101531516 |

| | | | | | | |
|---|---|---|---|---|---|---|
| T2DM | pericyte | *DCHS1* | 1.809498532 | 0.000898985 | 0.003037034 | 1 |
| T2DM | pericyte | *NETO1* | 1.657212459 | 0.00208793 | 0.00556221 | 1 |
| T2DM | pericyte | *USP54* | 2.142940887 | 0.000141162 | 0.000899173 | 0.887483264 |
| T2DM | pericyte | *C11orf74* | 1.683696369 | 0.00318547 | 0.007768443 | 1 |
| T2DM | pericyte | *RIC3* | 1.727837476 | 0.002876675 | 0.0071598 | 1 |
| T2DM | pericyte | *TRIM66* | 0.282176979 | 0.014750754 | 0.027092605 | 1 |
| T2DM | pericyte | *RPL27A* | 3.302027259 | 0.000624945 | 0.002316644 | 1 |
| T2DM | pericyte | *AKIP1* | 5.750668338 | 4.72E-05 | 0.000607238 | 0.296555911 |
| T2DM | pericyte | *C16orf46* | 8.415545611 | 0.017789423 | 0.03181852 | 1 |
| T2DM | pericyte | *TMEM41B* | 3.31234179 | 0.001737328 | 0.004889249 | 1 |
| T2DM | pericyte | *TMX3* | 2.550096146 | 0.000170359 | 0.000994477 | 1 |
| T2DM | pericyte | *CLEC4E* | 0.994006542 | 0.001349157 | 0.004041043 | 1 |
| T2DM | pericyte | *ZNF3* | 1.679883437 | 0.000906547 | 0.00304947 | 1 |
| T2DM | pericyte | *ZSCAN21* | 0.669640308 | 0.018683605 | 0.033125726 | 1 |
| T2DM | pericyte | *BEAN1* | 1.290018203 | 0.000963186 | 0.003197926 | 1 |
| T2DM | pericyte | *SEC11C* | 1.837221932 | 0.000891739 | 0.003028629 | 1 |
| T2DM | pericyte | *SPDYE6* | 1.311862191 | 0.005189467 | 0.011285644 | 1 |
| T2DM | pericyte | *ATF7IP2* | 1.636502004 | 0.004938948 | 0.010864649 | 1 |
| T2DM | pericyte | *ZNF592* | 1.814672136 | 0.004454432 | 0.010066505 | 1 |
| T2DM | pericyte | *AP1G1* | 3.059946757 | 0.000202615 | 0.001093325 | 1 |
| T2DM | pericyte | *SLFN5* | 1.993209329 | 0.001243571 | 0.003828762 | 1 |
| T2DM | pericyte | *C16orf45* | 3.109283309 | 0.000109427 | 0.000777368 | 0.687970329 |
| T2DM | pericyte | *FAM111A* | 1.916875846 | 0.001652587 | 0.004718355 | 1 |
| T2DM | pericyte | *ANKDD1A* | 1.282725063 | 0.006378543 | 0.01327438 | 1 |
| T2DM | pericyte | *STX3* | 2.018812654 | 0.001806352 | 0.00502724 | 1 |
| T2DM | pericyte | *MTMR10* | 2.298826998 | 7.56E-05 | 0.000656494 | 0.47530152 |
| T2DM | pericyte | *NYAP1* | -0.639400056 | 0.02547151 | 0.043146538 | 1 |
| T2DM | pericyte | *TSC22D4* | 4.260638868 | 1.26E-05 | 0.000458996 | 0.079363034 |
| T2DM | pericyte | *DIS3L* | 2.144281986 | 0.000569176 | 0.002174004 | 1 |
| T2DM | pericyte | *CCNDBP1* | 4.449126799 | 3.74E-05 | 0.000607238 | 0.235163514 |
| T2DM | pericyte | *SMAD3* | 2.354816 | 0.000459983 | 0.001874216 | 1 |
| T2DM | pericyte | *RCCD1* | 2.61449878 | 0.007609761 | 0.01535877 | 1 |
| T2DM | pericyte | *AKTIP* | 4.964145454 | 1.52E-06 | 0.000159759 | 0.009585549 |
| T2DM | pericyte | *EVA1C* | 1.543090878 | 0.000480714 | 0.00192612 | 1 |
| T2DM | pericyte | *NUDT21* | 2.769134118 | 0.000234327 | 0.001198741 | 1 |
| T2DM | pericyte | *TEF* | 1.167465699 | 0.001572241 | 0.004544679 | 1 |
| T2DM | pericyte | *PBX3* | 1.875703075 | 0.004516664 | 0.010159665 | 1 |
| T2DM | pericyte | *SNRPD1* | 2.878235767 | 0.000131085 | 0.000860073 | 0.824131142 |
| T2DM | pericyte | *SAMD14* | 1.507166199 | 0.021649611 | 0.037584749 | 1 |
| T2DM | pericyte | *URM1* | 1.278817731 | 0.023137543 | 0.039690514 | 1 |
| T2DM | pericyte | *ISLR2* | 0.835693985 | 0.025220881 | 0.042751059 | 1 |
| T2DM | pericyte | *COQ7* | 2.932224371 | 0.000153139 | 0.000938389 | 0.962787621 |
| T2DM | pericyte | *RNF214* | 3.806652616 | 6.46E-05 | 0.000632025 | 0.406391897 |
| T2DM | pericyte | *RBFOX3* | 1.56415364 | 0.001933421 | 0.005271214 | 1 |
| T2DM | pericyte | *MYO5B* | 0.601179682 | 0.019630979 | 0.034571418 | 1 |
| T2DM | pericyte | *FN3K* | 1.565650685 | 0.00093412 | 0.003123838 | 1 |
| T2DM | pericyte | *ZNF23* | 1.266884877 | 0.021045648 | 0.036702909 | 1 |
| T2DM | pericyte | *RAB8A* | 2.443992159 | 0.000474248 | 0.001905624 | 1 |
| T2DM | pericyte | *GATAD2A* | 4.50872325 | 4.22E-07 | 6.03E-05 | 0.002653024 |
| T2DM | pericyte | *ZNF641* | 1.586756139 | 6.33E-06 | 0.000343901 | 0.039770238 |

| | | | | | | |
|---|---|---|---|---|---|---|
| T2DM | pericyte | *CACNB3* | 0.847282596 | 0.013646276 | 0.025326807 | 1 |
| T2DM | pericyte | *TP53I13* | 1.60252665 | 0.000634345 | 0.002337355 | 1 |
| T2DM | pericyte | *ZNF701* | 1.715876617 | 0.008144553 | 0.016260656 | 1 |
| T2DM | pericyte | *TTYH1* | 3.351298006 | 0.007406882 | 0.015011949 | 1 |
| T2DM | pericyte | *LENG8* | 2.062803933 | 0.00514081 | 0.011222317 | 1 |
| T2DM | pericyte | *ZNF283* | 1.848444303 | 0.009162577 | 0.017962307 | 1 |
| T2DM | pericyte | *PSCA* | 1.725586732 | 4.79E-05 | 0.000607238 | 0.301047952 |
| T2DM | pericyte | *ZNF444* | 2.113599877 | 0.000569082 | 0.002174004 | 1 |
| T2DM | pericyte | *GLOD4* | 2.61398546 | 0.000252332 | 0.001255074 | 1 |
| T2DM | pericyte | *SERPINF2* | 1.098579743 | 0.00685928 | 0.014079103 | 1 |
| T2DM | pericyte | *CYB5D2* | 4.333600925 | 9.71E-05 | 0.000727674 | 0.610518315 |
| T2DM | pericyte | *C19orf48* | 0.64252314 | 0.014535988 | 0.026729381 | 1 |
| T2DM | pericyte | *ZNF83* | 2.059775092 | 0.002770927 | 0.006959976 | 1 |
| T2DM | pericyte | *CD320* | 6.293601404 | 7.04E-05 | 0.000639553 | 0.442340764 |
| T2DM | pericyte | *SPRYD3* | 4.25604702 | 0.00818434 | 0.016329718 | 1 |
| T2DM | pericyte | *SOAT2* | 0.816570129 | 0.025212774 | 0.042748843 | 1 |
| T2DM | pericyte | *ZNF558* | 0.95190992 | 0.008912311 | 0.017537308 | 1 |
| T2DM | pericyte | *NDUFV1* | 3.117294916 | 0.000796192 | 0.002774755 | 1 |
| T2DM | pericyte | *CDK2AP2* | 2.424788185 | 0.001993093 | 0.005384862 | 1 |
| T2DM | pericyte | *PRDX2* | 4.32571458 | 9.78E-05 | 0.000730927 | 0.614710004 |
| T2DM | pericyte | *MIS12* | 2.349878004 | 0.000615297 | 0.002292225 | 1 |
| T2DM | pericyte | *SRP68* | 2.252659958 | 0.005601126 | 0.011945142 | 1 |
| T2DM | pericyte | *TMC8* | 1.669819018 | 3.22E-05 | 0.000599418 | 0.202252254 |
| T2DM | pericyte | *TMEM68* | 2.152476074 | 0.005173308 | 0.011261976 | 1 |
| T2DM | pericyte | *CYP7A1* | 1.772470428 | 0.019657924 | 0.034609176 | 1 |
| T2DM | pericyte | *GSDMA* | 1.608192736 | 0.007015768 | 0.014339445 | 1 |
| T2DM | pericyte | *KRT24* | -0.552616832 | 0.007548434 | 0.015244781 | 1 |
| T2DM | pericyte | *MLST8* | 1.033462597 | 0.006645745 | 0.013734976 | 1 |
| T2DM | pericyte | *ABCA3* | 2.036402528 | 0.000880182 | 0.002999297 | 1 |
| T2DM | pericyte | *KCTD5* | 1.644888732 | 0.000734226 | 0.002610904 | 1 |
| T2DM | pericyte | *SDHAF2* | 5.753037327 | 6.43E-05 | 0.000632025 | 0.404054808 |
| T2DM | pericyte | *DDB1* | 4.010273075 | 0.000224658 | 0.00116826 | 1 |
| T2DM | pericyte | *VPS37C* | 1.922368883 | 0.000862311 | 0.002952804 | 1 |
| T2DM | pericyte | *VWCE* | 0.918463116 | 0.010091917 | 0.019558534 | 1 |
| T2DM | pericyte | *HRASLS5* | 0.304878008 | 0.016678441 | 0.030088195 | 1 |
| T2DM | pericyte | *TRANK1* | 2.506709341 | 0.001795901 | 0.005004801 | 1 |
| T2DM | pericyte | *TTC21A* | 2.883854987 | 0.000166838 | 0.000981205 | 1 |
| T2DM | pericyte | *RPSA* | 1.235629813 | 0.006246034 | 0.013041785 | 1 |
| T2DM | pericyte | *LTBP3* | 2.94297998 | 2.94E-08 | 7.39E-06 | 0.000184805 |
| T2DM | pericyte | *SCARA5* | 0.972399016 | 0.006022451 | 0.012675979 | 1 |
| T2DM | pericyte | *COPS6* | 1.926873979 | 0.018175614 | 0.032398663 | 1 |
| T2DM | pericyte | *NUDT16L1* | 2.028793741 | 0.003600643 | 0.008526835 | 1 |
| T2DM | pericyte | *RNF187* | 1.817912821 | 0.011223283 | 0.021486231 | 1 |
| T2DM | pericyte | *ZCCHC4* | 2.594592762 | 7.24E-05 | 0.00064687 | 0.455396725 |
| T2DM | pericyte | *GLYCTK* | 3.307674847 | 0.000323652 | 0.001453427 | 1 |
| T2DM | pericyte | *UBTD2* | 3.086306624 | 1.35E-05 | 0.000465154 | 0.085123137 |
| T2DM | pericyte | *POLR2J3* | 1.167969892 | 9.43E-05 | 0.000717456 | 0.593022385 |
| T2DM | pericyte | *COA6* | 5.741433428 | 4.42E-05 | 0.000607238 | 0.277825002 |
| T2DM | pericyte | *PXK* | 3.116900761 | 3.44E-08 | 8.33E-06 | 0.000216582 |
| T2DM | pericyte | *TAP1* | -2.372149001 | 0.000958253 | 0.003185399 | 1 |

| | | | | | | |
|---|---|---|---|---|---|---|
| T2DM | pericyte | *ING5* | 2.895193984 | 7.62E-05 | 0.000658 | 0.479023818 |
| T2DM | pericyte | *RFWD3* | 2.348536204 | 6.56E-05 | 0.000632701 | 0.412669425 |
| T2DM | pericyte | *RHOH* | 0.655255589 | 0.002994723 | 0.00739506 | 1 |
| T2DM | pericyte | *REEP4* | 2.58944809 | 0.002817041 | 0.007050453 | 1 |
| T2DM | pericyte | *BMP1* | 1.345389302 | 0.004689912 | 0.010459551 | 1 |
| T2DM | pericyte | *ATXN2L* | 2.831531085 | 0.000991446 | 0.003275472 | 1 |
| T2DM | pericyte | *FNTA* | 3.249897989 | 4.28E-05 | 0.000607238 | 0.26900902 |
| T2DM | pericyte | *TRAPPC11* | 3.355198768 | 8.38E-05 | 0.000681373 | 0.526805997 |
| T2DM | pericyte | *SLC20A2* | 2.866268239 | 2.24E-05 | 0.000547486 | 0.140704015 |
| T2DM | pericyte | *NBPF9* | 2.63165305 | 0.000398935 | 0.001682165 | 1 |
| T2DM | pericyte | *WFDC13* | 6.968261701 | 0.001411206 | 0.004182358 | 1 |
| T2DM | pericyte | *ZNF30* | 1.289795317 | 0.016950807 | 0.030552887 | 1 |
| T2DM | pericyte | *FAM84B* | 0.70551992 | 0.016432291 | 0.029746564 | 1 |
| T2DM | pericyte | *SLC16A4* | 1.459230519 | 0.000207095 | 0.001105269 | 1 |
| T2DM | pericyte | *TMEM208* | 1.157267276 | 0.014410158 | 0.026521272 | 1 |
| T2DM | pericyte | *DNAJC21* | 3.797015373 | 9.97E-05 | 0.000738177 | 0.626712309 |
| T2DM | pericyte | *GSTM4* | 1.077292453 | 0.001872482 | 0.005147484 | 1 |
| T2DM | pericyte | *ABHD15* | 0.856032414 | 0.000567383 | 0.002169792 | 1 |
| T2DM | pericyte | *SNTB2* | 3.307777852 | 4.09E-05 | 0.000607238 | 0.25724794 |
| T2DM | pericyte | *STX18* | 3.378048528 | 0.000539498 | 0.002097605 | 1 |
| T2DM | pericyte | *ZBTB49* | 1.489501925 | 0.001126649 | 0.003570184 | 1 |
| T2DM | pericyte | *INPP5D* | 2.757555611 | 5.88E-05 | 0.00062437 | 0.36969555 |
| T2DM | pericyte | *LETM1* | 2.318153456 | 0.000307336 | 0.001406275 | 1 |
| T2DM | pericyte | *CEP120* | 2.625217216 | 0.000778437 | 0.002737157 | 1 |
| T2DM | pericyte | *STXBP6* | 3.09955497 | 0.000243173 | 0.001224801 | 1 |
| T2DM | pericyte | *MFF* | 2.142232733 | 0.002260212 | 0.005940617 | 1 |
| T2DM | pericyte | *FEM1B* | 3.334270214 | 5.76E-06 | 0.000331374 | 0.036241994 |
| T2DM | pericyte | *IRS1* | -1.087166435 | 7.94E-15 | 1.66E-11 | 4.99E-11 |
| T2DM | pericyte | *UPF3A* | 0.971953809 | 0.027018184 | 0.04523657 | 1 |
| T2DM | pericyte | *C8orf46* | 0.191134998 | 0.015401218 | 0.028139337 | 1 |
| T2DM | pericyte | *HSPBAP1* | 1.340411372 | 0.003245946 | 0.007889627 | 1 |
| T2DM | pericyte | *ATF5* | 1.095123239 | 0.00417701 | 0.009563316 | 1 |
| T2DM | pericyte | *CPT1C* | 1.412123505 | 0.019693582 | 0.034652547 | 1 |
| T2DM | pericyte | *PCSK9* | -2.525515361 | 0.017127322 | 0.030791957 | 1 |
| T2DM | pericyte | *XPO6* | 3.754905055 | 1.05E-05 | 0.000413298 | 0.06625894 |
| T2DM | pericyte | *RAB24* | -1.092461259 | 0.013276799 | 0.024732218 | 1 |
| T2DM | pericyte | *GPRIN1* | 3.858149609 | 0.029497439 | 0.048674645 | 1 |
| T2DM | pericyte | *PGM2* | 1.990935412 | 0.000535524 | 0.002086022 | 1 |
| T2DM | pericyte | *SLC33A1* | 2.644466317 | 0.000104447 | 0.000757393 | 0.656659595 |
| T2DM | pericyte | *PTAFR* | 1.570501953 | 0.006601755 | 0.013666525 | 1 |
| T2DM | pericyte | *NPR1* | 4.566245419 | 3.07E-05 | 0.000599418 | 0.192749672 |
| T2DM | pericyte | *CLIC4* | 2.665094727 | 0.001140779 | 0.00360406 | 1 |
| T2DM | pericyte | *CCDC8* | 1.747887567 | 0.009538325 | 0.018606097 | 1 |
| T2DM | pericyte | *DTWD2* | 1.819566922 | 0.000291339 | 0.00136182 | 1 |
| T2DM | pericyte | *CLIC3* | 1.418688304 | 0.012167809 | 0.022965781 | 1 |
| T2DM | pericyte | *DFFB* | 2.4340186 | 0.001358332 | 0.004058896 | 1 |
| T2DM | pericyte | *FAM103A1* | -0.722184026 | 0.017013002 | 0.030621455 | 1 |
| T2DM | pericyte | *LUZP1* | 2.292733731 | 0.003668721 | 0.008644092 | 1 |
| T2DM | pericyte | *HEXDC* | 2.870391374 | 0.000100398 | 0.000740845 | 0.631199793 |
| T2DM | pericyte | *FASN* | 1.000303422 | 0.002078757 | 0.00554953 | 1 |

| | | | | | | |
|---|---|---|---|---|---|---|
| T2DM | pericyte | *DCXR* | 1.131818508 | 0.004160846 | 0.009533251 | 1 |
| T2DM | pericyte | *LIMS1* | 1.30972026 | 0.005866813 | 0.012389874 | 1 |
| T2DM | pericyte | *UGP2* | 1.392786355 | 0.000182245 | 0.001034031 | 1 |
| T2DM | pericyte | *CSGALNACT2* | 3.457375172 | 3.42E-05 | 0.000599418 | 0.21515895 |
| T2DM | pericyte | *AVEN* | 2.672844399 | 6.39E-05 | 0.000632025 | 0.401725265 |
| T2DM | pericyte | *GUSB* | 3.702251635 | 5.71E-05 | 0.00062437 | 0.359168966 |
| T2DM | pericyte | *BRD3* | 3.548195894 | 0.002984046 | 0.007374488 | 1 |
| T2DM | pericyte | *ZFPM2* | 2.307371668 | 5.67E-05 | 0.00062437 | 0.356439341 |
| T2DM | pericyte | *ZNF747* | 1.453552142 | 0.002518012 | 0.006458891 | 1 |
| T2DM | pericyte | *MAP3K2* | 3.048999075 | 0.000176205 | 0.001018057 | 1 |
| T2DM | pericyte | *MYO7B* | 0.902326343 | 0.020657678 | 0.036076339 | 1 |
| T2DM | pericyte | *NIPA1* | 4.256539836 | 0.000157365 | 0.000948564 | 0.989352037 |
| T2DM | pericyte | *RNF150* | 1.351766152 | 0.000935483 | 0.003125752 | 1 |
| T2DM | pericyte | *PDCD6IP* | 2.0042146 | 0.001037874 | 0.003365197 | 1 |
| T2DM | pericyte | *GLB1* | 1.521414792 | 0.002405072 | 0.006222505 | 1 |
| T2DM | pericyte | *FAXDC2* | 1.931587277 | 0.00031727 | 0.001432958 | 1 |
| T2DM | pericyte | *CRTAP* | 2.645623831 | 0.000184717 | 0.001035963 | 1 |
| T2DM | pericyte | *STX8* | 2.601158397 | 7.98E-05 | 0.000667567 | 0.50161792 |
| T2DM | pericyte | *NFRKB* | 1.620947345 | 0.000404557 | 0.001701304 | 1 |
| T2DM | pericyte | *SP7* | -1.520785293 | 0.00867484 | 0.017165966 | 1 |
| T2DM | pericyte | *DCLK2* | 0.966262695 | 0.02253992 | 0.038877497 | 1 |
| T2DM | pericyte | *GPRC5C* | 1.46614501 | 0.01745159 | 0.031285471 | 1 |
| T2DM | pericyte | *MGMT* | 1.431977704 | 0.011874808 | 0.022489668 | 1 |
| T2DM | pericyte | *METTL7B* | 3.179572613 | 0.003964739 | 0.009194509 | 1 |
| T2DM | pericyte | *NFXL1* | 3.240564558 | 0.002629246 | 0.006686921 | 1 |
| T2DM | pericyte | *CD14* | 1.214655246 | 0.00828837 | 0.016505854 | 1 |
| T2DM | pericyte | *SERPINB9* | 2.158748703 | 0.00175533 | 0.004926679 | 1 |
| T2DM | pericyte | *IRX2* | 0.933727327 | 0.008508917 | 0.016875571 | 1 |
| T2DM | pericyte | *STAT2* | 2.778569556 | 2.57E-05 | 0.000562456 | 0.161839115 |
| T2DM | pericyte | *NUDCD2* | 1.677203991 | 0.000719455 | 0.002572932 | 1 |
| T2DM | pericyte | *HSPA4* | 3.372580397 | 5.22E-05 | 0.000619691 | 0.328472004 |
| T2DM | pericyte | *SLC26A5* | 3.168206392 | 7.27E-06 | 0.000354405 | 0.045718305 |
| T2DM | pericyte | *GTSF1* | 2.892803268 | 0.004445231 | 0.010060176 | 1 |
| T2DM | pericyte | *ARMC10* | 1.74034837 | 0.007420509 | 0.015029877 | 1 |
| T2DM | pericyte | *ACYP2* | 2.733520518 | 9.69E-05 | 0.000726663 | 0.608943965 |
| T2DM | pericyte | *TRABD* | 2.304447467 | 0.000102993 | 0.00075118 | 0.647517401 |
| T2DM | pericyte | *KCNS3* | 0.656851583 | 0.000373034 | 0.00160305 | 1 |
| T2DM | pericyte | *AKAP13* | 2.652447688 | 2.49E-05 | 0.000558632 | 0.156694331 |
| T2DM | pericyte | *HTRA3* | 1.188172685 | 0.002292462 | 0.005992811 | 1 |
| T2DM | pericyte | *FOXN2* | 2.329566176 | 7.68E-05 | 0.000659789 | 0.482965697 |
| T2DM | pericyte | *MTSS1* | 2.304980679 | 6.88E-05 | 0.00063589 | 0.432405255 |
| T2DM | pericyte | *NDUFA3* | 3.513807181 | 0.002263967 | 0.005945514 | 1 |
| T2DM | pericyte | *PAQR8* | 1.007283656 | 0.023673165 | 0.040498827 | 1 |
| T2DM | pericyte | *NUDT6* | 1.752715475 | 0.000113775 | 0.000790392 | 0.715304712 |
| T2DM | pericyte | *ZNF160* | 2.015865428 | 0.000636852 | 0.002342824 | 1 |
| T2DM | pericyte | *TSNARE1* | 1.079104858 | 0.010155219 | 0.01964488 | 1 |
| T2DM | pericyte | *FEZ2* | 3.090591654 | 4.90E-05 | 0.000607238 | 0.308196736 |
| T2DM | pericyte | *C11orf24* | 1.48031524 | 0.002279298 | 0.005975306 | 1 |
| T2DM | pericyte | *ALK* | 0.625714758 | 0.012650509 | 0.023734333 | 1 |
| T2DM | pericyte | *TRMT61B* | 0.617854039 | 0.016073236 | 0.029189584 | 1 |

| | | | | | | |
|---|---|---|---|---|---|---|
| T2DM | pericyte | *MFN1* | 3.054917032 | 3.39E-05 | 0.000599418 | 0.213374516 |
| T2DM | pericyte | *KCNMB3* | 1.963146139 | 5.56E-05 | 0.00062437 | 0.349486752 |
| T2DM | pericyte | *ATP6V0E2* | 4.58631317 | 0.006133908 | 0.012850392 | 1 |
| T2DM | pericyte | *GRIK1* | 1.483035652 | 0.000586984 | 0.002216089 | 1 |
| T2DM | pericyte | *CLDN20* | 1.219260162 | 0.00318789 | 0.007771331 | 1 |
| T2DM | pericyte | *KCND3* | 3.006641599 | 2.47E-05 | 0.000558632 | 0.155049525 |
| T2DM | pericyte | *MCC* | 3.060754676 | 2.96E-05 | 0.000593226 | 0.186239237 |
| T2DM | pericyte | *LRRC8C* | 5.558181464 | 7.26E-05 | 0.000646895 | 0.456374699 |
| T2DM | pericyte | *RSL1D1* | 3.057981806 | 3.97E-05 | 0.000607238 | 0.249744334 |
| T2DM | pericyte | *PPID* | 3.327404332 | 0.001717534 | 0.004847304 | 1 |
| T2DM | pericyte | *TBCA* | 2.188526793 | 0.001828631 | 0.005073523 | 1 |
| T2DM | pericyte | *BCL2L1* | 2.81356396 | 0.000582851 | 0.00220613 | 1 |
| T2DM | pericyte | *PLRG1* | 2.659707386 | 0.000287435 | 0.001350599 | 1 |
| T2DM | pericyte | *NMUR1* | 5.398543355 | 5.68E-05 | 0.00062437 | 0.357171046 |
| T2DM | pericyte | *P2RY6* | 0.987840483 | 0.005272006 | 0.01142454 | 1 |
| T2DM | pericyte | *ZIK1* | 1.211411771 | 0.028742673 | 0.04761665 | 1 |
| T2DM | pericyte | *TCEA2* | 2.109851327 | 0.000598816 | 0.002247617 | 1 |
| T2DM | pericyte | *ANO5* | 0.862901457 | 0.003898612 | 0.00907463 | 1 |
| T2DM | pericyte | *VAT1L* | 1.272505588 | 0.006099722 | 0.012808601 | 1 |
| T2DM | pericyte | *CAMTA1* | 3.116049086 | 0.000110423 | 0.000780032 | 0.694228142 |
| T2DM | pericyte | *KNDC1* | -0.661855955 | 0.003335073 | 0.008049169 | 1 |
| T2DM | pericyte | *PWWP2B* | 0.960013979 | 0.00639941 | 0.013313398 | 1 |
| T2DM | pericyte | *ZNF570* | 0.821670334 | 0.016084817 | 0.02919955 | 1 |
| T2DM | pericyte | *NINJ2* | 2.479180246 | 0.000223768 | 0.00116598 | 1 |
| T2DM | pericyte | *TRAPPC12* | 2.550942089 | 0.003295502 | 0.007974913 | 1 |
| T2DM | pericyte | *AQP4* | 1.948679487 | 0.001636238 | 0.004681545 | 1 |
| T2DM | pericyte | *TLN2* | 0.73235581 | 0.005323775 | 0.011517747 | 1 |
| T2DM | pericyte | *TVP23B* | 2.867502587 | 0.000388788 | 0.001656035 | 1 |
| T2DM | pericyte | *SRGAP2C* | 2.992828683 | 0.001290338 | 0.003924699 | 1 |
| T2DM | pericyte | *ATPAF2* | 3.187840532 | 0.000140981 | 0.000898931 | 0.886346232 |
| T2DM | pericyte | *C20orf196* | 2.576130278 | 0.000344022 | 0.001511465 | 1 |
| T2DM | pericyte | *ANKRD20A4* | 1.870048887 | 0.00495088 | 0.01088328 | 1 |
| T2DM | pericyte | *SERF1A* | 4.007281185 | 0.005750558 | 0.012197625 | 1 |
| T2DM | pericyte | *MOB3A* | 1.418167764 | 0.018742397 | 0.033192521 | 1 |
| T2DM | pericyte | *NME6* | 0.915336544 | 0.005165088 | 0.011255775 | 1 |
| T2DM | pericyte | *MTBP* | 3.142780635 | 0.006680989 | 0.013785158 | 1 |
| T2DM | pericyte | *MALT1* | 2.511593505 | 0.000110118 | 0.000778752 | 0.692310362 |
| T2DM | pericyte | *ISG20* | 0.633844286 | 0.020854092 | 0.036399133 | 1 |
| T2DM | pericyte | *MBOAT1* | 2.753075732 | 0.000357573 | 0.001552529 | 1 |
| T2DM | pericyte | *CXCR6* | 1.109310548 | 0.014455908 | 0.026589904 | 1 |
| T2DM | pericyte | *C5orf34* | 1.668380923 | 0.004122541 | 0.009459276 | 1 |
| T2DM | pericyte | *C1QTNF4* | -0.493493427 | 1.55E-05 | 0.000488489 | 0.097697898 |
| T2DM | pericyte | *ZNF131* | 2.561201481 | 0.000168976 | 0.000990075 | 1 |
| T2DM | pericyte | *HINFP* | 1.685480557 | 0.000813452 | 0.002824311 | 1 |
| T2DM | pericyte | *SPTLC3* | 3.879393063 | 0.00039865 | 0.001682091 | 1 |
| T2DM | pericyte | *SUCLG2* | 2.743290975 | 0.000108224 | 0.000774167 | 0.680407425 |
| T2DM | pericyte | *CSDC2* | 1.677213434 | 0.016551796 | 0.029919821 | 1 |
| T2DM | pericyte | *RCAN2* | 3.503489073 | 0.000242064 | 0.001222377 | 1 |
| T2DM | pericyte | *GNB2* | 1.912725744 | 0.005483983 | 0.011769252 | 1 |
| T2DM | pericyte | *FGGY* | 2.937916861 | 2.73E-06 | 0.000229534 | 0.017185179 |

| | | | | | | |
|---|---|---|---|---|---|---|
| T2DM | pericyte | *ZNF24* | 2.725413261 | 6.36E-05 | 0.000632025 | 0.399603249 |
| T2DM | pericyte | *MANEA* | 2.323403139 | 0.000213461 | 0.001128705 | 1 |
| T2DM | pericyte | *CARNS1* | 2.076870527 | 0.007742132 | 0.015610899 | 1 |
| T2DM | pericyte | *CTSW* | -0.266764059 | 0.009335294 | 0.018249687 | 1 |
| T2DM | pericyte | *SNTG2* | 1.068348727 | 0.00388266 | 0.009047549 | 1 |
| T2DM | pericyte | *MRPL52* | 1.966773109 | 0.001015183 | 0.003325928 | 1 |
| T2DM | pericyte | *SMPDL3A* | 1.291862559 | 0.009332051 | 0.018249022 | 1 |
| T2DM | pericyte | *AGAP5* | 23.71258159 | 0.005872806 | 0.012398365 | 1 |
| T2DM | pericyte | *ZMAT3* | 4.970056626 | 0.000110048 | 0.000778752 | 0.691873227 |
| T2DM | pericyte | *ZFAND4* | 2.45013543 | 0.000382435 | 0.001635627 | 1 |
| T2DM | pericyte | *ZNF738* | 1.149442522 | 0.003094109 | 0.007586843 | 1 |
| T2DM | pericyte | *CCL19* | -1.076011725 | 0.027412578 | 0.045775001 | 1 |
| T2DM | pericyte | *CFL1* | 3.024517901 | 0.000131466 | 0.000860073 | 0.826529881 |
| T2DM | pericyte | *FAM192A* | 0.931522794 | 0.024029296 | 0.040974283 | 1 |
| T2DM | pericyte | *CBWD1* | 3.485413969 | 3.32E-05 | 0.000599418 | 0.20880205 |
| T2DM | pericyte | *RAB37* | 1.357571889 | 0.001687634 | 0.004790136 | 1 |
| T2DM | pericyte | *DCP2* | 2.970094351 | 8.39E-05 | 0.000681373 | 0.527382883 |
| T2DM | pericyte | *SNX32* | 1.9069647 | 2.07E-05 | 0.000527586 | 0.130334925 |
| T2DM | pericyte | *METAP1D* | 3.794244733 | 4.96E-05 | 0.000608237 | 0.312025462 |
| T2DM | pericyte | *NADSYN1* | 2.08259585 | 0.002046555 | 0.005482185 | 1 |
| T2DM | pericyte | *ANKRD13D* | 2.467395203 | 0.000260879 | 0.001281362 | 1 |
| T2DM | pericyte | *MRGPRF* | 2.583012345 | 0.000672087 | 0.002446108 | 1 |
| T2DM | pericyte | *GXYLT2* | 1.167876488 | 0.006200389 | 0.012959391 | 1 |
| T2DM | pericyte | *HPSE2* | 1.45069164 | 1.02E-05 | 0.000407741 | 0.063981244 |
| T2DM | pericyte | *HECTD4* | 2.798281384 | 3.95E-05 | 0.000607238 | 0.2483346 |
| T2DM | pericyte | *BNC2* | 1.543512445 | 0.00019588 | 0.001065597 | 1 |
| T2DM | pericyte | *TRMT112* | 1.358402556 | 0.028801327 | 0.04770125 | 1 |
| T2DM | pericyte | *NOC3L* | 1.971805104 | 0.000719126 | 0.002572932 | 1 |
| T2DM | pericyte | *VANGL1* | 1.120640371 | 0.001234674 | 0.003805096 | 1 |
| T2DM | pericyte | *GLRX* | 2.405311132 | 0.006718044 | 0.013843442 | 1 |
| T2DM | pericyte | *GOLGB1* | 2.986999816 | 2.60E-05 | 0.000562456 | 0.163516766 |
| T2DM | pericyte | *MAP3K11* | 4.959073474 | 0.000145793 | 0.000916603 | 0.916602599 |
| T2DM | pericyte | *C1QA* | 2.096951901 | 0.000233606 | 0.001198741 | 1 |
| T2DM | pericyte | *NDNF* | 2.978583737 | 0.016625243 | 0.030026689 | 1 |
| T2DM | pericyte | *OLR1* | 7.407514007 | 0.011700772 | 0.022258019 | 1 |
| T2DM | pericyte | *GLIPR1L1* | 2.078970277 | 0.000211267 | 0.00112039 | 1 |
| T2DM | pericyte | *ARV1* | 1.64730482 | 0.004020121 | 0.009281859 | 1 |
| T2DM | pericyte | *CCDC36* | 1.689394917 | 0.000163949 | 0.000970569 | 1 |
| T2DM | pericyte | *PPP1R14B* | 2.22831982 | 0.00167758 | 0.004772375 | 1 |
| T2DM | pericyte | *SMARCC1* | 3.237689371 | 2.28E-05 | 0.000548455 | 0.143642182 |
| T2DM | pericyte | *ZNF417* | 0.858021952 | 0.018471282 | 0.032832613 | 1 |
| T2DM | pericyte | *PEAK1* | 2.702270026 | 9.82E-05 | 0.000733178 | 0.617336188 |
| T2DM | pericyte | *TNFRSF10C* | 0.590395449 | 0.009908115 | 0.01925574 | 1 |
| T2DM | pericyte | *GMPPB* | 1.632529251 | 0.00324771 | 0.007889627 | 1 |
| T2DM | pericyte | *NUDT18* | 3.227288392 | 0.010917768 | 0.020958781 | 1 |
| T2DM | pericyte | *SULT1B1* | 1.5003304 | 0.009980685 | 0.01938479 | 1 |
| T2DM | pericyte | *NUDT4* | 1.376290397 | 0.027490111 | 0.045892281 | 1 |
| T2DM | pericyte | *SCAI* | 3.076352216 | 5.81E-05 | 0.00062437 | 0.365139707 |
| T2DM | pericyte | *AGFG1* | 1.705459833 | 0.000290351 | 0.001359225 | 1 |
| T2DM | pericyte | *RNF213* | 2.902507191 | 0.000217756 | 0.001144699 | 1 |

| | | | | | | |
|---|---|---|---|---|---|---|
| T2DM | pericyte | *PLK3* | 3.057909469 | 0.024131299 | 0.041114763 | 1 |
| T2DM | pericyte | *DPY19L1* | 3.576577116 | 0.001225726 | 0.003789498 | 1 |
| T2DM | pericyte | *TUBB8* | -1.337815368 | 0.014640052 | 0.026905001 | 1 |
| T2DM | pericyte | *GPR160* | 1.348119817 | 0.00169749 | 0.004807262 | 1 |
| T2DM | pericyte | *GOLIM4* | 1.753959247 | 0.015829761 | 0.028838513 | 1 |
| T2DM | pericyte | *USMG5* | 2.984398021 | 0.001899783 | 0.005202062 | 1 |
| T2DM | pericyte | *XXYLT1* | 2.016622435 | 0.000438953 | 0.001807948 | 1 |
| T2DM | pericyte | *CCS* | 2.280148855 | 7.27E-05 | 0.000646895 | 0.457354561 |
| T2DM | pericyte | *NRROS* | 0.626219798 | 0.011877177 | 0.022489668 | 1 |
| T2DM | pericyte | *SLC25A30* | 2.948550404 | 0.000221841 | 0.001158399 | 1 |
| T2DM | pericyte | *LEMD3* | 3.510993361 | 2.26E-05 | 0.000548455 | 0.141936998 |
| T2DM | pericyte | *C16orf91* | 3.335425163 | 6.75E-05 | 0.000632837 | 0.424628045 |
| T2DM | pericyte | *SOCS7* | 3.679684996 | 4.83E-05 | 0.000607238 | 0.303845626 |
| T2DM | pericyte | *TLR6* | 1.313484898 | 0.004015317 | 0.009274173 | 1 |
| T2DM | pericyte | *RGMB* | 2.139607584 | 0.002744294 | 0.006906875 | 1 |
| T2DM | pericyte | *FAM53A* | 0.854173854 | 0.01572742 | 0.028672552 | 1 |
| T2DM | pericyte | *CTU2* | 1.136970585 | 0.029363965 | 0.048530822 | 1 |
| T2DM | pericyte | *MGA* | 1.750059209 | 0.004263048 | 0.009731948 | 1 |
| T2DM | pericyte | *DDX23* | 2.811018831 | 0.000364158 | 0.001573515 | 1 |
| T2DM | pericyte | *RALGAPA1* | 1.809064276 | 0.002375819 | 0.006167125 | 1 |
| T2DM | pericyte | *LIG4* | 1.617483207 | 0.000444504 | 0.001824149 | 1 |
| T2DM | pericyte | *SNAPC5* | 2.05062509 | 0.003584958 | 0.008495527 | 1 |
| T2DM | pericyte | *LINGO2* | 0.958798944 | 0.001522985 | 0.004445221 | 1 |
| T2DM | pericyte | *ANKRD36C* | 2.61602071 | 0.001102096 | 0.003518983 | 1 |
| T2DM | pericyte | *TMEM81* | 4.702547495 | 0.000136539 | 0.000878628 | 0.858419173 |
| T2DM | pericyte | *IL20RB* | 1.327737414 | 3.51E-05 | 0.000600626 | 0.220429858 |
| T2DM | pericyte | *ANGEL2* | 3.509862392 | 4.04E-05 | 0.000607238 | 0.253786478 |
| T2DM | pericyte | *SH3PXD2B* | 1.405650379 | 0.000136511 | 0.000878628 | 0.858241593 |
| T2DM | pericyte | *LARP7* | 3.854019952 | 0.000105595 | 0.000763077 | 0.663877165 |
| T2DM | pericyte | *HRAS* | 1.630070232 | 0.013865013 | 0.025675799 | 1 |
| T2DM | pericyte | *BTC* | 0.875060088 | 0.002561897 | 0.006541705 | 1 |
| T2DM | pericyte | *NDUFA11* | 5.19705659 | 6.02E-05 | 0.00062437 | 0.378568092 |
| T2DM | pericyte | *FUT1* | 1.853416531 | 0.009956309 | 0.019343423 | 1 |
| T2DM | pericyte | *DHX36* | 2.233085248 | 0.008599941 | 0.017045344 | 1 |
| T2DM | pericyte | *FBXW8* | 3.631480274 | 4.15E-05 | 0.000607238 | 0.26063399 |
| T2DM | pericyte | *SLC22A1* | 1.107786334 | 0.029643558 | 0.048864459 | 1 |
| T2DM | pericyte | *ATR* | 2.997967157 | 1.83E-05 | 0.000512131 | 0.115077035 |
| T2DM | pericyte | *TVP23C* | 1.183022077 | 0.006814703 | 0.014003728 | 1 |
| T2DM | pericyte | *PACS1* | 2.154027414 | 0.001291877 | 0.003925772 | 1 |
| T2DM | pericyte | *SH3BP5L* | 3.618715555 | 3.48E-05 | 0.000599418 | 0.218719355 |
| T2DM | pericyte | *TP53I11* | 4.253889674 | 1.93E-05 | 0.000512152 | 0.121380051 |
| T2DM | pericyte | *PHYHD1* | 2.248003085 | 0.00104217 | 0.003375644 | 1 |
| T2DM | pericyte | *ZNF519* | 1.08149397 | 0.001566268 | 0.004533669 | 1 |
| T2DM | pericyte | *CHRNA7* | 1.380832798 | 0.00061514 | 0.002292225 | 1 |
| T2DM | pericyte | *TMEM9B* | 3.798900678 | 1.34E-06 | 0.00014989 | 0.008393832 |
| T2DM | pericyte | *SCUBE2* | 1.28796343 | 0.027696434 | 0.04618766 | 1 |
| T2DM | pericyte | *SMAD2* | 1.641069276 | 0.00132367 | 0.003995157 | 1 |
| T2DM | pericyte | *EIF3F* | 2.04686255 | 0.002703627 | 0.00683737 | 1 |
| T2DM | pericyte | *CLTB* | 1.572815655 | 0.000723036 | 0.002582631 | 1 |
| T2DM | pericyte | *PCSK1* | 1.979479969 | 0.000823817 | 0.002855045 | 1 |

| | | | | | | |
|---|---|---|---|---|---|---|
| T2DM | pericyte | *PPP2R2D* | 1.719187777 | 0.003213878 | 0.007822552 | 1 |
| T2DM | pericyte | *LRRC25* | 0.552593818 | 0.014940635 | 0.027401334 | 1 |
| T2DM | pericyte | *KCNE3* | 1.042788664 | 0.002210491 | 0.005824542 | 1 |
| T2DM | pericyte | *RAB6A* | 2.50403786 | 0.000468225 | 0.001894293 | 1 |
| T2DM | pericyte | *TOM1L2* | 3.082592953 | 3.12E-05 | 0.000599418 | 0.196113631 |
| T2DM | pericyte | *TEX26* | 1.815797498 | 0.004159565 | 0.009533251 | 1 |
| T2DM | pericyte | *GPR156* | 2.509296703 | 0.000313304 | 0.001424252 | 1 |
| T2DM | pericyte | *B3GNTL1* | 1.737229334 | 0.000843762 | 0.00290511 | 1 |
| T2DM | pericyte | *TTLL11* | 0.938721196 | 0.029408926 | 0.048575142 | 1 |
| T2DM | pericyte | *MSRA* | 2.424747554 | 0.002489671 | 0.006403521 | 1 |
| T2DM | pericyte | *CTDNEP1* | 4.568752723 | 2.04E-05 | 0.000524717 | 0.128555706 |
| T2DM | pericyte | *ETV4* | 2.028611243 | 0.000558215 | 0.002150427 | 1 |
| T2DM | pericyte | *ZDHHC21* | 2.287857306 | 0.001663858 | 0.004739771 | 1 |
| T2DM | pericyte | *LRRN1* | 2.245869399 | 0.000587244 | 0.002216089 | 1 |
| T2DM | pericyte | *ZNF613* | 1.508637761 | 0.009238091 | 0.018076526 | 1 |
| T2DM | pericyte | *IP6K1* | 2.417250674 | 0.003645278 | 0.008596348 | 1 |
| T2DM | pericyte | *SSNA1* | 5.512157982 | 9.11E-05 | 0.000704357 | 0.572642609 |
| T2DM | pericyte | *CHMP6* | 2.862740904 | 0.000143467 | 0.000908335 | 0.901976756 |
| T2DM | pericyte | *TMEM39A* | 2.458198837 | 0.000273175 | 0.001319089 | 1 |
| T2DM | pericyte | *TCP11L1* | 2.274037574 | 0.000225758 | 0.001172167 | 1 |
| T2DM | pericyte | *GPX2* | 2.381948083 | 0.00085524 | 0.002933386 | 1 |
| T2DM | pericyte | *CCDC57* | 1.252606453 | 0.029963052 | 0.049339368 | 1 |
| T2DM | pericyte | *SMIM19* | 1.025498256 | 0.007964023 | 0.015986531 | 1 |
| T2DM | pericyte | *SLC35G1* | 0.875751528 | 0.013791421 | 0.025562107 | 1 |
| T2DM | pericyte | *COX8A* | 3.746581916 | 0.000102365 | 0.000750264 | 0.643566881 |
| T2DM | pericyte | *B3GNT4* | 6.462125503 | 0.005071536 | 0.011090347 | 1 |
| T2DM | pericyte | *CRLF3* | 2.87866161 | 9.45E-05 | 0.000717456 | 0.594337258 |
| T2DM | pericyte | *RNPEP* | 2.386062426 | 0.00086223 | 0.002952804 | 1 |
| T2DM | pericyte | *WDR25* | 3.005993916 | 0.00016992 | 0.000993915 | 1 |
| T2DM | pericyte | *PLA2G16* | 0.471358422 | 0.006996324 | 0.014304354 | 1 |
| T2DM | pericyte | *RMDN1* | 2.559008642 | 3.13E-05 | 0.000599418 | 0.197096133 |
| T2DM | pericyte | *HORMAD2* | 1.136813916 | 7.92E-05 | 0.000667567 | 0.497707119 |
| T2DM | pericyte | *BOK* | 1.27581267 | 0.000771808 | 0.002722984 | 1 |
| T2DM | pericyte | *C8orf59* | 3.714004088 | 8.59E-06 | 0.000381879 | 0.054001021 |
| T2DM | pericyte | *RUFY1* | 1.222435148 | 0.029729375 | 0.048987533 | 1 |
| T2DM | pericyte | *LRRC37A3* | 2.493783263 | 0.0001551 | 0.000944434 | 0.975111135 |
| T2DM | pericyte | *IRX5* | 0.874124661 | 0.00516855 | 0.011259416 | 1 |
| T2DM | pericyte | *PNMA1* | 1.662292746 | 0.005355695 | 0.011570878 | 1 |
| T2DM | pericyte | *ANKLE2* | 4.367335272 | 3.23E-05 | 0.000599418 | 0.202963027 |
| T2DM | pericyte | *GCNT4* | 3.011552415 | 0.004377348 | 0.009938745 | 1 |
| T2DM | pericyte | *MUC20* | 1.509885778 | 0.004452203 | 0.010066505 | 1 |
| T2DM | pericyte | *THAP4* | 1.670529387 | 0.000508449 | 0.002006668 | 1 |
| T2DM | pericyte | *SHMT1* | 0.933317569 | 0.001298022 | 0.003940445 | 1 |
| T2DM | pericyte | *DPP7* | 4.268873584 | 0.000143989 | 0.000909584 | 0.905257299 |
| T2DM | pericyte | *MTHFR* | 2.001752875 | 0.001718569 | 0.004847304 | 1 |
| T2DM | pericyte | *C19orf18* | 0.815593225 | 0.011602469 | 0.022111163 | 1 |
| T2DM | pericyte | *WDR73* | 2.39065807 | 5.62E-05 | 0.00062437 | 0.353553485 |
| T2DM | pericyte | *POLE* | 4.152466386 | 1.33E-05 | 0.000462556 | 0.083480069 |
| T2DM | pericyte | *DSCAML1* | 3.342971397 | 0.000440759 | 0.00181233 | 1 |
| T2DM | pericyte | *TALDO1* | 1.541346449 | 0.000461154 | 0.001877771 | 1 |

| | | | | | | |
|---|---|---|---|---|---|---|
| T2DM | pericyte | *B3GNT8* | 4.841079917 | 0.001076432 | 0.003451061 | 1 |
| T2DM | pericyte | *AP006621.1* | 0.959142412 | 0.001023399 | 0.003343213 | 1 |
| T2DM | pericyte | *MAN1B1* | 2.948030496 | 0.00013139 | 0.000860073 | 0.826048675 |
| T2DM | pericyte | *TOP3A* | 2.417088544 | 0.000504258 | 0.001992628 | 1 |
| T2DM | pericyte | *ZBTB38* | 1.679258791 | 0.028380343 | 0.047103278 | 1 |
| T2DM | pericyte | *C8orf31* | 1.878583192 | 0.001683527 | 0.004782799 | 1 |
| T2DM | pericyte | *TIMM22* | 4.36362425 | 0.002082974 | 0.005553279 | 1 |
| T2DM | pericyte | *PPFIA3* | 0.904351172 | 0.016642128 | 0.030039924 | 1 |
| T2DM | pericyte | *MAGEF1* | 5.619442447 | 7.93E-05 | 0.000667567 | 0.498670775 |
| T2DM | pericyte | *NIM1K* | 1.245744402 | 0.002023451 | 0.005437849 | 1 |
| T2DM | pericyte | *ARIH2* | 1.43612073 | 0.003971544 | 0.0092035 | 1 |
| T2DM | pericyte | *ATOX1* | 2.917607166 | 6.35E-06 | 0.000343901 | 0.039947564 |
| T2DM | pericyte | *TBL1XR1* | 1.959986161 | 0.002741198 | 0.006905789 | 1 |
| T2DM | pericyte | *RPLP2* | 3.452219709 | 8.98E-05 | 0.000702125 | 0.564721909 |
| T2DM | pericyte | *C12orf54* | -2.153723423 | 0.002048603 | 0.005485334 | 1 |
| T2DM | pericyte | *IL17RA* | 1.43123364 | 0.003548417 | 0.008443942 | 1 |
| T2DM | pericyte | *THAP5* | 2.890085866 | 0.000295047 | 0.001375066 | 1 |
| T2DM | pericyte | *CD151* | 2.391230828 | 0.000796915 | 0.002775736 | 1 |
| T2DM | pericyte | *POLR2L* | 2.526381633 | 0.003258521 | 0.007906724 | 1 |
| T2DM | pericyte | *ZCCHC3* | 2.821086555 | 0.008877715 | 0.01748565 | 1 |
| T2DM | pericyte | *ZNF619* | 1.085337464 | 0.003709071 | 0.008717357 | 1 |
| T2DM | pericyte | *ZBTB41* | 2.846491474 | 0.001661346 | 0.004735303 | 1 |
| T2DM | pericyte | *BET1L* | 1.746285563 | 9.48E-05 | 0.000717456 | 0.596092126 |
| T2DM | pericyte | *IMP3* | 5.089813582 | 9.00E-05 | 0.000702125 | 0.565912476 |
| T2DM | pericyte | *ODF3B* | 1.121229513 | 0.003111792 | 0.007621283 | 1 |
| T2DM | pericyte | *DPY19L2* | -0.103019259 | 0.012778121 | 0.023959454 | 1 |
| T2DM | pericyte | *C2orf73* | 1.884270784 | 0.002022452 | 0.005437849 | 1 |
| T2DM | pericyte | *FAM26E* | 1.333848971 | 0.003669649 | 0.008644092 | 1 |
| T2DM | pericyte | *ALS2CL* | 1.307581725 | 0.002081873 | 0.005553279 | 1 |
| T2DM | pericyte | *MLF1* | 2.320137756 | 0.021424141 | 0.03724933 | 1 |
| T2DM | pericyte | *PRSS42* | 1.745376847 | 0.005454882 | 0.011724733 | 1 |
| T2DM | pericyte | *C2orf69* | 5.096905827 | 5.63E-05 | 0.00062437 | 0.354236062 |
| T2DM | pericyte | *GOLGA8Q* | 17.44482219 | 0.012682562 | 0.02378737 | 1 |
| T2DM | pericyte | *NDUFV2* | 2.739540965 | 0.00051167 | 0.002016848 | 1 |
| T2DM | pericyte | *DALRD3* | 2.167352504 | 0.000330459 | 0.001474519 | 1 |
| T2DM | pericyte | *ZNF114* | -0.694075225 | 0.02460112 | 0.041789672 | 1 |
| T2DM | pericyte | *LCORL* | 3.197425343 | 6.54E-05 | 0.00063262 | 0.411254768 |
| T2DM | pericyte | *ZNF454* | 0.777249609 | 0.007094706 | 0.014472556 | 1 |
| T2DM | pericyte | *ZC3H12D* | 0.839935622 | 0.004822461 | 0.010668126 | 1 |
| T2DM | pericyte | *KDELC2* | 1.670120323 | 0.007731481 | 0.015599429 | 1 |
| T2DM | pericyte | *PLEC* | 3.31668109 | 0.000275108 | 0.001323244 | 1 |
| T2DM | pericyte | *SH2D4B* | 2.853409113 | 0.002931422 | 0.00726729 | 1 |
| T2DM | pericyte | *RNF212* | 0.594746635 | 0.007741677 | 0.015610899 | 1 |
| T2DM | pericyte | *PRSS36* | 1.476100557 | 0.003609301 | 0.008537124 | 1 |
| T2DM | pericyte | *TMPRSS9* | 1.116927804 | 0.004505028 | 0.01014438 | 1 |
| T2DM | pericyte | *AQP11* | 1.457515551 | 0.001123948 | 0.003563421 | 1 |
| T2DM | pericyte | *TMEM11* | 1.817409357 | 0.0009236 | 0.003095242 | 1 |
| T2DM | pericyte | *ZNF223* | -0.450402125 | 0.013355886 | 0.024864807 | 1 |
| T2DM | pericyte | *BEND3* | 3.067795007 | 0.001024655 | 0.003343213 | 1 |
| T2DM | pericyte | *NT5DC1* | 2.60079257 | 0.000216017 | 0.001139346 | 1 |

| | | | | | | |
|---|---|---|---|---|---|---|
| T2DM | pericyte | *AC006111.1* | 4.175497867 | 0.002041325 | 0.005475175 | 1 |
| T2DM | pericyte | *LINC00843* | 2.097060787 | 0.000996347 | 0.003283509 | 1 |
| T2DM | pericyte | *COX14* | 1.274170718 | 0.001116437 | 0.003546762 | 1 |
| T2DM | pericyte | *P4HTM* | 1.670001279 | 0.001071818 | 0.003443289 | 1 |
| T2DM | pericyte | *ERBB4* | 1.158092445 | 0.000393714 | 0.001670229 | 1 |
| T2DM | pericyte | *PARP10* | 1.790989631 | 0.003251493 | 0.007895766 | 1 |
| T2DM | pericyte | *STX19* | 3.727184013 | 1.53E-08 | 5.05E-06 | 9.59E-05 |
| T2DM | pericyte | *FAM219B* | 1.860151793 | 0.000262338 | 0.001285515 | 1 |
| T2DM | pericyte | *GDPD4* | 1.904181189 | 0.004369895 | 0.009925409 | 1 |
| T2DM | pericyte | *MPI* | 2.298115252 | 0.00475112 | 0.010562338 | 1 |
| T2DM | pericyte | *EFCAB13* | 1.570475111 | 0.005389096 | 0.011615101 | 1 |
| T2DM | pericyte | *C17orf62* | 2.575503744 | 0.000496687 | 0.00197263 | 1 |
| T2DM | pericyte | *GAK* | 2.590707883 | 0.001556137 | 0.004517679 | 1 |
| T2DM | pericyte | *TUFM* | 1.296311008 | 0.00171214 | 0.004837854 | 1 |
| T2DM | pericyte | *RMI1* | 2.113085249 | 0.001028361 | 0.0033499 | 1 |
| T2DM | pericyte | *CTC1* | 1.348896967 | 0.000514682 | 0.002024911 | 1 |
| T2DM | pericyte | *SNX18* | 3.502231904 | 0.000122026 | 0.000824925 | 0.7671799 |
| T2DM | pericyte | *TRIML2* | -9.377899246 | 0.021338621 | 0.03712117 | 1 |
| T2DM | pericyte | *SPTY2D1* | 4.69518389 | 6.26E-05 | 0.000630549 | 0.393331514 |
| T2DM | pericyte | *FUCA1* | 1.664423299 | 0.010145557 | 0.019638275 | 1 |
| T2DM | pericyte | *ZNF664* | 3.867840902 | 0.000462072 | 0.00187827 | 1 |
| T2DM | pericyte | *CALR* | 4.839954786 | 2.14E-08 | 6.11E-06 | 0.000134332 |
| T2DM | pericyte | *HLA-DQB1* | -1.354489851 | 6.23E-13 | 5.60E-10 | 3.92E-09 |
| T2DM | pericyte | *FJX1* | -0.637684495 | 0.022829969 | 0.0392717 | 1 |
| T2DM | pericyte | *CIITA* | 1.151235864 | 0.000701225 | 0.002523526 | 1 |
| T2DM | pericyte | *PLD6* | 1.213045105 | 0.002597127 | 0.00661862 | 1 |
| T2DM | pericyte | *ZBTB42* | 2.514447778 | 0.009568025 | 0.018652456 | 1 |
| T2DM | pericyte | *LACC1* | 2.054625698 | 0.001624951 | 0.004656365 | 1 |
| T2DM | pericyte | *MAF1* | 2.089211727 | 0.000659999 | 0.002412817 | 1 |
| T2DM | pericyte | *FCER1A* | 1.01110779 | 0.015113027 | 0.027679893 | 1 |
| T2DM | pericyte | *PCED1B* | 1.347367272 | 0.001761846 | 0.004938667 | 1 |
| T2DM | pericyte | *SERTAD2* | 3.614987478 | 5.29E-05 | 0.000619691 | 0.33274996 |
| T2DM | pericyte | *PDXDC1* | 2.874430388 | 0.000138695 | 0.000890678 | 0.871973827 |
| T2DM | pericyte | *ZNF154* | 1.547918142 | 0.001071126 | 0.003442828 | 1 |
| T2DM | pericyte | *R3HDM2* | 2.922900527 | 3.20E-05 | 0.000599418 | 0.201032636 |
| T2DM | pericyte | *B3GNT3* | 4.906916775 | 1.91E-05 | 0.000512152 | 0.12037612 |
| T2DM | pericyte | *SEPT1* | 5.633010455 | 0.000469828 | 0.001897115 | 1 |
| T2DM | pericyte | *EXOC3* | 1.284710899 | 0.025908959 | 0.043764005 | 1 |
| T2DM | pericyte | *FGD6* | 1.412048739 | 0.000594275 | 0.002237251 | 1 |
| T2DM | pericyte | *HCLS1* | 2.86194908 | 6.66E-05 | 0.000632837 | 0.418728293 |
| T2DM | pericyte | *ZNF609* | 2.253774735 | 0.00063462 | 0.002337355 | 1 |
| T2DM | pericyte | *CCDC66* | 1.88675387 | 0.000526756 | 0.00206062 | 1 |
| T2DM | pericyte | *DEFB124* | -0.294207871 | 0.017666172 | 0.03163407 | 1 |
| T2DM | pericyte | *MCFD2* | 2.764323985 | 9.61E-05 | 0.000722468 | 0.603983259 |
| T2DM | pericyte | *CYP8B1* | 1.008741488 | 0.011828574 | 0.022440025 | 1 |
| T2DM | pericyte | *RNF182* | -0.536179399 | 0.003181776 | 0.007765461 | 1 |
| T2DM | pericyte | *MB21D2* | 1.960115344 | 0.022281157 | 0.038500272 | 1 |
| T2DM | pericyte | *SSTR2* | 1.426873131 | 0.004890063 | 0.010787308 | 1 |
| T2DM | pericyte | *ZNF594* | 2.664038624 | 0.014347781 | 0.026414201 | 1 |
| T2DM | pericyte | *PCGF5* | 3.139051376 | 0.00020277 | 0.001093325 | 1 |

| | | | | | | |
|---|---|---|---|---|---|---|
| T2DM | pericyte | *SHISA2* | 30.0718224 | 0.001837877 | 0.005086852 | 1 |
| T2DM | pericyte | *SRSF8* | 3.584852903 | 0.027665646 | 0.046160807 | 1 |
| T2DM | pericyte | *SLC36A4* | 1.511929598 | 0.030097364 | 0.049521625 | 1 |
| T2DM | pericyte | *ZDHHC20* | 3.048049122 | 4.26E-05 | 0.000607238 | 0.267956456 |
| T2DM | pericyte | *PSMG4* | 1.935807827 | 0.008213116 | 0.016366358 | 1 |
| T2DM | pericyte | *CAPS2* | 1.942407325 | 0.000111995 | 0.000783219 | 0.704113632 |
| T2DM | pericyte | *KCTD2* | 5.223980311 | 4.44E-05 | 0.000607238 | 0.279298227 |
| T2DM | pericyte | *D2HGDH* | 2.578624687 | 0.000154246 | 0.000942415 | 0.969744868 |
| T2DM | pericyte | *CMTR2* | 2.080879753 | 0.000284828 | 0.001344381 | 1 |
| T2DM | pericyte | *LRRC57* | 3.969953581 | 2.17E-05 | 0.000541196 | 0.136381279 |
| T2DM | pericyte | *BBS12* | 1.11826634 | 0.006416603 | 0.013331521 | 1 |
| T2DM | pericyte | *ZFP82* | 3.013022696 | 0.004729215 | 0.010532263 | 1 |
| T2DM | pericyte | *AEN* | 1.47083293 | 0.009254818 | 0.018103623 | 1 |
| T2DM | pericyte | *RPH3AL* | 1.915516585 | 0.001151413 | 0.003624363 | 1 |
| T2DM | pericyte | *SLC26A11* | 4.351236583 | 3.56E-05 | 0.00060096 | 0.223961121 |
| T2DM | pericyte | *POLR2A* | 2.301818792 | 0.010521852 | 0.020297908 | 1 |
| T2DM | pericyte | *TMEM132C* | 0.795434405 | 0.000545311 | 0.002113668 | 1 |
| T2DM | pericyte | *SLC25A41* | 3.898667529 | 0.011218217 | 0.021486231 | 1 |
| T2DM | pericyte | *TMEM136* | 0.956905495 | 0.002490315 | 0.006403521 | 1 |
| T2DM | pericyte | *DDX60L* | 2.715382003 | 0.000234484 | 0.001198741 | 1 |
| T2DM | pericyte | *ACBD4* | 1.043042791 | 0.001946256 | 0.00529329 | 1 |
| T2DM | pericyte | *SGSH* | 5.704631739 | 3.87E-05 | 0.000607238 | 0.243129854 |
| T2DM | pericyte | *SETD2* | 2.672059725 | 1.09E-05 | 0.000424012 | 0.068689873 |
| T2DM | pericyte | *MRPS23* | 2.192962124 | 0.000377882 | 0.001619456 | 1 |
| T2DM | pericyte | *GPR135* | 0.761695994 | 0.012941544 | 0.024254487 | 1 |
| T2DM | pericyte | *ATG9B* | 1.743970519 | 0.004768829 | 0.010593692 | 1 |
| T2DM | pericyte | *HKR1* | 2.553847034 | 0.000360742 | 0.001563051 | 1 |
| T2DM | pericyte | *ZBTB20* | 2.251739677 | 0.000118767 | 0.000808105 | 0.746688559 |
| T2DM | pericyte | *C3orf58* | 2.270070346 | 0.0018488 | 0.005106945 | 1 |
| T2DM | pericyte | *RELL1* | 3.266499168 | 3.39E-05 | 0.000599418 | 0.213148615 |
| T2DM | pericyte | *RFX7* | 2.616601288 | 4.62E-05 | 0.000607238 | 0.290503827 |
| T2DM | pericyte | *CLDN7* | -0.273865321 | 0.024021862 | 0.040972721 | 1 |
| T2DM | pericyte | *ADO* | 1.107745848 | 0.003925644 | 0.009117298 | 1 |
| T2DM | pericyte | *SNRPE* | 3.745125886 | 0.002433085 | 0.00628722 | 1 |
| T2DM | pericyte | *CHST15* | 2.641953031 | 0.001139038 | 0.003600367 | 1 |
| T2DM | pericyte | *IDH2* | 1.965483891 | 0.001877176 | 0.005158132 | 1 |
| T2DM | pericyte | *TNRC18* | 3.334944503 | 0.000117029 | 0.000799743 | 0.735763871 |
| T2DM | pericyte | *ZNF708* | 1.610926532 | 0.006110985 | 0.012819407 | 1 |
| T2DM | pericyte | *IST1* | 2.327065794 | 0.003486772 | 0.008335108 | 1 |
| T2DM | pericyte | *ERCC6L2* | 1.715691829 | 0.004770756 | 0.010593692 | 1 |
| T2DM | pericyte | *SYNM* | 2.676336975 | 0.000287134 | 0.001350194 | 1 |
| T2DM | pericyte | *KBTBD3* | 5.069931427 | 3.76E-05 | 0.000607238 | 0.236475019 |
| T2DM | pericyte | *CLN8* | 1.555888799 | 0.010528498 | 0.020304499 | 1 |
| T2DM | pericyte | *TRAPPC6B* | 1.771172146 | 0.002024814 | 0.005437849 | 1 |
| T2DM | pericyte | *CAPN12* | 4.246477723 | 3.07E-06 | 0.000244526 | 0.019317589 |
| T2DM | pericyte | *GLRX5* | 3.231699781 | 0.008088255 | 0.016173937 | 1 |
| T2DM | pericyte | *MFSD5* | 1.698988341 | 0.006323156 | 0.013192308 | 1 |
| T2DM | pericyte | *ADI1* | 3.268082198 | 0.00099649 | 0.003283509 | 1 |
| T2DM | pericyte | *CLEC4G* | 2.161503944 | 0.007271153 | 0.014771962 | 1 |
| T2DM | pericyte | *SATB1* | 2.986708544 | 0.001036543 | 0.003365197 | 1 |

| | | | | | | |
|---|---|---|---|---|---|---|
| T2DM | pericyte | *ACTL10* | -0.907251167 | 0.027213083 | 0.045514407 | 1 |
| T2DM | pericyte | *TRAK1* | 2.306448485 | 0.001293811 | 0.003929561 | 1 |
| T2DM | pericyte | *SKA2* | 4.074511339 | 4.24E-05 | 0.000607238 | 0.266551486 |
| T2DM | pericyte | *TTC3* | 0.725306606 | 6.81E-05 | 0.00063522 | 0.428138184 |
| T2DM | pericyte | *TSKU* | 2.707688941 | 0.000113498 | 0.000790347 | 0.713562002 |
| T2DM | pericyte | *GRID1* | -0.211650217 | 0.016045573 | 0.029164069 | 1 |
| T2DM | pericyte | *ACBD3* | 3.372069745 | 0.00030815 | 0.001407952 | 1 |
| T2DM | pericyte | *VMO1* | 0.610012139 | 0.02289453 | 0.039327298 | 1 |
| T2DM | pericyte | *ZNF721* | 1.985050175 | 0.00033818 | 0.001502572 | 1 |
| T2DM | pericyte | *HMGN4* | 1.362699397 | 0.014697163 | 0.027002065 | 1 |
| T2DM | pericyte | *ZNF320* | 0.704133686 | 6.00E-05 | 0.00062437 | 0.377039017 |
| T2DM | pericyte | *WBP2NL* | 3.922066352 | 0.002203666 | 0.005811429 | 1 |
| T2DM | pericyte | *BEGAIN* | 2.845719791 | 0.004044096 | 0.00933036 | 1 |
| T2DM | pericyte | *ARHGEF37* | 2.051548452 | 0.000185559 | 0.001039756 | 1 |
| T2DM | pericyte | *FAM43B* | 2.198497584 | 0.011845973 | 0.022452708 | 1 |
| T2DM | pericyte | *CSMD1* | 1.022429369 | 0.004305456 | 0.009804795 | 1 |
| T2DM | pericyte | *FANCF* | 4.204571161 | 0.000243236 | 0.001224801 | 1 |
| T2DM | pericyte | *RUVBL2* | 1.471238131 | 0.00077297 | 0.002724025 | 1 |
| T2DM | pericyte | *GDPGP1* | 1.364057584 | 0.001787841 | 0.004988689 | 1 |
| T2DM | pericyte | *PTTG1IP* | 1.626739437 | 0.004194809 | 0.009600569 | 1 |
| T2DM | pericyte | *CCBE1* | 0.865105233 | 0.011442448 | 0.021866468 | 1 |
| T2DM | pericyte | *ZNF623* | 2.14689557 | 0.020550783 | 0.035900121 | 1 |
| T2DM | pericyte | *CCDC125* | 1.710571514 | 0.000343244 | 0.00151137 | 1 |
| T2DM | pericyte | *FHL3* | -1.028483319 | 0.023864973 | 0.040782573 | 1 |
| T2DM | pericyte | *LRIT3* | 1.485026285 | 0.019809896 | 0.0348085 | 1 |
| T2DM | pericyte | *GPR132* | 0.985118866 | 0.005672933 | 0.01205738 | 1 |
| T2DM | pericyte | *PSMG1* | 0.471229026 | 0.017189593 | 0.030868601 | 1 |
| T2DM | pericyte | *ACSM5* | 0.683874582 | 0.001006915 | 0.003309186 | 1 |
| T2DM | pericyte | *SERHL2* | 5.221426151 | 6.86E-05 | 0.000635523 | 0.4314792 |
| T2DM | pericyte | *TANGO2* | 2.953881566 | 0.000320392 | 0.001443947 | 1 |
| T2DM | pericyte | *SFXN4* | 1.018061303 | 0.008716913 | 0.017228303 | 1 |
| T2DM | pericyte | *DGCR6* | -1.18459988 | 0.006971007 | 0.01426651 | 1 |
| T2DM | pericyte | *RP1L1* | 2.322226144 | 5.59E-06 | 0.000331374 | 0.035147545 |
| T2DM | pericyte | *NDUFB1* | 1.861387301 | 0.000638611 | 0.002346552 | 1 |
| T2DM | pericyte | *FAM19A1* | 1.782322467 | 0.000403303 | 0.001697168 | 1 |
| T2DM | pericyte | *BMP8A* | 3.129162275 | 0.000430771 | 0.001783951 | 1 |
| T2DM | pericyte | *CBX6* | 5.042426259 | 0.000299686 | 0.001384824 | 1 |
| T2DM | pericyte | *AIFM3* | 1.573855487 | 0.018607893 | 0.033028747 | 1 |
| T2DM | pericyte | *KCTD16* | 2.719938426 | 0.000425773 | 0.001768054 | 1 |
| T2DM | pericyte | *ZNF703* | -1.592509294 | 0.011946354 | 0.022602086 | 1 |
| T2DM | pericyte | *KCTD8* | 0.99306743 | 0.000527036 | 0.00206062 | 1 |
| T2DM | pericyte | *C9orf66* | 8.225643056 | 0.000260072 | 0.001279401 | 1 |
| T2DM | pericyte | *TUBA8* | 1.873920706 | 0.000195102 | 0.001065597 | 1 |
| T2DM | pericyte | *FAM162B* | -0.095097137 | 0.026448571 | 0.044531915 | 1 |
| T2DM | pericyte | *BTBD9* | 2.694871855 | 6.36E-05 | 0.000632025 | 0.400080018 |
| T2DM | pericyte | *ANKRD45* | 2.243508557 | 0.005115342 | 0.011174481 | 1 |
| T2DM | pericyte | *MAATS1* | 0.932358587 | 0.003955495 | 0.009176456 | 1 |
| T2DM | pericyte | *GPR39* | 1.252644913 | 0.003425713 | 0.008207872 | 1 |
| T2DM | pericyte | *TTC32* | 1.863187871 | 0.022202746 | 0.038422423 | 1 |
| T2DM | pericyte | *PP2D1* | 2.520284283 | 4.19E-05 | 0.000607238 | 0.263734416 |

| | | | | | | |
|---|---|---|---|---|---|---|
| T2DM | pericyte | *TMEM236* | 1.599886394 | 0.003181184 | 0.007765461 | 1 |
| T2DM | pericyte | *VPS33B* | 2.781630601 | 0.000343286 | 0.00151137 | 1 |
| T2DM | pericyte | *EIF3C* | 0.889864387 | 0.005818617 | 0.012320515 | 1 |
| T2DM | pericyte | *NIPSNAP1* | 1.46790049 | 0.003564329 | 0.008467163 | 1 |
| T2DM | pericyte | *CNTN2* | 1.536512432 | 0.003638771 | 0.008590671 | 1 |
| T2DM | pericyte | *CRELD2* | 3.915942865 | 0.000955335 | 0.003180036 | 1 |
| T2DM | pericyte | *TM2D3* | 2.937238564 | 0.001598596 | 0.004601818 | 1 |
| T2DM | pericyte | *CLECL1* | 0.686089347 | 0.002901758 | 0.007207962 | 1 |
| T2DM | pericyte | *MROH7* | 2.830457672 | 0.000231822 | 0.001193695 | 1 |
| T2DM | pericyte | *CSF1* | 1.496350866 | 0.000773943 | 0.002724402 | 1 |
| T2DM | pericyte | *PLA2G6* | 1.052801987 | 0.000506892 | 0.002001779 | 1 |
| T2DM | pericyte | *SS18L1* | 2.556134707 | 0.000196533 | 0.001067935 | 1 |
| T2DM | pericyte | *THAP7* | 3.349434248 | 0.000496492 | 0.00197263 | 1 |
| T2DM | pericyte | *WDR27* | 2.945563005 | 0.000638282 | 0.002346552 | 1 |
| T2DM | pericyte | *TXNRD2* | 1.702852599 | 0.000428213 | 0.001774672 | 1 |
| T2DM | pericyte | *PROS1* | 1.679282064 | 0.000669818 | 0.002441246 | 1 |
| T2DM | pericyte | *HDDC3* | 1.271415932 | 0.026767403 | 0.044912373 | 1 |
| T2DM | pericyte | *TMEM173* | 1.75478767 | 0.000598585 | 0.002247617 | 1 |
| T2DM | pericyte | *NELL2* | 0.638031961 | 0.006960402 | 0.014249446 | 1 |
| T2DM | pericyte | *ZNF93* | 5.415011958 | 0.001845815 | 0.005101215 | 1 |
| T2DM | pericyte | *SEPT5* | 2.762387348 | 0.004009959 | 0.0092652 | 1 |
| T2DM | pericyte | *ATL3* | 1.522941067 | 0.015544768 | 0.028368638 | 1 |
| T2DM | pericyte | *UBE2G2* | 1.893836034 | 0.004431803 | 0.010033398 | 1 |
| T2DM | pericyte | *OSBP2* | 2.216997329 | 0.000240967 | 0.00122084 | 1 |
| T2DM | pericyte | *TUSC5* | 0.907391298 | 0.000514344 | 0.002024845 | 1 |
| T2DM | pericyte | *TMED9* | 3.943830143 | 1.98E-05 | 0.000515685 | 0.12479573 |
| T2DM | pericyte | *H1FX* | 1.288401112 | 0.003484309 | 0.008334776 | 1 |
| T2DM | pericyte | *RBM43* | 2.700194357 | 0.016620205 | 0.030026215 | 1 |
| T2DM | pericyte | *ZFP90* | 4.398495167 | 7.06E-05 | 0.000639553 | 0.443661698 |
| T2DM | pericyte | *SORCS2* | 2.413347233 | 0.002067022 | 0.005527592 | 1 |
| T2DM | pericyte | *TMEM106A* | 2.860622307 | 1.51E-05 | 0.000481538 | 0.094862891 |
| T2DM | pericyte | *SIVA1* | 1.257553863 | 0.008431163 | 0.01674778 | 1 |
| T2DM | pericyte | *MAFF* | 1.2172817 | 0.005154259 | 0.01123607 | 1 |
| T2DM | pericyte | *BRF1* | 3.543629173 | 4.69E-05 | 0.000607238 | 0.294806157 |
| T2DM | pericyte | *ADSSL1* | 5.732429376 | 0.000143733 | 0.000909101 | 0.903646595 |
| T2DM | pericyte | *C6orf120* | 1.273818771 | 0.022253034 | 0.038469121 | 1 |
| T2DM | pericyte | *IFITM2* | 1.993811898 | 0.002327587 | 0.006061945 | 1 |
| T2DM | pericyte | *GP1BA* | 1.392838941 | 0.000782202 | 0.002742726 | 1 |
| T2DM | pericyte | *KIAA0825* | 2.343076914 | 0.006550711 | 0.01358322 | 1 |
| T2DM | pericyte | *KLHL33* | 0.767686015 | 0.026303098 | 0.044298842 | 1 |
| T2DM | pericyte | *TMEM105* | 9.522483633 | 0.026918194 | 0.045112808 | 1 |
| T2DM | pericyte | *ATP6V0A2* | 1.924569711 | 0.020225054 | 0.035448819 | 1 |
| T2DM | pericyte | *HS6ST3* | 2.912908887 | 0.008020486 | 0.01607421 | 1 |
| T2DM | pericyte | *TNFAIP8L1* | 3.043004313 | 0.005950916 | 0.012542209 | 1 |
| T2DM | pericyte | *SP140L* | 2.535503833 | 0.001135796 | 0.003593733 | 1 |
| T2DM | pericyte | *FAM174B* | 1.635411568 | 0.000562301 | 0.002159249 | 1 |
| T2DM | pericyte | *MUC1* | 1.148354783 | 0.001832636 | 0.005082393 | 1 |
| T2DM | pericyte | *SV2B* | 2.144926348 | 0.029513117 | 0.048687737 | 1 |
| T2DM | pericyte | *PDE6G* | 1.846155759 | 0.008031208 | 0.016090569 | 1 |
| T2DM | pericyte | *TLCD2* | 1.502765138 | 0.006620395 | 0.013691586 | 1 |

| | | | | | | |
|---|---|---|---|---|---|---|
| T2DM | pericyte | *AHNAK2* | 1.176692552 | 0.002368073 | 0.006149557 | 1 |
| T2DM | pericyte | *SP1* | 1.744230748 | 0.001816882 | 0.005049839 | 1 |
| T2DM | pericyte | *PDIA2* | 1.747581661 | 0.003747262 | 0.008790684 | 1 |
| T2DM | pericyte | *PCGF3* | 2.575269489 | 0.000189095 | 0.001048037 | 1 |
| T2DM | pericyte | *PSMD13* | 1.663803434 | 0.004778768 | 0.010605052 | 1 |
| T2DM | pericyte | *PBX1* | 1.938413667 | 0.000157752 | 0.000949988 | 0.991787027 |
| T2DM | pericyte | *UBE2L3* | 4.350957605 | 4.22E-06 | 0.000287028 | 0.026504282 |
| T2DM | pericyte | *C16orf52* | 3.28195635 | 0.000542214 | 0.002104259 | 1 |
| T2DM | pericyte | *ADARB2* | 3.74187847 | 0.000656897 | 0.002403905 | 1 |
| T2DM | pericyte | *SLC52A2* | 1.798040701 | 0.001232121 | 0.003802821 | 1 |
| T2DM | pericyte | *PIGP* | -0.316089955 | 0.022880073 | 0.0393251 | 1 |
| T2DM | pericyte | *ARL17A* | 1.205027246 | 0.004554384 | 0.010233528 | 1 |
| T2DM | pericyte | *NPIPB4* | 2.026209908 | 0.003268207 | 0.007927167 | 1 |
| T2DM | pericyte | *ZNF829* | 1.344130025 | 0.018174682 | 0.032398663 | 1 |
| T2DM | pericyte | *TRIM69* | 0.849885649 | 0.009492992 | 0.018523414 | 1 |
| T2DM | pericyte | *POMK* | 4.809173913 | 0.000254194 | 0.001258361 | 1 |
| T2DM | pericyte | *LINC00839* | 2.54690549 | 1.01E-05 | 0.000407741 | 0.063473081 |
| T2DM | pericyte | *RTN4RL1* | 0.93416762 | 0.002650287 | 0.006724115 | 1 |
| T2DM | pericyte | *ZNF566* | 1.613226099 | 0.027257197 | 0.045576063 | 1 |
| T2DM | pericyte | *ZNF529* | 1.597508879 | 0.022883384 | 0.0393251 | 1 |
| T2DM | pericyte | *ZNF284* | 3.480846108 | 0.001146469 | 0.0036184 | 1 |
| T2DM | pericyte | *GSAP* | 2.542165165 | 7.54E-06 | 0.000356314 | 0.04738975 |
| T2DM | pericyte | *WWOX* | 1.662205129 | 0.000155177 | 0.000944434 | 0.975600079 |
| T2DM | pericyte | *CYP4Z1* | 1.314425618 | 0.000903687 | 0.003044738 | 1 |
| T2DM | pericyte | *CCDC84* | 3.645287319 | 3.74E-05 | 0.000607238 | 0.235177516 |
| T2DM | pericyte | *POLR1D* | 1.92783826 | 0.000170638 | 0.000995155 | 1 |
| T2DM | pericyte | *KLHL32* | 0.848377538 | 0.010858733 | 0.020870944 | 1 |
| T2DM | pericyte | *NBPF12* | 1.733472186 | 0.001556844 | 0.004517679 | 1 |
| T2DM | pericyte | *TOR3A* | 5.244805479 | 0.000897826 | 0.00303638 | 1 |
| T2DM | pericyte | *CCDC30* | 2.150629083 | 0.002174232 | 0.005753114 | 1 |
| T2DM | pericyte | *GLDN* | 0.695703406 | 0.010177076 | 0.019681107 | 1 |
| T2DM | pericyte | *KPNA4* | 3.431508051 | 4.42E-05 | 0.000607238 | 0.277663061 |
| T2DM | pericyte | *ZNF197* | 1.223749826 | 0.013582423 | 0.025226793 | 1 |
| T2DM | pericyte | *PCLO* | 0.556742619 | 0.002152621 | 0.00570869 | 1 |
| T2DM | pericyte | *SEPT10* | 1.988074142 | 0.001186692 | 0.003706275 | 1 |
| T2DM | pericyte | *SMYD4* | 1.308965052 | 0.013966859 | 0.025826366 | 1 |
| T2DM | pericyte | *NF2* | 2.399366549 | 0.000530618 | 0.00207076 | 1 |
| T2DM | pericyte | *MIR22HG* | 1.040992869 | 0.004615634 | 0.010348962 | 1 |
| T2DM | pericyte | *C17orf58* | 0.774961078 | 0.001748682 | 0.00491671 | 1 |
| T2DM | pericyte | *BCR* | 1.493393453 | 0.00793513 | 0.015948901 | 1 |
| T2DM | pericyte | *FOXI2* | -0.823681496 | 0.025689262 | 0.043474668 | 1 |
| T2DM | pericyte | *HYAL3* | -0.643362016 | 0.030175317 | 0.049610935 | 1 |
| T2DM | pericyte | *ZSCAN30* | 4.023028446 | 4.12E-05 | 0.000607238 | 0.258790196 |
| T2DM | pericyte | *LILRB4* | 0.727591601 | 0.003582964 | 0.008494002 | 1 |
| T2DM | pericyte | *TMEM17* | 1.067224713 | 0.004414494 | 0.01000502 | 1 |
| T2DM | pericyte | *TNFRSF18* | 2.2909519 | 0.004675733 | 0.010446458 | 1 |
| T2DM | pericyte | *TMEM232* | 1.822990914 | 0.001369094 | 0.004081316 | 1 |
| T2DM | pericyte | *EFCAB6* | 3.661783868 | 1.02E-05 | 0.000407741 | 0.064362252 |
| T2DM | pericyte | *KANK3* | 4.024712261 | 0.001601732 | 0.004606629 | 1 |
| T2DM | pericyte | *EMID1* | 2.088524074 | 0.011715654 | 0.022272851 | 1 |

| | | | | | | |
|---|---|---|---|---|---|---|
| T2DM | pericyte | *CYP4A11* | 1.03436914 | 0.012023245 | 0.022733876 | 1 |
| T2DM | pericyte | *TMEM216* | 1.251899678 | 0.000341836 | 0.001508155 | 1 |
| T2DM | pericyte | *RPS19BP1* | 1.084760071 | 0.003271167 | 0.007929498 | 1 |
| T2DM | pericyte | *PLCD1* | 1.26743389 | 0.012562607 | 0.023597583 | 1 |
| T2DM | pericyte | *ENTPD5* | 4.123885604 | 7.03E-06 | 0.000348151 | 0.044215214 |
| T2DM | pericyte | *HEATR4* | 0.632693641 | 0.018767524 | 0.033227661 | 1 |
| T2DM | pericyte | *MRPS21* | 2.417832767 | 0.000396321 | 0.001673386 | 1 |
| T2DM | pericyte | *ANGPTL5* | 0.826545792 | 0.004003627 | 0.009253972 | 1 |
| T2DM | pericyte | *TSPYL4* | 1.701267079 | 0.003723206 | 0.008740776 | 1 |
| T2DM | pericyte | *MT1X* | 1.289203374 | 0.002309235 | 0.006026634 | 1 |
| T2DM | pericyte | *FNBP1* | 1.288115118 | 0.003395301 | 0.008147426 | 1 |
| T2DM | pericyte | *WDR86* | 0.999186146 | 0.020239911 | 0.035455091 | 1 |
| T2DM | pericyte | *FPR3* | 1.15258307 | 0.023793724 | 0.040671871 | 1 |
| T2DM | pericyte | *TLR5* | 4.262169154 | 0.000395369 | 0.001672514 | 1 |
| T2DM | pericyte | *ZNF385C* | 4.410977821 | 0.004705905 | 0.010491498 | 1 |
| T2DM | pericyte | *TET3* | 2.582908067 | 8.98E-05 | 0.000702125 | 0.564443994 |
| T2DM | pericyte | *TRPV2* | 2.302610728 | 0.00043102 | 0.001783951 | 1 |
| T2DM | pericyte | *C2orf88* | 1.877599057 | 0.000596803 | 0.002244078 | 1 |
| T2DM | pericyte | *SEMA4D* | 1.224639927 | 0.002034781 | 0.005462283 | 1 |
| T2DM | pericyte | *MCRS1* | 2.573998305 | 2.91E-06 | 0.000234693 | 0.018306051 |
| T2DM | pericyte | *TMEM220* | 3.298618271 | 0.000503704 | 0.00199169 | 1 |
| T2DM | pericyte | *CCDC157* | 2.162363128 | 0.003349474 | 0.008063243 | 1 |
| T2DM | pericyte | *GFRAL* | 1.264031477 | 0.001845914 | 0.005101215 | 1 |
| T2DM | pericyte | *C2orf66* | 3.991554754 | 0.004989044 | 0.010945263 | 1 |
| T2DM | pericyte | *C6orf132* | 1.141096143 | 0.007259109 | 0.014760031 | 1 |
| T2DM | pericyte | *FAM166A* | 3.753126416 | 0.005563122 | 0.011886436 | 1 |
| T2DM | pericyte | *ZNF626* | 1.52429595 | 0.0040857 | 0.009405637 | 1 |
| T2DM | pericyte | *SMTNL2* | -0.847901638 | 0.026268892 | 0.044259979 | 1 |
| T2DM | pericyte | *ZC3H6* | 2.658929952 | 0.000126397 | 0.00084164 | 0.794655797 |
| T2DM | pericyte | *PRKAR1B* | 1.203680072 | 0.014961391 | 0.027423401 | 1 |
| T2DM | pericyte | *NUTM2B* | -9.80731973 | 0.001064455 | 0.003424886 | 1 |
| T2DM | pericyte | *NCR3LG1* | 1.675888938 | 0.00443165 | 0.010033398 | 1 |
| T2DM | pericyte | *ZNF793* | 0.549444807 | 0.028943914 | 0.047899549 | 1 |
| T2DM | pericyte | *AGAP4* | 1.194021464 | 0.005639638 | 0.012018075 | 1 |
| T2DM | pericyte | *HYKK* | 1.238268057 | 0.001104104 | 0.003523606 | 1 |
| T2DM | pericyte | *RUFY4* | 1.937562513 | 0.005642971 | 0.012018075 | 1 |
| T2DM | pericyte | *PLSCR1* | 3.124189291 | 1.89E-05 | 0.000512131 | 0.118814467 |
| T2DM | pericyte | *JAKMIP3* | 0.802137792 | 0.007127414 | 0.014525139 | 1 |
| T2DM | pericyte | *CERKL* | 2.740852203 | 0.000832977 | 0.002875852 | 1 |
| T2DM | pericyte | *C19orf54* | 0.923589697 | 0.011820305 | 0.022434521 | 1 |
| T2DM | pericyte | *C22orf34* | 1.008475851 | 0.004824948 | 0.010669873 | 1 |
| T2DM | pericyte | *COL25A1* | -0.070907688 | 0.023604613 | 0.040404002 | 1 |
| T2DM | pericyte | *SRSF10* | 3.064488104 | 0.000431716 | 0.001785655 | 1 |
| T2DM | pericyte | *NBR1* | 3.914397903 | 8.34E-05 | 0.000681373 | 0.524143134 |
| T2DM | pericyte | *FAM72B* | 2.758598386 | 0.004626717 | 0.010362725 | 1 |
| T2DM | pericyte | *SUMO2* | 2.953836687 | 2.36E-05 | 0.000548455 | 0.148246533 |
| T2DM | pericyte | *GOLGA8M* | 3.175652625 | 9.08E-05 | 0.000704357 | 0.570792369 |
| T2DM | pericyte | *DPYD* | 2.420438038 | 0.00040863 | 0.001716137 | 1 |
| T2DM | pericyte | *S100A16* | 1.567024701 | 0.001242599 | 0.003827643 | 1 |
| T2DM | pericyte | *UROS* | 3.156348614 | 3.90E-05 | 0.000607238 | 0.245017593 |

| | | | | | | |
|---|---|---|---|---|---|---|
| T2DM | pericyte | *FAM221A* | 1.490499788 | 0.008456022 | 0.016781252 | 1 |
| T2DM | pericyte | *FSIP2* | 0.730120484 | 0.004622879 | 0.010361511 | 1 |
| T2DM | pericyte | *NOXA1* | 1.760082058 | 0.000950582 | 0.003167096 | 1 |
| T2DM | pericyte | *TMEM201* | 3.81715383 | 4.44E-06 | 0.000297085 | 0.027925957 |
| T2DM | pericyte | *LINC00910* | 1.14655792 | 0.02107421 | 0.036742528 | 1 |
| T2DM | pericyte | *BEND4* | 3.843691338 | 0.001270641 | 0.003881628 | 1 |
| T2DM | pericyte | *ZNF563* | 1.618339378 | 0.00420553 | 0.009621603 | 1 |
| T2DM | pericyte | *LRRK2* | 1.719756033 | 2.65E-05 | 0.000562456 | 0.166457816 |
| T2DM | pericyte | *FAM196A* | 2.225892816 | 0.013817806 | 0.025603462 | 1 |
| T2DM | pericyte | *C9orf152* | 2.037196874 | 0.005371889 | 0.01159484 | 1 |
| T2DM | pericyte | *SLC15A5* | 0.674404084 | 0.007415122 | 0.015023806 | 1 |
| T2DM | pericyte | *ADAT2* | 2.466104103 | 0.000501559 | 0.001985708 | 1 |
| T2DM | pericyte | *ZNF567* | 2.75966723 | 9.55E-05 | 0.000719108 | 0.600337253 |
| T2DM | pericyte | *ALKBH2* | 1.445495642 | 8.69E-05 | 0.000695771 | 0.546271034 |
| T2DM | pericyte | *LITAF* | 1.691617947 | 0.000500537 | 0.001982909 | 1 |
| T2DM | pericyte | *VSTM1* | 1.304020554 | 0.011305477 | 0.021630411 | 1 |
| T2DM | pericyte | *SF3B3* | 3.774655139 | 4.86E-05 | 0.000607238 | 0.30551376 |
| T2DM | pericyte | *SP6* | -6.716404781 | 0.008191263 | 0.016337938 | 1 |
| T2DM | pericyte | *ZNF573* | 3.681155678 | 0.000393548 | 0.001670229 | 1 |
| T2DM | pericyte | *FAM47E* | 0.565065211 | 0.009047045 | 0.017774617 | 1 |
| T2DM | pericyte | *S100A13* | 3.408532116 | 0.000901545 | 0.003040779 | 1 |
| T2DM | pericyte | *ZNF33A* | 2.392600867 | 0.000195872 | 0.001065597 | 1 |
| T2DM | pericyte | *ZNF600* | 0.893357291 | 0.007891642 | 0.015871643 | 1 |
| T2DM | pericyte | *C15orf61* | 4.542333902 | 4.64E-05 | 0.000607238 | 0.291628956 |
| T2DM | pericyte | *PNRC2* | 4.048070044 | 0.000380036 | 0.001626473 | 1 |
| T2DM | pericyte | *HLA-DRB1* | -2.391601831 | 3.03E-13 | 4.76E-10 | 1.91E-09 |
| T2DM | pericyte | *AKR1C3* | 1.548454368 | 0.001046181 | 0.003385334 | 1 |
| T2DM | pericyte | *WDSUB1* | 2.453319345 | 5.34E-05 | 0.000619691 | 0.335704117 |
| T2DM | pericyte | *SEMA4A* | 1.034842077 | 0.00468434 | 0.010454541 | 1 |
| T2DM | pericyte | *MPHOSPH8* | 2.678112442 | 0.000827989 | 0.002861776 | 1 |
| T2DM | pericyte | *SIRPB2* | 1.914090901 | 0.001059765 | 0.003417411 | 1 |
| T2DM | pericyte | *SRGAP3* | 0.985491504 | 0.000715571 | 0.002563417 | 1 |
| T2DM | pericyte | *SUPT5H* | 2.604918351 | 0.011152127 | 0.021376044 | 1 |
| T2DM | pericyte | *PPIA* | 3.449841294 | 0.000490069 | 0.001953752 | 1 |
| T2DM | pericyte | *ZNF471* | 1.300725351 | 0.008379479 | 0.016655638 | 1 |
| T2DM | pericyte | *GTF2IRD2* | 1.857016018 | 6.31E-06 | 0.000343901 | 0.039647035 |
| T2DM | pericyte | *GRM7* | 0.39528142 | 0.028003173 | 0.046612643 | 1 |
| T2DM | pericyte | *SUPT3H* | 3.083734367 | 0.00031311 | 0.001424252 | 1 |
| T2DM | pericyte | *GIMAP5* | 0.9823704 | 0.002070729 | 0.005535064 | 1 |
| T2DM | pericyte | *STK31* | 0.984063837 | 0.004724176 | 0.010527989 | 1 |
| T2DM | pericyte | *ZKSCAN7* | 3.525615473 | 0.013232242 | 0.024671146 | 1 |
| T2DM | pericyte | *ZNF565* | 1.784531465 | 0.002754498 | 0.00692701 | 1 |
| T2DM | pericyte | *NTNG2* | 2.346186023 | 0.001198654 | 0.003733971 | 1 |
| T2DM | pericyte | *WDR5* | 5.017998612 | 3.72E-06 | 0.000279804 | 0.023373811 |
| T2DM | pericyte | *LONP1* | 1.336009405 | 0.010884805 | 0.020914661 | 1 |
| T2DM | pericyte | *ZNF34* | 4.440743425 | 2.23E-05 | 0.000547486 | 0.140147554 |
| T2DM | pericyte | *INCA1* | 2.98084657 | 0.000181663 | 0.001034031 | 1 |
| T2DM | pericyte | *ZNF774* | 1.812068112 | 0.001835967 | 0.005086852 | 1 |
| T2DM | pericyte | *ZNF124* | 3.134667474 | 5.91E-05 | 0.00062437 | 0.371481176 |
| T2DM | pericyte | *ZNF569* | 1.653922077 | 0.005927209 | 0.012496433 | 1 |

| | | | | | | |
|---|---|---|---|---|---|---|
| T2DM | pericyte | *PIK3R4* | 1.074574949 | 0.02941396 | 0.048575142 | 1 |
| T2DM | pericyte | *ZNF605* | 2.280489352 | 0.000284045 | 0.001341693 | 1 |
| T2DM | pericyte | *RFX8* | 1.485197524 | 0.001260004 | 0.003865666 | 1 |
| T2DM | pericyte | *C20orf96* | 1.555784031 | 0.001588934 | 0.004578197 | 1 |
| T2DM | pericyte | *ANAPC7* | 1.937521444 | 0.000427564 | 0.001773148 | 1 |
| T2DM | pericyte | *TPK1* | 2.403326356 | 0.000152602 | 0.000937839 | 0.959408985 |
| T2DM | pericyte | *C1orf186* | 1.096372173 | 0.004771964 | 0.010593692 | 1 |
| T2DM | pericyte | *XRCC2* | 1.214191063 | 0.002970199 | 0.007351827 | 1 |
| T2DM | pericyte | *HDAC2* | 3.550050651 | 0.000779163 | 0.002738175 | 1 |
| T2DM | pericyte | *ADH1B* | 1.59318915 | 0.000104941 | 0.000759224 | 0.659765898 |
| T2DM | pericyte | *HRH1* | 3.536461732 | 6.74E-05 | 0.000632837 | 0.423574164 |
| T2DM | pericyte | *ZNF502* | 1.048461983 | 0.014286185 | 0.026316216 | 1 |
| T2DM | pericyte | *TECPR2* | 3.520000762 | 3.88E-05 | 0.000607238 | 0.24378385 |
| T2DM | pericyte | *ZFP62* | 4.519729119 | 0.000206302 | 0.001101974 | 1 |
| T2DM | pericyte | *ERI2* | 2.255112675 | 8.76E-05 | 0.000699239 | 0.550697649 |
| T2DM | pericyte | *TOMM7* | 2.531840984 | 0.023496797 | 0.040251869 | 1 |
| T2DM | pericyte | *TRPV1* | 1.559553118 | 4.37E-05 | 0.000607238 | 0.274688449 |
| T2DM | pericyte | *ZNF431* | 2.470186483 | 0.002324586 | 0.006059128 | 1 |
| T2DM | pericyte | *NF1* | 2.42060168 | 0.000305871 | 0.001401337 | 1 |
| T2DM | pericyte | *DAPK1* | 1.089828881 | 0.009192837 | 0.018013908 | 1 |
| T2DM | pericyte | *HLA-DQA1* | -0.681288128 | 0.006694128 | 0.013807737 | 1 |
| T2DM | pericyte | *GM2A* | 0.984678966 | 0.014598985 | 0.026837374 | 1 |
| T2DM | pericyte | *STRN3* | 2.778962856 | 0.000351398 | 0.001530521 | 1 |
| T2DM | pericyte | *MVB12B* | 3.168576374 | 5.35E-05 | 0.000619691 | 0.33610372 |
| T2DM | pericyte | *C6orf106* | 1.357556073 | 2.75E-05 | 0.000569342 | 0.173079937 |
| T2DM | pericyte | *PPTC7* | 1.95528812 | 0.000435913 | 0.001799464 | 1 |
| T2DM | pericyte | *NHLRC2* | 3.036923216 | 0.000275922 | 0.001324213 | 1 |
| T2DM | pericyte | *ZFP28* | 1.254145968 | 0.003001891 | 0.007409849 | 1 |
| T2DM | pericyte | *LAMB3* | 1.408909572 | 0.001854061 | 0.005116982 | 1 |
| T2DM | pericyte | *SRGAP1* | 3.070257817 | 0.000114466 | 0.000791693 | 0.719648505 |
| T2DM | pericyte | *SLC39A10* | 2.297937715 | 0.001348453 | 0.004041043 | 1 |
| T2DM | pericyte | *ZNF585A* | 1.191530023 | 0.000452345 | 0.00185029 | 1 |
| T2DM | pericyte | *FUT11* | 2.851337898 | 5.21E-06 | 0.000327358 | 0.032735816 |
| T2DM | pericyte | *ANXA4* | 2.668718815 | 4.56E-05 | 0.000607238 | 0.286593418 |
| T2DM | pericyte | *ZNF138* | 2.28222804 | 5.89E-05 | 0.00062437 | 0.370284195 |
| T2DM | pericyte | *ZNF429* | 1.679352305 | 0.002830627 | 0.007076004 | 1 |
| T2DM | pericyte | *ZNF470* | 1.157929598 | 0.023038593 | 0.03953156 | 1 |
| T2DM | pericyte | *GMFB* | 2.742306514 | 0.000162392 | 0.000963715 | 1 |
| T2DM | pericyte | *ZMYM1* | 3.969834862 | 0.000271864 | 0.001314776 | 1 |
| T2DM | pericyte | *ARRDC1* | 1.073926271 | 0.008150728 | 0.016267819 | 1 |
| T2DM | pericyte | *SLC25A29* | 1.934458316 | 7.98E-05 | 0.000667567 | 0.501894461 |
| T2DM | pericyte | *ZNF682* | 2.522626395 | 0.025594975 | 0.043338435 | 1 |
| T2DM | pericyte | *ZNF257* | 1.646258654 | 0.003888565 | 0.009057178 | 1 |
| T2DM | pericyte | *ADAM32* | 0.922818338 | 0.005452162 | 0.011722893 | 1 |
| T2DM | pericyte | *LRRC8B* | 1.610426645 | 0.002110492 | 0.005605687 | 1 |
| T2DM | pericyte | *SND1* | 1.651101047 | 0.002288203 | 0.005984165 | 1 |
| T2DM | pericyte | *SULT1A2* | 0.352792414 | 0.008083035 | 0.016172839 | 1 |
| T2DM | pericyte | *PIWIL2* | 1.127836763 | 0.001372039 | 0.004088155 | 1 |
| T2DM | pericyte | *ENTPD4* | 2.135920689 | 0.001246282 | 0.003833353 | 1 |
| T2DM | pericyte | *FAM110D* | 4.657253913 | 0.000882602 | 0.003004287 | 1 |

| | | | | | | |
|---|---|---|---|---|---|---|
| T2DM | pericyte | *KANK2* | 3.500716083 | 0.000583387 | 0.00220618 | 1 |
| T2DM | pericyte | *GTF2E2* | 3.503214296 | 4.69E-05 | 0.000607238 | 0.294717139 |
| T2DM | pericyte | *RAMP2-AS1* | 3.218209744 | 9.53E-06 | 0.000402119 | 0.059915685 |
| T2DM | pericyte | *DDI2* | 2.097900031 | 0.000416203 | 0.001738652 | 1 |
| T2DM | pericyte | *MRPL21* | 0.510419049 | 0.018707274 | 0.03315834 | 1 |
| T2DM | pericyte | *ZNF786* | 1.194558824 | 0.007980257 | 0.016008895 | 1 |
| T2DM | pericyte | *ZNF517* | 7.92372196 | 0.000301812 | 0.001391125 | 1 |
| T2DM | pericyte | *SLC22A5* | 0.683122008 | 0.011822104 | 0.022434521 | 1 |
| T2DM | pericyte | *C5AR1* | 3.831732181 | 5.80E-06 | 0.000331374 | 0.036451188 |
| T2DM | pericyte | *DCHS2* | -0.362496869 | 0.023604883 | 0.040404002 | 1 |
| T2DM | pericyte | *VEPH1* | 0.731787136 | 0.004883498 | 0.010780391 | 1 |
| T2DM | pericyte | *MAP3K5* | 1.955626852 | 0.000530129 | 0.002070139 | 1 |
| T2DM | pericyte | *STMN3* | 1.27601644 | 0.00333692 | 0.008049169 | 1 |
| T2DM | pericyte | *GYPE* | 1.245377485 | 0.000572505 | 0.00218539 | 1 |
| T2DM | pericyte | *PCDHB11* | -1.488066843 | 0.010539795 | 0.020320053 | 1 |
| T2DM | pericyte | *SLC2A10* | 2.762872762 | 7.94E-05 | 0.000667567 | 0.499475509 |
| T2DM | pericyte | *RPF2* | 2.381331178 | 0.000497345 | 0.00197275 | 1 |
| T2DM | pericyte | *MYO5A* | 1.689967913 | 0.000461885 | 0.00187827 | 1 |
| T2DM | pericyte | *ATG7* | 1.738252352 | 0.000563596 | 0.002159249 | 1 |
| T2DM | pericyte | *RAB40C* | 4.458184117 | 5.09E-05 | 0.000614643 | 0.32022897 |
| T2DM | pericyte | *ZNF624* | 4.486601314 | 0.002594316 | 0.006614137 | 1 |
| T2DM | pericyte | *BCO2* | 1.065830111 | 0.004120725 | 0.009459276 | 1 |
| T2DM | pericyte | *CCDC154* | 4.368027323 | 3.62E-05 | 0.00060096 | 0.227786714 |
| T2DM | pericyte | *C5orf42* | 1.161984403 | 0.007137551 | 0.014541082 | 1 |
| T2DM | pericyte | *ZNF841* | 1.174011459 | 0.008873747 | 0.01748565 | 1 |
| T2DM | pericyte | *MFAP5* | 2.187494013 | 6.76E-05 | 0.000632837 | 0.424756788 |
| T2DM | pericyte | *CDC42SE1* | 3.024545316 | 0.000521563 | 0.002043031 | 1 |
| T2DM | pericyte | *PDCD1LG2* | 0.85595942 | 0.01955302 | 0.03445343 | 1 |
| T2DM | pericyte | *NMB* | 5.348191771 | 1.80E-05 | 0.000512131 | 0.113315083 |
| T2DM | pericyte | *FAM114A1* | 2.254264098 | 7.02E-05 | 0.000639553 | 0.441365216 |
| T2DM | pericyte | *PHF2* | 2.238191166 | 0.00188508 | 0.005174823 | 1 |
| T2DM | pericyte | *C14orf178* | 0.651563018 | 0.019674224 | 0.034628177 | 1 |
| T2DM | pericyte | *PSAP* | 2.900827901 | 0.000116302 | 0.000798241 | 0.731188756 |
| T2DM | pericyte | *MAP1LC3C* | 0.853224809 | 0.022211076 | 0.038426262 | 1 |
| T2DM | pericyte | *TAF13* | 1.515668484 | 0.000771457 | 0.002722984 | 1 |
| T2DM | pericyte | *AC122129.1* | 4.749698879 | 0.001249294 | 0.003838863 | 1 |
| T2DM | pericyte | *ZNF181* | 2.785516429 | 0.000177577 | 0.001020443 | 1 |
| T2DM | pericyte | *SGTB* | 3.654304679 | 9.89E-05 | 0.00073561 | 0.621590294 |
| T2DM | pericyte | *ADH5* | 1.388267916 | 0.00977452 | 0.019031405 | 1 |
| T2DM | pericyte | *ZNF677* | 4.86542091 | 0.00132205 | 0.003994104 | 1 |
| T2DM | pericyte | *FCHSD1* | 2.373469672 | 0.013040732 | 0.024386402 | 1 |
| T2DM | pericyte | *RPL12* | 1.355692846 | 0.00267624 | 0.006784483 | 1 |
| T2DM | pericyte | *VPS13A* | 1.403658782 | 0.000275525 | 0.00132332 | 1 |
| T2DM | pericyte | *NOL8* | 2.990052105 | 0.003827327 | 0.008948459 | 1 |
| T2DM | pericyte | *ZNF335* | 1.508323469 | 0.002913955 | 0.007232543 | 1 |
| T2DM | pericyte | *AGAP9* | 1.014884254 | 0.003338078 | 0.008049169 | 1 |
| T2DM | pericyte | *ZNF273* | 1.829302391 | 0.000343186 | 0.00151137 | 1 |
| T2DM | pericyte | *ZNF84* | 2.579596666 | 0.001329075 | 0.004003783 | 1 |
| T2DM | pericyte | *SFI1* | 3.131270639 | 5.06E-05 | 0.000611501 | 0.317980276 |
| T2DM | pericyte | *HIBCH* | 1.34436638 | 0.01697506 | 0.030570668 | 1 |

| | | | | | | |
|---|---|---|---|---|---|---|
| T2DM | pericyte | *MIER1* | 3.776188922 | 1.01E-05 | 0.000407741 | 0.06332543 |
| T2DM | pericyte | *SVIP* | 2.041124215 | 0.00036748 | 0.001585434 | 1 |
| T2DM | pericyte | *ZNF251* | 3.293620137 | 0.000184346 | 0.001035729 | 1 |
| T2DM | pericyte | *DDRGK1* | 4.59502598 | 0.000765219 | 0.002704289 | 1 |
| T2DM | pericyte | *HSD17B11* | 2.476246507 | 6.55E-05 | 0.00063262 | 0.4118353 |
| T2DM | pericyte | *SULT1C2* | 0.653743869 | 0.001344431 | 0.004034577 | 1 |
| T2DM | pericyte | *RPS6KL1* | 0.76792354 | 0.004999195 | 0.010958836 | 1 |
| T2DM | pericyte | *DDX42* | 3.061913557 | 5.16E-05 | 0.000616617 | 0.324258298 |
| T2DM | pericyte | *SLC29A3* | 1.521541831 | 0.016253685 | 0.029457168 | 1 |
| T2DM | pericyte | *ANTXRL* | 28.98228008 | 0.007063419 | 0.014418089 | 1 |
| T2DM | pericyte | *STYX* | 2.960762447 | 6.44E-05 | 0.000632025 | 0.404810485 |
| T2DM | pericyte | *ZKSCAN8* | 2.662533463 | 0.001627756 | 0.004662279 | 1 |
| T2DM | pericyte | *ZNF813* | 1.951481658 | 0.00445739 | 0.010069568 | 1 |
| T2DM | pericyte | *UVRAG* | 1.255393634 | 0.00160798 | 0.004620369 | 1 |
| T2DM | pericyte | *MGEA5* | 1.759938718 | 0.002540713 | 0.006501205 | 1 |
| T2DM | pericyte | *MT1F* | -1.700056514 | 0.026922739 | 0.045112808 | 1 |
| T2DM | pericyte | *ZNF568* | 4.224862791 | 8.45E-08 | 1.83E-05 | 0.000531313 |
| T2DM | pericyte | *ZNF587* | 2.210520171 | 0.001654283 | 0.004721053 | 1 |
| T2DM | pericyte | *TMA16* | 3.637085297 | 1.49E-05 | 0.000478946 | 0.093619178 |
| T2DM | pericyte | *HLA-DRB5* | -0.752656124 | 3.13E-12 | 1.97E-09 | 1.97E-08 |
| T2DM | pericyte | *MAFK* | 1.370247382 | 0.001401383 | 0.004165718 | 1 |
| T2DM | pericyte | *C20orf203* | 1.423565421 | 0.023453104 | 0.040187971 | 1 |
| T2DM | pericyte | *ZNF627* | 3.147566938 | 7.77E-05 | 0.000663325 | 0.48848956 |
| T2DM | pericyte | *TLK1* | 2.775044727 | 4.17E-05 | 0.000607238 | 0.261903707 |
| T2DM | pericyte | *LRBA* | 2.706775502 | 0.000311908 | 0.001422709 | 1 |
| T2DM | pericyte | *MMP17* | 5.438549206 | 7.06E-05 | 0.000639553 | 0.443849706 |
| T2DM | pericyte | *CCDC69* | 1.165476916 | 0.001339738 | 0.004024334 | 1 |
| T2DM | pericyte | *MDM4* | 3.168329292 | 0.000174312 | 0.001009116 | 1 |
| T2DM | pericyte | *NCOA6* | 1.49952121 | 0.024014928 | 0.040972009 | 1 |
| T2DM | pericyte | *FAM19A2* | 2.423720943 | 0.000924708 | 0.003097305 | 1 |
| T2DM | pericyte | *TTC37* | 2.804700139 | 6.94E-05 | 0.000637766 | 0.436231715 |
| T2DM | pericyte | *TUSC1* | 1.378121732 | 0.003564909 | 0.008467163 | 1 |
| T2DM | pericyte | *PAPSS2* | 2.250972448 | 0.000683599 | 0.002474259 | 1 |
| T2DM | pericyte | *FAN1* | 3.409392983 | 0.00314335 | 0.007689588 | 1 |
| T2DM | pericyte | *CEP290* | 2.709648606 | 0.001615207 | 0.004636898 | 1 |
| T2DM | pericyte | *ECI2* | 2.292054204 | 0.000176504 | 0.001018057 | 1 |
| T2DM | pericyte | *ZNF652* | 3.447366437 | 0.000186953 | 0.001043394 | 1 |
| T2DM | pericyte | *APCDD1L* | 3.103021947 | 0.000244421 | 0.001229339 | 1 |
| T2DM | pericyte | *RASSF9* | 2.765110541 | 0.000148492 | 0.000922861 | 0.933568037 |
| T2DM | pericyte | *TMEM184B* | 4.052689668 | 7.33E-05 | 0.000646906 | 0.460659796 |
| T2DM | pericyte | *SFT2D1* | 3.823383869 | 0.000432899 | 0.001789371 | 1 |
| T2DM | pericyte | *ARHGAP11A* | 1.28355104 | 0.002691102 | 0.006811174 | 1 |
| T2DM | pericyte | *HMGN2* | 2.32751154 | 5.36E-05 | 0.000619691 | 0.336754266 |
| T2DM | pericyte | *DENND4B* | 1.230740242 | 0.024613831 | 0.041789672 | 1 |
| T2DM | pericyte | *CD3E* | 0.712371454 | 0.017586663 | 0.031509647 | 1 |
| T2DM | pericyte | *TSEN15* | 2.654227051 | 0.000121117 | 0.000821424 | 0.761460134 |
| T2DM | pericyte | *TYW1* | 1.121656277 | 0.001323342 | 0.003995157 | 1 |
| T2DM | pericyte | *SHISA4* | 1.627239657 | 0.012022004 | 0.022733876 | 1 |
| T2DM | pericyte | *CAPZA2* | 2.534673265 | 4.36E-05 | 0.000607238 | 0.274056114 |
| T2DM | pericyte | *MAP3K3* | 2.237959677 | 0.013918134 | 0.025765202 | 1 |

| | | | | | | |
|---|---|---|---|---|---|---|
| T2DM | pericyte | *C1orf174* | 1.788034728 | 0.020122067 | 0.035287988 | 1 |
| T2DM | pericyte | *KIAA0753* | 1.946210865 | 0.012349759 | 0.023274743 | 1 |
| T2DM | pericyte | *DCLRE1A* | 1.110787289 | 0.002561175 | 0.006541705 | 1 |
| T2DM | pericyte | *ZFP2* | 3.223979763 | 0.000246919 | 0.001237945 | 1 |
| T2DM | pericyte | *L3MBTL3* | 1.888719289 | 0.004490399 | 0.010118688 | 1 |
| T2DM | pericyte | *MFAP3L* | 0.993113769 | 0.003451906 | 0.00826433 | 1 |
| T2DM | pericyte | *NAGA* | 1.500303422 | 0.009838898 | 0.01913305 | 1 |
| T2DM | pericyte | *PJA2* | 2.068830794 | 0.005720607 | 0.012142288 | 1 |
| T2DM | pericyte | *AP001007.1* | 4.558868624 | 0.003848652 | 0.00899163 | 1 |
| T2DM | pericyte | *ZNF525* | 1.193040038 | 0.003111743 | 0.007621283 | 1 |
| T2DM | pericyte | *EFCAB2* | 2.649947635 | 0.000252025 | 0.001254535 | 1 |
| T2DM | pericyte | *CHML* | 1.522293248 | 0.000454995 | 0.001857501 | 1 |
| T2DM | pericyte | *CAPN8* | -0.296727228 | 0.019784954 | 0.034776823 | 1 |
| T2DM | pericyte | *ECT2L* | 1.271974918 | 0.019608754 | 0.034541955 | 1 |
| T2DM | pericyte | *FAM229B* | -0.676736505 | 0.02807225 | 0.046702895 | 1 |
| T2DM | pericyte | *DDO* | 1.892316135 | 0.001506113 | 0.004408255 | 1 |
| T2DM | pericyte | *HIST2H2BF* | 2.75687594 | 0.005144759 | 0.011223144 | 1 |
| T2DM | pericyte | *C6orf163* | 1.058479618 | 0.006648351 | 0.013735847 | 1 |
| T2DM | pericyte | *PCMTD2* | 3.18247202 | 4.12E-06 | 0.000287028 | 0.02589327 |
| T2DM | pericyte | *FOXO6* | -0.987175667 | 0.026745038 | 0.044912373 | 1 |
| T2DM | pericyte | *TRAF3IP1* | 1.981621291 | 0.002524233 | 0.00646957 | 1 |
| T2DM | pericyte | *GIGYF2* | 3.102536981 | 0.000128776 | 0.000851331 | 0.809616129 |
| T2DM | pericyte | *RUFY2* | 2.557229116 | 0.000172322 | 0.00100221 | 1 |
| T2DM | pericyte | *ASAH2B* | 1.314293704 | 0.001316831 | 0.003985998 | 1 |
| T2DM | pericyte | *AGAP6* | 0.970590583 | 0.000534523 | 0.002083415 | 1 |
| T2DM | pericyte | *C10orf128* | 1.316771951 | 0.003925192 | 0.009117298 | 1 |
| T2DM | pericyte | *SYT15* | 2.018931662 | 0.028111342 | 0.046743192 | 1 |
| T2DM | pericyte | *ZDBF2* | 2.263286331 | 8.05E-05 | 0.000670993 | 0.505928829 |
| T2DM | pericyte | *OXLD1* | 2.018776613 | 5.68E-06 | 0.000331374 | 0.035717527 |
| T2DM | pericyte | *HLA-DRA* | 2.778291066 | 5.39E-15 | 1.66E-11 | 3.39E-11 |
| T2DM | pericyte | *NOTCH4* | -0.237852097 | 1.40E-07 | 2.75E-05 | 0.000880153 |
| T2DM | pericyte | *AGER* | 2.002947543 | 0.00018937 | 0.001048037 | 1 |
| T2DM | pericyte | *PRRT1* | 1.255989391 | 0.001417947 | 0.004199074 | 1 |
| T2DM | pericyte | *DXO* | -0.949349408 | 0.001481071 | 0.004345073 | 1 |
| T2DM | pericyte | *NELFE* | 2.191256575 | 5.47E-07 | 7.17E-05 | 0.003442045 |
| T2DM | pericyte | *MSH5* | 1.071711094 | 0.001269996 | 0.003881608 | 1 |
| T2DM | pericyte | *BAG6* | 2.28502675 | 7.12E-06 | 0.000349569 | 0.044744821 |
| T2DM | pericyte | *NFKBIL1* | 1.818454775 | 1.80E-05 | 0.000512131 | 0.11339543 |
| T2DM | pericyte | *ZNF814* | 2.226329884 | 0.000261882 | 0.001284286 | 1 |
| T2DM | pericyte | *PSORS1C1* | 0.206459415 | 0.003950509 | 0.009168272 | 1 |
| T2DM | pericyte | *ABCF1* | 1.579504956 | 0.006787857 | 0.013963119 | 1 |
| T2DM | pericyte | *HLA-E* | 1.381258094 | 0.000878683 | 0.002995813 | 1 |
| T2DM | pericyte | *ZNF468* | 1.811951446 | 0.003705348 | 0.008711864 | 1 |
| T2DM | pericyte | *HLA-G* | 0.838622026 | 0.018793747 | 0.03326472 | 1 |
| T2DM | pericyte | *TBC1D8* | 2.624257803 | 0.000134014 | 0.000872202 | 0.842547329 |
| T2DM | pericyte | *FAM153C* | 2.035462579 | 0.027709013 | 0.046189572 | 1 |
| T2DM | pericyte | *ATXN2* | 2.265424132 | 0.000578125 | 0.002196177 | 1 |
| T2DM | pericyte | *FAM216A* | 2.077772842 | 0.000307642 | 0.001406649 | 1 |
| T2DM | pericyte | *ZNF155* | 1.397742656 | 0.012626234 | 0.023695861 | 1 |
| T2DM | pericyte | *ZNF783* | 2.095728879 | 0.003752704 | 0.008800168 | 1 |

| | | | | | | |
|---|---|---|---|---|---|---|
| T2DM | pericyte | *LRRC10B* | -0.979231332 | 0.02675433 | 0.044912373 | 1 |
| T2DM | pericyte | *PCDHA4* | -0.614952365 | 0.024719244 | 0.041945988 | 1 |
| T2DM | pericyte | *TRIM13* | 2.086680638 | 0.009087279 | 0.017842511 | 1 |
| T2DM | pericyte | *FAM71F2* | 5.23822008 | 0.000598484 | 0.002247617 | 1 |
| T2DM | pericyte | *LINC00654* | 1.606237793 | 0.005339487 | 0.011547765 | 1 |
| T2DM | pericyte | *SAP25* | -0.884536348 | 0.006950961 | 0.014234752 | 1 |
| T2DM | pericyte | *MT1M* | 4.831182504 | 0.000143027 | 0.000906847 | 0.899209221 |
| T2DM | pericyte | *SAMD9* | 2.082413883 | 0.000420025 | 0.00174765 | 1 |
| T2DM | pericyte | *EXOC3L4* | 1.986736677 | 0.019701635 | 0.034657016 | 1 |
| T2DM | pericyte | *ATP6AP1L* | 1.296383066 | 0.000182392 | 0.001034031 | 1 |
| T2DM | pericyte | *NAP1L4* | 1.501134253 | 0.021169473 | 0.036868672 | 1 |
| T2DM | pericyte | *DENND6B* | 2.282082093 | 0.000589955 | 0.002224983 | 1 |
| T2DM | pericyte | *EIF3CL* | 4.165263587 | 7.24E-05 | 0.00064687 | 0.455024361 |
| T2DM | pericyte | *LCMT1* | 2.647684107 | 0.000133591 | 0.000871003 | 0.839886736 |
| T2DM | pericyte | *LIN52* | 2.837262767 | 2.38E-08 | 6.50E-06 | 0.000149418 |
| T2DM | pericyte | *C1QTNF9B* | 75.03102057 | 0.002246226 | 0.005911269 | 1 |
| T2DM | pericyte | *RNPS1* | 2.126125949 | 0.003889674 | 0.009057178 | 1 |
| T2DM | pericyte | *NYNRIN* | 3.998563151 | 0.004480453 | 0.010103518 | 1 |
| T2DM | pericyte | *HCG27* | 1.659039481 | 7.49E-13 | 5.89E-10 | 4.71E-09 |
| T2DM | pericyte | *COLQ* | 1.642412491 | 0.004316274 | 0.00982383 | 1 |
| T2DM | pericyte | *METTL6* | 1.917764965 | 0.000103394 | 0.000753104 | 0.650035017 |
| T2DM | pericyte | *SACM1L* | 3.09940468 | 0.00031513 | 0.001426579 | 1 |
| T2DM | pericyte | *TAS2R14* | 1.247235192 | 0.00014634 | 0.000917289 | 0.920040493 |
| T2DM | pericyte | *ZRSR1* | 4.972492612 | 0.001114718 | 0.003544881 | 1 |
| T2DM | pericyte | *ZNF580* | 1.834643893 | 0.002156297 | 0.005715277 | 1 |
| T2DM | pericyte | *NUP62* | 2.288562702 | 0.000700123 | 0.002522449 | 1 |
| T2DM | pericyte | *FGFR1OP* | 4.101358191 | 8.94E-08 | 1.87E-05 | 0.000561852 |
| T2DM | pericyte | *ZNF254* | 3.430827174 | 9.50E-05 | 0.000717653 | 0.597087069 |
| T2DM | pericyte | *TCTEX1D2* | 1.428096892 | 0.003349959 | 0.008063243 | 1 |
| T2DM | pericyte | *ASIC3* | 0.892433465 | 0.021549934 | 0.037436982 | 1 |
| T2DM | pericyte | *GIMAP1* | 1.608764824 | 0.012999854 | 0.024324429 | 1 |
| T2DM | pericyte | *CCDC183* | 3.158972716 | 0.021778589 | 0.037782006 | 1 |
| T2DM | pericyte | *DNLZ* | 0.860409978 | 0.000291229 | 0.00136182 | 1 |
| T2DM | pericyte | *NOTCH2NL* | 0.878730285 | 0.015875879 | 0.028905778 | 1 |
| T2DM | pericyte | *SUPT4H1* | 1.955960981 | 0.029462692 | 0.048630072 | 1 |
| T2DM | pericyte | *GSTM2* | 0.80934271 | 0.02036271 | 0.035650336 | 1 |
| T2DM | pericyte | *ARHGAP19* | 2.48102319 | 0.000670829 | 0.002443511 | 1 |
| T2DM | pericyte | *SIPA1* | 2.570766978 | 0.000136496 | 0.000878628 | 0.858150966 |
| T2DM | pericyte | *ERV3-1* | 0.312304703 | 0.028359335 | 0.047080839 | 1 |
| T2DM | pericyte | *RBMXL1* | 2.415514347 | 0.001652461 | 0.004718355 | 1 |
| T2DM | pericyte | *HEXA* | 4.166846483 | 0.000289241 | 0.001357058 | 1 |
| T2DM | pericyte | *LEPROT* | 2.716378833 | 0.001473754 | 0.004327646 | 1 |
| T2DM | pericyte | *PPP1CB* | 2.235874902 | 0.000762913 | 0.002700345 | 1 |
| T2DM | pericyte | *NCKIPSD* | 2.611915521 | 0.000446552 | 0.001831358 | 1 |
| T2DM | pericyte | *ATF6B* | 1.99852099 | 9.53E-08 | 1.93E-05 | 0.000599329 |
| T2DM | pericyte | *SLC35F6* | 4.760110075 | 0.001059957 | 0.003417411 | 1 |
| T2DM | pericyte | *ATP6V1G2* | 0.479414867 | 0.015183519 | 0.027790039 | 1 |
| T2DM | pericyte | *PPM1N* | 2.319936562 | 0.000157113 | 0.000947957 | 0.987771 |
| T2DM | pericyte | *GALT* | 2.315290993 | 0.000233907 | 0.001198741 | 1 |
| T2DM | pericyte | *TAX1BP3* | 4.297386061 | 0.002042983 | 0.005477285 | 1 |

| | | | | | | |
|---|---|---|---|---|---|---|
| T2DM | pericyte | *AP1G2* | 3.309406466 | 5.02E-05 | 0.000610424 | 0.31532844 |
| T2DM | pericyte | *TM6SF2* | 12.71264837 | 0.003860045 | 0.009008206 | 1 |
| T2DM | pericyte | *REPIN1* | 1.439386608 | 0.008990864 | 0.01766976 | 1 |
| T2DM | pericyte | *SMIM7* | 2.453418379 | 4.39E-05 | 0.000607238 | 0.275945326 |
| T2DM | pericyte | *TSPAN4* | 3.297247336 | 0.000673487 | 0.002446108 | 1 |
| T2DM | pericyte | *AC079776.2* | 2.295547554 | 0.002090369 | 0.00556635 | 1 |
| T2DM | pericyte | *PAXIP1-AS2* | 3.010891677 | 0.000164378 | 0.000972196 | 1 |
| T2DM | pericyte | *LYRM4* | 2.065608322 | 0.024590512 | 0.041783933 | 1 |
| T2DM | pericyte | *STARD10* | 1.110466622 | 2.63E-07 | 4.46E-05 | 0.001651873 |
| T2DM | pericyte | *CPEB1* | 1.670337292 | 0.019786336 | 0.034776823 | 1 |
| T2DM | pericyte | *AC005082.1* | 3.24412934 | 0.008673445 | 0.017165966 | 1 |
| T2DM | pericyte | *TBC1D26* | -5.634432997 | 0.014226359 | 0.026229068 | 1 |
| T2DM | pericyte | *UBXN2B* | 1.562124507 | 0.002354406 | 0.006118532 | 1 |
| T2DM | pericyte | *UBE2QL1* | 1.251045829 | 0.005413096 | 0.011646864 | 1 |
| T2DM | pericyte | *GOLGA8B* | 3.847865948 | 2.27E-05 | 0.000548455 | 0.142840576 |
| T2DM | pericyte | *MYL5* | 4.896439771 | 0.000265692 | 0.001293785 | 1 |
| T2DM | pericyte | *TSTD1* | 1.448886445 | 0.001358107 | 0.004058896 | 1 |
| T2DM | pericyte | *LINC01138* | 3.932655107 | 4.76E-05 | 0.000607238 | 0.299025069 |
| T2DM | pericyte | *ATAD3C* | 2.145730389 | 0.001305053 | 0.003957968 | 1 |
| T2DM | pericyte | *CCDC7* | 2.015408428 | 0.006215644 | 0.012986957 | 1 |
| T2DM | pericyte | *RNASEK* | 2.00339793 | 0.000682115 | 0.002471734 | 1 |
| T2DM | pericyte | *NBPF1* | 1.50012025 | 0.012523798 | 0.023552832 | 1 |
| T2DM | pericyte | *FAM228B* | 0.544139327 | 0.008916828 | 0.017540706 | 1 |
| T2DM | pericyte | *RNU6ATAC35l* | 4.370294706 | 0.004913792 | 0.010828255 | 1 |
| T2DM | pericyte | *PPP3R1* | 1.373592578 | 0.002777473 | 0.006970848 | 1 |
| T2DM | pericyte | *AP4M1* | 0.641833926 | 0.017568744 | 0.031486514 | 1 |
| T2DM | pericyte | *APOL6* | 1.8854648 | 0.00019429 | 0.001065033 | 1 |
| T2DM | pericyte | *UBA52* | 2.811680946 | 2.37E-05 | 0.000548455 | 0.149060795 |
| T2DM | pericyte | *FAM185A* | 3.392910249 | 4.06E-05 | 0.000607238 | 0.25518908 |
| T2DM | pericyte | *HLA-DPB1* | 0.928863093 | 0.011590559 | 0.022101863 | 1 |
| T2DM | pericyte | *OSER1-AS1* | 1.064343134 | 0.002924171 | 0.007255038 | 1 |
| T2DM | pericyte | *LINC01133* | 2.178953938 | 0.000346822 | 0.00152055 | 1 |
| T2DM | pericyte | *AC069213.1* | -0.936898882 | 0.015740588 | 0.02868437 | 1 |
| T2DM | pericyte | *LINC00240* | 1.233497127 | 0.001099404 | 0.003513957 | 1 |
| T2DM | pericyte | *LINC00863* | 1.173301128 | 0.001364996 | 0.004072962 | 1 |
| T2DM | pericyte | *LINC00237* | -0.812283939 | 0.027337608 | 0.045686215 | 1 |
| T2DM | pericyte | *PLEKHM1* | 3.300912006 | 5.60E-06 | 0.000331374 | 0.035229362 |
| T2DM | pericyte | *SFTA1P* | -0.171991525 | 0.021002693 | 0.03663816 | 1 |
| T2DM | pericyte | *MIAT* | 0.710321741 | 0.01212831 | 0.022904982 | 1 |
| T2DM | pericyte | *LINC00626* | 18.07261582 | 0.002021862 | 0.005437849 | 1 |
| T2DM | pericyte | *ERCC6* | 3.787012945 | 0.000328502 | 0.001469959 | 1 |
| T2DM | pericyte | *LINC00115* | 0.859047346 | 0.006763965 | 0.013924377 | 1 |
| T2DM | pericyte | *NOL7* | 4.934391591 | 9.13E-06 | 0.000390594 | 0.057417304 |
| T2DM | pericyte | *ZNF503-AS1* | 5.679018865 | 4.15E-05 | 0.000607238 | 0.260920041 |
| T2DM | pericyte | *LINC00937* | 0.822177866 | 0.010491609 | 0.020251994 | 1 |
| T2DM | pericyte | *C14orf132* | 1.571304251 | 0.000895537 | 0.003035172 | 1 |
| T2DM | pericyte | *AC009299.3* | 2.382919399 | 0.002540654 | 0.006501205 | 1 |
| T2DM | pericyte | *LINC01013* | 1.474562532 | 0.002764206 | 0.006948647 | 1 |
| T2DM | pericyte | *PROB1* | -1.502222218 | 0.029543136 | 0.048724474 | 1 |
| T2DM | pericyte | *LINC00954* | 1.445762158 | 0.003563111 | 0.008467163 | 1 |

| | | | | | | |
|---|---|---|---|---|---|---|
| T2DM | pericyte | *RPL41* | 4.286207276 | 0.008676811 | 0.017165966 | 1 |
| T2DM | pericyte | *LINC00200* | 4.519533853 | 0.018253888 | 0.03251975 | 1 |
| T2DM | pericyte | *U91324.1* | 17.64370294 | 3.14E-05 | 0.000599418 | 0.197551412 |
| T2DM | pericyte | *ZNF688* | 3.761241245 | 8.99E-05 | 0.000702125 | 0.565287296 |
| T2DM | pericyte | *PSMG3-AS1* | 0.8412813 | 0.003291595 | 0.007971477 | 1 |
| T2DM | pericyte | *KLHL7-AS1* | 16.19080626 | 3.47E-05 | 0.000599418 | 0.218006723 |
| T2DM | pericyte | *AC092171.4* | 1.814426283 | 0.003571391 | 0.008479355 | 1 |
| T2DM | pericyte | *DIRC3* | 1.078120539 | 0.001502373 | 0.004399357 | 1 |
| T2DM | pericyte | *TAPBP* | 2.36753065 | 9.36E-05 | 0.000716959 | 0.588318822 |
| T2DM | pericyte | *AC107021.1* | 0.574876041 | 0.014248016 | 0.026261294 | 1 |
| T2DM | pericyte | *HLA-DQB2* | 0.652489484 | 1.23E-08 | 4.30E-06 | 7.74E-05 |
| T2DM | pericyte | *LINC00665* | 1.084219412 | 0.00318454 | 0.007768443 | 1 |
| T2DM | pericyte | *TNF* | 1.632241332 | 0.000123339 | 0.000831115 | 0.775429988 |
| T2DM | pericyte | *LYRM9* | 1.779606454 | 0.010850207 | 0.020860933 | 1 |
| T2DM | pericyte | *SNHG15* | 1.754451764 | 0.0080956 | 0.016183022 | 1 |
| T2DM | pericyte | *LINC00472* | 1.383770697 | 0.001840362 | 0.005090346 | 1 |
| T2DM | pericyte | *AC073133.1* | -0.889054835 | 0.011000055 | 0.021103859 | 1 |
| T2DM | pericyte | *RPS28* | 1.254458715 | 0.002110071 | 0.005605687 | 1 |
| T2DM | pericyte | *TRIM26* | 1.950306042 | 0.000255651 | 0.001263585 | 1 |
| T2DM | pericyte | *ZNF736* | 1.215030992 | 0.004463562 | 0.01007627 | 1 |
| T2DM | pericyte | *HLA-B* | 1.859292451 | 7.51E-08 | 1.69E-05 | 0.000472271 |
| T2DM | pericyte | *C19orf81* | 2.221256295 | 0.019304543 | 0.034072898 | 1 |
| T2DM | pericyte | *ZSCAN31* | 0.803851697 | 0.005202979 | 0.011306993 | 1 |
| T2DM | pericyte | *LINC00957* | 1.891299564 | 0.006241964 | 0.013037618 | 1 |
| T2DM | pericyte | *B3GALT4* | 2.246440096 | 0.000903267 | 0.003044738 | 1 |
| T2DM | pericyte | *LINC00941* | 3.09315064 | 0.007854575 | 0.015812268 | 1 |
| T2DM | pericyte | *ZBTB22* | 0.776757999 | 0.017678927 | 0.031647896 | 1 |
| T2DM | pericyte | *GS1-124K5.4* | 0.363466611 | 0.006375178 | 0.013271769 | 1 |
| T2DM | pericyte | *ZNF737* | 3.192863077 | 0.00011487 | 0.000793613 | 0.722188001 |
| T2DM | pericyte | *LINC00959* | 1.514648225 | 0.010011801 | 0.019438322 | 1 |
| T2DM | pericyte | *HLA-DQA2* | 1.902043435 | 8.28E-21 | 5.21E-17 | 5.21E-17 |
| T2DM | pericyte | *LRRC37A2* | 1.984338006 | 0.012505785 | 0.023525993 | 1 |
| T2DM | pericyte | *LY6G5B* | -0.505438049 | 7.41E-05 | 0.000649163 | 0.466098806 |
| T2DM | pericyte | *PSMB9* | 0.770776284 | 0.018825197 | 0.033311009 | 1 |
| T2DM | pericyte | *COX19* | 4.406544315 | 4.82E-05 | 0.000607238 | 0.303312858 |
| T2DM | pericyte | *AQP1* | 2.466508541 | 0.000285869 | 0.001346398 | 1 |
| T2DM | pericyte | *NSUN6* | 3.389977294 | 6.53E-05 | 0.00063262 | 0.41083829 |
| T2DM | pericyte | *CD302* | 3.334130492 | 8.83E-05 | 0.000701359 | 0.555261122 |
| T2DM | pericyte | *ATP5J2* | 2.655506912 | 0.003817839 | 0.008932919 | 1 |
| T2DM | pericyte | *ARHGAP8* | 1.336157566 | 0.015114534 | 0.027679893 | 1 |
| T2DM | pericyte | *ATP5O* | 1.583585274 | 0.002016546 | 0.00542956 | 1 |
| T2DM | pericyte | *PLEKHO2* | 2.106255049 | 0.004502039 | 0.010141282 | 1 |
| T2DM | pericyte | *PI4KA* | 2.391321009 | 0.001918798 | 0.005241694 | 1 |
| T2DM | pericyte | *AKAP2* | 2.887376681 | 0.004570853 | 0.010263196 | 1 |
| T2DM | pericyte | *C22orf39* | 1.062882922 | 0.001680457 | 0.004778397 | 1 |
| T2DM | pericyte | *EIF6* | 1.524269265 | 0.013465282 | 0.025038813 | 1 |
| T2DM | pericyte | *MRPL20* | 2.396117983 | 2.73E-05 | 0.000567661 | 0.171433684 |
| T2DM | pericyte | *AP5Z1* | 4.275487991 | 3.17E-05 | 0.000599418 | 0.19917639 |
| T2DM | pericyte | *C7orf73* | 3.600830417 | 4.27E-05 | 0.000607238 | 0.268584295 |
| T2DM | pericyte | *PALM2* | 1.63803198 | 5.95E-05 | 0.00062437 | 0.37383208 |

| | | | | | | |
|---|---|---|---|---|---|---|
| T2DM | pericyte | *NAT6* | 2.003277784 | 0.008122979 | 0.016222734 | 1 |
| T2DM | pericyte | *IL10RB* | 2.884306089 | 4.85E-05 | 0.000607238 | 0.304978035 |
| T2DM | pericyte | *NME1-NME2* | 2.276033629 | 0.000789523 | 0.002763771 | 1 |
| T2DM | pericyte | *PLA2G4B* | 1.973102576 | 1.88E-08 | 5.64E-06 | 0.000118417 |
| T2DM | pericyte | *TTC4* | 0.478652976 | 0.003658773 | 0.008624937 | 1 |
| T2DM | pericyte | *JMJD7* | 2.287718342 | 7.29E-10 | 3.53E-07 | 4.59E-06 |
| T2DM | pericyte | *TMEM199* | 1.885052741 | 0.001833466 | 0.005082451 | 1 |
| T2DM | pericyte | *LILRA6* | 0.566013974 | 0.020521104 | 0.035877693 | 1 |
| T2DM | pericyte | *APOBEC3C* | 7.077089249 | 0.001721163 | 0.004852445 | 1 |
| T2DM | pericyte | *N4BP2L2* | 2.338850123 | 0.000297624 | 0.001381518 | 1 |
| T2DM | pericyte | *LINC00847* | 0.930254706 | 0.00380645 | 0.008912906 | 1 |
| T2DM | pericyte | *NEAT1* | 1.678113102 | 0.000107376 | 0.000771465 | 0.675072244 |
| T2DM | pericyte | *ZNF585B* | 1.181439862 | 0.000198617 | 0.001076468 | 1 |
| T2DM | pericyte | *UBAP1L* | 1.346429334 | 0.025763886 | 0.043554061 | 1 |
| T2DM | pericyte | *DNAJC3-AS1* | 4.229000595 | 0.000135389 | 0.000877518 | 0.851192006 |
| T2DM | pericyte | *CDK11B* | 2.870943662 | 9.37E-05 | 0.000716959 | 0.589340277 |
| T2DM | pericyte | *ABHD14A* | 1.124385422 | 0.00116687 | 0.003664392 | 1 |
| T2DM | pericyte | *FMN1* | 0.989541742 | 0.00039278 | 0.00166915 | 1 |
| T2DM | pericyte | *PDCD6* | 2.405297625 | 6.49E-05 | 0.00063262 | 0.407934493 |
| T2DM | pericyte | *ZNF718* | 2.230742989 | 0.000172532 | 0.001002502 | 1 |
| T2DM | pericyte | *LINC00989* | -0.092431105 | 0.024380128 | 0.041471284 | 1 |
| T2DM | pericyte | *LINC01021* | 1.819546602 | 0.005152476 | 0.01123607 | 1 |
| T2DM | pericyte | *GLI4* | 0.945002002 | 0.003810882 | 0.008919962 | 1 |
| T2DM | pericyte | *ZNF550* | 1.788236689 | 0.001118329 | 0.003550571 | 1 |
| T2DM | pericyte | *CCDC71L* | 2.773537393 | 0.00110497 | 0.003524578 | 1 |
| T2DM | pericyte | *TMEM200B* | 1.234539938 | 0.004655223 | 0.010408033 | 1 |
| T2DM | pericyte | *PCDHGB6* | 1.34613622 | 0.021652929 | 0.037584749 | 1 |
| T2DM | pericyte | *TRNP1* | -0.674830352 | 0.026640801 | 0.044807575 | 1 |
| T2DM | pericyte | *ATXN7L3B* | 2.178758194 | 0.0002932 | 0.001367469 | 1 |
| T2DM | pericyte | *PRKDC* | 3.087572581 | 5.99E-05 | 0.00062437 | 0.376798365 |
| T2DM | pericyte | *CLDN23* | 1.819692175 | 0.003054767 | 0.007510479 | 1 |
| T2DM | pericyte | *ZNF260* | 4.900784641 | 4.70E-06 | 0.000304712 | 0.029557104 |
| T2DM | pericyte | *TAS2R20* | 1.062692468 | 0.000967508 | 0.00320988 | 1 |
| T2DM | pericyte | *ZNF432* | 1.789580648 | 0.015980115 | 0.029069408 | 1 |
| T2DM | pericyte | *HMBS* | 1.609004344 | 3.15E-07 | 5.08E-05 | 0.001981795 |
| T2DM | pericyte | *ZNF253* | 3.936508087 | 0.000153902 | 0.000941226 | 0.967580062 |
| T2DM | pericyte | *HP* | -1.378840406 | 0.029574149 | 0.04876283 | 1 |
| T2DM | pericyte | *KIAA1147* | 3.052935229 | 5.94E-05 | 0.00062437 | 0.373255724 |
| T2DM | pericyte | *ZBED6* | 1.922061754 | 0.005346778 | 0.011559557 | 1 |
| T2DM | pericyte | *OVCH1-AS1* | 2.572883221 | 0.000325539 | 0.001460857 | 1 |
| T2DM | pericyte | *CUX1* | 2.0475857 | 0.000147449 | 0.000921483 | 0.927012194 |
| T2DM | pericyte | *TEN1* | -1.627469602 | 0.02049293 | 0.035840788 | 1 |
| T2DM | pericyte | *RTEL1* | 1.707131086 | 0.009217168 | 0.018041201 | 1 |
| T2DM | pericyte | *ZNF578* | 0.70182512 | 0.013648357 | 0.025326807 | 1 |
| T2DM | pericyte | *CEP95* | 2.79133807 | 0.000913733 | 0.003067079 | 1 |
| T2DM | pericyte | *LINC00639* | 1.907283743 | 1.23E-05 | 0.000458996 | 0.077483066 |
| T2DM | pericyte | *LINC00648* | 1.30261143 | 0.003485307 | 0.008334776 | 1 |
| T2DM | pericyte | *IQCH-AS1* | 1.03330056 | 0.01306241 | 0.024419675 | 1 |
| T2DM | pericyte | *HOXB7* | 1.093500143 | 0.020118977 | 0.035287988 | 1 |
| T2DM | pericyte | *FRRS1L* | 2.300599718 | 0.00202056 | 0.005437849 | 1 |

| | | | | | | |
|---|---|---|---|---|---|---|
| T2DM | pericyte | *TMEM178B* | 2.805830113 | 0.000875534 | 0.002988317 | 1 |
| T2DM | pericyte | *HPR* | 1.61164076 | 0.008780718 | 0.017333855 | 1 |
| T2DM | pericyte | *LINC00662* | 3.272469996 | 8.21E-05 | 0.000678065 | 0.516007465 |
| T2DM | pericyte | *LINC00909* | 2.297118759 | 0.001595639 | 0.004595412 | 1 |
| T2DM | pericyte | *DYNLL2* | 1.112325741 | 0.011654219 | 0.022182887 | 1 |
| T2DM | pericyte | *MYO15B* | 1.972918948 | 0.006627558 | 0.013701893 | 1 |
| T2DM | pericyte | *ILF3-AS1* | 2.147871359 | 4.95E-05 | 0.000607238 | 0.310905927 |
| T2DM | pericyte | *LINC01140* | 2.578323356 | 7.95E-05 | 0.000667567 | 0.500005268 |
| T2DM | pericyte | *LINC00906* | 6.669129947 | 0.005517623 | 0.011823209 | 1 |
| T2DM | pericyte | *AL132989.1* | 16.54020795 | 1.55E-05 | 0.000488489 | 0.09767048 |
| T2DM | pericyte | *FBXO17* | 3.320496424 | 3.07E-05 | 0.000599418 | 0.19323029 |
| T2DM | pericyte | *SNHG8* | 1.559560828 | 0.024294856 | 0.041337419 | 1 |
| T2DM | pericyte | *LINC01050* | 14.184418 | 0.000313683 | 0.001424297 | 1 |
| T2DM | pericyte | *MILR1* | 0.464385734 | 0.019193672 | 0.033912286 | 1 |
| T2DM | pericyte | *GTF2H5* | 1.487532723 | 0.00209297 | 0.005570916 | 1 |
| T2DM | pericyte | *FGF14-AS2* | 1.675749712 | 0.011229611 | 0.0214918 | 1 |
| T2DM | pericyte | *POLR2J4* | 1.941843346 | 0.000851622 | 0.002925765 | 1 |
| T2DM | pericyte | *AC144652.1* | 0.951190034 | 0.001463429 | 0.004305372 | 1 |
| T2DM | SMC | *SEMA3F* | -0.829870168 | 7.05E-07 | 0.00016416 | 0.004432319 |
| T2DM | SMC | *RBM6* | -0.627372918 | 2.50E-07 | 7.87E-05 | 0.001573913 |
| T2DM | SMC | *CEP68* | 1.831910737 | 9.76E-09 | 5.11E-06 | 6.14E-05 |
| T2DM | SMC | *TUBG2* | -0.876081926 | 6.42E-08 | 2.69E-05 | 0.000403426 |
| T2DM | SMC | *STXBP2* | 1.36379222 | 0.000507397 | 0.044305623 | 1 |
| T2DM | SMC | *COBLL1* | -1.048814526 | 2.65E-05 | 0.004904684 | 0.166759265 |
| T2DM | SMC | *C3orf18* | 2.575746345 | 1.87E-07 | 6.19E-05 | 0.001176959 |
| T2DM | SMC | *PABPC4* | -1.883677685 | 6.10E-07 | 0.000147447 | 0.003833634 |
| T2DM | SMC | *RBL2* | 1.196009749 | 0.000231509 | 0.025094724 | 1 |
| T2DM | SMC | *EYA1* | 0.692664187 | 2.78E-05 | 0.004984877 | 0.174470696 |
| T2DM | SMC | *ATP13A1* | 1.773629344 | 2.73E-07 | 8.19E-05 | 0.001718891 |
| T2DM | SMC | *CPVL* | 1.366146931 | 0.00051679 | 0.044507688 | 1 |
| T2DM | SMC | *HSD17B1* | -0.633350764 | 3.35E-07 | 9.58E-05 | 0.00210697 |
| T2DM | SMC | *VAT1* | 1.507564416 | 0.000607815 | 0.048371341 | 1 |
| T2DM | SMC | *WFS1* | 4.781478117 | 3.85E-07 | 0.000105152 | 0.002418488 |
| T2DM | SMC | *CDK2AP1* | 0.654311189 | 5.92E-05 | 0.00907412 | 0.372038939 |
| T2DM | SMC | *BVES* | -0.690431061 | 0.000322415 | 0.031066738 | 1 |
| T2DM | SMC | *CD3EAP* | -1.390282723 | 7.97E-05 | 0.011386786 | 0.501018599 |
| T2DM | SMC | *TEX14* | 1.940156459 | 0.000214255 | 0.023824093 | 1 |
| T2DM | SMC | *GRK4* | 0.371683603 | 0.000623912 | 0.048426367 | 1 |
| T2DM | SMC | *OPRL1* | -1.144235073 | 0.000283571 | 0.028755003 | 1 |
| T2DM | SMC | *ZNF436* | 1.014798962 | 0.000238079 | 0.025369568 | 1 |
| T2DM | SMC | *PRR12* | 1.287726786 | 0.000215997 | 0.023824093 | 1 |
| T2DM | SMC | *C12orf65* | 2.027261306 | 0.000274209 | 0.028645086 | 1 |
| T2DM | SMC | *P2RX4* | 1.211533343 | 0.000650786 | 0.049295057 | 1 |
| T2DM | SMC | *SPTBN5* | 0.760586034 | 1.16E-05 | 0.002436837 | 0.073105118 |
| T2DM | SMC | *CARF* | 1.134740226 | 0.000373922 | 0.035087315 | 1 |
| T2DM | SMC | *HMG20A* | 2.044335592 | 0.000449844 | 0.039833329 | 1 |
| T2DM | SMC | *MAN2C1* | -0.387161065 | 0.00012735 | 0.017035131 | 0.800651155 |
| T2DM | SMC | *MYO19* | 2.11343314 | 0.000575759 | 0.046524961 | 1 |
| T2DM | SMC | *SIRT3* | 1.532979041 | 0.000301674 | 0.029634761 | 1 |
| T2DM | SMC | *WNT9A* | 1.251174452 | 0.000530545 | 0.045074844 | 1 |

| | | | | | | |
|---|---|---|---|---|---|---|
| T2DM | SMC | *HSD17B12* | -0.949163093 | 7.40E-08 | 2.91E-05 | 0.000465427 |
| T2DM | SMC | *SAR1B* | 1.689466213 | 0.000385064 | 0.035601465 | 1 |
| T2DM | SMC | *JAZF1* | -1.857171267 | 8.86E-05 | 0.012378156 | 0.557016998 |
| T2DM | SMC | *TSEN2* | 1.693129891 | 1.29E-06 | 0.00029027 | 0.008127553 |
| T2DM | SMC | *TIMP4* | 2.352066542 | 1.52E-05 | 0.002898322 | 0.095644619 |
| T2DM | SMC | *CLEC18A* | -2.006360578 | 9.46E-08 | 3.50E-05 | 0.000594795 |
| T2DM | SMC | *AP3S2* | 1.334345292 | 9.49E-11 | 7.46E-08 | 5.97E-07 |
| T2DM | SMC | *TRIM63* | 0.894739417 | 0.000277931 | 0.028645086 | 1 |
| T2DM | SMC | *CD1E* | 2.647359772 | 0.000180237 | 0.021733615 | 1 |
| T2DM | SMC | *LMX1A* | 0.628855026 | 0.000566025 | 0.046524961 | 1 |
| T2DM | SMC | *RFT1* | -1.687500839 | 0.000615575 | 0.048376496 | 1 |
| T2DM | SMC | *RNF123* | 1.150528551 | 5.26E-07 | 0.000135025 | 0.00330504 |
| T2DM | SMC | *TRMT61A* | -1.879975961 | 5.77E-06 | 0.001249945 | 0.036248398 |
| T2DM | SMC | *C2* | 3.41908608 | 5.12E-10 | 3.58E-07 | 3.22E-06 |
| T2DM | SMC | *LTBP3* | 1.569043481 | 1.35E-05 | 0.002661908 | 0.08518107 |
| T2DM | SMC | *PXK* | 1.495943138 | 0.00020846 | 0.023824093 | 1 |
| T2DM | SMC | *IRS1* | -1.10879418 | 1.09E-24 | 6.84E-21 | 6.84E-21 |
| T2DM | SMC | *RGS19* | -1.043312014 | 1.34E-05 | 0.002661908 | 0.084418711 |
| T2DM | SMC | *C1QTNF4* | -0.29079192 | 0.000567148 | 0.046524961 | 1 |
| T2DM | SMC | *HLA-DQB1* | -0.943276925 | 1.39E-13 | 2.19E-10 | 8.77E-10 |
| T2DM | SMC | *B3GNT3* | 3.037775025 | 0.000120636 | 0.016487828 | 0.758440086 |
| T2DM | SMC | *ZNF320* | 0.416352078 | 0.000640967 | 0.049143403 | 1 |
| T2DM | SMC | *RP1L1* | 1.608055936 | 6.65E-05 | 0.009956596 | 0.418177046 |
| T2DM | SMC | *BMP8A* | 3.619298902 | 4.17E-09 | 2.39E-06 | 2.62E-05 |
| T2DM | SMC | *ZNF703* | -1.714391425 | 7.91E-05 | 0.011386786 | 0.497295433 |
| T2DM | SMC | *TMEM201* | 2.42852763 | 0.000577214 | 0.046524961 | 1 |
| T2DM | SMC | *HLA-DRB1* | -1.786756807 | 1.24E-15 | 2.60E-12 | 7.80E-12 |
| T2DM | SMC | *HLA-DQA1* | -0.749228031 | 4.37E-05 | 0.007422193 | 0.274621139 |
| T2DM | SMC | *HLA-DRB5* | -0.517191837 | 2.90E-12 | 3.64E-09 | 1.82E-08 |
| T2DM | SMC | *OXLD1* | 1.469528159 | 0.000326134 | 0.031066738 | 1 |
| T2DM | SMC | *HLA-DRA* | 1.639927319 | 2.66E-11 | 2.79E-08 | 1.67E-07 |
| T2DM | SMC | *NOTCH4* | -0.170039803 | 1.08E-07 | 3.77E-05 | 0.000678164 |
| T2DM | SMC | *PRRT1* | 1.047429883 | 4.72E-05 | 0.007653393 | 0.297039905 |
| T2DM | SMC | *NELFE* | 1.20022913 | 0.000208469 | 0.023824093 | 1 |
| T2DM | SMC | *BAG6* | 1.17223766 | 0.000403185 | 0.036736599 | 1 |
| T2DM | SMC | *LIN52* | 1.405501983 | 0.000161483 | 0.020304892 | 1 |
| T2DM | SMC | *HCG27* | 1.041055815 | 5.20E-11 | 4.67E-08 | 3.27E-07 |
| T2DM | SMC | *FGFR1OP* | 2.106945 | 0.00017102 | 0.021082419 | 1 |
| T2DM | SMC | *DNLZ* | 0.624431988 | 0.000131253 | 0.017191433 | 0.825188781 |
| T2DM | SMC | *SIPA1* | 1.433970578 | 0.000423142 | 0.038004234 | 1 |
| T2DM | SMC | *ATF6B* | 1.21588642 | 4.87E-05 | 0.007653393 | 0.306135735 |
| T2DM | SMC | *STARD10* | 0.739727203 | 5.37E-07 | 0.000135025 | 0.003375636 |
| T2DM | SMC | *HLA-DQB2* | 0.424524529 | 1.87E-08 | 9.03E-06 | 0.000117409 |
| T2DM | SMC | *HLA-B* | 0.981568714 | 4.78E-05 | 0.007653393 | 0.300700643 |
| T2DM | SMC | *HLA-DQA2* | 1.254684637 | 4.36E-19 | 1.37E-15 | 2.74E-15 |
| T2DM | SMC | *NAT6* | 2.148294614 | 0.000141433 | 0.01814667 | 0.889186809 |
| T2DM | SMC | *PLA2G4B* | 1.336406878 | 2.19E-08 | 9.84E-06 | 0.000137766 |
| T2DM | SMC | *JMJD7* | 1.508327329 | 3.28E-09 | 2.06E-06 | 2.06E-05 |
| T2DM | SMC | *HMBS* | 0.915568938 | 2.99E-05 | 0.005217473 | 0.187829031 |
| T2DM | SMC | *LINC00639* | 0.942563732 | 0.000297569 | 0.029634761 | 1 |

| | | | | | | |
|---|---|---|---|---|---|---|
| T2DM | SMC | *AC009060.1* | -3.482500824 | 0.000183216 | 0.021733615 | 1 |
| T2DM | t_cell | *CFH* | 1.862765082 | 0.00242105 | 0.046690609 | 1 |
| T2DM | t_cell | *TMEM176A* | 1.497642865 | 0.000914859 | 0.032866956 | 1 |
| T2DM | t_cell | *RBM6* | -1.138652396 | 1.64E-06 | 0.000467868 | 0.010293093 |
| T2DM | t_cell | *PKD1* | 1.703227458 | 0.002828435 | 0.0481907 | 1 |
| T2DM | t_cell | *MLXIPL* | 0.564648857 | 0.000425382 | 0.024093458 | 1 |
| T2DM | t_cell | *ZNF200* | 2.306935014 | 0.001937914 | 0.043336413 | 1 |
| T2DM | t_cell | *SYT7* | 1.624186187 | 0.002872322 | 0.048413648 | 1 |
| T2DM | t_cell | *CEP68* | 3.58807905 | 7.41E-10 | 5.18E-07 | 4.66E-06 |
| T2DM | t_cell | *PSMC4* | 2.589675415 | 0.001340837 | 0.038668994 | 1 |
| T2DM | t_cell | *NR1H3* | 2.578248524 | 0.000101022 | 0.009866627 | 0.635123212 |
| T2DM | t_cell | *TUBG2* | -1.139635513 | 9.35E-05 | 0.009800546 | 0.588032747 |
| T2DM | t_cell | *RFC2* | 2.576367065 | 0.001786506 | 0.042869317 | 1 |
| T2DM | t_cell | *OPN3* | 1.321016307 | 0.00122426 | 0.036365626 | 1 |
| T2DM | t_cell | *ITIH4* | 1.004350323 | 2.86E-05 | 0.004083626 | 0.179679524 |
| T2DM | t_cell | *ATP2C2* | 3.026188345 | 0.001975237 | 0.043336413 | 1 |
| T2DM | t_cell | *SUGP2* | 2.357744239 | 0.00200556 | 0.043336413 | 1 |
| T2DM | t_cell | *CYBRD1* | 1.705466976 | 0.002186552 | 0.044670743 | 1 |
| T2DM | t_cell | *SDHA* | 2.5334595 | 0.000512805 | 0.026211401 | 1 |
| T2DM | t_cell | *LRCH4* | -2.018139362 | 9.98E-05 | 0.009866627 | 0.627558899 |
| T2DM | t_cell | *NAALAD2* | 1.145813368 | 0.00190613 | 0.043336413 | 1 |
| T2DM | t_cell | *MYH7B* | 2.248610981 | 0.002375111 | 0.046426982 | 1 |
| T2DM | t_cell | *CPNE3* | 1.989562058 | 0.000261306 | 0.018471093 | 1 |
| T2DM | t_cell | *EPDR1* | 1.276945891 | 0.000489199 | 0.025533014 | 1 |
| T2DM | t_cell | *C3orf18* | 5.292421711 | 4.09E-08 | 1.71E-05 | 0.000257045 |
| T2DM | t_cell | *ZNF343* | 1.942679983 | 0.002094939 | 0.044350959 | 1 |
| T2DM | t_cell | *PABPC4* | -3.099859957 | 1.20E-05 | 0.002158654 | 0.075552877 |
| T2DM | t_cell | *MYO15A* | 3.469868668 | 0.000485847 | 0.025533014 | 1 |
| T2DM | t_cell | *RPGRIP1* | 2.001601387 | 0.001487827 | 0.041026174 | 1 |
| T2DM | t_cell | *VNN3* | 3.41707729 | 0.002550145 | 0.046927985 | 1 |
| T2DM | t_cell | *TPSD1* | 9.875343387 | 0.001680406 | 0.042436657 | 1 |
| T2DM | t_cell | *PALMD* | 1.052646896 | 0.002765084 | 0.047627627 | 1 |
| T2DM | t_cell | *MARCH2* | 2.499572464 | 0.000406712 | 0.023458676 | 1 |
| T2DM | t_cell | *GGT5* | 2.527568114 | 0.002123372 | 0.044350959 | 1 |
| T2DM | t_cell | *TXN2* | 2.298584122 | 0.002227222 | 0.044755853 | 1 |
| T2DM | t_cell | *ISM2* | 3.820182083 | 0.000187882 | 0.014765181 | 1 |
| T2DM | t_cell | *PSMB5* | 2.250115268 | 0.000612189 | 0.027491664 | 1 |
| T2DM | t_cell | *PLTP* | 1.440270361 | 0.000267356 | 0.018471093 | 1 |
| T2DM | t_cell | *GSS* | 2.113718635 | 0.001815816 | 0.043036656 | 1 |
| T2DM | t_cell | *NINL* | 1.707243301 | 0.001253759 | 0.037006497 | 1 |
| T2DM | t_cell | *IFT52* | 2.247165129 | 0.001764145 | 0.042845976 | 1 |
| T2DM | t_cell | *GID8* | 2.909993969 | 0.002315651 | 0.045786221 | 1 |
| T2DM | t_cell | *MAP1LC3A* | 1.288137681 | 0.000287138 | 0.019370443 | 1 |
| T2DM | t_cell | *UGGT2* | 1.669148406 | 0.002019655 | 0.043336413 | 1 |
| T2DM | t_cell | *DTWD1* | 2.277467124 | 0.000485089 | 0.025533014 | 1 |
| T2DM | t_cell | *PDGFRL* | 2.237886191 | 0.001565151 | 0.042063823 | 1 |
| T2DM | t_cell | *EYA1* | 1.425406791 | 3.31E-06 | 0.000743782 | 0.020825889 |
| T2DM | t_cell | *NIPAL2* | 2.384013993 | 0.00060438 | 0.027491664 | 1 |
| T2DM | t_cell | *PLAT* | 1.748874881 | 0.003079038 | 0.049983077 | 1 |
| T2DM | t_cell | *RAB2A* | 1.526012894 | 0.003111811 | 0.049983077 | 1 |

| | | | | | | |
|---|---|---|---|---|---|---|
| T2DM | t_cell | *TLE6* | 2.356646578 | 0.001800658 | 0.043036656 | 1 |
| T2DM | t_cell | *POLR2I* | 2.476161119 | 0.002007817 | 0.043336413 | 1 |
| T2DM | t_cell | *CCDC9* | 3.188381288 | 0.000809342 | 0.03121675 | 1 |
| T2DM | t_cell | *PTPRS* | 0.994616903 | 0.003124609 | 0.049983077 | 1 |
| T2DM | t_cell | *TNPO2* | 2.536645617 | 0.002827902 | 0.0481907 | 1 |
| T2DM | t_cell | *GCDH* | -3.197061551 | 0.00066166 | 0.028492152 | 1 |
| T2DM | t_cell | *CPVL* | 2.563412121 | 5.04E-05 | 0.006099292 | 0.317163196 |
| T2DM | t_cell | *CRHR2* | 4.316505723 | 0.001102599 | 0.034834366 | 1 |
| T2DM | t_cell | *TMEM176B* | 1.156040864 | 0.000807373 | 0.03121675 | 1 |
| T2DM | t_cell | *MRPL32* | 1.718786062 | 0.001605595 | 0.042358851 | 1 |
| T2DM | t_cell | *SLC1A1* | 1.019350886 | 0.002329874 | 0.045918233 | 1 |
| T2DM | t_cell | *PSMB3* | 3.55242695 | 0.001820861 | 0.043036656 | 1 |
| T2DM | t_cell | *HSD17B1* | -1.05821755 | 1.54E-05 | 0.002622909 | 0.097047617 |
| T2DM | t_cell | *EZH1* | 2.164614972 | 0.002587207 | 0.046927985 | 1 |
| T2DM | t_cell | *VAT1* | 3.63209789 | 5.38E-05 | 0.006386111 | 0.338463862 |
| T2DM | t_cell | *CWH43* | 7.102838145 | 0.002575935 | 0.046927985 | 1 |
| T2DM | t_cell | *WFS1* | 2.718152839 | 2.31E-05 | 0.003549198 | 0.145517099 |
| T2DM | t_cell | *SLC2A9* | 3.313891952 | 0.000382267 | 0.023258718 | 1 |
| T2DM | t_cell | *FAM149A* | 1.099821741 | 0.000186291 | 0.014765181 | 1 |
| T2DM | t_cell | *PUS3* | 3.060524601 | 0.00228834 | 0.045672367 | 1 |
| T2DM | t_cell | *CARS* | 2.903131488 | 0.000325132 | 0.021073254 | 1 |
| T2DM | t_cell | *COQ5* | 0.640516407 | 0.000529501 | 0.02653421 | 1 |
| T2DM | t_cell | *WNT5B* | 2.509779224 | 0.002886904 | 0.048477534 | 1 |
| T2DM | t_cell | *PRR4* | 1.98014382 | 0.000520235 | 0.026376749 | 1 |
| T2DM | t_cell | *CREBL2* | 2.868280374 | 0.002698613 | 0.047259549 | 1 |
| T2DM | t_cell | *CDK2AP1* | 1.364882893 | 5.05E-06 | 0.001094005 | 0.031726152 |
| T2DM | t_cell | *MGP* | 1.614270221 | 0.001290891 | 0.037924442 | 1 |
| T2DM | t_cell | *ALDH5A1* | 1.93396675 | 0.000578183 | 0.027491664 | 1 |
| T2DM | t_cell | *GHR* | 0.790538948 | 0.000851648 | 0.032254892 | 1 |
| T2DM | t_cell | *PCDHB6* | 2.897210961 | 0.002718818 | 0.047401041 | 1 |
| T2DM | t_cell | *NPHP3* | 0.885234283 | 0.001121253 | 0.035012223 | 1 |
| T2DM | t_cell | *ZBTB47* | 2.156315169 | 0.000732528 | 0.029712291 | 1 |
| T2DM | t_cell | *CLCN2* | 3.542545022 | 0.002852929 | 0.048245511 | 1 |
| T2DM | t_cell | *LMAN2L* | 1.502810573 | 0.002167423 | 0.044670743 | 1 |
| T2DM | t_cell | *KCNIP3* | 2.805608592 | 0.002114212 | 0.044350959 | 1 |
| T2DM | t_cell | *CHMP3* | -2.526828456 | 0.001398843 | 0.03979424 | 1 |
| T2DM | t_cell | *GPX7* | -1.084526156 | 4.44E-05 | 0.005577165 | 0.278858243 |
| T2DM | t_cell | *MRPL37* | 2.160348527 | 0.000681183 | 0.028936464 | 1 |
| T2DM | t_cell | *TNNI3K* | 1.708549496 | 0.001955807 | 0.043336413 | 1 |
| T2DM | t_cell | *FAAH* | 0.587112658 | 0.001601343 | 0.042358851 | 1 |
| T2DM | t_cell | *TNFSF4* | -2.99144752 | 0.002672215 | 0.046927985 | 1 |
| T2DM | t_cell | *SERPINC1* | 1.61816948 | 0.000186549 | 0.014765181 | 1 |
| T2DM | t_cell | *MTFR1L* | 3.10959854 | 0.001850025 | 0.043078185 | 1 |
| T2DM | t_cell | *IRF2BPL* | 2.720661339 | 0.001702404 | 0.042452419 | 1 |
| T2DM | t_cell | *ADRA1A* | 0.539493408 | 2.11E-05 | 0.003402837 | 0.132710644 |
| T2DM | t_cell | *TEX14* | 3.422813057 | 0.000448169 | 0.024719054 | 1 |
| T2DM | t_cell | *PRB2* | 2.288967855 | 0.000985693 | 0.033337656 | 1 |
| T2DM | t_cell | *GPSM2* | 2.260066909 | 0.001871764 | 0.043336413 | 1 |
| T2DM | t_cell | *PMS2* | 4.365078628 | 0.00016858 | 0.014322474 | 1 |
| T2DM | t_cell | *SSPN* | 1.719677513 | 0.00022426 | 0.016795489 | 1 |

| | | | | | | |
|---|---|---|---|---|---|---|
| T2DM | t_cell | *EBPL* | 1.681925857 | 0.001168746 | 0.035326476 | 1 |
| T2DM | t_cell | *NLN* | 1.583615439 | 0.00080047 | 0.03121675 | 1 |
| T2DM | t_cell | *TSFM* | 2.287280341 | 0.002385226 | 0.046426982 | 1 |
| T2DM | t_cell | *ACVR1C* | 0.944935581 | 0.002562515 | 0.046927985 | 1 |
| T2DM | t_cell | *FAM210B* | 1.56390658 | 0.001350904 | 0.03878144 | 1 |
| T2DM | t_cell | *NEURL2* | 1.052396614 | 0.000884629 | 0.032866956 | 1 |
| T2DM | t_cell | *PEPD* | 2.142427035 | 0.000289617 | 0.019370443 | 1 |
| T2DM | t_cell | *HIF3A* | 1.281736065 | 0.003084639 | 0.049983077 | 1 |
| T2DM | t_cell | *HIST1H2BJ* | 1.82438292 | 0.002642687 | 0.046927985 | 1 |
| T2DM | t_cell | *GLO1* | 3.295985948 | 0.000339084 | 0.021533569 | 1 |
| T2DM | t_cell | *RPP40* | 1.674041913 | 0.001962294 | 0.043336413 | 1 |
| T2DM | t_cell | *GRK4* | 0.855966018 | 3.82E-05 | 0.005109178 | 0.240131379 |
| T2DM | t_cell | *ZNF436* | 2.078470635 | 2.05E-05 | 0.00339276 | 0.12892487 |
| T2DM | t_cell | *UROD* | 2.991515685 | 0.000568529 | 0.027491664 | 1 |
| T2DM | t_cell | *XRCC3* | 3.508543141 | 0.001438756 | 0.040562599 | 1 |
| T2DM | t_cell | *MCF2L* | 1.685862217 | 0.000646746 | 0.028312888 | 1 |
| T2DM | t_cell | *PRR12* | 2.633375025 | 9.92E-05 | 0.009866627 | 0.623576191 |
| T2DM | t_cell | *BAIAP2L2* | 2.219323559 | 0.00022878 | 0.016921624 | 1 |
| T2DM | t_cell | *RIBC2* | 1.657754428 | 0.001890108 | 0.043336413 | 1 |
| T2DM | t_cell | *EMC4* | 3.566125231 | 0.000559872 | 0.027286153 | 1 |
| T2DM | t_cell | *PLD2* | 2.65604096 | 0.000909599 | 0.032866956 | 1 |
| T2DM | t_cell | *SHC2* | 1.583339522 | 0.000821174 | 0.031289222 | 1 |
| T2DM | t_cell | *PDLIM4* | 2.626811824 | 0.000991742 | 0.033337656 | 1 |
| T2DM | t_cell | *ZNF132* | 1.411546702 | 0.001758726 | 0.042845976 | 1 |
| T2DM | t_cell | *FBXW9* | 2.540821644 | 0.001673526 | 0.042436657 | 1 |
| T2DM | t_cell | *SFTPD* | 0.648543562 | 0.001631396 | 0.042382577 | 1 |
| T2DM | t_cell | *ACP2* | 2.283911149 | 0.00170836 | 0.042452419 | 1 |
| T2DM | t_cell | *FADS2* | 2.343133744 | 0.002452342 | 0.046720821 | 1 |
| T2DM | t_cell | *CCDC146* | 2.153790822 | 0.002444408 | 0.046720821 | 1 |
| T2DM | t_cell | *NT5E* | 2.305005739 | 0.000182332 | 0.014765181 | 1 |
| T2DM | t_cell | *AOAH* | -0.950260817 | 0.001995623 | 0.043336413 | 1 |
| T2DM | t_cell | *TM6SF1* | 2.195203922 | 0.000772781 | 0.030735348 | 1 |
| T2DM | t_cell | *PRPF4* | 2.025439163 | 0.001487707 | 0.041026174 | 1 |
| T2DM | t_cell | *FLOT1* | 2.669125067 | 0.000483387 | 0.025533014 | 1 |
| T2DM | t_cell | *PRCP* | 2.770346981 | 0.001014855 | 0.033410211 | 1 |
| T2DM | t_cell | *RMDN3* | 2.46555582 | 0.000392577 | 0.023405167 | 1 |
| T2DM | t_cell | *SPTBN5* | 1.586427811 | 5.97E-06 | 0.00125128 | 0.037538385 |
| T2DM | t_cell | *IFT172* | 2.242945061 | 0.000484457 | 0.025533014 | 1 |
| T2DM | t_cell | *MYOF* | 1.724548284 | 0.000100978 | 0.009866627 | 0.634845714 |
| T2DM | t_cell | *TACC2* | 1.125436867 | 0.00293658 | 0.048927573 | 1 |
| T2DM | t_cell | *CARF* | 2.370364806 | 2.56E-05 | 0.003835652 | 0.161097376 |
| T2DM | t_cell | *FAM13A* | 1.848476329 | 0.000981876 | 0.033337656 | 1 |
| T2DM | t_cell | *PRDM5* | 1.99783926 | 0.002123337 | 0.044350959 | 1 |
| T2DM | t_cell | *SLCO1C1* | 1.862273374 | 0.001771903 | 0.042845976 | 1 |
| T2DM | t_cell | *ZCRB1* | 1.859869587 | 0.003074362 | 0.049983077 | 1 |
| T2DM | t_cell | *PRICKLE1* | 1.189564644 | 0.002203279 | 0.044670743 | 1 |
| T2DM | t_cell | *C1RL* | 2.308032008 | 0.000950218 | 0.033337656 | 1 |
| T2DM | t_cell | *DUOXA1* | 2.094853242 | 0.002850805 | 0.048245511 | 1 |
| T2DM | t_cell | *SRP14* | 2.783869823 | 0.001480125 | 0.041026174 | 1 |
| T2DM | t_cell | *HMG20A* | 3.497308095 | 8.93E-05 | 0.00951684 | 0.561493564 |

| | | | | | | |
|---|---|---|---|---|---|---|
| T2DM | t_cell | *MAN2C1* | -0.662194254 | 0.000817856 | 0.031289222 | 1 |
| T2DM | t_cell | *GCSH* | 0.884305633 | 0.002952923 | 0.048984235 | 1 |
| T2DM | t_cell | *TIGD7* | 1.870215224 | 0.000639124 | 0.028312888 | 1 |
| T2DM | t_cell | *MYO19* | 3.29364996 | 0.000992675 | 0.033337656 | 1 |
| T2DM | t_cell | *SPATA22* | 1.225385933 | 0.000420989 | 0.024061409 | 1 |
| T2DM | t_cell | *C18orf21* | 2.983973108 | 0.00097749 | 0.033337656 | 1 |
| T2DM | t_cell | *DUS3L* | 1.992453951 | 0.000788409 | 0.030979531 | 1 |
| T2DM | t_cell | *SIRT3* | 2.943943155 | 3.02E-05 | 0.004220372 | 0.18991672 |
| T2DM | t_cell | *TRPM2* | 1.057644881 | 0.000688703 | 0.02905957 | 1 |
| T2DM | t_cell | *CYP4B1* | 0.978937895 | 0.001952656 | 0.043336413 | 1 |
| T2DM | t_cell | *TARS2* | 3.654307795 | 0.000724092 | 0.029560822 | 1 |
| T2DM | t_cell | *SNAP47* | 2.49130251 | 0.000897833 | 0.032866956 | 1 |
| T2DM | t_cell | *WNT9A* | 2.348546846 | 5.97E-05 | 0.006951984 | 0.375407128 |
| T2DM | t_cell | *PLEKHA6* | 1.187563702 | 0.001649333 | 0.042436657 | 1 |
| T2DM | t_cell | *ABCG8* | 3.005801862 | 0.002420431 | 0.046690609 | 1 |
| T2DM | t_cell | *THSD7B* | 1.467555224 | 0.000127066 | 0.01139604 | 0.798865827 |
| T2DM | t_cell | *TMEFF2* | 1.458041562 | 0.001694392 | 0.042440811 | 1 |
| T2DM | t_cell | *NBEAL1* | 1.648487411 | 1.04E-05 | 0.001929425 | 0.065600439 |
| T2DM | t_cell | *ESPNL* | 8.335449103 | 0.001683156 | 0.042436657 | 1 |
| T2DM | t_cell | *OSBPL10* | 1.807164503 | 0.001156751 | 0.035303364 | 1 |
| T2DM | t_cell | *CISD2* | 3.016196047 | 0.002616741 | 0.046927985 | 1 |
| T2DM | t_cell | *SPATA9* | 1.072388612 | 0.001889448 | 0.043336413 | 1 |
| T2DM | t_cell | *PSPH* | 1.714712812 | 0.002519841 | 0.046927985 | 1 |
| T2DM | t_cell | *CCDC25* | 3.745751114 | 0.000334189 | 0.021439277 | 1 |
| T2DM | t_cell | *ZNF7* | 3.217337439 | 0.00018278 | 0.014765181 | 1 |
| T2DM | t_cell | *SEC16A* | 2.455976729 | 0.001834711 | 0.043040402 | 1 |
| T2DM | t_cell | *HSD17B12* | -1.122227544 | 0.000373545 | 0.023252228 | 1 |
| T2DM | t_cell | *APIP* | -0.541984873 | 0.001973504 | 0.043336413 | 1 |
| T2DM | t_cell | *GLYAT* | 0.914098249 | 0.000777306 | 0.030735348 | 1 |
| T2DM | t_cell | *CAPN5* | 2.281639839 | 0.001920104 | 0.043336413 | 1 |
| T2DM | t_cell | *P4HA3* | 2.151615891 | 0.000446013 | 0.024719054 | 1 |
| T2DM | t_cell | *PPP1R1C* | 1.629099397 | 0.000452154 | 0.024719054 | 1 |
| T2DM | t_cell | *SAR1B* | 3.198776573 | 0.000128697 | 0.01139604 | 0.809118813 |
| T2DM | t_cell | *PLA2R1* | 1.397970427 | 0.000611165 | 0.027491664 | 1 |
| T2DM | t_cell | *JAZF1* | -2.249387942 | 0.00154759 | 0.041938367 | 1 |
| T2DM | t_cell | *ANKRD29* | 1.814413056 | 0.000981821 | 0.033337656 | 1 |
| T2DM | t_cell | *ATP5G3* | 1.967828644 | 0.000541054 | 0.02657505 | 1 |
| T2DM | t_cell | *PDLIM3* | 1.799247461 | 0.00262379 | 0.046927985 | 1 |
| T2DM | t_cell | *TSEN2* | 3.263338963 | 2.13E-07 | 6.70E-05 | 0.001340878 |
| T2DM | t_cell | *FAM161B* | 4.705299374 | 0.000964705 | 0.033337656 | 1 |
| T2DM | t_cell | *SH3RF2* | 0.969238146 | 0.00307759 | 0.049983077 | 1 |
| T2DM | t_cell | *GDF6* | 2.677480018 | 0.001578982 | 0.042063823 | 1 |
| T2DM | t_cell | *TIMP4* | 2.299280952 | 2.83E-05 | 0.004083626 | 0.178048995 |
| T2DM | t_cell | *STEAP2* | 0.732877965 | 0.001012403 | 0.033410211 | 1 |
| T2DM | t_cell | *CLEC18A* | -4.115239022 | 2.16E-08 | 1.13E-05 | 0.000135909 |
| T2DM | t_cell | *KCNJ15* | -2.269679982 | 0.001051419 | 0.034250115 | 1 |
| T2DM | t_cell | *AP3S2* | 2.673461365 | 4.78E-11 | 3.76E-08 | 3.01E-07 |
| T2DM | t_cell | *C12orf43* | 2.9224102 | 0.000384747 | 0.023258718 | 1 |
| T2DM | t_cell | *TRIM63* | 2.005900922 | 0.000126666 | 0.01139604 | 0.796349884 |
| T2DM | t_cell | *HPD* | 1.502151796 | 0.00278733 | 0.04774917 | 1 |

| | | | | | | |
|---|---|---|---|---|---|---|
| T2DM | t_cell | *CD1E* | 4.634373155 | 2.27E-05 | 0.003549198 | 0.142737692 |
| T2DM | t_cell | *NPR2* | 2.451758526 | 0.001983655 | 0.043336413 | 1 |
| T2DM | t_cell | *DFFA* | 2.060962576 | 0.001732396 | 0.042777654 | 1 |
| T2DM | t_cell | *NR2F6* | 2.080238243 | 0.00121534 | 0.036365626 | 1 |
| T2DM | t_cell | *ZBTB7B* | 3.62344125 | 0.002781712 | 0.04774917 | 1 |
| T2DM | t_cell | *FDPS* | 3.037198171 | 0.00015404 | 0.013266401 | 0.968447252 |
| T2DM | t_cell | *VPS28* | 2.162184105 | 0.001613403 | 0.042358851 | 1 |
| T2DM | t_cell | *DVL3* | 2.121508026 | 0.001735057 | 0.042777654 | 1 |
| T2DM | t_cell | *AP2M1* | 2.729829533 | 0.001530139 | 0.041735408 | 1 |
| T2DM | t_cell | *TMEM143* | 1.695452874 | 0.002538294 | 0.046927985 | 1 |
| T2DM | t_cell | *JMJD8* | 2.003045727 | 0.00184765 | 0.043078185 | 1 |
| T2DM | t_cell | *TSSK3* | 1.226575919 | 0.003132389 | 0.049983077 | 1 |
| T2DM | t_cell | *KCNT2* | 1.762296279 | 0.003113945 | 0.049983077 | 1 |
| T2DM | t_cell | *LMX1A* | 1.444375961 | 4.04E-05 | 0.005294605 | 0.254141042 |
| T2DM | t_cell | *DNASE1L3* | 1.317777884 | 0.003085655 | 0.049983077 | 1 |
| T2DM | t_cell | *NMNAT3* | 2.05508907 | 0.000893051 | 0.032866956 | 1 |
| T2DM | t_cell | *DNALI1* | 1.754639811 | 0.002015633 | 0.043336413 | 1 |
| T2DM | t_cell | *RNF123* | 2.097062704 | 2.62E-06 | 0.000646003 | 0.016484952 |
| T2DM | t_cell | *MST1R* | 1.420733576 | 0.001657928 | 0.042436657 | 1 |
| T2DM | t_cell | *PRRC1* | 2.728456255 | 0.00201535 | 0.043336413 | 1 |
| T2DM | t_cell | *RHOBTB3* | 1.105964423 | 0.001128387 | 0.035012223 | 1 |
| T2DM | t_cell | *KLKB1* | 1.23284477 | 0.001468049 | 0.041026174 | 1 |
| T2DM | t_cell | *SUN1* | 2.008596446 | 0.000673556 | 0.028807114 | 1 |
| T2DM | t_cell | *SLC18A2* | 2.40983309 | 0.002149931 | 0.044670743 | 1 |
| T2DM | t_cell | *ZNF202* | 3.226645767 | 0.000710273 | 0.029378188 | 1 |
| T2DM | t_cell | *C2* | 5.956525917 | 8.65E-10 | 5.44E-07 | 5.44E-06 |
| T2DM | t_cell | *CLEC4E* | 1.187112835 | 0.002036639 | 0.043552214 | 1 |
| T2DM | t_cell | *CLEC4D* | 2.829608102 | 0.002981195 | 0.04911423 | 1 |
| T2DM | t_cell | *TEF* | 1.221096014 | 0.000267238 | 0.018471093 | 1 |
| T2DM | t_cell | *ZNF229* | 3.786912997 | 0.001572293 | 0.042063823 | 1 |
| T2DM | t_cell | *ZNF646* | 1.956141805 | 0.002299787 | 0.045755566 | 1 |
| T2DM | t_cell | *ZNF641* | 1.686080204 | 0.000743173 | 0.029950809 | 1 |
| T2DM | t_cell | *TTYH1* | 2.472822094 | 0.002109677 | 0.044350959 | 1 |
| T2DM | t_cell | *PSCA* | 3.273782638 | 0.00013775 | 0.012028281 | 0.866036236 |
| T2DM | t_cell | *PRDX2* | 4.390127753 | 0.000910669 | 0.032866956 | 1 |
| T2DM | t_cell | *KCTD5* | 4.136805996 | 0.002116764 | 0.044350959 | 1 |
| T2DM | t_cell | *LTBP3* | 3.001636557 | 2.67E-06 | 0.000646003 | 0.016796074 |
| T2DM | t_cell | *UBTD2* | 2.086694577 | 0.002589361 | 0.046927985 | 1 |
| T2DM | t_cell | *PXK* | 2.972738326 | 4.69E-05 | 0.005779331 | 0.294745856 |
| T2DM | t_cell | *TAP1* | -1.742502882 | 0.000508872 | 0.026211401 | 1 |
| T2DM | t_cell | *ZSWIM1* | 4.174158316 | 0.002235301 | 0.044755853 | 1 |
| T2DM | t_cell | *WFDC13* | 10.06035122 | 0.002204804 | 0.044670743 | 1 |
| T2DM | t_cell | *IRS1* | -1.618000545 | 9.21E-17 | 2.90E-13 | 5.79E-13 |
| T2DM | t_cell | *C8orf46* | 0.34246867 | 0.001682158 | 0.042436657 | 1 |
| T2DM | t_cell | *ZNF747* | 3.273499981 | 0.002378953 | 0.046426982 | 1 |
| T2DM | t_cell | *SEMA3E* | 1.481037555 | 0.002462567 | 0.046773889 | 1 |
| T2DM | t_cell | *EMB* | -0.52698321 | 0.000946281 | 0.033337656 | 1 |
| T2DM | t_cell | *SLC26A5* | 2.191563737 | 0.001431378 | 0.040536378 | 1 |
| T2DM | t_cell | *KCNS3* | 1.091156431 | 0.000904246 | 0.032866956 | 1 |
| T2DM | t_cell | *NUDT6* | 1.676473255 | 0.00162548 | 0.042382577 | 1 |

| | | | | | | |
|---|---|---|---|---|---|---|
| T2DM | t_cell | *MFN1* | 2.311991956 | 0.00312449 | 0.049983077 | 1 |
| T2DM | t_cell | *KCNMB3* | 1.849967312 | 0.000394616 | 0.023405167 | 1 |
| T2DM | t_cell | *KCND3* | 1.199678335 | 0.002596819 | 0.046927985 | 1 |
| T2DM | t_cell | *MRPL36* | 2.913229842 | 0.001759849 | 0.042845976 | 1 |
| T2DM | t_cell | *RGS19* | -1.442500546 | 1.93E-06 | 0.000521729 | 0.01211623 |
| T2DM | t_cell | *C1QTNF4* | -0.512692455 | 0.002172625 | 0.044670743 | 1 |
| T2DM | t_cell | *COPRS* | 1.849133056 | 0.002721771 | 0.047401041 | 1 |
| T2DM | t_cell | *FGGY* | 2.448153536 | 0.001944152 | 0.043336413 | 1 |
| T2DM | t_cell | *CTSW* | -0.359285713 | 0.000294678 | 0.019501485 | 1 |
| T2DM | t_cell | *PDE3A* | 2.082685432 | 0.000400094 | 0.023426917 | 1 |
| T2DM | t_cell | *SNX32* | 2.244519567 | 0.000534817 | 0.02653421 | 1 |
| T2DM | t_cell | *HPSE2* | 1.690745504 | 0.000120489 | 0.01130623 | 0.757517422 |
| T2DM | t_cell | *VANGL1* | 2.583925856 | 0.002537677 | 0.046927985 | 1 |
| T2DM | t_cell | *USMG5* | 2.805979442 | 0.001226263 | 0.036365626 | 1 |
| T2DM | t_cell | *C16orf91* | 4.424160501 | 0.000602003 | 0.027491664 | 1 |
| T2DM | t_cell | *LIG4* | 1.68869453 | 0.002669929 | 0.046927985 | 1 |
| T2DM | t_cell | *TMEM81* | 4.070766281 | 0.000892897 | 0.032866956 | 1 |
| T2DM | t_cell | *IL20RB* | 1.91929234 | 0.000263745 | 0.018471093 | 1 |
| T2DM | t_cell | *KLF14* | -31.22331699 | 3.70E-08 | 1.66E-05 | 0.000232312 |
| T2DM | t_cell | *SH3PXD2B* | 1.315006537 | 0.000382827 | 0.023258718 | 1 |
| T2DM | t_cell | *SH3BP5L* | 3.511941724 | 0.001765135 | 0.042845976 | 1 |
| T2DM | t_cell | *TP53I11* | 2.90902324 | 0.001120486 | 0.035012223 | 1 |
| T2DM | t_cell | *TMEM9B* | 3.631632729 | 0.00049141 | 0.025533014 | 1 |
| T2DM | t_cell | *ETV4* | 2.230525359 | 0.001015007 | 0.033410211 | 1 |
| T2DM | t_cell | *HORMAD2* | 1.660205852 | 0.00022056 | 0.016795489 | 1 |
| T2DM | t_cell | *PFN4* | 1.394940696 | 0.001576624 | 0.042063823 | 1 |
| T2DM | t_cell | *AP006621.1* | 1.037270682 | 0.002163021 | 0.044670743 | 1 |
| T2DM | t_cell | *TIMM22* | 3.116502185 | 0.001643697 | 0.042436657 | 1 |
| T2DM | t_cell | *IRX3* | 1.60640548 | 0.000760284 | 0.030445247 | 1 |
| T2DM | t_cell | *ATOX1* | 3.420927401 | 0.002668381 | 0.046927985 | 1 |
| T2DM | t_cell | *BET1L* | 2.952639255 | 0.000402435 | 0.023426917 | 1 |
| T2DM | t_cell | *IMP3* | 5.214412225 | 0.000602615 | 0.027491664 | 1 |
| T2DM | t_cell | *STX19* | 3.472423962 | 3.18E-06 | 0.000740482 | 0.019993022 |
| T2DM | t_cell | *CALR* | 3.499173954 | 0.00018386 | 0.014765181 | 1 |
| T2DM | t_cell | *HLA-DQB1* | -1.990221956 | 1.63E-14 | 3.41E-11 | 1.02E-10 |
| T2DM | t_cell | *TMEM132C* | 0.7371229 | 0.002541592 | 0.046927985 | 1 |
| T2DM | t_cell | *TTC3* | 0.79554224 | 0.001320013 | 0.038243876 | 1 |
| T2DM | t_cell | *ZNF320* | 0.897194947 | 0.000214148 | 0.016621566 | 1 |
| T2DM | t_cell | *ACSM5* | 1.139969503 | 0.001617007 | 0.042358851 | 1 |
| T2DM | t_cell | *RP1L1* | 3.753380697 | 6.69E-06 | 0.001356054 | 0.042037682 |
| T2DM | t_cell | *BMP8A* | 4.001113732 | 1.68E-07 | 5.55E-05 | 0.001055048 |
| T2DM | t_cell | *KCTD16* | 1.487035916 | 0.001832985 | 0.043040402 | 1 |
| T2DM | t_cell | *KCTD8* | 0.828012312 | 0.002572595 | 0.046927985 | 1 |
| T2DM | t_cell | *PP2D1* | 2.800948171 | 0.002482444 | 0.046927985 | 1 |
| T2DM | t_cell | *EIF3C* | 1.414591588 | 0.002010534 | 0.043336413 | 1 |
| T2DM | t_cell | *MROH7* | 4.221705204 | 0.001070623 | 0.034530246 | 1 |
| T2DM | t_cell | *CSF1* | 1.526178669 | 0.001388737 | 0.039686302 | 1 |
| T2DM | t_cell | *PLA2G6* | 1.236400001 | 0.002609397 | 0.046927985 | 1 |
| T2DM | t_cell | *LPAR5* | -0.810074026 | 0.002600408 | 0.046927985 | 1 |
| T2DM | t_cell | *TUSC5* | 0.853813018 | 0.00181669 | 0.043036656 | 1 |

| | | | | | | |
|---|---|---|---|---|---|---|
| T2DM | t_cell | *NDUFA6* | 0.75437371 | 0.002445314 | 0.046720821 | 1 |
| T2DM | t_cell | *TMEM106A* | 2.922833422 | 0.000996895 | 0.033337656 | 1 |
| T2DM | t_cell | *TNFAIP8L1* | 3.073971512 | 0.000653991 | 0.028356161 | 1 |
| T2DM | t_cell | *LINC00839* | 3.360071463 | 3.10E-05 | 0.004240348 | 0.195055993 |
| T2DM | t_cell | *CYP4Z1* | 1.620038547 | 0.002644849 | 0.046927985 | 1 |
| T2DM | t_cell | *CYP4X1* | 1.687092354 | 0.000890947 | 0.032866956 | 1 |
| T2DM | t_cell | *EFCAB6* | 1.639233582 | 0.001151725 | 0.035303364 | 1 |
| T2DM | t_cell | *C3orf70* | 1.66037247 | 0.001481792 | 0.041026174 | 1 |
| T2DM | t_cell | *EPOR* | 2.247538534 | 0.002057972 | 0.043859215 | 1 |
| T2DM | t_cell | *MCRS1* | 3.604906898 | 4.35E-05 | 0.005577165 | 0.273311299 |
| T2DM | t_cell | *GFRAL* | 1.466700884 | 0.002315893 | 0.045786221 | 1 |
| T2DM | t_cell | *UROS* | 2.488092444 | 0.002740529 | 0.047464748 | 1 |
| T2DM | t_cell | *TMEM201* | 5.123601502 | 0.00035026 | 0.022020866 | 1 |
| T2DM | t_cell | *BEND4* | 7.42111585 | 0.002345139 | 0.046074646 | 1 |
| T2DM | t_cell | *LRRK2* | 1.855690728 | 0.001176433 | 0.035388675 | 1 |
| T2DM | t_cell | *ALKBH2* | 2.636563632 | 0.000102009 | 0.009866627 | 0.641330783 |
| T2DM | t_cell | *S100A13* | 2.06263947 | 0.002531048 | 0.046927985 | 1 |
| T2DM | t_cell | *HLA-DRB1* | -3.267324608 | 1.27E-13 | 2.00E-10 | 8.01E-10 |
| T2DM | t_cell | *WDSUB1* | 2.103041561 | 0.001528303 | 0.041735408 | 1 |
| T2DM | t_cell | *GTF2IRD2* | 2.031220316 | 0.000642766 | 0.028312888 | 1 |
| T2DM | t_cell | *ZNF34* | 2.010891831 | 0.002731001 | 0.047430401 | 1 |
| T2DM | t_cell | *TRPV1* | 1.82030475 | 0.000252572 | 0.018464168 | 1 |
| T2DM | t_cell | *ZNF33B* | -1.589682153 | 0.000536002 | 0.02653421 | 1 |
| T2DM | t_cell | *HLA-DQA1* | -1.82042461 | 7.67E-06 | 0.001507759 | 0.048248304 |
| T2DM | t_cell | *C6orf106* | 1.34243698 | 0.002567749 | 0.046927985 | 1 |
| T2DM | t_cell | *ZNF585A* | 1.258541627 | 0.001095223 | 0.034776107 | 1 |
| T2DM | t_cell | *SLC25A29* | 2.220808082 | 0.001163945 | 0.035326476 | 1 |
| T2DM | t_cell | *RAMP2-AS1* | 1.183657593 | 0.000928964 | 0.033184081 | 1 |
| T2DM | t_cell | *C5AR1* | 2.291099052 | 0.001301404 | 0.038055472 | 1 |
| T2DM | t_cell | *GYPE* | 1.492227084 | 0.000979803 | 0.033337656 | 1 |
| T2DM | t_cell | *SLC2A10* | 2.337798582 | 0.000583664 | 0.027491664 | 1 |
| T2DM | t_cell | *NMB* | 3.510885484 | 0.002394524 | 0.046464117 | 1 |
| T2DM | t_cell | *PRIM1* | -2.581670011 | 0.002203578 | 0.044670743 | 1 |
| T2DM | t_cell | *SULT1C4* | 1.672595072 | 0.002433447 | 0.046720821 | 1 |
| T2DM | t_cell | *DDX42* | 2.390008823 | 0.00265949 | 0.046927985 | 1 |
| T2DM | t_cell | *TMEM116* | -1.000438756 | 0.000610339 | 0.027491664 | 1 |
| T2DM | t_cell | *ZNF568* | 3.219473599 | 0.000114792 | 0.010934838 | 0.72169931 |
| T2DM | t_cell | *HLA-DRB5* | -1.048256388 | 1.48E-12 | 1.86E-09 | 9.32E-09 |
| T2DM | t_cell | *HMGN2* | 2.158259287 | 0.001687476 | 0.042436657 | 1 |
| T2DM | t_cell | *CHML* | 1.774819996 | 0.001980335 | 0.043336413 | 1 |
| T2DM | t_cell | *CENPW* | -4.369197681 | 1.99E-06 | 0.000521729 | 0.012521489 |
| T2DM | t_cell | *AGAP6* | 1.19040111 | 0.002209735 | 0.044670743 | 1 |
| T2DM | t_cell | *SYT15* | 2.479303895 | 0.001130504 | 0.035012223 | 1 |
| T2DM | t_cell | *ZDBF2* | 2.579646625 | 0.001994681 | 0.043336413 | 1 |
| T2DM | t_cell | *OXLD1* | 2.997605762 | 0.00064849 | 0.028312888 | 1 |
| T2DM | t_cell | *HLA-DRA* | 2.869298171 | 2.73E-11 | 2.45E-08 | 1.72E-07 |
| T2DM | t_cell | *NOTCH4* | -0.36407073 | 5.23E-08 | 1.83E-05 | 0.000328843 |
| T2DM | t_cell | *PRRT1* | 2.211614184 | 1.02E-05 | 0.001929425 | 0.063921329 |
| T2DM | t_cell | *NELFE* | 2.271734865 | 0.00027528 | 0.018811805 | 1 |
| T2DM | t_cell | *BAG6* | 2.354224533 | 0.000224403 | 0.016795489 | 1 |

| | | | | | | |
|---|---|---|---|---|---|---|
| T2DM | t_cell | *NFKBIL1* | 2.521255474 | 8.67E-05 | 0.009395559 | 0.544942415 |
| T2DM | t_cell | *LINC01123* | -19.64381932 | 0.000309519 | 0.020270256 | 1 |
| T2DM | t_cell | *ATP6AP1L* | 1.492087043 | 0.001533463 | 0.041735408 | 1 |
| T2DM | t_cell | *LIN52* | 2.601801989 | 0.000127481 | 0.01139604 | 0.801473254 |
| T2DM | t_cell | *NYNRIN* | 1.964666644 | 0.002939073 | 0.048927573 | 1 |
| T2DM | t_cell | *HCG27* | 2.130302517 | 1.09E-11 | 1.14E-08 | 6.84E-08 |
| T2DM | t_cell | *METTL6* | 1.885836859 | 0.002611088 | 0.046927985 | 1 |
| T2DM | t_cell | *TAS2R14* | 1.576229821 | 0.000263739 | 0.018471093 | 1 |
| T2DM | t_cell | *ZRSR1* | 5.067526213 | 0.002993599 | 0.049140358 | 1 |
| T2DM | t_cell | *FGFR1OP* | 4.219190199 | 1.47E-05 | 0.002572711 | 0.09261761 |
| T2DM | t_cell | *DNLZ* | 1.296967403 | 7.07E-05 | 0.008078303 | 0.444306692 |
| T2DM | t_cell | *ATF6B* | 2.274023592 | 8.50E-05 | 0.009379654 | 0.534640269 |
| T2DM | t_cell | *PRCD* | 1.866227365 | 0.002601005 | 0.046927985 | 1 |
| T2DM | t_cell | *STARD10* | 1.588145343 | 5.20E-08 | 1.83E-05 | 0.0003272 |
| T2DM | t_cell | *LINC00240* | 0.928538696 | 0.000961727 | 0.033337656 | 1 |
| T2DM | t_cell | *PLEKHM1* | 2.747081931 | 0.002984195 | 0.04911423 | 1 |
| T2DM | t_cell | *SLC26A6* | 2.062476155 | 0.000579009 | 0.027491664 | 1 |
| T2DM | t_cell | *AC108463.2* | 2.90386521 | 0.001079982 | 0.034642079 | 1 |
| T2DM | t_cell | *HLA-DQB2* | 0.863661198 | 7.34E-09 | 4.19E-06 | 4.61E-05 |
| T2DM | t_cell | *ZSCAN31* | 0.925770881 | 0.002941725 | 0.048927573 | 1 |
| T2DM | t_cell | *GS1-124K5.4* | 0.592821134 | 0.001071003 | 0.034530246 | 1 |
| T2DM | t_cell | *HLA-DQA2* | 2.123566371 | 2.38E-18 | 1.50E-14 | 1.50E-14 |
| T2DM | t_cell | *ASB14* | 2.602798089 | 0.000450813 | 0.024719054 | 1 |
| T2DM | t_cell | *LY6G5B* | -0.589600669 | 0.002891534 | 0.048477534 | 1 |
| T2DM | t_cell | *C22orf39* | 5.061113073 | 0.001318339 | 0.038243876 | 1 |
| T2DM | t_cell | *PALM2* | 0.895663747 | 0.000695504 | 0.029144269 | 1 |
| T2DM | t_cell | *WDR92* | 3.108137834 | 0.001151959 | 0.035303364 | 1 |
| T2DM | t_cell | *NME1-NME2* | 3.059199222 | 0.001785711 | 0.042869317 | 1 |
| T2DM | t_cell | *PLA2G4B* | 2.628111167 | 4.93E-08 | 1.83E-05 | 0.000310208 |
| T2DM | t_cell | *JMJD7* | 2.881162697 | 3.36E-08 | 1.63E-05 | 0.000211282 |
| T2DM | t_cell | *ZNF585B* | 1.478467623 | 0.002644371 | 0.046927985 | 1 |
| T2DM | t_cell | *ZNF286B* | -3.357107649 | 0.001044886 | 0.034214561 | 1 |
| T2DM | t_cell | *TMEM200B* | 1.129271915 | 0.000699982 | 0.029144269 | 1 |
| T2DM | t_cell | *PCDHGA6* | 1.456161372 | 0.002751678 | 0.047526921 | 1 |
| T2DM | t_cell | *ZNF260* | 3.330870522 | 0.002854673 | 0.048245511 | 1 |
| T2DM | t_cell | *EBLN2* | 2.523743252 | 0.001091821 | 0.034776107 | 1 |
| T2DM | t_cell | *TAS2R20* | 1.49770919 | 0.000607052 | 0.027491664 | 1 |
| T2DM | t_cell | *HMBS* | 2.121915955 | 6.62E-07 | 0.000198204 | 0.004162283 |
| T2DM | t_cell | *RTEL1* | 2.285609721 | 0.002980409 | 0.04911423 | 1 |
| T2DM | t_cell | *LINC00639* | 1.361084691 | 7.81E-05 | 0.008770385 | 0.491141538 |
| T2DM | t_cell | *ILF3-AS1* | 2.19021856 | 0.001982814 | 0.043336413 | 1 |
| T2DM | t_cell | *LINC01140* | 1.270909535 | 0.002177217 | 0.044670743 | 1 |
| T2DM | t_cell | *LINC01050* | 6.737054256 | 0.00309685 | 0.049983077 | 1 |
| T2DM | t_cell | *LINC01023* | 1.965765278 | 0.00223526 | 0.044755853 | 1 |
| T2DM | t_cell | *AC144652.1* | 1.082675188 | 0.000722142 | 0.029560822 | 1 |
| HTN | adipocyte | *SEMA3F* | -0.371165 | 2.42E-06 | 0.000208841 | 0.015228882 |
| HTN | adipocyte | *RBM6* | -0.264003146 | 6.42E-06 | 0.000482329 | 0.040383732 |
| HTN | adipocyte | *COPZ2* | 0.446132491 | 0.000169951 | 0.005883099 | 1 |
| HTN | adipocyte | *GGNBP2* | -0.618015843 | 0.001114805 | 0.024134477 | 1 |
| HTN | adipocyte | *DLEC1* | -0.904626506 | 0.000241427 | 0.007823962 | 1 |

| | | | | | | |
|---|---|---|---|---|---|---|
| HTN | adipocyte | *HFE* | -0.826286125 | 3.99E-08 | 7.16E-06 | 0.000250708 |
| HTN | adipocyte | *CLCN6* | -1.827275439 | 7.90E-05 | 0.00344739 | 0.496424142 |
| HTN | adipocyte | *CEP68* | 0.879124785 | 5.30E-08 | 9.00E-06 | 0.000333007 |
| HTN | adipocyte | *NR1H3* | -1.870331563 | 1.81E-10 | 9.50E-08 | 1.14E-06 |
| HTN | adipocyte | *SNX1* | 0.806860591 | 0.000382919 | 0.011094061 | 1 |
| HTN | adipocyte | *LIMA1* | 0.293622966 | 0.000165348 | 0.005775251 | 1 |
| HTN | adipocyte | *ANAPC4* | 1.374808868 | 0.000654129 | 0.016190987 | 1 |
| HTN | adipocyte | *SDCCAG8* | 1.045897915 | 9.24E-10 | 3.87E-07 | 5.81E-06 |
| HTN | adipocyte | *ITIH4* | 0.210011297 | 0.000259202 | 0.008195801 | 1 |
| HTN | adipocyte | *PIGV* | -1.184392767 | 1.27E-07 | 1.85E-05 | 0.000798053 |
| HTN | adipocyte | *UHRF1BP1* | 0.422886923 | 1.02E-06 | 9.70E-05 | 0.006399114 |
| HTN | adipocyte | *TIE1* | -0.584495498 | 1.13E-05 | 0.000765002 | 0.071145204 |
| HTN | adipocyte | *POLR1A* | -0.318890984 | 0.000229066 | 0.007523257 | 1 |
| HTN | adipocyte | *ASNS* | 1.380525762 | 0.000625088 | 0.015782835 | 1 |
| HTN | adipocyte | *ACADVL* | -1.617776846 | 0.000595419 | 0.015341791 | 1 |
| HTN | adipocyte | *CLNS1A* | 0.632112676 | 1.65E-05 | 0.001018663 | 0.103903652 |
| HTN | adipocyte | *ATP2A3* | 1.213390877 | 1.88E-06 | 0.000168756 | 0.011812911 |
| HTN | adipocyte | *RGS11* | -0.282068751 | 0.002277757 | 0.040567302 | 1 |
| HTN | adipocyte | *SCGN* | 2.92008671 | 8.65E-05 | 0.003697824 | 0.543580185 |
| HTN | adipocyte | *DDX43* | -0.856912783 | 0.000664977 | 0.016330903 | 1 |
| HTN | adipocyte | *CCNT2* | -0.521292492 | 0.000657369 | 0.016207377 | 1 |
| HTN | adipocyte | *SCAMP1* | 1.115396488 | 0.00206293 | 0.038145995 | 1 |
| HTN | adipocyte | *MAST2* | 1.107489802 | 0.000851116 | 0.019966284 | 1 |
| HTN | adipocyte | *SNX10* | -0.195135414 | 6.50E-07 | 7.04E-05 | 0.004084847 |
| HTN | adipocyte | *AAMDC* | -0.385167191 | 1.77E-05 | 0.001061256 | 0.111431847 |
| HTN | adipocyte | *C3orf18* | 0.921637637 | 0.000263394 | 0.008279795 | 1 |
| HTN | adipocyte | *YPEL3* | 0.413291634 | 4.37E-05 | 0.002111325 | 0.274570418 |
| HTN | adipocyte | *ZNF268* | -1.90321191 | 0.000163063 | 0.005759414 | 1 |
| HTN | adipocyte | *PABPC4* | -0.79767069 | 2.36E-05 | 0.001346892 | 0.148158174 |
| HTN | adipocyte | *TGM1* | 0.378851281 | 0.001981992 | 0.037085661 | 1 |
| HTN | adipocyte | *WAC* | 1.106703676 | 0.001393606 | 0.02826323 | 1 |
| HTN | adipocyte | *TPSD1* | 5.224199129 | 0.001213152 | 0.025423629 | 1 |
| HTN | adipocyte | *MARCH2* | 1.591152954 | 1.29E-05 | 0.000844616 | 0.081083124 |
| HTN | adipocyte | *SLC7A4* | -1.087677179 | 0.000117034 | 0.004627615 | 0.7357908 |
| HTN | adipocyte | *CARD10* | 0.864078214 | 0.001220575 | 0.025494201 | 1 |
| HTN | adipocyte | *SLC17A9* | 1.17360308 | 0.002224394 | 0.039861457 | 1 |
| HTN | adipocyte | *DNTTIP1* | -0.525413309 | 0.000388732 | 0.011210817 | 1 |
| HTN | adipocyte | *POLI* | -0.181337333 | 6.14E-05 | 0.002851441 | 0.386024423 |
| HTN | adipocyte | *MGRN1* | 0.602063558 | 0.000580812 | 0.015151727 | 1 |
| HTN | adipocyte | *CRISPLD2* | 0.445069875 | 0.000829505 | 0.019679624 | 1 |
| HTN | adipocyte | *ZNF500* | -0.436301487 | 0.002139973 | 0.039114364 | 1 |
| HTN | adipocyte | *MTHFSD* | 0.297103374 | 0.001255734 | 0.026141718 | 1 |
| HTN | adipocyte | *KAT8* | 1.650191482 | 0.000126879 | 0.004834482 | 0.797689464 |
| HTN | adipocyte | *ZFAND1* | 0.593146884 | 0.002709115 | 0.045819587 | 1 |
| HTN | adipocyte | *EYA1* | 0.348480805 | 0.000398846 | 0.01139793 | 1 |
| HTN | adipocyte | *CCDC94* | 1.47031668 | 0.001109023 | 0.024126038 | 1 |
| HTN | adipocyte | *ICAM5* | -2.828477166 | 0.001719399 | 0.033663004 | 1 |
| HTN | adipocyte | *ATP13A1* | 0.859540711 | 7.54E-06 | 0.000538961 | 0.047428604 |
| HTN | adipocyte | *HOXA2* | -3.240670241 | 1.32E-07 | 1.85E-05 | 0.000831485 |
| HTN | adipocyte | *TAF6* | -0.862588943 | 0.000941996 | 0.021226993 | 1 |

| | | | | | | |
|---|---|---|---|---|---|---|
| HTN | adipocyte | *MEST* | -0.429776827 | 6.18E-06 | 0.000473724 | 0.038845379 |
| HTN | adipocyte | *ECM2* | -0.739437266 | 0.000552131 | 0.01470867 | 1 |
| HTN | adipocyte | *WNT3* | -1.641454466 | 5.20E-06 | 0.000408979 | 0.032718301 |
| HTN | adipocyte | *RAD51C* | -0.402164914 | 0.000130986 | 0.004872832 | 0.823508587 |
| HTN | adipocyte | *ICAM2* | 0.9360523 | 3.17E-05 | 0.001682963 | 0.199485783 |
| HTN | adipocyte | *SYNGR2* | -0.606537047 | 0.002517389 | 0.043600073 | 1 |
| HTN | adipocyte | *DDX5* | 0.578438084 | 9.46E-05 | 0.003964166 | 0.594624919 |
| HTN | adipocyte | *CYTH1* | 0.929516686 | 0.000111084 | 0.004476844 | 0.698387679 |
| HTN | adipocyte | *SLC2A9* | 1.187134819 | 0.000130216 | 0.004872832 | 0.818670604 |
| HTN | adipocyte | *MADD* | 1.38007528 | 0.00082426 | 0.019629242 | 1 |
| HTN | adipocyte | *KCTD10* | -0.51491149 | 0.000164007 | 0.005760393 | 1 |
| HTN | adipocyte | *ENDOU* | -0.507928248 | 0.002225445 | 0.039861457 | 1 |
| HTN | adipocyte | *CAND1* | -0.946888758 | 3.07E-05 | 0.001649698 | 0.193004893 |
| HTN | adipocyte | *CEP72* | 0.741949701 | 0.000934356 | 0.021226993 | 1 |
| HTN | adipocyte | *NNT* | -0.440077507 | 0.0009523 | 0.021382543 | 1 |
| HTN | adipocyte | *PCDHB6* | 0.779154082 | 0.001901067 | 0.035814658 | 1 |
| HTN | adipocyte | *NIT2* | -1.244973482 | 0.000838235 | 0.019811975 | 1 |
| HTN | adipocyte | *FAM162A* | -0.942882939 | 0.00189837 | 0.035814658 | 1 |
| HTN | adipocyte | *PCCB* | 1.409022686 | 0.000131802 | 0.004874332 | 0.828636397 |
| HTN | adipocyte | *TP53I3* | -1.27483906 | 0.000703124 | 0.017002088 | 1 |
| HTN | adipocyte | *DNAJC27* | -1.57816796 | 2.57E-07 | 3.37E-05 | 0.001618597 |
| HTN | adipocyte | *ITGA4* | 0.978196163 | 0.000367927 | 0.010859893 | 1 |
| HTN | adipocyte | *RTN4* | -0.600614949 | 0.00183124 | 0.035100624 | 1 |
| HTN | adipocyte | *ST3GAL5* | 1.012996906 | 0.002199171 | 0.039844928 | 1 |
| HTN | adipocyte | *PLCD4* | -1.655999131 | 0.001468189 | 0.029447438 | 1 |
| HTN | adipocyte | *ZNF142* | -0.876003896 | 0.000887006 | 0.020427126 | 1 |
| HTN | adipocyte | *CD207* | 14.85382031 | 0.000170432 | 0.005883099 | 1 |
| HTN | adipocyte | *PRDX1* | 1.102416073 | 0.000607247 | 0.015519368 | 1 |
| HTN | adipocyte | *FILIP1* | 0.574159997 | 0.000990298 | 0.022078036 | 1 |
| HTN | adipocyte | *TCF21* | 2.848051195 | 0.000266608 | 0.008297849 | 1 |
| HTN | adipocyte | *TRPM6* | 0.621158119 | 0.00025265 | 0.008104122 | 1 |
| HTN | adipocyte | *HDHD3* | -0.134405648 | 0.001814552 | 0.03488713 | 1 |
| HTN | adipocyte | *ZFP30* | 0.55237653 | 0.000575557 | 0.015077194 | 1 |
| HTN | adipocyte | *EPHX2* | -0.651797936 | 6.49E-06 | 0.000482329 | 0.04081558 |
| HTN | adipocyte | *TEX14* | 1.571033797 | 2.24E-09 | 7.29E-07 | 1.41E-05 |
| HTN | adipocyte | *TRIM6* | -0.667170454 | 7.62E-05 | 0.003397876 | 0.479100495 |
| HTN | adipocyte | *GJB6* | -6.287020442 | 0.00274553 | 0.046152795 | 1 |
| HTN | adipocyte | *CBX3* | -1.234994586 | 0.000719349 | 0.017327767 | 1 |
| HTN | adipocyte | *IKZF4* | -1.684499416 | 2.25E-05 | 0.00130932 | 0.141406515 |
| HTN | adipocyte | *FAM124B* | 0.644673749 | 1.50E-05 | 0.000950057 | 0.09405562 |
| HTN | adipocyte | *CDH26* | -1.803038973 | 0.001024794 | 0.022766353 | 1 |
| HTN | adipocyte | *BTN2A2* | 0.320032879 | 0.000616006 | 0.015616251 | 1 |
| HTN | adipocyte | *SNRPC* | -0.729741027 | 3.80E-07 | 4.51E-05 | 0.002389337 |
| HTN | adipocyte | *HIST1H1D* | 0.862795715 | 5.47E-08 | 9.06E-06 | 0.000344198 |
| HTN | adipocyte | *ZNF391* | -0.224833466 | 1.70E-05 | 0.001038659 | 0.106981918 |
| HTN | adipocyte | *GOT2* | 1.549417314 | 0.000588784 | 0.015285066 | 1 |
| HTN | adipocyte | *GRK4* | 0.171538267 | 0.00085106 | 0.019966284 | 1 |
| HTN | adipocyte | *OPRL1* | -0.8839841 | 2.66E-08 | 5.23E-06 | 0.000167378 |
| HTN | adipocyte | *TRAP1* | 0.299285793 | 5.13E-07 | 5.87E-05 | 0.003226904 |
| HTN | adipocyte | *APOL5* | -1.353470547 | 0.000108 | 0.004444097 | 0.678996805 |

| | | | | | | |
|---|---|---|---|---|---|---|
| HTN | adipocyte | *KLHDC10* | -1.537023226 | 1.81E-05 | 0.001072274 | 0.113661021 |
| HTN | adipocyte | *RIPK3* | 0.696396708 | 0.001311088 | 0.027025611 | 1 |
| HTN | adipocyte | *ADCY4* | 0.678230551 | 0.000258045 | 0.008195801 | 1 |
| HTN | adipocyte | *CDC16* | 0.794002416 | 4.40E-05 | 0.002111325 | 0.276583534 |
| HTN | adipocyte | *ATP8B3* | -0.292543455 | 0.00137466 | 0.027969225 | 1 |
| HTN | adipocyte | *DDA1* | -1.080983698 | 0.001489268 | 0.02972389 | 1 |
| HTN | adipocyte | *PGPEP1* | 0.681842054 | 0.000485713 | 0.013452329 | 1 |
| HTN | adipocyte | *CHMP1A* | 1.316488661 | 1.40E-06 | 0.000131075 | 0.008782027 |
| HTN | adipocyte | *HAUS8* | -1.990704725 | 3.39E-08 | 6.46E-06 | 0.000213074 |
| HTN | adipocyte | *PDLIM4* | 1.219023591 | 1.28E-07 | 1.85E-05 | 0.000804415 |
| HTN | adipocyte | *EXOC4* | 1.000881597 | 0.00068673 | 0.016734386 | 1 |
| HTN | adipocyte | *ANO1* | -1.71202109 | 0.000898298 | 0.020611682 | 1 |
| HTN | adipocyte | *KDM6B* | -2.003787906 | 5.66E-05 | 0.002675731 | 0.355872193 |
| HTN | adipocyte | *IMPA1* | 0.4518179 | 0.001153371 | 0.024664083 | 1 |
| HTN | adipocyte | *WNT2B* | -0.895823355 | 0.000210237 | 0.007106228 | 1 |
| HTN | adipocyte | *DDB2* | 2.09056312 | 1.47E-09 | 5.44E-07 | 9.26E-06 |
| HTN | adipocyte | *ACP2* | 2.41442145 | 1.35E-23 | 8.48E-20 | 8.48E-20 |
| HTN | adipocyte | *APLNR* | 1.369389999 | 0.000293341 | 0.008996254 | 1 |
| HTN | adipocyte | *TMEM165* | 1.224475649 | 6.81E-06 | 0.000498102 | 0.042836752 |
| HTN | adipocyte | *POM121C* | -1.001690609 | 0.000350919 | 0.010456051 | 1 |
| HTN | adipocyte | *AGT* | 0.22434996 | 0.000118993 | 0.004675684 | 0.748109503 |
| HTN | adipocyte | *USP37* | 1.177053013 | 0.0014588 | 0.02939575 | 1 |
| HTN | adipocyte | *GCC2* | 0.819738482 | 0.001593841 | 0.031510945 | 1 |
| HTN | adipocyte | *CIB2* | -0.458405228 | 0.000489761 | 0.013504948 | 1 |
| HTN | adipocyte | *NMT1* | 0.652303971 | 2.59E-09 | 7.76E-07 | 1.63E-05 |
| HTN | adipocyte | *BLK* | -0.813542839 | 0.003005385 | 0.049723311 | 1 |
| HTN | adipocyte | *FPGS* | -1.246697325 | 0.000684625 | 0.016734386 | 1 |
| HTN | adipocyte | *SULF1* | 0.639702471 | 0.002336146 | 0.041489684 | 1 |
| HTN | adipocyte | *RMDN3* | 1.312367191 | 3.19E-05 | 0.001682963 | 0.20027254 |
| HTN | adipocyte | *IFT172* | 1.358043829 | 0.000590786 | 0.015285066 | 1 |
| HTN | adipocyte | *DYNC2LI1* | 0.507277999 | 0.001192323 | 0.025116527 | 1 |
| HTN | adipocyte | *ARL3* | 1.448704243 | 8.92E-07 | 8.76E-05 | 0.00560515 |
| HTN | adipocyte | *TRPC3* | 1.031824654 | 0.002661615 | 0.045357084 | 1 |
| HTN | adipocyte | *G3BP2* | 1.156759055 | 0.00051768 | 0.014150657 | 1 |
| HTN | adipocyte | *FBN2* | 0.175149174 | 0.000128467 | 0.004865495 | 0.807672157 |
| HTN | adipocyte | *DEF8* | 0.870856122 | 3.37E-05 | 0.001751982 | 0.211989765 |
| HTN | adipocyte | *MYO19* | 1.202566193 | 0.002494696 | 0.043326389 | 1 |
| HTN | adipocyte | *RASL10B* | -0.797412341 | 0.000370826 | 0.010891356 | 1 |
| HTN | adipocyte | *SCRN2* | -0.189007123 | 6.17E-05 | 0.002851441 | 0.387795975 |
| HTN | adipocyte | *GREB1L* | -0.547804605 | 0.000145684 | 0.005279926 | 0.915912724 |
| HTN | adipocyte | *C18orf8* | -1.018187601 | 0.000171243 | 0.005883099 | 1 |
| HTN | adipocyte | *NPC1* | -1.063181013 | 0.001278866 | 0.026448124 | 1 |
| HTN | adipocyte | *TRIM65* | -0.635197278 | 0.002144186 | 0.039114364 | 1 |
| HTN | adipocyte | *RERE* | 0.867935337 | 0.0008677 | 0.020221516 | 1 |
| HTN | adipocyte | *RGS5* | -0.911439128 | 0.000309185 | 0.009390566 | 1 |
| HTN | adipocyte | *RAB13* | -1.744585274 | 0.001194503 | 0.025116527 | 1 |
| HTN | adipocyte | *CREB3L4* | -0.642719072 | 2.51E-05 | 0.001416564 | 0.158016016 |
| HTN | adipocyte | *CEP170* | 1.744096309 | 7.06E-09 | 1.71E-06 | 4.44E-05 |
| HTN | adipocyte | *GUK1* | 1.71536207 | 0.001521668 | 0.030274456 | 1 |
| HTN | adipocyte | *WNT9A* | 1.018857876 | 3.28E-06 | 0.000271715 | 0.020650351 |

| | | | | | | |
|---|---|---|---|---|---|---|
| HTN | adipocyte | *PLEKHA6* | 1.011017549 | 0.00037534 | 0.010924823 | 1 |
| HTN | adipocyte | *RABL2A* | -1.159845613 | 0.000197728 | 0.006756054 | 1 |
| HTN | adipocyte | *SLC10A4* | 1.541385336 | 9.03E-05 | 0.003808687 | 0.567494313 |
| HTN | adipocyte | *TSLP* | -0.395612566 | 0.000530295 | 0.014430171 | 1 |
| HTN | adipocyte | *ABT1* | 0.649967866 | 0.000571282 | 0.015027829 | 1 |
| HTN | adipocyte | *SLC18B1* | -0.218167995 | 0.000564749 | 0.014981345 | 1 |
| HTN | adipocyte | *CCDC25* | 1.186397223 | 0.001062062 | 0.023346802 | 1 |
| HTN | adipocyte | *ADHFE1* | -0.193766375 | 0.000642664 | 0.016097318 | 1 |
| HTN | adipocyte | *NRBF2* | -0.615034376 | 3.02E-06 | 0.000256383 | 0.018972358 |
| HTN | adipocyte | *PDCD11* | 2.910072638 | 7.11E-18 | 2.24E-14 | 4.47E-14 |
| HTN | adipocyte | *HSD17B12* | -0.479373172 | 1.33E-08 | 2.78E-06 | 8.35E-05 |
| HTN | adipocyte | *DUSP15* | -0.963359996 | 4.04E-05 | 0.001997333 | 0.25407154 |
| HTN | adipocyte | *DOC2A* | -1.35852729 | 7.56E-08 | 1.16E-05 | 0.000475605 |
| HTN | adipocyte | *ARID5B* | 0.847516796 | 0.001738634 | 0.033737003 | 1 |
| HTN | adipocyte | *KCTD14* | -2.102140115 | 0.000452707 | 0.012655433 | 1 |
| HTN | adipocyte | *ARL14EP* | -1.00409339 | 0.000598006 | 0.015345554 | 1 |
| HTN | adipocyte | *CWF19L2* | 1.716878994 | 9.36E-06 | 0.000653588 | 0.058822917 |
| HTN | adipocyte | *HS3ST3A1* | 0.507135717 | 0.001767965 | 0.034196007 | 1 |
| HTN | adipocyte | *UCHL1* | -1.227461106 | 0.000484341 | 0.013452329 | 1 |
| HTN | adipocyte | *DISP1* | 0.453515868 | 0.001724111 | 0.033663004 | 1 |
| HTN | adipocyte | *NEIL2* | 0.648685203 | 8.36E-07 | 8.35E-05 | 0.005258274 |
| HTN | adipocyte | *PDE1C* | 1.134241785 | 0.002772908 | 0.046488722 | 1 |
| HTN | adipocyte | *TTC39B* | 0.945651985 | 0.000286402 | 0.008826515 | 1 |
| HTN | adipocyte | *DCK* | 0.94566349 | 0.001061889 | 0.023346802 | 1 |
| HTN | adipocyte | *NDUFAF6* | -1.28650602 | 6.13E-07 | 6.77E-05 | 0.003856638 |
| HTN | adipocyte | *ANKRD9* | -1.458070271 | 6.77E-05 | 0.003039023 | 0.425463272 |
| HTN | adipocyte | *SFXN2* | 0.896589651 | 7.88E-05 | 0.00344739 | 0.495254762 |
| HTN | adipocyte | *VPS8* | 0.951200502 | 0.000110346 | 0.004475781 | 0.693746126 |
| HTN | adipocyte | *CLEC18A* | -0.801957708 | 5.80E-06 | 0.000450062 | 0.03645503 |
| HTN | adipocyte | *TRIM63* | 0.499075476 | 0.000109617 | 0.004475063 | 0.689159745 |
| HTN | adipocyte | *CLSTN2* | -0.6021393 | 0.002662122 | 0.045357084 | 1 |
| HTN | adipocyte | *SPATA2L* | 2.212382349 | 3.91E-05 | 0.001970224 | 0.246121044 |
| HTN | adipocyte | *ZNF276* | 1.651351898 | 2.64E-05 | 0.001467446 | 0.165821408 |
| HTN | adipocyte | *SLC5A11* | -0.29601881 | 5.97E-07 | 6.70E-05 | 0.003753067 |
| HTN | adipocyte | *MED8* | 0.691155286 | 6.49E-05 | 0.002957137 | 0.408084895 |
| HTN | adipocyte | *S100B* | -0.199080968 | 2.18E-05 | 0.001283591 | 0.137344194 |
| HTN | adipocyte | *FDPS* | 0.929011474 | 0.000974808 | 0.021810021 | 1 |
| HTN | adipocyte | *PAQR6* | -0.877342291 | 0.001158083 | 0.024680916 | 1 |
| HTN | adipocyte | *PMF1* | 0.797118225 | 0.001353345 | 0.027624934 | 1 |
| HTN | adipocyte | *ABCF3* | 0.579790277 | 4.53E-06 | 0.000364729 | 0.02844889 |
| HTN | adipocyte | *ACOX1* | -1.080293419 | 0.002218922 | 0.039861457 | 1 |
| HTN | adipocyte | *ASGR2* | 4.067384436 | 0.001878479 | 0.035814658 | 1 |
| HTN | adipocyte | *TNFSF13* | 1.236861043 | 2.81E-07 | 3.54E-05 | 0.001769147 |
| HTN | adipocyte | *TTLL10* | 2.004829251 | 0.000540855 | 0.014531429 | 1 |
| HTN | adipocyte | *LMOD1* | -0.805907614 | 2.31E-09 | 7.29E-07 | 1.45E-05 |
| HTN | adipocyte | *RFT1* | -0.941776756 | 0.000229754 | 0.007523257 | 1 |
| HTN | adipocyte | *FAM208A* | 1.15975485 | 1.09E-05 | 0.000746916 | 0.068716293 |
| HTN | adipocyte | *EXO5* | -0.826294076 | 2.52E-05 | 0.001416564 | 0.158655153 |
| HTN | adipocyte | *C1orf50* | -1.33958561 | 1.25E-05 | 0.000833131 | 0.078314287 |
| HTN | adipocyte | *BDH2* | -0.356563501 | 0.002368313 | 0.041942482 | 1 |

| | | | | | | |
|---|---|---|---|---|---|---|
| HTN | adipocyte | *CDC25A* | -0.776018053 | 5.89E-11 | 4.63E-08 | 3.70E-07 |
| HTN | adipocyte | *ZNF589* | -0.653886392 | 1.45E-10 | 8.62E-08 | 9.12E-07 |
| HTN | adipocyte | *ATRIP* | -4.606507039 | 0.000414057 | 0.011726019 | 1 |
| HTN | adipocyte | *APEH* | -0.933082754 | 5.15E-09 | 1.35E-06 | 3.24E-05 |
| HTN | adipocyte | *RNF123* | 0.741983227 | 1.52E-11 | 1.60E-08 | 9.58E-08 |
| HTN | adipocyte | *ERAP1* | -0.78168203 | 1.46E-07 | 1.99E-05 | 0.000916287 |
| HTN | adipocyte | *ERAP2* | 0.293544837 | 0.000874861 | 0.020221516 | 1 |
| HTN | adipocyte | *REEP3* | -1.161428512 | 2.88E-05 | 0.001573949 | 0.181004141 |
| HTN | adipocyte | *TRMT61A* | -1.008748899 | 3.09E-07 | 3.79E-05 | 0.001939866 |
| HTN | adipocyte | *XRRA1* | -0.343431626 | 0.001108159 | 0.024126038 | 1 |
| HTN | adipocyte | *TRIM66* | -0.159679251 | 3.48E-05 | 0.001792289 | 0.218659225 |
| HTN | adipocyte | *RIMKLB* | -0.508062201 | 7.25E-07 | 7.72E-05 | 0.004555948 |
| HTN | adipocyte | *ZNF646* | 1.012528124 | 0.000908363 | 0.020766832 | 1 |
| HTN | adipocyte | *TP53I13* | -0.550713378 | 0.002601892 | 0.044572459 | 1 |
| HTN | adipocyte | *KIR3DL1* | 0.782861872 | 6.43E-05 | 0.002949631 | 0.404099407 |
| HTN | adipocyte | *SRR* | 0.282573489 | 0.000224685 | 0.007434706 | 1 |
| HTN | adipocyte | *TMC8* | 0.88329917 | 0.000333021 | 0.010017707 | 1 |
| HTN | adipocyte | *CYP7A1* | 0.928039361 | 0.000313078 | 0.009463074 | 1 |
| HTN | adipocyte | *NUDT16L1* | -1.614383476 | 0.00269231 | 0.045747445 | 1 |
| HTN | adipocyte | *LRP1B* | -0.279196846 | 0.002817562 | 0.046986776 | 1 |
| HTN | adipocyte | *IRS1* | -0.319731954 | 3.30E-10 | 1.59E-07 | 2.07E-06 |
| HTN | adipocyte | *C8orf46* | 0.078945958 | 0.002122031 | 0.039009383 | 1 |
| HTN | adipocyte | *INO80E* | -1.305192822 | 4.98E-09 | 1.35E-06 | 3.13E-05 |
| HTN | adipocyte | *RGPD8* | -0.173101833 | 0.002736949 | 0.046131896 | 1 |
| HTN | adipocyte | *LIMS1* | 0.624102215 | 0.002419281 | 0.042358497 | 1 |
| HTN | adipocyte | *CALML6* | -1.009414967 | 3.06E-06 | 0.000256563 | 0.019242241 |
| HTN | adipocyte | *ELP5* | -1.960087567 | 3.62E-08 | 6.68E-06 | 0.000227277 |
| HTN | adipocyte | *TMEM133* | -2.912302491 | 0.00222014 | 0.039861457 | 1 |
| HTN | adipocyte | *TTLL6* | -8.198367257 | 0.000874312 | 0.020221516 | 1 |
| HTN | adipocyte | *PAQR8* | -1.167802671 | 0.000940474 | 0.021226993 | 1 |
| HTN | adipocyte | *SOX7* | 1.861941051 | 0.000115275 | 0.004592419 | 0.724733317 |
| HTN | adipocyte | *ALK* | -0.624399366 | 0.002544283 | 0.043900626 | 1 |
| HTN | adipocyte | *GIMAP8* | 0.535468162 | 0.000392208 | 0.011259411 | 1 |
| HTN | adipocyte | *ATP6V0E2* | -1.724525217 | 0.001164026 | 0.024723754 | 1 |
| HTN | adipocyte | *DLK2* | -1.480235972 | 4.07E-05 | 0.001997333 | 0.255658618 |
| HTN | adipocyte | *RGS19* | -0.838695553 | 1.41E-12 | 2.22E-09 | 8.88E-09 |
| HTN | adipocyte | *NME6* | 0.687912434 | 2.32E-09 | 7.29E-07 | 1.46E-05 |
| HTN | adipocyte | *C5orf34* | -1.082946153 | 0.000234366 | 0.007634514 | 1 |
| HTN | adipocyte | *FIBP* | 2.233966819 | 0.000158185 | 0.005650618 | 0.994508695 |
| HTN | adipocyte | *CTSW* | -0.123540449 | 0.000142514 | 0.005233458 | 0.895987018 |
| HTN | adipocyte | *SNTG2* | -0.804186714 | 0.002947962 | 0.049031318 | 1 |
| HTN | adipocyte | *ZNF738* | 0.446089849 | 0.000940721 | 0.021226993 | 1 |
| HTN | adipocyte | *SNX32* | 0.544121528 | 0.001169689 | 0.024760392 | 1 |
| HTN | adipocyte | *MYEOV* | -0.182262489 | 0.000259419 | 0.008195801 | 1 |
| HTN | adipocyte | *DCAKD* | -0.313383934 | 0.000283645 | 0.008784606 | 1 |
| HTN | adipocyte | *HECTD4* | 1.290109668 | 0.000761182 | 0.018196012 | 1 |
| HTN | adipocyte | *DMRT2* | -0.591733485 | 2.31E-05 | 0.001334442 | 0.145454164 |
| HTN | adipocyte | *OLR1* | 2.937896052 | 0.000532846 | 0.014430171 | 1 |
| HTN | adipocyte | *VEGFB* | -0.792684028 | 1.18E-09 | 4.63E-07 | 7.41E-06 |
| HTN | adipocyte | *MST1* | -0.427039893 | 7.60E-07 | 7.97E-05 | 0.004780326 |

| | | | | | | |
|---|---|---|---|---|---|---|
| HTN | adipocyte | *USMG5* | 1.318845338 | 0.002159812 | 0.039244907 | 1 |
| HTN | adipocyte | *NRROS* | 0.385402079 | 0.000372458 | 0.010891356 | 1 |
| HTN | adipocyte | *TLR6* | 1.116723108 | 3.14E-07 | 3.79E-05 | 0.001972382 |
| HTN | adipocyte | *CTU2* | -0.810530653 | 0.001345554 | 0.027555362 | 1 |
| HTN | adipocyte | *SNX31* | -0.776057071 | 4.24E-05 | 0.002066379 | 0.26656293 |
| HTN | adipocyte | *GLIS1* | -0.367418068 | 1.38E-05 | 0.000884872 | 0.086717465 |
| HTN | adipocyte | *KLF14* | -6.242040967 | 2.81E-09 | 8.04E-07 | 1.77E-05 |
| HTN | adipocyte | *RSRC1* | -0.8319811 | 1.51E-05 | 0.000951707 | 0.095170734 |
| HTN | adipocyte | *ARHGAP1* | 1.041688884 | 0.000202243 | 0.00687299 | 1 |
| HTN | adipocyte | *TP53I11* | 1.134178983 | 0.000870983 | 0.020221516 | 1 |
| HTN | adipocyte | *NRIP3* | -0.643412097 | 0.000100601 | 0.004188587 | 0.632476616 |
| HTN | adipocyte | *PPP2R2D* | -0.841806014 | 0.000108151 | 0.004444097 | 0.679946868 |
| HTN | adipocyte | *LRRC25* | 0.237373192 | 0.002100643 | 0.038729446 | 1 |
| HTN | adipocyte | *MSRA* | -1.281990023 | 0.001996342 | 0.037243324 | 1 |
| HTN | adipocyte | *CTDNEP1* | 2.57786684 | 3.72E-05 | 0.001900768 | 0.233794462 |
| HTN | adipocyte | *CRLF3* | 1.008003554 | 0.00107374 | 0.023521266 | 1 |
| HTN | adipocyte | *MTHFR* | -1.784034818 | 6.52E-06 | 0.000482329 | 0.040997974 |
| HTN | adipocyte | *TOP3A* | -1.390173755 | 0.000265187 | 0.008294678 | 1 |
| HTN | adipocyte | *ZBTB38* | 1.079455515 | 0.001647008 | 0.032460006 | 1 |
| HTN | adipocyte | *PPFIA3* | -0.544362516 | 0.001908368 | 0.035814658 | 1 |
| HTN | adipocyte | *ACOT4* | -0.422687976 | 0.002964063 | 0.049169034 | 1 |
| HTN | adipocyte | *PCDHB9* | 0.409124942 | 0.002256369 | 0.040300548 | 1 |
| HTN | adipocyte | *SH2B1* | -0.410678882 | 6.64E-05 | 0.003004652 | 0.417646602 |
| HTN | adipocyte | *COX14* | -1.432214937 | 2.17E-11 | 1.95E-08 | 1.37E-07 |
| HTN | adipocyte | *RPP25* | 0.78590431 | 5.80E-10 | 2.61E-07 | 3.65E-06 |
| HTN | adipocyte | *FAM219B* | -2.106725364 | 1.12E-08 | 2.52E-06 | 7.06E-05 |
| HTN | adipocyte | *MPI* | -2.309435822 | 1.20E-14 | 2.51E-11 | 7.54E-11 |
| HTN | adipocyte | *TRIM73* | -0.879347124 | 0.000650841 | 0.016190987 | 1 |
| HTN | adipocyte | *HYI* | -0.425165778 | 5.95E-08 | 9.36E-06 | 0.000374303 |
| HTN | adipocyte | *CTC1* | 0.695071663 | 0.000217942 | 0.007288293 | 1 |
| HTN | adipocyte | *DEFB124* | -1.570585914 | 0.000545003 | 0.014580561 | 1 |
| HTN | adipocyte | *ZNF443* | 0.602827332 | 0.000567303 | 0.014985863 | 1 |
| HTN | adipocyte | *ANKRD34A* | -1.05166503 | 0.000611618 | 0.015567786 | 1 |
| HTN | adipocyte | *SLC25A41* | -4.501162951 | 0.001773167 | 0.034196007 | 1 |
| HTN | adipocyte | *TMEM136* | 0.520674009 | 0.002425491 | 0.042358497 | 1 |
| HTN | adipocyte | *CLDN7* | -0.827636904 | 1.04E-08 | 2.42E-06 | 6.53E-05 |
| HTN | adipocyte | *ADO* | 2.639011411 | 0.00022033 | 0.007329179 | 1 |
| HTN | adipocyte | *CLN8* | 1.437761617 | 1.40E-08 | 2.84E-06 | 8.79E-05 |
| HTN | adipocyte | *COL18A1* | 0.661156271 | 0.000143177 | 0.005233458 | 0.900154821 |
| HTN | adipocyte | *ZNF320* | 0.203181495 | 0.000429197 | 0.01210029 | 1 |
| HTN | adipocyte | *RP1L1* | 1.119603678 | 1.75E-07 | 2.34E-05 | 0.001100668 |
| HTN | adipocyte | *BMP8A* | 1.295459269 | 0.000756443 | 0.018151745 | 1 |
| HTN | adipocyte | *KREMEN1* | -0.202231651 | 0.00040724 | 0.01158514 | 1 |
| HTN | adipocyte | *GPR39* | -1.171030405 | 0.000305036 | 0.009309522 | 1 |
| HTN | adipocyte | *EIF3C* | 0.521398199 | 1.76E-06 | 0.000160505 | 0.011074874 |
| HTN | adipocyte | *A3GALT2* | -0.439923353 | 1.55E-05 | 0.000962823 | 0.097245123 |
| HTN | adipocyte | *CCR10* | 2.518808901 | 2.81E-05 | 0.001547719 | 0.176439965 |
| HTN | adipocyte | *TMEM173* | 0.943454455 | 0.001541967 | 0.03058153 | 1 |
| HTN | adipocyte | *NUTM2A* | -0.774735393 | 0.001657966 | 0.032573854 | 1 |
| HTN | adipocyte | *DDX51* | 0.399621372 | 0.000639279 | 0.016076599 | 1 |

| | | | | | | |
|---|---|---|---|---|---|---|
| HTN | adipocyte | *CDK10* | -0.696041478 | 4.86E-06 | 0.000386873 | 0.030562938 |
| HTN | adipocyte | *TNFAIP8L1* | 1.258153382 | 0.00188919 | 0.035814658 | 1 |
| HTN | adipocyte | *TLCD2* | -0.704857326 | 0.002048828 | 0.037997005 | 1 |
| HTN | adipocyte | *LSAMP* | 0.885364762 | 4.86E-05 | 0.002316203 | 0.305738855 |
| HTN | adipocyte | *MYBL1* | -0.279785018 | 0.003016719 | 0.049779828 | 1 |
| HTN | adipocyte | *ARL17A* | -0.795092044 | 0.000146128 | 0.005279926 | 0.918707203 |
| HTN | adipocyte | *HYAL3* | -0.283964067 | 0.001905477 | 0.035814658 | 1 |
| HTN | adipocyte | *PLEKHN1* | 0.714371871 | 0.00024412 | 0.00787069 | 1 |
| HTN | adipocyte | *ADH1A* | -0.462534928 | 0.001398657 | 0.028274449 | 1 |
| HTN | adipocyte | *MCRS1* | 1.504057981 | 1.31E-07 | 1.85E-05 | 0.000822771 |
| HTN | adipocyte | *ZNF793* | -0.267499243 | 0.001117088 | 0.024134477 | 1 |
| HTN | adipocyte | *ZNF559* | -0.220373314 | 0.002548708 | 0.043900626 | 1 |
| HTN | adipocyte | *ZDHHC11* | 0.723687042 | 0.002416961 | 0.042358497 | 1 |
| HTN | adipocyte | *FBF1* | -0.368179436 | 2.63E-07 | 3.38E-05 | 0.001653792 |
| HTN | adipocyte | *NOC2L* | -0.344045101 | 0.000452914 | 0.012655433 | 1 |
| HTN | adipocyte | *HLA-DRB1* | -0.604326511 | 5.87E-08 | 9.36E-06 | 0.000369276 |
| HTN | adipocyte | *SULT1A1* | -0.443867845 | 3.22E-05 | 0.001688801 | 0.202656062 |
| HTN | adipocyte | *FAM180B* | -0.860551474 | 5.80E-09 | 1.46E-06 | 3.65E-05 |
| HTN | adipocyte | *C6orf106* | 0.5224355 | 7.11E-06 | 0.000513545 | 0.044678406 |
| HTN | adipocyte | *HIST1H3E* | 0.722014312 | 1.19E-08 | 2.59E-06 | 7.50E-05 |
| HTN | adipocyte | *WDR5B* | 0.771930755 | 0.001142554 | 0.024600133 | 1 |
| HTN | adipocyte | *ZNF429* | -0.967151209 | 0.002464626 | 0.042922721 | 1 |
| HTN | adipocyte | *ZNF100* | -0.56663112 | 7.71E-05 | 0.003411481 | 0.484430279 |
| HTN | adipocyte | *SULT1A2* | 0.186936751 | 5.87E-05 | 0.00275446 | 0.36909762 |
| HTN | adipocyte | *SLC22A5* | 0.437790823 | 4.78E-07 | 5.56E-05 | 0.003003362 |
| HTN | adipocyte | *RPS26* | -0.282663671 | 2.42E-06 | 0.000208841 | 0.015245419 |
| HTN | adipocyte | *TMEM116* | -0.597651417 | 1.05E-11 | 1.32E-08 | 6.62E-08 |
| HTN | adipocyte | *HLA-DRB5* | -0.129565199 | 0.00033578 | 0.010052613 | 1 |
| HTN | adipocyte | *DENND4B* | -1.054518847 | 0.000534791 | 0.014430171 | 1 |
| HTN | adipocyte | *AL590822.1* | -0.447570537 | 8.54E-05 | 0.003675719 | 0.536654963 |
| HTN | adipocyte | *CENPW* | -0.88300308 | 0.002581407 | 0.04434237 | 1 |
| HTN | adipocyte | *HSD17B8* | -0.679547294 | 9.30E-07 | 9.00E-05 | 0.005848726 |
| HTN | adipocyte | *HLA-DRA* | 0.393181138 | 0.001273103 | 0.026415846 | 1 |
| HTN | adipocyte | *PRRT1* | 0.533849871 | 8.74E-05 | 0.003714842 | 0.549796582 |
| HTN | adipocyte | *STK19* | 0.75138387 | 2.15E-06 | 0.000190471 | 0.013523468 |
| HTN | adipocyte | *C6orf48* | 0.975535016 | 1.73E-05 | 0.001048056 | 0.108997836 |
| HTN | adipocyte | *GPANK1* | 0.681197844 | 0.000153007 | 0.005496873 | 0.961952829 |
| HTN | adipocyte | *BAG6* | 0.849814996 | 8.37E-07 | 8.35E-05 | 0.005262431 |
| HTN | adipocyte | *CCHCR1* | -0.222070453 | 0.002146406 | 0.039114364 | 1 |
| HTN | adipocyte | *ATXN2* | -1.152914655 | 0.000213245 | 0.007169358 | 1 |
| HTN | adipocyte | *TRIM13* | 1.11354045 | 0.0023913 | 0.042230626 | 1 |
| HTN | adipocyte | *EIF3CL* | 2.189673882 | 0.000115413 | 0.004592419 | 0.725602189 |
| HTN | adipocyte | *LNP1* | -1.579293636 | 8.03E-05 | 0.003481546 | 0.504824117 |
| HTN | adipocyte | *SUPT4H1* | 1.379225179 | 3.92E-05 | 0.001970224 | 0.246626173 |
| HTN | adipocyte | *MXD3* | -0.499899307 | 0.000653702 | 0.016190987 | 1 |
| HTN | adipocyte | *SIPA1* | 1.431613625 | 1.10E-10 | 7.68E-08 | 6.91E-07 |
| HTN | adipocyte | *ATF6B* | 0.967587247 | 1.51E-10 | 8.62E-08 | 9.49E-07 |
| HTN | adipocyte | *CLIC1* | 1.84100294 | 3.85E-06 | 0.000314665 | 0.024229171 |
| HTN | adipocyte | *PPM1N* | 1.978393066 | 1.04E-05 | 0.000716946 | 0.065242058 |
| HTN | adipocyte | *LTB4R* | -0.545419533 | 8.96E-06 | 0.00063314 | 0.05634944 |

| | | | | | | |
|---|---|---|---|---|---|---|
| HTN | adipocyte | *LTB4R2* | -0.935174811 | 1.27E-05 | 0.000842462 | 0.080033879 |
| HTN | adipocyte | *REPIN1* | -0.965014992 | 0.000130933 | 0.004872832 | 0.823177416 |
| HTN | adipocyte | *KCNU1* | -0.529650652 | 0.001887933 | 0.035814658 | 1 |
| HTN | adipocyte | *AC016773.1* | 0.250830903 | 0.001030284 | 0.022807725 | 1 |
| HTN | adipocyte | *OR2A25* | -9.575939013 | 0.002032391 | 0.037803668 | 1 |
| HTN | adipocyte | *SLC12A8* | -0.651772227 | 0.000511575 | 0.01404485 | 1 |
| HTN | adipocyte | *EXOSC6* | -1.089988378 | 3.07E-05 | 0.001649698 | 0.193014688 |
| HTN | adipocyte | *HLA-DPB1* | 0.484386164 | 0.00012302 | 0.004722821 | 0.77342821 |
| HTN | adipocyte | *AL049840.1* | -0.512703202 | 0.000123198 | 0.004722821 | 0.774542715 |
| HTN | adipocyte | *SLC26A6* | 1.713105935 | 8.19E-07 | 8.35E-05 | 0.005151357 |
| HTN | adipocyte | *AC017074.2* | -2.17728303 | 0.001735413 | 0.033737003 | 1 |
| HTN | adipocyte | *SLC2A1-AS1* | -1.476521133 | 4.30E-08 | 7.50E-06 | 0.000270147 |
| HTN | adipocyte | *LINC01013* | 2.21596647 | 0.001470733 | 0.029447438 | 1 |
| HTN | adipocyte | *PROB1* | 2.010280495 | 0.000121572 | 0.004722821 | 0.764324418 |
| HTN | adipocyte | *HLA-B* | 0.409042216 | 0.000355027 | 0.010528551 | 1 |
| HTN | adipocyte | *ANKRD65* | 0.801463826 | 0.002408281 | 0.042358497 | 1 |
| HTN | adipocyte | *HLA-DQA2* | 0.285671997 | 1.34E-05 | 0.000867585 | 0.084155741 |
| HTN | adipocyte | *ARHGDIG* | -0.56663231 | 0.001148211 | 0.024637545 | 1 |
| HTN | adipocyte | *ZNF709* | -0.631425294 | 0.002711132 | 0.045819587 | 1 |
| HTN | adipocyte | *PLA2G4B* | 0.429482273 | 0.00016107 | 0.005721168 | 1 |
| HTN | adipocyte | *JMJD7* | 0.396039703 | 0.001317445 | 0.027067893 | 1 |
| HTN | adipocyte | *ZNF10* | -0.750193608 | 1.45E-06 | 0.000133629 | 0.009086747 |
| HTN | adipocyte | *HMBS* | 0.429672873 | 3.95E-05 | 0.001970224 | 0.248248263 |
| HTN | adipocyte | *UBE2Q2L* | -1.487796936 | 0.002801479 | 0.046842809 | 1 |
| HTN | adipocyte | *AC009060.1* | -1.800664894 | 0.000123072 | 0.004722821 | 0.773755907 |
| HTN | adipocyte | *AC144652.1* | 0.34895965 | 0.000699522 | 0.016980294 | 1 |
| HTN | ASPC | *SEMA3F* | -0.169767787 | 4.30E-07 | 3.71E-05 | 0.002705017 |
| HTN | ASPC | *M6PR* | -0.467490588 | 0.000382834 | 0.009987043 | 1 |
| HTN | ASPC | *RBM6* | -0.11248184 | 6.25E-06 | 0.00035702 | 0.039272194 |
| HTN | ASPC | *COPZ2* | 0.202207551 | 6.59E-05 | 0.002558591 | 0.414491788 |
| HTN | ASPC | *GGNBP2* | -0.298716849 | 0.000513616 | 0.012663146 | 1 |
| HTN | ASPC | *CD79B* | -0.185133278 | 0.003132184 | 0.04614018 | 1 |
| HTN | ASPC | *HFE* | -0.353186849 | 4.41E-08 | 6.02E-06 | 0.000277036 |
| HTN | ASPC | *CLCN6* | -1.713184989 | 3.90E-10 | 1.17E-07 | 2.45E-06 |
| HTN | ASPC | *CEP68* | 0.389125794 | 4.26E-08 | 5.95E-06 | 0.000267628 |
| HTN | ASPC | *NR1H3* | -0.94438032 | 1.19E-11 | 7.50E-09 | 7.50E-08 |
| HTN | ASPC | *SNX1* | 0.390262513 | 0.000230378 | 0.006583585 | 1 |
| HTN | ASPC | *YTHDC2* | 1.166879642 | 0.002881922 | 0.044084291 | 1 |
| HTN | ASPC | *LIMA1* | 0.135564487 | 5.26E-05 | 0.002100007 | 0.331008491 |
| HTN | ASPC | *ANAPC4* | 0.895250477 | 0.000149887 | 0.004665052 | 0.942340581 |
| HTN | ASPC | *SDCCAG8* | 0.500582457 | 1.83E-10 | 6.06E-08 | 1.15E-06 |
| HTN | ASPC | *ITIH4* | 0.087352938 | 0.000315732 | 0.008593104 | 1 |
| HTN | ASPC | *PITHD1* | 0.617874758 | 0.001637351 | 0.02916154 | 1 |
| HTN | ASPC | *PIGV* | -0.489454557 | 7.36E-07 | 5.61E-05 | 0.004629923 |
| HTN | ASPC | *HAGH* | 0.193332486 | 0.001090582 | 0.021835944 | 1 |
| HTN | ASPC | *UHRF1BP1* | 0.183284493 | 6.79E-07 | 5.43E-05 | 0.004270127 |
| HTN | ASPC | *TIE1* | -0.289543069 | 5.67E-07 | 4.76E-05 | 0.003567658 |
| HTN | ASPC | *POLR1A* | -0.140476771 | 0.000136432 | 0.00437627 | 0.857748825 |
| HTN | ASPC | *ASNS* | 0.530040079 | 0.002392889 | 0.038086307 | 1 |
| HTN | ASPC | *ACADVL* | -1.833714451 | 3.46E-07 | 3.20E-05 | 0.002178091 |

| | | | | | | |
|---|---|---|---|---|---|---|
| HTN | ASPC | *SMARCE1* | 0.556448729 | 0.003532786 | 0.049899903 | 1 |
| HTN | ASPC | *CLNS1A* | 0.280462421 | 1.20E-05 | 0.000641615 | 0.075710539 |
| HTN | ASPC | *ATP2A3* | 0.531487684 | 9.44E-06 | 0.000520407 | 0.05932643 |
| HTN | ASPC | *MARK3* | -1.145060424 | 4.29E-07 | 3.71E-05 | 0.002698286 |
| HTN | ASPC | *NT5C2* | 0.702703022 | 0.002946819 | 0.04464253 | 1 |
| HTN | ASPC | *FDFT1* | 1.628903934 | 0.001807886 | 0.031140209 | 1 |
| HTN | ASPC | *SCGN* | 1.245252105 | 8.07E-05 | 0.002967133 | 0.507379741 |
| HTN | ASPC | *DDX43* | -0.334624442 | 0.002202336 | 0.035870698 | 1 |
| HTN | ASPC | *CCNT2* | -0.230036768 | 0.000598612 | 0.01420178 | 1 |
| HTN | ASPC | *PLOD1* | 2.126813042 | 8.60E-05 | 0.003099174 | 0.540725784 |
| HTN | ASPC | *COL16A1* | -0.227163758 | 0.00315787 | 0.046307508 | 1 |
| HTN | ASPC | *SCAMP1* | 0.614835163 | 0.00336545 | 0.048307275 | 1 |
| HTN | ASPC | *MAST2* | 0.588842018 | 0.001900656 | 0.032383261 | 1 |
| HTN | ASPC | *B4GALT1* | 1.218200461 | 0.002333261 | 0.037517165 | 1 |
| HTN | ASPC | *SNX10* | -0.082714494 | 5.90E-07 | 4.88E-05 | 0.003711904 |
| HTN | ASPC | *AAMDC* | -0.161437418 | 2.72E-05 | 0.001302234 | 0.171313172 |
| HTN | ASPC | *C3orf18* | 0.420491228 | 0.000103084 | 0.003522226 | 0.648089639 |
| HTN | ASPC | *YPEL3* | 0.17013301 | 8.18E-05 | 0.002989674 | 0.514223877 |
| HTN | ASPC | *ZNF268* | -0.96702165 | 7.46E-05 | 0.002793529 | 0.469312917 |
| HTN | ASPC | *PABPC4* | -0.372208391 | 6.07E-06 | 0.000350366 | 0.038189925 |
| HTN | ASPC | *TGM1* | 0.197513025 | 0.000176228 | 0.0054046 | 1 |
| HTN | ASPC | *WAC* | 1.002887064 | 5.27E-05 | 0.002100007 | 0.33137484 |
| HTN | ASPC | *MARCH2* | 0.950087495 | 1.42E-07 | 1.57E-05 | 0.00089304 |
| HTN | ASPC | *SLC7A4* | -0.40531067 | 0.000912519 | 0.018905532 | 1 |
| HTN | ASPC | *GCAT* | 0.241167829 | 0.00186338 | 0.03183443 | 1 |
| HTN | ASPC | *CCDC134* | 1.436913831 | 0.001692412 | 0.029725527 | 1 |
| HTN | ASPC | *DNTTIP1* | -0.242272796 | 0.000145781 | 0.004578355 | 0.91652829 |
| HTN | ASPC | *POLI* | -0.074531953 | 0.000102143 | 0.003509156 | 0.642175551 |
| HTN | ASPC | *VWA8* | 0.567739745 | 0.000123181 | 0.004075995 | 0.77443904 |
| HTN | ASPC | *MGRN1* | 0.275253933 | 0.000359757 | 0.009463556 | 1 |
| HTN | ASPC | *CORO1A* | -0.34803629 | 0.000808663 | 0.017393541 | 1 |
| HTN | ASPC | *MTHFSD* | 0.133239351 | 0.000679072 | 0.01546858 | 1 |
| HTN | ASPC | *KAT8* | 1.091544767 | 7.16E-07 | 5.55E-05 | 0.004499048 |
| HTN | ASPC | *ZFAND1* | 0.282678987 | 0.001145593 | 0.022507325 | 1 |
| HTN | ASPC | *EYA1* | 0.173639192 | 4.51E-05 | 0.001851343 | 0.283255549 |
| HTN | ASPC | *SNX16* | 0.558382316 | 0.002545475 | 0.040108768 | 1 |
| HTN | ASPC | *PLEKHJ1* | 1.458628676 | 0.00196553 | 0.033218511 | 1 |
| HTN | ASPC | *CCDC94* | 0.723010347 | 0.000661515 | 0.015290237 | 1 |
| HTN | ASPC | *ICAM5* | -1.311054613 | 0.000649249 | 0.015153841 | 1 |
| HTN | ASPC | *ATP13A1* | 0.361279509 | 1.26E-05 | 0.000663349 | 0.078938498 |
| HTN | ASPC | *HBP1* | -1.180103856 | 0.001648492 | 0.029277026 | 1 |
| HTN | ASPC | *HOXA2* | -1.374479717 | 1.38E-06 | 9.70E-05 | 0.008681478 |
| HTN | ASPC | *EPHB6* | 0.325039232 | 0.001589465 | 0.028551325 | 1 |
| HTN | ASPC | *TAF6* | -0.360068689 | 0.001408962 | 0.026498517 | 1 |
| HTN | ASPC | *MEST* | -0.167736866 | 0.000107159 | 0.003641679 | 0.673710638 |
| HTN | ASPC | *ACTR3C* | 0.341921856 | 5.74E-05 | 0.002240615 | 0.360739064 |
| HTN | ASPC | *OGN* | -0.405897742 | 0.000910553 | 0.018905532 | 1 |
| HTN | ASPC | *ECM2* | -0.340805563 | 0.001161668 | 0.022681386 | 1 |
| HTN | ASPC | *WNT3* | -0.695417204 | 0.000146373 | 0.004578355 | 0.920249303 |
| HTN | ASPC | *RAD51C* | -0.175193971 | 9.27E-05 | 0.003275785 | 0.583089664 |

| | | | | | | |
|---|---|---|---|---|---|---|
| HTN | ASPC | *AKAP10* | 0.722862715 | 0.00191107 | 0.032385164 | 1 |
| HTN | ASPC | *ICAM2* | 0.395110555 | 0.00030572 | 0.00839328 | 1 |
| HTN | ASPC | *SYNGR2* | -0.278787111 | 0.00129873 | 0.025038153 | 1 |
| HTN | ASPC | *DDX5* | 0.286643289 | 2.11E-05 | 0.001028434 | 0.132668007 |
| HTN | ASPC | *CYTH1* | 0.468533127 | 0.000419997 | 0.010647251 | 1 |
| HTN | ASPC | *CWH43* | 2.119510248 | 0.001980124 | 0.03337545 | 1 |
| HTN | ASPC | *SLC2A9* | 0.581546159 | 0.00112998 | 0.022340194 | 1 |
| HTN | ASPC | *MADD* | 0.915731455 | 0.001522316 | 0.027822092 | 1 |
| HTN | ASPC | *PPFIBP1* | -0.425768118 | 0.002200157 | 0.035870698 | 1 |
| HTN | ASPC | *KCTD10* | -0.219925739 | 0.000175924 | 0.0054046 | 1 |
| HTN | ASPC | *ENDOU* | -0.205275274 | 0.003440112 | 0.049139247 | 1 |
| HTN | ASPC | *CAND1* | -0.515030891 | 9.63E-07 | 7.20E-05 | 0.006051806 |
| HTN | ASPC | *MAPK14* | -0.74341831 | 0.001028664 | 0.020929476 | 1 |
| HTN | ASPC | *GUCA1B* | 0.551648648 | 0.003114244 | 0.04614018 | 1 |
| HTN | ASPC | *CEP72* | 0.329370759 | 0.000624429 | 0.014758583 | 1 |
| HTN | ASPC | *NNT* | -0.190671475 | 0.001084724 | 0.021788051 | 1 |
| HTN | ASPC | *PCDHB6* | 0.329469313 | 0.001841488 | 0.031621293 | 1 |
| HTN | ASPC | *NPHP3* | 0.081755411 | 0.00344686 | 0.049139247 | 1 |
| HTN | ASPC | *NIT2* | -0.636370608 | 0.002938241 | 0.044620105 | 1 |
| HTN | ASPC | *FAM162A* | -0.435140601 | 0.001763693 | 0.030630774 | 1 |
| HTN | ASPC | *PCCB* | 1.048272356 | 3.56E-08 | 5.08E-06 | 0.000223681 |
| HTN | ASPC | *PDCD10* | 1.431837393 | 0.003069735 | 0.045832113 | 1 |
| HTN | ASPC | *HYAL1* | -0.138106393 | 0.003213944 | 0.046881818 | 1 |
| HTN | ASPC | *TP53I3* | -0.789326461 | 1.44E-05 | 0.000724049 | 0.090371456 |
| HTN | ASPC | *DNAJC27* | -0.77338154 | 5.76E-08 | 7.39E-06 | 0.000362091 |
| HTN | ASPC | *RTN4* | -0.307240577 | 0.00079676 | 0.017288947 | 1 |
| HTN | ASPC | *ZNF142* | -0.404915458 | 0.000568599 | 0.013787627 | 1 |
| HTN | ASPC | *PRKD3* | 0.709975415 | 0.002869766 | 0.044005407 | 1 |
| HTN | ASPC | *CD207* | 8.259072856 | 0.002049097 | 0.03426242 | 1 |
| HTN | ASPC | *C1orf21* | -0.200514362 | 0.000518684 | 0.01273814 | 1 |
| HTN | ASPC | *CD46* | 1.077912034 | 0.000683027 | 0.01550249 | 1 |
| HTN | ASPC | *NSUN4* | 0.417348725 | 0.001910659 | 0.032385164 | 1 |
| HTN | ASPC | *IRF6* | -0.108075261 | 0.003159841 | 0.046307508 | 1 |
| HTN | ASPC | *TCF21* | 1.105317124 | 0.000797486 | 0.017288947 | 1 |
| HTN | ASPC | *TRPM6* | 0.246062982 | 0.000851709 | 0.01796877 | 1 |
| HTN | ASPC | *ZFP30* | 0.237107948 | 0.000700435 | 0.015727276 | 1 |
| HTN | ASPC | *EPHX2* | -0.256038698 | 0.000102075 | 0.003509156 | 0.641743758 |
| HTN | ASPC | *TEX14* | 0.722516406 | 2.56E-10 | 8.05E-08 | 1.61E-06 |
| HTN | ASPC | *TRIM6* | -0.24764556 | 0.001024711 | 0.020916739 | 1 |
| HTN | ASPC | *CBX3* | -0.898901212 | 2.02E-06 | 0.000133794 | 0.01271045 |
| HTN | ASPC | *ATF1* | -0.667747209 | 0.001229833 | 0.023790653 | 1 |
| HTN | ASPC | *IKZF4* | -0.824628684 | 3.59E-05 | 0.001546706 | 0.22581914 |
| HTN | ASPC | *ACVR1C* | 0.560000903 | 2.16E-06 | 0.000141322 | 0.013566911 |
| HTN | ASPC | *FAM124B* | 0.25763427 | 4.81E-05 | 0.001952878 | 0.302696157 |
| HTN | ASPC | *CDH26* | -1.001973864 | 0.003256778 | 0.04728722 | 1 |
| HTN | ASPC | *PEPD* | 0.3464659 | 0.003364805 | 0.048307275 | 1 |
| HTN | ASPC | *BTN2A2* | 0.120951913 | 0.002319842 | 0.037397048 | 1 |
| HTN | ASPC | *SNRPC* | -0.312348517 | 3.66E-07 | 3.33E-05 | 0.002299333 |
| HTN | ASPC | *HIST1H1D* | 0.346989575 | 2.41E-07 | 2.44E-05 | 0.001514364 |
| HTN | ASPC | *ZNF391* | -0.090065298 | 4.30E-05 | 0.001788742 | 0.270282757 |

| | | | | | | |
|---|---|---|---|---|---|---|
| HTN | ASPC | *GRK4* | 0.076951171 | 0.000409393 | 0.010505526 | 1 |
| HTN | ASPC | *OPRL1* | -0.366203161 | 4.84E-08 | 6.48E-06 | 0.000304411 |
| HTN | ASPC | *KLC1* | 0.730436506 | 0.001107008 | 0.021955074 | 1 |
| HTN | ASPC | *XRCC3* | 0.66792022 | 0.000742944 | 0.016356655 | 1 |
| HTN | ASPC | *TRAP1* | 0.126097321 | 6.07E-07 | 4.96E-05 | 0.003815401 |
| HTN | ASPC | *RHOT1* | 0.324870291 | 0.001689438 | 0.029725527 | 1 |
| HTN | ASPC | *APOL5* | -0.51551143 | 0.00057019 | 0.013787627 | 1 |
| HTN | ASPC | *KLHDC10* | -1.269509416 | 5.68E-09 | 1.05E-06 | 3.57E-05 |
| HTN | ASPC | *SERGEF* | 1.34476138 | 0.002646846 | 0.041189899 | 1 |
| HTN | ASPC | *ADCY4* | 0.264676421 | 0.001557068 | 0.028161237 | 1 |
| HTN | ASPC | *CDC16* | 0.367380159 | 2.68E-05 | 0.00129397 | 0.168216115 |
| HTN | ASPC | *ATP8B3* | -0.130053368 | 0.000762311 | 0.016699133 | 1 |
| HTN | ASPC | *DDA1* | -0.490015436 | 0.001328988 | 0.025473626 | 1 |
| HTN | ASPC | *PGPEP1* | 0.302751186 | 0.00035251 | 0.009390811 | 1 |
| HTN | ASPC | *CHMP1A* | 0.595235502 | 1.06E-06 | 7.88E-05 | 0.006694794 |
| HTN | ASPC | *HAUS8* | -0.917182854 | 2.04E-08 | 3.21E-06 | 0.000128489 |
| HTN | ASPC | *PDLIM4* | 0.510379008 | 1.80E-07 | 1.92E-05 | 0.001131713 |
| HTN | ASPC | *EXOC4* | 0.823260118 | 1.06E-05 | 0.000579176 | 0.066605285 |
| HTN | ASPC | *ANO1* | -1.315979021 | 2.95E-06 | 0.000187502 | 0.018562663 |
| HTN | ASPC | *KDM6B* | -1.320504727 | 3.36E-07 | 3.15E-05 | 0.002110567 |
| HTN | ASPC | *IMPA1* | 0.191798111 | 0.001302287 | 0.025038153 | 1 |
| HTN | ASPC | *WNT2B* | -0.628893513 | 8.77E-08 | 1.06E-05 | 0.000551284 |
| HTN | ASPC | *DDB2* | 1.201870632 | 3.35E-11 | 1.62E-08 | 2.10E-07 |
| HTN | ASPC | *ACP2* | 1.05880279 | 1.39E-23 | 8.76E-20 | 8.76E-20 |
| HTN | ASPC | *APLNR* | 0.611664571 | 0.003072385 | 0.045832113 | 1 |
| HTN | ASPC | *TMEM165* | 0.752523555 | 9.65E-08 | 1.14E-05 | 0.000606845 |
| HTN | ASPC | *POM121C* | -0.507115302 | 0.000100476 | 0.003490004 | 0.631690724 |
| HTN | ASPC | *BLOC1S1* | 0.422995945 | 0.000819598 | 0.017526576 | 1 |
| HTN | ASPC | *AGT* | 0.093683184 | 0.0001388 | 0.004407254 | 0.872636278 |
| HTN | ASPC | *USP37* | 0.681994181 | 0.001036728 | 0.021025503 | 1 |
| HTN | ASPC | *GCC2* | 0.412316037 | 0.001727536 | 0.030253537 | 1 |
| HTN | ASPC | *CIB2* | -0.168249371 | 0.003117282 | 0.04614018 | 1 |
| HTN | ASPC | *NMT1* | 0.29297057 | 5.17E-10 | 1.48E-07 | 3.25E-06 |
| HTN | ASPC | *BLK* | -0.429908384 | 0.000202471 | 0.006061596 | 1 |
| HTN | ASPC | *SLC2A8* | 0.283174309 | 0.002127207 | 0.035380294 | 1 |
| HTN | ASPC | *FPGS* | -0.616838961 | 0.001064096 | 0.021442223 | 1 |
| HTN | ASPC | *MAP2K5* | 1.259868553 | 0.000250772 | 0.007101823 | 1 |
| HTN | ASPC | *PAQR5* | -0.680135015 | 0.001489018 | 0.027661438 | 1 |
| HTN | ASPC | *RMDN3* | 0.839612248 | 3.39E-08 | 4.95E-06 | 0.000212868 |
| HTN | ASPC | *IFT172* | 0.943900922 | 6.67E-06 | 0.000377521 | 0.041904794 |
| HTN | ASPC | *DYNC2LI1* | 0.242324666 | 0.000421879 | 0.010652019 | 1 |
| HTN | ASPC | *ARL3* | 0.677115751 | 6.90E-07 | 5.43E-05 | 0.004341 |
| HTN | ASPC | *FAM13A* | 0.630500256 | 0.00179254 | 0.03096072 | 1 |
| HTN | ASPC | *G3BP2* | 0.594345548 | 0.000401499 | 0.010387754 | 1 |
| HTN | ASPC | *FBN2* | 0.07119202 | 0.000229209 | 0.00658009 | 1 |
| HTN | ASPC | *MMAB* | 0.39589301 | 0.000232171 | 0.006604798 | 1 |
| HTN | ASPC | *GLTP* | -0.742701655 | 7.02E-05 | 0.002658615 | 0.441330022 |
| HTN | ASPC | *GPR84* | -1.011936225 | 0.002691032 | 0.041688068 | 1 |
| HTN | ASPC | *ULK3* | 4.106960992 | 1.14E-06 | 8.22E-05 | 0.007149628 |
| HTN | ASPC | *TICRR* | 0.663442177 | 0.000960368 | 0.019667215 | 1 |

| | | | | | | |
|---|---|---|---|---|---|---|
| HTN | ASPC | *DEF8* | 0.3793989 | 3.26E-05 | 0.001467405 | 0.20465925 |
| HTN | ASPC | *MYO19* | 0.834752717 | 0.000132399 | 0.004332191 | 0.832395048 |
| HTN | ASPC | *VPS53* | -0.358743778 | 0.00269212 | 0.041688068 | 1 |
| HTN | ASPC | *SCRN2* | -0.074130599 | 0.000224775 | 0.006482393 | 1 |
| HTN | ASPC | *GREB1L* | -0.23085736 | 0.000337788 | 0.009114473 | 1 |
| HTN | ASPC | *C18orf8* | -0.517897973 | 2.94E-05 | 0.001350185 | 0.184701076 |
| HTN | ASPC | *NPC1* | -0.753671765 | 1.20E-05 | 0.000641615 | 0.075225918 |
| HTN | ASPC | *TRIM65* | -0.323346206 | 0.00035175 | 0.009390811 | 1 |
| HTN | ASPC | *RERE* | 0.567421607 | 8.01E-05 | 0.002961318 | 0.503424026 |
| HTN | ASPC | *SYPL2* | 0.134071775 | 0.002439106 | 0.038626351 | 1 |
| HTN | ASPC | *NME7* | -0.660132207 | 0.002392251 | 0.038086307 | 1 |
| HTN | ASPC | *RGS5* | -0.461082033 | 4.32E-05 | 0.001788742 | 0.271888773 |
| HTN | ASPC | *ARNT* | -0.552412287 | 0.00034545 | 0.009281384 | 1 |
| HTN | ASPC | *RAB13* | -1.535162242 | 1.29E-07 | 1.45E-05 | 0.000808484 |
| HTN | ASPC | *CREB3L4* | -0.270113805 | 2.91E-05 | 0.001350185 | 0.18297666 |
| HTN | ASPC | *CEP170* | 1.107276141 | 7.56E-11 | 2.97E-08 | 4.75E-07 |
| HTN | ASPC | *GUK1* | 1.671381634 | 0.000132991 | 0.004332191 | 0.836112851 |
| HTN | ASPC | *WNT9A* | 0.361813735 | 0.000312219 | 0.008534434 | 1 |
| HTN | ASPC | *RABL2A* | -0.530702771 | 0.000206174 | 0.006114217 | 1 |
| HTN | ASPC | *CTDSPL* | -0.593907839 | 0.002553023 | 0.040125625 | 1 |
| HTN | ASPC | *AMT* | -0.089400637 | 0.002595573 | 0.040592953 | 1 |
| HTN | ASPC | *SLC10A4* | 0.691054355 | 3.34E-05 | 0.001467405 | 0.209838972 |
| HTN | ASPC | *ABT1* | 0.273464024 | 0.000667492 | 0.015371885 | 1 |
| HTN | ASPC | *IRAK1BP1* | 2.253775329 | 3.42E-05 | 0.001482764 | 0.215000826 |
| HTN | ASPC | *PHIP* | 1.699304799 | 8.64E-05 | 0.003099174 | 0.543456922 |
| HTN | ASPC | *SLC18B1* | -0.099515173 | 0.000214257 | 0.006265274 | 1 |
| HTN | ASPC | *CCDC25* | 0.595879057 | 0.000698678 | 0.015727276 | 1 |
| HTN | ASPC | *SLC25A37* | -1.731355807 | 1.44E-05 | 0.000724049 | 0.090506187 |
| HTN | ASPC | *ADHFE1* | -0.076160512 | 0.001754791 | 0.030630774 | 1 |
| HTN | ASPC | *NRBF2* | -0.257654935 | 5.32E-06 | 0.000312659 | 0.033454548 |
| HTN | ASPC | *PDCD11* | 1.557160667 | 1.69E-22 | 3.53E-19 | 1.06E-18 |
| HTN | ASPC | *HSD17B12* | -0.210504576 | 4.35E-09 | 8.29E-07 | 2.74E-05 |
| HTN | ASPC | *PTPRJ* | -0.551731908 | 0.000529067 | 0.012942582 | 1 |
| HTN | ASPC | *C11orf49* | -0.562513724 | 0.003488462 | 0.049406362 | 1 |
| HTN | ASPC | *DUSP15* | -0.411391317 | 3.29E-05 | 0.001467405 | 0.206748399 |
| HTN | ASPC | *DOC2A* | -0.567603703 | 1.06E-07 | 1.23E-05 | 0.000664056 |
| HTN | ASPC | *ARID5B* | 0.574603087 | 4.67E-05 | 0.001908504 | 0.29390964 |
| HTN | ASPC | *KCTD14* | -0.815257406 | 0.00145659 | 0.027173829 | 1 |
| HTN | ASPC | *ARL14EP* | -0.522104928 | 0.000136235 | 0.00437627 | 0.856507625 |
| HTN | ASPC | *CWF19L2* | 1.749220188 | 4.86E-11 | 2.18E-08 | 3.06E-07 |
| HTN | ASPC | *PRDM8* | -0.203916574 | 0.000578602 | 0.013884231 | 1 |
| HTN | ASPC | *MED21* | 2.064691713 | 0.001491527 | 0.027661438 | 1 |
| HTN | ASPC | *CAST* | -0.259936714 | 0.001511226 | 0.027719906 | 1 |
| HTN | ASPC | *PLA2R1* | 0.432038693 | 0.003340873 | 0.048285217 | 1 |
| HTN | ASPC | *JAZF1* | -0.529366821 | 0.003076372 | 0.045832113 | 1 |
| HTN | ASPC | *UCHL1* | -1.240783067 | 0.001631046 | 0.029131773 | 1 |
| HTN | ASPC | *NEIL2* | 0.324922979 | 1.19E-08 | 1.98E-06 | 7.49E-05 |
| HTN | ASPC | *PROM2* | 0.971588386 | 0.003005213 | 0.045092538 | 1 |
| HTN | ASPC | *AK9* | 0.466271921 | 0.000743688 | 0.016356655 | 1 |
| HTN | ASPC | *TTC39B* | 0.404333977 | 0.003133745 | 0.04614018 | 1 |

| | | | | | | |
|---|---|---|---|---|---|---|
| HTN | ASPC | *NDUFAF6* | -0.696936243 | 1.66E-08 | 2.67E-06 | 0.000104055 |
| HTN | ASPC | *ANKRD9* | -0.625819382 | 0.000894305 | 0.018741653 | 1 |
| HTN | ASPC | *SFXN2* | 0.430802271 | 1.32E-05 | 0.000681206 | 0.083107101 |
| HTN | ASPC | *VPS8* | 0.479699839 | 0.000153325 | 0.004748553 | 0.963956277 |
| HTN | ASPC | *CLEC18A* | -0.320918103 | 1.81E-05 | 0.000894793 | 0.113638763 |
| HTN | ASPC | *TRIM63* | 0.198784352 | 0.00027437 | 0.007632578 | 1 |
| HTN | ASPC | *SPATA2L* | 0.97768165 | 0.000114369 | 0.003836452 | 0.719039652 |
| HTN | ASPC | *ZNF276* | 0.890770515 | 7.07E-06 | 0.000396686 | 0.044428819 |
| HTN | ASPC | *SLC5A11* | -0.121642504 | 1.22E-06 | 8.68E-05 | 0.00763986 |
| HTN | ASPC | *UBE2Z* | -1.232003652 | 0.000209284 | 0.00617731 | 1 |
| HTN | ASPC | *MED8* | 0.351770511 | 3.52E-06 | 0.000218991 | 0.022118082 |
| HTN | ASPC | *S100B* | -0.079280343 | 6.81E-05 | 0.002611636 | 0.428308322 |
| HTN | ASPC | *FDPS* | 0.455607227 | 0.000586273 | 0.013961737 | 1 |
| HTN | ASPC | *PAQR6* | -0.352806642 | 0.002027048 | 0.033984141 | 1 |
| HTN | ASPC | *PMF1* | 0.361352923 | 0.000830233 | 0.017634047 | 1 |
| HTN | ASPC | *ABCF3* | 0.257391078 | 1.75E-06 | 0.000117173 | 0.011014292 |
| HTN | ASPC | *TNFSF13* | 0.529029792 | 1.92E-07 | 1.98E-05 | 0.001205218 |
| HTN | ASPC | *TTLL10* | 0.797951791 | 0.002156572 | 0.035565711 | 1 |
| HTN | ASPC | *LMOD1* | -0.358017461 | 7.07E-10 | 1.85E-07 | 4.44E-06 |
| HTN | ASPC | *RFT1* | -0.412343718 | 0.000272148 | 0.007604429 | 1 |
| HTN | ASPC | *GNL3* | -1.06138748 | 0.000907989 | 0.018905532 | 1 |
| HTN | ASPC | *FAM208A* | 0.597394059 | 3.62E-06 | 0.000223113 | 0.022757502 |
| HTN | ASPC | *EXO5* | -0.350623045 | 2.73E-05 | 0.001302234 | 0.171894828 |
| HTN | ASPC | *C1orf50* | -0.527695233 | 7.97E-05 | 0.002961318 | 0.500965893 |
| HTN | ASPC | *BDH2* | -0.167577385 | 0.00083445 | 0.017663928 | 1 |
| HTN | ASPC | *CDC25A* | -0.323215524 | 1.05E-10 | 3.65E-08 | 6.57E-07 |
| HTN | ASPC | *ZNF589* | -0.292919651 | 1.80E-11 | 1.03E-08 | 1.13E-07 |
| HTN | ASPC | *ATRIP* | -2.997094487 | 0.002921237 | 0.044469291 | 1 |
| HTN | ASPC | *APEH* | -0.403947904 | 2.72E-09 | 6.10E-07 | 1.71E-05 |
| HTN | ASPC | *RNF123* | 0.318807381 | 9.25E-12 | 6.46E-09 | 5.81E-08 |
| HTN | ASPC | *ERAP1* | -0.345847926 | 1.29E-07 | 1.45E-05 | 0.000810729 |
| HTN | ASPC | *ERAP2* | 0.118737518 | 0.001606671 | 0.028778186 | 1 |
| HTN | ASPC | *DCAF13* | 1.548654567 | 0.001653719 | 0.02928713 | 1 |
| HTN | ASPC | *ZNF367* | 0.260808367 | 0.001845875 | 0.031621293 | 1 |
| HTN | ASPC | *REEP3* | -0.701903901 | 2.92E-07 | 2.86E-05 | 0.001834888 |
| HTN | ASPC | *CEP57* | -0.412442041 | 0.001046476 | 0.021154969 | 1 |
| HTN | ASPC | *TRMT61A* | -0.411460027 | 1.55E-06 | 0.000105843 | 0.00973757 |
| HTN | ASPC | *XRRA1* | -0.153325235 | 0.000715662 | 0.015991305 | 1 |
| HTN | ASPC | *TRIM66* | -0.066867284 | 4.10E-05 | 0.001730402 | 0.257829875 |
| HTN | ASPC | *RIMKLB* | -0.226250686 | 2.83E-07 | 2.82E-05 | 0.001777005 |
| HTN | ASPC | *SMAD3* | -0.454281718 | 0.002612363 | 0.040754152 | 1 |
| HTN | ASPC | *ZNF646* | 0.45197727 | 0.000670665 | 0.015388582 | 1 |
| HTN | ASPC | *ZNF641* | 0.162808974 | 0.002373133 | 0.037964089 | 1 |
| HTN | ASPC | *KIR3DL1* | 0.324660744 | 8.43E-05 | 0.003063799 | 0.530037203 |
| HTN | ASPC | *SRR* | 0.124192999 | 0.000128046 | 0.004214785 | 0.805023932 |
| HTN | ASPC | *CYP7A1* | 0.383817726 | 0.000411142 | 0.010507527 | 1 |
| HTN | ASPC | *HRASLS5* | 0.061356402 | 0.000658678 | 0.015280839 | 1 |
| HTN | ASPC | *NUDT16L1* | -0.798856521 | 0.001370361 | 0.025950189 | 1 |
| HTN | ASPC | *PXK* | -0.353579136 | 0.002101939 | 0.035052753 | 1 |
| HTN | ASPC | *TMEM129* | 0.175912897 | 0.00346885 | 0.049340863 | 1 |

| | | | | | | |
|---|---|---|---|---|---|---|
| HTN | ASPC | *IRS1* | -0.129249444 | 3.40E-09 | 7.38E-07 | 2.14E-05 |
| HTN | ASPC | *C8orf46* | 0.036469875 | 0.000800652 | 0.017297933 | 1 |
| HTN | ASPC | *INO80E* | -0.580583235 | 3.66E-09 | 7.67E-07 | 2.30E-05 |
| HTN | ASPC | *LIMS1* | 0.386398239 | 9.42E-05 | 0.003309606 | 0.592419387 |
| HTN | ASPC | *CALML6* | -0.384515886 | 5.28E-05 | 0.002100007 | 0.331801079 |
| HTN | ASPC | *KCNAB3* | -0.190633094 | 0.003317407 | 0.048056537 | 1 |
| HTN | ASPC | *ELP5* | -0.911143952 | 5.35E-08 | 7.00E-06 | 0.000336228 |
| HTN | ASPC | *RNF34* | -0.502753332 | 0.000495933 | 0.012275314 | 1 |
| HTN | ASPC | *TMEM133* | -5.061745321 | 0.001369817 | 0.025950189 | 1 |
| HTN | ASPC | *PAQR8* | -0.859345413 | 3.88E-07 | 3.49E-05 | 0.002439952 |
| HTN | ASPC | *SOX7* | 0.850654346 | 0.00081061 | 0.017393541 | 1 |
| HTN | ASPC | *MRPL36* | 0.337876909 | 0.002181938 | 0.035816819 | 1 |
| HTN | ASPC | *DLK2* | -0.548069699 | 0.000939686 | 0.01930655 | 1 |
| HTN | ASPC | *RGS19* | -0.383233188 | 4.05E-14 | 4.24E-11 | 2.54E-10 |
| HTN | ASPC | *NME6* | 0.302031328 | 6.38E-10 | 1.74E-07 | 4.01E-06 |
| HTN | ASPC | *C5orf34* | -0.453161014 | 0.001105246 | 0.021955074 | 1 |
| HTN | ASPC | *FIBP* | 0.852023469 | 0.002301922 | 0.037203551 | 1 |
| HTN | ASPC | *CTSW* | -0.057522233 | 4.08E-05 | 0.001730402 | 0.256812878 |
| HTN | ASPC | *ZNF738* | 0.186356138 | 0.001222504 | 0.02372185 | 1 |
| HTN | ASPC | *HOXC5* | 0.260632737 | 0.001378281 | 0.026021784 | 1 |
| HTN | ASPC | *SNX32* | 0.204697937 | 0.003351832 | 0.048307275 | 1 |
| HTN | ASPC | *MYEOV* | -0.06612742 | 0.002559309 | 0.040125625 | 1 |
| HTN | ASPC | *DCAKD* | -0.130148814 | 0.000403574 | 0.010398651 | 1 |
| HTN | ASPC | *HECTD4* | 1.265478246 | 1.47E-06 | 0.00010183 | 0.009266566 |
| HTN | ASPC | *AHSA2* | 0.314666125 | 0.002518008 | 0.039775666 | 1 |
| HTN | ASPC | *DMRT2* | -0.229242998 | 0.000358918 | 0.009463556 | 1 |
| HTN | ASPC | *VEGFB* | -0.331163135 | 4.10E-09 | 8.20E-07 | 2.58E-05 |
| HTN | ASPC | *MST1* | -0.167864289 | 6.03E-06 | 0.000350366 | 0.037905922 |
| HTN | ASPC | *USMG5* | 0.934685343 | 3.88E-05 | 0.0016614 | 0.244225827 |
| HTN | ASPC | *CCS* | 1.016419378 | 8.07E-06 | 0.000448841 | 0.05071903 |
| HTN | ASPC | *NRROS* | 0.147733304 | 0.001512316 | 0.027719906 | 1 |
| HTN | ASPC | *TLR6* | 0.451051793 | 4.56E-06 | 0.000272982 | 0.028663099 |
| HTN | ASPC | *FAM53A* | 0.715909159 | 0.003132471 | 0.04614018 | 1 |
| HTN | ASPC | *CTU2* | -0.338479518 | 0.00176306 | 0.030630774 | 1 |
| HTN | ASPC | *SNX31* | -0.333264695 | 3.30E-05 | 0.001467405 | 0.207685391 |
| HTN | ASPC | *GLIS1* | -0.139047516 | 0.000285244 | 0.007865469 | 1 |
| HTN | ASPC | *KLF14* | -3.173969492 | 6.84E-08 | 8.60E-06 | 0.000430168 |
| HTN | ASPC | *RSRC1* | -0.452165003 | 3.95E-07 | 3.50E-05 | 0.002482072 |
| HTN | ASPC | *ARHGAP1* | 0.502876771 | 0.000138631 | 0.004407254 | 0.8715746 |
| HTN | ASPC | *NRIP3* | -0.287324615 | 4.31E-05 | 0.001788742 | 0.271077515 |
| HTN | ASPC | *PPP2R2D* | -0.41506114 | 2.94E-05 | 0.001350185 | 0.184975342 |
| HTN | ASPC | *LRRC25* | 0.09606169 | 0.003225016 | 0.04693444 | 1 |
| HTN | ASPC | *MSRA* | -1.114674117 | 1.32E-05 | 0.000681206 | 0.082720009 |
| HTN | ASPC | *CTDNEP1* | 1.839584534 | 1.91E-07 | 1.98E-05 | 0.001199269 |
| HTN | ASPC | *CRLF3* | 0.605294062 | 0.00013643 | 0.00437627 | 0.857737337 |
| HTN | ASPC | *MTHFR* | -1.844998611 | 2.31E-15 | 3.63E-12 | 1.45E-11 |
| HTN | ASPC | *TOP3A* | -0.807698228 | 3.13E-05 | 0.001424107 | 0.196526813 |
| HTN | ASPC | *ZBTB38* | 0.69770309 | 0.000437087 | 0.010948074 | 1 |
| HTN | ASPC | *PPFIA3* | -0.253716029 | 0.000692653 | 0.01566442 | 1 |
| HTN | ASPC | *CD151* | 0.473380454 | 0.002798049 | 0.043010589 | 1 |

| | | | | | | |
|---|---|---|---|---|---|---|
| HTN | ASPC | *PCDHB9* | 0.180918951 | 0.001413469 | 0.026498517 | 1 |
| HTN | ASPC | *SH2B1* | -0.165947863 | 0.000188025 | 0.005710687 | 1 |
| HTN | ASPC | *COX14* | -0.915887104 | 1.19E-12 | 1.07E-09 | 7.46E-09 |
| HTN | ASPC | *P4HTM* | -0.393943254 | 0.00318038 | 0.04650011 | 1 |
| HTN | ASPC | *RPP25* | 0.324910537 | 1.30E-09 | 3.14E-07 | 8.16E-06 |
| HTN | ASPC | *FAM219B* | -1.908357767 | 1.96E-12 | 1.54E-09 | 1.23E-08 |
| HTN | ASPC | *MPI* | -1.029986001 | 1.25E-14 | 1.57E-11 | 7.84E-11 |
| HTN | ASPC | *TRIM73* | -0.378909957 | 0.000494337 | 0.012275314 | 1 |
| HTN | ASPC | *HYI* | -0.171808095 | 3.09E-07 | 2.94E-05 | 0.001941024 |
| HTN | ASPC | *CTC1* | 0.302123757 | 0.000194028 | 0.005864678 | 1 |
| HTN | ASPC | *DEFB124* | -0.763588632 | 7.32E-05 | 0.002756516 | 0.460338145 |
| HTN | ASPC | *ZNF443* | 0.288252934 | 0.000110155 | 0.003723367 | 0.69254621 |
| HTN | ASPC | *CLDN7* | -0.375842127 | 1.11E-09 | 2.78E-07 | 6.96E-06 |
| HTN | ASPC | *ADO* | 1.311593673 | 0.000260284 | 0.007325255 | 1 |
| HTN | ASPC | *FAM89A* | 0.174108298 | 0.001692658 | 0.029725527 | 1 |
| HTN | ASPC | *CLN8* | 0.644838924 | 7.32E-08 | 9.03E-06 | 0.000460364 |
| HTN | ASPC | *FES* | -5.331319319 | 1.11E-22 | 3.50E-19 | 6.99E-19 |
| HTN | ASPC | *COL18A1* | 0.26303811 | 0.000717281 | 0.015991305 | 1 |
| HTN | ASPC | *ZNF320* | 0.088643328 | 0.000279286 | 0.007735113 | 1 |
| HTN | ASPC | *GAS6* | -0.488176875 | 0.001096097 | 0.021876703 | 1 |
| HTN | ASPC | *FAM43B* | -0.616499994 | 0.002225272 | 0.03615061 | 1 |
| HTN | ASPC | *RP1L1* | 0.506784704 | 2.54E-08 | 3.89E-06 | 0.000159516 |
| HTN | ASPC | *BMP8A* | 0.812913887 | 3.43E-06 | 0.000215845 | 0.021584483 |
| HTN | ASPC | *KREMEN1* | -0.089688683 | 0.000205677 | 0.006114217 | 1 |
| HTN | ASPC | *GPR39* | -0.653361023 | 0.003539901 | 0.049899903 | 1 |
| HTN | ASPC | *DENND5A* | -1.374816825 | 0.001510544 | 0.027719906 | 1 |
| HTN | ASPC | *EIF3C* | 0.228247333 | 7.41E-07 | 5.61E-05 | 0.004657528 |
| HTN | ASPC | *A3GALT2* | -0.186666706 | 1.38E-05 | 0.000706864 | 0.086944303 |
| HTN | ASPC | *CCR10* | 0.999586359 | 0.000210728 | 0.006190871 | 1 |
| HTN | ASPC | *LPAR5* | -0.181094841 | 0.001177799 | 0.022925141 | 1 |
| HTN | ASPC | *DDX51* | 0.171147212 | 0.00057617 | 0.013878859 | 1 |
| HTN | ASPC | *CDK10* | -0.309871653 | 2.29E-06 | 0.000148695 | 0.014423446 |
| HTN | ASPC | *TCN2* | -0.146414729 | 0.002157212 | 0.035565711 | 1 |
| HTN | ASPC | *LSAMP* | 0.341172383 | 0.001528145 | 0.027846639 | 1 |
| HTN | ASPC | *MYBL1* | -0.141945959 | 0.000483498 | 0.012062502 | 1 |
| HTN | ASPC | *ARL17A* | -0.368219793 | 5.60E-05 | 0.002199554 | 0.351928569 |
| HTN | ASPC | *HYAL3* | -0.114927964 | 0.002907845 | 0.044372867 | 1 |
| HTN | ASPC | *PLEKHN1* | 0.282561898 | 0.000629187 | 0.014815355 | 1 |
| HTN | ASPC | *MCRS1* | 0.697349494 | 2.93E-08 | 4.39E-06 | 0.00018447 |
| HTN | ASPC | *KLHL17* | 0.578537632 | 0.001578511 | 0.028435813 | 1 |
| HTN | ASPC | *ZC3H6* | 1.020255333 | 0.000583611 | 0.013951186 | 1 |
| HTN | ASPC | *ZNF793* | -0.116138517 | 0.000826362 | 0.017611315 | 1 |
| HTN | ASPC | *ZNF559* | -0.094868016 | 0.00213414 | 0.035401941 | 1 |
| HTN | ASPC | *GOLGA8M* | 0.551573343 | 0.003489172 | 0.049406362 | 1 |
| HTN | ASPC | *ZDHHC11* | 0.407068889 | 0.000114721 | 0.003836452 | 0.721252912 |
| HTN | ASPC | *FBF1* | -0.1531657 | 4.45E-07 | 3.78E-05 | 0.002800328 |
| HTN | ASPC | *NOC2L* | -0.134570788 | 0.00135779 | 0.025873058 | 1 |
| HTN | ASPC | *ZKSCAN3* | 0.585778889 | 0.000883441 | 0.018575891 | 1 |
| HTN | ASPC | *HLA-DRB1* | -0.272339358 | 4.17E-09 | 8.20E-07 | 2.62E-05 |
| HTN | ASPC | *ZNF34* | 1.102533186 | 0.00092513 | 0.019069808 | 1 |

| | | | | | | |
|---|---|---|---|---|---|---|
| HTN | ASPC | *SULT1A1* | -0.1719689 | 0.000202425 | 0.006061596 | 1 |
| HTN | ASPC | *MYO6* | -0.387897996 | 0.00226627 | 0.036721751 | 1 |
| HTN | ASPC | *FAM180B* | -0.41195405 | 5.81E-11 | 2.44E-08 | 3.65E-07 |
| HTN | ASPC | *ZNF431* | 0.697264488 | 0.001358058 | 0.025873058 | 1 |
| HTN | ASPC | *C6orf106* | 0.243571912 | 1.39E-06 | 9.70E-05 | 0.008728228 |
| HTN | ASPC | *ZFP28* | 1.184336998 | 0.000334807 | 0.009072983 | 1 |
| HTN | ASPC | *HIST1H3E* | 0.3066725 | 9.21E-09 | 1.65E-06 | 5.79E-05 |
| HTN | ASPC | *FUT11* | -0.610265227 | 0.00039362 | 0.01022599 | 1 |
| HTN | ASPC | *WDR5B* | 0.342776261 | 0.000744076 | 0.016356655 | 1 |
| HTN | ASPC | *ZNF429* | -0.508829368 | 0.003394004 | 0.048606163 | 1 |
| HTN | ASPC | *ZNF100* | -0.266608107 | 1.87E-05 | 0.000916483 | 0.117309859 |
| HTN | ASPC | *SULT1A2* | 0.084933922 | 1.64E-05 | 0.000820081 | 0.103330257 |
| HTN | ASPC | *DDI2* | 0.823729916 | 0.000432369 | 0.010873223 | 1 |
| HTN | ASPC | *SLC22A5* | 0.214774747 | 1.20E-08 | 1.98E-06 | 7.52E-05 |
| HTN | ASPC | *PCDHB11* | -0.436537628 | 0.002998637 | 0.045092538 | 1 |
| HTN | ASPC | *RPS26* | -0.117323894 | 3.72E-06 | 0.000227116 | 0.023392985 |
| HTN | ASPC | *HSD17B11* | 1.628291628 | 0.002703799 | 0.041766055 | 1 |
| HTN | ASPC | *RPS6KL1* | 0.161758884 | 0.002201646 | 0.035870698 | 1 |
| HTN | ASPC | *TMEM116* | -0.249957338 | 2.24E-11 | 1.17E-08 | 1.41E-07 |
| HTN | ASPC | *GFPT1* | 0.973999156 | 2.93E-06 | 0.000187502 | 0.018446711 |
| HTN | ASPC | *HLA-DRB5* | -0.055202613 | 0.000260992 | 0.007325255 | 1 |
| HTN | ASPC | *PNP* | -0.074969609 | 0.002999092 | 0.045092538 | 1 |
| HTN | ASPC | *DENND4B* | -0.781587688 | 2.96E-07 | 2.86E-05 | 0.001858938 |
| HTN | ASPC | *AL590822.1* | -0.17488478 | 0.000374048 | 0.00979851 | 1 |
| HTN | ASPC | *CENPW* | -0.372175308 | 0.002729848 | 0.042065091 | 1 |
| HTN | ASPC | *PCMTD2* | 0.959691894 | 0.000562406 | 0.013704821 | 1 |
| HTN | ASPC | *HSD17B8* | -0.271199371 | 4.52E-06 | 0.000272982 | 0.028439675 |
| HTN | ASPC | *PRRT1* | 0.227682885 | 6.96E-05 | 0.00265086 | 0.437391841 |
| HTN | ASPC | *MRPL38* | 0.219182084 | 0.001416176 | 0.026498517 | 1 |
| HTN | ASPC | *STK19* | 0.325054057 | 1.68E-06 | 0.0001139 | 0.010592695 |
| HTN | ASPC | *C6orf48* | 0.407075029 | 3.37E-05 | 0.001470603 | 0.211766775 |
| HTN | ASPC | *GPANK1* | 0.315627508 | 5.37E-05 | 0.002122933 | 0.337546355 |
| HTN | ASPC | *BAG6* | 0.371534903 | 6.91E-07 | 5.43E-05 | 0.004346879 |
| HTN | ASPC | *NFKBIL1* | 0.292180287 | 0.001143838 | 0.022507325 | 1 |
| HTN | ASPC | *HLA-C* | -0.13659273 | 0.002160983 | 0.035565711 | 1 |
| HTN | ASPC | *CCHCR1* | -0.093066967 | 0.002404461 | 0.038173852 | 1 |
| HTN | ASPC | *ATXN2* | -0.939919523 | 1.77E-07 | 1.92E-05 | 0.001113484 |
| HTN | ASPC | *TRIM13* | 0.829792382 | 0.000215443 | 0.006270786 | 1 |
| HTN | ASPC | *EIF3CL* | 0.949795167 | 8.79E-05 | 0.003120585 | 0.552343529 |
| HTN | ASPC | *ZDHHC11B* | 0.305331174 | 0.002978519 | 0.045014302 | 1 |
| HTN | ASPC | *LNP1* | -0.695389258 | 0.000636905 | 0.014941127 | 1 |
| HTN | ASPC | *SLC48A1* | -1.526490822 | 8.68E-05 | 0.003099174 | 0.545454609 |
| HTN | ASPC | *NOTCH2NL* | 0.155190865 | 0.002025946 | 0.033984141 | 1 |
| HTN | ASPC | *SUPT4H1* | 0.579606794 | 0.000141851 | 0.004481504 | 0.891819343 |
| HTN | ASPC | *MXD3* | -0.198586408 | 0.001507348 | 0.027719906 | 1 |
| HTN | ASPC | *SIPA1* | 0.599426956 | 2.09E-09 | 4.87E-07 | 1.31E-05 |
| HTN | ASPC | *ATF6B* | 0.428166907 | 8.50E-11 | 3.14E-08 | 5.34E-07 |
| HTN | ASPC | *CLIC1* | 0.69040037 | 0.000355586 | 0.009432782 | 1 |
| HTN | ASPC | *PPM1N* | 0.838269603 | 0.000122522 | 0.004075647 | 0.770297295 |
| HTN | ASPC | *LTB4R* | -0.227567638 | 1.27E-05 | 0.000665948 | 0.079913723 |

| | | | | | | |
|---|---|---|---|---|---|---|
| HTN | ASPC | *LTB4R2* | -0.379232901 | 2.80E-05 | 0.001324132 | 0.1761095 |
| HTN | ASPC | *GALT* | 0.495610484 | 0.00155879 | 0.028161237 | 1 |
| HTN | ASPC | *REPIN1* | -0.385855721 | 0.000650793 | 0.015153841 | 1 |
| HTN | ASPC | *AC016773.1* | 0.11412061 | 0.00041788 | 0.010636479 | 1 |
| HTN | ASPC | *SLC12A8* | -0.248745407 | 0.002339538 | 0.03752213 | 1 |
| HTN | ASPC | *EXOSC6* | -0.483097638 | 2.85E-05 | 0.001338661 | 0.179380617 |
| HTN | ASPC | *HLA-DPB1* | 0.197276911 | 0.000675586 | 0.015445127 | 1 |
| HTN | ASPC | *AL049840.1* | -0.22835907 | 6.79E-05 | 0.002611636 | 0.426716121 |
| HTN | ASPC | *SLC26A6* | 0.880826823 | 1.17E-08 | 1.98E-06 | 7.33E-05 |
| HTN | ASPC | *SLC2A1-AS1* | -0.676220928 | 1.10E-06 | 8.01E-05 | 0.006884604 |
| HTN | ASPC | *PROB1* | 0.859536283 | 9.96E-05 | 0.003479556 | 0.626320146 |
| HTN | ASPC | *CISD3* | 2.390331876 | 0.000914153 | 0.018905532 | 1 |
| HTN | ASPC | *HLA-B* | 0.16581429 | 0.001780963 | 0.030845493 | 1 |
| HTN | ASPC | *ANKRD65* | 0.354571229 | 0.001532517 | 0.027846639 | 1 |
| HTN | ASPC | *HLA-DQA2* | 0.115479582 | 3.32E-05 | 0.001467405 | 0.208642558 |
| HTN | ASPC | *PSG4* | 0.291203179 | 0.001156968 | 0.022659997 | 1 |
| HTN | ASPC | *PLA2G4B* | 0.179476339 | 0.000184565 | 0.005632821 | 1 |
| HTN | ASPC | *JMJD7* | 0.176093686 | 0.000723094 | 0.016063936 | 1 |
| HTN | ASPC | *ZNF10* | -0.306049106 | 4.65E-06 | 0.000275935 | 0.029249147 |
| HTN | ASPC | *HMBS* | 0.19463943 | 1.16E-05 | 0.000626301 | 0.07265086 |
| HTN | ASPC | *AC009060.1* | -0.727543864 | 0.00079259 | 0.017288947 | 1 |
| HTN | ASPC | *AC144652.1* | 0.161142137 | 0.000221074 | 0.006405027 | 1 |
| HTN | b_cell | *CFH* | -2.163707015 | 0.000469822 | 0.002180489 | 1 |
| HTN | b_cell | *STPG1* | 6.52885141 | 0.015024513 | 0.020525666 | 1 |
| HTN | b_cell | *NIPAL3* | -5.339299645 | 0.004572701 | 0.007703795 | 1 |
| HTN | b_cell | *SEMA3F* | -7.133059928 | 5.50E-09 | 3.85E-06 | 3.46E-05 |
| HTN | b_cell | *KRIT1* | -4.632152748 | 0.019682833 | 0.026002515 | 1 |
| HTN | b_cell | *RAD52* | -4.022695322 | 0.000720671 | 0.002388432 | 1 |
| HTN | b_cell | *BAD* | -6.579179747 | 0.0007498 | 0.002411911 | 1 |
| HTN | b_cell | *HS3ST1* | 4.842980246 | 0.000360532 | 0.002180344 | 1 |
| HTN | b_cell | *WNT16* | -15.14680636 | 0.005000538 | 0.008260216 | 1 |
| HTN | b_cell | *LASP1* | -4.86559656 | 0.005279791 | 0.008635287 | 1 |
| HTN | b_cell | *SNX11* | -5.633316833 | 0.000220232 | 0.002180344 | 1 |
| HTN | b_cell | *CYP26B1* | -5.522420151 | 0.000740404 | 0.002405521 | 1 |
| HTN | b_cell | *ICA1* | -2.586534788 | 0.00073192 | 0.002399172 | 1 |
| HTN | b_cell | *DBNDD1* | 5.106942757 | 0.000105862 | 0.002180344 | 0.665551642 |
| HTN | b_cell | *ALS2* | -4.113826656 | 0.000959894 | 0.002700157 | 1 |
| HTN | b_cell | *POLDIP2* | -4.32280524 | 0.000465495 | 0.002180489 | 1 |
| HTN | b_cell | *CD38* | 6.367379691 | 0.007654837 | 0.011574305 | 1 |
| HTN | b_cell | *RBM6* | -5.237157856 | 4.52E-07 | 6.93E-05 | 0.002841452 |
| HTN | b_cell | *ARHGAP33* | -8.347493495 | 5.22E-05 | 0.001785055 | 0.328450096 |
| HTN | b_cell | *PDK4* | -3.68499309 | 0.001562513 | 0.003569964 | 1 |
| HTN | b_cell | *SLC22A16* | -11.05945755 | 9.09E-05 | 0.002180344 | 0.57120335 |
| HTN | b_cell | *CDC27* | -3.270755427 | 0.031213213 | 0.039318267 | 1 |
| HTN | b_cell | *MEOX1* | -4.01526948 | 0.000917376 | 0.002637194 | 1 |
| HTN | b_cell | *LIG3* | -4.387533031 | 0.007075605 | 0.01084719 | 1 |
| HTN | b_cell | *CIAPIN1* | -6.48332231 | 0.002136738 | 0.00437294 | 1 |
| HTN | b_cell | *COPZ2* | -2.102733633 | 0.035378731 | 0.043975106 | 1 |
| HTN | b_cell | *GCFC2* | -3.915199426 | 0.001616078 | 0.003652902 | 1 |
| HTN | b_cell | *WDR54* | -6.651768571 | 0.000804322 | 0.002481486 | 1 |

| | | | | | | |
|---|---|---|---|---|---|---|
| HTN | b_cell | *ABCB4* | 3.546478837 | 0.000292705 | 0.002180344 | 1 |
| HTN | b_cell | *ZNF195* | -4.399551363 | 0.002587619 | 0.005038204 | 1 |
| HTN | b_cell | *PDK2* | -3.687852336 | 0.00072528 | 0.002388914 | 1 |
| HTN | b_cell | *GGNBP2* | -5.156723132 | 0.000117646 | 0.002180344 | 0.739643353 |
| HTN | b_cell | *TMEM98* | -4.711325076 | 0.000199785 | 0.002180344 | 1 |
| HTN | b_cell | *ABCC8* | -10.32097999 | 0.00104317 | 0.002813484 | 1 |
| HTN | b_cell | *CCL18* | 7.787114269 | 0.000377854 | 0.002180344 | 1 |
| HTN | b_cell | *SYNRG* | -4.331769466 | 0.003060763 | 0.00567808 | 1 |
| HTN | b_cell | *TMEM132A* | -11.97460804 | 0.000605033 | 0.002281385 | 1 |
| HTN | b_cell | *SPATA20* | -4.445155585 | 0.000652201 | 0.002323165 | 1 |
| HTN | b_cell | *TNFRSF12A* | 11.31053301 | 0.000424609 | 0.002180344 | 1 |
| HTN | b_cell | *MAP3K9* | 1.891322215 | 0.000285732 | 0.002180344 | 1 |
| HTN | b_cell | *RALA* | -4.931017662 | 0.006601572 | 0.010265664 | 1 |
| HTN | b_cell | *BAIAP2L1* | -3.966301203 | 0.000206186 | 0.002180344 | 1 |
| HTN | b_cell | *FARP2* | -4.25698893 | 0.010589923 | 0.015218022 | 1 |
| HTN | b_cell | *IFRD1* | -2.845081045 | 0.000830974 | 0.002520181 | 1 |
| HTN | b_cell | *GTF2IRD1* | -3.275398111 | 0.00092533 | 0.002648292 | 1 |
| HTN | b_cell | *VPS41* | -4.453029015 | 0.025942297 | 0.03329235 | 1 |
| HTN | b_cell | *ELAC2* | -4.75714695 | 0.00037147 | 0.002180344 | 1 |
| HTN | b_cell | *CCDC124* | -6.432628446 | 0.018036623 | 0.024085864 | 1 |
| HTN | b_cell | *CEACAM21* | 7.544585971 | 0.021259745 | 0.027857444 | 1 |
| HTN | b_cell | *NOS2* | -30.94564442 | 0.000427531 | 0.002180344 | 1 |
| HTN | b_cell | *TRAPPC6A* | -8.976958411 | 0.008246818 | 0.012274562 | 1 |
| HTN | b_cell | *CD79B* | 0.423301842 | 0.00033677 | 0.002180344 | 1 |
| HTN | b_cell | *ST7L* | -5.230012473 | 7.34E-05 | 0.002098808 | 0.46173769 |
| HTN | b_cell | *RPUSD1* | -9.129917884 | 0.000400445 | 0.002180344 | 1 |
| HTN | b_cell | *CDK11A* | -6.394935793 | 0.001964086 | 0.004148529 | 1 |
| HTN | b_cell | *DLEC1* | -4.629797856 | 0.000115784 | 0.002180344 | 0.727931843 |
| HTN | b_cell | *SS18L2* | -4.631621342 | 0.000463117 | 0.002180489 | 1 |
| HTN | b_cell | *MPND* | 5.769440945 | 5.98E-05 | 0.001949597 | 0.376272252 |
| HTN | b_cell | *MGST1* | -1.913501482 | 0.000412794 | 0.002180344 | 1 |
| HTN | b_cell | *NFIX* | -2.835536232 | 0.000545511 | 0.002214137 | 1 |
| HTN | b_cell | *MMP25* | -5.423718982 | 0.001053255 | 0.002826212 | 1 |
| HTN | b_cell | *IL32* | -5.211831784 | 0.000957466 | 0.002695741 | 1 |
| HTN | b_cell | *PKD1* | -3.615677276 | 0.003455102 | 0.006206645 | 1 |
| HTN | b_cell | *MAPK8IP2* | 3.063555095 | 0.000194325 | 0.002180344 | 1 |
| HTN | b_cell | *MED24* | -4.939523385 | 0.001090366 | 0.00287056 | 1 |
| HTN | b_cell | *RHOBTB2* | 7.809252304 | 0.003135712 | 0.005781297 | 1 |
| HTN | b_cell | *POMT2* | -6.870536234 | 0.002716472 | 0.005213204 | 1 |
| HTN | b_cell | *MLXIPL* | -2.416555413 | 4.62E-05 | 0.00167785 | 0.29029123 |
| HTN | b_cell | *METTL13* | -4.398690743 | 0.000358287 | 0.002180344 | 1 |
| HTN | b_cell | *DYRK4* | -5.522066899 | 0.001154878 | 0.002978146 | 1 |
| HTN | b_cell | *STARD3NL* | -4.132254139 | 0.001049561 | 0.002819912 | 1 |
| HTN | b_cell | *ZNF200* | -8.084173783 | 0.000888202 | 0.002596195 | 1 |
| HTN | b_cell | *CD4* | -3.393748069 | 0.000458792 | 0.002180489 | 1 |
| HTN | b_cell | *HFE* | -9.1391978 | 1.92E-07 | 4.32E-05 | 0.001209589 |
| HTN | b_cell | *SCMH1* | -3.667372849 | 0.01261433 | 0.017690451 | 1 |
| HTN | b_cell | *CLCN6* | -7.469412802 | 0.017599263 | 0.023551844 | 1 |
| HTN | b_cell | *MRC2* | -2.726476191 | 0.000455694 | 0.002180344 | 1 |
| HTN | b_cell | *TSPAN9* | -2.80408243 | 0.000655318 | 0.00232636 | 1 |

| | | | | | | |
|---|---|---|---|---|---|---|
| HTN | b_cell | *MKS1* | -7.973829555 | 0.008074926 | 0.012073023 | 1 |
| HTN | b_cell | *UTP18* | -4.358083131 | 0.000644474 | 0.002316643 | 1 |
| HTN | b_cell | *TTC19* | -4.016721924 | 0.000514314 | 0.002195202 | 1 |
| HTN | b_cell | *SYT7* | -12.13853361 | 9.77E-05 | 0.002180344 | 0.613942666 |
| HTN | b_cell | *LARS2* | -4.952402298 | 0.007387827 | 0.011240868 | 1 |
| HTN | b_cell | *PPP5C* | -5.413001345 | 0.002777831 | 0.005300219 | 1 |
| HTN | b_cell | *ZBTB32* | -5.604052817 | 0.000405489 | 0.002180344 | 1 |
| HTN | b_cell | *TMEM159* | -3.477042011 | 0.001464822 | 0.003432477 | 1 |
| HTN | b_cell | *ERCC1* | -3.30313039 | 0.000543561 | 0.002214137 | 1 |
| HTN | b_cell | *LTF* | -6.748880354 | 0.035668663 | 0.044317962 | 1 |
| HTN | b_cell | *ALOX5* | 3.116922083 | 0.000531899 | 0.002195202 | 1 |
| HTN | b_cell | *MAP4K5* | -4.115965583 | 0.001992734 | 0.004185234 | 1 |
| HTN | b_cell | *PSMC4* | -3.465275569 | 0.001156581 | 0.002978872 | 1 |
| HTN | b_cell | *MAN2B2* | -6.283601538 | 0.015075903 | 0.020567127 | 1 |
| HTN | b_cell | *CLDN11* | -2.920353824 | 0.000350902 | 0.002180344 | 1 |
| HTN | b_cell | *SLC25A39* | 8.792797158 | 0.0333403 | 0.041713526 | 1 |
| HTN | b_cell | *ANGEL1* | -5.897564809 | 0.001969906 | 0.00415458 | 1 |
| HTN | b_cell | *RNF14* | -4.032564216 | 0.000662783 | 0.002327958 | 1 |
| HTN | b_cell | *DDX11* | -6.003118582 | 0.000212598 | 0.002180344 | 1 |
| HTN | b_cell | *HEBP1* | -7.884206956 | 0.004296536 | 0.007332334 | 1 |
| HTN | b_cell | *TACC3* | -3.733187763 | 0.0016646 | 0.003726974 | 1 |
| HTN | b_cell | *MDH1* | -2.970900714 | 0.000554975 | 0.002220961 | 1 |
| HTN | b_cell | *COX15* | -5.802441853 | 0.026447599 | 0.033892388 | 1 |
| HTN | b_cell | *NPC1L1* | 10.66175718 | 2.85E-05 | 0.001261541 | 0.179069999 |
| HTN | b_cell | *CHDH* | -11.01604856 | 0.001661809 | 0.003724826 | 1 |
| HTN | b_cell | *IL20RA* | -12.50658595 | 0.000409832 | 0.002180344 | 1 |
| HTN | b_cell | *GLT8D1* | -3.494534752 | 0.000292671 | 0.002180344 | 1 |
| HTN | b_cell | *RUFY3* | -4.110211585 | 0.002844916 | 0.00538751 | 1 |
| HTN | b_cell | *WWTR1* | -3.31723077 | 0.000970971 | 0.002714314 | 1 |
| HTN | b_cell | *TTC27* | -4.322450958 | 0.016338198 | 0.02206148 | 1 |
| HTN | b_cell | *VSIG2* | -12.28632581 | 0.000768764 | 0.002432944 | 1 |
| HTN | b_cell | *PHLDB1* | -2.033541945 | 0.000864678 | 0.002559431 | 1 |
| HTN | b_cell | *MARCO* | -3.200978847 | 0.000576259 | 0.002241917 | 1 |
| HTN | b_cell | *MNAT1* | -3.392930004 | 0.000938194 | 0.002662292 | 1 |
| HTN | b_cell | *OSBPL5* | -4.337309269 | 0.000937631 | 0.002662292 | 1 |
| HTN | b_cell | *AQR* | -3.897805858 | 0.001066525 | 0.00284552 | 1 |
| HTN | b_cell | *CPS1* | -3.311513104 | 0.002015306 | 0.004214981 | 1 |
| HTN | b_cell | *NLRP2* | -6.465947088 | 0.00274008 | 0.005252099 | 1 |
| HTN | b_cell | *ZDHHC6* | -4.530095847 | 0.000940356 | 0.002665473 | 1 |
| HTN | b_cell | *RNH1* | -3.94394414 | 0.015289604 | 0.020819957 | 1 |
| HTN | b_cell | *NDUFS1* | -4.675099725 | 0.004807217 | 0.007993381 | 1 |
| HTN | b_cell | *BIRC3* | 1.984060379 | 0.00022376 | 0.002180344 | 1 |
| HTN | b_cell | *GLRX2* | -8.043405584 | 0.001569048 | 0.003577072 | 1 |
| HTN | b_cell | *SNAPC1* | -4.79698535 | 0.000813894 | 0.00249364 | 1 |
| HTN | b_cell | *GCLM* | -2.949947859 | 0.001737062 | 0.003831897 | 1 |
| HTN | b_cell | *UBR2* | -4.142853504 | 0.009245982 | 0.013537375 | 1 |
| HTN | b_cell | *NR1H3* | -3.855548687 | 2.42E-05 | 0.001143146 | 0.152038436 |
| HTN | b_cell | *TYMP* | -3.893214115 | 0.013622661 | 0.018877159 | 1 |
| HTN | b_cell | *TOMM34* | 6.441123668 | 0.000735525 | 0.00240209 | 1 |
| HTN | b_cell | *KPNA6* | -4.558949596 | 0.009549007 | 0.013887255 | 1 |

| | | | | | | |
|---|---|---|---|---|---|---|
| HTN | b_cell | *VIM* | -3.785711471 | 0.005665529 | 0.009126103 | 1 |
| HTN | b_cell | *FAS* | -3.592125489 | 0.001453484 | 0.003419708 | 1 |
| HTN | b_cell | *CD44* | -3.485478656 | 0.000639464 | 0.00231318 | 1 |
| HTN | b_cell | *BTN3A1* | -3.673615296 | 0.033882349 | 0.042324325 | 1 |
| HTN | b_cell | *MIPEP* | -4.401467284 | 0.001017961 | 0.002775525 | 1 |
| HTN | b_cell | *INSRR* | -11.46554517 | 0.010185468 | 0.014704027 | 1 |
| HTN | b_cell | *VRK2* | -4.581847815 | 0.01042663 | 0.01502802 | 1 |
| HTN | b_cell | *VEZT* | -4.579733511 | 0.002145095 | 0.004380258 | 1 |
| HTN | b_cell | *ARNTL2* | -6.881362402 | 0.007297579 | 0.011122395 | 1 |
| HTN | b_cell | *SLC39A9* | -5.391338967 | 0.003256287 | 0.005936589 | 1 |
| HTN | b_cell | *ANK1* | -5.464225963 | 0.000378488 | 0.002180344 | 1 |
| HTN | b_cell | *RABEP1* | -4.647857237 | 0.004682645 | 0.007843563 | 1 |
| HTN | b_cell | *BAK1* | -4.852493704 | 0.025508634 | 0.032775961 | 1 |
| HTN | b_cell | *CENPQ* | -5.404423824 | 0.000243766 | 0.002180344 | 1 |
| HTN | b_cell | *IFT88* | -4.55699486 | 0.001985775 | 0.00417265 | 1 |
| HTN | b_cell | *ZCCHC8* | -5.462960212 | 0.0062998 | 0.009913152 | 1 |
| HTN | b_cell | *FUT8* | -4.106482661 | 0.036201715 | 0.044927001 | 1 |
| HTN | b_cell | *UBA6* | -4.667663228 | 0.001993096 | 0.004185234 | 1 |
| HTN | b_cell | *PIAS1* | -3.981612083 | 0.010495413 | 0.015109838 | 1 |
| HTN | b_cell | *SLC4A7* | -4.31800557 | 0.017186471 | 0.023087894 | 1 |
| HTN | b_cell | *ASTE1* | -4.728327976 | 0.002317482 | 0.004647655 | 1 |
| HTN | b_cell | *MYOC* | -2.804148656 | 0.002619563 | 0.005079947 | 1 |
| HTN | b_cell | *SH3YL1* | -4.489509725 | 0.000365142 | 0.002180344 | 1 |
| HTN | b_cell | *VCL* | -4.146715781 | 0.003409824 | 0.00614609 | 1 |
| HTN | b_cell | *ADSS* | -5.59300824 | 0.023769543 | 0.030795984 | 1 |
| HTN | b_cell | *STAP1* | 0.460742928 | 0.000260907 | 0.002180344 | 1 |
| HTN | b_cell | *TBC1D23* | -4.389774923 | 0.002130219 | 0.00436386 | 1 |
| HTN | b_cell | *TUBG2* | -3.34945238 | 0.000306486 | 0.002180344 | 1 |
| HTN | b_cell | *NSUN2* | -5.931747031 | 0.009004077 | 0.013244724 | 1 |
| HTN | b_cell | *MRI1* | -3.736741393 | 0.002555317 | 0.00499387 | 1 |
| HTN | b_cell | *AGA* | -5.320727672 | 0.000613975 | 0.002286342 | 1 |
| HTN | b_cell | *MAT2B* | -6.565492599 | 0.00868598 | 0.012843075 | 1 |
| HTN | b_cell | *CLEC16A* | -4.624980399 | 0.038420865 | 0.047502848 | 1 |
| HTN | b_cell | *SKIV2L2* | -4.251064264 | 0.004746489 | 0.007922556 | 1 |
| HTN | b_cell | *C6* | -3.029363806 | 0.000316119 | 0.002180344 | 1 |
| HTN | b_cell | *RAI14* | -4.004800412 | 0.000197478 | 0.002180344 | 1 |
| HTN | b_cell | *PNKP* | -3.015231727 | 0.004716337 | 0.007881874 | 1 |
| HTN | b_cell | *PHLPP2* | -6.390645687 | 0.016774598 | 0.022597363 | 1 |
| HTN | b_cell | *SPDL1* | -5.823185665 | 0.00073307 | 0.002399172 | 1 |
| HTN | b_cell | *PQLC2* | -5.754738847 | 0.000257732 | 0.002180344 | 1 |
| HTN | b_cell | *CTNS* | -5.613851344 | 0.001174589 | 0.003006775 | 1 |
| HTN | b_cell | *INPP4A* | -5.069874587 | 0.024242805 | 0.031302165 | 1 |
| HTN | b_cell | *PSMA4* | -5.092389818 | 0.001194036 | 0.003030645 | 1 |
| HTN | b_cell | *LSG1* | -4.925242886 | 0.003529874 | 0.006306428 | 1 |
| HTN | b_cell | *THAP3* | -6.359156863 | 0.000739565 | 0.002405521 | 1 |
| HTN | b_cell | *SPATA7* | -4.766225418 | 0.001573515 | 0.003581136 | 1 |
| HTN | b_cell | *MED17* | -6.741982751 | 0.001164944 | 0.002995893 | 1 |
| HTN | b_cell | *RETSAT* | -2.24760902 | 0.00110358 | 0.002895745 | 1 |
| HTN | b_cell | *CAPG* | 7.340832244 | 0.000643234 | 0.002316643 | 1 |
| HTN | b_cell | *LCP2* | -3.022970967 | 0.000401275 | 0.002180344 | 1 |

| | | | | | | |
|---|---|---|---|---|---|---|
| HTN | b_cell | *CUL7* | -4.708488752 | 0.019061998 | 0.025272624 | 1 |
| HTN | b_cell | *CTNNA1* | -4.140514063 | 0.002702241 | 0.005190647 | 1 |
| HTN | b_cell | *CNTLN* | -4.065015526 | 0.002574513 | 0.005020459 | 1 |
| HTN | b_cell | *EPHA3* | -2.799925344 | 0.000415932 | 0.002180344 | 1 |
| HTN | b_cell | *DSG2* | -4.324499736 | 0.000672552 | 0.002334899 | 1 |
| HTN | b_cell | *WDR37* | -4.50702643 | 0.003043274 | 0.005650639 | 1 |
| HTN | b_cell | *YTHDC2* | -4.186084762 | 0.0258077 | 0.03312638 | 1 |
| HTN | b_cell | *ATP6V1H* | -4.311093513 | 0.001744766 | 0.003841901 | 1 |
| HTN | b_cell | *POLR2B* | -4.829766702 | 0.003183266 | 0.005841563 | 1 |
| HTN | b_cell | *CP* | -7.359788607 | 0.005762245 | 0.009243999 | 1 |
| HTN | b_cell | *ANO2* | -2.330861981 | 0.000963247 | 0.002705959 | 1 |
| HTN | b_cell | *GOPC* | -5.025385787 | 0.004184722 | 0.007192277 | 1 |
| HTN | b_cell | *TSPAN17* | -5.10481784 | 0.001005808 | 0.002758778 | 1 |
| HTN | b_cell | *CC2D2A* | -3.62984091 | 0.001044726 | 0.002814467 | 1 |
| HTN | b_cell | *RRM2B* | -6.743354607 | 0.002836704 | 0.005386395 | 1 |
| HTN | b_cell | *SNX29* | -4.147211481 | 0.013380102 | 0.018577893 | 1 |
| HTN | b_cell | *LMO3* | -2.538862786 | 0.000123158 | 0.002180344 | 0.774291359 |
| HTN | b_cell | *MRPS10* | -4.732581705 | 1.86E-05 | 0.000985596 | 0.116726785 |
| HTN | b_cell | *CELF2* | -3.212916194 | 0.003015098 | 0.005614905 | 1 |
| HTN | b_cell | *ERCC8* | -4.345085092 | 0.001947055 | 0.004124371 | 1 |
| HTN | b_cell | *ADAMTS6* | -5.247310674 | 0.007704441 | 0.01162275 | 1 |
| HTN | b_cell | *H6PD* | -3.626610082 | 0.010933625 | 0.015622659 | 1 |
| HTN | b_cell | *UTS2* | -5.198132878 | 0.005887859 | 0.009397555 | 1 |
| HTN | b_cell | *RFC2* | 8.147107848 | 0.003881819 | 0.006798049 | 1 |
| HTN | b_cell | *JKAMP* | -4.136354993 | 0.000622691 | 0.002291037 | 1 |
| HTN | b_cell | *DKK3* | -5.304501388 | 0.00109936 | 0.002885877 | 1 |
| HTN | b_cell | *ARHGEF5* | 6.33049157 | 0.000454423 | 0.002180344 | 1 |
| HTN | b_cell | *MCUR1* | -6.145680393 | 0.003083722 | 0.005714012 | 1 |
| HTN | b_cell | *LIMA1* | -1.198444987 | 0.013709968 | 0.018981407 | 1 |
| HTN | b_cell | *PTGER3* | -2.043709814 | 0.000504461 | 0.002195028 | 1 |
| HTN | b_cell | *MAPK9* | -6.099191222 | 0.007457006 | 0.011326937 | 1 |
| HTN | b_cell | *PIK3CB* | -4.036647039 | 0.00052784 | 0.002195202 | 1 |
| HTN | b_cell | *CYBA* | 5.137603618 | 0.033753854 | 0.042188962 | 1 |
| HTN | b_cell | *THOC3* | -9.638452065 | 0.024924786 | 0.032097937 | 1 |
| HTN | b_cell | *RRP12* | -5.182594065 | 0.001957955 | 0.004140486 | 1 |
| HTN | b_cell | *ALX4* | 7.904048127 | 0.000334761 | 0.002180344 | 1 |
| HTN | b_cell | *FOXN3* | -3.581055051 | 0.004847892 | 0.008048243 | 1 |
| HTN | b_cell | *AKR7A2* | -5.675907142 | 0.00169163 | 0.003759376 | 1 |
| HTN | b_cell | *MRTO4* | -7.874844087 | 0.006549196 | 0.01021707 | 1 |
| HTN | b_cell | *MCF2L2* | -5.520831654 | 0.001369882 | 0.003289051 | 1 |
| HTN | b_cell | *ANAPC4* | -4.359293122 | 0.009985024 | 0.014437867 | 1 |
| HTN | b_cell | *OPN3* | 5.639697868 | 0.03406231 | 0.042516005 | 1 |
| HTN | b_cell | *TBC1D22A* | -3.756571177 | 0.032525334 | 0.040799436 | 1 |
| HTN | b_cell | *PLEKHH1* | -3.747377223 | 0.005692734 | 0.009160536 | 1 |
| HTN | b_cell | *ATP9A* | -2.487297898 | 0.000353739 | 0.002180344 | 1 |
| HTN | b_cell | *GALC* | -5.570609242 | 0.034839022 | 0.043390041 | 1 |
| HTN | b_cell | *FAM114A2* | -5.032846681 | 0.002324748 | 0.004656162 | 1 |
| HTN | b_cell | *MRPL43* | -6.048956995 | 0.006200972 | 0.009795354 | 1 |
| HTN | b_cell | *TRAF1* | -9.394668517 | 0.001127236 | 0.002930908 | 1 |
| HTN | b_cell | *PCGF2* | -2.678945793 | 0.020307139 | 0.026715364 | 1 |

| | | | | | | |
|---|---|---|---|---|---|---|
| HTN | b_cell | *IL17RB* | -9.266551921 | 0.000423955 | 0.002180344 | 1 |
| HTN | b_cell | *TRAF3IP2* | -4.189807034 | 0.001051772 | 0.002823438 | 1 |
| HTN | b_cell | *DCBLD2* | -3.336850232 | 0.000621701 | 0.002291037 | 1 |
| HTN | b_cell | *TMCC3* | -6.238697679 | 0.009283617 | 0.013582988 | 1 |
| HTN | b_cell | *PITHD1* | -4.408429804 | 0.002510331 | 0.004923903 | 1 |
| HTN | b_cell | *ATP11B* | -3.541041094 | 0.019718837 | 0.026039137 | 1 |
| HTN | b_cell | *LAMC2* | -3.271772257 | 0.007138594 | 0.010929186 | 1 |
| HTN | b_cell | *ZC3H11A* | -8.739545456 | 0.001923427 | 0.004091873 | 1 |
| HTN | b_cell | *RIOK2* | -5.98649408 | 0.022461137 | 0.029254851 | 1 |
| HTN | b_cell | *FLYWCH1* | -4.45636706 | 0.000950462 | 0.002680823 | 1 |
| HTN | b_cell | *DNAJC25* | -4.790224542 | 0.000524976 | 0.002195202 | 1 |
| HTN | b_cell | *YBX3* | -4.858917491 | 0.002841166 | 0.00538751 | 1 |
| HTN | b_cell | *OGFR* | -4.999118128 | 0.003826469 | 0.006719836 | 1 |
| HTN | b_cell | *QSER1* | -4.546360753 | 0.001700982 | 0.003774824 | 1 |
| HTN | b_cell | *NCKAP1* | -2.968316138 | 0.000723363 | 0.002388914 | 1 |
| HTN | b_cell | *SFSWAP* | -4.445478539 | 0.002743693 | 0.005255819 | 1 |
| HTN | b_cell | *TNK2* | -4.935098786 | 0.028124797 | 0.035808141 | 1 |
| HTN | b_cell | *VMP1* | -3.99210583 | 0.002897426 | 0.005462323 | 1 |
| HTN | b_cell | *SLC6A16* | -4.697299334 | 0.000859605 | 0.002557769 | 1 |
| HTN | b_cell | *ISOC2* | -10.62461152 | 0.005161234 | 0.00847224 | 1 |
| HTN | b_cell | *EPN1* | -6.887204691 | 0.021613124 | 0.028267466 | 1 |
| HTN | b_cell | *ADCK1* | -3.856044609 | 0.000809135 | 0.002488487 | 1 |
| HTN | b_cell | *HAGH* | -2.614973706 | 0.032085809 | 0.040312446 | 1 |
| HTN | b_cell | *CASP8* | -4.903998875 | 0.022827434 | 0.029701175 | 1 |
| HTN | b_cell | *DMRT3* | -6.973815299 | 0.000551294 | 0.002219911 | 1 |
| HTN | b_cell | *ST3GAL6* | -2.772267486 | 0.000573632 | 0.002238625 | 1 |
| HTN | b_cell | *ATP2C2* | -7.892822533 | 0.000724458 | 0.002388914 | 1 |
| HTN | b_cell | *CDON* | -2.365615138 | 0.000348519 | 0.002180344 | 1 |
| HTN | b_cell | *TAF2* | -4.599227752 | 0.016346045 | 0.022067336 | 1 |
| HTN | b_cell | *TMEM161A* | -5.275389951 | 0.000620221 | 0.002291029 | 1 |
| HTN | b_cell | *LPAR2* | -5.033859261 | 0.000980382 | 0.002718863 | 1 |
| HTN | b_cell | *SUGP2* | -4.089990639 | 0.022929484 | 0.029827782 | 1 |
| HTN | b_cell | *ABCA7* | 6.441764576 | 0.002922421 | 0.005497604 | 1 |
| HTN | b_cell | *DDX20* | -6.386614257 | 0.000843222 | 0.002536213 | 1 |
| HTN | b_cell | *BTBD1* | -4.903875112 | 0.003159167 | 0.005812609 | 1 |
| HTN | b_cell | *CHI3L2* | 6.854891031 | 0.000254359 | 0.002180344 | 1 |
| HTN | b_cell | *PMS1* | -4.830002757 | 0.005321198 | 0.008678775 | 1 |
| HTN | b_cell | *TAF11* | -6.178618033 | 0.000487989 | 0.002188291 | 1 |
| HTN | b_cell | *ANKS1A* | -4.13888409 | 0.002834065 | 0.005383011 | 1 |
| HTN | b_cell | *IPO5* | -5.804137116 | 0.01339414 | 0.018593279 | 1 |
| HTN | b_cell | *WDR18* | -5.116081204 | 0.000414764 | 0.002180344 | 1 |
| HTN | b_cell | *MCM10* | 2.568762516 | 0.000112052 | 0.002180344 | 0.704468519 |
| HTN | b_cell | *NDUFB4* | -6.551229856 | 0.03847709 | 0.047553659 | 1 |
| HTN | b_cell | *ZC3H15* | -5.649309108 | 0.00522816 | 0.008568677 | 1 |
| HTN | b_cell | *TMEM206* | -5.140724145 | 0.001122231 | 0.002924292 | 1 |
| HTN | b_cell | *SEC61A2* | -3.869286433 | 0.000499589 | 0.002194994 | 1 |
| HTN | b_cell | *ASB1* | -5.977381576 | 0.000791737 | 0.002461747 | 1 |
| HTN | b_cell | *PPP2R5A* | -3.717404365 | 0.001519942 | 0.003510893 | 1 |
| HTN | b_cell | *TIE1* | -4.874848986 | 1.65E-06 | 0.000179109 | 0.010388328 |
| HTN | b_cell | *NFYC* | -5.621127163 | 0.006508138 | 0.010178275 | 1 |

| | | | | | | |
|---|---|---|---|---|---|---|
| HTN | b_cell | *ZMYND12* | -4.820455442 | 0.00626256 | 0.009872798 | 1 |
| HTN | b_cell | *SLC9A3* | -8.263939384 | 0.000897454 | 0.002599737 | 1 |
| HTN | b_cell | *NGEF* | -18.06525465 | 0.016061347 | 0.021739231 | 1 |
| HTN | b_cell | *CD84* | -3.442989244 | 0.000859642 | 0.002557769 | 1 |
| HTN | b_cell | *ATXN3* | -4.803471182 | 0.000335156 | 0.002180344 | 1 |
| HTN | b_cell | *GOLGA5* | -3.871341173 | 0.000662477 | 0.002327958 | 1 |
| HTN | b_cell | *EML1* | -2.601599236 | 0.001808953 | 0.003927449 | 1 |
| HTN | b_cell | *ZFAT* | 4.178215111 | 0.000420799 | 0.002180344 | 1 |
| HTN | b_cell | *MTFR1* | -5.43777885 | 0.002535401 | 0.004962661 | 1 |
| HTN | b_cell | *MYO9A* | -3.770266683 | 0.003533961 | 0.006309867 | 1 |
| HTN | b_cell | *STOML1* | 6.294447445 | 0.000368706 | 0.002180344 | 1 |
| HTN | b_cell | *METTL22* | -7.283982156 | 0.014437984 | 0.019832119 | 1 |
| HTN | b_cell | *TP53BP1* | -5.015481866 | 0.002960324 | 0.005545755 | 1 |
| HTN | b_cell | *DHX8* | -5.111560773 | 0.000257415 | 0.002180344 | 1 |
| HTN | b_cell | *SYT1* | -3.445737811 | 0.003531223 | 0.006307046 | 1 |
| HTN | b_cell | *ROGDI* | 6.994386264 | 0.000358653 | 0.002180344 | 1 |
| HTN | b_cell | *HDAC4* | -4.214623183 | 0.002398228 | 0.004760012 | 1 |
| HTN | b_cell | *RASSF1* | -6.48642908 | 0.005541968 | 0.008968431 | 1 |
| HTN | b_cell | *IFI35* | -3.897761983 | 0.011981958 | 0.01694726 | 1 |
| HTN | b_cell | *HEATR6* | -6.21186415 | 0.002623483 | 0.005082532 | 1 |
| HTN | b_cell | *POLR1A* | -5.732875337 | 1.48E-06 | 0.000166413 | 0.009319116 |
| HTN | b_cell | *STON1-GTF2A* | -1.335379648 | 0.030570005 | 0.038569862 | 1 |
| HTN | b_cell | *SRBD1* | -4.582455404 | 0.01768045 | 0.023645392 | 1 |
| HTN | b_cell | *CYFIP1* | -4.178014626 | 0.0016547 | 0.003713691 | 1 |
| HTN | b_cell | *PSME4* | -4.598964862 | 0.012719248 | 0.017822632 | 1 |
| HTN | b_cell | *PITX1* | -12.90507848 | 0.000405054 | 0.002180344 | 1 |
| HTN | b_cell | *SDK2* | 2.967513076 | 0.000267503 | 0.002180344 | 1 |
| HTN | b_cell | *NUCKS1* | -4.664915785 | 0.003737731 | 0.006595317 | 1 |
| HTN | b_cell | *DNAJA2* | -4.735369171 | 0.000680277 | 0.002339217 | 1 |
| HTN | b_cell | *KCNAB2* | -4.255892338 | 0.000566192 | 0.002232844 | 1 |
| HTN | b_cell | *GAL* | 4.00314607 | 0.000255196 | 0.002180344 | 1 |
| HTN | b_cell | *NEDD4* | -3.377025462 | 0.001769682 | 0.00386991 | 1 |
| HTN | b_cell | *PFN2* | -6.015177418 | 0.000431042 | 0.002180344 | 1 |
| HTN | b_cell | *DAPP1* | 6.705304351 | 0.000689381 | 0.002355324 | 1 |
| HTN | b_cell | *FGF10* | -2.56071836 | 0.002649298 | 0.005115521 | 1 |
| HTN | b_cell | *TMEM260* | -4.951826195 | 0.007207539 | 0.011011859 | 1 |
| HTN | b_cell | *CLTCL1* | -3.779053372 | 0.000267022 | 0.002180344 | 1 |
| HTN | b_cell | *DGCR2* | -5.246670958 | 0.002047041 | 0.004254461 | 1 |
| HTN | b_cell | *ZXDC* | -4.488705588 | 0.003539656 | 0.006311349 | 1 |
| HTN | b_cell | *WIPI1* | -3.842176081 | 0.00065129 | 0.002323165 | 1 |
| HTN | b_cell | *NDST1* | -5.547105822 | 0.000485002 | 0.002187427 | 1 |
| HTN | b_cell | *ASNS* | -6.91892237 | 0.008115463 | 0.0121221 | 1 |
| HTN | b_cell | *AP3M2* | -6.778527781 | 0.01107849 | 0.015790176 | 1 |
| HTN | b_cell | *ST6GALNAC2* | -4.548793205 | 0.001516985 | 0.003508935 | 1 |
| HTN | b_cell | *EIF2B3* | -3.125577194 | 0.000709383 | 0.002378257 | 1 |
| HTN | b_cell | *CAMK2A* | 8.632843607 | 0.000227562 | 0.002180344 | 1 |
| HTN | b_cell | *CDC42* | -4.948452023 | 0.031356942 | 0.039483496 | 1 |
| HTN | b_cell | *OSBPL3* | -4.267680899 | 0.001170088 | 0.003001724 | 1 |
| HTN | b_cell | *RAD18* | -5.469465415 | 0.001289116 | 0.003172074 | 1 |
| HTN | b_cell | *SNX13* | -4.440246042 | 0.005410765 | 0.008796865 | 1 |

| | | | | | | |
|---|---|---|---|---|---|---|
| HTN | b_cell | *VASH1* | -5.795364383 | 0.002638227 | 0.005101411 | 1 |
| HTN | b_cell | *TRIP13* | -22.15921602 | 0.002723845 | 0.00522576 | 1 |
| HTN | b_cell | *MBD3* | -6.552707164 | 0.004939243 | 0.008171848 | 1 |
| HTN | b_cell | *HLTF* | -5.000907406 | 0.001688422 | 0.00375889 | 1 |
| HTN | b_cell | *CPSF1* | -4.328929504 | 0.000314496 | 0.002180344 | 1 |
| HTN | b_cell | *CYBRD1* | -3.468226116 | 0.002348699 | 0.00469725 | 1 |
| HTN | b_cell | *EPN2* | -4.481110047 | 0.00092587 | 0.002648292 | 1 |
| HTN | b_cell | *LIMS2* | -2.822229426 | 0.00079215 | 0.002461813 | 1 |
| HTN | b_cell | *AFF4* | -4.034213786 | 0.009527097 | 0.013868223 | 1 |
| HTN | b_cell | *TRHDE* | -2.068615694 | 0.000412605 | 0.002180344 | 1 |
| HTN | b_cell | *TRNT1* | -4.374187345 | 0.001014766 | 0.002771431 | 1 |
| HTN | b_cell | *ACADVL* | -6.213659675 | 0.001678545 | 0.00374486 | 1 |
| HTN | b_cell | *CRMP1* | -3.550715338 | 0.000248121 | 0.002180344 | 1 |
| HTN | b_cell | *AP1M1* | -6.107765806 | 0.004484804 | 0.007579559 | 1 |
| HTN | b_cell | *PVR* | -5.896674242 | 0.000532128 | 0.002195202 | 1 |
| HTN | b_cell | *XRCC1* | -8.842881171 | 0.013162758 | 0.018336862 | 1 |
| HTN | b_cell | *MOV10L1* | -4.407048664 | 0.002768335 | 0.00528852 | 1 |
| HTN | b_cell | *TP63* | -3.198873563 | 0.00060717 | 0.002281385 | 1 |
| HTN | b_cell | *ALPK1* | -4.60719994 | 0.004682387 | 0.007843563 | 1 |
| HTN | b_cell | *LLGL2* | -6.034081419 | 0.000744296 | 0.002408333 | 1 |
| HTN | b_cell | *PDE8A* | -3.743626377 | 0.000917824 | 0.002637276 | 1 |
| HTN | b_cell | *NLE1* | 5.143146992 | 0.000344619 | 0.002180344 | 1 |
| HTN | b_cell | *SDHA* | -3.93567703 | 0.038601868 | 0.047698495 | 1 |
| HTN | b_cell | *SMARCE1* | -3.956746528 | 0.002103482 | 0.004335931 | 1 |
| HTN | b_cell | *GSDMB* | -2.866870363 | 0.000437641 | 0.002180344 | 1 |
| HTN | b_cell | *PICALM* | -3.531382233 | 0.003962583 | 0.006904866 | 1 |
| HTN | b_cell | *GLI2* | -2.707099198 | 0.004699619 | 0.007862295 | 1 |
| HTN | b_cell | *ATP2A3* | 9.117062251 | 3.76E-05 | 0.001482782 | 0.23636455 |
| HTN | b_cell | *CA12* | -3.692659554 | 0.001580037 | 0.003590058 | 1 |
| HTN | b_cell | *NTN4* | -2.685159785 | 0.00034421 | 0.002180344 | 1 |
| HTN | b_cell | *LMAN1* | -5.496358672 | 0.008140223 | 0.012150423 | 1 |
| HTN | b_cell | *ENO1* | -5.649647457 | 0.004090891 | 0.007081341 | 1 |
| HTN | b_cell | *SRI* | -4.79428421 | 0.0009263 | 0.002648315 | 1 |
| HTN | b_cell | *EIF4G3* | -3.574219217 | 0.019296159 | 0.025529241 | 1 |
| HTN | b_cell | *TTC38* | -2.977375384 | 0.000510306 | 0.002195202 | 1 |
| HTN | b_cell | *CELSR1* | -9.04711447 | 0.012481086 | 0.017523133 | 1 |
| HTN | b_cell | *ADD2* | 11.52201542 | 0.002745634 | 0.005256334 | 1 |
| HTN | b_cell | *VPS9D1* | -7.066545299 | 0.008854822 | 0.013052817 | 1 |
| HTN | b_cell | *MARK3* | -4.610483298 | 0.000682752 | 0.002340493 | 1 |
| HTN | b_cell | *SLC25A3* | -5.433417218 | 0.011755175 | 0.016663988 | 1 |
| HTN | b_cell | *FSCN1* | -5.531745597 | 0.000461344 | 0.002180489 | 1 |
| HTN | b_cell | *MOCOS* | 4.759708476 | 0.000124953 | 0.002180344 | 0.785581889 |
| HTN | b_cell | *PLD1* | -3.596570528 | 0.001215369 | 0.003062536 | 1 |
| HTN | b_cell | *BCAP29* | -4.948870611 | 0.002162487 | 0.004405559 | 1 |
| HTN | b_cell | *TUBA3D* | -14.56406181 | 0.002000019 | 0.004195568 | 1 |
| HTN | b_cell | *MKRN2* | -3.921349821 | 0.000668828 | 0.002333475 | 1 |
| HTN | b_cell | *MCM6* | 6.604713333 | 0.001764857 | 0.003863389 | 1 |
| HTN | b_cell | *RBMS2* | -3.061872947 | 0.000672409 | 0.002334899 | 1 |
| HTN | b_cell | *MLH1* | -4.688508367 | 0.00166456 | 0.003726974 | 1 |
| HTN | b_cell | *FMO4* | -4.577362812 | 0.000825288 | 0.002512631 | 1 |

| | | | | | | |
|---|---|---|---|---|---|---|
| HTN | b_cell | *RGS11* | -4.164963502 | 3.10E-05 | 0.001332022 | 0.195034772 |
| HTN | b_cell | *TPD52* | 1.94148513 | 0.00012899 | 0.002180344 | 0.810957933 |
| HTN | b_cell | *GPATCH1* | -4.344992751 | 0.000812313 | 0.00249056 | 1 |
| HTN | b_cell | *ICAM3* | 6.657456954 | 0.000466216 | 0.002180489 | 1 |
| HTN | b_cell | *NT5C2* | -3.619155727 | 0.038353612 | 0.047447689 | 1 |
| HTN | b_cell | *CAMSAP3* | -31.43005899 | 0.000416546 | 0.002180344 | 1 |
| HTN | b_cell | *XAB2* | -6.080741926 | 9.59E-05 | 0.002180344 | 0.603011944 |
| HTN | b_cell | *STXBP2* | -6.29937351 | 0.00101433 | 0.002771431 | 1 |
| HTN | b_cell | *MAP2K7* | -4.523174576 | 0.000657425 | 0.002327958 | 1 |
| HTN | b_cell | *CTTNBP2* | -3.946415803 | 0.000663269 | 0.002327958 | 1 |
| HTN | b_cell | *TM9SF3* | -4.433917946 | 0.007944222 | 0.011914438 | 1 |
| HTN | b_cell | *DYNC1I2* | -4.243576222 | 0.02978462 | 0.037692413 | 1 |
| HTN | b_cell | *APBB1IP* | -3.228081062 | 0.001679898 | 0.003745219 | 1 |
| HTN | b_cell | *CAPZB* | -4.397261973 | 0.003254702 | 0.005936266 | 1 |
| HTN | b_cell | *GPR137B* | -2.939582306 | 0.016304416 | 0.022030905 | 1 |
| HTN | b_cell | *NAALAD2* | -3.270312294 | 0.006838513 | 0.010579165 | 1 |
| HTN | b_cell | *FGFR1* | -3.404882914 | 0.000562804 | 0.002226547 | 1 |
| HTN | b_cell | *FBLN1* | -2.116011836 | 0.000556654 | 0.002226229 | 1 |
| HTN | b_cell | *PIAS2* | -4.04935773 | 0.00137367 | 0.003295025 | 1 |
| HTN | b_cell | *AMPH* | -2.382222718 | 0.006992088 | 0.010742731 | 1 |
| HTN | b_cell | *MCCC1* | -3.593302633 | 0.00184074 | 0.003968701 | 1 |
| HTN | b_cell | *FAP* | -2.717742047 | 0.001312047 | 0.003209666 | 1 |
| HTN | b_cell | *ACER3* | -2.867138185 | 0.000540026 | 0.002208942 | 1 |
| HTN | b_cell | *PIK3C3* | -4.7589378 | 0.01755966 | 0.02350385 | 1 |
| HTN | b_cell | *N4BP2* | -5.714703726 | 0.007355871 | 0.011197667 | 1 |
| HTN | b_cell | *TULP3* | -4.092824945 | 0.001316834 | 0.00321512 | 1 |
| HTN | b_cell | *SYNJ2* | -3.362895321 | 0.000479935 | 0.002183988 | 1 |
| HTN | b_cell | *ZCWPW1* | -4.16386741 | 0.005885972 | 0.009396929 | 1 |
| HTN | b_cell | *FGF20* | -42.88724647 | 0.000414349 | 0.002180344 | 1 |
| HTN | b_cell | *PCM1* | -4.129642326 | 0.008354567 | 0.012405566 | 1 |
| HTN | b_cell | *MYH7B* | -9.185006975 | 0.000545323 | 0.002214137 | 1 |
| HTN | b_cell | *FKBP7* | -4.356436314 | 0.000235846 | 0.002180344 | 1 |
| HTN | b_cell | *OSBPL6* | -5.203061818 | 0.001075067 | 0.002853079 | 1 |
| HTN | b_cell | *XRCC5* | -4.47935933 | 0.014225791 | 0.019586455 | 1 |
| HTN | b_cell | *LXN* | -4.436305098 | 0.000924912 | 0.002648292 | 1 |
| HTN | b_cell | *REXO1* | -7.008414519 | 0.003731389 | 0.006589675 | 1 |
| HTN | b_cell | *SAR1A* | -5.143503319 | 0.002024037 | 0.004224808 | 1 |
| HTN | b_cell | *RAPGEF3* | -1.94139647 | 0.027044055 | 0.034584254 | 1 |
| HTN | b_cell | *FDFT1* | -4.733408188 | 0.002640226 | 0.005102702 | 1 |
| HTN | b_cell | *DDX1* | -4.79845314 | 0.002143886 | 0.004380258 | 1 |
| HTN | b_cell | *DNM2* | -4.85127269 | 0.013188297 | 0.01835617 | 1 |
| HTN | b_cell | *STX7* | -4.1887451 | 0.019892987 | 0.026236041 | 1 |
| HTN | b_cell | *RABL2B* | -3.727932691 | 0.00034087 | 0.002180344 | 1 |
| HTN | b_cell | *DDX43* | -6.243527473 | 3.80E-05 | 0.001482782 | 0.238645947 |
| HTN | b_cell | *DCT* | -7.156648374 | 0.000490871 | 0.002190281 | 1 |
| HTN | b_cell | *SLC35C2* | -4.844993056 | 0.001058678 | 0.002833039 | 1 |
| HTN | b_cell | *SCTR* | -3.520883391 | 0.000379659 | 0.002180344 | 1 |
| HTN | b_cell | *RIF1* | -4.008931466 | 0.007517857 | 0.011402838 | 1 |
| HTN | b_cell | *KCNN2* | -9.972398691 | 0.000254226 | 0.002180344 | 1 |
| HTN | b_cell | *PSEN1* | -4.514899967 | 0.00911537 | 0.013386669 | 1 |

| | | | | | | |
|---|---|---|---|---|---|---|
| HTN | b_cell | *CPOX* | -4.834993449 | 0.000790488 | 0.002459174 | 1 |
| HTN | b_cell | *MOK* | -4.244572604 | 0.000689701 | 0.002355324 | 1 |
| HTN | b_cell | *RSBN1* | -5.703933978 | 0.003001341 | 0.005600763 | 1 |
| HTN | b_cell | *MAGI3* | -3.802780778 | 0.001529195 | 0.003525505 | 1 |
| HTN | b_cell | *COL4A4* | 1.534190424 | 0.00027696 | 0.002180344 | 1 |
| HTN | b_cell | *ZNF506* | -4.740467583 | 0.002843318 | 0.00538751 | 1 |
| HTN | b_cell | *KIAA0141* | -4.645087693 | 0.000425675 | 0.002180344 | 1 |
| HTN | b_cell | *PCDHA6* | -78.79295472 | 5.60E-05 | 0.001852286 | 0.351934361 |
| HTN | b_cell | *PCDHGA2* | 10.02235842 | 0.000322535 | 0.002180344 | 1 |
| HTN | b_cell | *HSPB11* | -4.154784123 | 0.001433457 | 0.003385825 | 1 |
| HTN | b_cell | *WDR70* | -3.732950136 | 0.004118461 | 0.007115352 | 1 |
| HTN | b_cell | *PGR* | -3.020251627 | 0.00071657 | 0.002385168 | 1 |
| HTN | b_cell | *CCNT2* | -6.927597945 | 0.000214249 | 0.002180344 | 1 |
| HTN | b_cell | *COBLL1* | -2.213351003 | 0.000209527 | 0.002180344 | 1 |
| HTN | b_cell | *GEMIN5* | -5.766412868 | 0.007848652 | 0.011796431 | 1 |
| HTN | b_cell | *GSK3B* | -3.981203555 | 0.004374375 | 0.007430882 | 1 |
| HTN | b_cell | *ERC1* | -3.75207128 | 0.017304542 | 0.023231616 | 1 |
| HTN | b_cell | *DOPEY1* | -3.403123962 | 0.021074423 | 0.027643417 | 1 |
| HTN | b_cell | *LYRM2* | -4.716967413 | 0.000102071 | 0.002180344 | 0.641720821 |
| HTN | b_cell | *ULK2* | -3.960186142 | 0.000402953 | 0.002180344 | 1 |
| HTN | b_cell | *PLOD1* | -5.281333388 | 0.001556299 | 0.003561868 | 1 |
| HTN | b_cell | *PIBF1* | -4.05302026 | 0.006562595 | 0.010222755 | 1 |
| HTN | b_cell | *TDRD3* | -4.206656352 | 0.02012093 | 0.026503308 | 1 |
| HTN | b_cell | *PDS5B* | -4.331828931 | 0.023397904 | 0.030336692 | 1 |
| HTN | b_cell | *OXCT1* | -4.331256962 | 0.000337826 | 0.002180344 | 1 |
| HTN | b_cell | *ZNF324* | -5.006200462 | 0.000505044 | 0.002195028 | 1 |
| HTN | b_cell | *ZNF586* | -4.876024996 | 0.003468574 | 0.006225725 | 1 |
| HTN | b_cell | *ZNF446* | 11.03008502 | 0.016829458 | 0.022656703 | 1 |
| HTN | b_cell | *CHMP2B* | -5.369609097 | 0.000454763 | 0.002180344 | 1 |
| HTN | b_cell | *PPIE* | -5.396607685 | 0.005103596 | 0.008392965 | 1 |
| HTN | b_cell | *ZMPSTE24* | -4.309364424 | 0.000646154 | 0.00232003 | 1 |
| HTN | b_cell | *NOA1* | 3.627653589 | 0.001979794 | 0.004161982 | 1 |
| HTN | b_cell | *GSTP1* | -4.288672678 | 0.000722391 | 0.002388914 | 1 |
| HTN | b_cell | *WBP11* | -5.663833693 | 0.001715396 | 0.003798764 | 1 |
| HTN | b_cell | *COL16A1* | -3.510369386 | 6.39E-05 | 0.001980182 | 0.401976857 |
| HTN | b_cell | *APOB* | -5.659088059 | 0.000329632 | 0.002180344 | 1 |
| HTN | b_cell | *NCOA1* | -3.786323457 | 0.010627026 | 0.015267851 | 1 |
| HTN | b_cell | *CD59* | -5.310712923 | 0.006464666 | 0.010117837 | 1 |
| HTN | b_cell | *TAF9* | -8.929306391 | 0.018610787 | 0.024752702 | 1 |
| HTN | b_cell | *FCN1* | -4.130192919 | 0.000347629 | 0.002180344 | 1 |
| HTN | b_cell | *SCAMP1* | -4.397131028 | 0.028952684 | 0.036772833 | 1 |
| HTN | b_cell | *OVGP1* | 7.865256253 | 0.000606123 | 0.002281385 | 1 |
| HTN | b_cell | *ABCB1* | -3.198488988 | 0.001037043 | 0.002801844 | 1 |
| HTN | b_cell | *ZNF213* | -3.998849129 | 0.000206221 | 0.002180344 | 1 |
| HTN | b_cell | *CPNE3* | -4.05893553 | 0.001453934 | 0.003419708 | 1 |
| HTN | b_cell | *RRN3* | -5.96762433 | 0.000707729 | 0.00237502 | 1 |
| HTN | b_cell | *CTTN* | -2.958867924 | 0.000513916 | 0.002195202 | 1 |
| HTN | b_cell | *EPS15* | -3.529405361 | 0.006043372 | 0.009587352 | 1 |
| HTN | b_cell | *USP40* | -3.654540627 | 0.003975334 | 0.006921331 | 1 |
| HTN | b_cell | *MAST2* | -4.034331417 | 0.009539749 | 0.013881653 | 1 |

| | | | | | | |
|---|---|---|---|---|---|---|
| HTN | b_cell | *B4GALT1* | -4.497148877 | 0.026968017 | 0.034496017 | 1 |
| HTN | b_cell | *EIF2AK1* | -4.697199352 | 0.003252217 | 0.005934319 | 1 |
| HTN | b_cell | *EPDR1* | -4.037517721 | 0.004996827 | 0.008258371 | 1 |
| HTN | b_cell | *SNX10* | -5.007204171 | 1.79E-07 | 4.18E-05 | 0.001127873 |
| HTN | b_cell | *SEPHS1* | -4.469185915 | 0.000592589 | 0.00226693 | 1 |
| HTN | b_cell | *ITPKC* | -3.133435512 | 0.000602752 | 0.002278713 | 1 |
| HTN | b_cell | *CEACAM6* | 38.89452675 | 0.000716157 | 0.002385168 | 1 |
| HTN | b_cell | *TMED2* | -6.198032941 | 0.014421731 | 0.019818454 | 1 |
| HTN | b_cell | *ACOX3* | -5.181121911 | 0.017402977 | 0.023328894 | 1 |
| HTN | b_cell | *HSD17B14* | -5.508239154 | 0.000494837 | 0.002192417 | 1 |
| HTN | b_cell | *TRIP6* | -5.027120477 | 0.008821765 | 0.013022408 | 1 |
| HTN | b_cell | *SRRT* | -5.536070556 | 0.002018107 | 0.00421905 | 1 |
| HTN | b_cell | *PIGS* | -4.920244567 | 0.000140079 | 0.002180344 | 0.880673869 |
| HTN | b_cell | *ATXN7L3* | 5.194222071 | 0.000293053 | 0.002180344 | 1 |
| HTN | b_cell | *PGS1* | -5.45007142 | 0.008828466 | 0.013028552 | 1 |
| HTN | b_cell | *PSMC5* | -3.684606675 | 0.001088344 | 0.002868262 | 1 |
| HTN | b_cell | *LPCAT2* | -3.694840098 | 0.001124208 | 0.002926641 | 1 |
| HTN | b_cell | *OGFOD1* | -6.859591347 | 0.000913957 | 0.002631803 | 1 |
| HTN | b_cell | *TXNDC16* | -5.793514406 | 0.012820651 | 0.017923823 | 1 |
| HTN | b_cell | *GMCL1* | -5.084003784 | 0.009313388 | 0.013620207 | 1 |
| HTN | b_cell | *KLHL42* | -6.548755962 | 0.021866628 | 0.028575242 | 1 |
| HTN | b_cell | *PTHLH* | -5.817002941 | 0.000302035 | 0.002180344 | 1 |
| HTN | b_cell | *PHACTR3* | -3.241478132 | 0.001776256 | 0.003880236 | 1 |
| HTN | b_cell | *ERGIC2* | -4.938883704 | 0.003012818 | 0.005612322 | 1 |
| HTN | b_cell | *AAMDC* | -4.364344749 | 1.86E-06 | 0.000186095 | 0.011723968 |
| HTN | b_cell | *METTL2A* | -6.643022886 | 0.004285134 | 0.007317488 | 1 |
| HTN | b_cell | *DDX18* | -4.232042833 | 0.001088465 | 0.002868262 | 1 |
| HTN | b_cell | *ASAP3* | -3.679553896 | 0.000628817 | 0.002295161 | 1 |
| HTN | b_cell | *EDEM2* | -5.952045714 | 0.013739242 | 0.019009378 | 1 |
| HTN | b_cell | *DOCK3* | -2.582203774 | 0.001705064 | 0.003778547 | 1 |
| HTN | b_cell | *C3orf18* | 5.146437397 | 1.83E-05 | 0.000981544 | 0.114840608 |
| HTN | b_cell | *FKBP1A* | -7.737237646 | 0.006853188 | 0.010588903 | 1 |
| HTN | b_cell | *NSFL1C* | -5.381734214 | 0.033169359 | 0.041516177 | 1 |
| HTN | b_cell | *ZNF343* | -5.042459207 | 0.000815249 | 0.002495359 | 1 |
| HTN | b_cell | *CPXM1* | -5.143959953 | 0.000156804 | 0.002180344 | 0.985828856 |
| HTN | b_cell | *MAVS* | -4.983964554 | 0.001823968 | 0.003947431 | 1 |
| HTN | b_cell | *LZTS3* | -12.58405511 | 0.00904664 | 0.013295051 | 1 |
| HTN | b_cell | *XRN2* | -4.767657439 | 0.003452221 | 0.006204721 | 1 |
| HTN | b_cell | *TESC* | -5.45147277 | 0.000419885 | 0.002180344 | 1 |
| HTN | b_cell | *ANAPC5* | -4.469710864 | 0.034771629 | 0.04334232 | 1 |
| HTN | b_cell | *SLC23A2* | -5.023921582 | 0.006281179 | 0.009894706 | 1 |
| HTN | b_cell | *TMEM230* | -5.24444498 | 0.014309139 | 0.019676632 | 1 |
| HTN | b_cell | *DZANK1* | -5.363145424 | 0.000550037 | 0.00221672 | 1 |
| HTN | b_cell | *TASP1* | -4.249763134 | 0.001803704 | 0.003925194 | 1 |
| HTN | b_cell | *KIF16B* | -4.030248992 | 0.001461096 | 0.003427611 | 1 |
| HTN | b_cell | *FUS* | -4.222456453 | 0.00769042 | 0.011611352 | 1 |
| HTN | b_cell | *FXYD5* | -7.17006189 | 0.002775429 | 0.005299736 | 1 |
| HTN | b_cell | *ZNF302* | -4.036941121 | 0.003722502 | 0.006575938 | 1 |
| HTN | b_cell | *FXYD3* | -20.64837576 | 0.000701941 | 0.002366275 | 1 |
| HTN | b_cell | *KCNH4* | -6.762963444 | 0.002923253 | 0.005497604 | 1 |

| | | | | | | |
|---|---|---|---|---|---|---|
| HTN | b_cell | *GANAB* | -5.392057536 | 0.002010164 | 0.004210907 | 1 |
| HTN | b_cell | *GMIP* | -7.583529014 | 0.001369772 | 0.003289051 | 1 |
| HTN | b_cell | *ANKRD24* | -4.932841815 | 0.002558917 | 0.004997799 | 1 |
| HTN | b_cell | *GPATCH2L* | -4.166341638 | 0.00503025 | 0.008298395 | 1 |
| HTN | b_cell | *SLC9A1* | -3.65719255 | 0.000455682 | 0.002180344 | 1 |
| HTN | b_cell | *SPTLC1* | -5.538016701 | 0.001362305 | 0.003280388 | 1 |
| HTN | b_cell | *RGS1* | -5.921587706 | 0.000417369 | 0.002180344 | 1 |
| HTN | b_cell | *MUL1* | -7.800038976 | 0.000159376 | 0.002180344 | 1 |
| HTN | b_cell | *GNPTG* | -5.661070379 | 0.00223568 | 0.00451372 | 1 |
| HTN | b_cell | *ZNF268* | -5.661137612 | 0.000102633 | 0.002180344 | 0.645256631 |
| HTN | b_cell | *PABPC4* | -7.163746948 | 0.019916303 | 0.026261282 | 1 |
| HTN | b_cell | *CERS4* | -6.562349341 | 0.010923095 | 0.015611161 | 1 |
| HTN | b_cell | *USP48* | -4.463825783 | 0.005073755 | 0.008353896 | 1 |
| HTN | b_cell | *EXOC1* | -4.28398369 | 0.003382306 | 0.006108868 | 1 |
| HTN | b_cell | *NLRC4* | -7.741653885 | 0.006929468 | 0.010664765 | 1 |
| HTN | b_cell | *LAMB1* | -2.828193099 | 0.000417647 | 0.002180344 | 1 |
| HTN | b_cell | *SLC26A4* | -7.748556084 | 0.000432681 | 0.002180344 | 1 |
| HTN | b_cell | *DLD* | -4.867748835 | 0.001427896 | 0.003381236 | 1 |
| HTN | b_cell | *TXNL1* | -4.989371101 | 0.003558441 | 0.006337653 | 1 |
| HTN | b_cell | *IL5RA* | -10.40675864 | 0.000399124 | 0.002180344 | 1 |
| HTN | b_cell | *ITGA6* | -2.777825069 | 0.000339258 | 0.002180344 | 1 |
| HTN | b_cell | *RAPGEF4* | -2.443594533 | 0.000511254 | 0.002195202 | 1 |
| HTN | b_cell | *SEL1L3* | 1.368553345 | 0.000222814 | 0.002180344 | 1 |
| HTN | b_cell | *TF* | -4.602982577 | 0.000375066 | 0.002180344 | 1 |
| HTN | b_cell | *MYO15A* | -6.470357157 | 0.000210749 | 0.002180344 | 1 |
| HTN | b_cell | *ALKBH5* | -3.746313895 | 0.018851679 | 0.025041307 | 1 |
| HTN | b_cell | *NLRP1* | -4.432956522 | 0.003786895 | 0.006665232 | 1 |
| HTN | b_cell | *SPAG7* | -4.846543098 | 0.006110899 | 0.009679824 | 1 |
| HTN | b_cell | *ZFHX4* | -2.288251542 | 0.000478619 | 0.002180489 | 1 |
| HTN | b_cell | *RGS17* | -4.439465235 | 0.004539974 | 0.007654282 | 1 |
| HTN | b_cell | *TMEM101* | -5.472477857 | 0.000387985 | 0.002180344 | 1 |
| HTN | b_cell | *CMA1* | -14.93050668 | 0.000977662 | 0.002717425 | 1 |
| HTN | b_cell | *PPP2R3C* | -4.510781067 | 0.006765214 | 0.010485809 | 1 |
| HTN | b_cell | *HAUS4* | 5.796259363 | 0.004336923 | 0.007387222 | 1 |
| HTN | b_cell | *OSGEP* | -6.039155218 | 0.0260618 | 0.033425242 | 1 |
| HTN | b_cell | *SLC22A17* | -7.170492784 | 0.000222039 | 0.002180344 | 1 |
| HTN | b_cell | *SCFD1* | -3.962147052 | 0.00698849 | 0.010739829 | 1 |
| HTN | b_cell | *RPGRIP1* | -5.566098575 | 0.003208709 | 0.005872825 | 1 |
| HTN | b_cell | *TOX4* | -5.37106126 | 0.005417314 | 0.008805235 | 1 |
| HTN | b_cell | *RFFL* | -6.254223119 | 0.000780746 | 0.002446934 | 1 |
| HTN | b_cell | *MFSD11* | -5.052246856 | 0.001530471 | 0.003526075 | 1 |
| HTN | b_cell | *DPYSL2* | -3.329706594 | 0.00102095 | 0.002777814 | 1 |
| HTN | b_cell | *COMT* | -5.049302489 | 0.002138697 | 0.004375525 | 1 |
| HTN | b_cell | *VNN3* | 3.870605423 | 0.000248179 | 0.002180344 | 1 |
| HTN | b_cell | *SEC22C* | -6.035134119 | 0.012949231 | 0.018087495 | 1 |
| HTN | b_cell | *FMO2* | -2.229974221 | 0.002799122 | 0.005334066 | 1 |
| HTN | b_cell | *PDE6C* | 8.744661152 | 0.000608558 | 0.002281385 | 1 |
| HTN | b_cell | *IKZF5* | -5.965244222 | 0.001228444 | 0.003088552 | 1 |
| HTN | b_cell | *BLNK* | 4.738394441 | 0.006030786 | 0.009577052 | 1 |
| HTN | b_cell | *WAC* | -3.481367904 | 0.023050353 | 0.029954025 | 1 |

| | | | | | | |
|---|---|---|---|---|---|---|
| HTN | b_cell | *CREM* | -4.291787466 | 0.001380374 | 0.00330666 | 1 |
| HTN | b_cell | *TPSD1* | -32.05819008 | 0.001190398 | 0.003027596 | 1 |
| HTN | b_cell | *ITPR3* | -3.244792248 | 0.000419768 | 0.002180344 | 1 |
| HTN | b_cell | *IFT74* | -3.527094558 | 0.008857463 | 0.01305365 | 1 |
| HTN | b_cell | *IL12RB1* | 6.188787788 | 0.004613577 | 0.007761216 | 1 |
| HTN | b_cell | *ACOT7* | -5.425063074 | 0.000334227 | 0.002180344 | 1 |
| HTN | b_cell | *CDC7* | -6.992989691 | 0.001063675 | 0.002842041 | 1 |
| HTN | b_cell | *PCSK5* | -2.34335084 | 0.000513742 | 0.002195202 | 1 |
| HTN | b_cell | *RAB18* | -4.596833889 | 0.000609388 | 0.002281385 | 1 |
| HTN | b_cell | *NRP1* | -3.315268786 | 0.002318283 | 0.004647655 | 1 |
| HTN | b_cell | *PRTFDC1* | -3.678268037 | 0.00034666 | 0.002180344 | 1 |
| HTN | b_cell | *PALMD* | -2.054861468 | 0.000278697 | 0.002180344 | 1 |
| HTN | b_cell | *TSPAN15* | -2.765632124 | 0.000373911 | 0.002180344 | 1 |
| HTN | b_cell | *OCEL1* | -6.514016289 | 0.032585997 | 0.040851079 | 1 |
| HTN | b_cell | *CATSPERG* | -6.454572561 | 0.017541843 | 0.023485001 | 1 |
| HTN | b_cell | *STX1B* | -8.938251825 | 4.11E-05 | 0.001539591 | 0.258651318 |
| HTN | b_cell | *HSD3B7* | -4.290344605 | 0.001257608 | 0.003131319 | 1 |
| HTN | b_cell | *MARCH2* | -3.801868515 | 0.010119251 | 0.014621864 | 1 |
| HTN | b_cell | *CDC34* | -4.420500212 | 0.000378872 | 0.002180344 | 1 |
| HTN | b_cell | *MTAP* | -5.842399148 | 0.001320444 | 0.003218935 | 1 |
| HTN | b_cell | *POLR2E* | -3.520318256 | 0.00035707 | 0.002180344 | 1 |
| HTN | b_cell | *MADCAM1* | -8.587334754 | 0.003031034 | 0.005637902 | 1 |
| HTN | b_cell | *TRMT2A* | -5.599992247 | 0.00041039 | 0.002180344 | 1 |
| HTN | b_cell | *ZDHHC8* | -3.786608845 | 0.001420843 | 0.003372154 | 1 |
| HTN | b_cell | *MED15* | -4.212650133 | 0.003089274 | 0.005715637 | 1 |
| HTN | b_cell | *LZTR1* | -5.952114674 | 0.001777995 | 0.003882687 | 1 |
| HTN | b_cell | *CECR2* | -2.895604367 | 0.001239673 | 0.003107584 | 1 |
| HTN | b_cell | *SMARCB1* | -4.585170028 | 0.01733021 | 0.023252106 | 1 |
| HTN | b_cell | *P2RX6* | -5.983967769 | 0.000327812 | 0.002180344 | 1 |
| HTN | b_cell | *SLC7A4* | -24.7798237 | 8.85E-07 | 0.000115006 | 0.005566379 |
| HTN | b_cell | *DDTL* | -5.46987244 | 0.00517239 | 0.008488336 | 1 |
| HTN | b_cell | *DDT* | -8.862016986 | 0.022018735 | 0.028756084 | 1 |
| HTN | b_cell | *GGT5* | -4.332769936 | 0.000326005 | 0.002180344 | 1 |
| HTN | b_cell | *SPECC1L* | -8.025379801 | 0.001524241 | 0.003516661 | 1 |
| HTN | b_cell | *PPIL2* | -4.05773621 | 0.030716707 | 0.038731636 | 1 |
| HTN | b_cell | *YPEL1* | -6.68006873 | 0.004447577 | 0.007526761 | 1 |
| HTN | b_cell | *TOP3B* | -4.665687773 | 0.000298533 | 0.002180344 | 1 |
| HTN | b_cell | *CRYBB3* | -14.04570403 | 0.000154962 | 0.002180344 | 0.974244797 |
| HTN | b_cell | *CYTH4* | -5.144427853 | 0.001106427 | 0.002900583 | 1 |
| HTN | b_cell | *CARD10* | -4.921829274 | 0.002764686 | 0.005284762 | 1 |
| HTN | b_cell | *LRP5L* | -8.641801363 | 0.001341006 | 0.003255175 | 1 |
| HTN | b_cell | *SLC25A1* | -4.456961969 | 0.00059189 | 0.00226693 | 1 |
| HTN | b_cell | *LGALS2* | 6.856567786 | 0.000437287 | 0.002180344 | 1 |
| HTN | b_cell | *GGA1* | -3.997779703 | 0.000546998 | 0.002214137 | 1 |
| HTN | b_cell | *SH3BP1* | -5.41968894 | 0.000457713 | 0.002180344 | 1 |
| HTN | b_cell | *HPS4* | -4.943690495 | 0.001939171 | 0.004111827 | 1 |
| HTN | b_cell | *TRIOBP* | -5.493792519 | 0.00014665 | 0.002180344 | 0.921987542 |
| HTN | b_cell | *CCDC134* | 10.59001484 | 0.001352449 | 0.003270974 | 1 |
| HTN | b_cell | *SLC16A8* | -8.97171749 | 0.001682432 | 0.003747346 | 1 |
| HTN | b_cell | *KDELR3* | -5.025430309 | 0.000132515 | 0.002180344 | 0.833124576 |

| | | | | | | |
|---|---|---|---|---|---|---|
| HTN | b_cell | *CYP2D6* | 5.635229836 | 0.006742609 | 0.010453954 | 1 |
| HTN | b_cell | *DDX17* | -3.233453993 | 0.014032095 | 0.01935918 | 1 |
| HTN | b_cell | *FBXO7* | -5.913640781 | 0.007901263 | 0.01186416 | 1 |
| HTN | b_cell | *POLDIP3* | -4.239937876 | 0.00055914 | 0.002226229 | 1 |
| HTN | b_cell | *TIMP3* | -2.706326466 | 0.000827437 | 0.002514184 | 1 |
| HTN | b_cell | *PPP6R2* | -4.693921147 | 0.004754017 | 0.007925883 | 1 |
| HTN | b_cell | *CYB5R3* | -4.63192584 | 0.001446821 | 0.003411913 | 1 |
| HTN | b_cell | *RHBDD3* | -3.834217869 | 0.000728507 | 0.002394922 | 1 |
| HTN | b_cell | *TTLL1* | -7.885869768 | 0.004435223 | 0.007509897 | 1 |
| HTN | b_cell | *RASL10A* | -6.899497061 | 0.000736315 | 0.00240229 | 1 |
| HTN | b_cell | *HMGXB4* | -5.721089962 | 0.003551197 | 0.006328104 | 1 |
| HTN | b_cell | *TOM1* | -2.769357908 | 0.003191937 | 0.005848938 | 1 |
| HTN | b_cell | *NEFH* | 7.930535891 | 0.000492763 | 0.002192417 | 1 |
| HTN | b_cell | *CHKB* | 6.213649486 | 0.037215983 | 0.046112906 | 1 |
| HTN | b_cell | *BIK* | -21.83240178 | 0.000385364 | 0.002180344 | 1 |
| HTN | b_cell | *MCAT* | -7.017908883 | 0.000110116 | 0.002180344 | 0.692297582 |
| HTN | b_cell | *ARSA* | -2.34342313 | 0.018983465 | 0.025184436 | 1 |
| HTN | b_cell | *TSPO* | -5.773901314 | 0.008652082 | 0.012798974 | 1 |
| HTN | b_cell | *RASD2* | -25.49204672 | 0.008851344 | 0.01305075 | 1 |
| HTN | b_cell | *TTLL12* | -5.586816218 | 0.000465575 | 0.002180489 | 1 |
| HTN | b_cell | *RBFOX2* | -3.159878454 | 0.000956695 | 0.002694776 | 1 |
| HTN | b_cell | *MTMR3* | -3.331967056 | 0.008095863 | 0.012098298 | 1 |
| HTN | b_cell | *APOL4* | -5.13373487 | 0.007142006 | 0.010929186 | 1 |
| HTN | b_cell | *SAMM50* | -3.461082898 | 0.000405689 | 0.002180344 | 1 |
| HTN | b_cell | *IFT27* | -4.709914122 | 0.000562643 | 0.002226547 | 1 |
| HTN | b_cell | *KIAA0930* | -4.961853682 | 0.003383343 | 0.006108868 | 1 |
| HTN | b_cell | *NCF4* | -3.637817985 | 0.00253007 | 0.004955312 | 1 |
| HTN | b_cell | *KCTD17* | -13.48919801 | 0.000775068 | 0.002438676 | 1 |
| HTN | b_cell | *EP300* | -4.415911975 | 0.017771747 | 0.023747285 | 1 |
| HTN | b_cell | *ACO2* | -4.903606744 | 0.000705906 | 0.00237262 | 1 |
| HTN | b_cell | *POLR3H* | -4.146225675 | 0.00326741 | 0.00594909 | 1 |
| HTN | b_cell | *DESI1* | -3.371022895 | 0.024091908 | 0.031140904 | 1 |
| HTN | b_cell | *ZBED4* | -5.837662466 | 0.002389759 | 0.004745691 | 1 |
| HTN | b_cell | *HDAC10* | -5.738968045 | 0.010641009 | 0.015284447 | 1 |
| HTN | b_cell | *SDR39U1* | -5.551872141 | 0.004013636 | 0.00697257 | 1 |
| HTN | b_cell | *RBM23* | -4.987849595 | 0.002106836 | 0.004337157 | 1 |
| HTN | b_cell | *VCPKMT* | -4.383779594 | 0.015841032 | 0.021491707 | 1 |
| HTN | b_cell | *SOS2* | -4.217947629 | 0.016061513 | 0.021739231 | 1 |
| HTN | b_cell | *PYGL* | -2.553697713 | 4.23E-05 | 0.00156551 | 0.266136625 |
| HTN | b_cell | *DDHD1* | -4.635637368 | 0.034773129 | 0.04334232 | 1 |
| HTN | b_cell | *CNIH1* | -5.483183614 | 0.00574918 | 0.009231316 | 1 |
| HTN | b_cell | *ATP6V1D* | -3.160926175 | 0.000649229 | 0.002323108 | 1 |
| HTN | b_cell | *GSTZ1* | -4.403129097 | 0.00089773 | 0.002599737 | 1 |
| HTN | b_cell | *DAAM1* | -2.903422705 | 0.000765395 | 0.002430321 | 1 |
| HTN | b_cell | *ISM2* | 12.54391885 | 0.000162167 | 0.002180344 | 1 |
| HTN | b_cell | *PPM1A* | -5.656399719 | 0.000658487 | 0.002327958 | 1 |
| HTN | b_cell | *SLC10A1* | -3.637399739 | 0.000668245 | 0.002332736 | 1 |
| HTN | b_cell | *DICER1* | -4.123912449 | 0.003413874 | 0.006150095 | 1 |
| HTN | b_cell | *ZFYVE21* | -5.934603433 | 0.01452863 | 0.019947914 | 1 |
| HTN | b_cell | *MTHFD1* | -2.82022925 | 0.000444453 | 0.002180344 | 1 |

| | | | | | | |
|---|---|---|---|---|---|---|
| HTN | b_cell | *VRK1* | -6.113866368 | 0.006329379 | 0.009943229 | 1 |
| HTN | b_cell | *PAPLN* | -6.047380157 | 0.032573146 | 0.040843114 | 1 |
| HTN | b_cell | *PSMB5* | -4.603190559 | 0.032451517 | 0.040723091 | 1 |
| HTN | b_cell | *CCNB1IP1* | -5.933086981 | 0.000345631 | 0.002180344 | 1 |
| HTN | b_cell | *TRIP11* | -4.139968619 | 0.001809113 | 0.003927449 | 1 |
| HTN | b_cell | *PCK2* | -8.81952108 | 0.000225292 | 0.002180344 | 1 |
| HTN | b_cell | *KIAA0391* | 14.71584083 | 0.00465173 | 0.007809192 | 1 |
| HTN | b_cell | *EMC9* | 6.155946354 | 0.027147133 | 0.034682793 | 1 |
| HTN | b_cell | *PLTP* | -3.68175345 | 0.00014409 | 0.002180344 | 0.905891147 |
| HTN | b_cell | *PYGB* | -3.902021585 | 0.000273678 | 0.002180344 | 1 |
| HTN | b_cell | *ABHD12* | -4.136361282 | 0.004045731 | 0.007020566 | 1 |
| HTN | b_cell | *PROCR* | -3.072047669 | 0.000892802 | 0.002599737 | 1 |
| HTN | b_cell | *NINL* | -3.48460229 | 0.000928927 | 0.002653415 | 1 |
| HTN | b_cell | *CD40* | 7.789641013 | 0.009605756 | 0.013950425 | 1 |
| HTN | b_cell | *UQCC1* | -4.795078765 | 0.004401081 | 0.007468177 | 1 |
| HTN | b_cell | *IFT52* | -5.621640646 | 0.001691576 | 0.003759376 | 1 |
| HTN | b_cell | *DOK5* | -4.10056112 | 0.000554645 | 0.002220961 | 1 |
| HTN | b_cell | *CSTF1* | -8.255898108 | 0.000531558 | 0.002195202 | 1 |
| HTN | b_cell | *BMP7* | -5.22940789 | 0.000416441 | 0.002180344 | 1 |
| HTN | b_cell | *TPD52L2* | -3.99059916 | 0.000745248 | 0.002408933 | 1 |
| HTN | b_cell | *NELFCD* | -5.64170866 | 0.002967869 | 0.005554926 | 1 |
| HTN | b_cell | *CTSZ* | -5.922025159 | 0.006300772 | 0.009913152 | 1 |
| HTN | b_cell | *PRPF6* | -4.985612884 | 0.000602686 | 0.002278713 | 1 |
| HTN | b_cell | *MTG2* | -3.709988356 | 0.020307416 | 0.026715364 | 1 |
| HTN | b_cell | *DIDO1* | -4.209345419 | 0.011044068 | 0.015755402 | 1 |
| HTN | b_cell | *SLC17A9* | 15.23254779 | 0.001810843 | 0.003929849 | 1 |
| HTN | b_cell | *BIRC7* | 4.220674625 | 0.00099459 | 0.002741337 | 1 |
| HTN | b_cell | *NKAIN4* | -13.83519654 | 0.000138777 | 0.002180344 | 0.872490747 |
| HTN | b_cell | *PTK6* | 2.648923239 | 0.000230526 | 0.002180344 | 1 |
| HTN | b_cell | *GMEB2* | -8.18958971 | 0.00718385 | 0.010981003 | 1 |
| HTN | b_cell | *RNF24* | -4.456006445 | 0.006105515 | 0.009678611 | 1 |
| HTN | b_cell | *NDUFAF5* | -3.193230549 | 0.000640097 | 0.002314142 | 1 |
| HTN | b_cell | *TRIB3* | -6.995719987 | 0.000548231 | 0.002215074 | 1 |
| HTN | b_cell | *ANGPT4* | -6.216585769 | 0.000301767 | 0.002180344 | 1 |
| HTN | b_cell | *RSPO4* | 7.010281184 | 0.000233937 | 0.002180344 | 1 |
| HTN | b_cell | *SIRPB1* | 1.815296834 | 0.001883325 | 0.004038359 | 1 |
| HTN | b_cell | *SEC23B* | -3.500731631 | 0.006786435 | 0.010508945 | 1 |
| HTN | b_cell | *CCM2L* | -4.932894086 | 0.000420672 | 0.002180344 | 1 |
| HTN | b_cell | *MYL9* | -3.922741339 | 0.013332607 | 0.018528853 | 1 |
| HTN | b_cell | *MROH8* | -7.100858015 | 0.00200967 | 0.004210907 | 1 |
| HTN | b_cell | *TTI1* | -6.287946923 | 0.021322436 | 0.027927949 | 1 |
| HTN | b_cell | *RPRD1B* | -4.366156851 | 0.000809441 | 0.002488487 | 1 |
| HTN | b_cell | *PXMP4* | -5.114536692 | 0.033476445 | 0.041867199 | 1 |
| HTN | b_cell | *BPI* | -11.71482234 | 0.032921554 | 0.041247073 | 1 |
| HTN | b_cell | *ACTR5* | -5.05790432 | 0.0004945 | 0.002192417 | 1 |
| HTN | b_cell | *WFDC2* | 11.51428884 | 0.000523363 | 0.002195202 | 1 |
| HTN | b_cell | *DHX35* | -5.943713583 | 0.004101062 | 0.007095362 | 1 |
| HTN | b_cell | *DNTTIP1* | -8.51949357 | 1.45E-05 | 0.000838228 | 0.091366894 |
| HTN | b_cell | *MAP1LC3A* | -4.399646217 | 0.006271813 | 0.009882429 | 1 |
| HTN | b_cell | *PIGU* | -5.381990069 | 0.016787145 | 0.022609421 | 1 |

| | | | | | | |
|---|---|---|---|---|---|---|
| HTN | b_cell | *APMAP* | -3.297579816 | 0.00055497 | 0.002220961 | 1 |
| HTN | b_cell | *CELF4* | -4.72143964 | 0.000289745 | 0.002180344 | 1 |
| HTN | b_cell | *METTL4* | -6.468665333 | 0.012722757 | 0.017822632 | 1 |
| HTN | b_cell | *LPIN2* | -4.937310837 | 0.00353854 | 0.00631115 | 1 |
| HTN | b_cell | *MYL12A* | -5.755253447 | 0.006650451 | 0.010331451 | 1 |
| HTN | b_cell | *CEP192* | -4.144721482 | 0.040333823 | 0.049624021 | 1 |
| HTN | b_cell | *LIPG* | -15.80824352 | 0.000561144 | 0.002226547 | 1 |
| HTN | b_cell | *POLI* | -4.262329471 | 2.54E-07 | 5.51E-05 | 0.001598685 |
| HTN | b_cell | *NALCN* | -4.432085328 | 0.000405094 | 0.002180344 | 1 |
| HTN | b_cell | *HTR2A* | -11.2220063 | 0.000412783 | 0.002180344 | 1 |
| HTN | b_cell | *KLF5* | -5.308925033 | 0.009491237 | 0.013827299 | 1 |
| HTN | b_cell | *STK24* | -5.026655147 | 0.005051586 | 0.008322672 | 1 |
| HTN | b_cell | *DNAJC3* | -5.014997796 | 0.016322763 | 0.022045373 | 1 |
| HTN | b_cell | *UGGT2* | -3.386216889 | 0.002351978 | 0.004700152 | 1 |
| HTN | b_cell | *ARHGEF7* | -3.72888993 | 0.002917065 | 0.005489252 | 1 |
| HTN | b_cell | *SGCG* | -2.775296142 | 0.000470505 | 0.002180489 | 1 |
| HTN | b_cell | *PARP4* | -4.43535686 | 0.002939258 | 0.005519448 | 1 |
| HTN | b_cell | *RGCC* | -2.335823472 | 0.000484714 | 0.002187427 | 1 |
| HTN | b_cell | *VWA8* | -3.147205483 | 0.009191994 | 0.013474019 | 1 |
| HTN | b_cell | *KATNAL1* | -3.994640816 | 0.002201339 | 0.004467654 | 1 |
| HTN | b_cell | *DHRS12* | -3.374951513 | 0.000386075 | 0.002180344 | 1 |
| HTN | b_cell | *MEDAG* | -3.125953567 | 0.000412049 | 0.002180344 | 1 |
| HTN | b_cell | *TSC22D1* | -3.950123272 | 0.003146317 | 0.00579575 | 1 |
| HTN | b_cell | *MGRN1* | -2.772964235 | 0.019373672 | 0.025612263 | 1 |
| HTN | b_cell | *CORO1A* | 4.545220294 | 0.036541225 | 0.0453215 | 1 |
| HTN | b_cell | *GDPD3* | -4.660375102 | 0.000940061 | 0.002665473 | 1 |
| HTN | b_cell | *NFAT5* | -3.38761423 | 0.002618878 | 0.005079947 | 1 |
| HTN | b_cell | *LONP2* | -4.369547599 | 0.006293499 | 0.009909149 | 1 |
| HTN | b_cell | *PLLP* | -7.761723857 | 0.000739429 | 0.002405521 | 1 |
| HTN | b_cell | *DHODH* | -2.613535491 | 0.017516057 | 0.023460471 | 1 |
| HTN | b_cell | *CTCF* | -5.252755534 | 0.021507704 | 0.028141298 | 1 |
| HTN | b_cell | *ACD* | -6.204890246 | 0.002316567 | 0.004647655 | 1 |
| HTN | b_cell | *USB1* | -9.438434543 | 0.004058655 | 0.007038502 | 1 |
| HTN | b_cell | *SETD6* | -4.112538298 | 0.000810284 | 0.002488644 | 1 |
| HTN | b_cell | *COG4* | -5.167489895 | 0.00016753 | 0.002180344 | 1 |
| HTN | b_cell | *SLC7A6OS* | -2.551118046 | 6.88E-05 | 0.002060872 | 0.43278306 |
| HTN | b_cell | *HCFC1R1* | -4.539225106 | 0.000419861 | 0.002180344 | 1 |
| HTN | b_cell | *NPRL3* | -3.936693868 | 0.001769158 | 0.00386991 | 1 |
| HTN | b_cell | *TAF1C* | 5.549569498 | 0.003089399 | 0.005715637 | 1 |
| HTN | b_cell | *NAGPA* | -6.666512915 | 0.000880465 | 0.002587702 | 1 |
| HTN | b_cell | *CRISPLD2* | -2.544527961 | 0.009426249 | 0.013743744 | 1 |
| HTN | b_cell | *ZNF500* | -8.887984098 | 0.000761358 | 0.00242729 | 1 |
| HTN | b_cell | *NME4* | -4.658774212 | 0.000586057 | 0.002261934 | 1 |
| HTN | b_cell | *NOMO3* | -6.510327283 | 0.000208615 | 0.002180344 | 1 |
| HTN | b_cell | *LMF1* | -5.114649447 | 0.004158925 | 0.007165569 | 1 |
| HTN | b_cell | *CLCN7* | -6.841142146 | 0.00700064 | 0.010750871 | 1 |
| HTN | b_cell | *HAGHL* | 15.45531497 | 0.000202257 | 0.002180344 | 1 |
| HTN | b_cell | *SLC7A5* | -17.80379337 | 0.001460259 | 0.003427611 | 1 |
| HTN | b_cell | *FBXO31* | -4.163326476 | 0.000679419 | 0.002339217 | 1 |
| HTN | b_cell | *STUB1* | -6.067962621 | 0.009380384 | 0.01369671 | 1 |

| | | | | | | |
|---|---|---|---|---|---|---|
| HTN | b_cell | *UBE2I* | -4.848312668 | 0.000376908 | 0.002180344 | 1 |
| HTN | b_cell | *EARS2* | -3.51108919 | 0.00163459 | 0.003683394 | 1 |
| HTN | b_cell | *BFAR* | -5.085339876 | 0.001165795 | 0.002996464 | 1 |
| HTN | b_cell | *RRN3P2* | -6.384974621 | 0.003233 | 0.00590525 | 1 |
| HTN | b_cell | *RBL2* | -3.686295442 | 0.010554893 | 0.015177126 | 1 |
| HTN | b_cell | *STX4* | 6.322844038 | 0.021113789 | 0.027683502 | 1 |
| HTN | b_cell | *NOMO1* | -4.976467059 | 0.000215581 | 0.002180344 | 1 |
| HTN | b_cell | *SYT17* | -3.894265513 | 0.000346909 | 0.002180344 | 1 |
| HTN | b_cell | *TMC5* | -5.883610654 | 0.000308729 | 0.002180344 | 1 |
| HTN | b_cell | *IQCH* | -4.127288168 | 0.000933579 | 0.002657043 | 1 |
| HTN | b_cell | *LACTB* | -5.252453462 | 0.00061695 | 0.002286342 | 1 |
| HTN | b_cell | *HERC1* | -3.815368116 | 0.008321972 | 0.012363006 | 1 |
| HTN | b_cell | *RASL12* | -6.258410209 | 0.000402726 | 0.002180344 | 1 |
| HTN | b_cell | *AP3B2* | 2.561050648 | 0.006913586 | 0.010652227 | 1 |
| HTN | b_cell | *TTC23* | -4.811225236 | 0.000914609 | 0.002631803 | 1 |
| HTN | b_cell | *FAH* | -3.255165559 | 0.000416146 | 0.002180344 | 1 |
| HTN | b_cell | *RPAP1* | -3.991547287 | 0.000337281 | 0.002180344 | 1 |
| HTN | b_cell | *EHD4* | -4.837275627 | 0.001133371 | 0.002943208 | 1 |
| HTN | b_cell | *ATP8B4* | -2.424353409 | 0.000505954 | 0.002195202 | 1 |
| HTN | b_cell | *DTWD1* | -3.37053798 | 0.001455931 | 0.003423125 | 1 |
| HTN | b_cell | *DNAJC17* | -5.053010512 | 0.00142718 | 0.003381236 | 1 |
| HTN | b_cell | *PDGFRL* | -3.061536152 | 0.00066824 | 0.002332736 | 1 |
| HTN | b_cell | *ZFAND1* | -3.057871669 | 0.007312544 | 0.011142502 | 1 |
| HTN | b_cell | *RIPK2* | -5.665475936 | 0.003273171 | 0.005956129 | 1 |
| HTN | b_cell | *NBN* | -4.894537264 | 0.002283742 | 0.004593054 | 1 |
| HTN | b_cell | *CPQ* | -3.168149113 | 0.002028401 | 0.004226899 | 1 |
| HTN | b_cell | *NIPAL2* | -3.540224498 | 0.00102108 | 0.002777814 | 1 |
| HTN | b_cell | *IKBKB* | -4.862220004 | 0.015073819 | 0.020567127 | 1 |
| HTN | b_cell | *PLAT* | -3.309491145 | 0.000467097 | 0.002180489 | 1 |
| HTN | b_cell | *RAB2A* | -3.139225203 | 0.007158485 | 0.010946781 | 1 |
| HTN | b_cell | *ZC2HC1A* | -4.817866166 | 0.007788316 | 0.01172537 | 1 |
| HTN | b_cell | *ARMC1* | -3.861686885 | 0.000184377 | 0.002180344 | 1 |
| HTN | b_cell | *SPAG1* | -4.823599709 | 0.001580698 | 0.003590263 | 1 |
| HTN | b_cell | *CHRAC1* | -5.703137504 | 0.000257208 | 0.002180344 | 1 |
| HTN | b_cell | *SNX16* | -4.292399774 | 0.004067372 | 0.007044509 | 1 |
| HTN | b_cell | *GSDMD* | -5.949522881 | 0.025104172 | 0.032295873 | 1 |
| HTN | b_cell | *TSTA3* | -9.56307812 | 0.001120621 | 0.002924292 | 1 |
| HTN | b_cell | *MTMR9* | -4.1899612 | 0.000972074 | 0.002716191 | 1 |
| HTN | b_cell | *LEPROTL1* | -5.820948629 | 0.002265193 | 0.004560252 | 1 |
| HTN | b_cell | *TNFRSF10A* | -3.320941914 | 0.002560598 | 0.004999527 | 1 |
| HTN | b_cell | *ERICH1* | -4.436487176 | 0.002464094 | 0.004856384 | 1 |
| HTN | b_cell | *TUSC3* | -3.079948458 | 0.000420385 | 0.002180344 | 1 |
| HTN | b_cell | *ARHGEF10* | -3.356374041 | 0.000347891 | 0.002180344 | 1 |
| HTN | b_cell | *KLHDC4* | -5.299020284 | 0.040578385 | 0.049905381 | 1 |
| HTN | b_cell | *MCM4* | -6.004648802 | 0.007983211 | 0.011960575 | 1 |
| HTN | b_cell | *MAN2B1* | -4.127415312 | 0.001509583 | 0.00349696 | 1 |
| HTN | b_cell | *TULP2* | 8.039993837 | 9.52E-05 | 0.002180344 | 0.598481738 |
| HTN | b_cell | *NUCB1* | -5.519476031 | 0.000667234 | 0.002331797 | 1 |
| HTN | b_cell | *TUBB4A* | -49.53940093 | 0.000786157 | 0.002452889 | 1 |
| HTN | b_cell | *SARS2* | -9.145362693 | 0.001955811 | 0.004138736 | 1 |

| | | | | | | |
|---|---|---|---|---|---|---|
| HTN | b_cell | *SNRNP70* | -5.297534679 | 0.004285508 | 0.007317488 | 1 |
| HTN | b_cell | *ERCC2* | -8.477121031 | 0.001169111 | 0.003001667 | 1 |
| HTN | b_cell | *PLEKHJ1* | -5.812491621 | 0.002127431 | 0.004361619 | 1 |
| HTN | b_cell | *CD37* | 1.301838357 | 0.0001965 | 0.002180344 | 1 |
| HTN | b_cell | *DKKL1* | -9.573388649 | 0.009388423 | 0.013698951 | 1 |
| HTN | b_cell | *STX10* | 8.961343601 | 0.006853228 | 0.010588903 | 1 |
| HTN | b_cell | *FCER2* | 1.408394833 | 0.000230789 | 0.002180344 | 1 |
| HTN | b_cell | *IL4I1* | -4.591063079 | 0.007140881 | 0.010929186 | 1 |
| HTN | b_cell | *TLE6* | -8.342428203 | 0.008965948 | 0.013194971 | 1 |
| HTN | b_cell | *AES* | -5.493350962 | 0.003002745 | 0.005600763 | 1 |
| HTN | b_cell | *C19orf53* | -7.95141035 | 0.027573008 | 0.03516971 | 1 |
| HTN | b_cell | *TIMM44* | -5.161951622 | 0.006820789 | 0.010556942 | 1 |
| HTN | b_cell | *OLFM2* | -2.733933613 | 0.00241181 | 0.004781787 | 1 |
| HTN | b_cell | *RASAL3* | -5.067778047 | 0.00066227 | 0.002327958 | 1 |
| HTN | b_cell | *SYDE1* | -6.329348249 | 0.000144196 | 0.002180344 | 0.906560624 |
| HTN | b_cell | *AURKC* | 2.806121905 | 0.000344757 | 0.002180344 | 1 |
| HTN | b_cell | *CCNE1* | -9.884741213 | 0.000335805 | 0.002180344 | 1 |
| HTN | b_cell | *PDCD5* | -6.144982781 | 0.014897631 | 0.020392208 | 1 |
| HTN | b_cell | *ANKRD27* | -3.526171021 | 0.000769081 | 0.002432944 | 1 |
| HTN | b_cell | *PRX* | 5.521533925 | 0.000384448 | 0.002180344 | 1 |
| HTN | b_cell | *CCDC94* | -6.257281827 | 0.016174703 | 0.021883012 | 1 |
| HTN | b_cell | *POLR2I* | -8.094409356 | 0.001201772 | 0.003041684 | 1 |
| HTN | b_cell | *CLIP3* | -4.687461931 | 0.000218832 | 0.002180344 | 1 |
| HTN | b_cell | *SLC1A5* | -3.844157463 | 0.000385603 | 0.002180344 | 1 |
| HTN | b_cell | *CACTIN* | -5.724862111 | 0.000423721 | 0.002180344 | 1 |
| HTN | b_cell | *CCDC9* | 6.527291185 | 0.000306782 | 0.002180344 | 1 |
| HTN | b_cell | *FZR1* | 9.444417345 | 0.014650862 | 0.020089415 | 1 |
| HTN | b_cell | *BBC3* | -11.71779604 | 0.02486475 | 0.032033747 | 1 |
| HTN | b_cell | *TGFB1* | -5.473546694 | 0.002010679 | 0.004210907 | 1 |
| HTN | b_cell | *PLIN3* | -4.098819331 | 0.000665191 | 0.002329372 | 1 |
| HTN | b_cell | *MRPL4* | -6.639301195 | 0.000529407 | 0.002195202 | 1 |
| HTN | b_cell | *ICAM5* | -45.46086909 | 2.86E-06 | 0.000264526 | 0.017987747 |
| HTN | b_cell | *CD33* | -5.63893288 | 0.000445355 | 0.002180344 | 1 |
| HTN | b_cell | *TYK2* | -6.017984947 | 0.004059425 | 0.007038502 | 1 |
| HTN | b_cell | *RABAC1* | -4.358073996 | 0.000541802 | 0.002213326 | 1 |
| HTN | b_cell | *PTPRS* | -2.265331734 | 0.002281459 | 0.004589931 | 1 |
| HTN | b_cell | *GRIN2D* | -5.56673154 | 0.000399728 | 0.002180344 | 1 |
| HTN | b_cell | *CARD8* | -5.181465668 | 0.000743575 | 0.002408333 | 1 |
| HTN | b_cell | *LIG1* | 3.59770679 | 0.000328198 | 0.002180344 | 1 |
| HTN | b_cell | *ZNF175* | -6.914608914 | 0.006315852 | 0.009929423 | 1 |
| HTN | b_cell | *PLA2G4C* | -4.99497488 | 0.000943829 | 0.0026705 | 1 |
| HTN | b_cell | *CAPS* | -5.815066521 | 0.006346094 | 0.009962021 | 1 |
| HTN | b_cell | *MIER2* | -8.166505103 | 0.005468879 | 0.008872991 | 1 |
| HTN | b_cell | *PPP2R1A* | -6.290583803 | 0.006221539 | 0.009815513 | 1 |
| HTN | b_cell | *TNPO2* | -6.843577958 | 0.003198934 | 0.005858345 | 1 |
| HTN | b_cell | *GCDH* | -3.854312954 | 0.000197364 | 0.002180344 | 1 |
| HTN | b_cell | *LILRB5* | -3.071500892 | 0.000420307 | 0.002180344 | 1 |
| HTN | b_cell | *MAST1* | -11.58172275 | 0.000406474 | 0.002180344 | 1 |
| HTN | b_cell | *PRPF31* | -6.265810216 | 0.00135747 | 0.003274908 | 1 |
| HTN | b_cell | *TFPT* | 9.82381499 | 0.003041522 | 0.005650639 | 1 |

| | | | | | | |
|---|---|---|---|---|---|---|
| HTN | b_cell | *PDE4C* | -9.487415744 | 0.018054549 | 0.024104683 | 1 |
| HTN | b_cell | *ELL* | -4.081715388 | 0.000680312 | 0.002339217 | 1 |
| HTN | b_cell | *UPK1A* | -18.34825262 | 0.002120183 | 0.004354653 | 1 |
| HTN | b_cell | *COPE* | 7.558454056 | 0.002653157 | 0.0051214 | 1 |
| HTN | b_cell | *MAG* | -12.23624735 | 0.039173563 | 0.048338409 | 1 |
| HTN | b_cell | *LSR* | -3.499615364 | 0.004246851 | 0.007265294 | 1 |
| HTN | b_cell | *KXD1* | -4.755394019 | 0.000621341 | 0.002291037 | 1 |
| HTN | b_cell | *HPN* | -5.617296376 | 0.000518906 | 0.002195202 | 1 |
| HTN | b_cell | *SIPA1L3* | 5.193026503 | 0.006933892 | 0.010668109 | 1 |
| HTN | b_cell | *SMG9* | -4.606037614 | 0.001187128 | 0.003022874 | 1 |
| HTN | b_cell | *GTPBP10* | -5.027973867 | 0.00085639 | 0.002551796 | 1 |
| HTN | b_cell | *PON3* | -5.363876414 | 0.000506731 | 0.002195202 | 1 |
| HTN | b_cell | *PON2* | -4.745102984 | 0.00181765 | 0.003939181 | 1 |
| HTN | b_cell | *HBP1* | -4.884313899 | 0.00135868 | 0.003275316 | 1 |
| HTN | b_cell | *WDR91* | -5.586344442 | 0.001838811 | 0.003965902 | 1 |
| HTN | b_cell | *DLX5* | 63.25229353 | 7.23E-05 | 0.002098808 | 0.454696183 |
| HTN | b_cell | *STEAP1B* | 10.63677372 | 0.000357706 | 0.002180344 | 1 |
| HTN | b_cell | *PTN* | -5.306697371 | 0.000397457 | 0.002180344 | 1 |
| HTN | b_cell | *MPP6* | -3.622551613 | 0.000628742 | 0.002295161 | 1 |
| HTN | b_cell | *DFNA5* | -3.424873063 | 0.003018986 | 0.005618817 | 1 |
| HTN | b_cell | *OGDH* | -4.748643124 | 0.007705364 | 0.01162275 | 1 |
| HTN | b_cell | *ADAP1* | -15.16255498 | 0.001287064 | 0.003170756 | 1 |
| HTN | b_cell | *H2AFV* | -5.790735451 | 0.004905324 | 0.008126423 | 1 |
| HTN | b_cell | *CAV1* | -2.216204036 | 0.00088624 | 0.002596195 | 1 |
| HTN | b_cell | *MET* | -3.133417706 | 0.000350692 | 0.002180344 | 1 |
| HTN | b_cell | *RNF32* | -3.609851102 | 0.001292233 | 0.003173543 | 1 |
| HTN | b_cell | *LMBR1* | -4.41723994 | 0.01627317 | 0.022006759 | 1 |
| HTN | b_cell | *HOXA2* | -18.5064947 | 6.48E-06 | 0.000472149 | 0.040713181 |
| HTN | b_cell | *HOXA5* | -3.070536267 | 0.002230381 | 0.004510262 | 1 |
| HTN | b_cell | *HOXA6* | -3.593399926 | 0.000920352 | 0.002642126 | 1 |
| HTN | b_cell | *CPED1* | -3.753235193 | 0.003259436 | 0.005936589 | 1 |
| HTN | b_cell | *HIBADH* | -3.882121674 | 0.001781987 | 0.003888703 | 1 |
| HTN | b_cell | *CPVL* | -3.232216538 | 0.000728843 | 0.002394922 | 1 |
| HTN | b_cell | *PLEKHA8* | -3.766460374 | 0.000568216 | 0.002232844 | 1 |
| HTN | b_cell | *NOD1* | -4.833370478 | 0.002681384 | 0.00515531 | 1 |
| HTN | b_cell | *NPTX2* | -191.3099911 | 0.000189471 | 0.002180344 | 1 |
| HTN | b_cell | *CYP3A5* | -5.111817777 | 0.001011939 | 0.002769536 | 1 |
| HTN | b_cell | *ZKSCAN1* | -4.332018995 | 0.003004311 | 0.005601453 | 1 |
| HTN | b_cell | *EIF3B* | -5.15161731 | 0.000855862 | 0.002551796 | 1 |
| HTN | b_cell | *SNX8* | -5.450012263 | 0.00249948 | 0.004913879 | 1 |
| HTN | b_cell | *TAF6* | -6.397676327 | 3.80E-05 | 0.001482782 | 0.238727925 |
| HTN | b_cell | *AIMP2* | -4.412559602 | 0.000622374 | 0.002291037 | 1 |
| HTN | b_cell | *RBM28* | -2.677840429 | 0.000327871 | 0.002180344 | 1 |
| HTN | b_cell | *USP42* | -4.595223277 | 0.001316061 | 0.003214865 | 1 |
| HTN | b_cell | *IMPDH1* | -7.323198264 | 0.000289054 | 0.002180344 | 1 |
| HTN | b_cell | *LSM5* | -5.7059449 | 0.001060584 | 0.002836194 | 1 |
| HTN | b_cell | *AP1S1* | -10.87435699 | 0.001314599 | 0.003213407 | 1 |
| HTN | b_cell | *PLOD3* | -6.495776207 | 0.002558459 | 0.004997799 | 1 |
| HTN | b_cell | *RPA3* | -5.231399658 | 0.004785104 | 0.00796293 | 1 |
| HTN | b_cell | *ZNHIT1* | -8.152989977 | 0.000171698 | 0.002180344 | 1 |

| | | | | | | |
|---|---|---|---|---|---|---|
| HTN | b_cell | *PHF14* | -4.069905627 | 0.008097604 | 0.012098298 | 1 |
| HTN | b_cell | *ZNF862* | -4.777297619 | 0.011907074 | 0.016848925 | 1 |
| HTN | b_cell | *MEST* | -3.099474813 | 9.16E-06 | 0.000585817 | 0.057610292 |
| HTN | b_cell | *POU6F2* | -3.645720836 | 0.000414613 | 0.002180344 | 1 |
| HTN | b_cell | *TSPAN13* | 4.325763112 | 0.000285575 | 0.002180344 | 1 |
| HTN | b_cell | *RARRES2* | -3.23661862 | 0.000322099 | 0.002180344 | 1 |
| HTN | b_cell | *MRPL32* | -5.198254604 | 0.005042224 | 0.008313786 | 1 |
| HTN | b_cell | *COA1* | -3.525783009 | 0.017169928 | 0.0230706 | 1 |
| HTN | b_cell | *BLVRA* | -4.640667967 | 0.003257762 | 0.005936589 | 1 |
| HTN | b_cell | *URGCP* | -4.284470644 | 0.000696178 | 0.002359622 | 1 |
| HTN | b_cell | *POLD2* | -7.1716998 | 0.005481562 | 0.00888747 | 1 |
| HTN | b_cell | *GCK* | -53.61149325 | 0.000398376 | 0.002180344 | 1 |
| HTN | b_cell | *BCL7B* | -4.966412922 | 0.026516417 | 0.033966731 | 1 |
| HTN | b_cell | *YKT6* | -4.602587812 | 0.000711782 | 0.002380505 | 1 |
| HTN | b_cell | *LIMK1* | -7.954354388 | 0.006209492 | 0.009803886 | 1 |
| HTN | b_cell | *SPATA6L* | -3.705458056 | 0.000388407 | 0.002180344 | 1 |
| HTN | b_cell | *SLC1A1* | -2.011851796 | 0.0155539 | 0.021124945 | 1 |
| HTN | b_cell | *FKTN* | -5.19872511 | 0.004214282 | 0.007227274 | 1 |
| HTN | b_cell | *NMRK1* | -3.732037581 | 0.000755532 | 0.002419498 | 1 |
| HTN | b_cell | *TMEM245* | -4.105596844 | 0.006593746 | 0.010261597 | 1 |
| HTN | b_cell | *MEGF9* | -3.989755275 | 0.034935169 | 0.043492555 | 1 |
| HTN | b_cell | *TRIM14* | -3.161023576 | 0.000498409 | 0.002194324 | 1 |
| HTN | b_cell | *CORO2A* | -4.396638239 | 0.000633514 | 0.002301586 | 1 |
| HTN | b_cell | *C5* | -3.607192021 | 0.000696214 | 0.002359622 | 1 |
| HTN | b_cell | *OGN* | -5.454598233 | 0.000155795 | 0.002180344 | 0.979485 |
| HTN | b_cell | *ECM2* | -2.686400737 | 0.000138263 | 0.002180344 | 0.869260406 |
| HTN | b_cell | *PTGR1* | -2.990001429 | 0.000776133 | 0.002438676 | 1 |
| HTN | b_cell | *SUSD1* | -5.10436801 | 0.001565692 | 0.003574258 | 1 |
| HTN | b_cell | *TNFSF8* | -4.315544017 | 0.000347998 | 0.002180344 | 1 |
| HTN | b_cell | *AK1* | 11.10969412 | 0.007323989 | 0.011157238 | 1 |
| HTN | b_cell | *PLGRKT* | -4.864735576 | 9.15E-05 | 0.002180344 | 0.57515718 |
| HTN | b_cell | *DOCK8* | -3.143796335 | 0.027933456 | 0.035578937 | 1 |
| HTN | b_cell | *ELAVL2* | -8.850414342 | 0.000369161 | 0.002180344 | 1 |
| HTN | b_cell | *FUBP3* | -4.734241708 | 0.001857082 | 0.003998439 | 1 |
| HTN | b_cell | *RGP1* | -8.131748432 | 0.039590237 | 0.048804671 | 1 |
| HTN | b_cell | *MPDZ* | -2.745371729 | 0.000533318 | 0.002197228 | 1 |
| HTN | b_cell | *PTGDS* | -9.279900171 | 0.001271838 | 0.003154259 | 1 |
| HTN | b_cell | *SHB* | -23.10012172 | 0.009619518 | 0.013963959 | 1 |
| HTN | b_cell | *DVL1* | -5.218083724 | 0.000935033 | 0.002659834 | 1 |
| HTN | b_cell | *PDLIM1* | -3.516766805 | 0.002876034 | 0.005430399 | 1 |
| HTN | b_cell | *CCNJ* | -8.177619501 | 0.002160072 | 0.004403494 | 1 |
| HTN | b_cell | *ATRNL1* | -4.008132115 | 0.000269829 | 0.002180344 | 1 |
| HTN | b_cell | *HPS1* | -6.013197986 | 0.014938497 | 0.020430352 | 1 |
| HTN | b_cell | *PHYH* | -3.738409736 | 0.000637509 | 0.002308767 | 1 |
| HTN | b_cell | *DNMBP* | -3.650583854 | 0.000786789 | 0.002453642 | 1 |
| HTN | b_cell | *TRDMT1* | -7.111952506 | 0.002198107 | 0.004463722 | 1 |
| HTN | b_cell | *DDX50* | -5.037573216 | 0.0041943 | 0.007201376 | 1 |
| HTN | b_cell | *MAPK8* | -4.060568139 | 0.004892447 | 0.008109364 | 1 |
| HTN | b_cell | *ATE1* | -3.501986745 | 0.012785183 | 0.017886169 | 1 |
| HTN | b_cell | *NSMCE4A* | -4.282544617 | 0.000770937 | 0.00243363 | 1 |

| | | | | | | |
|---|---|---|---|---|---|---|
| HTN | b_cell | *PLEKHA1* | -4.805123714 | 0.00309324 | 0.005719765 | 1 |
| HTN | b_cell | *SPOCK2* | -4.234752087 | 0.002432376 | 0.004813455 | 1 |
| HTN | b_cell | *MICU1* | -4.272489668 | 0.003733801 | 0.006590232 | 1 |
| HTN | b_cell | *CCSER2* | -4.145038517 | 0.003972484 | 0.006918284 | 1 |
| HTN | b_cell | *BMPR1A* | -2.436174789 | 0.000759556 | 0.002422796 | 1 |
| HTN | b_cell | *SFXN3* | -5.862363648 | 0.002140784 | 0.00437568 | 1 |
| HTN | b_cell | *KAZALD1* | -12.19619035 | 0.000153595 | 0.002180344 | 0.965651908 |
| HTN | b_cell | *ANKRD26* | -4.19216693 | 0.004665167 | 0.007825436 | 1 |
| HTN | b_cell | *ACBD5* | -5.152879208 | 0.006196851 | 0.009791306 | 1 |
| HTN | b_cell | *MTPAP* | -6.146879133 | 0.009875565 | 0.014296034 | 1 |
| HTN | b_cell | *PITRM1* | -6.587607254 | 0.003403575 | 0.006138346 | 1 |
| HTN | b_cell | *MAP3K8* | -4.647841164 | 0.001915793 | 0.004084296 | 1 |
| HTN | b_cell | *SHOC2* | -4.578831341 | 0.005765394 | 0.009246692 | 1 |
| HTN | b_cell | *TFAM* | -5.417857712 | 0.000949823 | 0.002680223 | 1 |
| HTN | b_cell | *DHRS11* | -4.859245571 | 0.000172417 | 0.002180344 | 1 |
| HTN | b_cell | *MLLT6* | -5.393453276 | 0.003992805 | 0.006942137 | 1 |
| HTN | b_cell | *CASC3* | -4.622167926 | 0.005264717 | 0.008613666 | 1 |
| HTN | b_cell | *RAPGEFL1* | -4.298307519 | 0.002378423 | 0.004732008 | 1 |
| HTN | b_cell | *RGS9* | -5.261660263 | 0.00047406 | 0.002180489 | 1 |
| HTN | b_cell | *WNT3* | -10.73801723 | 2.16E-05 | 0.001049416 | 0.135614268 |
| HTN | b_cell | *ASPA* | -2.718951238 | 0.000413971 | 0.002180344 | 1 |
| HTN | b_cell | *RAD51C* | -6.538909445 | 0.003011698 | 0.005611899 | 1 |
| HTN | b_cell | *RPS6KB1* | -4.859097264 | 0.002349375 | 0.00469725 | 1 |
| HTN | b_cell | *TRIM16L* | -4.762249371 | 0.000181974 | 0.002180344 | 1 |
| HTN | b_cell | *CDK5RAP3* | -5.760779953 | 0.000152208 | 0.002180344 | 0.956932956 |
| HTN | b_cell | *CBX1* | -4.245081437 | 0.002122974 | 0.004357537 | 1 |
| HTN | b_cell | *RECQL5* | -6.132238385 | 0.001690129 | 0.003759376 | 1 |
| HTN | b_cell | *PIGL* | -4.309319672 | 0.001998011 | 0.004192755 | 1 |
| HTN | b_cell | *GALK1* | -9.185141099 | 0.001268209 | 0.003148985 | 1 |
| HTN | b_cell | *CAMTA2* | -6.543568095 | 0.007780421 | 0.011716289 | 1 |
| HTN | b_cell | *MED13* | -4.185611155 | 0.012342826 | 0.017371692 | 1 |
| HTN | b_cell | *HOXB6* | -4.483767085 | 0.000123874 | 0.002180344 | 0.778798142 |
| HTN | b_cell | *C1QBP* | -4.975253555 | 0.00050723 | 0.002195202 | 1 |
| HTN | b_cell | *GOSR1* | -4.183985098 | 0.001548426 | 0.003551606 | 1 |
| HTN | b_cell | *DRG2* | -4.055004065 | 0.000432143 | 0.002180344 | 1 |
| HTN | b_cell | *AKAP10* | -3.849818194 | 0.033210673 | 0.041559614 | 1 |
| HTN | b_cell | *ALDH3A1* | 6.955646499 | 0.010360144 | 0.01494247 | 1 |
| HTN | b_cell | *SMARCD2* | -4.309703384 | 0.000513829 | 0.002195202 | 1 |
| HTN | b_cell | *ICAM2* | -3.084300526 | 0.003087686 | 0.005715637 | 1 |
| HTN | b_cell | *SYNGR2* | -6.94133936 | 2.67E-05 | 0.001226302 | 0.168003349 |
| HTN | b_cell | *C17orf75* | -9.386404275 | 0.0096531 | 0.014009474 | 1 |
| HTN | b_cell | *PEX12* | -10.44847011 | 4.52E-05 | 0.001652108 | 0.284162565 |
| HTN | b_cell | *DHX58* | -5.738987094 | 0.011657715 | 0.016540748 | 1 |
| HTN | b_cell | *RAB5C* | -5.26643142 | 0.000489998 | 0.002189495 | 1 |
| HTN | b_cell | *NAGLU* | -3.811215227 | 0.000486425 | 0.002188291 | 1 |
| HTN | b_cell | *EZH1* | -4.171262349 | 0.004063927 | 0.007042422 | 1 |
| HTN | b_cell | *DLX4* | -49.95428368 | 0.000405204 | 0.002180344 | 1 |
| HTN | b_cell | *SGCA* | 4.79818948 | 0.011787856 | 0.016702783 | 1 |
| HTN | b_cell | *VAT1* | -3.92833567 | 0.000513918 | 0.002195202 | 1 |
| HTN | b_cell | *LRRC59* | -8.842944411 | 0.004038939 | 0.007010714 | 1 |

| | | | | | | |
|---|---|---|---|---|---|---|
| HTN | b_cell | *ALOX12* | -8.691796596 | 0.001959036 | 0.004141379 | 1 |
| HTN | b_cell | *ABCC3* | -3.598444925 | 0.009874475 | 0.014296034 | 1 |
| HTN | b_cell | *MPP2* | -7.582105543 | 0.000424445 | 0.002180344 | 1 |
| HTN | b_cell | *EFNB3* | 19.64261776 | 0.001245664 | 0.003112676 | 1 |
| HTN | b_cell | *FAM20A* | -3.738076418 | 0.000903547 | 0.002608173 | 1 |
| HTN | b_cell | *YWHAE* | -4.400775265 | 0.003161062 | 0.005813975 | 1 |
| HTN | b_cell | *DPH1* | -6.811065543 | 0.000670222 | 0.002333871 | 1 |
| HTN | b_cell | *DHRS7B* | -4.213740269 | 0.001252627 | 0.003123021 | 1 |
| HTN | b_cell | *RCVRN* | -20.08626675 | 0.001837075 | 0.003963518 | 1 |
| HTN | b_cell | *SLC9A3R1* | -7.724411324 | 0.005860237 | 0.009372504 | 1 |
| HTN | b_cell | *MYH3* | -5.573170983 | 0.000602141 | 0.002278713 | 1 |
| HTN | b_cell | *TMEM104* | -6.119991221 | 0.00229311 | 0.00461042 | 1 |
| HTN | b_cell | *TMEM97* | 4.854859147 | 0.00063456 | 0.002303394 | 1 |
| HTN | b_cell | *RAB34* | -5.036963875 | 0.000255768 | 0.002180344 | 1 |
| HTN | b_cell | *SLAIN2* | -3.945212197 | 0.004774052 | 0.007950851 | 1 |
| HTN | b_cell | *OCIAD1* | -5.200924082 | 0.01337824 | 0.018577893 | 1 |
| HTN | b_cell | *CWH43* | -32.29760734 | 0.01466034 | 0.020098028 | 1 |
| HTN | b_cell | *KIAA1211* | 3.970524813 | 0.018965819 | 0.025166336 | 1 |
| HTN | b_cell | *AREG* | 3.828328425 | 0.000552493 | 0.002219911 | 1 |
| HTN | b_cell | *NDUFC1* | 6.998758507 | 0.039636609 | 0.048852256 | 1 |
| HTN | b_cell | *TBC1D9* | -4.435187595 | 0.004566563 | 0.007697046 | 1 |
| HTN | b_cell | *CPE* | -3.771894019 | 0.00069119 | 0.002356568 | 1 |
| HTN | b_cell | *WFS1* | -3.987150058 | 0.000429193 | 0.002180344 | 1 |
| HTN | b_cell | *GRPEL1* | -6.046699841 | 0.000872544 | 0.002573594 | 1 |
| HTN | b_cell | *AADAT* | -5.193781072 | 0.000355158 | 0.002180344 | 1 |
| HTN | b_cell | *CPZ* | -99.45782208 | 0.000415519 | 0.002180344 | 1 |
| HTN | b_cell | *TRIM2* | -3.113897111 | 0.000967557 | 0.002711104 | 1 |
| HTN | b_cell | *SLC2A9* | -3.703393488 | 0.0019299 | 0.004099081 | 1 |
| HTN | b_cell | *SH3D19* | -1.920947032 | 0.000403313 | 0.002180344 | 1 |
| HTN | b_cell | *RAPGEF2* | -3.873984299 | 0.002576784 | 0.005021774 | 1 |
| HTN | b_cell | *LRP2BP* | -5.128200207 | 0.025047451 | 0.032229498 | 1 |
| HTN | b_cell | *UFSP2* | -5.863909383 | 0.00987307 | 0.014296034 | 1 |
| HTN | b_cell | *KLF3* | -4.924706589 | 0.012333224 | 0.017365952 | 1 |
| HTN | b_cell | *KLHL5* | -3.15933142 | 0.006037825 | 0.00958581 | 1 |
| HTN | b_cell | *FAM149A* | -2.982359625 | 0.001006184 | 0.002758778 | 1 |
| HTN | b_cell | *UGDH* | -4.187901162 | 0.000537572 | 0.002207687 | 1 |
| HTN | b_cell | *DDX25* | -11.32881063 | 0.018964443 | 0.025166336 | 1 |
| HTN | b_cell | *HTATIP2* | -5.237722823 | 0.003580635 | 0.006369964 | 1 |
| HTN | b_cell | *CTSC* | -3.414207335 | 0.000304366 | 0.002180344 | 1 |
| HTN | b_cell | *ELP4* | -4.392386996 | 0.001065815 | 0.002845341 | 1 |
| HTN | b_cell | *C11orf63* | -3.65732076 | 0.000600363 | 0.002275156 | 1 |
| HTN | b_cell | *SNX15* | -8.742477173 | 6.10E-05 | 0.001965056 | 0.383697194 |
| HTN | b_cell | *DTX4* | -7.89539269 | 0.014176751 | 0.019537315 | 1 |
| HTN | b_cell | *PUS3* | -5.642047961 | 0.000387993 | 0.002180344 | 1 |
| HTN | b_cell | *DCPS* | 7.087310862 | 0.039556304 | 0.04878197 | 1 |
| HTN | b_cell | *FOXRED1* | -4.727742357 | 0.000367514 | 0.002180344 | 1 |
| HTN | b_cell | *NRXN2* | -14.40571292 | 0.029410725 | 0.037264254 | 1 |
| HTN | b_cell | *MS4A4A* | -3.016378763 | 0.000524754 | 0.002195202 | 1 |
| HTN | b_cell | *CCDC86* | -12.0692756 | 0.000407381 | 0.002180344 | 1 |
| HTN | b_cell | *PRPF19* | -5.55460442 | 0.000466645 | 0.002180489 | 1 |

| | | | | | | |
|---|---|---|---|---|---|---|
| HTN | b_cell | *HPX* | -7.711706895 | 0.003902209 | 0.006818562 | 1 |
| HTN | b_cell | *CHORDC1* | -5.407383141 | 0.005224288 | 0.008564562 | 1 |
| HTN | b_cell | *PANX1* | -5.369709639 | 0.000233549 | 0.002180344 | 1 |
| HTN | b_cell | *EIF4G2* | -5.069196065 | 0.014928059 | 0.02042052 | 1 |
| HTN | b_cell | *GALNT18* | -3.053946275 | 0.000190929 | 0.002180344 | 1 |
| HTN | b_cell | *CBL* | -3.820219909 | 0.00133529 | 0.003242552 | 1 |
| HTN | b_cell | *KIAA1549L* | -7.519150076 | 0.000409062 | 0.002180344 | 1 |
| HTN | b_cell | *MADD* | -5.121514042 | 0.010857898 | 0.015532106 | 1 |
| HTN | b_cell | *NAA40* | -4.464188819 | 0.00023081 | 0.002180344 | 1 |
| HTN | b_cell | *CARS* | -5.365408009 | 0.000346211 | 0.002180344 | 1 |
| HTN | b_cell | *ELMOD1* | -10.43235614 | 0.002323911 | 0.004655967 | 1 |
| HTN | b_cell | *NDUFS8* | -7.187980973 | 0.018158567 | 0.024222981 | 1 |
| HTN | b_cell | *TCIRG1* | -5.297905811 | 0.004125475 | 0.007119644 | 1 |
| HTN | b_cell | *GTF2H1* | -5.271894734 | 0.006766789 | 0.010485809 | 1 |
| HTN | b_cell | *PTPN5* | -4.898797827 | 0.010492845 | 0.0151096 | 1 |
| HTN | b_cell | *PPFIBP1* | -4.295518941 | 0.000327021 | 0.002180344 | 1 |
| HTN | b_cell | *PRDM4* | -5.013997549 | 0.007096231 | 0.010873509 | 1 |
| HTN | b_cell | *COQ5* | -2.209037175 | 0.003603295 | 0.00640484 | 1 |
| HTN | b_cell | *ASIC1* | -4.641563985 | 0.036347637 | 0.045090291 | 1 |
| HTN | b_cell | *CAPRIN2* | -6.342250275 | 0.004886744 | 0.008102046 | 1 |
| HTN | b_cell | *KCTD10* | -5.622634217 | 7.23E-06 | 0.000510628 | 0.045445868 |
| HTN | b_cell | *CYP27B1* | 5.269008322 | 0.000218633 | 0.002180344 | 1 |
| HTN | b_cell | *ACSS3* | -1.703849823 | 0.004111613 | 0.007105628 | 1 |
| HTN | b_cell | *GLI1* | -8.735789234 | 0.000415373 | 0.002180344 | 1 |
| HTN | b_cell | *LTA4H* | -5.45089023 | 0.005253185 | 0.00859991 | 1 |
| HTN | b_cell | *ITFG2* | -5.112870706 | 0.003912326 | 0.006830545 | 1 |
| HTN | b_cell | *PRR4* | -3.147653885 | 0.031490674 | 0.039628076 | 1 |
| HTN | b_cell | *PARP11* | -5.072115452 | 0.03905095 | 0.048196569 | 1 |
| HTN | b_cell | *GPN3* | -7.062450368 | 0.038273662 | 0.047358101 | 1 |
| HTN | b_cell | *VPS29* | -3.798456838 | 0.000704169 | 0.002372515 | 1 |
| HTN | b_cell | *KCNA1* | -13.20795567 | 0.000220402 | 0.002180344 | 1 |
| HTN | b_cell | *CREBL2* | -3.953102817 | 0.001087162 | 0.002868262 | 1 |
| HTN | b_cell | *LTBR* | -4.579874016 | 0.003497272 | 0.006260634 | 1 |
| HTN | b_cell | *MGP* | -3.03966529 | 0.000818188 | 0.002500703 | 1 |
| HTN | b_cell | *RASAL1* | 5.211168854 | 0.000190535 | 0.002180344 | 1 |
| HTN | b_cell | *EIF2B1* | -5.895190629 | 0.002897542 | 0.005462323 | 1 |
| HTN | b_cell | *DDX55* | -3.726104258 | 0.004817301 | 0.008005914 | 1 |
| HTN | b_cell | *ENDOU* | -12.09161362 | 4.40E-06 | 0.00036476 | 0.027687674 |
| HTN | b_cell | *STX2* | -4.571601719 | 0.003297647 | 0.005997197 | 1 |
| HTN | b_cell | *CAND1* | -5.133695876 | 3.62E-05 | 0.001457697 | 0.227400675 |
| HTN | b_cell | *NUP107* | -3.970486671 | 0.000812492 | 0.00249056 | 1 |
| HTN | b_cell | *CNOT2* | -4.438550123 | 0.006324324 | 0.009940257 | 1 |
| HTN | b_cell | *KRR1* | -5.466980565 | 0.007795941 | 0.011731231 | 1 |
| HTN | b_cell | *COPS7A* | -6.988129745 | 0.000119185 | 0.002180344 | 0.749313363 |
| HTN | b_cell | *GNB3* | -3.14358226 | 0.011161326 | 0.01589743 | 1 |
| HTN | b_cell | *CHPT1* | -2.680257431 | 0.00577108 | 0.009253451 | 1 |
| HTN | b_cell | *GNPTAB* | -4.505224503 | 0.001960585 | 0.00414326 | 1 |
| HTN | b_cell | *SPSB2* | -5.979243614 | 0.000242587 | 0.002180344 | 1 |
| HTN | b_cell | *ATN1* | -3.852277297 | 0.001185768 | 0.003022492 | 1 |
| HTN | b_cell | *NT5DC3* | -3.459239952 | 0.00092403 | 0.002648292 | 1 |

| | | | | | | |
|---|---|---|---|---|---|---|
| HTN | b_cell | *PRKAB1* | 7.007003117 | 9.80E-05 | 0.002180344 | 0.616019549 |
| HTN | b_cell | *ST8SIA1* | -3.00200677 | 0.000527286 | 0.002195202 | 1 |
| HTN | b_cell | *C2CD5* | -4.354192153 | 0.033535472 | 0.04193268 | 1 |
| HTN | b_cell | *COL12A1* | -2.197242739 | 0.00040955 | 0.002180344 | 1 |
| HTN | b_cell | *BTN3A3* | -5.88666926 | 0.001584316 | 0.003595883 | 1 |
| HTN | b_cell | *TDP2* | -5.937528412 | 0.001082344 | 0.002865134 | 1 |
| HTN | b_cell | *RWDD1* | -5.217848258 | 0.000230711 | 0.002180344 | 1 |
| HTN | b_cell | *SMIM8* | -3.161775447 | 0.001236517 | 0.003100909 | 1 |
| HTN | b_cell | *MCM9* | -4.615615593 | 0.004158427 | 0.007165569 | 1 |
| HTN | b_cell | *FAM184A* | -2.749923672 | 0.000816816 | 0.00249894 | 1 |
| HTN | b_cell | *GABRR2* | -6.003360934 | 0.001876567 | 0.004029364 | 1 |
| HTN | b_cell | *HDDC2* | -4.369784208 | 0.003439825 | 0.006187749 | 1 |
| HTN | b_cell | *TPD52L1* | -3.427641046 | 0.000416794 | 0.002180344 | 1 |
| HTN | b_cell | *UST* | -2.204208042 | 0.000517895 | 0.002195202 | 1 |
| HTN | b_cell | *TULP1* | -39.38969469 | 0.000723695 | 0.002388914 | 1 |
| HTN | b_cell | *SLC26A8* | -3.764001537 | 0.007144736 | 0.010929186 | 1 |
| HTN | b_cell | *MAPK14* | -4.612768714 | 0.000790519 | 0.002459174 | 1 |
| HTN | b_cell | *RNF8* | -3.49196758 | 0.000254421 | 0.002180344 | 1 |
| HTN | b_cell | *PHACTR1* | -4.739943956 | 0.002989255 | 0.005584167 | 1 |
| HTN | b_cell | *MDGA1* | 5.899899777 | 0.000722407 | 0.002388914 | 1 |
| HTN | b_cell | *CD83* | -3.971626141 | 0.03528983 | 0.043881954 | 1 |
| HTN | b_cell | *MDN1* | -4.229578729 | 0.008410181 | 0.012479303 | 1 |
| HTN | b_cell | *CAP2* | -2.730156565 | 0.000223971 | 0.002180344 | 1 |
| HTN | b_cell | *GPR63* | -5.674070486 | 0.008007867 | 0.011987015 | 1 |
| HTN | b_cell | *ASCC3* | -4.300191046 | 0.012393237 | 0.017427037 | 1 |
| HTN | b_cell | *BVES* | -7.68778485 | 0.002036584 | 0.004239268 | 1 |
| HTN | b_cell | *WASF1* | -3.082681367 | 0.000254748 | 0.002180344 | 1 |
| HTN | b_cell | *GPLD1* | -2.860374154 | 0.000843982 | 0.002536213 | 1 |
| HTN | b_cell | *ALDH5A1* | -4.22039248 | 0.013198842 | 0.018366781 | 1 |
| HTN | b_cell | *VNN1* | -3.564909686 | 0.007043881 | 0.010809099 | 1 |
| HTN | b_cell | *VNN2* | 5.836631064 | 0.000593152 | 0.00226693 | 1 |
| HTN | b_cell | *ACOT13* | -4.639519991 | 0.001048376 | 0.00281793 | 1 |
| HTN | b_cell | *C6orf62* | -4.287224841 | 0.002122732 | 0.004357537 | 1 |
| HTN | b_cell | *B3GAT2* | -8.855350875 | 0.0204215 | 0.026854208 | 1 |
| HTN | b_cell | *HBS1L* | -5.269850323 | 0.00461451 | 0.007761216 | 1 |
| HTN | b_cell | *PEX7* | -4.233833926 | 0.008202201 | 0.012219724 | 1 |
| HTN | b_cell | *SLC16A10* | -3.332510346 | 0.002234482 | 0.00451275 | 1 |
| HTN | b_cell | *PHACTR2* | -4.122113832 | 0.004017418 | 0.006977211 | 1 |
| HTN | b_cell | *PACRG* | -4.80119102 | 0.000594301 | 0.002268192 | 1 |
| HTN | b_cell | *TFEB* | -6.463181601 | 0.005975272 | 0.009505703 | 1 |
| HTN | b_cell | *TBP* | -6.2554277 | 0.02110898 | 0.02768297 | 1 |
| HTN | b_cell | *PRPH2* | -6.020537272 | 0.00073224 | 0.002399172 | 1 |
| HTN | b_cell | *DUSP22* | -4.778358313 | 0.001433604 | 0.003385825 | 1 |
| HTN | b_cell | *EXOC2* | -4.70104604 | 0.027608267 | 0.03520754 | 1 |
| HTN | b_cell | *TMEM30A* | -4.596555106 | 0.00387393 | 0.006789907 | 1 |
| HTN | b_cell | *SENP6* | -3.967611814 | 0.006501523 | 0.010170459 | 1 |
| HTN | b_cell | *TTK* | 2.640187134 | 0.000625974 | 0.002295161 | 1 |
| HTN | b_cell | *BTN2A1* | 8.323423921 | 0.00050984 | 0.002195202 | 1 |
| HTN | b_cell | *LAMA4* | -1.707691165 | 0.000450878 | 0.002180344 | 1 |
| HTN | b_cell | *CLIC5* | -3.679926832 | 0.000269326 | 0.002180344 | 1 |

| | | | | | | |
|---|---|---|---|---|---|---|
| HTN | b_cell | *LY86* | 3.561307359 | 0.000280421 | 0.002180344 | 1 |
| HTN | b_cell | *TBX18* | -3.387946617 | 0.000183583 | 0.002180344 | 1 |
| HTN | b_cell | *PCDHB2* | -5.478582599 | 0.006094987 | 0.009664359 | 1 |
| HTN | b_cell | *NUDT12* | -6.364835356 | 0.001719589 | 0.00380403 | 1 |
| HTN | b_cell | *CEP72* | 13.84628859 | 2.11E-05 | 0.001037194 | 0.132760805 |
| HTN | b_cell | *C7* | -3.560937276 | 0.000318289 | 0.002180344 | 1 |
| HTN | b_cell | *PAPD7* | -4.837783035 | 0.006130578 | 0.009706012 | 1 |
| HTN | b_cell | *GHR* | -1.538019319 | 0.001276848 | 0.003156737 | 1 |
| HTN | b_cell | *NME5* | -3.094792842 | 0.007219101 | 0.011026844 | 1 |
| HTN | b_cell | *NNT* | -5.558564408 | 7.18E-05 | 0.002098808 | 0.451554836 |
| HTN | b_cell | *PFDN1* | -3.915339782 | 0.002033618 | 0.004236366 | 1 |
| HTN | b_cell | *LOX* | -3.222330788 | 0.000415306 | 0.002180344 | 1 |
| HTN | b_cell | *APBB3* | 5.989216341 | 0.013066447 | 0.01821884 | 1 |
| HTN | b_cell | *TMCO6* | 3.740943995 | 0.000293345 | 0.002180344 | 1 |
| HTN | b_cell | *SPARC* | -2.791423919 | 0.000763225 | 0.002427608 | 1 |
| HTN | b_cell | *IK* | -4.82876762 | 0.021207967 | 0.027801186 | 1 |
| HTN | b_cell | *PDE8B* | -2.645237455 | 0.000519518 | 0.002195202 | 1 |
| HTN | b_cell | *CLK4* | -4.546176047 | 0.002372136 | 0.004723984 | 1 |
| HTN | b_cell | *PCDHB15* | -3.591503806 | 0.026733436 | 0.034216839 | 1 |
| HTN | b_cell | *HAVCR1* | 5.22146251 | 0.001211003 | 0.00305766 | 1 |
| HTN | b_cell | *RNF130* | -3.316872261 | 0.001119559 | 0.002924248 | 1 |
| HTN | b_cell | *ARSB* | -4.77429818 | 0.011459751 | 0.01629331 | 1 |
| HTN | b_cell | *CLINT1* | -4.155922488 | 0.001462315 | 0.003427879 | 1 |
| HTN | b_cell | *BTNL8* | -5.066806804 | 0.000265029 | 0.002180344 | 1 |
| HTN | b_cell | *TTC1* | -5.101228966 | 0.00942628 | 0.013743744 | 1 |
| HTN | b_cell | *MSH3* | -3.817332312 | 0.001749186 | 0.003844942 | 1 |
| HTN | b_cell | *RASGRF2* | -2.302302804 | 0.000404787 | 0.002180344 | 1 |
| HTN | b_cell | *POLR3G* | -3.854745128 | 0.000222678 | 0.002180344 | 1 |
| HTN | b_cell | *DROSHA* | -4.911925403 | 0.002523374 | 0.004945278 | 1 |
| HTN | b_cell | *CDH6* | -2.92656996 | 0.013133594 | 0.018304346 | 1 |
| HTN | b_cell | *SLC27A6* | -8.368153897 | 0.000477279 | 0.002180489 | 1 |
| HTN | b_cell | *TARS* | -4.526430381 | 0.000725756 | 0.002388914 | 1 |
| HTN | b_cell | *LNPEP* | -3.614767873 | 0.011839138 | 0.016760338 | 1 |
| HTN | b_cell | *SLC12A7* | -6.328564924 | 0.000929907 | 0.002653801 | 1 |
| HTN | b_cell | *IL4* | 6.799752636 | 0.001624326 | 0.003666835 | 1 |
| HTN | b_cell | *GNPDA1* | -6.211107496 | 0.00307871 | 0.005708006 | 1 |
| HTN | b_cell | *PPWD1* | -3.709524047 | 0.034943411 | 0.043494204 | 1 |
| HTN | b_cell | *TRIM23* | -5.491572044 | 0.006827747 | 0.010565111 | 1 |
| HTN | b_cell | *TRAPPC13* | -4.204626326 | 0.033009076 | 0.041348488 | 1 |
| HTN | b_cell | *TXNDC15* | -4.599018222 | 0.001275837 | 0.003156737 | 1 |
| HTN | b_cell | *SMAD5* | -4.735290249 | 0.003611628 | 0.006415909 | 1 |
| HTN | b_cell | *HMGXB3* | -5.931713097 | 0.003703024 | 0.006550623 | 1 |
| HTN | b_cell | *ATP6V0E1* | -5.31125772 | 0.001689857 | 0.003759376 | 1 |
| HTN | b_cell | *CNTN3* | -2.877622384 | 0.000413636 | 0.002180344 | 1 |
| HTN | b_cell | *ACTR8* | 8.373149015 | 0.018589117 | 0.024734345 | 1 |
| HTN | b_cell | *HGD* | 13.64838416 | 0.000616589 | 0.002286342 | 1 |
| HTN | b_cell | *NIT2* | -4.824037778 | 0.00022298 | 0.002180344 | 1 |
| HTN | b_cell | *FAM162A* | -4.743463727 | 0.000100923 | 0.002180344 | 0.634500092 |
| HTN | b_cell | *PCCB* | -3.749948543 | 0.007400121 | 0.01125685 | 1 |
| HTN | b_cell | *RBP1* | -10.27321441 | 0.000347631 | 0.002180344 | 1 |

| | | | | | | |
|---|---|---|---|---|---|---|
| HTN | b_cell | *RNF7* | -6.969800481 | 0.002206061 | 0.004474035 | 1 |
| HTN | b_cell | *BCHE* | -6.212335504 | 6.33E-05 | 0.001980182 | 0.397988048 |
| HTN | b_cell | *PDCD10* | -5.009202299 | 0.007967831 | 0.011944147 | 1 |
| HTN | b_cell | *WNT5A* | -8.774084571 | 0.003954091 | 0.006895061 | 1 |
| HTN | b_cell | *PRKAR2A* | -3.315600816 | 0.015061252 | 0.020562452 | 1 |
| HTN | b_cell | *HYAL1* | -5.543240622 | 9.12E-06 | 0.000585817 | 0.057354395 |
| HTN | b_cell | *CYB561D2* | -4.35559639 | 0.026009918 | 0.033365508 | 1 |
| HTN | b_cell | *IFT57* | 6.831488941 | 0.003328653 | 0.006039608 | 1 |
| HTN | b_cell | *IQCG* | -5.342255018 | 0.002839916 | 0.00538751 | 1 |
| HTN | b_cell | *GBE1* | -2.207184034 | 0.000524304 | 0.002195202 | 1 |
| HTN | b_cell | *UMPS* | -5.154144238 | 0.000758137 | 0.002419498 | 1 |
| HTN | b_cell | *NCBP2* | -5.93815 | 0.007671028 | 0.011595997 | 1 |
| HTN | b_cell | *SLC41A3* | -4.098818177 | 0.00037641 | 0.002180344 | 1 |
| HTN | b_cell | *PLXNA1* | -3.605495576 | 0.012009706 | 0.016982686 | 1 |
| HTN | b_cell | *ABTB1* | -6.348122064 | 0.006909975 | 0.010650408 | 1 |
| HTN | b_cell | *EFCC1* | -7.064343264 | 0.001002669 | 0.002756353 | 1 |
| HTN | b_cell | *NEK11* | -4.58642186 | 0.001501883 | 0.003481687 | 1 |
| HTN | b_cell | *PLSCR4* | -3.309689239 | 0.001678351 | 0.00374486 | 1 |
| HTN | b_cell | *HEMK1* | -6.557515975 | 0.006123701 | 0.00969766 | 1 |
| HTN | b_cell | *WDR48* | -4.899866371 | 0.02302401 | 0.029936049 | 1 |
| HTN | b_cell | *COMMD2* | -4.283628785 | 0.000868575 | 0.002568981 | 1 |
| HTN | b_cell | *GORASP1* | -7.744271025 | 0.031700328 | 0.039867965 | 1 |
| HTN | b_cell | *ABCC5* | -4.064749978 | 0.000315006 | 0.002180344 | 1 |
| HTN | b_cell | *ARHGEF26* | -3.302437488 | 0.000643725 | 0.002316643 | 1 |
| HTN | b_cell | *KLHL24* | -3.439400558 | 0.004127623 | 0.007121396 | 1 |
| HTN | b_cell | *PLCH1* | 11.35314297 | 0.000710729 | 0.002379315 | 1 |
| HTN | b_cell | *NKTR* | -3.905667823 | 0.011452338 | 0.016289785 | 1 |
| HTN | b_cell | *CLCN2* | -13.28161132 | 0.002058437 | 0.004270681 | 1 |
| HTN | b_cell | *MOB1A* | -5.790463166 | 0.004281997 | 0.007317454 | 1 |
| HTN | b_cell | *LMAN2L* | -5.018522069 | 0.007825557 | 0.011767346 | 1 |
| HTN | b_cell | *PIKFYVE* | -4.544264698 | 0.03513641 | 0.043717121 | 1 |
| HTN | b_cell | *KCNIP3* | -6.855103055 | 0.000725105 | 0.002388914 | 1 |
| HTN | b_cell | *FAHD2A* | 9.344339795 | 0.001020138 | 0.002777657 | 1 |
| HTN | b_cell | *NCL* | -5.219762612 | 0.029659646 | 0.037549375 | 1 |
| HTN | b_cell | *ACTR1B* | -5.897035687 | 0.00830131 | 0.012344319 | 1 |
| HTN | b_cell | *SLC35F5* | -5.69985624 | 0.001497323 | 0.003478813 | 1 |
| HTN | b_cell | *ACTR3* | -4.642727703 | 0.009360742 | 0.013673556 | 1 |
| HTN | b_cell | *STEAP3* | -3.862993737 | 0.004323481 | 0.007368318 | 1 |
| HTN | b_cell | *TFCP2L1* | -6.693785868 | 0.00069108 | 0.002356568 | 1 |
| HTN | b_cell | *TP53I3* | -4.3143818 | 9.99E-05 | 0.002180344 | 0.628334561 |
| HTN | b_cell | *DNAJC27* | -10.86022539 | 0.000184417 | 0.002180344 | 1 |
| HTN | b_cell | *STAM2* | -4.882900942 | 0.005284966 | 0.008641504 | 1 |
| HTN | b_cell | *GPD2* | -4.474075991 | 0.003452024 | 0.006204721 | 1 |
| HTN | b_cell | *CENPA* | -5.156175394 | 0.000395038 | 0.002180344 | 1 |
| HTN | b_cell | *TANC1* | -2.533359418 | 0.000212361 | 0.002180344 | 1 |
| HTN | b_cell | *PCSK4* | -8.47880574 | 0.000306957 | 0.002180344 | 1 |
| HTN | b_cell | *INO80B* | -4.788328186 | 0.00210472 | 0.004337061 | 1 |
| HTN | b_cell | *MOGS* | -2.971361407 | 0.000247021 | 0.002180344 | 1 |
| HTN | b_cell | *GRB14* | -3.78613521 | 0.000484701 | 0.002187427 | 1 |
| HTN | b_cell | *CLIP4* | -3.155498181 | 0.000682014 | 0.002339238 | 1 |

| | | | | | | |
|---|---|---|---|---|---|---|
| HTN | b_cell | *SPTBN1* | -2.141653096 | 0.000505202 | 0.002195028 | 1 |
| HTN | b_cell | *RTN4* | -3.879272229 | 0.000243031 | 0.002180344 | 1 |
| HTN | b_cell | *GALNT3* | 2.778328976 | 0.000199327 | 0.002180344 | 1 |
| HTN | b_cell | *POLE4* | -6.419106985 | 0.001298382 | 0.003186154 | 1 |
| HTN | b_cell | *MRPL19* | -4.032612552 | 0.000363931 | 0.002180344 | 1 |
| HTN | b_cell | *LANCL1* | -5.632303872 | 0.015397988 | 0.020952958 | 1 |
| HTN | b_cell | *WDR75* | -5.523677442 | 0.00644553 | 0.0100904 | 1 |
| HTN | b_cell | *EFEMP1* | -2.178274071 | 0.000354828 | 0.002180344 | 1 |
| HTN | b_cell | *GLS* | -3.939923197 | 0.008844066 | 0.013046138 | 1 |
| HTN | b_cell | *GNLY* | -3.131013346 | 0.000436623 | 0.002180344 | 1 |
| HTN | b_cell | *SF3B1* | -4.153374506 | 0.013950162 | 0.019271516 | 1 |
| HTN | b_cell | *ST3GAL5* | -2.638408658 | 0.000700604 | 0.002366204 | 1 |
| HTN | b_cell | *PLCD4* | -4.935227046 | 0.00026447 | 0.002180344 | 1 |
| HTN | b_cell | *CHMP3* | -8.483018594 | 0.00289899 | 0.005463414 | 1 |
| HTN | b_cell | *ZNF142* | -9.115528189 | 0.000120829 | 0.002180344 | 0.759650753 |
| HTN | b_cell | *WNT6* | -28.6921357 | 0.01302284 | 0.018182233 | 1 |
| HTN | b_cell | *MLPH* | -4.470340336 | 0.000415834 | 0.002180344 | 1 |
| HTN | b_cell | *HDLBP* | -4.275216787 | 0.010107127 | 0.014607702 | 1 |
| HTN | b_cell | *STK25* | -3.990021455 | 0.000471453 | 0.002180489 | 1 |
| HTN | b_cell | *TPO* | -2.934634629 | 0.000716651 | 0.002385168 | 1 |
| HTN | b_cell | *TAF1B* | -5.066729823 | 0.00695221 | 0.010689299 | 1 |
| HTN | b_cell | *STRN* | -4.700279222 | 0.004573018 | 0.007703795 | 1 |
| HTN | b_cell | *CEBPZ* | -3.926311054 | 0.006445513 | 0.0100904 | 1 |
| HTN | b_cell | *PRKD3* | -3.935911375 | 0.006618993 | 0.010285123 | 1 |
| HTN | b_cell | *SLC25A12* | -4.03804174 | 0.000966143 | 0.002709251 | 1 |
| HTN | b_cell | *SOS1* | -3.383476288 | 0.001806137 | 0.003927449 | 1 |
| HTN | b_cell | *WIPF1* | -3.870088677 | 0.001477817 | 0.003453917 | 1 |
| HTN | b_cell | *PNO1* | -4.056030942 | 0.000419491 | 0.002180344 | 1 |
| HTN | b_cell | *THADA* | -4.230689832 | 0.00922844 | 0.013514839 | 1 |
| HTN | b_cell | *ARID3A* | -5.246489047 | 0.001906561 | 0.004071517 | 1 |
| HTN | b_cell | *CD207* | -86.61575341 | 0.002371617 | 0.004723984 | 1 |
| HTN | b_cell | *VAX2* | -8.554763708 | 0.000587719 | 0.002263931 | 1 |
| HTN | b_cell | *MSH6* | -6.050586159 | 0.006369304 | 0.009983499 | 1 |
| HTN | b_cell | *ALMS1* | -4.133916662 | 0.000798556 | 0.002471947 | 1 |
| HTN | b_cell | *BCL9* | -6.272677515 | 0.012377905 | 0.017409371 | 1 |
| HTN | b_cell | *MORN1* | -4.479944926 | 0.000140728 | 0.002180344 | 0.884757243 |
| HTN | b_cell | *GPX7* | 4.428806128 | 0.000651966 | 0.002323165 | 1 |
| HTN | b_cell | *SCP2* | -3.844190906 | 0.001325466 | 0.003224925 | 1 |
| HTN | b_cell | *RALGPS2* | 0.764617511 | 0.000222852 | 0.002180344 | 1 |
| HTN | b_cell | *CEP104* | -3.572285836 | 0.001018914 | 0.002775525 | 1 |
| HTN | b_cell | *TMEM59* | -4.576074653 | 0.002510672 | 0.004923903 | 1 |
| HTN | b_cell | *WRAP73* | -6.469690685 | 0.001397198 | 0.003333657 | 1 |
| HTN | b_cell | *MRPL37* | -4.007990649 | 0.000630192 | 0.002297226 | 1 |
| HTN | b_cell | *ICMT* | -7.074931983 | 0.001294775 | 0.003178543 | 1 |
| HTN | b_cell | *QSOX1* | -6.481603225 | 0.012026989 | 0.016995657 | 1 |
| HTN | b_cell | *KCNC4* | -5.282900861 | 0.000775733 | 0.002438676 | 1 |
| HTN | b_cell | *EDEM3* | -5.094727657 | 0.00505062 | 0.008322672 | 1 |
| HTN | b_cell | *RAP1A* | -4.093455479 | 0.029366417 | 0.037223118 | 1 |
| HTN | b_cell | *CAPZA1* | -5.43030023 | 0.003908055 | 0.00682688 | 1 |
| HTN | b_cell | *DOCK7* | -3.979448872 | 0.002178898 | 0.004433246 | 1 |

| | | | | | | |
|---|---|---|---|---|---|---|
| HTN | b_cell | *FBXO6* | -7.711791691 | 0.000849429 | 0.002543028 | 1 |
| HTN | b_cell | *C1orf21* | -2.890389725 | 4.07E-05 | 0.001539591 | 0.256162912 |
| HTN | b_cell | *LEPR* | -4.534040707 | 0.017302362 | 0.023231616 | 1 |
| HTN | b_cell | *MFN2* | -4.803705055 | 0.000501056 | 0.002195028 | 1 |
| HTN | b_cell | *MIIP* | -7.742353185 | 0.000997607 | 0.002748073 | 1 |
| HTN | b_cell | *SMG7* | -4.649372234 | 0.006001951 | 0.009538489 | 1 |
| HTN | b_cell | *NCF2* | -3.500006202 | 0.000425736 | 0.002180344 | 1 |
| HTN | b_cell | *RGS2* | -8.685774342 | 0.002340767 | 0.004683769 | 1 |
| HTN | b_cell | *OLFML3* | -3.651319145 | 0.000465648 | 0.002180489 | 1 |
| HTN | b_cell | *TNNI3K* | -7.277095258 | 0.021058477 | 0.027628265 | 1 |
| HTN | b_cell | *CFHR3* | -5.85395415 | 0.023343264 | 0.030278338 | 1 |
| HTN | b_cell | *TTF2* | -5.541749824 | 0.002082177 | 0.004303303 | 1 |
| HTN | b_cell | *TMEM9* | -4.375211926 | 0.000156516 | 0.002180344 | 0.984018597 |
| HTN | b_cell | *WARS2* | -4.138505334 | 0.000558995 | 0.002226229 | 1 |
| HTN | b_cell | *OSCP1* | -4.069002201 | 0.00019465 | 0.002180344 | 1 |
| HTN | b_cell | *GNPAT* | -6.365499051 | 0.009557831 | 0.013896874 | 1 |
| HTN | b_cell | *C1orf109* | -4.862311497 | 0.000809142 | 0.002488487 | 1 |
| HTN | b_cell | *TBCE* | -5.712835253 | 0.003579653 | 0.006369964 | 1 |
| HTN | b_cell | *LGALS8* | -4.457371852 | 0.000331418 | 0.002180344 | 1 |
| HTN | b_cell | *MTR* | -4.300237192 | 0.003483746 | 0.006245312 | 1 |
| HTN | b_cell | *KCNQ4* | -6.342289731 | 0.000407567 | 0.002180344 | 1 |
| HTN | b_cell | *ACADM* | -4.782409531 | 0.003319363 | 0.006027971 | 1 |
| HTN | b_cell | *PADI2* | 6.542447449 | 0.000373639 | 0.002180344 | 1 |
| HTN | b_cell | *ZNHIT6* | -4.205428496 | 0.019208604 | 0.025440172 | 1 |
| HTN | b_cell | *PLA2G2D* | 7.45789784 | 0.000263503 | 0.002180344 | 1 |
| HTN | b_cell | *RBBP5* | -4.721553622 | 0.005299396 | 0.008658343 | 1 |
| HTN | b_cell | *GBP3* | -3.286440447 | 0.015090718 | 0.020580335 | 1 |
| HTN | b_cell | *KIF17* | -8.286105628 | 0.002943867 | 0.005524438 | 1 |
| HTN | b_cell | *CD46* | -3.326333426 | 0.002664438 | 0.005135292 | 1 |
| HTN | b_cell | *TMED5* | -4.341163347 | 0.001534438 | 0.003530383 | 1 |
| HTN | b_cell | *DR1* | -5.431051095 | 0.001326007 | 0.003224992 | 1 |
| HTN | b_cell | *CNN3* | -3.682731464 | 0.0006565 | 0.002327958 | 1 |
| HTN | b_cell | *PRRC2C* | -3.756730583 | 0.027993598 | 0.035648319 | 1 |
| HTN | b_cell | *ABCD3* | -5.148159707 | 0.012179589 | 0.017188002 | 1 |
| HTN | b_cell | *VAMP4* | -4.636492121 | 0.004458279 | 0.007542842 | 1 |
| HTN | b_cell | *PTBP2* | -3.786659628 | 0.008174994 | 0.012189374 | 1 |
| HTN | b_cell | *TNFSF4* | -6.806308595 | 0.000205975 | 0.002180344 | 1 |
| HTN | b_cell | *DARS2* | -6.515784316 | 0.001068242 | 0.002845984 | 1 |
| HTN | b_cell | *IRF6* | -8.710583068 | 2.00E-05 | 0.001016197 | 0.126008429 |
| HTN | b_cell | *DIEXF* | -7.835474049 | 0.000499957 | 0.002194994 | 1 |
| HTN | b_cell | *RCAN3* | -4.854618829 | 0.001472269 | 0.003446075 | 1 |
| HTN | b_cell | *RCOR3* | -4.422710482 | 0.004305133 | 0.007341028 | 1 |
| HTN | b_cell | *NSL1* | -4.782200443 | 0.005401223 | 0.008783761 | 1 |
| HTN | b_cell | *RPA2* | -6.491970281 | 0.008601527 | 0.012739175 | 1 |
| HTN | b_cell | *SLC5A9* | 10.59828371 | 0.000558381 | 0.002226229 | 1 |
| HTN | b_cell | *OSBPL9* | -4.274409783 | 0.011834 | 0.016756837 | 1 |
| HTN | b_cell | *TXNDC12* | -5.991793199 | 0.013046367 | 0.018202954 | 1 |
| HTN | b_cell | *ESYT2* | -3.476152042 | 0.005963502 | 0.009494185 | 1 |
| HTN | b_cell | *CD3EAP* | -4.837036748 | 4.85E-05 | 0.001724513 | 0.304732513 |
| HTN | b_cell | *COLEC11* | 9.610573836 | 0.016905985 | 0.022745117 | 1 |

| | | | | | | |
|---|---|---|---|---|---|---|
| HTN | b_cell | *A4GNT* | -5.75319019 | 0.013683232 | 0.018948564 | 1 |
| HTN | b_cell | *KMT2A* | -4.445488512 | 0.007508691 | 0.011394433 | 1 |
| HTN | b_cell | *IFT46* | -6.496297473 | 0.002028212 | 0.004226899 | 1 |
| HTN | b_cell | *CAMSAP2* | -4.292320672 | 0.012459113 | 0.017500099 | 1 |
| HTN | b_cell | *ATF6* | -4.468088169 | 0.004730641 | 0.007901578 | 1 |
| HTN | b_cell | *CASC1* | 5.504608018 | 0.039582764 | 0.048804671 | 1 |
| HTN | b_cell | *LRMP* | 2.684925075 | 0.000145158 | 0.002180344 | 0.912606397 |
| HTN | b_cell | *ATP10B* | -8.70341097 | 0.000374116 | 0.002180344 | 1 |
| HTN | b_cell | *FILIP1* | -2.489908915 | 0.00289259 | 0.00545626 | 1 |
| HTN | b_cell | *SGIP1* | -3.455142678 | 0.000525236 | 0.002195202 | 1 |
| HTN | b_cell | *AKAP7* | -4.208031929 | 0.000435318 | 0.002180344 | 1 |
| HTN | b_cell | *ALDH8A1* | -3.059121531 | 0.018797757 | 0.024974958 | 1 |
| HTN | b_cell | *MED28* | -6.321379473 | 0.0016698 | 0.00373449 | 1 |
| HTN | b_cell | *SLC16A7* | -3.774958162 | 0.002823973 | 0.005370332 | 1 |
| HTN | b_cell | *VAMP8* | -4.238826039 | 0.000794512 | 0.002466715 | 1 |
| HTN | b_cell | *FOXO3* | -3.906381726 | 0.004664808 | 0.007825436 | 1 |
| HTN | b_cell | *ABCG2* | -4.705447612 | 0.000477591 | 0.002180489 | 1 |
| HTN | b_cell | *SPP1* | 5.744977927 | 0.005839496 | 0.009346465 | 1 |
| HTN | b_cell | *FAM47E-STBL* | -4.070406808 | 0.001932132 | 0.004102437 | 1 |
| HTN | b_cell | *MFSD1* | -3.708567102 | 0.000693058 | 0.002357691 | 1 |
| HTN | b_cell | *RAB3GAP2* | -4.444993255 | 0.017811258 | 0.023795023 | 1 |
| HTN | b_cell | *UBN1* | -4.52183799 | 0.002908263 | 0.005475966 | 1 |
| HTN | b_cell | *UCHL3* | -5.79871756 | 0.000369728 | 0.002180344 | 1 |
| HTN | b_cell | *DNAH7* | -3.589290412 | 0.000141481 | 0.002180344 | 0.889489992 |
| HTN | b_cell | *NDUFB3* | -6.170790904 | 0.037850438 | 0.046862092 | 1 |
| HTN | b_cell | *TRPM6* | -3.342384139 | 0.019092973 | 0.025308353 | 1 |
| HTN | b_cell | *GDA* | 11.72613034 | 0.000986091 | 0.002724009 | 1 |
| HTN | b_cell | *C2orf40* | -5.789943321 | 0.000110551 | 0.002180344 | 0.695032987 |
| HTN | b_cell | *CPSF3* | -4.616423691 | 0.000842438 | 0.002536213 | 1 |
| HTN | b_cell | *PIGZ* | -2.477828599 | 0.0077235 | 0.011641727 | 1 |
| HTN | b_cell | *SENP5* | -4.498019264 | 0.003420754 | 0.006158728 | 1 |
| HTN | b_cell | *CCDC92* | -4.441210197 | 0.000487011 | 0.002188291 | 1 |
| HTN | b_cell | *FKBP15* | -4.896184815 | 0.004935416 | 0.008169816 | 1 |
| HTN | b_cell | *CTNNAL1* | -2.966209048 | 0.000177022 | 0.002180344 | 1 |
| HTN | b_cell | *WDR34* | 5.75379054 | 0.002196717 | 0.004463722 | 1 |
| HTN | b_cell | *TRIM32* | -8.946903355 | 0.000489525 | 0.002188936 | 1 |
| HTN | b_cell | *PHF19* | -5.035059297 | 0.000324379 | 0.002180344 | 1 |
| HTN | b_cell | *BSPRY* | 6.673423345 | 0.000240014 | 0.002180344 | 1 |
| HTN | b_cell | *PPP6C* | -3.934239838 | 0.001291767 | 0.003173543 | 1 |
| HTN | b_cell | *HDHD3* | -3.35222662 | 5.37E-06 | 0.000406808 | 0.033765026 |
| HTN | b_cell | *HSDL2* | -2.072759508 | 0.003811509 | 0.006702925 | 1 |
| HTN | b_cell | *INVS* | -4.084563809 | 0.005313764 | 0.008674814 | 1 |
| HTN | b_cell | *KDSR* | -3.882302484 | 0.000730095 | 0.002396922 | 1 |
| HTN | b_cell | *VSX2* | -8.470024666 | 0.00353481 | 0.006309867 | 1 |
| HTN | b_cell | *DNAL1* | -4.538863448 | 0.000746976 | 0.00240927 | 1 |
| HTN | b_cell | *TTLL5* | -4.720925434 | 0.023058169 | 0.029957989 | 1 |
| HTN | b_cell | *FLVCR2* | -3.759668637 | 0.000363331 | 0.002180344 | 1 |
| HTN | b_cell | *PPP4R4* | 5.111014099 | 0.000190463 | 0.002180344 | 1 |
| HTN | b_cell | *ZC2HC1C* | -4.746919281 | 0.000339135 | 0.002180344 | 1 |
| HTN | b_cell | *ALDH6A1* | -4.429090113 | 0.000662702 | 0.002327958 | 1 |

| | | | | | | |
|---|---|---|---|---|---|---|
| HTN | b_cell | *ESRRB* | -4.403666522 | 0.000435346 | 0.002180344 | 1 |
| HTN | b_cell | *COQ6* | 6.950252975 | 0.002300948 | 0.004621743 | 1 |
| HTN | b_cell | *GPR75* | -26.9864893 | 0.000203275 | 0.002180344 | 1 |
| HTN | b_cell | *KLHL29* | -3.679179518 | 0.000896032 | 0.002599737 | 1 |
| HTN | b_cell | *ATAD2B* | -4.219915283 | 0.004882637 | 0.008097614 | 1 |
| HTN | b_cell | *FKBP1B* | 9.71145049 | 9.94E-06 | 0.000618838 | 0.062502591 |
| HTN | b_cell | *ATL2* | -4.727566116 | 0.0036274 | 0.006436767 | 1 |
| HTN | b_cell | *CNRIP1* | -5.534640255 | 0.00197202 | 0.004156248 | 1 |
| HTN | b_cell | *CRIPT* | -7.939759298 | 0.001974545 | 0.004157389 | 1 |
| HTN | b_cell | *EPCAM* | -8.010289781 | 0.005265201 | 0.008613666 | 1 |
| HTN | b_cell | *SLC17A5* | -4.912701015 | 0.010894283 | 0.015580609 | 1 |
| HTN | b_cell | *OGFRL1* | -4.50541068 | 0.00375496 | 0.006622001 | 1 |
| HTN | b_cell | *CUTC* | -4.807136002 | 0.003185451 | 0.005842161 | 1 |
| HTN | b_cell | *PYROXD2* | -1.994521538 | 0.02031874 | 0.026724669 | 1 |
| HTN | b_cell | *MXI1* | -5.278659347 | 0.009166058 | 0.013449387 | 1 |
| HTN | b_cell | *C10orf88* | -4.569028391 | 0.000650867 | 0.002323165 | 1 |
| HTN | b_cell | *HELLS* | 3.996561597 | 0.003007636 | 0.00560599 | 1 |
| HTN | b_cell | *AVPI1* | -8.883017721 | 0.000538143 | 0.002208426 | 1 |
| HTN | b_cell | *WDR11* | -4.369873618 | 0.019375206 | 0.025612263 | 1 |
| HTN | b_cell | *C10orf76* | -4.116513397 | 0.002295552 | 0.004613854 | 1 |
| HTN | b_cell | *KCNIP2* | -2.528738707 | 0.00161503 | 0.003652902 | 1 |
| HTN | b_cell | *C10orf95* | 7.474152451 | 0.001674976 | 0.003742209 | 1 |
| HTN | b_cell | *KANSL1* | -3.871353883 | 0.001964473 | 0.004148529 | 1 |
| HTN | b_cell | *HOXB5* | -5.671014293 | 0.000144341 | 0.002180344 | 0.90747393 |
| HTN | b_cell | *HOXB3* | -4.371334952 | 0.001017924 | 0.002775525 | 1 |
| HTN | b_cell | *CD274* | -4.854873237 | 0.00083531 | 0.002525679 | 1 |
| HTN | b_cell | *NUP43* | -4.949181581 | 0.005060909 | 0.008335849 | 1 |
| HTN | b_cell | *LRP11* | -3.520861109 | 0.000253539 | 0.002180344 | 1 |
| HTN | b_cell | *CCDC170* | -2.496958118 | 0.001085255 | 0.002865833 | 1 |
| HTN | b_cell | *CYSTM1* | -4.14316013 | 0.002848057 | 0.005390046 | 1 |
| HTN | b_cell | *WDR55* | 9.319947576 | 0.015864123 | 0.021513749 | 1 |
| HTN | b_cell | *PCDHB10* | 3.175441651 | 2.87E-05 | 0.001261541 | 0.18031311 |
| HTN | b_cell | *PCDHB14* | -11.35824823 | 0.002625572 | 0.005082532 | 1 |
| HTN | b_cell | *SEC16B* | -8.540035883 | 0.004998108 | 0.008258371 | 1 |
| HTN | b_cell | *ACAT2* | -8.035394743 | 0.009660256 | 0.014016624 | 1 |
| HTN | b_cell | *SNX19* | -5.866645155 | 0.001875194 | 0.004027792 | 1 |
| HTN | b_cell | *MASTL* | -4.75449272 | 0.000598341 | 0.002274361 | 1 |
| HTN | b_cell | *MRC1* | -2.202295118 | 0.000750923 | 0.002413626 | 1 |
| HTN | b_cell | *IQSEC3* | -10.14231636 | 0.001921535 | 0.004091237 | 1 |
| HTN | b_cell | *CCDC77* | -7.827799798 | 0.006110527 | 0.009679824 | 1 |
| HTN | b_cell | *ENOX1* | -3.81219563 | 0.000633092 | 0.002301586 | 1 |
| HTN | b_cell | *WBP4* | -4.327015937 | 0.001324973 | 0.003224925 | 1 |
| HTN | b_cell | *SMAD9* | -3.569867595 | 0.000416228 | 0.002180344 | 1 |
| HTN | b_cell | *SIL1* | -4.547044823 | 0.008092522 | 0.012096455 | 1 |
| HTN | b_cell | *ARL1* | -4.663804434 | 0.00052948 | 0.002195202 | 1 |
| HTN | b_cell | *APAF1* | -4.845850195 | 0.002386715 | 0.004742502 | 1 |
| HTN | b_cell | *EPHX2* | -4.040042719 | 1.32E-05 | 0.000765656 | 0.082690876 |
| HTN | b_cell | *ZNF706* | -4.703173951 | 0.022281266 | 0.029050667 | 1 |
| HTN | b_cell | *CRISPLD1* | -5.389111785 | 0.000421408 | 0.002180344 | 1 |
| HTN | b_cell | *RDH10* | -3.539214891 | 0.000417247 | 0.002180344 | 1 |

| | | | | | | |
|---|---|---|---|---|---|---|
| HTN | b_cell | *TBX4* | -16.17061846 | 0.002355373 | 0.004701026 | 1 |
| HTN | b_cell | *TRIM6* | -7.815280974 | 8.06E-06 | 0.000545045 | 0.050689201 |
| HTN | b_cell | *CEP89* | -4.409308845 | 0.005316376 | 0.008674814 | 1 |
| HTN | b_cell | *ECHDC2* | -2.467942735 | 0.000408142 | 0.002180344 | 1 |
| HTN | b_cell | *PRB2* | -4.939975266 | 0.000819009 | 0.002501997 | 1 |
| HTN | b_cell | *ZSCAN18* | -6.468791933 | 0.002750125 | 0.005263329 | 1 |
| HTN | b_cell | *ZNF211* | -5.079647465 | 0.005628418 | 0.009072412 | 1 |
| HTN | b_cell | *PDZRN3* | -1.875370949 | 0.000416083 | 0.002180344 | 1 |
| HTN | b_cell | *LHX4* | 8.683232961 | 0.019115203 | 0.025332479 | 1 |
| HTN | b_cell | *SEC22A* | -3.868982396 | 0.032467397 | 0.040734889 | 1 |
| HTN | b_cell | *CSTA* | -3.439245356 | 0.008627659 | 0.01276915 | 1 |
| HTN | b_cell | *POPDC2* | 7.045172631 | 0.000410857 | 0.002180344 | 1 |
| HTN | b_cell | *B4GALT4* | 7.592823327 | 0.002364204 | 0.004711173 | 1 |
| HTN | b_cell | *CD80* | 4.82507985 | 0.000310818 | 0.002180344 | 1 |
| HTN | b_cell | *CAT* | -2.711201329 | 0.000508799 | 0.002195202 | 1 |
| HTN | b_cell | *GJB6* | -204.8972997 | 8.06E-05 | 0.002176137 | 0.506738945 |
| HTN | b_cell | *TMEM39B* | -5.043549853 | 0.000420819 | 0.002180344 | 1 |
| HTN | b_cell | *RNF115* | -4.577766674 | 0.010901783 | 0.015584245 | 1 |
| HTN | b_cell | *POLR3GL* | 8.99597559 | 0.002776994 | 0.005300219 | 1 |
| HTN | b_cell | *TNFSF10* | -4.576472039 | 0.001514266 | 0.003504366 | 1 |
| HTN | b_cell | *LIAS* | -3.19439327 | 0.000454396 | 0.002180344 | 1 |
| HTN | b_cell | *CLCC1* | -7.482060692 | 0.001759669 | 0.003854718 | 1 |
| HTN | b_cell | *GPSM2* | -3.836200347 | 0.002021841 | 0.004222363 | 1 |
| HTN | b_cell | *CXCR4* | 1.974021863 | 0.000304531 | 0.002180344 | 1 |
| HTN | b_cell | *ACVR2A* | -4.317703574 | 0.005638127 | 0.009084291 | 1 |
| HTN | b_cell | *POLK* | -4.407197757 | 0.007815438 | 0.011757755 | 1 |
| HTN | b_cell | *FLT3* | 4.941530761 | 0.000278768 | 0.002180344 | 1 |
| HTN | b_cell | *MTIF3* | -4.028963075 | 0.003415555 | 0.006151129 | 1 |
| HTN | b_cell | *RASL11A* | -7.467691635 | 0.001095551 | 0.002878282 | 1 |
| HTN | b_cell | *FMOD* | -3.713858968 | 0.000452045 | 0.002180344 | 1 |
| HTN | b_cell | *SERAC1* | -3.857640243 | 0.000339137 | 0.002180344 | 1 |
| HTN | b_cell | *LDB3* | -4.312751904 | 0.000242463 | 0.002180344 | 1 |
| HTN | b_cell | *FAM35A* | -5.428337317 | 0.004805253 | 0.007992229 | 1 |
| HTN | b_cell | *FAM213A* | -2.455606627 | 0.000468524 | 0.002180489 | 1 |
| HTN | b_cell | *NAA60* | -4.966521987 | 0.000289547 | 0.002180344 | 1 |
| HTN | b_cell | *ODF2L* | -4.998112917 | 0.00348194 | 0.006243855 | 1 |
| HTN | b_cell | *SPATA1* | -4.943927242 | 0.00634142 | 0.00995717 | 1 |
| HTN | b_cell | *LRRC39* | -2.544407762 | 0.034825921 | 0.043382319 | 1 |
| HTN | b_cell | *CCDC18* | -4.409368841 | 0.001593096 | 0.003611898 | 1 |
| HTN | b_cell | *RPAP2* | -4.190964068 | 0.002411679 | 0.004781787 | 1 |
| HTN | b_cell | *BBS9* | -4.003299003 | 0.005914437 | 0.009431708 | 1 |
| HTN | b_cell | *KIAA0087* | -14.16938534 | 0.000455958 | 0.002180344 | 1 |
| HTN | b_cell | *KLHL7* | -5.27554009 | 0.022091803 | 0.028839528 | 1 |
| HTN | b_cell | *HERPUD2* | -5.040015797 | 0.002456683 | 0.004847823 | 1 |
| HTN | b_cell | *CBX3* | -7.000477904 | 0.000262708 | 0.002180344 | 1 |
| HTN | b_cell | *FAM126A* | -4.249126125 | 0.014794592 | 0.0202644 | 1 |
| HTN | b_cell | *FKBP9* | -3.334853183 | 0.000607457 | 0.002281385 | 1 |
| HTN | b_cell | *CCZ1* | -5.331980856 | 0.015916603 | 0.021573588 | 1 |
| HTN | b_cell | *POLM* | -2.42102645 | 0.024099302 | 0.031143332 | 1 |
| HTN | b_cell | *SMU1* | -5.830528736 | 0.01711778 | 0.023010366 | 1 |

| | | | | | | |
|---|---|---|---|---|---|---|
| HTN | b_cell | *GLIPR2* | -5.818985173 | 0.005731361 | 0.009210907 | 1 |
| HTN | b_cell | *SLC25A51* | -4.962722145 | 1.97E-05 | 0.001016068 | 0.123548033 |
| HTN | b_cell | *DCAF10* | -4.166904433 | 0.002865533 | 0.005416598 | 1 |
| HTN | b_cell | *BICC1* | -2.440350487 | 0.000129792 | 0.002180344 | 0.816001224 |
| HTN | b_cell | *CISD1* | -3.721050635 | 0.00183181 | 0.003957685 | 1 |
| HTN | b_cell | *SLC25A16* | -2.312814614 | 0.00332163 | 0.006030347 | 1 |
| HTN | b_cell | *ACADS* | -3.633133818 | 0.000295027 | 0.002180344 | 1 |
| HTN | b_cell | *SSPN* | -4.259240706 | 0.000496897 | 0.002194324 | 1 |
| HTN | b_cell | *CCDC91* | -3.648303977 | 0.027504049 | 0.03508887 | 1 |
| HTN | b_cell | *PKN1* | -5.047269812 | 0.00466299 | 0.007825436 | 1 |
| HTN | b_cell | *EBPL* | -4.187419674 | 0.007077839 | 0.010847969 | 1 |
| HTN | b_cell | *ATP7B* | -2.086933133 | 0.015005019 | 0.020507947 | 1 |
| HTN | b_cell | *NLN* | -3.137889814 | 0.000205695 | 0.002180344 | 1 |
| HTN | b_cell | *CENPK* | 3.039808451 | 0.001320088 | 0.003218935 | 1 |
| HTN | b_cell | *OPTN* | -4.045989336 | 0.000446767 | 0.002180344 | 1 |
| HTN | b_cell | *ATF1* | -6.805304148 | 0.002938112 | 0.005518945 | 1 |
| HTN | b_cell | *TSFM* | -5.446620064 | 0.000644052 | 0.002316643 | 1 |
| HTN | b_cell | *ARHGAP9* | -8.350755511 | 0.001486165 | 0.003464107 | 1 |
| HTN | b_cell | *SPATS2* | -4.774186878 | 0.003998714 | 0.006950488 | 1 |
| HTN | b_cell | *PDE1B* | -6.329503418 | 0.008847041 | 0.013047466 | 1 |
| HTN | b_cell | *SMUG1* | -2.77241987 | 0.010704779 | 0.015365513 | 1 |
| HTN | b_cell | *SARDH* | -11.23358374 | 0.000416346 | 0.002180344 | 1 |
| HTN | b_cell | *DBH* | -24.75403735 | 0.016381823 | 0.022106143 | 1 |
| HTN | b_cell | *STIL* | -6.884350907 | 0.002595128 | 0.005048135 | 1 |
| HTN | b_cell | *TTC21B* | -4.66045048 | 0.012182239 | 0.017188002 | 1 |
| HTN | b_cell | *NMI* | -6.464481171 | 0.006359553 | 0.009975721 | 1 |
| HTN | b_cell | *ACVR1C* | -2.07181709 | 0.001922286 | 0.004091237 | 1 |
| HTN | b_cell | *PLA2G12A* | -5.600038203 | 0.035004384 | 0.043561473 | 1 |
| HTN | b_cell | *C4BPB* | -7.770897688 | 0.000137134 | 0.002180344 | 0.862163164 |
| HTN | b_cell | *RAB38* | -5.966746592 | 0.000431576 | 0.002180344 | 1 |
| HTN | b_cell | *AGO2* | -5.076193941 | 0.003910464 | 0.006829191 | 1 |
| HTN | b_cell | *CHPF* | -6.338678251 | 4.30E-05 | 0.001579949 | 0.270171269 |
| HTN | b_cell | *DNPEP* | -5.084803025 | 0.001127071 | 0.002930908 | 1 |
| HTN | b_cell | *MOGAT1* | -3.62429666 | 0.000504586 | 0.002195028 | 1 |
| HTN | b_cell | *FAM210B* | -4.936020204 | 0.000914663 | 0.002631803 | 1 |
| HTN | b_cell | *SNX21* | -2.790773922 | 0.000415316 | 0.002180344 | 1 |
| HTN | b_cell | *WFDC3* | -4.406060336 | 2.26E-05 | 0.001082615 | 0.141822524 |
| HTN | b_cell | *SDC4* | -4.805150201 | 0.000471946 | 0.002180489 | 1 |
| HTN | b_cell | *VAPB* | -4.709389117 | 0.004104545 | 0.007099112 | 1 |
| HTN | b_cell | *TOX2* | -4.172578246 | 8.20E-06 | 0.000548538 | 0.051562581 |
| HTN | b_cell | *ARFGEF2* | -4.411388566 | 0.006042202 | 0.009587352 | 1 |
| HTN | b_cell | *CDH26* | -4.895653872 | 0.000211422 | 0.002180344 | 1 |
| HTN | b_cell | *STX16* | -5.154263958 | 0.003323444 | 0.006031898 | 1 |
| HTN | b_cell | *RNF114* | -5.429321072 | 0.002900691 | 0.005464981 | 1 |
| HTN | b_cell | *KCNK15* | 4.380832132 | 0.00026888 | 0.002180344 | 1 |
| HTN | b_cell | *NEURL2* | -4.373432533 | 0.002219937 | 0.004497823 | 1 |
| HTN | b_cell | *PEPD* | -2.80852719 | 0.001880304 | 0.004034632 | 1 |
| HTN | b_cell | *CHST8* | -13.24389257 | 0.005804553 | 0.009297636 | 1 |
| HTN | b_cell | *STAMBP* | -5.945696677 | 0.009600458 | 0.013945952 | 1 |
| HTN | b_cell | *USP22* | -4.838914021 | 0.022337279 | 0.02911766 | 1 |

| | | | | | | |
|---|---|---|---|---|---|---|
| HTN | b_cell | *HIF3A* | -2.866103517 | 0.000643023 | 0.002316643 | 1 |
| HTN | b_cell | *BTN2A2* | 4.199714408 | 4.35E-06 | 0.00036476 | 0.027325023 |
| HTN | b_cell | *MRS2* | -5.682713665 | 0.001500324 | 0.003480291 | 1 |
| HTN | b_cell | *SNRPC* | -10.66160642 | 0.00025745 | 0.002180344 | 1 |
| HTN | b_cell | *SERPINB6* | -4.578021295 | 0.001524136 | 0.003516661 | 1 |
| HTN | b_cell | *PEX6* | -3.433712201 | 0.00039189 | 0.002180344 | 1 |
| HTN | b_cell | *NQO2* | -4.371777816 | 0.000968968 | 0.002711127 | 1 |
| HTN | b_cell | *ZNF391* | -4.85950053 | 3.16E-08 | 1.05E-05 | 0.000198702 |
| HTN | b_cell | *RPS10* | 5.751472152 | 0.000298535 | 0.002180344 | 1 |
| HTN | b_cell | *HIST1H2BJ* | 5.839243909 | 0.000190923 | 0.002180344 | 1 |
| HTN | b_cell | *MED20* | -5.57421911 | 0.000105781 | 0.002180344 | 0.665042471 |
| HTN | b_cell | *TBCC* | 10.08708412 | 0.001980012 | 0.004161982 | 1 |
| HTN | b_cell | *TCP11* | 12.58352899 | 0.00109295 | 0.00287385 | 1 |
| HTN | b_cell | *KLHDC3* | -4.201670352 | 0.000304635 | 0.002180344 | 1 |
| HTN | b_cell | *MEA1* | -5.549654099 | 0.003306164 | 0.006005737 | 1 |
| HTN | b_cell | *KLHL31* | -2.562619554 | 0.001789704 | 0.003900641 | 1 |
| HTN | b_cell | *COL21A1* | -3.51933837 | 0.000281582 | 0.002180344 | 1 |
| HTN | b_cell | *RPP40* | -7.171266452 | 0.001165096 | 0.002995893 | 1 |
| HTN | b_cell | *NUP153* | -4.710154175 | 0.031794194 | 0.03997802 | 1 |
| HTN | b_cell | *EEF1E1* | -6.19897629 | 0.000661782 | 0.002327958 | 1 |
| HTN | b_cell | *EREG* | 5.635480204 | 0.000237699 | 0.002180344 | 1 |
| HTN | b_cell | *CNOT1* | -4.302289544 | 0.028990664 | 0.0368062 | 1 |
| HTN | b_cell | *MT1G* | -33.96253593 | 0.000276608 | 0.002180344 | 1 |
| HTN | b_cell | *CLYBL* | -5.178584933 | 0.005568872 | 0.009005015 | 1 |
| HTN | b_cell | *TMTC4* | -3.775658979 | 0.000475858 | 0.002180489 | 1 |
| HTN | b_cell | *KIF25* | -10.22422936 | 0.001934951 | 0.004107035 | 1 |
| HTN | b_cell | *TEKT3* | -4.941099511 | 0.002665481 | 0.005135728 | 1 |
| HTN | b_cell | *MRPS7* | -6.091426527 | 0.038377391 | 0.04745843 | 1 |
| HTN | b_cell | *GGA3* | -5.146533311 | 0.001196353 | 0.003033854 | 1 |
| HTN | b_cell | *NUP85* | -5.474802486 | 0.000628129 | 0.002295161 | 1 |
| HTN | b_cell | *NT5C* | -6.885372318 | 0.024746706 | 0.031914367 | 1 |
| HTN | b_cell | *OPRL1* | -16.59392113 | 5.01E-10 | 9.64E-07 | 3.15E-06 |
| HTN | b_cell | *PPDPF* | -7.863827728 | 0.018154839 | 0.024222981 | 1 |
| HTN | b_cell | *CHCHD5* | -13.72879195 | 0.000779013 | 0.002445161 | 1 |
| HTN | b_cell | *PAX8* | -5.32459269 | 0.002570752 | 0.005014681 | 1 |
| HTN | b_cell | *POLR1B* | -3.777230622 | 0.002510901 | 0.004923903 | 1 |
| HTN | b_cell | *SLC25A23* | -5.549969139 | 0.002624282 | 0.005082532 | 1 |
| HTN | b_cell | *GTF2F1* | -7.339015257 | 0.002090261 | 0.004317172 | 1 |
| HTN | b_cell | *CLPP* | -4.264361021 | 0.000834531 | 0.002524879 | 1 |
| HTN | b_cell | *SH2D3A* | -5.966192778 | 0.000354273 | 0.002180344 | 1 |
| HTN | b_cell | *TRIP10* | -3.301337757 | 0.000615777 | 0.002286342 | 1 |
| HTN | b_cell | *GPR108* | -4.19330133 | 0.026485562 | 0.03393412 | 1 |
| HTN | b_cell | *RTN2* | -5.658471747 | 0.000344564 | 0.002180344 | 1 |
| HTN | b_cell | *SDCBP2* | -10.89674233 | 0.014664287 | 0.020099057 | 1 |
| HTN | b_cell | *PANK2* | -4.921121148 | 0.002382885 | 0.004736389 | 1 |
| HTN | b_cell | *PSMF1* | -4.897070106 | 0.000864309 | 0.002559431 | 1 |
| HTN | b_cell | *DTD1* | -5.270201472 | 0.032223835 | 0.040477772 | 1 |
| HTN | b_cell | *RRBP1* | -4.131131336 | 0.006541989 | 0.010210895 | 1 |
| HTN | b_cell | *BMP2* | -6.652673209 | 0.000377857 | 0.002180344 | 1 |
| HTN | b_cell | *DSTN* | -4.353410604 | 0.001771846 | 0.003873294 | 1 |

| | | | | | | |
|---|---|---|---|---|---|---|
| HTN | b_cell | *MGME1* | -3.402167996 | 0.000425805 | 0.002180344 | 1 |
| HTN | b_cell | *TMEM74B* | -9.472941069 | 0.019007606 | 0.025205826 | 1 |
| HTN | b_cell | *FAM110A* | -5.489787119 | 0.000433079 | 0.002180344 | 1 |
| HTN | b_cell | *MRPS26* | -2.753726299 | 0.031306227 | 0.039427534 | 1 |
| HTN | b_cell | *NCLN* | -6.649921067 | 0.002590624 | 0.005042493 | 1 |
| HTN | b_cell | *ZNF436* | -4.110251173 | 0.000617905 | 0.002286504 | 1 |
| HTN | b_cell | *GDF5* | -9.424574335 | 0.000178936 | 0.002180344 | 1 |
| HTN | b_cell | *MMP24* | -4.271770249 | 0.003566614 | 0.006350412 | 1 |
| HTN | b_cell | *UROD* | -8.427159179 | 0.001587525 | 0.003601866 | 1 |
| HTN | b_cell | *ST3GAL3* | -4.214363382 | 0.004372198 | 0.007430882 | 1 |
| HTN | b_cell | *KLC1* | -4.329085972 | 0.025441491 | 0.032703058 | 1 |
| HTN | b_cell | *XRCC3* | -4.168346128 | 0.001679861 | 0.003745219 | 1 |
| HTN | b_cell | *MCF2L* | -3.4046527 | 0.000568296 | 0.002232844 | 1 |
| HTN | b_cell | *F10* | -3.35400799 | 0.000416386 | 0.002180344 | 1 |
| HTN | b_cell | *PCID2* | -4.259869147 | 0.002851442 | 0.005394829 | 1 |
| HTN | b_cell | *PROZ* | 6.997048955 | 0.000247509 | 0.002180344 | 1 |
| HTN | b_cell | *IGFLR1* | 16.32000092 | 0.000315569 | 0.002180344 | 1 |
| HTN | b_cell | *PRR12* | -5.160454582 | 0.00459902 | 0.007739772 | 1 |
| HTN | b_cell | *TSKS* | -5.552794351 | 0.000433278 | 0.002180344 | 1 |
| HTN | b_cell | *FLRT1* | -6.140346015 | 4.99E-05 | 0.001750903 | 0.313515525 |
| HTN | b_cell | *DNAJC8* | -5.454275421 | 0.001195551 | 0.003033265 | 1 |
| HTN | b_cell | *ZNF384* | -7.242961001 | 0.012340949 | 0.017371692 | 1 |
| HTN | b_cell | *EMG1* | -7.384084491 | 0.008830071 | 0.013028552 | 1 |
| HTN | b_cell | *ATG14* | -4.025047459 | 0.001072803 | 0.002850682 | 1 |
| HTN | b_cell | *RHOJ* | -2.884071399 | 0.000412504 | 0.002180344 | 1 |
| HTN | b_cell | *L3HYPDH* | -2.542544483 | 0.002638745 | 0.005101411 | 1 |
| HTN | b_cell | *ZBTB1* | -4.027361237 | 0.003390157 | 0.006117913 | 1 |
| HTN | b_cell | *PZP* | -3.059828287 | 0.000645628 | 0.002319464 | 1 |
| HTN | b_cell | *RHOT1* | -3.161226116 | 0.01652585 | 0.022281368 | 1 |
| HTN | b_cell | *WDR60* | -3.507393522 | 0.002713861 | 0.005209784 | 1 |
| HTN | b_cell | *AIF1L* | -4.088476438 | 0.000419696 | 0.002180344 | 1 |
| HTN | b_cell | *MAP2K2* | -4.639184258 | 0.004261653 | 0.00728665 | 1 |
| HTN | b_cell | *CANX* | -4.93360625 | 0.018419069 | 0.024523653 | 1 |
| HTN | b_cell | *IPPK* | -5.320765879 | 0.000349304 | 0.002180344 | 1 |
| HTN | b_cell | *FGD3* | -4.655217049 | 0.000695163 | 0.002359111 | 1 |
| HTN | b_cell | *ABHD8* | -7.771223022 | 0.000219839 | 0.002180344 | 1 |
| HTN | b_cell | *MASP1* | -4.590421415 | 0.001871123 | 0.004022913 | 1 |
| HTN | b_cell | *ATP13A4* | -6.828788844 | 0.000181175 | 0.002180344 | 1 |
| HTN | b_cell | *HRASLS* | -8.329033602 | 0.001076786 | 0.002854029 | 1 |
| HTN | b_cell | *HELB* | -6.065860307 | 0.001242191 | 0.00310894 | 1 |
| HTN | b_cell | *RAP1B* | -3.98162194 | 0.01316041 | 0.018336862 | 1 |
| HTN | b_cell | *RAB3IP* | 7.351329112 | 0.023962562 | 0.03101763 | 1 |
| HTN | b_cell | *LRRC61* | -5.409025781 | 9.00E-05 | 0.002180344 | 0.565995727 |
| HTN | b_cell | *IDUA* | 7.181482947 | 0.000745959 | 0.002409022 | 1 |
| HTN | b_cell | *EMC1* | -6.96325341 | 0.001848279 | 0.00398086 | 1 |
| HTN | b_cell | *PLA2G5* | -5.333869121 | 0.000541494 | 0.002213326 | 1 |
| HTN | b_cell | *UBR4* | -4.100247904 | 0.003114311 | 0.005751959 | 1 |
| HTN | b_cell | *SIN3B* | -5.578967625 | 0.011803215 | 0.016717012 | 1 |
| HTN | b_cell | *WFIKKN1* | -43.23324694 | 0.001423213 | 0.003374109 | 1 |
| HTN | b_cell | *WDR24* | -9.466551934 | 0.001364178 | 0.003283533 | 1 |

| | | | | | | |
|---|---|---|---|---|---|---|
| HTN | b_cell | *CHTF18* | 11.19752572 | 0.013183919 | 0.018354141 | 1 |
| HTN | b_cell | *SMARCA4* | -4.886734303 | 0.004321898 | 0.007367617 | 1 |
| HTN | b_cell | *TICAM1* | -7.456813232 | 0.003164065 | 0.005814813 | 1 |
| HTN | b_cell | *AAMP* | -4.885282859 | 0.000714861 | 0.002383123 | 1 |
| HTN | b_cell | *PNKD* | -5.811539056 | 0.001797796 | 0.003916404 | 1 |
| HTN | b_cell | *ZNF835* | -6.244933231 | 0.002818567 | 0.005363296 | 1 |
| HTN | b_cell | *HIP1* | -3.603197365 | 0.00205892 | 0.004270681 | 1 |
| HTN | b_cell | *PTPN12* | -3.898264511 | 0.003663536 | 0.006491728 | 1 |
| HTN | b_cell | *POR* | -3.373159935 | 0.0006232 | 0.002291037 | 1 |
| HTN | b_cell | *FGL2* | -4.739699079 | 0.001140625 | 0.002953505 | 1 |
| HTN | b_cell | *STYXL1* | -6.912998133 | 0.000879068 | 0.002587407 | 1 |
| HTN | b_cell | *STEAP4* | -5.215092734 | 0.002841904 | 0.00538751 | 1 |
| HTN | b_cell | *GNAI1* | -2.318096397 | 0.000515185 | 0.002195202 | 1 |
| HTN | b_cell | *SGCE* | -3.334520107 | 0.000722726 | 0.002388914 | 1 |
| HTN | b_cell | *RBM48* | -3.516125279 | 0.00037253 | 0.002180344 | 1 |
| HTN | b_cell | *SRD5A3* | -5.44660941 | 0.000536804 | 0.002207251 | 1 |
| HTN | b_cell | *DGCR6L* | 7.405526695 | 9.58E-05 | 0.002180344 | 0.602255567 |
| HTN | b_cell | *VPREB3* | -73.7209434 | 0.000110632 | 0.002180344 | 0.69554413 |
| HTN | b_cell | *MGAT3* | -4.752144743 | 0.000393251 | 0.002180344 | 1 |
| HTN | b_cell | *ATF4* | -6.308991052 | 0.019276717 | 0.025508886 | 1 |
| HTN | b_cell | *A4GALT* | -4.117487211 | 0.000276413 | 0.002180344 | 1 |
| HTN | b_cell | *CDC42EP1* | -4.523561403 | 0.000557423 | 0.002226229 | 1 |
| HTN | b_cell | *TPST2* | -3.279186101 | 0.00053256 | 0.002195543 | 1 |
| HTN | b_cell | *TST* | -3.908276989 | 0.001284753 | 0.003168788 | 1 |
| HTN | b_cell | *APOL5* | -34.14532154 | 1.80E-06 | 0.000186095 | 0.011286064 |
| HTN | b_cell | *RIBC2* | 5.373293642 | 3.59E-05 | 0.001456416 | 0.225744547 |
| HTN | b_cell | *EMC4* | -6.017580333 | 0.000649113 | 0.002323108 | 1 |
| HTN | b_cell | *SPECC1* | -3.122491462 | 0.000659351 | 0.002327958 | 1 |
| HTN | b_cell | *POT1* | -5.562818294 | 0.016305042 | 0.022030905 | 1 |
| HTN | b_cell | *CDHR3* | -3.04749444 | 0.000342327 | 0.002180344 | 1 |
| HTN | b_cell | *PRKRIP1* | -4.814606089 | 0.001653112 | 0.003713691 | 1 |
| HTN | b_cell | *FOXP2* | -2.026315176 | 0.00052114 | 0.002195202 | 1 |
| HTN | b_cell | *FLNC* | -4.90487912 | 0.000941687 | 0.002668043 | 1 |
| HTN | b_cell | *IRF5* | -5.361091124 | 4.16E-05 | 0.001546827 | 0.261413789 |
| HTN | b_cell | *KLHDC10* | -4.509113209 | 0.000230934 | 0.002180344 | 1 |
| HTN | b_cell | *HOXD3* | -3.663626597 | 0.000224456 | 0.002180344 | 1 |
| HTN | b_cell | *PDE11A* | -4.781320722 | 0.009891046 | 0.014315148 | 1 |
| HTN | b_cell | *CHN1* | -2.805413503 | 0.000421575 | 0.002180344 | 1 |
| HTN | b_cell | *GAD1* | 5.683109046 | 0.000243174 | 0.002180344 | 1 |
| HTN | b_cell | *ORMDL1* | -2.560465999 | 0.019525929 | 0.025800655 | 1 |
| HTN | b_cell | *HOXD9* | -11.47075702 | 0.000258544 | 0.002180344 | 1 |
| HTN | b_cell | *PSMG2* | -4.136930831 | 0.002466192 | 0.004856384 | 1 |
| HTN | b_cell | *TWSG1* | -5.970753871 | 0.013429531 | 0.018634178 | 1 |
| HTN | b_cell | *WDFY4* | 2.358571151 | 0.001002434 | 0.002756353 | 1 |
| HTN | b_cell | *EIF2AK4* | -5.975665194 | 0.000599014 | 0.002274361 | 1 |
| HTN | b_cell | *TMOD2* | -4.795307225 | 0.001085275 | 0.002865833 | 1 |
| HTN | b_cell | *IVD* | -5.981455318 | 0.00756688 | 0.01146613 | 1 |
| HTN | b_cell | *KNSTRN* | -4.139319767 | 2.98E-05 | 0.001302192 | 0.187515603 |
| HTN | b_cell | *ARPP19* | -5.324762206 | 0.008523296 | 0.012629262 | 1 |
| HTN | b_cell | *VPS13C* | -3.746053126 | 0.01977877 | 0.026107313 | 1 |

| | | | | | | |
|---|---|---|---|---|---|---|
| HTN | b_cell | *CALML4* | -17.74646053 | 0.000990489 | 0.002732407 | 1 |
| HTN | b_cell | *ACKR4* | -5.636793911 | 0.001593774 | 0.003612133 | 1 |
| HTN | b_cell | *ANAPC13* | -4.290850477 | 0.000102195 | 0.002180344 | 0.642497208 |
| HTN | b_cell | *SUMF2* | -5.653674188 | 0.000894887 | 0.002599737 | 1 |
| HTN | b_cell | *SPCS3* | -5.869346138 | 0.023408073 | 0.03034362 | 1 |
| HTN | b_cell | *RPAIN* | -4.650590437 | 0.001177038 | 0.003008146 | 1 |
| HTN | b_cell | *PLD2* | 6.217464246 | 0.000617168 | 0.002286342 | 1 |
| HTN | b_cell | *ATP1B2* | -8.317177347 | 0.038469259 | 0.047553329 | 1 |
| HTN | b_cell | *CCL4* | -4.026787828 | 0.000411426 | 0.002180344 | 1 |
| HTN | b_cell | *PHF20L1* | -4.501567352 | 0.00335021 | 0.00606346 | 1 |
| HTN | b_cell | *PUS7L* | -5.733793875 | 0.017315916 | 0.023241923 | 1 |
| HTN | b_cell | *KRI1* | -5.672475765 | 0.017383369 | 0.02330758 | 1 |
| HTN | b_cell | *CDKN2D* | 3.657811271 | 0.00042162 | 0.002180344 | 1 |
| HTN | b_cell | *MTUS1* | -4.134780569 | 0.001197756 | 0.003035183 | 1 |
| HTN | b_cell | *NGDN* | -4.911957778 | 0.00079737 | 0.002471926 | 1 |
| HTN | b_cell | *RIPK3* | -4.983127706 | 0.007273289 | 0.011098827 | 1 |
| HTN | b_cell | *ADCY4* | -3.382728017 | 0.008627859 | 0.01276915 | 1 |
| HTN | b_cell | *RAB2B* | -4.280755198 | 0.001389381 | 0.003320045 | 1 |
| HTN | b_cell | *MIS18BP1* | -4.407150438 | 0.003512884 | 0.006281429 | 1 |
| HTN | b_cell | *RNASE1* | -4.199557623 | 0.03481898 | 0.043382268 | 1 |
| HTN | b_cell | *TEP1* | -6.069496764 | 0.029938707 | 0.037872164 | 1 |
| HTN | b_cell | *EPB41L4A* | -2.405319669 | 0.000461753 | 0.002180489 | 1 |
| HTN | b_cell | *CDO1* | -3.445615444 | 0.000417016 | 0.002180344 | 1 |
| HTN | b_cell | *REEP5* | -4.713918117 | 0.004991466 | 0.008251735 | 1 |
| HTN | b_cell | *QRICH2* | -4.794180437 | 0.000375098 | 0.002180344 | 1 |
| HTN | b_cell | *SEC14L1* | -3.605032285 | 0.001970765 | 0.004154996 | 1 |
| HTN | b_cell | *RHBDF2* | 5.081543443 | 7.58E-05 | 0.002135232 | 0.476430693 |
| HTN | b_cell | *AANAT* | 8.703893035 | 3.13E-05 | 0.001332022 | 0.196889514 |
| HTN | b_cell | *TMEM8A* | -6.05442814 | 0.00305672 | 0.005672254 | 1 |
| HTN | b_cell | *SHC2* | -4.110437127 | 0.00044033 | 0.002180344 | 1 |
| HTN | b_cell | *ERMARD* | -5.989513217 | 0.001236156 | 0.003100909 | 1 |
| HTN | b_cell | *PHF10* | -6.090853169 | 0.00918237 | 0.01346305 | 1 |
| HTN | b_cell | *GALNT8* | -6.377528849 | 3.24E-05 | 0.001350755 | 0.203856516 |
| HTN | b_cell | *DOCK6* | -2.734816807 | 0.001192273 | 0.003028615 | 1 |
| HTN | b_cell | *ECSIT* | -6.694271244 | 0.012522501 | 0.017573429 | 1 |
| HTN | b_cell | *ELOF1* | -6.612793499 | 0.000608461 | 0.002281385 | 1 |
| HTN | b_cell | *THEM6* | -5.56824892 | 0.00015137 | 0.002180344 | 0.951660831 |
| HTN | b_cell | *TOMM40* | -7.951688009 | 0.001412963 | 0.003357255 | 1 |
| HTN | b_cell | *DPP6* | -5.480260676 | 0.000355594 | 0.002180344 | 1 |
| HTN | b_cell | *FAM98C* | 8.319788351 | 0.000614454 | 0.002286342 | 1 |
| HTN | b_cell | *ATP8B3* | -8.371025087 | 1.42E-06 | 0.000162203 | 0.008921139 |
| HTN | b_cell | *PLVAP* | -2.056869611 | 0.010312382 | 0.014876995 | 1 |
| HTN | b_cell | *BST2* | -4.9522559 | 0.000762055 | 0.00242729 | 1 |
| HTN | b_cell | *DDA1* | -5.04637236 | 8.09E-05 | 0.002176137 | 0.508458854 |
| HTN | b_cell | *TULP4* | -4.364764717 | 0.003538321 | 0.00631115 | 1 |
| HTN | b_cell | *RTN4IP1* | -5.990852045 | 0.001384419 | 0.003311965 | 1 |
| HTN | b_cell | *QRSL1* | 5.180286867 | 0.003978695 | 0.006923348 | 1 |
| HTN | b_cell | *ACTN4* | -4.959488109 | 0.002421603 | 0.004794307 | 1 |
| HTN | b_cell | *NDUFA10* | -2.831179539 | 0.029743644 | 0.037648135 | 1 |
| HTN | b_cell | *PXDN* | -2.335828366 | 0.000415448 | 0.002180344 | 1 |

| | | | | | | |
|---|---|---|---|---|---|---|
| HTN | b_cell | *PGPEP1* | 10.58171212 | 4.10E-05 | 0.001539591 | 0.25798656 |
| HTN | b_cell | *KIAA1683* | -6.377413642 | 0.005147721 | 0.008454474 | 1 |
| HTN | b_cell | *HRC* | -8.614482744 | 0.001468906 | 0.003439483 | 1 |
| HTN | b_cell | *TRPM4* | -7.748031175 | 0.011030187 | 0.015742743 | 1 |
| HTN | b_cell | *CAMSAP1* | -5.125066496 | 0.001313819 | 0.003212751 | 1 |
| HTN | b_cell | *UBAC1* | -5.550409883 | 0.017304542 | 0.023231616 | 1 |
| HTN | b_cell | *TNNT3* | -4.558518382 | 0.003733663 | 0.006590232 | 1 |
| HTN | b_cell | *ATXN10* | -4.117389595 | 0.04010721 | 0.049403219 | 1 |
| HTN | b_cell | *TUBGCP2* | -4.086103778 | 0.001644864 | 0.003701815 | 1 |
| HTN | b_cell | *CYP2E1* | 7.528482003 | 0.032841917 | 0.041155498 | 1 |
| HTN | b_cell | *PAK4* | -4.740622255 | 0.000644336 | 0.002316643 | 1 |
| HTN | b_cell | *ZNF337* | -3.915160379 | 0.003366013 | 0.006089819 | 1 |
| HTN | b_cell | *POMT1* | -5.785893395 | 0.001908526 | 0.00407433 | 1 |
| HTN | b_cell | *UCK1* | 6.58823872 | 0.000217874 | 0.002180344 | 1 |
| HTN | b_cell | *MAP3K10* | -5.092356908 | 0.000178737 | 0.002180344 | 1 |
| HTN | b_cell | *SMPDL3B* | 9.067509282 | 0.000228319 | 0.002180344 | 1 |
| HTN | b_cell | *ATPIF1* | -6.44602471 | 0.002501982 | 0.004917149 | 1 |
| HTN | b_cell | *THEMIS2* | -3.265826209 | 0.00202777 | 0.004226899 | 1 |
| HTN | b_cell | *CLIP1* | -4.539854232 | 0.00592552 | 0.009445675 | 1 |
| HTN | b_cell | *ZNF317* | -3.961832418 | 0.00040764 | 0.002180344 | 1 |
| HTN | b_cell | *PPAN* | 11.20577678 | 0.000700016 | 0.002366173 | 1 |
| HTN | b_cell | *C12orf65* | -5.929482282 | 0.005794715 | 0.009284244 | 1 |
| HTN | b_cell | *PKDREJ* | -9.263242004 | 0.01839209 | 0.024498107 | 1 |
| HTN | b_cell | *HSD17B3* | -4.727905763 | 0.000795354 | 0.002467112 | 1 |
| HTN | b_cell | *HABP4* | -3.492073403 | 0.000494364 | 0.002192417 | 1 |
| HTN | b_cell | *EPS8L1* | 7.629074287 | 0.000233613 | 0.002180344 | 1 |
| HTN | b_cell | *LILRB2* | -5.022096394 | 0.000426543 | 0.002180344 | 1 |
| HTN | b_cell | *COX4I2* | 4.359072734 | 0.000316769 | 0.002180344 | 1 |
| HTN | b_cell | *ATP6V1E1* | -4.508978454 | 0.000775623 | 0.002438676 | 1 |
| HTN | b_cell | *CHMP1A* | 12.83532588 | 7.28E-05 | 0.002098808 | 0.457914931 |
| HTN | b_cell | *PPT1* | -3.831460952 | 0.000604247 | 0.002281385 | 1 |
| HTN | b_cell | *HAUS8* | -6.683552564 | 1.70E-05 | 0.000928737 | 0.106710675 |
| HTN | b_cell | *TBC1D5* | -3.623742498 | 0.009220732 | 0.01350946 | 1 |
| HTN | b_cell | *RFTN1* | -2.989064589 | 0.001004827 | 0.002758778 | 1 |
| HTN | b_cell | *KCNC3* | -9.074233307 | 6.85E-05 | 0.002060872 | 0.430655803 |
| HTN | b_cell | *PDLIM4* | 22.73856859 | 7.10E-06 | 0.00050748 | 0.044658259 |
| HTN | b_cell | *KIF3A* | -5.129392695 | 0.001831854 | 0.003957685 | 1 |
| HTN | b_cell | *AOC3* | -3.007522288 | 0.000549321 | 0.002216676 | 1 |
| HTN | b_cell | *ACLY* | -5.842666347 | 0.005594922 | 0.009033757 | 1 |
| HTN | b_cell | *AOC2* | -9.026891741 | 0.000287834 | 0.002180344 | 1 |
| HTN | b_cell | *DIAPH1* | -5.692571176 | 0.002374185 | 0.004726568 | 1 |
| HTN | b_cell | *EXOC4* | -2.952108175 | 0.018519293 | 0.024646655 | 1 |
| HTN | b_cell | *ACAP3* | -3.835072613 | 0.001365169 | 0.003284661 | 1 |
| HTN | b_cell | *C1orf159* | -5.463842957 | 0.001823097 | 0.003946904 | 1 |
| HTN | b_cell | *ANO1* | -9.539381936 | 0.004623151 | 0.007771591 | 1 |
| HTN | b_cell | *PPFIA1* | -3.636214432 | 0.007695242 | 0.011615843 | 1 |
| HTN | b_cell | *NINJ1* | -5.034364891 | 0.000337923 | 0.002180344 | 1 |
| HTN | b_cell | *NPHP4* | -4.314110394 | 0.000539357 | 0.002208907 | 1 |
| HTN | b_cell | *MAP1B* | -1.846777066 | 0.004748406 | 0.007922831 | 1 |
| HTN | b_cell | *CKMT2* | -3.638679825 | 0.000363938 | 0.002180344 | 1 |

| | | | | | | |
|---|---|---|---|---|---|---|
| HTN | b_cell | *ZCCHC9* | -5.721569866 | 0.013213691 | 0.018379308 | 1 |
| HTN | b_cell | *KHDRBS3* | -3.029561121 | 0.00075787 | 0.002419498 | 1 |
| HTN | b_cell | *CHD1L* | -6.285435466 | 0.00294442 | 0.005524438 | 1 |
| HTN | b_cell | *RBM8A* | -3.74251094 | 0.000174703 | 0.002180344 | 1 |
| HTN | b_cell | *ZNF304* | 8.0332401 | 0.018084305 | 0.024134159 | 1 |
| HTN | b_cell | *ZSCAN5A* | -5.708457522 | 0.007647483 | 0.011565968 | 1 |
| HTN | b_cell | *ZNF132* | -5.605874727 | 0.004402735 | 0.007468675 | 1 |
| HTN | b_cell | *RHPN2* | -6.069284789 | 0.00080063 | 0.002474709 | 1 |
| HTN | b_cell | *C19orf12* | -2.433701529 | 0.000747269 | 0.00240927 | 1 |
| HTN | b_cell | *ACTR10* | -5.09661037 | 0.019345496 | 0.02558913 | 1 |
| HTN | b_cell | *GCH1* | -4.044739052 | 0.001327193 | 0.003226629 | 1 |
| HTN | b_cell | *LGALS3* | -4.489162536 | 0.003131381 | 0.0057767 | 1 |
| HTN | b_cell | *DNAJB1* | -4.715547253 | 0.000140703 | 0.002180344 | 0.884599127 |
| HTN | b_cell | *FBXW9* | -6.754698273 | 0.000306374 | 0.002180344 | 1 |
| HTN | b_cell | *C19orf57* | -9.904696619 | 0.000516478 | 0.002195202 | 1 |
| HTN | b_cell | *MATN3* | -19.58962622 | 0.000689256 | 0.002355324 | 1 |
| HTN | b_cell | *CCT6B* | -5.73557153 | 0.003897747 | 0.006814343 | 1 |
| HTN | b_cell | *RAF1* | -4.69902244 | 0.018065224 | 0.024113814 | 1 |
| HTN | b_cell | *ENOSF1* | -3.448332824 | 0.000355366 | 0.002180344 | 1 |
| HTN | b_cell | *EMILIN2* | -3.766886603 | 0.000342308 | 0.002180344 | 1 |
| HTN | b_cell | *TRIM5* | -5.407087761 | 0.00858913 | 0.012723813 | 1 |
| HTN | b_cell | *CNGA4* | -8.000567401 | 0.000863872 | 0.002559431 | 1 |
| HTN | b_cell | *RRP8* | -7.033218833 | 0.03667778 | 0.04546396 | 1 |
| HTN | b_cell | *IMMT* | -4.752326788 | 0.000435615 | 0.002180344 | 1 |
| HTN | b_cell | *CARD6* | -6.731451358 | 0.008957585 | 0.013188838 | 1 |
| HTN | b_cell | *INPP5K* | -4.281296784 | 0.000171314 | 0.002180344 | 1 |
| HTN | b_cell | *MYBBP1A* | -4.505234622 | 0.000464102 | 0.002180489 | 1 |
| HTN | b_cell | *RPA1* | -4.285402595 | 0.00167711 | 0.00374486 | 1 |
| HTN | b_cell | *SERPINF1* | -3.897961878 | 0.000135446 | 0.002180344 | 0.851548197 |
| HTN | b_cell | *TMEM128* | -7.612977466 | 0.020670099 | 0.027169749 | 1 |
| HTN | b_cell | *SEC61G* | -4.762552593 | 0.000653698 | 0.002324546 | 1 |
| HTN | b_cell | *FIGNL1* | -7.108512463 | 0.003369398 | 0.00609419 | 1 |
| HTN | b_cell | *WBP2* | -5.433558327 | 0.001209302 | 0.00305766 | 1 |
| HTN | b_cell | *EIF5A* | -5.37686778 | 0.000130579 | 0.002180344 | 0.820951227 |
| HTN | b_cell | *KDM6B* | -10.08101173 | 0.000448831 | 0.002180344 | 1 |
| HTN | b_cell | *XAF1* | -3.052310411 | 0.000425097 | 0.002180344 | 1 |
| HTN | b_cell | *REEP2* | -8.537094698 | 7.00E-05 | 0.002084753 | 0.439882814 |
| HTN | b_cell | *BTBD3* | -3.456576465 | 0.000443328 | 0.002180344 | 1 |
| HTN | b_cell | *PCNA* | 12.74616591 | 0.001357293 | 0.003274908 | 1 |
| HTN | b_cell | *PTPRA* | -4.137911141 | 0.012433847 | 0.017472418 | 1 |
| HTN | b_cell | *IGHMBP2* | -5.932719059 | 0.00274555 | 0.005256334 | 1 |
| HTN | b_cell | *MMACHC* | -6.274742742 | 0.000223448 | 0.002180344 | 1 |
| HTN | b_cell | *CTNNBL1* | -5.379795506 | 0.009053264 | 0.013301676 | 1 |
| HTN | b_cell | *LPIN3* | -4.296288446 | 0.000552874 | 0.002219911 | 1 |
| HTN | b_cell | *SERINC3* | -4.601079473 | 0.001888005 | 0.004045632 | 1 |
| HTN | b_cell | *DMGDH* | -2.952818797 | 0.000388555 | 0.002180344 | 1 |
| HTN | b_cell | *BHMT2* | -2.390334343 | 9.46E-05 | 0.002180344 | 0.594679616 |
| HTN | b_cell | *KANK4* | -2.871306054 | 0.000827797 | 0.002514184 | 1 |
| HTN | b_cell | *RSG1* | -23.11725384 | 0.000103541 | 0.002180344 | 0.650960722 |
| HTN | b_cell | *ATP8A2* | -5.409730191 | 0.000418705 | 0.002180344 | 1 |

| | | | | | | |
|---|---|---|---|---|---|---|
| HTN | b_cell | *USPL1* | -5.241215568 | 0.018947555 | 0.025152719 | 1 |
| HTN | b_cell | *POMP* | -5.849897847 | 0.025011144 | 0.032195959 | 1 |
| HTN | b_cell | *WASF3* | -2.590548973 | 0.000422858 | 0.002180344 | 1 |
| HTN | b_cell | *RNF17* | 7.941828885 | 0.003161759 | 0.005813975 | 1 |
| HTN | b_cell | *PEMT* | -3.516929356 | 0.000984084 | 0.002723123 | 1 |
| HTN | b_cell | *SCO1* | -6.342355611 | 0.001728933 | 0.003822011 | 1 |
| HTN | b_cell | *TMCC2* | -10.31724905 | 1.87E-05 | 0.000985596 | 0.117564114 |
| HTN | b_cell | *COG6* | -5.344356883 | 0.006289459 | 0.009905267 | 1 |
| HTN | b_cell | *TRPC4* | -6.318179587 | 0.000475791 | 0.002180489 | 1 |
| HTN | b_cell | *RFC3* | -4.344992642 | 0.001416723 | 0.003364917 | 1 |
| HTN | b_cell | *PRAM1* | -5.645557345 | 4.86E-05 | 0.001724513 | 0.305238887 |
| HTN | b_cell | *ZNF414* | -4.83947234 | 0.000415439 | 0.002180344 | 1 |
| HTN | b_cell | *CNDP2* | -5.430962312 | 0.000690802 | 0.002356568 | 1 |
| HTN | b_cell | *RTN3* | -3.390876563 | 0.000960469 | 0.002700568 | 1 |
| HTN | b_cell | *C1QTNF6* | -4.53091476 | 0.000439424 | 0.002180344 | 1 |
| HTN | b_cell | *FAM83F* | -10.15417285 | 0.000335117 | 0.002180344 | 1 |
| HTN | b_cell | *GIMAP4* | -3.4113458 | 0.001574619 | 0.003581136 | 1 |
| HTN | b_cell | *AGAP3* | -4.544495269 | 0.001097166 | 0.002881321 | 1 |
| HTN | b_cell | *ACTR3B* | -6.265850959 | 0.008382963 | 0.012444791 | 1 |
| HTN | b_cell | *DYDC2* | -22.05763192 | 0.0011431 | 0.00295626 | 1 |
| HTN | b_cell | *TMTC1* | -2.282445981 | 0.000507971 | 0.002195202 | 1 |
| HTN | b_cell | *IPO8* | -5.626115847 | 0.008799337 | 0.012992351 | 1 |
| HTN | b_cell | *LARS* | -4.64839051 | 0.000775454 | 0.002438676 | 1 |
| HTN | b_cell | *SPINK5* | -8.701795348 | 0.00401918 | 0.006978345 | 1 |
| HTN | b_cell | *LRRCC1* | -5.507099354 | 0.000579963 | 0.002250503 | 1 |
| HTN | b_cell | *AMPD3* | -3.533040298 | 0.001567157 | 0.003576304 | 1 |
| HTN | b_cell | *SBF2* | -3.459827386 | 0.003684165 | 0.006522767 | 1 |
| HTN | b_cell | *MICAL2* | -6.191274064 | 0.0024672 | 0.004856384 | 1 |
| HTN | b_cell | *RRAS2* | -3.7224143 | 0.0065343 | 0.010206495 | 1 |
| HTN | b_cell | *HSD17B4* | -4.16891491 | 0.002957386 | 0.005543556 | 1 |
| HTN | b_cell | *DPF2* | -4.667253158 | 0.001034583 | 0.002800612 | 1 |
| HTN | b_cell | *GSC* | -12.63048688 | 0.000408716 | 0.002180344 | 1 |
| HTN | b_cell | *CATSPERB* | -3.076350838 | 0.000465861 | 0.002180489 | 1 |
| HTN | b_cell | *MED6* | -3.043997148 | 0.000899865 | 0.002604718 | 1 |
| HTN | b_cell | *EIF2S1* | -6.269467324 | 0.015750546 | 0.02137817 | 1 |
| HTN | b_cell | *ELP3* | -4.689913507 | 0.002237841 | 0.004516517 | 1 |
| HTN | b_cell | *ADAMDEC1* | -14.34572065 | 0.021454329 | 0.028077303 | 1 |
| HTN | b_cell | *MRO* | -6.469824117 | 0.002271998 | 0.004572359 | 1 |
| HTN | b_cell | *THUMPD3* | -5.611634558 | 0.000744247 | 0.002408333 | 1 |
| HTN | b_cell | *CNTN6* | -3.868973957 | 0.000525743 | 0.002195202 | 1 |
| HTN | b_cell | *KATNBL1* | -4.110435107 | 0.001832683 | 0.003958117 | 1 |
| HTN | b_cell | *GSTM5* | -5.815710035 | 6.49E-05 | 0.001999945 | 0.407988683 |
| HTN | b_cell | *GSTM3* | -3.410882072 | 0.000887382 | 0.002596195 | 1 |
| HTN | b_cell | *SYT6* | -14.64471937 | 0.000293646 | 0.002180344 | 1 |
| HTN | b_cell | *VAV3* | 4.874568743 | 0.0068476 | 0.010587292 | 1 |
| HTN | b_cell | *PSRC1* | 5.159699471 | 0.001280125 | 0.003161095 | 1 |
| HTN | b_cell | *WNT2B* | -5.517146244 | 2.47E-05 | 0.00115777 | 0.155141158 |
| HTN | b_cell | *CEPT1* | -3.839657782 | 0.001814311 | 0.003934658 | 1 |
| HTN | b_cell | *AP4B1* | -5.74640601 | 0.002360518 | 0.004705319 | 1 |
| HTN | b_cell | *NAPG* | -5.166020981 | 0.002398555 | 0.004760012 | 1 |

| | | | | | | |
|---|---|---|---|---|---|---|
| HTN | b_cell | *SPIRE1* | -3.936320454 | 0.000943304 | 0.002670217 | 1 |
| HTN | b_cell | *PPHLN1* | -4.173509253 | 0.023204449 | 0.030110706 | 1 |
| HTN | b_cell | *YWHAQ* | -5.23499494 | 0.018385018 | 0.024497496 | 1 |
| HTN | b_cell | *LPIN1* | -4.099688508 | 0.000235463 | 0.002180344 | 1 |
| HTN | b_cell | *LDHA* | -4.603135434 | 0.001085242 | 0.002865833 | 1 |
| HTN | b_cell | *ANO3* | -2.487711887 | 0.000503547 | 0.002195028 | 1 |
| HTN | b_cell | *FST* | -5.562408189 | 0.00010369 | 0.002180344 | 0.651902067 |
| HTN | b_cell | *NARS* | -4.806875563 | 0.001474226 | 0.003448441 | 1 |
| HTN | b_cell | *FBXO18* | -5.184326301 | 0.021241678 | 0.027839572 | 1 |
| HTN | b_cell | *ECHDC3* | -3.45044202 | 0.000455393 | 0.002180344 | 1 |
| HTN | b_cell | *IL15RA* | -3.797029376 | 0.000530291 | 0.002195202 | 1 |
| HTN | b_cell | *TMEM241* | -3.454616626 | 0.000868973 | 0.002568981 | 1 |
| HTN | b_cell | *EMP1* | -3.496737512 | 0.001211959 | 0.003058843 | 1 |
| HTN | b_cell | *KLRD1* | -3.345656139 | 0.000595543 | 0.002270576 | 1 |
| HTN | b_cell | *PRH2* | -13.61069874 | 8.82E-05 | 0.002180344 | 0.55476094 |
| HTN | b_cell | *ACP2* | 27.13584628 | 6.62E-13 | 4.16E-09 | 4.16E-09 |
| HTN | b_cell | *HOOK1* | 4.618922499 | 0.000248846 | 0.002180344 | 1 |
| HTN | b_cell | *ZCCHC11* | -3.933621107 | 0.027695874 | 0.035304939 | 1 |
| HTN | b_cell | *PRPF38A* | -6.47384378 | 0.014482187 | 0.01988849 | 1 |
| HTN | b_cell | *FHOD3* | -2.47003016 | 0.001047558 | 0.002816937 | 1 |
| HTN | b_cell | *TPGS2* | -4.081591835 | 0.001967714 | 0.004151348 | 1 |
| HTN | b_cell | *TIMM10* | -6.367019754 | 0.000267679 | 0.002180344 | 1 |
| HTN | b_cell | *DHX34* | -5.58008944 | 0.000847258 | 0.002540157 | 1 |
| HTN | b_cell | *APLNR* | -5.315082618 | 0.001596923 | 0.003617966 | 1 |
| HTN | b_cell | *FADS2* | -4.469419508 | 0.000277637 | 0.002180344 | 1 |
| HTN | b_cell | *TMEM165* | -2.857313964 | 0.0173309 | 0.023252106 | 1 |
| HTN | b_cell | *CLOCK* | -4.601348196 | 0.015534851 | 0.021108193 | 1 |
| HTN | b_cell | *CLDN10* | 3.64632502 | 0.00029686 | 0.002180344 | 1 |
| HTN | b_cell | *DZIP1* | -4.061286794 | 0.00022221 | 0.002180344 | 1 |
| HTN | b_cell | *BIVM* | -7.82999398 | 0.007105664 | 0.010882658 | 1 |
| HTN | b_cell | *ERCC5* | -11.72028609 | 0.00226334 | 0.004559315 | 1 |
| HTN | b_cell | *CARS2* | -4.273024743 | 0.000447342 | 0.002180344 | 1 |
| HTN | b_cell | *STT3A* | -5.016819601 | 0.000525261 | 0.002195202 | 1 |
| HTN | b_cell | *KLB* | -2.216546126 | 0.002080231 | 0.004303303 | 1 |
| HTN | b_cell | *WDR36* | -5.146099791 | 0.00171917 | 0.00380403 | 1 |
| HTN | b_cell | *NAA35* | -4.856362541 | 0.018993929 | 0.025193002 | 1 |
| HTN | b_cell | *TMEM2* | -3.361622965 | 0.000880815 | 0.002587702 | 1 |
| HTN | b_cell | *AGTPBP1* | -4.349546796 | 0.001173151 | 0.003004867 | 1 |
| HTN | b_cell | *GOLM1* | -6.015932634 | 0.000431045 | 0.002180344 | 1 |
| HTN | b_cell | *ADAM19* | 1.692809842 | 7.63E-05 | 0.002135232 | 0.479542214 |
| HTN | b_cell | *USP30* | -5.977749173 | 0.000467226 | 0.002180489 | 1 |
| HTN | b_cell | *HRK* | 4.814798778 | 0.000605801 | 0.002281385 | 1 |
| HTN | b_cell | *P2RX4* | -3.617604638 | 0.005128336 | 0.008427038 | 1 |
| HTN | b_cell | *DTX1* | -3.403845378 | 0.000173667 | 0.002180344 | 1 |
| HTN | b_cell | *DMTF1* | -4.503858803 | 0.008972566 | 0.013201621 | 1 |
| HTN | b_cell | *CCDC146* | -3.848637403 | 0.001693826 | 0.003762928 | 1 |
| HTN | b_cell | *TMEM60* | 11.91516936 | 0.000464221 | 0.002180489 | 1 |
| HTN | b_cell | *POM121C* | -5.832291727 | 9.18E-05 | 0.002180344 | 0.577270238 |
| HTN | b_cell | *SRPK2* | -3.887846368 | 0.014586367 | 0.020018443 | 1 |
| HTN | b_cell | *TES* | -5.40980307 | 0.005485776 | 0.008891229 | 1 |

| | | | | | | |
|---|---|---|---|---|---|---|
| HTN | b_cell | *MDFIC* | -2.972214713 | 0.001549645 | 0.00355181 | 1 |
| HTN | b_cell | *KHDC1* | -4.148055293 | 0.000869131 | 0.002568981 | 1 |
| HTN | b_cell | *SNX14* | -4.815572623 | 0.009513722 | 0.013855171 | 1 |
| HTN | b_cell | *NT5E* | -3.120565521 | 0.001057628 | 0.002833039 | 1 |
| HTN | b_cell | *PRR5L* | -3.324331702 | 0.001083273 | 0.002865833 | 1 |
| HTN | b_cell | *NAT10* | -5.45739516 | 0.000856822 | 0.002551796 | 1 |
| HTN | b_cell | *ITGA7* | -1.753918953 | 0.005682621 | 0.009148948 | 1 |
| HTN | b_cell | *RDH5* | -2.3987593 | 0.0007386 | 0.002405521 | 1 |
| HTN | b_cell | *BLOC1S1* | -4.665832707 | 0.030486914 | 0.038495928 | 1 |
| HTN | b_cell | *PPP1R1A* | -5.38560196 | 0.000478036 | 0.002180489 | 1 |
| HTN | b_cell | *B4GALNT1* | 7.88349277 | 0.000179718 | 0.002180344 | 1 |
| HTN | b_cell | *FAIM2* | -9.70678686 | 0.000464502 | 0.002180489 | 1 |
| HTN | b_cell | *KRT7* | 3.799741121 | 0.000238189 | 0.002180344 | 1 |
| HTN | b_cell | *SLC26A10* | -12.5612903 | 0.003436675 | 0.006183852 | 1 |
| HTN | b_cell | *MAP7* | -3.001461528 | 0.000504156 | 0.002195028 | 1 |
| HTN | b_cell | *AHI1* | -2.505585315 | 0.006133607 | 0.009706012 | 1 |
| HTN | b_cell | *PKIB* | -5.02130133 | 0.000285021 | 0.002180344 | 1 |
| HTN | b_cell | *SMPD2* | 4.534929482 | 0.025550159 | 0.032815904 | 1 |
| HTN | b_cell | *MICAL1* | -6.648239564 | 0.001912389 | 0.004079806 | 1 |
| HTN | b_cell | *PRADC1* | 8.205562279 | 0.000567919 | 0.002232844 | 1 |
| HTN | b_cell | *SMYD5* | -5.20766937 | 0.000611672 | 0.002284957 | 1 |
| HTN | b_cell | *DYSF* | -4.04083803 | 0.000233133 | 0.002180344 | 1 |
| HTN | b_cell | *KCNMB4* | 4.729137207 | 0.000260039 | 0.002180344 | 1 |
| HTN | b_cell | *USP15* | -3.949666349 | 0.019836727 | 0.026167332 | 1 |
| HTN | b_cell | *KLHL36* | -4.499830468 | 0.001120587 | 0.002924292 | 1 |
| HTN | b_cell | *MPHOSPH6* | 7.007642137 | 0.024803373 | 0.031974329 | 1 |
| HTN | b_cell | *FHOD1* | -3.802364548 | 0.000337829 | 0.002180344 | 1 |
| HTN | b_cell | *CCDC102A* | -5.94971925 | 0.001762221 | 0.003858964 | 1 |
| HTN | b_cell | *AGT* | 3.834620473 | 0.009387964 | 0.013698951 | 1 |
| HTN | b_cell | *ZNF670* | 5.792925962 | 0.000298316 | 0.002180344 | 1 |
| HTN | b_cell | *NTPCR* | -4.901966079 | 0.001672444 | 0.003737881 | 1 |
| HTN | b_cell | *RNASEL* | -4.59779994 | 0.00073276 | 0.002399172 | 1 |
| HTN | b_cell | *PIGC* | -5.179458897 | 0.00088016 | 0.002587702 | 1 |
| HTN | b_cell | *SP110* | 5.621150785 | 0.007281645 | 0.011106187 | 1 |
| HTN | b_cell | *SLC19A3* | -2.10722733 | 0.000526843 | 0.002195202 | 1 |
| HTN | b_cell | *TMBIM1* | -4.331804258 | 9.33E-05 | 0.002180344 | 0.586593191 |
| HTN | b_cell | *ARMC9* | -5.56883014 | 0.002092398 | 0.004320167 | 1 |
| HTN | b_cell | *TMEM127* | -5.496272934 | 0.001677547 | 0.00374486 | 1 |
| HTN | b_cell | *ANKRD36* | -4.483282857 | 0.004220469 | 0.00723394 | 1 |
| HTN | b_cell | *APPL2* | -4.192102061 | 0.003193681 | 0.005850429 | 1 |
| HTN | b_cell | *DRAM1* | -4.37449404 | 0.002022194 | 0.004222363 | 1 |
| HTN | b_cell | *SLC41A2* | -4.386726931 | 0.002416792 | 0.004788646 | 1 |
| HTN | b_cell | *NEK3* | -4.172136688 | 0.01305746 | 0.018210349 | 1 |
| HTN | b_cell | *VPS36* | -4.686911278 | 0.005513739 | 0.008929073 | 1 |
| HTN | b_cell | *CKAP2* | -3.389294151 | 0.012206457 | 0.017207725 | 1 |
| HTN | b_cell | *TBC1D4* | -3.84069343 | 0.001697569 | 0.003769909 | 1 |
| HTN | b_cell | *SUCLA2* | -3.582346206 | 0.003333161 | 0.006044299 | 1 |
| HTN | b_cell | *RCBTB1* | -5.533572351 | 0.000695313 | 0.002359111 | 1 |
| HTN | b_cell | *MED4* | -5.399888245 | 0.010808413 | 0.015485983 | 1 |
| HTN | b_cell | *RCBTB2* | -3.912453482 | 0.000965317 | 0.002708142 | 1 |

| | | | | | | |
|---|---|---|---|---|---|---|
| HTN | b_cell | *SETDB2* | -5.599902902 | 0.002198136 | 0.004463722 | 1 |
| HTN | b_cell | *SPDYE1* | -4.430358648 | 0.000735254 | 0.00240209 | 1 |
| HTN | b_cell | *CHST12* | -5.990012984 | 0.000458125 | 0.002180344 | 1 |
| HTN | b_cell | *GPNMB* | -3.054574455 | 0.000413006 | 0.002180344 | 1 |
| HTN | b_cell | *KDELR2* | -6.493808589 | 0.00840767 | 0.012478522 | 1 |
| HTN | b_cell | *NUPL2* | -5.264863654 | 0.000828831 | 0.002516107 | 1 |
| HTN | b_cell | *ZDHHC4* | -5.926123979 | 0.001406333 | 0.003346562 | 1 |
| HTN | b_cell | *AOAH* | -2.748727977 | 0.000351546 | 0.002180344 | 1 |
| HTN | b_cell | *TBRG4* | 6.920208714 | 0.000689264 | 0.002355324 | 1 |
| HTN | b_cell | *DDX56* | -7.636094374 | 0.00768582 | 0.011607195 | 1 |
| HTN | b_cell | *HUS1* | -3.789192924 | 0.001283245 | 0.003167554 | 1 |
| HTN | b_cell | *NACAD* | -23.22865656 | 2.06E-05 | 0.001028789 | 0.129404277 |
| HTN | b_cell | *DBNL* | 6.456025021 | 0.010776899 | 0.015444351 | 1 |
| HTN | b_cell | *TTC5* | -6.134582104 | 0.000512225 | 0.002195202 | 1 |
| HTN | b_cell | *ALPK3* | -3.180147782 | 0.000978831 | 0.002717425 | 1 |
| HTN | b_cell | *TM6SF1* | -3.739766463 | 0.000374781 | 0.002180344 | 1 |
| HTN | b_cell | *RSAD1* | -5.662878846 | 0.001325356 | 0.003224925 | 1 |
| HTN | b_cell | *MYCBPAP* | -4.300960183 | 0.008889863 | 0.013095259 | 1 |
| HTN | b_cell | *BRIP1* | 4.600883859 | 0.000352954 | 0.002180344 | 1 |
| HTN | b_cell | *RTP4* | -15.1245886 | 0.001926474 | 0.004094645 | 1 |
| HTN | b_cell | *MRPL47* | -7.239540215 | 0.01392447 | 0.019240252 | 1 |
| HTN | b_cell | *BLK* | 0.196065516 | 0.000258582 | 0.002180344 | 1 |
| HTN | b_cell | *EPRS* | -5.549528488 | 0.006244891 | 0.009847585 | 1 |
| HTN | b_cell | *RPS6KC1* | -4.149957754 | 0.002972567 | 0.005560407 | 1 |
| HTN | b_cell | *CBWD2* | -5.634271224 | 0.024283906 | 0.031343239 | 1 |
| HTN | b_cell | *IL1RN* | 4.968134885 | 0.000401836 | 0.002180344 | 1 |
| HTN | b_cell | *SAP130* | -6.104412716 | 0.007480934 | 0.011357796 | 1 |
| HTN | b_cell | *GYPC* | -4.452305276 | 0.000234473 | 0.002180344 | 1 |
| HTN | b_cell | *LRRC8A* | -3.926270814 | 0.002128228 | 0.004361619 | 1 |
| HTN | b_cell | *TXN* | -5.041583132 | 0.000718811 | 0.002385315 | 1 |
| HTN | b_cell | *ODF2* | -6.579524234 | 0.016487305 | 0.022234167 | 1 |
| HTN | b_cell | *TOR1B* | -3.784384324 | 0.004461943 | 0.007547009 | 1 |
| HTN | b_cell | *TOR1A* | -4.313378245 | 0.002661693 | 0.005131574 | 1 |
| HTN | b_cell | *SLC2A8* | -4.233348209 | 0.024163224 | 0.031219521 | 1 |
| HTN | b_cell | *ZFP37* | -4.608708293 | 0.000680892 | 0.002339217 | 1 |
| HTN | b_cell | *ZNF189* | -5.877963943 | 0.003204638 | 0.005867082 | 1 |
| HTN | b_cell | *ALDOB* | -8.984786511 | 0.000358731 | 0.002180344 | 1 |
| HTN | b_cell | *STX17* | -5.227278334 | 0.014126652 | 0.019476811 | 1 |
| HTN | b_cell | *PRPF4* | 7.949049573 | 0.003507847 | 0.006275992 | 1 |
| HTN | b_cell | *FPGS* | -5.003106669 | 0.00019111 | 0.002180344 | 1 |
| HTN | b_cell | *TEX10* | -4.246117216 | 0.000850352 | 0.00254458 | 1 |
| HTN | b_cell | *GARNL3* | -4.927155099 | 0.002193618 | 0.004458867 | 1 |
| HTN | b_cell | *GABBR2* | -9.424095543 | 0.000423348 | 0.002180344 | 1 |
| HTN | b_cell | *PSMB7* | -5.205018853 | 0.004301767 | 0.007339269 | 1 |
| HTN | b_cell | *RABEPK* | -4.012588346 | 0.000707935 | 0.00237502 | 1 |
| HTN | b_cell | *GOLGA1* | -3.400661996 | 0.000718522 | 0.002385315 | 1 |
| HTN | b_cell | *LMX1B* | -11.79699115 | 0.000431436 | 0.002180344 | 1 |
| HTN | b_cell | *DSCC1* | -2.471591449 | 0.003600632 | 0.006401916 | 1 |
| HTN | b_cell | *NOV* | -6.928120356 | 0.000977209 | 0.002717425 | 1 |
| HTN | b_cell | *POLR1E* | -6.082614829 | 0.000162264 | 0.002180344 | 1 |

| | | | | | | |
|---|---|---|---|---|---|---|
| HTN | b_cell | *IL11RA* | -5.690801741 | 0.000187962 | 0.002180344 | 1 |
| HTN | b_cell | *APTX* | -5.111782529 | 0.001012751 | 0.002769536 | 1 |
| HTN | b_cell | *TLN1* | -3.491629264 | 0.000925773 | 0.002648292 | 1 |
| HTN | b_cell | *SPAG8* | 12.77046307 | 0.000280691 | 0.002180344 | 1 |
| HTN | b_cell | *CD72* | -6.885727765 | 0.006304764 | 0.00991447 | 1 |
| HTN | b_cell | *TMEM8B* | 8.844968961 | 0.011012064 | 0.015724017 | 1 |
| HTN | b_cell | *GRHPR* | -5.448476486 | 0.001857974 | 0.003998439 | 1 |
| HTN | b_cell | *ARHGEF39* | 5.085285984 | 0.003838072 | 0.006734569 | 1 |
| HTN | b_cell | *DENND4C* | -3.823328908 | 0.005075502 | 0.008353896 | 1 |
| HTN | b_cell | *PPIL1* | 8.565834695 | 0.000364163 | 0.002180344 | 1 |
| HTN | b_cell | *KIF13A* | -3.266127886 | 0.001345623 | 0.003262603 | 1 |
| HTN | b_cell | *TMEM63B* | -4.090708527 | 0.000610025 | 0.002281515 | 1 |
| HTN | b_cell | *FRS3* | 9.335754036 | 0.009255288 | 0.013547845 | 1 |
| HTN | b_cell | *SLC22A23* | -3.232208621 | 0.000789343 | 0.002459167 | 1 |
| HTN | b_cell | *LRRC1* | -3.092720491 | 0.000944436 | 0.002671018 | 1 |
| HTN | b_cell | *FOXF2* | -11.99269413 | 0.020761756 | 0.027273122 | 1 |
| HTN | b_cell | *BPHL* | -2.79289672 | 0.000281014 | 0.002180344 | 1 |
| HTN | b_cell | *RIPK1* | -4.23203698 | 0.001482185 | 0.003458983 | 1 |
| HTN | b_cell | *TUBB2B* | -7.035893924 | 0.002985654 | 0.005579906 | 1 |
| HTN | b_cell | *TCF19* | -8.555488505 | 0.000604351 | 0.002281385 | 1 |
| HTN | b_cell | *FLOT1* | -4.751028943 | 0.000696987 | 0.002360969 | 1 |
| HTN | b_cell | *TPMT* | -7.76725067 | 0.023014563 | 0.029932263 | 1 |
| HTN | b_cell | *MTCH1* | -5.690040847 | 0.026202145 | 0.033584686 | 1 |
| HTN | b_cell | *FGFBP2* | 10.13060841 | 0.004263567 | 0.007287941 | 1 |
| HTN | b_cell | *FHDC1* | -5.096661701 | 0.000415584 | 0.002180344 | 1 |
| HTN | b_cell | *MYO7A* | -4.211789448 | 0.000833542 | 0.002524317 | 1 |
| HTN | b_cell | *ARRB1* | -4.392803513 | 0.001115893 | 0.002917097 | 1 |
| HTN | b_cell | *CCDC90B* | -4.632744623 | 0.000772595 | 0.002437181 | 1 |
| HTN | b_cell | *SYTL2* | -3.253196352 | 0.00024915 | 0.002180344 | 1 |
| HTN | b_cell | *PRCP* | -5.921192931 | 0.006613718 | 0.010279467 | 1 |
| HTN | b_cell | *PI15* | 6.827829211 | 0.000364962 | 0.002180344 | 1 |
| HTN | b_cell | *TTPA* | -3.011286288 | 0.000107339 | 0.002180344 | 0.67483736 |
| HTN | b_cell | *GGH* | -4.517643934 | 0.000235137 | 0.002180344 | 1 |
| HTN | b_cell | *SULF1* | -2.088633075 | 0.000876431 | 0.002582063 | 1 |
| HTN | b_cell | *TGS1* | -4.840786303 | 0.016746853 | 0.022564824 | 1 |
| HTN | b_cell | *NEK1* | -4.21174729 | 0.002435622 | 0.004818363 | 1 |
| HTN | b_cell | *DDX60* | -5.94772496 | 0.004757178 | 0.007928337 | 1 |
| HTN | b_cell | *DCUN1D5* | -2.924478514 | 0.000611219 | 0.002284621 | 1 |
| HTN | b_cell | *TRIM29* | -8.148544881 | 6.82E-05 | 0.002060872 | 0.428997859 |
| HTN | b_cell | *POU2F3* | 3.939706974 | 0.002592098 | 0.005043801 | 1 |
| HTN | b_cell | *PPP2R1B* | -2.165147347 | 0.00031186 | 0.002180344 | 1 |
| HTN | b_cell | *FDX1* | -4.436538446 | 0.004698249 | 0.007862295 | 1 |
| HTN | b_cell | *FXYD6* | -9.588040603 | 0.02886018 | 0.036670161 | 1 |
| HTN | b_cell | *TMPRSS13* | -11.9699569 | 0.025617818 | 0.032896083 | 1 |
| HTN | b_cell | *MAP2K5* | -3.758025805 | 0.027678513 | 0.035289964 | 1 |
| HTN | b_cell | *NDUFAF1* | -7.804141976 | 7.64E-05 | 0.002135232 | 0.480427239 |
| HTN | b_cell | *ITGA11* | -2.732667202 | 0.000360395 | 0.002180344 | 1 |
| HTN | b_cell | *PAQR5* | -6.975176684 | 0.000142639 | 0.002180344 | 0.896768533 |
| HTN | b_cell | *ITPKA* | 8.002327108 | 0.000225228 | 0.002180344 | 1 |
| HTN | b_cell | *UACA* | -3.421677061 | 0.001309584 | 0.003205463 | 1 |

| | | | | | | |
|---|---|---|---|---|---|---|
| HTN | b_cell | *PLCB2* | -3.951290859 | 8.41E-05 | 0.002180344 | 0.529027459 |
| HTN | b_cell | *RSL24D1* | -6.392345683 | 0.01366836 | 0.018932138 | 1 |
| HTN | b_cell | *RABGGTB* | -6.981720597 | 0.006359582 | 0.009975721 | 1 |
| HTN | b_cell | *DNASE2B* | -31.47547706 | 0.001521112 | 0.003512021 | 1 |
| HTN | b_cell | *DBT* | -5.473894652 | 0.006848817 | 0.010587292 | 1 |
| HTN | b_cell | *RTCA* | -4.713307701 | 0.007858298 | 0.011808107 | 1 |
| HTN | b_cell | *IFT172* | -4.151958742 | 0.009696272 | 0.014058218 | 1 |
| HTN | b_cell | *KHK* | -4.595667684 | 0.000249441 | 0.002180344 | 1 |
| HTN | b_cell | *ADCY3* | -5.114573609 | 0.00070524 | 0.00237262 | 1 |
| HTN | b_cell | *LHCGR* | -4.719850009 | 0.000336218 | 0.002180344 | 1 |
| HTN | b_cell | *THUMPD2* | -4.931412697 | 0.001753534 | 0.003846639 | 1 |
| HTN | b_cell | *PREB* | -3.496668092 | 0.00298971 | 0.005584167 | 1 |
| HTN | b_cell | *ABCG5* | -5.600061961 | 0.002873014 | 0.005428564 | 1 |
| HTN | b_cell | *ATRAID* | -7.811465959 | 0.001789939 | 0.003900641 | 1 |
| HTN | b_cell | *CENPO* | 10.38375993 | 0.029089351 | 0.036916583 | 1 |
| HTN | b_cell | *LRPPRC* | -4.584476187 | 0.004864207 | 0.008073197 | 1 |
| HTN | b_cell | *DTNB* | -3.970873205 | 0.001230489 | 0.003090725 | 1 |
| HTN | b_cell | *CYP2C9* | 4.014969748 | 0.006731159 | 0.010446506 | 1 |
| HTN | b_cell | *MYOF* | -3.479277771 | 0.004123096 | 0.007117492 | 1 |
| HTN | b_cell | *CH25H* | -5.06251354 | 0.001420467 | 0.003372154 | 1 |
| HTN | b_cell | *ATAD1* | -5.576565374 | 0.003215151 | 0.005881191 | 1 |
| HTN | b_cell | *BTBD16* | -7.773117587 | 0.007534963 | 0.011426028 | 1 |
| HTN | b_cell | *TACC2* | -2.529779215 | 0.000414975 | 0.002180344 | 1 |
| HTN | b_cell | *CALHM2* | -4.277118651 | 0.000180231 | 0.002180344 | 1 |
| HTN | b_cell | *ARL3* | -3.595236361 | 0.013732059 | 0.019003621 | 1 |
| HTN | b_cell | *PLCE1* | -3.000874984 | 0.000607635 | 0.002281385 | 1 |
| HTN | b_cell | *RBP4* | -3.616585005 | 0.000814803 | 0.002495209 | 1 |
| HTN | b_cell | *DNAJC13* | -4.744788148 | 0.006524501 | 0.010196256 | 1 |
| HTN | b_cell | *FAM149B1* | -3.777874064 | 0.020149921 | 0.026535935 | 1 |
| HTN | b_cell | *ATIC* | -4.204225371 | 0.028550096 | 0.036305513 | 1 |
| HTN | b_cell | *SMARCAL1* | -5.638979225 | 0.029165915 | 0.037006278 | 1 |
| HTN | b_cell | *BARD1* | -5.86453277 | 0.000836014 | 0.002525718 | 1 |
| HTN | b_cell | *STAT4* | -2.929437901 | 0.000834255 | 0.002524879 | 1 |
| HTN | b_cell | *CARF* | -4.659287502 | 0.000983171 | 0.002721796 | 1 |
| HTN | b_cell | *ASNSD1* | -4.095343279 | 0.000209017 | 0.002180344 | 1 |
| HTN | b_cell | *METTL5* | -6.554157709 | 0.006522514 | 0.010195684 | 1 |
| HTN | b_cell | *NAB1* | -5.461122475 | 0.008122917 | 0.012130351 | 1 |
| HTN | b_cell | *MDH1B* | 4.212132077 | 0.000281061 | 0.002180344 | 1 |
| HTN | b_cell | *ITGAV* | -4.259240144 | 0.002489164 | 0.004898083 | 1 |
| HTN | b_cell | *SLC35A5* | -3.881042539 | 0.00945364 | 0.013777245 | 1 |
| HTN | b_cell | *COX17* | 7.942207371 | 0.005344445 | 0.008709312 | 1 |
| HTN | b_cell | *USP8* | -3.551814093 | 0.0032596 | 0.005936589 | 1 |
| HTN | b_cell | *SPPL2A* | -4.762370547 | 0.017107861 | 0.023001951 | 1 |
| HTN | b_cell | *UBL7* | -7.236587044 | 7.97E-05 | 0.002176137 | 0.501246165 |
| HTN | b_cell | *ARHGAP24* | -3.451148011 | 0.005352853 | 0.008718866 | 1 |
| HTN | b_cell | *FAM13A* | -2.706508253 | 0.002128431 | 0.004361619 | 1 |
| HTN | b_cell | *HERC3* | -4.672644738 | 0.008143024 | 0.012151719 | 1 |
| HTN | b_cell | *AP1AR* | -6.217635157 | 0.005867897 | 0.009377597 | 1 |
| HTN | b_cell | *PRKG2* | -3.48796567 | 0.000585455 | 0.002261934 | 1 |
| HTN | b_cell | *BBS7* | -4.438139835 | 0.00077656 | 0.002438676 | 1 |

| | | | | | | |
|---|---|---|---|---|---|---|
| HTN | b_cell | *KIAA1109* | -3.989478771 | 0.023437791 | 0.030375879 | 1 |
| HTN | b_cell | *MMRN1* | -3.361998609 | 0.006843323 | 0.010584002 | 1 |
| HTN | b_cell | *PRDM5* | -3.191038769 | 0.000381674 | 0.002180344 | 1 |
| HTN | b_cell | *TRPC3* | -6.642393454 | 0.004444432 | 0.007523463 | 1 |
| HTN | b_cell | *NAAA* | -5.082610083 | 0.001135802 | 0.002947085 | 1 |
| HTN | b_cell | *NUP54* | -3.993366071 | 0.003673135 | 0.006505071 | 1 |
| HTN | b_cell | *BMP2K* | -3.719658427 | 0.003291398 | 0.005987564 | 1 |
| HTN | b_cell | *G3BP2* | -4.242163989 | 0.021649985 | 0.0283039 | 1 |
| HTN | b_cell | *SCARB2* | -4.729228561 | 0.00531005 | 0.008671243 | 1 |
| HTN | b_cell | *CNOT6L* | -4.763568742 | 0.019370405 | 0.025612263 | 1 |
| HTN | b_cell | *ANXA3* | -4.147907032 | 0.000421629 | 0.002180344 | 1 |
| HTN | b_cell | *PPA2* | -4.641505671 | 0.021979631 | 0.028710979 | 1 |
| HTN | b_cell | *ENPEP* | -4.259686274 | 0.000170858 | 0.002180344 | 1 |
| HTN | b_cell | *PAPSS1* | -3.938225433 | 0.003414002 | 0.006150095 | 1 |
| HTN | b_cell | *SLC39A8* | -4.380458187 | 0.020821255 | 0.027339856 | 1 |
| HTN | b_cell | *FBN2* | 2.546635399 | 0.022035694 | 0.028772255 | 1 |
| HTN | b_cell | *RNF185* | -4.980035318 | 0.004716097 | 0.007881874 | 1 |
| HTN | b_cell | *B4GALNT3* | -10.59075134 | 0.001228819 | 0.003088552 | 1 |
| HTN | b_cell | *ERP27* | 5.4867567 | 0.000672096 | 0.002334899 | 1 |
| HTN | b_cell | *ETV6* | -4.008884038 | 0.004957557 | 0.00819999 | 1 |
| HTN | b_cell | *KIF21A* | -2.548931905 | 0.00065739 | 0.002327958 | 1 |
| HTN | b_cell | *CPNE8* | -4.054294081 | 0.000679488 | 0.002339217 | 1 |
| HTN | b_cell | *ALG10* | -7.133353932 | 0.000609627 | 0.002281385 | 1 |
| HTN | b_cell | *SLCO1C1* | -5.032127856 | 0.001903072 | 0.004065448 | 1 |
| HTN | b_cell | *ZCRB1* | -5.378223092 | 0.029657425 | 0.037549375 | 1 |
| HTN | b_cell | *PRICKLE1* | -2.318317304 | 0.000515061 | 0.002195202 | 1 |
| HTN | b_cell | *C1RL* | -3.610828238 | 0.001277438 | 0.003156737 | 1 |
| HTN | b_cell | *CLSTN3* | -7.724999222 | 0.003299379 | 0.005998476 | 1 |
| HTN | b_cell | *KLRG1* | -4.040189055 | 0.001277414 | 0.003156737 | 1 |
| HTN | b_cell | *VAMP1* | -4.681538886 | 0.00660407 | 0.010267011 | 1 |
| HTN | b_cell | *CD27* | -5.477085994 | 0.002637384 | 0.005101411 | 1 |
| HTN | b_cell | *PEX5* | 5.170319475 | 0.00537127 | 0.008743961 | 1 |
| HTN | b_cell | *PPFIA2* | -2.375492506 | 0.000832943 | 0.002523717 | 1 |
| HTN | b_cell | *LGR5* | -6.750860263 | 0.000127671 | 0.002180344 | 0.802666407 |
| HTN | b_cell | *POC1B* | -5.770744891 | 0.007263811 | 0.011089748 | 1 |
| HTN | b_cell | *SNRPF* | -6.821190037 | 0.022674524 | 0.029508328 | 1 |
| HTN | b_cell | *AMDHD1* | -6.034764788 | 0.001699047 | 0.003771861 | 1 |
| HTN | b_cell | *GAS2L3* | -3.480622837 | 0.000418929 | 0.002180344 | 1 |
| HTN | b_cell | *TMEM132B* | -7.412687721 | 0.001488238 | 0.003464107 | 1 |
| HTN | b_cell | *TDG* | -4.384770642 | 0.004431721 | 0.007505989 | 1 |
| HTN | b_cell | *MMAB* | -3.534673526 | 0.006905583 | 0.010646248 | 1 |
| HTN | b_cell | *GLTP* | -5.2575812 | 8.81E-05 | 0.002180344 | 0.553664838 |
| HTN | b_cell | *MTMR6* | -3.731104632 | 0.001087989 | 0.002868262 | 1 |
| HTN | b_cell | *SLC46A3* | -3.808802562 | 0.001316218 | 0.003214865 | 1 |
| HTN | b_cell | *CCDC65* | -4.276791056 | 0.001205188 | 0.003049101 | 1 |
| HTN | b_cell | *ACVRL1* | -4.63092545 | 0.00423997 | 0.007258483 | 1 |
| HTN | b_cell | *GPR84* | -77.156426 | 4.45E-06 | 0.00036476 | 0.02799279 |
| HTN | b_cell | *KANSL2* | -2.955507984 | 0.000976633 | 0.002717425 | 1 |
| HTN | b_cell | *CERS5* | -4.331989472 | 0.014125659 | 0.019476811 | 1 |
| HTN | b_cell | *LMBR1L* | -7.534661294 | 0.00475074 | 0.007924623 | 1 |

| | | | | | | |
|---|---|---|---|---|---|---|
| HTN | b_cell | *ZNF740* | -4.778665395 | 0.000483196 | 0.002187427 | 1 |
| HTN | b_cell | *WDFY2* | -4.226266234 | 0.033904962 | 0.042344159 | 1 |
| HTN | b_cell | *HNRNPA1L2* | 12.19681519 | 0.001274858 | 0.003156737 | 1 |
| HTN | b_cell | *LPAR6* | -4.094224996 | 0.002055044 | 0.004266863 | 1 |
| HTN | b_cell | *VPS33A* | -6.600917801 | 0.000807074 | 0.002487292 | 1 |
| HTN | b_cell | *METTL21C* | 19.61101302 | 0.002012133 | 0.004211145 | 1 |
| HTN | b_cell | *GRTP1* | -2.768056707 | 0.009411659 | 0.013728794 | 1 |
| HTN | b_cell | *TSSK4* | -26.09052722 | 7.28E-05 | 0.002098808 | 0.457890813 |
| HTN | b_cell | *TMX1* | -5.473366288 | 0.001591985 | 0.003610683 | 1 |
| HTN | b_cell | *RTN1* | -2.377923742 | 0.000583797 | 0.002258667 | 1 |
| HTN | b_cell | *C14orf37* | -4.502852989 | 0.001613967 | 0.003652631 | 1 |
| HTN | b_cell | *RAB15* | -3.976962471 | 0.00102805 | 0.002792138 | 1 |
| HTN | b_cell | *STON2* | -3.220686576 | 0.000543667 | 0.002214137 | 1 |
| HTN | b_cell | *PTGR2* | -4.271774114 | 0.000409303 | 0.002180344 | 1 |
| HTN | b_cell | *AK7* | -7.775326146 | 0.000415716 | 0.002180344 | 1 |
| HTN | b_cell | *FBLN5* | -2.581369872 | 0.000562637 | 0.002226547 | 1 |
| HTN | b_cell | *C14orf79* | -5.083089702 | 0.000333873 | 0.002180344 | 1 |
| HTN | b_cell | *DUOXA1* | -6.469009357 | 0.007770464 | 0.011704098 | 1 |
| HTN | b_cell | *MFAP1* | -6.038307354 | 0.000985641 | 0.002724009 | 1 |
| HTN | b_cell | *TCF12* | -3.41176001 | 0.037844539 | 0.046862092 | 1 |
| HTN | b_cell | *SORD* | 13.5454152 | 0.000503806 | 0.002195028 | 1 |
| HTN | b_cell | *ZSCAN29* | -6.319891766 | 0.000741283 | 0.002405521 | 1 |
| HTN | b_cell | *LYSMD2* | -8.55660189 | 0.003184461 | 0.00584205 | 1 |
| HTN | b_cell | *SRP14* | -5.031388597 | 0.027048 | 0.034584254 | 1 |
| HTN | b_cell | *BAHD1* | -5.348460178 | 0.000900885 | 0.00260492 | 1 |
| HTN | b_cell | *DISP2* | 3.871808108 | 0.005971585 | 0.009502242 | 1 |
| HTN | b_cell | *ANP32A* | -4.218373563 | 0.000837131 | 0.002526663 | 1 |
| HTN | b_cell | *COMMD4* | -8.211551864 | 0.007480843 | 0.011357796 | 1 |
| HTN | b_cell | *UBE2Q2* | -4.832612273 | 0.002227306 | 0.004506943 | 1 |
| HTN | b_cell | *PSTPIP1* | -5.138109036 | 0.001430304 | 0.003383116 | 1 |
| HTN | b_cell | *HMG20A* | -4.18118706 | 0.002389829 | 0.004745691 | 1 |
| HTN | b_cell | *SCAPER* | -4.387260165 | 0.002977607 | 0.005566523 | 1 |
| HTN | b_cell | *NEIL1* | -3.179386926 | 0.000945095 | 0.002671678 | 1 |
| HTN | b_cell | *MAN2C1* | -2.177812439 | 0.010176753 | 0.01469482 | 1 |
| HTN | b_cell | *DNAJA4* | -4.727197571 | 0.000475442 | 0.002180489 | 1 |
| HTN | b_cell | *TPM1* | -4.063284927 | 0.002265257 | 0.004560252 | 1 |
| HTN | b_cell | *IGF1R* | -3.234566394 | 0.001191371 | 0.003028602 | 1 |
| HTN | b_cell | *CYP11A1* | -5.304371218 | 0.00091006 | 0.002624562 | 1 |
| HTN | b_cell | *BBS4* | -5.046000025 | 0.003719881 | 0.006575938 | 1 |
| HTN | b_cell | *ADAMTS17* | -3.069840746 | 0.000408277 | 0.002180344 | 1 |
| HTN | b_cell | *ULK3* | -7.175379255 | 0.020180165 | 0.026570198 | 1 |
| HTN | b_cell | *SCAMP2* | -4.817994812 | 0.006176311 | 0.009763759 | 1 |
| HTN | b_cell | *HAPLN3* | -5.933820394 | 8.07E-05 | 0.002176137 | 0.50738567 |
| HTN | b_cell | *POLG* | -6.082456413 | 0.00163349 | 0.003682235 | 1 |
| HTN | b_cell | *WDR93* | -2.885291792 | 0.001012322 | 0.002769536 | 1 |
| HTN | b_cell | *TICRR* | -7.000839793 | 0.004959582 | 0.008201182 | 1 |
| HTN | b_cell | *MFGE8* | -3.250326995 | 0.00035574 | 0.002180344 | 1 |
| HTN | b_cell | *UNC45A* | -6.87532396 | 0.000152938 | 0.002180344 | 0.961520962 |
| HTN | b_cell | *ST8SIA2* | -25.58953147 | 0.003969193 | 0.006914469 | 1 |
| HTN | b_cell | *IQGAP1* | -4.025691885 | 0.005188255 | 0.00851215 | 1 |

| | | | | | | |
|---|---|---|---|---|---|---|
| HTN | b_cell | *CRTC3* | -4.328951577 | 0.003404728 | 0.006138665 | 1 |
| HTN | b_cell | *SH3GL3* | 4.140537834 | 0.002056859 | 0.004269221 | 1 |
| HTN | b_cell | *SEC11A* | -4.377793759 | 0.001880948 | 0.004034636 | 1 |
| HTN | b_cell | *TGFB1I1* | -5.432401226 | 0.000302316 | 0.002180344 | 1 |
| HTN | b_cell | *PARN* | -4.007800042 | 0.004521 | 0.007628428 | 1 |
| HTN | b_cell | *FTO* | -3.87891503 | 0.003257039 | 0.005936589 | 1 |
| HTN | b_cell | *TXNL4B* | -6.724073208 | 0.001029113 | 0.002792956 | 1 |
| HTN | b_cell | *MARVELD3* | 3.873232718 | 0.000154155 | 0.002180344 | 0.969171884 |
| HTN | b_cell | *ZFHX3* | -3.449310328 | 0.001462108 | 0.003427879 | 1 |
| HTN | b_cell | *KIFC3* | -5.81599862 | 0.001943168 | 0.004118914 | 1 |
| HTN | b_cell | *ADAMTS18* | -3.460455006 | 0.000396168 | 0.002180344 | 1 |
| HTN | b_cell | *GCSH* | -5.210116134 | 0.000569045 | 0.002232844 | 1 |
| HTN | b_cell | *CMTM3* | -3.740693218 | 0.000444062 | 0.002180344 | 1 |
| HTN | b_cell | *MAP1LC3B* | -5.768008967 | 0.001118742 | 0.00292333 | 1 |
| HTN | b_cell | *TLDC1* | -6.840588615 | 0.02399761 | 0.031045817 | 1 |
| HTN | b_cell | *RPS2* | -6.509456168 | 0.001616411 | 0.003652902 | 1 |
| HTN | b_cell | *NDUFB10* | -7.6440063 | 0.005758239 | 0.00923993 | 1 |
| HTN | b_cell | *TIGD7* | -4.672090606 | 0.000193365 | 0.002180344 | 1 |
| HTN | b_cell | *DEF8* | -2.899670024 | 0.007419101 | 0.011280263 | 1 |
| HTN | b_cell | *GALNS* | -6.150909656 | 0.000789295 | 0.002459167 | 1 |
| HTN | b_cell | *COPS3* | -4.465262381 | 0.000627174 | 0.002295161 | 1 |
| HTN | b_cell | *MYO19* | -4.277970973 | 0.039441558 | 0.048659552 | 1 |
| HTN | b_cell | *DDX52* | -5.613135358 | 0.005587905 | 0.009031145 | 1 |
| HTN | b_cell | *RASL10B* | -18.73101921 | 7.89E-06 | 0.000539559 | 0.049586116 |
| HTN | b_cell | *UNC45B* | -17.1365372 | 0.014905707 | 0.02039882 | 1 |
| HTN | b_cell | *TOM1L1* | -3.547685755 | 0.000863178 | 0.002559431 | 1 |
| HTN | b_cell | *C17orf80* | -4.978732839 | 0.001655123 | 0.003713691 | 1 |
| HTN | b_cell | *VPS53* | -5.019654988 | 0.000377165 | 0.002180344 | 1 |
| HTN | b_cell | *SPATA22* | -2.875544292 | 0.000709657 | 0.002378257 | 1 |
| HTN | b_cell | *NPEPPS* | -4.249872317 | 0.002775892 | 0.005299736 | 1 |
| HTN | b_cell | *LRRC46* | -12.58723937 | 0.000229963 | 0.002180344 | 1 |
| HTN | b_cell | *SCRN2* | -3.887641314 | 4.36E-07 | 6.93E-05 | 0.002741787 |
| HTN | b_cell | *RHBDL3* | -11.47406508 | 0.000626288 | 0.002295161 | 1 |
| HTN | b_cell | *ARSG* | -4.963486347 | 0.003536744 | 0.00631115 | 1 |
| HTN | b_cell | *ABCA8* | -2.068693642 | 0.000518234 | 0.002195202 | 1 |
| HTN | b_cell | *G6PC3* | -10.15901865 | 0.005551091 | 0.008980882 | 1 |
| HTN | b_cell | *IMPA2* | -5.583766903 | 0.001361257 | 0.003280269 | 1 |
| HTN | b_cell | *GNAL* | -4.617874507 | 0.000635753 | 0.002305064 | 1 |
| HTN | b_cell | *RPRD1A* | -4.564445952 | 0.011063989 | 0.015773084 | 1 |
| HTN | b_cell | *C18orf21* | -6.276024694 | 0.000872738 | 0.002573594 | 1 |
| HTN | b_cell | *GALNT1* | -3.509309953 | 0.000718971 | 0.002385315 | 1 |
| HTN | b_cell | *ASXL3* | -4.652824376 | 0.000784776 | 0.002451011 | 1 |
| HTN | b_cell | *ADCYAP1* | 21.40077064 | 0.001452032 | 0.003419073 | 1 |
| HTN | b_cell | *ESCO1* | -4.819268285 | 0.038871652 | 0.048012982 | 1 |
| HTN | b_cell | *GREB1L* | -2.641322102 | 4.69E-05 | 0.001685537 | 0.294969013 |
| HTN | b_cell | *C18orf8* | -9.001625328 | 0.000266192 | 0.002180344 | 1 |
| HTN | b_cell | *NPC1* | -6.267924352 | 0.000469791 | 0.002180489 | 1 |
| HTN | b_cell | *SLC14A1* | 11.34725371 | 5.53E-05 | 0.001851584 | 0.347940435 |
| HTN | b_cell | *SLC13A5* | -33.32303423 | 0.001562452 | 0.003569964 | 1 |
| HTN | b_cell | *CCDC40* | -3.816491899 | 0.000146094 | 0.002180344 | 0.918492881 |

| | | | | | | |
|---|---|---|---|---|---|---|
| HTN | b_cell | *TMC6* | -3.945470239 | 0.015051596 | 0.020558197 | 1 |
| HTN | b_cell | *CARD14* | -4.478060308 | 0.00042632 | 0.002180344 | 1 |
| HTN | b_cell | *TBCD* | -4.38801251 | 0.002439396 | 0.004822793 | 1 |
| HTN | b_cell | *NARF* | -4.835210369 | 0.027476878 | 0.035061321 | 1 |
| HTN | b_cell | *TRIM65* | -8.263174194 | 0.014761547 | 0.020223544 | 1 |
| HTN | b_cell | *CBX8* | 2.18678471 | 0.00015215 | 0.002180344 | 0.956569772 |
| HTN | b_cell | *SECTM1* | -8.754189423 | 0.000400984 | 0.002180344 | 1 |
| HTN | b_cell | *RNF157* | -3.371798143 | 0.000144735 | 0.002180344 | 0.909949024 |
| HTN | b_cell | *RNF165* | -4.141823064 | 0.000266588 | 0.002180344 | 1 |
| HTN | b_cell | *MAPK4* | -6.519478053 | 0.010750877 | 0.015416241 | 1 |
| HTN | b_cell | *SMAD4* | -5.067860666 | 0.018702087 | 0.024858355 | 1 |
| HTN | b_cell | *NT5C3B* | -3.3534133 | 0.007745341 | 0.011669053 | 1 |
| HTN | b_cell | *PIP4K2B* | -4.437738382 | 0.001122565 | 0.002924292 | 1 |
| HTN | b_cell | *GRB7* | 5.523376796 | 0.000119038 | 0.002180344 | 0.748392749 |
| HTN | b_cell | *FKBP10* | -3.999855168 | 0.000821256 | 0.00250521 | 1 |
| HTN | b_cell | *TXNL4A* | -5.633446779 | 0.000106762 | 0.002180344 | 0.671215568 |
| HTN | b_cell | *NFIC* | -2.975476112 | 0.00045395 | 0.002180344 | 1 |
| HTN | b_cell | *TPGS1* | -4.432098454 | 0.001058954 | 0.002833039 | 1 |
| HTN | b_cell | *PRDM15* | -5.175793995 | 0.00086982 | 0.002569811 | 1 |
| HTN | b_cell | *PFKL* | -7.008910171 | 0.003084676 | 0.005714012 | 1 |
| HTN | b_cell | *MVB12A* | -5.314223585 | 0.003672847 | 0.006505071 | 1 |
| HTN | b_cell | *DUS3L* | 8.375624865 | 0.001018133 | 0.002775525 | 1 |
| HTN | b_cell | *DPP9* | 6.533594229 | 0.011528452 | 0.01637582 | 1 |
| HTN | b_cell | *SIRT3* | -3.878783753 | 0.004746981 | 0.007922556 | 1 |
| HTN | b_cell | *IFITM3* | -4.068915516 | 0.000689164 | 0.002355324 | 1 |
| HTN | b_cell | *COL6A1* | -2.157563537 | 0.000932497 | 0.002657043 | 1 |
| HTN | b_cell | *IFNAR1* | -4.958219413 | 0.002638411 | 0.005101411 | 1 |
| HTN | b_cell | *TRPM2* | -3.375595666 | 0.011189169 | 0.015933478 | 1 |
| HTN | b_cell | *TMEM50B* | -3.325101144 | 0.027180774 | 0.034718717 | 1 |
| HTN | b_cell | *URB1* | -4.731640059 | 0.0338434 | 0.042292307 | 1 |
| HTN | b_cell | *AKT1* | -5.078137958 | 0.000591404 | 0.00226693 | 1 |
| HTN | b_cell | *SAE1* | -4.061907197 | 0.004374121 | 0.007430882 | 1 |
| HTN | b_cell | *GEMIN7* | 6.167366305 | 0.001534681 | 0.003530383 | 1 |
| HTN | b_cell | *WTIP* | -3.25220367 | 0.000455987 | 0.002180344 | 1 |
| HTN | b_cell | *CAPN10* | -3.960655766 | 0.001787708 | 0.003898481 | 1 |
| HTN | b_cell | *MYO1F* | -2.810533383 | 0.000435891 | 0.002180344 | 1 |
| HTN | b_cell | *CACNG8* | -11.70166152 | 0.000421153 | 0.002180344 | 1 |
| HTN | b_cell | *FBN3* | -7.76157659 | 0.000973104 | 0.002717425 | 1 |
| HTN | b_cell | *EVI5L* | 9.507889025 | 0.00365289 | 0.006474688 | 1 |
| HTN | b_cell | *CTU1* | -6.206404518 | 0.0003102 | 0.002180344 | 1 |
| HTN | b_cell | *RCN3* | -3.511728805 | 0.000473181 | 0.002180489 | 1 |
| HTN | b_cell | *PEX14* | -3.286015923 | 0.00060856 | 0.002281385 | 1 |
| HTN | b_cell | *IL22RA1* | -8.292953391 | 0.012215198 | 0.017215187 | 1 |
| HTN | b_cell | *ZNF593* | -7.861645961 | 0.00870149 | 0.01285996 | 1 |
| HTN | b_cell | *C1orf216* | -5.387252575 | 0.000438734 | 0.002180344 | 1 |
| HTN | b_cell | *GPN2* | -3.828726901 | 0.000379419 | 0.002180344 | 1 |
| HTN | b_cell | *NBPF3* | -2.69464507 | 0.034062846 | 0.042516005 | 1 |
| HTN | b_cell | *HSPG2* | -2.343964692 | 0.008923821 | 0.013142203 | 1 |
| HTN | b_cell | *ITGB3BP* | -4.322548973 | 0.003923212 | 0.00684575 | 1 |
| HTN | b_cell | *SERBP1* | -5.448721189 | 0.01940982 | 0.025652625 | 1 |

| | | | | | | |
|---|---|---|---|---|---|---|
| HTN | b_cell | *PIGK* | -5.988372399 | 0.006861426 | 0.010596361 | 1 |
| HTN | b_cell | *RPS8* | -5.960902645 | 0.007187592 | 0.010984052 | 1 |
| HTN | b_cell | *MOB3C* | 5.05232685 | 0.000255892 | 0.002180344 | 1 |
| HTN | b_cell | *CYP4B1* | -2.398594863 | 0.01054297 | 0.015164414 | 1 |
| HTN | b_cell | *ZNF697* | -4.402331815 | 0.000780572 | 0.002446934 | 1 |
| HTN | b_cell | *CELSR2* | -12.42808409 | 0.002110591 | 0.004342044 | 1 |
| HTN | b_cell | *ATP1B1* | -3.593541904 | 0.000365046 | 0.002180344 | 1 |
| HTN | b_cell | *TIPRL* | -5.666115877 | 0.004669529 | 0.00782862 | 1 |
| HTN | b_cell | *NME7* | -5.887865313 | 0.002581691 | 0.005029777 | 1 |
| HTN | b_cell | *CREG1* | -5.559210348 | 0.002441919 | 0.004826265 | 1 |
| HTN | b_cell | *GPA33* | -14.2565152 | 0.000116198 | 0.002180344 | 0.730535171 |
| HTN | b_cell | *RXRG* | 24.67426193 | 0.005955735 | 0.009484221 | 1 |
| HTN | b_cell | *TMCO1* | -5.200032094 | 0.024855518 | 0.032028416 | 1 |
| HTN | b_cell | *MGST3* | -3.632279846 | 0.000901406 | 0.00260492 | 1 |
| HTN | b_cell | *ADCY10* | -4.875207212 | 0.000487568 | 0.002188291 | 1 |
| HTN | b_cell | *RFWD2* | -3.90536039 | 0.013031426 | 0.018186144 | 1 |
| HTN | b_cell | *PPOX* | -5.010725543 | 0.002909717 | 0.005477063 | 1 |
| HTN | b_cell | *RGS5* | -7.885727196 | 0.020079629 | 0.026459993 | 1 |
| HTN | b_cell | *RRNAD1* | -5.526670516 | 0.005482044 | 0.00888747 | 1 |
| HTN | b_cell | *PIGM* | -4.780476316 | 0.000714772 | 0.002383123 | 1 |
| HTN | b_cell | *RGL1* | -2.565990197 | 0.000656662 | 0.002327958 | 1 |
| HTN | b_cell | *TUFT1* | -3.594837016 | 0.006399539 | 0.010025891 | 1 |
| HTN | b_cell | *TARS2* | -4.416866749 | 0.001407463 | 0.003347484 | 1 |
| HTN | b_cell | *SNX27* | -5.687754616 | 0.012022805 | 0.016993565 | 1 |
| HTN | b_cell | *SELENBP1* | -2.912348311 | 0.001032144 | 0.002797022 | 1 |
| HTN | b_cell | *ENSA* | -4.824925046 | 0.039531723 | 0.04876122 | 1 |
| HTN | b_cell | *SEMA6C* | -8.1288348 | 0.00536477 | 0.00873564 | 1 |
| HTN | b_cell | *ARNT* | -5.691482779 | 0.000822328 | 0.002507264 | 1 |
| HTN | b_cell | *POGZ* | -4.504726056 | 0.002075587 | 0.004296746 | 1 |
| HTN | b_cell | *HORMAD1* | -1.910099029 | 0.014915439 | 0.020407696 | 1 |
| HTN | b_cell | *GABPB2* | -4.026523219 | 0.002087025 | 0.004311905 | 1 |
| HTN | b_cell | *DYRK3* | -6.378054203 | 0.000660573 | 0.002327958 | 1 |
| HTN | b_cell | *EIF2D* | -5.906992673 | 0.001553499 | 0.003556754 | 1 |
| HTN | b_cell | *TP53BP2* | -5.471404793 | 0.001799351 | 0.003918435 | 1 |
| HTN | b_cell | *RAB13* | -4.984500217 | 9.16E-05 | 0.002180344 | 0.576033117 |
| HTN | b_cell | *S100A8* | 4.28236741 | 0.000457055 | 0.002180344 | 1 |
| HTN | b_cell | *HAX1* | -5.853404284 | 0.018512982 | 0.024643472 | 1 |
| HTN | b_cell | *CREB3L4* | -9.254361069 | 3.79E-07 | 6.70E-05 | 0.002380901 |
| HTN | b_cell | *KCNN3* | -3.9835724 | 0.000619242 | 0.002288754 | 1 |
| HTN | b_cell | *GATAD2B* | -4.573369387 | 0.017453794 | 0.023382059 | 1 |
| HTN | b_cell | *RIT1* | -5.624591244 | 0.000942433 | 0.002668955 | 1 |
| HTN | b_cell | *FLG* | -4.749620024 | 0.000740335 | 0.002405521 | 1 |
| HTN | b_cell | *GALNT2* | -4.394939658 | 0.002081611 | 0.004303303 | 1 |
| HTN | b_cell | *SCCPDH* | -5.546902503 | 0.00120175 | 0.003041684 | 1 |
| HTN | b_cell | *NVL* | -4.75344036 | 0.002596511 | 0.005049261 | 1 |
| HTN | b_cell | *SDE2* | -9.210507539 | 0.012275546 | 0.017296359 | 1 |
| HTN | b_cell | *FBXO28* | -5.022322233 | 0.022449609 | 0.029245896 | 1 |
| HTN | b_cell | *GUK1* | -3.745246628 | 0.000924981 | 0.002648292 | 1 |
| HTN | b_cell | *CDC42BPA* | -3.113632456 | 0.000854547 | 0.002551796 | 1 |
| HTN | b_cell | *CNIH3* | -4.499526651 | 0.001607003 | 0.003638181 | 1 |

| | | | | | | |
|---|---|---|---|---|---|---|
| HTN | b_cell | *PYCR2* | 8.994731296 | 0.001646871 | 0.003703104 | 1 |
| HTN | b_cell | *WNT9A* | -3.377958754 | 0.007944167 | 0.011914438 | 1 |
| HTN | b_cell | *PLEKHA6* | -3.176335031 | 0.001871643 | 0.004022913 | 1 |
| HTN | b_cell | *ABCG8* | -21.76740111 | 0.000128585 | 0.002180344 | 0.808410864 |
| HTN | b_cell | *EML4* | -4.259490363 | 0.01258971 | 0.017659863 | 1 |
| HTN | b_cell | *CHAC2* | -9.859084979 | 0.001490001 | 0.003466926 | 1 |
| HTN | b_cell | *WDPCP* | -1.697034423 | 0.000226163 | 0.002180344 | 1 |
| HTN | b_cell | *ASXL2* | -4.550874117 | 0.012581058 | 0.017651665 | 1 |
| HTN | b_cell | *CIAO1* | -4.803448761 | 0.00917265 | 0.013455074 | 1 |
| HTN | b_cell | *ZNF514* | -2.411448812 | 0.022136461 | 0.028891826 | 1 |
| HTN | b_cell | *EXOC6B* | -3.662629929 | 0.00938095 | 0.01369671 | 1 |
| HTN | b_cell | *TEX261* | -9.475833357 | 0.021281255 | 0.027879819 | 1 |
| HTN | b_cell | *THNSL2* | -2.95535933 | 4.64E-05 | 0.00167785 | 0.291945951 |
| HTN | b_cell | *TMEM177* | -6.521232657 | 0.000530027 | 0.002195202 | 1 |
| HTN | b_cell | *RABL2A* | -8.820340848 | 0.000116252 | 0.002180344 | 0.730878015 |
| HTN | b_cell | *FBLN7* | -3.948932483 | 0.000997909 | 0.002748073 | 1 |
| HTN | b_cell | *ZC3H8* | -4.101965177 | 0.028396596 | 0.036124929 | 1 |
| HTN | b_cell | *FAHD2B* | -7.019125205 | 0.000564193 | 0.002229465 | 1 |
| HTN | b_cell | *LYG1* | -3.918435736 | 0.000989258 | 0.00273023 | 1 |
| HTN | b_cell | *AFF3* | 3.370328873 | 0.004414915 | 0.007481555 | 1 |
| HTN | b_cell | *UBXN4* | -4.359890128 | 0.006919411 | 0.010654503 | 1 |
| HTN | b_cell | *THSD7B* | -2.22873371 | 0.001994836 | 0.00418749 | 1 |
| HTN | b_cell | *POLR2D* | -6.928907827 | 0.000679552 | 0.002339217 | 1 |
| HTN | b_cell | *ZNF385B* | -2.892956 | 0.000349347 | 0.002180344 | 1 |
| HTN | b_cell | *TMEFF2* | -3.344430534 | 0.000514871 | 0.002195202 | 1 |
| HTN | b_cell | *PHOSPHO2* | -5.195914657 | 0.007070459 | 0.010841945 | 1 |
| HTN | b_cell | *GULP1* | -2.641048596 | 0.000407386 | 0.002180344 | 1 |
| HTN | b_cell | *FAM171B* | -4.21406735 | 0.001275158 | 0.003156737 | 1 |
| HTN | b_cell | *HSPD1* | -5.026282862 | 0.005327163 | 0.008683456 | 1 |
| HTN | b_cell | *PTH2R* | -3.331413325 | 0.000630303 | 0.002297226 | 1 |
| HTN | b_cell | *NBEAL1* | -3.106943496 | 0.000700791 | 0.002366204 | 1 |
| HTN | b_cell | *SUMF1* | -3.784490256 | 0.005497194 | 0.008905143 | 1 |
| HTN | b_cell | *ESPNL* | -45.71036706 | 0.002050543 | 0.004258924 | 1 |
| HTN | b_cell | *FANCD2* | 2.075222661 | 0.000190908 | 0.002180344 | 1 |
| HTN | b_cell | *OSBPL10* | 4.145813965 | 0.000549734 | 0.00221672 | 1 |
| HTN | b_cell | *SLC25A38* | 3.99507481 | 0.00042688 | 0.002180344 | 1 |
| HTN | b_cell | *CTDSPL* | -5.868868558 | 0.004165675 | 0.007171992 | 1 |
| HTN | b_cell | *STAC* | -3.684137405 | 0.000411793 | 0.002180344 | 1 |
| HTN | b_cell | *ARL6IP5* | -4.717775848 | 0.00213641 | 0.00437294 | 1 |
| HTN | b_cell | *TMF1* | -4.669286319 | 0.003474021 | 0.006231433 | 1 |
| HTN | b_cell | *COL8A1* | -2.263925082 | 0.00026898 | 0.002180344 | 1 |
| HTN | b_cell | *NXPE3* | -6.819378711 | 0.005614476 | 0.009060116 | 1 |
| HTN | b_cell | *PLA1A* | -7.98044379 | 0.002845019 | 0.00538751 | 1 |
| HTN | b_cell | *RABL3* | -6.177152967 | 0.002797245 | 0.005332408 | 1 |
| HTN | b_cell | *ADPRH* | -5.633425668 | 0.000483788 | 0.002187427 | 1 |
| HTN | b_cell | *IGSF11* | -9.506612443 | 0.000593505 | 0.00226693 | 1 |
| HTN | b_cell | *BOC* | -2.641470387 | 0.000901591 | 0.00260492 | 1 |
| HTN | b_cell | *AGTR1* | -3.192806562 | 0.000153897 | 0.002180344 | 0.967549236 |
| HTN | b_cell | *EIF2A* | -4.57029799 | 0.000440759 | 0.002180344 | 1 |
| HTN | b_cell | *AMT* | -6.391569188 | 2.08E-05 | 0.001030212 | 0.130836889 |

| | | | | | | |
|---|---|---|---|---|---|---|
| HTN | b_cell | *TCTA* | -5.388482554 | 0.000255852 | 0.002180344 | 1 |
| HTN | b_cell | *EAF2* | 2.820557314 | 0.000198329 | 0.002180344 | 1 |
| HTN | b_cell | *ILDR1* | 4.395015743 | 0.002671205 | 0.005142028 | 1 |
| HTN | b_cell | *MUC4* | -3.998700658 | 0.002421937 | 0.004794307 | 1 |
| HTN | b_cell | *DGKQ* | -4.660829309 | 0.001229133 | 0.003088552 | 1 |
| HTN | b_cell | *SLC26A1* | 7.010689387 | 0.00025308 | 0.002180344 | 1 |
| HTN | b_cell | *LYAR* | -4.289551238 | 0.000446415 | 0.002180344 | 1 |
| HTN | b_cell | *CENPC* | -5.808946887 | 0.001308422 | 0.003205335 | 1 |
| HTN | b_cell | *ATP10D* | -4.034068031 | 0.012022567 | 0.016993565 | 1 |
| HTN | b_cell | *SLC10A4* | 24.65496082 | 0.010132382 | 0.014637474 | 1 |
| HTN | b_cell | *PYURF* | -9.239044496 | 0.005111112 | 0.008400931 | 1 |
| HTN | b_cell | *CISD2* | -6.130699544 | 0.008311304 | 0.012350075 | 1 |
| HTN | b_cell | *CCNA2* | 6.312267954 | 0.004735695 | 0.007907919 | 1 |
| HTN | b_cell | *USP53* | -3.194051724 | 0.001067623 | 0.002845984 | 1 |
| HTN | b_cell | *MARCH1* | 3.603827321 | 0.027400133 | 0.034977591 | 1 |
| HTN | b_cell | *PDGFC* | -2.87663277 | 0.000504682 | 0.002195028 | 1 |
| HTN | b_cell | *CBR4* | -4.260090167 | 0.005809306 | 0.009302881 | 1 |
| HTN | b_cell | *CYP4V2* | -4.274148785 | 0.02021909 | 0.026611624 | 1 |
| HTN | b_cell | *NDUFS6* | -8.101783636 | 0.004896421 | 0.008113811 | 1 |
| HTN | b_cell | *FAM105A* | -4.049053712 | 0.001145139 | 0.0029591 | 1 |
| HTN | b_cell | *SSBP2* | -3.70014001 | 0.01511389 | 0.020607466 | 1 |
| HTN | b_cell | *BHMT* | -3.038088687 | 0.005523419 | 0.008940714 | 1 |
| HTN | b_cell | *CRHBP* | -4.776829807 | 0.000863542 | 0.002559431 | 1 |
| HTN | b_cell | *RASA1* | -3.359445998 | 0.006494716 | 0.010162339 | 1 |
| HTN | b_cell | *PPIP5K2* | -4.410282303 | 0.006820427 | 0.010556942 | 1 |
| HTN | b_cell | *SPATA9* | -2.916507209 | 0.000505578 | 0.002195143 | 1 |
| HTN | b_cell | *TSLP* | -8.900929358 | 4.94E-06 | 0.000398055 | 0.031048304 |
| HTN | b_cell | *FEM1C* | -6.316085602 | 0.006880423 | 0.010615269 | 1 |
| HTN | b_cell | *COMMD10* | -4.15722853 | 0.007227056 | 0.011036313 | 1 |
| HTN | b_cell | *YIPF5* | -5.171893589 | 0.00097807 | 0.002717425 | 1 |
| HTN | b_cell | *SLC25A48* | -7.129767009 | 0.003819848 | 0.006711958 | 1 |
| HTN | b_cell | *GABRB2* | -3.512644669 | 0.00121507 | 0.003062536 | 1 |
| HTN | b_cell | *MYLK4* | -8.218608069 | 0.001196749 | 0.003033854 | 1 |
| HTN | b_cell | *TBC1D7* | -6.999130975 | 0.00619239 | 0.009786716 | 1 |
| HTN | b_cell | *GFOD1* | -1.974924438 | 0.003758498 | 0.006622803 | 1 |
| HTN | b_cell | *PSD2* | 9.476187728 | 7.15E-05 | 0.002098808 | 0.449753208 |
| HTN | b_cell | *ZMAT2* | -3.375791388 | 0.002222731 | 0.004502033 | 1 |
| HTN | b_cell | *KLHL3* | -3.131836677 | 0.000335613 | 0.002180344 | 1 |
| HTN | b_cell | *TRIM7* | 5.295077567 | 0.00128382 | 0.003167729 | 1 |
| HTN | b_cell | *HIGD2A* | -7.107358941 | 0.000890545 | 0.002599737 | 1 |
| HTN | b_cell | *FAM193B* | -4.874002234 | 0.002080118 | 0.004303303 | 1 |
| HTN | b_cell | *MUT* | -5.41942551 | 0.001305084 | 0.003198854 | 1 |
| HTN | b_cell | *DOK3* | -5.941565007 | 0.000104324 | 0.002180344 | 0.655886835 |
| HTN | b_cell | *ABT1* | -3.46457256 | 0.020022179 | 0.026395353 | 1 |
| HTN | b_cell | *MLIP* | -6.705788233 | 0.005589659 | 0.009031388 | 1 |
| HTN | b_cell | *TPBG* | -3.87703721 | 0.000413566 | 0.002180344 | 1 |
| HTN | b_cell | *IRAK1BP1* | -4.364621133 | 0.002453729 | 0.004845036 | 1 |
| HTN | b_cell | *PHIP* | -3.782098878 | 0.012453969 | 0.017496783 | 1 |
| HTN | b_cell | *PM20D2* | -7.096313638 | 0.036607761 | 0.045395068 | 1 |
| HTN | b_cell | *RNF217* | -3.729211917 | 0.001198478 | 0.00303579 | 1 |

| | | | | | | |
|---|---|---|---|---|---|---|
| HTN | b_cell | *ABRACL* | 8.040228786 | 0.024764866 | 0.031931237 | 1 |
| HTN | b_cell | *SLC18B1* | -6.031187605 | 7.02E-07 | 9.59E-05 | 0.00441107 |
| HTN | b_cell | *AIG1* | -2.822242141 | 0.001069074 | 0.002845984 | 1 |
| HTN | b_cell | *DYNLT1* | -3.929414137 | 0.000355939 | 0.002180344 | 1 |
| HTN | b_cell | *PNLDC1* | -7.805152067 | 0.037734976 | 0.046737548 | 1 |
| HTN | b_cell | *SLC22A3* | -2.92114259 | 0.000414071 | 0.002180344 | 1 |
| HTN | b_cell | *GNA12* | -4.782438099 | 0.014227985 | 0.019586455 | 1 |
| HTN | b_cell | *C7orf50* | -4.477313288 | 0.000356081 | 0.002180344 | 1 |
| HTN | b_cell | *SDK1* | -1.997766582 | 0.001549072 | 0.003551792 | 1 |
| HTN | b_cell | *CCZ1B* | -6.796728169 | 0.0075177 | 0.011402838 | 1 |
| HTN | b_cell | *RBAK* | -8.943709187 | 0.009057372 | 0.013304602 | 1 |
| HTN | b_cell | *EGFR* | -2.046628338 | 0.000478543 | 0.002180489 | 1 |
| HTN | b_cell | *POMZP3* | -5.326153988 | 0.00957549 | 0.013916114 | 1 |
| HTN | b_cell | *CCT6A* | -6.348959323 | 0.003187549 | 0.005844305 | 1 |
| HTN | b_cell | *PSPH* | -3.898211408 | 0.002224232 | 0.004503622 | 1 |
| HTN | b_cell | *TMEM209* | -3.65904928 | 0.000402564 | 0.002180344 | 1 |
| HTN | b_cell | *AGBL3* | -3.290918896 | 0.000808748 | 0.002488487 | 1 |
| HTN | b_cell | *EPHA1* | -7.964826379 | 0.005555816 | 0.008986214 | 1 |
| HTN | b_cell | *NCAPG2* | -4.844364128 | 0.005041895 | 0.008313786 | 1 |
| HTN | b_cell | *FBXO25* | -4.570417813 | 0.000982423 | 0.002721721 | 1 |
| HTN | b_cell | *CSGALNACT1* | -4.274085071 | 0.022439889 | 0.029239292 | 1 |
| HTN | b_cell | *SLC25A37* | -4.658399828 | 0.000572382 | 0.002237915 | 1 |
| HTN | b_cell | *CHMP7* | 8.82228869 | 0.00867656 | 0.012832165 | 1 |
| HTN | b_cell | *DOCK5* | -5.088313669 | 0.006403125 | 0.010029011 | 1 |
| HTN | b_cell | *ST18* | -3.778347132 | 0.000839647 | 0.002531828 | 1 |
| HTN | b_cell | *RGS20* | -3.777946085 | 0.000892737 | 0.002599737 | 1 |
| HTN | b_cell | *TACC1* | -3.480113126 | 0.001919996 | 0.004090484 | 1 |
| HTN | b_cell | *ADHFE1* | -2.95988209 | 5.13E-06 | 0.000398561 | 0.032233874 |
| HTN | b_cell | *SLC26A7* | -2.741308781 | 0.006599339 | 0.010264731 | 1 |
| HTN | b_cell | *ATP6V0D2* | -6.586800599 | 0.000149226 | 0.002180344 | 0.938185406 |
| HTN | b_cell | *MTDH* | -4.35359582 | 0.016182146 | 0.021888372 | 1 |
| HTN | b_cell | *MAL2* | -3.996285826 | 0.000739402 | 0.002405521 | 1 |
| HTN | b_cell | *FAM135B* | 6.121018733 | 0.000248702 | 0.002180344 | 1 |
| HTN | b_cell | *ZNF7* | -4.594025937 | 0.009288356 | 0.01358676 | 1 |
| HTN | b_cell | *AK3* | -4.621454508 | 0.000722009 | 0.002388914 | 1 |
| HTN | b_cell | *CER1* | -8.92711741 | 0.032443678 | 0.040721382 | 1 |
| HTN | b_cell | *PLIN2* | -4.946417415 | 0.003172814 | 0.005825784 | 1 |
| HTN | b_cell | *HAUS6* | -6.174612305 | 0.00325269 | 0.005934319 | 1 |
| HTN | b_cell | *C9orf72* | -4.436339011 | 0.011112005 | 0.015834356 | 1 |
| HTN | b_cell | *SIGMAR1* | -6.353885863 | 0.000173973 | 0.002180344 | 1 |
| HTN | b_cell | *CBWD5* | -4.192519509 | 0.000489504 | 0.002188936 | 1 |
| HTN | b_cell | *ZNF462* | -3.78842958 | 0.000855571 | 0.002551796 | 1 |
| HTN | b_cell | *ALAD* | -2.98412219 | 0.021751842 | 0.02843115 | 1 |
| HTN | b_cell | *ASTN2* | -5.429697733 | 0.006008412 | 0.009543934 | 1 |
| HTN | b_cell | *WDR31* | -6.169308852 | 0.003103074 | 0.005736262 | 1 |
| HTN | b_cell | *POLE3* | 9.608781304 | 0.000315153 | 0.002180344 | 1 |
| HTN | b_cell | *SURF6* | 6.659067567 | 0.001190427 | 0.003027596 | 1 |
| HTN | b_cell | *ASB6* | -5.923533713 | 0.000378957 | 0.002180344 | 1 |
| HTN | b_cell | *NTMT1* | -6.876863137 | 0.004120821 | 0.007117472 | 1 |
| HTN | b_cell | *INPP5E* | -6.885456579 | 0.006147113 | 0.009722491 | 1 |

| | | | | | | |
|---|---|---|---|---|---|---|
| HTN | b_cell | *SEC16A* | -4.881812509 | 0.008045398 | 0.012031735 | 1 |
| HTN | b_cell | *DPH7* | -5.939146217 | 0.001703118 | 0.003778231 | 1 |
| HTN | b_cell | *SLC39A12* | -11.03767757 | 0.003300259 | 0.005998476 | 1 |
| HTN | b_cell | *RSU1* | -4.496937697 | 0.003569 | 0.00635286 | 1 |
| HTN | b_cell | *PARD3* | -2.249057929 | 0.000324303 | 0.002180344 | 1 |
| HTN | b_cell | *NRBF2* | -5.582758614 | 1.95E-06 | 0.00019128 | 0.012241904 |
| HTN | b_cell | *ADIRF* | -4.028421064 | 0.001261499 | 0.00313728 | 1 |
| HTN | b_cell | *HTR7* | -5.363309371 | 0.000449497 | 0.002180344 | 1 |
| HTN | b_cell | *RPP30* | -6.122604442 | 0.002252037 | 0.00454091 | 1 |
| HTN | b_cell | *DNAJB12* | -5.944311085 | 0.00135487 | 0.003272403 | 1 |
| HTN | b_cell | *PLEKHS1* | 12.77697801 | 1.08E-05 | 0.000667043 | 0.068038394 |
| HTN | b_cell | *FUOM* | -11.48917079 | 0.029489081 | 0.037356005 | 1 |
| HTN | b_cell | *LRRC27* | -4.112794012 | 0.016587145 | 0.022359215 | 1 |
| HTN | b_cell | *MTG1* | -2.797117402 | 0.031168827 | 0.039270223 | 1 |
| HTN | b_cell | *PAOX* | 4.373492288 | 0.000283437 | 0.002180344 | 1 |
| HTN | b_cell | *GSTO1* | -4.139363947 | 0.000964996 | 0.002708142 | 1 |
| HTN | b_cell | *PDCD11* | -2.74912167 | 0.030169113 | 0.038140602 | 1 |
| HTN | b_cell | *RGS10* | -3.861031846 | 0.000379835 | 0.002180344 | 1 |
| HTN | b_cell | *GAS2* | -6.612590097 | 0.001747554 | 0.003844252 | 1 |
| HTN | b_cell | *LIN7C* | -7.182265365 | 0.009389022 | 0.013698951 | 1 |
| HTN | b_cell | *ZNF215* | 3.966720456 | 5.78E-05 | 0.001902693 | 0.363414302 |
| HTN | b_cell | *HSD17B12* | -10.44526092 | 1.05E-08 | 5.08E-06 | 6.60E-05 |
| HTN | b_cell | *GLYAT* | -2.568009994 | 0.000827638 | 0.002514184 | 1 |
| HTN | b_cell | *PTPRJ* | -4.353138352 | 0.00032439 | 0.002180344 | 1 |
| HTN | b_cell | *C11orf49* | -5.587636857 | 0.011026762 | 0.015741429 | 1 |
| HTN | b_cell | *CCDC82* | -4.213425963 | 0.000981597 | 0.002721032 | 1 |
| HTN | b_cell | *SERPINH1* | -3.950384504 | 0.000273331 | 0.002180344 | 1 |
| HTN | b_cell | *CAPN5* | -4.884024877 | 0.000697875 | 0.002362702 | 1 |
| HTN | b_cell | *ZC3H12C* | -4.928309315 | 0.003833015 | 0.006727573 | 1 |
| HTN | b_cell | *DRD2* | -21.61906844 | 0.000666484 | 0.002330887 | 1 |
| HTN | b_cell | *C11orf52* | -36.70043371 | 0.000431119 | 0.002180344 | 1 |
| HTN | b_cell | *NPAT* | -5.022184107 | 0.013399533 | 0.018596658 | 1 |
| HTN | b_cell | *GRIK4* | -3.350361228 | 0.023771737 | 0.030795984 | 1 |
| HTN | b_cell | *ADAM33* | -3.456969728 | 0.000205439 | 0.002180344 | 1 |
| HTN | b_cell | *MTA2* | -10.52149978 | 0.004341461 | 0.007389282 | 1 |
| HTN | b_cell | *TMC2* | -14.23701323 | 0.024026229 | 0.03107445 | 1 |
| HTN | b_cell | *EML3* | -4.994502463 | 0.001150616 | 0.002969592 | 1 |
| HTN | b_cell | *INCENP* | -5.561361165 | 0.000454359 | 0.002180344 | 1 |
| HTN | b_cell | *MS4A3* | -94.88535265 | 0.000395719 | 0.002180344 | 1 |
| HTN | b_cell | *PLCH2* | -16.44102955 | 0.000456193 | 0.002180344 | 1 |
| HTN | b_cell | *CPSF7* | -5.422427754 | 0.008797845 | 0.012992351 | 1 |
| HTN | b_cell | *FEZ1* | -4.978434872 | 0.000433502 | 0.002180344 | 1 |
| HTN | b_cell | *COMMD7* | -5.113318088 | 0.001056188 | 0.002832872 | 1 |
| HTN | b_cell | *KIAA1755* | -3.448525262 | 0.009847656 | 0.014265486 | 1 |
| HTN | b_cell | *CNBD2* | 10.95862063 | 0.023027055 | 0.029936049 | 1 |
| HTN | b_cell | *CABLES2* | 4.570523983 | 0.004389884 | 0.007453199 | 1 |
| HTN | b_cell | *TRPT1* | -5.422596858 | 0.000271045 | 0.002180344 | 1 |
| HTN | b_cell | *DOC2A* | -19.2440483 | 9.17E-09 | 4.81E-06 | 5.77E-05 |
| HTN | b_cell | *TAOK2* | -6.889805265 | 0.0016234 | 0.003666061 | 1 |
| HTN | b_cell | *MPP7* | -3.846629485 | 0.00300305 | 0.005600763 | 1 |

| | | | | | | |
|---|---|---|---|---|---|---|
| HTN | b_cell | *CWC15* | -5.404997179 | 0.006557352 | 0.010222185 | 1 |
| HTN | b_cell | *TMCO3* | -5.673859931 | 0.013358766 | 0.018560567 | 1 |
| HTN | b_cell | *SAP18* | -6.100865784 | 0.005881162 | 0.009391635 | 1 |
| HTN | b_cell | *LYPD6B* | -7.019686038 | 0.000623469 | 0.002291037 | 1 |
| HTN | b_cell | *WDR17* | -8.217928317 | 0.01323963 | 0.018411315 | 1 |
| HTN | b_cell | *CCDC102B* | -3.104749288 | 0.000560134 | 0.002226547 | 1 |
| HTN | b_cell | *CNDP1* | -7.212410536 | 0.018600082 | 0.024743698 | 1 |
| HTN | b_cell | *FSIP1* | 13.53915804 | 0.008467452 | 0.012549474 | 1 |
| HTN | b_cell | *RGS18* | -5.089086193 | 0.000443016 | 0.002180344 | 1 |
| HTN | b_cell | *PRSS23* | -2.151710878 | 0.000734329 | 0.002400794 | 1 |
| HTN | b_cell | *MTMR12* | -4.094871399 | 0.002444749 | 0.004830338 | 1 |
| HTN | b_cell | *PPP1R1C* | 8.886735687 | 0.020737677 | 0.02725288 | 1 |
| HTN | b_cell | *C11orf53* | 38.49294501 | 0.000895924 | 0.002599737 | 1 |
| HTN | b_cell | *CCT5* | -4.268288353 | 0.001430096 | 0.003383116 | 1 |
| HTN | b_cell | *FAM173B* | -3.181551404 | 0.000663356 | 0.002327958 | 1 |
| HTN | b_cell | *DOCK1* | -3.562305982 | 0.001234487 | 0.003098291 | 1 |
| HTN | b_cell | *TEX12* | -32.63010082 | 0.000407402 | 0.002180344 | 1 |
| HTN | b_cell | *PIP4K2A* | -4.180231214 | 0.013713758 | 0.018982474 | 1 |
| HTN | b_cell | *CRIM1* | -2.031508546 | 0.000426052 | 0.002180344 | 1 |
| HTN | b_cell | *ABCB9* | -7.15666893 | 0.010363995 | 0.014944595 | 1 |
| HTN | b_cell | *RILPL2* | -4.47292483 | 0.003876245 | 0.006792071 | 1 |
| HTN | b_cell | *PRSS53* | -18.61522844 | 0.00023653 | 0.002180344 | 1 |
| HTN | b_cell | *GPR158* | -4.980675215 | 0.001121712 | 0.002924292 | 1 |
| HTN | b_cell | *THRB* | -2.901549634 | 0.001028122 | 0.002792138 | 1 |
| HTN | b_cell | *TMEM86A* | -6.935317546 | 0.000832774 | 0.002523717 | 1 |
| HTN | b_cell | *BTBD11* | -4.008292954 | 0.000428941 | 0.002180344 | 1 |
| HTN | b_cell | *ANK3* | -3.36128351 | 0.003756649 | 0.006622803 | 1 |
| HTN | b_cell | *IPMK* | -4.664446099 | 0.00151936 | 0.003510893 | 1 |
| HTN | b_cell | *PLBD2* | -7.230188277 | 0.00379776 | 0.006680615 | 1 |
| HTN | b_cell | *DLG5* | -3.098460724 | 0.001277329 | 0.003156737 | 1 |
| HTN | b_cell | *GXYLT1* | -5.647089084 | 0.034340707 | 0.042837306 | 1 |
| HTN | b_cell | *MIPOL1* | -2.826825528 | 0.000767017 | 0.002430563 | 1 |
| HTN | b_cell | *EXT2* | -4.33074747 | 0.016805854 | 0.022629771 | 1 |
| HTN | b_cell | *TMEM18* | -7.140699839 | 0.002205915 | 0.004474035 | 1 |
| HTN | b_cell | *KCTD14* | 12.52592539 | 0.013763617 | 0.019034725 | 1 |
| HTN | b_cell | *THRSP* | -3.234452039 | 0.001277858 | 0.003156737 | 1 |
| HTN | b_cell | *NDUFC2* | -9.253689955 | 0.003896872 | 0.006814343 | 1 |
| HTN | b_cell | *ME3* | -3.652979957 | 0.003490916 | 0.006252818 | 1 |
| HTN | b_cell | *ADAMTS12* | -2.109999393 | 0.00042292 | 0.002180344 | 1 |
| HTN | b_cell | *PTPRO* | -6.362196642 | 0.000739207 | 0.002405521 | 1 |
| HTN | b_cell | *VPS26B* | -5.79052999 | 0.017626608 | 0.023578401 | 1 |
| HTN | b_cell | *NCAPD3* | -4.820699365 | 0.001390686 | 0.003321901 | 1 |
| HTN | b_cell | *QDPR* | -2.54730589 | 0.008190802 | 0.012205634 | 1 |
| HTN | b_cell | *FAM160B1* | -3.810801212 | 0.000821051 | 0.00250521 | 1 |
| HTN | b_cell | *ANO4* | -3.731728849 | 0.000322846 | 0.002180344 | 1 |
| HTN | b_cell | *AKR1C2* | -2.76609056 | 0.000315598 | 0.002180344 | 1 |
| HTN | b_cell | *DPYSL4* | 4.596130859 | 0.005478328 | 0.00888603 | 1 |
| HTN | b_cell | *ADAM8* | -5.47139811 | 0.000547283 | 0.002214137 | 1 |
| HTN | b_cell | *ITIH2* | 2.432005605 | 0.000227102 | 0.002180344 | 1 |
| HTN | b_cell | *KIN* | -5.57756978 | 0.013212119 | 0.018379308 | 1 |

| | | | | | | |
|---|---|---|---|---|---|---|
| HTN | b_cell | *PIGF* | -5.181734492 | 0.003759304 | 0.006622803 | 1 |
| HTN | b_cell | *INPP1* | -4.519883417 | 0.000366591 | 0.002180344 | 1 |
| HTN | b_cell | *MFSD6* | -5.560406369 | 0.00392476 | 0.006846549 | 1 |
| HTN | b_cell | *RNF144A* | -2.504171845 | 0.000415772 | 0.002180344 | 1 |
| HTN | b_cell | *ASAP2* | -3.541203372 | 0.001447928 | 0.003411965 | 1 |
| HTN | b_cell | *ADAM17* | -4.607210697 | 0.012934903 | 0.018071496 | 1 |
| HTN | b_cell | *TMEM45B* | -11.95069724 | 0.002569852 | 0.005014482 | 1 |
| HTN | b_cell | *ACSL1* | -1.991046702 | 0.000748318 | 0.002411417 | 1 |
| HTN | b_cell | *CCDC122* | -2.544280008 | 0.030502773 | 0.03850822 | 1 |
| HTN | b_cell | *GABRA2* | -6.790490667 | 3.73E-05 | 0.001482782 | 0.234649842 |
| HTN | b_cell | *CENPJ* | -3.14966262 | 0.00076932 | 0.002432944 | 1 |
| HTN | b_cell | *PARP8* | -3.103576398 | 0.001977191 | 0.004160174 | 1 |
| HTN | b_cell | *CACUL1* | -4.889997101 | 0.005457127 | 0.008860209 | 1 |
| HTN | b_cell | *BEND6* | -4.319760485 | 0.000203421 | 0.002180344 | 1 |
| HTN | b_cell | *PTPN14* | -2.263021302 | 0.001018904 | 0.002775525 | 1 |
| HTN | b_cell | *ARL14EP* | -6.801068291 | 0.000148598 | 0.002180344 | 0.934232577 |
| HTN | b_cell | *EPG5* | -5.348497741 | 0.010556636 | 0.015177126 | 1 |
| HTN | b_cell | *G6PC2* | -232.2837481 | 0.000438611 | 0.002180344 | 1 |
| HTN | b_cell | *TGOLN2* | -5.529348318 | 0.023160335 | 0.030072083 | 1 |
| HTN | b_cell | *ATG10* | -2.561473629 | 0.005288616 | 0.008645223 | 1 |
| HTN | b_cell | *POC5* | -4.313847685 | 0.009524474 | 0.013867617 | 1 |
| HTN | b_cell | *TADA1* | -7.072961747 | 0.002799812 | 0.005334066 | 1 |
| HTN | b_cell | *GUCY1A2* | -2.578892158 | 0.001520065 | 0.003510893 | 1 |
| HTN | b_cell | *CWF19L2* | -3.397230932 | 0.014834333 | 0.020313247 | 1 |
| HTN | b_cell | *ZNF547* | 4.672515171 | 0.000313401 | 0.002180344 | 1 |
| HTN | b_cell | *ZNF773* | -6.712814927 | 0.007679912 | 0.011603846 | 1 |
| HTN | b_cell | *NMT2* | -3.563008755 | 0.000568814 | 0.002232844 | 1 |
| HTN | b_cell | *ZNF837* | 4.162085472 | 0.000718589 | 0.002385315 | 1 |
| HTN | b_cell | *ZFP36L2* | -4.2129983 | 0.005315641 | 0.008674814 | 1 |
| HTN | b_cell | *PLEKHH2* | -2.351989506 | 0.000187606 | 0.002180344 | 1 |
| HTN | b_cell | *TMEM123* | -5.376755881 | 0.005750207 | 0.009231316 | 1 |
| HTN | b_cell | *IGSF10* | -3.911592767 | 0.000214775 | 0.002180344 | 1 |
| HTN | b_cell | *SPEF2* | -3.827393164 | 0.00175039 | 0.003844942 | 1 |
| HTN | b_cell | *SPARCL1* | -2.151623393 | 0.000395289 | 0.002180344 | 1 |
| HTN | b_cell | *CCNO* | 31.0653187 | 0.002609781 | 0.005068796 | 1 |
| HTN | b_cell | *CLEC4F* | -13.40035741 | 0.000416101 | 0.002180344 | 1 |
| HTN | b_cell | *PELO* | -5.08637843 | 0.000415161 | 0.002180344 | 1 |
| HTN | b_cell | *SAR1B* | -5.548768551 | 0.008242788 | 0.012271467 | 1 |
| HTN | b_cell | *GPR180* | -3.961512324 | 0.00025085 | 0.002180344 | 1 |
| HTN | b_cell | *TCTEX1D1* | -3.975126094 | 0.035893034 | 0.044572242 | 1 |
| HTN | b_cell | *WDR78* | -3.947886307 | 0.003464891 | 0.006222157 | 1 |
| HTN | b_cell | *IFIT5* | -7.21768365 | 0.001243255 | 0.003110364 | 1 |
| HTN | b_cell | *PRDM8* | 2.669285868 | 0.012791932 | 0.01789163 | 1 |
| HTN | b_cell | *HHEX* | -5.427437957 | 0.000249681 | 0.002180344 | 1 |
| HTN | b_cell | *ZNF117* | -1.572429287 | 0.006302636 | 0.009913604 | 1 |
| HTN | b_cell | *RAB3C* | -3.306451011 | 0.001487415 | 0.003464107 | 1 |
| HTN | b_cell | *RAD17* | -5.838289557 | 0.000251966 | 0.002180344 | 1 |
| HTN | b_cell | *MED21* | -5.248744087 | 0.002541506 | 0.004971515 | 1 |
| HTN | b_cell | *SRP19* | -5.756639986 | 0.00191453 | 0.004082989 | 1 |
| HTN | b_cell | *CENPH* | -3.700655496 | 0.017448189 | 0.023379532 | 1 |

| | | | | | | |
|---|---|---|---|---|---|---|
| HTN | b_cell | *CARHSP1* | -4.428965653 | 0.000805758 | 0.002484455 | 1 |
| HTN | b_cell | *TEKT5* | -11.4647951 | 0.000753709 | 0.002416404 | 1 |
| HTN | b_cell | *BANK1* | 0.526186171 | 0.000230501 | 0.002180344 | 1 |
| HTN | b_cell | *CAST* | -4.991619702 | 0.000435539 | 0.002180344 | 1 |
| HTN | b_cell | *SCOC* | -4.053648817 | 0.000860794 | 0.002559246 | 1 |
| HTN | b_cell | *CLGN* | -32.00248495 | 0.000827416 | 0.002514184 | 1 |
| HTN | b_cell | *SYCP2L* | -4.144690846 | 0.000575995 | 0.002241917 | 1 |
| HTN | b_cell | *AHCTF1* | -4.332984901 | 0.002252823 | 0.004541039 | 1 |
| HTN | b_cell | *MERTK* | -2.213785175 | 0.000320579 | 0.002180344 | 1 |
| HTN | b_cell | *TMEM87B* | -4.273091003 | 0.009704919 | 0.014065198 | 1 |
| HTN | b_cell | *PTPRR* | -6.273811558 | 0.00026617 | 0.002180344 | 1 |
| HTN | b_cell | *PLA2R1* | -2.479271951 | 0.001166386 | 0.002996758 | 1 |
| HTN | b_cell | *SLC25A27* | -2.582697777 | 0.001190086 | 0.003027596 | 1 |
| HTN | b_cell | *FRMD1* | -5.458768076 | 0.000413986 | 0.002180344 | 1 |
| HTN | b_cell | *ASAP1* | -3.400539153 | 0.001374266 | 0.0032952 | 1 |
| HTN | b_cell | *INO80C* | -5.649365577 | 0.000444032 | 0.002180344 | 1 |
| HTN | b_cell | *LPCAT1* | -6.78189774 | 0.002230015 | 0.004510262 | 1 |
| HTN | b_cell | *C16orf89* | -3.640752471 | 0.010406676 | 0.0150027 | 1 |
| HTN | b_cell | *TEX29* | -5.652808867 | 0.001108027 | 0.002902569 | 1 |
| HTN | b_cell | *ADPRHL1* | -6.337194805 | 0.002040995 | 0.004245264 | 1 |
| HTN | b_cell | *TUBGCP5* | -5.7044864 | 0.01430254 | 0.019673727 | 1 |
| HTN | b_cell | *LURAP1L* | -2.401751929 | 0.000631784 | 0.002300923 | 1 |
| HTN | b_cell | *CFDP1* | -4.32508872 | 0.003512212 | 0.006281429 | 1 |
| HTN | b_cell | *ZDHHC7* | -4.176095091 | 0.001218445 | 0.003069057 | 1 |
| HTN | b_cell | *FAM92B* | 17.35120501 | 0.000235257 | 0.002180344 | 1 |
| HTN | b_cell | *JAZF1* | -4.188605233 | 0.040154304 | 0.049441855 | 1 |
| HTN | b_cell | *MCOLN2* | 3.00946596 | 0.000370321 | 0.002180344 | 1 |
| HTN | b_cell | *CACNA2D1* | -2.022587035 | 0.000448787 | 0.002180344 | 1 |
| HTN | b_cell | *ZUFSP* | -5.390662155 | 0.000938387 | 0.002662292 | 1 |
| HTN | b_cell | *PPP2R5E* | -3.980138802 | 0.016438645 | 0.022173302 | 1 |
| HTN | b_cell | *ANKRD29* | -3.363057257 | 0.00029205 | 0.002180344 | 1 |
| HTN | b_cell | *THY1* | -3.263519226 | 0.000411242 | 0.002180344 | 1 |
| HTN | b_cell | *TBCEL* | -3.655961354 | 0.000614103 | 0.002286342 | 1 |
| HTN | b_cell | *UBASH3B* | -4.063761865 | 0.000980309 | 0.002718863 | 1 |
| HTN | b_cell | *ROBO4* | -3.606770115 | 0.000323351 | 0.002180344 | 1 |
| HTN | b_cell | *ROBO3* | -3.894095371 | 0.000263377 | 0.002180344 | 1 |
| HTN | b_cell | *PRKCA* | -4.619528381 | 0.005719963 | 0.009194939 | 1 |
| HTN | b_cell | *LRRK1* | -5.062689326 | 0.005012792 | 0.008276109 | 1 |
| HTN | b_cell | *CEP112* | -2.512580099 | 0.001105102 | 0.00289853 | 1 |
| HTN | b_cell | *GAL3ST2* | -35.36069372 | 0.012767594 | 0.017869516 | 1 |
| HTN | b_cell | *ENPP3* | 7.021015069 | 0.034860614 | 0.043408334 | 1 |
| HTN | b_cell | *UCHL1* | -4.99626515 | 0.000209385 | 0.002180344 | 1 |
| HTN | b_cell | *MIA3* | -5.376625785 | 0.004781732 | 0.007959425 | 1 |
| HTN | b_cell | *DISP1* | 10.41367778 | 0.000121414 | 0.002180344 | 0.763330467 |
| HTN | b_cell | *FAM167A* | 5.407502458 | 0.000128325 | 0.002180344 | 0.806779445 |
| HTN | b_cell | *NEIL2* | 14.24602049 | 3.29E-09 | 3.30E-06 | 2.07E-05 |
| HTN | b_cell | *LONRF1* | -3.849620331 | 0.000397073 | 0.002180344 | 1 |
| HTN | b_cell | *CCSAP* | -7.75589139 | 0.001242189 | 0.00310894 | 1 |
| HTN | b_cell | *GBP5* | -4.87309608 | 0.000766623 | 0.002430538 | 1 |
| HTN | b_cell | *FAM69A* | -3.962716801 | 0.00033329 | 0.002180344 | 1 |

| | | | | | | |
|---|---|---|---|---|---|---|
| HTN | b_cell | *ATP5G3* | -4.721432558 | 0.017348858 | 0.023266269 | 1 |
| HTN | b_cell | *PDLIM3* | -3.930179491 | 0.000420099 | 0.002180344 | 1 |
| HTN | b_cell | *NCAM2* | -3.009786282 | 0.000371799 | 0.002180344 | 1 |
| HTN | b_cell | *L3MBTL4* | -4.051117609 | 0.00284586 | 0.00538751 | 1 |
| HTN | b_cell | *PDE1C* | -2.999228165 | 0.000729105 | 0.002394922 | 1 |
| HTN | b_cell | *RABGEF1* | -4.494367147 | 0.000852597 | 0.002550085 | 1 |
| HTN | b_cell | *MRPL39* | 5.027570849 | 0.024239355 | 0.031302165 | 1 |
| HTN | b_cell | *TSEN2* | -3.265979499 | 0.01715374 | 0.023053775 | 1 |
| HTN | b_cell | *FLCN* | -5.155004035 | 0.000585428 | 0.002261934 | 1 |
| HTN | b_cell | *CXXC1* | -6.498854257 | 0.000693891 | 0.002357691 | 1 |
| HTN | b_cell | *PPP4R1* | -4.563044335 | 0.001872695 | 0.004023799 | 1 |
| HTN | b_cell | *APCDD1* | -2.543332684 | 0.000586785 | 0.002263264 | 1 |
| HTN | b_cell | *MPPE1* | -5.279099602 | 0.00830154 | 0.012344319 | 1 |
| HTN | b_cell | *EPHB1* | -6.437672628 | 0.00381693 | 0.0067095 | 1 |
| HTN | b_cell | *ANKRD40* | -4.151200792 | 0.001140166 | 0.002953505 | 1 |
| HTN | b_cell | *CYP2U1* | -6.270128437 | 0.00146111 | 0.003427611 | 1 |
| HTN | b_cell | *FBXL18* | -5.263034185 | 0.001184223 | 0.003021596 | 1 |
| HTN | b_cell | *PROM2* | -15.04250664 | 0.013969822 | 0.019290198 | 1 |
| HTN | b_cell | *AK9* | -3.479062557 | 0.004161225 | 0.007167567 | 1 |
| HTN | b_cell | *CDK19* | -4.539501639 | 0.016881752 | 0.022717375 | 1 |
| HTN | b_cell | *GTF3C6* | 6.971120209 | 0.000542928 | 0.002214137 | 1 |
| HTN | b_cell | *AGPAT5* | 8.069222057 | 0.001751988 | 0.003844942 | 1 |
| HTN | b_cell | *MMS19* | -4.941162597 | 0.001499693 | 0.003480291 | 1 |
| HTN | b_cell | *GOLGA7B* | 4.250435772 | 0.00153142 | 0.003526754 | 1 |
| HTN | b_cell | *TRMT44* | -4.373755923 | 0.00033479 | 0.002180344 | 1 |
| HTN | b_cell | *C16orf87* | 7.033026555 | 0.000804401 | 0.002481486 | 1 |
| HTN | b_cell | *HEATR3* | -4.346191846 | 0.003372319 | 0.00609539 | 1 |
| HTN | b_cell | *CNOT8* | -5.952789845 | 0.007569043 | 0.011466644 | 1 |
| HTN | b_cell | *LRGUK* | -5.609334776 | 0.002153393 | 0.004395579 | 1 |
| HTN | b_cell | *SETD9* | -3.017752881 | 0.001183118 | 0.003020001 | 1 |
| HTN | b_cell | *ZKSCAN2* | -5.913801763 | 0.004189255 | 0.007198099 | 1 |
| HTN | b_cell | *FAM126B* | -4.223501518 | 0.002009632 | 0.004210907 | 1 |
| HTN | b_cell | *ALS2CR12* | -4.054251207 | 0.000176659 | 0.002180344 | 1 |
| HTN | b_cell | *SPAG17* | -2.472116904 | 0.00034502 | 0.002180344 | 1 |
| HTN | b_cell | *DEPTOR* | -4.56496688 | 0.000918288 | 0.002637404 | 1 |
| HTN | b_cell | *RRAGA* | -8.102917887 | 0.013369784 | 0.01857177 | 1 |
| HTN | b_cell | *TMBIM4* | -5.169798079 | 0.003039185 | 0.005650639 | 1 |
| HTN | b_cell | *FAM161B* | -5.47351975 | 0.001366796 | 0.003286059 | 1 |
| HTN | b_cell | *WIF1* | 10.23186579 | 0.001014235 | 0.002771431 | 1 |
| HTN | b_cell | *UGT2B4* | -7.122671742 | 0.000409373 | 0.002180344 | 1 |
| HTN | b_cell | *MMP16* | -2.721081932 | 0.000199746 | 0.002180344 | 1 |
| HTN | b_cell | *KCNMA1* | -2.446861029 | 0.000325909 | 0.002180344 | 1 |
| HTN | b_cell | *BATF* | 12.47927742 | 0.006923827 | 0.010658692 | 1 |
| HTN | b_cell | *DCK* | 6.511380273 | 7.04E-05 | 0.002086883 | 0.442419234 |
| HTN | b_cell | *DPY19L4* | -4.8929637 | 0.001115445 | 0.002917097 | 1 |
| HTN | b_cell | *NDUFAF6* | -6.115307745 | 5.10E-05 | 0.001756143 | 0.320861971 |
| HTN | b_cell | *DRAM2* | -4.321061495 | 0.004144865 | 0.007149182 | 1 |
| HTN | b_cell | *C8orf37* | -10.03613853 | 0.003954762 | 0.006895061 | 1 |
| HTN | b_cell | *ADAMTSL3* | -2.333941262 | 0.000594557 | 0.002268192 | 1 |
| HTN | b_cell | *WHAMM* | -4.392721662 | 0.006871248 | 0.010603715 | 1 |

| | | | | | | |
|---|---|---|---|---|---|---|
| HTN | b_cell | *CXCL13* | -8.257071378 | 0.000297472 | 0.002180344 | 1 |
| HTN | b_cell | *N6AMT1* | -4.061959317 | 0.001143037 | 0.00295626 | 1 |
| HTN | b_cell | *RWDD2B* | -3.894119131 | 0.0010302 | 0.002792956 | 1 |
| HTN | b_cell | *CCT8* | -4.823652975 | 0.001392986 | 0.00332613 | 1 |
| HTN | b_cell | *ANKRD9* | -6.215971014 | 7.99E-05 | 0.002176137 | 0.502462976 |
| HTN | b_cell | *SFXN2* | -3.180454264 | 0.007490291 | 0.011369257 | 1 |
| HTN | b_cell | *SH3RF2* | -3.278687092 | 0.000232451 | 0.002180344 | 1 |
| HTN | b_cell | *GDF6* | -11.83341654 | 0.000646565 | 0.00232018 | 1 |
| HTN | b_cell | *NODAL* | -6.240549303 | 0.000410489 | 0.002180344 | 1 |
| HTN | b_cell | *MED19* | -2.780949383 | 0.026961358 | 0.034494518 | 1 |
| HTN | b_cell | *WDYHV1* | -5.484688725 | 0.005629311 | 0.009072412 | 1 |
| HTN | b_cell | *ATAD2* | -6.094429745 | 0.015280622 | 0.020816743 | 1 |
| HTN | b_cell | *SASS6* | -5.354863793 | 0.0007645 | 0.002429935 | 1 |
| HTN | b_cell | *ITGAD* | -4.392545682 | 0.00629538 | 0.009909628 | 1 |
| HTN | b_cell | *VPS8* | -3.131950239 | 0.036214907 | 0.044934502 | 1 |
| HTN | b_cell | *RPUSD3* | -6.325593821 | 0.001383896 | 0.003311965 | 1 |
| HTN | b_cell | *GHRL* | -5.129218934 | 0.000599153 | 0.002274361 | 1 |
| HTN | b_cell | *EXOG* | -6.147686576 | 0.003350479 | 0.00606346 | 1 |
| HTN | b_cell | *SLC6A1* | -9.223685658 | 0.001193835 | 0.003030645 | 1 |
| HTN | b_cell | *FCHO2* | -3.93967487 | 0.002689372 | 0.005167507 | 1 |
| HTN | b_cell | *TIMP4* | -3.916779753 | 0.000612479 | 0.002285256 | 1 |
| HTN | b_cell | *CPT2* | 6.960574883 | 0.000780418 | 0.002446934 | 1 |
| HTN | b_cell | *LRP8* | 10.73812673 | 0.023031759 | 0.029936049 | 1 |
| HTN | b_cell | *STEAP2* | -4.148405228 | 9.11E-05 | 0.002180344 | 0.572612977 |
| HTN | b_cell | *GATAD1* | -3.797383951 | 0.001011519 | 0.002769536 | 1 |
| HTN | b_cell | *TMED6* | -20.67595015 | 0.000400953 | 0.002180344 | 1 |
| HTN | b_cell | *CLEC18A* | -22.02278599 | 1.23E-05 | 0.000723375 | 0.077401092 |
| HTN | b_cell | *IL34* | -9.648901928 | 0.025131722 | 0.0323247 | 1 |
| HTN | b_cell | *DHRS1* | -3.84955937 | 0.012811062 | 0.017914402 | 1 |
| HTN | b_cell | *AASDH* | -5.01874225 | 0.011359198 | 0.016164616 | 1 |
| HTN | b_cell | *RNF111* | -3.312019526 | 0.001380624 | 0.00330666 | 1 |
| HTN | b_cell | *DYRK1A* | -4.038768889 | 0.027410006 | 0.034983092 | 1 |
| HTN | b_cell | *KCNJ15* | -8.775775121 | 0.024247104 | 0.031302165 | 1 |
| HTN | b_cell | *CREB3L1* | -4.886504803 | 0.001160503 | 0.002986526 | 1 |
| HTN | b_cell | *C2CD2* | -2.526293571 | 0.002154098 | 0.004395591 | 1 |
| HTN | b_cell | *SLC38A10* | -5.78068662 | 0.007685119 | 0.011607195 | 1 |
| HTN | b_cell | *C9orf43* | -7.94951151 | 0.000333166 | 0.002180344 | 1 |
| HTN | b_cell | *WDR19* | -4.718437219 | 0.001808329 | 0.003927449 | 1 |
| HTN | b_cell | *SLC37A3* | -4.586171701 | 0.001488127 | 0.003464107 | 1 |
| HTN | b_cell | *AP3S2* | -2.828860216 | 0.013782241 | 0.01905629 | 1 |
| HTN | b_cell | *SPPL3* | -4.076642087 | 0.012474714 | 0.017518098 | 1 |
| HTN | b_cell | *FAM213B* | -6.449938873 | 0.006001311 | 0.009538489 | 1 |
| HTN | b_cell | *RADIL* | -3.575280459 | 0.000492433 | 0.002192417 | 1 |
| HTN | b_cell | *LDLRAP1* | -7.918479062 | 0.019179684 | 0.025407222 | 1 |
| HTN | b_cell | *PAFAH2* | -7.202306341 | 0.002106702 | 0.004337157 | 1 |
| HTN | b_cell | *TRIM63* | 8.181948116 | 6.23E-07 | 8.91E-05 | 0.003918672 |
| HTN | b_cell | *WDR66* | -3.205132248 | 0.009145118 | 0.013424086 | 1 |
| HTN | b_cell | *UBXN11* | -4.36005134 | 0.012367224 | 0.017401175 | 1 |
| HTN | b_cell | *GALNT14* | 13.67084203 | 0.001131327 | 0.002939114 | 1 |
| HTN | b_cell | *RHPN1* | -6.903398379 | 0.001535964 | 0.003532043 | 1 |

| | | | | | | |
|---|---|---|---|---|---|---|
| HTN | b_cell | *TPRG1L* | -7.311816213 | 0.00134323 | 0.003258057 | 1 |
| HTN | b_cell | *LRRC43* | -12.53510638 | 0.000421739 | 0.002180344 | 1 |
| HTN | b_cell | *XKR8* | -7.291686134 | 0.000367794 | 0.002180344 | 1 |
| HTN | b_cell | *EYA3* | -5.370652112 | 0.003981744 | 0.006926736 | 1 |
| HTN | b_cell | *MRAS* | -2.404623265 | 0.000798179 | 0.002471947 | 1 |
| HTN | b_cell | *ABHD3* | -3.706561801 | 0.001299642 | 0.003188002 | 1 |
| HTN | b_cell | *CLSTN2* | -2.292368713 | 0.000260499 | 0.002180344 | 1 |
| HTN | b_cell | *COLEC12* | -2.375307402 | 0.000457711 | 0.002180344 | 1 |
| HTN | b_cell | *RNF207* | -9.564547108 | 0.002349746 | 0.00469725 | 1 |
| HTN | b_cell | *AUTS2* | -2.975309975 | 0.001573751 | 0.003581136 | 1 |
| HTN | b_cell | *TSPAN33* | -5.169322701 | 0.022160598 | 0.028917327 | 1 |
| HTN | b_cell | *AHCYL2* | -4.378859407 | 0.008716053 | 0.012878454 | 1 |
| HTN | b_cell | *B4GALT5* | -3.408569603 | 0.000938151 | 0.002662292 | 1 |
| HTN | b_cell | *CD1D* | -10.97534765 | 0.012732915 | 0.017832888 | 1 |
| HTN | b_cell | *FAM86C1* | 18.95826307 | 0.004631111 | 0.007782892 | 1 |
| HTN | b_cell | *CD1E* | -19.28552661 | 0.002944551 | 0.005524438 | 1 |
| HTN | b_cell | *NCF1* | 3.738787153 | 0.000247294 | 0.002180344 | 1 |
| HTN | b_cell | *PPP1R9A* | -2.272152833 | 0.000383456 | 0.002180344 | 1 |
| HTN | b_cell | *ZFAND2B* | -7.953917187 | 0.000896511 | 0.002599737 | 1 |
| HTN | b_cell | *GDPD5* | -2.957346772 | 0.000418502 | 0.002180344 | 1 |
| HTN | b_cell | *DYNC1I1* | -5.183599723 | 0.001744578 | 0.003841901 | 1 |
| HTN | b_cell | *TMED4* | -5.167436594 | 0.001254475 | 0.003124756 | 1 |
| HTN | b_cell | *COPG2* | -2.516106648 | 0.000864032 | 0.002559431 | 1 |
| HTN | b_cell | *ELK4* | -5.875676502 | 0.009542947 | 0.013881653 | 1 |
| HTN | b_cell | *DUSP23* | -5.938880583 | 0.000469303 | 0.002180489 | 1 |
| HTN | b_cell | *RNF166* | -7.469783074 | 0.02223913 | 0.029002173 | 1 |
| HTN | b_cell | *F11R* | -3.987938468 | 0.000693735 | 0.002357691 | 1 |
| HTN | b_cell | *NIT1* | -3.132719929 | 0.003868635 | 0.006782518 | 1 |
| HTN | b_cell | *ZNF276* | -4.234107779 | 0.006348283 | 0.009962969 | 1 |
| HTN | b_cell | *FGF17* | -21.7015318 | 0.004195528 | 0.007201376 | 1 |
| HTN | b_cell | *CDA* | -7.724227944 | 0.00319167 | 0.005848938 | 1 |
| HTN | b_cell | *B4GALT3* | -5.425186613 | 0.002159605 | 0.004403494 | 1 |
| HTN | b_cell | *DMTN* | -3.955745448 | 0.000650497 | 0.002323165 | 1 |
| HTN | b_cell | *FAM160B2* | -4.737298295 | 0.000896495 | 0.002599737 | 1 |
| HTN | b_cell | *NDUFS2* | -4.735206983 | 0.008158233 | 0.012168638 | 1 |
| HTN | b_cell | *SLC5A11* | -6.274543285 | 3.67E-09 | 3.30E-06 | 2.31E-05 |
| HTN | b_cell | *CCAR2* | -6.142411998 | 0.002302496 | 0.004623377 | 1 |
| HTN | b_cell | *EPB41* | -2.843343833 | 0.005863799 | 0.009373431 | 1 |
| HTN | b_cell | *ALG8* | -6.423457369 | 0.004840603 | 0.008040388 | 1 |
| HTN | b_cell | *IFNAR2* | -4.951737774 | 0.006534271 | 0.010206495 | 1 |
| HTN | b_cell | *MRPL10* | -5.044085916 | 0.000537612 | 0.002207687 | 1 |
| HTN | b_cell | *SV2A* | -17.78220368 | 0.004202173 | 0.007208474 | 1 |
| HTN | b_cell | *RCAN1* | -4.709755809 | 0.000502562 | 0.002195028 | 1 |
| HTN | b_cell | *UBE2Z* | -6.985022011 | 0.002027607 | 0.004226899 | 1 |
| HTN | b_cell | *CLIC6* | -6.074203744 | 0.003977354 | 0.006922931 | 1 |
| HTN | b_cell | *SIM2* | 10.29918231 | 0.000154272 | 0.002180344 | 0.969910609 |
| HTN | b_cell | *HLCS* | -2.707519749 | 0.028935699 | 0.036758687 | 1 |
| HTN | b_cell | *ARHGAP27* | -4.487867493 | 0.000669218 | 0.00233354 | 1 |
| HTN | b_cell | *PADI4* | -12.34987768 | 0.000766492 | 0.002430538 | 1 |
| HTN | b_cell | *PSMD4* | -6.20776466 | 0.01044251 | 0.015044011 | 1 |

| | | | | | | |
|---|---|---|---|---|---|---|
| HTN | b_cell | *STARD9* | -2.76146273 | 0.0003132 | 0.002180344 | 1 |
| HTN | b_cell | *THEM4* | -3.675041196 | 0.002621241 | 0.005081636 | 1 |
| HTN | b_cell | *TCHH* | 5.100903629 | 0.000483897 | 0.002187427 | 1 |
| HTN | b_cell | *RSPRY1* | -5.221003502 | 0.021931307 | 0.028653809 | 1 |
| HTN | b_cell | *CCDC17* | -12.33825514 | 1.99E-05 | 0.001016068 | 0.124976316 |
| HTN | b_cell | *ACE* | -3.939172479 | 0.000308103 | 0.002180344 | 1 |
| HTN | b_cell | *TEPP* | 32.01112802 | 0.009949894 | 0.014393692 | 1 |
| HTN | b_cell | *EFCAB14* | -5.536330159 | 0.008310576 | 0.012350075 | 1 |
| HTN | b_cell | *LYPD5* | -10.96083281 | 0.001952568 | 0.004134656 | 1 |
| HTN | b_cell | *CCDC107* | -3.824093583 | 0.000827134 | 0.002514184 | 1 |
| HTN | b_cell | *NPR2* | -4.746736184 | 0.000389837 | 0.002180344 | 1 |
| HTN | b_cell | *ZNF235* | -4.515341417 | 0.002225154 | 0.004504038 | 1 |
| HTN | b_cell | *DFFA* | -4.91892122 | 0.003119726 | 0.005758578 | 1 |
| HTN | b_cell | *SSU72* | -4.334748656 | 0.000673505 | 0.002334899 | 1 |
| HTN | b_cell | *NR2F6* | -5.586178654 | 0.000566732 | 0.002232844 | 1 |
| HTN | b_cell | *ANKLE1* | -17.09594071 | 0.000184645 | 0.002180344 | 1 |
| HTN | b_cell | *CILP2* | -38.63605063 | 0.000548572 | 0.002215074 | 1 |
| HTN | b_cell | *HSF2BP* | -8.092009289 | 7.84E-05 | 0.002176137 | 0.492652078 |
| HTN | b_cell | *CSTB* | -4.014091074 | 0.000313806 | 0.002180344 | 1 |
| HTN | b_cell | *C21orf33* | -16.3426998 | 0.000619018 | 0.002288754 | 1 |
| HTN | b_cell | *ICOSLG* | 7.905026223 | 0.008421692 | 0.012488984 | 1 |
| HTN | b_cell | *LRRC3* | -8.844472348 | 0.001088691 | 0.002868262 | 1 |
| HTN | b_cell | *ITGB2* | -4.431018412 | 0.005001946 | 0.008260371 | 1 |
| HTN | b_cell | *FTCD* | 4.217357236 | 0.038892135 | 0.048028806 | 1 |
| HTN | b_cell | *LSS* | -3.738756919 | 0.000342487 | 0.002180344 | 1 |
| HTN | b_cell | *VAV2* | 6.216417441 | 0.017524552 | 0.02346685 | 1 |
| HTN | b_cell | *MCM3AP* | -3.761646194 | 0.018171559 | 0.02423517 | 1 |
| HTN | b_cell | *PCNT* | -4.711786464 | 0.001008871 | 0.00276494 | 1 |
| HTN | b_cell | *DIP2A* | -4.411500413 | 0.028716257 | 0.036502043 | 1 |
| HTN | b_cell | *S100B* | -5.381336626 | 3.22E-07 | 6.14E-05 | 0.002025994 |
| HTN | b_cell | *PRMT2* | -3.741634332 | 0.006583278 | 0.010252432 | 1 |
| HTN | b_cell | *ZNF208* | -3.241155806 | 0.000382209 | 0.002180344 | 1 |
| HTN | b_cell | *ADAMTS13* | -4.988057202 | 0.000344042 | 0.002180344 | 1 |
| HTN | b_cell | *ZNF714* | -6.30103542 | 0.004101248 | 0.007095362 | 1 |
| HTN | b_cell | *GPSM1* | -6.182640291 | 0.000235482 | 0.002180344 | 1 |
| HTN | b_cell | *C19orf47* | -6.017135983 | 0.000393626 | 0.002180344 | 1 |
| HTN | b_cell | *HIPK4* | 12.79654031 | 0.000226019 | 0.002180344 | 1 |
| HTN | b_cell | *RDH13* | -8.13507872 | 0.007354661 | 0.011197667 | 1 |
| HTN | b_cell | *MPZL3* | -5.682988798 | 0.00040601 | 0.002180344 | 1 |
| HTN | b_cell | *PCSK7* | -5.673322637 | 0.015846104 | 0.021493949 | 1 |
| HTN | b_cell | *S100A1* | 3.932524081 | 0.000183635 | 0.002180344 | 1 |
| HTN | b_cell | *ZBTB7B* | -8.248739701 | 0.003455266 | 0.006206645 | 1 |
| HTN | b_cell | *VPS11* | -6.344814745 | 0.000633138 | 0.002301586 | 1 |
| HTN | b_cell | *FDPS* | -3.666205981 | 0.007143369 | 0.010929186 | 1 |
| HTN | b_cell | *PAQR6* | 2.613645899 | 0.000925373 | 0.002648292 | 1 |
| HTN | b_cell | *PMF1* | 12.35915125 | 0.001661729 | 0.003724826 | 1 |
| HTN | b_cell | *SLC25A44* | -5.391500213 | 0.001035696 | 0.002800612 | 1 |
| HTN | b_cell | *NBEAL2* | -6.213582842 | 0.031876231 | 0.040073158 | 1 |
| HTN | b_cell | *FCRL3* | 0.46804483 | 0.00018412 | 0.002180344 | 1 |
| HTN | b_cell | *FGFR4* | 11.76288435 | 0.000209834 | 0.002180344 | 1 |

| | | | | | | |
|---|---|---|---|---|---|---|
| HTN | b_cell | *NACC1* | 5.607885377 | 0.000244749 | 0.002180344 | 1 |
| HTN | b_cell | *LY6K* | 17.02926333 | 0.001089 | 0.002868262 | 1 |
| HTN | b_cell | *TONSL* | -7.368171222 | 0.000613159 | 0.002286342 | 1 |
| HTN | b_cell | *MUM1* | -3.357234785 | 0.001574971 | 0.003581136 | 1 |
| HTN | b_cell | *PPP1R16A* | -2.979333188 | 0.00704681 | 0.010810955 | 1 |
| HTN | b_cell | *ORAI2* | 3.597473368 | 0.000283516 | 0.002180344 | 1 |
| HTN | b_cell | *FBXL13* | -4.397096742 | 0.001264444 | 0.003142117 | 1 |
| HTN | b_cell | *MFSD12* | -6.800869738 | 0.006537437 | 0.01020886 | 1 |
| HTN | b_cell | *YDJC* | 23.07654183 | 0.010855174 | 0.015531743 | 1 |
| HTN | b_cell | *CCDC116* | 7.824616106 | 0.000285705 | 0.002180344 | 1 |
| HTN | b_cell | *DVL3* | -6.278825388 | 0.006864431 | 0.010598399 | 1 |
| HTN | b_cell | *AP2M1* | -6.145894574 | 0.030114388 | 0.038079074 | 1 |
| HTN | b_cell | *ABCF3* | 6.86642894 | 0.010828068 | 0.015503544 | 1 |
| HTN | b_cell | *FBXO27* | -3.688908957 | 0.000414311 | 0.002180344 | 1 |
| HTN | b_cell | *DMKN* | 7.786852626 | 0.000552948 | 0.002219911 | 1 |
| HTN | b_cell | *BDH1* | -5.175276289 | 0.000416652 | 0.002180344 | 1 |
| HTN | b_cell | *COX7A1* | -5.032290093 | 0.000566056 | 0.002232844 | 1 |
| HTN | b_cell | *DUSP14* | -3.752257043 | 0.000447333 | 0.002180344 | 1 |
| HTN | b_cell | *LRRC56* | 5.724399531 | 0.000326673 | 0.002180344 | 1 |
| HTN | b_cell | *PGAP3* | -8.035425446 | 0.001290914 | 0.003173543 | 1 |
| HTN | b_cell | *IKZF3* | 0.959765453 | 0.00025562 | 0.002180344 | 1 |
| HTN | b_cell | *ACOX1* | -3.882326726 | 0.000359795 | 0.002180344 | 1 |
| HTN | b_cell | *PRPSAP1* | -4.599300858 | 0.002554979 | 0.00499387 | 1 |
| HTN | b_cell | *CYGB* | -5.751641729 | 0.000171186 | 0.002180344 | 1 |
| HTN | b_cell | *ZNF577* | -4.733825927 | 0.001736956 | 0.003831897 | 1 |
| HTN | b_cell | *CCL16* | -4.141582221 | 0.030216118 | 0.038192347 | 1 |
| HTN | b_cell | *ALDH16A1* | -6.535869114 | 0.004466181 | 0.007552147 | 1 |
| HTN | b_cell | *SIGLEC11* | -10.70809848 | 0.000137368 | 0.002180344 | 0.863633115 |
| HTN | b_cell | *MPP3* | -3.95867406 | 0.000581302 | 0.002253172 | 1 |
| HTN | b_cell | *ASB16* | 7.697578798 | 0.000781534 | 0.002448184 | 1 |
| HTN | b_cell | *PLCD3* | -4.684318694 | 0.000123157 | 0.002180344 | 0.774290635 |
| HTN | b_cell | *LARP4* | -3.563312598 | 0.028630175 | 0.036399981 | 1 |
| HTN | b_cell | *SCIMP* | 5.380256627 | 0.001145968 | 0.002960025 | 1 |
| HTN | b_cell | *ASGR2* | -24.21874646 | 0.002259073 | 0.004552178 | 1 |
| HTN | b_cell | *TNFSF13* | 18.42891987 | 7.33E-09 | 4.19E-06 | 4.61E-05 |
| HTN | b_cell | *SENP3* | 4.977580772 | 0.000773565 | 0.002438676 | 1 |
| HTN | b_cell | *SNRNP25* | -10.06220227 | 0.000128168 | 0.002180344 | 0.805789516 |
| HTN | b_cell | *JMJD8* | 7.552029776 | 0.003759628 | 0.006622803 | 1 |
| HTN | b_cell | *MEIOB* | -8.340168803 | 0.00242109 | 0.004794307 | 1 |
| HTN | b_cell | *ZG16B* | -71.00384526 | 0.000404531 | 0.002180344 | 1 |
| HTN | b_cell | *NXF1* | -4.668003238 | 0.000914206 | 0.002631803 | 1 |
| HTN | b_cell | *TPCN2* | -3.412247394 | 0.000268684 | 0.002180344 | 1 |
| HTN | b_cell | *CYP4A22* | 26.30714382 | 3.11E-05 | 0.001332022 | 0.19583421 |
| HTN | b_cell | *PDZK1IP1* | 91.52119803 | 0.002962587 | 0.005546689 | 1 |
| HTN | b_cell | *COA7* | 6.078017539 | 0.000308659 | 0.002180344 | 1 |
| HTN | b_cell | *C1orf123* | -6.106488326 | 0.00058829 | 0.002263931 | 1 |
| HTN | b_cell | *ACOT11* | -5.042973962 | 0.000333565 | 0.002180344 | 1 |
| HTN | b_cell | *USP24* | -3.035982011 | 0.030291825 | 0.038280342 | 1 |
| HTN | b_cell | *ZSWIM5* | -4.329783257 | 0.040153071 | 0.049441855 | 1 |
| HTN | b_cell | *RBP7* | -4.133808907 | 0.000499932 | 0.002194994 | 1 |

| | | | | | | |
|---|---|---|---|---|---|---|
| HTN | b_cell | *FBLIM1* | -4.523280971 | 0.000582613 | 0.002256862 | 1 |
| HTN | b_cell | *SLC25A34* | 15.05875291 | 0.004930136 | 0.008163225 | 1 |
| HTN | b_cell | *PDPN* | -5.001888433 | 0.000135057 | 0.002180344 | 0.84910082 |
| HTN | b_cell | *LRRC38* | -22.421751 | 0.005459591 | 0.008860209 | 1 |
| HTN | b_cell | *PEF1* | -4.385190634 | 0.000490703 | 0.002190281 | 1 |
| HTN | b_cell | *SYNC* | -5.082266269 | 0.000445996 | 0.002180344 | 1 |
| HTN | b_cell | *TSSK3* | -5.586089166 | 0.002466377 | 0.004856384 | 1 |
| HTN | b_cell | *CAMK2N1* | -6.091181712 | 0.000504864 | 0.002195028 | 1 |
| HTN | b_cell | *TTLL10* | -10.58084451 | 0.005156361 | 0.008466451 | 1 |
| HTN | b_cell | *NFIA* | -3.229437517 | 0.003145946 | 0.00579575 | 1 |
| HTN | b_cell | *FUBP1* | -4.704098161 | 0.021624561 | 0.028276543 | 1 |
| HTN | b_cell | *NEXN* | -3.40658822 | 0.000627721 | 0.002295161 | 1 |
| HTN | b_cell | *B3GALT2* | -6.369707505 | 0.000425354 | 0.002180344 | 1 |
| HTN | b_cell | *NTNG1* | -3.203984399 | 0.003914705 | 0.006832801 | 1 |
| HTN | b_cell | *HFM1* | -3.426089945 | 0.000403189 | 0.002180344 | 1 |
| HTN | b_cell | *KCNT2* | -2.573594279 | 0.000419628 | 0.002180344 | 1 |
| HTN | b_cell | *VCAM1* | -4.074455623 | 0.000946438 | 0.002674273 | 1 |
| HTN | b_cell | *EXTL2* | -4.93865725 | 0.00136235 | 0.003280388 | 1 |
| HTN | b_cell | *ZNF496* | -7.485443226 | 0.007920205 | 0.011886925 | 1 |
| HTN | b_cell | *TRIM58* | 2.702611703 | 0.000706087 | 0.00237262 | 1 |
| HTN | b_cell | *IGSF8* | -4.40615222 | 0.000417771 | 0.002180344 | 1 |
| HTN | b_cell | *VANGL2* | -8.499457814 | 0.009567185 | 0.013907259 | 1 |
| HTN | b_cell | *FCRLB* | -14.71291182 | 0.000485602 | 0.002187427 | 1 |
| HTN | b_cell | *KLHDC9* | 5.245005093 | 0.001824969 | 0.003948238 | 1 |
| HTN | b_cell | *LMX1A* | -2.92377335 | 0.005488521 | 0.008893385 | 1 |
| HTN | b_cell | *FLVCR1* | -3.771617869 | 0.001018765 | 0.002775525 | 1 |
| HTN | b_cell | *TDRD5* | -6.726414018 | 0.002962109 | 0.005546689 | 1 |
| HTN | b_cell | *SNED1* | -3.473604725 | 0.001652499 | 0.003713691 | 1 |
| HTN | b_cell | *C1orf115* | -4.109297687 | 0.005353082 | 0.008718866 | 1 |
| HTN | b_cell | *ACP6* | -7.898643626 | 0.015582412 | 0.02115453 | 1 |
| HTN | b_cell | *CNST* | -4.68442574 | 0.006089303 | 0.009657781 | 1 |
| HTN | b_cell | *PM20D1* | -7.184746214 | 0.000391113 | 0.002180344 | 1 |
| HTN | b_cell | *HAAO* | -7.304823864 | 0.01236931 | 0.017401175 | 1 |
| HTN | b_cell | *B3GALNT2* | -5.9833519 | 0.009030563 | 0.013274526 | 1 |
| HTN | b_cell | *PUS10* | -4.279629222 | 0.000882402 | 0.002588737 | 1 |
| HTN | b_cell | *RFTN2* | -3.830279505 | 0.000701751 | 0.002366275 | 1 |
| HTN | b_cell | *DISC1* | -4.960778342 | 0.002447346 | 0.00483395 | 1 |
| HTN | b_cell | *FAM84A* | -9.622555548 | 0.000413491 | 0.002180344 | 1 |
| HTN | b_cell | *NUP35* | -6.354135618 | 0.003042804 | 0.005650639 | 1 |
| HTN | b_cell | *FBXO41* | 13.99775398 | 0.001843753 | 0.003972472 | 1 |
| HTN | b_cell | *CCDC74A* | -21.41559235 | 0.000351273 | 0.002180344 | 1 |
| HTN | b_cell | *H3F3A* | -6.318334633 | 0.004241455 | 0.007258483 | 1 |
| HTN | b_cell | *NOSTRIN* | -3.238797713 | 0.000217847 | 0.002180344 | 1 |
| HTN | b_cell | *SGPP2* | 9.8707496 | 0.033124478 | 0.041468258 | 1 |
| HTN | b_cell | *BBS5* | -5.223766613 | 0.001151861 | 0.002971585 | 1 |
| HTN | b_cell | *HPGDS* | -4.585069291 | 0.006335653 | 0.0099506 | 1 |
| HTN | b_cell | *PDLIM5* | -3.934757938 | 0.00492413 | 0.008155427 | 1 |
| HTN | b_cell | *RPRD2* | -4.440611829 | 0.013980967 | 0.019301348 | 1 |
| HTN | b_cell | *CTSS* | -3.326786969 | 0.013251712 | 0.018424041 | 1 |
| HTN | b_cell | *PACRGL* | -4.669847039 | 0.015918542 | 0.021573588 | 1 |

| | | | | | | |
|---|---|---|---|---|---|---|
| HTN | b_cell | *C1QTNF7* | -2.415648804 | 0.000361171 | 0.002180344 | 1 |
| HTN | b_cell | *LYSMD1* | -5.572296557 | 0.000387835 | 0.002180344 | 1 |
| HTN | b_cell | *RNF149* | -4.450536546 | 0.007101576 | 0.010879048 | 1 |
| HTN | b_cell | *DHX57* | -4.262980528 | 0.001743201 | 0.003841397 | 1 |
| HTN | b_cell | *S100A9* | 6.627196254 | 0.000896701 | 0.002599737 | 1 |
| HTN | b_cell | *S100A12* | -48.81543967 | 0.000146248 | 0.002180344 | 0.919459627 |
| HTN | b_cell | *TDRD10* | -4.601607129 | 0.000769852 | 0.002432944 | 1 |
| HTN | b_cell | *DCAF16* | -4.565801492 | 0.000887515 | 0.002596195 | 1 |
| HTN | b_cell | *NIPAL1* | -5.02022476 | 0.000365343 | 0.002180344 | 1 |
| HTN | b_cell | *ANTXR2* | -3.566317629 | 0.001547936 | 0.003551606 | 1 |
| HTN | b_cell | *CGGBP1* | -2.432889769 | 0.008182963 | 0.012196843 | 1 |
| HTN | b_cell | *DCST2* | -7.38373131 | 0.000501584 | 0.002195028 | 1 |
| HTN | b_cell | *LMOD3* | -5.761352748 | 0.000413534 | 0.002180344 | 1 |
| HTN | b_cell | *NBPF10* | 4.011696081 | 0.000663819 | 0.002327958 | 1 |
| HTN | b_cell | *POGLUT1* | -5.936262433 | 0.007619204 | 0.011528745 | 1 |
| HTN | b_cell | *SLC22A15* | -3.357914992 | 0.000422028 | 0.002180344 | 1 |
| HTN | b_cell | *SLC15A2* | 3.101378903 | 0.001347305 | 0.003265422 | 1 |
| HTN | b_cell | *LRRC58* | -5.696029684 | 0.002297665 | 0.004616625 | 1 |
| HTN | b_cell | *FSTL1* | -2.548370279 | 0.000480273 | 0.002183988 | 1 |
| HTN | b_cell | *LMOD1* | -4.053273176 | 9.47E-07 | 0.000119113 | 0.005955669 |
| HTN | b_cell | *TMEM183A* | -6.080467754 | 0.000751514 | 0.002414293 | 1 |
| HTN | b_cell | *IGFBP7* | -3.719280452 | 0.000214366 | 0.002180344 | 1 |
| HTN | b_cell | *ARPC2* | -4.715686084 | 0.011786051 | 0.016702783 | 1 |
| HTN | b_cell | *GLB1L* | 5.842069667 | 0.005894547 | 0.009405841 | 1 |
| HTN | b_cell | *NUAK2* | 1.797484385 | 0.000323208 | 0.002180344 | 1 |
| HTN | b_cell | *PYHIN1* | -3.364173784 | 0.000571941 | 0.00223784 | 1 |
| HTN | b_cell | *AIM2* | 4.175670987 | 0.000275095 | 0.002180344 | 1 |
| HTN | b_cell | *ICA1L* | -4.018191307 | 0.002309438 | 0.004635836 | 1 |
| HTN | b_cell | *CD200R1* | -3.405809254 | 0.000664284 | 0.002327958 | 1 |
| HTN | b_cell | *GTPBP8* | -4.519718825 | 0.000654172 | 0.002324917 | 1 |
| HTN | b_cell | *SPICE1* | -5.315530836 | 0.000927347 | 0.002650105 | 1 |
| HTN | b_cell | *CADPS* | 7.010160043 | 0.000570642 | 0.002235498 | 1 |
| HTN | b_cell | *PPM1K* | 6.228033836 | 0.015435505 | 0.020995893 | 1 |
| HTN | b_cell | *GMPS* | -5.627384357 | 0.006916238 | 0.010652227 | 1 |
| HTN | b_cell | *DCLK3* | 7.284507806 | 0.000327891 | 0.002180344 | 1 |
| HTN | b_cell | *DNASE1L3* | -5.889497942 | 0.000449049 | 0.002180344 | 1 |
| HTN | b_cell | *CRELD1* | -3.6795728 | 0.005821321 | 0.009319746 | 1 |
| HTN | b_cell | *PCOLCE2* | -2.170572099 | 0.000441231 | 0.002180344 | 1 |
| HTN | b_cell | *TTC14* | -4.311421066 | 0.004084556 | 0.007072323 | 1 |
| HTN | b_cell | *CCDC158* | -4.94085592 | 0.000583011 | 0.002257015 | 1 |
| HTN | b_cell | *GYG1* | -5.904477257 | 0.003084044 | 0.005714012 | 1 |
| HTN | b_cell | *HPS3* | -5.746314212 | 0.031571628 | 0.039714051 | 1 |
| HTN | b_cell | *RYK* | -4.778215447 | 0.001750647 | 0.003844942 | 1 |
| HTN | b_cell | *PLB1* | -4.596038006 | 0.000421445 | 0.002180344 | 1 |
| HTN | b_cell | *CDCP1* | -4.687494007 | 0.00042108 | 0.002180344 | 1 |
| HTN | b_cell | *LZTFL1* | -3.765283279 | 0.000420118 | 0.002180344 | 1 |
| HTN | b_cell | *FYCO1* | -5.964152818 | 0.005390601 | 0.008770887 | 1 |
| HTN | b_cell | *LRRC2* | -3.51509285 | 0.00030696 | 0.002180344 | 1 |
| HTN | b_cell | *DTX3L* | -6.446553262 | 0.00215659 | 0.004399247 | 1 |
| HTN | b_cell | *NMNAT3* | -4.934757411 | 0.005108978 | 0.00839962 | 1 |

| | | | | | | |
|---|---|---|---|---|---|---|
| HTN | b_cell | *SMIM12* | -5.369053338 | 0.00144529 | 0.003409582 | 1 |
| HTN | b_cell | *YEATS2* | -5.167786627 | 0.010156263 | 0.014668602 | 1 |
| HTN | b_cell | *MEAF6* | -5.885710136 | 0.001646094 | 0.00370268 | 1 |
| HTN | b_cell | *DNALI1* | -8.274686626 | 0.000107244 | 0.002180344 | 0.674241678 |
| HTN | b_cell | *RPN1* | -5.817987841 | 0.024241577 | 0.031302165 | 1 |
| HTN | b_cell | *RFC4* | 3.563626062 | 0.00035926 | 0.002180344 | 1 |
| HTN | b_cell | *RFT1* | -8.218435544 | 3.58E-05 | 0.001456416 | 0.22531625 |
| HTN | b_cell | *GNL3* | -5.350232439 | 0.000641946 | 0.002316643 | 1 |
| HTN | b_cell | *UVSSA* | -4.44566844 | 0.030614119 | 0.03861777 | 1 |
| HTN | b_cell | *LRPAP1* | -5.876076465 | 0.017824046 | 0.023807048 | 1 |
| HTN | b_cell | *SLC51A* | -3.768503706 | 0.000625647 | 0.002295161 | 1 |
| HTN | b_cell | *PIGX* | -5.378289173 | 0.001411797 | 0.003355754 | 1 |
| HTN | b_cell | *ABLIM2* | -3.852748531 | 0.000476934 | 0.002180489 | 1 |
| HTN | b_cell | *C1orf50* | -13.45812619 | 0.036722367 | 0.045510254 | 1 |
| HTN | b_cell | *ERMAP* | -4.511526094 | 0.000977316 | 0.002717425 | 1 |
| HTN | b_cell | *METAP1* | -6.730255533 | 0.003523624 | 0.00629705 | 1 |
| HTN | b_cell | *SLC9B2* | -4.109166273 | 0.002683394 | 0.005157597 | 1 |
| HTN | b_cell | *BDH2* | -3.747556734 | 1.47E-05 | 0.000840482 | 0.092453063 |
| HTN | b_cell | *CDC25A* | -9.188272441 | 0.000429513 | 0.002180344 | 1 |
| HTN | b_cell | *ZNF589* | -12.00990229 | 1.40E-10 | 4.40E-07 | 8.80E-07 |
| HTN | b_cell | *ATRIP* | -13.40155685 | 0.00020397 | 0.002180344 | 1 |
| HTN | b_cell | *SHISA5* | -5.089635103 | 0.001747421 | 0.003844252 | 1 |
| HTN | b_cell | *APEH* | -16.69047653 | 1.76E-06 | 0.000186095 | 0.011066558 |
| HTN | b_cell | *RNF123* | 8.15830523 | 0.000241245 | 0.002180344 | 1 |
| HTN | b_cell | *RAD54L2* | -5.165361786 | 0.004422323 | 0.007492089 | 1 |
| HTN | b_cell | *TEX264* | 5.17632724 | 0.034364575 | 0.042858576 | 1 |
| HTN | b_cell | *GRM2* | 7.551773383 | 0.000241235 | 0.002180344 | 1 |
| HTN | b_cell | *POC1A* | 10.01098188 | 7.41E-05 | 0.002109335 | 0.466163073 |
| HTN | b_cell | *PPM1M* | -6.75842922 | 0.000127816 | 0.002180344 | 0.803580124 |
| HTN | b_cell | *C4orf3* | -6.504559397 | 0.000712842 | 0.002380505 | 1 |
| HTN | b_cell | *ANXA5* | -3.538511692 | 0.006043087 | 0.009587352 | 1 |
| HTN | b_cell | *MAP9* | -4.341821843 | 0.000608793 | 0.002281385 | 1 |
| HTN | b_cell | *CEP44* | -5.019155742 | 0.001755578 | 0.003848438 | 1 |
| HTN | b_cell | *C4orf45* | -3.592274713 | 0.000264687 | 0.002180344 | 1 |
| HTN | b_cell | *TMEM144* | -3.234819785 | 0.000512456 | 0.002195202 | 1 |
| HTN | b_cell | *NPY1R* | -2.435759351 | 0.000414177 | 0.002180344 | 1 |
| HTN | b_cell | *IL15* | -4.607598844 | 0.000440312 | 0.002180344 | 1 |
| HTN | b_cell | *ARFIP1* | -4.295298005 | 0.008700851 | 0.01285996 | 1 |
| HTN | b_cell | *OTUD4* | -5.502256712 | 0.010032067 | 0.014502553 | 1 |
| HTN | b_cell | *ITGA2* | -4.663718674 | 0.000385278 | 0.002180344 | 1 |
| HTN | b_cell | *NDUFAF2* | -3.759381624 | 0.002133712 | 0.004369592 | 1 |
| HTN | b_cell | *LMBRD2* | -5.834621925 | 0.00202591 | 0.004226899 | 1 |
| HTN | b_cell | *CMBL* | -4.215130483 | 0.000442923 | 0.002180344 | 1 |
| HTN | b_cell | *C5orf63* | -4.172400845 | 0.000422049 | 0.002180344 | 1 |
| HTN | b_cell | *PRRC1* | -4.541678919 | 0.001075013 | 0.002853079 | 1 |
| HTN | b_cell | *WDR41* | -3.820593004 | 0.000674193 | 0.002334899 | 1 |
| HTN | b_cell | *NDUFS4* | -4.579014545 | 0.002656622 | 0.005126513 | 1 |
| HTN | b_cell | *RHOBTB3* | -2.040444575 | 0.000577684 | 0.002246072 | 1 |
| HTN | b_cell | *TIGD6* | -6.936474715 | 0.000372712 | 0.002180344 | 1 |
| HTN | b_cell | *SERINC5* | -4.352657712 | 0.002742022 | 0.005254218 | 1 |

| | | | | | | |
|---|---|---|---|---|---|---|
| HTN | b_cell | *ENPP6* | -11.49941304 | 0.007284063 | 0.011107181 | 1 |
| HTN | b_cell | *PRIMPOL* | -4.929772879 | 0.002172074 | 0.004423657 | 1 |
| HTN | b_cell | *ERAP1* | -4.773130418 | 4.44E-06 | 0.00036476 | 0.027936311 |
| HTN | b_cell | *ERAP2* | 4.068111082 | 0.032355041 | 0.040626351 | 1 |
| HTN | b_cell | *RICTOR* | -4.29397769 | 0.012506816 | 0.017555337 | 1 |
| HTN | b_cell | *ANKRA2* | -4.291123202 | 0.000976233 | 0.002717425 | 1 |
| HTN | b_cell | *UBLCP1* | -4.814327183 | 0.004055038 | 0.007034775 | 1 |
| HTN | b_cell | *UTP15* | -5.526831797 | 0.000975274 | 0.002717425 | 1 |
| HTN | b_cell | *TLR3* | -4.485189801 | 0.000369539 | 0.002180344 | 1 |
| HTN | b_cell | *KLKB1* | -4.967132123 | 0.00417569 | 0.007183752 | 1 |
| HTN | b_cell | *CCDC127* | -6.068484211 | 0.000419196 | 0.002180344 | 1 |
| HTN | b_cell | *GDF9* | -11.54354352 | 0.015996307 | 0.021669637 | 1 |
| HTN | b_cell | *GRIK2* | -3.326731066 | 0.000471321 | 0.002180489 | 1 |
| HTN | b_cell | *MB21D1* | 7.340728372 | 0.004154955 | 0.007162655 | 1 |
| HTN | b_cell | *TXLNB* | -9.292802419 | 0.011505368 | 0.016350418 | 1 |
| HTN | b_cell | *DCBLD1* | -3.612007961 | 0.00033305 | 0.002180344 | 1 |
| HTN | b_cell | *TMEM200A* | -5.585524641 | 0.000472683 | 0.002180489 | 1 |
| HTN | b_cell | *PDSS2* | -4.187090383 | 0.02877745 | 0.036572433 | 1 |
| HTN | b_cell | *STXBP5* | -4.070494667 | 0.026152148 | 0.033534275 | 1 |
| HTN | b_cell | *PI16* | -2.543266033 | 0.000430817 | 0.002180344 | 1 |
| HTN | b_cell | *STK17A* | -4.881156981 | 0.005863023 | 0.009373431 | 1 |
| HTN | b_cell | *GALNT10* | -4.972350766 | 0.029311659 | 0.037168697 | 1 |
| HTN | b_cell | *SAP30L* | -6.172905487 | 0.004589939 | 0.00772989 | 1 |
| HTN | b_cell | *HCN1* | -6.301831382 | 0.000365723 | 0.002180344 | 1 |
| HTN | b_cell | *RP9* | -7.758554587 | 0.003271917 | 0.005955572 | 1 |
| HTN | b_cell | *KCNK5* | -7.739264401 | 0.000332176 | 0.002180344 | 1 |
| HTN | b_cell | *KIAA1324L* | -3.0810939 | 0.000642118 | 0.002316643 | 1 |
| HTN | b_cell | *USP49* | -4.437399371 | 0.001775844 | 0.003880236 | 1 |
| HTN | b_cell | *ZNF704* | -2.75683184 | 0.000835599 | 0.002525679 | 1 |
| HTN | b_cell | *FNDC1* | -3.523871521 | 0.000444659 | 0.002180344 | 1 |
| HTN | b_cell | *CHMP4C* | -8.2133787 | 5.01E-05 | 0.001750903 | 0.315162457 |
| HTN | b_cell | *LMTK2* | -4.687387822 | 0.006212416 | 0.009805739 | 1 |
| HTN | b_cell | *CTSB* | -4.639284849 | 8.81E-05 | 0.002180344 | 0.554038767 |
| HTN | b_cell | *PEX2* | -4.944769302 | 0.000456866 | 0.002180344 | 1 |
| HTN | b_cell | *TNFRSF11B* | -4.965153173 | 0.0005631 | 0.002226547 | 1 |
| HTN | b_cell | *DEFB1* | -7.262869361 | 0.000527207 | 0.002195202 | 1 |
| HTN | b_cell | *SUN1* | -4.10885827 | 0.001009977 | 0.002766764 | 1 |
| HTN | b_cell | *OXR1* | -3.75737881 | 0.010312003 | 0.014876995 | 1 |
| HTN | b_cell | *TMEM184A* | 2.989013206 | 0.0008451 | 0.002536213 | 1 |
| HTN | b_cell | *SLC4A2* | -7.959590265 | 0.00659406 | 0.010261597 | 1 |
| HTN | b_cell | *FASTK* | -3.949745499 | 0.000673902 | 0.002334899 | 1 |
| HTN | b_cell | *ALDH7A1* | -3.706473262 | 0.012712658 | 0.017820398 | 1 |
| HTN | b_cell | *COX6C* | -5.002375082 | 0.001519848 | 0.003510893 | 1 |
| HTN | b_cell | *YWHAZ* | -4.537561407 | 0.006962331 | 0.010702244 | 1 |
| HTN | b_cell | *SLC25A32* | -5.308731183 | 0.000758042 | 0.002419498 | 1 |
| HTN | b_cell | *DCAF13* | -6.514824233 | 0.010517716 | 0.015138481 | 1 |
| HTN | b_cell | *INTS8* | -4.561910219 | 0.001654456 | 0.003713691 | 1 |
| HTN | b_cell | *FREM1* | -2.263076318 | 0.000376442 | 0.002180344 | 1 |
| HTN | b_cell | *FAM219A* | -5.402299316 | 0.014613963 | 0.02005194 | 1 |
| HTN | b_cell | *TMEM65* | -5.167665941 | 0.016405862 | 0.022133832 | 1 |

| | | | | | | |
|---|---|---|---|---|---|---|
| HTN | b_cell | *CCDC171* | -4.309932574 | 0.001240352 | 0.003108048 | 1 |
| HTN | b_cell | *NIPSNAP3B* | -3.108725783 | 0.000498152 | 0.002194324 | 1 |
| HTN | b_cell | *TMC1* | -5.040704723 | 0.000334061 | 0.002180344 | 1 |
| HTN | b_cell | *C9orf64* | -3.554260188 | 0.005309622 | 0.008671243 | 1 |
| HTN | b_cell | *SVEP1* | -2.044185044 | 0.000434261 | 0.002180344 | 1 |
| HTN | b_cell | *ZNF367* | 9.572504011 | 1.20E-05 | 0.000717012 | 0.075286222 |
| HTN | b_cell | *NOL6* | -6.774714908 | 0.000721802 | 0.002388914 | 1 |
| HTN | b_cell | *AQP3* | -9.375743027 | 0.001291325 | 0.003173543 | 1 |
| HTN | b_cell | *TRMT10B* | -5.13264681 | 0.004391407 | 0.007453773 | 1 |
| HTN | b_cell | *VCP* | -4.71685758 | 0.01164771 | 0.016530283 | 1 |
| HTN | b_cell | *PIGO* | -6.973898871 | 0.001480854 | 0.003457158 | 1 |
| HTN | b_cell | *LRFN5* | -2.373675866 | 0.006328169 | 0.009943229 | 1 |
| HTN | b_cell | *WRN* | -4.320637002 | 0.002490218 | 0.004898624 | 1 |
| HTN | b_cell | *CFL2* | 8.21543849 | 0.004284359 | 0.007317488 | 1 |
| HTN | b_cell | *ZCCHC24* | -3.595585793 | 0.000782725 | 0.002448253 | 1 |
| HTN | b_cell | *PGM2L1* | 7.099008014 | 0.000599213 | 0.002274361 | 1 |
| HTN | b_cell | *GJB2* | 8.628905496 | 0.014281064 | 0.019650918 | 1 |
| HTN | b_cell | *CRYL1* | -4.321818553 | 0.025537357 | 0.032806163 | 1 |
| HTN | b_cell | *REEP3* | -4.985184314 | 8.05E-05 | 0.002176137 | 0.506126626 |
| HTN | b_cell | *HEPACAM* | -1.844925856 | 0.01162295 | 0.016498868 | 1 |
| HTN | b_cell | *LRR1* | -5.182494268 | 0.000380523 | 0.002180344 | 1 |
| HTN | b_cell | *RPL36AL* | -5.956072639 | 0.006368822 | 0.009983499 | 1 |
| HTN | b_cell | *C10orf25* | -4.760775244 | 0.019223847 | 0.025454997 | 1 |
| HTN | b_cell | *KLHDC2* | -4.055539094 | 0.005595268 | 0.009033757 | 1 |
| HTN | b_cell | *NOXRED1* | -8.930087773 | 0.015972038 | 0.021641423 | 1 |
| HTN | b_cell | *AKR1E2* | -3.941502928 | 0.000213431 | 0.002180344 | 1 |
| HTN | b_cell | *OXGR1* | -32.97212432 | 0.026618708 | 0.034083873 | 1 |
| HTN | b_cell | *ATP5C1* | -4.704340467 | 0.028438844 | 0.036171355 | 1 |
| HTN | b_cell | *TAF3* | -4.426479939 | 0.011726254 | 0.016626738 | 1 |
| HTN | b_cell | *VDAC2* | -4.137328291 | 0.000588866 | 0.002263931 | 1 |
| HTN | b_cell | *SLC18A2* | -5.070922104 | 0.000394718 | 0.002180344 | 1 |
| HTN | b_cell | *PDZD8* | -4.545756701 | 0.002062412 | 0.004276512 | 1 |
| HTN | b_cell | *QSOX2* | 4.152049631 | 0.000249868 | 0.002180344 | 1 |
| HTN | b_cell | *CLEC1B* | 2.035046771 | 0.033977918 | 0.042426847 | 1 |
| HTN | b_cell | *SNAPC4* | -5.455610477 | 0.003372414 | 0.00609539 | 1 |
| HTN | b_cell | *SDCCAG3* | -6.41464075 | 0.002113587 | 0.004345364 | 1 |
| HTN | b_cell | *AK8* | -7.12801882 | 0.000733926 | 0.002400726 | 1 |
| HTN | b_cell | *TSC1* | -5.370229514 | 0.007428199 | 0.011291365 | 1 |
| HTN | b_cell | *ZMYND19* | -11.58260806 | 0.001301749 | 0.003191925 | 1 |
| HTN | b_cell | *RET* | 3.663800469 | 0.000289762 | 0.002180344 | 1 |
| HTN | b_cell | *STK32C* | -4.962030132 | 0.000276575 | 0.002180344 | 1 |
| HTN | b_cell | *NSMF* | -4.985228048 | 0.000468375 | 0.002180489 | 1 |
| HTN | b_cell | *CASP7* | -6.971271648 | 0.004197155 | 0.007201832 | 1 |
| HTN | b_cell | *METTL3* | -5.588653832 | 0.001821927 | 0.00394573 | 1 |
| HTN | b_cell | *TRUB1* | -5.546761924 | 0.004177907 | 0.007184491 | 1 |
| HTN | b_cell | *HSPA12A* | -2.003215318 | 0.007454379 | 0.011325684 | 1 |
| HTN | b_cell | *UBTD1* | -4.804118372 | 0.000962635 | 0.002705447 | 1 |
| HTN | b_cell | *ARHGAP42* | -2.949938127 | 0.000599738 | 0.002274361 | 1 |
| HTN | b_cell | *TTC7B* | -3.477681739 | 0.002729468 | 0.00523495 | 1 |
| HTN | b_cell | *SLC39A13* | -6.450648067 | 0.000531032 | 0.002195202 | 1 |

| | | | | | | |
|---|---|---|---|---|---|---|
| HTN | b_cell | *PSMC3* | -4.165422375 | 0.003850578 | 0.006752742 | 1 |
| HTN | b_cell | *TC2N* | -2.577883349 | 0.000572007 | 0.00223784 | 1 |
| HTN | b_cell | *CPSF2* | -3.787927081 | 0.01474311 | 0.020202688 | 1 |
| HTN | b_cell | *CLMN* | -2.761519006 | 0.000366369 | 0.002180344 | 1 |
| HTN | b_cell | *PTER* | -4.574462216 | 0.001427666 | 0.003381236 | 1 |
| HTN | b_cell | *C1QL3* | -4.098102137 | 0.004227979 | 0.007244835 | 1 |
| HTN | b_cell | *ABTB2* | -4.449424669 | 0.000567411 | 0.002232844 | 1 |
| HTN | b_cell | *LIPC* | -4.411959291 | 0.000720198 | 0.002388124 | 1 |
| HTN | b_cell | *CEP57* | -6.277572213 | 0.000768534 | 0.002432944 | 1 |
| HTN | b_cell | *TCP11L2* | -5.254367916 | 0.003039848 | 0.005650639 | 1 |
| HTN | b_cell | *JAM3* | -5.53174267 | 0.001080158 | 0.002860555 | 1 |
| HTN | b_cell | *GPT2* | -2.894541563 | 0.001269523 | 0.003151003 | 1 |
| HTN | b_cell | *IKBIP* | -5.925466775 | 0.00462256 | 0.007771591 | 1 |
| HTN | b_cell | *HIF1AN* | -4.921130421 | 0.000888248 | 0.002596195 | 1 |
| HTN | b_cell | *NDUFB8* | -7.2627336 | 0.001751571 | 0.003844942 | 1 |
| HTN | b_cell | *AVPR1A* | -4.124708924 | 0.008648007 | 0.012795956 | 1 |
| HTN | b_cell | *TRMT61A* | -14.79769238 | 2.90E-07 | 5.89E-05 | 0.001825502 |
| HTN | b_cell | *BTRC* | -4.017333041 | 0.000976494 | 0.002717425 | 1 |
| HTN | b_cell | *LARP6* | -3.352043842 | 0.0015688 | 0.003577072 | 1 |
| HTN | b_cell | *ASPG* | -17.47556719 | 0.000361231 | 0.002180344 | 1 |
| HTN | b_cell | *GABRB3* | 9.309566727 | 0.000323434 | 0.002180344 | 1 |
| HTN | b_cell | *TBATA* | -24.5324364 | 0.029816851 | 0.037725607 | 1 |
| HTN | b_cell | *PCBD1* | -4.568699369 | 0.000460762 | 0.002180489 | 1 |
| HTN | b_cell | *COX11* | -3.755055478 | 0.000675218 | 0.002334899 | 1 |
| HTN | b_cell | *ZNF202* | -7.782850988 | 0.015471445 | 0.021037713 | 1 |
| HTN | b_cell | *FAM227B* | -3.423123829 | 0.000840497 | 0.002533175 | 1 |
| HTN | b_cell | *C2* | -5.157363673 | 0.014624398 | 0.020061879 | 1 |
| HTN | b_cell | *NUDT13* | -3.659695404 | 8.17E-05 | 0.002177448 | 0.513877797 |
| HTN | b_cell | *ILK* | -4.264744478 | 0.001138288 | 0.002949898 | 1 |
| HTN | b_cell | *TAF10* | -4.745161036 | 0.001076475 | 0.002854029 | 1 |
| HTN | b_cell | *DCHS1* | -4.70845049 | 0.000534856 | 0.00220212 | 1 |
| HTN | b_cell | *NETO1* | 1.183334642 | 0.000191293 | 0.002180344 | 1 |
| HTN | b_cell | *USP54* | -4.169759984 | 0.000798109 | 0.002471947 | 1 |
| HTN | b_cell | *C11orf74* | -3.523300262 | 0.000343931 | 0.002180344 | 1 |
| HTN | b_cell | *CYB5R2* | 4.699898158 | 0.000436777 | 0.002180344 | 1 |
| HTN | b_cell | *TUB* | -4.593211064 | 0.000349048 | 0.002180344 | 1 |
| HTN | b_cell | *RIC3* | -6.022519777 | 0.00077009 | 0.002432944 | 1 |
| HTN | b_cell | *LMO1* | -10.40952711 | 0.000771082 | 0.00243363 | 1 |
| HTN | b_cell | *XRRA1* | -4.873357019 | 1.70E-05 | 0.000928737 | 0.106804725 |
| HTN | b_cell | *TRIM66* | -4.266436882 | 2.70E-07 | 5.67E-05 | 0.001700544 |
| HTN | b_cell | *CDYL2* | -5.285642746 | 0.006139106 | 0.009712269 | 1 |
| HTN | b_cell | *PRTG* | -5.932812611 | 0.000929886 | 0.002653801 | 1 |
| HTN | b_cell | *AKIP1* | -8.39616235 | 0.006164772 | 0.009747968 | 1 |
| HTN | b_cell | *C16orf46* | -59.04089779 | 0.000673062 | 0.002334899 | 1 |
| HTN | b_cell | *TMEM41B* | -4.979679521 | 0.002323151 | 0.00465593 | 1 |
| HTN | b_cell | *TMX3* | -4.994551613 | 0.004536896 | 0.007653197 | 1 |
| HTN | b_cell | *CCDC68* | -4.509057471 | 0.001670333 | 0.00373449 | 1 |
| HTN | b_cell | *CLEC4E* | -8.570443451 | 0.000766241 | 0.002430538 | 1 |
| HTN | b_cell | *ZNF3* | -5.349468817 | 0.00164512 | 0.003701815 | 1 |
| HTN | b_cell | *CLEC4D* | -18.64827995 | 0.000529006 | 0.002195202 | 1 |

| | | | | | | |
|---|---|---|---|---|---|---|
| HTN | b_cell | *ZSCAN21* | -5.396734493 | 8.95E-05 | 0.002180344 | 0.562656708 |
| HTN | b_cell | *RIMKLB* | -5.68349625 | 3.83E-07 | 6.70E-05 | 0.002411042 |
| HTN | b_cell | *BEAN1* | -4.988845598 | 0.000982711 | 0.002721721 | 1 |
| HTN | b_cell | *TK2* | -3.424678677 | 0.001252785 | 0.003123021 | 1 |
| HTN | b_cell | *SEC11C* | 6.353535722 | 0.00049395 | 0.002192417 | 1 |
| HTN | b_cell | *GALR1* | -7.903464451 | 4.01E-05 | 0.001527848 | 0.25209486 |
| HTN | b_cell | *CENPV* | -3.761508584 | 0.001210881 | 0.00305766 | 1 |
| HTN | b_cell | *SPDYE6* | -8.405358944 | 0.003485698 | 0.00624703 | 1 |
| HTN | b_cell | *ATF7IP2* | -4.754696828 | 0.00704161 | 0.010808252 | 1 |
| HTN | b_cell | *ZNF592* | -4.059707924 | 0.001453535 | 0.003419708 | 1 |
| HTN | b_cell | *ACSM1* | 2.468022314 | 0.000283103 | 0.002180344 | 1 |
| HTN | b_cell | *AP1G1* | -4.786760346 | 0.004216594 | 0.007229268 | 1 |
| HTN | b_cell | *SLFN5* | -3.767973991 | 0.000684663 | 0.002345762 | 1 |
| HTN | b_cell | *CATSPER2* | -5.022802344 | 0.000383353 | 0.002180344 | 1 |
| HTN | b_cell | *C16orf45* | -4.479538612 | 0.006132442 | 0.009706012 | 1 |
| HTN | b_cell | *SAAL1* | -8.960236434 | 0.002659207 | 0.005128354 | 1 |
| HTN | b_cell | *YPEL4* | -10.71491402 | 0.015017919 | 0.020521116 | 1 |
| HTN | b_cell | *LDHC* | -3.383248139 | 0.02773244 | 0.035337222 | 1 |
| HTN | b_cell | *FAM111A* | -5.586176833 | 0.001241001 | 0.003108434 | 1 |
| HTN | b_cell | *SCNN1G* | -23.02548604 | 0.00430298 | 0.007339348 | 1 |
| HTN | b_cell | *ANKDD1A* | -5.053794692 | 0.001878859 | 0.004032908 | 1 |
| HTN | b_cell | *DCTN5* | -4.536656383 | 0.001329503 | 0.003229746 | 1 |
| HTN | b_cell | *STX3* | -4.297432182 | 0.00103356 | 0.002799652 | 1 |
| HTN | b_cell | *MTMR10* | -4.587671373 | 0.033106142 | 0.041453558 | 1 |
| HTN | b_cell | *TSC22D4* | -3.659293528 | 0.000637975 | 0.002309126 | 1 |
| HTN | b_cell | *MS4A6E* | -3.858462758 | 0.01418317 | 0.019541878 | 1 |
| HTN | b_cell | *MS4A7* | -2.784419933 | 0.000602533 | 0.002278713 | 1 |
| HTN | b_cell | *MS4A14* | -4.523039745 | 0.000101598 | 0.002180344 | 0.638746671 |
| HTN | b_cell | *DIS3L* | -3.421390735 | 0.000305988 | 0.002180344 | 1 |
| HTN | b_cell | *CCNDBP1* | -4.955398345 | 0.001245523 | 0.003112676 | 1 |
| HTN | b_cell | *SMAD3* | -4.09513232 | 0.000374636 | 0.002180344 | 1 |
| HTN | b_cell | *RCCD1* | -4.476483966 | 0.001232144 | 0.003093645 | 1 |
| HTN | b_cell | *AKTIP* | -6.392361249 | 0.001397807 | 0.003333844 | 1 |
| HTN | b_cell | *EVA1C* | -3.162658464 | 0.000652172 | 0.002323165 | 1 |
| HTN | b_cell | *CNPY4* | -4.218650427 | 0.007000806 | 0.010750871 | 1 |
| HTN | b_cell | *DUSP18* | -7.200721135 | 0.002140861 | 0.00437568 | 1 |
| HTN | b_cell | *TEF* | -2.852653119 | 0.001974289 | 0.004157389 | 1 |
| HTN | b_cell | *MEI1* | 5.158951284 | 0.002239221 | 0.004516517 | 1 |
| HTN | b_cell | *PBX3* | -3.990172196 | 0.013616975 | 0.01887344 | 1 |
| HTN | b_cell | *SNRPD1* | -4.266763641 | 0.00059783 | 0.002274361 | 1 |
| HTN | b_cell | *URM1* | -7.56234288 | 0.000401613 | 0.002180344 | 1 |
| HTN | b_cell | *CERCAM* | -3.352332271 | 0.002340054 | 0.004683769 | 1 |
| HTN | b_cell | *ENDOG* | 4.337403385 | 0.001244414 | 0.003112024 | 1 |
| HTN | b_cell | *ISLR2* | -8.519426159 | 3.35E-05 | 0.001383739 | 0.210328344 |
| HTN | b_cell | *SP2* | -6.659047061 | 0.000763382 | 0.002427608 | 1 |
| HTN | b_cell | *COQ7* | 7.765432883 | 0.017718335 | 0.023685981 | 1 |
| HTN | b_cell | *GPRC5B* | -4.643486671 | 0.00173532 | 0.003830743 | 1 |
| HTN | b_cell | *KATNAL2* | -2.482572128 | 0.001842378 | 0.003970871 | 1 |
| HTN | b_cell | *RNF214* | -4.32121666 | 0.001027744 | 0.002792138 | 1 |
| HTN | b_cell | *ENGASE* | -4.065656726 | 0.000554845 | 0.002220961 | 1 |

| | | | | | | |
|---|---|---|---|---|---|---|
| HTN | b_cell | *RBFOX3* | -7.015026593 | 0.000524995 | 0.002195202 | 1 |
| HTN | b_cell | *MYO5B* | -2.377135479 | 0.022224314 | 0.028994452 | 1 |
| HTN | b_cell | *RRM1* | -4.93809488 | 0.00596966 | 0.009501583 | 1 |
| HTN | b_cell | *TRIM68* | -7.175564149 | 0.000527755 | 0.002195202 | 1 |
| HTN | b_cell | *FN3K* | -4.229013554 | 0.000202863 | 0.002180344 | 1 |
| HTN | b_cell | *ZNF229* | -18.42616314 | 0.001066787 | 0.00284552 | 1 |
| HTN | b_cell | *VKORC1* | -4.851704337 | 3.47E-05 | 0.001426032 | 0.218182831 |
| HTN | b_cell | *SMG8* | -13.10594248 | 0.000492944 | 0.002192417 | 1 |
| HTN | b_cell | *RAB8A* | -7.58875351 | 0.008888285 | 0.013095259 | 1 |
| HTN | b_cell | *GATAD2A* | -4.76160303 | 0.017615992 | 0.023569215 | 1 |
| HTN | b_cell | *PROCA1* | -8.411692424 | 7.07E-05 | 0.002087068 | 0.444545542 |
| HTN | b_cell | *CACNB3* | 3.3771956 | 0.000201829 | 0.002180344 | 1 |
| HTN | b_cell | *TP53I13* | -4.497485112 | 0.000104134 | 0.002180344 | 0.654692257 |
| HTN | b_cell | *ZNF528* | -4.153813683 | 0.000694144 | 0.002357691 | 1 |
| HTN | b_cell | *ZNF701* | -6.374432235 | 0.010715471 | 0.01537735 | 1 |
| HTN | b_cell | *SERTAD3* | -10.36635407 | 0.001977176 | 0.004160174 | 1 |
| HTN | b_cell | *TMC4* | -6.04122398 | 0.00157298 | 0.003581136 | 1 |
| HTN | b_cell | *TTYH1* | -4.940574773 | 0.000845539 | 0.002536213 | 1 |
| HTN | b_cell | *LENG8* | -3.853200926 | 0.003878958 | 0.006794932 | 1 |
| HTN | b_cell | *CDC42EP5* | -4.782046841 | 0.001078725 | 0.002857963 | 1 |
| HTN | b_cell | *KIR3DL1* | 21.69834965 | 1.88E-05 | 0.000985596 | 0.118271504 |
| HTN | b_cell | *ZNF283* | 5.507532611 | 0.025182806 | 0.032383781 | 1 |
| HTN | b_cell | *PPP1R14A* | -9.329055226 | 0.010844952 | 0.015520649 | 1 |
| HTN | b_cell | *SPINT2* | -3.959835779 | 0.001146737 | 0.002960796 | 1 |
| HTN | b_cell | *PSCA* | -6.903024888 | 0.002809625 | 0.00534952 | 1 |
| HTN | b_cell | *CHAF1A* | -5.506163671 | 0.004599298 | 0.007739772 | 1 |
| HTN | b_cell | *UBXN6* | -5.922678271 | 0.000122337 | 0.002180344 | 0.769129734 |
| HTN | b_cell | *ZNF444* | -4.714254813 | 0.000518538 | 0.002195202 | 1 |
| HTN | b_cell | *GLOD4* | -6.713087915 | 0.007984497 | 0.011960575 | 1 |
| HTN | b_cell | *KIFC2* | -4.22773765 | 0.000559016 | 0.002226229 | 1 |
| HTN | b_cell | *SERPINF2* | 11.21801464 | 0.000817914 | 0.002500703 | 1 |
| HTN | b_cell | *CYB5D2* | -5.236744839 | 0.003169104 | 0.005822371 | 1 |
| HTN | b_cell | *ZNF83* | -3.689739179 | 0.00210618 | 0.004337157 | 1 |
| HTN | b_cell | *CD320* | -8.020035779 | 0.004478017 | 0.007570124 | 1 |
| HTN | b_cell | *SPRYD3* | -5.001249971 | 0.000511582 | 0.002195202 | 1 |
| HTN | b_cell | *SOAT2* | -7.66130313 | 0.000578504 | 0.002247872 | 1 |
| HTN | b_cell | *ZNF558* | -5.086994497 | 0.003763112 | 0.006627083 | 1 |
| HTN | b_cell | *NDUFV1* | -4.60812058 | 0.004753417 | 0.007925883 | 1 |
| HTN | b_cell | *CDK2AP2* | -7.081373562 | 0.000688987 | 0.002355324 | 1 |
| HTN | b_cell | *PRDX2* | -6.678660821 | 0.023196894 | 0.030107117 | 1 |
| HTN | b_cell | *MIS12* | -5.441993765 | 0.001271818 | 0.003154259 | 1 |
| HTN | b_cell | *HID1* | -5.806197689 | 0.001965718 | 0.004148529 | 1 |
| HTN | b_cell | *ATP5H* | -7.678207482 | 0.003520467 | 0.006293198 | 1 |
| HTN | b_cell | *SRP68* | 7.353310503 | 0.019276713 | 0.025508886 | 1 |
| HTN | b_cell | *MGAT5B* | -15.330525 | 0.005294806 | 0.008653092 | 1 |
| HTN | b_cell | *TMC8* | -3.69807264 | 0.003171664 | 0.005825374 | 1 |
| HTN | b_cell | *TMEM68* | -5.770501994 | 0.015544939 | 0.021117336 | 1 |
| HTN | b_cell | *GSDMA* | -10.08038695 | 0.000680454 | 0.002339217 | 1 |
| HTN | b_cell | *KRT24* | -9.777779499 | 0.005749509 | 0.009231316 | 1 |
| HTN | b_cell | *ZNF598* | -5.635474275 | 0.000181762 | 0.002180344 | 1 |

| | | | | | | |
|---|---|---|---|---|---|---|
| HTN | b_cell | *MLST8* | -6.598014311 | 0.000262047 | 0.002180344 | 1 |
| HTN | b_cell | *ECI1* | -7.9266191 | 0.000756301 | 0.002419498 | 1 |
| HTN | b_cell | *ABCA3* | -5.038531464 | 0.001036754 | 0.002801844 | 1 |
| HTN | b_cell | *KCTD5* | -6.052945301 | 0.001003924 | 0.002758597 | 1 |
| HTN | b_cell | *SDHAF2* | -9.024003794 | 0.001456514 | 0.003423215 | 1 |
| HTN | b_cell | *DDB1* | -5.364006951 | 0.002992193 | 0.005587146 | 1 |
| HTN | b_cell | *VPS37C* | -5.816608288 | 0.001626931 | 0.003671397 | 1 |
| HTN | b_cell | *TRANK1* | -4.856888201 | 0.004778781 | 0.00795662 | 1 |
| HTN | b_cell | *TTC21A* | -7.314784114 | 0.035238777 | 0.043827139 | 1 |
| HTN | b_cell | *ULK4* | -1.90885719 | 0.021371252 | 0.027984861 | 1 |
| HTN | b_cell | *LTBP3* | -3.239240923 | 0.0041935 | 0.007201376 | 1 |
| HTN | b_cell | *SCARA5* | -2.639280175 | 0.00036262 | 0.002180344 | 1 |
| HTN | b_cell | *COPS6* | -4.445401534 | 0.000732504 | 0.002399172 | 1 |
| HTN | b_cell | *NUDT16L1* | 4.705503525 | 0.000990916 | 0.002732407 | 1 |
| HTN | b_cell | *KIAA1586* | -4.124862334 | 0.000223727 | 0.002180344 | 1 |
| HTN | b_cell | *RNF187* | 8.15297508 | 0.002041887 | 0.004245264 | 1 |
| HTN | b_cell | *PTGDR* | -3.811547621 | 0.000414971 | 0.002180344 | 1 |
| HTN | b_cell | *GLYCTK* | -4.283470986 | 0.001500727 | 0.003480291 | 1 |
| HTN | b_cell | *UBTD2* | -3.777646709 | 0.00204194 | 0.004245264 | 1 |
| HTN | b_cell | *POLR2J3* | -5.237786215 | 0.003135166 | 0.005781297 | 1 |
| HTN | b_cell | *NT5DC2* | -8.751780327 | 3.93E-05 | 0.001519523 | 0.247366443 |
| HTN | b_cell | *COA6* | 10.10496806 | 0.002623773 | 0.005082532 | 1 |
| HTN | b_cell | *KIF5C* | -6.252720181 | 0.000294051 | 0.002180344 | 1 |
| HTN | b_cell | *PXK* | -5.742732439 | 0.000282264 | 0.002180344 | 1 |
| HTN | b_cell | *ACOX2* | -3.728478905 | 0.000232092 | 0.002180344 | 1 |
| HTN | b_cell | *FAM107A* | -4.60068431 | 0.002832937 | 0.005382495 | 1 |
| HTN | b_cell | *CX3CR1* | -4.779045735 | 0.000242182 | 0.002180344 | 1 |
| HTN | b_cell | *SCN11A* | -2.340716816 | 0.002876289 | 0.005430399 | 1 |
| HTN | b_cell | *SEPT2* | -3.229110524 | 0.004176334 | 0.007183752 | 1 |
| HTN | b_cell | *DTYMK* | 10.94160969 | 0.000693394 | 0.002357691 | 1 |
| HTN | b_cell | *TAP1* | -7.774315224 | 0.000162486 | 0.002180344 | 1 |
| HTN | b_cell | *ING5* | -4.491299298 | 0.001122831 | 0.002924292 | 1 |
| HTN | b_cell | *MLKL* | -2.375044151 | 0.023084852 | 0.029980266 | 1 |
| HTN | b_cell | *RHOH* | 2.332480565 | 0.000205708 | 0.002180344 | 1 |
| HTN | b_cell | *KLHL30* | -6.856389383 | 0.001974324 | 0.004157389 | 1 |
| HTN | b_cell | *REEP4* | 4.726446517 | 0.000178466 | 0.002180344 | 1 |
| HTN | b_cell | *BMP1* | -3.890813057 | 0.002354449 | 0.004701026 | 1 |
| HTN | b_cell | *ATXN2L* | -4.627406068 | 0.000527491 | 0.002195202 | 1 |
| HTN | b_cell | *SERINC2* | -8.412890359 | 0.000121589 | 0.002180344 | 0.764430805 |
| HTN | b_cell | *TRAPPC11* | -5.156283199 | 0.013452459 | 0.018661873 | 1 |
| HTN | b_cell | *TMEM223* | -5.676248559 | 0.000877366 | 0.002583606 | 1 |
| HTN | b_cell | *SLC20A2* | -4.193076013 | 0.002613314 | 0.005072525 | 1 |
| HTN | b_cell | *ZSWIM1* | 8.705478476 | 0.001899784 | 0.004061698 | 1 |
| HTN | b_cell | *NBPF9* | -6.250461331 | 0.000829772 | 0.002517751 | 1 |
| HTN | b_cell | *NDUFS5* | -7.203368233 | 0.001351239 | 0.003270974 | 1 |
| HTN | b_cell | *ZNF30* | -4.510714763 | 0.003233994 | 0.00590535 | 1 |
| HTN | b_cell | *FAM84B* | -6.824348837 | 0.000356115 | 0.002180344 | 1 |
| HTN | b_cell | *SLC16A4* | -4.057293547 | 0.002533915 | 0.004961297 | 1 |
| HTN | b_cell | *TMEM208* | -3.959709532 | 0.000252268 | 0.002180344 | 1 |
| HTN | b_cell | *LRP1B* | -2.311260399 | 4.92E-05 | 0.001739106 | 0.309560863 |

| | | | | | | |
|---|---|---|---|---|---|---|
| HTN | b_cell | *NPNT* | -5.854561955 | 0.00056207 | 0.002226547 | 1 |
| HTN | b_cell | *GSTM4* | -4.673993239 | 0.000223092 | 0.002180344 | 1 |
| HTN | b_cell | *ABHD15* | -3.797938503 | 0.002526597 | 0.004950052 | 1 |
| HTN | b_cell | *ADAL* | -3.120437743 | 0.002072139 | 0.00429102 | 1 |
| HTN | b_cell | *SNTB2* | -4.120927297 | 0.001901805 | 0.004064122 | 1 |
| HTN | b_cell | *ZNF507* | -5.061385284 | 0.001690747 | 0.003759376 | 1 |
| HTN | b_cell | *STX18* | -5.158790242 | 0.006553964 | 0.010219437 | 1 |
| HTN | b_cell | *ZBTB49* | -7.053667016 | 0.002111697 | 0.004342897 | 1 |
| HTN | b_cell | *TNIP2* | -8.278286879 | 3.24E-05 | 0.001350755 | 0.203964071 |
| HTN | b_cell | *ZNF608* | -2.760178386 | 0.000487896 | 0.002188291 | 1 |
| HTN | b_cell | *LETM1* | -3.602963159 | 0.000418437 | 0.002180344 | 1 |
| HTN | b_cell | *CEP120* | -5.647022308 | 0.001058501 | 0.002833039 | 1 |
| HTN | b_cell | *STXBP6* | -2.765023976 | 0.000453651 | 0.002180344 | 1 |
| HTN | b_cell | *MFF* | -5.139833872 | 0.009166634 | 0.013449387 | 1 |
| HTN | b_cell | *CPLX1* | -4.761592673 | 0.003961395 | 0.00690471 | 1 |
| HTN | b_cell | *FEM1B* | -5.026149793 | 0.014220367 | 0.019584544 | 1 |
| HTN | b_cell | *MFSD7* | 6.570020306 | 0.000978174 | 0.002717425 | 1 |
| HTN | b_cell | *COL4A3* | 5.7503134 | 0.001514453 | 0.003504366 | 1 |
| HTN | b_cell | *IRS1* | -4.156018461 | 2.55E-09 | 3.20E-06 | 1.60E-05 |
| HTN | b_cell | *UPF3A* | -2.995015509 | 0.007600253 | 0.011505608 | 1 |
| HTN | b_cell | *HSPBAP1* | -3.479817805 | 0.002680558 | 0.005155297 | 1 |
| HTN | b_cell | *PARM1* | -2.933001198 | 0.00155822 | 0.003563671 | 1 |
| HTN | b_cell | *FAM110B* | -2.975474723 | 0.000481335 | 0.002185085 | 1 |
| HTN | b_cell | *ATF5* | -4.97740548 | 0.000260316 | 0.002180344 | 1 |
| HTN | b_cell | *CPT1C* | -7.727531801 | 0.000964218 | 0.002707477 | 1 |
| HTN | b_cell | *PCSK9* | 10.89722852 | 0.000112589 | 0.002180344 | 0.707845787 |
| HTN | b_cell | *RGS14* | -5.792126798 | 0.000132894 | 0.002180344 | 0.835502937 |
| HTN | b_cell | *RAB24* | -8.044689478 | 0.000186681 | 0.002180344 | 1 |
| HTN | b_cell | *SH3TC2* | -6.375966829 | 0.000209266 | 0.002180344 | 1 |
| HTN | b_cell | *GPRIN1* | -24.8853678 | 0.033849991 | 0.042292307 | 1 |
| HTN | b_cell | *PGM2* | 9.856959581 | 0.012207166 | 0.017207725 | 1 |
| HTN | b_cell | *STK32A* | -3.720502137 | 5.27E-05 | 0.001789459 | 0.331049835 |
| HTN | b_cell | *P2RY12* | -2.979750776 | 0.002928008 | 0.005503255 | 1 |
| HTN | b_cell | *SLC33A1* | -5.489892155 | 0.003219068 | 0.005886643 | 1 |
| HTN | b_cell | *SNUPN* | -5.820261409 | 0.001069225 | 0.002845984 | 1 |
| HTN | b_cell | *PTK2* | -3.773891945 | 0.003106586 | 0.00573938 | 1 |
| HTN | b_cell | *PTAFR* | -4.33291353 | 9.34E-05 | 0.002180344 | 0.587220112 |
| HTN | b_cell | *RNASE6* | 2.720327539 | 0.000227936 | 0.002180344 | 1 |
| HTN | b_cell | *NPR1* | -3.330256963 | 0.000416917 | 0.002180344 | 1 |
| HTN | b_cell | *CLIC4* | -3.96547524 | 0.003651836 | 0.006474646 | 1 |
| HTN | b_cell | *SLC38A11* | 1.990662768 | 0.00025721 | 0.002180344 | 1 |
| HTN | b_cell | *CCDC8* | -11.58311085 | 0.000204248 | 0.002180344 | 1 |
| HTN | b_cell | *METTL15* | -4.316101125 | 0.003305124 | 0.006005583 | 1 |
| HTN | b_cell | *HINT1* | -6.499295573 | 0.004237344 | 0.007256927 | 1 |
| HTN | b_cell | *DTWD2* | -3.180347968 | 0.00052001 | 0.002195202 | 1 |
| HTN | b_cell | *CLIC3* | 12.5923305 | 0.000210117 | 0.002180344 | 1 |
| HTN | b_cell | *INO80E* | -12.01596979 | 1.40E-06 | 0.000162203 | 0.008771492 |
| HTN | b_cell | *C15orf40* | -4.928603943 | 0.013515961 | 0.018737563 | 1 |
| HTN | b_cell | *FAM103A1* | 2.381813403 | 0.002641036 | 0.005102702 | 1 |
| HTN | b_cell | *RGPD8* | -4.29345825 | 9.82E-06 | 0.000617661 | 0.061766071 |

| | | | | | | |
|---|---|---|---|---|---|---|
| HTN | b_cell | *LUZP1* | -4.777416509 | 0.004147887 | 0.007152431 | 1 |
| HTN | b_cell | *HEXDC* | -5.559841388 | 0.000623503 | 0.002291037 | 1 |
| HTN | b_cell | *LRRC45* | -5.856266917 | 0.000169794 | 0.002180344 | 1 |
| HTN | b_cell | *CHRNA5* | 7.929352003 | 0.001467699 | 0.003437936 | 1 |
| HTN | b_cell | *FASN* | -2.817000166 | 0.000854294 | 0.002551796 | 1 |
| HTN | b_cell | *MT1E* | -6.654061734 | 0.000471743 | 0.002180489 | 1 |
| HTN | b_cell | *DCXR* | -7.107123563 | 0.003748991 | 0.006613329 | 1 |
| HTN | b_cell | *LIMS1* | -2.452039073 | 0.005459027 | 0.008860209 | 1 |
| HTN | b_cell | *UGP2* | -2.665287564 | 0.00176658 | 0.003865816 | 1 |
| HTN | b_cell | *CSGALNACT2* | -4.46788935 | 0.001889228 | 0.004046874 | 1 |
| HTN | b_cell | *AVEN* | -4.614320021 | 0.001605065 | 0.003635102 | 1 |
| HTN | b_cell | *CALML6* | -26.53690368 | 9.66E-08 | 2.53E-05 | 0.000607605 |
| HTN | b_cell | *TPST1* | -2.388090125 | 0.000785299 | 0.002451427 | 1 |
| HTN | b_cell | *OTUD3* | -3.161534796 | 0.012399624 | 0.017432119 | 1 |
| HTN | b_cell | *BRD3* | -6.177690147 | 0.011061151 | 0.015773084 | 1 |
| HTN | b_cell | *ZFPM2* | -2.366428036 | 0.000647238 | 0.002320684 | 1 |
| HTN | b_cell | *ZNF747* | -8.32339329 | 0.000275297 | 0.002180344 | 1 |
| HTN | b_cell | *MAP3K2* | -4.381727028 | 0.016286657 | 0.022015574 | 1 |
| HTN | b_cell | *PUSL1* | -9.157924152 | 0.000879789 | 0.002587702 | 1 |
| HTN | b_cell | *MYO7B* | -4.7422138 | 0.002020862 | 0.004222363 | 1 |
| HTN | b_cell | *CNTROB* | -5.670751565 | 0.000441572 | 0.002180344 | 1 |
| HTN | b_cell | *KCNAB3* | -13.95558198 | 6.53E-06 | 0.000472149 | 0.041077005 |
| HTN | b_cell | *NIPA1* | -6.880580667 | 0.004842808 | 0.008041926 | 1 |
| HTN | b_cell | *RNF150* | -2.040864314 | 0.000497152 | 0.002194324 | 1 |
| HTN | b_cell | *HOXD4* | -15.07958765 | 5.11E-05 | 0.001756143 | 0.321098846 |
| HTN | b_cell | *CHRNB1* | 4.52474428 | 0.000255242 | 0.002180344 | 1 |
| HTN | b_cell | *ANKK1* | -6.167817529 | 0.017038566 | 0.022918585 | 1 |
| HTN | b_cell | *PDCD6IP* | -4.619662176 | 0.004718325 | 0.007883101 | 1 |
| HTN | b_cell | *ZNF212* | -6.867382 | 0.032755033 | 0.041054803 | 1 |
| HTN | b_cell | *GLB1* | -5.112993835 | 0.0012525 | 0.003123021 | 1 |
| HTN | b_cell | *FAXDC2* | -4.236923288 | 0.000819416 | 0.002502025 | 1 |
| HTN | b_cell | *CRTAP* | -5.834927177 | 0.004871427 | 0.008083046 | 1 |
| HTN | b_cell | *ELP5* | -11.40866842 | 2.17E-05 | 0.001049416 | 0.136424107 |
| HTN | b_cell | *STX8* | -4.349886996 | 0.006652603 | 0.010332242 | 1 |
| HTN | b_cell | *NFRKB* | -5.027028355 | 0.018733546 | 0.024894907 | 1 |
| HTN | b_cell | *SEMA3E* | -3.121508316 | 0.000474451 | 0.002180489 | 1 |
| HTN | b_cell | *DCLK2* | -3.578974449 | 0.001653662 | 0.003713691 | 1 |
| HTN | b_cell | *GPRC5C* | -6.951995332 | 0.000575053 | 0.00224084 | 1 |
| HTN | b_cell | *MGMT* | -4.213650066 | 0.003889054 | 0.006806927 | 1 |
| HTN | b_cell | *METTL7B* | -14.82965033 | 0.000485708 | 0.002187427 | 1 |
| HTN | b_cell | *CD14* | -3.471537153 | 0.002618191 | 0.005079947 | 1 |
| HTN | b_cell | *KRT72* | 11.94799888 | 0.000749384 | 0.002411911 | 1 |
| HTN | b_cell | *HSD17B13* | -5.112086173 | 0.003029627 | 0.005636952 | 1 |
| HTN | b_cell | *TMC7* | -4.188033905 | 0.001367887 | 0.003287426 | 1 |
| HTN | b_cell | *SERPINB9* | -6.064423749 | 0.030539618 | 0.038546995 | 1 |
| HTN | b_cell | *SMAGP* | -10.4144985 | 0.000124232 | 0.002180344 | 0.781046634 |
| HTN | b_cell | *IRX2* | -3.900609209 | 0.005590928 | 0.009031388 | 1 |
| HTN | b_cell | *EMB* | -3.013912885 | 0.000122985 | 0.002180344 | 0.773209385 |
| HTN | b_cell | *DLGAP1* | -3.04627403 | 0.000506823 | 0.002195202 | 1 |
| HTN | b_cell | *STAT2* | -4.785259209 | 0.039723424 | 0.04894966 | 1 |

| | | | | | | |
|---|---|---|---|---|---|---|
| HTN | b_cell | *NUDCD2* | -7.650079407 | 0.007542926 | 0.011432588 | 1 |
| HTN | b_cell | *HSPA4* | -5.130766973 | 0.001630943 | 0.003677812 | 1 |
| HTN | b_cell | *SLC26A5* | -6.062318923 | 0.016670492 | 0.022466748 | 1 |
| HTN | b_cell | *COMMD5* | -7.697320538 | 0.003222841 | 0.005890118 | 1 |
| HTN | b_cell | *GTSF1* | 11.39357996 | 0.000339951 | 0.002180344 | 1 |
| HTN | b_cell | *ARMC10* | -7.024011516 | 0.00149791 | 0.003478891 | 1 |
| HTN | b_cell | *RNF34* | -7.294991605 | 0.001422106 | 0.00337388 | 1 |
| HTN | b_cell | *ACYP2* | -3.636846492 | 0.003150321 | 0.005799727 | 1 |
| HTN | b_cell | *TRABD* | -6.859752746 | 0.001487802 | 0.003464107 | 1 |
| HTN | b_cell | *TMEM133* | -11.81723789 | 0.00029548 | 0.002180344 | 1 |
| HTN | b_cell | *TTLL6* | 10.33429922 | 0.000420598 | 0.002180344 | 1 |
| HTN | b_cell | *KCNS3* | -2.287642898 | 0.030629861 | 0.038629876 | 1 |
| HTN | b_cell | *AKAP13* | -3.635444233 | 0.035203811 | 0.043792315 | 1 |
| HTN | b_cell | *HTRA3* | -2.947698905 | 0.000405848 | 0.002180344 | 1 |
| HTN | b_cell | *FOXN2* | -4.078999242 | 0.001807005 | 0.003927449 | 1 |
| HTN | b_cell | *KBTBD2* | -5.341033489 | 0.001288811 | 0.003172074 | 1 |
| HTN | b_cell | *MTSS1* | -3.315251216 | 0.001899202 | 0.004061698 | 1 |
| HTN | b_cell | *TSEN34* | 8.740232988 | 0.000789815 | 0.002459174 | 1 |
| HTN | b_cell | *NDUFA3* | -4.811670697 | 0.000330647 | 0.002180344 | 1 |
| HTN | b_cell | *OSCAR* | -6.388222908 | 0.034248305 | 0.042730521 | 1 |
| HTN | b_cell | *PAQR8* | -19.81755431 | 0.003490836 | 0.006252818 | 1 |
| HTN | b_cell | *NUDT6* | -3.326373918 | 0.000633696 | 0.002301586 | 1 |
| HTN | b_cell | *ZNF160* | -5.824660831 | 0.000968632 | 0.002711127 | 1 |
| HTN | b_cell | *FPR1* | -3.453039641 | 0.000543956 | 0.002214137 | 1 |
| HTN | b_cell | *FEZ2* | -4.034528183 | 0.002667336 | 0.005136153 | 1 |
| HTN | b_cell | *SOX7* | -6.415014 | 0.001927581 | 0.004095539 | 1 |
| HTN | b_cell | *C11orf24* | -5.109010836 | 0.000397393 | 0.002180344 | 1 |
| HTN | b_cell | *ALK* | -3.558539093 | 0.000173914 | 0.002180344 | 1 |
| HTN | b_cell | *TRMT61B* | -3.783898214 | 0.007053213 | 0.010818139 | 1 |
| HTN | b_cell | *GIMAP8* | -2.632688045 | 0.003883165 | 0.006798512 | 1 |
| HTN | b_cell | *KCNMB3* | -2.915532846 | 0.001938104 | 0.004110966 | 1 |
| HTN | b_cell | *MORN4* | -4.04866985 | 0.000380476 | 0.002180344 | 1 |
| HTN | b_cell | *GRIK1* | -3.272213711 | 0.000753518 | 0.002416404 | 1 |
| HTN | b_cell | *CLDN20* | -4.346007983 | 0.004412112 | 0.007478822 | 1 |
| HTN | b_cell | *GAA* | 4.352502036 | 0.024092469 | 0.031140904 | 1 |
| HTN | b_cell | *CANT1* | -5.894159973 | 0.000765251 | 0.002430321 | 1 |
| HTN | b_cell | *CHST11* | -3.518790226 | 0.023068578 | 0.02996532 | 1 |
| HTN | b_cell | *PGAM1* | -4.490786317 | 0.00470791 | 0.00787407 | 1 |
| HTN | b_cell | *LURAP1* | -3.578223696 | 0.002118154 | 0.004351906 | 1 |
| HTN | b_cell | *KCND3* | -5.014274892 | 0.000228114 | 0.002180344 | 1 |
| HTN | b_cell | *MRPL36* | -5.099628835 | 0.024943416 | 0.03211535 | 1 |
| HTN | b_cell | *ZNF581* | -14.02524364 | 0.001542121 | 0.003543609 | 1 |
| HTN | b_cell | *MCC* | -3.934937469 | 0.023999152 | 0.031045817 | 1 |
| HTN | b_cell | *KRT27* | 7.253226008 | 0.000100056 | 0.002180344 | 0.629051148 |
| HTN | b_cell | *DLK2* | -23.0477024 | 2.01E-06 | 0.000194635 | 0.012651277 |
| HTN | b_cell | *HOPX* | 13.08236993 | 0.038040179 | 0.047087734 | 1 |
| HTN | b_cell | *RSL1D1* | -7.558508217 | 0.004810524 | 0.007996765 | 1 |
| HTN | b_cell | *COL24A1* | -1.814414675 | 0.01070094 | 0.015365513 | 1 |
| HTN | b_cell | *ETFDH* | -2.387296355 | 0.003502711 | 0.006268586 | 1 |
| HTN | b_cell | *TBCA* | -3.951842794 | 0.03570325 | 0.04435217 | 1 |

| | | | | | | |
|---|---|---|---|---|---|---|
| HTN | b_cell | *MAP6* | -5.541948547 | 0.001861679 | 0.004004235 | 1 |
| HTN | b_cell | *BCL2L1* | -4.223525768 | 0.002499536 | 0.004913879 | 1 |
| HTN | b_cell | *PLRG1* | -6.639854195 | 0.004357874 | 0.007412866 | 1 |
| HTN | b_cell | *ZNF584* | -3.198483764 | 0.011712846 | 0.016611474 | 1 |
| HTN | b_cell | *NMUR1* | -5.656737083 | 0.000484908 | 0.002187427 | 1 |
| HTN | b_cell | *P2RY6* | -4.752373505 | 0.000521076 | 0.002195202 | 1 |
| HTN | b_cell | *ZIK1* | 14.46073391 | 0.01075238 | 0.015416241 | 1 |
| HTN | b_cell | *TCEA2* | -4.382033076 | 0.000417447 | 0.002180344 | 1 |
| HTN | b_cell | *ANO5* | -4.181694966 | 0.000420234 | 0.002180344 | 1 |
| HTN | b_cell | *GPHN* | -2.700609418 | 0.002904633 | 0.005470769 | 1 |
| HTN | b_cell | *VAT1L* | -6.239856227 | 0.000200279 | 0.002180344 | 1 |
| HTN | b_cell | *CAMTA1* | -4.220540547 | 0.002932971 | 0.005510936 | 1 |
| HTN | b_cell | *LGALS4* | 3.952122787 | 9.09E-05 | 0.002180344 | 0.571194104 |
| HTN | b_cell | *LRRC34* | -2.928093984 | 0.001545542 | 0.003547581 | 1 |
| HTN | b_cell | *SPATA5L1* | -4.489299054 | 0.000887594 | 0.002596195 | 1 |
| HTN | b_cell | *SYCE1* | 4.027913168 | 0.000388078 | 0.002180344 | 1 |
| HTN | b_cell | *NXNL1* | -10.53658945 | 0.002838113 | 0.005387444 | 1 |
| HTN | b_cell | *BCL2* | -1.913505278 | 0.003336373 | 0.006048378 | 1 |
| HTN | b_cell | *RHNO1* | 2.912573801 | 0.000158597 | 0.002180344 | 0.997097351 |
| HTN | b_cell | *KNDC1* | 5.495090792 | 0.005859642 | 0.009372504 | 1 |
| HTN | b_cell | *METTL18* | -8.343039769 | 0.00135438 | 0.003272403 | 1 |
| HTN | b_cell | *PWWP2B* | -6.466679559 | 0.000663567 | 0.002327958 | 1 |
| HTN | b_cell | *ZNF570* | -9.947161865 | 0.006207606 | 0.00980337 | 1 |
| HTN | b_cell | *NINJ2* | -5.343323266 | 0.002666737 | 0.005136153 | 1 |
| HTN | b_cell | *FAM90A1* | -14.51639413 | 0.000414569 | 0.002180344 | 1 |
| HTN | b_cell | *TRAPPC12* | -5.007516141 | 0.003821021 | 0.006712142 | 1 |
| HTN | b_cell | *C3AR1* | -4.159205218 | 0.000192041 | 0.002180344 | 1 |
| HTN | b_cell | *AQP4* | -5.599441574 | 0.000413279 | 0.002180344 | 1 |
| HTN | b_cell | *TLN2* | -2.035853535 | 0.000356354 | 0.002180344 | 1 |
| HTN | b_cell | *TVP23B* | -5.091145099 | 0.000824194 | 0.002511734 | 1 |
| HTN | b_cell | *ATPAF2* | -4.47021891 | 0.000626892 | 0.002295161 | 1 |
| HTN | b_cell | *CYP4F22* | -12.66956434 | 0.000198296 | 0.002180344 | 1 |
| HTN | b_cell | *ZNF554* | -4.333501736 | 0.001172957 | 0.003004867 | 1 |
| HTN | b_cell | *ANKRD20A4* | 4.808946652 | 0.000241078 | 0.002180344 | 1 |
| HTN | b_cell | *LAMB2* | -3.429920919 | 0.000387037 | 0.002180344 | 1 |
| HTN | b_cell | *ORMDL3* | -3.006788156 | 0.00196509 | 0.004148529 | 1 |
| HTN | b_cell | *SERF1A* | -24.55442487 | 0.000355183 | 0.002180344 | 1 |
| HTN | b_cell | *LRRC15* | 3.957213623 | 0.000241377 | 0.002180344 | 1 |
| HTN | b_cell | *MOB3A* | -4.114370139 | 0.001479068 | 0.003455555 | 1 |
| HTN | b_cell | *SLFN12* | -3.752330328 | 0.019690671 | 0.026007406 | 1 |
| HTN | b_cell | *FRMD3* | -3.006708746 | 0.000364502 | 0.002180344 | 1 |
| HTN | b_cell | *MTBP* | -5.65221912 | 0.001923903 | 0.004091873 | 1 |
| HTN | b_cell | *ISG20* | 2.694443218 | 0.00082465 | 0.002511905 | 1 |
| HTN | b_cell | *MBOAT1* | -4.131067914 | 0.001050637 | 0.002821596 | 1 |
| HTN | b_cell | *CXCR6* | -5.589800345 | 0.019791603 | 0.026118767 | 1 |
| HTN | b_cell | *C5orf34* | -5.102748226 | 8.15E-05 | 0.002177448 | 0.512108097 |
| HTN | b_cell | *ZNF131* | -4.755183752 | 0.025947859 | 0.033292691 | 1 |
| HTN | b_cell | *HINFP* | -5.461118133 | 0.000845456 | 0.002536213 | 1 |
| HTN | b_cell | *SPTLC3* | -5.624533247 | 0.006591207 | 0.010261597 | 1 |
| HTN | b_cell | *COPRS* | -7.752744523 | 0.008256638 | 0.012286268 | 1 |

| | | | | | | |
|---|---|---|---|---|---|---|
| HTN | b_cell | *CLEC12A* | -1.772180808 | 0.006914902 | 0.010652227 | 1 |
| HTN | b_cell | *SUCLG2* | -3.863853092 | 0.029526409 | 0.037395756 | 1 |
| HTN | b_cell | *CSDC2* | -12.30449554 | 0.000158412 | 0.002180344 | 0.995933758 |
| HTN | b_cell | *RCAN2* | -2.638383031 | 0.000402152 | 0.002180344 | 1 |
| HTN | b_cell | *IL16* | -3.007326001 | 0.00114148 | 0.002954503 | 1 |
| HTN | b_cell | *PRSS27* | -24.82901319 | 0.000535903 | 0.002204987 | 1 |
| HTN | b_cell | *FGGY* | -4.010011622 | 0.003175562 | 0.005829126 | 1 |
| HTN | b_cell | *ZNF24* | -4.807021027 | 0.001289162 | 0.003172074 | 1 |
| HTN | b_cell | *FIBP* | -8.005492845 | 0.04060249 | 0.04992526 | 1 |
| HTN | b_cell | *CARNS1* | -14.21148635 | 0.000262699 | 0.002180344 | 1 |
| HTN | b_cell | *CTSW* | -3.80980033 | 8.55E-08 | 2.44E-05 | 0.000537518 |
| HTN | b_cell | *SNTG2* | -2.436944945 | 0.000182362 | 0.002180344 | 1 |
| HTN | b_cell | *PDE3A* | -2.631697978 | 0.001064352 | 0.002842642 | 1 |
| HTN | b_cell | *MRPL52* | -6.697716131 | 0.023928694 | 0.030980169 | 1 |
| HTN | b_cell | *EFEMP2* | -3.362084449 | 0.003594357 | 0.006392567 | 1 |
| HTN | b_cell | *AGAP5* | -142.2770633 | 0.000414858 | 0.002180344 | 1 |
| HTN | b_cell | *ZMAT3* | -6.853547811 | 0.013640449 | 0.018897643 | 1 |
| HTN | b_cell | *ZFAND4* | -5.190054409 | 0.000570697 | 0.002235498 | 1 |
| HTN | b_cell | *SLFN11* | -5.754609951 | 0.00017337 | 0.002180344 | 1 |
| HTN | b_cell | *CCL19* | -24.08208763 | 0.000253371 | 0.002180344 | 1 |
| HTN | b_cell | *EFCAB12* | -5.510829021 | 0.004644139 | 0.007800615 | 1 |
| HTN | b_cell | *FAM192A* | -4.791420374 | 0.00574631 | 0.009231316 | 1 |
| HTN | b_cell | *CBWD1* | -6.074520859 | 0.002863666 | 0.005414697 | 1 |
| HTN | b_cell | *HOXC5* | 12.62410347 | 1.82E-06 | 0.000186095 | 0.01144211 |
| HTN | b_cell | *DCP2* | -5.118184171 | 0.012705813 | 0.017814773 | 1 |
| HTN | b_cell | *SNX32* | 10.11708334 | 9.22E-06 | 0.000585817 | 0.057995915 |
| HTN | b_cell | *CYP7B1* | -2.655254628 | 0.000527495 | 0.002195202 | 1 |
| HTN | b_cell | *METAP1D* | -3.1730095 | 0.000501622 | 0.002195028 | 1 |
| HTN | b_cell | *NADSYN1* | -4.820555355 | 0.002233645 | 0.004512508 | 1 |
| HTN | b_cell | *MYEOV* | -4.075530782 | 3.93E-06 | 0.000342867 | 0.024686425 |
| HTN | b_cell | *ANKRD13D* | -5.761655427 | 0.01499679 | 0.020501157 | 1 |
| HTN | b_cell | *MRGPRF* | -9.928005159 | 0.006741406 | 0.010453954 | 1 |
| HTN | b_cell | *ADH6* | -5.036971742 | 6.33E-05 | 0.001980182 | 0.398234938 |
| HTN | b_cell | *GXYLT2* | -2.899252185 | 0.000530428 | 0.002195202 | 1 |
| HTN | b_cell | *HPSE2* | -2.729979025 | 0.009179246 | 0.01346161 | 1 |
| HTN | b_cell | *DCAKD* | -6.731594592 | 7.52E-06 | 0.000525319 | 0.047278704 |
| HTN | b_cell | *HECTD4* | -3.630492728 | 0.016036788 | 0.021715116 | 1 |
| HTN | b_cell | *BNC2* | -2.478083926 | 0.00168264 | 0.003747346 | 1 |
| HTN | b_cell | *TRMT112* | -5.098451889 | 0.001260677 | 0.003136478 | 1 |
| HTN | b_cell | *RHOD* | -5.145779006 | 0.000331867 | 0.002180344 | 1 |
| HTN | b_cell | *PARP15* | 6.9746215 | 0.000471565 | 0.002180489 | 1 |
| HTN | b_cell | *AHSA2* | -3.270413676 | 0.027710509 | 0.035316434 | 1 |
| HTN | b_cell | *ABLIM3* | -2.608252703 | 0.000448764 | 0.002180344 | 1 |
| HTN | b_cell | *VANGL1* | -4.218587413 | 0.000546652 | 0.002214137 | 1 |
| HTN | b_cell | *GLRX* | -2.402454791 | 0.000388321 | 0.002180344 | 1 |
| HTN | b_cell | *IQCB1* | -3.698951344 | 0.025030188 | 0.032213878 | 1 |
| HTN | b_cell | *GOLGB1* | -4.263025527 | 0.004666384 | 0.007825436 | 1 |
| HTN | b_cell | *DMRT2* | -4.926393351 | 1.10E-05 | 0.000671176 | 0.069131141 |
| HTN | b_cell | *ZNF483* | -3.58692688 | 0.001249458 | 0.003120831 | 1 |
| HTN | b_cell | *SLC2A14* | -5.564285706 | 0.005250909 | 0.008599236 | 1 |

| | | | | | | |
|---|---|---|---|---|---|---|
| HTN | b_cell | *MZT2A* | 5.146669808 | 0.000299251 | 0.002180344 | 1 |
| HTN | b_cell | *MAP3K11* | -6.247500001 | 0.004758011 | 0.007928337 | 1 |
| HTN | b_cell | *C1QA* | -5.287826154 | 0.001926509 | 0.004094645 | 1 |
| HTN | b_cell | *NDNF* | -5.26517694 | 0.000343756 | 0.002180344 | 1 |
| HTN | b_cell | *OLR1* | 9.14676994 | 0.000136401 | 0.002180344 | 0.857550149 |
| HTN | b_cell | *GLIPR1L1* | -3.121518006 | 0.001030039 | 0.002792956 | 1 |
| HTN | b_cell | *ARV1* | 8.309945495 | 0.001035337 | 0.002800612 | 1 |
| HTN | b_cell | *CCDC36* | -2.771614759 | 0.000562162 | 0.002226547 | 1 |
| HTN | b_cell | *THAP2* | 6.437534708 | 0.000978999 | 0.002717425 | 1 |
| HTN | b_cell | *PPP1R14B* | -10.0915728 | 0.019242266 | 0.025474021 | 1 |
| HTN | b_cell | *SMARCC1* | -4.472328535 | 0.030383356 | 0.038380583 | 1 |
| HTN | b_cell | *ZNF417* | -6.658107313 | 0.000191107 | 0.002180344 | 1 |
| HTN | b_cell | *VEGFB* | -9.498368779 | 3.14E-08 | 1.05E-05 | 0.000197721 |
| HTN | b_cell | *PEAK1* | -3.785246131 | 0.020755178 | 0.027270179 | 1 |
| HTN | b_cell | *TNFRSF10D* | -2.443732042 | 0.025342177 | 0.032582058 | 1 |
| HTN | b_cell | *MST1* | -8.909850141 | 1.62E-08 | 6.78E-06 | 0.000101751 |
| HTN | b_cell | *TNFRSF10C* | 3.97607157 | 0.021374789 | 0.027984861 | 1 |
| HTN | b_cell | *GMPPB* | -4.868565658 | 0.006392798 | 0.010017826 | 1 |
| HTN | b_cell | *CSPG4* | -3.920595288 | 0.000935405 | 0.002659834 | 1 |
| HTN | b_cell | *SNX33* | -5.555856889 | 0.000527439 | 0.002195202 | 1 |
| HTN | b_cell | *NUDT18* | -9.3751747 | 0.000897399 | 0.002599737 | 1 |
| HTN | b_cell | *SULT1B1* | -6.161866103 | 0.002378137 | 0.004732008 | 1 |
| HTN | b_cell | *NUDT4* | -3.998676709 | 0.000958566 | 0.00269763 | 1 |
| HTN | b_cell | *SCAI* | -4.713854355 | 0.009697867 | 0.014058218 | 1 |
| HTN | b_cell | *C1orf100* | -9.940648344 | 0.000554942 | 0.002220961 | 1 |
| HTN | b_cell | *AGFG1* | -3.434314965 | 0.000970176 | 0.0027133 | 1 |
| HTN | b_cell | *RNF213* | -4.175017832 | 0.004764635 | 0.007937271 | 1 |
| HTN | b_cell | *PLK3* | -4.488510535 | 0.000438676 | 0.002180344 | 1 |
| HTN | b_cell | *TUBB8* | -13.93957801 | 0.000542188 | 0.002213464 | 1 |
| HTN | b_cell | *GPR160* | -8.808959124 | 0.032560004 | 0.040834779 | 1 |
| HTN | b_cell | *GOLIM4* | -5.71683014 | 0.003605858 | 0.006407584 | 1 |
| HTN | b_cell | *USMG5* | -4.814973214 | 0.027295132 | 0.034857708 | 1 |
| HTN | b_cell | *HOXB2* | -6.485774764 | 0.001071137 | 0.002848663 | 1 |
| HTN | b_cell | *XXYLT1* | -4.234408565 | 0.000873232 | 0.002573845 | 1 |
| HTN | b_cell | *UBXN2A* | -4.215733855 | 0.024064058 | 0.031116975 | 1 |
| HTN | b_cell | *CEP19* | -5.442129233 | 0.000604706 | 0.002281385 | 1 |
| HTN | b_cell | *SLC25A30* | -7.70322152 | 0.002953569 | 0.005538053 | 1 |
| HTN | b_cell | *CTSF* | -3.722010597 | 0.000451736 | 0.002180344 | 1 |
| HTN | b_cell | *SOCS7* | -4.073966815 | 0.000663013 | 0.002327958 | 1 |
| HTN | b_cell | *TLR6* | -3.668774678 | 0.040284025 | 0.049591867 | 1 |
| HTN | b_cell | *FAM53A* | -4.896131418 | 0.001260285 | 0.003136478 | 1 |
| HTN | b_cell | *CTU2* | -8.25601324 | 1.66E-05 | 0.000924 | 0.104359129 |
| HTN | b_cell | *MGA* | -4.43536926 | 0.003382856 | 0.006108868 | 1 |
| HTN | b_cell | *SNX31* | -11.59669461 | 4.15E-07 | 6.93E-05 | 0.00260905 |
| HTN | b_cell | *DDX23* | -7.687139926 | 0.004062004 | 0.007041031 | 1 |
| HTN | b_cell | *GLIS1* | -5.517625774 | 1.21E-06 | 0.000143407 | 0.007600565 |
| HTN | b_cell | *RALGAPA1* | -4.551607212 | 0.004937065 | 0.008170395 | 1 |
| HTN | b_cell | *LIG4* | -4.978210892 | 0.000599792 | 0.002274361 | 1 |
| HTN | b_cell | *LINGO2* | -5.335128233 | 0.001728486 | 0.003822011 | 1 |
| HTN | b_cell | *BBS1* | 7.791545343 | 0.000449159 | 0.002180344 | 1 |

| | | | | | | |
|---|---|---|---|---|---|---|
| HTN | b_cell | *ANKRD36C* | -4.452242145 | 0.001742931 | 0.003841397 | 1 |
| HTN | b_cell | *TMEM81* | -29.44239473 | 0.004122011 | 0.007117492 | 1 |
| HTN | b_cell | *IL20RB* | 4.881679528 | 0.004509594 | 0.007613271 | 1 |
| HTN | b_cell | *ZNF497* | 3.766460433 | 0.000253112 | 0.002180344 | 1 |
| HTN | b_cell | *KLF14* | 5.852034006 | 0.000616611 | 0.002286342 | 1 |
| HTN | b_cell | *ANGEL2* | -4.381775406 | 0.001262447 | 0.003138396 | 1 |
| HTN | b_cell | *ZNF266* | -3.84141975 | 0.010577697 | 0.015203928 | 1 |
| HTN | b_cell | *BRSK2* | -10.18532938 | 0.000933555 | 0.002657043 | 1 |
| HTN | b_cell | *SH3PXD2B* | -2.411091939 | 0.000547192 | 0.002214137 | 1 |
| HTN | b_cell | *LARP7* | -4.178462165 | 0.001037906 | 0.002802971 | 1 |
| HTN | b_cell | *FGFBP3* | -7.527565776 | 0.000270833 | 0.002180344 | 1 |
| HTN | b_cell | *C4orf32* | -2.624293717 | 0.000407773 | 0.002180344 | 1 |
| HTN | b_cell | *HRAS* | -6.075411717 | 0.00480226 | 0.007989365 | 1 |
| HTN | b_cell | *C4orf26* | 12.45638462 | 0.000308569 | 0.002180344 | 1 |
| HTN | b_cell | *BTC* | -4.916983589 | 0.00046046 | 0.002180489 | 1 |
| HTN | b_cell | *RSRC1* | -6.168951501 | 8.10E-05 | 0.002176137 | 0.509216001 |
| HTN | b_cell | *PTDSS2* | -4.971738695 | 0.004166075 | 0.007171992 | 1 |
| HTN | b_cell | *FUT1* | -14.38298245 | 0.001121017 | 0.002924292 | 1 |
| HTN | b_cell | *DHX36* | -4.862698964 | 0.004241716 | 0.007258483 | 1 |
| HTN | b_cell | *FBXW8* | -4.646661751 | 0.001834426 | 0.00395916 | 1 |
| HTN | b_cell | *CHST2* | -10.0111066 | 0.000448919 | 0.002180344 | 1 |
| HTN | b_cell | *ATR* | -3.677400084 | 0.001029698 | 0.002792956 | 1 |
| HTN | b_cell | *SH3BP5L* | -6.300328996 | 0.000810746 | 0.002488849 | 1 |
| HTN | b_cell | *CADM2* | -2.897228388 | 8.60E-05 | 0.002180344 | 0.5404455 |
| HTN | b_cell | *PSMD2* | -3.559015337 | 0.000416886 | 0.002180344 | 1 |
| HTN | b_cell | *TP53I11* | -5.887758149 | 0.017346932 | 0.023266269 | 1 |
| HTN | b_cell | *PHYHD1* | -3.545482041 | 0.00105861 | 0.002833039 | 1 |
| HTN | b_cell | *ZNF519* | -5.878033968 | 0.002037033 | 0.004239268 | 1 |
| HTN | b_cell | *BANF1* | 5.72726851 | 7.52E-05 | 0.002130387 | 0.472945901 |
| HTN | b_cell | *CHRNA7* | -9.207440496 | 0.003390299 | 0.006117913 | 1 |
| HTN | b_cell | *NRIP3* | -8.833298298 | 8.96E-07 | 0.000115006 | 0.005635311 |
| HTN | b_cell | *SCUBE2* | -3.5814585 | 0.000459334 | 0.002180489 | 1 |
| HTN | b_cell | *SMAD2* | -3.812232369 | 0.003085662 | 0.005714155 | 1 |
| HTN | b_cell | *EIF3F* | -6.322521243 | 0.034101712 | 0.042556066 | 1 |
| HTN | b_cell | *ZNF25* | -4.772965599 | 0.00089129 | 0.002599737 | 1 |
| HTN | b_cell | *CLTB* | -4.473748681 | 0.007961924 | 0.011938139 | 1 |
| HTN | b_cell | *PCSK1* | -5.685322983 | 0.000327009 | 0.002180344 | 1 |
| HTN | b_cell | *LPL* | -2.073879925 | 0.00010066 | 0.002180344 | 0.632851896 |
| HTN | b_cell | *TBC1D10C* | 4.307734826 | 0.00034086 | 0.002180344 | 1 |
| HTN | b_cell | *PPP2R2D* | -5.566114042 | 6.13E-05 | 0.001965056 | 0.385150945 |
| HTN | b_cell | *MCTP1* | -2.024153102 | 0.000397689 | 0.002180344 | 1 |
| HTN | b_cell | *UBQLNL* | 3.143631196 | 0.000338277 | 0.002180344 | 1 |
| HTN | b_cell | *LIPT2* | -15.33367493 | 0.00157687 | 0.003584159 | 1 |
| HTN | b_cell | *KCNE3* | -4.148647009 | 0.000421636 | 0.002180344 | 1 |
| HTN | b_cell | *UCP3* | 3.267874407 | 0.000459922 | 0.002180489 | 1 |
| HTN | b_cell | *PAAF1* | -2.348811257 | 0.011880924 | 0.016815707 | 1 |
| HTN | b_cell | *MRPL48* | -5.69123983 | 0.010813778 | 0.015486612 | 1 |
| HTN | b_cell | *RAB6A* | -3.883813816 | 0.002116147 | 0.004349205 | 1 |
| HTN | b_cell | *RPS6KB2* | -5.758754827 | 0.001432786 | 0.003385825 | 1 |
| HTN | b_cell | *RMI2* | -7.162036049 | 0.003090098 | 0.005715637 | 1 |

| | | | | | | |
|---|---|---|---|---|---|---|
| HTN | b_cell | *TOM1L2* | -3.989106215 | 0.001508642 | 0.003496068 | 1 |
| HTN | b_cell | *TEX26* | -11.66738715 | 0.001349062 | 0.00326842 | 1 |
| HTN | b_cell | *GPR156* | -8.539143436 | 0.000706862 | 0.002373954 | 1 |
| HTN | b_cell | *TTLL11* | -4.283854953 | 0.001084219 | 0.002865833 | 1 |
| HTN | b_cell | *MSRA* | -5.788663252 | 0.007596813 | 0.01150317 | 1 |
| HTN | b_cell | *CTDNEP1* | -6.15428044 | 0.013172224 | 0.018345984 | 1 |
| HTN | b_cell | *ETV4* | 8.91729051 | 0.000616508 | 0.002286342 | 1 |
| HTN | b_cell | *SWI5* | -4.730668278 | 0.000679684 | 0.002339217 | 1 |
| HTN | b_cell | *ZDHHC21* | -4.88187337 | 0.003345659 | 0.00605997 | 1 |
| HTN | b_cell | *A2M* | -3.065126493 | 0.000756664 | 0.002419498 | 1 |
| HTN | b_cell | *LRRN1* | -4.812027048 | 0.000475823 | 0.002180489 | 1 |
| HTN | b_cell | *B3GALT6* | -8.63600785 | 0.000285353 | 0.002180344 | 1 |
| HTN | b_cell | *ZNF613* | -4.843971495 | 0.000586081 | 0.002261934 | 1 |
| HTN | b_cell | *IP6K1* | -4.982283595 | 0.011117292 | 0.015838299 | 1 |
| HTN | b_cell | *SSNA1* | -4.48268893 | 0.000351399 | 0.002180344 | 1 |
| HTN | b_cell | *CHMP6* | -7.198834332 | 0.026917197 | 0.034445027 | 1 |
| HTN | b_cell | *TMEM39A* | -5.041765633 | 0.002357629 | 0.004701051 | 1 |
| HTN | b_cell | *TCP11L1* | -4.024242339 | 0.023328852 | 0.030265887 | 1 |
| HTN | b_cell | *GPX2* | -7.637471436 | 0.001144525 | 0.002958728 | 1 |
| HTN | b_cell | *CCDC57* | -3.945608961 | 0.000931364 | 0.002656753 | 1 |
| HTN | b_cell | *CIDEA* | -2.006098793 | 0.001320424 | 0.003218935 | 1 |
| HTN | b_cell | *SMIM19* | -2.734122129 | 0.001681428 | 0.003747303 | 1 |
| HTN | b_cell | *ZNF404* | -3.672730889 | 0.003552072 | 0.006328104 | 1 |
| HTN | b_cell | *SLC35G1* | -4.524122769 | 0.014433045 | 0.019829666 | 1 |
| HTN | b_cell | *ZNF135* | -5.287700816 | 0.000383526 | 0.002180344 | 1 |
| HTN | b_cell | *COX8A* | -7.841269434 | 0.038899742 | 0.048028806 | 1 |
| HTN | b_cell | *B3GNT4* | 4.103625033 | 0.000230825 | 0.002180344 | 1 |
| HTN | b_cell | *CDC26* | -6.230440805 | 0.000712067 | 0.002380505 | 1 |
| HTN | b_cell | *RNPEP* | -4.24552245 | 0.000837094 | 0.002526663 | 1 |
| HTN | b_cell | *SPRYD4* | -3.705888006 | 0.018916253 | 0.02511647 | 1 |
| HTN | b_cell | *VPS37D* | -5.970511468 | 0.005870882 | 0.009379576 | 1 |
| HTN | b_cell | *SLCO3A1* | -3.621965099 | 0.001494065 | 0.003473811 | 1 |
| HTN | b_cell | *WDR25* | -6.164789018 | 0.00567409 | 0.009137552 | 1 |
| HTN | b_cell | *PLA2G16* | -1.67458333 | 0.004289908 | 0.007323012 | 1 |
| HTN | b_cell | *PHLDB3* | -7.339303975 | 0.035371581 | 0.043974912 | 1 |
| HTN | b_cell | *PRR15* | -18.27450992 | 0.011495823 | 0.016340547 | 1 |
| HTN | b_cell | *RMDN1* | -4.548314137 | 0.001178235 | 0.003009982 | 1 |
| HTN | b_cell | *HORMAD2* | -3.939118949 | 0.033078469 | 0.041427158 | 1 |
| HTN | b_cell | *RNF152* | -4.430830507 | 0.0005303 | 0.002195202 | 1 |
| HTN | b_cell | *ACSF3* | -5.790246605 | 0.008422664 | 0.012488984 | 1 |
| HTN | b_cell | *BOK* | -3.332423017 | 0.000494307 | 0.002192417 | 1 |
| HTN | b_cell | *ZNF843* | -7.915698807 | 0.000405621 | 0.002180344 | 1 |
| HTN | b_cell | *C8orf59* | -4.164307362 | 0.000872727 | 0.002573594 | 1 |
| HTN | b_cell | *PFN4* | -5.748933131 | 1.98E-05 | 0.001016068 | 0.124731376 |
| HTN | b_cell | *RUFY1* | -4.901110961 | 0.011793652 | 0.016707231 | 1 |
| HTN | b_cell | *BASP1* | -2.849854801 | 0.00055709 | 0.002226229 | 1 |
| HTN | b_cell | *VSIG10* | -2.684186898 | 0.00164063 | 0.003695679 | 1 |
| HTN | b_cell | *IRX5* | -4.974498491 | 0.001643238 | 0.003700229 | 1 |
| HTN | b_cell | *PNMA1* | -10.39371748 | 0.013029414 | 0.018186144 | 1 |
| HTN | b_cell | *ANKLE2* | -4.806588288 | 0.002160993 | 0.004403943 | 1 |

| | | | | | | |
|---|---|---|---|---|---|---|
| HTN | b_cell | *GCNT4* | 3.576080499 | 0.000231185 | 0.002180344 | 1 |
| HTN | b_cell | *MUC20* | 5.376645851 | 0.000317452 | 0.002180344 | 1 |
| HTN | b_cell | *SHMT1* | -2.399552525 | 0.004538211 | 0.007653362 | 1 |
| HTN | b_cell | *MTHFR* | -7.694232919 | 3.96E-05 | 0.001519523 | 0.249201847 |
| HTN | b_cell | *C19orf18* | -4.019484856 | 0.000225506 | 0.002180344 | 1 |
| HTN | b_cell | *TMEM80* | -4.012823867 | 0.000293587 | 0.002180344 | 1 |
| HTN | b_cell | *WDR73* | -6.211002659 | 0.000811316 | 0.002489381 | 1 |
| HTN | b_cell | *POLE* | -6.088256386 | 0.003468859 | 0.006225725 | 1 |
| HTN | b_cell | *DSCAML1* | -3.813180582 | 0.000910945 | 0.002625909 | 1 |
| HTN | b_cell | *EPS8L2* | -5.766973131 | 0.000388747 | 0.002180344 | 1 |
| HTN | b_cell | *TALDO1* | -4.428475322 | 0.000429312 | 0.002180344 | 1 |
| HTN | b_cell | *B3GNT8* | 2.192876269 | 0.000230507 | 0.002180344 | 1 |
| HTN | b_cell | *AP006621.1* | -3.854634208 | 0.001061693 | 0.002837951 | 1 |
| HTN | b_cell | *MAN1B1* | -6.898129343 | 0.005624395 | 0.009071465 | 1 |
| HTN | b_cell | *TOP3A* | -7.303401459 | 0.00042171 | 0.002180344 | 1 |
| HTN | b_cell | *C8orf31* | -5.746995231 | 0.000421095 | 0.002180344 | 1 |
| HTN | b_cell | *AC024940.1* | 1.359740285 | 0.000153298 | 0.002180344 | 0.96378725 |
| HTN | b_cell | *TIMM22* | -5.571082035 | 0.000422885 | 0.002180344 | 1 |
| HTN | b_cell | *PPFIA3* | -9.227469566 | 2.15E-06 | 0.00020503 | 0.013532004 |
| HTN | b_cell | *MAGEF1* | -5.30636503 | 0.0004139 | 0.002180344 | 1 |
| HTN | b_cell | *PAWR* | -5.656195258 | 0.005717026 | 0.009192568 | 1 |
| HTN | b_cell | *NAP1L5* | -6.964706808 | 0.001045203 | 0.002814467 | 1 |
| HTN | b_cell | *NIM1K* | -6.527119509 | 0.001351814 | 0.003270974 | 1 |
| HTN | b_cell | *ACOT4* | -7.737643038 | 5.11E-05 | 0.001756143 | 0.32137408 |
| HTN | b_cell | *ARIH2* | -4.552856475 | 0.000454066 | 0.002180344 | 1 |
| HTN | b_cell | *RBM44* | -6.010165183 | 0.000580255 | 0.002250503 | 1 |
| HTN | b_cell | *IRX3* | -5.821322102 | 0.000974034 | 0.002717425 | 1 |
| HTN | b_cell | *ATOX1* | 6.27136475 | 0.039865105 | 0.049114622 | 1 |
| HTN | b_cell | *TBL1XR1* | -3.77368681 | 0.006561202 | 0.010222755 | 1 |
| HTN | b_cell | *C18orf32* | 6.663427805 | 0.000676735 | 0.002336426 | 1 |
| HTN | b_cell | *C12orf54* | -22.60691751 | 0.000108829 | 0.002180344 | 0.684210163 |
| HTN | b_cell | *GBA* | -5.797729967 | 0.00016024 | 0.002180344 | 1 |
| HTN | b_cell | *ACAD9* | -2.972741853 | 0.000432211 | 0.002180344 | 1 |
| HTN | b_cell | *IL17RA* | -4.740478433 | 0.000732846 | 0.002399172 | 1 |
| HTN | b_cell | *CD163L1* | -2.585396601 | 0.00065179 | 0.002323165 | 1 |
| HTN | b_cell | *THAP5* | -8.726488747 | 0.008429248 | 0.012495799 | 1 |
| HTN | b_cell | *CD151* | -4.193197909 | 0.015072719 | 0.020567127 | 1 |
| HTN | b_cell | *POLR2L* | -5.839070489 | 0.000202392 | 0.002180344 | 1 |
| HTN | b_cell | *FAM87B* | -19.85067163 | 0.001159882 | 0.00298615 | 1 |
| HTN | b_cell | *ZBTB41* | -4.703698784 | 0.001134777 | 0.002945642 | 1 |
| HTN | b_cell | *ZNF354C* | -6.790340979 | 0.007997392 | 0.011976136 | 1 |
| HTN | b_cell | *CENPBD1* | -9.665163288 | 0.003349132 | 0.00606346 | 1 |
| HTN | b_cell | *RIC8A* | -7.886710622 | 0.000302231 | 0.002180344 | 1 |
| HTN | b_cell | *IMP3* | -4.307923194 | 0.00044354 | 0.002180344 | 1 |
| HTN | b_cell | *ODF3B* | -5.188820648 | 0.01419849 | 0.0195587 | 1 |
| HTN | b_cell | *DPY19L2* | -5.067083253 | 0.010488814 | 0.015107257 | 1 |
| HTN | b_cell | *C2orf73* | -6.179415367 | 0.000376229 | 0.002180344 | 1 |
| HTN | b_cell | *FAM26E* | -9.388885541 | 0.00449848 | 0.007596546 | 1 |
| HTN | b_cell | *ALS2CL* | -3.466137309 | 0.002219396 | 0.004497823 | 1 |
| HTN | b_cell | *MLF1* | -3.143554633 | 0.000676328 | 0.002336304 | 1 |

| | | | | | | |
|---|---|---|---|---|---|---|
| HTN | b_cell | *PRSS42* | 17.08976198 | 0.002318151 | 0.004647655 | 1 |
| HTN | b_cell | *C2orf69* | -5.532084156 | 0.001378633 | 0.003304409 | 1 |
| HTN | b_cell | *GRAMD1C* | -4.743431616 | 0.00043682 | 0.002180344 | 1 |
| HTN | b_cell | *TSSK6* | -15.78101695 | 0.00048506 | 0.002187427 | 1 |
| HTN | b_cell | *GOLGA8Q* | 87.01297504 | 0.000341901 | 0.002180344 | 1 |
| HTN | b_cell | *DALRD3* | -7.6779866 | 0.013991261 | 0.019311319 | 1 |
| HTN | b_cell | *ZNF114* | -9.638130328 | 0.000451713 | 0.002180344 | 1 |
| HTN | b_cell | *ZNF518B* | -6.809690893 | 0.000417071 | 0.002180344 | 1 |
| HTN | b_cell | *LCORL* | -4.374544123 | 0.015523277 | 0.021097026 | 1 |
| HTN | b_cell | *ZNF454* | -4.660324691 | 0.002853711 | 0.005397498 | 1 |
| HTN | b_cell | *SH2B1* | -3.774548161 | 7.90E-06 | 0.000539559 | 0.049639419 |
| HTN | b_cell | *ZC3H12D* | -9.820355621 | 0.007541029 | 0.011432469 | 1 |
| HTN | b_cell | *KDELC2* | -4.190067867 | 0.000239114 | 0.002180344 | 1 |
| HTN | b_cell | *PLEC* | -6.604412343 | 0.00569752 | 0.009165892 | 1 |
| HTN | b_cell | *SH2D4B* | 9.802975033 | 0.000291526 | 0.002180344 | 1 |
| HTN | b_cell | *RNF212* | -2.603967946 | 0.012205355 | 0.017207725 | 1 |
| HTN | b_cell | *PRSS36* | -6.907472649 | 0.000421472 | 0.002180344 | 1 |
| HTN | b_cell | *TMPRSS9* | -4.280205361 | 0.000275913 | 0.002180344 | 1 |
| HTN | b_cell | *AQP11* | -5.086886857 | 0.000746425 | 0.002409022 | 1 |
| HTN | b_cell | *ZFAND2A* | -11.43839466 | 0.000524808 | 0.002195202 | 1 |
| HTN | b_cell | *FAM220A* | -5.91151618 | 0.009022354 | 0.013265561 | 1 |
| HTN | b_cell | *BEND3* | -7.644834323 | 0.000672657 | 0.002334899 | 1 |
| HTN | b_cell | *NT5DC1* | -4.086935485 | 0.015373823 | 0.020925574 | 1 |
| HTN | b_cell | *AC006111.1* | -20.24603896 | 0.002355212 | 0.004701026 | 1 |
| HTN | b_cell | *COX14* | -3.749547544 | 1.66E-05 | 0.000924 | 0.104411984 |
| HTN | b_cell | *CTXN1* | 26.12211223 | 0.006768158 | 0.010485809 | 1 |
| HTN | b_cell | *CA8* | -3.895383793 | 0.002436986 | 0.004819545 | 1 |
| HTN | b_cell | *ERBB4* | -1.938917053 | 0.000415173 | 0.002180344 | 1 |
| HTN | b_cell | *GPR35* | -4.781876521 | 0.002948867 | 0.005530886 | 1 |
| HTN | b_cell | *CSRNP3* | -5.985875597 | 0.001191786 | 0.003028602 | 1 |
| HTN | b_cell | *ZNF713* | -6.042486669 | 0.010811544 | 0.015486612 | 1 |
| HTN | b_cell | *KCTD12* | -3.32697249 | 0.000774985 | 0.002438676 | 1 |
| HTN | b_cell | *RPP25* | 14.71524792 | 2.65E-06 | 0.000248264 | 0.016633679 |
| HTN | b_cell | *FAM219B* | -3.363276967 | 9.68E-05 | 0.002180344 | 0.608390382 |
| HTN | b_cell | *CPNE7* | 5.219838927 | 0.002927349 | 0.005503255 | 1 |
| HTN | b_cell | *GDPD4* | 5.176509871 | 0.000285967 | 0.002180344 | 1 |
| HTN | b_cell | *MPI* | -14.34786436 | 7.11E-08 | 2.13E-05 | 0.000446959 |
| HTN | b_cell | *TRIM73* | -28.34333906 | 3.11E-06 | 0.000279325 | 0.019552745 |
| HTN | b_cell | *EFCAB13* | -4.691189405 | 0.004341659 | 0.007389282 | 1 |
| HTN | b_cell | *HYI* | -5.743533097 | 2.65E-08 | 9.78E-06 | 0.000166326 |
| HTN | b_cell | *RMI1* | -9.250593719 | 0.002100869 | 0.004331966 | 1 |
| HTN | b_cell | *CTC1* | 10.33275706 | 1.23E-05 | 0.000723375 | 0.077209597 |
| HTN | b_cell | *SNX18* | -5.22537639 | 0.001779753 | 0.003885177 | 1 |
| HTN | b_cell | *C14orf39* | -3.29546252 | 0.000844364 | 0.002536213 | 1 |
| HTN | b_cell | *KLHDC7A* | -3.822352159 | 0.028395733 | 0.036124929 | 1 |
| HTN | b_cell | *TRIML2* | 12.82215783 | 0.000288582 | 0.002180344 | 1 |
| HTN | b_cell | *SPTY2D1* | -6.797869409 | 0.016286692 | 0.022015574 | 1 |
| HTN | b_cell | *TCAIM* | -3.702225887 | 0.001922302 | 0.004091237 | 1 |
| HTN | b_cell | *FUCA1* | -4.318498584 | 0.001286667 | 0.003170756 | 1 |
| HTN | b_cell | *ZNF664* | -6.571871223 | 0.00130982 | 0.003205463 | 1 |

| | | | | | | |
|---|---|---|---|---|---|---|
| HTN | b_cell | *CALR* | -5.175292336 | 0.005136313 | 0.008437941 | 1 |
| HTN | b_cell | *NSUN7* | -3.032272984 | 0.020220094 | 0.026611624 | 1 |
| HTN | b_cell | *ARID3B* | -7.280043623 | 0.007595076 | 0.01150317 | 1 |
| HTN | b_cell | *EGR3* | -10.28866205 | 0.001629328 | 0.003675488 | 1 |
| HTN | b_cell | *LINC00174* | -4.495930594 | 0.00191007 | 0.004076242 | 1 |
| HTN | b_cell | *GEMIN4* | 7.329482689 | 0.000498382 | 0.002194324 | 1 |
| HTN | b_cell | *FJX1* | -7.956941253 | 0.035635676 | 0.044285728 | 1 |
| HTN | b_cell | *CIITA* | 4.924376594 | 0.000267726 | 0.002180344 | 1 |
| HTN | b_cell | *PLD6* | -7.875969144 | 0.001569201 | 0.003577072 | 1 |
| HTN | b_cell | *ZBTB42* | -15.91152676 | 0.001532747 | 0.003528516 | 1 |
| HTN | b_cell | *LACC1* | -8.361324968 | 0.005014133 | 0.00827615 | 1 |
| HTN | b_cell | *MAF1* | -5.903871737 | 0.000658209 | 0.002327958 | 1 |
| HTN | b_cell | *FCER1A* | 7.59039844 | 0.012865189 | 0.017978094 | 1 |
| HTN | b_cell | *PCED1B* | -3.110008634 | 0.004111703 | 0.007105628 | 1 |
| HTN | b_cell | *FOXS1* | -13.69990823 | 0.001085343 | 0.002865833 | 1 |
| HTN | b_cell | *ATOH7* | -45.06490471 | 0.030383124 | 0.038380583 | 1 |
| HTN | b_cell | *CDH5* | -3.280630176 | 0.000265147 | 0.002180344 | 1 |
| HTN | b_cell | *GIPC3* | 4.457267146 | 0.000795425 | 0.002467112 | 1 |
| HTN | b_cell | *CITED4* | 8.418859384 | 0.001816247 | 0.003937497 | 1 |
| HTN | b_cell | *TIGD5* | -6.366386058 | 0.002050235 | 0.004258924 | 1 |
| HTN | b_cell | *PDXDC1* | -4.776350869 | 0.003618277 | 0.006424204 | 1 |
| HTN | b_cell | *ZNF154* | -4.058971355 | 0.001622693 | 0.003665783 | 1 |
| HTN | b_cell | *R3HDM2* | -3.923955675 | 0.002757732 | 0.005274677 | 1 |
| HTN | b_cell | *SSC5D* | -2.688018601 | 0.009541927 | 0.013881653 | 1 |
| HTN | b_cell | *ZADH2* | -5.808651842 | 0.009451765 | 0.013777245 | 1 |
| HTN | b_cell | *TMEM86B* | 6.894168117 | 0.028958857 | 0.036773245 | 1 |
| HTN | b_cell | *SEPT1* | -4.702180623 | 0.000367612 | 0.002180344 | 1 |
| HTN | b_cell | *EXOC3* | -5.063927998 | 0.007721607 | 0.011641727 | 1 |
| HTN | b_cell | *LYNX1* | -6.674022765 | 0.00093729 | 0.002662292 | 1 |
| HTN | b_cell | *TDRP* | -3.929228787 | 0.000391526 | 0.002180344 | 1 |
| HTN | b_cell | *ZNF816* | 6.404347261 | 0.000141012 | 0.002180344 | 0.886541686 |
| HTN | b_cell | *FGD6* | -4.272449093 | 0.001046836 | 0.002816937 | 1 |
| HTN | b_cell | *HCLS1* | -4.180198637 | 0.000843057 | 0.002536213 | 1 |
| HTN | b_cell | *ZNF609* | -4.059371442 | 0.005239967 | 0.008583552 | 1 |
| HTN | b_cell | *CCDC66* | -3.993220353 | 0.00862387 | 0.01276915 | 1 |
| HTN | b_cell | *DEFB124* | -29.69961135 | 4.66E-07 | 6.94E-05 | 0.002932408 |
| HTN | b_cell | *MCFD2* | -3.582705257 | 0.00080021 | 0.002474709 | 1 |
| HTN | b_cell | *CYP8B1* | -5.091352946 | 0.001212444 | 0.003058843 | 1 |
| HTN | b_cell | *GLIPR1L2* | -2.920191822 | 0.010702541 | 0.015365513 | 1 |
| HTN | b_cell | *RNF182* | -5.475995485 | 0.010938784 | 0.015626479 | 1 |
| HTN | b_cell | *MB21D2* | -4.617493012 | 0.003471067 | 0.006227911 | 1 |
| HTN | b_cell | *SSTR2* | 5.498106886 | 0.000259414 | 0.002180344 | 1 |
| HTN | b_cell | *ZNF594* | -4.031425847 | 0.000470836 | 0.002180489 | 1 |
| HTN | b_cell | *PCGF5* | -4.838698592 | 0.018628139 | 0.024770539 | 1 |
| HTN | b_cell | *SHISA2* | -119.2753822 | 0.000410284 | 0.002180344 | 1 |
| HTN | b_cell | *CHST13* | -26.5968318 | 2.66E-05 | 0.001226302 | 0.167434538 |
| HTN | b_cell | *SRSF8* | -6.922686265 | 0.000762123 | 0.00242729 | 1 |
| HTN | b_cell | *SLC36A4* | -5.551969813 | 0.029393504 | 0.037249942 | 1 |
| HTN | b_cell | *ZDHHC20* | -4.861516837 | 0.013999558 | 0.01931853 | 1 |
| HTN | b_cell | *OR51E1* | -5.628848347 | 0.006561323 | 0.010222755 | 1 |

| | | | | | | |
|---|---|---|---|---|---|---|
| HTN | b_cell | *PSMG4* | -3.112110871 | 0.00035808 | 0.002180344 | 1 |
| HTN | b_cell | *ZNF443* | 18.24561981 | 3.12E-07 | 6.12E-05 | 0.001959964 |
| HTN | b_cell | *CAPS2* | -4.63122786 | 0.001900025 | 0.004061698 | 1 |
| HTN | b_cell | *ZNF792* | -8.380541372 | 0.000510592 | 0.002195202 | 1 |
| HTN | b_cell | *KCTD2* | -5.583434059 | 0.00090124 | 0.00260492 | 1 |
| HTN | b_cell | *D2HGDH* | -3.573846051 | 0.002415467 | 0.00478753 | 1 |
| HTN | b_cell | *CMTR2* | -6.110016224 | 0.003817382 | 0.0067095 | 1 |
| HTN | b_cell | *LRRC57* | -7.626430277 | 0.004518667 | 0.00762654 | 1 |
| HTN | b_cell | *MRPL14* | -4.146636147 | 0.001530564 | 0.003526075 | 1 |
| HTN | b_cell | *BBS12* | -6.139094673 | 0.002333512 | 0.004672226 | 1 |
| HTN | b_cell | *ZFP82* | -7.167191923 | 0.002882517 | 0.005438891 | 1 |
| HTN | b_cell | *LSMEM1* | -8.255712004 | 0.001957517 | 0.004140486 | 1 |
| HTN | b_cell | *AEN* | -8.139101882 | 0.000168937 | 0.002180344 | 1 |
| HTN | b_cell | *FKRP* | -3.219008668 | 0.002606652 | 0.005065172 | 1 |
| HTN | b_cell | *RPH3AL* | -3.41872816 | 0.000390698 | 0.002180344 | 1 |
| HTN | b_cell | *ANKRD34A* | -11.19867621 | 2.82E-05 | 0.001261541 | 0.177368085 |
| HTN | b_cell | *SLC26A11* | -5.00486155 | 0.00061561 | 0.002286342 | 1 |
| HTN | b_cell | *HIGD1A* | -10.1059423 | 8.04E-05 | 0.002176137 | 0.50533313 |
| HTN | b_cell | *OR52N4* | -9.750581223 | 0.009137543 | 0.013416099 | 1 |
| HTN | b_cell | *ADIPOQ* | -1.803758761 | 0.001543947 | 0.00354651 | 1 |
| HTN | b_cell | *F2R* | -3.229185995 | 0.008213468 | 0.012233611 | 1 |
| HTN | b_cell | *POLR2A* | -4.557754132 | 0.003147542 | 0.005796309 | 1 |
| HTN | b_cell | *TMEM132C* | -1.967299774 | 0.000757937 | 0.002419498 | 1 |
| HTN | b_cell | *SLC25A41* | -27.04524457 | 0.000204879 | 0.002180344 | 1 |
| HTN | b_cell | *TMEM136* | -3.591438142 | 0.014533296 | 0.019949963 | 1 |
| HTN | b_cell | *TMEM132E* | -26.17674313 | 0.014836715 | 0.020313247 | 1 |
| HTN | b_cell | *ZNF322* | -3.887773756 | 0.00183377 | 0.003959104 | 1 |
| HTN | b_cell | *CCL13* | -7.248606994 | 0.000269496 | 0.002180344 | 1 |
| HTN | b_cell | *DDX60L* | -5.237668482 | 0.009273789 | 0.013571766 | 1 |
| HTN | b_cell | *OGFOD3* | 5.884306987 | 0.001138294 | 0.002949898 | 1 |
| HTN | b_cell | *ACBD4* | -5.671392153 | 0.005456006 | 0.008860209 | 1 |
| HTN | b_cell | *SGSH* | -9.173489944 | 0.003329619 | 0.006039618 | 1 |
| HTN | b_cell | *SETD2* | -3.979852531 | 0.002974039 | 0.005561505 | 1 |
| HTN | b_cell | *MRPS23* | -4.950010565 | 0.000632049 | 0.002300923 | 1 |
| HTN | b_cell | *GPR135* | -3.075231743 | 0.001091242 | 0.00287056 | 1 |
| HTN | b_cell | *ATG9B* | -10.32665919 | 0.000518786 | 0.002195202 | 1 |
| HTN | b_cell | *HKR1* | -6.081534621 | 0.007969828 | 0.011944293 | 1 |
| HTN | b_cell | *ZBTB20* | -3.007696089 | 0.00504401 | 0.00831455 | 1 |
| HTN | b_cell | *C3orf58* | -5.856139978 | 0.000523221 | 0.002195202 | 1 |
| HTN | b_cell | *ODF3L2* | -10.00227599 | 0.032303997 | 0.040570361 | 1 |
| HTN | b_cell | *RELL1* | -3.98593915 | 0.001124664 | 0.002926641 | 1 |
| HTN | b_cell | *CLDN7* | -9.758451715 | 4.41E-09 | 3.47E-06 | 2.77E-05 |
| HTN | b_cell | *ADO* | -5.918961113 | 0.001213069 | 0.003059192 | 1 |
| HTN | b_cell | *SNRPE* | 8.780400144 | 0.026604279 | 0.034072337 | 1 |
| HTN | b_cell | *CHST15* | -5.676193241 | 0.006271785 | 0.009882429 | 1 |
| HTN | b_cell | *WRB* | -3.517398598 | 0.030011805 | 0.037956994 | 1 |
| HTN | b_cell | *TNRC18* | -4.082615185 | 0.002208215 | 0.004476959 | 1 |
| HTN | b_cell | *FAM181B* | 3.310095541 | 0.000272733 | 0.002180344 | 1 |
| HTN | b_cell | *NOP10* | -5.196475682 | 0.00026188 | 0.002180344 | 1 |
| HTN | b_cell | *FAM89A* | -2.306841962 | 0.008962506 | 0.013192993 | 1 |

| | | | | | | |
|---|---|---|---|---|---|---|
| HTN | b_cell | *ZNF708* | -5.159146515 | 0.0013252 | 0.003224925 | 1 |
| HTN | b_cell | *IST1* | -4.777731382 | 0.002516271 | 0.004932896 | 1 |
| HTN | b_cell | *ERCC6L2* | -3.898943416 | 0.007676283 | 0.011601152 | 1 |
| HTN | b_cell | *MRPL41* | -5.638514494 | 0.000366279 | 0.002180344 | 1 |
| HTN | b_cell | *HHIPL1* | -11.65532368 | 0.011032919 | 0.015743069 | 1 |
| HTN | b_cell | *FAM153B* | -10.66616953 | 0.003164003 | 0.005814813 | 1 |
| HTN | b_cell | *BACE2* | -3.604937202 | 0.001351684 | 0.003270974 | 1 |
| HTN | b_cell | *SYNM* | -3.927566063 | 0.000416212 | 0.002180344 | 1 |
| HTN | b_cell | *IZUMO1* | 4.025140092 | 0.002625746 | 0.005082532 | 1 |
| HTN | b_cell | *C8orf33* | -4.402315608 | 0.001858349 | 0.003998439 | 1 |
| HTN | b_cell | *FBXL6* | -11.51524366 | 0.006245001 | 0.009847585 | 1 |
| HTN | b_cell | *KBTBD3* | 7.562031827 | 0.000783503 | 0.002448253 | 1 |
| HTN | b_cell | *CLN8* | 11.05253125 | 6.50E-06 | 0.000472149 | 0.040854522 |
| HTN | b_cell | *CACNB4* | -2.631344637 | 0.000477394 | 0.002180489 | 1 |
| HTN | b_cell | *TRAPPC6B* | -5.004670043 | 0.002613202 | 0.005072525 | 1 |
| HTN | b_cell | *NPLOC4* | -3.256994519 | 0.000693649 | 0.002357691 | 1 |
| HTN | b_cell | *TSHZ2* | -2.132350182 | 0.000670426 | 0.002333871 | 1 |
| HTN | b_cell | *CAPN12* | 6.097909501 | 0.000155409 | 0.002180344 | 0.977057692 |
| HTN | b_cell | *FES* | -10.33911322 | 0.000432297 | 0.002180344 | 1 |
| HTN | b_cell | *MXRA7* | -6.068452128 | 8.52E-05 | 0.002180344 | 0.535921744 |
| HTN | b_cell | *MFSD5* | -4.689374241 | 0.00287359 | 0.005428564 | 1 |
| HTN | b_cell | *ADI1* | -5.399340483 | 0.000717535 | 0.002385315 | 1 |
| HTN | b_cell | *SPNS3* | 2.245233655 | 0.000190443 | 0.002180344 | 1 |
| HTN | b_cell | *CLEC4G* | -13.92074367 | 0.000330386 | 0.002180344 | 1 |
| HTN | b_cell | *SATB1* | -6.138364371 | 0.000637118 | 0.002308682 | 1 |
| HTN | b_cell | *ACTL10* | -12.03910447 | 0.021385081 | 0.027992505 | 1 |
| HTN | b_cell | *TRAK1* | -3.816126219 | 0.029020641 | 0.03683682 | 1 |
| HTN | b_cell | *SKA2* | -6.093326069 | 0.004361295 | 0.007416679 | 1 |
| HTN | b_cell | *KCNB2* | -2.623015557 | 0.001253766 | 0.003124228 | 1 |
| HTN | b_cell | *BRICD5* | -6.919190694 | 0.009717965 | 0.014080859 | 1 |
| HTN | b_cell | *TSKU* | -3.438162762 | 0.000367146 | 0.002180344 | 1 |
| HTN | b_cell | *GRID1* | -4.100407256 | 0.000141159 | 0.002180344 | 0.887467746 |
| HTN | b_cell | *ACBD3* | -4.347250833 | 0.001474377 | 0.003448441 | 1 |
| HTN | b_cell | *PLCXD3* | -3.372231827 | 0.003072173 | 0.005697566 | 1 |
| HTN | b_cell | *GPIHBP1* | -5.288764142 | 5.89E-05 | 0.001928827 | 0.370334795 |
| HTN | b_cell | *VMO1* | -4.205830128 | 0.015400607 | 0.020952958 | 1 |
| HTN | b_cell | *SLC25A18* | -3.30512669 | 0.007607166 | 0.011513301 | 1 |
| HTN | b_cell | *ZNF721* | -3.537849906 | 0.010436913 | 0.015039392 | 1 |
| HTN | b_cell | *C11orf54* | -3.629280945 | 0.004066955 | 0.007044509 | 1 |
| HTN | b_cell | *CEP63* | -3.898177687 | 0.000289285 | 0.002180344 | 1 |
| HTN | b_cell | *HMGN4* | -8.697672798 | 0.001005539 | 0.002758778 | 1 |
| HTN | b_cell | *C12orf60* | -7.370205081 | 0.005950929 | 0.009483775 | 1 |
| HTN | b_cell | *LYSMD4* | -4.135181747 | 0.000178392 | 0.002180344 | 1 |
| HTN | b_cell | *WBP2NL* | -6.61007374 | 0.009614275 | 0.013959572 | 1 |
| HTN | b_cell | *GAS6* | -6.571112468 | 0.000528851 | 0.002195202 | 1 |
| HTN | b_cell | *BEGAIN* | -13.46867379 | 0.000407604 | 0.002180344 | 1 |
| HTN | b_cell | *ARHGEF37* | -5.032113745 | 0.000932325 | 0.002657043 | 1 |
| HTN | b_cell | *FAM43B* | -35.02651109 | 0.000750006 | 0.002411911 | 1 |
| HTN | b_cell | *CSMD1* | -2.620527942 | 0.000700029 | 0.002366173 | 1 |
| HTN | b_cell | *PTGDR2* | -99.17320081 | 0.024624419 | 0.031763176 | 1 |

| | | | | | | |
|---|---|---|---|---|---|---|
| HTN | b_cell | *GPR19* | -13.92884373 | 0.000222202 | 0.002180344 | 1 |
| HTN | b_cell | *FANCF* | 6.802473642 | 0.000281975 | 0.002180344 | 1 |
| HTN | b_cell | *C2CD4C* | -10.86911637 | 0.002464669 | 0.004856384 | 1 |
| HTN | b_cell | *RUVBL2* | -4.787090741 | 0.000776558 | 0.002438676 | 1 |
| HTN | b_cell | *GDPGP1* | 4.520352131 | 0.000628503 | 0.002295161 | 1 |
| HTN | b_cell | *CTNNA3* | -3.615522593 | 0.000745193 | 0.002408933 | 1 |
| HTN | b_cell | *PTTG1IP* | -4.517580215 | 0.02251853 | 0.029323529 | 1 |
| HTN | b_cell | *CCBE1* | -2.426193383 | 0.003152584 | 0.005802195 | 1 |
| HTN | b_cell | *ZNF623* | -6.204793517 | 0.001615531 | 0.003652902 | 1 |
| HTN | b_cell | *CCDC125* | -3.054025922 | 0.000319583 | 0.002180344 | 1 |
| HTN | b_cell | *FHL3* | -9.417941809 | 8.68E-05 | 0.002180344 | 0.545515422 |
| HTN | b_cell | *C19orf71* | -8.441923589 | 0.000895934 | 0.002599737 | 1 |
| HTN | b_cell | *LRIT3* | -13.90156418 | 6.08E-05 | 0.001965056 | 0.382421958 |
| HTN | b_cell | *NPIPA1* | 3.600716248 | 0.025723172 | 0.033024623 | 1 |
| HTN | b_cell | *SF3A3* | -5.820504929 | 0.001319365 | 0.003218935 | 1 |
| HTN | b_cell | *GRIN2A* | -5.637175273 | 0.000153096 | 0.002180344 | 0.962515562 |
| HTN | b_cell | *URAD* | -19.05203824 | 0.003989337 | 0.006938025 | 1 |
| HTN | b_cell | *MX2* | -6.63739121 | 0.012720022 | 0.017822632 | 1 |
| HTN | b_cell | *PSMG1* | -2.900000558 | 0.011325473 | 0.016123925 | 1 |
| HTN | b_cell | *KLRC4* | -12.51191136 | 0.000168777 | 0.002180344 | 1 |
| HTN | b_cell | *ACSM5* | -2.755808873 | 0.003644739 | 0.006463885 | 1 |
| HTN | b_cell | *AC131971.1* | -25.61797922 | 8.98E-05 | 0.002180344 | 0.564768216 |
| HTN | b_cell | *SERHL2* | 6.536951717 | 0.000717354 | 0.002385315 | 1 |
| HTN | b_cell | *PCBP3* | -1.328161763 | 0.007910176 | 0.011874708 | 1 |
| HTN | b_cell | *TANGO2* | -5.402343424 | 0.01465086 | 0.020089415 | 1 |
| HTN | b_cell | *SFXN4* | -5.515594727 | 0.000743385 | 0.002408333 | 1 |
| HTN | b_cell | *FAM167B* | 9.060955771 | 0.00039279 | 0.002180344 | 1 |
| HTN | b_cell | *MRPL54* | 6.20304218 | 0.006598372 | 0.010264731 | 1 |
| HTN | b_cell | *NDUFB1* | -5.329709429 | 0.006694091 | 0.010391544 | 1 |
| HTN | b_cell | *FAM19A1* | -3.081351816 | 0.000420277 | 0.002180344 | 1 |
| HTN | b_cell | *TRMT12* | 5.948686361 | 0.000276831 | 0.002180344 | 1 |
| HTN | b_cell | *CMTM4* | -6.520627169 | 0.001047129 | 0.002816937 | 1 |
| HTN | b_cell | *CBX6* | -7.154733733 | 0.034421053 | 0.0429205 | 1 |
| HTN | b_cell | *KREMEN1* | -4.021077531 | 6.54E-07 | 9.13E-05 | 0.004110645 |
| HTN | b_cell | *KCTD16* | -4.234412643 | 0.000477804 | 0.002180489 | 1 |
| HTN | b_cell | *B3GALT5* | 3.63933286 | 0.000381638 | 0.002180344 | 1 |
| HTN | b_cell | *ZNF703* | -8.053743352 | 0.000290178 | 0.002180344 | 1 |
| HTN | b_cell | *KCTD8* | -2.846509867 | 0.000440148 | 0.002180344 | 1 |
| HTN | b_cell | *C9orf66* | -13.47160441 | 0.000735872 | 0.00240209 | 1 |
| HTN | b_cell | *TUBA8* | -4.870928852 | 0.000475139 | 0.002180489 | 1 |
| HTN | b_cell | *FAM162B* | -5.515163342 | 0.000897536 | 0.002599737 | 1 |
| HTN | b_cell | *BTBD9* | -4.132700751 | 0.009006075 | 0.013244724 | 1 |
| HTN | b_cell | *NUDT14* | -5.735829483 | 0.027129296 | 0.03466705 | 1 |
| HTN | b_cell | *ANKRD45* | 26.78268296 | 0.005626059 | 0.009071821 | 1 |
| HTN | b_cell | *MAATS1* | -5.617233229 | 0.002125471 | 0.004361239 | 1 |
| HTN | b_cell | *GPR39* | -7.331842824 | 0.000261972 | 0.002180344 | 1 |
| HTN | b_cell | *FAM3B* | -2.662283052 | 0.012077198 | 0.017051279 | 1 |
| HTN | b_cell | *TTC32* | -4.003534379 | 0.001752149 | 0.003844942 | 1 |
| HTN | b_cell | *PP2D1* | -4.948609639 | 0.00203579 | 0.004239268 | 1 |
| HTN | b_cell | *DENND5A* | -5.057521159 | 0.004180821 | 0.007187537 | 1 |

| | | | | | | |
|---|---|---|---|---|---|---|
| HTN | b_cell | *TMEM236* | -6.484710474 | 0.000453221 | 0.002180344 | 1 |
| HTN | b_cell | *TBX1* | -5.814760509 | 0.000471658 | 0.002180489 | 1 |
| HTN | b_cell | *EIF3C* | 6.885588678 | 0.006781678 | 0.010504167 | 1 |
| HTN | b_cell | *CNTN2* | -8.744006007 | 0.000494383 | 0.002192417 | 1 |
| HTN | b_cell | *UBE2F* | -8.550765338 | 0.000259865 | 0.002180344 | 1 |
| HTN | b_cell | *GOLGA6L4* | 11.49579798 | 0.015286322 | 0.020819957 | 1 |
| HTN | b_cell | *PCDH9* | -1.631639308 | 0.000894715 | 0.002599737 | 1 |
| HTN | b_cell | *ACOT1* | -4.164271463 | 0.013178181 | 0.018350216 | 1 |
| HTN | b_cell | *OAF* | -2.738741836 | 0.001382253 | 0.003309301 | 1 |
| HTN | b_cell | *TM2D3* | -6.55831804 | 0.010743934 | 0.015414677 | 1 |
| HTN | b_cell | *TACSTD2* | -6.558756754 | 0.015189417 | 0.020705955 | 1 |
| HTN | b_cell | *CLECL1* | 3.122547585 | 0.000473362 | 0.002180489 | 1 |
| HTN | b_cell | *MROH7* | -12.51775254 | 0.005690177 | 0.009158767 | 1 |
| HTN | b_cell | *CSF1* | -3.655229262 | 0.000348291 | 0.002180344 | 1 |
| HTN | b_cell | *PLA2G6* | -3.353140847 | 0.000176602 | 0.002180344 | 1 |
| HTN | b_cell | *A3GALT2* | -12.62581031 | 1.73E-07 | 4.18E-05 | 0.001087934 |
| HTN | b_cell | *SS18L1* | -6.525757552 | 0.00349372 | 0.006256058 | 1 |
| HTN | b_cell | *TOP1MT* | -7.167952401 | 0.002607109 | 0.005065172 | 1 |
| HTN | b_cell | *THAP7* | -7.580089198 | 0.000881956 | 0.002588636 | 1 |
| HTN | b_cell | *CCR10* | -7.192246259 | 0.00337297 | 0.00609539 | 1 |
| HTN | b_cell | *WDR27* | -4.737455887 | 0.007825531 | 0.011767346 | 1 |
| HTN | b_cell | *TXNRD2* | -5.517487167 | 0.002409154 | 0.004779536 | 1 |
| HTN | b_cell | *PROS1* | -3.626269476 | 0.000712978 | 0.002380505 | 1 |
| HTN | b_cell | *HDDC3* | -5.31169069 | 0.000265987 | 0.002180344 | 1 |
| HTN | b_cell | *ZFP1* | -5.05209683 | 0.000399803 | 0.002180344 | 1 |
| HTN | b_cell | *SNN* | -11.35101245 | 0.011913979 | 0.016854902 | 1 |
| HTN | b_cell | *NELL2* | 3.753745139 | 0.017742989 | 0.023713897 | 1 |
| HTN | b_cell | *KRBA2* | -8.136251906 | 0.004234331 | 0.007253743 | 1 |
| HTN | b_cell | *ZNF93* | -5.499491098 | 0.000529805 | 0.002195202 | 1 |
| HTN | b_cell | *SEPT5* | -16.12951388 | 0.000419618 | 0.002180344 | 1 |
| HTN | b_cell | *ATL3* | -4.622179927 | 0.000398604 | 0.002180344 | 1 |
| HTN | b_cell | *NDUFA12* | -5.981883319 | 0.001663479 | 0.003726974 | 1 |
| HTN | b_cell | *UBE2G2* | -3.941214427 | 0.002465387 | 0.004856384 | 1 |
| HTN | b_cell | *OSBP2* | -4.4332697 | 0.001186498 | 0.003022492 | 1 |
| HTN | b_cell | *TUSC5* | -2.520102241 | 0.00389871 | 0.006814343 | 1 |
| HTN | b_cell | *TMED9* | -5.42950025 | 0.000590656 | 0.00226693 | 1 |
| HTN | b_cell | *BTBD6* | -3.958421919 | 0.002140426 | 0.00437568 | 1 |
| HTN | b_cell | *H1FX* | -6.65622039 | 0.000886913 | 0.002596195 | 1 |
| HTN | b_cell | *RBM43* | -6.790263479 | 0.000451664 | 0.002180344 | 1 |
| HTN | b_cell | *NUTM2A* | -23.29047587 | 2.87E-05 | 0.001261541 | 0.180400405 |
| HTN | b_cell | *ZFP90* | -5.51893859 | 0.035938788 | 0.044618317 | 1 |
| HTN | b_cell | *FAM227A* | -4.638560476 | 0.000933268 | 0.002657043 | 1 |
| HTN | b_cell | *NOC4L* | -5.085693825 | 0.003446921 | 0.00619874 | 1 |
| HTN | b_cell | *SORCS2* | -3.788347057 | 0.001440484 | 0.003400797 | 1 |
| HTN | b_cell | *TMEM106A* | -6.69537653 | 0.011352385 | 0.016158579 | 1 |
| HTN | b_cell | *SIVA1* | -5.750794503 | 0.001045297 | 0.002814467 | 1 |
| HTN | b_cell | *UBOX5* | -3.708408359 | 0.000763171 | 0.002427608 | 1 |
| HTN | b_cell | *MAFF* | -4.846766484 | 0.001395422 | 0.003330682 | 1 |
| HTN | b_cell | *BRF1* | -5.815016258 | 0.004489527 | 0.007583465 | 1 |
| HTN | b_cell | *FLRT2* | -2.100948961 | 0.000396665 | 0.002180344 | 1 |

| | | | | | | |
|---|---|---|---|---|---|---|
| HTN | b_cell | *ADSSL1* | -7.409200786 | 0.000496906 | 0.002194324 | 1 |
| HTN | b_cell | *C6orf120* | -9.089930143 | 0.003046441 | 0.005654849 | 1 |
| HTN | b_cell | *DDX51* | -2.374075379 | 0.032437371 | 0.040721382 | 1 |
| HTN | b_cell | *SIGIRR* | -6.037605098 | 0.018671678 | 0.024823185 | 1 |
| HTN | b_cell | *GP1BA* | -5.524080974 | 0.024542328 | 0.031663784 | 1 |
| HTN | b_cell | *KIAA0825* | -3.999980083 | 0.002357536 | 0.004701051 | 1 |
| HTN | b_cell | *CDK10* | -11.20993602 | 5.17E-06 | 0.000398561 | 0.032491372 |
| HTN | b_cell | *TMEM105* | -83.05955004 | 0.002509425 | 0.004923903 | 1 |
| HTN | b_cell | *TCN2* | -8.818987588 | 0.000114218 | 0.002180344 | 0.718087225 |
| HTN | b_cell | *GAS2L1* | -4.822691974 | 0.000185575 | 0.002180344 | 1 |
| HTN | b_cell | *ATP6V0A2* | -4.786360845 | 0.001111859 | 0.002911395 | 1 |
| HTN | b_cell | *HS6ST3* | 3.538076239 | 0.000299093 | 0.002180344 | 1 |
| HTN | b_cell | *TNFAIP8L1* | -4.82666357 | 0.001252715 | 0.003123021 | 1 |
| HTN | b_cell | *MAPK11* | -6.048081549 | 0.00040773 | 0.002180344 | 1 |
| HTN | b_cell | *SH3BGR* | -3.087120052 | 0.000561963 | 0.002226547 | 1 |
| HTN | b_cell | *FAM174B* | -6.052348727 | 0.000862896 | 0.002559431 | 1 |
| HTN | b_cell | *MUC1* | 8.304510761 | 0.009688555 | 0.014051199 | 1 |
| HTN | b_cell | *SV2B* | -6.070468895 | 0.00201452 | 0.004214738 | 1 |
| HTN | b_cell | *PDE6G* | 4.129087809 | 0.000158007 | 0.002180344 | 0.993390763 |
| HTN | b_cell | *TLCD2* | -6.119934823 | 0.00019413 | 0.002180344 | 1 |
| HTN | b_cell | *LSAMP* | -2.294857063 | 0.001716904 | 0.003800767 | 1 |
| HTN | b_cell | *AHNAK2* | -3.735961657 | 0.000446232 | 0.002180344 | 1 |
| HTN | b_cell | *PDIA2* | -9.124489137 | 0.003768561 | 0.006634821 | 1 |
| HTN | b_cell | *PCGF3* | -4.626322253 | 0.007159721 | 0.010946781 | 1 |
| HTN | b_cell | *PBX1* | -2.205524691 | 0.000539314 | 0.002208907 | 1 |
| HTN | b_cell | *PMEL* | 4.523817452 | 0.002081087 | 0.004303303 | 1 |
| HTN | b_cell | *SNAI3* | -4.930701129 | 0.000546619 | 0.002214137 | 1 |
| HTN | b_cell | *C6orf201* | -5.415745743 | 0.015567295 | 0.021138571 | 1 |
| HTN | b_cell | *MYBL1* | -4.556115049 | 3.06E-06 | 0.000278918 | 0.019245372 |
| HTN | b_cell | *C16orf52* | -3.970580241 | 0.001035128 | 0.002800612 | 1 |
| HTN | b_cell | *ADARB2* | -4.637719831 | 0.000345421 | 0.002180344 | 1 |
| HTN | b_cell | *SRL* | 6.653020208 | 0.000203675 | 0.002180344 | 1 |
| HTN | b_cell | *KCNIP4* | -2.230558176 | 0.00039138 | 0.002180344 | 1 |
| HTN | b_cell | *SLC52A2* | -4.796887536 | 0.000419802 | 0.002180344 | 1 |
| HTN | b_cell | *PIGP* | -2.758503047 | 0.011376479 | 0.016185545 | 1 |
| HTN | b_cell | *NPIPB4* | -4.776179905 | 0.000629006 | 0.002295161 | 1 |
| HTN | b_cell | *TRIM69* | 6.721919659 | 0.002466988 | 0.004856384 | 1 |
| HTN | b_cell | *LINC00839* | -8.648753204 | 0.040637057 | 0.049957993 | 1 |
| HTN | b_cell | *PTCH1* | -6.106210249 | 0.020789933 | 0.02730443 | 1 |
| HTN | b_cell | *RTN4RL1* | -3.565527739 | 0.000415971 | 0.002180344 | 1 |
| HTN | b_cell | *PLK5* | -9.348177396 | 0.018387734 | 0.024497496 | 1 |
| HTN | b_cell | *RASA3* | -4.845231793 | 0.000746303 | 0.002409022 | 1 |
| HTN | b_cell | *ZNF566* | -5.305473322 | 0.015896204 | 0.021552606 | 1 |
| HTN | b_cell | *ZNF284* | -8.900511079 | 0.000416971 | 0.002180344 | 1 |
| HTN | b_cell | *GSAP* | -3.738579143 | 0.001444692 | 0.003409452 | 1 |
| HTN | b_cell | *C2orf76* | -5.380486176 | 0.010536969 | 0.01515925 | 1 |
| HTN | b_cell | *WWOX* | -3.388591693 | 0.003699399 | 0.006547195 | 1 |
| HTN | b_cell | *CYP4Z1* | -4.363005288 | 0.000953778 | 0.00268897 | 1 |
| HTN | b_cell | *POLR1D* | -5.562646319 | 0.013958866 | 0.019279304 | 1 |
| HTN | b_cell | *FFAR4* | -2.412240307 | 0.01212705 | 0.017117819 | 1 |

| | | | | | | |
|---|---|---|---|---|---|---|
| HTN | b_cell | *BPIFB4* | -200.0972494 | 0.019124609 | 0.025339603 | 1 |
| HTN | b_cell | *CYP4F12* | -3.58407091 | 0.001484147 | 0.003462275 | 1 |
| HTN | b_cell | *ZNF749* | -5.615955905 | 0.007442777 | 0.011310791 | 1 |
| HTN | b_cell | *NBPF12* | -4.769540263 | 0.001068358 | 0.002845984 | 1 |
| HTN | b_cell | *GPAT2* | 7.625455097 | 0.000104411 | 0.002180344 | 0.65643101 |
| HTN | b_cell | *TOR3A* | 9.08225818 | 0.002584076 | 0.005032863 | 1 |
| HTN | b_cell | *CYP4X1* | -3.115826021 | 0.001458703 | 0.003427079 | 1 |
| HTN | b_cell | *CCDC30* | -5.154456452 | 0.003633698 | 0.006446123 | 1 |
| HTN | b_cell | *GLDN* | -3.000546388 | 0.000381226 | 0.002180344 | 1 |
| HTN | b_cell | *FCAR* | 4.477570742 | 0.000854418 | 0.002551796 | 1 |
| HTN | b_cell | *KPNA4* | -4.769585045 | 0.005086727 | 0.008369603 | 1 |
| HTN | b_cell | *ZNF501* | -4.92297461 | 0.000634979 | 0.002303584 | 1 |
| HTN | b_cell | *ZNF197* | -4.684677894 | 0.000902095 | 0.002605177 | 1 |
| HTN | b_cell | *PCLO* | -2.345245227 | 0.000444579 | 0.002180344 | 1 |
| HTN | b_cell | *RGS7BP* | -10.64376239 | 0.000612306 | 0.002285256 | 1 |
| HTN | b_cell | *ZNF396* | -8.442577561 | 3.14E-05 | 0.001332022 | 0.197139314 |
| HTN | b_cell | *TMEM222* | -4.376121432 | 0.00029388 | 0.002180344 | 1 |
| HTN | b_cell | *CLCNKA* | -15.28046795 | 0.004489056 | 0.007583465 | 1 |
| HTN | b_cell | *SEPT10* | -4.022365293 | 0.001073943 | 0.002852506 | 1 |
| HTN | b_cell | *SMYD4* | -4.892319179 | 0.000120074 | 0.002180344 | 0.754902928 |
| HTN | b_cell | *CEACAM19* | -4.645575483 | 0.000253333 | 0.002180344 | 1 |
| HTN | b_cell | *NF2* | -5.093288307 | 0.011518183 | 0.01636493 | 1 |
| HTN | b_cell | *MIR22HG* | -2.925828246 | 0.000257235 | 0.002180344 | 1 |
| HTN | b_cell | *PDE2A* | -3.497428302 | 0.001210271 | 0.00305766 | 1 |
| HTN | b_cell | *C17orf58* | -3.51136003 | 0.000335579 | 0.002180344 | 1 |
| HTN | b_cell | *CYP27C1* | -3.264609747 | 0.006739529 | 0.010453954 | 1 |
| HTN | b_cell | *BCR* | -4.109088845 | 0.000724946 | 0.002388914 | 1 |
| HTN | b_cell | *FOXI2* | -11.08658331 | 0.002828505 | 0.005375699 | 1 |
| HTN | b_cell | *HYAL3* | 2.048082739 | 0.013894689 | 0.019207544 | 1 |
| HTN | b_cell | *ZSCAN30* | -5.794425026 | 0.020074578 | 0.026458882 | 1 |
| HTN | b_cell | *LILRB4* | -3.662048246 | 0.002455324 | 0.004846663 | 1 |
| HTN | b_cell | *KRT16* | -9.432037765 | 0.000438648 | 0.002180344 | 1 |
| HTN | b_cell | *QRFPR* | -13.48916389 | 0.032057767 | 0.040285266 | 1 |
| HTN | b_cell | *TMEM17* | -6.21842547 | 0.00117692 | 0.003008146 | 1 |
| HTN | b_cell | *TNFRSF18* | -13.4453187 | 0.000447248 | 0.002180344 | 1 |
| HTN | b_cell | *RTN4RL2* | -14.43334686 | 0.000185094 | 0.002180344 | 1 |
| HTN | b_cell | *TMEM232* | -4.629716111 | 0.000503905 | 0.002195028 | 1 |
| HTN | b_cell | *EFCAB6* | -4.056241919 | 0.002752258 | 0.005265809 | 1 |
| HTN | b_cell | *KANK3* | -3.525169781 | 0.000463496 | 0.002180489 | 1 |
| HTN | b_cell | *EMID1* | -8.678318575 | 0.001428677 | 0.003381812 | 1 |
| HTN | b_cell | *CYP4A11* | -7.789892811 | 0.008013488 | 0.011992572 | 1 |
| HTN | b_cell | *TMEM216* | -4.736322818 | 0.001371186 | 0.003290323 | 1 |
| HTN | b_cell | *RPS19BP1* | -4.639433513 | 0.000204099 | 0.002180344 | 1 |
| HTN | b_cell | *C3orf70* | -4.629989587 | 0.031985236 | 0.040202154 | 1 |
| HTN | b_cell | *PLCD1* | -7.719332143 | 0.005373011 | 0.00874453 | 1 |
| HTN | b_cell | *ENTPD5* | -4.112713311 | 0.000712515 | 0.002380505 | 1 |
| HTN | b_cell | *MRPS21* | -5.684696909 | 0.001938111 | 0.004110966 | 1 |
| HTN | b_cell | *ANGPTL5* | -4.344345443 | 0.000433057 | 0.002180344 | 1 |
| HTN | b_cell | *TSPYL4* | -7.068397922 | 0.00147544 | 0.003449642 | 1 |
| HTN | b_cell | *MT1X* | -4.276541172 | 0.000441405 | 0.002180344 | 1 |

| | | | | | | |
|---|---|---|---|---|---|---|
| HTN | b_cell | *WDR86* | -3.429488096 | 0.002761095 | 0.005279503 | 1 |
| HTN | b_cell | *EPOR* | -5.030252038 | 0.000522945 | 0.002195202 | 1 |
| HTN | b_cell | *FPR3* | -2.87181652 | 0.000403675 | 0.002180344 | 1 |
| HTN | b_cell | *GJA4* | -7.117091771 | 0.000124632 | 0.002180344 | 0.78356057 |
| HTN | b_cell | *TLR5* | -3.844778871 | 0.000295768 | 0.002180344 | 1 |
| HTN | b_cell | *ZNF385C* | 9.067230269 | 0.000216003 | 0.002180344 | 1 |
| HTN | b_cell | *TET3* | -2.810696099 | 0.000435726 | 0.002180344 | 1 |
| HTN | b_cell | *ISG15* | -5.330241257 | 0.018417866 | 0.024523653 | 1 |
| HTN | b_cell | *DHRS4L2* | -7.029412928 | 0.001221448 | 0.003075388 | 1 |
| HTN | b_cell | *TRPV2* | -7.470960059 | 0.040305362 | 0.049605994 | 1 |
| HTN | b_cell | *C2orf88* | -3.281368353 | 0.000315078 | 0.002180344 | 1 |
| HTN | b_cell | *KBTBD12* | -6.120646051 | 0.005237455 | 0.008581673 | 1 |
| HTN | b_cell | *ADH1A* | -7.897948143 | 3.24E-05 | 0.001350755 | 0.20338806 |
| HTN | b_cell | *SEMA4D* | -4.443768849 | 0.000574477 | 0.002240531 | 1 |
| HTN | b_cell | *FAM69C* | -15.93954356 | 0.003246625 | 0.005926693 | 1 |
| HTN | b_cell | *DNAH17* | -4.814433866 | 0.000810118 | 0.002488644 | 1 |
| HTN | b_cell | *MCRS1* | 14.09265556 | 0.000597983 | 0.002274361 | 1 |
| HTN | b_cell | *ZFP69* | -6.109785828 | 0.000143456 | 0.002180344 | 0.901906124 |
| HTN | b_cell | *TMEM220* | -4.571821925 | 0.003345338 | 0.00605997 | 1 |
| HTN | b_cell | *CCDC157* | 2.822153392 | 0.000253123 | 0.002180344 | 1 |
| HTN | b_cell | *GFRAL* | 1.901577641 | 0.000245262 | 0.002180344 | 1 |
| HTN | b_cell | *LDLRAD2* | 6.721083831 | 0.029350565 | 0.037210527 | 1 |
| HTN | b_cell | *CPSF4L* | -13.0209592 | 0.001669395 | 0.00373449 | 1 |
| HTN | b_cell | *ZSCAN23* | -3.813081748 | 8.66E-05 | 0.002180344 | 0.544193414 |
| HTN | b_cell | *C6orf132* | -4.05080518 | 0.000802826 | 0.00248028 | 1 |
| HTN | b_cell | *MAPK12* | -10.10855453 | 0.000228738 | 0.002180344 | 1 |
| HTN | b_cell | *AGRN* | -8.399082204 | 0.000211094 | 0.002180344 | 1 |
| HTN | b_cell | *FAM166A* | -22.52092848 | 0.00041334 | 0.002180344 | 1 |
| HTN | b_cell | *ZNF626* | -4.691505803 | 0.000499465 | 0.002194994 | 1 |
| HTN | b_cell | *SMTNL2* | -32.42444773 | 0.000416361 | 0.002180344 | 1 |
| HTN | b_cell | *ZC3H6* | -4.053217002 | 0.010768574 | 0.015435938 | 1 |
| HTN | b_cell | *LINC00265* | -8.163909926 | 0.006886164 | 0.010618915 | 1 |
| HTN | b_cell | *PRKAR1B* | -3.576593473 | 0.000608905 | 0.002281385 | 1 |
| HTN | b_cell | *NUTM2B* | -97.77888959 | 0.021517957 | 0.028148854 | 1 |
| HTN | b_cell | *NCR3LG1* | -9.787174735 | 0.000432619 | 0.002180344 | 1 |
| HTN | b_cell | *ZNF793* | -6.775948006 | 0.000588633 | 0.002263931 | 1 |
| HTN | b_cell | *AGAP4* | -5.172720134 | 0.000292408 | 0.002180344 | 1 |
| HTN | b_cell | *HYKK* | -5.225075253 | 0.001540514 | 0.003541211 | 1 |
| HTN | b_cell | *RUFY4* | -11.59434404 | 0.000411252 | 0.002180344 | 1 |
| HTN | b_cell | *ZNF669* | -11.69123984 | 0.027392354 | 0.034974762 | 1 |
| HTN | b_cell | *CENPP* | -3.638704545 | 8.21E-05 | 0.002178401 | 0.516281014 |
| HTN | b_cell | *PLSCR1* | -5.190320678 | 0.011565658 | 0.016421249 | 1 |
| HTN | b_cell | *ZNF559* | -5.873078325 | 5.20E-06 | 0.000398561 | 0.03268204 |
| HTN | b_cell | *JAKMIP3* | -4.45198154 | 0.001005652 | 0.002758778 | 1 |
| HTN | b_cell | *PDCD1* | 16.16586525 | 0.000808717 | 0.002488487 | 1 |
| HTN | b_cell | *CERKL* | -9.989168744 | 0.005856057 | 0.009370585 | 1 |
| HTN | b_cell | *COL25A1* | -2.501709304 | 0.000527625 | 0.002195202 | 1 |
| HTN | b_cell | *NBR1* | -4.672536052 | 0.001354254 | 0.003272403 | 1 |
| HTN | b_cell | *NDOR1* | -8.51631923 | 0.007008961 | 0.010760766 | 1 |
| HTN | b_cell | *SUMO2* | -5.719773303 | 0.001407785 | 0.003347484 | 1 |

| | | | | | | |
|---|---|---|---|---|---|---|
| HTN | b_cell | *GOLGA8M* | -4.083479274 | 0.001228755 | 0.003088552 | 1 |
| HTN | b_cell | *DPYD* | -3.619876298 | 0.00786156 | 0.011810185 | 1 |
| HTN | b_cell | *S100A16* | -4.667086141 | 0.001328518 | 0.003228601 | 1 |
| HTN | b_cell | *RHCE* | -7.793770226 | 0.012286737 | 0.017308249 | 1 |
| HTN | b_cell | *PARVB* | -3.676034835 | 0.000800572 | 0.002474709 | 1 |
| HTN | b_cell | *SLC4A5* | 2.596312928 | 0.00020087 | 0.002180344 | 1 |
| HTN | b_cell | *UROS* | -5.191469503 | 0.002658517 | 0.005128354 | 1 |
| HTN | b_cell | *FAM221A* | -4.043399011 | 0.000321775 | 0.002180344 | 1 |
| HTN | b_cell | *FSIP2* | -2.374505718 | 0.002598606 | 0.005051773 | 1 |
| HTN | b_cell | *NOXA1* | -4.600230245 | 0.000477712 | 0.002180489 | 1 |
| HTN | b_cell | *SKOR1* | 2.279224061 | 0.000221782 | 0.002180344 | 1 |
| HTN | b_cell | *TMEM201* | -6.044373007 | 0.001562676 | 0.003569964 | 1 |
| HTN | b_cell | *ZDHHC11* | -4.114412402 | 0.013001776 | 0.018156856 | 1 |
| HTN | b_cell | *FAM26F* | 1.599292679 | 0.002547924 | 0.00498252 | 1 |
| HTN | b_cell | *BEND4* | 2.237190676 | 0.000241835 | 0.002180344 | 1 |
| HTN | b_cell | *ZNF563* | -5.750722304 | 0.00029463 | 0.002180344 | 1 |
| HTN | b_cell | *FBF1* | -5.129915965 | 2.64E-08 | 9.78E-06 | 0.000165765 |
| HTN | b_cell | *MSL1* | -6.327709904 | 0.000674841 | 0.002334899 | 1 |
| HTN | b_cell | *LRRK2* | -3.933728259 | 0.005448254 | 0.008853237 | 1 |
| HTN | b_cell | *FAM196A* | -2.98949555 | 0.000693771 | 0.002357691 | 1 |
| HTN | b_cell | *UTS2B* | -5.368808717 | 0.000285025 | 0.002180344 | 1 |
| HTN | b_cell | *C9orf152* | -12.31686 | 0.000412668 | 0.002180344 | 1 |
| HTN | b_cell | *SLC15A5* | -3.390279444 | 0.004371805 | 0.007430882 | 1 |
| HTN | b_cell | *LIPI* | -4.528480859 | 0.000454371 | 0.002180344 | 1 |
| HTN | b_cell | *ADAT2* | 6.388033837 | 0.027122495 | 0.034665405 | 1 |
| HTN | b_cell | *ZNF567* | -4.938662286 | 0.001173025 | 0.003004867 | 1 |
| HTN | b_cell | *ANKDD1B* | -4.228780324 | 0.007870059 | 0.01182013 | 1 |
| HTN | b_cell | *LITAF* | -3.779195652 | 0.000498401 | 0.002194324 | 1 |
| HTN | b_cell | *VSTM1* | -7.46123554 | 0.040462995 | 0.049773205 | 1 |
| HTN | b_cell | *SF3B3* | -5.490866131 | 0.005454556 | 0.008860209 | 1 |
| HTN | b_cell | *PLAC9* | -3.03760656 | 0.001176344 | 0.003008146 | 1 |
| HTN | b_cell | *ZNF527* | -6.710707029 | 0.000742502 | 0.002407482 | 1 |
| HTN | b_cell | *S100A13* | -3.252009408 | 0.000573105 | 0.002237955 | 1 |
| HTN | b_cell | *ZNF33A* | -4.133362704 | 0.015359712 | 0.020910894 | 1 |
| HTN | b_cell | *ZNF600* | -4.454849661 | 0.012774331 | 0.017874965 | 1 |
| HTN | b_cell | *PNRC2* | -6.51524872 | 0.00237962 | 0.004732892 | 1 |
| HTN | b_cell | *ZKSCAN3* | -4.666181019 | 0.004835825 | 0.008034575 | 1 |
| HTN | b_cell | *SLC35E2B* | -4.36467556 | 0.015055322 | 0.02055882 | 1 |
| HTN | b_cell | *BLOC1S2* | -5.015828008 | 0.003213103 | 0.005879156 | 1 |
| HTN | b_cell | *AKR1C3* | -3.096354625 | 0.000357305 | 0.002180344 | 1 |
| HTN | b_cell | *WDSUB1* | -4.760052888 | 0.002676606 | 0.005149272 | 1 |
| HTN | b_cell | *S100A4* | -2.801581337 | 0.009672798 | 0.014031583 | 1 |
| HTN | b_cell | *COLCA1* | -9.282738498 | 0.000560792 | 0.002226547 | 1 |
| HTN | b_cell | *ACADSB* | -4.125131887 | 0.001131309 | 0.002939114 | 1 |
| HTN | b_cell | *HRCT1* | -5.025676108 | 0.003018254 | 0.005618817 | 1 |
| HTN | b_cell | *MPHOSPH8* | -4.182364194 | 0.005207828 | 0.00854134 | 1 |
| HTN | b_cell | *SIRPB2* | -4.584003233 | 0.000316227 | 0.002180344 | 1 |
| HTN | b_cell | *SRGAP3* | -3.251149534 | 0.012063834 | 0.017039513 | 1 |
| HTN | b_cell | *XPNPEP3* | -3.389296365 | 0.001451742 | 0.003419073 | 1 |
| HTN | b_cell | *PPIA* | -4.862446229 | 0.001001302 | 0.002755316 | 1 |

| | | | | | | |
|---|---|---|---|---|---|---|
| HTN | b_cell | *ZNF471* | -5.15000913 | 0.000362622 | 0.002180344 | 1 |
| HTN | b_cell | *GRM7* | -2.727483595 | 0.000383661 | 0.002180344 | 1 |
| HTN | b_cell | *SUPT3H* | -4.456667011 | 0.010961955 | 0.015656023 | 1 |
| HTN | b_cell | *POM121* | -5.528094944 | 0.002357403 | 0.004701051 | 1 |
| HTN | b_cell | *GIMAP5* | -4.191279652 | 0.001669581 | 0.00373449 | 1 |
| HTN | b_cell | *STK31* | -5.827608948 | 0.000419503 | 0.002180344 | 1 |
| HTN | b_cell | *ZKSCAN7* | 8.052919316 | 0.000870838 | 0.002571612 | 1 |
| HTN | b_cell | *ZNF565* | -2.921776354 | 0.001075915 | 0.002854029 | 1 |
| HTN | b_cell | *WDR5* | -5.123480573 | 0.001661864 | 0.003724826 | 1 |
| HTN | b_cell | *LONP1* | -6.003034709 | 0.00135799 | 0.003274908 | 1 |
| HTN | b_cell | *SLC35F1* | -4.971624229 | 0.00059249 | 0.00226693 | 1 |
| HTN | b_cell | *ZNF34* | -6.82920355 | 0.003897953 | 0.006814343 | 1 |
| HTN | b_cell | *ZNF781* | -5.974729245 | 0.000280613 | 0.002180344 | 1 |
| HTN | b_cell | *INCA1* | -11.50862947 | 0.004715942 | 0.007881874 | 1 |
| HTN | b_cell | *ZNF774* | -11.87077775 | 0.001895585 | 0.004057726 | 1 |
| HTN | b_cell | *EVL* | -3.708917382 | 0.017406691 | 0.023328899 | 1 |
| HTN | b_cell | *ZNF124* | -5.228659982 | 0.002126726 | 0.004361619 | 1 |
| HTN | b_cell | *ZNF569* | -3.538979376 | 0.000480429 | 0.002183988 | 1 |
| HTN | b_cell | *PIK3R4* | -4.774401442 | 0.005785236 | 0.009271419 | 1 |
| HTN | b_cell | *ZNF775* | -7.323845222 | 0.005103473 | 0.008392965 | 1 |
| HTN | b_cell | *ZNF605* | -4.144721974 | 0.000548544 | 0.002215074 | 1 |
| HTN | b_cell | *RFX8* | -4.421616753 | 0.000345108 | 0.002180344 | 1 |
| HTN | b_cell | *MYL6B* | -4.519809227 | 0.000626864 | 0.002295161 | 1 |
| HTN | b_cell | *ZNF799* | -7.618264133 | 0.000653562 | 0.002324546 | 1 |
| HTN | b_cell | *C20orf96* | -4.771863194 | 0.00039434 | 0.002180344 | 1 |
| HTN | b_cell | *SULT1A1* | -4.354600383 | 5.62E-06 | 0.000420647 | 0.035334323 |
| HTN | b_cell | *ANAPC7* | -4.389641865 | 0.000956496 | 0.002694776 | 1 |
| HTN | b_cell | *TPK1* | -4.96510125 | 0.007935669 | 0.011907292 | 1 |
| HTN | b_cell | *C1orf186* | 0.59374995 | 0.000215132 | 0.002180344 | 1 |
| HTN | b_cell | *SPTSSB* | -15.74516475 | 0.000130984 | 0.002180344 | 0.823497906 |
| HTN | b_cell | *SULF2* | -4.260165919 | 0.000681895 | 0.002339238 | 1 |
| HTN | b_cell | *XRCC2* | -9.280421543 | 0.001039573 | 0.002805063 | 1 |
| HTN | b_cell | *MYO6* | -3.743882322 | 0.000200862 | 0.002180344 | 1 |
| HTN | b_cell | *HDAC2* | -4.894157171 | 0.000892917 | 0.002599737 | 1 |
| HTN | b_cell | *ADH1B* | -1.986729105 | 0.000405685 | 0.002180344 | 1 |
| HTN | b_cell | *ZNF502* | -3.841063324 | 0.002766779 | 0.005287155 | 1 |
| HTN | b_cell | *TECPR2* | -4.590401221 | 0.001461 | 0.003427611 | 1 |
| HTN | b_cell | *FAM180B* | -9.017694768 | 7.06E-09 | 4.19E-06 | 4.44E-05 |
| HTN | b_cell | *ZFP62* | -6.730864766 | 0.002461938 | 0.004856384 | 1 |
| HTN | b_cell | *ERI2* | -2.861609541 | 0.000395637 | 0.002180344 | 1 |
| HTN | b_cell | *TRPV1* | -5.562419303 | 0.002732656 | 0.005239465 | 1 |
| HTN | b_cell | *ZNF33B* | -4.861021937 | 0.006544265 | 0.010211912 | 1 |
| HTN | b_cell | *AMZ2* | -4.891809005 | 0.000210466 | 0.002180344 | 1 |
| HTN | b_cell | *ZNF431* | -3.907314395 | 0.005208763 | 0.00854134 | 1 |
| HTN | b_cell | *NF1* | -3.540703113 | 0.008030475 | 0.012012276 | 1 |
| HTN | b_cell | *DAPK1* | -3.023631329 | 0.000725551 | 0.002388914 | 1 |
| HTN | b_cell | *HLA-DQA1* | 2.298961705 | 0.003705771 | 0.006553638 | 1 |
| HTN | b_cell | *TLE1* | -2.591385821 | 0.000391586 | 0.002180344 | 1 |
| HTN | b_cell | *STRN3* | -3.890021729 | 0.005075852 | 0.008353896 | 1 |
| HTN | b_cell | *ZNF239* | -4.148815881 | 0.005573318 | 0.009009887 | 1 |

| | | | | | | |
|---|---|---|---|---|---|---|
| HTN | b_cell | *MVB12B* | -4.966410244 | 0.002880116 | 0.005435991 | 1 |
| HTN | b_cell | *PPTC7* | -5.528660026 | 0.012066154 | 0.017039513 | 1 |
| HTN | b_cell | *NHLRC2* | -4.172991085 | 0.003041459 | 0.005650639 | 1 |
| HTN | b_cell | *ZFP28* | -4.723659347 | 0.001114517 | 0.002915926 | 1 |
| HTN | b_cell | *LAMB3* | -5.606876762 | 0.000437415 | 0.002180344 | 1 |
| HTN | b_cell | *KPNA5* | -5.113095993 | 0.009968306 | 0.01441701 | 1 |
| HTN | b_cell | *ANKRD36B* | -7.031147751 | 0.001487688 | 0.003464107 | 1 |
| HTN | b_cell | *HCAR1* | -4.153722348 | 0.019804718 | 0.02613059 | 1 |
| HTN | b_cell | *SRGAP1* | -3.455507021 | 0.001917426 | 0.004086392 | 1 |
| HTN | b_cell | *AP2A1* | -7.410137572 | 0.005915258 | 0.009431708 | 1 |
| HTN | b_cell | *HIST1H3E* | 5.368121725 | 1.69E-07 | 4.18E-05 | 0.001062073 |
| HTN | b_cell | *ZNF585A* | -4.921349404 | 0.000193886 | 0.002180344 | 1 |
| HTN | b_cell | *FUT11* | -4.183110722 | 0.000113474 | 0.002180344 | 0.713409375 |
| HTN | b_cell | *ANXA4* | -4.347249283 | 0.007998695 | 0.011976136 | 1 |
| HTN | b_cell | *WDR5B* | 10.67523103 | 8.72E-06 | 0.000577032 | 0.05481804 |
| HTN | b_cell | *ZNF138* | -4.670787576 | 0.012407587 | 0.017439415 | 1 |
| HTN | b_cell | *ZNF429* | -5.541824337 | 0.000701925 | 0.002366275 | 1 |
| HTN | b_cell | *ZNF470* | -7.375313838 | 0.000265045 | 0.002180344 | 1 |
| HTN | b_cell | *ZNF100* | -8.599254683 | 8.65E-05 | 0.002180344 | 0.543517293 |
| HTN | b_cell | *GMFB* | -6.042027411 | 0.002641916 | 0.005102833 | 1 |
| HTN | b_cell | *ZNF763* | -5.490382588 | 0.000916953 | 0.002637183 | 1 |
| HTN | b_cell | *ARRDC1* | -4.785752576 | 0.000567947 | 0.002232844 | 1 |
| HTN | b_cell | *IGF2R* | -4.63516713 | 0.001113762 | 0.002915164 | 1 |
| HTN | b_cell | *GAL3ST4* | -8.816214243 | 0.000211353 | 0.002180344 | 1 |
| HTN | b_cell | *SLC25A29* | -4.530254873 | 0.002674452 | 0.005146703 | 1 |
| HTN | b_cell | *ZNF682* | -3.388264388 | 0.00043786 | 0.002180344 | 1 |
| HTN | b_cell | *ADAM32* | -3.53085708 | 0.000369675 | 0.002180344 | 1 |
| HTN | b_cell | *LRRC8B* | -2.954335261 | 0.000301074 | 0.002180344 | 1 |
| HTN | b_cell | *ABCB8* | -4.054067867 | 0.008325288 | 0.012365009 | 1 |
| HTN | b_cell | *SND1* | -3.611690446 | 0.016360916 | 0.02208267 | 1 |
| HTN | b_cell | *SULT1A2* | 2.670966164 | 0.015472942 | 0.021037713 | 1 |
| HTN | b_cell | *NEK5* | -4.327193703 | 0.001404214 | 0.003342783 | 1 |
| HTN | b_cell | *PIWIL2* | -4.163903941 | 0.000483947 | 0.002187427 | 1 |
| HTN | b_cell | *C1D* | -5.191229714 | 0.001388696 | 0.003320045 | 1 |
| HTN | b_cell | *TBC1D9B* | -5.250054843 | 0.00078278 | 0.002448253 | 1 |
| HTN | b_cell | *FAM110D* | -8.141271829 | 0.000426298 | 0.002180344 | 1 |
| HTN | b_cell | *KANK2* | -3.392574898 | 0.000487731 | 0.002188291 | 1 |
| HTN | b_cell | *C6orf141* | -7.02753425 | 1.76E-05 | 0.000953272 | 0.11057958 |
| HTN | b_cell | *CCL4L2* | -5.632105272 | 0.004694346 | 0.007859747 | 1 |
| HTN | b_cell | *GTF2E2* | -3.966594463 | 0.003700045 | 0.006547195 | 1 |
| HTN | b_cell | *ZNF165* | 14.27251314 | 1.64E-06 | 0.000179109 | 0.010301942 |
| HTN | b_cell | *RAMP2-AS1* | -3.545229808 | 0.000546104 | 0.002214137 | 1 |
| HTN | b_cell | *ZNF720* | -4.678873493 | 0.001181176 | 0.003016269 | 1 |
| HTN | b_cell | *DDI2* | -4.024832081 | 0.007287366 | 0.011109522 | 1 |
| HTN | b_cell | *MRPL21* | 3.866962597 | 0.022239458 | 0.029002173 | 1 |
| HTN | b_cell | *ZNF786* | -6.747621877 | 0.001267495 | 0.003148456 | 1 |
| HTN | b_cell | *ZNF517* | -5.73240955 | 0.000411784 | 0.002180344 | 1 |
| HTN | b_cell | *C5AR1* | -4.339869465 | 0.001249921 | 0.003120831 | 1 |
| HTN | b_cell | *DCHS2* | -3.258002287 | 0.002003174 | 0.004200786 | 1 |
| HTN | b_cell | *SHPK* | -5.495118835 | 0.000170651 | 0.002180344 | 1 |

| | | | | | | |
|---|---|---|---|---|---|---|
| HTN | b_cell | *MAP3K5* | -4.105565519 | 0.011543802 | 0.01639392 | 1 |
| HTN | b_cell | *GYPE* | 6.958907728 | 0.00056931 | 0.002232844 | 1 |
| HTN | b_cell | *PCDHB11* | -11.43248382 | 2.74E-05 | 0.001246309 | 0.17199066 |
| HTN | b_cell | *SLC2A10* | -3.439554907 | 0.00171007 | 0.003788305 | 1 |
| HTN | b_cell | *ZNF665* | -4.153373455 | 0.023355874 | 0.030288445 | 1 |
| HTN | b_cell | *RPF2* | -4.829674379 | 0.001800293 | 0.003919128 | 1 |
| HTN | b_cell | *MYO5A* | -3.873362792 | 0.000270681 | 0.002180344 | 1 |
| HTN | b_cell | *ATG7* | -2.936798656 | 0.00416885 | 0.007174804 | 1 |
| HTN | b_cell | *TTC30A* | -7.380319237 | 0.001286826 | 0.003170756 | 1 |
| HTN | b_cell | *RAB40C* | -4.691490041 | 0.000749637 | 0.002411911 | 1 |
| HTN | b_cell | *PIGN* | -4.329628669 | 0.022617679 | 0.029440445 | 1 |
| HTN | b_cell | *ZNF624* | 7.752166346 | 0.037398124 | 0.04632946 | 1 |
| HTN | b_cell | *HHLA3* | -7.559353713 | 0.017704994 | 0.023673181 | 1 |
| HTN | b_cell | *HOXA4* | -7.86456482 | 0.000881308 | 0.002587941 | 1 |
| HTN | b_cell | *BCO2* | -8.297591476 | 0.036192156 | 0.044924005 | 1 |
| HTN | b_cell | *ENTPD6* | -4.502093813 | 0.000551892 | 0.002219911 | 1 |
| HTN | b_cell | *CCDC154* | -10.6934076 | 0.012748439 | 0.017850654 | 1 |
| HTN | b_cell | *C5orf42* | -4.06487173 | 0.005955019 | 0.009484221 | 1 |
| HTN | b_cell | *MFAP5* | -2.084413657 | 0.000417283 | 0.002180344 | 1 |
| HTN | b_cell | *CDC42SE1* | -5.847357148 | 0.030389761 | 0.038380962 | 1 |
| HTN | b_cell | *PDCD1LG2* | -2.716532603 | 0.003831575 | 0.006726924 | 1 |
| HTN | b_cell | *NMB* | -11.14450232 | 0.011062993 | 0.015773084 | 1 |
| HTN | b_cell | *KLHL14* | 1.006571926 | 0.000161504 | 0.002180344 | 1 |
| HTN | b_cell | *FAM114A1* | -4.171461736 | 0.001738017 | 0.00383266 | 1 |
| HTN | b_cell | *PHF2* | -5.968325055 | 0.023994494 | 0.031045817 | 1 |
| HTN | b_cell | *RPS26* | -4.661168197 | 0.001447693 | 0.003411965 | 1 |
| HTN | b_cell | *C14orf178* | -6.403363584 | 0.005018267 | 0.0082808 | 1 |
| HTN | b_cell | *PSAP* | -4.178434701 | 0.018199603 | 0.024262278 | 1 |
| HTN | b_cell | *S100A10* | -4.775967948 | 0.000783402 | 0.002448253 | 1 |
| HTN | b_cell | *MAP1LC3C* | -5.581089348 | 0.008176013 | 0.012189374 | 1 |
| HTN | b_cell | *TAF13* | -3.855375174 | 0.000284911 | 0.002180344 | 1 |
| HTN | b_cell | *AC122129.1* | 16.1709853 | 0.000453997 | 0.002180344 | 1 |
| HTN | b_cell | *OCLN* | -5.269057605 | 0.000496446 | 0.002194324 | 1 |
| HTN | b_cell | *ZNF181* | 6.895596273 | 0.001571627 | 0.003581136 | 1 |
| HTN | b_cell | *SGTB* | -4.257362351 | 0.000503995 | 0.002195028 | 1 |
| HTN | b_cell | *ADH5* | -3.375599521 | 0.002959203 | 0.005545308 | 1 |
| HTN | b_cell | *ZNF677* | -4.069709171 | 0.000359023 | 0.002180344 | 1 |
| HTN | b_cell | *ZNF311* | -4.301982939 | 0.007795148 | 0.011731231 | 1 |
| HTN | b_cell | *ZNF71* | -4.022117077 | 0.00122859 | 0.003088552 | 1 |
| HTN | b_cell | *VPS13A* | -4.003424451 | 0.004882783 | 0.008097614 | 1 |
| HTN | b_cell | *LEKR1* | -3.665694714 | 0.000673974 | 0.002334899 | 1 |
| HTN | b_cell | *MRPL42* | -4.552774236 | 0.00440375 | 0.007468675 | 1 |
| HTN | b_cell | *ZNF335* | -4.542650013 | 0.018905176 | 0.025107064 | 1 |
| HTN | b_cell | *AGAP9* | 6.515250817 | 0.001043589 | 0.002813484 | 1 |
| HTN | b_cell | *ZNF84* | -5.02365042 | 0.00155024 | 0.00355188 | 1 |
| HTN | b_cell | *MAK16* | -4.109308587 | 0.000531118 | 0.002195202 | 1 |
| HTN | b_cell | *ZNF667* | -4.931497278 | 0.00139977 | 0.003335996 | 1 |
| HTN | b_cell | *PRIM1* | -6.241068053 | 0.003844971 | 0.006744792 | 1 |
| HTN | b_cell | *SULT1C4* | -5.599045593 | 0.000526197 | 0.002195202 | 1 |
| HTN | b_cell | *SFI1* | -5.964024744 | 0.023846158 | 0.030886031 | 1 |

| | | | | | | |
|---|---|---|---|---|---|---|
| HTN | b_cell | *ADH4* | -3.966276106 | 5.54E-05 | 0.001851584 | 0.348097748 |
| HTN | b_cell | *ZNF248* | -3.910474071 | 0.000453372 | 0.002180344 | 1 |
| HTN | b_cell | *HIBCH* | -3.246944907 | 0.003338448 | 0.006050395 | 1 |
| HTN | b_cell | *ZNF544* | -4.326771316 | 0.005613354 | 0.009060116 | 1 |
| HTN | b_cell | *SOWAHC* | -7.224069635 | 0.000326897 | 0.002180344 | 1 |
| HTN | b_cell | *MIER1* | -4.939576054 | 0.010898446 | 0.015583017 | 1 |
| HTN | b_cell | *SVIP* | -4.669613518 | 0.000666603 | 0.002330887 | 1 |
| HTN | b_cell | *ZNF251* | -4.959219919 | 0.005897231 | 0.009407737 | 1 |
| HTN | b_cell | *DDRGK1* | -6.341867147 | 0.001945347 | 0.004122143 | 1 |
| HTN | b_cell | *ZNF334* | -4.114538185 | 0.000520677 | 0.002195202 | 1 |
| HTN | b_cell | *HSD17B11* | -5.283623471 | 0.008029922 | 0.012012276 | 1 |
| HTN | b_cell | *DDX42* | -4.675148498 | 0.019778555 | 0.026107313 | 1 |
| HTN | b_cell | *SLC29A3* | -4.172807378 | 0.001495626 | 0.003476154 | 1 |
| HTN | b_cell | *ANTXRL* | -179.0875011 | 0.000358724 | 0.002180344 | 1 |
| HTN | b_cell | *STYX* | -6.406169336 | 0.004683432 | 0.007843563 | 1 |
| HTN | b_cell | *TMEM116* | -6.601034534 | 6.13E-10 | 9.64E-07 | 3.85E-06 |
| HTN | b_cell | *GFPT1* | -3.572094346 | 0.013508001 | 0.018730658 | 1 |
| HTN | b_cell | *UVRAG* | -3.52557473 | 0.001819892 | 0.00394268 | 1 |
| HTN | b_cell | *MGEA5* | -3.042707283 | 0.002063841 | 0.004278063 | 1 |
| HTN | b_cell | *MT1F* | -19.28131801 | 0.002068746 | 0.004285406 | 1 |
| HTN | b_cell | *ZNF587* | -5.301279302 | 0.002011471 | 0.004211145 | 1 |
| HTN | b_cell | *FLVCR1-AS1* | -4.05101434 | 0.000431142 | 0.002180344 | 1 |
| HTN | b_cell | *SH3BGRL2* | -3.004301574 | 0.000988448 | 0.002729192 | 1 |
| HTN | b_cell | *TMA16* | -5.820906406 | 0.006735371 | 0.010450463 | 1 |
| HTN | b_cell | *HLA-DRB5* | -2.443436061 | 0.007834795 | 0.011778421 | 1 |
| HTN | b_cell | *CNGA1* | -4.41398184 | 0.000252208 | 0.002180344 | 1 |
| HTN | b_cell | *MAFK* | -6.950355707 | 0.00238266 | 0.004736389 | 1 |
| HTN | b_cell | *HIST1H4E* | 16.58009236 | 0.000862791 | 0.002559431 | 1 |
| HTN | b_cell | *C20orf203* | 8.041110157 | 0.000336912 | 0.002180344 | 1 |
| HTN | b_cell | *ZNF627* | -4.431188802 | 0.0005928 | 0.00226693 | 1 |
| HTN | b_cell | *ARC* | -27.30058768 | 0.000432508 | 0.002180344 | 1 |
| HTN | b_cell | *MMP17* | -8.324197321 | 0.002352692 | 0.004700152 | 1 |
| HTN | b_cell | *CCDC69* | -3.703160417 | 0.000741514 | 0.002405521 | 1 |
| HTN | b_cell | *MDM4* | -4.392208637 | 0.013498466 | 0.018721566 | 1 |
| HTN | b_cell | *NCOA6* | -4.753249458 | 0.000489185 | 0.002188936 | 1 |
| HTN | b_cell | *FAM19A2* | -3.750379165 | 0.000559479 | 0.002226229 | 1 |
| HTN | b_cell | *TTC37* | -4.151047784 | 0.003722565 | 0.006575938 | 1 |
| HTN | b_cell | *PAPSS2* | -2.782948219 | 0.00154547 | 0.003547581 | 1 |
| HTN | b_cell | *FAN1* | -4.142777715 | 0.000704922 | 0.00237262 | 1 |
| HTN | b_cell | *CEP290* | -4.764548759 | 0.024484054 | 0.031595084 | 1 |
| HTN | b_cell | *ECI2* | -4.389133243 | 0.000675331 | 0.002334899 | 1 |
| HTN | b_cell | *ZNF652* | -4.609508767 | 0.030788757 | 0.038799141 | 1 |
| HTN | b_cell | *APCDD1L* | 9.112158824 | 0.002066665 | 0.004282506 | 1 |
| HTN | b_cell | *RASSF9* | -3.645790591 | 0.000436686 | 0.002180344 | 1 |
| HTN | b_cell | *TMEM184B* | -5.170336121 | 0.001090792 | 0.00287056 | 1 |
| HTN | b_cell | *ALPK2* | -5.10700051 | 0.001480746 | 0.003457158 | 1 |
| HTN | b_cell | *SFT2D1* | -5.112695967 | 0.001370133 | 0.003289051 | 1 |
| HTN | b_cell | *ARHGAP11A* | -7.44449768 | 0.000521016 | 0.002195202 | 1 |
| HTN | b_cell | *HMGN2* | -3.506699264 | 0.00121056 | 0.00305766 | 1 |
| HTN | b_cell | *DENND4B* | -8.083729707 | 6.88E-05 | 0.002060872 | 0.432512481 |

| | | | | | | |
|---|---|---|---|---|---|---|
| HTN | b_cell | *CD3E* | -4.721516353 | 0.003612585 | 0.006415909 | 1 |
| HTN | b_cell | *TSEN15* | -4.53814731 | 0.000863852 | 0.002559431 | 1 |
| HTN | b_cell | *TYW1* | -3.652595104 | 0.006950743 | 0.010689299 | 1 |
| HTN | b_cell | *ITPRIPL1* | 7.953559347 | 0.011459999 | 0.01629331 | 1 |
| HTN | b_cell | *PRMT6* | 7.065884695 | 0.000622627 | 0.002291037 | 1 |
| HTN | b_cell | *SHISA4* | -12.25299762 | 0.000153429 | 0.002180344 | 0.964607489 |
| HTN | b_cell | *CAPZA2* | -4.377683995 | 0.00342366 | 0.006162196 | 1 |
| HTN | b_cell | *MAP3K3* | -4.738112417 | 0.001759487 | 0.003854718 | 1 |
| HTN | b_cell | *C1orf174* | -5.417936531 | 0.001423276 | 0.003374109 | 1 |
| HTN | b_cell | *RASGEF1A* | -11.58111135 | 0.000448197 | 0.002180344 | 1 |
| HTN | b_cell | *KIAA0753* | -5.381867574 | 0.005514807 | 0.008929073 | 1 |
| HTN | b_cell | *DCLRE1A* | 12.66458389 | 0.008420465 | 0.012488984 | 1 |
| HTN | b_cell | *APRT* | 8.835587006 | 0.009338477 | 0.013650547 | 1 |
| HTN | b_cell | *TBKBP1* | -6.246538041 | 0.000168017 | 0.002180344 | 1 |
| HTN | b_cell | *ZFP2* | -5.518293075 | 0.001170228 | 0.003001724 | 1 |
| HTN | b_cell | *L3MBTL3* | 7.156636835 | 0.000658106 | 0.002327958 | 1 |
| HTN | b_cell | *MFAP3L* | -7.777887537 | 0.000888959 | 0.002597064 | 1 |
| HTN | b_cell | *NAGA* | -4.47827046 | 0.000847682 | 0.002540217 | 1 |
| HTN | b_cell | *SMG5* | -5.697913667 | 0.001750888 | 0.003844942 | 1 |
| HTN | b_cell | *PJA2* | -4.587202353 | 0.002567568 | 0.00501158 | 1 |
| HTN | b_cell | *AL590822.1* | -6.589898473 | 1.85E-06 | 0.000186095 | 0.011661411 |
| HTN | b_cell | *AP001007.1* | 4.258995532 | 0.000229257 | 0.002180344 | 1 |
| HTN | b_cell | *ZNF525* | -7.463573371 | 0.033611451 | 0.042019326 | 1 |
| HTN | b_cell | *AC105020.1* | 9.361035686 | 0.000197992 | 0.002180344 | 1 |
| HTN | b_cell | *EFCAB2* | -4.331542459 | 0.001806141 | 0.003927449 | 1 |
| HTN | b_cell | *CHML* | 5.60040802 | 0.000330937 | 0.002180344 | 1 |
| HTN | b_cell | *CAPN8* | -15.3394455 | 0.001276157 | 0.003156737 | 1 |
| HTN | b_cell | *CENPW* | -11.13691494 | 1.58E-05 | 0.000894532 | 0.099293084 |
| HTN | b_cell | *FAM229B* | -3.919272874 | 0.004405065 | 0.00746889 | 1 |
| HTN | b_cell | *DDO* | -6.65532581 | 0.000680527 | 0.002339217 | 1 |
| HTN | b_cell | *HIST2H2BF* | -17.30548241 | 0.000457813 | 0.002180344 | 1 |
| HTN | b_cell | *RBM20* | -5.20043281 | 0.00042621 | 0.002180344 | 1 |
| HTN | b_cell | *C6orf163* | -6.158552626 | 0.000221046 | 0.002180344 | 1 |
| HTN | b_cell | *PCMTD2* | -3.635553332 | 0.001704655 | 0.003778547 | 1 |
| HTN | b_cell | *OOEP* | 3.149701723 | 0.001980036 | 0.004161982 | 1 |
| HTN | b_cell | *FOXO6* | 10.47368094 | 0.002844717 | 0.00538751 | 1 |
| HTN | b_cell | *INPP5B* | -3.672710353 | 5.60E-05 | 0.001852286 | 0.351930905 |
| HTN | b_cell | *TRAF3IP1* | -4.281575493 | 0.000755482 | 0.002419498 | 1 |
| HTN | b_cell | *GIGYF2* | -4.167838034 | 0.004590967 | 0.00772989 | 1 |
| HTN | b_cell | *RUFY2* | -4.842250326 | 0.004681049 | 0.007843563 | 1 |
| HTN | b_cell | *PHACTR4* | -3.670979143 | 0.023167507 | 0.030075185 | 1 |
| HTN | b_cell | *ASAH2B* | -4.388132273 | 0.007404241 | 0.011260393 | 1 |
| HTN | b_cell | *C10orf128* | -3.00984728 | 0.001513282 | 0.003504236 | 1 |
| HTN | b_cell | *SYT15* | -12.0648708 | 0.003893185 | 0.006812261 | 1 |
| HTN | b_cell | *ZDBF2* | -5.365555066 | 0.002201494 | 0.004467654 | 1 |
| HTN | b_cell | *RING1* | -5.60931237 | 0.000195857 | 0.002180344 | 1 |
| HTN | b_cell | *HSD17B8* | -14.95787364 | 1.49E-08 | 6.68E-06 | 9.35E-05 |
| HTN | b_cell | *OXLD1* | -4.917506888 | 0.015077762 | 0.020567127 | 1 |
| HTN | b_cell | *HLA-DOA* | 1.315225302 | 0.001071124 | 0.002848663 | 1 |
| HTN | b_cell | *HLA-DRA* | 5.924352314 | 0.000210834 | 0.002180344 | 1 |

| | | | | | | |
|---|---|---|---|---|---|---|
| HTN | b_cell | *COL15A1* | -3.182173428 | 0.000890766 | 0.002599737 | 1 |
| HTN | b_cell | *NOTCH4* | -1.018897825 | 0.002804953 | 0.005342241 | 1 |
| HTN | b_cell | *AGER* | -5.970867082 | 0.038105932 | 0.047159841 | 1 |
| HTN | b_cell | *MRPL38* | 7.247438987 | 3.34E-06 | 0.000296063 | 0.021020451 |
| HTN | b_cell | *DXO* | -4.731630925 | 0.002233604 | 0.004512508 | 1 |
| HTN | b_cell | *SKIV2L* | -2.462055903 | 0.005327197 | 0.008683456 | 1 |
| HTN | b_cell | *LAYN* | -7.521464992 | 0.000249639 | 0.002180344 | 1 |
| HTN | b_cell | *C6orf48* | 13.19666169 | 0.000133144 | 0.002180344 | 0.837076924 |
| HTN | b_cell | *CARD16* | 6.866482363 | 0.006520978 | 0.010195684 | 1 |
| HTN | b_cell | *ABHD16A* | 3.596924268 | 0.000209401 | 0.002180344 | 1 |
| HTN | b_cell | *GPANK1* | 12.2480648 | 6.35E-05 | 0.001980182 | 0.399497861 |
| HTN | b_cell | *ZNF814* | -3.799784633 | 0.001500217 | 0.003480291 | 1 |
| HTN | b_cell | *MICB* | -2.896662393 | 0.020624798 | 0.027115873 | 1 |
| HTN | b_cell | *MICA* | -3.710441016 | 0.00600791 | 0.009543934 | 1 |
| HTN | b_cell | *HLA-C* | -6.22450374 | 9.04E-05 | 0.002180344 | 0.568545457 |
| HTN | b_cell | *POU5F1* | -2.762161009 | 0.016321124 | 0.022045373 | 1 |
| HTN | b_cell | *CCHCR1* | -5.55552448 | 1.17E-06 | 0.000141222 | 0.007343552 |
| HTN | b_cell | *ABCF1* | -5.707235771 | 0.001343144 | 0.003258057 | 1 |
| HTN | b_cell | *LILRB3* | -4.285584212 | 0.001289617 | 0.003172074 | 1 |
| HTN | b_cell | *LINC01123* | -184.4634279 | 0.000362611 | 0.002180344 | 1 |
| HTN | b_cell | *HLA-E* | -4.345690789 | 0.000804297 | 0.002481486 | 1 |
| HTN | b_cell | *ZNF468* | -7.050553616 | 0.000304404 | 0.002180344 | 1 |
| HTN | b_cell | *HCG9* | -5.984554336 | 0.000358705 | 0.002180344 | 1 |
| HTN | b_cell | *TBC1D8* | -4.551245674 | 0.006541561 | 0.010210895 | 1 |
| HTN | b_cell | *FAM153C* | 4.05279487 | 0.00019533 | 0.002180344 | 1 |
| HTN | b_cell | *TRIM27* | -3.669218451 | 0.005776648 | 0.009260017 | 1 |
| HTN | b_cell | *MALRD1* | -3.194577804 | 9.74E-05 | 0.002180344 | 0.612139902 |
| HTN | b_cell | *MROH6* | -5.720126124 | 0.010841162 | 0.015518758 | 1 |
| HTN | b_cell | *ATXN2* | -5.054325235 | 0.000607684 | 0.002281385 | 1 |
| HTN | b_cell | *ZNF155* | -5.506948256 | 0.001831016 | 0.003957685 | 1 |
| HTN | b_cell | *LRRC10B* | -42.51557259 | 0.005872119 | 0.009379576 | 1 |
| HTN | b_cell | *C12orf73* | -8.742440418 | 0.000579061 | 0.002248644 | 1 |
| HTN | b_cell | *PCDHA5* | -44.51539533 | 0.004106326 | 0.00710024 | 1 |
| HTN | b_cell | *PCDHA4* | -21.0678714 | 0.003543947 | 0.006317209 | 1 |
| HTN | b_cell | *TRIM13* | -3.889830294 | 0.020856169 | 0.027379982 | 1 |
| HTN | b_cell | *SPIRE2* | -3.737458674 | 0.014394634 | 0.019789868 | 1 |
| HTN | b_cell | *SLFN12L* | -3.263490409 | 0.000317912 | 0.002180344 | 1 |
| HTN | b_cell | *CLLU1OS* | -10.97162306 | 0.013906769 | 0.019220017 | 1 |
| HTN | b_cell | *SYCE1L* | 5.581144748 | 0.014303898 | 0.019673727 | 1 |
| HTN | b_cell | *TMEM231* | -4.502468594 | 0.008228061 | 0.012252444 | 1 |
| HTN | b_cell | *FAM71F2* | -4.693396209 | 0.000573039 | 0.002237955 | 1 |
| HTN | b_cell | *AC016586.1* | -39.65102051 | 0.030773091 | 0.038787174 | 1 |
| HTN | b_cell | *LINC00654* | -12.07786971 | 6.79E-05 | 0.002060872 | 0.426873516 |
| HTN | b_cell | *TMEM170B* | -1.749525306 | 0.028140591 | 0.035820995 | 1 |
| HTN | b_cell | *SAP25* | 5.395690362 | 0.013093096 | 0.01825195 | 1 |
| HTN | b_cell | *NT5M* | -4.564704585 | 0.002840903 | 0.00538751 | 1 |
| HTN | b_cell | *PRR13* | -9.122900057 | 0.00053898 | 0.002208907 | 1 |
| HTN | b_cell | *MT1A* | -12.75230869 | 0.000452381 | 0.002180344 | 1 |
| HTN | b_cell | *MT1M* | -6.058119507 | 0.000415015 | 0.002180344 | 1 |
| HTN | b_cell | *CFI* | -3.60604564 | 0.000643591 | 0.002316643 | 1 |

| | | | | | | |
|---|---|---|---|---|---|---|
| HTN | b_cell | *EXOC3L4* | -9.866318299 | 0.000317216 | 0.002180344 | 1 |
| HTN | b_cell | *TP53TG3D* | 7.648344305 | 0.000168762 | 0.002180344 | 1 |
| HTN | b_cell | *ATP6AP1L* | -3.700522068 | 0.004641335 | 0.007797989 | 1 |
| HTN | b_cell | *NAP1L4* | -4.229473607 | 0.001106808 | 0.002900583 | 1 |
| HTN | b_cell | *SMN2* | -5.752273668 | 0.00935783 | 0.013672479 | 1 |
| HTN | b_cell | *SERF1B* | -21.17193155 | 4.47E-06 | 0.00036476 | 0.028086487 |
| HTN | b_cell | *DENND6B* | -9.784525024 | 0.00921287 | 0.013501471 | 1 |
| HTN | b_cell | *EIF3CL* | -9.859653922 | 0.024904921 | 0.032078926 | 1 |
| HTN | b_cell | *LCMT1* | -4.883274674 | 0.001138053 | 0.002949898 | 1 |
| HTN | b_cell | *LIN52* | -3.517445578 | 0.00296925 | 0.005555855 | 1 |
| HTN | b_cell | *C17orf107* | 4.732184695 | 0.000368562 | 0.002180344 | 1 |
| HTN | b_cell | *LINC00487* | -21.10420865 | 0.000839395 | 0.002531828 | 1 |
| HTN | b_cell | *RFPL3S* | -8.083272821 | 0.005254057 | 0.00859991 | 1 |
| HTN | b_cell | *C1QTNF9B* | -64.51440623 | 0.000420735 | 0.002180344 | 1 |
| HTN | b_cell | *RNPS1* | -4.227237042 | 0.040311281 | 0.049605994 | 1 |
| HTN | b_cell | *NYNRIN* | -7.59044855 | 0.002145191 | 0.004380258 | 1 |
| HTN | b_cell | *ZDHHC11B* | -3.994311414 | 0.007723277 | 0.011641727 | 1 |
| HTN | b_cell | *GOLGA8O* | 21.74281482 | 0.000539666 | 0.002208907 | 1 |
| HTN | b_cell | *LNP1* | -6.158663363 | 8.60E-05 | 0.002180344 | 0.54083514 |
| HTN | b_cell | *VGLL3* | -2.288817151 | 0.000589119 | 0.002263931 | 1 |
| HTN | b_cell | *COLQ* | -4.460511317 | 0.000567744 | 0.002232844 | 1 |
| HTN | b_cell | *METTL6* | -4.543460444 | 0.007731077 | 0.011650354 | 1 |
| HTN | b_cell | *SACM1L* | -4.876588352 | 0.031388723 | 0.039515599 | 1 |
| HTN | b_cell | *SLC48A1* | -4.045856563 | 0.000197335 | 0.002180344 | 1 |
| HTN | b_cell | *TAS2R14* | 8.348858383 | 0.000152757 | 0.002180344 | 0.9603849 |
| HTN | b_cell | *ZRSR1* | -24.2919213 | 0.001169251 | 0.003001667 | 1 |
| HTN | b_cell | *AC084018.1* | -5.633784296 | 0.000663802 | 0.002327958 | 1 |
| HTN | b_cell | *C17orf51* | -6.286553937 | 0.000404104 | 0.002180344 | 1 |
| HTN | b_cell | *OR2A42* | -5.646798339 | 0.012189197 | 0.01719396 | 1 |
| HTN | b_cell | *RNF208* | -13.7237508 | 0.000406462 | 0.002180344 | 1 |
| HTN | b_cell | *MAP10* | 11.64617028 | 0.013054741 | 0.018210349 | 1 |
| HTN | b_cell | *NUP62* | -6.607713025 | 0.000756151 | 0.002419498 | 1 |
| HTN | b_cell | *FGFR1OP* | -5.283664532 | 0.004340698 | 0.007389282 | 1 |
| HTN | b_cell | *FAM106A* | -5.342398221 | 0.030568747 | 0.038569862 | 1 |
| HTN | b_cell | *ZNF254* | -4.562725665 | 0.002238548 | 0.004516517 | 1 |
| HTN | b_cell | *ASIC3* | -6.918833579 | 0.022371437 | 0.029156142 | 1 |
| HTN | b_cell | *NOTCH2NL* | 10.83815849 | 0.000495379 | 0.002193274 | 1 |
| HTN | b_cell | *MXD3* | -11.05026874 | 9.01E-06 | 0.000585817 | 0.056656417 |
| HTN | b_cell | *GSTM2* | -4.834357937 | 6.39E-05 | 0.001980182 | 0.401634509 |
| HTN | b_cell | *ARHGAP19* | 11.19893291 | 0.02549469 | 0.032764741 | 1 |
| HTN | b_cell | *LCAT* | -4.091540096 | 0.00034877 | 0.002180344 | 1 |
| HTN | b_cell | *SIPA1* | 14.65130131 | 9.48E-08 | 2.53E-05 | 0.000595979 |
| HTN | b_cell | *RBMXL1* | -5.625838312 | 0.000407408 | 0.002180344 | 1 |
| HTN | b_cell | *SRA1* | -4.511157413 | 0.001357138 | 0.003274908 | 1 |
| HTN | b_cell | *TMEM110* | -10.15364195 | 0.011711791 | 0.016611474 | 1 |
| HTN | b_cell | *HEXA* | -6.102371231 | 0.01407605 | 0.019415561 | 1 |
| HTN | b_cell | *LEPROT* | -4.903966802 | 0.00173189 | 0.003827204 | 1 |
| HTN | b_cell | *LBH* | 4.765122079 | 0.000326581 | 0.002180344 | 1 |
| HTN | b_cell | *PPP1CB* | -4.268404338 | 0.033373248 | 0.041746441 | 1 |
| HTN | b_cell | *NCKIPSD* | -5.887318799 | 0.000233038 | 0.002180344 | 1 |

| | | | | | | |
|---|---|---|---|---|---|---|
| HTN | b_cell | *SLC35F6* | -6.758552985 | 0.002826117 | 0.005372784 | 1 |
| HTN | b_cell | *ATP6V1G2* | 3.942811707 | 0.00015276 | 0.002180344 | 0.960402602 |
| HTN | b_cell | *EMP2* | -3.976083381 | 0.000253901 | 0.002180344 | 1 |
| HTN | b_cell | *LTB4R* | -12.74293283 | 3.43E-07 | 6.34E-05 | 0.002155157 |
| HTN | b_cell | *LTB4R2* | -18.18118096 | 5.68E-08 | 1.78E-05 | 0.00035684 |
| HTN | b_cell | *GALT* | -4.585726312 | 0.004354548 | 0.007409214 | 1 |
| HTN | b_cell | *ITGA1* | -2.244388964 | 0.000598533 | 0.002274361 | 1 |
| HTN | b_cell | *NUDT19* | -4.626535821 | 0.001038992 | 0.002804698 | 1 |
| HTN | b_cell | *ZNF726* | -5.481662073 | 0.000442948 | 0.002180344 | 1 |
| HTN | b_cell | *TAX1BP3* | -5.726380217 | 0.000575269 | 0.00224084 | 1 |
| HTN | b_cell | *AP1G2* | 7.077854393 | 0.001057908 | 0.002833039 | 1 |
| HTN | b_cell | *ZNF90* | 7.859072315 | 0.001432505 | 0.003385825 | 1 |
| HTN | b_cell | *TM6SF2* | -75.82349821 | 0.00036649 | 0.002180344 | 1 |
| HTN | b_cell | *REPIN1* | -4.92355216 | 5.30E-05 | 0.001792205 | 0.333350093 |
| HTN | b_cell | *SMIM7* | -5.532935967 | 0.000752302 | 0.002415588 | 1 |
| HTN | b_cell | *TSPAN4* | -4.056080636 | 0.000753463 | 0.002416404 | 1 |
| HTN | b_cell | *PAXIP1-AS2* | -5.714529525 | 0.001557271 | 0.003562795 | 1 |
| HTN | b_cell | *LYRM4* | -5.154976828 | 0.022560761 | 0.029372438 | 1 |
| HTN | b_cell | *PRCD* | -5.017520874 | 0.000793863 | 0.002465918 | 1 |
| HTN | b_cell | *C17orf67* | -3.805159775 | 0.000418855 | 0.002180344 | 1 |
| HTN | b_cell | *FIS1* | -7.503011023 | 0.000151055 | 0.002180344 | 0.94968014 |
| HTN | b_cell | *COLCA2* | -6.570827401 | 0.000592834 | 0.00226693 | 1 |
| HTN | b_cell | *NEURL1B* | -4.178482678 | 0.000374023 | 0.002180344 | 1 |
| HTN | b_cell | *VSTM5* | 4.232048469 | 0.003143014 | 0.005793061 | 1 |
| HTN | b_cell | *PPME1* | -6.493335442 | 0.000215752 | 0.002180344 | 1 |
| HTN | b_cell | *STARD10* | 5.215871773 | 0.002098814 | 0.004329147 | 1 |
| HTN | b_cell | *CPEB1* | -3.614671626 | 0.000651773 | 0.002323165 | 1 |
| HTN | b_cell | *C10orf105* | -9.602234815 | 0.000665429 | 0.002329372 | 1 |
| HTN | b_cell | *ARHGEF33* | -6.378915847 | 0.012302603 | 0.017326717 | 1 |
| HTN | b_cell | *AC005082.1* | 16.27572097 | 0.000348371 | 0.002180344 | 1 |
| HTN | b_cell | *ACSM4* | -3.593327321 | 0.016880895 | 0.022717375 | 1 |
| HTN | b_cell | *COL28A1* | -5.89446143 | 0.000710655 | 0.002379315 | 1 |
| HTN | b_cell | *GRID2IP* | -10.39623764 | 0.002180868 | 0.004434385 | 1 |
| HTN | b_cell | *UBE2QL1* | -7.493043015 | 0.000409139 | 0.002180344 | 1 |
| HTN | b_cell | *GOLGA8B* | -4.633208531 | 0.000740759 | 0.002405521 | 1 |
| HTN | b_cell | *KCNU1* | -6.847189564 | 0.000109251 | 0.002180344 | 0.686860174 |
| HTN | b_cell | *MYL5* | -7.486288316 | 0.00046121 | 0.002180489 | 1 |
| HTN | b_cell | *NPEPL1* | -6.400053293 | 0.000975479 | 0.002717425 | 1 |
| HTN | b_cell | *LINC01139* | -9.291303348 | 0.000843617 | 0.002536213 | 1 |
| HTN | b_cell | *TSTD1* | -5.178397425 | 0.000617498 | 0.002286342 | 1 |
| HTN | b_cell | *LINC01138* | -4.35245801 | 0.000967969 | 0.002711104 | 1 |
| HTN | b_cell | *CYB5RL* | 5.550973695 | 0.000782145 | 0.002448253 | 1 |
| HTN | b_cell | *ATAD3C* | 2.360169113 | 0.000243304 | 0.002180344 | 1 |
| HTN | b_cell | *CCDC7* | -4.0492786 | 0.001094298 | 0.002876193 | 1 |
| HTN | b_cell | *AC016773.1* | 7.069533238 | 4.31E-07 | 6.93E-05 | 0.002706902 |
| HTN | b_cell | *RNASEK* | -6.473098046 | 0.00118603 | 0.003022492 | 1 |
| HTN | b_cell | *FAM228B* | -4.024815817 | 0.000103553 | 0.002180344 | 0.651040813 |
| HTN | b_cell | *RNU6ATAC35I* | -25.79904969 | 0.000424415 | 0.002180344 | 1 |
| HTN | b_cell | *PPP3R1* | 6.713164026 | 0.000448678 | 0.002180344 | 1 |
| HTN | b_cell | *FANCG* | 2.167676802 | 0.000552735 | 0.002219911 | 1 |

| | | | | | | |
|---|---|---|---|---|---|---|
| HTN | b_cell | *AP4M1* | -6.871045397 | 0.000306314 | 0.002180344 | 1 |
| HTN | b_cell | *C19orf73* | -33.79313388 | 0.001352717 | 0.003270974 | 1 |
| HTN | b_cell | *ZNF880* | -3.580799005 | 0.035894586 | 0.044572242 | 1 |
| HTN | b_cell | *OR2A25* | -232.1098883 | 6.18E-05 | 0.001972109 | 0.388505388 |
| HTN | b_cell | *XKR9* | -4.674387794 | 0.005401308 | 0.008783761 | 1 |
| HTN | b_cell | *C1orf229* | -7.62403659 | 0.030759359 | 0.03877764 | 1 |
| HTN | b_cell | *SLC12A8* | -5.566294864 | 2.80E-05 | 0.001261541 | 0.176283192 |
| HTN | b_cell | *APOL6* | -2.49061702 | 0.000418065 | 0.002180344 | 1 |
| HTN | b_cell | *TIAF1* | -10.99728188 | 9.32E-05 | 0.002180344 | 0.585913974 |
| HTN | b_cell | *BTBD19* | -4.217999572 | 0.000557759 | 0.002226229 | 1 |
| HTN | b_cell | *FAM185A* | -6.350991296 | 0.0093367 | 0.013650547 | 1 |
| HTN | b_cell | *AC062017.1* | -4.185240031 | 0.000467738 | 0.002180489 | 1 |
| HTN | b_cell | *ADRA2B* | -38.78996607 | 0.006883073 | 0.010616752 | 1 |
| HTN | b_cell | *EXOSC6* | -6.096926212 | 2.28E-05 | 0.001085313 | 0.14326128 |
| HTN | b_cell | *TINCR* | 4.842978636 | 0.000727794 | 0.002394369 | 1 |
| HTN | b_cell | *LINC00205* | -6.537964436 | 0.000848786 | 0.002542314 | 1 |
| HTN | b_cell | *AC147651.3* | -11.80998152 | 0.001887486 | 0.004045632 | 1 |
| HTN | b_cell | *HLA-DPB1* | 6.4978561 | 3.96E-05 | 0.001519523 | 0.249016137 |
| HTN | b_cell | *OSER1-AS1* | -4.087066458 | 0.000922701 | 0.002647658 | 1 |
| HTN | b_cell | *LINC01133* | -3.562196335 | 0.000414319 | 0.002180344 | 1 |
| HTN | b_cell | *LINC01117* | -3.879883104 | 0.000439985 | 0.002180344 | 1 |
| HTN | b_cell | *RASAL2-AS1* | 3.151738058 | 0.000198947 | 0.002180344 | 1 |
| HTN | b_cell | *AC069213.1* | -23.3228405 | 0.002508708 | 0.004923903 | 1 |
| HTN | b_cell | *LINC00240* | -3.006680992 | 0.001155997 | 0.002978872 | 1 |
| HTN | b_cell | *LINC00863* | -4.992682654 | 0.010521899 | 0.015141034 | 1 |
| HTN | b_cell | *LINC00184* | -13.10155807 | 0.00019123 | 0.002180344 | 1 |
| HTN | b_cell | *TMEM233* | -2.975014067 | 0.005751402 | 0.009231316 | 1 |
| HTN | b_cell | *AL049840.1* | -5.585477619 | 5.05E-06 | 0.000398561 | 0.031734686 |
| HTN | b_cell | *LINC00237* | -24.97079837 | 0.000395127 | 0.002180344 | 1 |
| HTN | b_cell | *PLEKHM1* | -4.803486319 | 0.001891936 | 0.004051296 | 1 |
| HTN | b_cell | *LINC00092* | -10.46954118 | 0.000670226 | 0.002333871 | 1 |
| HTN | b_cell | *AP000350.6* | 4.948492067 | 0.000446874 | 0.002180344 | 1 |
| HTN | b_cell | *SFTA1P* | -18.44128289 | 0.000406807 | 0.002180344 | 1 |
| HTN | b_cell | *SLC26A6* | 14.47191551 | 9.84E-07 | 0.000121305 | 0.006186555 |
| HTN | b_cell | *MIAT* | -9.328956685 | 0.008136646 | 0.012147967 | 1 |
| HTN | b_cell | *LINC00626* | 130.7331362 | 0.007343267 | 0.011183895 | 1 |
| HTN | b_cell | *ERCC6* | -4.957181117 | 0.002539045 | 0.004968246 | 1 |
| HTN | b_cell | *LINC00115* | -7.824133669 | 0.029190569 | 0.037030086 | 1 |
| HTN | b_cell | *NOL7* | -6.066808218 | 0.009351935 | 0.013667042 | 1 |
| HTN | b_cell | *ELFN1* | -18.09935225 | 0.00067555 | 0.002334899 | 1 |
| HTN | b_cell | *ZNF503-AS1* | 8.007236472 | 0.000886355 | 0.002596195 | 1 |
| HTN | b_cell | *LINC00937* | -11.01327576 | 0.015242941 | 0.020774414 | 1 |
| HTN | b_cell | *FTCDNL1* | -2.476844616 | 0.002781936 | 0.005306442 | 1 |
| HTN | b_cell | *AC010536.1* | 11.37830753 | 0.017072769 | 0.022959679 | 1 |
| HTN | b_cell | *AC064836.3* | 6.930889845 | 0.000258658 | 0.002180344 | 1 |
| HTN | b_cell | *TMEM185B* | -8.101468408 | 0.001755458 | 0.003848438 | 1 |
| HTN | b_cell | *HSBP1L1* | -9.485536929 | 0.000843335 | 0.002536213 | 1 |
| HTN | b_cell | *TAS2R46* | 5.043133554 | 0.001156344 | 0.002978872 | 1 |
| HTN | b_cell | *AC093901.1* | 2.019964393 | 0.000266089 | 0.002180344 | 1 |
| HTN | b_cell | *C14orf132* | -4.037033803 | 0.000288085 | 0.002180344 | 1 |

| | | | | | | |
|---|---|---|---|---|---|---|
| HTN | b_cell | *AC017074.2* | -68.59198568 | 6.30E-05 | 0.001980182 | 0.396157365 |
| HTN | b_cell | *AC009299.3* | -6.664257918 | 0.000469437 | 0.002180489 | 1 |
| HTN | b_cell | *SLC2A1-AS1* | -18.09478913 | 0.001527855 | 0.003523706 | 1 |
| HTN | b_cell | *LINC01068* | -8.380642685 | 0.018183834 | 0.024246397 | 1 |
| HTN | b_cell | *AC108463.2* | -17.04740299 | 2.06E-05 | 0.001028789 | 0.12962746 |
| HTN | b_cell | *HCG20* | -14.65531909 | 0.002093996 | 0.004320628 | 1 |
| HTN | b_cell | *C19orf24* | -5.715796 | 0.002820269 | 0.005364911 | 1 |
| HTN | b_cell | *LINC01013* | 4.938877958 | 0.000203454 | 0.002180344 | 1 |
| HTN | b_cell | *AC073133.2* | -66.03629184 | 0.000518951 | 0.002195202 | 1 |
| HTN | b_cell | *ARL17B* | -3.580495121 | 0.000947363 | 0.002675683 | 1 |
| HTN | b_cell | *DHFR* | -3.243868641 | 0.014270142 | 0.019640189 | 1 |
| HTN | b_cell | *LINC00954* | -13.29458009 | 0.013760444 | 0.019034524 | 1 |
| HTN | b_cell | *RPL41* | 4.611899429 | 0.000230302 | 0.002180344 | 1 |
| HTN | b_cell | *PATL2* | 6.622101086 | 0.000663571 | 0.002327958 | 1 |
| HTN | b_cell | *ZNF492* | -6.389116194 | 0.006935034 | 0.010668109 | 1 |
| HTN | b_cell | *U91324.1* | -12.39960767 | 0.00043568 | 0.002180344 | 1 |
| HTN | b_cell | *ZNF688* | -11.40731316 | 0.006855758 | 0.010590209 | 1 |
| HTN | b_cell | *CISD3* | -5.735451339 | 0.000705672 | 0.00237262 | 1 |
| HTN | b_cell | *PSMG3-AS1* | -6.440062333 | 0.001735041 | 0.003830743 | 1 |
| HTN | b_cell | *AC112229.4* | 4.71417575 | 1.15E-05 | 0.000695457 | 0.072327518 |
| HTN | b_cell | *KLHL7-AS1* | 8.363752485 | 0.000258621 | 0.002180344 | 1 |
| HTN | b_cell | *AC092171.4* | -9.911583619 | 0.004001876 | 0.006954061 | 1 |
| HTN | b_cell | *HLA-DPA1* | 6.752155655 | 0.002018594 | 0.00421905 | 1 |
| HTN | b_cell | *DIRC3* | -1.874585361 | 0.00041604 | 0.002180344 | 1 |
| HTN | b_cell | *AC107021.1* | -2.488314517 | 0.000860952 | 0.002559246 | 1 |
| HTN | b_cell | *AC009495.3* | 13.75757252 | 0.000488434 | 0.002188728 | 1 |
| HTN | b_cell | *HLA-DQB2* | 1.738825854 | 0.028291293 | 0.036005538 | 1 |
| HTN | b_cell | *LINC00665* | -5.993980473 | 0.004650276 | 0.007808837 | 1 |
| HTN | b_cell | *TNF* | -6.345290263 | 0.003720732 | 0.006575938 | 1 |
| HTN | b_cell | *LINC00342* | -4.134661065 | 0.000514904 | 0.002195202 | 1 |
| HTN | b_cell | *LINC00702* | -5.119779868 | 0.000367221 | 0.002180344 | 1 |
| HTN | b_cell | *LINC00472* | -3.749511171 | 0.000319004 | 0.002180344 | 1 |
| HTN | b_cell | *RPS28* | -4.185770871 | 0.000466871 | 0.002180489 | 1 |
| HTN | b_cell | *AC123768.3* | -5.467273222 | 0.000589841 | 0.002265319 | 1 |
| HTN | b_cell | *TRIM26* | -5.386899278 | 0.008156353 | 0.012168638 | 1 |
| HTN | b_cell | *LINC01114* | -40.3020856 | 0.016007528 | 0.021680165 | 1 |
| HTN | b_cell | *ZNF736* | -3.850345074 | 0.021029312 | 0.02759576 | 1 |
| HTN | b_cell | *PINLYP* | -4.040100691 | 0.002708958 | 0.00520196 | 1 |
| HTN | b_cell | *C19orf81* | -15.05984198 | 0.015508616 | 0.021081658 | 1 |
| HTN | b_cell | *ZSCAN31* | -4.890436947 | 0.007281107 | 0.011106187 | 1 |
| HTN | b_cell | *LINC00957* | -12.78041482 | 0.000647444 | 0.002320684 | 1 |
| HTN | b_cell | *LINC00484* | -2.660528964 | 0.014136014 | 0.019485446 | 1 |
| HTN | b_cell | *KIAA0040* | 6.201469843 | 0.006107328 | 0.009679044 | 1 |
| HTN | b_cell | *B3GALT4* | 7.974022811 | 0.027103762 | 0.034648506 | 1 |
| HTN | b_cell | *LINC00941* | 3.281399168 | 0.000217941 | 0.002180344 | 1 |
| HTN | b_cell | *ZBTB22* | 2.794482268 | 9.51E-05 | 0.002180344 | 0.598133057 |
| HTN | b_cell | *B3GNT9* | -11.59769601 | 0.00015558 | 0.002180344 | 0.978134182 |
| HTN | b_cell | *CDKN2AIPNL* | -5.127959353 | 0.000181743 | 0.002180344 | 1 |
| HTN | b_cell | *GS1-124K5.4* | 2.380044972 | 0.009909283 | 0.014338242 | 1 |
| HTN | b_cell | *LINC00959* | -7.119115723 | 0.003118266 | 0.005757574 | 1 |

| | | | | | | |
|---|---|---|---|---|---|---|
| HTN | b_cell | *HLA-DQA2* | 3.758952055 | 4.44E-07 | 6.93E-05 | 0.002790378 |
| HTN | b_cell | *AC073257.2* | -50.56124963 | 0.000681264 | 0.002339217 | 1 |
| HTN | b_cell | *IRGM* | 2.197360625 | 0.000209974 | 0.002180344 | 1 |
| HTN | b_cell | *LRRC37A2* | -5.130451853 | 0.006367843 | 0.009983499 | 1 |
| HTN | b_cell | *TXNDC5* | -14.87178514 | 0.000892645 | 0.002599737 | 1 |
| HTN | b_cell | *ASB14* | -6.431058154 | 0.001784037 | 0.003891826 | 1 |
| HTN | b_cell | *NME1* | -3.117207959 | 0.024831022 | 0.032003411 | 1 |
| HTN | b_cell | *TNFSF12* | -8.637854717 | 0.00017242 | 0.002180344 | 1 |
| HTN | b_cell | *CDRT4* | -9.427707629 | 0.003392466 | 0.006120066 | 1 |
| HTN | b_cell | *LILRA4* | 29.26423375 | 0.000204265 | 0.002180344 | 1 |
| HTN | b_cell | *PSMB9* | -5.757924039 | 0.007365883 | 0.011210192 | 1 |
| HTN | b_cell | *COX19* | -6.037839352 | 0.004332705 | 0.007382036 | 1 |
| HTN | b_cell | *AQP1* | -4.214504853 | 0.000643568 | 0.002316643 | 1 |
| HTN | b_cell | *C1QTNF9* | -24.13468936 | 0.000394841 | 0.002180344 | 1 |
| HTN | b_cell | *ARHGEF25* | 11.01654598 | 0.00278623 | 0.00531302 | 1 |
| HTN | b_cell | *NSUN6* | -4.734966393 | 0.005622418 | 0.009070604 | 1 |
| HTN | b_cell | *HLA-DOB* | 0.952883419 | 0.000250402 | 0.002180344 | 1 |
| HTN | b_cell | *TDGF1* | -9.614727403 | 0.006551661 | 0.010218381 | 1 |
| HTN | b_cell | *CD302* | -6.566973552 | 0.005321565 | 0.008678775 | 1 |
| HTN | b_cell | *ATP5J2* | -4.595267094 | 0.001745263 | 0.003841901 | 1 |
| HTN | b_cell | *ARHGAP8* | -9.093564898 | 0.000544118 | 0.002214137 | 1 |
| HTN | b_cell | *ATP5O* | 5.201655682 | 0.001827523 | 0.003952403 | 1 |
| HTN | b_cell | *PLEKHO2* | -8.226807446 | 0.001366363 | 0.003286059 | 1 |
| HTN | b_cell | *PWP2* | -25.12348874 | 0.000195344 | 0.002180344 | 1 |
| HTN | b_cell | *PI4KA* | -4.915187667 | 0.002108125 | 0.004338389 | 1 |
| HTN | b_cell | *AKAP2* | -16.93930347 | 0.00042107 | 0.002180344 | 1 |
| HTN | b_cell | *LINC00969* | -4.048894286 | 0.003106128 | 0.00573938 | 1 |
| HTN | b_cell | *AMACR* | -2.297808098 | 0.036654147 | 0.045443624 | 1 |
| HTN | b_cell | *ARHGDIG* | -9.828985848 | 3.68E-05 | 0.001473442 | 0.231330434 |
| HTN | b_cell | *TCP10L* | -37.00214801 | 0.003624981 | 0.006434289 | 1 |
| HTN | b_cell | *ARFGAP3* | -5.329450721 | 0.00469842 | 0.007862295 | 1 |
| HTN | b_cell | *EIF6* | -4.738685129 | 0.000450911 | 0.002180344 | 1 |
| HTN | b_cell | *MRPL20* | -5.177139544 | 0.007697583 | 0.011616587 | 1 |
| HTN | b_cell | *LINC00960* | -5.320466452 | 0.000949745 | 0.002680223 | 1 |
| HTN | b_cell | *AP5Z1* | -4.013299479 | 0.000616416 | 0.002286342 | 1 |
| HTN | b_cell | *ZNF709* | -18.98637037 | 0.000655141 | 0.00232636 | 1 |
| HTN | b_cell | *STON1* | -3.849968511 | 0.001808943 | 0.003927449 | 1 |
| HTN | b_cell | *VSIG8* | -10.20451972 | 0.038630802 | 0.047724868 | 1 |
| HTN | b_cell | *C7orf73* | -5.421559781 | 0.008760019 | 0.012940376 | 1 |
| HTN | b_cell | *FGF14-IT1* | -14.57793395 | 0.000410609 | 0.002180344 | 1 |
| HTN | b_cell | *TICAM2* | -15.06450351 | 0.001897027 | 0.004059432 | 1 |
| HTN | b_cell | *PALM2* | -3.099396411 | 0.001027954 | 0.002792138 | 1 |
| HTN | b_cell | *NAT6* | -5.946396698 | 0.002996535 | 0.005593591 | 1 |
| HTN | b_cell | *UPK3B* | -9.163560006 | 0.013332994 | 0.018528853 | 1 |
| HTN | b_cell | *IL10RB* | -6.110946936 | 0.009596973 | 0.013944111 | 1 |
| HTN | b_cell | *WDR92* | -5.883592728 | 0.009492385 | 0.013827299 | 1 |
| HTN | b_cell | *PLA2G4B* | 7.981949365 | 0.0057082 | 0.009180724 | 1 |
| HTN | b_cell | *TTC4* | -2.544024895 | 0.026702827 | 0.034184621 | 1 |
| HTN | b_cell | *JMJD7* | 8.457014123 | 8.25E-07 | 0.000110372 | 0.005187481 |
| HTN | b_cell | *APOBEC3D* | 5.393321695 | 0.000265527 | 0.002180344 | 1 |

| | | | | | | |
|---|---|---|---|---|---|---|
| HTN | b_cell | *RN7SL832P* | -6.174201756 | 0.012042589 | 0.017013878 | 1 |
| HTN | b_cell | *LINC01011* | -7.502781421 | 0.001404097 | 0.003342783 | 1 |
| HTN | b_cell | *TMEM199* | -5.934446886 | 0.00021547 | 0.002180344 | 1 |
| HTN | b_cell | *P2RY11* | 5.843042387 | 0.001563716 | 0.003571044 | 1 |
| HTN | b_cell | *DBNDD2* | -11.35227152 | 0.031456111 | 0.039592506 | 1 |
| HTN | b_cell | *LILRA6* | -3.772451501 | 0.006669491 | 0.010355912 | 1 |
| HTN | b_cell | *APOBEC3C* | 9.417249179 | 0.000465272 | 0.002180489 | 1 |
| HTN | b_cell | *CCDC13* | -5.768160523 | 0.000127249 | 0.002180344 | 0.80001493 |
| HTN | b_cell | *ASPRV1* | -18.2493722 | 0.000274245 | 0.002180344 | 1 |
| HTN | b_cell | *OR2AE1* | 6.481531798 | 0.000318867 | 0.002180344 | 1 |
| HTN | b_cell | *CRYBB2* | -10.81166471 | 0.027917781 | 0.035566178 | 1 |
| HTN | b_cell | *N4BP2L2* | -3.691071013 | 0.005977586 | 0.009506977 | 1 |
| HTN | b_cell | *LINC00847* | 5.362244843 | 0.000451187 | 0.002180344 | 1 |
| HTN | b_cell | *NEAT1* | -2.508945002 | 0.001185638 | 0.003022492 | 1 |
| HTN | b_cell | *ZNF585B* | -5.368137229 | 0.000246226 | 0.002180344 | 1 |
| HTN | b_cell | *RAB30-AS1* | -5.248086031 | 0.001955249 | 0.004138736 | 1 |
| HTN | b_cell | *LINC00920* | 10.54879822 | 0.023861151 | 0.030899085 | 1 |
| HTN | b_cell | *UBAP1L* | -7.023960839 | 0.009222618 | 0.01350946 | 1 |
| HTN | b_cell | *PGAM5* | 12.69159708 | 0.002178556 | 0.004433246 | 1 |
| HTN | b_cell | *DNAJC3-AS1* | -6.033369828 | 0.001400355 | 0.003336125 | 1 |
| HTN | b_cell | *LINC00926* | 0.573450165 | 0.000226955 | 0.002180344 | 1 |
| HTN | b_cell | *BCKDHA* | -13.90125372 | 0.000968097 | 0.002711104 | 1 |
| HTN | b_cell | *CDK11B* | -6.195475985 | 0.014977818 | 0.020479674 | 1 |
| HTN | b_cell | *PCDHAC1* | 6.198756033 | 0.000481365 | 0.002185085 | 1 |
| HTN | b_cell | *ABHD14A* | -4.390986656 | 0.000397893 | 0.002180344 | 1 |
| HTN | b_cell | *SRP14-AS1* | -3.584640553 | 0.000474293 | 0.002180489 | 1 |
| HTN | b_cell | *CCDC153* | -8.287318445 | 6.62E-05 | 0.002030776 | 0.416309083 |
| HTN | b_cell | *C5orf17* | -4.003308616 | 0.000507894 | 0.002195202 | 1 |
| HTN | b_cell | *FMN1* | -2.514381805 | 0.000508246 | 0.002195202 | 1 |
| HTN | b_cell | *LINC01060* | -3.973148708 | 0.03152741 | 0.039666366 | 1 |
| HTN | b_cell | *NAIP* | -5.51011443 | 0.000714899 | 0.002383123 | 1 |
| HTN | b_cell | *ZNF286B* | 12.96824006 | 0.002818392 | 0.005363296 | 1 |
| HTN | b_cell | *LINC00942* | -15.44140083 | 0.012842894 | 0.017950929 | 1 |
| HTN | b_cell | *PDCD6* | -5.304376149 | 0.026165885 | 0.033545049 | 1 |
| HTN | b_cell | *LINC00603* | -6.025759918 | 0.001574905 | 0.003581136 | 1 |
| HTN | b_cell | *YJEFN3* | -5.285565856 | 0.000856451 | 0.002551796 | 1 |
| HTN | b_cell | *DNAH10OS* | -36.59262707 | 0.000344972 | 0.002180344 | 1 |
| HTN | b_cell | *PCDHA10* | -10.44572202 | 0.001389065 | 0.003320045 | 1 |
| HTN | b_cell | *ZNF718* | -5.389959578 | 0.007634771 | 0.01154952 | 1 |
| HTN | b_cell | *LINC00989* | -8.692223352 | 0.006866422 | 0.01059887 | 1 |
| HTN | b_cell | *LINC01021* | -10.86633266 | 0.000408978 | 0.002180344 | 1 |
| HTN | b_cell | *FAM218A* | -10.09743147 | 0.000413917 | 0.002180344 | 1 |
| HTN | b_cell | *GLI4* | -4.54601011 | 0.000526723 | 0.002195202 | 1 |
| HTN | b_cell | *SNHG18* | -5.806927346 | 0.00020403 | 0.002180344 | 1 |
| HTN | b_cell | *ZNF345* | -4.812180981 | 0.003796939 | 0.006680615 | 1 |
| HTN | b_cell | *SHANK3* | -2.856321266 | 0.000422341 | 0.002180344 | 1 |
| HTN | b_cell | *CCDC71L* | -3.90174876 | 0.000430725 | 0.002180344 | 1 |
| HTN | b_cell | *TMEM200B* | 6.756168658 | 0.004380201 | 0.007438769 | 1 |
| HTN | b_cell | *PCDHGB6* | -5.050316931 | 0.001493743 | 0.003473811 | 1 |
| HTN | b_cell | *TRNP1* | 3.950689985 | 0.000232047 | 0.002180344 | 1 |

| | | | | | | |
|---|---|---|---|---|---|---|
| HTN | b_cell | *PCDHGA5* | -5.692370363 | 0.000283885 | 0.002180344 | 1 |
| HTN | b_cell | *ATXN7L3B* | -5.141025672 | 0.000904538 | 0.002609834 | 1 |
| HTN | b_cell | *PRKDC* | -4.544246561 | 0.006213785 | 0.009805739 | 1 |
| HTN | b_cell | *PCDHGA6* | -4.14530547 | 0.007269542 | 0.011095802 | 1 |
| HTN | b_cell | *PCDHGA11* | -8.217521954 | 0.003127331 | 0.005770922 | 1 |
| HTN | b_cell | *CLDN23* | -8.842634446 | 0.004255517 | 0.007278139 | 1 |
| HTN | b_cell | *ZNF260* | -6.647174626 | 0.010749882 | 0.015416241 | 1 |
| HTN | b_cell | *SIGLEC14* | -4.402222382 | 0.03836248 | 0.047449323 | 1 |
| HTN | b_cell | *ARMS2* | -16.12635733 | 0.00217498 | 0.004428141 | 1 |
| HTN | b_cell | *FPGT* | -8.071902836 | 0.004195744 | 0.007201376 | 1 |
| HTN | b_cell | *MPV17L2* | -7.302477476 | 0.010227398 | 0.014761168 | 1 |
| HTN | b_cell | *DPP3* | -4.380058353 | 0.002233188 | 0.004512508 | 1 |
| HTN | b_cell | *EID3* | -4.915792183 | 0.000251769 | 0.002180344 | 1 |
| HTN | b_cell | *SNHG9* | -3.761164275 | 0.014402265 | 0.01979603 | 1 |
| HTN | b_cell | *EBLN2* | -7.314213078 | 0.00064914 | 0.002323108 | 1 |
| HTN | b_cell | *POLR2M* | -4.517822348 | 0.00045469 | 0.002180344 | 1 |
| HTN | b_cell | *TAS2R20* | 5.801718001 | 0.034793834 | 0.043359531 | 1 |
| HTN | b_cell | *LINC00346* | 11.61451616 | 0.000547134 | 0.002214137 | 1 |
| HTN | b_cell | *CTSO* | -6.500069832 | 0.016139911 | 0.021840642 | 1 |
| HTN | b_cell | *LINC00944* | -7.112874045 | 0.001173365 | 0.003004867 | 1 |
| HTN | b_cell | *ZNF10* | -13.83539416 | 4.75E-07 | 6.94E-05 | 0.002984306 |
| HTN | b_cell | *TAS2R31* | -9.206935399 | 0.00322196 | 0.005890118 | 1 |
| HTN | b_cell | *AC093668.2* | 4.957087528 | 0.000522149 | 0.002195202 | 1 |
| HTN | b_cell | *PGA5* | -66.62325097 | 0.002505467 | 0.004920922 | 1 |
| HTN | b_cell | *HP* | -20.81958179 | 2.51E-05 | 0.001169749 | 0.1579161 |
| HTN | b_cell | *KIAA1147* | -4.088429445 | 0.001071729 | 0.002849031 | 1 |
| HTN | b_cell | *ZBED6* | -11.49414727 | 0.000413111 | 0.002180344 | 1 |
| HTN | b_cell | *FNTB* | -6.197038807 | 0.000407697 | 0.002180344 | 1 |
| HTN | b_cell | *GALNT4* | -52.61929524 | 0.005952604 | 0.009484041 | 1 |
| HTN | b_cell | *OVCH1-AS1* | -4.966534936 | 0.000513641 | 0.002195202 | 1 |
| HTN | b_cell | *CUX1* | -3.492861275 | 0.018381134 | 0.024497496 | 1 |
| HTN | b_cell | *TEN1* | 10.32921931 | 0.000856378 | 0.002551796 | 1 |
| HTN | b_cell | *CHURC1* | -4.498145283 | 0.000977577 | 0.002717425 | 1 |
| HTN | b_cell | *C17orf49* | -18.64886315 | 0.001705058 | 0.003778547 | 1 |
| HTN | b_cell | *RTEL1* | 12.77425854 | 0.015822255 | 0.021470865 | 1 |
| HTN | b_cell | *ZNF578* | -6.383318512 | 0.000664204 | 0.002327958 | 1 |
| HTN | b_cell | *LINC00638* | -21.56472889 | 0.00017395 | 0.002180344 | 1 |
| HTN | b_cell | *CEP95* | -4.781491171 | 0.000986137 | 0.002724009 | 1 |
| HTN | b_cell | *LINC00639* | -2.918532891 | 0.000341028 | 0.002180344 | 1 |
| HTN | b_cell | *LINC00648* | -3.99244095 | 0.000396778 | 0.002180344 | 1 |
| HTN | b_cell | *LINC00924* | 3.615769716 | 0.000363759 | 0.002180344 | 1 |
| HTN | b_cell | *ITGB3* | -6.242542368 | 0.001001415 | 0.002755316 | 1 |
| HTN | b_cell | *UBE2Q2L* | 16.18786691 | 0.029307339 | 0.037168697 | 1 |
| HTN | b_cell | *IQCH-AS1* | -5.253868064 | 0.002505082 | 0.004920922 | 1 |
| HTN | b_cell | *AC083843.1* | -108.9259378 | 0.000172015 | 0.002180344 | 1 |
| HTN | b_cell | *SH3RF3-AS1* | -12.35118263 | 7.32E-05 | 0.002098808 | 0.460054915 |
| HTN | b_cell | *PWRN1* | -5.293401054 | 0.00037636 | 0.002180344 | 1 |
| HTN | b_cell | *HOXB7* | -5.591306421 | 0.000652952 | 0.002324524 | 1 |
| HTN | b_cell | *FRRS1L* | -12.23227332 | 0.000620615 | 0.002291037 | 1 |
| HTN | b_cell | *LINC00562* | -4.409914141 | 0.000698677 | 0.002364146 | 1 |

| | | | | | | |
|---|---|---|---|---|---|---|
| HTN | b_cell | *AC009120.5* | -8.187148759 | 0.00052677 | 0.002195202 | 1 |
| HTN | b_cell | *LINC00565* | -52.70286568 | 0.001551485 | 0.003553437 | 1 |
| HTN | b_cell | *TMEM178B* | -3.690194472 | 0.000333563 | 0.002180344 | 1 |
| HTN | b_cell | *C15orf65* | -5.570475278 | 0.002180734 | 0.004434385 | 1 |
| HTN | b_cell | *HPR* | -14.5904649 | 0.020093383 | 0.026472569 | 1 |
| HTN | b_cell | *AC123768.4* | 24.47281962 | 0.003228597 | 0.005898921 | 1 |
| HTN | b_cell | *LINC00667* | -3.299439241 | 0.008269428 | 0.012302388 | 1 |
| HTN | b_cell | *LINC00672* | -5.03382217 | 0.001582573 | 0.003593224 | 1 |
| HTN | b_cell | *LINC00909* | -5.122559258 | 0.001175333 | 0.003007455 | 1 |
| HTN | b_cell | *LINC00526* | -7.273727438 | 0.001308747 | 0.003205335 | 1 |
| HTN | b_cell | *LINC00668* | 40.50607016 | 0.001733945 | 0.003830398 | 1 |
| HTN | b_cell | *STRADA* | -4.664391978 | 0.000118822 | 0.002180344 | 0.747035696 |
| HTN | b_cell | *MYO15B* | -4.890725609 | 0.005980794 | 0.009509675 | 1 |
| HTN | b_cell | *LINC00663* | -2.070950987 | 0.020875671 | 0.027399863 | 1 |
| HTN | b_cell | *ILF3-AS1* | -4.42672489 | 0.015267894 | 0.020803912 | 1 |
| HTN | b_cell | *AC139100.2* | 13.80775324 | 0.000257819 | 0.002180344 | 1 |
| HTN | b_cell | *LINC01140* | -9.275156955 | 0.002093473 | 0.004320628 | 1 |
| HTN | b_cell | *LINC00906* | -40.08051066 | 0.000407447 | 0.002180344 | 1 |
| HTN | b_cell | *ZNF582-AS1* | -7.448598935 | 0.000308725 | 0.002180344 | 1 |
| HTN | b_cell | *AC137932.1* | -13.04021526 | 0.03901522 | 0.048161926 | 1 |
| HTN | b_cell | *AC003005.2* | -11.45665025 | 0.001814225 | 0.003934658 | 1 |
| HTN | b_cell | *AL132989.1* | 20.81973522 | 0.000985 | 0.002724009 | 1 |
| HTN | b_cell | *LINC00664* | -22.06111232 | 0.000100625 | 0.002180344 | 0.632626424 |
| HTN | b_cell | *FBXO17* | -3.041104759 | 0.000462811 | 0.002180489 | 1 |
| HTN | b_cell | *SNHG8* | -7.362154797 | 0.001277855 | 0.003156737 | 1 |
| HTN | b_cell | *HCG17* | -4.708291788 | 0.008306532 | 0.012348822 | 1 |
| HTN | b_cell | *YTHDF3-AS1* | 7.841888981 | 0.000191728 | 0.002180344 | 1 |
| HTN | b_cell | *LINC01050* | 6.062676562 | 0.000247268 | 0.002180344 | 1 |
| HTN | b_cell | *SRXN1* | -73.71852679 | 0.002575646 | 0.005021112 | 1 |
| HTN | b_cell | *GTF2H5* | -3.8766852 | 0.000491682 | 0.002192343 | 1 |
| HTN | b_cell | *FGF14-AS2* | -6.718874314 | 0.001398624 | 0.003334528 | 1 |
| HTN | b_cell | *CASC15* | -2.939114008 | 0.001321778 | 0.003220937 | 1 |
| HTN | b_cell | *LINC01023* | -6.378454179 | 0.000741106 | 0.002405521 | 1 |
| HTN | b_cell | *DOC2B* | -4.564862088 | 0.000288771 | 0.002180344 | 1 |
| HTN | b_cell | *POLR2J4* | -5.691701635 | 0.000589069 | 0.002263931 | 1 |
| HTN | b_cell | *PAXIP1-AS1* | -6.861369209 | 0.012758733 | 0.017861089 | 1 |
| HTN | dendritic_cell | *SEMA3F* | -1.945533384 | 8.38E-06 | 0.001646402 | 0.052684871 |
| HTN | dendritic_cell | *RBM6* | -2.06989231 | 2.28E-06 | 0.000596223 | 0.014309351 |
| HTN | dendritic_cell | *HFE* | -3.79444129 | 0.000164143 | 0.014726983 | 1 |
| HTN | dendritic_cell | *NR1H3* | -6.718104038 | 5.15E-06 | 0.001078743 | 0.032362299 |
| HTN | dendritic_cell | *ITIH4* | 1.697094507 | 0.000494962 | 0.033460528 | 1 |
| HTN | dendritic_cell | *PIGV* | -3.451778127 | 0.000349453 | 0.024778381 | 1 |
| HTN | dendritic_cell | *UHRF1BP1* | 2.129911402 | 0.000632506 | 0.040167306 | 1 |
| HTN | dendritic_cell | *TIE1* | -1.722668473 | 0.000698626 | 0.041436425 | 1 |
| HTN | dendritic_cell | *POLR1A* | -2.056516749 | 0.000160239 | 0.014600298 | 1 |
| HTN | dendritic_cell | *SNX10* | -1.629893132 | 7.18E-07 | 0.000265679 | 0.004516547 |
| HTN | dendritic_cell | *AAMDC* | -2.185483496 | 8.72E-05 | 0.009617671 | 0.548207237 |
| HTN | dendritic_cell | *C3orf18* | 6.944661385 | 0.000183608 | 0.015391226 | 1 |
| HTN | dendritic_cell | *DNTTIP1* | -3.633379241 | 0.000284579 | 0.021299366 | 1 |
| HTN | dendritic_cell | *POLI* | -1.353537768 | 5.41E-05 | 0.006803456 | 0.340172794 |

| | | | | | | |
|---|---|---|---|---|---|---|
| HTN | dendritic_cell | *ICAM5* | -23.20651316 | 0.000170751 | 0.014726983 | 1 |
| HTN | dendritic_cell | *RAD51C* | -3.272441804 | 6.98E-05 | 0.008441908 | 0.438979235 |
| HTN | dendritic_cell | *CAND1* | -3.877536753 | 0.000291246 | 0.021541947 | 1 |
| HTN | dendritic_cell | *DNAJC27* | -6.494523696 | 5.12E-05 | 0.006568202 | 0.3218419 |
| HTN | dendritic_cell | *EPHX2* | -4.391507134 | 0.000140965 | 0.013847621 | 0.886247719 |
| HTN | dendritic_cell | *SNRPC* | -5.414916997 | 6.00E-07 | 0.000235612 | 0.003769786 |
| HTN | dendritic_cell | *HIST1H1D* | 5.319601557 | 9.33E-07 | 0.000300403 | 0.005867712 |
| HTN | dendritic_cell | *ZNF391* | -1.608141985 | 2.10E-05 | 0.003158144 | 0.132190876 |
| HTN | dendritic_cell | *TRAP1* | 1.507699015 | 0.000459891 | 0.031772909 | 1 |
| HTN | dendritic_cell | *ATP8B3* | -2.051684173 | 0.000657273 | 0.040199503 | 1 |
| HTN | dendritic_cell | *PDLIM4* | 7.676076208 | 1.66E-05 | 0.002679611 | 0.104306914 |
| HTN | dendritic_cell | *WNT2B* | -5.675870477 | 0.000350768 | 0.024778381 | 1 |
| HTN | dendritic_cell | *ACP2* | 14.32073856 | 1.16E-16 | 7.26E-13 | 7.26E-13 |
| HTN | dendritic_cell | *AGT* | 1.625205041 | 0.000736301 | 0.043262817 | 1 |
| HTN | dendritic_cell | *NMT1* | 2.854807756 | 8.17E-05 | 0.009517665 | 0.513953899 |
| HTN | dendritic_cell | *FBN2* | 1.151123442 | 0.00021088 | 0.016997474 | 1 |
| HTN | dendritic_cell | *SCRN2* | -1.456419447 | 9.86E-05 | 0.010509169 | 0.620040998 |
| HTN | dendritic_cell | *C18orf8* | -5.518953114 | 0.000612873 | 0.039317699 | 1 |
| HTN | dendritic_cell | *RGS5* | -5.199403784 | 0.000264151 | 0.020008611 | 1 |
| HTN | dendritic_cell | *CREB3L4* | -2.238715273 | 0.000807973 | 0.044558977 | 1 |
| HTN | dendritic_cell | *SLC10A4* | 10.76792777 | 0.000658589 | 0.040199503 | 1 |
| HTN | dendritic_cell | *ABT1* | 4.240555736 | 0.00089048 | 0.048682149 | 1 |
| HTN | dendritic_cell | *SLC18B1* | -1.99464897 | 0.000152487 | 0.014203946 | 0.958683209 |
| HTN | dendritic_cell | *NRBF2* | -3.294567781 | 3.01E-05 | 0.004021492 | 0.189010133 |
| HTN | dendritic_cell | *PDCD11* | 5.713145581 | 0.000240867 | 0.018695452 | 1 |
| HTN | dendritic_cell | *HSD17B12* | -3.770140265 | 3.79E-08 | 2.38E-05 | 0.000238078 |
| HTN | dendritic_cell | *DUSP15* | -6.565312426 | 2.77E-05 | 0.003789261 | 0.174305988 |
| HTN | dendritic_cell | *KCTD14* | -16.93674602 | 0.000776233 | 0.044318466 | 1 |
| HTN | dendritic_cell | *NEIL2* | 2.952010379 | 0.000680508 | 0.040857162 | 1 |
| HTN | dendritic_cell | *NDUFAF6* | -3.440887602 | 0.00052844 | 0.035343652 | 1 |
| HTN | dendritic_cell | *SLC5A11* | -2.129009813 | 9.37E-07 | 0.000300403 | 0.005893055 |
| HTN | dendritic_cell | *S100B* | -1.518354968 | 2.17E-05 | 0.003158144 | 0.136728259 |
| HTN | dendritic_cell | *TNFSF13* | 8.050165948 | 1.97E-05 | 0.00310181 | 0.124072401 |
| HTN | dendritic_cell | *LMOD1* | -5.909625147 | 1.33E-08 | 1.19E-05 | 8.35E-05 |
| HTN | dendritic_cell | *RFT1* | -5.825585154 | 0.000796562 | 0.044318466 | 1 |
| HTN | dendritic_cell | *EXO5* | -3.458954357 | 0.000391747 | 0.027365696 | 1 |
| HTN | dendritic_cell | *CDC25A* | -5.455944645 | 3.52E-10 | 7.37E-07 | 2.21E-06 |
| HTN | dendritic_cell | *ZNF589* | -4.087038569 | 1.60E-09 | 2.51E-06 | 1.00E-05 |
| HTN | dendritic_cell | *APEH* | -5.292634178 | 8.34E-08 | 4.37E-05 | 0.00052435 |
| HTN | dendritic_cell | *RNF123* | 5.025967379 | 5.42E-09 | 5.68E-06 | 3.41E-05 |
| HTN | dendritic_cell | *ERAP1* | -4.326232893 | 4.54E-06 | 0.000985263 | 0.02857262 |
| HTN | dendritic_cell | *TRMT61A* | -7.425559326 | 3.72E-06 | 0.000934301 | 0.023357524 |
| HTN | dendritic_cell | *TRIM66* | -1.359178654 | 8.98E-06 | 0.001659866 | 0.056435446 |
| HTN | dendritic_cell | *RIMKLB* | -2.540188389 | 8.30E-06 | 0.001646402 | 0.052194998 |
| HTN | dendritic_cell | *KIR3DL1* | 6.44209618 | 8.57E-05 | 0.009617671 | 0.539096055 |
| HTN | dendritic_cell | *IRS1* | -1.718592881 | 1.89E-07 | 9.12E-05 | 0.001185892 |
| HTN | dendritic_cell | *INO80E* | -5.438043417 | 4.11E-06 | 0.000953582 | 0.025834437 |
| HTN | dendritic_cell | *CALML6* | -7.084359042 | 2.45E-05 | 0.00342847 | 0.154281138 |
| HTN | dendritic_cell | *ELP5* | -6.832522379 | 9.02E-05 | 0.009780172 | 0.567249995 |
| HTN | dendritic_cell | *NME6* | 5.279675495 | 2.20E-08 | 1.73E-05 | 0.000138231 |

| | | | | | | |
|---|---|---|---|---|---|---|
| HTN | dendritic_cell | *CTSW* | -1.073485222 | 3.52E-05 | 0.004606152 | 0.221095276 |
| HTN | dendritic_cell | *HOXC5* | 5.338659006 | 0.000657538 | 0.040199503 | 1 |
| HTN | dendritic_cell | *DCAKD* | -2.388121277 | 0.000221825 | 0.017432701 | 1 |
| HTN | dendritic_cell | *VEGFB* | -3.952770698 | 3.59E-07 | 0.000150402 | 0.002256033 |
| HTN | dendritic_cell | *MST1* | -3.141722285 | 1.76E-06 | 0.000480703 | 0.01105616 |
| HTN | dendritic_cell | *SNX31* | -6.490951923 | 1.66E-05 | 0.002679611 | 0.104504844 |
| HTN | dendritic_cell | *GLIS1* | -2.124723072 | 0.000216391 | 0.017220891 | 1 |
| HTN | dendritic_cell | *KLF14* | -17.89167647 | 0.000300438 | 0.021963414 | 1 |
| HTN | dendritic_cell | *RSRC1* | -3.820483571 | 0.000133815 | 0.013353917 | 0.841296782 |
| HTN | dendritic_cell | *NRIP3* | -3.943726831 | 8.65E-05 | 0.009617671 | 0.544055937 |
| HTN | dendritic_cell | *MTHFR* | -8.737387916 | 2.20E-05 | 0.003158144 | 0.138572109 |
| HTN | dendritic_cell | *PCDHB9* | 3.541856988 | 0.000612037 | 0.039317699 | 1 |
| HTN | dendritic_cell | *SH2B1* | -2.52301946 | 0.000197037 | 0.016228045 | 1 |
| HTN | dendritic_cell | *RPP25* | 5.283779088 | 3.90E-09 | 4.90E-06 | 2.45E-05 |
| HTN | dendritic_cell | *MPI* | -2.368974687 | 0.000746352 | 0.043447334 | 1 |
| HTN | dendritic_cell | *TRIM73* | -7.669858717 | 0.000148464 | 0.014142289 | 0.933391065 |
| HTN | dendritic_cell | *HYI* | -3.327788284 | 3.12E-07 | 0.000140016 | 0.001960224 |
| HTN | dendritic_cell | *CTC1* | 4.207752781 | 0.000767481 | 0.044267444 | 1 |
| HTN | dendritic_cell | *DEFB124* | -7.245736271 | 0.000788007 | 0.044318466 | 1 |
| HTN | dendritic_cell | *CLDN7* | -6.302130603 | 4.29E-08 | 2.45E-05 | 0.000270008 |
| HTN | dendritic_cell | *CLN8* | 6.682054833 | 0.000198753 | 0.016228045 | 1 |
| HTN | dendritic_cell | *ZNF320* | 1.819368434 | 0.000182303 | 0.015391226 | 1 |
| HTN | dendritic_cell | *KREMEN1* | -1.511027902 | 0.000153629 | 0.014203946 | 0.965868361 |
| HTN | dendritic_cell | *EIF3C* | 4.484011515 | 1.28E-06 | 0.000364731 | 0.008024077 |
| HTN | dendritic_cell | *A3GALT2* | -2.909636933 | 0.000341986 | 0.024713375 | 1 |
| HTN | dendritic_cell | *CDK10* | -4.183811849 | 1.25E-05 | 0.002175097 | 0.078303484 |
| HTN | dendritic_cell | *ARL17A* | -5.31541285 | 0.000170999 | 0.014726983 | 1 |
| HTN | dendritic_cell | *PLEKHN1* | 4.4519718 | 0.000682361 | 0.040857162 | 1 |
| HTN | dendritic_cell | *FBF1* | -2.63378873 | 1.14E-06 | 0.000342232 | 0.007186871 |
| HTN | dendritic_cell | *NOC2L* | -2.596489223 | 0.000566813 | 0.037120341 | 1 |
| HTN | dendritic_cell | *SULT1A1* | -3.040208833 | 0.000144756 | 0.014001256 | 0.910081632 |
| HTN | dendritic_cell | *FAM180B* | -3.136243793 | 4.25E-06 | 0.000953582 | 0.026700301 |
| HTN | dendritic_cell | *HIST1H3E* | 5.955089063 | 2.62E-08 | 1.83E-05 | 0.000164557 |
| HTN | dendritic_cell | *ZNF100* | -3.935480518 | 6.09E-05 | 0.007510357 | 0.383028191 |
| HTN | dendritic_cell | *SLC22A5* | 2.77524627 | 2.21E-05 | 0.003158144 | 0.138958326 |
| HTN | dendritic_cell | *RPS26* | -2.244625873 | 8.81E-06 | 0.001659866 | 0.055409359 |
| HTN | dendritic_cell | *TMEM116* | -3.902772015 | 2.14E-10 | 6.72E-07 | 1.34E-06 |
| HTN | dendritic_cell | *DENND4B* | -6.558886221 | 0.000467834 | 0.031970353 | 1 |
| HTN | dendritic_cell | *HSD17B8* | -5.433493824 | 9.56E-07 | 0.000300403 | 0.006008061 |
| HTN | dendritic_cell | *PRRT1* | 4.211743961 | 0.000122572 | 0.012633001 | 0.770613052 |
| HTN | dendritic_cell | *STK19* | 4.385180061 | 0.000246567 | 0.018904495 | 1 |
| HTN | dendritic_cell | *GPANK1* | 4.003399675 | 0.000536492 | 0.035504503 | 1 |
| HTN | dendritic_cell | *BAG6* | 5.318134593 | 8.17E-05 | 0.009517665 | 0.513616076 |
| HTN | dendritic_cell | *SIPA1* | 5.047750869 | 1.47E-05 | 0.002500517 | 0.092519127 |
| HTN | dendritic_cell | *ATF6B* | 5.093126283 | 4.20E-06 | 0.000953582 | 0.026411886 |
| HTN | dendritic_cell | *LTB4R* | -4.375490004 | 1.12E-05 | 0.002008489 | 0.070297128 |
| HTN | dendritic_cell | *LTB4R2* | -4.500107845 | 0.000167859 | 0.014726983 | 1 |
| HTN | dendritic_cell | *AL049840.1* | -4.007746407 | 0.000118489 | 0.012415631 | 0.744937849 |
| HTN | dendritic_cell | *PLA2G4B* | 3.301920964 | 0.000127738 | 0.012953095 | 0.803091916 |
| HTN | dendritic_cell | *ZNF10* | -3.825126051 | 0.000794242 | 0.044318466 | 1 |

| | | | | | | |
|---|---|---|---|---|---|---|
| HTN | dendritic_cell | *AC144652.1* | 2.017089691 | 0.000644155 | 0.040199503 | 1 |
| HTN | endothelial | *WNT16* | -0.957397166 | 0.001575762 | 0.045852374 | 1 |
| HTN | endothelial | *RBM6* | -0.288021887 | 0.001376473 | 0.04273724 | 1 |
| HTN | endothelial | *HFE* | -0.804511871 | 1.93E-05 | 0.002964758 | 0.121555064 |
| HTN | endothelial | *CEP68* | 0.636010953 | 9.51E-07 | 0.000259976 | 0.005979441 |
| HTN | endothelial | *LTF* | 0.37184504 | 0.001792022 | 0.049461154 | 1 |
| HTN | endothelial | *CCDC88C* | 0.454514766 | 0.001582625 | 0.045852374 | 1 |
| HTN | endothelial | *SDCCAG8* | 0.497589896 | 0.001361456 | 0.04273724 | 1 |
| HTN | endothelial | *PIGV* | -0.881845347 | 0.000102099 | 0.008843143 | 0.641897925 |
| HTN | endothelial | *HAGH* | 0.669876794 | 0.000332499 | 0.018177572 | 1 |
| HTN | endothelial | *UHRF1BP1* | 0.502779927 | 6.16E-05 | 0.006672015 | 0.386976853 |
| HTN | endothelial | *EML1* | 0.455928613 | 0.000541471 | 0.024977736 | 1 |
| HTN | endothelial | *FGFR3* | 0.216968116 | 0.001144691 | 0.039326095 | 1 |
| HTN | endothelial | *IFI35* | 0.675310753 | 0.000242739 | 0.015494955 | 1 |
| HTN | endothelial | *POLR1A* | -0.41993147 | 0.000211945 | 0.014012492 | 1 |
| HTN | endothelial | *NEDD4* | 0.357250472 | 0.001911993 | 0.049993244 | 1 |
| HTN | endothelial | *ST6GALNAC2* | 0.719951626 | 0.001027504 | 0.037909362 | 1 |
| HTN | endothelial | *CLNS1A* | 0.588501441 | 0.000827716 | 0.03232207 | 1 |
| HTN | endothelial | *ATP2A3* | 0.454371628 | 0.000164458 | 0.012164114 | 1 |
| HTN | endothelial | *TPD52* | 0.355904615 | 0.000823127 | 0.03232207 | 1 |
| HTN | endothelial | *SCAMP1* | 0.583775901 | 0.001369253 | 0.04273724 | 1 |
| HTN | endothelial | *SNX10* | -0.324621073 | 1.09E-07 | 4.70E-05 | 0.000685045 |
| HTN | endothelial | *SLC7A4* | -1.509861816 | 0.000108591 | 0.009102781 | 0.682708547 |
| HTN | endothelial | *FAM118A* | 0.551794291 | 0.000370387 | 0.019516802 | 1 |
| HTN | endothelial | *POLI* | -0.277791625 | 0.000256521 | 0.015967809 | 1 |
| HTN | endothelial | *MTHFSD* | 0.456957246 | 0.00180253 | 0.049461154 | 1 |
| HTN | endothelial | *ZFAND1* | 0.768743503 | 0.000862638 | 0.033272414 | 1 |
| HTN | endothelial | *LSR* | 0.307937315 | 0.001316546 | 0.042129838 | 1 |
| HTN | endothelial | *STEAP1B* | 0.28130504 | 0.001246842 | 0.041595028 | 1 |
| HTN | endothelial | *HOXA2* | -1.211692714 | 0.000602618 | 0.026366918 | 1 |
| HTN | endothelial | *CRHR2* | 2.952072326 | 9.71E-05 | 0.008665472 | 0.610331508 |
| HTN | endothelial | *MEST* | -0.362180944 | 0.000564974 | 0.025553888 | 1 |
| HTN | endothelial | *SLC1A1* | 0.36325522 | 0.000202516 | 0.013839302 | 1 |
| HTN | endothelial | *PDLIM1* | 0.357818271 | 0.001316172 | 0.042129838 | 1 |
| HTN | endothelial | *WNT3* | -1.234464795 | 0.00144124 | 0.043949851 | 1 |
| HTN | endothelial | *RAD51C* | -0.499256261 | 0.001152512 | 0.039379577 | 1 |
| HTN | endothelial | *ICAM2* | 0.193591299 | 0.001379936 | 0.04273724 | 1 |
| HTN | endothelial | *DDX5* | 0.462781786 | 0.001378243 | 0.04273724 | 1 |
| HTN | endothelial | *EFNB3* | -4.329359966 | 0.001184215 | 0.040027756 | 1 |
| HTN | endothelial | *KLHL5* | 0.465546574 | 0.000736759 | 0.030473704 | 1 |
| HTN | endothelial | *GUCA1B* | 1.317053168 | 0.001079797 | 0.038162287 | 1 |
| HTN | endothelial | *PCDHB6* | 1.491010774 | 0.000586993 | 0.026092832 | 1 |
| HTN | endothelial | *NPHP3* | 0.350745817 | 0.00116985 | 0.039755926 | 1 |
| HTN | endothelial | *FILIP1* | 0.299601104 | 0.00054429 | 0.024977736 | 1 |
| HTN | endothelial | *TRPM6* | 0.15714779 | 0.001464764 | 0.044019285 | 1 |
| HTN | endothelial | *SEC16B* | 1.113052063 | 0.001142198 | 0.039326095 | 1 |
| HTN | endothelial | *ZFP30* | 0.685129453 | 0.000585934 | 0.026092832 | 1 |
| HTN | endothelial | *TEX14* | 1.482008382 | 3.67E-06 | 0.000678119 | 0.023056054 |
| HTN | endothelial | *TRIM6* | -0.677750421 | 0.000524059 | 0.024405641 | 1 |
| HTN | endothelial | *NDUFAF4* | 0.521808176 | 0.001293206 | 0.042129838 | 1 |

| | | | | | | |
|---|---|---|---|---|---|---|
| HTN | endothelial | *FAM124B* | 0.196747429 | 0.000650399 | 0.027873805 | 1 |
| HTN | endothelial | *WFDC3* | -0.25621676 | 0.000559182 | 0.025475201 | 1 |
| HTN | endothelial | *BTN2A2* | 0.588376125 | 9.79E-05 | 0.008665472 | 0.61524851 |
| HTN | endothelial | *SNRPC* | -0.826639926 | 6.48E-05 | 0.006905808 | 0.407442688 |
| HTN | endothelial | *HIST1H1D* | 1.095843714 | 2.20E-05 | 0.003202294 | 0.138522536 |
| HTN | endothelial | *ZNF391* | -0.382490492 | 4.27E-06 | 0.00076768 | 0.026868806 |
| HTN | endothelial | *GRK4* | 0.317857621 | 0.000275648 | 0.016243271 | 1 |
| HTN | endothelial | *OPRL1* | -1.370463193 | 1.88E-08 | 1.18E-05 | 0.000117992 |
| HTN | endothelial | *GPR108* | 0.8480199 | 0.001073599 | 0.038162287 | 1 |
| HTN | endothelial | *XRCC3* | 0.854769452 | 0.000291923 | 0.016741209 | 1 |
| HTN | endothelial | *TRAP1* | 0.388787924 | 2.89E-05 | 0.003883897 | 0.181705452 |
| HTN | endothelial | *APOL5* | -2.000682068 | 6.69E-05 | 0.007011481 | 0.420688835 |
| HTN | endothelial | *RIPK3* | 0.786156037 | 0.000292911 | 0.016741209 | 1 |
| HTN | endothelial | *DOCK6* | 0.420564752 | 0.001064359 | 0.038162287 | 1 |
| HTN | endothelial | *ATP8B3* | -0.568503422 | 0.000112654 | 0.009277451 | 0.708256367 |
| HTN | endothelial | *STK33* | 0.206604459 | 0.001693084 | 0.047947822 | 1 |
| HTN | endothelial | *HAUS8* | -0.752103476 | 0.001080466 | 0.038162287 | 1 |
| HTN | endothelial | *PDLIM4* | 1.847435349 | 2.67E-07 | 8.85E-05 | 0.001681078 |
| HTN | endothelial | *MRPL35* | 0.5944846 | 0.00141566 | 0.04362871 | 1 |
| HTN | endothelial | *IMPA1* | 0.698282178 | 0.000252667 | 0.015885152 | 1 |
| HTN | endothelial | *ACP2* | 1.419421769 | 6.72E-08 | 3.25E-05 | 0.00042241 |
| HTN | endothelial | *AGT* | 0.314564434 | 0.001765666 | 0.049336644 | 1 |
| HTN | endothelial | *NACAD* | -1.434166528 | 0.001912875 | 0.049993244 | 1 |
| HTN | endothelial | *CIB2* | -0.695984755 | 0.001250431 | 0.041595028 | 1 |
| HTN | endothelial | *NMT1* | 0.751722913 | 5.54E-07 | 0.000173988 | 0.00348538 |
| HTN | endothelial | *BLK* | -1.387928454 | 5.80E-05 | 0.006392154 | 0.364352755 |
| HTN | endothelial | *LRRC8A* | 0.598596575 | 0.001020694 | 0.037909362 | 1 |
| HTN | endothelial | *TOR1B* | 0.774581168 | 0.001831462 | 0.049631055 | 1 |
| HTN | endothelial | *NRM* | 0.65919064 | 0.001357761 | 0.04273724 | 1 |
| HTN | endothelial | *SULF1* | 0.198699207 | 0.001924346 | 0.049993244 | 1 |
| HTN | endothelial | *ARL3* | 1.033380601 | 0.000219873 | 0.014250964 | 1 |
| HTN | endothelial | *TRPC3* | 1.574805967 | 0.000259756 | 0.016010657 | 1 |
| HTN | endothelial | *FBN2* | 0.331513496 | 2.24E-05 | 0.003202294 | 0.140900933 |
| HTN | endothelial | *ACVRL1* | 0.383508121 | 0.001447111 | 0.043949851 | 1 |
| HTN | endothelial | *GPR84* | -3.959143326 | 8.66E-05 | 0.008033078 | 0.544559919 |
| HTN | endothelial | *RASL10B* | -1.161259248 | 0.000771777 | 0.031304269 | 1 |
| HTN | endothelial | *SCRN2* | -0.286020762 | 0.00034275 | 0.018576486 | 1 |
| HTN | endothelial | *SLC14A1* | 0.830867143 | 0.000627025 | 0.027186953 | 1 |
| HTN | endothelial | *GUK1* | 0.681852457 | 0.001865426 | 0.049993244 | 1 |
| HTN | endothelial | *WNT9A* | 1.145989964 | 0.000195009 | 0.013558581 | 1 |
| HTN | endothelial | *GPR17* | 0.227585011 | 0.000436082 | 0.021745177 | 1 |
| HTN | endothelial | *GFOD1* | 0.263576487 | 0.000266442 | 0.01607007 | 1 |
| HTN | endothelial | *PSD2* | 2.863467993 | 0.000137976 | 0.010843176 | 0.867454096 |
| HTN | endothelial | *SLC18B1* | -0.343099293 | 0.001067121 | 0.038162287 | 1 |
| HTN | endothelial | *ADHFE1* | -0.266759661 | 0.000815606 | 0.03232207 | 1 |
| HTN | endothelial | *NRBF2* | -0.630705956 | 0.00010268 | 0.008843143 | 0.645549428 |
| HTN | endothelial | *PDCD11* | 1.327466287 | 3.02E-06 | 0.000593746 | 0.018999887 |
| HTN | endothelial | *GRIK4* | 0.239330707 | 0.001470344 | 0.044019285 | 1 |
| HTN | endothelial | *DOC2A* | -1.622400875 | 7.75E-05 | 0.007610188 | 0.487052047 |
| HTN | endothelial | *KCTD14* | -3.8605522 | 0.000124476 | 0.009994755 | 0.782581921 |

| | | | | | | |
|---|---|---|---|---|---|---|
| HTN | endothelial | *PTPN14* | 0.350216885 | 0.001064049 | 0.038162287 | 1 |
| HTN | endothelial | *PRDM8* | -0.52132136 | 0.000243996 | 0.015494955 | 1 |
| HTN | endothelial | *TMEM87B* | 0.495131811 | 0.001031096 | 0.037909362 | 1 |
| HTN | endothelial | *NEIL2* | 1.10411677 | 1.18E-07 | 4.70E-05 | 0.000739975 |
| HTN | endothelial | *AK9* | 0.687002836 | 0.000690458 | 0.028939413 | 1 |
| HTN | endothelial | *AGPAT5* | 0.624526174 | 0.001560485 | 0.045844722 | 1 |
| HTN | endothelial | *GOLGA7B* | -1.04717448 | 0.001721285 | 0.048311234 | 1 |
| HTN | endothelial | *ZNF276* | 0.965360802 | 0.000383084 | 0.019741385 | 1 |
| HTN | endothelial | *SLC5A11* | -0.473260101 | 3.14E-06 | 0.000598299 | 0.019743851 |
| HTN | endothelial | *S100B* | -0.316454312 | 4.46E-05 | 0.005195893 | 0.280578247 |
| HTN | endothelial | *FDPS* | 0.769296446 | 0.000651734 | 0.027873805 | 1 |
| HTN | endothelial | *PAQR6* | -1.008883091 | 0.000268388 | 0.01607007 | 1 |
| HTN | endothelial | *ABCF3* | 0.599342717 | 0.001298177 | 0.042129838 | 1 |
| HTN | endothelial | *TNFSF13* | 1.458089685 | 0.000153369 | 0.01177113 | 0.964233475 |
| HTN | endothelial | *VANGL2* | 1.071876547 | 0.000517787 | 0.024293506 | 1 |
| HTN | endothelial | *C1orf115* | 0.38918882 | 0.001311721 | 0.042129838 | 1 |
| HTN | endothelial | *LMOD1* | -0.626013078 | 2.68E-06 | 0.000543812 | 0.016858163 |
| HTN | endothelial | *EXO5* | -1.029674058 | 7.57E-05 | 0.007554854 | 0.475955803 |
| HTN | endothelial | *C1orf50* | -1.287900167 | 0.00015375 | 0.01177113 | 0.966628685 |
| HTN | endothelial | *SLC9B2* | 0.622174728 | 0.000752835 | 0.030935108 | 1 |
| HTN | endothelial | *CDC25A* | -1.189558979 | 1.89E-11 | 5.95E-08 | 1.19E-07 |
| HTN | endothelial | *ZNF589* | -0.830766706 | 1.74E-08 | 1.18E-05 | 0.000109654 |
| HTN | endothelial | *APEH* | -1.044469738 | 2.13E-07 | 7.43E-05 | 0.001337148 |
| HTN | endothelial | *RNF123* | 0.980692318 | 6.43E-09 | 5.78E-06 | 4.05E-05 |
| HTN | endothelial | *POC1A* | 1.222006202 | 0.001448078 | 0.043949851 | 1 |
| HTN | endothelial | *ENPP6* | 1.888836646 | 0.00106147 | 0.038162287 | 1 |
| HTN | endothelial | *ERAP2* | 0.506501155 | 0.000196251 | 0.013558581 | 1 |
| HTN | endothelial | *GPER1* | 0.553894378 | 0.000206114 | 0.01393375 | 1 |
| HTN | endothelial | *SPINT1* | -0.963930638 | 0.000730175 | 0.030401408 | 1 |
| HTN | endothelial | *TRMT61A* | -0.857501811 | 3.13E-05 | 0.004015831 | 0.196654092 |
| HTN | endothelial | *GABRB3* | 0.399395248 | 0.001488026 | 0.044128385 | 1 |
| HTN | endothelial | *TRIM66* | -0.259651313 | 3.43E-05 | 0.004224836 | 0.215466627 |
| HTN | endothelial | *NYAP1* | 0.560010035 | 0.000288143 | 0.016741209 | 1 |
| HTN | endothelial | *KIR3DL1* | 1.016828587 | 0.001880986 | 0.049993244 | 1 |
| HTN | endothelial | *SRR* | 0.523178692 | 4.45E-05 | 0.005195893 | 0.279827339 |
| HTN | endothelial | *MLKL* | 0.399099014 | 0.001916769 | 0.049993244 | 1 |
| HTN | endothelial | *BTNL3* | 0.150035352 | 0.001817279 | 0.049461154 | 1 |
| HTN | endothelial | *LGALS9* | 0.843428324 | 0.000771354 | 0.031304269 | 1 |
| HTN | endothelial | *IRS1* | -0.449398217 | 1.35E-08 | 1.06E-05 | 8.50E-05 |
| HTN | endothelial | *C8orf46* | 0.141865425 | 0.001570289 | 0.045852374 | 1 |
| HTN | endothelial | *CXCL10* | 0.503765438 | 0.001301604 | 0.042129838 | 1 |
| HTN | endothelial | *CALML6* | -1.435093393 | 2.60E-06 | 0.000543812 | 0.016343015 |
| HTN | endothelial | *GIMAP8* | 0.227446694 | 0.000821437 | 0.03232207 | 1 |
| HTN | endothelial | *KCNMB3* | 0.590975093 | 0.001320117 | 0.042129838 | 1 |
| HTN | endothelial | *DLK2* | -2.07068937 | 0.000603919 | 0.026366918 | 1 |
| HTN | endothelial | *RGS19* | -1.171616395 | 8.19E-12 | 5.15E-08 | 5.15E-08 |
| HTN | endothelial | *NME6* | 0.855469593 | 1.33E-06 | 0.000333792 | 0.008344788 |
| HTN | endothelial | *CTSW* | -0.241134995 | 9.75E-06 | 0.001612397 | 0.061271091 |
| HTN | endothelial | *ZNF738* | 0.747829474 | 8.69E-05 | 0.008033078 | 0.546249289 |
| HTN | endothelial | *SNX32* | 0.904689195 | 6.92E-05 | 0.007127618 | 0.434784727 |

| | | | | | | |
|---|---|---|---|---|---|---|
| HTN | endothelial | *MYEOV* | -0.275754892 | 0.000444714 | 0.021745177 | 1 |
| HTN | endothelial | *DCAKD* | -0.404629398 | 0.001105945 | 0.038771366 | 1 |
| HTN | endothelial | *DMRT2* | -0.421216738 | 0.000798318 | 0.032173239 | 1 |
| HTN | endothelial | *MAP3K11* | 0.606711885 | 0.001801013 | 0.049461154 | 1 |
| HTN | endothelial | *VEGFB* | -0.788355581 | 2.03E-06 | 0.000472358 | 0.012753668 |
| HTN | endothelial | *TNFRSF10D* | 0.27414377 | 0.000315811 | 0.017464815 | 1 |
| HTN | endothelial | *MST1* | -0.617956342 | 4.20E-05 | 0.005083195 | 0.264326117 |
| HTN | endothelial | *NRROS* | 0.664206071 | 0.000168948 | 0.012345191 | 1 |
| HTN | endothelial | *CTU2* | -0.908941405 | 0.001515005 | 0.044717544 | 1 |
| HTN | endothelial | *SNX31* | -1.015518795 | 2.17E-05 | 0.003202294 | 0.136638964 |
| HTN | endothelial | *GLIS1* | -0.437520186 | 7.36E-05 | 0.007464183 | 0.462779324 |
| HTN | endothelial | *KLF14* | -2.620828471 | 0.000155401 | 0.01177113 | 0.977003753 |
| HTN | endothelial | *TP53I11* | 0.421877258 | 0.001234645 | 0.041509158 | 1 |
| HTN | endothelial | *NRIP3* | -1.014501716 | 5.10E-05 | 0.005720357 | 0.320340004 |
| HTN | endothelial | *UFSP1* | -0.592326903 | 0.000901998 | 0.034161814 | 1 |
| HTN | endothelial | *RIMKLA* | 0.455341977 | 0.000662035 | 0.028123082 | 1 |
| HTN | endothelial | *PCDHB9* | 0.726076389 | 0.001454043 | 0.043949851 | 1 |
| HTN | endothelial | *COX14* | -0.563771247 | 0.000372517 | 0.019516802 | 1 |
| HTN | endothelial | *RPP25* | 1.385852568 | 1.74E-10 | 3.23E-07 | 1.09E-06 |
| HTN | endothelial | *MPI* | -1.436513303 | 6.64E-06 | 0.001149757 | 0.041768743 |
| HTN | endothelial | *TRIM73* | -1.571941919 | 0.000446179 | 0.021745177 | 1 |
| HTN | endothelial | *HYI* | -0.614056388 | 1.67E-06 | 0.000403861 | 0.010500394 |
| HTN | endothelial | *CTC1* | 0.967173707 | 0.000107099 | 0.009099092 | 0.673332801 |
| HTN | endothelial | *CIITA* | 0.56888195 | 0.001651674 | 0.047200337 | 1 |
| HTN | endothelial | *DEFB124* | -1.817607016 | 5.08E-05 | 0.005720357 | 0.319426094 |
| HTN | endothelial | *ZNF443* | 1.307536144 | 1.07E-05 | 0.001718497 | 0.067021364 |
| HTN | endothelial | *TMEM136* | 0.881447038 | 0.000465696 | 0.022521762 | 1 |
| HTN | endothelial | *CLDN7* | -0.94399846 | 5.31E-08 | 2.78E-05 | 0.000333728 |
| HTN | endothelial | *ADO* | 1.718072887 | 0.000974684 | 0.036481244 | 1 |
| HTN | endothelial | *FAM89A* | 0.665490559 | 0.000159016 | 0.01190157 | 0.999731883 |
| HTN | endothelial | *CLN8* | 1.02803236 | 0.000213965 | 0.014012492 | 1 |
| HTN | endothelial | *COL18A1* | 0.740169968 | 0.00018408 | 0.013151286 | 1 |
| HTN | endothelial | *ZNF320* | 0.380525393 | 0.000113626 | 0.009277451 | 0.7143637 |
| HTN | endothelial | *FAM43B* | -1.181421619 | 0.00130863 | 0.042129838 | 1 |
| HTN | endothelial | *RP1L1* | 0.735453013 | 2.23E-06 | 0.000484025 | 0.014036722 |
| HTN | endothelial | *KREMEN1* | -0.340860868 | 0.000310994 | 0.017457301 | 1 |
| HTN | endothelial | *EIF3C* | 0.989294806 | 1.74E-07 | 6.43E-05 | 0.00109319 |
| HTN | endothelial | *A3GALT2* | -0.781562058 | 2.14E-06 | 0.00048137 | 0.013478357 |
| HTN | endothelial | *LPAR5* | -0.691803173 | 0.000440424 | 0.021745177 | 1 |
| HTN | endothelial | *TMEM173* | 0.56951159 | 0.001139755 | 0.039326095 | 1 |
| HTN | endothelial | *NUTM2A* | -1.302250994 | 0.000517609 | 0.024293506 | 1 |
| HTN | endothelial | *TNFAIP8L1* | 0.539518116 | 0.001110044 | 0.038771366 | 1 |
| HTN | endothelial | *MYBL1* | -0.478680552 | 0.000896118 | 0.03414481 | 1 |
| HTN | endothelial | *SRL* | 0.332104595 | 0.000974845 | 0.036481244 | 1 |
| HTN | endothelial | *ZNF829* | 0.912496973 | 0.000821814 | 0.03232207 | 1 |
| HTN | endothelial | *ISG15* | 0.836791626 | 0.001898617 | 0.049993244 | 1 |
| HTN | endothelial | *MCRS1* | 1.122886422 | 0.000309592 | 0.017457301 | 1 |
| HTN | endothelial | *C22orf34* | 0.271706954 | 0.001900532 | 0.049993244 | 1 |
| HTN | endothelial | *GOLGA8M* | 0.760133201 | 0.00058934 | 0.026092832 | 1 |
| HTN | endothelial | *ZDHHC11* | 1.355845243 | 9.69E-05 | 0.008665472 | 0.609081325 |

| | | | | | | |
|---|---|---|---|---|---|---|
| HTN | endothelial | *FBF1* | -0.583214552 | 1.20E-07 | 4.70E-05 | 0.000752643 |
| HTN | endothelial | *HRCT1* | 0.745852028 | 0.001485962 | 0.044128385 | 1 |
| HTN | endothelial | *SULT1A1* | -0.421752607 | 0.000402904 | 0.020593957 | 1 |
| HTN | endothelial | *FAM180B* | -0.89844984 | 3.29E-08 | 1.88E-05 | 0.0002068 |
| HTN | endothelial | *C6orf106* | 0.531863048 | 0.000346079 | 0.018596578 | 1 |
| HTN | endothelial | *ZFP28* | 0.912629425 | 0.000833694 | 0.032354526 | 1 |
| HTN | endothelial | *KIAA1211L* | 0.195593386 | 0.000675786 | 0.02851454 | 1 |
| HTN | endothelial | *HIST1H3E* | 1.342476841 | 2.05E-10 | 3.23E-07 | 1.29E-06 |
| HTN | endothelial | *SLC25A29* | 0.435769196 | 0.001643514 | 0.047181613 | 1 |
| HTN | endothelial | *SLC22A5* | 0.479773244 | 0.000193617 | 0.013558581 | 1 |
| HTN | endothelial | *KLHL14* | 0.426085895 | 0.000887327 | 0.034016009 | 1 |
| HTN | endothelial | *RPS26* | -0.442612161 | 1.79E-05 | 0.002815603 | 0.11262413 |
| HTN | endothelial | *TMEM116* | -0.766610714 | 4.21E-09 | 5.29E-06 | 2.64E-05 |
| HTN | endothelial | *TGM2* | 0.188042244 | 0.000276448 | 0.016243271 | 1 |
| HTN | endothelial | *AL590822.1* | -0.449634949 | 0.00012559 | 0.009994755 | 0.789585678 |
| HTN | endothelial | *C10orf128* | 0.311185346 | 0.001711619 | 0.048255386 | 1 |
| HTN | endothelial | *HSD17B8* | -0.841730492 | 0.000316683 | 0.017464815 | 1 |
| HTN | endothelial | *HLA-DRA* | 0.525781941 | 0.000377379 | 0.019608129 | 1 |
| HTN | endothelial | *PRRT1* | 0.830804438 | 9.20E-07 | 0.000259976 | 0.005783972 |
| HTN | endothelial | *STK19* | 1.097478046 | 1.20E-06 | 0.000314334 | 0.007544024 |
| HTN | endothelial | *BAG6* | 0.796505856 | 0.000267172 | 0.01607007 | 1 |
| HTN | endothelial | *HLA-F* | 0.454445708 | 0.000503989 | 0.024004386 | 1 |
| HTN | endothelial | *ZDHHC11B* | 0.995979026 | 0.000444416 | 0.021745177 | 1 |
| HTN | endothelial | *SIPA1* | 1.491992288 | 5.81E-07 | 0.000173988 | 0.003653756 |
| HTN | endothelial | *ATF6B* | 1.125406053 | 6.21E-09 | 5.78E-06 | 3.90E-05 |
| HTN | endothelial | *LTB4R* | -0.797493595 | 3.37E-05 | 0.004224836 | 0.211585276 |
| HTN | endothelial | *LTB4R2* | -1.363479294 | 0.000170834 | 0.012345191 | 1 |
| HTN | endothelial | *AC016773.1* | 0.543219001 | 2.90E-05 | 0.003883897 | 0.182543181 |
| HTN | endothelial | *SLC12A8* | -0.555292601 | 0.001776354 | 0.04941564 | 1 |
| HTN | endothelial | *HLA-DPB1* | 0.625760048 | 8.00E-05 | 0.007735714 | 0.502821422 |
| HTN | endothelial | *TMEM233* | 0.420847629 | 0.000415736 | 0.021078479 | 1 |
| HTN | endothelial | *AL049840.1* | -0.548029648 | 0.001920567 | 0.049993244 | 1 |
| HTN | endothelial | *SLC26A6* | 1.767547144 | 3.13E-05 | 0.004015831 | 0.19677571 |
| HTN | endothelial | *SLC2A1-AS1* | -0.867424741 | 0.000212223 | 0.014012492 | 1 |
| HTN | endothelial | *AC108463.2* | -0.891882169 | 0.001681699 | 0.047840926 | 1 |
| HTN | endothelial | *PROB1* | 3.867334157 | 6.77E-06 | 0.001149757 | 0.04254102 |
| HTN | endothelial | *CISD3* | 0.902730668 | 0.001865833 | 0.049993244 | 1 |
| HTN | endothelial | *AC112229.4* | 0.483934785 | 0.000490017 | 0.023517082 | 1 |
| HTN | endothelial | *HLA-DPA1* | 0.547289686 | 0.001612849 | 0.046513664 | 1 |
| HTN | endothelial | *HLA-B* | 0.434888747 | 8.66E-05 | 0.008033078 | 0.544525765 |
| HTN | endothelial | *HLA-DQA2* | 0.384043452 | 0.00035499 | 0.018913742 | 1 |
| HTN | endothelial | *PSG4* | 1.317231146 | 2.65E-05 | 0.003703106 | 0.166639767 |
| HTN | endothelial | *ZNF10* | -0.740163308 | 0.001817326 | 0.049461154 | 1 |
| HTN | LEC | *SEMA3F* | -1.829341089 | 3.00E-07 | 0.000209412 | 0.00188471 |
| HTN | LEC | *WNT16* | -2.237252575 | 0.000342743 | 0.024892502 | 1 |
| HTN | LEC | *M6PR* | -2.881748164 | 0.000193438 | 0.01900229 | 1 |
| HTN | LEC | *RBM6* | -0.976748206 | 2.84E-05 | 0.005828192 | 0.178338765 |
| HTN | LEC | *CD79B* | -1.777287926 | 0.000121004 | 0.015650619 | 0.760754003 |
| HTN | LEC | *HFE* | -3.066718542 | 2.78E-07 | 0.000209412 | 0.001746811 |
| HTN | LEC | *NR1H3* | -2.157156841 | 0.000704666 | 0.034078726 | 1 |

| | | | | | | |
|---|---|---|---|---|---|---|
| HTN | LEC | *RCN1* | -2.695651728 | 0.000425352 | 0.027011981 | 1 |
| HTN | LEC | *ITIH4* | 0.974117214 | 0.000135363 | 0.015650619 | 0.851025676 |
| HTN | LEC | *UHRF1BP1* | 1.214793031 | 0.000172719 | 0.017898979 | 1 |
| HTN | LEC | *ATXN3* | -1.817205909 | 0.000199508 | 0.019297036 | 1 |
| HTN | LEC | *DHX8* | -2.479861578 | 0.000646476 | 0.032737455 | 1 |
| HTN | LEC | *GNB5* | -1.324626988 | 0.000949484 | 0.040358362 | 1 |
| HTN | LEC | *CPSF1* | -3.141768042 | 0.001255665 | 0.047556402 | 1 |
| HTN | LEC | *XAB2* | -3.211617756 | 0.000173666 | 0.017898979 | 1 |
| HTN | LEC | *KIAA0141* | -1.996523134 | 0.000816273 | 0.037187752 | 1 |
| HTN | LEC | *CCNT2* | -2.071561714 | 6.93E-05 | 0.010897467 | 0.435898674 |
| HTN | LEC | *NOA1* | -1.361623891 | 0.000900656 | 0.039256717 | 1 |
| HTN | LEC | *SNX10* | -0.499265759 | 0.00097857 | 0.041015144 | 1 |
| HTN | LEC | *AAMDC* | -1.618035842 | 7.72E-06 | 0.002655556 | 0.048515433 |
| HTN | LEC | *ZNF268* | -4.527204475 | 0.000234823 | 0.021396081 | 1 |
| HTN | LEC | *TGM1* | 1.789400834 | 0.000350696 | 0.024892502 | 1 |
| HTN | LEC | *TRIOBP* | -2.062013449 | 0.000465247 | 0.028125057 | 1 |
| HTN | LEC | *CCNB1IP1* | -2.682200656 | 0.000502617 | 0.029532252 | 1 |
| HTN | LEC | *DNTTIP1* | -2.213182542 | 8.73E-05 | 0.012760904 | 0.548718873 |
| HTN | LEC | *POLI* | -0.703797844 | 0.000392563 | 0.025979396 | 1 |
| HTN | LEC | *RNF40* | -2.69895445 | 0.00056355 | 0.030960839 | 1 |
| HTN | LEC | *TAF6* | -3.851537749 | 9.78E-05 | 0.013519342 | 0.615106714 |
| HTN | LEC | *WNT3* | -5.008991914 | 8.03E-06 | 0.002655556 | 0.05045556 |
| HTN | LEC | *SYNGR2* | -2.814987971 | 0.000155439 | 0.017152302 | 0.97724526 |
| HTN | LEC | *ENDOU* | -1.806644586 | 0.000314418 | 0.024892502 | 1 |
| HTN | LEC | *CAND1* | -2.28548922 | 0.000131622 | 0.015650619 | 0.827506334 |
| HTN | LEC | *RWDD1* | -2.217547689 | 0.000453274 | 0.028125057 | 1 |
| HTN | LEC | *PCDHB6* | 3.033714435 | 0.000566327 | 0.030960839 | 1 |
| HTN | LEC | *HYAL1* | -0.858483489 | 0.000542334 | 0.030960839 | 1 |
| HTN | LEC | *TP53I3* | -3.098315516 | 1.86E-05 | 0.004678128 | 0.116953196 |
| HTN | LEC | *DNAJC27* | -3.34056915 | 5.93E-07 | 0.000310753 | 0.003729041 |
| HTN | LEC | *CHMP3* | -2.426235537 | 0.000970699 | 0.040958286 | 1 |
| HTN | LEC | *C1orf21* | -1.468958203 | 1.42E-05 | 0.004045725 | 0.089005951 |
| HTN | LEC | *DIEXF* | -2.22969066 | 0.000470795 | 0.028183587 | 1 |
| HTN | LEC | *DDX59* | -1.906027361 | 0.000131652 | 0.015650619 | 0.827698265 |
| HTN | LEC | *TCF21* | 12.48848983 | 0.000377845 | 0.025802343 | 1 |
| HTN | LEC | *HOXB5* | -2.265266665 | 0.001023277 | 0.041643964 | 1 |
| HTN | LEC | *PCDHB12* | -1.09423094 | 0.000690504 | 0.033652722 | 1 |
| HTN | LEC | *CBX3* | -2.774738465 | 0.00033355 | 0.024892502 | 1 |
| HTN | LEC | *TOX2* | -0.777804068 | 0.000103484 | 0.013842654 | 0.650604746 |
| HTN | LEC | *SNRPC* | -3.345121654 | 1.01E-08 | 3.19E-05 | 6.37E-05 |
| HTN | LEC | *HIST1H1D* | 3.559414171 | 1.74E-06 | 0.000684908 | 0.01095852 |
| HTN | LEC | *OPRL1* | -0.664079008 | 0.000638851 | 0.032685822 | 1 |
| HTN | LEC | *CHCHD5* | -2.038086971 | 0.000889895 | 0.039124284 | 1 |
| HTN | LEC | *TRAP1* | 0.762088023 | 5.36E-05 | 0.009108635 | 0.337019491 |
| HTN | LEC | *OPN1SW* | -1.345607374 | 0.000938941 | 0.040358362 | 1 |
| HTN | LEC | *GALNT8* | -1.498132817 | 0.000639471 | 0.032685822 | 1 |
| HTN | LEC | *ANO1* | -2.930489211 | 7.49E-05 | 0.011215453 | 0.471049013 |
| HTN | LEC | *ENOSF1* | -1.572144626 | 0.000656887 | 0.032776552 | 1 |
| HTN | LEC | *REEP2* | -1.225411335 | 0.000950062 | 0.040358362 | 1 |
| HTN | LEC | *TMCC2* | -2.283940188 | 0.000356733 | 0.02491977 | 1 |

| | | | | | | |
|---|---|---|---|---|---|---|
| HTN | LEC | *ACP2* | 2.334087395 | 1.43E-06 | 0.000600151 | 0.009002267 |
| HTN | LEC | *FADS2* | -3.016409484 | 0.000561403 | 0.030960839 | 1 |
| HTN | LEC | *GOLM1* | -2.300480686 | 0.000734352 | 0.034704707 | 1 |
| HTN | LEC | *AGT* | 1.07600298 | 2.97E-05 | 0.005828192 | 0.186502159 |
| HTN | LEC | *CHST12* | -2.455347572 | 0.000303852 | 0.024491219 | 1 |
| HTN | LEC | *NACAD* | -6.007697134 | 0.000128201 | 0.015650619 | 0.805998027 |
| HTN | LEC | *CIB2* | -1.675827066 | 0.000155508 | 0.017152302 | 0.977681237 |
| HTN | LEC | *NMT1* | 1.571044747 | 2.39E-05 | 0.005359754 | 0.150007691 |
| HTN | LEC | *GYPC* | -2.31158639 | 0.000650896 | 0.032737455 | 1 |
| HTN | LEC | *VARS2* | 0.624858023 | 0.000802286 | 0.036817329 | 1 |
| HTN | LEC | *PAQR5* | -1.790607875 | 0.001013081 | 0.041643964 | 1 |
| HTN | LEC | *ENPEP* | -2.348521589 | 0.000462943 | 0.028125057 | 1 |
| HTN | LEC | *FBN2* | 0.726227874 | 0.000624926 | 0.032685822 | 1 |
| HTN | LEC | *GLTP* | -3.837641494 | 0.000134527 | 0.015650619 | 0.845773548 |
| HTN | LEC | *RPS2* | -2.282132196 | 0.001228433 | 0.047220168 | 1 |
| HTN | LEC | *TRIM65* | -2.324772214 | 0.001187217 | 0.046360455 | 1 |
| HTN | LEC | *GEMIN7* | -2.441191044 | 0.001266961 | 0.047696891 | 1 |
| HTN | LEC | *RGS5* | -3.32363181 | 1.84E-05 | 0.004678128 | 0.115864154 |
| HTN | LEC | *CREB3L4* | -3.152780515 | 2.85E-07 | 0.000209412 | 0.001789278 |
| HTN | LEC | *CTDSPL* | -1.313898064 | 0.000905396 | 0.039256717 | 1 |
| HTN | LEC | *SLC10A4* | 7.163481528 | 1.10E-05 | 0.003304415 | 0.069392713 |
| HTN | LEC | *PLEKHS1* | 4.060629696 | 0.001169994 | 0.045973461 | 1 |
| HTN | LEC | *PDCD11* | 1.853445945 | 0.001141645 | 0.045141665 | 1 |
| HTN | LEC | *HSD17B12* | -2.218972607 | 1.32E-09 | 8.30E-06 | 8.30E-06 |
| HTN | LEC | *DUSP15* | -3.779685337 | 0.000296697 | 0.024225119 | 1 |
| HTN | LEC | *PRDM8* | -1.608018503 | 6.34E-05 | 0.01021999 | 0.398579621 |
| HTN | LEC | *NEIL2* | 1.063941278 | 0.000210727 | 0.019773734 | 1 |
| HTN | LEC | *C8orf37* | -1.318322415 | 0.000876325 | 0.038798964 | 1 |
| HTN | LEC | *NTAN1* | -2.190742415 | 0.001114978 | 0.044648852 | 1 |
| HTN | LEC | *CLEC18A* | -3.805863044 | 2.48E-05 | 0.005382535 | 0.156093509 |
| HTN | LEC | *TRIM63* | 1.383725314 | 0.000320435 | 0.024892502 | 1 |
| HTN | LEC | *SLC5A11* | -1.501865321 | 3.27E-08 | 4.12E-05 | 0.000205886 |
| HTN | LEC | *CCDC17* | -3.005163213 | 0.000348566 | 0.024892502 | 1 |
| HTN | LEC | *ABCF3* | 1.740422254 | 0.001231765 | 0.047220168 | 1 |
| HTN | LEC | *CYGB* | -3.451802887 | 0.000463966 | 0.028125057 | 1 |
| HTN | LEC | *TNFSF13* | 4.933196351 | 2.08E-07 | 0.000209412 | 0.001309855 |
| HTN | LEC | *SNRNP25* | -3.59345867 | 0.000421545 | 0.027011981 | 1 |
| HTN | LEC | *C1orf50* | -2.189517986 | 0.000208214 | 0.019773734 | 1 |
| HTN | LEC | *CDC25A* | -1.476832957 | 0.000339096 | 0.024892502 | 1 |
| HTN | LEC | *APEH* | -1.906758022 | 0.000284892 | 0.023881524 | 1 |
| HTN | LEC | *RNF123* | 2.374469382 | 2.22E-08 | 3.49E-05 | 0.000139581 |
| HTN | LEC | *RPS14* | -2.104307493 | 0.001003083 | 0.041643964 | 1 |
| HTN | LEC | *REEP3* | -2.282107927 | 9.12E-06 | 0.002865755 | 0.057315099 |
| HTN | LEC | *CASP7* | -1.928204688 | 0.000277754 | 0.023597793 | 1 |
| HTN | LEC | *TRIM66* | -0.583351554 | 0.00039202 | 0.025979396 | 1 |
| HTN | LEC | *RIMKLB* | -1.548274106 | 2.39E-05 | 0.005359754 | 0.150073103 |
| HTN | LEC | *VKORC1* | -2.580800502 | 0.000131217 | 0.015650619 | 0.82496403 |
| HTN | LEC | *KIR3DL1* | 3.430514789 | 4.12E-05 | 0.007400271 | 0.259009475 |
| HTN | LEC | *UBXN6* | -3.227798476 | 0.000213937 | 0.01977976 | 1 |
| HTN | LEC | *SRR* | 1.158595287 | 0.000361011 | 0.02494152 | 1 |

| | | | | | | |
|---|---|---|---|---|---|---|
| HTN | LEC | *CYP7A1* | 4.138929073 | 0.000330609 | 0.024892502 | 1 |
| HTN | LEC | *HRASLS5* | 0.473012209 | 0.00058036 | 0.031185645 | 1 |
| HTN | LEC | *NUDT16L1* | -5.326985102 | 0.000136915 | 0.015650619 | 0.860784035 |
| HTN | LEC | *NT5DC2* | -3.101998752 | 0.000684832 | 0.033637024 | 1 |
| HTN | LEC | *TNIP2* | -1.683086066 | 0.000192618 | 0.01900229 | 1 |
| HTN | LEC | *IRS1* | -0.946562924 | 4.83E-06 | 0.001784521 | 0.030336853 |
| HTN | LEC | *C8orf46* | 0.408007654 | 0.000187586 | 0.01900229 | 1 |
| HTN | LEC | *CD52* | -2.050151087 | 0.000475181 | 0.028183587 | 1 |
| HTN | LEC | *INO80E* | -2.072444322 | 0.000603473 | 0.032152848 | 1 |
| HTN | LEC | *KCNAB3* | -1.767734666 | 0.000290891 | 0.02406361 | 1 |
| HTN | LEC | *KCND3* | -1.851633926 | 0.00074521 | 0.034704707 | 1 |
| HTN | LEC | *NME6* | 1.871077823 | 4.92E-07 | 0.000283966 | 0.003095987 |
| HTN | LEC | *VEGFB* | -1.448020974 | 0.000436969 | 0.027472222 | 1 |
| HTN | LEC | *MST1* | -1.262299781 | 0.000519695 | 0.030252989 | 1 |
| HTN | LEC | *SNX31* | -2.564358741 | 0.000834476 | 0.03720817 | 1 |
| HTN | LEC | *GLIS1* | -1.307935289 | 0.000743716 | 0.034704707 | 1 |
| HTN | LEC | *TOP3A* | -3.04750565 | 3.36E-05 | 0.006312766 | 0.211076955 |
| HTN | LEC | *PCDHB9* | 1.597692409 | 0.000774849 | 0.035819673 | 1 |
| HTN | LEC | *SH2B1* | -1.634702859 | 9.01E-05 | 0.012881157 | 0.5667709 |
| HTN | LEC | *COX14* | -1.938634181 | 0.001247484 | 0.047532932 | 1 |
| HTN | LEC | *CPNE7* | -4.412573524 | 0.000678644 | 0.033595546 | 1 |
| HTN | LEC | *TRIM73* | -2.310393017 | 0.00027768 | 0.023597793 | 1 |
| HTN | LEC | *ZNF443* | 2.0121814 | 0.000339832 | 0.024892502 | 1 |
| HTN | LEC | *ANKRD34A* | -2.732423653 | 0.000406316 | 0.026487546 | 1 |
| HTN | LEC | *MXRA7* | -1.269928854 | 0.000725527 | 0.034555985 | 1 |
| HTN | LEC | *ZNF320* | 0.907407999 | 0.000629938 | 0.032685822 | 1 |
| HTN | LEC | *CDK10* | -2.883456022 | 7.70E-07 | 0.00037262 | 0.004844059 |
| HTN | LEC | *FAM174B* | -1.884724155 | 0.000555077 | 0.030960839 | 1 |
| HTN | LEC | *KBTBD12* | -1.32963279 | 0.000822426 | 0.037198487 | 1 |
| HTN | LEC | *ZNF793* | -1.22314323 | 4.44E-05 | 0.007757437 | 0.279267744 |
| HTN | LEC | *NOC2L* | -1.644160713 | 0.000171293 | 0.017898979 | 1 |
| HTN | LEC | *MYO6* | -2.022985636 | 0.000832577 | 0.03720817 | 1 |
| HTN | LEC | *FAM3C* | -2.318956417 | 0.001221902 | 0.047220168 | 1 |
| HTN | LEC | *HIST1H3E* | 3.035261096 | 1.85E-08 | 3.49E-05 | 0.000116326 |
| HTN | LEC | *SULT1A2* | 0.544575647 | 7.46E-05 | 0.011215453 | 0.469149396 |
| HTN | LEC | *SLC22A5* | 1.766354225 | 4.97E-07 | 0.000283966 | 0.003123631 |
| HTN | LEC | *PCDHB11* | -5.516485576 | 2.92E-05 | 0.005828192 | 0.183814114 |
| HTN | LEC | *ZNF841* | -2.932210889 | 0.001026521 | 0.041643964 | 1 |
| HTN | LEC | *RPS26* | -1.354675447 | 9.52E-07 | 0.000427421 | 0.005983892 |
| HTN | LEC | *TMEM116* | -0.943244662 | 0.000381679 | 0.025802343 | 1 |
| HTN | LEC | *HLA-DRB5* | -0.624691871 | 0.000254708 | 0.02287645 | 1 |
| HTN | LEC | *CENPW* | -4.344525914 | 3.41E-05 | 0.006312766 | 0.214634045 |
| HTN | LEC | *C6orf163* | -3.903717928 | 0.000573243 | 0.031068758 | 1 |
| HTN | LEC | *STK19* | 2.132780333 | 0.000628025 | 0.032685822 | 1 |
| HTN | LEC | *GPANK1* | 2.057818132 | 0.001067855 | 0.043035942 | 1 |
| HTN | LEC | *LINC00654* | -4.553787964 | 0.001026692 | 0.041643964 | 1 |
| HTN | LEC | *LNP1* | -1.603783256 | 0.001322627 | 0.04949618 | 1 |
| HTN | LEC | *SIPA1* | 2.736231133 | 0.000263712 | 0.023351499 | 1 |
| HTN | LEC | *ATF6B* | 1.935099097 | 0.000352383 | 0.024892502 | 1 |
| HTN | LEC | *REPIN1* | -2.78405941 | 0.001128702 | 0.04491234 | 1 |

| | | | | | | |
|---|---|---|---|---|---|---|
| HTN | LEC | *AC016773.1* | 0.869779202 | 0.000408667 | 0.026487546 | 1 |
| HTN | LEC | *SLC2A1-AS1* | -2.516766145 | 1.99E-05 | 0.004823396 | 0.125408303 |
| HTN | LEC | *PROB1* | 5.166965136 | 0.000273142 | 0.023597793 | 1 |
| HTN | LEC | *HLA-DQA2* | 1.12624889 | 1.70E-05 | 0.00464906 | 0.106928373 |
| HTN | LEC | *TNFSF12* | -2.018311234 | 0.000552225 | 0.030960839 | 1 |
| HTN | LEC | *ATP5O* | -2.508065906 | 0.000532368 | 0.030706375 | 1 |
| HTN | LEC | *PLA2G4B* | 2.032667454 | 6.18E-05 | 0.01021999 | 0.388365914 |
| HTN | LEC | *JMJD7* | 1.900807855 | 0.000168805 | 0.017898979 | 1 |
| HTN | LEC | *HMBS* | 1.413070694 | 9.89E-05 | 0.013519342 | 0.621889745 |
| HTN | LEC | *SH3RF3-AS1* | -2.008800891 | 0.000722828 | 0.034555985 | 1 |
| HTN | LEC | *AC144652.1* | 1.588222252 | 0.000336833 | 0.024892502 | 1 |
| HTN | macrophage | *SEMA3F* | -0.440907419 | 1.28E-06 | 0.000149262 | 0.008057198 |
| HTN | macrophage | *M6PR* | -1.133716401 | 0.00016647 | 0.00742268 | 1 |
| HTN | macrophage | *RBM6* | -0.308894281 | 4.43E-06 | 0.000434706 | 0.027821158 |
| HTN | macrophage | *COPZ2* | 0.560927613 | 2.23E-05 | 0.001462082 | 0.140359911 |
| HTN | macrophage | *GGNBP2* | -0.657479715 | 0.001620839 | 0.040924567 | 1 |
| HTN | macrophage | *HFE* | -0.929125632 | 1.48E-08 | 3.86E-06 | 9.28E-05 |
| HTN | macrophage | *CLCN6* | -2.089814612 | 3.66E-06 | 0.000365549 | 0.023029583 |
| HTN | macrophage | *CEP68* | 0.925062437 | 3.66E-07 | 5.36E-05 | 0.002304025 |
| HTN | macrophage | *NR1H3* | -1.702885139 | 7.29E-10 | 4.30E-07 | 4.59E-06 |
| HTN | macrophage | *LIMA1* | 0.360145091 | 3.16E-05 | 0.001909593 | 0.198597622 |
| HTN | macrophage | *SDCCAG8* | 0.97794845 | 1.86E-07 | 3.16E-05 | 0.00116871 |
| HTN | macrophage | *ITIH4* | 0.242865174 | 0.000262471 | 0.01037835 | 1 |
| HTN | macrophage | *PIGV* | -1.139713403 | 8.96E-06 | 0.000731612 | 0.056334128 |
| HTN | macrophage | *UHRF1BP1* | 0.466974852 | 2.55E-06 | 0.000276155 | 0.016017008 |
| HTN | macrophage | *TIE1* | -0.659586228 | 1.14E-05 | 0.00088098 | 0.07135941 |
| HTN | macrophage | *POLR1A* | -0.361868086 | 0.000262071 | 0.01037835 | 1 |
| HTN | macrophage | *ASNS* | 1.48717766 | 0.000938994 | 0.026712464 | 1 |
| HTN | macrophage | *CLNS1A* | 0.670473627 | 5.06E-05 | 0.002815618 | 0.318164822 |
| HTN | macrophage | *ATP2A3* | 1.121041244 | 2.66E-05 | 0.001671681 | 0.167241523 |
| HTN | macrophage | *SCGN* | 2.902228111 | 0.000629244 | 0.019487961 | 1 |
| HTN | macrophage | *DDX43* | -0.940692909 | 0.001146796 | 0.03107717 | 1 |
| HTN | macrophage | *CCNT2* | -0.613198615 | 0.000363647 | 0.013292131 | 1 |
| HTN | macrophage | *SNX10* | -0.231507166 | 3.18E-07 | 4.76E-05 | 0.001998508 |
| HTN | macrophage | *AAMDC* | -0.408794267 | 6.83E-05 | 0.003578828 | 0.429650807 |
| HTN | macrophage | *C3orf18* | 1.129536632 | 3.28E-05 | 0.001961859 | 0.2059952 |
| HTN | macrophage | *YPEL3* | 0.449579951 | 0.000110463 | 0.005383551 | 0.694478052 |
| HTN | macrophage | *ZNF268* | -1.661253953 | 0.001978238 | 0.047260764 | 1 |
| HTN | macrophage | *PABPC4* | -0.991355625 | 1.28E-06 | 0.000149262 | 0.008060156 |
| HTN | macrophage | *TGM1* | 0.535453725 | 0.000205713 | 0.008679983 | 1 |
| HTN | macrophage | *MARCH2* | 1.355798675 | 0.000413003 | 0.014345578 | 1 |
| HTN | macrophage | *GCAT* | 0.693011072 | 0.000619885 | 0.019389146 | 1 |
| HTN | macrophage | *PCK2* | -1.269941789 | 0.001984546 | 0.047260764 | 1 |
| HTN | macrophage | *DNTTIP1* | -0.625208299 | 0.00021899 | 0.009117808 | 1 |
| HTN | macrophage | *POLI* | -0.204525613 | 9.65E-05 | 0.004777363 | 0.606725118 |
| HTN | macrophage | *MGRN1* | 0.608220395 | 0.001968851 | 0.047260764 | 1 |
| HTN | macrophage | *CORO1A* | -1.007903602 | 0.000238675 | 0.009680979 | 1 |
| HTN | macrophage | *CRISPLD2* | 0.475807946 | 0.001714878 | 0.042783477 | 1 |
| HTN | macrophage | *MTHFSD* | 0.360191779 | 0.000710059 | 0.021314448 | 1 |
| HTN | macrophage | *ZFAND1* | 0.685015986 | 0.001970284 | 0.047260764 | 1 |

| | | | | | | |
|---|---|---|---|---|---|---|
| HTN | macrophage | *EYA1* | 0.489204119 | 1.39E-05 | 0.001037687 | 0.087165681 |
| HTN | macrophage | *ICAM5* | -3.773466162 | 0.000330056 | 0.012134876 | 1 |
| HTN | macrophage | *ATP13A1* | 0.891708224 | 4.92E-05 | 0.002762695 | 0.30942189 |
| HTN | macrophage | *MEST* | -0.396971703 | 0.000469679 | 0.015790762 | 1 |
| HTN | macrophage | *ACTR3C* | 0.899584893 | 4.40E-05 | 0.002517582 | 0.276934066 |
| HTN | macrophage | *RAD51C* | -0.461849514 | 0.000129084 | 0.006148123 | 0.811552178 |
| HTN | macrophage | *ICAM2* | 0.919255189 | 0.000624906 | 0.019449422 | 1 |
| HTN | macrophage | *SYNGR2* | -0.810282238 | 0.000325368 | 0.012099761 | 1 |
| HTN | macrophage | *DDX5* | 0.58953062 | 0.000301551 | 0.011352417 | 1 |
| HTN | macrophage | *KCTD10* | -0.534566841 | 0.00055372 | 0.017761402 | 1 |
| HTN | macrophage | *CAND1* | -1.009626716 | 4.23E-05 | 0.002438786 | 0.265827636 |
| HTN | macrophage | *CEP72* | 0.822165694 | 0.001331924 | 0.034890869 | 1 |
| HTN | macrophage | *PCDHB6* | 0.966956931 | 0.000763894 | 0.022547415 | 1 |
| HTN | macrophage | *PCCB* | 1.642816473 | 3.34E-05 | 0.001981901 | 0.21008149 |
| HTN | macrophage | *TP53I3* | -1.349839776 | 0.000690117 | 0.020859447 | 1 |
| HTN | macrophage | *DNAJC27* | -1.400134248 | 1.67E-05 | 0.001179213 | 0.104949972 |
| HTN | macrophage | *RTN4* | -0.665349184 | 0.001339458 | 0.034942612 | 1 |
| HTN | macrophage | *ZNF142* | -0.955905107 | 0.001130013 | 0.030888671 | 1 |
| HTN | macrophage | *TCF21* | 3.211963317 | 0.000395433 | 0.014062863 | 1 |
| HTN | macrophage | *TRPM6* | 0.667377669 | 0.000371868 | 0.013514066 | 1 |
| HTN | macrophage | *ZFP30* | 0.591406515 | 0.00101536 | 0.028498081 | 1 |
| HTN | macrophage | *EPHX2* | -0.510641906 | 0.002024368 | 0.047626008 | 1 |
| HTN | macrophage | *TEX14* | 1.480567856 | 3.04E-07 | 4.66E-05 | 0.001911035 |
| HTN | macrophage | *CBX3* | -1.450613149 | 0.000158397 | 0.007216241 | 0.995841194 |
| HTN | macrophage | *ACVR1C* | 0.947907734 | 5.83E-05 | 0.003157556 | 0.366276536 |
| HTN | macrophage | *FAM124B* | 0.651504871 | 0.000161935 | 0.007324359 | 1 |
| HTN | macrophage | *SNRPC* | -0.848130537 | 2.66E-07 | 4.23E-05 | 0.001674598 |
| HTN | macrophage | *HIST1H1D* | 0.954951157 | 2.24E-07 | 3.71E-05 | 0.001410535 |
| HTN | macrophage | *ZNF391* | -0.22157954 | 0.000163696 | 0.007351121 | 1 |
| HTN | macrophage | *GRK4* | 0.215357878 | 0.000289089 | 0.011150335 | 1 |
| HTN | macrophage | *OPRL1* | -1.037062718 | 1.53E-08 | 3.86E-06 | 9.63E-05 |
| HTN | macrophage | *TRAP1* | 0.345209307 | 4.71E-07 | 6.74E-05 | 0.002963425 |
| HTN | macrophage | *APOL5* | -1.435072871 | 0.000474386 | 0.015864169 | 1 |
| HTN | macrophage | *KLHDC10* | -1.226556618 | 0.000450382 | 0.015388852 | 1 |
| HTN | macrophage | *EIF2AK4* | -0.623879872 | 0.001801981 | 0.044778881 | 1 |
| HTN | macrophage | *CDC16* | 0.804561586 | 0.000170552 | 0.007551148 | 1 |
| HTN | macrophage | *ATP8B3* | -0.365516895 | 0.000548047 | 0.017669593 | 1 |
| HTN | macrophage | *CHMP1A* | 1.34721729 | 8.28E-06 | 0.000703595 | 0.052066025 |
| HTN | macrophage | *HAUS8* | -2.166849804 | 1.65E-08 | 3.99E-06 | 0.000103647 |
| HTN | macrophage | *PDLIM4* | 1.432038843 | 3.51E-08 | 7.12E-06 | 0.000220864 |
| HTN | macrophage | *ANO1* | -1.621951587 | 0.001710256 | 0.042783477 | 1 |
| HTN | macrophage | *KDM6B* | -2.191163656 | 8.48E-06 | 0.000711248 | 0.05334362 |
| HTN | macrophage | *PRAM1* | -0.7286379 | 0.000654509 | 0.019975233 | 1 |
| HTN | macrophage | *WNT2B* | -1.406809741 | 1.69E-07 | 2.96E-05 | 0.001065235 |
| HTN | macrophage | *DDB2* | 1.662248228 | 6.49E-06 | 0.00056681 | 0.04081033 |
| HTN | macrophage | *ACP2* | 1.944144724 | 1.11E-14 | 2.33E-11 | 6.99E-11 |
| HTN | macrophage | *TMEM165* | 0.978154027 | 0.000515453 | 0.016790942 | 1 |
| HTN | macrophage | *POM121C* | -0.964987727 | 0.001315338 | 0.034600556 | 1 |
| HTN | macrophage | *BLOC1S1* | 1.026768574 | 0.000873964 | 0.024975513 | 1 |
| HTN | macrophage | *AGT* | 0.266997722 | 7.49E-05 | 0.003860155 | 0.47093888 |

| | | | | | | |
|---|---|---|---|---|---|---|
| HTN | macrophage | *NMT1* | 0.74293035 | 2.90E-09 | 1.22E-06 | 1.82E-05 |
| HTN | macrophage | *BLK* | -1.211951733 | 0.00015768 | 0.007216241 | 0.9913366 |
| HTN | macrophage | *PAQR5* | -0.999680332 | 0.000981839 | 0.027680826 | 1 |
| HTN | macrophage | *RMDN3* | 1.62078832 | 3.61E-06 | 0.000365549 | 0.022725969 |
| HTN | macrophage | *IFT172* | 1.294961817 | 0.000859972 | 0.024687881 | 1 |
| HTN | macrophage | *DYNC2LI1* | 0.601869809 | 0.000462746 | 0.015641314 | 1 |
| HTN | macrophage | *ARL3* | 1.375904666 | 2.34E-05 | 0.001500281 | 0.147027584 |
| HTN | macrophage | *FBN2* | 0.17644768 | 0.000797239 | 0.02331274 | 1 |
| HTN | macrophage | *MMAB* | 0.959791608 | 0.000324748 | 0.012099761 | 1 |
| HTN | macrophage | *GPR84* | -2.29846061 | 0.000427962 | 0.014783495 | 1 |
| HTN | macrophage | *SCRN2* | -0.19938354 | 0.000283982 | 0.01102096 | 1 |
| HTN | macrophage | *C18orf8* | -1.177611281 | 7.61E-05 | 0.003888671 | 0.478306584 |
| HTN | macrophage | *NPC1* | -1.303996794 | 0.00011252 | 0.00544164 | 0.707413156 |
| HTN | macrophage | *TRIM65* | -0.83220672 | 0.000403896 | 0.014185989 | 1 |
| HTN | macrophage | *SYPL2* | 0.385444766 | 0.001141237 | 0.031060407 | 1 |
| HTN | macrophage | *RGS5* | -0.830700775 | 0.00189136 | 0.046268406 | 1 |
| HTN | macrophage | *ARNT* | -0.97988178 | 0.001820213 | 0.044877169 | 1 |
| HTN | macrophage | *RAB13* | -2.525789351 | 1.76E-05 | 0.001215468 | 0.110607543 |
| HTN | macrophage | *CREB3L4* | -0.707887902 | 5.37E-05 | 0.002958856 | 0.337309582 |
| HTN | macrophage | *WNT9A* | 0.969294181 | 4.00E-05 | 0.002331165 | 0.251765872 |
| HTN | macrophage | *PLEKHA6* | 0.847251192 | 0.00118977 | 0.031966165 | 1 |
| HTN | macrophage | *SLC10A4* | 1.908834465 | 2.97E-05 | 0.001813474 | 0.186787814 |
| HTN | macrophage | *ABT1* | 0.740022125 | 0.000640686 | 0.019737376 | 1 |
| HTN | macrophage | *SLC18B1* | -0.288677527 | 2.72E-05 | 0.001678374 | 0.171194101 |
| HTN | macrophage | *NRBF2* | -0.678250716 | 7.42E-06 | 0.000639071 | 0.046652208 |
| HTN | macrophage | *PDCD11* | 2.88656458 | 2.72E-15 | 1.35E-11 | 1.71E-11 |
| HTN | macrophage | *HSD17B12* | -0.563734677 | 6.59E-09 | 2.07E-06 | 4.15E-05 |
| HTN | macrophage | *PTPRJ* | -0.780328878 | 0.000569935 | 0.018100963 | 1 |
| HTN | macrophage | *DUSP15* | -1.15461302 | 2.10E-05 | 0.00139127 | 0.132170617 |
| HTN | macrophage | *DOC2A* | -1.398151891 | 1.79E-08 | 4.17E-06 | 0.000112632 |
| HTN | macrophage | *ARL14EP* | -1.187066262 | 0.000151892 | 0.007126451 | 0.954944451 |
| HTN | macrophage | *CWF19L2* | 1.149132869 | 0.001993044 | 0.047284032 | 1 |
| HTN | macrophage | *HS3ST3A1* | 0.567109068 | 0.001885645 | 0.046268406 | 1 |
| HTN | macrophage | *NEIL2* | 0.859504642 | 2.51E-08 | 5.32E-06 | 0.000157743 |
| HTN | macrophage | *PROM2* | 2.658206503 | 0.000501902 | 0.016436202 | 1 |
| HTN | macrophage | *GOLGA7B* | -0.75734087 | 0.000753274 | 0.022338844 | 1 |
| HTN | macrophage | *NDUFAF6* | -1.25188497 | 4.58E-06 | 0.000441415 | 0.028815599 |
| HTN | macrophage | *ANKRD9* | -1.559085194 | 0.00080751 | 0.023503777 | 1 |
| HTN | macrophage | *CLEC18A* | -0.907632668 | 9.91E-06 | 0.000799066 | 0.062327151 |
| HTN | macrophage | *TRIM63* | 0.557637244 | 0.000193264 | 0.008322273 | 1 |
| HTN | macrophage | *ZNF276* | 1.350907588 | 0.00121496 | 0.032366336 | 1 |
| HTN | macrophage | *SLC5A11* | -0.334407315 | 1.13E-06 | 0.000138617 | 0.007104423 |
| HTN | macrophage | *MED8* | 0.902674714 | 4.77E-06 | 0.000441415 | 0.029966857 |
| HTN | macrophage | *S100B* | -0.217080714 | 6.80E-05 | 0.003578828 | 0.427743442 |
| HTN | macrophage | *PAQR6* | -0.982480778 | 0.001697025 | 0.042676793 | 1 |
| HTN | macrophage | *PMF1* | 0.866732268 | 0.002064408 | 0.048248832 | 1 |
| HTN | macrophage | *ABCF3* | 0.703983852 | 1.40E-06 | 0.000159752 | 0.008786376 |
| HTN | macrophage | *TNFSF13* | 1.160145769 | 1.30E-05 | 0.00098569 | 0.081812229 |
| HTN | macrophage | *TTLL10* | 1.958503058 | 0.001060029 | 0.029229829 | 1 |
| HTN | macrophage | *LMOD1* | -0.786722897 | 1.33E-07 | 2.54E-05 | 0.000837549 |

| | | | | | | |
|---|---|---|---|---|---|---|
| HTN | macrophage | *RFT1* | -1.022293456 | 0.000398153 | 0.014062863 | 1 |
| HTN | macrophage | *FAM208A* | 0.984852848 | 0.000499937 | 0.016436202 | 1 |
| HTN | macrophage | *EXO5* | -0.960238907 | 2.06E-05 | 0.001377814 | 0.129514557 |
| HTN | macrophage | *C1orf50* | -1.316576095 | 0.000233458 | 0.009550306 | 1 |
| HTN | macrophage | *CDC25A* | -0.910600052 | 2.52E-11 | 3.02E-08 | 1.59E-07 |
| HTN | macrophage | *ZNF589* | -0.779217557 | 3.27E-11 | 3.02E-08 | 2.06E-07 |
| HTN | macrophage | *APEH* | -1.077252526 | 4.56E-09 | 1.79E-06 | 2.87E-05 |
| HTN | macrophage | *RNF123* | 0.842742265 | 2.90E-11 | 3.02E-08 | 1.82E-07 |
| HTN | macrophage | *ERAP1* | -0.837424298 | 5.77E-07 | 7.77E-05 | 0.003629085 |
| HTN | macrophage | *CTSB* | -0.345954299 | 0.000858059 | 0.024687881 | 1 |
| HTN | macrophage | *REEP3* | -1.149297645 | 7.70E-05 | 0.00390359 | 0.48404516 |
| HTN | macrophage | *SPINT1* | -0.979089616 | 0.000570064 | 0.018100963 | 1 |
| HTN | macrophage | *TRMT61A* | -0.962716324 | 1.97E-05 | 0.001342328 | 0.123942461 |
| HTN | macrophage | *XRRA1* | -0.379774229 | 0.001481045 | 0.038161199 | 1 |
| HTN | macrophage | *TRIM66* | -0.181360268 | 4.70E-05 | 0.002661676 | 0.295446071 |
| HTN | macrophage | *RIMKLB* | -0.574844181 | 8.83E-07 | 0.000113291 | 0.005551259 |
| HTN | macrophage | *KIR3DL1* | 0.906896131 | 6.33E-05 | 0.003370513 | 0.397720482 |
| HTN | macrophage | *SRR* | 0.330277142 | 0.000192336 | 0.008322273 | 1 |
| HTN | macrophage | *CYP7A1* | 1.083244027 | 0.000274076 | 0.010702589 | 1 |
| HTN | macrophage | *HRASLS5* | 0.171326209 | 0.000501949 | 0.016436202 | 1 |
| HTN | macrophage | *IRS1* | -0.34775135 | 5.43E-09 | 1.80E-06 | 3.41E-05 |
| HTN | macrophage | *C8orf46* | 0.097748571 | 0.001048986 | 0.029052751 | 1 |
| HTN | macrophage | *INO80E* | -1.424572673 | 1.53E-08 | 3.86E-06 | 9.64E-05 |
| HTN | macrophage | *LIMS1* | 0.700814168 | 0.001551123 | 0.039650248 | 1 |
| HTN | macrophage | *CALML6* | -0.948990109 | 0.000226738 | 0.009378282 | 1 |
| HTN | macrophage | *ELP5* | -1.674052989 | 1.65E-05 | 0.001177727 | 0.103639958 |
| HTN | macrophage | *RNF34* | -1.039092719 | 0.001197845 | 0.032046166 | 1 |
| HTN | macrophage | *PAQR8* | -1.877569757 | 1.68E-07 | 2.96E-05 | 0.001055076 |
| HTN | macrophage | *SOX7* | 1.427743732 | 0.001232537 | 0.032696046 | 1 |
| HTN | macrophage | *MRPL36* | 0.938247015 | 0.001080486 | 0.029663811 | 1 |
| HTN | macrophage | *RGS19* | -1.049999457 | 4.28E-15 | 1.35E-11 | 2.69E-11 |
| HTN | macrophage | *NME6* | 0.809516191 | 1.27E-09 | 6.61E-07 | 7.99E-06 |
| HTN | macrophage | *CTSW* | -0.150066079 | 8.22E-05 | 0.004100792 | 0.516699748 |
| HTN | macrophage | *HOXC5* | 0.769421628 | 0.000397445 | 0.014062863 | 1 |
| HTN | macrophage | *DCAKD* | -0.344454021 | 0.000528113 | 0.017114673 | 1 |
| HTN | macrophage | *VEGFB* | -0.856697705 | 1.16E-08 | 3.33E-06 | 7.32E-05 |
| HTN | macrophage | *MST1* | -0.465266173 | 4.72E-06 | 0.000441415 | 0.029661955 |
| HTN | macrophage | *SNX31* | -0.80541707 | 0.000179877 | 0.007908299 | 1 |
| HTN | macrophage | *GLIS1* | -0.337706368 | 0.001035229 | 0.028892253 | 1 |
| HTN | macrophage | *KLF14* | -6.39836043 | 1.63E-05 | 0.001177151 | 0.102412101 |
| HTN | macrophage | *RSRC1* | -1.015834346 | 1.11E-06 | 0.000138617 | 0.007000537 |
| HTN | macrophage | *NRIP3* | -0.602939125 | 1.21E-05 | 0.000925973 | 0.075929762 |
| HTN | macrophage | *PPP2R2D* | -0.910884443 | 0.000135408 | 0.006400807 | 0.851307278 |
| HTN | macrophage | *MSRA* | -1.322714473 | 0.000643576 | 0.019737376 | 1 |
| HTN | macrophage | *MTHFR* | -2.426867216 | 7.52E-10 | 4.30E-07 | 4.73E-06 |
| HTN | macrophage | *TOP3A* | -1.371412217 | 0.000588786 | 0.018601505 | 1 |
| HTN | macrophage | *PPFIA3* | -0.620785608 | 0.001960498 | 0.047260764 | 1 |
| HTN | macrophage | *PCDHB9* | 0.535325593 | 0.000459805 | 0.015625912 | 1 |
| HTN | macrophage | *SH2B1* | -0.444790997 | 0.000192931 | 0.008322273 | 1 |
| HTN | macrophage | *COX14* | -1.487769212 | 1.65E-07 | 2.96E-05 | 0.001036442 |

| | | | | | | |
|---|---|---|---|---|---|---|
| HTN | macrophage | *RPP25* | 0.80832312 | 2.15E-08 | 4.82E-06 | 0.000134995 |
| HTN | macrophage | *FAM219B* | -1.894636023 | 2.51E-05 | 0.00159396 | 0.157802062 |
| HTN | macrophage | *MPI* | -2.181630773 | 3.36E-11 | 3.02E-08 | 2.11E-07 |
| HTN | macrophage | *TRIM73* | -1.050317447 | 0.000439086 | 0.015084873 | 1 |
| HTN | macrophage | *HYI* | -0.471234676 | 2.69E-07 | 4.23E-05 | 0.001693066 |
| HTN | macrophage | *CTC1* | 0.724544819 | 0.000711951 | 0.021314448 | 1 |
| HTN | macrophage | *CHST13* | -1.653216855 | 0.00145832 | 0.037730277 | 1 |
| HTN | macrophage | *ZNF443* | 0.758346831 | 0.000199667 | 0.008539508 | 1 |
| HTN | macrophage | *TMEM136* | 0.588076696 | 0.002030185 | 0.047626008 | 1 |
| HTN | macrophage | *CLDN7* | -0.752966636 | 2.76E-09 | 1.22E-06 | 1.74E-05 |
| HTN | macrophage | *IDH2* | -1.268171158 | 0.002080884 | 0.048453767 | 1 |
| HTN | macrophage | *FAM89A* | 0.463143254 | 0.001551449 | 0.039650248 | 1 |
| HTN | macrophage | *FES* | -2.523544345 | 5.06E-07 | 7.06E-05 | 0.003178552 |
| HTN | macrophage | *COL18A1* | 0.662930972 | 0.000613775 | 0.019294007 | 1 |
| HTN | macrophage | *ZNF320* | 0.241866835 | 0.000296631 | 0.011234454 | 1 |
| HTN | macrophage | *GAS6* | -0.958663242 | 0.000397308 | 0.014062863 | 1 |
| HTN | macrophage | *RP1L1* | 1.412857551 | 5.06E-09 | 1.80E-06 | 3.18E-05 |
| HTN | macrophage | *BMP8A* | 2.212840138 | 1.15E-06 | 0.000138617 | 0.007208102 |
| HTN | macrophage | *KREMEN1* | -0.24071806 | 0.000272579 | 0.010702589 | 1 |
| HTN | macrophage | *EIF3C* | 0.623507267 | 8.38E-07 | 0.00010973 | 0.00526706 |
| HTN | macrophage | *A3GALT2* | -0.498201058 | 2.29E-05 | 0.001482688 | 0.143820721 |
| HTN | macrophage | *CCR10* | 1.923080448 | 0.000156915 | 0.007216241 | 0.986522549 |
| HTN | macrophage | *LPAR5* | -0.539745375 | 0.00010204 | 0.005011929 | 0.641526908 |
| HTN | macrophage | *DDX51* | 0.472173265 | 0.000412934 | 0.014345578 | 1 |
| HTN | macrophage | *CDK10* | -0.815891347 | 2.69E-06 | 0.000284829 | 0.016882624 |
| HTN | macrophage | *TCN2* | -0.429641691 | 0.000829731 | 0.024039258 | 1 |
| HTN | macrophage | *MYBL1* | -0.356496164 | 0.001038595 | 0.028892253 | 1 |
| HTN | macrophage | *ARL17A* | -0.871217451 | 0.000207094 | 0.008680018 | 1 |
| HTN | macrophage | *PLEKHN1* | 0.811739172 | 0.000204044 | 0.008667744 | 1 |
| HTN | macrophage | *MCRS1* | 1.596353414 | 5.81E-07 | 7.77E-05 | 0.003649804 |
| HTN | macrophage | *ZDHHC11* | 1.083739408 | 5.69E-05 | 0.003110613 | 0.35772045 |
| HTN | macrophage | *FBF1* | -0.388093785 | 2.33E-06 | 0.000256475 | 0.014619092 |
| HTN | macrophage | *NOC2L* | -0.383118782 | 0.000773566 | 0.022726204 | 1 |
| HTN | macrophage | *HLA-DRB1* | -0.630844202 | 7.54E-09 | 2.26E-06 | 4.74E-05 |
| HTN | macrophage | *SULT1A1* | -0.493034872 | 7.90E-05 | 0.003974284 | 0.496785511 |
| HTN | macrophage | *FAM180B* | -1.12175122 | 1.02E-10 | 7.12E-08 | 6.40E-07 |
| HTN | macrophage | *C6orf106* | 0.58983735 | 8.73E-06 | 0.000721824 | 0.054858599 |
| HTN | macrophage | *HIST1H3E* | 0.802937239 | 3.94E-08 | 7.74E-06 | 0.000247718 |
| HTN | macrophage | *WDR5B* | 0.842262193 | 0.001925854 | 0.046929627 | 1 |
| HTN | macrophage | *ZNF100* | -0.707155521 | 1.42E-05 | 0.001052728 | 0.0894819 |
| HTN | macrophage | *SULT1A2* | 0.237306682 | 1.09E-05 | 0.000858652 | 0.068692139 |
| HTN | macrophage | *SLC22A5* | 0.594131653 | 5.24E-09 | 1.80E-06 | 3.30E-05 |
| HTN | macrophage | *MYO5A* | -0.52674535 | 0.001437492 | 0.037345085 | 1 |
| HTN | macrophage | *RPS26* | -0.3179819 | 4.77E-06 | 0.000441415 | 0.030016249 |
| HTN | macrophage | *RPS6KL1* | 0.44789109 | 0.001811447 | 0.044836875 | 1 |
| HTN | macrophage | *TMEM116* | -0.654629218 | 8.94E-11 | 7.02E-08 | 5.62E-07 |
| HTN | macrophage | *HLA-DRB5* | -0.141635639 | 0.000233935 | 0.009550306 | 1 |
| HTN | macrophage | *PNP* | -0.22114922 | 0.001305546 | 0.034487255 | 1 |
| HTN | macrophage | *DENND4B* | -1.226511423 | 2.23E-06 | 0.00025042 | 0.014023502 |
| HTN | macrophage | *HSD17B8* | -0.738346732 | 5.04E-06 | 0.000455101 | 0.031676974 |

| | | | | | | |
|---|---|---|---|---|---|---|
| HTN | macrophage | *HLA-DOA* | -0.426551418 | 0.001587418 | 0.040242337 | 1 |
| HTN | macrophage | *PRRT1* | 0.647654489 | 1.71E-05 | 0.001194575 | 0.10751175 |
| HTN | macrophage | *MRPL38* | 0.615944386 | 0.000952795 | 0.026982996 | 1 |
| HTN | macrophage | *STK19* | 0.849132115 | 2.72E-06 | 0.000284829 | 0.017089722 |
| HTN | macrophage | *C6orf48* | 0.922796429 | 0.000293796 | 0.011194517 | 1 |
| HTN | macrophage | *GPANK1* | 0.84113899 | 5.88E-05 | 0.003161079 | 0.369846197 |
| HTN | macrophage | *BAG6* | 0.89851457 | 5.07E-06 | 0.000455101 | 0.031857056 |
| HTN | macrophage | *HLA-C* | -0.36448376 | 0.001952635 | 0.047260764 | 1 |
| HTN | macrophage | *CCHCR1* | -0.257043942 | 0.002089085 | 0.04846522 | 1 |
| HTN | macrophage | *ATXN2* | -1.136639202 | 0.000250029 | 0.01007649 | 1 |
| HTN | macrophage | *EIF3CL* | 2.637498297 | 2.69E-05 | 0.001671681 | 0.168839774 |
| HTN | macrophage | *ZDHHC11B* | 0.791683815 | 0.002027403 | 0.047626008 | 1 |
| HTN | macrophage | *SUPT4H1* | 1.287049911 | 0.000681912 | 0.020711013 | 1 |
| HTN | macrophage | *MXD3* | -0.537501112 | 0.00155933 | 0.039690315 | 1 |
| HTN | macrophage | *SIPA1* | 1.113512651 | 5.62E-06 | 0.000497663 | 0.035334045 |
| HTN | macrophage | *ATF6B* | 1.037512205 | 1.37E-09 | 6.61E-07 | 8.59E-06 |
| HTN | macrophage | *LTB4R* | -0.648967935 | 3.57E-06 | 0.000365549 | 0.02246494 |
| HTN | macrophage | *LTB4R2* | -1.028312757 | 3.39E-05 | 0.001989556 | 0.212882533 |
| HTN | macrophage | *REPIN1* | -0.985287824 | 0.000739744 | 0.022041558 | 1 |
| HTN | macrophage | *AC016773.1* | 0.314575864 | 0.000388272 | 0.014029113 | 1 |
| HTN | macrophage | *EXOSC6* | -1.168904068 | 6.89E-05 | 0.003578828 | 0.433038133 |
| HTN | macrophage | *AL049840.1* | -0.520427734 | 0.000484664 | 0.016122131 | 1 |
| HTN | macrophage | *SLC26A6* | 2.130814761 | 2.54E-08 | 5.32E-06 | 0.000159743 |
| HTN | macrophage | *PROB1* | 2.484465196 | 1.99E-05 | 0.001342328 | 0.124836472 |
| HTN | macrophage | *ANKRD65* | 1.023288234 | 0.000327177 | 0.012099761 | 1 |
| HTN | macrophage | *HLA-DQA2* | 0.294345746 | 0.000120649 | 0.005790231 | 0.758520275 |
| HTN | macrophage | *PSG4* | 0.809999791 | 0.000293493 | 0.011194517 | 1 |
| HTN | macrophage | *PLA2G4B* | 0.498971779 | 0.00015389 | 0.007166694 | 0.967503725 |
| HTN | macrophage | *JMJD7* | 0.463085774 | 0.001183544 | 0.031935368 | 1 |
| HTN | macrophage | *ZNF10* | -0.77499243 | 1.48E-05 | 0.001082869 | 0.093126708 |
| HTN | macrophage | *HMBS* | 0.533717299 | 1.02E-05 | 0.000813233 | 0.064245388 |
| HTN | macrophage | *AC144652.1* | 0.436921291 | 0.000257992 | 0.010331188 | 1 |
| HTN | mast_cell | *RBM6* | -1.926552535 | 5.67E-06 | 0.001023171 | 0.035636281 |
| HTN | mast_cell | *HFE* | -4.334276837 | 2.07E-05 | 0.002711032 | 0.130129534 |
| HTN | mast_cell | *CEP68* | 4.064395894 | 0.000104922 | 0.008133045 | 0.659643915 |
| HTN | mast_cell | *LIMA1* | 1.712362052 | 0.00021385 | 0.014152399 | 1 |
| HTN | mast_cell | *SDCCAG8* | 3.992847692 | 3.57E-05 | 0.004158498 | 0.224558874 |
| HTN | mast_cell | *UHRF1BP1* | 2.821160543 | 3.12E-06 | 0.000785835 | 0.019645864 |
| HTN | mast_cell | *CLNS1A* | 3.398631208 | 0.000270998 | 0.01638236 | 1 |
| HTN | mast_cell | *DDX43* | -6.203197779 | 0.000212752 | 0.014152399 | 1 |
| HTN | mast_cell | *SNX10* | -1.154741982 | 4.09E-05 | 0.004285594 | 0.257135642 |
| HTN | mast_cell | *AAMDC* | -2.790045112 | 1.27E-05 | 0.002003974 | 0.080158976 |
| HTN | mast_cell | *YPEL3* | 2.498349116 | 9.86E-05 | 0.008050132 | 0.619860165 |
| HTN | mast_cell | *TGM1* | 3.089871366 | 8.33E-05 | 0.007177674 | 0.523970201 |
| HTN | mast_cell | *SLC7A4* | -3.949345399 | 0.000541482 | 0.027677197 | 1 |
| HTN | mast_cell | *DNTTIP1* | -3.29667069 | 0.001107754 | 0.048364241 | 1 |
| HTN | mast_cell | *POLI* | -1.195245606 | 0.000376407 | 0.021319544 | 1 |
| HTN | mast_cell | *CORO1A* | -6.225120261 | 0.000133114 | 0.009731278 | 0.836889883 |
| HTN | mast_cell | *MTHFSD* | 2.279710454 | 0.000682502 | 0.032506748 | 1 |
| HTN | mast_cell | *EYA1* | 1.875871505 | 0.000106454 | 0.008133045 | 0.669276709 |

| | | | | | | |
|---|---|---|---|---|---|---|
| HTN | mast_cell | *ICAM5* | -19.69061035 | 6.14E-05 | 0.005847017 | 0.38590312 |
| HTN | mast_cell | *ATP13A1* | 3.857158942 | 9.75E-05 | 0.008050132 | 0.613252423 |
| HTN | mast_cell | *SYNGR2* | -4.857476063 | 0.000231994 | 0.014732789 | 1 |
| HTN | mast_cell | *KCTD10* | -3.371564332 | 0.000416276 | 0.02295727 | 1 |
| HTN | mast_cell | *ENDOU* | -3.395566622 | 0.000449228 | 0.023934695 | 1 |
| HTN | mast_cell | *CAND1* | -4.548821383 | 0.00034377 | 0.019828273 | 1 |
| HTN | mast_cell | *PCDHB6* | 6.102011754 | 0.000234859 | 0.014765572 | 1 |
| HTN | mast_cell | *DNAJC27* | -5.658810853 | 0.000525261 | 0.027068169 | 1 |
| HTN | mast_cell | *C1orf21* | -3.138832442 | 0.000307318 | 0.018248021 | 1 |
| HTN | mast_cell | *IRF6* | -2.060167518 | 0.000387895 | 0.021581359 | 1 |
| HTN | mast_cell | *TCF21* | 20.26509671 | 0.000216385 | 0.014170942 | 1 |
| HTN | mast_cell | *TEX14* | 6.9167965 | 3.90E-05 | 0.004249342 | 0.245113002 |
| HTN | mast_cell | *TRIM6* | -4.606103037 | 0.000104755 | 0.008133045 | 0.658592147 |
| HTN | mast_cell | *FAM124B* | 4.580470654 | 3.85E-05 | 0.004249342 | 0.241990811 |
| HTN | mast_cell | *TOX2* | -1.253670347 | 0.000448683 | 0.023934695 | 1 |
| HTN | mast_cell | *SNRPC* | -4.338174019 | 1.24E-05 | 0.002000711 | 0.078027719 |
| HTN | mast_cell | *ZNF391* | -1.31511285 | 0.000327535 | 0.019066801 | 1 |
| HTN | mast_cell | *GRK4* | 1.343135843 | 0.000483079 | 0.025309319 | 1 |
| HTN | mast_cell | *OPRL1* | -6.623307063 | 3.13E-08 | 1.23E-05 | 0.000196508 |
| HTN | mast_cell | *TRAP1* | 2.059193763 | 6.46E-07 | 0.000169338 | 0.004064101 |
| HTN | mast_cell | *PDLIM4* | 4.61567562 | 5.86E-06 | 0.001023171 | 0.036834141 |
| HTN | mast_cell | *WNT2B* | -5.128862369 | 0.000265537 | 0.016208093 | 1 |
| HTN | mast_cell | *DDB2* | 4.213057065 | 0.000967365 | 0.043441607 | 1 |
| HTN | mast_cell | *ACP2* | 9.20177805 | 1.16E-13 | 7.30E-10 | 7.30E-10 |
| HTN | mast_cell | *AGT* | 1.788992152 | 3.33E-05 | 0.004100217 | 0.209111087 |
| HTN | mast_cell | *NMT1* | 4.092742299 | 2.22E-08 | 9.31E-06 | 0.000139649 |
| HTN | mast_cell | *BLK* | -7.772622458 | 0.000132283 | 0.009731278 | 0.831665094 |
| HTN | mast_cell | *RMDN3* | 4.143965567 | 0.000639266 | 0.030915904 | 1 |
| HTN | mast_cell | *FBN2* | 1.156832457 | 5.99E-05 | 0.00579236 | 0.376503416 |
| HTN | mast_cell | *GPR84* | -13.72151613 | 0.000635434 | 0.030915904 | 1 |
| HTN | mast_cell | *DEF8* | 2.396945065 | 0.00111589 | 0.04838347 | 1 |
| HTN | mast_cell | *SYPL2* | 1.992317188 | 0.000947796 | 0.042869011 | 1 |
| HTN | mast_cell | *CREB3L4* | -4.400188163 | 8.04E-05 | 0.007024111 | 0.505736011 |
| HTN | mast_cell | *WNT9A* | 2.95229838 | 0.001003094 | 0.044411651 | 1 |
| HTN | mast_cell | *SLC10A4* | 12.86671624 | 5.56E-06 | 0.001023171 | 0.034970869 |
| HTN | mast_cell | *TSLP* | -2.33571333 | 0.000626514 | 0.030842119 | 1 |
| HTN | mast_cell | *NRBF2* | -4.2778345 | 4.78E-06 | 0.00100245 | 0.030073499 |
| HTN | mast_cell | *PLEKHS1* | 8.156318066 | 0.000688956 | 0.032567412 | 1 |
| HTN | mast_cell | *PDCD11* | 8.001020425 | 3.42E-08 | 1.26E-05 | 0.000214866 |
| HTN | mast_cell | *HSD17B12* | -3.136110732 | 1.97E-07 | 6.20E-05 | 0.001240485 |
| HTN | mast_cell | *DUSP15* | -4.248568115 | 5.22E-05 | 0.005205414 | 0.327941098 |
| HTN | mast_cell | *DOC2A* | -5.206469075 | 0.000754561 | 0.035402442 | 1 |
| HTN | mast_cell | *NEIL2* | 2.578874679 | 9.51E-06 | 0.001574003 | 0.059812108 |
| HTN | mast_cell | *NDUFAF6* | -4.542085414 | 0.000507325 | 0.026359921 | 1 |
| HTN | mast_cell | *SFXN2* | 5.01779721 | 0.000165874 | 0.011459866 | 1 |
| HTN | mast_cell | *CLEC18A* | -3.697750584 | 1.73E-05 | 0.002411034 | 0.108496538 |
| HTN | mast_cell | *SLC5A11* | -1.230895374 | 0.001154588 | 0.049065736 | 1 |
| HTN | mast_cell | *MED8* | 4.292113792 | 3.53E-05 | 0.004158498 | 0.221838814 |
| HTN | mast_cell | *S100B* | -1.469658697 | 3.43E-05 | 0.004141871 | 0.215377303 |
| HTN | mast_cell | *PAQR6* | -7.076909019 | 0.000427937 | 0.023395113 | 1 |

| | | | | | | |
|---|---|---|---|---|---|---|
| HTN | mast_cell | *ABCF3* | 4.245435155 | 4.64E-06 | 0.00100245 | 0.029142462 |
| HTN | mast_cell | *LMOD1* | -3.868258475 | 5.73E-06 | 0.001023171 | 0.036045479 |
| HTN | mast_cell | *EXO5* | -3.119991212 | 0.000101627 | 0.008133045 | 0.638928091 |
| HTN | mast_cell | *C1orf50* | -8.50077691 | 1.41E-05 | 0.002162779 | 0.088673957 |
| HTN | mast_cell | *CDC25A* | -5.888437042 | 5.12E-11 | 8.04E-08 | 3.22E-07 |
| HTN | mast_cell | *ZNF589* | -4.914866967 | 2.36E-11 | 4.94E-08 | 1.48E-07 |
| HTN | mast_cell | *APEH* | -5.728319824 | 2.33E-09 | 1.33E-06 | 1.46E-05 |
| HTN | mast_cell | *RNF123* | 4.682579535 | 8.30E-11 | 1.04E-07 | 5.22E-07 |
| HTN | mast_cell | *ERAP1* | -4.740051522 | 4.44E-07 | 0.00012702 | 0.002794436 |
| HTN | mast_cell | *CHMP4C* | -2.073877508 | 0.000627929 | 0.030842119 | 1 |
| HTN | mast_cell | *TRMT61A* | -4.70656673 | 0.000361827 | 0.020680065 | 1 |
| HTN | mast_cell | *TRIM66* | -1.052190914 | 0.000187297 | 0.012661668 | 1 |
| HTN | mast_cell | *RIMKLB* | -2.918708234 | 2.22E-05 | 0.002850099 | 0.139654871 |
| HTN | mast_cell | *KIR3DL1* | 5.936525898 | 3.26E-05 | 0.004100217 | 0.205150126 |
| HTN | mast_cell | *SRR* | 2.19285067 | 8.65E-05 | 0.007345531 | 0.54356929 |
| HTN | mast_cell | *CYP7A1* | 6.837114808 | 0.000107371 | 0.008133045 | 0.675042743 |
| HTN | mast_cell | *HRASLS5* | 1.084747887 | 0.000107058 | 0.008133045 | 0.673072004 |
| HTN | mast_cell | *IRS1* | -2.325347235 | 1.54E-09 | 1.08E-06 | 9.69E-06 |
| HTN | mast_cell | *C8orf46* | 0.670363205 | 0.000556927 | 0.028237083 | 1 |
| HTN | mast_cell | *INO80E* | -7.882229893 | 1.66E-08 | 7.47E-06 | 0.000104563 |
| HTN | mast_cell | *PAQR8* | -9.770833782 | 9.72E-05 | 0.008050132 | 0.610898764 |
| HTN | mast_cell | *RGS19* | -6.065677653 | 2.35E-12 | 7.37E-09 | 1.47E-08 |
| HTN | mast_cell | *NME6* | 5.131401427 | 1.98E-09 | 1.25E-06 | 1.25E-05 |
| HTN | mast_cell | *HOXC5* | 4.160885598 | 0.000307665 | 0.018248021 | 1 |
| HTN | mast_cell | *VEGFB* | -2.95111006 | 0.000160034 | 0.011179233 | 1 |
| HTN | mast_cell | *NRROS* | 2.317326543 | 0.00061277 | 0.030575259 | 1 |
| HTN | mast_cell | *SNX31* | -5.509211925 | 7.87E-05 | 0.006968466 | 0.494761105 |
| HTN | mast_cell | *RSRC1* | -4.456316966 | 4.03E-05 | 0.004285594 | 0.253370209 |
| HTN | mast_cell | *LRRC25* | 1.6536757 | 0.000645289 | 0.030968939 | 1 |
| HTN | mast_cell | *PCDHB9* | 2.424194337 | 0.000456309 | 0.024107666 | 1 |
| HTN | mast_cell | *RPP25* | 5.082724555 | 6.71E-10 | 5.28E-07 | 4.22E-06 |
| HTN | mast_cell | *MPI* | -12.398567 | 1.07E-10 | 1.12E-07 | 6.73E-07 |
| HTN | mast_cell | *HYI* | -3.019261152 | 3.28E-07 | 9.83E-05 | 0.002064464 |
| HTN | mast_cell | *ANKRD34A* | -7.262753868 | 0.000917005 | 0.041776897 | 1 |
| HTN | mast_cell | *CLN8* | 5.620858986 | 6.35E-05 | 0.005895625 | 0.399046295 |
| HTN | mast_cell | *FES* | -9.518698445 | 0.000253301 | 0.015767353 | 1 |
| HTN | mast_cell | *ZNF320* | 1.666099798 | 5.33E-05 | 0.005232494 | 0.334879612 |
| HTN | mast_cell | *RP1L1* | 7.323402575 | 7.47E-08 | 2.61E-05 | 0.00046981 |
| HTN | mast_cell | *KREMEN1* | -0.90587608 | 0.000226242 | 0.014514104 | 1 |
| HTN | mast_cell | *EIF3C* | 3.498157218 | 1.56E-05 | 0.002280278 | 0.097777079 |
| HTN | mast_cell | *HYAL3* | -1.523017887 | 0.00078089 | 0.036098916 | 1 |
| HTN | mast_cell | *PLEKHN1* | 4.893279518 | 0.000222528 | 0.014423016 | 1 |
| HTN | mast_cell | *ZNF793* | -2.277439408 | 0.000149754 | 0.0106989 | 0.941503161 |
| HTN | mast_cell | *FBF1* | -2.464801243 | 3.62E-06 | 0.000876396 | 0.022786302 |
| HTN | mast_cell | *NOC2L* | -2.37639885 | 0.000181949 | 0.012433829 | 1 |
| HTN | mast_cell | *HLA-DRB1* | -4.105087314 | 4.29E-09 | 2.25E-06 | 2.69E-05 |
| HTN | mast_cell | *SULT1A1* | -2.872316826 | 0.000257977 | 0.015900998 | 1 |
| HTN | mast_cell | *C6orf106* | 3.012184468 | 0.000143629 | 0.010379254 | 0.902995061 |
| HTN | mast_cell | *HIST1H3E* | 3.950207569 | 8.21E-06 | 0.001395303 | 0.051626208 |
| HTN | mast_cell | *ZNF100* | -4.092293218 | 1.68E-05 | 0.002405649 | 0.105848555 |

| | | | | | | |
|---|---|---|---|---|---|---|
| HTN | mast_cell | *SULT1A2* | 1.428499863 | 5.13E-05 | 0.005200804 | 0.322449822 |
| HTN | mast_cell | *SLC22A5* | 3.430661176 | 9.26E-09 | 4.48E-06 | 5.82E-05 |
| HTN | mast_cell | *RPS26* | -1.917456794 | 2.03E-05 | 0.002709204 | 0.127332584 |
| HTN | mast_cell | *TMEM116* | -3.964208339 | 3.72E-10 | 3.34E-07 | 2.34E-06 |
| HTN | mast_cell | *HLA-DRB5* | -0.911571847 | 0.000384638 | 0.021581359 | 1 |
| HTN | mast_cell | *AL590822.1* | -3.22185433 | 6.57E-05 | 0.005985007 | 0.412965517 |
| HTN | mast_cell | *CENPW* | -6.891107355 | 0.001155039 | 0.049065736 | 1 |
| HTN | mast_cell | *HSD17B8* | -3.360877948 | 3.89E-06 | 0.000906073 | 0.02446398 |
| HTN | mast_cell | *PRRT1* | 4.07288998 | 7.74E-05 | 0.006949888 | 0.486492179 |
| HTN | mast_cell | *MRPL38* | 3.163387264 | 0.000611585 | 0.030575259 | 1 |
| HTN | mast_cell | *STK19* | 4.192155185 | 5.39E-06 | 0.001023171 | 0.033882372 |
| HTN | mast_cell | *C6orf48* | 5.056996061 | 0.000989277 | 0.044110529 | 1 |
| HTN | mast_cell | *GPANK1* | 5.517259093 | 4.31E-06 | 0.000968798 | 0.027126352 |
| HTN | mast_cell | *BAG6* | 4.160723108 | 1.88E-05 | 0.002572035 | 0.118313598 |
| HTN | mast_cell | *MICB* | -1.171404497 | 0.000880534 | 0.040408164 | 1 |
| HTN | mast_cell | *PSORS1C1* | 0.568059838 | 0.000764363 | 0.035596667 | 1 |
| HTN | mast_cell | *EIF3CL* | 9.391960669 | 0.000321525 | 0.018891829 | 1 |
| HTN | mast_cell | *SIPA1* | 6.626983305 | 4.71E-07 | 0.000128767 | 0.002961642 |
| HTN | mast_cell | *ATF6B* | 4.715104529 | 8.95E-08 | 2.96E-05 | 0.000562383 |
| HTN | mast_cell | *LTB4R* | -3.708211395 | 3.92E-05 | 0.004249342 | 0.246461854 |
| HTN | mast_cell | *LTB4R2* | -7.074407319 | 5.49E-06 | 0.001023171 | 0.034509814 |
| HTN | mast_cell | *AP4M1* | -2.08173765 | 0.000446554 | 0.023934695 | 1 |
| HTN | mast_cell | *SLC26A6* | 7.01274024 | 0.001132695 | 0.048775728 | 1 |
| HTN | mast_cell | *PROB1* | 12.67057293 | 3.85E-05 | 0.004249342 | 0.242355341 |
| HTN | mast_cell | *ANKRD65* | 4.192713998 | 0.001064626 | 0.046806338 | 1 |
| HTN | mast_cell | *HLA-DQA2* | 2.052427387 | 4.29E-05 | 0.00442658 | 0.270021389 |
| HTN | mast_cell | *PLA2G4B* | 3.404935001 | 6.38E-05 | 0.005895625 | 0.400902512 |
| HTN | mast_cell | *JMJD7* | 3.388984845 | 0.000122629 | 0.009178188 | 0.770967754 |
| HTN | mast_cell | *HMBS* | 2.612062723 | 1.56E-05 | 0.002280278 | 0.098051959 |
| HTN | mast_cell | *AC144652.1* | 2.098761674 | 0.000157424 | 0.01112048 | 0.989722742 |
| HTN | monocyte | *RBM6* | -1.694659934 | 0.000334091 | 0.044690059 | 1 |
| HTN | monocyte | *SDCCAG8* | 3.388972466 | 6.75E-05 | 0.013252232 | 0.42407143 |
| HTN | monocyte | *UHRF1BP1* | 3.380938073 | 1.64E-06 | 0.001106116 | 0.010291579 |
| HTN | monocyte | *SNX10* | -1.294127646 | 1.26E-05 | 0.00397217 | 0.079443403 |
| HTN | monocyte | *C3orf18* | 7.715071693 | 0.000184728 | 0.02765207 | 1 |
| HTN | monocyte | *YPEL3* | 2.968397212 | 0.000169925 | 0.026056497 | 1 |
| HTN | monocyte | *ATP13A1* | 6.457803925 | 2.68E-05 | 0.007021256 | 0.168510148 |
| HTN | monocyte | *HOXA2* | -18.06376227 | 0.000357905 | 0.046878119 | 1 |
| HTN | monocyte | *TEX14* | 10.84291097 | 3.87E-08 | 4.87E-05 | 0.000243353 |
| HTN | monocyte | *FAM124B* | 3.729402037 | 0.000253522 | 0.034649877 | 1 |
| HTN | monocyte | *SNRPC* | -5.129352722 | 1.70E-05 | 0.005085108 | 0.106787267 |
| HTN | monocyte | *TRAP1* | 2.317414228 | 1.32E-06 | 0.001039442 | 0.008315534 |
| HTN | monocyte | *CHMP1A* | 6.299560571 | 0.000146126 | 0.023556292 | 0.918695399 |
| HTN | monocyte | *DDB2* | 6.213113642 | 0.000234084 | 0.032704127 | 1 |
| HTN | monocyte | *ACP2* | 15.79156386 | 9.52E-18 | 5.99E-14 | 5.99E-14 |
| HTN | monocyte | *AGT* | 2.054900085 | 8.83E-05 | 0.016318807 | 0.554839449 |
| HTN | monocyte | *NMT1* | 3.802839797 | 3.42E-07 | 0.000307429 | 0.002152 |
| HTN | monocyte | *CREB3L4* | -5.198915693 | 8.74E-05 | 0.016318807 | 0.549289509 |
| HTN | monocyte | *SLC10A4* | 13.70832453 | 2.61E-05 | 0.007021256 | 0.16391349 |
| HTN | monocyte | *PDCD11* | 8.054286441 | 2.06E-07 | 0.000215597 | 0.001293585 |

| | | | | | | |
|---|---|---|---|---|---|---|
| HTN | monocyte | *HSD17B12* | -3.051804303 | 3.86E-06 | 0.001734706 | 0.024285879 |
| HTN | monocyte | *DUSP15* | -8.059996595 | 0.000117584 | 0.020171249 | 0.739252578 |
| HTN | monocyte | *CLEC18A* | -5.606569479 | 6.30E-05 | 0.012826885 | 0.396086993 |
| HTN | monocyte | *SLC5A11* | -1.719486502 | 0.000156182 | 0.024547946 | 0.981917847 |
| HTN | monocyte | *ABCF3* | 4.999555864 | 3.65E-06 | 0.001734706 | 0.022918619 |
| HTN | monocyte | *CDC25A* | -4.141719905 | 1.76E-06 | 0.001106116 | 0.011061162 |
| HTN | monocyte | *ZNF589* | -3.418016434 | 2.91E-06 | 0.001665243 | 0.018317678 |
| HTN | monocyte | *RNF123* | 4.391324588 | 7.88E-09 | 1.65E-05 | 4.95E-05 |
| HTN | monocyte | *KIR3DL1* | 6.348775816 | 0.000117106 | 0.020171249 | 0.736244078 |
| HTN | monocyte | *SRR* | 2.276332337 | 0.000124097 | 0.020531602 | 0.780200883 |
| HTN | monocyte | *NME6* | 5.384075871 | 4.87E-09 | 1.53E-05 | 3.06E-05 |
| HTN | monocyte | *HYI* | -2.979778001 | 7.43E-06 | 0.002691229 | 0.046699636 |
| HTN | monocyte | *CLDN7* | -3.90548422 | 6.32E-05 | 0.012826885 | 0.397633431 |
| HTN | monocyte | *ZNF320* | 1.893940473 | 0.000225878 | 0.032274928 | 1 |
| HTN | monocyte | *RP1L1* | 5.125282029 | 3.86E-05 | 0.009711728 | 0.242793198 |
| HTN | monocyte | *EIF3C* | 3.958019529 | 4.52E-06 | 0.00189518 | 0.028427698 |
| HTN | monocyte | *DDX51* | 3.391864317 | 0.000388192 | 0.049807362 | 1 |
| HTN | monocyte | *FBF1* | -2.846837202 | 5.16E-06 | 0.002028033 | 0.032448535 |
| HTN | monocyte | *C6orf106* | 3.307989783 | 4.44E-05 | 0.010696023 | 0.279298443 |
| HTN | monocyte | *HIST1H3E* | 3.936484941 | 3.23E-06 | 0.00169457 | 0.020334839 |
| HTN | monocyte | *SLC22A5* | 1.909672488 | 6.23E-05 | 0.012826885 | 0.391919435 |
| HTN | monocyte | *RPS26* | -2.356279196 | 1.07E-05 | 0.003549516 | 0.067440801 |
| HTN | monocyte | *TMEM116* | -2.069198643 | 0.000118711 | 0.020171249 | 0.746336227 |
| HTN | monocyte | *GPANK1* | 5.738742003 | 5.59E-05 | 0.012555171 | 0.351544794 |
| HTN | monocyte | *BAG6* | 4.538126791 | 4.59E-05 | 0.010696023 | 0.288792609 |
| HTN | monocyte | *SIPA1* | 7.199499502 | 7.71E-06 | 0.002691229 | 0.04844212 |
| HTN | monocyte | *ATF6B* | 6.056034282 | 3.61E-08 | 4.87E-05 | 0.000226719 |
| HTN | monocyte | *PLA2G4B* | 3.683110255 | 0.000193061 | 0.028227348 | 1 |
| HTN | monocyte | *HMBS* | 3.801517469 | 2.11E-05 | 0.006017876 | 0.132393281 |
| HTN | neutrophil | *PIGV* | -10.86554844 | 4.36E-05 | 0.021103524 | 0.274345812 |
| HTN | neutrophil | *POLR1A* | -6.193084679 | 1.38E-05 | 0.008698367 | 0.086983671 |
| HTN | neutrophil | *ZNF391* | -4.827386325 | 3.33E-06 | 0.007683136 | 0.020947563 |
| HTN | neutrophil | *WNT2B* | -9.623641691 | 3.50E-05 | 0.018352307 | 0.220227687 |
| HTN | neutrophil | *ACP2* | 13.70197566 | 2.05E-05 | 0.01171789 | 0.12889679 |
| HTN | neutrophil | *NRBF2* | -6.697017811 | 9.75E-05 | 0.043799608 | 0.613194513 |
| HTN | neutrophil | *HSD17B12* | -4.846692252 | 1.02E-05 | 0.008698367 | 0.064336894 |
| HTN | neutrophil | *CLEC18A* | -17.67035813 | 1.33E-05 | 0.008698367 | 0.08387271 |
| HTN | neutrophil | *S100B* | -4.196046104 | 9.78E-06 | 0.008698367 | 0.06149009 |
| HTN | neutrophil | *EXO5* | -15.48547035 | 4.09E-06 | 0.007683136 | 0.025744349 |
| HTN | neutrophil | *CDC25A* | -8.66800785 | 1.08E-06 | 0.006773933 | 0.006773933 |
| HTN | neutrophil | *APEH* | -9.386603765 | 4.89E-06 | 0.007683136 | 0.030732543 |
| HTN | neutrophil | *CTSW* | -2.859268376 | 1.30E-05 | 0.008698367 | 0.081907015 |
| HTN | neutrophil | *HSD17B8* | -11.75957511 | 6.32E-06 | 0.007950505 | 0.039752524 |
| HTN | nk_cell | *CFH* | -1.272892378 | 0.007838203 | 0.021632111 | 1 |
| HTN | nk_cell | *SEMA3F* | -1.872044868 | 1.31E-08 | 3.92E-06 | 8.23E-05 |
| HTN | nk_cell | *RAD52* | -1.916397275 | 0.018781546 | 0.041489664 | 1 |
| HTN | nk_cell | *BAD* | -2.691811271 | 0.002925264 | 0.011804324 | 1 |
| HTN | nk_cell | *HS3ST1* | -1.986839639 | 0.002319284 | 0.010460071 | 1 |
| HTN | nk_cell | *WNT16* | -3.261957995 | 0.00010527 | 0.003520371 | 0.661829693 |
| HTN | nk_cell | *SNX11* | -1.12614852 | 0.019196151 | 0.042173672 | 1 |

| | | | | | | |
|---|---|---|---|---|---|---|
| HTN | nk_cell | *TMEM176A* | 1.335945566 | 0.006284526 | 0.018663588 | 1 |
| HTN | nk_cell | *M6PR* | -2.940738911 | 0.00034153 | 0.006546344 | 1 |
| HTN | nk_cell | *CYP26B1* | -1.388181045 | 0.00213109 | 0.010104197 | 1 |
| HTN | nk_cell | *ICA1* | -1.128310409 | 0.014109505 | 0.033639157 | 1 |
| HTN | nk_cell | *ALS2* | -1.602880506 | 0.003122208 | 0.012264669 | 1 |
| HTN | nk_cell | *POLDIP2* | -1.724260646 | 0.001860222 | 0.009533293 | 1 |
| HTN | nk_cell | *CD38* | 0.339144547 | 0.002090336 | 0.010006758 | 1 |
| HTN | nk_cell | *RBM6* | -1.126219178 | 1.89E-06 | 0.000188514 | 0.011851575 |
| HTN | nk_cell | *PDK4* | -1.420200325 | 0.02134747 | 0.045744957 | 1 |
| HTN | nk_cell | *SLC22A16* | -1.864686073 | 0.000298519 | 0.006297947 | 1 |
| HTN | nk_cell | *GCFC2* | -1.424052888 | 0.00697555 | 0.019919637 | 1 |
| HTN | nk_cell | *WDR54* | 1.15026516 | 0.001429004 | 0.008851376 | 1 |
| HTN | nk_cell | *ABCB4* | -2.818132934 | 0.002225272 | 0.010252893 | 1 |
| HTN | nk_cell | *PDK2* | -1.236745974 | 0.004951517 | 0.015834277 | 1 |
| HTN | nk_cell | *GGNBP2* | -2.108615814 | 0.000103609 | 0.003490877 | 0.651392396 |
| HTN | nk_cell | *TMEM98* | -1.309180768 | 0.004771464 | 0.015470962 | 1 |
| HTN | nk_cell | *TMEM132A* | -1.850315621 | 0.001726118 | 0.009259473 | 1 |
| HTN | nk_cell | *TNFRSF12A* | 2.632662536 | 0.01309486 | 0.031811201 | 1 |
| HTN | nk_cell | *DLX6* | 2.024781705 | 0.001922451 | 0.009654896 | 1 |
| HTN | nk_cell | *MAP3K9* | -1.755110566 | 0.019831165 | 0.043176651 | 1 |
| HTN | nk_cell | *RALA* | -1.656538568 | 0.017786246 | 0.039709563 | 1 |
| HTN | nk_cell | *BAIAP2L1* | -1.981367001 | 0.001616452 | 0.009059476 | 1 |
| HTN | nk_cell | *GTF2IRD1* | -0.899705632 | 0.012196082 | 0.030262856 | 1 |
| HTN | nk_cell | *ARHGAP44* | -1.125931161 | 0.002779168 | 0.011427488 | 1 |
| HTN | nk_cell | *ELAC2* | -1.877841495 | 0.021838189 | 0.046503529 | 1 |
| HTN | nk_cell | *CEACAM21* | 0.236279116 | 0.001539657 | 0.008971107 | 1 |
| HTN | nk_cell | *NOS2* | -4.9642078 | 0.001162339 | 0.008849228 | 1 |
| HTN | nk_cell | *TRAPPC6A* | -2.66393467 | 0.001308006 | 0.00885079 | 1 |
| HTN | nk_cell | *ST7L* | -1.244279737 | 5.30E-05 | 0.002316216 | 0.333485144 |
| HTN | nk_cell | *RPUSD1* | 2.897098534 | 0.004600894 | 0.015167822 | 1 |
| HTN | nk_cell | *TSR3* | 2.212647207 | 0.022311117 | 0.047258931 | 1 |
| HTN | nk_cell | *CDK11A* | -1.203306078 | 0.008092607 | 0.022140218 | 1 |
| HTN | nk_cell | *DLEC1* | -3.064071846 | 6.05E-05 | 0.00256943 | 0.380275579 |
| HTN | nk_cell | *MGST1* | -1.002127242 | 0.002029895 | 0.009861903 | 1 |
| HTN | nk_cell | *NFIX* | -1.032061257 | 0.002235462 | 0.01026356 | 1 |
| HTN | nk_cell | *MMP25* | 0.47350213 | 0.001217394 | 0.00885079 | 1 |
| HTN | nk_cell | *IL32* | 0.357831285 | 0.001413779 | 0.00885079 | 1 |
| HTN | nk_cell | *MAPK8IP2* | 2.185670055 | 0.001057326 | 0.008846261 | 1 |
| HTN | nk_cell | *POMT2* | -1.873228149 | 0.006341084 | 0.018778331 | 1 |
| HTN | nk_cell | *MLXIPL* | -0.545851281 | 0.000198527 | 0.005142286 | 1 |
| HTN | nk_cell | *SPRTN* | -2.247914946 | 0.008585178 | 0.023135455 | 1 |
| HTN | nk_cell | *METTL13* | -2.462538204 | 0.003515107 | 0.013068882 | 1 |
| HTN | nk_cell | *DYRK4* | -1.165995525 | 0.021811539 | 0.046484456 | 1 |
| HTN | nk_cell | *STARD3NL* | 1.538570373 | 0.011064232 | 0.028082691 | 1 |
| HTN | nk_cell | *CD4* | -1.946383739 | 0.00509149 | 0.016150454 | 1 |
| HTN | nk_cell | *HFE* | -2.98068986 | 4.77E-10 | 2.72E-07 | 3.00E-06 |
| HTN | nk_cell | *CLCN6* | -3.374625492 | 2.51E-05 | 0.001349357 | 0.157874792 |
| HTN | nk_cell | *MRC2* | -1.072010057 | 0.005344885 | 0.016674626 | 1 |
| HTN | nk_cell | *TSPAN9* | -1.014711515 | 0.018765784 | 0.041469414 | 1 |
| HTN | nk_cell | *MKS1* | -1.970689552 | 0.00660529 | 0.019234451 | 1 |

| | | | | | | |
|---|---|---|---|---|---|---|
| HTN | nk_cell | *TTC19* | -1.639569155 | 0.009829293 | 0.025561939 | 1 |
| HTN | nk_cell | *SYT7* | -1.873312309 | 0.000330814 | 0.006479211 | 1 |
| HTN | nk_cell | *TMEM159* | -1.001420216 | 0.016033615 | 0.036742556 | 1 |
| HTN | nk_cell | *ERCC1* | -1.695709359 | 0.004579263 | 0.015120708 | 1 |
| HTN | nk_cell | *LTF* | 2.653201265 | 0.000421595 | 0.007214859 | 1 |
| HTN | nk_cell | *ALOX5* | -2.406865771 | 0.000536881 | 0.008086807 | 1 |
| HTN | nk_cell | *PSMC4* | 1.595792288 | 0.011549999 | 0.028964837 | 1 |
| HTN | nk_cell | *CLDN11* | -1.160078989 | 0.001775872 | 0.009341888 | 1 |
| HTN | nk_cell | *SLC25A39* | -1.460406037 | 0.007975815 | 0.021896922 | 1 |
| HTN | nk_cell | *HEBP1* | -2.446693907 | 0.003158713 | 0.012344206 | 1 |
| HTN | nk_cell | *MDH1* | -1.32723312 | 0.007498192 | 0.020970254 | 1 |
| HTN | nk_cell | *COX15* | -1.900592129 | 0.008972456 | 0.023922744 | 1 |
| HTN | nk_cell | *CCDC88C* | 0.397407492 | 0.000966717 | 0.008846261 | 1 |
| HTN | nk_cell | *IL20RA* | -1.974216998 | 0.001071947 | 0.008846261 | 1 |
| HTN | nk_cell | *GLT8D1* | -1.427794494 | 0.002414515 | 0.010645574 | 1 |
| HTN | nk_cell | *RUFY3* | -1.346181318 | 0.01505702 | 0.035217071 | 1 |
| HTN | nk_cell | *WWTR1* | -1.219377822 | 0.003460924 | 0.012974855 | 1 |
| HTN | nk_cell | *VSIG2* | -2.861614356 | 0.014105125 | 0.033639157 | 1 |
| HTN | nk_cell | *PHLDB1* | -0.482994791 | 0.020677216 | 0.04455026 | 1 |
| HTN | nk_cell | *MNAT1* | -1.495263039 | 0.014060338 | 0.033585618 | 1 |
| HTN | nk_cell | *OSBPL5* | 0.764144067 | 0.002932582 | 0.011811111 | 1 |
| HTN | nk_cell | *CPS1* | -0.628301778 | 0.008970666 | 0.023922744 | 1 |
| HTN | nk_cell | *NLRP2* | 0.298532802 | 0.001633261 | 0.009090004 | 1 |
| HTN | nk_cell | *GLRX2* | -1.78752899 | 0.001795819 | 0.009369553 | 1 |
| HTN | nk_cell | *SNAPC1* | -2.079654528 | 0.011830215 | 0.029573186 | 1 |
| HTN | nk_cell | *NR1H3* | -3.695254511 | 1.91E-08 | 5.46E-06 | 0.0001201 |
| HTN | nk_cell | *TOMM34* | -3.63951526 | 0.006614928 | 0.019244819 | 1 |
| HTN | nk_cell | *RNASET2* | -1.914964693 | 0.008230095 | 0.022389705 | 1 |
| HTN | nk_cell | *BTN3A1* | 0.598195033 | 0.003711955 | 0.013404401 | 1 |
| HTN | nk_cell | *INSRR* | -1.719308812 | 0.019993783 | 0.043421333 | 1 |
| HTN | nk_cell | *B4GALT7* | -2.400553221 | 0.00424779 | 0.014545674 | 1 |
| HTN | nk_cell | *VEZT* | -1.641113441 | 0.004844331 | 0.015613861 | 1 |
| HTN | nk_cell | *ARNTL2* | -1.58329597 | 0.001612801 | 0.009058261 | 1 |
| HTN | nk_cell | *ANK1* | -2.586222482 | 0.001806001 | 0.00938705 | 1 |
| HTN | nk_cell | *GRN* | -1.866736151 | 0.004361091 | 0.014746877 | 1 |
| HTN | nk_cell | *ZCCHC8* | -1.855914779 | 0.012074092 | 0.030003879 | 1 |
| HTN | nk_cell | *FUT8* | -1.431881869 | 0.01987345 | 0.043248315 | 1 |
| HTN | nk_cell | *SH3YL1* | -1.542059742 | 0.000938152 | 0.008846261 | 1 |
| HTN | nk_cell | *STAP1* | 0.900922215 | 0.011914043 | 0.029723645 | 1 |
| HTN | nk_cell | *MYOM2* | 0.13841052 | 0.001130408 | 0.008846261 | 1 |
| HTN | nk_cell | *DNAH5* | -1.389503512 | 0.002559069 | 0.010937366 | 1 |
| HTN | nk_cell | *C6* | -0.954338309 | 0.000730644 | 0.008846261 | 1 |
| HTN | nk_cell | *RAI14* | -1.751330499 | 0.001144181 | 0.008846261 | 1 |
| HTN | nk_cell | *PHLPP2* | -2.217649877 | 0.001759414 | 0.009314459 | 1 |
| HTN | nk_cell | *PQLC2* | -1.527695992 | 0.000546709 | 0.008118543 | 1 |
| HTN | nk_cell | *CTNS* | -1.013733895 | 0.000979616 | 0.008846261 | 1 |
| HTN | nk_cell | *RAB27B* | 0.56643154 | 0.001420647 | 0.00885079 | 1 |
| HTN | nk_cell | *SPATA7* | -1.827614132 | 0.004472843 | 0.014878713 | 1 |
| HTN | nk_cell | *RETSAT* | -0.691843049 | 0.021168563 | 0.045422101 | 1 |
| HTN | nk_cell | *CAPG* | -0.986661745 | 0.012970287 | 0.031569567 | 1 |

| | | | | | | |
|---|---|---|---|---|---|---|
| HTN | nk_cell | *LCP2* | 0.745055705 | 0.004694751 | 0.015371451 | 1 |
| HTN | nk_cell | *CTNNA1* | -1.452804799 | 0.004908224 | 0.015741663 | 1 |
| HTN | nk_cell | *CNTLN* | -1.44773381 | 0.020585278 | 0.044413055 | 1 |
| HTN | nk_cell | *EPHA3* | -1.846969249 | 0.014634369 | 0.034479556 | 1 |
| HTN | nk_cell | *DSG2* | -1.30268677 | 0.004219649 | 0.014465067 | 1 |
| HTN | nk_cell | *CC2D2A* | -1.42262895 | 0.003863521 | 0.013777629 | 1 |
| HTN | nk_cell | *RRM2B* | -2.082259842 | 0.002430294 | 0.010654267 | 1 |
| HTN | nk_cell | *LMO3* | -0.824116771 | 0.000202433 | 0.005142286 | 1 |
| HTN | nk_cell | *MRPS10* | -0.81553044 | 0.006822441 | 0.019666523 | 1 |
| HTN | nk_cell | *ADAMTS6* | -1.421958143 | 0.007084835 | 0.020105374 | 1 |
| HTN | nk_cell | *EPN3* | 6.344044325 | 8.95E-05 | 0.003215018 | 0.562628168 |
| HTN | nk_cell | *RCN1* | -2.206352718 | 0.000287692 | 0.006179456 | 1 |
| HTN | nk_cell | *RFC2* | -2.602931665 | 0.016039641 | 0.036742556 | 1 |
| HTN | nk_cell | *DKK3* | -1.774266163 | 0.008270414 | 0.022470223 | 1 |
| HTN | nk_cell | *ARHGEF5* | -2.301304879 | 0.000878461 | 0.008846261 | 1 |
| HTN | nk_cell | *PTGER3* | -0.73535536 | 0.001972218 | 0.009771592 | 1 |
| HTN | nk_cell | *HERPUD1* | -1.658847836 | 0.006347977 | 0.018781121 | 1 |
| HTN | nk_cell | *PIK3CB* | -1.77885908 | 0.003809702 | 0.013633851 | 1 |
| HTN | nk_cell | *CYBA* | 0.522856718 | 0.002564436 | 0.01095286 | 1 |
| HTN | nk_cell | *THOC3* | 3.175844139 | 0.014238801 | 0.03381816 | 1 |
| HTN | nk_cell | *RRP12* | -1.558906078 | 0.017460318 | 0.039194226 | 1 |
| HTN | nk_cell | *ALX4* | -1.554440546 | 0.001025616 | 0.008846261 | 1 |
| HTN | nk_cell | *MRTO4* | -1.823226766 | 0.002228543 | 0.010252893 | 1 |
| HTN | nk_cell | *MCF2L2* | -1.760321838 | 0.00450481 | 0.014962133 | 1 |
| HTN | nk_cell | *LY75* | 0.570962454 | 0.002059692 | 0.00993635 | 1 |
| HTN | nk_cell | *PLEKHH1* | -0.605246651 | 0.015309961 | 0.035506435 | 1 |
| HTN | nk_cell | *ATP9A* | -1.134609307 | 0.001017527 | 0.008846261 | 1 |
| HTN | nk_cell | *GINM1* | -1.716049912 | 0.009041945 | 0.02404683 | 1 |
| HTN | nk_cell | *MRPL43* | -1.289162664 | 0.014088925 | 0.033631019 | 1 |
| HTN | nk_cell | *ITIH4* | 0.802820105 | 0.001062803 | 0.008846261 | 1 |
| HTN | nk_cell | *IL17RB* | -2.958125321 | 0.001567401 | 0.008975583 | 1 |
| HTN | nk_cell | *TRAF3IP2* | -1.45132244 | 0.003528637 | 0.013080507 | 1 |
| HTN | nk_cell | *DCBLD2* | -1.404885587 | 0.002118019 | 0.010087867 | 1 |
| HTN | nk_cell | *MSH4* | -0.951940373 | 0.012637696 | 0.031000076 | 1 |
| HTN | nk_cell | *CDK14* | -1.23494276 | 0.003291176 | 0.012647692 | 1 |
| HTN | nk_cell | *ZC3H11A* | -1.895849927 | 0.011317215 | 0.028529003 | 1 |
| HTN | nk_cell | *DNAJC25* | -1.764443128 | 0.005932615 | 0.017931898 | 1 |
| HTN | nk_cell | *YBX3* | -1.464471806 | 0.001794431 | 0.009369553 | 1 |
| HTN | nk_cell | *PIGV* | -3.144195038 | 0.000199865 | 0.005142286 | 1 |
| HTN | nk_cell | *NCKAP1* | -1.103866453 | 0.003150731 | 0.012326475 | 1 |
| HTN | nk_cell | *ISOC2* | -1.374503953 | 0.001106715 | 0.008846261 | 1 |
| HTN | nk_cell | *CASP8* | 0.726466049 | 0.001801876 | 0.009385578 | 1 |
| HTN | nk_cell | *DMRT3* | -1.094369501 | 0.001443228 | 0.008873507 | 1 |
| HTN | nk_cell | *ATP2C2* | -2.591660274 | 0.013059594 | 0.031762348 | 1 |
| HTN | nk_cell | *CDON* | -0.747213144 | 0.001807949 | 0.00938705 | 1 |
| HTN | nk_cell | *TMEM161A* | -1.706795878 | 0.001740772 | 0.009286428 | 1 |
| HTN | nk_cell | *LPAR2* | 1.450389578 | 0.002292512 | 0.010399356 | 1 |
| HTN | nk_cell | *ABCA7* | 0.974058277 | 0.00910153 | 0.024154209 | 1 |
| HTN | nk_cell | *CHI3L2* | 1.195980437 | 0.002058981 | 0.00993635 | 1 |
| HTN | nk_cell | *TAF11* | -1.784056385 | 0.002109042 | 0.010060352 | 1 |

| | | | | | | |
|---|---|---|---|---|---|---|
| HTN | nk_cell | *UHRF1BP1* | 1.116926698 | 0.000791646 | 0.008846261 | 1 |
| HTN | nk_cell | *IPO5* | -1.837033232 | 0.013982538 | 0.033474803 | 1 |
| HTN | nk_cell | *ANKRD44* | 0.728340471 | 0.002525152 | 0.010838755 | 1 |
| HTN | nk_cell | *ZC3H15* | -1.944145095 | 0.012580958 | 0.030884998 | 1 |
| HTN | nk_cell | *SEC61A2* | -2.248435014 | 0.010271471 | 0.026465876 | 1 |
| HTN | nk_cell | *TIE1* | -2.693878304 | 6.09E-09 | 2.13E-06 | 3.83E-05 |
| HTN | nk_cell | *ZMYND12* | -0.752423755 | 0.012443637 | 0.030670335 | 1 |
| HTN | nk_cell | *SLC9A3* | -1.222040452 | 0.005653098 | 0.017328634 | 1 |
| HTN | nk_cell | *NGEF* | -3.725669347 | 0.001611016 | 0.009058261 | 1 |
| HTN | nk_cell | *CD84* | -1.117613474 | 0.017070292 | 0.038480074 | 1 |
| HTN | nk_cell | *ZNRD1* | 1.694966153 | 0.001162631 | 0.008849228 | 1 |
| HTN | nk_cell | *ATXN3* | -1.689864641 | 0.000316903 | 0.006421199 | 1 |
| HTN | nk_cell | *GOLGA5* | -1.885057802 | 0.006166579 | 0.018391499 | 1 |
| HTN | nk_cell | *EML1* | -0.621353389 | 0.016055365 | 0.036742556 | 1 |
| HTN | nk_cell | *STOML1* | -2.523698984 | 0.001197717 | 0.00885079 | 1 |
| HTN | nk_cell | *TP53BP1* | -1.554636265 | 0.003167044 | 0.012344206 | 1 |
| HTN | nk_cell | *DHX8* | -2.326935132 | 0.003682729 | 0.013360251 | 1 |
| HTN | nk_cell | *SYT1* | -0.778753158 | 0.01140211 | 0.028674026 | 1 |
| HTN | nk_cell | *RASSF1* | 0.593971576 | 0.004841762 | 0.015613861 | 1 |
| HTN | nk_cell | *IFI35* | 1.802742681 | 0.008880009 | 0.02372657 | 1 |
| HTN | nk_cell | *POLR1A* | -1.558820151 | 1.89E-06 | 0.000188514 | 0.011876381 |
| HTN | nk_cell | *CYFIP1* | -1.574035267 | 0.005678267 | 0.017397301 | 1 |
| HTN | nk_cell | *PITX1* | -2.070246763 | 0.001063907 | 0.008846261 | 1 |
| HTN | nk_cell | *SDK2* | -1.281025142 | 0.001043525 | 0.008846261 | 1 |
| HTN | nk_cell | *DNAJA2* | -2.036170057 | 0.002960433 | 0.011892808 | 1 |
| HTN | nk_cell | *KCNAB2* | 0.761547129 | 0.005594879 | 0.01721733 | 1 |
| HTN | nk_cell | *GAL* | -2.718613734 | 0.001007806 | 0.008846261 | 1 |
| HTN | nk_cell | *CLEC2D* | 0.719889599 | 0.003443874 | 0.01293636 | 1 |
| HTN | nk_cell | *PFN2* | -2.091917543 | 0.000868047 | 0.008846261 | 1 |
| HTN | nk_cell | *FGF10* | -0.532588208 | 0.022877794 | 0.048136778 | 1 |
| HTN | nk_cell | *CLTCL1* | -1.049293866 | 0.000565871 | 0.008121264 | 1 |
| HTN | nk_cell | *NDST1* | -1.54393534 | 0.000310961 | 0.006359244 | 1 |
| HTN | nk_cell | *ASNS* | -1.063777353 | 0.016623 | 0.037633705 | 1 |
| HTN | nk_cell | *AP3M2* | -1.754291934 | 0.000833881 | 0.008846261 | 1 |
| HTN | nk_cell | *ST6GALNAC2* | -1.166753562 | 0.016407588 | 0.037281986 | 1 |
| HTN | nk_cell | *TESK2* | -1.771038317 | 0.007054984 | 0.020042787 | 1 |
| HTN | nk_cell | *EIF2B3* | -1.519305172 | 0.016488537 | 0.037410116 | 1 |
| HTN | nk_cell | *CAMK2A* | -3.694401533 | 0.001256664 | 0.00885079 | 1 |
| HTN | nk_cell | *OSBPL3* | 0.944975573 | 0.007533176 | 0.021049367 | 1 |
| HTN | nk_cell | *VASH1* | -1.880296762 | 0.007704941 | 0.02138674 | 1 |
| HTN | nk_cell | *TRIP13* | -2.452469228 | 0.001082435 | 0.008846261 | 1 |
| HTN | nk_cell | *HLTF* | -1.425568554 | 0.009436764 | 0.024775264 | 1 |
| HTN | nk_cell | *EPN2* | -1.851320996 | 0.001696798 | 0.009212238 | 1 |
| HTN | nk_cell | *LIMS2* | -0.969788146 | 0.01570873 | 0.03614963 | 1 |
| HTN | nk_cell | *TRHDE* | -0.804818488 | 0.001287731 | 0.00885079 | 1 |
| HTN | nk_cell | *ACADVL* | -2.478220145 | 0.000469806 | 0.00757351 | 1 |
| HTN | nk_cell | *CRMP1* | -1.202804816 | 0.000525164 | 0.007994452 | 1 |
| HTN | nk_cell | *AP1M1* | -1.520979977 | 0.005182145 | 0.016306629 | 1 |
| HTN | nk_cell | *TP63* | -1.437159753 | 0.005885407 | 0.017857893 | 1 |
| HTN | nk_cell | *ALPK1* | -1.543113526 | 0.020514143 | 0.044335655 | 1 |

| | | | | | | |
|---|---|---|---|---|---|---|
| HTN | nk_cell | *LLGL2* | 0.290847848 | 0.002430163 | 0.010654267 | 1 |
| HTN | nk_cell | *PDE8A* | -1.379221238 | 0.009183892 | 0.024276248 | 1 |
| HTN | nk_cell | *GSDMB* | -1.244230292 | 0.010031414 | 0.025975083 | 1 |
| HTN | nk_cell | *ATP2A3* | 0.705003746 | 0.000203248 | 0.005142286 | 1 |
| HTN | nk_cell | *CA12* | -0.580742919 | 0.004451968 | 0.014861784 | 1 |
| HTN | nk_cell | *NTN4* | -1.075696054 | 0.000952265 | 0.008846261 | 1 |
| HTN | nk_cell | *LMAN1* | -1.855891459 | 0.008982159 | 0.023938464 | 1 |
| HTN | nk_cell | *TTC38* | 0.766838381 | 0.002550944 | 0.010917484 | 1 |
| HTN | nk_cell | *CELSR1* | -2.723586873 | 0.002410702 | 0.010645574 | 1 |
| HTN | nk_cell | *ADD2* | -1.619789329 | 0.000774466 | 0.008846261 | 1 |
| HTN | nk_cell | *MARK3* | -1.783344224 | 0.009721494 | 0.025371122 | 1 |
| HTN | nk_cell | *BCAP29* | -1.761312306 | 0.007364721 | 0.020652098 | 1 |
| HTN | nk_cell | *TUBA3D* | -2.300260329 | 0.003937089 | 0.013894565 | 1 |
| HTN | nk_cell | *RBMS2* | -1.373599785 | 0.005812082 | 0.017686622 | 1 |
| HTN | nk_cell | *FMO4* | -1.494392263 | 0.001692867 | 0.009212238 | 1 |
| HTN | nk_cell | *RGS11* | -1.049446623 | 4.46E-05 | 0.002078644 | 0.280186074 |
| HTN | nk_cell | *TPD52* | 0.814902796 | 0.000560685 | 0.008121264 | 1 |
| HTN | nk_cell | *ICAM3* | 0.383549527 | 0.00118328 | 0.00885079 | 1 |
| HTN | nk_cell | *CAMSAP3* | -4.978913029 | 0.001063796 | 0.008846261 | 1 |
| HTN | nk_cell | *XAB2* | -2.708774046 | 9.63E-05 | 0.003383954 | 0.60572773 |
| HTN | nk_cell | *NMRK2* | 2.617965032 | 0.006356963 | 0.018781121 | 1 |
| HTN | nk_cell | *CTTNBP2* | -1.570955253 | 0.009208173 | 0.024286608 | 1 |
| HTN | nk_cell | *FGFR1* | -1.262782723 | 0.000985378 | 0.008846261 | 1 |
| HTN | nk_cell | *FBLN1* | -0.876336757 | 0.016135557 | 0.036848619 | 1 |
| HTN | nk_cell | *FAP* | -0.594529497 | 0.009695667 | 0.025345803 | 1 |
| HTN | nk_cell | *ACER3* | -1.259918739 | 0.014095393 | 0.033631019 | 1 |
| HTN | nk_cell | *TULP3* | -1.319065591 | 0.00309368 | 0.012219765 | 1 |
| HTN | nk_cell | *FGF20* | -6.796155014 | 0.001057285 | 0.008846261 | 1 |
| HTN | nk_cell | *MYH7B* | -2.027714959 | 0.000364127 | 0.006654851 | 1 |
| HTN | nk_cell | *FKBP7* | -0.982774399 | 0.00234364 | 0.010532139 | 1 |
| HTN | nk_cell | *OSBPL6* | -1.708570178 | 0.001924222 | 0.009654896 | 1 |
| HTN | nk_cell | *LXN* | -1.528007244 | 0.003798196 | 0.013614173 | 1 |
| HTN | nk_cell | *RIMS1* | -1.404894705 | 0.001100706 | 0.008846261 | 1 |
| HTN | nk_cell | *RABL2B* | -2.552276399 | 0.010745482 | 0.027495662 | 1 |
| HTN | nk_cell | *DDX43* | -3.508289188 | 0.000157851 | 0.004430394 | 0.99240832 |
| HTN | nk_cell | *DCT* | -1.297654658 | 0.001323364 | 0.00885079 | 1 |
| HTN | nk_cell | *SCTR* | -1.581783164 | 0.001136778 | 0.008846261 | 1 |
| HTN | nk_cell | *KCNN2* | -5.770237527 | 0.000894375 | 0.008846261 | 1 |
| HTN | nk_cell | *CPOX* | -2.034066774 | 0.017109302 | 0.038526569 | 1 |
| HTN | nk_cell | *MOK* | -1.547408792 | 0.001858476 | 0.009533293 | 1 |
| HTN | nk_cell | *MAGI3* | -1.322278303 | 0.003331308 | 0.012697902 | 1 |
| HTN | nk_cell | *COL4A4* | -1.278499221 | 0.001589334 | 0.009018181 | 1 |
| HTN | nk_cell | *KIAA0141* | -2.366866433 | 0.010959745 | 0.027896324 | 1 |
| HTN | nk_cell | *PCDHA6* | -13.11831021 | 4.50E-05 | 0.002079562 | 0.282820406 |
| HTN | nk_cell | *PGR* | -0.659214258 | 0.002601813 | 0.01102956 | 1 |
| HTN | nk_cell | *C5orf22* | -1.994974282 | 0.003222984 | 0.012494105 | 1 |
| HTN | nk_cell | *CCNT2* | -1.507590386 | 0.005203959 | 0.016342136 | 1 |
| HTN | nk_cell | *COBLL1* | -1.063504087 | 0.000287988 | 0.006179456 | 1 |
| HTN | nk_cell | *GEMIN5* | -1.567509823 | 0.005206505 | 0.016342136 | 1 |
| HTN | nk_cell | *LYRM2* | -1.323906605 | 0.023389668 | 0.049033291 | 1 |

| | | | | | | |
|---|---|---|---|---|---|---|
| HTN | nk_cell | *ULK2* | -1.706191034 | 0.00139538 | 0.00885079 | 1 |
| HTN | nk_cell | *ZNF324* | -2.851181124 | 0.008084629 | 0.022128019 | 1 |
| HTN | nk_cell | *ZNF446* | -1.632557625 | 0.001295848 | 0.00885079 | 1 |
| HTN | nk_cell | *RPS5* | 1.387744936 | 0.020353074 | 0.044078461 | 1 |
| HTN | nk_cell | *CHMP2B* | -1.982994107 | 0.007054034 | 0.020042787 | 1 |
| HTN | nk_cell | *STARD7* | -1.978480099 | 0.007219556 | 0.020372239 | 1 |
| HTN | nk_cell | *NOA1* | -1.489510054 | 0.00010018 | 0.003441707 | 0.629832358 |
| HTN | nk_cell | *COL16A1* | -1.534788016 | 3.01E-05 | 0.001551557 | 0.189289963 |
| HTN | nk_cell | *APOB* | -0.892638198 | 0.000879309 | 0.008846261 | 1 |
| HTN | nk_cell | *CD59* | -1.655530207 | 0.010376938 | 0.026693866 | 1 |
| HTN | nk_cell | *TAF9* | -1.826045862 | 0.001361571 | 0.00885079 | 1 |
| HTN | nk_cell | *FCN1* | -0.646414959 | 0.001167073 | 0.00885079 | 1 |
| HTN | nk_cell | *OVGP1* | 1.628318802 | 0.010419814 | 0.0267713 | 1 |
| HTN | nk_cell | *ABCB1* | 0.67877605 | 0.002271765 | 0.010348636 | 1 |
| HTN | nk_cell | *ZNF213* | -1.784745887 | 0.002479872 | 0.010734818 | 1 |
| HTN | nk_cell | *CPNE3* | -1.125723247 | 0.006870992 | 0.019742515 | 1 |
| HTN | nk_cell | *RRN3* | -1.832653614 | 0.000738561 | 0.008846261 | 1 |
| HTN | nk_cell | *CTTN* | -1.135763805 | 0.001595765 | 0.009046506 | 1 |
| HTN | nk_cell | *MTIF2* | -1.540912324 | 0.021905596 | 0.046590151 | 1 |
| HTN | nk_cell | *ATG16L1* | 1.514112207 | 0.007802102 | 0.021551764 | 1 |
| HTN | nk_cell | *SNX10* | -0.866049715 | 1.38E-07 | 2.79E-05 | 0.000866416 |
| HTN | nk_cell | *SEPHS1* | -1.878395382 | 0.007276106 | 0.020467508 | 1 |
| HTN | nk_cell | *ACOX3* | -1.356929795 | 0.015080185 | 0.035258133 | 1 |
| HTN | nk_cell | *HSD17B14* | -0.915511057 | 0.001245545 | 0.00885079 | 1 |
| HTN | nk_cell | *ATXN7L3* | 1.435554972 | 0.003236317 | 0.012513362 | 1 |
| HTN | nk_cell | *OGFOD1* | -2.148428895 | 0.000820484 | 0.008846261 | 1 |
| HTN | nk_cell | *KLHL42* | -2.106748988 | 0.004860896 | 0.015631946 | 1 |
| HTN | nk_cell | *PTHLH* | -1.065455704 | 0.000888918 | 0.008846261 | 1 |
| HTN | nk_cell | *PHACTR3* | -0.620503076 | 0.008137334 | 0.022233558 | 1 |
| HTN | nk_cell | *AAMDC* | -1.61293464 | 2.86E-07 | 5.00E-05 | 0.00179949 |
| HTN | nk_cell | *GP6* | 1.083144512 | 0.001983544 | 0.00978483 | 1 |
| HTN | nk_cell | *ASAP3* | -0.805290086 | 0.001472027 | 0.008909666 | 1 |
| HTN | nk_cell | *FKBP1A* | -2.357035074 | 0.00242996 | 0.010654267 | 1 |
| HTN | nk_cell | *ZNF343* | -1.735399795 | 0.0083263 | 0.022553835 | 1 |
| HTN | nk_cell | *CPXM1* | -0.899776392 | 0.000466107 | 0.007546364 | 1 |
| HTN | nk_cell | *MAVS* | -1.675910826 | 0.010276474 | 0.026466606 | 1 |
| HTN | nk_cell | *LZTS3* | -1.103687321 | 0.001294126 | 0.00885079 | 1 |
| HTN | nk_cell | *TESC* | 1.151034275 | 0.002199157 | 0.010211299 | 1 |
| HTN | nk_cell | *RBBP9* | -1.642061366 | 0.000403055 | 0.007117989 | 1 |
| HTN | nk_cell | *DZANK1* | -2.157029831 | 0.001673639 | 0.009165654 | 1 |
| HTN | nk_cell | *FXYD5* | 0.891420995 | 0.003395734 | 0.012814513 | 1 |
| HTN | nk_cell | *FXYD3* | -3.22007211 | 0.001759697 | 0.009314459 | 1 |
| HTN | nk_cell | *GMIP* | 1.505461017 | 0.003821961 | 0.013659313 | 1 |
| HTN | nk_cell | *NECAP1* | -1.759154525 | 0.022699079 | 0.047840802 | 1 |
| HTN | nk_cell | *ANKRD24* | -0.790725069 | 0.006682968 | 0.019359449 | 1 |
| HTN | nk_cell | *RGS1* | -1.368081994 | 0.001073013 | 0.008846261 | 1 |
| HTN | nk_cell | *MRPS33* | -1.620784343 | 0.001857388 | 0.009533293 | 1 |
| HTN | nk_cell | *MUL1* | -3.042333416 | 0.002099329 | 0.010029243 | 1 |
| HTN | nk_cell | *GNPTG* | -2.053537415 | 0.008763838 | 0.023516111 | 1 |
| HTN | nk_cell | *PABPC4* | -3.039930631 | 6.90E-06 | 0.000522688 | 0.043383064 |

| | | | | | | |
|---|---|---|---|---|---|---|
| HTN | nk_cell | *NLRC4* | -1.409840472 | 0.00124983 | 0.00885079 | 1 |
| HTN | nk_cell | *LAMB1* | -0.853846389 | 0.001599737 | 0.009048265 | 1 |
| HTN | nk_cell | *SLC26A4* | -1.375495406 | 0.001031671 | 0.008846261 | 1 |
| HTN | nk_cell | *IL5RA* | 4.278945312 | 0.014273283 | 0.033849917 | 1 |
| HTN | nk_cell | *ITGA6* | -1.683523213 | 0.02044211 | 0.044195167 | 1 |
| HTN | nk_cell | *RAPGEF4* | -1.046480645 | 0.002511826 | 0.010816337 | 1 |
| HTN | nk_cell | *SEL1L3* | 1.387976518 | 0.004562189 | 0.015072246 | 1 |
| HTN | nk_cell | *TF* | -1.517404523 | 0.000936504 | 0.008846261 | 1 |
| HTN | nk_cell | *MYO15A* | -2.654716684 | 0.000365899 | 0.006667853 | 1 |
| HTN | nk_cell | *ZFHX4* | -0.752500303 | 0.001641517 | 0.009108692 | 1 |
| HTN | nk_cell | *TMEM101* | -2.928041108 | 0.008998588 | 0.023961932 | 1 |
| HTN | nk_cell | *CMA1* | -2.363906034 | 0.0020625 | 0.00993635 | 1 |
| HTN | nk_cell | *SLC22A17* | -1.52099713 | 0.000535832 | 0.008086807 | 1 |
| HTN | nk_cell | *MFSD11* | -1.534820895 | 0.011133326 | 0.028201136 | 1 |
| HTN | nk_cell | *DPYSL2* | -1.330221076 | 0.020643094 | 0.044491988 | 1 |
| HTN | nk_cell | *VNN3* | -3.19281131 | 0.001273727 | 0.00885079 | 1 |
| HTN | nk_cell | *PDE6C* | -1.48291408 | 0.013444542 | 0.032521354 | 1 |
| HTN | nk_cell | *IKZF5* | -1.957240578 | 0.001182222 | 0.00885079 | 1 |
| HTN | nk_cell | *BLNK* | -1.982137253 | 0.00133159 | 0.00885079 | 1 |
| HTN | nk_cell | *IL12RB1* | 0.34512492 | 0.002741058 | 0.011368608 | 1 |
| HTN | nk_cell | *PCSK5* | -0.974382617 | 0.001971782 | 0.009771592 | 1 |
| HTN | nk_cell | *NRP1* | -1.199452567 | 0.011152987 | 0.028239561 | 1 |
| HTN | nk_cell | *PALMD* | -0.667820072 | 0.000585924 | 0.008131793 | 1 |
| HTN | nk_cell | *TSPAN15* | -1.145889394 | 0.001121149 | 0.008846261 | 1 |
| HTN | nk_cell | *OCEL1* | -1.258047762 | 0.007408436 | 0.020756165 | 1 |
| HTN | nk_cell | *STX1B* | -2.970730639 | 0.000352712 | 0.006607413 | 1 |
| HTN | nk_cell | *HSD3B7* | -0.950409516 | 0.006984346 | 0.019919637 | 1 |
| HTN | nk_cell | *CDC34* | -2.973631658 | 0.015511457 | 0.035914868 | 1 |
| HTN | nk_cell | *MTAP* | -2.182279344 | 0.003688672 | 0.013360879 | 1 |
| HTN | nk_cell | *CEP170B* | -1.175503095 | 0.019634732 | 0.04293689 | 1 |
| HTN | nk_cell | *MADCAM1* | -1.200403442 | 0.000870226 | 0.008846261 | 1 |
| HTN | nk_cell | *TRMT2A* | -2.690723314 | 0.003873839 | 0.01379725 | 1 |
| HTN | nk_cell | *P2RX6* | -2.300986731 | 0.000916554 | 0.008846261 | 1 |
| HTN | nk_cell | *SLC7A4* | -3.964065789 | 2.58E-06 | 0.000245334 | 0.016192052 |
| HTN | nk_cell | *GGT5* | -2.993050116 | 0.004845454 | 0.015613861 | 1 |
| HTN | nk_cell | *SPECC1L* | -1.518952281 | 0.012545254 | 0.03082142 | 1 |
| HTN | nk_cell | *YPEL1* | 0.76317476 | 0.00238973 | 0.010625206 | 1 |
| HTN | nk_cell | *PPM1F* | -2.187368944 | 0.006812437 | 0.019650054 | 1 |
| HTN | nk_cell | *TOP3B* | -2.057568337 | 0.003031045 | 0.012060873 | 1 |
| HTN | nk_cell | *CYTH4* | 1.204889395 | 0.005617539 | 0.017270156 | 1 |
| HTN | nk_cell | *LRP5L* | -2.899517991 | 0.00061291 | 0.008358707 | 1 |
| HTN | nk_cell | *LGALS2* | -1.328400106 | 0.020032003 | 0.043487985 | 1 |
| HTN | nk_cell | *SH3BP1* | 0.488238721 | 0.001533237 | 0.008957475 | 1 |
| HTN | nk_cell | *TRIOBP* | -1.983093346 | 0.000121466 | 0.003780486 | 0.763658122 |
| HTN | nk_cell | *CCDC134* | 2.367489696 | 0.003220776 | 0.012494105 | 1 |
| HTN | nk_cell | *KDELR3* | -1.536902038 | 0.000502832 | 0.00788356 | 1 |
| HTN | nk_cell | *CYP2D6* | -1.181892328 | 0.018065519 | 0.040176129 | 1 |
| HTN | nk_cell | *FBXO7* | -1.872743858 | 0.015192502 | 0.035323691 | 1 |
| HTN | nk_cell | *TIMP3* | -1.23433422 | 0.011241207 | 0.028404792 | 1 |
| HTN | nk_cell | *CYB5R3* | -1.700858234 | 0.006681289 | 0.019359449 | 1 |

| | | | | | | |
|---|---|---|---|---|---|---|
| HTN | nk_cell | *RHBDD3* | -1.30440589 | 0.004908725 | 0.015741663 | 1 |
| HTN | nk_cell | *TTLL1* | -2.523201135 | 0.00997506 | 0.025864981 | 1 |
| HTN | nk_cell | *RASL10A* | -1.415930577 | 0.002441255 | 0.010654267 | 1 |
| HTN | nk_cell | *HMGXB4* | -2.008466241 | 0.0156123 | 0.03599515 | 1 |
| HTN | nk_cell | *BIK* | 5.854452705 | 0.013623066 | 0.03274759 | 1 |
| HTN | nk_cell | *MCAT* | -1.709322265 | 0.000254143 | 0.005756069 | 1 |
| HTN | nk_cell | *TSPO* | -1.644754476 | 0.015099417 | 0.035276861 | 1 |
| HTN | nk_cell | *TTLL12* | -1.9589064 | 0.001721145 | 0.009240684 | 1 |
| HTN | nk_cell | *RBFOX2* | -1.169344778 | 0.002731778 | 0.011343916 | 1 |
| HTN | nk_cell | *APOL4* | -0.855956879 | 0.00721943 | 0.020372239 | 1 |
| HTN | nk_cell | *SAMM50* | -2.285439724 | 0.00874912 | 0.023486643 | 1 |
| HTN | nk_cell | *FOXRED2* | -0.93586499 | 0.02099275 | 0.045106432 | 1 |
| HTN | nk_cell | *IFT27* | -2.125871409 | 0.002502085 | 0.010783493 | 1 |
| HTN | nk_cell | *KCTD17* | -1.474767671 | 0.001076876 | 0.008846261 | 1 |
| HTN | nk_cell | *ACO2* | -1.825308299 | 0.010563922 | 0.027064132 | 1 |
| HTN | nk_cell | *POLR3H* | -0.789183544 | 0.015025858 | 0.035183453 | 1 |
| HTN | nk_cell | *SDR39U1* | -1.111170633 | 0.006458744 | 0.018948262 | 1 |
| HTN | nk_cell | *PYGL* | -0.897656453 | 0.000119264 | 0.003730407 | 0.749811813 |
| HTN | nk_cell | *GSTZ1* | -1.080528401 | 0.009526873 | 0.024966841 | 1 |
| HTN | nk_cell | *DAAM1* | -1.107191663 | 0.003451495 | 0.012954956 | 1 |
| HTN | nk_cell | *ZFYVE21* | -1.3695657 | 0.006434556 | 0.018919244 | 1 |
| HTN | nk_cell | *MTHFD1* | -1.22014048 | 0.001947596 | 0.009717885 | 1 |
| HTN | nk_cell | *TELO2* | -2.997456168 | 0.000652531 | 0.00865499 | 1 |
| HTN | nk_cell | *CCNB1IP1* | -2.09998134 | 0.000217323 | 0.005316378 | 1 |
| HTN | nk_cell | *KIAA0391* | 2.409245626 | 0.004744521 | 0.015423878 | 1 |
| HTN | nk_cell | *PLTP* | -1.200230141 | 0.000826201 | 0.008846261 | 1 |
| HTN | nk_cell | *PYGB* | -1.39809939 | 0.019313969 | 0.042338537 | 1 |
| HTN | nk_cell | *NINL* | -1.328658962 | 0.015099124 | 0.035276861 | 1 |
| HTN | nk_cell | *CD40* | -1.336612815 | 0.001466851 | 0.008909666 | 1 |
| HTN | nk_cell | *UQCC1* | -1.307425209 | 0.017730881 | 0.039628171 | 1 |
| HTN | nk_cell | *IFT52* | -1.760540137 | 0.010142825 | 0.026209593 | 1 |
| HTN | nk_cell | *DOK5* | -0.966291741 | 0.001176418 | 0.00885079 | 1 |
| HTN | nk_cell | *BMP7* | -1.975386917 | 0.001337412 | 0.00885079 | 1 |
| HTN | nk_cell | *TPD52L2* | -1.675772221 | 0.012322682 | 0.030457938 | 1 |
| HTN | nk_cell | *PRPF6* | -2.234237435 | 0.001495549 | 0.008938486 | 1 |
| HTN | nk_cell | *TCFL5* | -1.072264755 | 0.011684995 | 0.029254947 | 1 |
| HTN | nk_cell | *NKAIN4* | -2.208802457 | 0.000323792 | 0.006479211 | 1 |
| HTN | nk_cell | *PTK6* | -5.56811527 | 0.001284586 | 0.00885079 | 1 |
| HTN | nk_cell | *CDC25B* | 1.474918649 | 0.004884163 | 0.015690718 | 1 |
| HTN | nk_cell | *ANGPT4* | -0.992914696 | 0.000709855 | 0.008846261 | 1 |
| HTN | nk_cell | *RSPO4* | 2.009931955 | 0.004254269 | 0.014559927 | 1 |
| HTN | nk_cell | *SIRPB1* | -1.038314451 | 0.003483565 | 0.013006455 | 1 |
| HTN | nk_cell | *SEC23B* | 1.470721497 | 0.003645524 | 0.013302035 | 1 |
| HTN | nk_cell | *CCM2L* | -2.093478138 | 0.002002878 | 0.009837575 | 1 |
| HTN | nk_cell | *MROH8* | -1.585440647 | 0.000639988 | 0.008562086 | 1 |
| HTN | nk_cell | *RPRD1B* | -1.880632617 | 0.002681181 | 0.011200387 | 1 |
| HTN | nk_cell | *BPI* | -1.763812343 | 0.003463384 | 0.012976336 | 1 |
| HTN | nk_cell | *DNTTIP1* | -2.058728338 | 2.26E-06 | 0.000222194 | 0.014220419 |
| HTN | nk_cell | *MAP1LC3A* | -0.792038899 | 0.010396528 | 0.026722432 | 1 |
| HTN | nk_cell | *APMAP* | 1.241909883 | 0.018126498 | 0.040254784 | 1 |

| | | | | | | |
|---|---|---|---|---|---|---|
| HTN | nk_cell | *CELF4* | -2.402698488 | 0.000657262 | 0.0086629 | 1 |
| HTN | nk_cell | *RBFA* | -1.151629884 | 0.007646232 | 0.021270736 | 1 |
| HTN | nk_cell | *MYL12A* | 1.00939054 | 0.00538777 | 0.016761499 | 1 |
| HTN | nk_cell | *POLI* | -0.842350408 | 4.42E-06 | 0.000389169 | 0.027786469 |
| HTN | nk_cell | *NALCN* | -2.33344077 | 0.002478296 | 0.010734818 | 1 |
| HTN | nk_cell | *HTR2A* | -1.763897741 | 0.001106502 | 0.008846261 | 1 |
| HTN | nk_cell | *STK24* | -1.70441456 | 0.016750326 | 0.037867422 | 1 |
| HTN | nk_cell | *UGGT2* | -1.047605577 | 0.01093884 | 0.027854388 | 1 |
| HTN | nk_cell | *SGCG* | -1.078920335 | 0.002607372 | 0.011031325 | 1 |
| HTN | nk_cell | *KATNAL1* | -1.240456636 | 0.005807631 | 0.017681635 | 1 |
| HTN | nk_cell | *DHRS12* | -1.143297007 | 0.00095878 | 0.008846261 | 1 |
| HTN | nk_cell | *MEDAG* | -0.771986621 | 0.001102402 | 0.008846261 | 1 |
| HTN | nk_cell | *CORO1A* | 0.244386087 | 0.003883636 | 0.013802386 | 1 |
| HTN | nk_cell | *LONP2* | -1.450622512 | 0.020621251 | 0.044463592 | 1 |
| HTN | nk_cell | *USB1* | 1.569581566 | 0.008082478 | 0.022128019 | 1 |
| HTN | nk_cell | *COG4* | -2.112528764 | 0.004380299 | 0.014766189 | 1 |
| HTN | nk_cell | *SLC7A6OS* | -1.093037708 | 0.00010801 | 0.003536779 | 0.679061584 |
| HTN | nk_cell | *HCFC1R1* | -2.463378767 | 0.002415586 | 0.010645574 | 1 |
| HTN | nk_cell | *NAGPA* | -2.02117526 | 0.001784344 | 0.009356274 | 1 |
| HTN | nk_cell | *COTL1* | -2.048872324 | 0.00100934 | 0.008846261 | 1 |
| HTN | nk_cell | *ZNF500* | -1.978143173 | 5.31E-05 | 0.002316216 | 0.333535147 |
| HTN | nk_cell | *NME4* | -1.488858272 | 0.002392113 | 0.010625206 | 1 |
| HTN | nk_cell | *NOMO3* | -2.416838833 | 0.008459534 | 0.022845829 | 1 |
| HTN | nk_cell | *LMF1* | -1.687208632 | 0.019700084 | 0.043019948 | 1 |
| HTN | nk_cell | *MTHFSD* | 1.063456324 | 0.004859588 | 0.015631946 | 1 |
| HTN | nk_cell | *HAGHL* | 3.688268261 | 0.011459754 | 0.028772952 | 1 |
| HTN | nk_cell | *SLC7A5* | 1.488528838 | 0.001901292 | 0.009616593 | 1 |
| HTN | nk_cell | *FBXO31* | -1.603310403 | 0.00378445 | 0.013588143 | 1 |
| HTN | nk_cell | *NUBP1* | -1.917434948 | 0.002436391 | 0.010654267 | 1 |
| HTN | nk_cell | *UBE2I* | -2.329539681 | 0.00821164 | 0.022349168 | 1 |
| HTN | nk_cell | *CPPED1* | -1.633150825 | 0.005690213 | 0.01742541 | 1 |
| HTN | nk_cell | *HMOX2* | 0.985013807 | 0.002437417 | 0.010654267 | 1 |
| HTN | nk_cell | *BFAR* | -2.070424318 | 0.014000259 | 0.03349301 | 1 |
| HTN | nk_cell | *STX4* | -1.83769028 | 0.004552561 | 0.015048343 | 1 |
| HTN | nk_cell | *NOMO1* | -2.530868961 | 0.000326101 | 0.006479211 | 1 |
| HTN | nk_cell | *SYT17* | -1.338491538 | 0.001154302 | 0.008846261 | 1 |
| HTN | nk_cell | *TMC5* | -1.197371582 | 0.000745398 | 0.008846261 | 1 |
| HTN | nk_cell | *RNF40* | -2.182825283 | 0.000407353 | 0.007133792 | 1 |
| HTN | nk_cell | *KNOP1* | -1.084153047 | 0.018175314 | 0.040334698 | 1 |
| HTN | nk_cell | *IQCH* | -1.494923818 | 0.004207657 | 0.014431826 | 1 |
| HTN | nk_cell | *LACTB* | -2.38323296 | 0.001367516 | 0.00885079 | 1 |
| HTN | nk_cell | *CSK* | 1.088347343 | 0.003261182 | 0.012570845 | 1 |
| HTN | nk_cell | *RASL12* | -0.987751873 | 0.001043444 | 0.008846261 | 1 |
| HTN | nk_cell | *TTC23* | -1.660136608 | 0.001386034 | 0.00885079 | 1 |
| HTN | nk_cell | *FAH* | -1.230696758 | 0.001256685 | 0.00885079 | 1 |
| HTN | nk_cell | *HOMER2* | -2.245242911 | 0.005891891 | 0.017862625 | 1 |
| HTN | nk_cell | *EHD4* | -1.660680907 | 0.00233085 | 0.010489659 | 1 |
| HTN | nk_cell | *ATP8B4* | 1.012964974 | 0.005368588 | 0.016717343 | 1 |
| HTN | nk_cell | *PDGFRL* | -1.004425463 | 0.004947416 | 0.015829213 | 1 |
| HTN | nk_cell | *NBN* | -1.768235212 | 0.017532801 | 0.039283223 | 1 |

| | | | | | | |
|---|---|---|---|---|---|---|
| HTN | nk_cell | *PLAT* | -1.152623888 | 0.001874324 | 0.009541601 | 1 |
| HTN | nk_cell | *GDAP1* | -2.335799195 | 0.004662743 | 0.015299929 | 1 |
| HTN | nk_cell | *ZC2HC1A* | -1.422402872 | 0.007159739 | 0.020248889 | 1 |
| HTN | nk_cell | *CHRAC1* | -3.745070116 | 0.003563931 | 0.013158269 | 1 |
| HTN | nk_cell | *LEPROTL1* | 1.310979076 | 0.014861675 | 0.034916049 | 1 |
| HTN | nk_cell | *TUSC3* | -1.685515809 | 0.003475149 | 0.012997181 | 1 |
| HTN | nk_cell | *ARHGEF10* | -0.887107935 | 0.001318408 | 0.00885079 | 1 |
| HTN | nk_cell | *KLHDC4* | 0.736585224 | 0.022107264 | 0.046892163 | 1 |
| HTN | nk_cell | *NUCB1* | -2.070295809 | 0.005132557 | 0.016231583 | 1 |
| HTN | nk_cell | *TUBB4A* | -7.557724044 | 0.001840683 | 0.009462282 | 1 |
| HTN | nk_cell | *SNRNP70* | -1.80584159 | 0.003619475 | 0.013268595 | 1 |
| HTN | nk_cell | *PPP1R37* | -1.736613694 | 0.018930684 | 0.04173114 | 1 |
| HTN | nk_cell | *CD37* | 0.953225938 | 0.001438208 | 0.008873507 | 1 |
| HTN | nk_cell | *AMH* | 1.538803448 | 0.004131862 | 0.014278329 | 1 |
| HTN | nk_cell | *DKKL1* | -1.439641287 | 0.018734782 | 0.041416789 | 1 |
| HTN | nk_cell | *STX10* | -2.04206587 | 0.008412796 | 0.022729372 | 1 |
| HTN | nk_cell | *FCER2* | -1.331787607 | 0.001174699 | 0.00885079 | 1 |
| HTN | nk_cell | *DMPK* | -1.306598451 | 0.007182721 | 0.02030475 | 1 |
| HTN | nk_cell | *C19orf53* | 1.793380733 | 0.002988543 | 0.011958531 | 1 |
| HTN | nk_cell | *CCDC61* | -2.369164103 | 0.003936836 | 0.013894565 | 1 |
| HTN | nk_cell | *IL27RA* | 0.656923612 | 0.001746079 | 0.009287307 | 1 |
| HTN | nk_cell | *C19orf44* | -0.78026022 | 0.022098615 | 0.046892163 | 1 |
| HTN | nk_cell | *RASAL3* | 0.22875141 | 0.001406826 | 0.00885079 | 1 |
| HTN | nk_cell | *SYDE1* | -2.498550548 | 0.000142959 | 0.004228148 | 0.898783805 |
| HTN | nk_cell | *AURKC* | 0.794756727 | 0.021518058 | 0.046014977 | 1 |
| HTN | nk_cell | *CCNE1* | -3.79553121 | 0.001007761 | 0.008846261 | 1 |
| HTN | nk_cell | *ANKRD27* | -1.609431463 | 0.019018971 | 0.041859912 | 1 |
| HTN | nk_cell | *RPS16* | 1.075327988 | 0.023200663 | 0.04873457 | 1 |
| HTN | nk_cell | *PRX* | -2.153802229 | 0.0226675 | 0.047790265 | 1 |
| HTN | nk_cell | *SLC1A5* | -1.816855979 | 0.013348932 | 0.032353407 | 1 |
| HTN | nk_cell | *PRKD2* | 1.137567245 | 0.003229008 | 0.012505129 | 1 |
| HTN | nk_cell | *CACTIN* | 1.754937097 | 0.003870716 | 0.013795462 | 1 |
| HTN | nk_cell | *FZR1* | 1.824354354 | 0.00775412 | 0.021466383 | 1 |
| HTN | nk_cell | *BBC3* | -1.581748765 | 0.001244781 | 0.00885079 | 1 |
| HTN | nk_cell | *CEACAM4* | -2.980390385 | 0.001006873 | 0.008846261 | 1 |
| HTN | nk_cell | *MRPL4* | -2.638351382 | 0.015290971 | 0.035487018 | 1 |
| HTN | nk_cell | *ICAM5* | -7.719634997 | 6.50E-06 | 0.000510895 | 0.04087163 |
| HTN | nk_cell | *CYTH2* | -2.034151072 | 0.006807353 | 0.019650054 | 1 |
| HTN | nk_cell | *GRWD1* | -1.19465541 | 0.000261047 | 0.005849812 | 1 |
| HTN | nk_cell | *GRIN2D* | -2.020110506 | 0.001138691 | 0.008846261 | 1 |
| HTN | nk_cell | *LIG1* | 1.425486828 | 0.007868353 | 0.021687127 | 1 |
| HTN | nk_cell | *ZNF175* | -1.33390746 | 0.001351604 | 0.00885079 | 1 |
| HTN | nk_cell | *PLA2G4C* | -1.326127186 | 0.011840079 | 0.029574326 | 1 |
| HTN | nk_cell | *BCAT2* | -2.17476338 | 0.003005807 | 0.011990806 | 1 |
| HTN | nk_cell | *MIER2* | -2.667117173 | 0.014696576 | 0.034592802 | 1 |
| HTN | nk_cell | *TNPO2* | -1.867216267 | 0.000406443 | 0.007133792 | 1 |
| HTN | nk_cell | *LILRB5* | -2.010122615 | 0.006336698 | 0.018774185 | 1 |
| HTN | nk_cell | *MAST1* | -1.814270671 | 0.001114003 | 0.008846261 | 1 |
| HTN | nk_cell | *PRPF31* | -1.857687633 | 0.012877929 | 0.031405563 | 1 |
| HTN | nk_cell | *TFPT* | -1.979814878 | 0.011437074 | 0.028738962 | 1 |

| | | | | | | |
|---|---|---|---|---|---|---|
| HTN | nk_cell | *RPL18A* | 1.218606653 | 0.00203261 | 0.009861903 | 1 |
| HTN | nk_cell | *PDE4C* | -1.443668258 | 0.001370761 | 0.00885079 | 1 |
| HTN | nk_cell | *COPE* | 1.481928929 | 0.009352737 | 0.024613083 | 1 |
| HTN | nk_cell | *KXD1* | -2.461995313 | 0.01966761 | 0.04297889 | 1 |
| HTN | nk_cell | *ZNF14* | -2.418669138 | 0.02382481 | 0.049762984 | 1 |
| HTN | nk_cell | *PON3* | -1.901258913 | 0.002163796 | 0.010159571 | 1 |
| HTN | nk_cell | *PON2* | -1.645128606 | 0.00376311 | 0.01352697 | 1 |
| HTN | nk_cell | *HBP1* | -1.88048272 | 0.001097576 | 0.008846261 | 1 |
| HTN | nk_cell | *WDR91* | -1.985373005 | 0.02293031 | 0.048217521 | 1 |
| HTN | nk_cell | *PTN* | -0.859845765 | 0.0010486 | 0.008846261 | 1 |
| HTN | nk_cell | *ADAP1* | 0.780570168 | 0.001562922 | 0.008975583 | 1 |
| HTN | nk_cell | *CAV1* | -0.797335069 | 0.014526622 | 0.034308367 | 1 |
| HTN | nk_cell | *MET* | -1.086080176 | 0.001087108 | 0.008846261 | 1 |
| HTN | nk_cell | *HOXA2* | -2.719361371 | 2.28E-05 | 0.00124716 | 0.143423362 |
| HTN | nk_cell | *HOXA5* | -0.709165842 | 0.021852389 | 0.046503529 | 1 |
| HTN | nk_cell | *HOXA6* | -0.72515237 | 0.003041689 | 0.012072664 | 1 |
| HTN | nk_cell | *CPED1* | -1.262857478 | 0.005927908 | 0.017931898 | 1 |
| HTN | nk_cell | *CPVL* | -1.401874099 | 0.007083143 | 0.020105374 | 1 |
| HTN | nk_cell | *NOD1* | -1.664584676 | 0.007739177 | 0.021443898 | 1 |
| HTN | nk_cell | *CRHR2* | 8.278675948 | 0.000424606 | 0.007214859 | 1 |
| HTN | nk_cell | *NPTX2* | -29.20545991 | 0.000579895 | 0.008131793 | 1 |
| HTN | nk_cell | *CYP3A5* | -1.296334722 | 0.001352468 | 0.00885079 | 1 |
| HTN | nk_cell | *EIF3B* | -1.902864883 | 0.023464724 | 0.049125115 | 1 |
| HTN | nk_cell | *TFR2* | 0.518682342 | 0.002769211 | 0.011416412 | 1 |
| HTN | nk_cell | *RBM28* | -1.288491043 | 0.020417894 | 0.044157998 | 1 |
| HTN | nk_cell | *LSM5* | -2.398582541 | 0.00725198 | 0.020436215 | 1 |
| HTN | nk_cell | *PLOD3* | -2.217800304 | 0.005764131 | 0.017583255 | 1 |
| HTN | nk_cell | *ZNHIT1* | -1.57668754 | 0.01124987 | 0.028404792 | 1 |
| HTN | nk_cell | *CEP41* | -1.105273321 | 0.018913755 | 0.041719711 | 1 |
| HTN | nk_cell | *MEST* | -1.036860114 | 1.77E-06 | 0.000188167 | 0.011101824 |
| HTN | nk_cell | *ACTR3C* | 1.659034661 | 1.22E-05 | 0.000807383 | 0.076701401 |
| HTN | nk_cell | *POU6F2* | -1.067416682 | 0.001109717 | 0.008846261 | 1 |
| HTN | nk_cell | *TSPAN13* | -2.639528315 | 0.003483835 | 0.013006455 | 1 |
| HTN | nk_cell | *RARRES2* | -1.870677253 | 0.0011567 | 0.008846261 | 1 |
| HTN | nk_cell | *URGCP* | -1.870670107 | 0.003557025 | 0.013146983 | 1 |
| HTN | nk_cell | *POLD2* | -1.400728781 | 0.002201021 | 0.010212412 | 1 |
| HTN | nk_cell | *GCK* | -8.496385147 | 0.001019809 | 0.008846261 | 1 |
| HTN | nk_cell | *CLIP2* | -2.126130845 | 0.020702258 | 0.044573662 | 1 |
| HTN | nk_cell | *SPATA6L* | -1.747645203 | 0.004472528 | 0.014878713 | 1 |
| HTN | nk_cell | *FKTN* | -1.738839522 | 0.0083199 | 0.022546211 | 1 |
| HTN | nk_cell | *TRIM14* | 1.151463238 | 0.006289817 | 0.018670482 | 1 |
| HTN | nk_cell | *C5* | -1.227096106 | 0.012054737 | 0.029976403 | 1 |
| HTN | nk_cell | *OGN* | -2.146951501 | 4.78E-05 | 0.002176757 | 0.30039243 |
| HTN | nk_cell | *ECM2* | -1.260538781 | 6.39E-05 | 0.002627011 | 0.401932744 |
| HTN | nk_cell | *PTGR1* | -0.83688838 | 0.009511635 | 0.024937301 | 1 |
| HTN | nk_cell | *TNFSF8* | -2.672945214 | 0.002263705 | 0.010320459 | 1 |
| HTN | nk_cell | *PLGRKT* | -1.503690829 | 0.002360834 | 0.010579162 | 1 |
| HTN | nk_cell | *ELAVL2* | -1.36249824 | 0.001113111 | 0.008846261 | 1 |
| HTN | nk_cell | *RGP1* | -2.665573954 | 0.019356873 | 0.042417796 | 1 |
| HTN | nk_cell | *MPDZ* | -0.993791257 | 0.001528117 | 0.008957475 | 1 |

| | | | | | | |
|---|---|---|---|---|---|---|
| HTN | nk_cell | *PTGDS* | 1.146900091 | 0.00219628 | 0.010211299 | 1 |
| HTN | nk_cell | *SHB* | -2.079188971 | 0.001242941 | 0.00885079 | 1 |
| HTN | nk_cell | *DVL1* | -1.65772794 | 0.005000931 | 0.015951725 | 1 |
| HTN | nk_cell | *ATRNL1* | -1.428523541 | 0.004267797 | 0.014577053 | 1 |
| HTN | nk_cell | *DNMBP* | -1.671323304 | 0.006608305 | 0.019234451 | 1 |
| HTN | nk_cell | *NSMCE4A* | -1.51047922 | 0.001762688 | 0.009314459 | 1 |
| HTN | nk_cell | *SPOCK2* | 0.479588199 | 0.001371205 | 0.00885079 | 1 |
| HTN | nk_cell | *KAZALD1* | -4.057996538 | 0.020536548 | 0.044364105 | 1 |
| HTN | nk_cell | *ACBD5* | 1.540855492 | 0.007708384 | 0.021386852 | 1 |
| HTN | nk_cell | *PITRM1* | -2.053598601 | 0.001989112 | 0.009795241 | 1 |
| HTN | nk_cell | *DHRS11* | -0.957819105 | 0.000804773 | 0.008846261 | 1 |
| HTN | nk_cell | *MLLT6* | 1.358500656 | 0.010849301 | 0.027715528 | 1 |
| HTN | nk_cell | *RGS9* | 0.40212366 | 0.001467777 | 0.008909666 | 1 |
| HTN | nk_cell | *WNT3* | -1.739343654 | 5.15E-05 | 0.002304845 | 0.324069188 |
| HTN | nk_cell | *ASPA* | -0.817743237 | 0.001142669 | 0.008846261 | 1 |
| HTN | nk_cell | *RAD51C* | -1.810488102 | 8.13E-07 | 0.000110176 | 0.00511177 |
| HTN | nk_cell | *SEPT4* | -1.058901726 | 0.000208166 | 0.005172882 | 1 |
| HTN | nk_cell | *RPS6KB1* | -1.75731046 | 0.019944031 | 0.043371886 | 1 |
| HTN | nk_cell | *TRIM16L* | -1.300216475 | 0.000687215 | 0.00874661 | 1 |
| HTN | nk_cell | *CDK5RAP3* | -2.427978317 | 0.00056708 | 0.008121264 | 1 |
| HTN | nk_cell | *GALK1* | -3.1405663 | 0.002654143 | 0.011139249 | 1 |
| HTN | nk_cell | *CAMTA2* | -1.701425373 | 0.022302169 | 0.047257747 | 1 |
| HTN | nk_cell | *HOXB6* | -1.109109429 | 0.000209875 | 0.005194824 | 1 |
| HTN | nk_cell | *C1QBP* | -2.335278099 | 0.003035325 | 0.012070265 | 1 |
| HTN | nk_cell | *SMARCD2* | -1.992316312 | 0.002155818 | 0.010155174 | 1 |
| HTN | nk_cell | *ICAM2* | 1.466392316 | 9.35E-05 | 0.003320848 | 0.587790022 |
| HTN | nk_cell | *SYNGR2* | -2.383161195 | 0.001546126 | 0.00897211 | 1 |
| HTN | nk_cell | *UTP6* | -1.853057284 | 0.020695722 | 0.044573662 | 1 |
| HTN | nk_cell | *CYTH1* | 1.377616626 | 0.000993073 | 0.008846261 | 1 |
| HTN | nk_cell | *PEX12* | -1.923368332 | 0.009186129 | 0.024276248 | 1 |
| HTN | nk_cell | *HSD17B1* | -0.605160194 | 0.016590141 | 0.037586385 | 1 |
| HTN | nk_cell | *DLX4* | -7.929434338 | 0.001026147 | 0.008846261 | 1 |
| HTN | nk_cell | *VAT1* | -1.642401853 | 0.008712599 | 0.023398594 | 1 |
| HTN | nk_cell | *LRRC59* | -2.159193663 | 0.001808148 | 0.00938705 | 1 |
| HTN | nk_cell | *ALOX12* | -2.193545322 | 0.000922394 | 0.008846261 | 1 |
| HTN | nk_cell | *MPP2* | -3.309003889 | 0.001871608 | 0.009540973 | 1 |
| HTN | nk_cell | *EFNB3* | -13.03851721 | 2.88E-05 | 0.001494252 | 0.180804466 |
| HTN | nk_cell | *DPH1* | -2.319042407 | 0.001808168 | 0.00938705 | 1 |
| HTN | nk_cell | *DHRS7B* | -1.72007675 | 0.015644234 | 0.036040783 | 1 |
| HTN | nk_cell | *RCVRN* | -3.13429742 | 0.003830032 | 0.013673714 | 1 |
| HTN | nk_cell | *SLC9A3R1* | 0.329974003 | 0.001392173 | 0.00885079 | 1 |
| HTN | nk_cell | *MYH3* | -1.832404538 | 0.005146385 | 0.016267131 | 1 |
| HTN | nk_cell | *TMEM104* | 1.674987979 | 0.015596026 | 0.03599515 | 1 |
| HTN | nk_cell | *TBC1D9* | -1.514621088 | 0.009677619 | 0.025319679 | 1 |
| HTN | nk_cell | *CPE* | -1.567802573 | 0.002455286 | 0.010682617 | 1 |
| HTN | nk_cell | *WFS1* | -2.382993226 | 0.005930525 | 0.017931898 | 1 |
| HTN | nk_cell | *AADAT* | -1.756255744 | 0.0010473 | 0.008846261 | 1 |
| HTN | nk_cell | *CPZ* | -15.62924838 | 0.001114803 | 0.008846261 | 1 |
| HTN | nk_cell | *TRIM2* | -0.927642441 | 0.013964824 | 0.033459164 | 1 |
| HTN | nk_cell | *SH3D19* | -0.796453155 | 0.001569776 | 0.008975583 | 1 |

| | | | | | | |
|---|---|---|---|---|---|---|
| HTN | nk_cell | *LRP2BP* | -1.059804862 | 0.019565471 | 0.042815216 | 1 |
| HTN | nk_cell | *FAM149A* | -0.644907546 | 0.016056055 | 0.036742556 | 1 |
| HTN | nk_cell | *UGDH* | -1.632493967 | 0.00141747 | 0.00885079 | 1 |
| HTN | nk_cell | *SC5D* | -1.890588979 | 0.002007354 | 0.009844174 | 1 |
| HTN | nk_cell | *C11orf63* | -1.922182308 | 0.014229007 | 0.03381816 | 1 |
| HTN | nk_cell | *SNX15* | -1.206651937 | 0.002301224 | 0.010422787 | 1 |
| HTN | nk_cell | *DTX4* | -1.237291806 | 0.000621994 | 0.008409632 | 1 |
| HTN | nk_cell | *PUS3* | -1.968753546 | 0.001355922 | 0.00885079 | 1 |
| HTN | nk_cell | *DCPS* | 1.331788466 | 0.016408343 | 0.037281986 | 1 |
| HTN | nk_cell | *FOXRED1* | -1.337242505 | 0.001208427 | 0.00885079 | 1 |
| HTN | nk_cell | *NRXN2* | -2.438198467 | 0.00104498 | 0.008846261 | 1 |
| HTN | nk_cell | *CCDC86* | -1.901036733 | 0.00108048 | 0.008846261 | 1 |
| HTN | nk_cell | *EIF4G2* | -1.671899417 | 0.023833291 | 0.049764164 | 1 |
| HTN | nk_cell | *GALNT18* | -1.169381867 | 0.003488371 | 0.013007939 | 1 |
| HTN | nk_cell | *KIAA1549L* | -1.212060561 | 0.000961027 | 0.008846261 | 1 |
| HTN | nk_cell | *CD5* | 0.807168683 | 0.00658371 | 0.019207324 | 1 |
| HTN | nk_cell | *CARS* | -2.370861361 | 0.000483191 | 0.007690692 | 1 |
| HTN | nk_cell | *SLC22A18* | 0.874629883 | 0.00463872 | 0.015244972 | 1 |
| HTN | nk_cell | *SLC35F2* | -1.611658404 | 0.000974634 | 0.008846261 | 1 |
| HTN | nk_cell | *C11orf21* | 0.236097416 | 0.002281225 | 0.010370254 | 1 |
| HTN | nk_cell | *ELMOD1* | 2.696801289 | 0.003428374 | 0.01289126 | 1 |
| HTN | nk_cell | *AIP* | 1.101267053 | 0.023248677 | 0.048802815 | 1 |
| HTN | nk_cell | *PTPN5* | -0.763494059 | 0.01559245 | 0.03599515 | 1 |
| HTN | nk_cell | *PPFIBP1* | -1.555438563 | 0.000143756 | 0.004228148 | 0.903793883 |
| HTN | nk_cell | *KCTD10* | -2.258495996 | 8.58E-06 | 0.000619721 | 0.05391577 |
| HTN | nk_cell | *CYP27B1* | -6.134953829 | 0.001682434 | 0.0091868 | 1 |
| HTN | nk_cell | *GLI1* | -2.419041908 | 0.001120132 | 0.008846261 | 1 |
| HTN | nk_cell | *WNT5B* | -2.770659977 | 0.008255057 | 0.0224479 | 1 |
| HTN | nk_cell | *TRPV4* | -1.375758026 | 0.002639226 | 0.011097616 | 1 |
| HTN | nk_cell | *KCNA1* | -3.409721114 | 0.0005779 | 0.008131793 | 1 |
| HTN | nk_cell | *NAA25* | -1.6697488 | 0.012197563 | 0.030262856 | 1 |
| HTN | nk_cell | *SCNN1A* | 1.873524307 | 0.006994131 | 0.019932957 | 1 |
| HTN | nk_cell | *ARHGDIB* | 0.520767989 | 0.004754881 | 0.015439713 | 1 |
| HTN | nk_cell | *CAND1* | -2.932466727 | 3.22E-06 | 0.000297919 | 0.020258521 |
| HTN | nk_cell | *COPS7A* | -2.66995549 | 5.17E-05 | 0.002304845 | 0.324983159 |
| HTN | nk_cell | *GNPTAB* | 0.885874383 | 0.008555513 | 0.023075294 | 1 |
| HTN | nk_cell | *SPSB2* | -2.024100254 | 0.000853919 | 0.008846261 | 1 |
| HTN | nk_cell | *NT5DC3* | -0.990163585 | 0.019153563 | 0.042117793 | 1 |
| HTN | nk_cell | *ST8SIA1* | -1.122717837 | 0.00237574 | 0.010598969 | 1 |
| HTN | nk_cell | *PHC1* | -2.131518627 | 0.00179208 | 0.009369553 | 1 |
| HTN | nk_cell | *COL12A1* | -0.839914662 | 0.001272904 | 0.00885079 | 1 |
| HTN | nk_cell | *RWDD1* | -1.88196701 | 0.015936882 | 0.036567583 | 1 |
| HTN | nk_cell | *FAM184A* | -0.834225519 | 0.007132491 | 0.020189992 | 1 |
| HTN | nk_cell | *TPD52L1* | -1.191645902 | 0.021642668 | 0.046187188 | 1 |
| HTN | nk_cell | *TULP1* | 4.495038825 | 0.001763036 | 0.009314459 | 1 |
| HTN | nk_cell | *RNF8* | -1.852933207 | 0.004362843 | 0.014746877 | 1 |
| HTN | nk_cell | *PHACTR1* | -1.639309263 | 0.007266059 | 0.020457553 | 1 |
| HTN | nk_cell | *SAYSD1* | -1.69410998 | 0.004117734 | 0.014255614 | 1 |
| HTN | nk_cell | *CAP2* | -0.929446758 | 0.000347755 | 0.006579952 | 1 |
| HTN | nk_cell | *BVES* | -1.229627339 | 0.005453194 | 0.016897109 | 1 |

| | | | | | | |
|---|---|---|---|---|---|---|
| HTN | nk_cell | *WASF1* | -0.780015013 | 0.002998128 | 0.011967765 | 1 |
| HTN | nk_cell | *GPLD1* | -0.635669987 | 0.00720924 | 0.020361406 | 1 |
| HTN | nk_cell | *VNN2* | 0.5762462 | 0.002098483 | 0.010029243 | 1 |
| HTN | nk_cell | *TBP* | -1.849556886 | 0.004470264 | 0.014878713 | 1 |
| HTN | nk_cell | *GUCA1B* | 3.245516195 | 0.007919387 | 0.021789579 | 1 |
| HTN | nk_cell | *PRPH2* | -1.170817778 | 0.003606529 | 0.013228851 | 1 |
| HTN | nk_cell | *DUSP22* | -1.894617048 | 0.014501539 | 0.034261998 | 1 |
| HTN | nk_cell | *TTK* | -2.18069032 | 0.000356277 | 0.006607413 | 1 |
| HTN | nk_cell | *BTN2A1* | 1.744270384 | 0.010026724 | 0.025975083 | 1 |
| HTN | nk_cell | *LAMA4* | -0.73724289 | 0.003964524 | 0.013928188 | 1 |
| HTN | nk_cell | *LY86* | -1.80698611 | 0.002281111 | 0.010370254 | 1 |
| HTN | nk_cell | *TBX18* | -1.13254338 | 0.000287022 | 0.006179456 | 1 |
| HTN | nk_cell | *PCDHB2* | -0.856167869 | 0.004732556 | 0.015416362 | 1 |
| HTN | nk_cell | *NUDT12* | -1.023872219 | 0.000955265 | 0.008846261 | 1 |
| HTN | nk_cell | *C7* | -2.131236789 | 0.001504626 | 0.008938486 | 1 |
| HTN | nk_cell | *NNT* | -1.587671159 | 0.000820006 | 0.008846261 | 1 |
| HTN | nk_cell | *PFDN1* | -1.155340277 | 0.017254424 | 0.038803448 | 1 |
| HTN | nk_cell | *LOX* | -1.552353317 | 0.001714174 | 0.009223501 | 1 |
| HTN | nk_cell | *APBB3* | -1.174910804 | 0.003324692 | 0.01269116 | 1 |
| HTN | nk_cell | *SPARC* | -1.208603482 | 0.00313602 | 0.012294167 | 1 |
| HTN | nk_cell | *PDE8B* | -1.148372286 | 0.007507962 | 0.020988243 | 1 |
| HTN | nk_cell | *CLK4* | -1.657215357 | 0.013987324 | 0.033474803 | 1 |
| HTN | nk_cell | *BTNL8* | -0.873536595 | 0.008291437 | 0.022504322 | 1 |
| HTN | nk_cell | *RASGRF2* | -1.016383625 | 0.001531527 | 0.008957475 | 1 |
| HTN | nk_cell | *POLR3G* | -1.282449251 | 0.00053187 | 0.008076978 | 1 |
| HTN | nk_cell | *CDH6* | -0.460018862 | 0.004631004 | 0.015227574 | 1 |
| HTN | nk_cell | *SLC27A6* | -4.580016404 | 0.007019521 | 0.019978148 | 1 |
| HTN | nk_cell | *IL4* | -2.665033491 | 0.007010672 | 0.019971043 | 1 |
| HTN | nk_cell | *TRIM23* | -1.670449495 | 0.014406705 | 0.034076356 | 1 |
| HTN | nk_cell | *C9* | -1.460591302 | 0.015990101 | 0.036662933 | 1 |
| HTN | nk_cell | *SMAD5* | -1.564885109 | 0.008209278 | 0.022349168 | 1 |
| HTN | nk_cell | *CNTN3* | -0.803589578 | 0.001127743 | 0.008846261 | 1 |
| HTN | nk_cell | *ACTR8* | 1.613254236 | 0.017711981 | 0.039627307 | 1 |
| HTN | nk_cell | *HGD* | -3.657413876 | 0.001449031 | 0.008873507 | 1 |
| HTN | nk_cell | *NIT2* | -2.289293976 | 0.000161055 | 0.004487306 | 1 |
| HTN | nk_cell | *FAM162A* | -3.104831752 | 7.85E-05 | 0.00292069 | 0.493596623 |
| HTN | nk_cell | *RBP1* | 1.019356805 | 0.004059204 | 0.014091781 | 1 |
| HTN | nk_cell | *BCHE* | -1.139060195 | 0.000132409 | 0.004002198 | 0.832457174 |
| HTN | nk_cell | *WNT5A* | -1.033883396 | 0.002607029 | 0.011031325 | 1 |
| HTN | nk_cell | *HYAL1* | -1.661990084 | 6.00E-05 | 0.002566137 | 0.377222078 |
| HTN | nk_cell | *CYB561D2* | 1.251948948 | 0.000698441 | 0.008747204 | 1 |
| HTN | nk_cell | *IFT57* | -1.767683743 | 0.002124183 | 0.01009268 | 1 |
| HTN | nk_cell | *IQCG* | -1.911821693 | 0.012314296 | 0.030456325 | 1 |
| HTN | nk_cell | *GBE1* | -1.14782527 | 0.003820263 | 0.013659313 | 1 |
| HTN | nk_cell | *UMPS* | -1.659992011 | 0.001957706 | 0.009759795 | 1 |
| HTN | nk_cell | *SLC41A3* | -1.855768498 | 0.002667331 | 0.011180705 | 1 |
| HTN | nk_cell | *EFCC1* | -0.858866956 | 0.000724761 | 0.008846261 | 1 |
| HTN | nk_cell | *NEK11* | -1.642759002 | 0.002756026 | 0.011391935 | 1 |
| HTN | nk_cell | *PLSCR4* | -0.970198684 | 0.006766688 | 0.019550629 | 1 |
| HTN | nk_cell | *HEMK1* | -1.460052814 | 0.023900835 | 0.049872072 | 1 |

| | | | | | | |
|---|---|---|---|---|---|---|
| HTN | nk_cell | *GORASP1* | -1.585597318 | 0.00092586 | 0.008846261 | 1 |
| HTN | nk_cell | *ABCC5* | -2.047328698 | 0.007609623 | 0.021215833 | 1 |
| HTN | nk_cell | *ARHGEF26* | -1.082379533 | 0.00276423 | 0.011410842 | 1 |
| HTN | nk_cell | *ZBTB47* | -1.251094043 | 0.01246629 | 0.030690284 | 1 |
| HTN | nk_cell | *CLCN2* | -3.867018342 | 0.001599542 | 0.009048265 | 1 |
| HTN | nk_cell | *NEK4* | -2.53034237 | 0.00291915 | 0.011794791 | 1 |
| HTN | nk_cell | *LMAN2L* | -1.129742244 | 0.00814754 | 0.022234893 | 1 |
| HTN | nk_cell | *KCNIP3* | -1.155468817 | 0.002292589 | 0.010399356 | 1 |
| HTN | nk_cell | *SLC35F5* | -1.996636253 | 0.004608485 | 0.015177343 | 1 |
| HTN | nk_cell | *TP53I3* | -3.56316851 | 4.46E-05 | 0.002078644 | 0.280616946 |
| HTN | nk_cell | *DNAJC27* | -4.239384516 | 7.39E-07 | 0.000108056 | 0.004646408 |
| HTN | nk_cell | *TANC1* | -1.349158741 | 0.000753238 | 0.008846261 | 1 |
| HTN | nk_cell | *ITGA4* | 0.53205642 | 0.000923824 | 0.008846261 | 1 |
| HTN | nk_cell | *PCSK4* | -1.299669724 | 0.001017707 | 0.008846261 | 1 |
| HTN | nk_cell | *MOGS* | -1.011304702 | 0.003542789 | 0.013117499 | 1 |
| HTN | nk_cell | *GRB14* | -0.757584775 | 0.001348776 | 0.00885079 | 1 |
| HTN | nk_cell | *SPTBN1* | -0.905213123 | 0.002645506 | 0.011110417 | 1 |
| HTN | nk_cell | *RTN4* | -1.70770939 | 0.001143842 | 0.008846261 | 1 |
| HTN | nk_cell | *HTRA2* | -2.407322316 | 0.003115187 | 0.012256059 | 1 |
| HTN | nk_cell | *GALNT3* | 0.490441719 | 0.001570898 | 0.008975583 | 1 |
| HTN | nk_cell | *MRPL19* | -1.893342232 | 0.000670834 | 0.00874661 | 1 |
| HTN | nk_cell | *LANCL1* | -1.889409549 | 0.010756464 | 0.027512568 | 1 |
| HTN | nk_cell | *EFEMP1* | -1.061618274 | 0.002391891 | 0.010625206 | 1 |
| HTN | nk_cell | *GNLY* | 0.091725287 | 0.001370689 | 0.00885079 | 1 |
| HTN | nk_cell | *PLCD4* | -2.147779849 | 0.000441847 | 0.007343364 | 1 |
| HTN | nk_cell | *CHMP3* | -2.059420664 | 0.000161449 | 0.004487306 | 1 |
| HTN | nk_cell | *ZNF142* | -3.12454512 | 0.000253955 | 0.005756069 | 1 |
| HTN | nk_cell | *MLPH* | -1.463824828 | 0.001399483 | 0.00885079 | 1 |
| HTN | nk_cell | *PASK* | -2.257052808 | 4.09E-05 | 0.00196317 | 0.257175335 |
| HTN | nk_cell | *STK25* | -2.001280863 | 0.003775698 | 0.013564464 | 1 |
| HTN | nk_cell | *TPO* | -0.71593652 | 0.014608547 | 0.034463015 | 1 |
| HTN | nk_cell | *ARID3A* | -1.984997662 | 0.008781142 | 0.023542448 | 1 |
| HTN | nk_cell | *VAX2* | -1.261182476 | 0.001213055 | 0.00885079 | 1 |
| HTN | nk_cell | *MSH6* | -1.980612805 | 0.008809268 | 0.023582574 | 1 |
| HTN | nk_cell | *PLEKHA3* | -1.966169607 | 0.006850024 | 0.01971231 | 1 |
| HTN | nk_cell | *BCL9* | -1.628064768 | 0.002958309 | 0.011891873 | 1 |
| HTN | nk_cell | *MORN1* | -2.03808923 | 0.00042275 | 0.007214859 | 1 |
| HTN | nk_cell | *GPX7* | 0.670604966 | 0.002295298 | 0.010404137 | 1 |
| HTN | nk_cell | *RALGPS2* | -1.389221482 | 0.005579064 | 0.017185485 | 1 |
| HTN | nk_cell | *FAM20B* | -2.033721491 | 0.015175941 | 0.035323691 | 1 |
| HTN | nk_cell | *WRAP73* | -2.439075369 | 0.013079188 | 0.031785411 | 1 |
| HTN | nk_cell | *ICMT* | 1.812541825 | 0.022254719 | 0.047173102 | 1 |
| HTN | nk_cell | *DOCK7* | -1.431751344 | 0.006393511 | 0.018844823 | 1 |
| HTN | nk_cell | *FBXO6* | 0.991050199 | 0.007726978 | 0.021424918 | 1 |
| HTN | nk_cell | *DNAJC6* | -3.267023549 | 8.16E-05 | 0.003006288 | 0.512858681 |
| HTN | nk_cell | *NCF2* | -1.633300779 | 0.001657861 | 0.009149879 | 1 |
| HTN | nk_cell | *RGS2* | -2.024888915 | 0.001297392 | 0.00885079 | 1 |
| HTN | nk_cell | *OLFML3* | -0.800945075 | 0.001321343 | 0.00885079 | 1 |
| HTN | nk_cell | *TMEM9* | -1.253211101 | 0.000610696 | 0.008358707 | 1 |
| HTN | nk_cell | *OSCP1* | -2.110980035 | 0.000225497 | 0.005431789 | 1 |

| | | | | | | |
|---|---|---|---|---|---|---|
| HTN | nk_cell | *GNPAT* | -2.040645909 | 0.00177266 | 0.009338615 | 1 |
| HTN | nk_cell | *KCNQ4* | -1.074696515 | 0.021460177 | 0.045922441 | 1 |
| HTN | nk_cell | *ACADM* | -1.570875321 | 0.018098186 | 0.040206111 | 1 |
| HTN | nk_cell | *CTBS* | -1.703390101 | 0.015112139 | 0.035280363 | 1 |
| HTN | nk_cell | *KIF17* | 1.541709798 | 0.004050006 | 0.014091781 | 1 |
| HTN | nk_cell | *TMED5* | -1.694413079 | 0.008541485 | 0.023047346 | 1 |
| HTN | nk_cell | *CNN3* | -1.110141949 | 0.001384299 | 0.00885079 | 1 |
| HTN | nk_cell | *TNFSF4* | -1.882686702 | 0.000632576 | 0.008516068 | 1 |
| HTN | nk_cell | *IRF6* | -1.338247243 | 6.16E-05 | 0.002580837 | 0.387125606 |
| HTN | nk_cell | *DIEXF* | -2.490071116 | 0.000185055 | 0.004993319 | 1 |
| HTN | nk_cell | *RCAN3* | -1.160977256 | 0.005633772 | 0.017286248 | 1 |
| HTN | nk_cell | *MTFR1L* | -1.523337898 | 0.00202247 | 0.009861903 | 1 |
| HTN | nk_cell | *SLC5A9* | -0.948911362 | 0.002002157 | 0.009837575 | 1 |
| HTN | nk_cell | *OSBPL9* | -1.421070159 | 0.019211811 | 0.042188144 | 1 |
| HTN | nk_cell | *CD3EAP* | -2.59633134 | 0.000106934 | 0.003536779 | 0.672296534 |
| HTN | nk_cell | *COLEC11* | -1.189015546 | 0.003899341 | 0.013830639 | 1 |
| HTN | nk_cell | *IFT46* | -1.897760687 | 0.022106605 | 0.046892163 | 1 |
| HTN | nk_cell | *DDX59* | -2.029217868 | 0.012324637 | 0.030457938 | 1 |
| HTN | nk_cell | *CAMSAP2* | -1.201138234 | 0.008537852 | 0.023047346 | 1 |
| HTN | nk_cell | *LRMP* | 0.93653272 | 0.002142828 | 0.010136915 | 1 |
| HTN | nk_cell | *SGIP1* | -1.648801429 | 0.006081084 | 0.018188286 | 1 |
| HTN | nk_cell | *ALDH8A1* | -0.532002398 | 0.014262427 | 0.033841117 | 1 |
| HTN | nk_cell | *MED28* | -2.07069215 | 0.004140885 | 0.014288553 | 1 |
| HTN | nk_cell | *ABCG2* | -2.421896332 | 0.003942724 | 0.013894565 | 1 |
| HTN | nk_cell | *FAM47E-STBL* | -0.641334862 | 0.003903138 | 0.013830639 | 1 |
| HTN | nk_cell | *MFSD1* | -1.762946328 | 0.012842813 | 0.031365595 | 1 |
| HTN | nk_cell | *UCHL3* | -2.628388474 | 0.001250881 | 0.00885079 | 1 |
| HTN | nk_cell | *DNAH7* | -0.870068068 | 0.002172512 | 0.010170205 | 1 |
| HTN | nk_cell | *GDA* | -0.881333834 | 0.003950073 | 0.013912665 | 1 |
| HTN | nk_cell | *C2orf40* | -1.750076954 | 0.000771094 | 0.008846261 | 1 |
| HTN | nk_cell | *CCDC92* | -1.504931029 | 0.002245216 | 0.010266299 | 1 |
| HTN | nk_cell | *CTNNAL1* | -0.791092306 | 0.002149401 | 0.010146672 | 1 |
| HTN | nk_cell | *WDR34* | -1.647926944 | 0.012594143 | 0.030905299 | 1 |
| HTN | nk_cell | *HDHD3* | -0.620123199 | 0.000625977 | 0.008445311 | 1 |
| HTN | nk_cell | *RBM18* | -2.243577508 | 0.010921318 | 0.027832317 | 1 |
| HTN | nk_cell | *KDSR* | -1.760619242 | 0.004783127 | 0.015492798 | 1 |
| HTN | nk_cell | *VSX2* | -1.38442324 | 0.003425514 | 0.012888215 | 1 |
| HTN | nk_cell | *DNAL1* | -1.598947138 | 0.018968311 | 0.041770147 | 1 |
| HTN | nk_cell | *IRF2BPL* | 2.317349984 | 0.004594834 | 0.015157026 | 1 |
| HTN | nk_cell | *ACOT2* | 0.381256114 | 0.022590749 | 0.047660416 | 1 |
| HTN | nk_cell | *FLVCR2* | -2.452387259 | 0.008128198 | 0.022218254 | 1 |
| HTN | nk_cell | *PPP4R4* | -1.081956965 | 0.015173583 | 0.035323691 | 1 |
| HTN | nk_cell | *ZC2HC1C* | -2.020856583 | 0.000877827 | 0.008846261 | 1 |
| HTN | nk_cell | *ALDH6A1* | -1.144635731 | 0.004056426 | 0.014091781 | 1 |
| HTN | nk_cell | *ESRRB* | -0.898637456 | 0.003731553 | 0.013454539 | 1 |
| HTN | nk_cell | *COQ6* | -2.13091839 | 0.002109041 | 0.010060352 | 1 |
| HTN | nk_cell | *GPR75* | -4.353990276 | 0.000440763 | 0.007343364 | 1 |
| HTN | nk_cell | *KLHL29* | -1.025711934 | 0.001546965 | 0.00897211 | 1 |
| HTN | nk_cell | *ATL2* | -1.523100537 | 0.003805365 | 0.013632098 | 1 |
| HTN | nk_cell | *CNRIP1* | -1.735637658 | 0.00289266 | 0.011702802 | 1 |

| | | | | | | |
|---|---|---|---|---|---|---|
| HTN | nk_cell | *CRIPT* | -2.452420531 | 0.001216667 | 0.00885079 | 1 |
| HTN | nk_cell | *CUTC* | -1.418527127 | 0.004048058 | 0.014091781 | 1 |
| HTN | nk_cell | *C10orf88* | -2.135859333 | 0.003977101 | 0.013953144 | 1 |
| HTN | nk_cell | *HELLS* | -2.907311772 | 0.000761626 | 0.008846261 | 1 |
| HTN | nk_cell | *TCTN3* | -1.732724196 | 0.011835456 | 0.029574326 | 1 |
| HTN | nk_cell | *AVPI1* | -2.031450523 | 0.000304104 | 0.006326876 | 1 |
| HTN | nk_cell | *C10orf95* | 0.524101508 | 0.00100031 | 0.008846261 | 1 |
| HTN | nk_cell | *HOXB5* | -2.190514327 | 0.000371305 | 0.006688814 | 1 |
| HTN | nk_cell | *HOXB3* | -0.995292595 | 0.001139315 | 0.008846261 | 1 |
| HTN | nk_cell | *CD274* | -1.567935612 | 0.015237428 | 0.035388883 | 1 |
| HTN | nk_cell | *LRP11* | -1.423480056 | 0.000435786 | 0.00732564 | 1 |
| HTN | nk_cell | *CYSTM1* | -1.427070167 | 0.006363293 | 0.018785762 | 1 |
| HTN | nk_cell | *PCDHB14* | -1.23710099 | 0.00111919 | 0.008846261 | 1 |
| HTN | nk_cell | *PCDHB12* | -1.384828574 | 2.81E-05 | 0.001471967 | 0.176636058 |
| HTN | nk_cell | *SEC16B* | -1.2852247 | 0.012700481 | 0.031105541 | 1 |
| HTN | nk_cell | *GORAB* | -2.102190446 | 0.008627952 | 0.023223444 | 1 |
| HTN | nk_cell | *SNX19* | -1.695959687 | 0.009395406 | 0.024704692 | 1 |
| HTN | nk_cell | *IQSEC3* | -1.253818375 | 0.000973331 | 0.008846261 | 1 |
| HTN | nk_cell | *CCDC77* | -2.438529636 | 0.004194052 | 0.014410776 | 1 |
| HTN | nk_cell | *ENOX1* | -1.609527876 | 0.001989585 | 0.009795241 | 1 |
| HTN | nk_cell | *WBP4* | -1.387380756 | 0.004819696 | 0.015579142 | 1 |
| HTN | nk_cell | *SMAD9* | -1.198539502 | 0.001265948 | 0.00885079 | 1 |
| HTN | nk_cell | *SIL1* | -1.493974882 | 0.010934362 | 0.027854268 | 1 |
| HTN | nk_cell | *ARL1* | -1.719928612 | 0.006813602 | 0.019650054 | 1 |
| HTN | nk_cell | *EPHX2* | -1.225466806 | 6.12E-06 | 0.000505677 | 0.038450983 |
| HTN | nk_cell | *CRISPLD1* | -1.601685022 | 0.001321704 | 0.00885079 | 1 |
| HTN | nk_cell | *RDH10* | -0.904052822 | 0.009707088 | 0.025365113 | 1 |
| HTN | nk_cell | *TBX4* | -2.451985037 | 0.005053497 | 0.016048648 | 1 |
| HTN | nk_cell | *TRIM6* | -1.747675147 | 5.02E-06 | 0.000431261 | 0.031578883 |
| HTN | nk_cell | *CEP89* | -0.835147383 | 0.02151127 | 0.046014977 | 1 |
| HTN | nk_cell | *ECHDC2* | -0.74519651 | 0.00612767 | 0.018298616 | 1 |
| HTN | nk_cell | *PRB2* | -1.440501915 | 0.005169494 | 0.016306629 | 1 |
| HTN | nk_cell | *ZSCAN18* | -1.727511231 | 0.001446934 | 0.008873507 | 1 |
| HTN | nk_cell | *PDZRN3* | -0.896950668 | 0.001705593 | 0.009223501 | 1 |
| HTN | nk_cell | *POPDC2* | -3.267383872 | 0.018083419 | 0.040187506 | 1 |
| HTN | nk_cell | *CD80* | -5.15195137 | 0.002878454 | 0.011652827 | 1 |
| HTN | nk_cell | *CAT* | -0.690772016 | 0.015037158 | 0.035196803 | 1 |
| HTN | nk_cell | *GJB6* | -31.34716513 | 0.00023023 | 0.005474297 | 1 |
| HTN | nk_cell | *TMEM39B* | -2.235995489 | 0.00115016 | 0.008846261 | 1 |
| HTN | nk_cell | *POLR3GL* | 1.728581885 | 0.016365992 | 0.03722612 | 1 |
| HTN | nk_cell | *TNFSF10* | -1.637437028 | 0.008768282 | 0.023517998 | 1 |
| HTN | nk_cell | *LIAS* | -1.789228337 | 0.007086609 | 0.020105374 | 1 |
| HTN | nk_cell | *CLCC1* | -2.493793786 | 0.001014762 | 0.008846261 | 1 |
| HTN | nk_cell | *CXCR4* | 1.272072159 | 0.01448814 | 0.03424321 | 1 |
| HTN | nk_cell | *POLK* | -1.449431042 | 0.015823455 | 0.036360403 | 1 |
| HTN | nk_cell | *FLT3* | -1.135402206 | 0.001126144 | 0.008846261 | 1 |
| HTN | nk_cell | *RASL11A* | -1.383345998 | 0.00331585 | 0.012680504 | 1 |
| HTN | nk_cell | *FMOD* | -1.842312829 | 0.004536839 | 0.015033969 | 1 |
| HTN | nk_cell | *LDB3* | -0.777761217 | 0.000557476 | 0.008121264 | 1 |
| HTN | nk_cell | *OPN4* | -3.207511808 | 0.001131175 | 0.008846261 | 1 |

| | | | | | | |
|---|---|---|---|---|---|---|
| HTN | nk_cell | *FAM35A* | -1.495351548 | 0.023776847 | 0.049679308 | 1 |
| HTN | nk_cell | *FAM213A* | -0.922208709 | 0.00240734 | 0.010643424 | 1 |
| HTN | nk_cell | *NAA60* | 1.663427088 | 0.017399837 | 0.039096775 | 1 |
| HTN | nk_cell | *ODF2L* | -1.598890394 | 0.003967772 | 0.013928188 | 1 |
| HTN | nk_cell | *RWDD3* | -3.185821763 | 0.001720656 | 0.009240684 | 1 |
| HTN | nk_cell | *CCDC18* | -1.695432334 | 0.007929123 | 0.021804049 | 1 |
| HTN | nk_cell | *KIAA0087* | -2.220202143 | 0.001154917 | 0.008846261 | 1 |
| HTN | nk_cell | *KLHL7* | -1.779618 | 0.0082018 | 0.022341732 | 1 |
| HTN | nk_cell | *CBX3* | -3.190005291 | 0.000146616 | 0.00426748 | 0.921775665 |
| HTN | nk_cell | *FAM126A* | -1.285809042 | 0.016046629 | 0.036742556 | 1 |
| HTN | nk_cell | *FKBP9* | -1.24916789 | 0.003493079 | 0.013010063 | 1 |
| HTN | nk_cell | *SLC25A51* | -0.878981793 | 7.63E-05 | 0.002917771 | 0.479736133 |
| HTN | nk_cell | *BICC1* | -0.748579904 | 0.000276943 | 0.006087905 | 1 |
| HTN | nk_cell | *ACADS* | -1.696137947 | 0.00064478 | 0.008600567 | 1 |
| HTN | nk_cell | *SSPN* | -1.104278401 | 0.001449878 | 0.008873507 | 1 |
| HTN | nk_cell | *PKN1* | 1.413505157 | 0.00258658 | 0.010988756 | 1 |
| HTN | nk_cell | *NLN* | -1.310091869 | 0.019575619 | 0.042822518 | 1 |
| HTN | nk_cell | *CENPK* | 0.470625283 | 0.021167387 | 0.045422101 | 1 |
| HTN | nk_cell | *OPTN* | -1.84230798 | 0.002583835 | 0.010988756 | 1 |
| HTN | nk_cell | *ATF1* | -2.249238709 | 0.010368169 | 0.026682227 | 1 |
| HTN | nk_cell | *TSFM* | -1.822808512 | 0.003204178 | 0.012465758 | 1 |
| HTN | nk_cell | *ARHGAP9* | 0.637260619 | 0.002865406 | 0.011629961 | 1 |
| HTN | nk_cell | *ORMDL2* | 1.107917618 | 0.002667577 | 0.011180705 | 1 |
| HTN | nk_cell | *IKZF4* | -2.472072607 | 3.04E-05 | 0.001552909 | 0.191007777 |
| HTN | nk_cell | *SARDH* | 3.359801006 | 0.003689337 | 0.013360879 | 1 |
| HTN | nk_cell | *STIL* | 1.752016318 | 0.002313616 | 0.010442 | 1 |
| HTN | nk_cell | *NDUFAF4* | 1.325293152 | 0.00094459 | 0.008846261 | 1 |
| HTN | nk_cell | *TTC21B* | -1.494546609 | 0.023486133 | 0.049153568 | 1 |
| HTN | nk_cell | *ACVR1C* | -0.534438707 | 0.021835071 | 0.046503529 | 1 |
| HTN | nk_cell | *PLA2G12A* | -1.910400802 | 0.014385321 | 0.034051398 | 1 |
| HTN | nk_cell | *C4BPB* | -1.361669045 | 0.00038292 | 0.006878338 | 1 |
| HTN | nk_cell | *RAB38* | 2.251429359 | 0.006593581 | 0.019209783 | 1 |
| HTN | nk_cell | *AGO2* | -1.781699017 | 0.012026974 | 0.02993412 | 1 |
| HTN | nk_cell | *CKS2* | 1.633877405 | 0.014747164 | 0.034698885 | 1 |
| HTN | nk_cell | *CHPF* | -1.524069665 | 0.001541708 | 0.00897211 | 1 |
| HTN | nk_cell | *MOGAT1* | -0.861288807 | 0.00141283 | 0.00885079 | 1 |
| HTN | nk_cell | *FAM210B* | -1.501869166 | 0.00602587 | 0.018126654 | 1 |
| HTN | nk_cell | *SNX21* | -1.056273222 | 0.001299723 | 0.00885079 | 1 |
| HTN | nk_cell | *WFDC3* | -0.70415599 | 0.000200775 | 0.005142286 | 1 |
| HTN | nk_cell | *SDC4* | -0.945713288 | 0.001232083 | 0.00885079 | 1 |
| HTN | nk_cell | *VAPB* | -0.939750946 | 0.02343068 | 0.049097432 | 1 |
| HTN | nk_cell | *TOX2* | -0.77516311 | 0.000544774 | 0.008118543 | 1 |
| HTN | nk_cell | *CDH26* | -3.85563484 | 0.000539607 | 0.008096683 | 1 |
| HTN | nk_cell | *STX16* | -1.801419973 | 0.019198513 | 0.042173672 | 1 |
| HTN | nk_cell | *RNF114* | -1.667085064 | 0.023233866 | 0.048788015 | 1 |
| HTN | nk_cell | *KCNK15* | -3.514255258 | 0.003538994 | 0.013111168 | 1 |
| HTN | nk_cell | *NEURL2* | -0.770110043 | 0.016337289 | 0.037212792 | 1 |
| HTN | nk_cell | *FASTKD3* | -1.306777938 | 0.002064162 | 0.009936744 | 1 |
| HTN | nk_cell | *ATP8A1* | 0.772546613 | 0.001080824 | 0.008846261 | 1 |
| HTN | nk_cell | *USP22* | -1.476423776 | 0.018365018 | 0.040698226 | 1 |

| | | | | | | |
|---|---|---|---|---|---|---|
| HTN | nk_cell | *HIF3A* | -0.768667017 | 0.004266467 | 0.014577053 | 1 |
| HTN | nk_cell | *ZNF45* | -2.390362734 | 0.002631373 | 0.011080672 | 1 |
| HTN | nk_cell | *BTN2A2* | 0.954688174 | 0.014649681 | 0.034495335 | 1 |
| HTN | nk_cell | *SNRPC* | -2.80010089 | 4.01E-09 | 1.58E-06 | 2.52E-05 |
| HTN | nk_cell | *HIST1H1D* | 0.711857628 | 1.68E-05 | 0.001013492 | 0.105630026 |
| HTN | nk_cell | *PEX6* | -0.744535276 | 0.01196475 | 0.02982648 | 1 |
| HTN | nk_cell | *NQO2* | -1.740856245 | 0.003586349 | 0.013193317 | 1 |
| HTN | nk_cell | *ZNF391* | -0.873662526 | 2.99E-07 | 5.07E-05 | 0.001876804 |
| HTN | nk_cell | *MED20* | -1.8450733 | 0.000324845 | 0.006479211 | 1 |
| HTN | nk_cell | *KLHDC3* | -2.440052859 | 0.01951957 | 0.042729644 | 1 |
| HTN | nk_cell | *KLHL31* | -0.494469315 | 0.015607101 | 0.03599515 | 1 |
| HTN | nk_cell | *COL21A1* | -1.254214732 | 0.000902237 | 0.008846261 | 1 |
| HTN | nk_cell | *CPNE5* | -1.86712211 | 0.000513747 | 0.007927918 | 1 |
| HTN | nk_cell | *EEF1E1* | 3.477656954 | 0.017468144 | 0.039194226 | 1 |
| HTN | nk_cell | *EREG* | -2.933353347 | 0.001019396 | 0.008846261 | 1 |
| HTN | nk_cell | *MYRF* | 0.618930575 | 0.002032006 | 0.009861903 | 1 |
| HTN | nk_cell | *MT1G* | -6.239361709 | 0.000676823 | 0.00874661 | 1 |
| HTN | nk_cell | *CLYBL* | -1.746393217 | 0.007841511 | 0.021632111 | 1 |
| HTN | nk_cell | *GRK4* | 0.685452635 | 0.001576116 | 0.008975583 | 1 |
| HTN | nk_cell | *MRPS7* | -1.379535975 | 0.021374202 | 0.045769622 | 1 |
| HTN | nk_cell | *OPRL1* | -2.701875939 | 1.45E-09 | 7.09E-07 | 9.11E-06 |
| HTN | nk_cell | *PSD4* | 0.389187287 | 0.001401273 | 0.00885079 | 1 |
| HTN | nk_cell | *SLC25A23* | -0.760732408 | 0.002423971 | 0.010654267 | 1 |
| HTN | nk_cell | *GTF2F1* | -1.969529712 | 0.004667505 | 0.015307565 | 1 |
| HTN | nk_cell | *TRIP10* | -1.31575404 | 0.003084468 | 0.012203935 | 1 |
| HTN | nk_cell | *RTN2* | -2.517367461 | 0.001120005 | 0.008846261 | 1 |
| HTN | nk_cell | *SDCBP2* | -1.35246541 | 0.004507441 | 0.014962133 | 1 |
| HTN | nk_cell | *PANK2* | -1.782649519 | 0.018509936 | 0.040976045 | 1 |
| HTN | nk_cell | *BMP2* | -1.692682555 | 0.000984435 | 0.008846261 | 1 |
| HTN | nk_cell | *PCSK2* | -1.925923382 | 0.000585522 | 0.008131793 | 1 |
| HTN | nk_cell | *MKKS* | -1.949179786 | 0.004028144 | 0.014077232 | 1 |
| HTN | nk_cell | *TMEM74B* | 2.288392015 | 0.007100072 | 0.020134484 | 1 |
| HTN | nk_cell | *NCLN* | -1.817464469 | 0.022373852 | 0.047346299 | 1 |
| HTN | nk_cell | *ZNF436* | -0.978285094 | 0.003139102 | 0.012296285 | 1 |
| HTN | nk_cell | *GDF5* | -1.604594502 | 0.002404006 | 0.010642983 | 1 |
| HTN | nk_cell | *UROD* | -2.15747333 | 0.001606488 | 0.009057712 | 1 |
| HTN | nk_cell | *MCF2L* | -1.088362921 | 0.015310632 | 0.035506435 | 1 |
| HTN | nk_cell | *F10* | -1.191709763 | 0.001282371 | 0.00885079 | 1 |
| HTN | nk_cell | *PROZ* | -4.790627046 | 0.000953263 | 0.008846261 | 1 |
| HTN | nk_cell | *IGFLR1* | 1.076039837 | 0.001498821 | 0.008938486 | 1 |
| HTN | nk_cell | *SCAF1* | -2.561943051 | 0.003324554 | 0.01269116 | 1 |
| HTN | nk_cell | *TSKS* | -0.87435887 | 0.001127542 | 0.008846261 | 1 |
| HTN | nk_cell | *FLRT1* | -1.456994415 | 0.000123395 | 0.003821606 | 0.775786088 |
| HTN | nk_cell | *TRAP1* | 0.788181118 | 0.000802418 | 0.008846261 | 1 |
| HTN | nk_cell | *ZNF384* | -1.952544191 | 0.0037344 | 0.013454539 | 1 |
| HTN | nk_cell | *EMG1* | -2.219742599 | 0.005168693 | 0.016306629 | 1 |
| HTN | nk_cell | *ATG14* | -1.647589827 | 0.00505429 | 0.016048648 | 1 |
| HTN | nk_cell | *RHOJ* | -1.260664398 | 0.001467505 | 0.008909666 | 1 |
| HTN | nk_cell | *HSPA2* | -1.743514867 | 0.000645692 | 0.008600567 | 1 |
| HTN | nk_cell | *PZP* | 0.534234267 | 0.001286208 | 0.00885079 | 1 |

| | | | | | | |
|---|---|---|---|---|---|---|
| HTN | nk_cell | *WDR60* | -0.964700047 | 0.019991772 | 0.043421333 | 1 |
| HTN | nk_cell | *AIF1L* | -1.335757096 | 0.001447079 | 0.008873507 | 1 |
| HTN | nk_cell | *MAP2K2* | 1.971457949 | 0.005875029 | 0.017843625 | 1 |
| HTN | nk_cell | *IPPK* | -2.910723685 | 0.002471882 | 0.010725134 | 1 |
| HTN | nk_cell | *FGD3* | 0.670769951 | 0.002197898 | 0.010211299 | 1 |
| HTN | nk_cell | *MASP1* | -0.959630544 | 0.010857857 | 0.027715528 | 1 |
| HTN | nk_cell | *HRASLS* | -1.634208128 | 0.003200731 | 0.012460059 | 1 |
| HTN | nk_cell | *LRRC61* | -1.106742122 | 0.004498235 | 0.014955263 | 1 |
| HTN | nk_cell | *IDUA* | -2.299146012 | 0.002138689 | 0.010132194 | 1 |
| HTN | nk_cell | *EMC1* | -2.158661138 | 0.002776674 | 0.011426483 | 1 |
| HTN | nk_cell | *PLA2G5* | -0.919940915 | 0.003147369 | 0.012320991 | 1 |
| HTN | nk_cell | *SIN3B* | -1.784869503 | 0.005249251 | 0.016451664 | 1 |
| HTN | nk_cell | *WDR24* | -2.82948731 | 0.008147965 | 0.022234893 | 1 |
| HTN | nk_cell | *TICAM1* | -1.996865386 | 0.003258206 | 0.01256708 | 1 |
| HTN | nk_cell | *AAMP* | -2.194849993 | 0.010027593 | 0.025975083 | 1 |
| HTN | nk_cell | *PNKD* | -2.120765985 | 0.004441841 | 0.014847412 | 1 |
| HTN | nk_cell | *POR* | -1.687107168 | 0.005709748 | 0.017451231 | 1 |
| HTN | nk_cell | *STYXL1* | -1.177237256 | 0.00770319 | 0.02138674 | 1 |
| HTN | nk_cell | *STEAP4* | -1.438486156 | 0.016465443 | 0.037384703 | 1 |
| HTN | nk_cell | *GNAI1* | -1.030608683 | 0.00524855 | 0.016451664 | 1 |
| HTN | nk_cell | *SGCE* | -1.098279517 | 0.002326013 | 0.010482898 | 1 |
| HTN | nk_cell | *SRD5A3* | -2.228293916 | 0.006685122 | 0.019359449 | 1 |
| HTN | nk_cell | *DGCR6L* | 1.964871274 | 0.001291873 | 0.00885079 | 1 |
| HTN | nk_cell | *VPREB3* | -13.40450154 | 0.000204481 | 0.005142286 | 1 |
| HTN | nk_cell | *MGAT3* | -1.064240683 | 0.001042171 | 0.008846261 | 1 |
| HTN | nk_cell | *ATF4* | -1.992383255 | 0.01850165 | 0.040972128 | 1 |
| HTN | nk_cell | *A4GALT* | -1.006124397 | 0.002010733 | 0.009853063 | 1 |
| HTN | nk_cell | *CDC42EP1* | -1.296923798 | 0.002798992 | 0.011473997 | 1 |
| HTN | nk_cell | *TPST2* | 0.983029146 | 0.00553551 | 0.017084806 | 1 |
| HTN | nk_cell | *BAIAP2L2* | -1.24956444 | 0.00223719 | 0.01026356 | 1 |
| HTN | nk_cell | *TST* | -0.605616159 | 0.003208529 | 0.012474969 | 1 |
| HTN | nk_cell | *APOL5* | -5.661309515 | 1.87E-06 | 0.000188514 | 0.011746458 |
| HTN | nk_cell | *EMC4* | -2.144821344 | 0.008305034 | 0.022525344 | 1 |
| HTN | nk_cell | *POT1* | -1.824890485 | 0.003583826 | 0.013191754 | 1 |
| HTN | nk_cell | *CDHR3* | -1.456500098 | 0.002480943 | 0.010734818 | 1 |
| HTN | nk_cell | *FOXP2* | -0.836346286 | 0.003223388 | 0.012494105 | 1 |
| HTN | nk_cell | *FLNC* | -1.059560269 | 0.009345246 | 0.02460367 | 1 |
| HTN | nk_cell | *IRF5* | -1.205230473 | 0.00102762 | 0.008846261 | 1 |
| HTN | nk_cell | *KLHDC10* | -2.47272717 | 8.34E-05 | 0.003049889 | 0.524580949 |
| HTN | nk_cell | *OPN1SW* | -1.259767171 | 0.000278204 | 0.006094325 | 1 |
| HTN | nk_cell | *HOXD1* | -4.718603552 | 0.00444218 | 0.014847412 | 1 |
| HTN | nk_cell | *HOXD3* | -1.272219792 | 0.000442922 | 0.007343364 | 1 |
| HTN | nk_cell | *CHN1* | -1.297330535 | 0.004357651 | 0.014745183 | 1 |
| HTN | nk_cell | *GAD1* | -2.060537387 | 0.000965034 | 0.008846261 | 1 |
| HTN | nk_cell | *HOXD9* | -1.802655423 | 0.000681565 | 0.00874661 | 1 |
| HTN | nk_cell | *TWSG1* | -1.804692699 | 0.010168073 | 0.026242477 | 1 |
| HTN | nk_cell | *WDFY4* | -1.272233185 | 0.000151092 | 0.004337502 | 0.949913017 |
| HTN | nk_cell | *EIF2AK4* | -1.890931316 | 0.000112132 | 0.003612201 | 0.704971848 |
| HTN | nk_cell | *IVD* | -1.333303053 | 0.018918864 | 0.041719711 | 1 |
| HTN | nk_cell | *KNSTRN* | -0.691791115 | 0.001262799 | 0.00885079 | 1 |

| | | | | | | |
|---|---|---|---|---|---|---|
| HTN | nk_cell | *CALML4* | -3.840603548 | 0.001424493 | 0.00885079 | 1 |
| HTN | nk_cell | *ACKR4* | -1.032225636 | 0.001750998 | 0.009297738 | 1 |
| HTN | nk_cell | *ANAPC13* | -0.826427765 | 0.018735382 | 0.041416789 | 1 |
| HTN | nk_cell | *SUMF2* | -2.203092133 | 0.01050372 | 0.026964837 | 1 |
| HTN | nk_cell | *RPAIN* | -1.469830341 | 0.014758054 | 0.034711517 | 1 |
| HTN | nk_cell | *CCL4* | 0.288945675 | 0.002230946 | 0.010252893 | 1 |
| HTN | nk_cell | *SLC44A2* | 1.316017503 | 0.008811113 | 0.023582574 | 1 |
| HTN | nk_cell | *CDKN2D* | 0.802840374 | 0.005193445 | 0.016325595 | 1 |
| HTN | nk_cell | *MTUS1* | -1.435980365 | 0.003164677 | 0.012344206 | 1 |
| HTN | nk_cell | *RIPK3* | 2.728112578 | 0.000885956 | 0.008846261 | 1 |
| HTN | nk_cell | *BCL2L2* | -1.697836841 | 0.001960652 | 0.009759795 | 1 |
| HTN | nk_cell | *EPB41L4A* | -1.312687129 | 0.003676052 | 0.013343729 | 1 |
| HTN | nk_cell | *CDO1* | -1.527832637 | 0.001523713 | 0.008957475 | 1 |
| HTN | nk_cell | *REEP5* | -1.609057316 | 0.013457362 | 0.032521354 | 1 |
| HTN | nk_cell | *RHBDF2* | 0.82402622 | 0.000353963 | 0.006607413 | 1 |
| HTN | nk_cell | *TMEM8A* | 2.021932029 | 0.00770088 | 0.02138674 | 1 |
| HTN | nk_cell | *SHC2* | -1.120531809 | 0.004686473 | 0.015353756 | 1 |
| HTN | nk_cell | *ERMARD* | -2.24214266 | 0.00510899 | 0.01618738 | 1 |
| HTN | nk_cell | *GALNT8* | -1.783962434 | 0.008399543 | 0.022703323 | 1 |
| HTN | nk_cell | *ECSIT* | -1.892749068 | 0.023152471 | 0.04866586 | 1 |
| HTN | nk_cell | *THEM6* | -0.923514832 | 0.000565731 | 0.008121264 | 1 |
| HTN | nk_cell | *DPP6* | -1.538794042 | 0.000460796 | 0.007524739 | 1 |
| HTN | nk_cell | *RPL36* | 1.359826143 | 0.014264189 | 0.033841117 | 1 |
| HTN | nk_cell | *ATP8B3* | -1.09663496 | 0.0044163 | 0.01480253 | 1 |
| HTN | nk_cell | *DDA1* | -3.815622493 | 0.000340252 | 0.006541794 | 1 |
| HTN | nk_cell | *LSM7* | 1.551309465 | 0.014615727 | 0.034467021 | 1 |
| HTN | nk_cell | *PXDN* | -0.888743067 | 0.001297647 | 0.00885079 | 1 |
| HTN | nk_cell | *PGPEP1* | 2.204095437 | 0.002172034 | 0.010170205 | 1 |
| HTN | nk_cell | *LSM4* | 1.314625667 | 0.001244924 | 0.00885079 | 1 |
| HTN | nk_cell | *HRC* | -1.291767077 | 0.004194975 | 0.014410776 | 1 |
| HTN | nk_cell | *TRPM4* | -1.610888792 | 0.000576019 | 0.008131793 | 1 |
| HTN | nk_cell | *CYP2E1* | -2.542543876 | 0.000234149 | 0.005519823 | 1 |
| HTN | nk_cell | *PAK4* | -2.114066454 | 0.003112824 | 0.012254431 | 1 |
| HTN | nk_cell | *YIPF2* | -1.393970761 | 0.001135339 | 0.008846261 | 1 |
| HTN | nk_cell | *MAP3K10* | -2.881819348 | 0.001565331 | 0.008975583 | 1 |
| HTN | nk_cell | *SMPDL3B* | -14.14624542 | 0.00224143 | 0.01026356 | 1 |
| HTN | nk_cell | *CLIP1* | -1.476529557 | 0.023496752 | 0.049159427 | 1 |
| HTN | nk_cell | *PPAN* | 3.220318902 | 0.01902239 | 0.041859912 | 1 |
| HTN | nk_cell | *HABP4* | -1.782407542 | 0.003797327 | 0.013614173 | 1 |
| HTN | nk_cell | *EPS8L1* | 1.983910527 | 0.001580996 | 0.008993168 | 1 |
| HTN | nk_cell | *LILRB2* | -1.581717171 | 0.001231236 | 0.00885079 | 1 |
| HTN | nk_cell | *CHMP1A* | 3.441108254 | 6.14E-05 | 0.002580837 | 0.385948417 |
| HTN | nk_cell | *HAUS8* | -5.661892095 | 1.90E-07 | 3.61E-05 | 0.001192858 |
| HTN | nk_cell | *KCNC3* | -1.811598087 | 0.000712509 | 0.008846261 | 1 |
| HTN | nk_cell | *KIF3A* | -1.549548175 | 0.01658317 | 0.037584134 | 1 |
| HTN | nk_cell | *AOC3* | -1.28798311 | 0.012313399 | 0.030456325 | 1 |
| HTN | nk_cell | *AOC2* | -1.428789726 | 0.000755071 | 0.008846261 | 1 |
| HTN | nk_cell | *DIAPH1* | 0.675864339 | 0.009171695 | 0.024276248 | 1 |
| HTN | nk_cell | *ANO1* | -3.808490018 | 0.000775238 | 0.008846261 | 1 |
| HTN | nk_cell | *NINJ1* | -1.392540125 | 0.000753554 | 0.008846261 | 1 |

| | | | | | | |
|---|---|---|---|---|---|---|
| HTN | nk_cell | *NPHP4* | -2.392361277 | 0.015157473 | 0.035323691 | 1 |
| HTN | nk_cell | *CKMT2* | -0.963645554 | 0.001188723 | 0.00885079 | 1 |
| HTN | nk_cell | *KHDRBS3* | -1.131055344 | 0.005953376 | 0.017968734 | 1 |
| HTN | nk_cell | *CHD1L* | -1.93134961 | 0.003554972 | 0.013146983 | 1 |
| HTN | nk_cell | *RHPN2* | -1.336388278 | 0.002399204 | 0.010637375 | 1 |
| HTN | nk_cell | *GCH1* | 1.110435564 | 0.001203901 | 0.00885079 | 1 |
| HTN | nk_cell | *LGALS3* | -1.524937313 | 0.0143809 | 0.034051398 | 1 |
| HTN | nk_cell | *DNAJB1* | -2.255569213 | 0.00064008 | 0.008562086 | 1 |
| HTN | nk_cell | *FBXW9* | -2.018683442 | 0.016120308 | 0.036827173 | 1 |
| HTN | nk_cell | *C19orf57* | -2.156588138 | 0.00135218 | 0.00885079 | 1 |
| HTN | nk_cell | *MATN3* | -6.089875552 | 0.001550119 | 0.008973847 | 1 |
| HTN | nk_cell | *CCT6B* | -1.33110789 | 0.022389771 | 0.047363556 | 1 |
| HTN | nk_cell | *ENOSF1* | -1.632731997 | 0.001712084 | 0.009223501 | 1 |
| HTN | nk_cell | *EMILIN2* | -1.358019498 | 0.001409321 | 0.00885079 | 1 |
| HTN | nk_cell | *CNGA4* | -2.161612338 | 0.008334131 | 0.022555611 | 1 |
| HTN | nk_cell | *RRP8* | -1.908777343 | 0.007749344 | 0.021462611 | 1 |
| HTN | nk_cell | *IMMT* | -1.9718987 | 0.002036454 | 0.009871385 | 1 |
| HTN | nk_cell | *MRPL35* | 1.382614874 | 0.004353831 | 0.014745183 | 1 |
| HTN | nk_cell | *INPP5K* | -1.531200189 | 0.000227396 | 0.005449811 | 1 |
| HTN | nk_cell | *SERPINF1* | -1.48688917 | 0.000101605 | 0.003471679 | 0.638788996 |
| HTN | nk_cell | *TMEM128* | -2.101149902 | 0.002126743 | 0.01009268 | 1 |
| HTN | nk_cell | *KDM6B* | -3.24479803 | 1.72E-05 | 0.001013492 | 0.108443681 |
| HTN | nk_cell | *REEP2* | -1.297134278 | 0.000237067 | 0.005540664 | 1 |
| HTN | nk_cell | *BTBD3* | -1.509121005 | 0.001466652 | 0.008909666 | 1 |
| HTN | nk_cell | *IGHMBP2* | -1.062020881 | 0.013626164 | 0.03274759 | 1 |
| HTN | nk_cell | *LPIN3* | -0.939784545 | 0.001129097 | 0.008846261 | 1 |
| HTN | nk_cell | *DMGDH* | -1.394377792 | 0.001266413 | 0.00885079 | 1 |
| HTN | nk_cell | *BHMT2* | -0.472459041 | 0.000451481 | 0.007450022 | 1 |
| HTN | nk_cell | *KANK4* | -0.652674858 | 0.011226345 | 0.028379587 | 1 |
| HTN | nk_cell | *RSG1* | -3.714833997 | 0.00025994 | 0.005849812 | 1 |
| HTN | nk_cell | *CASP9* | -1.340814745 | 0.001657496 | 0.009149879 | 1 |
| HTN | nk_cell | *ATP8A2* | 2.128858428 | 0.007274594 | 0.020467508 | 1 |
| HTN | nk_cell | *WASF3* | -0.890876469 | 0.001270132 | 0.00885079 | 1 |
| HTN | nk_cell | *RNF17* | -2.806071544 | 0.000330061 | 0.006479211 | 1 |
| HTN | nk_cell | *PEMT* | -0.965647529 | 0.004735725 | 0.015418697 | 1 |
| HTN | nk_cell | *COG6* | -1.731544134 | 0.005206031 | 0.016342136 | 1 |
| HTN | nk_cell | *TRPC4* | -0.89223411 | 0.001056045 | 0.008846261 | 1 |
| HTN | nk_cell | *PRAM1* | -1.060072476 | 0.000103408 | 0.003490877 | 0.650126343 |
| HTN | nk_cell | *ZNF414* | -2.860265615 | 0.00317142 | 0.012353606 | 1 |
| HTN | nk_cell | *RTN3* | -1.415019615 | 0.004641887 | 0.015245101 | 1 |
| HTN | nk_cell | *GSTT2B* | -0.860150152 | 0.004703836 | 0.015378584 | 1 |
| HTN | nk_cell | *FAM83F* | -3.40641352 | 0.000750266 | 0.008846261 | 1 |
| HTN | nk_cell | *GIMAP4* | 0.591622693 | 0.002016469 | 0.009858936 | 1 |
| HTN | nk_cell | *ACTR3B* | -1.861019891 | 0.014629171 | 0.034479556 | 1 |
| HTN | nk_cell | *DYDC2* | 5.247982849 | 0.020583251 | 0.044413055 | 1 |
| HTN | nk_cell | *TMTC1* | -1.053775787 | 0.002386018 | 0.010623862 | 1 |
| HTN | nk_cell | *SPINK5* | -1.382052454 | 0.006911222 | 0.0198044 | 1 |
| HTN | nk_cell | *IMPA1* | 1.570914869 | 0.003305929 | 0.012667763 | 1 |
| HTN | nk_cell | *MICAL2* | -1.534558256 | 0.004696764 | 0.015371451 | 1 |
| HTN | nk_cell | *DPF2* | -1.707719194 | 0.004965164 | 0.015869846 | 1 |

| | | | | | | |
|---|---|---|---|---|---|---|
| HTN | nk_cell | *GSC* | -8.341609394 | 0.006040368 | 0.018131247 | 1 |
| HTN | nk_cell | *COX16* | 1.416465041 | 0.001744436 | 0.009286428 | 1 |
| HTN | nk_cell | *MED6* | -0.976747594 | 0.012807843 | 0.03129534 | 1 |
| HTN | nk_cell | *MRO* | -1.146779634 | 0.008356393 | 0.022606128 | 1 |
| HTN | nk_cell | *THUMPD3* | -1.666621391 | 0.002046379 | 0.009904221 | 1 |
| HTN | nk_cell | *CNTN6* | -0.864845926 | 0.002021374 | 0.009861903 | 1 |
| HTN | nk_cell | *GSTM5* | -1.300082387 | 0.001472148 | 0.008909666 | 1 |
| HTN | nk_cell | *SYT6* | -5.504747694 | 0.001173154 | 0.00885079 | 1 |
| HTN | nk_cell | *VAV3* | 0.584375507 | 0.004731998 | 0.015416362 | 1 |
| HTN | nk_cell | *PSRC1* | -1.3313575 | 0.023463877 | 0.049125115 | 1 |
| HTN | nk_cell | *WNT2B* | -2.524437937 | 6.43E-07 | 9.62E-05 | 0.004040191 |
| HTN | nk_cell | *SPIRE1* | -1.397099241 | 0.007900306 | 0.021746597 | 1 |
| HTN | nk_cell | *FKBP11* | 1.09355046 | 0.00148621 | 0.008938486 | 1 |
| HTN | nk_cell | *LPIN1* | -1.469688423 | 0.003592552 | 0.01320145 | 1 |
| HTN | nk_cell | *LDHA* | -1.939390556 | 0.009863097 | 0.025613091 | 1 |
| HTN | nk_cell | *ANO3* | -0.706314928 | 0.002877449 | 0.011652827 | 1 |
| HTN | nk_cell | *FST* | -2.704374247 | 7.80E-05 | 0.002917771 | 0.490185499 |
| HTN | nk_cell | *NARS* | -1.801584185 | 0.009476941 | 0.024856708 | 1 |
| HTN | nk_cell | *ECHDC3* | -1.117909083 | 0.002610583 | 0.011037484 | 1 |
| HTN | nk_cell | *IL15RA* | -1.994796245 | 0.004893103 | 0.01571141 | 1 |
| HTN | nk_cell | *TMEM241* | -1.516095236 | 0.005946122 | 0.017955461 | 1 |
| HTN | nk_cell | *EMP1* | -1.146248981 | 0.008122866 | 0.022213336 | 1 |
| HTN | nk_cell | *KLRD1* | 0.309020185 | 0.001715011 | 0.009223501 | 1 |
| HTN | nk_cell | *PRH2* | -2.460376587 | 0.000198671 | 0.005142286 | 1 |
| HTN | nk_cell | *DDB2* | 2.856453237 | 0.000720349 | 0.008846261 | 1 |
| HTN | nk_cell | *ACP2* | 7.394435122 | 7.13E-17 | 4.48E-13 | 4.48E-13 |
| HTN | nk_cell | *HOOK1* | 1.399337104 | 0.002586824 | 0.010988756 | 1 |
| HTN | nk_cell | *PRPF38A* | -1.80459472 | 0.019750073 | 0.043114135 | 1 |
| HTN | nk_cell | *SLC43A3* | -1.665370137 | 0.013560792 | 0.032652892 | 1 |
| HTN | nk_cell | *APLNR* | -1.261385605 | 0.014118852 | 0.033644753 | 1 |
| HTN | nk_cell | *DZIP1* | -1.19548066 | 0.014233555 | 0.03381816 | 1 |
| HTN | nk_cell | *BIVM* | -2.586853555 | 0.009688239 | 0.02533692 | 1 |
| HTN | nk_cell | *KLB* | -0.478542283 | 0.012917189 | 0.031464691 | 1 |
| HTN | nk_cell | *WDR36* | -1.922102421 | 0.021792663 | 0.046475738 | 1 |
| HTN | nk_cell | *OSTF1* | 1.19161828 | 0.012387233 | 0.030576573 | 1 |
| HTN | nk_cell | *CTSL* | -1.713307135 | 0.020370647 | 0.044086147 | 1 |
| HTN | nk_cell | *HRK* | -3.398853092 | 0.000235809 | 0.005531824 | 1 |
| HTN | nk_cell | *DTX1* | -1.024274747 | 0.001035832 | 0.008846261 | 1 |
| HTN | nk_cell | *TES* | 0.879819253 | 0.009796933 | 0.025519983 | 1 |
| HTN | nk_cell | *KHDC1* | -0.778140139 | 0.022572989 | 0.047654931 | 1 |
| HTN | nk_cell | *PRR5L* | 0.656249977 | 0.001501869 | 0.008938486 | 1 |
| HTN | nk_cell | *NAT10* | -1.795070681 | 0.001880801 | 0.009566447 | 1 |
| HTN | nk_cell | *RDH5* | -0.621359723 | 0.003105784 | 0.012242047 | 1 |
| HTN | nk_cell | *BLOC1S1* | 2.64772515 | 0.005760886 | 0.017581888 | 1 |
| HTN | nk_cell | *PPP1R1A* | -2.164656865 | 0.001063275 | 0.008846261 | 1 |
| HTN | nk_cell | *FAIM2* | -1.515951176 | 0.001301717 | 0.00885079 | 1 |
| HTN | nk_cell | *SLC26A10* | -1.921415816 | 0.011947007 | 0.029794064 | 1 |
| HTN | nk_cell | *MAP7* | -1.401875253 | 0.01556625 | 0.035993017 | 1 |
| HTN | nk_cell | *PKIB* | -2.019422591 | 0.000489613 | 0.007734162 | 1 |
| HTN | nk_cell | *SMPD2* | -1.789493539 | 0.000778974 | 0.008846261 | 1 |

| | | | | | | |
|---|---|---|---|---|---|---|
| HTN | nk_cell | *PRADC1* | -1.350767743 | 0.001327547 | 0.00885079 | 1 |
| HTN | nk_cell | *KCNMB4* | -1.239989387 | 0.00626142 | 0.018612552 | 1 |
| HTN | nk_cell | *FHOD1* | -2.273298676 | 0.006022713 | 0.018126654 | 1 |
| HTN | nk_cell | *CCDC102A* | 1.651491141 | 0.009946772 | 0.025819717 | 1 |
| HTN | nk_cell | *AGT* | 0.705040639 | 0.003414986 | 0.012864002 | 1 |
| HTN | nk_cell | *NTPCR* | -1.321798321 | 0.003854051 | 0.013751655 | 1 |
| HTN | nk_cell | *PIGC* | -1.699461544 | 0.000899917 | 0.008846261 | 1 |
| HTN | nk_cell | *SLC19A3* | -0.51098565 | 0.007019499 | 0.019978148 | 1 |
| HTN | nk_cell | *TMBIM1* | -2.200920194 | 0.000326084 | 0.006479211 | 1 |
| HTN | nk_cell | *ARMC9* | -1.702243089 | 0.002378741 | 0.010598969 | 1 |
| HTN | nk_cell | *SLC41A2* | -1.083555461 | 0.002447124 | 0.010667096 | 1 |
| HTN | nk_cell | *VPS36* | -1.419020859 | 0.016317192 | 0.037195402 | 1 |
| HTN | nk_cell | *TBC1D4* | -1.459518428 | 0.00403263 | 0.014079207 | 1 |
| HTN | nk_cell | *SETDB2* | -1.724599154 | 0.01538084 | 0.035642956 | 1 |
| HTN | nk_cell | *CHST12* | 0.67410509 | 0.010224167 | 0.026354792 | 1 |
| HTN | nk_cell | *GPNMB* | -1.611327768 | 0.008628911 | 0.023223444 | 1 |
| HTN | nk_cell | *NUPL2* | -1.989802697 | 0.0156187 | 0.03599515 | 1 |
| HTN | nk_cell | *ZDHHC4* | 1.283023586 | 0.007332032 | 0.020584147 | 1 |
| HTN | nk_cell | *AOAH* | 0.308259979 | 0.002826394 | 0.011523696 | 1 |
| HTN | nk_cell | *DDX56* | -2.232831123 | 0.002928736 | 0.011809268 | 1 |
| HTN | nk_cell | *NACAD* | -3.803575463 | 3.86E-05 | 0.00190984 | 0.242549728 |
| HTN | nk_cell | *TTC5* | -2.326111286 | 0.005033428 | 0.016014758 | 1 |
| HTN | nk_cell | *ALPK3* | -0.813790963 | 0.004615533 | 0.015184955 | 1 |
| HTN | nk_cell | *TM6SF1* | -1.905248881 | 0.002450548 | 0.010669388 | 1 |
| HTN | nk_cell | *CIB2* | -2.014347358 | 2.08E-05 | 0.001159965 | 0.131076052 |
| HTN | nk_cell | *NMT1* | 2.072367707 | 8.24E-07 | 0.000110176 | 0.005178293 |
| HTN | nk_cell | *BRIP1* | 1.745289965 | 0.012671388 | 0.0310706 | 1 |
| HTN | nk_cell | *BLK* | -5.263277419 | 5.30E-07 | 8.32E-05 | 0.003329155 |
| HTN | nk_cell | *IL1RN* | -1.932221743 | 0.001713868 | 0.009223501 | 1 |
| HTN | nk_cell | *NIPSNAP3A* | -1.70957676 | 0.021851787 | 0.046503529 | 1 |
| HTN | nk_cell | *TOR1B* | 1.722365225 | 0.001327226 | 0.00885079 | 1 |
| HTN | nk_cell | *SLC2A8* | 2.653696311 | 0.000774001 | 0.008846261 | 1 |
| HTN | nk_cell | *ZNF189* | -1.825526917 | 0.015416594 | 0.035712647 | 1 |
| HTN | nk_cell | *ALDOB* | -2.031785686 | 0.004145927 | 0.014298103 | 1 |
| HTN | nk_cell | *FPGS* | -2.644444814 | 0.006560078 | 0.019169154 | 1 |
| HTN | nk_cell | *GARNL3* | -1.621735418 | 0.003132101 | 0.012291836 | 1 |
| HTN | nk_cell | *RABEPK* | -1.911160556 | 0.010108893 | 0.026143401 | 1 |
| HTN | nk_cell | *GOLGA1* | -1.642245869 | 0.012778344 | 0.031235399 | 1 |
| HTN | nk_cell | *DSCC1* | -0.401561541 | 0.011382726 | 0.028648197 | 1 |
| HTN | nk_cell | *DERL1* | 1.687958201 | 0.017726162 | 0.039628171 | 1 |
| HTN | nk_cell | *IL11RA* | -2.421841843 | 0.000583022 | 0.008131793 | 1 |
| HTN | nk_cell | *APTX* | -1.879944083 | 0.004187369 | 0.014410776 | 1 |
| HTN | nk_cell | *TMEM8B* | -1.462638437 | 0.001517208 | 0.0089561 | 1 |
| HTN | nk_cell | *PPIL1* | 1.127714668 | 0.001895306 | 0.009600838 | 1 |
| HTN | nk_cell | *KIF13A* | -1.253911875 | 0.005191619 | 0.016325595 | 1 |
| HTN | nk_cell | *BPHL* | -0.805480757 | 0.002673208 | 0.011190293 | 1 |
| HTN | nk_cell | *RIPK1* | -1.653335264 | 0.017955166 | 0.039987292 | 1 |
| HTN | nk_cell | *TUBB2B* | -1.092292192 | 0.005552369 | 0.01712003 | 1 |
| HTN | nk_cell | *UQCC2* | -1.716024078 | 0.001333614 | 0.00885079 | 1 |
| HTN | nk_cell | *FLOT1* | -1.872279248 | 0.006420355 | 0.018897364 | 1 |

| | | | | | | |
|---|---|---|---|---|---|---|
| HTN | nk_cell | *NRM* | 2.017544513 | 0.004674193 | 0.015321509 | 1 |
| HTN | nk_cell | *VARS2* | 0.393997335 | 0.021775587 | 0.046455078 | 1 |
| HTN | nk_cell | *FGFBP2* | 0.114973254 | 0.001223363 | 0.00885079 | 1 |
| HTN | nk_cell | *FHDC1* | -0.990954141 | 0.001114848 | 0.008846261 | 1 |
| HTN | nk_cell | *SYTL2* | 0.692806316 | 0.006721042 | 0.019427674 | 1 |
| HTN | nk_cell | *RAB30* | -1.218684637 | 0.002530138 | 0.010841214 | 1 |
| HTN | nk_cell | *PI15* | -8.21839017 | 0.015126892 | 0.035290029 | 1 |
| HTN | nk_cell | *TTPA* | -0.510902192 | 0.000687662 | 0.00874661 | 1 |
| HTN | nk_cell | *GGH* | -0.749164913 | 0.000792132 | 0.008846261 | 1 |
| HTN | nk_cell | *TGS1* | -1.184752527 | 0.023308887 | 0.048896554 | 1 |
| HTN | nk_cell | *DCUN1D5* | -1.001974727 | 0.00359276 | 0.01320145 | 1 |
| HTN | nk_cell | *TRIM29* | -1.313035339 | 0.000139462 | 0.004175231 | 0.8767985 |
| HTN | nk_cell | *PPP2R1B* | -0.933692883 | 0.00066002 | 0.008681056 | 1 |
| HTN | nk_cell | *FXYD6* | -2.187764433 | 0.014960913 | 0.035077441 | 1 |
| HTN | nk_cell | *TMPRSS13* | 2.191942896 | 0.000536455 | 0.008086807 | 1 |
| HTN | nk_cell | *NDUFAF1* | -2.443604147 | 0.000153427 | 0.004369177 | 0.964593539 |
| HTN | nk_cell | *ITGA11* | -1.001643642 | 0.001072295 | 0.008846261 | 1 |
| HTN | nk_cell | *PAQR5* | -2.271489156 | 7.65E-05 | 0.002917771 | 0.480914809 |
| HTN | nk_cell | *RMDN3* | 3.126417843 | 0.00091741 | 0.008846261 | 1 |
| HTN | nk_cell | *UACA* | -1.280297226 | 0.00413338 | 0.014278329 | 1 |
| HTN | nk_cell | *RABGGTB* | -2.279389909 | 0.01484208 | 0.03488305 | 1 |
| HTN | nk_cell | *SLC44A5* | -1.099516211 | 0.005182224 | 0.016306629 | 1 |
| HTN | nk_cell | *DNASE2B* | -3.987551782 | 0.002222556 | 0.010251805 | 1 |
| HTN | nk_cell | *DBT* | -1.534725143 | 0.023164492 | 0.048674854 | 1 |
| HTN | nk_cell | *KHK* | -0.65466514 | 0.003823825 | 0.013659313 | 1 |
| HTN | nk_cell | *ADCY3* | -2.146725551 | 0.001742371 | 0.009286428 | 1 |
| HTN | nk_cell | *LHCGR* | -0.7873788 | 0.000979192 | 0.008846261 | 1 |
| HTN | nk_cell | *THUMPD2* | -1.621297903 | 0.00169668 | 0.009212238 | 1 |
| HTN | nk_cell | *ATRAID* | -2.568641517 | 0.000472764 | 0.007601702 | 1 |
| HTN | nk_cell | *CYP2C8* | -1.389254439 | 0.005931161 | 0.017931898 | 1 |
| HTN | nk_cell | *CH25H* | -0.827462904 | 0.003810192 | 0.013633851 | 1 |
| HTN | nk_cell | *BTBD16* | -0.889228881 | 0.002078433 | 0.009982514 | 1 |
| HTN | nk_cell | *TACC2* | -0.936152071 | 0.001606934 | 0.009057712 | 1 |
| HTN | nk_cell | *CALHM2* | -1.913999071 | 0.012877195 | 0.031405563 | 1 |
| HTN | nk_cell | *PLCE1* | -1.074812218 | 0.002494066 | 0.010761973 | 1 |
| HTN | nk_cell | *ASCC1* | -1.823819983 | 0.010854607 | 0.027715528 | 1 |
| HTN | nk_cell | *STAT4* | 0.188527011 | 0.001191637 | 0.00885079 | 1 |
| HTN | nk_cell | *CARF* | -1.380489712 | 0.001631766 | 0.009090004 | 1 |
| HTN | nk_cell | *ASNSD1* | -1.940537507 | 0.005460609 | 0.016911749 | 1 |
| HTN | nk_cell | *METTL5* | -1.890056534 | 0.00267496 | 0.011190293 | 1 |
| HTN | nk_cell | *NAB1* | -1.646370981 | 0.003342062 | 0.012704157 | 1 |
| HTN | nk_cell | *MDH1B* | -2.292932822 | 0.000955777 | 0.008846261 | 1 |
| HTN | nk_cell | *ITGAV* | -1.38205468 | 0.015200937 | 0.035330236 | 1 |
| HTN | nk_cell | *UBL7* | -4.006398583 | 0.000559702 | 0.008121264 | 1 |
| HTN | nk_cell | *ARHGAP24* | -1.168319385 | 0.01883025 | 0.041582642 | 1 |
| HTN | nk_cell | *AP1AR* | -2.04694201 | 0.013490006 | 0.032532286 | 1 |
| HTN | nk_cell | *PRKG2* | -1.352660858 | 0.00345974 | 0.012974855 | 1 |
| HTN | nk_cell | *BBS7* | -1.284259148 | 0.001433575 | 0.008870952 | 1 |
| HTN | nk_cell | *MMRN1* | -0.546668443 | 0.001708116 | 0.009223501 | 1 |
| HTN | nk_cell | *PRDM5* | -1.392226444 | 0.001022405 | 0.008846261 | 1 |

| | | | | | | |
|---|---|---|---|---|---|---|
| HTN | nk_cell | *TRPC3* | 1.634740399 | 0.000297489 | 0.006297947 | 1 |
| HTN | nk_cell | *NAAA* | -1.571362023 | 0.021535682 | 0.046037005 | 1 |
| HTN | nk_cell | *G3BP2* | 2.024189517 | 0.009222704 | 0.024311589 | 1 |
| HTN | nk_cell | *SCARB2* | -1.614734247 | 0.016383994 | 0.037253587 | 1 |
| HTN | nk_cell | *ANXA3* | -2.2779003 | 0.004081183 | 0.014144651 | 1 |
| HTN | nk_cell | *ENPEP* | -1.859392354 | 0.000175303 | 0.004771114 | 1 |
| HTN | nk_cell | *RNF185* | -1.421149849 | 0.009143253 | 0.024254696 | 1 |
| HTN | nk_cell | *ERP27* | 0.403425642 | 0.002158614 | 0.010155174 | 1 |
| HTN | nk_cell | *ALG10* | -2.715979956 | 0.001618227 | 0.009059476 | 1 |
| HTN | nk_cell | *ZCRB1* | -1.285929057 | 0.014963845 | 0.035077441 | 1 |
| HTN | nk_cell | *PRICKLE1* | -0.911122689 | 0.002580328 | 0.010988756 | 1 |
| HTN | nk_cell | *KLRG1* | 0.563655962 | 0.010424119 | 0.026771421 | 1 |
| HTN | nk_cell | *VAMP1* | 1.176276518 | 0.006702275 | 0.019382337 | 1 |
| HTN | nk_cell | *CD27* | 3.619284043 | 0.000599182 | 0.008279245 | 1 |
| HTN | nk_cell | *PIANP* | 4.874023026 | 0.000762751 | 0.008846261 | 1 |
| HTN | nk_cell | *PPFIA2* | -0.722717356 | 0.01104431 | 0.028059143 | 1 |
| HTN | nk_cell | *GLIPR1* | 1.657656987 | 0.011068705 | 0.028082707 | 1 |
| HTN | nk_cell | *LGR5* | -1.076935717 | 0.000304362 | 0.006326876 | 1 |
| HTN | nk_cell | *GAS2L3* | -2.332365807 | 0.015797074 | 0.036331936 | 1 |
| HTN | nk_cell | *TMEM132B* | -0.94318838 | 0.0018689 | 0.009540973 | 1 |
| HTN | nk_cell | *ACVRL1* | -0.976870548 | 0.015263309 | 0.035435902 | 1 |
| HTN | nk_cell | *GPR84* | -12.07108126 | 1.53E-05 | 0.000952967 | 0.096249629 |
| HTN | nk_cell | *BRCA2* | -2.165288418 | 0.023876558 | 0.049837956 | 1 |
| HTN | nk_cell | *LMBR1L* | -2.655769087 | 0.000712494 | 0.008846261 | 1 |
| HTN | nk_cell | *METTL21C* | 2.434457557 | 0.00351732 | 0.013069378 | 1 |
| HTN | nk_cell | *TSSK4* | -4.215863129 | 0.000148925 | 0.004314717 | 0.936293507 |
| HTN | nk_cell | *C14orf37* | -1.595792846 | 0.004327175 | 0.014705378 | 1 |
| HTN | nk_cell | *STON2* | -1.498597948 | 0.002309919 | 0.010439478 | 1 |
| HTN | nk_cell | *PTGR2* | -0.763825967 | 0.006479652 | 0.018983026 | 1 |
| HTN | nk_cell | *AK7* | -2.069576282 | 0.001289154 | 0.00885079 | 1 |
| HTN | nk_cell | *SLC24A4* | -0.940499279 | 0.001089594 | 0.008846261 | 1 |
| HTN | nk_cell | *FBLN5* | -1.097675937 | 0.003942459 | 0.013894565 | 1 |
| HTN | nk_cell | *C14orf79* | -1.95073535 | 0.000764052 | 0.008846261 | 1 |
| HTN | nk_cell | *ZSCAN29* | -2.723363791 | 0.006294513 | 0.018675604 | 1 |
| HTN | nk_cell | *COMMD4* | -2.726202192 | 0.004339396 | 0.014723033 | 1 |
| HTN | nk_cell | *PSTPIP1* | 0.821130052 | 0.001710843 | 0.009223501 | 1 |
| HTN | nk_cell | *DNAJA4* | -1.829440947 | 0.001894756 | 0.009600838 | 1 |
| HTN | nk_cell | *TPM1* | -1.44114831 | 0.009199473 | 0.024280893 | 1 |
| HTN | nk_cell | *CYP11A1* | -0.850533151 | 0.002197307 | 0.010211299 | 1 |
| HTN | nk_cell | *BBS4* | -1.575264914 | 0.018274243 | 0.040511341 | 1 |
| HTN | nk_cell | *ADAMTS17* | -1.834380682 | 0.015515315 | 0.035914868 | 1 |
| HTN | nk_cell | *HAPLN3* | -3.052820371 | 0.000103832 | 0.003490877 | 0.652793932 |
| HTN | nk_cell | *WDR93* | -0.490319934 | 0.004430569 | 0.014832261 | 1 |
| HTN | nk_cell | *TICRR* | -1.056481547 | 0.012539127 | 0.03082142 | 1 |
| HTN | nk_cell | *MFGE8* | -1.625962411 | 0.001064126 | 0.008846261 | 1 |
| HTN | nk_cell | *UNC45A* | -1.71487533 | 0.000162482 | 0.004487306 | 1 |
| HTN | nk_cell | *ST8SIA2* | 4.419839685 | 0.000928836 | 0.008846261 | 1 |
| HTN | nk_cell | *SH3GL3* | -1.21855499 | 0.003221292 | 0.012494105 | 1 |
| HTN | nk_cell | *TGFB1I1* | -1.572626777 | 0.00126212 | 0.00885079 | 1 |
| HTN | nk_cell | *FTO* | -1.370342498 | 0.017934734 | 0.039984282 | 1 |

| | | | | | | |
|---|---|---|---|---|---|---|
| HTN | nk_cell | *TXNL4B* | -2.695022393 | 0.001511272 | 0.0089561 | 1 |
| HTN | nk_cell | *MARVELD3* | -1.456592374 | 0.005917023 | 0.017919231 | 1 |
| HTN | nk_cell | *ADAMTS18* | -1.395208982 | 0.004124833 | 0.014272329 | 1 |
| HTN | nk_cell | *GCSH* | -1.27989469 | 0.003655523 | 0.01330998 | 1 |
| HTN | nk_cell | *MAP1LC3B* | -2.063593649 | 0.008315575 | 0.022544208 | 1 |
| HTN | nk_cell | *TLDC1* | -2.040431167 | 0.003394075 | 0.012814513 | 1 |
| HTN | nk_cell | *NDUFB10* | 1.724293133 | 0.014781478 | 0.03475361 | 1 |
| HTN | nk_cell | *TIGD7* | -1.551510105 | 0.00099972 | 0.008846261 | 1 |
| HTN | nk_cell | *GALNS* | -2.297255136 | 0.002640692 | 0.011097616 | 1 |
| HTN | nk_cell | *DDX52* | -1.767723088 | 0.015133119 | 0.035290029 | 1 |
| HTN | nk_cell | *RASL10B* | -3.012974558 | 1.70E-05 | 0.001013492 | 0.1069466 |
| HTN | nk_cell | *TOM1L1* | -1.161104881 | 0.002528292 | 0.010841214 | 1 |
| HTN | nk_cell | *VPS53* | -1.915967441 | 0.004711293 | 0.015384215 | 1 |
| HTN | nk_cell | *SPATA22* | -0.676874081 | 0.00274839 | 0.011375332 | 1 |
| HTN | nk_cell | *LRRC46* | -2.178430928 | 0.002416293 | 0.010645574 | 1 |
| HTN | nk_cell | *SCRN2* | -0.878970922 | 4.18E-06 | 0.000375308 | 0.026271568 |
| HTN | nk_cell | *ARSG* | 0.786090019 | 0.001930896 | 0.009667839 | 1 |
| HTN | nk_cell | *ABCA8* | -0.772066707 | 0.004712899 | 0.015384215 | 1 |
| HTN | nk_cell | *G6PC3* | -1.928097091 | 0.00152365 | 0.008957475 | 1 |
| HTN | nk_cell | *IMPA2* | -1.650410254 | 0.002205245 | 0.01021693 | 1 |
| HTN | nk_cell | *ASXL3* | -1.692927303 | 0.005314949 | 0.016616154 | 1 |
| HTN | nk_cell | *GREB1L* | -1.279345112 | 8.88E-06 | 0.000634112 | 0.055801866 |
| HTN | nk_cell | *SLC14A1* | 0.406809651 | 0.000681817 | 0.00874661 | 1 |
| HTN | nk_cell | *CCDC40* | -1.537799077 | 0.000253042 | 0.005756069 | 1 |
| HTN | nk_cell | *TMC6* | 0.714572147 | 0.002823294 | 0.01152135 | 1 |
| HTN | nk_cell | *EIF4A3* | -1.916838997 | 0.004462888 | 0.014877081 | 1 |
| HTN | nk_cell | *CBX8* | -1.243943265 | 0.011768855 | 0.0294315 | 1 |
| HTN | nk_cell | *RNF165* | 0.428547019 | 0.002334692 | 0.010499432 | 1 |
| HTN | nk_cell | *MAPK4* | -0.873878388 | 0.007371132 | 0.02066086 | 1 |
| HTN | nk_cell | *FKBP10* | -0.769003559 | 0.02282139 | 0.048034175 | 1 |
| HTN | nk_cell | *NFIC* | -1.351256729 | 0.004744676 | 0.015423878 | 1 |
| HTN | nk_cell | *TPGS1* | 2.131538267 | 0.01931127 | 0.042338537 | 1 |
| HTN | nk_cell | *PRDM15* | -1.005419767 | 0.016943069 | 0.038261879 | 1 |
| HTN | nk_cell | *TMEM91* | -1.412584493 | 0.018597119 | 0.041154555 | 1 |
| HTN | nk_cell | *IFITM3* | -1.941100138 | 0.007224999 | 0.020378451 | 1 |
| HTN | nk_cell | *COL6A1* | -0.744585325 | 0.019241148 | 0.042237814 | 1 |
| HTN | nk_cell | *TRPM2* | 0.704177734 | 0.01746709 | 0.039194226 | 1 |
| HTN | nk_cell | *NTN5* | -1.485222579 | 0.00177837 | 0.009341888 | 1 |
| HTN | nk_cell | *GEMIN7* | -3.362193506 | 0.020714587 | 0.044584939 | 1 |
| HTN | nk_cell | *WTIP* | -1.191966658 | 0.001905123 | 0.009620491 | 1 |
| HTN | nk_cell | *MYO1F* | 0.520877072 | 0.002459894 | 0.010695267 | 1 |
| HTN | nk_cell | *CACNG8* | -2.452226757 | 0.001375203 | 0.00885079 | 1 |
| HTN | nk_cell | *EVI5L* | -1.374938243 | 0.003059687 | 0.012128785 | 1 |
| HTN | nk_cell | *RPS11* | 1.545702946 | 0.007931583 | 0.021804049 | 1 |
| HTN | nk_cell | *SH3BGRL3* | 1.290956439 | 0.014637496 | 0.034479556 | 1 |
| HTN | nk_cell | *IL22RA1* | -1.25458158 | 0.02197194 | 0.04671545 | 1 |
| HTN | nk_cell | *ZNF593* | -1.332192711 | 0.006901382 | 0.019785221 | 1 |
| HTN | nk_cell | *C1orf216* | -2.19803357 | 0.01588352 | 0.036471764 | 1 |
| HTN | nk_cell | *MAP3K6* | -2.362976394 | 0.000763321 | 0.008846261 | 1 |
| HTN | nk_cell | *MOB3C* | -1.530477245 | 0.002024469 | 0.009861903 | 1 |

| | | | | | | |
|---|---|---|---|---|---|---|
| HTN | nk_cell | *ZNF697* | -1.677794278 | 0.007643973 | 0.021270736 | 1 |
| HTN | nk_cell | *GPR161* | -1.807183974 | 0.005935617 | 0.017932352 | 1 |
| HTN | nk_cell | *ATP1B1* | -1.747354159 | 0.001659116 | 0.009149879 | 1 |
| HTN | nk_cell | *NME7* | -1.974717891 | 0.000432576 | 0.007291173 | 1 |
| HTN | nk_cell | *CREG1* | -1.739789365 | 0.005626394 | 0.017280478 | 1 |
| HTN | nk_cell | *GPA33* | 1.061304268 | 0.006981081 | 0.019919637 | 1 |
| HTN | nk_cell | *RXRG* | -2.089732516 | 0.001156835 | 0.008846261 | 1 |
| HTN | nk_cell | *XCL2* | 0.359203597 | 0.002717617 | 0.011305016 | 1 |
| HTN | nk_cell | *MGST3* | -1.545130535 | 0.004423504 | 0.014816499 | 1 |
| HTN | nk_cell | *ADCY10* | -2.166946916 | 0.003528382 | 0.013080507 | 1 |
| HTN | nk_cell | *RGS5* | -3.201048117 | 3.99E-06 | 0.000363517 | 0.025082654 |
| HTN | nk_cell | *SNX27* | -1.932808169 | 0.017953956 | 0.039987292 | 1 |
| HTN | nk_cell | *SELENBP1* | -0.788250639 | 0.010052892 | 0.026019979 | 1 |
| HTN | nk_cell | *CERS2* | -2.054578869 | 0.017897159 | 0.039914665 | 1 |
| HTN | nk_cell | *ARNT* | -2.313738422 | 0.000294653 | 0.006279604 | 1 |
| HTN | nk_cell | *POGZ* | -1.507008546 | 0.00978855 | 0.02551435 | 1 |
| HTN | nk_cell | *IKBKE* | 2.049954797 | 0.019972223 | 0.043418175 | 1 |
| HTN | nk_cell | *DYRK3* | -1.461088855 | 0.001152452 | 0.008846261 | 1 |
| HTN | nk_cell | *RAB13* | -1.854052191 | 5.44E-05 | 0.002341198 | 0.341814879 |
| HTN | nk_cell | *HAX1* | -1.377000561 | 0.014237804 | 0.03381816 | 1 |
| HTN | nk_cell | *CREB3L4* | -3.089101825 | 4.16E-07 | 6.89E-05 | 0.002617626 |
| HTN | nk_cell | *KCNN3* | -1.798273244 | 0.015107789 | 0.035280363 | 1 |
| HTN | nk_cell | *FLG* | -0.66893772 | 0.009835362 | 0.025562182 | 1 |
| HTN | nk_cell | *SCCPDH* | -1.609856 | 0.000859247 | 0.008846261 | 1 |
| HTN | nk_cell | *SDE2* | -2.920712988 | 0.019295514 | 0.042327598 | 1 |
| HTN | nk_cell | *CDC42BPA* | -1.216557196 | 0.003092439 | 0.012219765 | 1 |
| HTN | nk_cell | *CNIH3* | -1.585331262 | 0.023582437 | 0.049316509 | 1 |
| HTN | nk_cell | *PSEN2* | -2.16755854 | 0.001410301 | 0.00885079 | 1 |
| HTN | nk_cell | *PYCR2* | 1.603104036 | 0.00975113 | 0.025437906 | 1 |
| HTN | nk_cell | *PLEKHA6* | -0.751980221 | 0.013971376 | 0.03346211 | 1 |
| HTN | nk_cell | *ABCG8* | -4.265160265 | 0.000410066 | 0.007161351 | 1 |
| HTN | nk_cell | *CHAC2* | -1.202184754 | 0.000701096 | 0.008763009 | 1 |
| HTN | nk_cell | *WDPCP* | -0.783305617 | 0.002177479 | 0.010179792 | 1 |
| HTN | nk_cell | *EXOC6B* | -1.21546448 | 0.020408376 | 0.044152601 | 1 |
| HTN | nk_cell | *THNSL2* | -0.556561638 | 0.000550965 | 0.008121264 | 1 |
| HTN | nk_cell | *RALB* | -1.779240415 | 0.006670026 | 0.019342459 | 1 |
| HTN | nk_cell | *TMEM177* | 2.395657346 | 0.007330757 | 0.020584147 | 1 |
| HTN | nk_cell | *RABL2A* | -2.301874512 | 1.88E-05 | 0.001085979 | 0.118371701 |
| HTN | nk_cell | *SLC20A1* | 0.953067699 | 0.011526959 | 0.028918592 | 1 |
| HTN | nk_cell | *FBLN7* | -0.911462718 | 0.017025432 | 0.038420276 | 1 |
| HTN | nk_cell | *LIPT1* | -1.118242965 | 0.000694572 | 0.008747204 | 1 |
| HTN | nk_cell | *ZNF385B* | -0.80961351 | 0.001118953 | 0.008846261 | 1 |
| HTN | nk_cell | *TMEFF2* | -1.448736075 | 0.002585526 | 0.010988756 | 1 |
| HTN | nk_cell | *PHOSPHO2* | -0.803811787 | 0.015694552 | 0.036130228 | 1 |
| HTN | nk_cell | *GULP1* | -1.369550546 | 0.00259117 | 0.010999788 | 1 |
| HTN | nk_cell | *PTH2R* | -0.974868783 | 0.009209364 | 0.024286608 | 1 |
| HTN | nk_cell | *NBEAL1* | -0.970778554 | 0.004547354 | 0.015041515 | 1 |
| HTN | nk_cell | *ESPNL* | -9.488757558 | 0.01246747 | 0.030690284 | 1 |
| HTN | nk_cell | *FANCD2* | -2.081911996 | 0.014370602 | 0.03404219 | 1 |
| HTN | nk_cell | *OSBPL10* | -1.213193046 | 0.012739774 | 0.031168336 | 1 |

| | | | | | | |
|---|---|---|---|---|---|---|
| HTN | nk_cell | *FAM198A* | -1.154323155 | 0.002930246 | 0.011809268 | 1 |
| HTN | nk_cell | *SLC25A38* | 1.292021896 | 0.009088937 | 0.024130974 | 1 |
| HTN | nk_cell | *CTDSPL* | -2.014532297 | 0.000460379 | 0.007524739 | 1 |
| HTN | nk_cell | *STAC* | -1.168490859 | 0.001159427 | 0.008846261 | 1 |
| HTN | nk_cell | *COL8A1* | -0.72742749 | 0.000596829 | 0.008264905 | 1 |
| HTN | nk_cell | *ADPRH* | -1.092231776 | 0.005703138 | 0.017444961 | 1 |
| HTN | nk_cell | *IGSF11* | 0.864859563 | 0.022106226 | 0.046892163 | 1 |
| HTN | nk_cell | *BOC* | -0.886770855 | 0.010871432 | 0.027727665 | 1 |
| HTN | nk_cell | *AGTR1* | -2.011185313 | 0.000336414 | 0.006527878 | 1 |
| HTN | nk_cell | *EIF2A* | -0.949035931 | 0.016109693 | 0.0368163 | 1 |
| HTN | nk_cell | *AMT* | -1.068490119 | 0.000198428 | 0.005142286 | 1 |
| HTN | nk_cell | *TCTA* | -2.504041353 | 0.001399123 | 0.00885079 | 1 |
| HTN | nk_cell | *EAF2* | -1.211009006 | 0.017600913 | 0.039407743 | 1 |
| HTN | nk_cell | *LYAR* | 1.296353081 | 0.00401756 | 0.014048053 | 1 |
| HTN | nk_cell | *CENPC* | -1.886763303 | 0.000252992 | 0.005756069 | 1 |
| HTN | nk_cell | *SLC10A4* | 7.828125447 | 1.23E-06 | 0.000151855 | 0.007744627 |
| HTN | nk_cell | *TIFA* | 1.451846689 | 0.004777273 | 0.015481813 | 1 |
| HTN | nk_cell | *CCNA2* | -1.185848017 | 0.000245466 | 0.005679823 | 1 |
| HTN | nk_cell | *USP53* | -1.172831131 | 0.003650384 | 0.013309338 | 1 |
| HTN | nk_cell | *NDUFS6* | -2.954945729 | 0.003959449 | 0.01392229 | 1 |
| HTN | nk_cell | *LHFPL2* | -1.591339022 | 0.006850867 | 0.01971231 | 1 |
| HTN | nk_cell | *CRHBP* | -1.067665779 | 0.003671716 | 0.013343398 | 1 |
| HTN | nk_cell | *SPATA9* | -0.990209348 | 0.002302728 | 0.010422787 | 1 |
| HTN | nk_cell | *TSLP* | -1.395873214 | 1.90E-05 | 0.001086481 | 0.11951293 |
| HTN | nk_cell | *FEM1C* | -1.954512078 | 0.00435629 | 0.014745183 | 1 |
| HTN | nk_cell | *SLC25A48* | -1.021641172 | 0.016068891 | 0.036749771 | 1 |
| HTN | nk_cell | *GFOD1* | 1.160850917 | 0.005020953 | 0.015984969 | 1 |
| HTN | nk_cell | *PSD2* | 4.934306613 | 7.45E-05 | 0.002910639 | 0.468612912 |
| HTN | nk_cell | *KLHL3* | -1.59845311 | 0.006129614 | 0.018298616 | 1 |
| HTN | nk_cell | *TRIM7* | -1.327137221 | 0.000368313 | 0.006671285 | 1 |
| HTN | nk_cell | *MUT* | -1.946367506 | 0.003220005 | 0.012494105 | 1 |
| HTN | nk_cell | *DOK3* | -1.590492883 | 0.000309834 | 0.006359244 | 1 |
| HTN | nk_cell | *ABT1* | 2.636514185 | 2.20E-05 | 0.001213549 | 0.138344611 |
| HTN | nk_cell | *MLIP* | -1.234262433 | 0.005072387 | 0.01609798 | 1 |
| HTN | nk_cell | *FGD2* | -1.406375298 | 0.000921047 | 0.008846261 | 1 |
| HTN | nk_cell | *ANO7* | -0.992407254 | 0.00125044 | 0.00885079 | 1 |
| HTN | nk_cell | *RPL7L1* | 1.696532813 | 0.017081599 | 0.038491761 | 1 |
| HTN | nk_cell | *TPBG* | -1.586897118 | 0.001458479 | 0.008902387 | 1 |
| HTN | nk_cell | *RNF217* | -1.538528911 | 0.014605702 | 0.034463015 | 1 |
| HTN | nk_cell | *ABRACL* | 1.367825554 | 0.005695752 | 0.017433882 | 1 |
| HTN | nk_cell | *SLC18B1* | -1.145350276 | 7.76E-06 | 0.000574091 | 0.048797736 |
| HTN | nk_cell | *AIG1* | -1.05089632 | 0.018042523 | 0.040139187 | 1 |
| HTN | nk_cell | *PNLDC1* | 1.661784454 | 0.008645876 | 0.023259144 | 1 |
| HTN | nk_cell | *SLC22A3* | -0.78865661 | 0.00110305 | 0.008846261 | 1 |
| HTN | nk_cell | *GNA12* | -1.377913526 | 0.007775688 | 0.021497693 | 1 |
| HTN | nk_cell | *C7orf50* | -2.438408252 | 0.003470272 | 0.012986667 | 1 |
| HTN | nk_cell | *CCZ1B* | -1.715929285 | 0.004415303 | 0.01480253 | 1 |
| HTN | nk_cell | *EGFR* | -0.805825586 | 0.0030943 | 0.012219765 | 1 |
| HTN | nk_cell | *NOM1* | -2.269095583 | 0.015185556 | 0.035323691 | 1 |
| HTN | nk_cell | *FBXO25* | -1.924438559 | 0.021549056 | 0.046049937 | 1 |

| | | | | | | |
|---|---|---|---|---|---|---|
| HTN | nk_cell | *BIN3* | -1.782256477 | 0.006367498 | 0.018785762 | 1 |
| HTN | nk_cell | *SLC25A37* | -1.817590588 | 0.000443849 | 0.007343364 | 1 |
| HTN | nk_cell | *ST18* | -1.058017259 | 0.006974647 | 0.019919637 | 1 |
| HTN | nk_cell | *RGS20* | -0.786084537 | 0.00533108 | 0.016650026 | 1 |
| HTN | nk_cell | *TACC1* | -1.252611707 | 0.02394425 | 0.049946084 | 1 |
| HTN | nk_cell | *ADHFE1* | -0.80027198 | 9.86E-05 | 0.003441707 | 0.620122385 |
| HTN | nk_cell | *ATP6V0D2* | -1.892614594 | 0.003481041 | 0.013006455 | 1 |
| HTN | nk_cell | *MAL2* | -0.800869765 | 0.001980388 | 0.009780598 | 1 |
| HTN | nk_cell | *FAM135B* | -1.082749522 | 0.001472426 | 0.008909666 | 1 |
| HTN | nk_cell | *SLC39A4* | -2.112462781 | 0.00107515 | 0.008846261 | 1 |
| HTN | nk_cell | *PLIN2* | -1.712747898 | 0.006561479 | 0.019169154 | 1 |
| HTN | nk_cell | *CBWD5* | -1.833698209 | 0.001885291 | 0.009566447 | 1 |
| HTN | nk_cell | *ZNF462* | -1.282227407 | 0.001959329 | 0.009759795 | 1 |
| HTN | nk_cell | *ASTN2* | -1.782986183 | 0.002831515 | 0.011537095 | 1 |
| HTN | nk_cell | *WDR31* | -1.198287204 | 0.002873351 | 0.011647167 | 1 |
| HTN | nk_cell | *ASB6* | -1.675170427 | 0.001642102 | 0.009108692 | 1 |
| HTN | nk_cell | *SH3GLB2* | -1.686829743 | 0.011357824 | 0.02860843 | 1 |
| HTN | nk_cell | *DPH7* | -2.148221337 | 0.006665811 | 0.019342459 | 1 |
| HTN | nk_cell | *SLC39A12* | -1.340531011 | 0.002199065 | 0.010211299 | 1 |
| HTN | nk_cell | *PARD3* | -0.983364624 | 0.00079683 | 0.008846261 | 1 |
| HTN | nk_cell | *NRBF2* | -2.354309145 | 5.59E-08 | 1.35E-05 | 0.000351279 |
| HTN | nk_cell | *GLUD1* | -1.666280513 | 0.013615591 | 0.032747215 | 1 |
| HTN | nk_cell | *HTR7* | -1.685132221 | 0.001311115 | 0.00885079 | 1 |
| HTN | nk_cell | *RPP30* | -1.887469236 | 0.001121227 | 0.008846261 | 1 |
| HTN | nk_cell | *DNAJB12* | -2.318941941 | 0.005533047 | 0.017084806 | 1 |
| HTN | nk_cell | *FUOM* | -1.719008956 | 0.000688655 | 0.00874661 | 1 |
| HTN | nk_cell | *PAOX* | -2.078591857 | 0.001556379 | 0.008975583 | 1 |
| HTN | nk_cell | *GSTO1* | -1.731180349 | 0.011019943 | 0.028015521 | 1 |
| HTN | nk_cell | *PDCD11* | 2.868936556 | 0.000937909 | 0.008846261 | 1 |
| HTN | nk_cell | *GAS2* | -2.198184239 | 0.003018804 | 0.012027391 | 1 |
| HTN | nk_cell | *LIN7C* | -2.275718933 | 0.004730443 | 0.015416362 | 1 |
| HTN | nk_cell | *HSD17B12* | -2.085691321 | 2.00E-10 | 1.26E-07 | 1.26E-06 |
| HTN | nk_cell | *GLYAT* | -0.653033771 | 0.006126636 | 0.018298616 | 1 |
| HTN | nk_cell | *C11orf49* | -2.015215979 | 0.006049049 | 0.018131247 | 1 |
| HTN | nk_cell | *ENDOD1* | -1.749962084 | 0.001704881 | 0.009223501 | 1 |
| HTN | nk_cell | *SERPINH1* | -1.362689166 | 0.000493837 | 0.007781333 | 1 |
| HTN | nk_cell | *CAPN5* | 1.998475651 | 0.010152221 | 0.026223094 | 1 |
| HTN | nk_cell | *RPS3* | 0.772561157 | 0.006000619 | 0.018093953 | 1 |
| HTN | nk_cell | *ZC3H12C* | -1.676729857 | 0.016187431 | 0.03695366 | 1 |
| HTN | nk_cell | *DRD2* | -3.367892769 | 0.001815596 | 0.00938705 | 1 |
| HTN | nk_cell | *C11orf52* | 3.622001621 | 0.001666062 | 0.009156549 | 1 |
| HTN | nk_cell | *GLB1L2* | 1.95042328 | 0.00026958 | 0.005967776 | 1 |
| HTN | nk_cell | *GRIK4* | 0.476669635 | 0.000423944 | 0.007214859 | 1 |
| HTN | nk_cell | *ADAM33* | -0.934061052 | 0.000396898 | 0.007029004 | 1 |
| HTN | nk_cell | *TMC2* | -4.064674573 | 0.002217767 | 0.010237225 | 1 |
| HTN | nk_cell | *INCENP* | -3.427360879 | 0.016690216 | 0.037772277 | 1 |
| HTN | nk_cell | *MS4A3* | -15.07996723 | 0.000997189 | 0.008846261 | 1 |
| HTN | nk_cell | *PLCH2* | 0.296981564 | 0.001357933 | 0.00885079 | 1 |
| HTN | nk_cell | *CHEK1* | -1.348107666 | 0.001208341 | 0.00885079 | 1 |
| HTN | nk_cell | *DUSP15* | -3.965069055 | 1.26E-07 | 2.64E-05 | 0.000792468 |

| | | | | | | |
|---|---|---|---|---|---|---|
| HTN | nk_cell | *CNBD2* | -2.38436359 | 0.001455326 | 0.008891771 | 1 |
| HTN | nk_cell | *TRPT1* | -2.247982368 | 0.00089181 | 0.008846261 | 1 |
| HTN | nk_cell | *LYPD6B* | -1.107294544 | 0.001656309 | 0.009149879 | 1 |
| HTN | nk_cell | *WDR17* | -2.344723559 | 0.00167052 | 0.009156549 | 1 |
| HTN | nk_cell | *CCDC102B* | -1.601547947 | 0.004315493 | 0.014697456 | 1 |
| HTN | nk_cell | *CNDP1* | 1.034876632 | 0.023587521 | 0.049316509 | 1 |
| HTN | nk_cell | *FSIP1* | 3.130199306 | 0.00978668 | 0.02551435 | 1 |
| HTN | nk_cell | *RGS18* | -1.823610659 | 0.001505028 | 0.008938486 | 1 |
| HTN | nk_cell | *PRSS23* | -0.864440419 | 0.007987701 | 0.021919981 | 1 |
| HTN | nk_cell | *PPP1R1C* | -0.900425443 | 0.017775443 | 0.039699542 | 1 |
| HTN | nk_cell | *C11orf53* | -6.022343594 | 0.000805105 | 0.008846261 | 1 |
| HTN | nk_cell | *DOCK1* | -1.321695213 | 0.002743149 | 0.011368608 | 1 |
| HTN | nk_cell | *TEX12* | -5.143210726 | 0.001076065 | 0.008846261 | 1 |
| HTN | nk_cell | *PIP4K2A* | 0.725212698 | 0.003444483 | 0.01293636 | 1 |
| HTN | nk_cell | *CRIM1* | -1.214876071 | 0.004834527 | 0.015611027 | 1 |
| HTN | nk_cell | *ABCB9* | -1.57486872 | 0.018419635 | 0.040804879 | 1 |
| HTN | nk_cell | *DHX37* | -2.054087439 | 0.001862078 | 0.009533293 | 1 |
| HTN | nk_cell | *THRB* | -1.224373889 | 0.00682798 | 0.019673469 | 1 |
| HTN | nk_cell | *TMEM86A* | -1.104765589 | 0.001391705 | 0.00885079 | 1 |
| HTN | nk_cell | *ANK3* | 1.315475914 | 0.005038751 | 0.016015486 | 1 |
| HTN | nk_cell | *PLBD2* | -2.183411207 | 0.000669456 | 0.00874661 | 1 |
| HTN | nk_cell | *MIPOL1* | -1.069365458 | 0.007840285 | 0.021632111 | 1 |
| HTN | nk_cell | *KCTD14* | -10.04876594 | 7.06E-05 | 0.002810887 | 0.444120162 |
| HTN | nk_cell | *THRSP* | -0.625248168 | 0.006196691 | 0.018466505 | 1 |
| HTN | nk_cell | *ME3* | -0.970264694 | 0.01940499 | 0.042508422 | 1 |
| HTN | nk_cell | *ADAMTS12* | -0.974137603 | 0.00197369 | 0.009771592 | 1 |
| HTN | nk_cell | *VIPAS39* | -2.063544961 | 0.016266268 | 0.037106687 | 1 |
| HTN | nk_cell | *ANKRD50* | -1.913802367 | 0.007415269 | 0.020766057 | 1 |
| HTN | nk_cell | *PTPRO* | -1.740693713 | 0.000815103 | 0.008846261 | 1 |
| HTN | nk_cell | *FAM160B1* | -1.559417261 | 0.003233454 | 0.012509985 | 1 |
| HTN | nk_cell | *ANO4* | -0.844093563 | 0.000874343 | 0.008846261 | 1 |
| HTN | nk_cell | *TEX9* | -1.889396317 | 0.006402178 | 0.018861525 | 1 |
| HTN | nk_cell | *AKR1C2* | -1.073788632 | 0.000619398 | 0.008409632 | 1 |
| HTN | nk_cell | *ADAM8* | 0.377696653 | 0.001465171 | 0.008909666 | 1 |
| HTN | nk_cell | *KIN* | -1.756006225 | 0.005471762 | 0.016937947 | 1 |
| HTN | nk_cell | *INPP1* | -2.477073329 | 0.001765414 | 0.009319194 | 1 |
| HTN | nk_cell | *RNF144A* | -1.481204142 | 0.004033188 | 0.014079207 | 1 |
| HTN | nk_cell | *ASAP2* | -1.308529459 | 0.008173547 | 0.022284079 | 1 |
| HTN | nk_cell | *TMEM45B* | -1.816782947 | 0.006966761 | 0.019918158 | 1 |
| HTN | nk_cell | *GUF1* | -1.49538465 | 0.00532481 | 0.016638709 | 1 |
| HTN | nk_cell | *GABRA2* | -1.045565025 | 0.00015526 | 0.004377214 | 0.976118746 |
| HTN | nk_cell | *PARP8* | 0.76227461 | 0.004057058 | 0.014091781 | 1 |
| HTN | nk_cell | *BEND6* | -0.751034441 | 0.000927933 | 0.008846261 | 1 |
| HTN | nk_cell | *RABGAP1L* | 0.890365452 | 0.006636768 | 0.019290505 | 1 |
| HTN | nk_cell | *G6PC2* | 13.07896152 | 0.001422724 | 0.00885079 | 1 |
| HTN | nk_cell | *TADA1* | -2.373081285 | 0.004349079 | 0.014739978 | 1 |
| HTN | nk_cell | *ZNF773* | -1.398175405 | 0.007974099 | 0.021896922 | 1 |
| HTN | nk_cell | *NMT2* | -1.099812125 | 0.002936286 | 0.011818456 | 1 |
| HTN | nk_cell | *PLEKHH2* | -0.50724199 | 0.00303759 | 0.012070534 | 1 |
| HTN | nk_cell | *IGSF10* | -1.541269017 | 0.000655515 | 0.0086629 | 1 |

| | | | | | | |
|---|---|---|---|---|---|---|
| HTN | nk_cell | *CLEC4F* | -2.077080708 | 0.001210766 | 0.00885079 | 1 |
| HTN | nk_cell | *PELO* | -1.473386319 | 0.001206464 | 0.00885079 | 1 |
| HTN | nk_cell | *GPR180* | -1.441013196 | 0.005334659 | 0.016652929 | 1 |
| HTN | nk_cell | *HHEX* | -1.919502777 | 0.022770565 | 0.04795931 | 1 |
| HTN | nk_cell | *RAB3C* | -0.864339264 | 0.022317761 | 0.047258931 | 1 |
| HTN | nk_cell | *RAD17* | -1.587842034 | 0.002684441 | 0.011206561 | 1 |
| HTN | nk_cell | *CARHSP1* | -1.924553724 | 0.015957123 | 0.036600669 | 1 |
| HTN | nk_cell | *BANK1* | -1.635944014 | 0.004322533 | 0.014705378 | 1 |
| HTN | nk_cell | *CAST* | -1.318675395 | 0.014393981 | 0.034059074 | 1 |
| HTN | nk_cell | *SCOC* | -1.384095387 | 0.020144416 | 0.043716929 | 1 |
| HTN | nk_cell | *CLGN* | -4.176917426 | 0.001642955 | 0.009108692 | 1 |
| HTN | nk_cell | *SYCP2L* | -1.469909986 | 0.005355495 | 0.016684834 | 1 |
| HTN | nk_cell | *MERTK* | -0.344076894 | 0.016876169 | 0.038124497 | 1 |
| HTN | nk_cell | *PLA2R1* | -0.772899091 | 0.010666621 | 0.027304986 | 1 |
| HTN | nk_cell | *SLC25A27* | -0.569966851 | 0.013641078 | 0.032759341 | 1 |
| HTN | nk_cell | *FRMD1* | -0.860257616 | 0.001092793 | 0.008846261 | 1 |
| HTN | nk_cell | *INO80C* | -2.060245045 | 0.00188392 | 0.009566447 | 1 |
| HTN | nk_cell | *LPCAT1* | 0.439615103 | 0.00177355 | 0.009338615 | 1 |
| HTN | nk_cell | *NMRAL1* | 0.956132313 | 0.006436816 | 0.018919244 | 1 |
| HTN | nk_cell | *TEX29* | -1.430387423 | 0.018075294 | 0.040183653 | 1 |
| HTN | nk_cell | *LURAP1L* | -0.688397704 | 0.010867376 | 0.027727665 | 1 |
| HTN | nk_cell | *ZDHHC7* | -1.705333428 | 0.008788226 | 0.023551398 | 1 |
| HTN | nk_cell | *FAM92B* | -9.636729122 | 0.001064811 | 0.008846261 | 1 |
| HTN | nk_cell | *FBXO36* | -1.661225702 | 0.001207538 | 0.00885079 | 1 |
| HTN | nk_cell | *MCOLN2* | 0.792373636 | 0.005036906 | 0.016015486 | 1 |
| HTN | nk_cell | *CACNA2D1* | -0.944039344 | 0.00180976 | 0.00938705 | 1 |
| HTN | nk_cell | *ANKRD29* | -0.992083345 | 0.000789688 | 0.008846261 | 1 |
| HTN | nk_cell | *UBASH3B* | 1.646294237 | 0.013363303 | 0.032375757 | 1 |
| HTN | nk_cell | *ROBO4* | -1.028583978 | 0.000908175 | 0.008846261 | 1 |
| HTN | nk_cell | *ROBO3* | -1.705246641 | 0.002720634 | 0.011305105 | 1 |
| HTN | nk_cell | *LRRK1* | -1.721983179 | 0.016322907 | 0.037195402 | 1 |
| HTN | nk_cell | *CEP112* | -0.81197069 | 0.012523656 | 0.030804471 | 1 |
| HTN | nk_cell | *UCHL1* | -1.243592465 | 0.000404969 | 0.007131762 | 1 |
| HTN | nk_cell | *NEIL2* | 2.635019299 | 1.29E-06 | 0.000152832 | 0.00810012 |
| HTN | nk_cell | *LONRF1* | -1.811411657 | 0.00092626 | 0.008846261 | 1 |
| HTN | nk_cell | *CCSAP* | -2.01920176 | 0.000870775 | 0.008846261 | 1 |
| HTN | nk_cell | *GBP5* | 0.320604783 | 0.00128953 | 0.00885079 | 1 |
| HTN | nk_cell | *FAM69A* | -1.79956261 | 0.002479094 | 0.010734818 | 1 |
| HTN | nk_cell | *PDLIM3* | -1.411372835 | 0.008285475 | 0.022501418 | 1 |
| HTN | nk_cell | *NCAM2* | -0.8922034 | 0.001290095 | 0.00885079 | 1 |
| HTN | nk_cell | *PDE1C* | -1.00269589 | 0.004190582 | 0.014410776 | 1 |
| HTN | nk_cell | *RABGEF1* | -0.789002457 | 0.010160191 | 0.026232904 | 1 |
| HTN | nk_cell | *SLFN13* | 0.271429269 | 0.005388101 | 0.016761499 | 1 |
| HTN | nk_cell | *FLCN* | -1.726258142 | 0.006322791 | 0.018741813 | 1 |
| HTN | nk_cell | *PPP4R1* | -1.728773278 | 0.012240816 | 0.030322306 | 1 |
| HTN | nk_cell | *APCDD1* | -0.659834745 | 0.006365324 | 0.018785762 | 1 |
| HTN | nk_cell | *EPHB1* | -1.664718466 | 0.000909323 | 0.008846261 | 1 |
| HTN | nk_cell | *CYP2U1* | -2.276528291 | 0.004273361 | 0.014577656 | 1 |
| HTN | nk_cell | *TTC39B* | 1.649631018 | 0.00099256 | 0.008846261 | 1 |
| HTN | nk_cell | *MMS19* | -1.891467035 | 0.015652903 | 0.036047547 | 1 |

| | | | | | | |
|---|---|---|---|---|---|---|
| HTN | nk_cell | *TRMT44* | -1.903703457 | 0.000514119 | 0.007927918 | 1 |
| HTN | nk_cell | *LRGUK* | -1.085723965 | 0.003386614 | 0.012809409 | 1 |
| HTN | nk_cell | *SETD9* | -0.60990187 | 0.013067823 | 0.031770072 | 1 |
| HTN | nk_cell | *ZKSCAN2* | -0.935211247 | 0.020167845 | 0.043752672 | 1 |
| HTN | nk_cell | *KDM8* | 2.9304541 | 0.013537638 | 0.032622127 | 1 |
| HTN | nk_cell | *FAM126B* | -1.581957213 | 0.014927739 | 0.03504098 | 1 |
| HTN | nk_cell | *ALS2CR12* | -1.450965789 | 0.000298472 | 0.006297947 | 1 |
| HTN | nk_cell | *SPAG17* | -0.864133424 | 0.0008366 | 0.008846261 | 1 |
| HTN | nk_cell | *DEPTOR* | -1.372297567 | 0.001259399 | 0.00885079 | 1 |
| HTN | nk_cell | *WIF1* | -4.189713104 | 0.001942851 | 0.00970191 | 1 |
| HTN | nk_cell | *UGT2B4* | -1.1222759 | 0.001082918 | 0.008846261 | 1 |
| HTN | nk_cell | *MMP16* | -0.918483234 | 0.000811512 | 0.008846261 | 1 |
| HTN | nk_cell | *KCNMA1* | -1.339283127 | 0.002974923 | 0.011924071 | 1 |
| HTN | nk_cell | *BATF* | 0.425699035 | 0.001528177 | 0.008957475 | 1 |
| HTN | nk_cell | *DCK* | 2.348815226 | 0.022587783 | 0.047660416 | 1 |
| HTN | nk_cell | *DPY19L4* | -1.895201554 | 0.003656169 | 0.01330998 | 1 |
| HTN | nk_cell | *NDUFAF6* | -2.455375125 | 9.35E-07 | 0.000122453 | 0.005877767 |
| HTN | nk_cell | *C8orf37* | -1.893441726 | 5.28E-05 | 0.002316216 | 0.331707294 |
| HTN | nk_cell | *ADAMTSL3* | -0.710652905 | 0.018605165 | 0.041157871 | 1 |
| HTN | nk_cell | *CXCL13* | -1.838061116 | 0.00073744 | 0.008846261 | 1 |
| HTN | nk_cell | *RWDD2B* | -0.859672979 | 0.012006505 | 0.029895009 | 1 |
| HTN | nk_cell | *ANKRD9* | -1.107919432 | 0.000193199 | 0.005125085 | 1 |
| HTN | nk_cell | *TDRD9* | 0.637100812 | 0.007952252 | 0.021851315 | 1 |
| HTN | nk_cell | *SH3RF2* | -0.688858269 | 0.000949606 | 0.008846261 | 1 |
| HTN | nk_cell | *GDF6* | -1.865486803 | 0.001553388 | 0.008975583 | 1 |
| HTN | nk_cell | *NODAL* | -0.977727751 | 0.001123289 | 0.008846261 | 1 |
| HTN | nk_cell | *SASS6* | -2.118335287 | 0.007335906 | 0.020584147 | 1 |
| HTN | nk_cell | *ITGAD* | 0.319107287 | 0.001200172 | 0.00885079 | 1 |
| HTN | nk_cell | *GHRL* | 1.742817114 | 0.010200365 | 0.026304222 | 1 |
| HTN | nk_cell | *NTAN1* | -2.250923432 | 0.013467636 | 0.032521354 | 1 |
| HTN | nk_cell | *SLC6A1* | -1.336063642 | 0.005178282 | 0.016306629 | 1 |
| HTN | nk_cell | *TIMP4* | -1.249765619 | 0.002869329 | 0.011638368 | 1 |
| HTN | nk_cell | *CPT2* | -2.491307945 | 0.004054522 | 0.014091781 | 1 |
| HTN | nk_cell | *LRP8* | 1.609110618 | 0.004334809 | 0.014715413 | 1 |
| HTN | nk_cell | *STEAP2* | -0.745952397 | 0.000128789 | 0.003930579 | 0.809699247 |
| HTN | nk_cell | *GATAD1* | -1.223865523 | 0.009831202 | 0.025561939 | 1 |
| HTN | nk_cell | *TMED6* | -3.255844166 | 0.00106674 | 0.008846261 | 1 |
| HTN | nk_cell | *CLEC18A* | -3.732978871 | 5.14E-06 | 0.000431261 | 0.032344546 |
| HTN | nk_cell | *IL34* | -2.550566203 | 0.000418181 | 0.007214859 | 1 |
| HTN | nk_cell | *KCNJ15* | -2.426949822 | 7.17E-05 | 0.002833169 | 0.450473854 |
| HTN | nk_cell | *C9orf43* | -1.669500326 | 0.001629018 | 0.009090004 | 1 |
| HTN | nk_cell | *WDR19* | -1.363042795 | 0.001934344 | 0.009674796 | 1 |
| HTN | nk_cell | *SLC37A3* | -1.583738386 | 0.002975802 | 0.011924071 | 1 |
| HTN | nk_cell | *FAM213B* | -1.041939241 | 0.0134803 | 0.032521354 | 1 |
| HTN | nk_cell | *PEX10* | -1.453456378 | 0.000550865 | 0.008121264 | 1 |
| HTN | nk_cell | *RADIL* | -0.99007632 | 0.00141581 | 0.00885079 | 1 |
| HTN | nk_cell | *PAFAH2* | 1.140291938 | 0.015538288 | 0.03594158 | 1 |
| HTN | nk_cell | *GALNT14* | 2.98102456 | 0.006318022 | 0.018736512 | 1 |
| HTN | nk_cell | *HPD* | 1.362310958 | 0.020189687 | 0.043784946 | 1 |
| HTN | nk_cell | *RHPN1* | -1.096729223 | 0.002776593 | 0.011426483 | 1 |

| | | | | | | |
|---|---|---|---|---|---|---|
| HTN | nk_cell | *LRRC43* | 1.833077302 | 0.001973902 | 0.009771592 | 1 |
| HTN | nk_cell | *CLSTN2* | -0.782048137 | 0.000515905 | 0.007927918 | 1 |
| HTN | nk_cell | *COLEC12* | -1.469935754 | 0.021857472 | 0.046503529 | 1 |
| HTN | nk_cell | *RNF207* | -0.934474113 | 0.001107119 | 0.008846261 | 1 |
| HTN | nk_cell | *TSPAN33* | -1.139962783 | 0.000207004 | 0.005172882 | 1 |
| HTN | nk_cell | *PPP1R9A* | -0.955820867 | 0.001242621 | 0.00885079 | 1 |
| HTN | nk_cell | *ZFAND2B* | -2.41667698 | 0.001667702 | 0.009156549 | 1 |
| HTN | nk_cell | *GDPD5* | -1.344049067 | 0.00166847 | 0.009156549 | 1 |
| HTN | nk_cell | *DYNC1I1* | -1.184422066 | 0.001292276 | 0.00885079 | 1 |
| HTN | nk_cell | *COPG2* | -0.465874709 | 0.007122206 | 0.020181796 | 1 |
| HTN | nk_cell | *DUSP23* | -2.96449609 | 0.003376826 | 0.012789218 | 1 |
| HTN | nk_cell | *RNF166* | 0.719093499 | 0.002032929 | 0.009861903 | 1 |
| HTN | nk_cell | *ZNF276* | 1.002716701 | 0.000367666 | 0.006671285 | 1 |
| HTN | nk_cell | *B4GALT3* | 1.628073816 | 0.002525487 | 0.010838755 | 1 |
| HTN | nk_cell | *FAM160B2* | -1.452057023 | 0.002215624 | 0.01023485 | 1 |
| HTN | nk_cell | *SLC5A11* | -1.353168667 | 8.82E-09 | 2.77E-06 | 5.54E-05 |
| HTN | nk_cell | *FCER1G* | 1.000036607 | 0.00024261 | 0.005649223 | 1 |
| HTN | nk_cell | *CDC42SE2* | 0.915754206 | 0.013843811 | 0.033219862 | 1 |
| HTN | nk_cell | *ALG8* | -2.363127878 | 0.003487938 | 0.013007939 | 1 |
| HTN | nk_cell | *SV2A* | -2.769517064 | 0.009180231 | 0.024276248 | 1 |
| HTN | nk_cell | *RCAN1* | -1.819154533 | 0.020370629 | 0.044086147 | 1 |
| HTN | nk_cell | *UBE2Z* | -2.666144389 | 0.001237418 | 0.00885079 | 1 |
| HTN | nk_cell | *CLIC6* | -0.748157677 | 0.000826975 | 0.008846261 | 1 |
| HTN | nk_cell | *SIM2* | -1.011141117 | 0.020775379 | 0.04468519 | 1 |
| HTN | nk_cell | *PADI4* | 4.251690311 | 0.003967531 | 0.013928188 | 1 |
| HTN | nk_cell | *PSMD4* | -2.077763528 | 0.011659174 | 0.029203677 | 1 |
| HTN | nk_cell | *STARD9* | -1.687274832 | 0.005110856 | 0.01618738 | 1 |
| HTN | nk_cell | *TCHH* | -2.748229291 | 0.001059445 | 0.008846261 | 1 |
| HTN | nk_cell | *ACE* | -1.515393053 | 0.000719696 | 0.008846261 | 1 |
| HTN | nk_cell | *TEPP* | 3.29940119 | 0.008617437 | 0.023212437 | 1 |
| HTN | nk_cell | *ZFYVE28* | 0.909115495 | 0.009438008 | 0.024775264 | 1 |
| HTN | nk_cell | *LYPD5* | -1.842448812 | 0.000441205 | 0.007343364 | 1 |
| HTN | nk_cell | *NPR2* | -1.438383761 | 0.001120318 | 0.008846261 | 1 |
| HTN | nk_cell | *NR2F6* | -1.326879302 | 0.001574247 | 0.008975583 | 1 |
| HTN | nk_cell | *ANKLE1* | -2.814754614 | 0.00085197 | 0.008846261 | 1 |
| HTN | nk_cell | *CILP2* | -6.035474635 | 0.001533373 | 0.008957475 | 1 |
| HTN | nk_cell | *U2AF1* | -1.182506088 | 0.009239349 | 0.024345259 | 1 |
| HTN | nk_cell | *HSF2BP* | -1.792657118 | 0.000254523 | 0.005756069 | 1 |
| HTN | nk_cell | *C21orf33* | -2.945526826 | 0.00868835 | 0.023354457 | 1 |
| HTN | nk_cell | *LRRC3* | -1.344597785 | 0.003166199 | 0.012344206 | 1 |
| HTN | nk_cell | *ITGB2* | 0.447846912 | 0.005237992 | 0.016432761 | 1 |
| HTN | nk_cell | *LSS* | -1.516652098 | 0.003280833 | 0.012626477 | 1 |
| HTN | nk_cell | *VAV2* | -1.832302096 | 0.000662096 | 0.008690187 | 1 |
| HTN | nk_cell | *S100B* | -0.850091863 | 1.96E-05 | 0.001107396 | 0.122921008 |
| HTN | nk_cell | *ZNF208* | -1.108399753 | 0.001014032 | 0.008846261 | 1 |
| HTN | nk_cell | *ADAMTS13* | -1.536565452 | 0.013457706 | 0.032521354 | 1 |
| HTN | nk_cell | *GPSM1* | -2.166984322 | 0.001250204 | 0.00885079 | 1 |
| HTN | nk_cell | *RDH13* | 1.624702069 | 0.013385072 | 0.032416004 | 1 |
| HTN | nk_cell | *MPZL3* | 2.197238709 | 0.00648507 | 0.018989871 | 1 |
| HTN | nk_cell | *PCSK7* | 1.01967078 | 0.020575565 | 0.044413055 | 1 |

| | | | | | | |
|---|---|---|---|---|---|---|
| HTN | nk_cell | *S100A1* | -2.065386498 | 0.014339007 | 0.03398015 | 1 |
| HTN | nk_cell | *ZBTB7B* | -2.558619251 | 0.008363666 | 0.022616072 | 1 |
| HTN | nk_cell | *PAQR6* | -3.463211803 | 6.64E-06 | 0.000515704 | 0.041772045 |
| HTN | nk_cell | *SLC25A44* | 1.983498022 | 0.005153427 | 0.016281205 | 1 |
| HTN | nk_cell | *NBEAL2* | 1.189366758 | 0.002230433 | 0.010252893 | 1 |
| HTN | nk_cell | *FCRL3* | 0.220844602 | 0.000780031 | 0.008846261 | 1 |
| HTN | nk_cell | *LY6K* | -2.08436913 | 0.000804013 | 0.008846261 | 1 |
| HTN | nk_cell | *LY6E* | 0.96455148 | 0.012543566 | 0.03082142 | 1 |
| HTN | nk_cell | *TONSL* | -2.027643563 | 0.001909813 | 0.009636432 | 1 |
| HTN | nk_cell | *ORAI2* | 1.086029274 | 0.00284663 | 0.011578163 | 1 |
| HTN | nk_cell | *FBXL13* | -1.339574265 | 0.002370635 | 0.010592881 | 1 |
| HTN | nk_cell | *YDJC* | 0.762322609 | 0.001494324 | 0.008938486 | 1 |
| HTN | nk_cell | *DVL3* | -1.865922082 | 0.021560972 | 0.046059745 | 1 |
| HTN | nk_cell | *FBXO27* | -0.64597964 | 0.001312265 | 0.00885079 | 1 |
| HTN | nk_cell | *U2AF1L4* | -0.733221196 | 0.004299913 | 0.014652332 | 1 |
| HTN | nk_cell | *DUSP14* | -0.936909013 | 0.001603235 | 0.009056187 | 1 |
| HTN | nk_cell | *IKZF3* | 0.157775859 | 0.001572404 | 0.008975583 | 1 |
| HTN | nk_cell | *ACOX1* | -1.711094981 | 0.000423919 | 0.007214859 | 1 |
| HTN | nk_cell | *CYGB* | -1.386694611 | 0.000359015 | 0.006619138 | 1 |
| HTN | nk_cell | *ALDH16A1* | 2.646458694 | 0.012257774 | 0.030352352 | 1 |
| HTN | nk_cell | *SIGLEC11* | -2.329209747 | 0.000432355 | 0.007291173 | 1 |
| HTN | nk_cell | *MPP3* | -1.017496038 | 0.00221098 | 0.010227705 | 1 |
| HTN | nk_cell | *NAGS* | 6.209402681 | 0.000428794 | 0.007266376 | 1 |
| HTN | nk_cell | *PLCD3* | -1.057387194 | 0.000348473 | 0.006579952 | 1 |
| HTN | nk_cell | *RACGAP1* | -2.362853718 | 0.000330208 | 0.006479211 | 1 |
| HTN | nk_cell | *ASGR2* | -2.551771855 | 0.001923691 | 0.009654896 | 1 |
| HTN | nk_cell | *TNFSF13* | 5.18314948 | 4.56E-07 | 7.36E-05 | 0.002869198 |
| HTN | nk_cell | *SENP3* | -3.824449807 | 0.000142814 | 0.004228148 | 0.897874073 |
| HTN | nk_cell | *SNRNP25* | -2.527585456 | 0.000124846 | 0.003847593 | 0.784909053 |
| HTN | nk_cell | *MEIOB* | -1.092410522 | 0.00302181 | 0.012031743 | 1 |
| HTN | nk_cell | *FLYWCH2* | -1.276010228 | 0.003725724 | 0.013446397 | 1 |
| HTN | nk_cell | *ZG16B* | -11.27967927 | 0.00101917 | 0.008846261 | 1 |
| HTN | nk_cell | *TTC9C* | -2.172627591 | 0.015172789 | 0.035323691 | 1 |
| HTN | nk_cell | *RPL29* | 1.035893574 | 0.013585981 | 0.032688505 | 1 |
| HTN | nk_cell | *TPCN2* | -1.81115736 | 0.016486975 | 0.037410116 | 1 |
| HTN | nk_cell | *PDZK1IP1* | -19.39767717 | 0.002434184 | 0.010654267 | 1 |
| HTN | nk_cell | *COA7* | -2.112218427 | 0.002148553 | 0.010146672 | 1 |
| HTN | nk_cell | *SLC1A7* | 0.643960853 | 0.016275003 | 0.037113145 | 1 |
| HTN | nk_cell | *C1orf123* | -1.867337466 | 0.002181187 | 0.010183682 | 1 |
| HTN | nk_cell | *ACOT11* | -1.745449942 | 0.019159677 | 0.042117793 | 1 |
| HTN | nk_cell | *FBLIM1* | -1.591969258 | 0.020347456 | 0.044078461 | 1 |
| HTN | nk_cell | *PDPN* | -1.146872608 | 0.000340145 | 0.006541794 | 1 |
| HTN | nk_cell | *LAPTM5* | 0.91614084 | 0.011310073 | 0.028522435 | 1 |
| HTN | nk_cell | *SYNC* | -1.846465712 | 0.002273177 | 0.010348636 | 1 |
| HTN | nk_cell | *TSSK3* | -1.10468682 | 0.009446423 | 0.024787003 | 1 |
| HTN | nk_cell | *CAMK2N1* | -2.851525405 | 0.003901464 | 0.013830639 | 1 |
| HTN | nk_cell | *TTLL10* | -1.571424468 | 0.013330969 | 0.03232233 | 1 |
| HTN | nk_cell | *NFIA* | -1.113084166 | 0.009156882 | 0.024255774 | 1 |
| HTN | nk_cell | *NEXN* | -1.535863393 | 0.003039233 | 0.012070534 | 1 |
| HTN | nk_cell | *B3GALT2* | -2.886277285 | 0.001737095 | 0.009285921 | 1 |

| | | | | | | |
|---|---|---|---|---|---|---|
| HTN | nk_cell | *HFM1* | -1.026951548 | 0.001081741 | 0.008846261 | 1 |
| HTN | nk_cell | *KCNT2* | -1.476336366 | 0.003633342 | 0.013274443 | 1 |
| HTN | nk_cell | *VCAM1* | -1.114819918 | 0.009715412 | 0.025371122 | 1 |
| HTN | nk_cell | *EXTL2* | -1.079410044 | 0.004318456 | 0.014699583 | 1 |
| HTN | nk_cell | *ZNF496* | -2.145453621 | 0.003671196 | 0.013343398 | 1 |
| HTN | nk_cell | *TRIM58* | -1.520967678 | 0.00031866 | 0.006421199 | 1 |
| HTN | nk_cell | *VANGL2* | 1.825865924 | 0.00032822 | 0.006479211 | 1 |
| HTN | nk_cell | *FCRLB* | 1.528513394 | 0.006931547 | 0.019844553 | 1 |
| HTN | nk_cell | *TDRD5* | -1.045073457 | 0.00602588 | 0.018126654 | 1 |
| HTN | nk_cell | *SNED1* | -1.02933853 | 0.002006021 | 0.009844174 | 1 |
| HTN | nk_cell | *ACP6* | -2.2592547 | 0.003099774 | 0.012233696 | 1 |
| HTN | nk_cell | *PM20D1* | -2.048817077 | 0.00092482 | 0.008846261 | 1 |
| HTN | nk_cell | *HAAO* | -2.159995633 | 0.009159091 | 0.024255774 | 1 |
| HTN | nk_cell | *B3GALNT2* | -1.604520058 | 0.012689931 | 0.031091815 | 1 |
| HTN | nk_cell | *RFTN2* | -0.900590776 | 0.001213189 | 0.00885079 | 1 |
| HTN | nk_cell | *DISC1* | -1.8136168 | 0.012444731 | 0.030670335 | 1 |
| HTN | nk_cell | *FAM84A* | -4.658765638 | 0.001363175 | 0.00885079 | 1 |
| HTN | nk_cell | *FRZB* | -1.062170215 | 0.006465877 | 0.018960341 | 1 |
| HTN | nk_cell | *NUP35* | -2.210922062 | 0.015919274 | 0.036540516 | 1 |
| HTN | nk_cell | *CCDC74A* | -3.350378068 | 0.001000975 | 0.008846261 | 1 |
| HTN | nk_cell | *NOSTRIN* | -1.092366111 | 0.000582964 | 0.008131793 | 1 |
| HTN | nk_cell | *SGPP2* | -1.476922326 | 0.000455222 | 0.007492091 | 1 |
| HTN | nk_cell | *BBS5* | -1.086813942 | 0.006862493 | 0.019736138 | 1 |
| HTN | nk_cell | *HPGDS* | -0.902526762 | 0.006916728 | 0.019811148 | 1 |
| HTN | nk_cell | *PDLIM5* | -1.333835456 | 0.015814508 | 0.036353131 | 1 |
| HTN | nk_cell | *C1QTNF7* | -1.080391647 | 0.001283867 | 0.00885079 | 1 |
| HTN | nk_cell | *LYSMD1* | -2.505607019 | 0.014091942 | 0.033631019 | 1 |
| HTN | nk_cell | *DHX57* | -1.454031467 | 0.009192301 | 0.024280893 | 1 |
| HTN | nk_cell | *S100A12* | 2.600358296 | 0.003635769 | 0.013274443 | 1 |
| HTN | nk_cell | *TDRD10* | -2.024306951 | 0.004850305 | 0.015613861 | 1 |
| HTN | nk_cell | *NIPAL1* | -3.574468712 | 0.010637518 | 0.027241578 | 1 |
| HTN | nk_cell | *DCST2* | -3.269656377 | 0.003368643 | 0.012777125 | 1 |
| HTN | nk_cell | *LMOD3* | -2.384101427 | 0.00135193 | 0.00885079 | 1 |
| HTN | nk_cell | *NBPF10* | -2.144457409 | 0.002167009 | 0.010159571 | 1 |
| HTN | nk_cell | *SLC15A2* | -1.86774693 | 0.000768634 | 0.008846261 | 1 |
| HTN | nk_cell | *FSTL1* | -1.119910516 | 0.001863667 | 0.009533665 | 1 |
| HTN | nk_cell | *LMOD1* | -1.906209382 | 1.54E-09 | 7.09E-07 | 9.68E-06 |
| HTN | nk_cell | *IGFBP7* | -1.741400679 | 0.001794435 | 0.009369553 | 1 |
| HTN | nk_cell | *ARPC2* | 1.037103673 | 0.019786351 | 0.043148384 | 1 |
| HTN | nk_cell | *GLB1L* | -1.558555665 | 0.012736045 | 0.031168336 | 1 |
| HTN | nk_cell | *SERPINI1* | -1.641653919 | 0.01141234 | 0.028688277 | 1 |
| HTN | nk_cell | *PYHIN1* | 0.141600545 | 0.001376363 | 0.00885079 | 1 |
| HTN | nk_cell | *AIM2* | -1.408047586 | 0.001393707 | 0.00885079 | 1 |
| HTN | nk_cell | *SPICE1* | -1.776966514 | 0.018958253 | 0.041769319 | 1 |
| HTN | nk_cell | *TIPARP* | -1.795895049 | 0.015588248 | 0.03599515 | 1 |
| HTN | nk_cell | *RPP14* | -2.211223799 | 0.003707437 | 0.013397607 | 1 |
| HTN | nk_cell | *DNASE1L3* | -1.307959592 | 0.001144246 | 0.008846261 | 1 |
| HTN | nk_cell | *PCOLCE2* | -0.861783686 | 0.001481743 | 0.008938486 | 1 |
| HTN | nk_cell | *CCDC158* | -0.782852329 | 0.001386219 | 0.00885079 | 1 |
| HTN | nk_cell | *GYG1* | 1.758868773 | 0.00530873 | 0.016606623 | 1 |

| | | | | | | |
|---|---|---|---|---|---|---|
| HTN | nk_cell | *PLB1* | -1.769750354 | 0.001338029 | 0.00885079 | 1 |
| HTN | nk_cell | *CDCP1* | -1.853174169 | 0.001271173 | 0.00885079 | 1 |
| HTN | nk_cell | *LRRC2* | -0.868784569 | 0.00096825 | 0.008846261 | 1 |
| HTN | nk_cell | *NMNAT3* | -1.179139061 | 0.005178601 | 0.016306629 | 1 |
| HTN | nk_cell | *TPRA1* | -2.452911462 | 0.009802739 | 0.025519983 | 1 |
| HTN | nk_cell | *DNALI1* | -4.154597535 | 0.00015428 | 0.004369177 | 0.969957326 |
| HTN | nk_cell | *RFT1* | -3.446361826 | 1.72E-05 | 0.001013492 | 0.108161217 |
| HTN | nk_cell | *SFMBT1* | -2.207193244 | 0.003046359 | 0.012083569 | 1 |
| HTN | nk_cell | *GNL3* | -1.974289078 | 0.000415752 | 0.007214859 | 1 |
| HTN | nk_cell | *LRPAP1* | -1.719579985 | 0.016704976 | 0.037792079 | 1 |
| HTN | nk_cell | *EXO5* | -3.640696071 | 6.27E-07 | 9.62E-05 | 0.003944312 |
| HTN | nk_cell | *C1orf50* | -5.559826878 | 9.63E-07 | 0.000123557 | 0.006054272 |
| HTN | nk_cell | *BDH2* | -1.490676743 | 4.46E-06 | 0.000389169 | 0.02802019 |
| HTN | nk_cell | *CDC25A* | -3.222332931 | 1.72E-10 | 1.20E-07 | 1.08E-06 |
| HTN | nk_cell | *ZNF589* | -2.544612181 | 8.75E-13 | 2.75E-09 | 5.50E-09 |
| HTN | nk_cell | *ATRIP* | -2.110322144 | 0.000572597 | 0.008131793 | 1 |
| HTN | nk_cell | *APEH* | -1.749767915 | 0.001414374 | 0.00885079 | 1 |
| HTN | nk_cell | *RNF123* | 2.944651452 | 6.97E-12 | 1.09E-08 | 4.38E-08 |
| HTN | nk_cell | *POC1A* | 2.332414661 | 0.000361803 | 0.00663164 | 1 |
| HTN | nk_cell | *MAP9* | -1.490308115 | 0.00239308 | 0.010625206 | 1 |
| HTN | nk_cell | *C4orf45* | -1.300851706 | 0.000515605 | 0.007927918 | 1 |
| HTN | nk_cell | *TMEM144* | -1.752391376 | 0.004395498 | 0.014785711 | 1 |
| HTN | nk_cell | *NPY1R* | -0.887832249 | 0.001222031 | 0.00885079 | 1 |
| HTN | nk_cell | *IL15* | -2.381588695 | 0.003519407 | 0.013069412 | 1 |
| HTN | nk_cell | *OTUD4* | -1.775425001 | 0.00583761 | 0.017754044 | 1 |
| HTN | nk_cell | *ITGA2* | -1.748930601 | 0.001331848 | 0.00885079 | 1 |
| HTN | nk_cell | *CMBL* | -1.281342803 | 0.001201121 | 0.00885079 | 1 |
| HTN | nk_cell | *F2RL1* | 2.086603179 | 0.001870088 | 0.009540973 | 1 |
| HTN | nk_cell | *WDR41* | -1.42930101 | 0.017044784 | 0.038437392 | 1 |
| HTN | nk_cell | *NDUFS4* | -1.629614365 | 0.02054142 | 0.044364105 | 1 |
| HTN | nk_cell | *RHOBTB3* | -0.845438818 | 0.021630016 | 0.046187188 | 1 |
| HTN | nk_cell | *TIGD6* | -1.399309603 | 0.000955636 | 0.008846261 | 1 |
| HTN | nk_cell | *SERINC5* | -1.588740487 | 0.007664073 | 0.021310937 | 1 |
| HTN | nk_cell | *ENPP6* | -1.725916796 | 0.017256949 | 0.038803448 | 1 |
| HTN | nk_cell | *CASP3* | 1.682490648 | 0.014892085 | 0.034974425 | 1 |
| HTN | nk_cell | *ERAP1* | -2.45096958 | 2.39E-06 | 0.000231465 | 0.015045257 |
| HTN | nk_cell | *ERAP2* | 1.199974908 | 0.000612419 | 0.008358707 | 1 |
| HTN | nk_cell | *CCDC127* | -2.466590794 | 0.001491341 | 0.008938486 | 1 |
| HTN | nk_cell | *GRIK2* | -0.739741383 | 0.001656665 | 0.009149879 | 1 |
| HTN | nk_cell | *TXLNB* | 2.517577928 | 0.002531398 | 0.010841214 | 1 |
| HTN | nk_cell | *TMEM200A* | -1.636828878 | 0.001560909 | 0.008975583 | 1 |
| HTN | nk_cell | *PI16* | -1.307338974 | 0.002465705 | 0.010705724 | 1 |
| HTN | nk_cell | *DAGLB* | -1.862865947 | 0.000220256 | 0.005367235 | 1 |
| HTN | nk_cell | *STK17A* | 0.779118442 | 0.003283647 | 0.012626477 | 1 |
| HTN | nk_cell | *HCN1* | -1.630051986 | 0.000978623 | 0.008846261 | 1 |
| HTN | nk_cell | *RP9* | -1.588418705 | 0.000519857 | 0.007947617 | 1 |
| HTN | nk_cell | *KCNK5* | -1.214123771 | 0.00091785 | 0.008846261 | 1 |
| HTN | nk_cell | *ZNF704* | -1.090142627 | 0.004712824 | 0.015384215 | 1 |
| HTN | nk_cell | *FNDC1* | -1.095855314 | 0.003633746 | 0.013274443 | 1 |
| HTN | nk_cell | *CHMP4C* | -1.169763225 | 0.001202341 | 0.00885079 | 1 |

| | | | | | | |
|---|---|---|---|---|---|---|
| HTN | nk_cell | *CTSB* | -1.124822808 | 0.002376598 | 0.010598969 | 1 |
| HTN | nk_cell | *TNFRSF11B* | -0.777024572 | 0.00150321 | 0.008938486 | 1 |
| HTN | nk_cell | *DEFB1* | -1.316765312 | 0.002783043 | 0.011435944 | 1 |
| HTN | nk_cell | *SUN1* | -1.308107065 | 0.001477656 | 0.008931715 | 1 |
| HTN | nk_cell | *TMEM184A* | -1.57651238 | 8.18E-05 | 0.003006288 | 0.514075326 |
| HTN | nk_cell | *SLC25A32* | -2.246969823 | 0.009155716 | 0.024255774 | 1 |
| HTN | nk_cell | *DCAF13* | 2.116667056 | 0.012138009 | 0.030138888 | 1 |
| HTN | nk_cell | *FREM1* | -1.009083052 | 0.001167126 | 0.00885079 | 1 |
| HTN | nk_cell | *FAM219A* | -1.577796531 | 0.003102638 | 0.012237318 | 1 |
| HTN | nk_cell | *NUDT2* | -0.844456835 | 0.002823986 | 0.01152135 | 1 |
| HTN | nk_cell | *CCDC171* | -1.674573143 | 0.004330552 | 0.014708905 | 1 |
| HTN | nk_cell | *NIPSNAP3B* | -0.866346538 | 0.00429892 | 0.014652332 | 1 |
| HTN | nk_cell | *TMC1* | -1.280953386 | 0.000546821 | 0.008118543 | 1 |
| HTN | nk_cell | *SVEP1* | -0.702532794 | 0.001382424 | 0.00885079 | 1 |
| HTN | nk_cell | *ZNF367* | 1.758683919 | 4.72E-05 | 0.002166756 | 0.29684564 |
| HTN | nk_cell | *PIGO* | -2.258508155 | 0.023283261 | 0.0488591 | 1 |
| HTN | nk_cell | *WRN* | -1.533717939 | 0.015162757 | 0.035323691 | 1 |
| HTN | nk_cell | *ZCCHC24* | -1.397069555 | 0.002680979 | 0.011200387 | 1 |
| HTN | nk_cell | *PGM2L1* | -1.502088623 | 0.015130617 | 0.035290029 | 1 |
| HTN | nk_cell | *GJB2* | -4.525499239 | 0.000723382 | 0.008846261 | 1 |
| HTN | nk_cell | *REEP3* | -2.730646295 | 9.90E-06 | 0.000683746 | 0.062220862 |
| HTN | nk_cell | *LRR1* | -1.950899315 | 0.00110946 | 0.008846261 | 1 |
| HTN | nk_cell | *NOXRED1* | -1.383840708 | 0.001219967 | 0.00885079 | 1 |
| HTN | nk_cell | *AKR1E2* | -1.269145017 | 0.000688044 | 0.00874661 | 1 |
| HTN | nk_cell | *VDAC2* | -2.015386768 | 0.017376578 | 0.039058471 | 1 |
| HTN | nk_cell | *SLC18A2* | -2.035566708 | 0.000896781 | 0.008846261 | 1 |
| HTN | nk_cell | *QSOX2* | 1.259385258 | 0.002163935 | 0.010159571 | 1 |
| HTN | nk_cell | *SNAPC4* | -1.041018071 | 0.020767016 | 0.044682487 | 1 |
| HTN | nk_cell | *SDCCAG3* | -2.294481231 | 0.004505499 | 0.014962133 | 1 |
| HTN | nk_cell | *TSC1* | -1.629846133 | 0.011721122 | 0.029323794 | 1 |
| HTN | nk_cell | *RET* | -1.574839575 | 0.00074227 | 0.008846261 | 1 |
| HTN | nk_cell | *CASP7* | -1.745705647 | 0.000109216 | 0.003557714 | 0.68663874 |
| HTN | nk_cell | *UBTD1* | -1.876138532 | 0.002991 | 0.011958531 | 1 |
| HTN | nk_cell | *ARHGAP42* | -1.250276374 | 0.002149732 | 0.010146672 | 1 |
| HTN | nk_cell | *TTC7B* | -0.981458331 | 0.019267149 | 0.042280128 | 1 |
| HTN | nk_cell | *SLC39A13* | -3.381596085 | 0.006571486 | 0.019180563 | 1 |
| HTN | nk_cell | *IFI27L1* | -1.621749824 | 0.003245566 | 0.012541409 | 1 |
| HTN | nk_cell | *CLMN* | -1.173445458 | 0.001175212 | 0.00885079 | 1 |
| HTN | nk_cell | *ABTB2* | -1.344907523 | 0.001153623 | 0.008846261 | 1 |
| HTN | nk_cell | *LIPC* | 0.492776906 | 0.001646381 | 0.009119646 | 1 |
| HTN | nk_cell | *CEP57* | -2.112318069 | 0.003332518 | 0.012697902 | 1 |
| HTN | nk_cell | *TCP11L2* | -1.620923825 | 0.003011236 | 0.012004844 | 1 |
| HTN | nk_cell | *JAM3* | -1.859403532 | 0.002055146 | 0.00993635 | 1 |
| HTN | nk_cell | *GPT2* | -0.64101588 | 0.005522839 | 0.017062451 | 1 |
| HTN | nk_cell | *NDUFB8* | -2.39157984 | 0.006701553 | 0.019382337 | 1 |
| HTN | nk_cell | *TRMT61A* | -1.743840746 | 8.02E-07 | 0.000110176 | 0.005039064 |
| HTN | nk_cell | *BTRC* | -1.637263926 | 0.012350726 | 0.030510418 | 1 |
| HTN | nk_cell | *LARP6* | -0.672350226 | 0.006163373 | 0.018390662 | 1 |
| HTN | nk_cell | *ASPG* | -2.764535546 | 0.000923704 | 0.008846261 | 1 |
| HTN | nk_cell | *PCBD1* | -0.846571198 | 0.004462322 | 0.014877081 | 1 |

| | | | | | | |
|---|---|---|---|---|---|---|
| HTN | nk_cell | *COX11* | -1.38739596 | 0.004807649 | 0.015564205 | 1 |
| HTN | nk_cell | *FAM227B* | -1.251691195 | 0.012740993 | 0.031168336 | 1 |
| HTN | nk_cell | *NUDT13* | -0.535258188 | 0.009802888 | 0.025519983 | 1 |
| HTN | nk_cell | *TAF10* | 2.21169314 | 0.014932274 | 0.03504098 | 1 |
| HTN | nk_cell | *DCHS1* | -1.255484355 | 0.001993128 | 0.009805005 | 1 |
| HTN | nk_cell | *NETO1* | -1.053602297 | 0.006986289 | 0.019919637 | 1 |
| HTN | nk_cell | *C11orf74* | -1.341871967 | 0.000931925 | 0.008846261 | 1 |
| HTN | nk_cell | *CYB5R2* | -1.214244156 | 0.007123176 | 0.020181796 | 1 |
| HTN | nk_cell | *TUB* | -1.070325407 | 0.000814188 | 0.008846261 | 1 |
| HTN | nk_cell | *RIC3* | -1.810662989 | 0.001021407 | 0.008846261 | 1 |
| HTN | nk_cell | *LMO1* | -2.934566794 | 0.013472864 | 0.032521354 | 1 |
| HTN | nk_cell | *XRRA1* | -1.432613596 | 0.000334695 | 0.006514645 | 1 |
| HTN | nk_cell | *TRIM66* | -0.768521304 | 8.24E-07 | 0.000110176 | 0.005177789 |
| HTN | nk_cell | *CDYL2* | -1.114450927 | 0.009003921 | 0.023965983 | 1 |
| HTN | nk_cell | *PRTG* | -1.087041866 | 0.001228707 | 0.00885079 | 1 |
| HTN | nk_cell | *AKIP1* | -2.84540233 | 0.015190907 | 0.035323691 | 1 |
| HTN | nk_cell | *C16orf46* | -9.198465717 | 0.001830417 | 0.009432647 | 1 |
| HTN | nk_cell | *CLEC4E* | -1.093321576 | 0.000216943 | 0.005316378 | 1 |
| HTN | nk_cell | *CLEC4D* | -2.930683836 | 0.001290485 | 0.00885079 | 1 |
| HTN | nk_cell | *ZSCAN21* | -1.245759579 | 0.000512293 | 0.007927918 | 1 |
| HTN | nk_cell | *RIMKLB* | -2.03462161 | 4.30E-09 | 1.59E-06 | 2.71E-05 |
| HTN | nk_cell | *BEAN1* | -1.932225687 | 0.004074694 | 0.014129952 | 1 |
| HTN | nk_cell | *SEC11C* | 1.010550852 | 0.002353309 | 0.010568037 | 1 |
| HTN | nk_cell | *GALR1* | -1.219444872 | 0.000150666 | 0.004337502 | 0.947235003 |
| HTN | nk_cell | *SPDYE6* | 2.039010403 | 0.013553581 | 0.032648033 | 1 |
| HTN | nk_cell | *ACSM1* | -1.315568539 | 0.002070201 | 0.009950578 | 1 |
| HTN | nk_cell | *C16orf45* | -1.237740765 | 0.006977246 | 0.019919637 | 1 |
| HTN | nk_cell | *SAAL1* | -2.554125389 | 0.000369271 | 0.006671285 | 1 |
| HTN | nk_cell | *SCNN1G* | -3.621874501 | 0.007643725 | 0.021270736 | 1 |
| HTN | nk_cell | *ANKDD1A* | -1.509179796 | 0.003993913 | 0.013980919 | 1 |
| HTN | nk_cell | *MS4A14* | -1.137815492 | 0.001333121 | 0.00885079 | 1 |
| HTN | nk_cell | *DIS3L* | -1.966458856 | 0.002448325 | 0.010667096 | 1 |
| HTN | nk_cell | *SMAD3* | -1.912128601 | 0.00038805 | 0.006930874 | 1 |
| HTN | nk_cell | *DUSP18* | -1.928751441 | 0.001478911 | 0.008931715 | 1 |
| HTN | nk_cell | *MEI1* | 0.707219233 | 0.004094886 | 0.014184323 | 1 |
| HTN | nk_cell | *SNRPD1* | 1.721196917 | 0.008848896 | 0.02366355 | 1 |
| HTN | nk_cell | *ISLR2* | -2.268992613 | 9.99E-05 | 0.003441707 | 0.627979436 |
| HTN | nk_cell | *COQ7* | -1.968850111 | 0.00225805 | 0.01030215 | 1 |
| HTN | nk_cell | *GPRC5B* | -1.133910801 | 0.006214924 | 0.018509346 | 1 |
| HTN | nk_cell | *RNF214* | -1.702153794 | 0.003382958 | 0.012804729 | 1 |
| HTN | nk_cell | *RBFOX3* | 0.653339731 | 0.001414745 | 0.00885079 | 1 |
| HTN | nk_cell | *FN3K* | -2.99763862 | 0.001093296 | 0.008846261 | 1 |
| HTN | nk_cell | *ZNF229* | -2.900624495 | 0.002205092 | 0.01021693 | 1 |
| HTN | nk_cell | *VKORC1* | -1.776338955 | 0.007688937 | 0.021370623 | 1 |
| HTN | nk_cell | *RAB8A* | 1.469983156 | 0.003685604 | 0.013360879 | 1 |
| HTN | nk_cell | *PROCA1* | -2.541030655 | 0.000110113 | 0.003568466 | 0.692282336 |
| HTN | nk_cell | *ZNF641* | 1.603868693 | 0.000421531 | 0.007214859 | 1 |
| HTN | nk_cell | *TP53I13* | -2.149628228 | 0.000697888 | 0.008747204 | 1 |
| HTN | nk_cell | *ZNF701* | -2.045402903 | 0.006551074 | 0.019156559 | 1 |
| HTN | nk_cell | *SERTAD3* | -1.34394457 | 0.00113333 | 0.008846261 | 1 |

| | | | | | | |
|---|---|---|---|---|---|---|
| HTN | nk_cell | *TTYH1* | -1.848926751 | 0.014037554 | 0.033569457 | 1 |
| HTN | nk_cell | *CDC42EP5* | -0.917054889 | 0.003888875 | 0.013813196 | 1 |
| HTN | nk_cell | *KIR3DL1* | 0.206150597 | 0.000520824 | 0.007947617 | 1 |
| HTN | nk_cell | *ZNF283* | -2.069024961 | 0.000754125 | 0.008846261 | 1 |
| HTN | nk_cell | *PPP1R14A* | -2.540005633 | 0.00330329 | 0.012667763 | 1 |
| HTN | nk_cell | *CHAF1A* | -0.943732874 | 0.007242154 | 0.020417679 | 1 |
| HTN | nk_cell | *UBXN6* | -2.93550782 | 0.000137333 | 0.004131149 | 0.863410189 |
| HTN | nk_cell | *ZNF444* | -1.559884105 | 0.001333079 | 0.00885079 | 1 |
| HTN | nk_cell | *SRR* | 1.222790027 | 0.000130663 | 0.003968486 | 0.821476678 |
| HTN | nk_cell | *CYB5D2* | -1.548888135 | 0.006055753 | 0.018131247 | 1 |
| HTN | nk_cell | *CD320* | 1.521692354 | 0.004541038 | 0.015033969 | 1 |
| HTN | nk_cell | *SPRYD3* | -2.845825238 | 0.008833816 | 0.023633278 | 1 |
| HTN | nk_cell | *SOAT2* | -1.46146351 | 0.005018773 | 0.015984969 | 1 |
| HTN | nk_cell | *ZNF558* | -0.923086462 | 0.005648726 | 0.017323677 | 1 |
| HTN | nk_cell | *CDK2AP2* | 1.891280847 | 0.003508423 | 0.01305175 | 1 |
| HTN | nk_cell | *PRDX2* | -1.804809966 | 0.017089747 | 0.038496323 | 1 |
| HTN | nk_cell | *MIS12* | -2.03773549 | 0.005430915 | 0.016861315 | 1 |
| HTN | nk_cell | *MGAT5B* | -1.910973996 | 0.002486717 | 0.010752401 | 1 |
| HTN | nk_cell | *TMC8* | 0.386225743 | 0.000682186 | 0.00874661 | 1 |
| HTN | nk_cell | *TMEM99* | -1.05772844 | 0.011173806 | 0.028280886 | 1 |
| HTN | nk_cell | *MLST8* | -2.33560915 | 0.00051606 | 0.007927918 | 1 |
| HTN | nk_cell | *ECI1* | -2.064424169 | 0.001155568 | 0.008846261 | 1 |
| HTN | nk_cell | *KCTD5* | 2.374711684 | 0.004058736 | 0.014091781 | 1 |
| HTN | nk_cell | *SDHAF2* | -3.404179338 | 0.004819511 | 0.015579142 | 1 |
| HTN | nk_cell | *DDB1* | -1.886807329 | 0.015211265 | 0.035341175 | 1 |
| HTN | nk_cell | *TTC21A* | -2.20908552 | 0.003354839 | 0.012744336 | 1 |
| HTN | nk_cell | *RPSA* | 1.170822278 | 0.003914764 | 0.013850379 | 1 |
| HTN | nk_cell | *NAALADL1* | 0.749907691 | 0.0011593 | 0.008846261 | 1 |
| HTN | nk_cell | *SCARA5* | -1.35578207 | 0.001318992 | 0.00885079 | 1 |
| HTN | nk_cell | *KIAA1586* | -2.257014847 | 0.000560278 | 0.008121264 | 1 |
| HTN | nk_cell | *PTGDR* | 0.189756704 | 0.001545352 | 0.00897211 | 1 |
| HTN | nk_cell | *POLR2J3* | -1.02197983 | 0.018961291 | 0.041769319 | 1 |
| HTN | nk_cell | *COA6* | -1.767431739 | 0.001305849 | 0.00885079 | 1 |
| HTN | nk_cell | *PDHB* | -1.952284982 | 0.017943305 | 0.039987292 | 1 |
| HTN | nk_cell | *PXK* | -2.340426546 | 0.000202185 | 0.005142286 | 1 |
| HTN | nk_cell | *ACOX2* | -0.85173045 | 0.001631468 | 0.009090004 | 1 |
| HTN | nk_cell | *CX3CR1* | 0.133020084 | 0.00156271 | 0.008975583 | 1 |
| HTN | nk_cell | *DTYMK* | 2.769448571 | 0.01658246 | 0.037584134 | 1 |
| HTN | nk_cell | *TAP1* | 0.703576831 | 0.009606283 | 0.025153978 | 1 |
| HTN | nk_cell | *ING5* | -1.647736711 | 0.002921712 | 0.011797562 | 1 |
| HTN | nk_cell | *RHOH* | 0.300352404 | 0.001339827 | 0.00885079 | 1 |
| HTN | nk_cell | *KLHL30* | -1.193749754 | 0.006054423 | 0.018131247 | 1 |
| HTN | nk_cell | *BMP1* | -1.028859472 | 0.019653388 | 0.042962743 | 1 |
| HTN | nk_cell | *ATXN2L* | -2.188223815 | 0.012680068 | 0.031079761 | 1 |
| HTN | nk_cell | *NBPF9* | -1.813138647 | 0.0015127 | 0.0089561 | 1 |
| HTN | nk_cell | *WFDC13* | -4.889333058 | 0.018253479 | 0.040479585 | 1 |
| HTN | nk_cell | *LRP1B* | -0.90886388 | 4.82E-05 | 0.00217919 | 0.302907386 |
| HTN | nk_cell | *DNAJC21* | -2.063862117 | 0.014221719 | 0.03381816 | 1 |
| HTN | nk_cell | *NPNT* | -0.766962609 | 0.001054082 | 0.008846261 | 1 |
| HTN | nk_cell | *GSTM4* | -1.103832465 | 0.000928358 | 0.008846261 | 1 |

| | | | | | | |
|---|---|---|---|---|---|---|
| HTN | nk_cell | *TCTN2* | -0.855635952 | 0.000311539 | 0.006359244 | 1 |
| HTN | nk_cell | *ADAL* | -0.453692753 | 0.022544093 | 0.047627976 | 1 |
| HTN | nk_cell | *ZNF608* | -0.832947096 | 0.001607827 | 0.009057712 | 1 |
| HTN | nk_cell | *INPP5D* | 0.694446568 | 0.002864259 | 0.011629961 | 1 |
| HTN | nk_cell | *LETM1* | -2.130106488 | 0.003136606 | 0.012294167 | 1 |
| HTN | nk_cell | *STXBP6* | -1.062371388 | 0.00188286 | 0.009566447 | 1 |
| HTN | nk_cell | *MFF* | -1.677552608 | 0.013469119 | 0.032521354 | 1 |
| HTN | nk_cell | *LGALS9* | 1.792635379 | 0.001284417 | 0.00885079 | 1 |
| HTN | nk_cell | *CPLX1* | -0.774239998 | 0.022931508 | 0.048217521 | 1 |
| HTN | nk_cell | *SIGLEC7* | 0.609032661 | 0.000973698 | 0.008846261 | 1 |
| HTN | nk_cell | *COL4A3* | -1.332159267 | 0.001357113 | 0.00885079 | 1 |
| HTN | nk_cell | *IRS1* | -1.279737809 | 1.75E-11 | 2.20E-08 | 1.10E-07 |
| HTN | nk_cell | *C8orf46* | 0.298766863 | 0.006959158 | 0.019905472 | 1 |
| HTN | nk_cell | *FAM110B* | -1.274101171 | 0.001694207 | 0.009212238 | 1 |
| HTN | nk_cell | *ATF5* | -3.335575026 | 0.022374082 | 0.047346299 | 1 |
| HTN | nk_cell | *CPT1C* | -1.328454081 | 0.003791892 | 0.013607092 | 1 |
| HTN | nk_cell | *RAB24* | -1.37048388 | 0.000357373 | 0.006608242 | 1 |
| HTN | nk_cell | *SH3TC2* | -1.049084102 | 0.000545485 | 0.008118543 | 1 |
| HTN | nk_cell | *STK32A* | -1.112798233 | 7.77E-05 | 0.002917771 | 0.488303212 |
| HTN | nk_cell | *P2RY12* | 1.314729955 | 0.013533059 | 0.032622127 | 1 |
| HTN | nk_cell | *SNUPN* | -1.941402759 | 0.021319543 | 0.045726753 | 1 |
| HTN | nk_cell | *PTK2* | -1.309018249 | 0.006440237 | 0.018920453 | 1 |
| HTN | nk_cell | *PTAFR* | -2.64510101 | 0.000271639 | 0.005992272 | 1 |
| HTN | nk_cell | *RNASE6* | -2.483986341 | 0.001566138 | 0.008975583 | 1 |
| HTN | nk_cell | *NPR1* | -0.98755228 | 0.001230756 | 0.00885079 | 1 |
| HTN | nk_cell | *CD52* | 0.535440719 | 0.010811057 | 0.027629722 | 1 |
| HTN | nk_cell | *SLC38A11* | -1.164433352 | 0.000973629 | 0.008846261 | 1 |
| HTN | nk_cell | *CCDC8* | -1.805928973 | 0.000647848 | 0.008611034 | 1 |
| HTN | nk_cell | *DTWD2* | -1.48519925 | 0.004756915 | 0.015439713 | 1 |
| HTN | nk_cell | *CLIC3* | 0.182638834 | 0.001218243 | 0.00885079 | 1 |
| HTN | nk_cell | *INO80E* | -3.434729382 | 8.41E-06 | 0.000614574 | 0.052853401 |
| HTN | nk_cell | *FAM103A1* | -1.226157974 | 0.00703619 | 0.020016528 | 1 |
| HTN | nk_cell | *RGPD8* | -0.662907694 | 0.006666494 | 0.019342459 | 1 |
| HTN | nk_cell | *CHRNA5* | -1.92020787 | 0.002624251 | 0.011065505 | 1 |
| HTN | nk_cell | *MT1E* | -1.974315892 | 0.000854729 | 0.008846261 | 1 |
| HTN | nk_cell | *UGP2* | -0.877113704 | 0.02132509 | 0.045726753 | 1 |
| HTN | nk_cell | *CTNND2* | -1.099408986 | 0.000477613 | 0.007652021 | 1 |
| HTN | nk_cell | *CALML6* | -4.23362018 | 2.43E-07 | 4.50E-05 | 0.001530023 |
| HTN | nk_cell | *TPST1* | -0.736580359 | 0.015017801 | 0.035177688 | 1 |
| HTN | nk_cell | *GUSB* | -2.018909213 | 0.022784088 | 0.047971721 | 1 |
| HTN | nk_cell | *BRD3* | -1.762862264 | 0.018851395 | 0.041614719 | 1 |
| HTN | nk_cell | *ZFPM2* | -0.960996273 | 0.002123665 | 0.01009268 | 1 |
| HTN | nk_cell | *ZNF747* | -2.258269809 | 0.000571568 | 0.008131793 | 1 |
| HTN | nk_cell | *PUSL1* | -1.48633272 | 0.002441991 | 0.010654267 | 1 |
| HTN | nk_cell | *MYO7B* | -0.720703114 | 0.005541527 | 0.017094986 | 1 |
| HTN | nk_cell | *CNTROB* | -3.130916977 | 0.00243317 | 0.010654267 | 1 |
| HTN | nk_cell | *NIPA1* | -2.383338644 | 0.012861967 | 0.031390989 | 1 |
| HTN | nk_cell | *RNF150* | -0.899953458 | 0.011304874 | 0.028520763 | 1 |
| HTN | nk_cell | *FAM161A* | -1.538414554 | 0.001449 | 0.008873507 | 1 |
| HTN | nk_cell | *GLB1* | -2.078915961 | 0.002745542 | 0.011371029 | 1 |

| | | | | | | |
|---|---|---|---|---|---|---|
| HTN | nk_cell | *FAXDC2* | -1.573672554 | 0.001147706 | 0.008846261 | 1 |
| HTN | nk_cell | *CRTAP* | -1.997332369 | 0.007624176 | 0.021237571 | 1 |
| HTN | nk_cell | *ELP5* | -3.692412212 | 0.000808835 | 0.008846261 | 1 |
| HTN | nk_cell | *SP7* | -4.838389549 | 0.002808233 | 0.011494375 | 1 |
| HTN | nk_cell | *SEMA3E* | -0.905890836 | 0.001620154 | 0.009062195 | 1 |
| HTN | nk_cell | *DCLK2* | -0.962311386 | 0.009862004 | 0.025613091 | 1 |
| HTN | nk_cell | *GPRC5C* | -1.620755612 | 0.007591969 | 0.021185401 | 1 |
| HTN | nk_cell | *KRT72* | 0.528974888 | 0.001531503 | 0.008957475 | 1 |
| HTN | nk_cell | *ARL6IP1* | -2.149670625 | 8.74E-05 | 0.003158266 | 0.549538275 |
| HTN | nk_cell | *SERPINB9* | 1.352668201 | 0.020330582 | 0.044060107 | 1 |
| HTN | nk_cell | *DLGAP1* | -1.650151178 | 0.018867582 | 0.041635833 | 1 |
| HTN | nk_cell | *STAT2* | -1.573908955 | 0.019774563 | 0.04313764 | 1 |
| HTN | nk_cell | *SLC26A5* | -1.572517029 | 0.00323021 | 0.012505129 | 1 |
| HTN | nk_cell | *COMMD5* | -2.342421887 | 0.006037485 | 0.018131247 | 1 |
| HTN | nk_cell | *GTSF1* | 3.669069849 | 0.004416979 | 0.01480253 | 1 |
| HTN | nk_cell | *RNF34* | -1.897134882 | 0.020983705 | 0.04510241 | 1 |
| HTN | nk_cell | *TMEM133* | -2.398930353 | 0.000692491 | 0.008747204 | 1 |
| HTN | nk_cell | *TTLL6* | -5.534095157 | 0.000303895 | 0.006326876 | 1 |
| HTN | nk_cell | *HTRA3* | -1.062786465 | 0.006880607 | 0.019743667 | 1 |
| HTN | nk_cell | *OSCAR* | -0.970499786 | 0.005966334 | 0.017999204 | 1 |
| HTN | nk_cell | *PAQR8* | -2.071020457 | 6.34E-06 | 0.000510139 | 0.039865196 |
| HTN | nk_cell | *NUDT6* | -1.159893884 | 0.003161955 | 0.012344206 | 1 |
| HTN | nk_cell | *ZNF160* | -2.215467264 | 0.001490007 | 0.008938486 | 1 |
| HTN | nk_cell | *ZNF415* | -0.808599853 | 0.00057701 | 0.008131793 | 1 |
| HTN | nk_cell | *FEZ2* | -1.437392068 | 0.020312957 | 0.044037089 | 1 |
| HTN | nk_cell | *C11orf24* | -2.437998924 | 0.001360073 | 0.00885079 | 1 |
| HTN | nk_cell | *ALK* | -2.142880224 | 0.000463954 | 0.007546364 | 1 |
| HTN | nk_cell | *MORN4* | -0.659499854 | 0.001487826 | 0.008938486 | 1 |
| HTN | nk_cell | *CHST11* | 0.870471469 | 0.002836504 | 0.011549935 | 1 |
| HTN | nk_cell | *LURAP1* | -0.607548426 | 0.022565461 | 0.047654931 | 1 |
| HTN | nk_cell | *KCND3* | -1.323709882 | 0.000309743 | 0.006359244 | 1 |
| HTN | nk_cell | *MRPL36* | 3.238129483 | 0.000128722 | 0.003930579 | 0.809272656 |
| HTN | nk_cell | *ZNF581* | -1.409118415 | 0.001424569 | 0.00885079 | 1 |
| HTN | nk_cell | *MCC* | -0.920686602 | 0.004920592 | 0.015759429 | 1 |
| HTN | nk_cell | *DLK2* | -3.68753234 | 6.41E-06 | 0.000510139 | 0.040300971 |
| HTN | nk_cell | *HOPX* | 0.207830493 | 0.001502638 | 0.008938486 | 1 |
| HTN | nk_cell | *MAP6* | -1.267481071 | 0.011984684 | 0.029864332 | 1 |
| HTN | nk_cell | *NMUR1* | 0.285074048 | 0.001421455 | 0.00885079 | 1 |
| HTN | nk_cell | *ZIK1* | -1.342457669 | 0.001098204 | 0.008846261 | 1 |
| HTN | nk_cell | *RGS19* | -2.481876309 | 4.15E-08 | 1.09E-05 | 0.00026106 |
| HTN | nk_cell | *TCEA2* | -1.550462782 | 0.001338181 | 0.00885079 | 1 |
| HTN | nk_cell | *ANO5* | -1.611411069 | 0.00125114 | 0.00885079 | 1 |
| HTN | nk_cell | *VAT1L* | -2.225037961 | 0.000621179 | 0.008409632 | 1 |
| HTN | nk_cell | *SYCE1* | -1.0332469 | 0.013828115 | 0.033194868 | 1 |
| HTN | nk_cell | *METTL18* | 1.550350699 | 0.002239879 | 0.01026356 | 1 |
| HTN | nk_cell | *PWWP2B* | -2.434176723 | 0.005514392 | 0.017053115 | 1 |
| HTN | nk_cell | *NINJ2* | -1.580913502 | 0.003388269 | 0.012809409 | 1 |
| HTN | nk_cell | *FAM90A1* | 4.906883389 | 0.00476642 | 0.01546258 | 1 |
| HTN | nk_cell | *C3AR1* | -1.953143857 | 0.00060191 | 0.008298708 | 1 |
| HTN | nk_cell | *AQP4* | -0.891549458 | 0.001088216 | 0.008846261 | 1 |

| | | | | | | |
|---|---|---|---|---|---|---|
| HTN | nk_cell | *TLN2* | -0.649137254 | 0.001156994 | 0.008846261 | 1 |
| HTN | nk_cell | *SRGAP2C* | -1.687218815 | 0.004521727 | 0.014993723 | 1 |
| HTN | nk_cell | *CYP4F22* | -1.979197592 | 0.000561903 | 0.008121264 | 1 |
| HTN | nk_cell | *ANKRD20A4* | -1.744241525 | 0.001085415 | 0.008846261 | 1 |
| HTN | nk_cell | *LAMB2* | -1.402666013 | 0.001667764 | 0.009156549 | 1 |
| HTN | nk_cell | *SERF1A* | -5.166264489 | 0.000918103 | 0.008846261 | 1 |
| HTN | nk_cell | *MOB3A* | 1.454027013 | 0.022636604 | 0.047741136 | 1 |
| HTN | nk_cell | *NME6* | 2.442462963 | 2.52E-07 | 4.52E-05 | 0.001582561 |
| HTN | nk_cell | *FRMD3* | -1.691876309 | 0.001374575 | 0.00885079 | 1 |
| HTN | nk_cell | *ISG20* | 0.488717256 | 0.006667693 | 0.019342459 | 1 |
| HTN | nk_cell | *MBOAT1* | -1.754756961 | 0.009579249 | 0.02509364 | 1 |
| HTN | nk_cell | *C5orf34* | -3.130400773 | 3.10E-05 | 0.001573032 | 0.195055981 |
| HTN | nk_cell | *SPTLC3* | -1.401457657 | 0.001927039 | 0.009661321 | 1 |
| HTN | nk_cell | *COPRS* | -1.73767348 | 0.005178298 | 0.016306629 | 1 |
| HTN | nk_cell | *CSDC2* | -1.949518466 | 0.000415491 | 0.007214859 | 1 |
| HTN | nk_cell | *RCAN2* | -0.850972343 | 0.001131462 | 0.008846261 | 1 |
| HTN | nk_cell | *ZNF24* | -1.927804186 | 0.008148432 | 0.022234893 | 1 |
| HTN | nk_cell | *FIBP* | 1.886031139 | 0.00299412 | 0.011959359 | 1 |
| HTN | nk_cell | *CARNS1* | 1.144182112 | 0.002633499 | 0.011082203 | 1 |
| HTN | nk_cell | *CTSW* | 0.124011893 | 0.006865426 | 0.019736138 | 1 |
| HTN | nk_cell | *SNTG2* | -0.9166637 | 0.000208126 | 0.005172882 | 1 |
| HTN | nk_cell | *SMPDL3A* | -1.844033529 | 0.005704921 | 0.017444961 | 1 |
| HTN | nk_cell | *AGAP5* | -100.7364712 | 0.011051854 | 0.028062603 | 1 |
| HTN | nk_cell | *ZNF738* | 1.280838658 | 0.016778606 | 0.03791772 | 1 |
| HTN | nk_cell | *SLFN11* | -1.989669163 | 0.000199149 | 0.005142286 | 1 |
| HTN | nk_cell | *CCL19* | -3.803455277 | 0.000684315 | 0.00874661 | 1 |
| HTN | nk_cell | *CFL1* | 1.114614571 | 0.006433334 | 0.018919244 | 1 |
| HTN | nk_cell | *EFCAB12* | -0.865037381 | 0.00803749 | 0.022027767 | 1 |
| HTN | nk_cell | *RAB43* | 2.570268022 | 0.001343687 | 0.00885079 | 1 |
| HTN | nk_cell | *CBWD1* | -1.858292069 | 0.007770991 | 0.021494157 | 1 |
| HTN | nk_cell | *RAB37* | 0.449647513 | 0.001303206 | 0.00885079 | 1 |
| HTN | nk_cell | *CYP7B1* | -1.112328814 | 0.004819267 | 0.015579142 | 1 |
| HTN | nk_cell | *METAP1D* | -1.649792269 | 0.00350019 | 0.01302883 | 1 |
| HTN | nk_cell | *MYEOV* | -0.662129219 | 7.69E-06 | 0.000574091 | 0.048368935 |
| HTN | nk_cell | *MRGPRF* | -0.954377502 | 0.000583357 | 0.008131793 | 1 |
| HTN | nk_cell | *ADH6* | -0.796765555 | 0.00014606 | 0.00426748 | 0.918280734 |
| HTN | nk_cell | *GXYLT2* | -1.109079197 | 0.003633565 | 0.013274443 | 1 |
| HTN | nk_cell | *DCAKD* | -1.371760775 | 7.68E-05 | 0.002917771 | 0.482573984 |
| HTN | nk_cell | *NOC3L* | -2.167337645 | 0.008263739 | 0.022461794 | 1 |
| HTN | nk_cell | *RHOD* | -0.877452916 | 0.000245731 | 0.005679823 | 1 |
| HTN | nk_cell | *PARP15* | 0.432851638 | 0.000606126 | 0.008320337 | 1 |
| HTN | nk_cell | *ABLIM3* | -1.366989372 | 0.003564727 | 0.013158269 | 1 |
| HTN | nk_cell | *GLRX* | -1.44929257 | 0.002489694 | 0.010757873 | 1 |
| HTN | nk_cell | *DMRT2* | -0.859153875 | 2.68E-05 | 0.001427413 | 0.168434694 |
| HTN | nk_cell | *ZNF483* | -0.754800899 | 0.017890636 | 0.039914276 | 1 |
| HTN | nk_cell | *NDNF* | -1.141782851 | 0.000903759 | 0.008846261 | 1 |
| HTN | nk_cell | *OLR1* | 2.993576753 | 0.000693966 | 0.008747204 | 1 |
| HTN | nk_cell | *GLIPR1L1* | -1.05377164 | 0.022088395 | 0.046892163 | 1 |
| HTN | nk_cell | *CCDC36* | -0.769701352 | 0.002126619 | 0.01009268 | 1 |
| HTN | nk_cell | *THAP2* | -1.640506286 | 0.006051498 | 0.018131247 | 1 |

| | | | | | | |
|---|---|---|---|---|---|---|
| HTN | nk_cell | *ZNF417* | -1.210540789 | 0.000547521 | 0.008118543 | 1 |
| HTN | nk_cell | *VEGFB* | -3.028123342 | 5.43E-11 | 5.69E-08 | 3.41E-07 |
| HTN | nk_cell | *MST1* | -1.892024828 | 8.77E-08 | 1.90E-05 | 0.000551352 |
| HTN | nk_cell | *CSPG4* | -1.191068783 | 0.010083386 | 0.026088167 | 1 |
| HTN | nk_cell | *SNX33* | -1.456822518 | 0.000886099 | 0.008846261 | 1 |
| HTN | nk_cell | *NUDT18* | 2.894835206 | 0.002971802 | 0.011923243 | 1 |
| HTN | nk_cell | *SULT1B1* | 0.263004632 | 0.001237601 | 0.00885079 | 1 |
| HTN | nk_cell | *C1orf100* | -2.808892453 | 0.001139347 | 0.008846261 | 1 |
| HTN | nk_cell | *RNF213* | 0.932202274 | 0.021064369 | 0.045244855 | 1 |
| HTN | nk_cell | *PLK3* | -2.023870083 | 0.002044365 | 0.009902098 | 1 |
| HTN | nk_cell | *TUBB8* | 1.315296238 | 0.00216588 | 0.010159571 | 1 |
| HTN | nk_cell | *GPR160* | -2.481601572 | 0.006019195 | 0.018126654 | 1 |
| HTN | nk_cell | *XXYLT1* | -1.680076879 | 0.004131885 | 0.014278329 | 1 |
| HTN | nk_cell | *UBXN2A* | -1.104430939 | 0.013940384 | 0.033413341 | 1 |
| HTN | nk_cell | *NRROS* | 1.636708815 | 2.73E-05 | 0.001441455 | 0.171533101 |
| HTN | nk_cell | *CTSF* | -1.129656771 | 0.003691402 | 0.013360879 | 1 |
| HTN | nk_cell | *C16orf91* | -2.504258876 | 0.005592964 | 0.01721733 | 1 |
| HTN | nk_cell | *RGMB* | -1.077641895 | 0.009330673 | 0.024575593 | 1 |
| HTN | nk_cell | *FAM53A* | 3.53431789 | 0.002438775 | 0.010654267 | 1 |
| HTN | nk_cell | *CTU2* | -2.491011496 | 1.00E-05 | 0.000685781 | 0.063091822 |
| HTN | nk_cell | *GLIS1* | -1.144501284 | 1.39E-06 | 0.000162101 | 0.008753448 |
| HTN | nk_cell | *RALGAPA1* | -1.518941906 | 0.00997123 | 0.025864981 | 1 |
| HTN | nk_cell | *LIG4* | -1.548307383 | 0.006351277 | 0.018781121 | 1 |
| HTN | nk_cell | *SNAPC5* | -1.966203759 | 0.023361405 | 0.048990378 | 1 |
| HTN | nk_cell | *LINGO2* | 0.151204234 | 0.001222244 | 0.00885079 | 1 |
| HTN | nk_cell | *BBS1* | -2.062160247 | 0.00890043 | 0.02377103 | 1 |
| HTN | nk_cell | *TMEM81* | -4.292779755 | 0.002246926 | 0.010266299 | 1 |
| HTN | nk_cell | *ZNF497* | -4.058626391 | 0.003938463 | 0.013894565 | 1 |
| HTN | nk_cell | *BRSK2* | -1.618019321 | 0.001820614 | 0.009397539 | 1 |
| HTN | nk_cell | *SH3PXD2B* | -0.814483614 | 0.022433546 | 0.047424244 | 1 |
| HTN | nk_cell | *LARP7* | -1.784297576 | 0.016355347 | 0.03722612 | 1 |
| HTN | nk_cell | *FGFBP3* | -1.62847332 | 0.000605003 | 0.008320337 | 1 |
| HTN | nk_cell | *C4orf32* | -1.347014013 | 0.002598885 | 0.011025093 | 1 |
| HTN | nk_cell | *BTC* | -1.579048382 | 0.001903195 | 0.009618478 | 1 |
| HTN | nk_cell | *NDUFA11* | -2.690989443 | 0.015356408 | 0.03559946 | 1 |
| HTN | nk_cell | *RSRC1* | -2.401010085 | 2.39E-05 | 0.001296863 | 0.15043615 |
| HTN | nk_cell | *SLC22A1* | -2.344600721 | 0.022429597 | 0.047424244 | 1 |
| HTN | nk_cell | *CHST2* | 0.250960807 | 0.001390699 | 0.00885079 | 1 |
| HTN | nk_cell | *SH3BP5L* | -1.697662247 | 0.002256925 | 0.01030215 | 1 |
| HTN | nk_cell | *CADM2* | -1.119698126 | 0.000112612 | 0.003612201 | 0.707991384 |
| HTN | nk_cell | *TP53I11* | -1.327027728 | 0.022725937 | 0.047881355 | 1 |
| HTN | nk_cell | *PHYHD1* | -0.838148908 | 0.004643609 | 0.015245101 | 1 |
| HTN | nk_cell | *ZNF519* | -1.39852269 | 0.000564566 | 0.008121264 | 1 |
| HTN | nk_cell | *CHRNA7* | -1.795402946 | 0.017428757 | 0.039147766 | 1 |
| HTN | nk_cell | *NRIP3* | -1.419094665 | 2.96E-06 | 0.000277974 | 0.018624291 |
| HTN | nk_cell | *SCUBE2* | -1.339235406 | 0.002792875 | 0.011461362 | 1 |
| HTN | nk_cell | *ZNF25* | -1.112574202 | 0.01296328 | 0.031564733 | 1 |
| HTN | nk_cell | *PCSK1* | -2.713795033 | 0.001566523 | 0.008975583 | 1 |
| HTN | nk_cell | *LPL* | -0.753854428 | 0.000696625 | 0.008747204 | 1 |
| HTN | nk_cell | *TBC1D10C* | 0.255101035 | 0.001220191 | 0.00885079 | 1 |

| | | | | | | |
|---|---|---|---|---|---|---|
| HTN | nk_cell | *PPP2R2D* | -2.601670015 | 1.55E-05 | 0.00095645 | 0.097557895 |
| HTN | nk_cell | *UBQLNL* | -2.828104563 | 0.001402694 | 0.00885079 | 1 |
| HTN | nk_cell | *KCNE3* | -1.644823396 | 0.001424686 | 0.00885079 | 1 |
| HTN | nk_cell | *UCP2* | 1.529076495 | 0.017175401 | 0.038661563 | 1 |
| HTN | nk_cell | *RPS6KB2* | 2.211242353 | 0.009018431 | 0.023994445 | 1 |
| HTN | nk_cell | *GPR156* | 1.567271573 | 0.008989002 | 0.023946548 | 1 |
| HTN | nk_cell | *MSRA* | -2.386544878 | 0.000891928 | 0.008846261 | 1 |
| HTN | nk_cell | *ETV4* | -0.952380452 | 0.014122555 | 0.033644753 | 1 |
| HTN | nk_cell | *ARL4D* | -0.846603609 | 0.004741542 | 0.015423878 | 1 |
| HTN | nk_cell | *LRRN1* | -2.128158538 | 0.001761799 | 0.009314459 | 1 |
| HTN | nk_cell | *B3GALT6* | -2.246735082 | 0.000747169 | 0.008846261 | 1 |
| HTN | nk_cell | *ZNF613* | -1.317553451 | 0.011516896 | 0.028904881 | 1 |
| HTN | nk_cell | *UFSP1* | -1.32123445 | 0.00605913 | 0.018131247 | 1 |
| HTN | nk_cell | *ZNF135* | -1.660212945 | 0.001048264 | 0.008846261 | 1 |
| HTN | nk_cell | *COX8A* | 1.926208227 | 0.016342376 | 0.037212792 | 1 |
| HTN | nk_cell | *B3GNT4* | -6.037501225 | 0.001120926 | 0.008846261 | 1 |
| HTN | nk_cell | *CDC26* | -1.22888903 | 0.002091286 | 0.010006758 | 1 |
| HTN | nk_cell | *PHLDB3* | -1.83422998 | 0.014937144 | 0.03504098 | 1 |
| HTN | nk_cell | *RMDN1* | -1.572521329 | 0.021348132 | 0.045744957 | 1 |
| HTN | nk_cell | *RNF152* | -1.202891141 | 0.000880071 | 0.008846261 | 1 |
| HTN | nk_cell | *ACSF3* | -1.444597266 | 0.02254507 | 0.047627976 | 1 |
| HTN | nk_cell | *BOK* | -0.942160455 | 0.001872683 | 0.009540973 | 1 |
| HTN | nk_cell | *ZNF843* | -3.200820798 | 0.001495709 | 0.008938486 | 1 |
| HTN | nk_cell | *PFN4* | -1.202760561 | 4.19E-05 | 0.001982907 | 0.263140709 |
| HTN | nk_cell | *BASP1* | -1.109554963 | 0.011322676 | 0.028531328 | 1 |
| HTN | nk_cell | *LRRC37A3* | -2.177937447 | 0.001975546 | 0.009772037 | 1 |
| HTN | nk_cell | *IRX5* | -0.771392471 | 0.003990174 | 0.013975613 | 1 |
| HTN | nk_cell | *PNMA1* | -0.914140568 | 0.001229831 | 0.00885079 | 1 |
| HTN | nk_cell | *GCNT4* | -6.214322066 | 0.001445172 | 0.008873507 | 1 |
| HTN | nk_cell | *MTHFR* | -4.649484868 | 1.49E-06 | 0.000168827 | 0.00939668 |
| HTN | nk_cell | *C19orf18* | -1.832993803 | 0.001054731 | 0.008846261 | 1 |
| HTN | nk_cell | *TMEM80* | -1.125082718 | 0.005309259 | 0.016606623 | 1 |
| HTN | nk_cell | *WDR73* | -1.915762198 | 0.000330156 | 0.006479211 | 1 |
| HTN | nk_cell | *DSCAML1* | -1.397793899 | 0.003294358 | 0.01264854 | 1 |
| HTN | nk_cell | *EPS8L2* | -0.95968462 | 0.001038434 | 0.008846261 | 1 |
| HTN | nk_cell | *B3GNT8* | -2.441620067 | 0.001156922 | 0.008846261 | 1 |
| HTN | nk_cell | *AP006621.1* | -1.025493596 | 0.010196649 | 0.026304222 | 1 |
| HTN | nk_cell | *TOP3A* | -2.786062579 | 0.00331014 | 0.012674088 | 1 |
| HTN | nk_cell | *TIMM22* | -3.033902977 | 0.004058388 | 0.014091781 | 1 |
| HTN | nk_cell | *PPFIA3* | -2.554729148 | 0.000457231 | 0.007505506 | 1 |
| HTN | nk_cell | *MAGEF1* | -1.530243021 | 0.00110198 | 0.008846261 | 1 |
| HTN | nk_cell | *PAWR* | -1.630181172 | 0.002688383 | 0.011208507 | 1 |
| HTN | nk_cell | *NAP1L5* | -1.081534967 | 0.002675204 | 0.011190293 | 1 |
| HTN | nk_cell | *NIM1K* | -0.96201629 | 0.002712574 | 0.01129401 | 1 |
| HTN | nk_cell | *ACOT4* | -1.824761416 | 0.000106587 | 0.003536779 | 0.670114989 |
| HTN | nk_cell | *RBM44* | -2.395652298 | 0.011006925 | 0.027993745 | 1 |
| HTN | nk_cell | *IRX3* | -0.940334944 | 0.001676802 | 0.00917498 | 1 |
| HTN | nk_cell | *C12orf54* | -3.641947977 | 0.000234419 | 0.005519823 | 1 |
| HTN | nk_cell | *GBA* | -1.460699462 | 0.000230744 | 0.005474297 | 1 |
| HTN | nk_cell | *IL17RA* | -1.318540766 | 0.019770372 | 0.04313764 | 1 |

| | | | | | | |
|---|---|---|---|---|---|---|
| HTN | nk_cell | *POLR2L* | -2.805634042 | 0.001130884 | 0.008846261 | 1 |
| HTN | nk_cell | *FAM87B* | -2.636857098 | 0.000757053 | 0.008846261 | 1 |
| HTN | nk_cell | *ZCCHC3* | -3.075016961 | 0.012208059 | 0.030276949 | 1 |
| HTN | nk_cell | *ZNF619* | -2.483303863 | 0.001309203 | 0.00885079 | 1 |
| HTN | nk_cell | *ZBTB41* | -1.60802945 | 0.003627356 | 0.013274443 | 1 |
| HTN | nk_cell | *CENPBD1* | -1.697783556 | 0.019683286 | 0.042998201 | 1 |
| HTN | nk_cell | *BET1L* | -1.522352335 | 0.009404209 | 0.0247175 | 1 |
| HTN | nk_cell | *IMP3* | -2.466400493 | 0.006046292 | 0.018131247 | 1 |
| HTN | nk_cell | *DPY19L2* | -0.931245553 | 0.02180846 | 0.046484456 | 1 |
| HTN | nk_cell | *C2orf73* | -2.090754766 | 0.000963568 | 0.008846261 | 1 |
| HTN | nk_cell | *FAM26E* | -1.105920122 | 0.0017037 | 0.009223501 | 1 |
| HTN | nk_cell | *MLF1* | -1.378979734 | 0.010972037 | 0.027916308 | 1 |
| HTN | nk_cell | *STAP2* | -1.189220674 | 0.000853156 | 0.008846261 | 1 |
| HTN | nk_cell | *NDUFV2* | -1.391362362 | 0.001009962 | 0.008846261 | 1 |
| HTN | nk_cell | *DALRD3* | -1.769212657 | 0.000191756 | 0.005125085 | 1 |
| HTN | nk_cell | *ZNF114* | -1.487849583 | 0.001354214 | 0.00885079 | 1 |
| HTN | nk_cell | *ZNF518B* | -2.482428819 | 0.000918871 | 0.008846261 | 1 |
| HTN | nk_cell | *SH2B1* | -1.780631495 | 1.88E-06 | 0.000188514 | 0.011835231 |
| HTN | nk_cell | *ZC3H12D* | -3.330630419 | 0.012914093 | 0.031464691 | 1 |
| HTN | nk_cell | *PLEC* | -2.212432709 | 0.002366408 | 0.010587941 | 1 |
| HTN | nk_cell | *SH2D4B* | -3.656536838 | 0.001202641 | 0.00885079 | 1 |
| HTN | nk_cell | *PRSS36* | -2.108294944 | 0.00113835 | 0.008846261 | 1 |
| HTN | nk_cell | *GEN1* | -1.183820451 | 0.004054649 | 0.014091781 | 1 |
| HTN | nk_cell | *TMPRSS9* | -1.349459696 | 0.001109141 | 0.008846261 | 1 |
| HTN | nk_cell | *AQP11* | -1.913382428 | 0.002803918 | 0.011484192 | 1 |
| HTN | nk_cell | *TMEM11* | -1.999778084 | 0.001708876 | 0.009223501 | 1 |
| HTN | nk_cell | *ZFAND2A* | -3.277887638 | 0.000466921 | 0.007546364 | 1 |
| HTN | nk_cell | *BEND3* | -2.106313214 | 0.002753752 | 0.011390026 | 1 |
| HTN | nk_cell | *LINC00843* | -2.221140577 | 0.00151911 | 0.0089561 | 1 |
| HTN | nk_cell | *COX14* | -2.374921524 | 8.00E-08 | 1.80E-05 | 0.000503029 |
| HTN | nk_cell | *P4HTM* | -3.073020266 | 0.000572125 | 0.008131793 | 1 |
| HTN | nk_cell | *ERBB4* | -0.740701416 | 0.001301256 | 0.00885079 | 1 |
| HTN | nk_cell | *CSRNP3* | -1.047085836 | 0.001395946 | 0.00885079 | 1 |
| HTN | nk_cell | *RPP25* | 1.392046285 | 0.001737968 | 0.009285921 | 1 |
| HTN | nk_cell | *FAM219B* | -2.923435638 | 6.19E-06 | 0.000505677 | 0.03893716 |
| HTN | nk_cell | *CPNE7* | -4.304615088 | 4.19E-05 | 0.001982907 | 0.26372664 |
| HTN | nk_cell | *MPI* | -4.102927429 | 1.26E-10 | 9.92E-08 | 7.94E-07 |
| HTN | nk_cell | *TRIM73* | -4.504313741 | 6.75E-06 | 0.000517149 | 0.042406181 |
| HTN | nk_cell | *EFCAB13* | -1.21413347 | 0.00363585 | 0.013274443 | 1 |
| HTN | nk_cell | *HYI* | -1.810392569 | 2.33E-09 | 9.78E-07 | 1.47E-05 |
| HTN | nk_cell | *TUFM* | 1.662320214 | 0.004268547 | 0.014577053 | 1 |
| HTN | nk_cell | *CTC1* | 2.560387229 | 5.42E-05 | 0.002341198 | 0.340794075 |
| HTN | nk_cell | *C14orf39* | -0.626489364 | 0.00262839 | 0.011075529 | 1 |
| HTN | nk_cell | *TRIML2* | -18.25623994 | 0.000306392 | 0.006336471 | 1 |
| HTN | nk_cell | *SPTY2D1* | -2.2991164 | 0.006842231 | 0.019705501 | 1 |
| HTN | nk_cell | *ZNF664* | -2.607639926 | 0.007596604 | 0.02118893 | 1 |
| HTN | nk_cell | *CDH4* | -1.184016668 | 0.012846573 | 0.031365595 | 1 |
| HTN | nk_cell | *HLA-DQB1* | -0.612680171 | 0.018146846 | 0.040285741 | 1 |
| HTN | nk_cell | *EGR3* | -2.805524672 | 0.001575649 | 0.008975583 | 1 |
| HTN | nk_cell | *LINC00174* | -0.9487168 | 0.00111935 | 0.008846261 | 1 |

| | | | | | | |
|---|---|---|---|---|---|---|
| HTN | nk_cell | *ZBTB42* | -2.440830883 | 0.0040657 | 0.014106542 | 1 |
| HTN | nk_cell | *LACC1* | -1.819279059 | 0.003338226 | 0.012704157 | 1 |
| HTN | nk_cell | *MAF1* | -2.883078234 | 0.006692184 | 0.019370976 | 1 |
| HTN | nk_cell | *FCER1A* | -1.790470017 | 0.00090002 | 0.008846261 | 1 |
| HTN | nk_cell | *FOXS1* | -2.056272582 | 0.011046028 | 0.028059143 | 1 |
| HTN | nk_cell | *CDH5* | -0.991972007 | 0.004595084 | 0.015157026 | 1 |
| HTN | nk_cell | *CITED4* | -1.834663127 | 0.001310223 | 0.00885079 | 1 |
| HTN | nk_cell | *TIGD5* | -0.931602184 | 0.004548103 | 0.015041515 | 1 |
| HTN | nk_cell | *PDXDC1* | -1.684944576 | 0.003955857 | 0.01392229 | 1 |
| HTN | nk_cell | *SEPT1* | 0.670353283 | 0.00215961 | 0.010155174 | 1 |
| HTN | nk_cell | *TDRP* | -1.684040692 | 0.001633801 | 0.009090004 | 1 |
| HTN | nk_cell | *ZNF816* | 1.785516574 | 0.002367846 | 0.010587941 | 1 |
| HTN | nk_cell | *FGD6* | -1.77893712 | 0.006058649 | 0.018131247 | 1 |
| HTN | nk_cell | *HCLS1* | 1.12577693 | 0.004453571 | 0.014861784 | 1 |
| HTN | nk_cell | *DEFB124* | -4.750907983 | 1.73E-06 | 0.000187886 | 0.010897414 |
| HTN | nk_cell | *CYP8B1* | -0.8949293 | 0.004366073 | 0.014749867 | 1 |
| HTN | nk_cell | *SSTR2* | -1.331542235 | 0.001110848 | 0.008846261 | 1 |
| HTN | nk_cell | *SHISA2* | -18.65594241 | 0.001130745 | 0.008846261 | 1 |
| HTN | nk_cell | *CHST13* | -4.248844362 | 7.40E-05 | 0.002909489 | 0.465518221 |
| HTN | nk_cell | *PSMG4* | -1.605814641 | 0.001984358 | 0.00978483 | 1 |
| HTN | nk_cell | *CAPS2* | -1.67939321 | 0.003555586 | 0.013146983 | 1 |
| HTN | nk_cell | *ZNF792* | -1.548680884 | 0.002184826 | 0.010189912 | 1 |
| HTN | nk_cell | *KCTD2* | -2.478664522 | 0.009153 | 0.024255774 | 1 |
| HTN | nk_cell | *BBS12* | 3.692524131 | 0.001797908 | 0.009372676 | 1 |
| HTN | nk_cell | *LSMEM1* | -1.173091957 | 0.000301569 | 0.006326876 | 1 |
| HTN | nk_cell | *AEN* | 1.301907363 | 0.007481166 | 0.020931948 | 1 |
| HTN | nk_cell | *RPH3AL* | -1.5611735 | 0.014583865 | 0.034430628 | 1 |
| HTN | nk_cell | *ANKRD34A* | -4.551246969 | 7.76E-05 | 0.002917771 | 0.487623657 |
| HTN | nk_cell | *SLC26A11* | -2.063989774 | 0.001613687 | 0.009058261 | 1 |
| HTN | nk_cell | *HIGD1A* | -2.129205131 | 0.002613207 | 0.011041151 | 1 |
| HTN | nk_cell | *F2R* | 0.592048965 | 0.000466495 | 0.007546364 | 1 |
| HTN | nk_cell | *TMEM132C* | -0.573213976 | 0.004842803 | 0.015613861 | 1 |
| HTN | nk_cell | *SLC25A41* | -6.751848029 | 0.000339014 | 0.006541794 | 1 |
| HTN | nk_cell | *ACBD4* | -1.37965403 | 0.011453613 | 0.028769023 | 1 |
| HTN | nk_cell | *SGSH* | -2.843641777 | 0.002570401 | 0.010963442 | 1 |
| HTN | nk_cell | *MRPS23* | -2.246325244 | 0.00419541 | 0.014410776 | 1 |
| HTN | nk_cell | *GPR135* | -0.845244191 | 0.012518611 | 0.030804111 | 1 |
| HTN | nk_cell | *ATG9B* | -2.400724111 | 0.001941116 | 0.009700951 | 1 |
| HTN | nk_cell | *HKR1* | -2.118365906 | 0.004403106 | 0.014795471 | 1 |
| HTN | nk_cell | *RELL1* | -1.560659477 | 0.004345766 | 0.014736693 | 1 |
| HTN | nk_cell | *CLDN7* | -2.625633027 | 8.14E-11 | 7.31E-08 | 5.12E-07 |
| HTN | nk_cell | *ADO* | 4.534327247 | 0.011373438 | 0.028636286 | 1 |
| HTN | nk_cell | *SNRPE* | -2.304121492 | 0.023435883 | 0.049097432 | 1 |
| HTN | nk_cell | *CHST15* | -1.483699715 | 0.002907569 | 0.011755554 | 1 |
| HTN | nk_cell | *FAM181B* | -3.006192455 | 0.007449299 | 0.020852069 | 1 |
| HTN | nk_cell | *ZNF708* | -1.746977101 | 0.010544243 | 0.027046779 | 1 |
| HTN | nk_cell | *MRPL41* | -2.131803781 | 0.005796399 | 0.017664546 | 1 |
| HTN | nk_cell | *FAM153B* | -1.335726034 | 0.002029015 | 0.009861903 | 1 |
| HTN | nk_cell | *SYNM* | -2.348364043 | 0.002818343 | 0.011517757 | 1 |
| HTN | nk_cell | *CACNB4* | -0.772082598 | 0.014142462 | 0.033666663 | 1 |

| | | | | | | |
|---|---|---|---|---|---|---|
| HTN | nk_cell | *TSHZ2* | -0.865026456 | 0.004377981 | 0.014766189 | 1 |
| HTN | nk_cell | *CAPN12* | 0.679809697 | 0.000935302 | 0.008846261 | 1 |
| HTN | nk_cell | *FES* | -4.533337329 | 1.25E-05 | 0.000816648 | 0.078398201 |
| HTN | nk_cell | *GLRX5* | -2.30208086 | 0.002777106 | 0.011426483 | 1 |
| HTN | nk_cell | *MXRA7* | -1.18362359 | 0.00033384 | 0.006514645 | 1 |
| HTN | nk_cell | *MFSD5* | 2.416431432 | 0.000438986 | 0.007343364 | 1 |
| HTN | nk_cell | *ADI1* | -1.844835481 | 0.000797985 | 0.008846261 | 1 |
| HTN | nk_cell | *CLEC4G* | 2.601210993 | 0.005407264 | 0.016804482 | 1 |
| HTN | nk_cell | *KCNB2* | -0.528826273 | 0.006388174 | 0.018837921 | 1 |
| HTN | nk_cell | *TSKU* | -1.563668393 | 0.001094934 | 0.008846261 | 1 |
| HTN | nk_cell | *GRID1* | -2.364851325 | 0.000948821 | 0.008846261 | 1 |
| HTN | nk_cell | *GPIHBP1* | -1.029807019 | 0.000192928 | 0.005125085 | 1 |
| HTN | nk_cell | *LENG9* | -1.15968393 | 0.003306474 | 0.012667763 | 1 |
| HTN | nk_cell | *HMGN4* | -2.916386197 | 0.002615299 | 0.011042569 | 1 |
| HTN | nk_cell | *ZNF320* | 0.805746109 | 0.00094406 | 0.008846261 | 1 |
| HTN | nk_cell | *C12orf60* | -1.658125782 | 0.001753797 | 0.009304743 | 1 |
| HTN | nk_cell | *LYSMD4* | -1.108447759 | 0.0019798 | 0.009780598 | 1 |
| HTN | nk_cell | *WBP2NL* | -2.211396004 | 0.017185688 | 0.038670874 | 1 |
| HTN | nk_cell | *GAS6* | -2.347707074 | 0.000118103 | 0.003730407 | 0.742516405 |
| HTN | nk_cell | *BEGAIN* | -6.855813153 | 0.001624781 | 0.009079997 | 1 |
| HTN | nk_cell | *ARHGEF37* | -1.781489408 | 0.002984032 | 0.011949431 | 1 |
| HTN | nk_cell | *FAM43B* | -3.2826574 | 3.53E-05 | 0.001773256 | 0.221657036 |
| HTN | nk_cell | *CSMD1* | -1.065952226 | 0.011211261 | 0.028364265 | 1 |
| HTN | nk_cell | *GPR19* | 1.133941844 | 0.002515501 | 0.010818285 | 1 |
| HTN | nk_cell | *FANCF* | -1.925851552 | 0.001618054 | 0.009059476 | 1 |
| HTN | nk_cell | *C2CD4C* | -1.573374516 | 0.008293703 | 0.022504322 | 1 |
| HTN | nk_cell | *CTNNA3* | -0.828360235 | 0.001666996 | 0.009156549 | 1 |
| HTN | nk_cell | *CCDC125* | -2.068806203 | 0.005805215 | 0.017681635 | 1 |
| HTN | nk_cell | *FHL3* | 0.866800013 | 0.010529321 | 0.027019528 | 1 |
| HTN | nk_cell | *C19orf71* | 2.260353703 | 0.006475422 | 0.018979476 | 1 |
| HTN | nk_cell | *LRIT3* | -2.21455682 | 0.000163213 | 0.004487306 | 1 |
| HTN | nk_cell | *GRIN2A* | -1.230071157 | 0.00031853 | 0.006421199 | 1 |
| HTN | nk_cell | *URAD* | -2.973597933 | 0.008035306 | 0.022027767 | 1 |
| HTN | nk_cell | *GPR132* | 0.885951311 | 0.004700587 | 0.015375956 | 1 |
| HTN | nk_cell | *MX2* | -1.922569875 | 0.006456497 | 0.018948262 | 1 |
| HTN | nk_cell | *KLRC4* | 0.287969662 | 0.005172619 | 0.016306629 | 1 |
| HTN | nk_cell | *SERHL2* | -2.686326698 | 0.003064039 | 0.012138384 | 1 |
| HTN | nk_cell | *SFXN4* | -2.200265299 | 0.012394221 | 0.030581817 | 1 |
| HTN | nk_cell | *FAM167B* | -3.364710406 | 0.004927122 | 0.015772309 | 1 |
| HTN | nk_cell | *FAM19A1* | 0.746649193 | 0.002760246 | 0.011401882 | 1 |
| HTN | nk_cell | *BMP8A* | 5.92692651 | 0.000351527 | 0.006607413 | 1 |
| HTN | nk_cell | *CMTM4* | -1.863160645 | 0.001069726 | 0.008846261 | 1 |
| HTN | nk_cell | *KREMEN1* | -1.003303661 | 1.33E-05 | 0.000851753 | 0.083471807 |
| HTN | nk_cell | *KCTD16* | -0.87527723 | 0.001378716 | 0.00885079 | 1 |
| HTN | nk_cell | *B3GALT5* | -2.551434253 | 0.018027305 | 0.040119528 | 1 |
| HTN | nk_cell | *ZNF703* | -2.142237554 | 0.004171762 | 0.014371433 | 1 |
| HTN | nk_cell | *KCTD8* | -0.589803508 | 0.001353292 | 0.00885079 | 1 |
| HTN | nk_cell | *TUBA8* | -1.642919866 | 0.017489408 | 0.039227936 | 1 |
| HTN | nk_cell | *NPIPA5* | -0.541492992 | 0.003566358 | 0.013158269 | 1 |
| HTN | nk_cell | *ANKRD45* | -2.107194077 | 0.001227742 | 0.00885079 | 1 |

| | | | | | | |
|---|---|---|---|---|---|---|
| HTN | nk_cell | *MAATS1* | -1.046420563 | 0.010280174 | 0.026466606 | 1 |
| HTN | nk_cell | *GPR39* | -1.400269387 | 0.000204404 | 0.005142286 | 1 |
| HTN | nk_cell | *DENND5A* | -1.837124211 | 0.002289294 | 0.010399356 | 1 |
| HTN | nk_cell | *TMEM236* | -1.386421688 | 0.001211633 | 0.00885079 | 1 |
| HTN | nk_cell | *TBX1* | -1.969099432 | 0.000788979 | 0.008846261 | 1 |
| HTN | nk_cell | *EIF3C* | 2.43031566 | 1.64E-07 | 3.23E-05 | 0.001033545 |
| HTN | nk_cell | *CNTN2* | -2.236337008 | 0.00151916 | 0.0089561 | 1 |
| HTN | nk_cell | *GOLGA6L4* | -2.993394789 | 0.017717888 | 0.039627307 | 1 |
| HTN | nk_cell | *PCDH9* | -0.538440989 | 0.016463002 | 0.037384703 | 1 |
| HTN | nk_cell | *TM2D3* | -2.087516555 | 0.003758154 | 0.013516883 | 1 |
| HTN | nk_cell | *CSF1* | -1.116799444 | 0.001002992 | 0.008846261 | 1 |
| HTN | nk_cell | *PLA2G6* | -1.093715681 | 0.004385896 | 0.014776142 | 1 |
| HTN | nk_cell | *A3GALT2* | -2.010723376 | 5.80E-08 | 1.35E-05 | 0.000364661 |
| HTN | nk_cell | *SS18L1* | -1.802942111 | 0.001839523 | 0.009462282 | 1 |
| HTN | nk_cell | *THAP7* | -2.830397858 | 0.001972511 | 0.009771592 | 1 |
| HTN | nk_cell | *WDR27* | -1.493732515 | 0.015604715 | 0.03599515 | 1 |
| HTN | nk_cell | *PROS1* | -1.34811683 | 0.003393764 | 0.012814513 | 1 |
| HTN | nk_cell | *HDDC3* | -1.014387367 | 0.000656024 | 0.0086629 | 1 |
| HTN | nk_cell | *ZFP1* | -2.40097049 | 0.004992014 | 0.015931366 | 1 |
| HTN | nk_cell | *KRBA2* | -1.22427807 | 0.012401743 | 0.030588371 | 1 |
| HTN | nk_cell | *SEPT5* | -3.929603124 | 0.001200367 | 0.00885079 | 1 |
| HTN | nk_cell | *ATL3* | -1.97918957 | 0.000895901 | 0.008846261 | 1 |
| HTN | nk_cell | *NUTM2A* | -3.738185639 | 0.000107556 | 0.003536779 | 0.676207595 |
| HTN | nk_cell | *FAM227A* | -0.995352902 | 0.014298391 | 0.033896675 | 1 |
| HTN | nk_cell | *UBOX5* | -1.236117336 | 0.006412427 | 0.018882871 | 1 |
| HTN | nk_cell | *FLRT2* | -0.813761075 | 0.002307969 | 0.010438994 | 1 |
| HTN | nk_cell | *ADSSL1* | -1.165411883 | 0.0013131 | 0.00885079 | 1 |
| HTN | nk_cell | *SIGIRR* | 0.538068234 | 0.00232894 | 0.010488572 | 1 |
| HTN | nk_cell | *IFITM2* | 1.153530919 | 0.002570293 | 0.010963442 | 1 |
| HTN | nk_cell | *CDK10* | -2.721728947 | 4.87E-08 | 1.23E-05 | 0.000306366 |
| HTN | nk_cell | *TMEM105* | -13.2544174 | 0.003979551 | 0.013953953 | 1 |
| HTN | nk_cell | *TCN2* | -1.708676827 | 4.04E-05 | 0.00196317 | 0.254212993 |
| HTN | nk_cell | *GAS2L1* | -2.254049455 | 0.000199693 | 0.005142286 | 1 |
| HTN | nk_cell | *HS6ST3* | -1.507896093 | 0.001291311 | 0.00885079 | 1 |
| HTN | nk_cell | *MAPK11* | -1.370032831 | 0.000996014 | 0.008846261 | 1 |
| HTN | nk_cell | *SH3BGR* | -0.413308061 | 0.016078306 | 0.036757931 | 1 |
| HTN | nk_cell | *FAM174B* | -1.710179757 | 0.000384589 | 0.006888629 | 1 |
| HTN | nk_cell | *SV2B* | -1.495191788 | 0.007325997 | 0.020584147 | 1 |
| HTN | nk_cell | *PDE6G* | 2.150773259 | 0.013877592 | 0.033288219 | 1 |
| HTN | nk_cell | *TLCD2* | -1.338390404 | 0.000163447 | 0.004487306 | 1 |
| HTN | nk_cell | *AHNAK2* | -1.219235389 | 0.001448216 | 0.008873507 | 1 |
| HTN | nk_cell | *PBX1* | -0.826702685 | 0.001488535 | 0.008938486 | 1 |
| HTN | nk_cell | *PMEL* | -1.089192091 | 0.004849836 | 0.015613861 | 1 |
| HTN | nk_cell | *C6orf201* | -0.92521719 | 0.022000293 | 0.04675992 | 1 |
| HTN | nk_cell | *MYBL1* | 0.108904831 | 0.003398614 | 0.012817689 | 1 |
| HTN | nk_cell | *C16orf52* | -1.685053863 | 0.01636443 | 0.03722612 | 1 |
| HTN | nk_cell | *ADARB2* | -2.629967863 | 0.021154924 | 0.045422101 | 1 |
| HTN | nk_cell | *KCNIP4* | -0.755451119 | 0.008688735 | 0.023354457 | 1 |
| HTN | nk_cell | *ARL17A* | -2.854273235 | 1.50E-06 | 0.000168827 | 0.009454285 |
| HTN | nk_cell | *IFITM1* | 0.611548692 | 0.003605651 | 0.013228851 | 1 |

| | | | | | | |
|---|---|---|---|---|---|---|
| HTN | nk_cell | *PTCH1* | 0.898756206 | 0.013930405 | 0.033402158 | 1 |
| HTN | nk_cell | *RTN4RL1* | -1.656150399 | 0.001573997 | 0.008975583 | 1 |
| HTN | nk_cell | *RASA3* | 0.610470183 | 0.012889205 | 0.031420873 | 1 |
| HTN | nk_cell | *ZNF284* | -3.223197166 | 0.001270379 | 0.00885079 | 1 |
| HTN | nk_cell | *GSAP* | 0.78888967 | 0.003576743 | 0.013181114 | 1 |
| HTN | nk_cell | *CYP4Z1* | -1.200156772 | 0.006593719 | 0.019209783 | 1 |
| HTN | nk_cell | *BPIFB4* | 6.474121707 | 0.000929422 | 0.008846261 | 1 |
| HTN | nk_cell | *ZNF749* | -0.893908806 | 0.00209144 | 0.010006758 | 1 |
| HTN | nk_cell | *NBPF12* | -1.790036122 | 0.006233577 | 0.018547326 | 1 |
| HTN | nk_cell | *TOR3A* | -1.976772779 | 0.003651759 | 0.013309338 | 1 |
| HTN | nk_cell | *CCDC30* | -1.596037502 | 0.011581459 | 0.029032151 | 1 |
| HTN | nk_cell | *GLDN* | -1.125557496 | 0.002785515 | 0.011438624 | 1 |
| HTN | nk_cell | *FCAR* | -1.538350329 | 0.013461929 | 0.032521354 | 1 |
| HTN | nk_cell | *ZNF197* | -1.580400643 | 0.003698375 | 0.013378416 | 1 |
| HTN | nk_cell | *BTN3A2* | 0.348838664 | 0.014243755 | 0.03381816 | 1 |
| HTN | nk_cell | *PCLO* | -0.689364067 | 0.001582065 | 0.008993168 | 1 |
| HTN | nk_cell | *RGS7BP* | -1.366613735 | 0.000348517 | 0.006579952 | 1 |
| HTN | nk_cell | *ZNF396* | -1.412974437 | 8.60E-05 | 0.003125635 | 0.540734798 |
| HTN | nk_cell | *TMEM222* | -1.69626207 | 0.003491808 | 0.013010063 | 1 |
| HTN | nk_cell | *CLCNKA* | -2.263356078 | 0.01168895 | 0.029254947 | 1 |
| HTN | nk_cell | *SEPT10* | -1.356777378 | 0.005723041 | 0.017483362 | 1 |
| HTN | nk_cell | *SMYD4* | -2.176395558 | 0.001094457 | 0.008846261 | 1 |
| HTN | nk_cell | *MIR22HG* | -1.500478709 | 0.000957437 | 0.008846261 | 1 |
| HTN | nk_cell | *C17orf58* | -1.071103779 | 0.001567279 | 0.008975583 | 1 |
| HTN | nk_cell | *CYP27C1* | -0.505713386 | 0.014050902 | 0.033575834 | 1 |
| HTN | nk_cell | *FOXI2* | -1.754006628 | 0.003875607 | 0.01379725 | 1 |
| HTN | nk_cell | *HYAL3* | -1.358861972 | 0.000872937 | 0.008846261 | 1 |
| HTN | nk_cell | *ZSCAN30* | -1.850533297 | 0.004518431 | 0.0149907 | 1 |
| HTN | nk_cell | *KRT16* | -1.443938601 | 0.001222077 | 0.00885079 | 1 |
| HTN | nk_cell | *TMEM17* | -0.955530038 | 0.003745196 | 0.013477988 | 1 |
| HTN | nk_cell | *TNFRSF18* | 0.429724955 | 0.001382081 | 0.00885079 | 1 |
| HTN | nk_cell | *TMEM232* | -1.036938735 | 0.000479545 | 0.007652021 | 1 |
| HTN | nk_cell | *KANK3* | -1.73440248 | 0.003249422 | 0.012548599 | 1 |
| HTN | nk_cell | *EMID1* | -1.939635582 | 0.003334861 | 0.012699134 | 1 |
| HTN | nk_cell | *PLCD1* | -1.356665586 | 0.000557369 | 0.008121264 | 1 |
| HTN | nk_cell | *ENTPD5* | -1.814656854 | 0.004371717 | 0.014761003 | 1 |
| HTN | nk_cell | *HEATR4* | 0.880192062 | 0.016596243 | 0.037586665 | 1 |
| HTN | nk_cell | *ANGPTL5* | -1.506081313 | 0.001395414 | 0.00885079 | 1 |
| HTN | nk_cell | *WDR86* | -0.739464792 | 0.015524944 | 0.035923933 | 1 |
| HTN | nk_cell | *IFT140* | -1.317056429 | 0.003407271 | 0.012842634 | 1 |
| HTN | nk_cell | *TLR5* | -2.639980392 | 0.002082418 | 0.009986393 | 1 |
| HTN | nk_cell | *PLEKHN1* | 2.333102956 | 9.08E-06 | 0.000641657 | 0.057107441 |
| HTN | nk_cell | *TRPV2* | 1.265489952 | 0.003929378 | 0.013892416 | 1 |
| HTN | nk_cell | *C2orf88* | -1.271400321 | 0.000726555 | 0.008846261 | 1 |
| HTN | nk_cell | *KBTBD12* | -0.947375545 | 7.62E-05 | 0.002917771 | 0.479141818 |
| HTN | nk_cell | *FANCA* | 0.997545155 | 0.003982149 | 0.013955277 | 1 |
| HTN | nk_cell | *ADH1A* | -1.282627885 | 6.34E-05 | 0.002627011 | 0.39847097 |
| HTN | nk_cell | *SEMA4D* | 0.437055527 | 0.001968262 | 0.009771592 | 1 |
| HTN | nk_cell | *FAM69C* | -1.881650264 | 0.001058675 | 0.008846261 | 1 |
| HTN | nk_cell | *DNAH17* | 3.056240532 | 0.011249087 | 0.028404792 | 1 |

| | | | | | | |
|---|---|---|---|---|---|---|
| HTN | nk_cell | *TMEM220* | -1.36354961 | 0.005430413 | 0.016861315 | 1 |
| HTN | nk_cell | *CCDC157* | -1.651202154 | 0.000966759 | 0.008846261 | 1 |
| HTN | nk_cell | *GFRAL* | -1.104124977 | 0.005021523 | 0.015984969 | 1 |
| HTN | nk_cell | *LDLRAD2* | -0.697163659 | 0.005352544 | 0.016683909 | 1 |
| HTN | nk_cell | *ZSCAN23* | -0.572688155 | 0.001367675 | 0.00885079 | 1 |
| HTN | nk_cell | *C6orf132* | -0.843662312 | 0.002422236 | 0.010654267 | 1 |
| HTN | nk_cell | *MAPK12* | -2.104595873 | 0.000517011 | 0.007927918 | 1 |
| HTN | nk_cell | *AGRN* | -1.642445332 | 0.000582497 | 0.008131793 | 1 |
| HTN | nk_cell | *SMTNL2* | -5.080473913 | 0.001118312 | 0.008846261 | 1 |
| HTN | nk_cell | *PRKAR1B* | -1.326924411 | 0.021457747 | 0.045922441 | 1 |
| HTN | nk_cell | *NCR3LG1* | 2.384374451 | 0.002603446 | 0.01102956 | 1 |
| HTN | nk_cell | *ZNF793* | -1.121397458 | 0.001098108 | 0.008846261 | 1 |
| HTN | nk_cell | *AGAP4* | -3.322911422 | 0.001817166 | 0.009387445 | 1 |
| HTN | nk_cell | *HYKK* | -1.063699155 | 0.00388233 | 0.013802386 | 1 |
| HTN | nk_cell | *RUFY4* | -2.79980958 | 0.001061808 | 0.008846261 | 1 |
| HTN | nk_cell | *ZNF669* | -2.356464139 | 0.003603236 | 0.013228851 | 1 |
| HTN | nk_cell | *CENPP* | -1.077832424 | 0.001861629 | 0.009533293 | 1 |
| HTN | nk_cell | *ZNF559* | -1.021905423 | 0.000752854 | 0.008846261 | 1 |
| HTN | nk_cell | *ZP3* | -1.256563779 | 0.008568825 | 0.023101288 | 1 |
| HTN | nk_cell | *JAKMIP3* | -0.67233785 | 0.003283324 | 0.012626477 | 1 |
| HTN | nk_cell | *PDCD1* | 2.350528925 | 0.00363109 | 0.013274443 | 1 |
| HTN | nk_cell | *CERKL* | 1.417334954 | 0.000497992 | 0.007827185 | 1 |
| HTN | nk_cell | *C19orf54* | -1.500945435 | 0.004378624 | 0.014766189 | 1 |
| HTN | nk_cell | *C22orf34* | 1.214932036 | 0.004387952 | 0.014776142 | 1 |
| HTN | nk_cell | *NDOR1* | -2.359474198 | 0.002367406 | 0.010587941 | 1 |
| HTN | nk_cell | *FAM72B* | -3.253655374 | 0.001147155 | 0.008846261 | 1 |
| HTN | nk_cell | *GOLGA8M* | -0.999255794 | 0.007728919 | 0.021424918 | 1 |
| HTN | nk_cell | *S100A16* | -1.169838765 | 0.006349747 | 0.018781121 | 1 |
| HTN | nk_cell | *RHCE* | -1.118582315 | 0.00140803 | 0.00885079 | 1 |
| HTN | nk_cell | *SLC4A5* | -2.021595434 | 0.006644794 | 0.019304906 | 1 |
| HTN | nk_cell | *FAM221A* | -1.479899297 | 0.0013216 | 0.00885079 | 1 |
| HTN | nk_cell | *NOXA1* | -1.638433409 | 0.001339045 | 0.00885079 | 1 |
| HTN | nk_cell | *SKOR1* | 3.008518755 | 0.001688251 | 0.009197602 | 1 |
| HTN | nk_cell | *TMEM201* | -1.491457737 | 0.012556188 | 0.030836231 | 1 |
| HTN | nk_cell | *ZNF563* | -1.68562793 | 0.000754493 | 0.008846261 | 1 |
| HTN | nk_cell | *FBF1* | -1.583793503 | 6.58E-09 | 2.18E-06 | 4.14E-05 |
| HTN | nk_cell | *MSL1* | -1.414915348 | 0.000908889 | 0.008846261 | 1 |
| HTN | nk_cell | *FAM196A* | -1.133277219 | 0.005478686 | 0.016951033 | 1 |
| HTN | nk_cell | *UTS2B* | -1.043921142 | 0.000901187 | 0.008846261 | 1 |
| HTN | nk_cell | *C9orf152* | -5.381621563 | 0.001778629 | 0.009341888 | 1 |
| HTN | nk_cell | *NOC2L* | -1.114805815 | 0.006197602 | 0.018466505 | 1 |
| HTN | nk_cell | *SLC15A5* | -0.537911364 | 0.00726138 | 0.020453538 | 1 |
| HTN | nk_cell | *LIPI* | -0.869098389 | 0.002246631 | 0.010266299 | 1 |
| HTN | nk_cell | *ANKDD1B* | -0.695181987 | 0.005621111 | 0.017272691 | 1 |
| HTN | nk_cell | *LITAF* | 0.846656721 | 0.013030568 | 0.031704017 | 1 |
| HTN | nk_cell | *PLAC9* | -0.954100912 | 0.011889307 | 0.029673709 | 1 |
| HTN | nk_cell | *CLDN4* | -1.463927554 | 0.005450152 | 0.016896009 | 1 |
| HTN | nk_cell | *ZNF527* | -2.037056459 | 0.001931415 | 0.009667839 | 1 |
| HTN | nk_cell | *S100A13* | -1.701897712 | 0.017771134 | 0.039699542 | 1 |
| HTN | nk_cell | *ZNF600* | 0.842012471 | 0.001534468 | 0.008957475 | 1 |

| | | | | | | |
|---|---|---|---|---|---|---|
| HTN | nk_cell | *C15orf61* | -1.842533304 | 0.00074164 | 0.008846261 | 1 |
| HTN | nk_cell | *RRP7A* | -1.86374053 | 0.016552558 | 0.037541824 | 1 |
| HTN | nk_cell | *HLA-DRB1* | -1.80647662 | 9.80E-06 | 0.000683746 | 0.061586648 |
| HTN | nk_cell | *AKR1C3* | -1.733055221 | 0.002061974 | 0.00993635 | 1 |
| HTN | nk_cell | *SEMA4A* | -1.539987176 | 0.003581534 | 0.013191039 | 1 |
| HTN | nk_cell | *HRCT1* | -0.878200079 | 0.01404324 | 0.033570286 | 1 |
| HTN | nk_cell | *SIRPB2* | -1.934114482 | 0.000686102 | 0.00874661 | 1 |
| HTN | nk_cell | *ZNF471* | -1.692133516 | 0.00050884 | 0.007927918 | 1 |
| HTN | nk_cell | *GRM7* | -0.662789513 | 0.004910037 | 0.015741663 | 1 |
| HTN | nk_cell | *GIMAP5* | 1.987112681 | 0.00906256 | 0.024089096 | 1 |
| HTN | nk_cell | *STK31* | -2.672002162 | 0.001781156 | 0.009347351 | 1 |
| HTN | nk_cell | *ZKSCAN7* | 1.770849687 | 0.010138467 | 0.026209104 | 1 |
| HTN | nk_cell | *NTNG2* | 0.983194562 | 0.001670238 | 0.009156549 | 1 |
| HTN | nk_cell | *ASB13* | -2.051908121 | 0.002020124 | 0.009861903 | 1 |
| HTN | nk_cell | *ZNF781* | -1.067461791 | 0.001970905 | 0.009771592 | 1 |
| HTN | nk_cell | *INCA1* | 3.611320373 | 0.008183877 | 0.022302573 | 1 |
| HTN | nk_cell | *ZNF774* | -1.256369461 | 0.001428819 | 0.008851376 | 1 |
| HTN | nk_cell | *EVL* | 0.879029187 | 0.001238569 | 0.00885079 | 1 |
| HTN | nk_cell | *ZNF124* | -1.913962895 | 0.021369622 | 0.045769622 | 1 |
| HTN | nk_cell | *ZNF775* | -1.103916189 | 0.011389991 | 0.028655012 | 1 |
| HTN | nk_cell | *ZNF605* | -1.701073148 | 0.001915978 | 0.00964937 | 1 |
| HTN | nk_cell | *RFX8* | -1.376833463 | 0.002031418 | 0.009861903 | 1 |
| HTN | nk_cell | *C20orf96* | -3.120284128 | 0.006042522 | 0.018131247 | 1 |
| HTN | nk_cell | *SULT1A1* | -1.876531852 | 1.26E-06 | 0.000152275 | 0.007918316 |
| HTN | nk_cell | *C1orf186* | -1.41690482 | 0.00327949 | 0.012626477 | 1 |
| HTN | nk_cell | *SULF2* | -1.42441303 | 0.005840188 | 0.017754044 | 1 |
| HTN | nk_cell | *XRCC2* | -1.692987836 | 0.001600393 | 0.009048265 | 1 |
| HTN | nk_cell | *MYO6* | -1.950622711 | 0.000177619 | 0.004813319 | 1 |
| HTN | nk_cell | *ADH1B* | -0.824100525 | 0.001306609 | 0.00885079 | 1 |
| HTN | nk_cell | *HRH1* | -1.60141514 | 0.013108011 | 0.031828952 | 1 |
| HTN | nk_cell | *FAM180B* | -3.418966865 | 1.85E-12 | 3.87E-09 | 1.16E-08 |
| HTN | nk_cell | *ERI2* | -1.361216833 | 0.002081359 | 0.009986393 | 1 |
| HTN | nk_cell | *TRPV1* | -1.137679146 | 0.007337216 | 0.020584147 | 1 |
| HTN | nk_cell | *AMZ2* | -2.249087686 | 0.00648803 | 0.018989871 | 1 |
| HTN | nk_cell | *DAPK1* | -1.091218466 | 0.011999785 | 0.029890113 | 1 |
| HTN | nk_cell | *HLA-DQA1* | -1.206536311 | 0.00218187 | 0.010183682 | 1 |
| HTN | nk_cell | *TLE1* | -1.189547002 | 0.013479094 | 0.032521354 | 1 |
| HTN | nk_cell | *ZNF239* | -0.63407817 | 0.013641475 | 0.032759341 | 1 |
| HTN | nk_cell | *C6orf106* | 1.230098145 | 0.003742654 | 0.013476557 | 1 |
| HTN | nk_cell | *ZFP28* | -1.662939733 | 0.017518581 | 0.039270058 | 1 |
| HTN | nk_cell | *LAMB3* | -2.745308751 | 0.011297164 | 0.028512754 | 1 |
| HTN | nk_cell | *SRGAP1* | -1.217595779 | 0.011080047 | 0.028100144 | 1 |
| HTN | nk_cell | *FAM3C* | -2.230818149 | 0.005123014 | 0.016217719 | 1 |
| HTN | nk_cell | *AP2A1* | -2.244793445 | 0.00427329 | 0.014577656 | 1 |
| HTN | nk_cell | *ANXA4* | -1.455547937 | 0.015867017 | 0.036447183 | 1 |
| HTN | nk_cell | *ZNF429* | -1.900946589 | 0.00028036 | 0.006101112 | 1 |
| HTN | nk_cell | *ZNF470* | -2.674852921 | 0.000800782 | 0.008846261 | 1 |
| HTN | nk_cell | *ZNF100* | -2.28725711 | 5.11E-06 | 0.000431261 | 0.03213823 |
| HTN | nk_cell | *ZMYM1* | -1.869085166 | 0.019795181 | 0.04315267 | 1 |
| HTN | nk_cell | *GAL3ST4* | -1.384083212 | 0.000620529 | 0.008409632 | 1 |

| | | | | | | |
|---|---|---|---|---|---|---|
| HTN | nk_cell | *ZNF682* | -2.041132017 | 0.013112293 | 0.031828952 | 1 |
| HTN | nk_cell | *ZNF772* | -1.023221674 | 0.004401089 | 0.014795471 | 1 |
| HTN | nk_cell | *ZNF257* | -1.719422614 | 0.013151995 | 0.031913004 | 1 |
| HTN | nk_cell | *ADAM32* | -1.394256596 | 0.000956818 | 0.008846261 | 1 |
| HTN | nk_cell | *LRRC8B* | -1.804818766 | 0.002238566 | 0.01026356 | 1 |
| HTN | nk_cell | *SULT1A2* | 0.607793024 | 0.001353104 | 0.00885079 | 1 |
| HTN | nk_cell | *PIWIL2* | -1.080073654 | 0.002554593 | 0.010925664 | 1 |
| HTN | nk_cell | *ENTPD4* | -1.853699416 | 0.015055084 | 0.035217071 | 1 |
| HTN | nk_cell | *C1D* | -1.68965102 | 0.006880567 | 0.019743667 | 1 |
| HTN | nk_cell | *KANK2* | -1.624570069 | 0.001683609 | 0.0091868 | 1 |
| HTN | nk_cell | *C6orf141* | -1.693336086 | 6.66E-05 | 0.002685877 | 0.418996875 |
| HTN | nk_cell | *CCL4L2* | 0.419061411 | 0.009176193 | 0.024276248 | 1 |
| HTN | nk_cell | *RAMP2-AS1* | -0.750337445 | 0.001915926 | 0.00964937 | 1 |
| HTN | nk_cell | *ZNF517* | -1.658587236 | 0.001138637 | 0.008846261 | 1 |
| HTN | nk_cell | *SLC22A5* | 1.56388372 | 1.16E-05 | 0.000777309 | 0.073067021 |
| HTN | nk_cell | *C5AR1* | -1.419231896 | 0.005774014 | 0.017604862 | 1 |
| HTN | nk_cell | *SHPK* | -1.189266808 | 0.003959123 | 0.01392229 | 1 |
| HTN | nk_cell | *PCDHB11* | -2.146691909 | 3.97E-05 | 0.00194935 | 0.249516779 |
| HTN | nk_cell | *MYO5A* | -1.570223813 | 0.00649295 | 0.018995429 | 1 |
| HTN | nk_cell | *TTC30A* | -1.25648181 | 0.003676032 | 0.013343729 | 1 |
| HTN | nk_cell | *RAB40C* | -1.766935668 | 0.002062257 | 0.00993635 | 1 |
| HTN | nk_cell | *ZNF624* | -2.445827424 | 0.002434718 | 0.010654267 | 1 |
| HTN | nk_cell | *HHLA3* | -1.546888641 | 0.008872123 | 0.023715577 | 1 |
| HTN | nk_cell | *HOXA4* | -1.659207122 | 0.003370742 | 0.012777125 | 1 |
| HTN | nk_cell | *BCO2* | -2.167932137 | 0.000342644 | 0.006547732 | 1 |
| HTN | nk_cell | *ENTPD6* | -1.770499342 | 0.019994394 | 0.043421333 | 1 |
| HTN | nk_cell | *CCDC154* | -2.197903746 | 0.009381398 | 0.024678179 | 1 |
| HTN | nk_cell | *C5orf42* | -1.137720309 | 0.013577927 | 0.032681635 | 1 |
| HTN | nk_cell | *MFAP5* | -1.155595174 | 0.002502484 | 0.010783493 | 1 |
| HTN | nk_cell | *CDC42SE1* | 1.079347716 | 0.007129174 | 0.020189693 | 1 |
| HTN | nk_cell | *NMB* | 1.63038356 | 0.010560231 | 0.027064132 | 1 |
| HTN | nk_cell | *FAM114A1* | -1.325898045 | 0.002953763 | 0.011881194 | 1 |
| HTN | nk_cell | *PHF2* | -1.872425561 | 0.00584271 | 0.017754044 | 1 |
| HTN | nk_cell | *RPS26* | -0.803498968 | 0.000957792 | 0.008846261 | 1 |
| HTN | nk_cell | *C14orf178* | -0.904092937 | 0.001872516 | 0.009540973 | 1 |
| HTN | nk_cell | *S100A10* | -1.784691092 | 0.000583059 | 0.008131793 | 1 |
| HTN | nk_cell | *MAP1LC3C* | -0.829070348 | 0.019826628 | 0.043176651 | 1 |
| HTN | nk_cell | *TAF13* | -1.60038204 | 0.003466879 | 0.012981695 | 1 |
| HTN | nk_cell | *AC122129.1* | -1.9471894 | 0.001111607 | 0.008846261 | 1 |
| HTN | nk_cell | *OCLN* | -1.006796454 | 0.002177799 | 0.010179792 | 1 |
| HTN | nk_cell | *ZNF181* | -2.105343165 | 0.008331395 | 0.022555611 | 1 |
| HTN | nk_cell | *SGTB* | -2.059072533 | 0.007139779 | 0.020201527 | 1 |
| HTN | nk_cell | *ZNF677* | -2.197560577 | 0.001379278 | 0.00885079 | 1 |
| HTN | nk_cell | *ZNF71* | -0.9488295 | 0.009720458 | 0.025371122 | 1 |
| HTN | nk_cell | *LEKR1* | -1.216203643 | 0.00757295 | 0.021144291 | 1 |
| HTN | nk_cell | *AGAP9* | -3.82551306 | 0.00039358 | 0.00700973 | 1 |
| HTN | nk_cell | *MAK16* | -2.03188423 | 0.006225215 | 0.018531215 | 1 |
| HTN | nk_cell | *ZNF667* | -0.841606276 | 0.002140218 | 0.010132194 | 1 |
| HTN | nk_cell | *SULT1C4* | -1.907695168 | 0.010396543 | 0.026722432 | 1 |
| HTN | nk_cell | *SFI1* | 1.421736282 | 0.017989276 | 0.040049071 | 1 |

| | | | | | | |
|---|---|---|---|---|---|---|
| HTN | nk_cell | *ADH4* | -0.771964702 | 0.00093418 | 0.008846261 | 1 |
| HTN | nk_cell | *ZNF248* | -1.457772715 | 0.001054995 | 0.008846261 | 1 |
| HTN | nk_cell | *SOWAHC* | -2.166088322 | 0.000747543 | 0.008846261 | 1 |
| HTN | nk_cell | *ZNF334* | -0.885008952 | 0.002401375 | 0.010639496 | 1 |
| HTN | nk_cell | *ANTXRL* | -28.37945036 | 0.000924099 | 0.008846261 | 1 |
| HTN | nk_cell | *TMEM116* | -2.152404777 | 1.58E-09 | 7.09E-07 | 9.92E-06 |
| HTN | nk_cell | *ZNF813* | -3.2517964 | 0.000682667 | 0.00874661 | 1 |
| HTN | nk_cell | *UVRAG* | -1.079300022 | 0.00605441 | 0.018131247 | 1 |
| HTN | nk_cell | *MT1F* | 1.527504862 | 0.002111286 | 0.010063421 | 1 |
| HTN | nk_cell | *FLVCR1-AS1* | -0.642712057 | 0.017669012 | 0.03954613 | 1 |
| HTN | nk_cell | *SH3BGRL2* | -0.667107501 | 0.005565407 | 0.017151819 | 1 |
| HTN | nk_cell | *TMA16* | -1.903489326 | 0.009620751 | 0.025181374 | 1 |
| HTN | nk_cell | *HLA-DRB5* | -0.483399683 | 0.001439874 | 0.008873507 | 1 |
| HTN | nk_cell | *CNGA1* | -1.244019663 | 0.004003205 | 0.014005647 | 1 |
| HTN | nk_cell | *MAFK* | -2.162998772 | 0.002688473 | 0.011208507 | 1 |
| HTN | nk_cell | *HIST1H4E* | 2.709754123 | 0.004196935 | 0.014410776 | 1 |
| HTN | nk_cell | *C20orf203* | -1.749604719 | 0.016059893 | 0.036742556 | 1 |
| HTN | nk_cell | *ARC* | -4.278030996 | 0.001208047 | 0.00885079 | 1 |
| HTN | nk_cell | *MMP17* | -1.066666827 | 0.001088122 | 0.008846261 | 1 |
| HTN | nk_cell | *ZNF534* | -1.372638211 | 0.015617675 | 0.03599515 | 1 |
| HTN | nk_cell | *NCOA6* | -1.857245896 | 0.005607948 | 0.017249104 | 1 |
| HTN | nk_cell | *FAM19A2* | -1.632917761 | 0.001641373 | 0.009108692 | 1 |
| HTN | nk_cell | *TUSC1* | -1.01687573 | 0.001105821 | 0.008846261 | 1 |
| HTN | nk_cell | *PAPSS2* | -0.943814933 | 0.016253243 | 0.037090431 | 1 |
| HTN | nk_cell | *ANKRD13B* | 1.635726938 | 1.00E-04 | 0.003441707 | 0.628555589 |
| HTN | nk_cell | *ECI2* | -2.042626665 | 0.004394946 | 0.014785711 | 1 |
| HTN | nk_cell | *APCDD1L* | -3.013910879 | 0.010909294 | 0.027812949 | 1 |
| HTN | nk_cell | *RASSF9* | -1.381681637 | 0.001373929 | 0.00885079 | 1 |
| HTN | nk_cell | *FAM169A* | 0.233732635 | 0.001587377 | 0.009015212 | 1 |
| HTN | nk_cell | *TMEM184B* | -1.87529195 | 0.001570175 | 0.008975583 | 1 |
| HTN | nk_cell | *SCAMP5* | 1.326222591 | 0.004987244 | 0.015924228 | 1 |
| HTN | nk_cell | *PNP* | -0.771885397 | 0.002405548 | 0.010642983 | 1 |
| HTN | nk_cell | *ARHGAP11A* | -2.128373243 | 0.004151658 | 0.014310019 | 1 |
| HTN | nk_cell | *DENND4B* | -3.313878795 | 0.000216324 | 0.005316378 | 1 |
| HTN | nk_cell | *CD3E* | 0.232900065 | 0.001014446 | 0.008846261 | 1 |
| HTN | nk_cell | *ITPRIPL1* | 0.758231346 | 0.007622927 | 0.021237571 | 1 |
| HTN | nk_cell | *PRMT6* | -7.461212604 | 6.63E-05 | 0.002685877 | 0.416731948 |
| HTN | nk_cell | *SHISA4* | 1.755840027 | 0.012040154 | 0.029955064 | 1 |
| HTN | nk_cell | *MAP3K3* | -1.806763706 | 0.003295421 | 0.01264854 | 1 |
| HTN | nk_cell | *RASGEF1A* | 0.376704572 | 0.001389473 | 0.00885079 | 1 |
| HTN | nk_cell | *APRT* | 1.740333808 | 0.009198948 | 0.024280893 | 1 |
| HTN | nk_cell | *ZFP2* | -1.54979882 | 0.003164581 | 0.012344206 | 1 |
| HTN | nk_cell | *L3MBTL3* | 1.776249191 | 0.006242317 | 0.018564545 | 1 |
| HTN | nk_cell | *MFAP3L* | -1.25314718 | 0.001308207 | 0.00885079 | 1 |
| HTN | nk_cell | *AL590822.1* | -1.847248564 | 1.73E-06 | 0.000187886 | 0.010877776 |
| HTN | nk_cell | *AP001007.1* | -4.117727697 | 0.001176173 | 0.00885079 | 1 |
| HTN | nk_cell | *ZNF525* | -1.733409514 | 0.002819441 | 0.011517757 | 1 |
| HTN | nk_cell | *AC105020.1* | -1.978936862 | 0.002617155 | 0.01104299 | 1 |
| HTN | nk_cell | *EFCAB2* | -1.53918713 | 0.00299201 | 0.011958531 | 1 |
| HTN | nk_cell | *CAPN8* | 2.257985071 | 0.005100112 | 0.016169644 | 1 |

| | | | | | | |
|---|---|---|---|---|---|---|
| HTN | nk_cell | *CENPW* | -1.8022508 | 3.74E-05 | 0.001865555 | 0.235059883 |
| HTN | nk_cell | *FAM229B* | -0.516095706 | 0.003257783 | 0.01256708 | 1 |
| HTN | nk_cell | *HIST2H2BF* | -3.385555469 | 0.00107856 | 0.008846261 | 1 |
| HTN | nk_cell | *RBM20* | -1.957124671 | 0.002718814 | 0.011305016 | 1 |
| HTN | nk_cell | *FOXO6* | -0.76797458 | 0.014140439 | 0.033666663 | 1 |
| HTN | nk_cell | *INPP5B* | -0.628200946 | 0.002964936 | 0.01190329 | 1 |
| HTN | nk_cell | *TRAF3IP1* | -1.822088189 | 0.004433663 | 0.014834721 | 1 |
| HTN | nk_cell | *RUFY2* | -1.7069539 | 0.007762782 | 0.021480902 | 1 |
| HTN | nk_cell | *ZDBF2* | -1.037951135 | 0.002397962 | 0.010637375 | 1 |
| HTN | nk_cell | *RING1* | -1.740606773 | 0.000224914 | 0.005431789 | 1 |
| HTN | nk_cell | *HSD17B8* | -2.225423536 | 0.000117536 | 0.003730407 | 0.738950788 |
| HTN | nk_cell | *HLA-DOA* | -1.707181631 | 0.000253722 | 0.005756069 | 1 |
| HTN | nk_cell | *HLA-DMA* | -2.857486845 | 0.002768202 | 0.011416412 | 1 |
| HTN | nk_cell | *COL15A1* | -0.890079722 | 0.003312224 | 0.012674346 | 1 |
| HTN | nk_cell | *NOTCH4* | -0.174120159 | 0.007861067 | 0.021676548 | 1 |
| HTN | nk_cell | *AGER* | 1.659764199 | 0.001919652 | 0.009654896 | 1 |
| HTN | nk_cell | *MRPL38* | 1.696793479 | 2.00E-05 | 0.001121023 | 0.12555461 |
| HTN | nk_cell | *STK19* | 1.467170912 | 0.01108853 | 0.028110318 | 1 |
| HTN | nk_cell | *LAYN* | -1.946928221 | 0.001814205 | 0.00938705 | 1 |
| HTN | nk_cell | *C6orf48* | 3.057565921 | 1.30E-05 | 0.000844025 | 0.081870411 |
| HTN | nk_cell | *CARD16* | 0.946506819 | 0.019833652 | 0.043176651 | 1 |
| HTN | nk_cell | *LY6G5C* | 0.74431709 | 0.007285304 | 0.020484216 | 1 |
| HTN | nk_cell | *CCHCR1* | -1.036199069 | 0.000395438 | 0.007022931 | 1 |
| HTN | nk_cell | *LINC01123* | -28.48052546 | 0.000850838 | 0.008846261 | 1 |
| HTN | nk_cell | *HCG9* | -0.786297018 | 0.011219163 | 0.028372839 | 1 |
| HTN | nk_cell | *HLA-G* | 0.858416966 | 0.000675638 | 0.00874661 | 1 |
| HTN | nk_cell | *HLA-F* | 0.625289366 | 0.00056602 | 0.008121264 | 1 |
| HTN | nk_cell | *FAM153C* | -2.339719598 | 0.005913127 | 0.017916062 | 1 |
| HTN | nk_cell | *GABBR1* | -2.002395174 | 0.002357447 | 0.010571519 | 1 |
| HTN | nk_cell | *MALRD1* | -0.818593446 | 0.000479147 | 0.007652021 | 1 |
| HTN | nk_cell | *ATXN2* | -2.226915611 | 0.000265471 | 0.005897586 | 1 |
| HTN | nk_cell | *ZNF155* | -1.376698205 | 0.004832404 | 0.015611027 | 1 |
| HTN | nk_cell | *ZNF783* | -1.4082706 | 0.013203934 | 0.032026672 | 1 |
| HTN | nk_cell | *LRRC10B* | -6.421089089 | 0.012058267 | 0.029976403 | 1 |
| HTN | nk_cell | *PCDHA5* | -6.515253856 | 0.012267204 | 0.030363746 | 1 |
| HTN | nk_cell | *SLFN12L* | 0.188904412 | 0.001732422 | 0.009283228 | 1 |
| HTN | nk_cell | *SYCE1L* | -1.463940451 | 0.002696981 | 0.011236529 | 1 |
| HTN | nk_cell | *TMEM231* | -0.681323566 | 0.017585151 | 0.039386478 | 1 |
| HTN | nk_cell | *FAM71F2* | -1.230317691 | 0.001518383 | 0.0089561 | 1 |
| HTN | nk_cell | *LINC00654* | -1.946134564 | 0.000154153 | 0.004369177 | 0.969162406 |
| HTN | nk_cell | *NT5M* | -0.765419463 | 0.015799509 | 0.036331936 | 1 |
| HTN | nk_cell | *MT1A* | 1.748878001 | 0.001749159 | 0.009295824 | 1 |
| HTN | nk_cell | *MT1M* | -1.406704338 | 0.001135341 | 0.008846261 | 1 |
| HTN | nk_cell | *CFI* | -0.960923928 | 0.003734364 | 0.013454539 | 1 |
| HTN | nk_cell | *SAMD9* | 1.219915841 | 0.006570513 | 0.019180563 | 1 |
| HTN | nk_cell | *EXOC3L4* | -1.578790411 | 0.000942445 | 0.008846261 | 1 |
| HTN | nk_cell | *SMN2* | -1.258148159 | 0.000487453 | 0.007719437 | 1 |
| HTN | nk_cell | *SERF1B* | -3.353176418 | 1.36E-05 | 0.000860652 | 0.085204592 |
| HTN | nk_cell | *LCMT1* | -1.994588673 | 0.007192786 | 0.020324066 | 1 |
| HTN | nk_cell | *LINC00487* | -3.375895407 | 0.001165072 | 0.00885079 | 1 |

| | | | | | | |
|---|---|---|---|---|---|---|
| HTN | nk_cell | *RFPL3S* | -1.274896227 | 0.001130557 | 0.008846261 | 1 |
| HTN | nk_cell | *C1QTNF9B* | 9.264488596 | 0.001788471 | 0.009362295 | 1 |
| HTN | nk_cell | *NYNRIN* | -1.188694498 | 0.002068474 | 0.009949884 | 1 |
| HTN | nk_cell | *GOLGA8O* | 2.6604203 | 0.001680092 | 0.009184992 | 1 |
| HTN | nk_cell | *HLA-A* | 0.617563235 | 0.001811437 | 0.00938705 | 1 |
| HTN | nk_cell | *LNP1* | -1.956229296 | 6.47E-05 | 0.002639322 | 0.406455621 |
| HTN | nk_cell | *VGLL3* | -0.59491683 | 0.003707943 | 0.013397607 | 1 |
| HTN | nk_cell | *SLC48A1* | -1.284611302 | 0.000313464 | 0.006377819 | 1 |
| HTN | nk_cell | *TAS2R14* | 1.376176671 | 0.001120275 | 0.008846261 | 1 |
| HTN | nk_cell | *ZRSR1* | 7.759776414 | 0.001744103 | 0.009286428 | 1 |
| HTN | nk_cell | *AC084018.1* | -0.912895074 | 0.00088461 | 0.008846261 | 1 |
| HTN | nk_cell | *OR2A42* | -0.880092217 | 0.019104319 | 0.042025491 | 1 |
| HTN | nk_cell | *RNF208* | -2.168132544 | 0.001058516 | 0.008846261 | 1 |
| HTN | nk_cell | *NUP62* | -2.377891054 | 0.001124464 | 0.008846261 | 1 |
| HTN | nk_cell | *GIMAP1* | 0.615438391 | 0.019429002 | 0.042546198 | 1 |
| HTN | nk_cell | *CCDC183* | -3.635574794 | 0.007573874 | 0.021144291 | 1 |
| HTN | nk_cell | *SUPT4H1* | 2.244701029 | 6.74E-05 | 0.002700201 | 0.423931572 |
| HTN | nk_cell | *MXD3* | -2.303293958 | 9.21E-05 | 0.003289 | 0.578863941 |
| HTN | nk_cell | *GSTM2* | -1.587629386 | 9.54E-05 | 0.003369591 | 0.599787239 |
| HTN | nk_cell | *ARHGAP19* | -4.137429352 | 0.002229877 | 0.010252893 | 1 |
| HTN | nk_cell | *LCAT* | -0.951340093 | 0.001378273 | 0.00885079 | 1 |
| HTN | nk_cell | *SIPA1* | 1.106076117 | 1.03E-05 | 0.000693102 | 0.064458488 |
| HTN | nk_cell | *RBMXL1* | -2.585925322 | 0.001123156 | 0.008846261 | 1 |
| HTN | nk_cell | *SRA1* | -1.207052644 | 0.009410988 | 0.024724982 | 1 |
| HTN | nk_cell | *LEPROT* | -1.808465765 | 0.012755067 | 0.031190629 | 1 |
| HTN | nk_cell | *LBH* | 0.895463085 | 0.002016636 | 0.009858936 | 1 |
| HTN | nk_cell | *NCKIPSD* | -1.902178095 | 0.00026146 | 0.005849812 | 1 |
| HTN | nk_cell | *ATF6B* | 1.941299088 | 0.000225176 | 0.005431789 | 1 |
| HTN | nk_cell | *SLC35F6* | -2.132845431 | 0.015581276 | 0.03599515 | 1 |
| HTN | nk_cell | *CLIC1* | 1.981591796 | 0.001707184 | 0.009223501 | 1 |
| HTN | nk_cell | *KLRK1* | 0.50404998 | 0.000864513 | 0.008846261 | 1 |
| HTN | nk_cell | *EMP2* | -1.895654368 | 0.000537663 | 0.008086807 | 1 |
| HTN | nk_cell | *LTB4R* | -1.058500236 | 0.019822758 | 0.043176651 | 1 |
| HTN | nk_cell | *LTB4R2* | -3.875226463 | 1.83E-05 | 0.001066338 | 0.115164502 |
| HTN | nk_cell | *ITGA1* | -0.901280942 | 0.004137893 | 0.014286069 | 1 |
| HTN | nk_cell | *ZNF726* | -1.546547346 | 0.001182011 | 0.00885079 | 1 |
| HTN | nk_cell | *TAX1BP3* | -2.216209058 | 0.005520191 | 0.017062451 | 1 |
| HTN | nk_cell | *TM6SF2* | -11.81687753 | 0.001039154 | 0.008846261 | 1 |
| HTN | nk_cell | *REPIN1* | -3.251557097 | 0.000355706 | 0.006607413 | 1 |
| HTN | nk_cell | *SMIM7* | -2.083980217 | 0.014464179 | 0.034199434 | 1 |
| HTN | nk_cell | *TSPAN4* | -1.857064928 | 0.005126156 | 0.016219497 | 1 |
| HTN | nk_cell | *AC079776.2* | -1.695137272 | 0.004244847 | 0.014543516 | 1 |
| HTN | nk_cell | *PRCD* | -1.159232001 | 0.003931065 | 0.013892416 | 1 |
| HTN | nk_cell | *COLCA2* | -1.041471616 | 0.001285763 | 0.00885079 | 1 |
| HTN | nk_cell | *NEURL1B* | -1.086636096 | 0.001072093 | 0.008846261 | 1 |
| HTN | nk_cell | *VSTM5* | -0.882342483 | 0.006445676 | 0.018927587 | 1 |
| HTN | nk_cell | *PPME1* | -1.902238147 | 0.000199447 | 0.005142286 | 1 |
| HTN | nk_cell | *CPEB1* | -1.189501366 | 0.003523108 | 0.01307543 | 1 |
| HTN | nk_cell | *C10orf105* | -1.509660955 | 0.001501021 | 0.008938486 | 1 |
| HTN | nk_cell | *AC005082.1* | -3.381260523 | 0.001151268 | 0.008846261 | 1 |

| | | | | | | |
|---|---|---|---|---|---|---|
| HTN | nk_cell | *TBC1D26* | 5.504416624 | 0.012232761 | 0.030314295 | 1 |
| HTN | nk_cell | *COL28A1* | -1.680030375 | 0.00235667 | 0.010571519 | 1 |
| HTN | nk_cell | *UBXN2B* | 1.516861772 | 0.02212874 | 0.046921884 | 1 |
| HTN | nk_cell | *UBE2QL1* | -1.186038482 | 0.001052936 | 0.008846261 | 1 |
| HTN | nk_cell | *KCNU1* | -1.739272057 | 0.000169979 | 0.004646338 | 1 |
| HTN | nk_cell | *MYL5* | -4.427637176 | 0.004912994 | 0.015743117 | 1 |
| HTN | nk_cell | *NPEPL1* | -1.984057756 | 0.001017203 | 0.008846261 | 1 |
| HTN | nk_cell | *LINC01139* | -1.434414056 | 0.002154894 | 0.010155174 | 1 |
| HTN | nk_cell | *TSTD1* | 1.872065708 | 0.005736521 | 0.01751603 | 1 |
| HTN | nk_cell | *LINC01138* | 1.099314311 | 0.006631978 | 0.019285498 | 1 |
| HTN | nk_cell | *CYB5RL* | -1.231873203 | 0.000880956 | 0.008846261 | 1 |
| HTN | nk_cell | *ATAD3C* | -1.799065927 | 0.005629445 | 0.017281408 | 1 |
| HTN | nk_cell | *CCDC7* | -1.612147542 | 0.005892649 | 0.017862625 | 1 |
| HTN | nk_cell | *PAM16* | -1.387658188 | 0.000552837 | 0.008121264 | 1 |
| HTN | nk_cell | *RNASEK* | -2.614402245 | 0.003067585 | 0.012144776 | 1 |
| HTN | nk_cell | *NBPF1* | -1.45866028 | 0.008967901 | 0.023922744 | 1 |
| HTN | nk_cell | *FAM228B* | -0.762361148 | 0.006793285 | 0.019618456 | 1 |
| HTN | nk_cell | *RNU6ATAC35l* | -11.36937406 | 0.001410527 | 0.00885079 | 1 |
| HTN | nk_cell | *FANCG* | -0.918242827 | 0.00189665 | 0.009600838 | 1 |
| HTN | nk_cell | *AP4M1* | -1.296640022 | 0.000304922 | 0.006326876 | 1 |
| HTN | nk_cell | *C19orf73* | -5.168708564 | 0.003904785 | 0.013830639 | 1 |
| HTN | nk_cell | *ZNF880* | -0.726987296 | 0.00982689 | 0.025561939 | 1 |
| HTN | nk_cell | *OR2A25* | -37.85371303 | 0.000189709 | 0.005097002 | 1 |
| HTN | nk_cell | *SLC12A8* | -1.606813742 | 1.40E-05 | 0.000882274 | 0.088227379 |
| HTN | nk_cell | *UBA52* | 1.241886255 | 0.003369421 | 0.012777125 | 1 |
| HTN | nk_cell | *BTBD19* | -1.372288538 | 0.003666094 | 0.013338386 | 1 |
| HTN | nk_cell | *FAM185A* | -2.072769476 | 0.001733499 | 0.009283228 | 1 |
| HTN | nk_cell | *AC062017.1* | -1.070126133 | 0.00201334 | 0.009858153 | 1 |
| HTN | nk_cell | *ADRA2B* | -3.100472485 | 0.001052412 | 0.008846261 | 1 |
| HTN | nk_cell | *TINCR* | -3.3858775 | 0.008908407 | 0.023782231 | 1 |
| HTN | nk_cell | *LINC00205* | -1.541221685 | 0.019943985 | 0.043371886 | 1 |
| HTN | nk_cell | *HLA-DPB1* | 1.491218031 | 0.00251572 | 0.010818285 | 1 |
| HTN | nk_cell | *LINC01133* | -0.607524008 | 0.001130393 | 0.008846261 | 1 |
| HTN | nk_cell | *LINC01117* | -1.131156253 | 0.001543061 | 0.00897211 | 1 |
| HTN | nk_cell | *RASAL2-AS1* | -2.860484382 | 0.005274531 | 0.016522659 | 1 |
| HTN | nk_cell | *AC069213.1* | -3.787763891 | 0.003328395 | 0.012697584 | 1 |
| HTN | nk_cell | *AL049840.1* | -1.468080025 | 1.18E-06 | 0.000148329 | 0.007416448 |
| HTN | nk_cell | *LINC00237* | -3.950147937 | 0.001022651 | 0.008846261 | 1 |
| HTN | nk_cell | *SFTA1P* | -5.237887006 | 0.004446949 | 0.014855456 | 1 |
| HTN | nk_cell | *MIAT* | 0.36225271 | 0.002525652 | 0.010838755 | 1 |
| HTN | nk_cell | *LINC00626* | -16.39235501 | 0.001828093 | 0.009428402 | 1 |
| HTN | nk_cell | *ERCC6* | -1.62502719 | 0.007885105 | 0.021723774 | 1 |
| HTN | nk_cell | *LINC00115* | -1.641341715 | 0.006592573 | 0.019209783 | 1 |
| HTN | nk_cell | *ZNF503-AS1* | -1.116279589 | 0.001123679 | 0.008846261 | 1 |
| HTN | nk_cell | *LINC00937* | -2.051083345 | 0.000676226 | 0.00874661 | 1 |
| HTN | nk_cell | *AC064836.3* | -1.672051358 | 0.00119045 | 0.00885079 | 1 |
| HTN | nk_cell | *TMEM185B* | -1.276222532 | 0.003342242 | 0.012704157 | 1 |
| HTN | nk_cell | *HSBP1L1* | -3.209416383 | 0.000485187 | 0.007702956 | 1 |
| HTN | nk_cell | *TAS2R46* | -1.925378297 | 0.003878986 | 0.013801463 | 1 |
| HTN | nk_cell | *AC093901.1* | -1.587051315 | 0.001381878 | 0.00885079 | 1 |

| | | | | | | |
|---|---|---|---|---|---|---|
| HTN | nk_cell | *C14orf132* | -1.21674875 | 0.000665159 | 0.008712201 | 1 |
| HTN | nk_cell | *AC017074.2* | -8.052830733 | 0.00689227 | 0.019768113 | 1 |
| HTN | nk_cell | *SLC2A1-AS1* | -4.347955906 | 0.000227978 | 0.005449811 | 1 |
| HTN | nk_cell | *AC108463.2* | -2.785628773 | 4.07E-05 | 0.00196317 | 0.256128143 |
| HTN | nk_cell | *C19orf24* | 1.270378728 | 0.001317927 | 0.00885079 | 1 |
| HTN | nk_cell | *LINC01013* | -2.074276498 | 0.002377959 | 0.010598969 | 1 |
| HTN | nk_cell | *AC073133.2* | -10.05587782 | 0.001548923 | 0.008973847 | 1 |
| HTN | nk_cell | *ARL17B* | -1.132884889 | 0.023686834 | 0.049507688 | 1 |
| HTN | nk_cell | *LINC00954* | 1.288391499 | 0.002212451 | 0.010227705 | 1 |
| HTN | nk_cell | *AC067945.3* | 0.289192206 | 0.000558858 | 0.008121264 | 1 |
| HTN | nk_cell | *RPL41* | -2.283084693 | 0.007893132 | 0.021736366 | 1 |
| HTN | nk_cell | *PATL2* | 0.736789469 | 0.001684807 | 0.0091868 | 1 |
| HTN | nk_cell | *U91324.1* | 0.523078758 | 0.001438753 | 0.008873507 | 1 |
| HTN | nk_cell | *PSMG3-AS1* | -1.416470332 | 0.001505624 | 0.008938486 | 1 |
| HTN | nk_cell | *PPP5D1* | -1.804377272 | 0.017045244 | 0.038437392 | 1 |
| HTN | nk_cell | *AC092171.4* | -2.147107468 | 0.005015835 | 0.015984969 | 1 |
| HTN | nk_cell | *DIRC3* | -1.037019956 | 0.004047599 | 0.014091781 | 1 |
| HTN | nk_cell | *TAPBP* | 1.242387307 | 0.002586614 | 0.010988756 | 1 |
| HTN | nk_cell | *AC107021.1* | -0.52081881 | 0.00389184 | 0.013815921 | 1 |
| HTN | nk_cell | *AC009495.3* | 0.439378246 | 0.001282289 | 0.00885079 | 1 |
| HTN | nk_cell | *GOLGA8N* | 1.176063235 | 0.000361291 | 0.00663164 | 1 |
| HTN | nk_cell | *LINC00665* | -1.073751645 | 0.004470099 | 0.014878713 | 1 |
| HTN | nk_cell | *TNF* | 0.520500387 | 0.002157657 | 0.010155174 | 1 |
| HTN | nk_cell | *LYRM9* | -1.375446683 | 0.001370221 | 0.00885079 | 1 |
| HTN | nk_cell | *SNHG15* | -3.230540578 | 0.006098288 | 0.018231068 | 1 |
| HTN | nk_cell | *LINC00702* | -1.049745502 | 0.001048185 | 0.008846261 | 1 |
| HTN | nk_cell | *LINC00472* | -0.82609117 | 0.000894138 | 0.008846261 | 1 |
| HTN | nk_cell | *AC123768.3* | -1.754217514 | 0.003568971 | 0.013160187 | 1 |
| HTN | nk_cell | *HLA-B* | 0.781822785 | 0.000117082 | 0.003730407 | 0.736094391 |
| HTN | nk_cell | *ZSCAN31* | -0.774830591 | 0.005445852 | 0.016891008 | 1 |
| HTN | nk_cell | *LINC00957* | -3.047516949 | 0.013477422 | 0.032521354 | 1 |
| HTN | nk_cell | *B3GALT4* | 1.231494872 | 0.004728922 | 0.015416362 | 1 |
| HTN | nk_cell | *LINC00941* | -2.99081865 | 0.001769019 | 0.009330388 | 1 |
| HTN | nk_cell | *B3GNT9* | -2.071042425 | 0.000421766 | 0.007214859 | 1 |
| HTN | nk_cell | *CDKN2AIPNL* | -1.335441249 | 0.003123228 | 0.012264669 | 1 |
| HTN | nk_cell | *ZNF737* | -2.071165015 | 0.014992546 | 0.035131619 | 1 |
| HTN | nk_cell | *HLA-DQA2* | 0.737317433 | 0.006011178 | 0.018117103 | 1 |
| HTN | nk_cell | *AC073257.2* | -7.792867681 | 0.001916982 | 0.00964937 | 1 |
| HTN | nk_cell | *TXNDC5* | 3.563875244 | 0.001836659 | 0.009457064 | 1 |
| HTN | nk_cell | *ASB14* | -1.599252864 | 0.00249391 | 0.010761973 | 1 |
| HTN | nk_cell | *CDRT4* | -1.421227902 | 0.0090655 | 0.024089096 | 1 |
| HTN | nk_cell | *APOBEC3G* | 0.235094909 | 0.004871717 | 0.015658735 | 1 |
| HTN | nk_cell | *PSMB9* | 0.621098564 | 0.006873925 | 0.019742515 | 1 |
| HTN | nk_cell | *L1TD1* | -0.877970195 | 0.00284712 | 0.011578163 | 1 |
| HTN | nk_cell | *AQP1* | -1.68510215 | 0.00281466 | 0.011513186 | 1 |
| HTN | nk_cell | *C1QTNF9* | -3.637576671 | 0.001202739 | 0.00885079 | 1 |
| HTN | nk_cell | *LINC00886* | -1.404300738 | 0.005299207 | 0.016591691 | 1 |
| HTN | nk_cell | *CD302* | -1.827597722 | 0.004539925 | 0.015033969 | 1 |
| HTN | nk_cell | *ARHGAP8* | -1.426326582 | 0.001497801 | 0.008938486 | 1 |
| HTN | nk_cell | *ATP5O* | -2.699781117 | 0.009077461 | 0.024110688 | 1 |

| | | | | | | |
|---|---|---|---|---|---|---|
| HTN | nk_cell | *PWP2* | 1.536135801 | 0.002799604 | 0.011473997 | 1 |
| HTN | nk_cell | *AKAP2* | -7.444485905 | 0.00153069 | 0.008957475 | 1 |
| HTN | nk_cell | *LINC00969* | -1.098275888 | 0.011594052 | 0.029052136 | 1 |
| HTN | nk_cell | *ARHGDIG* | -2.640731518 | 0.000280455 | 0.006101112 | 1 |
| HTN | nk_cell | *TCP10L* | -5.660161939 | 0.007787675 | 0.021521367 | 1 |
| HTN | nk_cell | *C22orf39* | 1.564165615 | 0.002463066 | 0.010701656 | 1 |
| HTN | nk_cell | *LINC00960* | -0.81950131 | 0.001813426 | 0.00938705 | 1 |
| HTN | nk_cell | *DECR2* | -2.251897403 | 0.001096367 | 0.008846261 | 1 |
| HTN | nk_cell | *AP5Z1* | -2.016435508 | 0.01210544 | 0.030069893 | 1 |
| HTN | nk_cell | *ZNF709* | -3.104207477 | 0.000637051 | 0.008557988 | 1 |
| HTN | nk_cell | *VSIG8* | -1.42773584 | 0.002311397 | 0.010439478 | 1 |
| HTN | nk_cell | *C7orf73* | -1.743758108 | 0.005346914 | 0.016674626 | 1 |
| HTN | nk_cell | *FGF14-IT1* | -2.293948791 | 0.001093802 | 0.008846261 | 1 |
| HTN | nk_cell | *PALM2* | -0.474117651 | 0.003120875 | 0.012264669 | 1 |
| HTN | nk_cell | *NAT6* | -0.917206855 | 0.007047608 | 0.020039942 | 1 |
| HTN | nk_cell | *UPK3B* | -1.684502943 | 0.000293826 | 0.006279604 | 1 |
| HTN | nk_cell | *PLA2G4B* | 2.043921032 | 1.66E-05 | 0.001013492 | 0.104449243 |
| HTN | nk_cell | *JMJD7* | 1.324643906 | 0.009976552 | 0.025864981 | 1 |
| HTN | nk_cell | *APOBEC3D* | 1.215056881 | 0.001611556 | 0.009058261 | 1 |
| HTN | nk_cell | *RN7SL832P* | 1.564364419 | 0.008804658 | 0.023582574 | 1 |
| HTN | nk_cell | *TMEM199* | -2.199379743 | 0.000513546 | 0.007927918 | 1 |
| HTN | nk_cell | *P2RY11* | -1.964602805 | 0.000356098 | 0.006607413 | 1 |
| HTN | nk_cell | *APOBEC3C* | 0.464452821 | 0.001428558 | 0.008851376 | 1 |
| HTN | nk_cell | *ASPRV1* | -2.795807264 | 0.000976599 | 0.008846261 | 1 |
| HTN | nk_cell | *UBE2V1* | -3.292757559 | 0.01752068 | 0.039270058 | 1 |
| HTN | nk_cell | *LINC00847* | -3.578491819 | 0.008011551 | 0.021975838 | 1 |
| HTN | nk_cell | *NEAT1* | -1.029945832 | 0.020622846 | 0.044463592 | 1 |
| HTN | nk_cell | *ZNF585B* | -1.119579005 | 0.017793462 | 0.039711571 | 1 |
| HTN | nk_cell | *RAB30-AS1* | -1.813881041 | 0.004977993 | 0.015902765 | 1 |
| HTN | nk_cell | *LINC00920* | -2.047869795 | 0.000835804 | 0.008846261 | 1 |
| HTN | nk_cell | *UBAP1L* | -1.752279711 | 0.000879995 | 0.008846261 | 1 |
| HTN | nk_cell | *PGAM5* | -1.285686033 | 0.001098721 | 0.008846261 | 1 |
| HTN | nk_cell | *DNAJC3-AS1* | -1.932913379 | 0.006947072 | 0.019879945 | 1 |
| HTN | nk_cell | *BCKDHA* | -2.913797587 | 0.008172953 | 0.022284079 | 1 |
| HTN | nk_cell | *CDK11B* | 1.354463882 | 0.010801474 | 0.027616456 | 1 |
| HTN | nk_cell | *ABHD14A* | -2.482343273 | 0.014175597 | 0.033732769 | 1 |
| HTN | nk_cell | *SRP14-AS1* | -0.606342157 | 0.002853305 | 0.011595818 | 1 |
| HTN | nk_cell | *CCDC153* | -1.673919978 | 0.006275238 | 0.018644813 | 1 |
| HTN | nk_cell | *C5orf17* | -2.303159154 | 0.016962785 | 0.038292649 | 1 |
| HTN | nk_cell | *LINC01060* | -0.61998941 | 0.005401116 | 0.016793678 | 1 |
| HTN | nk_cell | *NAIP* | -1.661830811 | 0.004615627 | 0.015184955 | 1 |
| HTN | nk_cell | *ZNF286B* | 1.613405334 | 0.003424337 | 0.012888215 | 1 |
| HTN | nk_cell | *LINC00603* | -0.925348432 | 0.004201879 | 0.014419875 | 1 |
| HTN | nk_cell | *LINC00989* | -0.787435615 | 0.001026312 | 0.008846261 | 1 |
| HTN | nk_cell | *LINC01021* | -1.713104625 | 0.001078448 | 0.008846261 | 1 |
| HTN | nk_cell | *GLI4* | -1.510437572 | 0.001787729 | 0.009362295 | 1 |
| HTN | nk_cell | *SNHG18* | -1.766171771 | 0.000751309 | 0.008846261 | 1 |
| HTN | nk_cell | *ZNF345* | -1.452019065 | 0.016744158 | 0.037867094 | 1 |
| HTN | nk_cell | *SHANK3* | -0.847068459 | 0.004406751 | 0.014799809 | 1 |
| HTN | nk_cell | *ZNF550* | -2.13644441 | 0.004258918 | 0.014567909 | 1 |

| | | | | | | |
|---|---|---|---|---|---|---|
| HTN | nk_cell | *CCDC71L* | -1.812210492 | 0.002192485 | 0.010211299 | 1 |
| HTN | nk_cell | *TMEM200B* | -0.691058908 | 0.00432571 | 0.014705378 | 1 |
| HTN | nk_cell | *TRNP1* | -3.705998123 | 0.001100608 | 0.008846261 | 1 |
| HTN | nk_cell | *PCDHGA5* | -1.535929585 | 0.002127056 | 0.01009268 | 1 |
| HTN | nk_cell | *ZNF260* | -2.04395602 | 0.012458252 | 0.030690284 | 1 |
| HTN | nk_cell | *FPGT* | -2.650049363 | 0.004753588 | 0.015439713 | 1 |
| HTN | nk_cell | *EBLN2* | -2.684785069 | 0.002212128 | 0.010227705 | 1 |
| HTN | nk_cell | *TAS2R20* | 1.116089138 | 0.012222964 | 0.030301961 | 1 |
| HTN | nk_cell | *LINC00346* | -1.716329159 | 0.012369306 | 0.030544316 | 1 |
| HTN | nk_cell | *CTSO* | -1.992588512 | 0.003914237 | 0.013850379 | 1 |
| HTN | nk_cell | *LINC00944* | 0.269853028 | 0.001450925 | 0.008873507 | 1 |
| HTN | nk_cell | *ZNF10* | -2.942345005 | 3.49E-08 | 9.54E-06 | 0.000219408 |
| HTN | nk_cell | *HMBS* | 1.757628001 | 6.37E-05 | 0.002627011 | 0.400531118 |
| HTN | nk_cell | *TAS2R31* | -1.426907938 | 0.010549127 | 0.027048271 | 1 |
| HTN | nk_cell | *PGA5* | -10.03398373 | 0.006354635 | 0.018781121 | 1 |
| HTN | nk_cell | *ZBED6* | -4.455863919 | 0.001208805 | 0.00885079 | 1 |
| HTN | nk_cell | *FNTB* | -1.287188331 | 0.001077456 | 0.008846261 | 1 |
| HTN | nk_cell | *GALNT4* | -8.196307885 | 0.008693469 | 0.023357197 | 1 |
| HTN | nk_cell | *OVCH1-AS1* | -1.974437574 | 0.001073592 | 0.008846261 | 1 |
| HTN | nk_cell | *C17orf49* | -2.697901729 | 0.005878389 | 0.01784521 | 1 |
| HTN | nk_cell | *LINC00638* | -5.989018838 | 0.000828473 | 0.008846261 | 1 |
| HTN | nk_cell | *CEP95* | -1.892268245 | 0.011102268 | 0.0281338 | 1 |
| HTN | nk_cell | *LINC00639* | -0.683149657 | 0.00153772 | 0.008968132 | 1 |
| HTN | nk_cell | *LINC00648* | -1.321705292 | 0.00111798 | 0.008846261 | 1 |
| HTN | nk_cell | *LINC00924* | -0.991340071 | 0.000695105 | 0.008747204 | 1 |
| HTN | nk_cell | *ITGB3* | -1.026117812 | 0.021637415 | 0.046187188 | 1 |
| HTN | nk_cell | *UBE2Q2L* | -2.596896209 | 0.000262494 | 0.005852126 | 1 |
| HTN | nk_cell | *AC083843.1* | -16.91408146 | 0.000348368 | 0.006579952 | 1 |
| HTN | nk_cell | *SH3RF3-AS1* | -1.967846428 | 0.00014392 | 0.004228148 | 0.904823654 |
| HTN | nk_cell | *PWRN1* | -1.698853953 | 0.022103728 | 0.046892163 | 1 |
| HTN | nk_cell | *FRRS1L* | -5.133508736 | 0.00808335 | 0.022128019 | 1 |
| HTN | nk_cell | *LINC00562* | -0.787625287 | 0.018207389 | 0.040391621 | 1 |
| HTN | nk_cell | *AC009120.5* | -1.170803708 | 0.001050701 | 0.008846261 | 1 |
| HTN | nk_cell | *LINC00565* | -8.654637232 | 0.001519987 | 0.0089561 | 1 |
| HTN | nk_cell | *TMEM178B* | -1.090048293 | 0.000853978 | 0.008846261 | 1 |
| HTN | nk_cell | *C15orf65* | -0.867157499 | 0.004603182 | 0.015167822 | 1 |
| HTN | nk_cell | *HPR* | -2.023007635 | 0.004413243 | 0.01480253 | 1 |
| HTN | nk_cell | *LINC00672* | 1.175882917 | 0.020949533 | 0.045044362 | 1 |
| HTN | nk_cell | *DYNLL2* | -2.761562803 | 0.0033716 | 0.012777125 | 1 |
| HTN | nk_cell | *LINC00526* | -1.127509699 | 0.003317905 | 0.012680651 | 1 |
| HTN | nk_cell | *AC139100.2* | -9.004033933 | 0.001089393 | 0.008846261 | 1 |
| HTN | nk_cell | *LINC01140* | -1.136111165 | 0.001738433 | 0.009285921 | 1 |
| HTN | nk_cell | *LINC00906* | -6.329067714 | 0.001064114 | 0.008846261 | 1 |
| HTN | nk_cell | *ZNF582-AS1* | -1.181055477 | 0.000756928 | 0.008846261 | 1 |
| HTN | nk_cell | *AC003005.2* | -1.706095602 | 0.005440565 | 0.016882939 | 1 |
| HTN | nk_cell | *AL132989.1* | -1.828128633 | 0.001130355 | 0.008846261 | 1 |
| HTN | nk_cell | *LINC00664* | -3.464382859 | 0.000287154 | 0.006179456 | 1 |
| HTN | nk_cell | *FBXO17* | -0.919670851 | 0.001469122 | 0.008909666 | 1 |
| HTN | nk_cell | *AC009060.1* | -1.542259663 | 0.00011875 | 0.003730407 | 0.746578282 |
| HTN | nk_cell | *SNHG8* | -1.919449446 | 0.000908867 | 0.008846261 | 1 |

| | | | | | | |
|---|---|---|---|---|---|---|
| HTN | nk_cell | *LINC01050* | -7.089841548 | 0.001042359 | 0.008846261 | 1 |
| HTN | nk_cell | *GTF2H5* | -1.271521283 | 0.002438428 | 0.010654267 | 1 |
| HTN | nk_cell | *FGF14-AS2* | -1.427600037 | 0.011855521 | 0.029601136 | 1 |
| HTN | nk_cell | *LINC01023* | -1.525399803 | 0.00274296 | 0.011368608 | 1 |
| HTN | nk_cell | *DOC2B* | -0.761969435 | 0.002414432 | 0.010645574 | 1 |
| HTN | nk_cell | *POLR2J4* | -1.89207787 | 0.001519677 | 0.0089561 | 1 |
| HTN | nk_cell | *PAXIP1-AS1* | -1.550346326 | 0.003109713 | 0.012249854 | 1 |
| HTN | pericyte | *SEMA3F* | -1.999983448 | 9.12E-08 | 2.29E-05 | 0.00057307 |
| HTN | pericyte | *RBM6* | -1.254678633 | 3.78E-05 | 0.003011696 | 0.237923963 |
| HTN | pericyte | *COPZ2* | 2.290063151 | 6.36E-06 | 0.000741049 | 0.040016638 |
| HTN | pericyte | *MPND* | 4.523308409 | 0.000483721 | 0.017477881 | 1 |
| HTN | pericyte | *CEP68* | 4.427867105 | 1.37E-08 | 7.82E-06 | 8.60E-05 |
| HTN | pericyte | *TACC3* | 3.142366714 | 0.002258921 | 0.048141818 | 1 |
| HTN | pericyte | *NPC1L1* | 3.818480223 | 0.00185924 | 0.042816993 | 1 |
| HTN | pericyte | *NR1H3* | -4.968579794 | 2.32E-06 | 0.000338676 | 0.014563057 |
| HTN | pericyte | *SNX1* | 3.88214369 | 8.20E-05 | 0.004879994 | 0.515261928 |
| HTN | pericyte | *EPN3* | 8.596903257 | 0.001778034 | 0.041696949 | 1 |
| HTN | pericyte | *LIMA1* | 1.635722123 | 5.99E-06 | 0.000736734 | 0.037660523 |
| HTN | pericyte | *SDCCAG8* | 4.562388262 | 1.62E-09 | 1.27E-06 | 1.02E-05 |
| HTN | pericyte | *ITIH4* | 1.113737479 | 0.000261964 | 0.011517273 | 1 |
| HTN | pericyte | *PITHD1* | 3.637260338 | 0.000761127 | 0.023456887 | 1 |
| HTN | pericyte | *HAGH* | 2.50551809 | 0.000311501 | 0.012888322 | 1 |
| HTN | pericyte | *UHRF1BP1* | 2.262228508 | 5.18E-08 | 1.55E-05 | 0.000325486 |
| HTN | pericyte | *TIE1* | -3.139641428 | 7.29E-06 | 0.00083283 | 0.045805625 |
| HTN | pericyte | *ZNRD1* | 1.909367833 | 0.002082165 | 0.045946679 | 1 |
| HTN | pericyte | *ASNS* | 2.829886022 | 0.001609731 | 0.039074833 | 1 |
| HTN | pericyte | *SMARCE1* | 2.990690345 | 0.001713267 | 0.040646453 | 1 |
| HTN | pericyte | *CLNS1A* | 3.28103532 | 1.30E-06 | 0.000209131 | 0.00815609 |
| HTN | pericyte | *ATP2A3* | 1.48362207 | 0.001034132 | 0.027943661 | 1 |
| HTN | pericyte | *NAALAD2* | 2.151176598 | 0.002049638 | 0.045946679 | 1 |
| HTN | pericyte | *SCAMP1* | 4.16433226 | 0.001577303 | 0.038736341 | 1 |
| HTN | pericyte | *MAST2* | 2.931974347 | 0.001426189 | 0.036597765 | 1 |
| HTN | pericyte | *SNX10* | -0.819973188 | 4.00E-05 | 0.003027209 | 0.251258379 |
| HTN | pericyte | *AAMDC* | -1.643731724 | 0.00037001 | 0.01435959 | 1 |
| HTN | pericyte | *C3orf18* | 2.201043818 | 0.000353402 | 0.013973834 | 1 |
| HTN | pericyte | *YPEL3* | 2.201041928 | 2.67E-05 | 0.002295465 | 0.167568937 |
| HTN | pericyte | *GOLGA3* | 2.79713068 | 0.001694209 | 0.040346562 | 1 |
| HTN | pericyte | *ALKBH5* | 3.481952499 | 0.000395987 | 0.01508831 | 1 |
| HTN | pericyte | *NUP50* | 3.141965171 | 0.00095718 | 0.0269856 | 1 |
| HTN | pericyte | *FMO2* | 2.281516647 | 0.00100193 | 0.027627779 | 1 |
| HTN | pericyte | *WAC* | 3.665523192 | 0.002082838 | 0.045946679 | 1 |
| HTN | pericyte | *MARCH2* | 5.04070957 | 9.41E-06 | 0.001034772 | 0.059141657 |
| HTN | pericyte | *GCAT* | 2.835572231 | 0.000191514 | 0.009053012 | 1 |
| HTN | pericyte | *MTG2* | 2.262460539 | 0.001021066 | 0.027868916 | 1 |
| HTN | pericyte | *MANBAL* | 1.656775683 | 0.000941989 | 0.02667697 | 1 |
| HTN | pericyte | *PXMP4* | 3.789684789 | 0.000631376 | 0.020461148 | 1 |
| HTN | pericyte | *DNTTIP1* | -2.505571661 | 0.00089676 | 0.026321271 | 1 |
| HTN | pericyte | *POLI* | -0.849755733 | 0.000360595 | 0.014081128 | 1 |
| HTN | pericyte | *VWA8* | 3.918956687 | 0.000295842 | 0.012482925 | 1 |
| HTN | pericyte | *MGRN1* | 3.153580851 | 0.000234621 | 0.010536179 | 1 |

| | | | | | | |
|---|---|---|---|---|---|---|
| HTN | pericyte | *CRISPLD2* | 1.867035471 | 0.000150784 | 0.007553196 | 0.947978923 |
| HTN | pericyte | *NARFL* | 2.899003818 | 0.00209754 | 0.046109215 | 1 |
| HTN | pericyte | *MTHFSD* | 1.821740216 | 8.58E-05 | 0.004993087 | 0.539253383 |
| HTN | pericyte | *KAT8* | 7.468173306 | 1.46E-05 | 0.001459842 | 0.091837364 |
| HTN | pericyte | *TRIP4* | 2.9107781 | 0.000461949 | 0.016787719 | 1 |
| HTN | pericyte | *ZFAND1* | 3.259480746 | 0.000108269 | 0.005943943 | 0.680686382 |
| HTN | pericyte | *EYA1* | 1.312732971 | 4.98E-05 | 0.003554831 | 0.3128251 |
| HTN | pericyte | *SNX16* | 3.098584705 | 0.001273529 | 0.033361155 | 1 |
| HTN | pericyte | *PTPRS* | 2.23856442 | 0.000867891 | 0.025737876 | 1 |
| HTN | pericyte | *ATP13A1* | 4.09361861 | 2.87E-05 | 0.002439334 | 0.180510704 |
| HTN | pericyte | *EPHB6* | 4.301419432 | 0.000242798 | 0.010826038 | 1 |
| HTN | pericyte | *MEST* | -1.904797361 | 0.00017496 | 0.008445183 | 1 |
| HTN | pericyte | *ACTR3C* | 3.004439696 | 2.51E-05 | 0.002223309 | 0.157854936 |
| HTN | pericyte | *ICAM2* | 4.59291436 | 7.62E-05 | 0.004840533 | 0.479212737 |
| HTN | pericyte | *DDX5* | 2.954263442 | 1.47E-05 | 0.001459842 | 0.092107197 |
| HTN | pericyte | *CYTH1* | 3.540295272 | 0.000276692 | 0.011914807 | 1 |
| HTN | pericyte | *CWH43* | 14.36544943 | 0.001050614 | 0.028227395 | 1 |
| HTN | pericyte | *SLC2A9* | 5.19408603 | 0.000781577 | 0.023969646 | 1 |
| HTN | pericyte | *KCTD10* | -2.483373593 | 0.000323759 | 0.013227084 | 1 |
| HTN | pericyte | *PRR4* | 2.179296399 | 0.001463971 | 0.037112846 | 1 |
| HTN | pericyte | *ENDOU* | -2.29698837 | 0.000791122 | 0.024144573 | 1 |
| HTN | pericyte | *CAND1* | -3.697038081 | 0.000357448 | 0.014045463 | 1 |
| HTN | pericyte | *ALDH5A1* | 2.082083197 | 0.001946791 | 0.044345916 | 1 |
| HTN | pericyte | *CEP72* | 2.23586563 | 0.00040648 | 0.015394802 | 1 |
| HTN | pericyte | *PCDHB6* | 4.545344219 | 0.0005432 | 0.018560303 | 1 |
| HTN | pericyte | *NPHP3* | 1.10032844 | 0.001527319 | 0.038255992 | 1 |
| HTN | pericyte | *PCCB* | 4.136426846 | 2.45E-05 | 0.002200352 | 0.154024608 |
| HTN | pericyte | *PRKAR2A* | 2.328356521 | 0.002211716 | 0.047457543 | 1 |
| HTN | pericyte | *CEBPZ* | 3.42745241 | 0.00214641 | 0.046622834 | 1 |
| HTN | pericyte | *PRDX1* | 4.692101568 | 0.00054283 | 0.018560303 | 1 |
| HTN | pericyte | *NSUN4* | 4.504249114 | 0.000720754 | 0.022770741 | 1 |
| HTN | pericyte | *TCF21* | 15.22457304 | 0.000145036 | 0.007504753 | 0.91183963 |
| HTN | pericyte | *TRPM6* | 2.160326931 | 0.000219702 | 0.01015638 | 1 |
| HTN | pericyte | *HSDL2* | 1.703767629 | 0.00091101 | 0.026321271 | 1 |
| HTN | pericyte | *FKBP1B* | 1.972685335 | 0.001654649 | 0.039554294 | 1 |
| HTN | pericyte | *PCDHB10* | 1.970719817 | 0.001739934 | 0.041123924 | 1 |
| HTN | pericyte | *ZFP30* | 2.953474888 | 7.82E-05 | 0.004879994 | 0.491594768 |
| HTN | pericyte | *ACVR1C* | 1.718835029 | 0.000439093 | 0.016133988 | 1 |
| HTN | pericyte | *FAM124B* | 2.780861097 | 9.55E-06 | 0.001034772 | 0.060016792 |
| HTN | pericyte | *PEPD* | 2.749121554 | 0.000727372 | 0.022864944 | 1 |
| HTN | pericyte | *BTN2A2* | 1.369505603 | 0.000210513 | 0.009803669 | 1 |
| HTN | pericyte | *SNRPC* | -2.976600198 | 4.00E-05 | 0.003027209 | 0.251240131 |
| HTN | pericyte | *HIST1H1D* | 4.122867466 | 4.44E-08 | 1.51E-05 | 0.000279192 |
| HTN | pericyte | *ZNF391* | -0.96429556 | 0.000292066 | 0.01240687 | 1 |
| HTN | pericyte | *GRK4* | 1.022176709 | 7.94E-05 | 0.004879994 | 0.499399309 |
| HTN | pericyte | *MRPS7* | 3.732566775 | 0.001885861 | 0.04327157 | 1 |
| HTN | pericyte | *OPRL1* | -3.604121837 | 6.68E-08 | 1.82E-05 | 0.00041969 |
| HTN | pericyte | *XRCC3* | 3.970645137 | 0.000612656 | 0.020300292 | 1 |
| HTN | pericyte | *TRAP1* | 1.646242468 | 6.44E-08 | 1.82E-05 | 0.000404886 |
| HTN | pericyte | *RHOT1* | 2.970592771 | 0.000352318 | 0.013973834 | 1 |

| | | | | | | |
|---|---|---|---|---|---|---|
| HTN | pericyte | *YEATS4* | 2.034451241 | 0.000898817 | 0.026321271 | 1 |
| HTN | pericyte | *APOL5* | -5.839373762 | 0.001784075 | 0.041696949 | 1 |
| HTN | pericyte | *ORMDL1* | 1.786906782 | 0.002120488 | 0.046289965 | 1 |
| HTN | pericyte | *CDC16* | 3.809167908 | 3.87E-05 | 0.003027209 | 0.243544032 |
| HTN | pericyte | *ATP8B3* | -1.613669528 | 0.000827302 | 0.02476785 | 1 |
| HTN | pericyte | *PGPEP1* | 3.798295738 | 0.000112134 | 0.006077476 | 0.704987247 |
| HTN | pericyte | *CHMP1A* | 5.306369522 | 7.27E-07 | 0.000124636 | 0.004570773 |
| HTN | pericyte | *HAUS8* | -7.397590368 | 1.70E-05 | 0.001641505 | 0.106697849 |
| HTN | pericyte | *PDLIM4* | 4.016108175 | 4.92E-07 | 9.38E-05 | 0.003095094 |
| HTN | pericyte | *EXOC4* | 3.404847182 | 0.00062622 | 0.020461148 | 1 |
| HTN | pericyte | *MRPL35* | 1.839374728 | 0.00157131 | 0.038736341 | 1 |
| HTN | pericyte | *RSG1* | -3.743391741 | 0.002193822 | 0.047397104 | 1 |
| HTN | pericyte | *IMPA1* | 2.584653167 | 0.000108397 | 0.005943943 | 0.681494323 |
| HTN | pericyte | *WNT2B* | -4.497660541 | 0.000129815 | 0.006916495 | 0.816146369 |
| HTN | pericyte | *DDB2* | 5.202626425 | 7.34E-07 | 0.000124636 | 0.004611516 |
| HTN | pericyte | *ACP2* | 4.799166825 | 3.10E-12 | 9.76E-09 | 1.95E-08 |
| HTN | pericyte | *TMEM165* | 4.628764811 | 5.24E-06 | 0.000690371 | 0.032941842 |
| HTN | pericyte | *TMEM60* | 3.888214196 | 0.000820381 | 0.024678174 | 1 |
| HTN | pericyte | *BLOC1S1* | 4.916950447 | 0.000423598 | 0.015852132 | 1 |
| HTN | pericyte | *AGT* | 1.197925777 | 0.000108725 | 0.005943943 | 0.68355339 |
| HTN | pericyte | *USP37* | 4.452925551 | 0.002117823 | 0.046289965 | 1 |
| HTN | pericyte | *MRPS9* | 2.187571548 | 0.002081542 | 0.045946679 | 1 |
| HTN | pericyte | *NACAD* | -8.017270091 | 0.001441146 | 0.036682132 | 1 |
| HTN | pericyte | *NMT1* | 3.562380526 | 1.68E-10 | 1.91E-07 | 1.05E-06 |
| HTN | pericyte | *SLC2A8* | 3.836816188 | 0.000349997 | 0.013973834 | 1 |
| HTN | pericyte | *VARS2* | 0.678265155 | 0.001166235 | 0.030937222 | 1 |
| HTN | pericyte | *FDX1* | 4.772312015 | 0.002049912 | 0.045946679 | 1 |
| HTN | pericyte | *RMDN3* | 7.458197935 | 2.13E-07 | 4.62E-05 | 0.001339365 |
| HTN | pericyte | *IFT172* | 5.984631282 | 8.28E-05 | 0.004879994 | 0.520534372 |
| HTN | pericyte | *DYNC2LI1* | 2.879844797 | 0.000209591 | 0.009803669 | 1 |
| HTN | pericyte | *ARL3* | 6.006802542 | 1.29E-05 | 0.001333052 | 0.081316152 |
| HTN | pericyte | *G3BP2* | 3.693798896 | 0.000377151 | 0.014458213 | 1 |
| HTN | pericyte | *FBN2* | 0.921785688 | 0.000146565 | 0.007504753 | 0.921454455 |
| HTN | pericyte | *CD27* | 3.007121848 | 0.002252656 | 0.048141818 | 1 |
| HTN | pericyte | *MMAB* | 3.599622363 | 5.95E-05 | 0.004056445 | 0.374053976 |
| HTN | pericyte | *TICRR* | 2.640625227 | 0.002351928 | 0.049453412 | 1 |
| HTN | pericyte | *DEF8* | 3.82336536 | 6.09E-06 | 0.000736734 | 0.038310172 |
| HTN | pericyte | *MYO19* | 5.006590832 | 0.000554874 | 0.018856704 | 1 |
| HTN | pericyte | *SCRN2* | -0.861911775 | 0.000576144 | 0.01937016 | 1 |
| HTN | pericyte | *RERE* | 3.459847581 | 0.000441394 | 0.016133988 | 1 |
| HTN | pericyte | *CREB3L4* | -2.388148657 | 0.001546624 | 0.038433308 | 1 |
| HTN | pericyte | *CEP170* | 6.130977981 | 6.68E-07 | 0.000119978 | 0.004199234 |
| HTN | pericyte | *WNT9A* | 4.281400875 | 1.06E-05 | 0.001132328 | 0.066807375 |
| HTN | pericyte | *PLEKHA6* | 4.226785705 | 0.000227387 | 0.010308814 | 1 |
| HTN | pericyte | *ZC3H8* | 3.18713709 | 0.001560534 | 0.038626284 | 1 |
| HTN | pericyte | *SLC10A4* | 8.37146338 | 6.13E-05 | 0.004056445 | 0.385362293 |
| HTN | pericyte | *PSD2* | 9.119510491 | 0.001023973 | 0.027868916 | 1 |
| HTN | pericyte | *ZMAT2* | 2.32040089 | 0.001336629 | 0.034724748 | 1 |
| HTN | pericyte | *RPL7L1* | 2.889206472 | 0.001071295 | 0.028660565 | 1 |
| HTN | pericyte | *CCDC25* | 4.433440092 | 0.000282897 | 0.012099131 | 1 |

| | | | | | | |
|---|---|---|---|---|---|---|
| HTN | pericyte | *NRBF2* | -2.632960709 | 0.000104099 | 0.005896146 | 0.654472176 |
| HTN | pericyte | *PDCD11* | 11.09552295 | 9.84E-16 | 6.18E-12 | 6.18E-12 |
| HTN | pericyte | *HSD17B12* | -2.449571704 | 3.05E-08 | 1.28E-05 | 0.000191665 |
| HTN | pericyte | *TTC12* | 1.315423944 | 0.001916375 | 0.043811813 | 1 |
| HTN | pericyte | *ARID5B* | 3.795518219 | 0.000151376 | 0.007553196 | 0.951702707 |
| HTN | pericyte | *CWF19L2* | 5.310006514 | 8.25E-05 | 0.004879994 | 0.518703766 |
| HTN | pericyte | *NEIL2* | 3.818291069 | 4.30E-09 | 3.01E-06 | 2.71E-05 |
| HTN | pericyte | *PROM2* | 6.655696518 | 0.001008754 | 0.027694493 | 1 |
| HTN | pericyte | *AK9* | 4.315573475 | 0.000505915 | 0.017912053 | 1 |
| HTN | pericyte | *NDUFAF6* | -4.165152474 | 0.000150473 | 0.007553196 | 0.946025948 |
| HTN | pericyte | *SFXN2* | 1.891397107 | 0.000252348 | 0.011172608 | 1 |
| HTN | pericyte | *VPS8* | 3.819067626 | 0.000168046 | 0.008253971 | 1 |
| HTN | pericyte | *CLEC18A* | -4.253691874 | 3.12E-06 | 0.000446157 | 0.019630894 |
| HTN | pericyte | *TRIM63* | 2.555808054 | 3.05E-05 | 0.002522785 | 0.191731622 |
| HTN | pericyte | *SPATA2L* | 5.079565245 | 0.000325828 | 0.013227084 | 1 |
| HTN | pericyte | *ZNF276* | 4.473142664 | 9.30E-05 | 0.005315644 | 0.58472088 |
| HTN | pericyte | *SLC5A11* | -1.292042946 | 2.33E-05 | 0.002126746 | 0.146745481 |
| HTN | pericyte | *MED8* | 3.543962527 | 9.50E-07 | 0.000157215 | 0.005974153 |
| HTN | pericyte | *ATAD3B* | 4.526913013 | 0.000804784 | 0.024325361 | 1 |
| HTN | pericyte | *ZNF66* | 4.320778134 | 0.001546508 | 0.038433308 | 1 |
| HTN | pericyte | *S100B* | -0.936751229 | 0.000166737 | 0.008253971 | 1 |
| HTN | pericyte | *FDPS* | 4.435592716 | 0.000266189 | 0.011621748 | 1 |
| HTN | pericyte | *PMF1* | 4.453533616 | 8.31E-05 | 0.004879994 | 0.522159406 |
| HTN | pericyte | *ABCF3* | 2.569760425 | 4.58E-05 | 0.003349015 | 0.288015282 |
| HTN | pericyte | *TNFSF13* | 6.906286555 | 2.17E-08 | 1.14E-05 | 0.000136258 |
| HTN | pericyte | *LMOD1* | -1.70222282 | 5.82E-06 | 0.000731937 | 0.03659685 |
| HTN | pericyte | *FAM208A* | 4.833952677 | 2.60E-05 | 0.002272832 | 0.163643923 |
| HTN | pericyte | *EXO5* | -4.096210597 | 4.95E-05 | 0.003554831 | 0.311367351 |
| HTN | pericyte | *C1orf50* | -3.632776181 | 0.00027113 | 0.011755833 | 1 |
| HTN | pericyte | *CDC25A* | -3.898878604 | 4.90E-10 | 4.40E-07 | 3.08E-06 |
| HTN | pericyte | *ZNF589* | -2.835868763 | 3.13E-08 | 1.28E-05 | 0.000197005 |
| HTN | pericyte | *APEH* | -3.548258863 | 3.47E-08 | 1.28E-05 | 0.000218056 |
| HTN | pericyte | *RNF123* | 3.915744524 | 1.00E-11 | 2.10E-08 | 6.31E-08 |
| HTN | pericyte | *ERAP1* | -2.943167256 | 5.83E-05 | 0.004030906 | 0.366812467 |
| HTN | pericyte | *ERAP2* | 1.542456662 | 0.000758857 | 0.023456887 | 1 |
| HTN | pericyte | *CHMP4C* | -1.230658481 | 0.000644469 | 0.020756381 | 1 |
| HTN | pericyte | *BRI3* | 3.657196053 | 0.002064352 | 0.045946679 | 1 |
| HTN | pericyte | *C9orf64* | 1.79930648 | 0.00118426 | 0.03128338 | 1 |
| HTN | pericyte | *ZNF367* | 3.498049077 | 0.000227919 | 0.010308814 | 1 |
| HTN | pericyte | *LARP6* | 3.058140835 | 0.00141847 | 0.036548859 | 1 |
| HTN | pericyte | *C2* | 2.902254228 | 0.001595287 | 0.038874312 | 1 |
| HTN | pericyte | *TRIM66* | -0.698877228 | 0.000438827 | 0.016133988 | 1 |
| HTN | pericyte | *RIMKLB* | -1.923918754 | 0.000136658 | 0.007219927 | 0.859171271 |
| HTN | pericyte | *ZNF641* | 2.254180353 | 0.000188585 | 0.008982077 | 1 |
| HTN | pericyte | *SRR* | 1.64899608 | 1.25E-05 | 0.001304558 | 0.078273459 |
| HTN | pericyte | *TMC8* | 2.18719619 | 0.001524582 | 0.038255992 | 1 |
| HTN | pericyte | *CYP7A1* | 5.168629101 | 7.50E-05 | 0.004811001 | 0.471478129 |
| HTN | pericyte | *HRASLS5* | 0.848594332 | 0.000105151 | 0.005902564 | 0.661087186 |
| HTN | pericyte | *TMEM129* | 2.445911689 | 0.000629504 | 0.020461148 | 1 |
| HTN | pericyte | *IRS1* | -0.98842915 | 3.93E-05 | 0.003027209 | 0.247339073 |

| | | | | | | |
|---|---|---|---|---|---|---|
| HTN | pericyte | *C8orf46* | 0.413534067 | 0.002278165 | 0.048387913 | 1 |
| HTN | pericyte | *INO80E* | -5.011535094 | 4.75E-06 | 0.000649584 | 0.029880846 |
| HTN | pericyte | *LIMS1* | 3.2927206 | 5.53E-05 | 0.003863004 | 0.347670384 |
| HTN | pericyte | *CALML6* | -3.551790735 | 0.00163595 | 0.039406965 | 1 |
| HTN | pericyte | *GIMAP8* | 2.630526794 | 0.000528817 | 0.018284581 | 1 |
| HTN | pericyte | *RGS19* | -1.272490913 | 0.001520817 | 0.038255992 | 1 |
| HTN | pericyte | *NME6* | 3.674735963 | 6.34E-11 | 9.97E-08 | 3.99E-07 |
| HTN | pericyte | *CTSW* | -0.594205908 | 0.000750069 | 0.023453415 | 1 |
| HTN | pericyte | *ZNF738* | 2.506199266 | 0.000173622 | 0.008445183 | 1 |
| HTN | pericyte | *HOXC5* | 3.456288363 | 0.000143121 | 0.007498336 | 0.899800355 |
| HTN | pericyte | *SNX32* | 2.568311173 | 0.000845963 | 0.025206491 | 1 |
| HTN | pericyte | *HPSE2* | 1.863111573 | 0.000968916 | 0.027194528 | 1 |
| HTN | pericyte | *HECTD4* | 4.071169947 | 0.000499441 | 0.017840836 | 1 |
| HTN | pericyte | *AHSA2* | 3.367904803 | 0.000529313 | 0.018284581 | 1 |
| HTN | pericyte | *VEGFB* | -2.605718572 | 3.35E-05 | 0.002715193 | 0.210712956 |
| HTN | pericyte | *MST1* | -1.510274518 | 0.000527173 | 0.018284581 | 1 |
| HTN | pericyte | *GMPPB* | 3.073860623 | 0.001255659 | 0.03303066 | 1 |
| HTN | pericyte | *CCS* | 3.48390318 | 0.000418063 | 0.015738702 | 1 |
| HTN | pericyte | *NRROS* | 1.578894448 | 0.000222206 | 0.010197163 | 1 |
| HTN | pericyte | *TLR6* | 3.56531401 | 5.47E-06 | 0.000702185 | 0.034407085 |
| HTN | pericyte | *RGMB* | 3.795312756 | 0.001984558 | 0.045043016 | 1 |
| HTN | pericyte | *TP53I11* | 5.202805208 | 0.002350091 | 0.049453412 | 1 |
| HTN | pericyte | *LRRC25* | 1.340022244 | 0.000591351 | 0.019775652 | 1 |
| HTN | pericyte | *CTDNEP1* | 7.458140764 | 5.17E-05 | 0.003649463 | 0.324802242 |
| HTN | pericyte | *CRLF3* | 4.174865469 | 0.000980768 | 0.027372518 | 1 |
| HTN | pericyte | *ZBTB38* | 4.028252889 | 0.002207208 | 0.047457543 | 1 |
| HTN | pericyte | *CD151* | 4.126825135 | 0.000753553 | 0.023453415 | 1 |
| HTN | pericyte | *PCDHB9* | 2.458137443 | 0.000516762 | 0.018150186 | 1 |
| HTN | pericyte | *PRSS42* | 3.322634012 | 0.002074333 | 0.045946679 | 1 |
| HTN | pericyte | *RPP25* | 3.556027929 | 4.56E-08 | 1.51E-05 | 0.000286974 |
| HTN | pericyte | *MPI* | -7.476842778 | 8.93E-08 | 2.29E-05 | 0.000561404 |
| HTN | pericyte | *HYI* | -1.624953812 | 4.52E-05 | 0.003349015 | 0.28400456 |
| HTN | pericyte | *TUFM* | 2.431268224 | 0.000617406 | 0.020322694 | 1 |
| HTN | pericyte | *CTC1* | 2.652108992 | 7.04E-05 | 0.004564725 | 0.442778328 |
| HTN | pericyte | *ZNF443* | 3.23480947 | 1.90E-05 | 0.001756688 | 0.11945476 |
| HTN | pericyte | *TMEM136* | 1.783092327 | 0.001035609 | 0.027943661 | 1 |
| HTN | pericyte | *CLDN7* | -0.718889513 | 0.000563925 | 0.019061255 | 1 |
| HTN | pericyte | *FAM89A* | 2.321823236 | 0.000311599 | 0.012888322 | 1 |
| HTN | pericyte | *CLN8* | 5.534625821 | 1.18E-07 | 2.85E-05 | 0.000741019 |
| HTN | pericyte | *FES* | -6.128483424 | 0.001078762 | 0.028738029 | 1 |
| HTN | pericyte | *MFSD5* | 3.334885882 | 0.001809136 | 0.042015652 | 1 |
| HTN | pericyte | *ZNF320* | 1.20466912 | 6.43E-05 | 0.004210828 | 0.404239515 |
| HTN | pericyte | *RP1L1* | 4.854235824 | 3.28E-08 | 1.28E-05 | 0.000206111 |
| HTN | pericyte | *EIF3C* | 2.316738282 | 2.92E-05 | 0.002448973 | 0.18367295 |
| HTN | pericyte | *A3GALT2* | -2.311811559 | 1.86E-05 | 0.001756688 | 0.117016125 |
| HTN | pericyte | *DDX51* | 2.249963875 | 8.11E-05 | 0.004879994 | 0.510178067 |
| HTN | pericyte | *CDK10* | -2.682358697 | 0.000376098 | 0.014458213 | 1 |
| HTN | pericyte | *SP1* | 3.004113402 | 0.001768111 | 0.041633385 | 1 |
| HTN | pericyte | *ZNF829* | 3.238240916 | 0.000924612 | 0.026321271 | 1 |
| HTN | pericyte | *WDR86* | 2.362126269 | 0.001397367 | 0.036153266 | 1 |

| | | | | | | |
|---|---|---|---|---|---|---|
| HTN | pericyte | *PLEKHN1* | 3.339158179 | 8.13E-05 | 0.004879994 | 0.511272608 |
| HTN | pericyte | *MCRS1* | 4.763497781 | 4.70E-07 | 9.23E-05 | 0.002953135 |
| HTN | pericyte | *ZNF793* | -1.423914199 | 0.000983965 | 0.027372518 | 1 |
| HTN | pericyte | *GOLGA8M* | 4.622664695 | 0.000911577 | 0.026321271 | 1 |
| HTN | pericyte | *ZDHHC11* | 3.495779473 | 4.53E-05 | 0.003349015 | 0.284762638 |
| HTN | pericyte | *FBF1* | -1.71275533 | 5.27E-06 | 0.000690371 | 0.033137816 |
| HTN | pericyte | *FAM47E* | 1.224717963 | 0.00099034 | 0.027428495 | 1 |
| HTN | pericyte | *HLA-DRB1* | -2.974170245 | 1.30E-07 | 3.02E-05 | 0.000814986 |
| HTN | pericyte | *ZNF34* | 5.958823911 | 0.000925243 | 0.026321271 | 1 |
| HTN | pericyte | *C6orf106* | 2.861191676 | 2.69E-07 | 5.45E-05 | 0.001689197 |
| HTN | pericyte | *HIST1H3E* | 3.628042461 | 5.03E-08 | 1.55E-05 | 0.000316089 |
| HTN | pericyte | *SULT1A2* | 1.086599392 | 2.05E-06 | 0.000314038 | 0.012875572 |
| HTN | pericyte | *ZNF165* | 3.172957209 | 0.000613497 | 0.020300292 | 1 |
| HTN | pericyte | *DDI2* | 3.280255102 | 0.00131568 | 0.034322326 | 1 |
| HTN | pericyte | *SLC22A5* | 2.656205709 | 1.21E-08 | 7.62E-06 | 7.62E-05 |
| HTN | pericyte | *RPS26* | -1.373294443 | 1.49E-05 | 0.001459842 | 0.093429896 |
| HTN | pericyte | *RPS6KL1* | 1.679403723 | 0.000352223 | 0.013973834 | 1 |
| HTN | pericyte | *TMEM116* | -2.211917167 | 2.06E-07 | 4.62E-05 | 0.001296085 |
| HTN | pericyte | *HLA-DRB5* | -0.670243874 | 0.000301686 | 0.012644654 | 1 |
| HTN | pericyte | *ANKRD13B* | 1.941599162 | 0.000647089 | 0.020756381 | 1 |
| HTN | pericyte | *TGM2* | 0.930753718 | 0.001811077 | 0.042015652 | 1 |
| HTN | pericyte | *PCMTD2* | 3.737188847 | 0.001643774 | 0.039444312 | 1 |
| HTN | pericyte | *HSD17B8* | -3.374667965 | 3.82E-06 | 0.000533061 | 0.023987747 |
| HTN | pericyte | *HLA-DRA* | 2.025505261 | 0.00090818 | 0.026321271 | 1 |
| HTN | pericyte | *PRRT1* | 2.267177345 | 0.000799612 | 0.024285799 | 1 |
| HTN | pericyte | *MRPL38* | 2.791771965 | 0.000175969 | 0.008445183 | 1 |
| HTN | pericyte | *STK19* | 3.408645698 | 1.89E-05 | 0.001756688 | 0.119119588 |
| HTN | pericyte | *C6orf48* | 5.136641227 | 1.94E-06 | 0.000305218 | 0.012208734 |
| HTN | pericyte | *GPANK1* | 4.385855679 | 6.05E-07 | 0.000111925 | 0.003805455 |
| HTN | pericyte | *BAG6* | 4.508605768 | 2.51E-07 | 5.25E-05 | 0.001575805 |
| HTN | pericyte | *NFKBIL1* | 2.564240237 | 0.000432087 | 0.016074154 | 1 |
| HTN | pericyte | *POU5F1* | -0.687035462 | 0.001433157 | 0.036627058 | 1 |
| HTN | pericyte | *PSORS1C1* | 0.388620453 | 0.001589039 | 0.038872727 | 1 |
| HTN | pericyte | *EIF3CL* | 7.234015319 | 6.02E-05 | 0.004056445 | 0.378720686 |
| HTN | pericyte | *FGFR1OP* | 4.069160971 | 0.00201428 | 0.045553167 | 1 |
| HTN | pericyte | *NOTCH2NL* | 2.132724933 | 0.000655992 | 0.020935148 | 1 |
| HTN | pericyte | *MXD3* | -2.126633761 | 0.000488032 | 0.01753288 | 1 |
| HTN | pericyte | *SIPA1* | 6.439742223 | 2.67E-08 | 1.28E-05 | 0.000167552 |
| HTN | pericyte | *ATF6B* | 4.102435066 | 1.83E-10 | 1.91E-07 | 1.15E-06 |
| HTN | pericyte | *CLIC1* | 4.94565762 | 0.001819104 | 0.042046711 | 1 |
| HTN | pericyte | *PPM1N* | 3.572510731 | 0.000704147 | 0.022358433 | 1 |
| HTN | pericyte | *LTB4R* | -2.036826237 | 0.000918297 | 0.026321271 | 1 |
| HTN | pericyte | *GALT* | 3.758746771 | 0.000507133 | 0.017912053 | 1 |
| HTN | pericyte | *WEE2* | 2.390248244 | 0.000924261 | 0.026321271 | 1 |
| HTN | pericyte | *UBXN2B* | 2.707540507 | 0.002150568 | 0.046622834 | 1 |
| HTN | pericyte | *AC016773.1* | 1.490011323 | 6.12E-05 | 0.004056445 | 0.384817402 |
| HTN | pericyte | *HLA-DPB1* | 2.423753836 | 0.00012589 | 0.006764683 | 0.791467869 |
| HTN | pericyte | *HLA-B* | 2.1341453 | 0.000326101 | 0.013227084 | 1 |
| HTN | pericyte | *HLA-DQA2* | 1.57651855 | 6.36E-06 | 0.000741049 | 0.04000849 |
| HTN | pericyte | *LINC00960* | -0.932164352 | 0.001631255 | 0.039406965 | 1 |

| | | | | | | |
|---|---|---|---|---|---|---|
| HTN | pericyte | *PLA2G4B* | 2.356931327 | 9.25E-05 | 0.005315644 | 0.581607085 |
| HTN | pericyte | *JMJD7* | 2.42229161 | 0.000146824 | 0.007504753 | 0.923084559 |
| HTN | pericyte | *ZNF10* | -3.561286959 | 8.70E-06 | 0.000977093 | 0.054717207 |
| HTN | pericyte | *HMBS* | 2.558912135 | 2.20E-06 | 0.000328804 | 0.013809755 |
| HTN | pericyte | *CUX1* | 2.821048093 | 0.002336003 | 0.049449325 | 1 |
| HTN | pericyte | *AC144652.1* | 2.129706902 | 3.37E-05 | 0.002715193 | 0.211785041 |
| HTN | SMC | *SEMA3F* | -1.56063817 | 5.65E-08 | 1.37E-05 | 0.00035492 |
| HTN | SMC | *M6PR* | -3.518848804 | 5.95E-05 | 0.00328279 | 0.374238107 |
| HTN | SMC | *RBM6* | -1.040376603 | 6.41E-07 | 8.22E-05 | 0.004028184 |
| HTN | SMC | *COPZ2* | 1.224060543 | 0.001230568 | 0.031577893 | 1 |
| HTN | SMC | *GGNBP2* | -2.244005829 | 0.000123103 | 0.005690788 | 0.773947174 |
| HTN | SMC | *CD79B* | -1.643375773 | 0.001893133 | 0.04020989 | 1 |
| HTN | SMC | *ST7L* | -1.051858188 | 0.001241383 | 0.031597481 | 1 |
| HTN | SMC | *DLEC1* | -3.282051231 | 0.001040956 | 0.027924153 | 1 |
| HTN | SMC | *HFE* | -2.769741548 | 4.74E-08 | 1.24E-05 | 0.000298012 |
| HTN | SMC | *CLCN6* | -6.407813026 | 1.06E-07 | 2.30E-05 | 0.000668193 |
| HTN | SMC | *TTC19* | -2.413610256 | 0.001369212 | 0.033108611 | 1 |
| HTN | SMC | *CEP68* | 2.116176212 | 0.000115144 | 0.005402296 | 0.723907616 |
| HTN | SMC | *NR1H3* | -3.488037024 | 9.25E-07 | 0.000107651 | 0.005813168 |
| HTN | SMC | *PHLPP2* | -2.767912472 | 0.001902447 | 0.040271665 | 1 |
| HTN | SMC | *PQLC2* | -6.40765946 | 0.001858082 | 0.039869484 | 1 |
| HTN | SMC | *SDCCAG8* | 1.920140419 | 0.000201002 | 0.008342799 | 1 |
| HTN | SMC | *PIGV* | -2.059272639 | 0.000391416 | 0.013747672 | 1 |
| HTN | SMC | *UHRF1BP1* | 1.289488662 | 2.40E-05 | 0.00167753 | 0.150977686 |
| HTN | SMC | *TIE1* | -2.458368794 | 1.52E-07 | 2.90E-05 | 0.000956113 |
| HTN | SMC | *ATXN3* | -1.834575599 | 0.001032974 | 0.027924153 | 1 |
| HTN | SMC | *POLR1A* | -1.195583237 | 6.52E-05 | 0.003535323 | 0.410097446 |
| HTN | SMC | *NDST1* | -4.018336274 | 0.000468158 | 0.015739633 | 1 |
| HTN | SMC | *EPN2* | -2.837326514 | 0.001687352 | 0.037259806 | 1 |
| HTN | SMC | *ACADVL* | -3.967334858 | 0.000255957 | 0.009872392 | 1 |
| HTN | SMC | *DERL2* | -4.497834222 | 0.001571205 | 0.035661241 | 1 |
| HTN | SMC | *MARK3* | -3.591202652 | 2.51E-05 | 0.001730766 | 0.157499716 |
| HTN | SMC | *DDX43* | -2.94973577 | 0.000505208 | 0.01645723 | 1 |
| HTN | SMC | *CCNT2* | -2.16669609 | 3.06E-05 | 0.002022425 | 0.192130386 |
| HTN | SMC | *COL16A1* | -2.208683709 | 0.000282972 | 0.010652975 | 1 |
| HTN | SMC | *SNX10* | -0.697458836 | 5.80E-07 | 7.75E-05 | 0.003643434 |
| HTN | SMC | *AAMDC* | -1.442695431 | 4.89E-06 | 0.000452221 | 0.030751055 |
| HTN | SMC | *C3orf18* | 2.602233402 | 0.002177601 | 0.043880066 | 1 |
| HTN | SMC | *YPEL3* | 1.152226565 | 0.001251756 | 0.031665961 | 1 |
| HTN | SMC | *MUL1* | -3.10725407 | 0.001780557 | 0.038734821 | 1 |
| HTN | SMC | *PABPC4* | -3.329220353 | 2.87E-07 | 4.47E-05 | 0.001803941 |
| HTN | SMC | *TGM1* | 1.356846611 | 0.000952662 | 0.026384958 | 1 |
| HTN | SMC | *SLC7A4* | -3.73968309 | 0.00029269 | 0.010888399 | 1 |
| HTN | SMC | *TRIOBP* | -1.808076363 | 0.001779836 | 0.038734821 | 1 |
| HTN | SMC | *PPM1A* | -2.768632351 | 0.000645796 | 0.019805447 | 1 |
| HTN | SMC | *PRPF6* | -3.379743394 | 0.00249351 | 0.048384866 | 1 |
| HTN | SMC | *DNTTIP1* | -1.876786501 | 0.000119435 | 0.00556211 | 0.750884906 |
| HTN | SMC | *POLI* | -0.668856884 | 4.46E-05 | 0.002654735 | 0.280361406 |
| HTN | SMC | *CORO1A* | -2.250819355 | 0.001668564 | 0.037259806 | 1 |
| HTN | SMC | *ZNF500* | -1.693295293 | 0.000790886 | 0.023565392 | 1 |

| | | | | | | |
|---|---|---|---|---|---|---|
| HTN | SMC | *UBE2I* | -3.601839374 | 0.001097815 | 0.029122196 | 1 |
| HTN | SMC | *RNF40* | -3.754048354 | 0.001526436 | 0.035543348 | 1 |
| HTN | SMC | *EYA1* | 0.904284426 | 0.001447216 | 0.034334527 | 1 |
| HTN | SMC | *CARD8* | -2.917454029 | 0.000951396 | 0.026384958 | 1 |
| HTN | SMC | *TNPO2* | -3.282377986 | 0.002121644 | 0.043075535 | 1 |
| HTN | SMC | *ATP13A1* | 1.907964825 | 0.001285558 | 0.032072628 | 1 |
| HTN | SMC | *HOXA2* | -3.692717368 | 0.001379583 | 0.033231554 | 1 |
| HTN | SMC | *TAF6* | -3.070727312 | 0.000422138 | 0.014537432 | 1 |
| HTN | SMC | *ZNHIT1* | -1.680995503 | 0.002549544 | 0.049018304 | 1 |
| HTN | SMC | *MEST* | -0.981999757 | 0.000833962 | 0.024050991 | 1 |
| HTN | SMC | *OGN* | -3.44381017 | 4.68E-05 | 0.002725577 | 0.294362332 |
| HTN | SMC | *ECM2* | -2.678514341 | 7.40E-05 | 0.003846861 | 0.465470233 |
| HTN | SMC | *TRDMT1* | -3.651672374 | 0.001182331 | 0.030843622 | 1 |
| HTN | SMC | *RPL28* | -2.059737924 | 0.000460085 | 0.015551367 | 1 |
| HTN | SMC | *WNT3* | -5.368804896 | 0.000100532 | 0.004861871 | 0.632043193 |
| HTN | SMC | *RAD51C* | -1.48782211 | 4.20E-05 | 0.002573544 | 0.264047849 |
| HTN | SMC | *SYNGR2* | -2.782818552 | 6.74E-05 | 0.003588458 | 0.423438028 |
| HTN | SMC | *C17orf75* | -4.058506376 | 0.002045898 | 0.042311061 | 1 |
| HTN | SMC | *ELP4* | -2.999629014 | 0.001550228 | 0.035661241 | 1 |
| HTN | SMC | *PPFIBP1* | -2.718413399 | 0.000526813 | 0.017072557 | 1 |
| HTN | SMC | *KCTD10* | -1.767357767 | 0.000106451 | 0.005108837 | 0.66925759 |
| HTN | SMC | *ENDOU* | -1.903669211 | 0.001547506 | 0.035661241 | 1 |
| HTN | SMC | *CAND1* | -3.730730491 | 1.75E-07 | 3.09E-05 | 0.001102049 |
| HTN | SMC | *COPS7A* | -2.702296579 | 0.001453263 | 0.034348366 | 1 |
| HTN | SMC | *RWDD1* | -2.426465991 | 0.002044429 | 0.042311061 | 1 |
| HTN | SMC | *MAPK14* | -3.022233601 | 0.000718865 | 0.021728372 | 1 |
| HTN | SMC | *NNT* | -1.526046721 | 0.00037398 | 0.013209069 | 1 |
| HTN | SMC | *PCDHB6* | 2.606499384 | 0.002407657 | 0.047155579 | 1 |
| HTN | SMC | *NIT2* | -4.459859044 | 0.000158646 | 0.006878663 | 0.997406188 |
| HTN | SMC | *FAM162A* | -3.513964525 | 0.000202275 | 0.008342799 | 1 |
| HTN | SMC | *HYAL1* | -1.250042735 | 0.001561519 | 0.035661241 | 1 |
| HTN | SMC | *ABCC5* | -2.82533319 | 0.00186612 | 0.039905776 | 1 |
| HTN | SMC | *SLC35F5* | -2.988805023 | 0.000337658 | 0.012087948 | 1 |
| HTN | SMC | *TP53I3* | -4.59817593 | 1.67E-05 | 0.001296318 | 0.105001756 |
| HTN | SMC | *DNAJC27* | -2.421977017 | 5.39E-05 | 0.003001157 | 0.339130716 |
| HTN | SMC | *RTN4* | -2.043011344 | 0.000219494 | 0.008845906 | 1 |
| HTN | SMC | *MRPL19* | -2.743751816 | 0.002123973 | 0.043075535 | 1 |
| HTN | SMC | *ZNF142* | -3.628787731 | 3.05E-05 | 0.002022425 | 0.191949778 |
| HTN | SMC | *CAPZA1* | -2.828892778 | 0.001187713 | 0.030855988 | 1 |
| HTN | SMC | *C1orf21* | -1.854231462 | 4.47E-05 | 0.002654735 | 0.28113487 |
| HTN | SMC | *IRF6* | -0.966715719 | 0.002189746 | 0.043983814 | 1 |
| HTN | SMC | *CD3EAP* | -1.955262407 | 0.001238457 | 0.031597481 | 1 |
| HTN | SMC | *DDX59* | -2.867193537 | 0.00036702 | 0.013036481 | 1 |
| HTN | SMC | *HDHD3* | -0.487454987 | 0.001827291 | 0.039478275 | 1 |
| HTN | SMC | *HELLS* | -3.039827851 | 0.000538587 | 0.017301877 | 1 |
| HTN | SMC | *EPHX2* | -2.263144041 | 1.41E-05 | 0.001153664 | 0.088832146 |
| HTN | SMC | *TEX14* | 4.198832344 | 3.11E-06 | 0.000301143 | 0.019574321 |
| HTN | SMC | *TRIM6* | -1.741013215 | 0.002224608 | 0.044541746 | 1 |
| HTN | SMC | *CBX3* | -4.178034017 | 1.77E-05 | 0.001358045 | 0.111359705 |
| HTN | SMC | *OPTN* | -2.468140691 | 0.002538654 | 0.048958645 | 1 |

| | | | | | | |
|---|---|---|---|---|---|---|
| HTN | SMC | *ATF1* | -3.688032224 | 0.000338393 | 0.012087948 | 1 |
| HTN | SMC | *IKZF4* | -5.743453627 | 3.57E-06 | 0.000339956 | 0.022437068 |
| HTN | SMC | *FAM124B* | 1.458552882 | 0.000802939 | 0.023589149 | 1 |
| HTN | SMC | *TOX2* | -0.620708414 | 0.001418317 | 0.033893511 | 1 |
| HTN | SMC | *SNRPC* | -2.690554896 | 8.16E-08 | 1.83E-05 | 0.000512727 |
| HTN | SMC | *HIST1H1D* | 2.861832092 | 7.26E-07 | 9.13E-05 | 0.004564424 |
| HTN | SMC | *ZNF391* | -0.687181037 | 9.09E-05 | 0.00453381 | 0.571260106 |
| HTN | SMC | *GRK4* | 0.598807465 | 0.001335455 | 0.032542663 | 1 |
| HTN | SMC | *OPRL1* | -2.764393234 | 3.31E-07 | 4.96E-05 | 0.002081834 |
| HTN | SMC | *PRDX5* | -5.209085131 | 0.000588806 | 0.018235571 | 1 |
| HTN | SMC | *TRAP1* | 0.96268477 | 5.43E-06 | 0.000494307 | 0.034107192 |
| HTN | SMC | *KLHDC10* | -3.82365313 | 8.93E-06 | 0.000779625 | 0.056132986 |
| HTN | SMC | *EIF2AK4* | -1.933213637 | 0.000808386 | 0.023638713 | 1 |
| HTN | SMC | *ATP8B3* | -1.122297406 | 0.000688374 | 0.02090728 | 1 |
| HTN | SMC | *DDA1* | -4.179869398 | 0.00012882 | 0.005884661 | 0.809889803 |
| HTN | SMC | *MAP3K10* | -3.186040604 | 0.001677856 | 0.037259806 | 1 |
| HTN | SMC | *HAUS8* | -6.140254583 | 3.85E-08 | 1.10E-05 | 0.000241746 |
| HTN | SMC | *PDLIM4* | 3.562125711 | 4.30E-06 | 0.000403163 | 0.027011941 |
| HTN | SMC | *IMMT* | -2.327526484 | 0.002618444 | 0.049885322 | 1 |
| HTN | SMC | *KDM6B* | -5.214207197 | 2.36E-05 | 0.001665888 | 0.148264074 |
| HTN | SMC | *DSTYK* | -3.036605494 | 0.00168211 | 0.037259806 | 1 |
| HTN | SMC | *WNT2B* | -4.042346631 | 1.63E-07 | 3.02E-05 | 0.001025611 |
| HTN | SMC | *FST* | -3.206712089 | 0.001300027 | 0.032178235 | 1 |
| HTN | SMC | *ACP2* | 6.309582403 | 1.61E-15 | 1.01E-11 | 1.01E-11 |
| HTN | SMC | *FADS2* | -4.626314757 | 0.001043769 | 0.027924153 | 1 |
| HTN | SMC | *POM121C* | -3.690178706 | 1.50E-05 | 0.001179142 | 0.094331393 |
| HTN | SMC | *AGT* | 0.783977648 | 0.000206886 | 0.008435387 | 1 |
| HTN | SMC | *NMT1* | 1.953437137 | 2.86E-07 | 4.47E-05 | 0.001798014 |
| HTN | SMC | *FPGS* | -4.37706031 | 7.28E-05 | 0.003816574 | 0.457988825 |
| HTN | SMC | *NDUFAF1* | -2.091103707 | 0.001537247 | 0.035661241 | 1 |
| HTN | SMC | *ATRAID* | -3.456220222 | 0.001323488 | 0.032432167 | 1 |
| HTN | SMC | *ASCC1* | -2.355314361 | 0.001462078 | 0.034411501 | 1 |
| HTN | SMC | *FBN2* | 0.584556007 | 0.000437357 | 0.014943836 | 1 |
| HTN | SMC | *GLTP* | -5.430073702 | 9.27E-06 | 0.000787459 | 0.058271956 |
| HTN | SMC | *LMBR1L* | -3.997364473 | 0.000557618 | 0.017795657 | 1 |
| HTN | SMC | *VPS53* | -2.832206421 | 0.000165738 | 0.007136963 | 1 |
| HTN | SMC | *SCRN2* | -0.6512564 | 0.000147489 | 0.006576335 | 0.927263245 |
| HTN | SMC | *C18orf8* | -4.049489797 | 2.32E-06 | 0.000235645 | 0.01460997 |
| HTN | SMC | *NPC1* | -4.076587053 | 8.42E-06 | 0.000745368 | 0.05292114 |
| HTN | SMC | *TRIM65* | -1.916276779 | 0.00148765 | 0.034768972 | 1 |
| HTN | SMC | *RPS8* | -2.521205747 | 0.00207223 | 0.042575529 | 1 |
| HTN | SMC | *NME7* | -3.259065234 | 0.000822267 | 0.023862417 | 1 |
| HTN | SMC | *ARNT* | -2.93052388 | 0.000267937 | 0.010271447 | 1 |
| HTN | SMC | *RAB13* | -8.061816851 | 1.88E-07 | 3.10E-05 | 0.001179347 |
| HTN | SMC | *CREB3L4* | -2.304687776 | 1.44E-05 | 0.001162678 | 0.090718926 |
| HTN | SMC | *RABL2A* | -3.022245215 | 0.000500252 | 0.016380637 | 1 |
| HTN | SMC | *CTDSPL* | -2.7026919 | 0.000840741 | 0.024057181 | 1 |
| HTN | SMC | *AMT* | -0.818104689 | 0.001156703 | 0.030300789 | 1 |
| HTN | SMC | *CENPC* | -2.252235972 | 0.000313454 | 0.011524463 | 1 |
| HTN | SMC | *SLC10A4* | 5.753063585 | 4.12E-05 | 0.002564372 | 0.259001567 |

| | | | | | | |
|---|---|---|---|---|---|---|
| HTN | SMC | *SLC18B1* | -0.889721675 | 8.68E-05 | 0.00436322 | 0.545402438 |
| HTN | SMC | *C7orf50* | -3.703904867 | 0.001325762 | 0.032432167 | 1 |
| HTN | SMC | *SLC25A37* | -3.533757667 | 0.001036041 | 0.027924153 | 1 |
| HTN | SMC | *ADHFE1* | -0.667870775 | 0.000870204 | 0.024533512 | 1 |
| HTN | SMC | *NRBF2* | -1.711502385 | 3.03E-05 | 0.002022425 | 0.190578246 |
| HTN | SMC | *PDCD11* | 5.9583651 | 6.95E-09 | 2.73E-06 | 4.37E-05 |
| HTN | SMC | *HSD17B12* | -1.880860059 | 5.40E-10 | 4.24E-07 | 3.39E-06 |
| HTN | SMC | *EIF3M* | -2.633688852 | 0.002030021 | 0.042260744 | 1 |
| HTN | SMC | *PTPRJ* | -2.478270377 | 0.000823627 | 0.023862417 | 1 |
| HTN | SMC | *C11orf49* | -2.912684221 | 0.001213731 | 0.031402162 | 1 |
| HTN | SMC | *DUSP15* | -3.174009453 | 9.42E-05 | 0.004627214 | 0.592283367 |
| HTN | SMC | *DOC2A* | -4.117071185 | 8.17E-07 | 9.69E-05 | 0.005136921 |
| HTN | SMC | *KCTD14* | -7.083480311 | 0.00125924 | 0.031667373 | 1 |
| HTN | SMC | *ARL14EP* | -3.480437788 | 4.88E-05 | 0.002790835 | 0.306543063 |
| HTN | SMC | *CAST* | -2.097299075 | 0.000152886 | 0.006704619 | 0.961194928 |
| HTN | SMC | *JAZF1* | -2.837833382 | 0.000489131 | 0.01610036 | 1 |
| HTN | SMC | *NEIL2* | 2.294982924 | 7.66E-07 | 9.44E-05 | 0.004814673 |
| HTN | SMC | *FAM69A* | -2.359379116 | 0.001254147 | 0.031665961 | 1 |
| HTN | SMC | *NDUFAF6* | -4.216113004 | 3.57E-08 | 1.07E-05 | 0.000224724 |
| HTN | SMC | *CLEC18A* | -2.758741872 | 1.94E-05 | 0.001470662 | 0.12206494 |
| HTN | SMC | *TRIM63* | 1.375914519 | 0.001138276 | 0.030068666 | 1 |
| HTN | SMC | *SLC5A11* | -1.103412098 | 2.91E-07 | 4.47E-05 | 0.001831747 |
| HTN | SMC | *UBE2Z* | -4.379417306 | 0.000423151 | 0.014537432 | 1 |
| HTN | SMC | *MED8* | 2.102081568 | 0.000480557 | 0.016070537 | 1 |
| HTN | SMC | *S100B* | -0.67715784 | 5.05E-05 | 0.00286181 | 0.317660936 |
| HTN | SMC | *ABCF3* | 1.911826792 | 2.36E-05 | 0.001665888 | 0.148097434 |
| HTN | SMC | *TNFSF13* | 4.283463597 | 4.34E-07 | 6.06E-05 | 0.002728519 |
| HTN | SMC | *SENP3* | -5.056164537 | 0.001284818 | 0.032072628 | 1 |
| HTN | SMC | *ACP6* | -2.178461564 | 0.001886727 | 0.04020966 | 1 |
| HTN | SMC | *RFT1* | -3.492869439 | 3.69E-05 | 0.002389616 | 0.231792775 |
| HTN | SMC | *GNL3* | -3.87138988 | 0.000801012 | 0.023589149 | 1 |
| HTN | SMC | *EXO5* | -2.436235992 | 0.000107319 | 0.005111493 | 0.674717054 |
| HTN | SMC | *C1orf50* | -4.93004358 | 1.04E-05 | 0.000870194 | 0.065264584 |
| HTN | SMC | *BDH2* | -1.589493527 | 0.000135614 | 0.006090021 | 0.852602945 |
| HTN | SMC | *CDC25A* | -2.422992591 | 2.14E-09 | 1.12E-06 | 1.34E-05 |
| HTN | SMC | *ZNF589* | -2.322577494 | 3.16E-11 | 4.96E-08 | 1.98E-07 |
| HTN | SMC | *ATRIP* | -17.99424562 | 0.000238139 | 0.009357384 | 1 |
| HTN | SMC | *APEH* | -2.584571576 | 1.78E-07 | 3.09E-05 | 0.001121947 |
| HTN | SMC | *RNF123* | 2.433739861 | 6.52E-10 | 4.56E-07 | 4.10E-06 |
| HTN | SMC | *C4orf3* | -3.916775444 | 0.001694974 | 0.037259806 | 1 |
| HTN | SMC | *ERAP1* | -2.913226708 | 5.22E-09 | 2.19E-06 | 3.28E-05 |
| HTN | SMC | *RPS14* | -3.420099471 | 0.00058337 | 0.018235571 | 1 |
| HTN | SMC | *REEP3* | -4.252105708 | 1.25E-07 | 2.54E-05 | 0.00078679 |
| HTN | SMC | *STK32C* | -4.012999722 | 0.002000078 | 0.041914975 | 1 |
| HTN | SMC | *CEP57* | -3.280299936 | 3.80E-05 | 0.002437385 | 0.238863709 |
| HTN | SMC | *TRMT61A* | -3.770044095 | 1.20E-07 | 2.51E-05 | 0.000753757 |
| HTN | SMC | *XRRA1* | -1.461159382 | 7.20E-05 | 0.003802303 | 0.452474042 |
| HTN | SMC | *TRIM66* | -0.588207154 | 2.21E-05 | 0.001619126 | 0.139235132 |
| HTN | SMC | *RIMKLB* | -2.04953251 | 1.31E-08 | 4.33E-06 | 8.22E-05 |
| HTN | SMC | *SAAL1* | -4.117425754 | 0.001570636 | 0.035661241 | 1 |

| | | | | | | |
|---|---|---|---|---|---|---|
| HTN | SMC | *SMAD3* | -2.244986933 | 0.002306904 | 0.045897173 | 1 |
| HTN | SMC | *SP2* | -3.886307661 | 0.002568019 | 0.049144116 | 1 |
| HTN | SMC | *KIR3DL1* | 2.705694447 | 0.000111923 | 0.005290654 | 0.703657006 |
| HTN | SMC | *UBXN6* | -3.003255063 | 0.001041398 | 0.027924153 | 1 |
| HTN | SMC | *SRR* | 0.965168707 | 0.000486929 | 0.01610036 | 1 |
| HTN | SMC | *CYP7A1* | 2.85193196 | 0.001680532 | 0.037259806 | 1 |
| HTN | SMC | *HRASLS5* | 0.485096986 | 0.001151397 | 0.030288009 | 1 |
| HTN | SMC | *PXK* | -2.175360098 | 0.00169408 | 0.037259806 | 1 |
| HTN | SMC | *ATXN2L* | -3.624809266 | 0.001772229 | 0.038734821 | 1 |
| HTN | SMC | *TNIP2* | -1.97400253 | 0.001413921 | 0.033893511 | 1 |
| HTN | SMC | *CEP120* | -3.231681995 | 0.002086754 | 0.042734278 | 1 |
| HTN | SMC | *IRS1* | -1.117666033 | 1.76E-09 | 1.00E-06 | 1.10E-05 |
| HTN | SMC | *C8orf46* | 0.312311215 | 0.000841829 | 0.024057181 | 1 |
| HTN | SMC | *PTAFR* | -2.450132393 | 0.002399817 | 0.047148911 | 1 |
| HTN | SMC | *INO80E* | -4.811366495 | 1.71E-10 | 1.79E-07 | 1.08E-06 |
| HTN | SMC | *CALML6* | -2.861716004 | 0.000223252 | 0.008883447 | 1 |
| HTN | SMC | *ELP5* | -6.728361501 | 8.27E-09 | 3.06E-06 | 5.20E-05 |
| HTN | SMC | *ARL6IP1* | -2.497263846 | 0.002524773 | 0.048840759 | 1 |
| HTN | SMC | *RNF34* | -3.816950854 | 2.23E-05 | 0.001619126 | 0.140486832 |
| HTN | SMC | *TTLL6* | -24.23138601 | 0.000149387 | 0.006614067 | 0.939197471 |
| HTN | SMC | *PAQR8* | -6.17898119 | 7.87E-07 | 9.52E-05 | 0.004950247 |
| HTN | SMC | *RGS19* | -3.025954265 | 1.99E-13 | 6.26E-10 | 1.25E-09 |
| HTN | SMC | *NME6* | 2.266846831 | 1.41E-08 | 4.42E-06 | 8.85E-05 |
| HTN | SMC | *CTSW* | -0.490284644 | 4.08E-05 | 0.002563748 | 0.256374838 |
| HTN | SMC | *SLFN11* | -2.180797616 | 0.002335558 | 0.046320681 | 1 |
| HTN | SMC | *HOXC5* | 1.912142105 | 0.002449909 | 0.047834087 | 1 |
| HTN | SMC | *DCAKD* | -1.092019412 | 0.000232787 | 0.009204604 | 1 |
| HTN | SMC | *VEGFB* | -2.506993135 | 9.84E-09 | 3.44E-06 | 6.18E-05 |
| HTN | SMC | *MST1* | -1.524375726 | 1.73E-06 | 0.000190908 | 0.010881758 |
| HTN | SMC | *TLR6* | 1.945525353 | 0.001794862 | 0.038911379 | 1 |
| HTN | SMC | *CTU2* | -2.322111726 | 0.0010405 | 0.027924153 | 1 |
| HTN | SMC | *SNX31* | -1.175991466 | 0.002353756 | 0.046534787 | 1 |
| HTN | SMC | *GLIS1* | -1.041137837 | 0.000584157 | 0.018235571 | 1 |
| HTN | SMC | *RALGAPA1* | -2.232592207 | 0.002238618 | 0.044679977 | 1 |
| HTN | SMC | *RSRC1* | -3.332340959 | 4.20E-08 | 1.15E-05 | 0.000264268 |
| HTN | SMC | *NRIP3* | -2.495903297 | 2.20E-05 | 0.001619126 | 0.138383009 |
| HTN | SMC | *PPP2R2D* | -3.223487865 | 2.50E-06 | 0.000249193 | 0.015699131 |
| HTN | SMC | *MSRA* | -3.883840909 | 7.81E-05 | 0.003991232 | 0.49092158 |
| HTN | SMC | *MTHFR* | -7.187774987 | 1.29E-10 | 1.62E-07 | 8.08E-07 |
| HTN | SMC | *WDR73* | -2.362548303 | 0.001360298 | 0.033020062 | 1 |
| HTN | SMC | *TOP3A* | -3.531777529 | 0.000207966 | 0.008435387 | 1 |
| HTN | SMC | *PPFIA3* | -2.149812912 | 0.000418623 | 0.014537432 | 1 |
| HTN | SMC | *SH2B1* | -1.52193728 | 3.30E-05 | 0.002161778 | 0.207530723 |
| HTN | SMC | *COX14* | -2.623890615 | 1.13E-05 | 0.000938317 | 0.071312109 |
| HTN | SMC | *P4HTM* | -2.910216532 | 0.001067259 | 0.02843159 | 1 |
| HTN | SMC | *RPP25* | 2.675383295 | 2.58E-09 | 1.16E-06 | 1.62E-05 |
| HTN | SMC | *FAM219B* | -3.175825563 | 0.000221907 | 0.008883447 | 1 |
| HTN | SMC | *MPI* | -4.810954329 | 2.52E-09 | 1.16E-06 | 1.59E-05 |
| HTN | SMC | *TRIM73* | -3.051950342 | 0.000775542 | 0.023329355 | 1 |
| HTN | SMC | *HYI* | -1.540904019 | 6.84E-08 | 1.59E-05 | 0.000430114 |

| | | | | | | |
|---|---|---|---|---|---|---|
| HTN | SMC | *DEFB124* | -6.62976222 | 4.88E-05 | 0.002790835 | 0.306991805 |
| HTN | SMC | *CHST13* | -8.364269904 | 0.001466881 | 0.034411501 | 1 |
| HTN | SMC | *ZNF443* | 2.182554498 | 0.000416374 | 0.014537432 | 1 |
| HTN | SMC | *CLDN7* | -3.043341543 | 1.30E-09 | 8.18E-07 | 8.18E-06 |
| HTN | SMC | *IDH2* | -3.886603674 | 0.000642276 | 0.019794078 | 1 |
| HTN | SMC | *CLN8* | 2.217620828 | 0.002056032 | 0.042381232 | 1 |
| HTN | SMC | *FES* | -4.174932213 | 0.000335301 | 0.012087948 | 1 |
| HTN | SMC | *ZNF320* | 0.72013863 | 0.000588237 | 0.018235571 | 1 |
| HTN | SMC | *GAS6* | -3.634977418 | 6.20E-05 | 0.003391088 | 0.389975142 |
| HTN | SMC | *RP1L1* | 3.374119867 | 1.11E-06 | 0.000124335 | 0.006962756 |
| HTN | SMC | *BMP8A* | 3.479994456 | 0.001003943 | 0.02768328 | 1 |
| HTN | SMC | *KREMEN1* | -0.785399057 | 0.000129169 | 0.005884661 | 0.812083246 |
| HTN | SMC | *EIF3C* | 1.876413289 | 1.97E-06 | 0.000210303 | 0.012407868 |
| HTN | SMC | *A3GALT2* | -1.63993286 | 9.12E-06 | 0.000785861 | 0.057367851 |
| HTN | SMC | *LPAR5* | -1.470332384 | 0.001291679 | 0.032097958 | 1 |
| HTN | SMC | *ATL3* | -2.499266977 | 0.001955634 | 0.041120631 | 1 |
| HTN | SMC | *CDK10* | -2.607492566 | 5.39E-07 | 7.37E-05 | 0.003389373 |
| HTN | SMC | *TCN2* | -1.233293293 | 0.001423236 | 0.033893511 | 1 |
| HTN | SMC | *MYBL1* | -1.128984601 | 0.000446292 | 0.015166686 | 1 |
| HTN | SMC | *ARL17A* | -2.405259686 | 0.000131205 | 0.005934435 | 0.824886464 |
| HTN | SMC | *SMYD4* | -2.394586603 | 0.000649291 | 0.019815994 | 1 |
| HTN | SMC | *HYAL3* | -1.053420995 | 0.002571722 | 0.049144116 | 1 |
| HTN | SMC | *PLEKHN1* | 2.126707352 | 0.001562716 | 0.035661241 | 1 |
| HTN | SMC | *MCRS1* | 3.581689308 | 0.000153565 | 0.006704619 | 0.965465108 |
| HTN | SMC | *ZNF793* | -1.03760553 | 0.000288497 | 0.010796319 | 1 |
| HTN | SMC | *CENPP* | -0.988280602 | 0.002394474 | 0.047148911 | 1 |
| HTN | SMC | *ZNF559* | -0.870373658 | 0.000951425 | 0.026384958 | 1 |
| HTN | SMC | *FBF1* | -1.223775561 | 9.68E-07 | 0.000110707 | 0.006088878 |
| HTN | SMC | *NOC2L* | -1.265966841 | 0.000271869 | 0.010359025 | 1 |
| HTN | SMC | *HLA-DRB1* | -2.432565484 | 2.30E-10 | 2.06E-07 | 1.44E-06 |
| HTN | SMC | *SULT1A1* | -1.496543752 | 7.71E-05 | 0.003971534 | 0.484527174 |
| HTN | SMC | *MYO6* | -2.880382474 | 0.000188959 | 0.008026935 | 1 |
| HTN | SMC | *FAM180B* | -2.341127514 | 1.42E-07 | 2.79E-05 | 0.000892964 |
| HTN | SMC | *C6orf106* | 1.425315055 | 0.000328537 | 0.011939386 | 1 |
| HTN | SMC | *ANKRD36B* | -2.348126766 | 0.001229307 | 0.031577893 | 1 |
| HTN | SMC | *AP2A1* | -2.828150672 | 0.000539394 | 0.017301877 | 1 |
| HTN | SMC | *HIST1H3E* | 2.3842378 | 5.06E-08 | 1.27E-05 | 0.000318181 |
| HTN | SMC | *FUT11* | -4.524810639 | 4.59E-05 | 0.002698836 | 0.288775442 |
| HTN | SMC | *ZNF100* | -2.284375598 | 2.83E-06 | 0.000278051 | 0.017795286 |
| HTN | SMC | *IGF2R* | -2.904335879 | 0.00193633 | 0.040851366 | 1 |
| HTN | SMC | *SULT1A2* | 0.676177372 | 4.22E-05 | 0.002573544 | 0.265075077 |
| HTN | SMC | *SLC22A5* | 1.66476932 | 1.82E-07 | 3.09E-05 | 0.001144929 |
| HTN | SMC | *PCDHB11* | -4.223134312 | 0.000584219 | 0.018235571 | 1 |
| HTN | SMC | *MYO5A* | -1.954548539 | 0.001314021 | 0.032397056 | 1 |
| HTN | SMC | *RPS26* | -1.030998253 | 2.20E-06 | 0.000226557 | 0.013819997 |
| HTN | SMC | *TMEM116* | -2.238943619 | 1.27E-12 | 2.67E-09 | 8.01E-09 |
| HTN | SMC | *HLA-DRB5* | -0.466969751 | 0.000242658 | 0.009475709 | 1 |
| HTN | SMC | *NCOA6* | -2.848442948 | 0.000275863 | 0.010447905 | 1 |
| HTN | SMC | *PNP* | -0.674997838 | 0.001840068 | 0.039618183 | 1 |
| HTN | SMC | *DENND4B* | -4.713447453 | 3.89E-07 | 5.68E-05 | 0.002443964 |

| | | | | | | |
|---|---|---|---|---|---|---|
| HTN | SMC | *AL590822.1* | -1.537294486 | 0.000191236 | 0.008069141 | 1 |
| HTN | SMC | *HSD17B8* | -2.408636354 | 2.08E-06 | 0.000217753 | 0.013065156 |
| HTN | SMC | *HLA-DMA* | -3.319971128 | 0.001609745 | 0.036324175 | 1 |
| HTN | SMC | *PRRT1* | 1.542984508 | 0.000484724 | 0.01610036 | 1 |
| HTN | SMC | *STK19* | 2.206099497 | 8.34E-05 | 0.004226082 | 0.524034166 |
| HTN | SMC | *GPANK1* | 2.113396668 | 0.000879125 | 0.024674361 | 1 |
| HTN | SMC | *BAG6* | 2.11466399 | 0.00020303 | 0.008342799 | 1 |
| HTN | SMC | *HLA-C* | -1.358250296 | 0.0001802 | 0.007706914 | 1 |
| HTN | SMC | *CCHCR1* | -0.769373625 | 0.002022845 | 0.042251247 | 1 |
| HTN | SMC | *ATXN2* | -3.865702708 | 1.84E-06 | 0.000199177 | 0.01155227 |
| HTN | SMC | *LNP1* | -5.243492767 | 6.60E-05 | 0.003545416 | 0.414813632 |
| HTN | SMC | *SLC48A1* | -4.273439961 | 0.001611968 | 0.036324175 | 1 |
| HTN | SMC | *NOTCH2NL* | 1.295992748 | 0.002493327 | 0.048384866 | 1 |
| HTN | SMC | *MXD3* | -1.899938494 | 0.000302924 | 0.011202825 | 1 |
| HTN | SMC | *SIPA1* | 3.028935803 | 1.46E-05 | 0.001162678 | 0.091851544 |
| HTN | SMC | *ATF6B* | 2.603324485 | 4.14E-07 | 5.91E-05 | 0.002602549 |
| HTN | SMC | *LTB4R* | -1.748888495 | 2.56E-05 | 0.001747942 | 0.160810626 |
| HTN | SMC | *LTB4R2* | -2.532995871 | 0.000244881 | 0.009503514 | 1 |
| HTN | SMC | *REPIN1* | -3.370410889 | 9.19E-05 | 0.004547753 | 0.577564637 |
| HTN | SMC | *PPME1* | -1.876338876 | 0.000321038 | 0.011734685 | 1 |
| HTN | SMC | *AC016773.1* | 0.920708622 | 0.000856431 | 0.02425398 | 1 |
| HTN | SMC | *AP4M1* | -1.055039606 | 0.001021493 | 0.027924153 | 1 |
| HTN | SMC | *EXOSC6* | -3.262779106 | 4.48E-05 | 0.002654735 | 0.281401924 |
| HTN | SMC | *AL049840.1* | -2.106876457 | 6.24E-06 | 0.000560205 | 0.039214377 |
| HTN | SMC | *SLC26A6* | 2.883439865 | 0.000785775 | 0.023524621 | 1 |
| HTN | SMC | *SLC2A1-AS1* | -3.961417333 | 2.24E-05 | 0.001619126 | 0.14086398 |
| HTN | SMC | *PROB1* | 5.698342362 | 0.000569421 | 0.018080569 | 1 |
| HTN | SMC | *SNHG15* | -3.807985916 | 0.002095295 | 0.042769878 | 1 |
| HTN | SMC | *HLA-DQA2* | 0.975071116 | 5.38E-05 | 0.003001157 | 0.338209666 |
| HTN | SMC | *PLA2G4B* | 1.524231712 | 0.000202911 | 0.008342799 | 1 |
| HTN | SMC | *JMJD7* | 1.469028194 | 0.000794778 | 0.023569654 | 1 |
| HTN | SMC | *ZNF10* | -2.686992312 | 6.21E-07 | 8.14E-05 | 0.003906683 |
| HTN | SMC | *HMBS* | 1.466358581 | 9.95E-05 | 0.004850355 | 0.62569576 |
| HTN | SMC | *CEP95* | -3.031236854 | 0.002151517 | 0.043493847 | 1 |
| HTN | SMC | *AC009060.1* | -6.578402987 | 3.89E-05 | 0.002472867 | 0.244813814 |
| HTN | SMC | *AC144652.1* | 1.173467956 | 0.000856167 | 0.02425398 | 1 |
| HTN | t_cell | *C1orf112* | -3.188799633 | 0.008593598 | 0.033247969 | 1 |
| HTN | t_cell | *NIPAL3* | -1.430254027 | 0.002939103 | 0.01807841 | 1 |
| HTN | t_cell | *SEMA3F* | -1.193094606 | 0.01312506 | 0.043870233 | 1 |
| HTN | t_cell | *HECW1* | 2.046880046 | 0.00162003 | 0.014238085 | 1 |
| HTN | t_cell | *TMEM176A* | 2.586727277 | 0.000857846 | 0.013695656 | 1 |
| HTN | t_cell | *M6PR* | -3.892883942 | 0.013932391 | 0.045645096 | 1 |
| HTN | t_cell | *CYP26B1* | 3.314823323 | 0.002740362 | 0.017634242 | 1 |
| HTN | t_cell | *ICA1* | 3.455869674 | 0.008623345 | 0.033313811 | 1 |
| HTN | t_cell | *RBM6* | -1.850129041 | 5.79E-06 | 0.000559733 | 0.036382613 |
| HTN | t_cell | *CIAPIN1* | 4.169283408 | 0.007906605 | 0.031601289 | 1 |
| HTN | t_cell | *FAM214B* | 3.946057356 | 0.006275055 | 0.027301917 | 1 |
| HTN | t_cell | *COPZ2* | 3.330352513 | 9.18E-07 | 0.000134263 | 0.00577333 |
| HTN | t_cell | *WDR54* | -2.576369126 | 0.003049822 | 0.018353849 | 1 |
| HTN | t_cell | *PDK2* | 3.571117193 | 0.002762246 | 0.01766657 | 1 |

| | | | | | | |
|---|---|---|---|---|---|---|
| HTN | t_cell | *GGNBP2* | -3.053909387 | 0.00493484 | 0.023746745 | 1 |
| HTN | t_cell | *ABCC8* | 9.76808123 | 0.002065904 | 0.015441262 | 1 |
| HTN | t_cell | *CCL18* | 2.224206633 | 0.001888779 | 0.015146366 | 1 |
| HTN | t_cell | *SYNRG* | -2.534970235 | 0.006395828 | 0.027525449 | 1 |
| HTN | t_cell | *SPATA20* | 2.980508036 | 0.004936694 | 0.023746745 | 1 |
| HTN | t_cell | *TNFRSF12A* | 4.90196802 | 0.003574717 | 0.019909231 | 1 |
| HTN | t_cell | *FARP2* | 2.635448526 | 0.004847443 | 0.023537205 | 1 |
| HTN | t_cell | *IFRD1* | 1.901264934 | 0.015019731 | 0.048128379 | 1 |
| HTN | t_cell | *ARHGAP44* | 3.573746761 | 0.005880266 | 0.026471702 | 1 |
| HTN | t_cell | *CEACAM21* | -0.524700537 | 0.001016414 | 0.013965754 | 1 |
| HTN | t_cell | *NOS2* | 13.51174998 | 0.003066746 | 0.018432728 | 1 |
| HTN | t_cell | *TRAPPC6A* | -3.667167363 | 0.000862947 | 0.013695656 | 1 |
| HTN | t_cell | *CD79B* | -3.4852829 | 0.001056175 | 0.013965754 | 1 |
| HTN | t_cell | *PAX6* | 2.15059894 | 0.010427108 | 0.037545949 | 1 |
| HTN | t_cell | *RPUSD1* | -8.20853748 | 0.002283641 | 0.016194559 | 1 |
| HTN | t_cell | *BAIAP3* | -4.948624878 | 0.001134843 | 0.013965754 | 1 |
| HTN | t_cell | *PIGQ* | 3.553213183 | 0.000592734 | 0.012118368 | 1 |
| HTN | t_cell | *MPND* | 5.959347038 | 0.000212524 | 0.006800219 | 1 |
| HTN | t_cell | *MGST1* | 1.616872275 | 0.013899066 | 0.045559661 | 1 |
| HTN | t_cell | *MMP25* | -1.275248701 | 0.002289178 | 0.016195448 | 1 |
| HTN | t_cell | *IL32* | -0.820658215 | 0.001442584 | 0.014126985 | 1 |
| HTN | t_cell | *PKD1* | 3.036005471 | 0.001947848 | 0.015192726 | 1 |
| HTN | t_cell | *MAPK8IP2* | 7.170796198 | 0.007613155 | 0.031016532 | 1 |
| HTN | t_cell | *POMT2* | 2.177682674 | 0.010259935 | 0.037347798 | 1 |
| HTN | t_cell | *CD4* | -1.859547326 | 0.001583825 | 0.014185269 | 1 |
| HTN | t_cell | *MKS1* | 5.876388773 | 0.009390616 | 0.035226014 | 1 |
| HTN | t_cell | *DPF1* | 4.508152954 | 0.002442372 | 0.016594315 | 1 |
| HTN | t_cell | *CEP68* | 4.946164179 | 7.79E-07 | 0.000119438 | 0.004896963 |
| HTN | t_cell | *TMEM159* | 2.885242685 | 0.001229797 | 0.013965754 | 1 |
| HTN | t_cell | *LTF* | 3.707863101 | 0.000181941 | 0.006250611 | 1 |
| HTN | t_cell | *MTMR11* | 1.176046226 | 0.010757703 | 0.038297665 | 1 |
| HTN | t_cell | *CCDC88C* | -0.763519028 | 0.003178747 | 0.018851478 | 1 |
| HTN | t_cell | *NPC1L1* | 6.186680423 | 9.42E-05 | 0.003973597 | 0.592066023 |
| HTN | t_cell | *CHDH* | 11.2355468 | 0.002540234 | 0.016935789 | 1 |
| HTN | t_cell | *IL20RA* | 3.517120007 | 0.001342099 | 0.013965754 | 1 |
| HTN | t_cell | *WWTR1* | 2.393969915 | 0.014538197 | 0.047089975 | 1 |
| HTN | t_cell | *CPS1* | 1.563520153 | 0.004011548 | 0.021084486 | 1 |
| HTN | t_cell | *NLRP2* | -1.458698601 | 0.002689005 | 0.017428634 | 1 |
| HTN | t_cell | *BIRC3* | -0.973091303 | 0.001404634 | 0.01399841 | 1 |
| HTN | t_cell | *TYMP* | 3.501772901 | 0.005721357 | 0.026005629 | 1 |
| HTN | t_cell | *RNASET2* | -3.522616481 | 0.002756133 | 0.017645427 | 1 |
| HTN | t_cell | *CD44* | -1.91643977 | 0.000806741 | 0.013695656 | 1 |
| HTN | t_cell | *BTN3A1* | -1.079878376 | 0.007577975 | 0.030916761 | 1 |
| HTN | t_cell | *SNX1* | 3.142309113 | 0.010125999 | 0.037022875 | 1 |
| HTN | t_cell | *ARNTL2* | 3.577760993 | 0.01177515 | 0.040837464 | 1 |
| HTN | t_cell | *ANK1* | -4.25045847 | 0.001545487 | 0.014185269 | 1 |
| HTN | t_cell | *FUT8* | -2.511242752 | 0.011471642 | 0.040157134 | 1 |
| HTN | t_cell | *SLC4A7* | -2.401811195 | 0.010196462 | 0.037183965 | 1 |
| HTN | t_cell | *MYOC* | 1.587145634 | 0.011094965 | 0.039209692 | 1 |
| HTN | t_cell | *ADSS* | -3.305636083 | 0.008424325 | 0.032896729 | 1 |

| | | | | | | |
|---|---|---|---|---|---|---|
| HTN | t_cell | *STAP1* | -4.643239987 | 0.00011471 | 0.004593514 | 0.721181665 |
| HTN | t_cell | *AGA* | -4.129153362 | 0.004000327 | 0.021081356 | 1 |
| HTN | t_cell | *DNAH5* | 3.870672599 | 0.003257439 | 0.019003909 | 1 |
| HTN | t_cell | *C6* | 1.586889279 | 0.00420691 | 0.021679382 | 1 |
| HTN | t_cell | *PQLC2* | 2.31332739 | 0.003638922 | 0.020086785 | 1 |
| HTN | t_cell | *INPP4A* | -2.203349076 | 0.000277027 | 0.007952839 | 1 |
| HTN | t_cell | *RAB27B* | -1.131440026 | 0.006855405 | 0.02882113 | 1 |
| HTN | t_cell | *TDP1* | -4.023404789 | 0.007809849 | 0.031354101 | 1 |
| HTN | t_cell | *RETSAT* | 1.758626757 | 0.002592935 | 0.017105755 | 1 |
| HTN | t_cell | *AP2S1* | 3.407692171 | 0.005666388 | 0.025834063 | 1 |
| HTN | t_cell | *LCP2* | -1.074379292 | 0.001024517 | 0.013965754 | 1 |
| HTN | t_cell | *DSG2* | 3.141345526 | 0.000947658 | 0.013965754 | 1 |
| HTN | t_cell | *ANO2* | 1.585439987 | 0.013241234 | 0.044069688 | 1 |
| HTN | t_cell | *H6PD* | 2.453268516 | 0.005468069 | 0.025304138 | 1 |
| HTN | t_cell | *UTS2* | 2.849758192 | 0.003497882 | 0.019705361 | 1 |
| HTN | t_cell | *RFC2* | 3.421116284 | 0.015759337 | 0.049613898 | 1 |
| HTN | t_cell | *ARHGEF5* | 4.394554425 | 0.007637446 | 0.031058618 | 1 |
| HTN | t_cell | *NFE2L3* | -2.967953517 | 0.00372724 | 0.020378085 | 1 |
| HTN | t_cell | *LIMA1* | 1.817910906 | 2.25E-06 | 0.000252267 | 0.014126945 |
| HTN | t_cell | *PTGER3* | 1.566277355 | 0.001861281 | 0.015083316 | 1 |
| HTN | t_cell | *CYBA* | -3.02893066 | 0.00518975 | 0.024403857 | 1 |
| HTN | t_cell | *THOC3* | -0.796978977 | 0.001452317 | 0.014156149 | 1 |
| HTN | t_cell | *ALX4* | 6.063769562 | 0.003627463 | 0.020082835 | 1 |
| HTN | t_cell | *AKR7A2* | 3.893402536 | 0.004196667 | 0.021679382 | 1 |
| HTN | t_cell | *MCF2L2* | -1.829073784 | 0.001626805 | 0.014244742 | 1 |
| HTN | t_cell | *LY75* | -2.319851024 | 0.003041007 | 0.018351697 | 1 |
| HTN | t_cell | *OPN3* | 2.272724227 | 0.001203979 | 0.013965754 | 1 |
| HTN | t_cell | *SDCCAG8* | 4.683359368 | 1.37E-07 | 3.19E-05 | 0.000861938 |
| HTN | t_cell | *PLEKHH1* | 2.844094976 | 0.000151245 | 0.005580753 | 0.950875199 |
| HTN | t_cell | *ATP9A* | 1.690043254 | 0.004378534 | 0.022253712 | 1 |
| HTN | t_cell | *ITIH4* | 1.284682062 | 0.001835174 | 0.015003561 | 1 |
| HTN | t_cell | *TRAF1* | -1.335994062 | 0.000338356 | 0.00869179 | 1 |
| HTN | t_cell | *PCGF2* | 1.185307949 | 0.015449804 | 0.049125217 | 1 |
| HTN | t_cell | *IL17RB* | 7.312428342 | 0.015681963 | 0.049519087 | 1 |
| HTN | t_cell | *TRAF3IP2* | 3.365267179 | 0.011515853 | 0.040289466 | 1 |
| HTN | t_cell | *DCBLD2* | 2.451543941 | 0.014305504 | 0.04651904 | 1 |
| HTN | t_cell | *ZC3H11A* | 8.684689564 | 0.001264797 | 0.013965754 | 1 |
| HTN | t_cell | *RIOK2* | 4.269817031 | 0.015500703 | 0.049218648 | 1 |
| HTN | t_cell | *DNAJC25* | 2.668930064 | 0.00424802 | 0.021837532 | 1 |
| HTN | t_cell | *PIGV* | -6.820203252 | 8.46E-06 | 0.000718963 | 0.053203242 |
| HTN | t_cell | *NCKAP1* | 2.201176115 | 0.013125482 | 0.043870233 | 1 |
| HTN | t_cell | *ISOC2* | 5.584457109 | 0.014555173 | 0.047120687 | 1 |
| HTN | t_cell | *ADCK1* | 3.262405687 | 0.002634344 | 0.017252209 | 1 |
| HTN | t_cell | *CASP8* | -1.306843706 | 0.002596741 | 0.017112907 | 1 |
| HTN | t_cell | *DMRT3* | 3.295762187 | 0.001435949 | 0.014126985 | 1 |
| HTN | t_cell | *ATP2C2* | 5.181415484 | 0.002044807 | 0.015375679 | 1 |
| HTN | t_cell | *ABCA7* | -1.849775576 | 0.000510154 | 0.011059782 | 1 |
| HTN | t_cell | *UHRF1BP1* | 3.098754697 | 4.42E-08 | 1.39E-05 | 0.000278196 |
| HTN | t_cell | *ANKRD44* | -1.054540053 | 0.002308905 | 0.016247738 | 1 |
| HTN | t_cell | *CD84* | -1.612538001 | 0.006441507 | 0.027678129 | 1 |

| | | | | | | |
|---|---|---|---|---|---|---|
| HTN | t_cell | *EML1* | 2.892569983 | 0.000527005 | 0.011231463 | 1 |
| HTN | t_cell | *MTFR1* | -3.321865096 | 0.001756781 | 0.014724258 | 1 |
| HTN | t_cell | *RRP15* | 1.885536151 | 0.003418739 | 0.019478401 | 1 |
| HTN | t_cell | *SYT1* | 2.110105284 | 0.011686008 | 0.040726125 | 1 |
| HTN | t_cell | *ROGDI* | 5.626901585 | 0.005790737 | 0.026210483 | 1 |
| HTN | t_cell | *RASSF1* | -3.375058474 | 0.001415057 | 0.01399841 | 1 |
| HTN | t_cell | *FGFR3* | 0.985988228 | 0.008704134 | 0.033490142 | 1 |
| HTN | t_cell | *POLR1A* | -1.616753313 | 0.005818966 | 0.026281493 | 1 |
| HTN | t_cell | *PITX1* | 6.844126729 | 0.00195927 | 0.015192726 | 1 |
| HTN | t_cell | *SDK2* | -5.547347326 | 0.006306235 | 0.02737997 | 1 |
| HTN | t_cell | *KCNAB2* | -1.913232638 | 0.001227039 | 0.013965754 | 1 |
| HTN | t_cell | *CLEC2D* | -1.180019387 | 0.002280728 | 0.016194559 | 1 |
| HTN | t_cell | *GNB5* | -3.155968144 | 0.000237594 | 0.007211618 | 1 |
| HTN | t_cell | *FGF10* | 1.790218444 | 0.002958913 | 0.01807841 | 1 |
| HTN | t_cell | *AP3M2* | -3.579866788 | 0.001638129 | 0.014280696 | 1 |
| HTN | t_cell | *CAMK2A* | 15.37030868 | 0.001925165 | 0.015190044 | 1 |
| HTN | t_cell | *OSBPL3* | -1.568613333 | 0.001552823 | 0.014185269 | 1 |
| HTN | t_cell | *VASH1* | 3.563420205 | 0.008687579 | 0.033446913 | 1 |
| HTN | t_cell | *TRIP13* | 4.404036514 | 0.00146654 | 0.014185269 | 1 |
| HTN | t_cell | *LIMS2* | 2.8524077 | 0.001269661 | 0.013965754 | 1 |
| HTN | t_cell | *TRHDE* | 1.504326028 | 0.002923707 | 0.018049857 | 1 |
| HTN | t_cell | *NFATC3* | -2.869385069 | 0.001761196 | 0.014724258 | 1 |
| HTN | t_cell | *TP63* | 1.916458991 | 0.001208654 | 0.013965754 | 1 |
| HTN | t_cell | *LLGL2* | -1.451295765 | 0.000989277 | 0.013965754 | 1 |
| HTN | t_cell | *GLI2* | 1.557381322 | 0.002810262 | 0.017736088 | 1 |
| HTN | t_cell | *CLNS1A* | 3.859983247 | 2.33E-05 | 0.001500094 | 0.1462035 |
| HTN | t_cell | *CA12* | 1.734399019 | 0.002284806 | 0.016194559 | 1 |
| HTN | t_cell | *TIPIN* | 1.479125314 | 0.000827003 | 0.013695656 | 1 |
| HTN | t_cell | *TTC38* | 2.50082972 | 0.003681969 | 0.020236593 | 1 |
| HTN | t_cell | *CELSR1* | 4.07980576 | 0.007726636 | 0.031266496 | 1 |
| HTN | t_cell | *ADD2* | 3.755482695 | 0.00473087 | 0.023218561 | 1 |
| HTN | t_cell | *FSCN1* | 2.940460593 | 0.004669376 | 0.02313741 | 1 |
| HTN | t_cell | *MOCOS* | 1.903976828 | 0.002192006 | 0.015895207 | 1 |
| HTN | t_cell | *TUBA3D* | 5.043087688 | 0.002090271 | 0.015533729 | 1 |
| HTN | t_cell | *MCM6* | -4.17772251 | 0.012561402 | 0.042782049 | 1 |
| HTN | t_cell | *RBMS2* | 2.626737126 | 0.010444278 | 0.037579299 | 1 |
| HTN | t_cell | *FMO4* | 3.258222583 | 0.013174614 | 0.043962605 | 1 |
| HTN | t_cell | *TPD52* | -0.917719739 | 0.006858026 | 0.02882113 | 1 |
| HTN | t_cell | *ICAM3* | -1.142560822 | 0.003737225 | 0.020378085 | 1 |
| HTN | t_cell | *CAMSAP3* | -6.103372693 | 0.001084125 | 0.013965754 | 1 |
| HTN | t_cell | *XAB2* | -5.553176599 | 0.005920646 | 0.026531078 | 1 |
| HTN | t_cell | *STXBP2* | -2.111756025 | 0.007779603 | 0.031318911 | 1 |
| HTN | t_cell | *SIRT6* | 4.657912663 | 0.00499435 | 0.023855009 | 1 |
| HTN | t_cell | *NAALAD2* | 2.335510519 | 0.000230063 | 0.007125155 | 1 |
| HTN | t_cell | *FAP* | 1.791044966 | 0.009072139 | 0.034463166 | 1 |
| HTN | t_cell | *TULP3* | 3.004196806 | 0.009650181 | 0.0358665 | 1 |
| HTN | t_cell | *ZCWPW1* | 3.524640546 | 0.001621518 | 0.014238085 | 1 |
| HTN | t_cell | *FGF20* | 12.07989961 | 0.001334325 | 0.013965754 | 1 |
| HTN | t_cell | *OSBPL6* | 3.087596 | 0.007338938 | 0.030216047 | 1 |
| HTN | t_cell | *CDC14A* | -0.966691957 | 0.001457756 | 0.014185269 | 1 |

| | | | | | | |
|---|---|---|---|---|---|---|
| HTN | t_cell | *RIMS1* | 2.86683116 | 0.003620106 | 0.020070197 | 1 |
| HTN | t_cell | *DCT* | 2.92283364 | 0.001274742 | 0.013965754 | 1 |
| HTN | t_cell | *SCTR* | 2.64854597 | 0.004341651 | 0.022119904 | 1 |
| HTN | t_cell | *KCNN2* | 3.443282695 | 0.003181393 | 0.018851478 | 1 |
| HTN | t_cell | *MOK* | 2.731300826 | 0.004460801 | 0.022508072 | 1 |
| HTN | t_cell | *COL4A4* | 2.027218123 | 0.015040507 | 0.04814647 | 1 |
| HTN | t_cell | *STK17B* | -1.340050845 | 0.001576494 | 0.014185269 | 1 |
| HTN | t_cell | *PCDHA6* | -16.18658931 | 0.007554061 | 0.030859249 | 1 |
| HTN | t_cell | *PCDHGA2* | 4.181678822 | 0.003260113 | 0.019003909 | 1 |
| HTN | t_cell | *PGR* | 1.695909006 | 0.003532772 | 0.019830839 | 1 |
| HTN | t_cell | *CCNT2* | -3.48492992 | 0.000166127 | 0.005968241 | 1 |
| HTN | t_cell | *PLOD1* | 3.975868533 | 0.001177949 | 0.013965754 | 1 |
| HTN | t_cell | *EPYC* | 12.22163701 | 0.001098212 | 0.013965754 | 1 |
| HTN | t_cell | *CYLD* | -2.113916803 | 0.001264546 | 0.013965754 | 1 |
| HTN | t_cell | *RPS5* | -2.711958023 | 0.002854453 | 0.01787445 | 1 |
| HTN | t_cell | *APOB* | 2.362470707 | 0.004961419 | 0.023811023 | 1 |
| HTN | t_cell | *CD82* | -1.328287837 | 0.012276169 | 0.042116263 | 1 |
| HTN | t_cell | *CTTN* | 2.12864992 | 0.003968092 | 0.020966245 | 1 |
| HTN | t_cell | *ATG16L1* | -2.911253423 | 0.008057107 | 0.031979187 | 1 |
| HTN | t_cell | *USP40* | 2.93630136 | 0.004676045 | 0.02313741 | 1 |
| HTN | t_cell | *MAST2* | 3.721675415 | 0.006776398 | 0.028631191 | 1 |
| HTN | t_cell | *SNX10* | -1.251445855 | 7.37E-06 | 0.00066222 | 0.046355424 |
| HTN | t_cell | *ITPKC* | 3.030021085 | 0.002306815 | 0.016247738 | 1 |
| HTN | t_cell | *TRIP6* | 3.747856047 | 0.001584218 | 0.014185269 | 1 |
| HTN | t_cell | *LPCAT2* | 2.714362639 | 0.006995535 | 0.029165071 | 1 |
| HTN | t_cell | *PTHLH* | 1.914385779 | 0.004293643 | 0.021964306 | 1 |
| HTN | t_cell | *PHACTR3* | 1.986827094 | 0.012691676 | 0.043004354 | 1 |
| HTN | t_cell | *AAMDC* | -1.465880184 | 0.010591509 | 0.037813069 | 1 |
| HTN | t_cell | *ALG6* | -4.117848487 | 0.002728688 | 0.017579998 | 1 |
| HTN | t_cell | *GP6* | -1.835092878 | 0.002965537 | 0.01809613 | 1 |
| HTN | t_cell | *ASAP3* | 2.19187811 | 0.002506867 | 0.016874383 | 1 |
| HTN | t_cell | *C3orf18* | 7.022691832 | 2.25E-05 | 0.001475584 | 0.141656044 |
| HTN | t_cell | *ZNF343* | 3.142804283 | 0.003766199 | 0.020414857 | 1 |
| HTN | t_cell | *LZTS3* | 3.7635562 | 0.002302894 | 0.016247738 | 1 |
| HTN | t_cell | *TESC* | 5.717105493 | 0.012894857 | 0.043352924 | 1 |
| HTN | t_cell | *FXYD5* | -1.922414506 | 0.001120058 | 0.013965754 | 1 |
| HTN | t_cell | *GMIP* | -2.237707859 | 0.00137011 | 0.013965754 | 1 |
| HTN | t_cell | *MLF2* | 3.749238922 | 0.010326422 | 0.037440723 | 1 |
| HTN | t_cell | *RGS1* | -1.748639698 | 0.001120634 | 0.013965754 | 1 |
| HTN | t_cell | *YPEL3* | 2.544615483 | 0.000298208 | 0.008171005 | 1 |
| HTN | t_cell | *GOLGA3* | 3.410276685 | 0.008681122 | 0.033442532 | 1 |
| HTN | t_cell | *PABPC4* | -5.165014146 | 2.18E-05 | 0.001443057 | 0.13709041 |
| HTN | t_cell | *CERS4* | -4.06633673 | 0.005670103 | 0.025834063 | 1 |
| HTN | t_cell | *NLRC4* | 3.307100006 | 0.004196626 | 0.021679382 | 1 |
| HTN | t_cell | *SLC26A4* | 3.38643713 | 0.001607187 | 0.014197745 | 1 |
| HTN | t_cell | *IL5RA* | -1.639265636 | 0.001056834 | 0.013965754 | 1 |
| HTN | t_cell | *ITGA6* | -2.900901632 | 0.00339338 | 0.019412359 | 1 |
| HTN | t_cell | *RAPGEF4* | 2.306432951 | 0.003101325 | 0.018587253 | 1 |
| HTN | t_cell | *SEL1L3* | -1.323051204 | 0.001541451 | 0.014185269 | 1 |
| HTN | t_cell | *TF* | 2.728785788 | 0.004703739 | 0.023211958 | 1 |

| | | | | | | |
|---|---|---|---|---|---|---|
| HTN | t_cell | *NLRP1* | -2.691208927 | 0.011839378 | 0.041010561 | 1 |
| HTN | t_cell | *ZFHX4* | 1.943532562 | 0.00293363 | 0.01807841 | 1 |
| HTN | t_cell | *RGS17* | 2.780382591 | 0.001053849 | 0.013965754 | 1 |
| HTN | t_cell | *RPGRIP1* | 3.649933614 | 0.000745292 | 0.013311501 | 1 |
| HTN | t_cell | *TGM1* | 3.460906254 | 0.000123467 | 0.004863616 | 0.776237235 |
| HTN | t_cell | *RFFL* | -4.243629393 | 0.003314132 | 0.019133102 | 1 |
| HTN | t_cell | *VNN3* | 5.864697762 | 0.002574064 | 0.017027838 | 1 |
| HTN | t_cell | *FMO2* | 2.360674986 | 0.000110529 | 0.004541789 | 0.694893715 |
| HTN | t_cell | *TMEM38B* | -3.68935726 | 0.002552056 | 0.016987039 | 1 |
| HTN | t_cell | *PDE6C* | 2.585290823 | 0.001392845 | 0.013966213 | 1 |
| HTN | t_cell | *TPSD1* | 18.87666974 | 0.00047298 | 0.010658152 | 1 |
| HTN | t_cell | *IL12RB1* | -1.462269568 | 0.000415448 | 0.009759197 | 1 |
| HTN | t_cell | *CDC7* | -1.77195382 | 0.000411481 | 0.009725493 | 1 |
| HTN | t_cell | *PRTFDC1* | -3.824585163 | 0.00317779 | 0.018851478 | 1 |
| HTN | t_cell | *TSPAN15* | 2.579385738 | 0.015719612 | 0.049524434 | 1 |
| HTN | t_cell | *OCEL1* | 1.695732197 | 0.009821925 | 0.036174836 | 1 |
| HTN | t_cell | *MARCH2* | 5.162647899 | 2.13E-05 | 0.00142385 | 0.133841924 |
| HTN | t_cell | *MADCAM1* | 2.434548366 | 0.003306324 | 0.019112361 | 1 |
| HTN | t_cell | *ZDHHC8* | 3.232887597 | 0.01026515 | 0.037347798 | 1 |
| HTN | t_cell | *CECR2* | 1.466552625 | 0.001897467 | 0.015177294 | 1 |
| HTN | t_cell | *P2RX6* | 1.864191706 | 0.002121209 | 0.015666072 | 1 |
| HTN | t_cell | *UPB1* | -1.400649019 | 0.007798027 | 0.031326641 | 1 |
| HTN | t_cell | *CYTH4* | -2.862771284 | 0.002567644 | 0.017027838 | 1 |
| HTN | t_cell | *CARD10* | 3.096932856 | 0.000153157 | 0.005580753 | 0.962899203 |
| HTN | t_cell | *SLC25A1* | 3.277657924 | 0.00232692 | 0.016291034 | 1 |
| HTN | t_cell | *LGALS2* | 2.620646701 | 0.003685477 | 0.020236593 | 1 |
| HTN | t_cell | *SH3BP1* | -1.57606403 | 0.001257818 | 0.013965754 | 1 |
| HTN | t_cell | *PIK3IP1* | -1.638819489 | 0.01317722 | 0.043962605 | 1 |
| HTN | t_cell | *GCAT* | 4.764532615 | 2.06E-05 | 0.001393711 | 0.129615139 |
| HTN | t_cell | *ANKRD54* | 2.035171832 | 0.015584936 | 0.049361458 | 1 |
| HTN | t_cell | *CCDC134* | -3.166118142 | 0.009303086 | 0.035002095 | 1 |
| HTN | t_cell | *SLC16A8* | 4.502695568 | 0.006393589 | 0.027525449 | 1 |
| HTN | t_cell | *RHBDD3* | 2.874004803 | 0.000976469 | 0.013965754 | 1 |
| HTN | t_cell | *TOM1* | 2.211111085 | 0.003472357 | 0.01961645 | 1 |
| HTN | t_cell | *BIK* | -13.40957643 | 0.001313941 | 0.013965754 | 1 |
| HTN | t_cell | *RASD2* | -7.013350532 | 0.000167151 | 0.005970884 | 1 |
| HTN | t_cell | *TXN2* | 3.922169763 | 0.002385091 | 0.016478098 | 1 |
| HTN | t_cell | *KIAA0930* | 2.633065502 | 0.009858617 | 0.036246273 | 1 |
| HTN | t_cell | *DDHD1* | -3.057567424 | 0.009843439 | 0.036232845 | 1 |
| HTN | t_cell | *ISM2* | 4.992748647 | 0.004492692 | 0.022614535 | 1 |
| HTN | t_cell | *SLC10A1* | 3.769837477 | 0.015602044 | 0.049390761 | 1 |
| HTN | t_cell | *ZFYVE21* | 3.386409972 | 0.002407628 | 0.016542906 | 1 |
| HTN | t_cell | *MTHFD1* | 2.941800972 | 0.009045694 | 0.03440522 | 1 |
| HTN | t_cell | *TELO2* | -6.336381698 | 0.004946643 | 0.023776411 | 1 |
| HTN | t_cell | *VRK1* | -3.609294905 | 0.013371341 | 0.044338409 | 1 |
| HTN | t_cell | *PAPLN* | 2.971764885 | 0.010739488 | 0.038254482 | 1 |
| HTN | t_cell | *NINL* | 2.525290445 | 0.005469751 | 0.025304138 | 1 |
| HTN | t_cell | *CD40* | 2.650927998 | 0.003831755 | 0.020625209 | 1 |
| HTN | t_cell | *DOK5* | 2.937718769 | 0.004693684 | 0.023180827 | 1 |
| HTN | t_cell | *BMP7* | 2.867423115 | 0.001909881 | 0.015190044 | 1 |

| | | | | | | |
|---|---|---|---|---|---|---|
| HTN | t_cell | *MTG2* | 3.454567568 | 0.006396482 | 0.027525449 | 1 |
| HTN | t_cell | *SLC17A9* | 4.353175734 | 0.000363374 | 0.009061689 | 1 |
| HTN | t_cell | *PTK6* | 12.5587974 | 0.008108068 | 0.03212062 | 1 |
| HTN | t_cell | *GMEB2* | -4.897863818 | 0.007285552 | 0.030094786 | 1 |
| HTN | t_cell | *SLC52A3* | 3.458332894 | 0.009006922 | 0.034298316 | 1 |
| HTN | t_cell | *RSPO4* | 3.765326116 | 0.014877232 | 0.047867531 | 1 |
| HTN | t_cell | *CCM2L* | 3.162611494 | 0.002015025 | 0.015300078 | 1 |
| HTN | t_cell | *MYL9* | 3.670640806 | 0.001042904 | 0.013965754 | 1 |
| HTN | t_cell | *MANBAL* | 2.402180057 | 0.001382283 | 0.013965754 | 1 |
| HTN | t_cell | *PXMP4* | 4.298090938 | 0.013061553 | 0.043726295 | 1 |
| HTN | t_cell | *BPI* | 2.577885511 | 0.003277364 | 0.019032949 | 1 |
| HTN | t_cell | *MAP1LC3A* | 1.802081146 | 0.003143341 | 0.018722726 | 1 |
| HTN | t_cell | *CELF4* | 2.149316791 | 0.003730391 | 0.020378085 | 1 |
| HTN | t_cell | *RBFA* | 3.107677838 | 0.001045994 | 0.013965754 | 1 |
| HTN | t_cell | *LPIN2* | -2.60567245 | 0.004673765 | 0.02313741 | 1 |
| HTN | t_cell | *SMCHD1* | -1.948949513 | 0.004737998 | 0.02323541 | 1 |
| HTN | t_cell | *MYL12A* | -3.196611545 | 0.008493669 | 0.033064829 | 1 |
| HTN | t_cell | *LIPG* | -6.331007245 | 0.004405379 | 0.02235401 | 1 |
| HTN | t_cell | *POLI* | -1.096297256 | 0.000694691 | 0.012921657 | 1 |
| HTN | t_cell | *NALCN* | 3.071987277 | 0.004269934 | 0.021914345 | 1 |
| HTN | t_cell | *HTR2A* | 11.05611464 | 0.009685981 | 0.035926705 | 1 |
| HTN | t_cell | *KLF5* | 3.505913827 | 0.015511221 | 0.049227181 | 1 |
| HTN | t_cell | *UGGT2* | 2.300211499 | 0.01326398 | 0.044119901 | 1 |
| HTN | t_cell | *SGCG* | 1.944450428 | 0.00274632 | 0.017638397 | 1 |
| HTN | t_cell | *MGRN1* | 3.729237023 | 0.000287389 | 0.008066144 | 1 |
| HTN | t_cell | *CORO1A* | -1.125351149 | 0.00014537 | 0.005440117 | 0.913938451 |
| HTN | t_cell | *GDPD3* | 3.739441815 | 0.010354856 | 0.037457412 | 1 |
| HTN | t_cell | *SETD6* | 4.296155333 | 0.000991496 | 0.013965754 | 1 |
| HTN | t_cell | *COG4* | -3.754597757 | 0.004594184 | 0.022916271 | 1 |
| HTN | t_cell | *COTL1* | -4.066507868 | 0.005406663 | 0.025141785 | 1 |
| HTN | t_cell | *CRISPLD2* | 3.252856936 | 0.000227165 | 0.00707022 | 1 |
| HTN | t_cell | *NME4* | 4.881290181 | 0.004677533 | 0.02313741 | 1 |
| HTN | t_cell | *NARFL* | 5.18526927 | 0.002075662 | 0.015461717 | 1 |
| HTN | t_cell | *MTHFSD* | 2.208724218 | 0.000701242 | 0.012928761 | 1 |
| HTN | t_cell | *SLC7A5* | -1.530746853 | 0.001081643 | 0.013965754 | 1 |
| HTN | t_cell | *KAT8* | 6.125257304 | 0.001938238 | 0.015190044 | 1 |
| HTN | t_cell | *TMC5* | 2.008595988 | 0.002789628 | 0.017682732 | 1 |
| HTN | t_cell | *CSK* | -3.220329266 | 0.002326905 | 0.016291034 | 1 |
| HTN | t_cell | *TRIP4* | 2.848455029 | 0.010480867 | 0.037634027 | 1 |
| HTN | t_cell | *FAH* | 2.649618773 | 0.003748351 | 0.020396571 | 1 |
| HTN | t_cell | *ATP8B4* | 2.784375741 | 0.015173512 | 0.048522822 | 1 |
| HTN | t_cell | *PDGFRL* | 3.018978484 | 0.013017157 | 0.043647394 | 1 |
| HTN | t_cell | *ZFAND1* | 3.932532391 | 0.00097762 | 0.013965754 | 1 |
| HTN | t_cell | *EYA1* | 2.633572467 | 5.69E-07 | 0.000101821 | 0.003576204 |
| HTN | t_cell | *PLAT* | 3.017965517 | 0.002948058 | 0.01807841 | 1 |
| HTN | t_cell | *GDAP1* | -3.878865922 | 0.002774046 | 0.017670137 | 1 |
| HTN | t_cell | *SNX16* | 4.360836193 | 0.000950233 | 0.013965754 | 1 |
| HTN | t_cell | *LEPROTL1* | -1.298408275 | 0.001079596 | 0.013965754 | 1 |
| HTN | t_cell | *TNFRSF10A* | -0.964222067 | 0.013046872 | 0.043723711 | 1 |
| HTN | t_cell | *TUSC3* | 2.853750764 | 0.005044836 | 0.02399159 | 1 |

| | | | | | | |
|---|---|---|---|---|---|---|
| HTN | t_cell | *ARHGEF10* | 1.479908758 | 0.012671898 | 0.042994185 | 1 |
| HTN | t_cell | *KLHDC4* | -2.460719826 | 0.012883344 | 0.043346584 | 1 |
| HTN | t_cell | *MCM4* | 3.419863736 | 0.001240315 | 0.013965754 | 1 |
| HTN | t_cell | *TULP2* | 3.697304388 | 0.002611677 | 0.017160523 | 1 |
| HTN | t_cell | *NFKBIB* | -4.390270826 | 0.006175357 | 0.027131006 | 1 |
| HTN | t_cell | *FCGRT* | 2.472475199 | 0.009195721 | 0.034806442 | 1 |
| HTN | t_cell | *PEX11G* | 3.573717076 | 5.57E-05 | 0.002822215 | 0.349954703 |
| HTN | t_cell | *PLEKHJ1* | 5.009478995 | 0.003259164 | 0.019003909 | 1 |
| HTN | t_cell | *CD37* | -1.667095289 | 0.004312236 | 0.02202635 | 1 |
| HTN | t_cell | *FCER2* | 5.090337464 | 0.002981815 | 0.018165378 | 1 |
| HTN | t_cell | *TIMM44* | 3.477423314 | 0.014292525 | 0.04650989 | 1 |
| HTN | t_cell | *IL27RA* | -0.919169018 | 0.001312381 | 0.013965754 | 1 |
| HTN | t_cell | *C19orf44* | 1.492179708 | 0.014030186 | 0.045917636 | 1 |
| HTN | t_cell | *RASAL3* | -0.560419519 | 0.001166142 | 0.013965754 | 1 |
| HTN | t_cell | *CCNE1* | -5.097972119 | 0.000985932 | 0.013965754 | 1 |
| HTN | t_cell | *RPS16* | -2.646405339 | 0.003555094 | 0.019909231 | 1 |
| HTN | t_cell | *CACTIN* | 5.755809675 | 0.009104433 | 0.034523262 | 1 |
| HTN | t_cell | *CCDC9* | 4.653200257 | 0.00443289 | 0.022457356 | 1 |
| HTN | t_cell | *TGFB1* | -2.945570855 | 0.00132957 | 0.013965754 | 1 |
| HTN | t_cell | *CEACAM4* | 6.628853543 | 0.001560269 | 0.014185269 | 1 |
| HTN | t_cell | *ICAM5* | -25.47027213 | 5.52E-05 | 0.00281998 | 0.346857488 |
| HTN | t_cell | *PTPRS* | 2.128336092 | 0.000232253 | 0.00715771 | 1 |
| HTN | t_cell | *CYTH2* | 3.570512543 | 0.003724871 | 0.020378085 | 1 |
| HTN | t_cell | *GRIN2D* | 4.890196609 | 0.006450333 | 0.027678129 | 1 |
| HTN | t_cell | *CARD8* | -3.832015711 | 0.000963756 | 0.013965754 | 1 |
| HTN | t_cell | *LIG1* | -4.843648563 | 0.006931678 | 0.029003866 | 1 |
| HTN | t_cell | *GCDH* | -3.989290743 | 0.013390519 | 0.044342537 | 1 |
| HTN | t_cell | *MAST1* | 4.812830232 | 0.00155617 | 0.014185269 | 1 |
| HTN | t_cell | *UPK1A* | 5.53929878 | 0.002201065 | 0.015906408 | 1 |
| HTN | t_cell | *PON3* | 2.447025289 | 0.00117263 | 0.013965754 | 1 |
| HTN | t_cell | *WDR91* | 3.589021471 | 0.005100864 | 0.024174482 | 1 |
| HTN | t_cell | *PTN* | 3.771951039 | 0.004535313 | 0.022712185 | 1 |
| HTN | t_cell | *DFNA5* | 3.294018811 | 0.001175147 | 0.013965754 | 1 |
| HTN | t_cell | *ADAP1* | -3.085366177 | 0.001168056 | 0.013965754 | 1 |
| HTN | t_cell | *CAV1* | 2.239661969 | 0.003939551 | 0.020880704 | 1 |
| HTN | t_cell | *MET* | 2.233140204 | 0.006933802 | 0.029003866 | 1 |
| HTN | t_cell | *HOXA5* | 2.495535194 | 0.000178743 | 0.006208591 | 1 |
| HTN | t_cell | *HOXA6* | 2.068766071 | 0.000913992 | 0.013965754 | 1 |
| HTN | t_cell | *CPED1* | 2.136430606 | 0.012404288 | 0.042371471 | 1 |
| HTN | t_cell | *CRHR2* | 9.188875042 | 5.27E-05 | 0.002715239 | 0.331259134 |
| HTN | t_cell | *CCL24* | 2.851351814 | 0.01209058 | 0.041674054 | 1 |
| HTN | t_cell | *TAF6* | -6.331020632 | 3.80E-05 | 0.002130724 | 0.238641108 |
| HTN | t_cell | *TFR2* | -2.50836603 | 0.000406262 | 0.009638365 | 1 |
| HTN | t_cell | *AP1S1* | -6.756759182 | 0.00445766 | 0.022508072 | 1 |
| HTN | t_cell | *POU6F2* | 2.66620696 | 0.00292553 | 0.018049857 | 1 |
| HTN | t_cell | *TSPAN13* | 5.009365334 | 0.008382752 | 0.032754729 | 1 |
| HTN | t_cell | *TMEM176B* | 1.593288258 | 0.007192088 | 0.02984598 | 1 |
| HTN | t_cell | *GCK* | 15.00907702 | 0.001389941 | 0.013965754 | 1 |
| HTN | t_cell | *LIMK1* | 3.719910142 | 0.005510407 | 0.025373449 | 1 |
| HTN | t_cell | *SLC1A1* | 1.712753867 | 0.002902742 | 0.018049857 | 1 |

| | | | | | | |
|---|---|---|---|---|---|---|
| HTN | t_cell | *TRIM14* | -2.177719419 | 0.003756235 | 0.020410933 | 1 |
| HTN | t_cell | *CORO2A* | -1.823508903 | 0.002566655 | 0.017027838 | 1 |
| HTN | t_cell | *C5* | 1.886023578 | 0.009592394 | 0.035734315 | 1 |
| HTN | t_cell | *TNFSF8* | -0.733494828 | 0.000977793 | 0.013965754 | 1 |
| HTN | t_cell | *PLGRKT* | -2.429286518 | 0.004876517 | 0.023562138 | 1 |
| HTN | t_cell | *ELAVL2* | -3.436958526 | 0.004200344 | 0.021679382 | 1 |
| HTN | t_cell | *MPDZ* | 2.392304421 | 0.013636997 | 0.044934906 | 1 |
| HTN | t_cell | *PTGDS* | -6.122242052 | 0.005916994 | 0.026531078 | 1 |
| HTN | t_cell | *DVL1* | 4.809991126 | 0.008651087 | 0.033388202 | 1 |
| HTN | t_cell | *PDLIM1* | 2.99361951 | 0.001660508 | 0.014360746 | 1 |
| HTN | t_cell | *DNMBP* | 2.635339774 | 0.012320393 | 0.042152138 | 1 |
| HTN | t_cell | *TRDMT1* | -4.735273306 | 0.004537379 | 0.022712185 | 1 |
| HTN | t_cell | *NSMCE4A* | 2.932786437 | 0.014975437 | 0.048098218 | 1 |
| HTN | t_cell | *SPOCK2* | -0.400171095 | 0.000918101 | 0.013965754 | 1 |
| HTN | t_cell | *CCSER2* | -2.356926037 | 0.010321121 | 0.037440723 | 1 |
| HTN | t_cell | *RPL28* | -2.398063951 | 7.44E-05 | 0.003388133 | 0.467562301 |
| HTN | t_cell | *PBLD* | 1.170584786 | 0.003984605 | 0.021016119 | 1 |
| HTN | t_cell | *RAPGEFL1* | 2.44879878 | 0.003936548 | 0.020880704 | 1 |
| HTN | t_cell | *RGS9* | -1.989094288 | 0.001376869 | 0.013965754 | 1 |
| HTN | t_cell | *ASPA* | 2.113258678 | 0.003361825 | 0.019313893 | 1 |
| HTN | t_cell | *RAD51C* | -2.058995318 | 0.003908911 | 0.020844209 | 1 |
| HTN | t_cell | *GALK1* | 4.78123026 | 0.004127395 | 0.021445397 | 1 |
| HTN | t_cell | *RNF167* | -4.23806246 | 0.006820309 | 0.028739465 | 1 |
| HTN | t_cell | *SYNGR2* | -3.943056005 | 0.002920229 | 0.018049857 | 1 |
| HTN | t_cell | *DDX5* | 2.384234785 | 0.004790288 | 0.023346157 | 1 |
| HTN | t_cell | *NAGLU* | 3.487869856 | 0.00873354 | 0.033540599 | 1 |
| HTN | t_cell | *DLX4* | 14.02136244 | 0.001382155 | 0.013965754 | 1 |
| HTN | t_cell | *SGCA* | 1.324820332 | 0.015777102 | 0.04962013 | 1 |
| HTN | t_cell | *MPP2* | 2.582226813 | 0.001335195 | 0.013965754 | 1 |
| HTN | t_cell | *EFNB3* | -19.78996629 | 3.75E-05 | 0.002123491 | 0.235707522 |
| HTN | t_cell | *FAM20A* | 2.447511901 | 0.007244866 | 0.029985826 | 1 |
| HTN | t_cell | *SLC9A3R1* | -0.920998159 | 0.001449615 | 0.014156149 | 1 |
| HTN | t_cell | *CWH43* | 16.24952467 | 5.97E-05 | 0.00288513 | 0.375066965 |
| HTN | t_cell | *WFS1* | 3.196471272 | 0.003766699 | 0.020414857 | 1 |
| HTN | t_cell | *GRPEL1* | 5.33941713 | 0.004015863 | 0.021084486 | 1 |
| HTN | t_cell | *AADAT* | 3.116219246 | 0.010498119 | 0.037634027 | 1 |
| HTN | t_cell | *CPZ* | 27.93545455 | 0.001354384 | 0.013965754 | 1 |
| HTN | t_cell | *SLC2A9* | 4.984607236 | 0.001870419 | 0.015083316 | 1 |
| HTN | t_cell | *STIM2* | -3.177883315 | 0.003571958 | 0.019909231 | 1 |
| HTN | t_cell | *DTX4* | 2.733808584 | 0.014087566 | 0.046057477 | 1 |
| HTN | t_cell | *NRXN2* | 4.73512556 | 0.002539853 | 0.016935789 | 1 |
| HTN | t_cell | *CCDC86* | 4.765003027 | 0.001535898 | 0.014185269 | 1 |
| HTN | t_cell | *CD5* | -0.495583284 | 0.00099993 | 0.013965754 | 1 |
| HTN | t_cell | *SLC35F2* | -2.713889568 | 0.001634136 | 0.014269189 | 1 |
| HTN | t_cell | *C11orf21* | -0.843781173 | 0.000513977 | 0.011103937 | 1 |
| HTN | t_cell | *ELMOD1* | 5.538990421 | 0.002773257 | 0.017670137 | 1 |
| HTN | t_cell | *AIP* | -4.250011882 | 0.002290083 | 0.016195448 | 1 |
| HTN | t_cell | *ASIC1* | 4.210385257 | 0.00021693 | 0.006819187 | 1 |
| HTN | t_cell | *KCTD10* | -2.21479898 | 0.012760267 | 0.043107898 | 1 |
| HTN | t_cell | *CYP27B1* | 11.38172633 | 0.001276267 | 0.013965754 | 1 |

| | | | | | | |
|---|---|---|---|---|---|---|
| HTN | t_cell | *ACSS3* | 1.063604049 | 0.005140227 | 0.024246359 | 1 |
| HTN | t_cell | *GLI1* | 8.012020117 | 0.006107057 | 0.027019752 | 1 |
| HTN | t_cell | *TRPV4* | 2.446999491 | 0.002416023 | 0.016563558 | 1 |
| HTN | t_cell | *GPN3* | 4.265177509 | 0.00378235 | 0.020429237 | 1 |
| HTN | t_cell | *KCNA1* | 3.0379689 | 0.005666794 | 0.025834063 | 1 |
| HTN | t_cell | *LTBR* | 2.779051841 | 0.006959364 | 0.029067341 | 1 |
| HTN | t_cell | *ARHGDIB* | -1.183654459 | 0.000495337 | 0.010888745 | 1 |
| HTN | t_cell | *CAND1* | -4.293150825 | 0.000667967 | 0.012876096 | 1 |
| HTN | t_cell | *ATN1* | 3.005082925 | 0.005416957 | 0.025171035 | 1 |
| HTN | t_cell | *C12orf57* | -2.955447084 | 0.005042037 | 0.02399159 | 1 |
| HTN | t_cell | *ST8SIA1* | -1.573559328 | 0.001914267 | 0.015190044 | 1 |
| HTN | t_cell | *COL12A1* | 2.199188404 | 0.011753658 | 0.040837464 | 1 |
| HTN | t_cell | *BTN3A3* | -2.000965304 | 0.000364835 | 0.009061689 | 1 |
| HTN | t_cell | *SMIM8* | 3.297687172 | 0.005878918 | 0.026471702 | 1 |
| HTN | t_cell | *RNGTT* | -2.667843259 | 0.014975278 | 0.048098218 | 1 |
| HTN | t_cell | *TULP1* | 11.66265012 | 0.001338618 | 0.013965754 | 1 |
| HTN | t_cell | *MDGA1* | -3.422742709 | 0.000760942 | 0.013406921 | 1 |
| HTN | t_cell | *CD83* | 2.626919154 | 0.005898907 | 0.026504502 | 1 |
| HTN | t_cell | *SAYSD1* | 5.324787673 | 0.004079867 | 0.021283851 | 1 |
| HTN | t_cell | *ALDH5A1* | 3.391772688 | 0.000442166 | 0.010182786 | 1 |
| HTN | t_cell | *VNN2* | -1.763712111 | 0.001333441 | 0.013965754 | 1 |
| HTN | t_cell | *SLC16A10* | -0.921836041 | 0.006751282 | 0.028544258 | 1 |
| HTN | t_cell | *PHACTR2* | -2.215135729 | 0.00450488 | 0.022657744 | 1 |
| HTN | t_cell | *TTK* | -3.861779663 | 0.000591953 | 0.012118368 | 1 |
| HTN | t_cell | *CLIC5* | -2.485051696 | 0.006701191 | 0.028432724 | 1 |
| HTN | t_cell | *LY86* | 3.300453318 | 0.003561045 | 0.019909231 | 1 |
| HTN | t_cell | *NUDT12* | 2.761438874 | 0.006120728 | 0.027024952 | 1 |
| HTN | t_cell | *C7* | 2.496965625 | 0.011187184 | 0.039424791 | 1 |
| HTN | t_cell | *GHR* | 1.068603954 | 0.008677549 | 0.033442532 | 1 |
| HTN | t_cell | *NME5* | 2.451550422 | 5.25E-05 | 0.002715239 | 0.329762362 |
| HTN | t_cell | *PCDHB6* | 6.952596793 | 2.82E-05 | 0.001703056 | 0.17711781 |
| HTN | t_cell | *PDE8B* | 1.602363933 | 0.002944311 | 0.01807841 | 1 |
| HTN | t_cell | *RASGRF2* | -2.638581266 | 0.009744853 | 0.036081209 | 1 |
| HTN | t_cell | *SLC27A6* | 8.607301818 | 0.008043746 | 0.031966519 | 1 |
| HTN | t_cell | *GNPDA1* | 3.508140403 | 0.009629249 | 0.035836682 | 1 |
| HTN | t_cell | *BNIP1* | 1.714710221 | 0.003840303 | 0.020638868 | 1 |
| HTN | t_cell | *CNTN3* | 1.682321089 | 0.002055642 | 0.015403841 | 1 |
| HTN | t_cell | *HGD* | 4.241951248 | 0.001606696 | 0.014197745 | 1 |
| HTN | t_cell | *NPHP3* | 1.54926802 | 0.000902438 | 0.013965754 | 1 |
| HTN | t_cell | *PCCB* | 6.614064596 | 8.39E-05 | 0.003637376 | 0.527419541 |
| HTN | t_cell | *WNT5A* | 2.98846599 | 0.00153872 | 0.014185269 | 1 |
| HTN | t_cell | *PLXNA1* | 3.380349984 | 0.00076337 | 0.013406921 | 1 |
| HTN | t_cell | *ZBTB47* | 3.536687811 | 0.001262973 | 0.013965754 | 1 |
| HTN | t_cell | *CLCN2* | 5.121210747 | 0.012061184 | 0.041618368 | 1 |
| HTN | t_cell | *LMAN2L* | 2.353463144 | 0.005186467 | 0.024403857 | 1 |
| HTN | t_cell | *KCNIP3* | 4.867731152 | 0.001907344 | 0.015190044 | 1 |
| HTN | t_cell | *FAHD2A* | 4.016910167 | 0.004156595 | 0.021578936 | 1 |
| HTN | t_cell | *CENPA* | 3.504962457 | 0.01471149 | 0.047504436 | 1 |
| HTN | t_cell | *ITGA4* | -0.51743246 | 0.001808029 | 0.014955249 | 1 |
| HTN | t_cell | *GRB14* | 3.297385274 | 0.012882426 | 0.043346584 | 1 |

| | | | | | | |
|---|---|---|---|---|---|---|
| HTN | t_cell | *GNLY* | -1.362334187 | 0.001588903 | 0.014185269 | 1 |
| HTN | t_cell | *PLCD4* | 1.783616299 | 0.002850002 | 0.01787445 | 1 |
| HTN | t_cell | *CHMP3* | -3.324264086 | 0.014418096 | 0.046797403 | 1 |
| HTN | t_cell | *MLPH* | 4.373948105 | 0.006677516 | 0.028402239 | 1 |
| HTN | t_cell | *PASK* | -2.051263822 | 0.000123776 | 0.004863616 | 0.778178521 |
| HTN | t_cell | *SDC1* | -5.157826885 | 0.010903865 | 0.038686569 | 1 |
| HTN | t_cell | *WIPF1* | -2.30215297 | 0.002767535 | 0.017670137 | 1 |
| HTN | t_cell | *CD207* | 31.64336131 | 0.000236905 | 0.007211618 | 1 |
| HTN | t_cell | *VAX2* | 7.461610786 | 0.01554696 | 0.049315709 | 1 |
| HTN | t_cell | *MRPL37* | 3.203305735 | 0.003366952 | 0.019313893 | 1 |
| HTN | t_cell | *ICMT* | 3.642796941 | 0.011776415 | 0.040837464 | 1 |
| HTN | t_cell | *CAPZA1* | -3.624622687 | 0.005042861 | 0.02399159 | 1 |
| HTN | t_cell | *C1orf21* | -2.374925396 | 0.005061161 | 0.024043138 | 1 |
| HTN | t_cell | *DNAJC6* | -6.889177994 | 0.000691187 | 0.012921657 | 1 |
| HTN | t_cell | *TNNI3K* | 3.424756197 | 0.000302109 | 0.008186882 | 1 |
| HTN | t_cell | *OSCP1* | 1.730702554 | 0.015804649 | 0.049681915 | 1 |
| HTN | t_cell | *C1orf109* | 3.793342666 | 0.006221612 | 0.027220094 | 1 |
| HTN | t_cell | *GBP3* | -1.203067834 | 0.000824635 | 0.013695656 | 1 |
| HTN | t_cell | *CD46* | 2.704682655 | 0.015378132 | 0.049010075 | 1 |
| HTN | t_cell | *FAAH* | 0.915209824 | 0.00419044 | 0.021679382 | 1 |
| HTN | t_cell | *NSUN4* | 4.800984182 | 0.000668047 | 0.012876096 | 1 |
| HTN | t_cell | *CNN3* | 2.656953721 | 0.007014563 | 0.02922502 | 1 |
| HTN | t_cell | *SERPINC1* | 2.301343546 | 0.001979016 | 0.015266349 | 1 |
| HTN | t_cell | *RCAN3* | -0.625363241 | 0.001353616 | 0.013965754 | 1 |
| HTN | t_cell | *SLC5A9* | 2.434980135 | 0.000762151 | 0.013406921 | 1 |
| HTN | t_cell | *IFT46* | 2.837069441 | 0.015192534 | 0.048557886 | 1 |
| HTN | t_cell | *CAMSAP2* | 2.559505802 | 0.009206381 | 0.034825822 | 1 |
| HTN | t_cell | *ATP10B* | -7.38852537 | 0.006344835 | 0.027453531 | 1 |
| HTN | t_cell | *FILIP1* | 3.389881947 | 0.000445107 | 0.0102131 | 1 |
| HTN | t_cell | *FAM47E-STBL* | 1.357260889 | 0.004370803 | 0.022232394 | 1 |
| HTN | t_cell | *UCHL3* | 4.626407191 | 0.007416642 | 0.030396627 | 1 |
| HTN | t_cell | *TRPM6* | 4.263139313 | 1.30E-05 | 0.00102461 | 0.081968823 |
| HTN | t_cell | *GDA* | 4.156276423 | 0.011738529 | 0.040837464 | 1 |
| HTN | t_cell | *C2orf40* | -3.209718808 | 0.005510506 | 0.025373449 | 1 |
| HTN | t_cell | *CNTRL* | -2.682776949 | 0.009286428 | 0.034960343 | 1 |
| HTN | t_cell | *PHF19* | -2.23759291 | 0.00080251 | 0.013695656 | 1 |
| HTN | t_cell | *BSPRY* | -4.244284805 | 0.001043715 | 0.013965754 | 1 |
| HTN | t_cell | *HDHD3* | -0.794312589 | 0.010372642 | 0.037478621 | 1 |
| HTN | t_cell | *HSDL2* | 2.224496934 | 0.000360784 | 0.009036853 | 1 |
| HTN | t_cell | *IRF2BPL* | 4.62381121 | 0.001908034 | 0.015190044 | 1 |
| HTN | t_cell | *ACOT2* | 0.697569877 | 0.014615955 | 0.047293107 | 1 |
| HTN | t_cell | *PPP4R4* | -1.835699627 | 0.014237718 | 0.046403595 | 1 |
| HTN | t_cell | *ZC2HC1C* | 2.855339621 | 0.004989037 | 0.023855009 | 1 |
| HTN | t_cell | *EIF2B2* | 2.375646079 | 0.011391528 | 0.039960965 | 1 |
| HTN | t_cell | *KLHL29* | 2.174750719 | 0.007633997 | 0.031058618 | 1 |
| HTN | t_cell | *FKBP1B* | 3.148239325 | 7.13E-05 | 0.003294966 | 0.448115372 |
| HTN | t_cell | *HELLS* | -4.540203214 | 7.81E-05 | 0.003478747 | 0.491269878 |
| HTN | t_cell | *KCNIP2* | 1.123940854 | 0.007533835 | 0.030796632 | 1 |
| HTN | t_cell | *HOXB5* | 1.602038149 | 0.013851872 | 0.045470226 | 1 |
| HTN | t_cell | *CCDC170* | 1.391819905 | 0.005375237 | 0.025032679 | 1 |

| | | | | | | |
|---|---|---|---|---|---|---|
| HTN | t_cell | *WDR55* | 4.963209971 | 0.006572927 | 0.028120771 | 1 |
| HTN | t_cell | *PCDHB10* | 2.805630682 | 0.002023816 | 0.015311349 | 1 |
| HTN | t_cell | *PCDHB14* | 2.698295739 | 0.001629147 | 0.014245408 | 1 |
| HTN | t_cell | *SEC16B* | 5.879512743 | 0.002465284 | 0.016701769 | 1 |
| HTN | t_cell | *MRC1* | 2.66241744 | 0.004330962 | 0.022083339 | 1 |
| HTN | t_cell | *IQSEC3* | 3.577411605 | 0.004004735 | 0.021084486 | 1 |
| HTN | t_cell | *ENOX1* | 2.291448826 | 0.011849291 | 0.041022299 | 1 |
| HTN | t_cell | *SMAD9* | 3.561220906 | 0.008711058 | 0.033496282 | 1 |
| HTN | t_cell | *ZFP30* | 3.636485716 | 0.000153897 | 0.005580753 | 0.96754942 |
| HTN | t_cell | *ADRA1A* | 0.66754541 | 0.002190004 | 0.015895207 | 1 |
| HTN | t_cell | *TEX14* | 7.95979818 | 2.02E-06 | 0.000230873 | 0.012698039 |
| HTN | t_cell | *PRB2* | 4.011059458 | 0.000777644 | 0.013547949 | 1 |
| HTN | t_cell | *A1BG* | -4.575221579 | 0.007561941 | 0.030871379 | 1 |
| HTN | t_cell | *PDZRN3* | 1.645747702 | 0.004707372 | 0.023211958 | 1 |
| HTN | t_cell | *CSTA* | 1.993798869 | 0.000141099 | 0.005409072 | 0.887087886 |
| HTN | t_cell | *CXCR4* | -0.605579511 | 0.000848493 | 0.013695656 | 1 |
| HTN | t_cell | *FLT3* | 1.883764002 | 0.001288117 | 0.013965754 | 1 |
| HTN | t_cell | *RASL11A* | 6.461869626 | 0.001008106 | 0.013965754 | 1 |
| HTN | t_cell | *FMOD* | 3.442715123 | 0.011396534 | 0.039960965 | 1 |
| HTN | t_cell | *LRRC39* | 1.710639707 | 0.010490145 | 0.037634027 | 1 |
| HTN | t_cell | *PMS2* | 5.059923156 | 0.011134768 | 0.039284112 | 1 |
| HTN | t_cell | *KIAA0087* | 4.509214512 | 0.008519528 | 0.0330955 | 1 |
| HTN | t_cell | *HERPUD2* | -3.188636203 | 0.010287205 | 0.037384772 | 1 |
| HTN | t_cell | *CBX3* | -5.232342482 | 0.002221626 | 0.016006305 | 1 |
| HTN | t_cell | *FKBP9* | 2.373039092 | 0.006809714 | 0.028714066 | 1 |
| HTN | t_cell | *SLC25A16* | 1.984826731 | 0.004316282 | 0.02202635 | 1 |
| HTN | t_cell | *ZWINT* | 2.630208132 | 0.001556222 | 0.014185269 | 1 |
| HTN | t_cell | *CENPK* | -0.696512633 | 0.009984754 | 0.036603002 | 1 |
| HTN | t_cell | *ATF1* | -4.42310476 | 0.005644124 | 0.02582577 | 1 |
| HTN | t_cell | *ARHGAP9* | -1.487387606 | 0.00080275 | 0.013695656 | 1 |
| HTN | t_cell | *ORMDL2* | -3.406097102 | 0.010223156 | 0.037238114 | 1 |
| HTN | t_cell | *PDE1B* | 3.639251474 | 0.004442 | 0.022485392 | 1 |
| HTN | t_cell | *IKZF4* | -7.256964899 | 0.000627139 | 0.012503451 | 1 |
| HTN | t_cell | *SARDH* | -6.246177168 | 0.001356113 | 0.013965754 | 1 |
| HTN | t_cell | *STIL* | 3.823083477 | 0.001166218 | 0.013965754 | 1 |
| HTN | t_cell | *NDUFAF4* | 1.706100379 | 0.014761292 | 0.047603702 | 1 |
| HTN | t_cell | *ACVR1C* | 2.203735087 | 4.24E-05 | 0.002335689 | 0.266307634 |
| HTN | t_cell | *MOGAT1* | 1.536941722 | 0.001810452 | 0.014955249 | 1 |
| HTN | t_cell | *FAM124B* | 4.717874604 | 2.56E-07 | 5.54E-05 | 0.001606652 |
| HTN | t_cell | *FAM210B* | 2.205442399 | 0.008515709 | 0.0330955 | 1 |
| HTN | t_cell | *SNX21* | 2.302837965 | 0.003881196 | 0.02075047 | 1 |
| HTN | t_cell | *TOX2* | -1.355700729 | 0.00029944 | 0.008171005 | 1 |
| HTN | t_cell | *KCNK15* | 3.791376822 | 0.002585981 | 0.0170778 | 1 |
| HTN | t_cell | *PEPD* | 3.532653491 | 0.000503594 | 0.010955345 | 1 |
| HTN | t_cell | *CHST8* | 5.400007354 | 0.006453974 | 0.027678129 | 1 |
| HTN | t_cell | *HIF3A* | 1.810711892 | 0.014955734 | 0.048095497 | 1 |
| HTN | t_cell | *BTN2A2* | 1.676624262 | 0.011929748 | 0.041255404 | 1 |
| HTN | t_cell | *SNRPC* | -4.841584584 | 1.01E-06 | 0.000142956 | 0.006375642 |
| HTN | t_cell | *NQO2* | 2.700266256 | 0.011340756 | 0.039809788 | 1 |
| HTN | t_cell | *RPS10* | -4.714008629 | 0.000847076 | 0.013695656 | 1 |

| | | | | | | |
|---|---|---|---|---|---|---|
| HTN | t_cell | *HIST1H2BJ* | 2.827308726 | 0.00668398 | 0.028402239 | 1 |
| HTN | t_cell | *KLHL31* | 1.036153917 | 0.006331027 | 0.027425133 | 1 |
| HTN | t_cell | *EREG* | 8.07170769 | 0.001867322 | 0.015083316 | 1 |
| HTN | t_cell | *MT1G* | 12.03236023 | 0.007966657 | 0.03174941 | 1 |
| HTN | t_cell | *CLYBL* | -2.910879625 | 0.006189408 | 0.027154784 | 1 |
| HTN | t_cell | *GRK4* | 1.546428137 | 1.49E-05 | 0.001102931 | 0.093372424 |
| HTN | t_cell | *MRPS7* | 5.14559945 | 0.001279816 | 0.013965754 | 1 |
| HTN | t_cell | *MIF4GD* | 5.548876723 | 0.012611099 | 0.042834132 | 1 |
| HTN | t_cell | *CHCHD5* | -5.555003474 | 0.000574294 | 0.01191612 | 1 |
| HTN | t_cell | *INSIG2* | -3.653242322 | 0.01091314 | 0.038697639 | 1 |
| HTN | t_cell | *PSD4* | -1.098277518 | 0.001264974 | 0.013965754 | 1 |
| HTN | t_cell | *SH2D3A* | -1.233064983 | 0.000938207 | 0.013965754 | 1 |
| HTN | t_cell | *TRIP10* | 2.666608269 | 0.005220918 | 0.024532073 | 1 |
| HTN | t_cell | *SNRPD2* | -5.098132051 | 0.002249413 | 0.01610713 | 1 |
| HTN | t_cell | *RTN2* | 3.308768352 | 0.004453629 | 0.022508072 | 1 |
| HTN | t_cell | *SDCBP2* | 3.058895752 | 0.000763429 | 0.013406921 | 1 |
| HTN | t_cell | *RRBP1* | 2.83966259 | 0.012604971 | 0.042834132 | 1 |
| HTN | t_cell | *BMP2* | 4.075993818 | 0.002993261 | 0.018217455 | 1 |
| HTN | t_cell | *ZNF436* | 2.212696895 | 0.008303752 | 0.032558321 | 1 |
| HTN | t_cell | *XRCC3* | 6.363662373 | 0.000766587 | 0.013424879 | 1 |
| HTN | t_cell | *F10* | 3.192244446 | 0.006863283 | 0.028823956 | 1 |
| HTN | t_cell | *PROZ* | -15.90588222 | 0.001408757 | 0.01399841 | 1 |
| HTN | t_cell | *IGFLR1* | -6.936139935 | 0.001288306 | 0.013965754 | 1 |
| HTN | t_cell | *TSKS* | 3.941472175 | 0.003790337 | 0.020454806 | 1 |
| HTN | t_cell | *TRAP1* | 2.146764184 | 2.74E-07 | 5.73E-05 | 0.00171974 |
| HTN | t_cell | *PZP* | -0.671628394 | 0.001547636 | 0.014185269 | 1 |
| HTN | t_cell | *RHOT1* | 3.021492544 | 0.009677846 | 0.035917722 | 1 |
| HTN | t_cell | *AIF1L* | 2.841496846 | 0.002371863 | 0.016422799 | 1 |
| HTN | t_cell | *FGD3* | -0.938410235 | 0.001129105 | 0.013965754 | 1 |
| HTN | t_cell | *MASP1* | 2.991171109 | 0.003118937 | 0.018622965 | 1 |
| HTN | t_cell | *ATP13A4* | -2.751550112 | 0.000674808 | 0.012876096 | 1 |
| HTN | t_cell | *HRASLS* | 2.77528407 | 0.001872645 | 0.015083316 | 1 |
| HTN | t_cell | *HELB* | -1.128180312 | 0.000747582 | 0.013314577 | 1 |
| HTN | t_cell | *RAB3IP* | -2.1247139 | 0.000867837 | 0.013695656 | 1 |
| HTN | t_cell | *TAS2R4* | 2.67591728 | 0.006684055 | 0.028402239 | 1 |
| HTN | t_cell | *WDR24* | 3.279451357 | 0.00092565 | 0.013965754 | 1 |
| HTN | t_cell | *TICAM1* | 4.278223035 | 0.006113074 | 0.027024952 | 1 |
| HTN | t_cell | *GNAI1* | 2.251131662 | 0.00551702 | 0.025373449 | 1 |
| HTN | t_cell | *SGCE* | 2.760026024 | 0.005733068 | 0.026005629 | 1 |
| HTN | t_cell | *MGAT3* | 1.98395211 | 0.001669549 | 0.014378705 | 1 |
| HTN | t_cell | *CDC42EP1* | 3.060214089 | 0.005784416 | 0.026200737 | 1 |
| HTN | t_cell | *BAIAP2L2* | 2.667137373 | 0.009942657 | 0.036491235 | 1 |
| HTN | t_cell | *TST* | 1.813408599 | 0.006887434 | 0.028893444 | 1 |
| HTN | t_cell | *APOL5* | -7.693534647 | 0.002441862 | 0.016594315 | 1 |
| HTN | t_cell | *FOXP2* | 1.744872274 | 0.007721884 | 0.031266496 | 1 |
| HTN | t_cell | *STRIP2* | 1.986334314 | 0.014359308 | 0.046630667 | 1 |
| HTN | t_cell | *HOXD1* | 8.517291992 | 0.00415996 | 0.021578936 | 1 |
| HTN | t_cell | *GAD1* | 4.166808688 | 0.001464552 | 0.014185269 | 1 |
| HTN | t_cell | *CALML4* | 3.042423364 | 0.001930359 | 0.015190044 | 1 |
| HTN | t_cell | *ACKR4* | 2.510034066 | 0.01428522 | 0.04650989 | 1 |

| | | | | | | |
|---|---|---|---|---|---|---|
| HTN | t_cell | *CCL4* | -1.085128767 | 0.000866454 | 0.013695656 | 1 |
| HTN | t_cell | *KRI1* | -2.217216419 | 0.002562026 | 0.017026912 | 1 |
| HTN | t_cell | *SLC44A2* | -2.915573098 | 0.007876918 | 0.031542745 | 1 |
| HTN | t_cell | *CDKN2D* | -3.335407607 | 0.000697079 | 0.012927825 | 1 |
| HTN | t_cell | *ADCY4* | 4.400107243 | 5.27E-05 | 0.002715239 | 0.331088698 |
| HTN | t_cell | *BCL2L2* | 2.936405914 | 0.004729673 | 0.023218561 | 1 |
| HTN | t_cell | *PARP2* | 2.728049727 | 0.001597344 | 0.014197745 | 1 |
| HTN | t_cell | *CDO1* | 2.308654283 | 0.00311849 | 0.018622965 | 1 |
| HTN | t_cell | *GALNT8* | -3.28830524 | 0.003685526 | 0.020236593 | 1 |
| HTN | t_cell | *DOCK6* | 2.659010679 | 0.001887148 | 0.015146366 | 1 |
| HTN | t_cell | *CDC16* | 3.973524842 | 0.000278999 | 0.007956921 | 1 |
| HTN | t_cell | *RPL36* | -2.367014444 | 0.006234044 | 0.027236577 | 1 |
| HTN | t_cell | *ATP8B3* | -1.790046026 | 0.006142328 | 0.027048346 | 1 |
| HTN | t_cell | *GTPBP3* | -4.320712797 | 0.007306852 | 0.030143163 | 1 |
| HTN | t_cell | *DDA1* | -6.725956068 | 0.000305349 | 0.008215297 | 1 |
| HTN | t_cell | *MRPL34* | 1.450329404 | 0.0139506 | 0.04568095 | 1 |
| HTN | t_cell | *STK33* | 2.619354239 | 0.00113604 | 0.013965754 | 1 |
| HTN | t_cell | *PXDN* | 1.731504022 | 0.002950421 | 0.01807841 | 1 |
| HTN | t_cell | *PGPEP1* | 4.730766958 | 6.02E-05 | 0.002888581 | 0.378404123 |
| HTN | t_cell | *UBAC1* | 3.134898399 | 0.015455651 | 0.049125217 | 1 |
| HTN | t_cell | *TNNT3* | 2.621166067 | 0.011580752 | 0.040426533 | 1 |
| HTN | t_cell | *TMEM160* | -5.812984824 | 0.015018339 | 0.048128379 | 1 |
| HTN | t_cell | *ARHGEF16* | 1.869834625 | 0.010941557 | 0.038754688 | 1 |
| HTN | t_cell | *SMPDL3B* | 9.85446349 | 0.001186998 | 0.013965754 | 1 |
| HTN | t_cell | *PPAN* | -7.309464697 | 0.005395326 | 0.025107633 | 1 |
| HTN | t_cell | *EPS8L1* | -9.516987018 | 0.001689085 | 0.014467678 | 1 |
| HTN | t_cell | *LILRB2* | 3.195239035 | 0.00214207 | 0.015732705 | 1 |
| HTN | t_cell | *CHMP1A* | 6.805395259 | 2.39E-06 | 0.000263124 | 0.014998042 |
| HTN | t_cell | *HAUS8* | -7.514702041 | 0.000291072 | 0.008097215 | 1 |
| HTN | t_cell | *PDLIM4* | 8.150624908 | 2.72E-09 | 1.66E-06 | 1.71E-05 |
| HTN | t_cell | *AOC2* | 3.641307201 | 0.003868702 | 0.020717656 | 1 |
| HTN | t_cell | *DIAPH1* | -1.895092717 | 0.000593292 | 0.012118368 | 1 |
| HTN | t_cell | *NPHP4* | 3.667929826 | 0.01061816 | 0.037886704 | 1 |
| HTN | t_cell | *KHDRBS3* | 2.303756077 | 0.001230923 | 0.013965754 | 1 |
| HTN | t_cell | *ZNF132* | 2.28199286 | 0.003241502 | 0.019003909 | 1 |
| HTN | t_cell | *C19orf57* | 6.081823429 | 0.008626513 | 0.033313811 | 1 |
| HTN | t_cell | *MATN3* | 4.340101501 | 0.0013357 | 0.013965754 | 1 |
| HTN | t_cell | *CCT6B* | 2.063178862 | 0.012725442 | 0.0430365 | 1 |
| HTN | t_cell | *CNGA4* | 3.210661667 | 0.001202701 | 0.013965754 | 1 |
| HTN | t_cell | *TRIM22* | -2.589750909 | 0.006386974 | 0.027525449 | 1 |
| HTN | t_cell | *CARD6* | 3.392232576 | 0.006703558 | 0.028432724 | 1 |
| HTN | t_cell | *KDM6B* | -7.8188004 | 0.000290725 | 0.008097215 | 1 |
| HTN | t_cell | *REEP2* | -2.068620337 | 0.001240625 | 0.013965754 | 1 |
| HTN | t_cell | *BTBD3* | 2.861185467 | 0.009722366 | 0.036019161 | 1 |
| HTN | t_cell | *LPIN3* | 2.719652675 | 0.003332379 | 0.019206707 | 1 |
| HTN | t_cell | *KANK4* | 0.898774623 | 0.008287347 | 0.032555655 | 1 |
| HTN | t_cell | *WASF3* | 2.149093905 | 0.003628774 | 0.020082835 | 1 |
| HTN | t_cell | *PEMT* | 2.367635552 | 0.002903199 | 0.018049857 | 1 |
| HTN | t_cell | *CHI3L1* | 1.107945137 | 0.002660987 | 0.017318452 | 1 |
| HTN | t_cell | *TRPC4* | 2.696685849 | 0.001785081 | 0.014844048 | 1 |

| | | | | | | |
|---|---|---|---|---|---|---|
| HTN | t_cell | *PRAM1* | -4.761127534 | 0.002045748 | 0.015375679 | 1 |
| HTN | t_cell | *SLC2A11* | 4.390925716 | 0.01057212 | 0.037801145 | 1 |
| HTN | t_cell | *FAM83F* | 3.22630116 | 0.001784188 | 0.014844048 | 1 |
| HTN | t_cell | *GIMAP4* | -1.082028564 | 0.009964731 | 0.036550914 | 1 |
| HTN | t_cell | *DYDC2* | 6.417491254 | 0.001949847 | 0.015192726 | 1 |
| HTN | t_cell | *SPINK5* | 4.094691503 | 0.009578874 | 0.035727231 | 1 |
| HTN | t_cell | *GSC* | 6.797101495 | 0.001961529 | 0.015192726 | 1 |
| HTN | t_cell | *CATSPERB* | -1.501506189 | 0.00074126 | 0.013302814 | 1 |
| HTN | t_cell | *MRO* | 2.469814896 | 0.00186704 | 0.015083316 | 1 |
| HTN | t_cell | *EDEM1* | -3.189201352 | 0.007320898 | 0.030161524 | 1 |
| HTN | t_cell | *CNTN6* | 1.647396405 | 0.002341877 | 0.016323038 | 1 |
| HTN | t_cell | *SYT6* | 6.839829468 | 0.004881644 | 0.023562138 | 1 |
| HTN | t_cell | *VAV3* | -2.57554684 | 0.001352799 | 0.013965754 | 1 |
| HTN | t_cell | *ECHDC3* | 1.534819577 | 0.00368309 | 0.020236593 | 1 |
| HTN | t_cell | *TMEM241* | 2.399698568 | 0.012238925 | 0.042047063 | 1 |
| HTN | t_cell | *KLRD1* | -3.002680246 | 0.014232871 | 0.046403595 | 1 |
| HTN | t_cell | *ACP2* | 10.95046229 | 2.04E-18 | 1.28E-14 | 1.28E-14 |
| HTN | t_cell | *HOOK1* | -1.293793275 | 0.001160563 | 0.013965754 | 1 |
| HTN | t_cell | *TPGS2* | 3.570949229 | 0.012593454 | 0.042834132 | 1 |
| HTN | t_cell | *APLNR* | 3.243454039 | 0.000203263 | 0.006698301 | 1 |
| HTN | t_cell | *TMEM165* | 4.033037155 | 0.000818898 | 0.013695656 | 1 |
| HTN | t_cell | *CLDN10* | 3.191560729 | 0.005011 | 0.023884881 | 1 |
| HTN | t_cell | *KLB* | 1.392008645 | 0.001274781 | 0.013965754 | 1 |
| HTN | t_cell | *AGTPBP1* | -2.676497056 | 0.002749031 | 0.017638397 | 1 |
| HTN | t_cell | *POM121C* | -4.21898447 | 0.00548527 | 0.025357272 | 1 |
| HTN | t_cell | *TES* | -2.351169723 | 0.01219093 | 0.041927997 | 1 |
| HTN | t_cell | *NT5E* | 2.937358286 | 0.00550256 | 0.025373449 | 1 |
| HTN | t_cell | *ITGA7* | 1.23389927 | 0.001611327 | 0.014208155 | 1 |
| HTN | t_cell | *RDH5* | 1.577698975 | 0.000819023 | 0.013695656 | 1 |
| HTN | t_cell | *BLOC1S1* | 5.823679338 | 9.05E-05 | 0.003897841 | 0.56908485 |
| HTN | t_cell | *PPP1R1A* | 2.373878395 | 0.007444042 | 0.030469199 | 1 |
| HTN | t_cell | *FAIM2* | 2.768872413 | 0.001229225 | 0.013965754 | 1 |
| HTN | t_cell | *SLC26A10* | 7.365441332 | 0.006889021 | 0.028893444 | 1 |
| HTN | t_cell | *MICAL1* | -2.915129319 | 0.005893974 | 0.026504502 | 1 |
| HTN | t_cell | *PRADC1* | 4.310528082 | 0.003286531 | 0.019043707 | 1 |
| HTN | t_cell | *KCNMB4* | 3.532444544 | 0.007358898 | 0.03025216 | 1 |
| HTN | t_cell | *AGT* | 1.718522231 | 4.78E-05 | 0.002570449 | 0.300742542 |
| HTN | t_cell | *SP110* | -2.423230832 | 0.003495093 | 0.019705361 | 1 |
| HTN | t_cell | *SLC41A2* | 3.026886677 | 0.012852426 | 0.043326115 | 1 |
| HTN | t_cell | *RNASEH2B* | -3.542943006 | 0.005514146 | 0.025373449 | 1 |
| HTN | t_cell | *SPDYE1* | 2.856031099 | 0.009347243 | 0.035105206 | 1 |
| HTN | t_cell | *CHST12* | -4.617958523 | 0.002645539 | 0.017271549 | 1 |
| HTN | t_cell | *AOAH* | -1.700461495 | 0.001283486 | 0.013965754 | 1 |
| HTN | t_cell | *ALPK3* | 1.915269417 | 0.000419915 | 0.009814148 | 1 |
| HTN | t_cell | *NMT1* | 4.317866066 | 2.90E-09 | 1.66E-06 | 1.82E-05 |
| HTN | t_cell | *BRIP1* | -2.977823946 | 0.002237123 | 0.016055701 | 1 |
| HTN | t_cell | *RTP4* | -3.009031752 | 4.12E-05 | 0.002289688 | 0.258734707 |
| HTN | t_cell | *BLK* | -2.178544205 | 2.80E-05 | 0.001703056 | 0.176028156 |
| HTN | t_cell | *SAP130* | -3.38162894 | 0.010980018 | 0.038869017 | 1 |
| HTN | t_cell | *SLC2A8* | 4.949243351 | 3.48E-05 | 0.001991638 | 0.219080155 |

| | | | | | | |
|---|---|---|---|---|---|---|
| HTN | t_cell | *GABBR2* | 7.626330727 | 0.00336658 | 0.019313893 | 1 |
| HTN | t_cell | *LMX1B* | 7.630291767 | 0.003214872 | 0.018960508 | 1 |
| HTN | t_cell | *IL11RA* | -4.325758536 | 0.002293904 | 0.016204243 | 1 |
| HTN | t_cell | *SPAG8* | 3.804535507 | 0.000816888 | 0.013695656 | 1 |
| HTN | t_cell | *TMEM8B* | 3.699340106 | 0.01421213 | 0.046368274 | 1 |
| HTN | t_cell | *PPIL1* | -3.482967748 | 0.001336542 | 0.013965754 | 1 |
| HTN | t_cell | *ZSCAN9* | 2.840407834 | 0.00056682 | 0.01191612 | 1 |
| HTN | t_cell | *FRS3* | 3.676548263 | 0.010348308 | 0.037457412 | 1 |
| HTN | t_cell | *UQCC2* | 3.34489988 | 0.015207647 | 0.048557886 | 1 |
| HTN | t_cell | *VARS2* | 0.970807625 | 0.000628454 | 0.012503451 | 1 |
| HTN | t_cell | *FHDC1* | 2.997147125 | 0.002031603 | 0.015333355 | 1 |
| HTN | t_cell | *SYTL2* | -1.749783468 | 0.00048899 | 0.010824941 | 1 |
| HTN | t_cell | *RAB30* | 2.7090109 | 0.012282401 | 0.042116263 | 1 |
| HTN | t_cell | *SULF1* | 2.750577343 | 0.001760131 | 0.014724258 | 1 |
| HTN | t_cell | *DDX60* | -3.403529686 | 0.007881923 | 0.031542745 | 1 |
| HTN | t_cell | *PPP2R1B* | 1.608010881 | 0.01035223 | 0.037457412 | 1 |
| HTN | t_cell | *FDX1* | 4.379366707 | 0.003642617 | 0.020088714 | 1 |
| HTN | t_cell | *FXYD6* | 3.393248729 | 0.005499934 | 0.025373449 | 1 |
| HTN | t_cell | *TMPRSS13* | -2.590444507 | 0.01051201 | 0.037657554 | 1 |
| HTN | t_cell | *CASP1* | -3.724708795 | 0.003899207 | 0.020827297 | 1 |
| HTN | t_cell | *PAQR5* | 1.845095578 | 0.014345105 | 0.046608617 | 1 |
| HTN | t_cell | *RMDN3* | 5.712558672 | 1.74E-06 | 0.00020668 | 0.010954033 |
| HTN | t_cell | *PLCB2* | -1.485128156 | 0.001567416 | 0.014185269 | 1 |
| HTN | t_cell | *SLC44A5* | 2.562370471 | 0.015576827 | 0.049360641 | 1 |
| HTN | t_cell | *DNASE2B* | 8.135710908 | 0.000798291 | 0.013695656 | 1 |
| HTN | t_cell | *IFT172* | 4.194864331 | 0.000145493 | 0.005440117 | 0.914716087 |
| HTN | t_cell | *KHK* | -1.122651459 | 0.003584681 | 0.019944152 | 1 |
| HTN | t_cell | *DYNC2LI1* | 3.096241169 | 1.76E-05 | 0.001257422 | 0.110653164 |
| HTN | t_cell | *LHCGR* | 1.279160988 | 0.001996212 | 0.015283681 | 1 |
| HTN | t_cell | *PREB* | 2.608913054 | 0.005063323 | 0.024043138 | 1 |
| HTN | t_cell | *ABCG5* | 2.60505474 | 0.00066169 | 0.012858576 | 1 |
| HTN | t_cell | *DTNB* | -2.11637692 | 0.009278942 | 0.034953091 | 1 |
| HTN | t_cell | *ATAD1* | -3.668165022 | 0.01305855 | 0.043726295 | 1 |
| HTN | t_cell | *BTBD16* | 2.251006318 | 0.005445679 | 0.025248513 | 1 |
| HTN | t_cell | *TACC2* | 1.798918809 | 0.005650041 | 0.025834044 | 1 |
| HTN | t_cell | *ARL3* | 6.449391784 | 6.94E-05 | 0.003253998 | 0.436035753 |
| HTN | t_cell | *PLCE1* | 2.461878935 | 0.007241285 | 0.029985826 | 1 |
| HTN | t_cell | *RBP4* | 2.859197267 | 0.008522605 | 0.0330955 | 1 |
| HTN | t_cell | *FAM149B1* | 3.075788191 | 0.008306593 | 0.032558321 | 1 |
| HTN | t_cell | *RPS24* | -2.871644829 | 0.007715028 | 0.031266496 | 1 |
| HTN | t_cell | *BARD1* | -4.202929776 | 0.003044215 | 0.018351697 | 1 |
| HTN | t_cell | *STAT4* | -0.490337599 | 0.001557729 | 0.014185269 | 1 |
| HTN | t_cell | *MDH1B* | 3.27413576 | 0.00265337 | 0.017304706 | 1 |
| HTN | t_cell | *SENP7* | -2.839270228 | 0.015119189 | 0.048373711 | 1 |
| HTN | t_cell | *FAM13A* | 2.507575085 | 0.009255731 | 0.03489313 | 1 |
| HTN | t_cell | *CNOT6L* | -1.859416816 | 0.000805929 | 0.013695656 | 1 |
| HTN | t_cell | *ANXA3* | 2.595510042 | 0.001960653 | 0.015192726 | 1 |
| HTN | t_cell | *FBN2* | 1.398342196 | 1.13E-05 | 0.000925454 | 0.071259971 |
| HTN | t_cell | *PARVG* | -2.686120586 | 0.000376036 | 0.009198979 | 1 |
| HTN | t_cell | *B4GALNT3* | -3.30863407 | 0.001539322 | 0.014185269 | 1 |

| | | | | | | |
|---|---|---|---|---|---|---|
| HTN | t_cell | *ERP27* | -1.07258066 | 0.000670236 | 0.012876096 | 1 |
| HTN | t_cell | *ALG10* | -5.907876022 | 0.004658007 | 0.023113569 | 1 |
| HTN | t_cell | *SLCO1C1* | 3.753429813 | 0.000244864 | 0.007330748 | 1 |
| HTN | t_cell | *PRICKLE1* | 1.994253078 | 0.00281261 | 0.017736088 | 1 |
| HTN | t_cell | *CLSTN3* | -2.886622838 | 0.002005274 | 0.015283681 | 1 |
| HTN | t_cell | *KLRG1* | -0.940116583 | 0.002000086 | 0.015283681 | 1 |
| HTN | t_cell | *CD27* | -0.684308701 | 0.002335738 | 0.016323038 | 1 |
| HTN | t_cell | *POC1B* | -3.37253761 | 0.014064962 | 0.0460075 | 1 |
| HTN | t_cell | *GAS2L3* | 3.48333907 | 0.009405687 | 0.035261509 | 1 |
| HTN | t_cell | *MMAB* | 4.778093429 | 1.65E-05 | 0.001208179 | 0.103903395 |
| HTN | t_cell | *ACVRL1* | 2.675432936 | 0.001104384 | 0.013965754 | 1 |
| HTN | t_cell | *LMBR1L* | -4.953818785 | 0.001559738 | 0.014185269 | 1 |
| HTN | t_cell | *HNRNPA1L2* | -6.800165598 | 0.003465084 | 0.019608443 | 1 |
| HTN | t_cell | *RHOF* | -3.542325373 | 0.001918771 | 0.015190044 | 1 |
| HTN | t_cell | *GRTP1* | 1.468896472 | 0.002523857 | 0.016925325 | 1 |
| HTN | t_cell | *RTN1* | 2.636730208 | 0.005961737 | 0.026620342 | 1 |
| HTN | t_cell | *EFCAB11* | 2.566806337 | 0.000952574 | 0.013965754 | 1 |
| HTN | t_cell | *AK7* | 6.612583437 | 0.003827576 | 0.020620371 | 1 |
| HTN | t_cell | *FBLN5* | 2.903016045 | 0.002620942 | 0.017200276 | 1 |
| HTN | t_cell | *DUOXA1* | 3.669172825 | 0.002351976 | 0.016336366 | 1 |
| HTN | t_cell | *SORD* | 5.613087005 | 0.01479971 | 0.047691178 | 1 |
| HTN | t_cell | *LYSMD2* | -3.092329395 | 0.002381309 | 0.016470068 | 1 |
| HTN | t_cell | *PSTPIP1* | -1.270771775 | 0.002418538 | 0.016563558 | 1 |
| HTN | t_cell | *NEIL1* | 3.371942813 | 0.003143962 | 0.018722726 | 1 |
| HTN | t_cell | *CYP11A1* | 1.260528472 | 0.009900366 | 0.036378494 | 1 |
| HTN | t_cell | *HAPLN3* | -2.389781216 | 0.000135525 | 0.005227285 | 0.852047407 |
| HTN | t_cell | *TICRR* | 6.241245536 | 2.96E-05 | 0.00175547 | 0.186079831 |
| HTN | t_cell | *MARVELD3* | 4.312292523 | 0.000988598 | 0.013965754 | 1 |
| HTN | t_cell | *KIFC3* | 2.70724704 | 0.00621066 | 0.027210047 | 1 |
| HTN | t_cell | *NUDT7* | 2.788470079 | 0.0077844 | 0.031318911 | 1 |
| HTN | t_cell | *GCSH* | 1.480753106 | 0.00376196 | 0.020414857 | 1 |
| HTN | t_cell | *RPS2* | -2.384487262 | 0.0006931 | 0.012921657 | 1 |
| HTN | t_cell | *DEF8* | 5.301364402 | 7.07E-05 | 0.00329156 | 0.44436056 |
| HTN | t_cell | *SCRN2* | -0.904588399 | 0.006447674 | 0.027678129 | 1 |
| HTN | t_cell | *ARSG* | -2.16209887 | 0.007349626 | 0.030240246 | 1 |
| HTN | t_cell | *IMPA2* | 3.323371298 | 0.005328149 | 0.024885186 | 1 |
| HTN | t_cell | *C18orf21* | 3.841375491 | 0.013488833 | 0.044516691 | 1 |
| HTN | t_cell | *ASXL3* | 4.256583094 | 0.00263928 | 0.017264821 | 1 |
| HTN | t_cell | *C18orf8* | -5.235769131 | 0.000596897 | 0.012144635 | 1 |
| HTN | t_cell | *NPC1* | -5.062786096 | 0.000195025 | 0.006553493 | 1 |
| HTN | t_cell | *SLC14A1* | -1.04016473 | 0.006253787 | 0.027260919 | 1 |
| HTN | t_cell | *TMC6* | -1.089496413 | 0.010372594 | 0.037478621 | 1 |
| HTN | t_cell | *CBX8* | 5.020331014 | 0.000703805 | 0.012938078 | 1 |
| HTN | t_cell | *RNF157* | -0.923384995 | 0.000331141 | 0.00863852 | 1 |
| HTN | t_cell | *GRB7* | 3.766386734 | 0.00088921 | 0.013906632 | 1 |
| HTN | t_cell | *TXNL4A* | -2.536947689 | 0.014107314 | 0.046081546 | 1 |
| HTN | t_cell | *CCDC97* | -4.776440124 | 0.013466838 | 0.044490809 | 1 |
| HTN | t_cell | *SIRT3* | 3.737664206 | 0.002047768 | 0.015375679 | 1 |
| HTN | t_cell | *IFITM3* | 3.171984091 | 0.015302492 | 0.048815976 | 1 |
| HTN | t_cell | *WTIP* | 2.847171414 | 0.012230081 | 0.04203965 | 1 |

| | | | | | | |
|---|---|---|---|---|---|---|
| HTN | t_cell | *MYO1F* | -1.402925968 | 0.001356657 | 0.013965754 | 1 |
| HTN | t_cell | *CACNG8* | 3.446627935 | 0.001222222 | 0.013965754 | 1 |
| HTN | t_cell | *RCN3* | 2.869195473 | 0.003818827 | 0.020590881 | 1 |
| HTN | t_cell | *SH3BGRL3* | -3.559111591 | 0.008227509 | 0.032409993 | 1 |
| HTN | t_cell | *MAP3K6* | 2.789801351 | 0.009047674 | 0.03440522 | 1 |
| HTN | t_cell | *RPS8* | -2.815830507 | 0.000519822 | 0.011154004 | 1 |
| HTN | t_cell | *MOB3C* | 5.058804308 | 0.004593597 | 0.022916271 | 1 |
| HTN | t_cell | *CYP4B1* | 2.027684632 | 0.000188061 | 0.006391036 | 1 |
| HTN | t_cell | *SYPL2* | 2.519845511 | 1.48E-05 | 0.001102931 | 0.093346272 |
| HTN | t_cell | *ZNF697* | 3.618759655 | 0.001025487 | 0.013965754 | 1 |
| HTN | t_cell | *GPR161* | 3.536396445 | 0.002852075 | 0.01787445 | 1 |
| HTN | t_cell | *GPA33* | -1.951791886 | 0.000416012 | 0.009759197 | 1 |
| HTN | t_cell | *RXRG* | 3.948351476 | 0.001796801 | 0.014903014 | 1 |
| HTN | t_cell | *XCL2* | -2.981526778 | 0.006493852 | 0.02783016 | 1 |
| HTN | t_cell | *ADCY10* | 2.622348822 | 0.001824771 | 0.014996514 | 1 |
| HTN | t_cell | *TUFT1* | 2.068367101 | 0.000876981 | 0.013783948 | 1 |
| HTN | t_cell | *SELENBP1* | 2.358381085 | 0.001169155 | 0.013965754 | 1 |
| HTN | t_cell | *SEMA6C* | 3.305209331 | 0.000924832 | 0.013965754 | 1 |
| HTN | t_cell | *IKBKE* | -1.524005706 | 0.001232552 | 0.013965754 | 1 |
| HTN | t_cell | *DYRK3* | 2.252839483 | 0.003270715 | 0.019032949 | 1 |
| HTN | t_cell | *HHIPL2* | 4.446375365 | 0.001043614 | 0.013965754 | 1 |
| HTN | t_cell | *CREB3L4* | -2.573420152 | 0.010940503 | 0.038754688 | 1 |
| HTN | t_cell | *KCNN3* | 3.896154445 | 0.002168777 | 0.01581798 | 1 |
| HTN | t_cell | *SCCPDH* | 2.380773257 | 0.012721412 | 0.0430365 | 1 |
| HTN | t_cell | *CEP170* | 4.97133338 | 2.61E-05 | 0.001623963 | 0.164020238 |
| HTN | t_cell | *SNAP47* | 4.355461131 | 0.000726383 | 0.013156092 | 1 |
| HTN | t_cell | *WNT9A* | 4.573664834 | 5.36E-06 | 0.000534608 | 0.033680292 |
| HTN | t_cell | *PLEKHA6* | 2.461705476 | 0.000146235 | 0.005440117 | 0.919379787 |
| HTN | t_cell | *EML4* | -1.705940735 | 0.002392579 | 0.016483077 | 1 |
| HTN | t_cell | *TMEM177* | 4.566133749 | 0.005299065 | 0.024788113 | 1 |
| HTN | t_cell | *SLC20A1* | -3.362854033 | 0.001095702 | 0.013965754 | 1 |
| HTN | t_cell | *THSD7B* | 2.390415627 | 0.000279701 | 0.007956921 | 1 |
| HTN | t_cell | *GPR17* | 1.221915106 | 0.000206627 | 0.006718648 | 1 |
| HTN | t_cell | *ZNF385B* | 1.820581705 | 0.009257474 | 0.03489313 | 1 |
| HTN | t_cell | *TMEFF2* | 2.37154486 | 0.002956961 | 0.01807841 | 1 |
| HTN | t_cell | *ESPNL* | 14.61142425 | 0.001349979 | 0.013965754 | 1 |
| HTN | t_cell | *FANCD2* | -2.507149565 | 0.004315749 | 0.02202635 | 1 |
| HTN | t_cell | *OSBPL10* | 2.558794325 | 0.007403268 | 0.030370454 | 1 |
| HTN | t_cell | *SLC25A38* | -5.171569635 | 0.001562049 | 0.014185269 | 1 |
| HTN | t_cell | *NXPE3* | -3.481446988 | 0.011447083 | 0.040093487 | 1 |
| HTN | t_cell | *BOC* | 2.269314438 | 0.000852588 | 0.013695656 | 1 |
| HTN | t_cell | *EAF2* | 3.194522545 | 0.007719497 | 0.031266496 | 1 |
| HTN | t_cell | *LYAR* | -4.461091917 | 0.006227548 | 0.027227116 | 1 |
| HTN | t_cell | *ATP10D* | 2.613301913 | 0.013857317 | 0.045470226 | 1 |
| HTN | t_cell | *SLC10A4* | 13.37478295 | 7.22E-07 | 0.000113436 | 0.004537452 |
| HTN | t_cell | *LHFPL2* | 2.713379112 | 0.006255448 | 0.027260919 | 1 |
| HTN | t_cell | *GTF2H2* | 3.31023102 | 0.007101363 | 0.029527954 | 1 |
| HTN | t_cell | *SPATA9* | 1.737276406 | 0.003389404 | 0.019407271 | 1 |
| HTN | t_cell | *MYLK4* | 3.136905919 | 0.002908548 | 0.018049857 | 1 |
| HTN | t_cell | *GFOD1* | 2.316273035 | 0.001420704 | 0.014021922 | 1 |

| | | | | | | |
|---|---|---|---|---|---|---|
| HTN | t_cell | *ABT1* | 4.417948646 | 4.55E-06 | 0.000461793 | 0.028631157 |
| HTN | t_cell | *MLIP* | 2.375178007 | 0.001423359 | 0.014026106 | 1 |
| HTN | t_cell | *FGD2* | 2.505641544 | 0.013188711 | 0.043964702 | 1 |
| HTN | t_cell | *TPBG* | 3.623611399 | 0.00633391 | 0.027425133 | 1 |
| HTN | t_cell | *IRAK1BP1* | 3.520425494 | 0.005846699 | 0.026387793 | 1 |
| HTN | t_cell | *MMS22L* | -3.797421072 | 0.009807286 | 0.036174836 | 1 |
| HTN | t_cell | *ABRACL* | -1.909660255 | 0.004560606 | 0.022792156 | 1 |
| HTN | t_cell | *SLC18B1* | -1.592923038 | 0.00045489 | 0.010320043 | 1 |
| HTN | t_cell | *AIG1* | 2.403238464 | 0.006584026 | 0.028120771 | 1 |
| HTN | t_cell | *DYNLT1* | -2.817812966 | 0.000781124 | 0.013566092 | 1 |
| HTN | t_cell | *SLC22A3* | 2.826229312 | 0.007767149 | 0.031318911 | 1 |
| HTN | t_cell | *EGFR* | 1.725161157 | 0.010179685 | 0.0371602 | 1 |
| HTN | t_cell | *EPHA1* | -2.139445326 | 0.009704605 | 0.035974558 | 1 |
| HTN | t_cell | *CCDC25* | 4.50736217 | 0.011980315 | 0.041384748 | 1 |
| HTN | t_cell | *ST18* | 2.699252388 | 0.001304968 | 0.013965754 | 1 |
| HTN | t_cell | *SLC26A7* | 1.986050859 | 0.001012096 | 0.013965754 | 1 |
| HTN | t_cell | *MAL2* | 1.517723778 | 0.002482091 | 0.016797534 | 1 |
| HTN | t_cell | *FAM135B* | 2.476176062 | 0.001048796 | 0.013965754 | 1 |
| HTN | t_cell | *ZNF7* | 3.76096078 | 0.010894221 | 0.038674176 | 1 |
| HTN | t_cell | *CER1* | 3.589282978 | 0.003549945 | 0.019909231 | 1 |
| HTN | t_cell | *ZNF462* | 2.630961505 | 0.010350695 | 0.037457412 | 1 |
| HTN | t_cell | *WDR31* | 2.268203154 | 0.000855789 | 0.013695656 | 1 |
| HTN | t_cell | *SURF1* | 2.888542777 | 0.000681794 | 0.01291096 | 1 |
| HTN | t_cell | *SLC39A12* | 3.103638136 | 0.015428451 | 0.049125217 | 1 |
| HTN | t_cell | *HTR7* | 4.943724058 | 0.004086147 | 0.021283851 | 1 |
| HTN | t_cell | *PLEKHS1* | 9.01654682 | 0.000648743 | 0.012745772 | 1 |
| HTN | t_cell | *FUOM* | 2.722224105 | 0.01572033 | 0.049524434 | 1 |
| HTN | t_cell | *MTG1* | 2.113047186 | 0.004596405 | 0.022916271 | 1 |
| HTN | t_cell | *PDCD11* | 10.50335101 | 4.07E-09 | 1.72E-06 | 2.56E-05 |
| HTN | t_cell | *RGS10* | -3.168533226 | 0.002012291 | 0.015300078 | 1 |
| HTN | t_cell | *ZNF215* | 1.437861338 | 0.002612155 | 0.017160523 | 1 |
| HTN | t_cell | *HSD17B12* | -2.073740759 | 0.00012955 | 0.005058897 | 0.814482351 |
| HTN | t_cell | *EIF3M* | -3.952548604 | 0.013093947 | 0.043811411 | 1 |
| HTN | t_cell | *GLYAT* | 1.3755215 | 0.003241837 | 0.019003909 | 1 |
| HTN | t_cell | *PTPRJ* | -3.472568216 | 0.000204239 | 0.006698301 | 1 |
| HTN | t_cell | *CAPN5* | 3.644624626 | 0.003918458 | 0.020877409 | 1 |
| HTN | t_cell | *RPS3* | -1.717007175 | 0.002657097 | 0.017311056 | 1 |
| HTN | t_cell | *ZC3H12C* | 2.78839923 | 0.010418982 | 0.03753819 | 1 |
| HTN | t_cell | *DRD2* | 11.16634181 | 0.006519885 | 0.027922694 | 1 |
| HTN | t_cell | *C11orf52* | 10.38344548 | 0.001299409 | 0.013965754 | 1 |
| HTN | t_cell | *GLB1L2* | -1.963455739 | 0.012869371 | 0.043346584 | 1 |
| HTN | t_cell | *P4HA3* | 3.463741286 | 0.001000602 | 0.013965754 | 1 |
| HTN | t_cell | *MTA2* | -3.62169847 | 0.01385241 | 0.045470226 | 1 |
| HTN | t_cell | *TMC2* | -8.631927097 | 0.015706936 | 0.049524434 | 1 |
| HTN | t_cell | *INCENP* | 4.620412485 | 0.004863183 | 0.023537205 | 1 |
| HTN | t_cell | *MS4A3* | 26.54657304 | 0.001409947 | 0.01399841 | 1 |
| HTN | t_cell | *PLCH2* | -1.009146586 | 0.001092676 | 0.013965754 | 1 |
| HTN | t_cell | *KIAA1755* | 2.267981941 | 0.006181 | 0.027136836 | 1 |
| HTN | t_cell | *CNBD2* | 3.729943732 | 0.002793075 | 0.017683851 | 1 |
| HTN | t_cell | *TBX6* | 2.093700805 | 0.003939233 | 0.020880704 | 1 |

| | | | | | | |
|---|---|---|---|---|---|---|
| HTN | t_cell | *MPP7* | -1.281265457 | 0.014994832 | 0.048098218 | 1 |
| HTN | t_cell | *SAP18* | 4.313895162 | 0.009594325 | 0.035734315 | 1 |
| HTN | t_cell | *PRSS23* | 1.803055284 | 0.005437225 | 0.025227921 | 1 |
| HTN | t_cell | *PPP1R1C* | 2.376305831 | 0.002898639 | 0.018049857 | 1 |
| HTN | t_cell | *C11orf53* | 9.705416007 | 0.002831989 | 0.017822536 | 1 |
| HTN | t_cell | *CCT5* | 3.598819926 | 0.015723043 | 0.049524434 | 1 |
| HTN | t_cell | *FAM173B* | 3.443426104 | 0.006310694 | 0.02737997 | 1 |
| HTN | t_cell | *TEX12* | 9.163435948 | 0.001340123 | 0.013965754 | 1 |
| HTN | t_cell | *PIP4K2A* | -1.510095621 | 0.002696714 | 0.017460596 | 1 |
| HTN | t_cell | *GPR158* | 2.159993026 | 0.010871571 | 0.038632973 | 1 |
| HTN | t_cell | *BTBD11* | -0.93195972 | 0.001127504 | 0.013965754 | 1 |
| HTN | t_cell | *ANK3* | -0.476024707 | 0.00176084 | 0.014724258 | 1 |
| HTN | t_cell | *MIPOL1* | 2.164225201 | 0.006091279 | 0.027009862 | 1 |
| HTN | t_cell | *ME3* | 2.457683349 | 0.00428891 | 0.021964306 | 1 |
| HTN | t_cell | *ADAMTS12* | 1.640062062 | 0.003857949 | 0.02069343 | 1 |
| HTN | t_cell | *ANKRD50* | 3.004108805 | 0.011122201 | 0.039283865 | 1 |
| HTN | t_cell | *AKR1C2* | 1.712851759 | 0.006258872 | 0.027260919 | 1 |
| HTN | t_cell | *ADAM8* | -1.223816197 | 0.001384868 | 0.013965754 | 1 |
| HTN | t_cell | *ITIH2* | 4.821305241 | 0.002392154 | 0.016483077 | 1 |
| HTN | t_cell | *MFSD6* | -3.377712094 | 0.007954621 | 0.031732678 | 1 |
| HTN | t_cell | *ASAP2* | 2.506792534 | 0.011748643 | 0.040837464 | 1 |
| HTN | t_cell | *TMEM45B* | -0.928106144 | 0.001662629 | 0.014360746 | 1 |
| HTN | t_cell | *ACSL1* | 1.380252708 | 0.012695416 | 0.043004354 | 1 |
| HTN | t_cell | *CENPJ* | 3.176969833 | 0.005723077 | 0.026005629 | 1 |
| HTN | t_cell | *PARP8* | -0.998980031 | 0.003240791 | 0.019003909 | 1 |
| HTN | t_cell | *RABGAP1L* | -1.765172866 | 0.012637678 | 0.042901233 | 1 |
| HTN | t_cell | *PTPN14* | 2.850577166 | 0.001159443 | 0.013965754 | 1 |
| HTN | t_cell | *G6PC2* | -30.47185056 | 0.001116597 | 0.013965754 | 1 |
| HTN | t_cell | *TGOLN2* | -3.528439547 | 0.009218246 | 0.034828794 | 1 |
| HTN | t_cell | *GUCY1A2* | 2.578312274 | 0.000607854 | 0.012248652 | 1 |
| HTN | t_cell | *TMEM123* | -2.500845528 | 0.003180379 | 0.018851478 | 1 |
| HTN | t_cell | *CLEC4F* | 6.060374482 | 0.001586181 | 0.014185269 | 1 |
| HTN | t_cell | *PELO* | 4.13464237 | 0.003575238 | 0.019909231 | 1 |
| HTN | t_cell | *WDR78* | 3.028929345 | 0.00060594 | 0.012248652 | 1 |
| HTN | t_cell | *IFIT5* | -2.629938932 | 0.010582156 | 0.037801145 | 1 |
| HTN | t_cell | *PRDM8* | -2.15007288 | 1.21E-05 | 0.000978456 | 0.076319579 |
| HTN | t_cell | *RAB3C* | 2.787080317 | 0.000476933 | 0.010708859 | 1 |
| HTN | t_cell | *CENPH* | 2.37256395 | 0.012456151 | 0.042491491 | 1 |
| HTN | t_cell | *TEKT5* | -5.272486468 | 0.003942325 | 0.020880704 | 1 |
| HTN | t_cell | *BANK1* | 2.715738951 | 0.008531149 | 0.03309951 | 1 |
| HTN | t_cell | *SCOC* | 3.224404696 | 0.004192141 | 0.021679382 | 1 |
| HTN | t_cell | *CLGN* | 7.970629001 | 0.000906614 | 0.013965754 | 1 |
| HTN | t_cell | *SYCP2L* | 2.807543098 | 0.002913329 | 0.018049857 | 1 |
| HTN | t_cell | *PTPRR* | 2.215905467 | 0.003426382 | 0.01949243 | 1 |
| HTN | t_cell | *PLA2R1* | 2.491389272 | 0.000378327 | 0.009219153 | 1 |
| HTN | t_cell | *FRMD1* | 1.536438581 | 0.001313917 | 0.013965754 | 1 |
| HTN | t_cell | *INO80C* | 3.926497318 | 0.012550075 | 0.042782049 | 1 |
| HTN | t_cell | *TEX29* | 2.765482409 | 0.014994795 | 0.048098218 | 1 |
| HTN | t_cell | *FAM92B* | 17.12939205 | 0.001340373 | 0.013965754 | 1 |
| HTN | t_cell | *JAZF1* | -3.039217176 | 0.012783405 | 0.043162871 | 1 |

| | | | | | | |
|---|---|---|---|---|---|---|
| HTN | t_cell | *MCOLN2* | -0.807252017 | 0.001227366 | 0.013965754 | 1 |
| HTN | t_cell | *CACNA2D1* | 1.652943516 | 0.007038566 | 0.029305605 | 1 |
| HTN | t_cell | *HS3ST3A1* | 3.339202591 | 3.07E-05 | 0.001803661 | 0.192991774 |
| HTN | t_cell | *UBASH3B* | -1.448111408 | 0.002099399 | 0.015564764 | 1 |
| HTN | t_cell | *PRKCA* | -1.459808655 | 0.001547136 | 0.014185269 | 1 |
| HTN | t_cell | *CEP112* | 2.127502353 | 0.006154468 | 0.027077075 | 1 |
| HTN | t_cell | *ENPP3* | 2.854148111 | 0.011204859 | 0.039464956 | 1 |
| HTN | t_cell | *UCHL1* | 1.794213131 | 0.004508798 | 0.022659323 | 1 |
| HTN | t_cell | *DISP1* | 3.26884589 | 9.15E-05 | 0.003914598 | 0.575445877 |
| HTN | t_cell | *NEIL2* | 5.433413162 | 4.09E-09 | 1.72E-06 | 2.57E-05 |
| HTN | t_cell | *CCSAP* | -3.565718136 | 0.001190904 | 0.013965754 | 1 |
| HTN | t_cell | *GBP5* | -0.522329267 | 0.001315927 | 0.013965754 | 1 |
| HTN | t_cell | *FAM69A* | -3.038524651 | 0.007713672 | 0.031266496 | 1 |
| HTN | t_cell | *PDE1C* | 4.053364726 | 0.003696158 | 0.020277262 | 1 |
| HTN | t_cell | *TSEN2* | 3.088231302 | 0.004269353 | 0.021914345 | 1 |
| HTN | t_cell | *SLFN13* | -1.343262642 | 0.000837212 | 0.013695656 | 1 |
| HTN | t_cell | *CYP2U1* | 3.561294142 | 0.006247588 | 0.027260919 | 1 |
| HTN | t_cell | *GOLGA7B* | -4.103837533 | 3.24E-05 | 0.001888736 | 0.203983455 |
| HTN | t_cell | *KDM8* | -4.182405371 | 0.008580572 | 0.033218015 | 1 |
| HTN | t_cell | *SPAG17* | 1.757792581 | 0.008056813 | 0.031979187 | 1 |
| HTN | t_cell | *RRAGA* | 5.716893438 | 0.012592858 | 0.042834132 | 1 |
| HTN | t_cell | *UGT2B4* | 6.32849683 | 0.005371434 | 0.025032679 | 1 |
| HTN | t_cell | *BATF* | -0.764870081 | 0.001089066 | 0.013965754 | 1 |
| HTN | t_cell | *NDUFAF6* | -3.776703788 | 0.003524696 | 0.019803187 | 1 |
| HTN | t_cell | *CXCL13* | 5.076397369 | 0.011728508 | 0.040837464 | 1 |
| HTN | t_cell | *SFXN2* | 6.065053854 | 2.64E-05 | 0.001628023 | 0.166058357 |
| HTN | t_cell | *GDF6* | 4.514049798 | 0.001928957 | 0.015190044 | 1 |
| HTN | t_cell | *NODAL* | 3.553659851 | 0.001986956 | 0.015283681 | 1 |
| HTN | t_cell | *MED19* | 2.052874101 | 0.011287135 | 0.039665856 | 1 |
| HTN | t_cell | *ATAD2* | -3.402959534 | 0.008809375 | 0.033688892 | 1 |
| HTN | t_cell | *MPV17L* | 2.400677017 | 0.000143652 | 0.005440117 | 0.903140673 |
| HTN | t_cell | *GHRL* | -4.714767989 | 0.015771349 | 0.04962013 | 1 |
| HTN | t_cell | *TIMP4* | 2.77730193 | 0.003243469 | 0.019003909 | 1 |
| HTN | t_cell | *NECAP2* | -4.376013464 | 0.007263667 | 0.030043863 | 1 |
| HTN | t_cell | *LRP8* | -2.58302168 | 0.001296876 | 0.013965754 | 1 |
| HTN | t_cell | *SUSD3* | -0.708644826 | 0.005005633 | 0.023877401 | 1 |
| HTN | t_cell | *TMED6* | 7.237107078 | 0.00148351 | 0.014185269 | 1 |
| HTN | t_cell | *CLEC18A* | -6.086690151 | 1.43E-06 | 0.000184061 | 0.009018989 |
| HTN | t_cell | *IL34* | 2.329450659 | 0.005592553 | 0.025683261 | 1 |
| HTN | t_cell | *KCNJ15* | -3.417391418 | 0.004083722 | 0.021283851 | 1 |
| HTN | t_cell | *CREB3L1* | 2.878878872 | 0.013792535 | 0.045352337 | 1 |
| HTN | t_cell | *WDR19* | 2.58871082 | 0.008521547 | 0.0330955 | 1 |
| HTN | t_cell | *RADIL* | 2.743192979 | 0.002896208 | 0.018049857 | 1 |
| HTN | t_cell | *LDLRAP1* | -2.226635787 | 0.001601735 | 0.014197745 | 1 |
| HTN | t_cell | *TRIM63* | 3.860713653 | 1.75E-05 | 0.001257422 | 0.110225117 |
| HTN | t_cell | *WDR66* | 1.761665509 | 0.000717693 | 0.013040852 | 1 |
| HTN | t_cell | *HPD* | 2.680470135 | 0.001896025 | 0.015177294 | 1 |
| HTN | t_cell | *LRRC43* | -13.37104876 | 0.004534038 | 0.022712185 | 1 |
| HTN | t_cell | *MRAS* | 1.342560293 | 0.012080996 | 0.041663864 | 1 |
| HTN | t_cell | *CLSTN2* | 1.250745471 | 0.008077626 | 0.032040401 | 1 |

| | | | | | | |
|---|---|---|---|---|---|---|
| HTN | t_cell | *RNF207* | 5.139342347 | 0.008677512 | 0.033442532 | 1 |
| HTN | t_cell | *FAM86C1* | 3.273389422 | 0.001929148 | 0.015190044 | 1 |
| HTN | t_cell | *NCF1* | 4.045292647 | 0.002092905 | 0.015534937 | 1 |
| HTN | t_cell | *ZFAND2B* | -4.204089674 | 0.005260025 | 0.024671565 | 1 |
| HTN | t_cell | *DYNC1I1* | 2.627049494 | 0.004982632 | 0.023855009 | 1 |
| HTN | t_cell | *DUSP23* | 3.802072273 | 0.002227695 | 0.016006305 | 1 |
| HTN | t_cell | *RNF166* | -1.987781637 | 0.001697705 | 0.014482324 | 1 |
| HTN | t_cell | *SLC5A11* | -1.505253165 | 0.000250056 | 0.007450726 | 1 |
| HTN | t_cell | *FCER1G* | 2.482905614 | 0.001930182 | 0.015190044 | 1 |
| HTN | t_cell | *CDC42SE2* | -1.266772163 | 0.001105874 | 0.013965754 | 1 |
| HTN | t_cell | *RAPGEF6* | -4.458410469 | 0.002449614 | 0.016613506 | 1 |
| HTN | t_cell | *IFNAR2* | -2.065978913 | 0.004997144 | 0.023855009 | 1 |
| HTN | t_cell | *SV2A* | -4.492496696 | 0.007211718 | 0.029887984 | 1 |
| HTN | t_cell | *SIM2* | 2.40820714 | 0.000569485 | 0.01191612 | 1 |
| HTN | t_cell | *TCHH* | 3.04500547 | 0.010304028 | 0.037424276 | 1 |
| HTN | t_cell | *MED8* | 4.975140307 | 4.07E-07 | 7.75E-05 | 0.002557239 |
| HTN | t_cell | *TEPP* | -6.602094461 | 0.000486533 | 0.010824941 | 1 |
| HTN | t_cell | *ZFYVE28* | -1.810800537 | 0.001346629 | 0.013965754 | 1 |
| HTN | t_cell | *C16orf86* | 4.30577332 | 0.00023859 | 0.007211618 | 1 |
| HTN | t_cell | *NPR2* | 3.525569337 | 0.00963321 | 0.035836682 | 1 |
| HTN | t_cell | *DFFA* | 3.092140266 | 0.006220889 | 0.027220094 | 1 |
| HTN | t_cell | *NR2F6* | 2.776569902 | 0.011948918 | 0.041298982 | 1 |
| HTN | t_cell | *CILP2* | 11.7412149 | 0.00138356 | 0.013965754 | 1 |
| HTN | t_cell | *SLC37A1* | -4.114372223 | 0.014696772 | 0.047481296 | 1 |
| HTN | t_cell | *ITGB2* | -1.48126234 | 0.006083999 | 0.027009862 | 1 |
| HTN | t_cell | *FTCD* | 1.545831943 | 0.004204755 | 0.021679382 | 1 |
| HTN | t_cell | *S100B* | -0.971088572 | 0.003108993 | 0.018615466 | 1 |
| HTN | t_cell | *FCN2* | 1.986190833 | 0.004855901 | 0.023537205 | 1 |
| HTN | t_cell | *HIPK4* | 5.227177648 | 0.006082656 | 0.027009862 | 1 |
| HTN | t_cell | *MPZL3* | -0.558277648 | 0.001082929 | 0.013965754 | 1 |
| HTN | t_cell | *PCSK7* | -2.819111266 | 0.001492076 | 0.014185269 | 1 |
| HTN | t_cell | *S100A1* | 5.139889947 | 0.01294058 | 0.043460163 | 1 |
| HTN | t_cell | *FDPS* | 4.452240233 | 0.001240865 | 0.013965754 | 1 |
| HTN | t_cell | *PAQR6* | -6.427524135 | 0.000965066 | 0.013965754 | 1 |
| HTN | t_cell | *NBEAL2* | -2.423519776 | 0.008290378 | 0.032555655 | 1 |
| HTN | t_cell | *FCRL3* | -0.74782131 | 0.012965904 | 0.043521965 | 1 |
| HTN | t_cell | *LY6K* | 4.978673501 | 0.003119132 | 0.018622965 | 1 |
| HTN | t_cell | *TONSL* | 7.668689116 | 0.005933833 | 0.026565948 | 1 |
| HTN | t_cell | *ZNF333* | 3.769643351 | 0.006829426 | 0.02875861 | 1 |
| HTN | t_cell | *PPP1R16A* | 1.805974032 | 0.002611457 | 0.017160523 | 1 |
| HTN | t_cell | *ORAI2* | -3.705863334 | 0.012599318 | 0.042834132 | 1 |
| HTN | t_cell | *RPL8* | -3.345856979 | 0.003748855 | 0.020396571 | 1 |
| HTN | t_cell | *SCGB3A1* | -2.778960379 | 0.004883325 | 0.023562138 | 1 |
| HTN | t_cell | *YDJC* | -2.158319987 | 0.001123679 | 0.013965754 | 1 |
| HTN | t_cell | *DVL3* | 3.045608144 | 0.008858891 | 0.033857657 | 1 |
| HTN | t_cell | *ABCF3* | 4.281336091 | 1.49E-08 | 5.52E-06 | 9.39E-05 |
| HTN | t_cell | *DMKN* | 2.133963597 | 0.004058574 | 0.021228168 | 1 |
| HTN | t_cell | *BDH1* | -2.427157578 | 0.001263095 | 0.013965754 | 1 |
| HTN | t_cell | *DUSP14* | 1.973597757 | 0.004818552 | 0.023465715 | 1 |
| HTN | t_cell | *PGAP3* | -5.580968064 | 0.009320483 | 0.035025628 | 1 |

| | | | | | | |
|---|---|---|---|---|---|---|
| HTN | t_cell | *IKZF3* | -0.442562539 | 0.000925439 | 0.013965754 | 1 |
| HTN | t_cell | *ALDH16A1* | 5.699351097 | 0.006095382 | 0.027009862 | 1 |
| HTN | t_cell | *SIGLEC11* | -4.572259721 | 0.005880934 | 0.026471702 | 1 |
| HTN | t_cell | *MPP3* | 2.135096659 | 0.001253615 | 0.013965754 | 1 |
| HTN | t_cell | *NAGS* | 10.3041965 | 0.008490015 | 0.033064829 | 1 |
| HTN | t_cell | *ASGR2* | -10.7874513 | 0.008147153 | 0.03219431 | 1 |
| HTN | t_cell | *TNFSF13* | 7.713154086 | 8.30E-06 | 0.000714561 | 0.052162923 |
| HTN | t_cell | *SENP3* | -7.583090161 | 0.004954517 | 0.023796066 | 1 |
| HTN | t_cell | *WDR90* | 2.423486551 | 0.001352648 | 0.013965754 | 1 |
| HTN | t_cell | *MEIOB* | 3.308319465 | 0.000806708 | 0.013695656 | 1 |
| HTN | t_cell | *FLYWCH2* | 4.251379293 | 0.001509309 | 0.014185269 | 1 |
| HTN | t_cell | *ZG16B* | 19.93618982 | 0.001390054 | 0.013965754 | 1 |
| HTN | t_cell | *RPL29* | -2.564106443 | 0.002575708 | 0.017027838 | 1 |
| HTN | t_cell | *CYP4A22* | 14.58839447 | 0.000626986 | 0.012503451 | 1 |
| HTN | t_cell | *COA7* | -2.577731945 | 0.001546539 | 0.014185269 | 1 |
| HTN | t_cell | *SLC1A7* | 1.255899469 | 0.004631157 | 0.023016667 | 1 |
| HTN | t_cell | *FBLIM1* | 2.567473005 | 0.004226457 | 0.021762274 | 1 |
| HTN | t_cell | *SLC25A34* | 6.181781987 | 0.002005573 | 0.015283681 | 1 |
| HTN | t_cell | *LRRC38* | 14.13532254 | 0.002340052 | 0.016323038 | 1 |
| HTN | t_cell | *LAPTM5* | -2.248508982 | 0.00120682 | 0.013965754 | 1 |
| HTN | t_cell | *TSSK3* | 1.930665589 | 0.006789364 | 0.028647472 | 1 |
| HTN | t_cell | *CAMK2N1* | 6.808957177 | 0.005515037 | 0.025373449 | 1 |
| HTN | t_cell | *B3GALT2* | -6.832857446 | 0.011561229 | 0.040380805 | 1 |
| HTN | t_cell | *HFM1* | 2.641382839 | 0.004124719 | 0.021445397 | 1 |
| HTN | t_cell | *KCNT2* | 2.498840343 | 0.014684226 | 0.047465155 | 1 |
| HTN | t_cell | *VCAM1* | 2.510103551 | 0.000572787 | 0.01191612 | 1 |
| HTN | t_cell | *LMX1A* | 1.856535616 | 0.00213034 | 0.015682578 | 1 |
| HTN | t_cell | *C1orf115* | 2.715264647 | 0.001300425 | 0.013965754 | 1 |
| HTN | t_cell | *ACP6* | -4.026595889 | 0.002422961 | 0.016573967 | 1 |
| HTN | t_cell | *PM20D1* | -5.922501597 | 0.001693339 | 0.014482324 | 1 |
| HTN | t_cell | *HAAO* | 2.432715064 | 0.005753329 | 0.026078715 | 1 |
| HTN | t_cell | *RFTN2* | 2.742078423 | 0.008122616 | 0.032133155 | 1 |
| HTN | t_cell | *FAM84A* | 4.894217039 | 0.001844798 | 0.015062654 | 1 |
| HTN | t_cell | *FRZB* | 4.667585558 | 0.003416444 | 0.019478401 | 1 |
| HTN | t_cell | *FBXO41* | -4.288264458 | 0.002749424 | 0.017638397 | 1 |
| HTN | t_cell | *CCDC74A* | 15.2684983 | 0.004996182 | 0.023855009 | 1 |
| HTN | t_cell | *SGPP2* | -1.014543638 | 0.000693377 | 0.012921657 | 1 |
| HTN | t_cell | *BBS5* | 2.466977169 | 0.001540913 | 0.014185269 | 1 |
| HTN | t_cell | *HPGDS* | 2.914995441 | 0.006206687 | 0.027210047 | 1 |
| HTN | t_cell | *C1QTNF7* | 1.670752254 | 0.00498561 | 0.023855009 | 1 |
| HTN | t_cell | *TDRD10* | 3.259510698 | 0.008539431 | 0.03309951 | 1 |
| HTN | t_cell | *PBXIP1* | -1.476006533 | 0.001729037 | 0.014591212 | 1 |
| HTN | t_cell | *DCST2* | -3.317382143 | 0.001812613 | 0.014955249 | 1 |
| HTN | t_cell | *LMOD1* | -4.820539522 | 8.13E-08 | 2.13E-05 | 0.000511228 |
| HTN | t_cell | *PYHIN1* | -0.541963111 | 0.001161395 | 0.013965754 | 1 |
| HTN | t_cell | *CD200R1* | -1.492421101 | 0.003639072 | 0.020086785 | 1 |
| HTN | t_cell | *CADPS* | 4.194636507 | 0.004862369 | 0.023537205 | 1 |
| HTN | t_cell | *PPM1K* | -3.117853137 | 0.005732266 | 0.026005629 | 1 |
| HTN | t_cell | *DNASE1L3* | 2.273893076 | 0.002956273 | 0.01807841 | 1 |
| HTN | t_cell | *CRELD1* | 2.599400197 | 0.002225837 | 0.016006305 | 1 |

| | | | | | | |
|---|---|---|---|---|---|---|
| HTN | t_cell | *CCDC158* | 1.746933998 | 0.001713096 | 0.014532983 | 1 |
| HTN | t_cell | *PLB1* | 4.638630796 | 0.009671004 | 0.035913527 | 1 |
| HTN | t_cell | *CDCP1* | 2.692501241 | 0.00197781 | 0.015266349 | 1 |
| HTN | t_cell | *NMNAT3* | 3.010241036 | 0.004615767 | 0.022969232 | 1 |
| HTN | t_cell | *TPRA1* | 3.608959499 | 0.013871175 | 0.045491956 | 1 |
| HTN | t_cell | *SFMBT1* | -2.93803321 | 0.001508846 | 0.014185269 | 1 |
| HTN | t_cell | *EXO5* | -3.839882206 | 0.003455852 | 0.019578594 | 1 |
| HTN | t_cell | *CDC25A* | -4.893125602 | 7.47E-09 | 2.94E-06 | 4.70E-05 |
| HTN | t_cell | *ZNF589* | -4.232872009 | 1.89E-09 | 1.48E-06 | 1.19E-05 |
| HTN | t_cell | *ATRIP* | 7.149014071 | 0.013270357 | 0.044119901 | 1 |
| HTN | t_cell | *APEH* | -6.742996634 | 7.12E-11 | 8.96E-08 | 4.48E-07 |
| HTN | t_cell | *RNF123* | 4.591849031 | 2.12E-09 | 1.48E-06 | 1.33E-05 |
| HTN | t_cell | *MST1R* | 2.816390063 | 0.000283715 | 0.008034755 | 1 |
| HTN | t_cell | *GRM2* | -4.433036583 | 0.001035504 | 0.013965754 | 1 |
| HTN | t_cell | *POC1A* | 4.808892125 | 0.000213081 | 0.006800219 | 1 |
| HTN | t_cell | *PPM1M* | -2.585236992 | 0.000242587 | 0.007297348 | 1 |
| HTN | t_cell | *TMEM144* | 2.321082401 | 0.00393535 | 0.020880704 | 1 |
| HTN | t_cell | *NPY1R* | 1.852884965 | 0.003193343 | 0.018882638 | 1 |
| HTN | t_cell | *CMBL* | 2.76907955 | 0.002444144 | 0.016594315 | 1 |
| HTN | t_cell | *RHOBTB3* | 1.446613507 | 0.013160006 | 0.043962252 | 1 |
| HTN | t_cell | *TIGD6* | 3.333130998 | 0.002354183 | 0.016336366 | 1 |
| HTN | t_cell | *SERINC5* | -2.279827472 | 0.002553325 | 0.016987039 | 1 |
| HTN | t_cell | *ENPP6* | 5.559865322 | 0.0002166 | 0.006819187 | 1 |
| HTN | t_cell | *ERAP1* | -4.360061461 | 2.63E-06 | 0.000280502 | 0.016549618 |
| HTN | t_cell | *TLR3* | 3.358094466 | 0.00674784 | 0.028544258 | 1 |
| HTN | t_cell | *CCDC127* | -4.653156032 | 0.004029147 | 0.021109371 | 1 |
| HTN | t_cell | *MB21D1* | -3.719316839 | 0.005622995 | 0.025766596 | 1 |
| HTN | t_cell | *TMEM200A* | -1.131298613 | 0.00115827 | 0.013965754 | 1 |
| HTN | t_cell | *STK17A* | -1.966108832 | 0.002914814 | 0.018049857 | 1 |
| HTN | t_cell | *GALNT10* | -2.987294051 | 0.007950609 | 0.031732678 | 1 |
| HTN | t_cell | *RPS14* | -2.627704445 | 0.000498554 | 0.010921292 | 1 |
| HTN | t_cell | *KCNK5* | 3.218199174 | 0.002150058 | 0.015772949 | 1 |
| HTN | t_cell | *USP49* | 3.224905919 | 0.005907335 | 0.026509219 | 1 |
| HTN | t_cell | *ZNF704* | 2.263493166 | 0.00831808 | 0.032583032 | 1 |
| HTN | t_cell | *TNFRSF11B* | 3.267749884 | 0.001696165 | 0.014482324 | 1 |
| HTN | t_cell | *GPER1* | 2.594452793 | 0.000515723 | 0.011103937 | 1 |
| HTN | t_cell | *ALDH7A1* | 2.119353906 | 0.001852889 | 0.015083316 | 1 |
| HTN | t_cell | *FREM1* | 1.800643476 | 0.006295262 | 0.027370891 | 1 |
| HTN | t_cell | *TMEM65* | -3.00177966 | 0.003438216 | 0.019509081 | 1 |
| HTN | t_cell | *TMC1* | 2.137112625 | 0.005276464 | 0.024717026 | 1 |
| HTN | t_cell | *C9orf64* | 3.600391383 | 4.87E-05 | 0.002572845 | 0.306168528 |
| HTN | t_cell | *SVEP1* | 1.699040711 | 0.003332991 | 0.019206707 | 1 |
| HTN | t_cell | *AQP3* | -0.675840343 | 0.001079822 | 0.013965754 | 1 |
| HTN | t_cell | *LRFN5* | 1.911588837 | 0.000276757 | 0.007952839 | 1 |
| HTN | t_cell | *CFL2* | 3.586306244 | 0.00295443 | 0.01807841 | 1 |
| HTN | t_cell | *ZCCHC24* | 2.570300359 | 0.006962846 | 0.029067341 | 1 |
| HTN | t_cell | *REEP3* | -3.803434653 | 0.005102578 | 0.024174482 | 1 |
| HTN | t_cell | *AK8* | 2.497281066 | 0.003618018 | 0.020070197 | 1 |
| HTN | t_cell | *ZMYND19* | -5.638639374 | 0.002072191 | 0.01545417 | 1 |
| HTN | t_cell | *STOX1* | 0.936470534 | 0.001223917 | 0.013965754 | 1 |

| | | | | | | |
|---|---|---|---|---|---|---|
| HTN | t_cell | *RET* | 2.449299312 | 0.002132751 | 0.015682578 | 1 |
| HTN | t_cell | *VWA2* | 3.410176093 | 0.00407937 | 0.021283851 | 1 |
| HTN | t_cell | *HSPA12A* | 1.668825661 | 0.000202954 | 0.006698301 | 1 |
| HTN | t_cell | *UBTD1* | 2.492892821 | 0.003420416 | 0.019478401 | 1 |
| HTN | t_cell | *ARHGAP42* | 2.002155638 | 0.004600021 | 0.022916271 | 1 |
| HTN | t_cell | *TTC7B* | 2.389130523 | 0.003402065 | 0.019433479 | 1 |
| HTN | t_cell | *PSMC3* | 3.258417147 | 0.003513578 | 0.019762162 | 1 |
| HTN | t_cell | *TC2N* | -0.608814882 | 0.000389697 | 0.00938708 | 1 |
| HTN | t_cell | *IFI27L1* | 2.301055301 | 0.001940131 | 0.015190044 | 1 |
| HTN | t_cell | *ABTB2* | 3.159899444 | 0.007786139 | 0.031318911 | 1 |
| HTN | t_cell | *LIPC* | -1.474506485 | 0.002270848 | 0.016174171 | 1 |
| HTN | t_cell | *GPT2* | 1.632651065 | 0.002505072 | 0.016874383 | 1 |
| HTN | t_cell | *SPINT1* | -4.876060195 | 1.86E-05 | 0.001315114 | 0.117045116 |
| HTN | t_cell | *AVPR1A* | 1.611062907 | 0.003307499 | 0.019112361 | 1 |
| HTN | t_cell | *TRMT61A* | -4.029855286 | 0.001950907 | 0.015192726 | 1 |
| HTN | t_cell | *LARP6* | 3.702625028 | 0.000319305 | 0.008399457 | 1 |
| HTN | t_cell | *ASPG* | 4.946756069 | 0.00193169 | 0.015190044 | 1 |
| HTN | t_cell | *GABRB3* | 3.252177133 | 0.011179313 | 0.039419148 | 1 |
| HTN | t_cell | *ZNF202* | 5.04322221 | 0.002068004 | 0.015441262 | 1 |
| HTN | t_cell | *C2* | 4.311687479 | 0.00976605 | 0.03613841 | 1 |
| HTN | t_cell | *APBB1* | -2.290079835 | 0.009780389 | 0.036148917 | 1 |
| HTN | t_cell | *DCHS1* | 4.32074438 | 0.00404417 | 0.021170439 | 1 |
| HTN | t_cell | *NETO1* | 4.754077621 | 0.001715202 | 0.014532983 | 1 |
| HTN | t_cell | *C11orf74* | 2.519490971 | 0.007846485 | 0.031481079 | 1 |
| HTN | t_cell | *TUB* | 1.867881276 | 0.002493602 | 0.01683918 | 1 |
| HTN | t_cell | *RIC3* | -4.119429334 | 0.004105394 | 0.021366402 | 1 |
| HTN | t_cell | *LMO1* | 3.008458009 | 0.001526808 | 0.014185269 | 1 |
| HTN | t_cell | *XRRA1* | -2.002657745 | 0.004290952 | 0.021964306 | 1 |
| HTN | t_cell | *TRIM66* | -0.879712789 | 0.001184115 | 0.013965754 | 1 |
| HTN | t_cell | *RPL27A* | -2.54317414 | 0.002999959 | 0.018222937 | 1 |
| HTN | t_cell | *PRTG* | 2.877750713 | 0.002438141 | 0.016594315 | 1 |
| HTN | t_cell | *AKIP1* | 3.731793542 | 0.004491413 | 0.022614535 | 1 |
| HTN | t_cell | *C16orf46* | 20.24684067 | 0.001248341 | 0.013965754 | 1 |
| HTN | t_cell | *CCDC68* | 1.806492882 | 0.006143636 | 0.027048346 | 1 |
| HTN | t_cell | *CLEC4D* | 4.506613605 | 0.005902068 | 0.026504502 | 1 |
| HTN | t_cell | *RIMKLB* | -1.701185526 | 0.006169162 | 0.027122744 | 1 |
| HTN | t_cell | *BEAN1* | 2.604571819 | 0.003382402 | 0.01938483 | 1 |
| HTN | t_cell | *TK2* | 1.869958119 | 0.011769488 | 0.040837464 | 1 |
| HTN | t_cell | *SEC11C* | -2.464254821 | 0.001467809 | 0.014185269 | 1 |
| HTN | t_cell | *CENPV* | 4.656402942 | 0.007756225 | 0.031318911 | 1 |
| HTN | t_cell | *SPDYE6* | 3.10434927 | 0.001580223 | 0.014185269 | 1 |
| HTN | t_cell | *ATF7IP2* | -2.236749478 | 0.003565672 | 0.019909231 | 1 |
| HTN | t_cell | *SLFN5* | -2.676065866 | 0.001713891 | 0.014532983 | 1 |
| HTN | t_cell | *NYAP1* | 2.2780534 | 0.001072269 | 0.013965754 | 1 |
| HTN | t_cell | *TSC22D4* | -2.982246767 | 0.012185962 | 0.041927997 | 1 |
| HTN | t_cell | *MS4A6E* | 1.402840434 | 0.014838105 | 0.047766086 | 1 |
| HTN | t_cell | *SMAD3* | -3.539170499 | 0.002998749 | 0.018222937 | 1 |
| HTN | t_cell | *RCCD1* | 4.857484666 | 0.000662666 | 0.012858576 | 1 |
| HTN | t_cell | *AKTIP* | -3.199004504 | 0.002922853 | 0.018049857 | 1 |
| HTN | t_cell | *TEF* | 1.697211528 | 0.003186148 | 0.018861878 | 1 |

| | | | | | | |
|---|---|---|---|---|---|---|
| HTN | t_cell | *MEI1* | -1.108497602 | 0.009441365 | 0.035311041 | 1 |
| HTN | t_cell | *SAMD14* | 3.494032429 | 0.002729136 | 0.017579998 | 1 |
| HTN | t_cell | *COQ7* | 3.168791386 | 0.004083389 | 0.021283851 | 1 |
| HTN | t_cell | *GPRC5B* | 2.828245828 | 0.005310772 | 0.024824404 | 1 |
| HTN | t_cell | *RBFOX3* | 4.788836286 | 0.001481819 | 0.014185269 | 1 |
| HTN | t_cell | *MYO5B* | 1.70928995 | 0.006356081 | 0.027483273 | 1 |
| HTN | t_cell | *TRIM68* | 1.698726376 | 0.014183273 | 0.046298149 | 1 |
| HTN | t_cell | *ZNF646* | 4.714014916 | 1.90E-05 | 0.001323962 | 0.119396363 |
| HTN | t_cell | *SMG8* | 5.887938306 | 0.001190157 | 0.013965754 | 1 |
| HTN | t_cell | *PROCA1* | -3.975287714 | 0.008538163 | 0.03309951 | 1 |
| HTN | t_cell | *RPL13* | -1.966772004 | 0.001639997 | 0.014280696 | 1 |
| HTN | t_cell | *ZNF641* | 2.262052693 | 0.008432055 | 0.032906474 | 1 |
| HTN | t_cell | *CACNB3* | 4.051356995 | 0.015380448 | 0.049010075 | 1 |
| HTN | t_cell | *SERTAD3* | 3.278599879 | 0.001936066 | 0.015190044 | 1 |
| HTN | t_cell | *TMC4* | 7.239429867 | 0.001569023 | 0.014185269 | 1 |
| HTN | t_cell | *TTYH1* | 4.548261848 | 0.000996206 | 0.013965754 | 1 |
| HTN | t_cell | *CDC42EP5* | 4.612873194 | 0.00157108 | 0.014185269 | 1 |
| HTN | t_cell | *KIR3DL1* | 4.770765416 | 0.000715254 | 0.013040852 | 1 |
| HTN | t_cell | *PPP1R14A* | 4.977652863 | 0.012329852 | 0.042152138 | 1 |
| HTN | t_cell | *SERPINF2* | 3.171301331 | 0.000552698 | 0.011699694 | 1 |
| HTN | t_cell | *SRR* | 2.098331657 | 0.000134195 | 0.005207942 | 0.84368663 |
| HTN | t_cell | *CDK2AP2* | -3.467454439 | 0.008337075 | 0.032637105 | 1 |
| HTN | t_cell | *NUDT8* | 2.166424416 | 0.001935443 | 0.015190044 | 1 |
| HTN | t_cell | *HID1* | 2.399295456 | 0.008126565 | 0.032133155 | 1 |
| HTN | t_cell | *MGAT5B* | 3.66447766 | 0.00181503 | 0.014955563 | 1 |
| HTN | t_cell | *TMC8* | -0.537029498 | 0.002535325 | 0.016935789 | 1 |
| HTN | t_cell | *CYP7A1* | 7.258591777 | 7.63E-05 | 0.003452106 | 0.479842782 |
| HTN | t_cell | *GSDMA* | 4.64410283 | 0.004617959 | 0.022969232 | 1 |
| HTN | t_cell | *ZNF598* | -4.435677615 | 0.000312755 | 0.008296576 | 1 |
| HTN | t_cell | *KCTD5* | 6.237914468 | 0.006983022 | 0.029132221 | 1 |
| HTN | t_cell | *VWCE* | 2.518231766 | 0.004535053 | 0.022712185 | 1 |
| HTN | t_cell | *HRASLS5* | 1.229040898 | 9.01E-06 | 0.000755211 | 0.056640838 |
| HTN | t_cell | *TTC21A* | 3.648423044 | 0.009051413 | 0.03440522 | 1 |
| HTN | t_cell | *NAALADL1* | 2.490673258 | 0.000902779 | 0.013965754 | 1 |
| HTN | t_cell | *NUDT16L1* | -8.841943308 | 0.000112065 | 0.004575014 | 0.704552159 |
| HTN | t_cell | *RNF187* | 2.974611541 | 0.012326349 | 0.042152138 | 1 |
| HTN | t_cell | *PTGDR* | -1.095398524 | 0.00126322 | 0.013965754 | 1 |
| HTN | t_cell | *UBTD2* | 3.271269245 | 0.005973179 | 0.026652503 | 1 |
| HTN | t_cell | *COA6* | 5.152040034 | 0.012844974 | 0.043324222 | 1 |
| HTN | t_cell | *KIF5C* | -0.957157093 | 0.000863726 | 0.013695656 | 1 |
| HTN | t_cell | *FAM107A* | 2.284613641 | 0.00234518 | 0.016327958 | 1 |
| HTN | t_cell | *CX3CR1* | -1.26620974 | 0.001005527 | 0.013965754 | 1 |
| HTN | t_cell | *DTYMK* | 5.621817138 | 0.012908733 | 0.043376378 | 1 |
| HTN | t_cell | *TAP1* | -3.388840804 | 8.31E-05 | 0.003629567 | 0.522657605 |
| HTN | t_cell | *RHOH* | -0.381132552 | 0.001256369 | 0.013965754 | 1 |
| HTN | t_cell | *KLHL30* | 1.941737901 | 0.014450456 | 0.046854057 | 1 |
| HTN | t_cell | *REEP4* | 4.902234407 | 0.002022523 | 0.015311349 | 1 |
| HTN | t_cell | *BMP1* | 2.392991125 | 0.000847686 | 0.013695656 | 1 |
| HTN | t_cell | *NBPF9* | 3.866203897 | 0.004758826 | 0.023249344 | 1 |
| HTN | t_cell | *WFDC13* | 18.05133579 | 0.00138718 | 0.013965754 | 1 |

| | | | | | | |
|---|---|---|---|---|---|---|
| HTN | t_cell | *FAM84B* | -1.907789722 | 0.009808886 | 0.036174836 | 1 |
| HTN | t_cell | *CNNM3* | -4.360412375 | 0.010992785 | 0.038892313 | 1 |
| HTN | t_cell | *TNIP2* | -2.939081243 | 0.015642707 | 0.049469668 | 1 |
| HTN | t_cell | *BTNL3* | 0.917282466 | 0.004411964 | 0.022369369 | 1 |
| HTN | t_cell | *ZNF608* | 1.867740916 | 0.001681845 | 0.014464786 | 1 |
| HTN | t_cell | *INPP5D* | -1.565316833 | 0.001499207 | 0.014185269 | 1 |
| HTN | t_cell | *LETM1* | 3.72590113 | 0.012886077 | 0.043346584 | 1 |
| HTN | t_cell | *TMEM129* | 3.394353835 | 0.000436852 | 0.010097392 | 1 |
| HTN | t_cell | *CEP120* | -4.021777095 | 0.004720063 | 0.023213281 | 1 |
| HTN | t_cell | *STXBP6* | 2.74681432 | 0.00981921 | 0.036174836 | 1 |
| HTN | t_cell | *SIGLEC7* | 3.073940929 | 0.00761723 | 0.031016532 | 1 |
| HTN | t_cell | *MFSD7* | 2.336666688 | 0.005944521 | 0.026581226 | 1 |
| HTN | t_cell | *IRS1* | -1.284367525 | 0.000122121 | 0.004859338 | 0.767775389 |
| HTN | t_cell | *UPF3A* | 2.46590272 | 0.00604937 | 0.026935121 | 1 |
| HTN | t_cell | *C8orf46* | 0.731975868 | 9.30E-05 | 0.003948945 | 0.584443914 |
| HTN | t_cell | *FAM110B* | 2.676911729 | 0.010448324 | 0.037579299 | 1 |
| HTN | t_cell | *CPT1C* | 3.062889539 | 0.013840242 | 0.045470226 | 1 |
| HTN | t_cell | *XPO6* | -2.780650955 | 0.005619167 | 0.025766596 | 1 |
| HTN | t_cell | *RGS14* | -1.30770077 | 0.00026818 | 0.007813354 | 1 |
| HTN | t_cell | *CXCL10* | 2.127479976 | 0.001492549 | 0.014185269 | 1 |
| HTN | t_cell | *P2RY12* | 2.623118314 | 0.00111967 | 0.013965754 | 1 |
| HTN | t_cell | *RNASE6* | 7.574197318 | 0.006959318 | 0.029067341 | 1 |
| HTN | t_cell | *NPR1* | 2.401611527 | 0.002679122 | 0.017407175 | 1 |
| HTN | t_cell | *CD52* | -0.981170603 | 0.000401589 | 0.009599953 | 1 |
| HTN | t_cell | *CLIC4* | 3.098631712 | 0.008304204 | 0.032558321 | 1 |
| HTN | t_cell | *CCDC8* | 2.847714605 | 0.003025382 | 0.018324256 | 1 |
| HTN | t_cell | *INO80E* | -3.183151295 | 0.008174758 | 0.032280832 | 1 |
| HTN | t_cell | *RGPD8* | -1.013359338 | 0.01462564 | 0.047300102 | 1 |
| HTN | t_cell | *CHRNA5* | 2.095571241 | 0.007604019 | 0.031002897 | 1 |
| HTN | t_cell | *FASN* | 1.547844511 | 0.009095133 | 0.034508812 | 1 |
| HTN | t_cell | *MT1E* | 3.155667148 | 0.014310136 | 0.04651904 | 1 |
| HTN | t_cell | *DCXR* | 4.612835506 | 0.002348891 | 0.016335702 | 1 |
| HTN | t_cell | *LIMS1* | 3.348090667 | 0.000176331 | 0.006193265 | 1 |
| HTN | t_cell | *UGP2* | 2.213667906 | 0.008744397 | 0.033540599 | 1 |
| HTN | t_cell | *CALML6* | -5.807254315 | 3.73E-07 | 7.33E-05 | 0.002346711 |
| HTN | t_cell | *OTUD3* | 2.125671742 | 0.00777203 | 0.031318911 | 1 |
| HTN | t_cell | *ZNF747* | 4.64804364 | 0.012029785 | 0.041532818 | 1 |
| HTN | t_cell | *MYO7B* | 3.259991894 | 0.001277922 | 0.013965754 | 1 |
| HTN | t_cell | *KCNAB3* | -3.323152083 | 1.49E-05 | 0.001102931 | 0.093749113 |
| HTN | t_cell | *SEMA3E* | 2.557345583 | 0.002338782 | 0.016323038 | 1 |
| HTN | t_cell | *DCLK2* | 2.479069358 | 0.011538486 | 0.040341854 | 1 |
| HTN | t_cell | *METTL7B* | 4.994063076 | 0.001290741 | 0.013965754 | 1 |
| HTN | t_cell | *KRT72* | -4.951362764 | 0.008777102 | 0.033626836 | 1 |
| HTN | t_cell | *SERPINB9* | -1.929786334 | 0.00198215 | 0.015271785 | 1 |
| HTN | t_cell | *IRX2* | 2.493660772 | 0.002165311 | 0.015811049 | 1 |
| HTN | t_cell | *EMB* | -1.013254693 | 0.00021539 | 0.006819187 | 1 |
| HTN | t_cell | *SLC26A5* | 3.236478948 | 0.006129647 | 0.027024952 | 1 |
| HTN | t_cell | *GTSF1* | -2.428720231 | 0.000911375 | 0.013965754 | 1 |
| HTN | t_cell | *RNF34* | -4.726595939 | 0.000153071 | 0.005580753 | 0.962360107 |
| HTN | t_cell | *TRABD* | -3.221958184 | 0.001255016 | 0.013965754 | 1 |

| | | | | | | |
|---|---|---|---|---|---|---|
| HTN | t_cell | *TMEM133* | 6.336410911 | 0.004291612 | 0.021964306 | 1 |
| HTN | t_cell | *TTLL6* | 6.658024254 | 0.00514083 | 0.024246359 | 1 |
| HTN | t_cell | *KCNS3* | 1.887306111 | 0.000833532 | 0.013695656 | 1 |
| HTN | t_cell | *KBTBD2* | -3.624707756 | 0.009501687 | 0.035494419 | 1 |
| HTN | t_cell | *OSCAR* | 1.927794835 | 0.002225187 | 0.016006305 | 1 |
| HTN | t_cell | *PAQR8* | -1.629906285 | 7.81E-05 | 0.003478747 | 0.490821498 |
| HTN | t_cell | *NUDT6* | 2.704073812 | 0.00308862 | 0.018530928 | 1 |
| HTN | t_cell | *SOX7* | 7.714471423 | 6.84E-05 | 0.003234325 | 0.430165223 |
| HTN | t_cell | *C11orf24* | 2.883092794 | 0.003403254 | 0.019433479 | 1 |
| HTN | t_cell | *KCNMB3* | 2.712362663 | 0.002491432 | 0.01683918 | 1 |
| HTN | t_cell | *ATP6V0E2* | -4.328098611 | 0.000256848 | 0.00758123 | 1 |
| HTN | t_cell | *CHST11* | -1.170193128 | 0.008934284 | 0.034088258 | 1 |
| HTN | t_cell | *KCND3* | 1.654438558 | 0.015615601 | 0.049408798 | 1 |
| HTN | t_cell | *MRPL36* | 6.01083004 | 0.000172014 | 0.006075584 | 1 |
| HTN | t_cell | *ZNF581* | 5.707227734 | 0.002681046 | 0.017407175 | 1 |
| HTN | t_cell | *MCC* | 2.856879165 | 0.01124819 | 0.039564767 | 1 |
| HTN | t_cell | *HOPX* | -1.966522509 | 0.000837033 | 0.013695656 | 1 |
| HTN | t_cell | *LRRC8C* | -3.916319453 | 0.004862969 | 0.023537205 | 1 |
| HTN | t_cell | *RSL1D1* | -4.472865299 | 0.009094056 | 0.034508812 | 1 |
| HTN | t_cell | *MAP6* | 2.585587973 | 0.000677905 | 0.012876096 | 1 |
| HTN | t_cell | *NMUR1* | 5.621735404 | 0.013559583 | 0.044703249 | 1 |
| HTN | t_cell | *P2RY6* | 2.971074999 | 0.001490522 | 0.014185269 | 1 |
| HTN | t_cell | *RGS19* | -3.722067416 | 8.62E-13 | 1.81E-09 | 5.42E-09 |
| HTN | t_cell | *ANO5* | 2.136178982 | 0.001757512 | 0.014724258 | 1 |
| HTN | t_cell | *LGALS4* | 2.826378434 | 0.005808867 | 0.026273633 | 1 |
| HTN | t_cell | *PWWP2B* | 2.991215246 | 0.003708078 | 0.02032492 | 1 |
| HTN | t_cell | *AQP4* | 2.237823843 | 0.001517417 | 0.014185269 | 1 |
| HTN | t_cell | *CYP4F22* | -3.357327976 | 0.000657521 | 0.012837993 | 1 |
| HTN | t_cell | *C20orf196* | -2.906453799 | 0.005729465 | 0.026005629 | 1 |
| HTN | t_cell | *ZNF554* | 2.270264446 | 0.004543899 | 0.022726723 | 1 |
| HTN | t_cell | *ANKRD20A4* | 4.197840549 | 0.001467882 | 0.014185269 | 1 |
| HTN | t_cell | *SERF1A* | -10.8520914 | 0.000948608 | 0.013965754 | 1 |
| HTN | t_cell | *NME6* | 4.99583089 | 1.47E-09 | 1.32E-06 | 9.27E-06 |
| HTN | t_cell | *ISG20* | -0.785105314 | 0.000677408 | 0.012876096 | 1 |
| HTN | t_cell | *CXCR6* | -1.179117904 | 0.00798072 | 0.031776306 | 1 |
| HTN | t_cell | *SPTLC3* | 2.896870619 | 0.010738995 | 0.038254482 | 1 |
| HTN | t_cell | *COPRS* | 3.39033376 | 0.001378157 | 0.013965754 | 1 |
| HTN | t_cell | *CSDC2* | -5.114350346 | 0.0150272 | 0.048128379 | 1 |
| HTN | t_cell | *PRSS27* | -10.10725502 | 0.00475933 | 0.023249344 | 1 |
| HTN | t_cell | *CTSW* | -0.784623163 | 4.19E-06 | 0.000431752 | 0.026336886 |
| HTN | t_cell | *PDE3A* | 2.623980608 | 0.009425301 | 0.035286598 | 1 |
| HTN | t_cell | *RAD9A* | 1.465283367 | 0.008800633 | 0.033675947 | 1 |
| HTN | t_cell | *EFEMP2* | 2.805516982 | 0.000674401 | 0.012876096 | 1 |
| HTN | t_cell | *AGAP5* | 40.0959398 | 0.001353075 | 0.013965754 | 1 |
| HTN | t_cell | *ZNF738* | 2.988401037 | 0.001275739 | 0.013965754 | 1 |
| HTN | t_cell | *CCL19* | 6.217076812 | 0.002244333 | 0.016089078 | 1 |
| HTN | t_cell | *HOXC5* | 4.890430831 | 5.46E-06 | 0.000536447 | 0.034332583 |
| HTN | t_cell | *RAB37* | -1.332557907 | 0.00162378 | 0.014238085 | 1 |
| HTN | t_cell | *SNX32* | 3.15257387 | 0.004638809 | 0.023036485 | 1 |
| HTN | t_cell | *METAP1D* | 3.049377243 | 0.011296426 | 0.039676331 | 1 |

| | | | | | | |
|---|---|---|---|---|---|---|
| HTN | t_cell | *ANKRD13D* | -3.060441142 | 0.013197233 | 0.043969795 | 1 |
| HTN | t_cell | *LCLAT1* | -2.849920033 | 0.009775372 | 0.036148917 | 1 |
| HTN | t_cell | *GXYLT2* | 2.468632355 | 0.002827138 | 0.017809834 | 1 |
| HTN | t_cell | *HPSE2* | 2.708442737 | 0.000336395 | 0.00869179 | 1 |
| HTN | t_cell | *DCAKD* | -2.05615446 | 0.000688302 | 0.012921657 | 1 |
| HTN | t_cell | *BNC2* | 2.117672512 | 0.001547359 | 0.014185269 | 1 |
| HTN | t_cell | *PARP15* | -0.875948478 | 0.010768454 | 0.038314244 | 1 |
| HTN | t_cell | *AHSA2* | 2.973278069 | 0.013401164 | 0.044342537 | 1 |
| HTN | t_cell | *ABLIM3* | 2.625301309 | 0.006105777 | 0.027019752 | 1 |
| HTN | t_cell | *VANGL1* | 3.700987941 | 0.011837553 | 0.041010561 | 1 |
| HTN | t_cell | *GLRX* | 2.134220115 | 0.007366957 | 0.03025216 | 1 |
| HTN | t_cell | *MAP3K11* | 4.101015438 | 0.002110651 | 0.015629757 | 1 |
| HTN | t_cell | *NDNF* | 3.266613328 | 0.005596657 | 0.025683343 | 1 |
| HTN | t_cell | *OLR1* | 7.170406875 | 0.000309709 | 0.008250608 | 1 |
| HTN | t_cell | *GLIPR1L1* | 2.717689251 | 0.004536262 | 0.022712185 | 1 |
| HTN | t_cell | *CCDC36* | 1.910332759 | 0.001832535 | 0.015001497 | 1 |
| HTN | t_cell | *PPP1R14B* | 4.406685952 | 0.010664386 | 0.038030058 | 1 |
| HTN | t_cell | *ZNF417* | -5.197793743 | 0.007782656 | 0.031318911 | 1 |
| HTN | t_cell | *VEGFB* | -2.463017409 | 0.001590681 | 0.014185269 | 1 |
| HTN | t_cell | *MST1* | -1.767424257 | 0.002846754 | 0.01787445 | 1 |
| HTN | t_cell | *CSPG4* | 2.677001438 | 0.007110512 | 0.029546456 | 1 |
| HTN | t_cell | *SNX33* | 1.984449563 | 0.013386901 | 0.044342537 | 1 |
| HTN | t_cell | *NUDT18* | 5.033418325 | 0.001483351 | 0.014185269 | 1 |
| HTN | t_cell | *SULT1B1* | -1.271049708 | 0.003234525 | 0.019003909 | 1 |
| HTN | t_cell | *RNF213* | -2.479759311 | 0.006927321 | 0.029003866 | 1 |
| HTN | t_cell | *PLK3* | -4.275518824 | 0.005992102 | 0.026699039 | 1 |
| HTN | t_cell | *USMG5* | 5.189028795 | 0.000501569 | 0.010949181 | 1 |
| HTN | t_cell | *UBXN2A* | 2.633158219 | 0.013792467 | 0.045352337 | 1 |
| HTN | t_cell | *CCS* | 4.745004175 | 0.00048881 | 0.010824941 | 1 |
| HTN | t_cell | *NRROS* | 2.750853305 | 0.000308801 | 0.008250608 | 1 |
| HTN | t_cell | *C16orf91* | 6.582030794 | 0.00296757 | 0.01809613 | 1 |
| HTN | t_cell | *SOCS7* | 3.191816717 | 0.014986545 | 0.048098218 | 1 |
| HTN | t_cell | *TLR6* | 6.144859017 | 1.12E-07 | 2.72E-05 | 0.000706881 |
| HTN | t_cell | *RGMB* | 3.33927251 | 0.000295843 | 0.008157738 | 1 |
| HTN | t_cell | *FAM53A* | 2.81301145 | 0.000323221 | 0.008467033 | 1 |
| HTN | t_cell | *DDX23* | -4.803917361 | 0.014274961 | 0.04650087 | 1 |
| HTN | t_cell | *BBS1* | 4.236144152 | 0.010574113 | 0.037801145 | 1 |
| HTN | t_cell | *TMEM81* | 6.595570322 | 0.00172811 | 0.014591212 | 1 |
| HTN | t_cell | *KLF14* | -57.64263257 | 3.30E-09 | 1.69E-06 | 2.08E-05 |
| HTN | t_cell | *FGFBP3* | -7.313866804 | 0.001488446 | 0.014185269 | 1 |
| HTN | t_cell | *BTC* | 2.183019248 | 0.002532603 | 0.016935789 | 1 |
| HTN | t_cell | *NLRP6* | -2.446758499 | 0.000454645 | 0.010320043 | 1 |
| HTN | t_cell | *RSRC1* | -4.082765327 | 0.000113835 | 0.004587677 | 0.715677539 |
| HTN | t_cell | *CHST2* | 4.771447881 | 0.001873719 | 0.015083316 | 1 |
| HTN | t_cell | *PACS1* | -2.030654338 | 0.004011592 | 0.021084486 | 1 |
| HTN | t_cell | *ARHGAP1* | 4.246260836 | 0.004386274 | 0.022275043 | 1 |
| HTN | t_cell | *TP53I11* | 4.966307006 | 0.00120329 | 0.013965754 | 1 |
| HTN | t_cell | *PHYHD1* | 2.185590855 | 0.000944039 | 0.013965754 | 1 |
| HTN | t_cell | *NRIP3* | -4.731823008 | 5.79E-05 | 0.002863966 | 0.364279127 |
| HTN | t_cell | *SCUBE2* | 2.280238763 | 0.005109504 | 0.024174482 | 1 |

| | | | | | | |
|---|---|---|---|---|---|---|
| HTN | t_cell | *EIF3F* | -3.19591653 | 0.010876469 | 0.038632973 | 1 |
| HTN | t_cell | *PCSK1* | 1.803544401 | 0.00219972 | 0.015906408 | 1 |
| HTN | t_cell | *TBC1D10C* | -0.618058046 | 0.001647877 | 0.014328546 | 1 |
| HTN | t_cell | *LRRC25* | 2.03344215 | 0.000106569 | 0.00440788 | 0.669997711 |
| HTN | t_cell | *KCNE3* | 2.756337782 | 0.002768665 | 0.017670137 | 1 |
| HTN | t_cell | *UCP2* | -2.249152303 | 0.002400159 | 0.016509626 | 1 |
| HTN | t_cell | *TEX26* | 3.113061832 | 0.009856026 | 0.036246273 | 1 |
| HTN | t_cell | *GPR156* | 2.671523689 | 0.003971815 | 0.020966245 | 1 |
| HTN | t_cell | *ETV4* | 3.0300577 | 0.009357774 | 0.035123777 | 1 |
| HTN | t_cell | *A2M* | -2.2561515 | 0.004775647 | 0.023300746 | 1 |
| HTN | t_cell | *ARL4D* | 3.856760269 | 0.001324341 | 0.013965754 | 1 |
| HTN | t_cell | *LRRN1* | 3.134197408 | 0.002776891 | 0.017670355 | 1 |
| HTN | t_cell | *B3GALT6* | -7.533983472 | 0.00981754 | 0.036174836 | 1 |
| HTN | t_cell | *GPX2* | 4.9627237 | 0.003300182 | 0.019105197 | 1 |
| HTN | t_cell | *SLC35G1* | 2.568355004 | 0.005462931 | 0.025304138 | 1 |
| HTN | t_cell | *B3GNT4* | 10.81392596 | 0.001290313 | 0.013965754 | 1 |
| HTN | t_cell | *PLA2G16* | 0.989937867 | 0.006631315 | 0.028284312 | 1 |
| HTN | t_cell | *PHLDB3* | -3.110945695 | 0.009484477 | 0.035451192 | 1 |
| HTN | t_cell | *BOK* | 1.686268502 | 0.001556633 | 0.014185269 | 1 |
| HTN | t_cell | *ZNF843* | -5.907218298 | 0.00180495 | 0.014950884 | 1 |
| HTN | t_cell | *IRX5* | 2.196317646 | 0.000868819 | 0.013695656 | 1 |
| HTN | t_cell | *GCNT4* | -4.827228732 | 0.00115989 | 0.013965754 | 1 |
| HTN | t_cell | *MUC20* | 3.730566006 | 0.004692812 | 0.023180827 | 1 |
| HTN | t_cell | *SHMT1* | 2.263014494 | 0.002310399 | 0.016247738 | 1 |
| HTN | t_cell | *MTHFR* | -4.864208477 | 0.007317599 | 0.030161524 | 1 |
| HTN | t_cell | *RIMKLA* | 2.578116616 | 0.010173263 | 0.0371602 | 1 |
| HTN | t_cell | *AP006621.1* | 1.684795876 | 0.003735128 | 0.020378085 | 1 |
| HTN | t_cell | *C8orf31* | 4.221891664 | 0.002844853 | 0.01787445 | 1 |
| HTN | t_cell | *AC024940.1* | 4.455385648 | 0.000357267 | 0.008984542 | 1 |
| HTN | t_cell | *MAGEF1* | 4.985655275 | 0.006678283 | 0.028402239 | 1 |
| HTN | t_cell | *RBM44* | 4.629373767 | 0.015477308 | 0.049169193 | 1 |
| HTN | t_cell | *SAMD12* | -1.651732492 | 0.003860886 | 0.02069343 | 1 |
| HTN | t_cell | *RPLP2* | -2.735489881 | 0.012189051 | 0.041927997 | 1 |
| HTN | t_cell | *THAP5* | -5.39061006 | 0.015449911 | 0.049125217 | 1 |
| HTN | t_cell | *CD151* | 3.985816305 | 0.001057015 | 0.013965754 | 1 |
| HTN | t_cell | *FAM87B* | 3.400111804 | 0.010279834 | 0.037379592 | 1 |
| HTN | t_cell | *PCDHB9* | 3.158738387 | 7.70E-06 | 0.000681454 | 0.048383221 |
| HTN | t_cell | *CENPBD1* | 2.000307079 | 0.006619167 | 0.028251666 | 1 |
| HTN | t_cell | *BET1L* | 3.54836795 | 0.013310834 | 0.044207721 | 1 |
| HTN | t_cell | *ODF3B* | 4.546849145 | 0.000222444 | 0.006957723 | 1 |
| HTN | t_cell | *C2orf73* | 2.238606397 | 0.00165808 | 0.014360746 | 1 |
| HTN | t_cell | *FAM26E* | 2.829599389 | 0.012272661 | 0.042116263 | 1 |
| HTN | t_cell | *MLF1* | 3.960895404 | 0.003197929 | 0.018882638 | 1 |
| HTN | t_cell | *PRSS42* | 5.172677051 | 0.000113568 | 0.004587677 | 0.714001841 |
| HTN | t_cell | *GRAMD1C* | 4.7808289 | 0.009147807 | 0.034645943 | 1 |
| HTN | t_cell | *STAP2* | -4.211719126 | 0.015555976 | 0.049319424 | 1 |
| HTN | t_cell | *TSSK6* | -3.289813641 | 0.000859879 | 0.013695656 | 1 |
| HTN | t_cell | *ZNF114* | 2.765301114 | 0.001855141 | 0.015083316 | 1 |
| HTN | t_cell | *ZNF454* | 2.24672534 | 0.004750238 | 0.023249344 | 1 |
| HTN | t_cell | *SH2B1* | -1.77045369 | 0.010540772 | 0.037739086 | 1 |

| | | | | | | |
|---|---|---|---|---|---|---|
| HTN | t_cell | *ZC3H12D* | -0.984581451 | 0.00109385 | 0.013965754 | 1 |
| HTN | t_cell | *PLEC* | -3.897655418 | 0.002175004 | 0.015838902 | 1 |
| HTN | t_cell | *RNF212* | 1.306684437 | 0.011252076 | 0.039564767 | 1 |
| HTN | t_cell | *PRSS36* | 4.872082934 | 0.002641762 | 0.017264821 | 1 |
| HTN | t_cell | *AQP11* | 3.067175693 | 0.008369881 | 0.032724778 | 1 |
| HTN | t_cell | *AC006111.1* | 8.462731794 | 0.000252502 | 0.007488119 | 1 |
| HTN | t_cell | *LINC00843* | 4.786216288 | 0.006782669 | 0.028638443 | 1 |
| HTN | t_cell | *P4HTM* | -4.57489028 | 0.007968915 | 0.03174941 | 1 |
| HTN | t_cell | *ERBB4* | 1.793254501 | 0.005936879 | 0.026565948 | 1 |
| HTN | t_cell | *CSRNP3* | 3.156009435 | 0.006706812 | 0.028432724 | 1 |
| HTN | t_cell | *ZNF713* | 2.167896392 | 0.001296124 | 0.013965754 | 1 |
| HTN | t_cell | *RPP25* | 5.448611126 | 5.66E-12 | 8.90E-09 | 3.56E-08 |
| HTN | t_cell | *STX19* | 3.381425008 | 0.008270018 | 0.032516326 | 1 |
| HTN | t_cell | *TRIM73* | -5.554005454 | 0.002718735 | 0.017558598 | 1 |
| HTN | t_cell | *HYI* | -1.983688667 | 0.000238463 | 0.007211618 | 1 |
| HTN | t_cell | *C17orf62* | -3.483337468 | 0.010463848 | 0.037613614 | 1 |
| HTN | t_cell | *TRIML2* | 23.09245283 | 0.011419914 | 0.040020625 | 1 |
| HTN | t_cell | *ARID3B* | -3.673804096 | 0.001717889 | 0.014536166 | 1 |
| HTN | t_cell | *GEMIN4* | 2.290316654 | 0.008118391 | 0.032133155 | 1 |
| HTN | t_cell | *CIITA* | 3.554506655 | 0.012443198 | 0.042470349 | 1 |
| HTN | t_cell | *ZBTB42* | -4.637547587 | 0.004713184 | 0.023212384 | 1 |
| HTN | t_cell | *MAF1* | 4.458411984 | 0.004861576 | 0.023537205 | 1 |
| HTN | t_cell | *CITED4* | 2.732151472 | 0.012742455 | 0.043070868 | 1 |
| HTN | t_cell | *TIGD5* | 2.642799625 | 0.001572982 | 0.014185269 | 1 |
| HTN | t_cell | *ZNF154* | 3.467515336 | 0.002947343 | 0.01807841 | 1 |
| HTN | t_cell | *SEPT1* | -1.48437328 | 0.000986223 | 0.013965754 | 1 |
| HTN | t_cell | *TDRP* | 2.869858645 | 0.007899224 | 0.031591873 | 1 |
| HTN | t_cell | *FGD6* | 2.830082066 | 0.008935494 | 0.034088258 | 1 |
| HTN | t_cell | *HCLS1* | -1.943875901 | 0.001994951 | 0.015283681 | 1 |
| HTN | t_cell | *CYP8B1* | 2.536545086 | 0.002123031 | 0.015666072 | 1 |
| HTN | t_cell | *SSTR2* | 4.868513492 | 0.001968112 | 0.015219586 | 1 |
| HTN | t_cell | *PCGF5* | -2.935114537 | 0.008008052 | 0.031853166 | 1 |
| HTN | t_cell | *SHISA2* | -6.104728078 | 0.001093978 | 0.013965754 | 1 |
| HTN | t_cell | *SRSF8* | -3.918793794 | 0.001950859 | 0.015192726 | 1 |
| HTN | t_cell | *ZDHHC20* | -2.850831979 | 0.010312481 | 0.037433352 | 1 |
| HTN | t_cell | *ZNF443* | 4.764582963 | 5.83E-07 | 0.000101821 | 0.003665545 |
| HTN | t_cell | *ZNF792* | -2.939816582 | 0.003018364 | 0.018299376 | 1 |
| HTN | t_cell | *CMTR2* | 3.175873796 | 0.008740205 | 0.033540599 | 1 |
| HTN | t_cell | *MRPL14* | 4.152400992 | 0.009125063 | 0.034580632 | 1 |
| HTN | t_cell | *BBS12* | 7.358639797 | 0.000750754 | 0.013333299 | 1 |
| HTN | t_cell | *AEN* | -4.153632454 | 0.000318472 | 0.008399457 | 1 |
| HTN | t_cell | *TRAPPC5* | 4.362551183 | 0.003261527 | 0.019003909 | 1 |
| HTN | t_cell | *TMEM132C* | 1.393668181 | 0.000893974 | 0.013946442 | 1 |
| HTN | t_cell | *SLC25A41* | 7.14241322 | 0.003242176 | 0.019003909 | 1 |
| HTN | t_cell | *TMEM136* | 3.964700876 | 0.000208388 | 0.006718648 | 1 |
| HTN | t_cell | *ZNF322* | 2.435923988 | 0.013349158 | 0.044288208 | 1 |
| HTN | t_cell | *ACBD4* | 4.866712597 | 0.007432061 | 0.030439981 | 1 |
| HTN | t_cell | *MRPS23* | -4.272734169 | 0.014109588 | 0.046081546 | 1 |
| HTN | t_cell | *GPR135* | 1.53590134 | 0.013775242 | 0.045342904 | 1 |
| HTN | t_cell | *ZFP41* | 2.473886071 | 0.002260509 | 0.016149794 | 1 |

| | | | | | | |
|---|---|---|---|---|---|---|
| HTN | t_cell | *ATG9B* | 3.549696368 | 0.000995786 | 0.013965754 | 1 |
| HTN | t_cell | *CLDN7* | -2.125528064 | 0.00821035 | 0.032382981 | 1 |
| HTN | t_cell | *CHST15* | 2.633236723 | 0.011761744 | 0.040837464 | 1 |
| HTN | t_cell | *FAM89A* | 2.602279418 | 2.38E-05 | 0.00151218 | 0.149705832 |
| HTN | t_cell | *HHIPL1* | 2.815723383 | 0.002709127 | 0.017522924 | 1 |
| HTN | t_cell | *FAM153B* | -1.003024493 | 0.001069949 | 0.013965754 | 1 |
| HTN | t_cell | *SYNM* | 3.347365064 | 0.004345821 | 0.02212322 | 1 |
| HTN | t_cell | *C8orf33* | 4.159255713 | 0.002755653 | 0.017645427 | 1 |
| HTN | t_cell | *CLN8* | 7.225500102 | 1.08E-06 | 0.000142956 | 0.006788555 |
| HTN | t_cell | *MFSD5* | 4.360884544 | 0.000181028 | 0.006250611 | 1 |
| HTN | t_cell | *CLEC4G* | 3.627346637 | 0.00251552 | 0.01691452 | 1 |
| HTN | t_cell | *SATB1* | -2.961764269 | 0.00060713 | 0.012248652 | 1 |
| HTN | t_cell | *TSKU* | 2.096314252 | 0.003050703 | 0.018353849 | 1 |
| HTN | t_cell | *PLCXD3* | 2.377244258 | 0.003198665 | 0.018882638 | 1 |
| HTN | t_cell | *COL18A1* | 3.902374136 | 6.29E-05 | 0.002995098 | 0.39535288 |
| HTN | t_cell | *SLC25A18* | 1.996513588 | 0.00069255 | 0.012921657 | 1 |
| HTN | t_cell | *HMGN4* | -5.564082062 | 0.000717003 | 0.013040852 | 1 |
| HTN | t_cell | *ZNF320* | 1.621514233 | 9.84E-05 | 0.004126322 | 0.618948261 |
| HTN | t_cell | *C12orf60* | 2.932759825 | 0.009652635 | 0.0358665 | 1 |
| HTN | t_cell | *BEGAIN* | 5.775267795 | 0.001618039 | 0.014238085 | 1 |
| HTN | t_cell | *CSMD1* | 2.587256538 | 0.001709889 | 0.014532983 | 1 |
| HTN | t_cell | *PTGDR2* | -20.00272377 | 0.005435607 | 0.025227921 | 1 |
| HTN | t_cell | *GPR19* | -1.03213969 | 0.00089977 | 0.013965754 | 1 |
| HTN | t_cell | *C2CD4C* | 2.737471858 | 0.00980235 | 0.036174836 | 1 |
| HTN | t_cell | *CCBE1* | 2.038541337 | 0.000940972 | 0.013965754 | 1 |
| HTN | t_cell | *LRIT3* | -4.634960034 | 0.000168789 | 0.005995334 | 1 |
| HTN | t_cell | *NPIPA1* | 1.137628682 | 0.013407659 | 0.044342537 | 1 |
| HTN | t_cell | *URAD* | 7.126928476 | 0.000838338 | 0.013695656 | 1 |
| HTN | t_cell | *GPR132* | -2.040971673 | 0.000615044 | 0.012353944 | 1 |
| HTN | t_cell | *MX2* | -3.276179771 | 0.008614872 | 0.03330978 | 1 |
| HTN | t_cell | *KLRC4* | -1.546575598 | 0.000100634 | 0.004189973 | 0.632685958 |
| HTN | t_cell | *ACSM5* | 2.269668335 | 0.000258167 | 0.00758456 | 1 |
| HTN | t_cell | *RP1L1* | 8.955542826 | 3.98E-10 | 4.17E-07 | 2.50E-06 |
| HTN | t_cell | *FAM19A1* | -0.692568305 | 0.001095202 | 0.013965754 | 1 |
| HTN | t_cell | *BMP8A* | 6.519128248 | 6.97E-07 | 0.000112416 | 0.004384238 |
| HTN | t_cell | *KREMEN1* | -1.284177239 | 0.001509535 | 0.014185269 | 1 |
| HTN | t_cell | *KCTD16* | 2.205008995 | 0.007153112 | 0.029703841 | 1 |
| HTN | t_cell | *KCTD8* | 1.297523104 | 0.005961082 | 0.026620342 | 1 |
| HTN | t_cell | *C9orf66* | 4.149308932 | 0.000789068 | 0.013628763 | 1 |
| HTN | t_cell | *ANKRD45* | 3.801015999 | 0.001324124 | 0.013965754 | 1 |
| HTN | t_cell | *GPR39* | 1.750237441 | 0.010781997 | 0.038340733 | 1 |
| HTN | t_cell | *TBX1* | 3.977558599 | 0.009234858 | 0.034870603 | 1 |
| HTN | t_cell | *EIF3C* | 3.456945955 | 1.11E-05 | 0.00092157 | 0.07003931 |
| HTN | t_cell | *CNTN2* | 3.861675618 | 0.001350821 | 0.013965754 | 1 |
| HTN | t_cell | *GOLGA6L4* | 2.526096044 | 0.004903063 | 0.023621115 | 1 |
| HTN | t_cell | *PCDH9* | 1.433307244 | 0.006261296 | 0.027260919 | 1 |
| HTN | t_cell | *OAF* | 2.021904047 | 0.015207315 | 0.048557886 | 1 |
| HTN | t_cell | *A3GALT2* | -2.404037867 | 0.000733287 | 0.013209674 | 1 |
| HTN | t_cell | *CCR10* | 9.432686354 | 1.24E-05 | 0.000990415 | 0.078242791 |
| HTN | t_cell | *PROS1* | 2.323464282 | 0.011543661 | 0.040341854 | 1 |

| | | | | | | |
|---|---|---|---|---|---|---|
| HTN | t_cell | *LPAR5* | -2.077737853 | 6.92E-06 | 0.000639792 | 0.043505839 |
| HTN | t_cell | *NELL2* | -0.447662363 | 0.000372778 | 0.009190797 | 1 |
| HTN | t_cell | *KRBA2* | 3.112432615 | 0.008010157 | 0.031853166 | 1 |
| HTN | t_cell | *SEPT5* | 4.518921499 | 0.001354539 | 0.013965754 | 1 |
| HTN | t_cell | *TUSC5* | 1.71383314 | 0.00026844 | 0.007813354 | 1 |
| HTN | t_cell | *BTBD6* | 1.859855343 | 0.015304006 | 0.048815976 | 1 |
| HTN | t_cell | *H1FX* | -4.524652583 | 0.006853218 | 0.02882113 | 1 |
| HTN | t_cell | *NUTM2A* | -5.938352984 | 0.000824075 | 0.013695656 | 1 |
| HTN | t_cell | *SORCS2* | 3.547939855 | 0.000347484 | 0.00884467 | 1 |
| HTN | t_cell | *MAFF* | 3.346173226 | 0.003231613 | 0.019003909 | 1 |
| HTN | t_cell | *ADSSL1* | 3.94626203 | 0.001781189 | 0.014844048 | 1 |
| HTN | t_cell | *C6orf120* | -4.087064931 | 0.013294139 | 0.04417561 | 1 |
| HTN | t_cell | *DDX51* | 3.257710971 | 2.34E-05 | 0.001500094 | 0.147009238 |
| HTN | t_cell | *SIGIRR* | -1.630111837 | 0.002785757 | 0.017682732 | 1 |
| HTN | t_cell | *KLHL33* | 1.774696501 | 0.010576915 | 0.037801145 | 1 |
| HTN | t_cell | *CDK10* | -4.899678255 | 8.87E-07 | 0.000132818 | 0.005578361 |
| HTN | t_cell | *GAS2L1* | 2.110294989 | 0.011611669 | 0.040499614 | 1 |
| HTN | t_cell | *TNFAIP8L1* | 5.504836976 | 0.000381185 | 0.009252934 | 1 |
| HTN | t_cell | *MAPK11* | 3.236505403 | 0.007733323 | 0.031266496 | 1 |
| HTN | t_cell | *SP140L* | -2.544652766 | 0.002525204 | 0.016925325 | 1 |
| HTN | t_cell | *SV2B* | 3.742772608 | 0.003734041 | 0.020378085 | 1 |
| HTN | t_cell | *LSAMP* | 3.057852489 | 7.91E-05 | 0.003478747 | 0.497460774 |
| HTN | t_cell | *AHNAK2* | 3.798079277 | 0.013407852 | 0.044342537 | 1 |
| HTN | t_cell | *SP1* | 3.423949021 | 0.001587934 | 0.014185269 | 1 |
| HTN | t_cell | *PCGF3* | 3.29009607 | 0.011775043 | 0.040837464 | 1 |
| HTN | t_cell | *PBX1* | 1.633706815 | 0.008945638 | 0.034106263 | 1 |
| HTN | t_cell | *PMEL* | -1.969293283 | 0.013319734 | 0.044213922 | 1 |
| HTN | t_cell | *MYBL1* | -0.599139142 | 5.78E-05 | 0.002863966 | 0.36307936 |
| HTN | t_cell | *ADARB2* | 3.221630076 | 0.008219538 | 0.032398895 | 1 |
| HTN | t_cell | *SRL* | 1.811680363 | 0.003750348 | 0.020396571 | 1 |
| HTN | t_cell | *ARL17A* | -5.516529561 | 3.56E-06 | 0.000373026 | 0.022381578 |
| HTN | t_cell | *RTN4RL1* | 2.223057417 | 0.002201141 | 0.015906408 | 1 |
| HTN | t_cell | *RASA3* | -2.088351623 | 0.000341195 | 0.008719901 | 1 |
| HTN | t_cell | *ZNF284* | 6.43472968 | 0.002804387 | 0.017719777 | 1 |
| HTN | t_cell | *CYP4Z1* | 3.232539818 | 0.000479812 | 0.010735157 | 1 |
| HTN | t_cell | *FFAR4* | 1.530171138 | 0.000422672 | 0.009841989 | 1 |
| HTN | t_cell | *BPIFB4* | 131.3222549 | 0.003279945 | 0.019032949 | 1 |
| HTN | t_cell | *KLHL32* | -2.240263118 | 0.000195969 | 0.006553493 | 1 |
| HTN | t_cell | *CYP4X1* | 3.235509386 | 0.000207636 | 0.006718648 | 1 |
| HTN | t_cell | *ZNF501* | 2.799463698 | 0.00342908 | 0.01949243 | 1 |
| HTN | t_cell | *RPS23* | -2.488706741 | 0.002785754 | 0.017682732 | 1 |
| HTN | t_cell | *PCLO* | 1.231865712 | 0.006366642 | 0.027510019 | 1 |
| HTN | t_cell | *CYP27C1* | 1.033976322 | 0.007290946 | 0.030097292 | 1 |
| HTN | t_cell | *HYAL3* | -2.138938135 | 0.001307138 | 0.013965754 | 1 |
| HTN | t_cell | *ZSCAN30* | 2.79714116 | 0.006582324 | 0.028120771 | 1 |
| HTN | t_cell | *TRABD2A* | -0.781753013 | 0.005678108 | 0.025849575 | 1 |
| HTN | t_cell | *TMEM17* | 2.568954807 | 0.002062747 | 0.015438678 | 1 |
| HTN | t_cell | *TNFRSF18* | -0.609761296 | 0.001098815 | 0.013965754 | 1 |
| HTN | t_cell | *RTN4RL2* | -6.354871212 | 0.000353589 | 0.008960886 | 1 |
| HTN | t_cell | *EFCAB6* | 3.036593919 | 0.000456334 | 0.010320043 | 1 |

| | | | | | | |
|---|---|---|---|---|---|---|
| HTN | t_cell | *EMID1* | 4.256173196 | 0.003144775 | 0.018722726 | 1 |
| HTN | t_cell | *HEATR4* | 1.752100259 | 0.004753167 | 0.023249344 | 1 |
| HTN | t_cell | *ANGPTL5* | 3.270544673 | 0.002803045 | 0.017719777 | 1 |
| HTN | t_cell | *MT1X* | 2.09768797 | 0.008987438 | 0.034244863 | 1 |
| HTN | t_cell | *FNBP1* | -1.933085377 | 0.000294252 | 0.008149603 | 1 |
| HTN | t_cell | *WDR86* | 4.104308735 | 0.000385515 | 0.009322041 | 1 |
| HTN | t_cell | *EPOR* | 3.282340823 | 0.008797391 | 0.033675947 | 1 |
| HTN | t_cell | *FPR3* | 1.617028278 | 0.013415933 | 0.044345936 | 1 |
| HTN | t_cell | *TLR5* | 2.266666829 | 0.005670591 | 0.025834063 | 1 |
| HTN | t_cell | *ZNF385C* | 5.191341165 | 0.000828863 | 0.013695656 | 1 |
| HTN | t_cell | *TRPV2* | -2.451483289 | 0.001511335 | 0.014185269 | 1 |
| HTN | t_cell | *SEMA4D* | -0.7852808 | 0.001013959 | 0.013965754 | 1 |
| HTN | t_cell | *FAM69C* | 2.709978226 | 0.012692476 | 0.043004354 | 1 |
| HTN | t_cell | *MCRS1* | 8.178688604 | 6.70E-08 | 1.85E-05 | 0.000420927 |
| HTN | t_cell | *TMEM220* | 2.884694367 | 0.005254123 | 0.024669658 | 1 |
| HTN | t_cell | *CCDC157* | -5.850589647 | 0.002020613 | 0.015311349 | 1 |
| HTN | t_cell | *GFRAL* | 2.255506806 | 0.006390406 | 0.027525449 | 1 |
| HTN | t_cell | *LDLRAD2* | 2.103373714 | 0.010410903 | 0.037530589 | 1 |
| HTN | t_cell | *C2orf66* | 9.08036028 | 0.001407183 | 0.01399841 | 1 |
| HTN | t_cell | *MORN2* | 2.093556229 | 0.004238754 | 0.021807728 | 1 |
| HTN | t_cell | *C6orf132* | 2.091299984 | 0.002049438 | 0.015375679 | 1 |
| HTN | t_cell | *FAM166A* | 8.931215813 | 0.001531355 | 0.014185269 | 1 |
| HTN | t_cell | *ZNF626* | 2.742901928 | 0.01007924 | 0.036906336 | 1 |
| HTN | t_cell | *SMTNL2* | 9.100576973 | 0.001296985 | 0.013965754 | 1 |
| HTN | t_cell | *ZNF793* | -1.905655073 | 0.00117463 | 0.013965754 | 1 |
| HTN | t_cell | *HYKK* | 2.845124071 | 0.003847413 | 0.020656433 | 1 |
| HTN | t_cell | *RUFY4* | 4.983821282 | 0.001650051 | 0.014328546 | 1 |
| HTN | t_cell | *ZNF669* | 3.830545827 | 0.009429217 | 0.035286598 | 1 |
| HTN | t_cell | *ZNF559* | -1.68298693 | 0.001313894 | 0.013965754 | 1 |
| HTN | t_cell | *PDCD1* | -1.942724585 | 0.001605371 | 0.014197745 | 1 |
| HTN | t_cell | *CERKL* | -1.171796118 | 0.00250061 | 0.016868387 | 1 |
| HTN | t_cell | *FAM72B* | 4.568025342 | 0.001280798 | 0.013965754 | 1 |
| HTN | t_cell | *GOLGA8M* | 3.714156997 | 0.000276489 | 0.007952839 | 1 |
| HTN | t_cell | *S100A16* | 4.469576714 | 0.002271639 | 0.016174171 | 1 |
| HTN | t_cell | *SLC4A5* | 5.314924551 | 0.00242533 | 0.016573967 | 1 |
| HTN | t_cell | *UROS* | 3.669236062 | 0.010128733 | 0.037022875 | 1 |
| HTN | t_cell | *FSIP2* | 1.506512815 | 0.005654885 | 0.025834063 | 1 |
| HTN | t_cell | *SKOR1* | 6.243118287 | 0.001537865 | 0.014185269 | 1 |
| HTN | t_cell | *ZDHHC11* | 5.838519246 | 1.73E-06 | 0.00020668 | 0.010852426 |
| HTN | t_cell | *FAM26F* | -1.440285914 | 0.009914658 | 0.036409728 | 1 |
| HTN | t_cell | *LINC00910* | 3.811176674 | 0.010062466 | 0.036866388 | 1 |
| HTN | t_cell | *FBF1* | -2.285386139 | 7.06E-06 | 0.000643721 | 0.044416729 |
| HTN | t_cell | *FAM196A* | 2.860264575 | 0.002539114 | 0.016935789 | 1 |
| HTN | t_cell | *C9orf152* | 3.425800915 | 0.001451874 | 0.014156149 | 1 |
| HTN | t_cell | *NOC2L* | -2.70995796 | 4.83E-05 | 0.002572845 | 0.303659805 |
| HTN | t_cell | *ALKBH2* | 3.45401307 | 0.003044508 | 0.018351697 | 1 |
| HTN | t_cell | *LITAF* | -3.10538539 | 0.013488833 | 0.044516691 | 1 |
| HTN | t_cell | *FAM47E* | 2.166958577 | 7.32E-05 | 0.003359644 | 0.460271248 |
| HTN | t_cell | *S100A13* | 3.281633536 | 0.005165606 | 0.024344949 | 1 |
| HTN | t_cell | *C15orf61* | -6.027045677 | 0.006127802 | 0.027024952 | 1 |

| | | | | | | |
|---|---|---|---|---|---|---|
| HTN | t_cell | *ZKSCAN3* | 6.018567901 | 7.88E-05 | 0.003478747 | 0.495673343 |
| HTN | t_cell | *HLA-DRB1* | -4.057492571 | 8.52E-08 | 2.14E-05 | 0.000535886 |
| HTN | t_cell | *AKR1C3* | 1.851166886 | 0.014734801 | 0.047555284 | 1 |
| HTN | t_cell | *COLCA1* | 2.36846755 | 0.004840279 | 0.023535059 | 1 |
| HTN | t_cell | *HRCT1* | 2.829100322 | 0.000453499 | 0.010320043 | 1 |
| HTN | t_cell | *SUPT3H* | -2.56993469 | 0.014480916 | 0.046928619 | 1 |
| HTN | t_cell | *GIMAP5* | -1.177327526 | 0.005099773 | 0.024174482 | 1 |
| HTN | t_cell | *STK31* | -4.429042831 | 0.002003178 | 0.015283681 | 1 |
| HTN | t_cell | *ZKSCAN7* | 4.347501182 | 0.015674244 | 0.049519087 | 1 |
| HTN | t_cell | *NTNG2* | -3.600049245 | 0.002211964 | 0.015966264 | 1 |
| HTN | t_cell | *ASB13* | 3.684210901 | 0.005986071 | 0.026691083 | 1 |
| HTN | t_cell | *ZNF34* | 4.133010104 | 0.000337025 | 0.00869179 | 1 |
| HTN | t_cell | *INCA1* | 3.017892567 | 0.000707297 | 0.012964364 | 1 |
| HTN | t_cell | *ZNF774* | 3.513298745 | 0.001962223 | 0.015192726 | 1 |
| HTN | t_cell | *EVL* | -1.183931226 | 0.010184034 | 0.0371602 | 1 |
| HTN | t_cell | *ZNF605* | 2.882627009 | 0.003472739 | 0.01961645 | 1 |
| HTN | t_cell | *MYL6B* | 4.218721763 | 0.002176684 | 0.015838902 | 1 |
| HTN | t_cell | *C20orf96* | 4.364397807 | 0.008482349 | 0.033061704 | 1 |
| HTN | t_cell | *SULT1A1* | -2.76088045 | 0.000285778 | 0.008056901 | 1 |
| HTN | t_cell | *TPK1* | -2.240541091 | 0.00388143 | 0.02075047 | 1 |
| HTN | t_cell | *C1orf186* | 4.380332709 | 0.001335374 | 0.013965754 | 1 |
| HTN | t_cell | *ADH1B* | 1.44539091 | 0.003088979 | 0.018530928 | 1 |
| HTN | t_cell | *HRH1* | 2.399173403 | 0.00794014 | 0.031715158 | 1 |
| HTN | t_cell | *ERI2* | 2.083774103 | 0.004474298 | 0.022558071 | 1 |
| HTN | t_cell | *TRPV1* | 2.343483497 | 0.006096229 | 0.027009862 | 1 |
| HTN | t_cell | *C6orf106* | 3.457998966 | 6.15E-06 | 0.000576901 | 0.038652399 |
| HTN | t_cell | *ZFP28* | 3.671308045 | 0.000375165 | 0.009198979 | 1 |
| HTN | t_cell | *KIAA1211L* | 1.738212887 | 0.002878238 | 0.017969694 | 1 |
| HTN | t_cell | *LAMB3* | 2.670895904 | 0.002255569 | 0.016132836 | 1 |
| HTN | t_cell | *ANKRD36B* | -3.752036963 | 0.001952766 | 0.015192726 | 1 |
| HTN | t_cell | *HCAR1* | 2.938189787 | 0.000204561 | 0.006698301 | 1 |
| HTN | t_cell | *SRGAP1* | 2.419727039 | 0.006721842 | 0.028477238 | 1 |
| HTN | t_cell | *SLC39A10* | -2.880182559 | 0.003608109 | 0.02003903 | 1 |
| HTN | t_cell | *PCDHB16* | 2.331459638 | 0.000391821 | 0.009402202 | 1 |
| HTN | t_cell | *HIST1H3E* | 5.116474007 | 2.51E-08 | 8.77E-06 | 0.000157913 |
| HTN | t_cell | *FUT11* | -6.263174661 | 0.000535638 | 0.01137687 | 1 |
| HTN | t_cell | *WDR5B* | 4.775462937 | 0.003037776 | 0.018351697 | 1 |
| HTN | t_cell | *ZNF100* | -2.337496882 | 0.008262166 | 0.032505781 | 1 |
| HTN | t_cell | *GAL3ST4* | -4.82344459 | 0.000974477 | 0.013965754 | 1 |
| HTN | t_cell | *SLC25A29* | 3.388355322 | 0.003923079 | 0.020880704 | 1 |
| HTN | t_cell | *SULT1A2* | 1.561818056 | 2.55E-06 | 0.000276425 | 0.016032654 |
| HTN | t_cell | *PIWIL2* | 2.037213997 | 0.011018519 | 0.038961435 | 1 |
| HTN | t_cell | *ENTPD4* | -3.260952817 | 0.010402294 | 0.037530589 | 1 |
| HTN | t_cell | *FAM110D* | 4.353245274 | 0.004870176 | 0.02355292 | 1 |
| HTN | t_cell | *KANK2* | 2.536562562 | 0.015445754 | 0.049125217 | 1 |
| HTN | t_cell | *CCL4L2* | -1.638874339 | 0.001290439 | 0.013965754 | 1 |
| HTN | t_cell | *ZNF165* | 4.814024939 | 5.83E-05 | 0.002863966 | 0.36658759 |
| HTN | t_cell | *RAMP2-AS1* | 1.926380471 | 0.001707354 | 0.014532983 | 1 |
| HTN | t_cell | *DDI2* | 3.573820227 | 0.012837748 | 0.043323094 | 1 |
| HTN | t_cell | *ZNF517* | 4.33479549 | 0.003164896 | 0.018824691 | 1 |

| | | | | | | |
|---|---|---|---|---|---|---|
| HTN | t_cell | *SLC22A5* | 3.667409678 | 3.50E-09 | 1.69E-06 | 2.20E-05 |
| HTN | t_cell | *C5AR1* | 3.388167654 | 0.00563713 | 0.025812554 | 1 |
| HTN | t_cell | *VEPH1* | 2.99036761 | 0.005818763 | 0.026281493 | 1 |
| HTN | t_cell | *STMN3* | -1.059950248 | 0.001991558 | 0.015283681 | 1 |
| HTN | t_cell | *GYPE* | 2.244398012 | 0.003902427 | 0.020827297 | 1 |
| HTN | t_cell | *SLC2A10* | 3.901529209 | 0.000834848 | 0.013695656 | 1 |
| HTN | t_cell | *HOXA4* | 3.260477563 | 0.000700031 | 0.012928761 | 1 |
| HTN | t_cell | *CDC42SE1* | -2.228825135 | 0.001539832 | 0.014185269 | 1 |
| HTN | t_cell | *PDCD1LG2* | 2.367519285 | 0.001242161 | 0.013965754 | 1 |
| HTN | t_cell | *KLHL14* | 4.66882188 | 0.001416095 | 0.01399841 | 1 |
| HTN | t_cell | *FAM114A1* | 2.210631107 | 0.004990048 | 0.023855009 | 1 |
| HTN | t_cell | *RPS26* | -2.114978613 | 1.03E-06 | 0.000142956 | 0.006484044 |
| HTN | t_cell | *MAP1LC3C* | 2.173798721 | 0.000657383 | 0.012837993 | 1 |
| HTN | t_cell | *AC122129.1* | 3.484259305 | 0.001312521 | 0.013965754 | 1 |
| HTN | t_cell | *SGTB* | -2.305004364 | 0.001771159 | 0.014787883 | 1 |
| HTN | t_cell | *ADH5* | 3.198334196 | 0.002432336 | 0.016594315 | 1 |
| HTN | t_cell | *ZNF311* | 1.844102149 | 0.012715144 | 0.0430365 | 1 |
| HTN | t_cell | *FCHSD1* | -5.413910931 | 0.006432569 | 0.027661807 | 1 |
| HTN | t_cell | *RPL12* | -2.302656354 | 0.005528217 | 0.025406358 | 1 |
| HTN | t_cell | *ZNF273* | 3.38254321 | 0.014441404 | 0.046848868 | 1 |
| HTN | t_cell | *ZNF667* | 1.918016514 | 0.015831593 | 0.049741742 | 1 |
| HTN | t_cell | *SULT1C4* | 2.538340113 | 0.007405425 | 0.030370454 | 1 |
| HTN | t_cell | *SFI1* | -1.756145447 | 0.001439187 | 0.014126985 | 1 |
| HTN | t_cell | *SOWAHC* | 3.304157324 | 0.002720228 | 0.017558598 | 1 |
| HTN | t_cell | *AC135178.1* | 11.68610616 | 0.001314953 | 0.013965754 | 1 |
| HTN | t_cell | *RPS6KL1* | 3.191870621 | 1.92E-05 | 0.001323962 | 0.120480544 |
| HTN | t_cell | *SLC29A3* | 5.096184117 | 0.003274518 | 0.019032949 | 1 |
| HTN | t_cell | *ANTXRL* | -2.92606676 | 0.001085475 | 0.013965754 | 1 |
| HTN | t_cell | *STYX* | -3.023178276 | 0.002357431 | 0.016340867 | 1 |
| HTN | t_cell | *TMEM116* | -3.605136136 | 6.54E-13 | 1.81E-09 | 4.11E-09 |
| HTN | t_cell | *GFPT1* | 3.973603943 | 0.000992707 | 0.013965754 | 1 |
| HTN | t_cell | *MT1F* | -2.5206327 | 0.001491569 | 0.014185269 | 1 |
| HTN | t_cell | *HLA-DRB5* | -1.041453964 | 4.27E-05 | 0.002335689 | 0.268604225 |
| HTN | t_cell | *HIST1H4E* | 4.770520491 | 0.006080629 | 0.027009862 | 1 |
| HTN | t_cell | *C20orf203* | 4.06082275 | 0.005110209 | 0.024174482 | 1 |
| HTN | t_cell | *ARC* | -20.20980127 | 0.002862389 | 0.017906305 | 1 |
| HTN | t_cell | *LRBA* | -2.16300046 | 0.012407488 | 0.042371471 | 1 |
| HTN | t_cell | *MMP17* | 3.108068078 | 0.002624901 | 0.017208292 | 1 |
| HTN | t_cell | *TUSC1* | 5.759056916 | 0.005262378 | 0.024671565 | 1 |
| HTN | t_cell | *PAPSS2* | 2.806226352 | 0.00363468 | 0.020086785 | 1 |
| HTN | t_cell | *ANKRD13B* | 2.105825057 | 4.74E-05 | 0.002567791 | 0.297863735 |
| HTN | t_cell | *RPL10A* | -2.332152866 | 0.010113836 | 0.037011459 | 1 |
| HTN | t_cell | *APCDD1L* | 3.773030181 | 0.00365644 | 0.020147271 | 1 |
| HTN | t_cell | *RCSD1* | -1.375224109 | 0.000300223 | 0.008171005 | 1 |
| HTN | t_cell | *RASSF9* | 2.924333651 | 0.003773965 | 0.020419033 | 1 |
| HTN | t_cell | *FAM169A* | -1.078807211 | 0.001142351 | 0.013965754 | 1 |
| HTN | t_cell | *SCAMP5* | 2.366322618 | 0.00078803 | 0.013628763 | 1 |
| HTN | t_cell | *PNP* | -1.58010791 | 0.000178193 | 0.006208591 | 1 |
| HTN | t_cell | *HMGN2* | 3.107917148 | 0.008479534 | 0.033061704 | 1 |
| HTN | t_cell | *DENND4B* | -6.726462539 | 2.56E-05 | 0.001612016 | 0.161201591 |

| | | | | | | |
|---|---|---|---|---|---|---|
| HTN | t_cell | *CD3E* | -0.393398004 | 0.001067107 | 0.013965754 | 1 |
| HTN | t_cell | *ITPRIPL1* | -3.111741054 | 0.000432799 | 0.010040615 | 1 |
| HTN | t_cell | *PRMT6* | -14.78476583 | 0.007652023 | 0.031097782 | 1 |
| HTN | t_cell | *RASGEF1A* | -1.316498637 | 0.001099736 | 0.013965754 | 1 |
| HTN | t_cell | *MFAP3L* | 4.060824761 | 0.001881668 | 0.01512794 | 1 |
| HTN | t_cell | *TGM2* | 1.432455698 | 0.000824763 | 0.013695656 | 1 |
| HTN | t_cell | *AP001007.1* | 7.98752164 | 0.001263229 | 0.013965754 | 1 |
| HTN | t_cell | *AC105020.1* | 4.507766561 | 0.001005327 | 0.013965754 | 1 |
| HTN | t_cell | *CENPW* | -6.734768872 | 1.99E-05 | 0.001360891 | 0.125201995 |
| HTN | t_cell | *DDO* | 4.320974046 | 0.002323752 | 0.016291034 | 1 |
| HTN | t_cell | *RBM20* | 3.837963929 | 0.009578315 | 0.035727231 | 1 |
| HTN | t_cell | *FOXO6* | 2.929402064 | 0.001385067 | 0.013965754 | 1 |
| HTN | t_cell | *SYT15* | 3.429624353 | 0.008749257 | 0.033540599 | 1 |
| HTN | t_cell | *ZDBF2* | 3.884663656 | 0.006737541 | 0.028524525 | 1 |
| HTN | t_cell | *HSD17B8* | -4.716956429 | 3.93E-08 | 1.30E-05 | 0.000247137 |
| HTN | t_cell | *HLA-DRA* | 2.426081822 | 0.001045149 | 0.013965754 | 1 |
| HTN | t_cell | *COL15A1* | 1.768824014 | 0.011133933 | 0.039284112 | 1 |
| HTN | t_cell | *NOTCH4* | -0.330975135 | 0.003970449 | 0.020966245 | 1 |
| HTN | t_cell | *PRRT1* | 4.196002832 | 1.09E-06 | 0.000142956 | 0.006861871 |
| HTN | t_cell | *MRPL38* | 3.909593447 | 0.000856246 | 0.013695656 | 1 |
| HTN | t_cell | *STK19* | 5.343551623 | 1.98E-07 | 4.43E-05 | 0.001241734 |
| HTN | t_cell | *LAYN* | -3.422558534 | 0.003281647 | 0.019032949 | 1 |
| HTN | t_cell | *CARD16* | -1.61837672 | 0.000523522 | 0.011195174 | 1 |
| HTN | t_cell | *MSH5* | 2.257582591 | 0.002184019 | 0.015873903 | 1 |
| HTN | t_cell | *LY6G5C* | 1.22414377 | 0.009253024 | 0.03489313 | 1 |
| HTN | t_cell | *GPANK1* | 4.932540592 | 3.46E-05 | 0.001991638 | 0.217738864 |
| HTN | t_cell | *BAG6* | 5.257288717 | 1.61E-06 | 0.000199046 | 0.010151328 |
| HTN | t_cell | *NFKBIL1* | 4.433155798 | 5.88E-05 | 0.00286689 | 0.36982877 |
| HTN | t_cell | *MICB* | -1.045289188 | 0.002000511 | 0.015283681 | 1 |
| HTN | t_cell | *HLA-C* | -2.502002813 | 1.32E-05 | 0.001024785 | 0.083007624 |
| HTN | t_cell | *CCHCR1* | -1.524113781 | 0.003246675 | 0.019003909 | 1 |
| HTN | t_cell | *PSORS1C1* | 0.558180484 | 0.000632923 | 0.012544984 | 1 |
| HTN | t_cell | *LILRB3* | 1.923300197 | 0.015719412 | 0.049524434 | 1 |
| HTN | t_cell | *LINC01123* | -33.77573706 | 0.000305771 | 0.008215297 | 1 |
| HTN | t_cell | *FAM153C* | -1.28602776 | 0.001604462 | 0.014197745 | 1 |
| HTN | t_cell | *PCDHGA1* | 2.614333589 | 0.006373504 | 0.027520757 | 1 |
| HTN | t_cell | *PCDHA5* | 11.82524268 | 0.010218038 | 0.037238114 | 1 |
| HTN | t_cell | *PCDHA4* | 7.490314606 | 0.000634532 | 0.012544984 | 1 |
| HTN | t_cell | *SLFN12L* | -0.471024431 | 0.000881199 | 0.01381571 | 1 |
| HTN | t_cell | *SYCE1L* | -2.471803527 | 0.015668809 | 0.049519087 | 1 |
| HTN | t_cell | *FAM71F2* | 2.730822052 | 0.00168652 | 0.014465416 | 1 |
| HTN | t_cell | *TMEM170B* | 1.349478743 | 0.001245027 | 0.013965754 | 1 |
| HTN | t_cell | *PRR13* | -4.698098434 | 0.001193216 | 0.013965754 | 1 |
| HTN | t_cell | *MT1A* | 5.08917801 | 0.001390578 | 0.013965754 | 1 |
| HTN | t_cell | *MT1M* | 4.09655665 | 0.002441434 | 0.016594315 | 1 |
| HTN | t_cell | *CFI* | 2.010841781 | 0.004777264 | 0.023300746 | 1 |
| HTN | t_cell | *SAMD9* | -2.295770913 | 0.001500776 | 0.014185269 | 1 |
| HTN | t_cell | *EXOC3L4* | 3.364382262 | 0.003039679 | 0.018351697 | 1 |
| HTN | t_cell | *ATP6AP1L* | 2.315975579 | 0.004201529 | 0.021679382 | 1 |
| HTN | t_cell | *NAP1L4* | -2.743232776 | 0.007875143 | 0.031542745 | 1 |

| | | | | | | |
|---|---|---|---|---|---|---|
| HTN | t_cell | *C1QTNF9B* | 18.12548235 | 0.001265244 | 0.013965754 | 1 |
| HTN | t_cell | *NYNRIN* | 3.621864973 | 0.001415466 | 0.01399841 | 1 |
| HTN | t_cell | *ZDHHC11B* | 4.825788187 | 0.000646382 | 0.012739203 | 1 |
| HTN | t_cell | *GOLGA8O* | 6.779779673 | 0.00360006 | 0.020012004 | 1 |
| HTN | t_cell | *LNP1* | 1.735249065 | 0.013508295 | 0.044557528 | 1 |
| HTN | t_cell | *METTL6* | 3.202741691 | 0.002921905 | 0.018049857 | 1 |
| HTN | t_cell | *SLC48A1* | 2.014159839 | 0.012123558 | 0.041764828 | 1 |
| HTN | t_cell | *TAS2R14* | 2.377382129 | 0.001358579 | 0.013965754 | 1 |
| HTN | t_cell | *ZRSR1* | 10.09947423 | 0.000573791 | 0.01191612 | 1 |
| HTN | t_cell | *C17orf51* | 2.431613333 | 0.0087307 | 0.033540599 | 1 |
| HTN | t_cell | *RNF208* | 6.411192619 | 0.001745515 | 0.014702092 | 1 |
| HTN | t_cell | *MAP10* | 2.949755267 | 0.000977675 | 0.013965754 | 1 |
| HTN | t_cell | *FGFR1OP* | 5.043685914 | 0.002570618 | 0.017027838 | 1 |
| HTN | t_cell | *GIMAP1* | -2.692436852 | 5.80E-05 | 0.002863966 | 0.364460203 |
| HTN | t_cell | *CCDC183* | 7.49694851 | 0.000573954 | 0.01191612 | 1 |
| HTN | t_cell | *SUPT4H1* | 5.928899264 | 0.003361443 | 0.019313893 | 1 |
| HTN | t_cell | *MXD3* | -2.842831633 | 0.006060761 | 0.026966739 | 1 |
| HTN | t_cell | *ARHGAP19* | -6.068617342 | 0.000833232 | 0.013695656 | 1 |
| HTN | t_cell | *ERV3-1* | 0.839398778 | 0.007686079 | 0.031216007 | 1 |
| HTN | t_cell | *RBMXL1* | -5.801225962 | 0.005331692 | 0.024885186 | 1 |
| HTN | t_cell | *TMEM110* | 7.170521743 | 0.000985485 | 0.013965754 | 1 |
| HTN | t_cell | *LBH* | -1.161244503 | 0.001271924 | 0.013965754 | 1 |
| HTN | t_cell | *ATF6B* | 5.426317848 | 4.80E-08 | 1.44E-05 | 0.000301776 |
| HTN | t_cell | *SLC35F6* | 5.222805646 | 0.01049027 | 0.037634027 | 1 |
| HTN | t_cell | *KLRK1* | -0.98354668 | 0.003016489 | 0.018299376 | 1 |
| HTN | t_cell | *LTB4R* | -4.021695164 | 6.76E-08 | 1.85E-05 | 0.000425159 |
| HTN | t_cell | *LTB4R2* | -7.409068621 | 6.77E-07 | 0.000112062 | 0.004258355 |
| HTN | t_cell | *ZNF726* | 4.332303681 | 0.00401769 | 0.021084486 | 1 |
| HTN | t_cell | *AP1G2* | -3.637191137 | 0.001267664 | 0.013965754 | 1 |
| HTN | t_cell | *TM6SF2* | 20.38508214 | 0.001857919 | 0.015083316 | 1 |
| HTN | t_cell | *AC079776.2* | 5.168041626 | 0.0003661 | 0.009061689 | 1 |
| HTN | t_cell | *WEE2* | 3.699058293 | 0.000338713 | 0.00869179 | 1 |
| HTN | t_cell | *PRCD* | 3.077823273 | 0.003840858 | 0.020638868 | 1 |
| HTN | t_cell | *COLCA2* | 1.808355385 | 0.011245796 | 0.039564767 | 1 |
| HTN | t_cell | *MBLAC1* | 1.411900486 | 0.007277895 | 0.030082922 | 1 |
| HTN | t_cell | *STARD10* | 1.25419244 | 0.012326176 | 0.042152138 | 1 |
| HTN | t_cell | *C10orf105* | 2.856869458 | 0.008179317 | 0.032280832 | 1 |
| HTN | t_cell | *AC005082.1* | 7.897458212 | 0.003456695 | 0.019578594 | 1 |
| HTN | t_cell | *COL28A1* | 3.496911916 | 0.002325097 | 0.016291034 | 1 |
| HTN | t_cell | *UBE2QL1* | 3.068375008 | 0.001582452 | 0.014185269 | 1 |
| HTN | t_cell | *GOLGA8B* | -2.518690958 | 0.001339334 | 0.013965754 | 1 |
| HTN | t_cell | *MYL5* | 6.995420205 | 0.010378929 | 0.037479796 | 1 |
| HTN | t_cell | *LINC01138* | -2.161929946 | 0.001899878 | 0.015177294 | 1 |
| HTN | t_cell | *AC016773.1* | 1.68467789 | 0.00226906 | 0.016174171 | 1 |
| HTN | t_cell | *AP000350.4* | 0.806193712 | 0.009418862 | 0.035286598 | 1 |
| HTN | t_cell | *RNU6ATAC35I* | 11.49628014 | 0.001549997 | 0.014185269 | 1 |
| HTN | t_cell | *C1orf229* | 3.199087605 | 0.000491354 | 0.010839102 | 1 |
| HTN | t_cell | *TIAF1* | -3.090334456 | 0.007792297 | 0.031323637 | 1 |
| HTN | t_cell | *AC062017.1* | 1.969881393 | 0.014764947 | 0.047603702 | 1 |
| HTN | t_cell | *ADRA2B* | 5.9745575 | 0.001376004 | 0.013965754 | 1 |

| | | | | | | |
|---|---|---|---|---|---|---|
| HTN | t_cell | *EXOSC6* | -6.811076676 | 6.01E-06 | 0.000572526 | 0.037786735 |
| HTN | t_cell | *TINCR* | -5.757759037 | 0.009217124 | 0.034828794 | 1 |
| HTN | t_cell | *AC147651.3* | -2.105260673 | 0.002163553 | 0.015811049 | 1 |
| HTN | t_cell | *HLA-DPB1* | 3.033627373 | 0.0001907 | 0.006445865 | 1 |
| HTN | t_cell | *OSER1-AS1* | 4.355115904 | 0.00072822 | 0.013156092 | 1 |
| HTN | t_cell | *LINC01133* | 2.850676422 | 0.003570823 | 0.019909231 | 1 |
| HTN | t_cell | *LINC01117* | 1.886939913 | 0.002028959 | 0.015331808 | 1 |
| HTN | t_cell | *RASAL2-AS1* | 9.016899534 | 0.003771294 | 0.020419033 | 1 |
| HTN | t_cell | *AC069213.1* | -5.786772777 | 0.00132618 | 0.013965754 | 1 |
| HTN | t_cell | *LINC00240* | 1.474278604 | 0.002278218 | 0.016194559 | 1 |
| HTN | t_cell | *LINC00863* | 3.417272742 | 0.006680883 | 0.028402239 | 1 |
| HTN | t_cell | *LINC00184* | 2.909764417 | 0.007047145 | 0.029321907 | 1 |
| HTN | t_cell | *TMEM233* | 1.966052343 | 0.000154454 | 0.005580753 | 0.971050998 |
| HTN | t_cell | *LINC00237* | -4.644531964 | 0.001040254 | 0.013965754 | 1 |
| HTN | t_cell | *LINC00092* | -2.693569829 | 0.002523814 | 0.016925325 | 1 |
| HTN | t_cell | *SFTA1P* | 6.155305053 | 0.004830903 | 0.023507652 | 1 |
| HTN | t_cell | *SLC26A6* | 5.25227413 | 3.37E-07 | 6.84E-05 | 0.002120797 |
| HTN | t_cell | *MIAT* | -0.639314253 | 0.000742689 | 0.013302814 | 1 |
| HTN | t_cell | *LINC00626* | 27.30546173 | 0.003954123 | 0.020925564 | 1 |
| HTN | t_cell | *LINC00115* | -2.762678899 | 0.008898068 | 0.033986728 | 1 |
| HTN | t_cell | *ELFN1* | -14.46280862 | 0.013211729 | 0.043994779 | 1 |
| HTN | t_cell | *ZNF503-AS1* | 2.548549864 | 0.001856031 | 0.015083316 | 1 |
| HTN | t_cell | *AC064836.3* | 5.182945407 | 0.001607888 | 0.014197745 | 1 |
| HTN | t_cell | *TAS2R46* | -3.287928287 | 0.007785913 | 0.031318911 | 1 |
| HTN | t_cell | *AC093901.1* | 2.726896432 | 0.00631477 | 0.02737997 | 1 |
| HTN | t_cell | *C14orf132* | 2.027037468 | 0.007873063 | 0.031542745 | 1 |
| HTN | t_cell | *ZNF717* | 0.859586302 | 0.000405648 | 0.009638365 | 1 |
| HTN | t_cell | *SLC2A1-AS1* | -6.485554915 | 0.00137504 | 0.013965754 | 1 |
| HTN | t_cell | *LINC01013* | 3.163108715 | 0.00085758 | 0.013695656 | 1 |
| HTN | t_cell | *PROB1* | 13.73663976 | 6.17E-07 | 0.000104843 | 0.00387919 |
| HTN | t_cell | *ARL17B* | 2.061820754 | 0.012285864 | 0.042116263 | 1 |
| HTN | t_cell | *LINC00954* | -1.899106135 | 0.001131367 | 0.013965754 | 1 |
| HTN | t_cell | *AC067945.3* | -0.668360052 | 0.005129373 | 0.024234763 | 1 |
| HTN | t_cell | *RPL41* | -3.867641644 | 0.009581058 | 0.035727231 | 1 |
| HTN | t_cell | *PATL2* | -1.724295497 | 0.002129682 | 0.015682578 | 1 |
| HTN | t_cell | *U91324.1* | 6.740221355 | 0.001864608 | 0.015083316 | 1 |
| HTN | t_cell | *CISD3* | 4.297316126 | 0.000931316 | 0.013965754 | 1 |
| HTN | t_cell | *PPP5D1* | 2.857334802 | 0.005347428 | 0.02494012 | 1 |
| HTN | t_cell | *AC112229.4* | 2.033345746 | 0.000999205 | 0.013965754 | 1 |
| HTN | t_cell | *KLHL7-AS1* | 4.697111613 | 0.001413248 | 0.01399841 | 1 |
| HTN | t_cell | *RPS18* | -2.158184858 | 0.007204837 | 0.029879162 | 1 |
| HTN | t_cell | *DIRC3* | 1.57436354 | 0.006300334 | 0.027374015 | 1 |
| HTN | t_cell | *AC107021.1* | 1.476495167 | 0.00178733 | 0.014844048 | 1 |
| HTN | t_cell | *LINC00865* | 2.946525447 | 0.00343226 | 0.01949288 | 1 |
| HTN | t_cell | *AC009495.3* | -1.868807801 | 0.002041799 | 0.015375679 | 1 |
| HTN | t_cell | *GOLGA8N* | 2.19025039 | 0.002417038 | 0.016563558 | 1 |
| HTN | t_cell | *TNF* | -0.778650666 | 0.00166518 | 0.014360746 | 1 |
| HTN | t_cell | *LINC00702* | 2.547292009 | 0.008091646 | 0.032075774 | 1 |
| HTN | t_cell | *RPS28* | -4.310057168 | 0.008352813 | 0.032678368 | 1 |
| HTN | t_cell | *AC123768.3* | 2.94264211 | 0.001178712 | 0.013965754 | 1 |

| | | | | | | |
|---|---|---|---|---|---|---|
| HTN | t_cell | *C19orf81* | 4.702266214 | 0.010499457 | 0.037634027 | 1 |
| HTN | t_cell | *ANKRD65* | 4.149574135 | 1.49E-05 | 0.001102931 | 0.093502878 |
| HTN | t_cell | *LINC00957* | 3.483599541 | 0.012561741 | 0.042782049 | 1 |
| HTN | t_cell | *LINC00941* | 6.90428603 | 0.001045295 | 0.013965754 | 1 |
| HTN | t_cell | *LINC00545* | -6.576419722 | 0.002872894 | 0.01795416 | 1 |
| HTN | t_cell | *HLA-DQA2* | 2.090895472 | 5.51E-07 | 0.00010182 | 0.003461868 |
| HTN | t_cell | *AC073257.2* | -11.83482218 | 0.001282888 | 0.013965754 | 1 |
| HTN | t_cell | *TXNDC5* | 4.530363086 | 0.000992201 | 0.013965754 | 1 |
| HTN | t_cell | *CDRT4* | 4.685537095 | 0.007365745 | 0.03025216 | 1 |
| HTN | t_cell | *APOBEC3G* | -1.026331672 | 0.001513305 | 0.014185269 | 1 |
| HTN | t_cell | *ADSL* | -2.735052133 | 0.001210845 | 0.013965754 | 1 |
| HTN | t_cell | *LILRA4* | -10.11227906 | 0.00182014 | 0.014978034 | 1 |
| HTN | t_cell | *PSMB9* | -1.943937573 | 0.002165131 | 0.015811049 | 1 |
| HTN | t_cell | *AQP1* | 2.461929484 | 0.013711391 | 0.045156373 | 1 |
| HTN | t_cell | *ARHGEF25* | 2.84114593 | 0.004722409 | 0.023213281 | 1 |
| HTN | t_cell | *ATP5J2* | 4.304906855 | 0.001384269 | 0.013965754 | 1 |
| HTN | t_cell | *ARHGAP8* | 5.441035017 | 0.00193797 | 0.015190044 | 1 |
| HTN | t_cell | *PLEKHO2* | 2.622175248 | 0.010407083 | 0.037530589 | 1 |
| HTN | t_cell | *PWP2* | 6.134338878 | 0.008257904 | 0.032505781 | 1 |
| HTN | t_cell | *AKAP2* | 4.774476589 | 0.001276822 | 0.013965754 | 1 |
| HTN | t_cell | *LINC00960* | -1.337084366 | 0.003514251 | 0.019762162 | 1 |
| HTN | t_cell | *DECR2* | 2.307343903 | 0.001662899 | 0.014360746 | 1 |
| HTN | t_cell | *ZNF709* | -3.057611918 | 2.86E-05 | 0.00171397 | 0.179966848 |
| HTN | t_cell | *PSG4* | 4.253128186 | 7.99E-06 | 0.000697853 | 0.050245422 |
| HTN | t_cell | *STON1* | 1.88733919 | 0.003941307 | 0.020880704 | 1 |
| HTN | t_cell | *VSIG8* | 2.978312582 | 0.003358767 | 0.019313893 | 1 |
| HTN | t_cell | *FGF14-IT1* | 5.488161139 | 0.001511219 | 0.014185269 | 1 |
| HTN | t_cell | *PALM2* | 1.437938568 | 0.001527866 | 0.014185269 | 1 |
| HTN | t_cell | *WDR92* | 4.071887026 | 0.01318109 | 0.043962605 | 1 |
| HTN | t_cell | *PLA2G4B* | 3.147238103 | 0.000143733 | 0.005440117 | 0.903646827 |
| HTN | t_cell | *LEFTY1* | 14.44577164 | 0.000580452 | 0.012004292 | 1 |
| HTN | t_cell | *JMJD7* | 2.794096608 | 0.001827323 | 0.014997888 | 1 |
| HTN | t_cell | *APOBEC3D* | -2.556524774 | 0.002079745 | 0.015473797 | 1 |
| HTN | t_cell | *RN7SL832P* | 2.769080552 | 0.002682925 | 0.017407175 | 1 |
| HTN | t_cell | *P2RY11* | -4.230020188 | 0.000593679 | 0.012118368 | 1 |
| HTN | t_cell | *APOBEC3C* | -5.291850724 | 0.004890847 | 0.023580332 | 1 |
| HTN | t_cell | *LINC00920* | -4.442316183 | 0.005279937 | 0.024717026 | 1 |
| HTN | t_cell | *BCKDHA* | 3.68330322 | 0.004459383 | 0.022508072 | 1 |
| HTN | t_cell | *C5orf17* | 3.526159481 | 0.012297384 | 0.04213278 | 1 |
| HTN | t_cell | *LINC01060* | 1.962685665 | 0.003257714 | 0.019003909 | 1 |
| HTN | t_cell | *ZNF286B* | -4.845552803 | 0.005882133 | 0.026471702 | 1 |
| HTN | t_cell | *YJEFN3* | 4.205485294 | 0.009312836 | 0.035017823 | 1 |
| HTN | t_cell | *PCDHA10* | 3.358517208 | 0.000777924 | 0.013547949 | 1 |
| HTN | t_cell | *LINC00989* | 4.216356408 | 0.012994094 | 0.043593314 | 1 |
| HTN | t_cell | *LINC01021* | 5.03877731 | 0.001685746 | 0.014465416 | 1 |
| HTN | t_cell | *FAM218A* | 3.784957705 | 0.00148538 | 0.014185269 | 1 |
| HTN | t_cell | *TMEM200B* | 1.617362646 | 0.004714842 | 0.023212384 | 1 |
| HTN | t_cell | *PCDHGB6* | 2.436944015 | 0.006576915 | 0.028120771 | 1 |
| HTN | t_cell | *PCDHGA6* | 2.108414985 | 0.01161457 | 0.040499614 | 1 |
| HTN | t_cell | *CLDN23* | 9.864620537 | 0.000869185 | 0.013695656 | 1 |

| | | | | | | |
|---|---|---|---|---|---|---|
| HTN | t_cell | *FPGT* | 3.742685482 | 0.005130662 | 0.024234763 | 1 |
| HTN | t_cell | *MPV17L2* | 3.504459325 | 0.002393677 | 0.016483077 | 1 |
| HTN | t_cell | *POLR2M* | 4.035219707 | 0.006686069 | 0.028402239 | 1 |
| HTN | t_cell | *TAS2R20* | 1.998945553 | 0.007730664 | 0.031266496 | 1 |
| HTN | t_cell | *LINC00346* | 3.409734251 | 0.005187989 | 0.024403857 | 1 |
| HTN | t_cell | *LINC00944* | -0.573608057 | 0.001160943 | 0.013965754 | 1 |
| HTN | t_cell | *ZNF10* | -5.154016047 | 1.97E-06 | 0.000229392 | 0.012387167 |
| HTN | t_cell | *HMBS* | 3.576189605 | 1.07E-06 | 0.000142956 | 0.006738113 |
| HTN | t_cell | *ZBED6* | 3.828121375 | 0.001408599 | 0.01399841 | 1 |
| HTN | t_cell | *FNTB* | 4.431654061 | 0.002790086 | 0.017682732 | 1 |
| HTN | t_cell | *RTEL1* | 4.72208716 | 0.000354901 | 0.008960886 | 1 |
| HTN | t_cell | *LINC00648* | 2.949352226 | 0.004027607 | 0.021109371 | 1 |
| HTN | t_cell | *PWRN1* | 1.957589391 | 0.011927255 | 0.041255404 | 1 |
| HTN | t_cell | *HOXB7* | 3.715177292 | 0.002164065 | 0.015811049 | 1 |
| HTN | t_cell | *FRRS1L* | 3.647293284 | 0.000925852 | 0.013965754 | 1 |
| HTN | t_cell | *AC009120.5* | 3.241840232 | 0.001746853 | 0.014702092 | 1 |
| HTN | t_cell | *HPR* | 5.852246214 | 0.003479152 | 0.019635032 | 1 |
| HTN | t_cell | *GOLGA8H* | 1.480958219 | 0.006129725 | 0.027024952 | 1 |
| HTN | t_cell | *DYNLL2* | -3.866009666 | 0.001831309 | 0.015001497 | 1 |
| HTN | t_cell | *LINC00526* | 3.579440325 | 0.00474606 | 0.023249344 | 1 |
| HTN | t_cell | *LINC00668* | -46.27800323 | 0.006913705 | 0.028977644 | 1 |
| HTN | t_cell | *ILF3-AS1* | 3.198933637 | 0.008551572 | 0.033126143 | 1 |
| HTN | t_cell | *AC139100.2* | 6.111174911 | 0.001341148 | 0.013965754 | 1 |
| HTN | t_cell | *LINC01140* | 2.189274439 | 0.002116926 | 0.015657783 | 1 |
| HTN | t_cell | *LINC00906* | 11.65639292 | 0.001371103 | 0.013965754 | 1 |
| HTN | t_cell | *AC137932.1* | 7.088338051 | 0.000184731 | 0.006311992 | 1 |
| HTN | t_cell | *AL132989.1* | 4.414764712 | 0.001376398 | 0.013965754 | 1 |
| HTN | t_cell | *FBXO17* | 2.252208987 | 0.002014966 | 0.015300078 | 1 |
| HTN | t_cell | *YTHDF3-AS1* | 3.853526892 | 0.001167767 | 0.013965754 | 1 |
| HTN | t_cell | *LINC01050* | 11.40006954 | 0.003572805 | 0.019909231 | 1 |
| HTN | t_cell | *MILR1* | 1.280345654 | 0.003778941 | 0.020428376 | 1 |
| HTN | t_cell | *FGF14-AS2* | 3.124445801 | 0.001100384 | 0.013965754 | 1 |
| HTN | t_cell | *CASC15* | 3.227287158 | 0.001441274 | 0.014126985 | 1 |
| HTN | t_cell | *LINC01023* | 3.545907967 | 0.001327512 | 0.013965754 | 1 |
| HTN | t_cell | *AP000347.2* | 1.714079942 | 0.007460297 | 0.030515868 | 1 |
| HTN | t_cell | *POLR2J4* | 4.746728068 | 0.01480725 | 0.047691178 | 1 |
| HTN | t_cell | *AC144652.1* | 2.639964994 | 1.59E-06 | 0.000199046 | 0.01002381 |
| Obesity | adipocyte | *C1orf112* | 1.262873035 | 0.01397912 | 0.038614555 | 1 |
| Obesity | adipocyte | *CFH* | 1.144080227 | 1.11E-06 | 4.24E-05 | 0.006993093 |
| Obesity | adipocyte | *NIPAL3* | 0.983592006 | 0.000173422 | 0.001259013 | 1 |
| Obesity | adipocyte | *SEMA3F* | -0.630108791 | 6.05E-20 | 3.80E-17 | 3.80E-16 |
| Obesity | adipocyte | *KRIT1* | 0.948897742 | 0.011682089 | 0.03346027 | 1 |
| Obesity | adipocyte | *HS3ST1* | 1.334301154 | 6.46E-05 | 0.000613065 | 0.406163828 |
| Obesity | adipocyte | *WNT16* | 0.61201517 | 0.001216019 | 0.005604921 | 1 |
| Obesity | adipocyte | *LASP1* | 1.151992491 | 0.006974746 | 0.022464257 | 1 |
| Obesity | adipocyte | *ICA1* | 1.252979077 | 1.07E-05 | 0.000165792 | 0.067311627 |
| Obesity | adipocyte | *ALS2* | 1.160895522 | 0.009491957 | 0.028498537 | 1 |
| Obesity | adipocyte | *RBM6* | -0.506196295 | 4.94E-23 | 4.43E-20 | 3.10E-19 |
| Obesity | adipocyte | *RECQL* | 1.997046254 | 4.07E-05 | 0.000437222 | 0.255774654 |
| Obesity | adipocyte | *ARHGAP33* | 0.499756243 | 0.003301958 | 0.012520755 | 1 |

| | | | | | | |
|---|---|---|---|---|---|---|
| Obesity | adipocyte | *PDK4* | 1.35731861 | 0.000563997 | 0.003110396 | 1 |
| Obesity | adipocyte | *CDC27* | 0.711946396 | 2.27E-05 | 0.000282607 | 0.142716593 |
| Obesity | adipocyte | *FAM214B* | 1.521388335 | 0.000169505 | 0.001233424 | 1 |
| Obesity | adipocyte | *COPZ2* | 0.299375739 | 0.003966654 | 0.014381982 | 1 |
| Obesity | adipocyte | *GCFC2* | 1.131341726 | 0.007180176 | 0.022984607 | 1 |
| Obesity | adipocyte | *WDR54* | 1.164873384 | 0.001015172 | 0.004899217 | 1 |
| Obesity | adipocyte | *ABCB4* | 1.939672101 | 3.54E-05 | 0.000393241 | 0.222574658 |
| Obesity | adipocyte | *GGNBP2* | -0.817839922 | 8.32E-07 | 3.44E-05 | 0.005228778 |
| Obesity | adipocyte | *TMEM98* | 0.380888867 | 0.000974048 | 0.004743485 | 1 |
| Obesity | adipocyte | *ABCC8* | 1.618607746 | 0.009727622 | 0.029039677 | 1 |
| Obesity | adipocyte | *SYNRG* | 1.43530059 | 0.000667652 | 0.003489218 | 1 |
| Obesity | adipocyte | *TMEM132A* | 2.641633877 | 1.71E-05 | 0.000234277 | 0.107299001 |
| Obesity | adipocyte | *ZNF263* | 1.960258986 | 0.000467162 | 0.002660371 | 1 |
| Obesity | adipocyte | *RALA* | 1.715536974 | 0.000101985 | 0.000847459 | 0.641177435 |
| Obesity | adipocyte | *ARHGAP44* | 1.33014743 | 0.000112546 | 0.000917735 | 0.707573796 |
| Obesity | adipocyte | *NOS2* | 5.50734828 | 0.008451372 | 0.026113702 | 1 |
| Obesity | adipocyte | *RPUSD1* | 6.359785718 | 6.53E-05 | 0.000617597 | 0.410702075 |
| Obesity | adipocyte | *TSR3* | 1.529920992 | 0.014106998 | 0.038916497 | 1 |
| Obesity | adipocyte | *MMP25* | 0.717151524 | 0.002840202 | 0.011090901 | 1 |
| Obesity | adipocyte | *PKD1* | 0.692455963 | 0.000350786 | 0.002102373 | 1 |
| Obesity | adipocyte | *MAPK8IP2* | 1.195644165 | 0.003689954 | 0.013609479 | 1 |
| Obesity | adipocyte | *SEC62* | 1.531868752 | 0.000270157 | 0.001738461 | 1 |
| Obesity | adipocyte | *RPS20* | 1.277601967 | 0.000888901 | 0.004414315 | 1 |
| Obesity | adipocyte | *SPRTN* | 1.854734765 | 0.003391486 | 0.012752554 | 1 |
| Obesity | adipocyte | *STARD3NL* | 2.01276619 | 2.74E-07 | 1.61E-05 | 0.001721312 |
| Obesity | adipocyte | *CD4* | 1.802001843 | 2.57E-06 | 6.50E-05 | 0.016162925 |
| Obesity | adipocyte | *LRRC23* | 0.435319849 | 0.011430586 | 0.032965182 | 1 |
| Obesity | adipocyte | *HFE* | -0.71198694 | 6.46E-08 | 5.07E-06 | 0.000405984 |
| Obesity | adipocyte | *SCMH1* | 0.67299894 | 0.003312719 | 0.012542775 | 1 |
| Obesity | adipocyte | *CLCN6* | 1.929395989 | 1.93E-06 | 5.81E-05 | 0.01211217 |
| Obesity | adipocyte | *TSPAN9* | 0.374747313 | 0.008275615 | 0.025695067 | 1 |
| Obesity | adipocyte | *UTP18* | 2.151659906 | 0.000350562 | 0.002102373 | 1 |
| Obesity | adipocyte | *TTC19* | 1.195092886 | 1.75E-07 | 1.11E-05 | 0.001097938 |
| Obesity | adipocyte | *SYT7* | 0.687747432 | 0.015533043 | 0.042158204 | 1 |
| Obesity | adipocyte | *LARS2* | 0.748913468 | 0.010433308 | 0.030737678 | 1 |
| Obesity | adipocyte | *PPP5C* | 0.764968106 | 0.009629283 | 0.028841974 | 1 |
| Obesity | adipocyte | *ERCC1* | 1.117692351 | 0.003933997 | 0.014296553 | 1 |
| Obesity | adipocyte | *ALOX5* | 0.777506256 | 0.011495818 | 0.033107744 | 1 |
| Obesity | adipocyte | *MAP4K5* | 1.202507207 | 0.000551437 | 0.003049149 | 1 |
| Obesity | adipocyte | *PSMC4* | 0.759955892 | 0.000128891 | 0.001012094 | 0.810337779 |
| Obesity | adipocyte | *MAN2B2* | 1.268194528 | 0.00136577 | 0.006213167 | 1 |
| Obesity | adipocyte | *CLDN11* | 0.790314137 | 5.62E-05 | 0.000553957 | 0.353033844 |
| Obesity | adipocyte | *ANGEL1* | 1.840816389 | 0.000239345 | 0.001597411 | 1 |
| Obesity | adipocyte | *RNF14* | 0.87393867 | 0.014650282 | 0.040230121 | 1 |
| Obesity | adipocyte | *SLC30A9* | 1.092340845 | 0.000313412 | 0.001947056 | 1 |
| Obesity | adipocyte | *MTMR11* | 0.179248954 | 0.003609875 | 0.013373768 | 1 |
| Obesity | adipocyte | *NUDCD3* | 0.428826991 | 0.001016815 | 0.004899217 | 1 |
| Obesity | adipocyte | *IL20RA* | 7.090070484 | 4.76E-06 | 9.50E-05 | 0.029934024 |
| Obesity | adipocyte | *RUFY3* | 0.547137835 | 0.007883537 | 0.024720099 | 1 |
| Obesity | adipocyte | *WWTR1* | 1.067926348 | 0.001139501 | 0.00533039 | 1 |

| | | | | | | |
|---|---|---|---|---|---|---|
| Obesity | adipocyte | *VSIG2* | 0.774085106 | 0.013081425 | 0.036715589 | 1 |
| Obesity | adipocyte | *OSBPL5* | 2.106195046 | 9.53E-06 | 0.000153475 | 0.059887475 |
| Obesity | adipocyte | *AQR* | 1.040019355 | 0.002798561 | 0.010982868 | 1 |
| Obesity | adipocyte | *ZDHHC6* | 1.166120167 | 0.000575412 | 0.003151231 | 1 |
| Obesity | adipocyte | *RNH1* | 0.453064099 | 0.008778102 | 0.026881602 | 1 |
| Obesity | adipocyte | *NDUFS1* | 0.974394497 | 0.000681455 | 0.003546613 | 1 |
| Obesity | adipocyte | *AKAP11* | 1.729019694 | 0.000159671 | 0.001175468 | 1 |
| Obesity | adipocyte | *UBR2* | 1.216243869 | 0.00116702 | 0.005434857 | 1 |
| Obesity | adipocyte | *HSF2* | 1.441421917 | 0.007925649 | 0.024815017 | 1 |
| Obesity | adipocyte | *NR1H3* | 0.962273236 | 0.000178433 | 0.001285002 | 1 |
| Obesity | adipocyte | *TYMP* | 0.621831027 | 0.003305985 | 0.012528467 | 1 |
| Obesity | adipocyte | *TOMM34* | 2.405838455 | 0.000273203 | 0.001752683 | 1 |
| Obesity | adipocyte | *KPNA6* | 1.053986236 | 0.00039678 | 0.002342306 | 1 |
| Obesity | adipocyte | *VIM* | 1.366840847 | 4.21E-06 | 8.80E-05 | 0.026489744 |
| Obesity | adipocyte | *CD44* | 0.99695707 | 1.75E-05 | 0.000238414 | 0.109908695 |
| Obesity | adipocyte | *MIPEP* | 1.558442912 | 0.002912248 | 0.011302039 | 1 |
| Obesity | adipocyte | *SNX1* | 0.627386794 | 0.001608158 | 0.007100062 | 1 |
| Obesity | adipocyte | *ANK1* | 1.399167008 | 0.005038856 | 0.017377557 | 1 |
| Obesity | adipocyte | *RABEP1* | 1.103897409 | 2.58E-08 | 2.46E-06 | 0.000162372 |
| Obesity | adipocyte | *GRN* | 1.38371438 | 0.00013754 | 0.001054529 | 0.864713494 |
| Obesity | adipocyte | *ARID4A* | 1.667373402 | 4.01E-05 | 0.000431649 | 0.252083285 |
| Obesity | adipocyte | *IFT88* | 0.648316324 | 0.012994603 | 0.03653715 | 1 |
| Obesity | adipocyte | *ZCCHC8* | 2.693881983 | 1.13E-09 | 1.85E-07 | 7.08E-06 |
| Obesity | adipocyte | *FUT8* | 1.527135254 | 6.12E-05 | 0.000590248 | 0.384841375 |
| Obesity | adipocyte | *PIAS1* | 1.41348442 | 3.90E-05 | 0.000423188 | 0.245166968 |
| Obesity | adipocyte | *SLC4A7* | 1.407547999 | 1.20E-05 | 0.00018131 | 0.075424882 |
| Obesity | adipocyte | *SH3YL1* | -0.484761368 | 0.00065732 | 0.00344882 | 1 |
| Obesity | adipocyte | *ADSS* | 1.90714573 | 9.01E-06 | 0.000147085 | 0.056627624 |
| Obesity | adipocyte | *STAP1* | -0.85700325 | 0.001861982 | 0.007925713 | 1 |
| Obesity | adipocyte | *CUL3* | 1.066996226 | 0.002582517 | 0.010243714 | 1 |
| Obesity | adipocyte | *NSUN2* | 1.30286805 | 0.00043811 | 0.00252697 | 1 |
| Obesity | adipocyte | *BOD1L1* | 0.666748784 | 0.006273558 | 0.020661005 | 1 |
| Obesity | adipocyte | *MAT2B* | 1.993866635 | 1.29E-05 | 0.0001921 | 0.081342103 |
| Obesity | adipocyte | *CLEC16A* | 1.335069982 | 0.001790257 | 0.007682829 | 1 |
| Obesity | adipocyte | *SKIV2L2* | 1.163808473 | 0.000651216 | 0.00343761 | 1 |
| Obesity | adipocyte | *DNAH5* | 1.341244474 | 0.000258873 | 0.001693585 | 1 |
| Obesity | adipocyte | *RAI14* | 0.478840243 | 0.002436701 | 0.009788843 | 1 |
| Obesity | adipocyte | *INPP4A* | 0.394458262 | 0.012231769 | 0.034796892 | 1 |
| Obesity | adipocyte | *LSG1* | 0.366505681 | 0.000201286 | 0.001406097 | 1 |
| Obesity | adipocyte | *TDP1* | 2.069538024 | 0.000752315 | 0.003845369 | 1 |
| Obesity | adipocyte | *SPATA7* | 1.220043529 | 0.00667376 | 0.021683686 | 1 |
| Obesity | adipocyte | *MED17* | 0.777924763 | 0.01603305 | 0.043225987 | 1 |
| Obesity | adipocyte | *LCP2* | 0.607549194 | 0.01072458 | 0.031331522 | 1 |
| Obesity | adipocyte | *CTNNA1* | 1.292251882 | 1.30E-05 | 0.0001921 | 0.081450574 |
| Obesity | adipocyte | *CNTLN* | 1.439543178 | 2.91E-06 | 6.99E-05 | 0.018303581 |
| Obesity | adipocyte | *EPHA3* | 1.446029677 | 1.94E-06 | 5.81E-05 | 0.012176739 |
| Obesity | adipocyte | *DSG2* | 1.34050052 | 0.000408809 | 0.002399799 | 1 |
| Obesity | adipocyte | *YTHDC2* | 1.518702932 | 8.95E-05 | 0.000772236 | 0.562959748 |
| Obesity | adipocyte | *ATP6V1H* | 1.616092532 | 0.000309524 | 0.001932321 | 1 |
| Obesity | adipocyte | *POLR2B* | 1.681589017 | 0.000133555 | 0.001032791 | 0.83965896 |

| | | | | | | |
|---|---|---|---|---|---|---|
| Obesity | adipocyte | *CP* | 2.326892893 | 1.40E-06 | 5.05E-05 | 0.008784351 |
| Obesity | adipocyte | *DTNBP1* | 1.837012779 | 7.84E-06 | 0.000136183 | 0.049298407 |
| Obesity | adipocyte | *ANO2* | 0.351749505 | 0.000518454 | 0.002902509 | 1 |
| Obesity | adipocyte | *GOPC* | 1.062572521 | 0.003343859 | 0.012621094 | 1 |
| Obesity | adipocyte | *TSPAN17* | 0.47210528 | 0.001327897 | 0.006058409 | 1 |
| Obesity | adipocyte | *SNX29* | 0.88882847 | 0.002667727 | 0.010555065 | 1 |
| Obesity | adipocyte | *CELF2* | 0.986135129 | 0.001759201 | 0.007608682 | 1 |
| Obesity | adipocyte | *ADAMTS6* | 0.368881282 | 0.013619546 | 0.03790442 | 1 |
| Obesity | adipocyte | *UTS2* | -0.378699112 | 0.011986103 | 0.034175342 | 1 |
| Obesity | adipocyte | *RFC2* | 1.016194246 | 0.011910036 | 0.034004721 | 1 |
| Obesity | adipocyte | *JKAMP* | 0.62835207 | 0.00030511 | 0.001918225 | 1 |
| Obesity | adipocyte | *DKK3* | 1.383335207 | 2.31E-06 | 6.25E-05 | 0.014543849 |
| Obesity | adipocyte | *NFE2L3* | 2.187977418 | 1.80E-05 | 0.000241715 | 0.113246457 |
| Obesity | adipocyte | *HERPUD1* | 1.62888102 | 0.00026688 | 0.00173155 | 1 |
| Obesity | adipocyte | *CYBA* | 1.282126621 | 0.000120404 | 0.000966769 | 0.756979753 |
| Obesity | adipocyte | *ALX4* | 3.676728146 | 0.001984017 | 0.008310136 | 1 |
| Obesity | adipocyte | *FOXN3* | 1.0611929 | 0.000776838 | 0.003938693 | 1 |
| Obesity | adipocyte | *ANAPC4* | 1.869923373 | 1.19E-07 | 8.30E-06 | 0.000746774 |
| Obesity | adipocyte | *OPN3* | 0.267752859 | 0.011351072 | 0.03278098 | 1 |
| Obesity | adipocyte | *SDCCAG8* | 0.724816586 | 1.26E-06 | 4.65E-05 | 0.007906639 |
| Obesity | adipocyte | *TBC1D22A* | 1.350197185 | 3.20E-05 | 0.000362662 | 0.200914922 |
| Obesity | adipocyte | *GINM1* | 1.157724419 | 0.00343176 | 0.012880883 | 1 |
| Obesity | adipocyte | *ITIH4* | 0.326688406 | 8.50E-11 | 1.84E-08 | 5.35E-07 |
| Obesity | adipocyte | *TRAF3IP2* | 1.248917505 | 0.006438487 | 0.021082692 | 1 |
| Obesity | adipocyte | *DCBLD2* | 1.073238885 | 0.013172834 | 0.036922696 | 1 |
| Obesity | adipocyte | *TMCC3* | 1.023846769 | 0.001930929 | 0.008147951 | 1 |
| Obesity | adipocyte | *ATP11B* | 0.525364262 | 6.25E-05 | 0.000599545 | 0.392701671 |
| Obesity | adipocyte | *CDK14* | 1.423966165 | 0.000123729 | 0.00098716 | 0.777881769 |
| Obesity | adipocyte | *CROCC* | 0.242402154 | 0.013488743 | 0.037657073 | 1 |
| Obesity | adipocyte | *RIOK2* | 1.14580537 | 0.01476984 | 0.040496287 | 1 |
| Obesity | adipocyte | *YBX3* | 1.691177103 | 1.53E-06 | 5.27E-05 | 0.00958893 |
| Obesity | adipocyte | *QSER1* | 1.866174004 | 2.71E-05 | 0.000321149 | 0.170208721 |
| Obesity | adipocyte | *NCKAP1* | 0.783510527 | 0.008703747 | 0.026705934 | 1 |
| Obesity | adipocyte | *SFSWAP* | 0.972665321 | 0.008878397 | 0.027162277 | 1 |
| Obesity | adipocyte | *VMP1* | 1.344202433 | 0.000184823 | 0.001314461 | 1 |
| Obesity | adipocyte | *EIF4B* | 0.378470953 | 0.013420636 | 0.03750024 | 1 |
| Obesity | adipocyte | *EPN1* | 2.348572221 | 5.34E-06 | 0.000101392 | 0.033560656 |
| Obesity | adipocyte | *TM7SF3* | 1.256225274 | 1.98E-05 | 0.000256719 | 0.124310214 |
| Obesity | adipocyte | *TAF2* | 0.752765208 | 0.003196506 | 0.012214139 | 1 |
| Obesity | adipocyte | *LPAR2* | 1.149147075 | 6.41E-06 | 0.000115994 | 0.040311849 |
| Obesity | adipocyte | *BTBD1* | 1.828971385 | 7.55E-06 | 0.000132196 | 0.047458512 |
| Obesity | adipocyte | *PMS1* | 1.645901946 | 0.000288754 | 0.001835585 | 1 |
| Obesity | adipocyte | *ANKS1A* | 1.581862387 | 1.14E-05 | 0.000173732 | 0.071403692 |
| Obesity | adipocyte | *UHRF1BP1* | 0.775206604 | 1.38E-24 | 1.74E-21 | 8.71E-21 |
| Obesity | adipocyte | *IPO5* | 1.204118788 | 0.009178229 | 0.027829061 | 1 |
| Obesity | adipocyte | *ANKRD44* | 1.320857077 | 9.62E-07 | 3.83E-05 | 0.006049418 |
| Obesity | adipocyte | *NDUFB4* | 1.872782939 | 0.000124968 | 0.00099201 | 0.785672075 |
| Obesity | adipocyte | *MAP2K4* | 1.474805906 | 0.000257746 | 0.001687968 | 1 |
| Obesity | adipocyte | *TBC1D1* | 1.640979037 | 2.57E-06 | 6.50E-05 | 0.016180289 |
| Obesity | adipocyte | *NGEF* | 6.177935317 | 1.77E-06 | 5.67E-05 | 0.011113347 |

| | | | | | | |
|---|---|---|---|---|---|---|
| Obesity | adipocyte | *MYO9A* | 1.012535744 | 2.62E-05 | 0.000312547 | 0.164513504 |
| Obesity | adipocyte | *SP100* | 1.462456831 | 9.93E-06 | 0.000157261 | 0.062432598 |
| Obesity | adipocyte | *TP53BP1* | 1.152793183 | 4.98E-05 | 0.000508417 | 0.312975102 |
| Obesity | adipocyte | *SYT1* | 0.357595904 | 0.006734114 | 0.021823388 | 1 |
| Obesity | adipocyte | *ROGDI* | 3.885199531 | 0.000761456 | 0.003876333 | 1 |
| Obesity | adipocyte | *HDAC4* | 1.591798548 | 9.56E-05 | 0.000812388 | 0.600869154 |
| Obesity | adipocyte | *RASSF1* | 0.699264314 | 0.004954134 | 0.017153523 | 1 |
| Obesity | adipocyte | *IFI35* | 0.403164208 | 0.011257011 | 0.032569181 | 1 |
| Obesity | adipocyte | *HEATR6* | 1.711059275 | 0.001634072 | 0.007189232 | 1 |
| Obesity | adipocyte | *SRBD1* | 1.521790241 | 0.000316374 | 0.001955872 | 1 |
| Obesity | adipocyte | *CYFIP1* | 1.737004436 | 1.21E-05 | 0.000181367 | 0.075811595 |
| Obesity | adipocyte | *PITX1* | -1.993666656 | 0.008687814 | 0.02668309 | 1 |
| Obesity | adipocyte | *SDK2* | 1.408071773 | 0.00386206 | 0.014083974 | 1 |
| Obesity | adipocyte | *NUCKS1* | 1.401485268 | 8.90E-05 | 0.000769285 | 0.559269921 |
| Obesity | adipocyte | *GAL* | 3.099772935 | 0.000140713 | 0.001068434 | 0.88466341 |
| Obesity | adipocyte | *NEDD4* | 0.471620444 | 0.012693817 | 0.035819581 | 1 |
| Obesity | adipocyte | *TMEM260* | 1.560970167 | 0.000648143 | 0.003436811 | 1 |
| Obesity | adipocyte | *DGCR2* | 1.374708437 | 0.001620246 | 0.007138395 | 1 |
| Obesity | adipocyte | *ZXDC* | 1.375869084 | 0.000131667 | 0.001022252 | 0.827793505 |
| Obesity | adipocyte | *ASNS* | 1.048672368 | 0.002997834 | 0.011569909 | 1 |
| Obesity | adipocyte | *TESK2* | 1.300302059 | 0.000650518 | 0.003436811 | 1 |
| Obesity | adipocyte | *CAMK2A* | 7.085123933 | 0.001509343 | 0.006742043 | 1 |
| Obesity | adipocyte | *CDC42* | 1.655480179 | 0.000219602 | 0.001500692 | 1 |
| Obesity | adipocyte | *OSBPL3* | 1.099154943 | 1.65E-05 | 0.000229145 | 0.104032015 |
| Obesity | adipocyte | *RAD18* | 1.164170757 | 0.006934194 | 0.022356553 | 1 |
| Obesity | adipocyte | *SNX13* | 1.582843664 | 6.28E-05 | 0.000602007 | 0.394916405 |
| Obesity | adipocyte | *VASH1* | 1.398667074 | 0.000316387 | 0.001955872 | 1 |
| Obesity | adipocyte | *TCF3* | 1.306072185 | 0.002456894 | 0.0098448 | 1 |
| Obesity | adipocyte | *MBD3* | 0.754304155 | 0.000815663 | 0.004099178 | 1 |
| Obesity | adipocyte | *CYBRD1* | 0.527705075 | 0.004720788 | 0.016488664 | 1 |
| Obesity | adipocyte | *EPN2* | 1.489582994 | 1.43E-05 | 0.000205963 | 0.09000574 |
| Obesity | adipocyte | *LIMS2* | 0.717680738 | 0.000418424 | 0.002435768 | 1 |
| Obesity | adipocyte | *AFF4* | 0.946015773 | 0.000203058 | 0.00141376 | 1 |
| Obesity | adipocyte | *TRNT1* | 1.782018356 | 0.000335286 | 0.002040602 | 1 |
| Obesity | adipocyte | *NDE1* | 1.113191564 | 4.57E-05 | 0.000478648 | 0.287322515 |
| Obesity | adipocyte | *PVR* | 0.67386139 | 0.00066879 | 0.003492263 | 1 |
| Obesity | adipocyte | *ALPK1* | 1.575959572 | 1.78E-05 | 0.000240148 | 0.111695826 |
| Obesity | adipocyte | *PDE8A* | 0.55054315 | 0.005410104 | 0.018336023 | 1 |
| Obesity | adipocyte | *SDHA* | 0.57471004 | 0.002002693 | 0.008377201 | 1 |
| Obesity | adipocyte | *SMARCE1* | 1.226744843 | 0.000227855 | 0.001537428 | 1 |
| Obesity | adipocyte | *PICALM* | 1.119873936 | 0.00089595 | 0.004442302 | 1 |
| Obesity | adipocyte | *CLNS1A* | 0.337925386 | 0.008535085 | 0.026329773 | 1 |
| Obesity | adipocyte | *ATP2A3* | 0.630248468 | 0.004696569 | 0.016440607 | 1 |
| Obesity | adipocyte | *NTN4* | 1.047589311 | 0.000178101 | 0.00128408 | 1 |
| Obesity | adipocyte | *LMAN1* | 1.212604502 | 0.007974542 | 0.024943255 | 1 |
| Obesity | adipocyte | *ENO1* | 1.489273625 | 0.001603002 | 0.007082271 | 1 |
| Obesity | adipocyte | *TIPIN* | 0.239602344 | 2.11E-05 | 0.000268503 | 0.132909025 |
| Obesity | adipocyte | *SRI* | 2.164690622 | 4.86E-05 | 0.00050334 | 0.305527269 |
| Obesity | adipocyte | *EIF4G3* | 1.195833576 | 6.10E-05 | 0.000590248 | 0.383801225 |
| Obesity | adipocyte | *CELSR1* | 2.567455981 | 3.02E-06 | 7.03E-05 | 0.018963487 |

| | | | | | | |
|---|---|---|---|---|---|---|
| Obesity | adipocyte | *ADD2* | 2.054110857 | 6.71E-05 | 0.000629487 | 0.421756245 |
| Obesity | adipocyte | *SLC25A3* | 1.26868364 | 8.22E-05 | 0.000727931 | 0.516831106 |
| Obesity | adipocyte | *PLD1* | 1.646768022 | 4.77E-06 | 9.50E-05 | 0.030018584 |
| Obesity | adipocyte | *BCAP29* | 1.025514918 | 0.002492756 | 0.009969439 | 1 |
| Obesity | adipocyte | *MKRN2* | 1.494657896 | 0.000863313 | 0.004304242 | 1 |
| Obesity | adipocyte | *RBMS2* | 1.238274338 | 0.001195728 | 0.005527604 | 1 |
| Obesity | adipocyte | *FMO4* | 1.825234903 | 0.001591379 | 0.007050741 | 1 |
| Obesity | adipocyte | *GPATCH1* | 0.693286829 | 0.010451423 | 0.030766502 | 1 |
| Obesity | adipocyte | *NT5C2* | 1.826707089 | 1.71E-09 | 2.62E-07 | 1.07E-05 |
| Obesity | adipocyte | *CAMSAP3* | 13.8615468 | 0.016315947 | 0.043836906 | 1 |
| Obesity | adipocyte | *MAP2K7* | 1.635438662 | 0.008999356 | 0.02746551 | 1 |
| Obesity | adipocyte | *TM9SF3* | 1.217996557 | 0.000154397 | 0.001145894 | 0.970693497 |
| Obesity | adipocyte | *DNAJC10* | 1.986607503 | 1.35E-05 | 0.000198148 | 0.084807555 |
| Obesity | adipocyte | *DYNC1I2* | 2.004106265 | 2.40E-09 | 3.51E-07 | 1.51E-05 |
| Obesity | adipocyte | *SIRT6* | 3.326436726 | 0.000414983 | 0.002424589 | 1 |
| Obesity | adipocyte | *CAPZB* | 1.141762737 | 0.000414882 | 0.002424589 | 1 |
| Obesity | adipocyte | *FGFR1* | 0.704557672 | 0.006263969 | 0.02065106 | 1 |
| Obesity | adipocyte | *FBLN1* | 0.41521847 | 0.00110299 | 0.005217833 | 1 |
| Obesity | adipocyte | *MCCC1* | 0.365448824 | 0.01884916 | 0.049562807 | 1 |
| Obesity | adipocyte | *ACER3* | 0.556558338 | 0.004807266 | 0.016725666 | 1 |
| Obesity | adipocyte | *TULP3* | 1.130357928 | 0.013882916 | 0.038416325 | 1 |
| Obesity | adipocyte | *SYNJ2* | 0.671168277 | 0.005271435 | 0.017972621 | 1 |
| Obesity | adipocyte | *FGF20* | 9.008856501 | 2.10E-05 | 0.000268001 | 0.1321243 |
| Obesity | adipocyte | *MYH7B* | -0.748123748 | 0.00210229 | 0.008712654 | 1 |
| Obesity | adipocyte | *OSBPL6* | 1.415723939 | 0.0133129 | 0.03726545 | 1 |
| Obesity | adipocyte | *XRCC5* | 1.516595403 | 7.63E-05 | 0.000691526 | 0.479413418 |
| Obesity | adipocyte | *DDX1* | 1.402913086 | 0.004068405 | 0.014647266 | 1 |
| Obesity | adipocyte | *DNM2* | 1.404061829 | 0.000809664 | 0.004072287 | 1 |
| Obesity | adipocyte | *DCT* | -0.701990941 | 0.007584021 | 0.023960171 | 1 |
| Obesity | adipocyte | *SLC35C2* | 1.779137397 | 0.001788582 | 0.007682414 | 1 |
| Obesity | adipocyte | *PSEN1* | 0.632080738 | 0.01376134 | 0.038197591 | 1 |
| Obesity | adipocyte | *MAGI3* | 1.551631088 | 7.39E-05 | 0.000675074 | 0.464451001 |
| Obesity | adipocyte | *STK17B* | 1.566718353 | 3.19E-05 | 0.000362662 | 0.200771916 |
| Obesity | adipocyte | *KIAA0141* | 1.167799651 | 0.002442774 | 0.009800714 | 1 |
| Obesity | adipocyte | *PCDHGA2* | -1.141128734 | 0.000782589 | 0.003959442 | 1 |
| Obesity | adipocyte | *HSPB11* | 1.287986436 | 0.002795967 | 0.010979539 | 1 |
| Obesity | adipocyte | *WDR70* | 1.123148081 | 0.000241464 | 0.001608137 | 1 |
| Obesity | adipocyte | *C5orf22* | 1.258341498 | 0.016033856 | 0.043225987 | 1 |
| Obesity | adipocyte | *CCNT2* | -0.418721424 | 0.001777395 | 0.007653755 | 1 |
| Obesity | adipocyte | *COBLL1* | 0.387178188 | 0.000473904 | 0.002691451 | 1 |
| Obesity | adipocyte | *GSK3B* | 1.128871574 | 0.000965269 | 0.004722681 | 1 |
| Obesity | adipocyte | *ERC1* | 1.311313628 | 9.74E-05 | 0.000820815 | 0.612328069 |
| Obesity | adipocyte | *DOPEY1* | 0.389214545 | 0.000324205 | 0.001990125 | 1 |
| Obesity | adipocyte | *PLOD1* | 2.123082299 | 0.000636502 | 0.003399903 | 1 |
| Obesity | adipocyte | *PIBF1* | 0.737571337 | 0.007979939 | 0.024947724 | 1 |
| Obesity | adipocyte | *TDRD3* | 1.346555954 | 0.000579997 | 0.003165315 | 1 |
| Obesity | adipocyte | *PDS5B* | 1.432082861 | 4.24E-05 | 0.000450057 | 0.26643364 |
| Obesity | adipocyte | *EPYC* | -1.156119993 | 0.018101129 | 0.04787623 | 1 |
| Obesity | adipocyte | *CYLD* | 1.20325558 | 0.003317472 | 0.012549306 | 1 |
| Obesity | adipocyte | *RPS5* | 1.844441835 | 2.44E-05 | 0.000298201 | 0.153275252 |

| | | | | | | |
|---|---|---|---|---|---|---|
| Obesity | adipocyte | *PPIE* | 0.659037838 | 0.001672895 | 0.007303811 | 1 |
| Obesity | adipocyte | *ZMPSTE24* | 1.644127697 | 0.002347372 | 0.00952739 | 1 |
| Obesity | adipocyte | *STARD7* | 1.679417614 | 0.000259333 | 0.001694832 | 1 |
| Obesity | adipocyte | *GSTP1* | 2.369085894 | 1.27E-07 | 8.76E-06 | 0.000797119 |
| Obesity | adipocyte | *WBP11* | 1.946979257 | 0.001123709 | 0.005280087 | 1 |
| Obesity | adipocyte | *COL16A1* | -0.544412337 | 0.000520441 | 0.002906785 | 1 |
| Obesity | adipocyte | *APOB* | -0.674229038 | 0.000809611 | 0.004072287 | 1 |
| Obesity | adipocyte | *NCOA1* | 1.353070949 | 3.07E-05 | 0.000352365 | 0.192896483 |
| Obesity | adipocyte | *CD59* | 0.96151586 | 0.000317637 | 0.001961671 | 1 |
| Obesity | adipocyte | *CD82* | 0.510073397 | 0.005777911 | 0.019301662 | 1 |
| Obesity | adipocyte | *SCAMP1* | 1.813763982 | 1.04E-08 | 1.24E-06 | 6.56E-05 |
| Obesity | adipocyte | *HACE1* | 0.888322815 | 0.010011762 | 0.029746667 | 1 |
| Obesity | adipocyte | *CTTN* | 1.250458261 | 0.006876997 | 0.022194907 | 1 |
| Obesity | adipocyte | *ATG16L1* | 1.238247135 | 0.009675171 | 0.02891055 | 1 |
| Obesity | adipocyte | *MAST2* | 1.326668875 | 5.00E-06 | 9.72E-05 | 0.031424335 |
| Obesity | adipocyte | *B4GALT1* | 1.529058781 | 6.94E-05 | 0.000645118 | 0.436099673 |
| Obesity | adipocyte | *CHMP5* | 1.137423813 | 0.003843248 | 0.014045825 | 1 |
| Obesity | adipocyte | *ITPKC* | 0.945246066 | 0.012848309 | 0.036158155 | 1 |
| Obesity | adipocyte | *TMED2* | 1.995241827 | 0.000262467 | 0.001709977 | 1 |
| Obesity | adipocyte | *ACHE* | -0.202984032 | 0.005333695 | 0.01811612 | 1 |
| Obesity | adipocyte | *SRRT* | 0.497942983 | 0.005761901 | 0.019268653 | 1 |
| Obesity | adipocyte | *ATXN7L3* | 1.275807896 | 0.006126105 | 0.020270959 | 1 |
| Obesity | adipocyte | *LPCAT2* | 0.624685085 | 0.003643913 | 0.013476046 | 1 |
| Obesity | adipocyte | *OGFOD1* | -0.860544943 | 0.000184032 | 0.001310318 | 1 |
| Obesity | adipocyte | *GMCL1* | 1.05657724 | 0.00426988 | 0.015209481 | 1 |
| Obesity | adipocyte | *KLHL42* | 0.885550845 | 0.002311488 | 0.009418227 | 1 |
| Obesity | adipocyte | *PTHLH* | 1.14392792 | 4.92E-06 | 9.70E-05 | 0.030913589 |
| Obesity | adipocyte | *PHACTR3* | -0.27504698 | 0.011560905 | 0.033234298 | 1 |
| Obesity | adipocyte | *ERGIC2* | 1.363919005 | 0.001070633 | 0.00511091 | 1 |
| Obesity | adipocyte | *ALG6* | 0.820947959 | 0.01898862 | 0.049855734 | 1 |
| Obesity | adipocyte | *DDX18* | 0.904896253 | 0.003844897 | 0.014045825 | 1 |
| Obesity | adipocyte | *ASAP3* | 1.459469323 | 0.000115782 | 0.000940373 | 0.727923548 |
| Obesity | adipocyte | *DOCK3* | 0.303587259 | 0.007066633 | 0.02269046 | 1 |
| Obesity | adipocyte | *C3orf18* | 1.379938835 | 4.40E-10 | 7.69E-08 | 2.77E-06 |
| Obesity | adipocyte | *FKBP1A* | 2.019286889 | 7.87E-06 | 0.000136344 | 0.049493047 |
| Obesity | adipocyte | *NSFL1C* | 1.336521015 | 0.000545157 | 0.003025067 | 1 |
| Obesity | adipocyte | *XRN2* | 1.807812188 | 9.59E-06 | 0.000153475 | 0.060313985 |
| Obesity | adipocyte | *TESC* | 2.479897599 | 1.72E-06 | 5.58E-05 | 0.010788765 |
| Obesity | adipocyte | *SLC23A2* | 1.819324576 | 1.61E-05 | 0.000226277 | 0.101372077 |
| Obesity | adipocyte | *TASP1* | 0.850384615 | 0.000468058 | 0.002663061 | 1 |
| Obesity | adipocyte | *OAS1* | 1.84657438 | 0.000177222 | 0.001280682 | 1 |
| Obesity | adipocyte | *KIF16B* | 1.565258224 | 0.000299291 | 0.001887301 | 1 |
| Obesity | adipocyte | *FUS* | 1.656194482 | 7.81E-06 | 0.000135968 | 0.049084599 |
| Obesity | adipocyte | *FXYD5* | 1.597116144 | 3.72E-06 | 7.99E-05 | 0.023410061 |
| Obesity | adipocyte | *GANAB* | 1.439369236 | 0.000441292 | 0.002538336 | 1 |
| Obesity | adipocyte | *GMIP* | 2.48131349 | 1.33E-06 | 4.83E-05 | 0.008357815 |
| Obesity | adipocyte | *MLF2* | 1.534251694 | 0.011713854 | 0.033535974 | 1 |
| Obesity | adipocyte | *PAPOLA* | 1.541809843 | 2.47E-05 | 0.00029974 | 0.155447763 |
| Obesity | adipocyte | *RGS1* | 2.529416695 | 5.11E-06 | 9.86E-05 | 0.032151963 |
| Obesity | adipocyte | *YPEL3* | 0.562360634 | 2.12E-10 | 4.16E-08 | 1.33E-06 |

| | | | | | | |
|---|---|---|---|---|---|---|
| Obesity | adipocyte | *LYZ* | 0.842455361 | 4.89E-05 | 0.000504605 | 0.307304746 |
| Obesity | adipocyte | *GNPTG* | 1.690812068 | 0.00341366 | 0.012820597 | 1 |
| Obesity | adipocyte | *PABPC4* | -0.857092999 | 2.10E-07 | 1.28E-05 | 0.001319319 |
| Obesity | adipocyte | *PDPR* | 1.184624729 | 0.001431041 | 0.00645869 | 1 |
| Obesity | adipocyte | *EXOC1* | 1.563408595 | 9.51E-05 | 0.000812388 | 0.597588333 |
| Obesity | adipocyte | *LAMB1* | 0.376434299 | 0.004666689 | 0.01634511 | 1 |
| Obesity | adipocyte | *TXNL1* | 1.565409271 | 0.000499182 | 0.002809631 | 1 |
| Obesity | adipocyte | *IL5RA* | 6.410695112 | 5.90E-05 | 0.000575842 | 0.370842447 |
| Obesity | adipocyte | *ITGA6* | 1.090001052 | 2.49E-05 | 0.000300831 | 0.156733129 |
| Obesity | adipocyte | *SEL1L3* | 1.697768864 | 5.57E-07 | 2.57E-05 | 0.003500106 |
| Obesity | adipocyte | *TF* | -0.617099112 | 0.01623466 | 0.043655819 | 1 |
| Obesity | adipocyte | *ALKBH5* | 0.569088855 | 0.002010656 | 0.00839933 | 1 |
| Obesity | adipocyte | *NLRP1* | 1.023477564 | 0.001602498 | 0.007082271 | 1 |
| Obesity | adipocyte | *SPAG7* | 0.325570287 | 0.008764882 | 0.026854197 | 1 |
| Obesity | adipocyte | *RGS17* | -0.666694545 | 0.001227995 | 0.005655973 | 1 |
| Obesity | adipocyte | *PPP2R3C* | 1.5058458 | 1.73E-05 | 0.000237172 | 0.108861917 |
| Obesity | adipocyte | *OSGEP* | 1.893728853 | 0.00017143 | 0.001245987 | 1 |
| Obesity | adipocyte | *TOX4* | 1.499775424 | 0.000146214 | 0.00110354 | 0.919248584 |
| Obesity | adipocyte | *AGO1* | 1.111503077 | 0.004273026 | 0.015212069 | 1 |
| Obesity | adipocyte | *RFFL* | 2.013705165 | 0.011602233 | 0.033328892 | 1 |
| Obesity | adipocyte | *DPYSL2* | 1.148917349 | 3.03E-05 | 0.000351403 | 0.190444718 |
| Obesity | adipocyte | *COMT* | 1.869674157 | 0.000130205 | 0.001016439 | 0.818597535 |
| Obesity | adipocyte | *VNN3* | 2.2570296 | 0.004481612 | 0.015793661 | 1 |
| Obesity | adipocyte | *SUCO* | 1.679818799 | 0.000157299 | 0.001162091 | 0.988939285 |
| Obesity | adipocyte | *MSH2* | 2.29865348 | 1.12E-05 | 0.000172633 | 0.070606724 |
| Obesity | adipocyte | *MAP3K1* | 0.694444303 | 0.010800579 | 0.031509624 | 1 |
| Obesity | adipocyte | *TMEM38B* | 2.404474216 | 0.000268618 | 0.001733879 | 1 |
| Obesity | adipocyte | *PDE6C* | 0.632020163 | 0.011606547 | 0.033328892 | 1 |
| Obesity | adipocyte | *BLNK* | 0.49692654 | 0.005836653 | 0.019465497 | 1 |
| Obesity | adipocyte | *WAC* | 1.227408763 | 5.15E-05 | 0.000521472 | 0.323833822 |
| Obesity | adipocyte | *CREM* | 1.654191956 | 0.000215236 | 0.001479877 | 1 |
| Obesity | adipocyte | *TPSD1* | 3.579624335 | 0.011318069 | 0.032715725 | 1 |
| Obesity | adipocyte | *ITPR3* | 1.605555806 | 1.50E-06 | 5.24E-05 | 0.009423476 |
| Obesity | adipocyte | *IFT74* | 0.380904847 | 0.00435318 | 0.015436232 | 1 |
| Obesity | adipocyte | *PCSK5* | 1.330752481 | 0.000227912 | 0.001537428 | 1 |
| Obesity | adipocyte | *NRP1* | 1.305796098 | 1.80E-05 | 0.000241715 | 0.11310473 |
| Obesity | adipocyte | *PRTFDC1* | 1.474715364 | 0.000336053 | 0.002043294 | 1 |
| Obesity | adipocyte | *OCEL1* | -0.402552208 | 0.005222017 | 0.017852539 | 1 |
| Obesity | adipocyte | *STX1B* | -0.931027668 | 2.34E-06 | 6.25E-05 | 0.014705761 |
| Obesity | adipocyte | *HSD3B7* | 0.557946968 | 0.006219927 | 0.020548306 | 1 |
| Obesity | adipocyte | *ATP5D* | 1.642660212 | 0.003905631 | 0.014218126 | 1 |
| Obesity | adipocyte | *MTAP* | 1.542447261 | 0.013881507 | 0.038416325 | 1 |
| Obesity | adipocyte | *LZTR1* | 1.582547756 | 0.009012091 | 0.027491032 | 1 |
| Obesity | adipocyte | *SMARCB1* | 0.579357104 | 0.000106792 | 0.000879952 | 0.671403537 |
| Obesity | adipocyte | *P2RX6* | -0.762481977 | 0.000423747 | 0.002462195 | 1 |
| Obesity | adipocyte | *GGT5* | 1.296458896 | 0.0004165 | 0.002429066 | 1 |
| Obesity | adipocyte | *YPEL1* | 1.330649366 | 0.009162581 | 0.027801713 | 1 |
| Obesity | adipocyte | *PPM1F* | 1.041889322 | 0.005812829 | 0.019397695 | 1 |
| Obesity | adipocyte | *CYTH4* | 1.755148459 | 1.91E-06 | 5.81E-05 | 0.011978186 |
| Obesity | adipocyte | *SH3BP1* | 2.331509071 | 1.01E-05 | 0.000160299 | 0.063799167 |

| | | | | | | |
|---|---|---|---|---|---|---|
| Obesity | adipocyte | *HPS4* | 1.29922751 | 0.00760191 | 0.023980537 | 1 |
| Obesity | adipocyte | *TFIP11* | 0.491700935 | 0.001098131 | 0.005198755 | 1 |
| Obesity | adipocyte | *ANKRD54* | -0.453290701 | 0.000147115 | 0.001107681 | 0.924913306 |
| Obesity | adipocyte | *CCDC134* | 2.497564503 | 0.000143892 | 0.001088926 | 0.904648246 |
| Obesity | adipocyte | *FBXO7* | 1.564957734 | 6.95E-08 | 5.26E-06 | 0.000436727 |
| Obesity | adipocyte | *POLDIP3* | 1.881339051 | 0.003122007 | 0.011982942 | 1 |
| Obesity | adipocyte | *TIMP3* | 1.321615842 | 5.79E-05 | 0.000566037 | 0.363961602 |
| Obesity | adipocyte | *CYB5R3* | 1.296994243 | 0.000500208 | 0.00281289 | 1 |
| Obesity | adipocyte | *TTLL1* | 2.024055547 | 0.001082967 | 0.005154138 | 1 |
| Obesity | adipocyte | *AP1B1* | 0.973517372 | 0.000931066 | 0.004585154 | 1 |
| Obesity | adipocyte | *HMGXB4* | 1.217416458 | 0.010272656 | 0.030341529 | 1 |
| Obesity | adipocyte | *TSPO* | 0.955166255 | 0.000280428 | 0.001786271 | 1 |
| Obesity | adipocyte | *RBFOX2* | 1.395696346 | 0.000168236 | 0.001227027 | 1 |
| Obesity | adipocyte | *KCTD17* | 3.336092074 | 8.73E-06 | 0.000143664 | 0.05487979 |
| Obesity | adipocyte | *EP300* | 1.360113665 | 2.61E-05 | 0.000311977 | 0.163788051 |
| Obesity | adipocyte | *ACO2* | 0.890711001 | 0.000264496 | 0.001721414 | 1 |
| Obesity | adipocyte | *DESI1* | 0.507671549 | 7.40E-06 | 0.000129939 | 0.046518218 |
| Obesity | adipocyte | *ZBED4* | 1.917855982 | 0.001077442 | 0.005131725 | 1 |
| Obesity | adipocyte | *SOS2* | 1.35526009 | 0.000482345 | 0.002729523 | 1 |
| Obesity | adipocyte | *DICER1* | 1.311065124 | 4.98E-08 | 4.17E-06 | 0.000313023 |
| Obesity | adipocyte | *ZFYVE21* | 1.48226069 | 1.44E-05 | 0.000206073 | 0.090259878 |
| Obesity | adipocyte | *GSKIP* | 1.103120207 | 0.001769141 | 0.007629186 | 1 |
| Obesity | adipocyte | *VRK1* | 1.974694602 | 0.000297297 | 0.001878502 | 1 |
| Obesity | adipocyte | *PSMB5* | 0.472589883 | 0.00477507 | 0.016641278 | 1 |
| Obesity | adipocyte | *TRIP11* | 1.408191522 | 0.001154144 | 0.005386864 | 1 |
| Obesity | adipocyte | *PLTP* | 0.336407273 | 0.000589967 | 0.00321136 | 1 |
| Obesity | adipocyte | *ABHD12* | 1.085910849 | 3.05E-08 | 2.78E-06 | 0.000192009 |
| Obesity | adipocyte | *NDRG3* | 2.102684838 | 4.73E-07 | 2.34E-05 | 0.002970899 |
| Obesity | adipocyte | *BMP7* | 2.288191354 | 2.56E-06 | 6.50E-05 | 0.01607166 |
| Obesity | adipocyte | *TPD52L2* | 0.825128563 | 0.006441936 | 0.021083005 | 1 |
| Obesity | adipocyte | *NELFCD* | 1.548108488 | 0.004142025 | 0.014863532 | 1 |
| Obesity | adipocyte | *CTSZ* | 0.636945512 | 0.003982239 | 0.014417956 | 1 |
| Obesity | adipocyte | *DIDO1* | 0.898880525 | 0.000408122 | 0.002398005 | 1 |
| Obesity | adipocyte | *SLC17A9* | 1.19529179 | 0.000373233 | 0.002219976 | 1 |
| Obesity | adipocyte | *PTK6* | 6.947233204 | 0.002881385 | 0.01123131 | 1 |
| Obesity | adipocyte | *CDC25B* | 0.630678306 | 0.007021692 | 0.022603881 | 1 |
| Obesity | adipocyte | *RNF24* | 1.051903494 | 7.21E-05 | 0.000664998 | 0.453407512 |
| Obesity | adipocyte | *SLC52A3* | 0.578573132 | 0.006257325 | 0.020639979 | 1 |
| Obesity | adipocyte | *ANGPT4* | -1.358556183 | 0.000348643 | 0.002097529 | 1 |
| Obesity | adipocyte | *CCM2L* | 2.01106825 | 1.24E-06 | 4.62E-05 | 0.007813135 |
| Obesity | adipocyte | *MYL9* | 0.673701747 | 0.001884588 | 0.008000272 | 1 |
| Obesity | adipocyte | *MANBAL* | 0.311791802 | 0.001473484 | 0.006607554 | 1 |
| Obesity | adipocyte | *MAPRE1* | 1.287178865 | 0.003823546 | 0.013984081 | 1 |
| Obesity | adipocyte | *TTI1* | 1.666436873 | 0.00333725 | 0.012616529 | 1 |
| Obesity | adipocyte | *PXMP4* | 0.726515068 | 0.002814276 | 0.011023896 | 1 |
| Obesity | adipocyte | *PIGU* | 0.897565608 | 0.001391534 | 0.00629847 | 1 |
| Obesity | adipocyte | *METTL4* | 2.334931656 | 2.10E-05 | 0.000268001 | 0.131801251 |
| Obesity | adipocyte | *LPIN2* | 1.334074458 | 0.002833031 | 0.011069772 | 1 |
| Obesity | adipocyte | *SMCHD1* | 1.46386573 | 1.18E-05 | 0.000178905 | 0.07424577 |
| Obesity | adipocyte | *MYL12A* | 1.495571522 | 0.000599466 | 0.003245577 | 1 |

| | | | | | | |
|---|---|---|---|---|---|---|
| Obesity | adipocyte | *HTR2A* | 4.268908474 | 4.25E-06 | 8.80E-05 | 0.026745689 |
| Obesity | adipocyte | *DNAJC3* | 1.182728587 | 0.003902478 | 0.014214877 | 1 |
| Obesity | adipocyte | *ARHGEF7* | 0.960454312 | 0.004362078 | 0.015459067 | 1 |
| Obesity | adipocyte | *SGCG* | -0.427642901 | 0.018651956 | 0.049105882 | 1 |
| Obesity | adipocyte | *PARP4* | 1.676597741 | 1.68E-05 | 0.000232712 | 0.10588398 |
| Obesity | adipocyte | *MEDAG* | 1.80772547 | 3.54E-06 | 7.79E-05 | 0.022282328 |
| Obesity | adipocyte | *TSC22D1* | 1.169234892 | 4.95E-05 | 0.000508417 | 0.311437158 |
| Obesity | adipocyte | *CORO1A* | -1.112644675 | 3.37E-08 | 3.02E-06 | 0.00021164 |
| Obesity | adipocyte | *ELMO3* | 0.266854679 | 0.001076673 | 0.005131725 | 1 |
| Obesity | adipocyte | *NFAT5* | 1.075122793 | 0.001109808 | 0.005238261 | 1 |
| Obesity | adipocyte | *CTCF* | 1.067968709 | 0.001158329 | 0.005402387 | 1 |
| Obesity | adipocyte | *COG4* | -0.785284365 | 0.000111936 | 0.000913947 | 0.703739494 |
| Obesity | adipocyte | *HCFC1R1* | 2.979040227 | 2.95E-06 | 6.99E-05 | 0.018521679 |
| Obesity | adipocyte | *NAGPA* | 1.129251959 | 0.000191541 | 0.001353055 | 1 |
| Obesity | adipocyte | *LMF1* | 1.753296283 | 2.10E-05 | 0.000268001 | 0.132096283 |
| Obesity | adipocyte | *CLCN7* | 1.641948224 | 8.71E-05 | 0.000757209 | 0.54746238 |
| Obesity | adipocyte | *SLC7A5* | 6.842775566 | 5.56E-06 | 0.000104394 | 0.034971856 |
| Obesity | adipocyte | *RHBDL1* | -0.34405969 | 0.008078555 | 0.025217737 | 1 |
| Obesity | adipocyte | *NUBP1* | 1.849004469 | 0.001958126 | 0.008240117 | 1 |
| Obesity | adipocyte | *UBE2I* | 1.302204023 | 0.000210923 | 0.001457222 | 1 |
| Obesity | adipocyte | *CLUAP1* | 0.402416101 | 0.000151915 | 0.001132963 | 0.955087499 |
| Obesity | adipocyte | *CPPED1* | 1.263251313 | 0.00094293 | 0.004638653 | 1 |
| Obesity | adipocyte | *BFAR* | 1.20409694 | 0.018214376 | 0.048135258 | 1 |
| Obesity | adipocyte | *STX4* | 0.983600388 | 0.002207048 | 0.009069094 | 1 |
| Obesity | adipocyte | *KAT8* | 2.82603087 | 6.48E-14 | 2.71E-11 | 4.07E-10 |
| Obesity | adipocyte | *TMC5* | -0.802419371 | 0.005693542 | 0.019101013 | 1 |
| Obesity | adipocyte | *KNOP1* | -0.629665433 | 0.00912806 | 0.027710339 | 1 |
| Obesity | adipocyte | *IQCH* | 1.807157787 | 8.59E-05 | 0.000752918 | 0.539842327 |
| Obesity | adipocyte | *LACTB* | 1.107517116 | 0.011157769 | 0.032356502 | 1 |
| Obesity | adipocyte | *CSK* | 2.423173097 | 2.39E-06 | 6.25E-05 | 0.015039523 |
| Obesity | adipocyte | *HERC1* | 1.311847798 | 0.000119385 | 0.000961831 | 0.750573489 |
| Obesity | adipocyte | *TRIP4* | 0.5707693 | 0.000539581 | 0.003004732 | 1 |
| Obesity | adipocyte | *RASL12* | 1.964249866 | 2.38E-05 | 0.000293854 | 0.149571711 |
| Obesity | adipocyte | *EHD4* | 1.450538172 | 1.42E-05 | 0.000205406 | 0.089494977 |
| Obesity | adipocyte | *ATP8B4* | 1.163284576 | 0.001115861 | 0.00526288 | 1 |
| Obesity | adipocyte | *DTWD1* | 0.659430988 | 0.008280294 | 0.025695067 | 1 |
| Obesity | adipocyte | *DNAJC17* | 1.700250465 | 0.002172316 | 0.008943911 | 1 |
| Obesity | adipocyte | *PDGFRL* | 0.723989538 | 0.00110419 | 0.005219582 | 1 |
| Obesity | adipocyte | *ZDHHC2* | -0.498373125 | 0.012622172 | 0.035681474 | 1 |
| Obesity | adipocyte | *RIPK2* | 0.976714373 | 0.002994023 | 0.011562298 | 1 |
| Obesity | adipocyte | *CPQ* | 0.512947181 | 0.000910189 | 0.00450225 | 1 |
| Obesity | adipocyte | *NIPAL2* | 0.671258699 | 0.003478053 | 0.013002031 | 1 |
| Obesity | adipocyte | *IKBKB* | 1.878371894 | 1.32E-06 | 4.83E-05 | 0.008320885 |
| Obesity | adipocyte | *PLAT* | 1.78672442 | 2.78E-07 | 1.62E-05 | 0.001750718 |
| Obesity | adipocyte | *GDAP1* | 1.219816721 | 2.29E-05 | 0.000283956 | 0.143681813 |
| Obesity | adipocyte | *ZC2HC1A* | 0.84118705 | 0.003951923 | 0.014353402 | 1 |
| Obesity | adipocyte | *ERI1* | 1.53635168 | 0.004937668 | 0.017122516 | 1 |
| Obesity | adipocyte | *MTMR9* | 2.015156082 | 4.81E-05 | 0.000499361 | 0.302612725 |
| Obesity | adipocyte | *LEPROTL1* | 2.09448089 | 1.49E-05 | 0.000212312 | 0.09362951 |
| Obesity | adipocyte | *ERICH1* | 1.718328613 | 1.27E-05 | 0.000188607 | 0.079592193 |

| | | | | | | |
|---|---|---|---|---|---|---|
| Obesity | adipocyte | *TUSC3* | 1.500666543 | 1.03E-06 | 3.97E-05 | 0.006474247 |
| Obesity | adipocyte | *MCM4* | 0.684604049 | 0.000654759 | 0.003447018 | 1 |
| Obesity | adipocyte | *PPP1R37* | 1.089214891 | 3.78E-05 | 0.000415113 | 0.237859717 |
| Obesity | adipocyte | *FCGRT* | 0.377393339 | 0.015615007 | 0.042314375 | 1 |
| Obesity | adipocyte | *CD37* | 0.812126663 | 0.001636251 | 0.007193783 | 1 |
| Obesity | adipocyte | *FCER2* | 2.319249405 | 0.000129045 | 0.001012094 | 0.811304178 |
| Obesity | adipocyte | *AES* | 1.981261146 | 1.49E-05 | 0.000212312 | 0.093458467 |
| Obesity | adipocyte | *SGTA* | 1.411705849 | 0.000649589 | 0.003436811 | 1 |
| Obesity | adipocyte | *C19orf53* | 1.104751078 | 0.017882701 | 0.047418194 | 1 |
| Obesity | adipocyte | *IL27RA* | 1.239318274 | 8.10E-06 | 0.000137554 | 0.050894947 |
| Obesity | adipocyte | *RASAL3* | 0.595558115 | 0.003089474 | 0.011879831 | 1 |
| Obesity | adipocyte | *CCNE1* | -1.862519002 | 0.000303239 | 0.001908374 | 1 |
| Obesity | adipocyte | *ANKRD27* | 0.868896013 | 0.015516172 | 0.042153084 | 1 |
| Obesity | adipocyte | *CCDC94* | 1.07614735 | 0.00642533 | 0.021050572 | 1 |
| Obesity | adipocyte | *FZR1* | 2.917021121 | 2.77E-05 | 0.000327993 | 0.174164204 |
| Obesity | adipocyte | *BBC3* | 2.99884564 | 0.000175004 | 0.001269029 | 1 |
| Obesity | adipocyte | *TGFB1* | 0.951826496 | 0.00424708 | 0.01514631 | 1 |
| Obesity | adipocyte | *CEACAM4* | 6.010091545 | 2.41E-05 | 0.000297116 | 0.151826375 |
| Obesity | adipocyte | *PLIN3* | 1.184180653 | 0.002087446 | 0.008674009 | 1 |
| Obesity | adipocyte | *MRPL4* | 1.000723197 | 0.006362975 | 0.020909152 | 1 |
| Obesity | adipocyte | *ICAM5* | -2.074509597 | 0.008627586 | 0.026549992 | 1 |
| Obesity | adipocyte | *CD33* | 0.51647021 | 0.010667148 | 0.031207239 | 1 |
| Obesity | adipocyte | *BABAM1* | 2.552034534 | 0.001561179 | 0.006941656 | 1 |
| Obesity | adipocyte | *TYK2* | 0.960188205 | 0.002827921 | 0.011067494 | 1 |
| Obesity | adipocyte | *RABAC1* | 0.891660929 | 0.011784528 | 0.033692281 | 1 |
| Obesity | adipocyte | *CYTH2* | 1.825291741 | 6.47E-05 | 0.000613065 | 0.406574492 |
| Obesity | adipocyte | *GRIN2D* | 2.981848306 | 0.000118771 | 0.000958553 | 0.746712564 |
| Obesity | adipocyte | *PLA2G4C* | 0.430900028 | 0.003244124 | 0.012361095 | 1 |
| Obesity | adipocyte | *BCAT2* | 1.666046054 | 0.014325088 | 0.039431624 | 1 |
| Obesity | adipocyte | *PPP2R1A* | 1.791946158 | 0.001189049 | 0.005508881 | 1 |
| Obesity | adipocyte | *LILRB5* | 1.797103151 | 2.05E-06 | 5.99E-05 | 0.012915572 |
| Obesity | adipocyte | *UPK1A* | -1.406541498 | 0.001575958 | 0.006997209 | 1 |
| Obesity | adipocyte | *COPE* | 1.327694264 | 0.006522856 | 0.021290682 | 1 |
| Obesity | adipocyte | *KXD1* | 2.031231536 | 0.003342982 | 0.012621094 | 1 |
| Obesity | adipocyte | *ZNF14* | 1.905724589 | 0.004016282 | 0.014495042 | 1 |
| Obesity | adipocyte | *ATP13A1* | -0.57477921 | 0.000626396 | 0.003354472 | 1 |
| Obesity | adipocyte | *SIPA1L3* | 0.944642428 | 0.006222996 | 0.020548306 | 1 |
| Obesity | adipocyte | *PON3* | 2.802210789 | 8.25E-06 | 0.000139123 | 0.051892699 |
| Obesity | adipocyte | *PON2* | 1.501849135 | 0.002509045 | 0.010021833 | 1 |
| Obesity | adipocyte | *OGDH* | 1.691797649 | 2.88E-05 | 0.00033687 | 0.180899048 |
| Obesity | adipocyte | *ADAP1* | 4.960281209 | 1.81E-06 | 5.73E-05 | 0.011404863 |
| Obesity | adipocyte | *MET* | 1.085628166 | 0.000215443 | 0.001479877 | 1 |
| Obesity | adipocyte | *LMBR1* | 0.828813181 | 0.005681658 | 0.019081509 | 1 |
| Obesity | adipocyte | *HOXA2* | -1.90635056 | 0.000392995 | 0.002326518 | 1 |
| Obesity | adipocyte | *HIBADH* | 1.084823313 | 0.003455031 | 0.012945043 | 1 |
| Obesity | adipocyte | *CPVL* | 0.535288738 | 0.017447439 | 0.046401036 | 1 |
| Obesity | adipocyte | *PLEKHA8* | 1.76490943 | 3.29E-05 | 0.000370365 | 0.206916596 |
| Obesity | adipocyte | *NOD1* | 1.518447309 | 0.000129525 | 0.001012841 | 0.814323881 |
| Obesity | adipocyte | *SNX8* | 1.985994315 | 0.000198022 | 0.001389468 | 1 |
| Obesity | adipocyte | *TAF6* | -0.544221405 | 0.017144672 | 0.045750659 | 1 |

| | | | | | | |
|---|---|---|---|---|---|---|
| Obesity | adipocyte | *USP42* | 1.430440787 | 0.00197584 | 0.008292135 | 1 |
| Obesity | adipocyte | *C1GALT1* | 1.323162733 | 0.000856662 | 0.004277869 | 1 |
| Obesity | adipocyte | *RPA3* | 1.456445856 | 0.001466026 | 0.006578804 | 1 |
| Obesity | adipocyte | *PHF14* | 1.445904361 | 4.58E-05 | 0.000478648 | 0.287667704 |
| Obesity | adipocyte | *ZNF862* | 0.718953589 | 0.005875611 | 0.019576029 | 1 |
| Obesity | adipocyte | *MEST* | -0.332062586 | 6.61E-05 | 0.000622148 | 0.415542425 |
| Obesity | adipocyte | *POU6F2* | -0.570804322 | 0.015156163 | 0.041356806 | 1 |
| Obesity | adipocyte | *TSPAN13* | 3.504886405 | 6.46E-07 | 2.86E-05 | 0.004063114 |
| Obesity | adipocyte | *BLVRA* | 0.363943196 | 0.009880726 | 0.029426872 | 1 |
| Obesity | adipocyte | *GCK* | -9.591075124 | 0.012681374 | 0.035800537 | 1 |
| Obesity | adipocyte | *CLIP2* | 2.145587461 | 4.64E-06 | 9.38E-05 | 0.029159862 |
| Obesity | adipocyte | *LIMK1* | 2.699162481 | 3.85E-06 | 8.20E-05 | 0.024180036 |
| Obesity | adipocyte | *FKTN* | 1.233503791 | 0.005656512 | 0.01902787 | 1 |
| Obesity | adipocyte | *TRIM14* | 0.807928231 | 0.001109193 | 0.005238261 | 1 |
| Obesity | adipocyte | *CORO2A* | 0.993620161 | 0.002250794 | 0.009212722 | 1 |
| Obesity | adipocyte | *OGN* | 1.323638566 | 2.63E-09 | 3.76E-07 | 1.65E-05 |
| Obesity | adipocyte | *TNFSF8* | 1.648068686 | 1.64E-05 | 0.000227909 | 0.103242824 |
| Obesity | adipocyte | *KDM4C* | 1.388725034 | 0.000225895 | 0.001527101 | 1 |
| Obesity | adipocyte | *FUBP3* | 1.404101377 | 0.004525714 | 0.015913403 | 1 |
| Obesity | adipocyte | *RGP1* | 2.217069392 | 7.99E-06 | 0.000136462 | 0.050218162 |
| Obesity | adipocyte | *MPDZ* | 1.506293553 | 0.000273588 | 0.001753362 | 1 |
| Obesity | adipocyte | *PTGDS* | 4.125897944 | 2.77E-06 | 6.83E-05 | 0.017415262 |
| Obesity | adipocyte | *PDLIM1* | 0.448639417 | 0.012274789 | 0.034887703 | 1 |
| Obesity | adipocyte | *HPS1* | 1.296541175 | 0.001260736 | 0.005798278 | 1 |
| Obesity | adipocyte | *DNMBP* | 1.812470174 | 1.63E-05 | 0.000227175 | 0.102455814 |
| Obesity | adipocyte | *TRDMT1* | 1.406985605 | 0.000740536 | 0.003794416 | 1 |
| Obesity | adipocyte | *DDX50* | 1.664248745 | 0.000449312 | 0.002579746 | 1 |
| Obesity | adipocyte | *MAPK8* | 1.342330512 | 0.00017894 | 0.001285708 | 1 |
| Obesity | adipocyte | *ATE1* | 0.624210826 | 0.004621623 | 0.016208471 | 1 |
| Obesity | adipocyte | *PLEKHA1* | 1.160421705 | 0.004192215 | 0.01500083 | 1 |
| Obesity | adipocyte | *MICU1* | 1.779361916 | 1.05E-06 | 4.02E-05 | 0.006594007 |
| Obesity | adipocyte | *CCSER2* | 1.781734176 | 1.75E-06 | 5.64E-05 | 0.010997225 |
| Obesity | adipocyte | *LIPA* | 0.555252918 | 0.001918724 | 0.008128717 | 1 |
| Obesity | adipocyte | *SFXN3* | 2.026209621 | 4.65E-08 | 3.95E-06 | 0.000292516 |
| Obesity | adipocyte | *TNKS2* | 1.088824771 | 0.006589797 | 0.021433032 | 1 |
| Obesity | adipocyte | *ACBD5* | 1.643045541 | 2.15E-06 | 6.10E-05 | 0.013536065 |
| Obesity | adipocyte | *MTPAP* | 1.897402073 | 0.000106484 | 0.000878562 | 0.66946421 |
| Obesity | adipocyte | *PITRM1* | 1.565881202 | 0.003786887 | 0.01388231 | 1 |
| Obesity | adipocyte | *MAP3K8* | 1.08515349 | 0.002702085 | 0.010684281 | 1 |
| Obesity | adipocyte | *SHOC2* | 1.15441891 | 0.001859981 | 0.00792256 | 1 |
| Obesity | adipocyte | *RPL28* | 0.401048147 | 0.007450303 | 0.023656594 | 1 |
| Obesity | adipocyte | *MLLT6* | 1.767236842 | 4.17E-05 | 0.000444689 | 0.261921788 |
| Obesity | adipocyte | *RGS9* | 3.814395457 | 5.82E-06 | 0.000108265 | 0.036593447 |
| Obesity | adipocyte | *RAD51C* | -0.277264279 | 0.002595495 | 0.010282217 | 1 |
| Obesity | adipocyte | *TRIM37* | 1.37778056 | 0.000890493 | 0.004418728 | 1 |
| Obesity | adipocyte | *RPS6KB1* | 1.440405174 | 0.003225754 | 0.012298553 | 1 |
| Obesity | adipocyte | *CBX1* | 0.856881175 | 0.000134599 | 0.001037493 | 0.846223803 |
| Obesity | adipocyte | *PIGL* | 1.197680967 | 0.006896049 | 0.022244977 | 1 |
| Obesity | adipocyte | *GALK1* | 3.865244383 | 4.77E-06 | 9.50E-05 | 0.029981591 |
| Obesity | adipocyte | *HOXB6* | 0.775939576 | 0.000362674 | 0.002167423 | 1 |

| | | | | | | |
|---|---|---|---|---|---|---|
| Obesity | adipocyte | *RNF167* | 0.963676816 | 0.009238279 | 0.027977389 | 1 |
| Obesity | adipocyte | *CHRNE* | 0.250356158 | 0.000753872 | 0.003849565 | 1 |
| Obesity | adipocyte | *C1QBP* | 2.918682564 | 2.17E-07 | 1.31E-05 | 0.001367333 |
| Obesity | adipocyte | *GOSR1* | 1.631970693 | 0.000339626 | 0.002060983 | 1 |
| Obesity | adipocyte | *DRG2* | 1.918106943 | 0.009347368 | 0.028224104 | 1 |
| Obesity | adipocyte | *AKAP10* | 0.795587628 | 0.012780116 | 0.035998472 | 1 |
| Obesity | adipocyte | *ICAM2* | 0.550370657 | 0.005197079 | 0.01777695 | 1 |
| Obesity | adipocyte | *UTP6* | 1.879913282 | 0.000181426 | 0.001298735 | 1 |
| Obesity | adipocyte | *CYTH1* | 0.693524641 | 0.000986976 | 0.0047953 | 1 |
| Obesity | adipocyte | *EZH1* | 0.690064312 | 0.016520727 | 0.044330266 | 1 |
| Obesity | adipocyte | *DLX4* | -5.037255255 | 0.001364944 | 0.006213167 | 1 |
| Obesity | adipocyte | *VAT1* | 0.848857258 | 0.000415346 | 0.002424589 | 1 |
| Obesity | adipocyte | *LRRC59* | 2.22374901 | 0.007367051 | 0.023451467 | 1 |
| Obesity | adipocyte | *MPP2* | 2.755468542 | 2.25E-06 | 6.20E-05 | 0.014168638 |
| Obesity | adipocyte | *EFTUD2* | 1.272869928 | 0.000125973 | 0.000997474 | 0.791994091 |
| Obesity | adipocyte | *EFNB3* | 3.914059764 | 0.001794266 | 0.007694781 | 1 |
| Obesity | adipocyte | *FAM20A* | 1.398133762 | 1.72E-06 | 5.58E-05 | 0.010825284 |
| Obesity | adipocyte | *YWHAE* | 1.427508126 | 0.000843116 | 0.004216921 | 1 |
| Obesity | adipocyte | *SLAIN2* | 0.800966583 | 0.002887692 | 0.011231541 | 1 |
| Obesity | adipocyte | *KIAA1211* | 0.173756207 | 0.010732759 | 0.031340854 | 1 |
| Obesity | adipocyte | *UBE2D3* | 1.642282919 | 8.60E-06 | 0.000142266 | 0.05406096 |
| Obesity | adipocyte | *NDUFC1* | 1.871253072 | 0.001374857 | 0.006244393 | 1 |
| Obesity | adipocyte | *TBC1D9* | 1.348015615 | 0.001030128 | 0.004947607 | 1 |
| Obesity | adipocyte | *CPE* | 0.921840877 | 0.005154647 | 0.017660637 | 1 |
| Obesity | adipocyte | *CPZ* | 23.90178547 | 2.46E-05 | 0.000299601 | 0.154594115 |
| Obesity | adipocyte | *SLC2A9* | 0.917266225 | 0.000733265 | 0.003766372 | 1 |
| Obesity | adipocyte | *STIM2* | 1.55647643 | 1.91E-06 | 5.81E-05 | 0.012009468 |
| Obesity | adipocyte | *RAPGEF2* | 0.90081293 | 0.002214702 | 0.009088661 | 1 |
| Obesity | adipocyte | *UFSP2* | 1.692232893 | 0.000515252 | 0.002889731 | 1 |
| Obesity | adipocyte | *KLF3* | 1.765885247 | 1.35E-05 | 0.000198119 | 0.084597018 |
| Obesity | adipocyte | *UGDH* | 1.407247267 | 0.00098942 | 0.004803463 | 1 |
| Obesity | adipocyte | *ELP4* | 0.852963367 | 0.009389818 | 0.028291901 | 1 |
| Obesity | adipocyte | *FOXRED1* | 0.427104696 | 0.003054435 | 0.01175948 | 1 |
| Obesity | adipocyte | *NRXN2* | 3.863691768 | 4.58E-06 | 9.31E-05 | 0.028767756 |
| Obesity | adipocyte | *EIF4G2* | 1.785342277 | 6.22E-05 | 0.00059814 | 0.391183527 |
| Obesity | adipocyte | *CBL* | 0.739233136 | 0.00078282 | 0.003959442 | 1 |
| Obesity | adipocyte | *CD5* | 0.389170024 | 0.011177028 | 0.032382477 | 1 |
| Obesity | adipocyte | *MADD* | 2.068966278 | 1.02E-08 | 1.23E-06 | 6.42E-05 |
| Obesity | adipocyte | *AIP* | 0.61029691 | 0.015396012 | 0.041902479 | 1 |
| Obesity | adipocyte | *TCIRG1* | 1.530735935 | 3.87E-07 | 2.07E-05 | 0.002433966 |
| Obesity | adipocyte | *GTF2H1* | 1.030654898 | 8.95E-05 | 0.000772236 | 0.5625437 |
| Obesity | adipocyte | *PRDM4* | 1.500394291 | 9.56E-05 | 0.000812388 | 0.601260195 |
| Obesity | adipocyte | *KCTD10* | -0.315339453 | 0.008378118 | 0.025917351 | 1 |
| Obesity | adipocyte | *GLI1* | 3.017979532 | 5.75E-06 | 0.000107188 | 0.0361222 |
| Obesity | adipocyte | *LTA4H* | 1.820605074 | 8.32E-05 | 0.000733728 | 0.523148293 |
| Obesity | adipocyte | *WNT5B* | 1.223496153 | 0.002545475 | 0.010122327 | 1 |
| Obesity | adipocyte | *PARP11* | 1.29092104 | 0.004467802 | 0.015762667 | 1 |
| Obesity | adipocyte | *ARPC3* | 1.38382952 | 0.002980019 | 0.011515751 | 1 |
| Obesity | adipocyte | *VPS29* | 1.968931556 | 9.35E-06 | 0.000151431 | 0.05875512 |
| Obesity | adipocyte | *NAA25* | 1.310441259 | 0.000342245 | 0.002068937 | 1 |

| | | | | | | |
|---|---|---|---|---|---|---|
| Obesity | adipocyte | *LTBR* | 0.48808354 | 0.015446515 | 0.04202174 | 1 |
| Obesity | adipocyte | *ARHGDIB* | 0.476468708 | 0.019033487 | 0.049918989 | 1 |
| Obesity | adipocyte | *EIF2B1* | 1.52100333 | 0.008380782 | 0.025917351 | 1 |
| Obesity | adipocyte | *DDX55* | 0.513971402 | 1.83E-05 | 0.000244141 | 0.114990289 |
| Obesity | adipocyte | *NUP107* | 1.01265758 | 0.001857131 | 0.007915783 | 1 |
| Obesity | adipocyte | *CNOT2* | 1.178337485 | 0.001762087 | 0.007608682 | 1 |
| Obesity | adipocyte | *GNPTAB* | 1.494592843 | 5.41E-05 | 0.000539517 | 0.339895854 |
| Obesity | adipocyte | *ST8SIA1* | 0.881230202 | 0.000418154 | 0.002435768 | 1 |
| Obesity | adipocyte | *C2CD5* | 1.618427529 | 4.93E-06 | 9.70E-05 | 0.03100495 |
| Obesity | adipocyte | *PHC1* | 2.479294479 | 0.000217874 | 0.001492127 | 1 |
| Obesity | adipocyte | *BTN3A3* | 0.264477552 | 0.001159902 | 0.005405709 | 1 |
| Obesity | adipocyte | *TDP2* | 1.021473015 | 0.01735246 | 0.046167971 | 1 |
| Obesity | adipocyte | *SMIM8* | 0.821901886 | 0.004175774 | 0.014950507 | 1 |
| Obesity | adipocyte | *MCM9* | 1.537314328 | 0.000435587 | 0.002514722 | 1 |
| Obesity | adipocyte | *RNGTT* | 1.674982478 | 2.42E-05 | 0.000297225 | 0.15217938 |
| Obesity | adipocyte | *GABRR2* | 0.56303719 | 0.008356869 | 0.025881594 | 1 |
| Obesity | adipocyte | *UST* | 0.632647934 | 0.00064365 | 0.003423543 | 1 |
| Obesity | adipocyte | *SLC26A8* | -0.251004652 | 0.005690943 | 0.019101013 | 1 |
| Obesity | adipocyte | *PHACTR1* | 1.016582441 | 0.00011592 | 0.000940373 | 0.728788707 |
| Obesity | adipocyte | *MDGA1* | 0.85980005 | 0.005098123 | 0.017505134 | 1 |
| Obesity | adipocyte | *CAP2* | -0.577597315 | 7.83E-05 | 0.000704086 | 0.492156321 |
| Obesity | adipocyte | *ASCC3* | 1.359275712 | 0.00010451 | 0.000866669 | 0.657052739 |
| Obesity | adipocyte | *VNN2* | 0.592955587 | 0.011219408 | 0.03247533 | 1 |
| Obesity | adipocyte | *C6orf62* | 0.760294929 | 0.003270765 | 0.012424955 | 1 |
| Obesity | adipocyte | *B3GAT2* | -0.621269381 | 0.017245904 | 0.045962271 | 1 |
| Obesity | adipocyte | *PHACTR2* | 1.422039284 | 5.38E-05 | 0.000537967 | 0.338381212 |
| Obesity | adipocyte | *PACRG* | 1.700052442 | 0.006369718 | 0.020911968 | 1 |
| Obesity | adipocyte | *TFEB* | 1.480549937 | 0.001331868 | 0.006072121 | 1 |
| Obesity | adipocyte | *DUSP22* | 1.832139469 | 0.000320005 | 0.001974014 | 1 |
| Obesity | adipocyte | *EXOC2* | 1.529282791 | 0.00036758 | 0.002190495 | 1 |
| Obesity | adipocyte | *SENP6* | 1.426638444 | 5.79E-05 | 0.000566037 | 0.363776413 |
| Obesity | adipocyte | *LAMA4* | 0.318814522 | 0.010522508 | 0.03093516 | 1 |
| Obesity | adipocyte | *FBRSL1* | 1.159086973 | 0.00213381 | 0.00882003 | 1 |
| Obesity | adipocyte | *LY86* | 2.145576962 | 2.78E-05 | 0.000328374 | 0.175023092 |
| Obesity | adipocyte | *PRSS16* | 1.374774583 | 2.37E-08 | 2.33E-06 | 0.000148823 |
| Obesity | adipocyte | *C7* | 1.006292459 | 0.00994491 | 0.029604002 | 1 |
| Obesity | adipocyte | *DAP* | 1.5381453 | 8.80E-06 | 0.000144447 | 0.055323191 |
| Obesity | adipocyte | *NME5* | 0.35432489 | 0.006741168 | 0.021834994 | 1 |
| Obesity | adipocyte | *PFDN1* | 0.566895619 | 0.002588502 | 0.010260981 | 1 |
| Obesity | adipocyte | *LOX* | 2.255620619 | 4.00E-05 | 0.000431649 | 0.251754683 |
| Obesity | adipocyte | *PDE8B* | -0.46227946 | 0.00367248 | 0.013557769 | 1 |
| Obesity | adipocyte | *CLK4* | 1.07639746 | 0.016984993 | 0.045382342 | 1 |
| Obesity | adipocyte | *RNF130* | 1.410384724 | 0.000175996 | 0.00127329 | 1 |
| Obesity | adipocyte | *CLINT1* | 0.915878975 | 0.008763134 | 0.026854197 | 1 |
| Obesity | adipocyte | *TTC1* | 1.301192848 | 0.005022651 | 0.017342079 | 1 |
| Obesity | adipocyte | *LNPEP* | 0.59468195 | 0.011374557 | 0.032833718 | 1 |
| Obesity | adipocyte | *SLC12A7* | 0.931966544 | 0.009876779 | 0.029426872 | 1 |
| Obesity | adipocyte | *GNPDA1* | 2.047312502 | 0.000907967 | 0.004494792 | 1 |
| Obesity | adipocyte | *SKP1* | 1.786201035 | 3.24E-05 | 0.000366706 | 0.203888636 |
| Obesity | adipocyte | *TXNDC15* | 1.389253972 | 0.000972283 | 0.004738561 | 1 |

| | | | | | | |
|---|---|---|---|---|---|---|
| Obesity | adipocyte | *SMAD5* | 1.062213168 | 0.00583934 | 0.019465497 | 1 |
| Obesity | adipocyte | *HMGXB3* | 1.159705391 | 0.007833088 | 0.024574163 | 1 |
| Obesity | adipocyte | *BNIP1* | -0.224740298 | 0.002098768 | 0.008703796 | 1 |
| Obesity | adipocyte | *PDCD10* | 1.384379694 | 0.002326457 | 0.009466947 | 1 |
| Obesity | adipocyte | *PRKAR2A* | 0.535161553 | 0.000943756 | 0.004639087 | 1 |
| Obesity | adipocyte | *HYAL1* | -0.295365778 | 0.002267467 | 0.009268897 | 1 |
| Obesity | adipocyte | *IFT57* | 2.109081966 | 0.000320263 | 0.001974014 | 1 |
| Obesity | adipocyte | *NEK11* | 1.301994466 | 0.002776128 | 0.010928939 | 1 |
| Obesity | adipocyte | *WDR48* | 0.623361087 | 0.007115476 | 0.022812338 | 1 |
| Obesity | adipocyte | *ABCC5* | 0.688795246 | 0.009958824 | 0.02963139 | 1 |
| Obesity | adipocyte | *ARHGEF26* | 0.624567825 | 0.005148377 | 0.01764877 | 1 |
| Obesity | adipocyte | *KLHL24* | 0.782661559 | 6.99E-05 | 0.000648798 | 0.439236549 |
| Obesity | adipocyte | *PLCH1* | 1.459254652 | 0.000315658 | 0.001955216 | 1 |
| Obesity | adipocyte | *NKTR* | 1.249836972 | 8.39E-06 | 0.000139725 | 0.052779272 |
| Obesity | adipocyte | *MOB1A* | 2.098386263 | 6.27E-06 | 0.000114593 | 0.03942014 |
| Obesity | adipocyte | *KCNIP3* | 1.274617484 | 0.001303132 | 0.005967073 | 1 |
| Obesity | adipocyte | *FAHD2A* | 0.888776143 | 0.001509909 | 0.006742043 | 1 |
| Obesity | adipocyte | *NCL* | 1.216662393 | 0.002916212 | 0.01131044 | 1 |
| Obesity | adipocyte | *ACTR1B* | 2.284317453 | 1.01E-08 | 1.23E-06 | 6.35E-05 |
| Obesity | adipocyte | *ACTR3* | 1.650088729 | 4.23E-06 | 8.80E-05 | 0.026580304 |
| Obesity | adipocyte | *TFCP2L1* | 0.977764416 | 0.00016213 | 0.001190786 | 1 |
| Obesity | adipocyte | *DNAJC27* | 0.856600454 | 0.001402843 | 0.006345089 | 1 |
| Obesity | adipocyte | *STAM2* | 1.72340151 | 9.57E-05 | 0.000812388 | 0.601979208 |
| Obesity | adipocyte | *GPD2* | 1.132131158 | 0.004247327 | 0.01514631 | 1 |
| Obesity | adipocyte | *ITGB6* | -0.339421554 | 0.005330844 | 0.01811612 | 1 |
| Obesity | adipocyte | *ITGA4* | 1.000543296 | 3.18E-05 | 0.000362662 | 0.200087058 |
| Obesity | adipocyte | *POLE4* | 2.824750886 | 6.40E-06 | 0.000115994 | 0.040216062 |
| Obesity | adipocyte | *WDR75* | 2.004684012 | 5.33E-05 | 0.000534324 | 0.335021283 |
| Obesity | adipocyte | *GLS* | 1.398655533 | 5.02E-05 | 0.000509485 | 0.315371296 |
| Obesity | adipocyte | *GGCX* | 0.373483913 | 0.003545606 | 0.013166702 | 1 |
| Obesity | adipocyte | *GNLY* | 1.177339586 | 0.00040949 | 0.002401553 | 1 |
| Obesity | adipocyte | *SF3B1* | 1.182067973 | 0.000675009 | 0.003518889 | 1 |
| Obesity | adipocyte | *ST3GAL5* | 1.212677962 | 2.83E-05 | 0.000332421 | 0.177845297 |
| Obesity | adipocyte | *PLCD4* | -1.2710263 | 0.005285916 | 0.017992722 | 1 |
| Obesity | adipocyte | *CHMP3* | -0.652756502 | 0.000520604 | 0.002906785 | 1 |
| Obesity | adipocyte | *ZNF142* | 0.543246302 | 0.018553303 | 0.048866617 | 1 |
| Obesity | adipocyte | *MLPH* | 1.786306906 | 2.11E-06 | 6.01E-05 | 0.013240586 |
| Obesity | adipocyte | *STK25* | 0.997628329 | 0.008698363 | 0.026702446 | 1 |
| Obesity | adipocyte | *STRN* | 1.703209442 | 5.23E-05 | 0.000527228 | 0.328990041 |
| Obesity | adipocyte | *RMDN2* | 1.371396851 | 0.007505571 | 0.02377003 | 1 |
| Obesity | adipocyte | *WIPF1* | 1.116790659 | 5.96E-05 | 0.000579949 | 0.374502467 |
| Obesity | adipocyte | *COX7A2L* | 1.558862834 | 0.004514168 | 0.015890579 | 1 |
| Obesity | adipocyte | *ORC4* | 0.826314109 | 0.003120044 | 0.011982721 | 1 |
| Obesity | adipocyte | *THADA* | 1.428409484 | 0.000187156 | 0.001328051 | 1 |
| Obesity | adipocyte | *CD207* | 19.2720403 | 2.53E-08 | 2.45E-06 | 0.000159118 |
| Obesity | adipocyte | *PLEKHA3* | 1.932290007 | 0.000927427 | 0.004576716 | 1 |
| Obesity | adipocyte | *SCP2* | 0.800198351 | 0.006082218 | 0.020146946 | 1 |
| Obesity | adipocyte | *RALGPS2* | 0.691021835 | 0.008452582 | 0.026113702 | 1 |
| Obesity | adipocyte | *FAM20B* | 1.7390052 | 0.001093236 | 0.005183391 | 1 |
| Obesity | adipocyte | *TMEM59* | 1.498546336 | 9.25E-05 | 0.000793731 | 0.581804575 |

| | | | | | | |
|---|---|---|---|---|---|---|
| Obesity | adipocyte | *QSOX1* | 1.124080392 | 0.001785439 | 0.007677877 | 1 |
| Obesity | adipocyte | *RAP1A* | 1.294739433 | 7.63E-05 | 0.000691526 | 0.479595396 |
| Obesity | adipocyte | *DOCK7* | 1.026892564 | 0.011660925 | 0.033414874 | 1 |
| Obesity | adipocyte | *C1orf21* | 0.386580076 | 0.000622417 | 0.003341704 | 1 |
| Obesity | adipocyte | *LEPR* | 0.932147752 | 0.000309057 | 0.001932321 | 1 |
| Obesity | adipocyte | *SMG7* | 1.487582403 | 0.000130464 | 0.001016439 | 0.820228261 |
| Obesity | adipocyte | *NCF2* | 1.938346979 | 3.21E-06 | 7.31E-05 | 0.020187647 |
| Obesity | adipocyte | *RGS2* | 1.539863524 | 0.006546464 | 0.021347313 | 1 |
| Obesity | adipocyte | *OLFML3* | 1.423168858 | 1.87E-05 | 0.000247605 | 0.117860197 |
| Obesity | adipocyte | *WARS2* | 0.460939608 | 0.00145051 | 0.006518482 | 1 |
| Obesity | adipocyte | *GNPAT* | 0.640387239 | 0.010610041 | 0.031123891 | 1 |
| Obesity | adipocyte | *ACADM* | 0.775151825 | 0.012627971 | 0.035681822 | 1 |
| Obesity | adipocyte | *MFAP2* | -0.310676872 | 0.001373089 | 0.006241944 | 1 |
| Obesity | adipocyte | *PRDX1* | 1.080106526 | 0.000124675 | 0.000990936 | 0.783830061 |
| Obesity | adipocyte | *PRRC2C* | 1.622540964 | 1.91E-07 | 1.18E-05 | 0.001200822 |
| Obesity | adipocyte | *ABCD3* | 1.505568871 | 0.001525736 | 0.006803051 | 1 |
| Obesity | adipocyte | *VAMP4* | 0.63726632 | 0.013781661 | 0.038203397 | 1 |
| Obesity | adipocyte | *PTBP2* | 0.601069759 | 0.000119483 | 0.000961831 | 0.751190345 |
| Obesity | adipocyte | *TNFSF4* | -0.737551576 | 0.000630811 | 0.003372369 | 1 |
| Obesity | adipocyte | *DARS2* | -1.339617626 | 0.000166331 | 0.001215955 | 1 |
| Obesity | adipocyte | *SERPINC1* | 0.599730695 | 3.99E-09 | 5.34E-07 | 2.51E-05 |
| Obesity | adipocyte | *RCAN3* | 1.045883672 | 1.98E-05 | 0.000256719 | 0.124279925 |
| Obesity | adipocyte | *RCOR3* | 1.437442034 | 0.000576759 | 0.00315586 | 1 |
| Obesity | adipocyte | *RPA2* | 1.812139227 | 0.000570978 | 0.003135143 | 1 |
| Obesity | adipocyte | *SLC5A9* | 2.096813323 | 0.002046664 | 0.008515803 | 1 |
| Obesity | adipocyte | *OSBPL9* | 1.283570345 | 0.001019273 | 0.004902962 | 1 |
| Obesity | adipocyte | *TXNDC12* | 1.236104685 | 0.011609776 | 0.033328892 | 1 |
| Obesity | adipocyte | *COLEC11* | -0.558272646 | 0.003257707 | 0.012393985 | 1 |
| Obesity | adipocyte | *KMT2A* | 1.60364604 | 1.89E-05 | 0.000248405 | 0.118737426 |
| Obesity | adipocyte | *CAMSAP2* | 0.873762952 | 0.003255654 | 0.012393985 | 1 |
| Obesity | adipocyte | *ATF6* | 1.302647991 | 0.000233858 | 0.001569121 | 1 |
| Obesity | adipocyte | *LRMP* | 0.391992653 | 0.001597656 | 0.007068588 | 1 |
| Obesity | adipocyte | *ATP10B* | 1.827239547 | 0.00266518 | 0.010551631 | 1 |
| Obesity | adipocyte | *SPCS2* | 1.17035386 | 0.002414182 | 0.009729461 | 1 |
| Obesity | adipocyte | *SGIP1* | 1.109645329 | 1.58E-05 | 0.000223295 | 0.099589362 |
| Obesity | adipocyte | *VAMP8* | 0.578904257 | 0.004350434 | 0.015435204 | 1 |
| Obesity | adipocyte | *FOXO3* | 1.386016313 | 9.81E-05 | 0.000825832 | 0.616896749 |
| Obesity | adipocyte | *ABCG2* | 2.081081355 | 1.63E-05 | 0.000227305 | 0.10274177 |
| Obesity | adipocyte | *SPP1* | 1.264018145 | 0.000115075 | 0.000937147 | 0.723477681 |
| Obesity | adipocyte | *MFSD1* | 1.672696351 | 1.39E-05 | 0.000203849 | 0.087655213 |
| Obesity | adipocyte | *RAB3GAP2* | 1.522293083 | 0.000215525 | 0.001479877 | 1 |
| Obesity | adipocyte | *WDR35* | 1.621348763 | 0.000688572 | 0.003574776 | 1 |
| Obesity | adipocyte | *SENP5* | 1.183568164 | 0.002237639 | 0.009170821 | 1 |
| Obesity | adipocyte | *CCDC92* | 1.271522757 | 2.00E-13 | 6.98E-11 | 1.26E-09 |
| Obesity | adipocyte | *PTBP3* | 1.571231293 | 2.35E-06 | 6.25E-05 | 0.014777461 |
| Obesity | adipocyte | *FKBP15* | 1.545517433 | 3.67E-06 | 7.91E-05 | 0.02309711 |
| Obesity | adipocyte | *CNTRL* | 1.602381981 | 3.83E-05 | 0.000418541 | 0.240660882 |
| Obesity | adipocyte | *TRIM32* | 1.916035392 | 0.000309433 | 0.001932321 | 1 |
| Obesity | adipocyte | *BSPRY* | 10.85283978 | 0.000248859 | 0.00164346 | 1 |
| Obesity | adipocyte | *HDHD3* | -0.093058505 | 0.013642628 | 0.037951858 | 1 |

| | | | | | | |
|---|---|---|---|---|---|---|
| Obesity | adipocyte | *INVS* | 1.581011254 | 6.65E-08 | 5.16E-06 | 0.000417956 |
| Obesity | adipocyte | *KDSR* | 1.853736094 | 0.000225817 | 0.001527101 | 1 |
| Obesity | adipocyte | *DCAF4* | 0.72350452 | 4.39E-05 | 0.00046362 | 0.275853646 |
| Obesity | adipocyte | *TTLL5* | 1.707296814 | 4.98E-05 | 0.000508417 | 0.313184766 |
| Obesity | adipocyte | *FLVCR2* | 1.171848143 | 0.010184117 | 0.030158992 | 1 |
| Obesity | adipocyte | *ATAD2B* | 1.500615772 | 8.56E-05 | 0.00075182 | 0.538303182 |
| Obesity | adipocyte | *MXI1* | 1.523300124 | 0.001116843 | 0.005263563 | 1 |
| Obesity | adipocyte | *C10orf88* | 1.608958275 | 0.010912568 | 0.031792083 | 1 |
| Obesity | adipocyte | *WDR11* | 1.023319229 | 0.00294746 | 0.011410517 | 1 |
| Obesity | adipocyte | *C10orf76* | 1.575964973 | 1.42E-05 | 0.000205406 | 0.089557012 |
| Obesity | adipocyte | *KANSL1* | 0.571140714 | 0.009765232 | 0.029124295 | 1 |
| Obesity | adipocyte | *HOXB5* | 0.871968593 | 0.000145963 | 0.001102969 | 0.917670265 |
| Obesity | adipocyte | *HOXB3* | 0.860066762 | 9.12E-08 | 6.66E-06 | 0.000573082 |
| Obesity | adipocyte | *CYSTM1* | 1.511239934 | 0.000132031 | 0.001023525 | 0.830078877 |
| Obesity | adipocyte | *PCDHB14* | 3.221293186 | 0.003266952 | 0.012417974 | 1 |
| Obesity | adipocyte | *GORAB* | 1.099585315 | 0.007903625 | 0.024758391 | 1 |
| Obesity | adipocyte | *SNX19* | -0.566301716 | 0.003356065 | 0.012657218 | 1 |
| Obesity | adipocyte | *MRC1* | 0.516083815 | 0.010274712 | 0.030341529 | 1 |
| Obesity | adipocyte | *ENOX1* | 0.985444233 | 0.013579423 | 0.037859791 | 1 |
| Obesity | adipocyte | *DNAJC15* | 1.781353075 | 0.000193004 | 0.001360329 | 1 |
| Obesity | adipocyte | *SMAD9* | 1.948804758 | 1.81E-06 | 5.73E-05 | 0.011371644 |
| Obesity | adipocyte | *APAF1* | 0.847526892 | 0.000931325 | 0.004585154 | 1 |
| Obesity | adipocyte | *ADRA1A* | 0.096132288 | 0.000344775 | 0.002080228 | 1 |
| Obesity | adipocyte | *ZNF706* | 0.761032559 | 0.006681092 | 0.021685095 | 1 |
| Obesity | adipocyte | *CRISPLD1* | 2.160729638 | 1.84E-06 | 5.80E-05 | 0.011593897 |
| Obesity | adipocyte | *TEX14* | 0.844179957 | 0.000242341 | 0.00160938 | 1 |
| Obesity | adipocyte | *PYROXD1* | 0.563840151 | 0.002147835 | 0.008860525 | 1 |
| Obesity | adipocyte | *A1BG* | 2.124668474 | 2.87E-06 | 6.96E-05 | 0.018013755 |
| Obesity | adipocyte | *ZSCAN18* | 0.948693858 | 0.005732402 | 0.019180209 | 1 |
| Obesity | adipocyte | *POPDC2* | 1.428467969 | 0.006798706 | 0.021976074 | 1 |
| Obesity | adipocyte | *B4GALT4* | 1.624486993 | 0.01005084 | 0.029848668 | 1 |
| Obesity | adipocyte | *GJB6* | -6.652397665 | 0.000294221 | 0.001865246 | 1 |
| Obesity | adipocyte | *RNF115* | 1.615014083 | 1.20E-05 | 0.000181341 | 0.075619391 |
| Obesity | adipocyte | *TNFSF10* | 1.294803122 | 0.000129108 | 0.001012094 | 0.811699493 |
| Obesity | adipocyte | *ZNF639* | 1.075561366 | 0.008258136 | 0.025664311 | 1 |
| Obesity | adipocyte | *GPSM2* | 1.186487932 | 5.23E-06 | 0.00010026 | 0.03288532 |
| Obesity | adipocyte | *ACVR2A* | 1.570934013 | 2.96E-05 | 0.000344651 | 0.186111708 |
| Obesity | adipocyte | *FLT3* | 2.825907774 | 1.23E-05 | 0.000184851 | 0.077452606 |
| Obesity | adipocyte | *ODF2L* | 0.621664527 | 0.002122661 | 0.008780734 | 1 |
| Obesity | adipocyte | *SPATA1* | 1.350326888 | 0.001927939 | 0.008147951 | 1 |
| Obesity | adipocyte | *RPAP2* | 1.012358535 | 0.015205247 | 0.041455069 | 1 |
| Obesity | adipocyte | *BBS9* | 0.732150175 | 0.009446873 | 0.028397009 | 1 |
| Obesity | adipocyte | *KLHL7* | 1.712834739 | 0.000469335 | 0.00266791 | 1 |
| Obesity | adipocyte | *HERPUD2* | 2.012173289 | 1.53E-05 | 0.000217012 | 0.096136259 |
| Obesity | adipocyte | *FAM126A* | 0.874323578 | 0.001614919 | 0.00712491 | 1 |
| Obesity | adipocyte | *CCZ1* | 0.636981165 | 0.000646078 | 0.003433553 | 1 |
| Obesity | adipocyte | *SMU1* | 1.772287065 | 0.000138711 | 0.001057037 | 0.872078222 |
| Obesity | adipocyte | *DCAF10* | 1.020079765 | 0.012195467 | 0.034725046 | 1 |
| Obesity | adipocyte | *TRIM24* | 0.992278697 | 0.005646845 | 0.019025569 | 1 |
| Obesity | adipocyte | *RBM19* | 1.149222681 | 0.010700798 | 0.031291124 | 1 |

| | | | | | | |
|---|---|---|---|---|---|---|
| Obesity | adipocyte | *CCDC91* | 0.882173801 | 0.001944064 | 0.008191909 | 1 |
| Obesity | adipocyte | *PKN1* | 1.157711789 | 0.001146332 | 0.005354377 | 1 |
| Obesity | adipocyte | *IKZF4* | -0.976103348 | 0.005023052 | 0.017342079 | 1 |
| Obesity | adipocyte | *SMUG1* | 0.135552079 | 0.009660861 | 0.02889526 | 1 |
| Obesity | adipocyte | *SARDH* | 6.22435534 | 2.17E-06 | 6.11E-05 | 0.013616873 |
| Obesity | adipocyte | *TTC21B* | 1.294717711 | 0.002791044 | 0.010967057 | 1 |
| Obesity | adipocyte | *NMI* | 0.902587992 | 0.015246797 | 0.041550332 | 1 |
| Obesity | adipocyte | *ACVR1C* | -0.432139654 | 0.004600028 | 0.016147613 | 1 |
| Obesity | adipocyte | *PLA2G12A* | 1.49136954 | 0.00241124 | 0.009729461 | 1 |
| Obesity | adipocyte | *RAB38* | 1.971799366 | 0.00325867 | 0.012393985 | 1 |
| Obesity | adipocyte | *AGO2* | 1.639466384 | 0.000268228 | 0.00173349 | 1 |
| Obesity | adipocyte | *DNPEP* | 1.74756591 | 0.001977081 | 0.008292135 | 1 |
| Obesity | adipocyte | *MOGAT1* | -0.41234746 | 0.009180447 | 0.027829061 | 1 |
| Obesity | adipocyte | *ARFGEF2* | 1.642868642 | 6.35E-05 | 0.000606894 | 0.399336129 |
| Obesity | adipocyte | *CSE1L* | 0.85851628 | 0.003114383 | 0.011968294 | 1 |
| Obesity | adipocyte | *NEURL2* | 0.279348736 | 0.002897535 | 0.01125884 | 1 |
| Obesity | adipocyte | *PEPD* | 0.544537896 | 0.013517554 | 0.037712654 | 1 |
| Obesity | adipocyte | *CHST8* | 1.690081433 | 0.001761371 | 0.007608682 | 1 |
| Obesity | adipocyte | *STAMBP* | 1.099009392 | 0.011442353 | 0.032983986 | 1 |
| Obesity | adipocyte | *ATP8A1* | 0.519959342 | 0.000222438 | 0.001515135 | 1 |
| Obesity | adipocyte | *HIF3A* | 0.510109473 | 0.000124171 | 0.000988183 | 0.780664544 |
| Obesity | adipocyte | *SNRPC* | -1.204458993 | 1.02E-21 | 7.58E-19 | 6.39E-18 |
| Obesity | adipocyte | *SERPINB6* | 1.860863316 | 9.55E-05 | 0.000812388 | 0.600580063 |
| Obesity | adipocyte | *HIST1H2BJ* | 0.894239297 | 1.13E-08 | 1.32E-06 | 7.10E-05 |
| Obesity | adipocyte | *GLO1* | 1.476532534 | 9.57E-05 | 0.000812388 | 0.601423774 |
| Obesity | adipocyte | *NUP153* | 1.879539765 | 5.43E-07 | 2.55E-05 | 0.003415507 |
| Obesity | adipocyte | *EEF1E1* | 3.506491103 | 1.91E-06 | 5.81E-05 | 0.011996678 |
| Obesity | adipocyte | *EREG* | 7.249524276 | 0.00012524 | 0.000992914 | 0.787380996 |
| Obesity | adipocyte | *CNOT1* | 1.158027503 | 0.000239275 | 0.001597411 | 1 |
| Obesity | adipocyte | *BBS2* | 0.462836209 | 0.000300667 | 0.001894083 | 1 |
| Obesity | adipocyte | *MT1G* | 3.041012403 | 0.011406729 | 0.032911475 | 1 |
| Obesity | adipocyte | *GOT2* | 1.626992504 | 3.75E-05 | 0.000413382 | 0.235627631 |
| Obesity | adipocyte | *CLYBL* | 1.712480212 | 0.000292945 | 0.001860347 | 1 |
| Obesity | adipocyte | *TMTC4* | 1.932444452 | 0.002122907 | 0.008780734 | 1 |
| Obesity | adipocyte | *GRK4* | 0.181691205 | 5.44E-05 | 0.000541627 | 0.341766426 |
| Obesity | adipocyte | *GGA3* | 2.037240286 | 0.000131148 | 0.001020451 | 0.824524556 |
| Obesity | adipocyte | *MIF4GD* | 2.238510227 | 0.016155695 | 0.043499294 | 1 |
| Obesity | adipocyte | *PPP1R12C* | 0.956184843 | 0.005034163 | 0.017370901 | 1 |
| Obesity | adipocyte | *OPRL1* | 0.446687656 | 0.001325086 | 0.006049974 | 1 |
| Obesity | adipocyte | *INSIG2* | 2.188602972 | 1.76E-05 | 0.000238689 | 0.110437228 |
| Obesity | adipocyte | *PSD4* | 1.193085032 | 0.000459064 | 0.002628537 | 1 |
| Obesity | adipocyte | *TRIP10* | 1.227107836 | 0.001631522 | 0.007183037 | 1 |
| Obesity | adipocyte | *RTN2* | 1.857209192 | 9.23E-05 | 0.00079315 | 0.580586098 |
| Obesity | adipocyte | *SYMPK* | 1.364955958 | 0.000441746 | 0.002538626 | 1 |
| Obesity | adipocyte | *GPCPD1* | 1.585505951 | 5.36E-05 | 0.000536256 | 0.336768824 |
| Obesity | adipocyte | *PANK2* | 1.894665246 | 7.29E-05 | 0.000669682 | 0.458062542 |
| Obesity | adipocyte | *DTD1* | 0.915299022 | 0.014929096 | 0.040879454 | 1 |
| Obesity | adipocyte | *RRBP1* | 0.615135838 | 0.017349927 | 0.046167971 | 1 |
| Obesity | adipocyte | *PCSK2* | 1.960000239 | 3.05E-05 | 0.000351733 | 0.191694729 |
| Obesity | adipocyte | *DSTN* | 1.741362962 | 3.67E-06 | 7.91E-05 | 0.023072269 |

| | | | | | | |
|---|---|---|---|---|---|---|
| Obesity | adipocyte | *ITPA* | 2.013190428 | 0.000515764 | 0.002890023 | 1 |
| Obesity | adipocyte | *FAM110A* | 2.444915955 | 0.003490796 | 0.013024708 | 1 |
| Obesity | adipocyte | *UROD* | 1.015965635 | 0.001928764 | 0.008147951 | 1 |
| Obesity | adipocyte | *KLC1* | 1.935321932 | 1.49E-08 | 1.63E-06 | 9.35E-05 |
| Obesity | adipocyte | *XRCC3* | 1.998686043 | 6.23E-08 | 4.96E-06 | 0.000391821 |
| Obesity | adipocyte | *MCF2L* | 0.381615504 | 0.009405818 | 0.028307505 | 1 |
| Obesity | adipocyte | *F10* | 1.871525489 | 2.02E-06 | 5.99E-05 | 0.0126928 |
| Obesity | adipocyte | *COX6B1* | 2.343685819 | 0.000104629 | 0.000866669 | 0.65780155 |
| Obesity | adipocyte | *PRDX5* | 1.446283522 | 0.001967792 | 0.008275258 | 1 |
| Obesity | adipocyte | *BECN1* | 1.609635471 | 0.001523865 | 0.006799532 | 1 |
| Obesity | adipocyte | *TRAP1* | 0.159722653 | 0.002203935 | 0.009062224 | 1 |
| Obesity | adipocyte | *ZNF384* | 1.320007045 | 0.001441886 | 0.006484363 | 1 |
| Obesity | adipocyte | *RHOJ* | 1.433014646 | 2.90E-06 | 6.99E-05 | 0.018206782 |
| Obesity | adipocyte | *ZBTB1* | 0.962712726 | 0.00243389 | 0.009783802 | 1 |
| Obesity | adipocyte | *PZP* | 1.22733451 | 0.001063465 | 0.005088283 | 1 |
| Obesity | adipocyte | *RHOT1* | 0.577853344 | 0.002888723 | 0.011231541 | 1 |
| Obesity | adipocyte | *AIF1L* | 1.728434024 | 1.44E-06 | 5.18E-05 | 0.009069754 |
| Obesity | adipocyte | *CANX* | 1.986446612 | 3.93E-07 | 2.07E-05 | 0.002469242 |
| Obesity | adipocyte | *IPPK* | 1.585397457 | 0.000118044 | 0.000953909 | 0.742141245 |
| Obesity | adipocyte | *FGD3* | 1.904149371 | 1.95E-06 | 5.81E-05 | 0.012264628 |
| Obesity | adipocyte | *HELB* | 1.127132122 | 0.001588229 | 0.007041745 | 1 |
| Obesity | adipocyte | *RAP1B* | 1.342168358 | 7.55E-05 | 0.000689004 | 0.474723573 |
| Obesity | adipocyte | *YEATS4* | 0.434543356 | 0.000307472 | 0.001927297 | 1 |
| Obesity | adipocyte | *SIN3B* | 1.517523569 | 0.00322288 | 0.012295053 | 1 |
| Obesity | adipocyte | *HIP1* | 1.130108445 | 0.002166948 | 0.008927657 | 1 |
| Obesity | adipocyte | *PTPN12* | 1.430063279 | 2.16E-05 | 0.0002729 | 0.135631356 |
| Obesity | adipocyte | *FGL2* | 0.516927282 | 0.003451333 | 0.012943383 | 1 |
| Obesity | adipocyte | *CASD1* | 1.666488468 | 0.00015637 | 0.00115659 | 0.983101084 |
| Obesity | adipocyte | *VPREB3* | -2.97341734 | 0.005664086 | 0.019032662 | 1 |
| Obesity | adipocyte | *MGAT3* | -0.713523753 | 0.015521614 | 0.042153084 | 1 |
| Obesity | adipocyte | *TPST2* | 1.21723295 | 0.00374135 | 0.013779652 | 1 |
| Obesity | adipocyte | *RIBC2* | 0.431645806 | 0.000101494 | 0.000846278 | 0.63809325 |
| Obesity | adipocyte | *EMC4* | 0.970069836 | 0.000320721 | 0.001974899 | 1 |
| Obesity | adipocyte | *SPECC1* | 0.977758961 | 0.018208424 | 0.048135258 | 1 |
| Obesity | adipocyte | *GAD1* | 6.199238599 | 0.007204629 | 0.023027708 | 1 |
| Obesity | adipocyte | *TWSG1* | 1.161787673 | 0.009189594 | 0.027843361 | 1 |
| Obesity | adipocyte | *EIF2AK4* | 0.524606909 | 0.000958682 | 0.004697767 | 1 |
| Obesity | adipocyte | *ARPP19* | 1.161110106 | 0.00528171 | 0.017992722 | 1 |
| Obesity | adipocyte | *VPS13C* | 1.124105707 | 5.47E-05 | 0.000542409 | 0.343887551 |
| Obesity | adipocyte | *SUMF2* | 0.865941124 | 0.012311882 | 0.034977317 | 1 |
| Obesity | adipocyte | *SPCS3* | 1.312809032 | 0.001296868 | 0.00594272 | 1 |
| Obesity | adipocyte | *SERGEF* | 1.975497597 | 7.97E-06 | 0.000136462 | 0.050134354 |
| Obesity | adipocyte | *RPAIN* | -0.961529443 | 9.72E-06 | 0.000154742 | 0.061122973 |
| Obesity | adipocyte | *PLD2* | 0.950533843 | 6.84E-05 | 0.000638629 | 0.42979747 |
| Obesity | adipocyte | *CCL4* | 0.352860459 | 0.013598514 | 0.037895138 | 1 |
| Obesity | adipocyte | *PHF20L1* | 1.425430826 | 0.000201811 | 0.001408199 | 1 |
| Obesity | adipocyte | *SLC44A2* | 1.839670746 | 1.41E-05 | 0.000204947 | 0.088847059 |
| Obesity | adipocyte | *CDKN2D* | 0.945008201 | 0.000425219 | 0.002468469 | 1 |
| Obesity | adipocyte | *MTUS1* | 0.739312192 | 0.005015791 | 0.017342079 | 1 |
| Obesity | adipocyte | *NGDN* | 1.945978132 | 0.001174005 | 0.005455262 | 1 |

| | | | | | | |
|---|---|---|---|---|---|---|
| Obesity | adipocyte | *RIPK3* | 0.575524913 | 0.002433319 | 0.009783802 | 1 |
| Obesity | adipocyte | *RAB2B* | 1.200712723 | 0.008141928 | 0.025340743 | 1 |
| Obesity | adipocyte | *MIS18BP1* | 0.303976125 | 0.017973045 | 0.047617587 | 1 |
| Obesity | adipocyte | *EPB41L4A* | 1.576509277 | 4.60E-05 | 0.000480798 | 0.289440492 |
| Obesity | adipocyte | *REEP5* | 1.599473855 | 0.000233562 | 0.001569121 | 1 |
| Obesity | adipocyte | *SHC2* | 0.502986828 | 0.000923109 | 0.004558985 | 1 |
| Obesity | adipocyte | *PHF10* | 1.698854198 | 0.002240656 | 0.009177203 | 1 |
| Obesity | adipocyte | *ELOF1* | 2.353446419 | 0.005095494 | 0.017505134 | 1 |
| Obesity | adipocyte | *DPP6* | 0.848480914 | 0.015842328 | 0.0428206 | 1 |
| Obesity | adipocyte | *RPL36* | 1.399040941 | 2.68E-05 | 0.000318494 | 0.168483325 |
| Obesity | adipocyte | *ATP8B3* | -0.42239712 | 1.31E-07 | 8.80E-06 | 0.000822159 |
| Obesity | adipocyte | *BST2* | 1.761160534 | 4.16E-06 | 8.73E-05 | 0.026184425 |
| Obesity | adipocyte | *ACTN4* | 1.051577962 | 0.004422774 | 0.015621339 | 1 |
| Obesity | adipocyte | *PGPEP1* | 0.633272145 | 0.000214528 | 0.001479877 | 1 |
| Obesity | adipocyte | *LSM4* | 0.428495962 | 0.000594527 | 0.003227798 | 1 |
| Obesity | adipocyte | *CAMSAP1* | 1.755490338 | 0.002144638 | 0.008853143 | 1 |
| Obesity | adipocyte | *ATXN10* | 1.261745987 | 0.000600497 | 0.003246193 | 1 |
| Obesity | adipocyte | *TUBGCP2* | 0.801049997 | 0.005456991 | 0.018455138 | 1 |
| Obesity | adipocyte | *CYP2E1* | -0.943204488 | 0.000332446 | 0.002029214 | 1 |
| Obesity | adipocyte | *PAK4* | 2.374135783 | 0.000115854 | 0.000940373 | 0.728375136 |
| Obesity | adipocyte | *TMEM160* | 3.651744908 | 3.42E-06 | 7.63E-05 | 0.021502745 |
| Obesity | adipocyte | *THEMIS2* | 0.369329022 | 0.007751159 | 0.024377956 | 1 |
| Obesity | adipocyte | *CLIP1* | 2.14394672 | 1.75E-10 | 3.55E-08 | 1.10E-06 |
| Obesity | adipocyte | *PKDREJ* | -0.450108556 | 0.000639603 | 0.003410367 | 1 |
| Obesity | adipocyte | *LILRB2* | 2.310895976 | 6.24E-06 | 0.000114438 | 0.039252231 |
| Obesity | adipocyte | *PPT1* | 1.356368367 | 0.00015868 | 0.001169541 | 0.997618894 |
| Obesity | adipocyte | *TBC1D5* | 1.34000186 | 1.62E-05 | 0.000226767 | 0.101818384 |
| Obesity | adipocyte | *RFTN1* | 0.748417733 | 0.005166366 | 0.017682008 | 1 |
| Obesity | adipocyte | *KIF3A* | 0.851803752 | 0.000108252 | 0.000889651 | 0.680583296 |
| Obesity | adipocyte | *AOC2* | -0.858806081 | 0.017220998 | 0.045915359 | 1 |
| Obesity | adipocyte | *DIAPH1* | 1.034688843 | 4.26E-05 | 0.000451214 | 0.267569891 |
| Obesity | adipocyte | *EXOC4* | 1.43271905 | 2.83E-08 | 2.65E-06 | 0.000177672 |
| Obesity | adipocyte | *ANO1* | 1.560136898 | 0.000545686 | 0.003025331 | 1 |
| Obesity | adipocyte | *NINJ1* | 1.92750411 | 1.63E-05 | 0.00022712 | 0.10220404 |
| Obesity | adipocyte | *NPHP4* | 2.474908456 | 9.99E-05 | 0.000837425 | 0.62806905 |
| Obesity | adipocyte | *ZSCAN5A* | 1.080695341 | 0.012423986 | 0.035232116 | 1 |
| Obesity | adipocyte | *GCH1* | 0.614223579 | 0.00751165 | 0.02377003 | 1 |
| Obesity | adipocyte | *LGALS3* | 1.481758396 | 8.35E-06 | 0.000139617 | 0.0524961 |
| Obesity | adipocyte | *MATN3* | -2.032473173 | 0.004070109 | 0.014647266 | 1 |
| Obesity | adipocyte | *RAF1* | 1.472141148 | 0.00032446 | 0.001990125 | 1 |
| Obesity | adipocyte | *EMILIN2* | 0.944767591 | 3.16E-11 | 7.96E-09 | 1.99E-07 |
| Obesity | adipocyte | *TRIM5* | 1.503536258 | 1.87E-05 | 0.000247605 | 0.117695057 |
| Obesity | adipocyte | *TRIM22* | 1.09450916 | 0.00025332 | 0.001665923 | 1 |
| Obesity | adipocyte | *IMMT* | 0.564813992 | 0.009382119 | 0.028291901 | 1 |
| Obesity | adipocyte | *CARD6* | 1.989970789 | 6.80E-07 | 2.95E-05 | 0.004277155 |
| Obesity | adipocyte | *RPA1* | 0.667545839 | 0.00516651 | 0.017682008 | 1 |
| Obesity | adipocyte | *SERPINF1* | 0.414146236 | 0.012669329 | 0.035782603 | 1 |
| Obesity | adipocyte | *REEP2* | -0.18634881 | 0.01777547 | 0.047213511 | 1 |
| Obesity | adipocyte | *TERF2* | 1.189690393 | 0.005234068 | 0.0178743 | 1 |
| Obesity | adipocyte | *BTBD3* | 1.344921854 | 0.00720419 | 0.023027708 | 1 |

| | | | | | | |
|---|---|---|---|---|---|---|
| Obesity | adipocyte | *PTPRA* | 1.374215579 | 7.76E-05 | 0.000700305 | 0.488112446 |
| Obesity | adipocyte | *IGHMBP2* | 0.487227418 | 2.08E-06 | 5.99E-05 | 0.013051374 |
| Obesity | adipocyte | *MMACHC* | -0.401458937 | 0.017117944 | 0.045698733 | 1 |
| Obesity | adipocyte | *MUTYH* | 1.960000828 | 0.000191414 | 0.001353055 | 1 |
| Obesity | adipocyte | *CTNNBL1* | 1.554689745 | 0.000261172 | 0.001703308 | 1 |
| Obesity | adipocyte | *USPL1* | 1.199187542 | 0.00903277 | 0.027540748 | 1 |
| Obesity | adipocyte | *POMP* | 1.60508052 | 0.000969541 | 0.004732534 | 1 |
| Obesity | adipocyte | *RNF17* | -0.774552756 | 0.003786576 | 0.01388231 | 1 |
| Obesity | adipocyte | *PEMT* | -0.585022715 | 0.001066405 | 0.005097592 | 1 |
| Obesity | adipocyte | *CHI3L1* | 0.121887135 | 0.010209359 | 0.030191082 | 1 |
| Obesity | adipocyte | *TRPC4* | -0.897820511 | 0.010068228 | 0.02988619 | 1 |
| Obesity | adipocyte | *CNDP2* | 2.350191639 | 3.90E-05 | 0.000423188 | 0.24514792 |
| Obesity | adipocyte | *FAM83F* | -1.439138781 | 0.008292841 | 0.025708625 | 1 |
| Obesity | adipocyte | *GIMAP4* | 0.242891104 | 0.013684791 | 0.038052313 | 1 |
| Obesity | adipocyte | *TMTC1* | 1.361188809 | 9.69E-05 | 0.000818743 | 0.608966721 |
| Obesity | adipocyte | *IPO8* | 1.70102962 | 0.000222741 | 0.001515558 | 1 |
| Obesity | adipocyte | *LARS* | -0.250788198 | 0.002883605 | 0.011231541 | 1 |
| Obesity | adipocyte | *SWAP70* | 1.642946152 | 1.80E-06 | 5.73E-05 | 0.011342419 |
| Obesity | adipocyte | *SBF2* | 1.262901023 | 7.90E-05 | 0.000709524 | 0.496666883 |
| Obesity | adipocyte | *ELP3* | 1.24760576 | 0.007463948 | 0.023687957 | 1 |
| Obesity | adipocyte | *MRO* | -0.449226418 | 0.010807464 | 0.031515086 | 1 |
| Obesity | adipocyte | *EDEM1* | 1.779218613 | 0.000519144 | 0.00290379 | 1 |
| Obesity | adipocyte | *CNTN6* | -0.469196878 | 0.00363158 | 0.013438341 | 1 |
| Obesity | adipocyte | *KATNBL1* | 1.534875674 | 3.36E-05 | 0.000376993 | 0.211115854 |
| Obesity | adipocyte | *GSTM5* | 0.317766185 | 0.001685074 | 0.007346783 | 1 |
| Obesity | adipocyte | *SYT6* | 2.636001002 | 0.000968994 | 0.004732534 | 1 |
| Obesity | adipocyte | *VAV3* | 0.753858478 | 0.001643492 | 0.007215525 | 1 |
| Obesity | adipocyte | *WNT2B* | 0.730683799 | 0.000552832 | 0.003051495 | 1 |
| Obesity | adipocyte | *NAPG* | 1.181464764 | 0.000884687 | 0.004396859 | 1 |
| Obesity | adipocyte | *PPHLN1* | 0.866653431 | 0.000434725 | 0.002512053 | 1 |
| Obesity | adipocyte | *FST* | 0.962804586 | 0.000306026 | 0.001920414 | 1 |
| Obesity | adipocyte | *NARS* | 1.518151507 | 0.003796323 | 0.013900688 | 1 |
| Obesity | adipocyte | *KIAA1468* | 0.882166875 | 0.013335072 | 0.037310903 | 1 |
| Obesity | adipocyte | *FBXO18* | 1.260064491 | 0.007071939 | 0.022695907 | 1 |
| Obesity | adipocyte | *ECHDC3* | -0.37219519 | 0.010312655 | 0.030424994 | 1 |
| Obesity | adipocyte | *IL15RA* | 2.224513802 | 4.26E-06 | 8.80E-05 | 0.026766147 |
| Obesity | adipocyte | *KLRD1* | 1.58982397 | 0.000267652 | 0.00173349 | 1 |
| Obesity | adipocyte | *DDB2* | 1.351627766 | 7.96E-06 | 0.000136462 | 0.050049864 |
| Obesity | adipocyte | *ACP2* | 1.073092995 | 3.72E-07 | 2.03E-05 | 0.002337483 |
| Obesity | adipocyte | *ZCCHC11* | 1.13765405 | 8.15E-05 | 0.000724371 | 0.512671162 |
| Obesity | adipocyte | *PRPF38A* | 1.197894607 | 0.002105231 | 0.008719096 | 1 |
| Obesity | adipocyte | *APLNR* | 1.255239831 | 0.000149839 | 0.001124589 | 0.942034728 |
| Obesity | adipocyte | *CLOCK* | 1.459023813 | 0.000584394 | 0.003183784 | 1 |
| Obesity | adipocyte | *DZIP1* | 0.344277217 | 0.00253829 | 0.010105344 | 1 |
| Obesity | adipocyte | *STT3A* | 0.906957863 | 0.002014867 | 0.008405753 | 1 |
| Obesity | adipocyte | *WDR36* | 1.698916831 | 0.00027011 | 0.001738461 | 1 |
| Obesity | adipocyte | *OSTF1* | 1.932622584 | 3.65E-06 | 7.91E-05 | 0.022935352 |
| Obesity | adipocyte | *NAA35* | 1.567404308 | 0.000491387 | 0.002773207 | 1 |
| Obesity | adipocyte | *CTSL* | 1.399300664 | 0.000349355 | 0.002099806 | 1 |
| Obesity | adipocyte | *TMEM2* | 1.221727556 | 0.002980138 | 0.011515751 | 1 |

| | | | | | | |
|---|---|---|---|---|---|---|
| Obesity | adipocyte | *AGTPBP1* | 1.050940406 | 5.20E-05 | 0.000525734 | 0.327006747 |
| Obesity | adipocyte | *P2RX4* | 0.720098802 | 3.77E-06 | 8.06E-05 | 0.023689676 |
| Obesity | adipocyte | *TMEM60* | -0.711354032 | 0.000971262 | 0.004737255 | 1 |
| Obesity | adipocyte | *POM121C* | -1.672975368 | 9.06E-12 | 2.48E-09 | 5.70E-08 |
| Obesity | adipocyte | *SRPK2* | 1.431629542 | 3.48E-05 | 0.000387378 | 0.218481445 |
| Obesity | adipocyte | *MTO1* | 0.534849644 | 0.009674549 | 0.02891055 | 1 |
| Obesity | adipocyte | *ANKRD6* | 1.401138562 | 0.000252459 | 0.001661999 | 1 |
| Obesity | adipocyte | *SNX14* | 0.811732499 | 0.017732625 | 0.047119617 | 1 |
| Obesity | adipocyte | *EPHA7* | 0.111458007 | 0.003379877 | 0.012724125 | 1 |
| Obesity | adipocyte | *ATP5G2* | 1.795483189 | 0.000329106 | 0.002012732 | 1 |
| Obesity | adipocyte | *RDH5* | -0.428995422 | 0.003486754 | 0.013017353 | 1 |
| Obesity | adipocyte | *FAIM2* | 3.084315673 | 8.65E-05 | 0.000756031 | 0.543586292 |
| Obesity | adipocyte | *HNRNPA1* | 1.811211391 | 0.000119842 | 0.000963487 | 0.753447118 |
| Obesity | adipocyte | *SLC26A10* | 0.939776048 | 0.012526057 | 0.035457595 | 1 |
| Obesity | adipocyte | *MICAL1* | 0.967703266 | 0.000732982 | 0.003766372 | 1 |
| Obesity | adipocyte | *USP15* | 1.066120872 | 0.000640088 | 0.003410367 | 1 |
| Obesity | adipocyte | *NTPCR* | 0.92692456 | 0.001454866 | 0.006533387 | 1 |
| Obesity | adipocyte | *PIGC* | 0.710663468 | 0.002828927 | 0.011067494 | 1 |
| Obesity | adipocyte | *SP110* | 1.336334255 | 2.03E-06 | 5.99E-05 | 0.012772786 |
| Obesity | adipocyte | *USP37* | 1.836143826 | 1.41E-08 | 1.60E-06 | 8.88E-05 |
| Obesity | adipocyte | *ARMC9* | 1.756433716 | 9.10E-05 | 0.00078262 | 0.572095547 |
| Obesity | adipocyte | *MRPS9* | 0.511457551 | 0.000340809 | 0.002062241 | 1 |
| Obesity | adipocyte | *ANKRD36* | 1.466478769 | 2.09E-05 | 0.000268001 | 0.13152734 |
| Obesity | adipocyte | *APPL2* | 1.438194244 | 0.000221427 | 0.001509879 | 1 |
| Obesity | adipocyte | *SLC41A2* | 1.632739629 | 9.71E-05 | 0.000819547 | 0.610562416 |
| Obesity | adipocyte | *NEK3* | 0.283895477 | 0.016382072 | 0.043995764 | 1 |
| Obesity | adipocyte | *VPS36* | 0.896680117 | 0.000205667 | 0.001429692 | 1 |
| Obesity | adipocyte | *TBC1D4* | 0.901126267 | 0.005342331 | 0.01812587 | 1 |
| Obesity | adipocyte | *LRCH1* | 1.288128375 | 1.57E-05 | 0.000222046 | 0.098810553 |
| Obesity | adipocyte | *MED4* | 0.789404021 | 0.004407865 | 0.015586192 | 1 |
| Obesity | adipocyte | *RCBTB2* | 1.248642269 | 0.002355904 | 0.009543538 | 1 |
| Obesity | adipocyte | *GPNMB* | 0.577117277 | 0.00565541 | 0.01902787 | 1 |
| Obesity | adipocyte | *KDELR2* | 2.319967724 | 3.23E-05 | 0.000365792 | 0.20301444 |
| Obesity | adipocyte | *HUS1* | 0.714287217 | 0.015254217 | 0.041552539 | 1 |
| Obesity | adipocyte | *TM6SF1* | 1.165186536 | 7.56E-07 | 3.22E-05 | 0.004750927 |
| Obesity | adipocyte | *RSAD1* | 1.389214828 | 0.006107923 | 0.020221439 | 1 |
| Obesity | adipocyte | *NMT1* | 0.298983336 | 0.001817783 | 0.007779716 | 1 |
| Obesity | adipocyte | *BLK* | -0.657343421 | 0.006170853 | 0.020397558 | 1 |
| Obesity | adipocyte | *EPRS* | 1.822231318 | 0.000314742 | 0.001953388 | 1 |
| Obesity | adipocyte | *RPS6KC1* | 1.114985032 | 0.002291652 | 0.009349524 | 1 |
| Obesity | adipocyte | *IL1RN* | 2.27136673 | 2.50E-06 | 6.46E-05 | 0.015697441 |
| Obesity | adipocyte | *UGGT1* | 1.730947732 | 0.000295613 | 0.00186974 | 1 |
| Obesity | adipocyte | *GYPC* | 0.677653802 | 0.005921829 | 0.019719564 | 1 |
| Obesity | adipocyte | *ODF2* | 1.8733303 | 0.000237617 | 0.001589256 | 1 |
| Obesity | adipocyte | *STX17* | 1.873304992 | 6.61E-05 | 0.000622148 | 0.415594981 |
| Obesity | adipocyte | *PRPF4* | 0.362301778 | 0.011013786 | 0.032057255 | 1 |
| Obesity | adipocyte | *TEX10* | 1.497193599 | 0.00470462 | 0.016459623 | 1 |
| Obesity | adipocyte | *GARNL3* | 1.774412169 | 7.06E-06 | 0.000126154 | 0.04440636 |
| Obesity | adipocyte | *DSCC1* | 0.111662319 | 0.003278406 | 0.01244646 | 1 |
| Obesity | adipocyte | *NOV* | 0.689793844 | 0.003853739 | 0.014061786 | 1 |

| | | | | | | |
|---|---|---|---|---|---|---|
| Obesity | adipocyte | *TLN1* | 0.901801123 | 0.003621354 | 0.013408393 | 1 |
| Obesity | adipocyte | *CD72* | 0.767223603 | 0.008292043 | 0.025708625 | 1 |
| Obesity | adipocyte | *DENND4C* | 1.404840168 | 2.22E-05 | 0.000278827 | 0.139692459 |
| Obesity | adipocyte | *PPIL1* | 5.420071357 | 2.14E-05 | 0.000271105 | 0.134467845 |
| Obesity | adipocyte | *KIF13A* | 0.936194405 | 0.00927452 | 0.028073619 | 1 |
| Obesity | adipocyte | *ZSCAN9* | 0.698690236 | 1.54E-07 | 1.00E-05 | 0.000969942 |
| Obesity | adipocyte | *SLC22A23* | 0.980424925 | 0.016814744 | 0.045023125 | 1 |
| Obesity | adipocyte | *LRRC1* | 0.485224931 | 0.006403633 | 0.021001378 | 1 |
| Obesity | adipocyte | *RIPK1* | 1.393855676 | 0.002518356 | 0.010039888 | 1 |
| Obesity | adipocyte | *TCF19* | 0.871650567 | 6.02E-07 | 2.68E-05 | 0.003784726 |
| Obesity | adipocyte | *FLOT1* | 1.103900265 | 1.05E-05 | 0.000164341 | 0.066229345 |
| Obesity | adipocyte | *NRM* | 0.592458015 | 8.83E-05 | 0.000764481 | 0.555013293 |
| Obesity | adipocyte | *MTCH1* | 2.036883516 | 2.05E-05 | 0.00026474 | 0.129193011 |
| Obesity | adipocyte | *FAM8A1* | 2.36398941 | 0.000656547 | 0.00344882 | 1 |
| Obesity | adipocyte | *FHDC1* | 3.063547795 | 2.77E-06 | 6.83E-05 | 0.017430301 |
| Obesity | adipocyte | *ARRB1* | 1.237463954 | 0.000322598 | 0.001982574 | 1 |
| Obesity | adipocyte | *PRCP* | 1.130954773 | 4.23E-05 | 0.000450046 | 0.265977443 |
| Obesity | adipocyte | *NARS2* | 1.275093628 | 0.007361664 | 0.02344619 | 1 |
| Obesity | adipocyte | *TTPA* | -0.101365664 | 0.008168689 | 0.025411453 | 1 |
| Obesity | adipocyte | *SULF1* | 0.724671001 | 8.18E-05 | 0.000725583 | 0.514438382 |
| Obesity | adipocyte | *NEK1* | 0.860667502 | 0.007279815 | 0.023247877 | 1 |
| Obesity | adipocyte | *DDX60* | 1.670210281 | 1.56E-06 | 5.34E-05 | 0.009833913 |
| Obesity | adipocyte | *DCUN1D5* | 0.419708057 | 0.017006264 | 0.045419874 | 1 |
| Obesity | adipocyte | *SLC37A4* | 1.706021611 | 0.015983888 | 0.043166111 | 1 |
| Obesity | adipocyte | *FDX1* | 1.001484004 | 0.002480382 | 0.009926263 | 1 |
| Obesity | adipocyte | *FXYD6* | 0.805838541 | 0.004954795 | 0.017153523 | 1 |
| Obesity | adipocyte | *CASP1* | 1.762843375 | 0.000505465 | 0.002837375 | 1 |
| Obesity | adipocyte | *MAP2K5* | 2.159602329 | 1.15E-09 | 1.85E-07 | 7.22E-06 |
| Obesity | adipocyte | *ITGA11* | 0.635134635 | 0.009635236 | 0.028846061 | 1 |
| Obesity | adipocyte | *PAQR5* | 0.853406072 | 0.014987741 | 0.040986485 | 1 |
| Obesity | adipocyte | *UACA* | 1.523116324 | 2.93E-05 | 0.000342653 | 0.184347513 |
| Obesity | adipocyte | *SLC44A5* | 0.682819908 | 0.000270842 | 0.001739311 | 1 |
| Obesity | adipocyte | *RTCA* | 0.442086924 | 0.012265937 | 0.034878312 | 1 |
| Obesity | adipocyte | *IFT172* | 1.823622371 | 1.36E-07 | 9.02E-06 | 0.000856966 |
| Obesity | adipocyte | *ADCY3* | 1.907349224 | 3.30E-06 | 7.45E-05 | 0.02074894 |
| Obesity | adipocyte | *LHCGR* | -0.539504458 | 0.003377861 | 0.012724125 | 1 |
| Obesity | adipocyte | *THUMPD2* | 1.181520799 | 0.002752113 | 0.010847985 | 1 |
| Obesity | adipocyte | *CENPO* | 3.898373674 | 5.92E-06 | 0.000109736 | 0.037201375 |
| Obesity | adipocyte | *LRPPRC* | 1.370537153 | 0.001404221 | 0.006346754 | 1 |
| Obesity | adipocyte | *DTNB* | 0.619447664 | 0.000534767 | 0.002980565 | 1 |
| Obesity | adipocyte | *CALHM2* | 0.436670123 | 0.015483433 | 0.042103955 | 1 |
| Obesity | adipocyte | *DNAJC13* | 1.703585348 | 2.62E-05 | 0.000312547 | 0.164712267 |
| Obesity | adipocyte | *RPS24* | 1.578677873 | 3.70E-05 | 0.000408476 | 0.23242297 |
| Obesity | adipocyte | *SMARCAL1* | 1.377207443 | 0.007599051 | 0.023980537 | 1 |
| Obesity | adipocyte | *BARD1* | 1.623033237 | 0.000309811 | 0.001932321 | 1 |
| Obesity | adipocyte | *STAT4* | 0.399153246 | 0.01451313 | 0.039879392 | 1 |
| Obesity | adipocyte | *CARF* | 0.723082732 | 4.10E-07 | 2.15E-05 | 0.002578178 |
| Obesity | adipocyte | *NAB1* | 1.20502906 | 0.016034842 | 0.043225987 | 1 |
| Obesity | adipocyte | *FAM117B* | 2.024265419 | 4.17E-07 | 2.16E-05 | 0.00262411 |
| Obesity | adipocyte | *SENP7* | 1.220286049 | 0.000175708 | 0.001272669 | 1 |

| | | | | | | |
|---|---|---|---|---|---|---|
| Obesity | adipocyte | *COX17* | 2.175611178 | 1.98E-05 | 0.000256719 | 0.124508524 |
| Obesity | adipocyte | *USP8* | 0.688476859 | 0.00884475 | 0.027072512 | 1 |
| Obesity | adipocyte | *SPPL2A* | 1.058972859 | 0.00202625 | 0.008447635 | 1 |
| Obesity | adipocyte | *UBL7* | 1.279435746 | 4.08E-05 | 0.000438168 | 0.256766427 |
| Obesity | adipocyte | *ARHGAP24* | 1.122556365 | 0.000462424 | 0.002640562 | 1 |
| Obesity | adipocyte | *HERC3* | 1.628333257 | 8.69E-05 | 0.000757 | 0.546554161 |
| Obesity | adipocyte | *PRKG2* | 1.186564833 | 4.13E-05 | 0.000441502 | 0.259394457 |
| Obesity | adipocyte | *BBS7* | 2.162333843 | 6.45E-05 | 0.000613065 | 0.405377499 |
| Obesity | adipocyte | *KIAA1109* | 1.353363142 | 5.00E-05 | 0.000508503 | 0.314254669 |
| Obesity | adipocyte | *PRDM5* | 0.796242745 | 0.008988991 | 0.027447201 | 1 |
| Obesity | adipocyte | *NAAA* | 0.606683853 | 0.005041833 | 0.01737829 | 1 |
| Obesity | adipocyte | *G3BP2* | 0.993195348 | 0.000662532 | 0.003468173 | 1 |
| Obesity | adipocyte | *SCARB2* | 1.449083876 | 0.00054833 | 0.003036273 | 1 |
| Obesity | adipocyte | *ANXA3* | 2.009477593 | 7.75E-07 | 3.27E-05 | 0.00487435 |
| Obesity | adipocyte | *GSTCD* | 0.893373018 | 0.003526264 | 0.013110362 | 1 |
| Obesity | adipocyte | *ENPEP* | 0.840138994 | 0.000639548 | 0.003410367 | 1 |
| Obesity | adipocyte | *ERP27* | 1.351183132 | 0.000464144 | 0.002645577 | 1 |
| Obesity | adipocyte | *ETV6* | 1.461223721 | 8.85E-06 | 0.000144887 | 0.055636644 |
| Obesity | adipocyte | *PRICKLE1* | 0.697683921 | 0.008342528 | 0.025849914 | 1 |
| Obesity | adipocyte | *CLSTN3* | 0.515412472 | 0.015773848 | 0.042690565 | 1 |
| Obesity | adipocyte | *KLRG1* | 0.397419604 | 0.000205801 | 0.001429692 | 1 |
| Obesity | adipocyte | *CD27* | 0.887349049 | 0.006731422 | 0.021823388 | 1 |
| Obesity | adipocyte | *GLIPR1* | 0.679593288 | 0.002372875 | 0.009587574 | 1 |
| Obesity | adipocyte | *POC1B* | 1.997330836 | 9.98E-05 | 0.000837425 | 0.627430423 |
| Obesity | adipocyte | *GAS2L3* | 2.219210615 | 2.93E-06 | 6.99E-05 | 0.018423255 |
| Obesity | adipocyte | *RITA1* | -0.280736868 | 0.004311399 | 0.01532265 | 1 |
| Obesity | adipocyte | *MTMR6* | 0.586137127 | 0.017861929 | 0.047383099 | 1 |
| Obesity | adipocyte | *BRCA2* | 1.850591742 | 6.88E-05 | 0.000641238 | 0.432835646 |
| Obesity | adipocyte | *CERS5* | 0.923094615 | 0.003137109 | 0.012033561 | 1 |
| Obesity | adipocyte | *HNRNPA1L2* | 1.021892822 | 0.013520649 | 0.037712654 | 1 |
| Obesity | adipocyte | *LPAR6* | 1.450777028 | 4.96E-05 | 0.000508417 | 0.311806067 |
| Obesity | adipocyte | *RHOF* | -0.846093896 | 4.44E-05 | 0.000468253 | 0.279078513 |
| Obesity | adipocyte | *TMX1* | 2.079785223 | 7.70E-05 | 0.000696989 | 0.484407301 |
| Obesity | adipocyte | *RTN1* | 0.786230877 | 0.013007867 | 0.036558095 | 1 |
| Obesity | adipocyte | *STON2* | 1.124540116 | 3.04E-05 | 0.000351403 | 0.191163239 |
| Obesity | adipocyte | *PTGR2* | -0.192283593 | 0.00197193 | 0.008281581 | 1 |
| Obesity | adipocyte | *AK7* | 5.554737343 | 3.05E-06 | 7.08E-05 | 0.019198263 |
| Obesity | adipocyte | *FBLN5* | 1.852739533 | 7.59E-07 | 3.22E-05 | 0.004771611 |
| Obesity | adipocyte | *NIPA2* | 1.070552485 | 0.009984779 | 0.029680521 | 1 |
| Obesity | adipocyte | *TCF12* | 1.296410527 | 2.84E-05 | 0.000333654 | 0.178838655 |
| Obesity | adipocyte | *SORD* | 1.042163838 | 0.001644699 | 0.007215789 | 1 |
| Obesity | adipocyte | *SRP14* | 0.625281394 | 0.016034477 | 0.043225987 | 1 |
| Obesity | adipocyte | *UBE2Q2* | 1.343607675 | 0.000476317 | 0.002700273 | 1 |
| Obesity | adipocyte | *HMG20A* | 1.12457511 | 0.000599769 | 0.003245577 | 1 |
| Obesity | adipocyte | *TPM1* | 1.152391825 | 5.08E-05 | 0.000514858 | 0.319211935 |
| Obesity | adipocyte | *IGF1R* | 1.259807852 | 2.48E-06 | 6.45E-05 | 0.015601967 |
| Obesity | adipocyte | *BBS4* | 1.646823731 | 2.64E-05 | 0.000314796 | 0.166212452 |
| Obesity | adipocyte | *SCAMP2* | 0.80245891 | 0.000608541 | 0.003279449 | 1 |
| Obesity | adipocyte | *POLG* | 1.092991795 | 0.01179186 | 0.033697919 | 1 |
| Obesity | adipocyte | *WDR93* | -0.244733837 | 5.45E-05 | 0.000541676 | 0.34270666 |

| | | | | | | |
|---|---|---|---|---|---|---|
| Obesity | adipocyte | *IQGAP1* | 0.510728641 | 0.007315707 | 0.023323452 | 1 |
| Obesity | adipocyte | *SH3GL3* | -0.457235565 | 9.52E-05 | 0.000812388 | 0.59840218 |
| Obesity | adipocyte | *SEC11A* | 1.084472261 | 0.012972944 | 0.036492572 | 1 |
| Obesity | adipocyte | *FTO* | 0.928810191 | 0.008110558 | 0.025275119 | 1 |
| Obesity | adipocyte | *ZFHX3* | 1.451232062 | 4.96E-05 | 0.000508417 | 0.311653511 |
| Obesity | adipocyte | *CMTM3* | 2.694125887 | 1.48E-06 | 5.22E-05 | 0.009284559 |
| Obesity | adipocyte | *TLDC1* | 1.473675789 | 0.003777042 | 0.013870509 | 1 |
| Obesity | adipocyte | *NDUFB10* | 2.142289916 | 2.53E-05 | 0.000304601 | 0.159001497 |
| Obesity | adipocyte | *MYO19* | 2.571859965 | 1.49E-13 | 5.51E-11 | 9.36E-10 |
| Obesity | adipocyte | *VPS53* | 0.566592982 | 0.009590258 | 0.028738777 | 1 |
| Obesity | adipocyte | *NPEPPS* | 1.545433635 | 0.000123029 | 0.000984074 | 0.773481779 |
| Obesity | adipocyte | *ABCA8* | 0.364431177 | 0.007432617 | 0.023613269 | 1 |
| Obesity | adipocyte | *IMPA2* | 2.215303642 | 0.000252093 | 0.001661329 | 1 |
| Obesity | adipocyte | *GNAL* | 0.993169744 | 0.013692047 | 0.03805566 | 1 |
| Obesity | adipocyte | *GALNT1* | 1.046715291 | 0.001182524 | 0.005486735 | 1 |
| Obesity | adipocyte | *ASXL3* | 1.347639683 | 0.005963347 | 0.019836807 | 1 |
| Obesity | adipocyte | *ESCO1* | 1.346989359 | 0.001008489 | 0.004880963 | 1 |
| Obesity | adipocyte | *C18orf8* | -1.265689024 | 9.49E-08 | 6.86E-06 | 0.000596599 |
| Obesity | adipocyte | *NPC1* | -0.995790238 | 0.000569494 | 0.003129728 | 1 |
| Obesity | adipocyte | *CARD14* | 2.466077687 | 0.001008479 | 0.004880963 | 1 |
| Obesity | adipocyte | *TRIM65* | 0.962780088 | 1.08E-07 | 7.61E-06 | 0.000677465 |
| Obesity | adipocyte | *SECTM1* | 0.873092355 | 0.015621388 | 0.042314375 | 1 |
| Obesity | adipocyte | *RNF165* | 1.33232944 | 5.66E-05 | 0.000555602 | 0.355585408 |
| Obesity | adipocyte | *PIP4K2B* | 1.049908881 | 0.005064426 | 0.017437047 | 1 |
| Obesity | adipocyte | *MVB12A* | 0.483850307 | 0.013861227 | 0.038390103 | 1 |
| Obesity | adipocyte | *CCDC97* | 1.905571985 | 0.000339946 | 0.002060983 | 1 |
| Obesity | adipocyte | *SIRT3* | 0.503435106 | 0.011947752 | 0.03408145 | 1 |
| Obesity | adipocyte | *IFITM3* | 2.105207082 | 4.64E-06 | 9.38E-05 | 0.029170456 |
| Obesity | adipocyte | *DOPEY2* | 0.679894504 | 0.010601208 | 0.031115683 | 1 |
| Obesity | adipocyte | *AKT1* | 2.406863265 | 0.001177307 | 0.005466564 | 1 |
| Obesity | adipocyte | *MYO1F* | 0.929637455 | 7.74E-05 | 0.000699321 | 0.486727265 |
| Obesity | adipocyte | *CACNG8* | 5.597506305 | 5.78E-07 | 2.62E-05 | 0.003635591 |
| Obesity | adipocyte | *RPS11* | 1.519044094 | 7.47E-08 | 5.59E-06 | 0.000469604 |
| Obesity | adipocyte | *RCN3* | 2.066922206 | 1.69E-06 | 5.56E-05 | 0.010627273 |
| Obesity | adipocyte | *RERE* | 0.73909407 | 0.001196756 | 0.005528293 | 1 |
| Obesity | adipocyte | *SH3BGRL3* | 1.795629154 | 3.10E-06 | 7.16E-05 | 0.019469043 |
| Obesity | adipocyte | *SERBP1* | 1.593445172 | 0.000430857 | 0.002494286 | 1 |
| Obesity | adipocyte | *PIGK* | 1.407620346 | 0.004008722 | 0.014489576 | 1 |
| Obesity | adipocyte | *RPS8* | 0.918539839 | 5.25E-05 | 0.000528075 | 0.330047135 |
| Obesity | adipocyte | *MOB3C* | 1.333209316 | 0.018516995 | 0.048811885 | 1 |
| Obesity | adipocyte | *GPR161* | 1.665860945 | 3.60E-06 | 7.87E-05 | 0.022631993 |
| Obesity | adipocyte | *TIPRL* | 2.207165964 | 1.56E-06 | 5.34E-05 | 0.009823182 |
| Obesity | adipocyte | *TMCO1* | 1.648933674 | 0.000650499 | 0.003436811 | 1 |
| Obesity | adipocyte | *MGST3* | 0.999198583 | 0.011167774 | 0.032370585 | 1 |
| Obesity | adipocyte | *RFWD2* | 1.073802133 | 0.000488877 | 0.002761519 | 1 |
| Obesity | adipocyte | *RGS5* | 0.575127136 | 0.009308047 | 0.028148 | 1 |
| Obesity | adipocyte | *RGL1* | 1.36565398 | 1.60E-05 | 0.000224391 | 0.100302613 |
| Obesity | adipocyte | *TUFT1* | -0.392965544 | 0.000968234 | 0.004732534 | 1 |
| Obesity | adipocyte | *SNX27* | 1.491966069 | 0.000738468 | 0.00378696 | 1 |
| Obesity | adipocyte | *ENSA* | 1.827675858 | 2.16E-05 | 0.000273167 | 0.13603717 |

| | | | | | | |
|---|---|---|---|---|---|---|
| Obesity | adipocyte | *SEMA6C* | 1.927680447 | 7.77E-08 | 5.75E-06 | 0.000488787 |
| Obesity | adipocyte | *ARNT* | 0.680209885 | 0.010116771 | 0.030001951 | 1 |
| Obesity | adipocyte | *POGZ* | 0.835200309 | 0.000105348 | 0.000871474 | 0.66232056 |
| Obesity | adipocyte | *IKBKE* | 1.349055628 | 0.004262107 | 0.015190401 | 1 |
| Obesity | adipocyte | *TP53BP2* | 2.052191288 | 9.59E-06 | 0.000153475 | 0.060315631 |
| Obesity | adipocyte | *HAX1* | 1.134365471 | 3.95E-05 | 0.00042694 | 0.248479304 |
| Obesity | adipocyte | *KCNN3* | 1.510508644 | 3.86E-06 | 8.20E-05 | 0.024260673 |
| Obesity | adipocyte | *RIT1* | 1.694679651 | 0.000137806 | 0.001055284 | 0.866388112 |
| Obesity | adipocyte | *GALNT2* | 1.235965319 | 0.006064565 | 0.020109664 | 1 |
| Obesity | adipocyte | *CEP170* | 1.472780093 | 2.34E-08 | 2.33E-06 | 0.000147109 |
| Obesity | adipocyte | *NVL* | 1.411779755 | 0.002888001 | 0.011231541 | 1 |
| Obesity | adipocyte | *FBXO28* | 1.54866986 | 0.001055161 | 0.005056246 | 1 |
| Obesity | adipocyte | *CNIH4* | 1.573387448 | 0.003181591 | 0.012174475 | 1 |
| Obesity | adipocyte | *GUK1* | 2.732190267 | 7.99E-09 | 1.02E-06 | 5.02E-05 |
| Obesity | adipocyte | *CDC42BPA* | 1.314717691 | 4.19E-07 | 2.16E-05 | 0.00263289 |
| Obesity | adipocyte | *CNIH3* | 0.899535589 | 0.002581245 | 0.010243714 | 1 |
| Obesity | adipocyte | *WNT9A* | 0.522051763 | 0.006466077 | 0.021140003 | 1 |
| Obesity | adipocyte | *PLEKHA6* | 0.854053313 | 0.000597644 | 0.003241921 | 1 |
| Obesity | adipocyte | *EML4* | 1.546827005 | 5.65E-05 | 0.000555602 | 0.355455638 |
| Obesity | adipocyte | *ASXL2* | 0.899638908 | 0.012721327 | 0.035881106 | 1 |
| Obesity | adipocyte | *EXOC6B* | 1.25170522 | 0.000129477 | 0.001012841 | 0.814022592 |
| Obesity | adipocyte | *TEX261* | 3.297266993 | 0.000139494 | 0.001060461 | 0.87700155 |
| Obesity | adipocyte | *RALB* | 0.647417351 | 0.018837221 | 0.049552138 | 1 |
| Obesity | adipocyte | *SLC20A1* | 1.180193399 | 0.001856083 | 0.007915783 | 1 |
| Obesity | adipocyte | *ZC3H8* | 0.76793895 | 0.000688184 | 0.003574776 | 1 |
| Obesity | adipocyte | *FAHD2B* | -0.529199863 | 0.004326635 | 0.015368109 | 1 |
| Obesity | adipocyte | *AFF3* | -0.333104412 | 0.003956285 | 0.01435434 | 1 |
| Obesity | adipocyte | *UBXN4* | 1.399191977 | 7.63E-05 | 0.000691526 | 0.479919098 |
| Obesity | adipocyte | *ZNF385B* | 0.714271393 | 0.000147865 | 0.001111996 | 0.92962892 |
| Obesity | adipocyte | *PHOSPHO2* | -0.162552501 | 0.017915137 | 0.047484178 | 1 |
| Obesity | adipocyte | *GULP1* | 1.021786856 | 4.01E-06 | 8.48E-05 | 0.025196611 |
| Obesity | adipocyte | *FAM171B* | 0.50333891 | 0.001650835 | 0.007232613 | 1 |
| Obesity | adipocyte | *HSPD1* | 2.010499945 | 3.01E-06 | 7.03E-05 | 0.018949345 |
| Obesity | adipocyte | *METTL21A* | 2.091782427 | 1.41E-05 | 0.000204947 | 0.088946801 |
| Obesity | adipocyte | *NBEAL1* | 0.467679331 | 2.61E-06 | 6.56E-05 | 0.016394307 |
| Obesity | adipocyte | *SUMF1* | 1.026220058 | 0.000491929 | 0.002773773 | 1 |
| Obesity | adipocyte | *FANCD2* | 2.392710334 | 1.37E-05 | 0.000201001 | 0.086229556 |
| Obesity | adipocyte | *TAMM41* | 1.415608981 | 7.97E-05 | 0.000713463 | 0.500851296 |
| Obesity | adipocyte | *OSBPL10* | 0.82269351 | 7.05E-05 | 0.000653897 | 0.443342325 |
| Obesity | adipocyte | *CTDSPL* | 1.013984068 | 0.000452206 | 0.002591628 | 1 |
| Obesity | adipocyte | *STAC* | 2.521402985 | 1.33E-05 | 0.000196414 | 0.083504106 |
| Obesity | adipocyte | *ARL6IP5* | 1.345991194 | 0.002223976 | 0.009120767 | 1 |
| Obesity | adipocyte | *TMF1* | 0.995068982 | 0.017796863 | 0.047250372 | 1 |
| Obesity | adipocyte | *LIMD1* | 1.451020517 | 1.03E-05 | 0.000161981 | 0.064792544 |
| Obesity | adipocyte | *NXPE3* | 1.180427803 | 0.000232229 | 0.001563192 | 1 |
| Obesity | adipocyte | *RABL3* | 1.62512877 | 0.001664176 | 0.007275853 | 1 |
| Obesity | adipocyte | *BOC* | 0.56408652 | 0.006068649 | 0.020112597 | 1 |
| Obesity | adipocyte | *AGTR1* | 0.50760096 | 0.009045441 | 0.027566014 | 1 |
| Obesity | adipocyte | *AMT* | -0.176958958 | 0.003990783 | 0.014436165 | 1 |
| Obesity | adipocyte | *LYAR* | 2.896389498 | 5.50E-05 | 0.000544919 | 0.346023359 |

| | | | | | | |
|---|---|---|---|---|---|---|
| Obesity | adipocyte | *CENPC* | -0.466371263 | 0.006704913 | 0.02175118 | 1 |
| Obesity | adipocyte | *USP53* | 1.370529832 | 8.80E-05 | 0.000763508 | 0.552962258 |
| Obesity | adipocyte | *PDGFC* | 0.785058924 | 0.012346584 | 0.035028418 | 1 |
| Obesity | adipocyte | *CBR4* | 1.231955415 | 0.00164115 | 0.00721028 | 1 |
| Obesity | adipocyte | *LHFPL2* | 1.68958593 | 1.56E-05 | 0.000220887 | 0.098073754 |
| Obesity | adipocyte | *SSBP2* | 1.335987673 | 2.55E-05 | 0.00030701 | 0.160566027 |
| Obesity | adipocyte | *TSLP* | -0.249387911 | 0.012597544 | 0.035627871 | 1 |
| Obesity | adipocyte | *FEM1C* | 1.742626474 | 4.87E-05 | 0.000503368 | 0.306047769 |
| Obesity | adipocyte | *YIPF5* | 1.7110658 | 0.003028719 | 0.011681935 | 1 |
| Obesity | adipocyte | *SLC25A48* | -0.281061884 | 0.001969318 | 0.008276139 | 1 |
| Obesity | adipocyte | *PCYOX1L* | -0.41296418 | 0.007284606 | 0.023247877 | 1 |
| Obesity | adipocyte | *TBC1D7* | 1.400007149 | 0.002013234 | 0.008404518 | 1 |
| Obesity | adipocyte | *GFOD1* | 0.301951143 | 0.002537368 | 0.010105344 | 1 |
| Obesity | adipocyte | *KLHL3* | 0.520217615 | 0.008055139 | 0.025170309 | 1 |
| Obesity | adipocyte | *ABT1* | 0.634942061 | 0.000121056 | 0.000969526 | 0.761077887 |
| Obesity | adipocyte | *FGD2* | 1.137877599 | 0.000111193 | 0.000909061 | 0.699067731 |
| Obesity | adipocyte | *RPL7L1* | 0.57820069 | 0.001333731 | 0.006076207 | 1 |
| Obesity | adipocyte | *TPBG* | 1.962465478 | 2.37E-06 | 6.25E-05 | 0.014925632 |
| Obesity | adipocyte | *IRAK1BP1* | 1.511429635 | 0.001557113 | 0.006933124 | 1 |
| Obesity | adipocyte | *PHIP* | 1.543628178 | 6.78E-06 | 0.000121491 | 0.04264346 |
| Obesity | adipocyte | *PM20D2* | 1.543272226 | 0.005715962 | 0.019155787 | 1 |
| Obesity | adipocyte | *RNF217* | 1.03627235 | 0.014120491 | 0.038936635 | 1 |
| Obesity | adipocyte | *AIG1* | 0.54787017 | 0.013610183 | 0.037895138 | 1 |
| Obesity | adipocyte | *DYNLT1* | 0.959855972 | 0.003197785 | 0.012214139 | 1 |
| Obesity | adipocyte | *GIGYF1* | 0.772785078 | 0.012446184 | 0.035279153 | 1 |
| Obesity | adipocyte | *NOM1* | 1.509654303 | 0.007282138 | 0.023247877 | 1 |
| Obesity | adipocyte | *FBXO25* | 1.238753992 | 0.009584383 | 0.028734868 | 1 |
| Obesity | adipocyte | *CSGALNACT1* | 0.670317025 | 0.000310359 | 0.001933258 | 1 |
| Obesity | adipocyte | *SLC25A37* | 1.301333933 | 0.002368134 | 0.009574572 | 1 |
| Obesity | adipocyte | *DOCK5* | 1.631121164 | 5.01E-06 | 9.72E-05 | 0.03148026 |
| Obesity | adipocyte | *TACC1* | 1.169017513 | 5.27E-05 | 0.000529324 | 0.331356802 |
| Obesity | adipocyte | *ADHFE1* | -0.159967241 | 0.001288508 | 0.005908718 | 1 |
| Obesity | adipocyte | *ATP6V0D2* | -0.504445515 | 0.000854673 | 0.004271327 | 1 |
| Obesity | adipocyte | *MTDH* | 1.496813886 | 0.000135845 | 0.001045361 | 0.854060038 |
| Obesity | adipocyte | *FAM135B* | 2.207350115 | 0.004404796 | 0.015584105 | 1 |
| Obesity | adipocyte | *PLIN2* | 1.125062152 | 0.018285685 | 0.048267098 | 1 |
| Obesity | adipocyte | *HAUS6* | 1.823136337 | 0.002430288 | 0.009783802 | 1 |
| Obesity | adipocyte | *ZNF462* | 1.751570872 | 0.000137333 | 0.001054227 | 0.863412094 |
| Obesity | adipocyte | *ASTN2* | 1.520271614 | 0.000250952 | 0.001655548 | 1 |
| Obesity | adipocyte | *CIZ1* | 0.911393317 | 0.001016936 | 0.004899217 | 1 |
| Obesity | adipocyte | *SH3GLB2* | 1.558099939 | 2.27E-05 | 0.000282578 | 0.142419206 |
| Obesity | adipocyte | *LRSAM1* | 0.918594804 | 0.012747059 | 0.035937561 | 1 |
| Obesity | adipocyte | *DPH7* | 1.682444923 | 0.000523682 | 0.002921374 | 1 |
| Obesity | adipocyte | *RSU1* | 1.406995267 | 6.15E-05 | 0.00059232 | 0.386784834 |
| Obesity | adipocyte | *PARD3* | 0.535849983 | 0.004708022 | 0.016462367 | 1 |
| Obesity | adipocyte | *NRBF2* | -0.373112954 | 0.001209299 | 0.005578036 | 1 |
| Obesity | adipocyte | *ADIRF* | 1.381851827 | 0.000994805 | 0.004825878 | 1 |
| Obesity | adipocyte | *GLUD1* | 1.472189881 | 1.74E-05 | 0.000237294 | 0.109155354 |
| Obesity | adipocyte | *HTR7* | 2.527469772 | 5.64E-06 | 0.000105498 | 0.035447271 |
| Obesity | adipocyte | *MTG1* | 0.342577719 | 0.000426183 | 0.002469507 | 1 |

| | | | | | | |
|---|---|---|---|---|---|---|
| Obesity | adipocyte | *PAOX* | 2.683073269 | 0.002812652 | 0.011023896 | 1 |
| Obesity | adipocyte | *GSTO1* | 1.074043566 | 0.00375159 | 0.013809276 | 1 |
| Obesity | adipocyte | *RGS10* | 1.242709874 | 0.000382126 | 0.002267667 | 1 |
| Obesity | adipocyte | *GAS2* | 2.160214328 | 0.00180598 | 0.007734464 | 1 |
| Obesity | adipocyte | *HSD17B12* | -0.466846052 | 2.60E-10 | 4.66E-08 | 1.63E-06 |
| Obesity | adipocyte | *PTPRJ* | 0.665195881 | 0.008097276 | 0.025251774 | 1 |
| Obesity | adipocyte | *C11orf49* | 0.842062141 | 0.004795566 | 0.016694199 | 1 |
| Obesity | adipocyte | *ENDOD1* | 2.048969136 | 1.93E-06 | 5.81E-05 | 0.012140825 |
| Obesity | adipocyte | *CAPN5* | 1.058471252 | 0.000131704 | 0.001022252 | 0.828024436 |
| Obesity | adipocyte | *ZC3H12C* | 1.865072991 | 5.93E-06 | 0.000109736 | 0.037310122 |
| Obesity | adipocyte | *TTC12* | 0.300226406 | 2.21E-05 | 0.000277513 | 0.138655424 |
| Obesity | adipocyte | *C11orf52* | -3.273923299 | 0.001552258 | 0.006916406 | 1 |
| Obesity | adipocyte | *INCENP* | 2.966567471 | 0.001795935 | 0.007696689 | 1 |
| Obesity | adipocyte | *PLCH2* | 13.97134 | 7.31E-09 | 9.57E-07 | 4.59E-05 |
| Obesity | adipocyte | *CPSF7* | 1.324036919 | 0.008082351 | 0.025217737 | 1 |
| Obesity | adipocyte | *CCDC15* | -0.505591479 | 3.86E-05 | 0.000421087 | 0.242940825 |
| Obesity | adipocyte | *FEZ1* | 1.863106725 | 9.69E-05 | 0.000818743 | 0.609145135 |
| Obesity | adipocyte | *DUSP15* | -0.508211009 | 0.013366117 | 0.037364506 | 1 |
| Obesity | adipocyte | *COMMD7* | 1.708353606 | 0.004473868 | 0.015775215 | 1 |
| Obesity | adipocyte | *TRPT1* | -0.953048852 | 0.000315215 | 0.001954398 | 1 |
| Obesity | adipocyte | *TBX6* | 0.449287352 | 5.33E-07 | 2.55E-05 | 0.003352038 |
| Obesity | adipocyte | *DOC2A* | -1.74477121 | 3.04E-15 | 1.47E-12 | 1.91E-11 |
| Obesity | adipocyte | *TAOK2* | 1.398472224 | 0.000352736 | 0.00211205 | 1 |
| Obesity | adipocyte | *ARID5B* | 0.675961959 | 0.004338428 | 0.015401296 | 1 |
| Obesity | adipocyte | *TMCO3* | 2.02790688 | 8.28E-06 | 0.000139179 | 0.052052947 |
| Obesity | adipocyte | *SAP18* | 1.591784716 | 0.001851541 | 0.007902674 | 1 |
| Obesity | adipocyte | *LYPD6B* | -1.00798093 | 0.005659634 | 0.01902787 | 1 |
| Obesity | adipocyte | *PDCD4* | 1.637346926 | 1.18E-05 | 0.000178905 | 0.074119422 |
| Obesity | adipocyte | *WDR17* | 0.849719752 | 0.015099631 | 0.041238653 | 1 |
| Obesity | adipocyte | *CCDC102B* | 1.550175577 | 6.74E-05 | 0.000631485 | 0.423726409 |
| Obesity | adipocyte | *FSIP1* | 1.6056455 | 0.001284305 | 0.005893739 | 1 |
| Obesity | adipocyte | *RGS18* | 2.220864043 | 1.12E-06 | 4.24E-05 | 0.007043085 |
| Obesity | adipocyte | *DOCK1* | 0.703634405 | 0.012846397 | 0.036158155 | 1 |
| Obesity | adipocyte | *PIP4K2A* | 0.744892761 | 0.00505319 | 0.017407894 | 1 |
| Obesity | adipocyte | *RILPL2* | 0.525359784 | 0.001087135 | 0.005170057 | 1 |
| Obesity | adipocyte | *DHX37* | 2.628379259 | 0.000552224 | 0.003050819 | 1 |
| Obesity | adipocyte | *PRSS53* | -0.948003493 | 9.34E-07 | 3.74E-05 | 0.005873698 |
| Obesity | adipocyte | *UBE3B* | 0.446269814 | 0.003515636 | 0.01307858 | 1 |
| Obesity | adipocyte | *IPMK* | 1.872626076 | 4.41E-06 | 9.06E-05 | 0.027724719 |
| Obesity | adipocyte | *PLBD2* | 0.501061884 | 0.015268812 | 0.041574283 | 1 |
| Obesity | adipocyte | *GXYLT1* | 1.682799597 | 0.000183951 | 0.001310318 | 1 |
| Obesity | adipocyte | *FAM177A1* | 2.086028613 | 5.05E-07 | 2.46E-05 | 0.003175307 |
| Obesity | adipocyte | *MIPOL1* | 0.737550299 | 0.00065667 | 0.00344882 | 1 |
| Obesity | adipocyte | *TMEM18* | 1.725722202 | 0.002028527 | 0.008451522 | 1 |
| Obesity | adipocyte | *ANKRD50* | 2.139086101 | 1.87E-05 | 0.000247605 | 0.117333728 |
| Obesity | adipocyte | *PTPRO* | 1.173190516 | 0.000150804 | 0.001127351 | 0.948102343 |
| Obesity | adipocyte | *VPS26B* | 1.120815903 | 0.00932927 | 0.028185066 | 1 |
| Obesity | adipocyte | *VTI1A* | 1.074655059 | 8.29E-05 | 0.000731757 | 0.521010873 |
| Obesity | adipocyte | *ANO4* | 0.815898365 | 0.000414236 | 0.002424589 | 1 |
| Obesity | adipocyte | *DPYSL4* | 0.212804294 | 0.000298047 | 0.001881346 | 1 |

| | | | | | | |
|---|---|---|---|---|---|---|
| Obesity | adipocyte | *INPP1* | 1.171442461 | 0.000138876 | 0.001057037 | 0.873112893 |
| Obesity | adipocyte | *MFSD6* | 1.782734018 | 3.56E-05 | 0.000394699 | 0.223794319 |
| Obesity | adipocyte | *ASAP2* | 1.326421035 | 9.96E-05 | 0.000837425 | 0.626407078 |
| Obesity | adipocyte | *ADAM17* | 1.486101928 | 0.000425945 | 0.002469507 | 1 |
| Obesity | adipocyte | *BICD1* | 1.34115317 | 0.000123557 | 0.000987041 | 0.776801445 |
| Obesity | adipocyte | *CACUL1* | 1.271502123 | 0.002433615 | 0.009783802 | 1 |
| Obesity | adipocyte | *BEND6* | -0.207647255 | 0.008564609 | 0.026394951 | 1 |
| Obesity | adipocyte | *RABGAP1L* | 1.32908113 | 2.20E-08 | 2.30E-06 | 0.000138282 |
| Obesity | adipocyte | *PTPN14* | 0.596749526 | 0.003150636 | 0.01207808 | 1 |
| Obesity | adipocyte | *RNF219* | 1.276029091 | 0.009118936 | 0.027696015 | 1 |
| Obesity | adipocyte | *ARL14EP* | -0.64211699 | 0.012152345 | 0.034617939 | 1 |
| Obesity | adipocyte | *G6PC2* | 45.57644682 | 1.06E-05 | 0.000165348 | 0.066800771 |
| Obesity | adipocyte | *TGOLN2* | 1.081911581 | 0.00392705 | 0.014279562 | 1 |
| Obesity | adipocyte | *POC5* | -0.7080478 | 3.29E-05 | 0.000370365 | 0.206863808 |
| Obesity | adipocyte | *TADA1* | 1.731523206 | 0.012573926 | 0.035577081 | 1 |
| Obesity | adipocyte | *CWF19L2* | 1.626892705 | 1.61E-06 | 5.39E-05 | 0.010125911 |
| Obesity | adipocyte | *ZFP36L2* | 1.624120036 | 1.01E-06 | 3.93E-05 | 0.006368747 |
| Obesity | adipocyte | *TMEM123* | 1.776837193 | 9.10E-06 | 0.00014821 | 0.057208901 |
| Obesity | adipocyte | *SPARCL1* | 0.430781878 | 0.001129986 | 0.005297703 | 1 |
| Obesity | adipocyte | *MBNL1* | 1.014142654 | 0.000150409 | 0.001125743 | 0.945624387 |
| Obesity | adipocyte | *CLEC4F* | 3.20711978 | 1.78E-05 | 0.000240148 | 0.111908846 |
| Obesity | adipocyte | *RASGRP3* | 0.240844318 | 0.00996454 | 0.029634372 | 1 |
| Obesity | adipocyte | *TCTEX1D1* | 0.297796444 | 0.000962265 | 0.004711651 | 1 |
| Obesity | adipocyte | *HHEX* | 0.500054888 | 0.00935117 | 0.028224104 | 1 |
| Obesity | adipocyte | *RAB3C* | 0.501745456 | 0.003169141 | 0.01213422 | 1 |
| Obesity | adipocyte | *MED21* | 1.555762251 | 0.0047323 | 0.016510528 | 1 |
| Obesity | adipocyte | *CARHSP1* | 1.035737442 | 0.011042535 | 0.032111202 | 1 |
| Obesity | adipocyte | *CLGN* | 7.346408156 | 0.016983916 | 0.045382342 | 1 |
| Obesity | adipocyte | *AHCTF1* | 1.441930939 | 0.000998864 | 0.004841832 | 1 |
| Obesity | adipocyte | *PTPRR* | 1.542824232 | 8.01E-05 | 0.000715194 | 0.503307866 |
| Obesity | adipocyte | *FRMD1* | -0.67027581 | 0.003461801 | 0.012962681 | 1 |
| Obesity | adipocyte | *ASAP1* | 0.955487431 | 0.000605337 | 0.003266744 | 1 |
| Obesity | adipocyte | *LPCAT1* | 1.980132517 | 3.45E-06 | 7.67E-05 | 0.021695779 |
| Obesity | adipocyte | *TUBGCP5* | 1.471740594 | 0.005301664 | 0.018026804 | 1 |
| Obesity | adipocyte | *CFDP1* | 1.501327263 | 0.000275354 | 0.00176288 | 1 |
| Obesity | adipocyte | *ZDHHC7* | 1.377842388 | 0.000154339 | 0.001145894 | 0.970328731 |
| Obesity | adipocyte | *FAM92B* | -9.494075523 | 0.006764684 | 0.02188861 | 1 |
| Obesity | adipocyte | *JAZF1* | 1.549684559 | 3.80E-09 | 5.19E-07 | 2.39E-05 |
| Obesity | adipocyte | *PPP2R5E* | 0.695539742 | 0.001139412 | 0.00533039 | 1 |
| Obesity | adipocyte | *ANKRD29* | 0.62674177 | 0.004717503 | 0.016486349 | 1 |
| Obesity | adipocyte | *THY1* | 1.014490237 | 0.001118587 | 0.005264294 | 1 |
| Obesity | adipocyte | *UBASH3B* | 1.481645318 | 5.45E-05 | 0.000541676 | 0.34288092 |
| Obesity | adipocyte | *ROBO3* | 0.614210741 | 0.00235294 | 0.009540644 | 1 |
| Obesity | adipocyte | *CC2D1B* | 0.344780324 | 0.006234677 | 0.020576069 | 1 |
| Obesity | adipocyte | *PRKCA* | 1.581038544 | 3.04E-07 | 1.72E-05 | 0.001911577 |
| Obesity | adipocyte | *LRRK1* | 1.587244949 | 2.40E-06 | 6.25E-05 | 0.015065625 |
| Obesity | adipocyte | *CEP112* | 0.731587722 | 3.04E-05 | 0.000351403 | 0.190890686 |
| Obesity | adipocyte | *FAM167A* | 0.92485423 | 0.004496283 | 0.015836489 | 1 |
| Obesity | adipocyte | *NEIL2* | 0.392872309 | 0.000653629 | 0.003444566 | 1 |
| Obesity | adipocyte | *GBP5* | 1.182931242 | 4.53E-06 | 9.25E-05 | 0.028493068 |

| | | | | | | |
|---|---|---|---|---|---|---|
| Obesity | adipocyte | *PDLIM3* | 0.58130726 | 0.000671099 | 0.003501409 | 1 |
| Obesity | adipocyte | *L3MBTL4* | 1.609775933 | 1.41E-05 | 0.000204947 | 0.088577313 |
| Obesity | adipocyte | *PDE1C* | 1.65873405 | 5.77E-07 | 2.62E-05 | 0.003627966 |
| Obesity | adipocyte | *RABGEF1* | -0.179631691 | 0.00765675 | 0.024117228 | 1 |
| Obesity | adipocyte | *FLCN* | 0.538034791 | 0.006679673 | 0.021685095 | 1 |
| Obesity | adipocyte | *DPH3* | 2.173110683 | 0.007724469 | 0.024306176 | 1 |
| Obesity | adipocyte | *PPP4R1* | 1.881275199 | 1.04E-05 | 0.000163505 | 0.065565668 |
| Obesity | adipocyte | *PROM2* | -1.547249605 | 0.014980172 | 0.040983612 | 1 |
| Obesity | adipocyte | *AK9* | 0.669074281 | 0.007891867 | 0.024733883 | 1 |
| Obesity | adipocyte | *CDK19* | 1.359435684 | 0.000186105 | 0.001322079 | 1 |
| Obesity | adipocyte | *AGPAT5* | 0.701031769 | 0.000602766 | 0.003255661 | 1 |
| Obesity | adipocyte | *MMS19* | 1.43673243 | 0.000747311 | 0.003826016 | 1 |
| Obesity | adipocyte | *PDIA4* | 2.319842938 | 2.34E-06 | 6.25E-05 | 0.014689625 |
| Obesity | adipocyte | *DEPTOR* | 1.545128609 | 9.20E-07 | 3.71E-05 | 0.00578148 |
| Obesity | adipocyte | *UGT2B4* | -0.721034902 | 0.001822811 | 0.007790628 | 1 |
| Obesity | adipocyte | *KCNMA1* | 0.504425182 | 0.003313751 | 0.012542775 | 1 |
| Obesity | adipocyte | *BATF* | 2.83990494 | 2.91E-06 | 6.99E-05 | 0.018312328 |
| Obesity | adipocyte | *DCK* | 0.633729633 | 0.012222137 | 0.034785231 | 1 |
| Obesity | adipocyte | *ADAMTSL3* | 0.297653135 | 0.001067032 | 0.005097592 | 1 |
| Obesity | adipocyte | *ATAD2* | 1.335229781 | 0.018082054 | 0.047866053 | 1 |
| Obesity | adipocyte | *VPS8* | 0.737527596 | 0.000616631 | 0.003313858 | 1 |
| Obesity | adipocyte | *NTAN1* | 1.048890864 | 0.001091295 | 0.00517809 | 1 |
| Obesity | adipocyte | *FCHO2* | 1.506810775 | 4.68E-06 | 9.42E-05 | 0.029394814 |
| Obesity | adipocyte | *TIMP4* | -0.74962892 | 0.00377705 | 0.013870509 | 1 |
| Obesity | adipocyte | *NECAP2* | 1.650937562 | 0.000234905 | 0.00157279 | 1 |
| Obesity | adipocyte | *STEAP2* | -0.214134591 | 0.000138395 | 0.001056221 | 0.870087584 |
| Obesity | adipocyte | *SUSD3* | 0.226814746 | 0.015571474 | 0.042233761 | 1 |
| Obesity | adipocyte | *TMED6* | -2.805967962 | 0.003818929 | 0.013975326 | 1 |
| Obesity | adipocyte | *CLEC18A* | -1.305473832 | 3.39E-17 | 1.94E-14 | 2.13E-13 |
| Obesity | adipocyte | *DYRK1A* | 1.243355877 | 0.000100727 | 0.00084206 | 0.633272902 |
| Obesity | adipocyte | *CREB3L1* | 0.525012509 | 0.003978462 | 0.014416479 | 1 |
| Obesity | adipocyte | *SLC37A3* | 1.701870978 | 0.000200222 | 0.001401779 | 1 |
| Obesity | adipocyte | *SPPL3* | 0.935555094 | 0.000615641 | 0.003313814 | 1 |
| Obesity | adipocyte | *RADIL* | 1.166642891 | 0.002346457 | 0.00952739 | 1 |
| Obesity | adipocyte | *GALNT14* | 3.095700576 | 4.77E-05 | 0.000496044 | 0.300106902 |
| Obesity | adipocyte | *HPD* | 0.416229503 | 0.000484414 | 0.002738769 | 1 |
| Obesity | adipocyte | *LRRC43* | -1.782357775 | 0.006868454 | 0.022178721 | 1 |
| Obesity | adipocyte | *EYA3* | 0.832178485 | 0.001769412 | 0.007629186 | 1 |
| Obesity | adipocyte | *COLEC12* | 1.25637994 | 2.66E-06 | 6.67E-05 | 0.016748719 |
| Obesity | adipocyte | *AUTS2* | 1.05055173 | 0.000146717 | 0.001106006 | 0.922408765 |
| Obesity | adipocyte | *AHCYL2* | 0.755757709 | 0.013768006 | 0.038199228 | 1 |
| Obesity | adipocyte | *CD1E* | 1.134016512 | 0.00016373 | 0.001199735 | 1 |
| Obesity | adipocyte | *NCF1* | 2.780469609 | 4.06E-06 | 8.58E-05 | 0.025553812 |
| Obesity | adipocyte | *TMED4* | 1.718090655 | 0.003191693 | 0.012205702 | 1 |
| Obesity | adipocyte | *COPG2* | -0.127751553 | 0.004622521 | 0.016208471 | 1 |
| Obesity | adipocyte | *ELK4* | 1.924309584 | 8.44E-06 | 0.000139977 | 0.053051352 |
| Obesity | adipocyte | *DUSP23* | 2.200016845 | 0.00929245 | 0.028114357 | 1 |
| Obesity | adipocyte | *SLC5A11* | -0.274068516 | 1.29E-07 | 8.80E-06 | 0.000812409 |
| Obesity | adipocyte | *FCER1G* | 0.353770624 | 0.000980673 | 0.00476836 | 1 |
| Obesity | adipocyte | *CDC42SE2* | 1.534123203 | 9.14E-06 | 0.000148547 | 0.057487735 |

| | | | | | | |
|---|---|---|---|---|---|---|
| Obesity | adipocyte | *IFNAR2* | 0.302025609 | 0.01314121 | 0.036866928 | 1 |
| Obesity | adipocyte | *ARHGAP27* | 2.36096262 | 8.66E-06 | 0.000142901 | 0.054445408 |
| Obesity | adipocyte | *STARD9* | 0.771724028 | 0.000806387 | 0.004063111 | 1 |
| Obesity | adipocyte | *ACE* | 1.154999407 | 0.00029431 | 0.001865246 | 1 |
| Obesity | adipocyte | *EFCAB14* | 1.98384649 | 3.16E-05 | 0.000361509 | 0.198829693 |
| Obesity | adipocyte | *ZFYVE28* | 0.86470032 | 0.000608735 | 0.003279449 | 1 |
| Obesity | adipocyte | *CCDC107* | -0.564531071 | 0.006558256 | 0.021374678 | 1 |
| Obesity | adipocyte | *DFFA* | 0.908049758 | 8.80E-05 | 0.000763508 | 0.553543553 |
| Obesity | adipocyte | *ANKLE1* | -0.495996586 | 0.010190064 | 0.030162398 | 1 |
| Obesity | adipocyte | *CILP2* | -5.470422365 | 0.005415732 | 0.018343994 | 1 |
| Obesity | adipocyte | *SLC37A1* | 1.74717146 | 2.58E-06 | 6.50E-05 | 0.016194529 |
| Obesity | adipocyte | *C21orf33* | -0.829963986 | 0.002141503 | 0.008846013 | 1 |
| Obesity | adipocyte | *ZNF66* | 0.778591858 | 0.005658982 | 0.01902787 | 1 |
| Obesity | adipocyte | *FAM207A* | 0.513498121 | 0.012480314 | 0.035359953 | 1 |
| Obesity | adipocyte | *LSS* | 0.448475121 | 0.006578243 | 0.021418058 | 1 |
| Obesity | adipocyte | *VAV2* | 1.127127772 | 0.001887196 | 0.008005937 | 1 |
| Obesity | adipocyte | *DIP2A* | 0.551397531 | 0.004848543 | 0.016850629 | 1 |
| Obesity | adipocyte | *PRMT2* | 0.473186692 | 0.005285297 | 0.017992722 | 1 |
| Obesity | adipocyte | *C19orf47* | 1.056617307 | 0.001903226 | 0.008068498 | 1 |
| Obesity | adipocyte | *MPZL3* | 2.285855772 | 3.23E-06 | 7.33E-05 | 0.020293571 |
| Obesity | adipocyte | *PCSK7* | 0.953592073 | 0.008553007 | 0.026372122 | 1 |
| Obesity | adipocyte | *ZBTB7B* | -1.659079109 | 3.94E-05 | 0.000426583 | 0.247844729 |
| Obesity | adipocyte | *PAQR6* | -0.690969243 | 0.003468062 | 0.012978397 | 1 |
| Obesity | adipocyte | *PMF1* | 0.742369635 | 0.000651784 | 0.003437721 | 1 |
| Obesity | adipocyte | *NBEAL2* | 1.315412486 | 1.88E-05 | 0.000247687 | 0.118146886 |
| Obesity | adipocyte | *ZNF333* | 0.895598559 | 0.010723633 | 0.031331522 | 1 |
| Obesity | adipocyte | *ORAI2* | 1.693695 | 1.85E-07 | 1.16E-05 | 0.00116048 |
| Obesity | adipocyte | *RPL8* | 1.737524646 | 5.36E-06 | 0.000101483 | 0.033692477 |
| Obesity | adipocyte | *FBXL13* | 1.175751359 | 0.005418291 | 0.018343994 | 1 |
| Obesity | adipocyte | *MFSD12* | 2.295673643 | 5.18E-06 | 9.96E-05 | 0.032576105 |
| Obesity | adipocyte | *YDJC* | 5.618741241 | 7.20E-07 | 3.10E-05 | 0.004527612 |
| Obesity | adipocyte | *AP2M1* | 1.227413744 | 0.000648403 | 0.003436811 | 1 |
| Obesity | adipocyte | *DUSP14* | -0.626003338 | 0.001438028 | 0.006476592 | 1 |
| Obesity | adipocyte | *PGAP3* | 3.214494915 | 0.000286302 | 0.001821841 | 1 |
| Obesity | adipocyte | *IKZF3* | 1.14665895 | 0.000136513 | 0.001049213 | 0.85825652 |
| Obesity | adipocyte | *CYGB* | 0.841319986 | 0.014947147 | 0.040911062 | 1 |
| Obesity | adipocyte | *ALDH16A1* | 0.972618178 | 0.012338332 | 0.035020811 | 1 |
| Obesity | adipocyte | *ASGR2* | 4.173734373 | 0.000268282 | 0.00173349 | 1 |
| Obesity | adipocyte | *JMJD8* | 0.422044173 | 0.004063777 | 0.014641239 | 1 |
| Obesity | adipocyte | *FLYWCH2* | 1.690624418 | 0.000914867 | 0.004521832 | 1 |
| Obesity | adipocyte | *ZG16B* | 37.79417695 | 1.26E-05 | 0.00018782 | 0.079072363 |
| Obesity | adipocyte | *RPL29* | 0.782579419 | 0.003478514 | 0.013002031 | 1 |
| Obesity | adipocyte | *TPCN2* | 0.753767342 | 0.000954786 | 0.004684547 | 1 |
| Obesity | adipocyte | *NOL9* | 1.340944152 | 0.000100854 | 0.00084206 | 0.634070901 |
| Obesity | adipocyte | *ZSWIM5* | 0.562038145 | 0.00939684 | 0.028294031 | 1 |
| Obesity | adipocyte | *RBP7* | 2.312230352 | 8.63E-07 | 3.54E-05 | 0.005423701 |
| Obesity | adipocyte | *SLC25A34* | 2.593629894 | 0.000766267 | 0.003897672 | 1 |
| Obesity | adipocyte | *LAPTM5* | 0.734846844 | 0.001596625 | 0.007068588 | 1 |
| Obesity | adipocyte | *PEF1* | 3.596966457 | 1.48E-06 | 5.22E-05 | 0.009297283 |
| Obesity | adipocyte | *RBBP4* | 1.128979677 | 0.001264653 | 0.005812043 | 1 |

| | | | | | | |
|---|---|---|---|---|---|---|
| Obesity | adipocyte | *CAMK2N1* | 2.715929676 | 1.45E-07 | 9.53E-06 | 0.000914749 |
| Obesity | adipocyte | *NFIA* | 1.239805278 | 3.19E-05 | 0.000362662 | 0.200828131 |
| Obesity | adipocyte | *TM2D1* | 1.557839896 | 0.000394828 | 0.002332971 | 1 |
| Obesity | adipocyte | *KCNT2* | 1.149422073 | 2.11E-06 | 6.01E-05 | 0.013286473 |
| Obesity | adipocyte | *VCAM1* | 0.665554106 | 0.003774995 | 0.013870509 | 1 |
| Obesity | adipocyte | *ZNF496* | 1.799754063 | 3.30E-06 | 7.45E-05 | 0.020777345 |
| Obesity | adipocyte | *SNED1* | 1.172762271 | 0.001433076 | 0.006463236 | 1 |
| Obesity | adipocyte | *ACP6* | -0.640995659 | 0.000706484 | 0.00365213 | 1 |
| Obesity | adipocyte | *CNST* | 1.336126129 | 0.002005857 | 0.008384855 | 1 |
| Obesity | adipocyte | *HAAO* | 1.398462836 | 7.17E-05 | 0.000662504 | 0.45050281 |
| Obesity | adipocyte | *RFTN2* | 1.744967954 | 0.000578952 | 0.003162355 | 1 |
| Obesity | adipocyte | *DISC1* | 1.098785793 | 5.22E-05 | 0.000526711 | 0.328140817 |
| Obesity | adipocyte | *FAM84A* | -1.246449273 | 0.003699941 | 0.013635127 | 1 |
| Obesity | adipocyte | *CLHC1* | 0.13793301 | 0.002081664 | 0.008655704 | 1 |
| Obesity | adipocyte | *FBXO41* | 1.450408679 | 0.000629451 | 0.003367966 | 1 |
| Obesity | adipocyte | *SMC6* | 1.307028254 | 0.001568817 | 0.006970426 | 1 |
| Obesity | adipocyte | *CCDC74A* | 7.569582297 | 9.71E-06 | 0.000154742 | 0.061075612 |
| Obesity | adipocyte | *NOSTRIN* | 0.582509296 | 0.001952781 | 0.008223132 | 1 |
| Obesity | adipocyte | *HPGDS* | 0.622140233 | 0.000495276 | 0.002790144 | 1 |
| Obesity | adipocyte | *PDLIM5* | 1.448498243 | 9.59E-06 | 0.000153475 | 0.060277399 |
| Obesity | adipocyte | *RPRD2* | 0.97489709 | 0.000143932 | 0.001088926 | 0.904897756 |
| Obesity | adipocyte | *PACRGL* | 0.563125522 | 0.007642393 | 0.024084073 | 1 |
| Obesity | adipocyte | *RNF149* | 1.547394108 | 3.06E-05 | 0.000352365 | 0.192632238 |
| Obesity | adipocyte | *DHX57* | 1.179596358 | 0.001501886 | 0.00671576 | 1 |
| Obesity | adipocyte | *TDRD10* | 1.598608846 | 1.66E-06 | 5.50E-05 | 0.010449438 |
| Obesity | adipocyte | *ANTXR2* | 0.993226325 | 1.65E-07 | 1.06E-05 | 0.001036606 |
| Obesity | adipocyte | *PBXIP1* | 2.854650286 | 2.25E-06 | 6.20E-05 | 0.014153236 |
| Obesity | adipocyte | *SLC22A15* | 1.175637498 | 6.78E-06 | 0.000121491 | 0.042622034 |
| Obesity | adipocyte | *LRRC58* | 2.06841066 | 0.000259848 | 0.001696432 | 1 |
| Obesity | adipocyte | *LMOD1* | -0.878429471 | 1.02E-13 | 3.99E-11 | 6.38E-10 |
| Obesity | adipocyte | *TMEM183A* | 2.363799448 | 0.002938738 | 0.011383763 | 1 |
| Obesity | adipocyte | *ARPC2* | 1.582883182 | 2.23E-05 | 0.00027944 | 0.140278733 |
| Obesity | adipocyte | *SERPINI1* | 1.364423795 | 0.002554793 | 0.010152962 | 1 |
| Obesity | adipocyte | *AIM2* | 1.828181995 | 2.30E-06 | 6.25E-05 | 0.014466131 |
| Obesity | adipocyte | *GTPBP8* | 1.561759453 | 0.004108579 | 0.014760363 | 1 |
| Obesity | adipocyte | *CADPS* | 2.840590153 | 2.78E-06 | 6.83E-05 | 0.017493213 |
| Obesity | adipocyte | *PPM1K* | 1.058003878 | 0.004880463 | 0.016942834 | 1 |
| Obesity | adipocyte | *GMPS* | 1.862012634 | 0.000215614 | 0.001479877 | 1 |
| Obesity | adipocyte | *TIPARP* | 1.020151769 | 0.000780309 | 0.003953103 | 1 |
| Obesity | adipocyte | *DNASE1L3* | 1.062244037 | 0.000231183 | 0.001557822 | 1 |
| Obesity | adipocyte | *GYG1* | 2.114692858 | 2.29E-08 | 2.32E-06 | 0.000143829 |
| Obesity | adipocyte | *HPS3* | 0.899470214 | 0.003576429 | 0.013273324 | 1 |
| Obesity | adipocyte | *RYK* | 1.215157636 | 0.0009764 | 0.00475126 | 1 |
| Obesity | adipocyte | *PLB1* | 2.401923104 | 2.93E-06 | 6.99E-05 | 0.018448339 |
| Obesity | adipocyte | *CDCP1* | 2.181215185 | 3.61E-06 | 7.87E-05 | 0.022708532 |
| Obesity | adipocyte | *FYCO1* | 1.104585792 | 0.013055932 | 0.036676784 | 1 |
| Obesity | adipocyte | *SMIM12* | 1.70674903 | 0.002441838 | 0.009800714 | 1 |
| Obesity | adipocyte | *TPRA1* | 1.290098378 | 0.007432903 | 0.023613269 | 1 |
| Obesity | adipocyte | *YEATS2* | 1.741249855 | 2.78E-05 | 0.000328374 | 0.174881365 |
| Obesity | adipocyte | *MEAF6* | 1.539006727 | 0.003502223 | 0.013044119 | 1 |

| | | | | | | |
|---|---|---|---|---|---|---|
| Obesity | adipocyte | *DNALI1* | -1.617233868 | 1.07E-05 | 0.000165792 | 0.067246533 |
| Obesity | adipocyte | *RPN1* | 1.901330583 | 4.51E-05 | 0.000473844 | 0.283358878 |
| Obesity | adipocyte | *RFT1* | -0.956180563 | 1.94E-05 | 0.000252965 | 0.121676387 |
| Obesity | adipocyte | *SFMBT1* | 1.499243158 | 0.003793117 | 0.013897043 | 1 |
| Obesity | adipocyte | *FAM208A* | 0.608585828 | 0.008377128 | 0.025917351 | 1 |
| Obesity | adipocyte | *LRPAP1* | 0.990897371 | 0.006313605 | 0.020782006 | 1 |
| Obesity | adipocyte | *SLC51A* | -0.931305295 | 0.001758226 | 0.007608682 | 1 |
| Obesity | adipocyte | *ABLIM2* | 1.538476295 | 0.016943255 | 0.045309335 | 1 |
| Obesity | adipocyte | *C1orf50* | -0.741518281 | 0.005725868 | 0.019178759 | 1 |
| Obesity | adipocyte | *DNAJB14* | 1.111800334 | 6.03E-05 | 0.0005852 | 0.37920961 |
| Obesity | adipocyte | *SLC9B2* | 0.907929352 | 0.000308905 | 0.001932321 | 1 |
| Obesity | adipocyte | *ATRIP* | -3.713785472 | 0.001145777 | 0.005354377 | 1 |
| Obesity | adipocyte | *SHISA5* | 2.456090802 | 5.09E-08 | 4.21E-06 | 0.000319998 |
| Obesity | adipocyte | *APEH* | -0.91575533 | 5.75E-11 | 1.39E-08 | 3.62E-07 |
| Obesity | adipocyte | *RNF123* | 0.921375513 | 1.09E-21 | 7.58E-19 | 6.83E-18 |
| Obesity | adipocyte | *MST1R* | 0.593536584 | 4.92E-07 | 2.42E-05 | 0.003092275 |
| Obesity | adipocyte | *TEX264* | -0.46575745 | 0.002211403 | 0.00908105 | 1 |
| Obesity | adipocyte | *GRM2* | -9.220821092 | 0.010380675 | 0.030596954 | 1 |
| Obesity | adipocyte | *POC1A* | 0.802161647 | 0.001708024 | 0.007436529 | 1 |
| Obesity | adipocyte | *PPM1M* | -0.409941373 | 0.002315377 | 0.009427963 | 1 |
| Obesity | adipocyte | *C4orf3* | 1.163315583 | 0.002788673 | 0.010964596 | 1 |
| Obesity | adipocyte | *CEP44* | 1.47059057 | 0.005356536 | 0.018164262 | 1 |
| Obesity | adipocyte | *IL15* | 1.337564393 | 3.29E-05 | 0.000370365 | 0.207034295 |
| Obesity | adipocyte | *ARFIP1* | 0.792500523 | 0.003786603 | 0.01388231 | 1 |
| Obesity | adipocyte | *OTUD4* | 1.78891935 | 0.000348025 | 0.002097529 | 1 |
| Obesity | adipocyte | *ITGA2* | 2.189813278 | 4.35E-07 | 2.20E-05 | 0.002732177 |
| Obesity | adipocyte | *PGGT1B* | 1.801551109 | 0.000280002 | 0.001786271 | 1 |
| Obesity | adipocyte | *C5orf63* | -1.024012551 | 0.014659966 | 0.040230121 | 1 |
| Obesity | adipocyte | *PRRC1* | 1.362223938 | 0.00161895 | 0.007137686 | 1 |
| Obesity | adipocyte | *NDUFS4* | 1.515389607 | 0.000266604 | 0.001731546 | 1 |
| Obesity | adipocyte | *SERINC5* | 1.068708384 | 0.007594271 | 0.023980504 | 1 |
| Obesity | adipocyte | *CASP3* | 1.616220734 | 0.000928787 | 0.00457983 | 1 |
| Obesity | adipocyte | *ERAP1* | -0.413770906 | 0.001479066 | 0.006623139 | 1 |
| Obesity | adipocyte | *RICTOR* | 1.50002939 | 7.37E-05 | 0.000674035 | 0.463062281 |
| Obesity | adipocyte | *ANKRA2* | 1.519244101 | 0.002901923 | 0.011268925 | 1 |
| Obesity | adipocyte | *TLR3* | 1.660598698 | 0.001719253 | 0.007475066 | 1 |
| Obesity | adipocyte | *MB21D1* | 2.154196705 | 1.89E-06 | 5.81E-05 | 0.011898969 |
| Obesity | adipocyte | *TXLNB* | 0.878555071 | 0.006042508 | 0.020057681 | 1 |
| Obesity | adipocyte | *DCBLD1* | 0.985688436 | 0.000269499 | 0.001737784 | 1 |
| Obesity | adipocyte | *TMEM200A* | 1.381719392 | 4.47E-05 | 0.000470847 | 0.281095776 |
| Obesity | adipocyte | *PDSS2* | 1.474743893 | 0.00015456 | 0.001145894 | 0.971718258 |
| Obesity | adipocyte | *STXBP5* | 1.454192402 | 4.97E-05 | 0.000508417 | 0.312229107 |
| Obesity | adipocyte | *PI16* | 1.407332144 | 3.62E-06 | 7.87E-05 | 0.022743604 |
| Obesity | adipocyte | *SAP30L* | 0.977983983 | 0.01190881 | 0.034004721 | 1 |
| Obesity | adipocyte | *RPS14* | 1.243970278 | 4.18E-05 | 0.00044565 | 0.26293344 |
| Obesity | adipocyte | *SLU7* | 0.883341142 | 0.004730053 | 0.016510528 | 1 |
| Obesity | adipocyte | *CAMLG* | 0.567688217 | 0.00605682 | 0.020094578 | 1 |
| Obesity | adipocyte | *FNDC1* | 0.369683171 | 0.007566316 | 0.023916255 | 1 |
| Obesity | adipocyte | *BRI3* | 0.73738878 | 0.003647247 | 0.013480449 | 1 |
| Obesity | adipocyte | *LMTK2* | 1.286178208 | 0.000233651 | 0.001569121 | 1 |

| | | | | | | |
|---|---|---|---|---|---|---|
| Obesity | adipocyte | *SUN1* | 0.579639826 | 0.009092692 | 0.027652248 | 1 |
| Obesity | adipocyte | *OXR1* | 0.955270708 | 0.000568661 | 0.003129728 | 1 |
| Obesity | adipocyte | *INTS1* | 2.343967157 | 0.000408118 | 0.002398005 | 1 |
| Obesity | adipocyte | *SLC4A2* | 2.414941127 | 0.001090534 | 0.00517809 | 1 |
| Obesity | adipocyte | *COX6C* | 2.031705922 | 0.000105565 | 0.000872124 | 0.66368601 |
| Obesity | adipocyte | *YWHAZ* | -0.363102486 | 0.002516556 | 0.010039078 | 1 |
| Obesity | adipocyte | *DCAF13* | 2.456549353 | 1.40E-05 | 0.000204119 | 0.087975257 |
| Obesity | adipocyte | *FAM219A* | 1.525815206 | 0.002264934 | 0.009264568 | 1 |
| Obesity | adipocyte | *TMEM65* | 1.378669291 | 7.60E-05 | 0.000691526 | 0.478036983 |
| Obesity | adipocyte | *SYK* | 1.155289255 | 3.04E-05 | 0.000351403 | 0.190942453 |
| Obesity | adipocyte | *HNRNPK* | 1.42066368 | 0.00075436 | 0.003849565 | 1 |
| Obesity | adipocyte | *TRMT10B* | 1.352079179 | 0.005472521 | 0.018477841 | 1 |
| Obesity | adipocyte | *LRFN5* | 0.39437602 | 2.02E-05 | 0.000260852 | 0.126774104 |
| Obesity | adipocyte | *WRN* | 1.182215235 | 0.000623641 | 0.003345419 | 1 |
| Obesity | adipocyte | *MARCH8* | 1.807861875 | 9.46E-05 | 0.000810152 | 0.594651304 |
| Obesity | adipocyte | *ZCCHC24* | 1.355453032 | 0.003371325 | 0.012707145 | 1 |
| Obesity | adipocyte | *PGM2L1* | 0.691302864 | 0.011200613 | 0.032435862 | 1 |
| Obesity | adipocyte | *TAF3* | 1.648535413 | 1.91E-06 | 5.81E-05 | 0.012029905 |
| Obesity | adipocyte | *QSOX2* | 3.0780632 | 1.80E-05 | 0.000241715 | 0.113364312 |
| Obesity | adipocyte | *SDCCAG3* | 1.885306706 | 0.001770476 | 0.007629186 | 1 |
| Obesity | adipocyte | *TSC1* | 1.173152261 | 0.000329929 | 0.002015808 | 1 |
| Obesity | adipocyte | *METTL3* | 1.288859239 | 0.003763528 | 0.013845114 | 1 |
| Obesity | adipocyte | *SLC39A13* | 4.39820025 | 3.13E-11 | 7.96E-09 | 1.97E-07 |
| Obesity | adipocyte | *PSMC3* | 0.629106238 | 0.000460861 | 0.002634028 | 1 |
| Obesity | adipocyte | *TCP11L2* | 1.755326481 | 0.000575285 | 0.003151231 | 1 |
| Obesity | adipocyte | *JAM3* | 0.701496012 | 0.008671085 | 0.026644729 | 1 |
| Obesity | adipocyte | *NDUFB8* | 1.581608255 | 0.003596685 | 0.013332759 | 1 |
| Obesity | adipocyte | *TRMT61A* | -0.827820636 | 1.61E-06 | 5.39E-05 | 0.010091658 |
| Obesity | adipocyte | *BTRC* | 0.836029388 | 0.01104823 | 0.032112909 | 1 |
| Obesity | adipocyte | *ZNF202* | 1.740512007 | 0.002093957 | 0.008689575 | 1 |
| Obesity | adipocyte | *FAM227B* | 0.527218377 | 0.005076808 | 0.017464919 | 1 |
| Obesity | adipocyte | *APBB1* | 0.986788396 | 0.004924462 | 0.017086143 | 1 |
| Obesity | adipocyte | *ILK* | 1.488728378 | 0.000234277 | 0.001570257 | 1 |
| Obesity | adipocyte | *DCHS1* | 1.579498765 | 5.28E-06 | 0.000100646 | 0.033213042 |
| Obesity | adipocyte | *USP54* | 0.638985133 | 0.008076243 | 0.025217737 | 1 |
| Obesity | adipocyte | *LMO1* | 0.941628373 | 0.013979072 | 0.038614555 | 1 |
| Obesity | adipocyte | *RPL27A* | 1.314763299 | 0.000163432 | 0.001198948 | 1 |
| Obesity | adipocyte | *PRTG* | 2.316838897 | 0.001312874 | 0.005998576 | 1 |
| Obesity | adipocyte | *AKIP1* | 2.92103829 | 0.000111023 | 0.000908855 | 0.698000702 |
| Obesity | adipocyte | *TMEM41B* | 1.41106115 | 0.005018456 | 0.017342079 | 1 |
| Obesity | adipocyte | *TMX3* | 1.469347767 | 0.000590701 | 0.003212574 | 1 |
| Obesity | adipocyte | *CCDC68* | 0.303863558 | 0.015162627 | 0.041356806 | 1 |
| Obesity | adipocyte | *ZSCAN21* | -0.250336491 | 0.003581062 | 0.013282678 | 1 |
| Obesity | adipocyte | *SEC11C* | 2.913405057 | 1.00E-06 | 3.92E-05 | 0.006304108 |
| Obesity | adipocyte | *AP1G1* | 1.105835954 | 0.010472546 | 0.030809966 | 1 |
| Obesity | adipocyte | *CATSPER2* | 0.977157047 | 6.58E-05 | 0.00062133 | 0.413805886 |
| Obesity | adipocyte | *C16orf45* | 1.387893881 | 0.000153852 | 0.001145894 | 0.967265391 |
| Obesity | adipocyte | *SAAL1* | 2.763770044 | 2.95E-07 | 1.69E-05 | 0.001855816 |
| Obesity | adipocyte | *YPEL4* | 0.822625048 | 0.000257617 | 0.001687968 | 1 |
| Obesity | adipocyte | *MTMR10* | 0.649241205 | 0.002871801 | 0.011207334 | 1 |

| | | | | | | |
|---|---|---|---|---|---|---|
| Obesity | adipocyte | *TSC22D4* | 1.670238398 | 0.001783203 | 0.00767351 | 1 |
| Obesity | adipocyte | *CCNDBP1* | 2.069632247 | 0.000392415 | 0.002325271 | 1 |
| Obesity | adipocyte | *SNRPD1* | 2.065496319 | 0.000871505 | 0.004334773 | 1 |
| Obesity | adipocyte | *URM1* | -0.636138245 | 0.01008172 | 0.029912116 | 1 |
| Obesity | adipocyte | *ISLR2* | 0.515746577 | 0.00241303 | 0.009729461 | 1 |
| Obesity | adipocyte | *SP2* | 2.00928836 | 1.46E-06 | 5.22E-05 | 0.009202852 |
| Obesity | adipocyte | *GPRC5B* | 1.012666999 | 2.25E-07 | 1.33E-05 | 0.001412853 |
| Obesity | adipocyte | *ZNF646* | 1.735181332 | 8.32E-11 | 1.84E-08 | 5.23E-07 |
| Obesity | adipocyte | *VKORC1* | -0.628326338 | 1.87E-05 | 0.000247605 | 0.117859281 |
| Obesity | adipocyte | *RAB8A* | 1.035415915 | 0.001996407 | 0.008356464 | 1 |
| Obesity | adipocyte | *GATAD2A* | 0.998230307 | 0.005980084 | 0.019871452 | 1 |
| Obesity | adipocyte | *RPL13* | 1.242146754 | 0.000280191 | 0.001786271 | 1 |
| Obesity | adipocyte | *LENG8* | 0.623472846 | 0.016277282 | 0.043751719 | 1 |
| Obesity | adipocyte | *CDC42EP5* | 0.745499535 | 0.007762213 | 0.024400517 | 1 |
| Obesity | adipocyte | *PSCA* | 0.500930063 | 0.006425003 | 0.021050572 | 1 |
| Obesity | adipocyte | *UBXN6* | -0.712695885 | 0.003382304 | 0.012725641 | 1 |
| Obesity | adipocyte | *FAM57A* | 0.308393494 | 0.009449108 | 0.028397009 | 1 |
| Obesity | adipocyte | *SRR* | 0.302492276 | 6.42E-06 | 0.000115994 | 0.040365752 |
| Obesity | adipocyte | *ZNF83* | 0.683673062 | 0.009739696 | 0.029061923 | 1 |
| Obesity | adipocyte | *CD320* | 2.584300115 | 6.00E-07 | 2.68E-05 | 0.003770313 |
| Obesity | adipocyte | *SPRYD3* | 2.998307024 | 5.96E-05 | 0.000579949 | 0.374839133 |
| Obesity | adipocyte | *NDUFV1* | 0.870679334 | 0.011339267 | 0.032761935 | 1 |
| Obesity | adipocyte | *CDK2AP2* | 0.905418317 | 0.009841786 | 0.029338695 | 1 |
| Obesity | adipocyte | *NUDT8* | -0.365996322 | 0.001312068 | 0.005998576 | 1 |
| Obesity | adipocyte | *PRDX2* | 1.415590841 | 0.001012772 | 0.004896445 | 1 |
| Obesity | adipocyte | *TMC8* | 1.099088275 | 3.41E-07 | 1.89E-05 | 0.002141214 |
| Obesity | adipocyte | *MLST8* | -1.585317841 | 0.018932524 | 0.049740401 | 1 |
| Obesity | adipocyte | *SDHAF2* | 2.981293634 | 0.002444355 | 0.009800804 | 1 |
| Obesity | adipocyte | *DDB1* | 1.240830972 | 0.014675244 | 0.040254477 | 1 |
| Obesity | adipocyte | *VWCE* | 1.220787 | 2.58E-05 | 0.000309507 | 0.16218145 |
| Obesity | adipocyte | *BSCL2* | -1.249658383 | 0.010311334 | 0.030424994 | 1 |
| Obesity | adipocyte | *TRANK1* | 1.476555528 | 9.08E-07 | 3.68E-05 | 0.005708678 |
| Obesity | adipocyte | *TTC21A* | 1.878597788 | 0.0001544 | 0.001145894 | 0.970714046 |
| Obesity | adipocyte | *LTBP3* | 0.421205596 | 0.010452879 | 0.030766502 | 1 |
| Obesity | adipocyte | *SCARA5* | 0.860934617 | 0.000758616 | 0.003865007 | 1 |
| Obesity | adipocyte | *ZCCHC4* | 1.08192742 | 0.002933299 | 0.011369698 | 1 |
| Obesity | adipocyte | *UBTD2* | 1.085688093 | 0.000100401 | 0.000840512 | 0.631224142 |
| Obesity | adipocyte | *COA6* | 2.675491821 | 0.003290112 | 0.012483364 | 1 |
| Obesity | adipocyte | *KIF5C* | 1.055982657 | 0.00454018 | 0.01594643 | 1 |
| Obesity | adipocyte | *SEPT2* | 0.172709737 | 0.015949246 | 0.043091066 | 1 |
| Obesity | adipocyte | *TAP1* | 0.900413184 | 0.000355294 | 0.00212534 | 1 |
| Obesity | adipocyte | *RFWD3* | 0.619107502 | 0.000826004 | 0.004139136 | 1 |
| Obesity | adipocyte | *RHOH* | 0.63884461 | 0.005470095 | 0.018477841 | 1 |
| Obesity | adipocyte | *FNTA* | 1.488266989 | 0.000120697 | 0.000967883 | 0.758820375 |
| Obesity | adipocyte | *TRAPPC11* | 1.671084973 | 0.000178703 | 0.001285477 | 1 |
| Obesity | adipocyte | *SLC20A2* | 1.119360085 | 0.003536423 | 0.01314036 | 1 |
| Obesity | adipocyte | *ZSWIM1* | 0.964911789 | 0.002394373 | 0.009668221 | 1 |
| Obesity | adipocyte | *NBPF9* | 2.226229561 | 0.003983456 | 0.014417956 | 1 |
| Obesity | adipocyte | *NDUFS5* | 0.709096195 | 0.01102646 | 0.032079294 | 1 |
| Obesity | adipocyte | *FAM84B* | 0.257705929 | 0.010363147 | 0.030559619 | 1 |

| | | | | | | |
|---|---|---|---|---|---|---|
| Obesity | adipocyte | *DNAJC21* | 1.785137533 | 0.000270498 | 0.001738878 | 1 |
| Obesity | adipocyte | *NPNT* | 1.069563663 | 0.001649419 | 0.007231448 | 1 |
| Obesity | adipocyte | *CNNM3* | 1.409170214 | 0.000864418 | 0.004306338 | 1 |
| Obesity | adipocyte | *SNTB2* | 1.374081847 | 0.001726792 | 0.007502655 | 1 |
| Obesity | adipocyte | *STX18* | 1.400576275 | 0.001710853 | 0.007443689 | 1 |
| Obesity | adipocyte | *BTNL3* | 0.113608478 | 0.008211644 | 0.025532445 | 1 |
| Obesity | adipocyte | *INPP5D* | 1.459362056 | 1.60E-06 | 5.39E-05 | 0.010073105 |
| Obesity | adipocyte | *CEP120* | 0.937546888 | 0.005460736 | 0.018457876 | 1 |
| Obesity | adipocyte | *STXBP6* | 1.026149965 | 6.75E-05 | 0.000631925 | 0.424653406 |
| Obesity | adipocyte | *CPLX1* | 0.302488342 | 0.006361725 | 0.020909152 | 1 |
| Obesity | adipocyte | *FEM1B* | 1.008623066 | 0.000128675 | 0.001012094 | 0.808980066 |
| Obesity | adipocyte | *COL4A3* | 0.578603067 | 0.011615015 | 0.033328892 | 1 |
| Obesity | adipocyte | *C8orf46* | 0.073297745 | 0.001119716 | 0.005265261 | 1 |
| Obesity | adipocyte | *FAM110B* | 1.826773573 | 7.36E-05 | 0.000674035 | 0.463014332 |
| Obesity | adipocyte | *XPO6* | 2.041452034 | 2.39E-06 | 6.25E-05 | 0.015057289 |
| Obesity | adipocyte | *RAB24* | -0.848150771 | 7.27E-05 | 0.000669214 | 0.45707326 |
| Obesity | adipocyte | *PGM2* | 2.106400629 | 2.47E-05 | 0.00029974 | 0.155565108 |
| Obesity | adipocyte | *SLC33A1* | 1.891979799 | 0.000334779 | 0.002039492 | 1 |
| Obesity | adipocyte | *PTK2* | 1.157941318 | 8.03E-05 | 0.000715468 | 0.505120575 |
| Obesity | adipocyte | *RNASE6* | 3.219715706 | 6.05E-06 | 0.000111519 | 0.038027873 |
| Obesity | adipocyte | *NPR1* | 1.446062882 | 0.015498503 | 0.042126714 | 1 |
| Obesity | adipocyte | *CD52* | 0.449242727 | 0.01419495 | 0.039107647 | 1 |
| Obesity | adipocyte | *CLIC4* | 1.081358877 | 0.000787292 | 0.003975667 | 1 |
| Obesity | adipocyte | *CCDC8* | 1.541646038 | 0.00731429 | 0.023323452 | 1 |
| Obesity | adipocyte | *METTL15* | 0.663831409 | 0.000181313 | 0.001298735 | 1 |
| Obesity | adipocyte | *INO80E* | -1.350801642 | 4.72E-12 | 1.48E-09 | 2.97E-08 |
| Obesity | adipocyte | *FAM103A1* | -0.352021521 | 0.000194863 | 0.001370363 | 1 |
| Obesity | adipocyte | *RGPD8* | -0.127777625 | 0.011527151 | 0.033167597 | 1 |
| Obesity | adipocyte | *LUZP1* | 1.072073833 | 0.002091048 | 0.008683235 | 1 |
| Obesity | adipocyte | *CSGALNACT2* | 1.632988618 | 2.48E-05 | 0.000300332 | 0.156172806 |
| Obesity | adipocyte | *AVEN* | 1.239113723 | 0.011056518 | 0.032122148 | 1 |
| Obesity | adipocyte | *GUSB* | 1.826150079 | 0.000277703 | 0.001775359 | 1 |
| Obesity | adipocyte | *BRD3* | 1.358134054 | 0.006365535 | 0.020909152 | 1 |
| Obesity | adipocyte | *ZFPM2* | 1.14317391 | 0.000306069 | 0.001920414 | 1 |
| Obesity | adipocyte | *MAP3K2* | 1.533926337 | 7.58E-05 | 0.000690514 | 0.476454413 |
| Obesity | adipocyte | *MYO7B* | 0.50343327 | 0.01437622 | 0.039537748 | 1 |
| Obesity | adipocyte | *CNTROB* | 2.090430114 | 0.002580386 | 0.010243714 | 1 |
| Obesity | adipocyte | *NIPA1* | 1.63126876 | 0.006041599 | 0.020057681 | 1 |
| Obesity | adipocyte | *HOXD4* | -0.406794127 | 0.011526134 | 0.033167597 | 1 |
| Obesity | adipocyte | *GLB1* | 0.922846978 | 0.007035353 | 0.022636265 | 1 |
| Obesity | adipocyte | *CRTAP* | 1.752199167 | 0.000657728 | 0.00344882 | 1 |
| Obesity | adipocyte | *ELP5* | -0.799783339 | 0.010235761 | 0.030254927 | 1 |
| Obesity | adipocyte | *STX8* | 1.019996893 | 0.003165573 | 0.012127946 | 1 |
| Obesity | adipocyte | *SEMA3E* | 1.410057677 | 1.28E-06 | 4.71E-05 | 0.008058303 |
| Obesity | adipocyte | *ADORA2B* | -0.376019808 | 2.84E-06 | 6.93E-05 | 0.017873093 |
| Obesity | adipocyte | *MGMT* | 0.485889521 | 0.011278849 | 0.032617352 | 1 |
| Obesity | adipocyte | *METTL7B* | -1.402946665 | 0.001283107 | 0.005892546 | 1 |
| Obesity | adipocyte | *NFXL1* | 1.788232129 | 0.000101736 | 0.000847175 | 0.63961711 |
| Obesity | adipocyte | *DLGAP1* | 1.238682767 | 0.00010907 | 0.000895199 | 0.685722564 |
| Obesity | adipocyte | *HSPA4* | 2.019950353 | 0.000155104 | 0.001148575 | 0.975140106 |

| | | | | | | |
|---|---|---|---|---|---|---|
| Obesity | adipocyte | *SLC26A5* | 1.330542775 | 0.005526737 | 0.018640877 | 1 |
| Obesity | adipocyte | *COMMD5* | 1.060887548 | 0.007512494 | 0.02377003 | 1 |
| Obesity | adipocyte | *GTSF1* | 4.974110007 | 3.38E-05 | 0.000378405 | 0.212285008 |
| Obesity | adipocyte | *AKAP13* | 1.024684646 | 7.37E-06 | 0.000129817 | 0.046344725 |
| Obesity | adipocyte | *KBTBD2* | 0.951635425 | 0.007542391 | 0.023852623 | 1 |
| Obesity | adipocyte | *MTSS1* | 1.252879439 | 2.07E-06 | 5.99E-05 | 0.013010321 |
| Obesity | adipocyte | *RPS9* | 0.670564331 | 0.000757831 | 0.003864141 | 1 |
| Obesity | adipocyte | *OSCAR* | 0.441339471 | 0.008595914 | 0.02646548 | 1 |
| Obesity | adipocyte | *TSNARE1* | 0.408207603 | 0.001194648 | 0.005526676 | 1 |
| Obesity | adipocyte | *FEZ2* | 1.221699139 | 0.002353672 | 0.009540644 | 1 |
| Obesity | adipocyte | *SOX7* | 2.429940909 | 9.02E-09 | 1.13E-06 | 5.67E-05 |
| Obesity | adipocyte | *TRMT61B* | 0.339977964 | 7.18E-06 | 0.000127589 | 0.045166374 |
| Obesity | adipocyte | *MFN1* | 0.730071316 | 0.005342315 | 0.01812587 | 1 |
| Obesity | adipocyte | *GIMAP8* | 0.410001115 | 0.001929822 | 0.008147951 | 1 |
| Obesity | adipocyte | *ATP6V0E2* | -1.0996276 | 0.018012025 | 0.047700758 | 1 |
| Obesity | adipocyte | *CHST11* | 0.323861947 | 0.018750388 | 0.049344365 | 1 |
| Obesity | adipocyte | *MCC* | 1.316769168 | 3.90E-05 | 0.000423188 | 0.245448925 |
| Obesity | adipocyte | *DLK2* | -0.99307609 | 0.00166074 | 0.007270941 | 1 |
| Obesity | adipocyte | *LRRC8C* | 2.589215487 | 3.20E-06 | 7.31E-05 | 0.020127344 |
| Obesity | adipocyte | *RSL1D1* | 1.740901818 | 0.000245839 | 0.001626938 | 1 |
| Obesity | adipocyte | *BCL2L1* | 1.312358416 | 4.98E-06 | 9.72E-05 | 0.031283286 |
| Obesity | adipocyte | *PLRG1* | 1.931978797 | 0.000577443 | 0.003156855 | 1 |
| Obesity | adipocyte | *P2RY6* | 1.859155962 | 4.71E-05 | 0.000490248 | 0.296109815 |
| Obesity | adipocyte | *RGS19* | 0.370587994 | 0.000350539 | 0.002102373 | 1 |
| Obesity | adipocyte | *TCEA2* | 3.104265847 | 3.80E-05 | 0.000416162 | 0.238876943 |
| Obesity | adipocyte | *CAMTA1* | 1.806558884 | 1.64E-06 | 5.46E-05 | 0.010321345 |
| Obesity | adipocyte | *NXNL1* | -0.485841617 | 0.001016766 | 0.004899217 | 1 |
| Obesity | adipocyte | *NINJ2* | 1.465279782 | 0.000126953 | 0.001002708 | 0.798155358 |
| Obesity | adipocyte | *FAM90A1* | 9.624337101 | 0.000200032 | 0.001401779 | 1 |
| Obesity | adipocyte | *TRAPPC12* | 1.073757489 | 0.005121185 | 0.017574723 | 1 |
| Obesity | adipocyte | *AQP4* | -0.676476684 | 0.00304617 | 0.011734849 | 1 |
| Obesity | adipocyte | *SRGAP2C* | 1.18931128 | 6.40E-05 | 0.000610395 | 0.402250621 |
| Obesity | adipocyte | *ATPAF2* | 2.059361631 | 0.000860811 | 0.004295172 | 1 |
| Obesity | adipocyte | *C20orf196* | 1.17768189 | 0.004538746 | 0.01594643 | 1 |
| Obesity | adipocyte | *ZNF554* | -0.397332363 | 0.016934934 | 0.045306354 | 1 |
| Obesity | adipocyte | *FRMD3* | 1.932299409 | 7.16E-10 | 1.22E-07 | 4.50E-06 |
| Obesity | adipocyte | *MTBP* | 1.356543779 | 0.005699078 | 0.019109387 | 1 |
| Obesity | adipocyte | *MALT1* | 1.766675919 | 3.16E-06 | 7.28E-05 | 0.019885376 |
| Obesity | adipocyte | *MBOAT1* | 1.800954108 | 4.50E-06 | 9.22E-05 | 0.028317645 |
| Obesity | adipocyte | *CXCR6* | 0.351183928 | 0.009094598 | 0.027652248 | 1 |
| Obesity | adipocyte | *C1QTNF4* | -0.278472858 | 4.63E-15 | 2.08E-12 | 2.91E-11 |
| Obesity | adipocyte | *ZNF131* | 1.204849542 | 0.000796525 | 0.004019063 | 1 |
| Obesity | adipocyte | *SPTLC3* | 1.966338174 | 6.30E-05 | 0.000602955 | 0.396141159 |
| Obesity | adipocyte | *SUCLG2* | 0.741560665 | 0.00529207 | 0.01800392 | 1 |
| Obesity | adipocyte | *RCAN2* | 1.472539964 | 4.73E-06 | 9.49E-05 | 0.02971288 |
| Obesity | adipocyte | *ZNF24* | 1.808667794 | 0.000647757 | 0.003436811 | 1 |
| Obesity | adipocyte | *FIBP* | 3.737361161 | 5.18E-13 | 1.71E-10 | 3.26E-09 |
| Obesity | adipocyte | *CARNS1* | 1.755188937 | 0.00149481 | 0.006688875 | 1 |
| Obesity | adipocyte | *CTSW* | -0.142402218 | 5.49E-07 | 2.56E-05 | 0.003450242 |
| Obesity | adipocyte | *SNTG2* | -0.806996693 | 0.000655191 | 0.003447018 | 1 |

| | | | | | | |
|---|---|---|---|---|---|---|
| Obesity | adipocyte | *SMPDL3A* | 0.762345176 | 0.002035795 | 0.008476187 | 1 |
| Obesity | adipocyte | *EFEMP2* | 0.891931957 | 1.32E-07 | 8.80E-06 | 0.000826968 |
| Obesity | adipocyte | *AGAP5* | -24.23359374 | 0.018992283 | 0.049855734 | 1 |
| Obesity | adipocyte | *ZMAT3* | 2.112795125 | 0.000818718 | 0.004111247 | 1 |
| Obesity | adipocyte | *SLFN11* | 0.463435446 | 0.005440727 | 0.018410036 | 1 |
| Obesity | adipocyte | *FUT10* | -0.351850754 | 0.000340506 | 0.002062241 | 1 |
| Obesity | adipocyte | *CFL1* | 1.734958379 | 8.49E-05 | 0.000746756 | 0.533930322 |
| Obesity | adipocyte | *RAB37* | 1.491775605 | 0.00012871 | 0.001012094 | 0.809198813 |
| Obesity | adipocyte | *DCP2* | 1.589947402 | 8.61E-05 | 0.000753994 | 0.541368006 |
| Obesity | adipocyte | *SNX32* | 0.830592697 | 1.50E-08 | 1.63E-06 | 9.45E-05 |
| Obesity | adipocyte | *NADSYN1* | 1.008074792 | 0.000773659 | 0.003932088 | 1 |
| Obesity | adipocyte | *ANKRD13D* | 0.817804678 | 0.010974876 | 0.031958799 | 1 |
| Obesity | adipocyte | *LCLAT1* | 1.270117222 | 0.000195403 | 0.001372625 | 1 |
| Obesity | adipocyte | *HPSE2* | 0.845094405 | 2.59E-10 | 4.66E-08 | 1.63E-06 |
| Obesity | adipocyte | *BNC2* | 0.595267711 | 0.005800409 | 0.019366528 | 1 |
| Obesity | adipocyte | *NOC3L* | 1.279138689 | 0.001018892 | 0.004902962 | 1 |
| Obesity | adipocyte | *PARP14* | 0.167138245 | 0.018326508 | 0.048350297 | 1 |
| Obesity | adipocyte | *ABLIM3* | 1.370031836 | 1.51E-06 | 5.24E-05 | 0.009481106 |
| Obesity | adipocyte | *GOLGB1* | 1.333284344 | 0.000450062 | 0.002581697 | 1 |
| Obesity | adipocyte | *MAP3K11* | 2.069683658 | 5.67E-05 | 0.000555811 | 0.356275164 |
| Obesity | adipocyte | *C1QA* | 1.909610122 | 2.38E-06 | 6.25E-05 | 0.014954741 |
| Obesity | adipocyte | *NDNF* | 1.869701207 | 3.49E-07 | 1.92E-05 | 0.002194234 |
| Obesity | adipocyte | *OLR1* | 3.05582233 | 3.86E-05 | 0.000421087 | 0.242967352 |
| Obesity | adipocyte | *ARV1* | 0.771185047 | 0.013210457 | 0.037011651 | 1 |
| Obesity | adipocyte | *PPP1R14B* | 2.14995606 | 0.00193104 | 0.008147951 | 1 |
| Obesity | adipocyte | *SMARCC1* | 1.462897586 | 0.000225684 | 0.001527101 | 1 |
| Obesity | adipocyte | *ZNF417* | -0.765111098 | 0.011491428 | 0.033107744 | 1 |
| Obesity | adipocyte | *PEAK1* | 1.307900527 | 8.02E-05 | 0.000715194 | 0.504211449 |
| Obesity | adipocyte | *MST1* | -0.367230244 | 1.18E-06 | 4.44E-05 | 0.00741996 |
| Obesity | adipocyte | *GMPPB* | 1.018271944 | 5.40E-07 | 2.55E-05 | 0.003397303 |
| Obesity | adipocyte | *C1orf100* | 4.001809664 | 0.000321768 | 0.00197941 | 1 |
| Obesity | adipocyte | *RNF213* | 1.348794074 | 7.80E-05 | 0.000702269 | 0.490183951 |
| Obesity | adipocyte | *PLK3* | 1.870532612 | 2.74E-06 | 6.81E-05 | 0.017239166 |
| Obesity | adipocyte | *GOLIM4* | 0.623680918 | 0.009567312 | 0.028697373 | 1 |
| Obesity | adipocyte | *HOXB2* | 0.585786084 | 0.000460365 | 0.00263359 | 1 |
| Obesity | adipocyte | *CCS* | 0.958975469 | 0.007322732 | 0.023334017 | 1 |
| Obesity | adipocyte | *SLC25A30* | 2.218012467 | 2.83E-05 | 0.000332421 | 0.177750501 |
| Obesity | adipocyte | *LEMD3* | 1.095906886 | 0.007504304 | 0.02377003 | 1 |
| Obesity | adipocyte | *TLR6* | 0.648668416 | 0.000689748 | 0.003577927 | 1 |
| Obesity | adipocyte | *DDX23* | 1.167670673 | 0.013348007 | 0.037330481 | 1 |
| Obesity | adipocyte | *RALGAPA1* | 0.55862451 | 0.012001197 | 0.034202866 | 1 |
| Obesity | adipocyte | *LINGO2* | 2.113610068 | 1.02E-07 | 7.29E-06 | 0.000641684 |
| Obesity | adipocyte | *BBS1* | 1.173845304 | 0.000398362 | 0.002349439 | 1 |
| Obesity | adipocyte | *ANGEL2* | 1.59987216 | 0.001171946 | 0.005449723 | 1 |
| Obesity | adipocyte | *SH3PXD2B* | 0.316927812 | 0.003032822 | 0.01169059 | 1 |
| Obesity | adipocyte | *LARP7* | 1.378036474 | 0.00509323 | 0.017505134 | 1 |
| Obesity | adipocyte | *C4orf32* | 1.423704958 | 1.71E-05 | 0.000234277 | 0.107194227 |
| Obesity | adipocyte | *HRAS* | 1.1779041 | 6.88E-05 | 0.000641238 | 0.432589097 |
| Obesity | adipocyte | *SRP72* | 0.523444093 | 0.000244856 | 0.001623853 | 1 |
| Obesity | adipocyte | *FUT1* | 0.661375118 | 0.012328658 | 0.035009157 | 1 |

| | | | | | | |
|---|---|---|---|---|---|---|
| Obesity | adipocyte | *FBXW8* | 1.721128987 | 0.000400033 | 0.002357081 | 1 |
| Obesity | adipocyte | *CHST2* | 3.340879372 | 2.21E-05 | 0.000277513 | 0.138756396 |
| Obesity | adipocyte | *ATR* | 0.932178477 | 0.007931812 | 0.024821951 | 1 |
| Obesity | adipocyte | *CADM2* | 0.534222914 | 1.94E-06 | 5.81E-05 | 0.012213048 |
| Obesity | adipocyte | *PSMD2* | 0.443247331 | 0.001788938 | 0.007682414 | 1 |
| Obesity | adipocyte | *GOLGA8A* | 0.662818301 | 0.000949088 | 0.004661651 | 1 |
| Obesity | adipocyte | *TP53I11* | 1.174086078 | 8.26E-05 | 0.000730748 | 0.519562021 |
| Obesity | adipocyte | *ZNF519* | 1.148649892 | 0.002479181 | 0.009926263 | 1 |
| Obesity | adipocyte | *TMEM9B* | 2.77176303 | 1.31E-10 | 2.75E-08 | 8.24E-07 |
| Obesity | adipocyte | *PCSK1* | -0.997094003 | 0.00015101 | 0.001127554 | 0.949400264 |
| Obesity | adipocyte | *PPP2R2D* | 0.465402025 | 0.014480655 | 0.039807556 | 1 |
| Obesity | adipocyte | *MCTP1* | 0.381465264 | 0.004657306 | 0.016321341 | 1 |
| Obesity | adipocyte | *LIPT2* | -1.121183311 | 0.002283973 | 0.009324248 | 1 |
| Obesity | adipocyte | *RAB6A* | 0.777257185 | 0.010613905 | 0.031123891 | 1 |
| Obesity | adipocyte | *TOM1L2* | 1.037083465 | 0.014037867 | 0.038759801 | 1 |
| Obesity | adipocyte | *B3GNTL1* | 0.777987026 | 0.014912911 | 0.040852929 | 1 |
| Obesity | adipocyte | *CTDNEP1* | 2.22908039 | 4.62E-05 | 0.000482158 | 0.290741306 |
| Obesity | adipocyte | *A2M* | 1.362456898 | 1.61E-06 | 5.39E-05 | 0.010132554 |
| Obesity | adipocyte | *IP6K1* | 1.52513953 | 6.55E-07 | 2.88E-05 | 0.004118979 |
| Obesity | adipocyte | *SSNA1* | 2.810415824 | 0.000381806 | 0.002267667 | 1 |
| Obesity | adipocyte | *TCP11L1* | 0.649166733 | 0.000806547 | 0.004063111 | 1 |
| Obesity | adipocyte | *SMIM19* | 0.30249017 | 0.007381578 | 0.023473941 | 1 |
| Obesity | adipocyte | *COX8A* | 3.024090325 | 1.11E-05 | 0.000171449 | 0.069951118 |
| Obesity | adipocyte | *B3GNT4* | -5.240807555 | 0.004752058 | 0.016570265 | 1 |
| Obesity | adipocyte | *SLCO3A1* | 1.336886892 | 1.50E-06 | 5.24E-05 | 0.009417115 |
| Obesity | adipocyte | *RMDN1* | 0.81710079 | 0.001380063 | 0.006255557 | 1 |
| Obesity | adipocyte | *RNF152* | 1.06585693 | 0.009548652 | 0.028655072 | 1 |
| Obesity | adipocyte | *BOK* | -0.470091876 | 0.015820583 | 0.042780218 | 1 |
| Obesity | adipocyte | *C8orf59* | 1.115841617 | 0.002277646 | 0.00930446 | 1 |
| Obesity | adipocyte | *RUFY1* | 0.926804277 | 2.92E-07 | 1.68E-05 | 0.00183577 |
| Obesity | adipocyte | *LRRC37A3* | 1.525631718 | 0.006268263 | 0.020654387 | 1 |
| Obesity | adipocyte | *ANKLE2* | 2.480039385 | 1.88E-07 | 1.17E-05 | 0.001182807 |
| Obesity | adipocyte | *GCNT4* | 5.721923371 | 5.27E-06 | 0.000100618 | 0.033103367 |
| Obesity | adipocyte | *DPP7* | 1.898093243 | 0.000431508 | 0.00249576 | 1 |
| Obesity | adipocyte | *MTHFR* | 1.366195518 | 8.01E-05 | 0.000715194 | 0.503656658 |
| Obesity | adipocyte | *C19orf18* | -0.490635534 | 0.011624327 | 0.033340393 | 1 |
| Obesity | adipocyte | *POLE* | 1.711244711 | 0.00230675 | 0.009405016 | 1 |
| Obesity | adipocyte | *DSCAML1* | 1.20859192 | 8.05E-06 | 0.000137212 | 0.05063131 |
| Obesity | adipocyte | *TALDO1* | -0.51354054 | 0.003500013 | 0.013044119 | 1 |
| Obesity | adipocyte | *B3GNT8* | 5.609885023 | 3.02E-06 | 7.03E-05 | 0.0189912 |
| Obesity | adipocyte | *MAN1B1* | 2.374406214 | 0.000157631 | 0.001163175 | 0.991025424 |
| Obesity | adipocyte | *ZBTB38* | 1.102381721 | 0.000241152 | 0.001607767 | 1 |
| Obesity | adipocyte | *C8orf31* | 4.136457456 | 1.08E-05 | 0.000167313 | 0.068096356 |
| Obesity | adipocyte | *PPFIA3* | -0.440684037 | 0.004100558 | 0.014748401 | 1 |
| Obesity | adipocyte | *PAWR* | 1.896369953 | 0.000221351 | 0.001509879 | 1 |
| Obesity | adipocyte | *NAP1L5* | 0.954346885 | 0.011083057 | 0.032183538 | 1 |
| Obesity | adipocyte | *ATOX1* | 1.412824219 | 0.000480691 | 0.002722614 | 1 |
| Obesity | adipocyte | *RPLP2* | 1.57906926 | 0.000215055 | 0.001479877 | 1 |
| Obesity | adipocyte | *CD151* | 1.324696964 | 2.57E-06 | 6.50E-05 | 0.016129479 |
| Obesity | adipocyte | *FAM87B* | -1.306436033 | 4.57E-05 | 0.000478648 | 0.287620549 |

| | | | | | | |
|---|---|---|---|---|---|---|
| Obesity | adipocyte | *ZNF354C* | 2.127771846 | 0.000685039 | 0.003562319 | 1 |
| Obesity | adipocyte | *ODF3B* | 0.663341918 | 0.008638885 | 0.026569195 | 1 |
| Obesity | adipocyte | *C2orf73* | -0.94644257 | 0.01172779 | 0.033560588 | 1 |
| Obesity | adipocyte | *PRSS42* | -0.598346505 | 0.001742519 | 0.007565759 | 1 |
| Obesity | adipocyte | *GRAMD1C* | 1.805458658 | 8.38E-05 | 0.000737701 | 0.526718782 |
| Obesity | adipocyte | *TSSK6* | -0.666493095 | 0.01720463 | 0.045891179 | 1 |
| Obesity | adipocyte | *LCORL* | 1.534264709 | 2.36E-05 | 0.000292667 | 0.148381963 |
| Obesity | adipocyte | *SH2B1* | -1.023873416 | 6.50E-30 | 2.04E-26 | 4.09E-26 |
| Obesity | adipocyte | *ZC3H12D* | 3.139081588 | 3.55E-06 | 7.79E-05 | 0.022293268 |
| Obesity | adipocyte | *PLEC* | 1.070735303 | 0.00120561 | 0.005565101 | 1 |
| Obesity | adipocyte | *SH2D4B* | 7.168451977 | 4.99E-06 | 9.72E-05 | 0.031393537 |
| Obesity | adipocyte | *PRSS36* | 2.980956202 | 3.32E-06 | 7.45E-05 | 0.020859958 |
| Obesity | adipocyte | *P4HTM* | -0.644902298 | 0.013074731 | 0.03671319 | 1 |
| Obesity | adipocyte | *CSRNP3* | 0.771385873 | 0.017853581 | 0.047380946 | 1 |
| Obesity | adipocyte | *RPP25* | -0.363107838 | 0.001074022 | 0.005123199 | 1 |
| Obesity | adipocyte | *STX19* | 1.545554702 | 1.59E-09 | 2.50E-07 | 1.00E-05 |
| Obesity | adipocyte | *GDPD4* | 2.784649617 | 0.007150056 | 0.022899848 | 1 |
| Obesity | adipocyte | *MPI* | 0.765181761 | 0.003500454 | 0.013044119 | 1 |
| Obesity | adipocyte | *TRIM73* | -1.554985874 | 5.73E-12 | 1.72E-09 | 3.60E-08 |
| Obesity | adipocyte | *C17orf62* | 1.63670734 | 1.13E-05 | 0.000173684 | 0.071210276 |
| Obesity | adipocyte | *GAK* | 1.073985635 | 0.005270326 | 0.017972621 | 1 |
| Obesity | adipocyte | *TUFM* | 1.360010409 | 3.38E-24 | 3.54E-21 | 2.12E-20 |
| Obesity | adipocyte | *CTC1* | 0.536071593 | 0.001127323 | 0.005290235 | 1 |
| Obesity | adipocyte | *FUCA1* | 0.453799365 | 0.018550625 | 0.048866617 | 1 |
| Obesity | adipocyte | *CALR* | 1.430849614 | 5.37E-07 | 2.55E-05 | 0.003377671 |
| Obesity | adipocyte | *EGR3* | 2.427571895 | 4.94E-06 | 9.70E-05 | 0.031050729 |
| Obesity | adipocyte | *LACC1* | 1.248596244 | 0.000569423 | 0.003129728 | 1 |
| Obesity | adipocyte | *MAF1* | 2.224739139 | 0.007090317 | 0.022743276 | 1 |
| Obesity | adipocyte | *CDH5* | 0.359210881 | 0.000412869 | 0.002419112 | 1 |
| Obesity | adipocyte | *SERTAD2* | 1.794799709 | 7.58E-06 | 0.000132393 | 0.047661321 |
| Obesity | adipocyte | *PDXDC1* | 0.935604831 | 0.003483954 | 0.013014626 | 1 |
| Obesity | adipocyte | *R3HDM2* | 1.343503218 | 0.000310576 | 0.001933258 | 1 |
| Obesity | adipocyte | *B3GNT3* | 1.319043816 | 0.000183489 | 0.001309413 | 1 |
| Obesity | adipocyte | *FGD6* | 1.738702461 | 0.000624323 | 0.003346221 | 1 |
| Obesity | adipocyte | *HCLS1* | 1.907412696 | 9.74E-07 | 3.83E-05 | 0.00612345 |
| Obesity | adipocyte | *ZNF609* | 0.681127923 | 0.012755865 | 0.035946268 | 1 |
| Obesity | adipocyte | *MB21D2* | 1.029327694 | 0.000836041 | 0.004184864 | 1 |
| Obesity | adipocyte | *SSTR2* | -1.082141318 | 0.004146067 | 0.014863876 | 1 |
| Obesity | adipocyte | *PCGF5* | 1.257282367 | 0.001170103 | 0.005445178 | 1 |
| Obesity | adipocyte | *SHISA2* | 22.48761095 | 2.38E-05 | 0.000293854 | 0.149342591 |
| Obesity | adipocyte | *CHST13* | 1.546477349 | 0.013480677 | 0.037651273 | 1 |
| Obesity | adipocyte | *SRSF8* | 2.051084847 | 0.004520366 | 0.015903492 | 1 |
| Obesity | adipocyte | *SLC36A4* | 0.774194094 | 0.00906525 | 0.027599626 | 1 |
| Obesity | adipocyte | *ZDHHC20* | 1.661696577 | 3.07E-05 | 0.000352365 | 0.193096146 |
| Obesity | adipocyte | *KCTD2* | 1.770749175 | 0.00850124 | 0.026238241 | 1 |
| Obesity | adipocyte | *ZFP82* | 1.116074429 | 0.006474135 | 0.021155346 | 1 |
| Obesity | adipocyte | *FKRP* | 0.206260088 | 0.013834854 | 0.038333947 | 1 |
| Obesity | adipocyte | *RPH3AL* | 0.71327266 | 0.000703231 | 0.003638859 | 1 |
| Obesity | adipocyte | *ANKRD34A* | -0.728074788 | 0.00675925 | 0.021882289 | 1 |
| Obesity | adipocyte | *SLC26A11* | 1.013143578 | 0.019022941 | 0.049915372 | 1 |

| | | | | | | |
|---|---|---|---|---|---|---|
| Obesity | adipocyte | *POLR2A* | 1.591810424 | 8.31E-06 | 0.000139378 | 0.052266636 |
| Obesity | adipocyte | *SLC25A41* | -4.135025085 | 0.001039663 | 0.004985784 | 1 |
| Obesity | adipocyte | *ZNF322* | -0.830829606 | 4.62E-07 | 2.30E-05 | 0.00290387 |
| Obesity | adipocyte | *DDX60L* | 1.204106629 | 1.90E-05 | 0.000248876 | 0.119211604 |
| Obesity | adipocyte | *SGSH* | 2.934493876 | 3.11E-05 | 0.000356539 | 0.195739654 |
| Obesity | adipocyte | *MRPS23* | 2.24574869 | 0.001062818 | 0.005088283 | 1 |
| Obesity | adipocyte | *ATG9B* | 2.96235001 | 7.93E-06 | 0.000136462 | 0.049858467 |
| Obesity | adipocyte | *ZBTB20* | 0.996256121 | 0.000242418 | 0.00160938 | 1 |
| Obesity | adipocyte | *AMIGO1* | 0.188786521 | 0.017558676 | 0.046677124 | 1 |
| Obesity | adipocyte | *RELL1* | 1.531603653 | 0.000463318 | 0.002643267 | 1 |
| Obesity | adipocyte | *RFX7* | 1.334407036 | 0.000332942 | 0.002030265 | 1 |
| Obesity | adipocyte | *ADO* | 1.945233199 | 0.001867199 | 0.007937173 | 1 |
| Obesity | adipocyte | *SNRPE* | 1.376057323 | 0.007785621 | 0.024449648 | 1 |
| Obesity | adipocyte | *CHST15* | 1.015727228 | 0.000110511 | 0.000905843 | 0.694781492 |
| Obesity | adipocyte | *TNRC18* | 1.554373996 | 6.06E-05 | 0.000586722 | 0.380782317 |
| Obesity | adipocyte | *ERCC6L2* | 0.54995923 | 0.007608208 | 0.023988368 | 1 |
| Obesity | adipocyte | *FBXL6* | -1.131124366 | 0.005078077 | 0.017464919 | 1 |
| Obesity | adipocyte | *KBTBD3* | 1.74548414 | 0.009484317 | 0.028489203 | 1 |
| Obesity | adipocyte | *CLN8* | 0.728413574 | 0.001026068 | 0.004931873 | 1 |
| Obesity | adipocyte | *FES* | 2.352107275 | 2.51E-06 | 6.46E-05 | 0.015755102 |
| Obesity | adipocyte | *CAV3* | -0.237408142 | 0.00103307 | 0.004957949 | 1 |
| Obesity | adipocyte | *MFSD5* | 0.583153213 | 0.016782916 | 0.044957049 | 1 |
| Obesity | adipocyte | *SPNS3* | 1.417069544 | 0.003848801 | 0.014051922 | 1 |
| Obesity | adipocyte | *CLEC4G* | 2.035346484 | 0.001678123 | 0.007321555 | 1 |
| Obesity | adipocyte | *SKA2* | 1.505296891 | 0.008937153 | 0.027302176 | 1 |
| Obesity | adipocyte | *BRICD5* | -0.337196572 | 0.002337005 | 0.009497575 | 1 |
| Obesity | adipocyte | *ACBD3* | 1.018949965 | 0.016040437 | 0.043225987 | 1 |
| Obesity | adipocyte | *COL18A1* | 0.50286538 | 0.000955239 | 0.004684547 | 1 |
| Obesity | adipocyte | *LYSMD4* | 0.308469439 | 0.001243123 | 0.005721459 | 1 |
| Obesity | adipocyte | *WBP2NL* | 1.4629481 | 0.015781968 | 0.042694161 | 1 |
| Obesity | adipocyte | *GAS6* | 0.895736427 | 0.00013047 | 0.001016439 | 0.820266506 |
| Obesity | adipocyte | *BEGAIN* | -2.213916527 | 0.013255245 | 0.03712059 | 1 |
| Obesity | adipocyte | *ARHGEF37* | 1.55843952 | 0.008755977 | 0.026853085 | 1 |
| Obesity | adipocyte | *GPR19* | 2.109231709 | 0.000565136 | 0.003113944 | 1 |
| Obesity | adipocyte | *FANCF* | 3.500163964 | 0.001439103 | 0.006476592 | 1 |
| Obesity | adipocyte | *C2CD4C* | 0.721694327 | 0.000776631 | 0.003938693 | 1 |
| Obesity | adipocyte | *CCBE1* | 0.373015241 | 0.00022473 | 0.001527101 | 1 |
| Obesity | adipocyte | *C19orf71* | 1.553128378 | 0.001671409 | 0.007302394 | 1 |
| Obesity | adipocyte | *LRIT3* | -0.822792533 | 0.004880053 | 0.016942834 | 1 |
| Obesity | adipocyte | *URAD* | -1.106653744 | 0.01314851 | 0.036870956 | 1 |
| Obesity | adipocyte | *GPR132* | 0.971904726 | 0.007054014 | 0.022672544 | 1 |
| Obesity | adipocyte | *PSMG1* | 0.253206612 | 4.13E-05 | 0.000441502 | 0.25960346 |
| Obesity | adipocyte | *TANGO2* | 1.145061589 | 0.000191027 | 0.001352466 | 1 |
| Obesity | adipocyte | *RP1L1* | 1.028920641 | 4.16E-08 | 3.64E-06 | 0.000261841 |
| Obesity | adipocyte | *BMP8A* | 1.549784413 | 4.16E-06 | 8.73E-05 | 0.026165695 |
| Obesity | adipocyte | *CBX6* | 2.827571107 | 6.73E-08 | 5.16E-06 | 0.000422906 |
| Obesity | adipocyte | *KCTD16* | 0.940314982 | 7.31E-05 | 0.000671154 | 0.459740505 |
| Obesity | adipocyte | *C9orf66* | -2.22403234 | 0.007057447 | 0.022672544 | 1 |
| Obesity | adipocyte | *BTBD9* | 0.793993437 | 0.005267801 | 0.017972621 | 1 |
| Obesity | adipocyte | *MAATS1* | 0.464174425 | 0.000326835 | 0.002000792 | 1 |

| | | | | | | |
|---|---|---|---|---|---|---|
| Obesity | adipocyte | *GPR39* | -0.747327687 | 0.008472623 | 0.02616276 | 1 |
| Obesity | adipocyte | *DENND5A* | 1.289923587 | 0.001094447 | 0.005185219 | 1 |
| Obesity | adipocyte | *TBX1* | 2.156299862 | 2.99E-06 | 7.03E-05 | 0.018785291 |
| Obesity | adipocyte | *UQCR10* | 2.193403234 | 2.95E-05 | 0.000344032 | 0.185433383 |
| Obesity | adipocyte | *EIF3C* | 1.020741742 | 1.14E-26 | 1.79E-23 | 7.15E-23 |
| Obesity | adipocyte | *CRELD2* | 1.266788754 | 0.003909473 | 0.014223876 | 1 |
| Obesity | adipocyte | *PLA2G6* | 0.396980423 | 5.00E-05 | 0.000508503 | 0.314107757 |
| Obesity | adipocyte | *WDR27* | 1.294948439 | 0.000188279 | 0.001334512 | 1 |
| Obesity | adipocyte | *TXNRD2* | 1.226113377 | 0.009051726 | 0.027571803 | 1 |
| Obesity | adipocyte | *PROS1* | 0.571022867 | 0.008642278 | 0.026569195 | 1 |
| Obesity | adipocyte | *SNN* | 2.249589408 | 2.04E-06 | 5.99E-05 | 0.012824092 |
| Obesity | adipocyte | *ZNF93* | 2.84808669 | 0.000167512 | 0.001223165 | 1 |
| Obesity | adipocyte | *SEPT5* | 3.881071233 | 1.83E-05 | 0.000244265 | 0.115293218 |
| Obesity | adipocyte | *ATL3* | 0.689814893 | 0.00367133 | 0.013557769 | 1 |
| Obesity | adipocyte | *NDUFA12* | 2.284694374 | 0.00026593 | 0.001728956 | 1 |
| Obesity | adipocyte | *OSBP2* | 1.53344435 | 6.44E-05 | 0.000613015 | 0.404590083 |
| Obesity | adipocyte | *TMED9* | 1.255394601 | 0.000325984 | 0.001997525 | 1 |
| Obesity | adipocyte | *H1FX* | 1.788032902 | 1.88E-06 | 5.81E-05 | 0.01183829 |
| Obesity | adipocyte | *ZFP90* | 2.018167903 | 1.33E-05 | 0.000196414 | 0.083672337 |
| Obesity | adipocyte | *NOC4L* | -0.295418391 | 0.013890881 | 0.038421455 | 1 |
| Obesity | adipocyte | *SORCS2* | 0.973538168 | 0.000181579 | 0.001298735 | 1 |
| Obesity | adipocyte | *TMEM106A* | 1.409956765 | 5.39E-06 | 0.000101756 | 0.033884744 |
| Obesity | adipocyte | *MAFF* | -0.92781668 | 0.000267756 | 0.00173349 | 1 |
| Obesity | adipocyte | *IFITM2* | 0.65642066 | 0.006594762 | 0.02143809 | 1 |
| Obesity | adipocyte | *GP1BA* | 0.570106795 | 0.000138433 | 0.001056221 | 0.870325996 |
| Obesity | adipocyte | *KLHL33* | 0.323805571 | 0.003690816 | 0.013609479 | 1 |
| Obesity | adipocyte | *HS6ST3* | 1.451261736 | 8.16E-05 | 0.000724371 | 0.512854802 |
| Obesity | adipocyte | *TNFAIP8L1* | 0.874697446 | 0.013609484 | 0.037895138 | 1 |
| Obesity | adipocyte | *SP140L* | 0.980211901 | 0.002783862 | 0.010952529 | 1 |
| Obesity | adipocyte | *FAM174B* | 0.717086728 | 0.014784667 | 0.040519269 | 1 |
| Obesity | adipocyte | *SV2B* | 0.757457889 | 0.004146849 | 0.014863876 | 1 |
| Obesity | adipocyte | *TLCD2* | -0.550392837 | 0.005958694 | 0.01983182 | 1 |
| Obesity | adipocyte | *LSAMP* | 0.518636897 | 0.006581786 | 0.021418058 | 1 |
| Obesity | adipocyte | *AHNAK2* | 1.613129079 | 9.72E-07 | 3.83E-05 | 0.006109387 |
| Obesity | adipocyte | *SP1* | 0.649499808 | 0.001983429 | 0.008310136 | 1 |
| Obesity | adipocyte | *PDIA2* | -0.435096399 | 0.009391633 | 0.028291901 | 1 |
| Obesity | adipocyte | *PCGF3* | 1.265589092 | 2.11E-05 | 0.000268503 | 0.132681897 |
| Obesity | adipocyte | *PMEL* | 0.316819629 | 0.009422292 | 0.028343516 | 1 |
| Obesity | adipocyte | *C16orf52* | 1.604528461 | 0.000142633 | 0.001081708 | 0.896735837 |
| Obesity | adipocyte | *SRL* | 0.246734554 | 0.002539596 | 0.010105344 | 1 |
| Obesity | adipocyte | *SLC52A2* | 2.679382718 | 0.002360725 | 0.009550757 | 1 |
| Obesity | adipocyte | *POMK* | 1.468013329 | 0.002514046 | 0.010035435 | 1 |
| Obesity | adipocyte | *WWOX* | 0.514088339 | 0.004238659 | 0.015132566 | 1 |
| Obesity | adipocyte | *CCDC84* | 1.289671441 | 0.004941777 | 0.017127316 | 1 |
| Obesity | adipocyte | *POLR1D* | 1.308379953 | 0.005967792 | 0.019841092 | 1 |
| Obesity | adipocyte | *FFAR4* | -0.17618981 | 0.014058212 | 0.038798936 | 1 |
| Obesity | adipocyte | *BPIFB4* | 18.39954569 | 0.001429554 | 0.006456612 | 1 |
| Obesity | adipocyte | *TOR3A* | 2.41510349 | 0.00082454 | 0.004137177 | 1 |
| Obesity | adipocyte | *KPNA4* | 1.6220207 | 0.000210125 | 0.00145331 | 1 |
| Obesity | adipocyte | *RPS23* | 0.568632304 | 0.006331002 | 0.020828367 | 1 |

| | | | | | | |
|---|---|---|---|---|---|---|
| Obesity | adipocyte | *BTN3A2* | -0.267031879 | 2.49E-10 | 4.66E-08 | 1.57E-06 |
| Obesity | adipocyte | *ZNF396* | -0.43342944 | 0.010888456 | 0.031736544 | 1 |
| Obesity | adipocyte | *BCR* | 0.624020827 | 0.00638759 | 0.020959695 | 1 |
| Obesity | adipocyte | *HYAL3* | -0.349690753 | 1.26E-05 | 0.00018782 | 0.078997216 |
| Obesity | adipocyte | *LILRB4* | 0.258301989 | 0.017318956 | 0.046117865 | 1 |
| Obesity | adipocyte | *TRABD2A* | 0.210254833 | 0.002832748 | 0.011069772 | 1 |
| Obesity | adipocyte | *TNFRSF18* | 4.100024679 | 2.33E-06 | 6.25E-05 | 0.014679079 |
| Obesity | adipocyte | *KANK3* | 1.799079467 | 0.000706958 | 0.00365213 | 1 |
| Obesity | adipocyte | *EMID1* | 1.477256015 | 0.000134658 | 0.001037493 | 0.846594519 |
| Obesity | adipocyte | *MRPS21* | 1.004861654 | 0.000256566 | 0.00168551 | 1 |
| Obesity | adipocyte | *ANGPTL5* | 2.678410425 | 0.000225433 | 0.001527101 | 1 |
| Obesity | adipocyte | *FNBP1* | 0.42716401 | 0.001561238 | 0.006941656 | 1 |
| Obesity | adipocyte | *EPOR* | 1.179967734 | 7.21E-05 | 0.000664998 | 0.453528637 |
| Obesity | adipocyte | *GJA4* | 0.683215884 | 0.007195891 | 0.02302319 | 1 |
| Obesity | adipocyte | *IFT140* | 1.525412128 | 0.001013245 | 0.004896445 | 1 |
| Obesity | adipocyte | *TLR5* | 1.315031222 | 0.005226449 | 0.017857979 | 1 |
| Obesity | adipocyte | *TET3* | 1.739191732 | 2.72E-06 | 6.77E-05 | 0.017069273 |
| Obesity | adipocyte | *ISG15* | 0.493091315 | 0.013907928 | 0.038451689 | 1 |
| Obesity | adipocyte | *TRPV2* | 2.015961345 | 2.06E-06 | 5.99E-05 | 0.012959088 |
| Obesity | adipocyte | *KBTBD12* | 0.219962308 | 0.009313965 | 0.028152355 | 1 |
| Obesity | adipocyte | *ADH1A* | -0.510694305 | 5.59E-05 | 0.000552854 | 0.351615258 |
| Obesity | adipocyte | *SEMA4D* | 1.620817017 | 3.39E-05 | 0.000379484 | 0.21326975 |
| Obesity | adipocyte | *ZSCAN23* | 0.135627401 | 0.001937015 | 0.00816768 | 1 |
| Obesity | adipocyte | *FAM166A* | -1.854342502 | 0.001090699 | 0.00517809 | 1 |
| Obesity | adipocyte | *ZC3H6* | 1.084477802 | 0.002881518 | 0.01123131 | 1 |
| Obesity | adipocyte | *NCR3LG1* | 4.118702606 | 2.25E-06 | 6.20E-05 | 0.014140915 |
| Obesity | adipocyte | *HYKK* | 0.628052583 | 0.006784149 | 0.021940302 | 1 |
| Obesity | adipocyte | *CENPP* | 0.442748478 | 5.30E-08 | 4.33E-06 | 0.000333402 |
| Obesity | adipocyte | *PLSCR1* | 1.625555072 | 3.82E-07 | 2.07E-05 | 0.002403252 |
| Obesity | adipocyte | *JAKMIP3* | -0.33193546 | 0.000542692 | 0.003016714 | 1 |
| Obesity | adipocyte | *CERKL* | 1.118534176 | 0.004140839 | 0.014863532 | 1 |
| Obesity | adipocyte | *C22orf34* | 0.477368937 | 0.000776761 | 0.003938693 | 1 |
| Obesity | adipocyte | *SRSF10* | 1.470163201 | 3.77E-05 | 0.000414535 | 0.237113898 |
| Obesity | adipocyte | *NBR1* | 1.891735867 | 0.000149897 | 0.001124589 | 0.942405527 |
| Obesity | adipocyte | *FAM72B* | 5.221784288 | 5.10E-06 | 9.86E-05 | 0.032070156 |
| Obesity | adipocyte | *SUMO2* | 1.717243026 | 0.000716998 | 0.003694893 | 1 |
| Obesity | adipocyte | *GOLGA8M* | -0.70157494 | 0.004990131 | 0.017266348 | 1 |
| Obesity | adipocyte | *DPYD* | 1.210091092 | 9.84E-06 | 0.000156185 | 0.061849453 |
| Obesity | adipocyte | *S100A16* | 1.005940057 | 0.002711775 | 0.010709125 | 1 |
| Obesity | adipocyte | *BEND4* | 2.881172487 | 0.000133219 | 0.001031461 | 0.837546428 |
| Obesity | adipocyte | *FBF1* | 0.218788098 | 0.00047457 | 0.002692801 | 1 |
| Obesity | adipocyte | *FAM196A* | 1.27207341 | 9.62E-05 | 0.000815381 | 0.605012418 |
| Obesity | adipocyte | *C9orf152* | -1.206103187 | 0.01677133 | 0.044945163 | 1 |
| Obesity | adipocyte | *NELFB* | 2.288119383 | 0.000548625 | 0.003036273 | 1 |
| Obesity | adipocyte | *ADAT2* | 0.624417073 | 0.005144023 | 0.017643464 | 1 |
| Obesity | adipocyte | *LITAF* | 0.926333672 | 0.001375614 | 0.006244393 | 1 |
| Obesity | adipocyte | *SF3B3* | 1.809446457 | 0.000438738 | 0.002528273 | 1 |
| Obesity | adipocyte | *SP6* | -6.448384782 | 0.001580604 | 0.007012885 | 1 |
| Obesity | adipocyte | *PLAC9* | 0.520261046 | 0.000826247 | 0.004139136 | 1 |
| Obesity | adipocyte | *ZNF573* | 1.202939841 | 0.008112803 | 0.025275119 | 1 |

| | | | | | | |
|---|---|---|---|---|---|---|
| Obesity | adipocyte | *S100A13* | 1.683844561 | 4.32E-06 | 8.90E-05 | 0.027155603 |
| Obesity | adipocyte | *ZNF33A* | 0.827735727 | 0.004454727 | 0.015725362 | 1 |
| Obesity | adipocyte | *ZKSCAN3* | -0.82716991 | 0.01014811 | 0.0300807 | 1 |
| Obesity | adipocyte | *SLC35E2B* | 0.266054739 | 0.008574001 | 0.026410949 | 1 |
| Obesity | adipocyte | *SEMA4A* | 0.639009733 | 0.000599866 | 0.003245577 | 1 |
| Obesity | adipocyte | *MPHOSPH8* | 1.085183046 | 0.001745846 | 0.007574972 | 1 |
| Obesity | adipocyte | *SIRPB2* | 1.641191136 | 3.76E-05 | 0.000413665 | 0.236202492 |
| Obesity | adipocyte | *SUPT5H* | 1.229136898 | 0.003406469 | 0.012801239 | 1 |
| Obesity | adipocyte | *PPIA* | 1.820478329 | 6.67E-05 | 0.000626644 | 0.419224721 |
| Obesity | adipocyte | *SUPT3H* | 1.453183291 | 0.000128079 | 0.001010333 | 0.805235716 |
| Obesity | adipocyte | *ZNF565* | 0.580761205 | 0.01173824 | 0.03357521 | 1 |
| Obesity | adipocyte | *NTNG2* | 2.248618897 | 9.02E-05 | 0.00077681 | 0.56707129 |
| Obesity | adipocyte | *LONP1* | 0.484271255 | 0.006525711 | 0.021290682 | 1 |
| Obesity | adipocyte | *ASB13* | 2.647921926 | 0.000182717 | 0.001305388 | 1 |
| Obesity | adipocyte | *ZNF124* | 1.86128297 | 0.000348463 | 0.002097529 | 1 |
| Obesity | adipocyte | *RFX8* | 0.556117903 | 0.004060022 | 0.014636099 | 1 |
| Obesity | adipocyte | *SULT1A1* | -0.518646064 | 2.92E-08 | 2.70E-06 | 0.000183279 |
| Obesity | adipocyte | *TPK1* | 0.855570209 | 1.70E-05 | 0.000234144 | 0.106769655 |
| Obesity | adipocyte | *C1orf186* | 0.856869624 | 0.016132909 | 0.043456556 | 1 |
| Obesity | adipocyte | *SULF2* | 0.522971981 | 0.004198937 | 0.015016336 | 1 |
| Obesity | adipocyte | *HDAC2* | 1.608271715 | 0.000225474 | 0.001527101 | 1 |
| Obesity | adipocyte | *HRH1* | 1.296808186 | 1.70E-06 | 5.57E-05 | 0.010699786 |
| Obesity | adipocyte | *TECPR2* | 1.712257764 | 0.000574001 | 0.003148991 | 1 |
| Obesity | adipocyte | *ZFP62* | 1.935910112 | 0.000698427 | 0.003619752 | 1 |
| Obesity | adipocyte | *TOMM7* | 2.18964438 | 7.34E-06 | 0.000129676 | 0.046164774 |
| Obesity | adipocyte | *ZNF431* | 1.026674078 | 0.001844859 | 0.007879503 | 1 |
| Obesity | adipocyte | *NF1* | 0.966135714 | 0.000206474 | 0.001431205 | 1 |
| Obesity | adipocyte | *GM2A* | 0.44854409 | 0.005732373 | 0.019180209 | 1 |
| Obesity | adipocyte | *STRN3* | 1.048966613 | 0.004037107 | 0.014561844 | 1 |
| Obesity | adipocyte | *MVB12B* | 1.558176422 | 0.000181508 | 0.001298735 | 1 |
| Obesity | adipocyte | *C6orf106* | 0.610177428 | 2.09E-09 | 3.13E-07 | 1.32E-05 |
| Obesity | adipocyte | *KIAA1211L* | 0.190360987 | 0.009710024 | 0.029000912 | 1 |
| Obesity | adipocyte | *KPNA5* | 1.334544591 | 0.003866921 | 0.014093524 | 1 |
| Obesity | adipocyte | *SRGAP1* | 1.668787025 | 4.50E-07 | 2.26E-05 | 0.002827428 |
| Obesity | adipocyte | *FAM3C* | 1.507294645 | 8.86E-07 | 3.62E-05 | 0.00556991 |
| Obesity | adipocyte | *SLC39A10* | 1.333601707 | 0.000866771 | 0.004314641 | 1 |
| Obesity | adipocyte | *ZNF585A* | 0.333586205 | 0.014995925 | 0.040991034 | 1 |
| Obesity | adipocyte | *FUT11* | 1.657938423 | 3.60E-08 | 3.19E-06 | 0.000226512 |
| Obesity | adipocyte | *ANXA4* | 1.549915173 | 1.05E-05 | 0.000164341 | 0.066213653 |
| Obesity | adipocyte | *ZNF138* | 1.000475344 | 0.000373147 | 0.002219976 | 1 |
| Obesity | adipocyte | *ZMYM1* | 1.776361947 | 0.000580726 | 0.003166546 | 1 |
| Obesity | adipocyte | *ARRDC1* | 1.232637514 | 0.003452529 | 0.012943383 | 1 |
| Obesity | adipocyte | *IGF2R* | 0.726559839 | 0.018100877 | 0.04787623 | 1 |
| Obesity | adipocyte | *GAL3ST4* | 1.234895441 | 0.000709261 | 0.003661019 | 1 |
| Obesity | adipocyte | *SLC25A29* | 1.056146964 | 8.13E-06 | 0.000137794 | 0.05112158 |
| Obesity | adipocyte | *SND1* | 0.823570576 | 0.001190052 | 0.005509466 | 1 |
| Obesity | adipocyte | *SULT1A2* | 0.445237663 | 8.01E-28 | 1.68E-24 | 5.04E-24 |
| Obesity | adipocyte | *ENTPD4* | 0.660979667 | 0.014165556 | 0.039043775 | 1 |
| Obesity | adipocyte | *FAM110D* | 1.532186784 | 0.000169503 | 0.001233424 | 1 |
| Obesity | adipocyte | *KANK2* | 1.81363617 | 8.68E-05 | 0.000757 | 0.545815179 |

| | | | | | | |
|---|---|---|---|---|---|---|
| Obesity | adipocyte | *CCL4L2* | -0.302862229 | 0.000738478 | 0.00378696 | 1 |
| Obesity | adipocyte | *GTF2E2* | 1.092192737 | 0.0006493 | 0.003436811 | 1 |
| Obesity | adipocyte | *ZNF165* | -0.613043077 | 7.11E-05 | 0.000658191 | 0.446911901 |
| Obesity | adipocyte | *MRPL21* | -0.225602301 | 0.000366338 | 0.00218517 | 1 |
| Obesity | adipocyte | *PCDHB11* | -0.867967696 | 0.0042896 | 0.015253799 | 1 |
| Obesity | adipocyte | *MYO5A* | 0.506269387 | 0.003511332 | 0.013070304 | 1 |
| Obesity | adipocyte | *ZNF624* | 1.535590252 | 0.018287296 | 0.048267098 | 1 |
| Obesity | adipocyte | *MFAP5* | 1.460812557 | 8.40E-06 | 0.000139725 | 0.052816168 |
| Obesity | adipocyte | *CDC42SE1* | 1.362107917 | 0.000440192 | 0.002534329 | 1 |
| Obesity | adipocyte | *FAM114A1* | 1.208188303 | 0.000723418 | 0.003724921 | 1 |
| Obesity | adipocyte | *PHF2* | 1.545060069 | 0.000543614 | 0.00301917 | 1 |
| Obesity | adipocyte | *RPS26* | -0.210685275 | 5.97E-05 | 0.000579949 | 0.375227309 |
| Obesity | adipocyte | *PSAP* | 1.491148922 | 4.90E-06 | 9.70E-05 | 0.030833536 |
| Obesity | adipocyte | *ZNF780A* | -0.265226305 | 2.09E-05 | 0.000268001 | 0.131521189 |
| Obesity | adipocyte | *AC122129.1* | -1.805645236 | 0.008279828 | 0.025695067 | 1 |
| Obesity | adipocyte | *ZNF677* | 1.745390923 | 0.010201822 | 0.030182992 | 1 |
| Obesity | adipocyte | *FCHSD1* | 1.401158088 | 5.62E-05 | 0.000553957 | 0.353424604 |
| Obesity | adipocyte | *LEKR1* | -0.472618303 | 0.010656748 | 0.031191329 | 1 |
| Obesity | adipocyte | *SFI1* | 2.187468959 | 1.15E-05 | 0.000175893 | 0.07246789 |
| Obesity | adipocyte | *ADH4* | -0.233246843 | 9.47E-06 | 0.000153028 | 0.059527855 |
| Obesity | adipocyte | *SOWAHC* | 3.389503782 | 0.000503102 | 0.002826635 | 1 |
| Obesity | adipocyte | *MIER1* | 1.671436288 | 6.65E-06 | 0.000119827 | 0.041819562 |
| Obesity | adipocyte | *SVIP* | 1.833721947 | 0.00420688 | 0.015036048 | 1 |
| Obesity | adipocyte | *DDRGK1* | 1.744964778 | 0.004785262 | 0.016667557 | 1 |
| Obesity | adipocyte | *HSD17B11* | 1.961419741 | 3.13E-07 | 1.76E-05 | 0.001965847 |
| Obesity | adipocyte | *QRICH1* | 0.361871995 | 0.002336006 | 0.009497575 | 1 |
| Obesity | adipocyte | *DDX42* | 0.870295409 | 0.001118673 | 0.005264294 | 1 |
| Obesity | adipocyte | *SLC29A3* | 0.691808499 | 0.012499733 | 0.035399019 | 1 |
| Obesity | adipocyte | *ANTXRL* | 80.88161419 | 0.00016062 | 0.001181073 | 1 |
| Obesity | adipocyte | *STYX* | 2.193597428 | 0.000177572 | 0.001281741 | 1 |
| Obesity | adipocyte | *ZKSCAN8* | 1.538754973 | 5.54E-08 | 4.47E-06 | 0.000348406 |
| Obesity | adipocyte | *ZNF813* | 1.186889379 | 0.016178315 | 0.043539268 | 1 |
| Obesity | adipocyte | *GFPT1* | 1.116009924 | 0.000257318 | 0.001687968 | 1 |
| Obesity | adipocyte | *SH3BGRL2* | 0.336319863 | 0.010646594 | 0.031176123 | 1 |
| Obesity | adipocyte | *TMA16* | 1.277548884 | 0.005522246 | 0.018635727 | 1 |
| Obesity | adipocyte | *MAFK* | 2.670736818 | 7.92E-06 | 0.000136462 | 0.049799519 |
| Obesity | adipocyte | *TLK1* | 1.349861534 | 0.000748675 | 0.003829876 | 1 |
| Obesity | adipocyte | *LRBA* | 1.478374694 | 1.50E-05 | 0.000212911 | 0.094106625 |
| Obesity | adipocyte | *MDM4* | 1.464386883 | 0.000241934 | 0.00160938 | 1 |
| Obesity | adipocyte | *TTC37* | 0.960175441 | 0.004158712 | 0.014897903 | 1 |
| Obesity | adipocyte | *TUSC1* | 3.657038203 | 0.001923159 | 0.008142022 | 1 |
| Obesity | adipocyte | *PAPSS2* | 0.791409357 | 0.001821262 | 0.007789301 | 1 |
| Obesity | adipocyte | *CEP290* | 1.00059065 | 0.006581716 | 0.021418058 | 1 |
| Obesity | adipocyte | *ANKRD13B* | 0.347028993 | 0.001187988 | 0.005508024 | 1 |
| Obesity | adipocyte | *ECI2* | 1.672702158 | 0.002195848 | 0.009034881 | 1 |
| Obesity | adipocyte | *ZNF652* | 1.553741314 | 0.000394035 | 0.002330479 | 1 |
| Obesity | adipocyte | *RPL10A* | 0.613402273 | 0.001663976 | 0.007275853 | 1 |
| Obesity | adipocyte | *APCDD1L* | 3.130463828 | 0.00011767 | 0.000952111 | 0.739790519 |
| Obesity | adipocyte | *RASSF9* | 2.18619256 | 0.000616703 | 0.003313858 | 1 |
| Obesity | adipocyte | *FAM169A* | 0.422372866 | 0.013778691 | 0.038203397 | 1 |

| | | | | | | |
|---|---|---|---|---|---|---|
| Obesity | adipocyte | *SFT2D1* | 1.465633822 | 0.00216336 | 0.008918717 | 1 |
| Obesity | adipocyte | *CAPZA2* | 1.69002995 | 2.43E-05 | 0.000297672 | 0.152705837 |
| Obesity | adipocyte | *MAP3K3* | 0.92266218 | 0.010524941 | 0.03093516 | 1 |
| Obesity | adipocyte | *RASGEF1A* | 8.436342068 | 5.26E-07 | 2.54E-05 | 0.003308454 |
| Obesity | adipocyte | *APRT* | 2.976070012 | 2.03E-05 | 0.000261778 | 0.127485712 |
| Obesity | adipocyte | *L3MBTL3* | 1.061914039 | 0.000339342 | 0.002060983 | 1 |
| Obesity | adipocyte | *NAGA* | 0.834854512 | 0.001758076 | 0.007608682 | 1 |
| Obesity | adipocyte | *TGM2* | 0.136846003 | 0.010576601 | 0.031063577 | 1 |
| Obesity | adipocyte | *ZNF525* | 1.050636532 | 0.010766477 | 0.031424717 | 1 |
| Obesity | adipocyte | *EFCAB2* | 1.07164488 | 0.001377649 | 0.006249118 | 1 |
| Obesity | adipocyte | *CHML* | 0.298884769 | 0.016760786 | 0.044936059 | 1 |
| Obesity | adipocyte | *CENPW* | 0.887142539 | 0.000541932 | 0.003015156 | 1 |
| Obesity | adipocyte | *DDO* | 1.025797275 | 0.015043786 | 0.041103992 | 1 |
| Obesity | adipocyte | *PCMTD2* | 1.333967848 | 0.000663074 | 0.003468173 | 1 |
| Obesity | adipocyte | *SYS1* | 3.344496941 | 3.19E-06 | 7.31E-05 | 0.020074554 |
| Obesity | adipocyte | *GIGYF2* | 1.39294356 | 0.000344391 | 0.00207991 | 1 |
| Obesity | adipocyte | *C10orf128* | 0.514282186 | 0.000218455 | 0.001494477 | 1 |
| Obesity | adipocyte | *SYT15* | 0.828278309 | 0.001439128 | 0.006476592 | 1 |
| Obesity | adipocyte | *HLA-DMA* | 0.870046019 | 0.006836916 | 0.022088228 | 1 |
| Obesity | adipocyte | *NOTCH4* | -0.074859551 | 4.49E-08 | 3.87E-06 | 0.000282205 |
| Obesity | adipocyte | *AGER* | 0.591349887 | 4.96E-05 | 0.000508417 | 0.311792114 |
| Obesity | adipocyte | *MRPL38* | -0.524414708 | 0.000206057 | 0.001429887 | 1 |
| Obesity | adipocyte | *SKIV2L* | -0.230455021 | 4.28E-07 | 2.19E-05 | 0.002688924 |
| Obesity | adipocyte | *NELFE* | 0.708470679 | 2.24E-07 | 1.33E-05 | 0.00140754 |
| Obesity | adipocyte | *GPANK1* | -0.443729282 | 0.00483629 | 0.016817343 | 1 |
| Obesity | adipocyte | *BAG6* | 0.363135002 | 0.016184392 | 0.043539268 | 1 |
| Obesity | adipocyte | *HLA-C* | 0.319197452 | 0.000200531 | 0.001402376 | 1 |
| Obesity | adipocyte | *POU5F1* | 0.189489805 | 6.78E-07 | 2.95E-05 | 0.004263708 |
| Obesity | adipocyte | *CCHCR1* | 0.353979781 | 2.28E-08 | 2.32E-06 | 0.000143645 |
| Obesity | adipocyte | *ABCF1* | 0.956636214 | 5.66E-07 | 2.60E-05 | 0.003557102 |
| Obesity | adipocyte | *HLA-E* | 1.027223833 | 9.02E-17 | 4.73E-14 | 5.67E-13 |
| Obesity | adipocyte | *TBC1D8* | 0.902336904 | 0.002963054 | 0.011463826 | 1 |
| Obesity | adipocyte | *ZFP57* | 0.272897437 | 1.80E-08 | 1.92E-06 | 0.000113183 |
| Obesity | adipocyte | *ATXN2* | 1.164683682 | 1.93E-05 | 0.000252859 | 0.121372462 |
| Obesity | adipocyte | *PCDHGA1* | -0.35455598 | 0.011087887 | 0.032183538 | 1 |
| Obesity | adipocyte | *MT1A* | 5.066544461 | 0.014353516 | 0.039492583 | 1 |
| Obesity | adipocyte | *MT1M* | 3.298943763 | 2.25E-06 | 6.20E-05 | 0.014158326 |
| Obesity | adipocyte | *SAMD9* | 1.140906376 | 0.000382333 | 0.002267667 | 1 |
| Obesity | adipocyte | *SMN2* | -0.222650691 | 0.004010354 | 0.014489576 | 1 |
| Obesity | adipocyte | *SERF1B* | -0.631102898 | 0.005622844 | 0.018954862 | 1 |
| Obesity | adipocyte | *DENND6B* | 3.006307094 | 3.63E-05 | 0.00040203 | 0.228352786 |
| Obesity | adipocyte | *EIF3CL* | 5.848382977 | 5.89E-32 | 3.70E-28 | 3.70E-28 |
| Obesity | adipocyte | *C17orf107* | 0.3931305 | 0.000150085 | 0.001124653 | 0.943583738 |
| Obesity | adipocyte | *NYNRIN* | 1.556094955 | 1.03E-05 | 0.000161515 | 0.064444684 |
| Obesity | adipocyte | *METTL6* | 0.78959068 | 0.001310711 | 0.005997409 | 1 |
| Obesity | adipocyte | *SACM1L* | 0.87235466 | 0.013714177 | 0.038100324 | 1 |
| Obesity | adipocyte | *SLC48A1* | 1.626792945 | 0.000784179 | 0.003963128 | 1 |
| Obesity | adipocyte | *AC084018.1* | -0.281117503 | 0.00010204 | 0.000847459 | 0.641526166 |
| Obesity | adipocyte | *RNF208* | -2.257947212 | 0.014242486 | 0.039221423 | 1 |
| Obesity | adipocyte | *ZNF254* | 1.260543554 | 0.002715002 | 0.010714419 | 1 |

| | | | | | | |
|---|---|---|---|---|---|---|
| Obesity | adipocyte | *AC022498.1* | -0.450276248 | 0.006452958 | 0.02110809 | 1 |
| Obesity | adipocyte | *ASIC3* | 0.424596994 | 0.000202571 | 0.001411935 | 1 |
| Obesity | adipocyte | *NOTCH2NL* | 0.316437701 | 0.002360308 | 0.009550757 | 1 |
| Obesity | adipocyte | *SUPT4H1* | 0.99354919 | 0.000714379 | 0.003684414 | 1 |
| Obesity | adipocyte | *ARHGAP19* | 1.439851949 | 0.000640944 | 0.003412035 | 1 |
| Obesity | adipocyte | *SIPA1* | 0.902187818 | 3.42E-06 | 7.63E-05 | 0.021483174 |
| Obesity | adipocyte | *TMEM110* | 1.47111852 | 6.08E-06 | 0.000111809 | 0.038238607 |
| Obesity | adipocyte | *HEXA* | 2.197086903 | 2.82E-06 | 6.91E-05 | 0.017749269 |
| Obesity | adipocyte | *LBH* | 1.864410445 | 4.36E-05 | 0.00046131 | 0.274018429 |
| Obesity | adipocyte | *SLC35F6* | 2.91691736 | 1.43E-08 | 1.60E-06 | 8.96E-05 |
| Obesity | adipocyte | *EMP2* | 0.643230418 | 0.001383545 | 0.006266821 | 1 |
| Obesity | adipocyte | *PPM1N* | 1.058506929 | 0.007042018 | 0.022646121 | 1 |
| Obesity | adipocyte | *GALT* | 0.992567568 | 0.000727411 | 0.003742418 | 1 |
| Obesity | adipocyte | *TAX1BP3* | 1.603652013 | 0.000192402 | 0.001357607 | 1 |
| Obesity | adipocyte | *AP1G2* | 1.826170156 | 0.000246132 | 0.001627163 | 1 |
| Obesity | adipocyte | *ZNF90* | 0.735882938 | 0.006954368 | 0.022410103 | 1 |
| Obesity | adipocyte | *TM6SF2* | 12.20940697 | 2.41E-05 | 0.000297116 | 0.151579971 |
| Obesity | adipocyte | *TSPAN4* | 1.903478124 | 3.51E-05 | 0.000390424 | 0.220589736 |
| Obesity | adipocyte | *PAXIP1-AS2* | 2.540305699 | 8.19E-06 | 0.000138375 | 0.051475662 |
| Obesity | adipocyte | *LYRM4* | 1.378471346 | 0.000313399 | 0.001947056 | 1 |
| Obesity | adipocyte | *NEURL1B* | 1.5583805 | 1.77E-05 | 0.000240105 | 0.111408633 |
| Obesity | adipocyte | *UBE2QL1* | -0.901513644 | 0.002754192 | 0.010849376 | 1 |
| Obesity | adipocyte | *LINC01138* | 1.221161664 | 0.018897065 | 0.049667996 | 1 |
| Obesity | adipocyte | *ATAD3C* | 0.703316141 | 0.009095729 | 0.027652248 | 1 |
| Obesity | adipocyte | *CCDC7* | 0.800872222 | 0.00576907 | 0.019282372 | 1 |
| Obesity | adipocyte | *RNU6ATAC35I* | -2.156286986 | 0.001136077 | 0.005322293 | 1 |
| Obesity | adipocyte | *AP4M1* | -0.188475271 | 0.016547057 | 0.044381973 | 1 |
| Obesity | adipocyte | *C19orf73* | -2.007394603 | 0.009654918 | 0.028891229 | 1 |
| Obesity | adipocyte | *OR2A25* | -6.630491439 | 0.014659108 | 0.040230121 | 1 |
| Obesity | adipocyte | *SLC12A8* | -0.630195096 | 0.000124004 | 0.000988104 | 0.779613787 |
| Obesity | adipocyte | *UBA52* | 0.697888259 | 0.002716524 | 0.010714419 | 1 |
| Obesity | adipocyte | *BTBD19* | 0.725594918 | 0.00334448 | 0.012621094 | 1 |
| Obesity | adipocyte | *FAM185A* | 0.93237905 | 0.010627107 | 0.031141982 | 1 |
| Obesity | adipocyte | *ADRA2B* | 5.554646957 | 1.47E-05 | 0.000210625 | 0.092464557 |
| Obesity | adipocyte | *EXOSC6* | -1.566613217 | 7.67E-12 | 2.19E-09 | 4.82E-08 |
| Obesity | adipocyte | *HLA-DPB1* | 0.287939614 | 0.009115902 | 0.027696015 | 1 |
| Obesity | adipocyte | *OSER1-AS1* | 0.9299807 | 0.015536911 | 0.042158204 | 1 |
| Obesity | adipocyte | *LINC01133* | 1.783845184 | 2.26E-06 | 6.20E-05 | 0.014193048 |
| Obesity | adipocyte | *LINC01117* | 1.001635311 | 1.97E-05 | 0.000256719 | 0.123876502 |
| Obesity | adipocyte | *LINC00240* | 0.679662413 | 2.92E-09 | 4.09E-07 | 1.84E-05 |
| Obesity | adipocyte | *LINC00184* | -1.034994197 | 6.12E-05 | 0.000590248 | 0.384522156 |
| Obesity | adipocyte | *TMEM233* | 0.319183734 | 0.007129596 | 0.02284596 | 1 |
| Obesity | adipocyte | *AL049840.1* | -0.432454786 | 0.000216024 | 0.001481069 | 1 |
| Obesity | adipocyte | *LINC00237* | 5.834438852 | 2.47E-05 | 0.00029974 | 0.155106156 |
| Obesity | adipocyte | *PLEKHM1* | 1.121962364 | 0.000164312 | 0.001202596 | 1 |
| Obesity | adipocyte | *ERCC6* | 1.438451839 | 0.001753452 | 0.007602725 | 1 |
| Obesity | adipocyte | *NOL7* | 1.820237649 | 0.00029471 | 0.001865901 | 1 |
| Obesity | adipocyte | *ELFN1* | 3.784514062 | 2.37E-06 | 6.25E-05 | 0.014905876 |
| Obesity | adipocyte | *C14orf132* | 0.882847469 | 0.000423378 | 0.002462195 | 1 |
| Obesity | adipocyte | *SLC2A1-AS1* | -0.733961395 | 0.001866541 | 0.007937173 | 1 |

| | | | | | | |
|---|---|---|---|---|---|---|
| Obesity | adipocyte | *HCG20* | -2.509982253 | 2.24E-05 | 0.000279971 | 0.140825455 |
| Obesity | adipocyte | *LINC01013* | 2.673777239 | 1.17E-05 | 0.000177737 | 0.073405352 |
| Obesity | adipocyte | *LINC00954* | 4.056506861 | 2.21E-06 | 6.20E-05 | 0.013911381 |
| Obesity | adipocyte | *PATL2* | 1.997938731 | 5.52E-06 | 0.000103995 | 0.03473449 |
| Obesity | adipocyte | *U91324.1* | -1.770854279 | 0.013756199 | 0.038197591 | 1 |
| Obesity | adipocyte | *CISD3* | 3.075524143 | 0.000194183 | 0.001367112 | 1 |
| Obesity | adipocyte | *KLHL7-AS1* | -1.558465259 | 0.004103977 | 0.014752259 | 1 |
| Obesity | adipocyte | *HLA-DPA1* | 0.64781185 | 0.000404272 | 0.002379828 | 1 |
| Obesity | adipocyte | *RPS18* | 0.890930653 | 8.06E-07 | 3.36E-05 | 0.005066902 |
| Obesity | adipocyte | *TAPBP* | 0.788806727 | 3.43E-05 | 0.000383242 | 0.21576504 |
| Obesity | adipocyte | *TNF* | -0.463275739 | 0.006167252 | 0.020396377 | 1 |
| Obesity | adipocyte | *LINC00342* | 1.023011853 | 0.017281604 | 0.0460379 | 1 |
| Obesity | adipocyte | *SNHG15* | 1.309493909 | 0.000138387 | 0.001056221 | 0.870040955 |
| Obesity | adipocyte | *LINC00702* | 0.724529564 | 0.004386984 | 0.015529823 | 1 |
| Obesity | adipocyte | *RPS28* | 2.382659995 | 0.001698358 | 0.007399569 | 1 |
| Obesity | adipocyte | *HLA-B* | 0.395602418 | 7.95E-05 | 0.000712998 | 0.499811277 |
| Obesity | adipocyte | *ZSCAN31* | 0.363296305 | 2.45E-05 | 0.000298874 | 0.153920208 |
| Obesity | adipocyte | *LINC00484* | -0.130268078 | 0.006504079 | 0.021242154 | 1 |
| Obesity | adipocyte | *ZNF737* | 1.367824036 | 0.009385081 | 0.028291901 | 1 |
| Obesity | adipocyte | *PSMB9* | 0.576085915 | 1.19E-06 | 4.44E-05 | 0.007455379 |
| Obesity | adipocyte | *COX19* | 1.530477284 | 0.007480764 | 0.023717381 | 1 |
| Obesity | adipocyte | *AQP1* | 0.925538581 | 0.002707435 | 0.010698706 | 1 |
| Obesity | adipocyte | *NSUN6* | 1.283512274 | 0.003956747 | 0.01435434 | 1 |
| Obesity | adipocyte | *ATP5J2* | 0.834013292 | 0.018244606 | 0.048194889 | 1 |
| Obesity | adipocyte | *ATP5O* | 0.712962582 | 0.016396849 | 0.044016649 | 1 |
| Obesity | adipocyte | *PI4KA* | 1.457585429 | 3.46E-06 | 7.67E-05 | 0.021775248 |
| Obesity | adipocyte | *ARFGAP3* | 1.82862247 | 0.000206909 | 0.001432643 | 1 |
| Obesity | adipocyte | *C22orf39* | 1.7823349 | 0.00770046 | 0.024242759 | 1 |
| Obesity | adipocyte | *MRPL20* | 1.279784365 | 0.001475388 | 0.00661138 | 1 |
| Obesity | adipocyte | *AP5Z1* | 1.529006023 | 0.00089781 | 0.004448015 | 1 |
| Obesity | adipocyte | *FGF14-IT1* | 4.658922883 | 7.13E-06 | 0.00012694 | 0.044809975 |
| Obesity | adipocyte | *NAT6* | 0.996668469 | 8.67E-05 | 0.000756837 | 0.544922374 |
| Obesity | adipocyte | *LINC01011* | 0.521144482 | 0.007479094 | 0.023717381 | 1 |
| Obesity | adipocyte | *APOBEC3C* | 3.146966492 | 7.89E-07 | 3.31E-05 | 0.004959605 |
| Obesity | adipocyte | *ZNF585B* | 0.256228547 | 0.016875947 | 0.045167763 | 1 |
| Obesity | adipocyte | *CDK11B* | 1.873395862 | 0.001127551 | 0.005290235 | 1 |
| Obesity | adipocyte | *PCDHAC1* | -1.114175888 | 0.000698963 | 0.003619752 | 1 |
| Obesity | adipocyte | *ABHD14A* | 0.948213043 | 0.0190402 | 0.049918989 | 1 |
| Obesity | adipocyte | *SRP14-AS1* | 0.114164012 | 0.011930507 | 0.034047707 | 1 |
| Obesity | adipocyte | *ZNF286B* | 1.208474365 | 0.004371175 | 0.015482578 | 1 |
| Obesity | adipocyte | *PDCD6* | 1.056642532 | 0.008906646 | 0.027235448 | 1 |
| Obesity | adipocyte | *DNAH10OS* | 2.951485746 | 0.008125466 | 0.025302032 | 1 |
| Obesity | adipocyte | *PCDHA10* | 1.071553793 | 0.015114619 | 0.041261662 | 1 |
| Obesity | adipocyte | *ZNF718* | 1.63224354 | 0.000245192 | 0.001624367 | 1 |
| Obesity | adipocyte | *LINC00989* | 2.027752951 | 0.000363934 | 0.002172892 | 1 |
| Obesity | adipocyte | *LINC01021* | -1.096270751 | 0.001760661 | 0.007608682 | 1 |
| Obesity | adipocyte | *FAM218A* | -1.195713819 | 0.002806079 | 0.0110055 | 1 |
| Obesity | adipocyte | *GLI4* | -0.625633612 | 0.011141599 | 0.032324519 | 1 |
| Obesity | adipocyte | *SNHG18* | -0.641095018 | 0.000277868 | 0.001775359 | 1 |
| Obesity | adipocyte | *TRNP1* | 6.812002877 | 1.83E-05 | 0.000244141 | 0.114749899 |

| | | | | | | |
|---|---|---|---|---|---|---|
| Obesity | adipocyte | *ATXN7L3B* | 1.652900705 | 0.007379097 | 0.023473941 | 1 |
| Obesity | adipocyte | *PRKDC* | 1.544443565 | 0.000126137 | 0.000997517 | 0.793026271 |
| Obesity | adipocyte | *PCDHGA6* | 0.321888764 | 0.00893473 | 0.027302176 | 1 |
| Obesity | adipocyte | *CLDN23* | 1.00977666 | 0.015588161 | 0.042260789 | 1 |
| Obesity | adipocyte | *ARMS2* | -0.619162788 | 0.018477308 | 0.048727699 | 1 |
| Obesity | adipocyte | *FPGT* | 2.18269766 | 0.004413151 | 0.015596111 | 1 |
| Obesity | adipocyte | *MPV17L2* | 0.982488001 | 2.11E-06 | 6.01E-05 | 0.013255979 |
| Obesity | adipocyte | *DPP3* | 0.294024699 | 0.00249679 | 0.009979224 | 1 |
| Obesity | adipocyte | *CTSO* | 2.148700013 | 2.96E-06 | 6.99E-05 | 0.018581179 |
| Obesity | adipocyte | *HMBS* | 0.59743606 | 6.62E-11 | 1.54E-08 | 4.16E-07 |
| Obesity | adipocyte | *AC093668.2* | -0.495996261 | 0.003059963 | 0.011773556 | 1 |
| Obesity | adipocyte | *PGA5* | -3.236481595 | 0.010578514 | 0.031063577 | 1 |
| Obesity | adipocyte | *HP* | -0.893918449 | 0.000117123 | 0.000948911 | 0.73635469 |
| Obesity | adipocyte | *KIAA1147* | 1.174279184 | 6.47E-05 | 0.000613065 | 0.407075343 |
| Obesity | adipocyte | *ZBED6* | -1.780566825 | 0.011659543 | 0.033414874 | 1 |
| Obesity | adipocyte | *DUXA* | -0.295560018 | 0.010629982 | 0.031141982 | 1 |
| Obesity | adipocyte | *CEP95* | 0.916563788 | 0.007815601 | 0.024531544 | 1 |
| Obesity | adipocyte | *LINC00639* | 0.355458973 | 0.003221463 | 0.012295053 | 1 |
| Obesity | adipocyte | *LINC00648* | 1.834914305 | 0.001872096 | 0.007952611 | 1 |
| Obesity | adipocyte | *LINC00924* | 0.76992208 | 0.011542421 | 0.033196342 | 1 |
| Obesity | adipocyte | *UBE2Q2L* | -1.688176092 | 0.000107134 | 0.00088161 | 0.673550114 |
| Obesity | adipocyte | *AC083843.1* | -6.682216971 | 7.24E-06 | 0.000128284 | 0.045540709 |
| Obesity | adipocyte | *SH3RF3-AS1* | -0.414712841 | 0.000677255 | 0.003527675 | 1 |
| Obesity | adipocyte | *HOXB7* | 1.623556537 | 6.33E-06 | 0.00011529 | 0.039775084 |
| Obesity | adipocyte | *LINC00562* | -0.275290383 | 0.000592969 | 0.003222125 | 1 |
| Obesity | adipocyte | *AC009120.5* | -0.943260022 | 0.004209233 | 0.015036048 | 1 |
| Obesity | adipocyte | *TMEM178B* | 1.566668384 | 0.00065974 | 0.003456489 | 1 |
| Obesity | adipocyte | *GOLGA8H* | 0.224949692 | 0.015684805 | 0.042467859 | 1 |
| Obesity | adipocyte | *LINC00662* | 1.199223923 | 0.007779504 | 0.024442648 | 1 |
| Obesity | adipocyte | *STRADA* | -0.593538368 | 0.010162148 | 0.030108116 | 1 |
| Obesity | adipocyte | *MYO15B* | 0.871726513 | 0.000133949 | 0.001034566 | 0.842136891 |
| Obesity | adipocyte | *LINC00906* | -5.4686382 | 0.004284708 | 0.015245024 | 1 |
| Obesity | adipocyte | *AC009060.1* | -2.082240628 | 3.92E-07 | 2.07E-05 | 0.002466961 |
| Obesity | adipocyte | *LINC01050* | 3.958399627 | 0.004012463 | 0.014489576 | 1 |
| Obesity | adipocyte | *GTF2H5* | 0.804870495 | 1.76E-05 | 0.000238689 | 0.110513207 |
| Obesity | ASPC | *SEMA3F* | -0.295310036 | 9.58E-24 | 8.61E-21 | 6.02E-20 |
| Obesity | ASPC | *WNT16* | 0.262754874 | 0.001015746 | 0.020145098 | 1 |
| Obesity | ASPC | *RBM6* | -0.227804582 | 1.42E-25 | 1.79E-22 | 8.93E-22 |
| Obesity | ASPC | *GGNBP2* | -0.495839615 | 4.50E-11 | 8.84E-09 | 2.83E-07 |
| Obesity | ASPC | *HFE* | -0.336647923 | 2.52E-09 | 4.06E-07 | 1.58E-05 |
| Obesity | ASPC | *TTC19* | 0.439164034 | 0.00011339 | 0.003674653 | 0.712882634 |
| Obesity | ASPC | *PSMC4* | 0.284586248 | 0.001061588 | 0.020856886 | 1 |
| Obesity | ASPC | *NR1H3* | 0.365224372 | 0.002735504 | 0.043984951 | 1 |
| Obesity | ASPC | *BTN3A1* | -0.141060906 | 0.001256484 | 0.023865613 | 1 |
| Obesity | ASPC | *RABEP1* | 0.391257894 | 3.61E-05 | 0.001532209 | 0.226766951 |
| Obesity | ASPC | *ZCCHC8* | 2.079117205 | 5.98E-09 | 8.54E-07 | 3.76E-05 |
| Obesity | ASPC | *SH3YL1* | -0.270913658 | 1.38E-05 | 0.000676454 | 0.086586083 |
| Obesity | ASPC | *VCL* | -0.421594045 | 0.000227146 | 0.006307324 | 1 |
| Obesity | ASPC | *STAP1* | -0.514647389 | 1.24E-05 | 0.000623349 | 0.077918676 |
| Obesity | ASPC | *LSG1* | 0.125844632 | 0.002907451 | 0.045727107 | 1 |

| | | | | | | |
|---|---|---|---|---|---|---|
| Obesity | ASPC | *AP2S1* | -1.72642852 | 0.000404257 | 0.009927979 | 1 |
| Obesity | ASPC | *RCN1* | -0.414934294 | 0.001902407 | 0.03303987 | 1 |
| Obesity | ASPC | *ANAPC4* | 0.668584874 | 0.001218685 | 0.023430795 | 1 |
| Obesity | ASPC | *SDCCAG8* | 0.246071532 | 0.000344162 | 0.008904318 | 1 |
| Obesity | ASPC | *ITIH4* | 0.135385898 | 1.80E-10 | 3.33E-08 | 1.13E-06 |
| Obesity | ASPC | *TAF11* | -0.209073369 | 0.001941916 | 0.033540726 | 1 |
| Obesity | ASPC | *UHRF1BP1* | 0.317226219 | 9.28E-23 | 6.48E-20 | 5.83E-19 |
| Obesity | ASPC | *NFYC* | -0.858049469 | 0.000468648 | 0.011245765 | 1 |
| Obesity | ASPC | *TIPIN* | 0.099805984 | 2.95E-05 | 0.00133519 | 0.18559145 |
| Obesity | ASPC | *MARK3* | -1.040090483 | 1.55E-07 | 1.40E-05 | 0.000976949 |
| Obesity | ASPC | *NT5C2* | 0.833120578 | 5.64E-05 | 0.002215759 | 0.354521513 |
| Obesity | ASPC | *DYNC1I2* | 0.908436026 | 2.14E-05 | 0.001019172 | 0.134530681 |
| Obesity | ASPC | *PIK3C3* | -1.097904608 | 0.001553823 | 0.028315604 | 1 |
| Obesity | ASPC | *RSBN1* | -0.784467234 | 0.000722483 | 0.015771697 | 1 |
| Obesity | ASPC | *PCDHGA2* | -0.486189982 | 0.000720799 | 0.015771697 | 1 |
| Obesity | ASPC | *CCNT2* | -0.232921362 | 7.23E-05 | 0.002614147 | 0.454861571 |
| Obesity | ASPC | *COBLL1* | 0.181901807 | 0.000324867 | 0.008474844 | 1 |
| Obesity | ASPC | *COL16A1* | -0.306321938 | 5.47E-06 | 0.000337041 | 0.034378176 |
| Obesity | ASPC | *SCAMP1* | 0.66706001 | 0.000278846 | 0.007397074 | 1 |
| Obesity | ASPC | *OGFOD1* | -0.517283747 | 5.33E-07 | 4.29E-05 | 0.003350012 |
| Obesity | ASPC | *PTHLH* | 0.319203927 | 0.003302878 | 0.049440944 | 1 |
| Obesity | ASPC | *C3orf18* | 0.580105344 | 9.49E-10 | 1.61E-07 | 5.97E-06 |
| Obesity | ASPC | *YPEL3* | 0.216670657 | 1.00E-08 | 1.36E-06 | 6.30E-05 |
| Obesity | ASPC | *PABPC4* | -0.462271516 | 1.38E-10 | 2.62E-08 | 8.65E-07 |
| Obesity | ASPC | *RAPGEF4* | -0.86394697 | 0.001363623 | 0.025439455 | 1 |
| Obesity | ASPC | *RGS17* | -0.340427665 | 0.000663912 | 0.014840523 | 1 |
| Obesity | ASPC | *PDE6C* | 0.352692434 | 0.002023371 | 0.034851874 | 1 |
| Obesity | ASPC | *STX1B* | -0.374278039 | 2.23E-05 | 0.001056054 | 0.140455125 |
| Obesity | ASPC | *SMARCB1* | 0.203763672 | 0.001669013 | 0.030152551 | 1 |
| Obesity | ASPC | *ANKRD54* | -0.203560377 | 6.89E-05 | 0.00253453 | 0.433404585 |
| Obesity | ASPC | *FBXO7* | 0.502719353 | 0.00099646 | 0.019825146 | 1 |
| Obesity | ASPC | *SAMM50* | -1.300481477 | 0.003061481 | 0.047291231 | 1 |
| Obesity | ASPC | *ACO2* | 0.379916064 | 0.00126183 | 0.02389496 | 1 |
| Obesity | ASPC | *DESI1* | 0.20245331 | 2.87E-05 | 0.001306673 | 0.180320858 |
| Obesity | ASPC | *DICER1* | 0.434103286 | 0.00071474 | 0.015711785 | 1 |
| Obesity | ASPC | *ZFYVE21* | 0.595373063 | 0.000416214 | 0.010103238 | 1 |
| Obesity | ASPC | *ABHD12* | 0.370089033 | 3.70E-05 | 0.001563236 | 0.2329222 |
| Obesity | ASPC | *ANGPT4* | -0.714174303 | 0.000159184 | 0.004811474 | 1 |
| Obesity | ASPC | *UGGT2* | -0.429774313 | 0.00129292 | 0.024337085 | 1 |
| Obesity | ASPC | *CORO1A* | -0.725815847 | 1.46E-15 | 6.13E-13 | 9.19E-12 |
| Obesity | ASPC | *ELMO3* | 0.105837235 | 0.002119557 | 0.036309682 | 1 |
| Obesity | ASPC | *COG4* | -0.442490264 | 1.59E-06 | 0.000110797 | 0.009971694 |
| Obesity | ASPC | *NAGPA* | 0.425986911 | 0.001938746 | 0.033540726 | 1 |
| Obesity | ASPC | *CLUAP1* | 0.160238569 | 0.000414496 | 0.010100522 | 1 |
| Obesity | ASPC | *KAT8* | 1.398887062 | 3.96E-13 | 1.13E-10 | 2.49E-09 |
| Obesity | ASPC | *RNF40* | -1.132988033 | 5.74E-06 | 0.000350258 | 0.036076603 |
| Obesity | ASPC | *KNOP1* | -0.517382965 | 3.78E-06 | 0.00024278 | 0.023792441 |
| Obesity | ASPC | *ZDHHC2* | -0.401341828 | 9.57E-06 | 0.000519016 | 0.060151593 |
| Obesity | ASPC | *MCM4* | 0.252426929 | 0.003337359 | 0.049838419 | 1 |
| Obesity | ASPC | *SNRNP70* | -0.489513361 | 5.36E-05 | 0.002117487 | 0.336680399 |

| | | | | | | |
|---|---|---|---|---|---|---|
| Obesity | ASPC | *PPP1R37* | 0.387324531 | 0.000819552 | 0.017209809 | 1 |
| Obesity | ASPC | *CCNE1* | -1.222853584 | 0.001200546 | 0.023192646 | 1 |
| Obesity | ASPC | *ICAM5* | -1.24057726 | 0.000227734 | 0.006307324 | 1 |
| Obesity | ASPC | *ELL* | -0.515696168 | 0.000542458 | 0.012678202 | 1 |
| Obesity | ASPC | *ATP13A1* | -0.320551833 | 9.59E-06 | 0.000519016 | 0.060268635 |
| Obesity | ASPC | *MEST* | -0.135196215 | 0.000361168 | 0.009170156 | 1 |
| Obesity | ASPC | *OGN* | 0.453134964 | 2.34E-05 | 0.001086368 | 0.146945028 |
| Obesity | ASPC | *RGP1* | 0.960343315 | 0.000210914 | 0.006027357 | 1 |
| Obesity | ASPC | *MICU1* | 0.913885818 | 0.002170306 | 0.036977549 | 1 |
| Obesity | ASPC | *ACBD5* | 0.615828449 | 0.000647084 | 0.014529356 | 1 |
| Obesity | ASPC | *RAD51C* | -0.139935063 | 0.00036173 | 0.009170156 | 1 |
| Obesity | ASPC | *HOXB6* | 0.36925048 | 0.000158027 | 0.004811474 | 0.993517691 |
| Obesity | ASPC | *CHRNE* | 0.101482405 | 0.001202609 | 0.023192646 | 1 |
| Obesity | ASPC | *NUP88* | -0.801006821 | 1.21E-06 | 8.74E-05 | 0.007600511 |
| Obesity | ASPC | *C1QBP* | 1.099714342 | 0.000584954 | 0.01347108 | 1 |
| Obesity | ASPC | *CARS* | -0.388498821 | 0.002237745 | 0.037616854 | 1 |
| Obesity | ASPC | *KCTD10* | -0.161064976 | 0.001701024 | 0.0306428 | 1 |
| Obesity | ASPC | *TRPV4* | -0.440491319 | 6.81E-05 | 0.002518193 | 0.428092842 |
| Obesity | ASPC | *DDX55* | 0.199712836 | 0.000111458 | 0.003630767 | 0.700738072 |
| Obesity | ASPC | *SLC26A8* | -0.117556015 | 0.002190423 | 0.037219425 | 1 |
| Obesity | ASPC | *CAP2* | -0.286931313 | 0.00012072 | 0.003852614 | 0.758965055 |
| Obesity | ASPC | *PRSS16* | 0.593660953 | 1.07E-08 | 1.41E-06 | 6.74E-05 |
| Obesity | ASPC | *NUDT12* | -1.342893081 | 0.000275042 | 0.00732707 | 1 |
| Obesity | ASPC | *NME5* | 0.218189434 | 0.00024212 | 0.006676345 | 1 |
| Obesity | ASPC | *IK* | -0.399564829 | 0.000204709 | 0.005876741 | 1 |
| Obesity | ASPC | *BNIP1* | -0.096755555 | 0.00177221 | 0.031563413 | 1 |
| Obesity | ASPC | *HYAL1* | -0.145103179 | 0.000405838 | 0.009928035 | 1 |
| Obesity | ASPC | *KLHL24* | 0.304362103 | 0.001028229 | 0.020328535 | 1 |
| Obesity | ASPC | *PLCH1* | 0.627571859 | 0.000603485 | 0.01380583 | 1 |
| Obesity | ASPC | *ACTR1B* | 0.845279461 | 3.53E-05 | 0.001518877 | 0.221755999 |
| Obesity | ASPC | *PLCD4* | -2.029728682 | 3.31E-05 | 0.001454427 | 0.207983007 |
| Obesity | ASPC | *CHMP3* | -0.440086547 | 1.36E-07 | 1.24E-05 | 0.000857063 |
| Obesity | ASPC | *CD207* | 8.318845852 | 0.000389602 | 0.009643416 | 1 |
| Obesity | ASPC | *DNAJC16* | -0.366150824 | 0.000746396 | 0.016237342 | 1 |
| Obesity | ASPC | *EDEM3* | -1.304341826 | 0.002665719 | 0.042972761 | 1 |
| Obesity | ASPC | *MFAP2* | -0.135519075 | 0.000930824 | 0.01881701 | 1 |
| Obesity | ASPC | *PRRC2C* | 0.667165842 | 0.002767685 | 0.044275913 | 1 |
| Obesity | ASPC | *TNFSF4* | -0.283802138 | 0.002487932 | 0.04127078 | 1 |
| Obesity | ASPC | *DARS2* | -0.978908365 | 7.23E-08 | 7.21E-06 | 0.000454238 |
| Obesity | ASPC | *SERPINC1* | 0.268586562 | 5.46E-10 | 9.80E-08 | 3.43E-06 |
| Obesity | ASPC | *CCDC92* | 0.52251367 | 1.57E-11 | 3.41E-09 | 9.89E-08 |
| Obesity | ASPC | *INVS* | 0.696115133 | 8.07E-05 | 0.002820869 | 0.507410982 |
| Obesity | ASPC | *DCAF4* | 0.300483344 | 8.42E-05 | 0.002875892 | 0.529164105 |
| Obesity | ASPC | *CNRIP1* | -0.435107042 | 0.001321195 | 0.024795083 | 1 |
| Obesity | ASPC | *CRIPT* | -0.433239647 | 0.000883281 | 0.018207176 | 1 |
| Obesity | ASPC | *HOXB5* | 0.422483492 | 3.30E-05 | 0.001454427 | 0.207538533 |
| Obesity | ASPC | *HOXB3* | 0.321001208 | 7.03E-06 | 0.000401666 | 0.044183302 |
| Obesity | ASPC | *WDR55* | -0.539866991 | 0.000159142 | 0.004811474 | 1 |
| Obesity | ASPC | *SNX19* | -0.364095324 | 3.00E-05 | 0.001345465 | 0.18836511 |
| Obesity | ASPC | *ADRA1A* | 0.045050253 | 8.02E-05 | 0.002820869 | 0.504058176 |

| | | | | | | |
|---|---|---|---|---|---|---|
| Obesity | ASPC | *TEX14* | 0.301230631 | 0.002600059 | 0.042397856 | 1 |
| Obesity | ASPC | *GJB6* | -2.777152337 | 0.000340563 | 0.00884759 | 1 |
| Obesity | ASPC | *GPSM2* | 0.381151971 | 0.003145749 | 0.047771311 | 1 |
| Obesity | ASPC | *ARHGAP9* | -0.870199546 | 7.57E-06 | 0.000425178 | 0.047619953 |
| Obesity | ASPC | *SPATS2* | -0.768124015 | 0.001499446 | 0.027484021 | 1 |
| Obesity | ASPC | *IKZF4* | -0.638423777 | 0.000257641 | 0.006951896 | 1 |
| Obesity | ASPC | *SNRPC* | -0.543274075 | 5.25E-24 | 5.50E-21 | 3.30E-20 |
| Obesity | ASPC | *HIST1H2BJ* | 0.406074957 | 5.87E-10 | 1.03E-07 | 3.69E-06 |
| Obesity | ASPC | *NUP153* | 0.892134696 | 0.000688634 | 0.015298384 | 1 |
| Obesity | ASPC | *EEF1E1* | 1.624413837 | 3.91E-05 | 0.00162858 | 0.245915543 |
| Obesity | ASPC | *GRK4* | 0.07899047 | 3.41E-05 | 0.001479342 | 0.214504624 |
| Obesity | ASPC | *OPRL1* | 0.183015746 | 0.001838242 | 0.032372631 | 1 |
| Obesity | ASPC | *PCSK2* | 1.020355181 | 1.13E-06 | 8.44E-05 | 0.007093384 |
| Obesity | ASPC | *KLC1* | 0.829195296 | 2.33E-05 | 0.001086368 | 0.146570495 |
| Obesity | ASPC | *XRCC3* | 0.795081604 | 4.49E-06 | 0.000279566 | 0.028236179 |
| Obesity | ASPC | *YEATS4* | 0.159408771 | 0.001896401 | 0.033026791 | 1 |
| Obesity | ASPC | *RIBC2* | 0.166635216 | 0.000382213 | 0.009573588 | 1 |
| Obesity | ASPC | *GAD1* | -4.270603896 | 0.003133923 | 0.04770696 | 1 |
| Obesity | ASPC | *RPAIN* | -0.556546546 | 2.96E-08 | 3.38E-06 | 0.000185841 |
| Obesity | ASPC | *ATP8B3* | -0.190465729 | 1.79E-08 | 2.16E-06 | 0.000112499 |
| Obesity | ASPC | *PGPEP1* | 0.227622395 | 0.002147302 | 0.036685012 | 1 |
| Obesity | ASPC | *CYP2E1* | -0.441507207 | 0.000110921 | 0.003630767 | 0.697362525 |
| Obesity | ASPC | *CLIP1* | 1.265910927 | 7.00E-08 | 7.09E-06 | 0.000439857 |
| Obesity | ASPC | *PKDREJ* | -0.180804128 | 0.00116378 | 0.022722628 | 1 |
| Obesity | ASPC | *EXOC4* | 0.623365902 | 0.000138798 | 0.004363122 | 0.872624355 |
| Obesity | ASPC | *EMILIN2* | 0.337861682 | 5.87E-07 | 4.68E-05 | 0.003693338 |
| Obesity | ASPC | *IGHMBP2* | 0.189628608 | 1.45E-05 | 0.00069626 | 0.091210062 |
| Obesity | ASPC | *BTBD2* | -0.405977848 | 0.00230859 | 0.038593411 | 1 |
| Obesity | ASPC | *LARS* | -0.134304666 | 0.000197381 | 0.005718597 | 1 |
| Obesity | ASPC | *GSTM5* | 0.127397546 | 0.002750099 | 0.044106822 | 1 |
| Obesity | ASPC | *YWHAQ* | -0.931813513 | 0.000301553 | 0.007965811 | 1 |
| Obesity | ASPC | *ACP2* | 0.370042898 | 6.45E-05 | 0.002415134 | 0.405742478 |
| Obesity | ASPC | *P2RX4* | 0.214005131 | 0.002209778 | 0.037346444 | 1 |
| Obesity | ASPC | *TMEM60* | -0.405839733 | 1.42E-05 | 0.000684941 | 0.089042334 |
| Obesity | ASPC | *POM121C* | -1.024676142 | 2.72E-19 | 1.56E-16 | 1.71E-15 |
| Obesity | ASPC | *USP37* | 0.671147233 | 0.00022608 | 0.006307324 | 1 |
| Obesity | ASPC | *BLK* | -0.441659254 | 1.30E-05 | 0.00064738 | 0.081569845 |
| Obesity | ASPC | *DSCC1* | 0.047030419 | 0.003346926 | 0.049862854 | 1 |
| Obesity | ASPC | *ZSCAN9* | 0.271141697 | 2.12E-06 | 0.00014365 | 0.013359441 |
| Obesity | ASPC | *TCF19* | 0.334077265 | 3.60E-05 | 0.001532209 | 0.226626287 |
| Obesity | ASPC | *NRM* | 0.205233859 | 0.00157905 | 0.028617558 | 1 |
| Obesity | ASPC | *MAP2K5* | 1.709047508 | 1.38E-08 | 1.74E-06 | 8.71E-05 |
| Obesity | ASPC | *IFT172* | 0.870411119 | 2.09E-06 | 0.000142946 | 0.013151055 |
| Obesity | ASPC | *CARF* | 0.284487957 | 5.81E-06 | 0.000351085 | 0.036512856 |
| Obesity | ASPC | *UBL7* | 0.45742067 | 0.000824889 | 0.017229498 | 1 |
| Obesity | ASPC | *RITA1* | -0.123666623 | 0.002960223 | 0.046295825 | 1 |
| Obesity | ASPC | *RHOF* | -0.391044403 | 8.12E-05 | 0.002820869 | 0.510577279 |
| Obesity | ASPC | *WDR93* | -0.091462021 | 0.000786023 | 0.016923728 | 1 |
| Obesity | ASPC | *SH3GL3* | -0.18455267 | 0.000192458 | 0.005654141 | 1 |
| Obesity | ASPC | *MYO19* | 1.294862759 | 1.26E-11 | 2.83E-09 | 7.91E-08 |

| | | | | | | |
|---|---|---|---|---|---|---|
| Obesity | ASPC | *C18orf8* | -0.795799795 | 2.24E-13 | 6.70E-11 | 1.41E-09 |
| Obesity | ASPC | *NPC1* | -1.08705894 | 5.44E-13 | 1.42E-10 | 3.42E-09 |
| Obesity | ASPC | *TRIM65* | 0.344225806 | 1.39E-05 | 0.000679839 | 0.08769927 |
| Obesity | ASPC | *NTN5* | -0.259117102 | 0.002298519 | 0.038535443 | 1 |
| Obesity | ASPC | *RPS11* | 0.479090152 | 0.002889333 | 0.045641302 | 1 |
| Obesity | ASPC | *GPN2* | -1.130485894 | 0.00294394 | 0.046155994 | 1 |
| Obesity | ASPC | *TUFT1* | -0.167228941 | 0.00099336 | 0.019825146 | 1 |
| Obesity | ASPC | *SEMA6C* | 0.620480097 | 0.000555808 | 0.012846927 | 1 |
| Obesity | ASPC | *S100A8* | -0.481267453 | 0.000507595 | 0.011952241 | 1 |
| Obesity | ASPC | *HAX1* | 0.421204943 | 0.000855074 | 0.017771388 | 1 |
| Obesity | ASPC | *CEP170* | 0.498220664 | 0.000820806 | 0.017209809 | 1 |
| Obesity | ASPC | *GUK1* | 1.369414375 | 0.00034823 | 0.008972641 | 1 |
| Obesity | ASPC | *AFF3* | -0.180792458 | 0.000547587 | 0.012750664 | 1 |
| Obesity | ASPC | *HSPD1* | 1.009086268 | 0.000391709 | 0.009657536 | 1 |
| Obesity | ASPC | *NBEAL1* | 0.176180212 | 5.01E-05 | 0.0020136 | 0.314973914 |
| Obesity | ASPC | *AMT* | -0.083956745 | 0.001231874 | 0.023540403 | 1 |
| Obesity | ASPC | *EAF2* | -0.380623011 | 0.000665664 | 0.014840523 | 1 |
| Obesity | ASPC | *CENPC* | -0.346961245 | 1.00E-05 | 0.000524288 | 0.062914541 |
| Obesity | ASPC | *MARCH1* | -0.180353396 | 0.000985775 | 0.019800536 | 1 |
| Obesity | ASPC | *SLC25A48* | -0.123978478 | 0.001173472 | 0.022840928 | 1 |
| Obesity | ASPC | *PCYOX1L* | -0.246522035 | 0.000194762 | 0.005695196 | 1 |
| Obesity | ASPC | *ABT1* | 0.236046633 | 0.000794246 | 0.016970865 | 1 |
| Obesity | ASPC | *ADHFE1* | -0.068192287 | 0.001373777 | 0.025477681 | 1 |
| Obesity | ASPC | *ATP6V0D2* | -0.294450693 | 6.01E-06 | 0.00035668 | 0.037808048 |
| Obesity | ASPC | *NRBF2* | -0.178635363 | 0.000311422 | 0.008192091 | 1 |
| Obesity | ASPC | *MTG1* | 0.134889269 | 0.001082975 | 0.02121079 | 1 |
| Obesity | ASPC | *HSD17B12* | -0.217605807 | 4.16E-12 | 9.69E-10 | 2.62E-08 |
| Obesity | ASPC | *TTC12* | 0.12032927 | 6.17E-05 | 0.002349144 | 0.387608709 |
| Obesity | ASPC | *CCDC15* | -0.231533903 | 1.01E-05 | 0.00052646 | 0.0637016 |
| Obesity | ASPC | *TRPT1* | -0.460756393 | 0.000110862 | 0.003630767 | 0.696987601 |
| Obesity | ASPC | *TBX6* | 0.203774719 | 8.82E-08 | 8.67E-06 | 0.000554723 |
| Obesity | ASPC | *DOC2A* | -0.768221024 | 2.03E-16 | 9.81E-14 | 1.28E-12 |
| Obesity | ASPC | *PRSS53* | -0.393402892 | 1.42E-06 | 0.000100299 | 0.008926571 |
| Obesity | ASPC | *DPYSL4* | 0.094339368 | 0.000145952 | 0.004498037 | 0.917599517 |
| Obesity | ASPC | *BEND6* | -0.097818363 | 0.003189047 | 0.048195999 | 1 |
| Obesity | ASPC | *ARL14EP* | -0.529816254 | 9.75E-06 | 0.000519016 | 0.061270165 |
| Obesity | ASPC | *POC5* | -0.411497023 | 4.05E-08 | 4.32E-06 | 0.000254612 |
| Obesity | ASPC | *JAZF1* | 0.61096894 | 9.49E-05 | 0.003175031 | 0.596905915 |
| Obesity | ASPC | *CEP112* | 0.244810641 | 0.003026096 | 0.046975471 | 1 |
| Obesity | ASPC | *NEIL2* | 0.193017944 | 0.000109593 | 0.003626389 | 0.689013921 |
| Obesity | ASPC | *RABGEF1* | -0.084555461 | 0.003087893 | 0.04746598 | 1 |
| Obesity | ASPC | *EPHB1* | -1.238327231 | 0.000197153 | 0.005718597 | 1 |
| Obesity | ASPC | *EXOG* | -1.0339369 | 0.002787118 | 0.044361047 | 1 |
| Obesity | ASPC | *TIMP4* | -0.385195442 | 0.002485213 | 0.04127078 | 1 |
| Obesity | ASPC | *STEAP2* | -0.088559814 | 0.000223927 | 0.006284945 | 1 |
| Obesity | ASPC | *CLEC18A* | -0.533018288 | 4.08E-16 | 1.83E-13 | 2.56E-12 |
| Obesity | ASPC | *HPD* | 0.238599679 | 6.80E-06 | 0.000395572 | 0.042721773 |
| Obesity | ASPC | *COPG2* | -0.0626751 | 0.001033887 | 0.020376323 | 1 |
| Obesity | ASPC | *SLC5A11* | -0.11613774 | 1.20E-07 | 1.11E-05 | 0.000756359 |
| Obesity | ASPC | *RAPGEF6* | -1.159659759 | 0.000991504 | 0.019825146 | 1 |

| | | | | | | |
|---|---|---|---|---|---|---|
| Obesity | ASPC | *UBE2Z* | -1.095857558 | 0.000164718 | 0.004931344 | 1 |
| Obesity | ASPC | *CCDC107* | -0.3340877 | 0.000464114 | 0.011179635 | 1 |
| Obesity | ASPC | *C19orf47* | 0.500461642 | 0.001223847 | 0.023458309 | 1 |
| Obesity | ASPC | *ZBTB7B* | -1.181427723 | 1.09E-07 | 1.03E-05 | 0.000687467 |
| Obesity | ASPC | *PEF1* | 2.365594545 | 0.000200067 | 0.00576982 | 1 |
| Obesity | ASPC | *FUBP1* | -1.666224526 | 1.99E-14 | 6.58E-12 | 1.25E-10 |
| Obesity | ASPC | *ACP6* | -0.314224531 | 0.000217201 | 0.006178937 | 1 |
| Obesity | ASPC | *LYSMD1* | 0.432786974 | 0.003084717 | 0.04746598 | 1 |
| Obesity | ASPC | *ANTXR2* | 0.358018434 | 7.21E-05 | 0.002614147 | 0.453021819 |
| Obesity | ASPC | *SLC15A2* | -0.211924809 | 0.001757293 | 0.031386654 | 1 |
| Obesity | ASPC | *LMOD1* | -0.419478443 | 1.57E-16 | 8.25E-14 | 9.90E-13 |
| Obesity | ASPC | *ICA1L* | -0.355137953 | 7.52E-05 | 0.002671575 | 0.472868851 |
| Obesity | ASPC | *GYG1* | 0.734015991 | 0.000126069 | 0.004003001 | 0.792594278 |
| Obesity | ASPC | *DNALI1* | -0.71117043 | 6.94E-06 | 0.000400372 | 0.0436406 |
| Obesity | ASPC | *RFT1* | -0.507879733 | 3.04E-07 | 2.54E-05 | 0.001908386 |
| Obesity | ASPC | *GNL3* | -1.540305794 | 3.79E-08 | 4.18E-06 | 0.000238532 |
| Obesity | ASPC | *SLC51A* | -0.634598864 | 9.82E-06 | 0.000519016 | 0.061762937 |
| Obesity | ASPC | *ATRIP* | -3.51902268 | 6.55E-05 | 0.00243808 | 0.412035541 |
| Obesity | ASPC | *SHISA5* | 0.998169931 | 0.002610428 | 0.042407652 | 1 |
| Obesity | ASPC | *APEH* | -0.425051484 | 8.70E-13 | 2.19E-10 | 5.47E-09 |
| Obesity | ASPC | *RNF123* | 0.3821736 | 1.00E-20 | 6.31E-18 | 6.31E-17 |
| Obesity | ASPC | *MST1R* | 0.245489447 | 8.35E-07 | 6.48E-05 | 0.005251824 |
| Obesity | ASPC | *TEX264* | -0.233099042 | 0.000357636 | 0.00914008 | 1 |
| Obesity | ASPC | *POC1A* | 0.377381954 | 0.000753032 | 0.016325209 | 1 |
| Obesity | ASPC | *PPM1M* | -0.236670812 | 4.10E-05 | 0.001697225 | 0.25797825 |
| Obesity | ASPC | *C5orf63* | -1.186966875 | 0.000796311 | 0.016970865 | 1 |
| Obesity | ASPC | *ERAP1* | -0.231812155 | 5.29E-05 | 0.002104954 | 0.332582707 |
| Obesity | ASPC | *TMEM184A* | -0.341482398 | 0.002535884 | 0.041845417 | 1 |
| Obesity | ASPC | *YWHAZ* | -0.207623818 | 8.62E-05 | 0.002930408 | 0.542125511 |
| Obesity | ASPC | *CCDC171* | -1.15400196 | 0.000161328 | 0.004852948 | 1 |
| Obesity | ASPC | *LRFN5* | 0.12523549 | 0.001887993 | 0.032971706 | 1 |
| Obesity | ASPC | *REEP3* | -0.430991339 | 0.000320779 | 0.008403063 | 1 |
| Obesity | ASPC | *SLC39A13* | 2.38350487 | 2.81E-07 | 2.39E-05 | 0.001765594 |
| Obesity | ASPC | *TRMT61A* | -0.348887293 | 3.26E-06 | 0.000213434 | 0.020489674 |
| Obesity | ASPC | *RIMKLB* | -0.114057994 | 0.003110228 | 0.047468779 | 1 |
| Obesity | ASPC | *SAAL1* | 1.262672576 | 6.99E-05 | 0.002555956 | 0.439624428 |
| Obesity | ASPC | *YWHAB* | -0.617330639 | 0.002909312 | 0.045727107 | 1 |
| Obesity | ASPC | *PBX3* | -0.484585263 | 0.001266668 | 0.023914545 | 1 |
| Obesity | ASPC | *URM1* | -0.411366423 | 0.000222071 | 0.006260809 | 1 |
| Obesity | ASPC | *SP2* | 0.770874243 | 0.000511598 | 0.012001559 | 1 |
| Obesity | ASPC | *GPRC5B* | 0.310005735 | 0.002196738 | 0.037226124 | 1 |
| Obesity | ASPC | *ZNF646* | 0.677775772 | 5.66E-09 | 8.48E-07 | 3.56E-05 |
| Obesity | ASPC | *VKORC1* | -0.376538695 | 5.85E-09 | 8.54E-07 | 3.68E-05 |
| Obesity | ASPC | *PSCA* | 0.321711038 | 0.000114565 | 0.003693694 | 0.720270345 |
| Obesity | ASPC | *UBXN6* | -0.424270084 | 8.12E-05 | 0.002820869 | 0.510468099 |
| Obesity | ASPC | *SRR* | 0.125524083 | 9.75E-06 | 0.000519016 | 0.061327095 |
| Obesity | ASPC | *MLST8* | -1.429016859 | 0.000248462 | 0.006821019 | 1 |
| Obesity | ASPC | *COPS6* | -0.420160567 | 0.001579496 | 0.028617558 | 1 |
| Obesity | ASPC | *ATXN2L* | -1.95313821 | 4.81E-13 | 1.32E-10 | 3.03E-09 |
| Obesity | ASPC | *C8orf46* | 0.033951245 | 0.000363826 | 0.009186236 | 1 |

| | | | | | | |
|---|---|---|---|---|---|---|
| Obesity | ASPC | *RAB24* | -0.368365516 | 5.93E-05 | 0.002301587 | 0.37285702 |
| Obesity | ASPC | *INO80E* | -0.683477195 | 2.14E-15 | 8.41E-13 | 1.35E-11 |
| Obesity | ASPC | *FAM103A1* | -0.151371534 | 0.000144376 | 0.004471404 | 0.907694958 |
| Obesity | ASPC | *ELP5* | -0.485763283 | 0.000923787 | 0.018795636 | 1 |
| Obesity | ASPC | *ADORA2B* | -0.164614409 | 1.20E-06 | 8.74E-05 | 0.007533758 |
| Obesity | ASPC | *TTLL6* | -5.415300196 | 6.97E-07 | 5.48E-05 | 0.004384461 |
| Obesity | ASPC | *SOX7* | 0.794001404 | 0.000355965 | 0.009134495 | 1 |
| Obesity | ASPC | *TRMT61B* | 0.133649234 | 3.39E-05 | 0.001478236 | 0.212866015 |
| Obesity | ASPC | *FRMD3* | 0.82837794 | 0.001461118 | 0.026859799 | 1 |
| Obesity | ASPC | *C1QTNF4* | -0.117594795 | 4.10E-15 | 1.43E-12 | 2.58E-11 |
| Obesity | ASPC | *GNB2* | -0.622280014 | 0.003108015 | 0.047468779 | 1 |
| Obesity | ASPC | *FIBP* | 1.267830562 | 2.19E-07 | 1.91E-05 | 0.001378685 |
| Obesity | ASPC | *CTSW* | -0.068783171 | 2.10E-08 | 2.49E-06 | 0.000131791 |
| Obesity | ASPC | *SNTG2* | -0.641946072 | 9.73E-08 | 9.27E-06 | 0.000611494 |
| Obesity | ASPC | *EFEMP2* | 0.380050219 | 3.07E-07 | 2.54E-05 | 0.001932758 |
| Obesity | ASPC | *CCL19* | -7.517998808 | 4.49E-05 | 0.001820209 | 0.28213241 |
| Obesity | ASPC | *FUT10* | -0.168775222 | 6.25E-05 | 0.002367138 | 0.392944932 |
| Obesity | ASPC | *SNX32* | 0.298790689 | 9.98E-07 | 7.56E-05 | 0.006276639 |
| Obesity | ASPC | *HPSE2* | 0.34855717 | 2.51E-09 | 4.06E-07 | 1.58E-05 |
| Obesity | ASPC | *IQCB1* | -0.209664657 | 0.000708535 | 0.015630035 | 1 |
| Obesity | ASPC | *MST1* | -0.144993163 | 7.97E-06 | 0.000443291 | 0.050091903 |
| Obesity | ASPC | *GMPPB* | 0.47051015 | 9.46E-08 | 9.15E-06 | 0.000594981 |
| Obesity | ASPC | *GPR160* | -0.768673211 | 0.001723531 | 0.030898538 | 1 |
| Obesity | ASPC | *HOXB2* | 0.21682559 | 0.002644983 | 0.042858271 | 1 |
| Obesity | ASPC | *LINGO2* | 0.62784896 | 0.0025308 | 0.041845417 | 1 |
| Obesity | ASPC | *BBS1* | 0.472069774 | 0.00136806 | 0.02544673 | 1 |
| Obesity | ASPC | *KLF14* | 1.711538185 | 0.000889614 | 0.018238212 | 1 |
| Obesity | ASPC | *HRAS* | 0.408232632 | 0.00172505 | 0.030898538 | 1 |
| Obesity | ASPC | *RSRC1* | -0.247161555 | 0.001541606 | 0.028174637 | 1 |
| Obesity | ASPC | *DHX36* | -1.282734907 | 1.12E-05 | 0.000579619 | 0.070713467 |
| Obesity | ASPC | *CADM2* | 0.175172437 | 0.000387823 | 0.009637316 | 1 |
| Obesity | ASPC | *TMEM9B* | 1.142073305 | 2.42E-06 | 0.000161524 | 0.015183256 |
| Obesity | ASPC | *PCSK1* | -0.719394788 | 0.000808873 | 0.017122502 | 1 |
| Obesity | ASPC | *MSRA* | -1.239776591 | 3.08E-08 | 3.46E-06 | 0.000193515 |
| Obesity | ASPC | *IP6K1* | 0.555961384 | 0.000502167 | 0.011868876 | 1 |
| Obesity | ASPC | *CCDC57* | -0.321713779 | 0.000249536 | 0.006821019 | 1 |
| Obesity | ASPC | *RNF152* | -1.120460502 | 0.000890589 | 0.018238212 | 1 |
| Obesity | ASPC | *RUFY1* | 0.325919993 | 7.37E-05 | 0.002636241 | 0.463231775 |
| Obesity | ASPC | *ANKLE2* | 1.635396496 | 0.000383926 | 0.009578336 | 1 |
| Obesity | ASPC | *TALDO1* | -0.262175996 | 0.000856486 | 0.017771388 | 1 |
| Obesity | ASPC | *AP006621.1* | 0.191750438 | 0.001245403 | 0.023726818 | 1 |
| Obesity | ASPC | *PPFIA3* | -0.244163015 | 0.000192247 | 0.005654141 | 1 |
| Obesity | ASPC | *CD151* | 0.445059788 | 0.0013276 | 0.024841127 | 1 |
| Obesity | ASPC | *POLR2L* | -0.426333424 | 0.001837891 | 0.032372631 | 1 |
| Obesity | ASPC | *SH2B1* | -0.453940742 | 1.78E-31 | 1.12E-27 | 1.12E-27 |
| Obesity | ASPC | *P4HTM* | -0.458751171 | 8.70E-05 | 0.002939787 | 0.546800317 |
| Obesity | ASPC | *RPP25* | -0.163563511 | 0.000483511 | 0.011558304 | 1 |
| Obesity | ASPC | *STX19* | 0.587783033 | 2.71E-07 | 2.33E-05 | 0.001701725 |
| Obesity | ASPC | *TRIM73* | -0.667896154 | 2.32E-12 | 5.61E-10 | 1.46E-08 |
| Obesity | ASPC | *TUFM* | 0.564740578 | 8.91E-23 | 6.48E-20 | 5.60E-19 |

| | | | | | | |
|---|---|---|---|---|---|---|
| Obesity | ASPC | *FARSA* | -0.416100933 | 0.000803632 | 0.017069039 | 1 |
| Obesity | ASPC | *B3GNT3* | 0.518239017 | 0.000490308 | 0.011632331 | 1 |
| Obesity | ASPC | *ZNF322* | -0.413959503 | 8.73E-09 | 1.22E-06 | 5.49E-05 |
| Obesity | ASPC | *CLDN7* | -0.162672888 | 0.002591778 | 0.042397856 | 1 |
| Obesity | ASPC | *FBXL6* | -0.632550426 | 0.000821209 | 0.017209809 | 1 |
| Obesity | ASPC | *CAV3* | -0.098509349 | 0.001388402 | 0.025673188 | 1 |
| Obesity | ASPC | *BRICD5* | -0.146284054 | 0.001833904 | 0.032372631 | 1 |
| Obesity | ASPC | *GRID1* | -0.372353899 | 7.38E-05 | 0.002636241 | 0.463978378 |
| Obesity | ASPC | *LYSMD4* | 0.119695168 | 0.003272945 | 0.049109803 | 1 |
| Obesity | ASPC | *CCBE1* | 0.132214485 | 0.003025173 | 0.046975471 | 1 |
| Obesity | ASPC | *PSMG1* | 0.107702211 | 3.75E-05 | 0.001572477 | 0.235871511 |
| Obesity | ASPC | *RP1L1* | 0.454147966 | 1.18E-08 | 1.52E-06 | 7.44E-05 |
| Obesity | ASPC | *BMP8A* | 0.907014478 | 3.26E-09 | 5.00E-07 | 2.05E-05 |
| Obesity | ASPC | *EIF3C* | 0.439844809 | 1.16E-27 | 1.82E-24 | 7.28E-24 |
| Obesity | ASPC | *PLA2G6* | 0.214653345 | 9.60E-07 | 7.36E-05 | 0.006032413 |
| Obesity | ASPC | *SNN* | 0.65719701 | 0.002776334 | 0.04430155 | 1 |
| Obesity | ASPC | *H1FX* | 0.56681547 | 0.002664191 | 0.042972761 | 1 |
| Obesity | ASPC | *MAFF* | -0.504177417 | 1.16E-05 | 0.000594608 | 0.073136825 |
| Obesity | ASPC | *GP1BA* | 0.287897343 | 9.15E-06 | 0.000504408 | 0.057502546 |
| Obesity | ASPC | *BTN3A2* | -0.118894136 | 1.95E-11 | 4.09E-09 | 1.23E-07 |
| Obesity | ASPC | *HYAL3* | -0.142889509 | 2.35E-05 | 0.001086368 | 0.147746034 |
| Obesity | ASPC | *ADH1A* | -0.180465099 | 0.002045433 | 0.035135618 | 1 |
| Obesity | ASPC | *ZSCAN23* | 0.055598994 | 0.00256963 | 0.042291273 | 1 |
| Obesity | ASPC | *HYKK* | 0.321911804 | 0.0022168 | 0.037364666 | 1 |
| Obesity | ASPC | *CENPP* | 0.169034183 | 1.38E-06 | 9.85E-05 | 0.008670969 |
| Obesity | ASPC | *FBF1* | 0.091697766 | 0.000555319 | 0.012846927 | 1 |
| Obesity | ASPC | *FAM47E* | 0.12836672 | 0.000623562 | 0.014152839 | 1 |
| Obesity | ASPC | *ZKSCAN3* | -0.547755441 | 0.000382123 | 0.009573588 | 1 |
| Obesity | ASPC | *INCA1* | 0.741649101 | 0.000173577 | 0.005171934 | 1 |
| Obesity | ASPC | *SULT1A1* | -0.221107372 | 4.89E-08 | 5.12E-06 | 0.000307276 |
| Obesity | ASPC | *C6orf106* | 0.250140373 | 1.50E-08 | 1.85E-06 | 9.42E-05 |
| Obesity | ASPC | *HCAR1* | 0.220154029 | 0.000259907 | 0.006983065 | 1 |
| Obesity | ASPC | *FUT11* | 0.530623559 | 0.000431217 | 0.010427151 | 1 |
| Obesity | ASPC | *SULT1A2* | 0.19208012 | 8.77E-29 | 1.84E-25 | 5.52E-25 |
| Obesity | ASPC | *CCL4L2* | -0.178515113 | 4.01E-06 | 0.000252385 | 0.025238488 |
| Obesity | ASPC | *GTF2E2* | 0.597911581 | 0.001406641 | 0.025934167 | 1 |
| Obesity | ASPC | *ZNF165* | -0.257867224 | 8.25E-05 | 0.002850029 | 0.518705225 |
| Obesity | ASPC | *MRPL21* | -0.097959927 | 0.000251967 | 0.006857656 | 1 |
| Obesity | ASPC | *PCDHB11* | -0.439874276 | 0.000636575 | 0.014396209 | 1 |
| Obesity | ASPC | *RPS26* | -0.08898184 | 6.12E-05 | 0.002349144 | 0.384537668 |
| Obesity | ASPC | *ZNF780A* | -0.118548194 | 6.70E-06 | 0.000393851 | 0.042142017 |
| Obesity | ASPC | *NOL8* | -1.276352753 | 2.75E-05 | 0.001261203 | 0.172784825 |
| Obesity | ASPC | *MAK16* | -1.054855663 | 0.000611469 | 0.013928645 | 1 |
| Obesity | ASPC | *ADH4* | -0.090645433 | 6.16E-05 | 0.002349144 | 0.38705985 |
| Obesity | ASPC | *ZKSCAN8* | 0.598579033 | 7.14E-06 | 0.000404451 | 0.044894059 |
| Obesity | ASPC | *GFPT1* | 0.603778531 | 0.000929789 | 0.01881701 | 1 |
| Obesity | ASPC | *MGEA5* | -0.479201731 | 0.002314254 | 0.038593411 | 1 |
| Obesity | ASPC | *PRMT6* | -1.361809164 | 8.32E-05 | 0.002859518 | 0.523291873 |
| Obesity | ASPC | *MAP3K3* | -1.461157649 | 4.13E-05 | 0.001697669 | 0.259743358 |
| Obesity | ASPC | *CENPW* | 0.365947967 | 0.000762252 | 0.016468309 | 1 |

| | | | | | | |
|---|---|---|---|---|---|---|
| Obesity | ASPC | *HLA-DRA* | -0.139158773 | 0.002603081 | 0.042397856 | 1 |
| Obesity | ASPC | *NOTCH4* | -0.031728832 | 4.00E-08 | 4.32E-06 | 0.000251344 |
| Obesity | ASPC | *AGER* | 0.220741287 | 0.000896515 | 0.01829996 | 1 |
| Obesity | ASPC | *MRPL38* | -0.206235221 | 0.000603882 | 0.01380583 | 1 |
| Obesity | ASPC | *SKIV2L* | -0.099668745 | 2.16E-07 | 1.91E-05 | 0.001360857 |
| Obesity | ASPC | *NELFE* | 0.273003643 | 3.09E-06 | 0.000204529 | 0.019430289 |
| Obesity | ASPC | *GPANK1* | -0.229439494 | 0.000795371 | 0.016970865 | 1 |
| Obesity | ASPC | *POU5F1* | 0.078631508 | 1.19E-06 | 8.74E-05 | 0.007473952 |
| Obesity | ASPC | *CCHCR1* | 0.145136195 | 6.40E-08 | 6.60E-06 | 0.000402538 |
| Obesity | ASPC | *ABCF1* | 0.35200172 | 5.03E-05 | 0.0020136 | 0.31613524 |
| Obesity | ASPC | *HLA-E* | 0.465967167 | 6.89E-14 | 2.17E-11 | 4.33E-10 |
| Obesity | ASPC | *ZFP57* | 0.114881071 | 2.55E-08 | 2.97E-06 | 0.000160285 |
| Obesity | ASPC | *FAM216A* | -0.599230597 | 0.001870259 | 0.032752975 | 1 |
| Obesity | ASPC | *SMN2* | -0.096713436 | 0.003110726 | 0.047468779 | 1 |
| Obesity | ASPC | *EIF3CL* | 2.450823117 | 6.43E-31 | 2.02E-27 | 4.04E-27 |
| Obesity | ASPC | *C17orf107* | 0.159881833 | 0.000256978 | 0.006951896 | 1 |
| Obesity | ASPC | *COLQ* | -1.460416271 | 1.37E-05 | 0.000676454 | 0.086214734 |
| Obesity | ASPC | *AC084018.1* | -0.115857786 | 0.000152228 | 0.004668588 | 0.957060513 |
| Obesity | ASPC | *ASIC3* | 0.193567331 | 6.40E-05 | 0.002407963 | 0.402129897 |
| Obesity | ASPC | *NOTCH2NL* | 0.149163733 | 0.00070644 | 0.015630035 | 1 |
| Obesity | ASPC | *SIPA1* | 0.261208723 | 0.002870964 | 0.045543615 | 1 |
| Obesity | ASPC | *TMEM110* | 0.604017131 | 1.22E-05 | 0.000618875 | 0.076740553 |
| Obesity | ASPC | *PPP1CB* | -0.759536697 | 3.11E-05 | 0.001384563 | 0.195223444 |
| Obesity | ASPC | *SLC35F6* | 1.173860122 | 0.001180306 | 0.022903038 | 1 |
| Obesity | ASPC | *CLIC1* | -0.803745018 | 2.04E-06 | 0.000140908 | 0.012822649 |
| Obesity | ASPC | *GALT* | 0.426630986 | 0.001867807 | 0.032752975 | 1 |
| Obesity | ASPC | *SLC12A8* | -0.279727224 | 9.23E-05 | 0.003104197 | 0.580484889 |
| Obesity | ASPC | *EXOSC6* | -0.796333997 | 3.25E-15 | 1.20E-12 | 2.04E-11 |
| Obesity | ASPC | *LINC00240* | 0.291396912 | 2.93E-09 | 4.61E-07 | 1.84E-05 |
| Obesity | ASPC | *AL049840.1* | -0.227263151 | 5.94E-06 | 0.000355683 | 0.037346698 |
| Obesity | ASPC | *PLEKHM1* | 0.436894719 | 0.002875905 | 0.045543615 | 1 |
| Obesity | ASPC | *HCG20* | -1.191466567 | 3.51E-06 | 0.000227421 | 0.022059857 |
| Obesity | ASPC | *RPL41* | -1.491560218 | 0.000137907 | 0.004356883 | 0.867019706 |
| Obesity | ASPC | *RPS18* | 0.261682269 | 0.001785984 | 0.031718875 | 1 |
| Obesity | ASPC | *TNF* | -0.282025681 | 5.75E-05 | 0.002245049 | 0.361452881 |
| Obesity | ASPC | *ZSCAN31* | 0.141103266 | 0.000119524 | 0.003833905 | 0.751445356 |
| Obesity | ASPC | *PSMB9* | 0.203400444 | 0.000220124 | 0.006233884 | 1 |
| Obesity | ASPC | *NAT6* | 0.398036337 | 0.000263028 | 0.007036843 | 1 |
| Obesity | ASPC | *N4BP2L2* | -0.665919739 | 0.002997383 | 0.046760657 | 1 |
| Obesity | ASPC | *PCDHAC1* | -0.458527385 | 0.00094066 | 0.018954893 | 1 |
| Obesity | ASPC | *DNAH10OS* | 2.965019049 | 3.68E-07 | 3.00E-05 | 0.002312762 |
| Obesity | ASPC | *LINC00989* | -2.386156876 | 0.000141895 | 0.004438267 | 0.892091685 |
| Obesity | ASPC | *SNHG18* | -0.243035337 | 0.003055808 | 0.047291231 | 1 |
| Obesity | ASPC | *MPV17L2* | 0.405887891 | 4.00E-06 | 0.000252385 | 0.025131318 |
| Obesity | ASPC | *DPP3* | 0.121576924 | 0.003206241 | 0.048339655 | 1 |
| Obesity | ASPC | *HMBS* | 0.25735289 | 3.49E-11 | 7.07E-09 | 2.19E-07 |
| Obesity | ASPC | *AC093668.2* | -0.211579741 | 0.003188282 | 0.048195999 | 1 |
| Obesity | ASPC | *HP* | -0.368091131 | 0.000182827 | 0.005421843 | 1 |
| Obesity | ASPC | *AC083843.1* | -2.581720233 | 0.000646673 | 0.014529356 | 1 |
| Obesity | ASPC | *SH3RF3-AS1* | -0.196203197 | 0.000143181 | 0.004456341 | 0.900180943 |

| | | | | | | |
|---|---|---|---|---|---|---|
| Obesity | ASPC | *HOXB7* | 0.659089909 | 4.20E-05 | 0.001713494 | 0.263878105 |
| Obesity | ASPC | *LINC00562* | -0.118388197 | 0.000490081 | 0.011632331 | 1 |
| Obesity | ASPC | *STRADA* | -0.406334895 | 9.99E-05 | 0.003324034 | 0.62824245 |
| Obesity | ASPC | *AC139100.2* | -10.57206477 | 0.003268917 | 0.049109803 | 1 |
| Obesity | ASPC | *LINC01140* | 0.382670867 | 0.002578636 | 0.042328673 | 1 |
| Obesity | ASPC | *AC009060.1* | -1.087230099 | 1.02E-08 | 1.36E-06 | 6.39E-05 |
| Obesity | ASPC | *GTF2H5* | 0.274467063 | 0.00087374 | 0.018069746 | 1 |
| Obesity | b_cell | *STPG1* | 7.292360995 | 0.001901559 | 0.03507628 | 1 |
| Obesity | b_cell | *SEMA3F* | -8.237783247 | 1.43E-14 | 1.50E-11 | 8.98E-11 |
| Obesity | b_cell | *HS3ST1* | 3.89135635 | 0.001052964 | 0.034102799 | 1 |
| Obesity | b_cell | *WNT16* | 14.80711539 | 0.001719813 | 0.03507628 | 1 |
| Obesity | b_cell | *DBNDD1* | 3.516186027 | 0.002290961 | 0.037063572 | 1 |
| Obesity | b_cell | *RBM6* | -7.06635667 | 7.31E-15 | 9.19E-12 | 4.60E-11 |
| Obesity | b_cell | *SLC22A16* | -8.317444081 | 0.00077178 | 0.032503045 | 1 |
| Obesity | b_cell | *ABCB4* | 2.64859495 | 0.001995445 | 0.035765074 | 1 |
| Obesity | b_cell | *GGNBP2* | -3.617700868 | 0.002021302 | 0.035898098 | 1 |
| Obesity | b_cell | *MAP3K9* | 1.350673474 | 0.00307228 | 0.040573336 | 1 |
| Obesity | b_cell | *MPND* | 4.180113884 | 0.000889998 | 0.034102799 | 1 |
| Obesity | b_cell | *MAPK8IP2* | 2.161816767 | 0.002660721 | 0.038366866 | 1 |
| Obesity | b_cell | *LRRC23* | 7.982695918 | 0.000262115 | 0.020859717 | 1 |
| Obesity | b_cell | *HFE* | -6.268327267 | 4.50E-05 | 0.005661961 | 0.283098065 |
| Obesity | b_cell | *ALOX5* | 2.369342368 | 0.002617247 | 0.038001458 | 1 |
| Obesity | b_cell | *DDX11* | -4.006372814 | 0.004741845 | 0.0486292 | 1 |
| Obesity | b_cell | *NPC1L1* | 6.738164672 | 0.002505786 | 0.037294603 | 1 |
| Obesity | b_cell | *BIRC3* | 1.378726668 | 0.003382837 | 0.04270662 | 1 |
| Obesity | b_cell | *TOMM34* | 5.567483937 | 0.000851599 | 0.034013039 | 1 |
| Obesity | b_cell | *BTN3A1* | -4.288964464 | 0.004652146 | 0.048264092 | 1 |
| Obesity | b_cell | *CENPQ* | -4.00351211 | 0.001902499 | 0.03507628 | 1 |
| Obesity | b_cell | *SH3YL1* | -3.332673158 | 0.0025042 | 0.037294603 | 1 |
| Obesity | b_cell | *TUBG2* | -2.484161344 | 0.002224375 | 0.037063572 | 1 |
| Obesity | b_cell | *CAPG* | 5.512207966 | 0.00339664 | 0.042794937 | 1 |
| Obesity | b_cell | *UTS2* | -5.021419747 | 0.002372202 | 0.037063572 | 1 |
| Obesity | b_cell | *RFC2* | 8.056986998 | 0.001093596 | 0.034102799 | 1 |
| Obesity | b_cell | *ARHGEF5* | 4.849942582 | 0.002140023 | 0.036745583 | 1 |
| Obesity | b_cell | *ALX4* | 5.918066693 | 0.002146277 | 0.036745583 | 1 |
| Obesity | b_cell | *ITIH4* | 6.323717516 | 2.82E-07 | 7.71E-05 | 0.001774159 |
| Obesity | b_cell | *PIGV* | 4.658009826 | 0.003569574 | 0.043263022 | 1 |
| Obesity | b_cell | *ABCA7* | 5.72312468 | 0.002509244 | 0.037294603 | 1 |
| Obesity | b_cell | *CHI3L2* | 4.980468046 | 0.002391482 | 0.037063572 | 1 |
| Obesity | b_cell | *ZFAT* | 3.135454488 | 0.002485796 | 0.037294603 | 1 |
| Obesity | b_cell | *STOML1* | 4.83295913 | 0.001775958 | 0.03507628 | 1 |
| Obesity | b_cell | *ROGDI* | 5.411577493 | 0.001601142 | 0.034403393 | 1 |
| Obesity | b_cell | *SDK2* | 2.163561768 | 0.002391987 | 0.037063572 | 1 |
| Obesity | b_cell | *GAL* | 2.841240677 | 0.003017154 | 0.040292366 | 1 |
| Obesity | b_cell | *DAPP1* | 5.143758665 | 0.002921927 | 0.039848499 | 1 |
| Obesity | b_cell | *CAMK2A* | 6.047784191 | 0.003168467 | 0.041157333 | 1 |
| Obesity | b_cell | *NLE1* | 3.810049962 | 0.002445646 | 0.037063572 | 1 |
| Obesity | b_cell | *ATP2A3* | 6.260576607 | 0.001211718 | 0.034102799 | 1 |
| Obesity | b_cell | *ADD2* | 10.67333194 | 0.001514534 | 0.034251993 | 1 |
| Obesity | b_cell | *MOCOS* | 3.541522623 | 0.001107165 | 0.034102799 | 1 |

| | | | | | | |
|---|---|---|---|---|---|---|
| Obesity | b_cell | *MCM6* | 6.022255603 | 0.00111594 | 0.034102799 | 1 |
| Obesity | b_cell | *TPD52* | 1.301862799 | 0.003351524 | 0.04256681 | 1 |
| Obesity | b_cell | *ICAM3* | 5.38091756 | 0.001226422 | 0.034102799 | 1 |
| Obesity | b_cell | *XAB2* | -3.857583147 | 0.004695274 | 0.048392116 | 1 |
| Obesity | b_cell | *COL4A4* | 1.133100895 | 0.002150847 | 0.036745583 | 1 |
| Obesity | b_cell | *PCDHA6* | -54.26040305 | 0.001525459 | 0.034251993 | 1 |
| Obesity | b_cell | *EPYC* | -39.48474157 | 0.000671131 | 0.032011478 | 1 |
| Obesity | b_cell | *ZNF446* | 12.04729169 | 0.002821677 | 0.039195703 | 1 |
| Obesity | b_cell | *COL16A1* | -2.443430114 | 0.001476492 | 0.034128696 | 1 |
| Obesity | b_cell | *ACHE* | -5.737839465 | 0.00211911 | 0.03650095 | 1 |
| Obesity | b_cell | *ATXN7L3* | 4.099897138 | 0.001087427 | 0.034102799 | 1 |
| Obesity | b_cell | *C3orf18* | 4.308432697 | 4.14E-05 | 0.005441085 | 0.260512957 |
| Obesity | b_cell | *CPXM1* | -3.755346594 | 0.001617032 | 0.034403393 | 1 |
| Obesity | b_cell | *KCNH4* | -5.580540351 | 0.005029588 | 0.04956273 | 1 |
| Obesity | b_cell | *SEL1L3* | 0.970582186 | 0.002774194 | 0.039029882 | 1 |
| Obesity | b_cell | *VNN3* | 2.712070289 | 0.003350365 | 0.04256681 | 1 |
| Obesity | b_cell | *PDE6C* | 9.147591337 | 4.15E-05 | 0.005441085 | 0.261172098 |
| Obesity | b_cell | *BLNK* | 5.009592956 | 0.000903723 | 0.034102799 | 1 |
| Obesity | b_cell | *STX1B* | -6.895276369 | 0.000300946 | 0.022141385 | 1 |
| Obesity | b_cell | *LGALS2* | 6.15315643 | 0.000310183 | 0.022141385 | 1 |
| Obesity | b_cell | *TFIP11* | 9.103413147 | 0.001077788 | 0.034102799 | 1 |
| Obesity | b_cell | *CCDC134* | 9.969983845 | 0.000560253 | 0.030042288 | 1 |
| Obesity | b_cell | *PIGH* | 9.153464004 | 0.002739445 | 0.038879503 | 1 |
| Obesity | b_cell | *ISM2* | 8.670901134 | 0.002893568 | 0.039738693 | 1 |
| Obesity | b_cell | *KIAA0391* | 13.13200437 | 0.003873478 | 0.044373001 | 1 |
| Obesity | b_cell | *GSS* | 8.619122934 | 0.00263364 | 0.038151367 | 1 |
| Obesity | b_cell | *CD40* | 7.52933537 | 0.004200211 | 0.045990468 | 1 |
| Obesity | b_cell | *SLC17A9* | 13.89176971 | 0.001140432 | 0.034102799 | 1 |
| Obesity | b_cell | *PTK6* | 1.82062363 | 0.003822826 | 0.044344465 | 1 |
| Obesity | b_cell | *RSPO4* | 4.785592527 | 0.004100063 | 0.045382214 | 1 |
| Obesity | b_cell | *SIRPB1* | 1.527939913 | 0.002798858 | 0.039103157 | 1 |
| Obesity | b_cell | *WFDC2* | 8.31795327 | 0.004193255 | 0.045990468 | 1 |
| Obesity | b_cell | *ELMO3* | 7.711368697 | 2.41E-05 | 0.003892803 | 0.151819326 |
| Obesity | b_cell | *ACD* | -5.431837538 | 0.002313804 | 0.037063572 | 1 |
| Obesity | b_cell | *COG4* | -3.432044912 | 0.00429009 | 0.046251976 | 1 |
| Obesity | b_cell | *HAGHL* | 11.47094214 | 0.001616127 | 0.034403393 | 1 |
| Obesity | b_cell | *RHBDL1* | -8.74041443 | 0.005078815 | 0.049782492 | 1 |
| Obesity | b_cell | *CLUAP1* | 6.654506955 | 0.001871406 | 0.03507628 | 1 |
| Obesity | b_cell | *STX4* | 7.463324698 | 0.001851019 | 0.03507628 | 1 |
| Obesity | b_cell | *TULP2* | 5.063567331 | 0.004975397 | 0.049260352 | 1 |
| Obesity | b_cell | *TUBB4A* | -41.14512357 | 0.001445596 | 0.034102799 | 1 |
| Obesity | b_cell | *PPP1R37* | 14.41077827 | 0.000302558 | 0.022141385 | 1 |
| Obesity | b_cell | *CD37* | 0.911668197 | 0.002886931 | 0.039738693 | 1 |
| Obesity | b_cell | *STX10* | 9.160413526 | 0.001572435 | 0.034403393 | 1 |
| Obesity | b_cell | *FCER2* | 0.970010882 | 0.003753203 | 0.044074841 | 1 |
| Obesity | b_cell | *AURKC* | 2.055584625 | 0.002737403 | 0.038879503 | 1 |
| Obesity | b_cell | *PRX* | 4.290101592 | 0.001618523 | 0.034403393 | 1 |
| Obesity | b_cell | *CCDC9* | 4.956272864 | 0.001731271 | 0.03507628 | 1 |
| Obesity | b_cell | *FZR1* | 10.7965385 | 0.001414367 | 0.034102799 | 1 |
| Obesity | b_cell | *ICAM5* | -30.34894062 | 0.000356857 | 0.023129496 | 1 |

| | | | | | | |
|---|---|---|---|---|---|---|
| Obesity | b_cell | *CYTH2* | 11.45639102 | 0.001543329 | 0.034403393 | 1 |
| Obesity | b_cell | *LIG1* | 2.655573871 | 0.002445758 | 0.037063572 | 1 |
| Obesity | b_cell | *TFPT* | 8.787097324 | 0.002437998 | 0.037063572 | 1 |
| Obesity | b_cell | *UPK1A* | -16.11169354 | 0.002051872 | 0.035933485 | 1 |
| Obesity | b_cell | *COPE* | 6.770433609 | 0.002082869 | 0.036274231 | 1 |
| Obesity | b_cell | *ZNF14* | 8.654403964 | 0.004881921 | 0.048848715 | 1 |
| Obesity | b_cell | *ATP13A1* | -8.066565526 | 0.002162468 | 0.036843998 | 1 |
| Obesity | b_cell | *SIPA1L3* | 4.977422037 | 0.003078333 | 0.040573336 | 1 |
| Obesity | b_cell | *DLX5* | 46.41687841 | 0.000876428 | 0.034102799 | 1 |
| Obesity | b_cell | *HOXA2* | -10.44452182 | 0.003637512 | 0.04355457 | 1 |
| Obesity | b_cell | *TAF6* | -3.887754433 | 0.004234854 | 0.04605545 | 1 |
| Obesity | b_cell | *C1GALT1* | 6.044766041 | 0.004562063 | 0.047802818 | 1 |
| Obesity | b_cell | *MEST* | -1.904694611 | 0.001842336 | 0.03507628 | 1 |
| Obesity | b_cell | *TSPAN13* | 3.302659882 | 0.001547728 | 0.034403393 | 1 |
| Obesity | b_cell | *AK1* | 11.17235551 | 0.002046695 | 0.035933485 | 1 |
| Obesity | b_cell | *PLGRKT* | -3.061509621 | 0.004907642 | 0.048897535 | 1 |
| Obesity | b_cell | *ELAVL2* | -6.292957631 | 0.003815141 | 0.044344465 | 1 |
| Obesity | b_cell | *DHRS11* | -3.237959271 | 0.004241467 | 0.04605545 | 1 |
| Obesity | b_cell | *CHRNE* | 6.104299986 | 0.001517209 | 0.034251993 | 1 |
| Obesity | b_cell | *EFNB3* | 16.7162516 | 0.001693596 | 0.034796197 | 1 |
| Obesity | b_cell | *TMEM97* | 4.198731161 | 0.000734992 | 0.032503045 | 1 |
| Obesity | b_cell | *KIAA1211* | 5.341368584 | 0.000310986 | 0.022141385 | 1 |
| Obesity | b_cell | *NDUFC1* | 8.370446121 | 0.004877169 | 0.048848715 | 1 |
| Obesity | b_cell | *SNX15* | -6.193083309 | 0.00117984 | 0.034102799 | 1 |
| Obesity | b_cell | *CD5* | 10.66379615 | 0.000629572 | 0.031842914 | 1 |
| Obesity | b_cell | *ASIC1* | -6.356879544 | 0.001056996 | 0.034102799 | 1 |
| Obesity | b_cell | *KCTD10* | -3.230621598 | 0.003226707 | 0.041485287 | 1 |
| Obesity | b_cell | *CYP27B1* | 3.59854895 | 0.003918755 | 0.044434752 | 1 |
| Obesity | b_cell | *KCNA1* | -8.76145966 | 0.005113506 | 0.049992378 | 1 |
| Obesity | b_cell | *PRKAB1* | 5.485833138 | 0.000490418 | 0.029691623 | 1 |
| Obesity | b_cell | *SLC26A8* | -4.4069045 | 0.000320631 | 0.022151694 | 1 |
| Obesity | b_cell | *MDGA1* | 4.892420356 | 0.001354183 | 0.034102799 | 1 |
| Obesity | b_cell | *CAP2* | -1.824159376 | 0.004846167 | 0.048848715 | 1 |
| Obesity | b_cell | *VNN2* | 4.946190596 | 0.000879777 | 0.034102799 | 1 |
| Obesity | b_cell | *TTK* | 1.916869137 | 0.00454942 | 0.047802818 | 1 |
| Obesity | b_cell | *LY86* | 2.696717069 | 0.001668105 | 0.034726413 | 1 |
| Obesity | b_cell | *PRSS16* | 33.70955643 | 1.40E-07 | 4.61E-05 | 0.000879022 |
| Obesity | b_cell | *CEP72* | 8.211190131 | 0.003937365 | 0.044434752 | 1 |
| Obesity | b_cell | *TMCO6* | 2.615576017 | 0.003812478 | 0.044344465 | 1 |
| Obesity | b_cell | *BTNL8* | -3.71088245 | 0.002273641 | 0.037063572 | 1 |
| Obesity | b_cell | *ACTR8* | 9.038392578 | 0.003661271 | 0.043568476 | 1 |
| Obesity | b_cell | *HGD* | 11.42003198 | 0.001055796 | 0.034102799 | 1 |
| Obesity | b_cell | *BCHE* | -3.979599094 | 0.003418453 | 0.042795618 | 1 |
| Obesity | b_cell | *HYAL1* | -3.869887547 | 0.000401975 | 0.025527444 | 1 |
| Obesity | b_cell | *IFT57* | 6.485640049 | 0.001437219 | 0.034102799 | 1 |
| Obesity | b_cell | *PLCH1* | 11.89644758 | 5.00E-05 | 0.00616674 | 0.314503764 |
| Obesity | b_cell | *FAHD2A* | 9.800310196 | 8.18E-05 | 0.009186662 | 0.514453093 |
| Obesity | b_cell | *ITGB6* | -6.130950836 | 0.004613498 | 0.048119676 | 1 |
| Obesity | b_cell | *GALNT3* | 1.906993225 | 0.003526759 | 0.043053853 | 1 |
| Obesity | b_cell | *CHMP3* | -7.122132565 | 0.004263799 | 0.046138564 | 1 |

| | | | | | | |
|---|---|---|---|---|---|---|
| Obesity | b_cell | *RALGPS2* | 0.527063223 | 0.003640971 | 0.04355457 | 1 |
| Obesity | b_cell | *PADI2* | 4.6200898 | 0.004087354 | 0.045321336 | 1 |
| Obesity | b_cell | *TNFSF4* | -4.971532336 | 0.001952746 | 0.035482412 | 1 |
| Obesity | b_cell | *SERPINC1* | 9.001500691 | 8.42E-05 | 0.009281941 | 0.529070658 |
| Obesity | b_cell | *SLC5A9* | 9.02404989 | 0.000780604 | 0.032503045 | 1 |
| Obesity | b_cell | *CD3EAP* | -2.934360657 | 0.004862425 | 0.048848715 | 1 |
| Obesity | b_cell | *LRMP* | 2.051596525 | 0.000908253 | 0.034102799 | 1 |
| Obesity | b_cell | *SPP1* | 5.494352707 | 0.00256713 | 0.037783333 | 1 |
| Obesity | b_cell | *DNAH7* | -2.521323718 | 0.002257563 | 0.037063572 | 1 |
| Obesity | b_cell | *GDA* | 9.85083389 | 0.001556657 | 0.034403393 | 1 |
| Obesity | b_cell | *WDR34* | 4.62724523 | 0.004883225 | 0.048848715 | 1 |
| Obesity | b_cell | *BSPRY* | 4.622774868 | 0.003648004 | 0.04355457 | 1 |
| Obesity | b_cell | *HDHD3* | -2.061234096 | 0.001392855 | 0.034102799 | 1 |
| Obesity | b_cell | *DCAF4* | 13.92018084 | 3.33E-05 | 0.004696291 | 0.209246317 |
| Obesity | b_cell | *COQ6* | 6.265776491 | 0.001676498 | 0.034785943 | 1 |
| Obesity | b_cell | *HELLS* | 3.805998795 | 0.001240318 | 0.034102799 | 1 |
| Obesity | b_cell | *C10orf95* | 6.778718044 | 0.001126505 | 0.034102799 | 1 |
| Obesity | b_cell | *MASTL* | -3.603921971 | 0.00295497 | 0.040001035 | 1 |
| Obesity | b_cell | *A1BG* | 29.30436125 | 0.000899417 | 0.034102799 | 1 |
| Obesity | b_cell | *POPDC2* | 5.561279569 | 0.001435357 | 0.034102799 | 1 |
| Obesity | b_cell | *B4GALT4* | 7.075846134 | 0.001196311 | 0.034102799 | 1 |
| Obesity | b_cell | *CD80* | 3.583226822 | 0.002210959 | 0.037063572 | 1 |
| Obesity | b_cell | *GJB6* | -183.439779 | 5.51E-05 | 0.006540292 | 0.346635452 |
| Obesity | b_cell | *POLR3GL* | 8.489834647 | 0.00124764 | 0.034102799 | 1 |
| Obesity | b_cell | *CXCR4* | 1.354692058 | 0.004626163 | 0.048153458 | 1 |
| Obesity | b_cell | *FLT3* | 3.632695929 | 0.002263809 | 0.037063572 | 1 |
| Obesity | b_cell | *CENPK* | 2.552518757 | 0.002058618 | 0.035951481 | 1 |
| Obesity | b_cell | *KCNK15* | 3.039454591 | 0.003870201 | 0.044373001 | 1 |
| Obesity | b_cell | *ZNF45* | 8.836043023 | 0.004673058 | 0.048391578 | 1 |
| Obesity | b_cell | *BTN2A2* | 2.613662976 | 0.001086683 | 0.034102799 | 1 |
| Obesity | b_cell | *SNRPC* | -16.33752213 | 1.54E-10 | 1.07E-07 | 9.67E-07 |
| Obesity | b_cell | *RPS10* | 4.34051716 | 0.001815113 | 0.03507628 | 1 |
| Obesity | b_cell | *HIST1H2BJ* | 6.540985615 | 1.80E-06 | 0.000389641 | 0.011299598 |
| Obesity | b_cell | *EREG* | 3.910499916 | 0.003567612 | 0.043263022 | 1 |
| Obesity | b_cell | *RBCK1* | 7.420398251 | 0.002585425 | 0.037801319 | 1 |
| Obesity | b_cell | *PCSK2* | 38.74142562 | 2.16E-07 | 6.17E-05 | 0.001356358 |
| Obesity | b_cell | *PROZ* | 4.886508629 | 0.003450156 | 0.04295274 | 1 |
| Obesity | b_cell | *IGFLR1* | 12.15400779 | 0.002169867 | 0.036870141 | 1 |
| Obesity | b_cell | *RAB3IP* | 9.308406872 | 0.001078116 | 0.034102799 | 1 |
| Obesity | b_cell | *LRRC61* | -3.404145549 | 0.004867881 | 0.048848715 | 1 |
| Obesity | b_cell | *IDUA* | 6.053294622 | 0.001156869 | 0.034102799 | 1 |
| Obesity | b_cell | *CHTF18* | 12.75941823 | 0.001235892 | 0.034102799 | 1 |
| Obesity | b_cell | *DGCR6L* | 5.058443277 | 0.002325929 | 0.037063572 | 1 |
| Obesity | b_cell | *RIBC2* | 4.656983937 | 4.27E-05 | 0.005482944 | 0.268664273 |
| Obesity | b_cell | *GAD1* | 4.004427857 | 0.003132275 | 0.040993234 | 1 |
| Obesity | b_cell | *RPAIN* | -3.647423827 | 0.003643774 | 0.04355457 | 1 |
| Obesity | b_cell | *PLD2* | 5.790214902 | 0.000267475 | 0.021020222 | 1 |
| Obesity | b_cell | *CDKN2D* | 2.988275806 | 0.000994982 | 0.034102799 | 1 |
| Obesity | b_cell | *RHBDF2* | 3.353679435 | 0.002836654 | 0.039195703 | 1 |
| Obesity | b_cell | *FAM98C* | 6.461172485 | 0.002360378 | 0.037063572 | 1 |

| | | | | | | |
|---|---|---|---|---|---|---|
| Obesity | b_cell | *ATP8B3* | -8.518779093 | 2.07E-08 | 8.66E-06 | 0.000129955 |
| Obesity | b_cell | *PGPEP1* | 9.556820719 | 2.28E-05 | 0.003772914 | 0.14337072 |
| Obesity | b_cell | *HRC* | -8.351161301 | 0.000427534 | 0.026612914 | 1 |
| Obesity | b_cell | *UCK1* | 4.74783521 | 0.002326031 | 0.037063572 | 1 |
| Obesity | b_cell | *SMPDL3B* | 6.303397213 | 0.003417327 | 0.042795618 | 1 |
| Obesity | b_cell | *PPAN* | 9.388433944 | 0.001168714 | 0.034102799 | 1 |
| Obesity | b_cell | *PKDREJ* | -12.86346446 | 0.000183883 | 0.015836622 | 1 |
| Obesity | b_cell | *EPS8L1* | 5.274463996 | 0.003650908 | 0.04355457 | 1 |
| Obesity | b_cell | *COX4I2* | 3.064917751 | 0.003811539 | 0.044344465 | 1 |
| Obesity | b_cell | *RHPN2* | -4.450437386 | 0.004974667 | 0.049260352 | 1 |
| Obesity | b_cell | *REEP2* | -5.457926897 | 0.003683763 | 0.043697767 | 1 |
| Obesity | b_cell | *PCNA* | 11.78345089 | 0.000707745 | 0.032011478 | 1 |
| Obesity | b_cell | *MMACHC* | -4.442000615 | 0.002834299 | 0.039195703 | 1 |
| Obesity | b_cell | *LARS* | -3.817384164 | 0.001607901 | 0.034403393 | 1 |
| Obesity | b_cell | *VAV3* | 4.758686502 | 0.002537872 | 0.037615155 | 1 |
| Obesity | b_cell | *ACP2* | 14.95579693 | 5.92E-06 | 0.001162752 | 0.037208059 |
| Obesity | b_cell | *HOOK1* | 3.223686889 | 0.003481958 | 0.043053853 | 1 |
| Obesity | b_cell | *POM121C* | -4.422664816 | 0.000701553 | 0.032011478 | 1 |
| Obesity | b_cell | *PRADC1* | 7.058364482 | 0.000701956 | 0.032011478 | 1 |
| Obesity | b_cell | *KCNMB4* | 3.238093068 | 0.004261386 | 0.046138564 | 1 |
| Obesity | b_cell | *MPHOSPH6* | 8.175854674 | 0.002741004 | 0.038879503 | 1 |
| Obesity | b_cell | *ZNF670* | 5.013744246 | 0.000347816 | 0.022778303 | 1 |
| Obesity | b_cell | *SP110* | 6.120221568 | 0.000832621 | 0.033555694 | 1 |
| Obesity | b_cell | *TBRG4* | 5.140938296 | 0.003950824 | 0.044434752 | 1 |
| Obesity | b_cell | *DBNL* | 6.9206313 | 0.001770907 | 0.03507628 | 1 |
| Obesity | b_cell | *BRIP1* | 3.562778849 | 0.001565779 | 0.034403393 | 1 |
| Obesity | b_cell | *BLK* | 0.131611452 | 0.005074231 | 0.049782492 | 1 |
| Obesity | b_cell | *IL1RN* | 3.911863017 | 0.001448298 | 0.034102799 | 1 |
| Obesity | b_cell | *PRPF4* | 8.260504339 | 0.000523347 | 0.030042288 | 1 |
| Obesity | b_cell | *SPAG8* | 10.40243616 | 0.000718949 | 0.032285949 | 1 |
| Obesity | b_cell | *TMEM8B* | 9.215557492 | 0.002449945 | 0.037063572 | 1 |
| Obesity | b_cell | *PPIL1* | 6.710350896 | 0.00141403 | 0.034102799 | 1 |
| Obesity | b_cell | *ZSCAN9* | 10.55001459 | 6.36E-05 | 0.007408943 | 0.400082944 |
| Obesity | b_cell | *FRS3* | 10.06116757 | 0.001338886 | 0.034102799 | 1 |
| Obesity | b_cell | *PI15* | 5.012607299 | 0.002790635 | 0.039075109 | 1 |
| Obesity | b_cell | *TTPA* | -2.243137826 | 0.000980315 | 0.034102799 | 1 |
| Obesity | b_cell | *TRIM29* | -5.151192885 | 0.004024456 | 0.044861264 | 1 |
| Obesity | b_cell | *CENPO* | 12.49743707 | 0.002659505 | 0.038366866 | 1 |
| Obesity | b_cell | *CYP2C9* | 3.64847922 | 0.004897428 | 0.048873227 | 1 |
| Obesity | b_cell | *MDH1B* | 2.996436682 | 0.003152224 | 0.041039897 | 1 |
| Obesity | b_cell | *COX17* | 7.919650569 | 0.001489413 | 0.034251459 | 1 |
| Obesity | b_cell | *MMRN1* | -3.425452201 | 0.001643111 | 0.034665241 | 1 |
| Obesity | b_cell | *ERP27* | 4.432875908 | 0.001689768 | 0.034796197 | 1 |
| Obesity | b_cell | *PEX5* | 5.225120146 | 0.001300852 | 0.034102799 | 1 |
| Obesity | b_cell | *AMDHD1* | -4.788596019 | 0.004437706 | 0.047071491 | 1 |
| Obesity | b_cell | *RITA1* | -6.975502577 | 0.001393292 | 0.034102799 | 1 |
| Obesity | b_cell | *GPR84* | -50.98703764 | 0.00053164 | 0.030042288 | 1 |
| Obesity | b_cell | *HNRNPA1L2* | 10.75392463 | 0.001168554 | 0.034102799 | 1 |
| Obesity | b_cell | *PTGR2* | -3.675011337 | 0.000514371 | 0.030042288 | 1 |
| Obesity | b_cell | *DUOXA1* | -6.125408806 | 0.003983824 | 0.044645819 | 1 |

| | | | | | | |
|---|---|---|---|---|---|---|
| Obesity | b_cell | *SORD* | 11.80552708 | 0.00052744 | 0.030042288 | 1 |
| Obesity | b_cell | *WDR93* | -2.756891654 | 0.00033262 | 0.022246626 | 1 |
| Obesity | b_cell | *MARVELD3* | 2.523173792 | 0.004848153 | 0.048848715 | 1 |
| Obesity | b_cell | *PRPSAP2* | 7.026616439 | 0.001663773 | 0.034726413 | 1 |
| Obesity | b_cell | *C18orf8* | -6.380655597 | 0.003136273 | 0.040993234 | 1 |
| Obesity | b_cell | *SLC14A1* | 8.413736144 | 0.00063441 | 0.031842914 | 1 |
| Obesity | b_cell | *TRIM65* | 10.67504015 | 0.00031696 | 0.022141385 | 1 |
| Obesity | b_cell | *CBX8* | 1.541970599 | 0.002275652 | 0.037063572 | 1 |
| Obesity | b_cell | *GRB7* | 3.985798958 | 0.001508673 | 0.034251993 | 1 |
| Obesity | b_cell | *DPP9* | 6.696710356 | 0.003061676 | 0.040573336 | 1 |
| Obesity | b_cell | *GEMIN7* | 4.992977257 | 0.003376098 | 0.04270662 | 1 |
| Obesity | b_cell | *EVI5L* | 8.719683438 | 0.002298119 | 0.037063572 | 1 |
| Obesity | b_cell | *MOB3C* | 3.852418644 | 0.001441233 | 0.034102799 | 1 |
| Obesity | b_cell | *RXRG* | 26.16328221 | 0.000854789 | 0.034013039 | 1 |
| Obesity | b_cell | *TUFT1* | -3.881536712 | 0.000769198 | 0.032503045 | 1 |
| Obesity | b_cell | *PSEN2* | 11.91437835 | 0.000305845 | 0.022141385 | 1 |
| Obesity | b_cell | *PYCR2* | 7.080544382 | 0.004615264 | 0.048119676 | 1 |
| Obesity | b_cell | *FBLN7* | -3.182634406 | 0.002442278 | 0.037063572 | 1 |
| Obesity | b_cell | *FAHD2B* | -5.39569589 | 0.002457598 | 0.037063572 | 1 |
| Obesity | b_cell | *FANCD2* | 1.466123787 | 0.002596698 | 0.037878051 | 1 |
| Obesity | b_cell | *OSBPL10* | 3.480995572 | 0.000911605 | 0.034102799 | 1 |
| Obesity | b_cell | *SLC25A38* | 3.006098076 | 0.002450826 | 0.037063572 | 1 |
| Obesity | b_cell | *AMT* | -4.567776646 | 0.000510825 | 0.030042288 | 1 |
| Obesity | b_cell | *SLC26A1* | 5.639272323 | 0.000771468 | 0.032503045 | 1 |
| Obesity | b_cell | *CCNA2* | 6.677199072 | 0.000638175 | 0.031842914 | 1 |
| Obesity | b_cell | *TSLP* | -5.10323337 | 0.002774989 | 0.039029882 | 1 |
| Obesity | b_cell | *SLC25A48* | -8.383943224 | 0.000102196 | 0.010889975 | 0.642508523 |
| Obesity | b_cell | *PSD2* | 5.890866913 | 0.004790832 | 0.048659065 | 1 |
| Obesity | b_cell | *TRIM7* | 4.449485494 | 0.001990224 | 0.035765074 | 1 |
| Obesity | b_cell | *SDK1* | -1.624360074 | 0.003277422 | 0.041795437 | 1 |
| Obesity | b_cell | *ADHFE1* | -1.93179272 | 0.000674248 | 0.032011478 | 1 |
| Obesity | b_cell | *SLC26A7* | -2.493894443 | 0.0047527 | 0.0486292 | 1 |
| Obesity | b_cell | *ATP6V0D2* | -5.774072258 | 0.000145841 | 0.013985015 | 0.916902378 |
| Obesity | b_cell | *FAM135B* | 4.452275967 | 0.002323536 | 0.037063572 | 1 |
| Obesity | b_cell | *POLE3* | 6.560936772 | 0.004924469 | 0.04891746 | 1 |
| Obesity | b_cell | *SURF6* | 5.813233548 | 0.001217224 | 0.034102799 | 1 |
| Obesity | b_cell | *NRBF2* | -3.279367279 | 0.001403905 | 0.034102799 | 1 |
| Obesity | b_cell | *PLEKHS1* | 7.996993417 | 0.001650971 | 0.034714562 | 1 |
| Obesity | b_cell | *PAOX* | 3.278089413 | 0.001874696 | 0.03507628 | 1 |
| Obesity | b_cell | *HSD17B12* | -8.923874884 | 2.32E-08 | 9.11E-06 | 0.00014569 |
| Obesity | b_cell | *CHEK1* | 6.555142906 | 0.002465821 | 0.037087602 | 1 |
| Obesity | b_cell | *CNBD2* | 13.08887053 | 0.001899547 | 0.03507628 | 1 |
| Obesity | b_cell | *TRPT1* | -3.834154863 | 0.003262995 | 0.041780955 | 1 |
| Obesity | b_cell | *DOC2A* | -18.07157885 | 7.08E-10 | 4.05E-07 | 4.45E-06 |
| Obesity | b_cell | *FSIP1* | 13.71356023 | 0.002287048 | 0.037063572 | 1 |
| Obesity | b_cell | *C11orf53* | 30.1872753 | 0.002912522 | 0.039806572 | 1 |
| Obesity | b_cell | *PRSS53* | -23.25628251 | 1.54E-07 | 4.61E-05 | 0.000967998 |
| Obesity | b_cell | *DPYSL4* | 6.402012155 | 9.90E-06 | 0.00188633 | 0.062248886 |
| Obesity | b_cell | *ITIH2* | 1.690421832 | 0.003409148 | 0.042795618 | 1 |
| Obesity | b_cell | *BEND6* | -3.566294553 | 0.000458573 | 0.028265183 | 1 |

| | | | | | | |
|---|---|---|---|---|---|---|
| Obesity | b_cell | *POC5* | -4.793838152 | 0.000994047 | 0.034102799 | 1 |
| Obesity | b_cell | *ZNF547* | 3.509746422 | 0.001978908 | 0.035751135 | 1 |
| Obesity | b_cell | *BANK1* | 0.362524214 | 0.003738571 | 0.044015719 | 1 |
| Obesity | b_cell | *FAM92B* | 12.0624994 | 0.003482894 | 0.043053853 | 1 |
| Obesity | b_cell | *MCOLN2* | 2.232524537 | 0.002542777 | 0.037615155 | 1 |
| Obesity | b_cell | *FAM167A* | 3.984471909 | 0.001262552 | 0.034102799 | 1 |
| Obesity | b_cell | *NEIL2* | 10.35455856 | 8.88E-07 | 0.000206807 | 0.005583795 |
| Obesity | b_cell | *RABGEF1* | -3.687865868 | 0.001764977 | 0.03507628 | 1 |
| Obesity | b_cell | *GTF3C6* | 5.873696145 | 0.000864605 | 0.034102799 | 1 |
| Obesity | b_cell | *AGPAT5* | 7.826147713 | 0.000519508 | 0.030042288 | 1 |
| Obesity | b_cell | *GOLGA7B* | 3.309311946 | 0.00481489 | 0.048824537 | 1 |
| Obesity | b_cell | *C16orf87* | 5.871574823 | 0.001380666 | 0.034102799 | 1 |
| Obesity | b_cell | *SETD9* | -2.470528322 | 0.002418874 | 0.037063572 | 1 |
| Obesity | b_cell | *WIF1* | 7.760335633 | 0.004396635 | 0.04685024 | 1 |
| Obesity | b_cell | *BATF* | 12.51791237 | 0.001947278 | 0.035482412 | 1 |
| Obesity | b_cell | *DCK* | 4.646794996 | 0.00118568 | 0.034102799 | 1 |
| Obesity | b_cell | *SH3RF2* | -2.230163653 | 0.004228818 | 0.04605545 | 1 |
| Obesity | b_cell | *CPT2* | 5.816168128 | 0.001331217 | 0.034102799 | 1 |
| Obesity | b_cell | *STEAP2* | -3.544550734 | 0.000133522 | 0.013539606 | 0.839455554 |
| Obesity | b_cell | *CLEC18A* | -35.57430668 | 7.06E-16 | 1.11E-12 | 4.44E-12 |
| Obesity | b_cell | *PEX10* | 9.350631403 | 0.001948038 | 0.035482412 | 1 |
| Obesity | b_cell | *TRIM63* | 4.571028329 | 0.001464879 | 0.034128696 | 1 |
| Obesity | b_cell | *WDR66* | -3.308841275 | 0.002110531 | 0.036453042 | 1 |
| Obesity | b_cell | *GALNT14* | 11.47778141 | 0.001777203 | 0.03507628 | 1 |
| Obesity | b_cell | *RHPN1* | -5.655831893 | 0.003024972 | 0.040292366 | 1 |
| Obesity | b_cell | *CD1D* | -11.42568401 | 0.003047773 | 0.040510256 | 1 |
| Obesity | b_cell | *FAM86C1* | 18.8699528 | 0.001269893 | 0.034102799 | 1 |
| Obesity | b_cell | *NCF1* | 2.69124639 | 0.002572175 | 0.037783333 | 1 |
| Obesity | b_cell | *COPG2* | -2.271424946 | 0.000591076 | 0.030967447 | 1 |
| Obesity | b_cell | *SLC5A11* | -4.939602511 | 1.13E-07 | 4.17E-05 | 0.000709012 |
| Obesity | b_cell | *ANKLE1* | -13.46134435 | 0.00076968 | 0.032503045 | 1 |
| Obesity | b_cell | *C21orf33* | -13.41712152 | 0.001325237 | 0.034102799 | 1 |
| Obesity | b_cell | *VAV2* | 6.678691578 | 0.003512957 | 0.043053853 | 1 |
| Obesity | b_cell | *HIPK4* | 9.003478801 | 0.003019533 | 0.040292366 | 1 |
| Obesity | b_cell | *S100A1* | 2.78640517 | 0.002458328 | 0.037063572 | 1 |
| Obesity | b_cell | *PMF1* | 12.39076313 | 0.000311711 | 0.022141385 | 1 |
| Obesity | b_cell | *FCRL3* | 0.317719701 | 0.003722381 | 0.043917606 | 1 |
| Obesity | b_cell | *FGFR4* | 8.478926651 | 0.002265363 | 0.037063572 | 1 |
| Obesity | b_cell | *NACC1* | 4.262848335 | 0.001440776 | 0.034102799 | 1 |
| Obesity | b_cell | *LY6K* | 13.33411545 | 0.003461098 | 0.043003799 | 1 |
| Obesity | b_cell | *ORAI2* | 2.863354674 | 0.000960833 | 0.034102799 | 1 |
| Obesity | b_cell | *RPL8* | 6.565565145 | 0.005128851 | 0.049992378 | 1 |
| Obesity | b_cell | *YDJC* | 24.72949785 | 0.001794035 | 0.03507628 | 1 |
| Obesity | b_cell | *CCDC116* | 5.439042162 | 0.003957923 | 0.044434752 | 1 |
| Obesity | b_cell | *LRRC56* | 4.773110003 | 0.000619109 | 0.031842914 | 1 |
| Obesity | b_cell | *IKZF3* | 0.672184084 | 0.003430729 | 0.042795618 | 1 |
| Obesity | b_cell | *TNFSF13* | 9.461189993 | 0.00069357 | 0.032011478 | 1 |
| Obesity | b_cell | *SENP3* | 3.916635131 | 0.002500335 | 0.037294603 | 1 |
| Obesity | b_cell | *JMJD8* | 8.950182258 | 8.66E-05 | 0.009387418 | 0.544470222 |
| Obesity | b_cell | *CYP4A22* | 15.7186318 | 0.004460906 | 0.04721501 | 1 |

| | | | | | | |
|---|---|---|---|---|---|---|
| Obesity | b_cell | *COA7* | 4.650647591 | 0.001605067 | 0.034403393 | 1 |
| Obesity | b_cell | *SLC25A34* | 13.92339781 | 0.002946607 | 0.040001035 | 1 |
| Obesity | b_cell | *LAPTM5* | 6.725395077 | 0.003509062 | 0.043053853 | 1 |
| Obesity | b_cell | *FBXO41* | 12.3921023 | 0.001619756 | 0.034403393 | 1 |
| Obesity | b_cell | *SGPP2* | 12.07179394 | 0.002895285 | 0.039738693 | 1 |
| Obesity | b_cell | *NBPF10* | 3.009505579 | 0.003521419 | 0.043053853 | 1 |
| Obesity | b_cell | *LMOD1* | -3.006555507 | 3.26E-05 | 0.004696291 | 0.204882114 |
| Obesity | b_cell | *NUAK2* | 1.275276997 | 0.003555222 | 0.043263022 | 1 |
| Obesity | b_cell | *AIM2* | 3.124650275 | 0.001865821 | 0.03507628 | 1 |
| Obesity | b_cell | *CADPS* | 5.738883167 | 0.001264244 | 0.034102799 | 1 |
| Obesity | b_cell | *PPM1K* | 6.999441147 | 0.001850055 | 0.03507628 | 1 |
| Obesity | b_cell | *DCLK3* | 5.725248505 | 0.001254972 | 0.034102799 | 1 |
| Obesity | b_cell | *DNALI1* | -6.00034356 | 0.001330731 | 0.034102799 | 1 |
| Obesity | b_cell | *RFC4* | 2.714302169 | 0.001896794 | 0.03507628 | 1 |
| Obesity | b_cell | *RFT1* | -5.450696837 | 0.00173765 | 0.03507628 | 1 |
| Obesity | b_cell | *SFMBT1* | 7.664333181 | 0.005083563 | 0.049782492 | 1 |
| Obesity | b_cell | *BDH2* | -2.232502439 | 0.003186024 | 0.041215094 | 1 |
| Obesity | b_cell | *APEH* | -14.92468623 | 1.04E-06 | 0.000233044 | 0.00652522 |
| Obesity | b_cell | *RNF123* | 13.11348224 | 1.55E-11 | 1.22E-08 | 9.74E-08 |
| Obesity | b_cell | *MST1R* | 13.88421495 | 2.15E-06 | 0.000450433 | 0.013513002 |
| Obesity | b_cell | *GRM2* | 5.079288756 | 0.004779905 | 0.0486292 | 1 |
| Obesity | b_cell | *POC1A* | 9.168083366 | 3.36E-05 | 0.004696291 | 0.211333106 |
| Obesity | b_cell | *PPM1M* | -5.565279896 | 0.000313944 | 0.022141385 | 1 |
| Obesity | b_cell | *ERAP1* | -2.637779387 | 0.003757613 | 0.044074841 | 1 |
| Obesity | b_cell | *NSA2* | 4.142833632 | 0.003942146 | 0.044434752 | 1 |
| Obesity | b_cell | *MB21D1* | 7.174866812 | 0.001356788 | 0.034102799 | 1 |
| Obesity | b_cell | *SLU7* | 8.199055341 | 0.002450556 | 0.037063572 | 1 |
| Obesity | b_cell | *ZNF367* | 6.503216537 | 0.00067565 | 0.032011478 | 1 |
| Obesity | b_cell | *CFL2* | 8.033030256 | 0.001401855 | 0.034102799 | 1 |
| Obesity | b_cell | *PGM2L1* | 5.580966842 | 0.002038746 | 0.035903639 | 1 |
| Obesity | b_cell | *QSOX2* | 3.064722971 | 0.002005781 | 0.035824849 | 1 |
| Obesity | b_cell | *RET* | 2.55571953 | 0.003865428 | 0.044373001 | 1 |
| Obesity | b_cell | *TRMT61A* | -9.403093503 | 0.000195812 | 0.016636049 | 1 |
| Obesity | b_cell | *NETO1* | 0.792295927 | 0.004319955 | 0.046347363 | 1 |
| Obesity | b_cell | *CYB5R2* | 3.38286613 | 0.00382292 | 0.044344465 | 1 |
| Obesity | b_cell | *ZSCAN21* | -3.871495412 | 0.001323649 | 0.034102799 | 1 |
| Obesity | b_cell | *RIMKLB* | -2.768998376 | 0.004712094 | 0.04840676 | 1 |
| Obesity | b_cell | *SEC11C* | 5.171237158 | 0.00118971 | 0.034102799 | 1 |
| Obesity | b_cell | *MS4A6E* | -4.550709817 | 0.000952256 | 0.034102799 | 1 |
| Obesity | b_cell | *MS4A14* | -3.14273535 | 0.002032784 | 0.035903639 | 1 |
| Obesity | b_cell | *MEI1* | 5.601616604 | 0.000149037 | 0.013985015 | 0.93699598 |
| Obesity | b_cell | *ENDOG* | 4.057098511 | 0.000558539 | 0.030042288 | 1 |
| Obesity | b_cell | *COQ7* | 8.685922643 | 0.002411762 | 0.037063572 | 1 |
| Obesity | b_cell | *VKORC1* | -3.809038052 | 0.000204239 | 0.017120643 | 1 |
| Obesity | b_cell | *CACNB3* | 2.388016498 | 0.002671369 | 0.038432253 | 1 |
| Obesity | b_cell | *ZNF283* | 6.560941307 | 0.002294158 | 0.037063572 | 1 |
| Obesity | b_cell | *UBXN6* | -3.827738456 | 0.004560223 | 0.047802818 | 1 |
| Obesity | b_cell | *SERPINF2* | 10.15377152 | 0.000535652 | 0.030042288 | 1 |
| Obesity | b_cell | *NUDT8* | -6.271812326 | 0.000729754 | 0.032503045 | 1 |
| Obesity | b_cell | *KRT24* | -10.41154055 | 0.000780534 | 0.032503045 | 1 |

| | | | | | | |
|---|---|---|---|---|---|---|
| Obesity | b_cell | *NUDT16L1* | 3.968048683 | 0.00150524 | 0.034251993 | 1 |
| Obesity | b_cell | *NT5DC2* | -5.957304212 | 0.001387262 | 0.034102799 | 1 |
| Obesity | b_cell | *COA6* | 9.479928567 | 0.001248211 | 0.034102799 | 1 |
| Obesity | b_cell | *DTYMK* | 9.329578625 | 0.000940811 | 0.034102799 | 1 |
| Obesity | b_cell | *RHOH* | 1.67113076 | 0.002373056 | 0.037063572 | 1 |
| Obesity | b_cell | *REEP4* | 3.09289925 | 0.005069306 | 0.049782492 | 1 |
| Obesity | b_cell | *ZSWIM1* | 9.26680639 | 0.000158296 | 0.014291176 | 0.995209363 |
| Obesity | b_cell | *MFSD7* | 5.761489387 | 0.00095223 | 0.034102799 | 1 |
| Obesity | b_cell | *COL4A3* | 6.006717613 | 0.00015212 | 0.014064382 | 0.956377988 |
| Obesity | b_cell | *RAB24* | -6.500113213 | 0.000561959 | 0.030042288 | 1 |
| Obesity | b_cell | *PGM2* | 10.60750485 | 0.002035758 | 0.035903639 | 1 |
| Obesity | b_cell | *RNASE6* | 1.908179881 | 0.003133183 | 0.040993234 | 1 |
| Obesity | b_cell | *SLC38A11* | 1.401666319 | 0.003273844 | 0.041795437 | 1 |
| Obesity | b_cell | *INO80E* | -8.962711639 | 3.89E-05 | 0.005321128 | 0.244771866 |
| Obesity | b_cell | *RGPD8* | -2.782932372 | 0.001060199 | 0.034102799 | 1 |
| Obesity | b_cell | *LRRC45* | -3.936818555 | 0.003881339 | 0.044373001 | 1 |
| Obesity | b_cell | *HOXD4* | -10.30248651 | 0.001571673 | 0.034403393 | 1 |
| Obesity | b_cell | *ADORA2B* | -7.74792342 | 4.12E-07 | 0.000103512 | 0.002587803 |
| Obesity | b_cell | *GTSF1* | 8.687906388 | 0.001796464 | 0.03507628 | 1 |
| Obesity | b_cell | *TTLL6* | 7.471119784 | 0.003571419 | 0.043263022 | 1 |
| Obesity | b_cell | *TSEN34* | 7.50586925 | 0.000984092 | 0.034102799 | 1 |
| Obesity | b_cell | *LURAP1* | -3.133950957 | 0.002104581 | 0.036453042 | 1 |
| Obesity | b_cell | *DLK2* | -12.93639951 | 0.002307086 | 0.037063572 | 1 |
| Obesity | b_cell | *HOPX* | 15.87126469 | 0.00400194 | 0.044769032 | 1 |
| Obesity | b_cell | *ZIK1* | 15.59629566 | 0.001658452 | 0.034726413 | 1 |
| Obesity | b_cell | *RGS19* | 3.36939344 | 0.00017094 | 0.014926376 | 1 |
| Obesity | b_cell | *LGALS4* | 2.850872408 | 0.001253837 | 0.034102799 | 1 |
| Obesity | b_cell | *NXNL1* | -9.997934261 | 0.00121814 | 0.034102799 | 1 |
| Obesity | b_cell | *CYP4F22* | -9.499928986 | 0.001434277 | 0.034102799 | 1 |
| Obesity | b_cell | *ZNF554* | -3.3462166 | 0.004195825 | 0.045990468 | 1 |
| Obesity | b_cell | *ANKRD20A4* | 3.405263203 | 0.002969596 | 0.040064056 | 1 |
| Obesity | b_cell | *C1QTNF4* | -7.220293765 | 1.67E-16 | 3.50E-13 | 1.05E-12 |
| Obesity | b_cell | *CTSW* | -3.606818541 | 6.96E-09 | 3.36E-06 | 4.37E-05 |
| Obesity | b_cell | *FUT10* | -4.693615251 | 0.004570402 | 0.047810511 | 1 |
| Obesity | b_cell | *HOXC5* | 6.62647519 | 0.004206222 | 0.045990468 | 1 |
| Obesity | b_cell | *SNX32* | 10.48790818 | 1.49E-07 | 4.61E-05 | 0.000934322 |
| Obesity | b_cell | *PARP15* | 5.212132185 | 0.002817323 | 0.039195703 | 1 |
| Obesity | b_cell | *MZT2A* | 3.992312995 | 0.00134715 | 0.034102799 | 1 |
| Obesity | b_cell | *OLR1* | 6.552529721 | 0.001791447 | 0.03507628 | 1 |
| Obesity | b_cell | *ARV1* | 7.858075659 | 0.000388939 | 0.024951613 | 1 |
| Obesity | b_cell | *THAP2* | 5.491893387 | 0.001306095 | 0.034102799 | 1 |
| Obesity | b_cell | *MST1* | -6.389294651 | 3.69E-06 | 0.000748602 | 0.023206648 |
| Obesity | b_cell | *BBS1* | 7.3836593 | 0.000144008 | 0.013985015 | 0.905379003 |
| Obesity | b_cell | *IL20RB* | 4.333010091 | 0.003956836 | 0.044434752 | 1 |
| Obesity | b_cell | *ZNF497* | 2.667320293 | 0.003067764 | 0.040573336 | 1 |
| Obesity | b_cell | *KLF14* | 4.56134783 | 0.002291001 | 0.037063572 | 1 |
| Obesity | b_cell | *C4orf26* | 9.578278349 | 0.001523933 | 0.034251993 | 1 |
| Obesity | b_cell | *NDUFA11* | 8.820605059 | 0.004925203 | 0.04891746 | 1 |
| Obesity | b_cell | *TMEM9B* | 10.25204102 | 0.002392695 | 0.037063572 | 1 |
| Obesity | b_cell | *TBC1D10C* | 3.314176392 | 0.001635376 | 0.034618213 | 1 |

| | | | | | | |
|---|---|---|---|---|---|---|
| Obesity | b_cell | *LIPT2* | -12.7968036 | 0.002584522 | 0.037801319 | 1 |
| Obesity | b_cell | *ETV4* | 8.740884888 | 0.000124737 | 0.013070342 | 0.78422054 |
| Obesity | b_cell | *UFSP1* | 4.179556241 | 0.000821445 | 0.033535241 | 1 |
| Obesity | b_cell | *B3GNT4* | 2.794053763 | 0.004169526 | 0.045989136 | 1 |
| Obesity | b_cell | *GCNT4* | 2.465392047 | 0.003723252 | 0.043917606 | 1 |
| Obesity | b_cell | *MUC20* | 3.758268809 | 0.004024412 | 0.044861264 | 1 |
| Obesity | b_cell | *B3GNT8* | 1.506240643 | 0.003844848 | 0.044373001 | 1 |
| Obesity | b_cell | *AC024940.1* | 0.920034908 | 0.00342484 | 0.042795618 | 1 |
| Obesity | b_cell | *PPFIA3* | -6.119114777 | 0.000330902 | 0.022246626 | 1 |
| Obesity | b_cell | *ATOX1* | 8.305659946 | 0.001848855 | 0.03507628 | 1 |
| Obesity | b_cell | *C18orf32* | 5.359823869 | 0.001779182 | 0.03507628 | 1 |
| Obesity | b_cell | *FAM87B* | -17.01604249 | 0.001470032 | 0.034128696 | 1 |
| Obesity | b_cell | *TSSK6* | -12.12805267 | 0.002189964 | 0.037063572 | 1 |
| Obesity | b_cell | *GOLGA8Q* | 70.20238502 | 0.000962416 | 0.034102799 | 1 |
| Obesity | b_cell | *SH2B1* | -4.198549645 | 1.36E-08 | 6.11E-06 | 8.55E-05 |
| Obesity | b_cell | *SH2D4B* | 7.334166731 | 0.001951764 | 0.035482412 | 1 |
| Obesity | b_cell | *RPP25* | -9.982557075 | 0.000272591 | 0.021157797 | 1 |
| Obesity | b_cell | *STX19* | 10.5424472 | 0.001756726 | 0.03507628 | 1 |
| Obesity | b_cell | *GDPD4* | 3.71462084 | 0.002932293 | 0.039903301 | 1 |
| Obesity | b_cell | *TRIM73* | -39.25716134 | 1.61E-13 | 1.44E-10 | 1.01E-09 |
| Obesity | b_cell | *HYI* | -2.739682447 | 0.002432116 | 0.037063572 | 1 |
| Obesity | b_cell | *C17orf62* | 9.316577578 | 0.001776843 | 0.03507628 | 1 |
| Obesity | b_cell | *TUFM* | 4.492332325 | 0.004773882 | 0.0486292 | 1 |
| Obesity | b_cell | *CTC1* | 8.352065177 | 5.34E-05 | 0.006451248 | 0.335464888 |
| Obesity | b_cell | *TRIML2* | 8.685179882 | 0.005017932 | 0.049525497 | 1 |
| Obesity | b_cell | *CIITA* | 3.448903901 | 0.003524612 | 0.043053853 | 1 |
| Obesity | b_cell | *FOXS1* | -10.35470819 | 0.004780157 | 0.0486292 | 1 |
| Obesity | b_cell | *CITED4* | 7.168752956 | 0.002402451 | 0.037063572 | 1 |
| Obesity | b_cell | *SSTR2* | 4.01138227 | 0.002322899 | 0.037063572 | 1 |
| Obesity | b_cell | *ZNF443* | 8.897310261 | 0.004360899 | 0.046627501 | 1 |
| Obesity | b_cell | *ANKRD34A* | -6.639613866 | 0.00456147 | 0.047802818 | 1 |
| Obesity | b_cell | *ZNF322* | -3.768415055 | 0.000559229 | 0.030042288 | 1 |
| Obesity | b_cell | *OGFOD3* | 4.834757665 | 0.002248156 | 0.037063572 | 1 |
| Obesity | b_cell | *CLDN7* | -4.710837014 | 0.001211188 | 0.034102799 | 1 |
| Obesity | b_cell | *SNRPE* | 10.92078304 | 0.00160695 | 0.034403393 | 1 |
| Obesity | b_cell | *IZUMO1* | 4.205054598 | 0.000329817 | 0.022246626 | 1 |
| Obesity | b_cell | *KBTBD3* | 6.402057776 | 0.001151056 | 0.034102799 | 1 |
| Obesity | b_cell | *CLN8* | 7.306220837 | 0.000653151 | 0.032011478 | 1 |
| Obesity | b_cell | *CAPN12* | 4.028322109 | 0.00429635 | 0.046251976 | 1 |
| Obesity | b_cell | *SPNS3* | 1.554741292 | 0.003152898 | 0.041039897 | 1 |
| Obesity | b_cell | *BRICD5* | -8.266402815 | 0.000416755 | 0.026201401 | 1 |
| Obesity | b_cell | *GRID1* | -2.797663528 | 0.003007677 | 0.040292366 | 1 |
| Obesity | b_cell | *FANCF* | 5.169102709 | 0.001612375 | 0.034403393 | 1 |
| Obesity | b_cell | *GDPGP1* | 3.87981342 | 0.000798706 | 0.032820022 | 1 |
| Obesity | b_cell | *LRIT3* | -9.372571656 | 0.002014131 | 0.03587207 | 1 |
| Obesity | b_cell | *SERHL2* | 5.7964883 | 0.000606169 | 0.031495721 | 1 |
| Obesity | b_cell | *FAM167B* | 6.506032814 | 0.003632381 | 0.04355457 | 1 |
| Obesity | b_cell | *TRMT12* | 4.180285465 | 0.003497856 | 0.043053853 | 1 |
| Obesity | b_cell | *B3GALT5* | 2.530643659 | 0.004763604 | 0.0486292 | 1 |
| Obesity | b_cell | *ANKRD45* | 26.37307534 | 0.00182231 | 0.03507628 | 1 |

| | | | | | | |
|---|---|---|---|---|---|---|
| Obesity | b_cell | *EIF3C* | 18.86240343 | 2.37E-17 | 7.45E-14 | 1.49E-13 |
| Obesity | b_cell | *CLECL1* | 2.313315795 | 0.003087643 | 0.040610897 | 1 |
| Obesity | b_cell | *NELL2* | 4.59755433 | 0.000902578 | 0.034102799 | 1 |
| Obesity | b_cell | *KRBA2* | -7.43558372 | 0.002825733 | 0.039195703 | 1 |
| Obesity | b_cell | *NUTM2A* | -14.0046756 | 0.004058849 | 0.045164578 | 1 |
| Obesity | b_cell | *NOC4L* | -5.001308945 | 0.001017035 | 0.034102799 | 1 |
| Obesity | b_cell | *MAFF* | -3.834855435 | 0.003867142 | 0.044373001 | 1 |
| Obesity | b_cell | *HS6ST3* | 2.719253887 | 0.001492747 | 0.034251459 | 1 |
| Obesity | b_cell | *PDE6G* | 2.956136524 | 0.001996746 | 0.035765074 | 1 |
| Obesity | b_cell | *PMEL* | 4.740669871 | 0.00022735 | 0.01879469 | 1 |
| Obesity | b_cell | *MYBL1* | -2.67420414 | 0.001751251 | 0.03507628 | 1 |
| Obesity | b_cell | *SRL* | 5.909910325 | 0.000162974 | 0.014431256 | 1 |
| Obesity | b_cell | *FFAR4* | -2.379865178 | 0.00469137 | 0.048392116 | 1 |
| Obesity | b_cell | *TOR3A* | 8.388278089 | 0.001459304 | 0.034128696 | 1 |
| Obesity | b_cell | *FCAR* | 3.612622657 | 0.002109657 | 0.036453042 | 1 |
| Obesity | b_cell | *BTN3A2* | -5.863307977 | 2.08E-10 | 1.31E-07 | 1.31E-06 |
| Obesity | b_cell | *ZNF396* | -5.321834487 | 0.002713976 | 0.038814065 | 1 |
| Obesity | b_cell | *ZNF385C* | 7.12811171 | 0.000887565 | 0.034102799 | 1 |
| Obesity | b_cell | *ADH1A* | -5.672681884 | 0.000648666 | 0.032011478 | 1 |
| Obesity | b_cell | *CCDC157* | 2.010063365 | 0.002901234 | 0.039738693 | 1 |
| Obesity | b_cell | *GFRAL* | 1.347637968 | 0.002981738 | 0.040141723 | 1 |
| Obesity | b_cell | *JAKMIP3* | -3.868830801 | 0.00109349 | 0.034102799 | 1 |
| Obesity | b_cell | *PDCD1* | 12.55319994 | 0.002958562 | 0.040001035 | 1 |
| Obesity | b_cell | *SLC4A5* | 1.779444755 | 0.003594779 | 0.043409901 | 1 |
| Obesity | b_cell | *SKOR1* | 1.542596692 | 0.004292975 | 0.046251976 | 1 |
| Obesity | b_cell | *FAM26F* | 1.47462216 | 0.00147654 | 0.034128696 | 1 |
| Obesity | b_cell | *BEND4* | 1.549795215 | 0.003665933 | 0.043568476 | 1 |
| Obesity | b_cell | *ADAT2* | 8.710373113 | 0.000568639 | 0.030042288 | 1 |
| Obesity | b_cell | *BLOC1S2* | -4.181796738 | 0.00500161 | 0.049442017 | 1 |
| Obesity | b_cell | *ZKSCAN7* | 6.902936991 | 0.001104465 | 0.034102799 | 1 |
| Obesity | b_cell | *NTNG2* | 17.08824953 | 0.004439859 | 0.047071491 | 1 |
| Obesity | b_cell | *SULT1A1* | -3.345337555 | 6.76E-05 | 0.007723568 | 0.424796214 |
| Obesity | b_cell | *C1orf186* | 0.401929391 | 0.004205882 | 0.045990468 | 1 |
| Obesity | b_cell | *FAM180B* | -4.502201081 | 0.000959559 | 0.034102799 | 1 |
| Obesity | b_cell | *HLA-DQA1* | 2.232892782 | 0.001275573 | 0.034102799 | 1 |
| Obesity | b_cell | *HIST1H3E* | 2.780974861 | 0.001960053 | 0.035512542 | 1 |
| Obesity | b_cell | *WDR5B* | 6.285689106 | 0.002765335 | 0.039029882 | 1 |
| Obesity | b_cell | *SULT1A2* | 8.908498825 | 2.81E-20 | 1.77E-16 | 1.77E-16 |
| Obesity | b_cell | *CCL4L2* | -7.470486592 | 1.83E-05 | 0.003117221 | 0.115337192 |
| Obesity | b_cell | *PCDHB11* | -7.836200342 | 0.001021542 | 0.034102799 | 1 |
| Obesity | b_cell | *ZNF624* | 9.21774243 | 0.004638132 | 0.048198239 | 1 |
| Obesity | b_cell | *KLHL14* | 0.65715745 | 0.004887202 | 0.048848715 | 1 |
| Obesity | b_cell | *C14orf178* | -5.767720035 | 0.003886421 | 0.044373001 | 1 |
| Obesity | b_cell | *ZNF780A* | -5.209753548 | 0.000780652 | 0.032503045 | 1 |
| Obesity | b_cell | *AC122129.1* | 12.91087215 | 0.00137453 | 0.034102799 | 1 |
| Obesity | b_cell | *OCLN* | -4.251830272 | 0.001323265 | 0.034102799 | 1 |
| Obesity | b_cell | *ZNF181* | 6.132175561 | 0.001310243 | 0.034102799 | 1 |
| Obesity | b_cell | *AGAP9* | 5.414773048 | 0.001844354 | 0.03507628 | 1 |
| Obesity | b_cell | *ADH4* | -3.606892878 | 2.81E-05 | 0.004200417 | 0.176417506 |
| Obesity | b_cell | *APCDD1L* | 8.42052645 | 0.001133895 | 0.034102799 | 1 |

| | | | | | | |
|---|---|---|---|---|---|---|
| Obesity | b_cell | *RCSD1* | 4.592074383 | 0.00434531 | 0.046539968 | 1 |
| Obesity | b_cell | *FAM169A* | 9.708332984 | 0.001872258 | 0.03507628 | 1 |
| Obesity | b_cell | *APRT* | 9.250663614 | 0.001853279 | 0.03507628 | 1 |
| Obesity | b_cell | *L3MBTL3* | 6.343275893 | 0.000555837 | 0.030042288 | 1 |
| Obesity | b_cell | *AP001007.1* | 2.920117447 | 0.003895959 | 0.044373001 | 1 |
| Obesity | b_cell | *AC105020.1* | 7.018508856 | 0.001429729 | 0.034102799 | 1 |
| Obesity | b_cell | *CHML* | 4.722848651 | 0.000540362 | 0.030042288 | 1 |
| Obesity | b_cell | *ECT2L* | 8.944023076 | 0.001418959 | 0.034102799 | 1 |
| Obesity | b_cell | *HLA-DOA* | 1.004416789 | 0.004310962 | 0.046329948 | 1 |
| Obesity | b_cell | *HLA-DMA* | 7.774780788 | 0.004228047 | 0.04605545 | 1 |
| Obesity | b_cell | *NOTCH4* | -1.576074663 | 1.29E-07 | 4.49E-05 | 0.000808494 |
| Obesity | b_cell | *SKIV2L* | -3.939976119 | 3.50E-07 | 9.17E-05 | 0.002201209 |
| Obesity | b_cell | *CARD16* | 6.508788386 | 0.003206838 | 0.041399158 | 1 |
| Obesity | b_cell | *ABHD16A* | 2.704682658 | 0.001445733 | 0.034102799 | 1 |
| Obesity | b_cell | *POU5F1* | 5.054593227 | 5.17E-07 | 0.000125001 | 0.00325002 |
| Obesity | b_cell | *HLA-G* | -6.488434209 | 0.004423682 | 0.047058699 | 1 |
| Obesity | b_cell | *ZFP57* | 6.17862495 | 1.69E-09 | 8.88E-07 | 1.07E-05 |
| Obesity | b_cell | *FAM153C* | 2.772281232 | 0.003597353 | 0.043409901 | 1 |
| Obesity | b_cell | *PCDHGA1* | -8.67457981 | 0.001060207 | 0.034102799 | 1 |
| Obesity | b_cell | *PCDHA4* | -19.16367569 | 0.0024417 | 0.037063572 | 1 |
| Obesity | b_cell | *TP53TG3D* | 5.13644109 | 0.003895093 | 0.044373001 | 1 |
| Obesity | b_cell | *SERF1B* | -14.78007067 | 0.000252351 | 0.020340175 | 1 |
| Obesity | b_cell | *C17orf107* | 4.873020826 | 2.78E-05 | 0.004200417 | 0.174943838 |
| Obesity | b_cell | *VGLL3* | -1.657103507 | 0.004474382 | 0.047278054 | 1 |
| Obesity | b_cell | *AC084018.1* | -6.299231151 | 1.37E-05 | 0.002463655 | 0.086227914 |
| Obesity | b_cell | *LINGO4* | -60.46439074 | 0.003185561 | 0.041215094 | 1 |
| Obesity | b_cell | *NOTCH2NL* | 10.29079311 | 0.000159119 | 0.014291176 | 1 |
| Obesity | b_cell | *ARHGAP19* | 14.9284464 | 0.000661985 | 0.032011478 | 1 |
| Obesity | b_cell | *SIPA1* | 10.10455729 | 2.58E-05 | 0.004060983 | 0.162439318 |
| Obesity | b_cell | *LBH* | 3.627567399 | 0.001769677 | 0.03507628 | 1 |
| Obesity | b_cell | *AP1G2* | 6.04730513 | 0.001381971 | 0.034102799 | 1 |
| Obesity | b_cell | *ZNF90* | 6.808826996 | 0.001590576 | 0.034403393 | 1 |
| Obesity | b_cell | *ARL16* | -2.09360942 | 0.004709177 | 0.04840676 | 1 |
| Obesity | b_cell | *AC005082.1* | 12.38541653 | 0.001869956 | 0.03507628 | 1 |
| Obesity | b_cell | *CYB5RL* | 4.861256831 | 0.000774462 | 0.032503045 | 1 |
| Obesity | b_cell | *ATAD3C* | 1.744042038 | 0.001947791 | 0.035482412 | 1 |
| Obesity | b_cell | *AC016773.1* | 3.669484416 | 0.002716429 | 0.038814065 | 1 |
| Obesity | b_cell | *PPP3R1* | 5.09118571 | 0.002345986 | 0.037063572 | 1 |
| Obesity | b_cell | *FANCG* | 1.560563524 | 0.004494386 | 0.04740974 | 1 |
| Obesity | b_cell | *SLC12A8* | -3.713745519 | 0.001408064 | 0.034102799 | 1 |
| Obesity | b_cell | *EXOSC6* | -4.640597369 | 0.000230188 | 0.01879469 | 1 |
| Obesity | b_cell | *TINCR* | 3.547876224 | 0.004679828 | 0.048391578 | 1 |
| Obesity | b_cell | *HLA-DPB1* | 4.474394327 | 0.001214286 | 0.034102799 | 1 |
| Obesity | b_cell | *RASAL2-AS1* | 2.089219734 | 0.004833955 | 0.048848715 | 1 |
| Obesity | b_cell | *AC069213.1* | -21.02802118 | 0.001854289 | 0.03507628 | 1 |
| Obesity | b_cell | *LINC00184* | -8.813359002 | 0.004142467 | 0.045770984 | 1 |
| Obesity | b_cell | *AL049840.1* | -3.658875842 | 0.000637777 | 0.031842914 | 1 |
| Obesity | b_cell | *AP000350.6* | 4.050683819 | 0.001023176 | 0.034102799 | 1 |
| Obesity | b_cell | *SLC26A6* | 7.73501192 | 0.002784426 | 0.039075109 | 1 |
| Obesity | b_cell | *LINC00626* | 128.6740543 | 0.002563798 | 0.037783333 | 1 |

| | | | | | | |
|---|---|---|---|---|---|---|
| Obesity | b_cell | *ZNF503-AS1* | 6.863086306 | 0.001124457 | 0.034102799 | 1 |
| Obesity | b_cell | *FTCDNL1* | -2.028833821 | 0.005128508 | 0.049992378 | 1 |
| Obesity | b_cell | *AC064836.3* | 5.079773579 | 0.002213346 | 0.037063572 | 1 |
| Obesity | b_cell | *AC093901.1* | 1.421935837 | 0.003358221 | 0.04256681 | 1 |
| Obesity | b_cell | *AC108463.2* | -10.63500172 | 0.002406243 | 0.037063572 | 1 |
| Obesity | b_cell | *HCG20* | -14.16109112 | 0.000682934 | 0.032011478 | 1 |
| Obesity | b_cell | *LINC01013* | 3.529984676 | 0.002413446 | 0.037063572 | 1 |
| Obesity | b_cell | *RPL41* | 3.228068644 | 0.003215998 | 0.041432331 | 1 |
| Obesity | b_cell | *PATL2* | 5.709579926 | 0.000792107 | 0.032762994 | 1 |
| Obesity | b_cell | *KLHL7-AS1* | 6.113860891 | 0.002273067 | 0.037063572 | 1 |
| Obesity | b_cell | *HLA-DPA1* | 6.447413425 | 0.000747978 | 0.032503045 | 1 |
| Obesity | b_cell | *RPS18* | 7.697567743 | 0.000286204 | 0.021943452 | 1 |
| Obesity | b_cell | *AC009495.3* | 10.50561523 | 0.002335349 | 0.037063572 | 1 |
| Obesity | b_cell | *HLA-DQB2* | 2.009045027 | 0.003791548 | 0.044344465 | 1 |
| Obesity | b_cell | *TNF* | -7.264685197 | 0.000147916 | 0.013985015 | 0.929948868 |
| Obesity | b_cell | *SNHG15* | 12.97533795 | 0.00105023 | 0.034102799 | 1 |
| Obesity | b_cell | *LINC00484* | -2.981194961 | 0.001684038 | 0.034796197 | 1 |
| Obesity | b_cell | *KIAA0040* | 6.893978686 | 0.000491161 | 0.029691623 | 1 |
| Obesity | b_cell | *LINC00941* | 2.254528152 | 0.003700976 | 0.043819272 | 1 |
| Obesity | b_cell | *LINC00545* | -61.53035901 | 0.001148817 | 0.034102799 | 1 |
| Obesity | b_cell | *ZBTB22* | 2.000039691 | 0.001414648 | 0.034102799 | 1 |
| Obesity | b_cell | *GS1-124K5.4* | 2.890164781 | 0.000346271 | 0.022778303 | 1 |
| Obesity | b_cell | *IRGM* | 1.527628623 | 0.00323395 | 0.041493562 | 1 |
| Obesity | b_cell | *LILRA4* | 19.87804747 | 0.003947027 | 0.044434752 | 1 |
| Obesity | b_cell | *ARHGEF25* | 10.01009906 | 0.001896553 | 0.03507628 | 1 |
| Obesity | b_cell | *HLA-DOB* | 0.68204365 | 0.002745745 | 0.038879503 | 1 |
| Obesity | b_cell | *ATP5O* | 4.829381097 | 0.000940933 | 0.034102799 | 1 |
| Obesity | b_cell | *PLA2G4B* | -7.203520772 | 0.004378141 | 0.046732379 | 1 |
| Obesity | b_cell | *APOBEC3C* | 7.72334429 | 0.001033608 | 0.034102799 | 1 |
| Obesity | b_cell | *LINC00847* | 4.277216142 | 0.001382721 | 0.034102799 | 1 |
| Obesity | b_cell | *PGAM5* | 11.65639931 | 0.001288411 | 0.034102799 | 1 |
| Obesity | b_cell | *LINC00926* | 0.390944929 | 0.004078256 | 0.045300351 | 1 |
| Obesity | b_cell | *ZNF286B* | 12.69783541 | 0.000827492 | 0.033555694 | 1 |
| Obesity | b_cell | *SNHG18* | -4.160351043 | 0.002365329 | 0.037063572 | 1 |
| Obesity | b_cell | *TMEM200B* | 7.028736588 | 0.00070413 | 0.032011478 | 1 |
| Obesity | b_cell | *TRNP1* | 2.706683361 | 0.003953212 | 0.044434752 | 1 |
| Obesity | b_cell | *ARMS2* | -13.66586826 | 0.002998254 | 0.040277828 | 1 |
| Obesity | b_cell | *LINC00346* | 9.504303815 | 0.001230411 | 0.034102799 | 1 |
| Obesity | b_cell | *PGA5* | -61.47807173 | 0.001438592 | 0.034102799 | 1 |
| Obesity | b_cell | *HP* | -18.80550492 | 1.37E-05 | 0.002463655 | 0.086200365 |
| Obesity | b_cell | *DUXA* | -9.400126729 | 0.001262952 | 0.034102799 | 1 |
| Obesity | b_cell | *LINC00924* | 2.663035023 | 0.002695373 | 0.038689069 | 1 |
| Obesity | b_cell | *ITGB3* | -5.725464377 | 0.000566154 | 0.030042288 | 1 |
| Obesity | b_cell | *AC083843.1* | -96.37381195 | 0.000146198 | 0.013985015 | 0.919148422 |
| Obesity | b_cell | *SH3RF3-AS1* | -11.81839959 | 1.46E-05 | 0.002548229 | 0.091736237 |
| Obesity | b_cell | *LINC00562* | -4.356154149 | 0.00013051 | 0.013451127 | 0.820518767 |
| Obesity | b_cell | *C15orf65* | -4.64607769 | 0.00350246 | 0.043053853 | 1 |
| Obesity | b_cell | *LINC00668* | 35.16824858 | 0.001887493 | 0.03507628 | 1 |
| Obesity | b_cell | *AC139100.2* | 9.953150189 | 0.002610275 | 0.037987959 | 1 |
| Obesity | b_cell | *AL132989.1* | 17.80279127 | 0.001277505 | 0.034102799 | 1 |

| | | | | | | |
|---|---|---|---|---|---|---|
| Obesity | b_cell | *YTHDF3-AS1* | 6.250333496 | 0.000680918 | 0.032011478 | 1 |
| Obesity | b_cell | *LINC01050* | 4.18579869 | 0.003835672 | 0.044373001 | 1 |
| Obesity | dendritic_cell | *SEMA3F* | -1.961787909 | 2.87E-07 | 6.69E-05 | 0.001806397 |
| Obesity | dendritic_cell | *WNT16* | 3.988197611 | 0.000420194 | 0.02751835 | 1 |
| Obesity | dendritic_cell | *RBM6* | -3.150293556 | 2.06E-16 | 2.16E-13 | 1.29E-12 |
| Obesity | dendritic_cell | *HFE* | -4.497703499 | 3.33E-07 | 7.48E-05 | 0.00209543 |
| Obesity | dendritic_cell | *PSMC4* | 3.8943254 | 0.000635688 | 0.03842513 | 1 |
| Obesity | dendritic_cell | *LSG1* | 2.548002605 | 0.000518115 | 0.032573902 | 1 |
| Obesity | dendritic_cell | *SDCCAG8* | 2.602705817 | 0.000666349 | 0.039522013 | 1 |
| Obesity | dendritic_cell | *ITIH4* | 2.771162888 | 8.15E-11 | 4.27E-08 | 5.13E-07 |
| Obesity | dendritic_cell | *UHRF1BP1* | 5.088064292 | 1.11E-20 | 2.33E-17 | 6.99E-17 |
| Obesity | dendritic_cell | *TIPIN* | 1.755235363 | 9.29E-06 | 0.001243312 | 0.058435671 |
| Obesity | dendritic_cell | *C3orf18* | 8.022066546 | 8.02E-07 | 0.0001616 | 0.005039078 |
| Obesity | dendritic_cell | *YPEL3* | 3.46946455 | 1.36E-08 | 5.03E-06 | 8.55E-05 |
| Obesity | dendritic_cell | *PABPC4* | -5.278165559 | 4.70E-07 | 0.000101912 | 0.002955459 |
| Obesity | dendritic_cell | *DESI1* | 2.384375089 | 7.20E-05 | 0.006289724 | 0.452860112 |
| Obesity | dendritic_cell | *ABHD12* | 4.805315763 | 9.06E-05 | 0.007597071 | 0.56978029 |
| Obesity | dendritic_cell | *CORO1A* | -1.771776617 | 0.000430776 | 0.027920475 | 1 |
| Obesity | dendritic_cell | *CLUAP1* | 3.269483282 | 0.000106471 | 0.008693325 | 0.669386047 |
| Obesity | dendritic_cell | *KAT8* | 4.153126625 | 0.000159853 | 0.011964232 | 1 |
| Obesity | dendritic_cell | *CHRNE* | 2.121974877 | 0.000736069 | 0.042848746 | 1 |
| Obesity | dendritic_cell | *DDX55* | 3.090974152 | 5.96E-05 | 0.005273934 | 0.374449294 |
| Obesity | dendritic_cell | *PRSS16* | 7.985005198 | 1.65E-05 | 0.001852665 | 0.103933669 |
| Obesity | dendritic_cell | *SERPINC1* | 5.085120815 | 2.91E-09 | 1.22E-06 | 1.83E-05 |
| Obesity | dendritic_cell | *CCDC92* | 3.157934802 | 3.94E-06 | 0.000590452 | 0.024799004 |
| Obesity | dendritic_cell | *DCAF4* | 4.54107815 | 5.25E-05 | 0.004855985 | 0.330206971 |
| Obesity | dendritic_cell | *ADRA1A* | 0.955635281 | 1.82E-05 | 0.001971937 | 0.114372361 |
| Obesity | dendritic_cell | *SNRPC* | -7.763306563 | 2.97E-16 | 2.67E-13 | 1.87E-12 |
| Obesity | dendritic_cell | *HIST1H2BJ* | 5.338612642 | 3.02E-06 | 0.000463596 | 0.019007439 |
| Obesity | dendritic_cell | *GRK4* | 1.340925051 | 1.68E-05 | 0.001852665 | 0.105601894 |
| Obesity | dendritic_cell | *XRCC3* | 4.508650721 | 0.000635472 | 0.03842513 | 1 |
| Obesity | dendritic_cell | *ATP8B3* | -1.766894226 | 0.000804058 | 0.046377207 | 1 |
| Obesity | dendritic_cell | *IGHMBP2* | 3.787287551 | 4.52E-06 | 0.000660913 | 0.028419252 |
| Obesity | dendritic_cell | *MRPS9* | 3.140944365 | 0.000837807 | 0.047884468 | 1 |
| Obesity | dendritic_cell | *ZSCAN9* | 4.696239242 | 7.49E-06 | 0.001047053 | 0.047117382 |
| Obesity | dendritic_cell | *TCF19* | 5.717718184 | 1.34E-05 | 0.00161661 | 0.084063721 |
| Obesity | dendritic_cell | *CARF* | 3.513944876 | 1.39E-05 | 0.001651635 | 0.087536659 |
| Obesity | dendritic_cell | *KLRG1* | 2.447175672 | 0.000679057 | 0.039899377 | 1 |
| Obesity | dendritic_cell | *MYO19* | 4.851865525 | 0.000122831 | 0.009533844 | 0.772241336 |
| Obesity | dendritic_cell | *C18orf8* | -4.90526617 | 0.000503042 | 0.031945721 | 1 |
| Obesity | dendritic_cell | *TRIM65* | 5.376828501 | 1.24E-05 | 0.00153196 | 0.078129936 |
| Obesity | dendritic_cell | *TUFT1* | -2.866269274 | 0.000885714 | 0.049278635 | 1 |
| Obesity | dendritic_cell | *NBEAL1* | 1.778693651 | 0.000173308 | 0.01252399 | 1 |
| Obesity | dendritic_cell | *ATP6V0D2* | -4.742387618 | 1.50E-05 | 0.001748593 | 0.09442402 |
| Obesity | dendritic_cell | *MTG1* | 1.750821355 | 0.000641743 | 0.03842513 | 1 |
| Obesity | dendritic_cell | *HSD17B12* | -3.933440986 | 5.54E-11 | 3.17E-08 | 3.48E-07 |
| Obesity | dendritic_cell | *TTC12* | 2.467818525 | 2.03E-05 | 0.002161809 | 0.12754675 |
| Obesity | dendritic_cell | *TBX6* | 3.983881645 | 1.58E-07 | 3.82E-05 | 0.000994418 |
| Obesity | dendritic_cell | *DOC2A* | -5.093234493 | 8.16E-08 | 2.23E-05 | 0.000513045 |
| Obesity | dendritic_cell | *PRSS53* | -5.516828857 | 0.00020658 | 0.014592924 | 1 |

| | | | | | | |
|---|---|---|---|---|---|---|
| Obesity | dendritic_cell | *DPYSL4* | 1.921720129 | 0.000112839 | 0.008979991 | 0.709419311 |
| Obesity | dendritic_cell | *NEIL2* | 3.061219139 | 5.73E-05 | 0.005142398 | 0.359967877 |
| Obesity | dendritic_cell | *CLEC18A* | -2.690325088 | 1.10E-06 | 0.000192652 | 0.00693547 |
| Obesity | dendritic_cell | *HPD* | 2.488650417 | 4.42E-05 | 0.00422073 | 0.277580649 |
| Obesity | dendritic_cell | *SLC5A11* | -1.393308493 | 0.00024636 | 0.017020508 | 1 |
| Obesity | dendritic_cell | *LMOD1* | -6.227063931 | 7.90E-12 | 4.96E-09 | 4.96E-08 |
| Obesity | dendritic_cell | *APEH* | -3.401999708 | 8.31E-05 | 0.007061341 | 0.522539214 |
| Obesity | dendritic_cell | *RNF123* | 6.930300395 | 3.96E-20 | 6.22E-17 | 2.49E-16 |
| Obesity | dendritic_cell | *MST1R* | 4.717935847 | 1.68E-06 | 0.000270903 | 0.010565228 |
| Obesity | dendritic_cell | *TRMT61A* | -5.657699077 | 5.65E-05 | 0.005142398 | 0.355180034 |
| Obesity | dendritic_cell | *ZNF646* | 10.15219069 | 2.51E-08 | 8.30E-06 | 0.00015777 |
| Obesity | dendritic_cell | *VKORC1* | -3.457313503 | 0.000544011 | 0.033863365 | 1 |
| Obesity | dendritic_cell | *SRR* | 2.564537016 | 1.02E-06 | 0.000188324 | 0.00642756 |
| Obesity | dendritic_cell | *C8orf46* | 0.719515451 | 0.000142431 | 0.010820042 | 0.895465907 |
| Obesity | dendritic_cell | *TRMT61B* | 1.855159646 | 3.64E-05 | 0.003636553 | 0.229102833 |
| Obesity | dendritic_cell | *C1QTNF4* | -2.227962643 | 2.09E-15 | 1.64E-12 | 1.31E-11 |
| Obesity | dendritic_cell | *CTSW* | -1.022348173 | 6.78E-06 | 0.00096824 | 0.042602572 |
| Obesity | dendritic_cell | *EFEMP2* | 3.395510068 | 2.48E-05 | 0.002601026 | 0.156061584 |
| Obesity | dendritic_cell | *SNX32* | 5.928861762 | 1.11E-07 | 2.92E-05 | 0.000700372 |
| Obesity | dendritic_cell | *HPSE2* | 2.406326044 | 1.63E-05 | 0.001852665 | 0.102700051 |
| Obesity | dendritic_cell | *GMPPB* | 1.970941981 | 0.000872211 | 0.048960613 | 1 |
| Obesity | dendritic_cell | *RUFY1* | 3.730734592 | 0.000182905 | 0.013067347 | 1 |
| Obesity | dendritic_cell | *SH2B1* | -4.51794665 | 2.63E-14 | 1.84E-11 | 1.66E-10 |
| Obesity | dendritic_cell | *RPP25* | -2.894278708 | 0.000228656 | 0.015972925 | 1 |
| Obesity | dendritic_cell | *STX19* | 7.639856216 | 2.63E-06 | 0.000413921 | 0.016556846 |
| Obesity | dendritic_cell | *TRIM73* | -10.55026988 | 2.52E-09 | 1.13E-06 | 1.59E-05 |
| Obesity | dendritic_cell | *TUFM* | 9.061575951 | 1.53E-18 | 1.92E-15 | 9.59E-15 |
| Obesity | dendritic_cell | *LYSMD4* | 2.527681941 | 0.000440901 | 0.028285175 | 1 |
| Obesity | dendritic_cell | *PSMG1* | 2.211943301 | 9.08E-06 | 0.001241411 | 0.0571049 |
| Obesity | dendritic_cell | *RP1L1* | 4.779504116 | 1.57E-06 | 0.000259212 | 0.009850057 |
| Obesity | dendritic_cell | *EIF3C* | 8.7907482 | 2.02E-27 | 1.27E-23 | 1.27E-23 |
| Obesity | dendritic_cell | *PLA2G6* | 2.070435883 | 2.86E-05 | 0.002902926 | 0.179981399 |
| Obesity | dendritic_cell | *GP1BA* | 3.090870477 | 7.39E-05 | 0.006368348 | 0.46488942 |
| Obesity | dendritic_cell | *BTN3A2* | -2.003078437 | 5.06E-09 | 1.99E-06 | 3.18E-05 |
| Obesity | dendritic_cell | *HYAL3* | -2.703914911 | 1.05E-05 | 0.001349511 | 0.065738193 |
| Obesity | dendritic_cell | *ZSCAN23* | 1.203376041 | 0.000562376 | 0.034663297 | 1 |
| Obesity | dendritic_cell | *CENPP* | 2.773774321 | 9.66E-07 | 0.000184131 | 0.006076315 |
| Obesity | dendritic_cell | *FBF1* | 1.833321398 | 0.000109463 | 0.008823021 | 0.688195636 |
| Obesity | dendritic_cell | *FAM47E* | 1.398109959 | 0.00086203 | 0.048825042 | 1 |
| Obesity | dendritic_cell | *SULT1A1* | -2.562693547 | 0.000254561 | 0.017395901 | 1 |
| Obesity | dendritic_cell | *C6orf106* | 2.653158114 | 1.05E-06 | 0.000188324 | 0.006591334 |
| Obesity | dendritic_cell | *SULT1A2* | 2.782720411 | 2.28E-23 | 7.17E-20 | 1.43E-19 |
| Obesity | dendritic_cell | *ZNF165* | -5.281850726 | 4.17E-05 | 0.004093421 | 0.261978974 |
| Obesity | dendritic_cell | *RPS26* | -1.72851548 | 9.24E-05 | 0.007646921 | 0.581165975 |
| Obesity | dendritic_cell | *ZNF780A* | -2.276725997 | 1.07E-05 | 0.001351121 | 0.067556044 |
| Obesity | dendritic_cell | *ZKSCAN8* | 4.219062398 | 0.00030972 | 0.020496964 | 1 |
| Obesity | dendritic_cell | *NOTCH4* | -0.610659869 | 1.34E-07 | 3.37E-05 | 0.00084351 |
| Obesity | dendritic_cell | *SKIV2L* | -1.90045755 | 8.23E-07 | 0.0001616 | 0.005171198 |
| Obesity | dendritic_cell | *NELFE* | 2.790018783 | 4.43E-05 | 0.00422073 | 0.27856816 |
| Obesity | dendritic_cell | *POU5F1* | 1.560053115 | 1.44E-06 | 0.000245309 | 0.009076434 |

| | | | | | | |
|---|---|---|---|---|---|---|
| Obesity | dendritic_cell | *CCHCR1* | 2.698258856 | 5.12E-08 | 1.53E-05 | 0.000321613 |
| Obesity | dendritic_cell | *ABCF1* | 3.935722704 | 0.000170622 | 0.012473269 | 1 |
| Obesity | dendritic_cell | *HLA-E* | 3.50590509 | 8.14E-08 | 2.23E-05 | 0.000511876 |
| Obesity | dendritic_cell | *ZFP57* | 2.296787893 | 1.47E-08 | 5.12E-06 | 9.22E-05 |
| Obesity | dendritic_cell | *EIF3CL* | 7.888196312 | 4.97E-08 | 1.53E-05 | 0.000312731 |
| Obesity | dendritic_cell | *ASIC3* | 3.904745368 | 1.05E-05 | 0.001349511 | 0.066126044 |
| Obesity | dendritic_cell | *NOTCH2NL* | 3.266630527 | 0.000142845 | 0.010820042 | 0.89806352 |
| Obesity | dendritic_cell | *LINC00240* | 1.690931576 | 4.96E-05 | 0.004655043 | 0.311887869 |
| Obesity | dendritic_cell | *AL049840.1* | -3.330501263 | 0.000258003 | 0.017441571 | 1 |
| Obesity | dendritic_cell | *ZSCAN31* | 2.811509518 | 2.61E-05 | 0.002692754 | 0.164257987 |
| Obesity | dendritic_cell | *PSMB9* | 3.763243718 | 0.000121572 | 0.009533844 | 0.764321691 |
| Obesity | dendritic_cell | *MPV17L2* | 8.053949821 | 7.46E-07 | 0.000156402 | 0.004692049 |
| Obesity | dendritic_cell | *HMBS* | 4.127505945 | 1.51E-10 | 7.30E-08 | 9.49E-07 |
| Obesity | dendritic_cell | *SH3RF3-AS1* | -2.913920707 | 0.000165713 | 0.012256878 | 1 |
| Obesity | dendritic_cell | *GTF2H5* | 4.098942216 | 0.000306413 | 0.020493808 | 1 |
| Obesity | endothelial | *SCYL3* | -0.593345226 | 0.000581351 | 0.001179018 | 1 |
| Obesity | endothelial | *C1orf112* | -0.971237819 | 2.41E-06 | 1.16E-05 | 0.015172001 |
| Obesity | endothelial | *NFYA* | -1.015861446 | 1.11E-05 | 4.01E-05 | 0.069493214 |
| Obesity | endothelial | *NIPAL3* | 0.48273627 | 0.002316608 | 0.004020489 | 1 |
| Obesity | endothelial | *ENPP4* | 0.348615951 | 0.016666072 | 0.023846233 | 1 |
| Obesity | endothelial | *KRIT1* | -0.67009486 | 0.000422827 | 0.000890856 | 1 |
| Obesity | endothelial | *RAD52* | -0.928063541 | 2.52E-05 | 7.92E-05 | 0.158730293 |
| Obesity | endothelial | *BAD* | -1.592283918 | 3.45E-07 | 2.44E-06 | 0.002167215 |
| Obesity | endothelial | *HS3ST1* | -0.980630734 | 0.002707322 | 0.004606477 | 1 |
| Obesity | endothelial | *HECW1* | -0.513857387 | 3.59E-08 | 4.81E-07 | 0.000225397 |
| Obesity | endothelial | *MAD1L1* | -0.930943393 | 3.98E-06 | 1.75E-05 | 0.025009275 |
| Obesity | endothelial | *LASP1* | -0.890978939 | 1.88E-06 | 9.39E-06 | 0.011790236 |
| Obesity | endothelial | *SNX11* | -0.651825097 | 1.37E-05 | 4.77E-05 | 0.086182461 |
| Obesity | endothelial | *TMEM176A* | -0.407428867 | 0.004493938 | 0.007259349 | 1 |
| Obesity | endothelial | *M6PR* | -0.650113149 | 0.004731144 | 0.007611234 | 1 |
| Obesity | endothelial | *CYP26B1* | -1.070492336 | 3.49E-07 | 2.47E-06 | 0.002196535 |
| Obesity | endothelial | *ICA1* | 0.280079148 | 2.00E-05 | 6.55E-05 | 0.125510101 |
| Obesity | endothelial | *ALS2* | -0.65336775 | 0.002478618 | 0.004265828 | 1 |
| Obesity | endothelial | *POLDIP2* | -0.82259473 | 0.011904584 | 0.017492889 | 1 |
| Obesity | endothelial | *CD38* | -0.776678149 | 4.56E-09 | 2.02E-07 | 2.87E-05 |
| Obesity | endothelial | *RBM6* | -0.827679333 | 8.27E-26 | 1.30E-22 | 5.20E-22 |
| Obesity | endothelial | *RECQL* | -0.892198554 | 0.000102413 | 0.000257651 | 0.643869597 |
| Obesity | endothelial | *PDK4* | -0.674618506 | 7.65E-05 | 0.000200111 | 0.480667032 |
| Obesity | endothelial | *SLC22A16* | -0.980505745 | 5.38E-06 | 2.23E-05 | 0.0338173 |
| Obesity | endothelial | *MEOX1* | 0.149375403 | 8.04E-06 | 3.08E-05 | 0.050532196 |
| Obesity | endothelial | *CIAPIN1* | -1.258883335 | 1.44E-05 | 4.98E-05 | 0.09059215 |
| Obesity | endothelial | *GCFC2* | -0.625705074 | 0.001330186 | 0.002449584 | 1 |
| Obesity | endothelial | *WDR54* | 0.590457668 | 2.19E-05 | 7.04E-05 | 0.13740781 |
| Obesity | endothelial | *ABCB4* | -1.400403522 | 1.47E-07 | 1.27E-06 | 0.000924577 |
| Obesity | endothelial | *ZNF195* | -0.595013514 | 0.001115384 | 0.00209201 | 1 |
| Obesity | endothelial | *PDK2* | -1.1126089 | 4.38E-07 | 2.95E-06 | 0.002752586 |
| Obesity | endothelial | *GGNBP2* | -1.071754698 | 5.85E-12 | 1.36E-09 | 3.68E-08 |
| Obesity | endothelial | *TMEM98* | -0.407536321 | 0.002412914 | 0.004171017 | 1 |
| Obesity | endothelial | *ABCC8* | -1.104736765 | 8.48E-08 | 8.38E-07 | 0.000533361 |
| Obesity | endothelial | *CCL18* | -0.386481273 | 0.010632629 | 0.015844355 | 1 |

| | | | | | | |
|---|---|---|---|---|---|---|
| Obesity | endothelial | *SYNRG* | -0.722170132 | 7.01E-05 | 0.000186655 | 0.440506689 |
| Obesity | endothelial | *TMEM132A* | 1.001834753 | 7.58E-06 | 2.94E-05 | 0.047668702 |
| Obesity | endothelial | *TAC1* | 1.535571081 | 0.000612254 | 0.001231366 | 1 |
| Obesity | endothelial | *ZNF263* | -0.864460642 | 0.000839118 | 0.001636333 | 1 |
| Obesity | endothelial | *SPATA20* | -0.948772391 | 0.000313945 | 0.000685101 | 1 |
| Obesity | endothelial | *TNFRSF12A* | -1.929619654 | 2.88E-08 | 4.36E-07 | 0.00018103 |
| Obesity | endothelial | *MAP3K9* | 0.60441371 | 0.019772399 | 0.027884493 | 1 |
| Obesity | endothelial | *RALA* | -0.669397575 | 0.001055828 | 0.001993391 | 1 |
| Obesity | endothelial | *BAIAP2L1* | 0.264342325 | 5.50E-05 | 0.000152136 | 0.345804675 |
| Obesity | endothelial | *FARP2* | -0.589864117 | 0.001540204 | 0.002789761 | 1 |
| Obesity | endothelial | *IFRD1* | -0.689534809 | 7.36E-07 | 4.45E-06 | 0.004624798 |
| Obesity | endothelial | *GTF2IRD1* | -0.341945121 | 0.023057927 | 0.03194473 | 1 |
| Obesity | endothelial | *VPS41* | -0.566048186 | 0.00132288 | 0.002439702 | 1 |
| Obesity | endothelial | *ARHGAP44* | -0.989065986 | 3.01E-05 | 9.21E-05 | 0.189535515 |
| Obesity | endothelial | *ELAC2* | -1.094627419 | 1.35E-06 | 7.22E-06 | 0.008482132 |
| Obesity | endothelial | *CCDC124* | -0.727357445 | 0.000361184 | 0.000775004 | 1 |
| Obesity | endothelial | *CEACAM21* | -0.746005136 | 0.005712999 | 0.009013664 | 1 |
| Obesity | endothelial | *NOS2* | -6.804820423 | 1.27E-06 | 6.85E-06 | 0.007957257 |
| Obesity | endothelial | *TRAPPC6A* | -1.134697747 | 0.000257804 | 0.000575777 | 1 |
| Obesity | endothelial | *ST7L* | -0.316306638 | 0.006720887 | 0.010448619 | 1 |
| Obesity | endothelial | *RPUSD1* | -2.922593181 | 1.92E-07 | 1.57E-06 | 0.001210193 |
| Obesity | endothelial | *BAIAP3* | 0.887136028 | 1.33E-05 | 4.65E-05 | 0.083694489 |
| Obesity | endothelial | *TSR3* | -1.635328074 | 1.45E-06 | 7.62E-06 | 0.009089934 |
| Obesity | endothelial | *PIGQ* | -0.907365815 | 0.000601259 | 0.001213519 | 1 |
| Obesity | endothelial | *PSMB1* | -1.003379442 | 1.35E-05 | 4.71E-05 | 0.085114235 |
| Obesity | endothelial | *CDK11A* | -1.133731352 | 3.23E-06 | 1.48E-05 | 0.020302635 |
| Obesity | endothelial | *DLEC1* | -0.844133896 | 3.73E-08 | 4.91E-07 | 0.000234383 |
| Obesity | endothelial | *SS18L2* | -0.708514214 | 0.018197968 | 0.025832157 | 1 |
| Obesity | endothelial | *MGST1* | -0.554800177 | 3.18E-08 | 4.54E-07 | 0.000200113 |
| Obesity | endothelial | *NFIX* | -0.741619225 | 1.64E-07 | 1.39E-06 | 0.001032218 |
| Obesity | endothelial | *PKD1* | -0.510044418 | 0.006323564 | 0.0098783 | 1 |
| Obesity | endothelial | *MED24* | -0.685598716 | 0.001216272 | 0.002266361 | 1 |
| Obesity | endothelial | *SEC62* | -0.564166711 | 0.004058272 | 0.006630549 | 1 |
| Obesity | endothelial | *POMT2* | -1.130284839 | 4.45E-08 | 5.43E-07 | 0.000280085 |
| Obesity | endothelial | *MLXIPL* | -0.173431495 | 0.002439523 | 0.004210069 | 1 |
| Obesity | endothelial | *SPRTN* | -0.674647289 | 0.019368413 | 0.027376172 | 1 |
| Obesity | endothelial | *METTL13* | -1.222179533 | 0.000116573 | 0.00028786 | 0.732892674 |
| Obesity | endothelial | *DYRK4* | -0.632100904 | 0.003419251 | 0.00567648 | 1 |
| Obesity | endothelial | *STARD3NL* | 0.384076377 | 0.031211842 | 0.042127275 | 1 |
| Obesity | endothelial | *NCAPD2* | -0.380065914 | 0.002330309 | 0.004040446 | 1 |
| Obesity | endothelial | *CD4* | -0.987170072 | 5.28E-08 | 6.01E-07 | 0.000331907 |
| Obesity | endothelial | *LRRC23* | -0.431619453 | 0.035038049 | 0.046829128 | 1 |
| Obesity | endothelial | *HFE* | -1.202987339 | 2.92E-13 | 1.15E-10 | 1.84E-09 |
| Obesity | endothelial | *SCMH1* | -0.373173159 | 0.025751929 | 0.035311315 | 1 |
| Obesity | endothelial | *CLCN6* | -0.843483126 | 0.000163564 | 0.000388047 | 1 |
| Obesity | endothelial | *MRC2* | -0.745323157 | 3.84E-06 | 1.70E-05 | 0.024166096 |
| Obesity | endothelial | *TSPAN9* | 0.281119242 | 4.49E-05 | 0.000127635 | 0.282072083 |
| Obesity | endothelial | *MKS1* | -1.243252783 | 0.000513967 | 0.001055984 | 1 |
| Obesity | endothelial | *UTP18* | -1.113976764 | 6.95E-06 | 2.74E-05 | 0.043723825 |
| Obesity | endothelial | *SYT7* | 0.311067939 | 7.66E-07 | 4.60E-06 | 0.004817381 |

| | | | | | | |
|---|---|---|---|---|---|---|
| Obesity | endothelial | *LARS2* | -0.480851417 | 0.028914094 | 0.03925349 | 1 |
| Obesity | endothelial | *CEP68* | 0.306031629 | 0.007060779 | 0.010931081 | 1 |
| Obesity | endothelial | *ZBTB32* | -1.787825677 | 3.08E-05 | 9.36E-05 | 0.193421138 |
| Obesity | endothelial | *TMEM159* | -0.869690932 | 3.45E-07 | 2.44E-06 | 0.002167239 |
| Obesity | endothelial | *ERCC1* | -0.791594102 | 4.88E-07 | 3.21E-06 | 0.003067696 |
| Obesity | endothelial | *LTF* | 0.519840813 | 6.13E-07 | 3.85E-06 | 0.003853752 |
| Obesity | endothelial | *ALOX5* | -1.176224323 | 4.82E-06 | 2.04E-05 | 0.030320261 |
| Obesity | endothelial | *MAP4K5* | -0.467331707 | 0.009799216 | 0.014710524 | 1 |
| Obesity | endothelial | *MAN2B2* | -1.000927188 | 3.89E-05 | 0.000113541 | 0.244795457 |
| Obesity | endothelial | *CLDN11* | 0.375963925 | 0.004953504 | 0.007928381 | 1 |
| Obesity | endothelial | *SLC25A39* | -0.948717323 | 0.00269257 | 0.004583858 | 1 |
| Obesity | endothelial | *HEBP1* | -0.731945983 | 0.00726936 | 0.01121533 | 1 |
| Obesity | endothelial | *TACC3* | -0.633030831 | 0.019879397 | 0.028022819 | 1 |
| Obesity | endothelial | *ZC3H3* | -0.535352614 | 0.011997743 | 0.017599116 | 1 |
| Obesity | endothelial | *MDH1* | -0.646408186 | 3.05E-05 | 9.30E-05 | 0.191823301 |
| Obesity | endothelial | *SLC30A9* | -0.464727594 | 0.013092195 | 0.019053387 | 1 |
| Obesity | endothelial | *COX15* | -0.979169585 | 7.76E-06 | 2.99E-05 | 0.048781437 |
| Obesity | endothelial | *CCDC88C* | 0.275213094 | 0.028889748 | 0.039228908 | 1 |
| Obesity | endothelial | *NPC1L1* | -1.098172333 | 1.66E-06 | 8.52E-06 | 0.010437274 |
| Obesity | endothelial | *CHDH* | -2.605413401 | 0.000352384 | 0.000757673 | 1 |
| Obesity | endothelial | *IL20RA* | -3.89777783 | 5.83E-08 | 6.41E-07 | 0.000366801 |
| Obesity | endothelial | *GLT8D1* | -0.884677761 | 1.92E-05 | 6.32E-05 | 0.120487736 |
| Obesity | endothelial | *RUFY3* | -0.51157024 | 0.001488629 | 0.002713486 | 1 |
| Obesity | endothelial | *TTC27* | -0.618604279 | 0.000682663 | 0.001358197 | 1 |
| Obesity | endothelial | *VSIG2* | 0.336783051 | 8.74E-07 | 5.10E-06 | 0.005496723 |
| Obesity | endothelial | *PHLDB1* | -0.296283214 | 0.001283231 | 0.002376339 | 1 |
| Obesity | endothelial | *MARCO* | -0.487066202 | 1.21E-05 | 4.31E-05 | 0.07629362 |
| Obesity | endothelial | *MNAT1* | -0.69444149 | 8.36E-05 | 0.000216561 | 0.525811135 |
| Obesity | endothelial | *AQR* | -0.545796912 | 0.005758197 | 0.009077679 | 1 |
| Obesity | endothelial | *CPS1* | -0.352896196 | 0.013098718 | 0.019058468 | 1 |
| Obesity | endothelial | *NLRP2* | -0.68952396 | 0.000229973 | 0.000520647 | 1 |
| Obesity | endothelial | *RNH1* | -0.532190989 | 0.002300106 | 0.003995791 | 1 |
| Obesity | endothelial | *NDUFS1* | -0.572740123 | 0.002830987 | 0.004794831 | 1 |
| Obesity | endothelial | *BIRC3* | -0.810549256 | 0.001460823 | 0.002669824 | 1 |
| Obesity | endothelial | *GLRX2* | -0.88260964 | 3.13E-05 | 9.50E-05 | 0.197049442 |
| Obesity | endothelial | *SNAPC1* | -1.17466801 | 4.53E-08 | 5.46E-07 | 0.000284614 |
| Obesity | endothelial | *GCLM* | -0.491681633 | 0.004952798 | 0.007928381 | 1 |
| Obesity | endothelial | *UBR2* | -0.564145266 | 0.000965351 | 0.001845851 | 1 |
| Obesity | endothelial | *HSF2* | -1.122158099 | 1.84E-05 | 6.12E-05 | 0.115837921 |
| Obesity | endothelial | *NR1H3* | -0.584187716 | 0.004299061 | 0.006980423 | 1 |
| Obesity | endothelial | *TYMP* | -0.709515223 | 8.03E-05 | 0.000208591 | 0.504789447 |
| Obesity | endothelial | *KPNA6* | -0.592716619 | 0.003110577 | 0.00520527 | 1 |
| Obesity | endothelial | *FAS* | -0.539690283 | 0.000135388 | 0.000327757 | 0.851186099 |
| Obesity | endothelial | *RNASET2* | -0.858095502 | 2.37E-07 | 1.84E-06 | 0.001487272 |
| Obesity | endothelial | *CD44* | -0.680047982 | 1.66E-07 | 1.40E-06 | 0.001046742 |
| Obesity | endothelial | *MIPEP* | -0.962441247 | 4.81E-06 | 2.04E-05 | 0.030238714 |
| Obesity | endothelial | *INSRR* | -0.921793675 | 7.19E-05 | 0.000190681 | 0.452104128 |
| Obesity | endothelial | *VRK2* | -0.396589595 | 0.031093353 | 0.041985376 | 1 |
| Obesity | endothelial | *SNX1* | -0.45577111 | 0.008073067 | 0.012328242 | 1 |
| Obesity | endothelial | *SLC39A9* | -0.807389616 | 5.97E-05 | 0.000163358 | 0.3751305 |

| | | | | | | |
|---|---|---|---|---|---|---|
| Obesity | endothelial | *ANK1* | 0.458065678 | 6.50E-05 | 0.000175163 | 0.408582886 |
| Obesity | endothelial | *BAK1* | 0.355003225 | 0.034704404 | 0.046452328 | 1 |
| Obesity | endothelial | *GRN* | -0.924172528 | 1.30E-05 | 4.57E-05 | 0.08182907 |
| Obesity | endothelial | *CENPQ* | -0.743171195 | 1.95E-05 | 6.42E-05 | 0.122822689 |
| Obesity | endothelial | *ZCCHC8* | -0.609226047 | 0.004301281 | 0.006982224 | 1 |
| Obesity | endothelial | *UBA6* | -0.611301383 | 0.000968439 | 0.001850632 | 1 |
| Obesity | endothelial | *PIAS1* | -0.572036055 | 0.00043276 | 0.000907525 | 1 |
| Obesity | endothelial | *SLC4A7* | -0.723924146 | 9.89E-08 | 9.37E-07 | 0.000621874 |
| Obesity | endothelial | *ASTE1* | -0.720288856 | 0.000120714 | 0.00029692 | 0.758928131 |
| Obesity | endothelial | *MYOC* | -0.432945654 | 6.83E-05 | 0.000182541 | 0.4291532 |
| Obesity | endothelial | *SH3YL1* | -1.000293237 | 1.80E-10 | 1.92E-08 | 1.13E-06 |
| Obesity | endothelial | *VCL* | -0.815744584 | 3.02E-06 | 1.40E-05 | 0.018996951 |
| Obesity | endothelial | *STAP1* | -2.636111319 | 9.73E-12 | 2.04E-09 | 6.12E-08 |
| Obesity | endothelial | *TBC1D23* | -0.818207969 | 8.88E-06 | 3.35E-05 | 0.055826862 |
| Obesity | endothelial | *CUL3* | -0.570769869 | 0.001595209 | 0.002876959 | 1 |
| Obesity | endothelial | *MYOM2* | -0.559237856 | 0.018833712 | 0.026662362 | 1 |
| Obesity | endothelial | *TUBG2* | -0.411365623 | 0.000167268 | 0.000395641 | 1 |
| Obesity | endothelial | *MRI1* | -0.530303503 | 0.002784952 | 0.004723225 | 1 |
| Obesity | endothelial | *AGA* | -1.115229903 | 0.00130098 | 0.002404251 | 1 |
| Obesity | endothelial | *BOD1L1* | -0.496867894 | 0.007419194 | 0.011432469 | 1 |
| Obesity | endothelial | *MAT2B* | -0.694113625 | 0.016502175 | 0.023638454 | 1 |
| Obesity | endothelial | *CLEC16A* | -0.657811183 | 0.000924803 | 0.001779687 | 1 |
| Obesity | endothelial | *SKIV2L2* | -0.516581535 | 0.005055993 | 0.008067774 | 1 |
| Obesity | endothelial | *DNAH5* | -0.821194122 | 3.75E-07 | 2.61E-06 | 0.002358852 |
| Obesity | endothelial | *C6* | -0.687475735 | 1.28E-08 | 3.34E-07 | 8.07E-05 |
| Obesity | endothelial | *RAI14* | 0.342881903 | 0.000141536 | 0.000341325 | 0.889834332 |
| Obesity | endothelial | *PHLPP2* | -0.824011683 | 5.05E-05 | 0.000140765 | 0.31728471 |
| Obesity | endothelial | *PQLC2* | -1.513292854 | 1.79E-08 | 3.45E-07 | 0.000112347 |
| Obesity | endothelial | *CTNS* | -0.436718335 | 0.001271612 | 0.002358387 | 1 |
| Obesity | endothelial | *INPP4A* | -0.436187977 | 0.007968156 | 0.012183062 | 1 |
| Obesity | endothelial | *RAB27B* | -0.438297127 | 0.009994663 | 0.014971753 | 1 |
| Obesity | endothelial | *PSMA4* | -1.014652341 | 4.30E-05 | 0.000123263 | 0.270070285 |
| Obesity | endothelial | *LSG1* | 0.324342411 | 0.020536367 | 0.028832546 | 1 |
| Obesity | endothelial | *THAP3* | -1.320805538 | 2.10E-08 | 3.66E-07 | 0.000132138 |
| Obesity | endothelial | *TDP1* | -1.069503137 | 7.21E-05 | 0.000190978 | 0.45319111 |
| Obesity | endothelial | *SPATA7* | -0.880228342 | 9.27E-06 | 3.47E-05 | 0.05825176 |
| Obesity | endothelial | *MED17* | -0.71073226 | 0.01285537 | 0.018739093 | 1 |
| Obesity | endothelial | *RETSAT* | -0.477089166 | 1.07E-05 | 3.89E-05 | 0.067047918 |
| Obesity | endothelial | *CAPG* | -0.818808157 | 3.29E-07 | 2.36E-06 | 0.002071078 |
| Obesity | endothelial | *AP2S1* | -1.265702112 | 1.15E-05 | 4.14E-05 | 0.072119096 |
| Obesity | endothelial | *LCP2* | -0.896288783 | 1.25E-05 | 4.43E-05 | 0.078766577 |
| Obesity | endothelial | *TRIT1* | -0.477744181 | 0.008571241 | 0.013009994 | 1 |
| Obesity | endothelial | *CUL7* | -0.805162159 | 8.24E-05 | 0.000213647 | 0.517880356 |
| Obesity | endothelial | *EPHA3* | -0.52539086 | 0.022767169 | 0.03159064 | 1 |
| Obesity | endothelial | *DSG2* | 0.215457854 | 2.40E-06 | 1.16E-05 | 0.015072024 |
| Obesity | endothelial | *WDR37* | -0.641475925 | 0.001232202 | 0.002291969 | 1 |
| Obesity | endothelial | *YTHDC2* | -0.572693221 | 0.002336569 | 0.004050182 | 1 |
| Obesity | endothelial | *ATP6V1H* | -0.80679121 | 2.39E-05 | 7.56E-05 | 0.150240036 |
| Obesity | endothelial | *CP* | 0.44062842 | 0.027410765 | 0.037438948 | 1 |
| Obesity | endothelial | *ANO2* | 0.134443124 | 1.11E-06 | 6.20E-06 | 0.006999009 |

| | | | | | | |
|---|---|---|---|---|---|---|
| Obesity | endothelial | *GOPC* | -0.666716445 | 0.001091095 | 0.002051964 | 1 |
| Obesity | endothelial | *CC2D2A* | -0.635400708 | 0.000824114 | 0.00161057 | 1 |
| Obesity | endothelial | *RRM2B* | -1.116021168 | 5.22E-07 | 3.39E-06 | 0.003278799 |
| Obesity | endothelial | *SNX29* | -0.708760343 | 7.18E-07 | 4.37E-06 | 0.004512968 |
| Obesity | endothelial | *LMO3* | -0.483875424 | 9.83E-09 | 3.16E-07 | 6.18E-05 |
| Obesity | endothelial | *MRPS10* | -0.352837984 | 0.00051787 | 0.001062961 | 1 |
| Obesity | endothelial | *CELF2* | -0.582411617 | 5.33E-07 | 3.44E-06 | 0.003350363 |
| Obesity | endothelial | *ERCC8* | -0.654822021 | 0.001440659 | 0.002636804 | 1 |
| Obesity | endothelial | *ADAMTS6* | 0.259655748 | 0.029922267 | 0.040491023 | 1 |
| Obesity | endothelial | *H6PD* | -0.434954099 | 0.012729614 | 0.018598904 | 1 |
| Obesity | endothelial | *UTS2* | -0.652988346 | 0.007586371 | 0.011670055 | 1 |
| Obesity | endothelial | *EPN3* | -2.506570389 | 5.97E-05 | 0.000163448 | 0.375603883 |
| Obesity | endothelial | *RFC2* | -1.136784165 | 0.000317581 | 0.000692074 | 1 |
| Obesity | endothelial | *JKAMP* | -0.429845869 | 0.012804512 | 0.018682285 | 1 |
| Obesity | endothelial | *DKK3* | -0.790227369 | 0.011401231 | 0.016826183 | 1 |
| Obesity | endothelial | *ARHGEF5* | -1.873718002 | 2.13E-08 | 3.68E-07 | 0.000133638 |
| Obesity | endothelial | *LIMA1* | -0.272258559 | 0.001590285 | 0.002869726 | 1 |
| Obesity | endothelial | *PTGER3* | -0.516596534 | 4.73E-08 | 5.61E-07 | 0.00029763 |
| Obesity | endothelial | *MAPK9* | -0.828244528 | 0.000254249 | 0.000568849 | 1 |
| Obesity | endothelial | *FAM160A2* | -0.490079399 | 0.005645797 | 0.008927345 | 1 |
| Obesity | endothelial | *HERPUD1* | -0.627252475 | 0.0050239 | 0.008022585 | 1 |
| Obesity | endothelial | *CYBA* | -0.914429632 | 2.69E-07 | 2.03E-06 | 0.001694069 |
| Obesity | endothelial | *THOC3* | -1.730866785 | 8.07E-08 | 8.07E-07 | 0.000507304 |
| Obesity | endothelial | *RRP12* | -0.919196157 | 4.17E-05 | 0.000120398 | 0.262226669 |
| Obesity | endothelial | *ALX4* | -1.715197708 | 6.81E-09 | 2.55E-07 | 4.28E-05 |
| Obesity | endothelial | *FOXN3* | -0.608934947 | 6.14E-05 | 0.000167149 | 0.386281942 |
| Obesity | endothelial | *AKR7A2* | -1.113294419 | 2.16E-06 | 1.06E-05 | 0.013578572 |
| Obesity | endothelial | *MRTO4* | -1.276039705 | 2.61E-05 | 8.17E-05 | 0.164132871 |
| Obesity | endothelial | *OPN3* | -0.314392532 | 0.035952495 | 0.047949372 | 1 |
| Obesity | endothelial | *PLEKHH1* | -0.701358434 | 1.54E-07 | 1.32E-06 | 0.000965694 |
| Obesity | endothelial | *ATP9A* | -0.853432047 | 4.60E-08 | 5.53E-07 | 0.000289378 |
| Obesity | endothelial | *CHRDL2* | -0.968049531 | 4.33E-05 | 0.000124156 | 0.272397697 |
| Obesity | endothelial | *GALC* | -0.981633934 | 4.23E-06 | 1.84E-05 | 0.026583472 |
| Obesity | endothelial | *MRPL43* | -0.909564683 | 1.51E-05 | 5.17E-05 | 0.094843554 |
| Obesity | endothelial | *ITIH4* | 0.389104739 | 4.18E-06 | 1.82E-05 | 0.026255446 |
| Obesity | endothelial | *TRAF1* | -0.829276863 | 4.99E-08 | 5.77E-07 | 0.000313646 |
| Obesity | endothelial | *PCGF2* | 0.230633984 | 0.002795138 | 0.004739222 | 1 |
| Obesity | endothelial | *IL17RB* | -2.485500628 | 3.27E-05 | 9.84E-05 | 0.205284418 |
| Obesity | endothelial | *TRAF3IP2* | -0.800963289 | 0.000758541 | 0.001494032 | 1 |
| Obesity | endothelial | *DCBLD2* | -0.790173855 | 1.14E-07 | 1.05E-06 | 0.000716245 |
| Obesity | endothelial | *F7* | -4.491656815 | 0.00101358 | 0.001921706 | 1 |
| Obesity | endothelial | *GDI2* | -0.583951391 | 0.003269823 | 0.005452885 | 1 |
| Obesity | endothelial | *TMCC3* | 0.362763951 | 0.000214597 | 0.000491143 | 1 |
| Obesity | endothelial | *PITHD1* | -1.03866507 | 8.44E-05 | 0.000217738 | 0.53062864 |
| Obesity | endothelial | *ATP11B* | -0.288296029 | 0.032029816 | 0.0431387 | 1 |
| Obesity | endothelial | *CDK14* | -0.618433766 | 0.000240241 | 0.000541277 | 1 |
| Obesity | endothelial | *ZC3H11A* | -1.571730827 | 1.86E-06 | 9.31E-06 | 0.011678972 |
| Obesity | endothelial | *RIOK2* | -1.033767428 | 3.93E-05 | 0.000114471 | 0.247371319 |
| Obesity | endothelial | *DNAJC25* | -0.970156937 | 6.00E-07 | 3.80E-06 | 0.003773128 |
| Obesity | endothelial | *PIGV* | -0.982452941 | 7.61E-07 | 4.57E-06 | 0.004782668 |

| | | | | | | |
|---|---|---|---|---|---|---|
| Obesity | endothelial | *QSER1* | -0.650584641 | 0.00258321 | 0.004417064 | 1 |
| Obesity | endothelial | *SFSWAP* | -0.526118429 | 0.007904935 | 0.012094993 | 1 |
| Obesity | endothelial | *TNK2* | -0.691232246 | 8.72E-05 | 0.000224194 | 0.548379168 |
| Obesity | endothelial | *VMP1* | -0.700824366 | 1.65E-05 | 5.60E-05 | 0.103874221 |
| Obesity | endothelial | *SPHK2* | -1.160919825 | 1.81E-08 | 3.45E-07 | 0.000113922 |
| Obesity | endothelial | *ISOC2* | -1.303002792 | 1.41E-08 | 3.34E-07 | 8.89E-05 |
| Obesity | endothelial | *ADCK1* | -0.964482187 | 1.57E-07 | 1.34E-06 | 0.000985283 |
| Obesity | endothelial | *CASP8* | -0.968650517 | 6.71E-08 | 7.12E-07 | 0.000422135 |
| Obesity | endothelial | *DMRT3* | -0.889356341 | 3.88E-08 | 5.03E-07 | 0.000243692 |
| Obesity | endothelial | *ST3GAL6* | -0.644209461 | 5.32E-07 | 3.44E-06 | 0.0033443 |
| Obesity | endothelial | *CDON* | -0.335780725 | 0.018329614 | 0.025996061 | 1 |
| Obesity | endothelial | *TMEM161A* | -1.104874186 | 2.89E-05 | 8.90E-05 | 0.181445239 |
| Obesity | endothelial | *LPAR2* | -0.831900539 | 4.18E-05 | 0.000120583 | 0.26287024 |
| Obesity | endothelial | *SUGP2* | -0.753982778 | 4.40E-05 | 0.000125604 | 0.276704893 |
| Obesity | endothelial | *ABCA7* | -1.08108155 | 0.000695924 | 0.001383268 | 1 |
| Obesity | endothelial | *DDX20* | -1.310477057 | 0.000309071 | 0.000675401 | 1 |
| Obesity | endothelial | *CHI3L2* | -0.595687818 | 0.000779035 | 0.001531996 | 1 |
| Obesity | endothelial | *PMS1* | -0.541165884 | 0.009526848 | 0.014332447 | 1 |
| Obesity | endothelial | *TAF11* | -0.709793952 | 5.87E-05 | 0.000161408 | 0.369300869 |
| Obesity | endothelial | *NTHL1* | 0.479249998 | 0.000702595 | 0.001395644 | 1 |
| Obesity | endothelial | *UHRF1BP1* | 0.589599272 | 8.03E-08 | 8.05E-07 | 0.000504897 |
| Obesity | endothelial | *IPO5* | -0.94990668 | 3.82E-05 | 0.000111821 | 0.240079018 |
| Obesity | endothelial | *WDR18* | -0.498553404 | 0.003469163 | 0.005745686 | 1 |
| Obesity | endothelial | *ANKRD44* | -0.632819656 | 9.15E-05 | 0.000233808 | 0.575401516 |
| Obesity | endothelial | *ADAT1* | -0.437837405 | 0.008613281 | 0.013067495 | 1 |
| Obesity | endothelial | *NDUFB4* | -0.683214347 | 0.019394815 | 0.027401169 | 1 |
| Obesity | endothelial | *ZC3H15* | -0.674673587 | 0.001747524 | 0.003122992 | 1 |
| Obesity | endothelial | *MAP2K4* | -0.514569115 | 0.007782293 | 0.011924757 | 1 |
| Obesity | endothelial | *TMEM206* | -0.956758417 | 0.000289394 | 0.000636383 | 1 |
| Obesity | endothelial | *SEC61A2* | -1.078224801 | 4.51E-05 | 0.000127947 | 0.283244656 |
| Obesity | endothelial | *ASB1* | -0.899325451 | 3.83E-09 | 1.84E-07 | 2.41E-05 |
| Obesity | endothelial | *PPP2R5A* | -0.434181895 | 0.016326553 | 0.023429591 | 1 |
| Obesity | endothelial | *TIE1* | 0.159340418 | 1.83E-05 | 6.07E-05 | 0.114752558 |
| Obesity | endothelial | *NFYC* | -0.879207534 | 2.12E-05 | 6.86E-05 | 0.133161495 |
| Obesity | endothelial | *ZMYND12* | -0.465838573 | 0.004473023 | 0.007231138 | 1 |
| Obesity | endothelial | *SLC9A3* | -0.381251905 | 0.008237051 | 0.012539066 | 1 |
| Obesity | endothelial | *NGEF* | 1.141580289 | 0.000932894 | 0.001791418 | 1 |
| Obesity | endothelial | *CD84* | -0.847851359 | 8.55E-07 | 5.00E-06 | 0.005377312 |
| Obesity | endothelial | *ZNRD1* | -0.782923536 | 0.000413398 | 0.000873919 | 1 |
| Obesity | endothelial | *GOLGA5* | -0.833939042 | 0.000388731 | 0.000826496 | 1 |
| Obesity | endothelial | *EML1* | 0.260601315 | 0.023899223 | 0.03300119 | 1 |
| Obesity | endothelial | *ZFAT* | -1.032398221 | 1.64E-07 | 1.39E-06 | 0.001030525 |
| Obesity | endothelial | *MTFR1* | -0.906555094 | 1.98E-06 | 9.87E-06 | 0.012470011 |
| Obesity | endothelial | *STAG3* | -1.165051767 | 3.93E-08 | 5.06E-07 | 0.000247159 |
| Obesity | endothelial | *STOML1* | -2.014837444 | 1.32E-05 | 4.61E-05 | 0.082894462 |
| Obesity | endothelial | *METTL22* | -1.255518771 | 7.32E-07 | 4.44E-06 | 0.004603441 |
| Obesity | endothelial | *TP53BP1* | -0.646792031 | 0.000511681 | 0.001051975 | 1 |
| Obesity | endothelial | *RRP15* | -0.402560175 | 0.000887482 | 0.001715744 | 1 |
| Obesity | endothelial | *SYT1* | 0.313740753 | 0.002178039 | 0.003810053 | 1 |
| Obesity | endothelial | *ROGDI* | -2.328466688 | 1.06E-06 | 5.97E-06 | 0.006655206 |

| | | | | | | |
|---|---|---|---|---|---|---|
| Obesity | endothelial | *HDAC4* | -0.727896959 | 6.64E-05 | 0.00017833 | 0.417598323 |
| Obesity | endothelial | *RASSF1* | -0.667203367 | 0.003337607 | 0.005551661 | 1 |
| Obesity | endothelial | *FGFR3* | 0.165143979 | 0.004685915 | 0.00754813 | 1 |
| Obesity | endothelial | *IFI35* | 0.511026929 | 0.001512426 | 0.002748951 | 1 |
| Obesity | endothelial | *HEATR6* | -1.182446861 | 5.56E-06 | 2.29E-05 | 0.034986743 |
| Obesity | endothelial | *POLR1A* | -0.449783266 | 5.84E-06 | 2.37E-05 | 0.036745368 |
| Obesity | endothelial | *SRBD1* | -0.704024969 | 0.000160502 | 0.000381648 | 1 |
| Obesity | endothelial | *CYFIP1* | -0.46565676 | 0.018397692 | 0.026086217 | 1 |
| Obesity | endothelial | *PSME4* | -0.737195682 | 1.20E-05 | 4.27E-05 | 0.075239328 |
| Obesity | endothelial | *PITX1* | -1.513975988 | 2.79E-08 | 4.27E-07 | 0.000175217 |
| Obesity | endothelial | *NUCKS1* | -0.458033466 | 0.018329973 | 0.025996061 | 1 |
| Obesity | endothelial | *DNAJA2* | -1.083623322 | 7.50E-05 | 0.000196925 | 0.471240592 |
| Obesity | endothelial | *KCNAB2* | -0.954975626 | 6.10E-08 | 6.64E-07 | 0.000383556 |
| Obesity | endothelial | *GAL* | -1.253941418 | 1.17E-08 | 3.34E-07 | 7.34E-05 |
| Obesity | endothelial | *CLEC2D* | -0.520295129 | 0.003471892 | 0.005748692 | 1 |
| Obesity | endothelial | *NEDD4* | 0.305066677 | 0.002466298 | 0.004250443 | 1 |
| Obesity | endothelial | *PIGB* | -0.864879553 | 3.00E-05 | 9.18E-05 | 0.188385774 |
| Obesity | endothelial | *DAPP1* | -0.760739443 | 2.41E-06 | 1.16E-05 | 0.015137921 |
| Obesity | endothelial | *FGF10* | -0.411595729 | 1.59E-05 | 5.41E-05 | 0.099910733 |
| Obesity | endothelial | *TMEM260* | -0.662651524 | 0.001414617 | 0.002592159 | 1 |
| Obesity | endothelial | *CLTCL1* | -0.682419094 | 0.000124339 | 0.000304763 | 0.781717262 |
| Obesity | endothelial | *DGCR2* | -0.802232456 | 0.000218041 | 0.000497578 | 1 |
| Obesity | endothelial | *MNT* | -1.112020892 | 8.62E-08 | 8.47E-07 | 0.00054214 |
| Obesity | endothelial | *ZXDC* | -0.468738637 | 0.017724466 | 0.025216953 | 1 |
| Obesity | endothelial | *WIPI1* | -0.87098102 | 3.51E-08 | 4.77E-07 | 0.000220956 |
| Obesity | endothelial | *ASNS* | -1.049122067 | 3.55E-06 | 1.59E-05 | 0.02233093 |
| Obesity | endothelial | *ST6GALNAC2* | 0.696935542 | 0.000283047 | 0.000623516 | 1 |
| Obesity | endothelial | *TESK2* | -0.888051275 | 0.000101415 | 0.000255528 | 0.637596225 |
| Obesity | endothelial | *EIF2B3* | -0.756269691 | 1.80E-07 | 1.49E-06 | 0.001129462 |
| Obesity | endothelial | *CAMK2A* | -2.22488421 | 4.47E-08 | 5.43E-07 | 0.000281336 |
| Obesity | endothelial | *CDC42* | -0.522582591 | 0.015154997 | 0.021853088 | 1 |
| Obesity | endothelial | *OSBPL3* | -0.82235177 | 1.19E-06 | 6.51E-06 | 0.00746681 |
| Obesity | endothelial | *RAD18* | -0.969833496 | 1.39E-05 | 4.83E-05 | 0.087547944 |
| Obesity | endothelial | *SNX13* | -0.583119265 | 0.001748367 | 0.00312361 | 1 |
| Obesity | endothelial | *VASH1* | 0.385771534 | 0.022516495 | 0.031304998 | 1 |
| Obesity | endothelial | *TRIP13* | -1.875930857 | 1.75E-08 | 3.42E-07 | 0.000109965 |
| Obesity | endothelial | *MBD3* | -0.660083648 | 0.005269177 | 0.008370959 | 1 |
| Obesity | endothelial | *HLTF* | -0.918060736 | 9.74E-08 | 9.25E-07 | 0.000612627 |
| Obesity | endothelial | *CPSF1* | -1.528402785 | 2.81E-07 | 2.10E-06 | 0.001768592 |
| Obesity | endothelial | *CYBRD1* | -0.548878173 | 0.000450903 | 0.000940553 | 1 |
| Obesity | endothelial | *LIMS2* | 0.260505956 | 1.95E-05 | 6.42E-05 | 0.122838166 |
| Obesity | endothelial | *TRHDE* | -0.521648825 | 1.54E-08 | 3.34E-07 | 9.70E-05 |
| Obesity | endothelial | *NFATC3* | -0.67960481 | 5.27E-05 | 0.000146267 | 0.331279075 |
| Obesity | endothelial | *ACADVL* | -0.949231019 | 1.70E-07 | 1.42E-06 | 0.00106861 |
| Obesity | endothelial | *CRMP1* | -0.703633005 | 1.84E-08 | 3.45E-07 | 0.000115497 |
| Obesity | endothelial | *DERL2* | -1.089874825 | 8.93E-05 | 0.000228805 | 0.561258029 |
| Obesity | endothelial | *AP1M1* | -0.732474165 | 4.79E-05 | 0.000134443 | 0.300834724 |
| Obesity | endothelial | *PVR* | 0.658777141 | 0.000583077 | 0.001181756 | 1 |
| Obesity | endothelial | *XRCC1* | -1.091878124 | 0.001368185 | 0.00251514 | 1 |
| Obesity | endothelial | *TP63* | -0.823250193 | 1.53E-08 | 3.34E-07 | 9.59E-05 |

| | | | | | | |
|---|---|---|---|---|---|---|
| Obesity | endothelial | *LLGL2* | 0.392016146 | 0.012303693 | 0.018039486 | 1 |
| Obesity | endothelial | *PDE8A* | -0.443757471 | 0.004105029 | 0.006697836 | 1 |
| Obesity | endothelial | *NLE1* | -1.454238931 | 1.23E-06 | 6.67E-06 | 0.007702949 |
| Obesity | endothelial | *SDHA* | -0.475924047 | 0.009768996 | 0.01466866 | 1 |
| Obesity | endothelial | *GSDMB* | -0.75271673 | 1.54E-08 | 3.34E-07 | 9.68E-05 |
| Obesity | endothelial | *PICALM* | -0.544475265 | 0.000235006 | 0.000530515 | 1 |
| Obesity | endothelial | *GLI2* | -0.456045064 | 6.92E-07 | 4.24E-06 | 0.004351355 |
| Obesity | endothelial | *ATP2A3* | 0.390286925 | 0.000217683 | 0.00049694 | 1 |
| Obesity | endothelial | *CA12* | -0.533417847 | 2.08E-06 | 1.03E-05 | 0.013056994 |
| Obesity | endothelial | *LMAN1* | -0.631858652 | 0.004901643 | 0.007865397 | 1 |
| Obesity | endothelial | *ENO1* | -1.007508193 | 9.57E-06 | 3.56E-05 | 0.06017 |
| Obesity | endothelial | *TTC38* | -0.872957866 | 5.76E-08 | 6.38E-07 | 0.000362253 |
| Obesity | endothelial | *GRAMD4* | -0.901375261 | 0.000424028 | 0.000892523 | 1 |
| Obesity | endothelial | *CELSR1* | 0.424921257 | 3.56E-05 | 0.000105715 | 0.223692366 |
| Obesity | endothelial | *ADD2* | 0.393500006 | 6.10E-06 | 2.45E-05 | 0.038330563 |
| Obesity | endothelial | *VPS9D1* | -1.212064865 | 2.14E-05 | 6.93E-05 | 0.134802861 |
| Obesity | endothelial | *MARK3* | -0.696263182 | 3.09E-05 | 9.39E-05 | 0.194301786 |
| Obesity | endothelial | *SLC25A3* | -0.721869349 | 0.000965842 | 0.001846229 | 1 |
| Obesity | endothelial | *TUBA3D* | -1.816303387 | 7.14E-06 | 2.80E-05 | 0.044885146 |
| Obesity | endothelial | *MKRN2* | -0.634170255 | 0.025479124 | 0.034983021 | 1 |
| Obesity | endothelial | *MCM6* | -1.131218145 | 4.51E-06 | 1.93E-05 | 0.028352002 |
| Obesity | endothelial | *MLH1* | -0.723418225 | 0.000303677 | 0.000664538 | 1 |
| Obesity | endothelial | *FMO4* | -1.068852898 | 4.24E-07 | 2.87E-06 | 0.002664286 |
| Obesity | endothelial | *RGS11* | -0.409844211 | 0.000872009 | 0.001687907 | 1 |
| Obesity | endothelial | *TPD52* | 0.32482571 | 0.000487526 | 0.001007452 | 1 |
| Obesity | endothelial | *GPATCH1* | -0.822459566 | 9.56E-05 | 0.000242655 | 0.600813399 |
| Obesity | endothelial | *ICAM3* | -0.654242441 | 0.000443292 | 0.000927209 | 1 |
| Obesity | endothelial | *XAB2* | -1.716910325 | 1.28E-07 | 1.15E-06 | 0.000803174 |
| Obesity | endothelial | *STXBP2* | -0.922323678 | 1.60E-08 | 3.37E-07 | 0.000100284 |
| Obesity | endothelial | *MAP2K7* | -1.142832199 | 9.40E-06 | 3.50E-05 | 0.059083434 |
| Obesity | endothelial | *CTTNBP2* | -0.787103071 | 3.81E-05 | 0.000111791 | 0.239697235 |
| Obesity | endothelial | *TM9SF3* | -0.419284141 | 0.029382519 | 0.039854993 | 1 |
| Obesity | endothelial | *APBB1IP* | -0.612078447 | 1.96E-08 | 3.58E-07 | 0.000123362 |
| Obesity | endothelial | *LRCH4* | -0.777935764 | 4.34E-05 | 0.000124206 | 0.272880341 |
| Obesity | endothelial | *SIRT6* | -1.6018321 | 4.93E-08 | 5.72E-07 | 0.000309645 |
| Obesity | endothelial | *CAPZB* | -0.670471716 | 0.000156042 | 0.000372876 | 0.981037378 |
| Obesity | endothelial | *GPR137B* | -0.540801418 | 0.000351134 | 0.000755245 | 1 |
| Obesity | endothelial | *NAALAD2* | -0.567494065 | 3.77E-06 | 1.68E-05 | 0.02367729 |
| Obesity | endothelial | *FGFR1* | -0.686047758 | 3.08E-06 | 1.42E-05 | 0.01934072 |
| Obesity | endothelial | *FBLN1* | -0.487950682 | 5.12E-06 | 2.14E-05 | 0.032163589 |
| Obesity | endothelial | *PIAS2* | -0.709416901 | 0.000153111 | 0.00036629 | 0.96261137 |
| Obesity | endothelial | *AMPH* | -0.361522216 | 5.26E-05 | 0.000146051 | 0.330512974 |
| Obesity | endothelial | *MCCC1* | -0.4975622 | 0.000272766 | 0.000604257 | 1 |
| Obesity | endothelial | *FAP* | -0.529380277 | 0.000307637 | 0.000672735 | 1 |
| Obesity | endothelial | *PIK3C3* | -0.510690985 | 0.009337621 | 0.014074712 | 1 |
| Obesity | endothelial | *N4BP2* | -0.979617136 | 2.10E-07 | 1.68E-06 | 0.00131843 |
| Obesity | endothelial | *TULP3* | -0.928265617 | 3.91E-07 | 2.69E-06 | 0.002460662 |
| Obesity | endothelial | *SYNJ2* | 0.270299627 | 2.46E-05 | 7.74E-05 | 0.154865168 |
| Obesity | endothelial | *ZCWPW1* | -0.657245654 | 0.000459158 | 0.000955553 | 1 |
| Obesity | endothelial | *FGF20* | -11.22851544 | 1.66E-08 | 3.39E-07 | 0.000104177 |

| | | | | | | |
|---|---|---|---|---|---|---|
| Obesity | endothelial | *PCM1* | -0.513421037 | 0.001705532 | 0.003055765 | 1 |
| Obesity | endothelial | *MYH7B* | -1.516222869 | 6.72E-07 | 4.14E-06 | 0.004227086 |
| Obesity | endothelial | *FKBP7* | -0.332926346 | 0.019409642 | 0.027415956 | 1 |
| Obesity | endothelial | *OSBPL6* | -1.129592849 | 3.06E-07 | 2.22E-06 | 0.001926154 |
| Obesity | endothelial | *REXO1* | -1.050421764 | 0.000378828 | 0.000808175 | 1 |
| Obesity | endothelial | *SAR1A* | -0.859661211 | 1.02E-06 | 5.82E-06 | 0.006421523 |
| Obesity | endothelial | *RAPGEF3* | 0.164672663 | 2.02E-06 | 1.00E-05 | 0.012694211 |
| Obesity | endothelial | *FDFT1* | -0.887300528 | 1.88E-05 | 6.21E-05 | 0.118241768 |
| Obesity | endothelial | *SCGN* | -2.501342353 | 0.000621661 | 0.001248685 | 1 |
| Obesity | endothelial | *DDX1* | -0.78254398 | 0.000266391 | 0.00059222 | 1 |
| Obesity | endothelial | *DNM2* | -0.643682479 | 0.001578722 | 0.002850496 | 1 |
| Obesity | endothelial | *RIMS1* | -1.133761269 | 1.81E-08 | 3.45E-07 | 0.000113839 |
| Obesity | endothelial | *STX7* | -0.58769329 | 0.000321497 | 0.000699638 | 1 |
| Obesity | endothelial | *RABL2B* | -1.226633092 | 3.09E-08 | 4.48E-07 | 0.000194554 |
| Obesity | endothelial | *DCT* | -0.848319449 | 5.32E-08 | 6.04E-07 | 0.000334729 |
| Obesity | endothelial | *SLC35C2* | -0.946218748 | 0.000133966 | 0.000325192 | 0.842246115 |
| Obesity | endothelial | *SCTR* | -0.664631265 | 9.91E-09 | 3.16E-07 | 6.23E-05 |
| Obesity | endothelial | *KCNN2* | 0.346294168 | 2.50E-06 | 1.20E-05 | 0.015717922 |
| Obesity | endothelial | *PSEN1* | -0.653076966 | 0.000245086 | 0.000550502 | 1 |
| Obesity | endothelial | *CPOX* | -1.117441924 | 2.67E-06 | 1.26E-05 | 0.016793531 |
| Obesity | endothelial | *MOK* | -1.055776955 | 6.15E-06 | 2.47E-05 | 0.038676626 |
| Obesity | endothelial | *RSBN1* | -0.876306912 | 6.85E-05 | 0.000183035 | 0.43049771 |
| Obesity | endothelial | *COL4A4* | -0.672764619 | 0.001235266 | 0.002296989 | 1 |
| Obesity | endothelial | *PKP1* | 0.281355243 | 8.84E-06 | 3.34E-05 | 0.055570367 |
| Obesity | endothelial | *STK17B* | -0.814871951 | 2.91E-07 | 2.15E-06 | 0.001830568 |
| Obesity | endothelial | *ZNF506* | -0.891469984 | 0.000178044 | 0.00041705 | 1 |
| Obesity | endothelial | *PCDHA6* | -4.43419729 | 0.000394704 | 0.00083778 | 1 |
| Obesity | endothelial | *PCDHGA2* | -2.109307446 | 7.84E-11 | 1.04E-08 | 4.93E-07 |
| Obesity | endothelial | *HSPB11* | -0.856782518 | 0.000115046 | 0.000284985 | 0.723291411 |
| Obesity | endothelial | *WDR70* | -0.538316258 | 0.000957575 | 0.001832099 | 1 |
| Obesity | endothelial | *PGR* | -0.475361833 | 1.25E-06 | 6.76E-06 | 0.00783159 |
| Obesity | endothelial | *CCNT2* | -0.593745604 | 0.000173705 | 0.000409172 | 1 |
| Obesity | endothelial | *GEMIN5* | -0.849344304 | 1.79E-06 | 9.05E-06 | 0.011247117 |
| Obesity | endothelial | *GSK3B* | -0.384892799 | 0.022152637 | 0.030867382 | 1 |
| Obesity | endothelial | *ULK2* | -0.698967164 | 9.78E-06 | 3.62E-05 | 0.061460531 |
| Obesity | endothelial | *PLOD1* | -1.204382661 | 1.05E-06 | 5.94E-06 | 0.006606308 |
| Obesity | endothelial | *TDRD3* | -0.415955592 | 0.018905922 | 0.026758561 | 1 |
| Obesity | endothelial | *OXCT1* | -0.639219409 | 1.72E-05 | 5.78E-05 | 0.107999957 |
| Obesity | endothelial | *EPYC* | -3.63990036 | 6.74E-07 | 4.14E-06 | 0.004237194 |
| Obesity | endothelial | *CYLD* | -0.692815808 | 0.001489029 | 0.002713486 | 1 |
| Obesity | endothelial | *ZNF324* | -1.386199674 | 3.91E-05 | 0.000113979 | 0.246081107 |
| Obesity | endothelial | *ZNF586* | -0.483424444 | 0.010199661 | 0.015249766 | 1 |
| Obesity | endothelial | *ZNF446* | -1.696438868 | 1.32E-07 | 1.17E-06 | 0.000827753 |
| Obesity | endothelial | *ZNF264* | -0.559873867 | 0.024410831 | 0.033611672 | 1 |
| Obesity | endothelial | *RPS5* | -0.689292007 | 0.005489445 | 0.008697616 | 1 |
| Obesity | endothelial | *CHMP2B* | -0.709612434 | 0.001869066 | 0.003318503 | 1 |
| Obesity | endothelial | *PPIE* | -0.591653448 | 0.006099307 | 0.009559059 | 1 |
| Obesity | endothelial | *ZMPSTE24* | -1.039840956 | 3.87E-06 | 1.71E-05 | 0.024317169 |
| Obesity | endothelial | *STARD7* | -0.748441915 | 0.003051437 | 0.005115836 | 1 |
| Obesity | endothelial | *NOA1* | -1.060894575 | 1.61E-05 | 5.49E-05 | 0.101489425 |

| | | | | | | |
|---|---|---|---|---|---|---|
| Obesity | endothelial | *GSTP1* | -0.829793438 | 0.000977455 | 0.0018622 | 1 |
| Obesity | endothelial | *WBP11* | -0.580768791 | 0.030676527 | 0.041484905 | 1 |
| Obesity | endothelial | *COL16A1* | -0.754646729 | 1.25E-12 | 3.74E-10 | 7.84E-09 |
| Obesity | endothelial | *APOB* | -0.814370169 | 6.02E-09 | 2.37E-07 | 3.79E-05 |
| Obesity | endothelial | *NCOA1* | -0.494471145 | 0.001553168 | 0.002810815 | 1 |
| Obesity | endothelial | *CD59* | 0.328077977 | 0.002233546 | 0.003891991 | 1 |
| Obesity | endothelial | *TAF9* | -1.366552014 | 0.000165835 | 0.0003924 | 1 |
| Obesity | endothelial | *FCN1* | -0.805102905 | 4.46E-08 | 5.43E-07 | 0.000280391 |
| Obesity | endothelial | *SCAMP1* | 0.360382428 | 0.023995639 | 0.033108118 | 1 |
| Obesity | endothelial | *SEH1L* | -0.880876306 | 0.000323298 | 0.000702826 | 1 |
| Obesity | endothelial | *SLC25A24* | -0.696470986 | 0.000194294 | 0.000451748 | 1 |
| Obesity | endothelial | *ABCB1* | 0.203150754 | 3.53E-06 | 1.59E-05 | 0.02220883 |
| Obesity | endothelial | *ZNF213* | -0.708601091 | 0.000205419 | 0.000473586 | 1 |
| Obesity | endothelial | *CPNE3* | -0.648246559 | 0.00034262 | 0.000740479 | 1 |
| Obesity | endothelial | *RRN3* | -0.890053081 | 0.000170991 | 0.000403687 | 1 |
| Obesity | endothelial | *MTIF2* | -1.086177476 | 1.61E-07 | 1.37E-06 | 0.001012487 |
| Obesity | endothelial | *EPS15* | -0.59287977 | 3.42E-05 | 0.000102569 | 0.215292308 |
| Obesity | endothelial | *ATG16L1* | -0.849025209 | 0.001271661 | 0.002358387 | 1 |
| Obesity | endothelial | *USP40* | -0.695020769 | 3.97E-05 | 0.000115371 | 0.249662314 |
| Obesity | endothelial | *MAST2* | -0.590181674 | 0.002898014 | 0.004891225 | 1 |
| Obesity | endothelial | *B4GALT1* | -0.664478738 | 0.001068218 | 0.002013156 | 1 |
| Obesity | endothelial | *EIF2AK1* | -0.856528452 | 5.99E-05 | 0.00016378 | 0.376857442 |
| Obesity | endothelial | *EPDR1* | -0.530470305 | 2.00E-05 | 6.55E-05 | 0.125648067 |
| Obesity | endothelial | *SNX10* | -0.243936187 | 5.15E-06 | 2.15E-05 | 0.032375276 |
| Obesity | endothelial | *TMED2* | -1.017347942 | 6.02E-05 | 0.000164351 | 0.37833513 |
| Obesity | endothelial | *ACOX3* | -0.623952733 | 0.004204406 | 0.006839095 | 1 |
| Obesity | endothelial | *HSD17B14* | -1.194576921 | 2.46E-07 | 1.89E-06 | 0.001548451 |
| Obesity | endothelial | *TRIP6* | -1.171491775 | 1.22E-08 | 3.34E-07 | 7.66E-05 |
| Obesity | endothelial | *ACHE* | -0.517829379 | 2.30E-05 | 7.33E-05 | 0.14467903 |
| Obesity | endothelial | *PIGS* | -0.374853225 | 0.01992678 | 0.028083314 | 1 |
| Obesity | endothelial | *PGS1* | -0.763373728 | 0.001717431 | 0.003072748 | 1 |
| Obesity | endothelial | *PSMC5* | -0.636787397 | 0.005610576 | 0.008873885 | 1 |
| Obesity | endothelial | *LPCAT2* | 0.308122805 | 0.006514267 | 0.010157539 | 1 |
| Obesity | endothelial | *OGFOD1* | -1.116688758 | 3.29E-06 | 1.50E-05 | 0.020665384 |
| Obesity | endothelial | *TXNDC16* | -0.766602299 | 0.000107178 | 0.00026803 | 0.673828464 |
| Obesity | endothelial | *GMCL1* | -0.786810587 | 0.000326914 | 0.00070995 | 1 |
| Obesity | endothelial | *KLHL42* | -0.696723554 | 0.003533504 | 0.005839942 | 1 |
| Obesity | endothelial | *PTHLH* | -0.735339814 | 6.77E-06 | 2.68E-05 | 0.042564578 |
| Obesity | endothelial | *PHACTR3* | -0.707446505 | 3.15E-07 | 2.27E-06 | 0.001982295 |
| Obesity | endothelial | *AAMDC* | -0.351117782 | 0.002289646 | 0.003978718 | 1 |
| Obesity | endothelial | *METTL2A* | -0.683003193 | 0.006768797 | 0.010515302 | 1 |
| Obesity | endothelial | *ALG6* | -0.561897835 | 0.024441517 | 0.033646555 | 1 |
| Obesity | endothelial | *GP6* | -1.281580455 | 3.43E-05 | 0.000102731 | 0.215837136 |
| Obesity | endothelial | *DDX18* | -0.545576171 | 0.008685247 | 0.01317032 | 1 |
| Obesity | endothelial | *EDEM2* | -0.779842529 | 8.43E-05 | 0.000217738 | 0.530240379 |
| Obesity | endothelial | *TPX2* | -0.518751872 | 8.70E-05 | 0.00022375 | 0.546845697 |
| Obesity | endothelial | *DOCK3* | -0.416190447 | 0.000317513 | 0.000692074 | 1 |
| Obesity | endothelial | *FKBP1A* | 0.476915519 | 0.005123591 | 0.008165277 | 1 |
| Obesity | endothelial | *NSFL1C* | -0.85613758 | 0.000123535 | 0.000303029 | 0.776662276 |
| Obesity | endothelial | *CPXM1* | -0.641602756 | 7.43E-08 | 7.62E-07 | 0.000467103 |

| | | | | | | |
|---|---|---|---|---|---|---|
| Obesity | endothelial | *MAVS* | -0.781573264 | 7.57E-05 | 0.000198584 | 0.476205273 |
| Obesity | endothelial | *LZTS3* | -0.979205358 | 1.40E-08 | 3.34E-07 | 8.77E-05 |
| Obesity | endothelial | *TESC* | 0.357886907 | 6.43E-06 | 2.57E-05 | 0.040442766 |
| Obesity | endothelial | *RBBP9* | -1.480164063 | 0.000132254 | 0.000321407 | 0.831479772 |
| Obesity | endothelial | *ANAPC5* | -0.639532073 | 0.001114489 | 0.00209158 | 1 |
| Obesity | endothelial | *SLC23A2* | -0.675498385 | 0.001299548 | 0.002402391 | 1 |
| Obesity | endothelial | *TMEM230* | -0.785694441 | 3.48E-05 | 0.000103868 | 0.218641551 |
| Obesity | endothelial | *DZANK1* | -1.3599665 | 2.28E-07 | 1.79E-06 | 0.001434144 |
| Obesity | endothelial | *TASP1* | -0.592671705 | 0.000703284 | 0.001396572 | 1 |
| Obesity | endothelial | *OAS1* | -1.031247696 | 0.001237718 | 0.002300188 | 1 |
| Obesity | endothelial | *KIF16B* | -0.645619594 | 0.001218727 | 0.00226959 | 1 |
| Obesity | endothelial | *FUS* | -0.553790307 | 0.001811322 | 0.003222349 | 1 |
| Obesity | endothelial | *FXYD5* | 0.49607955 | 0.002209005 | 0.00385671 | 1 |
| Obesity | endothelial | *ZNF302* | -0.452225246 | 0.000930764 | 0.00178842 | 1 |
| Obesity | endothelial | *FXYD3* | -1.506230527 | 1.05E-05 | 3.86E-05 | 0.066298519 |
| Obesity | endothelial | *KCNH4* | 0.339432357 | 0.005271294 | 0.008370959 | 1 |
| Obesity | endothelial | *GANAB* | -0.835316242 | 0.000252813 | 0.000566643 | 1 |
| Obesity | endothelial | *GMIP* | -1.463997493 | 1.00E-07 | 9.44E-07 | 0.000630361 |
| Obesity | endothelial | *MLF2* | -1.131059973 | 1.64E-05 | 5.56E-05 | 0.102910429 |
| Obesity | endothelial | *NECAP1* | -1.060906956 | 3.57E-05 | 0.000105993 | 0.224515399 |
| Obesity | endothelial | *ANKRD24* | -0.762761865 | 1.03E-05 | 3.77E-05 | 0.064484707 |
| Obesity | endothelial | *GPATCH2L* | -0.681202406 | 7.37E-05 | 0.000194245 | 0.463468333 |
| Obesity | endothelial | *SLC9A1* | -1.1827811 | 2.39E-07 | 1.85E-06 | 0.001500525 |
| Obesity | endothelial | *SPTLC1* | -0.605220851 | 0.002284722 | 0.003971259 | 1 |
| Obesity | endothelial | *RGS1* | -0.881433671 | 1.85E-08 | 3.45E-07 | 0.000116095 |
| Obesity | endothelial | *YPEL3* | 0.269476864 | 0.035995259 | 0.04799597 | 1 |
| Obesity | endothelial | *MRPS33* | -1.218062848 | 4.52E-07 | 3.03E-06 | 0.00284442 |
| Obesity | endothelial | *MAEA* | -0.591411804 | 0.001595701 | 0.00287702 | 1 |
| Obesity | endothelial | *STRN4* | -1.317963289 | 5.42E-06 | 2.24E-05 | 0.034072986 |
| Obesity | endothelial | *LYZ* | -0.64470259 | 0.00053268 | 0.001090511 | 1 |
| Obesity | endothelial | *MUL1* | -1.128903086 | 1.77E-05 | 5.92E-05 | 0.111001297 |
| Obesity | endothelial | *SPG21* | -0.950445511 | 8.47E-06 | 3.23E-05 | 0.053254838 |
| Obesity | endothelial | *GNPTG* | -0.899025322 | 0.000429607 | 0.000902118 | 1 |
| Obesity | endothelial | *ZNF268* | -1.331713002 | 0.00027214 | 0.000603082 | 1 |
| Obesity | endothelial | *GOLGA3* | -0.750186377 | 2.16E-05 | 6.98E-05 | 0.135817668 |
| Obesity | endothelial | *PABPC4* | -1.491083487 | 1.55E-14 | 8.88E-12 | 9.77E-11 |
| Obesity | endothelial | *CERS4* | -0.811082001 | 4.14E-09 | 1.90E-07 | 2.60E-05 |
| Obesity | endothelial | *USP48* | -0.543849695 | 0.001613388 | 0.002905578 | 1 |
| Obesity | endothelial | *PDPR* | -0.809219309 | 0.000301646 | 0.000660782 | 1 |
| Obesity | endothelial | *NLRC4* | -1.120498684 | 2.51E-08 | 4.05E-07 | 0.000157869 |
| Obesity | endothelial | *LAMB1* | 0.324334714 | 0.01650127 | 0.023638454 | 1 |
| Obesity | endothelial | *SLC26A4* | -1.850171083 | 1.50E-08 | 3.34E-07 | 9.41E-05 |
| Obesity | endothelial | *DLD* | -0.789006296 | 0.00042196 | 0.000889625 | 1 |
| Obesity | endothelial | *TXNL1* | -0.681276747 | 0.002566569 | 0.004398181 | 1 |
| Obesity | endothelial | *IL5RA* | -2.090516217 | 1.22E-08 | 3.34E-07 | 7.65E-05 |
| Obesity | endothelial | *ITGA6* | 0.209699056 | 7.19E-06 | 2.82E-05 | 0.045216913 |
| Obesity | endothelial | *SEL1L3* | 0.403633173 | 0.000266003 | 0.000591777 | 1 |
| Obesity | endothelial | *TF* | -1.084985526 | 1.35E-08 | 3.34E-07 | 8.48E-05 |
| Obesity | endothelial | *MYO15A* | -1.583184463 | 8.91E-08 | 8.69E-07 | 0.000559869 |
| Obesity | endothelial | *ALKBH5* | -0.450348793 | 0.026540748 | 0.036345389 | 1 |

| | | | | | | |
|---|---|---|---|---|---|---|
| Obesity | endothelial | *ZFHX4* | -0.608527979 | 1.16E-08 | 3.34E-07 | 7.27E-05 |
| Obesity | endothelial | *RGS17* | -0.881072175 | 9.76E-11 | 1.23E-08 | 6.14E-07 |
| Obesity | endothelial | *TMEM101* | -1.756650047 | 2.51E-07 | 1.91E-06 | 0.001580519 |
| Obesity | endothelial | *CMA1* | -1.228346374 | 0.015800877 | 0.022716696 | 1 |
| Obesity | endothelial | *SLC22A17* | -1.308648445 | 7.27E-05 | 0.000192086 | 0.456781417 |
| Obesity | endothelial | *SCFD1* | -0.533846002 | 0.000936103 | 0.001797032 | 1 |
| Obesity | endothelial | *RPGRIP1* | -0.89960182 | 2.50E-07 | 1.91E-06 | 0.001573845 |
| Obesity | endothelial | *TOX4* | -0.526337421 | 0.019931387 | 0.028083512 | 1 |
| Obesity | endothelial | *TGM1* | -0.422465866 | 0.008293138 | 0.012621389 | 1 |
| Obesity | endothelial | *AGO1* | -0.980219533 | 2.08E-05 | 6.77E-05 | 0.130750667 |
| Obesity | endothelial | *RFFL* | -1.436583328 | 2.42E-05 | 7.64E-05 | 0.15209593 |
| Obesity | endothelial | *MFSD11* | -0.705680079 | 1.64E-05 | 5.57E-05 | 0.103244182 |
| Obesity | endothelial | *COMT* | -0.957368711 | 1.72E-06 | 8.78E-06 | 0.010825625 |
| Obesity | endothelial | *VNN3* | -2.367588811 | 0.029394871 | 0.039863148 | 1 |
| Obesity | endothelial | *SEC22C* | -0.655610914 | 0.007381076 | 0.01137652 | 1 |
| Obesity | endothelial | *SUCO* | -0.520579404 | 0.016479616 | 0.023616901 | 1 |
| Obesity | endothelial | *TMEM38B* | -1.229483661 | 3.24E-08 | 4.54E-07 | 0.000203464 |
| Obesity | endothelial | *CWF19L1* | -0.406050336 | 0.003131651 | 0.005237747 | 1 |
| Obesity | endothelial | *IKZF5* | -0.96672063 | 0.000105019 | 0.000263155 | 0.660256505 |
| Obesity | endothelial | *BLNK* | -0.713271639 | 2.09E-06 | 1.03E-05 | 0.013159561 |
| Obesity | endothelial | *WAC* | -0.439481509 | 0.008774714 | 0.013299573 | 1 |
| Obesity | endothelial | *CREM* | -0.863210397 | 7.89E-06 | 3.04E-05 | 0.049624194 |
| Obesity | endothelial | *TPSD1* | -9.031996601 | 5.77E-08 | 6.38E-07 | 0.000362754 |
| Obesity | endothelial | *ITPR3* | 0.387187767 | 0.022117081 | 0.030831505 | 1 |
| Obesity | endothelial | *DSP* | 0.347741134 | 1.20E-05 | 4.27E-05 | 0.075230218 |
| Obesity | endothelial | *IL12RB1* | -1.307250916 | 1.28E-07 | 1.15E-06 | 0.000801925 |
| Obesity | endothelial | *ACOT7* | -1.159299397 | 2.00E-06 | 9.96E-06 | 0.012595045 |
| Obesity | endothelial | *CDC7* | -0.433163256 | 0.007446682 | 0.011469203 | 1 |
| Obesity | endothelial | *PCSK5* | 0.24691134 | 0.000630944 | 0.001265713 | 1 |
| Obesity | endothelial | *RAB18* | -0.522525242 | 0.003838843 | 0.006304808 | 1 |
| Obesity | endothelial | *PRTFDC1* | -0.698222917 | 3.04E-08 | 4.48E-07 | 0.00019094 |
| Obesity | endothelial | *PALMD* | 0.25089187 | 0.004306577 | 0.006985411 | 1 |
| Obesity | endothelial | *OCEL1* | -0.932567188 | 9.29E-10 | 7.04E-08 | 5.84E-06 |
| Obesity | endothelial | *CATSPERG* | -0.842678127 | 2.36E-05 | 7.50E-05 | 0.14866164 |
| Obesity | endothelial | *STX1B* | -1.417662543 | 2.71E-12 | 7.09E-10 | 1.70E-08 |
| Obesity | endothelial | *HSD3B7* | -0.810807656 | 4.04E-05 | 0.0001171 | 0.253873789 |
| Obesity | endothelial | *ATP5D* | -1.235143472 | 1.11E-05 | 4.02E-05 | 0.069767864 |
| Obesity | endothelial | *MARCH2* | -0.614150101 | 0.011009201 | 0.016312715 | 1 |
| Obesity | endothelial | *CDC34* | -1.351679586 | 3.63E-05 | 0.000107433 | 0.228188059 |
| Obesity | endothelial | *MTAP* | -1.185746016 | 8.35E-07 | 4.91E-06 | 0.005252329 |
| Obesity | endothelial | *POLR2E* | -1.219164596 | 2.90E-07 | 2.14E-06 | 0.001822653 |
| Obesity | endothelial | *MADCAM1* | -1.013816504 | 3.54E-08 | 4.79E-07 | 0.00022241 |
| Obesity | endothelial | *TRMT2A* | -1.205593628 | 1.71E-08 | 3.41E-07 | 0.000107729 |
| Obesity | endothelial | *ZDHHC8* | -0.573601707 | 0.016578413 | 0.023742251 | 1 |
| Obesity | endothelial | *MED15* | -0.756613494 | 6.51E-05 | 0.000175367 | 0.409306686 |
| Obesity | endothelial | *SERPIND1* | -0.825685361 | 6.74E-05 | 0.000180593 | 0.423904309 |
| Obesity | endothelial | *SNAP29* | -0.631196359 | 3.38E-05 | 0.000101331 | 0.2123908 |
| Obesity | endothelial | *LZTR1* | -1.074398145 | 0.00013529 | 0.000327646 | 0.850567926 |
| Obesity | endothelial | *CECR2* | -0.614755511 | 8.95E-06 | 3.37E-05 | 0.056268636 |
| Obesity | endothelial | *P2RX6* | -0.781577045 | 7.49E-09 | 2.69E-07 | 4.71E-05 |

| | | | | | | |
|---|---|---|---|---|---|---|
| Obesity | endothelial | *SLC7A4* | -1.017107063 | 0.00289229 | 0.004882875 | 1 |
| Obesity | endothelial | *DDTL* | -0.363547537 | 0.005709889 | 0.009012822 | 1 |
| Obesity | endothelial | *DDT* | -1.335761011 | 1.82E-05 | 6.07E-05 | 0.114573945 |
| Obesity | endothelial | *GGT5* | -1.369516952 | 3.12E-08 | 4.49E-07 | 0.00019615 |
| Obesity | endothelial | *SPECC1L* | -0.750809112 | 5.97E-05 | 0.000163358 | 0.375233433 |
| Obesity | endothelial | *PPIL2* | -0.536112062 | 5.01E-05 | 0.000139761 | 0.314745616 |
| Obesity | endothelial | *UPB1* | 0.271761349 | 0.015763317 | 0.022678254 | 1 |
| Obesity | endothelial | *YPEL1* | -0.936185236 | 0.000807778 | 0.001581594 | 1 |
| Obesity | endothelial | *PPM1F* | 0.347314158 | 0.028743706 | 0.039047467 | 1 |
| Obesity | endothelial | *CRYBB3* | 0.536089251 | 2.06E-06 | 1.02E-05 | 0.012979284 |
| Obesity | endothelial | *CYTH4* | -0.971327879 | 3.57E-08 | 4.80E-07 | 0.000224694 |
| Obesity | endothelial | *CARD10* | 0.196384256 | 2.31E-06 | 1.12E-05 | 0.014504474 |
| Obesity | endothelial | *LRP5L* | -0.727937573 | 0.014458056 | 0.020900851 | 1 |
| Obesity | endothelial | *SLC25A1* | -1.129027009 | 3.67E-07 | 2.57E-06 | 0.002304969 |
| Obesity | endothelial | *GGA1* | -0.729900724 | 5.12E-05 | 0.000142579 | 0.321657728 |
| Obesity | endothelial | *SH3BP1* | -1.179246951 | 1.97E-08 | 3.58E-07 | 0.000124033 |
| Obesity | endothelial | *HPS4* | -0.950594721 | 1.54E-05 | 5.27E-05 | 0.096944706 |
| Obesity | endothelial | *TRIOBP* | -0.566845722 | 0.001547607 | 0.002801557 | 1 |
| Obesity | endothelial | *ANKRD54* | -1.043367644 | 7.56E-11 | 1.04E-08 | 4.75E-07 |
| Obesity | endothelial | *EIF3L* | -0.7183574 | 0.002209004 | 0.00385671 | 1 |
| Obesity | endothelial | *CCDC134* | -1.35117997 | 1.60E-06 | 8.26E-06 | 0.010046112 |
| Obesity | endothelial | *SLC16A8* | -1.627388782 | 9.91E-05 | 0.000250632 | 0.622820892 |
| Obesity | endothelial | *SLC5A4* | 0.061919068 | 0.006229985 | 0.009744627 | 1 |
| Obesity | endothelial | *CYP2D6* | -0.672582844 | 0.000781611 | 0.001536582 | 1 |
| Obesity | endothelial | *DDX17* | -0.371411017 | 0.009983364 | 0.014958391 | 1 |
| Obesity | endothelial | *CBY1* | -0.354206934 | 0.001800281 | 0.003207244 | 1 |
| Obesity | endothelial | *POLDIP3* | -1.179841293 | 2.10E-06 | 1.04E-05 | 0.01323287 |
| Obesity | endothelial | *PPP6R2* | -0.675259496 | 0.000102827 | 0.000258487 | 0.646475012 |
| Obesity | endothelial | *RHBDD3* | -1.055857447 | 3.40E-06 | 1.54E-05 | 0.021376315 |
| Obesity | endothelial | *TTLL1* | -1.08572939 | 0.001489666 | 0.002713861 | 1 |
| Obesity | endothelial | *RASL10A* | 0.657301023 | 0.001051289 | 0.001987806 | 1 |
| Obesity | endothelial | *AP1B1* | -0.759445979 | 0.000457706 | 0.000953163 | 1 |
| Obesity | endothelial | *HMGXB4* | -0.68958605 | 0.003798719 | 0.006243804 | 1 |
| Obesity | endothelial | *TOM1* | -0.603977823 | 2.41E-06 | 1.16E-05 | 0.015130718 |
| Obesity | endothelial | *NEFH* | -1.319535775 | 0.002755846 | 0.004680174 | 1 |
| Obesity | endothelial | *CHKB* | -0.59871389 | 0.000207101 | 0.00047694 | 1 |
| Obesity | endothelial | *BIK* | -4.122577911 | 7.13E-08 | 7.41E-07 | 0.00044808 |
| Obesity | endothelial | *MCAT* | -1.191145943 | 1.25E-05 | 4.43E-05 | 0.078727179 |
| Obesity | endothelial | *THOC5* | -1.003693797 | 4.29E-06 | 1.85E-05 | 0.02695286 |
| Obesity | endothelial | *TSPO* | -0.688609365 | 0.00122649 | 0.00228202 | 1 |
| Obesity | endothelial | *RASD2* | -1.713243854 | 0.002468442 | 0.004251821 | 1 |
| Obesity | endothelial | *TTLL12* | -1.737655013 | 2.45E-08 | 3.99E-07 | 0.000153979 |
| Obesity | endothelial | *MTMR3* | -0.511586136 | 0.000488454 | 0.001008509 | 1 |
| Obesity | endothelial | *SAMM50* | -1.024166961 | 0.000256956 | 0.000574292 | 1 |
| Obesity | endothelial | *TXN2* | -0.816029103 | 0.000175185 | 0.000411496 | 1 |
| Obesity | endothelial | *FOXRED2* | -0.580787207 | 2.92E-05 | 9.00E-05 | 0.183732115 |
| Obesity | endothelial | *IFT27* | -1.276159266 | 1.08E-05 | 3.92E-05 | 0.067688565 |
| Obesity | endothelial | *KIAA0930* | -0.837529828 | 1.32E-07 | 1.17E-06 | 0.000831072 |
| Obesity | endothelial | *NCF4* | -0.629590938 | 5.13E-05 | 0.000142986 | 0.322720062 |
| Obesity | endothelial | *SLC25A17* | -0.900140834 | 1.27E-05 | 4.47E-05 | 0.079816196 |

| | | | | | | |
|---|---|---|---|---|---|---|
| Obesity | endothelial | *KCTD17* | -3.083288585 | 1.15E-07 | 1.05E-06 | 0.000721068 |
| Obesity | endothelial | *EP300* | -0.626011155 | 0.000307624 | 0.000672735 | 1 |
| Obesity | endothelial | *ACO2* | -0.603042904 | 0.000947484 | 0.001815553 | 1 |
| Obesity | endothelial | *POLR3H* | -0.644804125 | 1.10E-05 | 3.99E-05 | 0.069137811 |
| Obesity | endothelial | *ZBED4* | -0.939770931 | 0.000264156 | 0.000588291 | 1 |
| Obesity | endothelial | *SDR39U1* | -0.485777613 | 0.000129959 | 0.000316688 | 0.817054798 |
| Obesity | endothelial | *RBM23* | -0.846502074 | 0.000142325 | 0.000342966 | 0.894797296 |
| Obesity | endothelial | *VCPKMT* | -0.785763927 | 0.000160057 | 0.000380967 | 1 |
| Obesity | endothelial | *SOS2* | -0.652429546 | 0.000125019 | 0.000306072 | 0.785992098 |
| Obesity | endothelial | *PYGL* | -0.379014801 | 1.94E-06 | 9.67E-06 | 0.012168149 |
| Obesity | endothelial | *TRIM9* | -0.433483201 | 0.004148019 | 0.006759615 | 1 |
| Obesity | endothelial | *DDHD1* | -0.832049087 | 4.73E-07 | 3.14E-06 | 0.002973447 |
| Obesity | endothelial | *ATP6V1D* | -0.486300851 | 0.002408287 | 0.004164164 | 1 |
| Obesity | endothelial | *PIGH* | -0.725912487 | 0.000636391 | 0.001275418 | 1 |
| Obesity | endothelial | *VTI1B* | -1.117209978 | 3.31E-06 | 1.51E-05 | 0.020833903 |
| Obesity | endothelial | *GSTZ1* | -0.761565036 | 0.000151658 | 0.000363229 | 0.953476348 |
| Obesity | endothelial | *TMED8* | -0.870729562 | 5.20E-05 | 0.000144741 | 0.327114201 |
| Obesity | endothelial | *DAAM1* | -0.473376163 | 0.002571741 | 0.004404395 | 1 |
| Obesity | endothelial | *PPM1A* | -0.772838475 | 2.19E-05 | 7.04E-05 | 0.137572702 |
| Obesity | endothelial | *MTHFD1* | -0.88901016 | 4.57E-07 | 3.04E-06 | 0.002870633 |
| Obesity | endothelial | *TELO2* | -1.488250767 | 5.84E-06 | 2.37E-05 | 0.036717103 |
| Obesity | endothelial | *PAPLN* | -0.803866547 | 1.48E-08 | 3.34E-07 | 9.29E-05 |
| Obesity | endothelial | *CCNB1IP1* | -0.86274015 | 6.12E-05 | 0.000166624 | 0.384734937 |
| Obesity | endothelial | *TRIP11* | -0.618843702 | 0.001407504 | 0.002580628 | 1 |
| Obesity | endothelial | *PCK2* | -1.068045913 | 1.22E-08 | 3.34E-07 | 7.66E-05 |
| Obesity | endothelial | *KIAA0391* | -1.546299572 | 8.04E-09 | 2.76E-07 | 5.05E-05 |
| Obesity | endothelial | *PLTP* | -0.340491839 | 0.001372314 | 0.002521992 | 1 |
| Obesity | endothelial | *GSS* | -0.832777043 | 2.68E-06 | 1.26E-05 | 0.016859047 |
| Obesity | endothelial | *PYGB* | -0.541167534 | 0.000929005 | 0.001785839 | 1 |
| Obesity | endothelial | *ABHD12* | -0.416700507 | 0.013586561 | 0.019727185 | 1 |
| Obesity | endothelial | *PROCR* | -0.419580905 | 0.002759047 | 0.004684344 | 1 |
| Obesity | endothelial | *NINL* | -0.66584229 | 3.34E-08 | 4.63E-07 | 0.000209768 |
| Obesity | endothelial | *CD40* | 0.442938766 | 0.00821215 | 0.012504187 | 1 |
| Obesity | endothelial | *UQCC1* | -0.610880101 | 0.000367824 | 0.000787371 | 1 |
| Obesity | endothelial | *IFT52* | -0.497634267 | 0.027274571 | 0.037261023 | 1 |
| Obesity | endothelial | *NDRG3* | -0.697304384 | 0.006642611 | 0.010337152 | 1 |
| Obesity | endothelial | *DOK5* | -1.131777926 | 5.28E-06 | 2.19E-05 | 0.033226644 |
| Obesity | endothelial | *BMP7* | -0.851968966 | 1.72E-08 | 3.41E-07 | 0.000107985 |
| Obesity | endothelial | *TPD52L2* | -0.878246938 | 9.68E-05 | 0.000245598 | 0.608837181 |
| Obesity | endothelial | *NELFCD* | -0.846002554 | 0.000859896 | 0.0016696 | 1 |
| Obesity | endothelial | *CTSZ* | -0.77130835 | 6.74E-05 | 0.000180593 | 0.424032872 |
| Obesity | endothelial | *PRPF6* | -0.782217832 | 0.000604573 | 0.001219256 | 1 |
| Obesity | endothelial | *MTG2* | -0.621063664 | 0.00153824 | 0.00278781 | 1 |
| Obesity | endothelial | *TCFL5* | -0.570891668 | 0.011712786 | 0.01724954 | 1 |
| Obesity | endothelial | *DIDO1* | -0.438572731 | 0.021819156 | 0.030449952 | 1 |
| Obesity | endothelial | *GID8* | -1.294693728 | 6.05E-07 | 3.82E-06 | 0.003802227 |
| Obesity | endothelial | *SLC17A9* | -1.174073011 | 4.08E-07 | 2.79E-06 | 0.002562297 |
| Obesity | endothelial | *BIRC7* | -0.679127159 | 0.009363495 | 0.014110329 | 1 |
| Obesity | endothelial | *NKAIN4* | -1.55625838 | 2.45E-08 | 3.99E-07 | 0.000153999 |
| Obesity | endothelial | *PTK6* | -2.05128983 | 1.71E-08 | 3.41E-07 | 0.000107589 |

| | | | | | | |
|---|---|---|---|---|---|---|
| Obesity | endothelial | *GMEB2* | -1.037839736 | 0.000546835 | 0.001114409 | 1 |
| Obesity | endothelial | *CDC25B* | 0.432706561 | 0.013029141 | 0.018974799 | 1 |
| Obesity | endothelial | *RNF24* | -0.698366802 | 2.74E-07 | 2.05E-06 | 0.0017209 |
| Obesity | endothelial | *NDUFAF5* | -0.186017498 | 0.007826539 | 0.011983793 | 1 |
| Obesity | endothelial | *SLC52A3* | 0.569388264 | 9.10E-08 | 8.81E-07 | 0.0005719 |
| Obesity | endothelial | *ANGPT4* | -1.235947989 | 3.02E-10 | 2.93E-08 | 1.90E-06 |
| Obesity | endothelial | *RSPO4* | -1.096621053 | 2.38E-07 | 1.84E-06 | 0.001497055 |
| Obesity | endothelial | *SEC23B* | -0.536734651 | 0.005089036 | 0.008112265 | 1 |
| Obesity | endothelial | *CCM2L* | 0.17043249 | 1.55E-06 | 8.07E-06 | 0.009744667 |
| Obesity | endothelial | *MYL9* | -0.57559117 | 0.001940615 | 0.003428111 | 1 |
| Obesity | endothelial | *MROH8* | -0.81138144 | 5.28E-08 | 6.01E-07 | 0.000331904 |
| Obesity | endothelial | *MANBAL* | -0.283849976 | 0.02411226 | 0.033236803 | 1 |
| Obesity | endothelial | *IDH3B* | -1.16624162 | 0.000408046 | 0.000862894 | 1 |
| Obesity | endothelial | *MAPRE1* | -0.898531762 | 0.00199187 | 0.003507526 | 1 |
| Obesity | endothelial | *TTI1* | -0.873841897 | 0.000969218 | 0.001850827 | 1 |
| Obesity | endothelial | *RPRD1B* | -0.867495618 | 3.45E-05 | 0.000103102 | 0.216824274 |
| Obesity | endothelial | *PXMP4* | -0.625791635 | 0.020053857 | 0.028230766 | 1 |
| Obesity | endothelial | *ACTR5* | -0.907370779 | 0.009210758 | 0.013918774 | 1 |
| Obesity | endothelial | *DHX35* | -1.098955777 | 5.61E-07 | 3.59E-06 | 0.003527573 |
| Obesity | endothelial | *DNTTIP1* | -0.671709308 | 4.18E-05 | 0.000120605 | 0.263039522 |
| Obesity | endothelial | *MAP1LC3A* | -0.552858096 | 0.000450611 | 0.000940553 | 1 |
| Obesity | endothelial | *APMAP* | -0.760340151 | 1.13E-06 | 6.26E-06 | 0.007095157 |
| Obesity | endothelial | *CELF4* | -1.267613378 | 2.40E-08 | 3.95E-07 | 0.000150759 |
| Obesity | endothelial | *RBFA* | -1.154680207 | 5.76E-06 | 2.35E-05 | 0.036242985 |
| Obesity | endothelial | *LPIN2* | -0.915613023 | 1.94E-07 | 1.57E-06 | 0.001220503 |
| Obesity | endothelial | *CEP76* | -0.642952835 | 0.005486271 | 0.008694778 | 1 |
| Obesity | endothelial | *CEP192* | -0.46455527 | 0.001529156 | 0.002772945 | 1 |
| Obesity | endothelial | *LIPG* | -2.450014684 | 3.78E-05 | 0.000111117 | 0.237789372 |
| Obesity | endothelial | *NALCN* | -1.162602452 | 2.59E-07 | 1.96E-06 | 0.00162936 |
| Obesity | endothelial | *HTR2A* | -1.057348159 | 4.24E-08 | 5.30E-07 | 0.000266496 |
| Obesity | endothelial | *KLF5* | -0.703414904 | 0.010208783 | 0.015259776 | 1 |
| Obesity | endothelial | *STK24* | -0.777715399 | 6.52E-05 | 0.000175642 | 0.410123798 |
| Obesity | endothelial | *DNAJC3* | -0.735896513 | 0.000281909 | 0.000621663 | 1 |
| Obesity | endothelial | *UGGT2* | -0.709582491 | 1.21E-05 | 4.31E-05 | 0.076198969 |
| Obesity | endothelial | *ARHGEF7* | -0.556467639 | 0.00130811 | 0.00241388 | 1 |
| Obesity | endothelial | *SGCG* | -0.63032366 | 6.85E-09 | 2.55E-07 | 4.31E-05 |
| Obesity | endothelial | *PARP4* | -0.422602194 | 0.028668486 | 0.038953701 | 1 |
| Obesity | endothelial | *RGCC* | -0.192175714 | 0.014618507 | 0.021118233 | 1 |
| Obesity | endothelial | *VWA8* | -0.609314852 | 0.0002281 | 0.000517153 | 1 |
| Obesity | endothelial | *KATNAL1* | -0.773069655 | 1.24E-05 | 4.40E-05 | 0.078250928 |
| Obesity | endothelial | *DHRS12* | -0.939966874 | 0.00010603 | 0.000265582 | 0.666611547 |
| Obesity | endothelial | *MEDAG* | -1.047484623 | 1.09E-06 | 6.11E-06 | 0.006865284 |
| Obesity | endothelial | *CLN5* | -0.443971058 | 0.023160717 | 0.032073002 | 1 |
| Obesity | endothelial | *MGRN1* | -0.606129678 | 8.42E-05 | 0.000217622 | 0.529282364 |
| Obesity | endothelial | *CORO1A* | -1.875345511 | 3.44E-19 | 3.09E-16 | 2.17E-15 |
| Obesity | endothelial | *GDPD3* | 0.466405493 | 0.013325336 | 0.019370262 | 1 |
| Obesity | endothelial | *ELMO3* | 0.331954142 | 0.017987767 | 0.025562634 | 1 |
| Obesity | endothelial | *LONP2* | -0.545928499 | 0.001930752 | 0.003413565 | 1 |
| Obesity | endothelial | *PLLP* | 0.313301007 | 6.08E-06 | 2.45E-05 | 0.038245279 |
| Obesity | endothelial | *CTCF* | -0.477472027 | 0.02573759 | 0.035299351 | 1 |

| | | | | | | |
|---|---|---|---|---|---|---|
| Obesity | endothelial | *ACD* | -1.109338309 | 2.25E-06 | 1.10E-05 | 0.014166505 |
| Obesity | endothelial | *USB1* | -1.363794705 | 8.98E-06 | 3.38E-05 | 0.05646814 |
| Obesity | endothelial | *SETD6* | -0.861031014 | 0.007634744 | 0.011730117 | 1 |
| Obesity | endothelial | *COG4* | -1.046314707 | 7.59E-07 | 4.57E-06 | 0.004774953 |
| Obesity | endothelial | *SLC7A6OS* | -0.257457201 | 0.017814134 | 0.02533306 | 1 |
| Obesity | endothelial | *MON1B* | -0.976796724 | 0.000365414 | 0.000782746 | 1 |
| Obesity | endothelial | *HCFC1R1* | -1.524710671 | 6.89E-06 | 2.71E-05 | 0.043292734 |
| Obesity | endothelial | *NPRL3* | -0.520538321 | 0.001874349 | 0.003324128 | 1 |
| Obesity | endothelial | *NAGPA* | -0.781763953 | 0.001665451 | 0.002989916 | 1 |
| Obesity | endothelial | *COTL1* | -0.885851096 | 4.29E-08 | 5.34E-07 | 0.000269468 |
| Obesity | endothelial | *CRISPLD2* | -0.534402254 | 7.41E-05 | 0.000195016 | 0.465698892 |
| Obesity | endothelial | *ZNF500* | -0.398960748 | 0.027008296 | 0.036937384 | 1 |
| Obesity | endothelial | *NME4* | -1.246644529 | 3.40E-09 | 1.74E-07 | 2.14E-05 |
| Obesity | endothelial | *NOMO3* | -0.919003059 | 0.002047622 | 0.003598938 | 1 |
| Obesity | endothelial | *LMF1* | -0.669690851 | 0.001523861 | 0.00276494 | 1 |
| Obesity | endothelial | *NARFL* | -1.057287078 | 7.04E-05 | 0.000187519 | 0.442733313 |
| Obesity | endothelial | *CLCN7* | -1.009028424 | 7.97E-06 | 3.07E-05 | 0.050137651 |
| Obesity | endothelial | *HAGHL* | -1.088546319 | 7.44E-08 | 7.62E-07 | 0.000467859 |
| Obesity | endothelial | *SLC7A5* | -2.563222328 | 1.68E-08 | 3.40E-07 | 0.000105356 |
| Obesity | endothelial | *STUB1* | -1.311840215 | 7.24E-08 | 7.48E-07 | 0.000455056 |
| Obesity | endothelial | *RHBDL1* | -0.837525819 | 2.05E-08 | 3.63E-07 | 0.000128574 |
| Obesity | endothelial | *NUBP1* | -1.083064546 | 0.000266133 | 0.000591856 | 1 |
| Obesity | endothelial | *UBE2I* | -0.723016334 | 0.001569859 | 0.002838568 | 1 |
| Obesity | endothelial | *EARS2* | -0.488444688 | 0.000114795 | 0.000284476 | 0.721715426 |
| Obesity | endothelial | *CPPED1* | -0.875803615 | 2.25E-05 | 7.20E-05 | 0.141501943 |
| Obesity | endothelial | *BFAR* | -0.874830596 | 0.000466886 | 0.00096907 | 1 |
| Obesity | endothelial | *RBL2* | -0.337420824 | 0.033617652 | 0.045112951 | 1 |
| Obesity | endothelial | *STX4* | -0.961422079 | 2.71E-05 | 8.42E-05 | 0.170206964 |
| Obesity | endothelial | *KAT8* | -0.566648132 | 0.017096184 | 0.024405928 | 1 |
| Obesity | endothelial | *NOMO1* | -1.343053697 | 0.000115499 | 0.00028577 | 0.72614194 |
| Obesity | endothelial | *SYT17* | -0.716935327 | 1.14E-08 | 3.34E-07 | 7.18E-05 |
| Obesity | endothelial | *TMC5* | -1.162585108 | 6.72E-09 | 2.54E-07 | 4.23E-05 |
| Obesity | endothelial | *RNF40* | -1.225982495 | 1.11E-05 | 4.03E-05 | 0.069979738 |
| Obesity | endothelial | *KNOP1* | -0.86187447 | 9.20E-05 | 0.000234909 | 0.578345352 |
| Obesity | endothelial | *IQCH* | -0.754193178 | 0.002088263 | 0.003666268 | 1 |
| Obesity | endothelial | *LACTB* | -1.031060145 | 1.56E-06 | 8.12E-06 | 0.009813352 |
| Obesity | endothelial | *CSK* | -1.244374455 | 3.00E-05 | 9.19E-05 | 0.188847851 |
| Obesity | endothelial | *HERC1* | -0.373215281 | 0.022814041 | 0.031646375 | 1 |
| Obesity | endothelial | *RASL12* | -1.00547003 | 1.67E-08 | 3.39E-07 | 0.000104878 |
| Obesity | endothelial | *AP3B2* | -0.266197276 | 0.009323939 | 0.01405746 | 1 |
| Obesity | endothelial | *TTC23* | -0.576029318 | 0.006738806 | 0.010473887 | 1 |
| Obesity | endothelial | *FAH* | -0.742856258 | 1.40E-08 | 3.34E-07 | 8.77E-05 |
| Obesity | endothelial | *RPAP1* | -1.371023039 | 2.40E-07 | 1.85E-06 | 0.001508978 |
| Obesity | endothelial | *HOMER2* | -0.713630941 | 0.001616807 | 0.002910902 | 1 |
| Obesity | endothelial | *EHD4* | 0.36006709 | 0.014038471 | 0.020359831 | 1 |
| Obesity | endothelial | *ATP8B4* | -0.709097305 | 5.50E-08 | 6.18E-07 | 0.00034592 |
| Obesity | endothelial | *DTWD1* | -0.676134956 | 3.52E-05 | 0.000104807 | 0.221247495 |
| Obesity | endothelial | *PDGFRL* | -0.733446017 | 1.32E-07 | 1.17E-06 | 0.000830758 |
| Obesity | endothelial | *RIPK2* | -0.878887305 | 0.000100711 | 0.000253877 | 0.633168407 |
| Obesity | endothelial | *EYA1* | -0.410500113 | 1.22E-05 | 4.33E-05 | 0.076730407 |

| | | | | | | |
|---|---|---|---|---|---|---|
| Obesity | endothelial | *NBN* | -0.837046549 | 2.69E-05 | 8.37E-05 | 0.169177702 |
| Obesity | endothelial | *CPQ* | -0.500337499 | 6.46E-05 | 0.000174146 | 0.40593368 |
| Obesity | endothelial | *NIPAL2* | -0.639478124 | 0.001028757 | 0.001948131 | 1 |
| Obesity | endothelial | *PLAT* | 0.258032996 | 1.02E-05 | 3.77E-05 | 0.064319459 |
| Obesity | endothelial | *GDAP1* | 0.452378873 | 0.00224005 | 0.003901162 | 1 |
| Obesity | endothelial | *RAB2A* | -0.673347036 | 5.23E-07 | 3.40E-06 | 0.003290146 |
| Obesity | endothelial | *ZC2HC1A* | -0.574425374 | 0.004482517 | 0.007242762 | 1 |
| Obesity | endothelial | *ARMC1* | -0.386251236 | 0.006922542 | 0.010743525 | 1 |
| Obesity | endothelial | *CHRAC1* | -1.73555761 | 3.37E-06 | 1.53E-05 | 0.021196192 |
| Obesity | endothelial | *SNX16* | -0.907205985 | 4.55E-05 | 0.000129089 | 0.286319581 |
| Obesity | endothelial | *ERI1* | -0.98633476 | 2.20E-05 | 7.06E-05 | 0.138165477 |
| Obesity | endothelial | *MTMR9* | -0.56078062 | 0.015699917 | 0.022592212 | 1 |
| Obesity | endothelial | *TNFRSF10A* | 0.31534454 | 5.80E-06 | 2.36E-05 | 0.03647554 |
| Obesity | endothelial | *TUSC3* | 0.205260809 | 6.72E-06 | 2.67E-05 | 0.042253858 |
| Obesity | endothelial | *KLHDC4* | -0.467592424 | 0.014101165 | 0.020436612 | 1 |
| Obesity | endothelial | *MAN2B1* | -0.881853375 | 5.54E-07 | 3.55E-06 | 0.003482871 |
| Obesity | endothelial | *NFKBIB* | -1.618459693 | 6.22E-07 | 3.90E-06 | 0.003911567 |
| Obesity | endothelial | *TUBB4A* | -4.066495975 | 1.02E-06 | 5.81E-06 | 0.006403243 |
| Obesity | endothelial | *SARS2* | -0.606827046 | 0.000430298 | 0.000903267 | 1 |
| Obesity | endothelial | *SNRNP70* | -0.925701244 | 4.66E-08 | 5.58E-07 | 0.000293082 |
| Obesity | endothelial | *PPP1R37* | -0.709326121 | 0.005812327 | 0.00915154 | 1 |
| Obesity | endothelial | *FCGRT* | -0.450067298 | 0.007015982 | 0.010869758 | 1 |
| Obesity | endothelial | *PEX11G* | -1.140484296 | 1.51E-06 | 7.90E-06 | 0.009488866 |
| Obesity | endothelial | *ERCC2* | -1.452711238 | 7.11E-08 | 7.40E-07 | 0.000446984 |
| Obesity | endothelial | *PLEKHJ1* | -1.321037301 | 2.44E-07 | 1.87E-06 | 0.001531284 |
| Obesity | endothelial | *CD37* | -0.931533566 | 8.85E-05 | 0.000227128 | 0.55623649 |
| Obesity | endothelial | *AMH* | -0.873555416 | 0.001448969 | 0.002650472 | 1 |
| Obesity | endothelial | *STX10* | -1.167521271 | 2.28E-07 | 1.79E-06 | 0.00143303 |
| Obesity | endothelial | *FCER2* | -1.368040327 | 1.83E-08 | 3.45E-07 | 0.000115303 |
| Obesity | endothelial | *DMPK* | -0.900677837 | 0.00053981 | 0.001103018 | 1 |
| Obesity | endothelial | *IL4I1* | -0.762288539 | 0.000103876 | 0.000260707 | 0.653071281 |
| Obesity | endothelial | *TLE6* | -0.798029797 | 0.002902624 | 0.004896378 | 1 |
| Obesity | endothelial | *SGTA* | -0.785929013 | 0.012502481 | 0.018305332 | 1 |
| Obesity | endothelial | *C19orf53* | -1.377331201 | 4.59E-05 | 0.00012985 | 0.288396412 |
| Obesity | endothelial | *TIMM44* | -0.909943793 | 3.57E-05 | 0.000105993 | 0.224599221 |
| Obesity | endothelial | *CCDC61* | -0.934942703 | 0.001050839 | 0.001987553 | 1 |
| Obesity | endothelial | *IL27RA* | 0.669570076 | 0.000164404 | 0.000389599 | 1 |
| Obesity | endothelial | *C19orf44* | -0.406635751 | 0.003439093 | 0.005706407 | 1 |
| Obesity | endothelial | *RASAL3* | -0.755771679 | 0.001870493 | 0.003320099 | 1 |
| Obesity | endothelial | *CCNE1* | -1.472959962 | 2.48E-09 | 1.39E-07 | 1.56E-05 |
| Obesity | endothelial | *PDCD5* | -0.571605927 | 0.022144707 | 0.030863173 | 1 |
| Obesity | endothelial | *ANKRD27* | -0.736057332 | 0.000805281 | 0.00157818 | 1 |
| Obesity | endothelial | *RPS16* | -0.631754048 | 0.002772371 | 0.004705695 | 1 |
| Obesity | endothelial | *PRX* | -0.778990194 | 0.034592907 | 0.046322813 | 1 |
| Obesity | endothelial | *CCDC94* | -0.995185959 | 0.002822701 | 0.004782086 | 1 |
| Obesity | endothelial | *POLR2I* | -0.993213389 | 0.001952765 | 0.003446668 | 1 |
| Obesity | endothelial | *SLC1A5* | -1.073771263 | 1.84E-07 | 1.52E-06 | 0.001157839 |
| Obesity | endothelial | *PRKD2* | 0.283981191 | 0.032169001 | 0.043307604 | 1 |
| Obesity | endothelial | *CACTIN* | -1.93950525 | 2.36E-06 | 1.14E-05 | 0.014806189 |
| Obesity | endothelial | *FZR1* | -1.413636245 | 1.87E-05 | 6.18E-05 | 0.117363696 |

| | | | | | | |
|---|---|---|---|---|---|---|
| Obesity | endothelial | *BBC3* | -1.898387998 | 3.49E-05 | 0.00010403 | 0.219399501 |
| Obesity | endothelial | *TGFB1* | -0.534399342 | 0.014344871 | 0.020751542 | 1 |
| Obesity | endothelial | *CEACAM4* | -2.01813555 | 3.37E-08 | 4.64E-07 | 0.00021194 |
| Obesity | endothelial | *PLIN3* | -1.076169329 | 4.65E-06 | 1.98E-05 | 0.029219966 |
| Obesity | endothelial | *MRPL4* | -0.846721792 | 0.004324591 | 0.007005592 | 1 |
| Obesity | endothelial | *ICAM5* | 1.160564028 | 0.00011781 | 0.000290459 | 0.740670944 |
| Obesity | endothelial | *CD33* | -0.777733963 | 1.74E-06 | 8.82E-06 | 0.010914451 |
| Obesity | endothelial | *BABAM1* | -1.491857142 | 3.65E-05 | 0.000108029 | 0.229777799 |
| Obesity | endothelial | *TYK2* | -0.837173517 | 0.000315647 | 0.000688574 | 1 |
| Obesity | endothelial | *RABAC1* | -0.589384211 | 0.022593811 | 0.031405546 | 1 |
| Obesity | endothelial | *PTPRS* | -0.477544141 | 3.46E-06 | 1.56E-05 | 0.021727457 |
| Obesity | endothelial | *GRWD1* | -0.864273262 | 5.63E-09 | 2.31E-07 | 3.54E-05 |
| Obesity | endothelial | *GRIN2D* | -1.151494088 | 8.25E-09 | 2.82E-07 | 5.19E-05 |
| Obesity | endothelial | *ZNF175* | -0.69434849 | 0.001113326 | 0.002090022 | 1 |
| Obesity | endothelial | *CAPS* | -0.823045256 | 2.14E-05 | 6.92E-05 | 0.134437086 |
| Obesity | endothelial | *BCAT2* | -0.951287519 | 0.002014162 | 0.003545084 | 1 |
| Obesity | endothelial | *PPP2R1A* | -0.941907159 | 0.000229404 | 0.000519923 | 1 |
| Obesity | endothelial | *TNPO2* | -1.043566685 | 7.38E-07 | 4.46E-06 | 0.00464027 |
| Obesity | endothelial | *GCDH* | -1.249133737 | 2.82E-09 | 1.52E-07 | 1.77E-05 |
| Obesity | endothelial | *LILRB5* | -0.881517848 | 2.76E-08 | 4.26E-07 | 0.000173387 |
| Obesity | endothelial | *MAST1* | -3.925954323 | 5.06E-06 | 2.12E-05 | 0.031781058 |
| Obesity | endothelial | *PRPF31* | -0.955295185 | 1.81E-05 | 6.05E-05 | 0.114075158 |
| Obesity | endothelial | *PDE4C* | -1.143836578 | 1.83E-05 | 6.10E-05 | 0.115348463 |
| Obesity | endothelial | *ELL* | -1.131297137 | 1.11E-07 | 1.03E-06 | 0.000698948 |
| Obesity | endothelial | *UPK1A* | -2.263609271 | 5.15E-09 | 2.19E-07 | 3.24E-05 |
| Obesity | endothelial | *COPE* | -1.30512427 | 3.55E-06 | 1.59E-05 | 0.022295712 |
| Obesity | endothelial | *LSR* | 0.300505957 | 0.000342192 | 0.000739808 | 1 |
| Obesity | endothelial | *KXD1* | -1.210256084 | 4.51E-05 | 0.000127947 | 0.283274065 |
| Obesity | endothelial | *HPN* | -1.443640018 | 3.70E-06 | 1.65E-05 | 0.023290069 |
| Obesity | endothelial | *ZNF14* | -1.059281687 | 0.000325744 | 0.000707654 | 1 |
| Obesity | endothelial | *ATP13A1* | -1.403806318 | 1.59E-11 | 3.12E-09 | 9.98E-08 |
| Obesity | endothelial | *SIPA1L3* | -0.742574785 | 0.000102412 | 0.000257651 | 0.643867163 |
| Obesity | endothelial | *ETHE1* | -0.67791472 | 0.007614584 | 0.011704863 | 1 |
| Obesity | endothelial | *SMG9* | -0.998246632 | 0.000432421 | 0.000907117 | 1 |
| Obesity | endothelial | *GTPBP10* | -0.532636909 | 0.000823703 | 0.001610267 | 1 |
| Obesity | endothelial | *RASA4* | 0.361009177 | 4.27E-07 | 2.89E-06 | 0.002687181 |
| Obesity | endothelial | *PON3* | -1.470856375 | 0.000612451 | 0.001231366 | 1 |
| Obesity | endothelial | *HBP1* | -0.722127671 | 0.000258118 | 0.000576275 | 1 |
| Obesity | endothelial | *WDR91* | -1.181146341 | 6.46E-08 | 6.95E-07 | 0.000405996 |
| Obesity | endothelial | *STEAP1B* | 0.335491634 | 1.09E-05 | 3.97E-05 | 0.068756574 |
| Obesity | endothelial | *PTN* | -1.21546021 | 3.83E-08 | 5.00E-07 | 0.000240923 |
| Obesity | endothelial | *MPP6* | -0.684011359 | 1.57E-08 | 3.35E-07 | 9.90E-05 |
| Obesity | endothelial | *DFNA5* | -0.60207349 | 6.38E-06 | 2.55E-05 | 0.040116878 |
| Obesity | endothelial | *OGDH* | -0.78472296 | 2.01E-05 | 6.58E-05 | 0.126183252 |
| Obesity | endothelial | *ADAP1* | -1.304002714 | 2.49E-08 | 4.04E-07 | 0.000156746 |
| Obesity | endothelial | *H2AFV* | -0.889346001 | 0.000111323 | 0.000276635 | 0.699885308 |
| Obesity | endothelial | *MET* | 0.152814563 | 3.39E-06 | 1.54E-05 | 0.021340204 |
| Obesity | endothelial | *RNF32* | 0.317937237 | 0.002650729 | 0.004518746 | 1 |
| Obesity | endothelial | *HOXA2* | -1.921593407 | 5.14E-10 | 4.62E-08 | 3.23E-06 |
| Obesity | endothelial | *HOXA5* | -0.805338815 | 2.71E-07 | 2.03E-06 | 0.001704392 |

| | | | | | | |
|---|---|---|---|---|---|---|
| Obesity | endothelial | *HOXA6* | -1.002649851 | 5.07E-08 | 5.82E-07 | 0.000319038 |
| Obesity | endothelial | *BRAT1* | -0.96629411 | 6.49E-08 | 6.95E-07 | 0.000408115 |
| Obesity | endothelial | *CPED1* | -0.64242478 | 6.24E-08 | 6.75E-07 | 0.000392117 |
| Obesity | endothelial | *HIBADH* | -0.726671392 | 7.68E-06 | 2.97E-05 | 0.04829751 |
| Obesity | endothelial | *CPVL* | -0.874595998 | 4.38E-07 | 2.95E-06 | 0.002754559 |
| Obesity | endothelial | *PLEKHA8* | -0.817195522 | 0.000420461 | 0.000886761 | 1 |
| Obesity | endothelial | *CRHR2* | 2.225148852 | 0.000790193 | 0.001551997 | 1 |
| Obesity | endothelial | *EPHB6* | -0.567794441 | 1.85E-06 | 9.27E-06 | 0.011626941 |
| Obesity | endothelial | *CHCHD2* | -0.477664507 | 0.02359158 | 0.032633721 | 1 |
| Obesity | endothelial | *NPTX2* | 7.653215266 | 0.020482901 | 0.02877033 | 1 |
| Obesity | endothelial | *CYP3A5* | -1.104696012 | 2.29E-07 | 1.79E-06 | 0.001441679 |
| Obesity | endothelial | *ZKSCAN1* | -0.68952661 | 0.000705401 | 0.001400334 | 1 |
| Obesity | endothelial | *EIF3B* | -0.664738871 | 0.005252194 | 0.008351174 | 1 |
| Obesity | endothelial | *SNX8* | -1.012332266 | 2.25E-06 | 1.10E-05 | 0.014135262 |
| Obesity | endothelial | *TAF6* | -1.527689191 | 1.46E-07 | 1.27E-06 | 0.000918104 |
| Obesity | endothelial | *AIMP2* | -1.200322022 | 2.39E-06 | 1.16E-05 | 0.015040977 |
| Obesity | endothelial | *TFR2* | -1.637373009 | 8.98E-06 | 3.38E-05 | 0.056480581 |
| Obesity | endothelial | *RBM28* | -0.700071938 | 1.30E-05 | 4.57E-05 | 0.082001249 |
| Obesity | endothelial | *USP42* | -0.513062664 | 0.032691513 | 0.043951588 | 1 |
| Obesity | endothelial | *LSM5* | -1.139051572 | 7.66E-05 | 0.000200298 | 0.481316346 |
| Obesity | endothelial | *AP1S1* | -1.966480904 | 0.000216296 | 0.000494312 | 1 |
| Obesity | endothelial | *C1GALT1* | -0.819097303 | 1.05E-05 | 3.85E-05 | 0.066032248 |
| Obesity | endothelial | *RPA3* | -0.470288099 | 0.035396816 | 0.047278475 | 1 |
| Obesity | endothelial | *ZNF862* | -0.596430313 | 0.002919763 | 0.004920008 | 1 |
| Obesity | endothelial | *MEST* | -0.598864196 | 7.37E-11 | 1.04E-08 | 4.63E-07 |
| Obesity | endothelial | *ACTR3C* | -0.751965346 | 3.89E-06 | 1.72E-05 | 0.024430849 |
| Obesity | endothelial | *POU6F2* | -0.690585261 | 1.47E-08 | 3.34E-07 | 9.26E-05 |
| Obesity | endothelial | *TSPAN13* | 0.278954609 | 3.36E-06 | 1.53E-05 | 0.021113434 |
| Obesity | endothelial | *RARRES2* | -0.870829324 | 4.09E-08 | 5.16E-07 | 0.000257083 |
| Obesity | endothelial | *TMEM176B* | -0.29288366 | 0.010385004 | 0.015512122 | 1 |
| Obesity | endothelial | *MRPL32* | -0.563951926 | 0.000465274 | 0.000966361 | 1 |
| Obesity | endothelial | *BLVRA* | -0.458206529 | 0.004584571 | 0.007396253 | 1 |
| Obesity | endothelial | *URGCP* | -0.948062727 | 1.22E-05 | 4.32E-05 | 0.076527475 |
| Obesity | endothelial | *POLD2* | -1.135624378 | 1.85E-07 | 1.52E-06 | 0.001160104 |
| Obesity | endothelial | *GCK* | -6.036426337 | 2.88E-08 | 4.36E-07 | 0.000181213 |
| Obesity | endothelial | *BCL7B* | -0.543221255 | 0.004320379 | 0.007003365 | 1 |
| Obesity | endothelial | *YKT6* | -0.994874601 | 0.005264391 | 0.008368451 | 1 |
| Obesity | endothelial | *CLIP2* | -0.87378532 | 0.000882948 | 0.001707504 | 1 |
| Obesity | endothelial | *LIMK1* | -1.195649803 | 3.35E-08 | 4.64E-07 | 0.000210894 |
| Obesity | endothelial | *SPATA6L* | 0.26074233 | 6.72E-05 | 0.000179999 | 0.422278125 |
| Obesity | endothelial | *SLC1A1* | 0.297465546 | 0.000509179 | 0.001047516 | 1 |
| Obesity | endothelial | *FKTN* | -0.619163731 | 0.004923613 | 0.007892595 | 1 |
| Obesity | endothelial | *MEGF9* | -0.749392271 | 1.50E-07 | 1.29E-06 | 0.000944979 |
| Obesity | endothelial | *CORO2A* | 0.351191266 | 1.50E-05 | 5.14E-05 | 0.094098302 |
| Obesity | endothelial | *C5* | -0.641375546 | 1.13E-06 | 6.28E-06 | 0.007130501 |
| Obesity | endothelial | *OGN* | -0.571329987 | 0.000980738 | 0.001867498 | 1 |
| Obesity | endothelial | *ECM2* | -0.566537297 | 3.15E-07 | 2.27E-06 | 0.001979711 |
| Obesity | endothelial | *PTGR1* | -0.285451891 | 0.033082989 | 0.044433402 | 1 |
| Obesity | endothelial | *SUSD1* | -0.449434187 | 0.003527442 | 0.005834524 | 1 |
| Obesity | endothelial | *TNFSF8* | -1.078296351 | 3.23E-08 | 4.54E-07 | 0.000203179 |

| | | | | | | |
|---|---|---|---|---|---|---|
| Obesity | endothelial | *AK1* | -0.798628524 | 0.003779929 | 0.006214543 | 1 |
| Obesity | endothelial | *PLGRKT* | -0.757175636 | 1.55E-06 | 8.07E-06 | 0.009730271 |
| Obesity | endothelial | *KDM4C* | -0.509988002 | 0.003480084 | 0.005760739 | 1 |
| Obesity | endothelial | *DOCK8* | -0.541055645 | 3.99E-06 | 1.76E-05 | 0.025093945 |
| Obesity | endothelial | *ELAVL2* | 0.484035063 | 0.003584143 | 0.005917413 | 1 |
| Obesity | endothelial | *FUBP3* | -0.76012269 | 0.000507495 | 0.001044735 | 1 |
| Obesity | endothelial | *PTGDS* | -1.184355555 | 1.54E-08 | 3.34E-07 | 9.70E-05 |
| Obesity | endothelial | *SHB* | 1.076713003 | 8.14E-06 | 3.12E-05 | 0.051184938 |
| Obesity | endothelial | *DVL1* | -1.326133044 | 4.20E-05 | 0.000120896 | 0.263795417 |
| Obesity | endothelial | *PDLIM1* | 0.367322235 | 0.00016503 | 0.000390935 | 1 |
| Obesity | endothelial | *CCNJ* | -0.864766219 | 0.002129202 | 0.003733202 | 1 |
| Obesity | endothelial | *ATRNL1* | -0.776372504 | 3.03E-07 | 2.21E-06 | 0.001905228 |
| Obesity | endothelial | *HPS1* | -1.015690322 | 7.52E-07 | 4.53E-06 | 0.004725841 |
| Obesity | endothelial | *DNMBP* | -0.796517241 | 8.44E-05 | 0.000217738 | 0.530449676 |
| Obesity | endothelial | *DDX50* | -0.753878814 | 0.000420302 | 0.000886724 | 1 |
| Obesity | endothelial | *MAPK8* | -0.405348427 | 0.021194165 | 0.029669943 | 1 |
| Obesity | endothelial | *ATE1* | -0.458517519 | 0.005586132 | 0.008837446 | 1 |
| Obesity | endothelial | *NSMCE4A* | -0.579964955 | 0.016466146 | 0.023602977 | 1 |
| Obesity | endothelial | *PLEKHA1* | -0.685216065 | 0.000974646 | 0.001857975 | 1 |
| Obesity | endothelial | *SPOCK2* | -0.457993822 | 0.013792344 | 0.020016728 | 1 |
| Obesity | endothelial | *MICU1* | -0.533752796 | 0.0029556 | 0.004976394 | 1 |
| Obesity | endothelial | *BMPR1A* | -0.429285187 | 1.08E-05 | 3.93E-05 | 0.067985595 |
| Obesity | endothelial | *LIPA* | -0.417707613 | 0.02023337 | 0.028457985 | 1 |
| Obesity | endothelial | *SFXN3* | -0.615998462 | 0.029536277 | 0.040011759 | 1 |
| Obesity | endothelial | *KAZALD1* | 0.94641441 | 0.000929136 | 0.001785839 | 1 |
| Obesity | endothelial | *TNKS2* | -0.654077929 | 0.001141752 | 0.002137641 | 1 |
| Obesity | endothelial | *ACBD5* | -0.728018922 | 0.000710158 | 0.001408887 | 1 |
| Obesity | endothelial | *MTPAP* | -1.007171838 | 0.000111479 | 0.000276914 | 0.700869655 |
| Obesity | endothelial | *PITRM1* | -0.767503422 | 0.004556187 | 0.007354235 | 1 |
| Obesity | endothelial | *MAP3K8* | -0.660159154 | 0.000879529 | 0.001701415 | 1 |
| Obesity | endothelial | *SHOC2* | -0.556913467 | 0.002357061 | 0.004082325 | 1 |
| Obesity | endothelial | *TFAM* | -1.167369725 | 3.39E-05 | 0.000101625 | 0.213209438 |
| Obesity | endothelial | *TADA2A* | -0.257092185 | 0.021043628 | 0.02947233 | 1 |
| Obesity | endothelial | *DHRS11* | -0.56665954 | 2.65E-05 | 8.26E-05 | 0.166497652 |
| Obesity | endothelial | *PSMB3* | -1.069578076 | 1.47E-05 | 5.06E-05 | 0.092399063 |
| Obesity | endothelial | *CASC3* | -0.7253525 | 0.001007262 | 0.001911701 | 1 |
| Obesity | endothelial | *RAPGEFL1* | -0.846564323 | 0.000131853 | 0.000320809 | 0.828958213 |
| Obesity | endothelial | *RGS9* | -1.343374655 | 1.98E-08 | 3.59E-07 | 0.000124487 |
| Obesity | endothelial | *WNT3* | -1.554893023 | 4.60E-06 | 1.97E-05 | 0.028937539 |
| Obesity | endothelial | *ASPA* | -0.617667732 | 1.38E-08 | 3.34E-07 | 8.66E-05 |
| Obesity | endothelial | *RAD51C* | -0.756199923 | 1.86E-08 | 3.45E-07 | 0.000116726 |
| Obesity | endothelial | *SEPT4* | -1.381838758 | 9.18E-08 | 8.82E-07 | 0.000576852 |
| Obesity | endothelial | *TRIM37* | -0.796923064 | 0.000233452 | 0.000527576 | 1 |
| Obesity | endothelial | *PNPO* | -0.722899801 | 0.000402377 | 0.00085234 | 1 |
| Obesity | endothelial | *RPS6KB1* | -0.836927097 | 7.89E-05 | 0.000205359 | 0.496352463 |
| Obesity | endothelial | *TRIM16L* | -0.799363541 | 8.23E-08 | 8.20E-07 | 0.000517441 |
| Obesity | endothelial | *CDK5RAP3* | -0.668135106 | 0.020134921 | 0.028332195 | 1 |
| Obesity | endothelial | *RECQL5* | -1.00475469 | 2.05E-06 | 1.01E-05 | 0.012864383 |
| Obesity | endothelial | *PIGL* | -0.717174757 | 0.000229508 | 0.000519969 | 1 |
| Obesity | endothelial | *CAMTA2* | -0.807567509 | 0.000464944 | 0.000965996 | 1 |

| | | | | | | |
|---|---|---|---|---|---|---|
| Obesity | endothelial | *MED13* | -0.702353931 | 1.42E-05 | 4.91E-05 | 0.089052512 |
| Obesity | endothelial | *RNF167* | -0.893661474 | 0.003266964 | 0.005449564 | 1 |
| Obesity | endothelial | *CHRNE* | 0.409233944 | 0.001605518 | 0.00289389 | 1 |
| Obesity | endothelial | *NUP88* | -0.94483488 | 6.85E-06 | 2.70E-05 | 0.043082204 |
| Obesity | endothelial | *GOSR1* | -0.664309159 | 0.001127979 | 0.002113742 | 1 |
| Obesity | endothelial | *DRG2* | -1.273774674 | 6.42E-05 | 0.000173362 | 0.403413477 |
| Obesity | endothelial | *FTSJ3* | -0.411840299 | 5.15E-06 | 2.15E-05 | 0.032378982 |
| Obesity | endothelial | *AKAP10* | -0.729359453 | 3.65E-05 | 0.000107888 | 0.229369681 |
| Obesity | endothelial | *ALDH3A1* | 0.616129284 | 0.035743377 | 0.047690707 | 1 |
| Obesity | endothelial | *SMARCD2* | -1.121515751 | 3.12E-08 | 4.49E-07 | 0.000196066 |
| Obesity | endothelial | *ICAM2* | 0.240509471 | 5.63E-06 | 2.31E-05 | 0.035372088 |
| Obesity | endothelial | *UTP6* | -0.872039354 | 6.13E-05 | 0.000166899 | 0.385536507 |
| Obesity | endothelial | *DDX5* | -0.292207025 | 0.021033488 | 0.029464692 | 1 |
| Obesity | endothelial | *C17orf75* | -0.654724415 | 0.035622951 | 0.047550211 | 1 |
| Obesity | endothelial | *RAB5C* | -0.619835381 | 0.033335215 | 0.044753042 | 1 |
| Obesity | endothelial | *NAGLU* | -1.082147845 | 7.35E-08 | 7.57E-07 | 0.000461929 |
| Obesity | endothelial | *HSD17B1* | -0.206759188 | 0.021716566 | 0.030333714 | 1 |
| Obesity | endothelial | *EZH1* | -0.477148361 | 0.010802473 | 0.016059387 | 1 |
| Obesity | endothelial | *DLX4* | -3.479584923 | 6.78E-08 | 7.15E-07 | 0.000426416 |
| Obesity | endothelial | *LRRC59* | -1.566217383 | 8.67E-07 | 5.06E-06 | 0.005451008 |
| Obesity | endothelial | *ALOX12* | -0.944404742 | 0.004963471 | 0.007940291 | 1 |
| Obesity | endothelial | *ABCC3* | -0.277786997 | 0.011979357 | 0.01758035 | 1 |
| Obesity | endothelial | *MPP2* | 0.969386834 | 0.002429424 | 0.004195121 | 1 |
| Obesity | endothelial | *EFTUD2* | -0.482708345 | 0.036255195 | 0.048301846 | 1 |
| Obesity | endothelial | *EFNB3* | -4.629095053 | 7.48E-05 | 0.000196569 | 0.470193058 |
| Obesity | endothelial | *FAM20A* | -0.751345656 | 0.000193299 | 0.000449602 | 1 |
| Obesity | endothelial | *YWHAE* | -0.628440669 | 0.001008533 | 0.001913292 | 1 |
| Obesity | endothelial | *DPH1* | -1.677250035 | 3.95E-07 | 2.71E-06 | 0.002486254 |
| Obesity | endothelial | *DHRS7B* | -0.700129662 | 0.000742825 | 0.001465833 | 1 |
| Obesity | endothelial | *RCVRN* | -1.919018298 | 2.65E-07 | 2.00E-06 | 0.001663347 |
| Obesity | endothelial | *SLC9A3R1* | -1.093437511 | 0.000113839 | 0.00028244 | 0.715703292 |
| Obesity | endothelial | *MYH3* | -1.299108706 | 1.05E-06 | 5.94E-06 | 0.006590609 |
| Obesity | endothelial | *TMEM104* | -0.999418319 | 7.40E-08 | 7.60E-07 | 0.000465006 |
| Obesity | endothelial | *RAB34* | -1.037027406 | 3.28E-05 | 9.87E-05 | 0.20613432 |
| Obesity | endothelial | *SLAIN2* | -0.514194255 | 0.007744112 | 0.011874935 | 1 |
| Obesity | endothelial | *OCIAD1* | -0.568290739 | 0.004184772 | 0.006814209 | 1 |
| Obesity | endothelial | *CWH43* | -3.375550849 | 3.94E-08 | 5.06E-07 | 0.000247527 |
| Obesity | endothelial | *KIAA1211* | 0.293890608 | 0.009228572 | 0.013940421 | 1 |
| Obesity | endothelial | *AREG* | 0.340411422 | 0.000100354 | 0.000253181 | 0.630927056 |
| Obesity | endothelial | *UBE2D3* | -0.451835704 | 0.009595333 | 0.014421674 | 1 |
| Obesity | endothelial | *NDUFC1* | -0.862120606 | 0.003857058 | 0.006333069 | 1 |
| Obesity | endothelial | *TBC1D9* | -0.408719564 | 0.030833817 | 0.041688647 | 1 |
| Obesity | endothelial | *CPE* | -0.837276866 | 2.39E-05 | 7.56E-05 | 0.150104395 |
| Obesity | endothelial | *WFS1* | -0.919022479 | 0.003858708 | 0.006334125 | 1 |
| Obesity | endothelial | *GRPEL1* | -1.381621185 | 0.000212925 | 0.000488741 | 1 |
| Obesity | endothelial | *AADAT* | -0.784189521 | 3.34E-08 | 4.63E-07 | 0.000210186 |
| Obesity | endothelial | *CPZ* | -6.947250804 | 6.54E-08 | 6.99E-07 | 0.000411038 |
| Obesity | endothelial | *SLC2A9* | -1.003061619 | 8.32E-07 | 4.91E-06 | 0.005229895 |
| Obesity | endothelial | *SH3D19* | -0.514260408 | 1.50E-05 | 5.15E-05 | 0.094328357 |
| Obesity | endothelial | *LRP2BP* | 0.395172087 | 0.011834981 | 0.017417257 | 1 |

| | | | | | | |
|---|---|---|---|---|---|---|
| Obesity | endothelial | *UFSP2* | -0.642824599 | 0.008295353 | 0.012621705 | 1 |
| Obesity | endothelial | *UGDH* | -1.00165269 | 5.64E-08 | 6.28E-07 | 0.000354795 |
| Obesity | endothelial | *DDX25* | 0.805993018 | 5.97E-06 | 2.41E-05 | 0.037516747 |
| Obesity | endothelial | *HTATIP2* | -0.41500562 | 0.016819555 | 0.024038314 | 1 |
| Obesity | endothelial | *CTSC* | -0.452432697 | 0.01421434 | 0.020591141 | 1 |
| Obesity | endothelial | *ELP4* | -0.45006314 | 0.014090124 | 0.020425319 | 1 |
| Obesity | endothelial | *SC5D* | -1.218243913 | 2.15E-07 | 1.71E-06 | 0.001351361 |
| Obesity | endothelial | *C11orf63* | -1.003863472 | 8.53E-07 | 5.00E-06 | 0.005365917 |
| Obesity | endothelial | *SNX15* | -0.584821791 | 4.32E-05 | 0.000124007 | 0.27185922 |
| Obesity | endothelial | *DCPS* | -1.319180834 | 4.48E-06 | 1.92E-05 | 0.02813815 |
| Obesity | endothelial | *FOXRED1* | -0.431293015 | 0.004087928 | 0.0066738 | 1 |
| Obesity | endothelial | *NRXN2* | -1.104503629 | 2.00E-08 | 3.61E-07 | 0.000125784 |
| Obesity | endothelial | *MS4A4A* | -0.421241766 | 0.000547312 | 0.001115019 | 1 |
| Obesity | endothelial | *PRPF19* | -1.416608406 | 1.54E-07 | 1.32E-06 | 0.00096682 |
| Obesity | endothelial | *HPX* | 0.55113228 | 0.010437282 | 0.015582806 | 1 |
| Obesity | endothelial | *CHORDC1* | -0.88458551 | 0.000170614 | 0.000402948 | 1 |
| Obesity | endothelial | *PANX1* | -1.175459355 | 9.17E-08 | 8.82E-07 | 0.000576223 |
| Obesity | endothelial | *EIF4G2* | -0.640810805 | 0.002340197 | 0.004054236 | 1 |
| Obesity | endothelial | *GALNT18* | 0.162282049 | 2.84E-05 | 8.77E-05 | 0.178338392 |
| Obesity | endothelial | *CBL* | -0.459578692 | 0.010042147 | 0.015028559 | 1 |
| Obesity | endothelial | *MADD* | -0.799564008 | 0.000514929 | 0.001057614 | 1 |
| Obesity | endothelial | *NAA40* | -1.096797689 | 0.000147787 | 0.000354632 | 0.929135903 |
| Obesity | endothelial | *CARS* | -0.765193902 | 0.00029278 | 0.000642929 | 1 |
| Obesity | endothelial | *C11orf21* | -0.668744321 | 0.006467324 | 0.010086844 | 1 |
| Obesity | endothelial | *RPS13* | -0.484397098 | 0.021974089 | 0.030645763 | 1 |
| Obesity | endothelial | *AIP* | -0.877254695 | 0.004394162 | 0.007110964 | 1 |
| Obesity | endothelial | *NDUFS8* | -1.214417942 | 1.13E-05 | 4.10E-05 | 0.071296029 |
| Obesity | endothelial | *TCIRG1* | -0.652340488 | 0.011205569 | 0.016571826 | 1 |
| Obesity | endothelial | *GTF2H1* | -0.51556454 | 0.008629732 | 0.013089294 | 1 |
| Obesity | endothelial | *PTPN5* | 0.320219375 | 9.01E-06 | 3.38E-05 | 0.056621794 |
| Obesity | endothelial | *PRDM4* | -0.578825235 | 0.018740455 | 0.026536316 | 1 |
| Obesity | endothelial | *ASIC1* | -0.822697821 | 1.65E-09 | 1.09E-07 | 1.04E-05 |
| Obesity | endothelial | *CAPRIN2* | -1.002807567 | 1.31E-05 | 4.60E-05 | 0.082588734 |
| Obesity | endothelial | *KCTD10* | -0.438668456 | 0.007683232 | 0.011790207 | 1 |
| Obesity | endothelial | *CYP27B1* | -2.766929785 | 1.50E-07 | 1.29E-06 | 0.000940707 |
| Obesity | endothelial | *ACSS3* | -0.26385974 | 0.000224074 | 0.000509495 | 1 |
| Obesity | endothelial | *GLI1* | -1.036006802 | 2.86E-08 | 4.35E-07 | 0.000179986 |
| Obesity | endothelial | *METAP2* | -0.713771111 | 0.000469347 | 0.000973214 | 1 |
| Obesity | endothelial | *WNT5B* | -0.640304817 | 0.021575468 | 0.030150027 | 1 |
| Obesity | endothelial | *TRPV4* | -1.850286488 | 1.78E-11 | 3.40E-09 | 1.12E-07 |
| Obesity | endothelial | *ITFG2* | -0.886725355 | 8.33E-07 | 4.91E-06 | 0.005235772 |
| Obesity | endothelial | *PARP11* | -0.875321167 | 4.61E-05 | 0.000130327 | 0.289846205 |
| Obesity | endothelial | *ARPC3* | -0.836268658 | 0.001304083 | 0.002409277 | 1 |
| Obesity | endothelial | *GPN3* | -1.365614586 | 8.49E-08 | 8.38E-07 | 0.000533618 |
| Obesity | endothelial | *VPS29* | -0.685081427 | 0.000869034 | 0.001683184 | 1 |
| Obesity | endothelial | *AKAP3* | -0.531092958 | 9.93E-05 | 0.000251003 | 0.624341631 |
| Obesity | endothelial | *KCNA1* | -1.532213531 | 2.26E-09 | 1.33E-07 | 1.42E-05 |
| Obesity | endothelial | *CREBL2* | -0.942439096 | 2.42E-06 | 1.16E-05 | 0.015222854 |
| Obesity | endothelial | *NAA25* | -0.685716913 | 0.000453718 | 0.000945797 | 1 |
| Obesity | endothelial | *CDK2AP1* | 0.268244476 | 0.023326819 | 0.032295907 | 1 |

| | | | | | | |
|---|---|---|---|---|---|---|
| Obesity | endothelial | *MGP* | -0.340194394 | 0.015977764 | 0.022960503 | 1 |
| Obesity | endothelial | *RASAL1* | -2.400323496 | 1.93E-07 | 1.57E-06 | 0.001210608 |
| Obesity | endothelial | *ARHGDIB* | -0.615625254 | 0.008083165 | 0.012334401 | 1 |
| Obesity | endothelial | *EIF2B1* | -0.969484345 | 0.00011467 | 0.000284278 | 0.720929419 |
| Obesity | endothelial | *ENDOU* | 0.420528416 | 0.023738602 | 0.03280822 | 1 |
| Obesity | endothelial | *STX2* | -0.561305605 | 0.001217581 | 0.002268129 | 1 |
| Obesity | endothelial | *CAND1* | -0.609746116 | 0.000764552 | 0.001504457 | 1 |
| Obesity | endothelial | *MDM1* | -0.838436774 | 3.80E-05 | 0.000111405 | 0.23863045 |
| Obesity | endothelial | *NUP107* | -0.50357464 | 0.011523832 | 0.016991166 | 1 |
| Obesity | endothelial | *CNOT2* | -0.561779739 | 0.001741543 | 0.003113187 | 1 |
| Obesity | endothelial | *KRR1* | -0.887437529 | 6.76E-05 | 0.000180914 | 0.425148487 |
| Obesity | endothelial | *COPS7A* | -0.989489769 | 2.88E-05 | 8.89E-05 | 0.181009526 |
| Obesity | endothelial | *CHPT1* | -0.349305558 | 5.35E-05 | 0.0001482 | 0.33656333 |
| Obesity | endothelial | *GNPTAB* | -0.73783613 | 8.90E-05 | 0.000228175 | 0.559485701 |
| Obesity | endothelial | *C12orf57* | -0.814203394 | 0.007137704 | 0.011042014 | 1 |
| Obesity | endothelial | *NT5DC3* | -0.766204468 | 3.81E-07 | 2.64E-06 | 0.002393126 |
| Obesity | endothelial | *ST8SIA1* | -0.784401672 | 2.05E-08 | 3.63E-07 | 0.000129095 |
| Obesity | endothelial | *C2CD5* | -0.551072033 | 0.003654519 | 0.006020953 | 1 |
| Obesity | endothelial | *COL12A1* | -0.581265397 | 1.44E-08 | 3.34E-07 | 9.08E-05 |
| Obesity | endothelial | *BTN3A3* | 0.345992821 | 0.00588295 | 0.009251152 | 1 |
| Obesity | endothelial | *TDP2* | -0.904777425 | 0.000360338 | 0.000773453 | 1 |
| Obesity | endothelial | *SMIM8* | -0.612159785 | 0.001768384 | 0.003156682 | 1 |
| Obesity | endothelial | *MCM9* | -0.524058397 | 0.009710374 | 0.014584118 | 1 |
| Obesity | endothelial | *FAM184A* | 0.346641695 | 0.024753746 | 0.034054005 | 1 |
| Obesity | endothelial | *RNGTT* | -0.576854338 | 0.002863347 | 0.004844323 | 1 |
| Obesity | endothelial | *TPD52L1* | 0.23417947 | 8.35E-06 | 3.19E-05 | 0.05252073 |
| Obesity | endothelial | *UST* | -0.549263447 | 4.06E-08 | 5.14E-07 | 0.00025519 |
| Obesity | endothelial | *TULP1* | 2.659014297 | 2.40E-05 | 7.57E-05 | 0.150613495 |
| Obesity | endothelial | *MAPK14* | -0.490155909 | 0.007481544 | 0.011514434 | 1 |
| Obesity | endothelial | *MRPL18* | -0.49735804 | 0.031273064 | 0.042200849 | 1 |
| Obesity | endothelial | *PHACTR1* | 0.283568507 | 0.022364444 | 0.031114242 | 1 |
| Obesity | endothelial | *MDGA1* | -1.20819484 | 4.10E-07 | 2.80E-06 | 0.002578664 |
| Obesity | endothelial | *CD83* | -0.790762524 | 1.38E-07 | 1.21E-06 | 0.000864637 |
| Obesity | endothelial | *MDN1* | -0.448985075 | 0.020983525 | 0.029420924 | 1 |
| Obesity | endothelial | *CAP2* | -0.915165363 | 7.93E-11 | 1.04E-08 | 4.99E-07 |
| Obesity | endothelial | *ASCC3* | -0.391182103 | 0.029596984 | 0.040085359 | 1 |
| Obesity | endothelial | *BVES* | -0.592917798 | 1.02E-05 | 3.76E-05 | 0.064062344 |
| Obesity | endothelial | *WASF1* | -0.296398567 | 0.001007391 | 0.001911701 | 1 |
| Obesity | endothelial | *GPLD1* | -0.355976231 | 0.000454064 | 0.000946008 | 1 |
| Obesity | endothelial | *ALDH5A1* | -0.777764528 | 4.83E-05 | 0.000135559 | 0.303787977 |
| Obesity | endothelial | *VNN1* | -0.447477934 | 8.54E-05 | 0.000219854 | 0.536662735 |
| Obesity | endothelial | *VNN2* | -0.912445353 | 0.000783112 | 0.001539051 | 1 |
| Obesity | endothelial | *ACOT13* | -0.832946563 | 0.00058856 | 0.001191332 | 1 |
| Obesity | endothelial | *C6orf62* | -0.703431794 | 1.05E-05 | 3.85E-05 | 0.066068334 |
| Obesity | endothelial | *B3GAT2* | -0.967737179 | 0.00249557 | 0.004289134 | 1 |
| Obesity | endothelial | *PERP* | 0.369539462 | 0.001101981 | 0.002070578 | 1 |
| Obesity | endothelial | *SLC16A10* | -0.619917046 | 5.50E-06 | 2.27E-05 | 0.034599135 |
| Obesity | endothelial | *TBP* | -0.688018215 | 0.008120303 | 0.012376326 | 1 |
| Obesity | endothelial | *PRPH2* | -0.966465558 | 1.08E-06 | 6.08E-06 | 0.006818788 |
| Obesity | endothelial | *DNPH1* | -0.588167039 | 4.55E-05 | 0.000128889 | 0.285746049 |

| | | | | | | |
|---|---|---|---|---|---|---|
| Obesity | endothelial | *DUSP22* | -0.59301575 | 0.01163394 | 0.017141453 | 1 |
| Obesity | endothelial | *TMEM30A* | -0.744372002 | 0.000893651 | 0.001726609 | 1 |
| Obesity | endothelial | *SENP6* | -0.435613855 | 0.0091431 | 0.013827921 | 1 |
| Obesity | endothelial | *LAMA4* | -0.3226837 | 0.004365549 | 0.00707012 | 1 |
| Obesity | endothelial | *CLIC5* | 0.178967148 | 5.32E-06 | 2.21E-05 | 0.033462771 |
| Obesity | endothelial | *FBRSL1* | -0.880457027 | 0.000423422 | 0.00089181 | 1 |
| Obesity | endothelial | *LY86* | -0.914228105 | 3.10E-08 | 4.48E-07 | 0.00019501 |
| Obesity | endothelial | *PRSS16* | 1.978435498 | 1.06E-05 | 3.88E-05 | 0.066657251 |
| Obesity | endothelial | *TBX18* | -0.638286335 | 3.75E-09 | 1.84E-07 | 2.35E-05 |
| Obesity | endothelial | *PCDHB2* | -0.318035352 | 0.032946393 | 0.044268855 | 1 |
| Obesity | endothelial | *NUDT12* | -1.297837293 | 5.89E-07 | 3.74E-06 | 0.003701699 |
| Obesity | endothelial | *CEP72* | -0.573993042 | 0.03405609 | 0.045642856 | 1 |
| Obesity | endothelial | *C7* | -1.192381043 | 9.91E-07 | 5.66E-06 | 0.006229744 |
| Obesity | endothelial | *PAPD7* | -0.925331552 | 5.38E-07 | 3.46E-06 | 0.003379848 |
| Obesity | endothelial | *GHR* | -0.242966086 | 0.006944863 | 0.010770191 | 1 |
| Obesity | endothelial | *NNT* | -0.438507089 | 0.000747114 | 0.001473372 | 1 |
| Obesity | endothelial | *LOX* | -1.019072154 | 6.74E-08 | 7.12E-07 | 0.000423794 |
| Obesity | endothelial | *APBB3* | -0.874524593 | 2.67E-05 | 8.32E-05 | 0.167958931 |
| Obesity | endothelial | *TMCO6* | -1.553703798 | 3.57E-06 | 1.60E-05 | 0.022429697 |
| Obesity | endothelial | *SPARC* | -0.661980051 | 8.22E-07 | 4.86E-06 | 0.005166312 |
| Obesity | endothelial | *IK* | -0.987675403 | 2.37E-05 | 7.53E-05 | 0.149248833 |
| Obesity | endothelial | *PCDHB6* | 0.880334541 | 0.020427625 | 0.028711933 | 1 |
| Obesity | endothelial | *PDE8B* | -0.67454598 | 7.49E-10 | 5.95E-08 | 4.71E-06 |
| Obesity | endothelial | *CLK4* | -0.793135405 | 5.19E-05 | 0.000144327 | 0.326034128 |
| Obesity | endothelial | *HAVCR1* | -0.598656977 | 1.05E-06 | 5.94E-06 | 0.006597053 |
| Obesity | endothelial | *RNF130* | -0.700517667 | 9.70E-06 | 3.60E-05 | 0.061007705 |
| Obesity | endothelial | *ARSB* | -0.766571684 | 5.11E-06 | 2.14E-05 | 0.032148336 |
| Obesity | endothelial | *CLINT1* | -0.683468214 | 0.000162945 | 0.000386726 | 1 |
| Obesity | endothelial | *BTNL8* | 0.157282948 | 6.20E-05 | 0.000168584 | 0.389934255 |
| Obesity | endothelial | *TTC1* | -0.814520957 | 6.54E-05 | 0.00017586 | 0.411073565 |
| Obesity | endothelial | *MSH3* | -0.575497323 | 4.89E-05 | 0.000136908 | 0.307357478 |
| Obesity | endothelial | *RASGRF2* | 0.284245273 | 0.00038232 | 0.000814407 | 1 |
| Obesity | endothelial | *POLR3G* | -1.042741244 | 6.17E-07 | 3.87E-06 | 0.003876265 |
| Obesity | endothelial | *DROSHA* | -0.462550455 | 0.015158954 | 0.021853782 | 1 |
| Obesity | endothelial | *CDH6* | -0.196077187 | 0.005012738 | 0.008008916 | 1 |
| Obesity | endothelial | *SLC27A6* | 0.80907169 | 5.90E-05 | 0.000161912 | 0.370939811 |
| Obesity | endothelial | *TARS* | -1.077106413 | 1.98E-05 | 6.50E-05 | 0.124481372 |
| Obesity | endothelial | *LNPEP* | -0.567575601 | 0.000340971 | 0.000737422 | 1 |
| Obesity | endothelial | *IL4* | -1.070045922 | 0.007896544 | 0.012085096 | 1 |
| Obesity | endothelial | *GNPDA1* | -1.123619873 | 4.46E-06 | 1.91E-05 | 0.028033066 |
| Obesity | endothelial | *SKP1* | -0.597880588 | 0.00857768 | 0.013016624 | 1 |
| Obesity | endothelial | *TRIM23* | -0.498864787 | 0.031671706 | 0.042692971 | 1 |
| Obesity | endothelial | *TRAPPC13* | -0.708933152 | 0.000437229 | 0.000915676 | 1 |
| Obesity | endothelial | *C9* | -0.885839949 | 1.44E-06 | 7.60E-06 | 0.009062462 |
| Obesity | endothelial | *SMAD5* | -0.457840441 | 0.025263475 | 0.034709673 | 1 |
| Obesity | endothelial | *ATP6V0E1* | -0.825042721 | 3.72E-05 | 0.000109683 | 0.234064571 |
| Obesity | endothelial | *BNIP1* | -0.522910066 | 1.35E-05 | 4.71E-05 | 0.085021904 |
| Obesity | endothelial | *CNTN3* | -0.690186218 | 1.43E-08 | 3.34E-07 | 9.01E-05 |
| Obesity | endothelial | *ACTR8* | -1.41126389 | 2.46E-05 | 7.73E-05 | 0.154548161 |
| Obesity | endothelial | *HGD* | -2.108514301 | 2.66E-08 | 4.15E-07 | 0.000167019 |

| | | | | | | |
|---|---|---|---|---|---|---|
| Obesity | endothelial | *NIT2* | -0.951253011 | 7.80E-08 | 7.89E-07 | 0.000490141 |
| Obesity | endothelial | *FAM162A* | -0.764093134 | 0.002580578 | 0.004417064 | 1 |
| Obesity | endothelial | *PCCB* | -0.925836862 | 2.68E-06 | 1.26E-05 | 0.016847141 |
| Obesity | endothelial | *BCHE* | -0.488366512 | 0.016417628 | 0.023549538 | 1 |
| Obesity | endothelial | *WNT5A* | -1.084650401 | 1.31E-08 | 3.34E-07 | 8.25E-05 |
| Obesity | endothelial | *CYB561D2* | -0.669850638 | 0.00014199 | 0.000342289 | 0.892690556 |
| Obesity | endothelial | *IFT57* | -0.600146009 | 0.028134959 | 0.038344783 | 1 |
| Obesity | endothelial | *IQCG* | -0.916708135 | 4.27E-05 | 0.000122703 | 0.268474552 |
| Obesity | endothelial | *GBE1* | -0.61530163 | 1.68E-05 | 5.69E-05 | 0.105813225 |
| Obesity | endothelial | *UMPS* | -1.003766152 | 7.72E-05 | 0.000201624 | 0.485309759 |
| Obesity | endothelial | *NCBP2* | -0.899711467 | 0.000822171 | 0.001607773 | 1 |
| Obesity | endothelial | *SLC41A3* | 0.38898242 | 0.029404501 | 0.039863905 | 1 |
| Obesity | endothelial | *PLXNA1* | -0.541950347 | 0.000144993 | 0.000348727 | 0.911573554 |
| Obesity | endothelial | *ABTB1* | -1.122534586 | 4.94E-05 | 0.000138141 | 0.31067872 |
| Obesity | endothelial | *EFCC1* | 0.330389882 | 0.000836091 | 0.001632064 | 1 |
| Obesity | endothelial | *NEK11* | -0.852115341 | 1.84E-06 | 9.25E-06 | 0.011580955 |
| Obesity | endothelial | *HEMK1* | -0.649691602 | 0.000864146 | 0.001676816 | 1 |
| Obesity | endothelial | *COMMD2* | -0.634465838 | 0.006520754 | 0.010165133 | 1 |
| Obesity | endothelial | *ABCC5* | -0.864321769 | 2.36E-07 | 1.84E-06 | 0.001484743 |
| Obesity | endothelial | *ARHGEF26* | -0.632793785 | 7.99E-06 | 3.07E-05 | 0.050259565 |
| Obesity | endothelial | *KLHL24* | -0.40851135 | 0.011985464 | 0.017585207 | 1 |
| Obesity | endothelial | *PLCH1* | -1.081332386 | 5.70E-05 | 0.000157033 | 0.358192184 |
| Obesity | endothelial | *ZBTB47* | -0.945801239 | 0.000197664 | 0.00045806 | 1 |
| Obesity | endothelial | *CLCN2* | -1.911889896 | 9.88E-05 | 0.000250075 | 0.621185891 |
| Obesity | endothelial | *NEK4* | -0.96210148 | 0.000231844 | 0.000524318 | 1 |
| Obesity | endothelial | *MOB1A* | -0.817735851 | 0.000848131 | 0.00165057 | 1 |
| Obesity | endothelial | *LMAN2L* | -0.810091568 | 2.39E-05 | 7.57E-05 | 0.150506329 |
| Obesity | endothelial | *PIKFYVE* | -0.525797548 | 0.005652598 | 0.008933606 | 1 |
| Obesity | endothelial | *KCNIP3* | -1.327992091 | 5.70E-07 | 3.63E-06 | 0.003583335 |
| Obesity | endothelial | *NCL* | -0.652338841 | 0.002878848 | 0.004865409 | 1 |
| Obesity | endothelial | *SLC35F5* | -0.449373725 | 0.016465196 | 0.023602977 | 1 |
| Obesity | endothelial | *ACTR3* | -0.5271773 | 0.00639537 | 0.009982048 | 1 |
| Obesity | endothelial | *STEAP3* | 0.235419351 | 0.02375183 | 0.032819286 | 1 |
| Obesity | endothelial | *TFCP2L1* | -0.820243919 | 0.000509141 | 0.001047516 | 1 |
| Obesity | endothelial | *TP53I3* | -0.996151397 | 4.17E-05 | 0.00012035 | 0.26200146 |
| Obesity | endothelial | *DNAJC27* | -0.71052663 | 0.001156095 | 0.002161919 | 1 |
| Obesity | endothelial | *STAM2* | -0.720865421 | 0.00034509 | 0.000744538 | 1 |
| Obesity | endothelial | *GPD2* | -0.427267775 | 0.024352384 | 0.033538921 | 1 |
| Obesity | endothelial | *CENPA* | -1.568616767 | 5.27E-06 | 2.19E-05 | 0.033126764 |
| Obesity | endothelial | *TANC1* | 0.272637766 | 0.000439279 | 0.000919663 | 1 |
| Obesity | endothelial | *NRBP1* | -0.649249907 | 0.003298074 | 0.005492713 | 1 |
| Obesity | endothelial | *ITGB6* | -0.949232501 | 1.83E-08 | 3.45E-07 | 0.000114933 |
| Obesity | endothelial | *ITGA4* | -0.836566675 | 1.38E-06 | 7.34E-06 | 0.008673819 |
| Obesity | endothelial | *PPM1G* | -0.7292243 | 0.003142561 | 0.0052532 | 1 |
| Obesity | endothelial | *PCSK4* | -0.70476075 | 0.000197143 | 0.00045702 | 1 |
| Obesity | endothelial | *INO80B* | -1.112922757 | 5.59E-06 | 2.30E-05 | 0.035123521 |
| Obesity | endothelial | *MOGS* | -0.519869648 | 1.09E-05 | 3.97E-05 | 0.068785817 |
| Obesity | endothelial | *GRB14* | 0.324419749 | 2.65E-05 | 8.27E-05 | 0.166879615 |
| Obesity | endothelial | *CLIP4* | -0.598869395 | 2.63E-08 | 4.14E-07 | 0.000165079 |
| Obesity | endothelial | *RTN4* | -0.678911562 | 3.06E-07 | 2.22E-06 | 0.001923433 |

| | | | | | | |
|---|---|---|---|---|---|---|
| Obesity | endothelial | *HTRA2* | -1.262715434 | 0.000581781 | 0.001179508 | 1 |
| Obesity | endothelial | *GALNT3* | -0.668357672 | 0.001863522 | 0.003310912 | 1 |
| Obesity | endothelial | *POLE4* | 0.489516209 | 0.000942627 | 0.001808452 | 1 |
| Obesity | endothelial | *MRPL19* | -0.781195555 | 0.000116326 | 0.000287478 | 0.731343525 |
| Obesity | endothelial | *LANCL1* | -0.753116885 | 0.001072736 | 0.002020458 | 1 |
| Obesity | endothelial | *WDR75* | -0.49408729 | 0.03725647 | 0.0495623 | 1 |
| Obesity | endothelial | *EFEMP1* | -0.574036329 | 2.22E-07 | 1.75E-06 | 0.001393841 |
| Obesity | endothelial | *GNLY* | -0.547724351 | 0.027658601 | 0.037736464 | 1 |
| Obesity | endothelial | *SF3B1* | -0.637319401 | 0.000174959 | 0.000411496 | 1 |
| Obesity | endothelial | *ST3GAL5* | -0.789667232 | 1.05E-07 | 9.84E-07 | 0.000662205 |
| Obesity | endothelial | *PLCD4* | -1.733689637 | 2.64E-06 | 1.25E-05 | 0.01660703 |
| Obesity | endothelial | *CHMP3* | -1.074779912 | 1.68E-06 | 8.61E-06 | 0.010587181 |
| Obesity | endothelial | *ZNF142* | -0.550367593 | 0.022287985 | 0.03103534 | 1 |
| Obesity | endothelial | *WNT6* | -2.430713344 | 3.89E-05 | 0.000113539 | 0.244677462 |
| Obesity | endothelial | *MLPH* | 0.213584013 | 2.78E-06 | 1.30E-05 | 0.017493495 |
| Obesity | endothelial | *HDLBP* | -0.606075457 | 0.000233958 | 0.000528338 | 1 |
| Obesity | endothelial | *STK25* | -0.820436711 | 0.00111507 | 0.00209201 | 1 |
| Obesity | endothelial | *TPO* | 0.117511385 | 6.48E-06 | 2.58E-05 | 0.040745368 |
| Obesity | endothelial | *TAF1B* | -0.925965296 | 1.09E-06 | 6.10E-06 | 0.00685551 |
| Obesity | endothelial | *SLC25A12* | -0.632581459 | 0.000692687 | 0.001377269 | 1 |
| Obesity | endothelial | *RMDN2* | -1.00550041 | 1.08E-06 | 6.05E-06 | 0.006768769 |
| Obesity | endothelial | *SDC1* | 1.340222964 | 0.000198066 | 0.000458766 | 1 |
| Obesity | endothelial | *SOS1* | -0.459894743 | 0.001067934 | 0.002013156 | 1 |
| Obesity | endothelial | *WIPF1* | -0.679389984 | 7.44E-05 | 0.000195738 | 0.467813244 |
| Obesity | endothelial | *THADA* | -0.485152119 | 0.005826454 | 0.009169191 | 1 |
| Obesity | endothelial | *ARID3A* | -0.84776995 | 2.62E-08 | 4.14E-07 | 0.000164526 |
| Obesity | endothelial | *CD207* | 1.695866435 | 4.56E-07 | 3.04E-06 | 0.002868002 |
| Obesity | endothelial | *VAX2* | 0.800569662 | 0.000160094 | 0.000380967 | 1 |
| Obesity | endothelial | *MSH6* | -0.690756866 | 0.005680545 | 0.008971008 | 1 |
| Obesity | endothelial | *PLEKHA3* | -0.706412549 | 0.007602829 | 0.011692511 | 1 |
| Obesity | endothelial | *ALMS1* | -0.539430385 | 0.005785405 | 0.009116001 | 1 |
| Obesity | endothelial | *BCL9* | -1.022747359 | 5.68E-06 | 2.32E-05 | 0.035681516 |
| Obesity | endothelial | *DHCR24* | -1.247579865 | 7.68E-09 | 2.70E-07 | 4.83E-05 |
| Obesity | endothelial | *DNAJC16* | -1.001827704 | 1.26E-06 | 6.83E-06 | 0.007923835 |
| Obesity | endothelial | *MORN1* | -1.075605881 | 1.93E-06 | 9.67E-06 | 0.012152463 |
| Obesity | endothelial | *SCP2* | -0.713727826 | 2.62E-05 | 8.18E-05 | 0.164428599 |
| Obesity | endothelial | *RALGPS2* | -0.853549985 | 2.05E-08 | 3.63E-07 | 0.000129166 |
| Obesity | endothelial | *CEP104* | -0.707083186 | 0.000672264 | 0.001339686 | 1 |
| Obesity | endothelial | *FAM20B* | -0.70718818 | 0.003912835 | 0.006412928 | 1 |
| Obesity | endothelial | *TMEM59* | -0.561542657 | 0.00571329 | 0.009013664 | 1 |
| Obesity | endothelial | *WRAP73* | -1.270133123 | 0.000195749 | 0.00045429 | 1 |
| Obesity | endothelial | *MRPL37* | -1.149251597 | 1.09E-05 | 3.96E-05 | 0.068529272 |
| Obesity | endothelial | *ICMT* | -1.050331232 | 0.002020475 | 0.003555198 | 1 |
| Obesity | endothelial | *PARK7* | -1.011358452 | 7.27E-06 | 2.84E-05 | 0.045722265 |
| Obesity | endothelial | *KCNC4* | 0.347289099 | 0.001802509 | 0.003209179 | 1 |
| Obesity | endothelial | *EDEM3* | -0.835338688 | 1.69E-05 | 5.71E-05 | 0.10621942 |
| Obesity | endothelial | *RAP1A* | -0.519829137 | 0.003453416 | 0.005727149 | 1 |
| Obesity | endothelial | *CAPZA1* | -0.689518314 | 0.000148937 | 0.000357119 | 0.936364812 |
| Obesity | endothelial | *DOCK7* | -0.712065873 | 6.58E-08 | 7.02E-07 | 0.000413887 |
| Obesity | endothelial | *FBXO6* | -1.252152423 | 2.20E-05 | 7.07E-05 | 0.138440154 |

| | | | | | | |
|---|---|---|---|---|---|---|
| Obesity | endothelial | *C1orf21* | 0.30062762 | 0.005007459 | 0.008002515 | 1 |
| Obesity | endothelial | *DNAJC6* | 0.452876014 | 0.001957073 | 0.003453302 | 1 |
| Obesity | endothelial | *MFN2* | -0.978873967 | 0.001053269 | 0.00199095 | 1 |
| Obesity | endothelial | *MIIP* | -1.156696359 | 7.73E-08 | 7.84E-07 | 0.000486297 |
| Obesity | endothelial | *NCF2* | -1.075946107 | 5.56E-08 | 6.24E-07 | 0.000349792 |
| Obesity | endothelial | *RGS2* | -0.998646808 | 5.28E-09 | 2.21E-07 | 3.32E-05 |
| Obesity | endothelial | *OLFML3* | -0.870593719 | 1.27E-07 | 1.15E-06 | 0.000799429 |
| Obesity | endothelial | *TNNI3K* | -1.145057409 | 2.21E-06 | 1.09E-05 | 0.013924119 |
| Obesity | endothelial | *CRYZ* | -0.426011377 | 0.014666055 | 0.021177191 | 1 |
| Obesity | endothelial | *TTF2* | -1.090480669 | 1.22E-06 | 6.65E-06 | 0.007669714 |
| Obesity | endothelial | *TMEM9* | -0.742658888 | 7.72E-06 | 2.98E-05 | 0.048560408 |
| Obesity | endothelial | *WARS2* | -0.376088751 | 0.019759351 | 0.027872344 | 1 |
| Obesity | endothelial | *OSCP1* | -0.858986234 | 9.13E-09 | 3.02E-07 | 5.74E-05 |
| Obesity | endothelial | *C1orf109* | -0.968642706 | 0.000362614 | 0.000777542 | 1 |
| Obesity | endothelial | *TBCE* | -0.922846145 | 2.18E-05 | 7.03E-05 | 0.136969614 |
| Obesity | endothelial | *LGALS8* | -0.390391998 | 0.017046259 | 0.024345714 | 1 |
| Obesity | endothelial | *MTR* | -0.701026745 | 0.000111048 | 0.000276062 | 0.698160143 |
| Obesity | endothelial | *KCNQ4* | -0.643528007 | 3.22E-05 | 9.70E-05 | 0.202339216 |
| Obesity | endothelial | *ACADM* | -0.457326203 | 0.024007894 | 0.033114881 | 1 |
| Obesity | endothelial | *PADI2* | -1.379037961 | 3.97E-09 | 1.84E-07 | 2.50E-05 |
| Obesity | endothelial | *MFAP2* | -0.752656079 | 5.62E-06 | 2.30E-05 | 0.035330754 |
| Obesity | endothelial | *CTBS* | -0.652991265 | 0.003373894 | 0.005607103 | 1 |
| Obesity | endothelial | *KLHL12* | -0.924649555 | 0.00027381 | 0.000606141 | 1 |
| Obesity | endothelial | *ZNHIT6* | -0.397850342 | 0.02855146 | 0.038819859 | 1 |
| Obesity | endothelial | *PLA2G2D* | -5.415471861 | 2.55E-06 | 1.21E-05 | 0.016041436 |
| Obesity | endothelial | *RBBP5* | -0.7175777 | 0.000406078 | 0.000859022 | 1 |
| Obesity | endothelial | *KIF17* | 0.519634567 | 7.32E-05 | 0.000193104 | 0.459974133 |
| Obesity | endothelial | *CD46* | -0.487685858 | 0.004158806 | 0.006775206 | 1 |
| Obesity | endothelial | *FAAH* | -0.14053189 | 0.037551289 | 0.049913959 | 1 |
| Obesity | endothelial | *NSUN4* | -1.044780818 | 9.76E-06 | 3.62E-05 | 0.061347316 |
| Obesity | endothelial | *TMED5* | -0.85979606 | 3.42E-07 | 2.43E-06 | 0.00214815 |
| Obesity | endothelial | *DR1* | -0.826061543 | 0.001935871 | 0.003421653 | 1 |
| Obesity | endothelial | *ABCD3* | -0.624225088 | 0.003461333 | 0.005737253 | 1 |
| Obesity | endothelial | *TNFSF4* | -1.199029115 | 7.92E-11 | 1.04E-08 | 4.98E-07 |
| Obesity | endothelial | *IRF6* | 0.256070684 | 2.65E-07 | 2.00E-06 | 0.001668168 |
| Obesity | endothelial | *DIEXF* | -0.623832197 | 0.028400065 | 0.038664185 | 1 |
| Obesity | endothelial | *SERPINC1* | 0.63580687 | 0.000178718 | 0.000418201 | 1 |
| Obesity | endothelial | *RCOR3* | -0.657685067 | 0.000390311 | 0.000829296 | 1 |
| Obesity | endothelial | *MTFR1L* | -1.196586957 | 7.33E-07 | 4.44E-06 | 0.004610399 |
| Obesity | endothelial | *NSL1* | -0.527656491 | 0.011106514 | 0.016437537 | 1 |
| Obesity | endothelial | *RPA2* | -1.081464942 | 5.47E-06 | 2.26E-05 | 0.034389365 |
| Obesity | endothelial | *SLC5A9* | -1.638250424 | 1.71E-06 | 8.74E-06 | 0.010772136 |
| Obesity | endothelial | *OSBPL9* | -0.661329624 | 0.00011357 | 0.000281885 | 0.714015575 |
| Obesity | endothelial | *TXNDC12* | -0.80191695 | 0.000994374 | 0.001890422 | 1 |
| Obesity | endothelial | *ESYT2* | -0.578711776 | 0.000535788 | 0.001096516 | 1 |
| Obesity | endothelial | *CD3EAP* | -0.993207995 | 3.07E-05 | 9.34E-05 | 0.192866579 |
| Obesity | endothelial | *A4GNT* | -0.353197639 | 0.006219406 | 0.009733981 | 1 |
| Obesity | endothelial | *IFT46* | -0.921667037 | 0.000353892 | 0.000760655 | 1 |
| Obesity | endothelial | *ATF6* | -0.534742154 | 0.003994492 | 0.006533135 | 1 |
| Obesity | endothelial | *CASC1* | -0.970508586 | 8.33E-05 | 0.000215944 | 0.523857233 |

| | | | | | | |
|---|---|---|---|---|---|---|
| Obesity | endothelial | *LRMP* | 0.403692584 | 0.003668298 | 0.006038908 | 1 |
| Obesity | endothelial | *ATP10B* | -1.988246212 | 0.000463819 | 0.000964296 | 1 |
| Obesity | endothelial | *SPCS2* | -0.757740426 | 0.000928082 | 0.001784905 | 1 |
| Obesity | endothelial | *ELOVL4* | -0.25635818 | 0.037552573 | 0.049913959 | 1 |
| Obesity | endothelial | *FILIP1* | 0.28493225 | 0.00017218 | 0.000406189 | 1 |
| Obesity | endothelial | *AKAP7* | -0.421013182 | 0.00246845 | 0.004251821 | 1 |
| Obesity | endothelial | *TCF21* | -4.175084439 | 3.35E-05 | 0.00010066 | 0.210782374 |
| Obesity | endothelial | *MED28* | -0.824997425 | 0.000171341 | 0.000404363 | 1 |
| Obesity | endothelial | *SLC16A7* | -0.712170938 | 4.21E-07 | 2.86E-06 | 0.002646681 |
| Obesity | endothelial | *VAMP8* | -0.665048348 | 0.000298998 | 0.000655439 | 1 |
| Obesity | endothelial | *FOXO3* | -0.674904468 | 7.65E-06 | 2.96E-05 | 0.048122628 |
| Obesity | endothelial | *TGIF2* | -1.354948167 | 0.000490575 | 0.001012224 | 1 |
| Obesity | endothelial | *ABCG2* | 0.422198695 | 2.92E-05 | 9.00E-05 | 0.183769023 |
| Obesity | endothelial | *SPP1* | -0.876277605 | 5.00E-06 | 2.10E-05 | 0.031464346 |
| Obesity | endothelial | *FAM47E-STBL* | -0.566469967 | 2.64E-05 | 8.23E-05 | 0.165695683 |
| Obesity | endothelial | *MFSD1* | -0.86633116 | 2.54E-06 | 1.21E-05 | 0.015949868 |
| Obesity | endothelial | *RAB3GAP2* | -0.544423689 | 0.003740465 | 0.006151269 | 1 |
| Obesity | endothelial | *UBN1* | -0.475664103 | 0.017701763 | 0.025196058 | 1 |
| Obesity | endothelial | *UCHL3* | -1.422949951 | 2.70E-08 | 4.21E-07 | 0.000169817 |
| Obesity | endothelial | *WDR35* | -0.988392987 | 5.32E-07 | 3.44E-06 | 0.003345903 |
| Obesity | endothelial | *DNAH7* | -0.468328244 | 2.26E-05 | 7.23E-05 | 0.142219597 |
| Obesity | endothelial | *TRPM6* | 0.192171163 | 8.80E-06 | 3.33E-05 | 0.055321375 |
| Obesity | endothelial | *GDA* | -0.793525181 | 5.68E-07 | 3.62E-06 | 0.003572561 |
| Obesity | endothelial | *C2orf40* | 0.384415799 | 0.000195686 | 0.00045429 | 1 |
| Obesity | endothelial | *CPSF3* | -1.029807383 | 2.32E-05 | 7.39E-05 | 0.146003797 |
| Obesity | endothelial | *FKBP15* | -0.782804265 | 1.46E-05 | 5.03E-05 | 0.091612621 |
| Obesity | endothelial | *CTNNAL1* | 0.241001946 | 0.018522729 | 0.026251667 | 1 |
| Obesity | endothelial | *WDR34* | -0.850964811 | 0.000355479 | 0.000763806 | 1 |
| Obesity | endothelial | *CNTRL* | -0.597464342 | 0.002957704 | 0.004978604 | 1 |
| Obesity | endothelial | *TRIM32* | -1.074893293 | 2.94E-07 | 2.16E-06 | 0.001848627 |
| Obesity | endothelial | *PHF19* | -1.115571197 | 2.06E-09 | 1.25E-07 | 1.30E-05 |
| Obesity | endothelial | *BSPRY* | -3.836937201 | 1.03E-08 | 3.23E-07 | 6.45E-05 |
| Obesity | endothelial | *PPP6C* | -0.837817212 | 1.54E-05 | 5.27E-05 | 0.097079722 |
| Obesity | endothelial | *HDHD3* | -0.256363991 | 4.26E-05 | 0.000122483 | 0.267625686 |
| Obesity | endothelial | *HSDL2* | -0.514927993 | 3.75E-06 | 1.67E-05 | 0.023568104 |
| Obesity | endothelial | *INVS* | -0.38529385 | 0.023536031 | 0.032571214 | 1 |
| Obesity | endothelial | *KDSR* | -0.925569314 | 1.46E-05 | 5.04E-05 | 0.091805416 |
| Obesity | endothelial | *DCAF4* | 0.539469555 | 0.029408111 | 0.039863905 | 1 |
| Obesity | endothelial | *VSX2* | 0.28303635 | 0.002231757 | 0.003889951 | 1 |
| Obesity | endothelial | *IFI27L2* | -0.514992837 | 0.02567958 | 0.035237405 | 1 |
| Obesity | endothelial | *DNAL1* | -0.681336878 | 0.002689518 | 0.004579903 | 1 |
| Obesity | endothelial | *IRF2BPL* | -1.602630946 | 4.76E-08 | 5.62E-07 | 0.00029953 |
| Obesity | endothelial | *FLVCR2* | -1.13195634 | 4.97E-08 | 5.76E-07 | 0.000312759 |
| Obesity | endothelial | *PPP4R4* | -0.919438626 | 2.48E-07 | 1.90E-06 | 0.001558271 |
| Obesity | endothelial | *ALDH6A1* | -0.509365064 | 0.001511272 | 0.002747648 | 1 |
| Obesity | endothelial | *ESRRB* | -0.329108706 | 0.008088365 | 0.01233662 | 1 |
| Obesity | endothelial | *COQ6* | -1.022339859 | 0.000538376 | 0.001101096 | 1 |
| Obesity | endothelial | *GPR75* | -3.223158845 | 3.32E-05 | 9.98E-05 | 0.208871187 |
| Obesity | endothelial | *KLHL29* | -0.790637004 | 7.25E-07 | 4.41E-06 | 0.004555922 |
| Obesity | endothelial | *ATAD2B* | -0.553739373 | 0.001888017 | 0.00334271 | 1 |

| | | | | | | |
|---|---|---|---|---|---|---|
| Obesity | endothelial | *CNRIP1* | -0.487376301 | 0.025028266 | 0.034416603 | 1 |
| Obesity | endothelial | *CRIPT* | -1.272298865 | 3.14E-07 | 2.27E-06 | 0.001974523 |
| Obesity | endothelial | *SLC17A5* | -0.649996186 | 4.56E-05 | 0.000129287 | 0.286886892 |
| Obesity | endothelial | *OGFRL1* | -0.793291037 | 1.57E-05 | 5.36E-05 | 0.098752638 |
| Obesity | endothelial | *CUTC* | -0.859100001 | 3.38E-06 | 1.53E-05 | 0.021247907 |
| Obesity | endothelial | *MXI1* | -0.656180188 | 0.004779255 | 0.007682734 | 1 |
| Obesity | endothelial | *C10orf88* | -1.054509678 | 0.000131997 | 0.000321032 | 0.829868187 |
| Obesity | endothelial | *HELLS* | -0.870902509 | 0.000242525 | 0.000545725 | 1 |
| Obesity | endothelial | *TCTN3* | -0.919488717 | 7.80E-05 | 0.000203392 | 0.490378445 |
| Obesity | endothelial | *AVPI1* | -0.689689879 | 8.51E-07 | 4.99E-06 | 0.005351765 |
| Obesity | endothelial | *WDR11* | -0.725575501 | 5.11E-05 | 0.000142565 | 0.321483956 |
| Obesity | endothelial | *C10orf76* | -0.639895527 | 0.000246343 | 0.000553127 | 1 |
| Obesity | endothelial | *KCNIP2* | -0.328922898 | 0.000334874 | 0.000725983 | 1 |
| Obesity | endothelial | *KANSL1* | -0.471342989 | 0.00140107 | 0.002570331 | 1 |
| Obesity | endothelial | *HOXB3* | 0.463718683 | 0.002633436 | 0.004492623 | 1 |
| Obesity | endothelial | *CAAP1* | -0.704579311 | 0.000864717 | 0.00167689 | 1 |
| Obesity | endothelial | *CD274* | -1.345441777 | 1.17E-05 | 4.20E-05 | 0.073406645 |
| Obesity | endothelial | *NUP43* | -0.435374778 | 0.03101643 | 0.041890503 | 1 |
| Obesity | endothelial | *LRP11* | -0.867476092 | 4.83E-05 | 0.000135564 | 0.303933542 |
| Obesity | endothelial | *CCDC170* | -0.430084116 | 5.33E-07 | 3.44E-06 | 0.003351759 |
| Obesity | endothelial | *WDR55* | -1.260342655 | 2.06E-05 | 6.71E-05 | 0.12920119 |
| Obesity | endothelial | *PCDHB14* | 0.688399098 | 0.000209818 | 0.000482666 | 1 |
| Obesity | endothelial | *SEC16B* | 0.681286728 | 0.022945035 | 0.031802345 | 1 |
| Obesity | endothelial | *GORAB* | -0.802335011 | 0.003957698 | 0.006476326 | 1 |
| Obesity | endothelial | *ACAT2* | -1.102171389 | 4.88E-07 | 3.21E-06 | 0.00306771 |
| Obesity | endothelial | *SNX19* | -1.009713633 | 1.22E-07 | 1.11E-06 | 0.000766603 |
| Obesity | endothelial | *MASTL* | -0.616703729 | 4.26E-08 | 5.32E-07 | 0.000267968 |
| Obesity | endothelial | *MRC1* | -0.694049185 | 4.93E-08 | 5.72E-07 | 0.000310251 |
| Obesity | endothelial | *CCDC77* | -1.026806335 | 1.21E-06 | 6.62E-06 | 0.007618946 |
| Obesity | endothelial | *ENOX1* | -0.776684288 | 5.63E-08 | 6.28E-07 | 0.000353948 |
| Obesity | endothelial | *DNAJC15* | -0.481337442 | 0.034980644 | 0.046762345 | 1 |
| Obesity | endothelial | *WBP4* | -0.825623015 | 3.64E-05 | 0.000107699 | 0.228861366 |
| Obesity | endothelial | *SMAD9* | 0.298069628 | 1.72E-05 | 5.80E-05 | 0.108383222 |
| Obesity | endothelial | *SIL1* | -0.785456722 | 2.79E-06 | 1.30E-05 | 0.017517284 |
| Obesity | endothelial | *ARL1* | -0.818186663 | 8.35E-05 | 0.000216407 | 0.52521987 |
| Obesity | endothelial | *APAF1* | -0.575408969 | 0.007550968 | 0.011618437 | 1 |
| Obesity | endothelial | *EPHX2* | -0.420626632 | 0.020737959 | 0.029096083 | 1 |
| Obesity | endothelial | *ZNF706* | -0.688379886 | 0.000307774 | 0.000672802 | 1 |
| Obesity | endothelial | *CRISPLD1* | 0.670148573 | 0.010880367 | 0.016141354 | 1 |
| Obesity | endothelial | *RDH10* | -0.471500282 | 0.000158724 | 0.000378421 | 0.997895451 |
| Obesity | endothelial | *SCPEP1* | -0.225570489 | 0.012652073 | 0.018511423 | 1 |
| Obesity | endothelial | *TBX4* | -0.811656478 | 0.012701671 | 0.018562391 | 1 |
| Obesity | endothelial | *TRIM6* | -0.605550907 | 0.000400721 | 0.000849405 | 1 |
| Obesity | endothelial | *ECHDC2* | -0.469122646 | 2.03E-07 | 1.64E-06 | 0.001274172 |
| Obesity | endothelial | *PRB2* | -0.887893608 | 0.022701041 | 0.031512794 | 1 |
| Obesity | endothelial | *A1BG* | -1.486195368 | 5.99E-05 | 0.000163729 | 0.376413498 |
| Obesity | endothelial | *PDZRN3* | -0.639944104 | 1.37E-06 | 7.30E-06 | 0.008596269 |
| Obesity | endothelial | *LHX4* | -1.389084051 | 2.09E-05 | 6.78E-05 | 0.131279929 |
| Obesity | endothelial | *SEC22A* | -0.731181494 | 1.35E-05 | 4.69E-05 | 0.084635139 |
| Obesity | endothelial | *CSTA* | -0.476234575 | 1.52E-05 | 5.20E-05 | 0.09556417 |

| | | | | | | |
|---|---|---|---|---|---|---|
| Obesity | endothelial | *POPDC2* | 0.750357685 | 0.001305837 | 0.002410392 | 1 |
| Obesity | endothelial | *B4GALT4* | -1.246172932 | 2.64E-06 | 1.25E-05 | 0.016624964 |
| Obesity | endothelial | *CD80* | -1.860979957 | 3.69E-08 | 4.89E-07 | 0.000232088 |
| Obesity | endothelial | *CAT* | -0.274277103 | 0.004254521 | 0.006913459 | 1 |
| Obesity | endothelial | *TMEM39B* | -0.729226272 | 0.012646822 | 0.018508609 | 1 |
| Obesity | endothelial | *POLR3GL* | -1.534940528 | 8.22E-07 | 4.86E-06 | 0.005167198 |
| Obesity | endothelial | *TNFSF10* | 0.324710588 | 0.000331818 | 0.000719657 | 1 |
| Obesity | endothelial | *ZNF639* | -0.919406708 | 0.001169533 | 0.002185748 | 1 |
| Obesity | endothelial | *LIAS* | -0.935151568 | 0.000101635 | 0.000255898 | 0.638976512 |
| Obesity | endothelial | *CLCC1* | -1.055224224 | 2.63E-05 | 8.22E-05 | 0.165348941 |
| Obesity | endothelial | *CXCR4* | -1.704949822 | 1.41E-06 | 7.49E-06 | 0.008892334 |
| Obesity | endothelial | *ACVR2A* | -0.693235821 | 0.000217047 | 0.000495668 | 1 |
| Obesity | endothelial | *POLK* | -0.457417669 | 0.012905808 | 0.018805539 | 1 |
| Obesity | endothelial | *FLT3* | -1.237496205 | 1.19E-08 | 3.34E-07 | 7.50E-05 |
| Obesity | endothelial | *MTIF3* | -0.526785523 | 0.009263224 | 0.013982687 | 1 |
| Obesity | endothelial | *RASL11A* | -2.512117898 | 5.09E-09 | 2.18E-07 | 3.20E-05 |
| Obesity | endothelial | *FMOD* | -1.001494576 | 3.61E-08 | 4.82E-07 | 0.000227082 |
| Obesity | endothelial | *SERAC1* | -1.203524806 | 1.10E-05 | 3.99E-05 | 0.069097499 |
| Obesity | endothelial | *LDB3* | -0.512755893 | 1.88E-05 | 6.22E-05 | 0.118377857 |
| Obesity | endothelial | *OPN4* | -0.993678604 | 0.001513365 | 0.002749862 | 1 |
| Obesity | endothelial | *FAM35A* | -0.594732767 | 0.000524394 | 0.001074947 | 1 |
| Obesity | endothelial | *NAA60* | -1.617019234 | 1.63E-05 | 5.55E-05 | 0.102665549 |
| Obesity | endothelial | *RWDD3* | -2.059415415 | 2.82E-07 | 2.10E-06 | 0.001771762 |
| Obesity | endothelial | *CCDC18* | -0.743009678 | 1.58E-07 | 1.34E-06 | 0.000990739 |
| Obesity | endothelial | *RPAP2* | -0.603628291 | 0.001326089 | 0.002444188 | 1 |
| Obesity | endothelial | *BBS9* | -0.51603288 | 0.002633609 | 0.004492623 | 1 |
| Obesity | endothelial | *KIAA0087* | 0.842886088 | 0.000159241 | 0.000379246 | 1 |
| Obesity | endothelial | *KLHL7* | -0.736106592 | 0.0008481 | 0.00165057 | 1 |
| Obesity | endothelial | *HERPUD2* | -0.643364898 | 0.003886775 | 0.006376868 | 1 |
| Obesity | endothelial | *CBX3* | -0.754742899 | 0.001011274 | 0.001917913 | 1 |
| Obesity | endothelial | *FAM126A* | -0.692896729 | 1.70E-05 | 5.73E-05 | 0.106824379 |
| Obesity | endothelial | *POLM* | -0.219442255 | 0.01047267 | 0.015628217 | 1 |
| Obesity | endothelial | *SMU1* | -0.690925668 | 0.003457258 | 0.00573201 | 1 |
| Obesity | endothelial | *GLIPR2* | -0.823278116 | 1.16E-08 | 3.34E-07 | 7.28E-05 |
| Obesity | endothelial | *DCAF10* | -0.618191832 | 0.000683011 | 0.00135846 | 1 |
| Obesity | endothelial | *BICC1* | -0.387627135 | 1.06E-05 | 3.86E-05 | 0.066423448 |
| Obesity | endothelial | *CISD1* | -0.404257289 | 0.016427677 | 0.023558577 | 1 |
| Obesity | endothelial | *SLC25A16* | -0.372286387 | 0.001891922 | 0.003347738 | 1 |
| Obesity | endothelial | *ZWINT* | -0.406573285 | 0.006603025 | 0.010285732 | 1 |
| Obesity | endothelial | *RBM19* | -0.876825676 | 0.000268115 | 0.000595632 | 1 |
| Obesity | endothelial | *ACADS* | -0.98959737 | 1.74E-08 | 3.42E-07 | 0.000109559 |
| Obesity | endothelial | *SSPN* | -0.587576525 | 3.20E-05 | 9.65E-05 | 0.201096992 |
| Obesity | endothelial | *CCDC91* | -0.611864662 | 3.93E-05 | 0.000114255 | 0.246791868 |
| Obesity | endothelial | *EBPL* | 0.370892633 | 0.007020402 | 0.010873926 | 1 |
| Obesity | endothelial | *ATP7B* | 0.211632473 | 0.001521342 | 0.002761165 | 1 |
| Obesity | endothelial | *NLN* | -0.579897433 | 0.000837007 | 0.001633228 | 1 |
| Obesity | endothelial | *OPTN* | -0.856066169 | 6.64E-07 | 4.09E-06 | 0.004172761 |
| Obesity | endothelial | *ATF1* | -0.83931776 | 0.000100246 | 0.00025301 | 0.630247249 |
| Obesity | endothelial | *TSFM* | -0.928523113 | 0.000653593 | 0.001307394 | 1 |
| Obesity | endothelial | *ARHGAP9* | -1.404856639 | 1.19E-10 | 1.41E-08 | 7.46E-07 |

| | | | | | | |
|---|---|---|---|---|---|---|
| Obesity | endothelial | *SPATS2* | -0.425002137 | 0.034537125 | 0.046257969 | 1 |
| Obesity | endothelial | *PDE1B* | -0.724251808 | 5.46E-08 | 6.14E-07 | 0.000342975 |
| Obesity | endothelial | *IKZF4* | -1.528981353 | 1.47E-07 | 1.27E-06 | 0.000922761 |
| Obesity | endothelial | *SARDH* | -1.642445344 | 2.04E-08 | 3.63E-07 | 0.000127986 |
| Obesity | endothelial | *DBH* | -1.844244301 | 0.001579376 | 0.002850858 | 1 |
| Obesity | endothelial | *STIL* | -1.186773441 | 0.000732153 | 0.001447955 | 1 |
| Obesity | endothelial | *TTC21B* | -0.671082017 | 0.000636839 | 0.001275911 | 1 |
| Obesity | endothelial | *NMI* | -0.876768738 | 0.0007075 | 0.001404057 | 1 |
| Obesity | endothelial | *ACVR1C* | -0.60884633 | 3.21E-09 | 1.69E-07 | 2.02E-05 |
| Obesity | endothelial | *SLC36A1* | -0.294260075 | 0.007127101 | 0.011028325 | 1 |
| Obesity | endothelial | *PLA2G12A* | -0.676505573 | 0.006016645 | 0.009442498 | 1 |
| Obesity | endothelial | *C4BPB* | -0.527688339 | 0.000137682 | 0.000332541 | 0.865604757 |
| Obesity | endothelial | *RAB38* | -1.76301874 | 5.71E-09 | 2.33E-07 | 3.59E-05 |
| Obesity | endothelial | *AGO2* | -0.737476901 | 0.000451442 | 0.000941366 | 1 |
| Obesity | endothelial | *CHPF* | -0.631438629 | 9.18E-07 | 5.31E-06 | 0.005770575 |
| Obesity | endothelial | *DNPEP* | -0.641208501 | 0.027219441 | 0.037193789 | 1 |
| Obesity | endothelial | *MOGAT1* | -0.672400642 | 1.74E-08 | 3.42E-07 | 0.000109149 |
| Obesity | endothelial | *FAM124B* | 0.238854245 | 2.26E-06 | 1.10E-05 | 0.014192248 |
| Obesity | endothelial | *FAM210B* | -0.856890017 | 7.54E-06 | 2.92E-05 | 0.047384506 |
| Obesity | endothelial | *SNX21* | -0.742816622 | 1.83E-08 | 3.45E-07 | 0.000115088 |
| Obesity | endothelial | *WFDC3* | -0.193661541 | 0.002892063 | 0.004882875 | 1 |
| Obesity | endothelial | *SDC4* | -1.228657162 | 1.10E-08 | 3.34E-07 | 6.92E-05 |
| Obesity | endothelial | *TOX2* | 0.150829889 | 0.001062839 | 0.00200422 | 1 |
| Obesity | endothelial | *CSE1L* | -0.562679041 | 0.014600181 | 0.021096607 | 1 |
| Obesity | endothelial | *CDH26* | -1.383337528 | 1.50E-06 | 7.89E-06 | 0.009457311 |
| Obesity | endothelial | *STX16* | -0.911753859 | 8.34E-06 | 3.18E-05 | 0.052427116 |
| Obesity | endothelial | *RNF114* | -0.886039834 | 2.49E-05 | 7.82E-05 | 0.156643698 |
| Obesity | endothelial | *KCNK15* | -1.866823128 | 3.32E-07 | 2.37E-06 | 0.002087315 |
| Obesity | endothelial | *FASTKD3* | -0.773682614 | 0.032508024 | 0.043735918 | 1 |
| Obesity | endothelial | *PEPD* | -0.652100799 | 2.24E-05 | 7.18E-05 | 0.140918204 |
| Obesity | endothelial | *CHST8* | -0.743224455 | 6.46E-07 | 4.02E-06 | 0.004059693 |
| Obesity | endothelial | *STAMBP* | -0.871165689 | 9.84E-05 | 0.000249141 | 0.618367852 |
| Obesity | endothelial | *ATP8A1* | 0.268828869 | 1.20E-05 | 4.29E-05 | 0.075658398 |
| Obesity | endothelial | *USP22* | -0.74211961 | 0.000340009 | 0.000735594 | 1 |
| Obesity | endothelial | *HIF3A* | 0.245228881 | 2.28E-06 | 1.11E-05 | 0.014335661 |
| Obesity | endothelial | *ZNF45* | -0.834490049 | 0.006162318 | 0.009649438 | 1 |
| Obesity | endothelial | *MRS2* | -0.69142967 | 0.000373665 | 0.000798788 | 1 |
| Obesity | endothelial | *SNRPC* | -1.904206709 | 7.65E-26 | 1.30E-22 | 4.81E-22 |
| Obesity | endothelial | *PEX6* | -0.269458522 | 0.011864951 | 0.017453193 | 1 |
| Obesity | endothelial | *NQO2* | -0.92304896 | 2.89E-07 | 2.14E-06 | 0.00181567 |
| Obesity | endothelial | *ZNF391* | -0.153945946 | 0.034532 | 0.046257969 | 1 |
| Obesity | endothelial | *RPS10* | -2.581252068 | 0.000187373 | 0.000436464 | 1 |
| Obesity | endothelial | *HIST1H2BJ* | 1.612763213 | 2.98E-10 | 2.93E-08 | 1.87E-06 |
| Obesity | endothelial | *MED20* | -0.60448979 | 0.011678275 | 0.017202744 | 1 |
| Obesity | endothelial | *TBCC* | -0.902475574 | 0.000325149 | 0.000706607 | 1 |
| Obesity | endothelial | *KLHDC3* | -0.991191467 | 0.000175517 | 0.000412052 | 1 |
| Obesity | endothelial | *KLHL31* | -0.264458138 | 0.001797729 | 0.003203606 | 1 |
| Obesity | endothelial | *COL21A1* | 0.247443007 | 5.31E-05 | 0.000146958 | 0.333574242 |
| Obesity | endothelial | *GLO1* | -0.620689884 | 0.01140891 | 0.016833565 | 1 |
| Obesity | endothelial | *CPNE5* | 0.277551482 | 4.71E-05 | 0.000132928 | 0.296295953 |

| | | | | | | |
|---|---|---|---|---|---|---|
| Obesity | endothelial | *RIOK1* | -0.541218463 | 0.022872245 | 0.031708447 | 1 |
| Obesity | endothelial | *RPP40* | -0.637454839 | 0.010663389 | 0.015882665 | 1 |
| Obesity | endothelial | *NUP153* | -0.622042383 | 0.001140745 | 0.002137027 | 1 |
| Obesity | endothelial | *EEF1E1* | -1.151183294 | 0.016169955 | 0.02322607 | 1 |
| Obesity | endothelial | *EREG* | -2.694151467 | 1.77E-08 | 3.43E-07 | 0.000111457 |
| Obesity | endothelial | *MYRF* | -0.619126571 | 0.003900756 | 0.006396467 | 1 |
| Obesity | endothelial | *CNOT1* | -0.535753584 | 0.004931737 | 0.007899574 | 1 |
| Obesity | endothelial | *MT1G* | -6.721665588 | 3.45E-07 | 2.44E-06 | 0.002168763 |
| Obesity | endothelial | *CLYBL* | -0.654299992 | 0.002551617 | 0.004377085 | 1 |
| Obesity | endothelial | *KIF25* | 0.504577635 | 2.42E-06 | 1.16E-05 | 0.015188611 |
| Obesity | endothelial | *GRK4* | 0.234628116 | 0.002162692 | 0.003786367 | 1 |
| Obesity | endothelial | *MRPS7* | -1.126239106 | 3.69E-05 | 0.000108772 | 0.231901188 |
| Obesity | endothelial | *GGA3* | -1.02818359 | 7.89E-06 | 3.04E-05 | 0.049628423 |
| Obesity | endothelial | *NUP85* | -0.645072748 | 0.005809419 | 0.009149252 | 1 |
| Obesity | endothelial | *MIF4GD* | -1.65353241 | 9.55E-05 | 0.000242655 | 0.600669711 |
| Obesity | endothelial | *NT5C* | -0.87778263 | 7.79E-09 | 2.71E-07 | 4.90E-05 |
| Obesity | endothelial | *DDX31* | -0.890396224 | 2.02E-05 | 6.61E-05 | 0.12691677 |
| Obesity | endothelial | *PPP1R12C* | -1.004890179 | 1.07E-05 | 3.92E-05 | 0.067559743 |
| Obesity | endothelial | *PPDPF* | -1.056238295 | 0.000833426 | 0.001628262 | 1 |
| Obesity | endothelial | *PLGLB2* | -3.876220274 | 0.005733058 | 0.009042582 | 1 |
| Obesity | endothelial | *CHCHD5* | -1.142692259 | 0.000667601 | 0.001332024 | 1 |
| Obesity | endothelial | *PAX8* | -0.676925668 | 1.17E-05 | 4.20E-05 | 0.073542575 |
| Obesity | endothelial | *INSIG2* | -1.004519631 | 0.000158595 | 0.000378258 | 0.997087474 |
| Obesity | endothelial | *PSD4* | -1.141059178 | 3.04E-07 | 2.21E-06 | 0.001913022 |
| Obesity | endothelial | *SLC25A23* | -0.588913275 | 7.69E-05 | 0.000201001 | 0.483418503 |
| Obesity | endothelial | *CLPP* | -0.74003564 | 0.007216806 | 0.011145187 | 1 |
| Obesity | endothelial | *SH2D3A* | -1.930989046 | 3.72E-07 | 2.60E-06 | 0.002339939 |
| Obesity | endothelial | *TRIP10* | -0.883676994 | 5.60E-06 | 2.30E-05 | 0.035237113 |
| Obesity | endothelial | *SNRPD2* | -1.28041034 | 4.65E-05 | 0.00013144 | 0.292453572 |
| Obesity | endothelial | *RTN2* | -1.588912589 | 0.000126971 | 0.000310367 | 0.798263954 |
| Obesity | endothelial | *SYMPK* | -0.805094456 | 0.000106267 | 0.00026607 | 0.668102006 |
| Obesity | endothelial | *GPCPD1* | -0.66346645 | 0.000269006 | 0.000597114 | 1 |
| Obesity | endothelial | *SDCBP2* | 0.728107244 | 4.04E-05 | 0.000117131 | 0.254056768 |
| Obesity | endothelial | *PANK2* | -0.792878077 | 0.000243513 | 0.000547359 | 1 |
| Obesity | endothelial | *RRBP1* | -0.438359989 | 0.022383946 | 0.031127597 | 1 |
| Obesity | endothelial | *BMP2* | -1.872473451 | 1.64E-07 | 1.39E-06 | 0.001031955 |
| Obesity | endothelial | *MKKS* | -0.808896862 | 0.000728238 | 0.001442026 | 1 |
| Obesity | endothelial | *DSTN* | -0.612917744 | 0.001916231 | 0.003389799 | 1 |
| Obesity | endothelial | *MGME1* | -0.866938225 | 0.000108482 | 0.000270753 | 0.682027593 |
| Obesity | endothelial | *ITPA* | -1.028732697 | 8.06E-08 | 8.06E-07 | 0.000506482 |
| Obesity | endothelial | *TMEM74B* | 1.122136604 | 5.60E-05 | 0.000154734 | 0.352329784 |
| Obesity | endothelial | *FAM110A* | -1.477787019 | 9.91E-09 | 3.16E-07 | 6.23E-05 |
| Obesity | endothelial | *MRPS26* | -0.340484588 | 0.001305236 | 0.002409991 | 1 |
| Obesity | endothelial | *NCLN* | -0.60078608 | 0.024117432 | 0.033236803 | 1 |
| Obesity | endothelial | *ZNF436* | -0.719246513 | 0.000198496 | 0.000459479 | 1 |
| Obesity | endothelial | *MMP24* | -0.696053325 | 0.000601527 | 0.001213672 | 1 |
| Obesity | endothelial | *UROD* | -0.948798983 | 0.000369592 | 0.000790618 | 1 |
| Obesity | endothelial | *ST3GAL3* | -0.571498321 | 0.001667837 | 0.002992491 | 1 |
| Obesity | endothelial | *XRCC3* | 0.764445942 | 0.00021506 | 0.000492023 | 1 |
| Obesity | endothelial | *MCF2L* | 0.196381655 | 4.03E-06 | 1.77E-05 | 0.025329889 |

| | | | | | | |
|---|---|---|---|---|---|---|
| Obesity | endothelial | *F10* | -0.811506245 | 1.48E-08 | 3.34E-07 | 9.30E-05 |
| Obesity | endothelial | *PCID2* | -0.843443305 | 6.82E-07 | 4.18E-06 | 0.004289473 |
| Obesity | endothelial | *PROZ* | -3.539012283 | 1.25E-07 | 1.13E-06 | 0.000787139 |
| Obesity | endothelial | *IGFLR1* | -2.464821099 | 1.21E-08 | 3.34E-07 | 7.59E-05 |
| Obesity | endothelial | *COX6B1* | -1.062778993 | 7.12E-05 | 0.000189351 | 0.447624784 |
| Obesity | endothelial | *PRDX5* | -0.862241832 | 0.007032763 | 0.01089039 | 1 |
| Obesity | endothelial | *SCAF1* | -1.472512865 | 2.54E-06 | 1.21E-05 | 0.015962373 |
| Obesity | endothelial | *PRR12* | -0.510853335 | 0.006972033 | 0.010806995 | 1 |
| Obesity | endothelial | *TSKS* | -0.887782968 | 2.33E-08 | 3.88E-07 | 0.000146797 |
| Obesity | endothelial | *SBDS* | -1.002781698 | 7.67E-05 | 0.000200468 | 0.481925656 |
| Obesity | endothelial | *BECN1* | -0.919113517 | 0.000553837 | 0.001126852 | 1 |
| Obesity | endothelial | *DNAJC8* | -0.87245451 | 0.000172928 | 0.000407649 | 1 |
| Obesity | endothelial | *ATG14* | -0.836712012 | 6.03E-06 | 2.43E-05 | 0.037940363 |
| Obesity | endothelial | *RHOJ* | 0.293879806 | 6.22E-05 | 0.00016895 | 0.391120094 |
| Obesity | endothelial | *L3HYPDH* | -0.424564011 | 0.000403249 | 0.0008539 | 1 |
| Obesity | endothelial | *HSPA2* | 0.88066603 | 0.000280079 | 0.000618279 | 1 |
| Obesity | endothelial | *ZBTB1* | -0.727854833 | 1.30E-05 | 4.57E-05 | 0.082029683 |
| Obesity | endothelial | *RHOT1* | -0.442809578 | 0.00848407 | 0.012893243 | 1 |
| Obesity | endothelial | *WDR60* | -0.461612075 | 0.004703131 | 0.007571981 | 1 |
| Obesity | endothelial | *AIF1L* | 0.311756171 | 1.13E-05 | 4.10E-05 | 0.071213989 |
| Obesity | endothelial | *MAP2K2* | -1.02943217 | 1.82E-05 | 6.07E-05 | 0.114526339 |
| Obesity | endothelial | *CANX* | -0.483688644 | 0.02371117 | 0.03278472 | 1 |
| Obesity | endothelial | *IPPK* | -0.909570966 | 0.000515712 | 0.001058877 | 1 |
| Obesity | endothelial | *FGD3* | -1.150760304 | 2.16E-07 | 1.71E-06 | 0.001355493 |
| Obesity | endothelial | *ABHD8* | -0.620475314 | 0.003279143 | 0.005465528 | 1 |
| Obesity | endothelial | *MASP1* | -0.66926906 | 2.05E-07 | 1.65E-06 | 0.001291314 |
| Obesity | endothelial | *ATP13A4* | -0.887767585 | 5.42E-07 | 3.49E-06 | 0.003410117 |
| Obesity | endothelial | *HRASLS* | -0.735572589 | 1.21E-06 | 6.60E-06 | 0.007585898 |
| Obesity | endothelial | *HELB* | -0.668080137 | 0.006263793 | 0.009791264 | 1 |
| Obesity | endothelial | *RAP1B* | -0.449604857 | 0.006654549 | 0.010353167 | 1 |
| Obesity | endothelial | *RAB3IP* | -1.043926686 | 3.99E-05 | 0.000115707 | 0.250620879 |
| Obesity | endothelial | *LRRC61* | -0.490135583 | 2.53E-07 | 1.92E-06 | 0.001591679 |
| Obesity | endothelial | *IDUA* | -1.495782796 | 1.75E-06 | 8.87E-06 | 0.010976613 |
| Obesity | endothelial | *TMEM175* | -0.910668181 | 1.64E-05 | 5.56E-05 | 0.103069863 |
| Obesity | endothelial | *EMC1* | -0.64726727 | 0.006025257 | 0.009453653 | 1 |
| Obesity | endothelial | *PLA2G5* | -0.569441622 | 0.000587118 | 0.001188795 | 1 |
| Obesity | endothelial | *UBR4* | -0.499525795 | 0.000953893 | 0.001826165 | 1 |
| Obesity | endothelial | *SIN3B* | -0.567833879 | 0.014258153 | 0.020645097 | 1 |
| Obesity | endothelial | *WFIKKN1* | -3.293501958 | 4.68E-06 | 1.99E-05 | 0.029432037 |
| Obesity | endothelial | *WDR24* | -2.726299793 | 1.27E-08 | 3.34E-07 | 8.00E-05 |
| Obesity | endothelial | *CHTF18* | -1.00067113 | 0.008079436 | 0.012334401 | 1 |
| Obesity | endothelial | *TICAM1* | -1.324242966 | 6.08E-05 | 0.000165895 | 0.382387705 |
| Obesity | endothelial | *AAMP* | -1.086679257 | 6.07E-05 | 0.000165686 | 0.38174048 |
| Obesity | endothelial | *PNKD* | -0.952248515 | 2.89E-05 | 8.92E-05 | 0.181801444 |
| Obesity | endothelial | *ZNF835* | -0.741534809 | 0.00020231 | 0.000467446 | 1 |
| Obesity | endothelial | *POR* | -0.891724886 | 1.38E-06 | 7.36E-06 | 0.008695453 |
| Obesity | endothelial | *FGL2* | -0.653855683 | 0.000280651 | 0.000619324 | 1 |
| Obesity | endothelial | *STEAP4* | -1.035201562 | 2.69E-07 | 2.03E-06 | 0.001694302 |
| Obesity | endothelial | *SGCE* | -0.932120379 | 2.55E-06 | 1.21E-05 | 0.016032489 |
| Obesity | endothelial | *RBM48* | -1.188025513 | 5.46E-07 | 3.51E-06 | 0.003433719 |

| | | | | | | |
|---|---|---|---|---|---|---|
| Obesity | endothelial | *SRD5A3* | -0.835597997 | 0.001545712 | 0.002798932 | 1 |
| Obesity | endothelial | *VPREB3* | -5.505097021 | 3.40E-08 | 4.64E-07 | 0.000213608 |
| Obesity | endothelial | *ATF4* | -1.085969325 | 1.77E-06 | 8.96E-06 | 0.011106129 |
| Obesity | endothelial | *BAIAP2L2* | -0.809069024 | 1.41E-07 | 1.23E-06 | 0.00088442 |
| Obesity | endothelial | *TST* | -0.572156948 | 0.000159093 | 0.000379157 | 1 |
| Obesity | endothelial | *APOL5* | -1.927580179 | 1.14E-05 | 4.13E-05 | 0.071843759 |
| Obesity | endothelial | *RIBC2* | 0.435593456 | 0.022357883 | 0.031111998 | 1 |
| Obesity | endothelial | *POT1* | -0.56597749 | 0.007209085 | 0.011135999 | 1 |
| Obesity | endothelial | *CDHR3* | -0.65564512 | 1.59E-06 | 8.22E-06 | 0.009984104 |
| Obesity | endothelial | *PRKRIP1* | -0.702135465 | 9.27E-05 | 0.000236123 | 0.582986576 |
| Obesity | endothelial | *FOXP2* | -0.618360919 | 2.65E-08 | 4.15E-07 | 0.000166814 |
| Obesity | endothelial | *STRIP2* | -0.56552277 | 0.004197859 | 0.006830212 | 1 |
| Obesity | endothelial | *IRF5* | -0.418363665 | 0.00021846 | 0.000498171 | 1 |
| Obesity | endothelial | *KLHDC10* | -0.617489086 | 0.000597896 | 0.00120864 | 1 |
| Obesity | endothelial | *OPN1SW* | -0.449823307 | 0.002189924 | 0.00382826 | 1 |
| Obesity | endothelial | *HOXD1* | 0.600837758 | 6.60E-07 | 4.09E-06 | 0.004146554 |
| Obesity | endothelial | *HOXD3* | 0.363431114 | 8.38E-05 | 0.000216881 | 0.526804939 |
| Obesity | endothelial | *PDE11A* | -0.503609847 | 0.0004261 | 0.000895949 | 1 |
| Obesity | endothelial | *CHN1* | 0.214352286 | 1.55E-05 | 5.27E-05 | 0.097149286 |
| Obesity | endothelial | *GAD1* | -1.510654607 | 3.06E-08 | 4.48E-07 | 0.00019218 |
| Obesity | endothelial | *ORMDL1* | -0.400607685 | 0.003465882 | 0.005743279 | 1 |
| Obesity | endothelial | *PSMG2* | -0.692981329 | 0.000136736 | 0.000330639 | 0.859660103 |
| Obesity | endothelial | *TWSG1* | -1.067912348 | 5.35E-07 | 3.45E-06 | 0.003365535 |
| Obesity | endothelial | *WDFY4* | -0.785144115 | 1.99E-09 | 1.23E-07 | 1.25E-05 |
| Obesity | endothelial | *TMOD2* | -0.736342882 | 0.000807758 | 0.001581594 | 1 |
| Obesity | endothelial | *KNSTRN* | -0.233360983 | 0.002300922 | 0.003996104 | 1 |
| Obesity | endothelial | *ARPP19* | -0.569552307 | 0.016867278 | 0.02410104 | 1 |
| Obesity | endothelial | *VPS13C* | -0.457265861 | 0.004733791 | 0.007613544 | 1 |
| Obesity | endothelial | *ACKR4* | -0.608701604 | 0.010807737 | 0.016063415 | 1 |
| Obesity | endothelial | *ANAPC13* | -0.40840046 | 0.000543529 | 0.001108749 | 1 |
| Obesity | endothelial | *SUMF2* | -0.906159285 | 5.20E-06 | 2.17E-05 | 0.032687838 |
| Obesity | endothelial | *SERGEF* | -0.612284734 | 0.005861903 | 0.009220362 | 1 |
| Obesity | endothelial | *DCTD* | -0.600173782 | 0.000860916 | 0.001671065 | 1 |
| Obesity | endothelial | *RPAIN* | -1.300985555 | 1.33E-10 | 1.55E-08 | 8.39E-07 |
| Obesity | endothelial | *PLD2* | 0.611967674 | 0.001454539 | 0.002659111 | 1 |
| Obesity | endothelial | *ATP1B2* | -1.366184603 | 3.37E-05 | 0.000101181 | 0.211974012 |
| Obesity | endothelial | *CCL4* | -0.598754154 | 0.000107944 | 0.000269519 | 0.678640919 |
| Obesity | endothelial | *PHF20L1* | -0.605167038 | 0.001237105 | 0.002299727 | 1 |
| Obesity | endothelial | *PUS7L* | -0.618274699 | 0.003733575 | 0.006141545 | 1 |
| Obesity | endothelial | *SLC44A2* | 0.358633383 | 0.021238541 | 0.029718831 | 1 |
| Obesity | endothelial | *MTUS1* | 0.303934317 | 0.003637329 | 0.005995775 | 1 |
| Obesity | endothelial | *RIPK3* | 0.725037839 | 0.000136482 | 0.000330277 | 0.858059255 |
| Obesity | endothelial | *ADCY4* | 0.169026092 | 1.82E-06 | 9.14E-06 | 0.011421129 |
| Obesity | endothelial | *RAB2B* | -0.563018788 | 0.023906106 | 0.033003445 | 1 |
| Obesity | endothelial | *BCL2L2* | -1.112164123 | 2.29E-07 | 1.79E-06 | 0.001438177 |
| Obesity | endothelial | *DTD2* | -0.502037582 | 0.00014843 | 0.000356039 | 0.933177367 |
| Obesity | endothelial | *PARP2* | -0.862083648 | 2.44E-05 | 7.68E-05 | 0.153153423 |
| Obesity | endothelial | *DAD1* | -0.524684602 | 0.021837149 | 0.030468299 | 1 |
| Obesity | endothelial | *TEP1* | -1.037713308 | 1.06E-05 | 3.89E-05 | 0.066922894 |
| Obesity | endothelial | *EPB41L4A* | 0.274225581 | 0.000673054 | 0.001340776 | 1 |

| | | | | | | |
|---|---|---|---|---|---|---|
| Obesity | endothelial | *CDO1* | -0.746455292 | 1.23E-07 | 1.11E-06 | 0.000771394 |
| Obesity | endothelial | *REEP5* | -0.759809724 | 6.69E-05 | 0.00017955 | 0.420864053 |
| Obesity | endothelial | *SEC14L1* | 0.278122824 | 0.002404265 | 0.004158354 | 1 |
| Obesity | endothelial | *TMEM8A* | -1.139726324 | 0.000129451 | 0.000315694 | 0.81385855 |
| Obesity | endothelial | *SHC2* | 0.55306187 | 0.000897156 | 0.001732315 | 1 |
| Obesity | endothelial | *ERMARD* | -1.021979605 | 8.94E-06 | 3.37E-05 | 0.056215979 |
| Obesity | endothelial | *PHF10* | -0.84935141 | 0.000659696 | 0.001317925 | 1 |
| Obesity | endothelial | *DOCK6* | 0.286255472 | 0.010937319 | 0.016217671 | 1 |
| Obesity | endothelial | *ECSIT* | -0.972444984 | 0.000388535 | 0.00082636 | 1 |
| Obesity | endothelial | *ELOF1* | -1.805458219 | 1.34E-06 | 7.18E-06 | 0.008412158 |
| Obesity | endothelial | *CDC16* | -0.684669787 | 7.21E-05 | 0.000190978 | 0.453063913 |
| Obesity | endothelial | *TOMM40* | -1.099639021 | 1.54E-05 | 5.27E-05 | 0.097028787 |
| Obesity | endothelial | *DPP6* | -0.951258655 | 4.21E-08 | 5.27E-07 | 0.000264551 |
| Obesity | endothelial | *FAM98C* | -0.915857802 | 0.000160658 | 0.000381875 | 1 |
| Obesity | endothelial | *ATP8B3* | -0.938268345 | 3.33E-13 | 1.16E-10 | 2.10E-09 |
| Obesity | endothelial | *GTPBP3* | -0.740257532 | 0.008458667 | 0.012858929 | 1 |
| Obesity | endothelial | *PLVAP* | 0.143456474 | 2.45E-05 | 7.71E-05 | 0.153930944 |
| Obesity | endothelial | *BST2* | 0.317233054 | 3.43E-05 | 0.000102731 | 0.215769186 |
| Obesity | endothelial | *DDA1* | -1.164243966 | 2.36E-05 | 7.50E-05 | 0.148571414 |
| Obesity | endothelial | *MRPL34* | -0.254988553 | 0.022060357 | 0.030759252 | 1 |
| Obesity | endothelial | *LSM7* | -0.690200912 | 0.000835808 | 0.001632064 | 1 |
| Obesity | endothelial | *RTN4IP1* | -0.831209856 | 0.000250688 | 0.000562483 | 1 |
| Obesity | endothelial | *STK33* | 0.269066073 | 3.00E-06 | 1.39E-05 | 0.018878234 |
| Obesity | endothelial | *CACNG6* | 1.023369135 | 0.002749903 | 0.004671343 | 1 |
| Obesity | endothelial | *PXDN* | -0.795615388 | 2.26E-06 | 1.10E-05 | 0.014238013 |
| Obesity | endothelial | *PGPEP1* | -0.442079946 | 0.018146918 | 0.025771329 | 1 |
| Obesity | endothelial | *KIAA1683* | -0.3689518 | 0.027200034 | 0.037175351 | 1 |
| Obesity | endothelial | *HRC* | -0.958901385 | 1.62E-07 | 1.38E-06 | 0.001021245 |
| Obesity | endothelial | *TRPM4* | 0.371281124 | 0.021085874 | 0.02952492 | 1 |
| Obesity | endothelial | *CAMSAP1* | -0.921137706 | 0.000230831 | 0.000522214 | 1 |
| Obesity | endothelial | *UBAC1* | -0.924380639 | 2.10E-05 | 6.81E-05 | 0.131827275 |
| Obesity | endothelial | *TNNT3* | -0.601414987 | 0.003144421 | 0.005254911 | 1 |
| Obesity | endothelial | *ATXN10* | -0.648042046 | 0.000337108 | 0.000730071 | 1 |
| Obesity | endothelial | *TUBGCP2* | -0.742649679 | 0.000217009 | 0.000495668 | 1 |
| Obesity | endothelial | *CYP2E1* | -1.935861695 | 4.38E-08 | 5.40E-07 | 0.000275534 |
| Obesity | endothelial | *ZNF337* | -0.517509066 | 0.00581936 | 0.009160319 | 1 |
| Obesity | endothelial | *CEP85* | -0.536839041 | 0.00280489 | 0.004753192 | 1 |
| Obesity | endothelial | *POMT1* | -1.159007624 | 2.51E-06 | 1.20E-05 | 0.01577732 |
| Obesity | endothelial | *UCK1* | -0.489415316 | 0.023990471 | 0.033108118 | 1 |
| Obesity | endothelial | *YIPF2* | -1.193761737 | 1.38E-06 | 7.33E-06 | 0.008648385 |
| Obesity | endothelial | *TMEM160* | -1.256611838 | 0.002489728 | 0.004282604 | 1 |
| Obesity | endothelial | *MAP3K10* | -0.800692556 | 0.002614081 | 0.004467173 | 1 |
| Obesity | endothelial | *ARHGEF16* | -0.413610565 | 0.004123288 | 0.006724543 | 1 |
| Obesity | endothelial | *SMPDL3B* | 2.203954329 | 7.44E-06 | 2.89E-05 | 0.046758174 |
| Obesity | endothelial | *ATPIF1* | -0.919707805 | 2.21E-05 | 7.10E-05 | 0.13906434 |
| Obesity | endothelial | *THEMIS2* | -0.570015866 | 0.000230104 | 0.000520757 | 1 |
| Obesity | endothelial | *CLIP1* | -0.570768172 | 0.000618802 | 0.001243737 | 1 |
| Obesity | endothelial | *ZNF317* | -1.150781679 | 4.86E-05 | 0.000136101 | 0.305411657 |
| Obesity | endothelial | *PPAN* | -2.324101061 | 3.20E-07 | 2.30E-06 | 0.002010092 |
| Obesity | endothelial | *ZNF236* | -0.751462095 | 0.000108849 | 0.000271405 | 0.684331034 |

| | | | | | | |
|---|---|---|---|---|---|---|
| Obesity | endothelial | *C12orf65* | -0.684055318 | 0.00885342 | 0.013415631 | 1 |
| Obesity | endothelial | *PKDREJ* | -0.808800042 | 0.000463463 | 0.000963875 | 1 |
| Obesity | endothelial | *EPS8L1* | 3.071182494 | 0.01319094 | 0.019179334 | 1 |
| Obesity | endothelial | *LILRB2* | -1.088290211 | 1.32E-08 | 3.34E-07 | 8.28E-05 |
| Obesity | endothelial | *COX4I2* | -3.249548862 | 4.32E-09 | 1.96E-07 | 2.71E-05 |
| Obesity | endothelial | *ATP6V1E1* | -0.797337492 | 0.000656798 | 0.00131297 | 1 |
| Obesity | endothelial | *CHMP1A* | -0.970978031 | 2.54E-05 | 7.95E-05 | 0.159650869 |
| Obesity | endothelial | *PPT1* | -0.960556341 | 9.92E-05 | 0.00025099 | 0.623961485 |
| Obesity | endothelial | *HAUS8* | -1.045084608 | 2.12E-07 | 1.69E-06 | 0.001332591 |
| Obesity | endothelial | *TBC1D5* | -0.384593951 | 0.009968581 | 0.014939802 | 1 |
| Obesity | endothelial | *RFTN1* | -0.623343929 | 1.71E-05 | 5.77E-05 | 0.107748497 |
| Obesity | endothelial | *KCNC3* | -0.753294428 | 0.000134381 | 0.000326071 | 0.844850539 |
| Obesity | endothelial | *KIF3A* | -0.606516676 | 0.00292453 | 0.004926721 | 1 |
| Obesity | endothelial | *AOC3* | -0.816090073 | 8.55E-05 | 0.000220123 | 0.53776131 |
| Obesity | endothelial | *ACLY* | -0.873466145 | 0.000178493 | 0.000417946 | 1 |
| Obesity | endothelial | *AOC2* | -1.404715632 | 4.58E-08 | 5.52E-07 | 0.000288169 |
| Obesity | endothelial | *ACAP3* | -0.916687444 | 2.28E-07 | 1.79E-06 | 0.001433086 |
| Obesity | endothelial | *C1orf159* | -0.793790266 | 0.002519621 | 0.004325739 | 1 |
| Obesity | endothelial | *ANO1* | -1.097139523 | 9.50E-06 | 3.54E-05 | 0.05973434 |
| Obesity | endothelial | *PPFIA1* | -0.623495703 | 0.000204191 | 0.0004711 | 1 |
| Obesity | endothelial | *NINJ1* | -1.17488531 | 1.02E-06 | 5.82E-06 | 0.006433676 |
| Obesity | endothelial | *NPHP4* | -0.869731613 | 0.003387149 | 0.005627644 | 1 |
| Obesity | endothelial | *MAP1B* | -0.318434998 | 0.000368201 | 0.000787909 | 1 |
| Obesity | endothelial | *CKMT2* | -0.864464067 | 4.90E-05 | 0.0001372 | 0.308150909 |
| Obesity | endothelial | *ZCCHC9* | -0.960556244 | 3.53E-05 | 0.000105124 | 0.222021324 |
| Obesity | endothelial | *KHDRBS3* | -0.808148453 | 1.44E-08 | 3.34E-07 | 9.05E-05 |
| Obesity | endothelial | *CHD1L* | -0.97315806 | 3.74E-07 | 2.60E-06 | 0.002349559 |
| Obesity | endothelial | *RBM8A* | -0.964948715 | 8.14E-07 | 4.83E-06 | 0.005119528 |
| Obesity | endothelial | *ZNF304* | -0.907251884 | 0.001182949 | 0.002209506 | 1 |
| Obesity | endothelial | *ZSCAN5A* | -0.916232549 | 3.01E-05 | 9.21E-05 | 0.189378427 |
| Obesity | endothelial | *RHPN2* | -1.37083879 | 6.74E-09 | 2.54E-07 | 4.24E-05 |
| Obesity | endothelial | *C19orf12* | -0.598637152 | 1.05E-07 | 9.84E-07 | 0.000661 |
| Obesity | endothelial | *ACTR10* | -0.587212712 | 0.005034719 | 0.008035866 | 1 |
| Obesity | endothelial | *GCH1* | 0.374037175 | 6.23E-05 | 0.000169178 | 0.391815487 |
| Obesity | endothelial | *DNAJB1* | -1.024947233 | 3.33E-08 | 4.63E-07 | 0.000209164 |
| Obesity | endothelial | *FBXW9* | -1.020162888 | 7.91E-07 | 4.73E-06 | 0.004973647 |
| Obesity | endothelial | *MATN3* | -1.63227798 | 3.21E-08 | 4.54E-07 | 0.000201581 |
| Obesity | endothelial | *GAS2L2* | -0.203614563 | 0.024331224 | 0.033524086 | 1 |
| Obesity | endothelial | *CCT6B* | -0.819190197 | 3.11E-05 | 9.44E-05 | 0.195654677 |
| Obesity | endothelial | *RAF1* | -0.603597668 | 0.002140299 | 0.003751341 | 1 |
| Obesity | endothelial | *ENOSF1* | -0.649727243 | 2.16E-05 | 6.98E-05 | 0.13594126 |
| Obesity | endothelial | *EMILIN2* | -0.345053813 | 0.004927119 | 0.007895975 | 1 |
| Obesity | endothelial | *RRP8* | -0.810813235 | 0.00451265 | 0.007287703 | 1 |
| Obesity | endothelial | *IMMT* | -0.466486969 | 0.015583554 | 0.022445316 | 1 |
| Obesity | endothelial | *RPA1* | -0.639009156 | 0.001717455 | 0.003072748 | 1 |
| Obesity | endothelial | *SERPINF1* | -0.568120832 | 4.05E-06 | 1.77E-05 | 0.025444397 |
| Obesity | endothelial | *TMEM128* | -1.130923192 | 0.000208831 | 0.000480571 | 1 |
| Obesity | endothelial | *SEC61G* | -0.698306123 | 0.001701756 | 0.003051608 | 1 |
| Obesity | endothelial | *FIGNL1* | -1.403009863 | 3.47E-07 | 2.45E-06 | 0.002183688 |
| Obesity | endothelial | *WBP2* | -0.867182093 | 0.001773056 | 0.003164123 | 1 |

| | | | | | | |
|---|---|---|---|---|---|---|
| Obesity | endothelial | *KDM6B* | -1.175132129 | 2.98E-05 | 9.15E-05 | 0.187437411 |
| Obesity | endothelial | *XAF1* | 0.234880404 | 0.000539842 | 0.001103018 | 1 |
| Obesity | endothelial | *REEP2* | -0.313273062 | 0.022983566 | 0.031848728 | 1 |
| Obesity | endothelial | *PRMT7* | -0.562718648 | 0.000175215 | 0.000411496 | 1 |
| Obesity | endothelial | *TERF2* | -0.849796887 | 0.000497889 | 0.001025967 | 1 |
| Obesity | endothelial | *PCNA* | -1.108979323 | 2.03E-05 | 6.64E-05 | 0.127872994 |
| Obesity | endothelial | *PTPRA* | -0.565225507 | 0.001088391 | 0.002047491 | 1 |
| Obesity | endothelial | *MMACHC* | -1.218020061 | 3.33E-07 | 2.38E-06 | 0.002095829 |
| Obesity | endothelial | *MUTYH* | -0.919043873 | 0.007443777 | 0.011467539 | 1 |
| Obesity | endothelial | *CTNNBL1* | -0.708886139 | 0.001405559 | 0.002577816 | 1 |
| Obesity | endothelial | *LPIN3* | -0.96816623 | 2.06E-08 | 3.63E-07 | 0.000129605 |
| Obesity | endothelial | *DMGDH* | -0.8354401 | 9.83E-05 | 0.000249141 | 0.618223132 |
| Obesity | endothelial | *BHMT2* | -0.233831344 | 8.86E-06 | 3.34E-05 | 0.055673408 |
| Obesity | endothelial | *KANK4* | -0.283677556 | 0.001628901 | 0.002930996 | 1 |
| Obesity | endothelial | *CASP9* | -0.655950035 | 9.30E-06 | 3.48E-05 | 0.058487851 |
| Obesity | endothelial | *ATP8A2* | -1.022760773 | 0.021797101 | 0.030425927 | 1 |
| Obesity | endothelial | *USPL1* | -0.787207224 | 0.000175673 | 0.000412111 | 1 |
| Obesity | endothelial | *POMP* | -0.725285498 | 0.003945475 | 0.006459687 | 1 |
| Obesity | endothelial | *WASF3* | -0.850820445 | 1.07E-07 | 1.00E-06 | 0.000675193 |
| Obesity | endothelial | *RNF17* | -1.065831123 | 0.021842117 | 0.03046847 | 1 |
| Obesity | endothelial | *PEMT* | -1.10177272 | 2.63E-08 | 4.14E-07 | 0.000165105 |
| Obesity | endothelial | *SCO1* | -0.908490037 | 0.000392864 | 0.000834438 | 1 |
| Obesity | endothelial | *DSTYK* | -0.827986225 | 7.87E-05 | 0.000204907 | 0.495056345 |
| Obesity | endothelial | *TMCC2* | -0.848339146 | 0.000868319 | 0.001682319 | 1 |
| Obesity | endothelial | *COG6* | -0.521481282 | 0.004105847 | 0.006697836 | 1 |
| Obesity | endothelial | *TRPC4* | -1.039110992 | 1.37E-08 | 3.34E-07 | 8.63E-05 |
| Obesity | endothelial | *BTBD2* | -1.142345338 | 4.96E-07 | 3.25E-06 | 0.003116007 |
| Obesity | endothelial | *PRAM1* | -1.046478704 | 3.70E-09 | 1.83E-07 | 2.33E-05 |
| Obesity | endothelial | *ZNF414* | -1.647787934 | 2.55E-06 | 1.21E-05 | 0.016009239 |
| Obesity | endothelial | *RTN3* | -0.688749825 | 4.30E-07 | 2.91E-06 | 0.002704358 |
| Obesity | endothelial | *SLC2A11* | -1.21086231 | 1.95E-08 | 3.57E-07 | 0.000122842 |
| Obesity | endothelial | *C1QTNF6* | -1.434646685 | 9.19E-08 | 8.82E-07 | 0.000577665 |
| Obesity | endothelial | *FAM83F* | 1.143302724 | 0.031812397 | 0.042873427 | 1 |
| Obesity | endothelial | *GIMAP4* | 0.34201007 | 1.18E-05 | 4.23E-05 | 0.074449419 |
| Obesity | endothelial | *ADCK2* | -0.350027155 | 0.008060522 | 0.012312075 | 1 |
| Obesity | endothelial | *AGAP3* | -1.064064876 | 1.27E-07 | 1.15E-06 | 0.000800985 |
| Obesity | endothelial | *ACTR3B* | -0.775693681 | 0.002744704 | 0.004665032 | 1 |
| Obesity | endothelial | *SFTPD* | 0.18888266 | 0.015817081 | 0.022734793 | 1 |
| Obesity | endothelial | *DYDC2* | -2.51752788 | 4.16E-07 | 2.83E-06 | 0.002617569 |
| Obesity | endothelial | *TMTC1* | 0.251526499 | 0.002579752 | 0.004416912 | 1 |
| Obesity | endothelial | *IPO8* | -0.524841069 | 0.021245208 | 0.029721546 | 1 |
| Obesity | endothelial | *LARS* | -0.614875016 | 2.94E-07 | 2.16E-06 | 0.00184959 |
| Obesity | endothelial | *SPINK5* | 0.851695779 | 0.003069526 | 0.005143419 | 1 |
| Obesity | endothelial | *SWAP70* | 0.299285499 | 0.017230343 | 0.02458197 | 1 |
| Obesity | endothelial | *AMPD3* | -0.584451147 | 3.56E-07 | 2.50E-06 | 0.002236651 |
| Obesity | endothelial | *SBF2* | -0.429269482 | 0.004099309 | 0.006690642 | 1 |
| Obesity | endothelial | *RRAS2* | -0.253328116 | 0.01965817 | 0.027735843 | 1 |
| Obesity | endothelial | *HSD17B4* | -0.779996341 | 8.07E-06 | 3.09E-05 | 0.050743544 |
| Obesity | endothelial | *DPF2* | -0.855849836 | 4.13E-05 | 0.000119507 | 0.259927936 |
| Obesity | endothelial | *GSC* | -1.261897943 | 3.74E-08 | 4.91E-07 | 0.000234917 |

| | | | | | | |
|---|---|---|---|---|---|---|
| Obesity | endothelial | *CATSPERB* | -0.505006742 | 3.48E-05 | 0.000103882 | 0.218774937 |
| Obesity | endothelial | *MED6* | -0.389566894 | 0.032597193 | 0.043846503 | 1 |
| Obesity | endothelial | *EIF2S1* | -0.621790448 | 0.013479348 | 0.01958056 | 1 |
| Obesity | endothelial | *ELP3* | -0.815505866 | 0.000103262 | 0.000259373 | 0.649210549 |
| Obesity | endothelial | *MRO* | -0.826399411 | 1.25E-08 | 3.34E-07 | 7.83E-05 |
| Obesity | endothelial | *THUMPD3* | -1.038228668 | 1.33E-06 | 7.17E-06 | 0.008386871 |
| Obesity | endothelial | *EDEM1* | -1.09565411 | 5.73E-07 | 3.64E-06 | 0.003604392 |
| Obesity | endothelial | *CNTN6* | -0.625193605 | 3.82E-09 | 1.84E-07 | 2.40E-05 |
| Obesity | endothelial | *GSTM5* | -0.256932986 | 0.037406531 | 0.049751399 | 1 |
| Obesity | endothelial | *SYT6* | 0.335016887 | 1.09E-06 | 6.11E-06 | 0.00688191 |
| Obesity | endothelial | *VAV3* | -0.811185013 | 1.20E-05 | 4.29E-05 | 0.075739293 |
| Obesity | endothelial | *PSRC1* | -0.709902333 | 0.000253827 | 0.000568135 | 1 |
| Obesity | endothelial | *WNT2B* | 0.377102197 | 0.00220734 | 0.003855945 | 1 |
| Obesity | endothelial | *CEPT1* | -0.763073359 | 7.75E-07 | 4.64E-06 | 0.00487232 |
| Obesity | endothelial | *AP4B1* | -1.016194776 | 0.000536552 | 0.001097722 | 1 |
| Obesity | endothelial | *NAPG* | -0.735139944 | 0.000670853 | 0.001337663 | 1 |
| Obesity | endothelial | *SPIRE1* | -0.786248461 | 7.39E-07 | 4.46E-06 | 0.004648345 |
| Obesity | endothelial | *PPHLN1* | -0.478589955 | 0.006347168 | 0.009911734 | 1 |
| Obesity | endothelial | *FKBP11* | -1.082005552 | 8.59E-06 | 3.27E-05 | 0.054033956 |
| Obesity | endothelial | *YWHAQ* | -0.861777346 | 3.62E-05 | 0.000107126 | 0.227429248 |
| Obesity | endothelial | *LPIN1* | -0.62730285 | 1.43E-06 | 7.56E-06 | 0.008980728 |
| Obesity | endothelial | *LDHA* | -0.933644665 | 2.28E-05 | 7.28E-05 | 0.143401685 |
| Obesity | endothelial | *ANO3* | -0.485194193 | 4.86E-08 | 5.69E-07 | 0.00030542 |
| Obesity | endothelial | *FST* | -0.763207156 | 5.15E-07 | 3.35E-06 | 0.003238088 |
| Obesity | endothelial | *TIMM17A* | -1.017016843 | 3.14E-06 | 1.45E-05 | 0.019756128 |
| Obesity | endothelial | *NARS* | -0.887075928 | 0.000236392 | 0.000533259 | 1 |
| Obesity | endothelial | *KIAA1468* | -0.452768599 | 0.017618048 | 0.02508826 | 1 |
| Obesity | endothelial | *FBXO18* | -0.537530814 | 0.015208013 | 0.021919481 | 1 |
| Obesity | endothelial | *ECHDC3* | -0.728695942 | 9.63E-09 | 3.14E-07 | 6.05E-05 |
| Obesity | endothelial | *TMEM241* | -0.757375162 | 2.99E-05 | 9.16E-05 | 0.188026961 |
| Obesity | endothelial | *EMP1* | -0.432668806 | 0.00163133 | 0.002933688 | 1 |
| Obesity | endothelial | *DDB2* | -0.770131406 | 2.98E-05 | 9.15E-05 | 0.187663485 |
| Obesity | endothelial | *HOOK1* | -1.977140398 | 4.88E-07 | 3.21E-06 | 0.003067735 |
| Obesity | endothelial | *ZCCHC11* | -0.437070927 | 0.010669149 | 0.015887479 | 1 |
| Obesity | endothelial | *FHOD3* | -0.464548154 | 2.16E-07 | 1.71E-06 | 0.001359689 |
| Obesity | endothelial | *TPGS2* | -0.842872093 | 0.000108963 | 0.000271413 | 0.685047439 |
| Obesity | endothelial | *SLC43A3* | -1.169854824 | 5.17E-08 | 5.91E-07 | 0.000324802 |
| Obesity | endothelial | *TIMM10* | -0.724818616 | 0.007678537 | 0.01178588 | 1 |
| Obesity | endothelial | *DHX34* | -1.029798784 | 8.41E-08 | 8.34E-07 | 0.000528596 |
| Obesity | endothelial | *APLNR* | 0.214901343 | 1.34E-06 | 7.18E-06 | 0.008414548 |
| Obesity | endothelial | *FADS2* | -1.068308333 | 1.37E-07 | 1.21E-06 | 0.000861568 |
| Obesity | endothelial | *TMEM165* | -0.622623978 | 0.000160439 | 0.000381643 | 1 |
| Obesity | endothelial | *CLOCK* | -0.429906435 | 0.024984078 | 0.034363355 | 1 |
| Obesity | endothelial | *CLDN10* | 0.412678655 | 3.55E-05 | 0.000105544 | 0.223186594 |
| Obesity | endothelial | *BIVM* | -1.353454592 | 2.37E-06 | 1.15E-05 | 0.014909112 |
| Obesity | endothelial | *ERCC5* | -2.009492265 | 7.14E-08 | 7.41E-07 | 0.000448808 |
| Obesity | endothelial | *CARS2* | -0.701850874 | 0.000108881 | 0.000271405 | 0.684533738 |
| Obesity | endothelial | *STT3A* | -0.6509412 | 0.007158862 | 0.011069323 | 1 |
| Obesity | endothelial | *KLB* | -0.322811009 | 0.000496395 | 0.001023438 | 1 |
| Obesity | endothelial | *WDR36* | -0.861159328 | 7.62E-05 | 0.000199589 | 0.479013904 |

| | | | | | | |
|---|---|---|---|---|---|---|
| Obesity | endothelial | *UBQLN1* | -0.788503722 | 6.54E-05 | 0.00017586 | 0.411161044 |
| Obesity | endothelial | *NAA35* | -0.573848095 | 0.004918122 | 0.007887814 | 1 |
| Obesity | endothelial | *CTSL* | -0.844199195 | 6.50E-07 | 4.04E-06 | 0.004087799 |
| Obesity | endothelial | *GOLM1* | -0.70103513 | 0.003211071 | 0.00536202 | 1 |
| Obesity | endothelial | *ADAM19* | -0.340256644 | 5.73E-05 | 0.000157654 | 0.360082576 |
| Obesity | endothelial | *USP30* | -0.583106429 | 0.000175082 | 0.000411496 | 1 |
| Obesity | endothelial | *HRK* | 0.818155571 | 1.94E-05 | 6.39E-05 | 0.122072957 |
| Obesity | endothelial | *P2RX4* | -0.370652457 | 0.034781362 | 0.046535523 | 1 |
| Obesity | endothelial | *DTX1* | -0.469752835 | 8.43E-07 | 4.95E-06 | 0.005297868 |
| Obesity | endothelial | *DMTF1* | -0.409927257 | 0.011217441 | 0.016574396 | 1 |
| Obesity | endothelial | *CCDC146* | -0.80891877 | 1.95E-07 | 1.58E-06 | 0.001225072 |
| Obesity | endothelial | *TMEM60* | -1.55321971 | 1.47E-08 | 3.34E-07 | 9.21E-05 |
| Obesity | endothelial | *POM121C* | -1.568266947 | 2.21E-15 | 1.54E-12 | 1.39E-11 |
| Obesity | endothelial | *SRPK2* | -0.559277771 | 0.000483923 | 0.001001126 | 1 |
| Obesity | endothelial | *MDFIC* | -0.611091374 | 1.29E-05 | 4.55E-05 | 0.081311045 |
| Obesity | endothelial | *MTO1* | -0.661713717 | 0.00042435 | 0.000892867 | 1 |
| Obesity | endothelial | *ANKRD6* | -0.723229227 | 0.001053788 | 0.001991333 | 1 |
| Obesity | endothelial | *KHDC1* | -0.39001645 | 4.54E-05 | 0.000128889 | 0.285741348 |
| Obesity | endothelial | *SNX14* | -0.541535479 | 0.004992997 | 0.007981432 | 1 |
| Obesity | endothelial | *NT5E* | -0.712062963 | 0.000969473 | 0.001850827 | 1 |
| Obesity | endothelial | *EPHA7* | 0.199411014 | 1.65E-08 | 3.39E-07 | 0.000103853 |
| Obesity | endothelial | *PRR5L* | -0.793526834 | 2.62E-06 | 1.24E-05 | 0.016486139 |
| Obesity | endothelial | *NAT10* | -0.798670071 | 0.000842429 | 0.00164177 | 1 |
| Obesity | endothelial | *ATP5G2* | -0.701197408 | 0.004632553 | 0.007465999 | 1 |
| Obesity | endothelial | *ITGA7* | -0.333454844 | 2.25E-05 | 7.20E-05 | 0.141483567 |
| Obesity | endothelial | *RDH5* | -0.628819743 | 4.34E-09 | 1.96E-07 | 2.73E-05 |
| Obesity | endothelial | *BLOC1S1* | -1.040362558 | 7.18E-07 | 4.37E-06 | 0.004511871 |
| Obesity | endothelial | *PPP1R1A* | -1.176341883 | 1.32E-06 | 7.12E-06 | 0.008320469 |
| Obesity | endothelial | *B4GALNT1* | -1.609721353 | 3.56E-07 | 2.50E-06 | 0.002236294 |
| Obesity | endothelial | *FAIM2* | -1.181357923 | 3.21E-08 | 4.54E-07 | 0.000202003 |
| Obesity | endothelial | *HNRNPA1* | -0.951799292 | 0.000225188 | 0.000511289 | 1 |
| Obesity | endothelial | *SLC26A10* | -1.636249622 | 1.23E-05 | 4.35E-05 | 0.077037654 |
| Obesity | endothelial | *MAP7* | -0.866735104 | 7.26E-08 | 7.50E-07 | 0.000456633 |
| Obesity | endothelial | *AHI1* | -0.338402234 | 0.000645415 | 0.001291857 | 1 |
| Obesity | endothelial | *PKIB* | -1.000692878 | 3.98E-06 | 1.75E-05 | 0.024999821 |
| Obesity | endothelial | *SMPD2* | -0.865121951 | 2.46E-06 | 1.18E-05 | 0.015479486 |
| Obesity | endothelial | *MICAL1* | -1.086855177 | 4.65E-07 | 3.09E-06 | 0.002921892 |
| Obesity | endothelial | *PRADC1* | -1.313005283 | 0.00195983 | 0.003456227 | 1 |
| Obesity | endothelial | *SMYD5* | -1.127116959 | 0.000175584 | 0.000412055 | 1 |
| Obesity | endothelial | *DYSF* | 0.23225302 | 3.96E-05 | 0.000115216 | 0.249213163 |
| Obesity | endothelial | *USP15* | -0.517541527 | 0.001887364 | 0.003342495 | 1 |
| Obesity | endothelial | *KLHL36* | -0.747812482 | 0.001326946 | 0.00244505 | 1 |
| Obesity | endothelial | *MPHOSPH6* | -1.039082057 | 6.39E-05 | 0.000172956 | 0.401949741 |
| Obesity | endothelial | *FHOD1* | -1.062840053 | 5.88E-06 | 2.38E-05 | 0.036936294 |
| Obesity | endothelial | *RNASEL* | -0.846239495 | 3.95E-05 | 0.000114899 | 0.248412376 |
| Obesity | endothelial | *SLC19A3* | -0.298891315 | 5.59E-05 | 0.000154461 | 0.351552192 |
| Obesity | endothelial | *TMBIM1* | -0.61853811 | 0.001529723 | 0.002773174 | 1 |
| Obesity | endothelial | *ARMC9* | -0.909397296 | 0.000253202 | 0.000567314 | 1 |
| Obesity | endothelial | *TMEM127* | -0.988823935 | 0.000494161 | 0.001019288 | 1 |
| Obesity | endothelial | *GCC2* | -0.588933076 | 0.000598072 | 0.00120864 | 1 |

| | | | | | | |
|---|---|---|---|---|---|---|
| Obesity | endothelial | *ANKRD36* | -0.438518224 | 0.023819461 | 0.032905504 | 1 |
| Obesity | endothelial | *USP44* | -0.415336205 | 0.011023233 | 0.016325811 | 1 |
| Obesity | endothelial | *APPL2* | -0.740609068 | 4.40E-05 | 0.000125604 | 0.276701857 |
| Obesity | endothelial | *DRAM1* | -0.456234012 | 0.019967464 | 0.02812174 | 1 |
| Obesity | endothelial | *SLC41A2* | -0.870769161 | 4.57E-07 | 3.04E-06 | 0.002870712 |
| Obesity | endothelial | *RNASEH2B* | -0.790248917 | 3.81E-05 | 0.000111791 | 0.23979166 |
| Obesity | endothelial | *CKAP2* | -0.305293681 | 0.003623429 | 0.005974428 | 1 |
| Obesity | endothelial | *TBC1D4* | -0.662130369 | 3.27E-06 | 1.50E-05 | 0.020548227 |
| Obesity | endothelial | *SUCLA2* | -0.364742659 | 0.020461135 | 0.02874618 | 1 |
| Obesity | endothelial | *RCBTB1* | -0.444062549 | 0.028190258 | 0.038411823 | 1 |
| Obesity | endothelial | *RCBTB2* | -0.494356112 | 0.027651308 | 0.037734703 | 1 |
| Obesity | endothelial | *SETDB2* | -0.523444438 | 0.011247165 | 0.01661441 | 1 |
| Obesity | endothelial | *C7orf25* | -0.352935417 | 0.021401574 | 0.029920323 | 1 |
| Obesity | endothelial | *SPDYE1* | -1.044873655 | 0.000109708 | 0.000273161 | 0.689732052 |
| Obesity | endothelial | *CHST12* | -0.828786762 | 0.000467319 | 0.000969648 | 1 |
| Obesity | endothelial | *GPNMB* | -0.729383879 | 6.22E-08 | 6.75E-07 | 0.000391214 |
| Obesity | endothelial | *KDELR2* | -1.06998933 | 2.15E-05 | 6.93E-05 | 0.134893556 |
| Obesity | endothelial | *NUPL2* | -0.949398061 | 3.79E-06 | 1.68E-05 | 0.023820127 |
| Obesity | endothelial | *ZDHHC4* | -1.124571109 | 4.48E-08 | 5.43E-07 | 0.000281477 |
| Obesity | endothelial | *AOAH* | -0.639656025 | 4.68E-06 | 1.99E-05 | 0.029438876 |
| Obesity | endothelial | *TBRG4* | -1.484545631 | 3.49E-08 | 4.76E-07 | 0.000219361 |
| Obesity | endothelial | *DDX56* | -1.292751472 | 4.51E-07 | 3.02E-06 | 0.002835155 |
| Obesity | endothelial | *NACAD* | -2.224035531 | 3.84E-08 | 5.00E-07 | 0.000241534 |
| Obesity | endothelial | *DBNL* | -0.65072346 | 0.004469504 | 0.007227308 | 1 |
| Obesity | endothelial | *ALPK3* | -0.986901968 | 4.17E-07 | 2.83E-06 | 0.002619939 |
| Obesity | endothelial | *TM6SF1* | 0.323296434 | 1.46E-05 | 5.03E-05 | 0.091493337 |
| Obesity | endothelial | *CIB2* | -0.559629104 | 0.003042628 | 0.005103789 | 1 |
| Obesity | endothelial | *RSAD1* | -1.223862748 | 1.33E-05 | 4.65E-05 | 0.083717762 |
| Obesity | endothelial | *MYCBPAP* | -0.416057129 | 0.001989639 | 0.003504864 | 1 |
| Obesity | endothelial | *BRIP1* | -0.803879428 | 1.85E-08 | 3.45E-07 | 0.000116086 |
| Obesity | endothelial | *MRPL47* | -1.151460727 | 4.67E-05 | 0.0001318 | 0.293649948 |
| Obesity | endothelial | *BLK* | -2.208235595 | 2.70E-13 | 1.13E-10 | 1.70E-09 |
| Obesity | endothelial | *EPRS* | -0.700144398 | 0.002498241 | 0.004291378 | 1 |
| Obesity | endothelial | *RPS6KC1* | -0.690835496 | 6.50E-05 | 0.000175163 | 0.40865477 |
| Obesity | endothelial | *CBWD2* | -1.075166776 | 1.16E-05 | 4.18E-05 | 0.073091369 |
| Obesity | endothelial | *IL1RN* | -1.196764556 | 3.24E-08 | 4.54E-07 | 0.000203813 |
| Obesity | endothelial | *SAP130* | -0.49775043 | 0.029989702 | 0.040573543 | 1 |
| Obesity | endothelial | *UGGT1* | -0.812808324 | 0.000144263 | 0.000347236 | 0.906980262 |
| Obesity | endothelial | *GYPC* | -0.751203701 | 3.75E-06 | 1.67E-05 | 0.023573017 |
| Obesity | endothelial | *NIPSNAP3A* | -0.840506796 | 0.000600213 | 0.001212188 | 1 |
| Obesity | endothelial | *LRRC8A* | 0.41074607 | 0.010031247 | 0.015015821 | 1 |
| Obesity | endothelial | *TXN* | -1.100957013 | 5.22E-06 | 2.17E-05 | 0.032801792 |
| Obesity | endothelial | *TOR1A* | -0.864240026 | 5.63E-06 | 2.31E-05 | 0.035393942 |
| Obesity | endothelial | *ZNF189* | -0.973611386 | 2.35E-06 | 1.14E-05 | 0.014776016 |
| Obesity | endothelial | *ALDOB* | -1.106462782 | 2.44E-05 | 7.69E-05 | 0.153446681 |
| Obesity | endothelial | *STX17* | -0.530733687 | 0.016212065 | 0.023281236 | 1 |
| Obesity | endothelial | *FPGS* | -0.878217082 | 0.000374744 | 0.000800821 | 1 |
| Obesity | endothelial | *TEX10* | -0.858824902 | 0.000265789 | 0.000591511 | 1 |
| Obesity | endothelial | *GARNL3* | -0.911203937 | 2.49E-07 | 1.90E-06 | 0.001566662 |
| Obesity | endothelial | *GABBR2* | -3.141862082 | 9.69E-06 | 3.60E-05 | 0.060951197 |

| | | | | | | |
|---|---|---|---|---|---|---|
| Obesity | endothelial | *PSMB7* | -0.865047787 | 2.19E-05 | 7.05E-05 | 0.137909985 |
| Obesity | endothelial | *RABEPK* | -1.040400856 | 4.43E-06 | 1.91E-05 | 0.027873826 |
| Obesity | endothelial | *GOLGA1* | -0.818670929 | 7.85E-05 | 0.000204461 | 0.493568618 |
| Obesity | endothelial | *LMX1B* | -1.470553322 | 9.44E-09 | 3.09E-07 | 5.94E-05 |
| Obesity | endothelial | *DERL1* | -0.951737485 | 0.000143764 | 0.000346301 | 0.903846604 |
| Obesity | endothelial | *NOV* | -0.853225711 | 0.006401963 | 0.009989858 | 1 |
| Obesity | endothelial | *IL11RA* | -1.015698578 | 6.09E-05 | 0.000166166 | 0.383178829 |
| Obesity | endothelial | *APTX* | -0.807421014 | 4.75E-05 | 0.000133691 | 0.298611774 |
| Obesity | endothelial | *TLN1* | -0.773549565 | 7.16E-05 | 0.000190103 | 0.450163801 |
| Obesity | endothelial | *SPAG8* | -0.882970493 | 5.27E-06 | 2.19E-05 | 0.033116993 |
| Obesity | endothelial | *CD72* | -0.827309288 | 7.28E-06 | 2.84E-05 | 0.045783417 |
| Obesity | endothelial | *TMEM8B* | -1.310722467 | 1.17E-07 | 1.07E-06 | 0.000738491 |
| Obesity | endothelial | *GRHPR* | -0.566628212 | 0.003567589 | 0.00589163 | 1 |
| Obesity | endothelial | *ARHGEF39* | -0.582164012 | 3.66E-05 | 0.000108154 | 0.230259494 |
| Obesity | endothelial | *DENND4C* | -0.587402542 | 0.000212143 | 0.000487302 | 1 |
| Obesity | endothelial | *PPIL1* | -2.264085044 | 9.50E-08 | 9.08E-07 | 0.000597493 |
| Obesity | endothelial | *KIF13A* | -0.503067859 | 0.001880921 | 0.003333902 | 1 |
| Obesity | endothelial | *ZSCAN9* | 0.445437634 | 0.02435266 | 0.033538921 | 1 |
| Obesity | endothelial | *TMEM63B* | -1.153262663 | 6.31E-10 | 5.29E-08 | 3.97E-06 |
| Obesity | endothelial | *FRS3* | 0.560583785 | 0.024097946 | 0.033224515 | 1 |
| Obesity | endothelial | *SLC22A23* | -0.716571975 | 0.000238683 | 0.000538041 | 1 |
| Obesity | endothelial | *LRRC1* | 0.275467501 | 1.18E-05 | 4.23E-05 | 0.074233983 |
| Obesity | endothelial | *BPHL* | -0.437228028 | 2.83E-06 | 1.32E-05 | 0.017781226 |
| Obesity | endothelial | *RIPK1* | -0.59372994 | 0.004275547 | 0.006944036 | 1 |
| Obesity | endothelial | *TUBB2B* | -0.634859 | 1.19E-05 | 4.25E-05 | 0.074737725 |
| Obesity | endothelial | *UQCC2* | -1.270392887 | 2.42E-08 | 3.96E-07 | 0.000152033 |
| Obesity | endothelial | *TCF19* | 0.856639222 | 0.000358085 | 0.000768879 | 1 |
| Obesity | endothelial | *FLOT1* | 0.400905497 | 0.007166275 | 0.011078035 | 1 |
| Obesity | endothelial | *TPMT* | -0.803243519 | 0.015616614 | 0.022487781 | 1 |
| Obesity | endothelial | *NRM* | 0.906987813 | 4.76E-07 | 3.16E-06 | 0.00299432 |
| Obesity | endothelial | *MTCH1* | -0.933988056 | 4.60E-05 | 0.000130186 | 0.289402784 |
| Obesity | endothelial | *VARS2* | -0.159874138 | 0.003350296 | 0.005570831 | 1 |
| Obesity | endothelial | *FAM8A1* | -1.292766866 | 2.77E-06 | 1.30E-05 | 0.017433528 |
| Obesity | endothelial | *FGFBP2* | -1.182286486 | 1.08E-05 | 3.94E-05 | 0.068059692 |
| Obesity | endothelial | *FHDC1* | -1.336824839 | 0.001376505 | 0.002528956 | 1 |
| Obesity | endothelial | *CCDC90B* | -0.903069469 | 0.000203648 | 0.000470193 | 1 |
| Obesity | endothelial | *SYTL2* | -1.007113688 | 9.33E-09 | 3.07E-07 | 5.86E-05 |
| Obesity | endothelial | *RAB30* | -0.658398656 | 0.002557292 | 0.004384427 | 1 |
| Obesity | endothelial | *NARS2* | -0.863177154 | 1.25E-05 | 4.43E-05 | 0.078815174 |
| Obesity | endothelial | *PI15* | -2.421711293 | 0.028468738 | 0.038724136 | 1 |
| Obesity | endothelial | *TTPA* | -0.283389237 | 3.24E-07 | 2.32E-06 | 0.00203893 |
| Obesity | endothelial | *SULF1* | 0.229882048 | 4.14E-05 | 0.000119687 | 0.260438391 |
| Obesity | endothelial | *TGS1* | -0.664224263 | 0.000229836 | 0.000520525 | 1 |
| Obesity | endothelial | *NEK1* | -0.551986711 | 0.002583354 | 0.004417064 | 1 |
| Obesity | endothelial | *DDX60* | 0.37304275 | 0.000522373 | 0.001071503 | 1 |
| Obesity | endothelial | *DCUN1D5* | -0.609332829 | 4.70E-06 | 2.00E-05 | 0.029534768 |
| Obesity | endothelial | *TRIM29* | -0.802391055 | 3.24E-08 | 4.54E-07 | 0.000203876 |
| Obesity | endothelial | *POU2F3* | -0.73500825 | 2.24E-05 | 7.17E-05 | 0.1406589 |
| Obesity | endothelial | *PPP2R1B* | -0.679922922 | 1.29E-07 | 1.15E-06 | 0.000809241 |
| Obesity | endothelial | *FDX1* | -0.918756063 | 3.24E-06 | 1.48E-05 | 0.020358397 |

| | | | | | | |
|---|---|---|---|---|---|---|
| Obesity | endothelial | *FXYD6* | 0.703996644 | 0.014428421 | 0.020867606 | 1 |
| Obesity | endothelial | *TMPRSS13* | -1.803786426 | 1.40E-05 | 4.87E-05 | 0.088316673 |
| Obesity | endothelial | *MAP2K5* | -0.364393396 | 0.036030367 | 0.048022666 | 1 |
| Obesity | endothelial | *NDUFAF1* | -0.575100298 | 0.005650833 | 0.008933062 | 1 |
| Obesity | endothelial | *ITGA11* | -0.580215271 | 1.71E-08 | 3.41E-07 | 0.000107772 |
| Obesity | endothelial | *PAQR5* | -0.903831486 | 1.45E-08 | 3.34E-07 | 9.09E-05 |
| Obesity | endothelial | *RMDN3* | -0.820442563 | 0.000549497 | 0.001119109 | 1 |
| Obesity | endothelial | *ITPKA* | -2.738501425 | 1.30E-05 | 4.57E-05 | 0.081973316 |
| Obesity | endothelial | *PLCB2* | -0.255011965 | 0.008459445 | 0.012858929 | 1 |
| Obesity | endothelial | *RSL24D1* | -0.926004095 | 2.33E-05 | 7.40E-05 | 0.146304535 |
| Obesity | endothelial | *RABGGTB* | -1.032767141 | 0.000104162 | 0.000261214 | 0.654864303 |
| Obesity | endothelial | *SLC44A5* | 0.406496369 | 1.51E-06 | 7.90E-06 | 0.009482359 |
| Obesity | endothelial | *DNASE2B* | -3.563979993 | 8.98E-09 | 2.99E-07 | 5.64E-05 |
| Obesity | endothelial | *DBT* | -0.731753076 | 0.000401401 | 0.000850558 | 1 |
| Obesity | endothelial | *KHK* | -0.206224181 | 0.011087916 | 0.016413875 | 1 |
| Obesity | endothelial | *ADCY3* | -0.896219356 | 6.58E-08 | 7.02E-07 | 0.00041386 |
| Obesity | endothelial | *DYNC2LI1* | -0.477135496 | 0.001273853 | 0.002361756 | 1 |
| Obesity | endothelial | *LHCGR* | -0.709802455 | 1.34E-08 | 3.34E-07 | 8.40E-05 |
| Obesity | endothelial | *THUMPD2* | -0.582786598 | 0.006639267 | 0.010337066 | 1 |
| Obesity | endothelial | *PREB* | -0.833073346 | 1.15E-06 | 6.33E-06 | 0.007206653 |
| Obesity | endothelial | *ATRAID* | -0.818558162 | 0.001346477 | 0.002476682 | 1 |
| Obesity | endothelial | *CENPO* | -1.64678002 | 1.72E-07 | 1.43E-06 | 0.00107981 |
| Obesity | endothelial | *LRPPRC* | -0.559177999 | 0.004304858 | 0.006984424 | 1 |
| Obesity | endothelial | *STAMBPL1* | -0.673999564 | 1.20E-07 | 1.09E-06 | 0.000755436 |
| Obesity | endothelial | *ATAD1* | -0.87637603 | 2.86E-05 | 8.83E-05 | 0.1795356 |
| Obesity | endothelial | *BTBD16* | -0.861755999 | 1.21E-05 | 4.29E-05 | 0.075846136 |
| Obesity | endothelial | *TACC2* | -0.621977221 | 6.33E-05 | 0.000171482 | 0.397838069 |
| Obesity | endothelial | *CALHM2* | -0.535257831 | 0.028366335 | 0.038626629 | 1 |
| Obesity | endothelial | *PLCE1* | -0.740364738 | 9.51E-05 | 0.000241554 | 0.5976046 |
| Obesity | endothelial | *RBP4* | -0.864808955 | 3.53E-06 | 1.59E-05 | 0.022209581 |
| Obesity | endothelial | *DNAJC13* | -0.691128585 | 0.000389973 | 0.000828857 | 1 |
| Obesity | endothelial | *FAM149B1* | -0.585532336 | 0.001516208 | 0.002753437 | 1 |
| Obesity | endothelial | *ASCC1* | -0.781803656 | 3.17E-05 | 9.61E-05 | 0.199564216 |
| Obesity | endothelial | *DNA2* | -0.18541859 | 0.017662792 | 0.025146279 | 1 |
| Obesity | endothelial | *ATIC* | -0.779438974 | 8.28E-08 | 8.22E-07 | 0.000520518 |
| Obesity | endothelial | *SMARCAL1* | -0.892359702 | 0.000107683 | 0.000269186 | 0.677001723 |
| Obesity | endothelial | *STAT4* | -0.678972901 | 2.71E-05 | 8.43E-05 | 0.170620681 |
| Obesity | endothelial | *ASNSD1* | -0.714841796 | 0.001320842 | 0.002436658 | 1 |
| Obesity | endothelial | *METTL5* | -0.969121865 | 0.000297975 | 0.000653425 | 1 |
| Obesity | endothelial | *NAB1* | -0.745138157 | 0.000907708 | 0.001750004 | 1 |
| Obesity | endothelial | *FASTKD1* | -0.506206453 | 0.01919788 | 0.027153447 | 1 |
| Obesity | endothelial | *MDH1B* | 1.240938236 | 0.000662126 | 0.001322358 | 1 |
| Obesity | endothelial | *SLC35A5* | -0.463330736 | 0.000380616 | 0.000811714 | 1 |
| Obesity | endothelial | *USP8* | -0.468010199 | 0.010023563 | 0.015007893 | 1 |
| Obesity | endothelial | *SPPL2A* | -0.726462456 | 0.000102485 | 0.000257729 | 0.644322412 |
| Obesity | endothelial | *PPCDC* | -0.498063498 | 0.002154292 | 0.003772712 | 1 |
| Obesity | endothelial | *ARHGAP24* | -0.595134374 | 3.35E-07 | 2.38E-06 | 0.002105183 |
| Obesity | endothelial | *FAM13A* | -0.658338452 | 9.46E-07 | 5.45E-06 | 0.005944824 |
| Obesity | endothelial | *HERC3* | -0.663205948 | 0.000527859 | 0.001080993 | 1 |
| Obesity | endothelial | *AP1AR* | -0.588892225 | 0.022371874 | 0.031117693 | 1 |

| | | | | | | |
|---|---|---|---|---|---|---|
| Obesity | endothelial | *PRKG2* | -0.737484568 | 3.39E-08 | 4.64E-07 | 0.00021328 |
| Obesity | endothelial | *BBS7* | -0.759303149 | 0.003124021 | 0.005226375 | 1 |
| Obesity | endothelial | *PRDM5* | 0.304160281 | 0.025394774 | 0.034874824 | 1 |
| Obesity | endothelial | *NAAA* | -0.78070073 | 2.91E-05 | 8.96E-05 | 0.182783471 |
| Obesity | endothelial | *NUP54* | -0.596201954 | 0.002084015 | 0.003660856 | 1 |
| Obesity | endothelial | *BMP2K* | -0.646262907 | 8.19E-06 | 3.13E-05 | 0.051487988 |
| Obesity | endothelial | *SCARB2* | -0.722521179 | 0.000161195 | 0.00038286 | 1 |
| Obesity | endothelial | *ANXA3* | 0.177906892 | 2.20E-06 | 1.08E-05 | 0.01380575 |
| Obesity | endothelial | *GSTCD* | -0.497115083 | 0.024094735 | 0.033224515 | 1 |
| Obesity | endothelial | *ENPEP* | -0.788296055 | 5.65E-07 | 3.61E-06 | 0.003552951 |
| Obesity | endothelial | *PAPSS1* | -0.755911225 | 4.91E-06 | 2.07E-05 | 0.030852738 |
| Obesity | endothelial | *RNF185* | -0.808547118 | 7.33E-05 | 0.000193339 | 0.460726371 |
| Obesity | endothelial | *PARVG* | -1.085031907 | 4.41E-06 | 1.90E-05 | 0.027703142 |
| Obesity | endothelial | *B4GALNT3* | -1.772534561 | 1.40E-08 | 3.34E-07 | 8.78E-05 |
| Obesity | endothelial | *ERP27* | -1.670338506 | 4.05E-05 | 0.000117352 | 0.25477139 |
| Obesity | endothelial | *KIF21A* | -0.579789473 | 5.36E-07 | 3.45E-06 | 0.003370307 |
| Obesity | endothelial | *CPNE8* | 0.238821176 | 0.009644846 | 0.014489163 | 1 |
| Obesity | endothelial | *ALG10* | -1.848502632 | 2.05E-06 | 1.02E-05 | 0.012907425 |
| Obesity | endothelial | *SLCO1C1* | -0.805331661 | 1.48E-06 | 7.75E-06 | 0.00928728 |
| Obesity | endothelial | *ZCRB1* | -0.466344124 | 0.027642445 | 0.037730797 | 1 |
| Obesity | endothelial | *PRICKLE1* | -0.766379625 | 9.02E-08 | 8.80E-07 | 0.000567333 |
| Obesity | endothelial | *C1RL* | -0.629767659 | 0.000350233 | 0.000753823 | 1 |
| Obesity | endothelial | *CLSTN3* | -0.590905317 | 0.027862237 | 0.037989565 | 1 |
| Obesity | endothelial | *VAMP1* | -0.559740168 | 0.00336748 | 0.005597923 | 1 |
| Obesity | endothelial | *CD27* | -1.1640571 | 7.10E-05 | 0.000188838 | 0.446224656 |
| Obesity | endothelial | *PEX5* | -0.802287853 | 0.000718179 | 0.00142435 | 1 |
| Obesity | endothelial | *PPFIA2* | -0.459839341 | 2.78E-06 | 1.30E-05 | 0.017462615 |
| Obesity | endothelial | *GLIPR1* | -0.839438735 | 6.26E-06 | 2.51E-05 | 0.039368993 |
| Obesity | endothelial | *LGR5* | -0.832308882 | 2.81E-05 | 8.70E-05 | 0.176534385 |
| Obesity | endothelial | *POC1B* | -0.679945965 | 0.004855151 | 0.007794773 | 1 |
| Obesity | endothelial | *GAS2L3* | -1.130158745 | 1.11E-07 | 1.03E-06 | 0.000698328 |
| Obesity | endothelial | *TMEM132B* | -1.055028623 | 0.000213564 | 0.000489314 | 1 |
| Obesity | endothelial | *TDG* | -0.853254974 | 0.000103233 | 0.000259373 | 0.649024303 |
| Obesity | endothelial | *RITA1* | -0.514996998 | 0.001702537 | 0.00305178 | 1 |
| Obesity | endothelial | *MMAB* | -1.047624388 | 5.57E-06 | 2.29E-05 | 0.035026073 |
| Obesity | endothelial | *GLTP* | -1.178138866 | 5.55E-05 | 0.000153261 | 0.348669426 |
| Obesity | endothelial | *SLC46A3* | -0.634953282 | 7.18E-05 | 0.000190513 | 0.45132541 |
| Obesity | endothelial | *SUOX* | -0.693469651 | 0.000343542 | 0.000741706 | 1 |
| Obesity | endothelial | *CCDC65* | -0.857977437 | 8.70E-06 | 3.30E-05 | 0.054718834 |
| Obesity | endothelial | *ACVRL1* | 0.320936393 | 0.002328398 | 0.004038245 | 1 |
| Obesity | endothelial | *GPR84* | -4.819343077 | 4.81E-08 | 5.66E-07 | 0.00030235 |
| Obesity | endothelial | *BRCA2* | -1.256428337 | 1.43E-06 | 7.56E-06 | 0.008994742 |
| Obesity | endothelial | *KANSL2* | -0.759455706 | 1.89E-05 | 6.25E-05 | 0.119112855 |
| Obesity | endothelial | *CERS5* | -0.549762189 | 0.009127614 | 0.01381447 | 1 |
| Obesity | endothelial | *ZNF740* | -0.96107358 | 2.56E-05 | 8.02E-05 | 0.16101749 |
| Obesity | endothelial | *WDFY2* | -0.807691775 | 3.72E-07 | 2.60E-06 | 0.002341346 |
| Obesity | endothelial | *HNRNPA1L2* | 1.141716883 | 0.000127918 | 0.000312197 | 0.804219441 |
| Obesity | endothelial | *VPS33A* | -1.537360962 | 0.000173685 | 0.000409172 | 1 |
| Obesity | endothelial | *RHOF* | -1.071046603 | 2.60E-11 | 4.41E-09 | 1.63E-07 |
| Obesity | endothelial | *METTL21C* | -2.123687292 | 0.000133641 | 0.000324526 | 0.840199037 |

| | | | | | | |
|---|---|---|---|---|---|---|
| Obesity | endothelial | *GRTP1* | -0.469258096 | 7.11E-06 | 2.79E-05 | 0.044722265 |
| Obesity | endothelial | *CBLN3* | -0.181324375 | 0.000351008 | 0.000755231 | 1 |
| Obesity | endothelial | *TSSK4* | -3.548904661 | 2.97E-07 | 2.17E-06 | 0.001866267 |
| Obesity | endothelial | *TMX1* | -0.9526547 | 0.000154178 | 0.000368701 | 0.969315579 |
| Obesity | endothelial | *RTN1* | -0.647512744 | 7.83E-08 | 7.89E-07 | 0.000492315 |
| Obesity | endothelial | *RAB15* | -0.855388663 | 9.53E-07 | 5.48E-06 | 0.005991541 |
| Obesity | endothelial | *STON2* | -0.871398379 | 7.98E-07 | 4.76E-06 | 0.005018034 |
| Obesity | endothelial | *PTGR2* | -0.260057226 | 0.014313913 | 0.020716291 | 1 |
| Obesity | endothelial | *AK7* | 0.444342822 | 4.25E-06 | 1.84E-05 | 0.026689762 |
| Obesity | endothelial | *SLC24A4* | -0.695507018 | 2.45E-07 | 1.88E-06 | 0.001539614 |
| Obesity | endothelial | *FBLN5* | -0.661498787 | 0.001887038 | 0.003342495 | 1 |
| Obesity | endothelial | *C14orf79* | -1.261018469 | 0.001593472 | 0.002874651 | 1 |
| Obesity | endothelial | *NIPA2* | -0.674965779 | 0.008106171 | 0.012357784 | 1 |
| Obesity | endothelial | *DUOXA1* | -0.867410924 | 5.87E-09 | 2.35E-07 | 3.69E-05 |
| Obesity | endothelial | *MFAP1* | -0.560585524 | 0.026219412 | 0.035928824 | 1 |
| Obesity | endothelial | *TCF12* | -0.331057653 | 0.027491822 | 0.037533352 | 1 |
| Obesity | endothelial | *SORD* | -1.282043 | 4.29E-05 | 0.000123111 | 0.269612644 |
| Obesity | endothelial | *ZSCAN29* | -1.062164248 | 3.81E-07 | 2.64E-06 | 0.002396848 |
| Obesity | endothelial | *SRP14* | 0.35747847 | 0.009626185 | 0.014464585 | 1 |
| Obesity | endothelial | *ANP32A* | -0.873624085 | 3.49E-05 | 0.000104007 | 0.219245865 |
| Obesity | endothelial | *COMMD4* | -1.144957609 | 0.000279528 | 0.000617279 | 1 |
| Obesity | endothelial | *PSTPIP1* | -0.982457677 | 1.13E-07 | 1.04E-06 | 0.000710481 |
| Obesity | endothelial | *HMG20A* | -0.459764387 | 0.028801258 | 0.039117197 | 1 |
| Obesity | endothelial | *MAN2C1* | -0.171616515 | 0.012837411 | 0.018717255 | 1 |
| Obesity | endothelial | *DNAJA4* | -1.147624636 | 4.05E-08 | 5.14E-07 | 0.000254535 |
| Obesity | endothelial | *TPM1* | -0.530317937 | 0.005173464 | 0.00824058 | 1 |
| Obesity | endothelial | *BBS4* | -0.790097165 | 0.000282761 | 0.000623106 | 1 |
| Obesity | endothelial | *ADAMTS17* | -0.947611338 | 1.57E-07 | 1.34E-06 | 0.000984593 |
| Obesity | endothelial | *ULK3* | -1.359813913 | 9.34E-06 | 3.49E-05 | 0.05869382 |
| Obesity | endothelial | *SCAMP2* | -0.61590178 | 0.004211248 | 0.006848453 | 1 |
| Obesity | endothelial | *HAPLN3* | 0.440208164 | 5.77E-05 | 0.000158662 | 0.362543146 |
| Obesity | endothelial | *POLG* | -0.782479401 | 0.001987166 | 0.003501488 | 1 |
| Obesity | endothelial | *WDR93* | -0.482242373 | 1.48E-10 | 1.70E-08 | 9.32E-07 |
| Obesity | endothelial | *TICRR* | -1.728461038 | 6.86E-08 | 7.19E-07 | 0.000431548 |
| Obesity | endothelial | *MFGE8* | -0.98092209 | 1.44E-08 | 3.34E-07 | 9.04E-05 |
| Obesity | endothelial | *UNC45A* | -0.460765013 | 0.010880716 | 0.016141354 | 1 |
| Obesity | endothelial | *ST8SIA2* | -3.594058395 | 8.85E-06 | 3.34E-05 | 0.055662034 |
| Obesity | endothelial | *IQGAP1* | -0.448187956 | 0.003532212 | 0.005839343 | 1 |
| Obesity | endothelial | *CRTC3* | -0.746436746 | 2.13E-05 | 6.90E-05 | 0.134083411 |
| Obesity | endothelial | *SH3GL3* | -0.990480701 | 1.00E-10 | 1.24E-08 | 6.31E-07 |
| Obesity | endothelial | *SEC11A* | -0.80042731 | 4.50E-05 | 0.000127828 | 0.282627486 |
| Obesity | endothelial | *TGFB1I1* | -0.666869573 | 0.017843459 | 0.025363289 | 1 |
| Obesity | endothelial | *PARN* | -0.603686775 | 0.00073657 | 0.00145486 | 1 |
| Obesity | endothelial | *FTO* | -0.555147547 | 0.000672293 | 0.001339686 | 1 |
| Obesity | endothelial | *TXNL4B* | -1.187012138 | 2.03E-05 | 6.62E-05 | 0.127355895 |
| Obesity | endothelial | *MARVELD3* | -1.609045192 | 8.50E-08 | 8.38E-07 | 0.000534338 |
| Obesity | endothelial | *ZFHX3* | -0.681102389 | 4.95E-07 | 3.25E-06 | 0.003111754 |
| Obesity | endothelial | *ADAMTS18* | 0.176246162 | 4.28E-06 | 1.85E-05 | 0.026898836 |
| Obesity | endothelial | *GCSH* | -0.761421433 | 3.08E-07 | 2.24E-06 | 0.001937763 |
| Obesity | endothelial | *CMTM3* | -0.898405405 | 4.71E-08 | 5.59E-07 | 0.000296344 |

| | | | | | | |
|---|---|---|---|---|---|---|
| Obesity | endothelial | *MAP1LC3B* | -0.683212693 | 0.003185432 | 0.005322033 | 1 |
| Obesity | endothelial | *TLDC1* | -1.034343217 | 0.000152414 | 0.0003649 | 0.958227264 |
| Obesity | endothelial | *NDUFB10* | -1.141999512 | 0.000136845 | 0.000330775 | 0.860346458 |
| Obesity | endothelial | *TIGD7* | -0.434417409 | 0.020824393 | 0.029210835 | 1 |
| Obesity | endothelial | *DEF8* | -0.914456568 | 3.10E-06 | 1.43E-05 | 0.019464477 |
| Obesity | endothelial | *GALNS* | -1.11977307 | 1.06E-08 | 3.30E-07 | 6.66E-05 |
| Obesity | endothelial | *COPS3* | -1.176183075 | 1.52E-06 | 7.96E-06 | 0.009571234 |
| Obesity | endothelial | *RANBP10* | -0.695305778 | 0.002618753 | 0.004472725 | 1 |
| Obesity | endothelial | *PRPSAP2* | -0.633085407 | 0.003071531 | 0.005145407 | 1 |
| Obesity | endothelial | *DDX52* | -0.650741616 | 0.004065235 | 0.0066402 | 1 |
| Obesity | endothelial | *RASL10B* | -1.214217192 | 5.90E-05 | 0.00016193 | 0.371143491 |
| Obesity | endothelial | *UNC45B* | -1.579367419 | 0.012752767 | 0.01861975 | 1 |
| Obesity | endothelial | *TOM1L1* | -0.583745315 | 0.007470956 | 0.011500956 | 1 |
| Obesity | endothelial | *VPS53* | -0.476347159 | 0.00694052 | 0.010766112 | 1 |
| Obesity | endothelial | *SPATA22* | -0.462078291 | 4.86E-06 | 2.06E-05 | 0.030544696 |
| Obesity | endothelial | *SGSM2* | -0.83493027 | 0.000597124 | 0.001207501 | 1 |
| Obesity | endothelial | *NPEPPS* | -0.615004399 | 0.000845781 | 0.001647791 | 1 |
| Obesity | endothelial | *RHBDL3* | 0.556088039 | 0.00045412 | 0.000946008 | 1 |
| Obesity | endothelial | *ARSG* | -0.958342597 | 8.91E-06 | 3.36E-05 | 0.056010809 |
| Obesity | endothelial | *ABCA8* | -0.436128796 | 2.72E-06 | 1.28E-05 | 0.017086009 |
| Obesity | endothelial | *IMPA2* | -0.919397945 | 3.09E-08 | 4.48E-07 | 0.000194368 |
| Obesity | endothelial | *RPRD1A* | -0.531233677 | 0.004845342 | 0.007783001 | 1 |
| Obesity | endothelial | *C18orf21* | -1.141527555 | 8.02E-06 | 3.08E-05 | 0.050419707 |
| Obesity | endothelial | *GALNT1* | -0.46549651 | 0.016648334 | 0.023831529 | 1 |
| Obesity | endothelial | *ASXL3* | 0.364491967 | 4.47E-05 | 0.000127253 | 0.28084835 |
| Obesity | endothelial | *ADCYAP1* | -1.889723602 | 8.17E-06 | 3.13E-05 | 0.051384823 |
| Obesity | endothelial | *ESCO1* | -0.708707385 | 0.000417956 | 0.000882366 | 1 |
| Obesity | endothelial | *GREB1L* | -0.527251576 | 2.06E-07 | 1.65E-06 | 0.001295368 |
| Obesity | endothelial | *C18orf8* | -1.554395991 | 1.56E-13 | 7.53E-11 | 9.78E-10 |
| Obesity | endothelial | *NPC1* | -1.173264095 | 7.57E-10 | 5.95E-08 | 4.76E-06 |
| Obesity | endothelial | *SLC14A1* | 0.765086235 | 0.000321942 | 0.000700122 | 1 |
| Obesity | endothelial | *SLC13A5* | 1.412184896 | 0.002980458 | 0.005007734 | 1 |
| Obesity | endothelial | *CCDC40* | -0.89351576 | 9.63E-07 | 5.53E-06 | 0.006053698 |
| Obesity | endothelial | *CARD14* | -1.260105077 | 0.000464563 | 0.000965523 | 1 |
| Obesity | endothelial | *EIF4A3* | -1.08881683 | 1.75E-07 | 1.46E-06 | 0.001102742 |
| Obesity | endothelial | *TBCD* | 0.291985979 | 0.016800514 | 0.02401656 | 1 |
| Obesity | endothelial | *NARF* | -0.727633867 | 0.000848257 | 0.00165057 | 1 |
| Obesity | endothelial | *WDR45B* | -0.802836367 | 0.000208317 | 0.000479564 | 1 |
| Obesity | endothelial | *RNF165* | 0.371258217 | 1.66E-05 | 5.61E-05 | 0.10417882 |
| Obesity | endothelial | *MAPK4* | -0.702757901 | 0.000172916 | 0.000407649 | 1 |
| Obesity | endothelial | *SMAD4* | -0.772674116 | 0.000417702 | 0.000882127 | 1 |
| Obesity | endothelial | *GRB7* | 0.92649963 | 6.44E-05 | 0.00017389 | 0.405163835 |
| Obesity | endothelial | *NFIC* | -0.631155619 | 5.54E-06 | 2.28E-05 | 0.034822972 |
| Obesity | endothelial | *TPGS1* | -0.926402831 | 0.000948384 | 0.001816725 | 1 |
| Obesity | endothelial | *PRDM15* | -0.519564443 | 0.000742169 | 0.001464997 | 1 |
| Obesity | endothelial | *PFKL* | -0.55627362 | 0.009272485 | 0.013989949 | 1 |
| Obesity | endothelial | *CCDC97* | -0.63249985 | 0.033404465 | 0.044836438 | 1 |
| Obesity | endothelial | *SIRT3* | -0.610518636 | 0.001958458 | 0.003454778 | 1 |
| Obesity | endothelial | *COL6A1* | -0.49841238 | 6.38E-06 | 2.55E-05 | 0.040099586 |
| Obesity | endothelial | *TRPM2* | -0.42581431 | 1.01E-05 | 3.74E-05 | 0.06373164 |

| | | | | | | |
|---|---|---|---|---|---|---|
| Obesity | endothelial | *DOPEY2* | -0.824405685 | 2.07E-05 | 6.75E-05 | 0.130127158 |
| Obesity | endothelial | *AKT1* | -1.326637198 | 1.86E-05 | 6.18E-05 | 0.117196682 |
| Obesity | endothelial | *SAE1* | -0.735278317 | 3.04E-05 | 9.28E-05 | 0.191152063 |
| Obesity | endothelial | *NTN5* | -1.237522514 | 0.000180516 | 0.00042174 | 1 |
| Obesity | endothelial | *GEMIN7* | -1.46892958 | 2.63E-08 | 4.14E-07 | 0.000165547 |
| Obesity | endothelial | *WTIP* | -0.956698606 | 3.44E-05 | 0.000103016 | 0.216539627 |
| Obesity | endothelial | *MYO1F* | -0.753103907 | 4.33E-05 | 0.000124162 | 0.272534539 |
| Obesity | endothelial | *CACNG8* | 0.348028093 | 1.30E-06 | 7.02E-06 | 0.008176472 |
| Obesity | endothelial | *EVI5L* | -1.059187462 | 0.001104522 | 0.002074733 | 1 |
| Obesity | endothelial | *CTU1* | -1.830954402 | 3.19E-08 | 4.54E-07 | 0.000200459 |
| Obesity | endothelial | *RCN3* | -0.986176911 | 4.90E-08 | 5.71E-07 | 0.000307988 |
| Obesity | endothelial | *RERE* | -0.371931793 | 0.010723129 | 0.015960302 | 1 |
| Obesity | endothelial | *PEX14* | -0.809282406 | 3.55E-06 | 1.59E-05 | 0.022290173 |
| Obesity | endothelial | *SH3BGRL3* | -0.909473231 | 0.000165544 | 0.000391857 | 1 |
| Obesity | endothelial | *ZNF593* | -0.744455801 | 1.60E-05 | 5.45E-05 | 0.100708534 |
| Obesity | endothelial | *C1orf216* | -1.07680942 | 9.93E-05 | 0.000251003 | 0.624496276 |
| Obesity | endothelial | *GPN2* | -1.265798157 | 2.36E-05 | 7.48E-05 | 0.148124158 |
| Obesity | endothelial | *PIGK* | -0.752014488 | 0.001702822 | 0.00305178 | 1 |
| Obesity | endothelial | *MOB3C* | -1.49726008 | 8.20E-07 | 4.86E-06 | 0.005156052 |
| Obesity | endothelial | *CYP4B1* | -0.42833155 | 0.00086638 | 0.001679078 | 1 |
| Obesity | endothelial | *SYPL2* | -0.338641257 | 0.000377975 | 0.000807178 | 1 |
| Obesity | endothelial | *ZNF697* | -0.988141518 | 1.82E-08 | 3.45E-07 | 0.000114485 |
| Obesity | endothelial | *CELSR2* | -1.349838104 | 6.17E-05 | 0.000167646 | 0.387598261 |
| Obesity | endothelial | *GPR161* | 0.558784591 | 0.012758586 | 0.018623922 | 1 |
| Obesity | endothelial | *ATP1B1* | -1.210074961 | 1.28E-08 | 3.34E-07 | 8.03E-05 |
| Obesity | endothelial | *NME7* | -0.514504434 | 0.011875656 | 0.017460769 | 1 |
| Obesity | endothelial | *CREG1* | -0.844145796 | 1.27E-06 | 6.85E-06 | 0.007965223 |
| Obesity | endothelial | *GPA33* | -1.789733195 | 1.59E-07 | 1.35E-06 | 0.000996796 |
| Obesity | endothelial | *RXRG* | -3.06493167 | 0.000792931 | 0.001556888 | 1 |
| Obesity | endothelial | *TMCO1* | -0.727447925 | 0.00096323 | 0.001842358 | 1 |
| Obesity | endothelial | *XCL2* | -0.848116511 | 1.80E-05 | 6.01E-05 | 0.112903958 |
| Obesity | endothelial | *MGST3* | -0.57226685 | 0.002567123 | 0.004398181 | 1 |
| Obesity | endothelial | *ADCY10* | -1.276184221 | 0.00055162 | 0.001123069 | 1 |
| Obesity | endothelial | *RFWD2* | -0.408571262 | 0.012391489 | 0.018163976 | 1 |
| Obesity | endothelial | *RGS5* | -0.504601182 | 0.029733328 | 0.040252677 | 1 |
| Obesity | endothelial | *RRNAD1* | -0.624852519 | 0.001431161 | 0.002621709 | 1 |
| Obesity | endothelial | *PIGM* | -0.915957075 | 0.002279312 | 0.003964048 | 1 |
| Obesity | endothelial | *RGL1* | -0.663704449 | 9.97E-08 | 9.41E-07 | 0.000626651 |
| Obesity | endothelial | *TUFT1* | -0.905122458 | 3.75E-11 | 6.05E-09 | 2.36E-07 |
| Obesity | endothelial | *TARS2* | -0.707207485 | 0.011852047 | 0.017438291 | 1 |
| Obesity | endothelial | *SNX27* | -0.94171678 | 6.27E-06 | 2.51E-05 | 0.039430402 |
| Obesity | endothelial | *SELENBP1* | -0.631238703 | 1.81E-06 | 9.12E-06 | 0.011384134 |
| Obesity | endothelial | *CERS2* | -0.734960908 | 0.006939947 | 0.010766112 | 1 |
| Obesity | endothelial | *SEMA6C* | 0.554267621 | 4.26E-06 | 1.85E-05 | 0.026773302 |
| Obesity | endothelial | *ARNT* | -0.693061084 | 8.40E-05 | 0.000217134 | 0.527852573 |
| Obesity | endothelial | *HORMAD1* | -0.198116157 | 9.56E-05 | 0.00024269 | 0.601142299 |
| Obesity | endothelial | *GABPB2* | -0.677312009 | 1.92E-07 | 1.57E-06 | 0.001205763 |
| Obesity | endothelial | *IKBKE* | -1.089516125 | 1.56E-08 | 3.35E-07 | 9.83E-05 |
| Obesity | endothelial | *DYRK3* | 0.539829332 | 0.000431883 | 0.000906291 | 1 |
| Obesity | endothelial | *EIF2D* | -0.732243513 | 0.000648564 | 0.001297747 | 1 |

| | | | | | | |
|---|---|---|---|---|---|---|
| Obesity | endothelial | *HHIPL2* | -1.205935907 | 1.10E-06 | 6.12E-06 | 0.006898537 |
| Obesity | endothelial | *TP53BP2* | -0.665132355 | 0.009320507 | 0.014055656 | 1 |
| Obesity | endothelial | *RAB13* | 0.461650742 | 0.032696291 | 0.043951588 | 1 |
| Obesity | endothelial | *S100A8* | -1.811630569 | 4.14E-11 | 6.34E-09 | 2.60E-07 |
| Obesity | endothelial | *KCNN3* | 0.219793368 | 2.26E-06 | 1.10E-05 | 0.014194999 |
| Obesity | endothelial | *GATAD2B* | -0.452638697 | 0.009367315 | 0.014112703 | 1 |
| Obesity | endothelial | *FLG* | -0.335689868 | 0.000297589 | 0.000652805 | 1 |
| Obesity | endothelial | *GALNT2* | -0.711758292 | 0.000336194 | 0.000728577 | 1 |
| Obesity | endothelial | *SCCPDH* | -0.974254813 | 1.06E-06 | 5.96E-06 | 0.006637525 |
| Obesity | endothelial | *CEP170* | -0.496420627 | 0.004711211 | 0.007581106 | 1 |
| Obesity | endothelial | *SNAP47* | -0.74711439 | 0.002858592 | 0.004838978 | 1 |
| Obesity | endothelial | *NVL* | -0.662833983 | 0.001735579 | 0.003103409 | 1 |
| Obesity | endothelial | *SDE2* | -0.927863525 | 0.005200984 | 0.008280219 | 1 |
| Obesity | endothelial | *FBXO28* | -0.734895465 | 0.000436412 | 0.000914574 | 1 |
| Obesity | endothelial | *CNIH4* | -1.034529439 | 1.97E-05 | 6.46E-05 | 0.123665187 |
| Obesity | endothelial | *GUK1* | 0.401119426 | 0.036577827 | 0.04870072 | 1 |
| Obesity | endothelial | *CDC42BPA* | 0.25407751 | 0.02265985 | 0.031476465 | 1 |
| Obesity | endothelial | *CNIH3* | -0.763694837 | 8.51E-05 | 0.000219228 | 0.534915534 |
| Obesity | endothelial | *PSEN2* | -0.906038347 | 7.18E-05 | 0.000190588 | 0.45169281 |
| Obesity | endothelial | *PYCR2* | -1.170036035 | 6.86E-06 | 2.71E-05 | 0.043158902 |
| Obesity | endothelial | *PLEKHA6* | -0.874053026 | 3.05E-05 | 9.29E-05 | 0.191609296 |
| Obesity | endothelial | *ABCG8* | -1.341135975 | 0.017384244 | 0.02477777 | 1 |
| Obesity | endothelial | *EML4* | -0.601015173 | 0.000638412 | 0.001278654 | 1 |
| Obesity | endothelial | *WDPCP* | -0.235676584 | 0.010892717 | 0.016155346 | 1 |
| Obesity | endothelial | *ASXL2* | -0.677069198 | 0.000289762 | 0.000636971 | 1 |
| Obesity | endothelial | *CIAO1* | -0.747595414 | 0.000541886 | 0.001106476 | 1 |
| Obesity | endothelial | *ZNF514* | -0.231606621 | 0.011213066 | 0.016571826 | 1 |
| Obesity | endothelial | *TEX261* | -1.648425749 | 1.66E-06 | 8.51E-06 | 0.010414308 |
| Obesity | endothelial | *THNSL2* | -0.158453786 | 0.003208756 | 0.005359578 | 1 |
| Obesity | endothelial | *RALB* | 0.343279184 | 0.023733336 | 0.032808154 | 1 |
| Obesity | endothelial | *TMEM177* | -1.382467768 | 0.001727226 | 0.003089351 | 1 |
| Obesity | endothelial | *RABL2A* | -0.853272966 | 0.001469626 | 0.002684352 | 1 |
| Obesity | endothelial | *FBLN7* | -0.887511056 | 1.63E-09 | 1.09E-07 | 1.03E-05 |
| Obesity | endothelial | *ZC3H8* | -0.528413429 | 0.007649846 | 0.011749334 | 1 |
| Obesity | endothelial | *LIPT1* | -0.589455276 | 0.001709005 | 0.003060242 | 1 |
| Obesity | endothelial | *FAHD2B* | -1.409936461 | 1.59E-08 | 3.36E-07 | 9.99E-05 |
| Obesity | endothelial | *AFF3* | -0.597620411 | 1.51E-10 | 1.70E-08 | 9.52E-07 |
| Obesity | endothelial | *UBXN4* | -0.517291962 | 0.006178763 | 0.00967278 | 1 |
| Obesity | endothelial | *THSD7B* | -0.408912601 | 0.000157319 | 0.000375642 | 0.989064546 |
| Obesity | endothelial | *GPR17* | 0.210876684 | 0.000196619 | 0.000456142 | 1 |
| Obesity | endothelial | *POLR2D* | -1.566911265 | 5.66E-08 | 6.29E-07 | 0.000355931 |
| Obesity | endothelial | *ZNF385B* | -0.713627806 | 9.84E-06 | 3.64E-05 | 0.061860598 |
| Obesity | endothelial | *TMEFF2* | 0.398120332 | 0.002448027 | 0.004223585 | 1 |
| Obesity | endothelial | *PHOSPHO2* | -0.372818677 | 0.00147037 | 0.00268493 | 1 |
| Obesity | endothelial | *GULP1* | 0.314182256 | 0.005270105 | 0.008370959 | 1 |
| Obesity | endothelial | *FAM171B* | 0.485462924 | 0.020362538 | 0.028633253 | 1 |
| Obesity | endothelial | *HSPD1* | -0.721259772 | 0.001207032 | 0.002250477 | 1 |
| Obesity | endothelial | *PTH2R* | -0.55136852 | 3.83E-07 | 2.65E-06 | 0.002407393 |
| Obesity | endothelial | *SUMF1* | -0.600289081 | 4.76E-05 | 0.000133834 | 0.299118616 |
| Obesity | endothelial | *ESPNL* | -6.134283435 | 0.000110611 | 0.000275186 | 0.695411175 |

| | | | | | | |
|---|---|---|---|---|---|---|
| Obesity | endothelial | *FANCD2* | -1.344463951 | 0.000283181 | 0.000623593 | 1 |
| Obesity | endothelial | *TAMM41* | -0.517686457 | 0.018637677 | 0.026402676 | 1 |
| Obesity | endothelial | *OSBPL10* | 0.323676898 | 0.001620281 | 0.002916321 | 1 |
| Obesity | endothelial | *FAM198A* | -0.737598409 | 4.31E-08 | 5.35E-07 | 0.000270727 |
| Obesity | endothelial | *CSRNP1* | -0.718066151 | 0.019250247 | 0.02721527 | 1 |
| Obesity | endothelial | *SLC25A38* | -1.6067269 | 1.10E-06 | 6.11E-06 | 0.006884446 |
| Obesity | endothelial | *CTDSPL* | -0.703832956 | 0.000273565 | 0.000605813 | 1 |
| Obesity | endothelial | *STAC* | -0.787270472 | 1.34E-08 | 3.34E-07 | 8.42E-05 |
| Obesity | endothelial | *ARL6IP5* | -0.853499336 | 7.25E-06 | 2.83E-05 | 0.045594053 |
| Obesity | endothelial | *TMF1* | -0.886378014 | 7.00E-06 | 2.75E-05 | 0.044002061 |
| Obesity | endothelial | *LIMD1* | -0.817026823 | 3.19E-06 | 1.47E-05 | 0.020040332 |
| Obesity | endothelial | *NXPE3* | 0.438528495 | 0.000461536 | 0.000960185 | 1 |
| Obesity | endothelial | *PLA1A* | 0.286294303 | 1.76E-05 | 5.91E-05 | 0.110839583 |
| Obesity | endothelial | *BOC* | -0.799238034 | 1.03E-05 | 3.79E-05 | 0.064887976 |
| Obesity | endothelial | *AGTR1* | -0.729567198 | 7.08E-08 | 7.38E-07 | 0.000444925 |
| Obesity | endothelial | *EIF2A* | -0.607339631 | 2.95E-07 | 2.16E-06 | 0.001852002 |
| Obesity | endothelial | *AMT* | -0.470633599 | 4.04E-06 | 1.77E-05 | 0.025382423 |
| Obesity | endothelial | *TCTA* | -1.161933188 | 1.73E-07 | 1.44E-06 | 0.001087333 |
| Obesity | endothelial | *EAF2* | -1.138309815 | 3.75E-10 | 3.47E-08 | 2.36E-06 |
| Obesity | endothelial | *MUC4* | -0.321057974 | 0.002152828 | 0.003771198 | 1 |
| Obesity | endothelial | *DGKQ* | -0.705931624 | 3.39E-05 | 0.0001016 | 0.213056007 |
| Obesity | endothelial | *LYAR* | -1.09109094 | 0.001120393 | 0.002100778 | 1 |
| Obesity | endothelial | *CENPC* | -0.709459183 | 1.72E-05 | 5.80E-05 | 0.108380105 |
| Obesity | endothelial | *PYURF* | -1.273374284 | 1.49E-05 | 5.12E-05 | 0.09351532 |
| Obesity | endothelial | *CISD2* | -1.048523148 | 3.14E-07 | 2.27E-06 | 0.001972119 |
| Obesity | endothelial | *USP53* | -0.651481309 | 5.99E-05 | 0.00016378 | 0.376742699 |
| Obesity | endothelial | *MARCH1* | -0.736414704 | 7.66E-07 | 4.60E-06 | 0.004813956 |
| Obesity | endothelial | *PDGFC* | -0.640643959 | 3.21E-07 | 2.30E-06 | 0.002015147 |
| Obesity | endothelial | *CBR4* | -0.565215812 | 0.001287418 | 0.002382689 | 1 |
| Obesity | endothelial | *CYP4V2* | -0.317796673 | 0.016777231 | 0.023988731 | 1 |
| Obesity | endothelial | *NDUFS6* | -1.059725236 | 0.000121819 | 0.000299286 | 0.765872968 |
| Obesity | endothelial | *NKD2* | -2.13859963 | 0.010735561 | 0.015975024 | 1 |
| Obesity | endothelial | *FAM105A* | -0.713972583 | 0.001539996 | 0.002789761 | 1 |
| Obesity | endothelial | *BHMT* | -0.247231238 | 0.007968308 | 0.012183062 | 1 |
| Obesity | endothelial | *CRHBP* | 0.372731018 | 0.000343779 | 0.000741963 | 1 |
| Obesity | endothelial | *RASA1* | -0.449045045 | 0.001802898 | 0.003209179 | 1 |
| Obesity | endothelial | *PPIP5K2* | -0.48911476 | 0.00775155 | 0.011883442 | 1 |
| Obesity | endothelial | *GTF2H2* | -0.910724687 | 0.000103584 | 0.000260078 | 0.65123538 |
| Obesity | endothelial | *SPATA9* | -0.581613809 | 7.03E-08 | 7.35E-07 | 0.00044223 |
| Obesity | endothelial | *TSLP* | -0.513713836 | 0.003468275 | 0.005745686 | 1 |
| Obesity | endothelial | *FEM1C* | -0.681924912 | 0.008129869 | 0.012387903 | 1 |
| Obesity | endothelial | *YIPF5* | -0.983217962 | 0.00044525 | 0.00093079 | 1 |
| Obesity | endothelial | *SLC25A48* | -0.754230869 | 3.37E-08 | 4.64E-07 | 0.000211581 |
| Obesity | endothelial | *GABRB2* | -0.593673781 | 2.51E-07 | 1.91E-06 | 0.001578348 |
| Obesity | endothelial | *PCYOX1L* | -1.278608446 | 3.04E-08 | 4.48E-07 | 0.000191334 |
| Obesity | endothelial | *GFOD1* | 0.28315327 | 7.67E-06 | 2.96E-05 | 0.048207172 |
| Obesity | endothelial | *PSD2* | 1.998201181 | 0.002375057 | 0.004109168 | 1 |
| Obesity | endothelial | *ZMAT2* | -1.176436594 | 6.34E-07 | 3.96E-06 | 0.003985088 |
| Obesity | endothelial | *KLHL3* | 0.332239976 | 2.24E-05 | 7.18E-05 | 0.140835215 |
| Obesity | endothelial | *TRIM7* | 0.759178953 | 0.004701664 | 0.007571557 | 1 |

| | | | | | | |
|---|---|---|---|---|---|---|
| Obesity | endothelial | *HIGD2A* | -1.178589733 | 1.71E-07 | 1.43E-06 | 0.001076475 |
| Obesity | endothelial | *FAM193B* | -0.730742534 | 0.000774912 | 0.001524366 | 1 |
| Obesity | endothelial | *MUT* | -0.943855305 | 0.000400203 | 0.00084888 | 1 |
| Obesity | endothelial | *DOK3* | -0.673055769 | 0.000488379 | 0.001008509 | 1 |
| Obesity | endothelial | *MLIP* | -0.849964124 | 0.002882434 | 0.004870159 | 1 |
| Obesity | endothelial | *FGD2* | -0.831237067 | 4.87E-08 | 5.69E-07 | 0.000306282 |
| Obesity | endothelial | *PHIP* | -0.414472765 | 0.014960531 | 0.021587527 | 1 |
| Obesity | endothelial | *MMS22L* | -0.63796543 | 0.003703558 | 0.006095358 | 1 |
| Obesity | endothelial | *PM20D2* | -1.237741533 | 3.88E-05 | 0.000113265 | 0.243860545 |
| Obesity | endothelial | *RNF217* | -0.712479102 | 0.000124665 | 0.000305444 | 0.783768396 |
| Obesity | endothelial | *ABRACL* | -0.842108355 | 0.002163661 | 0.003787009 | 1 |
| Obesity | endothelial | *SLC18B1* | -0.283846709 | 0.001983822 | 0.003496577 | 1 |
| Obesity | endothelial | *AIG1* | -0.53706272 | 0.00073549 | 0.001453832 | 1 |
| Obesity | endothelial | *DYNLT1* | -0.796266821 | 0.000373527 | 0.000798764 | 1 |
| Obesity | endothelial | *WTAP* | -0.454870093 | 0.007228489 | 0.011160488 | 1 |
| Obesity | endothelial | *VIP* | 0.27401194 | 1.16E-06 | 6.39E-06 | 0.007314598 |
| Obesity | endothelial | *SLC22A3* | -0.672562547 | 1.42E-08 | 3.34E-07 | 8.91E-05 |
| Obesity | endothelial | *GNA12* | -0.807455203 | 8.93E-06 | 3.36E-05 | 0.056113326 |
| Obesity | endothelial | *SDK1* | -0.51008894 | 9.14E-08 | 8.82E-07 | 0.00057461 |
| Obesity | endothelial | *CCZ1B* | -0.927574805 | 9.46E-07 | 5.45E-06 | 0.005949758 |
| Obesity | endothelial | *RBAK* | -1.462270261 | 9.98E-06 | 3.69E-05 | 0.062742267 |
| Obesity | endothelial | *EGFR* | -0.584275256 | 5.48E-06 | 2.26E-05 | 0.03443069 |
| Obesity | endothelial | *CCT6A* | -0.576939982 | 0.022604197 | 0.031413039 | 1 |
| Obesity | endothelial | *PSPH* | -0.713370245 | 2.83E-06 | 1.32E-05 | 0.01778787 |
| Obesity | endothelial | *GIGYF1* | -0.62498972 | 0.010983586 | 0.016282434 | 1 |
| Obesity | endothelial | *TRIM4* | -0.346115833 | 0.00034322 | 0.000741266 | 1 |
| Obesity | endothelial | *AGBL3* | -0.484183323 | 0.019961082 | 0.02811905 | 1 |
| Obesity | endothelial | *NOM1* | -0.798043379 | 0.003304376 | 0.005501751 | 1 |
| Obesity | endothelial | *NCAPG2* | -0.767722199 | 0.001281575 | 0.002373973 | 1 |
| Obesity | endothelial | *CSGALNACT1* | 0.300158771 | 0.004320988 | 0.007003365 | 1 |
| Obesity | endothelial | *CCDC25* | -0.532836338 | 0.036647419 | 0.048772718 | 1 |
| Obesity | endothelial | *BIN3* | -0.491866008 | 0.033240805 | 0.044635827 | 1 |
| Obesity | endothelial | *SLC25A37* | -0.477654731 | 0.025731639 | 0.03529889 | 1 |
| Obesity | endothelial | *CHMP7* | -0.861292581 | 3.69E-06 | 1.65E-05 | 0.023213795 |
| Obesity | endothelial | *DOCK5* | -0.77524001 | 0.000182799 | 0.0004266 | 1 |
| Obesity | endothelial | *ST18* | -0.837679796 | 3.04E-08 | 4.48E-07 | 0.000191367 |
| Obesity | endothelial | *RGS20* | -0.657151925 | 0.000422318 | 0.000890082 | 1 |
| Obesity | endothelial | *ADHFE1* | -0.408729417 | 4.66E-09 | 2.03E-07 | 2.93E-05 |
| Obesity | endothelial | *SLC26A7* | -0.66874559 | 1.60E-06 | 8.29E-06 | 0.010087376 |
| Obesity | endothelial | *ATP6V0D2* | -1.192042632 | 9.26E-13 | 2.91E-10 | 5.82E-09 |
| Obesity | endothelial | *MTDH* | -0.61232031 | 0.000640547 | 0.001282522 | 1 |
| Obesity | endothelial | *MAL2* | -0.538728983 | 5.98E-06 | 2.41E-05 | 0.037575851 |
| Obesity | endothelial | *UTP23* | -1.114894358 | 2.76E-06 | 1.29E-05 | 0.017324485 |
| Obesity | endothelial | *ZNF7* | -0.721132443 | 0.001476451 | 0.00269447 | 1 |
| Obesity | endothelial | *SLC39A4* | -1.198338733 | 4.64E-06 | 1.98E-05 | 0.029166117 |
| Obesity | endothelial | *CER1* | -0.8807372 | 0.000194795 | 0.000452745 | 1 |
| Obesity | endothelial | *PLIN2* | -0.847225077 | 9.19E-08 | 8.82E-07 | 0.000577484 |
| Obesity | endothelial | *HAUS6* | -1.01809745 | 8.44E-05 | 0.000217738 | 0.530363155 |
| Obesity | endothelial | *C9orf72* | -0.671885614 | 0.000204733 | 0.000472177 | 1 |
| Obesity | endothelial | *SIGMAR1* | -0.800191304 | 0.00048391 | 0.001001126 | 1 |

| | | | | | | |
|---|---|---|---|---|---|---|
| Obesity | endothelial | *ALAD* | -0.269139768 | 0.010571909 | 0.015761345 | 1 |
| Obesity | endothelial | *ASTN2* | -0.847565654 | 1.05E-06 | 5.94E-06 | 0.00661013 |
| Obesity | endothelial | *WDR31* | -1.122191502 | 1.67E-06 | 8.57E-06 | 0.010511003 |
| Obesity | endothelial | *POLE3* | -0.73554197 | 0.022696827 | 0.031512794 | 1 |
| Obesity | endothelial | *SURF6* | -1.30447136 | 1.62E-06 | 8.34E-06 | 0.010159647 |
| Obesity | endothelial | *NTMT1* | -0.637460952 | 0.017058025 | 0.024356984 | 1 |
| Obesity | endothelial | *CIZ1* | -0.771393489 | 3.82E-05 | 0.000111887 | 0.240445397 |
| Obesity | endothelial | *SH3GLB2* | -0.994066946 | 3.30E-05 | 9.92E-05 | 0.207422033 |
| Obesity | endothelial | *LRSAM1* | -0.776536934 | 0.001084552 | 0.00204088 | 1 |
| Obesity | endothelial | *INPP5E* | -1.239718268 | 6.34E-05 | 0.000171844 | 0.398849321 |
| Obesity | endothelial | *SEC16A* | -0.88086098 | 3.01E-05 | 9.21E-05 | 0.18927016 |
| Obesity | endothelial | *DPH7* | -0.990932589 | 0.000174968 | 0.000411496 | 1 |
| Obesity | endothelial | *SLC39A12* | -1.158132873 | 1.72E-06 | 8.78E-06 | 0.01083841 |
| Obesity | endothelial | *RSU1* | -0.414246683 | 0.028490539 | 0.038745408 | 1 |
| Obesity | endothelial | *PARD3* | -0.470529352 | 0.000558725 | 0.001135694 | 1 |
| Obesity | endothelial | *NRBF2* | -0.909417283 | 1.57E-10 | 1.73E-08 | 9.85E-07 |
| Obesity | endothelial | *GLUD1* | -0.763884612 | 2.40E-05 | 7.57E-05 | 0.150659857 |
| Obesity | endothelial | *HTR7* | -0.848196715 | 1.70E-08 | 3.41E-07 | 0.000106702 |
| Obesity | endothelial | *RPP30* | -0.881622665 | 0.000213346 | 0.000489299 | 1 |
| Obesity | endothelial | *FRA10AC1* | -0.433847481 | 0.014690608 | 0.021207774 | 1 |
| Obesity | endothelial | *DNAJB12* | -0.856553822 | 0.00105437 | 0.001991835 | 1 |
| Obesity | endothelial | *FUOM* | -1.40551166 | 1.54E-08 | 3.34E-07 | 9.68E-05 |
| Obesity | endothelial | *LRRC27* | -0.4190085 | 0.000799465 | 0.001567759 | 1 |
| Obesity | endothelial | *PAOX* | -1.708658819 | 6.34E-07 | 3.96E-06 | 0.003983034 |
| Obesity | endothelial | *GSTO1* | -0.784427593 | 0.002582517 | 0.004417064 | 1 |
| Obesity | endothelial | *PDCD11* | -1.126056075 | 6.07E-06 | 2.44E-05 | 0.03813587 |
| Obesity | endothelial | *RGS10* | -1.087042777 | 2.53E-08 | 4.06E-07 | 0.000159287 |
| Obesity | endothelial | *GAS2* | 0.465580987 | 0.014584537 | 0.021078847 | 1 |
| Obesity | endothelial | *LIN7C* | -1.220574033 | 4.90E-06 | 2.07E-05 | 0.030802704 |
| Obesity | endothelial | *ZNF215* | -0.269633941 | 0.004626327 | 0.007457877 | 1 |
| Obesity | endothelial | *HSD17B12* | -0.422668922 | 4.66E-05 | 0.000131483 | 0.29268223 |
| Obesity | endothelial | *EIF3M* | -0.588215556 | 0.008503996 | 0.012917281 | 1 |
| Obesity | endothelial | *GLYAT* | -0.408519064 | 1.81E-05 | 6.04E-05 | 0.113662129 |
| Obesity | endothelial | *PTPRJ* | -0.734909343 | 4.50E-06 | 1.93E-05 | 0.028293125 |
| Obesity | endothelial | *C11orf49* | -0.581935504 | 0.002658568 | 0.004529652 | 1 |
| Obesity | endothelial | *ENDOD1* | 0.462669544 | 0.00033751 | 0.000730691 | 1 |
| Obesity | endothelial | *CCDC82* | -0.432827812 | 0.002229332 | 0.003888235 | 1 |
| Obesity | endothelial | *SERPINH1* | -0.828222537 | 0.000106684 | 0.000267008 | 0.670723904 |
| Obesity | endothelial | *INTS4* | -0.516145805 | 0.005434965 | 0.008619986 | 1 |
| Obesity | endothelial | *TTC12* | 0.278922831 | 0.014340087 | 0.020749397 | 1 |
| Obesity | endothelial | *DRD2* | -2.906444317 | 1.98E-06 | 9.84E-06 | 0.012424134 |
| Obesity | endothelial | *C11orf52* | -2.55909546 | 7.47E-08 | 7.64E-07 | 0.000469694 |
| Obesity | endothelial | *NPAT* | -0.505137075 | 0.021020265 | 0.029452732 | 1 |
| Obesity | endothelial | *GLB1L2* | 0.907014563 | 0.028656279 | 0.038945531 | 1 |
| Obesity | endothelial | *GRIK4* | 0.252096753 | 0.000129752 | 0.000316304 | 0.815747684 |
| Obesity | endothelial | *ADAM33* | -0.599309304 | 4.85E-09 | 2.10E-07 | 3.05E-05 |
| Obesity | endothelial | *MTA2* | 0.626502591 | 0.004037558 | 0.006600137 | 1 |
| Obesity | endothelial | *TMC2* | -1.403231624 | 1.46E-08 | 3.34E-07 | 9.20E-05 |
| Obesity | endothelial | *ROM1* | 0.30852065 | 0.001380891 | 0.002535532 | 1 |
| Obesity | endothelial | *EML3* | -0.759886872 | 0.006622352 | 0.010313284 | 1 |

| | | | | | | |
|---|---|---|---|---|---|---|
| Obesity | endothelial | *INCENP* | -1.376721004 | 0.001188423 | 0.002218271 | 1 |
| Obesity | endothelial | *MS4A3* | -6.866712535 | 6.11E-08 | 6.64E-07 | 0.000383978 |
| Obesity | endothelial | *PLCH2* | -1.960700808 | 6.05E-08 | 6.61E-07 | 0.000380071 |
| Obesity | endothelial | *CPSF7* | -0.900224213 | 1.56E-05 | 5.33E-05 | 0.098170758 |
| Obesity | endothelial | *CCDC15* | -1.107618868 | 2.27E-11 | 3.97E-09 | 1.43E-07 |
| Obesity | endothelial | *CHEK1* | -0.329474309 | 0.027433598 | 0.037461996 | 1 |
| Obesity | endothelial | *FEZ1* | -1.365705193 | 3.13E-08 | 4.50E-07 | 0.000197066 |
| Obesity | endothelial | *CNBD2* | -1.300961022 | 3.06E-08 | 4.48E-07 | 0.000192255 |
| Obesity | endothelial | *CABLES2* | -1.013771313 | 2.65E-06 | 1.25E-05 | 0.016686575 |
| Obesity | endothelial | *TRPT1* | -1.825368184 | 4.39E-09 | 1.97E-07 | 2.76E-05 |
| Obesity | endothelial | *TBX6* | 0.471802346 | 0.001525625 | 0.002767341 | 1 |
| Obesity | endothelial | *DOC2A* | -2.628638991 | 2.59E-13 | 1.13E-10 | 1.63E-09 |
| Obesity | endothelial | *TAOK2* | 0.44839712 | 0.000149399 | 0.00035809 | 0.939268975 |
| Obesity | endothelial | *MPP7* | -0.428611106 | 0.000918402 | 0.001768451 | 1 |
| Obesity | endothelial | *ARID5B* | -0.574423603 | 0.000115364 | 0.000285548 | 0.725292461 |
| Obesity | endothelial | *SAP18* | -0.677086202 | 0.016753897 | 0.023960817 | 1 |
| Obesity | endothelial | *LYPD6B* | 0.768357333 | 0.017272747 | 0.024635608 | 1 |
| Obesity | endothelial | *WDR17* | -1.102382597 | 3.23E-08 | 4.54E-07 | 0.000203261 |
| Obesity | endothelial | *CCDC102B* | -0.842048168 | 1.86E-07 | 1.53E-06 | 0.001166328 |
| Obesity | endothelial | *RGS18* | -0.996390489 | 4.42E-08 | 5.42E-07 | 0.000278184 |
| Obesity | endothelial | *MTMR12* | -0.69261834 | 0.000866202 | 0.001679078 | 1 |
| Obesity | endothelial | *PPP1R1C* | -0.792125583 | 6.41E-06 | 2.56E-05 | 0.040285665 |
| Obesity | endothelial | *C11orf53* | 3.574255995 | 0.000666651 | 0.00133055 | 1 |
| Obesity | endothelial | *CCT5* | -0.925305899 | 2.80E-05 | 8.69E-05 | 0.176282099 |
| Obesity | endothelial | *FAM173B* | -1.024900136 | 1.02E-06 | 5.82E-06 | 0.006430405 |
| Obesity | endothelial | *TEX12* | -3.883001177 | 2.71E-08 | 4.21E-07 | 0.000170392 |
| Obesity | endothelial | *PIP4K2A* | -0.612012111 | 0.000546223 | 0.001113523 | 1 |
| Obesity | endothelial | *ABCB9* | 0.340039015 | 1.44E-05 | 4.98E-05 | 0.090375718 |
| Obesity | endothelial | *DHX37* | -0.956956511 | 0.005676714 | 0.008967211 | 1 |
| Obesity | endothelial | *PRSS53* | -1.749929046 | 2.07E-07 | 1.66E-06 | 0.00129875 |
| Obesity | endothelial | *GPR158* | -0.742921114 | 6.46E-07 | 4.02E-06 | 0.004061687 |
| Obesity | endothelial | *THRB* | -0.637519376 | 8.19E-07 | 4.86E-06 | 0.005148271 |
| Obesity | endothelial | *TMEM86A* | -0.634950639 | 1.20E-05 | 4.27E-05 | 0.07517902 |
| Obesity | endothelial | *BTBD11* | -1.107474694 | 2.08E-08 | 3.64E-07 | 0.000130617 |
| Obesity | endothelial | *ANK3* | -0.639389252 | 1.86E-05 | 6.17E-05 | 0.116868333 |
| Obesity | endothelial | *IPMK* | -0.938688712 | 1.13E-06 | 6.26E-06 | 0.007074579 |
| Obesity | endothelial | *PLBD2* | -0.698580065 | 2.38E-05 | 7.53E-05 | 0.149357563 |
| Obesity | endothelial | *DLG5* | -0.661133265 | 4.90E-06 | 2.07E-05 | 0.030789808 |
| Obesity | endothelial | *TMEM18* | -1.271693158 | 6.76E-05 | 0.000180904 | 0.424944158 |
| Obesity | endothelial | *KCTD14* | -2.283715545 | 0.009530769 | 0.014334916 | 1 |
| Obesity | endothelial | *THRSP* | -0.388450978 | 0.002028555 | 0.003567419 | 1 |
| Obesity | endothelial | *NDUFC2* | -0.98327736 | 0.002494642 | 0.00428871 | 1 |
| Obesity | endothelial | *ME3* | -0.48495675 | 0.006569621 | 0.010236235 | 1 |
| Obesity | endothelial | *ADAMTS12* | -0.554304164 | 8.77E-09 | 2.95E-07 | 5.51E-05 |
| Obesity | endothelial | *VIPAS39* | -1.066300369 | 4.04E-06 | 1.77E-05 | 0.025387366 |
| Obesity | endothelial | *ANKRD50* | -0.634545902 | 0.009212028 | 0.013918774 | 1 |
| Obesity | endothelial | *PTPRO* | -0.864691744 | 3.73E-08 | 4.91E-07 | 0.000234198 |
| Obesity | endothelial | *VPS26B* | -0.94095055 | 0.000219369 | 0.000500063 | 1 |
| Obesity | endothelial | *NCAPD3* | -0.888008301 | 4.38E-05 | 0.000125145 | 0.275193172 |
| Obesity | endothelial | *VTI1A* | -0.376759939 | 0.016611552 | 0.023784292 | 1 |

| | | | | | | |
|---|---|---|---|---|---|---|
| Obesity | endothelial | *QDPR* | -0.227464714 | 0.005923062 | 0.009307246 | 1 |
| Obesity | endothelial | *FAM160B1* | -0.720211302 | 0.000790181 | 0.001551997 | 1 |
| Obesity | endothelial | *ANO4* | -0.592400529 | 1.67E-07 | 1.40E-06 | 0.001048669 |
| Obesity | endothelial | *AKR1C2* | -0.816401168 | 2.81E-08 | 4.30E-07 | 0.000176902 |
| Obesity | endothelial | *DPYSL4* | 0.288782987 | 0.00476029 | 0.007654206 | 1 |
| Obesity | endothelial | *ADAM8* | -0.939960932 | 1.03E-06 | 5.83E-06 | 0.006455007 |
| Obesity | endothelial | *KIN* | -0.587930559 | 0.003414115 | 0.00566945 | 1 |
| Obesity | endothelial | *PIGF* | -0.862154657 | 6.87E-05 | 0.000183554 | 0.431902547 |
| Obesity | endothelial | *INPP1* | 0.399840406 | 0.00025348 | 0.000567734 | 1 |
| Obesity | endothelial | *ASAP2* | -0.396557797 | 0.030883414 | 0.041746726 | 1 |
| Obesity | endothelial | *ADAM17* | -0.727722819 | 6.99E-05 | 0.00018637 | 0.439646571 |
| Obesity | endothelial | *TMEM45B* | -2.525236716 | 1.27E-06 | 6.86E-06 | 0.00798021 |
| Obesity | endothelial | *ACSL1* | -0.559700218 | 2.03E-08 | 3.63E-07 | 0.000127767 |
| Obesity | endothelial | *CCDC122* | -0.288242989 | 0.004375402 | 0.007082429 | 1 |
| Obesity | endothelial | *GUF1* | -1.00958699 | 2.52E-05 | 7.90E-05 | 0.158266751 |
| Obesity | endothelial | *GABRA2* | 0.236805814 | 2.56E-06 | 1.22E-05 | 0.016066912 |
| Obesity | endothelial | *CENPJ* | -0.879982781 | 3.91E-05 | 0.000113833 | 0.245652645 |
| Obesity | endothelial | *PARP8* | -0.687553679 | 3.81E-07 | 2.64E-06 | 0.002396816 |
| Obesity | endothelial | *CACUL1* | -0.747236405 | 4.96E-05 | 0.000138546 | 0.311727913 |
| Obesity | endothelial | *BEND6* | -0.636854654 | 8.69E-10 | 6.66E-08 | 5.46E-06 |
| Obesity | endothelial | *PTPN14* | 0.320118364 | 0.00063161 | 0.001266644 | 1 |
| Obesity | endothelial | *ARL14EP* | -1.21996694 | 7.70E-09 | 2.70E-07 | 4.84E-05 |
| Obesity | endothelial | *EPG5* | -0.592146487 | 0.001032776 | 0.001954564 | 1 |
| Obesity | endothelial | *SPC25* | 0.142335578 | 4.57E-06 | 1.95E-05 | 0.028705695 |
| Obesity | endothelial | *G6PC2* | 9.83592074 | 1.96E-06 | 9.77E-06 | 0.012318626 |
| Obesity | endothelial | *TGOLN2* | -0.464771303 | 0.034041547 | 0.045633093 | 1 |
| Obesity | endothelial | *POC5* | -1.248957176 | 2.14E-10 | 2.24E-08 | 1.34E-06 |
| Obesity | endothelial | *TADA1* | -0.941296844 | 0.00264807 | 0.004515436 | 1 |
| Obesity | endothelial | *GUCY1A2* | -0.647564763 | 7.99E-08 | 8.04E-07 | 0.000502409 |
| Obesity | endothelial | *ZNF547* | -1.175485154 | 0.00193918 | 0.003426538 | 1 |
| Obesity | endothelial | *ZNF773* | -0.775998763 | 4.05E-05 | 0.000117275 | 0.254485755 |
| Obesity | endothelial | *NMT2* | -0.691812602 | 9.69E-06 | 3.60E-05 | 0.060895335 |
| Obesity | endothelial | *ZFP36L2* | -0.462689683 | 0.012427202 | 0.018212079 | 1 |
| Obesity | endothelial | *PLEKHH2* | -0.193792025 | 0.001606025 | 0.002893975 | 1 |
| Obesity | endothelial | *TMEM123* | -0.568933589 | 0.012481142 | 0.018282605 | 1 |
| Obesity | endothelial | *IGSF10* | -0.721865997 | 6.24E-08 | 6.75E-07 | 0.00039227 |
| Obesity | endothelial | *SPEF2* | -0.745504635 | 0.000156323 | 0.000373405 | 0.982802891 |
| Obesity | endothelial | *SPARCL1* | 0.204271945 | 2.35E-05 | 7.48E-05 | 0.148050646 |
| Obesity | endothelial | *MBNL1* | -0.369321122 | 0.005574091 | 0.008820616 | 1 |
| Obesity | endothelial | *CCNO* | 2.173117654 | 0.000680347 | 0.001354445 | 1 |
| Obesity | endothelial | *CLEC4F* | 0.840198825 | 5.93E-06 | 2.40E-05 | 0.037312762 |
| Obesity | endothelial | *RASGRP3* | 0.313493231 | 0.004159741 | 0.006775206 | 1 |
| Obesity | endothelial | *SAR1B* | -0.734524139 | 0.00013527 | 0.000327646 | 0.850441016 |
| Obesity | endothelial | *GPR180* | -0.702265074 | 2.80E-06 | 1.31E-05 | 0.017590967 |
| Obesity | endothelial | *TCTEX1D1* | 0.36152783 | 3.51E-08 | 4.77E-07 | 0.000220512 |
| Obesity | endothelial | *PRDM8* | -0.668914676 | 7.59E-08 | 7.75E-07 | 0.000477377 |
| Obesity | endothelial | *HHEX* | 0.372188156 | 7.05E-06 | 2.77E-05 | 0.044314436 |
| Obesity | endothelial | *RAB3C* | 0.142493816 | 1.64E-06 | 8.47E-06 | 0.010324756 |
| Obesity | endothelial | *RAD17* | -0.616640819 | 0.000220327 | 0.000501701 | 1 |
| Obesity | endothelial | *SRP19* | -1.181564522 | 0.000164117 | 0.000389212 | 1 |

| | | | | | | |
|---|---|---|---|---|---|---|
| Obesity | endothelial | *BANK1* | -0.771550193 | 2.62E-08 | 4.14E-07 | 0.000164727 |
| Obesity | endothelial | *CAST* | -0.482764433 | 0.000847419 | 0.001650471 | 1 |
| Obesity | endothelial | *SCOC* | -0.568976949 | 0.032763289 | 0.044032235 | 1 |
| Obesity | endothelial | *CLGN* | 1.522915936 | 2.97E-06 | 1.38E-05 | 0.018658379 |
| Obesity | endothelial | *SYCP2L* | -0.744183333 | 0.011421804 | 0.016848636 | 1 |
| Obesity | endothelial | *AHCTF1* | -0.497713749 | 0.013479258 | 0.01958056 | 1 |
| Obesity | endothelial | *TMEM87B* | 0.276954595 | 0.036002703 | 0.04799597 | 1 |
| Obesity | endothelial | *PTPRR* | 0.197530495 | 1.18E-06 | 6.50E-06 | 0.007447189 |
| Obesity | endothelial | *PLA2R1* | -0.650673747 | 0.000465844 | 0.000967225 | 1 |
| Obesity | endothelial | *FRMD1* | -1.546972307 | 2.40E-08 | 3.95E-07 | 0.000150584 |
| Obesity | endothelial | *INO80C* | -1.140636574 | 0.002090786 | 0.003669673 | 1 |
| Obesity | endothelial | *LPCAT1* | -0.899182075 | 0.002804617 | 0.004753192 | 1 |
| Obesity | endothelial | *C16orf89* | -0.504036407 | 0.000896939 | 0.001732315 | 1 |
| Obesity | endothelial | *TEX29* | -0.76154428 | 0.000224992 | 0.000511027 | 1 |
| Obesity | endothelial | *TUBGCP5* | -0.925695826 | 3.55E-05 | 0.000105544 | 0.223225738 |
| Obesity | endothelial | *LURAP1L* | -0.317377247 | 0.002721804 | 0.004627362 | 1 |
| Obesity | endothelial | *CFDP1* | -0.609021647 | 0.001155787 | 0.002161919 | 1 |
| Obesity | endothelial | *ZDHHC7* | -0.812460498 | 1.30E-07 | 1.16E-06 | 0.000814635 |
| Obesity | endothelial | *FAM92B* | -7.307576821 | 2.52E-08 | 4.05E-07 | 0.000158519 |
| Obesity | endothelial | *JAZF1* | -0.502072711 | 0.001220956 | 0.002272395 | 1 |
| Obesity | endothelial | *FBXO36* | -1.108205549 | 0.000127396 | 0.000311271 | 0.800941217 |
| Obesity | endothelial | *MCOLN2* | -0.804461582 | 7.23E-05 | 0.000191161 | 0.454390592 |
| Obesity | endothelial | *CACNA2D1* | -0.617652661 | 7.04E-07 | 4.30E-06 | 0.004423675 |
| Obesity | endothelial | *ZUFSP* | -0.993209342 | 1.69E-05 | 5.71E-05 | 0.106254897 |
| Obesity | endothelial | *PPP2R5E* | -0.405798586 | 0.018239058 | 0.02588464 | 1 |
| Obesity | endothelial | *ANKRD29* | 0.184996317 | 5.37E-06 | 2.23E-05 | 0.033780002 |
| Obesity | endothelial | *THY1* | -0.986674199 | 5.20E-09 | 2.19E-07 | 3.27E-05 |
| Obesity | endothelial | *TBCEL* | -0.678436134 | 0.003529016 | 0.005835592 | 1 |
| Obesity | endothelial | *UBASH3B* | -0.961599832 | 1.91E-07 | 1.56E-06 | 0.001200008 |
| Obesity | endothelial | *ROBO4* | 0.228500873 | 1.47E-05 | 5.05E-05 | 0.092141907 |
| Obesity | endothelial | *PRKCA* | -0.740847816 | 1.63E-05 | 5.54E-05 | 0.102397663 |
| Obesity | endothelial | *LRRK1* | -0.797077933 | 0.000117287 | 0.000289337 | 0.737385191 |
| Obesity | endothelial | *CEP112* | 0.30771509 | 0.003589784 | 0.00592517 | 1 |
| Obesity | endothelial | *ENPP3* | -1.159790275 | 1.84E-05 | 6.11E-05 | 0.115695082 |
| Obesity | endothelial | *UCHL1* | -0.814791404 | 1.41E-08 | 3.34E-07 | 8.88E-05 |
| Obesity | endothelial | *MIA3* | -0.788844023 | 7.22E-05 | 0.000191089 | 0.453859035 |
| Obesity | endothelial | *DISP1* | -0.563450911 | 0.001796656 | 0.003202602 | 1 |
| Obesity | endothelial | *LONRF1* | -0.835287675 | 8.18E-06 | 3.13E-05 | 0.051401783 |
| Obesity | endothelial | *CCSAP* | -1.285001778 | 4.32E-08 | 5.36E-07 | 0.000271695 |
| Obesity | endothelial | *GBP5* | -0.686659655 | 0.031585322 | 0.042594792 | 1 |
| Obesity | endothelial | *FAM69A* | -0.761461596 | 8.03E-09 | 2.76E-07 | 5.05E-05 |
| Obesity | endothelial | *ATP5G3* | -0.466500137 | 0.01715879 | 0.024489742 | 1 |
| Obesity | endothelial | *PDLIM3* | 0.528157755 | 0.000195392 | 0.000453797 | 1 |
| Obesity | endothelial | *NCAM2* | -0.359755153 | 0.029618965 | 0.04010649 | 1 |
| Obesity | endothelial | *PDE1C* | -0.705312666 | 2.09E-07 | 1.67E-06 | 0.001315534 |
| Obesity | endothelial | *RABGEF1* | -0.532012631 | 6.45E-08 | 6.95E-07 | 0.000405472 |
| Obesity | endothelial | *TSEN2* | -0.669380005 | 0.000215474 | 0.000492792 | 1 |
| Obesity | endothelial | *FLCN* | -0.644337925 | 8.75E-06 | 3.32E-05 | 0.055003078 |
| Obesity | endothelial | *DPH3* | -1.523458309 | 1.65E-06 | 8.49E-06 | 0.01037864 |
| Obesity | endothelial | *CXXC1* | -1.363465686 | 1.30E-05 | 4.55E-05 | 0.081434719 |

| | | | | | | |
|---|---|---|---|---|---|---|
| Obesity | endothelial | *APCDD1* | -0.402021994 | 1.18E-05 | 4.22E-05 | 0.073929116 |
| Obesity | endothelial | *EPHB1* | -0.81099391 | 0.001289977 | 0.002386723 | 1 |
| Obesity | endothelial | *ANKRD40* | -0.998570507 | 2.86E-05 | 8.83E-05 | 0.17981009 |
| Obesity | endothelial | *CYP2U1* | -1.096377607 | 3.57E-08 | 4.80E-07 | 0.000224722 |
| Obesity | endothelial | *FBXL18* | -1.10984867 | 1.14E-06 | 6.28E-06 | 0.007146215 |
| Obesity | endothelial | *PROM2* | -4.146528704 | 3.04E-08 | 4.48E-07 | 0.000190937 |
| Obesity | endothelial | *CDK19* | -0.435687649 | 0.019518723 | 0.027545278 | 1 |
| Obesity | endothelial | *GTF3C6* | -0.790881856 | 0.002889161 | 0.004880214 | 1 |
| Obesity | endothelial | *TTC39B* | -0.547057711 | 0.010593414 | 0.015789661 | 1 |
| Obesity | endothelial | *AGPAT5* | 0.458704364 | 0.007958979 | 0.01217472 | 1 |
| Obesity | endothelial | *MMS19* | -0.51783749 | 0.019216171 | 0.027173205 | 1 |
| Obesity | endothelial | *GOLGA7B* | -1.551110427 | 1.13E-07 | 1.04E-06 | 0.000712604 |
| Obesity | endothelial | *TRMT44* | -0.868670352 | 0.000841726 | 0.001640908 | 1 |
| Obesity | endothelial | *C16orf87* | -1.004501628 | 8.06E-07 | 4.79E-06 | 0.005065739 |
| Obesity | endothelial | *HEATR3* | -0.384565751 | 0.001378928 | 0.002532668 | 1 |
| Obesity | endothelial | *CNOT8* | -0.883645301 | 3.08E-05 | 9.36E-05 | 0.193558615 |
| Obesity | endothelial | *LRGUK* | -0.515457941 | 0.00016219 | 0.000385078 | 1 |
| Obesity | endothelial | *SETD9* | -0.545062964 | 4.49E-07 | 3.01E-06 | 0.002824536 |
| Obesity | endothelial | *PDIA4* | -0.89506083 | 0.002914432 | 0.004914977 | 1 |
| Obesity | endothelial | *KDM8* | -1.6340686 | 0.000118394 | 0.000291784 | 0.744340257 |
| Obesity | endothelial | *FAM126B* | -0.704895506 | 0.000122825 | 0.000301524 | 0.772202323 |
| Obesity | endothelial | *ALS2CR12* | -0.656073641 | 0.001951819 | 0.003445966 | 1 |
| Obesity | endothelial | *SPAG17* | -0.597379164 | 1.70E-08 | 3.41E-07 | 0.000106877 |
| Obesity | endothelial | *DEPTOR* | -0.780588588 | 0.000624878 | 0.001254345 | 1 |
| Obesity | endothelial | *RRAGA* | -1.558936149 | 5.83E-06 | 2.37E-05 | 0.036645215 |
| Obesity | endothelial | *WIF1* | -1.797419502 | 0.00105824 | 0.001996745 | 1 |
| Obesity | endothelial | *UGT2B4* | -1.102371055 | 1.77E-08 | 3.43E-07 | 0.000111539 |
| Obesity | endothelial | *MMP16* | 0.257689487 | 0.004652004 | 0.007495425 | 1 |
| Obesity | endothelial | *KCNMA1* | -0.642194497 | 6.38E-07 | 3.98E-06 | 0.004008207 |
| Obesity | endothelial | *BATF* | -1.668635573 | 2.51E-08 | 4.05E-07 | 0.000158047 |
| Obesity | endothelial | *DCK* | -0.926533184 | 0.001299592 | 0.002402391 | 1 |
| Obesity | endothelial | *DPY19L4* | -0.568638723 | 0.01065093 | 0.015867867 | 1 |
| Obesity | endothelial | *DRAM2* | -0.753218983 | 1.47E-06 | 7.71E-06 | 0.009218771 |
| Obesity | endothelial | *C8orf37* | -0.673895985 | 0.003664889 | 0.006034876 | 1 |
| Obesity | endothelial | *ADAMTSL3* | -0.3100008 | 0.005163111 | 0.008226173 | 1 |
| Obesity | endothelial | *WHAMM* | -0.550296568 | 0.00017695 | 0.000414797 | 1 |
| Obesity | endothelial | *RWDD2B* | -0.742595439 | 3.04E-05 | 9.27E-05 | 0.19089645 |
| Obesity | endothelial | *CCT8* | -0.398724221 | 0.002022198 | 0.003557235 | 1 |
| Obesity | endothelial | *ANKRD9* | -0.935780168 | 7.75E-09 | 2.71E-07 | 4.88E-05 |
| Obesity | endothelial | *SFXN2* | -0.820021747 | 0.000330872 | 0.000718051 | 1 |
| Obesity | endothelial | *C14orf2* | -0.701427344 | 8.13E-05 | 0.000211186 | 0.511280642 |
| Obesity | endothelial | *TDRD9* | -0.804615521 | 3.53E-09 | 1.78E-07 | 2.22E-05 |
| Obesity | endothelial | *SH3RF2* | 0.193535036 | 0.000482544 | 0.000998931 | 1 |
| Obesity | endothelial | *GDF6* | -1.235605247 | 2.95E-08 | 4.42E-07 | 0.000185303 |
| Obesity | endothelial | *MED19* | -0.599557028 | 8.55E-05 | 0.000220079 | 0.537433505 |
| Obesity | endothelial | *ATAD2* | -0.966568717 | 4.87E-08 | 5.69E-07 | 0.000306123 |
| Obesity | endothelial | *SASS6* | -1.151833553 | 7.51E-06 | 2.92E-05 | 0.047229506 |
| Obesity | endothelial | *ITGAD* | -0.5979186 | 6.84E-06 | 2.70E-05 | 0.042998689 |
| Obesity | endothelial | *VPS8* | -0.545574345 | 0.000735842 | 0.001453878 | 1 |
| Obesity | endothelial | *MPV17L* | -0.369078179 | 0.006100039 | 0.009559059 | 1 |

| | | | | | | |
|---|---|---|---|---|---|---|
| Obesity | endothelial | *RPUSD3* | -1.314986712 | 2.38E-07 | 1.84E-06 | 0.001497011 |
| Obesity | endothelial | *GHRL* | -1.333120666 | 1.83E-05 | 6.09E-05 | 0.115155831 |
| Obesity | endothelial | *EXOG* | -0.694095519 | 0.005189104 | 0.008263399 | 1 |
| Obesity | endothelial | *TIMP4* | -0.623777877 | 0.022680837 | 0.031498657 | 1 |
| Obesity | endothelial | *CPT2* | -1.224097889 | 2.97E-08 | 4.43E-07 | 0.000186778 |
| Obesity | endothelial | *NECAP2* | -0.658814968 | 0.014806841 | 0.021370663 | 1 |
| Obesity | endothelial | *LRP8* | -1.084410074 | 0.011052978 | 0.016366009 | 1 |
| Obesity | endothelial | *STEAP2* | -0.504927353 | 1.70E-10 | 1.85E-08 | 1.07E-06 |
| Obesity | endothelial | *TMED6* | -6.737800571 | 9.04E-08 | 8.80E-07 | 0.000568649 |
| Obesity | endothelial | *CLEC18A* | -1.553657376 | 1.95E-09 | 1.22E-07 | 1.23E-05 |
| Obesity | endothelial | *IL34* | -1.131627637 | 3.98E-09 | 1.84E-07 | 2.50E-05 |
| Obesity | endothelial | *DHRS1* | -0.37826631 | 0.010835075 | 0.016096435 | 1 |
| Obesity | endothelial | *AASDH* | -0.457741101 | 0.028319952 | 0.038571824 | 1 |
| Obesity | endothelial | *RNF111* | -0.635697446 | 4.26E-05 | 0.000122576 | 0.267950598 |
| Obesity | endothelial | *MX1* | 0.197964858 | 0.000496498 | 0.001023438 | 1 |
| Obesity | endothelial | *CREB3L1* | -0.735990022 | 0.002229535 | 0.003888235 | 1 |
| Obesity | endothelial | *C2CD2* | -0.289096579 | 0.007359052 | 0.011348138 | 1 |
| Obesity | endothelial | *SLC38A10* | -0.930617675 | 6.75E-06 | 2.68E-05 | 0.042462573 |
| Obesity | endothelial | *C9orf43* | -0.969895242 | 5.24E-06 | 2.18E-05 | 0.032939228 |
| Obesity | endothelial | *WDR19* | -0.920566122 | 5.11E-07 | 3.33E-06 | 0.003211805 |
| Obesity | endothelial | *SLC37A3* | -0.93510242 | 1.58E-06 | 8.16E-06 | 0.009907732 |
| Obesity | endothelial | *AP3S2* | -0.308713794 | 0.025857361 | 0.035445705 | 1 |
| Obesity | endothelial | *SPPL3* | -0.462489454 | 0.007234208 | 0.011166577 | 1 |
| Obesity | endothelial | *FAM213B* | 0.403933432 | 0.001438336 | 0.002634087 | 1 |
| Obesity | endothelial | *PEX10* | -0.993097623 | 0.001100586 | 0.002068576 | 1 |
| Obesity | endothelial | *RADIL* | -0.837921106 | 3.09E-08 | 4.48E-07 | 0.00019396 |
| Obesity | endothelial | *LDLRAP1* | -1.024090172 | 0.002375173 | 0.004109168 | 1 |
| Obesity | endothelial | *PAFAH2* | -0.686319793 | 0.0119442 | 0.017541039 | 1 |
| Obesity | endothelial | *TRIM63* | -0.388685383 | 0.012647206 | 0.018508609 | 1 |
| Obesity | endothelial | *WDR66* | -0.678834019 | 5.82E-08 | 6.41E-07 | 0.000365613 |
| Obesity | endothelial | *UBXN11* | -0.782637133 | 3.96E-06 | 1.75E-05 | 0.024912372 |
| Obesity | endothelial | *RHPN1* | 0.347697482 | 0.033908072 | 0.045483262 | 1 |
| Obesity | endothelial | *TPRG1L* | -1.32844401 | 0.000798781 | 0.001566907 | 1 |
| Obesity | endothelial | *LRRC43* | -1.059238273 | 5.41E-08 | 6.10E-07 | 0.00034012 |
| Obesity | endothelial | *XKR8* | -0.859090444 | 0.009245137 | 0.013958736 | 1 |
| Obesity | endothelial | *MRAS* | -0.405081159 | 0.001150021 | 0.00215184 | 1 |
| Obesity | endothelial | *ABHD3* | -0.648694832 | 1.93E-05 | 6.36E-05 | 0.121326494 |
| Obesity | endothelial | *CLSTN2* | -0.537981932 | 6.92E-09 | 2.56E-07 | 4.35E-05 |
| Obesity | endothelial | *COLEC12* | -0.662598668 | 3.98E-08 | 5.07E-07 | 0.000250267 |
| Obesity | endothelial | *RNF207* | -1.790916666 | 1.37E-06 | 7.30E-06 | 0.008590351 |
| Obesity | endothelial | *AUTS2* | -0.60008177 | 4.33E-07 | 2.93E-06 | 0.002723336 |
| Obesity | endothelial | *TSPAN33* | -0.377017372 | 0.001060778 | 0.002000934 | 1 |
| Obesity | endothelial | *AHCYL2* | -0.631980231 | 0.000338462 | 0.000732498 | 1 |
| Obesity | endothelial | *B4GALT5* | -0.602024159 | 0.001676303 | 0.003006823 | 1 |
| Obesity | endothelial | *CD1D* | -1.458892105 | 1.73E-09 | 1.13E-07 | 1.09E-05 |
| Obesity | endothelial | *FAM86C1* | -3.071311249 | 5.03E-06 | 2.11E-05 | 0.031648944 |
| Obesity | endothelial | *NCF1* | -1.220570241 | 1.52E-08 | 3.34E-07 | 9.58E-05 |
| Obesity | endothelial | *ZFAND2B* | -1.144527377 | 1.15E-05 | 4.14E-05 | 0.072199588 |
| Obesity | endothelial | *GDPD5* | -0.886633943 | 2.08E-07 | 1.66E-06 | 0.001304648 |
| Obesity | endothelial | *DYNC1I1* | -0.973902008 | 1.40E-07 | 1.23E-06 | 0.00088049 |

| | | | | | | |
|---|---|---|---|---|---|---|
| Obesity | endothelial | *TMED4* | -1.025329765 | 3.08E-05 | 9.37E-05 | 0.193782054 |
| Obesity | endothelial | *COPG2* | -0.368572723 | 1.74E-09 | 1.13E-07 | 1.09E-05 |
| Obesity | endothelial | *DUSP23* | 0.687414192 | 0.000872465 | 0.00168827 | 1 |
| Obesity | endothelial | *RNF166* | -1.111216367 | 2.96E-08 | 4.42E-07 | 0.000185846 |
| Obesity | endothelial | *SPATA2L* | -1.817401668 | 6.27E-09 | 2.40E-07 | 3.94E-05 |
| Obesity | endothelial | *FAM160B2* | -0.801355119 | 0.001018141 | 0.001929772 | 1 |
| Obesity | endothelial | *NDUFS2* | -0.418997825 | 0.034628296 | 0.046360327 | 1 |
| Obesity | endothelial | *SLC5A11* | -0.541338092 | 1.12E-09 | 8.21E-08 | 7.06E-06 |
| Obesity | endothelial | *CDC42SE2* | -0.527652083 | 0.004402697 | 0.007122944 | 1 |
| Obesity | endothelial | *RAPGEF6* | -1.927808297 | 2.86E-08 | 4.35E-07 | 0.000179793 |
| Obesity | endothelial | *ALG8* | -0.934027108 | 4.85E-05 | 0.000136075 | 0.305216547 |
| Obesity | endothelial | *IFNAR2* | 0.265868455 | 0.022755353 | 0.031581215 | 1 |
| Obesity | endothelial | *MRPL10* | -0.841176549 | 4.57E-05 | 0.000129318 | 0.287085857 |
| Obesity | endothelial | *SV2A* | -3.524777784 | 0.000157414 | 0.000375727 | 0.989664766 |
| Obesity | endothelial | *UBE2Z* | -0.747322503 | 0.002428684 | 0.004195121 | 1 |
| Obesity | endothelial | *CLIC6* | -0.377335666 | 7.14E-05 | 0.000189682 | 0.448788763 |
| Obesity | endothelial | *CBR1* | -0.442707834 | 0.000281595 | 0.00062119 | 1 |
| Obesity | endothelial | *SIM2* | -1.049108954 | 2.38E-09 | 1.35E-07 | 1.50E-05 |
| Obesity | endothelial | *HLCS* | -0.343871139 | 0.002779979 | 0.004716063 | 1 |
| Obesity | endothelial | *PADI4* | -2.071915579 | 1.08E-07 | 1.00E-06 | 0.000676251 |
| Obesity | endothelial | *PSMD4* | -1.066654325 | 2.68E-06 | 1.26E-05 | 0.016848393 |
| Obesity | endothelial | *STARD9* | 0.274343127 | 0.010074566 | 0.015069901 | 1 |
| Obesity | endothelial | *THEM4* | -0.828726647 | 4.77E-06 | 2.03E-05 | 0.03001469 |
| Obesity | endothelial | *TCHH* | -2.092872376 | 5.94E-09 | 2.36E-07 | 3.73E-05 |
| Obesity | endothelial | *MED8* | -0.623411817 | 0.000127439 | 0.000311271 | 0.801210375 |
| Obesity | endothelial | *RSPRY1* | -0.540023037 | 0.013804103 | 0.02002917 | 1 |
| Obesity | endothelial | *ACE* | 0.424609619 | 0.000428797 | 0.000900716 | 1 |
| Obesity | endothelial | *TEPP* | -5.977517459 | 3.91E-07 | 2.69E-06 | 0.002458665 |
| Obesity | endothelial | *ATP6V0D1* | -1.122954626 | 1.43E-08 | 3.34E-07 | 8.97E-05 |
| Obesity | endothelial | *C16orf86* | -0.963362428 | 9.17E-06 | 3.44E-05 | 0.057670342 |
| Obesity | endothelial | *CCDC117* | -1.316019197 | 2.66E-07 | 2.01E-06 | 0.001674511 |
| Obesity | endothelial | *CCDC107* | -1.000100656 | 3.54E-10 | 3.37E-08 | 2.22E-06 |
| Obesity | endothelial | *NPR2* | -1.474691603 | 8.75E-07 | 5.10E-06 | 0.005503943 |
| Obesity | endothelial | *CALM3* | -1.349293321 | 3.93E-07 | 2.70E-06 | 0.002473865 |
| Obesity | endothelial | *DFFA* | -0.541475836 | 0.013074755 | 0.019032412 | 1 |
| Obesity | endothelial | *ATAD3B* | -0.934089007 | 0.000449593 | 0.000938754 | 1 |
| Obesity | endothelial | *SSU72* | -1.059753224 | 3.83E-05 | 0.000112138 | 0.241095936 |
| Obesity | endothelial | *NR2F6* | -1.048772997 | 2.73E-06 | 1.28E-05 | 0.017134979 |
| Obesity | endothelial | *ANKLE1* | -1.3092811 | 4.43E-08 | 5.42E-07 | 0.000278521 |
| Obesity | endothelial | *CILP2* | -2.883282689 | 1.90E-08 | 3.50E-07 | 0.000119548 |
| Obesity | endothelial | *ABCG1* | 0.346178207 | 0.00025384 | 0.000568135 | 1 |
| Obesity | endothelial | *SLC37A1* | 0.491135657 | 0.000864414 | 0.001676819 | 1 |
| Obesity | endothelial | *U2AF1* | -0.614651195 | 0.023997724 | 0.033108118 | 1 |
| Obesity | endothelial | *HSF2BP* | -0.629162499 | 0.007702893 | 0.011817494 | 1 |
| Obesity | endothelial | *CSTB* | -0.667827896 | 0.000290003 | 0.000637275 | 1 |
| Obesity | endothelial | *C21orf33* | -1.853835497 | 4.50E-08 | 5.45E-07 | 0.000283227 |
| Obesity | endothelial | *C21orf2* | -0.307474973 | 0.008021005 | 0.012254693 | 1 |
| Obesity | endothelial | *LRRC3* | -0.938858296 | 0.00016418 | 0.000389215 | 1 |
| Obesity | endothelial | *ITGB2* | -0.516235278 | 3.78E-05 | 0.000111089 | 0.237596884 |
| Obesity | endothelial | *FAM207A* | -0.677301164 | 0.00155623 | 0.002815544 | 1 |

| | | | | | | |
|---|---|---|---|---|---|---|
| Obesity | endothelial | *FTCD* | 0.248425027 | 0.027708235 | 0.03779598 | 1 |
| Obesity | endothelial | *LSS* | -0.525654791 | 0.000136594 | 0.000330421 | 0.858763441 |
| Obesity | endothelial | *VAV2* | -0.821533255 | 0.002103256 | 0.0036895 | 1 |
| Obesity | endothelial | *MCM3AP* | -0.526298893 | 3.32E-05 | 9.98E-05 | 0.208776126 |
| Obesity | endothelial | *PCNT* | -0.357373933 | 0.004706632 | 0.007575678 | 1 |
| Obesity | endothelial | *S100B* | -0.251224637 | 0.00021424 | 0.000490505 | 1 |
| Obesity | endothelial | *ADAMTS13* | -0.772325767 | 0.000240344 | 0.000541277 | 1 |
| Obesity | endothelial | *ZNF714* | -0.88515081 | 3.78E-05 | 0.000111089 | 0.237619843 |
| Obesity | endothelial | *GPSM1* | -0.668127238 | 0.014249886 | 0.020637879 | 1 |
| Obesity | endothelial | *RDH13* | -0.969189031 | 0.002964101 | 0.004986701 | 1 |
| Obesity | endothelial | *MPZL3* | 0.436079325 | 1.26E-05 | 4.43E-05 | 0.079085919 |
| Obesity | endothelial | *PCSK7* | -0.713780264 | 0.001031041 | 0.001951869 | 1 |
| Obesity | endothelial | *S100A1* | -1.202828853 | 0.00437185 | 0.007078501 | 1 |
| Obesity | endothelial | *ZBTB7B* | -1.656833742 | 4.10E-11 | 6.34E-09 | 2.58E-07 |
| Obesity | endothelial | *VPS11* | -1.284635712 | 5.93E-08 | 6.50E-07 | 0.000372632 |
| Obesity | endothelial | *PAQR6* | -1.452145929 | 2.08E-09 | 1.25E-07 | 1.31E-05 |
| Obesity | endothelial | *PMF1* | -0.661919584 | 0.002556233 | 0.004383807 | 1 |
| Obesity | endothelial | *SLC25A44* | -1.008354821 | 0.003253734 | 0.005430376 | 1 |
| Obesity | endothelial | *NACC1* | -1.56244771 | 5.05E-06 | 2.12E-05 | 0.031727035 |
| Obesity | endothelial | *LY6K* | 1.347245518 | 0.004829976 | 0.007760301 | 1 |
| Obesity | endothelial | *LY6E* | 0.363475705 | 0.02843414 | 0.038695304 | 1 |
| Obesity | endothelial | *TONSL* | -1.934134984 | 0.001072363 | 0.002020361 | 1 |
| Obesity | endothelial | *MUM1* | -0.808954978 | 1.17E-05 | 4.21E-05 | 0.073814667 |
| Obesity | endothelial | *ZNF333* | -0.832062538 | 0.000398784 | 0.000846155 | 1 |
| Obesity | endothelial | *ORAI2* | 0.68157446 | 0.000475514 | 0.000985027 | 1 |
| Obesity | endothelial | *FBXL13* | -0.763011192 | 0.000467934 | 0.000970604 | 1 |
| Obesity | endothelial | *MFSD12* | -1.052001582 | 0.000127515 | 0.000311335 | 0.801686352 |
| Obesity | endothelial | *YDJC* | -3.546927145 | 9.30E-06 | 3.48E-05 | 0.058468225 |
| Obesity | endothelial | *DVL3* | -0.948523533 | 4.94E-05 | 0.000138141 | 0.31063432 |
| Obesity | endothelial | *AP2M1* | -0.926629312 | 8.39E-05 | 0.000217088 | 0.527523981 |
| Obesity | endothelial | *FBXO27* | -0.234263158 | 0.01004593 | 0.015030643 | 1 |
| Obesity | endothelial | *DMKN* | 0.359889615 | 5.72E-05 | 0.000157573 | 0.359738851 |
| Obesity | endothelial | *U2AF1L4* | -0.275549006 | 0.002190274 | 0.00382826 | 1 |
| Obesity | endothelial | *BDH1* | -1.446845383 | 2.26E-08 | 3.81E-07 | 0.000141855 |
| Obesity | endothelial | *COX7A1* | -0.558077355 | 0.003016819 | 0.005063198 | 1 |
| Obesity | endothelial | *DUSP14* | -0.867720275 | 6.28E-10 | 5.29E-08 | 3.95E-06 |
| Obesity | endothelial | *IKZF3* | -1.101611254 | 7.36E-07 | 4.45E-06 | 0.004628729 |
| Obesity | endothelial | *ACOX1* | -0.839940779 | 1.12E-06 | 6.24E-06 | 0.007049551 |
| Obesity | endothelial | *ZNF577* | -0.375316041 | 0.025681214 | 0.035237405 | 1 |
| Obesity | endothelial | *TMEM143* | -0.641200785 | 7.40E-05 | 0.000194843 | 0.46508909 |
| Obesity | endothelial | *CCL16* | 0.253519266 | 3.16E-05 | 9.56E-05 | 0.198520237 |
| Obesity | endothelial | *SIGLEC11* | -1.125845175 | 1.43E-07 | 1.25E-06 | 0.000897989 |
| Obesity | endothelial | *ASB16* | -1.026072951 | 0.000201732 | 0.000466283 | 1 |
| Obesity | endothelial | *PLCD3* | -0.519355358 | 0.009879419 | 0.01481889 | 1 |
| Obesity | endothelial | *RACGAP1* | -1.589122834 | 1.51E-08 | 3.34E-07 | 9.48E-05 |
| Obesity | endothelial | *LARP4* | -0.557045079 | 0.000915838 | 0.001764595 | 1 |
| Obesity | endothelial | *SPC24* | -2.32838601 | 3.19E-05 | 9.65E-05 | 0.200844061 |
| Obesity | endothelial | *LEMD2* | -0.968727755 | 0.000146836 | 0.00035262 | 0.923159937 |
| Obesity | endothelial | *SCIMP* | -0.995101173 | 1.80E-09 | 1.14E-07 | 1.13E-05 |
| Obesity | endothelial | *ASGR2* | -1.581850708 | 4.75E-08 | 5.61E-07 | 0.000298454 |

| | | | | | | |
|---|---|---|---|---|---|---|
| Obesity | endothelial | *SENP3* | -1.805635184 | 1.07E-06 | 6.03E-06 | 0.006737764 |
| Obesity | endothelial | *SNRNP25* | -1.164153679 | 0.004621646 | 0.007452241 | 1 |
| Obesity | endothelial | *WDR90* | -0.701440937 | 8.33E-07 | 4.91E-06 | 0.005234158 |
| Obesity | endothelial | *ZG16B* | -4.94773028 | 6.66E-08 | 7.07E-07 | 0.000418688 |
| Obesity | endothelial | *TTC9C* | -0.80428612 | 0.000820843 | 0.001605675 | 1 |
| Obesity | endothelial | *NXF1* | -0.983793598 | 4.24E-06 | 1.84E-05 | 0.026650674 |
| Obesity | endothelial | *TPCN2* | -0.671239659 | 0.00022189 | 0.000504893 | 1 |
| Obesity | endothelial | *CYP4A22* | -2.197040098 | 8.38E-07 | 4.92E-06 | 0.005268618 |
| Obesity | endothelial | *PDZK1IP1* | 6.951132498 | 0.02361341 | 0.032656733 | 1 |
| Obesity | endothelial | *COA7* | -1.800170382 | 1.26E-05 | 4.43E-05 | 0.079098845 |
| Obesity | endothelial | *SLC1A7* | -0.295438316 | 0.000386036 | 0.000821601 | 1 |
| Obesity | endothelial | *C1orf123* | -1.002205163 | 0.000199116 | 0.000460745 | 1 |
| Obesity | endothelial | *ACOT11* | -0.783198894 | 0.000180309 | 0.000421414 | 1 |
| Obesity | endothelial | *USP24* | -0.543596895 | 0.000174364 | 0.000410417 | 1 |
| Obesity | endothelial | *RBP7* | 0.296406602 | 1.47E-05 | 5.06E-05 | 0.09246569 |
| Obesity | endothelial | *FBLIM1* | 0.436381936 | 6.42E-05 | 0.000173458 | 0.403809568 |
| Obesity | endothelial | *SLC25A34* | -2.008137934 | 2.97E-08 | 4.43E-07 | 0.000187 |
| Obesity | endothelial | *PDPN* | -0.605731277 | 1.51E-08 | 3.34E-07 | 9.46E-05 |
| Obesity | endothelial | *LRRC38* | -2.490414665 | 1.15E-07 | 1.05E-06 | 0.000723298 |
| Obesity | endothelial | *LAPTM5* | -0.826580636 | 0.000100583 | 0.000253656 | 0.632363198 |
| Obesity | endothelial | *SYNC* | -0.803394203 | 0.000116567 | 0.00028786 | 0.73285568 |
| Obesity | endothelial | *TSSK3* | -0.734348643 | 0.000119346 | 0.000293903 | 0.750328795 |
| Obesity | endothelial | *CAMK2N1* | -0.954876101 | 4.09E-08 | 5.16E-07 | 0.000257309 |
| Obesity | endothelial | *TTLL10* | 0.415560678 | 1.35E-06 | 7.22E-06 | 0.008470176 |
| Obesity | endothelial | *OMA1* | -0.413857696 | 0.00946418 | 0.014248395 | 1 |
| Obesity | endothelial | *TM2D1* | -0.65966729 | 0.000852915 | 0.001658093 | 1 |
| Obesity | endothelial | *FUBP1* | -0.703448764 | 0.000204125 | 0.0004711 | 1 |
| Obesity | endothelial | *NEXN* | -0.898780212 | 6.35E-07 | 3.96E-06 | 0.003991141 |
| Obesity | endothelial | *HFM1* | -1.126579713 | 9.36E-06 | 3.49E-05 | 0.058855609 |
| Obesity | endothelial | *VCAM1* | 0.201384346 | 3.16E-06 | 1.46E-05 | 0.019893731 |
| Obesity | endothelial | *EXTL2* | -0.647619973 | 2.95E-06 | 1.37E-05 | 0.018574669 |
| Obesity | endothelial | *ARPC5* | -0.841746764 | 4.96E-06 | 2.09E-05 | 0.03118689 |
| Obesity | endothelial | *IGSF8* | -1.31122231 | 3.64E-08 | 4.85E-07 | 0.000228867 |
| Obesity | endothelial | *VANGL2* | 0.974997189 | 0.000309648 | 0.000676426 | 1 |
| Obesity | endothelial | *FCRLB* | -1.100404432 | 5.58E-06 | 2.29E-05 | 0.03507862 |
| Obesity | endothelial | *LMX1A* | -0.500111624 | 2.35E-07 | 1.83E-06 | 0.001475566 |
| Obesity | endothelial | *FLVCR1* | -0.553195699 | 0.035302635 | 0.0471627 | 1 |
| Obesity | endothelial | *TDRD5* | -0.609796392 | 0.001367666 | 0.002514922 | 1 |
| Obesity | endothelial | *SNED1* | -0.547061693 | 0.000859102 | 0.001668575 | 1 |
| Obesity | endothelial | *C1orf115* | 0.501498087 | 2.24E-06 | 1.10E-05 | 0.014105706 |
| Obesity | endothelial | *ACP6* | -1.289562906 | 5.25E-10 | 4.65E-08 | 3.30E-06 |
| Obesity | endothelial | *HAAO* | -0.913248432 | 7.18E-06 | 2.81E-05 | 0.045171526 |
| Obesity | endothelial | *B3GALNT2* | -0.622180694 | 0.000290104 | 0.000637275 | 1 |
| Obesity | endothelial | *FAM84A* | -1.801236481 | 1.56E-08 | 3.35E-07 | 9.82E-05 |
| Obesity | endothelial | *FRZB* | -1.269540743 | 1.47E-07 | 1.27E-06 | 0.000924145 |
| Obesity | endothelial | *NUP35* | -0.843792524 | 0.002151118 | 0.003769253 | 1 |
| Obesity | endothelial | *FBXO41* | -1.153161469 | 3.65E-08 | 4.85E-07 | 0.000229333 |
| Obesity | endothelial | *CCDC74A* | -2.012930157 | 3.95E-08 | 5.06E-07 | 0.000248599 |
| Obesity | endothelial | *NOSTRIN* | 0.178588183 | 4.03E-06 | 1.77E-05 | 0.025311594 |
| Obesity | endothelial | *SGPP2* | -1.334204054 | 9.91E-07 | 5.66E-06 | 0.006230296 |

| | | | | | | |
|---|---|---|---|---|---|---|
| Obesity | endothelial | *BBS5* | -0.91001195 | 3.82E-05 | 0.000111821 | 0.239983914 |
| Obesity | endothelial | *HPGDS* | -0.697225314 | 7.58E-06 | 2.94E-05 | 0.047646275 |
| Obesity | endothelial | *CTSS* | -0.309270188 | 0.003000505 | 0.005039854 | 1 |
| Obesity | endothelial | *C1QTNF7* | -0.710509599 | 5.96E-08 | 6.53E-07 | 0.000374806 |
| Obesity | endothelial | *LYSMD1* | -0.781285611 | 0.019486245 | 0.027508588 | 1 |
| Obesity | endothelial | *ERCC3* | -0.54074203 | 0.000446362 | 0.000932628 | 1 |
| Obesity | endothelial | *RNF149* | -0.765693264 | 4.76E-06 | 2.02E-05 | 0.029924086 |
| Obesity | endothelial | *DHX57* | -0.700482644 | 0.001252839 | 0.002326912 | 1 |
| Obesity | endothelial | *S100A9* | -1.660533083 | 3.37E-07 | 2.40E-06 | 0.002121779 |
| Obesity | endothelial | *S100A12* | -3.870656801 | 0.011015122 | 0.016317642 | 1 |
| Obesity | endothelial | *TDRD10* | 0.416893183 | 0.002453969 | 0.004232675 | 1 |
| Obesity | endothelial | *DCAF16* | -0.852457326 | 0.00072351 | 0.001433566 | 1 |
| Obesity | endothelial | *NIPAL1* | -1.206185689 | 1.88E-08 | 3.47E-07 | 0.000118478 |
| Obesity | endothelial | *ANTXR2* | -0.463368264 | 5.62E-05 | 0.000155169 | 0.353631139 |
| Obesity | endothelial | *CGGBP1* | -0.245362455 | 0.008868698 | 0.013435543 | 1 |
| Obesity | endothelial | *PBXIP1* | -1.153114051 | 5.83E-06 | 2.37E-05 | 0.036680427 |
| Obesity | endothelial | *DCST2* | 0.916947251 | 9.25E-05 | 0.000235862 | 0.58163663 |
| Obesity | endothelial | *LMOD3* | -1.337505281 | 1.40E-08 | 3.34E-07 | 8.78E-05 |
| Obesity | endothelial | *NBPF10* | -0.759194382 | 2.24E-07 | 1.77E-06 | 0.001409264 |
| Obesity | endothelial | *SLC22A15* | -0.721649317 | 1.33E-06 | 7.14E-06 | 0.008347718 |
| Obesity | endothelial | *SLC15A2* | -1.097111258 | 2.31E-07 | 1.80E-06 | 0.001449206 |
| Obesity | endothelial | *LRRC58* | -1.062332981 | 4.38E-06 | 1.89E-05 | 0.027567553 |
| Obesity | endothelial | *FSTL1* | -0.64170583 | 9.60E-07 | 5.52E-06 | 0.006034565 |
| Obesity | endothelial | *LMOD1* | -1.079011777 | 2.41E-20 | 3.03E-17 | 1.52E-16 |
| Obesity | endothelial | *TMEM183A* | -0.932111831 | 0.009148684 | 0.013833038 | 1 |
| Obesity | endothelial | *ARPC2* | -0.596689327 | 0.001863737 | 0.003310912 | 1 |
| Obesity | endothelial | *CCT3* | -0.464771922 | 0.021572175 | 0.030150027 | 1 |
| Obesity | endothelial | *GLB1L* | -0.852383974 | 0.000363254 | 0.000778649 | 1 |
| Obesity | endothelial | *NUAK2* | -0.440061013 | 0.011583468 | 0.017071089 | 1 |
| Obesity | endothelial | *PYHIN1* | -1.006949385 | 6.28E-10 | 5.29E-08 | 3.95E-06 |
| Obesity | endothelial | *AIM2* | 0.238236353 | 2.49E-06 | 1.19E-05 | 0.015628439 |
| Obesity | endothelial | *CD200R1* | -0.782853516 | 7.84E-07 | 4.69E-06 | 0.004926561 |
| Obesity | endothelial | *GTPBP8* | -0.735183869 | 0.009482369 | 0.014272361 | 1 |
| Obesity | endothelial | *SPICE1* | -0.511783242 | 0.017287711 | 0.024651358 | 1 |
| Obesity | endothelial | *CADPS* | -1.144527838 | 1.43E-08 | 3.34E-07 | 8.97E-05 |
| Obesity | endothelial | *PPM1K* | -0.916239517 | 9.05E-05 | 0.000231452 | 0.568909982 |
| Obesity | endothelial | *TIPARP* | -0.763017913 | 0.000209947 | 0.00048268 | 1 |
| Obesity | endothelial | *RPP14* | -1.423415092 | 9.66E-07 | 5.54E-06 | 0.006074284 |
| Obesity | endothelial | *DNASE1L3* | 0.318346812 | 3.92E-06 | 1.73E-05 | 0.024614498 |
| Obesity | endothelial | *CRELD1* | -0.566306546 | 0.000405717 | 0.000858547 | 1 |
| Obesity | endothelial | *PCOLCE2* | -0.586936901 | 1.17E-08 | 3.34E-07 | 7.37E-05 |
| Obesity | endothelial | *TTC14* | -0.495285093 | 0.001578411 | 0.002850496 | 1 |
| Obesity | endothelial | *RCHY1* | -0.851028614 | 8.70E-05 | 0.000223752 | 0.547074186 |
| Obesity | endothelial | *CCDC158* | -0.8178658 | 6.60E-07 | 4.09E-06 | 0.00415252 |
| Obesity | endothelial | *GYG1* | -0.663077044 | 0.019154068 | 0.027097576 | 1 |
| Obesity | endothelial | *HPS3* | -0.775252257 | 0.0007254 | 0.001436859 | 1 |
| Obesity | endothelial | *RYK* | -0.408890245 | 0.031500664 | 0.042489739 | 1 |
| Obesity | endothelial | *PLB1* | -1.26466808 | 2.07E-08 | 3.63E-07 | 0.000130086 |
| Obesity | endothelial | *CDCP1* | -0.911307414 | 1.42E-08 | 3.34E-07 | 8.94E-05 |
| Obesity | endothelial | *LZTFL1* | -1.278414719 | 2.65E-06 | 1.25E-05 | 0.016652304 |

| | | | | | | |
|---|---|---|---|---|---|---|
| Obesity | endothelial | *FYCO1* | -0.964049788 | 3.07E-05 | 9.34E-05 | 0.192784279 |
| Obesity | endothelial | *LRRC2* | -0.677134967 | 1.19E-06 | 6.54E-06 | 0.007502928 |
| Obesity | endothelial | *TPRA1* | -1.218929191 | 3.41E-06 | 1.54E-05 | 0.021410987 |
| Obesity | endothelial | *MEAF6* | -1.000806058 | 0.000132726 | 0.000322431 | 0.834450835 |
| Obesity | endothelial | *DNALI1* | -1.982895279 | 8.48E-13 | 2.81E-10 | 5.33E-09 |
| Obesity | endothelial | *RFC4* | -0.895418121 | 0.007465163 | 0.011494852 | 1 |
| Obesity | endothelial | *RFT1* | -1.573227853 | 3.74E-10 | 3.47E-08 | 2.35E-06 |
| Obesity | endothelial | *SFMBT1* | -1.069755725 | 4.81E-07 | 3.18E-06 | 0.003025541 |
| Obesity | endothelial | *GNL3* | -1.102003155 | 4.17E-07 | 2.83E-06 | 0.002619308 |
| Obesity | endothelial | *UVSSA* | -0.428548602 | 0.013131787 | 0.019102161 | 1 |
| Obesity | endothelial | *FAM208A* | -0.503654743 | 0.005951397 | 0.009349434 | 1 |
| Obesity | endothelial | *LRPAP1* | -0.794808828 | 0.001491509 | 0.00271643 | 1 |
| Obesity | endothelial | *SLC51A* | -1.360614805 | 6.48E-08 | 6.95E-07 | 0.000407701 |
| Obesity | endothelial | *PIGX* | -0.966810393 | 2.12E-05 | 6.87E-05 | 0.133392523 |
| Obesity | endothelial | *ABLIM2* | 0.402192986 | 5.61E-05 | 0.000154863 | 0.352777143 |
| Obesity | endothelial | *EXO5* | -0.994824778 | 1.26E-05 | 4.43E-05 | 0.078952705 |
| Obesity | endothelial | *C1orf50* | -1.770909601 | 2.75E-09 | 1.49E-07 | 1.73E-05 |
| Obesity | endothelial | *ERMAP* | -0.633212965 | 6.92E-05 | 0.000184593 | 0.435085536 |
| Obesity | endothelial | *SLC9B2* | 0.482010601 | 0.002864061 | 0.004844323 | 1 |
| Obesity | endothelial | *BDH2* | -0.655584165 | 9.68E-06 | 3.60E-05 | 0.060853057 |
| Obesity | endothelial | *CDC25A* | -0.484498917 | 0.001785262 | 0.003184098 | 1 |
| Obesity | endothelial | *ZNF589* | -0.396983537 | 0.002098196 | 0.003681652 | 1 |
| Obesity | endothelial | *ATRIP* | -3.629262387 | 0.000348506 | 0.000750876 | 1 |
| Obesity | endothelial | *SHISA5* | 0.468271968 | 0.003007058 | 0.005048163 | 1 |
| Obesity | endothelial | *APEH* | -1.585369098 | 2.33E-19 | 2.45E-16 | 1.47E-15 |
| Obesity | endothelial | *RNF123* | 0.875602258 | 3.22E-09 | 1.69E-07 | 2.02E-05 |
| Obesity | endothelial | *MST1R* | 0.677195078 | 0.000655449 | 0.00131069 | 1 |
| Obesity | endothelial | *RAD54L2* | -0.657366072 | 0.000974282 | 0.001857843 | 1 |
| Obesity | endothelial | *TEX264* | -0.780195962 | 0.000257222 | 0.000574681 | 1 |
| Obesity | endothelial | *GRM2* | 13.11471372 | 7.84E-07 | 4.69E-06 | 0.004930927 |
| Obesity | endothelial | *POC1A* | 1.056871641 | 0.001652963 | 0.002969194 | 1 |
| Obesity | endothelial | *PPM1M* | -1.080362627 | 2.25E-12 | 6.15E-10 | 1.41E-08 |
| Obesity | endothelial | *C4orf3* | -0.674954576 | 0.013168841 | 0.019151631 | 1 |
| Obesity | endothelial | *ANXA5* | -0.742429419 | 2.05E-06 | 1.01E-05 | 0.012859972 |
| Obesity | endothelial | *MAP9* | -1.042978831 | 1.08E-08 | 3.32E-07 | 6.81E-05 |
| Obesity | endothelial | *CEP44* | -0.914062871 | 5.21E-05 | 0.000144899 | 0.327616172 |
| Obesity | endothelial | *TMEM144* | -0.917346324 | 1.59E-07 | 1.35E-06 | 0.000996995 |
| Obesity | endothelial | *NPY1R* | -0.614756601 | 1.61E-08 | 3.37E-07 | 0.000101381 |
| Obesity | endothelial | *IL15* | 0.352805543 | 0.000426636 | 0.000896776 | 1 |
| Obesity | endothelial | *ARFIP1* | -0.483152668 | 0.007180452 | 0.011097222 | 1 |
| Obesity | endothelial | *ITGA2* | 0.319466874 | 4.13E-06 | 1.80E-05 | 0.025939482 |
| Obesity | endothelial | *NDUFAF2* | -0.659857696 | 0.00026839 | 0.000596033 | 1 |
| Obesity | endothelial | *LMBRD2* | -1.021357184 | 2.23E-05 | 7.14E-05 | 0.139951136 |
| Obesity | endothelial | *PGGT1B* | -0.767283739 | 0.000802124 | 0.001572484 | 1 |
| Obesity | endothelial | *CMBL* | -0.945871609 | 1.18E-08 | 3.34E-07 | 7.40E-05 |
| Obesity | endothelial | *C5orf63* | -1.544634836 | 7.71E-08 | 7.84E-07 | 0.000484925 |
| Obesity | endothelial | *PRRC1* | -0.957631147 | 2.41E-05 | 7.61E-05 | 0.151437082 |
| Obesity | endothelial | *F2RL1* | -0.7113448 | 0.005840698 | 0.009189307 | 1 |
| Obesity | endothelial | *WDR41* | -0.668235487 | 2.99E-05 | 9.16E-05 | 0.187966466 |
| Obesity | endothelial | *NDUFS4* | -0.673041851 | 0.000630277 | 0.001264778 | 1 |

| | | | | | | |
|---|---|---|---|---|---|---|
| Obesity | endothelial | *RHOBTB3* | -0.503033288 | 6.92E-07 | 4.24E-06 | 0.004351775 |
| Obesity | endothelial | *TIGD6* | -1.628864118 | 3.25E-08 | 4.54E-07 | 0.000204413 |
| Obesity | endothelial | *SERINC5* | -0.658891035 | 0.000318745 | 0.000694129 | 1 |
| Obesity | endothelial | *ENPP6* | 1.393548121 | 0.005795746 | 0.009130006 | 1 |
| Obesity | endothelial | *PRIMPOL* | -0.669054443 | 0.000424044 | 0.000892523 | 1 |
| Obesity | endothelial | *ERAP1* | -0.583044198 | 0.000101447 | 0.000255528 | 0.637798368 |
| Obesity | endothelial | *ANKRA2* | -0.998429632 | 3.57E-05 | 0.000106011 | 0.224742662 |
| Obesity | endothelial | *UBLCP1* | -0.705992953 | 0.000219603 | 0.000500415 | 1 |
| Obesity | endothelial | *UTP15* | -1.326773671 | 3.47E-05 | 0.000103771 | 0.218335104 |
| Obesity | endothelial | *TLR3* | 0.555794983 | 0.014089448 | 0.020425319 | 1 |
| Obesity | endothelial | *CCDC127* | -1.350428358 | 1.32E-05 | 4.61E-05 | 0.082802483 |
| Obesity | endothelial | *GDF9* | -1.245938311 | 0.000146613 | 0.000352219 | 0.921757296 |
| Obesity | endothelial | *GRIK2* | -0.637597526 | 0.000400657 | 0.000849405 | 1 |
| Obesity | endothelial | *TXLNB* | -1.193026958 | 2.17E-07 | 1.71E-06 | 0.001362585 |
| Obesity | endothelial | *TMEM200A* | -1.479905683 | 2.67E-06 | 1.26E-05 | 0.016793901 |
| Obesity | endothelial | *PDSS2* | -0.568848246 | 0.001494191 | 0.002720527 | 1 |
| Obesity | endothelial | *STXBP5* | -0.581106709 | 0.000940699 | 0.001805304 | 1 |
| Obesity | endothelial | *PI16* | -0.791276222 | 5.65E-08 | 6.28E-07 | 0.000355023 |
| Obesity | endothelial | *DAGLB* | -0.676916855 | 0.000294942 | 0.000647224 | 1 |
| Obesity | endothelial | *STK17A* | -0.567273964 | 0.006129821 | 0.009600942 | 1 |
| Obesity | endothelial | *SAP30L* | -1.089100926 | 3.80E-05 | 0.00011161 | 0.239179336 |
| Obesity | endothelial | *HCN1* | -1.264046845 | 3.88E-09 | 1.84E-07 | 2.44E-05 |
| Obesity | endothelial | *SLU7* | -0.80856337 | 0.00073067 | 0.00144593 | 1 |
| Obesity | endothelial | *RP9* | -1.198987849 | 6.09E-07 | 3.84E-06 | 0.003829122 |
| Obesity | endothelial | *CAMLG* | -0.571202463 | 0.009142172 | 0.013827921 | 1 |
| Obesity | endothelial | *KCNK5* | 0.918280437 | 0.002581305 | 0.004417064 | 1 |
| Obesity | endothelial | *USP49* | -1.002743969 | 1.53E-07 | 1.32E-06 | 0.000964943 |
| Obesity | endothelial | *ZNF704* | -0.623402684 | 0.000126945 | 0.000310367 | 0.798103982 |
| Obesity | endothelial | *FNDC1* | -0.560890738 | 2.08E-05 | 6.77E-05 | 0.13063515 |
| Obesity | endothelial | *CHMP4C* | 0.288359571 | 0.002129356 | 0.003733202 | 1 |
| Obesity | endothelial | *CTSB* | -0.53468671 | 1.27E-06 | 6.86E-06 | 0.007976959 |
| Obesity | endothelial | *PEX2* | -1.04310797 | 1.08E-05 | 3.92E-05 | 0.06769133 |
| Obesity | endothelial | *DEFB1* | -0.748532134 | 6.62E-07 | 4.09E-06 | 0.004161056 |
| Obesity | endothelial | *OXR1* | -0.440329747 | 0.00903613 | 0.013682599 | 1 |
| Obesity | endothelial | *GPER1* | 0.485404331 | 0.000202951 | 0.000468756 | 1 |
| Obesity | endothelial | *TMEM184A* | -1.536069024 | 2.29E-05 | 7.30E-05 | 0.144010565 |
| Obesity | endothelial | *INTS1* | -1.235279867 | 8.98E-06 | 3.38E-05 | 0.056439365 |
| Obesity | endothelial | *SLC4A2* | -1.375832615 | 8.35E-07 | 4.91E-06 | 0.005247496 |
| Obesity | endothelial | *FASTK* | -1.242322412 | 6.12E-07 | 3.85E-06 | 0.003847403 |
| Obesity | endothelial | *ALDH7A1* | -0.385029893 | 0.022311248 | 0.031060854 | 1 |
| Obesity | endothelial | *COX6C* | -0.949233961 | 3.90E-05 | 0.000113609 | 0.24505417 |
| Obesity | endothelial | *YWHAZ* | -0.737917478 | 7.81E-08 | 7.89E-07 | 0.000490801 |
| Obesity | endothelial | *SLC25A32* | -0.762986125 | 0.00094674 | 0.001814681 | 1 |
| Obesity | endothelial | *INTS8* | -0.658828371 | 0.001754121 | 0.003133 | 1 |
| Obesity | endothelial | *FREM1* | -0.582493708 | 8.58E-09 | 2.92E-07 | 5.39E-05 |
| Obesity | endothelial | *RPP25L* | -1.470672754 | 1.82E-05 | 6.05E-05 | 0.114153139 |
| Obesity | endothelial | *TMEM65* | -0.480321916 | 0.019380433 | 0.027387004 | 1 |
| Obesity | endothelial | *SYK* | -0.828650541 | 9.96E-08 | 9.41E-07 | 0.000626047 |
| Obesity | endothelial | *TMC1* | 0.506050406 | 0.005750871 | 0.009068403 | 1 |
| Obesity | endothelial | *HGSNAT* | -0.461215787 | 0.011905849 | 0.017492889 | 1 |

| | | | | | | |
|---|---|---|---|---|---|---|
| Obesity | endothelial | *C9orf64* | -0.949495613 | 5.54E-07 | 3.55E-06 | 0.003481636 |
| Obesity | endothelial | *HNRNPK* | -0.687001423 | 0.001486649 | 0.002711505 | 1 |
| Obesity | endothelial | *SVEP1* | -0.516908028 | 3.26E-08 | 4.55E-07 | 0.000205169 |
| Obesity | endothelial | *ZNF367* | -0.694565199 | 0.000971898 | 0.001853859 | 1 |
| Obesity | endothelial | *NOL6* | -1.460492749 | 8.02E-05 | 0.000208405 | 0.50413084 |
| Obesity | endothelial | *AQP3* | -0.77956623 | 0.029536015 | 0.040011759 | 1 |
| Obesity | endothelial | *TRMT10B* | -0.78125992 | 0.000255249 | 0.000570883 | 1 |
| Obesity | endothelial | *VCP* | -0.820466156 | 5.24E-06 | 2.18E-05 | 0.03292046 |
| Obesity | endothelial | *PIGO* | -1.252637238 | 0.002867689 | 0.004849156 | 1 |
| Obesity | endothelial | *LRFN5* | -0.297289114 | 0.002484448 | 0.004274691 | 1 |
| Obesity | endothelial | *WRN* | -0.402045539 | 0.032608856 | 0.043852808 | 1 |
| Obesity | endothelial | *CFL2* | -1.014137783 | 5.16E-06 | 2.15E-05 | 0.032440028 |
| Obesity | endothelial | *ZCCHC24* | -0.877174108 | 4.92E-06 | 2.07E-05 | 0.030928866 |
| Obesity | endothelial | *GJB2* | 0.553014323 | 1.23E-05 | 4.35E-05 | 0.077033377 |
| Obesity | endothelial | *REEP3* | -0.752299115 | 5.27E-05 | 0.000146267 | 0.331586496 |
| Obesity | endothelial | *LRR1* | -1.255193855 | 4.18E-06 | 1.82E-05 | 0.026286977 |
| Obesity | endothelial | *RPL36AL* | -0.702998677 | 0.000519857 | 0.001066691 | 1 |
| Obesity | endothelial | *KLHDC2* | -0.461970965 | 0.022838883 | 0.031669179 | 1 |
| Obesity | endothelial | *OXGR1* | 2.252296984 | 0.003324707 | 0.005532671 | 1 |
| Obesity | endothelial | *ATP5C1* | -0.723224284 | 0.000487496 | 0.001007452 | 1 |
| Obesity | endothelial | *TAF3* | -0.484145853 | 0.011208772 | 0.016571826 | 1 |
| Obesity | endothelial | *VDAC2* | -1.171492731 | 4.13E-06 | 1.80E-05 | 0.025935698 |
| Obesity | endothelial | *SLC18A2* | -1.131814276 | 2.54E-05 | 7.95E-05 | 0.159421829 |
| Obesity | endothelial | *PDZD8* | -0.397887453 | 0.029902717 | 0.040473279 | 1 |
| Obesity | endothelial | *SNAPC4* | -0.628686045 | 9.79E-05 | 0.000248254 | 0.615669123 |
| Obesity | endothelial | *SDCCAG3* | -1.165160239 | 7.96E-07 | 4.75E-06 | 0.005002466 |
| Obesity | endothelial | *AK8* | -1.257994028 | 9.31E-06 | 3.48E-05 | 0.058548765 |
| Obesity | endothelial | *TSC1* | -0.607990235 | 0.005233159 | 0.008323013 | 1 |
| Obesity | endothelial | *ZMYND19* | -1.375338831 | 1.47E-08 | 3.34E-07 | 9.27E-05 |
| Obesity | endothelial | *RET* | -0.764402149 | 1.36E-08 | 3.34E-07 | 8.55E-05 |
| Obesity | endothelial | *STK32C* | -1.288461107 | 6.66E-07 | 4.10E-06 | 0.004186794 |
| Obesity | endothelial | *NSMF* | -1.553775803 | 8.00E-07 | 4.77E-06 | 0.005029472 |
| Obesity | endothelial | *VWA2* | 0.375455144 | 2.82E-05 | 8.74E-05 | 0.177429284 |
| Obesity | endothelial | *METTL3* | -0.741064683 | 0.005783627 | 0.009115484 | 1 |
| Obesity | endothelial | *TRUB1* | -0.518102406 | 0.002720843 | 0.004626978 | 1 |
| Obesity | endothelial | *HSPA12A* | -0.477389354 | 1.45E-07 | 1.27E-06 | 0.000913522 |
| Obesity | endothelial | *UBTD1* | -1.046039272 | 1.89E-07 | 1.55E-06 | 0.001188217 |
| Obesity | endothelial | *ARHGAP42* | -0.630467491 | 2.02E-08 | 3.63E-07 | 0.000126819 |
| Obesity | endothelial | *OTOGL* | -1.375285596 | 0.000149888 | 0.000359126 | 0.942347276 |
| Obesity | endothelial | *TTC7B* | -0.69626515 | 5.17E-05 | 0.000144032 | 0.325223862 |
| Obesity | endothelial | *SLC39A13* | -1.293610722 | 9.87E-05 | 0.000249862 | 0.620407504 |
| Obesity | endothelial | *TC2N* | -0.209816849 | 0.035899392 | 0.047888708 | 1 |
| Obesity | endothelial | *CPSF2* | -0.343455033 | 0.005570187 | 0.008816659 | 1 |
| Obesity | endothelial | *IFI27L1* | -1.005411159 | 0.000428664 | 0.000900716 | 1 |
| Obesity | endothelial | *CLMN* | -0.8189355 | 7.50E-07 | 4.52E-06 | 0.004712919 |
| Obesity | endothelial | *PTER* | -0.998343724 | 2.55E-06 | 1.21E-05 | 0.016001417 |
| Obesity | endothelial | *ABTB2* | -0.904455426 | 1.35E-08 | 3.34E-07 | 8.51E-05 |
| Obesity | endothelial | *LIPC* | -0.823199079 | 1.88E-07 | 1.55E-06 | 0.001184567 |
| Obesity | endothelial | *GPT2* | -0.534632176 | 6.79E-06 | 2.68E-05 | 0.042672597 |
| Obesity | endothelial | *HIF1AN* | -1.158495314 | 3.14E-06 | 1.45E-05 | 0.019772453 |

| | | | | | | |
|---|---|---|---|---|---|---|
| Obesity | endothelial | *NDUFB8* | -1.337651255 | 4.48E-05 | 0.000127479 | 0.281473622 |
| Obesity | endothelial | *SPINT1* | -1.133078583 | 5.72E-06 | 2.34E-05 | 0.035978465 |
| Obesity | endothelial | *AVPR1A* | -0.351622152 | 0.008890755 | 0.013465714 | 1 |
| Obesity | endothelial | *TRMT61A* | -1.313567599 | 3.17E-13 | 1.16E-10 | 1.99E-09 |
| Obesity | endothelial | *BTRC* | -0.622678247 | 0.000470483 | 0.000975248 | 1 |
| Obesity | endothelial | *LARP6* | -0.545705837 | 0.02925351 | 0.039688567 | 1 |
| Obesity | endothelial | *ASPG* | -2.030483569 | 3.02E-07 | 2.21E-06 | 0.001899988 |
| Obesity | endothelial | *GABRB3* | 0.36715491 | 0.000848884 | 0.001651279 | 1 |
| Obesity | endothelial | *PCBD1* | -0.265696602 | 0.021602163 | 0.030180622 | 1 |
| Obesity | endothelial | *COX11* | -0.493428626 | 0.023585857 | 0.032632985 | 1 |
| Obesity | endothelial | *FAM227B* | -0.596613782 | 0.00027802 | 0.000614164 | 1 |
| Obesity | endothelial | *STXBP4* | -0.751993878 | 0.000175196 | 0.000411496 | 1 |
| Obesity | endothelial | *C2* | -1.007375409 | 1.13E-08 | 3.34E-07 | 7.11E-05 |
| Obesity | endothelial | *TAF10* | -0.95002078 | 0.002429527 | 0.004195121 | 1 |
| Obesity | endothelial | *DCHS1* | 0.546820704 | 8.83E-05 | 0.000226772 | 0.555039133 |
| Obesity | endothelial | *NETO1* | 0.62188856 | 0.00026036 | 0.000580455 | 1 |
| Obesity | endothelial | *USP54* | 0.338446338 | 0.013591174 | 0.019729326 | 1 |
| Obesity | endothelial | *C11orf74* | -0.946728995 | 0.000277934 | 0.000614164 | 1 |
| Obesity | endothelial | *WDR88* | -1.040745654 | 2.26E-08 | 3.81E-07 | 0.000142043 |
| Obesity | endothelial | *CYB5R2* | -0.725270414 | 8.51E-06 | 3.24E-05 | 0.053474427 |
| Obesity | endothelial | *TUB* | -0.646735003 | 1.57E-08 | 3.35E-07 | 9.87E-05 |
| Obesity | endothelial | *CDYL2* | -0.621111777 | 1.76E-05 | 5.90E-05 | 0.11041751 |
| Obesity | endothelial | *AKIP1* | -1.444507592 | 4.23E-06 | 1.84E-05 | 0.026603216 |
| Obesity | endothelial | *C16orf46* | -5.360969968 | 4.11E-07 | 2.80E-06 | 0.002584999 |
| Obesity | endothelial | *TMEM41B* | -0.880177922 | 3.68E-05 | 0.000108713 | 0.231668378 |
| Obesity | endothelial | *ZNF143* | -1.027751776 | 1.77E-05 | 5.94E-05 | 0.111447243 |
| Obesity | endothelial | *TMX3* | -0.661849103 | 0.001860914 | 0.003307766 | 1 |
| Obesity | endothelial | *CCDC68* | 0.166363822 | 9.12E-07 | 5.28E-06 | 0.005730626 |
| Obesity | endothelial | *CLEC4E* | -0.629941991 | 3.33E-06 | 1.52E-05 | 0.020939937 |
| Obesity | endothelial | *ZNF3* | -0.934535663 | 0.000442028 | 0.000925111 | 1 |
| Obesity | endothelial | *CLEC4D* | -1.501664137 | 2.92E-06 | 1.36E-05 | 0.01835035 |
| Obesity | endothelial | *ZSCAN21* | -0.598156457 | 1.16E-05 | 4.17E-05 | 0.072777664 |
| Obesity | endothelial | *BEAN1* | -1.003236539 | 5.71E-07 | 3.64E-06 | 0.0035926 |
| Obesity | endothelial | *TK2* | -0.650222359 | 1.23E-05 | 4.37E-05 | 0.07760378 |
| Obesity | endothelial | *SEC11C* | 0.555057432 | 0.00061927 | 0.00124428 | 1 |
| Obesity | endothelial | *GALR1* | 0.295363637 | 1.35E-05 | 4.70E-05 | 0.084781749 |
| Obesity | endothelial | *CENPV* | -1.139931634 | 5.78E-06 | 2.36E-05 | 0.036365724 |
| Obesity | endothelial | *SPDYE6* | -0.902390431 | 0.000525993 | 0.001077874 | 1 |
| Obesity | endothelial | *ZNF592* | -0.899898083 | 5.86E-06 | 2.38E-05 | 0.036872985 |
| Obesity | endothelial | *AP1G1* | -0.752434879 | 0.000107945 | 0.000269519 | 0.678647674 |
| Obesity | endothelial | *YPEL4* | -0.703839584 | 7.76E-05 | 0.00020249 | 0.487597084 |
| Obesity | endothelial | *LDHC* | -0.326122088 | 0.007366163 | 0.011356319 | 1 |
| Obesity | endothelial | *FAM111A* | -0.606361685 | 0.002980586 | 0.005007734 | 1 |
| Obesity | endothelial | *ANKDD1A* | -0.780919845 | 7.57E-05 | 0.000198558 | 0.475745271 |
| Obesity | endothelial | *MTMR10* | -0.440894941 | 0.020094941 | 0.028282269 | 1 |
| Obesity | endothelial | *YWHAB* | -0.748020176 | 0.000510745 | 0.001050395 | 1 |
| Obesity | endothelial | *NYAP1* | 0.297519415 | 0.02777119 | 0.037873638 | 1 |
| Obesity | endothelial | *TSC22D4* | -1.071374624 | 8.88E-05 | 0.000227751 | 0.55821697 |
| Obesity | endothelial | *MS4A6E* | -0.604291316 | 5.86E-09 | 2.35E-07 | 3.69E-05 |
| Obesity | endothelial | *MS4A7* | -0.665406827 | 2.11E-07 | 1.69E-06 | 0.001328298 |

| | | | | | | |
|---|---|---|---|---|---|---|
| Obesity | endothelial | *MS4A14* | -0.58673235 | 2.05E-08 | 3.63E-07 | 0.000129038 |
| Obesity | endothelial | *DIS3L* | -1.066532255 | 2.37E-08 | 3.93E-07 | 0.000148898 |
| Obesity | endothelial | *CCNDBP1* | -1.074608278 | 6.70E-06 | 2.66E-05 | 0.042120694 |
| Obesity | endothelial | *SMAD3* | -0.780268337 | 3.84E-06 | 1.70E-05 | 0.024150148 |
| Obesity | endothelial | *RCCD1* | -0.945450923 | 4.50E-09 | 2.01E-07 | 2.83E-05 |
| Obesity | endothelial | *EVA1C* | 0.267607312 | 0.000191154 | 0.000444941 | 1 |
| Obesity | endothelial | *CNPY4* | -0.438737334 | 0.016666192 | 0.023846233 | 1 |
| Obesity | endothelial | *NKX3-1* | 2.8837083 | 0.010023341 | 0.015007893 | 1 |
| Obesity | endothelial | *TEF* | -0.369005806 | 0.007822114 | 0.011979934 | 1 |
| Obesity | endothelial | *PBX3* | -0.773589408 | 9.07E-08 | 8.81E-07 | 0.000570228 |
| Obesity | endothelial | *SNRPD1* | -1.22340355 | 1.54E-05 | 5.25E-05 | 0.096570333 |
| Obesity | endothelial | *SAMD14* | 1.196088666 | 0.002301806 | 0.003996535 | 1 |
| Obesity | endothelial | *PIP5KL1* | 0.389557541 | 0.000557465 | 0.0011335 | 1 |
| Obesity | endothelial | *URM1* | -1.075367977 | 2.18E-05 | 7.03E-05 | 0.137059225 |
| Obesity | endothelial | *ENDOG* | -0.45566129 | 0.019430916 | 0.02743984 | 1 |
| Obesity | endothelial | *ISLR2* | 0.432225031 | 2.48E-07 | 1.90E-06 | 0.001559391 |
| Obesity | endothelial | *SP2* | -0.805392461 | 0.001992266 | 0.003507526 | 1 |
| Obesity | endothelial | *COQ7* | -0.999163094 | 0.001019975 | 0.001932666 | 1 |
| Obesity | endothelial | *GPRC5B* | 0.460406146 | 1.37E-05 | 4.78E-05 | 0.086435303 |
| Obesity | endothelial | *KATNAL2* | -0.279756185 | 0.02448156 | 0.033686927 | 1 |
| Obesity | endothelial | *HDHD2* | -0.326875723 | 0.015691154 | 0.022584772 | 1 |
| Obesity | endothelial | *CCL23* | 0.230446088 | 0.011115914 | 0.016447576 | 1 |
| Obesity | endothelial | *RNF214* | -0.793725897 | 0.000331841 | 0.000719657 | 1 |
| Obesity | endothelial | *ENGASE* | -0.587510332 | 0.000932845 | 0.001791418 | 1 |
| Obesity | endothelial | *MYO5B* | -0.340227115 | 0.001267441 | 0.002351948 | 1 |
| Obesity | endothelial | *RRM1* | -0.366799209 | 0.00700788 | 0.010859882 | 1 |
| Obesity | endothelial | *OR51E2* | -0.545381534 | 0.00765099 | 0.011749334 | 1 |
| Obesity | endothelial | *TRIM68* | -0.764746213 | 0.008441281 | 0.012837527 | 1 |
| Obesity | endothelial | *FN3K* | -1.056504949 | 0.000348277 | 0.00075064 | 1 |
| Obesity | endothelial | *ZNF23* | -1.575929131 | 6.04E-07 | 3.82E-06 | 0.003798728 |
| Obesity | endothelial | *VKORC1* | -1.457980961 | 3.12E-12 | 7.85E-10 | 1.96E-08 |
| Obesity | endothelial | *RAB8A* | -0.876463656 | 0.004961974 | 0.007939916 | 1 |
| Obesity | endothelial | *GATAD2A* | -0.629850943 | 0.002220874 | 0.003875281 | 1 |
| Obesity | endothelial | *PROCA1* | -0.804812209 | 2.65E-05 | 8.27E-05 | 0.1667284 |
| Obesity | endothelial | *RPL13* | -0.492687951 | 0.01265862 | 0.018513113 | 1 |
| Obesity | endothelial | *ZNF641* | -0.536586786 | 0.000167919 | 0.00039703 | 1 |
| Obesity | endothelial | *TP53I13* | -0.764780758 | 2.17E-07 | 1.71E-06 | 0.001364128 |
| Obesity | endothelial | *ZNF701* | -0.724441678 | 0.00228095 | 0.0039658 | 1 |
| Obesity | endothelial | *SERTAD3* | -1.992551634 | 1.57E-06 | 8.14E-06 | 0.009856152 |
| Obesity | endothelial | *TMC4* | -1.705876514 | 8.63E-08 | 8.47E-07 | 0.000542725 |
| Obesity | endothelial | *TTYH1* | -1.170110003 | 7.19E-07 | 4.37E-06 | 0.004518036 |
| Obesity | endothelial | *LENG8* | -0.673132297 | 0.000114618 | 0.000284263 | 0.720606217 |
| Obesity | endothelial | *CDC42EP5* | 0.413902155 | 1.79E-05 | 5.99E-05 | 0.112526942 |
| Obesity | endothelial | *ZNF283* | -0.711148925 | 0.004082164 | 0.00666612 | 1 |
| Obesity | endothelial | *PPP1R14A* | 0.521903876 | 0.037476563 | 0.049834 | 1 |
| Obesity | endothelial | *C19orf33* | 0.615527362 | 1.78E-05 | 5.96E-05 | 0.111914576 |
| Obesity | endothelial | *DNAAF3* | 1.57754582 | 8.42E-05 | 0.000217622 | 0.529474147 |
| Obesity | endothelial | *PSCA* | -0.47488128 | 0.00083615 | 0.001632064 | 1 |
| Obesity | endothelial | *CHAF1A* | -0.351586693 | 0.003731748 | 0.006140147 | 1 |
| Obesity | endothelial | *UBXN6* | -1.395164752 | 3.58E-11 | 5.92E-09 | 2.25E-07 |

| | | | | | | |
|---|---|---|---|---|---|---|
| Obesity | endothelial | *ZNF444* | -1.090425891 | 0.000871794 | 0.001687907 | 1 |
| Obesity | endothelial | *GLOD4* | -0.837769043 | 0.002278916 | 0.003964048 | 1 |
| Obesity | endothelial | *KIFC2* | -0.514347922 | 0.00010402 | 0.000260963 | 0.653972636 |
| Obesity | endothelial | *SERPINF2* | -0.63696935 | 0.00205987 | 0.003619454 | 1 |
| Obesity | endothelial | *SRR* | 0.356757434 | 0.001470885 | 0.002685091 | 1 |
| Obesity | endothelial | *CYB5D2* | -0.78527375 | 0.001466044 | 0.002678586 | 1 |
| Obesity | endothelial | *CD320* | 0.285939759 | 7.23E-06 | 2.82E-05 | 0.045424703 |
| Obesity | endothelial | *SPRYD3* | -1.462044931 | 6.08E-07 | 3.83E-06 | 0.003823014 |
| Obesity | endothelial | *SOAT2* | -0.569598351 | 0.000317904 | 0.000692537 | 1 |
| Obesity | endothelial | *ZNF558* | -0.602428187 | 0.0001078 | 0.000269372 | 0.677739756 |
| Obesity | endothelial | *NDUFV1* | -0.690349512 | 0.002976217 | 0.005004168 | 1 |
| Obesity | endothelial | *NUDT8* | -0.90313821 | 1.24E-08 | 3.34E-07 | 7.77E-05 |
| Obesity | endothelial | *PRDX2* | -0.866533323 | 0.003102697 | 0.005194848 | 1 |
| Obesity | endothelial | *ZNF232* | -0.284457856 | 0.034882592 | 0.046651108 | 1 |
| Obesity | endothelial | *MIS12* | -1.206237094 | 1.66E-07 | 1.39E-06 | 0.001042007 |
| Obesity | endothelial | *CD300C* | -0.662638571 | 0.008084948 | 0.012334401 | 1 |
| Obesity | endothelial | *HID1* | 0.208504539 | 1.31E-06 | 7.06E-06 | 0.008235396 |
| Obesity | endothelial | *ATP5H* | -1.130247377 | 6.61E-05 | 0.000177785 | 0.415838575 |
| Obesity | endothelial | *SRP68* | -1.253462783 | 3.89E-07 | 2.68E-06 | 0.002446178 |
| Obesity | endothelial | *MGAT5B* | 1.323234906 | 0.000487622 | 0.001007452 | 1 |
| Obesity | endothelial | *TMC8* | -0.661233707 | 0.000920812 | 0.001772549 | 1 |
| Obesity | endothelial | *TMEM68* | -0.611642058 | 0.012659118 | 0.018513113 | 1 |
| Obesity | endothelial | *GSDMA* | -1.19520424 | 1.81E-07 | 1.50E-06 | 0.001136936 |
| Obesity | endothelial | *KRT24* | -1.114137399 | 4.91E-08 | 5.72E-07 | 0.000308804 |
| Obesity | endothelial | *MLST8* | -2.230387038 | 5.77E-06 | 2.35E-05 | 0.03628779 |
| Obesity | endothelial | *ECI1* | -0.695298395 | 0.001005779 | 0.001909796 | 1 |
| Obesity | endothelial | *KCTD5* | -1.57744851 | 5.62E-08 | 6.28E-07 | 0.000353303 |
| Obesity | endothelial | *SDHAF2* | -1.769434802 | 1.44E-05 | 4.98E-05 | 0.090442399 |
| Obesity | endothelial | *DDB1* | -0.76483169 | 0.001188699 | 0.002218271 | 1 |
| Obesity | endothelial | *VPS37C* | -0.991644738 | 0.000320575 | 0.000697872 | 1 |
| Obesity | endothelial | *VWCE* | 0.862906324 | 0.000115123 | 0.000285064 | 0.723777349 |
| Obesity | endothelial | *BSCL2* | 2.082803229 | 0.003132522 | 0.005237811 | 1 |
| Obesity | endothelial | *HRASLS5* | -0.133639978 | 0.020997046 | 0.029433317 | 1 |
| Obesity | endothelial | *TRANK1* | -0.547274647 | 0.008514301 | 0.012929809 | 1 |
| Obesity | endothelial | *TTC21A* | -0.925904705 | 0.003294113 | 0.005487569 | 1 |
| Obesity | endothelial | *LTBP3* | -0.418451395 | 0.022646891 | 0.031465416 | 1 |
| Obesity | endothelial | *SCARA5* | -0.692228059 | 1.03E-08 | 3.23E-07 | 6.49E-05 |
| Obesity | endothelial | *COPS6* | -1.003250067 | 1.88E-05 | 6.20E-05 | 0.117927221 |
| Obesity | endothelial | *NUDT16L1* | -1.507727907 | 8.80E-07 | 5.12E-06 | 0.005531796 |
| Obesity | endothelial | *KIAA1586* | -1.051312509 | 1.78E-06 | 9.01E-06 | 0.011176903 |
| Obesity | endothelial | *RNF187* | -1.030953378 | 0.000271987 | 0.000602955 | 1 |
| Obesity | endothelial | *ZCCHC4* | -0.807216049 | 0.00095269 | 0.001824417 | 1 |
| Obesity | endothelial | *PTGDR* | -0.760180618 | 0.000489123 | 0.001009559 | 1 |
| Obesity | endothelial | *GLYCTK* | -0.676271071 | 0.025348681 | 0.03481913 | 1 |
| Obesity | endothelial | *UBTD2* | -0.519784413 | 0.011949163 | 0.017544228 | 1 |
| Obesity | endothelial | *POLR2J3* | -0.491583046 | 0.002776311 | 0.004711112 | 1 |
| Obesity | endothelial | *NT5DC2* | -1.48220938 | 9.98E-08 | 9.41E-07 | 0.000627506 |
| Obesity | endothelial | *COA6* | -1.700155489 | 9.17E-07 | 5.31E-06 | 0.005767778 |
| Obesity | endothelial | *KIF5C* | 0.679882833 | 0.005556273 | 0.008799065 | 1 |
| Obesity | endothelial | *PXK* | -0.87389561 | 2.99E-06 | 1.39E-05 | 0.018801522 |

| | | | | | | |
|---|---|---|---|---|---|---|
| Obesity | endothelial | *ACOX2* | -0.449005466 | 0.000131855 | 0.000320809 | 0.82897174 |
| Obesity | endothelial | *FAM107A* | 0.257379776 | 1.57E-06 | 8.14E-06 | 0.009856631 |
| Obesity | endothelial | *CX3CR1* | -0.639751235 | 0.000668125 | 0.001332647 | 1 |
| Obesity | endothelial | *SCN11A* | 0.143804661 | 0.002716852 | 0.004621441 | 1 |
| Obesity | endothelial | *DTYMK* | -1.261414083 | 2.01E-05 | 6.58E-05 | 0.126304124 |
| Obesity | endothelial | *TAP1* | 0.502333847 | 0.000899978 | 0.001737232 | 1 |
| Obesity | endothelial | *ING5* | -0.947468546 | 2.89E-07 | 2.14E-06 | 0.001816459 |
| Obesity | endothelial | *RHOH* | -1.004862266 | 1.44E-06 | 7.59E-06 | 0.009042089 |
| Obesity | endothelial | *KLHL30* | -0.633515833 | 5.28E-05 | 0.000146429 | 0.332099976 |
| Obesity | endothelial | *REEP4* | -1.738329001 | 1.41E-05 | 4.89E-05 | 0.088746119 |
| Obesity | endothelial | *BMP1* | -0.834154125 | 3.01E-06 | 1.39E-05 | 0.018955005 |
| Obesity | endothelial | *ATXN2L* | -1.062720003 | 2.30E-06 | 1.12E-05 | 0.01448589 |
| Obesity | endothelial | *TRAPPC11* | -0.698954614 | 0.001054837 | 0.001992117 | 1 |
| Obesity | endothelial | *SLC20A2* | -0.7379297 | 2.98E-05 | 9.15E-05 | 0.187474223 |
| Obesity | endothelial | *ZSWIM1* | -0.831129607 | 0.036366276 | 0.048439571 | 1 |
| Obesity | endothelial | *NBPF9* | -1.119927397 | 1.60E-08 | 3.37E-07 | 0.000100651 |
| Obesity | endothelial | *WFDC13* | -2.83089574 | 1.15E-06 | 6.34E-06 | 0.007238344 |
| Obesity | endothelial | *NDUFS5* | -0.772733388 | 0.000991701 | 0.001885912 | 1 |
| Obesity | endothelial | *ZNF30* | 0.414020175 | 0.003947612 | 0.006461504 | 1 |
| Obesity | endothelial | *FAM84B* | 0.276848445 | 2.84E-07 | 2.11E-06 | 0.001783933 |
| Obesity | endothelial | *TMEM208* | -0.924007882 | 0.000364553 | 0.000781166 | 1 |
| Obesity | endothelial | *LRP1B* | -0.298301765 | 0.006253208 | 0.009777149 | 1 |
| Obesity | endothelial | *DNAJC21* | -0.596768803 | 0.016317163 | 0.023421462 | 1 |
| Obesity | endothelial | *NPNT* | 0.34444088 | 1.47E-05 | 5.05E-05 | 0.092252459 |
| Obesity | endothelial | *CNNM3* | -1.273938138 | 1.70E-05 | 5.75E-05 | 0.107089841 |
| Obesity | endothelial | *GSTM4* | -0.451509729 | 0.001872447 | 0.003322629 | 1 |
| Obesity | endothelial | *TCTN2* | -0.453148585 | 1.87E-05 | 6.18E-05 | 0.117402926 |
| Obesity | endothelial | *ABHD15* | -0.886094833 | 1.37E-06 | 7.30E-06 | 0.008597433 |
| Obesity | endothelial | *SNTB2* | -0.714898948 | 0.00018418 | 0.000429503 | 1 |
| Obesity | endothelial | *ZNF507* | -0.756032356 | 0.00027002 | 0.000599017 | 1 |
| Obesity | endothelial | *STX18* | -0.708226238 | 0.000663465 | 0.001324612 | 1 |
| Obesity | endothelial | *ZBTB49* | -0.801094233 | 0.013972433 | 0.020268732 | 1 |
| Obesity | endothelial | *TNIP2* | -0.52804958 | 0.015664805 | 0.022552011 | 1 |
| Obesity | endothelial | *BTNL3* | 0.221157206 | 1.51E-07 | 1.30E-06 | 0.00094824 |
| Obesity | endothelial | *ZNF608* | 0.343219254 | 0.01723123 | 0.02458197 | 1 |
| Obesity | endothelial | *LETM1* | -1.193895499 | 1.27E-05 | 4.47E-05 | 0.079748156 |
| Obesity | endothelial | *TMEM129* | -0.502308475 | 0.00296516 | 0.004987149 | 1 |
| Obesity | endothelial | *MFF* | -0.553373142 | 0.006814252 | 0.010580687 | 1 |
| Obesity | endothelial | *CPLX1* | 0.221434116 | 2.94E-07 | 2.16E-06 | 0.001845824 |
| Obesity | endothelial | *SIGLEC7* | -0.831011376 | 2.97E-07 | 2.17E-06 | 0.001868382 |
| Obesity | endothelial | *FEM1B* | -0.707752875 | 0.001873241 | 0.0033231 | 1 |
| Obesity | endothelial | *MFSD7* | -1.220906312 | 2.26E-06 | 1.10E-05 | 0.014220847 |
| Obesity | endothelial | *COL4A3* | -0.701619766 | 0.001327985 | 0.002446246 | 1 |
| Obesity | endothelial | *C8orf46* | 0.108617377 | 0.00569688 | 0.008994547 | 1 |
| Obesity | endothelial | *HSPBAP1* | -0.439637191 | 0.011397409 | 0.016824491 | 1 |
| Obesity | endothelial | *PARM1* | 0.273772528 | 2.42E-05 | 7.64E-05 | 0.152267286 |
| Obesity | endothelial | *FAM110B* | -0.80045602 | 3.79E-08 | 4.98E-07 | 0.000238457 |
| Obesity | endothelial | *ATF5* | -1.462575351 | 7.17E-09 | 2.61E-07 | 4.51E-05 |
| Obesity | endothelial | *CPT1C* | 0.708322054 | 0.000250038 | 0.000561224 | 1 |
| Obesity | endothelial | *PCSK9* | -3.871748439 | 6.59E-06 | 2.62E-05 | 0.041459695 |

| | | | | | | |
|---|---|---|---|---|---|---|
| Obesity | endothelial | *XPO6* | -0.81042737 | 7.53E-05 | 0.000197758 | 0.473432649 |
| Obesity | endothelial | *RGS14* | -0.310589315 | 0.025143353 | 0.034559742 | 1 |
| Obesity | endothelial | *RAB24* | -1.486133725 | 7.14E-06 | 2.80E-05 | 0.044870347 |
| Obesity | endothelial | *CXCL10* | 0.392178393 | 0.00424149 | 0.006895849 | 1 |
| Obesity | endothelial | *SH3TC2* | 0.321627545 | 0.000256422 | 0.000573303 | 1 |
| Obesity | endothelial | *GPRIN1* | -2.803733764 | 0.000679376 | 0.001352942 | 1 |
| Obesity | endothelial | *STK32A* | -0.448464426 | 6.96E-09 | 2.56E-07 | 4.37E-05 |
| Obesity | endothelial | *P2RY12* | -0.498941849 | 6.78E-06 | 2.68E-05 | 0.042601652 |
| Obesity | endothelial | *SLC33A1* | -0.890906349 | 0.000122548 | 0.00030096 | 0.770458423 |
| Obesity | endothelial | *SNUPN* | -0.938478872 | 5.59E-06 | 2.30E-05 | 0.035136592 |
| Obesity | endothelial | *RNASE6* | -1.289422984 | 1.40E-08 | 3.34E-07 | 8.83E-05 |
| Obesity | endothelial | *NPR1* | 0.41901572 | 0.00685435 | 0.010640321 | 1 |
| Obesity | endothelial | *CD52* | -0.782811035 | 1.66E-05 | 5.61E-05 | 0.104215105 |
| Obesity | endothelial | *CLIC4* | 0.28545506 | 0.015772792 | 0.022683644 | 1 |
| Obesity | endothelial | *SLC38A11* | -1.140064467 | 6.27E-09 | 2.40E-07 | 3.94E-05 |
| Obesity | endothelial | *CCDC8* | -1.27954723 | 6.08E-08 | 6.64E-07 | 0.00038239 |
| Obesity | endothelial | *HINT1* | -0.805449651 | 0.000853635 | 0.00165898 | 1 |
| Obesity | endothelial | *DTWD2* | -0.794977477 | 0.00017584 | 0.000412348 | 1 |
| Obesity | endothelial | *INO80E* | -1.487134263 | 5.84E-11 | 8.53E-09 | 3.67E-07 |
| Obesity | endothelial | *DFFB* | -1.033963385 | 0.001125391 | 0.002109521 | 1 |
| Obesity | endothelial | *C15orf40* | -0.493959744 | 0.004991472 | 0.007981023 | 1 |
| Obesity | endothelial | *FAM103A1* | -0.655124379 | 6.71E-05 | 0.000179967 | 0.42202213 |
| Obesity | endothelial | *RGPD8* | -0.305329899 | 0.00037643 | 0.00080415 | 1 |
| Obesity | endothelial | *HEXDC* | -0.878084407 | 0.000237614 | 0.000535824 | 1 |
| Obesity | endothelial | *LRRC45* | -0.726159569 | 5.41E-08 | 6.10E-07 | 0.000340179 |
| Obesity | endothelial | *CHRNA5* | -1.324618056 | 1.71E-05 | 5.75E-05 | 0.107329815 |
| Obesity | endothelial | *FASN* | -0.364208726 | 0.001413606 | 0.002591061 | 1 |
| Obesity | endothelial | *DCXR* | -1.172190953 | 1.48E-07 | 1.28E-06 | 0.000927876 |
| Obesity | endothelial | *LIMS1* | -0.574918594 | 5.23E-05 | 0.000145266 | 0.328592715 |
| Obesity | endothelial | *UGP2* | -0.541386697 | 2.27E-05 | 7.24E-05 | 0.142549214 |
| Obesity | endothelial | *CSGALNACT2* | -0.889108673 | 7.90E-06 | 3.04E-05 | 0.049671542 |
| Obesity | endothelial | *CALML6* | -1.017841673 | 0.000140195 | 0.000338221 | 0.881405222 |
| Obesity | endothelial | *OTUD3* | -0.320009733 | 0.034798073 | 0.046547976 | 1 |
| Obesity | endothelial | *GUSB* | -0.994451963 | 2.07E-05 | 6.74E-05 | 0.130027599 |
| Obesity | endothelial | *BRD3* | -1.090674192 | 2.71E-05 | 8.42E-05 | 0.170299509 |
| Obesity | endothelial | *ZNF747* | -2.789524589 | 1.79E-09 | 1.14E-07 | 1.13E-05 |
| Obesity | endothelial | *MAP3K2* | -0.568500564 | 0.001507331 | 0.002741275 | 1 |
| Obesity | endothelial | *PUSL1* | -2.072277014 | 3.69E-08 | 4.89E-07 | 0.000232111 |
| Obesity | endothelial | *MYO7B* | 0.533888383 | 6.83E-06 | 2.70E-05 | 0.042953308 |
| Obesity | endothelial | *CNTROB* | -1.32187786 | 7.84E-05 | 0.000204139 | 0.492588505 |
| Obesity | endothelial | *NIPA1* | -0.704615214 | 0.012816427 | 0.018690994 | 1 |
| Obesity | endothelial | *RNF150* | -0.569163688 | 3.31E-09 | 1.72E-07 | 2.08E-05 |
| Obesity | endothelial | *CHRNB1* | -0.501681007 | 0.005886847 | 0.009253283 | 1 |
| Obesity | endothelial | *PDCD6IP* | -0.475316423 | 0.005084927 | 0.00810777 | 1 |
| Obesity | endothelial | *ZNF212* | -1.210670077 | 4.11E-06 | 1.80E-05 | 0.025865645 |
| Obesity | endothelial | *GLB1* | -0.791092178 | 1.87E-05 | 6.20E-05 | 0.117849763 |
| Obesity | endothelial | *FAXDC2* | -0.790024828 | 1.19E-05 | 4.25E-05 | 0.074820466 |
| Obesity | endothelial | *CRTAP* | -0.957395388 | 1.27E-05 | 4.46E-05 | 0.079611622 |
| Obesity | endothelial | *ELP5* | -1.298068422 | 2.12E-05 | 6.87E-05 | 0.133340028 |
| Obesity | endothelial | *STX8* | -0.522159161 | 0.003212797 | 0.005363477 | 1 |

| | | | | | | |
|---|---|---|---|---|---|---|
| Obesity | endothelial | *NFRKB* | -0.938989293 | 1.22E-08 | 3.34E-07 | 7.68E-05 |
| Obesity | endothelial | *SEMA3E* | 0.314164142 | 4.59E-05 | 0.000129851 | 0.288529263 |
| Obesity | endothelial | *GPRC5C* | -0.61155478 | 0.015117285 | 0.02180371 | 1 |
| Obesity | endothelial | *ADORA2B* | -0.784248923 | 1.02E-08 | 3.23E-07 | 6.44E-05 |
| Obesity | endothelial | *METTL7B* | -1.500274164 | 3.57E-08 | 4.80E-07 | 0.000224723 |
| Obesity | endothelial | *NFXL1* | -0.753633877 | 0.001575035 | 0.002845474 | 1 |
| Obesity | endothelial | *CD14* | -0.964262171 | 8.66E-09 | 2.93E-07 | 5.44E-05 |
| Obesity | endothelial | *HSD17B13* | 0.246663524 | 0.000138427 | 0.000334214 | 0.870292225 |
| Obesity | endothelial | *TMC7* | 0.195614317 | 2.12E-06 | 1.04E-05 | 0.013300226 |
| Obesity | endothelial | *ARL6IP1* | 0.396358148 | 0.011724302 | 0.017262455 | 1 |
| Obesity | endothelial | *SERPINB9* | -0.792806461 | 0.001713674 | 0.003067731 | 1 |
| Obesity | endothelial | *SMAGP* | 0.319024102 | 0.000124767 | 0.000305575 | 0.784409887 |
| Obesity | endothelial | *IRX2* | -0.517892292 | 1.09E-06 | 6.10E-06 | 0.006839701 |
| Obesity | endothelial | *EMB* | -0.40700578 | 0.000983529 | 0.001871503 | 1 |
| Obesity | endothelial | *DLGAP1* | 0.29995323 | 7.22E-05 | 0.000191089 | 0.454027771 |
| Obesity | endothelial | *STAT2* | -0.640671506 | 0.00094314 | 0.001808883 | 1 |
| Obesity | endothelial | *NUDCD2* | -1.289036976 | 7.55E-06 | 2.93E-05 | 0.047490511 |
| Obesity | endothelial | *HSPA4* | -0.843568038 | 0.000503343 | 0.001036868 | 1 |
| Obesity | endothelial | *COMMD5* | -1.168098525 | 4.35E-06 | 1.88E-05 | 0.027358556 |
| Obesity | endothelial | *GTSF1* | 1.750050497 | 3.72E-05 | 0.00010958 | 0.233733087 |
| Obesity | endothelial | *RNF34* | -0.657010928 | 0.001840927 | 0.003273165 | 1 |
| Obesity | endothelial | *ACYP2* | -0.663024411 | 2.22E-06 | 1.09E-05 | 0.013959166 |
| Obesity | endothelial | *TRABD* | -0.872764265 | 5.75E-06 | 2.35E-05 | 0.036180871 |
| Obesity | endothelial | *TMEM133* | -1.072333233 | 4.49E-08 | 5.44E-07 | 0.000282374 |
| Obesity | endothelial | *TTLL6* | 1.485544768 | 0.000378815 | 0.000808175 | 1 |
| Obesity | endothelial | *KCNS3* | -0.293407589 | 0.016237764 | 0.023312817 | 1 |
| Obesity | endothelial | *GPR37* | -0.503366948 | 0.008495617 | 0.012907671 | 1 |
| Obesity | endothelial | *AKAP13* | -0.407810666 | 0.007261073 | 0.011205294 | 1 |
| Obesity | endothelial | *HTRA3* | -0.556670717 | 2.83E-05 | 8.75E-05 | 0.177800974 |
| Obesity | endothelial | *FOXN2* | -0.786930783 | 2.91E-06 | 1.35E-05 | 0.018293023 |
| Obesity | endothelial | *MTSS1* | -0.64925052 | 1.73E-05 | 5.80E-05 | 0.108590954 |
| Obesity | endothelial | *TSEN34* | -1.581328162 | 5.84E-07 | 3.71E-06 | 0.003673665 |
| Obesity | endothelial | *NDUFA3* | -1.308801881 | 7.47E-06 | 2.90E-05 | 0.046974517 |
| Obesity | endothelial | *OSCAR* | -0.693110501 | 4.17E-05 | 0.000120423 | 0.262401999 |
| Obesity | endothelial | *PAQR8* | -0.939265344 | 0.035468967 | 0.047364783 | 1 |
| Obesity | endothelial | *FPR1* | -0.507671287 | 2.08E-05 | 6.77E-05 | 0.130979811 |
| Obesity | endothelial | *SOX7* | 0.254839341 | 5.97E-07 | 3.78E-06 | 0.003754687 |
| Obesity | endothelial | *C11orf24* | -1.073360272 | 1.88E-08 | 3.47E-07 | 0.000118494 |
| Obesity | endothelial | *ALK* | -0.60874571 | 3.83E-07 | 2.65E-06 | 0.002404861 |
| Obesity | endothelial | *MFN1* | -0.553109956 | 0.004546484 | 0.007340458 | 1 |
| Obesity | endothelial | *GIMAP8* | 0.266856076 | 7.33E-06 | 2.86E-05 | 0.046062594 |
| Obesity | endothelial | *ATP6V0E2* | -1.044454094 | 4.58E-09 | 2.02E-07 | 2.88E-05 |
| Obesity | endothelial | *MORN4* | -0.194767172 | 0.036095359 | 0.048099093 | 1 |
| Obesity | endothelial | *GRIK1* | -1.163512691 | 2.30E-07 | 1.80E-06 | 0.001445758 |
| Obesity | endothelial | *CLDN20* | -0.558879102 | 0.031828384 | 0.042885674 | 1 |
| Obesity | endothelial | *GAA* | -0.406918122 | 0.002497205 | 0.004290771 | 1 |
| Obesity | endothelial | *PGAM1* | -0.312596273 | 0.0190251 | 0.026921181 | 1 |
| Obesity | endothelial | *KCND3* | -0.803865573 | 0.000838567 | 0.001635765 | 1 |
| Obesity | endothelial | *ZNF581* | -1.872775105 | 0.004921771 | 0.007891654 | 1 |
| Obesity | endothelial | *DLK2* | -2.378651083 | 6.77E-06 | 2.68E-05 | 0.042553488 |

| | | | | | | |
|---|---|---|---|---|---|---|
| Obesity | endothelial | *ZNF561* | -0.439959288 | 0.013377747 | 0.019441955 | 1 |
| Obesity | endothelial | *HOPX* | -1.543675608 | 6.86E-08 | 7.19E-07 | 0.00043111 |
| Obesity | endothelial | *LRRC8C* | 0.479602469 | 0.003870954 | 0.006352567 | 1 |
| Obesity | endothelial | *RSL1D1* | -1.064574806 | 4.91E-05 | 0.00013734 | 0.308602395 |
| Obesity | endothelial | *PPID* | -0.86669321 | 0.00143971 | 0.002635835 | 1 |
| Obesity | endothelial | *COL24A1* | -0.143407033 | 0.017831111 | 0.025351469 | 1 |
| Obesity | endothelial | *ETFDH* | -0.229657802 | 0.005464883 | 0.008663066 | 1 |
| Obesity | endothelial | *TBCA* | -0.71393953 | 6.88E-05 | 0.000183715 | 0.432464734 |
| Obesity | endothelial | *MAP6* | -1.162215682 | 1.94E-06 | 9.70E-06 | 0.012213013 |
| Obesity | endothelial | *PLRG1* | -1.039685096 | 7.13E-05 | 0.00018943 | 0.448001777 |
| Obesity | endothelial | *NMUR1* | -1.06288496 | 2.62E-08 | 4.14E-07 | 0.000164458 |
| Obesity | endothelial | *P2RY6* | -0.970012673 | 1.14E-08 | 3.34E-07 | 7.17E-05 |
| Obesity | endothelial | *ZIK1* | -1.484540846 | 1.94E-07 | 1.57E-06 | 0.001220049 |
| Obesity | endothelial | *TCEA2* | 0.493562624 | 0.000165279 | 0.000391379 | 1 |
| Obesity | endothelial | *ANO5* | -1.053197853 | 1.63E-08 | 3.39E-07 | 0.000102532 |
| Obesity | endothelial | *VAT1L* | 0.529739071 | 4.33E-05 | 0.000124007 | 0.271947103 |
| Obesity | endothelial | *LRRC34* | -0.29917367 | 0.009505621 | 0.014303935 | 1 |
| Obesity | endothelial | *SYCE1* | -0.750159961 | 1.98E-06 | 9.87E-06 | 0.012478953 |
| Obesity | endothelial | *NXNL1* | -1.148662875 | 2.04E-07 | 1.65E-06 | 0.001285362 |
| Obesity | endothelial | *BCL2* | -0.320903034 | 1.21E-05 | 4.31E-05 | 0.076277891 |
| Obesity | endothelial | *RHNO1* | -1.003818752 | 0.001804186 | 0.003210563 | 1 |
| Obesity | endothelial | *KNDC1* | -0.555608697 | 0.000580113 | 0.001176886 | 1 |
| Obesity | endothelial | *METTL18* | -0.932190799 | 0.018087918 | 0.025693344 | 1 |
| Obesity | endothelial | *PWWP2B* | -1.293039895 | 0.000147701 | 0.000354562 | 0.928597909 |
| Obesity | endothelial | *ZNF570* | -0.790715701 | 0.021326752 | 0.029825634 | 1 |
| Obesity | endothelial | *FAM90A1* | -4.843581864 | 9.31E-06 | 3.48E-05 | 0.05853382 |
| Obesity | endothelial | *TRAPPC12* | -0.716746692 | 0.000383361 | 0.000816183 | 1 |
| Obesity | endothelial | *C3AR1* | -0.902520635 | 2.09E-08 | 3.64E-07 | 0.000131126 |
| Obesity | endothelial | *AQP4* | -0.840640025 | 1.85E-08 | 3.45E-07 | 0.000116606 |
| Obesity | endothelial | *TLN2* | -0.52172384 | 5.67E-07 | 3.62E-06 | 0.003563349 |
| Obesity | endothelial | *TVP23B* | -1.167923099 | 2.62E-06 | 1.24E-05 | 0.016480485 |
| Obesity | endothelial | *SRGAP2C* | -0.925499879 | 1.68E-05 | 5.68E-05 | 0.105681439 |
| Obesity | endothelial | *ATPAF2* | -1.131594943 | 0.000144662 | 0.000348064 | 0.909490802 |
| Obesity | endothelial | *CYP4F22* | -1.525031116 | 0.000126506 | 0.000309471 | 0.795340898 |
| Obesity | endothelial | *ZNF554* | -1.027658571 | 2.16E-09 | 1.28E-07 | 1.36E-05 |
| Obesity | endothelial | *ANKRD20A4* | -3.223541934 | 0.000293975 | 0.000645328 | 1 |
| Obesity | endothelial | *LAMB2* | -0.666241841 | 0.000179484 | 0.000419799 | 1 |
| Obesity | endothelial | *ORMDL3* | -0.308838119 | 0.002959255 | 0.004979882 | 1 |
| Obesity | endothelial | *SERF1A* | 3.026084118 | 0.00149521 | 0.002721593 | 1 |
| Obesity | endothelial | *LRRC15* | -1.305542125 | 1.50E-05 | 5.15E-05 | 0.094443762 |
| Obesity | endothelial | *MOB3A* | -0.91771333 | 1.67E-07 | 1.40E-06 | 0.001052516 |
| Obesity | endothelial | *NME6* | -0.419475028 | 0.00675624 | 0.010498388 | 1 |
| Obesity | endothelial | *FRMD3* | 0.376403191 | 0.000971257 | 0.0018532 | 1 |
| Obesity | endothelial | *MTBP* | -1.01801731 | 3.79E-05 | 0.000111396 | 0.238499137 |
| Obesity | endothelial | *MBOAT1* | -0.900415022 | 1.03E-06 | 5.83E-06 | 0.006456652 |
| Obesity | endothelial | *C5orf34* | -0.718393109 | 0.006970571 | 0.010806995 | 1 |
| Obesity | endothelial | *C1QTNF4* | -0.521820981 | 2.46E-17 | 1.94E-14 | 1.55E-13 |
| Obesity | endothelial | *ZNF131* | -0.551970642 | 0.003408637 | 0.00566185 | 1 |
| Obesity | endothelial | *HINFP* | -0.997580638 | 0.001141383 | 0.002137586 | 1 |
| Obesity | endothelial | *COPRS* | -1.135743406 | 0.000806397 | 0.001579874 | 1 |

| | | | | | | |
|---|---|---|---|---|---|---|
| Obesity | endothelial | *CLEC12A* | -0.114076196 | 0.014182469 | 0.020549708 | 1 |
| Obesity | endothelial | *SUCLG2* | -0.498438267 | 0.004448061 | 0.007194484 | 1 |
| Obesity | endothelial | *CSDC2* | -0.766999933 | 1.33E-05 | 4.65E-05 | 0.083912388 |
| Obesity | endothelial | *RCAN2* | -0.865465719 | 2.42E-07 | 1.86E-06 | 0.001523274 |
| Obesity | endothelial | *IL16* | -0.57939849 | 4.45E-07 | 2.98E-06 | 0.002795367 |
| Obesity | endothelial | *GNB2* | -1.087494697 | 0.000121708 | 0.000299132 | 0.765180933 |
| Obesity | endothelial | *ZNF24* | -0.912438122 | 6.92E-05 | 0.000184586 | 0.4348842 |
| Obesity | endothelial | *MANEA* | -1.096175941 | 6.64E-05 | 0.000178282 | 0.417179406 |
| Obesity | endothelial | *FIBP* | -0.929210684 | 0.016744284 | 0.023952518 | 1 |
| Obesity | endothelial | *CARNS1* | 1.465763502 | 4.22E-06 | 1.84E-05 | 0.026554346 |
| Obesity | endothelial | *CTSW* | -0.338366208 | 1.35E-12 | 3.85E-10 | 8.47E-09 |
| Obesity | endothelial | *SNTG2* | 0.204206449 | 0.001220036 | 0.002271355 | 1 |
| Obesity | endothelial | *PDE3A* | -0.732217086 | 1.47E-08 | 3.34E-07 | 9.21E-05 |
| Obesity | endothelial | *SMPDL3A* | -0.913935912 | 6.02E-06 | 2.43E-05 | 0.037855938 |
| Obesity | endothelial | *AGAP5* | -45.18003264 | 9.64E-08 | 9.18E-07 | 0.000605967 |
| Obesity | endothelial | *ZMAT3* | -1.060439114 | 4.63E-08 | 5.55E-07 | 0.000290987 |
| Obesity | endothelial | *ZFAND4* | -0.742518331 | 0.001450825 | 0.002653095 | 1 |
| Obesity | endothelial | *CCL19* | -4.041920535 | 2.52E-09 | 1.39E-07 | 1.58E-05 |
| Obesity | endothelial | *FUT10* | -0.711897968 | 3.16E-07 | 2.27E-06 | 0.001985326 |
| Obesity | endothelial | *CFL1* | -0.771003173 | 0.002274463 | 0.003957806 | 1 |
| Obesity | endothelial | *EFCAB12* | -0.638572507 | 2.49E-06 | 1.19E-05 | 0.015640358 |
| Obesity | endothelial | *FAM192A* | -0.629192264 | 8.48E-06 | 3.23E-05 | 0.053286791 |
| Obesity | endothelial | *RAB43* | 1.932582948 | 3.84E-07 | 2.65E-06 | 0.002412957 |
| Obesity | endothelial | *CBWD1* | -1.161565716 | 1.79E-06 | 9.05E-06 | 0.011250851 |
| Obesity | endothelial | *HOXC5* | -0.715428544 | 0.000154699 | 0.000369807 | 0.972593063 |
| Obesity | endothelial | *RAB37* | -1.362829517 | 2.98E-05 | 9.14E-05 | 0.187142771 |
| Obesity | endothelial | *DCP2* | -0.532872175 | 0.010875733 | 0.016141354 | 1 |
| Obesity | endothelial | *SNX32* | 1.021155933 | 2.88E-07 | 2.14E-06 | 0.001812548 |
| Obesity | endothelial | *CYP7B1* | -0.670795833 | 1.04E-07 | 9.78E-07 | 0.000654145 |
| Obesity | endothelial | *METAP1D* | -0.928441543 | 1.15E-06 | 6.34E-06 | 0.007232597 |
| Obesity | endothelial | *NADSYN1* | -0.650962065 | 0.004271151 | 0.006938689 | 1 |
| Obesity | endothelial | *RNASEH2C* | -0.514525109 | 0.022200346 | 0.030927006 | 1 |
| Obesity | endothelial | *MYEOV* | -0.212046615 | 0.002037568 | 0.003582267 | 1 |
| Obesity | endothelial | *ANKRD13D* | -0.91466412 | 3.51E-05 | 0.000104675 | 0.220863989 |
| Obesity | endothelial | *MRGPRF* | -0.990146295 | 1.76E-07 | 1.46E-06 | 0.00110519 |
| Obesity | endothelial | *ADH6* | -0.322421684 | 0.000608551 | 0.001225483 | 1 |
| Obesity | endothelial | *GXYLT2* | -0.688728469 | 6.52E-09 | 2.49E-07 | 4.10E-05 |
| Obesity | endothelial | *HPSE2* | -0.275310221 | 0.006074087 | 0.009523139 | 1 |
| Obesity | endothelial | *DCAKD* | -0.396260268 | 0.00026324 | 0.00058646 | 1 |
| Obesity | endothelial | *HECTD4* | -0.648745943 | 0.000285953 | 0.000629478 | 1 |
| Obesity | endothelial | *BNC2* | -0.529995805 | 7.05E-05 | 0.000187531 | 0.442948539 |
| Obesity | endothelial | *TRMT112* | -0.530308937 | 0.036528458 | 0.048645291 | 1 |
| Obesity | endothelial | *NOC3L* | -0.757646028 | 0.001921086 | 0.003397432 | 1 |
| Obesity | endothelial | *PARP15* | -0.575058035 | 3.29E-05 | 9.90E-05 | 0.206816278 |
| Obesity | endothelial | *AHSA2* | -0.523934306 | 0.002652826 | 0.004521094 | 1 |
| Obesity | endothelial | *ABLIM3* | 0.197180013 | 1.05E-05 | 3.84E-05 | 0.065849823 |
| Obesity | endothelial | *GLRX* | -0.642950166 | 2.86E-08 | 4.35E-07 | 0.000180057 |
| Obesity | endothelial | *IQCB1* | -0.772197008 | 6.08E-07 | 3.83E-06 | 0.003821605 |
| Obesity | endothelial | *GOLGB1* | -0.539350353 | 0.002235721 | 0.003894702 | 1 |
| Obesity | endothelial | *DMRT2* | -0.598012553 | 5.38E-08 | 6.10E-07 | 0.00033852 |

| | | | | | | |
|---|---|---|---|---|---|---|
| Obesity | endothelial | *SLC2A14* | -0.456020107 | 0.012492523 | 0.018295014 | 1 |
| Obesity | endothelial | *MZT2A* | -0.701121795 | 0.000606188 | 0.001221507 | 1 |
| Obesity | endothelial | *MAP3K11* | 0.475195359 | 0.005229218 | 0.00831885 | 1 |
| Obesity | endothelial | *C1QA* | -1.014472487 | 4.85E-07 | 3.20E-06 | 0.003047626 |
| Obesity | endothelial | *NDNF* | 0.421436114 | 4.98E-06 | 2.10E-05 | 0.031323811 |
| Obesity | endothelial | *OLR1* | -1.319446519 | 2.00E-07 | 1.62E-06 | 0.001259179 |
| Obesity | endothelial | *GLIPR1L1* | -0.575369892 | 0.005060008 | 0.008072132 | 1 |
| Obesity | endothelial | *ARV1* | -0.717378611 | 0.008081308 | 0.012334401 | 1 |
| Obesity | endothelial | *CCDC36* | -0.707349136 | 0.000132051 | 0.000321038 | 0.830203807 |
| Obesity | endothelial | *THAP2* | -0.975330495 | 8.19E-05 | 0.00021256 | 0.515032519 |
| Obesity | endothelial | *PPP1R14B* | -1.703380117 | 7.20E-06 | 2.82E-05 | 0.045242009 |
| Obesity | endothelial | *SMARCC1* | -0.514078013 | 0.006710264 | 0.010434685 | 1 |
| Obesity | endothelial | *ZNF417* | -1.804443727 | 1.46E-09 | 9.95E-08 | 9.15E-06 |
| Obesity | endothelial | *VEGFB* | -0.528267897 | 0.000277019 | 0.000612383 | 1 |
| Obesity | endothelial | *PEAK1* | -0.446536597 | 0.004885393 | 0.007841324 | 1 |
| Obesity | endothelial | *TNFRSF10D* | 0.26373913 | 7.55E-05 | 0.000198249 | 0.474806741 |
| Obesity | endothelial | *MST1* | -0.640756532 | 1.22E-06 | 6.65E-06 | 0.007663525 |
| Obesity | endothelial | *CSPG4* | -0.880670782 | 3.40E-09 | 1.74E-07 | 2.14E-05 |
| Obesity | endothelial | *SNX33* | -0.943755453 | 1.13E-06 | 6.26E-06 | 0.007084778 |
| Obesity | endothelial | *NUDT18* | -1.903951778 | 8.35E-07 | 4.91E-06 | 0.005246648 |
| Obesity | endothelial | *SULT1B1* | 0.592481844 | 0.004126772 | 0.006728479 | 1 |
| Obesity | endothelial | *NUDT4* | -0.670581925 | 0.001488197 | 0.002713486 | 1 |
| Obesity | endothelial | *SCAI* | -0.788373092 | 1.32E-05 | 4.62E-05 | 0.083060126 |
| Obesity | endothelial | *C1orf100* | 1.005467695 | 0.003829485 | 0.006291083 | 1 |
| Obesity | endothelial | *AGFG1* | -0.623628116 | 6.42E-06 | 2.56E-05 | 0.040364473 |
| Obesity | endothelial | *RNF213* | -0.492887381 | 0.004478612 | 0.007238312 | 1 |
| Obesity | endothelial | *PLK3* | -0.801034882 | 0.02445231 | 0.033654044 | 1 |
| Obesity | endothelial | *TUBB8* | 0.941897256 | 0.000177787 | 0.000416603 | 1 |
| Obesity | endothelial | *GPR160* | 0.246124897 | 8.00E-06 | 3.07E-05 | 0.050274625 |
| Obesity | endothelial | *USMG5* | -0.905756819 | 0.000252404 | 0.000566131 | 1 |
| Obesity | endothelial | *XXYLT1* | -0.956001638 | 5.19E-06 | 2.16E-05 | 0.032624914 |
| Obesity | endothelial | *UBXN2A* | -0.772590346 | 4.40E-06 | 1.90E-05 | 0.027656957 |
| Obesity | endothelial | *CCS* | -0.867387719 | 3.96E-06 | 1.75E-05 | 0.024897205 |
| Obesity | endothelial | *CEP19* | -0.860420763 | 2.96E-05 | 9.09E-05 | 0.186023391 |
| Obesity | endothelial | *CTSF* | -0.694279091 | 2.65E-06 | 1.25E-05 | 0.016666049 |
| Obesity | endothelial | *LEMD3* | -0.772924339 | 0.000232663 | 0.00052598 | 1 |
| Obesity | endothelial | *C16orf91* | -2.082063868 | 6.16E-06 | 2.47E-05 | 0.038723325 |
| Obesity | endothelial | *SOCS7* | -1.027986397 | 2.96E-07 | 2.17E-06 | 0.001858283 |
| Obesity | endothelial | *TLR6* | -0.750060572 | 0.000161074 | 0.000382718 | 1 |
| Obesity | endothelial | *RGMB* | -0.962153805 | 0.001338919 | 0.002464468 | 1 |
| Obesity | endothelial | *FAM53A* | -1.312306047 | 1.07E-06 | 6.02E-06 | 0.006717194 |
| Obesity | endothelial | *CTU2* | -0.779636184 | 0.001884948 | 0.003340099 | 1 |
| Obesity | endothelial | *MGA* | -0.747483389 | 3.08E-06 | 1.42E-05 | 0.019385746 |
| Obesity | endothelial | *SNX31* | -0.851240227 | 4.79E-05 | 0.000134443 | 0.301017715 |
| Obesity | endothelial | *GLIS1* | -0.396431367 | 4.08E-05 | 0.000118155 | 0.256751421 |
| Obesity | endothelial | *LIG4* | -0.761535999 | 1.02E-06 | 5.80E-06 | 0.006388892 |
| Obesity | endothelial | *SNAPC5* | -1.247854555 | 3.48E-05 | 0.00010398 | 0.21908677 |
| Obesity | endothelial | *LINGO2* | -1.059040068 | 3.82E-05 | 0.000111873 | 0.240303009 |
| Obesity | endothelial | *ANKRD36C* | -0.748193678 | 6.21E-05 | 0.000168666 | 0.390293647 |
| Obesity | endothelial | *TMEM81* | -4.202718741 | 1.32E-07 | 1.17E-06 | 0.000829505 |

| | | | | | | |
|---|---|---|---|---|---|---|
| Obesity | endothelial | *ZNF497* | -1.818344938 | 0.000258521 | 0.000576968 | 1 |
| Obesity | endothelial | *KLF14* | -2.523351714 | 3.18E-05 | 9.61E-05 | 0.199826322 |
| Obesity | endothelial | *ANGEL2* | -0.600911651 | 0.006802161 | 0.010564522 | 1 |
| Obesity | endothelial | *BRSK2* | 0.659197655 | 0.00021355 | 0.000489314 | 1 |
| Obesity | endothelial | *LARP7* | -0.825134389 | 0.000135198 | 0.000327646 | 0.849988865 |
| Obesity | endothelial | *FGFBP3* | -1.47045643 | 9.47E-08 | 9.06E-07 | 0.00059527 |
| Obesity | endothelial | *C4orf32* | 0.263418729 | 4.39E-05 | 0.000125357 | 0.27586045 |
| Obesity | endothelial | *BTC* | 0.56822406 | 0.011142328 | 0.016482781 | 1 |
| Obesity | endothelial | *NLRP6* | -3.527907891 | 2.40E-07 | 1.85E-06 | 0.001508698 |
| Obesity | endothelial | *NDUFA11* | -1.298851744 | 1.04E-06 | 5.88E-06 | 0.006509658 |
| Obesity | endothelial | *RSRC1* | -0.66253999 | 1.46E-05 | 5.04E-05 | 0.091769566 |
| Obesity | endothelial | *FUT1* | 0.524940754 | 2.32E-07 | 1.80E-06 | 0.001455597 |
| Obesity | endothelial | *DHX36* | -0.749522482 | 9.44E-05 | 0.000240107 | 0.593496665 |
| Obesity | endothelial | *SLC22A1* | -1.176114422 | 5.29E-08 | 6.01E-07 | 0.000332534 |
| Obesity | endothelial | *ATR* | -0.597559274 | 0.000633116 | 0.001269261 | 1 |
| Obesity | endothelial | *PACS1* | -0.626060483 | 0.00018697 | 0.000435687 | 1 |
| Obesity | endothelial | *SH3BP5L* | -0.83797332 | 0.014265712 | 0.020651286 | 1 |
| Obesity | endothelial | *CADM2* | 0.170531919 | 3.03E-07 | 2.21E-06 | 0.001907276 |
| Obesity | endothelial | *ARHGAP1* | -0.854281149 | 7.26E-07 | 4.41E-06 | 0.004564126 |
| Obesity | endothelial | *CHST1* | 0.598574272 | 2.76E-05 | 8.57E-05 | 0.173613885 |
| Obesity | endothelial | *TP53I11* | 0.451942589 | 7.69E-05 | 0.000201001 | 0.483609249 |
| Obesity | endothelial | *PHYHD1* | -0.906597137 | 5.85E-06 | 2.37E-05 | 0.036785595 |
| Obesity | endothelial | *ZNF519* | -0.888265801 | 1.04E-05 | 3.83E-05 | 0.065521423 |
| Obesity | endothelial | *CHRNA7* | 0.462183501 | 0.007758533 | 0.011891247 | 1 |
| Obesity | endothelial | *NRIP3* | -0.723162152 | 0.000969103 | 0.001850827 | 1 |
| Obesity | endothelial | *SCUBE2* | 0.406060003 | 0.00076131 | 0.001498546 | 1 |
| Obesity | endothelial | *ZNF25* | -0.790257691 | 2.17E-07 | 1.71E-06 | 0.001364375 |
| Obesity | endothelial | *CLTB* | -0.805152104 | 4.22E-05 | 0.000121506 | 0.265368201 |
| Obesity | endothelial | *PCSK1* | -1.088699381 | 8.43E-10 | 6.54E-08 | 5.30E-06 |
| Obesity | endothelial | *LPL* | -0.319789961 | 9.95E-05 | 0.000251369 | 0.625658031 |
| Obesity | endothelial | *TBC1D10C* | -0.61154555 | 7.82E-05 | 0.000203925 | 0.491866739 |
| Obesity | endothelial | *PPP2R2D* | -0.473184359 | 0.007625109 | 0.011718177 | 1 |
| Obesity | endothelial | *MCTP1* | 0.203031247 | 6.25E-05 | 0.00016947 | 0.392661465 |
| Obesity | endothelial | *LRRC25* | -0.31823078 | 0.005014994 | 0.008010485 | 1 |
| Obesity | endothelial | *LIPT2* | -2.738280431 | 7.72E-08 | 7.84E-07 | 0.000485209 |
| Obesity | endothelial | *KCNE3* | -1.214773225 | 0.000224232 | 0.000509671 | 1 |
| Obesity | endothelial | *UCP3* | -1.335113956 | 0.000382397 | 0.000814407 | 1 |
| Obesity | endothelial | *UCP2* | -0.918308791 | 1.56E-08 | 3.35E-07 | 9.78E-05 |
| Obesity | endothelial | *PAAF1* | -0.168540135 | 0.029156453 | 0.039565426 | 1 |
| Obesity | endothelial | *MRPL48* | -0.713009482 | 1.67E-05 | 5.67E-05 | 0.105281487 |
| Obesity | endothelial | *RPS6KB2* | -1.302233938 | 1.98E-05 | 6.50E-05 | 0.124375745 |
| Obesity | endothelial | *TOM1L2* | -0.643888185 | 0.001057627 | 0.001996187 | 1 |
| Obesity | endothelial | *TEX26* | -0.899634369 | 4.53E-05 | 0.000128484 | 0.284592521 |
| Obesity | endothelial | *GPR156* | -1.238869127 | 1.33E-07 | 1.18E-06 | 0.000836779 |
| Obesity | endothelial | *B3GNTL1* | -0.959851524 | 8.81E-07 | 5.12E-06 | 0.005539977 |
| Obesity | endothelial | *TTLL11* | -0.647567573 | 6.65E-05 | 0.00017833 | 0.417828011 |
| Obesity | endothelial | *LINC01106* | 3.892179298 | 0.00020052 | 0.000463652 | 1 |
| Obesity | endothelial | *CTDNEP1* | -1.203792006 | 2.26E-06 | 1.10E-05 | 0.014239816 |
| Obesity | endothelial | *ETV4* | -0.646203748 | 0.00070114 | 0.001393195 | 1 |
| Obesity | endothelial | *ZDHHC21* | -0.482765452 | 0.008209451 | 0.012503106 | 1 |

| | | | | | | |
|---|---|---|---|---|---|---|
| Obesity | endothelial | *A2M* | 0.264612607 | 0.00044853 | 0.000936846 | 1 |
| Obesity | endothelial | *ARL4D* | -1.092709238 | 5.07E-08 | 5.82E-07 | 0.000319058 |
| Obesity | endothelial | *LRRN1* | -1.313425754 | 4.06E-08 | 5.14E-07 | 0.000255339 |
| Obesity | endothelial | *B3GALT6* | -1.396010974 | 0.012675516 | 0.018528474 | 1 |
| Obesity | endothelial | *SSNA1* | -1.349426528 | 0.000108916 | 0.000271405 | 0.684755827 |
| Obesity | endothelial | *CHMP6* | -1.344766723 | 7.34E-06 | 2.86E-05 | 0.046149885 |
| Obesity | endothelial | *TCP11L1* | -0.53221948 | 0.004848012 | 0.007785301 | 1 |
| Obesity | endothelial | *CCDC57* | -1.045279234 | 1.88E-08 | 3.47E-07 | 0.0001181 |
| Obesity | endothelial | *CIDEA* | -0.198815027 | 0.000213053 | 0.000488855 | 1 |
| Obesity | endothelial | *SLC35G1* | -0.760705966 | 9.08E-08 | 8.81E-07 | 0.000570852 |
| Obesity | endothelial | *ZNF135* | -0.77500337 | 0.000955659 | 0.001828989 | 1 |
| Obesity | endothelial | *COX8A* | -0.904660906 | 0.009190146 | 0.013892389 | 1 |
| Obesity | endothelial | *B3GNT4* | -3.18154314 | 5.12E-08 | 5.86E-07 | 0.000321633 |
| Obesity | endothelial | *CDC26* | -0.784680829 | 7.28E-06 | 2.84E-05 | 0.045794179 |
| Obesity | endothelial | *CRLF3* | -0.724424156 | 6.64E-05 | 0.00017833 | 0.417709305 |
| Obesity | endothelial | *RNPEP* | -0.829827377 | 0.00056416 | 0.00114563 | 1 |
| Obesity | endothelial | *SPRYD4* | -0.541899856 | 7.64E-06 | 2.96E-05 | 0.048031113 |
| Obesity | endothelial | *WDR25* | -0.610886303 | 0.014661169 | 0.021174998 | 1 |
| Obesity | endothelial | *PHLDB3* | -0.785103582 | 0.001323382 | 0.002439913 | 1 |
| Obesity | endothelial | *PRR15* | 1.087908156 | 4.36E-06 | 1.88E-05 | 0.027428907 |
| Obesity | endothelial | *HORMAD2* | -0.398929407 | 0.005668624 | 0.008956683 | 1 |
| Obesity | endothelial | *LRRC37A* | -0.796440773 | 8.79E-06 | 3.33E-05 | 0.05526947 |
| Obesity | endothelial | *ACSF3* | -0.926952279 | 2.67E-06 | 1.26E-05 | 0.016816693 |
| Obesity | endothelial | *BOK* | -1.027795313 | 4.71E-08 | 5.59E-07 | 0.000296306 |
| Obesity | endothelial | *ZNF843* | -2.497885246 | 8.60E-07 | 5.03E-06 | 0.005407312 |
| Obesity | endothelial | *C8orf59* | -0.748231899 | 0.000624003 | 0.001252988 | 1 |
| Obesity | endothelial | *PFN4* | -0.552164794 | 0.000793662 | 0.001557837 | 1 |
| Obesity | endothelial | *RUFY1* | -0.34170974 | 0.036639678 | 0.048772718 | 1 |
| Obesity | endothelial | *BASP1* | -0.685748147 | 1.16E-06 | 6.38E-06 | 0.007284129 |
| Obesity | endothelial | *LRRC37A3* | -1.082032173 | 4.79E-05 | 0.000134443 | 0.300955272 |
| Obesity | endothelial | *IRX5* | -0.673962352 | 4.34E-06 | 1.88E-05 | 0.027308394 |
| Obesity | endothelial | *PNMA1* | -1.592503038 | 2.33E-09 | 1.34E-07 | 1.47E-05 |
| Obesity | endothelial | *ANKLE2* | -0.579941246 | 0.011482049 | 0.016933531 | 1 |
| Obesity | endothelial | *GCNT4* | 1.519610641 | 2.19E-05 | 7.04E-05 | 0.137451909 |
| Obesity | endothelial | *MUC20* | -1.692263113 | 6.03E-09 | 2.37E-07 | 3.79E-05 |
| Obesity | endothelial | *THAP4* | -1.083825981 | 5.00E-06 | 2.10E-05 | 0.031462503 |
| Obesity | endothelial | *SHMT1* | -0.430935255 | 0.001485822 | 0.002710785 | 1 |
| Obesity | endothelial | *DPP7* | -1.012961944 | 1.38E-06 | 7.33E-06 | 0.008651711 |
| Obesity | endothelial | *SEC24C* | -1.080467033 | 1.49E-05 | 5.13E-05 | 0.09377893 |
| Obesity | endothelial | *C19orf18* | -0.711114929 | 0.004323227 | 0.007005187 | 1 |
| Obesity | endothelial | *TMEM80* | -0.778466592 | 7.32E-07 | 4.44E-06 | 0.004601519 |
| Obesity | endothelial | *WDR73* | -0.620136772 | 0.001286708 | 0.002382077 | 1 |
| Obesity | endothelial | *POLE* | -1.108503878 | 1.04E-05 | 3.83E-05 | 0.065574954 |
| Obesity | endothelial | *DSCAML1* | -0.796596724 | 5.41E-08 | 6.10E-07 | 0.000340258 |
| Obesity | endothelial | *EPS8L2* | -0.705543274 | 3.68E-07 | 2.57E-06 | 0.002311025 |
| Obesity | endothelial | *TALDO1* | -1.1007196 | 1.31E-09 | 9.25E-08 | 8.24E-06 |
| Obesity | endothelial | *RIMKLA* | 0.512927062 | 1.18E-05 | 4.22E-05 | 0.074043757 |
| Obesity | endothelial | *B3GNT8* | -4.272316896 | 2.28E-07 | 1.79E-06 | 0.001435456 |
| Obesity | endothelial | *MAN1B1* | -1.193092688 | 3.71E-06 | 1.65E-05 | 0.023314253 |
| Obesity | endothelial | *FBXO39* | 0.296184754 | 0.000812247 | 0.001589354 | 1 |

| | | | | | | |
|---|---|---|---|---|---|---|
| Obesity | endothelial | *TOP3A* | -0.994408498 | 1.04E-05 | 3.83E-05 | 0.065540147 |
| Obesity | endothelial | *C8orf31* | -0.956190209 | 1.75E-08 | 3.42E-07 | 0.000110269 |
| Obesity | endothelial | *PPFIA3* | 0.287296229 | 0.002946163 | 0.004961834 | 1 |
| Obesity | endothelial | *MAGEF1* | -1.089705407 | 1.34E-08 | 3.34E-07 | 8.41E-05 |
| Obesity | endothelial | *NIM1K* | -1.531325707 | 4.89E-06 | 2.07E-05 | 0.030725958 |
| Obesity | endothelial | *ACOT4* | -0.615407326 | 3.18E-05 | 9.61E-05 | 0.199765027 |
| Obesity | endothelial | *ARIH2* | -0.696964978 | 3.46E-06 | 1.56E-05 | 0.021779845 |
| Obesity | endothelial | *RBM44* | 0.398862571 | 2.11E-05 | 6.86E-05 | 0.132831442 |
| Obesity | endothelial | *IRX3* | -0.80063298 | 1.75E-08 | 3.42E-07 | 0.000109743 |
| Obesity | endothelial | *ATOX1* | -0.794889108 | 0.002086162 | 0.003663604 | 1 |
| Obesity | endothelial | *TBL1XR1* | -0.648951907 | 1.80E-05 | 6.03E-05 | 0.113391551 |
| Obesity | endothelial | *SAMD12* | 0.210087351 | 0.000275998 | 0.00061034 | 1 |
| Obesity | endothelial | *C18orf32* | -1.254713037 | 5.72E-05 | 0.000157541 | 0.359509483 |
| Obesity | endothelial | *RPLP2* | -0.550248081 | 0.004911853 | 0.007879771 | 1 |
| Obesity | endothelial | *GBA* | -0.889015614 | 0.002976871 | 0.005004168 | 1 |
| Obesity | endothelial | *IL17RA* | -0.726343915 | 1.03E-05 | 3.77E-05 | 0.064509885 |
| Obesity | endothelial | *CD163L1* | -0.455312605 | 0.000117309 | 0.000289337 | 0.73752043 |
| Obesity | endothelial | *THAP5* | -0.716208504 | 0.026549114 | 0.036348928 | 1 |
| Obesity | endothelial | *POLR2L* | -0.993345638 | 6.11E-05 | 0.000166509 | 0.384135538 |
| Obesity | endothelial | *FAM87B* | -1.6142324 | 1.20E-09 | 8.57E-08 | 7.54E-06 |
| Obesity | endothelial | *ZCCHC3* | -1.750805165 | 8.08E-07 | 4.80E-06 | 0.005080366 |
| Obesity | endothelial | *ZBTB41* | -0.937966088 | 6.44E-06 | 2.57E-05 | 0.040489648 |
| Obesity | endothelial | *CENPBD1* | -1.075596917 | 4.26E-06 | 1.85E-05 | 0.026784188 |
| Obesity | endothelial | *BET1L* | -0.983737531 | 3.20E-05 | 9.65E-05 | 0.200934725 |
| Obesity | endothelial | *RIC8A* | -1.318647862 | 0.000120853 | 0.000297147 | 0.759804655 |
| Obesity | endothelial | *IMP3* | -1.360888361 | 3.42E-06 | 1.55E-05 | 0.021532736 |
| Obesity | endothelial | *C2orf73* | -1.717896579 | 2.07E-08 | 3.63E-07 | 0.000129947 |
| Obesity | endothelial | *FAM26E* | 0.603680404 | 4.34E-05 | 0.000124206 | 0.272792094 |
| Obesity | endothelial | *ALS2CL* | 0.343828171 | 8.69E-06 | 3.30E-05 | 0.054657056 |
| Obesity | endothelial | *MLF1* | -1.080796294 | 8.99E-07 | 5.22E-06 | 0.005655063 |
| Obesity | endothelial | *PRSS42* | -1.461096857 | 1.04E-07 | 9.78E-07 | 0.000654965 |
| Obesity | endothelial | *C2orf69* | -0.796232125 | 0.005204955 | 0.008284443 | 1 |
| Obesity | endothelial | *GRAMD1C* | 0.23334271 | 3.62E-06 | 1.62E-05 | 0.022771522 |
| Obesity | endothelial | *STAP2* | -1.340846629 | 6.89E-05 | 0.000184045 | 0.433425705 |
| Obesity | endothelial | *TSSK6* | -1.920636278 | 3.44E-06 | 1.56E-05 | 0.021655754 |
| Obesity | endothelial | *NDUFV2* | -1.102827701 | 0.00389707 | 0.00639209 | 1 |
| Obesity | endothelial | *DALRD3* | -1.067817887 | 9.89E-07 | 5.66E-06 | 0.006219525 |
| Obesity | endothelial | *ZNF114* | 0.645031698 | 3.54E-05 | 0.000105392 | 0.222693526 |
| Obesity | endothelial | *ZNF518B* | -1.3481424 | 1.00E-05 | 3.70E-05 | 0.062954517 |
| Obesity | endothelial | *ZNF454* | 0.465003689 | 4.35E-06 | 1.88E-05 | 0.027361527 |
| Obesity | endothelial | *SH2B1* | -1.529725843 | 1.09E-33 | 6.87E-30 | 6.87E-30 |
| Obesity | endothelial | *ZC3H12D* | -1.036220218 | 1.35E-08 | 3.34E-07 | 8.48E-05 |
| Obesity | endothelial | *KDELC2* | -1.043804424 | 1.69E-06 | 8.63E-06 | 0.010609612 |
| Obesity | endothelial | *SH2D4B* | 1.433387318 | 0.004928234 | 0.007895975 | 1 |
| Obesity | endothelial | *RNF212* | -0.287676103 | 0.004579 | 0.007389162 | 1 |
| Obesity | endothelial | *PRSS36* | -1.15377198 | 1.67E-08 | 3.39E-07 | 0.000104886 |
| Obesity | endothelial | *TMPRSS9* | -0.885911898 | 7.37E-06 | 2.87E-05 | 0.046366114 |
| Obesity | endothelial | *AQP11* | -0.850612261 | 0.002250298 | 0.003917923 | 1 |
| Obesity | endothelial | *TMEM11* | -1.338554271 | 4.74E-05 | 0.000133541 | 0.298063412 |
| Obesity | endothelial | *KCNG2* | -0.327900794 | 0.010852715 | 0.016118833 | 1 |

| | | | | | | |
|---|---|---|---|---|---|---|
| Obesity | endothelial | *FAM220A* | -0.770868552 | 0.000315797 | 0.000688664 | 1 |
| Obesity | endothelial | *BEND3* | -1.978261723 | 0.001241017 | 0.002305636 | 1 |
| Obesity | endothelial | *NT5DC1* | -0.625308089 | 0.000855547 | 0.001662182 | 1 |
| Obesity | endothelial | *AC006111.1* | -4.880371832 | 9.47E-05 | 0.000240861 | 0.595649994 |
| Obesity | endothelial | *LINC00843* | -1.656515423 | 1.56E-07 | 1.34E-06 | 0.000982408 |
| Obesity | endothelial | *COX14* | -0.705169325 | 3.68E-07 | 2.57E-06 | 0.002310786 |
| Obesity | endothelial | *P4HTM* | -1.369738859 | 9.74E-11 | 1.23E-08 | 6.13E-07 |
| Obesity | endothelial | *CTXN1* | -1.426115144 | 0.009808555 | 0.014717514 | 1 |
| Obesity | endothelial | *CA8* | 0.221535539 | 4.44E-06 | 1.91E-05 | 0.027883174 |
| Obesity | endothelial | *ERBB4* | -0.658544225 | 5.47E-07 | 3.51E-06 | 0.003436902 |
| Obesity | endothelial | *CSRNP3* | -0.951844294 | 0.00166461 | 0.002989261 | 1 |
| Obesity | endothelial | *PARP10* | -0.805693338 | 0.000174217 | 0.000410226 | 1 |
| Obesity | endothelial | *KCTD12* | 0.294514493 | 8.46E-05 | 0.00021823 | 0.53204566 |
| Obesity | endothelial | *RPP25* | -0.546848591 | 0.004010469 | 0.00655756 | 1 |
| Obesity | endothelial | *STX19* | 0.797092317 | 0.00073804 | 0.001457305 | 1 |
| Obesity | endothelial | *FAM219B* | -0.856105069 | 2.15E-06 | 1.06E-05 | 0.013545587 |
| Obesity | endothelial | *CPNE7* | -1.210406971 | 0.007084157 | 0.010964573 | 1 |
| Obesity | endothelial | *GDPD4* | -2.331932482 | 0.002524252 | 0.004331466 | 1 |
| Obesity | endothelial | *MPI* | -0.646790754 | 0.020507168 | 0.028797982 | 1 |
| Obesity | endothelial | *TRIM73* | -2.98478514 | 2.65E-14 | 1.39E-11 | 1.67E-10 |
| Obesity | endothelial | *HYI* | -0.438890416 | 9.23E-05 | 0.000235407 | 0.580277699 |
| Obesity | endothelial | *C17orf62* | -0.867661607 | 0.001186422 | 0.002215336 | 1 |
| Obesity | endothelial | *GAK* | -0.660078511 | 0.002916957 | 0.004917915 | 1 |
| Obesity | endothelial | *TUFM* | 1.002739883 | 8.68E-08 | 8.50E-07 | 0.000546004 |
| Obesity | endothelial | *RMI1* | -0.494841971 | 0.035632415 | 0.047552748 | 1 |
| Obesity | endothelial | *SNX18* | -0.846456865 | 0.000591449 | 0.00119641 | 1 |
| Obesity | endothelial | *C14orf39* | -0.370017146 | 6.59E-07 | 4.09E-06 | 0.004145436 |
| Obesity | endothelial | *KLHDC7A* | -0.317909325 | 0.001564237 | 0.002829216 | 1 |
| Obesity | endothelial | *TRIML2* | -7.881856943 | 1.08E-08 | 3.32E-07 | 6.80E-05 |
| Obesity | endothelial | *FARSA* | -1.478993058 | 2.04E-07 | 1.65E-06 | 0.001284688 |
| Obesity | endothelial | *SPTY2D1* | -1.110800629 | 6.94E-06 | 2.73E-05 | 0.043605461 |
| Obesity | endothelial | *TCAIM* | -0.438541775 | 0.009806828 | 0.014717514 | 1 |
| Obesity | endothelial | *FUCA1* | -0.780892134 | 9.11E-05 | 0.000232969 | 0.572869886 |
| Obesity | endothelial | *ZNF664* | -1.25518871 | 3.89E-08 | 5.04E-07 | 0.000244687 |
| Obesity | endothelial | *CDH4* | -0.790623265 | 0.000131464 | 0.00032023 | 0.826513916 |
| Obesity | endothelial | *ARID3B* | -0.847100725 | 0.000600013 | 0.001212172 | 1 |
| Obesity | endothelial | *EGR3* | -1.448007419 | 0.003073504 | 0.005147341 | 1 |
| Obesity | endothelial | *LINC00174* | -0.465472272 | 7.69E-06 | 2.97E-05 | 0.048326403 |
| Obesity | endothelial | *GEMIN4* | -0.712513855 | 2.71E-07 | 2.03E-06 | 0.001705104 |
| Obesity | endothelial | *PLD6* | -0.824162699 | 1.38E-05 | 4.78E-05 | 0.086583608 |
| Obesity | endothelial | *MAF1* | -1.607023175 | 2.92E-07 | 2.15E-06 | 0.001837919 |
| Obesity | endothelial | *FCER1A* | -1.060893122 | 6.24E-07 | 3.91E-06 | 0.003925652 |
| Obesity | endothelial | *RPRML* | 1.73316152 | 1.35E-07 | 1.19E-06 | 0.000847282 |
| Obesity | endothelial | *PCED1B* | -0.552529339 | 0.00040366 | 0.000854482 | 1 |
| Obesity | endothelial | *PIPOX* | -1.284363688 | 3.57E-09 | 1.78E-07 | 2.25E-05 |
| Obesity | endothelial | *FOXS1* | -0.994165283 | 0.002368333 | 0.00409959 | 1 |
| Obesity | endothelial | *ATOH7* | -2.908127014 | 0.016357403 | 0.023468505 | 1 |
| Obesity | endothelial | *CDH5* | 0.17078849 | 4.96E-07 | 3.25E-06 | 0.003117316 |
| Obesity | endothelial | *GIPC3* | 0.409785613 | 0.003657924 | 0.006024985 | 1 |
| Obesity | endothelial | *CITED4* | -1.333230743 | 5.96E-06 | 2.41E-05 | 0.037492586 |

| | | | | | | |
|---|---|---|---|---|---|---|
| Obesity | endothelial | *TIGD5* | -1.058553763 | 5.07E-08 | 5.82E-07 | 0.000318518 |
| Obesity | endothelial | *PDXDC1* | -0.502488184 | 0.005956518 | 0.009355141 | 1 |
| Obesity | endothelial | *R3HDM2* | -0.62932666 | 0.000123092 | 0.000302061 | 0.77388035 |
| Obesity | endothelial | *B3GNT3* | 2.482533924 | 1.19E-08 | 3.34E-07 | 7.50E-05 |
| Obesity | endothelial | *SEPT1* | -1.063570501 | 1.47E-08 | 3.34E-07 | 9.24E-05 |
| Obesity | endothelial | *EXOC3* | -0.574431249 | 0.006230857 | 0.009744627 | 1 |
| Obesity | endothelial | *TDRP* | 0.35620107 | 2.86E-05 | 8.83E-05 | 0.17981551 |
| Obesity | endothelial | *ZNF816* | -0.840691895 | 0.006324186 | 0.0098783 | 1 |
| Obesity | endothelial | *FGD6* | -0.774376797 | 0.000604683 | 0.001219256 | 1 |
| Obesity | endothelial | *HCLS1* | -1.017330926 | 1.68E-06 | 8.61E-06 | 0.010573153 |
| Obesity | endothelial | *ZNF609* | -0.609707802 | 6.40E-05 | 0.000173065 | 0.402549404 |
| Obesity | endothelial | *CCDC66* | -0.431515309 | 0.00929551 | 0.014021322 | 1 |
| Obesity | endothelial | *DEFB124* | -1.908968441 | 1.17E-06 | 6.40E-06 | 0.007333601 |
| Obesity | endothelial | *MCFD2* | -0.627077106 | 0.011921905 | 0.017512387 | 1 |
| Obesity | endothelial | *CYP8B1* | -0.875491243 | 1.35E-07 | 1.19E-06 | 0.000848805 |
| Obesity | endothelial | *GLIPR1L2* | -0.303497475 | 0.028435232 | 0.038695304 | 1 |
| Obesity | endothelial | *RNF182* | -0.678142269 | 9.72E-08 | 9.24E-07 | 0.000610806 |
| Obesity | endothelial | *MB21D2* | -0.737351152 | 0.000486509 | 0.001006146 | 1 |
| Obesity | endothelial | *SSTR2* | -1.393596332 | 1.64E-08 | 3.39E-07 | 0.000103018 |
| Obesity | endothelial | *ZNF594* | -1.187818854 | 2.71E-08 | 4.21E-07 | 0.000170636 |
| Obesity | endothelial | *SHISA2* | 3.770382624 | 1.43E-06 | 7.56E-06 | 0.008987521 |
| Obesity | endothelial | *CHST13* | 2.241050972 | 0.002213573 | 0.003863613 | 1 |
| Obesity | endothelial | *SLC36A4* | -0.479078277 | 0.030936095 | 0.041799963 | 1 |
| Obesity | endothelial | *OR51E1* | 0.323305332 | 0.000888624 | 0.001717423 | 1 |
| Obesity | endothelial | *CAPS2* | -0.81811787 | 3.40E-06 | 1.54E-05 | 0.021349224 |
| Obesity | endothelial | *ZNF792* | 0.420171619 | 7.28E-05 | 0.000192262 | 0.457425922 |
| Obesity | endothelial | *KCTD2* | -0.699729456 | 0.021235963 | 0.029718831 | 1 |
| Obesity | endothelial | *D2HGDH* | -0.893997796 | 1.41E-07 | 1.24E-06 | 0.000887148 |
| Obesity | endothelial | *CMTR2* | -1.083051993 | 3.28E-06 | 1.50E-05 | 0.020640116 |
| Obesity | endothelial | *MRPL14* | -0.739949248 | 0.006548643 | 0.010206078 | 1 |
| Obesity | endothelial | *BBS12* | -1.337991843 | 0.000458124 | 0.000953717 | 1 |
| Obesity | endothelial | *ZFP82* | -0.762043691 | 0.006227875 | 0.009744627 | 1 |
| Obesity | endothelial | *LSMEM1* | -0.669815583 | 2.18E-06 | 1.07E-05 | 0.013685082 |
| Obesity | endothelial | *AEN* | -1.761138775 | 0.000609085 | 0.001226167 | 1 |
| Obesity | endothelial | *ANKRD34A* | -1.521685972 | 5.76E-09 | 2.34E-07 | 3.62E-05 |
| Obesity | endothelial | *SLC26A11* | -1.079450873 | 6.38E-05 | 0.000172593 | 0.400934237 |
| Obesity | endothelial | *HIGD1A* | -0.901463524 | 7.45E-05 | 0.00019586 | 0.468301317 |
| Obesity | endothelial | *ADIPOQ* | -0.176994597 | 0.01036761 | 0.01548982 | 1 |
| Obesity | endothelial | *HIST3H2A* | -1.158111595 | 3.81E-06 | 1.69E-05 | 0.023968105 |
| Obesity | endothelial | *TMEM132C* | -0.550792924 | 2.29E-08 | 3.82E-07 | 0.000143731 |
| Obesity | endothelial | *SLC25A41* | -2.357189098 | 1.76E-08 | 3.42E-07 | 0.000110589 |
| Obesity | endothelial | *TMEM132E* | 1.382384782 | 9.42E-06 | 3.51E-05 | 0.059211741 |
| Obesity | endothelial | *ZNF322* | -1.283242042 | 2.38E-10 | 2.41E-08 | 1.49E-06 |
| Obesity | endothelial | *CCL13* | -0.936277156 | 4.09E-07 | 2.79E-06 | 0.002572862 |
| Obesity | endothelial | *ACBD4* | -1.108286241 | 5.10E-07 | 3.33E-06 | 0.00320435 |
| Obesity | endothelial | *SGSH* | -1.521923247 | 2.41E-06 | 1.16E-05 | 0.015153402 |
| Obesity | endothelial | *SETD2* | -0.595141277 | 0.000265296 | 0.000590621 | 1 |
| Obesity | endothelial | *MRPS23* | -1.290173997 | 3.86E-05 | 0.000112707 | 0.242545008 |
| Obesity | endothelial | *GPR135* | -0.464103501 | 8.86E-05 | 0.000227283 | 0.556843388 |
| Obesity | endothelial | *ATG9B* | 0.260247623 | 1.08E-06 | 6.08E-06 | 0.006811654 |

| | | | | | | |
|---|---|---|---|---|---|---|
| Obesity | endothelial | *HKR1* | -0.621186563 | 0.005270835 | 0.008370959 | 1 |
| Obesity | endothelial | *ZBTB20* | -0.292340706 | 0.021792381 | 0.030425927 | 1 |
| Obesity | endothelial | *RELL1* | -0.50362578 | 0.015460582 | 0.022273299 | 1 |
| Obesity | endothelial | *CLDN7* | -0.941043603 | 5.80E-10 | 5.06E-08 | 3.64E-06 |
| Obesity | endothelial | *SNRPE* | -1.247255615 | 6.99E-05 | 0.000186357 | 0.439429009 |
| Obesity | endothelial | *IDH2* | -1.072426584 | 6.47E-08 | 6.95E-07 | 0.000406772 |
| Obesity | endothelial | *TNRC18* | -0.593879466 | 0.001781524 | 0.003178332 | 1 |
| Obesity | endothelial | *FAM181B* | 1.067890675 | 0.005452485 | 0.008645593 | 1 |
| Obesity | endothelial | *NOP10* | -1.060621539 | 7.29E-05 | 0.000192523 | 0.458398329 |
| Obesity | endothelial | *TDRKH* | -0.616398256 | 0.012041592 | 0.017659317 | 1 |
| Obesity | endothelial | *ZNF708* | -0.765802549 | 2.08E-05 | 6.77E-05 | 0.130808938 |
| Obesity | endothelial | *IST1* | -0.773594094 | 0.000157648 | 0.000376141 | 0.991132714 |
| Obesity | endothelial | *ERCC6L2* | -0.541483288 | 0.00166668 | 0.002991269 | 1 |
| Obesity | endothelial | *MRPL41* | -0.947253192 | 1.33E-05 | 4.65E-05 | 0.083669584 |
| Obesity | endothelial | *HHIPL1* | -1.705916492 | 1.28E-08 | 3.34E-07 | 8.02E-05 |
| Obesity | endothelial | *FAM153B* | -0.881861012 | 0.011871796 | 0.017459176 | 1 |
| Obesity | endothelial | *BACE2* | 0.2753472 | 0.00261785 | 0.004472398 | 1 |
| Obesity | endothelial | *SYNM* | -1.187093243 | 4.34E-08 | 5.38E-07 | 0.000273076 |
| Obesity | endothelial | *IZUMO1* | 0.507642589 | 1.43E-07 | 1.25E-06 | 0.000897351 |
| Obesity | endothelial | *FBXL6* | -2.193441189 | 3.98E-09 | 1.84E-07 | 2.50E-05 |
| Obesity | endothelial | *CACNB4* | -0.411712298 | 1.72E-05 | 5.79E-05 | 0.10810606 |
| Obesity | endothelial | *TRAPPC6B* | -0.878823019 | 3.25E-06 | 1.49E-05 | 0.020428371 |
| Obesity | endothelial | *NPLOC4* | -0.644903211 | 3.04E-05 | 9.28E-05 | 0.191230073 |
| Obesity | endothelial | *CAPN12* | 0.465320965 | 0.000227928 | 0.000517136 | 1 |
| Obesity | endothelial | *FES* | 0.484663956 | 0.016045577 | 0.023052684 | 1 |
| Obesity | endothelial | *GLRX5* | -1.182509733 | 9.04E-05 | 0.000231315 | 0.568340269 |
| Obesity | endothelial | *CAV3* | -0.617995599 | 3.81E-09 | 1.84E-07 | 2.40E-05 |
| Obesity | endothelial | *MFSD5* | -0.846123814 | 0.002362866 | 0.004091252 | 1 |
| Obesity | endothelial | *ADI1* | -1.064462162 | 2.48E-08 | 4.03E-07 | 0.00015597 |
| Obesity | endothelial | *SPNS3* | 0.59298904 | 3.21E-06 | 1.48E-05 | 0.020189551 |
| Obesity | endothelial | *CLEC4G* | -1.13055804 | 8.44E-08 | 8.36E-07 | 0.000530632 |
| Obesity | endothelial | *SATB1* | -1.169758803 | 5.80E-06 | 2.36E-05 | 0.036457184 |
| Obesity | endothelial | *ACTL10* | -1.222276508 | 4.91E-06 | 2.07E-05 | 0.030883723 |
| Obesity | endothelial | *TRAK1* | -0.525576556 | 0.002625083 | 0.004481101 | 1 |
| Obesity | endothelial | *SKA2* | -1.075179835 | 1.65E-06 | 8.49E-06 | 0.010386299 |
| Obesity | endothelial | *TTC3* | -0.205915552 | 0.007158878 | 0.011069323 | 1 |
| Obesity | endothelial | *KCNB2* | -0.30645548 | 3.03E-05 | 9.25E-05 | 0.190319093 |
| Obesity | endothelial | *BRICD5* | -0.816296939 | 1.02E-05 | 3.75E-05 | 0.063983331 |
| Obesity | endothelial | *TSKU* | -0.911299204 | 3.95E-08 | 5.06E-07 | 0.000248612 |
| Obesity | endothelial | *GRID1* | -1.070475726 | 7.52E-12 | 1.63E-09 | 4.73E-08 |
| Obesity | endothelial | *ACBD3* | -0.686539168 | 0.000904595 | 0.001744537 | 1 |
| Obesity | endothelial | *PLCXD3* | 0.184640126 | 1.56E-06 | 8.10E-06 | 0.009786236 |
| Obesity | endothelial | *GPIHBP1* | 0.204192077 | 1.53E-06 | 7.97E-06 | 0.009590578 |
| Obesity | endothelial | *SLC25A18* | -0.517090962 | 2.45E-05 | 7.70E-05 | 0.153791125 |
| Obesity | endothelial | *ZNF721* | -0.360102887 | 0.029139869 | 0.039551458 | 1 |
| Obesity | endothelial | *LENG9* | -0.641174317 | 0.004142297 | 0.00675204 | 1 |
| Obesity | endothelial | *CEP63* | -0.483073408 | 4.66E-05 | 0.000131647 | 0.293176903 |
| Obesity | endothelial | *HMGN4* | -1.138581108 | 6.61E-07 | 4.09E-06 | 0.004156292 |
| Obesity | endothelial | *C12orf60* | -0.74888602 | 0.009391258 | 0.014145386 | 1 |
| Obesity | endothelial | *WBP2NL* | -1.120499692 | 5.78E-06 | 2.36E-05 | 0.036360031 |

| | | | | | | |
|---|---|---|---|---|---|---|
| Obesity | endothelial | *BEGAIN* | -2.991433981 | 1.28E-08 | 3.34E-07 | 8.03E-05 |
| Obesity | endothelial | *ARHGEF37* | 0.40558056 | 0.028649384 | 0.038944578 | 1 |
| Obesity | endothelial | *FAM43B* | -1.934561405 | 1.83E-09 | 1.15E-07 | 1.15E-05 |
| Obesity | endothelial | *PTGDR2* | -8.259251743 | 0.000195032 | 0.000453129 | 1 |
| Obesity | endothelial | *C2CD4C* | 0.337091745 | 1.89E-07 | 1.55E-06 | 0.001190807 |
| Obesity | endothelial | *CTNNA3* | -0.6208727 | 2.89E-05 | 8.92E-05 | 0.181989657 |
| Obesity | endothelial | *CCBE1* | -0.314440864 | 9.26E-05 | 0.00023594 | 0.582063098 |
| Obesity | endothelial | *ZNF623* | -1.034447902 | 1.15E-05 | 4.14E-05 | 0.072223934 |
| Obesity | endothelial | *CCDC125* | -0.88015947 | 5.31E-07 | 3.44E-06 | 0.003335377 |
| Obesity | endothelial | *C10orf107* | 0.147971556 | 0.004303043 | 0.006983281 | 1 |
| Obesity | endothelial | *FHL3* | -1.373920391 | 0.000214155 | 0.000490489 | 1 |
| Obesity | endothelial | *LRIT3* | -2.186488924 | 1.45E-07 | 1.27E-06 | 0.00091423 |
| Obesity | endothelial | *NPIPA1* | -0.247488989 | 0.00958138 | 0.014404146 | 1 |
| Obesity | endothelial | *SF3A3* | -0.790764977 | 0.000721359 | 0.001429756 | 1 |
| Obesity | endothelial | *GRIN2A* | -0.920688414 | 2.90E-06 | 1.35E-05 | 0.018234465 |
| Obesity | endothelial | *URAD* | -1.996727676 | 1.84E-07 | 1.52E-06 | 0.001157437 |
| Obesity | endothelial | *GPR132* | -1.20899385 | 1.28E-08 | 3.34E-07 | 8.08E-05 |
| Obesity | endothelial | *MX2* | -0.718734109 | 0.001021658 | 0.00193527 | 1 |
| Obesity | endothelial | *PSMG1* | 0.333954722 | 0.001503082 | 0.00273434 | 1 |
| Obesity | endothelial | *KLRC4* | -0.523048357 | 0.021005048 | 0.02943797 | 1 |
| Obesity | endothelial | *ACSM5* | -0.50522395 | 4.21E-06 | 1.83E-05 | 0.026456576 |
| Obesity | endothelial | *AC131971.1* | -2.54102742 | 1.57E-06 | 8.16E-06 | 0.009899206 |
| Obesity | endothelial | *SERHL2* | -0.955888489 | 0.006447757 | 0.010058821 | 1 |
| Obesity | endothelial | *PCBP3* | -0.130120898 | 0.000605801 | 0.00122112 | 1 |
| Obesity | endothelial | *TANGO2* | -0.815516639 | 1.51E-05 | 5.17E-05 | 0.094900637 |
| Obesity | endothelial | *SFXN4* | -0.996372978 | 3.99E-05 | 0.000115707 | 0.250563546 |
| Obesity | endothelial | *FAM167B* | 0.776244461 | 2.05E-06 | 1.01E-05 | 0.012889881 |
| Obesity | endothelial | *MRPL54* | -0.636331667 | 7.43E-05 | 0.000195608 | 0.467306413 |
| Obesity | endothelial | *RP1L1* | 0.896089239 | 4.54E-11 | 6.79E-09 | 2.85E-07 |
| Obesity | endothelial | *NDUFB1* | -0.630709006 | 0.001891058 | 0.003347152 | 1 |
| Obesity | endothelial | *FAM19A1* | -0.83554037 | 2.26E-08 | 3.81E-07 | 0.000141922 |
| Obesity | endothelial | *TRMT12* | -1.238068849 | 8.00E-05 | 0.000208105 | 0.503198832 |
| Obesity | endothelial | *FIGLA* | 1.252629063 | 4.49E-05 | 0.000127635 | 0.282073077 |
| Obesity | endothelial | *AIFM3* | -1.209621599 | 1.07E-08 | 3.32E-07 | 6.74E-05 |
| Obesity | endothelial | *KCTD16* | -0.760299334 | 0.000526882 | 0.001079344 | 1 |
| Obesity | endothelial | *B3GALT5* | -1.250534281 | 3.18E-05 | 9.61E-05 | 0.199695559 |
| Obesity | endothelial | *ZNF703* | -0.815098351 | 6.72E-06 | 2.67E-05 | 0.042269044 |
| Obesity | endothelial | *KCTD8* | -0.496677757 | 1.19E-07 | 1.08E-06 | 0.000747629 |
| Obesity | endothelial | *C9orf66* | -1.124903207 | 3.83E-08 | 5.00E-07 | 0.000240701 |
| Obesity | endothelial | *FAM162B* | -0.516824464 | 0.003979774 | 0.006510758 | 1 |
| Obesity | endothelial | *BTBD9* | -0.576246343 | 0.000211864 | 0.000486838 | 1 |
| Obesity | endothelial | *NUDT14* | 0.415314518 | 0.001975676 | 0.003483196 | 1 |
| Obesity | endothelial | *ANKRD45* | 1.54698356 | 0.017717977 | 0.025213427 | 1 |
| Obesity | endothelial | *MAATS1* | 0.606613817 | 1.18E-05 | 4.22E-05 | 0.074047603 |
| Obesity | endothelial | *GPR39* | -0.735696211 | 5.30E-09 | 2.21E-07 | 3.33E-05 |
| Obesity | endothelial | *FAM3B* | -0.166152426 | 0.026916991 | 0.036828535 | 1 |
| Obesity | endothelial | *NPIPA7* | 1.592119129 | 7.22E-05 | 0.000191089 | 0.453860786 |
| Obesity | endothelial | *DENND5A* | -0.650195879 | 0.000681561 | 0.001356433 | 1 |
| Obesity | endothelial | *TMEM236* | -1.267241486 | 7.22E-09 | 2.61E-07 | 4.54E-05 |
| Obesity | endothelial | *VPS33B* | -0.664563175 | 0.022284167 | 0.03103534 | 1 |

| | | | | | | |
|---|---|---|---|---|---|---|
| Obesity | endothelial | *TBX1* | 0.531068879 | 0.000119863 | 0.000295022 | 0.753576729 |
| Obesity | endothelial | *UQCR10* | -1.213363611 | 9.08E-06 | 3.41E-05 | 0.057101999 |
| Obesity | endothelial | *EIF3C* | 1.847738685 | 7.11E-29 | 2.23E-25 | 4.47E-25 |
| Obesity | endothelial | *NIPSNAP1* | -0.889083513 | 0.001258317 | 0.002336397 | 1 |
| Obesity | endothelial | *CNTN2* | -1.894776439 | 3.55E-08 | 4.80E-07 | 0.000223209 |
| Obesity | endothelial | *CRELD2* | -1.237895658 | 1.51E-05 | 5.16E-05 | 0.094706614 |
| Obesity | endothelial | *UBE2F* | -1.285434525 | 1.15E-07 | 1.06E-06 | 0.00072535 |
| Obesity | endothelial | *GOLGA6L4* | -1.699611699 | 3.06E-08 | 4.48E-07 | 0.000192661 |
| Obesity | endothelial | *PCDH9* | -0.429007945 | 3.74E-06 | 1.67E-05 | 0.02353684 |
| Obesity | endothelial | *OAF* | -0.821869724 | 4.43E-08 | 5.42E-07 | 0.000278739 |
| Obesity | endothelial | *TACSTD2* | 0.404098417 | 0.000244969 | 0.000550435 | 1 |
| Obesity | endothelial | *CLECL1* | -0.330774757 | 0.006357252 | 0.009925017 | 1 |
| Obesity | endothelial | *MROH7* | -1.330191373 | 0.00098083 | 0.001867498 | 1 |
| Obesity | endothelial | *GDF3* | 1.096650568 | 1.85E-06 | 9.26E-06 | 0.011608469 |
| Obesity | endothelial | *PKP3* | -0.708186237 | 0.007987214 | 0.012208999 | 1 |
| Obesity | endothelial | *CSF1* | -0.475951577 | 0.003106417 | 0.005199691 | 1 |
| Obesity | endothelial | *SS18L1* | -0.925947204 | 0.000131706 | 0.000320696 | 0.828036577 |
| Obesity | endothelial | *THAP7* | -1.602590671 | 1.09E-07 | 1.01E-06 | 0.000686896 |
| Obesity | endothelial | *CCR10* | -1.491474087 | 4.37E-08 | 5.40E-07 | 0.000274625 |
| Obesity | endothelial | *WDR27* | -0.478090205 | 0.017768319 | 0.025273625 | 1 |
| Obesity | endothelial | *TXNRD2* | -1.126314799 | 4.72E-06 | 2.01E-05 | 0.029665009 |
| Obesity | endothelial | *ZFP1* | -0.749503964 | 0.030250507 | 0.040917586 | 1 |
| Obesity | endothelial | *LPAR5* | -0.535908252 | 0.001868501 | 0.003318437 | 1 |
| Obesity | endothelial | *TMEM173* | 0.390621923 | 0.010800779 | 0.016059387 | 1 |
| Obesity | endothelial | *NELL2* | -0.38422495 | 0.020648133 | 0.02897652 | 1 |
| Obesity | endothelial | *ZNF93* | -1.519815993 | 1.89E-07 | 1.55E-06 | 0.001188058 |
| Obesity | endothelial | *SEPT5* | -3.81519034 | 2.39E-08 | 3.95E-07 | 0.000150365 |
| Obesity | endothelial | *ATL3* | -0.448300795 | 0.020622129 | 0.028946489 | 1 |
| Obesity | endothelial | *UBE2G2* | -0.773283331 | 1.88E-05 | 6.20E-05 | 0.117899184 |
| Obesity | endothelial | *TUSC5* | -0.572858164 | 1.76E-06 | 8.95E-06 | 0.011091754 |
| Obesity | endothelial | *BTBD6* | -0.764416317 | 2.67E-05 | 8.32E-05 | 0.167905936 |
| Obesity | endothelial | *H1FX* | -0.775440784 | 0.004823768 | 0.007752308 | 1 |
| Obesity | endothelial | *NUTM2A* | -1.421706928 | 1.50E-05 | 5.15E-05 | 0.094240306 |
| Obesity | endothelial | *ZFP90* | -0.569629144 | 0.020891201 | 0.029298011 | 1 |
| Obesity | endothelial | *FAM227A* | -0.472225867 | 0.003006122 | 0.00504794 | 1 |
| Obesity | endothelial | *NOC4L* | -0.946894469 | 2.41E-07 | 1.86E-06 | 0.001517034 |
| Obesity | endothelial | *SORCS2* | -0.777710281 | 1.83E-06 | 9.20E-06 | 0.011515115 |
| Obesity | endothelial | *TMEM106A* | -0.981741615 | 3.11E-05 | 9.43E-05 | 0.195443657 |
| Obesity | endothelial | *UBOX5* | -1.114026964 | 4.34E-07 | 2.93E-06 | 0.002726745 |
| Obesity | endothelial | *MAFF* | -1.284629167 | 4.00E-06 | 1.76E-05 | 0.025155651 |
| Obesity | endothelial | *BRF1* | -0.789512341 | 0.001339052 | 0.002464468 | 1 |
| Obesity | endothelial | *CIB1* | -0.62769965 | 0.001265381 | 0.00234882 | 1 |
| Obesity | endothelial | *FLRT2* | -0.476804923 | 9.96E-05 | 0.000251494 | 0.626221111 |
| Obesity | endothelial | *ADSSL1* | -1.268382487 | 8.20E-08 | 8.18E-07 | 0.000515565 |
| Obesity | endothelial | *C6orf120* | -1.484814793 | 7.16E-08 | 7.41E-07 | 0.000449949 |
| Obesity | endothelial | *KIAA0825* | -0.77166207 | 1.83E-05 | 6.09E-05 | 0.115094354 |
| Obesity | endothelial | *RBM11* | 0.863052009 | 0.008299463 | 0.012624903 | 1 |
| Obesity | endothelial | *CCDC137* | -0.449759154 | 0.033641968 | 0.045135948 | 1 |
| Obesity | endothelial | *TMEM105* | -6.620314021 | 1.07E-06 | 6.04E-06 | 0.006752712 |
| Obesity | endothelial | *TCN2* | 0.238864538 | 0.01125285 | 0.016618902 | 1 |

| | | | | | | |
|---|---|---|---|---|---|---|
| Obesity | endothelial | *ATP6V0A2* | -0.90137346 | 4.77E-05 | 0.000134233 | 0.300144448 |
| Obesity | endothelial | *TNFAIP8L1* | 0.549998277 | 0.000146072 | 0.000351053 | 0.918355801 |
| Obesity | endothelial | *MAPK11* | 0.359488567 | 5.24E-06 | 2.18E-05 | 0.03295614 |
| Obesity | endothelial | *SH3BGR* | -0.143295357 | 0.020458526 | 0.02874618 | 1 |
| Obesity | endothelial | *MUC1* | -0.444413953 | 0.031396764 | 0.042358682 | 1 |
| Obesity | endothelial | *SV2B* | 0.294820648 | 1.15E-05 | 4.16E-05 | 0.07259228 |
| Obesity | endothelial | *TLCD2* | -1.18537176 | 7.68E-09 | 2.70E-07 | 4.83E-05 |
| Obesity | endothelial | *LSAMP* | -0.614723712 | 4.71E-08 | 5.59E-07 | 0.00029617 |
| Obesity | endothelial | *SP1* | -0.575526544 | 0.000976231 | 0.001860433 | 1 |
| Obesity | endothelial | *PDIA2* | -1.000172421 | 4.02E-07 | 2.75E-06 | 0.00252561 |
| Obesity | endothelial | *PSMD13* | -0.805923191 | 0.00360764 | 0.005949956 | 1 |
| Obesity | endothelial | *PBX1* | -0.511651802 | 0.000266575 | 0.00059242 | 1 |
| Obesity | endothelial | *UBE2L3* | -1.006081112 | 3.48E-06 | 1.57E-05 | 0.021878564 |
| Obesity | endothelial | *PMEL* | 0.576754648 | 2.26E-05 | 7.24E-05 | 0.14234095 |
| Obesity | endothelial | *SNAI3* | -1.088994444 | 4.91E-07 | 3.23E-06 | 0.003085871 |
| Obesity | endothelial | *MYBL1* | -0.592147896 | 2.69E-06 | 1.26E-05 | 0.016908732 |
| Obesity | endothelial | *ADARB2* | 0.18934548 | 3.93E-06 | 1.74E-05 | 0.02468621 |
| Obesity | endothelial | *SRL* | 0.416866345 | 2.26E-06 | 1.10E-05 | 0.014179445 |
| Obesity | endothelial | *KCNIP4* | -0.473360314 | 1.79E-06 | 9.05E-06 | 0.01126908 |
| Obesity | endothelial | *SLC52A2* | -0.996150828 | 0.011900558 | 0.017492889 | 1 |
| Obesity | endothelial | *ARL17A* | -0.883050241 | 2.12E-05 | 6.86E-05 | 0.133027533 |
| Obesity | endothelial | *IFITM1* | 0.881099662 | 4.50E-05 | 0.000127853 | 0.282810902 |
| Obesity | endothelial | *POMK* | -0.7027371 | 0.033928923 | 0.045501523 | 1 |
| Obesity | endothelial | *LINC00839* | 0.845586991 | 0.000226672 | 0.000514471 | 1 |
| Obesity | endothelial | *PTCH1* | -0.598972535 | 0.007608768 | 0.011698784 | 1 |
| Obesity | endothelial | *RTN4RL1* | -1.142309491 | 8.63E-08 | 8.47E-07 | 0.000542492 |
| Obesity | endothelial | *PLK5* | -0.61586185 | 0.020425715 | 0.028711933 | 1 |
| Obesity | endothelial | *RASA3* | -0.681938951 | 3.24E-07 | 2.32E-06 | 0.002037042 |
| Obesity | endothelial | *ZNF566* | -0.858082778 | 2.68E-05 | 8.33E-05 | 0.168298427 |
| Obesity | endothelial | *ZNF284* | -1.487215139 | 1.65E-08 | 3.39E-07 | 0.000103865 |
| Obesity | endothelial | *GSAP* | -0.778066282 | 3.22E-05 | 9.70E-05 | 0.20216669 |
| Obesity | endothelial | *C2orf76* | -0.53125315 | 0.01274965 | 0.018619524 | 1 |
| Obesity | endothelial | *WWOX* | -0.433420216 | 0.000583598 | 0.001182431 | 1 |
| Obesity | endothelial | *CYP4Z1* | -0.910468947 | 2.42E-08 | 3.96E-07 | 0.000152152 |
| Obesity | endothelial | *CCDC84* | -0.760545646 | 0.000180798 | 0.000422243 | 1 |
| Obesity | endothelial | *POLR1D* | -0.686872899 | 0.003422744 | 0.005680778 | 1 |
| Obesity | endothelial | *FFAR4* | -0.488320118 | 1.29E-07 | 1.15E-06 | 0.000807982 |
| Obesity | endothelial | *BPIFB4* | 8.749448733 | 3.65E-07 | 2.57E-06 | 0.002297143 |
| Obesity | endothelial | *CYP4F12* | 0.356280705 | 0.002696132 | 0.004588679 | 1 |
| Obesity | endothelial | *KLHL32* | -0.78644971 | 0.000228045 | 0.000517153 | 1 |
| Obesity | endothelial | *NBPF12* | -0.876111633 | 7.61E-05 | 0.000199541 | 0.478699646 |
| Obesity | endothelial | *CYP4X1* | -0.897608694 | 4.42E-07 | 2.97E-06 | 0.002780136 |
| Obesity | endothelial | *CCDC30* | -0.681437116 | 0.001637665 | 0.002944237 | 1 |
| Obesity | endothelial | *FCAR* | -0.577077579 | 0.01302127 | 0.018967731 | 1 |
| Obesity | endothelial | *KPNA4* | -0.602135737 | 0.002861921 | 0.00484331 | 1 |
| Obesity | endothelial | *ZNF197* | -0.442858506 | 0.031835126 | 0.042885674 | 1 |
| Obesity | endothelial | *BTN3A2* | -0.350290279 | 2.56E-06 | 1.22E-05 | 0.016098329 |
| Obesity | endothelial | *PCLO* | -0.508650963 | 1.16E-06 | 6.38E-06 | 0.007295599 |
| Obesity | endothelial | *ZNF396* | -1.198781756 | 2.29E-08 | 3.82E-07 | 0.000144125 |
| Obesity | endothelial | *TMEM222* | -0.811433569 | 0.000213402 | 0.000489299 | 1 |

| | | | | | | |
|---|---|---|---|---|---|---|
| Obesity | endothelial | *CLCNKA* | -1.139338768 | 4.72E-05 | 0.00013294 | 0.296456793 |
| Obesity | endothelial | *SMYD4* | -0.639784583 | 0.00025275 | 0.000566643 | 1 |
| Obesity | endothelial | *NF2* | -0.932594672 | 6.51E-06 | 2.59E-05 | 0.040917847 |
| Obesity | endothelial | *MIR22HG* | -0.686987405 | 3.81E-07 | 2.64E-06 | 0.002398411 |
| Obesity | endothelial | *PDE2A* | 0.223603796 | 1.57E-05 | 5.37E-05 | 0.099008284 |
| Obesity | endothelial | *C17orf58* | -0.591879134 | 2.95E-05 | 9.08E-05 | 0.185597619 |
| Obesity | endothelial | *CYP27C1* | -0.305080306 | 0.000215952 | 0.000493707 | 1 |
| Obesity | endothelial | *BCR* | 0.380744806 | 0.006069204 | 0.009517856 | 1 |
| Obesity | endothelial | *HYAL3* | -0.479823857 | 0.000560381 | 0.001138693 | 1 |
| Obesity | endothelial | *ZSCAN30* | -0.711268248 | 0.003503506 | 0.005797985 | 1 |
| Obesity | endothelial | *LILRB4* | -0.435584636 | 0.000414407 | 0.000875757 | 1 |
| Obesity | endothelial | *KRT16* | -0.743785042 | 1.05E-06 | 5.94E-06 | 0.006588439 |
| Obesity | endothelial | *QRFPR* | -1.269793506 | 0.002746417 | 0.004666683 | 1 |
| Obesity | endothelial | *RTN4RL2* | -2.618631784 | 5.82E-08 | 6.41E-07 | 0.000365822 |
| Obesity | endothelial | *TMEM232* | -0.81008257 | 2.45E-06 | 1.18E-05 | 0.015393835 |
| Obesity | endothelial | *EFCAB6* | -0.89814528 | 2.80E-07 | 2.09E-06 | 0.00176322 |
| Obesity | endothelial | *KANK3* | 0.252716384 | 9.71E-06 | 3.60E-05 | 0.061021042 |
| Obesity | endothelial | *EMID1* | -1.3962838 | 6.53E-06 | 2.60E-05 | 0.04107878 |
| Obesity | endothelial | *CYP4A11* | -0.587346923 | 0.003802844 | 0.006248949 | 1 |
| Obesity | endothelial | *PLCD1* | -0.66508117 | 0.002524334 | 0.004331466 | 1 |
| Obesity | endothelial | *ENTPD5* | -1.023844794 | 8.60E-06 | 3.27E-05 | 0.054089668 |
| Obesity | endothelial | *MRPS21* | -0.709061507 | 0.000504888 | 0.001039709 | 1 |
| Obesity | endothelial | *ANGPTL5* | -0.777699124 | 1.31E-08 | 3.34E-07 | 8.24E-05 |
| Obesity | endothelial | *MT1X* | -0.923218662 | 2.17E-06 | 1.06E-05 | 0.013631368 |
| Obesity | endothelial | *WDR86* | -0.820469807 | 1.13E-08 | 3.34E-07 | 7.13E-05 |
| Obesity | endothelial | *EPOR* | -0.877166082 | 7.15E-06 | 2.80E-05 | 0.044950385 |
| Obesity | endothelial | *FPR3* | -0.691488087 | 7.35E-08 | 7.57E-07 | 0.000462324 |
| Obesity | endothelial | *GJA4* | 0.296010128 | 2.12E-06 | 1.04E-05 | 0.013323103 |
| Obesity | endothelial | *TLR5* | -0.914396711 | 1.53E-08 | 3.34E-07 | 9.61E-05 |
| Obesity | endothelial | *ZNF385C* | -2.011860701 | 3.35E-05 | 0.000100557 | 0.210465061 |
| Obesity | endothelial | *TET3* | -0.811125491 | 0.000209978 | 0.00048268 | 1 |
| Obesity | endothelial | *DHRS4L2* | -0.583964816 | 9.61E-05 | 0.000243845 | 0.604246949 |
| Obesity | endothelial | *TRPV2* | -1.193724796 | 3.61E-08 | 4.82E-07 | 0.000227182 |
| Obesity | endothelial | *C2orf88* | -0.760619512 | 4.07E-05 | 0.000117867 | 0.256006607 |
| Obesity | endothelial | *FANCA* | -0.908299858 | 1.39E-06 | 7.40E-06 | 0.00876532 |
| Obesity | endothelial | *ADH1A* | -1.009418786 | 1.00E-09 | 7.51E-08 | 6.31E-06 |
| Obesity | endothelial | *FAM69C* | 0.59597625 | 9.02E-05 | 0.000231057 | 0.567245099 |
| Obesity | endothelial | *MCRS1* | -0.686081508 | 0.011821139 | 0.017400961 | 1 |
| Obesity | endothelial | *ZFP69* | -0.977075721 | 0.000137303 | 0.000331754 | 0.863224367 |
| Obesity | endothelial | *TMEM220* | -0.890363608 | 8.55E-06 | 3.25E-05 | 0.053728831 |
| Obesity | endothelial | *GFRAL* | 0.664276357 | 0.001574319 | 0.002844996 | 1 |
| Obesity | endothelial | *CPSF4L* | 1.107617402 | 6.53E-05 | 0.000175698 | 0.410431575 |
| Obesity | endothelial | *KLHL17* | -1.043039886 | 4.76E-07 | 3.16E-06 | 0.002995079 |
| Obesity | endothelial | *OR7D2* | -0.946147977 | 0.0126746 | 0.018528474 | 1 |
| Obesity | endothelial | *C6orf132* | -0.747701206 | 2.17E-08 | 3.74E-07 | 0.000136351 |
| Obesity | endothelial | *MAPK12* | 0.43317982 | 2.02E-06 | 1.00E-05 | 0.012714699 |
| Obesity | endothelial | *AGRN* | 0.330891558 | 3.35E-06 | 1.52E-05 | 0.021047139 |
| Obesity | endothelial | *FAM166A* | -2.216201531 | 3.99E-08 | 5.07E-07 | 0.000250665 |
| Obesity | endothelial | *SMTNL2* | -7.600523481 | 0.004242774 | 0.006896153 | 1 |
| Obesity | endothelial | *ZC3H6* | -0.676939252 | 0.000189739 | 0.00044181 | 1 |

| | | | | | | |
|---|---|---|---|---|---|---|
| Obesity | endothelial | *LINC00265* | -1.121592581 | 1.59E-08 | 3.36E-07 | 9.99E-05 |
| Obesity | endothelial | *LAMTOR4* | -0.91747505 | 7.64E-05 | 0.000199932 | 0.480036212 |
| Obesity | endothelial | *PRKAR1B* | 0.382307784 | 0.000310225 | 0.000677451 | 1 |
| Obesity | endothelial | *NUTM2B* | -9.569719258 | 9.91E-06 | 3.67E-05 | 0.062309587 |
| Obesity | endothelial | *NCR3LG1* | -2.811302249 | 0.000381807 | 0.000813702 | 1 |
| Obesity | endothelial | *ZNF793* | 0.264511821 | 0.001206754 | 0.002250477 | 1 |
| Obesity | endothelial | *AGAP4* | -1.500888058 | 4.72E-05 | 0.000133018 | 0.296763519 |
| Obesity | endothelial | *RUFY4* | -1.37157675 | 2.78E-08 | 4.27E-07 | 0.000174468 |
| Obesity | endothelial | *PLSCR1* | 0.330605521 | 0.00053916 | 0.001102341 | 1 |
| Obesity | endothelial | *ZP3* | -0.366758749 | 0.025861213 | 0.035445705 | 1 |
| Obesity | endothelial | *JAKMIP3* | -0.679520967 | 3.56E-09 | 1.78E-07 | 2.24E-05 |
| Obesity | endothelial | *PDCD1* | -1.370603219 | 0.005025122 | 0.008022585 | 1 |
| Obesity | endothelial | *CERKL* | -1.387243375 | 3.09E-05 | 9.39E-05 | 0.194388078 |
| Obesity | endothelial | *C19orf54* | -1.027035958 | 4.71E-06 | 2.00E-05 | 0.029612552 |
| Obesity | endothelial | *C22orf34* | 0.301045445 | 8.48E-05 | 0.00021848 | 0.532873222 |
| Obesity | endothelial | *COL25A1* | -0.707141107 | 3.18E-05 | 9.62E-05 | 0.200173979 |
| Obesity | endothelial | *NBR1* | -0.746445142 | 0.000988418 | 0.001880238 | 1 |
| Obesity | endothelial | *NDOR1* | -1.456801855 | 3.69E-07 | 2.58E-06 | 0.002316992 |
| Obesity | endothelial | *FAM72B* | 2.066992307 | 0.003535045 | 0.005840954 | 1 |
| Obesity | endothelial | *SUMO2* | -1.034442879 | 3.60E-05 | 0.000106603 | 0.226211306 |
| Obesity | endothelial | *GOLGA8M* | 0.433644271 | 0.025128858 | 0.034547371 | 1 |
| Obesity | endothelial | *DPYD* | -0.490271687 | 0.000735588 | 0.001453832 | 1 |
| Obesity | endothelial | *UROS* | -1.016118526 | 8.28E-06 | 3.16E-05 | 0.052027625 |
| Obesity | endothelial | *NOXA1* | -1.379443291 | 9.84E-06 | 3.64E-05 | 0.061864527 |
| Obesity | endothelial | *SKOR1* | -3.701932192 | 0.001039715 | 0.001967104 | 1 |
| Obesity | endothelial | *LINC00910* | -1.226675457 | 3.28E-06 | 1.50E-05 | 0.020628568 |
| Obesity | endothelial | *BEND4* | 0.569710944 | 6.73E-07 | 4.14E-06 | 0.004232296 |
| Obesity | endothelial | *ZNF563* | -0.91306536 | 3.51E-06 | 1.58E-05 | 0.022072854 |
| Obesity | endothelial | *MSL1* | -0.418388662 | 0.003903461 | 0.006399233 | 1 |
| Obesity | endothelial | *LRRK2* | -0.606427901 | 0.000728508 | 0.001442106 | 1 |
| Obesity | endothelial | *FAM196A* | 0.366362142 | 0.000328497 | 0.000713142 | 1 |
| Obesity | endothelial | *UTS2B* | -0.5833787 | 1.65E-06 | 8.48E-06 | 0.010357925 |
| Obesity | endothelial | *C9orf152* | -2.906480182 | 0.001707965 | 0.003059252 | 1 |
| Obesity | endothelial | *NELFB* | -1.142977881 | 6.36E-05 | 0.000172089 | 0.399589876 |
| Obesity | endothelial | *SLC15A5* | -0.357099166 | 0.000995757 | 0.001892481 | 1 |
| Obesity | endothelial | *ADAT2* | -0.702759654 | 0.000275032 | 0.000608419 | 1 |
| Obesity | endothelial | *ZNF567* | -0.586555445 | 0.032131871 | 0.043266882 | 1 |
| Obesity | endothelial | *ANKDD1B* | -0.492927858 | 0.000287154 | 0.000631678 | 1 |
| Obesity | endothelial | *LITAF* | -0.562317768 | 0.005386757 | 0.008549998 | 1 |
| Obesity | endothelial | *SF3B3* | -0.830261159 | 0.000259644 | 0.000579065 | 1 |
| Obesity | endothelial | *SP6* | 4.9536838 | 1.90E-05 | 6.26E-05 | 0.119250532 |
| Obesity | endothelial | *PLAC9* | -0.452001481 | 0.000945968 | 0.001813755 | 1 |
| Obesity | endothelial | *ZNF573* | -0.657365467 | 0.009136836 | 0.0138251 | 1 |
| Obesity | endothelial | *ZNF527* | -1.381944501 | 1.15E-05 | 4.15E-05 | 0.072345006 |
| Obesity | endothelial | *S100A13* | -0.588119467 | 0.020032303 | 0.028206738 | 1 |
| Obesity | endothelial | *ZNF600* | -0.702118461 | 0.00028209 | 0.000621845 | 1 |
| Obesity | endothelial | *LINC00994* | -0.571409725 | 0.02701978 | 0.036945054 | 1 |
| Obesity | endothelial | *C15orf61* | -1.457116007 | 1.05E-07 | 9.79E-07 | 0.000657128 |
| Obesity | endothelial | *PNRC2* | -1.245594227 | 7.14E-07 | 4.36E-06 | 0.004488278 |
| Obesity | endothelial | *ZKSCAN3* | -1.412845668 | 1.20E-06 | 6.56E-06 | 0.00753234 |

| | | | | | | |
|---|---|---|---|---|---|---|
| Obesity | endothelial | *BLOC1S2* | -0.568245766 | 8.85E-06 | 3.34E-05 | 0.055647715 |
| Obesity | endothelial | *HLA-DRB1* | 0.217797017 | 0.02175748 | 0.030384113 | 1 |
| Obesity | endothelial | *AKR1C3* | -0.814429264 | 6.14E-06 | 2.47E-05 | 0.038591162 |
| Obesity | endothelial | *ZNF79* | -0.453248373 | 0.022355859 | 0.031111998 | 1 |
| Obesity | endothelial | *S100A4* | -0.33781366 | 0.00176717 | 0.00315541 | 1 |
| Obesity | endothelial | *COLCA1* | -0.910799218 | 1.52E-06 | 7.94E-06 | 0.009540097 |
| Obesity | endothelial | *ACADSB* | -0.697557425 | 0.000387823 | 0.000825125 | 1 |
| Obesity | endothelial | *SEMA4A* | -0.529830723 | 0.026408946 | 0.036172776 | 1 |
| Obesity | endothelial | *HRCT1* | 0.598281701 | 0.003598491 | 0.005937983 | 1 |
| Obesity | endothelial | *MPHOSPH8* | -0.556382421 | 0.001651201 | 0.002966877 | 1 |
| Obesity | endothelial | *SIRPB2* | -1.095603063 | 1.70E-08 | 3.41E-07 | 0.00010708 |
| Obesity | endothelial | *SRGAP3* | -0.39257536 | 0.000579943 | 0.001176886 | 1 |
| Obesity | endothelial | *SUPT5H* | -0.832689575 | 0.000302701 | 0.000662634 | 1 |
| Obesity | endothelial | *XPNPEP3* | -0.402406823 | 0.000926679 | 0.001782752 | 1 |
| Obesity | endothelial | *PPIA* | -0.599905213 | 0.026309487 | 0.036044399 | 1 |
| Obesity | endothelial | *GTF2IRD2* | -0.604243318 | 0.000589759 | 0.001193375 | 1 |
| Obesity | endothelial | *GRM7* | -0.373742982 | 1.37E-06 | 7.30E-06 | 0.00858833 |
| Obesity | endothelial | *SUPT3H* | -0.467339832 | 0.011966208 | 0.017565153 | 1 |
| Obesity | endothelial | *POM121* | -0.625023305 | 0.002316881 | 0.004020489 | 1 |
| Obesity | endothelial | *GIMAP5* | 0.331663285 | 5.90E-05 | 0.000161912 | 0.37078737 |
| Obesity | endothelial | *STK31* | 0.738712073 | 0.001793486 | 0.003197857 | 1 |
| Obesity | endothelial | *ZKSCAN7* | -0.992263129 | 2.76E-08 | 4.26E-07 | 0.000173695 |
| Obesity | endothelial | *ZNF565* | -0.47097481 | 0.010988821 | 0.016286356 | 1 |
| Obesity | endothelial | *NTNG2* | -1.737495378 | 9.60E-08 | 9.15E-07 | 0.000603272 |
| Obesity | endothelial | *WDR5* | -1.177872677 | 4.68E-07 | 3.11E-06 | 0.002943883 |
| Obesity | endothelial | *ASB13* | -1.35383857 | 1.32E-06 | 7.09E-06 | 0.008278052 |
| Obesity | endothelial | *SLC35F1* | -1.260457162 | 3.45E-06 | 1.56E-05 | 0.021665748 |
| Obesity | endothelial | *ZNF34* | -1.314366233 | 0.000608069 | 0.001224905 | 1 |
| Obesity | endothelial | *INCA1* | -1.133162472 | 0.009231094 | 0.01394088 | 1 |
| Obesity | endothelial | *ZNF774* | -2.015318304 | 0.000134933 | 0.000327158 | 0.848320668 |
| Obesity | endothelial | *EVL* | -0.497840201 | 0.003915681 | 0.006415921 | 1 |
| Obesity | endothelial | *ZNF124* | -0.840430242 | 0.000350122 | 0.000753823 | 1 |
| Obesity | endothelial | *ZNF569* | -0.505952022 | 0.031634051 | 0.042651357 | 1 |
| Obesity | endothelial | *PIK3R4* | -0.736659876 | 5.66E-05 | 0.000155984 | 0.355643464 |
| Obesity | endothelial | *ZNF605* | -1.171215672 | 4.64E-07 | 3.09E-06 | 0.002916912 |
| Obesity | endothelial | *RFX8* | -0.927363044 | 1.60E-05 | 5.46E-05 | 0.100857183 |
| Obesity | endothelial | *MYL6B* | -1.34796117 | 8.96E-05 | 0.000229663 | 0.563593822 |
| Obesity | endothelial | *C20orf96* | -1.201089162 | 0.001941823 | 0.003429281 | 1 |
| Obesity | endothelial | *SULT1A1* | -0.81020668 | 8.28E-15 | 5.20E-12 | 5.20E-11 |
| Obesity | endothelial | *ANAPC7* | -1.084495456 | 5.09E-06 | 2.13E-05 | 0.032014744 |
| Obesity | endothelial | *C1orf186* | -1.474948901 | 4.79E-07 | 3.17E-06 | 0.003010863 |
| Obesity | endothelial | *MYO18A* | 0.331833005 | 0.017391135 | 0.024781974 | 1 |
| Obesity | endothelial | *SPTSSB* | 1.202251884 | 4.45E-06 | 1.91E-05 | 0.027971731 |
| Obesity | endothelial | *SULF2* | 0.273237379 | 3.60E-05 | 0.000106603 | 0.226121254 |
| Obesity | endothelial | *XRCC2* | -1.65402503 | 9.90E-09 | 3.16E-07 | 6.22E-05 |
| Obesity | endothelial | *HDAC2* | -0.754058083 | 0.001801434 | 0.003208389 | 1 |
| Obesity | endothelial | *ADH1B* | -0.544289765 | 3.89E-08 | 5.04E-07 | 0.000244828 |
| Obesity | endothelial | *FAM180B* | -0.89127513 | 3.81E-10 | 3.47E-08 | 2.40E-06 |
| Obesity | endothelial | *ERI2* | -0.843047184 | 8.83E-06 | 3.34E-05 | 0.055510144 |
| Obesity | endothelial | *TOMM7* | -0.551582314 | 0.030914327 | 0.04177953 | 1 |

| | | | | | | |
|---|---|---|---|---|---|---|
| Obesity | endothelial | *TRPV1* | -0.645474738 | 2.77E-05 | 8.59E-05 | 0.174046532 |
| Obesity | endothelial | *ZNF33B* | 0.261983721 | 0.033016077 | 0.044353008 | 1 |
| Obesity | endothelial | *AMZ2* | -0.70940206 | 0.006120974 | 0.009589475 | 1 |
| Obesity | endothelial | *ZNF431* | -0.708702226 | 0.000145428 | 0.00034964 | 0.914307295 |
| Obesity | endothelial | *NF1* | -0.409921341 | 0.010866062 | 0.016134844 | 1 |
| Obesity | endothelial | *DAPK1* | -0.500014029 | 0.001147548 | 0.002147852 | 1 |
| Obesity | endothelial | *STRN3* | -0.59391456 | 0.000436791 | 0.000915063 | 1 |
| Obesity | endothelial | *ZNF239* | -0.353492896 | 0.0213291 | 0.029825634 | 1 |
| Obesity | endothelial | *PPTC7* | -0.98013734 | 2.24E-08 | 3.81E-07 | 0.000140657 |
| Obesity | endothelial | *NHLRC2* | -0.744120769 | 0.000134829 | 0.000327033 | 0.847668922 |
| Obesity | endothelial | *ZFP28* | 0.590707897 | 0.013494663 | 0.019598278 | 1 |
| Obesity | endothelial | *KIAA1211L* | 0.259059986 | 2.71E-07 | 2.03E-06 | 0.001701133 |
| Obesity | endothelial | *LAMB3* | -0.881027137 | 3.09E-08 | 4.48E-07 | 0.000194497 |
| Obesity | endothelial | *KPNA5* | -0.722442248 | 0.000445334 | 0.00093079 | 1 |
| Obesity | endothelial | *SRGAP1* | -0.6407371 | 9.21E-05 | 0.000235117 | 0.579328082 |
| Obesity | endothelial | *FAM3C* | -0.510669495 | 0.029417191 | 0.039867618 | 1 |
| Obesity | endothelial | *SLC39A10* | -0.536696698 | 0.010792716 | 0.016056272 | 1 |
| Obesity | endothelial | *AP2A1* | -0.584891148 | 0.007291035 | 0.01124601 | 1 |
| Obesity | endothelial | *ANXA4* | -0.615744621 | 0.000544269 | 0.001109899 | 1 |
| Obesity | endothelial | *WDR5B* | -0.974665994 | 0.0002338 | 0.000528171 | 1 |
| Obesity | endothelial | *ZNF138* | -0.63398578 | 0.001277686 | 0.002368163 | 1 |
| Obesity | endothelial | *ZNF429* | -0.901651478 | 1.53E-06 | 8.01E-06 | 0.009648114 |
| Obesity | endothelial | *ZNF100* | -0.635370977 | 7.15E-05 | 0.000189901 | 0.449496061 |
| Obesity | endothelial | *GMFB* | -0.934212395 | 2.14E-08 | 3.69E-07 | 0.000134282 |
| Obesity | endothelial | *ZMYM1* | -0.894263338 | 6.12E-05 | 0.000166624 | 0.384624354 |
| Obesity | endothelial | *IGF2R* | -0.739802656 | 6.04E-05 | 0.000164769 | 0.379461967 |
| Obesity | endothelial | *GAL3ST4* | -1.009718323 | 4.37E-07 | 2.95E-06 | 0.002749883 |
| Obesity | endothelial | *SLC25A29* | 0.489921756 | 5.27E-05 | 0.000146267 | 0.331572593 |
| Obesity | endothelial | *ZNF682* | -1.052707467 | 3.75E-05 | 0.00011049 | 0.235895738 |
| Obesity | endothelial | *ZNF772* | -0.464500515 | 0.000224991 | 0.000511027 | 1 |
| Obesity | endothelial | *ZNF257* | -0.708771575 | 0.013041373 | 0.018988215 | 1 |
| Obesity | endothelial | *ADAM32* | -0.924248165 | 7.28E-05 | 0.000192262 | 0.457584631 |
| Obesity | endothelial | *LRRC8B* | -0.844151035 | 8.55E-06 | 3.25E-05 | 0.05373477 |
| Obesity | endothelial | *SND1* | -0.560099496 | 0.00061228 | 0.001231366 | 1 |
| Obesity | endothelial | *SULT1A2* | 0.354138936 | 2.32E-09 | 1.34E-07 | 1.46E-05 |
| Obesity | endothelial | *PIWIL2* | -0.774965452 | 0.000541534 | 0.001106115 | 1 |
| Obesity | endothelial | *ENTPD4* | -0.65864918 | 0.002633976 | 0.004492623 | 1 |
| Obesity | endothelial | *C1D* | -0.581480323 | 0.010796472 | 0.016058061 | 1 |
| Obesity | endothelial | *TBC1D9B* | -0.474495817 | 0.009883189 | 0.01481889 | 1 |
| Obesity | endothelial | *FAM110D* | 0.221392438 | 9.70E-07 | 5.56E-06 | 0.006099094 |
| Obesity | endothelial | *KANK2* | -0.823934475 | 4.97E-07 | 3.25E-06 | 0.003122481 |
| Obesity | endothelial | *C6orf141* | 0.172172929 | 7.86E-06 | 3.03E-05 | 0.04940231 |
| Obesity | endothelial | *CCL4L2* | -0.933465242 | 1.45E-11 | 2.93E-09 | 9.09E-08 |
| Obesity | endothelial | *GTF2E2* | -0.689545368 | 0.000221102 | 0.000503283 | 1 |
| Obesity | endothelial | *ZNF165* | -1.02282272 | 0.000185229 | 0.000431789 | 1 |
| Obesity | endothelial | *ZNF720* | -0.612286178 | 0.003064852 | 0.005136956 | 1 |
| Obesity | endothelial | *DDI2* | -0.807532098 | 2.77E-07 | 2.07E-06 | 0.001744579 |
| Obesity | endothelial | *MRPL21* | -0.458957642 | 1.86E-05 | 6.17E-05 | 0.116939865 |
| Obesity | endothelial | *ZNF786* | -0.744790406 | 0.010319145 | 0.015421075 | 1 |
| Obesity | endothelial | *ZNF517* | -1.055679002 | 0.025168436 | 0.034586657 | 1 |

| | | | | | | |
|---|---|---|---|---|---|---|
| Obesity | endothelial | *SLC22A5* | -0.288593267 | 0.010436193 | 0.015582806 | 1 |
| Obesity | endothelial | *C5AR1* | -0.87960505 | 6.09E-09 | 2.38E-07 | 3.83E-05 |
| Obesity | endothelial | *DCHS2* | -0.347951812 | 1.81E-05 | 6.04E-05 | 0.113820615 |
| Obesity | endothelial | *VEPH1* | -0.992803176 | 1.67E-06 | 8.57E-06 | 0.010502204 |
| Obesity | endothelial | *SHPK* | -0.410342887 | 0.000811505 | 0.001588398 | 1 |
| Obesity | endothelial | *MAP3K5* | -0.647080287 | 0.000108616 | 0.00027098 | 0.682869587 |
| Obesity | endothelial | *STMN3* | -1.091827802 | 0.000415475 | 0.000877718 | 1 |
| Obesity | endothelial | *GYPE* | -0.575327455 | 0.002182265 | 0.003816384 | 1 |
| Obesity | endothelial | *PCDHB11* | -2.350516149 | 1.97E-11 | 3.54E-09 | 1.24E-07 |
| Obesity | endothelial | *SLC2A10* | -0.795564062 | 1.80E-06 | 9.09E-06 | 0.011329758 |
| Obesity | endothelial | *ZNF665* | -0.548907703 | 0.002164507 | 0.003787435 | 1 |
| Obesity | endothelial | *RPF2* | -0.792161532 | 0.000794279 | 0.001558561 | 1 |
| Obesity | endothelial | *MYO5A* | -0.53461043 | 0.000222444 | 0.000505972 | 1 |
| Obesity | endothelial | *ATG7* | -0.616578535 | 2.46E-05 | 7.73E-05 | 0.154678399 |
| Obesity | endothelial | *TTC30A* | -0.732188587 | 0.001644436 | 0.002955565 | 1 |
| Obesity | endothelial | *RAB40C* | -1.12879556 | 3.31E-07 | 2.37E-06 | 0.002083732 |
| Obesity | endothelial | *PIGN* | -0.485856605 | 0.000523768 | 0.001074014 | 1 |
| Obesity | endothelial | *ZNF624* | -1.390005873 | 1.47E-06 | 7.71E-06 | 0.009220956 |
| Obesity | endothelial | *HHLA3* | -1.183370873 | 9.39E-07 | 5.42E-06 | 0.00590398 |
| Obesity | endothelial | *HOXA4* | -1.37046634 | 7.72E-08 | 7.84E-07 | 0.00048508 |
| Obesity | endothelial | *BCO2* | -1.166325876 | 3.98E-08 | 5.07E-07 | 0.000250155 |
| Obesity | endothelial | *ENTPD6* | -1.15715045 | 1.27E-07 | 1.14E-06 | 0.000796251 |
| Obesity | endothelial | *CCDC154* | -1.424459595 | 2.51E-09 | 1.39E-07 | 1.58E-05 |
| Obesity | endothelial | *C5orf42* | -0.538602595 | 0.000349577 | 0.000752927 | 1 |
| Obesity | endothelial | *ZNF841* | -0.871895418 | 0.005887249 | 0.009253283 | 1 |
| Obesity | endothelial | *MFAP5* | -0.677487848 | 1.07E-07 | 1.00E-06 | 0.000675839 |
| Obesity | endothelial | *CDC42SE1* | -0.644785842 | 0.008759919 | 0.01328035 | 1 |
| Obesity | endothelial | *MPEG1* | -0.357553561 | 0.000381346 | 0.000812995 | 1 |
| Obesity | endothelial | *PDCD1LG2* | -0.497659954 | 0.000900355 | 0.001737425 | 1 |
| Obesity | endothelial | *NMB* | -1.743310163 | 1.41E-06 | 7.45E-06 | 0.008833722 |
| Obesity | endothelial | *KLHL14* | 0.439925226 | 8.83E-05 | 0.000226772 | 0.555137358 |
| Obesity | endothelial | *FAM114A1* | -0.780993361 | 1.32E-05 | 4.62E-05 | 0.083139821 |
| Obesity | endothelial | *RPS26* | -0.381089859 | 2.46E-05 | 7.73E-05 | 0.154475219 |
| Obesity | endothelial | *C14orf178* | -0.729812783 | 4.11E-08 | 5.17E-07 | 0.000258348 |
| Obesity | endothelial | *PSAP* | -0.663014101 | 9.44E-05 | 0.000240107 | 0.593543617 |
| Obesity | endothelial | *S100A10* | -0.541311144 | 0.003275187 | 0.005460382 | 1 |
| Obesity | endothelial | *MAP1LC3C* | -0.659869777 | 1.11E-06 | 6.20E-06 | 0.007003866 |
| Obesity | endothelial | *TAF13* | -0.714813899 | 0.000915274 | 0.001764049 | 1 |
| Obesity | endothelial | *ZNF780A* | -0.594880513 | 1.54E-08 | 3.34E-07 | 9.67E-05 |
| Obesity | endothelial | *AC122129.1* | -2.873792906 | 1.30E-08 | 3.34E-07 | 8.17E-05 |
| Obesity | endothelial | *CCDC180* | -0.785977853 | 0.000443326 | 0.000927209 | 1 |
| Obesity | endothelial | *OCLN* | 0.225732798 | 0.004976246 | 0.007958702 | 1 |
| Obesity | endothelial | *SGTB* | -1.018325446 | 0.000433043 | 0.000907816 | 1 |
| Obesity | endothelial | *ADH5* | -0.826705674 | 2.05E-07 | 1.65E-06 | 0.001286974 |
| Obesity | endothelial | *RPL12* | -0.525954857 | 0.002322887 | 0.0040298 | 1 |
| Obesity | endothelial | *LEKR1* | -0.997107907 | 6.48E-10 | 5.33E-08 | 4.08E-06 |
| Obesity | endothelial | *NOL8* | -1.062831393 | 1.13E-06 | 6.27E-06 | 0.007112685 |
| Obesity | endothelial | *MRPL42* | -0.772567678 | 3.05E-05 | 9.30E-05 | 0.191850209 |
| Obesity | endothelial | *ZNF335* | -0.654283607 | 0.002836202 | 0.004802371 | 1 |
| Obesity | endothelial | *AGAP9* | -1.734529239 | 4.92E-07 | 3.23E-06 | 0.003092805 |

| | | | | | | |
|---|---|---|---|---|---|---|
| Obesity | endothelial | *ZNF273* | -0.943544877 | 7.76E-05 | 0.000202538 | 0.487914793 |
| Obesity | endothelial | *ZNF84* | -0.761075291 | 0.000425015 | 0.000893969 | 1 |
| Obesity | endothelial | *MAK16* | -0.699806041 | 0.032296603 | 0.043460775 | 1 |
| Obesity | endothelial | *ZNF667* | -0.657213739 | 0.000969719 | 0.001850827 | 1 |
| Obesity | endothelial | *PRIM1* | -0.731780896 | 0.010480063 | 0.015635538 | 1 |
| Obesity | endothelial | *SULT1C4* | 0.384838704 | 7.35E-06 | 2.86E-05 | 0.046222615 |
| Obesity | endothelial | *ZNF649* | -1.153748124 | 2.28E-05 | 7.27E-05 | 0.143219797 |
| Obesity | endothelial | *ADH4* | -0.469407441 | 1.32E-07 | 1.17E-06 | 0.000831798 |
| Obesity | endothelial | *ZNF248* | -0.762584009 | 1.39E-06 | 7.37E-06 | 0.008722584 |
| Obesity | endothelial | *HIBCH* | -0.576237459 | 0.000302122 | 0.000661596 | 1 |
| Obesity | endothelial | *SOWAHC* | -1.84748033 | 1.53E-08 | 3.34E-07 | 9.63E-05 |
| Obesity | endothelial | *AC135178.1* | 0.650462582 | 1.24E-06 | 6.75E-06 | 0.007814864 |
| Obesity | endothelial | *SVIP* | 0.385707656 | 0.000124091 | 0.000304274 | 0.780157453 |
| Obesity | endothelial | *DDRGK1* | -1.0577828 | 0.000263136 | 0.000586435 | 1 |
| Obesity | endothelial | *ZNF334* | -0.677743764 | 3.43E-06 | 1.55E-05 | 0.021575046 |
| Obesity | endothelial | *SULT1C2* | 0.282498374 | 0.006275797 | 0.00980759 | 1 |
| Obesity | endothelial | *RPS6KL1* | -0.532041319 | 0.001004261 | 0.001907489 | 1 |
| Obesity | endothelial | *DDX42* | -0.568597186 | 0.003606673 | 0.005949923 | 1 |
| Obesity | endothelial | *SLC29A3* | -1.09543112 | 3.33E-06 | 1.52E-05 | 0.020955004 |
| Obesity | endothelial | *ANTXRL* | -20.83907837 | 2.22E-08 | 3.81E-07 | 0.000139298 |
| Obesity | endothelial | *STYX* | -0.857010031 | 0.00111018 | 0.002084738 | 1 |
| Obesity | endothelial | *MYL4* | 0.798447885 | 9.34E-07 | 5.40E-06 | 0.005873522 |
| Obesity | endothelial | *GFPT1* | -0.661345661 | 1.17E-05 | 4.20E-05 | 0.073531901 |
| Obesity | endothelial | *UVRAG* | -0.5118135 | 1.94E-05 | 6.40E-05 | 0.122280717 |
| Obesity | endothelial | *MGEA5* | -0.674544344 | 5.94E-06 | 2.40E-05 | 0.037326569 |
| Obesity | endothelial | *MT1F* | -2.451631999 | 1.47E-06 | 7.72E-06 | 0.009234858 |
| Obesity | endothelial | *ZNF568* | -0.594745563 | 0.011212238 | 0.016571826 | 1 |
| Obesity | endothelial | *ZNF587* | -0.887362573 | 0.000218258 | 0.00049789 | 1 |
| Obesity | endothelial | *SH3BGRL2* | 0.284521082 | 4.11E-06 | 1.80E-05 | 0.02582691 |
| Obesity | endothelial | *ZNF808* | -0.663006968 | 0.005417261 | 0.008594075 | 1 |
| Obesity | endothelial | *TMA16* | -0.728704531 | 0.002510718 | 0.004311631 | 1 |
| Obesity | endothelial | *CNGA1* | -0.570032809 | 2.29E-05 | 7.30E-05 | 0.143841684 |
| Obesity | endothelial | *MAFK* | -1.301754778 | 1.40E-06 | 7.40E-06 | 0.008774582 |
| Obesity | endothelial | *HIST1H4E* | 0.917596301 | 0.010521098 | 0.015691721 | 1 |
| Obesity | endothelial | *C20orf203* | 0.485832298 | 0.000321626 | 0.000699676 | 1 |
| Obesity | endothelial | *ZNF627* | -1.097929662 | 1.09E-07 | 1.01E-06 | 0.000684817 |
| Obesity | endothelial | *ARC* | -5.043587358 | 3.13E-09 | 1.67E-07 | 1.97E-05 |
| Obesity | endothelial | *TLK1* | -0.573692584 | 0.001400509 | 0.002570053 | 1 |
| Obesity | endothelial | *LRBA* | -0.438306389 | 0.006642145 | 0.010337152 | 1 |
| Obesity | endothelial | *MMP17* | -0.754965484 | 1.61E-08 | 3.37E-07 | 0.000101125 |
| Obesity | endothelial | *MDM4* | -0.622082504 | 0.000542722 | 0.001107638 | 1 |
| Obesity | endothelial | *ZNF534* | -0.90400858 | 0.026847839 | 0.036741917 | 1 |
| Obesity | endothelial | *NCOA6* | -0.643146588 | 0.000347503 | 0.00074923 | 1 |
| Obesity | endothelial | *FAM19A2* | -0.79414832 | 5.87E-05 | 0.000161403 | 0.369129709 |
| Obesity | endothelial | *TTC37* | -0.595452334 | 0.000851618 | 0.001656086 | 1 |
| Obesity | endothelial | *TUSC1* | -2.213249773 | 6.39E-07 | 3.98E-06 | 0.004019201 |
| Obesity | endothelial | *PAPSS2* | -0.674405953 | 2.02E-05 | 6.61E-05 | 0.127026224 |
| Obesity | endothelial | *FAN1* | -0.822520613 | 0.000903095 | 0.001742179 | 1 |
| Obesity | endothelial | *ZNF652* | -0.713692023 | 0.000240853 | 0.000542156 | 1 |
| Obesity | endothelial | *APCDD1L* | -0.798284836 | 0.034089519 | 0.045677922 | 1 |

| | | | | | | |
|---|---|---|---|---|---|---|
| Obesity | endothelial | *RCSD1* | -0.45843433 | 0.002901907 | 0.004896378 | 1 |
| Obesity | endothelial | *RASSF9* | 0.340862425 | 4.27E-05 | 0.000122643 | 0.268221163 |
| Obesity | endothelial | *FAM169A* | 0.559354359 | 0.008100229 | 0.012351719 | 1 |
| Obesity | endothelial | *SCAMP5* | -0.75013116 | 2.88E-06 | 1.34E-05 | 0.018130638 |
| Obesity | endothelial | *ALPK2* | -0.632540584 | 5.42E-09 | 2.24E-07 | 3.41E-05 |
| Obesity | endothelial | *PNP* | 0.169773213 | 0.027954746 | 0.038107434 | 1 |
| Obesity | endothelial | *SFT2D1* | -0.952832301 | 1.95E-06 | 9.73E-06 | 0.012264699 |
| Obesity | endothelial | *ARHGAP11A* | -1.300904674 | 1.23E-06 | 6.69E-06 | 0.007730055 |
| Obesity | endothelial | *HMGN2* | -0.955256109 | 1.14E-06 | 6.28E-06 | 0.007141271 |
| Obesity | endothelial | *DENND4B* | -0.930367548 | 4.45E-05 | 0.000126977 | 0.279984462 |
| Obesity | endothelial | *CD3E* | -0.496723992 | 0.011358392 | 0.016770833 | 1 |
| Obesity | endothelial | *TSEN15* | -0.964285151 | 7.86E-05 | 0.000204691 | 0.494328073 |
| Obesity | endothelial | *TYW1* | -0.637215161 | 2.77E-06 | 1.30E-05 | 0.01744049 |
| Obesity | endothelial | *ITPRIPL1* | -1.083084421 | 0.001571098 | 0.002839992 | 1 |
| Obesity | endothelial | *PRMT6* | -4.143457701 | 4.82E-12 | 1.17E-09 | 3.03E-08 |
| Obesity | endothelial | *C1orf174* | -0.953176601 | 0.000110424 | 0.000274836 | 0.694234733 |
| Obesity | endothelial | *RASGEF1A* | -2.111476871 | 2.12E-08 | 3.68E-07 | 0.000133056 |
| Obesity | endothelial | *KIAA0753* | -0.536753257 | 0.019488349 | 0.027508588 | 1 |
| Obesity | endothelial | *DCLRE1A* | -1.267410689 | 6.61E-08 | 7.03E-07 | 0.000415261 |
| Obesity | endothelial | *APRT* | -1.348554678 | 2.33E-05 | 7.42E-05 | 0.146753444 |
| Obesity | endothelial | *TBKBP1* | 0.512504091 | 0.000181801 | 0.000424429 | 1 |
| Obesity | endothelial | *ZFP2* | 0.408791439 | 0.032249373 | 0.04340651 | 1 |
| Obesity | endothelial | *L3MBTL3* | -0.667075864 | 0.007658041 | 0.011757291 | 1 |
| Obesity | endothelial | *NAGA* | -1.028940167 | 1.76E-05 | 5.91E-05 | 0.110870858 |
| Obesity | endothelial | *SMG5* | -0.88154397 | 0.000220274 | 0.000501701 | 1 |
| Obesity | endothelial | *TGM2* | 0.227270791 | 5.15E-07 | 3.35E-06 | 0.003240332 |
| Obesity | endothelial | *PJA2* | -0.655018328 | 0.000378257 | 0.000807504 | 1 |
| Obesity | endothelial | *AL590822.1* | -0.473428756 | 3.97E-06 | 1.75E-05 | 0.024980793 |
| Obesity | endothelial | *AP001007.1* | -4.172181168 | 1.83E-08 | 3.45E-07 | 0.000115055 |
| Obesity | endothelial | *EFCAB2* | -0.557884456 | 0.002621662 | 0.004476477 | 1 |
| Obesity | endothelial | *DDO* | -1.378905728 | 3.23E-06 | 1.48E-05 | 0.02029266 |
| Obesity | endothelial | *HIST2H2BF* | -2.149871565 | 2.27E-08 | 3.82E-07 | 0.00014287 |
| Obesity | endothelial | *RBM20* | -1.05915753 | 0.00431888 | 0.007003365 | 1 |
| Obesity | endothelial | *C6orf163* | -1.933577365 | 8.06E-07 | 4.79E-06 | 0.005066321 |
| Obesity | endothelial | *PCMTD2* | -0.706034368 | 0.001161402 | 0.002171196 | 1 |
| Obesity | endothelial | *OOEP* | -0.683637107 | 3.88E-05 | 0.000113281 | 0.24400748 |
| Obesity | endothelial | *FOXO6* | -1.059716501 | 5.69E-06 | 2.33E-05 | 0.035793005 |
| Obesity | endothelial | *SYS1* | -0.964061249 | 0.018274218 | 0.025928687 | 1 |
| Obesity | endothelial | *INPP5B* | -0.265999736 | 0.000243174 | 0.000546988 | 1 |
| Obesity | endothelial | *NEU4* | -1.725672766 | 0.012786694 | 0.018660618 | 1 |
| Obesity | endothelial | *TRAF3IP1* | -0.962737317 | 5.31E-06 | 2.20E-05 | 0.033399689 |
| Obesity | endothelial | *GIGYF2* | -0.526789714 | 0.00291867 | 0.004919484 | 1 |
| Obesity | endothelial | *RUFY2* | -0.682354551 | 0.000342792 | 0.000740596 | 1 |
| Obesity | endothelial | *C10orf128* | 0.382487427 | 1.07E-05 | 3.89E-05 | 0.067006883 |
| Obesity | endothelial | *SYT15* | 0.403867281 | 2.59E-07 | 1.96E-06 | 0.001625895 |
| Obesity | endothelial | *ZDBF2* | -0.798710161 | 0.003646919 | 0.006010008 | 1 |
| Obesity | endothelial | *RING1* | -0.846377747 | 2.32E-05 | 7.39E-05 | 0.145824403 |
| Obesity | endothelial | *OXLD1* | -0.829733258 | 0.000394379 | 0.000837374 | 1 |
| Obesity | endothelial | *AGER* | 0.601479155 | 0.003337884 | 0.005551661 | 1 |
| Obesity | endothelial | *PRRT1* | 0.410873419 | 0.005551119 | 0.008793118 | 1 |

| | | | | | | |
|---|---|---|---|---|---|---|
| Obesity | endothelial | *MRPL38* | -1.171049347 | 8.44E-07 | 4.95E-06 | 0.005308709 |
| Obesity | endothelial | *SKIV2L* | -0.445659461 | 1.18E-09 | 8.55E-08 | 7.44E-06 |
| Obesity | endothelial | *LAYN* | 0.584061011 | 2.12E-05 | 6.86E-05 | 0.133035148 |
| Obesity | endothelial | *C6orf48* | -0.788289538 | 0.001182825 | 0.002209506 | 1 |
| Obesity | endothelial | *MSH5* | -0.554266998 | 0.000235091 | 0.000530517 | 1 |
| Obesity | endothelial | *GPANK1* | -1.021575538 | 5.70E-08 | 6.32E-07 | 0.000358427 |
| Obesity | endothelial | *NFKBIL1* | -0.794105171 | 9.21E-05 | 0.000235117 | 0.579272354 |
| Obesity | endothelial | *ZNF814* | -0.548838423 | 0.012906935 | 0.018805539 | 1 |
| Obesity | endothelial | *MICA* | -0.264414681 | 0.0052092 | 0.008289102 | 1 |
| Obesity | endothelial | *HLA-C* | 0.27823721 | 0.001082411 | 0.00203746 | 1 |
| Obesity | endothelial | *POU5F1* | 0.331246844 | 1.29E-07 | 1.15E-06 | 0.000811129 |
| Obesity | endothelial | *CCHCR1* | 0.476705979 | 1.39E-05 | 4.83E-05 | 0.08745739 |
| Obesity | endothelial | *LILRB3* | -0.552122533 | 0.001383623 | 0.002539807 | 1 |
| Obesity | endothelial | *LINC01123* | 3.409817724 | 3.54E-06 | 1.59E-05 | 0.022269232 |
| Obesity | endothelial | *HLA-E* | 0.407776153 | 3.98E-07 | 2.73E-06 | 0.002501372 |
| Obesity | endothelial | *ZNF468* | -1.656309193 | 0.000169921 | 0.000401464 | 1 |
| Obesity | endothelial | *TBC1D8* | -0.462011345 | 0.017566045 | 0.025025543 | 1 |
| Obesity | endothelial | *HLA-F* | 0.363145874 | 0.001497166 | 0.002724366 | 1 |
| Obesity | endothelial | *ZFP57* | 0.421827564 | 4.52E-06 | 1.93E-05 | 0.02840524 |
| Obesity | endothelial | *FAM153C* | -1.67359996 | 0.007718278 | 0.011838208 | 1 |
| Obesity | endothelial | *GABBR1* | -0.996196527 | 2.34E-05 | 7.45E-05 | 0.147421918 |
| Obesity | endothelial | *TRIM27* | -0.690912751 | 0.000367225 | 0.000786358 | 1 |
| Obesity | endothelial | *MALRD1* | -0.352422875 | 0.000139846 | 0.000337509 | 0.879210353 |
| Obesity | endothelial | *ATXN2* | -0.405618784 | 0.01273974 | 0.018609374 | 1 |
| Obesity | endothelial | *FAM216A* | -1.273771776 | 1.65E-06 | 8.48E-06 | 0.010352261 |
| Obesity | endothelial | *ZNF783* | -0.981013045 | 1.80E-05 | 6.01E-05 | 0.112988045 |
| Obesity | endothelial | *ZNF425* | -0.682527326 | 0.000119347 | 0.000293903 | 0.750334514 |
| Obesity | endothelial | *LRRC10B* | -3.35909663 | 1.99E-05 | 6.53E-05 | 0.125150601 |
| Obesity | endothelial | *C12orf73* | -0.933986887 | 4.10E-05 | 0.000118618 | 0.25787531 |
| Obesity | endothelial | *PCDHGA1* | -0.875750901 | 0.00021257 | 0.000488105 | 1 |
| Obesity | endothelial | *PCDHA5* | -3.573058041 | 0.000744198 | 0.001468081 | 1 |
| Obesity | endothelial | *SPIRE2* | -0.357964043 | 0.00800885 | 0.012239096 | 1 |
| Obesity | endothelial | *CLLU1OS* | -0.798323984 | 0.003930385 | 0.006436658 | 1 |
| Obesity | endothelial | *TMEM231* | 0.504470081 | 8.18E-05 | 0.000212316 | 0.514228786 |
| Obesity | endothelial | *FAM71F2* | -0.986960634 | 1.53E-08 | 3.34E-07 | 9.62E-05 |
| Obesity | endothelial | *AC016586.1* | -3.849242123 | 4.28E-05 | 0.000123061 | 0.269380516 |
| Obesity | endothelial | *LINC00654* | -1.08783891 | 3.34E-07 | 2.38E-06 | 0.002098353 |
| Obesity | endothelial | *TMEM170B* | -0.29100114 | 0.000556318 | 0.001131535 | 1 |
| Obesity | endothelial | *NT5M* | -0.567808705 | 0.001340769 | 0.002466906 | 1 |
| Obesity | endothelial | *PRR13* | -1.434039707 | 5.43E-06 | 2.24E-05 | 0.034148142 |
| Obesity | endothelial | *MT1A* | -1.361935489 | 4.70E-08 | 5.59E-07 | 0.000295523 |
| Obesity | endothelial | *MT1M* | -2.050302344 | 4.41E-06 | 1.90E-05 | 0.027716659 |
| Obesity | endothelial | *CFI* | 0.203678113 | 5.93E-06 | 2.40E-05 | 0.037292518 |
| Obesity | endothelial | *EXOC3L4* | -1.033006086 | 7.16E-09 | 2.61E-07 | 4.50E-05 |
| Obesity | endothelial | *ATP6AP1L* | -0.697963903 | 4.79E-05 | 0.000134518 | 0.301320861 |
| Obesity | endothelial | *NAP1L4* | -0.751980533 | 2.19E-05 | 7.05E-05 | 0.13780038 |
| Obesity | endothelial | *SMN2* | -0.589408972 | 3.70E-06 | 1.65E-05 | 0.023251417 |
| Obesity | endothelial | *SERF1B* | -1.417015731 | 0.000287096 | 0.000631678 | 1 |
| Obesity | endothelial | *EIF3CL* | 2.590175778 | 3.13E-05 | 9.50E-05 | 0.196943567 |
| Obesity | endothelial | *LCMT1* | -0.80497978 | 0.000562405 | 0.001142437 | 1 |

| | | | | | | |
|---|---|---|---|---|---|---|
| Obesity | endothelial | *CDPF1* | -0.578358431 | 0.00352237 | 0.005827669 | 1 |
| Obesity | endothelial | *C17orf107* | 0.442813099 | 0.012812362 | 0.018689402 | 1 |
| Obesity | endothelial | *LINC00487* | 0.740147842 | 5.27E-05 | 0.000146267 | 0.331492466 |
| Obesity | endothelial | *RFPL3S* | -0.421463245 | 0.0092682 | 0.01398684 | 1 |
| Obesity | endothelial | *C1QTNF9B* | 5.768276238 | 2.51E-05 | 7.87E-05 | 0.15766453 |
| Obesity | endothelial | *RNPS1* | -0.766527794 | 0.00034651 | 0.000747344 | 1 |
| Obesity | endothelial | *NYNRIN* | 0.518085528 | 3.76E-05 | 0.000110776 | 0.236661541 |
| Obesity | endothelial | *GOLGA8O* | -2.037094257 | 2.12E-07 | 1.69E-06 | 0.001329958 |
| Obesity | endothelial | *HCG27* | -0.23751551 | 0.017372514 | 0.024766665 | 1 |
| Obesity | endothelial | *LNP1* | -0.968419849 | 8.99E-09 | 2.99E-07 | 5.65E-05 |
| Obesity | endothelial | *VGLL3* | -0.452481254 | 1.53E-09 | 1.03E-07 | 9.60E-06 |
| Obesity | endothelial | *COLQ* | -1.068425794 | 2.60E-09 | 1.42E-07 | 1.63E-05 |
| Obesity | endothelial | *METTL6* | -0.693640068 | 0.000300811 | 0.000659185 | 1 |
| Obesity | endothelial | *SACM1L* | -0.60166851 | 0.004586749 | 0.007397869 | 1 |
| Obesity | endothelial | *SLC48A1* | 0.291463209 | 1.45E-05 | 4.99E-05 | 0.090847769 |
| Obesity | endothelial | *ZRSR1* | -5.967446284 | 2.61E-05 | 8.16E-05 | 0.163996233 |
| Obesity | endothelial | *AC084018.1* | -0.570386099 | 4.83E-06 | 2.05E-05 | 0.030384051 |
| Obesity | endothelial | *C17orf51* | -0.758314211 | 1.34E-07 | 1.18E-06 | 0.000841233 |
| Obesity | endothelial | *OR2A42* | -0.544359829 | 0.00303047 | 0.005084752 | 1 |
| Obesity | endothelial | *RNF208* | -1.703813076 | 2.54E-08 | 4.06E-07 | 0.000159677 |
| Obesity | endothelial | *ZNF580* | -0.77564932 | 2.08E-05 | 6.77E-05 | 0.130611129 |
| Obesity | endothelial | *NUP62* | -1.379149043 | 1.70E-06 | 8.67E-06 | 0.010669587 |
| Obesity | endothelial | *FGFR1OP* | -1.150041891 | 3.84E-05 | 0.000112226 | 0.241397621 |
| Obesity | endothelial | *ZNF254* | -0.744912479 | 0.000159251 | 0.000379246 | 1 |
| Obesity | endothelial | *LINGO4* | -6.190429114 | 6.69E-07 | 4.12E-06 | 0.004203431 |
| Obesity | endothelial | *GIMAP1* | 0.347105387 | 2.72E-05 | 8.43E-05 | 0.170797783 |
| Obesity | endothelial | *CCDC183* | -2.058117133 | 1.50E-07 | 1.29E-06 | 0.000941566 |
| Obesity | endothelial | *ARHGEF35* | -1.332542318 | 2.22E-05 | 7.12E-05 | 0.139414412 |
| Obesity | endothelial | *NOTCH2NL* | 0.459834495 | 0.01155163 | 0.017028159 | 1 |
| Obesity | endothelial | *SUPT4H1* | -0.771759467 | 0.010442084 | 0.015586272 | 1 |
| Obesity | endothelial | *GSTM2* | -0.610779577 | 1.81E-06 | 9.14E-06 | 0.011409652 |
| Obesity | endothelial | *LCAT* | -0.584663447 | 0.000757591 | 0.001492627 | 1 |
| Obesity | endothelial | *CCL14* | 0.130013498 | 0.000542808 | 0.001107638 | 1 |
| Obesity | endothelial | *RBMXL1* | -1.346153159 | 0.000206095 | 0.000474796 | 1 |
| Obesity | endothelial | *SRA1* | -0.693357423 | 0.000116209 | 0.000287302 | 0.730608142 |
| Obesity | endothelial | *HEXA* | -0.76867657 | 0.002420163 | 0.004182398 | 1 |
| Obesity | endothelial | *LBH* | -1.334753162 | 4.43E-08 | 5.42E-07 | 0.000278356 |
| Obesity | endothelial | *PPP1CB* | -0.830934476 | 1.75E-05 | 5.87E-05 | 0.109839935 |
| Obesity | endothelial | *NCKIPSD* | -1.047031629 | 0.000273915 | 0.00060616 | 1 |
| Obesity | endothelial | *SLC35F6* | -0.964663079 | 0.003316797 | 0.005520969 | 1 |
| Obesity | endothelial | *CLIC1* | -0.982079057 | 5.89E-05 | 0.000161877 | 0.370536795 |
| Obesity | endothelial | *ATP6V1G2* | -0.318636273 | 0.000730986 | 0.0014461 | 1 |
| Obesity | endothelial | *KLRK1* | -0.787125011 | 0.00419079 | 0.006820475 | 1 |
| Obesity | endothelial | *EMP2* | 0.361902466 | 0.000756753 | 0.001491445 | 1 |
| Obesity | endothelial | *PPM1N* | -1.496353391 | 1.92E-07 | 1.57E-06 | 0.001209574 |
| Obesity | endothelial | *LTB4R* | -0.380333307 | 0.023860984 | 0.032955625 | 1 |
| Obesity | endothelial | *GALT* | -0.595053276 | 0.026966287 | 0.036887956 | 1 |
| Obesity | endothelial | *ITGA1* | 0.273298955 | 0.018000202 | 0.025574525 | 1 |
| Obesity | endothelial | *NUDT19* | -0.670094948 | 0.000513453 | 0.001055272 | 1 |
| Obesity | endothelial | *ZNF726* | -1.740811999 | 5.03E-08 | 5.80E-07 | 0.000316071 |

| | | | | | | |
|---|---|---|---|---|---|---|
| Obesity | endothelial | *TAX1BP3* | -0.908473019 | 0.030999988 | 0.041877294 | 1 |
| Obesity | endothelial | *ZNF90* | -1.051517963 | 0.000269067 | 0.000597114 | 1 |
| Obesity | endothelial | *TM6SF2* | -18.22393324 | 1.24E-06 | 6.72E-06 | 0.007776285 |
| Obesity | endothelial | *REPIN1* | -1.184111657 | 3.00E-08 | 4.46E-07 | 0.000188466 |
| Obesity | endothelial | *SMIM7* | -0.826000893 | 0.000183057 | 0.000427042 | 1 |
| Obesity | endothelial | *TSPAN4* | -0.82480017 | 0.000178735 | 0.000418201 | 1 |
| Obesity | endothelial | *CPNE1* | -0.415130966 | 5.91E-05 | 0.000161986 | 0.371433618 |
| Obesity | endothelial | *ARL16* | -0.216288108 | 0.002230571 | 0.003888963 | 1 |
| Obesity | endothelial | *AC079776.2* | -1.178741135 | 9.14E-08 | 8.82E-07 | 0.00057445 |
| Obesity | endothelial | *LYRM4* | -0.682144731 | 0.001608726 | 0.002898012 | 1 |
| Obesity | endothelial | *PRCD* | 0.499267313 | 0.005983614 | 0.009393003 | 1 |
| Obesity | endothelial | *C17orf67* | 0.249310552 | 0.010879817 | 0.016141354 | 1 |
| Obesity | endothelial | *COLCA2* | -0.696334497 | 3.77E-07 | 2.62E-06 | 0.002368389 |
| Obesity | endothelial | *MBLAC1* | -0.35088124 | 0.002469687 | 0.004252786 | 1 |
| Obesity | endothelial | *NEURL1B* | 0.257830307 | 6.25E-06 | 2.51E-05 | 0.039308726 |
| Obesity | endothelial | *PPME1* | -0.412417287 | 0.009537153 | 0.014341087 | 1 |
| Obesity | endothelial | *STARD10* | -0.314946101 | 0.001079416 | 0.002032432 | 1 |
| Obesity | endothelial | *CPEB1* | -0.587137742 | 3.22E-08 | 4.54E-07 | 0.00020273 |
| Obesity | endothelial | *C10orf105* | 0.733964009 | 5.01E-05 | 0.000139761 | 0.314882494 |
| Obesity | endothelial | *ARHGEF33* | -0.684041704 | 8.30E-06 | 3.17E-05 | 0.052176755 |
| Obesity | endothelial | *AC005082.1* | -2.283137215 | 1.76E-07 | 1.46E-06 | 0.001108527 |
| Obesity | endothelial | *ACSM4* | -0.358183717 | 0.000760689 | 0.001497792 | 1 |
| Obesity | endothelial | *COL28A1* | -1.009229103 | 0.004946272 | 0.007920839 | 1 |
| Obesity | endothelial | *PHB2* | -1.280643541 | 3.67E-05 | 0.000108298 | 0.230674913 |
| Obesity | endothelial | *GRID2IP* | 0.637536069 | 8.33E-05 | 0.000215944 | 0.523879124 |
| Obesity | endothelial | *UBXN2B* | -0.765623127 | 5.95E-05 | 0.000162866 | 0.373777232 |
| Obesity | endothelial | *UBE2QL1* | -0.874913387 | 2.92E-08 | 4.40E-07 | 0.000183863 |
| Obesity | endothelial | *KCNU1* | -0.769392585 | 4.25E-06 | 1.85E-05 | 0.026735129 |
| Obesity | endothelial | *MYL5* | -1.628322809 | 1.41E-08 | 3.34E-07 | 8.86E-05 |
| Obesity | endothelial | *NPEPL1* | -1.106492803 | 4.70E-08 | 5.59E-07 | 0.000295392 |
| Obesity | endothelial | *LINC01139* | -0.711208794 | 0.000126423 | 0.00030939 | 0.794822627 |
| Obesity | endothelial | *TSTD1* | -1.090250338 | 2.93E-08 | 4.40E-07 | 0.000184051 |
| Obesity | endothelial | *LINC01138* | -0.787996559 | 2.25E-08 | 3.81E-07 | 0.000141375 |
| Obesity | endothelial | *CYB5RL* | -0.98530076 | 0.000111984 | 0.000278058 | 0.704043052 |
| Obesity | endothelial | *ATAD3C* | 1.28555202 | 2.83E-07 | 2.10E-06 | 0.001776224 |
| Obesity | endothelial | *CCDC7* | -0.420383509 | 0.018563491 | 0.026303509 | 1 |
| Obesity | endothelial | *RNASEK* | -1.003747216 | 0.000552805 | 0.001125117 | 1 |
| Obesity | endothelial | *NBPF1* | -0.825811734 | 2.95E-05 | 9.08E-05 | 0.18569156 |
| Obesity | endothelial | *RNU6ATAC35I* | -4.987873736 | 1.41E-08 | 3.34E-07 | 8.89E-05 |
| Obesity | endothelial | *C6orf226* | -1.093529229 | 1.00E-05 | 3.69E-05 | 0.062866101 |
| Obesity | endothelial | *PPP3R1* | -1.062304883 | 2.03E-05 | 6.62E-05 | 0.127347163 |
| Obesity | endothelial | *FANCG* | -0.438526911 | 0.007784472 | 0.011925189 | 1 |
| Obesity | endothelial | *AP4M1* | -0.295326488 | 0.018170847 | 0.025799484 | 1 |
| Obesity | endothelial | *C19orf73* | -2.65372288 | 6.83E-08 | 7.18E-07 | 0.000429369 |
| Obesity | endothelial | *ZNF880* | -0.294364357 | 0.005394656 | 0.008560375 | 1 |
| Obesity | endothelial | *OR2A25* | -13.45181115 | 1.24E-06 | 6.72E-06 | 0.007777941 |
| Obesity | endothelial | *TIGD1* | -0.447733804 | 0.004188382 | 0.006818322 | 1 |
| Obesity | endothelial | *XKR9* | -0.302714815 | 0.001519938 | 0.002759414 | 1 |
| Obesity | endothelial | *C1orf229* | -0.956859176 | 2.49E-07 | 1.90E-06 | 0.001562994 |
| Obesity | endothelial | *SLC12A8* | -1.064982926 | 7.54E-12 | 1.63E-09 | 4.74E-08 |

| | | | | | | |
|---|---|---|---|---|---|---|
| Obesity | endothelial | *APOL6* | -0.792901075 | 7.83E-08 | 7.89E-07 | 0.000492334 |
| Obesity | endothelial | *TIAF1* | 0.528796779 | 0.002260705 | 0.003934953 | 1 |
| Obesity | endothelial | *FAM185A* | -0.877343794 | 2.81E-05 | 8.71E-05 | 0.176880428 |
| Obesity | endothelial | *AC062017.1* | -0.703486143 | 6.98E-07 | 4.27E-06 | 0.00438833 |
| Obesity | endothelial | *ADRA2B* | -1.95677531 | 3.81E-08 | 4.99E-07 | 0.000239366 |
| Obesity | endothelial | *EXOSC6* | -1.776732828 | 1.90E-11 | 3.52E-09 | 1.20E-07 |
| Obesity | endothelial | *VPS52* | -0.858172752 | 0.001292616 | 0.002390901 | 1 |
| Obesity | endothelial | *LINC00205* | -1.079291335 | 1.08E-05 | 3.92E-05 | 0.067639051 |
| Obesity | endothelial | *OSER1-AS1* | -1.070072128 | 0.000152809 | 0.000365706 | 0.960710124 |
| Obesity | endothelial | *LINC01133* | -0.756650499 | 1.30E-08 | 3.34E-07 | 8.16E-05 |
| Obesity | endothelial | *LINC01117* | 0.391490684 | 3.56E-05 | 0.000105826 | 0.224034258 |
| Obesity | endothelial | *RASAL2-AS1* | -3.059760938 | 4.14E-08 | 5.19E-07 | 0.000260089 |
| Obesity | endothelial | *AC069213.1* | -2.435611062 | 3.82E-06 | 1.69E-05 | 0.023993967 |
| Obesity | endothelial | *LINC00863* | -0.606591727 | 0.002196823 | 0.00383864 | 1 |
| Obesity | endothelial | *LINC00184* | -1.303851839 | 2.36E-09 | 1.35E-07 | 1.48E-05 |
| Obesity | endothelial | *TMEM233* | 0.399735041 | 0.000128117 | 0.000312562 | 0.805471966 |
| Obesity | endothelial | *AL049840.1* | -0.996388568 | 1.17E-10 | 1.41E-08 | 7.37E-07 |
| Obesity | endothelial | *LINC00237* | -2.847499355 | 2.74E-08 | 4.25E-07 | 0.000172465 |
| Obesity | endothelial | *PLEKHM1* | -0.694752551 | 0.000982181 | 0.001869503 | 1 |
| Obesity | endothelial | *LINC00092* | -0.494358013 | 0.008524022 | 0.012941446 | 1 |
| Obesity | endothelial | *SFTA1P* | -3.215265011 | 1.63E-08 | 3.39E-07 | 0.000102591 |
| Obesity | endothelial | *FAM225B* | -0.594937212 | 0.00120525 | 0.002248489 | 1 |
| Obesity | endothelial | *SLC26A6* | -1.125527897 | 0.002455447 | 0.004234064 | 1 |
| Obesity | endothelial | *LINC00626* | -9.113935715 | 8.85E-07 | 5.14E-06 | 0.005561958 |
| Obesity | endothelial | *ERCC6* | -0.903633312 | 4.44E-05 | 0.000126563 | 0.278945462 |
| Obesity | endothelial | *LINC00115* | -0.569727901 | 0.018496069 | 0.026219793 | 1 |
| Obesity | endothelial | *NOL7* | -0.530265319 | 0.034962804 | 0.046748436 | 1 |
| Obesity | endothelial | *ELFN1* | 0.363510275 | 1.13E-06 | 6.26E-06 | 0.007100972 |
| Obesity | endothelial | *ZNF503-AS1* | -1.517266 | 9.45E-08 | 9.06E-07 | 0.000594424 |
| Obesity | endothelial | *LINC00937* | -1.293690773 | 3.99E-09 | 1.84E-07 | 2.51E-05 |
| Obesity | endothelial | *FTCDNL1* | -0.247515772 | 0.005558375 | 0.008800178 | 1 |
| Obesity | endothelial | *AC010536.1* | -1.240937428 | 2.40E-05 | 7.57E-05 | 0.150634153 |
| Obesity | endothelial | *AC064836.3* | 1.108629848 | 7.35E-05 | 0.000193827 | 0.462084134 |
| Obesity | endothelial | *HSBP1L1* | -0.839777444 | 0.016881881 | 0.024116424 | 1 |
| Obesity | endothelial | *AC093901.1* | -1.318675207 | 0.00016839 | 0.000397995 | 1 |
| Obesity | endothelial | *AC009299.3* | -1.219004035 | 0.000104375 | 0.000261645 | 0.656205695 |
| Obesity | endothelial | *SLC2A1-AS1* | -1.241190424 | 1.41E-09 | 9.82E-08 | 8.84E-06 |
| Obesity | endothelial | *LINC01068* | -0.808970002 | 5.31E-05 | 0.000146958 | 0.333593934 |
| Obesity | endothelial | *AC108463.2* | -1.452691423 | 5.07E-09 | 2.18E-07 | 3.19E-05 |
| Obesity | endothelial | *C19orf24* | -1.047526616 | 9.15E-05 | 0.000233808 | 0.575202993 |
| Obesity | endothelial | *LINC01013* | 0.734446904 | 2.64E-05 | 8.25E-05 | 0.166165269 |
| Obesity | endothelial | *AC073133.2* | 2.914272154 | 0.000196954 | 0.000456751 | 1 |
| Obesity | endothelial | *ARL17B* | -0.683095115 | 1.25E-05 | 4.43E-05 | 0.078820099 |
| Obesity | endothelial | *DHFR* | -0.375597427 | 5.86E-05 | 0.000161074 | 0.368215846 |
| Obesity | endothelial | *LINC00954* | -1.307562362 | 1.51E-08 | 3.34E-07 | 9.49E-05 |
| Obesity | endothelial | *AC067945.3* | -0.851272933 | 0.000419192 | 0.000884679 | 1 |
| Obesity | endothelial | *RPL41* | -0.989239073 | 0.010522713 | 0.015691721 | 1 |
| Obesity | endothelial | *LINC00200* | -3.396414856 | 9.31E-05 | 0.000236958 | 0.58528509 |
| Obesity | endothelial | *PATL2* | -1.133118732 | 9.35E-07 | 5.40E-06 | 0.005879259 |
| Obesity | endothelial | *U91324.1* | -1.185455022 | 4.82E-08 | 5.66E-07 | 0.000302961 |

| | | | | | | |
|---|---|---|---|---|---|---|
| Obesity | endothelial | *ZNF688* | -1.772741195 | 5.13E-06 | 2.15E-05 | 0.032258697 |
| Obesity | endothelial | *CISD3* | 0.689082148 | 0.006677489 | 0.010386287 | 1 |
| Obesity | endothelial | *PSMG3-AS1* | -0.924037407 | 6.53E-07 | 4.05E-06 | 0.004103102 |
| Obesity | endothelial | *PPP5D1* | -0.944034879 | 2.89E-06 | 1.35E-05 | 0.018189025 |
| Obesity | endothelial | *KLHL7-AS1* | -2.670540501 | 1.38E-08 | 3.34E-07 | 8.65E-05 |
| Obesity | endothelial | *AC092171.4* | -1.148897448 | 1.17E-07 | 1.07E-06 | 0.000736664 |
| Obesity | endothelial | *HLA-DPA1* | 0.345889285 | 0.022817404 | 0.031646375 | 1 |
| Obesity | endothelial | *DIRC3* | -0.564062742 | 2.29E-08 | 3.82E-07 | 0.000143754 |
| Obesity | endothelial | *AC107021.1* | -0.424682583 | 0.008141402 | 0.012402471 | 1 |
| Obesity | endothelial | *LINC00865* | -0.919063494 | 3.22E-06 | 1.48E-05 | 0.020220992 |
| Obesity | endothelial | *AC009495.3* | -1.993240766 | 6.26E-05 | 0.000169839 | 0.393687572 |
| Obesity | endothelial | *TNF* | -1.066035644 | 1.42E-09 | 9.83E-08 | 8.95E-06 |
| Obesity | endothelial | *LYRM9* | -1.441965195 | 1.58E-06 | 8.16E-06 | 0.009908006 |
| Obesity | endothelial | *LINC00342* | -1.406611497 | 3.42E-07 | 2.43E-06 | 0.002148799 |
| Obesity | endothelial | *SNHG15* | -0.75780553 | 0.017588256 | 0.02505151 | 1 |
| Obesity | endothelial | *LINC00702* | -0.689262849 | 8.72E-08 | 8.52E-07 | 0.000547925 |
| Obesity | endothelial | *AC123768.3* | -1.477616081 | 1.73E-06 | 8.79E-06 | 0.010856969 |
| Obesity | endothelial | *PINLYP* | -0.45704301 | 0.000116816 | 0.000288347 | 0.73442038 |
| Obesity | endothelial | *HLA-B* | 0.28513137 | 0.003284954 | 0.005473762 | 1 |
| Obesity | endothelial | *C19orf81* | -1.253720465 | 5.41E-05 | 0.000149754 | 0.340241212 |
| Obesity | endothelial | *ZSCAN31* | 0.466128668 | 4.18E-06 | 1.82E-05 | 0.02630925 |
| Obesity | endothelial | *LINC00957* | 1.312067127 | 0.000198115 | 0.000458766 | 1 |
| Obesity | endothelial | *LINC00484* | -0.217650769 | 0.009425482 | 0.014193535 | 1 |
| Obesity | endothelial | *KIAA0040* | -0.792121023 | 0.001210082 | 0.002255494 | 1 |
| Obesity | endothelial | *LINC00941* | -3.792288764 | 4.99E-07 | 3.26E-06 | 0.003134558 |
| Obesity | endothelial | *LINC00545* | -5.972715417 | 5.14E-07 | 3.35E-06 | 0.003228922 |
| Obesity | endothelial | *B3GNT9* | -1.100446141 | 6.41E-06 | 2.56E-05 | 0.040298003 |
| Obesity | endothelial | *ZNF737* | -0.813307178 | 0.00163105 | 0.002933688 | 1 |
| Obesity | endothelial | *AC073257.2* | -4.745394891 | 6.40E-05 | 0.000173004 | 0.402233819 |
| Obesity | endothelial | *IRGM* | -0.504001687 | 0.001831569 | 0.003257447 | 1 |
| Obesity | endothelial | *LRRC37A2* | -0.992809204 | 6.17E-06 | 2.48E-05 | 0.038801909 |
| Obesity | endothelial | *TXNDC5* | 1.265157712 | 0.009881296 | 0.01481889 | 1 |
| Obesity | endothelial | *CDRT4* | -1.171974116 | 0.000575503 | 0.001168288 | 1 |
| Obesity | endothelial | *ADSL* | -0.925921406 | 0.00019286 | 0.000448745 | 1 |
| Obesity | endothelial | *LILRA4* | -8.792370242 | 0.002611289 | 0.004463614 | 1 |
| Obesity | endothelial | *LILRA2* | -0.231490958 | 0.034712623 | 0.046453439 | 1 |
| Obesity | endothelial | *LY6G5B* | -0.149551738 | 0.03395593 | 0.045528029 | 1 |
| Obesity | endothelial | *PSMB9* | 0.433991023 | 1.30E-05 | 4.55E-05 | 0.081435055 |
| Obesity | endothelial | *COX19* | -1.066814512 | 1.46E-06 | 7.68E-06 | 0.009174705 |
| Obesity | endothelial | *L1TD1* | -0.381405355 | 0.000916556 | 0.001765438 | 1 |
| Obesity | endothelial | *AQP1* | 0.262715402 | 4.46E-05 | 0.000127123 | 0.280434222 |
| Obesity | endothelial | *C1QTNF9* | 0.484573764 | 1.73E-06 | 8.79E-06 | 0.010860853 |
| Obesity | endothelial | *ARHGEF25* | -1.015424489 | 2.48E-06 | 1.19E-05 | 0.015606669 |
| Obesity | endothelial | *LINC00886* | -0.777909456 | 0.000259181 | 0.000578236 | 1 |
| Obesity | endothelial | *NSUN6* | -0.612157588 | 0.002464011 | 0.004247667 | 1 |
| Obesity | endothelial | *CD302* | -1.286028364 | 2.84E-07 | 2.11E-06 | 0.001782772 |
| Obesity | endothelial | *ATP5J2* | -0.986902141 | 0.000271554 | 0.000602209 | 1 |
| Obesity | endothelial | *ARHGAP8* | -1.7546686 | 3.40E-08 | 4.64E-07 | 0.000213575 |
| Obesity | endothelial | *ATP5O* | -0.645935949 | 0.031192549 | 0.042110276 | 1 |
| Obesity | endothelial | *PLEKHO2* | -1.107527297 | 0.000205816 | 0.000474326 | 1 |

| | | | | | | |
|---|---|---|---|---|---|---|
| Obesity | endothelial | *PWP2* | -2.638841728 | 0.019808455 | 0.027929077 | 1 |
| Obesity | endothelial | *PI4KA* | -0.476770186 | 0.015075664 | 0.021748669 | 1 |
| Obesity | endothelial | *AKAP2* | 1.014842838 | 4.68E-06 | 1.99E-05 | 0.029406894 |
| Obesity | endothelial | *LINC00969* | -0.52872889 | 0.000107126 | 0.000268008 | 0.673503235 |
| Obesity | endothelial | *AMACR* | -0.177950716 | 0.01017307 | 0.015213627 | 1 |
| Obesity | endothelial | *ARHGDIG* | -0.836540181 | 4.03E-05 | 0.00011696 | 0.253451599 |
| Obesity | endothelial | *ARFGAP3* | -0.673832532 | 0.002869866 | 0.004851532 | 1 |
| Obesity | endothelial | *C22orf39* | -1.427202602 | 0.000243435 | 0.000547359 | 1 |
| Obesity | endothelial | *EIF6* | -1.336804162 | 1.17E-07 | 1.07E-06 | 0.000737101 |
| Obesity | endothelial | *MRPL20* | -0.728178295 | 0.003044585 | 0.00510571 | 1 |
| Obesity | endothelial | *DECR2* | -1.299174474 | 1.34E-08 | 3.34E-07 | 8.43E-05 |
| Obesity | endothelial | *AP5Z1* | -0.988807008 | 4.39E-05 | 0.000125357 | 0.275910373 |
| Obesity | endothelial | *ZNF709* | 0.620643109 | 0.00786886 | 0.012045659 | 1 |
| Obesity | endothelial | *STON1* | 0.411332078 | 0.012453488 | 0.018246348 | 1 |
| Obesity | endothelial | *VSIG8* | -1.283316011 | 3.29E-06 | 1.50E-05 | 0.020662965 |
| Obesity | endothelial | *C7orf73* | -0.92949089 | 2.34E-06 | 1.13E-05 | 0.014710388 |
| Obesity | endothelial | *FGF14-IT1* | -1.054885308 | 6.45E-08 | 6.95E-07 | 0.000405656 |
| Obesity | endothelial | *TICAM2* | -1.375270267 | 9.28E-06 | 3.47E-05 | 0.058339292 |
| Obesity | endothelial | *PALM2* | -0.488058031 | 1.11E-08 | 3.34E-07 | 7.00E-05 |
| Obesity | endothelial | *NAT6* | -0.666281599 | 0.000450818 | 0.000940553 | 1 |
| Obesity | endothelial | *UPK3B* | -0.592577917 | 0.003259206 | 0.005438065 | 1 |
| Obesity | endothelial | *IL10RB* | -0.479309316 | 0.035584779 | 0.047509346 | 1 |
| Obesity | endothelial | *NME1-NME2* | -1.133972246 | 7.36E-05 | 0.000194042 | 0.462788999 |
| Obesity | endothelial | *PLA2G4B* | -0.64794032 | 9.04E-05 | 0.000231315 | 0.568323452 |
| Obesity | endothelial | *APOBEC3D* | -2.279979389 | 7.57E-05 | 0.000198584 | 0.476081032 |
| Obesity | endothelial | *TMEM199* | -1.131607841 | 6.30E-05 | 0.000170888 | 0.396290315 |
| Obesity | endothelial | *P2RY11* | -1.050066017 | 0.003553059 | 0.005869175 | 1 |
| Obesity | endothelial | *LILRA6* | -0.373701461 | 0.000750574 | 0.001479729 | 1 |
| Obesity | endothelial | *APOBEC3C* | -1.213223037 | 2.22E-08 | 3.81E-07 | 0.00013987 |
| Obesity | endothelial | *CCDC13* | -0.36016451 | 0.010822845 | 0.016082067 | 1 |
| Obesity | endothelial | *OR2AE1* | 1.801763295 | 0.001875668 | 0.003325529 | 1 |
| Obesity | endothelial | *UBE2V1* | -1.299305025 | 0.001305224 | 0.002409991 | 1 |
| Obesity | endothelial | *N4BP2L2* | -0.443906944 | 0.002491945 | 0.004285246 | 1 |
| Obesity | endothelial | *LINC00847* | -1.456065548 | 8.26E-08 | 8.22E-07 | 0.000519301 |
| Obesity | endothelial | *NEAT1* | -0.465955635 | 2.07E-05 | 6.74E-05 | 0.129885911 |
| Obesity | endothelial | *RAB30-AS1* | -0.789425465 | 1.05E-05 | 3.86E-05 | 0.06621283 |
| Obesity | endothelial | *LINC00920* | -1.599004267 | 7.03E-06 | 2.77E-05 | 0.044227389 |
| Obesity | endothelial | *UBAP1L* | -1.12029286 | 3.59E-07 | 2.52E-06 | 0.00225617 |
| Obesity | endothelial | *DNAJC3-AS1* | -1.507823391 | 6.27E-07 | 3.92E-06 | 0.003940338 |
| Obesity | endothelial | *LINC00926* | -1.359281726 | 0.000119895 | 0.000295022 | 0.753781866 |
| Obesity | endothelial | *BCKDHA* | -1.925228775 | 3.66E-05 | 0.000108149 | 0.230140967 |
| Obesity | endothelial | *CDK11B* | -0.887267361 | 0.000480301 | 0.000994616 | 1 |
| Obesity | endothelial | *PCDHAC1* | -2.825281433 | 2.06E-09 | 1.25E-07 | 1.29E-05 |
| Obesity | endothelial | *ABHD14A* | -1.065831331 | 1.79E-06 | 9.05E-06 | 0.011260115 |
| Obesity | endothelial | *C5orf17* | -1.142649383 | 1.23E-05 | 4.37E-05 | 0.077513069 |
| Obesity | endothelial | *FMN1* | -0.567449523 | 9.07E-07 | 5.26E-06 | 0.005702157 |
| Obesity | endothelial | *NAIP* | -1.182912773 | 9.13E-06 | 3.42E-05 | 0.057378237 |
| Obesity | endothelial | *ZNF286B* | -1.648435806 | 0.000355904 | 0.000764459 | 1 |
| Obesity | endothelial | *LINC00942* | -1.418129618 | 9.26E-05 | 0.000236008 | 0.582467465 |
| Obesity | endothelial | *PDCD6* | -0.74516872 | 0.001281292 | 0.002373973 | 1 |

| | | | | | | |
|---|---|---|---|---|---|---|
| Obesity | endothelial | *LINC00603* | -0.814484446 | 1.65E-07 | 1.39E-06 | 0.001035423 |
| Obesity | endothelial | *YJEFN3* | -0.968774403 | 1.03E-08 | 3.23E-07 | 6.46E-05 |
| Obesity | endothelial | *DNAH10OS* | -2.353884961 | 4.97E-05 | 0.000138843 | 0.312535537 |
| Obesity | endothelial | *LINC00989* | -1.019382838 | 6.16E-09 | 2.39E-07 | 3.87E-05 |
| Obesity | endothelial | *LINC01021* | -1.926867961 | 1.48E-08 | 3.34E-07 | 9.28E-05 |
| Obesity | endothelial | *FAM218A* | -0.996647818 | 3.95E-08 | 5.06E-07 | 0.000248386 |
| Obesity | endothelial | *ATP6V1E2* | -0.346407067 | 0.002439182 | 0.004210069 | 1 |
| Obesity | endothelial | *GLI4* | -1.087674769 | 1.06E-09 | 7.84E-08 | 6.66E-06 |
| Obesity | endothelial | *SNHG18* | -0.847015496 | 2.37E-10 | 2.41E-08 | 1.49E-06 |
| Obesity | endothelial | *SHANK3* | 0.149805496 | 1.23E-05 | 4.35E-05 | 0.077092751 |
| Obesity | endothelial | *ZNF550* | -1.011915856 | 0.000313732 | 0.000684872 | 1 |
| Obesity | endothelial | *CCDC71L* | -1.155159604 | 6.55E-06 | 2.61E-05 | 0.041190152 |
| Obesity | endothelial | *TMEM200B* | -0.498656457 | 0.000144225 | 0.000347236 | 0.906745156 |
| Obesity | endothelial | *TRNP1* | -2.527182612 | 3.25E-08 | 4.54E-07 | 0.00020448 |
| Obesity | endothelial | *ALG11* | -0.46163309 | 0.007203139 | 0.01112955 | 1 |
| Obesity | endothelial | *ATXN7L3B* | -0.945608786 | 0.000609284 | 0.001226175 | 1 |
| Obesity | endothelial | *PRKDC* | -0.552264582 | 0.003921598 | 0.006423942 | 1 |
| Obesity | endothelial | *ZNF260* | -0.936869634 | 0.000336302 | 0.000728577 | 1 |
| Obesity | endothelial | *SIGLEC14* | -0.377335444 | 0.002339724 | 0.004054236 | 1 |
| Obesity | endothelial | *ARMS2* | -1.752909748 | 2.65E-07 | 2.00E-06 | 0.001665398 |
| Obesity | endothelial | *FPGT* | -1.403819931 | 3.15E-07 | 2.27E-06 | 0.001982611 |
| Obesity | endothelial | *MPV17L2* | 1.175684102 | 0.000362225 | 0.000776974 | 1 |
| Obesity | endothelial | *EID3* | -0.279637811 | 0.035220517 | 0.047062994 | 1 |
| Obesity | endothelial | *SNHG9* | -0.279971456 | 0.015774279 | 0.022683644 | 1 |
| Obesity | endothelial | *EBLN2* | -1.336601246 | 4.80E-06 | 2.04E-05 | 0.030162031 |
| Obesity | endothelial | *POLR2M* | -1.047442774 | 0.002567413 | 0.004398181 | 1 |
| Obesity | endothelial | *LINC00346* | 0.984401764 | 0.005082179 | 0.008105444 | 1 |
| Obesity | endothelial | *CTSO* | -0.616049164 | 0.023078697 | 0.031966462 | 1 |
| Obesity | endothelial | *LINC00944* | -0.685532077 | 0.00266742 | 0.004543502 | 1 |
| Obesity | endothelial | *ZNF10* | -0.470591401 | 0.023528811 | 0.032568391 | 1 |
| Obesity | endothelial | *HMBS* | 0.290943861 | 0.028233499 | 0.038462406 | 1 |
| Obesity | endothelial | *TAS2R31* | -0.509403307 | 0.01443722 | 0.020875529 | 1 |
| Obesity | endothelial | *AC093668.2* | -0.916882878 | 0.000473656 | 0.0009815 | 1 |
| Obesity | endothelial | *CLEC12B* | -0.135442909 | 0.027084287 | 0.037025204 | 1 |
| Obesity | endothelial | *PGA5* | 3.063766162 | 0.010682802 | 0.015904044 | 1 |
| Obesity | endothelial | *ZNF253* | -0.990655751 | 1.18E-05 | 4.22E-05 | 0.073942502 |
| Obesity | endothelial | *HP* | -1.541907607 | 0.000116165 | 0.000287302 | 0.730328704 |
| Obesity | endothelial | *GATC* | -0.827359499 | 0.002476063 | 0.004262597 | 1 |
| Obesity | endothelial | *ZBED6* | -2.299708339 | 1.41E-08 | 3.34E-07 | 8.84E-05 |
| Obesity | endothelial | *FNTB* | -1.962732305 | 2.60E-08 | 4.14E-07 | 0.000163705 |
| Obesity | endothelial | *GALNT4* | -2.562147359 | 0.018688618 | 0.026468876 | 1 |
| Obesity | endothelial | *CUX1* | -0.603290018 | 0.000199328 | 0.000461064 | 1 |
| Obesity | endothelial | *TEN1* | -1.290460923 | 0.001441504 | 0.002637582 | 1 |
| Obesity | endothelial | *C17orf49* | -0.943961322 | 0.009955611 | 0.014923922 | 1 |
| Obesity | endothelial | *RTEL1* | -0.676928425 | 0.025566286 | 0.035095031 | 1 |
| Obesity | endothelial | *ZNF578* | 0.39844687 | 0.004052939 | 0.006623557 | 1 |
| Obesity | endothelial | *LINC00640* | -0.34972308 | 0.00058658 | 0.001188089 | 1 |
| Obesity | endothelial | *LINC00638* | -1.441531823 | 1.11E-07 | 1.02E-06 | 0.00069585 |
| Obesity | endothelial | *DUXA* | -0.744653217 | 0.000179907 | 0.000420631 | 1 |
| Obesity | endothelial | *CEP95* | -0.758286691 | 4.38E-05 | 0.000125145 | 0.275157755 |

| | | | | | | |
|---|---|---|---|---|---|---|
| Obesity | endothelial | *LINC00639* | 0.136259125 | 1.80E-06 | 9.07E-06 | 0.011306532 |
| Obesity | endothelial | *LINC00648* | -0.620261231 | 2.24E-08 | 3.81E-07 | 0.000140836 |
| Obesity | endothelial | *LINC00924* | -0.843629932 | 6.80E-08 | 7.16E-07 | 0.000427363 |
| Obesity | endothelial | *UBE2Q2L* | -2.45703823 | 6.53E-10 | 5.33E-08 | 4.11E-06 |
| Obesity | endothelial | *IQCH-AS1* | -0.551844428 | 0.000600658 | 0.001212697 | 1 |
| Obesity | endothelial | *AC083843.1* | -10.40822069 | 2.54E-10 | 2.53E-08 | 1.59E-06 |
| Obesity | endothelial | *PWRN1* | -0.606590483 | 0.026673575 | 0.03651138 | 1 |
| Obesity | endothelial | *HOXB7* | 0.856529279 | 0.015357922 | 0.022130473 | 1 |
| Obesity | endothelial | *LINC00562* | -0.683752519 | 6.67E-10 | 5.38E-08 | 4.19E-06 |
| Obesity | endothelial | *AC009120.5* | -1.247196251 | 1.40E-08 | 3.34E-07 | 8.82E-05 |
| Obesity | endothelial | *TMEM178B* | 0.421794666 | 0.000240376 | 0.000541277 | 1 |
| Obesity | endothelial | *LINC01003* | -0.51649669 | 0.02844478 | 0.03869992 | 1 |
| Obesity | endothelial | *HPR* | -1.445404423 | 0.001094202 | 0.002057191 | 1 |
| Obesity | endothelial | *LINC00662* | -0.858359896 | 4.75E-05 | 0.000133691 | 0.298665403 |
| Obesity | endothelial | *AC123768.4* | -2.508909843 | 0.001004159 | 0.001907489 | 1 |
| Obesity | endothelial | *LINC00667* | -0.515003668 | 5.54E-05 | 0.000153035 | 0.348001722 |
| Obesity | endothelial | *LINC00672* | -0.654560549 | 0.000659449 | 0.00131785 | 1 |
| Obesity | endothelial | *LINC00909* | -0.936953185 | 4.18E-06 | 1.82E-05 | 0.026251012 |
| Obesity | endothelial | *DYNLL2* | -1.6594506 | 9.83E-08 | 9.33E-07 | 0.000618266 |
| Obesity | endothelial | *LINC00526* | -1.477911784 | 3.99E-07 | 2.73E-06 | 0.002507016 |
| Obesity | endothelial | *LINC00668* | 12.55588616 | 4.20E-05 | 0.000120919 | 0.263967146 |
| Obesity | endothelial | *STRADA* | -0.893331846 | 0.000110652 | 0.000275186 | 0.695670138 |
| Obesity | endothelial | *ILF3-AS1* | -0.872342396 | 2.96E-06 | 1.37E-05 | 0.018610756 |
| Obesity | endothelial | *AC139100.2* | -1.505820859 | 6.92E-08 | 7.23E-07 | 0.000434792 |
| Obesity | endothelial | *LINC01140* | -0.684691756 | 9.88E-06 | 3.66E-05 | 0.062137611 |
| Obesity | endothelial | *LINC00906* | -2.795828099 | 6.74E-08 | 7.12E-07 | 0.000423587 |
| Obesity | endothelial | *AC137932.1* | -1.927943358 | 0.006068912 | 0.009517856 | 1 |
| Obesity | endothelial | *AC003005.2* | -0.821017585 | 0.005312665 | 0.008434526 | 1 |
| Obesity | endothelial | *AL132989.1* | -3.069947519 | 1.93E-08 | 3.53E-07 | 0.000121102 |
| Obesity | endothelial | *LINC00664* | -2.275388234 | 5.93E-05 | 0.00016256 | 0.372912421 |
| Obesity | endothelial | *AC026740.1* | 1.558746064 | 0.036890537 | 0.049085885 | 1 |
| Obesity | endothelial | *FBXO17* | -0.687645224 | 7.55E-09 | 2.70E-07 | 4.75E-05 |
| Obesity | endothelial | *AC009060.1* | -1.824602474 | 3.77E-05 | 0.000110776 | 0.236728032 |
| Obesity | endothelial | *SNHG8* | -0.658916115 | 0.005973078 | 0.009378806 | 1 |
| Obesity | endothelial | *HCG17* | -0.784027085 | 6.43E-05 | 0.000173476 | 0.404025455 |
| Obesity | endothelial | *YTHDF3-AS1* | -1.135765921 | 1.15E-05 | 4.16E-05 | 0.07251243 |
| Obesity | endothelial | *LINC01050* | -3.246327898 | 1.82E-07 | 1.51E-06 | 0.001144155 |
| Obesity | endothelial | *SRXN1* | -4.752747782 | 0.000720442 | 0.001428389 | 1 |
| Obesity | endothelial | *MILR1* | -0.287462402 | 0.001514522 | 0.002751169 | 1 |
| Obesity | endothelial | *GTF2H5* | -0.442241015 | 0.020214583 | 0.028437925 | 1 |
| Obesity | endothelial | *FGF14-AS2* | -0.88131717 | 0.009125723 | 0.01381447 | 1 |
| Obesity | endothelial | *LINC01023* | -1.19850536 | 8.03E-08 | 8.05E-07 | 0.0005048 |
| Obesity | endothelial | *DOC2B* | 0.149113742 | 5.41E-06 | 2.24E-05 | 0.033992619 |
| Obesity | endothelial | *AC144652.1* | -0.406251429 | 0.00345076 | 0.005724255 | 1 |
| Obesity | endothelial | *PAXIP1-AS1* | -0.727833156 | 7.78E-05 | 0.000202978 | 0.48917741 |
| Obesity | LEC | *SCYL3* | 1.657931011 | 0.000443636 | 0.000805877 | 1 |
| Obesity | LEC | *C1orf112* | 1.644794542 | 1.30E-05 | 0.000105108 | 0.081832078 |
| Obesity | LEC | *CFH* | 1.405232074 | 1.72E-05 | 0.000105108 | 0.108335157 |
| Obesity | LEC | *NFYA* | 2.209744372 | 0.002018508 | 0.003108859 | 1 |
| Obesity | LEC | *NIPAL3* | -0.861655198 | 0.029584238 | 0.037243914 | 1 |

| | | | | | | |
|---|---|---|---|---|---|---|
| Obesity | LEC | *ENPP4* | 1.393346788 | 0.000201762 | 0.000419888 | 1 |
| Obesity | LEC | *SEMA3F* | -2.538407918 | 4.60E-16 | 4.13E-13 | 2.89E-12 |
| Obesity | LEC | *KRIT1* | 1.547705007 | 0.005105521 | 0.007252239 | 1 |
| Obesity | LEC | *RAD52* | 1.454811692 | 1.80E-05 | 0.000105108 | 0.113426579 |
| Obesity | LEC | *BAD* | 0.89508576 | 2.36E-05 | 0.000109118 | 0.148295098 |
| Obesity | LEC | *HS3ST1* | -0.221283923 | 0.000124891 | 0.000288248 | 0.785186804 |
| Obesity | LEC | *HECW1* | 0.560823704 | 0.002117255 | 0.00324942 | 1 |
| Obesity | LEC | *MAD1L1* | 1.712993 | 0.000132654 | 0.000302087 | 0.833998117 |
| Obesity | LEC | *LASP1* | 1.92033323 | 0.000121485 | 0.000282462 | 0.763778238 |
| Obesity | LEC | *SNX11* | 0.751522651 | 0.00129412 | 0.002075014 | 1 |
| Obesity | LEC | *TMEM176A* | 0.741253593 | 2.11E-05 | 0.000106989 | 0.132733056 |
| Obesity | LEC | *CYP26B1* | 1.052975158 | 1.33E-05 | 0.000105108 | 0.083715009 |
| Obesity | LEC | *ICA1* | -0.709020066 | 0.020887327 | 0.026910447 | 1 |
| Obesity | LEC | *DBNDD1* | 1.204762768 | 0.00574625 | 0.008100152 | 1 |
| Obesity | LEC | *ALS2* | 1.85607091 | 0.000109327 | 0.000259961 | 0.687335746 |
| Obesity | LEC | *CD38* | 1.490553297 | 0.005514164 | 0.007799224 | 1 |
| Obesity | LEC | *RBM6* | -1.319399727 | 1.05E-10 | 3.31E-08 | 6.61E-07 |
| Obesity | LEC | *RECQL* | 1.829534145 | 1.95E-05 | 0.000105217 | 0.122466966 |
| Obesity | LEC | *ARHGAP33* | 1.376417521 | 1.35E-06 | 5.03E-05 | 0.008501776 |
| Obesity | LEC | *PDK4* | 1.185358926 | 1.66E-05 | 0.000105108 | 0.104130946 |
| Obesity | LEC | *CDC27* | 1.818565427 | 2.17E-05 | 0.000107915 | 0.136404044 |
| Obesity | LEC | *MEOX1* | 0.719565492 | 0.00132666 | 0.002120699 | 1 |
| Obesity | LEC | *LIG3* | 1.067588197 | 0.009750638 | 0.013206001 | 1 |
| Obesity | LEC | *FAM214B* | 2.497301536 | 2.75E-06 | 7.03E-05 | 0.017300095 |
| Obesity | LEC | *COPZ2* | 1.438869335 | 4.94E-07 | 2.77E-05 | 0.003107262 |
| Obesity | LEC | *GCFC2* | 1.815084111 | 0.000216223 | 0.000444245 | 1 |
| Obesity | LEC | *ABCB4* | 1.317078855 | 1.57E-05 | 0.000105108 | 0.098818628 |
| Obesity | LEC | *PDK2* | 2.005186236 | 6.38E-05 | 0.000178168 | 0.400877939 |
| Obesity | LEC | *ASB4* | 1.28083395 | 0.024410006 | 0.03110371 | 1 |
| Obesity | LEC | *TMEM98* | 1.279289976 | 0.011008859 | 0.014808022 | 1 |
| Obesity | LEC | *ABCC8* | -2.131236916 | 0.000926423 | 0.001536382 | 1 |
| Obesity | LEC | *CCL18* | 0.748395658 | 0.000110042 | 0.000261197 | 0.69183447 |
| Obesity | LEC | *SYNRG* | 1.750300731 | 0.000202725 | 0.000421611 | 1 |
| Obesity | LEC | *TMEM132A* | 1.76222221 | 1.70E-05 | 0.000105108 | 0.107038943 |
| Obesity | LEC | *ZNF263* | 2.242467965 | 0.002140273 | 0.003281126 | 1 |
| Obesity | LEC | *TNFRSF12A* | 0.878185384 | 0.000125737 | 0.000289988 | 0.790507394 |
| Obesity | LEC | *BAIAP2L1* | 0.84261932 | 0.000154059 | 0.000339312 | 0.968570734 |
| Obesity | LEC | *FARP2* | 1.567313912 | 0.000106005 | 0.00025435 | 0.666455923 |
| Obesity | LEC | *IFRD1* | 1.102441671 | 0.000137166 | 0.000309868 | 0.862362174 |
| Obesity | LEC | *GTF2IRD1* | 1.246750862 | 5.71E-06 | 9.65E-05 | 0.035909649 |
| Obesity | LEC | *VPS41* | 1.462463774 | 0.001088899 | 0.00177755 | 1 |
| Obesity | LEC | *ARHGAP44* | -0.934570785 | 0.008943335 | 0.012180838 | 1 |
| Obesity | LEC | *ELAC2* | 1.452203602 | 0.011292344 | 0.015160146 | 1 |
| Obesity | LEC | *CCDC124* | 1.797863727 | 0.000257245 | 0.000513429 | 1 |
| Obesity | LEC | *CEACAM21* | 0.763754614 | 4.25E-05 | 0.000137166 | 0.267416794 |
| Obesity | LEC | *NOS2* | -2.313703996 | 0.000170806 | 0.000367006 | 1 |
| Obesity | LEC | *CD79B* | -1.663012402 | 3.97E-05 | 0.000131291 | 0.249583319 |
| Obesity | LEC | *ST7L* | 0.648536535 | 0.021952557 | 0.028160727 | 1 |
| Obesity | LEC | *PAX6* | -0.598640073 | 0.001205179 | 0.001947804 | 1 |
| Obesity | LEC | *RPUSD1* | 2.600495228 | 1.51E-05 | 0.000105108 | 0.094846681 |

| | | | | | | |
|---|---|---|---|---|---|---|
| Obesity | LEC | *BAIAP3* | -1.877384384 | 0.000144138 | 0.000322145 | 0.906193215 |
| Obesity | LEC | *TSR3* | 1.51176379 | 1.72E-05 | 0.000105108 | 0.107930815 |
| Obesity | LEC | *PIGQ* | 1.289691224 | 2.37E-05 | 0.000109118 | 0.14891552 |
| Obesity | LEC | *PSMB1* | 1.717784634 | 7.64E-05 | 0.000201692 | 0.480073154 |
| Obesity | LEC | *CDK11A* | 2.373037035 | 0.000878381 | 0.00146278 | 1 |
| Obesity | LEC | *DLEC1* | 0.944710886 | 2.67E-05 | 0.000112975 | 0.167679722 |
| Obesity | LEC | *SS18L2* | -1.181141931 | 0.002838804 | 0.004230282 | 1 |
| Obesity | LEC | *MPND* | 3.083878691 | 7.76E-06 | 0.000105108 | 0.04881729 |
| Obesity | LEC | *MGST1* | 0.675399127 | 2.33E-05 | 0.000109118 | 0.146651178 |
| Obesity | LEC | *MMP25* | 0.879333461 | 8.25E-06 | 0.000105108 | 0.051888832 |
| Obesity | LEC | *IL32* | 1.245279234 | 0.000772165 | 0.001303949 | 1 |
| Obesity | LEC | *PKD1* | 2.012555816 | 1.95E-06 | 5.93E-05 | 0.012273648 |
| Obesity | LEC | *MAPK8IP2* | -1.701296323 | 0.002749105 | 0.004108301 | 1 |
| Obesity | LEC | *MED24* | 2.102750787 | 0.000163508 | 0.000355209 | 1 |
| Obesity | LEC | *RHOBTB2* | 1.983954275 | 0.011433597 | 0.015333409 | 1 |
| Obesity | LEC | *SEC62* | 1.760064514 | 0.000168151 | 0.000362913 | 1 |
| Obesity | LEC | *RPS20* | 1.579076101 | 0.014854075 | 0.019541237 | 1 |
| Obesity | LEC | *MLXIPL* | 0.545305673 | 7.84E-07 | 3.74E-05 | 0.004930534 |
| Obesity | LEC | *METTL13* | 2.000695585 | 7.68E-05 | 0.00020244 | 0.482962416 |
| Obesity | LEC | *DYRK4* | 0.856244639 | 3.40E-05 | 0.000120775 | 0.213965635 |
| Obesity | LEC | *NCAPD2* | 0.998299787 | 0.000138579 | 0.000312492 | 0.871246476 |
| Obesity | LEC | *ZNF200* | -1.039767618 | 0.004728258 | 0.006757572 | 1 |
| Obesity | LEC | *CD4* | 0.630709077 | 2.30E-05 | 0.000109118 | 0.14475005 |
| Obesity | LEC | *LRRC23* | 1.032195362 | 6.59E-06 | 0.000102597 | 0.04144923 |
| Obesity | LEC | *HFE* | -1.640128148 | 0.001694777 | 0.002648537 | 1 |
| Obesity | LEC | *SCMH1* | 1.584448788 | 0.000606804 | 0.001057072 | 1 |
| Obesity | LEC | *CLCN6* | 2.471244866 | 8.55E-06 | 0.000105108 | 0.053739841 |
| Obesity | LEC | *MRC2* | 0.660681384 | 7.72E-05 | 0.000202909 | 0.485093089 |
| Obesity | LEC | *TSPAN9* | -0.524892305 | 0.011428393 | 0.015329701 | 1 |
| Obesity | LEC | *MKS1* | 2.320981987 | 6.84E-06 | 0.000104405 | 0.04300611 |
| Obesity | LEC | *UTP18* | 2.417183 | 0.001223284 | 0.001971997 | 1 |
| Obesity | LEC | *TTC19* | 2.136189547 | 2.05E-06 | 6.09E-05 | 0.012916233 |
| Obesity | LEC | *DPF1* | -0.980929985 | 7.87E-05 | 0.000205977 | 0.494963563 |
| Obesity | LEC | *LARS2* | 1.902556224 | 0.002239154 | 0.003416055 | 1 |
| Obesity | LEC | *ZBTB32* | 1.866056044 | 1.49E-05 | 0.000105108 | 0.093712213 |
| Obesity | LEC | *TMEM159* | 0.877168831 | 3.37E-05 | 0.000120167 | 0.211734529 |
| Obesity | LEC | *ERCC1* | 0.769103891 | 1.21E-05 | 0.000105108 | 0.075952598 |
| Obesity | LEC | *LTF* | -0.717533706 | 0.000361984 | 0.000678631 | 1 |
| Obesity | LEC | *ALOX5* | 1.430181152 | 5.05E-05 | 0.000153071 | 0.317775872 |
| Obesity | LEC | *PSMC4* | 2.825105723 | 1.77E-05 | 0.000105108 | 0.111477704 |
| Obesity | LEC | *MAN2B2* | 1.543630406 | 1.10E-05 | 0.000105108 | 0.069393776 |
| Obesity | LEC | *CLDN11* | -0.272030672 | 0.000258058 | 0.000514235 | 1 |
| Obesity | LEC | *SLC25A39* | 1.36738223 | 5.42E-05 | 0.000160434 | 0.340601533 |
| Obesity | LEC | *ANGEL1* | 1.431469012 | 1.23E-05 | 0.000105108 | 0.077375418 |
| Obesity | LEC | *DDX11* | 0.738575467 | 0.006998115 | 0.009705967 | 1 |
| Obesity | LEC | *TACC3* | 1.535051094 | 4.69E-05 | 0.000145969 | 0.294711347 |
| Obesity | LEC | *ZC3H3* | 1.609128478 | 0.004848527 | 0.006909041 | 1 |
| Obesity | LEC | *MDH1* | 1.677301239 | 2.24E-06 | 6.43E-05 | 0.014075197 |
| Obesity | LEC | *SLC30A9* | 1.80682476 | 2.10E-05 | 0.000106989 | 0.132000015 |
| Obesity | LEC | *MTMR11* | 1.018310474 | 0.000213988 | 0.000439798 | 1 |

| | | | | | | |
|---|---|---|---|---|---|---|
| Obesity | LEC | *COX15* | 1.533180026 | 0.000212648 | 0.000438046 | 1 |
| Obesity | LEC | *CCDC88C* | 0.943552687 | 0.00019259 | 0.000404684 | 1 |
| Obesity | LEC | *NUDCD3* | 1.630805703 | 7.65E-07 | 3.73E-05 | 0.004810406 |
| Obesity | LEC | *CHDH* | -1.091595367 | 0.000128213 | 0.00029408 | 0.806074076 |
| Obesity | LEC | *IL20RA* | -0.576881968 | 0.000100186 | 0.000244515 | 0.629870034 |
| Obesity | LEC | *RUFY3* | 1.395866247 | 0.001366994 | 0.002180739 | 1 |
| Obesity | LEC | *WWTR1* | -0.586511543 | 0.001509682 | 0.002385965 | 1 |
| Obesity | LEC | *TTC27* | 1.527758235 | 0.000314508 | 0.000603759 | 1 |
| Obesity | LEC | *VSIG2* | 1.822969379 | 2.82E-05 | 0.000114165 | 0.177194834 |
| Obesity | LEC | *PHLDB1* | 0.750692474 | 2.42E-05 | 0.000109613 | 0.152362614 |
| Obesity | LEC | *MARCO* | 0.59196805 | 0.001662944 | 0.002603319 | 1 |
| Obesity | LEC | *MNAT1* | 1.601595884 | 4.76E-05 | 0.000147275 | 0.299263255 |
| Obesity | LEC | *AQR* | 1.753674261 | 0.000198719 | 0.000414928 | 1 |
| Obesity | LEC | *CPS1* | 1.632155533 | 8.35E-05 | 0.000214782 | 0.524949819 |
| Obesity | LEC | *NLRP2* | 1.022152513 | 0.000151482 | 0.000335222 | 0.952366974 |
| Obesity | LEC | *ZDHHC6* | 2.193818937 | 0.00025749 | 0.000513592 | 1 |
| Obesity | LEC | *RNH1* | 1.5092501 | 5.03E-06 | 9.23E-05 | 0.031629339 |
| Obesity | LEC | *NDUFS1* | 1.896819867 | 8.31E-06 | 0.000105108 | 0.052215506 |
| Obesity | LEC | *BIRC3* | 1.712993359 | 0.000403588 | 0.000744311 | 1 |
| Obesity | LEC | *AKAP11* | 1.899584724 | 0.000111402 | 0.000263552 | 0.700383005 |
| Obesity | LEC | *GLRX2* | 1.019209079 | 0.002814374 | 0.004197857 | 1 |
| Obesity | LEC | *SNAPC1* | 0.815469234 | 4.98E-05 | 0.000151436 | 0.312867632 |
| Obesity | LEC | *GCLM* | 1.587886007 | 5.11E-06 | 9.29E-05 | 0.032155688 |
| Obesity | LEC | *UBR2* | 1.533111965 | 0.000436601 | 0.000795165 | 1 |
| Obesity | LEC | *HSF2* | 2.324933596 | 2.59E-05 | 0.000112124 | 0.162855694 |
| Obesity | LEC | *NR1H3* | 2.597221891 | 3.17E-06 | 7.47E-05 | 0.019937105 |
| Obesity | LEC | *TYMP* | 1.300087399 | 9.84E-06 | 0.000105108 | 0.061892188 |
| Obesity | LEC | *KPNA6* | 1.836936391 | 8.32E-06 | 0.000105108 | 0.052326338 |
| Obesity | LEC | *VIM* | 1.448457097 | 0.000541031 | 0.000955466 | 1 |
| Obesity | LEC | *FAS* | 1.12519712 | 1.49E-05 | 0.000105108 | 0.093697343 |
| Obesity | LEC | *RNASET2* | 1.142842382 | 6.95E-05 | 0.000188556 | 0.43707231 |
| Obesity | LEC | *CD44* | 0.917998021 | 1.44E-05 | 0.000105108 | 0.090694165 |
| Obesity | LEC | *MIPEP* | 1.557469291 | 1.64E-05 | 0.000105108 | 0.102823968 |
| Obesity | LEC | *INSRR* | 2.020435241 | 0.000939019 | 0.001553992 | 1 |
| Obesity | LEC | *B4GALT7* | -1.091228734 | 0.004642058 | 0.006641925 | 1 |
| Obesity | LEC | *VRK2* | 1.431813892 | 0.003820566 | 0.005557589 | 1 |
| Obesity | LEC | *SNX1* | 1.559739978 | 7.10E-06 | 0.000105108 | 0.044645226 |
| Obesity | LEC | *ARNTL2* | -1.071982975 | 0.00576827 | 0.008127546 | 1 |
| Obesity | LEC | *SLC39A9* | 1.274511295 | 0.000233838 | 0.00047424 | 1 |
| Obesity | LEC | *ANK1* | 0.709658925 | 3.99E-05 | 0.000131515 | 0.250963265 |
| Obesity | LEC | *RABEP1* | 1.891333629 | 1.37E-07 | 1.33E-05 | 0.000861464 |
| Obesity | LEC | *BAK1* | 1.207964548 | 8.27E-06 | 0.000105108 | 0.051970113 |
| Obesity | LEC | *GRN* | 1.303648846 | 2.58E-05 | 0.000112124 | 0.162282868 |
| Obesity | LEC | *ARID4A* | 1.754201436 | 0.000109902 | 0.000261034 | 0.690956534 |
| Obesity | LEC | *ZCCHC8* | 1.721454711 | 0.008721662 | 0.011891799 | 1 |
| Obesity | LEC | *ABCF2* | 1.488246308 | 7.04E-05 | 0.000190166 | 0.44254744 |
| Obesity | LEC | *FUT8* | 1.335487371 | 2.13E-05 | 0.00010722 | 0.133846924 |
| Obesity | LEC | *PIAS1* | 1.479652922 | 0.000102813 | 0.000249503 | 0.646386868 |
| Obesity | LEC | *SLC4A7* | 0.952855525 | 1.58E-05 | 0.000105108 | 0.099256401 |
| Obesity | LEC | *MYOC* | 0.706420037 | 3.32E-05 | 0.000119352 | 0.208431543 |

| | | | | | | |
|---|---|---|---|---|---|---|
| Obesity | LEC | *VCL* | 1.070807832 | 0.002511411 | 0.003784573 | 1 |
| Obesity | LEC | *ADSS* | 2.021633462 | 5.19E-05 | 0.000155484 | 0.326049213 |
| Obesity | LEC | *STAP1* | 1.235864161 | 0.025805154 | 0.0327884 | 1 |
| Obesity | LEC | *TBC1D23* | 1.763536746 | 0.000148182 | 0.000329777 | 0.931620213 |
| Obesity | LEC | *CUL3* | 1.543434495 | 0.000140217 | 0.00031509 | 0.881541168 |
| Obesity | LEC | *MYOM2* | 0.935873492 | 0.000345247 | 0.000652144 | 1 |
| Obesity | LEC | *NSUN2* | 2.043158873 | 7.29E-06 | 0.000105108 | 0.045851068 |
| Obesity | LEC | *MRI1* | 1.294923637 | 0.011972392 | 0.015989732 | 1 |
| Obesity | LEC | *AGA* | 2.36581668 | 1.99E-05 | 0.000105667 | 0.12532124 |
| Obesity | LEC | *BOD1L1* | 1.726136876 | 0.000126723 | 0.000291097 | 0.796707921 |
| Obesity | LEC | *MAT2B* | 2.44029322 | 8.61E-06 | 0.000105108 | 0.05415926 |
| Obesity | LEC | *CLEC16A* | 1.4355468 | 2.85E-05 | 0.000114878 | 0.178980035 |
| Obesity | LEC | *SKIV2L2* | 1.704590235 | 3.87E-05 | 0.000129293 | 0.243457866 |
| Obesity | LEC | *DNAH5* | 3.19776385 | 4.33E-06 | 8.75E-05 | 0.027214703 |
| Obesity | LEC | *C6* | 0.584687907 | 2.57E-05 | 0.000112124 | 0.161619206 |
| Obesity | LEC | *RAI14* | -0.332015286 | 0.000277777 | 0.000546846 | 1 |
| Obesity | LEC | *PNKP* | 1.448926494 | 0.000393689 | 0.000729049 | 1 |
| Obesity | LEC | *PHLPP2* | 1.827757997 | 9.22E-05 | 0.000230354 | 0.579401504 |
| Obesity | LEC | *PQLC2* | 2.468690214 | 2.78E-05 | 0.00011384 | 0.174591177 |
| Obesity | LEC | *INPP4A* | 1.538148989 | 8.06E-05 | 0.000209468 | 0.506912745 |
| Obesity | LEC | *RAB27B* | 1.03974661 | 0.000164102 | 0.00035613 | 1 |
| Obesity | LEC | *PSMA4* | 1.862877381 | 8.61E-05 | 0.000219223 | 0.541261648 |
| Obesity | LEC | *THAP3* | 1.305885395 | 2.09E-05 | 0.00010698 | 0.131691931 |
| Obesity | LEC | *TDP1* | 2.594672758 | 5.92E-05 | 0.00017021 | 0.371908402 |
| Obesity | LEC | *SPATA7* | 1.594655753 | 2.26E-05 | 0.000109111 | 0.142070532 |
| Obesity | LEC | *MED17* | 2.389838444 | 1.66E-05 | 0.000105108 | 0.104300943 |
| Obesity | LEC | *RETSAT* | 0.859619 | 8.41E-06 | 0.000105108 | 0.052863627 |
| Obesity | LEC | *CAPG* | 0.887029277 | 3.71E-05 | 0.000126017 | 0.233004544 |
| Obesity | LEC | *AP2S1* | 2.267062919 | 0.000193599 | 0.000405989 | 1 |
| Obesity | LEC | *LCP2* | 0.79137452 | 4.18E-05 | 0.000135461 | 0.262794145 |
| Obesity | LEC | *CUL7* | 0.882276055 | 0.000422098 | 0.000773733 | 1 |
| Obesity | LEC | *CTNNA1* | 1.50524797 | 0.002295355 | 0.003492473 | 1 |
| Obesity | LEC | *CNTLN* | 1.547756798 | 0.002487007 | 0.003753196 | 1 |
| Obesity | LEC | *EPHA3* | 1.370091348 | 1.44E-05 | 0.000105108 | 0.090824887 |
| Obesity | LEC | *DSG2* | 0.846479288 | 1.73E-05 | 0.000105108 | 0.108622372 |
| Obesity | LEC | *WDR37* | 1.406520998 | 6.27E-05 | 0.000176459 | 0.394329121 |
| Obesity | LEC | *YTHDC2* | 1.744760112 | 0.00078784 | 0.0013265 | 1 |
| Obesity | LEC | *ATP6V1H* | 1.797966677 | 4.62E-05 | 0.000145061 | 0.29059555 |
| Obesity | LEC | *POLR2B* | 1.819060102 | 0.001496232 | 0.00236828 | 1 |
| Obesity | LEC | *CP* | 0.894141297 | 1.91E-05 | 0.000105211 | 0.120287211 |
| Obesity | LEC | *DTNBP1* | 1.557435898 | 0.013319396 | 0.017636698 | 1 |
| Obesity | LEC | *ANO2* | -0.325230425 | 0.001089092 | 0.00177755 | 1 |
| Obesity | LEC | *GOPC* | 1.40329107 | 0.02121278 | 0.027289697 | 1 |
| Obesity | LEC | *TSPAN17* | 0.948635626 | 1.73E-06 | 5.62E-05 | 0.01087275 |
| Obesity | LEC | *CC2D2A* | 1.59776884 | 3.95E-05 | 0.000130742 | 0.248278346 |
| Obesity | LEC | *RRM2B* | 1.969482525 | 0.000447002 | 0.000810462 | 1 |
| Obesity | LEC | *SNX29* | 1.01086697 | 3.01E-05 | 0.000115831 | 0.18943606 |
| Obesity | LEC | *LMO3* | 0.652911927 | 0.000168836 | 0.000364017 | 1 |
| Obesity | LEC | *CELF2* | 0.952381712 | 1.68E-05 | 0.000105108 | 0.10571799 |
| Obesity | LEC | *H6PD* | 1.306663357 | 0.006880158 | 0.009557126 | 1 |

| | | | | | | |
|---|---|---|---|---|---|---|
| Obesity | LEC | *UTS2* | 0.743758859 | 0.001257689 | 0.002021841 | 1 |
| Obesity | LEC | *EPN3* | 4.673546568 | 0.004733224 | 0.006760058 | 1 |
| Obesity | LEC | *RFC2* | 1.12996133 | 1.56E-05 | 0.000105108 | 0.098242869 |
| Obesity | LEC | *JKAMP* | 1.756644999 | 4.44E-07 | 2.59E-05 | 0.002794351 |
| Obesity | LEC | *DKK3* | -0.160213586 | 0.000111424 | 0.000263552 | 0.700520771 |
| Obesity | LEC | *ARHGEF5* | 3.890065334 | 0.001119231 | 0.001820597 | 1 |
| Obesity | LEC | *NFE2L3* | 2.628011733 | 3.15E-05 | 0.000116847 | 0.197727058 |
| Obesity | LEC | *MCUR1* | 1.680202918 | 3.56E-05 | 0.000123048 | 0.22408094 |
| Obesity | LEC | *LIMA1* | 0.660900809 | 0.000200843 | 0.00041839 | 1 |
| Obesity | LEC | *PTGER3* | 0.885451985 | 9.27E-06 | 0.000105108 | 0.05828615 |
| Obesity | LEC | *MAPK9* | 1.474329164 | 0.000362037 | 0.000678631 | 1 |
| Obesity | LEC | *FAM160A2* | 0.947185639 | 0.038351884 | 0.04749132 | 1 |
| Obesity | LEC | *HERPUD1* | 1.730291209 | 2.40E-05 | 0.000109361 | 0.150612685 |
| Obesity | LEC | *PIK3CB* | 1.309238249 | 0.003439988 | 0.005051905 | 1 |
| Obesity | LEC | *CYBA* | 1.355932184 | 2.64E-05 | 0.000112582 | 0.166105278 |
| Obesity | LEC | *THOC3* | 1.244096496 | 3.11E-05 | 0.000116544 | 0.195328397 |
| Obesity | LEC | *RRP12* | 0.952951392 | 3.18E-05 | 0.000117138 | 0.199954269 |
| Obesity | LEC | *ALX4* | 4.690023415 | 4.78E-05 | 0.000147775 | 0.300425893 |
| Obesity | LEC | *FOXN3* | 1.230423324 | 1.93E-05 | 0.000105211 | 0.12121349 |
| Obesity | LEC | *AKR7A2* | 2.533320043 | 3.34E-05 | 0.000119779 | 0.210093211 |
| Obesity | LEC | *MRTO4* | 2.645105505 | 0.000255856 | 0.00051111 | 1 |
| Obesity | LEC | *MCF2L2* | 2.171504339 | 0.000185084 | 0.000392056 | 1 |
| Obesity | LEC | *ANAPC4* | 2.39458021 | 9.47E-06 | 0.000105108 | 0.059550899 |
| Obesity | LEC | *LY75* | 1.955519823 | 3.49E-05 | 0.000122312 | 0.219671992 |
| Obesity | LEC | *OPN3* | 1.549039917 | 0.000343157 | 0.000649675 | 1 |
| Obesity | LEC | *SDCCAG8* | 1.539538111 | 2.31E-07 | 1.84E-05 | 0.001453037 |
| Obesity | LEC | *TBC1D22A* | 1.534212631 | 0.000179667 | 0.000383396 | 1 |
| Obesity | LEC | *PLEKHH1* | 1.315541606 | 0.000784067 | 0.001321563 | 1 |
| Obesity | LEC | *ATP9A* | 1.208515176 | 0.04015702 | 0.04955195 | 1 |
| Obesity | LEC | *CHRDL2* | -0.732899714 | 6.88E-05 | 0.000187286 | 0.432469131 |
| Obesity | LEC | *GALC* | 1.48402202 | 0.000110462 | 0.000262066 | 0.694474804 |
| Obesity | LEC | *GINM1* | 2.073587198 | 0.000104398 | 0.000251766 | 0.656352896 |
| Obesity | LEC | *MRPL43* | 1.466058599 | 0.001854099 | 0.002872528 | 1 |
| Obesity | LEC | *ITIH4* | 1.634712126 | 2.54E-13 | 1.33E-10 | 1.60E-09 |
| Obesity | LEC | *TRAF1* | 1.631085976 | 2.54E-05 | 0.000111691 | 0.159784899 |
| Obesity | LEC | *IL17RB* | -2.02594474 | 0.001542892 | 0.002433558 | 1 |
| Obesity | LEC | *TRAF3IP2* | 2.317361637 | 0.000132098 | 0.000301233 | 0.830500096 |
| Obesity | LEC | *DCBLD2* | 1.850295166 | 7.84E-05 | 0.000205585 | 0.492897795 |
| Obesity | LEC | *MSH4* | 1.481533543 | 0.002301635 | 0.003500334 | 1 |
| Obesity | LEC | *GDI2* | 1.486187275 | 0.002185578 | 0.003345685 | 1 |
| Obesity | LEC | *TMCC3* | 2.166088162 | 0.00016377 | 0.000355585 | 1 |
| Obesity | LEC | *PITHD1* | 1.256625243 | 1.03E-05 | 0.000105108 | 0.064886258 |
| Obesity | LEC | *ATP11B* | 1.764500872 | 3.52E-06 | 7.79E-05 | 0.022129725 |
| Obesity | LEC | *CDK14* | 1.186260639 | 1.77E-05 | 0.000105108 | 0.111141952 |
| Obesity | LEC | *CROCC* | 1.408870564 | 5.75E-06 | 9.67E-05 | 0.036179192 |
| Obesity | LEC | *ZC3H11A* | 3.623482262 | 2.43E-06 | 6.70E-05 | 0.01527088 |
| Obesity | LEC | *RIOK2* | 2.510917926 | 0.000311855 | 0.000599284 | 1 |
| Obesity | LEC | *FLYWCH1* | 0.931843326 | 0.006465537 | 0.009031067 | 1 |
| Obesity | LEC | *DNAJC25* | -1.051241692 | 0.002828561 | 0.004216018 | 1 |
| Obesity | LEC | *OGFR* | 1.631983006 | 0.013555181 | 0.017937576 | 1 |

| | | | | | | |
|---|---|---|---|---|---|---|
| Obesity | LEC | *PIGV* | 1.505534095 | 0.000259904 | 0.000517585 | 1 |
| Obesity | LEC | *QSER1* | 1.457496233 | 1.67E-05 | 0.000105108 | 0.105123362 |
| Obesity | LEC | *SFSWAP* | 1.730495925 | 0.00010929 | 0.000259961 | 0.687103555 |
| Obesity | LEC | *TNK2* | 1.345903427 | 0.007710388 | 0.010597991 | 1 |
| Obesity | LEC | *VMP1* | 1.335807197 | 3.59E-05 | 0.000123286 | 0.225855644 |
| Obesity | LEC | *EIF4B* | 1.80922682 | 3.53E-06 | 7.79E-05 | 0.022213051 |
| Obesity | LEC | *SLC6A16* | 1.424862653 | 0.019777515 | 0.025574092 | 1 |
| Obesity | LEC | *SPHK2* | 1.203900729 | 5.02E-05 | 0.000152246 | 0.315301216 |
| Obesity | LEC | *ISOC2* | 2.543621078 | 1.51E-05 | 0.000105108 | 0.09504582 |
| Obesity | LEC | *EPN1* | 2.158237913 | 8.84E-06 | 0.000105108 | 0.055594529 |
| Obesity | LEC | *ADCK1* | 1.346994739 | 1.37E-05 | 0.000105108 | 0.086137262 |
| Obesity | LEC | *HAGH* | 1.286937005 | 0.003607444 | 0.005270079 | 1 |
| Obesity | LEC | *CASP8* | 1.346158869 | 7.99E-05 | 0.000208081 | 0.502307313 |
| Obesity | LEC | *TM7SF3* | 2.026585182 | 0.000385891 | 0.000716508 | 1 |
| Obesity | LEC | *DMRT3* | 1.000365978 | 2.75E-05 | 0.00011384 | 0.172733324 |
| Obesity | LEC | *ST3GAL6* | 0.755843532 | 0.037218449 | 0.046197905 | 1 |
| Obesity | LEC | *ATP2C2* | -1.497411081 | 0.000465618 | 0.000839881 | 1 |
| Obesity | LEC | *TAF2* | 1.858380492 | 0.000186742 | 0.000394639 | 1 |
| Obesity | LEC | *TMEM161A* | 1.606996788 | 5.24E-06 | 9.34E-05 | 0.032953174 |
| Obesity | LEC | *LPAR2* | 0.821037832 | 6.03E-06 | 9.93E-05 | 0.037927559 |
| Obesity | LEC | *SUGP2* | 1.351656386 | 0.003340052 | 0.004917776 | 1 |
| Obesity | LEC | *ABCA7* | 1.120001556 | 3.92E-05 | 0.000130236 | 0.246220423 |
| Obesity | LEC | *BTBD1* | 2.125322626 | 5.03E-05 | 0.000152613 | 0.316214654 |
| Obesity | LEC | *CHI3L2* | 1.067136657 | 3.53E-05 | 0.000122918 | 0.222236266 |
| Obesity | LEC | *PMS1* | 1.669889976 | 2.52E-05 | 0.000111345 | 0.158232572 |
| Obesity | LEC | *ANKS1A* | 1.274413423 | 0.016223963 | 0.021241161 | 1 |
| Obesity | LEC | *NTHL1* | -1.221057118 | 0.000297093 | 0.0005756 | 1 |
| Obesity | LEC | *UHRF1BP1* | 2.89877888 | 1.34E-24 | 8.45E-21 | 8.45E-21 |
| Obesity | LEC | *IPO5* | 2.111229142 | 5.44E-05 | 0.000160812 | 0.341725757 |
| Obesity | LEC | *ANKRD44* | 1.206711041 | 5.87E-06 | 9.81E-05 | 0.03688865 |
| Obesity | LEC | *NDUFB4* | 2.606744503 | 0.001785434 | 0.002777784 | 1 |
| Obesity | LEC | *ZC3H15* | 1.49398351 | 5.39E-05 | 0.000159836 | 0.338852171 |
| Obesity | LEC | *MAP2K4* | 1.744522387 | 8.77E-05 | 0.000222231 | 0.551391285 |
| Obesity | LEC | *TMEM206* | 2.045668481 | 1.94E-05 | 0.000105217 | 0.122049808 |
| Obesity | LEC | *SEC61A2* | 2.066336922 | 2.14E-05 | 0.000107417 | 0.134701477 |
| Obesity | LEC | *ASB1* | 1.181533883 | 8.24E-05 | 0.000212956 | 0.518335316 |
| Obesity | LEC | *TBC1D1* | 1.944549162 | 0.000337029 | 0.000640151 | 1 |
| Obesity | LEC | *PPP2R5A* | 1.495893656 | 0.003611748 | 0.005274578 | 1 |
| Obesity | LEC | *TIE1* | -0.324723474 | 6.60E-05 | 0.000182779 | 0.414976949 |
| Obesity | LEC | *NFYC* | 1.595736448 | 0.001668238 | 0.002610956 | 1 |
| Obesity | LEC | *ZMYND12* | 0.731098789 | 0.001129097 | 0.001834746 | 1 |
| Obesity | LEC | *SLC9A3* | 0.905628302 | 0.012358384 | 0.016440364 | 1 |
| Obesity | LEC | *NGEF* | 1.96863929 | 2.07E-05 | 0.000106696 | 0.130382807 |
| Obesity | LEC | *CD84* | 0.853053813 | 0.000117448 | 0.000275315 | 0.738393591 |
| Obesity | LEC | *ZNRD1* | 1.134468198 | 0.011760121 | 0.015731039 | 1 |
| Obesity | LEC | *GOLGA5* | 1.949270478 | 0.003942831 | 0.005716923 | 1 |
| Obesity | LEC | *EML1* | 0.990581604 | 0.000146077 | 0.000325439 | 0.918388229 |
| Obesity | LEC | *ZFAT* | 0.761865108 | 4.29E-05 | 0.000137948 | 0.269857544 |
| Obesity | LEC | *MTFR1* | 1.443339197 | 0.000424482 | 0.00077692 | 1 |
| Obesity | LEC | *STAG3* | 1.900905494 | 0.000395038 | 0.000731156 | 1 |

| | | | | | | |
|---|---|---|---|---|---|---|
| Obesity | LEC | *MYO9A* | 1.580774647 | 7.97E-05 | 0.000207712 | 0.501000249 |
| Obesity | LEC | *SP100* | 1.556607258 | 0.001053258 | 0.001726683 | 1 |
| Obesity | LEC | *STOML1* | 2.916730625 | 2.02E-05 | 0.000106047 | 0.127259079 |
| Obesity | LEC | *METTL22* | 1.807831382 | 0.036763723 | 0.045642481 | 1 |
| Obesity | LEC | *TP53BP1* | 2.122670739 | 2.64E-05 | 0.000112582 | 0.165841202 |
| Obesity | LEC | *RRP15* | 0.773358563 | 0.004866369 | 0.006929753 | 1 |
| Obesity | LEC | *SYT1* | -0.344640689 | 0.000375826 | 0.000700094 | 1 |
| Obesity | LEC | *ROGDI* | 0.961368021 | 2.93E-05 | 0.000115648 | 0.18430199 |
| Obesity | LEC | *RASSF1* | 1.889285404 | 0.001727777 | 0.00269475 | 1 |
| Obesity | LEC | *FGFR3* | -0.39218296 | 0.000107914 | 0.000257479 | 0.678455849 |
| Obesity | LEC | *IFI35* | 1.251373479 | 7.59E-06 | 0.000105108 | 0.047739267 |
| Obesity | LEC | *HEATR6* | 1.590643925 | 1.12E-05 | 0.000105108 | 0.070436286 |
| Obesity | LEC | *POLR1A* | 0.611452899 | 0.0038429 | 0.005587491 | 1 |
| Obesity | LEC | *SRBD1* | 1.594238122 | 3.55E-05 | 0.000123048 | 0.223331809 |
| Obesity | LEC | *CYFIP1* | 1.809693167 | 0.000495354 | 0.000886 | 1 |
| Obesity | LEC | *PSME4* | 1.579050132 | 0.000268174 | 0.000531026 | 1 |
| Obesity | LEC | *PITX1* | 4.118094121 | 1.74E-05 | 0.000105108 | 0.10926705 |
| Obesity | LEC | *SDK2* | -1.168926746 | 0.000919973 | 0.001526491 | 1 |
| Obesity | LEC | *NUCKS1* | 1.825674786 | 0.000583303 | 0.001020943 | 1 |
| Obesity | LEC | *DNAJA2* | 1.81979644 | 1.35E-05 | 0.000105108 | 0.084674426 |
| Obesity | LEC | *KCNAB2* | 0.930613292 | 8.32E-05 | 0.000214185 | 0.522825997 |
| Obesity | LEC | *GAL* | -0.903972057 | 0.000156047 | 0.000342227 | 0.981068197 |
| Obesity | LEC | *CLEC2D* | 1.251262164 | 0.000318442 | 0.000609451 | 1 |
| Obesity | LEC | *NEDD4* | -0.798165716 | 0.02081642 | 0.026834701 | 1 |
| Obesity | LEC | *PIGB* | 1.798394305 | 0.000126727 | 0.000291097 | 0.796732125 |
| Obesity | LEC | *GNB5* | -0.882982632 | 0.011839149 | 0.015823284 | 1 |
| Obesity | LEC | *PFN2* | 1.901823759 | 4.68E-05 | 0.000145967 | 0.294270298 |
| Obesity | LEC | *DAPP1* | 1.074823787 | 3.49E-05 | 0.000122228 | 0.219277475 |
| Obesity | LEC | *FGF10* | 1.175323862 | 8.71E-05 | 0.000220912 | 0.547419693 |
| Obesity | LEC | *TMEM260* | 1.426434983 | 1.83E-05 | 0.000105108 | 0.115091456 |
| Obesity | LEC | *CLTCL1* | 1.875013528 | 0.000375422 | 0.000699739 | 1 |
| Obesity | LEC | *DGCR2* | 1.551034683 | 0.019123408 | 0.024809919 | 1 |
| Obesity | LEC | *MNT* | 0.594393223 | 0.000141697 | 0.000317678 | 0.890847547 |
| Obesity | LEC | *ZXDC* | 1.984967466 | 0.000126485 | 0.000290753 | 0.795210616 |
| Obesity | LEC | *POLB* | -1.057569597 | 0.008517347 | 0.011635693 | 1 |
| Obesity | LEC | *WIPI1* | 1.554358053 | 1.61E-05 | 0.000105108 | 0.101474222 |
| Obesity | LEC | *NDST1* | 2.131321406 | 0.000151688 | 0.0003355 | 0.953661479 |
| Obesity | LEC | *ASNS* | 1.799416905 | 4.89E-06 | 9.11E-05 | 0.030739712 |
| Obesity | LEC | *AP3M2* | 1.630656562 | 0.031415276 | 0.039391272 | 1 |
| Obesity | LEC | *ST6GALNAC2* | 0.77866343 | 1.13E-05 | 0.000105108 | 0.071164407 |
| Obesity | LEC | *EIF2B3* | 1.153436795 | 1.51E-05 | 0.000105108 | 0.094815037 |
| Obesity | LEC | *CAMK2A* | 3.319541104 | 3.02E-05 | 0.000115831 | 0.190085001 |
| Obesity | LEC | *OSBPL3* | 1.736925149 | 2.27E-05 | 0.000109111 | 0.142854296 |
| Obesity | LEC | *RAD18* | 2.291838519 | 0.000134501 | 0.000304944 | 0.84560987 |
| Obesity | LEC | *SNX13* | 1.689787169 | 0.000134683 | 0.000305246 | 0.846752245 |
| Obesity | LEC | *VASH1* | 1.485545172 | 2.69E-05 | 0.000113184 | 0.168932991 |
| Obesity | LEC | *TRIP13* | 1.656979751 | 3.03E-05 | 0.000115831 | 0.190727785 |
| Obesity | LEC | *MBD3* | 3.160867728 | 3.43E-06 | 7.73E-05 | 0.021534246 |
| Obesity | LEC | *HLTF* | 1.026073355 | 0.002783897 | 0.004155356 | 1 |
| Obesity | LEC | *CYBRD1* | 1.29551551 | 1.38E-05 | 0.000105108 | 0.08656158 |

| | | | | | | |
|---|---|---|---|---|---|---|
| Obesity | LEC | *EPN2* | 1.246797956 | 0.031794225 | 0.039779163 | 1 |
| Obesity | LEC | *LIMS2* | 1.771428312 | 0.003394461 | 0.004990873 | 1 |
| Obesity | LEC | *AFF4* | 1.719569042 | 3.50E-05 | 0.000122546 | 0.22033688 |
| Obesity | LEC | *TRHDE* | 0.556372851 | 1.84E-05 | 0.000105108 | 0.115927506 |
| Obesity | LEC | *NFATC3* | 1.215770121 | 0.009206726 | 0.012512483 | 1 |
| Obesity | LEC | *TRNT1* | 1.997091767 | 4.42E-05 | 0.000140994 | 0.278151527 |
| Obesity | LEC | *ACADVL* | 1.061364967 | 2.76E-05 | 0.00011384 | 0.173513706 |
| Obesity | LEC | *CRMP1* | 1.522073964 | 0.019436759 | 0.025159338 | 1 |
| Obesity | LEC | *DERL2* | 2.068230933 | 0.009337898 | 0.012677039 | 1 |
| Obesity | LEC | *NDE1* | 1.84211148 | 1.02E-05 | 0.000105108 | 0.064191054 |
| Obesity | LEC | *AP1M1* | 1.29114422 | 0.003262001 | 0.004813002 | 1 |
| Obesity | LEC | *PVR* | 1.651114988 | 0.021737201 | 0.027895852 | 1 |
| Obesity | LEC | *XRCC1* | 1.090969826 | 3.02E-05 | 0.000115831 | 0.189900237 |
| Obesity | LEC | *MOV10L1* | -0.260876426 | 0.000171759 | 0.000368675 | 1 |
| Obesity | LEC | *TP63* | -1.087190225 | 0.008102593 | 0.011098257 | 1 |
| Obesity | LEC | *ALPK1* | 1.281035889 | 9.80E-06 | 0.000105108 | 0.061626134 |
| Obesity | LEC | *LLGL2* | 0.904438786 | 2.55E-05 | 0.000111795 | 0.160426195 |
| Obesity | LEC | *PDE8A* | 1.48600204 | 1.34E-05 | 0.000105108 | 0.084480549 |
| Obesity | LEC | *NLE1* | 1.018538686 | 4.09E-05 | 0.000133262 | 0.256992506 |
| Obesity | LEC | *SDHA* | 2.127322567 | 5.55E-06 | 9.59E-05 | 0.034907614 |
| Obesity | LEC | *GSDMB* | 0.717574884 | 0.000152819 | 0.000337113 | 0.960771099 |
| Obesity | LEC | *ADAM11* | 0.758088158 | 0.01920378 | 0.024903911 | 1 |
| Obesity | LEC | *PICALM* | 1.245669288 | 3.49E-05 | 0.000122308 | 0.219541997 |
| Obesity | LEC | *GLI2* | 0.537366617 | 0.000465566 | 0.000839881 | 1 |
| Obesity | LEC | *CLNS1A* | 1.566954858 | 2.32E-05 | 0.000109118 | 0.145580252 |
| Obesity | LEC | *ATP2A3* | 0.802077804 | 1.54E-05 | 0.000105108 | 0.097027795 |
| Obesity | LEC | *CA12* | 0.585608444 | 2.88E-05 | 0.000115038 | 0.180810896 |
| Obesity | LEC | *NTN4* | -0.87531916 | 0.008458629 | 0.01156074 | 1 |
| Obesity | LEC | *LMAN1* | 1.904748547 | 0.001354041 | 0.002161171 | 1 |
| Obesity | LEC | *ENO1* | 1.993135862 | 0.004575655 | 0.006558027 | 1 |
| Obesity | LEC | *TIPIN* | 1.24791943 | 7.39E-07 | 3.69E-05 | 0.004643588 |
| Obesity | LEC | *SRI* | 1.74681124 | 1.40E-05 | 0.000105108 | 0.088213521 |
| Obesity | LEC | *EIF4G3* | 1.258804404 | 0.00674926 | 0.009393978 | 1 |
| Obesity | LEC | *TTC38* | 1.637728456 | 4.82E-05 | 0.000148518 | 0.303124879 |
| Obesity | LEC | *GRAMD4* | 1.63689447 | 2.52E-05 | 0.000111345 | 0.158203883 |
| Obesity | LEC | *CELSR1* | -0.190076517 | 9.79E-05 | 0.000240947 | 0.615618403 |
| Obesity | LEC | *ADD2* | -1.50154196 | 0.000698356 | 0.001195036 | 1 |
| Obesity | LEC | *VPS9D1* | 2.424508535 | 0.003031366 | 0.004499102 | 1 |
| Obesity | LEC | *MARK3* | 1.124476683 | 0.001039137 | 0.00170665 | 1 |
| Obesity | LEC | *SLC25A3* | 2.158660348 | 1.75E-05 | 0.000105108 | 0.109991818 |
| Obesity | LEC | *FSCN1* | -0.563505293 | 7.64E-05 | 0.000201692 | 0.480228066 |
| Obesity | LEC | *MOCOS* | 0.984843425 | 1.62E-05 | 0.000105108 | 0.102150972 |
| Obesity | LEC | *SEC31B* | 0.780323684 | 9.88E-05 | 0.00024246 | 0.620939127 |
| Obesity | LEC | *TUBA3D* | 2.421362863 | 2.33E-05 | 0.000109118 | 0.146276025 |
| Obesity | LEC | *MKRN2* | 2.878858407 | 9.25E-06 | 0.000105108 | 0.058153239 |
| Obesity | LEC | *MCM6* | 1.802428344 | 1.66E-05 | 0.000105108 | 0.104138577 |
| Obesity | LEC | *RBMS2* | 1.291286393 | 0.030066912 | 0.037828833 | 1 |
| Obesity | LEC | *MLH1* | 1.567175197 | 0.003534898 | 0.00517919 | 1 |
| Obesity | LEC | *FMO4* | 2.045021505 | 2.85E-05 | 0.000114887 | 0.179388103 |
| Obesity | LEC | *RGS11* | 0.728350785 | 0.000946627 | 0.001565345 | 1 |

| | | | | | | |
|---|---|---|---|---|---|---|
| Obesity | LEC | *TPD52* | 1.417054439 | 0.001798806 | 0.002796511 | 1 |
| Obesity | LEC | *GPATCH1* | 2.011385829 | 4.57E-06 | 8.97E-05 | 0.028751011 |
| Obesity | LEC | *ICAM3* | 1.563537669 | 0.000105862 | 0.000254223 | 0.665557079 |
| Obesity | LEC | *NT5C2* | 1.917471458 | 2.00E-05 | 0.000105734 | 0.125505826 |
| Obesity | LEC | *CAMSAP3* | -1.791838372 | 0.000107965 | 0.000257502 | 0.678774515 |
| Obesity | LEC | *XAB2* | -2.4282256 | 0.001183359 | 0.001915495 | 1 |
| Obesity | LEC | *STXBP2* | 1.546099036 | 0.006474698 | 0.009041853 | 1 |
| Obesity | LEC | *MAP2K7* | 1.872444676 | 1.59E-05 | 0.000105108 | 0.10004231 |
| Obesity | LEC | *NMRK2* | 4.029506435 | 0.000120741 | 0.000281252 | 0.759099766 |
| Obesity | LEC | *CTTNBP2* | -0.916466181 | 0.022306442 | 0.028573879 | 1 |
| Obesity | LEC | *TM9SF3* | 1.825178901 | 0.000173714 | 0.000372489 | 1 |
| Obesity | LEC | *DNAJC10* | 2.043053757 | 0.000697924 | 0.001194622 | 1 |
| Obesity | LEC | *DYNC1I2* | 1.657426509 | 0.004729543 | 0.006757872 | 1 |
| Obesity | LEC | *APBB1IP* | 0.824545509 | 2.55E-05 | 0.000111732 | 0.160222975 |
| Obesity | LEC | *LRCH4* | 0.929980235 | 0.000208356 | 0.000430616 | 1 |
| Obesity | LEC | *SIRT6* | 3.756454241 | 7.32E-05 | 0.000195742 | 0.460213213 |
| Obesity | LEC | *CAPZB* | 1.641015305 | 0.000209226 | 0.000431845 | 1 |
| Obesity | LEC | *GPR137B* | 1.051370924 | 0.000566133 | 0.000995046 | 1 |
| Obesity | LEC | *NAALAD2* | 1.533716184 | 2.79E-05 | 0.000113898 | 0.175630942 |
| Obesity | LEC | *FGFR1* | 1.63353008 | 0.000446364 | 0.000809663 | 1 |
| Obesity | LEC | *FBLN1* | 0.784312626 | 9.67E-06 | 0.000105108 | 0.06078479 |
| Obesity | LEC | *PIAS2* | 1.506466759 | 0.00026613 | 0.000527477 | 1 |
| Obesity | LEC | *AMPH* | 0.82943219 | 7.10E-05 | 0.000191374 | 0.4466664 |
| Obesity | LEC | *MCCC1* | 1.079986548 | 2.11E-06 | 6.24E-05 | 0.013295549 |
| Obesity | LEC | *FAP* | 0.461533706 | 6.22E-05 | 0.000175786 | 0.391291739 |
| Obesity | LEC | *ACER3* | 1.434874993 | 2.14E-06 | 6.25E-05 | 0.013439415 |
| Obesity | LEC | *PIK3C3* | 1.266783144 | 0.026104678 | 0.033128807 | 1 |
| Obesity | LEC | *N4BP2* | 1.483763169 | 0.000185474 | 0.000392619 | 1 |
| Obesity | LEC | *SYNJ2* | 1.591788287 | 5.21E-05 | 0.000155937 | 0.327622675 |
| Obesity | LEC | *ZCWPW1* | 1.14842541 | 2.65E-06 | 6.98E-05 | 0.016674693 |
| Obesity | LEC | *FGF20* | -6.532662334 | 0.000271993 | 0.000537236 | 1 |
| Obesity | LEC | *PCM1* | 1.374113209 | 0.000213402 | 0.00043906 | 1 |
| Obesity | LEC | *FKBP7* | 0.73364359 | 0.019396125 | 0.025117084 | 1 |
| Obesity | LEC | *OSBPL6* | -1.01072124 | 0.003639288 | 0.005311096 | 1 |
| Obesity | LEC | *XRCC5* | 1.68807295 | 0.000287911 | 0.000563015 | 1 |
| Obesity | LEC | *REXO1* | 2.140160756 | 1.95E-05 | 0.000105217 | 0.122675225 |
| Obesity | LEC | *SAR1A* | 1.251530848 | 1.16E-05 | 0.000105108 | 0.072617088 |
| Obesity | LEC | *RAPGEF3* | 0.571430278 | 0.006267473 | 0.008775858 | 1 |
| Obesity | LEC | *FDFT1* | 1.779047404 | 8.40E-06 | 0.000105108 | 0.052829471 |
| Obesity | LEC | *SCGN* | 5.895058588 | 3.36E-06 | 7.66E-05 | 0.02111141 |
| Obesity | LEC | *DDX1* | 1.915870614 | 6.73E-05 | 0.000184822 | 0.423243373 |
| Obesity | LEC | *DNM2* | 1.65456899 | 6.56E-05 | 0.000182036 | 0.412492725 |
| Obesity | LEC | *RIMS1* | 1.469262474 | 3.15E-05 | 0.000116847 | 0.19797205 |
| Obesity | LEC | *STX7* | 1.145734974 | 0.017871532 | 0.02327705 | 1 |
| Obesity | LEC | *DDX43* | 1.289537317 | 7.62E-05 | 0.000201393 | 0.479115113 |
| Obesity | LEC | *DCT* | 1.024621423 | 2.35E-05 | 0.000109118 | 0.14744901 |
| Obesity | LEC | *SLC35C2* | 1.573172889 | 1.53E-05 | 0.000105108 | 0.096024309 |
| Obesity | LEC | *SCTR* | 0.660815571 | 2.79E-05 | 0.000113866 | 0.1754679 |
| Obesity | LEC | *RIF1* | 1.558072387 | 7.55E-05 | 0.000200408 | 0.474966116 |
| Obesity | LEC | *KCNN2* | -2.982544635 | 0.001211801 | 0.001956999 | 1 |

| | | | | | | |
|---|---|---|---|---|---|---|
| Obesity | LEC | *PSEN1* | 1.521098914 | 6.91E-05 | 0.000187781 | 0.434150729 |
| Obesity | LEC | *CPOX* | 2.167052421 | 6.79E-05 | 0.000186128 | 0.427162752 |
| Obesity | LEC | *MOK* | 2.0076233 | 2.58E-05 | 0.000112124 | 0.162481971 |
| Obesity | LEC | *RSBN1* | 1.601294541 | 0.000519269 | 0.00092138 | 1 |
| Obesity | LEC | *MAGI3* | 1.581562108 | 0.002093787 | 0.003215997 | 1 |
| Obesity | LEC | *COL4A4* | 0.630312353 | 1.27E-05 | 0.000105108 | 0.080032807 |
| Obesity | LEC | *STK17B* | 1.037048208 | 1.53E-05 | 0.000105108 | 0.096323146 |
| Obesity | LEC | *ZNF506* | -0.974516937 | 0.031312765 | 0.039278402 | 1 |
| Obesity | LEC | *PCDHGA2* | 1.589000622 | 0.016052911 | 0.021025969 | 1 |
| Obesity | LEC | *HSPB11* | 1.617963171 | 0.016741898 | 0.021873715 | 1 |
| Obesity | LEC | *WDR70* | 1.46919455 | 3.51E-05 | 0.000122587 | 0.220656291 |
| Obesity | LEC | *PGR* | 1.640644233 | 6.37E-05 | 0.000178168 | 0.400402865 |
| Obesity | LEC | *C5orf22* | 2.623747014 | 0.000351466 | 0.000661578 | 1 |
| Obesity | LEC | *COBLL1* | 1.498614109 | 1.54E-07 | 1.43E-05 | 0.000969181 |
| Obesity | LEC | *GEMIN5* | 1.370900503 | 0.002323215 | 0.00353059 | 1 |
| Obesity | LEC | *GSK3B* | 1.113503214 | 0.025577538 | 0.032525481 | 1 |
| Obesity | LEC | *ERC1* | 1.30471155 | 3.83E-05 | 0.000128424 | 0.240926177 |
| Obesity | LEC | *DOPEY1* | 1.595872937 | 1.13E-05 | 0.000105108 | 0.071269102 |
| Obesity | LEC | *LYRM2* | 1.52278828 | 0.002442352 | 0.003696453 | 1 |
| Obesity | LEC | *ULK2* | 1.735946511 | 0.000442286 | 0.000804048 | 1 |
| Obesity | LEC | *PLOD1* | 1.790842126 | 7.98E-06 | 0.000105108 | 0.050190224 |
| Obesity | LEC | *PIBF1* | 1.331346724 | 0.005099626 | 0.007245502 | 1 |
| Obesity | LEC | *TDRD3* | 1.579610476 | 0.000934897 | 0.001548393 | 1 |
| Obesity | LEC | *PDS5B* | 1.623338397 | 0.000588747 | 0.001028958 | 1 |
| Obesity | LEC | *CYLD* | 1.599431542 | 5.16E-05 | 0.000155003 | 0.324420878 |
| Obesity | LEC | *ZNF324* | 1.526915062 | 1.47E-05 | 0.000105108 | 0.092675924 |
| Obesity | LEC | *ZNF586* | 1.734604407 | 6.93E-05 | 0.000188201 | 0.435872663 |
| Obesity | LEC | *ZNF264* | 1.660597813 | 0.022019523 | 0.028235109 | 1 |
| Obesity | LEC | *RPS5* | 2.249139053 | 0.000145902 | 0.000325164 | 0.917286274 |
| Obesity | LEC | *CHMP2B* | 1.611378725 | 2.90E-05 | 0.000115266 | 0.182596497 |
| Obesity | LEC | *PPIE* | 2.237486052 | 7.82E-07 | 3.74E-05 | 0.004919509 |
| Obesity | LEC | *ZMPSTE24* | 1.802528823 | 0.015361555 | 0.02018351 | 1 |
| Obesity | LEC | *NOA1* | -1.25461714 | 0.000474952 | 0.00085437 | 1 |
| Obesity | LEC | *GSTP1* | 2.651173602 | 3.89E-05 | 0.000129587 | 0.244406907 |
| Obesity | LEC | *WBP11* | 1.484480307 | 1.39E-05 | 0.000105108 | 0.087675888 |
| Obesity | LEC | *COL16A1* | 0.582980376 | 0.004336697 | 0.006253397 | 1 |
| Obesity | LEC | *APOB* | 1.057143303 | 9.02E-05 | 0.000226751 | 0.567330153 |
| Obesity | LEC | *NCOA1* | 1.410627005 | 0.000107839 | 0.000257479 | 0.677982438 |
| Obesity | LEC | *CD59* | -0.770597126 | 0.004951315 | 0.007042741 | 1 |
| Obesity | LEC | *CD82* | 1.901030781 | 1.45E-06 | 5.18E-05 | 0.009109518 |
| Obesity | LEC | *TAF9* | -1.406169384 | 0.005956162 | 0.008366039 | 1 |
| Obesity | LEC | *FCN1* | 0.817399022 | 0.0001193 | 0.000278619 | 0.750042109 |
| Obesity | LEC | *SCAMP1* | 2.141833806 | 1.37E-06 | 5.05E-05 | 0.008627343 |
| Obesity | LEC | *HACE1* | 1.621756619 | 1.20E-05 | 0.000105108 | 0.07545248 |
| Obesity | LEC | *SEH1L* | 1.866468831 | 1.80E-05 | 0.000105108 | 0.113360296 |
| Obesity | LEC | *OVGP1* | -1.808269542 | 0.000131716 | 0.000300471 | 0.828097779 |
| Obesity | LEC | *SLC25A24* | 1.744012131 | 0.000574464 | 0.001008561 | 1 |
| Obesity | LEC | *PILRA* | 0.475900984 | 3.04E-05 | 0.000115831 | 0.191235999 |
| Obesity | LEC | *ABCB1* | 0.721400017 | 2.11E-05 | 0.000106989 | 0.132366899 |
| Obesity | LEC | *ZNF213* | 1.0349038 | 0.022147037 | 0.028387038 | 1 |

| | | | | | | |
|---|---|---|---|---|---|---|
| Obesity | LEC | *RRN3* | 2.124081621 | 0.000663191 | 0.001139203 | 1 |
| Obesity | LEC | *EPS15* | 0.865964664 | 0.018183599 | 0.023663898 | 1 |
| Obesity | LEC | *ATG16L1* | 1.704853624 | 1.82E-05 | 0.000105108 | 0.11468523 |
| Obesity | LEC | *USP40* | 1.678408728 | 3.30E-05 | 0.000119154 | 0.207661067 |
| Obesity | LEC | *MAST2* | 1.792983881 | 6.24E-06 | 9.96E-05 | 0.039208175 |
| Obesity | LEC | *B4GALT1* | 1.477276849 | 2.12E-05 | 0.00010722 | 0.133558661 |
| Obesity | LEC | *CHMP5* | 1.964604112 | 1.14E-05 | 0.000105108 | 0.071479251 |
| Obesity | LEC | *FOLH1* | 0.799877537 | 0.000345392 | 0.000652144 | 1 |
| Obesity | LEC | *EIF2AK1* | 1.562842148 | 2.36E-05 | 0.000109118 | 0.148280593 |
| Obesity | LEC | *SEPHS1* | -1.179933108 | 0.0131481 | 0.017424558 | 1 |
| Obesity | LEC | *ITPKC* | 0.975957967 | 1.23E-05 | 0.000105108 | 0.077549825 |
| Obesity | LEC | *CEACAM6* | -6.343443398 | 0.001827807 | 0.002837388 | 1 |
| Obesity | LEC | *TMED2* | 1.844901848 | 1.39E-05 | 0.000105108 | 0.087513768 |
| Obesity | LEC | *ACOX3* | 1.951158027 | 0.001145449 | 0.001859875 | 1 |
| Obesity | LEC | *HSD17B14* | 0.770437111 | 4.01E-05 | 0.000131769 | 0.25203375 |
| Obesity | LEC | *TRIP6* | 0.761566346 | 0.000130976 | 0.000299435 | 0.82344571 |
| Obesity | LEC | *ACHE* | -1.286555614 | 2.07E-05 | 0.000106696 | 0.130265535 |
| Obesity | LEC | *SRRT* | 2.161776167 | 1.37E-05 | 0.000105108 | 0.085872966 |
| Obesity | LEC | *PIGS* | 1.170510909 | 0.000306662 | 0.000591225 | 1 |
| Obesity | LEC | *ATXN7L3* | 1.170537991 | 1.05E-05 | 0.000105108 | 0.066270983 |
| Obesity | LEC | *PGS1* | 1.328664191 | 2.42E-05 | 0.000109361 | 0.151902633 |
| Obesity | LEC | *PSMC5* | 1.487535085 | 0.004081186 | 0.005903916 | 1 |
| Obesity | LEC | *LPCAT2* | 0.952283628 | 3.80E-05 | 0.000127833 | 0.238919832 |
| Obesity | LEC | *OGFOD1* | -1.619219258 | 0.016602179 | 0.021718248 | 1 |
| Obesity | LEC | *TXNDC16* | 1.609398513 | 0.000142645 | 0.000319376 | 0.896806806 |
| Obesity | LEC | *GMCL1* | 2.175826304 | 0.000247611 | 0.000497995 | 1 |
| Obesity | LEC | *KLHL42* | 1.882650229 | 0.008585912 | 0.011716872 | 1 |
| Obesity | LEC | *PTHLH* | 2.763146378 | 4.44E-07 | 2.59E-05 | 0.002791608 |
| Obesity | LEC | *PHACTR3* | 0.534030131 | 0.00022449 | 0.000458236 | 1 |
| Obesity | LEC | *METTL2A* | 0.910362081 | 9.39E-05 | 0.000233799 | 0.590577426 |
| Obesity | LEC | *GP6* | 1.4657737 | 5.68E-05 | 0.000165989 | 0.357373446 |
| Obesity | LEC | *DDX18* | 1.952082007 | 0.0001037 | 0.000250928 | 0.651961113 |
| Obesity | LEC | *EDEM2* | 1.180983189 | 0.030505262 | 0.038326655 | 1 |
| Obesity | LEC | *TPX2* | -0.864536424 | 0.023664993 | 0.030197242 | 1 |
| Obesity | LEC | *DOCK3* | 1.23376302 | 3.58E-06 | 7.81E-05 | 0.022504781 |
| Obesity | LEC | *C3orf18* | 1.164069026 | 1.03E-07 | 1.09E-05 | 0.000646736 |
| Obesity | LEC | *NSFL1C* | 1.255234489 | 4.61E-06 | 9.00E-05 | 0.029012587 |
| Obesity | LEC | *CPXM1* | 0.477983155 | 0.004991344 | 0.007094863 | 1 |
| Obesity | LEC | *MAVS* | 1.946178572 | 2.37E-05 | 0.000109118 | 0.149073673 |
| Obesity | LEC | *LZTS3* | 2.473474484 | 7.85E-05 | 0.000205645 | 0.493547682 |
| Obesity | LEC | *XRN2* | 1.846127944 | 0.000849369 | 0.0014187 | 1 |
| Obesity | LEC | *TESC* | 2.159069184 | 1.33E-05 | 0.000105108 | 0.083567304 |
| Obesity | LEC | *RBBP9* | 2.677579999 | 0.000885123 | 0.00147255 | 1 |
| Obesity | LEC | *ANAPC5* | 1.529896059 | 0.002946574 | 0.004383054 | 1 |
| Obesity | LEC | *SLC23A2* | 1.62578173 | 2.04E-05 | 0.000106095 | 0.128268499 |
| Obesity | LEC | *TMEM230* | 1.493658491 | 4.65E-05 | 0.000145571 | 0.292503083 |
| Obesity | LEC | *TASP1* | 1.723656363 | 3.21E-05 | 0.00011749 | 0.201620798 |
| Obesity | LEC | *OAS1* | 1.298854921 | 2.33E-05 | 0.000109118 | 0.146781648 |
| Obesity | LEC | *KIF16B* | 1.681090174 | 3.02E-05 | 0.000115831 | 0.189618478 |
| Obesity | LEC | *FUS* | 1.673445915 | 0.00024838 | 0.000499061 | 1 |

| | | | | | | |
|---|---|---|---|---|---|---|
| Obesity | LEC | *ZNF302* | 0.738747948 | 0.026643849 | 0.033772153 | 1 |
| Obesity | LEC | *FXYD3* | 3.243650665 | 1.80E-05 | 0.000105108 | 0.113280384 |
| Obesity | LEC | *GANAB* | 2.269372304 | 3.64E-05 | 0.000124476 | 0.228537434 |
| Obesity | LEC | *GMIP* | 0.985344102 | 2.02E-05 | 0.000106047 | 0.127275706 |
| Obesity | LEC | *NECAP1* | 1.262001506 | 0.000104052 | 0.000251238 | 0.654173746 |
| Obesity | LEC | *ANKRD24* | 0.852038925 | 1.33E-05 | 0.000105108 | 0.083550018 |
| Obesity | LEC | *GPATCH2L* | 1.470296563 | 0.000562446 | 0.000988842 | 1 |
| Obesity | LEC | *SLC9A1* | 1.366096698 | 1.51E-05 | 0.000105108 | 0.095200973 |
| Obesity | LEC | *SPTLC1* | 1.597088614 | 1.43E-05 | 0.000105108 | 0.089830752 |
| Obesity | LEC | *PAPOLA* | 1.237806594 | 0.02764929 | 0.034954975 | 1 |
| Obesity | LEC | *RGS1* | 0.825972626 | 3.10E-05 | 0.000116539 | 0.194630623 |
| Obesity | LEC | *YPEL3* | 2.311333511 | 7.21E-12 | 3.24E-09 | 4.53E-08 |
| Obesity | LEC | *MRPS33* | 1.13153663 | 2.51E-05 | 0.000111345 | 0.158084395 |
| Obesity | LEC | *MAEA* | 1.140073969 | 0.031975715 | 0.039982363 | 1 |
| Obesity | LEC | *STRN4* | 2.063614492 | 0.000154744 | 0.000340286 | 0.972876411 |
| Obesity | LEC | *LYZ* | 0.672054496 | 8.19E-06 | 0.000105108 | 0.051486229 |
| Obesity | LEC | *MUL1* | 1.665023945 | 5.99E-05 | 0.000171669 | 0.376522368 |
| Obesity | LEC | *GNPTG* | 1.177196873 | 1.41E-05 | 0.000105108 | 0.088641629 |
| Obesity | LEC | *GOLGA3* | 1.428798519 | 1.63E-05 | 0.000105108 | 0.102422137 |
| Obesity | LEC | *CERS4* | 2.003576284 | 0.000757472 | 0.001283619 | 1 |
| Obesity | LEC | *USP48* | 1.181297032 | 0.014272788 | 0.018815898 | 1 |
| Obesity | LEC | *PDPR* | 1.568177648 | 1.21E-05 | 0.000105108 | 0.076033178 |
| Obesity | LEC | *EXOC1* | 1.705834125 | 0.000223449 | 0.000457151 | 1 |
| Obesity | LEC | *NLRC4* | 0.908479379 | 2.07E-05 | 0.000106696 | 0.12987263 |
| Obesity | LEC | *SLC26A4* | -0.154931433 | 8.03E-05 | 0.000208625 | 0.504663697 |
| Obesity | LEC | *DLD* | 1.674760879 | 1.07E-05 | 0.000105108 | 0.067389465 |
| Obesity | LEC | *TXNL1* | 1.792683966 | 1.22E-05 | 0.000105108 | 0.076894628 |
| Obesity | LEC | *IL5RA* | -1.630739058 | 0.000276596 | 0.000545299 | 1 |
| Obesity | LEC | *ITGA6* | -0.423568258 | 0.000466969 | 0.000841696 | 1 |
| Obesity | LEC | *RAPGEF4* | -0.30854215 | 0.000156718 | 0.000343501 | 0.985288788 |
| Obesity | LEC | *SEL1L3* | 2.583899914 | 5.72E-05 | 0.000166529 | 0.359451556 |
| Obesity | LEC | *TF* | 2.204773843 | 0.000106593 | 0.000255389 | 0.670150376 |
| Obesity | LEC | *MYO15A* | 2.165481415 | 0.001778096 | 0.002767052 | 1 |
| Obesity | LEC | *ALKBH5* | 1.394179283 | 1.08E-06 | 4.46E-05 | 0.006786337 |
| Obesity | LEC | *NLRP1* | 1.123135293 | 3.16E-05 | 0.000116919 | 0.198669328 |
| Obesity | LEC | *SPAG7* | 1.407299115 | 2.38E-06 | 6.66E-05 | 0.014933967 |
| Obesity | LEC | *ZFHX4* | 1.230840065 | 0.025173589 | 0.032031239 | 1 |
| Obesity | LEC | *TMEM101* | 2.887393685 | 0.000750115 | 0.001273211 | 1 |
| Obesity | LEC | *CMA1* | -1.286993976 | 5.94E-05 | 0.00017072 | 0.373194044 |
| Obesity | LEC | *PPP2R3C* | 1.830118448 | 9.86E-06 | 0.000105108 | 0.062018869 |
| Obesity | LEC | *OSGEP* | 2.057457531 | 4.05E-05 | 0.000132652 | 0.254691071 |
| Obesity | LEC | *SLC22A17* | 1.696441649 | 0.000715391 | 0.00122219 | 1 |
| Obesity | LEC | *SCFD1* | 1.275824756 | 4.45E-05 | 0.000141409 | 0.279579573 |
| Obesity | LEC | *RPGRIP1* | -1.18525454 | 0.000608553 | 0.001059533 | 1 |
| Obesity | LEC | *TOX4* | 1.729104347 | 6.38E-06 | 0.00010078 | 0.040110279 |
| Obesity | LEC | *TGM1* | 1.13742914 | 0.009443868 | 0.012809838 | 1 |
| Obesity | LEC | *AGO1* | 1.08256454 | 1.56E-05 | 0.000105108 | 0.09779816 |
| Obesity | LEC | *RFFL* | 0.999534006 | 2.02E-05 | 0.000106047 | 0.127282282 |
| Obesity | LEC | *MFSD11* | 1.277795464 | 8.53E-05 | 0.000217791 | 0.535984643 |
| Obesity | LEC | *DPYSL2* | 1.365995947 | 0.018705137 | 0.024302376 | 1 |

| | | | | | | |
|---|---|---|---|---|---|---|
| Obesity | LEC | *NUP50* | 1.621862787 | 0.000165471 | 0.000358358 | 1 |
| Obesity | LEC | *COMT* | 1.653237482 | 1.96E-05 | 0.00010526 | 0.123154374 |
| Obesity | LEC | *VNN3* | 2.999629277 | 1.77E-05 | 0.000105108 | 0.111265252 |
| Obesity | LEC | *SEC22C* | 2.191824786 | 0.000587346 | 0.001027162 | 1 |
| Obesity | LEC | *FMO2* | 1.311026708 | 3.04E-06 | 7.33E-05 | 0.019112658 |
| Obesity | LEC | *SUCO* | 1.679592988 | 3.73E-05 | 0.000126137 | 0.234396297 |
| Obesity | LEC | *MSH2* | 2.294554368 | 6.18E-05 | 0.000175234 | 0.388367546 |
| Obesity | LEC | *MAP3K1* | -0.607116571 | 0.000643052 | 0.001107938 | 1 |
| Obesity | LEC | *TMEM38B* | 2.856699531 | 0.000141511 | 0.000317516 | 0.889678452 |
| Obesity | LEC | *IKZF5* | 1.495606639 | 1.69E-05 | 0.000105108 | 0.106540181 |
| Obesity | LEC | *BLNK* | 0.725821523 | 1.36E-05 | 0.000105108 | 0.085513624 |
| Obesity | LEC | *WAC* | 1.570668744 | 0.000440567 | 0.000801692 | 1 |
| Obesity | LEC | *CREM* | 1.701913895 | 3.29E-05 | 0.000119012 | 0.206843485 |
| Obesity | LEC | *TPSD1* | 4.74692704 | 5.98E-05 | 0.000171484 | 0.375744523 |
| Obesity | LEC | *ITPR3* | 2.447532169 | 0.000255355 | 0.000510305 | 1 |
| Obesity | LEC | *DSP* | -0.104419568 | 9.07E-05 | 0.000227792 | 0.570390435 |
| Obesity | LEC | *IFT74* | 1.267356707 | 0.00669785 | 0.009328911 | 1 |
| Obesity | LEC | *IL12RB1* | 0.869156766 | 0.00016468 | 0.000357015 | 1 |
| Obesity | LEC | *CDC7* | 0.816254193 | 0.007324886 | 0.010123447 | 1 |
| Obesity | LEC | *PCSK5* | 1.045430383 | 1.49E-05 | 0.000105108 | 0.09344271 |
| Obesity | LEC | *NRP1* | 0.901300365 | 1.49E-05 | 0.000105108 | 0.093399777 |
| Obesity | LEC | *PRTFDC1* | 0.885195644 | 1.79E-05 | 0.000105108 | 0.112227077 |
| Obesity | LEC | *PALMD* | 1.28883169 | 3.57E-05 | 0.000123048 | 0.224314991 |
| Obesity | LEC | *TSPAN15* | 1.598706646 | 1.12E-05 | 0.000105108 | 0.070292079 |
| Obesity | LEC | *OCEL1* | 0.822759564 | 0.00212056 | 0.003253285 | 1 |
| Obesity | LEC | *CATSPERG* | 0.95475452 | 0.001311922 | 0.00210088 | 1 |
| Obesity | LEC | *STX1B* | 0.762688615 | 0.00385255 | 0.005600227 | 1 |
| Obesity | LEC | *HSD3B7* | 1.068939342 | 9.97E-06 | 0.000105108 | 0.062707925 |
| Obesity | LEC | *ATP5D* | 2.71186174 | 0.000469365 | 0.000845045 | 1 |
| Obesity | LEC | *MARCH2* | 2.05515859 | 2.13E-05 | 0.00010722 | 0.134120068 |
| Obesity | LEC | *CDC34* | 3.100166929 | 0.00195367 | 0.003016386 | 1 |
| Obesity | LEC | *MTAP* | 1.519085689 | 1.46E-05 | 0.000105108 | 0.091692044 |
| Obesity | LEC | *CEP170B* | 0.769764508 | 3.32E-05 | 0.000119352 | 0.208645792 |
| Obesity | LEC | *POLR2E* | 1.65073791 | 2.76E-05 | 0.00011384 | 0.173207316 |
| Obesity | LEC | *MADCAM1* | 1.573659126 | 1.18E-05 | 0.000105108 | 0.074366066 |
| Obesity | LEC | *TRMT2A* | 1.057911095 | 5.15E-05 | 0.000154924 | 0.323790546 |
| Obesity | LEC | *MED15* | 1.550896713 | 3.44E-05 | 0.000121524 | 0.216433896 |
| Obesity | LEC | *LZTR1* | 2.03198134 | 2.21E-05 | 0.000108445 | 0.138980887 |
| Obesity | LEC | *CECR2* | 1.1212842 | 9.38E-05 | 0.000233645 | 0.58972014 |
| Obesity | LEC | *P2RX6* | 0.785216049 | 6.72E-05 | 0.000184604 | 0.422190118 |
| Obesity | LEC | *SLC7A4* | 2.608894479 | 8.31E-05 | 0.000214185 | 0.522656225 |
| Obesity | LEC | *DDTL* | 0.593399315 | 0.012191258 | 0.016242093 | 1 |
| Obesity | LEC | *DDT* | 2.300872033 | 0.019674892 | 0.025457099 | 1 |
| Obesity | LEC | *GGT5* | -0.921431689 | 0.000533736 | 0.000944175 | 1 |
| Obesity | LEC | *PPIL2* | 0.85838141 | 0.000391571 | 0.000725769 | 1 |
| Obesity | LEC | *UPB1* | 0.648278645 | 0.026808168 | 0.033973584 | 1 |
| Obesity | LEC | *YPEL1* | -1.060436563 | 0.009387633 | 0.012741806 | 1 |
| Obesity | LEC | *PRODH* | 4.548542964 | 0.035704674 | 0.044415371 | 1 |
| Obesity | LEC | *PPM1F* | -0.82266676 | 0.000847816 | 0.001416481 | 1 |
| Obesity | LEC | *TOP3B* | 1.161448643 | 1.18E-05 | 0.000105108 | 0.073954971 |

| | | | | | | |
|---|---|---|---|---|---|---|
| Obesity | LEC | *CRYBB3* | -0.403411767 | 9.90E-05 | 0.000242553 | 0.62239138 |
| Obesity | LEC | *CYTH4* | 0.482567419 | 2.85E-05 | 0.000114887 | 0.179453304 |
| Obesity | LEC | *CARD10* | -0.245459873 | 8.79E-05 | 0.000222231 | 0.552345898 |
| Obesity | LEC | *LRP5L* | 2.067945715 | 0.00027607 | 0.000544433 | 1 |
| Obesity | LEC | *SLC25A1* | 1.363269769 | 1.09E-05 | 0.000105108 | 0.068650183 |
| Obesity | LEC | *LGALS2* | 1.172235135 | 3.96E-06 | 8.35E-05 | 0.024886103 |
| Obesity | LEC | *GGA1* | 1.804832179 | 0.000101429 | 0.000246974 | 0.637686691 |
| Obesity | LEC | *SH3BP1* | 1.337449824 | 1.85E-05 | 0.000105108 | 0.11600452 |
| Obesity | LEC | *HPS4* | 1.644902041 | 1.47E-05 | 0.000105108 | 0.092247677 |
| Obesity | LEC | *PIK3IP1* | 1.520541881 | 0.000105817 | 0.00025421 | 0.665268724 |
| Obesity | LEC | *TFIP11* | 1.983220193 | 4.17E-07 | 2.56E-05 | 0.002620099 |
| Obesity | LEC | *GCAT* | 1.152162067 | 1.75E-06 | 5.62E-05 | 0.011005466 |
| Obesity | LEC | *EIF3L* | 1.730285483 | 0.012354738 | 0.016438992 | 1 |
| Obesity | LEC | *CCDC134* | -1.243908738 | 0.008983334 | 0.012232666 | 1 |
| Obesity | LEC | *SLC5A4* | -0.166986459 | 0.01200531 | 0.016021521 | 1 |
| Obesity | LEC | *DDX17* | 1.079971573 | 0.008882116 | 0.012100079 | 1 |
| Obesity | LEC | *FBXO7* | 2.015215754 | 1.17E-06 | 4.64E-05 | 0.007332262 |
| Obesity | LEC | *POLDIP3* | 2.543191763 | 0.000937484 | 0.001551859 | 1 |
| Obesity | LEC | *TIMP3* | 1.330037634 | 0.002264797 | 0.003451825 | 1 |
| Obesity | LEC | *PPP6R2* | 1.43470538 | 5.37E-05 | 0.000159567 | 0.337802932 |
| Obesity | LEC | *RHBDD3* | 1.957244861 | 6.24E-06 | 9.96E-05 | 0.039243692 |
| Obesity | LEC | *TTLL1* | -0.953910889 | 0.0010595 | 0.001735107 | 1 |
| Obesity | LEC | *AP1B1* | 1.406921057 | 1.13E-05 | 0.000105108 | 0.071199012 |
| Obesity | LEC | *HMGXB4* | 1.193507273 | 1.58E-05 | 0.000105108 | 0.099064937 |
| Obesity | LEC | *TOM1* | 0.969654996 | 0.000185767 | 0.000392823 | 1 |
| Obesity | LEC | *NEFH* | 2.408796557 | 0.020168668 | 0.026063497 | 1 |
| Obesity | LEC | *CHKB* | -0.98263404 | 0.009253491 | 0.012567876 | 1 |
| Obesity | LEC | *BIK* | -0.376468125 | 7.99E-05 | 0.000208108 | 0.50258063 |
| Obesity | LEC | *MCAT* | -1.240372349 | 0.002319001 | 0.003525038 | 1 |
| Obesity | LEC | *THOC5* | 1.547540341 | 0.000200594 | 0.00041801 | 1 |
| Obesity | LEC | *TSPO* | 1.864673728 | 0.005778034 | 0.008137656 | 1 |
| Obesity | LEC | *RASD2* | -3.798487925 | 1.48E-05 | 0.000105108 | 0.093098099 |
| Obesity | LEC | *MTMR3* | 1.118173053 | 0.001248355 | 0.002008807 | 1 |
| Obesity | LEC | *APOL4* | 0.929973274 | 6.52E-06 | 0.000102185 | 0.040976212 |
| Obesity | LEC | *SAMM50* | 1.193045131 | 2.86E-05 | 0.000114981 | 0.179883298 |
| Obesity | LEC | *TXN2* | 1.702561758 | 0.000611805 | 0.001064604 | 1 |
| Obesity | LEC | *FOXRED2* | 0.656027733 | 7.39E-05 | 0.000196872 | 0.464531784 |
| Obesity | LEC | *IFT27* | 1.935370264 | 1.49E-05 | 0.000105108 | 0.093920878 |
| Obesity | LEC | *KIAA0930* | 0.992361514 | 3.13E-05 | 0.000116803 | 0.196813093 |
| Obesity | LEC | *NCF4* | 0.611120123 | 2.34E-05 | 0.000109118 | 0.146838786 |
| Obesity | LEC | *SLC25A17* | 2.018603207 | 0.000290255 | 0.000565986 | 1 |
| Obesity | LEC | *KCTD17* | -1.063766399 | 0.000182527 | 0.000387685 | 1 |
| Obesity | LEC | *EP300* | 1.782093984 | 3.23E-05 | 0.000117812 | 0.203226062 |
| Obesity | LEC | *ACO2* | 1.958600979 | 4.26E-07 | 2.56E-05 | 0.002680472 |
| Obesity | LEC | *DESI1* | 2.397536652 | 1.86E-07 | 1.54E-05 | 0.001169761 |
| Obesity | LEC | *HDAC10* | 2.136180678 | 0.004086975 | 0.005909571 | 1 |
| Obesity | LEC | *SDR39U1* | 0.861635373 | 0.003402759 | 0.005001797 | 1 |
| Obesity | LEC | *RBM23* | 1.765845432 | 0.007675725 | 0.010559581 | 1 |
| Obesity | LEC | *VCPKMT* | 1.323345584 | 0.003769716 | 0.005487429 | 1 |
| Obesity | LEC | *SOS2* | 1.56154764 | 8.42E-05 | 0.000215705 | 0.529554656 |

| | | | | | | |
|---|---|---|---|---|---|---|
| Obesity | LEC | *PYGL* | 0.486879276 | 0.003190668 | 0.004717717 | 1 |
| Obesity | LEC | *DDHD1* | 0.840389012 | 2.75E-05 | 0.00011384 | 0.172773159 |
| Obesity | LEC | *CNIH1* | 1.663814808 | 7.98E-05 | 0.000207817 | 0.501462376 |
| Obesity | LEC | *ATP6V1D* | 1.095505837 | 0.000502078 | 0.000895988 | 1 |
| Obesity | LEC | *PIGH* | 1.146497912 | 6.49E-06 | 0.000101993 | 0.040797342 |
| Obesity | LEC | *VTI1B* | 2.193618115 | 2.26E-05 | 0.000109111 | 0.141832633 |
| Obesity | LEC | *GSTZ1* | 1.638609826 | 0.006893 | 0.009570735 | 1 |
| Obesity | LEC | *TMED8* | 1.891379399 | 0.000801417 | 0.001348277 | 1 |
| Obesity | LEC | *DAAM1* | 1.316756955 | 0.002062039 | 0.003168918 | 1 |
| Obesity | LEC | *ISM2* | 3.052450076 | 0.000364298 | 0.000682259 | 1 |
| Obesity | LEC | *PPM1A* | 1.489927988 | 0.002540761 | 0.003822389 | 1 |
| Obesity | LEC | *SLC10A1* | 1.656349444 | 3.73E-06 | 7.98E-05 | 0.023452844 |
| Obesity | LEC | *DICER1* | 1.789909066 | 1.29E-06 | 4.98E-05 | 0.008114662 |
| Obesity | LEC | *ZFYVE21* | 1.674739571 | 0.028324982 | 0.035787613 | 1 |
| Obesity | LEC | *MTHFD1* | 1.046270391 | 1.05E-05 | 0.000105108 | 0.066063584 |
| Obesity | LEC | *TELO2* | 1.867312177 | 3.03E-05 | 0.000115831 | 0.190414798 |
| Obesity | LEC | *VRK1* | 2.311680202 | 0.000409056 | 0.00075285 | 1 |
| Obesity | LEC | *PSMB5* | 2.355133496 | 2.63E-05 | 0.000112582 | 0.165507483 |
| Obesity | LEC | *TRIP11* | 1.460378922 | 0.010524127 | 0.014189402 | 1 |
| Obesity | LEC | *PCK2* | 1.275380738 | 4.23E-05 | 0.00013685 | 0.266035446 |
| Obesity | LEC | *KIAA0391* | -1.596157428 | 0.000519531 | 0.00092138 | 1 |
| Obesity | LEC | *PLTP* | 0.802366132 | 1.85E-06 | 5.72E-05 | 0.011606983 |
| Obesity | LEC | *GSS* | 2.35497716 | 0.000124103 | 0.000287273 | 0.780232723 |
| Obesity | LEC | *PYGB* | 1.293992239 | 3.57E-06 | 7.81E-05 | 0.022456596 |
| Obesity | LEC | *ABHD12* | 2.175990679 | 4.42E-08 | 5.56E-06 | 0.000277881 |
| Obesity | LEC | *PROCR* | 0.900839127 | 0.023472934 | 0.029976506 | 1 |
| Obesity | LEC | *UQCC1* | 1.525522427 | 0.000117878 | 0.000276015 | 0.741101496 |
| Obesity | LEC | *IFT52* | 1.604825465 | 2.69E-05 | 0.000113184 | 0.169293283 |
| Obesity | LEC | *DOK5* | 1.014885597 | 1.65E-05 | 0.000105108 | 0.10376296 |
| Obesity | LEC | *CSTF1* | 1.829087711 | 2.43E-05 | 0.000109618 | 0.152521806 |
| Obesity | LEC | *BMP7* | 1.629949466 | 1.63E-05 | 0.000105108 | 0.102239046 |
| Obesity | LEC | *TPD52L2* | 2.045887792 | 3.91E-05 | 0.000130057 | 0.245546937 |
| Obesity | LEC | *NELFCD* | 1.869573244 | 3.24E-05 | 0.000117999 | 0.203784858 |
| Obesity | LEC | *CTSZ* | 1.732985058 | 3.81E-05 | 0.000127862 | 0.239357113 |
| Obesity | LEC | *PRPF6* | 1.544054856 | 0.020174658 | 0.026063497 | 1 |
| Obesity | LEC | *MTG2* | 1.165311969 | 1.01E-05 | 0.000105108 | 0.063798597 |
| Obesity | LEC | *TCFL5* | -0.831896299 | 0.000618548 | 0.001073368 | 1 |
| Obesity | LEC | *DIDO1* | 1.494093749 | 0.008047775 | 0.011030382 | 1 |
| Obesity | LEC | *GID8* | 1.856975977 | 0.006884973 | 0.009561702 | 1 |
| Obesity | LEC | *SLC17A9* | 0.917146189 | 2.11E-05 | 0.000106989 | 0.132608531 |
| Obesity | LEC | *BIRC7* | 1.429713369 | 0.03779538 | 0.046860206 | 1 |
| Obesity | LEC | *NKAIN4* | 1.599901872 | 0.000311891 | 0.000599284 | 1 |
| Obesity | LEC | *PTK6* | -0.773820709 | 0.000110995 | 0.000262933 | 0.697823287 |
| Obesity | LEC | *GMEB2* | 1.220185454 | 1.72E-05 | 0.000105108 | 0.107925237 |
| Obesity | LEC | *CDC25B* | -1.014779021 | 0.007698808 | 0.010586702 | 1 |
| Obesity | LEC | *RNF24* | 1.220083772 | 6.88E-06 | 0.000104435 | 0.043236281 |
| Obesity | LEC | *NDUFAF5* | 0.354131403 | 0.03295262 | 0.041146599 | 1 |
| Obesity | LEC | *TRIB3* | 2.688760165 | 5.66E-05 | 0.000165507 | 0.355701749 |
| Obesity | LEC | *SLC52A3* | 2.806005895 | 3.53E-06 | 7.79E-05 | 0.022199111 |
| Obesity | LEC | *ANGPT4* | 2.149523743 | 0.002978481 | 0.004426882 | 1 |

| | | | | | | |
|---|---|---|---|---|---|---|
| Obesity | LEC | *RSPO4* | 1.017286328 | 6.66E-05 | 0.000183725 | 0.418991497 |
| Obesity | LEC | *SIRPB1* | 0.898660636 | 0.005880929 | 0.008275157 | 1 |
| Obesity | LEC | *SEC23B* | 1.192101531 | 0.000217463 | 0.000446503 | 1 |
| Obesity | LEC | *CCM2L* | -0.402448253 | 0.000144784 | 0.000323473 | 0.910254227 |
| Obesity | LEC | *MYL9* | 1.461646533 | 1.84E-06 | 5.72E-05 | 0.011573617 |
| Obesity | LEC | *MROH8* | 1.02450556 | 0.000619658 | 0.001074997 | 1 |
| Obesity | LEC | *MANBAL* | 0.790626915 | 8.77E-07 | 3.91E-05 | 0.005512006 |
| Obesity | LEC | *IDH3B* | 2.019651482 | 1.07E-05 | 0.000105108 | 0.067387555 |
| Obesity | LEC | *TTI1* | 1.973173367 | 1.78E-05 | 0.000105108 | 0.111944649 |
| Obesity | LEC | *RPRD1B* | 1.911472742 | 0.000163398 | 0.000355093 | 1 |
| Obesity | LEC | *PXMP4* | 2.147488941 | 1.34E-06 | 5.03E-05 | 0.008434876 |
| Obesity | LEC | *BPI* | 2.451732654 | 0.020782664 | 0.026802176 | 1 |
| Obesity | LEC | *WFDC2* | 3.405593979 | 0.040292924 | 0.049680646 | 1 |
| Obesity | LEC | *MAP1LC3A* | 1.625540486 | 8.80E-05 | 0.000222432 | 0.553406242 |
| Obesity | LEC | *PIGU* | 1.462818249 | 5.68E-06 | 9.65E-05 | 0.035686791 |
| Obesity | LEC | *APMAP* | 1.162132641 | 3.99E-06 | 8.37E-05 | 0.025078539 |
| Obesity | LEC | *RBFA* | 1.591980347 | 6.88E-05 | 0.000187286 | 0.432466938 |
| Obesity | LEC | *LPIN2* | 1.883762349 | 0.000254944 | 0.000509645 | 1 |
| Obesity | LEC | *SMCHD1* | 1.554758751 | 0.00029548 | 0.000573183 | 1 |
| Obesity | LEC | *CEP76* | 1.734306861 | 0.014293159 | 0.018834855 | 1 |
| Obesity | LEC | *LIPG* | 2.229839525 | 5.23E-05 | 0.000156459 | 0.329097172 |
| Obesity | LEC | *POLI* | 0.563514494 | 0.001184499 | 0.001916846 | 1 |
| Obesity | LEC | *NALCN* | -0.08552832 | 8.34E-05 | 0.000214516 | 0.524062562 |
| Obesity | LEC | *HTR2A* | 4.161541355 | 1.54E-05 | 0.000105108 | 0.096608014 |
| Obesity | LEC | *KLF5* | 2.03490003 | 0.000290953 | 0.000566322 | 1 |
| Obesity | LEC | *DNAJC3* | 1.70214975 | 0.000188088 | 0.000397082 | 1 |
| Obesity | LEC | *ARHGEF7* | 1.456228256 | 0.003817239 | 0.005554035 | 1 |
| Obesity | LEC | *SGCG* | 0.570059059 | 3.68E-05 | 0.000125526 | 0.231344603 |
| Obesity | LEC | *PARP4* | 1.735191142 | 0.000100426 | 0.000245005 | 0.631376773 |
| Obesity | LEC | *VWA8* | 1.287221926 | 0.000110732 | 0.000262608 | 0.696173878 |
| Obesity | LEC | *KATNAL1* | 1.338458532 | 0.000148366 | 0.000330071 | 0.932779566 |
| Obesity | LEC | *TSC22D1* | 1.654144452 | 0.000989083 | 0.001629978 | 1 |
| Obesity | LEC | *CLN5* | 1.436616186 | 0.001118191 | 0.001819846 | 1 |
| Obesity | LEC | *MGRN1* | 1.185639302 | 7.91E-05 | 0.000206694 | 0.497512459 |
| Obesity | LEC | *GDPD3* | 1.830980655 | 0.000690292 | 0.001182847 | 1 |
| Obesity | LEC | *LONP2* | 1.497365202 | 0.001063554 | 0.001739933 | 1 |
| Obesity | LEC | *PLLP* | 0.8655562 | 3.36E-05 | 0.00011999 | 0.211062699 |
| Obesity | LEC | *DHODH* | 1.029697589 | 0.000693423 | 0.001187889 | 1 |
| Obesity | LEC | *CTCF* | 1.96309523 | 0.000648919 | 0.001116212 | 1 |
| Obesity | LEC | *ACD* | -1.88657244 | 0.002268049 | 0.00345427 | 1 |
| Obesity | LEC | *USB1* | 3.367699187 | 1.19E-05 | 0.000105108 | 0.074571782 |
| Obesity | LEC | *SETD6* | 2.936076872 | 0.001368775 | 0.002183026 | 1 |
| Obesity | LEC | *SLC7A6OS* | 0.895121403 | 3.64E-06 | 7.87E-05 | 0.022856415 |
| Obesity | LEC | *MON1B* | 1.872888665 | 0.00019747 | 0.00041273 | 1 |
| Obesity | LEC | *HCFC1R1* | 2.477956092 | 1.63E-05 | 0.000105108 | 0.102702497 |
| Obesity | LEC | *NPRL3* | 1.337185047 | 4.36E-05 | 0.000139798 | 0.274284416 |
| Obesity | LEC | *NAGPA* | 2.572466772 | 4.64E-07 | 2.67E-05 | 0.002914902 |
| Obesity | LEC | *COTL1* | 1.288258339 | 1.72E-05 | 0.000105108 | 0.108349043 |
| Obesity | LEC | *CRISPLD2* | 1.140597613 | 2.64E-05 | 0.000112582 | 0.166171244 |
| Obesity | LEC | *ZNF500* | 1.428485701 | 0.0002894 | 0.000565225 | 1 |

| | | | | | | |
|---|---|---|---|---|---|---|
| Obesity | LEC | *NME4* | 2.733380675 | 0.002181719 | 0.003341405 | 1 |
| Obesity | LEC | *NOMO3* | 1.247575923 | 2.30E-05 | 0.000109118 | 0.144530981 |
| Obesity | LEC | *LMF1* | 1.880491204 | 9.35E-06 | 0.000105108 | 0.058781926 |
| Obesity | LEC | *NARFL* | 2.267845803 | 1.38E-05 | 0.000105108 | 0.086687648 |
| Obesity | LEC | *MTHFSD* | 0.753827872 | 0.000155167 | 0.000340976 | 0.975531967 |
| Obesity | LEC | *CLCN7* | 2.496210334 | 1.13E-05 | 0.000105108 | 0.07116309 |
| Obesity | LEC | *HAGHL* | 1.043327108 | 2.97E-05 | 0.000115831 | 0.18653179 |
| Obesity | LEC | *SLC7A5* | -0.98942953 | 0.000152788 | 0.000337113 | 0.960577367 |
| Obesity | LEC | *FBXO31* | 1.692110736 | 2.28E-05 | 0.000109111 | 0.143288097 |
| Obesity | LEC | *NUBP1* | 2.65640265 | 0.001570411 | 0.002472002 | 1 |
| Obesity | LEC | *UBE2I* | 2.006550599 | 0.003016049 | 0.004479542 | 1 |
| Obesity | LEC | *CLUAP1* | 1.614183625 | 2.69E-08 | 3.84E-06 | 0.000169113 |
| Obesity | LEC | *EARS2* | 0.795239161 | 0.000342376 | 0.000648935 | 1 |
| Obesity | LEC | *CPPED1* | 1.722297207 | 2.46E-05 | 0.000110453 | 0.15474519 |
| Obesity | LEC | *BFAR* | 2.25333731 | 0.000124201 | 0.000287273 | 0.78084974 |
| Obesity | LEC | *RRN3P2* | 1.54401374 | 2.89E-05 | 0.000115115 | 0.181519364 |
| Obesity | LEC | *RBL2* | 1.275908433 | 0.000184129 | 0.00039056 | 1 |
| Obesity | LEC | *STX4* | 1.806570498 | 1.53E-05 | 0.000105108 | 0.096036334 |
| Obesity | LEC | *KAT8* | 2.431718359 | 1.33E-08 | 2.46E-06 | 8.38E-05 |
| Obesity | LEC | *NOMO1* | 1.888072806 | 1.68E-05 | 0.000105108 | 0.105516291 |
| Obesity | LEC | *SYT17* | -1.19785399 | 0.005359849 | 0.007592918 | 1 |
| Obesity | LEC | *TMC5* | 0.771041581 | 6.35E-05 | 0.000178091 | 0.399458754 |
| Obesity | LEC | *RNF40* | -1.411112476 | 0.03942303 | 0.048713167 | 1 |
| Obesity | LEC | *IQCH* | 1.166573478 | 7.01E-06 | 0.000104897 | 0.044056811 |
| Obesity | LEC | *LACTB* | 1.538808618 | 2.03E-05 | 0.000106065 | 0.127711973 |
| Obesity | LEC | *CSK* | 2.821353389 | 7.96E-05 | 0.000207606 | 0.500538885 |
| Obesity | LEC | *HERC1* | 1.412348614 | 6.31E-05 | 0.00017717 | 0.39685977 |
| Obesity | LEC | *TRIP4* | 1.956371116 | 2.80E-05 | 0.000114038 | 0.176313837 |
| Obesity | LEC | *RASL12* | -1.246844817 | 0.000537572 | 0.000949892 | 1 |
| Obesity | LEC | *AP3B2* | 0.543558076 | 0.002438686 | 0.003691793 | 1 |
| Obesity | LEC | *TTC23* | 1.561416386 | 0.00018806 | 0.000397082 | 1 |
| Obesity | LEC | *FAH* | 1.001973542 | 1.67E-05 | 0.000105108 | 0.104980491 |
| Obesity | LEC | *HOMER2* | 1.143680146 | 6.86E-06 | 0.000104405 | 0.043119158 |
| Obesity | LEC | *EHD4* | 1.939364821 | 9.86E-06 | 0.000105108 | 0.062007183 |
| Obesity | LEC | *ATP8B4* | 0.791236756 | 1.50E-05 | 0.000105108 | 0.094596236 |
| Obesity | LEC | *DTWD1* | 1.744370091 | 2.69E-05 | 0.000113184 | 0.169316761 |
| Obesity | LEC | *DNAJC17* | 1.786714416 | 2.06E-05 | 0.000106611 | 0.129425747 |
| Obesity | LEC | *PDGFRL* | 0.728152498 | 1.90E-05 | 0.000105211 | 0.119234381 |
| Obesity | LEC | *RIPK2* | 0.969672668 | 1.62E-05 | 0.000105108 | 0.101619931 |
| Obesity | LEC | *EYA1* | -0.170598796 | 0.000133443 | 0.000303202 | 0.8389591 |
| Obesity | LEC | *NBN* | 1.598283491 | 3.69E-05 | 0.000125724 | 0.232156856 |
| Obesity | LEC | *CPQ* | 1.081062377 | 1.10E-05 | 0.000105108 | 0.06929079 |
| Obesity | LEC | *IKBKB* | 1.990319799 | 0.000125989 | 0.000290463 | 0.792093335 |
| Obesity | LEC | *PLAT* | 1.803827313 | 9.95E-06 | 0.000105108 | 0.062578228 |
| Obesity | LEC | *GDAP1* | -0.714307842 | 0.001421367 | 0.002261111 | 1 |
| Obesity | LEC | *RAB2A* | 0.75751773 | 0.017052528 | 0.022270303 | 1 |
| Obesity | LEC | *ARMC1* | 0.982679721 | 0.009206735 | 0.012512483 | 1 |
| Obesity | LEC | *SPAG1* | 1.542204531 | 4.97E-05 | 0.000151436 | 0.312487546 |
| Obesity | LEC | *CHRAC1* | 2.038982789 | 7.45E-05 | 0.000198233 | 0.468623022 |
| Obesity | LEC | *GSDMD* | 1.211639926 | 0.007594274 | 0.010452102 | 1 |

| | | | | | | |
|---|---|---|---|---|---|---|
| Obesity | LEC | *TSTA3* | -1.204690325 | 0.002738856 | 0.004094932 | 1 |
| Obesity | LEC | *ERI1* | 1.438548935 | 1.43E-05 | 0.000105108 | 0.08975459 |
| Obesity | LEC | *MTMR9* | 1.807454094 | 2.20E-05 | 0.000108376 | 0.138147027 |
| Obesity | LEC | *LEPROTL1* | 1.882650288 | 1.96E-05 | 0.000105317 | 0.123325927 |
| Obesity | LEC | *TNFRSF10A* | 1.411528694 | 7.59E-05 | 0.00020093 | 0.477008335 |
| Obesity | LEC | *TUSC3* | 1.211550474 | 1.29E-05 | 0.000105108 | 0.081271108 |
| Obesity | LEC | *ARHGEF10* | 1.516131773 | 0.004082859 | 0.005904976 | 1 |
| Obesity | LEC | *KLHDC4* | 1.785466994 | 1.37E-05 | 0.000105108 | 0.086240717 |
| Obesity | LEC | *MCM4* | 1.968553405 | 2.81E-07 | 2.10E-05 | 0.001764113 |
| Obesity | LEC | *MAN2B1* | 1.124373103 | 0.00055684 | 0.000980357 | 1 |
| Obesity | LEC | *TULP2* | 1.342670696 | 0.00019594 | 0.000410352 | 1 |
| Obesity | LEC | *NUCB1* | 1.570098608 | 3.21E-05 | 0.000117552 | 0.202071654 |
| Obesity | LEC | *NFKBIB* | 0.98144989 | 2.48E-05 | 0.000110994 | 0.155922145 |
| Obesity | LEC | *SARS2* | -0.881165648 | 0.000354921 | 0.000667081 | 1 |
| Obesity | LEC | *SNRNP70* | 1.06972308 | 0.007550086 | 0.010398114 | 1 |
| Obesity | LEC | *PPP1R37* | 3.344306746 | 0.000196261 | 0.000410749 | 1 |
| Obesity | LEC | *FCGRT* | 1.225118176 | 3.87E-05 | 0.00012912 | 0.2430037 |
| Obesity | LEC | *PEX11G* | 0.822979403 | 6.88E-05 | 0.000187286 | 0.432318891 |
| Obesity | LEC | *CD37* | 2.33756517 | 2.89E-05 | 0.000115266 | 0.181918341 |
| Obesity | LEC | *AMH* | -1.498973322 | 0.004523076 | 0.006493852 | 1 |
| Obesity | LEC | *DKKL1* | 1.542088496 | 0.001849353 | 0.002866588 | 1 |
| Obesity | LEC | *STX10* | 1.612438484 | 1.49E-05 | 0.000105108 | 0.09396233 |
| Obesity | LEC | *FCER2* | 0.996110022 | 3.31E-05 | 0.000119154 | 0.207804849 |
| Obesity | LEC | *DMPK* | 2.084548934 | 0.00134425 | 0.002146089 | 1 |
| Obesity | LEC | *IL4I1* | 1.162831778 | 5.73E-05 | 0.000166529 | 0.359974689 |
| Obesity | LEC | *AES* | 1.828019378 | 1.61E-05 | 0.000105108 | 0.101102038 |
| Obesity | LEC | *SGTA* | 2.791260807 | 8.99E-06 | 0.000105108 | 0.05652893 |
| Obesity | LEC | *C19orf53* | -1.408466583 | 0.025922593 | 0.032904369 | 1 |
| Obesity | LEC | *TIMM44* | 1.372765558 | 1.58E-05 | 0.000105108 | 0.099334748 |
| Obesity | LEC | *CCDC61* | -1.491381441 | 0.018576993 | 0.024150859 | 1 |
| Obesity | LEC | *IL27RA* | 2.441141979 | 1.30E-06 | 4.99E-05 | 0.008191483 |
| Obesity | LEC | *C19orf44* | 1.18205852 | 0.00254141 | 0.003822451 | 1 |
| Obesity | LEC | *OLFM2* | 0.734603242 | 2.45E-06 | 6.73E-05 | 0.015415697 |
| Obesity | LEC | *RASAL3* | 1.026906199 | 1.24E-05 | 0.000105108 | 0.077656069 |
| Obesity | LEC | *SYDE1* | -1.896169449 | 0.000393167 | 0.000728297 | 1 |
| Obesity | LEC | *AURKC* | 1.319487173 | 0.000312776 | 0.000600641 | 1 |
| Obesity | LEC | *CCNE1* | 1.287169423 | 7.49E-05 | 0.000199064 | 0.47098498 |
| Obesity | LEC | *PDCD5* | 2.043912935 | 2.22E-05 | 0.000108559 | 0.139877716 |
| Obesity | LEC | *ANKRD27* | 1.640048936 | 3.31E-05 | 0.000119217 | 0.208033683 |
| Obesity | LEC | *RPS16* | 1.275977087 | 0.033665264 | 0.041994745 | 1 |
| Obesity | LEC | *PRX* | 1.044721317 | 2.36E-05 | 0.000109118 | 0.148438507 |
| Obesity | LEC | *CCDC94* | 1.147012734 | 1.18E-05 | 0.000105108 | 0.074060969 |
| Obesity | LEC | *TBCB* | 1.376840038 | 3.21E-05 | 0.00011749 | 0.201553024 |
| Obesity | LEC | *POLR2I* | 2.469345747 | 0.001103189 | 0.001799156 | 1 |
| Obesity | LEC | *CLIP3* | 1.480227447 | 0.000250255 | 0.000502348 | 1 |
| Obesity | LEC | *SLC1A5* | 0.997968239 | 6.93E-05 | 0.000188201 | 0.435779508 |
| Obesity | LEC | *CACTIN* | -1.067929605 | 0.000456293 | 0.000826005 | 1 |
| Obesity | LEC | *CCDC9* | 1.493495092 | 3.11E-05 | 0.000116624 | 0.195836068 |
| Obesity | LEC | *FZR1* | 1.555857473 | 1.04E-05 | 0.000105108 | 0.065627436 |
| Obesity | LEC | *TGFB1* | 1.736644325 | 0.003411786 | 0.005012829 | 1 |

| | | | | | | |
|---|---|---|---|---|---|---|
| Obesity | LEC | *CEACAM4* | -2.256403501 | 0.000193065 | 0.000405275 | 1 |
| Obesity | LEC | *PLIN3* | 1.295320652 | 9.48E-06 | 0.000105108 | 0.059625796 |
| Obesity | LEC | *MRPL4* | 3.108253171 | 0.000170524 | 0.00036661 | 1 |
| Obesity | LEC | *ICAM5* | 3.427091226 | 0.003492796 | 0.005123474 | 1 |
| Obesity | LEC | *CD33* | 0.762105891 | 1.82E-05 | 0.000105108 | 0.114598719 |
| Obesity | LEC | *BABAM1* | 1.87889787 | 1.28E-05 | 0.000105108 | 0.080482358 |
| Obesity | LEC | *TYK2* | 1.295164424 | 7.79E-06 | 0.000105108 | 0.049005419 |
| Obesity | LEC | *RABAC1* | 1.790449714 | 0.012117583 | 0.016161062 | 1 |
| Obesity | LEC | *PTPRS* | 1.110993628 | 2.96E-06 | 7.23E-05 | 0.018578528 |
| Obesity | LEC | *GRWD1* | 0.649739157 | 0.004712773 | 0.006736972 | 1 |
| Obesity | LEC | *GRIN2D* | 2.131584008 | 2.20E-05 | 0.000108376 | 0.138309099 |
| Obesity | LEC | *CARD8* | 1.495754259 | 0.000915762 | 0.001520306 | 1 |
| Obesity | LEC | *LIG1* | 2.37433455 | 6.49E-05 | 0.000180539 | 0.408198832 |
| Obesity | LEC | *PLA2G4C* | -0.628034481 | 0.023495629 | 0.029993304 | 1 |
| Obesity | LEC | *CAPS* | 1.459957034 | 1.27E-05 | 0.000105108 | 0.079640548 |
| Obesity | LEC | *BCAT2* | 2.531689424 | 3.50E-05 | 0.000122515 | 0.220158992 |
| Obesity | LEC | *MIER2* | 2.463444611 | 0.000269415 | 0.000532812 | 1 |
| Obesity | LEC | *TNPO2* | 1.633456132 | 0.000150641 | 0.000334135 | 0.947081089 |
| Obesity | LEC | *GCDH* | 1.361449236 | 0.035702491 | 0.044415371 | 1 |
| Obesity | LEC | *LILRB5* | 0.587711778 | 2.37E-05 | 0.000109118 | 0.148911223 |
| Obesity | LEC | *MAST1* | 1.606385838 | 3.05E-05 | 0.000115831 | 0.191566712 |
| Obesity | LEC | *TFPT* | 2.878437386 | 0.00034318 | 0.000649675 | 1 |
| Obesity | LEC | *RPL18A* | 1.344179022 | 0.023826309 | 0.03039075 | 1 |
| Obesity | LEC | *ELL* | 1.449162242 | 0.00350696 | 0.005141851 | 1 |
| Obesity | LEC | *UPK1A* | 2.366016933 | 0.00125677 | 0.002021313 | 1 |
| Obesity | LEC | *COPE* | 2.487442621 | 0.000169949 | 0.000365665 | 1 |
| Obesity | LEC | *MAG* | -1.611499687 | 0.022000056 | 0.028215902 | 1 |
| Obesity | LEC | *LSR* | 1.114240149 | 0.005673518 | 0.008011997 | 1 |
| Obesity | LEC | *KXD1* | 2.18241743 | 0.018903245 | 0.02453948 | 1 |
| Obesity | LEC | *HPN* | 0.772381561 | 5.70E-05 | 0.000166253 | 0.358109016 |
| Obesity | LEC | *ZNF14* | 2.176590986 | 2.13E-05 | 0.00010722 | 0.134047907 |
| Obesity | LEC | *SIPA1L3* | 1.330197801 | 0.019287317 | 0.02500193 | 1 |
| Obesity | LEC | *ZNF85* | 2.411399058 | 0.001343548 | 0.002145514 | 1 |
| Obesity | LEC | *SMG9* | 1.015260102 | 3.70E-05 | 0.000125857 | 0.232584173 |
| Obesity | LEC | *RASA4* | 1.502224893 | 0.000495578 | 0.000886149 | 1 |
| Obesity | LEC | *PON3* | 1.024729718 | 2.01E-05 | 0.000105931 | 0.126270193 |
| Obesity | LEC | *HBP1* | 1.501643706 | 0.009016857 | 0.012275457 | 1 |
| Obesity | LEC | *WDR91* | 1.441364006 | 2.81E-05 | 0.000114038 | 0.176872637 |
| Obesity | LEC | *DLX5* | 21.07593014 | 0.028983772 | 0.036546525 | 1 |
| Obesity | LEC | *STEAP1B* | 1.67420765 | 0.002795854 | 0.004171223 | 1 |
| Obesity | LEC | *PTN* | 0.738039855 | 2.94E-05 | 0.000115648 | 0.184573921 |
| Obesity | LEC | *MPP6* | 0.851737655 | 1.63E-05 | 0.000105108 | 0.102285012 |
| Obesity | LEC | *DFNA5* | 0.963108146 | 1.86E-05 | 0.000105108 | 0.116654475 |
| Obesity | LEC | *OGDH* | 1.751903042 | 8.78E-05 | 0.000222231 | 0.552312415 |
| Obesity | LEC | *ADAP1* | 1.728958905 | 2.38E-05 | 0.000109261 | 0.149775404 |
| Obesity | LEC | *H2AFV* | 1.875395224 | 0.00041108 | 0.000756132 | 1 |
| Obesity | LEC | *CAV1* | 1.174551808 | 0.005941781 | 0.008351436 | 1 |
| Obesity | LEC | *MET* | -0.715889026 | 0.000956468 | 0.001579957 | 1 |
| Obesity | LEC | *RNF32* | -0.999533896 | 0.00195179 | 0.003014223 | 1 |
| Obesity | LEC | *HOXA2* | 3.161102551 | 0.018643842 | 0.024227746 | 1 |

| | | | | | | |
|---|---|---|---|---|---|---|
| Obesity | LEC | *HOXA5* | 0.737956196 | 7.08E-05 | 0.000190901 | 0.445166588 |
| Obesity | LEC | *CPED1* | 0.949612772 | 1.59E-05 | 0.000105108 | 0.100044805 |
| Obesity | LEC | *HIBADH* | 1.402826654 | 1.98E-05 | 0.000105667 | 0.124553459 |
| Obesity | LEC | *CPVL* | 0.856061837 | 5.06E-05 | 0.0001531 | 0.318406445 |
| Obesity | LEC | *ABHD11* | -0.664640358 | 0.026162363 | 0.033195313 | 1 |
| Obesity | LEC | *PLEKHA8* | 1.706398438 | 5.26E-06 | 9.34E-05 | 0.033071825 |
| Obesity | LEC | *NOD1* | 1.555981148 | 0.011995844 | 0.016012286 | 1 |
| Obesity | LEC | *EPHB6* | 1.076362643 | 5.23E-06 | 9.34E-05 | 0.032864997 |
| Obesity | LEC | *CHCHD2* | 1.683991584 | 0.000631756 | 0.001092669 | 1 |
| Obesity | LEC | *CCL24* | -1.496701961 | 0.000292606 | 0.00056845 | 1 |
| Obesity | LEC | *NPTX2* | 22.79892033 | 0.001123001 | 0.001825784 | 1 |
| Obesity | LEC | *CYP3A5* | 1.919133289 | 1.92E-05 | 0.000105211 | 0.120673335 |
| Obesity | LEC | *ZKSCAN1* | 1.717233556 | 0.000578586 | 0.00101438 | 1 |
| Obesity | LEC | *EIF3B* | 1.562222601 | 0.017705852 | 0.0230756 | 1 |
| Obesity | LEC | *SNX8* | 1.871180814 | 2.43E-05 | 0.000109689 | 0.152905826 |
| Obesity | LEC | *AIMP2* | 2.492009285 | 0.000484877 | 0.000869718 | 1 |
| Obesity | LEC | *TFR2* | 2.558899489 | 0.000227695 | 0.000463575 | 1 |
| Obesity | LEC | *RBM28* | 1.288278275 | 0.000630538 | 0.001091162 | 1 |
| Obesity | LEC | *IMPDH1* | 1.670579379 | 0.012824663 | 0.017017445 | 1 |
| Obesity | LEC | *AP1S1* | 0.921191509 | 2.93E-05 | 0.000115648 | 0.184314254 |
| Obesity | LEC | *C1GALT1* | 1.749245818 | 3.74E-05 | 0.000126288 | 0.235395583 |
| Obesity | LEC | *PLOD3* | 2.090764842 | 0.003164544 | 0.00468019 | 1 |
| Obesity | LEC | *RPA3* | 1.41027756 | 7.83E-06 | 0.000105108 | 0.049252742 |
| Obesity | LEC | *PHF14* | 1.366048403 | 0.004933605 | 0.007020727 | 1 |
| Obesity | LEC | *CEP41* | 1.828197443 | 0.003122726 | 0.004622693 | 1 |
| Obesity | LEC | *ZNF862* | 2.032798667 | 7.58E-06 | 0.000105108 | 0.047632628 |
| Obesity | LEC | *MEST* | 0.40334268 | 0.004403737 | 0.006339888 | 1 |
| Obesity | LEC | *ACTR3C* | 1.438046225 | 7.04E-05 | 0.000190166 | 0.442602412 |
| Obesity | LEC | *POU6F2* | 1.49378233 | 1.46E-05 | 0.000105108 | 0.091567363 |
| Obesity | LEC | *TSPAN13* | 1.643219812 | 1.05E-05 | 0.000105108 | 0.066007594 |
| Obesity | LEC | *RARRES2* | 1.3387053 | 8.12E-05 | 0.00021039 | 0.510615895 |
| Obesity | LEC | *TMEM176B* | 0.617529854 | 3.05E-05 | 0.000115831 | 0.191648451 |
| Obesity | LEC | *MRPL32* | 1.693836149 | 0.00013367 | 0.000303606 | 0.840382283 |
| Obesity | LEC | *COA1* | 1.195511021 | 0.002393931 | 0.003629285 | 1 |
| Obesity | LEC | *BLVRA* | 1.42311928 | 2.59E-06 | 6.85E-05 | 0.016305838 |
| Obesity | LEC | *URGCP* | 1.518364953 | 1.93E-05 | 0.000105211 | 0.121504119 |
| Obesity | LEC | *POLD2* | 1.518198423 | 3.45E-05 | 0.000121733 | 0.217172351 |
| Obesity | LEC | *GCK* | -11.76932712 | 0.000613381 | 0.001066757 | 1 |
| Obesity | LEC | *BCL7B* | 1.0120087 | 0.000918393 | 0.001524271 | 1 |
| Obesity | LEC | *TBL2* | 1.106703365 | 1.07E-05 | 0.000105108 | 0.067265481 |
| Obesity | LEC | *CLIP2* | -1.033503443 | 0.013135786 | 0.01741191 | 1 |
| Obesity | LEC | *LIMK1* | 1.607395526 | 1.44E-05 | 0.000105108 | 0.090228795 |
| Obesity | LEC | *SPATA6L* | -0.479233197 | 7.72E-05 | 0.000202909 | 0.485330592 |
| Obesity | LEC | *FKTN* | 1.967261758 | 0.000960689 | 0.001586096 | 1 |
| Obesity | LEC | *TMEM245* | 1.55007385 | 0.000178777 | 0.000382043 | 1 |
| Obesity | LEC | *MEGF9* | 0.93139516 | 0.00013405 | 0.000304249 | 0.842770383 |
| Obesity | LEC | *TRIM14* | 1.032875911 | 1.19E-05 | 0.000105108 | 0.075104024 |
| Obesity | LEC | *CORO2A* | 2.253868333 | 4.05E-05 | 0.000132669 | 0.254856243 |
| Obesity | LEC | *C5* | 0.829999302 | 1.82E-05 | 0.000105108 | 0.114308344 |
| Obesity | LEC | *OGN* | 2.096104822 | 9.14E-08 | 1.02E-05 | 0.000574688 |

| | | | | | | |
|---|---|---|---|---|---|---|
| Obesity | LEC | *ECM2* | 0.802777047 | 1.26E-05 | 0.000105108 | 0.079507015 |
| Obesity | LEC | *PTGR1* | 1.005306766 | 1.79E-05 | 0.000105108 | 0.112672637 |
| Obesity | LEC | *SUSD1* | 1.293677071 | 3.16E-05 | 0.000116919 | 0.198807009 |
| Obesity | LEC | *TNFSF8* | 0.686090812 | 2.51E-05 | 0.000111345 | 0.157943523 |
| Obesity | LEC | *AK1* | 2.230971958 | 1.57E-05 | 0.000105108 | 0.098467367 |
| Obesity | LEC | *KDM4C* | 1.517368721 | 6.81E-05 | 0.00018633 | 0.428186453 |
| Obesity | LEC | *DOCK8* | 1.04864634 | 7.85E-05 | 0.000205645 | 0.4933929 |
| Obesity | LEC | *ELAVL2* | 1.323418536 | 0.012298095 | 0.016370554 | 1 |
| Obesity | LEC | *FUBP3* | 2.014184888 | 0.000523201 | 0.000927103 | 1 |
| Obesity | LEC | *RGP1* | 2.365567932 | 7.21E-07 | 3.63E-05 | 0.004533783 |
| Obesity | LEC | *MPDZ* | -0.636801187 | 0.002207774 | 0.003374733 | 1 |
| Obesity | LEC | *PTGDS* | 1.435004218 | 1.83E-05 | 0.000105108 | 0.1152676 |
| Obesity | LEC | *SHB* | 1.844744637 | 2.03E-05 | 0.000106065 | 0.127914949 |
| Obesity | LEC | *DVL1* | 2.731566287 | 0.00072122 | 0.001230144 | 1 |
| Obesity | LEC | *CCNJ* | -1.043641341 | 0.00180477 | 0.00280509 | 1 |
| Obesity | LEC | *ATRNL1* | 0.998761451 | 0.000237519 | 0.000480837 | 1 |
| Obesity | LEC | *HPS1* | 1.575979227 | 1.36E-05 | 0.000105108 | 0.085420336 |
| Obesity | LEC | *PHYH* | 0.987658913 | 0.00153833 | 0.00242758 | 1 |
| Obesity | LEC | *TRDMT1* | 2.453417297 | 1.25E-05 | 0.000105108 | 0.078861937 |
| Obesity | LEC | *DDX50* | 1.512484209 | 1.35E-05 | 0.000105108 | 0.085016916 |
| Obesity | LEC | *MAPK8* | 1.592308188 | 0.000765 | 0.001293587 | 1 |
| Obesity | LEC | *ATE1* | 1.493369307 | 2.06E-05 | 0.00010658 | 0.129281571 |
| Obesity | LEC | *PLEKHA1* | 1.581523158 | 8.31E-05 | 0.000214185 | 0.522374866 |
| Obesity | LEC | *SPOCK2* | 0.723001537 | 2.95E-05 | 0.000115831 | 0.18564377 |
| Obesity | LEC | *MICU1* | 1.816596108 | 3.42E-05 | 0.000121059 | 0.214818219 |
| Obesity | LEC | *CCSER2* | 1.794708129 | 0.000139412 | 0.000313703 | 0.876485052 |
| Obesity | LEC | *BMPR1A* | 0.843413887 | 3.63E-05 | 0.000124419 | 0.228308573 |
| Obesity | LEC | *LIPA* | 0.99517744 | 7.79E-06 | 0.000105108 | 0.048946134 |
| Obesity | LEC | *KAZALD1* | -2.771866895 | 0.001843708 | 0.002858543 | 1 |
| Obesity | LEC | *TNKS2* | 1.503513386 | 0.008261117 | 0.011306386 | 1 |
| Obesity | LEC | *ANKRD26* | 1.596547013 | 5.68E-05 | 0.000165809 | 0.356821852 |
| Obesity | LEC | *ACBD5* | 1.243604992 | 2.00E-06 | 6.04E-05 | 0.012586173 |
| Obesity | LEC | *MTPAP* | 2.067739218 | 5.52E-06 | 9.57E-05 | 0.034721372 |
| Obesity | LEC | *MAP3K8* | 1.533732739 | 2.41E-05 | 0.000109361 | 0.151211357 |
| Obesity | LEC | *SHOC2* | 1.670002331 | 0.000188909 | 0.000398413 | 1 |
| Obesity | LEC | *TFAM* | 2.644333527 | 0.00015484 | 0.000340378 | 0.97348085 |
| Obesity | LEC | *RPL28* | 1.202875569 | 0.015404197 | 0.020235308 | 1 |
| Obesity | LEC | *PBLD* | 0.598523313 | 0.009099126 | 0.012376938 | 1 |
| Obesity | LEC | *DHRS11* | 0.605680912 | 0.014415245 | 0.018987774 | 1 |
| Obesity | LEC | *MLLT6* | 2.105374775 | 0.000932529 | 0.001544877 | 1 |
| Obesity | LEC | *PSMB3* | 2.425511129 | 0.00031537 | 0.000604676 | 1 |
| Obesity | LEC | *CASC3* | 1.673179661 | 0.000253269 | 0.000507103 | 1 |
| Obesity | LEC | *RAPGEFL1* | 0.893836892 | 0.000385564 | 0.000716112 | 1 |
| Obesity | LEC | *RGS9* | -1.685345888 | 0.025094216 | 0.031936707 | 1 |
| Obesity | LEC | *ASPA* | 1.680982035 | 2.39E-05 | 0.000109302 | 0.149962178 |
| Obesity | LEC | *TRIM37* | 2.043935402 | 0.000179962 | 0.000383532 | 1 |
| Obesity | LEC | *PNPO* | -1.140345627 | 0.004567578 | 0.006548771 | 1 |
| Obesity | LEC | *RPS6KB1* | 1.793806892 | 0.004466206 | 0.006419532 | 1 |
| Obesity | LEC | *CDK5RAP3* | 2.630079359 | 0.000813304 | 0.001364259 | 1 |
| Obesity | LEC | *CBX1* | 2.670271362 | 1.79E-05 | 0.000105108 | 0.112372525 |

| | | | | | | |
|---|---|---|---|---|---|---|
| Obesity | LEC | *PIGL* | 1.575686468 | 2.77E-05 | 0.00011384 | 0.17390929 |
| Obesity | LEC | *GALK1* | -1.542172821 | 0.002123151 | 0.003256465 | 1 |
| Obesity | LEC | *MED13* | 1.10247352 | 0.010565326 | 0.014241896 | 1 |
| Obesity | LEC | *HOXB6* | 1.753392173 | 1.73E-07 | 1.51E-05 | 0.00108726 |
| Obesity | LEC | *RNF167* | 2.776533072 | 0.000131084 | 0.000299464 | 0.824125019 |
| Obesity | LEC | *C1QBP* | 1.243532393 | 5.98E-06 | 9.92E-05 | 0.037618053 |
| Obesity | LEC | *GOSR1* | 1.918825976 | 0.000475941 | 0.000855904 | 1 |
| Obesity | LEC | *AKAP10* | 1.462392182 | 6.71E-05 | 0.000184604 | 0.421792498 |
| Obesity | LEC | *SMARCD2* | 1.777291191 | 4.69E-05 | 0.000145969 | 0.294581639 |
| Obesity | LEC | *ICAM2* | 0.994897207 | 2.03E-05 | 0.000106047 | 0.127362245 |
| Obesity | LEC | *UTP6* | 2.064036604 | 2.61E-05 | 0.000112507 | 0.164260011 |
| Obesity | LEC | *DDX5* | 1.030789248 | 0.001303881 | 0.002089066 | 1 |
| Obesity | LEC | *C17orf75* | 1.980328253 | 1.38E-05 | 0.000105108 | 0.086971493 |
| Obesity | LEC | *CYTH1* | 1.31370856 | 3.34E-05 | 0.000119779 | 0.210074859 |
| Obesity | LEC | *DHX58* | 1.769774725 | 4.61E-05 | 0.000144768 | 0.289824751 |
| Obesity | LEC | *RAB5C* | 1.294479034 | 0.000140998 | 0.000316704 | 0.88645504 |
| Obesity | LEC | *NAGLU* | 1.366569139 | 1.56E-05 | 0.000105108 | 0.097950931 |
| Obesity | LEC | *EZH1* | 1.723101735 | 4.09E-05 | 0.000133262 | 0.257061891 |
| Obesity | LEC | *DLX4* | 6.967513982 | 3.15E-05 | 0.000116847 | 0.197996208 |
| Obesity | LEC | *VAT1* | 2.647748469 | 3.73E-05 | 0.000126137 | 0.23443693 |
| Obesity | LEC | *ALOX12* | 2.245590229 | 0.005204808 | 0.007383264 | 1 |
| Obesity | LEC | *ABCC3* | 0.575457919 | 9.78E-06 | 0.000105108 | 0.061505084 |
| Obesity | LEC | *MPP2* | 1.896323943 | 1.78E-05 | 0.000105108 | 0.111638976 |
| Obesity | LEC | *EFTUD2* | 1.856620374 | 4.85E-06 | 9.10E-05 | 0.030473583 |
| Obesity | LEC | *EFNB3* | 10.30215377 | 7.64E-06 | 0.000105108 | 0.048026169 |
| Obesity | LEC | *FAM20A* | 0.582280798 | 2.00E-05 | 0.000105931 | 0.125959745 |
| Obesity | LEC | *YWHAE* | 1.570033069 | 0.004463986 | 0.006417809 | 1 |
| Obesity | LEC | *DPH1* | 1.971288616 | 1.21E-05 | 0.000105108 | 0.075902456 |
| Obesity | LEC | *DHRS7B* | 1.588867494 | 2.20E-05 | 0.000108376 | 0.138615175 |
| Obesity | LEC | *RCVRN* | -1.732853749 | 6.15E-05 | 0.000174839 | 0.386918324 |
| Obesity | LEC | *SLC9A3R1* | 2.075316395 | 4.42E-05 | 0.000140994 | 0.278180662 |
| Obesity | LEC | *TMEM104* | 2.241258533 | 4.87E-05 | 0.000149292 | 0.305899521 |
| Obesity | LEC | *TMEM97* | 1.639247972 | 2.91E-05 | 0.000115266 | 0.182926549 |
| Obesity | LEC | *RAB34* | 1.512268819 | 0.000204188 | 0.000424092 | 1 |
| Obesity | LEC | *SLAIN2* | 1.505447652 | 0.008082513 | 0.011073166 | 1 |
| Obesity | LEC | *OCIAD1* | 1.633491611 | 0.001174963 | 0.001904355 | 1 |
| Obesity | LEC | *CWH43* | 6.054387494 | 0.000152792 | 0.000337113 | 0.960600569 |
| Obesity | LEC | *KIAA1211* | 1.018948048 | 9.80E-06 | 0.000105108 | 0.061601011 |
| Obesity | LEC | *AREG* | 0.821014291 | 0.02064943 | 0.026641282 | 1 |
| Obesity | LEC | *UBE2D3* | 1.547404781 | 2.93E-05 | 0.000115648 | 0.183964481 |
| Obesity | LEC | *NDUFC1* | 1.912943869 | 2.25E-05 | 0.000109111 | 0.141634722 |
| Obesity | LEC | *TBC1D9* | 1.36128636 | 1.77E-05 | 0.000105108 | 0.111518802 |
| Obesity | LEC | *CPE* | 1.68276228 | 0.002929403 | 0.004359091 | 1 |
| Obesity | LEC | *WFS1* | -0.560803629 | 0.000367094 | 0.000686881 | 1 |
| Obesity | LEC | *GRPEL1* | 3.766710943 | 0.000124653 | 0.000287806 | 0.783695583 |
| Obesity | LEC | *AADAT* | 2.972571075 | 0.000368993 | 0.000689962 | 1 |
| Obesity | LEC | *CPZ* | 13.85100339 | 3.21E-05 | 0.00011749 | 0.201730852 |
| Obesity | LEC | *SLC2A9* | 0.719942393 | 2.28E-05 | 0.000109118 | 0.143631317 |
| Obesity | LEC | *SH3D19* | 0.794004852 | 0.037522381 | 0.046538412 | 1 |
| Obesity | LEC | *STIM2* | 1.830828436 | 0.000751253 | 0.001274454 | 1 |

| | | | | | | |
|---|---|---|---|---|---|---|
| Obesity | LEC | *RAPGEF2* | 1.440738256 | 0.002418246 | 0.003661733 | 1 |
| Obesity | LEC | *UFSP2* | 1.989348347 | 2.33E-05 | 0.000109118 | 0.14641018 |
| Obesity | LEC | *KLF3* | 1.254752804 | 0.039791728 | 0.049139774 | 1 |
| Obesity | LEC | *FAM149A* | 0.732930869 | 0.001954263 | 0.00301656 | 1 |
| Obesity | LEC | *UGDH* | 0.953010934 | 2.15E-05 | 0.000107557 | 0.13523656 |
| Obesity | LEC | *DDX25* | 2.394293683 | 0.000247585 | 0.000497995 | 1 |
| Obesity | LEC | *CTSC* | 1.060159996 | 9.13E-05 | 0.000228894 | 0.573837221 |
| Obesity | LEC | *ELP4* | 1.5710763 | 0.000331958 | 0.000632048 | 1 |
| Obesity | LEC | *SC5D* | 2.976807675 | 0.000280684 | 0.000551112 | 1 |
| Obesity | LEC | *C11orf63* | 0.996500677 | 1.30E-05 | 0.000105108 | 0.081594467 |
| Obesity | LEC | *DTX4* | -0.49457139 | 0.00019971 | 0.000416442 | 1 |
| Obesity | LEC | *PUS3* | -0.745519152 | 0.000322629 | 0.000616712 | 1 |
| Obesity | LEC | *DCPS* | 1.7170045 | 0.00017519 | 0.000375015 | 1 |
| Obesity | LEC | *FOXRED1* | 1.301939063 | 6.24E-07 | 3.24E-05 | 0.003922929 |
| Obesity | LEC | *MS4A4A* | 0.516635839 | 1.82E-05 | 0.000105108 | 0.114360471 |
| Obesity | LEC | *CCDC86* | -1.180034717 | 0.000158888 | 0.000347453 | 0.998928595 |
| Obesity | LEC | *HPX* | -0.786612014 | 0.002444004 | 0.003697173 | 1 |
| Obesity | LEC | *CHORDC1* | 1.551115771 | 4.38E-05 | 0.000140225 | 0.275402392 |
| Obesity | LEC | *FOLR3* | 0.705694189 | 0.014057996 | 0.018552187 | 1 |
| Obesity | LEC | *EIF4G2* | 1.883009043 | 0.000927465 | 0.001537704 | 1 |
| Obesity | LEC | *GALNT18* | -0.320538642 | 1.00E-04 | 0.00024418 | 0.62851844 |
| Obesity | LEC | *CBL* | 1.804440905 | 1.33E-05 | 0.000105108 | 0.08380694 |
| Obesity | LEC | *KIAA1549L* | 1.035325285 | 6.06E-05 | 0.000173168 | 0.381250836 |
| Obesity | LEC | *CD5* | 1.07664095 | 5.35E-06 | 9.39E-05 | 0.033621035 |
| Obesity | LEC | *ACCS* | 0.864910747 | 0.000126473 | 0.000290753 | 0.795138859 |
| Obesity | LEC | *NAA40* | 2.342981618 | 5.74E-05 | 0.000166792 | 0.360771162 |
| Obesity | LEC | *CARS* | 1.176139144 | 0.000469042 | 0.000844706 | 1 |
| Obesity | LEC | *SLC22A18* | 0.706817959 | 5.67E-06 | 9.65E-05 | 0.035616425 |
| Obesity | LEC | *SLC35F2* | 0.731367174 | 0.000295303 | 0.000573015 | 1 |
| Obesity | LEC | *C11orf21* | 1.06688819 | 0.001922362 | 0.002972361 | 1 |
| Obesity | LEC | *ELMOD1* | -1.901072065 | 0.001274054 | 0.002045448 | 1 |
| Obesity | LEC | *AIP* | 2.220422861 | 1.60E-05 | 0.000105108 | 0.100658144 |
| Obesity | LEC | *NDUFS8* | 2.73829043 | 0.000161406 | 0.000351736 | 1 |
| Obesity | LEC | *TCIRG1* | 1.594803459 | 3.23E-06 | 7.50E-05 | 0.020312615 |
| Obesity | LEC | *GTF2H1* | 1.711284195 | 2.98E-06 | 7.26E-05 | 0.018724398 |
| Obesity | LEC | *PTPN5* | 0.908257584 | 0.000606292 | 0.001056474 | 1 |
| Obesity | LEC | *PPFIBP1* | -0.186542785 | 0.000112518 | 0.000265819 | 0.70739755 |
| Obesity | LEC | *PRDM4* | 2.376552149 | 0.000465583 | 0.000839881 | 1 |
| Obesity | LEC | *ASIC1* | 0.784644331 | 0.004078308 | 0.005901111 | 1 |
| Obesity | LEC | *CAPRIN2* | 1.666443171 | 5.17E-05 | 0.000155257 | 0.325108009 |
| Obesity | LEC | *CYP27B1* | 5.768010279 | 2.40E-05 | 0.000109361 | 0.150812345 |
| Obesity | LEC | *ACSS3* | 0.657311215 | 0.000577938 | 0.001013527 | 1 |
| Obesity | LEC | *METAP2* | 1.520765244 | 3.99E-05 | 0.000131515 | 0.250740678 |
| Obesity | LEC | *LTA4H* | 0.850130862 | 1.84E-05 | 0.000105108 | 0.115784473 |
| Obesity | LEC | *WNT5B* | -1.053893508 | 0.005496263 | 0.007777404 | 1 |
| Obesity | LEC | *TRPV4* | 1.178484422 | 0.032256367 | 0.040325269 | 1 |
| Obesity | LEC | *ITFG2* | 1.362782843 | 2.64E-05 | 0.000112582 | 0.165784107 |
| Obesity | LEC | *PRR4* | 1.32917072 | 4.97E-05 | 0.000151436 | 0.312771211 |
| Obesity | LEC | *GPN3* | 2.355127272 | 5.35E-05 | 0.00015899 | 0.33632356 |
| Obesity | LEC | *VPS29* | 2.085400799 | 6.65E-05 | 0.000183512 | 0.417856611 |

| | | | | | | |
|---|---|---|---|---|---|---|
| Obesity | LEC | *AKAP3* | 0.959895406 | 5.34E-05 | 0.000158981 | 0.335926468 |
| Obesity | LEC | *KCNA1* | 1.551612988 | 0.00026607 | 0.000527477 | 1 |
| Obesity | LEC | *CREBL2* | 1.816791847 | 0.001613032 | 0.002531485 | 1 |
| Obesity | LEC | *NAA25* | 1.816372619 | 0.002073588 | 0.003185886 | 1 |
| Obesity | LEC | *SCNN1A* | 1.051152135 | 2.78E-05 | 0.00011384 | 0.174869963 |
| Obesity | LEC | *LTBR* | 1.762971119 | 5.21E-06 | 9.34E-05 | 0.032766176 |
| Obesity | LEC | *CDK2AP1* | 0.908526699 | 0.003559472 | 0.005212764 | 1 |
| Obesity | LEC | *MGP* | 0.938722806 | 6.37E-05 | 0.000178168 | 0.400638125 |
| Obesity | LEC | *RASAL1* | 2.317375484 | 0.017205208 | 0.022451047 | 1 |
| Obesity | LEC | *ARHGDIB* | 1.415172713 | 5.38E-05 | 0.000159684 | 0.338209752 |
| Obesity | LEC | *EIF2B1* | 1.488515891 | 1.41E-05 | 0.000105108 | 0.088792842 |
| Obesity | LEC | *DDX55* | 2.161308402 | 2.01E-08 | 3.33E-06 | 0.00012642 |
| Obesity | LEC | *ENDOU* | -1.409423555 | 0.001322272 | 0.002115838 | 1 |
| Obesity | LEC | *STX2* | 1.349969962 | 0.005799693 | 0.008164503 | 1 |
| Obesity | LEC | *CAND1* | 1.202120785 | 0.021606797 | 0.027756831 | 1 |
| Obesity | LEC | *NUP107* | 1.974854861 | 4.43E-05 | 0.000141063 | 0.278459056 |
| Obesity | LEC | *CNOT2* | 1.536849041 | 0.000132973 | 0.000302461 | 0.836002268 |
| Obesity | LEC | *KRR1* | 1.797273097 | 0.000810513 | 0.001361758 | 1 |
| Obesity | LEC | *COPS7A* | 1.453083144 | 9.22E-05 | 0.000230354 | 0.579571641 |
| Obesity | LEC | *CHPT1* | 0.62752217 | 0.000247461 | 0.000497995 | 1 |
| Obesity | LEC | *GNPTAB* | 1.558634642 | 1.69E-05 | 0.000105108 | 0.105986451 |
| Obesity | LEC | *ATN1* | 1.482532292 | 1.57E-05 | 0.000105108 | 0.098913701 |
| Obesity | LEC | *NT5DC3* | 1.190647017 | 0.010308567 | 0.013916677 | 1 |
| Obesity | LEC | *ST8SIA1* | 0.808963101 | 2.29E-05 | 0.000109118 | 0.143970047 |
| Obesity | LEC | *C2CD5* | 1.615334647 | 4.52E-05 | 0.000143 | 0.284427933 |
| Obesity | LEC | *PHC1* | 2.743003723 | 2.52E-05 | 0.000111345 | 0.158554523 |
| Obesity | LEC | *COL12A1* | 0.941165047 | 1.64E-05 | 0.000105108 | 0.102858184 |
| Obesity | LEC | *BTN3A3* | 1.382109349 | 2.72E-06 | 7.03E-05 | 0.01711327 |
| Obesity | LEC | *RSPH4A* | -0.688979228 | 0.000615498 | 0.001069846 | 1 |
| Obesity | LEC | *SMIM8* | 1.29762209 | 4.70E-06 | 9.00E-05 | 0.029544035 |
| Obesity | LEC | *MCM9* | 1.863720688 | 0.00011943 | 0.000278818 | 0.75085819 |
| Obesity | LEC | *FAM184A* | 1.921079982 | 0.000145608 | 0.000324766 | 0.915437732 |
| Obesity | LEC | *RNGTT* | 1.707549476 | 5.17E-05 | 0.000155274 | 0.325298316 |
| Obesity | LEC | *GABRR2* | 0.989342039 | 3.66E-06 | 7.87E-05 | 0.022982618 |
| Obesity | LEC | *HDDC2* | 1.125261168 | 3.04E-05 | 0.000115831 | 0.19101367 |
| Obesity | LEC | *TPD52L1* | 1.464747857 | 0.000816696 | 0.001369218 | 1 |
| Obesity | LEC | *UST* | 0.642107217 | 1.91E-05 | 0.000105211 | 0.120188374 |
| Obesity | LEC | *MTRF1L* | 1.906922577 | 0.000297946 | 0.000576897 | 1 |
| Obesity | LEC | *TULP1* | 9.282508399 | 1.40E-05 | 0.000105108 | 0.087736377 |
| Obesity | LEC | *SLC26A8* | 0.469309822 | 0.027127887 | 0.034351062 | 1 |
| Obesity | LEC | *MAPK14* | 1.366973618 | 0.006148331 | 0.00861863 | 1 |
| Obesity | LEC | *RNF8* | 1.853255333 | 4.27E-05 | 0.000137557 | 0.268762611 |
| Obesity | LEC | *PHACTR1* | 1.144495141 | 2.48E-05 | 0.000110998 | 0.156185988 |
| Obesity | LEC | *CD83* | 0.703070209 | 6.02E-05 | 0.00017227 | 0.378476644 |
| Obesity | LEC | *MDN1* | 1.444709166 | 0.005606246 | 0.007925899 | 1 |
| Obesity | LEC | *SAYSD1* | 1.728826727 | 1.81E-05 | 0.000105108 | 0.113946015 |
| Obesity | LEC | *CAP2* | 0.681225328 | 0.000835431 | 0.001397645 | 1 |
| Obesity | LEC | *GPR63* | -0.542386316 | 0.000661071 | 0.001136493 | 1 |
| Obesity | LEC | *ASCC3* | 1.636494306 | 0.000483636 | 0.000868253 | 1 |
| Obesity | LEC | *BVES* | 0.648926147 | 0.010137003 | 0.013693885 | 1 |

| | | | | | | |
|---|---|---|---|---|---|---|
| Obesity | LEC | *WASF1* | 0.887911668 | 0.000607171 | 0.00105742 | 1 |
| Obesity | LEC | *GPLD1* | 0.868886163 | 0.000278345 | 0.000547546 | 1 |
| Obesity | LEC | *ALDH5A1* | 0.798805138 | 1.74E-05 | 0.000105108 | 0.10968786 |
| Obesity | LEC | *VNN1* | 0.628768559 | 0.000143233 | 0.000320464 | 0.90050405 |
| Obesity | LEC | *VNN2* | 2.984506144 | 0.000633919 | 0.001095505 | 1 |
| Obesity | LEC | *ACOT13* | 1.892193281 | 2.45E-05 | 0.000110187 | 0.154151757 |
| Obesity | LEC | *C6orf62* | 1.574068909 | 2.69E-05 | 0.000113184 | 0.169423147 |
| Obesity | LEC | *B3GAT2* | 1.569369546 | 0.001315528 | 0.002106117 | 1 |
| Obesity | LEC | *HBS1L* | 1.181101447 | 0.01702448 | 0.022238293 | 1 |
| Obesity | LEC | *PERP* | -0.907840676 | 0.002465818 | 0.003728379 | 1 |
| Obesity | LEC | *SLC16A10* | 0.543650039 | 3.05E-05 | 0.000115831 | 0.191821331 |
| Obesity | LEC | *PHACTR2* | 1.306120771 | 2.52E-05 | 0.000111345 | 0.15841973 |
| Obesity | LEC | *PACRG* | 3.221747505 | 0.000311698 | 0.00059928 | 1 |
| Obesity | LEC | *TFEB* | -1.000321584 | 0.004935187 | 0.00702139 | 1 |
| Obesity | LEC | *FAM120B* | 1.475939779 | 0.000213489 | 0.00043906 | 1 |
| Obesity | LEC | *TBP* | 2.199548208 | 0.000754668 | 0.001279901 | 1 |
| Obesity | LEC | *DNPH1* | 0.913040608 | 0.004125856 | 0.005960307 | 1 |
| Obesity | LEC | *DUSP22* | 2.079358021 | 0.000102908 | 0.000249503 | 0.646985659 |
| Obesity | LEC | *EXOC2* | 1.764928555 | 0.000966892 | 0.001595499 | 1 |
| Obesity | LEC | *TMEM30A* | 1.441941671 | 0.025920329 | 0.032904369 | 1 |
| Obesity | LEC | *SENP6* | 1.524097229 | 0.00016949 | 0.000364927 | 1 |
| Obesity | LEC | *TTK* | 1.640441388 | 0.000463401 | 0.000836945 | 1 |
| Obesity | LEC | *CLIC5* | 0.562820177 | 4.47E-05 | 0.000141942 | 0.281046515 |
| Obesity | LEC | *FBRSL1* | 1.580334259 | 8.97E-06 | 0.000105108 | 0.056373474 |
| Obesity | LEC | *LY86* | 0.746134999 | 2.10E-05 | 0.000106989 | 0.132090889 |
| Obesity | LEC | *PRSS16* | 5.514102445 | 1.61E-06 | 5.41E-05 | 0.01010711 |
| Obesity | LEC | *TBX18* | 1.195719214 | 0.001750381 | 0.002726622 | 1 |
| Obesity | LEC | *PCDHB2* | -1.100043345 | 0.000448053 | 0.000812024 | 1 |
| Obesity | LEC | *NUDT12* | 1.37129094 | 5.07E-05 | 0.0001531 | 0.318561737 |
| Obesity | LEC | *C7* | 1.645689645 | 5.06E-05 | 0.0001531 | 0.318312627 |
| Obesity | LEC | *PAPD7* | 1.562379151 | 0.009987423 | 0.013503425 | 1 |
| Obesity | LEC | *GHR* | 0.760618709 | 3.73E-05 | 0.000126137 | 0.234286705 |
| Obesity | LEC | *DAP* | 2.228820508 | 2.19E-05 | 0.000108369 | 0.13756303 |
| Obesity | LEC | *NME5* | 1.461990505 | 0.006201472 | 0.008685624 | 1 |
| Obesity | LEC | *NNT* | 1.218271707 | 0.00034713 | 0.000654788 | 1 |
| Obesity | LEC | *PFDN1* | 1.929365392 | 0.000109085 | 0.000259584 | 0.685819674 |
| Obesity | LEC | *APBB3* | 1.122691335 | 0.001191242 | 0.001927261 | 1 |
| Obesity | LEC | *TMCO6* | -1.830972208 | 0.009943997 | 0.013450497 | 1 |
| Obesity | LEC | *SPARC* | 1.103769882 | 3.73E-05 | 0.000126146 | 0.234632267 |
| Obesity | LEC | *PCDHB6* | 3.384715696 | 1.11E-05 | 0.000105108 | 0.070043171 |
| Obesity | LEC | *PDE8B* | 1.165272489 | 0.001323075 | 0.002116529 | 1 |
| Obesity | LEC | *CLK4* | 1.706781325 | 3.94E-05 | 0.000130647 | 0.247752832 |
| Obesity | LEC | *PCDHB15* | -0.622850969 | 0.001622322 | 0.00254416 | 1 |
| Obesity | LEC | *HAVCR1* | 0.783256682 | 0.000801974 | 0.001348492 | 1 |
| Obesity | LEC | *RNF130* | 1.05067201 | 1.50E-05 | 0.000105108 | 0.094072278 |
| Obesity | LEC | *ARSB* | 1.155378047 | 0.000318434 | 0.000609451 | 1 |
| Obesity | LEC | *CLINT1* | 1.317227286 | 1.65E-05 | 0.000105108 | 0.103686179 |
| Obesity | LEC | *BTNL8* | 0.461461652 | 0.009043694 | 0.012306862 | 1 |
| Obesity | LEC | *MSH3* | 0.789751435 | 0.02495287 | 0.031763251 | 1 |
| Obesity | LEC | *RASGRF2* | 0.918602959 | 1.01E-05 | 0.000105108 | 0.063759889 |

| | | | | | | |
|---|---|---|---|---|---|---|
| Obesity | LEC | *POLR3G* | 0.797743923 | 6.33E-05 | 0.000177525 | 0.398011118 |
| Obesity | LEC | *DROSHA* | 1.596184021 | 2.59E-05 | 0.000112124 | 0.163023195 |
| Obesity | LEC | *CDH6* | 0.442295858 | 0.019966798 | 0.025813543 | 1 |
| Obesity | LEC | *SLC27A6* | 1.162992777 | 3.47E-05 | 0.000121951 | 0.218101272 |
| Obesity | LEC | *TARS* | 2.427513914 | 0.000106632 | 0.000255389 | 0.670395328 |
| Obesity | LEC | *LNPEP* | 1.686475322 | 1.94E-05 | 0.000105217 | 0.121900913 |
| Obesity | LEC | *SLC12A7* | 1.944211201 | 0.014300843 | 0.018841031 | 1 |
| Obesity | LEC | *GNPDA1* | 2.580020494 | 0.00029042 | 0.000565986 | 1 |
| Obesity | LEC | *SKP1* | 2.057698131 | 9.50E-06 | 0.000105108 | 0.059696976 |
| Obesity | LEC | *PPWD1* | 1.081682627 | 0.003334565 | 0.004911079 | 1 |
| Obesity | LEC | *TRIM23* | 1.763998657 | 0.008050383 | 0.011031552 | 1 |
| Obesity | LEC | *TRAPPC13* | 1.381278691 | 0.001486327 | 0.002353789 | 1 |
| Obesity | LEC | *C9* | 1.1002644 | 0.00069772 | 0.001194599 | 1 |
| Obesity | LEC | *HMGXB3* | 1.643377389 | 0.012194731 | 0.016243278 | 1 |
| Obesity | LEC | *ATP6V0E1* | 1.759070507 | 0.000730643 | 0.001244528 | 1 |
| Obesity | LEC | *BNIP1* | -0.844210834 | 0.010705507 | 0.014418493 | 1 |
| Obesity | LEC | *CNTN3* | 0.848132935 | 1.71E-05 | 0.000105108 | 0.107601236 |
| Obesity | LEC | *ACTR8* | 1.50674986 | 2.52E-05 | 0.000111345 | 0.158635942 |
| Obesity | LEC | *HGD* | -1.386434056 | 0.000353471 | 0.000664554 | 1 |
| Obesity | LEC | *NIT2* | 0.814987707 | 1.91E-05 | 0.000105211 | 0.120331598 |
| Obesity | LEC | *FAM162A* | 2.097781033 | 0.000332196 | 0.000632309 | 1 |
| Obesity | LEC | *PCCB* | 2.264216509 | 1.11E-05 | 0.000105108 | 0.069903516 |
| Obesity | LEC | *RBP1* | 2.561861865 | 1.83E-05 | 0.000105108 | 0.115135582 |
| Obesity | LEC | *RNF7* | 1.672528033 | 0.000238509 | 0.000482311 | 1 |
| Obesity | LEC | *BCHE* | -0.342100977 | 3.99E-05 | 0.000131515 | 0.251062035 |
| Obesity | LEC | *PDCD10* | 1.918102839 | 0.000177924 | 0.000380349 | 1 |
| Obesity | LEC | *WNT5A* | -1.219667744 | 0.000185001 | 0.000392012 | 1 |
| Obesity | LEC | *PRKAR2A* | 1.665473646 | 3.42E-07 | 2.39E-05 | 0.0021497 |
| Obesity | LEC | *HYAL1* | -1.046985834 | 1.44E-06 | 5.18E-05 | 0.009070629 |
| Obesity | LEC | *CYB561D2* | 1.334049642 | 1.07E-05 | 0.000105108 | 0.067395841 |
| Obesity | LEC | *C3orf14* | 0.652631017 | 0.001597312 | 0.002509321 | 1 |
| Obesity | LEC | *IFT57* | 2.401788996 | 0.000115316 | 0.000271366 | 0.724990878 |
| Obesity | LEC | *IQCG* | 1.425477029 | 1.47E-05 | 0.000105108 | 0.092641914 |
| Obesity | LEC | *GBE1* | 1.013949209 | 1.65E-05 | 0.000105108 | 0.103470925 |
| Obesity | LEC | *UMPS* | 1.798698507 | 0.000307882 | 0.000593032 | 1 |
| Obesity | LEC | *NCBP2* | 2.318462581 | 2.23E-05 | 0.000108559 | 0.140149241 |
| Obesity | LEC | *SLC41A3* | -0.68890127 | 0.00022167 | 0.000454102 | 1 |
| Obesity | LEC | *PLXNA1* | 0.925358212 | 4.58E-06 | 8.97E-05 | 0.028779308 |
| Obesity | LEC | *ABTB1* | 2.537751506 | 0.000169455 | 0.000364927 | 1 |
| Obesity | LEC | *EFCC1* | -0.195620023 | 5.38E-05 | 0.000159748 | 0.338506073 |
| Obesity | LEC | *NEK11* | 1.970370469 | 0.000146708 | 0.000326728 | 0.922352462 |
| Obesity | LEC | *PLSCR4* | -0.50549232 | 0.000809442 | 0.001360321 | 1 |
| Obesity | LEC | *HEMK1* | 0.978522447 | 7.35E-06 | 0.000105108 | 0.046213627 |
| Obesity | LEC | *WDR48* | 1.815120636 | 0.000986105 | 0.001625496 | 1 |
| Obesity | LEC | *COMMD2* | 1.765748853 | 8.62E-06 | 0.000105108 | 0.054184863 |
| Obesity | LEC | *ABCC5* | 1.376510418 | 6.86E-05 | 0.000187191 | 0.431100753 |
| Obesity | LEC | *AADAC* | 0.233347689 | 0.013619199 | 0.018010918 | 1 |
| Obesity | LEC | *ARHGEF26* | 0.783726748 | 4.91E-06 | 9.11E-05 | 0.030876226 |
| Obesity | LEC | *KLHL24* | 1.586411555 | 3.99E-07 | 2.51E-05 | 0.002508683 |
| Obesity | LEC | *PLCH1* | 0.978481076 | 6.85E-06 | 0.000104405 | 0.043054181 |

| | | | | | | |
|---|---|---|---|---|---|---|
| Obesity | LEC | *NKTR* | 1.034597666 | 0.031873651 | 0.039870602 | 1 |
| Obesity | LEC | *CLCN2* | 4.342996179 | 8.34E-06 | 0.000105108 | 0.052431583 |
| Obesity | LEC | *NEK4* | 2.011763895 | 0.000155324 | 0.000341202 | 0.976519822 |
| Obesity | LEC | *MOB1A* | 2.135900249 | 0.001978127 | 0.003052014 | 1 |
| Obesity | LEC | *LMAN2L* | 1.593951407 | 0.001410274 | 0.002244656 | 1 |
| Obesity | LEC | *PIKFYVE* | 1.315600427 | 3.48E-05 | 0.000122161 | 0.218789704 |
| Obesity | LEC | *KCNIP3* | 3.338701969 | 1.13E-05 | 0.000105108 | 0.071307594 |
| Obesity | LEC | *FAHD2A* | 3.033294893 | 3.82E-07 | 2.50E-05 | 0.002401202 |
| Obesity | LEC | *NCL* | 1.884764947 | 7.72E-05 | 0.000202909 | 0.485358097 |
| Obesity | LEC | *ACTR1B* | 2.774278307 | 4.22E-07 | 2.56E-05 | 0.002650286 |
| Obesity | LEC | *SLC35F5* | 1.423456973 | 0.005408377 | 0.007658213 | 1 |
| Obesity | LEC | *ACTR3* | 1.769925245 | 0.000440844 | 0.000801731 | 1 |
| Obesity | LEC | *STEAP3* | 0.663391993 | 9.82E-05 | 0.000241277 | 0.617186038 |
| Obesity | LEC | *TFCP2L1* | 1.289864373 | 2.84E-06 | 7.08E-05 | 0.017836278 |
| Obesity | LEC | *STAM2* | 1.824023542 | 7.59E-05 | 0.000200935 | 0.477277668 |
| Obesity | LEC | *GPD2* | 1.305232345 | 3.30E-05 | 0.000119154 | 0.207604421 |
| Obesity | LEC | *CENPA* | -0.851632049 | 0.000220323 | 0.000451784 | 1 |
| Obesity | LEC | *TANC1* | -0.592090657 | 0.002471095 | 0.003734458 | 1 |
| Obesity | LEC | *ITGB6* | 0.691327784 | 0.007264736 | 0.010042523 | 1 |
| Obesity | LEC | *ITGA4* | 1.322575965 | 7.92E-06 | 0.000105108 | 0.049767992 |
| Obesity | LEC | *PPM1G* | 1.620367592 | 5.44E-05 | 0.000160966 | 0.342213693 |
| Obesity | LEC | *PCSK4* | 1.184870383 | 4.85E-05 | 0.000149091 | 0.305189166 |
| Obesity | LEC | *MOGS* | 0.946716923 | 0.004314833 | 0.006223298 | 1 |
| Obesity | LEC | *GRB14* | 0.640706137 | 2.96E-05 | 0.000115831 | 0.185905376 |
| Obesity | LEC | *CLIP4* | 0.738183659 | 1.34E-05 | 0.000105108 | 0.084052551 |
| Obesity | LEC | *RTN4* | 0.906504073 | 0.003334722 | 0.004911079 | 1 |
| Obesity | LEC | *HTRA2* | -1.49967433 | 0.031214474 | 0.039162921 | 1 |
| Obesity | LEC | *GALNT3* | 1.129101395 | 0.000153688 | 0.000338674 | 0.966235532 |
| Obesity | LEC | *POLE4* | 2.35096648 | 1.90E-05 | 0.000105211 | 0.119167721 |
| Obesity | LEC | *MRPL19* | 1.467995816 | 0.001855721 | 0.002874334 | 1 |
| Obesity | LEC | *LANCL1* | 1.940380979 | 2.13E-05 | 0.00010722 | 0.134131934 |
| Obesity | LEC | *WDR75* | 2.061238853 | 3.34E-05 | 0.000119779 | 0.21004887 |
| Obesity | LEC | *EFEMP1* | -0.533841999 | 0.002561657 | 0.0038483 | 1 |
| Obesity | LEC | *DNAH6* | 0.314049851 | 0.006766627 | 0.009409817 | 1 |
| Obesity | LEC | *GGCX* | 0.98774854 | 1.35E-06 | 5.03E-05 | 0.008497527 |
| Obesity | LEC | *GNLY* | 0.578970672 | 2.66E-05 | 0.000112975 | 0.167475399 |
| Obesity | LEC | *SF3B1* | 1.55933639 | 0.000122489 | 0.000283851 | 0.770087324 |
| Obesity | LEC | *ST3GAL5* | 1.133764442 | 1.85E-05 | 0.000105108 | 0.116357445 |
| Obesity | LEC | *PLCD4* | 2.903681676 | 0.002218168 | 0.003388867 | 1 |
| Obesity | LEC | *CHMP3* | -1.711630591 | 0.007858461 | 0.010785012 | 1 |
| Obesity | LEC | *ZNF142* | 1.643244572 | 4.52E-06 | 8.91E-05 | 0.028409651 |
| Obesity | LEC | *WNT6* | 4.74771977 | 0.002461974 | 0.003723462 | 1 |
| Obesity | LEC | *HDLBP* | 1.591599255 | 0.000365274 | 0.000683882 | 1 |
| Obesity | LEC | *PASK* | 2.005788654 | 0.026938027 | 0.034117522 | 1 |
| Obesity | LEC | *TPO* | 0.471767074 | 0.000377815 | 0.000703383 | 1 |
| Obesity | LEC | *TAF1B* | 1.561413543 | 8.77E-05 | 0.000222231 | 0.551594322 |
| Obesity | LEC | *CEBPZ* | 1.643124994 | 0.00204045 | 0.00313728 | 1 |
| Obesity | LEC | *PRKD3* | 1.549579357 | 0.000635207 | 0.001096827 | 1 |
| Obesity | LEC | *SLC25A12* | 1.211354192 | 2.51E-05 | 0.000111345 | 0.15769083 |
| Obesity | LEC | *RMDN2* | 2.081947477 | 0.002340522 | 0.003554314 | 1 |

| | | | | | | |
|---|---|---|---|---|---|---|
| Obesity | LEC | *SDC1* | 3.21258544 | 0.002158024 | 0.003306726 | 1 |
| Obesity | LEC | *SOS1* | 1.086394931 | 1.71E-05 | 0.000105108 | 0.107572365 |
| Obesity | LEC | *WIPF1* | 1.473150384 | 6.22E-05 | 0.000175786 | 0.391196276 |
| Obesity | LEC | *COX7A2L* | -0.95314618 | 0.003489705 | 0.005120135 | 1 |
| Obesity | LEC | *THADA* | 1.557737692 | 8.08E-05 | 0.000209699 | 0.50794598 |
| Obesity | LEC | *ARID3A* | 1.146421661 | 1.47E-05 | 0.000105108 | 0.092414583 |
| Obesity | LEC | *CD207* | 15.03767775 | 4.81E-06 | 9.10E-05 | 0.030253238 |
| Obesity | LEC | *GRIN3B* | 2.015963188 | 0.001574187 | 0.002476705 | 1 |
| Obesity | LEC | *VAX2* | 0.968693643 | 2.70E-05 | 0.000113262 | 0.169764001 |
| Obesity | LEC | *MSH6* | 1.940707116 | 4.32E-05 | 0.000138608 | 0.271532348 |
| Obesity | LEC | *PLEKHA3* | 2.136831998 | 2.84E-05 | 0.000114842 | 0.178644374 |
| Obesity | LEC | *ALMS1* | 1.547764332 | 7.91E-05 | 0.000206694 | 0.497384355 |
| Obesity | LEC | *BCL9* | 1.254480097 | 3.36E-05 | 0.000120044 | 0.21139795 |
| Obesity | LEC | *DHCR24* | 1.490281453 | 5.30E-05 | 0.000157883 | 0.333291362 |
| Obesity | LEC | *GPX7* | 0.606625054 | 0.000462142 | 0.000835151 | 1 |
| Obesity | LEC | *SCP2* | 1.626267518 | 1.86E-05 | 0.000105108 | 0.116656506 |
| Obesity | LEC | *FAM20B* | 2.242378909 | 0.000228879 | 0.000465683 | 1 |
| Obesity | LEC | *TMEM59* | 1.858121534 | 0.000151181 | 0.000334675 | 0.950477904 |
| Obesity | LEC | *WRAP73* | 2.833824887 | 0.00128929 | 0.002068854 | 1 |
| Obesity | LEC | *MRPL37* | -1.307946341 | 0.003381197 | 0.004973698 | 1 |
| Obesity | LEC | *ICMT* | 1.505284019 | 1.99E-05 | 0.000105667 | 0.125128346 |
| Obesity | LEC | *KCNC4* | -1.09605961 | 0.022716605 | 0.02907559 | 1 |
| Obesity | LEC | *EDEM3* | 1.609737434 | 0.000111581 | 0.000263825 | 0.701511556 |
| Obesity | LEC | *CAPZA1* | 1.513828442 | 0.000116568 | 0.000273661 | 0.732862984 |
| Obesity | LEC | *DOCK7* | 1.268078761 | 2.02E-05 | 0.000106047 | 0.126986848 |
| Obesity | LEC | *FBXO6* | 0.950403486 | 0.000115664 | 0.000272047 | 0.727181221 |
| Obesity | LEC | *DNAJC6* | 0.931746665 | 6.67E-05 | 0.000183765 | 0.419351107 |
| Obesity | LEC | *LEPR* | 1.769113367 | 0.000131532 | 0.000300161 | 0.826943744 |
| Obesity | LEC | *MIIP* | 2.003037205 | 5.63E-05 | 0.000165104 | 0.353817525 |
| Obesity | LEC | *SMG7* | 1.705009459 | 0.001742528 | 0.002715061 | 1 |
| Obesity | LEC | *NCF2* | 0.936243572 | 1.83E-05 | 0.000105108 | 0.115231359 |
| Obesity | LEC | *RGS2* | 1.289418122 | 2.88E-05 | 0.000115038 | 0.181069492 |
| Obesity | LEC | *OLFML3* | 1.032771403 | 1.16E-05 | 0.000105108 | 0.07311331 |
| Obesity | LEC | *CFHR3* | -0.778921735 | 6.88E-05 | 0.000187286 | 0.432659514 |
| Obesity | LEC | *CRYZ* | 1.239050479 | 0.013727068 | 0.018142122 | 1 |
| Obesity | LEC | *TTF2* | 0.983244603 | 1.65E-05 | 0.000105108 | 0.103953578 |
| Obesity | LEC | *WARS2* | 1.79021709 | 3.00E-06 | 7.28E-05 | 0.018864605 |
| Obesity | LEC | *OSCP1* | 1.838440699 | 0.000163045 | 0.000354572 | 1 |
| Obesity | LEC | *GNPAT* | 1.454433824 | 0.023576331 | 0.030090214 | 1 |
| Obesity | LEC | *C1orf109* | 0.948352843 | 6.69E-06 | 0.000103165 | 0.042091381 |
| Obesity | LEC | *TBCE* | 1.241593278 | 2.34E-05 | 0.000109118 | 0.147336402 |
| Obesity | LEC | *MTR* | 1.416466613 | 3.86E-05 | 0.000128947 | 0.242549756 |
| Obesity | LEC | *KCNQ4* | -0.736259011 | 9.41E-05 | 0.000233893 | 0.591378803 |
| Obesity | LEC | *ACADM* | 1.925929771 | 0.000230574 | 0.000468525 | 1 |
| Obesity | LEC | *PADI2* | 1.230525377 | 6.60E-05 | 0.000182779 | 0.415035362 |
| Obesity | LEC | *MFAP2* | -1.419074743 | 0.001916954 | 0.002965524 | 1 |
| Obesity | LEC | *CTBS* | 1.664563425 | 0.002240423 | 0.003417161 | 1 |
| Obesity | LEC | *KLHL12* | 1.148631755 | 6.29E-05 | 0.000176718 | 0.395141794 |
| Obesity | LEC | *ZNHIT6* | 1.178782804 | 0.02088802 | 0.026910447 | 1 |
| Obesity | LEC | *PLA2G2D* | -3.802987012 | 3.05E-05 | 0.000115831 | 0.191472721 |

| | | | | | | |
|---|---|---|---|---|---|---|
| Obesity | LEC | *KIF17* | 1.217548467 | 0.000335802 | 0.000638206 | 1 |
| Obesity | LEC | *CD46* | 1.333318441 | 2.32E-05 | 0.000109118 | 0.145839197 |
| Obesity | LEC | *PRDX1* | 1.721793807 | 0.016671094 | 0.021799328 | 1 |
| Obesity | LEC | *FAAH* | 0.54703613 | 0.000830289 | 0.001389783 | 1 |
| Obesity | LEC | *NSUN4* | 1.32281375 | 0.038514396 | 0.047673828 | 1 |
| Obesity | LEC | *TMED5* | 1.859428075 | 8.44E-05 | 0.000215965 | 0.530409892 |
| Obesity | LEC | *PRRC2C* | 1.757582763 | 2.14E-05 | 0.000107284 | 0.134319044 |
| Obesity | LEC | *ABCD3* | 1.886898618 | 7.69E-05 | 0.00020244 | 0.483223327 |
| Obesity | LEC | *VAMP4* | 2.04926358 | 0.000104668 | 0.000252318 | 0.6580465 |
| Obesity | LEC | *PTBP2* | 1.581796905 | 8.82E-05 | 0.000222743 | 0.554408116 |
| Obesity | LEC | *TNFSF4* | 0.749487169 | 0.000631122 | 0.001091873 | 1 |
| Obesity | LEC | *DARS2* | -1.923438049 | 0.001921211 | 0.002971378 | 1 |
| Obesity | LEC | *IRF6* | 1.066419758 | 0.00031702 | 0.000607468 | 1 |
| Obesity | LEC | *DIEXF* | -1.554952353 | 0.00533962 | 0.007565966 | 1 |
| Obesity | LEC | *SERPINC1* | 3.01764367 | 2.60E-14 | 1.49E-11 | 1.64E-10 |
| Obesity | LEC | *RCAN3* | 1.220841237 | 4.11E-06 | 8.49E-05 | 0.025814546 |
| Obesity | LEC | *RCOR3* | 1.500077997 | 2.87E-05 | 0.000115038 | 0.180619956 |
| Obesity | LEC | *MTFR1L* | 1.659622905 | 1.78E-05 | 0.000105108 | 0.111900138 |
| Obesity | LEC | *NSL1* | 1.656838308 | 0.002117524 | 0.00324942 | 1 |
| Obesity | LEC | *RPA2* | 2.274800383 | 4.04E-05 | 0.00013241 | 0.253764863 |
| Obesity | LEC | *OSBPL9* | 1.200955684 | 0.022573333 | 0.028903981 | 1 |
| Obesity | LEC | *TXNDC12* | 1.863007448 | 1.99E-05 | 0.000105667 | 0.124845577 |
| Obesity | LEC | *ESYT2* | 1.199678214 | 0.000517881 | 0.00092001 | 1 |
| Obesity | LEC | *A4GNT* | 0.748175206 | 0.029387421 | 0.037010961 | 1 |
| Obesity | LEC | *KMT2A* | 1.686288751 | 0.000415817 | 0.000763505 | 1 |
| Obesity | LEC | *DDX59* | -0.913125896 | 0.036404753 | 0.045241487 | 1 |
| Obesity | LEC | *ATF6* | 1.69171583 | 0.000340563 | 0.000645693 | 1 |
| Obesity | LEC | *CASC1* | 0.713989257 | 0.000117551 | 0.000275351 | 0.739043105 |
| Obesity | LEC | *LRMP* | 1.82610285 | 2.70E-05 | 0.000113262 | 0.169779511 |
| Obesity | LEC | *ATP10B* | -1.606259094 | 0.001114469 | 0.001814728 | 1 |
| Obesity | LEC | *SPCS2* | 1.986896798 | 4.24E-05 | 0.000136912 | 0.266392047 |
| Obesity | LEC | *ELOVL4* | 1.139202588 | 1.88E-05 | 0.000105211 | 0.118462572 |
| Obesity | LEC | *FILIP1* | -0.678517142 | 0.036169785 | 0.044967261 | 1 |
| Obesity | LEC | *SGIP1* | -0.208087024 | 0.000152327 | 0.000336381 | 0.95767745 |
| Obesity | LEC | *AKAP7* | 0.942985039 | 0.000126062 | 0.000290524 | 0.792549547 |
| Obesity | LEC | *ALDH8A1* | 0.410468221 | 0.040288784 | 0.049680646 | 1 |
| Obesity | LEC | *SLC16A7* | 1.147466601 | 2.37E-05 | 0.000109118 | 0.149097973 |
| Obesity | LEC | *VAMP8* | 1.314846509 | 1.48E-06 | 5.25E-05 | 0.009285915 |
| Obesity | LEC | *FOXO3* | 1.192869222 | 1.97E-05 | 0.00010532 | 0.123646033 |
| Obesity | LEC | *TGIF2* | 2.061201538 | 1.92E-05 | 0.000105211 | 0.120932149 |
| Obesity | LEC | *ABCG2* | -1.203662993 | 0.001106387 | 0.001803904 | 1 |
| Obesity | LEC | *FAM47E-STBL* | 0.994149696 | 0.016694812 | 0.021821264 | 1 |
| Obesity | LEC | *MFSD1* | 1.037052461 | 1.15E-05 | 0.000105108 | 0.072290455 |
| Obesity | LEC | *RAB3GAP2* | 1.702988547 | 5.99E-05 | 0.000171669 | 0.376641745 |
| Obesity | LEC | *UCHL3* | 4.078813542 | 0.000233532 | 0.000473771 | 1 |
| Obesity | LEC | *WDR35* | 2.23220041 | 0.000120956 | 0.000281648 | 0.760449533 |
| Obesity | LEC | *NDUFB3* | 1.889084352 | 0.000206758 | 0.00042844 | 1 |
| Obesity | LEC | *TRPM6* | 1.089462306 | 0.020731634 | 0.026741852 | 1 |
| Obesity | LEC | *GDA* | 1.425189417 | 2.49E-05 | 0.000111016 | 0.15656797 |
| Obesity | LEC | *C2orf40* | 1.145945418 | 0.000264195 | 0.0005248 | 1 |

| | | | | | | |
|---|---|---|---|---|---|---|
| Obesity | LEC | *CPSF3* | 1.984448581 | 1.93E-05 | 0.000105211 | 0.121509696 |
| Obesity | LEC | *PIGZ* | 0.779884871 | 0.000138675 | 0.000312492 | 0.871851367 |
| Obesity | LEC | *SENP5* | 1.753544604 | 9.00E-05 | 0.000226393 | 0.565756478 |
| Obesity | LEC | *CCDC92* | 2.922862093 | 2.61E-08 | 3.84E-06 | 0.00016436 |
| Obesity | LEC | *PTBP3* | 1.297413957 | 0.028417115 | 0.035882386 | 1 |
| Obesity | LEC | *FKBP15* | 1.500414829 | 5.05E-06 | 9.23E-05 | 0.031752285 |
| Obesity | LEC | *CNTRL* | 1.623322146 | 3.27E-05 | 0.000118624 | 0.205486656 |
| Obesity | LEC | *TRIM32* | 4.197518477 | 3.47E-05 | 0.000121951 | 0.21816392 |
| Obesity | LEC | *PHF19* | 0.802468799 | 4.55E-05 | 0.000143383 | 0.285888205 |
| Obesity | LEC | *BSPRY* | 2.97568469 | 3.48E-05 | 0.000122199 | 0.219008767 |
| Obesity | LEC | *PPP6C* | 1.780624443 | 3.07E-05 | 0.000115968 | 0.193203158 |
| Obesity | LEC | *RBM18* | 1.803150189 | 0.040515576 | 0.049925798 | 1 |
| Obesity | LEC | *HSDL2* | 0.881074839 | 0.000104119 | 0.000251284 | 0.654594151 |
| Obesity | LEC | *INVS* | 1.922008047 | 1.22E-05 | 0.000105108 | 0.076561315 |
| Obesity | LEC | *KDSR* | 1.272774316 | 1.42E-05 | 0.000105108 | 0.089146599 |
| Obesity | LEC | *DCAF4* | 2.460647713 | 1.69E-08 | 2.96E-06 | 0.000106406 |
| Obesity | LEC | *IFI27L2* | 1.583001135 | 0.000500921 | 0.000894176 | 1 |
| Obesity | LEC | *DNAL1* | 1.449622858 | 0.029360254 | 0.036984154 | 1 |
| Obesity | LEC | *IRF2BPL* | -1.455486681 | 0.001498933 | 0.002370765 | 1 |
| Obesity | LEC | *TTLL5* | 1.627818123 | 0.004518548 | 0.006488833 | 1 |
| Obesity | LEC | *PPP4R4* | 0.957939782 | 0.000212798 | 0.000438212 | 1 |
| Obesity | LEC | *ZC2HC1C* | 2.257533632 | 7.63E-06 | 0.000105108 | 0.047972009 |
| Obesity | LEC | *ALDH6A1* | 0.818104597 | 7.29E-05 | 0.000195433 | 0.458441882 |
| Obesity | LEC | *ESRRB* | 1.058036209 | 0.001717813 | 0.002681204 | 1 |
| Obesity | LEC | *COQ6* | 1.910040665 | 3.36E-05 | 0.000120044 | 0.211362414 |
| Obesity | LEC | *GPR75* | -1.244336437 | 0.000179288 | 0.00038287 | 1 |
| Obesity | LEC | *KLHL29* | 1.749300896 | 0.000733868 | 0.001248668 | 1 |
| Obesity | LEC | *ATAD2B* | 1.61453714 | 0.000126155 | 0.000290574 | 0.793136768 |
| Obesity | LEC | *FKBP1B* | 1.241897473 | 0.001217214 | 0.001963931 | 1 |
| Obesity | LEC | *ATL2* | 1.579726484 | 5.11E-05 | 0.000154003 | 0.321250335 |
| Obesity | LEC | *CNRIP1* | 1.282186147 | 0.021157457 | 0.027224095 | 1 |
| Obesity | LEC | *CRIPT* | 1.379881591 | 0.001330876 | 0.002126357 | 1 |
| Obesity | LEC | *EPCAM* | -0.881188714 | 0.000525389 | 0.000930456 | 1 |
| Obesity | LEC | *SLC17A5* | 1.464071946 | 3.38E-05 | 0.000120302 | 0.212213338 |
| Obesity | LEC | *OGFRL1* | 1.452379959 | 0.000162193 | 0.000353083 | 1 |
| Obesity | LEC | *CUTC* | 1.753975411 | 0.000124021 | 0.000287189 | 0.779718577 |
| Obesity | LEC | *PYROXD2* | 0.833150624 | 0.000508302 | 0.000904783 | 1 |
| Obesity | LEC | *MXI1* | 2.060730598 | 9.51E-05 | 0.000235634 | 0.598039145 |
| Obesity | LEC | *C10orf88* | 2.483671309 | 0.001599355 | 0.002511902 | 1 |
| Obesity | LEC | *HELLS* | 1.467581995 | 3.68E-05 | 0.000125546 | 0.23150734 |
| Obesity | LEC | *TCTN3* | -0.668203338 | 0.000654206 | 0.001124997 | 1 |
| Obesity | LEC | *AVPI1* | 1.311185607 | 1.56E-05 | 0.000105108 | 0.097767309 |
| Obesity | LEC | *WDR11* | 1.715676215 | 0.000131282 | 0.000299806 | 0.825366891 |
| Obesity | LEC | *C10orf76* | 1.725096654 | 1.56E-05 | 0.000105108 | 0.098286925 |
| Obesity | LEC | *KCNIP2* | 0.516453235 | 2.32E-05 | 0.000109118 | 0.145747296 |
| Obesity | LEC | *C10orf95* | 1.238967849 | 1.80E-05 | 0.000105108 | 0.112905845 |
| Obesity | LEC | *KANSL1* | 1.336474323 | 0.000196396 | 0.000410895 | 1 |
| Obesity | LEC | *HOXB3* | 2.304123011 | 1.66E-08 | 2.96E-06 | 0.000104663 |
| Obesity | LEC | *CD274* | -1.477584129 | 0.029865254 | 0.037590161 | 1 |
| Obesity | LEC | *LRP11* | 1.087336352 | 0.000233362 | 0.0004736 | 1 |

| | | | | | | |
|---|---|---|---|---|---|---|
| Obesity | LEC | *CCDC170* | 0.506710915 | 3.35E-05 | 0.000119881 | 0.21068916 |
| Obesity | LEC | *CYSTM1* | 1.429929703 | 9.73E-06 | 0.000105108 | 0.061145726 |
| Obesity | LEC | *PCDHB14* | -1.221560492 | 0.000534876 | 0.00094566 | 1 |
| Obesity | LEC | *PCDHB12* | -0.846820154 | 0.002700174 | 0.004042866 | 1 |
| Obesity | LEC | *SEC16B* | 1.348780017 | 0.000233199 | 0.000473555 | 1 |
| Obesity | LEC | *GORAB* | 2.193254222 | 0.000121669 | 0.000282576 | 0.764932653 |
| Obesity | LEC | *ACAT2* | 2.542017008 | 0.000256898 | 0.000512899 | 1 |
| Obesity | LEC | *SNX19* | 0.883199574 | 0.009642218 | 0.01306479 | 1 |
| Obesity | LEC | *MRC1* | 0.724140342 | 3.47E-05 | 0.000122027 | 0.218428301 |
| Obesity | LEC | *IQSEC3* | 4.171136853 | 0.000336147 | 0.000638669 | 1 |
| Obesity | LEC | *CCDC77* | 1.22223047 | 1.90E-05 | 0.000105211 | 0.119250062 |
| Obesity | LEC | *ENOX1* | 1.553093727 | 1.89E-05 | 0.000105211 | 0.118710467 |
| Obesity | LEC | *DNAJC15* | 1.98046442 | 6.22E-05 | 0.000175786 | 0.390899728 |
| Obesity | LEC | *WBP4* | 1.852671372 | 0.001179666 | 0.001911407 | 1 |
| Obesity | LEC | *SMAD9* | -0.247017423 | 0.000124438 | 0.000287507 | 0.782343122 |
| Obesity | LEC | *SIL1* | 1.320947246 | 6.05E-05 | 0.000172864 | 0.380128374 |
| Obesity | LEC | *ZFP30* | 1.588017868 | 0.000162841 | 0.000354251 | 1 |
| Obesity | LEC | *APAF1* | 1.980819821 | 6.83E-05 | 0.000186748 | 0.429333624 |
| Obesity | LEC | *ADRA1A* | 0.597194467 | 1.14E-06 | 4.64E-05 | 0.007187324 |
| Obesity | LEC | *EPHX2* | 1.083944642 | 0.002252307 | 0.003434235 | 1 |
| Obesity | LEC | *ZNF706* | 1.838675535 | 0.000466637 | 0.000841339 | 1 |
| Obesity | LEC | *CRISPLD1* | 0.748552187 | 2.89E-05 | 0.000115115 | 0.181535867 |
| Obesity | LEC | *RDH10* | 1.20696166 | 0.000101889 | 0.000247805 | 0.640576443 |
| Obesity | LEC | *SCPEP1* | 0.671771958 | 6.99E-06 | 0.000104897 | 0.04393804 |
| Obesity | LEC | *TEX14* | 1.312106386 | 1.16E-06 | 4.64E-05 | 0.007300515 |
| Obesity | LEC | *TRIM6* | 1.929695822 | 5.61E-05 | 0.000164778 | 0.352789897 |
| Obesity | LEC | *CEP89* | 0.877308453 | 0.0123754 | 0.016459517 | 1 |
| Obesity | LEC | *PRB2* | 2.489560247 | 1.55E-05 | 0.000105108 | 0.097570639 |
| Obesity | LEC | *PYROXD1* | 1.710448584 | 1.93E-06 | 5.90E-05 | 0.012147647 |
| Obesity | LEC | *A1BG* | 2.070748005 | 3.15E-06 | 7.45E-05 | 0.019809179 |
| Obesity | LEC | *ZNF211* | 1.380898598 | 0.000212439 | 0.00043776 | 1 |
| Obesity | LEC | *PDZRN3* | 1.184125647 | 2.49E-05 | 0.000110998 | 0.156284937 |
| Obesity | LEC | *LHX4* | 2.242309268 | 0.002660134 | 0.003985763 | 1 |
| Obesity | LEC | *SEC22A* | 0.922928177 | 0.001320068 | 0.002112848 | 1 |
| Obesity | LEC | *CSTA* | 0.751926459 | 1.19E-05 | 0.000105108 | 0.074701091 |
| Obesity | LEC | *POPDC2* | 3.961495132 | 0.000635593 | 0.001097191 | 1 |
| Obesity | LEC | *B4GALT4* | 2.362601864 | 0.007521317 | 0.010360762 | 1 |
| Obesity | LEC | *CAT* | 0.687849658 | 9.05E-05 | 0.000227269 | 0.568854138 |
| Obesity | LEC | *PILRB* | 0.433883239 | 0.003901337 | 0.005661982 | 1 |
| Obesity | LEC | *RNF115* | 1.526865824 | 0.004989257 | 0.007093501 | 1 |
| Obesity | LEC | *POLR3GL* | -1.512517201 | 0.035792231 | 0.044515481 | 1 |
| Obesity | LEC | *TNFSF10* | 1.684832008 | 2.11E-05 | 0.000106989 | 0.132420435 |
| Obesity | LEC | *ZNF639* | 2.6887579 | 9.76E-06 | 0.000105108 | 0.061369461 |
| Obesity | LEC | *LIAS* | 1.795992784 | 1.78E-05 | 0.000105108 | 0.112168061 |
| Obesity | LEC | *CLCC1* | 2.134465885 | 8.70E-05 | 0.000220912 | 0.547177514 |
| Obesity | LEC | *GPSM2* | 2.183310651 | 1.28E-05 | 0.000105108 | 0.080374916 |
| Obesity | LEC | *CXCR4* | 0.913219635 | 0.00011098 | 0.000262933 | 0.697732209 |
| Obesity | LEC | *ACVR2A* | 1.556274498 | 3.44E-05 | 0.000121483 | 0.216118751 |
| Obesity | LEC | *POLK* | 1.571847146 | 0.000281995 | 0.000553341 | 1 |
| Obesity | LEC | *FLT3* | -1.196271127 | 0.000530537 | 0.000939045 | 1 |

| | | | | | | |
|---|---|---|---|---|---|---|
| Obesity | LEC | *MTIF3* | 1.755412058 | 0.001050482 | 0.001724352 | 1 |
| Obesity | LEC | *RASL11A* | 1.314275288 | 0.00013422 | 0.000304415 | 0.843838366 |
| Obesity | LEC | *SERAC1* | 1.221033257 | 1.70E-05 | 0.000105108 | 0.107067718 |
| Obesity | LEC | *LDB3* | 0.76039587 | 2.19E-05 | 0.000108376 | 0.137894428 |
| Obesity | LEC | *OPN4* | 3.449854899 | 9.95E-06 | 0.000105108 | 0.062539188 |
| Obesity | LEC | *FAM35A* | 1.419954171 | 6.27E-05 | 0.000176459 | 0.394386725 |
| Obesity | LEC | *FAM213A* | 1.411926642 | 0.00263101 | 0.003944946 | 1 |
| Obesity | LEC | *NAA60* | 2.544957135 | 6.36E-05 | 0.000178132 | 0.400070392 |
| Obesity | LEC | *ODF2L* | 1.472881698 | 0.003476663 | 0.005103381 | 1 |
| Obesity | LEC | *SPATA1* | 2.000115865 | 0.000352007 | 0.000662398 | 1 |
| Obesity | LEC | *LRRC39* | 0.900794741 | 0.000128085 | 0.000293895 | 0.805273321 |
| Obesity | LEC | *CCDC18* | 1.215270009 | 1.55E-05 | 0.000105108 | 0.097258764 |
| Obesity | LEC | *RPAP2* | 1.724303908 | 0.000380182 | 0.000707161 | 1 |
| Obesity | LEC | *PMS2* | 1.628857073 | 4.22E-06 | 8.64E-05 | 0.026510613 |
| Obesity | LEC | *KIAA0087* | 1.742102864 | 0.000290917 | 0.000566322 | 1 |
| Obesity | LEC | *KLHL7* | 1.722729465 | 0.007909379 | 0.010850156 | 1 |
| Obesity | LEC | *HERPUD2* | 1.731589429 | 1.61E-05 | 0.000105108 | 0.10153413 |
| Obesity | LEC | *FKBP9* | 1.631351974 | 6.18E-05 | 0.000175234 | 0.388269915 |
| Obesity | LEC | *CCZ1* | 2.1078529 | 5.92E-07 | 3.13E-05 | 0.003722643 |
| Obesity | LEC | *POLM* | 0.418646666 | 0.03368769 | 0.042014383 | 1 |
| Obesity | LEC | *SMU1* | 1.184441528 | 9.79E-06 | 0.000105108 | 0.061544831 |
| Obesity | LEC | *GLIPR2* | 1.606836214 | 0.000112551 | 0.000265819 | 0.707611036 |
| Obesity | LEC | *SLC25A51* | 0.73573358 | 1.40E-06 | 5.10E-05 | 0.008819934 |
| Obesity | LEC | *DCAF10* | 1.658636639 | 0.000717936 | 0.001225872 | 1 |
| Obesity | LEC | *TRIM24* | -0.847649079 | 0.009494942 | 0.01287356 | 1 |
| Obesity | LEC | *BICC1* | 0.593546576 | 7.38E-06 | 0.000105108 | 0.046397521 |
| Obesity | LEC | *CISD1* | 1.044393726 | 0.006201649 | 0.008685624 | 1 |
| Obesity | LEC | *SLC25A16* | 1.128953688 | 3.10E-06 | 7.35E-05 | 0.019474755 |
| Obesity | LEC | *RBM19* | 2.199641708 | 9.48E-05 | 0.000235119 | 0.595791127 |
| Obesity | LEC | *ACADS* | 0.955917916 | 2.99E-05 | 0.000115831 | 0.187685551 |
| Obesity | LEC | *SSPN* | 1.028349616 | 2.01E-05 | 0.000106011 | 0.126471421 |
| Obesity | LEC | *CCDC91* | 1.045594883 | 8.74E-06 | 0.000105108 | 0.054967545 |
| Obesity | LEC | *PKN1* | 2.160244883 | 0.000472135 | 0.000849546 | 1 |
| Obesity | LEC | *EBPL* | -0.566440032 | 0.001885107 | 0.002917693 | 1 |
| Obesity | LEC | *NLN* | 1.169875124 | 0.007020701 | 0.009732999 | 1 |
| Obesity | LEC | *CENPK* | 0.587966777 | 3.44E-05 | 0.000121488 | 0.216249292 |
| Obesity | LEC | *OPTN* | 1.292027246 | 0.001526376 | 0.002409321 | 1 |
| Obesity | LEC | *ATF1* | 1.701429619 | 0.000108243 | 0.000258067 | 0.680522557 |
| Obesity | LEC | *TSFM* | 1.535919713 | 0.000118165 | 0.00027648 | 0.742901784 |
| Obesity | LEC | *ARHGAP9* | 0.665077051 | 0.000107674 | 0.000257468 | 0.676943849 |
| Obesity | LEC | *ORMDL2* | 1.656597282 | 4.05E-05 | 0.000132638 | 0.254532604 |
| Obesity | LEC | *PDE1B* | 0.819125427 | 2.02E-05 | 0.000106047 | 0.127283264 |
| Obesity | LEC | *IKZF4* | 1.266826018 | 0.000290905 | 0.000566322 | 1 |
| Obesity | LEC | *SMUG1* | 0.961728293 | 7.69E-06 | 0.000105108 | 0.048331949 |
| Obesity | LEC | *SARDH* | 1.566065525 | 2.98E-05 | 0.000115831 | 0.187506198 |
| Obesity | LEC | *STIL* | 0.863215314 | 2.72E-05 | 0.000113713 | 0.171031197 |
| Obesity | LEC | *TTC21B* | 1.650401199 | 0.003333364 | 0.004911079 | 1 |
| Obesity | LEC | *ACVR1C* | 0.855850937 | 9.35E-05 | 0.000233128 | 0.587716228 |
| Obesity | LEC | *SLC36A1* | 0.951289301 | 4.55E-05 | 0.000143462 | 0.286206188 |
| Obesity | LEC | *PLA2G12A* | 1.991490872 | 2.08E-05 | 0.000106878 | 0.130906467 |

| | | | | | | |
|---|---|---|---|---|---|---|
| Obesity | LEC | *C4BPB* | 1.044229477 | 4.05E-05 | 0.000132638 | 0.254514599 |
| Obesity | LEC | *RAB38* | 1.090952416 | 3.85E-05 | 0.00012891 | 0.242287228 |
| Obesity | LEC | *CKS2* | 2.174126578 | 0.000204534 | 0.000424531 | 1 |
| Obesity | LEC | *CHPF* | -1.059962287 | 0.006752494 | 0.009396399 | 1 |
| Obesity | LEC | *DNPEP* | 2.651025621 | 0.001068765 | 0.001747549 | 1 |
| Obesity | LEC | *MOGAT1* | 1.491330241 | 7.57E-05 | 0.000200562 | 0.47593329 |
| Obesity | LEC | *FAM210B* | 1.904802591 | 0.000166373 | 0.000359816 | 1 |
| Obesity | LEC | *SNX21* | 0.760247862 | 1.80E-05 | 0.000105108 | 0.113154419 |
| Obesity | LEC | *SDC4* | -1.503554613 | 0.002443553 | 0.003697173 | 1 |
| Obesity | LEC | *VAPB* | 0.937509411 | 0.011168108 | 0.015002969 | 1 |
| Obesity | LEC | *TOX2* | -0.510944853 | 0.003578532 | 0.005234581 | 1 |
| Obesity | LEC | *ARFGEF2* | 1.849790009 | 7.70E-05 | 0.000202634 | 0.483888943 |
| Obesity | LEC | *CSE1L* | 1.926045338 | 2.24E-05 | 0.000108932 | 0.140957982 |
| Obesity | LEC | *CDH26* | 2.700333753 | 1.30E-05 | 0.000105108 | 0.081832995 |
| Obesity | LEC | *STX16* | 1.765143449 | 2.75E-05 | 0.00011384 | 0.172660748 |
| Obesity | LEC | *RNF114* | 1.828232648 | 2.97E-05 | 0.000115831 | 0.18659789 |
| Obesity | LEC | *KCNK15* | 1.953971497 | 7.86E-05 | 0.000205818 | 0.494373871 |
| Obesity | LEC | *NEURL2* | 1.470488505 | 0.000484398 | 0.000869123 | 1 |
| Obesity | LEC | *FASTKD3* | 2.141719411 | 6.20E-05 | 0.000175678 | 0.38982861 |
| Obesity | LEC | *PEPD* | 1.196242339 | 0.037929471 | 0.046996962 | 1 |
| Obesity | LEC | *CHST8* | -0.663524115 | 0.000395059 | 0.000731156 | 1 |
| Obesity | LEC | *STAMBP* | 2.106045762 | 9.16E-05 | 0.000229542 | 0.576149854 |
| Obesity | LEC | *ATP8A1* | 1.345198528 | 3.22E-06 | 7.50E-05 | 0.020265219 |
| Obesity | LEC | *HIF3A* | -0.335157649 | 0.000636966 | 0.00109926 | 1 |
| Obesity | LEC | *ZNF45* | 2.186851381 | 2.13E-05 | 0.00010722 | 0.134004001 |
| Obesity | LEC | *MRS2* | 1.489120631 | 0.007845302 | 0.010774009 | 1 |
| Obesity | LEC | *SNRPC* | -4.208779654 | 1.83E-16 | 2.30E-13 | 1.15E-12 |
| Obesity | LEC | *SERPINB6* | 1.941949916 | 6.39E-05 | 0.000178506 | 0.401816378 |
| Obesity | LEC | *PEX6* | 0.653024161 | 0.026613284 | 0.033740213 | 1 |
| Obesity | LEC | *NQO2* | 1.46167292 | 1.62E-05 | 0.000105108 | 0.101602581 |
| Obesity | LEC | *ZNF391* | 0.560096896 | 0.00063314 | 0.001094461 | 1 |
| Obesity | LEC | *RPS10* | -2.225670962 | 0.004543438 | 0.006521596 | 1 |
| Obesity | LEC | *HIST1H2BJ* | 3.050779103 | 3.11E-11 | 1.15E-08 | 1.96E-07 |
| Obesity | LEC | *MED20* | 2.006074778 | 5.98E-05 | 0.000171484 | 0.375663499 |
| Obesity | LEC | *TBCC* | 0.975904712 | 0.000120095 | 0.000280266 | 0.755037899 |
| Obesity | LEC | *TCP11* | -1.204773923 | 0.006934576 | 0.009626337 | 1 |
| Obesity | LEC | *KLHDC3* | 2.413571228 | 0.000621461 | 0.001077234 | 1 |
| Obesity | LEC | *MEA1* | -1.123110137 | 0.002702931 | 0.004046031 | 1 |
| Obesity | LEC | *KLHL31* | 1.011240504 | 0.000223096 | 0.000456577 | 1 |
| Obesity | LEC | *COL21A1* | 0.913417078 | 3.93E-05 | 0.000130538 | 0.247254629 |
| Obesity | LEC | *GLO1* | 2.490239246 | 1.80E-05 | 0.000105108 | 0.113442866 |
| Obesity | LEC | *CPNE5* | -0.476358194 | 6.07E-05 | 0.000173168 | 0.381315529 |
| Obesity | LEC | *RIOK1* | 1.752668029 | 9.19E-05 | 0.000230093 | 0.577763517 |
| Obesity | LEC | *NUP153* | 2.022753476 | 1.05E-05 | 0.000105108 | 0.065988108 |
| Obesity | LEC | *EREG* | -0.989195099 | 0.000102248 | 0.000248318 | 0.64283201 |
| Obesity | LEC | *CNOT1* | 1.832823634 | 9.15E-05 | 0.000229207 | 0.575081186 |
| Obesity | LEC | *BBS2* | 1.85318782 | 1.77E-06 | 5.64E-05 | 0.011102284 |
| Obesity | LEC | *MT1G* | 4.546443279 | 4.45E-05 | 0.000141409 | 0.27984809 |
| Obesity | LEC | *GOT2* | 2.593248024 | 9.63E-06 | 0.000105108 | 0.060569132 |
| Obesity | LEC | *CLYBL* | 1.40396889 | 0.030185966 | 0.037955834 | 1 |

| | | | | | | |
|---|---|---|---|---|---|---|
| Obesity | LEC | *TMTC4* | -1.082809829 | 0.031537011 | 0.039517342 | 1 |
| Obesity | LEC | *KIF25* | -0.95448539 | 0.000169355 | 0.000364927 | 1 |
| Obesity | LEC | *GRK4* | 0.904593606 | 1.28E-05 | 0.000105108 | 0.080240259 |
| Obesity | LEC | *TEKT3* | 1.080338928 | 6.24E-06 | 9.96E-05 | 0.03923459 |
| Obesity | LEC | *MRPS7* | 2.943726233 | 4.71E-05 | 0.000146395 | 0.296011363 |
| Obesity | LEC | *NUP85* | 1.921729872 | 0.001217344 | 0.001963931 | 1 |
| Obesity | LEC | *MIF4GD* | -1.660066598 | 0.006381388 | 0.008919472 | 1 |
| Obesity | LEC | *NT5C* | 0.991307245 | 2.42E-05 | 0.000109361 | 0.151835082 |
| Obesity | LEC | *DDX31* | 1.115174445 | 0.017228959 | 0.022477375 | 1 |
| Obesity | LEC | *PPP1R12C* | 2.35602056 | 0.000521705 | 0.000924714 | 1 |
| Obesity | LEC | *OPRL1* | -0.501197548 | 0.00324046 | 0.004783464 | 1 |
| Obesity | LEC | *PPDPF* | 2.573265321 | 0.000347748 | 0.000655757 | 1 |
| Obesity | LEC | *CHCHD5* | -1.773745811 | 0.0009535 | 0.001575467 | 1 |
| Obesity | LEC | *PAX8* | 0.796949037 | 0.000149827 | 0.000332967 | 0.941964267 |
| Obesity | LEC | *INSIG2* | 2.495106494 | 0.000723485 | 0.001233338 | 1 |
| Obesity | LEC | *POLR1B* | 1.096867009 | 0.013652186 | 0.018050746 | 1 |
| Obesity | LEC | *PSD4* | 1.186426259 | 1.66E-05 | 0.000105108 | 0.104452484 |
| Obesity | LEC | *CLPP* | 2.002202121 | 0.007758527 | 0.010659498 | 1 |
| Obesity | LEC | *SH2D3A* | 1.486042744 | 4.21E-05 | 0.000136137 | 0.264377584 |
| Obesity | LEC | *TRIP10* | 2.030377281 | 3.56E-05 | 0.000123048 | 0.223542494 |
| Obesity | LEC | *GPR108* | 2.066971664 | 0.00019315 | 0.000405318 | 1 |
| Obesity | LEC | *RTN2* | 1.861548137 | 1.58E-05 | 0.000105108 | 0.099124962 |
| Obesity | LEC | *SYMPK* | 1.995457197 | 1.84E-05 | 0.000105108 | 0.115783384 |
| Obesity | LEC | *GPCPD1* | 1.450690388 | 2.59E-05 | 0.000112124 | 0.162920166 |
| Obesity | LEC | *SDCBP2* | 1.85938615 | 3.57E-05 | 0.000123048 | 0.224680465 |
| Obesity | LEC | *PANK2* | 1.688767042 | 1.95E-05 | 0.000105217 | 0.122300552 |
| Obesity | LEC | *PSMF1* | 1.583858163 | 0.00036673 | 0.000686403 | 1 |
| Obesity | LEC | *RBCK1* | 2.040825622 | 7.39E-05 | 0.000196872 | 0.464616887 |
| Obesity | LEC | *RRBP1* | 1.587457335 | 0.001709282 | 0.002668552 | 1 |
| Obesity | LEC | *MKKS* | 1.910587066 | 4.25E-05 | 0.000137166 | 0.267473122 |
| Obesity | LEC | *DSTN* | 1.839324365 | 0.000211356 | 0.000435814 | 1 |
| Obesity | LEC | *MGME1* | 1.645486468 | 2.43E-05 | 0.000109618 | 0.152588285 |
| Obesity | LEC | *ITPA* | 1.767610095 | 1.61E-05 | 0.000105108 | 0.101517087 |
| Obesity | LEC | *TMEM74B* | -0.775370389 | 0.0005116 | 0.000910138 | 1 |
| Obesity | LEC | *FAM110A* | 0.761967483 | 3.46E-05 | 0.000121788 | 0.21739126 |
| Obesity | LEC | *NCLN* | 2.534516051 | 0.00054548 | 0.000962513 | 1 |
| Obesity | LEC | *ZNF436* | 0.674908825 | 1.85E-05 | 0.000105108 | 0.11658662 |
| Obesity | LEC | *GDF5* | 0.967777762 | 0.003533266 | 0.005178005 | 1 |
| Obesity | LEC | *MMP24* | 1.553039984 | 4.88E-05 | 0.000149615 | 0.306711451 |
| Obesity | LEC | *ST3GAL3* | 1.414499435 | 0.000878552 | 0.00146278 | 1 |
| Obesity | LEC | *KLC1* | 2.345843221 | 2.41E-06 | 6.68E-05 | 0.015152869 |
| Obesity | LEC | *XRCC3* | 2.633282454 | 0.004059703 | 0.005878248 | 1 |
| Obesity | LEC | *MCF2L* | 1.702065675 | 0.001117441 | 0.001819096 | 1 |
| Obesity | LEC | *F10* | 0.929193637 | 1.76E-05 | 0.000105108 | 0.110736992 |
| Obesity | LEC | *PROZ* | -0.766516548 | 9.47E-05 | 0.000235119 | 0.595674006 |
| Obesity | LEC | *IGFLR1* | 2.24560163 | 3.58E-05 | 0.000123048 | 0.224808015 |
| Obesity | LEC | *CAPNS1* | -1.022549837 | 0.002342626 | 0.003555792 | 1 |
| Obesity | LEC | *PRDX5* | 2.569671423 | 5.01E-05 | 0.000152019 | 0.314679723 |
| Obesity | LEC | *SCAF1* | 1.894119828 | 1.41E-05 | 0.000105108 | 0.08850014 |
| Obesity | LEC | *PRR12* | 1.808741391 | 0.000121116 | 0.000281916 | 0.761455171 |

| | | | | | | |
|---|---|---|---|---|---|---|
| Obesity | LEC | *TSKS* | 0.990206911 | 2.27E-05 | 0.000109111 | 0.142893052 |
| Obesity | LEC | *FLRT1* | 1.153192665 | 6.13E-06 | 9.96E-05 | 0.03852533 |
| Obesity | LEC | *SBDS* | 2.647396384 | 0.000199868 | 0.000416634 | 1 |
| Obesity | LEC | *BECN1* | 1.871605948 | 0.020428816 | 0.026383724 | 1 |
| Obesity | LEC | *TRAP1* | 0.807184591 | 1.02E-06 | 4.34E-05 | 0.006429287 |
| Obesity | LEC | *DNAJC8* | 1.302565137 | 0.000103907 | 0.000251061 | 0.653261984 |
| Obesity | LEC | *ZNF384* | 2.816328044 | 0.000182011 | 0.000387111 | 1 |
| Obesity | LEC | *ATG14* | 1.835781361 | 0.000627625 | 0.001086719 | 1 |
| Obesity | LEC | *RHOJ* | -0.150412828 | 0.000106009 | 0.00025435 | 0.666481492 |
| Obesity | LEC | *HSPA2* | 3.762428 | 0.000213448 | 0.00043906 | 1 |
| Obesity | LEC | *ZBTB1* | 1.785449843 | 0.000201363 | 0.000419196 | 1 |
| Obesity | LEC | *PZP* | 0.777314134 | 1.75E-05 | 0.000105108 | 0.109790397 |
| Obesity | LEC | *RHOT1* | 1.189041384 | 2.25E-06 | 6.43E-05 | 0.01415312 |
| Obesity | LEC | *WDR60* | 1.151020752 | 0.000614544 | 0.001068483 | 1 |
| Obesity | LEC | *AIF1L* | 0.698827644 | 2.44E-05 | 0.000109899 | 0.153601049 |
| Obesity | LEC | *MAP2K2* | 1.738550509 | 0.002342618 | 0.003555792 | 1 |
| Obesity | LEC | *CANX* | 2.059036116 | 2.27E-05 | 0.000109111 | 0.142732382 |
| Obesity | LEC | *IPPK* | 2.644666321 | 0.000726443 | 0.001237709 | 1 |
| Obesity | LEC | *FGD3* | 0.993838737 | 1.70E-05 | 0.000105108 | 0.106820122 |
| Obesity | LEC | *ABHD8* | 1.960605825 | 0.000126483 | 0.000290753 | 0.79520169 |
| Obesity | LEC | *MASP1* | 0.712473664 | 5.40E-05 | 0.000159889 | 0.339285184 |
| Obesity | LEC | *ATP13A4* | 0.791689124 | 0.000410067 | 0.00075449 | 1 |
| Obesity | LEC | *HRASLS* | 3.617796541 | 0.003084096 | 0.004570889 | 1 |
| Obesity | LEC | *HELB* | 1.850500689 | 1.42E-05 | 0.000105108 | 0.089467456 |
| Obesity | LEC | *RAP1B* | 1.498797034 | 0.000459674 | 0.000831646 | 1 |
| Obesity | LEC | *RAB3IP* | 1.626153437 | 1.19E-05 | 0.000105108 | 0.074782641 |
| Obesity | LEC | *TAS2R4* | -0.711067794 | 0.004425642 | 0.006365595 | 1 |
| Obesity | LEC | *IDUA* | 3.052953448 | 0.002724298 | 0.004075103 | 1 |
| Obesity | LEC | *TMEM175* | 1.328577799 | 2.30E-05 | 0.000109118 | 0.144329278 |
| Obesity | LEC | *EMC1* | 1.663119954 | 0.003769263 | 0.005487429 | 1 |
| Obesity | LEC | *UBR4* | 0.960318069 | 0.00225271 | 0.003434235 | 1 |
| Obesity | LEC | *SIN3B* | 2.098819514 | 0.000502907 | 0.000897212 | 1 |
| Obesity | LEC | *WFIKKN1* | -6.202897927 | 0.008440322 | 0.011538806 | 1 |
| Obesity | LEC | *WDR24* | 1.001506516 | 7.43E-05 | 0.000197765 | 0.467298222 |
| Obesity | LEC | *CHTF18* | 0.802735737 | 1.94E-05 | 0.000105217 | 0.12196323 |
| Obesity | LEC | *SMARCA4* | 1.807493443 | 0.00018495 | 0.000392012 | 1 |
| Obesity | LEC | *TICAM1* | 1.77248567 | 2.72E-05 | 0.000113713 | 0.171076988 |
| Obesity | LEC | *AAMP* | 2.773310098 | 6.78E-05 | 0.000185932 | 0.426528288 |
| Obesity | LEC | *PNKD* | 1.760687784 | 7.77E-05 | 0.000203839 | 0.488194738 |
| Obesity | LEC | *ZNF835* | 1.00952637 | 0.000101971 | 0.000247909 | 0.641092107 |
| Obesity | LEC | *PTPN12* | 1.514094754 | 0.001677807 | 0.002623973 | 1 |
| Obesity | LEC | *POR* | 1.965408843 | 0.000434394 | 0.000792523 | 1 |
| Obesity | LEC | *STYXL1* | 1.272784613 | 0.000268761 | 0.000531686 | 1 |
| Obesity | LEC | *STEAP4* | 1.454906572 | 0.000121893 | 0.000282991 | 0.766338543 |
| Obesity | LEC | *GNAI1* | 1.499402355 | 4.19E-05 | 0.000135866 | 0.263716529 |
| Obesity | LEC | *RBM48* | 1.371288173 | 2.63E-05 | 0.000112582 | 0.165257868 |
| Obesity | LEC | *CASD1* | 1.621451877 | 0.007518556 | 0.010359229 | 1 |
| Obesity | LEC | *SRD5A3* | -0.995112591 | 0.017969648 | 0.023399995 | 1 |
| Obesity | LEC | *VPREB3* | 7.58711083 | 0.001700035 | 0.002655433 | 1 |
| Obesity | LEC | *MGAT3* | 1.002660896 | 2.15E-05 | 0.000107557 | 0.135414809 |

| | | | | | | |
|---|---|---|---|---|---|---|
| Obesity | LEC | *ATF4* | 1.991450795 | 0.00014562 | 0.000324766 | 0.915514475 |
| Obesity | LEC | *A4GALT* | -0.597676563 | 0.036598794 | 0.045455674 | 1 |
| Obesity | LEC | *TPST2* | 1.98154034 | 0.002004535 | 0.003090366 | 1 |
| Obesity | LEC | *BAIAP2L2* | 1.058515385 | 1.80E-05 | 0.000105108 | 0.113357399 |
| Obesity | LEC | *TST* | 0.759366448 | 2.76E-05 | 0.00011384 | 0.173270944 |
| Obesity | LEC | *RIBC2* | 2.254223352 | 1.17E-07 | 1.19E-05 | 0.000737694 |
| Obesity | LEC | *EMC4* | 2.626151063 | 8.14E-07 | 3.79E-05 | 0.005115561 |
| Obesity | LEC | *SPECC1* | 1.331829857 | 2.12E-05 | 0.00010722 | 0.13333085 |
| Obesity | LEC | *CDHR3* | 1.13320715 | 2.53E-05 | 0.000111345 | 0.158888981 |
| Obesity | LEC | *PRKRIP1* | 1.286748755 | 0.000640935 | 0.001105503 | 1 |
| Obesity | LEC | *FOXP2* | -0.319561332 | 0.000208062 | 0.000430314 | 1 |
| Obesity | LEC | *FLNC* | -0.515553275 | 0.000132713 | 0.000302087 | 0.834363683 |
| Obesity | LEC | *IRF5* | 0.61282492 | 0.000379214 | 0.000705778 | 1 |
| Obesity | LEC | *KLHDC10* | 1.588384881 | 0.000131029 | 0.000299447 | 0.823777699 |
| Obesity | LEC | *OPN1SW* | -0.922461228 | 0.009569962 | 0.012969682 | 1 |
| Obesity | LEC | *HOXD1* | -4.4024809 | 0.001120905 | 0.001822847 | 1 |
| Obesity | LEC | *HOXD3* | -0.555061356 | 0.000373452 | 0.000697118 | 1 |
| Obesity | LEC | *CHN1* | -0.841275034 | 0.011983321 | 0.015998968 | 1 |
| Obesity | LEC | *GAD1* | -0.29262861 | 8.22E-05 | 0.00021261 | 0.51706792 |
| Obesity | LEC | *ORMDL1* | 0.883484016 | 0.006633954 | 0.009247819 | 1 |
| Obesity | LEC | *HOXD9* | -0.146129444 | 6.61E-05 | 0.000182807 | 0.415336706 |
| Obesity | LEC | *PSMG2* | 1.140008374 | 0.005173047 | 0.007341523 | 1 |
| Obesity | LEC | *WDFY4* | 0.983218384 | 0.001402756 | 0.002233821 | 1 |
| Obesity | LEC | *EIF2AK4* | 1.427360777 | 2.14E-06 | 6.25E-05 | 0.013431903 |
| Obesity | LEC | *TMOD2* | -1.067433306 | 0.022508457 | 0.028826782 | 1 |
| Obesity | LEC | *IVD* | 1.409383467 | 0.006720997 | 0.009356711 | 1 |
| Obesity | LEC | *KNSTRN* | 0.355396299 | 0.013564272 | 0.017945829 | 1 |
| Obesity | LEC | *ARPP19* | 2.073824223 | 9.37E-05 | 0.000233549 | 0.589243189 |
| Obesity | LEC | *VPS13C* | 1.489983062 | 0.000315032 | 0.000604212 | 1 |
| Obesity | LEC | *ANAPC13* | 0.601466408 | 0.003881102 | 0.005635216 | 1 |
| Obesity | LEC | *SUMF2* | 1.99928491 | 0.001303561 | 0.002089066 | 1 |
| Obesity | LEC | *SPCS3* | 1.70327859 | 0.01404361 | 0.018537093 | 1 |
| Obesity | LEC | *SERGEF* | 2.143844304 | 0.000227655 | 0.000463575 | 1 |
| Obesity | LEC | *RPAIN* | 0.84222434 | 0.030237942 | 0.038013586 | 1 |
| Obesity | LEC | *PLD2* | -0.792188598 | 0.005144818 | 0.007304399 | 1 |
| Obesity | LEC | *ATP1B2* | 1.843790767 | 0.000292679 | 0.00056845 | 1 |
| Obesity | LEC | *CCL4* | 0.586486549 | 1.75E-05 | 0.000105108 | 0.109971434 |
| Obesity | LEC | *PHF20L1* | 1.73383252 | 0.000869101 | 0.001448579 | 1 |
| Obesity | LEC | *PUS7L* | 1.801503077 | 0.0003266 | 0.000623544 | 1 |
| Obesity | LEC | *KRI1* | 1.062852924 | 0.001128416 | 0.001834113 | 1 |
| Obesity | LEC | *CDKN2D* | 3.584444457 | 2.82E-06 | 7.08E-05 | 0.017742383 |
| Obesity | LEC | *MTUS1* | -0.806632216 | 0.01942503 | 0.025149334 | 1 |
| Obesity | LEC | *NGDN* | 1.716149364 | 0.04015123 | 0.04955195 | 1 |
| Obesity | LEC | *RIPK3* | -0.876014057 | 0.00404908 | 0.005864217 | 1 |
| Obesity | LEC | *ADCY4* | -0.514300273 | 0.000244617 | 0.000492919 | 1 |
| Obesity | LEC | *BCL2L2* | 1.511781663 | 1.44E-05 | 0.000105108 | 0.090358495 |
| Obesity | LEC | *DTD2* | 0.855922104 | 0.003314036 | 0.004886337 | 1 |
| Obesity | LEC | *PARP2* | 1.191063072 | 3.72E-05 | 0.000126137 | 0.233846846 |
| Obesity | LEC | *MIS18BP1* | 1.47092711 | 0.000140229 | 0.00031509 | 0.881620812 |
| Obesity | LEC | *RNASE1* | 0.931950791 | 9.93E-05 | 0.000242855 | 0.624138156 |

| | | | | | | |
|---|---|---|---|---|---|---|
| Obesity | LEC | *DAD1* | 1.718867124 | 0.000243892 | 0.000491616 | 1 |
| Obesity | LEC | *CDO1* | 1.444032873 | 5.19E-06 | 9.32E-05 | 0.032612596 |
| Obesity | LEC | *REEP5* | 1.749830004 | 7.10E-05 | 0.000191335 | 0.446384213 |
| Obesity | LEC | *QRICH2* | -1.095034724 | 0.000460996 | 0.000833798 | 1 |
| Obesity | LEC | *SEC14L1* | 1.222036098 | 0.009234079 | 0.012546932 | 1 |
| Obesity | LEC | *RHBDF2* | 1.005588927 | 0.000154648 | 0.000340193 | 0.972270499 |
| Obesity | LEC | *AANAT* | -0.857429596 | 0.000358387 | 0.00067259 | 1 |
| Obesity | LEC | *TMEM8A* | 2.233703177 | 0.000626831 | 0.001085644 | 1 |
| Obesity | LEC | *SHC2* | -0.348471489 | 0.000588084 | 0.001028166 | 1 |
| Obesity | LEC | *ERMARD* | 1.103783921 | 4.16E-05 | 0.000135062 | 0.261749539 |
| Obesity | LEC | *PHF10* | 2.249666202 | 0.000144086 | 0.000322143 | 0.905866164 |
| Obesity | LEC | *DOCK6* | -0.857891187 | 0.003297312 | 0.00486282 | 1 |
| Obesity | LEC | *ECSIT* | 1.46460748 | 1.70E-05 | 0.000105108 | 0.107048814 |
| Obesity | LEC | *ELOF1* | 3.658528531 | 1.55E-05 | 0.000105108 | 0.097427046 |
| Obesity | LEC | *THEM6* | -0.81836523 | 0.000194735 | 0.000408236 | 1 |
| Obesity | LEC | *TOMM40* | 2.096134753 | 3.56E-05 | 0.000123048 | 0.22354944 |
| Obesity | LEC | *DPP6* | 0.641042238 | 2.86E-05 | 0.000114981 | 0.179944635 |
| Obesity | LEC | *FAM98C* | 1.460644627 | 3.99E-05 | 0.000131515 | 0.250589686 |
| Obesity | LEC | *GTPBP3* | 2.021470635 | 2.78E-05 | 0.00011384 | 0.174778997 |
| Obesity | LEC | *PLVAP* | 0.442397839 | 0.013274268 | 0.017584349 | 1 |
| Obesity | LEC | *BST2* | 1.352192764 | 1.60E-05 | 0.000105108 | 0.100484869 |
| Obesity | LEC | *DDA1* | 1.721883241 | 2.75E-05 | 0.00011384 | 0.172950069 |
| Obesity | LEC | *MRPL34* | 0.931596925 | 0.002416351 | 0.003659745 | 1 |
| Obesity | LEC | *LSM7* | 1.884407996 | 0.000125609 | 0.0002898 | 0.789706125 |
| Obesity | LEC | *TULP4* | 1.811588781 | 6.37E-05 | 0.000178168 | 0.400765862 |
| Obesity | LEC | *RTN4IP1* | 2.399037877 | 1.98E-05 | 0.000105667 | 0.124649161 |
| Obesity | LEC | *RSPH3* | 1.086699722 | 0.000200983 | 0.000418543 | 1 |
| Obesity | LEC | *STK33* | -0.441890681 | 0.000749654 | 0.001272816 | 1 |
| Obesity | LEC | *NDUFA10* | 0.808151895 | 0.006762986 | 0.009408916 | 1 |
| Obesity | LEC | *CACNG6* | 2.563751846 | 0.001031704 | 0.001695327 | 1 |
| Obesity | LEC | *PXDN* | 1.582797553 | 0.003887492 | 0.005643192 | 1 |
| Obesity | LEC | *PGPEP1* | 2.743539158 | 1.14E-06 | 4.64E-05 | 0.007151269 |
| Obesity | LEC | *HRC* | -1.764142988 | 2.88E-06 | 7.13E-05 | 0.018116881 |
| Obesity | LEC | *TRPM4* | 1.837157412 | 0.000301733 | 0.000583153 | 1 |
| Obesity | LEC | *CAMSAP1* | 1.727964875 | 1.69E-05 | 0.000105108 | 0.106358607 |
| Obesity | LEC | *UBAC1* | 1.905235928 | 0.000137265 | 0.000309981 | 0.862986612 |
| Obesity | LEC | *TNNT3* | 1.547671401 | 5.61E-05 | 0.000164778 | 0.35263799 |
| Obesity | LEC | *ATXN10* | 1.655498332 | 0.00015066 | 0.000334135 | 0.947202491 |
| Obesity | LEC | *TUBGCP2* | 1.647249453 | 7.00E-06 | 0.000104897 | 0.044011373 |
| Obesity | LEC | *CYP2E1* | 0.846762099 | 0.000812139 | 0.001363033 | 1 |
| Obesity | LEC | *HBZ* | 4.862263823 | 0.02883295 | 0.036370939 | 1 |
| Obesity | LEC | *PAK4* | -1.067997496 | 0.003601615 | 0.005263449 | 1 |
| Obesity | LEC | *POMT1* | 1.053493505 | 0.000105578 | 0.000253961 | 0.663770803 |
| Obesity | LEC | *UCK1* | 1.369888017 | 0.019684974 | 0.025464904 | 1 |
| Obesity | LEC | *MAP3K10* | 0.934200011 | 2.09E-05 | 0.00010698 | 0.131408456 |
| Obesity | LEC | *SMPDL3B* | 4.516863859 | 2.91E-05 | 0.000115266 | 0.182829989 |
| Obesity | LEC | *ATPIF1* | 1.216062449 | 0.033859126 | 0.042219818 | 1 |
| Obesity | LEC | *THEMIS2* | 0.681303997 | 9.22E-06 | 0.000105108 | 0.057960886 |
| Obesity | LEC | *CLIP1* | 1.59891877 | 1.00E-06 | 4.29E-05 | 0.006305939 |
| Obesity | LEC | *PPAN* | 4.392408787 | 2.36E-05 | 0.000109118 | 0.148065422 |

| | | | | | | |
|---|---|---|---|---|---|---|
| Obesity | LEC | *ZNF236* | 1.321580904 | 2.30E-05 | 0.000109118 | 0.144888542 |
| Obesity | LEC | *C12orf65* | 2.32103287 | 0.001806811 | 0.002806874 | 1 |
| Obesity | LEC | *PKDREJ* | -2.604091538 | 3.90E-06 | 8.25E-05 | 0.024505527 |
| Obesity | LEC | *HSD17B3* | 1.179583748 | 0.004045786 | 0.005860797 | 1 |
| Obesity | LEC | *HABP4* | 2.002667132 | 0.000468462 | 0.000843903 | 1 |
| Obesity | LEC | *EPS8L1* | -1.535992286 | 0.000116841 | 0.000274198 | 0.734576687 |
| Obesity | LEC | *LILRB2* | 0.701324379 | 3.07E-05 | 0.000115897 | 0.192967731 |
| Obesity | LEC | *COX4I2* | 1.651963561 | 6.10E-05 | 0.000173812 | 0.383255011 |
| Obesity | LEC | *ATP6V1E1* | 1.686225863 | 0.000145436 | 0.000324701 | 0.914358703 |
| Obesity | LEC | *PPT1* | 2.407149325 | 0.000836562 | 0.001399167 | 1 |
| Obesity | LEC | *HAUS8* | 2.313096662 | 1.49E-05 | 0.000105108 | 0.093459839 |
| Obesity | LEC | *TBC1D5* | 1.279098358 | 0.002539952 | 0.003822087 | 1 |
| Obesity | LEC | *RFTN1* | 1.281938109 | 7.22E-06 | 0.000105108 | 0.045364631 |
| Obesity | LEC | *PDLIM4* | -0.607427492 | 0.000506093 | 0.000901617 | 1 |
| Obesity | LEC | *KIF3A* | 2.010343732 | 3.88E-06 | 8.24E-05 | 0.024378404 |
| Obesity | LEC | *AOC3* | 1.154987761 | 0.000163794 | 0.000355585 | 1 |
| Obesity | LEC | *ACLY* | 1.506807404 | 0.025855271 | 0.03283858 | 1 |
| Obesity | LEC | *AOC2* | 1.853479088 | 1.82E-05 | 0.000105108 | 0.114669089 |
| Obesity | LEC | *DIAPH1* | 1.730056804 | 0.002593332 | 0.003891236 | 1 |
| Obesity | LEC | *EXOC4* | 1.50537103 | 0.000373932 | 0.000697627 | 1 |
| Obesity | LEC | *ACAP3* | 1.383248506 | 1.84E-05 | 0.000105108 | 0.115719489 |
| Obesity | LEC | *C1orf159* | 1.312144451 | 2.33E-05 | 0.000109118 | 0.146465084 |
| Obesity | LEC | *PPFIA1* | 1.193040483 | 1.92E-05 | 0.000105211 | 0.12100898 |
| Obesity | LEC | *NINJ1* | 2.452218999 | 3.23E-06 | 7.50E-05 | 0.020304316 |
| Obesity | LEC | *NPHP4* | 2.235518107 | 2.27E-05 | 0.000109111 | 0.142869339 |
| Obesity | LEC | *ZCCHC9* | 1.48387367 | 1.23E-05 | 0.000105108 | 0.077616491 |
| Obesity | LEC | *KHDRBS3* | -0.822261938 | 0.001925192 | 0.00297534 | 1 |
| Obesity | LEC | *RBM8A* | 1.119080047 | 0.003104475 | 0.004597841 | 1 |
| Obesity | LEC | *ZNF304* | 1.072885417 | 0.000416226 | 0.000764032 | 1 |
| Obesity | LEC | *ZSCAN5A* | 1.955803141 | 8.78E-05 | 0.000222231 | 0.551782556 |
| Obesity | LEC | *ZNF132* | -1.244356928 | 0.007413687 | 0.010232679 | 1 |
| Obesity | LEC | *RHPN2* | -0.404209627 | 5.25E-05 | 0.000156802 | 0.330068497 |
| Obesity | LEC | *C19orf12* | 0.67007814 | 0.021043014 | 0.027093473 | 1 |
| Obesity | LEC | *ACTR10* | 1.770457915 | 0.00041498 | 0.000762191 | 1 |
| Obesity | LEC | *GCH1* | 1.337674545 | 1.95E-05 | 0.000105217 | 0.122732212 |
| Obesity | LEC | *LGALS3* | 1.784114158 | 0.000774506 | 0.001306848 | 1 |
| Obesity | LEC | *DNAJB1* | 1.090508313 | 0.000291891 | 0.00056727 | 1 |
| Obesity | LEC | *FBXW9* | 0.914596642 | 4.55E-05 | 0.000143383 | 0.285905867 |
| Obesity | LEC | *C19orf57* | 1.140675614 | 3.16E-05 | 0.000116919 | 0.198878852 |
| Obesity | LEC | *MATN3* | -4.4994204 | 0.002218638 | 0.003388867 | 1 |
| Obesity | LEC | *CCT6B* | 1.734595908 | 0.000211591 | 0.000436154 | 1 |
| Obesity | LEC | *RAF1* | 1.557632687 | 3.57E-05 | 0.000123048 | 0.224225151 |
| Obesity | LEC | *EMILIN2* | 1.127052145 | 4.15E-08 | 5.32E-06 | 0.000260814 |
| Obesity | LEC | *TRIM5* | 1.937773366 | 5.89E-05 | 0.000169777 | 0.370085148 |
| Obesity | LEC | *CNGA4* | 1.204356778 | 2.32E-05 | 0.000109118 | 0.146074288 |
| Obesity | LEC | *TRIM22* | 1.665309894 | 4.30E-05 | 0.000138063 | 0.270327549 |
| Obesity | LEC | *IMMT* | 1.847133087 | 1.87E-05 | 0.000105211 | 0.117822232 |
| Obesity | LEC | *INPP5K* | 1.345806272 | 0.001159571 | 0.001881834 | 1 |
| Obesity | LEC | *MYBBP1A* | 2.424158278 | 0.000760761 | 0.001287458 | 1 |
| Obesity | LEC | *RPA1* | 1.943860571 | 4.10E-06 | 8.49E-05 | 0.025782832 |

| | | | | | | |
|---|---|---|---|---|---|---|
| Obesity | LEC | *SERPINF1* | 0.987608453 | 7.28E-06 | 0.000105108 | 0.045761955 |
| Obesity | LEC | *TMEM128* | -1.079547191 | 0.001070674 | 0.001750216 | 1 |
| Obesity | LEC | *SEC61G* | 1.640307992 | 0.003006188 | 0.004465951 | 1 |
| Obesity | LEC | *FIGNL1* | 2.544058593 | 0.000290543 | 0.000566051 | 1 |
| Obesity | LEC | *WBP2* | 2.474957681 | 0.001056988 | 0.001732347 | 1 |
| Obesity | LEC | *KDM6B* | 1.857261485 | 2.90E-05 | 0.000115266 | 0.182579459 |
| Obesity | LEC | *XAF1* | 0.923065723 | 0.000323011 | 0.000617254 | 1 |
| Obesity | LEC | *REEP2* | -1.150009525 | 0.000395651 | 0.00073182 | 1 |
| Obesity | LEC | *PRMT7* | 1.025951453 | 0.000491383 | 0.000879399 | 1 |
| Obesity | LEC | *TERF2* | 2.201010212 | 8.86E-05 | 0.000223412 | 0.556743664 |
| Obesity | LEC | *BTBD3* | -1.005813962 | 0.00488827 | 0.006959365 | 1 |
| Obesity | LEC | *PCNA* | 1.60416865 | 8.33E-06 | 0.000105108 | 0.052399372 |
| Obesity | LEC | *PTPRA* | 1.577741087 | 0.000426296 | 0.000779558 | 1 |
| Obesity | LEC | *IGHMBP2* | 1.731028967 | 6.33E-10 | 1.59E-07 | 3.98E-06 |
| Obesity | LEC | *MMACHC* | 0.718362652 | 0.000534438 | 0.000945151 | 1 |
| Obesity | LEC | *CTNNBL1* | 1.635347879 | 1.80E-05 | 0.000105108 | 0.11303911 |
| Obesity | LEC | *LPIN3* | 0.934668933 | 1.84E-05 | 0.000105108 | 0.115749606 |
| Obesity | LEC | *SERINC3* | 1.133899991 | 0.000822801 | 0.001377663 | 1 |
| Obesity | LEC | *KANK4* | 0.998806936 | 1.27E-05 | 0.000105108 | 0.079777732 |
| Obesity | LEC | *RSG1* | -0.778790635 | 0.000107342 | 0.000256892 | 0.674856093 |
| Obesity | LEC | *CASP9* | 0.683625204 | 0.002572394 | 0.003861662 | 1 |
| Obesity | LEC | *ATP8A2* | 3.737544522 | 3.99E-05 | 0.000131515 | 0.251016002 |
| Obesity | LEC | *USPL1* | 1.842292617 | 4.95E-05 | 0.000151155 | 0.311476764 |
| Obesity | LEC | *POMP* | 2.115964366 | 0.002729991 | 0.004082648 | 1 |
| Obesity | LEC | *RNF17* | 1.647576917 | 0.005145726 | 0.007304399 | 1 |
| Obesity | LEC | *PEMT* | 0.857641121 | 0.0004363 | 0.000794847 | 1 |
| Obesity | LEC | *SCO1* | 1.869521361 | 4.69E-05 | 0.000145969 | 0.294707243 |
| Obesity | LEC | *DSTYK* | 1.704173233 | 0.003358833 | 0.004941957 | 1 |
| Obesity | LEC | *TMCC2* | -1.497471836 | 0.007497188 | 0.010336583 | 1 |
| Obesity | LEC | *COG6* | 1.485392696 | 0.000101625 | 0.000247258 | 0.638915897 |
| Obesity | LEC | *TRPC4* | -1.508648734 | 0.001654838 | 0.002592566 | 1 |
| Obesity | LEC | *RFC3* | 1.699006013 | 0.000289982 | 0.000565868 | 1 |
| Obesity | LEC | *BTBD2* | 1.421563165 | 0.001293153 | 0.002073993 | 1 |
| Obesity | LEC | *PRAM1* | 0.810247176 | 0.000179319 | 0.00038287 | 1 |
| Obesity | LEC | *ZNF414* | 1.462351218 | 1.66E-05 | 0.000105108 | 0.104294129 |
| Obesity | LEC | *CNDP2* | 2.306734803 | 2.64E-05 | 0.000112582 | 0.165836561 |
| Obesity | LEC | *RTN3* | 1.31574781 | 1.59E-05 | 0.000105108 | 0.099749771 |
| Obesity | LEC | *SLC2A11* | 1.833919536 | 1.65E-05 | 0.000105108 | 0.103582372 |
| Obesity | LEC | *C1QTNF6* | 1.581890006 | 1.45E-05 | 0.000105108 | 0.0911565 |
| Obesity | LEC | *FAM83F* | -0.96197519 | 0.000156994 | 0.000343724 | 0.987021205 |
| Obesity | LEC | *ADCK2* | 0.682120412 | 0.011436633 | 0.01533421 | 1 |
| Obesity | LEC | *AGAP3* | 1.533751637 | 0.000208858 | 0.000431228 | 1 |
| Obesity | LEC | *ACTR3B* | 1.652964457 | 1.53E-05 | 0.000105108 | 0.096311141 |
| Obesity | LEC | *SFTPD* | 0.623978896 | 0.003428868 | 0.00503675 | 1 |
| Obesity | LEC | *DYDC2* | 3.276773907 | 6.42E-05 | 0.000178992 | 0.403626417 |
| Obesity | LEC | *IPO8* | 2.032369895 | 0.000230565 | 0.000468525 | 1 |
| Obesity | LEC | *SPINK5* | -0.85326458 | 0.000237551 | 0.000480837 | 1 |
| Obesity | LEC | *LRRCC1* | 1.36185438 | 5.12E-05 | 0.00015431 | 0.322044214 |
| Obesity | LEC | *AMPD3* | 0.661773434 | 1.70E-05 | 0.000105108 | 0.106627203 |
| Obesity | LEC | *MICAL2* | -0.57255237 | 0.000733372 | 0.001248163 | 1 |

| | | | | | | |
|---|---|---|---|---|---|---|
| Obesity | LEC | *RRAS2* | 1.163432846 | 0.00019923 | 0.000415719 | 1 |
| Obesity | LEC | *HSD17B4* | 1.2086448 | 4.66E-05 | 0.000145679 | 0.293106658 |
| Obesity | LEC | *GSC* | 1.757023981 | 3.05E-05 | 0.000115831 | 0.191972194 |
| Obesity | LEC | *CATSPERB* | -0.415674782 | 0.000317238 | 0.000607516 | 1 |
| Obesity | LEC | *MED6* | 1.597350055 | 1.09E-05 | 0.000105108 | 0.068223665 |
| Obesity | LEC | *ADAMDEC1* | 2.880171936 | 0.00147656 | 0.002338909 | 1 |
| Obesity | LEC | *MRO* | 0.934330003 | 0.000287859 | 0.000563015 | 1 |
| Obesity | LEC | *THUMPD3* | 1.389863131 | 0.005609519 | 0.007928742 | 1 |
| Obesity | LEC | *EDEM1* | 2.523881536 | 0.000250726 | 0.000503044 | 1 |
| Obesity | LEC | *CNTN6* | 0.848832671 | 0.000122044 | 0.000283212 | 0.767292406 |
| Obesity | LEC | *KATNBL1* | 1.605444397 | 9.90E-06 | 0.000105108 | 0.062267806 |
| Obesity | LEC | *GSTM5* | 0.890350209 | 8.28E-07 | 3.83E-05 | 0.005203583 |
| Obesity | LEC | *GSTM3* | 0.611541801 | 0.003658861 | 0.005338422 | 1 |
| Obesity | LEC | *SYT6* | 1.932886551 | 3.57E-05 | 0.000123048 | 0.224305473 |
| Obesity | LEC | *VAV3* | -0.309200795 | 0.000180134 | 0.00038377 | 1 |
| Obesity | LEC | *PSRC1* | 1.437739458 | 0.001053166 | 0.001726683 | 1 |
| Obesity | LEC | *WNT2B* | 1.774609125 | 1.49E-05 | 0.000105108 | 0.093809658 |
| Obesity | LEC | *CEPT1* | 0.950081428 | 2.09E-05 | 0.000106951 | 0.131203809 |
| Obesity | LEC | *AP4B1* | 2.304175114 | 0.001837594 | 0.002851173 | 1 |
| Obesity | LEC | *NAPG* | 2.058465185 | 2.73E-05 | 0.000113713 | 0.171441584 |
| Obesity | LEC | *SPIRE1* | 0.868227567 | 6.99E-05 | 0.000189301 | 0.439366879 |
| Obesity | LEC | *PPHLN1* | 1.621812343 | 1.07E-05 | 0.000105108 | 0.067001173 |
| Obesity | LEC | *YWHAQ* | 1.4375304 | 0.011921336 | 0.015929743 | 1 |
| Obesity | LEC | *LPIN1* | 1.167125987 | 0.000295301 | 0.000573015 | 1 |
| Obesity | LEC | *LDHA* | 1.572131856 | 1.41E-05 | 0.000105108 | 0.088833896 |
| Obesity | LEC | *ANO3* | 0.606257283 | 0.000254535 | 0.000508989 | 1 |
| Obesity | LEC | *FST* | 2.432880389 | 4.25E-06 | 8.66E-05 | 0.026745742 |
| Obesity | LEC | *NARS* | 1.787871593 | 1.57E-05 | 0.000105108 | 0.09887725 |
| Obesity | LEC | *KIAA1468* | 1.590301424 | 0.00034422 | 0.000650861 | 1 |
| Obesity | LEC | *FBXO18* | 1.966915198 | 0.000503807 | 0.000898053 | 1 |
| Obesity | LEC | *ECHDC3* | 1.470294877 | 0.00102952 | 0.00169218 | 1 |
| Obesity | LEC | *IL15RA* | 2.267414063 | 5.18E-05 | 0.000155338 | 0.325588758 |
| Obesity | LEC | *TMEM241* | 1.707847957 | 0.000103837 | 0.00025099 | 0.652824124 |
| Obesity | LEC | *EMP1* | 0.68532349 | 4.37E-05 | 0.000139975 | 0.274770217 |
| Obesity | LEC | *KLRD1* | 1.97724211 | 3.67E-05 | 0.000125385 | 0.230958274 |
| Obesity | LEC | *PRH2* | -1.931812967 | 0.000641629 | 0.001106397 | 1 |
| Obesity | LEC | *DDB2* | 2.317691645 | 0.000117031 | 0.00027444 | 0.73577291 |
| Obesity | LEC | *ACP2* | 2.370650721 | 2.23E-08 | 3.42E-06 | 0.000140355 |
| Obesity | LEC | *ZCCHC11* | 1.665507023 | 6.25E-05 | 0.00017606 | 0.39314093 |
| Obesity | LEC | *PRPF38A* | -0.974805059 | 0.030935251 | 0.038820344 | 1 |
| Obesity | LEC | *FHOD3* | 0.840578583 | 0.000161137 | 0.000351393 | 1 |
| Obesity | LEC | *TIMM10* | -1.076634887 | 0.031925825 | 0.039927921 | 1 |
| Obesity | LEC | *DHX34* | 0.899008583 | 0.000351009 | 0.000661017 | 1 |
| Obesity | LEC | *APLNR* | 1.2227894 | 1.65E-05 | 0.000105108 | 0.103452511 |
| Obesity | LEC | *TMEM165* | 1.149026555 | 0.003132772 | 0.004636473 | 1 |
| Obesity | LEC | *CLOCK* | 1.705260621 | 8.39E-05 | 0.00021504 | 0.527559019 |
| Obesity | LEC | *CLDN10* | -0.448899223 | 5.59E-05 | 0.000164486 | 0.35145188 |
| Obesity | LEC | *DZIP1* | 0.949908381 | 2.78E-06 | 7.05E-05 | 0.017457948 |
| Obesity | LEC | *BIVM* | 1.956939453 | 1.34E-05 | 0.000105108 | 0.084095496 |
| Obesity | LEC | *ERCC5* | 2.03564293 | 1.83E-05 | 0.000105108 | 0.115272609 |

| | | | | | | |
|---|---|---|---|---|---|---|
| Obesity | LEC | *CARS2* | 1.406610636 | 0.006592885 | 0.009192607 | 1 |
| Obesity | LEC | *STT3A* | 2.149938369 | 0.001590194 | 0.002499388 | 1 |
| Obesity | LEC | *KLB* | 0.820158911 | 1.32E-06 | 5.03E-05 | 0.008317489 |
| Obesity | LEC | *WDR36* | 1.940805684 | 0.002299463 | 0.003497876 | 1 |
| Obesity | LEC | *OSTF1* | 1.82335312 | 2.94E-05 | 0.000115699 | 0.185078386 |
| Obesity | LEC | *UBQLN1* | 1.834540029 | 0.000546145 | 0.000963145 | 1 |
| Obesity | LEC | *NAA35* | 1.679056875 | 0.003876907 | 0.005630425 | 1 |
| Obesity | LEC | *CTSL* | 1.52559653 | 1.59E-05 | 0.000105108 | 0.09989293 |
| Obesity | LEC | *TMEM2* | 1.119512034 | 1.30E-05 | 0.000105108 | 0.0818544 |
| Obesity | LEC | *AGTPBP1* | 1.6802824 | 2.03E-05 | 0.000106065 | 0.12781705 |
| Obesity | LEC | *ADAM19* | -0.370447582 | 4.00E-05 | 0.000131529 | 0.251220588 |
| Obesity | LEC | *USP30* | 1.131189195 | 1.83E-05 | 0.000105108 | 0.114846459 |
| Obesity | LEC | *HRK* | -2.311582027 | 6.54E-05 | 0.000181705 | 0.411341406 |
| Obesity | LEC | *P2RX4* | 2.239274317 | 2.06E-07 | 1.66E-05 | 0.001296878 |
| Obesity | LEC | *DTX1* | -0.438536644 | 0.000431209 | 0.00078717 | 1 |
| Obesity | LEC | *DMTF1* | 1.480115701 | 0.000426004 | 0.000779479 | 1 |
| Obesity | LEC | *CCDC146* | 1.425980084 | 5.07E-05 | 0.0001531 | 0.318600265 |
| Obesity | LEC | *SRPK2* | 1.50190567 | 0.000136301 | 0.000308357 | 0.856926323 |
| Obesity | LEC | *MTO1* | 1.924414797 | 0.000270876 | 0.000535365 | 1 |
| Obesity | LEC | *ANKRD6* | -0.886447093 | 0.012294015 | 0.016368589 | 1 |
| Obesity | LEC | *KHDC1* | 0.770557739 | 0.000408397 | 0.000752297 | 1 |
| Obesity | LEC | *NT5E* | -0.974262865 | 0.02472816 | 0.031489962 | 1 |
| Obesity | LEC | *NAT10* | 2.034714604 | 0.000535702 | 0.000946854 | 1 |
| Obesity | LEC | *ATP5G2* | 2.173186411 | 0.001514828 | 0.002393497 | 1 |
| Obesity | LEC | *ITGA7* | 0.571830027 | 0.001457634 | 0.00231301 | 1 |
| Obesity | LEC | *RDH5* | 0.641527947 | 6.15E-05 | 0.000174839 | 0.386865152 |
| Obesity | LEC | *BLOC1S1* | 2.118335873 | 0.002024487 | 0.003116899 | 1 |
| Obesity | LEC | *PPP1R1A* | 1.131590365 | 1.52E-05 | 0.000105108 | 0.095467584 |
| Obesity | LEC | *B4GALNT1* | 2.439581737 | 0.001423763 | 0.002263833 | 1 |
| Obesity | LEC | *FAIM2* | 1.363015973 | 3.02E-05 | 0.000115831 | 0.189904085 |
| Obesity | LEC | *KRT7* | -1.345831665 | 4.41E-05 | 0.000140779 | 0.277333693 |
| Obesity | LEC | *HNRNPA1* | 2.578288177 | 0.000759775 | 0.00128676 | 1 |
| Obesity | LEC | *SLC26A10* | 2.217231438 | 2.07E-05 | 0.000106696 | 0.130333206 |
| Obesity | LEC | *MAP7* | 0.638609875 | 4.90E-05 | 0.000149938 | 0.307955452 |
| Obesity | LEC | *PKIB* | 2.139779958 | 1.36E-05 | 0.000105108 | 0.085767725 |
| Obesity | LEC | *MICAL1* | 1.355149446 | 1.46E-05 | 0.000105108 | 0.091764923 |
| Obesity | LEC | *PRADC1* | 4.214250977 | 1.27E-05 | 0.000105108 | 0.079691404 |
| Obesity | LEC | *SMYD5* | 1.956528215 | 7.98E-06 | 0.000105108 | 0.050164156 |
| Obesity | LEC | *DYSF* | -0.228482014 | 7.93E-05 | 0.000206997 | 0.498863133 |
| Obesity | LEC | *KCNMB4* | 1.245618787 | 0.000205008 | 0.000425375 | 1 |
| Obesity | LEC | *USP15* | 1.455925341 | 0.000320644 | 0.000613291 | 1 |
| Obesity | LEC | *KLHL36* | 2.350581581 | 9.49E-05 | 0.000235176 | 0.596406716 |
| Obesity | LEC | *MPHOSPH6* | 1.972342419 | 1.98E-05 | 0.000105598 | 0.124183349 |
| Obesity | LEC | *FHOD1* | 1.990522148 | 1.14E-05 | 0.000105108 | 0.071526735 |
| Obesity | LEC | *CCDC102A* | -0.598240389 | 0.000329237 | 0.000628008 | 1 |
| Obesity | LEC | *NTPCR* | 2.535934936 | 3.04E-05 | 0.000115831 | 0.190818084 |
| Obesity | LEC | *RNASEL* | 1.718093389 | 3.15E-05 | 0.000116847 | 0.197819418 |
| Obesity | LEC | *PIGC* | 1.86381422 | 6.31E-05 | 0.000177084 | 0.396491891 |
| Obesity | LEC | *SP110* | 2.147333397 | 7.22E-05 | 0.00019394 | 0.45420685 |
| Obesity | LEC | *USP37* | 2.211013871 | 0.000408552 | 0.000752363 | 1 |

| | | | | | | |
|---|---|---|---|---|---|---|
| Obesity | LEC | *SLC19A3* | 0.517951846 | 0.000191874 | 0.000403583 | 1 |
| Obesity | LEC | *TMBIM1* | 1.376338879 | 0.003056709 | 0.004531367 | 1 |
| Obesity | LEC | *ARMC9* | 1.15502498 | 1.79E-05 | 0.000105108 | 0.112259308 |
| Obesity | LEC | *TMEM127* | 1.110401145 | 3.83E-05 | 0.000128384 | 0.240592139 |
| Obesity | LEC | *GCC2* | 1.273293293 | 0.005731337 | 0.008080941 | 1 |
| Obesity | LEC | *MRPS9* | 2.038028832 | 2.94E-07 | 2.15E-05 | 0.001848582 |
| Obesity | LEC | *ANKRD36* | 1.727609594 | 0.001081007 | 0.00176527 | 1 |
| Obesity | LEC | *USP44* | -0.682125351 | 0.012251889 | 0.016315955 | 1 |
| Obesity | LEC | *APPL2* | 1.29812619 | 1.89E-05 | 0.000105211 | 0.119072826 |
| Obesity | LEC | *DRAM1* | 1.553311906 | 0.000300433 | 0.000580818 | 1 |
| Obesity | LEC | *SLC41A2* | 1.115263444 | 1.39E-05 | 0.000105108 | 0.087507659 |
| Obesity | LEC | *NEK3* | 1.437899371 | 8.37E-05 | 0.000214842 | 0.525934016 |
| Obesity | LEC | *VPS36* | 1.24310868 | 0.021844455 | 0.028027774 | 1 |
| Obesity | LEC | *RNASEH2B* | 1.318748097 | 6.88E-05 | 0.000187286 | 0.432684348 |
| Obesity | LEC | *TBC1D4* | 1.497302302 | 2.41E-05 | 0.000109361 | 0.15151615 |
| Obesity | LEC | *LRCH1* | 1.660094132 | 0.000155922 | 0.000342157 | 0.980280262 |
| Obesity | LEC | *SUCLA2* | 1.141139994 | 0.003730312 | 0.00543926 | 1 |
| Obesity | LEC | *RCBTB2* | 1.90294593 | 0.001839864 | 0.002853992 | 1 |
| Obesity | LEC | *SETDB2* | 1.295473404 | 2.95E-05 | 0.000115811 | 0.185413077 |
| Obesity | LEC | *GPNMB* | 0.573288637 | 3.16E-05 | 0.000116919 | 0.198581046 |
| Obesity | LEC | *KDELR2* | 2.415775948 | 0.000852479 | 0.001423235 | 1 |
| Obesity | LEC | *ZDHHC4* | 1.270461516 | 0.003099427 | 0.004591447 | 1 |
| Obesity | LEC | *AOAH* | 0.721341271 | 3.39E-05 | 0.000120512 | 0.213112188 |
| Obesity | LEC | *TBRG4* | 2.064902239 | 0.013691058 | 0.018098335 | 1 |
| Obesity | LEC | *DDX56* | 1.122626418 | 1.60E-05 | 0.000105108 | 0.100415178 |
| Obesity | LEC | *HUS1* | 1.306047 | 1.44E-05 | 0.000105108 | 0.09055326 |
| Obesity | LEC | *NACAD* | -4.576464992 | 0.000859246 | 0.001433672 | 1 |
| Obesity | LEC | *DBNL* | 2.093366105 | 0.00063255 | 0.001093741 | 1 |
| Obesity | LEC | *TTC5* | 2.018735359 | 0.000602397 | 0.001050561 | 1 |
| Obesity | LEC | *MTHFS* | 0.540637491 | 0.003864768 | 0.005615391 | 1 |
| Obesity | LEC | *ALPK3* | -1.006758604 | 0.00474718 | 0.006778452 | 1 |
| Obesity | LEC | *CIB2* | -1.19835429 | 0.002005952 | 0.003091793 | 1 |
| Obesity | LEC | *NMT1* | 1.528160608 | 2.66E-06 | 6.98E-05 | 0.016752019 |
| Obesity | LEC | *BRIP1* | 0.71768761 | 2.59E-05 | 0.000112124 | 0.1630286 |
| Obesity | LEC | *RTP4* | 1.085038818 | 0.031547805 | 0.039518042 | 1 |
| Obesity | LEC | *MRPL47* | 2.472700601 | 3.80E-05 | 0.000127862 | 0.23914971 |
| Obesity | LEC | *BLK* | -1.07635877 | 7.82E-06 | 0.000105108 | 0.049166238 |
| Obesity | LEC | *EPRS* | 1.879992959 | 0.004697386 | 0.006716504 | 1 |
| Obesity | LEC | *RPS6KC1* | 1.546054531 | 0.000303487 | 0.000585642 | 1 |
| Obesity | LEC | *CBWD2* | 1.578821478 | 2.60E-05 | 0.000112153 | 0.163519123 |
| Obesity | LEC | *IL1RN* | 2.367235172 | 1.99E-05 | 0.000105667 | 0.125020243 |
| Obesity | LEC | *SAP130* | 1.833593637 | 0.000442373 | 0.000804048 | 1 |
| Obesity | LEC | *UGGT1* | 1.925944392 | 7.19E-05 | 0.000193256 | 0.452219141 |
| Obesity | LEC | *NIPSNAP3A* | 1.350904298 | 0.001722628 | 0.002687386 | 1 |
| Obesity | LEC | *ODF2* | 2.243371218 | 0.00467546 | 0.006686673 | 1 |
| Obesity | LEC | *TOR1B* | 1.429825222 | 0.002562867 | 0.003849198 | 1 |
| Obesity | LEC | *TOR1A* | 1.259387543 | 0.000499001 | 0.000891255 | 1 |
| Obesity | LEC | *SLC2A8* | 2.260381103 | 7.85E-06 | 0.000105108 | 0.049324798 |
| Obesity | LEC | *ZFP37* | 1.404690362 | 0.00061742 | 0.001071923 | 1 |
| Obesity | LEC | *ZNF189* | 1.027726801 | 2.57E-05 | 0.000112124 | 0.161690119 |

| | | | | | | |
|---|---|---|---|---|---|---|
| Obesity | LEC | *ALDOB* | -1.112503356 | 0.000158862 | 0.000347453 | 0.9987659 |
| Obesity | LEC | *STX17* | 1.398925406 | 8.08E-06 | 0.000105108 | 0.050813251 |
| Obesity | LEC | *PRPF4* | 1.73335557 | 4.43E-06 | 8.83E-05 | 0.027823484 |
| Obesity | LEC | *FPGS* | 1.88750869 | 0.000284057 | 0.000556172 | 1 |
| Obesity | LEC | *TEX10* | 2.158504382 | 0.000503797 | 0.000898053 | 1 |
| Obesity | LEC | *GARNL3* | 1.106569118 | 1.48E-05 | 0.000105108 | 0.093311248 |
| Obesity | LEC | *GABBR2* | 1.792245673 | 2.32E-05 | 0.000109118 | 0.145903341 |
| Obesity | LEC | *PSMB7* | 1.702984597 | 3.40E-05 | 0.000120775 | 0.214012419 |
| Obesity | LEC | *RABEPK* | 1.185591164 | 1.34E-05 | 0.000105108 | 0.084513178 |
| Obesity | LEC | *GOLGA1* | 1.759373589 | 2.52E-05 | 0.000111345 | 0.158673265 |
| Obesity | LEC | *DSCC1* | 0.68801215 | 5.71E-05 | 0.000166393 | 0.3587429 |
| Obesity | LEC | *NOV* | 1.135249956 | 1.95E-05 | 0.000105217 | 0.122594234 |
| Obesity | LEC | *IL11RA* | 1.008566043 | 0.000274278 | 0.000541579 | 1 |
| Obesity | LEC | *APTX* | 1.324020302 | 6.14E-05 | 0.000174617 | 0.385903795 |
| Obesity | LEC | *TLN1* | 1.997353192 | 0.000158331 | 0.000346477 | 0.995429612 |
| Obesity | LEC | *SPAG8* | 2.017263731 | 8.82E-06 | 0.000105108 | 0.055432959 |
| Obesity | LEC | *CD72* | 1.276438142 | 3.07E-06 | 7.33E-05 | 0.019273059 |
| Obesity | LEC | *TMEM8B* | 0.843464151 | 2.58E-05 | 0.000112124 | 0.162410999 |
| Obesity | LEC | *ARHGEF39* | 0.958810557 | 0.00045918 | 0.000830992 | 1 |
| Obesity | LEC | *DENND4C* | 1.367392891 | 2.60E-05 | 0.000112153 | 0.163514289 |
| Obesity | LEC | *PPIL1* | 3.31556613 | 1.16E-05 | 0.000105108 | 0.072849255 |
| Obesity | LEC | *KIF13A* | 1.403712258 | 5.74E-05 | 0.000166843 | 0.361048994 |
| Obesity | LEC | *ZSCAN9* | 1.837541928 | 3.38E-09 | 7.07E-07 | 2.12E-05 |
| Obesity | LEC | *SLC22A23* | -0.435181247 | 0.000381714 | 0.00070959 | 1 |
| Obesity | LEC | *LRRC1* | -0.264973857 | 0.000189805 | 0.000399887 | 1 |
| Obesity | LEC | *FOXF2* | 2.078226329 | 0.003384642 | 0.004977601 | 1 |
| Obesity | LEC | *BPHL* | 0.652202305 | 0.000719131 | 0.001227247 | 1 |
| Obesity | LEC | *RIPK1* | 1.901012504 | 0.000190477 | 0.000401048 | 1 |
| Obesity | LEC | *TUBB2B* | 0.861099752 | 0.006505552 | 0.009080908 | 1 |
| Obesity | LEC | *UQCC2* | 2.063046354 | 0.000127674 | 0.000293059 | 0.802688385 |
| Obesity | LEC | *TCF19* | 3.557280455 | 1.80E-07 | 1.53E-05 | 0.001130591 |
| Obesity | LEC | *FLOT1* | 2.242351208 | 1.75E-05 | 0.000105108 | 0.110099494 |
| Obesity | LEC | *TPMT* | 1.981626239 | 0.000105592 | 0.000253961 | 0.663854799 |
| Obesity | LEC | *NRM* | 1.513131194 | 3.65E-07 | 2.44E-05 | 0.002293597 |
| Obesity | LEC | *MTCH1* | 2.120784819 | 4.51E-05 | 0.000142544 | 0.283235433 |
| Obesity | LEC | *FAM8A1* | 1.800730005 | 1.41E-05 | 0.000105108 | 0.088584775 |
| Obesity | LEC | *FGFBP2* | 0.79520218 | 0.000240278 | 0.000485201 | 1 |
| Obesity | LEC | *FHDC1* | -1.067892671 | 0.000617412 | 0.001071923 | 1 |
| Obesity | LEC | *MYO7A* | 0.910600878 | 0.013091185 | 0.017356449 | 1 |
| Obesity | LEC | *ARRB1* | 1.526118627 | 0.005729416 | 0.008080044 | 1 |
| Obesity | LEC | *CCDC90B* | 2.172308212 | 0.000364171 | 0.000682224 | 1 |
| Obesity | LEC | *SYTL2* | 1.173356569 | 5.87E-05 | 0.000169536 | 0.369248687 |
| Obesity | LEC | *RAB30* | 1.567863853 | 4.40E-05 | 0.000140597 | 0.27683495 |
| Obesity | LEC | *NARS2* | 1.170097568 | 1.41E-05 | 0.000105108 | 0.088673182 |
| Obesity | LEC | *PI15* | 4.229264325 | 1.65E-05 | 0.000105108 | 0.103433251 |
| Obesity | LEC | *GGH* | 0.511019908 | 0.022729468 | 0.029086132 | 1 |
| Obesity | LEC | *SULF1* | 1.409116187 | 1.22E-05 | 0.000105108 | 0.076474802 |
| Obesity | LEC | *TGS1* | 1.094470884 | 0.039329116 | 0.048606675 | 1 |
| Obesity | LEC | *NEK1* | 1.412529574 | 2.90E-05 | 0.000115266 | 0.182500666 |
| Obesity | LEC | *DDX60* | -0.972738894 | 0.023989309 | 0.030580046 | 1 |

| | | | | | | |
|---|---|---|---|---|---|---|
| Obesity | LEC | *MMP27* | 0.693520519 | 7.86E-05 | 0.000205732 | 0.493962669 |
| Obesity | LEC | *DCUN1D5* | 1.51142161 | 1.18E-05 | 0.000105108 | 0.074400531 |
| Obesity | LEC | *TRIM29* | -0.446277933 | 3.93E-05 | 0.000130538 | 0.247281451 |
| Obesity | LEC | *SLC37A4* | 1.706886658 | 1.94E-05 | 0.000105217 | 0.122176445 |
| Obesity | LEC | *POU2F3* | -1.27748617 | 7.67E-05 | 0.00020244 | 0.48231778 |
| Obesity | LEC | *PPP2R1B* | 0.884399301 | 1.50E-05 | 0.000105108 | 0.094358791 |
| Obesity | LEC | *FXYD6* | -0.440347308 | 0.000192739 | 0.000404862 | 1 |
| Obesity | LEC | *TMPRSS13* | 2.432497418 | 0.000358135 | 0.000672319 | 1 |
| Obesity | LEC | *CASP1* | 1.9923029 | 2.39E-05 | 0.000109361 | 0.150512803 |
| Obesity | LEC | *MAP2K5* | 1.825569698 | 0.000249923 | 0.000501841 | 1 |
| Obesity | LEC | *NDUFAF1* | 1.670014102 | 0.006768883 | 0.009410873 | 1 |
| Obesity | LEC | *ITGA11* | 0.510709717 | 2.78E-05 | 0.00011384 | 0.17486708 |
| Obesity | LEC | *PAQR5* | -1.404323253 | 0.003231032 | 0.004770666 | 1 |
| Obesity | LEC | *RMDN3* | 1.665632109 | 4.51E-06 | 8.91E-05 | 0.028354475 |
| Obesity | LEC | *ITPKA* | -2.383433392 | 8.97E-05 | 0.000225697 | 0.563790084 |
| Obesity | LEC | *PLCB2* | 0.685313396 | 0.000115233 | 0.000271338 | 0.724472855 |
| Obesity | LEC | *RSL24D1* | 1.17642414 | 0.029226722 | 0.036838092 | 1 |
| Obesity | LEC | *SPTBN5* | 0.825293507 | 0.002774438 | 0.004144189 | 1 |
| Obesity | LEC | *RABGGTB* | 2.176360707 | 4.11E-05 | 0.00013378 | 0.258329749 |
| Obesity | LEC | *SLC44A5* | -0.226858988 | 0.000210858 | 0.00043493 | 1 |
| Obesity | LEC | *DNASE2B* | 3.583682416 | 8.09E-05 | 0.000209834 | 0.508637157 |
| Obesity | LEC | *DBT* | 1.296532167 | 3.26E-05 | 0.000118274 | 0.204732954 |
| Obesity | LEC | *RTCA* | 1.819184383 | 0.001268169 | 0.00203652 | 1 |
| Obesity | LEC | *IFT172* | 2.393994022 | 3.61E-07 | 2.44E-05 | 0.002272478 |
| Obesity | LEC | *ADCY3* | 1.38125065 | 9.50E-06 | 0.000105108 | 0.05975171 |
| Obesity | LEC | *DYNC2LI1* | 1.269686356 | 0.004414497 | 0.00635247 | 1 |
| Obesity | LEC | *LHCGR* | 0.784847262 | 5.00E-05 | 0.000151905 | 0.314291406 |
| Obesity | LEC | *THUMPD2* | 1.616455623 | 0.009838714 | 0.013316684 | 1 |
| Obesity | LEC | *PREB* | 1.038280687 | 0.001589248 | 0.002498525 | 1 |
| Obesity | LEC | *ABCG5* | 1.816413533 | 6.19E-06 | 9.96E-05 | 0.038903787 |
| Obesity | LEC | *ATRAID* | 1.970046597 | 0.00602432 | 0.008452332 | 1 |
| Obesity | LEC | *CENPO* | 1.807505113 | 1.21E-05 | 0.000105108 | 0.076126757 |
| Obesity | LEC | *LRPPRC* | 1.561343012 | 3.16E-05 | 0.000116919 | 0.198731203 |
| Obesity | LEC | *DTNB* | 1.876171148 | 3.47E-05 | 0.000121951 | 0.218170108 |
| Obesity | LEC | *CYP2C9* | 0.809678229 | 0.002986805 | 0.004438204 | 1 |
| Obesity | LEC | *CYP2C8* | -0.776896952 | 0.000328267 | 0.000626538 | 1 |
| Obesity | LEC | *MYOF* | 0.894276779 | 0.006997488 | 0.009705967 | 1 |
| Obesity | LEC | *STAMBPL1* | 1.812653287 | 0.000124385 | 0.000287502 | 0.78200602 |
| Obesity | LEC | *ATAD1* | 1.888467656 | 0.00106158 | 0.001737608 | 1 |
| Obesity | LEC | *BTBD16* | 1.02429593 | 2.91E-05 | 0.000115266 | 0.182919422 |
| Obesity | LEC | *TACC2* | 1.628336201 | 1.66E-05 | 0.000105108 | 0.104481696 |
| Obesity | LEC | *CALHM2* | 1.678647025 | 3.74E-05 | 0.000126288 | 0.235400854 |
| Obesity | LEC | *ARL3* | 1.808750484 | 2.15E-05 | 0.000107557 | 0.135335572 |
| Obesity | LEC | *PLCE1* | -0.536472531 | 0.000722602 | 0.001232167 | 1 |
| Obesity | LEC | *RBP4* | 1.843073614 | 1.10E-05 | 0.000105108 | 0.069254524 |
| Obesity | LEC | *DNAJC13* | 1.762230273 | 0.001047826 | 0.001720471 | 1 |
| Obesity | LEC | *FAM149B1* | 1.201998341 | 2.07E-05 | 0.000106696 | 0.130065292 |
| Obesity | LEC | *ASCC1* | 1.371911876 | 0.001243793 | 0.002002436 | 1 |
| Obesity | LEC | *RPS24* | 1.342158903 | 0.031353563 | 0.039321734 | 1 |
| Obesity | LEC | *DNA2* | 0.417898282 | 0.01154529 | 0.015469999 | 1 |

| | | | | | | |
|---|---|---|---|---|---|---|
| Obesity | LEC | *ATIC* | 0.847550366 | 0.003481633 | 0.005109483 | 1 |
| Obesity | LEC | *SMARCAL1* | 1.785768152 | 3.21E-05 | 0.00011749 | 0.201716744 |
| Obesity | LEC | *BARD1* | 1.618870446 | 1.83E-05 | 0.000105108 | 0.115225375 |
| Obesity | LEC | *STAT4* | 0.689898398 | 1.85E-05 | 0.000105108 | 0.116567607 |
| Obesity | LEC | *CARF* | 2.459229931 | 1.63E-07 | 1.45E-05 | 0.001027774 |
| Obesity | LEC | *ASNSD1* | 1.655322646 | 9.71E-05 | 0.000239182 | 0.610153979 |
| Obesity | LEC | *FASTKD1* | 1.845663753 | 9.20E-05 | 0.000230222 | 0.578547639 |
| Obesity | LEC | *MDH1B* | 1.682587489 | 3.33E-05 | 0.00011952 | 0.209159392 |
| Obesity | LEC | *FAM117B* | 1.852346645 | 0.003745565 | 0.005458592 | 1 |
| Obesity | LEC | *ITGAV* | 1.503148685 | 7.35E-05 | 0.000196142 | 0.462307128 |
| Obesity | LEC | *COX17* | 1.72016035 | 1.22E-05 | 0.000105108 | 0.077006384 |
| Obesity | LEC | *USP8* | 1.640100968 | 0.000412587 | 0.000758461 | 1 |
| Obesity | LEC | *SPPL2A* | 1.661579949 | 0.000662243 | 0.001137885 | 1 |
| Obesity | LEC | *UBL7* | 3.310042522 | 9.02E-08 | 1.02E-05 | 0.0005669 |
| Obesity | LEC | *ARHGAP24* | 0.790227306 | 1.43E-05 | 0.000105108 | 0.090070617 |
| Obesity | LEC | *FAM13A* | 0.800054421 | 3.59E-05 | 0.000123286 | 0.225860614 |
| Obesity | LEC | *HERC3* | 1.360008196 | 1.82E-05 | 0.000105108 | 0.11446872 |
| Obesity | LEC | *AP1AR* | 2.143538761 | 0.00246751 | 0.00373004 | 1 |
| Obesity | LEC | *PRKG2* | 0.507767922 | 2.67E-05 | 0.000112975 | 0.167725359 |
| Obesity | LEC | *BBS7* | 2.132075334 | 0.003466914 | 0.005090259 | 1 |
| Obesity | LEC | *KIAA1109* | 1.368702291 | 0.004473532 | 0.006428594 | 1 |
| Obesity | LEC | *MMRN1* | -0.039687536 | 3.06E-05 | 0.000115834 | 0.192284115 |
| Obesity | LEC | *PRDM5* | -0.921240759 | 0.011636187 | 0.015585153 | 1 |
| Obesity | LEC | *TRPC3* | 1.092073224 | 8.64E-05 | 0.000219867 | 0.54350999 |
| Obesity | LEC | *NAAA* | 1.922238955 | 0.000958591 | 0.001583047 | 1 |
| Obesity | LEC | *BMP2K* | -0.755160629 | 0.03674926 | 0.045633537 | 1 |
| Obesity | LEC | *G3BP2* | 2.057596667 | 2.13E-05 | 0.00010722 | 0.133675445 |
| Obesity | LEC | *SCARB2* | 1.742266814 | 0.000647894 | 0.001115058 | 1 |
| Obesity | LEC | *CNOT6L* | 1.099572292 | 0.004170595 | 0.006023554 | 1 |
| Obesity | LEC | *PPA2* | 1.418170826 | 0.00247674 | 0.003739497 | 1 |
| Obesity | LEC | *GSTCD* | 1.694094666 | 1.68E-05 | 0.000105108 | 0.105593624 |
| Obesity | LEC | *PAPSS1* | 1.112629631 | 0.000384473 | 0.000714508 | 1 |
| Obesity | LEC | *SLC39A8* | 1.591141896 | 0.001734479 | 0.002704531 | 1 |
| Obesity | LEC | *RNF185* | 1.65917682 | 2.97E-05 | 0.000115831 | 0.186897141 |
| Obesity | LEC | *PARVG* | 0.994950115 | 3.30E-05 | 0.000119154 | 0.207744945 |
| Obesity | LEC | *ERP27* | 1.901604345 | 1.37E-05 | 0.000105108 | 0.086317746 |
| Obesity | LEC | *ETV6* | 1.537578147 | 0.000103315 | 0.000250113 | 0.6495436 |
| Obesity | LEC | *KIF21A* | 0.858346347 | 0.000223951 | 0.00045788 | 1 |
| Obesity | LEC | *ALG10* | -1.611410372 | 0.003629032 | 0.005297359 | 1 |
| Obesity | LEC | *SLCO1C1* | 0.838759963 | 1.31E-05 | 0.000105108 | 0.082234654 |
| Obesity | LEC | *ZCRB1* | 1.263283705 | 0.037857764 | 0.046917358 | 1 |
| Obesity | LEC | *PRICKLE1* | 1.125647016 | 3.56E-05 | 0.000123048 | 0.223545693 |
| Obesity | LEC | *C1RL* | 1.721136386 | 0.00012424 | 0.000287273 | 0.781095132 |
| Obesity | LEC | *CLSTN3* | 1.25347204 | 1.28E-05 | 0.000105108 | 0.080760164 |
| Obesity | LEC | *KLRG1* | 1.567151636 | 1.52E-07 | 1.43E-05 | 0.000954841 |
| Obesity | LEC | *VAMP1* | 0.751704808 | 0.000188156 | 0.000397091 | 1 |
| Obesity | LEC | *CD27* | 1.934522157 | 3.65E-06 | 7.87E-05 | 0.022939314 |
| Obesity | LEC | *PEX5* | 0.924785458 | 2.35E-05 | 0.000109118 | 0.148041064 |
| Obesity | LEC | *PIANP* | -2.435185872 | 0.002908747 | 0.004330404 | 1 |
| Obesity | LEC | *PPFIA2* | 1.097233764 | 0.000307259 | 0.000592075 | 1 |

| | | | | | | |
|---|---|---|---|---|---|---|
| Obesity | LEC | *GLIPR1* | 0.948180721 | 1.33E-05 | 0.000105108 | 0.083918884 |
| Obesity | LEC | *LGR5* | 0.901788756 | 4.82E-05 | 0.000148518 | 0.302870961 |
| Obesity | LEC | *GAS2L3* | 1.013627565 | 1.72E-05 | 0.000105108 | 0.108049904 |
| Obesity | LEC | *TMEM132B* | 1.534988915 | 8.54E-05 | 0.000217903 | 0.536696015 |
| Obesity | LEC | *TDG* | 1.877491729 | 0.001449537 | 0.002301905 | 1 |
| Obesity | LEC | *RITA1* | -1.54464793 | 3.74E-05 | 0.000126288 | 0.235388039 |
| Obesity | LEC | *MMAB* | 1.465516571 | 0.001522175 | 0.002403898 | 1 |
| Obesity | LEC | *SLC46A3* | 1.14104327 | 4.01E-05 | 0.000131769 | 0.251902848 |
| Obesity | LEC | *SUOX* | 0.860153966 | 0.000406325 | 0.00074892 | 1 |
| Obesity | LEC | *CCDC65* | 1.421626607 | 0.000418354 | 0.000767715 | 1 |
| Obesity | LEC | *ACVRL1* | -1.389533007 | 0.015192345 | 0.019965358 | 1 |
| Obesity | LEC | *BRCA2* | 2.038488416 | 1.17E-05 | 0.000105108 | 0.073793521 |
| Obesity | LEC | *CERS5* | 1.784075812 | 3.56E-05 | 0.000123048 | 0.224102748 |
| Obesity | LEC | *LMBR1L* | 2.094586399 | 0.000164626 | 0.000357015 | 1 |
| Obesity | LEC | *ZNF740* | 2.277716846 | 0.000617545 | 0.001071923 | 1 |
| Obesity | LEC | *WDFY2* | 1.208765319 | 0.000106037 | 0.00025435 | 0.666651626 |
| Obesity | LEC | *HNRNPA1L2* | -2.079704364 | 0.003495645 | 0.005126457 | 1 |
| Obesity | LEC | *LPAR6* | 1.744568367 | 6.85E-05 | 0.000187169 | 0.43086374 |
| Obesity | LEC | *VPS33A* | -1.803960377 | 0.017761904 | 0.023143852 | 1 |
| Obesity | LEC | *RHOF* | 1.094441139 | 0.023985194 | 0.030580046 | 1 |
| Obesity | LEC | *METTL21C* | -2.969339389 | 8.85E-05 | 0.000223403 | 0.556495927 |
| Obesity | LEC | *GRTP1* | 0.683826389 | 0.004839607 | 0.006897894 | 1 |
| Obesity | LEC | *TSSK4* | 2.845061659 | 0.000371585 | 0.000694045 | 1 |
| Obesity | LEC | *TMX1* | 2.23050364 | 6.62E-05 | 0.000182861 | 0.416008424 |
| Obesity | LEC | *RTN1* | 0.611206183 | 1.55E-05 | 0.000105108 | 0.097323321 |
| Obesity | LEC | *C14orf37* | 1.478492058 | 0.00022428 | 0.000458137 | 1 |
| Obesity | LEC | *RAB15* | 0.987310911 | 0.003730569 | 0.00543926 | 1 |
| Obesity | LEC | *STON2* | -0.24487296 | 0.000151821 | 0.000335618 | 0.954497679 |
| Obesity | LEC | *EFCAB11* | 1.803733418 | 6.25E-06 | 9.96E-05 | 0.039323576 |
| Obesity | LEC | *AK7* | 5.665444734 | 6.88E-05 | 0.000187286 | 0.432599779 |
| Obesity | LEC | *FBLN5* | 1.045325613 | 7.86E-06 | 0.000105108 | 0.049427884 |
| Obesity | LEC | *C14orf79* | 1.000276738 | 2.78E-05 | 0.00011384 | 0.174547861 |
| Obesity | LEC | *NIPA2* | 1.644190104 | 1.92E-05 | 0.000105211 | 0.120618046 |
| Obesity | LEC | *DUOXA1* | 1.001525805 | 0.00074968 | 0.001272816 | 1 |
| Obesity | LEC | *MFAP1* | 2.30454072 | 0.000330314 | 0.000629679 | 1 |
| Obesity | LEC | *TCF12* | 1.322104832 | 0.00133297 | 0.002129162 | 1 |
| Obesity | LEC | *SORD* | 0.863498116 | 1.79E-05 | 0.000105108 | 0.112770352 |
| Obesity | LEC | *ZSCAN29* | 1.734621536 | 5.65E-06 | 9.65E-05 | 0.03552561 |
| Obesity | LEC | *SRP14* | 1.971697232 | 0.000635055 | 0.001096827 | 1 |
| Obesity | LEC | *BAHD1* | 1.805602667 | 1.34E-05 | 0.000105108 | 0.084554099 |
| Obesity | LEC | *DISP2* | 0.526695711 | 0.037826079 | 0.046887334 | 1 |
| Obesity | LEC | *ANP32A* | 1.894421887 | 4.67E-05 | 0.000145712 | 0.293318251 |
| Obesity | LEC | *COMMD4* | 0.976353144 | 2.81E-05 | 0.000114038 | 0.176824195 |
| Obesity | LEC | *UBE2Q2* | 1.340849619 | 2.60E-05 | 0.000112153 | 0.163306297 |
| Obesity | LEC | *PSTPIP1* | 1.147240754 | 5.84E-05 | 0.000168887 | 0.367160426 |
| Obesity | LEC | *HMG20A* | 1.955270687 | 1.05E-05 | 0.000105108 | 0.065968935 |
| Obesity | LEC | *SCAPER* | 1.214819813 | 0.003607829 | 0.005270079 | 1 |
| Obesity | LEC | *DNAJA4* | 1.931961442 | 1.12E-05 | 0.000105108 | 0.070131824 |
| Obesity | LEC | *TPM1* | 1.535508021 | 0.000103754 | 0.000250928 | 0.652300546 |
| Obesity | LEC | *CYP11A1* | 1.930954715 | 1.52E-07 | 1.43E-05 | 0.00095829 |

| | | | | | | |
|---|---|---|---|---|---|---|
| Obesity | LEC | *BBS4* | 1.482951265 | 2.75E-06 | 7.03E-05 | 0.017281795 |
| Obesity | LEC | *ADAMTS17* | 0.544516542 | 3.20E-05 | 0.000117412 | 0.200892676 |
| Obesity | LEC | *ULK3* | 2.521961557 | 5.76E-05 | 0.000167083 | 0.362402356 |
| Obesity | LEC | *SCAMP2* | 2.252837443 | 3.40E-05 | 0.000120731 | 0.213694556 |
| Obesity | LEC | *HAPLN3* | 2.171163145 | 0.000174261 | 0.000373408 | 1 |
| Obesity | LEC | *POLG* | 1.902835727 | 1.43E-05 | 0.000105108 | 0.09015318 |
| Obesity | LEC | *WDR93* | 0.3006151 | 0.005605935 | 0.007925899 | 1 |
| Obesity | LEC | *TICRR* | 1.198526366 | 5.19E-05 | 0.000155604 | 0.326457541 |
| Obesity | LEC | *MFGE8* | 1.005420001 | 4.08E-05 | 0.000133256 | 0.256760263 |
| Obesity | LEC | *UNC45A* | 1.685249097 | 0.000159407 | 0.000348308 | 1 |
| Obesity | LEC | *ST8SIA2* | 5.227497867 | 3.73E-05 | 0.000126137 | 0.234428726 |
| Obesity | LEC | *IQGAP1* | 1.288623275 | 8.35E-05 | 0.000214782 | 0.525207317 |
| Obesity | LEC | *CRTC3* | 1.619038869 | 5.39E-05 | 0.000159887 | 0.339119937 |
| Obesity | LEC | *SH3GL3* | -2.287469607 | 1.21E-05 | 0.000105108 | 0.075992735 |
| Obesity | LEC | *SEC11A* | 1.822698427 | 0.0005486 | 0.000966662 | 1 |
| Obesity | LEC | *TGFB1I1* | -1.002187765 | 0.000168402 | 0.000363236 | 1 |
| Obesity | LEC | *PARN* | 1.552129991 | 6.88E-05 | 0.000187286 | 0.432818193 |
| Obesity | LEC | *FTO* | 1.390686488 | 0.000241498 | 0.000487101 | 1 |
| Obesity | LEC | *MARVELD3* | -0.920907924 | 8.39E-05 | 0.00021504 | 0.527268452 |
| Obesity | LEC | *ZFHX3* | 1.435043853 | 0.003087774 | 0.004575262 | 1 |
| Obesity | LEC | *KIFC3* | 1.483795363 | 1.95E-05 | 0.000105217 | 0.122614545 |
| Obesity | LEC | *CMTM3* | 1.289014093 | 9.82E-06 | 0.000105108 | 0.061725671 |
| Obesity | LEC | *MAP1LC3B* | 1.753133075 | 0.000132163 | 0.000301272 | 0.830909182 |
| Obesity | LEC | *TLDC1* | 1.53517277 | 8.71E-06 | 0.000105108 | 0.054787269 |
| Obesity | LEC | *NDUFB10* | 3.042571931 | 6.83E-05 | 0.000186798 | 0.429635115 |
| Obesity | LEC | *TIGD7* | 1.488006746 | 1.04E-06 | 4.37E-05 | 0.006556089 |
| Obesity | LEC | *DEF8* | 1.457412026 | 0.000342916 | 0.000649568 | 1 |
| Obesity | LEC | *GALNS* | 1.732315853 | 0.000683988 | 0.001172365 | 1 |
| Obesity | LEC | *COPS3* | 1.822174398 | 1.55E-05 | 0.000105108 | 0.097667674 |
| Obesity | LEC | *MYO19* | 3.061881645 | 1.84E-07 | 1.54E-05 | 0.001154871 |
| Obesity | LEC | *DDX52* | 1.925009309 | 0.001411026 | 0.002245285 | 1 |
| Obesity | LEC | *RASL10B* | 1.677486008 | 0.002190985 | 0.003352329 | 1 |
| Obesity | LEC | *UNC45B* | 3.070554898 | 0.000629674 | 0.001089967 | 1 |
| Obesity | LEC | *TOM1L1* | 1.451091153 | 5.05E-06 | 9.23E-05 | 0.03176192 |
| Obesity | LEC | *C17orf80* | 1.359976855 | 1.11E-05 | 0.000105108 | 0.069596891 |
| Obesity | LEC | *VPS53* | 1.404610973 | 1.30E-05 | 0.000105108 | 0.081635077 |
| Obesity | LEC | *SPATA22* | 1.113802363 | 0.000331386 | 0.000631148 | 1 |
| Obesity | LEC | *SGSM2* | 1.636821481 | 8.92E-05 | 0.000224703 | 0.56085846 |
| Obesity | LEC | *NPEPPS* | 1.307520382 | 0.018634363 | 0.024220434 | 1 |
| Obesity | LEC | *LRRC46* | 1.630990193 | 0.000249469 | 0.00050109 | 1 |
| Obesity | LEC | *SCRN2* | 0.555458155 | 0.001071078 | 0.00175042 | 1 |
| Obesity | LEC | *RHBDL3* | 1.098858694 | 4.65E-05 | 0.000145508 | 0.292180969 |
| Obesity | LEC | *ARSG* | 1.754718344 | 0.000896371 | 0.001490475 | 1 |
| Obesity | LEC | *ABCA8* | 0.788927962 | 4.63E-06 | 9.00E-05 | 0.029124627 |
| Obesity | LEC | *G6PC3* | -1.412260754 | 0.022944942 | 0.029337981 | 1 |
| Obesity | LEC | *IMPA2* | 0.877062759 | 1.67E-05 | 0.000105108 | 0.10493809 |
| Obesity | LEC | *GNAL* | 1.502170402 | 7.69E-06 | 0.000105108 | 0.048348461 |
| Obesity | LEC | *RPRD1A* | 1.542637572 | 0.001577655 | 0.00248154 | 1 |
| Obesity | LEC | *C18orf21* | 3.519667563 | 2.77E-05 | 0.00011384 | 0.173957993 |
| Obesity | LEC | *ASXL3* | 2.28445067 | 4.84E-05 | 0.0001487 | 0.304240843 |

| | | | | | | |
|---|---|---|---|---|---|---|
| Obesity | LEC | *ESCO1* | 1.665634263 | 2.61E-05 | 0.000112528 | 0.164403309 |
| Obesity | LEC | *GREB1L* | 0.786509959 | 4.97E-05 | 0.000151436 | 0.312575389 |
| Obesity | LEC | *C18orf8* | 0.722968304 | 0.007400318 | 0.010218712 | 1 |
| Obesity | LEC | *NPC1* | 0.985366629 | 0.009189953 | 0.012495076 | 1 |
| Obesity | LEC | *SLC14A1* | 1.99229475 | 0.019561433 | 0.025315506 | 1 |
| Obesity | LEC | *SLC13A5* | 3.425310678 | 0.014741133 | 0.019404838 | 1 |
| Obesity | LEC | *CCDC40* | 1.043709275 | 0.012112492 | 0.016157699 | 1 |
| Obesity | LEC | *TMC6* | 1.270519203 | 2.65E-05 | 0.000112661 | 0.166701802 |
| Obesity | LEC | *CARD14* | 3.370955576 | 0.000253882 | 0.000508006 | 1 |
| Obesity | LEC | *EIF4A3* | 1.706897893 | 0.000732651 | 0.001247272 | 1 |
| Obesity | LEC | *NARF* | 1.359803839 | 0.038358665 | 0.04749132 | 1 |
| Obesity | LEC | *TRIM65* | 3.169681517 | 4.44E-07 | 2.59E-05 | 0.002793595 |
| Obesity | LEC | *SECTM1* | 3.446677382 | 8.90E-05 | 0.000224335 | 0.559716702 |
| Obesity | LEC | *RNF157* | 0.85791572 | 2.85E-05 | 0.000114887 | 0.179359345 |
| Obesity | LEC | *WDR45B* | 1.496873728 | 9.12E-05 | 0.000228778 | 0.57331852 |
| Obesity | LEC | *RNF165* | -1.050727817 | 0.004625778 | 0.006623153 | 1 |
| Obesity | LEC | *MAPK4* | 0.8615379 | 0.000358833 | 0.000673227 | 1 |
| Obesity | LEC | *SMAD4* | 1.803699173 | 0.000220624 | 0.000452253 | 1 |
| Obesity | LEC | *PIP4K2B* | 2.086057067 | 2.06E-05 | 0.000106628 | 0.129553488 |
| Obesity | LEC | *GRB7* | 1.868827896 | 1.29E-05 | 0.000105108 | 0.080958968 |
| Obesity | LEC | *TXNL4A* | 1.175821996 | 0.020915539 | 0.02694038 | 1 |
| Obesity | LEC | *NFIC* | 1.364651598 | 0.000345299 | 0.000652144 | 1 |
| Obesity | LEC | *TPGS1* | 1.905301895 | 6.64E-06 | 0.000102808 | 0.04174005 |
| Obesity | LEC | *PFKL* | 1.161485538 | 0.000104284 | 0.000251585 | 0.655631298 |
| Obesity | LEC | *MVB12A* | 1.527931539 | 2.93E-06 | 7.22E-05 | 0.018399547 |
| Obesity | LEC | *DUS3L* | 0.847839684 | 9.62E-05 | 0.000237845 | 0.605077994 |
| Obesity | LEC | *TMEM91* | -0.970309746 | 0.015090489 | 0.019839795 | 1 |
| Obesity | LEC | *SIRT3* | 1.359500008 | 2.05E-06 | 6.09E-05 | 0.012867929 |
| Obesity | LEC | *IFITM3* | 1.459840074 | 1.42E-05 | 0.000105108 | 0.089146914 |
| Obesity | LEC | *COL6A1* | 1.085793735 | 1.79E-05 | 0.000105108 | 0.112586719 |
| Obesity | LEC | *IFNAR1* | 1.032058282 | 0.001326547 | 0.002120699 | 1 |
| Obesity | LEC | *TRPM2* | 0.4324744 | 6.43E-05 | 0.000179062 | 0.404143115 |
| Obesity | LEC | *TMEM50B* | 0.642997074 | 0.03174694 | 0.039735818 | 1 |
| Obesity | LEC | *DOPEY2* | 1.264676409 | 0.03779674 | 0.046860206 | 1 |
| Obesity | LEC | *AKT1* | 2.457273005 | 2.10E-05 | 0.000106989 | 0.131959962 |
| Obesity | LEC | *SAE1* | 1.075577005 | 1.57E-05 | 0.000105108 | 0.098442411 |
| Obesity | LEC | *NTN5* | 0.962027712 | 0.005714878 | 0.00806316 | 1 |
| Obesity | LEC | *GEMIN7* | -2.015181349 | 0.002367638 | 0.003592022 | 1 |
| Obesity | LEC | *CAPN10* | 1.404735055 | 0.000167186 | 0.000361077 | 1 |
| Obesity | LEC | *MYO1F* | 0.827798284 | 1.23E-05 | 0.000105108 | 0.077422273 |
| Obesity | LEC | *CACNG8* | 1.611346574 | 2.78E-05 | 0.00011384 | 0.174799947 |
| Obesity | LEC | *EVI5L* | 2.859504497 | 0.000291305 | 0.000566831 | 1 |
| Obesity | LEC | *RPS11* | 1.835558122 | 0.003055941 | 0.004531298 | 1 |
| Obesity | LEC | *CTU1* | -2.176826937 | 0.006518199 | 0.009094522 | 1 |
| Obesity | LEC | *RCN3* | 0.935838799 | 1.64E-05 | 0.000105108 | 0.103168112 |
| Obesity | LEC | *RERE* | 1.338001473 | 0.001053139 | 0.001726683 | 1 |
| Obesity | LEC | *ARHGEF19* | -0.456554671 | 0.002061417 | 0.003168735 | 1 |
| Obesity | LEC | *PEX14* | 1.624572104 | 5.13E-05 | 0.000154319 | 0.322218547 |
| Obesity | LEC | *SH3BGRL3* | 2.249286387 | 0.000699385 | 0.001196472 | 1 |
| Obesity | LEC | *IL22RA1* | 1.555689792 | 0.000557638 | 0.000980937 | 1 |

| | | | | | | |
|---|---|---|---|---|---|---|
| Obesity | LEC | *ZNF593* | 1.648287878 | 0.000154504 | 0.000340103 | 0.97136531 |
| Obesity | LEC | *C1orf216* | 2.972167412 | 4.82E-05 | 0.000148518 | 0.303000238 |
| Obesity | LEC | *MAP3K6* | 2.477053329 | 8.08E-05 | 0.000209699 | 0.508099527 |
| Obesity | LEC | *GPN2* | 2.149549351 | 0.012513348 | 0.016622095 | 1 |
| Obesity | LEC | *NBPF3* | 0.876977413 | 6.93E-05 | 0.000188201 | 0.435771943 |
| Obesity | LEC | *ITGB3BP* | 1.231788578 | 0.000314801 | 0.000604138 | 1 |
| Obesity | LEC | *SERBP1* | 1.881766062 | 0.003765799 | 0.005484267 | 1 |
| Obesity | LEC | *PIGK* | 1.600802811 | 2.41E-05 | 0.000109361 | 0.151600491 |
| Obesity | LEC | *RPS8* | 1.209417078 | 0.03650142 | 0.045343692 | 1 |
| Obesity | LEC | *CYP4B1* | 0.847564861 | 0.000257402 | 0.000513579 | 1 |
| Obesity | LEC | *SYPL2* | 0.758376886 | 1.07E-05 | 0.000105108 | 0.067383345 |
| Obesity | LEC | *ZNF697* | 1.143106525 | 2.43E-05 | 0.000109663 | 0.152761136 |
| Obesity | LEC | *CELSR2* | 1.897109438 | 0.000172726 | 0.000370497 | 1 |
| Obesity | LEC | *GPR161* | 3.073699149 | 0.001050738 | 0.001724352 | 1 |
| Obesity | LEC | *ATP1B1* | 1.092961238 | 5.75E-05 | 0.000166878 | 0.361457559 |
| Obesity | LEC | *TIPRL* | 1.937275289 | 6.54E-06 | 0.000102291 | 0.041120926 |
| Obesity | LEC | *CREG1* | 1.073937309 | 1.13E-05 | 0.000105108 | 0.071297835 |
| Obesity | LEC | *GPA33* | -1.246827247 | 0.000124478 | 0.000287507 | 0.782593962 |
| Obesity | LEC | *RXRG* | 6.61899522 | 1.26E-05 | 0.000105108 | 0.0794429 |
| Obesity | LEC | *TMCO1* | 2.066173987 | 0.00020773 | 0.000430029 | 1 |
| Obesity | LEC | *MGST3* | 1.665569978 | 3.79E-05 | 0.000127514 | 0.238068706 |
| Obesity | LEC | *ADCY10* | 1.241495826 | 1.72E-05 | 0.000105108 | 0.107865001 |
| Obesity | LEC | *RFWD2* | 1.435724145 | 0.00089872 | 0.001493986 | 1 |
| Obesity | LEC | *PPOX* | 2.221779732 | 0.000224311 | 0.000458137 | 1 |
| Obesity | LEC | *RRNAD1* | 1.715752727 | 0.000105255 | 0.000253345 | 0.661738122 |
| Obesity | LEC | *PIGM* | 2.461711252 | 0.004618634 | 0.006614431 | 1 |
| Obesity | LEC | *RGL1* | 0.990300044 | 1.32E-05 | 0.000105108 | 0.083188846 |
| Obesity | LEC | *TUFT1* | 0.595431566 | 0.028749291 | 0.036272686 | 1 |
| Obesity | LEC | *SNX27* | 1.845381858 | 7.68E-05 | 0.00020244 | 0.482606438 |
| Obesity | LEC | *SELENBP1* | 1.357419816 | 5.32E-06 | 9.39E-05 | 0.033416646 |
| Obesity | LEC | *CERS2* | 1.592762643 | 6.55E-05 | 0.000181705 | 0.411562605 |
| Obesity | LEC | *ENSA* | 1.856581613 | 5.61E-05 | 0.000164758 | 0.352417038 |
| Obesity | LEC | *SEMA6C* | 4.406699601 | 1.36E-06 | 5.03E-05 | 0.008537463 |
| Obesity | LEC | *ARNT* | 1.382988309 | 1.99E-05 | 0.000105667 | 0.125204957 |
| Obesity | LEC | *POGZ* | 1.688599097 | 1.12E-05 | 0.000105108 | 0.070302371 |
| Obesity | LEC | *GABPB2* | 0.943713494 | 0.000811928 | 0.001363033 | 1 |
| Obesity | LEC | *IKBKE* | 2.505020206 | 2.81E-05 | 0.000114038 | 0.17671294 |
| Obesity | LEC | *DYRK3* | 1.358397784 | 2.11E-05 | 0.000106989 | 0.132661872 |
| Obesity | LEC | *EIF2D* | 1.302125675 | 0.000305734 | 0.000589617 | 1 |
| Obesity | LEC | *HHIPL2* | 2.024584929 | 0.009304559 | 0.012634506 | 1 |
| Obesity | LEC | *TP53BP2* | 2.133888119 | 4.73E-05 | 0.000146745 | 0.297451376 |
| Obesity | LEC | *RAB13* | 1.077258072 | 4.02E-05 | 0.000132041 | 0.252857804 |
| Obesity | LEC | *S100A8* | -3.022728509 | 0.000198292 | 0.000414174 | 1 |
| Obesity | LEC | *HAX1* | 1.76780967 | 3.92E-07 | 2.51E-05 | 0.0024619 |
| Obesity | LEC | *KCNN3* | -0.191468377 | 0.000119162 | 0.000278398 | 0.749168837 |
| Obesity | LEC | *GATAD2B* | 1.351656293 | 0.002946902 | 0.004383054 | 1 |
| Obesity | LEC | *RIT1* | 2.003215502 | 1.03E-05 | 0.000105108 | 0.064861859 |
| Obesity | LEC | *GALNT2* | 1.585692972 | 2.65E-05 | 0.000112661 | 0.166675765 |
| Obesity | LEC | *CEP170* | 2.052360987 | 1.22E-05 | 0.000105108 | 0.07679546 |
| Obesity | LEC | *NVL* | 1.657727859 | 0.007233601 | 0.010001682 | 1 |

| | | | | | | |
|---|---|---|---|---|---|---|
| Obesity | LEC | *SDE2* | 3.086540015 | 0.0001169 | 0.000274234 | 0.734947506 |
| Obesity | LEC | *FBXO28* | 1.543344573 | 0.013742728 | 0.018155187 | 1 |
| Obesity | LEC | *CNIH4* | 1.599205256 | 0.038013444 | 0.047091728 | 1 |
| Obesity | LEC | *CDC42BPA* | 0.988646358 | 0.034523927 | 0.043014651 | 1 |
| Obesity | LEC | *PSEN2* | 2.915322673 | 3.55E-06 | 7.80E-05 | 0.0223164 |
| Obesity | LEC | *PYCR2* | -1.221769974 | 0.001773469 | 0.002760534 | 1 |
| Obesity | LEC | *WNT9A* | -0.87912525 | 0.002703903 | 0.004046522 | 1 |
| Obesity | LEC | *PLEKHA6* | 1.650545871 | 3.99E-06 | 8.37E-05 | 0.025100141 |
| Obesity | LEC | *ABCG8* | 2.891854558 | 4.86E-05 | 0.000149219 | 0.305600918 |
| Obesity | LEC | *EML4* | 1.440654222 | 1.90E-05 | 0.000105211 | 0.119531551 |
| Obesity | LEC | *CHAC2* | -1.243892755 | 0.004858816 | 0.006920566 | 1 |
| Obesity | LEC | *WDPCP* | 0.87494007 | 8.64E-06 | 0.000105108 | 0.054342317 |
| Obesity | LEC | *ASXL2* | 1.587719299 | 0.000179347 | 0.00038287 | 1 |
| Obesity | LEC | *CIAO1* | 1.877463115 | 3.28E-05 | 0.000118709 | 0.206078322 |
| Obesity | LEC | *ZNF514* | 0.63081568 | 0.002779333 | 0.004150274 | 1 |
| Obesity | LEC | *EXOC6B* | 1.268248698 | 3.53E-05 | 0.000122918 | 0.222098495 |
| Obesity | LEC | *TEX261* | 1.393502317 | 1.63E-05 | 0.000105108 | 0.10270068 |
| Obesity | LEC | *THNSL2* | 0.482175567 | 5.72E-05 | 0.000166529 | 0.359656532 |
| Obesity | LEC | *TMEM177* | 3.888619044 | 0.003225889 | 0.004764191 | 1 |
| Obesity | LEC | *RABL2A* | 2.086481214 | 0.001102302 | 0.001798177 | 1 |
| Obesity | LEC | *SLC20A1* | 1.257194231 | 2.70E-05 | 0.000113184 | 0.169436336 |
| Obesity | LEC | *FBLN7* | -1.17620078 | 3.55E-05 | 0.000123048 | 0.222938759 |
| Obesity | LEC | *ZC3H8* | 1.40226588 | 0.019295144 | 0.025006921 | 1 |
| Obesity | LEC | *FAHD2B* | -1.223762681 | 2.38E-05 | 0.000109261 | 0.149797461 |
| Obesity | LEC | *LYG1* | 1.566948192 | 0.000349465 | 0.000658599 | 1 |
| Obesity | LEC | *AFF3* | 0.540544021 | 0.002012041 | 0.003099658 | 1 |
| Obesity | LEC | *UBXN4* | 1.671569599 | 2.05E-05 | 0.000106439 | 0.128897972 |
| Obesity | LEC | *THSD7B* | 0.927437565 | 8.07E-07 | 3.79E-05 | 0.005075957 |
| Obesity | LEC | *GPR17* | 0.559687569 | 0.001651585 | 0.002588115 | 1 |
| Obesity | LEC | *POLR2D* | 0.716860182 | 5.64E-05 | 0.000165243 | 0.354280437 |
| Obesity | LEC | *ZNF385B* | 0.403266795 | 2.29E-05 | 0.000109118 | 0.143827004 |
| Obesity | LEC | *TMEFF2* | 2.166360408 | 0.001179922 | 0.001911407 | 1 |
| Obesity | LEC | *GULP1* | 1.123956789 | 0.036447098 | 0.045285159 | 1 |
| Obesity | LEC | *FAM171B* | -0.302294802 | 0.000265117 | 0.000526133 | 1 |
| Obesity | LEC | *HSPD1* | 1.878177245 | 2.58E-06 | 6.85E-05 | 0.016235136 |
| Obesity | LEC | *METTL21A* | 1.413305473 | 1.14E-05 | 0.000105108 | 0.071689926 |
| Obesity | LEC | *PTH2R* | 0.526392658 | 5.81E-05 | 0.000168116 | 0.365025196 |
| Obesity | LEC | *NBEAL1* | 1.687103396 | 4.70E-07 | 2.69E-05 | 0.002956741 |
| Obesity | LEC | *SUMF1* | 1.342371555 | 1.76E-05 | 0.000105108 | 0.110507659 |
| Obesity | LEC | *ESPNL* | -1.164618879 | 9.56E-05 | 0.000236646 | 0.601081496 |
| Obesity | LEC | *TAMM41* | 2.16018865 | 1.44E-05 | 0.000105108 | 0.090807741 |
| Obesity | LEC | *CSRNP1* | 1.263903171 | 0.000152204 | 0.000336228 | 0.956905084 |
| Obesity | LEC | *SLC25A38* | 1.042483375 | 3.23E-05 | 0.000117692 | 0.202814199 |
| Obesity | LEC | *CTDSPL* | -0.764547671 | 0.027393779 | 0.034666806 | 1 |
| Obesity | LEC | *STAC* | 2.665251908 | 8.39E-05 | 0.00021504 | 0.527708721 |
| Obesity | LEC | *ARL6IP5* | 1.637543678 | 5.72E-05 | 0.000166529 | 0.359533363 |
| Obesity | LEC | *TMF1* | 1.778695067 | 0.000151714 | 0.0003355 | 0.953826343 |
| Obesity | LEC | *LIMD1* | 1.691866068 | 1.11E-05 | 0.000105108 | 0.069774533 |
| Obesity | LEC | *COL8A1* | 0.707534629 | 0.000324763 | 0.000620227 | 1 |
| Obesity | LEC | *PLA1A* | -0.382811151 | 4.91E-05 | 0.000149999 | 0.30869798 |

| | | | | | | |
|---|---|---|---|---|---|---|
| Obesity | LEC | *RABL3* | 2.476551113 | 2.77E-05 | 0.00011384 | 0.174187295 |
| Obesity | LEC | *IGSF11* | 0.789120271 | 2.02E-05 | 0.000106047 | 0.127308712 |
| Obesity | LEC | *BOC* | 0.726714704 | 1.84E-05 | 0.000105108 | 0.115801175 |
| Obesity | LEC | *AGTR1* | 0.828975225 | 3.57E-05 | 0.000123048 | 0.224496583 |
| Obesity | LEC | *TCTA* | 1.536621441 | 0.006160308 | 0.008633495 | 1 |
| Obesity | LEC | *SLC26A1* | 1.291886909 | 0.000822829 | 0.001377663 | 1 |
| Obesity | LEC | *ATP10D* | 1.254688554 | 0.006764836 | 0.009409408 | 1 |
| Obesity | LEC | *SLC10A4* | 4.268750726 | 0.00277305 | 0.0041431 | 1 |
| Obesity | LEC | *PYURF* | 2.768206679 | 6.23E-05 | 0.000175889 | 0.391742418 |
| Obesity | LEC | *CISD2* | 1.732604114 | 1.77E-05 | 0.000105108 | 0.111414515 |
| Obesity | LEC | *TIFA* | 0.941539271 | 4.58E-05 | 0.00014419 | 0.288136063 |
| Obesity | LEC | *CCNA2* | 1.307452822 | 1.13E-05 | 0.000105108 | 0.070898401 |
| Obesity | LEC | *USP53* | 1.465855502 | 2.44E-05 | 0.000109899 | 0.15347772 |
| Obesity | LEC | *MARCH1* | 0.611263978 | 0.002881197 | 0.004291421 | 1 |
| Obesity | LEC | *PDGFC* | 0.935552687 | 8.54E-06 | 0.000105108 | 0.053663826 |
| Obesity | LEC | *CBR4* | 1.615138458 | 0.000221964 | 0.000454557 | 1 |
| Obesity | LEC | *FAM105A* | 0.837539965 | 5.50E-05 | 0.000162502 | 0.345966266 |
| Obesity | LEC | *LHFPL2* | 1.782891008 | 2.49E-05 | 0.000110998 | 0.15626639 |
| Obesity | LEC | *SSBP2* | 1.409706425 | 0.00027694 | 0.000545752 | 1 |
| Obesity | LEC | *CRHBP* | 0.615145311 | 8.31E-05 | 0.000214185 | 0.522148556 |
| Obesity | LEC | *RASA1* | 1.107595123 | 0.001382437 | 0.002203698 | 1 |
| Obesity | LEC | *PPIP5K2* | 1.159556643 | 0.02519503 | 0.032052035 | 1 |
| Obesity | LEC | *GTF2H2* | 1.723535874 | 0.000145249 | 0.000324398 | 0.913180098 |
| Obesity | LEC | *SPATA9* | 0.414431776 | 2.87E-05 | 0.000115038 | 0.180702024 |
| Obesity | LEC | *FEM1C* | 2.496604505 | 0.000263003 | 0.000523095 | 1 |
| Obesity | LEC | *GABRB2* | 0.496406112 | 0.00015056 | 0.000334135 | 0.946569777 |
| Obesity | LEC | *PCYOX1L* | -2.197257855 | 0.000429813 | 0.000785078 | 1 |
| Obesity | LEC | *MYLK4* | 2.827683025 | 2.64E-05 | 0.000112582 | 0.165731216 |
| Obesity | LEC | *GFOD1* | 1.220469879 | 4.64E-06 | 9.00E-05 | 0.029156308 |
| Obesity | LEC | *PSD2* | -3.649240131 | 0.000255921 | 0.00051111 | 1 |
| Obesity | LEC | *ZMAT2* | 1.221390402 | 0.01972746 | 0.025514614 | 1 |
| Obesity | LEC | *KLHL3* | -0.309115826 | 0.000290375 | 0.000565986 | 1 |
| Obesity | LEC | *TRIM7* | 1.624479383 | 3.00E-05 | 0.000115831 | 0.188697386 |
| Obesity | LEC | *HIGD2A* | 1.411667684 | 4.24E-05 | 0.000136912 | 0.266431291 |
| Obesity | LEC | *FAM193B* | 1.888571941 | 0.000634597 | 0.001096376 | 1 |
| Obesity | LEC | *MUT* | 2.47264681 | 0.000375524 | 0.000699739 | 1 |
| Obesity | LEC | *DOK3* | 1.007365949 | 0.019181382 | 0.024879998 | 1 |
| Obesity | LEC | *FGD2* | 0.615029539 | 2.28E-05 | 0.000109111 | 0.143217197 |
| Obesity | LEC | *ANO7* | 1.027510199 | 0.001982118 | 0.003057306 | 1 |
| Obesity | LEC | *RPL7L1* | 2.184131137 | 2.58E-06 | 6.85E-05 | 0.016192827 |
| Obesity | LEC | *PHIP* | 1.258230825 | 0.012381089 | 0.016463602 | 1 |
| Obesity | LEC | *MMS22L* | 1.513083177 | 6.60E-05 | 0.000182779 | 0.415090718 |
| Obesity | LEC | *PM20D2* | 2.234254004 | 0.011307289 | 0.015176969 | 1 |
| Obesity | LEC | *RNF217* | 1.327259614 | 1.75E-05 | 0.000105108 | 0.109858303 |
| Obesity | LEC | *ABRACL* | 1.710035472 | 0.004123351 | 0.005958058 | 1 |
| Obesity | LEC | *AIG1* | 1.319273845 | 1.71E-06 | 5.60E-05 | 0.010744204 |
| Obesity | LEC | *DYNLT1* | 1.534257515 | 1.10E-05 | 0.000105108 | 0.069397994 |
| Obesity | LEC | *WTAP* | 1.543137736 | 0.00019659 | 0.000411165 | 1 |
| Obesity | LEC | *VIP* | -0.44755908 | 0.000175096 | 0.000374941 | 1 |
| Obesity | LEC | *SLC22A3* | 1.657959532 | 1.84E-05 | 0.000105108 | 0.115774951 |

| | | | | | | |
|---|---|---|---|---|---|---|
| Obesity | LEC | *GNA12* | 1.482365039 | 9.57E-05 | 0.000236748 | 0.601577929 |
| Obesity | LEC | *C7orf50* | 2.061107849 | 1.89E-05 | 0.000105211 | 0.118893507 |
| Obesity | LEC | *EGFR* | 1.203963027 | 7.25E-05 | 0.000194526 | 0.455968696 |
| Obesity | LEC | *POMZP3* | 1.3843686 | 8.56E-05 | 0.000218223 | 0.538138738 |
| Obesity | LEC | *PSPH* | 1.017544013 | 0.030827325 | 0.038700358 | 1 |
| Obesity | LEC | *GIGYF1* | 2.135025595 | 3.00E-05 | 0.000115831 | 0.188322635 |
| Obesity | LEC | *TMEM209* | 2.359402345 | 1.54E-05 | 0.000105108 | 0.096695236 |
| Obesity | LEC | *AGBL3* | 1.300247591 | 0.034735119 | 0.043252069 | 1 |
| Obesity | LEC | *EPHA1* | -1.277885781 | 0.003902684 | 0.005662629 | 1 |
| Obesity | LEC | *NOM1* | 1.872372004 | 0.020174251 | 0.026063497 | 1 |
| Obesity | LEC | *NCAPG2* | 1.94414038 | 0.002864398 | 0.00426741 | 1 |
| Obesity | LEC | *FBXO25* | -0.946772074 | 0.025796505 | 0.032784036 | 1 |
| Obesity | LEC | *CSGALNACT1* | -0.38194497 | 0.000462739 | 0.000835989 | 1 |
| Obesity | LEC | *CCDC25* | 2.193786923 | 0.000861859 | 0.001437652 | 1 |
| Obesity | LEC | *BIN3* | 1.333529521 | 7.90E-05 | 0.000206614 | 0.496905681 |
| Obesity | LEC | *SLC25A37* | 2.071925243 | 0.000139093 | 0.000313096 | 0.874476755 |
| Obesity | LEC | *CHMP7* | 1.188206751 | 1.84E-05 | 0.000105108 | 0.115701044 |
| Obesity | LEC | *DOCK5* | -0.305077278 | 0.000195789 | 0.000410171 | 1 |
| Obesity | LEC | *ST18* | 0.539523631 | 5.81E-05 | 0.000168116 | 0.365149008 |
| Obesity | LEC | *RGS20* | -0.854744113 | 0.000519044 | 0.000921296 | 1 |
| Obesity | LEC | *TACC1* | 1.406663403 | 0.001395335 | 0.00222313 | 1 |
| Obesity | LEC | *ATP6V0D2* | -2.821857545 | 5.32E-06 | 9.39E-05 | 0.033462059 |
| Obesity | LEC | *MTDH* | 1.642087949 | 0.000612878 | 0.001066178 | 1 |
| Obesity | LEC | *MAL2* | 1.604847115 | 0.008635067 | 0.011778838 | 1 |
| Obesity | LEC | *UTP23* | 2.135462081 | 0.000241328 | 0.000486916 | 1 |
| Obesity | LEC | *FAM135B* | -2.257492093 | 0.003583919 | 0.005241241 | 1 |
| Obesity | LEC | *ZNF7* | 1.876575827 | 0.000267752 | 0.000530525 | 1 |
| Obesity | LEC | *SLC39A4* | -1.77915562 | 0.001621413 | 0.002543369 | 1 |
| Obesity | LEC | *AK3* | 1.635534132 | 0.005257283 | 0.007450977 | 1 |
| Obesity | LEC | *CER1* | 2.429494691 | 9.68E-05 | 0.000238821 | 0.60875523 |
| Obesity | LEC | *PLIN2* | 1.102521941 | 1.44E-05 | 0.000105108 | 0.090669731 |
| Obesity | LEC | *HAUS6* | 1.727905311 | 1.55E-05 | 0.000105108 | 0.097393657 |
| Obesity | LEC | *C9orf72* | 1.089890702 | 1.45E-05 | 0.000105108 | 0.091077091 |
| Obesity | LEC | *SIGMAR1* | 1.156317913 | 0.000185222 | 0.000392215 | 1 |
| Obesity | LEC | *CBWD5* | 1.525520389 | 1.42E-05 | 0.000105108 | 0.089206028 |
| Obesity | LEC | *ALAD* | 0.945739954 | 0.001736194 | 0.002706534 | 1 |
| Obesity | LEC | *ASTN2* | 1.818052381 | 1.39E-05 | 0.000105108 | 0.087091965 |
| Obesity | LEC | *WDR31* | 2.743686103 | 4.71E-06 | 9.00E-05 | 0.029595198 |
| Obesity | LEC | *POLE3* | 1.908110824 | 0.00041389 | 0.000760411 | 1 |
| Obesity | LEC | *SURF1* | 1.57942224 | 0.000337923 | 0.00064131 | 1 |
| Obesity | LEC | *SURF6* | 2.756259094 | 0.000126176 | 0.000290574 | 0.793266661 |
| Obesity | LEC | *ASB6* | 1.838298352 | 1.10E-05 | 0.000105108 | 0.068979519 |
| Obesity | LEC | *NTMT1* | 1.39211966 | 7.32E-05 | 0.000195742 | 0.459991205 |
| Obesity | LEC | *CIZ1* | 2.094175335 | 3.08E-06 | 7.33E-05 | 0.019349575 |
| Obesity | LEC | *LRSAM1* | 1.290942525 | 1.63E-05 | 0.000105108 | 0.10235801 |
| Obesity | LEC | *INPP5E* | 1.857487511 | 9.89E-05 | 0.000242506 | 0.622028912 |
| Obesity | LEC | *SEC16A* | 0.952161388 | 0.000310269 | 0.000596898 | 1 |
| Obesity | LEC | *SLC39A12* | 1.275647958 | 4.53E-05 | 0.000143138 | 0.284988104 |
| Obesity | LEC | *RSU1* | 1.473524292 | 0.007509611 | 0.010349172 | 1 |
| Obesity | LEC | *ADIRF* | 1.660652311 | 0.004669273 | 0.006679345 | 1 |

| | | | | | | |
|---|---|---|---|---|---|---|
| Obesity | LEC | *GLUD1* | 1.236303534 | 1.02E-05 | 0.000105108 | 0.064199912 |
| Obesity | LEC | *HTR7* | 0.752353729 | 3.07E-05 | 0.000115897 | 0.192935207 |
| Obesity | LEC | *RPP30* | 1.839529688 | 0.007182215 | 0.009941565 | 1 |
| Obesity | LEC | *FRA10AC1* | 1.426506539 | 0.000339583 | 0.00064403 | 1 |
| Obesity | LEC | *DNAJB12* | 1.83640945 | 3.52E-05 | 0.000122857 | 0.221604244 |
| Obesity | LEC | *PLEKHS1* | 4.150813528 | 0.000150311 | 0.000333807 | 0.945007232 |
| Obesity | LEC | *MTG1* | 1.935888023 | 3.95E-07 | 2.51E-05 | 0.002481676 |
| Obesity | LEC | *PAOX* | 1.173337718 | 1.93E-05 | 0.000105211 | 0.121041311 |
| Obesity | LEC | *GSTO1* | 2.039352298 | 9.66E-05 | 0.000238637 | 0.607569437 |
| Obesity | LEC | *PDCD11* | 1.529766429 | 0.002161066 | 0.00331058 | 1 |
| Obesity | LEC | *RGS10* | 1.100472183 | 2.36E-05 | 0.000109118 | 0.148289593 |
| Obesity | LEC | *GAS2* | -1.185089497 | 0.032518811 | 0.040629126 | 1 |
| Obesity | LEC | *LIN7C* | 2.388742934 | 3.13E-05 | 0.000116821 | 0.196959926 |
| Obesity | LEC | *ZNF215* | 0.59892542 | 0.000668859 | 0.001148312 | 1 |
| Obesity | LEC | *HSD17B12* | -1.722904702 | 7.49E-08 | 8.72E-06 | 0.000470935 |
| Obesity | LEC | *EIF3M* | 1.804772795 | 4.29E-05 | 0.000137948 | 0.269964169 |
| Obesity | LEC | *GLYAT* | 0.506234955 | 7.78E-06 | 0.000105108 | 0.048897096 |
| Obesity | LEC | *PTPRJ* | 1.161069838 | 3.32E-05 | 0.000119352 | 0.208746066 |
| Obesity | LEC | *C11orf49* | 1.671548072 | 0.000124182 | 0.000287273 | 0.780729244 |
| Obesity | LEC | *ENDOD1* | -1.12727865 | 0.003109967 | 0.00460489 | 1 |
| Obesity | LEC | *CAPN5* | 0.726434521 | 1.87E-05 | 0.000105211 | 0.117470879 |
| Obesity | LEC | *INTS4* | 1.74180888 | 0.000208097 | 0.000430314 | 1 |
| Obesity | LEC | *RPS3* | 1.317046696 | 0.007958244 | 0.010912428 | 1 |
| Obesity | LEC | *ZC3H12C* | 1.789942406 | 2.21E-05 | 0.000108376 | 0.138676839 |
| Obesity | LEC | *TTC12* | 1.625790295 | 2.75E-08 | 3.84E-06 | 0.000172759 |
| Obesity | LEC | *DRD2* | 3.321767199 | 1.24E-05 | 0.000105108 | 0.077885183 |
| Obesity | LEC | *C11orf52* | 5.161683173 | 2.89E-05 | 0.000115115 | 0.18148268 |
| Obesity | LEC | *NPAT* | 1.550447068 | 0.011723429 | 0.015691973 | 1 |
| Obesity | LEC | *GLB1L2* | 2.392249623 | 0.040101444 | 0.049502803 | 1 |
| Obesity | LEC | *GRIK4* | 0.754572336 | 0.034216911 | 0.04265749 | 1 |
| Obesity | LEC | *ADAM33* | 0.607891798 | 4.95E-05 | 0.000151155 | 0.311281903 |
| Obesity | LEC | *TMC2* | 5.379724802 | 0.000983334 | 0.001621354 | 1 |
| Obesity | LEC | *INCENP* | 1.339140708 | 1.85E-05 | 0.000105108 | 0.116235901 |
| Obesity | LEC | *MS4A3* | 13.21013454 | 3.28E-05 | 0.00011885 | 0.206442788 |
| Obesity | LEC | *PLCH2* | 2.36136031 | 2.16E-05 | 0.000107764 | 0.136106327 |
| Obesity | LEC | *CPSF7* | 2.012871615 | 0.000101488 | 0.00024702 | 0.638052469 |
| Obesity | LEC | *CHEK1* | 1.447352198 | 0.000122168 | 0.000283212 | 0.768071756 |
| Obesity | LEC | *FEZ1* | 1.246548284 | 1.99E-05 | 0.000105667 | 0.124918712 |
| Obesity | LEC | *KIAA1755* | 1.126725578 | 3.85E-05 | 0.00012891 | 0.242347819 |
| Obesity | LEC | *CNBD2* | 1.556318488 | 1.32E-05 | 0.000105108 | 0.082701588 |
| Obesity | LEC | *TBX6* | 1.960065118 | 2.05E-11 | 8.07E-09 | 1.29E-07 |
| Obesity | LEC | *TAOK2* | -1.133704198 | 0.021450293 | 0.027572682 | 1 |
| Obesity | LEC | *MPP7* | -0.164151441 | 7.76E-05 | 0.000203672 | 0.487590753 |
| Obesity | LEC | *CWC15* | 1.961364082 | 6.63E-05 | 0.000183069 | 0.416665881 |
| Obesity | LEC | *ARID5B* | 1.428357543 | 9.59E-06 | 0.000105108 | 0.060310377 |
| Obesity | LEC | *LYPD6B* | -0.560194436 | 0.00011386 | 0.000268407 | 0.715840867 |
| Obesity | LEC | *PDCD4* | 1.357179377 | 1.81E-05 | 0.000105108 | 0.113594576 |
| Obesity | LEC | *WDR17* | 0.994006295 | 2.74E-05 | 0.000113825 | 0.171989989 |
| Obesity | LEC | *CCDC102B* | -0.6888806 | 0.001805257 | 0.002805153 | 1 |
| Obesity | LEC | *FSIP1* | 2.256184461 | 1.11E-05 | 0.000105108 | 0.069872145 |

| | | | | | | |
|---|---|---|---|---|---|---|
| Obesity | LEC | *RGS18* | 1.31204207 | 1.24E-05 | 0.000105108 | 0.07784802 |
| Obesity | LEC | *PRSS23* | 1.082979575 | 0.009251895 | 0.012567876 | 1 |
| Obesity | LEC | *MTMR12* | 1.815712016 | 0.001244077 | 0.002002436 | 1 |
| Obesity | LEC | *PPP1R1C* | -0.697936545 | 0.000226 | 0.000460869 | 1 |
| Obesity | LEC | *C11orf53* | -1.471367058 | 8.70E-05 | 0.000220912 | 0.547255203 |
| Obesity | LEC | *CCT5* | 2.20026817 | 9.84E-05 | 0.000241679 | 0.618698157 |
| Obesity | LEC | *FAM173B* | 1.946487553 | 0.003403481 | 0.005001797 | 1 |
| Obesity | LEC | *DOCK1* | 1.059665749 | 0.026836044 | 0.034002058 | 1 |
| Obesity | LEC | *TEX12* | 4.537256136 | 3.09E-05 | 0.000116378 | 0.194118254 |
| Obesity | LEC | *PIP4K2A* | 1.369484355 | 0.000277819 | 0.000546846 | 1 |
| Obesity | LEC | *ABCB9* | 1.628450201 | 3.75E-05 | 0.000126557 | 0.236028458 |
| Obesity | LEC | *RILPL2* | 1.101524775 | 1.16E-06 | 4.64E-05 | 0.007308461 |
| Obesity | LEC | *DHX37* | 2.01605422 | 1.47E-05 | 0.000105108 | 0.092321934 |
| Obesity | LEC | *PRSS53* | -2.550876747 | 1.20E-08 | 2.29E-06 | 7.55E-05 |
| Obesity | LEC | *GPR158* | -0.761591177 | 0.000291722 | 0.00056727 | 1 |
| Obesity | LEC | *THRB* | 1.07313471 | 1.83E-05 | 0.000105108 | 0.115223142 |
| Obesity | LEC | *TMEM86A* | 0.668213178 | 0.015944046 | 0.020892084 | 1 |
| Obesity | LEC | *BTBD11* | 1.082817231 | 1.84E-05 | 0.000105108 | 0.115735643 |
| Obesity | LEC | *UBE3B* | 1.999993487 | 8.13E-06 | 0.000105108 | 0.051127156 |
| Obesity | LEC | *ANK3* | 0.765263984 | 1.08E-05 | 0.000105108 | 0.068033649 |
| Obesity | LEC | *IPMK* | 1.209488347 | 1.41E-05 | 0.000105108 | 0.088804477 |
| Obesity | LEC | *PLBD2* | 1.933437254 | 8.32E-06 | 0.000105108 | 0.052332243 |
| Obesity | LEC | *DLG5* | 1.157893785 | 0.000283835 | 0.000556082 | 1 |
| Obesity | LEC | *GXYLT1* | 2.021497515 | 4.79E-05 | 0.000147994 | 0.301167393 |
| Obesity | LEC | *FAM177A1* | 2.240165198 | 4.15E-05 | 0.000134786 | 0.260675821 |
| Obesity | LEC | *EXT2* | 1.249051401 | 0.00013736 | 0.000310083 | 0.863581807 |
| Obesity | LEC | *THRSP* | 1.257641647 | 0.000543518 | 0.00095959 | 1 |
| Obesity | LEC | *NDUFC2* | 2.343573075 | 0.000479179 | 0.000861059 | 1 |
| Obesity | LEC | *ME3* | 1.690353963 | 5.61E-05 | 0.000164758 | 0.352394569 |
| Obesity | LEC | *VIPAS39* | 1.529247902 | 3.51E-05 | 0.000122587 | 0.220624542 |
| Obesity | LEC | *ANKRD50* | 2.063036501 | 0.003018361 | 0.004481916 | 1 |
| Obesity | LEC | *C4orf33* | 1.436651089 | 1.10E-05 | 0.000105108 | 0.068918705 |
| Obesity | LEC | *PTPRO* | 0.669762356 | 2.20E-05 | 0.000108376 | 0.138118629 |
| Obesity | LEC | *VPS26B* | 2.029956782 | 1.15E-05 | 0.000105108 | 0.072553189 |
| Obesity | LEC | *NCAPD3* | 1.324426588 | 1.01E-05 | 0.000105108 | 0.063641783 |
| Obesity | LEC | *VTI1A* | 1.480442728 | 5.67E-05 | 0.000165809 | 0.356704343 |
| Obesity | LEC | *FAM160B1* | 1.508811995 | 0.018753909 | 0.024360708 | 1 |
| Obesity | LEC | *ANO4* | 1.249520286 | 6.79E-06 | 0.000104088 | 0.042676246 |
| Obesity | LEC | *TEX9* | 1.926517549 | 0.007934638 | 0.010882432 | 1 |
| Obesity | LEC | *AKR1C2* | 1.376064194 | 3.21E-05 | 0.00011749 | 0.201760864 |
| Obesity | LEC | *DPYSL4* | 1.184309001 | 1.81E-05 | 0.000105108 | 0.113667689 |
| Obesity | LEC | *ADAM8* | 1.162136872 | 7.67E-06 | 0.000105108 | 0.048212235 |
| Obesity | LEC | *ITIH2* | -2.108801108 | 0.002559869 | 0.003846533 | 1 |
| Obesity | LEC | *KIN* | 1.838608639 | 0.000295205 | 0.000573015 | 1 |
| Obesity | LEC | *PIGF* | 1.793086199 | 0.0001327 | 0.000302087 | 0.83428764 |
| Obesity | LEC | *INPP1* | 2.26154648 | 0.002409903 | 0.003650858 | 1 |
| Obesity | LEC | *RNF144A* | 1.728547542 | 0.000395529 | 0.00073181 | 1 |
| Obesity | LEC | *ASAP2* | 1.577433343 | 5.20E-05 | 0.000155665 | 0.326740059 |
| Obesity | LEC | *ADAM17* | 1.399212825 | 2.04E-05 | 0.000106214 | 0.128519143 |
| Obesity | LEC | *TMEM45B* | 3.200776947 | 0.000156753 | 0.000343501 | 0.985505481 |

| | | | | | | |
|---|---|---|---|---|---|---|
| Obesity | LEC | *CENPU* | 1.208420199 | 5.86E-05 | 0.000169213 | 0.368207817 |
| Obesity | LEC | *ACSL1* | 0.60277227 | 0.000619922 | 0.001075159 | 1 |
| Obesity | LEC | *GUF1* | 1.492845783 | 0.021242617 | 0.02732249 | 1 |
| Obesity | LEC | *GABRA2* | 1.545560579 | 7.04E-05 | 0.000190166 | 0.442897574 |
| Obesity | LEC | *CENPJ* | 1.794087728 | 0.000199187 | 0.000415719 | 1 |
| Obesity | LEC | *PARP8* | 0.99295486 | 0.000400585 | 0.000739207 | 1 |
| Obesity | LEC | *CACUL1* | 1.447405193 | 2.42E-05 | 0.000109361 | 0.151859237 |
| Obesity | LEC | *BEND6* | 0.389907836 | 0.007858447 | 0.010785012 | 1 |
| Obesity | LEC | *RABGAP1L* | 1.249646208 | 0.005905441 | 0.008304073 | 1 |
| Obesity | LEC | *PTPN14* | -0.444443066 | 0.001212336 | 0.001957359 | 1 |
| Obesity | LEC | *RNF219* | 1.587804148 | 3.01E-05 | 0.000115831 | 0.189295546 |
| Obesity | LEC | *ARL14EP* | 1.052109203 | 0.001458495 | 0.002313792 | 1 |
| Obesity | LEC | *EPG5* | 1.541021765 | 0.001644168 | 0.002577777 | 1 |
| Obesity | LEC | *SPC25* | 0.501484446 | 0.01175541 | 0.015728083 | 1 |
| Obesity | LEC | *G6PC2* | -29.84359607 | 0.000252491 | 0.000506028 | 1 |
| Obesity | LEC | *TGOLN2* | 1.692976866 | 7.53E-05 | 0.000199955 | 0.473692607 |
| Obesity | LEC | *POC5* | -1.988868903 | 8.36E-05 | 0.000214829 | 0.525685402 |
| Obesity | LEC | *GUCY1A2* | 1.342318302 | 0.004637934 | 0.006637535 | 1 |
| Obesity | LEC | *CWF19L2* | 1.888726893 | 9.72E-05 | 0.000239567 | 0.611373957 |
| Obesity | LEC | *ZNF547* | 2.273713731 | 1.62E-05 | 0.000105108 | 0.102083676 |
| Obesity | LEC | *NMT2* | 0.778477785 | 0.04025141 | 0.049648934 | 1 |
| Obesity | LEC | *ZNF837* | -1.219150326 | 0.000196015 | 0.000410372 | 1 |
| Obesity | LEC | *CCDC50* | 1.294690984 | 0.000157019 | 0.000343724 | 0.987175361 |
| Obesity | LEC | *ZFP36L2* | 1.596448881 | 2.09E-05 | 0.00010698 | 0.131488943 |
| Obesity | LEC | *PLEKHH2* | 0.399025265 | 0.013166239 | 0.01744492 | 1 |
| Obesity | LEC | *IGSF10* | 0.517710549 | 3.58E-05 | 0.000123124 | 0.225070807 |
| Obesity | LEC | *SPEF2* | 1.600489963 | 5.16E-05 | 0.000155003 | 0.324293788 |
| Obesity | LEC | *SPARCL1* | 0.917615586 | 1.02E-05 | 0.000105108 | 0.064260724 |
| Obesity | LEC | *MBNL1* | 1.200020339 | 0.000164943 | 0.000357462 | 1 |
| Obesity | LEC | *CLEC4F* | 1.856791265 | 3.01E-05 | 0.000115831 | 0.189505959 |
| Obesity | LEC | *PELO* | 0.969315023 | 2.34E-05 | 0.000109118 | 0.147017426 |
| Obesity | LEC | *RASGRP3* | -0.149528737 | 0.000179715 | 0.000383396 | 1 |
| Obesity | LEC | *SAR1B* | 1.848435999 | 2.56E-05 | 0.000111859 | 0.160853694 |
| Obesity | LEC | *GPR180* | 1.245179666 | 0.001014768 | 0.001669679 | 1 |
| Obesity | LEC | *WDR78* | 1.168516707 | 6.21E-05 | 0.00017578 | 0.390240275 |
| Obesity | LEC | *IFIT5* | 0.736543174 | 6.81E-05 | 0.00018633 | 0.42804688 |
| Obesity | LEC | *PRDM8* | -1.175487118 | 0.000838379 | 0.001401459 | 1 |
| Obesity | LEC | *HHEX* | -0.424155818 | 0.000208774 | 0.000431196 | 1 |
| Obesity | LEC | *ZNF117* | 0.6823494 | 8.48E-06 | 0.000105108 | 0.05329183 |
| Obesity | LEC | *RAB3C* | 1.852894213 | 6.41E-05 | 0.000178795 | 0.402825976 |
| Obesity | LEC | *MED21* | 1.674309403 | 1.45E-05 | 0.000105108 | 0.090978776 |
| Obesity | LEC | *SRP19* | 1.006422402 | 2.27E-05 | 0.000109111 | 0.142997594 |
| Obesity | LEC | *CARHSP1* | 2.001743778 | 0.000170564 | 0.00036661 | 1 |
| Obesity | LEC | *TEKT5* | 1.696228166 | 2.35E-05 | 0.000109118 | 0.147837689 |
| Obesity | LEC | *BANK1* | 1.071537675 | 1.41E-05 | 0.000105108 | 0.088604853 |
| Obesity | LEC | *CAST* | 0.979723718 | 0.005945464 | 0.008354746 | 1 |
| Obesity | LEC | *SCOC* | 1.872454501 | 0.000157006 | 0.000343724 | 0.987097695 |
| Obesity | LEC | *CLGN* | 3.89308581 | 2.29E-05 | 0.000109118 | 0.143901204 |
| Obesity | LEC | *AHCTF1* | 1.634539665 | 2.01E-05 | 0.000105931 | 0.126173475 |
| Obesity | LEC | *MERTK* | 0.351503257 | 0.011292031 | 0.015160146 | 1 |

| | | | | | | |
|---|---|---|---|---|---|---|
| Obesity | LEC | *TMEM87B* | 1.241676805 | 0.037292943 | 0.0462721 | 1 |
| Obesity | LEC | *PTPRR* | 1.543743505 | 1.20E-05 | 0.000105108 | 0.075478559 |
| Obesity | LEC | *PLA2R1* | -0.780692216 | 0.030086156 | 0.03784547 | 1 |
| Obesity | LEC | *INO80C* | -0.308202747 | 0.000150144 | 0.000333552 | 0.943952237 |
| Obesity | LEC | *LPCAT1* | 1.317833038 | 1.27E-05 | 0.000105108 | 0.079898452 |
| Obesity | LEC | *NMRAL1* | 0.877495401 | 0.039953722 | 0.049330136 | 1 |
| Obesity | LEC | *C16orf89* | 0.754363014 | 1.15E-05 | 0.000105108 | 0.072492879 |
| Obesity | LEC | *TEX29* | 2.245460112 | 0.004511716 | 0.006480502 | 1 |
| Obesity | LEC | *ADPRHL1* | 0.911113742 | 0.000426179 | 0.000779558 | 1 |
| Obesity | LEC | *TUBGCP5* | 2.152782481 | 0.000507251 | 0.000903168 | 1 |
| Obesity | LEC | *LURAP1L* | 0.920233904 | 6.31E-06 | 0.000100256 | 0.039701423 |
| Obesity | LEC | *ZDHHC7* | 1.368016359 | 1.36E-05 | 0.000105108 | 0.085471998 |
| Obesity | LEC | *FAM92B* | 8.485584543 | 3.17E-05 | 0.000117047 | 0.199539568 |
| Obesity | LEC | *C7orf31* | -0.874899905 | 0.000648749 | 0.001116212 | 1 |
| Obesity | LEC | *JAZF1* | 1.381010676 | 1.03E-06 | 4.34E-05 | 0.00646919 |
| Obesity | LEC | *FBXO36* | 2.089229455 | 0.000291874 | 0.00056727 | 1 |
| Obesity | LEC | *CACNA2D1* | 1.30332894 | 7.33E-05 | 0.00019586 | 0.460859658 |
| Obesity | LEC | *ZUFSP* | 2.130435949 | 1.82E-05 | 0.000105108 | 0.114241358 |
| Obesity | LEC | *HS3ST3A1* | 1.017902948 | 3.83E-05 | 0.000128424 | 0.240925805 |
| Obesity | LEC | *PPP2R5E* | 1.648148724 | 0.00050084 | 0.000894176 | 1 |
| Obesity | LEC | *ANKRD29* | 1.145820534 | 3.42E-05 | 0.000121139 | 0.21514365 |
| Obesity | LEC | *THY1* | 0.725714206 | 3.57E-05 | 0.000123048 | 0.224667475 |
| Obesity | LEC | *UBASH3B* | 0.957660417 | 1.39E-05 | 0.000105108 | 0.087647323 |
| Obesity | LEC | *ROBO4* | -0.724977278 | 0.000333524 | 0.000634453 | 1 |
| Obesity | LEC | *CC2D1B* | 1.884566994 | 3.21E-05 | 0.00011749 | 0.201721678 |
| Obesity | LEC | *PRKCA* | 1.640951331 | 1.15E-05 | 0.000105108 | 0.072009729 |
| Obesity | LEC | *LRRK1* | 1.714626336 | 3.02E-05 | 0.000115831 | 0.190027941 |
| Obesity | LEC | *GAL3ST2* | 6.075368204 | 0.004564848 | 0.00654635 | 1 |
| Obesity | LEC | *ENPP3* | -0.641457205 | 0.000343444 | 0.00064995 | 1 |
| Obesity | LEC | *UCHL1* | 0.756538967 | 2.94E-05 | 0.000115648 | 0.184537054 |
| Obesity | LEC | *MIA3* | 1.500516326 | 3.01E-05 | 0.000115831 | 0.189167111 |
| Obesity | LEC | *DISP1* | 1.281173078 | 0.006060472 | 0.008499261 | 1 |
| Obesity | LEC | *FAM167A* | 4.796113795 | 1.54E-06 | 5.35E-05 | 0.009678623 |
| Obesity | LEC | *NEIL2* | 1.384141511 | 3.64E-08 | 4.77E-06 | 0.000228921 |
| Obesity | LEC | *LONRF1* | 1.680333019 | 0.004799228 | 0.006844243 | 1 |
| Obesity | LEC | *CCSAP* | 0.815665124 | 4.80E-05 | 0.000148174 | 0.301681347 |
| Obesity | LEC | *GBP5* | 3.516812409 | 0.000169406 | 0.000364927 | 1 |
| Obesity | LEC | *FAM69A* | 0.858535672 | 0.000119044 | 0.000278227 | 0.748430331 |
| Obesity | LEC | *ATP5G3* | 1.904764296 | 3.43E-05 | 0.000121366 | 0.215667228 |
| Obesity | LEC | *PDLIM3* | -0.586126226 | 0.001517475 | 0.002397077 | 1 |
| Obesity | LEC | *NCAM2* | 1.200685839 | 3.21E-05 | 0.00011749 | 0.20184857 |
| Obesity | LEC | *L3MBTL4* | -0.680448059 | 0.039036543 | 0.048273553 | 1 |
| Obesity | LEC | *PDE1C* | -0.829804933 | 0.014525949 | 0.019129586 | 1 |
| Obesity | LEC | *MRPL39* | 1.004865992 | 0.004288643 | 0.006188363 | 1 |
| Obesity | LEC | *FLCN* | 1.936790146 | 4.04E-06 | 8.45E-05 | 0.0254195 |
| Obesity | LEC | *DPH3* | 1.182251231 | 1.78E-05 | 0.000105108 | 0.112216636 |
| Obesity | LEC | *APCDD1* | 0.796503277 | 0.007003767 | 0.009711663 | 1 |
| Obesity | LEC | *MPPE1* | 0.936229571 | 0.010469615 | 0.014121962 | 1 |
| Obesity | LEC | *EPHB1* | -1.072150866 | 0.003617398 | 0.005281603 | 1 |
| Obesity | LEC | *ANKRD40* | 1.832887868 | 6.46E-05 | 0.000179856 | 0.40629574 |

| | | | | | | |
|---|---|---|---|---|---|---|
| Obesity | LEC | *CYP2U1* | 1.773541883 | 1.30E-05 | 0.000105108 | 0.081739094 |
| Obesity | LEC | *FBXL18* | 1.261877558 | 1.16E-05 | 0.000105108 | 0.072871826 |
| Obesity | LEC | *PROM2* | 2.673649726 | 0.000268492 | 0.000531322 | 1 |
| Obesity | LEC | *CDK19* | 1.644428147 | 0.000122126 | 0.000283212 | 0.767808965 |
| Obesity | LEC | *GTF3C6* | 2.027395857 | 1.35E-05 | 0.000105108 | 0.085169593 |
| Obesity | LEC | *AGPAT5* | 2.171802018 | 0.000641904 | 0.001106567 | 1 |
| Obesity | LEC | *GOLGA7B* | 1.161163985 | 0.000108601 | 0.000258788 | 0.682774908 |
| Obesity | LEC | *TRMT44* | 2.686317717 | 4.70E-05 | 0.000146133 | 0.295335015 |
| Obesity | LEC | *HEATR3* | 0.896517605 | 0.000412739 | 0.000758519 | 1 |
| Obesity | LEC | *CNOT8* | 1.325190574 | 5.10E-05 | 0.000153894 | 0.320869406 |
| Obesity | LEC | *PDIA4* | 2.647872559 | 7.06E-05 | 0.000190427 | 0.443694215 |
| Obesity | LEC | *FAM126B* | 1.637937221 | 9.48E-05 | 0.000235176 | 0.596226671 |
| Obesity | LEC | *ALS2CR12* | 1.334356819 | 4.82E-06 | 9.10E-05 | 0.030326479 |
| Obesity | LEC | *SPAG17* | 0.844945852 | 1.93E-05 | 0.000105211 | 0.121518726 |
| Obesity | LEC | *DEPTOR* | 1.644126498 | 1.19E-05 | 0.000105108 | 0.075061197 |
| Obesity | LEC | *TMBIM4* | 1.498245309 | 0.001400907 | 0.002231443 | 1 |
| Obesity | LEC | *WIF1* | -3.462942034 | 0.003947928 | 0.005721674 | 1 |
| Obesity | LEC | *UGT2B4* | 1.116877111 | 2.81E-05 | 0.000114038 | 0.17636833 |
| Obesity | LEC | *KCNMA1* | 1.047870019 | 1.76E-05 | 0.000105108 | 0.110942846 |
| Obesity | LEC | *BATF* | 0.814015216 | 3.03E-05 | 0.000115831 | 0.190496426 |
| Obesity | LEC | *DCK* | 2.513315834 | 0.001072178 | 0.001751762 | 1 |
| Obesity | LEC | *NDUFAF6* | 1.653455735 | 0.000107836 | 0.000257479 | 0.677967581 |
| Obesity | LEC | *DRAM2* | 1.279350168 | 2.63E-05 | 0.000112582 | 0.165183718 |
| Obesity | LEC | *C8orf37* | -1.045179295 | 0.002581606 | 0.003874566 | 1 |
| Obesity | LEC | *ADAMTSL3* | 1.24856301 | 2.04E-05 | 0.000106095 | 0.128114641 |
| Obesity | LEC | *WHAMM* | 0.960143631 | 0.00031718 | 0.000607516 | 1 |
| Obesity | LEC | *CXCL13* | -1.887495788 | 0.001206535 | 0.001948994 | 1 |
| Obesity | LEC | *N6AMT1* | 0.773253738 | 0.000302873 | 0.000584637 | 1 |
| Obesity | LEC | *ANKRD9* | 0.753047152 | 5.35E-05 | 0.00015899 | 0.336175986 |
| Obesity | LEC | *SFXN2* | 1.76368074 | 4.21E-05 | 0.000136342 | 0.264913142 |
| Obesity | LEC | *C14orf2* | 1.34362481 | 0.00022746 | 0.000463397 | 1 |
| Obesity | LEC | *NODAL* | 2.975985977 | 1.49E-05 | 0.000105108 | 0.093537612 |
| Obesity | LEC | *MED19* | 0.598250488 | 0.001354714 | 0.002161698 | 1 |
| Obesity | LEC | *ATAD2* | 1.195229101 | 1.51E-05 | 0.000105108 | 0.094729842 |
| Obesity | LEC | *SASS6* | 1.632289723 | 5.75E-05 | 0.000166916 | 0.361706904 |
| Obesity | LEC | *ITGAD* | 0.726315312 | 0.000289204 | 0.000565018 | 1 |
| Obesity | LEC | *VPS8* | 1.354447949 | 1.52E-05 | 0.000105108 | 0.095629871 |
| Obesity | LEC | *MPV17L* | 1.098082504 | 0.000345417 | 0.000652144 | 1 |
| Obesity | LEC | *RPUSD3* | 2.978596909 | 0.000373287 | 0.000697016 | 1 |
| Obesity | LEC | *GHRL* | 0.913931128 | 2.03E-05 | 0.000106065 | 0.127544598 |
| Obesity | LEC | *EXOG* | 1.685642776 | 9.01E-05 | 0.0002266 | 0.566727175 |
| Obesity | LEC | *SLC6A1* | 1.432522286 | 4.11E-05 | 0.00013378 | 0.258226071 |
| Obesity | LEC | *FCHO2* | 1.331652556 | 2.16E-05 | 0.000107661 | 0.135694718 |
| Obesity | LEC | *CPT2* | 1.362575997 | 1.91E-05 | 0.000105211 | 0.119901582 |
| Obesity | LEC | *LRP8* | 2.366790111 | 4.58E-05 | 0.000144184 | 0.287936354 |
| Obesity | LEC | *GATAD1* | 1.630914213 | 0.000169865 | 0.000365609 | 1 |
| Obesity | LEC | *SUSD3* | 1.384977144 | 9.42E-06 | 0.000105108 | 0.059215635 |
| Obesity | LEC | *TMED6* | -6.682322918 | 0.009921357 | 0.01342565 | 1 |
| Obesity | LEC | *CLEC18A* | -6.20706318 | 3.93E-15 | 2.74E-12 | 2.47E-11 |
| Obesity | LEC | *IL34* | -1.284319351 | 0.00106429 | 0.001740684 | 1 |

| | | | | | | |
|---|---|---|---|---|---|---|
| Obesity | LEC | *DHRS1* | 0.730286453 | 0.02283409 | 0.029208123 | 1 |
| Obesity | LEC | *AASDH* | 1.962474878 | 9.68E-05 | 0.000238821 | 0.608664944 |
| Obesity | LEC | *RNF111* | 1.384223272 | 4.42E-05 | 0.000140884 | 0.277683198 |
| Obesity | LEC | *DYRK1A* | 1.421675961 | 0.003217478 | 0.004752886 | 1 |
| Obesity | LEC | *KCNJ15* | 1.078572585 | 0.000268136 | 0.000531026 | 1 |
| Obesity | LEC | *LCA5L* | 0.634423379 | 0.03155544 | 0.039519731 | 1 |
| Obesity | LEC | *MX1* | 0.559916914 | 0.00095223 | 0.001574196 | 1 |
| Obesity | LEC | *CREB3L1* | 0.953046973 | 1.17E-05 | 0.000105108 | 0.073257688 |
| Obesity | LEC | *C2CD2* | 0.739364272 | 4.98E-05 | 0.000151488 | 0.313124917 |
| Obesity | LEC | *SLC38A10* | 1.721940278 | 3.10E-05 | 0.000116544 | 0.195201595 |
| Obesity | LEC | *C9orf43* | 1.055791329 | 9.81E-05 | 0.000241238 | 0.616831155 |
| Obesity | LEC | *WDR19* | 1.546594718 | 2.38E-05 | 0.000109162 | 0.149333393 |
| Obesity | LEC | *SLC37A3* | 1.998306195 | 0.000756044 | 0.001281545 | 1 |
| Obesity | LEC | *SPPL3* | 1.614870815 | 0.000176685 | 0.000377947 | 1 |
| Obesity | LEC | *PEX10* | 2.722421784 | 0.001118762 | 0.001820304 | 1 |
| Obesity | LEC | *RADIL* | -0.228361754 | 0.0001144 | 0.000269579 | 0.719235554 |
| Obesity | LEC | *LDLRAP1* | 2.294916403 | 2.27E-05 | 0.000109111 | 0.142574457 |
| Obesity | LEC | *PAFAH2* | 1.614677163 | 0.017443119 | 0.022742615 | 1 |
| Obesity | LEC | *TRIM63* | 1.35739568 | 5.52E-05 | 0.000162792 | 0.346747442 |
| Obesity | LEC | *UBXN11* | 0.939925556 | 0.000242212 | 0.000488385 | 1 |
| Obesity | LEC | *GALNT14* | -2.382742224 | 0.038571871 | 0.0477177 | 1 |
| Obesity | LEC | *HPD* | 1.157228146 | 0.018985837 | 0.024636524 | 1 |
| Obesity | LEC | *RHPN1* | -1.446443461 | 3.82E-05 | 0.000128354 | 0.240406149 |
| Obesity | LEC | *TPRG1L* | -1.367420742 | 0.000878061 | 0.001462738 | 1 |
| Obesity | LEC | *LRRC43* | 9.269899178 | 0.000976427 | 0.001610387 | 1 |
| Obesity | LEC | *XKR8* | 1.127057363 | 1.32E-05 | 0.000105108 | 0.082726167 |
| Obesity | LEC | *EYA3* | 1.732822774 | 3.53E-05 | 0.000122918 | 0.222232323 |
| Obesity | LEC | *MRAS* | 1.081158151 | 0.000268364 | 0.000531235 | 1 |
| Obesity | LEC | *ABHD3* | 1.165652832 | 2.46E-05 | 0.000110453 | 0.154726804 |
| Obesity | LEC | *CLSTN2* | 0.924141887 | 4.80E-05 | 0.000148264 | 0.302012831 |
| Obesity | LEC | *COLEC12* | -0.446822145 | 0.000549365 | 0.000967739 | 1 |
| Obesity | LEC | *RNF207* | 3.725307135 | 6.78E-05 | 0.000185861 | 0.426180004 |
| Obesity | LEC | *AUTS2* | 0.961815864 | 0.021589133 | 0.027739807 | 1 |
| Obesity | LEC | *MITD1* | 1.953642101 | 6.09E-05 | 0.000173812 | 0.383081035 |
| Obesity | LEC | *AHCYL2* | 1.543340058 | 0.000118033 | 0.000276274 | 0.742072051 |
| Obesity | LEC | *B4GALT5* | 1.360781368 | 0.015884623 | 0.020827242 | 1 |
| Obesity | LEC | *CD1D* | 1.112550966 | 0.001004088 | 0.001653406 | 1 |
| Obesity | LEC | *FAM86C1* | 6.91042528 | 0.000668402 | 0.001147841 | 1 |
| Obesity | LEC | *CD1E* | 4.401650773 | 3.11E-07 | 2.22E-05 | 0.001954113 |
| Obesity | LEC | *NCF1* | 0.937154944 | 3.01E-05 | 0.000115831 | 0.189152535 |
| Obesity | LEC | *PPP1R9A* | -0.531335307 | 0.000674892 | 0.001157721 | 1 |
| Obesity | LEC | *ZFAND2B* | 1.559194195 | 0.02619582 | 0.033224353 | 1 |
| Obesity | LEC | *GDPD5* | 1.577272164 | 0.026899434 | 0.034075507 | 1 |
| Obesity | LEC | *DYNC1I1* | -0.501467455 | 0.000350515 | 0.000660379 | 1 |
| Obesity | LEC | *TMED4* | 1.996097628 | 2.37E-05 | 0.000109118 | 0.149067537 |
| Obesity | LEC | *DUSP23* | 2.210728104 | 1.46E-05 | 0.000105108 | 0.092042142 |
| Obesity | LEC | *RNF166* | 1.717414069 | 3.22E-05 | 0.000117652 | 0.202472763 |
| Obesity | LEC | *F11R* | 1.306989796 | 0.02017672 | 0.026063497 | 1 |
| Obesity | LEC | *SPATA2L* | 1.463392713 | 8.30E-05 | 0.000214119 | 0.521594826 |
| Obesity | LEC | *NIT1* | 1.381460052 | 2.16E-06 | 6.29E-05 | 0.013584808 |

| | | | | | | |
|---|---|---|---|---|---|---|
| Obesity | LEC | *ZNF276* | 1.341941792 | 8.83E-05 | 0.000222863 | 0.554927817 |
| Obesity | LEC | *FGF17* | -2.415149713 | 8.10E-05 | 0.000209934 | 0.509300956 |
| Obesity | LEC | *CDA* | 0.708254986 | 0.000263529 | 0.000523809 | 1 |
| Obesity | LEC | *B4GALT3* | 2.688788865 | 6.00E-05 | 0.000171896 | 0.377483475 |
| Obesity | LEC | *DMTN* | -0.348263999 | 0.000773898 | 0.001306173 | 1 |
| Obesity | LEC | *FAM160B2* | 2.05234355 | 0.000160938 | 0.000351102 | 1 |
| Obesity | LEC | *NDUFS2* | 1.281342789 | 1.22E-05 | 0.000105108 | 0.076976238 |
| Obesity | LEC | *SLC5A11* | -1.285446934 | 6.55E-08 | 7.92E-06 | 0.00041174 |
| Obesity | LEC | *FCER1G* | 1.519628682 | 1.06E-06 | 4.43E-05 | 0.006684168 |
| Obesity | LEC | *CCAR2* | 2.302392774 | 0.000112745 | 0.000266173 | 0.70882989 |
| Obesity | LEC | *CDC42SE2* | 1.698547436 | 0.000208752 | 0.000431196 | 1 |
| Obesity | LEC | *EPB41* | 1.196060063 | 2.83E-06 | 7.08E-05 | 0.017792431 |
| Obesity | LEC | *ALG8* | 1.55890543 | 2.24E-05 | 0.000108832 | 0.140641719 |
| Obesity | LEC | *IFNAR2* | 1.257894507 | 3.38E-05 | 0.000120325 | 0.212494701 |
| Obesity | LEC | *MRPL10* | 1.325578133 | 0.000593334 | 0.001035903 | 1 |
| Obesity | LEC | *SV2A* | -0.640579361 | 9.39E-05 | 0.000233693 | 0.590075695 |
| Obesity | LEC | *RCAN1* | -1.152863642 | 0.026563847 | 0.03368433 | 1 |
| Obesity | LEC | *UBE2Z* | 1.805425712 | 0.000812534 | 0.001363332 | 1 |
| Obesity | LEC | *CLIC6* | 1.025895893 | 1.50E-06 | 5.27E-05 | 0.009434806 |
| Obesity | LEC | *CBR1* | 0.606626544 | 0.004559859 | 0.006540688 | 1 |
| Obesity | LEC | *CBR3* | 1.430629068 | 0.0005872 | 0.001027162 | 1 |
| Obesity | LEC | *SIM2* | 1.212257591 | 0.014267766 | 0.018813222 | 1 |
| Obesity | LEC | *HLCS* | 0.802480788 | 0.00228324 | 0.003476564 | 1 |
| Obesity | LEC | *ARHGAP27* | -0.89368682 | 0.001683719 | 0.002632565 | 1 |
| Obesity | LEC | *PADI4* | 5.573646315 | 5.21E-05 | 0.000155829 | 0.327241492 |
| Obesity | LEC | *PSMD4* | 1.902965303 | 2.31E-05 | 0.000109118 | 0.145148848 |
| Obesity | LEC | *STARD9* | -0.729909052 | 0.015925221 | 0.020876118 | 1 |
| Obesity | LEC | *TCHH* | 1.603098495 | 0.000784299 | 0.001321599 | 1 |
| Obesity | LEC | *MED8* | 1.368323872 | 1.85E-05 | 0.000105108 | 0.116368392 |
| Obesity | LEC | *ACE* | 0.528210202 | 3.66E-05 | 0.000124867 | 0.22988034 |
| Obesity | LEC | *TEPP* | -2.197167432 | 3.91E-05 | 0.000130236 | 0.246076742 |
| Obesity | LEC | *ATP6V0D1* | 1.070033356 | 0.000264644 | 0.000525495 | 1 |
| Obesity | LEC | *ZFYVE28* | 1.647645686 | 4.18E-06 | 8.58E-05 | 0.026265507 |
| Obesity | LEC | *C16orf86* | 1.882277213 | 0.000335494 | 0.000637814 | 1 |
| Obesity | LEC | *LYPD5* | -0.558783115 | 5.65E-05 | 0.00016541 | 0.354970126 |
| Obesity | LEC | *CCDC117* | 2.724790621 | 0.000331186 | 0.000631148 | 1 |
| Obesity | LEC | *CCDC107* | 0.761648278 | 0.000514686 | 0.000914852 | 1 |
| Obesity | LEC | *NPR2* | 1.978080387 | 3.12E-05 | 0.000116657 | 0.196175562 |
| Obesity | LEC | *ZNF235* | 1.649632187 | 6.93E-05 | 0.000188201 | 0.435466373 |
| Obesity | LEC | *CALM3* | 2.075010004 | 0.000303766 | 0.000586001 | 1 |
| Obesity | LEC | *DFFA* | 2.643520149 | 1.82E-06 | 5.72E-05 | 0.011452963 |
| Obesity | LEC | *ATAD3B* | 2.378122581 | 9.89E-05 | 0.000242506 | 0.6219632 |
| Obesity | LEC | *SSU72* | 1.631112954 | 1.60E-05 | 0.000105108 | 0.100345837 |
| Obesity | LEC | *NR2F6* | 0.936525766 | 2.48E-05 | 0.000110994 | 0.155946958 |
| Obesity | LEC | *CILP2* | 5.155837268 | 0.000103183 | 0.000249985 | 0.648710798 |
| Obesity | LEC | *ABCG1* | -0.562413256 | 0.000760756 | 0.001287458 | 1 |
| Obesity | LEC | *RSPH1* | -0.877931841 | 4.84E-05 | 0.0001487 | 0.304165358 |
| Obesity | LEC | *NDUFV3* | 0.573649971 | 0.000759939 | 0.00128676 | 1 |
| Obesity | LEC | *U2AF1* | 0.890180316 | 0.00021732 | 0.000446354 | 1 |
| Obesity | LEC | *HSF2BP* | 1.460115557 | 1.86E-05 | 0.000105129 | 0.116924107 |

| | | | | | | |
|---|---|---|---|---|---|---|
| Obesity | LEC | *CSTB* | 1.572047962 | 0.001508267 | 0.002384329 | 1 |
| Obesity | LEC | *C21orf33* | 1.496551247 | 0.013973708 | 0.018448699 | 1 |
| Obesity | LEC | *ZNF66* | 2.313931774 | 2.85E-05 | 0.000114887 | 0.179244641 |
| Obesity | LEC | *LRRC3* | 1.244492728 | 0.000238738 | 0.00048262 | 1 |
| Obesity | LEC | *ITGB2* | 0.827458944 | 0.000135682 | 0.000307179 | 0.853035226 |
| Obesity | LEC | *FAM207A* | 2.497146823 | 2.19E-06 | 6.31E-05 | 0.013739864 |
| Obesity | LEC | *LSS* | 0.838569011 | 2.59E-06 | 6.85E-05 | 0.016293111 |
| Obesity | LEC | *PCNT* | 1.082239826 | 3.18E-06 | 7.47E-05 | 0.020020882 |
| Obesity | LEC | *DIP2A* | 1.218102887 | 0.021681822 | 0.027841833 | 1 |
| Obesity | LEC | *PRMT2* | 1.617018131 | 8.37E-05 | 0.000214895 | 0.526278104 |
| Obesity | LEC | *ADAMTS13* | 1.587106393 | 0.007385861 | 0.010203232 | 1 |
| Obesity | LEC | *ZNF714* | 1.853058749 | 0.001080492 | 0.001764888 | 1 |
| Obesity | LEC | *GPSM1* | -0.581236076 | 0.000446322 | 0.000809663 | 1 |
| Obesity | LEC | *C19orf47* | 1.511206296 | 1.65E-06 | 5.47E-05 | 0.010367358 |
| Obesity | LEC | *HIPK4* | 1.347446586 | 4.90E-05 | 0.000149956 | 0.308189503 |
| Obesity | LEC | *RDH13* | 1.427672991 | 1.74E-05 | 0.000105108 | 0.10939987 |
| Obesity | LEC | *MPZL3* | 0.892667189 | 2.78E-05 | 0.00011384 | 0.174722099 |
| Obesity | LEC | *PCSK7* | 1.858604808 | 9.89E-05 | 0.000242506 | 0.622009738 |
| Obesity | LEC | *S100A1* | -1.03306425 | 0.000427557 | 0.000781637 | 1 |
| Obesity | LEC | *ZBTB7B* | 1.004622735 | 0.000206237 | 0.000427501 | 1 |
| Obesity | LEC | *VPS11* | 1.704683072 | 0.002035466 | 0.003131907 | 1 |
| Obesity | LEC | *PMF1* | 1.759440137 | 5.12E-07 | 2.85E-05 | 0.003219242 |
| Obesity | LEC | *SLC25A44* | -1.699732541 | 0.002291219 | 0.003487867 | 1 |
| Obesity | LEC | *NBEAL2* | 1.945847299 | 1.74E-06 | 5.62E-05 | 0.010953887 |
| Obesity | LEC | *FCRL3* | 0.978803685 | 4.58E-05 | 0.00014419 | 0.288235807 |
| Obesity | LEC | *NACC1* | 2.585761333 | 0.031540961 | 0.039517342 | 1 |
| Obesity | LEC | *LY6K* | 4.632192189 | 9.45E-05 | 0.0002347 | 0.594259948 |
| Obesity | LEC | *TONSL* | -1.015029867 | 0.000151124 | 0.000334666 | 0.950118067 |
| Obesity | LEC | *MUM1* | 1.349088038 | 0.000755797 | 0.001281471 | 1 |
| Obesity | LEC | *ZNF333* | 2.143584321 | 2.52E-05 | 0.000111345 | 0.158476361 |
| Obesity | LEC | *ORAI2* | 2.053459386 | 1.19E-06 | 4.70E-05 | 0.007467551 |
| Obesity | LEC | *RPL8* | 1.406137419 | 0.038562857 | 0.047715938 | 1 |
| Obesity | LEC | *LRWD1* | -0.995980272 | 0.000113036 | 0.000266562 | 0.710655074 |
| Obesity | LEC | *FBXL13* | 1.798137491 | 3.23E-05 | 0.000117692 | 0.202900255 |
| Obesity | LEC | *SCGB3A1* | -0.915761907 | 0.000106817 | 0.000255735 | 0.671560909 |
| Obesity | LEC | *MFSD12* | 1.323467143 | 1.39E-05 | 0.000105108 | 0.087365183 |
| Obesity | LEC | *YDJC* | 2.069380455 | 2.18E-05 | 0.00010816 | 0.13682249 |
| Obesity | LEC | *CCDC116* | 2.191436875 | 0.032328021 | 0.040406812 | 1 |
| Obesity | LEC | *DVL3* | 2.028937277 | 1.72E-05 | 0.000105108 | 0.108392459 |
| Obesity | LEC | *AP2M1* | 1.77563611 | 8.76E-06 | 0.000105108 | 0.055089326 |
| Obesity | LEC | *FBXO27* | 0.986958997 | 1.39E-06 | 5.08E-05 | 0.008742311 |
| Obesity | LEC | *DMKN* | -0.607525362 | 0.000166785 | 0.000360583 | 1 |
| Obesity | LEC | *BDH1* | 0.836104882 | 2.68E-05 | 0.000113184 | 0.168733629 |
| Obesity | LEC | *DUSP14* | 0.857799342 | 0.000241256 | 0.000486916 | 1 |
| Obesity | LEC | *LRRC56* | 1.690250644 | 4.08E-05 | 0.000133178 | 0.256215748 |
| Obesity | LEC | *IKZF3* | 1.31287675 | 1.31E-05 | 0.000105108 | 0.082495098 |
| Obesity | LEC | *ACOX1* | 1.091618607 | 1.24E-05 | 0.000105108 | 0.077892407 |
| Obesity | LEC | *ZNF577* | 1.634818926 | 0.000422249 | 0.000773733 | 1 |
| Obesity | LEC | *TMEM143* | 1.527905812 | 3.36E-06 | 7.66E-05 | 0.021149831 |
| Obesity | LEC | *ALDH16A1* | 1.661341299 | 1.54E-05 | 0.000105108 | 0.096520453 |

| | | | | | | |
|---|---|---|---|---|---|---|
| Obesity | LEC | *SIGLEC11* | 1.22871256 | 0.000369585 | 0.000690514 | 1 |
| Obesity | LEC | *MPP3* | 1.094315471 | 3.25E-05 | 0.000118187 | 0.204346008 |
| Obesity | LEC | *NAGS* | -2.575995652 | 0.015844289 | 0.020778691 | 1 |
| Obesity | LEC | *ASB16* | -1.442362744 | 0.001228528 | 0.001978927 | 1 |
| Obesity | LEC | *PLCD3* | -1.195831724 | 0.001025491 | 0.001685999 | 1 |
| Obesity | LEC | *RACGAP1* | 1.020613131 | 4.64E-05 | 0.000145361 | 0.291594498 |
| Obesity | LEC | *LARP4* | 1.15143258 | 2.71E-05 | 0.000113653 | 0.170593599 |
| Obesity | LEC | *SPC24* | 2.089849899 | 1.22E-05 | 0.000105108 | 0.076387804 |
| Obesity | LEC | *IP6K3* | -0.513802615 | 8.38E-05 | 0.00021504 | 0.527123156 |
| Obesity | LEC | *LEMD2* | 1.38626585 | 0.028688783 | 0.036203609 | 1 |
| Obesity | LEC | *SCIMP* | 1.244120889 | 0.000811046 | 0.001361925 | 1 |
| Obesity | LEC | *ASGR2* | 2.380081821 | 3.24E-05 | 0.000117939 | 0.203563478 |
| Obesity | LEC | *TNFSF13* | 3.221613701 | 0.000107592 | 0.000257393 | 0.676429061 |
| Obesity | LEC | *SENP3* | 3.253244572 | 0.000261309 | 0.000520054 | 1 |
| Obesity | LEC | *JMJD8* | 1.913317076 | 1.52E-06 | 5.31E-05 | 0.009566618 |
| Obesity | LEC | *MEIOB* | 1.781243749 | 1.17E-05 | 0.000105108 | 0.073456019 |
| Obesity | LEC | *FLYWCH2* | 1.879661577 | 7.38E-06 | 0.000105108 | 0.046401256 |
| Obesity | LEC | *ZG16B* | 9.900330316 | 3.22E-05 | 0.000117657 | 0.202605555 |
| Obesity | LEC | *TTC9C* | 1.95489159 | 0.002550898 | 0.003833969 | 1 |
| Obesity | LEC | *NXF1* | 1.717383066 | 0.018444172 | 0.023988107 | 1 |
| Obesity | LEC | *RPL29* | 1.857490422 | 0.000112786 | 0.000266173 | 0.709085111 |
| Obesity | LEC | *TPCN2* | 2.084366078 | 5.35E-07 | 2.92E-05 | 0.003362777 |
| Obesity | LEC | *CYP4A22* | 4.278694306 | 0.000102885 | 0.000249503 | 0.646836719 |
| Obesity | LEC | *PDZK1IP1* | 16.97851132 | 0.000479218 | 0.000861059 | 1 |
| Obesity | LEC | *COA7* | 4.119319511 | 7.62E-05 | 0.000201393 | 0.479027212 |
| Obesity | LEC | *SLC1A7* | 0.557285646 | 0.003319062 | 0.0048926 | 1 |
| Obesity | LEC | *ACOT11* | 1.871885919 | 0.00069593 | 0.001191859 | 1 |
| Obesity | LEC | *USP24* | 1.268404623 | 0.000647794 | 0.001115058 | 1 |
| Obesity | LEC | *ZSWIM5* | -0.678262321 | 0.00463447 | 0.006634088 | 1 |
| Obesity | LEC | *RBP7* | 0.863367363 | 1.87E-05 | 0.000105211 | 0.117352484 |
| Obesity | LEC | *FBLIM1* | 1.657197934 | 4.08E-05 | 0.000133244 | 0.256493813 |
| Obesity | LEC | *SLC25A34* | 4.521632336 | 0.000769248 | 0.001299722 | 1 |
| Obesity | LEC | *PDPN* | -0.221490806 | 5.73E-05 | 0.000166529 | 0.359975362 |
| Obesity | LEC | *LRRC38* | 5.462977688 | 4.45E-05 | 0.000141409 | 0.279753981 |
| Obesity | LEC | *PEF1* | 3.129347656 | 0.003592379 | 0.005251171 | 1 |
| Obesity | LEC | *SYNC* | 1.235934639 | 0.007850581 | 0.010778904 | 1 |
| Obesity | LEC | *RBBP4* | 2.095710983 | 1.59E-05 | 0.000105108 | 0.100179743 |
| Obesity | LEC | *CAMK2N1* | 0.875512228 | 2.19E-05 | 0.000108369 | 0.137628501 |
| Obesity | LEC | *TTLL10* | 1.857023338 | 2.85E-05 | 0.000114878 | 0.178876493 |
| Obesity | LEC | *OMA1* | 1.263276026 | 0.000312788 | 0.000600641 | 1 |
| Obesity | LEC | *TM2D1* | 1.681213286 | 4.25E-05 | 0.000137166 | 0.267473215 |
| Obesity | LEC | *NEXN* | 1.928392939 | 8.61E-05 | 0.000219223 | 0.541077962 |
| Obesity | LEC | *B3GALT2* | -1.259447946 | 0.00042219 | 0.000773733 | 1 |
| Obesity | LEC | *NTNG1* | 0.48213244 | 0.000435217 | 0.000793564 | 1 |
| Obesity | LEC | *HFM1* | -0.752095514 | 0.000720129 | 0.001228617 | 1 |
| Obesity | LEC | *KCNT2* | -0.748517683 | 0.008150913 | 0.011159579 | 1 |
| Obesity | LEC | *VCAM1* | 0.823813531 | 2.94E-05 | 0.000115666 | 0.184718668 |
| Obesity | LEC | *EXTL2* | 0.932856738 | 3.94E-05 | 0.000130647 | 0.247836726 |
| Obesity | LEC | *TRIM58* | -1.674392613 | 0.014211464 | 0.018742915 | 1 |
| Obesity | LEC | *IGSF8* | 2.716400535 | 2.21E-05 | 0.000108376 | 0.138721589 |

| | | | | | | |
|---|---|---|---|---|---|---|
| Obesity | LEC | *VANGL2* | 1.595505465 | 5.25E-05 | 0.000156831 | 0.330285262 |
| Obesity | LEC | *FCRLB* | -1.937373731 | 0.011788628 | 0.015763331 | 1 |
| Obesity | LEC | *KLHDC9* | -1.207865692 | 0.02113361 | 0.027198978 | 1 |
| Obesity | LEC | *LMX1A* | 0.631254744 | 0.000189412 | 0.000399207 | 1 |
| Obesity | LEC | *C1orf115* | 1.874068952 | 0.001843387 | 0.002858543 | 1 |
| Obesity | LEC | *BROX* | 1.165570361 | 0.002025566 | 0.003117437 | 1 |
| Obesity | LEC | *ACP6* | 1.002319856 | 0.012894183 | 0.017106083 | 1 |
| Obesity | LEC | *PM20D1* | -1.323346238 | 0.000597274 | 0.001041915 | 1 |
| Obesity | LEC | *HAAO* | 1.206973177 | 4.43E-06 | 8.83E-05 | 0.027858328 |
| Obesity | LEC | *B3GALNT2* | 1.528701871 | 4.54E-05 | 0.000143383 | 0.285727905 |
| Obesity | LEC | *PUS10* | 2.184208623 | 0.000582781 | 0.001020598 | 1 |
| Obesity | LEC | *RFTN2* | -0.891340266 | 0.011060522 | 0.014871151 | 1 |
| Obesity | LEC | *DISC1* | 1.387832324 | 3.24E-05 | 0.000118 | 0.203904797 |
| Obesity | LEC | *FAM84A* | -0.539388647 | 0.000108744 | 0.000258868 | 0.683671101 |
| Obesity | LEC | *CLHC1* | 0.773233903 | 0.000121343 | 0.000282237 | 0.762885696 |
| Obesity | LEC | *FRZB* | 2.212347831 | 0.000461999 | 0.000835131 | 1 |
| Obesity | LEC | *NUP35* | 1.974481344 | 6.24E-05 | 0.000175889 | 0.39240863 |
| Obesity | LEC | *FBXO41* | -1.227915259 | 0.000447063 | 0.000810462 | 1 |
| Obesity | LEC | *SMC6* | 1.433994636 | 3.72E-05 | 0.000126137 | 0.233914771 |
| Obesity | LEC | *CCDC74A* | 2.938027064 | 3.09E-05 | 0.000116537 | 0.19450065 |
| Obesity | LEC | *NOSTRIN* | 0.831970786 | 1.53E-05 | 0.000105108 | 0.09624199 |
| Obesity | LEC | *SGPP2* | -1.505746196 | 0.001550337 | 0.00244285 | 1 |
| Obesity | LEC | *BBS5* | 0.918855574 | 1.50E-05 | 0.000105108 | 0.094463309 |
| Obesity | LEC | *HPGDS* | 1.284024106 | 5.63E-06 | 9.64E-05 | 0.035374069 |
| Obesity | LEC | *PDLIM5* | 1.424901623 | 4.71E-05 | 0.000146463 | 0.296317542 |
| Obesity | LEC | *RPRD2* | 1.398622526 | 0.007114565 | 0.009854433 | 1 |
| Obesity | LEC | *CTSS* | 0.549819129 | 0.000912926 | 0.001515997 | 1 |
| Obesity | LEC | *PACRGL* | 1.71439468 | 1.21E-05 | 0.000105108 | 0.076130868 |
| Obesity | LEC | *C1QTNF7* | 1.152426862 | 2.26E-05 | 0.000109111 | 0.142182306 |
| Obesity | LEC | *LYSMD1* | 2.377463005 | 6.33E-07 | 3.26E-05 | 0.003979292 |
| Obesity | LEC | *ERCC3* | 1.030308636 | 1.86E-05 | 0.000105108 | 0.116669849 |
| Obesity | LEC | *RNF149* | 1.398689891 | 2.56E-05 | 0.000111859 | 0.160705962 |
| Obesity | LEC | *DHX57* | 2.05057653 | 1.34E-05 | 0.000105108 | 0.084521428 |
| Obesity | LEC | *S100A12* | 6.276032523 | 7.29E-05 | 0.000195433 | 0.458486004 |
| Obesity | LEC | *TDRD10* | -1.004071083 | 0.006334511 | 0.00885592 | 1 |
| Obesity | LEC | *GNPDA2* | 1.444126187 | 6.22E-05 | 0.000175786 | 0.39130043 |
| Obesity | LEC | *NIPAL1* | 1.303280663 | 1.88E-05 | 0.000105211 | 0.118475658 |
| Obesity | LEC | *ANTXR2* | 1.83595058 | 2.40E-07 | 1.85E-05 | 0.001507996 |
| Obesity | LEC | *CGGBP1* | 0.461228732 | 0.035619303 | 0.044326714 | 1 |
| Obesity | LEC | *PBXIP1* | 2.100678141 | 7.28E-06 | 0.000105108 | 0.045767665 |
| Obesity | LEC | *DCST2* | 2.497971019 | 6.23E-06 | 9.96E-05 | 0.039150146 |
| Obesity | LEC | *LMOD3* | 2.25860415 | 1.48E-05 | 0.000105108 | 0.093336927 |
| Obesity | LEC | *NBPF10* | 1.03909686 | 7.53E-05 | 0.000199955 | 0.473521352 |
| Obesity | LEC | *POGLUT1* | 1.119564713 | 0.003789216 | 0.005514537 | 1 |
| Obesity | LEC | *SLC22A15* | 0.695826309 | 9.63E-06 | 0.000105108 | 0.060572272 |
| Obesity | LEC | *SLC15A2* | -1.38626468 | 0.020791861 | 0.026808538 | 1 |
| Obesity | LEC | *LRRC58* | 2.156912893 | 3.98E-05 | 0.000131395 | 0.25003167 |
| Obesity | LEC | *FSTL1* | 0.980938698 | 0.00034465 | 0.000651478 | 1 |
| Obesity | LEC | *IGFBP7* | -0.517566342 | 0.000290231 | 0.000565986 | 1 |
| Obesity | LEC | *ARPC2* | 1.73356625 | 0.000110054 | 0.000261197 | 0.691911328 |

| | | | | | | |
|---|---|---|---|---|---|---|
| Obesity | LEC | *CCT3* | 1.592429807 | 1.37E-05 | 0.000105108 | 0.085927274 |
| Obesity | LEC | *NUAK2* | 1.205006869 | 0.002471621 | 0.003734458 | 1 |
| Obesity | LEC | *SPTA1* | 0.374796074 | 0.03931623 | 0.048600303 | 1 |
| Obesity | LEC | *PYHIN1* | 0.513937945 | 0.000120538 | 0.000281091 | 0.757820559 |
| Obesity | LEC | *AIM2* | 3.959451361 | 0.000223665 | 0.000457444 | 1 |
| Obesity | LEC | *EFHB* | 0.381297216 | 0.007035061 | 0.009750756 | 1 |
| Obesity | LEC | *ICA1L* | 0.916604358 | 0.016022219 | 0.020990142 | 1 |
| Obesity | LEC | *CD200R1* | 0.756279084 | 4.60E-05 | 0.000144505 | 0.289044833 |
| Obesity | LEC | *GTPBP8* | 2.556868982 | 0.000328464 | 0.000626723 | 1 |
| Obesity | LEC | *SPICE1* | 1.833569745 | 6.10E-05 | 0.000173884 | 0.383588378 |
| Obesity | LEC | *CADPS* | 1.190438058 | 1.93E-05 | 0.000105211 | 0.12149768 |
| Obesity | LEC | *PPM1K* | 1.431451807 | 1.33E-05 | 0.000105108 | 0.083423021 |
| Obesity | LEC | *GMPS* | 1.736421086 | 0.012098708 | 0.016142737 | 1 |
| Obesity | LEC | *TIPARP* | 1.300818807 | 4.45E-06 | 8.83E-05 | 0.027988229 |
| Obesity | LEC | *DCLK3* | -1.817861815 | 0.030573976 | 0.038405312 | 1 |
| Obesity | LEC | *RPL9* | -0.536237105 | 0.004395745 | 0.006331282 | 1 |
| Obesity | LEC | *RPP14* | 1.542921053 | 2.93E-05 | 0.000115648 | 0.18400965 |
| Obesity | LEC | *DNASE1L3* | 1.246326101 | 1.67E-05 | 0.000105108 | 0.105127634 |
| Obesity | LEC | *CRELD1* | 1.970545972 | 1.35E-05 | 0.000105108 | 0.084605119 |
| Obesity | LEC | *PCOLCE2* | 0.858262681 | 4.73E-05 | 0.000146731 | 0.297276447 |
| Obesity | LEC | *TTC14* | 1.392764661 | 0.000310048 | 0.000596656 | 1 |
| Obesity | LEC | *RCHY1* | 1.534581354 | 0.000150789 | 0.000334277 | 0.948008356 |
| Obesity | LEC | *CCDC158* | 1.367459771 | 8.76E-06 | 0.000105108 | 0.055101451 |
| Obesity | LEC | *GYG1* | 1.386688436 | 1.74E-06 | 5.62E-05 | 0.010936511 |
| Obesity | LEC | *HPS3* | 1.73458266 | 7.16E-05 | 0.000192536 | 0.449957077 |
| Obesity | LEC | *RYK* | 1.791533949 | 9.01E-05 | 0.0002266 | 0.566635292 |
| Obesity | LEC | *PLB1* | 0.641287036 | 3.01E-05 | 0.000115831 | 0.189236321 |
| Obesity | LEC | *KIAA1143* | 0.805049126 | 0.007391496 | 0.010208772 | 1 |
| Obesity | LEC | *CDCP1* | 1.556233085 | 1.63E-05 | 0.000105108 | 0.102694053 |
| Obesity | LEC | *FYCO1* | 1.448842986 | 1.30E-05 | 0.000105108 | 0.081933496 |
| Obesity | LEC | *LRRC2* | 0.578139778 | 6.66E-05 | 0.000183725 | 0.418989721 |
| Obesity | LEC | *DTX3L* | 1.418927399 | 3.14E-05 | 0.000116847 | 0.19765792 |
| Obesity | LEC | *NMNAT3* | 1.632724868 | 1.24E-05 | 0.000105108 | 0.078015307 |
| Obesity | LEC | *TPRA1* | 2.921979488 | 0.000385551 | 0.000716112 | 1 |
| Obesity | LEC | *YEATS2* | 1.753463479 | 0.004398779 | 0.006334202 | 1 |
| Obesity | LEC | *MEAF6* | 2.234665921 | 0.00078202 | 0.001318466 | 1 |
| Obesity | LEC | *DNALI1* | 1.144631394 | 0.001552638 | 0.002445861 | 1 |
| Obesity | LEC | *RPN1* | 1.975850297 | 3.06E-05 | 0.000115831 | 0.192164202 |
| Obesity | LEC | *RFC4* | 2.036037324 | 1.95E-05 | 0.000105217 | 0.122906909 |
| Obesity | LEC | *SFMBT1* | 2.031960216 | 2.67E-05 | 0.000113078 | 0.168146272 |
| Obesity | LEC | *GNL3* | 1.505504751 | 0.010292139 | 0.013900468 | 1 |
| Obesity | LEC | *UVSSA* | 1.594442862 | 6.24E-05 | 0.000175889 | 0.392281433 |
| Obesity | LEC | *FAM208A* | 1.717071834 | 0.000130343 | 0.000298206 | 0.819468781 |
| Obesity | LEC | *SLC51A* | 1.159067439 | 0.000762721 | 0.001290427 | 1 |
| Obesity | LEC | *PIGX* | 1.274824211 | 4.82E-05 | 0.000148518 | 0.303047328 |
| Obesity | LEC | *ABLIM2* | -1.124023769 | 0.004765089 | 0.006800934 | 1 |
| Obesity | LEC | *EXO5* | 1.272308606 | 0.000567741 | 0.000997315 | 1 |
| Obesity | LEC | *C1orf50* | -2.166017718 | 2.77E-05 | 0.00011384 | 0.174232785 |
| Obesity | LEC | *ERMAP* | 0.853447508 | 2.82E-05 | 0.000114165 | 0.177298157 |
| Obesity | LEC | *DNAJB14* | 1.908084665 | 5.92E-05 | 0.00017021 | 0.371878282 |

| | | | | | | |
|---|---|---|---|---|---|---|
| Obesity | LEC | *SLC9B2* | 2.495311593 | 4.91E-06 | 9.11E-05 | 0.030871747 |
| Obesity | LEC | *BDH2* | 0.59319597 | 0.003571755 | 0.005228123 | 1 |
| Obesity | LEC | *CDC25A* | 1.007812902 | 0.005214571 | 0.007395445 | 1 |
| Obesity | LEC | *ZNF589* | 0.8511147 | 0.000544519 | 0.000961087 | 1 |
| Obesity | LEC | *ATRIP* | 1.698133815 | 6.22E-05 | 0.000175786 | 0.391222113 |
| Obesity | LEC | *SHISA5* | 2.79497811 | 6.12E-05 | 0.00017432 | 0.385072179 |
| Obesity | LEC | *RNF123* | 3.678660272 | 4.14E-23 | 8.68E-20 | 2.60E-19 |
| Obesity | LEC | *MST1R* | 2.160941411 | 3.23E-10 | 8.82E-08 | 2.03E-06 |
| Obesity | LEC | *POC1A* | 1.144033239 | 1.79E-06 | 5.67E-05 | 0.011223321 |
| Obesity | LEC | *PPM1M* | 0.581121966 | 0.006995062 | 0.009705967 | 1 |
| Obesity | LEC | *C4orf3* | 2.493338685 | 5.08E-05 | 0.000153502 | 0.319591419 |
| Obesity | LEC | *ANXA5* | 1.419830533 | 5.27E-05 | 0.000157313 | 0.331614812 |
| Obesity | LEC | *MAP9* | 2.201491295 | 4.48E-05 | 0.000142019 | 0.281765715 |
| Obesity | LEC | *CEP44* | 1.838375811 | 2.54E-05 | 0.000111668 | 0.159461615 |
| Obesity | LEC | *C4orf45* | 1.434559931 | 4.44E-06 | 8.83E-05 | 0.02794515 |
| Obesity | LEC | *TMEM144* | 1.030897774 | 1.47E-05 | 0.000105108 | 0.092709644 |
| Obesity | LEC | *NPY1R* | 1.823619985 | 0.000227331 | 0.000463283 | 1 |
| Obesity | LEC | *IL15* | 2.117219845 | 6.16E-05 | 0.00017491 | 0.387251293 |
| Obesity | LEC | *ARFIP1* | 1.635346302 | 0.000591122 | 0.00103233 | 1 |
| Obesity | LEC | *OTUD4* | 1.691680492 | 0.011392293 | 0.015284538 | 1 |
| Obesity | LEC | *ITGA2* | -0.436475956 | 0.000222382 | 0.000455263 | 1 |
| Obesity | LEC | *NDUFAF2* | 1.434300993 | 0.000107903 | 0.000257479 | 0.678386634 |
| Obesity | LEC | *PGGT1B* | 2.095206279 | 0.000166117 | 0.00035951 | 1 |
| Obesity | LEC | *CMBL* | 2.570388615 | 3.32E-05 | 0.000119352 | 0.208727055 |
| Obesity | LEC | *C5orf63* | -1.405991955 | 0.001660684 | 0.002600428 | 1 |
| Obesity | LEC | *PRRC1* | 1.583949441 | 1.11E-05 | 0.000105108 | 0.069594036 |
| Obesity | LEC | *F2RL1* | 1.651847019 | 0.016346189 | 0.02139673 | 1 |
| Obesity | LEC | *WDR41* | 1.132755299 | 1.58E-05 | 0.000105108 | 0.099244986 |
| Obesity | LEC | *NDUFS4* | 1.621528991 | 0.004799788 | 0.006844243 | 1 |
| Obesity | LEC | *RHOBTB3* | 0.706478803 | 2.65E-05 | 0.000112661 | 0.166737718 |
| Obesity | LEC | *TIGD6* | -1.915551456 | 0.002966899 | 0.004410711 | 1 |
| Obesity | LEC | *SERINC5* | 1.608852925 | 0.000162169 | 0.000353083 | 1 |
| Obesity | LEC | *ENPP6* | -1.567949071 | 0.000763052 | 0.00129064 | 1 |
| Obesity | LEC | *CASP3* | 2.171809807 | 0.006308192 | 0.008828941 | 1 |
| Obesity | LEC | *PRIMPOL* | 1.421706034 | 0.010301305 | 0.013909859 | 1 |
| Obesity | LEC | *ERAP2* | 0.856845167 | 0.00877669 | 0.011959049 | 1 |
| Obesity | LEC | *RICTOR* | 1.556608108 | 0.001588257 | 0.002497592 | 1 |
| Obesity | LEC | *ANKRA2* | 2.411500319 | 0.000161887 | 0.000352662 | 1 |
| Obesity | LEC | *UTP15* | 1.354049877 | 1.92E-05 | 0.000105211 | 0.120832244 |
| Obesity | LEC | *TLR3* | 3.285274001 | 5.75E-05 | 0.000166843 | 0.361214933 |
| Obesity | LEC | *NSA2* | 0.815808101 | 0.006282275 | 0.008794625 | 1 |
| Obesity | LEC | *CCDC127* | 1.46664044 | 1.46E-05 | 0.000105108 | 0.091841406 |
| Obesity | LEC | *MB21D1* | 1.279267758 | 1.44E-05 | 0.000105108 | 0.09060185 |
| Obesity | LEC | *TXLNB* | -0.790540788 | 0.000248309 | 0.000499061 | 1 |
| Obesity | LEC | *DCBLD1* | 1.73658762 | 0.01133617 | 0.015212487 | 1 |
| Obesity | LEC | *PDSS2* | 1.626499613 | 0.000621744 | 0.001077427 | 1 |
| Obesity | LEC | *STXBP5* | 1.474604997 | 2.73E-05 | 0.000113803 | 0.171841972 |
| Obesity | LEC | *PI16* | 0.619816212 | 1.92E-05 | 0.000105211 | 0.120852884 |
| Obesity | LEC | *DAGLB* | 1.353259917 | 0.000316309 | 0.000606292 | 1 |
| Obesity | LEC | *STK17A* | 1.578540225 | 2.23E-05 | 0.000108559 | 0.140115362 |

| | | | | | | |
|---|---|---|---|---|---|---|
| Obesity | LEC | *SAP30L* | 2.435513883 | 0.00010788 | 0.000257479 | 0.678242529 |
| Obesity | LEC | *HCN1* | 2.103095969 | 0.000373947 | 0.000697627 | 1 |
| Obesity | LEC | *SLU7* | 2.358523477 | 2.26E-05 | 0.000109111 | 0.142261346 |
| Obesity | LEC | *CAMLG* | 2.375623691 | 7.93E-05 | 0.000206997 | 0.49878169 |
| Obesity | LEC | *KCNK5* | -2.235122406 | 0.001819586 | 0.002825324 | 1 |
| Obesity | LEC | *KIAA1324L* | -0.678031281 | 0.001978202 | 0.003052014 | 1 |
| Obesity | LEC | *USP49* | 1.72038099 | 7.61E-05 | 0.000201309 | 0.478511858 |
| Obesity | LEC | *ZNF704* | 1.409648172 | 5.59E-05 | 0.000164486 | 0.351507194 |
| Obesity | LEC | *FNDC1* | 0.693620758 | 1.08E-05 | 0.000105108 | 0.068056835 |
| Obesity | LEC | *CHMP4C* | -0.533306242 | 0.000195693 | 0.000410106 | 1 |
| Obesity | LEC | *BRI3* | 1.734797683 | 2.08E-05 | 0.000106878 | 0.13092575 |
| Obesity | LEC | *LMTK2* | 2.14636725 | 2.79E-05 | 0.00011384 | 0.175299634 |
| Obesity | LEC | *CTSB* | 0.582273476 | 0.003549786 | 0.005199791 | 1 |
| Obesity | LEC | *TNFRSF11B* | 3.740294849 | 1.85E-05 | 0.000105108 | 0.116121138 |
| Obesity | LEC | *OXR1* | 1.590928313 | 0.00017569 | 0.000375958 | 1 |
| Obesity | LEC | *GPER1* | 1.382297279 | 5.35E-06 | 9.39E-05 | 0.03363323 |
| Obesity | LEC | *TMEM184A* | 0.803348713 | 0.000363463 | 0.0006811 | 1 |
| Obesity | LEC | *INTS1* | 1.994667517 | 1.45E-05 | 0.000105108 | 0.090968265 |
| Obesity | LEC | *SLC4A2* | 1.247607 | 1.61E-05 | 0.000105108 | 0.101295557 |
| Obesity | LEC | *FASTK* | 1.885507854 | 2.26E-05 | 0.000109111 | 0.1419499 |
| Obesity | LEC | *ALDH7A1* | 1.077198513 | 0.028375908 | 0.035844753 | 1 |
| Obesity | LEC | *DCAF13* | 2.145354879 | 0.015718219 | 0.02061766 | 1 |
| Obesity | LEC | *FREM1* | 0.851672597 | 2.39E-05 | 0.000109361 | 0.150481505 |
| Obesity | LEC | *RPP25L* | 2.068452757 | 9.84E-05 | 0.000241679 | 0.61868745 |
| Obesity | LEC | *FAM219A* | 1.703417405 | 2.31E-05 | 0.000109118 | 0.145036022 |
| Obesity | LEC | *CCDC171* | 1.471888421 | 0.0002105 | 0.000434333 | 1 |
| Obesity | LEC | *SYK* | 0.502215034 | 2.40E-05 | 0.000109361 | 0.150860399 |
| Obesity | LEC | *NIPSNAP3B* | 0.689822337 | 0.000400834 | 0.00073945 | 1 |
| Obesity | LEC | *METTL2B* | 0.73548357 | 9.29E-05 | 0.000231771 | 0.583832312 |
| Obesity | LEC | *TMC1* | 1.211956603 | 1.49E-05 | 0.000105108 | 0.093856391 |
| Obesity | LEC | *HGSNAT* | 1.497147386 | 0.001644925 | 0.00257832 | 1 |
| Obesity | LEC | *C9orf64* | 1.399997097 | 0.000605043 | 0.00105459 | 1 |
| Obesity | LEC | *HNRNPK* | 1.80279662 | 4.08E-05 | 0.000133178 | 0.256233627 |
| Obesity | LEC | *SVEP1* | 1.049155316 | 1.02E-05 | 0.000105108 | 0.064070043 |
| Obesity | LEC | *ZNF367* | 1.348808789 | 8.63E-06 | 0.000105108 | 0.054272443 |
| Obesity | LEC | *NOL6* | 0.908508259 | 4.90E-05 | 0.000149938 | 0.307972079 |
| Obesity | LEC | *TRMT10B* | 1.909823936 | 6.43E-05 | 0.000179062 | 0.404137089 |
| Obesity | LEC | *VCP* | 1.509088334 | 0.000369421 | 0.000690412 | 1 |
| Obesity | LEC | *PIGO* | 1.286890354 | 3.38E-05 | 0.000120325 | 0.212384161 |
| Obesity | LEC | *LRFN5* | 0.832417747 | 4.28E-07 | 2.56E-05 | 0.002691662 |
| Obesity | LEC | *WRN* | 1.734006188 | 0.000777742 | 0.001311604 | 1 |
| Obesity | LEC | *MARCH8* | 2.053376622 | 0.000156063 | 0.000342227 | 0.981165308 |
| Obesity | LEC | *CFL2* | 2.361667915 | 4.28E-05 | 0.000137578 | 0.268965576 |
| Obesity | LEC | *ZCCHC24* | 1.66895575 | 3.02E-05 | 0.000115831 | 0.190112895 |
| Obesity | LEC | *PGM2L1* | -0.562716828 | 0.000238209 | 0.00048186 | 1 |
| Obesity | LEC | *GJB2* | -2.872024222 | 0.039114095 | 0.048359944 | 1 |
| Obesity | LEC | *CRYL1* | 1.27121554 | 1.43E-05 | 0.000105108 | 0.090099075 |
| Obesity | LEC | *REEP3* | -0.983242035 | 0.028968747 | 0.036534907 | 1 |
| Obesity | LEC | *HEPACAM* | 0.347779834 | 0.001214929 | 0.001961042 | 1 |
| Obesity | LEC | *LRR1* | -1.249137912 | 0.007221302 | 0.00998907 | 1 |

| | | | | | | |
|---|---|---|---|---|---|---|
| Obesity | LEC | *C10orf25* | 0.940567859 | 0.001004881 | 0.001654277 | 1 |
| Obesity | LEC | *KLHDC2* | 1.687998905 | 0.00049587 | 0.000886419 | 1 |
| Obesity | LEC | *NOXRED1* | -1.398453614 | 0.004547618 | 0.006524617 | 1 |
| Obesity | LEC | *AKR1E2* | 1.43848606 | 0.000140002 | 0.000314805 | 0.880195111 |
| Obesity | LEC | *OXGR1* | -0.845497485 | 8.28E-05 | 0.000213751 | 0.520482555 |
| Obesity | LEC | *TAF3* | 1.495270479 | 8.34E-06 | 0.000105108 | 0.052438946 |
| Obesity | LEC | *SLC18A2* | 1.510790149 | 1.79E-05 | 0.000105108 | 0.112439217 |
| Obesity | LEC | *PDZD8* | 1.382091213 | 4.16E-05 | 0.000134903 | 0.261306941 |
| Obesity | LEC | *QSOX2* | 2.962472315 | 0.011589343 | 0.01552572 | 1 |
| Obesity | LEC | *SNAPC4* | 0.975210625 | 0.012775988 | 0.016956436 | 1 |
| Obesity | LEC | *SDCCAG3* | 2.294528365 | 2.77E-05 | 0.00011384 | 0.174174704 |
| Obesity | LEC | *AK8* | 0.721642845 | 2.51E-05 | 0.000111345 | 0.157821986 |
| Obesity | LEC | *TSC1* | 2.289845962 | 5.23E-05 | 0.000156286 | 0.328513269 |
| Obesity | LEC | *ZMYND19* | 0.986782027 | 2.96E-05 | 0.000115831 | 0.185974401 |
| Obesity | LEC | *STOX1* | 0.606929203 | 4.72E-05 | 0.000146638 | 0.296941187 |
| Obesity | LEC | *RET* | -2.185688522 | 0.0005195 | 0.00092138 | 1 |
| Obesity | LEC | *STK32C* | 1.89475948 | 0.00018917 | 0.00039883 | 1 |
| Obesity | LEC | *NSMF* | -1.419433945 | 0.00632596 | 0.008845932 | 1 |
| Obesity | LEC | *VWA2* | 1.087094073 | 0.001174311 | 0.001903789 | 1 |
| Obesity | LEC | *METTL3* | 2.02499471 | 0.010353708 | 0.013971616 | 1 |
| Obesity | LEC | *TRUB1* | 1.577831641 | 3.35E-05 | 0.000119881 | 0.210750275 |
| Obesity | LEC | *HSPA12A* | 0.601593028 | 0.006854338 | 0.009523364 | 1 |
| Obesity | LEC | *UBTD1* | -0.952165101 | 0.004296715 | 0.006198588 | 1 |
| Obesity | LEC | *ARHGAP42* | 1.665583508 | 7.35E-05 | 0.000196122 | 0.462063885 |
| Obesity | LEC | *OTOGL* | -1.986801938 | 0.002231747 | 0.003407235 | 1 |
| Obesity | LEC | *PACSIN3* | 0.931753679 | 0.000726089 | 0.001237442 | 1 |
| Obesity | LEC | *TTC7B* | 1.356589945 | 0.000339332 | 0.000643748 | 1 |
| Obesity | LEC | *SLC39A13* | 1.715155577 | 3.49E-06 | 7.79E-05 | 0.021955803 |
| Obesity | LEC | *PSMC3* | 1.351298521 | 6.11E-07 | 3.20E-05 | 0.003840948 |
| Obesity | LEC | *CPSF2* | 0.667237181 | 0.034569468 | 0.043062858 | 1 |
| Obesity | LEC | *IFI27L1* | 2.21946569 | 0.002600231 | 0.003900657 | 1 |
| Obesity | LEC | *CLMN* | 1.116382553 | 0.000143396 | 0.000320716 | 0.901532963 |
| Obesity | LEC | *PTER* | 1.508718561 | 4.84E-05 | 0.0001487 | 0.303992568 |
| Obesity | LEC | *C1QL3* | 0.712496321 | 5.45E-06 | 9.50E-05 | 0.034270282 |
| Obesity | LEC | *ABTB2* | 1.000343604 | 1.92E-05 | 0.000105211 | 0.120569604 |
| Obesity | LEC | *LIPC* | -0.668734567 | 0.000357704 | 0.00067171 | 1 |
| Obesity | LEC | *CEP57* | 1.378200646 | 0.007457667 | 0.01028435 | 1 |
| Obesity | LEC | *JAM3* | 0.747049874 | 3.15E-05 | 0.000116847 | 0.198056238 |
| Obesity | LEC | *GPT2* | 0.786399349 | 1.84E-05 | 0.000105108 | 0.115402018 |
| Obesity | LEC | *IKBIP* | 1.212345697 | 1.78E-05 | 0.000105108 | 0.112182374 |
| Obesity | LEC | *HIF1AN* | 2.453995178 | 0.000108669 | 0.000258788 | 0.68320007 |
| Obesity | LEC | *NDUFB8* | 2.565385717 | 1.29E-05 | 0.000105108 | 0.081104861 |
| Obesity | LEC | *SPINT1* | 1.71152887 | 7.42E-05 | 0.000197667 | 0.466691393 |
| Obesity | LEC | *AVPR1A* | -0.531822678 | 0.002267367 | 0.003454067 | 1 |
| Obesity | LEC | *BTRC* | 1.636334632 | 0.000152116 | 0.000336153 | 0.956356118 |
| Obesity | LEC | *LARP6* | 1.84036778 | 1.38E-05 | 0.000105108 | 0.086571704 |
| Obesity | LEC | *ASPG* | 2.512244423 | 1.46E-05 | 0.000105108 | 0.092090955 |
| Obesity | LEC | *TBATA* | 6.412331133 | 6.73E-05 | 0.000184723 | 0.422830963 |
| Obesity | LEC | *COX11* | 1.944726831 | 0.000351063 | 0.000661017 | 1 |
| Obesity | LEC | *ZNF202* | 3.176896878 | 0.002111478 | 0.003241725 | 1 |

| | | | | | | |
|---|---|---|---|---|---|---|
| Obesity | LEC | *FAM227B* | 1.614764382 | 1.15E-05 | 0.000105108 | 0.072249855 |
| Obesity | LEC | *STXBP4* | 1.747705846 | 0.000843414 | 0.001409501 | 1 |
| Obesity | LEC | *C2* | 1.28303833 | 7.32E-05 | 0.000195742 | 0.460349193 |
| Obesity | LEC | *APBB1* | 0.981942256 | 1.48E-05 | 0.000105108 | 0.092840128 |
| Obesity | LEC | *ILK* | 1.999543566 | 8.10E-05 | 0.000209893 | 0.508990631 |
| Obesity | LEC | *TAF10* | 1.655685634 | 1.52E-05 | 0.000105108 | 0.095510606 |
| Obesity | LEC | *DCHS1* | 3.332667736 | 2.36E-05 | 0.000109118 | 0.148176719 |
| Obesity | LEC | *NETO1* | 1.05300591 | 6.70E-05 | 0.000184604 | 0.421535222 |
| Obesity | LEC | *USP54* | 1.875650968 | 8.23E-05 | 0.000212766 | 0.517658787 |
| Obesity | LEC | *WDR88* | 2.623952797 | 2.81E-05 | 0.000114038 | 0.176702441 |
| Obesity | LEC | *CYB5R2* | 1.250362212 | 0.000147079 | 0.000327438 | 0.924684227 |
| Obesity | LEC | *TUB* | 2.468512692 | 0.000284049 | 0.000556172 | 1 |
| Obesity | LEC | *RIC3* | 1.64436562 | 6.66E-06 | 0.000102922 | 0.041889052 |
| Obesity | LEC | *LMO1* | 2.311034792 | 3.17E-05 | 0.000117011 | 0.199197814 |
| Obesity | LEC | *XRRA1* | 0.721746238 | 0.018948021 | 0.024592528 | 1 |
| Obesity | LEC | *TRIM66* | 0.451437478 | 0.001720796 | 0.002685193 | 1 |
| Obesity | LEC | *PRTG* | -1.205727803 | 0.004778114 | 0.006816429 | 1 |
| Obesity | LEC | *C16orf46* | 9.084029976 | 1.09E-05 | 0.000105108 | 0.068315764 |
| Obesity | LEC | *TMEM41B* | 1.987543928 | 7.25E-05 | 0.000194493 | 0.455696342 |
| Obesity | LEC | *ZNF143* | 2.011249536 | 8.50E-05 | 0.000217214 | 0.534346818 |
| Obesity | LEC | *TMX3* | 1.889045198 | 0.000285518 | 0.000558857 | 1 |
| Obesity | LEC | *CCDC68* | 1.449285069 | 0.006835407 | 0.009499161 | 1 |
| Obesity | LEC | *CLEC4E* | 0.811440051 | 0.000229338 | 0.000466316 | 1 |
| Obesity | LEC | *ZNF3* | 1.536019934 | 0.029331194 | 0.036954953 | 1 |
| Obesity | LEC | *CLEC4D* | 2.854438693 | 1.56E-05 | 0.000105108 | 0.097871915 |
| Obesity | LEC | *BEAN1* | 1.976130314 | 7.09E-06 | 0.000105108 | 0.044549099 |
| Obesity | LEC | *TK2* | 1.139147024 | 4.90E-05 | 0.000149938 | 0.307777895 |
| Obesity | LEC | *SEC11C* | 2.392220723 | 1.39E-05 | 0.000105108 | 0.087215981 |
| Obesity | LEC | *GALR1* | 0.862131995 | 0.000239934 | 0.000484881 | 1 |
| Obesity | LEC | *CENPV* | 1.435913961 | 6.30E-05 | 0.000176855 | 0.395801432 |
| Obesity | LEC | *SPDYE6* | 1.667848712 | 6.02E-06 | 9.93E-05 | 0.037830293 |
| Obesity | LEC | *ATF7IP2* | 1.52148242 | 0.000106453 | 0.000255252 | 0.669271189 |
| Obesity | LEC | *ZNF592* | 1.385425775 | 0.000199304 | 0.000415734 | 1 |
| Obesity | LEC | *ACSM1* | -0.467588623 | 5.31E-05 | 0.000157954 | 0.333597905 |
| Obesity | LEC | *AP1G1* | 1.599078727 | 4.91E-05 | 0.000149956 | 0.30844392 |
| Obesity | LEC | *SLFN5* | 1.400397654 | 0.00011895 | 0.000278111 | 0.747840982 |
| Obesity | LEC | *CATSPER2* | 2.623801636 | 2.53E-06 | 6.83E-05 | 0.0159068 |
| Obesity | LEC | *C16orf45* | 1.735508612 | 2.20E-05 | 0.000108376 | 0.138375057 |
| Obesity | LEC | *SAAL1* | 3.380597792 | 0.000276999 | 0.000545752 | 1 |
| Obesity | LEC | *YPEL4* | 1.497362316 | 3.32E-06 | 7.65E-05 | 0.020893875 |
| Obesity | LEC | *FAM111A* | 1.408216431 | 0.000484029 | 0.000868709 | 1 |
| Obesity | LEC | *SCNN1G* | -1.574553639 | 6.71E-05 | 0.000184604 | 0.422130018 |
| Obesity | LEC | *ANKDD1A* | -0.873967588 | 0.013732652 | 0.018145688 | 1 |
| Obesity | LEC | *DCTN5* | 1.382235708 | 1.42E-05 | 0.000105108 | 0.089049077 |
| Obesity | LEC | *STX3* | -1.053612905 | 0.001495282 | 0.002367373 | 1 |
| Obesity | LEC | *MTMR10* | 2.074128436 | 1.53E-05 | 0.000105108 | 0.095884119 |
| Obesity | LEC | *YWHAB* | 1.577016054 | 0.000816022 | 0.001368453 | 1 |
| Obesity | LEC | *NYAP1* | -0.420441451 | 9.98E-05 | 0.000243896 | 0.627545293 |
| Obesity | LEC | *MS4A6E* | 0.542856582 | 0.016678284 | 0.021804195 | 1 |
| Obesity | LEC | *MS4A7* | 0.394924641 | 4.47E-05 | 0.000141942 | 0.281187278 |

| | | | | | | |
|---|---|---|---|---|---|---|
| Obesity | LEC | *MS4A14* | 0.455169069 | 0.001504951 | 0.002379685 | 1 |
| Obesity | LEC | *DIS3L* | 1.405178326 | 0.001062866 | 0.001739261 | 1 |
| Obesity | LEC | *CCNDBP1* | 2.490162106 | 0.000170882 | 0.000367044 | 1 |
| Obesity | LEC | *SMAD3* | 1.054905073 | 0.000185716 | 0.000392823 | 1 |
| Obesity | LEC | *AKTIP* | 0.96496746 | 1.97E-05 | 0.000105385 | 0.123827661 |
| Obesity | LEC | *CNPY4* | 0.954387956 | 0.014587196 | 0.019206221 | 1 |
| Obesity | LEC | *NKX3-1* | 5.820896401 | 2.41E-05 | 0.000109361 | 0.15148006 |
| Obesity | LEC | *DUSP18* | -1.254722883 | 0.012634445 | 0.016772119 | 1 |
| Obesity | LEC | *TEF* | 1.209594568 | 1.12E-05 | 0.000105108 | 0.070687781 |
| Obesity | LEC | *MEI1* | 1.535631136 | 2.90E-05 | 0.000115266 | 0.18250756 |
| Obesity | LEC | *PBX3* | 0.929851657 | 0.000122139 | 0.000283212 | 0.767886399 |
| Obesity | LEC | *SNRPD1* | 2.732500647 | 9.41E-05 | 0.000233893 | 0.591516213 |
| Obesity | LEC | *URM1* | 1.189874861 | 0.019319943 | 0.025028741 | 1 |
| Obesity | LEC | *CERCAM* | 1.234738058 | 0.000661364 | 0.001136685 | 1 |
| Obesity | LEC | *ENDOG* | 1.258527095 | 7.16E-06 | 0.000105108 | 0.045010908 |
| Obesity | LEC | *ISLR2* | -0.285298056 | 0.000259577 | 0.000517098 | 1 |
| Obesity | LEC | *SP2* | 2.218806975 | 9.23E-07 | 4.06E-05 | 0.00580127 |
| Obesity | LEC | *COQ7* | 1.454772377 | 1.41E-05 | 0.000105108 | 0.088836476 |
| Obesity | LEC | *GPRC5B* | -0.666745384 | 0.014829209 | 0.019512608 | 1 |
| Obesity | LEC | *KATNAL2* | 0.839101922 | 6.36E-05 | 0.000178132 | 0.400083562 |
| Obesity | LEC | *ENGASE* | 2.136555891 | 0.000128798 | 0.000295314 | 0.809751429 |
| Obesity | LEC | *RBFOX3* | -1.948720625 | 0.035942709 | 0.044693792 | 1 |
| Obesity | LEC | *TRIM68* | -1.301299341 | 0.000145521 | 0.000324766 | 0.914893439 |
| Obesity | LEC | *FN3K* | 1.115059478 | 6.85E-05 | 0.000187169 | 0.430785167 |
| Obesity | LEC | *ZNF23* | -2.242341294 | 0.000465695 | 0.000839881 | 1 |
| Obesity | LEC | *ZNF229* | 3.337804953 | 2.01E-06 | 6.04E-05 | 0.012615648 |
| Obesity | LEC | *VKORC1* | -2.402737016 | 4.76E-05 | 0.000147215 | 0.298994209 |
| Obesity | LEC | *SMG8* | 1.843865951 | 2.98E-05 | 0.000115831 | 0.187172648 |
| Obesity | LEC | *RAB8A* | 1.328569794 | 9.44E-06 | 0.000105108 | 0.059338095 |
| Obesity | LEC | *GATAD2A* | 1.58252982 | 5.15E-05 | 0.000154892 | 0.323568747 |
| Obesity | LEC | *PROCA1* | 0.942183472 | 2.51E-05 | 0.000111345 | 0.157507419 |
| Obesity | LEC | *ZNF641* | 0.83487451 | 0.000159445 | 0.000348308 | 1 |
| Obesity | LEC | *CACNB3* | -1.517601791 | 0.010112402 | 0.013663588 | 1 |
| Obesity | LEC | *TP53I13* | 0.882499824 | 4.50E-05 | 0.000142544 | 0.283167091 |
| Obesity | LEC | *ZNF528* | 2.289008038 | 6.97E-05 | 0.000188832 | 0.437900886 |
| Obesity | LEC | *ZNF701* | 1.986286925 | 0.001898626 | 0.002937894 | 1 |
| Obesity | LEC | *SERTAD3* | 1.040804367 | 2.50E-05 | 0.000111345 | 0.157466621 |
| Obesity | LEC | *TMC4* | 2.975476367 | 8.07E-05 | 0.000209586 | 0.507407858 |
| Obesity | LEC | *TTYH1* | 2.655240706 | 6.56E-06 | 0.000102375 | 0.041256972 |
| Obesity | LEC | *LENG8* | 1.907391973 | 0.000246789 | 0.000496817 | 1 |
| Obesity | LEC | *CDC42EP5* | -0.891527966 | 0.000510409 | 0.000908277 | 1 |
| Obesity | LEC | *KIR3DL1* | 1.72857028 | 0.018253032 | 0.02374934 | 1 |
| Obesity | LEC | *ZNF283* | 1.919473532 | 0.003906181 | 0.005666396 | 1 |
| Obesity | LEC | *PPP1R14A* | 1.467160103 | 1.99E-05 | 0.000105667 | 0.125261038 |
| Obesity | LEC | *C19orf33* | 1.459702422 | 0.005638427 | 0.00796524 | 1 |
| Obesity | LEC | *PSCA* | 2.114036016 | 0.008582006 | 0.011714085 | 1 |
| Obesity | LEC | *CHAF1A* | 0.542448517 | 0.001010926 | 0.001663794 | 1 |
| Obesity | LEC | *FAM57A* | 1.2679812 | 0.018314746 | 0.023824707 | 1 |
| Obesity | LEC | *KIFC2* | 0.745481749 | 0.000540479 | 0.00095476 | 1 |
| Obesity | LEC | *SERPINF2* | 1.330229758 | 5.74E-06 | 9.67E-05 | 0.036075798 |

| | | | | | | |
|---|---|---|---|---|---|---|
| Obesity | LEC | *SRR* | 1.696633144 | 2.41E-09 | 5.22E-07 | 1.51E-05 |
| Obesity | LEC | *CYB5D2* | 1.261577225 | 1.11E-05 | 0.000105108 | 0.070010544 |
| Obesity | LEC | *C19orf48* | 1.119680603 | 4.31E-06 | 8.74E-05 | 0.027095947 |
| Obesity | LEC | *ZNF83* | 1.5900213 | 0.000852537 | 0.001423235 | 1 |
| Obesity | LEC | *CD320* | 2.419039652 | 1.50E-05 | 0.000105108 | 0.094042524 |
| Obesity | LEC | *SPRYD3* | 3.22962773 | 0.000274389 | 0.000541628 | 1 |
| Obesity | LEC | *SOAT2* | 1.66806249 | 0.000299064 | 0.000578705 | 1 |
| Obesity | LEC | *ZNF558* | 1.252714887 | 0.001947825 | 0.00300884 | 1 |
| Obesity | LEC | *NDUFV1* | 2.136230696 | 6.03E-05 | 0.000172507 | 0.379170086 |
| Obesity | LEC | *NUDT8* | -0.948341849 | 8.28E-06 | 0.000105108 | 0.052081836 |
| Obesity | LEC | *PRDX2* | 1.861295133 | 2.81E-06 | 7.08E-05 | 0.01766847 |
| Obesity | LEC | *MIS12* | 1.202769074 | 2.81E-05 | 0.000114038 | 0.17655881 |
| Obesity | LEC | *CD300C* | 1.281651138 | 0.021083224 | 0.027139687 | 1 |
| Obesity | LEC | *HID1* | -0.252705993 | 0.000111272 | 0.00026349 | 0.699566436 |
| Obesity | LEC | *ATP5H* | 1.759557806 | 3.42E-05 | 0.000121059 | 0.21487888 |
| Obesity | LEC | *SRP68* | 1.383448014 | 9.61E-05 | 0.000237527 | 0.604031114 |
| Obesity | LEC | *MGAT5B* | 1.679913129 | 1.25E-05 | 0.000105108 | 0.078507963 |
| Obesity | LEC | *TMC8* | 1.249369615 | 1.26E-06 | 4.90E-05 | 0.007938142 |
| Obesity | LEC | *TMEM68* | 1.639669287 | 2.86E-05 | 0.000114981 | 0.179839153 |
| Obesity | LEC | *CYP7A1* | 3.281237757 | 0.001150724 | 0.001867957 | 1 |
| Obesity | LEC | *GSDMA* | -1.985946966 | 0.001808944 | 0.002809494 | 1 |
| Obesity | LEC | *TMEM99* | 1.134926046 | 0.010496451 | 0.014155124 | 1 |
| Obesity | LEC | *ZNF598* | 1.344135131 | 1.39E-05 | 0.000105108 | 0.087600944 |
| Obesity | LEC | *MLST8* | -2.554024425 | 0.002549025 | 0.003832071 | 1 |
| Obesity | LEC | *ECI1* | 1.366087024 | 2.63E-05 | 0.000112582 | 0.16562331 |
| Obesity | LEC | *ABCA3* | -1.134655885 | 0.001524264 | 0.002406592 | 1 |
| Obesity | LEC | *KCTD5* | -1.368155581 | 0.001675756 | 0.002621417 | 1 |
| Obesity | LEC | *SDHAF2* | -1.685603539 | 0.037454571 | 0.046463475 | 1 |
| Obesity | LEC | *HRASLS5* | 0.428119326 | 0.000375482 | 0.000699739 | 1 |
| Obesity | LEC | *TRANK1* | 1.785390411 | 7.14E-06 | 0.000105108 | 0.044918701 |
| Obesity | LEC | *TTC21A* | 2.355690967 | 1.50E-05 | 0.000105108 | 0.09431277 |
| Obesity | LEC | *RPSA* | 1.502896391 | 4.78E-05 | 0.000147799 | 0.300623517 |
| Obesity | LEC | *ULK4* | 0.585157293 | 0.012475592 | 0.016582251 | 1 |
| Obesity | LEC | *LTBP3* | 1.762075628 | 0.000154553 | 0.000340103 | 0.97167371 |
| Obesity | LEC | *NAALADL1* | -1.11720816 | 0.000423347 | 0.000775294 | 1 |
| Obesity | LEC | *SCARA5* | 0.627592581 | 2.59E-05 | 0.000112124 | 0.162845244 |
| Obesity | LEC | *RNF187* | 1.331591246 | 0.000862239 | 0.001437904 | 1 |
| Obesity | LEC | *PTGDR* | 0.587437452 | 5.30E-05 | 0.000157883 | 0.333162605 |
| Obesity | LEC | *GLYCTK* | 2.577706206 | 1.49E-06 | 5.27E-05 | 0.009372235 |
| Obesity | LEC | *POLR2J3* | 1.12547217 | 0.002779832 | 0.004150274 | 1 |
| Obesity | LEC | *NT5DC2* | -2.246846006 | 0.004959104 | 0.007052226 | 1 |
| Obesity | LEC | *COA6* | 0.913419666 | 2.44E-05 | 0.000109899 | 0.153517769 |
| Obesity | LEC | *PDHB* | 2.220163181 | 0.000183167 | 0.000388781 | 1 |
| Obesity | LEC | *ACOX2* | 0.650908263 | 0.000380565 | 0.000707663 | 1 |
| Obesity | LEC | *FAM107A* | 0.796896307 | 1.59E-05 | 0.000105108 | 0.099794879 |
| Obesity | LEC | *CX3CR1* | 0.60173592 | 0.000121263 | 0.000282154 | 0.76238089 |
| Obesity | LEC | *SEPT2* | 1.03617016 | 9.81E-05 | 0.000241238 | 0.616846803 |
| Obesity | LEC | *DTYMK* | 1.193868462 | 1.45E-05 | 0.000105108 | 0.091006825 |
| Obesity | LEC | *TAP1* | 1.610325771 | 9.21E-06 | 0.000105108 | 0.057920284 |
| Obesity | LEC | *ING5* | 1.787573555 | 2.54E-05 | 0.000111691 | 0.159853801 |

| | | | | | | |
|---|---|---|---|---|---|---|
| Obesity | LEC | *RFWD3* | 1.981131175 | 9.60E-07 | 4.17E-05 | 0.006037699 |
| Obesity | LEC | *RHOH* | 0.617594075 | 2.79E-05 | 0.00011384 | 0.175313468 |
| Obesity | LEC | *KLHL30* | 1.155000657 | 2.79E-05 | 0.00011384 | 0.175273782 |
| Obesity | LEC | *REEP4* | 1.724047934 | 0.000388348 | 0.000720431 | 1 |
| Obesity | LEC | *BMP1* | 1.779187408 | 0.000719053 | 0.001227247 | 1 |
| Obesity | LEC | *ATXN2L* | 1.401197153 | 0.00238318 | 0.003613858 | 1 |
| Obesity | LEC | *FNTA* | 2.511119537 | 0.000513237 | 0.000912534 | 1 |
| Obesity | LEC | *SERINC2* | 2.064589764 | 0.011199503 | 0.015041931 | 1 |
| Obesity | LEC | *TRAPPC11* | 1.954580373 | 0.000103772 | 0.000250928 | 0.652411743 |
| Obesity | LEC | *SLC20A2* | 1.698078896 | 2.21E-05 | 0.000108376 | 0.138697387 |
| Obesity | LEC | *ZSWIM1* | 1.655759036 | 3.05E-06 | 7.33E-05 | 0.019151754 |
| Obesity | LEC | *NBPF9* | 0.822431024 | 2.42E-05 | 0.000109361 | 0.151861053 |
| Obesity | LEC | *WFDC13* | 10.06815464 | 0.007198992 | 0.009962593 | 1 |
| Obesity | LEC | *NDUFS5* | 1.858952363 | 0.010893119 | 0.014661751 | 1 |
| Obesity | LEC | *ZNF30* | 1.667535105 | 0.002953169 | 0.004391337 | 1 |
| Obesity | LEC | *FAM84B* | 1.174787663 | 0.011739504 | 0.015710145 | 1 |
| Obesity | LEC | *SLC16A4* | 1.247816022 | 0.005135066 | 0.007292559 | 1 |
| Obesity | LEC | *LRP1B* | 0.659189807 | 0.000174834 | 0.000374507 | 1 |
| Obesity | LEC | *NPNT* | 0.77020351 | 2.66E-05 | 0.000112975 | 0.167489576 |
| Obesity | LEC | *CNNM3* | 2.118811719 | 5.03E-06 | 9.23E-05 | 0.031631156 |
| Obesity | LEC | *GSTM4* | 1.026611929 | 0.000105719 | 0.000254073 | 0.664654336 |
| Obesity | LEC | *ABHD15* | 0.780888121 | 7.22E-05 | 0.00019394 | 0.454090264 |
| Obesity | LEC | *ADAL* | 0.594708914 | 0.001595029 | 0.00250636 | 1 |
| Obesity | LEC | *ZNF507* | 1.117873981 | 0.00077307 | 0.001305126 | 1 |
| Obesity | LEC | *STX18* | 1.862899266 | 0.000396795 | 0.000733505 | 1 |
| Obesity | LEC | *ZBTB49* | 2.627990074 | 3.52E-05 | 0.000122857 | 0.221388221 |
| Obesity | LEC | *GFM1* | 1.480717012 | 0.007230493 | 0.009999585 | 1 |
| Obesity | LEC | *TNIP2* | -0.9816612 | 0.012985016 | 0.01721932 | 1 |
| Obesity | LEC | *BTNL3* | 0.701593108 | 1.92E-05 | 0.000105211 | 0.120725104 |
| Obesity | LEC | *INPP5D* | 1.67408764 | 2.23E-05 | 0.000108559 | 0.140079096 |
| Obesity | LEC | *LETM1* | 2.367196945 | 2.97E-05 | 0.000115831 | 0.186928665 |
| Obesity | LEC | *TMEM129* | 1.154052881 | 3.57E-05 | 0.000123048 | 0.224318537 |
| Obesity | LEC | *CEP120* | 1.963858178 | 0.00016334 | 0.00035509 | 1 |
| Obesity | LEC | *STXBP6* | -0.258894548 | 0.00015539 | 0.000341228 | 0.97693554 |
| Obesity | LEC | *MFF* | 1.736365251 | 3.85E-05 | 0.00012891 | 0.242350137 |
| Obesity | LEC | *LGALS9* | 1.550078219 | 0.022805785 | 0.029177853 | 1 |
| Obesity | LEC | *SIGLEC7* | 1.024289385 | 0.000253728 | 0.00050786 | 1 |
| Obesity | LEC | *FEM1B* | 2.107899849 | 5.11E-06 | 9.29E-05 | 0.032130177 |
| Obesity | LEC | *MFSD7* | 2.378377475 | 0.023291686 | 0.029763177 | 1 |
| Obesity | LEC | *COL4A3* | 0.861172193 | 4.16E-06 | 8.57E-05 | 0.026144842 |
| Obesity | LEC | *IRS1* | 0.705187173 | 9.98E-05 | 0.000243865 | 0.627221785 |
| Obesity | LEC | *UPF3A* | 0.989542802 | 0.014115788 | 0.018624546 | 1 |
| Obesity | LEC | *C8orf46* | 0.497086639 | 2.01E-07 | 1.64E-05 | 0.001264588 |
| Obesity | LEC | *FAM110B* | 1.563162484 | 0.015436847 | 0.020269728 | 1 |
| Obesity | LEC | *ATF5* | 0.657316284 | 4.45E-05 | 0.000141409 | 0.279600874 |
| Obesity | LEC | *CPT1C* | -0.637438779 | 0.000179783 | 0.000383411 | 1 |
| Obesity | LEC | *PCSK9* | 5.771765532 | 0.001091931 | 0.00178172 | 1 |
| Obesity | LEC | *XPO6* | 1.920879842 | 2.99E-05 | 0.000115831 | 0.188001721 |
| Obesity | LEC | *RGS14* | -0.699508214 | 0.017629178 | 0.022980435 | 1 |
| Obesity | LEC | *RAB24* | 0.907369082 | 0.015700906 | 0.020599248 | 1 |

| | | | | | | |
|---|---|---|---|---|---|---|
| Obesity | LEC | *CXCL10* | 1.032366398 | 7.02E-05 | 0.000189971 | 0.441174712 |
| Obesity | LEC | *SH3TC2* | -1.476270811 | 0.004266154 | 0.006157325 | 1 |
| Obesity | LEC | *GPRIN1* | -1.904676087 | 0.00010406 | 0.000251238 | 0.654224382 |
| Obesity | LEC | *STK32A* | 0.457283887 | 0.000419013 | 0.0007687 | 1 |
| Obesity | LEC | *P2RY12* | 0.50932395 | 2.08E-05 | 0.000106878 | 0.130751383 |
| Obesity | LEC | *SLC33A1* | 2.190916847 | 0.00022551 | 0.000460068 | 1 |
| Obesity | LEC | *SNUPN* | 1.710800473 | 8.70E-06 | 0.000105108 | 0.054685851 |
| Obesity | LEC | *PTAFR* | 0.686012541 | 3.37E-05 | 0.000120302 | 0.212116381 |
| Obesity | LEC | *RNASE6* | 1.000234026 | 3.05E-05 | 0.000115831 | 0.191688732 |
| Obesity | LEC | *NPR1* | 1.225838211 | 1.44E-05 | 0.000105108 | 0.090505189 |
| Obesity | LEC | *CLIC4* | 1.521193055 | 0.005898749 | 0.008296518 | 1 |
| Obesity | LEC | *SLC38A11* | 0.764277995 | 3.80E-05 | 0.000127833 | 0.238846725 |
| Obesity | LEC | *CCDC8* | 1.539574275 | 2.93E-05 | 0.000115648 | 0.184248203 |
| Obesity | LEC | *METTL15* | 1.731854649 | 6.40E-05 | 0.000178795 | 0.402663157 |
| Obesity | LEC | *DTWD2* | 1.974623552 | 0.001705258 | 0.00266293 | 1 |
| Obesity | LEC | *CLIC3* | 1.316511017 | 9.91E-05 | 0.000242611 | 0.623024892 |
| Obesity | LEC | *DFFB* | 1.429551309 | 1.74E-05 | 0.000105108 | 0.109325825 |
| Obesity | LEC | *C15orf40* | 1.136172909 | 1.25E-05 | 0.000105108 | 0.078447965 |
| Obesity | LEC | *LUZP1* | 1.535761172 | 0.007507119 | 0.010348007 | 1 |
| Obesity | LEC | *HEXDC* | 1.913313362 | 0.00017674 | 0.000377947 | 1 |
| Obesity | LEC | *LRRC45* | -1.170665499 | 0.013550358 | 0.017934969 | 1 |
| Obesity | LEC | *FASN* | 1.410804291 | 1.70E-06 | 5.58E-05 | 0.010665244 |
| Obesity | LEC | *MT1E* | 0.917025711 | 5.04E-05 | 0.000152943 | 0.317110636 |
| Obesity | LEC | *DCXR* | 3.28351983 | 4.15E-05 | 0.000134809 | 0.260949704 |
| Obesity | LEC | *LIMS1* | 1.215505518 | 0.001878013 | 0.002907429 | 1 |
| Obesity | LEC | *UGP2* | 0.981117883 | 0.02861104 | 0.036120002 | 1 |
| Obesity | LEC | *CSGALNACT2* | 1.48033602 | 1.66E-05 | 0.000105108 | 0.104070805 |
| Obesity | LEC | *AVEN* | 1.844619271 | 0.005724385 | 0.008074761 | 1 |
| Obesity | LEC | *CTNND2* | 0.622124282 | 0.002816962 | 0.004200721 | 1 |
| Obesity | LEC | *CALML6* | 1.885299187 | 0.007583681 | 0.010442094 | 1 |
| Obesity | LEC | *TPST1* | 0.739666724 | 0.024549362 | 0.031268606 | 1 |
| Obesity | LEC | *BRD3* | 2.135244281 | 1.83E-05 | 0.000105108 | 0.114945834 |
| Obesity | LEC | *ZFPM2* | -0.553207842 | 0.008026002 | 0.011002939 | 1 |
| Obesity | LEC | *ZNF747* | -1.432827256 | 0.000142546 | 0.000319268 | 0.896185155 |
| Obesity | LEC | *MAP3K2* | 1.690500543 | 0.000173999 | 0.000372973 | 1 |
| Obesity | LEC | *PUSL1* | 1.310479413 | 6.06E-05 | 0.000173168 | 0.381049378 |
| Obesity | LEC | *MYO7B* | 1.138271277 | 4.35E-06 | 8.77E-05 | 0.027352171 |
| Obesity | LEC | *CNTROB* | 2.93379866 | 6.59E-05 | 0.000182575 | 0.414080776 |
| Obesity | LEC | *KCNAB3* | -1.374840857 | 0.001286748 | 0.002065301 | 1 |
| Obesity | LEC | *NIPA1* | 1.341491133 | 1.47E-05 | 0.000105108 | 0.092626103 |
| Obesity | LEC | *RNF150* | 0.713786633 | 0.000337231 | 0.000640341 | 1 |
| Obesity | LEC | *CHRNB1* | 0.721962044 | 0.000455365 | 0.000824561 | 1 |
| Obesity | LEC | *PDCD6IP* | 1.402935577 | 0.002039089 | 0.003136715 | 1 |
| Obesity | LEC | *ZNF212* | 2.338391888 | 0.000589028 | 0.001028958 | 1 |
| Obesity | LEC | *FAM161A* | 1.722756362 | 0.000308909 | 0.000594645 | 1 |
| Obesity | LEC | *GLB1* | 1.412901944 | 1.19E-05 | 0.000105108 | 0.075034614 |
| Obesity | LEC | *FAXDC2* | 1.492406565 | 0.002474692 | 0.003737455 | 1 |
| Obesity | LEC | *CRTAP* | 2.298768505 | 0.000185606 | 0.000392765 | 1 |
| Obesity | LEC | *ELP5* | 1.516773281 | 0.000945472 | 0.001564259 | 1 |
| Obesity | LEC | *STX8* | 1.379422772 | 3.74E-05 | 0.000126288 | 0.235264872 |

| | | | | | | |
|---|---|---|---|---|---|---|
| Obesity | LEC | *NFRKB* | 1.290865376 | 0.000142065 | 0.000318305 | 0.893164648 |
| Obesity | LEC | *SP7* | 4.128417233 | 0.001261527 | 0.002026372 | 1 |
| Obesity | LEC | *SEMA3E* | 1.509496384 | 1.22E-05 | 0.000105108 | 0.07665262 |
| Obesity | LEC | *DCLK2* | 1.325615017 | 0.008702273 | 0.011867937 | 1 |
| Obesity | LEC | *MGMT* | 1.431797399 | 2.28E-05 | 0.000109111 | 0.143288885 |
| Obesity | LEC | *METTL7B* | 2.869025284 | 2.67E-05 | 0.000113078 | 0.168101072 |
| Obesity | LEC | *NFXL1* | 2.369650568 | 0.000287204 | 0.000561982 | 1 |
| Obesity | LEC | *CD14* | 0.743275456 | 0.000346107 | 0.000653053 | 1 |
| Obesity | LEC | *KRT72* | 3.136191831 | 0.023481928 | 0.029981902 | 1 |
| Obesity | LEC | *HSD17B13* | 0.655845842 | 0.011647476 | 0.015596951 | 1 |
| Obesity | LEC | *ARL6IP1* | -0.938851385 | 0.002333927 | 0.003545155 | 1 |
| Obesity | LEC | *SERPINB9* | -0.865018222 | 0.000801777 | 0.001348492 | 1 |
| Obesity | LEC | *SMAGP* | -0.41541086 | 5.90E-05 | 0.000170005 | 0.371120195 |
| Obesity | LEC | *IRX2* | 0.71228898 | 5.52E-05 | 0.000162907 | 0.347154834 |
| Obesity | LEC | *EMB* | 0.79328417 | 0.000100799 | 0.000245535 | 0.633725504 |
| Obesity | LEC | *STAT2* | 1.314461485 | 1.67E-05 | 0.000105108 | 0.104685555 |
| Obesity | LEC | *NUDCD2* | 0.85193556 | 3.06E-05 | 0.000115867 | 0.192455502 |
| Obesity | LEC | *HSPA4* | 2.191569682 | 8.80E-05 | 0.000222432 | 0.553410184 |
| Obesity | LEC | *COMMD5* | 1.456167493 | 8.81E-06 | 0.000105108 | 0.055359504 |
| Obesity | LEC | *GTSF1* | 2.438939736 | 2.09E-05 | 0.00010698 | 0.131626607 |
| Obesity | LEC | *ARMC10* | 1.857120317 | 0.000741178 | 0.001259402 | 1 |
| Obesity | LEC | *RNF34* | 1.613093277 | 0.007403759 | 0.01022122 | 1 |
| Obesity | LEC | *ACYP2* | 1.193526902 | 1.99E-05 | 0.000105667 | 0.124840061 |
| Obesity | LEC | *TMEM133* | 1.608601671 | 3.02E-05 | 0.000115831 | 0.189618126 |
| Obesity | LEC | *TTLL6* | -0.62571556 | 8.01E-05 | 0.000208554 | 0.503866492 |
| Obesity | LEC | *KCNS3* | 0.715191076 | 0.01709363 | 0.022319346 | 1 |
| Obesity | LEC | *GPR37* | -0.6754722 | 0.000130068 | 0.000297792 | 0.81773643 |
| Obesity | LEC | *AKAP13* | 1.212369605 | 1.66E-05 | 0.000105108 | 0.104135166 |
| Obesity | LEC | *HTRA3* | 0.8121449 | 2.84E-05 | 0.000114842 | 0.178694004 |
| Obesity | LEC | *FOXN2* | 1.424098816 | 2.22E-05 | 0.000108497 | 0.13941906 |
| Obesity | LEC | *GPR27* | 3.957831374 | 0.000283774 | 0.000556082 | 1 |
| Obesity | LEC | *KBTBD2* | 1.476781055 | 0.023968511 | 0.030565929 | 1 |
| Obesity | LEC | *MTSS1* | 0.871660593 | 8.55E-06 | 0.000105108 | 0.053733321 |
| Obesity | LEC | *RPS9* | 1.88120631 | 6.57E-05 | 0.000182176 | 0.412992289 |
| Obesity | LEC | *NDUFA3* | 1.892312773 | 2.44E-05 | 0.000109899 | 0.153638689 |
| Obesity | LEC | *OSCAR* | 0.828355527 | 1.28E-05 | 0.000105108 | 0.080763201 |
| Obesity | LEC | *PAQR8* | 1.735252357 | 0.000186827 | 0.000394685 | 1 |
| Obesity | LEC | *NUDT6* | 1.347451913 | 2.76E-05 | 0.00011384 | 0.173328259 |
| Obesity | LEC | *ZNF160* | 1.490143382 | 1.36E-05 | 0.000105108 | 0.085506847 |
| Obesity | LEC | *ZNF415* | -0.597182556 | 1.95E-05 | 0.000105217 | 0.122653796 |
| Obesity | LEC | *TSNARE1* | 1.699878586 | 3.53E-07 | 2.44E-05 | 0.002221282 |
| Obesity | LEC | *FPR1* | 0.554015039 | 1.52E-05 | 0.000105108 | 0.095382701 |
| Obesity | LEC | *FEZ2* | 1.322687795 | 0.0125135 | 0.016622095 | 1 |
| Obesity | LEC | *SOX7* | 6.229947859 | 7.09E-06 | 0.000105108 | 0.044592651 |
| Obesity | LEC | *C11orf24* | -1.435623757 | 0.030909945 | 0.038796331 | 1 |
| Obesity | LEC | *ALK* | 0.802643543 | 1.05E-05 | 0.000105108 | 0.06572446 |
| Obesity | LEC | *TRMT61B* | 1.591998472 | 7.60E-07 | 3.73E-05 | 0.004779802 |
| Obesity | LEC | *MFN1* | 1.822795375 | 4.01E-05 | 0.000131769 | 0.252074855 |
| Obesity | LEC | *GIMAP8* | -0.567538912 | 0.003353322 | 0.004935004 | 1 |
| Obesity | LEC | *ATP6V0E2* | 0.858911549 | 5.76E-05 | 0.000166947 | 0.361941929 |

| | | | | | | |
|---|---|---|---|---|---|---|
| Obesity | LEC | *GRIK1* | -1.115235035 | 0.001467931 | 0.002327 | 1 |
| Obesity | LEC | *CANT1* | -1.304443927 | 0.031761706 | 0.039746387 | 1 |
| Obesity | LEC | *CHST11* | 1.239800139 | 0.00012073 | 0.000281252 | 0.759027155 |
| Obesity | LEC | *LURAP1* | -0.979166887 | 0.000172165 | 0.00036942 | 1 |
| Obesity | LEC | *MRPL36* | 1.775916311 | 3.30E-06 | 7.62E-05 | 0.020719035 |
| Obesity | LEC | *ZNF581* | 3.53889457 | 1.26E-05 | 0.000105108 | 0.07919193 |
| Obesity | LEC | *MCC* | 0.982665352 | 8.34E-06 | 0.000105108 | 0.052464343 |
| Obesity | LEC | *DLK2* | 1.863937416 | 0.001933943 | 0.002988129 | 1 |
| Obesity | LEC | *ZNF561* | 1.569878965 | 0.001193788 | 0.001930883 | 1 |
| Obesity | LEC | *HOPX* | 1.110720143 | 3.13E-05 | 0.000116784 | 0.196547241 |
| Obesity | LEC | *RSL1D1* | 2.30891897 | 5.81E-06 | 9.74E-05 | 0.036519889 |
| Obesity | LEC | *PPID* | 1.681333942 | 2.18E-05 | 0.000108281 | 0.137145639 |
| Obesity | LEC | *COL24A1* | 0.413916086 | 0.002228962 | 0.00340381 | 1 |
| Obesity | LEC | *TBCA* | 1.204075742 | 7.84E-05 | 0.000205585 | 0.492994014 |
| Obesity | LEC | *MAP6* | -0.469054382 | 0.000123148 | 0.000285274 | 0.774232617 |
| Obesity | LEC | *BCL2L1* | 1.509719982 | 0.016499293 | 0.021592643 | 1 |
| Obesity | LEC | *ZNF584* | 0.668225938 | 0.000141645 | 0.000317678 | 0.890520577 |
| Obesity | LEC | *NMUR1* | 0.802644761 | 2.73E-05 | 0.000113713 | 0.171478466 |
| Obesity | LEC | *P2RY6* | 0.670720503 | 3.35E-05 | 0.00011987 | 0.21048117 |
| Obesity | LEC | *ZIK1* | -1.982324525 | 0.006319038 | 0.008838219 | 1 |
| Obesity | LEC | *RGS19* | 1.241018113 | 5.25E-07 | 2.90E-05 | 0.003301056 |
| Obesity | LEC | *TCEA2* | -1.163991869 | 0.001788842 | 0.002782398 | 1 |
| Obesity | LEC | *GPHN* | 1.094753663 | 0.002545882 | 0.003828261 | 1 |
| Obesity | LEC | *VAT1L* | 0.828583402 | 3.02E-05 | 0.000115831 | 0.189871023 |
| Obesity | LEC | *CAMTA1* | 1.418719578 | 0.015677249 | 0.020572503 | 1 |
| Obesity | LEC | *LRRC34* | 0.831422435 | 0.000820299 | 0.001374525 | 1 |
| Obesity | LEC | *SPATA5L1* | 1.752126525 | 1.29E-05 | 0.000105108 | 0.080884284 |
| Obesity | LEC | *SYCE1* | 1.040208111 | 0.001163164 | 0.00188669 | 1 |
| Obesity | LEC | *BCL2* | 0.520714326 | 0.000248147 | 0.000498912 | 1 |
| Obesity | LEC | *RHNO1* | 1.311929444 | 0.000716153 | 0.00122316 | 1 |
| Obesity | LEC | *PWWP2B* | 2.006765161 | 2.02E-05 | 0.000106047 | 0.126787475 |
| Obesity | LEC | *NINJ2* | 1.049738492 | 1.00E-05 | 0.000105108 | 0.063084752 |
| Obesity | LEC | *FAM90A1* | -2.681207712 | 0.000440824 | 0.000801731 | 1 |
| Obesity | LEC | *TRAPPC12* | 1.857004722 | 0.000219504 | 0.000450398 | 1 |
| Obesity | LEC | *C3AR1* | 0.852856378 | 9.42E-05 | 0.000234031 | 0.592099313 |
| Obesity | LEC | *AQP4* | 0.851256358 | 2.88E-05 | 0.000115038 | 0.181027207 |
| Obesity | LEC | *TLN2* | 0.542960198 | 0.001546222 | 0.002437587 | 1 |
| Obesity | LEC | *TVP23B* | 1.699638574 | 0.031633697 | 0.039601962 | 1 |
| Obesity | LEC | *SRGAP2C* | 1.930603102 | 1.55E-05 | 0.000105108 | 0.097416772 |
| Obesity | LEC | *ATPAF2* | 2.940349157 | 6.61E-05 | 0.000182825 | 0.415744534 |
| Obesity | LEC | *CYP4F22* | -3.185869924 | 0.00169598 | 0.002649758 | 1 |
| Obesity | LEC | *C20orf196* | 1.740852593 | 2.94E-05 | 0.000115699 | 0.18511876 |
| Obesity | LEC | *ANKRD20A4* | -0.952512707 | 0.000141429 | 0.000317445 | 0.889163083 |
| Obesity | LEC | *LAMB2* | 1.386748867 | 0.000148802 | 0.000330806 | 0.935519401 |
| Obesity | LEC | *ORMDL3* | 0.938509432 | 4.39E-05 | 0.000140462 | 0.276287881 |
| Obesity | LEC | *SERF1A* | -2.013876373 | 0.000136635 | 0.000308779 | 0.859023954 |
| Obesity | LEC | *LRRC15* | -0.209002357 | 7.34E-05 | 0.000195952 | 0.461465795 |
| Obesity | LEC | *MOB3A* | 1.788512565 | 0.00373053 | 0.00543926 | 1 |
| Obesity | LEC | *NME6* | 0.965247032 | 0.003038173 | 0.004507078 | 1 |
| Obesity | LEC | *FRMD3* | 1.873563564 | 2.73E-06 | 7.03E-05 | 0.017177313 |

| | | | | | | |
|---|---|---|---|---|---|---|
| Obesity | LEC | *MTBP* | 1.568412643 | 1.58E-05 | 0.000105108 | 0.099287168 |
| Obesity | LEC | *MALT1* | 1.994840923 | 6.00E-05 | 0.000171891 | 0.377299895 |
| Obesity | LEC | *MBOAT1* | -0.839205273 | 0.010326637 | 0.013938078 | 1 |
| Obesity | LEC | *C5orf34* | 1.743708455 | 1.14E-05 | 0.000105108 | 0.071665174 |
| Obesity | LEC | *C1QTNF4* | -0.801723067 | 1.04E-07 | 1.09E-05 | 0.000655371 |
| Obesity | LEC | *ZNF131* | 1.681119192 | 0.000120449 | 0.000280989 | 0.757264209 |
| Obesity | LEC | *HINFP* | 2.824191564 | 0.000225534 | 0.000460068 | 1 |
| Obesity | LEC | *SPTLC3* | 0.723856138 | 1.86E-05 | 0.000105129 | 0.116942538 |
| Obesity | LEC | *COPRS* | 3.208597792 | 0.000785999 | 0.001323755 | 1 |
| Obesity | LEC | *CLEC12A* | 0.247250199 | 0.016102151 | 0.02108607 | 1 |
| Obesity | LEC | *SUCLG2* | 1.453566999 | 4.83E-05 | 0.000148656 | 0.3037036 |
| Obesity | LEC | *CSDC2* | -1.343408806 | 0.037283578 | 0.046269613 | 1 |
| Obesity | LEC | *RCAN2* | 1.720425852 | 2.90E-05 | 0.000115266 | 0.182260732 |
| Obesity | LEC | *IL16* | 0.647030618 | 1.59E-05 | 0.000105108 | 0.099891801 |
| Obesity | LEC | *GNB2* | 1.533698471 | 0.000192969 | 0.000405209 | 1 |
| Obesity | LEC | *PRSS27* | 7.635243236 | 0.000277903 | 0.000546846 | 1 |
| Obesity | LEC | *FGGY* | 1.721013542 | 3.92E-05 | 0.000130236 | 0.246323071 |
| Obesity | LEC | *ZNF24* | 2.206256615 | 0.000810782 | 0.001361845 | 1 |
| Obesity | LEC | *MANEA* | 1.8078128 | 3.52E-05 | 0.00012278 | 0.221127019 |
| Obesity | LEC | *FIBP* | 1.165584385 | 3.43E-06 | 7.73E-05 | 0.021555583 |
| Obesity | LEC | *CARNS1* | 7.282261697 | 4.45E-06 | 8.83E-05 | 0.027996095 |
| Obesity | LEC | *NIPAL4* | -1.055084644 | 3.39E-05 | 0.000120512 | 0.213155477 |
| Obesity | LEC | *SNTG2* | -0.233443077 | 6.71E-05 | 0.000184604 | 0.42214846 |
| Obesity | LEC | *PDE3A* | 1.219897565 | 0.003570843 | 0.005228123 | 1 |
| Obesity | LEC | *MRPL52* | 2.307308275 | 1.25E-05 | 0.000105108 | 0.078597608 |
| Obesity | LEC | *SMPDL3A* | 1.895065319 | 5.48E-06 | 9.52E-05 | 0.03447336 |
| Obesity | LEC | *RAD9A* | 0.918700517 | 0.001109757 | 0.001808929 | 1 |
| Obesity | LEC | *EFEMP2* | 3.290295281 | 1.01E-07 | 1.09E-05 | 0.000633867 |
| Obesity | LEC | *AGAP5* | -6.710058118 | 0.000100501 | 0.000245094 | 0.631852576 |
| Obesity | LEC | *ZMAT3* | 2.582130013 | 0.000171735 | 0.000368675 | 1 |
| Obesity | LEC | *ZFAND4* | 1.636131618 | 0.006666369 | 0.009288887 | 1 |
| Obesity | LEC | *ZNF738* | 1.418893944 | 1.42E-05 | 0.000105108 | 0.088986546 |
| Obesity | LEC | *SLFN11* | 1.455221932 | 3.72E-05 | 0.000126137 | 0.233585792 |
| Obesity | LEC | *CCL19* | 4.582690891 | 5.98E-05 | 0.000171484 | 0.375893265 |
| Obesity | LEC | *CFL1* | 2.19951064 | 0.001015244 | 0.001670026 | 1 |
| Obesity | LEC | *FAM192A* | 1.16731878 | 0.000109826 | 0.000260967 | 0.690473529 |
| Obesity | LEC | *CBWD1* | 2.442727111 | 0.002204274 | 0.003370202 | 1 |
| Obesity | LEC | *RAB37* | 1.621561666 | 7.40E-06 | 0.000105108 | 0.046552346 |
| Obesity | LEC | *DCP2* | 1.88408233 | 0.000315027 | 0.000604212 | 1 |
| Obesity | LEC | *SNX32* | 2.89436821 | 2.89E-10 | 8.26E-08 | 1.82E-06 |
| Obesity | LEC | *CYP7B1* | 1.36699331 | 0.00648016 | 0.009047471 | 1 |
| Obesity | LEC | *METAP1D* | 1.902095363 | 1.09E-05 | 0.000105108 | 0.068300337 |
| Obesity | LEC | *NADSYN1* | 2.282320686 | 0.000480259 | 0.000862683 | 1 |
| Obesity | LEC | *RNASEH2C* | 1.585557539 | 0.006984943 | 0.009694114 | 1 |
| Obesity | LEC | *MYEOV* | 0.446207073 | 0.001542486 | 0.002433528 | 1 |
| Obesity | LEC | *ANKRD13D* | 1.541069391 | 5.26E-05 | 0.000157018 | 0.33083651 |
| Obesity | LEC | *MRGPRF* | 2.244111433 | 0.018839987 | 0.024467465 | 1 |
| Obesity | LEC | *ADH6* | 0.46325494 | 0.008436638 | 0.011538209 | 1 |
| Obesity | LEC | *GXYLT2* | 0.772982115 | 3.76E-05 | 0.000126834 | 0.236672122 |
| Obesity | LEC | *HPSE2* | 2.592947773 | 4.54E-11 | 1.59E-08 | 2.86E-07 |

| | | | | | | |
|---|---|---|---|---|---|---|
| Obesity | LEC | *DCAKD* | 0.672407929 | 0.009479782 | 0.012855779 | 1 |
| Obesity | LEC | *HECTD4* | 1.434499124 | 0.000375364 | 0.000699739 | 1 |
| Obesity | LEC | *BNC2* | 1.066563768 | 6.93E-06 | 0.000104897 | 0.043579889 |
| Obesity | LEC | *NOC3L* | 2.385650879 | 7.19E-05 | 0.000193228 | 0.451961168 |
| Obesity | LEC | *RHOD* | -0.735051765 | 0.003031153 | 0.004499102 | 1 |
| Obesity | LEC | *PARP14* | 0.745502762 | 0.009928648 | 0.013432626 | 1 |
| Obesity | LEC | *PARP15* | 1.325708881 | 0.000240087 | 0.000485034 | 1 |
| Obesity | LEC | *AHSA2* | 1.690972828 | 0.000297605 | 0.000576415 | 1 |
| Obesity | LEC | *ABLIM3* | 1.594410957 | 0.011091121 | 0.014909103 | 1 |
| Obesity | LEC | *GLRX* | 0.929451813 | 1.30E-05 | 0.000105108 | 0.081620837 |
| Obesity | LEC | *GOLGB1* | 1.439618756 | 1.59E-05 | 0.000105108 | 0.099794603 |
| Obesity | LEC | *DMRT2* | 0.542066859 | 0.000138158 | 0.00031166 | 0.868596983 |
| Obesity | LEC | *ZNF483* | 0.84011488 | 0.012482 | 0.016587262 | 1 |
| Obesity | LEC | *C1QA* | 1.012626737 | 1.53E-05 | 0.000105108 | 0.095988564 |
| Obesity | LEC | *NDNF* | -1.061647486 | 0.002521284 | 0.003796722 | 1 |
| Obesity | LEC | *OLR1* | 6.1096879 | 0.030398965 | 0.038208375 | 1 |
| Obesity | LEC | *GLIPR1L1* | 1.650398268 | 6.62E-06 | 0.000102716 | 0.04159999 |
| Obesity | LEC | *ARV1* | 1.927567285 | 2.35E-06 | 6.63E-05 | 0.014794107 |
| Obesity | LEC | *CCDC36* | 1.613719216 | 1.55E-06 | 5.36E-05 | 0.009758766 |
| Obesity | LEC | *THAP2* | 1.461162119 | 1.91E-05 | 0.000105211 | 0.120239971 |
| Obesity | LEC | *PPP1R14B* | 1.782762531 | 2.41E-05 | 0.000109361 | 0.151424451 |
| Obesity | LEC | *SMARCC1* | 1.391737788 | 0.012148184 | 0.016195003 | 1 |
| Obesity | LEC | *ZNF417* | 1.34471679 | 0.000435554 | 0.000793948 | 1 |
| Obesity | LEC | *VEGFB* | 1.198530478 | 0.000884328 | 0.001471617 | 1 |
| Obesity | LEC | *PEAK1* | 1.278322915 | 3.65E-05 | 0.000124801 | 0.229509888 |
| Obesity | LEC | *TNFRSF10C* | 0.733697338 | 0.015420526 | 0.020252527 | 1 |
| Obesity | LEC | *GMPPB* | 1.509634847 | 0.029510349 | 0.037158335 | 1 |
| Obesity | LEC | *CSPG4* | 1.369125636 | 0.005986788 | 0.008403424 | 1 |
| Obesity | LEC | *NUDT18* | -1.926283039 | 0.002293422 | 0.003490377 | 1 |
| Obesity | LEC | *SULT1B1* | 1.588596485 | 9.59E-05 | 0.000237111 | 0.602736421 |
| Obesity | LEC | *NUDT4* | -0.894811979 | 0.00458768 | 0.006573096 | 1 |
| Obesity | LEC | *SCAI* | 1.363809317 | 1.66E-05 | 0.000105108 | 0.104367498 |
| Obesity | LEC | *C1orf100* | -1.99761172 | 0.001430464 | 0.002273338 | 1 |
| Obesity | LEC | *AGFG1* | 1.21034922 | 3.18E-05 | 0.000117047 | 0.199681697 |
| Obesity | LEC | *HAP1* | -2.100469028 | 0.001604102 | 0.0025181 | 1 |
| Obesity | LEC | *RNF213* | 1.533758731 | 0.000204533 | 0.000424531 | 1 |
| Obesity | LEC | *PLK3* | 1.532120735 | 9.51E-06 | 0.000105108 | 0.059815529 |
| Obesity | LEC | *TUBB8* | 2.134719016 | 0.000224359 | 0.000458137 | 1 |
| Obesity | LEC | *GPR160* | 2.09845362 | 0.00028876 | 0.000564325 | 1 |
| Obesity | LEC | *GOLIM4* | 1.778461292 | 0.000420249 | 0.000770743 | 1 |
| Obesity | LEC | *HOXB2* | 1.034467113 | 1.36E-06 | 5.03E-05 | 0.008558053 |
| Obesity | LEC | *UBXN2A* | 1.42886172 | 0.000159815 | 0.000348996 | 1 |
| Obesity | LEC | *CCS* | 1.641464012 | 4.68E-06 | 9.00E-05 | 0.029427305 |
| Obesity | LEC | *NRROS* | 0.882206745 | 0.000180465 | 0.000384344 | 1 |
| Obesity | LEC | *SLC25A30* | 2.464742823 | 9.55E-06 | 0.000105108 | 0.060026191 |
| Obesity | LEC | *CTSF* | 0.794347906 | 0.003330919 | 0.004908929 | 1 |
| Obesity | LEC | *C16orf91* | 1.694206017 | 2.40E-05 | 0.000109361 | 0.150875424 |
| Obesity | LEC | *SOCS7* | 1.843018215 | 2.05E-05 | 0.000106468 | 0.129039797 |
| Obesity | LEC | *TLR6* | 1.5735684 | 2.88E-06 | 7.13E-05 | 0.01808625 |
| Obesity | LEC | *RGMB* | 1.994538245 | 0.012351369 | 0.016437988 | 1 |

| | | | | | | |
|---|---|---|---|---|---|---|
| Obesity | LEC | *FAM53A* | 0.839862585 | 1.91E-05 | 0.000105211 | 0.119892251 |
| Obesity | LEC | *CTU2* | 3.312694151 | 7.93E-05 | 0.000206961 | 0.498361647 |
| Obesity | LEC | *MGA* | 1.147483041 | 0.017440479 | 0.022742615 | 1 |
| Obesity | LEC | *SNX31* | 1.464350277 | 0.029297719 | 0.036920176 | 1 |
| Obesity | LEC | *RALGAPA1* | 1.390630563 | 0.000671666 | 0.001152815 | 1 |
| Obesity | LEC | *LIG4* | 1.384823251 | 4.67E-05 | 0.000145875 | 0.293792906 |
| Obesity | LEC | *LINGO2* | 0.730664718 | 1.09E-05 | 0.000105108 | 0.068801532 |
| Obesity | LEC | *BBS1* | 1.84357319 | 8.76E-07 | 3.91E-05 | 0.005505169 |
| Obesity | LEC | *ANKRD36C* | 1.46648913 | 0.000557916 | 0.000981151 | 1 |
| Obesity | LEC | *TMEM81* | 2.821419995 | 3.81E-05 | 0.000127862 | 0.239347278 |
| Obesity | LEC | *IL20RB* | 1.147449139 | 0.000184494 | 0.000391201 | 1 |
| Obesity | LEC | *ZNF497* | 4.634377364 | 1.21E-05 | 0.000105108 | 0.076051416 |
| Obesity | LEC | *KLF14* | -1.04567973 | 0.000182199 | 0.000387381 | 1 |
| Obesity | LEC | *ANGEL2* | 1.828374986 | 0.003830251 | 0.005570389 | 1 |
| Obesity | LEC | *ZNF266* | 1.238396537 | 3.62E-05 | 0.000124065 | 0.227410853 |
| Obesity | LEC | *BRSK2* | -1.158707485 | 0.000130311 | 0.000298206 | 0.819265701 |
| Obesity | LEC | *SH3PXD2B* | 0.975055625 | 6.72E-06 | 0.000103236 | 0.042223433 |
| Obesity | LEC | *LARP7* | 1.853700821 | 6.29E-05 | 0.000176749 | 0.395387329 |
| Obesity | LEC | *FGFBP3* | -1.447391476 | 0.000758045 | 0.001284243 | 1 |
| Obesity | LEC | *HRAS* | 1.555407208 | 1.20E-06 | 4.71E-05 | 0.007531838 |
| Obesity | LEC | *SRP72* | 1.950338163 | 1.62E-06 | 5.43E-05 | 0.010215919 |
| Obesity | LEC | *C4orf26* | 3.92293872 | 6.21E-05 | 0.00017578 | 0.390407251 |
| Obesity | LEC | *BTC* | 1.220444765 | 2.36E-05 | 0.000109118 | 0.148112577 |
| Obesity | LEC | *NLRP6* | -4.880719344 | 3.13E-05 | 0.000116803 | 0.196755048 |
| Obesity | LEC | *NDUFA11* | 2.759098354 | 0.000581731 | 0.001019043 | 1 |
| Obesity | LEC | *PTDSS2* | 2.203131464 | 2.58E-05 | 0.000112124 | 0.162277463 |
| Obesity | LEC | *FUT1* | -1.406679823 | 0.008619728 | 0.011760467 | 1 |
| Obesity | LEC | *DHX36* | 1.25585565 | 0.020529775 | 0.026497781 | 1 |
| Obesity | LEC | *FBXW8* | 1.720822723 | 2.31E-05 | 0.000109118 | 0.144940351 |
| Obesity | LEC | *SLC22A1* | 1.738992152 | 0.025822843 | 0.032804246 | 1 |
| Obesity | LEC | *CHST2* | -2.375106872 | 0.001617527 | 0.002537907 | 1 |
| Obesity | LEC | *ATR* | 1.663753983 | 6.18E-05 | 0.000175234 | 0.388570224 |
| Obesity | LEC | *TVP23C* | 0.947542999 | 2.49E-05 | 0.000111016 | 0.156574649 |
| Obesity | LEC | *PACS1* | 1.416856041 | 0.000204183 | 0.000424092 | 1 |
| Obesity | LEC | *SH3BP5L* | 1.107944996 | 1.17E-05 | 0.000105108 | 0.073312603 |
| Obesity | LEC | *CADM2* | 1.789931129 | 2.65E-08 | 3.84E-06 | 0.000166916 |
| Obesity | LEC | *PSMD2* | 1.770947237 | 0.001181595 | 0.001913336 | 1 |
| Obesity | LEC | *ARHGAP1* | 1.059736932 | 2.37E-05 | 0.000109118 | 0.149019859 |
| Obesity | LEC | *CHST1* | -0.527324498 | 4.88E-05 | 0.000149724 | 0.307083949 |
| Obesity | LEC | *GOLGA8A* | 1.562190648 | 9.63E-07 | 4.17E-05 | 0.006051452 |
| Obesity | LEC | *TP53I11* | 2.464869044 | 0.00373888 | 0.005450113 | 1 |
| Obesity | LEC | *PHYHD1* | 1.454363748 | 1.51E-05 | 0.000105108 | 0.095060288 |
| Obesity | LEC | *ZNF519* | 1.664376448 | 1.15E-05 | 0.000105108 | 0.072522295 |
| Obesity | LEC | *BANF1* | -0.910103249 | 0.007208206 | 0.00997315 | 1 |
| Obesity | LEC | *CHRNA7* | -1.824988549 | 6.51E-05 | 0.000181 | 0.409420915 |
| Obesity | LEC | *TMEM9B* | 2.165965351 | 4.90E-07 | 2.77E-05 | 0.003081434 |
| Obesity | LEC | *NRIP3* | 1.168902844 | 0.00111166 | 0.001811043 | 1 |
| Obesity | LEC | *SCUBE2* | 2.028904136 | 0.001034849 | 0.001700051 | 1 |
| Obesity | LEC | *SMAD2* | 1.383121466 | 0.000355943 | 0.000668602 | 1 |
| Obesity | LEC | *EIF3F* | 1.682027451 | 8.02E-05 | 0.000208562 | 0.50430301 |

| | | | | | | |
|---|---|---|---|---|---|---|
| Obesity | LEC | *CLTB* | 1.933292079 | 0.00013143 | 0.000300036 | 0.826300368 |
| Obesity | LEC | *PCSK1* | 0.739228649 | 7.78E-05 | 0.000204061 | 0.488930888 |
| Obesity | LEC | *LPL* | 0.534237026 | 0.004177313 | 0.006031872 | 1 |
| Obesity | LEC | *PPP2R2D* | 1.793005157 | 2.64E-05 | 0.000112582 | 0.166025184 |
| Obesity | LEC | *MCTP1* | 1.096260788 | 5.90E-05 | 0.000169976 | 0.370717842 |
| Obesity | LEC | *LRRC25* | 0.490943741 | 0.023150294 | 0.029588514 | 1 |
| Obesity | LEC | *UBQLNL* | -3.185112304 | 0.000899041 | 0.001494123 | 1 |
| Obesity | LEC | *LIPT2* | 1.906970107 | 0.001298971 | 0.002082262 | 1 |
| Obesity | LEC | *KCNE3* | 1.056788732 | 2.90E-05 | 0.000115266 | 0.182014631 |
| Obesity | LEC | *UCP3* | 1.575089031 | 0.000485006 | 0.000869718 | 1 |
| Obesity | LEC | *UCP2* | 1.771717843 | 7.34E-05 | 0.000195935 | 0.461231175 |
| Obesity | LEC | *RAB6A* | 1.553720356 | 0.001059095 | 0.001734896 | 1 |
| Obesity | LEC | *TOM1L2* | 1.741843262 | 4.83E-05 | 0.000148656 | 0.30361957 |
| Obesity | LEC | *TEX26* | 2.044956873 | 5.99E-06 | 9.92E-05 | 0.03767753 |
| Obesity | LEC | *GPR156* | 1.205780394 | 2.49E-05 | 0.000111016 | 0.156642873 |
| Obesity | LEC | *B3GNTL1* | 1.230858158 | 8.35E-06 | 0.000105108 | 0.052522843 |
| Obesity | LEC | *LINC01106* | -3.432176305 | 0.000443793 | 0.000805929 | 1 |
| Obesity | LEC | *MSRA* | 1.284056264 | 0.000583297 | 0.001020943 | 1 |
| Obesity | LEC | *CTDNEP1* | 1.53252745 | 5.97E-06 | 9.92E-05 | 0.0375608 |
| Obesity | LEC | *ETV4* | 1.400612719 | 2.19E-06 | 6.31E-05 | 0.013747418 |
| Obesity | LEC | *SWI5* | 1.894421717 | 2.87E-05 | 0.000115038 | 0.180658383 |
| Obesity | LEC | *A2M* | 1.195692028 | 1.56E-05 | 0.000105108 | 0.098155908 |
| Obesity | LEC | *ARL4D* | 1.024858774 | 9.33E-05 | 0.000232753 | 0.586537257 |
| Obesity | LEC | *LRRN1* | 2.756663266 | 2.54E-05 | 0.000111691 | 0.159941811 |
| Obesity | LEC | *B3GALT6* | -1.201962801 | 0.000262252 | 0.000521766 | 1 |
| Obesity | LEC | *ZNF613* | 1.190820677 | 6.24E-06 | 9.96E-05 | 0.03921012 |
| Obesity | LEC | *IP6K1* | 2.445059661 | 1.55E-05 | 0.000105108 | 0.09717944 |
| Obesity | LEC | *SSNA1* | 3.330974344 | 0.000608801 | 0.001059672 | 1 |
| Obesity | LEC | *CHMP6* | 3.027374243 | 0.00028265 | 0.00055428 | 1 |
| Obesity | LEC | *UFSP1* | 1.397738298 | 0.00079067 | 0.001330909 | 1 |
| Obesity | LEC | *TMEM39A* | 1.572954635 | 0.007432978 | 0.010257053 | 1 |
| Obesity | LEC | *TCP11L1* | 1.779606077 | 5.15E-06 | 9.31E-05 | 0.032405747 |
| Obesity | LEC | *CIDEA* | 0.306804968 | 0.008507049 | 0.01162439 | 1 |
| Obesity | LEC | *SMIM19* | 1.553868384 | 5.92E-07 | 3.13E-05 | 0.003719345 |
| Obesity | LEC | *SLC35G1* | 1.227430374 | 0.000797697 | 0.001342377 | 1 |
| Obesity | LEC | *COX8A* | 2.594176316 | 0.010666816 | 0.014369461 | 1 |
| Obesity | LEC | *B3GNT4* | 5.347172302 | 3.05E-05 | 0.000115831 | 0.192034862 |
| Obesity | LEC | *CDC26* | -1.159811537 | 0.000182502 | 0.000387685 | 1 |
| Obesity | LEC | *CRLF3* | 1.560544367 | 0.000160846 | 0.000351102 | 1 |
| Obesity | LEC | *RNPEP* | 1.409175772 | 0.000132578 | 0.000302087 | 0.833519256 |
| Obesity | LEC | *SPRYD4* | 1.231009722 | 0.000397381 | 0.000734372 | 1 |
| Obesity | LEC | *SLCO3A1* | 1.712939505 | 0.000133397 | 0.000303202 | 0.838663979 |
| Obesity | LEC | *WDR25* | 1.711228754 | 3.89E-05 | 0.000129587 | 0.244531005 |
| Obesity | LEC | *PLA2G16* | 0.790368242 | 2.25E-05 | 0.000109111 | 0.14161722 |
| Obesity | LEC | *PHLDB3* | -0.6292765 | 0.000455037 | 0.000824206 | 1 |
| Obesity | LEC | *RMDN1* | 1.807846402 | 2.62E-05 | 0.000112582 | 0.164713489 |
| Obesity | LEC | *HORMAD2* | 1.098476371 | 0.004118643 | 0.005952622 | 1 |
| Obesity | LEC | *RNF152* | -0.248387023 | 0.000104864 | 0.000252502 | 0.659281908 |
| Obesity | LEC | *LRRC37A* | 1.046362073 | 0.000480541 | 0.000862942 | 1 |
| Obesity | LEC | *ACSF3* | 1.228007428 | 0.015470039 | 0.02030907 | 1 |

| | | | | | | |
|---|---|---|---|---|---|---|
| Obesity | LEC | *BOK* | 1.20905102 | 5.66E-05 | 0.000165507 | 0.355839834 |
| Obesity | LEC | *ZNF843* | 1.097201233 | 3.22E-05 | 0.000117652 | 0.202478744 |
| Obesity | LEC | *C8orf59* | 2.161047316 | 1.61E-05 | 0.000105108 | 0.101025605 |
| Obesity | LEC | *RUFY1* | 2.149272199 | 7.01E-08 | 8.31E-06 | 0.000440527 |
| Obesity | LEC | *BASP1* | 1.189126439 | 5.70E-05 | 0.000166389 | 0.35856893 |
| Obesity | LEC | *LRRC37A3* | 1.384014577 | 1.63E-05 | 0.000105108 | 0.102648953 |
| Obesity | LEC | *VSIG10* | 0.94543728 | 1.73E-05 | 0.000105108 | 0.108763459 |
| Obesity | LEC | *IRX5* | 1.04675434 | 9.50E-06 | 0.000105108 | 0.059749869 |
| Obesity | LEC | *PNMA1* | 2.631556607 | 0.000263423 | 0.000523763 | 1 |
| Obesity | LEC | *ANKLE2* | 2.245999908 | 1.88E-05 | 0.000105211 | 0.118000282 |
| Obesity | LEC | *GCNT4* | 13.18161402 | 0.000300081 | 0.000580316 | 1 |
| Obesity | LEC | *MUC20* | 2.854612239 | 0.000936409 | 0.001550489 | 1 |
| Obesity | LEC | *THAP4* | 1.431574875 | 7.57E-05 | 0.000200562 | 0.475621713 |
| Obesity | LEC | *SHMT1* | 0.931510477 | 2.83E-05 | 0.000114535 | 0.177987961 |
| Obesity | LEC | *DPP7* | 2.158496657 | 7.69E-05 | 0.00020244 | 0.483202941 |
| Obesity | LEC | *SEC24C* | 2.224041553 | 0.000193267 | 0.000405429 | 1 |
| Obesity | LEC | *MTHFR* | 2.217165385 | 2.54E-05 | 0.000111691 | 0.159635632 |
| Obesity | LEC | *C19orf18* | -1.456718018 | 0.007352704 | 0.010159659 | 1 |
| Obesity | LEC | *WDR73* | 1.559795779 | 9.50E-05 | 0.000235521 | 0.597517128 |
| Obesity | LEC | *POLE* | 2.200532656 | 1.96E-05 | 0.00010532 | 0.123453796 |
| Obesity | LEC | *DSCAML1* | 1.972479225 | 0.000100677 | 0.000245333 | 0.632957997 |
| Obesity | LEC | *TALDO1* | 0.867307583 | 0.001659493 | 0.002599211 | 1 |
| Obesity | LEC | *RIMKLA* | -0.88483797 | 0.005470975 | 0.007743364 | 1 |
| Obesity | LEC | *B3GNT8* | -3.595494846 | 0.002509066 | 0.003782853 | 1 |
| Obesity | LEC | *AP006621.1* | 1.113632994 | 2.67E-07 | 2.02E-05 | 0.001677687 |
| Obesity | LEC | *MAN1B1* | 2.394171745 | 3.66E-05 | 0.000124867 | 0.229836379 |
| Obesity | LEC | *FBXO39* | -0.550390086 | 0.00015368 | 0.000338674 | 0.966188745 |
| Obesity | LEC | *ZBTB38* | 1.812922711 | 9.92E-05 | 0.000242787 | 0.623719468 |
| Obesity | LEC | *C8orf31* | 1.17278354 | 2.20E-05 | 0.000108376 | 0.138336671 |
| Obesity | LEC | *AC024940.1* | -0.719035327 | 0.000379796 | 0.000706652 | 1 |
| Obesity | LEC | *TIMM22* | 1.945288018 | 2.37E-05 | 0.000109118 | 0.14916374 |
| Obesity | LEC | *NAP1L5* | 1.235189191 | 1.01E-05 | 0.000105108 | 0.063434539 |
| Obesity | LEC | *NIM1K* | -0.778371049 | 0.00020767 | 0.000430029 | 1 |
| Obesity | LEC | *ACOT4* | -0.940202669 | 0.002794606 | 0.004170351 | 1 |
| Obesity | LEC | *ARIH2* | 1.013652553 | 0.01715536 | 0.022395296 | 1 |
| Obesity | LEC | *IRX3* | -0.760892788 | 6.36E-05 | 0.000178132 | 0.400067138 |
| Obesity | LEC | *ATOX1* | 2.242639121 | 1.35E-05 | 0.000105108 | 0.085159555 |
| Obesity | LEC | *TBL1XR1* | 1.263105889 | 0.000735868 | 0.001251733 | 1 |
| Obesity | LEC | *C18orf32* | 1.286852923 | 3.53E-05 | 0.000122857 | 0.22175629 |
| Obesity | LEC | *RPLP2* | 1.78215766 | 0.000189862 | 0.000399887 | 1 |
| Obesity | LEC | *C12orf54* | -1.257052225 | 0.0001365 | 0.000308585 | 0.858175721 |
| Obesity | LEC | *GBA* | 3.176783749 | 0.000165307 | 0.000358128 | 1 |
| Obesity | LEC | *ACAD9* | 0.501472862 | 0.032388928 | 0.040474894 | 1 |
| Obesity | LEC | *IL17RA* | 1.183184603 | 0.00282196 | 0.004207177 | 1 |
| Obesity | LEC | *CD163L1* | 0.458428621 | 1.89E-05 | 0.000105211 | 0.118743386 |
| Obesity | LEC | *THAP5* | 1.540712081 | 3.17E-05 | 0.000117047 | 0.199571923 |
| Obesity | LEC | *CD151* | 2.448444136 | 3.71E-06 | 7.97E-05 | 0.023343005 |
| Obesity | LEC | *POLR2L* | 1.479028332 | 0.003198403 | 0.00472693 | 1 |
| Obesity | LEC | *FAM87B* | 1.680794298 | 0.002237447 | 0.003415107 | 1 |
| Obesity | LEC | *PCDHB9* | 1.186238206 | 0.004355939 | 0.006278265 | 1 |

| | | | | | | |
|---|---|---|---|---|---|---|
| Obesity | LEC | *ZNF619* | 3.797595475 | 0.001111173 | 0.001810768 | 1 |
| Obesity | LEC | *ZBTB41* | 1.151493931 | 1.84E-05 | 0.000105108 | 0.115762715 |
| Obesity | LEC | *ZNF354C* | 2.692535672 | 0.000512395 | 0.000911295 | 1 |
| Obesity | LEC | *CENPBD1* | -0.569268901 | 0.000109833 | 0.000260967 | 0.690517936 |
| Obesity | LEC | *BET1L* | 1.927778172 | 7.56E-06 | 0.000105108 | 0.047541786 |
| Obesity | LEC | *RIC8A* | 1.354954946 | 0.000126315 | 0.000290753 | 0.794139948 |
| Obesity | LEC | *IMP3* | 2.582855661 | 1.89E-05 | 0.000105211 | 0.1187428 |
| Obesity | LEC | *ODF3B* | 2.771105858 | 1.56E-05 | 0.000105108 | 0.098333871 |
| Obesity | LEC | *DPY19L2* | -0.652346051 | 0.000583979 | 0.001021842 | 1 |
| Obesity | LEC | *FAM26E* | 0.933973878 | 2.77E-05 | 0.00011384 | 0.173928482 |
| Obesity | LEC | *MLF1* | 1.147019131 | 2.31E-05 | 0.000109118 | 0.145215047 |
| Obesity | LEC | *GRAMD1C* | 1.365046047 | 2.32E-05 | 0.000109118 | 0.14580502 |
| Obesity | LEC | *STAP2* | -0.975045693 | 0.000228605 | 0.000465276 | 1 |
| Obesity | LEC | *GOLGA8Q* | 28.62316466 | 0.001257741 | 0.002021841 | 1 |
| Obesity | LEC | *ZNF114* | 1.284572703 | 7.04E-05 | 0.000190166 | 0.442750296 |
| Obesity | LEC | *ZNF518B* | -0.909591675 | 0.000345557 | 0.000652212 | 1 |
| Obesity | LEC | *LCORL* | 1.676872716 | 0.000769682 | 0.001300104 | 1 |
| Obesity | LEC | *ZNF454* | -0.741408032 | 0.031502439 | 0.039484816 | 1 |
| Obesity | LEC | *SH2B1* | -2.909132537 | 1.71E-15 | 1.34E-12 | 1.08E-11 |
| Obesity | LEC | *ZC3H12D* | 0.79266172 | 2.75E-05 | 0.00011384 | 0.172701947 |
| Obesity | LEC | *KDELC2* | 1.50638741 | 2.11E-05 | 0.000106989 | 0.132880325 |
| Obesity | LEC | *PLEC* | -0.973716041 | 0.04047344 | 0.049883654 | 1 |
| Obesity | LEC | *SH2D4B* | -1.708054702 | 0.000280615 | 0.000551112 | 1 |
| Obesity | LEC | *RNF212* | 0.831543757 | 0.000731805 | 0.001246169 | 1 |
| Obesity | LEC | *PRSS36* | 0.96444114 | 3.03E-05 | 0.000115831 | 0.190338267 |
| Obesity | LEC | *GEN1* | 0.945324135 | 0.000672503 | 0.001153937 | 1 |
| Obesity | LEC | *AQP11* | 1.691313599 | 8.83E-06 | 0.000105108 | 0.05551177 |
| Obesity | LEC | *KCNG2* | 0.682351478 | 0.00715422 | 0.009904995 | 1 |
| Obesity | LEC | *ZFAND2A* | -1.128665999 | 0.002508013 | 0.003782853 | 1 |
| Obesity | LEC | *FAM220A* | 1.033627568 | 0.014197249 | 0.018728096 | 1 |
| Obesity | LEC | *BEND3* | 1.869270566 | 1.84E-05 | 0.000105108 | 0.115698035 |
| Obesity | LEC | *NT5DC1* | 1.37277594 | 6.61E-05 | 0.000182814 | 0.41553695 |
| Obesity | LEC | *AC006111.1* | 3.314045845 | 2.64E-05 | 0.000112582 | 0.166018634 |
| Obesity | LEC | *LINC00843* | 3.416648777 | 3.83E-05 | 0.000128424 | 0.241051176 |
| Obesity | LEC | *COX14* | 2.212152524 | 2.58E-05 | 0.000112124 | 0.162264144 |
| Obesity | LEC | *P4HTM* | 0.606664206 | 0.000237348 | 0.000480737 | 1 |
| Obesity | LEC | *CA8* | -0.612269577 | 0.015634442 | 0.020520613 | 1 |
| Obesity | LEC | *ERBB4* | 1.112241003 | 1.85E-05 | 0.000105108 | 0.116193177 |
| Obesity | LEC | *CSRNP3* | -0.535549571 | 0.000207859 | 0.000430155 | 1 |
| Obesity | LEC | *ZNF713* | -0.96982852 | 0.000295936 | 0.000573889 | 1 |
| Obesity | LEC | *PARP10* | 1.146164925 | 4.45E-05 | 0.000141409 | 0.279553878 |
| Obesity | LEC | *KCTD12* | 1.049884903 | 0.021728731 | 0.027890676 | 1 |
| Obesity | LEC | *RPP25* | -1.362913909 | 4.20E-07 | 2.56E-05 | 0.002638456 |
| Obesity | LEC | *STX19* | 1.160952567 | 3.36E-07 | 2.37E-05 | 0.002113737 |
| Obesity | LEC | *FAM219B* | 1.757662138 | 2.78E-06 | 7.05E-05 | 0.01748432 |
| Obesity | LEC | *CPNE7* | -3.009565468 | 0.008105165 | 0.011099362 | 1 |
| Obesity | LEC | *GDPD4* | 6.282131985 | 0.000643022 | 0.001107938 | 1 |
| Obesity | LEC | *MPI* | 1.812863277 | 7.76E-07 | 3.74E-05 | 0.004880043 |
| Obesity | LEC | *TRIM73* | -3.884646668 | 2.90E-12 | 1.40E-09 | 1.82E-08 |
| Obesity | LEC | *EFCAB13* | 1.040757059 | 0.002741956 | 0.004098592 | 1 |

| | | | | | | |
|---|---|---|---|---|---|---|
| Obesity | LEC | *C17orf62* | 1.710768144 | 4.25E-06 | 8.66E-05 | 0.026717323 |
| Obesity | LEC | *GAK* | 1.850580735 | 0.002011958 | 0.003099658 | 1 |
| Obesity | LEC | *TUFM* | 4.164342745 | 5.60E-22 | 8.81E-19 | 3.52E-18 |
| Obesity | LEC | *RMI1* | 1.786221115 | 0.000265795 | 0.000527146 | 1 |
| Obesity | LEC | *CTC1* | 1.710418938 | 0.005771265 | 0.008129944 | 1 |
| Obesity | LEC | *SNX18* | 1.327334462 | 3.16E-05 | 0.000116919 | 0.198802613 |
| Obesity | LEC | *C14orf39* | 0.549799479 | 0.0002056 | 0.000426463 | 1 |
| Obesity | LEC | *TRIML2* | 13.12071463 | 0.000236771 | 0.000479722 | 1 |
| Obesity | LEC | *FARSA* | 1.385030891 | 0.001770083 | 0.002755946 | 1 |
| Obesity | LEC | *SPTY2D1* | 2.460175911 | 0.001199708 | 0.00193996 | 1 |
| Obesity | LEC | *TCAIM* | 1.453289679 | 0.000376788 | 0.000701679 | 1 |
| Obesity | LEC | *FUCA1* | 1.545073323 | 1.42E-05 | 0.000105108 | 0.089418273 |
| Obesity | LEC | *ZNF664* | 0.885846336 | 2.69E-05 | 0.000113184 | 0.169212639 |
| Obesity | LEC | *CALR* | 2.327262138 | 2.36E-05 | 0.000109118 | 0.148598101 |
| Obesity | LEC | *CDH4* | 0.966264119 | 5.45E-05 | 0.000161076 | 0.34262594 |
| Obesity | LEC | *ARID3B* | 1.911688432 | 3.69E-05 | 0.000125724 | 0.232213052 |
| Obesity | LEC | *EGR3* | -1.913055272 | 0.00975894 | 0.013214399 | 1 |
| Obesity | LEC | *GEMIN4* | 0.943260862 | 0.004854015 | 0.006915293 | 1 |
| Obesity | LEC | *FJX1* | 2.235885772 | 1.22E-05 | 0.000105108 | 0.076468194 |
| Obesity | LEC | *CIITA* | 0.990271272 | 8.48E-05 | 0.000216794 | 0.532833549 |
| Obesity | LEC | *PLD6* | -1.073749446 | 0.000264712 | 0.000525495 | 1 |
| Obesity | LEC | *ZBTB42* | 2.748477718 | 4.91E-06 | 9.11E-05 | 0.030851538 |
| Obesity | LEC | *LACC1* | 3.449836511 | 6.12E-05 | 0.000174262 | 0.384770442 |
| Obesity | LEC | *FCER1A* | 1.039341279 | 0.000120717 | 0.000281252 | 0.758947099 |
| Obesity | LEC | *PCED1B* | 1.132090229 | 1.80E-05 | 0.000105108 | 0.112959612 |
| Obesity | LEC | *PIPOX* | 1.124002255 | 0.000387386 | 0.00071886 | 1 |
| Obesity | LEC | *FOXS1* | 1.195951548 | 0.033464774 | 0.041761221 | 1 |
| Obesity | LEC | *CDH5* | -0.420582586 | 0.003945368 | 0.005719283 | 1 |
| Obesity | LEC | *GIPC3* | -1.611609076 | 8.27E-06 | 0.000105108 | 0.05200829 |
| Obesity | LEC | *CITED4* | 1.04532469 | 3.65E-05 | 0.000124736 | 0.229265305 |
| Obesity | LEC | *TIGD5* | -0.774376379 | 9.11E-05 | 0.000228566 | 0.572557009 |
| Obesity | LEC | *PDXDC1* | 1.630367351 | 0.000929887 | 0.001540907 | 1 |
| Obesity | LEC | *R3HDM2* | 1.565496527 | 0.000107705 | 0.000257468 | 0.677141377 |
| Obesity | LEC | *B3GNT3* | 6.531886606 | 2.23E-08 | 3.42E-06 | 0.000140063 |
| Obesity | LEC | *SSC5D* | 0.657198311 | 0.005711978 | 0.008060877 | 1 |
| Obesity | LEC | *ZADH2* | 1.954180167 | 0.000150672 | 0.000334135 | 0.94727243 |
| Obesity | LEC | *TMEM86B* | 1.070136597 | 3.25E-05 | 0.000118253 | 0.204578188 |
| Obesity | LEC | *SEPT1* | 1.285214196 | 1.85E-05 | 0.000105108 | 0.116047528 |
| Obesity | LEC | *EXOC3* | 1.44402113 | 0.015190484 | 0.019965358 | 1 |
| Obesity | LEC | *LYNX1* | -0.749510588 | 0.019309146 | 0.02501991 | 1 |
| Obesity | LEC | *TDRP* | -0.459747234 | 0.000203956 | 0.000423891 | 1 |
| Obesity | LEC | *ZNF816* | 1.432411993 | 0.002360721 | 0.003582393 | 1 |
| Obesity | LEC | *FGD6* | -0.724980216 | 0.001564845 | 0.002463857 | 1 |
| Obesity | LEC | *HCLS1* | 1.202032512 | 1.21E-05 | 0.000105108 | 0.075885274 |
| Obesity | LEC | *ZNF609* | 1.356711585 | 9.20E-05 | 0.000230166 | 0.578176001 |
| Obesity | LEC | *CCDC66* | 1.451328605 | 6.18E-05 | 0.000175234 | 0.388668049 |
| Obesity | LEC | *DEFB124* | 2.738145131 | 0.000203722 | 0.000423545 | 1 |
| Obesity | LEC | *CYP8B1* | -0.216520898 | 8.48E-05 | 0.000216794 | 0.532879351 |
| Obesity | LEC | *GLIPR1L2* | 0.700899151 | 0.001323377 | 0.002116529 | 1 |
| Obesity | LEC | *RNF182* | 0.822020059 | 0.002651698 | 0.00397407 | 1 |

| | | | | | | |
|---|---|---|---|---|---|---|
| Obesity | LEC | *MB21D2* | 1.453705733 | 6.19E-06 | 9.96E-05 | 0.038892235 |
| Obesity | LEC | *SSTR2* | 1.178603592 | 3.01E-05 | 0.000115831 | 0.189493107 |
| Obesity | LEC | *ZNF594* | 2.019403761 | 0.002397042 | 0.003633126 | 1 |
| Obesity | LEC | *PCGF5* | 1.61323568 | 5.66E-05 | 0.000165507 | 0.355760072 |
| Obesity | LEC | *SHISA2* | -2.455734649 | 8.16E-05 | 0.000211307 | 0.513265585 |
| Obesity | LEC | *CHST13* | 3.267377976 | 1.80E-05 | 0.000105108 | 0.113083564 |
| Obesity | LEC | *SRSF8* | -1.437887677 | 0.002214287 | 0.003383866 | 1 |
| Obesity | LEC | *SLC36A4* | 1.766255446 | 0.00013045 | 0.000298341 | 0.820140092 |
| Obesity | LEC | *ZDHHC20* | 1.723061195 | 0.001983749 | 0.00305907 | 1 |
| Obesity | LEC | *OR51E1* | 0.735654239 | 0.008519022 | 0.011635693 | 1 |
| Obesity | LEC | *PSMG4* | -0.944914893 | 0.006081835 | 0.008527319 | 1 |
| Obesity | LEC | *ZNF443* | 1.840349972 | 0.000181544 | 0.00038651 | 1 |
| Obesity | LEC | *CAPS2* | 1.081004761 | 1.28E-05 | 0.000105108 | 0.080322481 |
| Obesity | LEC | *ZNF792* | 1.342956237 | 0.002510141 | 0.003783567 | 1 |
| Obesity | LEC | *D2HGDH* | 1.155829592 | 6.67E-05 | 0.000183725 | 0.419077779 |
| Obesity | LEC | *CMTR2* | 1.812998027 | 1.75E-05 | 0.000105108 | 0.109837534 |
| Obesity | LEC | *LRRC57* | 1.805275768 | 0.031610241 | 0.039580479 | 1 |
| Obesity | LEC | *MRPL14* | -0.996289722 | 0.008440746 | 0.011538806 | 1 |
| Obesity | LEC | *BBS12* | 1.412589424 | 1.20E-05 | 0.000105108 | 0.075363939 |
| Obesity | LEC | *ZFP82* | 2.051983503 | 0.012433196 | 0.016529394 | 1 |
| Obesity | LEC | *LSMEM1* | 1.629177008 | 0.000307291 | 0.000592075 | 1 |
| Obesity | LEC | *AEN* | 4.102637978 | 0.000409022 | 0.00075285 | 1 |
| Obesity | LEC | *FKRP* | 1.441389812 | 3.59E-06 | 7.81E-05 | 0.022573344 |
| Obesity | LEC | *TRAPPC5* | 1.74008217 | 0.000129597 | 0.000297038 | 0.814775158 |
| Obesity | LEC | *RPH3AL* | 1.870872009 | 0.001255924 | 0.00202047 | 1 |
| Obesity | LEC | *ANKRD34A* | -2.585157635 | 0.000132907 | 0.000302421 | 0.835588533 |
| Obesity | LEC | *SLC26A11* | 1.388156943 | 3.45E-05 | 0.000121618 | 0.216723516 |
| Obesity | LEC | *HIGD1A* | 0.856232033 | 0.000100608 | 0.000245258 | 0.632519608 |
| Obesity | LEC | *OR52N4* | 1.382892648 | 0.014815892 | 0.019499165 | 1 |
| Obesity | LEC | *ADIPOQ* | 0.45412387 | 0.003757848 | 0.005475223 | 1 |
| Obesity | LEC | *HIST3H2A* | 1.133163601 | 0.006699544 | 0.009328911 | 1 |
| Obesity | LEC | *POLR2A* | 1.946947745 | 7.20E-06 | 0.000105108 | 0.045292064 |
| Obesity | LEC | *TMEM132C* | 0.664610557 | 5.86E-05 | 0.000169303 | 0.368573085 |
| Obesity | LEC | *SLC25A41* | -1.594085337 | 8.66E-05 | 0.00021996 | 0.544286649 |
| Obesity | LEC | *TMEM132E* | -3.151164045 | 0.000138808 | 0.000312678 | 0.872683585 |
| Obesity | LEC | *ZNF322* | -1.854386947 | 1.65E-06 | 5.47E-05 | 0.010400841 |
| Obesity | LEC | *CCL13* | 1.132005245 | 0.000291426 | 0.000566892 | 1 |
| Obesity | LEC | *OGFOD3* | 1.020402468 | 7.50E-05 | 0.000199334 | 0.4718229 |
| Obesity | LEC | *ACBD4* | 1.044865447 | 1.71E-05 | 0.000105108 | 0.107578667 |
| Obesity | LEC | *SGSH* | 1.721639556 | 9.41E-06 | 0.000105108 | 0.059154853 |
| Obesity | LEC | *SETD2* | 1.296170251 | 0.000213921 | 0.000439798 | 1 |
| Obesity | LEC | *MRPS23* | 3.263467701 | 8.02E-05 | 0.000208562 | 0.504144537 |
| Obesity | LEC | *GPR135* | 1.053133826 | 6.24E-06 | 9.96E-05 | 0.039239058 |
| Obesity | LEC | *ATG9B* | -1.658097515 | 0.000342502 | 0.000648978 | 1 |
| Obesity | LEC | *HKR1* | 1.780740599 | 0.00454706 | 0.006524617 | 1 |
| Obesity | LEC | *ZBTB20* | 1.039550101 | 0.004111893 | 0.005944233 | 1 |
| Obesity | LEC | *C3orf58* | 1.687900491 | 3.69E-05 | 0.000125724 | 0.232152483 |
| Obesity | LEC | *AMIGO1* | 1.27140556 | 1.64E-05 | 0.000105108 | 0.102842821 |
| Obesity | LEC | *ODF3L2* | 1.458394094 | 0.022192277 | 0.028439226 | 1 |
| Obesity | LEC | *RELL1* | 1.297620657 | 1.32E-05 | 0.000105108 | 0.082928835 |

| | | | | | | |
|---|---|---|---|---|---|---|
| Obesity | LEC | *RFX7* | 1.546565726 | 0.003573286 | 0.005228123 | 1 |
| Obesity | LEC | *CLDN7* | 0.723694534 | 0.028630605 | 0.036137444 | 1 |
| Obesity | LEC | *ADO* | 1.905993685 | 1.21E-05 | 0.000105108 | 0.076072848 |
| Obesity | LEC | *SNRPE* | 1.358565027 | 8.74E-06 | 0.000105108 | 0.05497817 |
| Obesity | LEC | *CHST15* | -0.589787571 | 0.00057765 | 0.001013527 | 1 |
| Obesity | LEC | *IDH2* | 1.398073306 | 3.59E-05 | 0.000123286 | 0.225729 |
| Obesity | LEC | *PTCHD3* | -0.624701018 | 0.000746526 | 0.001268146 | 1 |
| Obesity | LEC | *WRB* | -0.694210515 | 0.010382713 | 0.014007751 | 1 |
| Obesity | LEC | *TNRC18* | 1.699818202 | 7.57E-05 | 0.000200562 | 0.47584108 |
| Obesity | LEC | *FAM181B* | 2.674050186 | 0.002265485 | 0.003452037 | 1 |
| Obesity | LEC | *NOP10* | 0.913661502 | 0.000252297 | 0.0005058 | 1 |
| Obesity | LEC | *FAM89A* | 1.287905111 | 0.004062904 | 0.005881529 | 1 |
| Obesity | LEC | *TDRKH* | 0.948644126 | 0.000344034 | 0.000650705 | 1 |
| Obesity | LEC | *ZNF708* | 1.486790352 | 0.000192054 | 0.000403826 | 1 |
| Obesity | LEC | *IST1* | 1.570792961 | 5.85E-05 | 0.000169202 | 0.368014347 |
| Obesity | LEC | *ERCC6L2* | 1.583726916 | 3.28E-05 | 0.000118709 | 0.205965703 |
| Obesity | LEC | *FAM153B* | 1.209769328 | 9.91E-05 | 0.000242611 | 0.622959298 |
| Obesity | LEC | *BACE2* | -0.457757611 | 0.000496863 | 0.000887942 | 1 |
| Obesity | LEC | *SYNM* | -0.581593399 | 0.000275263 | 0.000543182 | 1 |
| Obesity | LEC | *IZUMO1* | -0.509577312 | 0.01260389 | 0.01673509 | 1 |
| Obesity | LEC | *C8orf33* | 1.829207832 | 0.00769483 | 0.010583547 | 1 |
| Obesity | LEC | *FBXL6* | 2.142757966 | 0.027532592 | 0.034814443 | 1 |
| Obesity | LEC | *KBTBD3* | 0.956289834 | 2.25E-05 | 0.000109111 | 0.141566805 |
| Obesity | LEC | *CLN8* | 1.424653548 | 6.36E-06 | 0.000100677 | 0.039968719 |
| Obesity | LEC | *CACNB4* | 0.628324077 | 4.28E-05 | 0.000137557 | 0.268786132 |
| Obesity | LEC | *TRAPPC6B* | 1.684551048 | 0.000680266 | 0.001166622 | 1 |
| Obesity | LEC | *NPLOC4* | 0.98600184 | 0.002902611 | 0.004322291 | 1 |
| Obesity | LEC | *TSHZ2* | -0.606283495 | 0.03014787 | 0.037915515 | 1 |
| Obesity | LEC | *CAPN12* | 1.434840041 | 9.96E-05 | 0.00024367 | 0.626475916 |
| Obesity | LEC | *FES* | 1.687680672 | 1.14E-05 | 0.000105108 | 0.071365791 |
| Obesity | LEC | *GLRX5* | 2.780866921 | 0.000680675 | 0.001167004 | 1 |
| Obesity | LEC | *MFSD5* | 2.463416604 | 0.005917706 | 0.008319458 | 1 |
| Obesity | LEC | *ADI1* | 1.575724077 | 3.93E-05 | 0.000130538 | 0.247369913 |
| Obesity | LEC | *CLEC4G* | 2.9601757 | 2.10E-05 | 0.000106989 | 0.132268559 |
| Obesity | LEC | *SATB1* | 1.69150765 | 2.59E-05 | 0.000112124 | 0.162721052 |
| Obesity | LEC | *ACTL10* | -0.629432333 | 6.72E-05 | 0.000184641 | 0.42245798 |
| Obesity | LEC | *TRAK1* | 1.467852647 | 0.000515952 | 0.000916844 | 1 |
| Obesity | LEC | *SKA2* | 1.421650663 | 1.41E-05 | 0.000105108 | 0.088364145 |
| Obesity | LEC | *TTC3* | 0.573290569 | 0.00232495 | 0.003532373 | 1 |
| Obesity | LEC | *KCNB2* | 0.411822997 | 2.92E-05 | 0.000115567 | 0.183520501 |
| Obesity | LEC | *BRICD5* | -1.601275208 | 0.001022149 | 0.001680945 | 1 |
| Obesity | LEC | *TSKU* | 2.210617171 | 1.66E-05 | 0.000105108 | 0.104516645 |
| Obesity | LEC | *ACBD3* | 1.620038921 | 5.77E-05 | 0.000167226 | 0.362880156 |
| Obesity | LEC | *PLCXD3* | -0.606229858 | 0.002671341 | 0.004000648 | 1 |
| Obesity | LEC | *VMO1* | 1.010891557 | 1.23E-05 | 0.000105108 | 0.077196094 |
| Obesity | LEC | *COL18A1* | -0.579103036 | 0.025439387 | 0.032356347 | 1 |
| Obesity | LEC | *SLC25A18* | 0.603111194 | 6.47E-05 | 0.000180055 | 0.40692485 |
| Obesity | LEC | *ZNF721* | 1.222975799 | 0.00857168 | 0.01170253 | 1 |
| Obesity | LEC | *LENG9* | 1.323795004 | 3.00E-05 | 0.000115831 | 0.188646692 |
| Obesity | LEC | *CEP63* | 0.764750976 | 0.00175116 | 0.002727159 | 1 |

| | | | | | | |
|---|---|---|---|---|---|---|
| Obesity | LEC | *HMGN4* | 2.010408195 | 0.000617009 | 0.001071923 | 1 |
| Obesity | LEC | *C12orf60* | 1.748043885 | 0.033281892 | 0.041541246 | 1 |
| Obesity | LEC | *LYSMD4* | 1.510273098 | 1.26E-07 | 1.24E-05 | 0.000791105 |
| Obesity | LEC | *WBP2NL* | 2.183372914 | 3.04E-05 | 0.000115831 | 0.191050336 |
| Obesity | LEC | *GAS6* | 1.928843768 | 0.001452845 | 0.002305992 | 1 |
| Obesity | LEC | *BEGAIN* | 7.738827328 | 2.15E-05 | 0.000107557 | 0.135121281 |
| Obesity | LEC | *ARHGEF37* | 1.750954622 | 0.04024445 | 0.049648934 | 1 |
| Obesity | LEC | *FAM43B* | 1.863304457 | 0.001060004 | 0.00173548 | 1 |
| Obesity | LEC | *CSMD1* | 1.246181274 | 1.79E-05 | 0.000105108 | 0.112798773 |
| Obesity | LEC | *PTGDR2* | -1.530511554 | 8.54E-05 | 0.000217903 | 0.536666068 |
| Obesity | LEC | *GPR19* | 1.908420099 | 1.96E-05 | 0.000105217 | 0.122999183 |
| Obesity | LEC | *FANCF* | -1.884727894 | 0.003147726 | 0.004657508 | 1 |
| Obesity | LEC | *C2CD4C* | 2.511550811 | 1.81E-06 | 5.72E-05 | 0.011386816 |
| Obesity | LEC | *RUVBL2* | 2.282153503 | 5.97E-05 | 0.000171484 | 0.375144402 |
| Obesity | LEC | *GDPGP1* | -0.947485735 | 0.030658645 | 0.038503975 | 1 |
| Obesity | LEC | *CTNNA3* | 1.662963452 | 0.000168416 | 0.000363236 | 1 |
| Obesity | LEC | *PLGLB1* | 2.337079232 | 6.44E-05 | 0.000179424 | 0.405139556 |
| Obesity | LEC | *CCBE1* | 1.194722207 | 2.97E-07 | 2.15E-05 | 0.001869261 |
| Obesity | LEC | *ZNF623* | 1.449087216 | 9.61E-06 | 0.000105108 | 0.060409941 |
| Obesity | LEC | *CCDC125* | 1.252375688 | 1.76E-05 | 0.000105108 | 0.110486999 |
| Obesity | LEC | *C10orf107* | 0.453719416 | 0.000580032 | 0.00101635 | 1 |
| Obesity | LEC | *FHL3* | 1.138326357 | 4.48E-05 | 0.000141991 | 0.28142957 |
| Obesity | LEC | *C19orf71* | 1.962376714 | 1.35E-05 | 0.000105108 | 0.084851265 |
| Obesity | LEC | *LRIT3* | 1.495935585 | 0.002193771 | 0.003355776 | 1 |
| Obesity | LEC | *SF3A3* | 2.170816504 | 0.000213082 | 0.000438654 | 1 |
| Obesity | LEC | *GRIN2A* | 1.363769621 | 0.000427859 | 0.000781963 | 1 |
| Obesity | LEC | *URAD* | 3.1176024 | 0.000110982 | 0.000262933 | 0.697746636 |
| Obesity | LEC | *GPR132* | 1.475523512 | 3.55E-05 | 0.000123048 | 0.223389987 |
| Obesity | LEC | *MX2* | 0.982181226 | 0.000191096 | 0.000402217 | 1 |
| Obesity | LEC | *PSMG1* | 1.469863179 | 2.91E-08 | 3.97E-06 | 0.000182687 |
| Obesity | LEC | *ACSM5* | 1.054975306 | 4.91E-05 | 0.000149956 | 0.308458578 |
| Obesity | LEC | *AC131971.1* | -1.706360085 | 0.00014571 | 0.000324851 | 0.916080631 |
| Obesity | LEC | *SERHL2* | 1.522869027 | 6.97E-06 | 0.000104897 | 0.043803485 |
| Obesity | LEC | *PCBP3* | 0.21844044 | 0.005953499 | 0.008364167 | 1 |
| Obesity | LEC | *TANGO2* | 1.385495299 | 3.50E-06 | 7.79E-05 | 0.021997375 |
| Obesity | LEC | *SFXN4* | 1.868015982 | 1.26E-05 | 0.000105108 | 0.079472179 |
| Obesity | LEC | *FAM167B* | 3.488506912 | 0.000115791 | 0.000272243 | 0.727979094 |
| Obesity | LEC | *MRPL54* | -0.941490052 | 0.001736948 | 0.002707039 | 1 |
| Obesity | LEC | *RP1L1* | 2.288840555 | 0.002183527 | 0.00334336 | 1 |
| Obesity | LEC | *NDUFB1* | 1.743551661 | 6.75E-05 | 0.000185198 | 0.424473448 |
| Obesity | LEC | *FAM19A1* | 0.643768362 | 2.21E-05 | 0.000108445 | 0.139177234 |
| Obesity | LEC | *TRMT12* | 1.139014557 | 8.36E-05 | 0.000214782 | 0.525357103 |
| Obesity | LEC | *BMP8A* | 2.601829619 | 1.84E-08 | 3.12E-06 | 0.000115494 |
| Obesity | LEC | *CMTM4* | 1.985193044 | 2.18E-05 | 0.000108281 | 0.137199172 |
| Obesity | LEC | *CBX6* | 2.846014208 | 1.40E-05 | 0.000105108 | 0.087793516 |
| Obesity | LEC | *KREMEN1* | 0.632757206 | 0.001497274 | 0.002369334 | 1 |
| Obesity | LEC | *KCTD16* | 2.946384899 | 1.85E-05 | 0.000105108 | 0.116409715 |
| Obesity | LEC | *B3GALT5* | 2.302719596 | 0.00105868 | 0.001734669 | 1 |
| Obesity | LEC | *ZNF703* | 1.164186688 | 3.44E-05 | 0.000121483 | 0.215998988 |
| Obesity | LEC | *KCTD8* | 0.463176709 | 4.35E-05 | 0.000139427 | 0.273416114 |

| | | | | | | |
|---|---|---|---|---|---|---|
| Obesity | LEC | *C9orf66* | 2.26678709 | 4.13E-05 | 0.000134397 | 0.259654706 |
| Obesity | LEC | *TUBA8* | 1.183515608 | 2.69E-05 | 0.000113184 | 0.168913395 |
| Obesity | LEC | *NPIPA5* | 0.64322251 | 0.000992092 | 0.00163408 | 1 |
| Obesity | LEC | *FAM162B* | -0.468440518 | 0.000323832 | 0.000618637 | 1 |
| Obesity | LEC | *BTBD9* | 1.196721667 | 2.57E-05 | 0.000112124 | 0.161692622 |
| Obesity | LEC | *NUDT14* | 1.27568824 | 0.001549607 | 0.00244231 | 1 |
| Obesity | LEC | *ANKRD45* | 1.876553474 | 3.19E-05 | 0.000117214 | 0.200436409 |
| Obesity | LEC | *MAATS1* | 0.898614141 | 2.38E-06 | 6.66E-05 | 0.014975876 |
| Obesity | LEC | *GPR39* | 0.838957676 | 5.16E-05 | 0.000155003 | 0.324242078 |
| Obesity | LEC | *NPIPA7* | -1.80000442 | 0.016717942 | 0.021846956 | 1 |
| Obesity | LEC | *PP2D1* | 1.503281798 | 7.74E-06 | 0.000105108 | 0.048680739 |
| Obesity | LEC | *DENND5A* | 1.695591062 | 0.000188329 | 0.000397323 | 1 |
| Obesity | LEC | *TMEM236* | 0.954489794 | 3.73E-05 | 0.000126137 | 0.234488864 |
| Obesity | LEC | *VPS33B* | 2.26886727 | 0.000161335 | 0.000351703 | 1 |
| Obesity | LEC | *TBX1* | -0.095726505 | 8.22E-05 | 0.00021261 | 0.517044036 |
| Obesity | LEC | *UQCR10* | 3.00832261 | 0.000709562 | 0.001212892 | 1 |
| Obesity | LEC | *CNTN2* | 1.251677146 | 2.38E-05 | 0.000109181 | 0.149468868 |
| Obesity | LEC | *CRELD2* | 1.214981507 | 1.07E-05 | 0.000105108 | 0.067142089 |
| Obesity | LEC | *UBE2F* | 1.835824132 | 0.001235757 | 0.001990063 | 1 |
| Obesity | LEC | *GOLGA6L4* | -1.030619405 | 9.40E-05 | 0.000233893 | 0.591049988 |
| Obesity | LEC | *PCDH9* | 0.669942568 | 0.000126892 | 0.000291369 | 0.797767653 |
| Obesity | LEC | *OAF* | -0.866908801 | 8.66E-05 | 0.00021996 | 0.544385165 |
| Obesity | LEC | *CLECL1* | 0.923711796 | 0.002094209 | 0.003215997 | 1 |
| Obesity | LEC | *MROH7* | 2.569631274 | 5.97E-05 | 0.000171484 | 0.375234676 |
| Obesity | LEC | *GDF3* | -0.657806499 | 0.000302748 | 0.000584575 | 1 |
| Obesity | LEC | *CSF1* | 1.120183464 | 1.44E-05 | 0.000105108 | 0.090245878 |
| Obesity | LEC | *PLA2G6* | 1.864830884 | 1.35E-10 | 4.04E-08 | 8.49E-07 |
| Obesity | LEC | *A3GALT2* | 1.25738923 | 0.000446114 | 0.000809663 | 1 |
| Obesity | LEC | *SS18L1* | 1.087980017 | 3.15E-05 | 0.000116847 | 0.19803366 |
| Obesity | LEC | *TOP1MT* | -1.212502644 | 0.002474793 | 0.003737455 | 1 |
| Obesity | LEC | *THAP7* | 2.125004538 | 2.16E-05 | 0.000107661 | 0.135842468 |
| Obesity | LEC | *CCR10* | 1.20240455 | 3.39E-05 | 0.000120512 | 0.213186179 |
| Obesity | LEC | *WDR27* | 1.67434683 | 1.33E-05 | 0.000105108 | 0.083903421 |
| Obesity | LEC | *TXNRD2* | -1.085045797 | 0.018074843 | 0.023532106 | 1 |
| Obesity | LEC | *PROS1* | 1.573395699 | 0.002513318 | 0.003785632 | 1 |
| Obesity | LEC | *HDDC3* | 2.082664649 | 0.004576119 | 0.006558027 | 1 |
| Obesity | LEC | *ZFP1* | 3.018515715 | 7.08E-05 | 0.000190901 | 0.445180963 |
| Obesity | LEC | *LPAR5* | 0.783426266 | 0.000389966 | 0.000723007 | 1 |
| Obesity | LEC | *TMEM173* | -1.349757801 | 0.005434702 | 0.007693756 | 1 |
| Obesity | LEC | *SNN* | 2.285077887 | 1.60E-06 | 5.41E-05 | 0.010082921 |
| Obesity | LEC | *NELL2* | 0.522197276 | 1.71E-05 | 0.000105108 | 0.107481447 |
| Obesity | LEC | *KRBA2* | -1.995422846 | 5.98E-05 | 0.000171484 | 0.375726363 |
| Obesity | LEC | *SEPT5* | 2.863290538 | 2.37E-05 | 0.000109118 | 0.148907235 |
| Obesity | LEC | *ATL3* | 1.387399171 | 7.37E-06 | 0.000105108 | 0.046321501 |
| Obesity | LEC | *NDUFA12* | 2.057689764 | 1.88E-05 | 0.000105211 | 0.11842079 |
| Obesity | LEC | *UBE2G2* | 1.423206671 | 6.74E-05 | 0.000184971 | 0.423769402 |
| Obesity | LEC | *OSBP2* | -0.279843678 | 0.000167181 | 0.000361077 | 1 |
| Obesity | LEC | *TUSC5* | 0.738599103 | 0.000139772 | 0.000314399 | 0.878745166 |
| Obesity | LEC | *TMED9* | 2.252156602 | 1.91E-05 | 0.000105211 | 0.119832693 |
| Obesity | LEC | *BTBD6* | 0.817878005 | 0.025860316 | 0.03283858 | 1 |

| | | | | | | |
|---|---|---|---|---|---|---|
| Obesity | LEC | *H1FX* | 1.473875575 | 2.46E-06 | 6.73E-05 | 0.015480068 |
| Obesity | LEC | *RBM43* | 1.778012307 | 3.33E-05 | 0.000119713 | 0.209617266 |
| Obesity | LEC | *FAM227A* | 1.682563565 | 0.001308978 | 0.002096699 | 1 |
| Obesity | LEC | *NOC4L* | -1.269504403 | 0.017774153 | 0.023155015 | 1 |
| Obesity | LEC | *NDUFA6* | 0.541674988 | 0.024882367 | 0.03167992 | 1 |
| Obesity | LEC | *SORCS2* | 1.865624737 | 2.51E-06 | 6.81E-05 | 0.015801415 |
| Obesity | LEC | *TMEM106A* | 1.428714843 | 5.36E-06 | 9.39E-05 | 0.033723231 |
| Obesity | LEC | *SIVA1* | 2.306658431 | 0.001603115 | 0.00251718 | 1 |
| Obesity | LEC | *UBOX5* | 1.384685012 | 0.038937471 | 0.048160512 | 1 |
| Obesity | LEC | *MAFF* | -1.891830479 | 0.01327874 | 0.017586568 | 1 |
| Obesity | LEC | *BRF1* | 1.602191849 | 1.56E-05 | 0.000105108 | 0.098176442 |
| Obesity | LEC | *CIB1* | 1.650824662 | 0.000269909 | 0.000533623 | 1 |
| Obesity | LEC | *FLRT2* | -0.35276957 | 0.000464126 | 0.000838012 | 1 |
| Obesity | LEC | *ADSSL1* | 4.169573109 | 1.35E-05 | 0.000105108 | 0.084994633 |
| Obesity | LEC | *C6orf120* | 1.021146726 | 0.000102944 | 0.000249503 | 0.647211821 |
| Obesity | LEC | *SIGIRR* | 1.616345062 | 0.003287068 | 0.00484885 | 1 |
| Obesity | LEC | *IFITM2* | 1.937991961 | 0.00013635 | 0.000308357 | 0.857232797 |
| Obesity | LEC | *GP1BA* | 3.191007203 | 5.86E-10 | 1.54E-07 | 3.68E-06 |
| Obesity | LEC | *KLHL33* | 1.778764871 | 0.000461614 | 0.000834676 | 1 |
| Obesity | LEC | *CCDC137* | 1.51027139 | 4.40E-05 | 0.000140597 | 0.27681418 |
| Obesity | LEC | *TMEM105* | 11.06503503 | 0.001789673 | 0.002783002 | 1 |
| Obesity | LEC | *GAS2L1* | -1.225397947 | 0.00303801 | 0.004507078 | 1 |
| Obesity | LEC | *ATP6V0A2* | 1.599467377 | 0.006648384 | 0.009265881 | 1 |
| Obesity | LEC | *HS6ST3* | -0.812939342 | 0.000891657 | 0.001483028 | 1 |
| Obesity | LEC | *TNFAIP8L1* | -0.554123452 | 0.000191524 | 0.000402982 | 1 |
| Obesity | LEC | *MAPK11* | 1.163639586 | 3.19E-05 | 0.000117214 | 0.200341076 |
| Obesity | LEC | *SP140L* | 1.951006588 | 0.000989641 | 0.00163047 | 1 |
| Obesity | LEC | *SH3BGR* | -0.464436321 | 0.002601548 | 0.003901701 | 1 |
| Obesity | LEC | *MUC1* | 1.112399743 | 0.000289999 | 0.000565868 | 1 |
| Obesity | LEC | *SV2B* | -0.301960773 | 0.000112927 | 0.000266407 | 0.709974177 |
| Obesity | LEC | *PDE6G* | 2.701956434 | 2.11E-05 | 0.000106989 | 0.132851215 |
| Obesity | LEC | *TLCD2* | 1.430334147 | 0.00142498 | 0.002265196 | 1 |
| Obesity | LEC | *LSAMP* | 1.179458396 | 2.59E-05 | 0.000112124 | 0.162536745 |
| Obesity | LEC | *AHNAK2* | 1.925430332 | 7.71E-06 | 0.000105108 | 0.048496751 |
| Obesity | LEC | *SP1* | 1.450596754 | 1.09E-06 | 4.50E-05 | 0.006883659 |
| Obesity | LEC | *PDIA2* | 1.296100853 | 0.001134966 | 0.00184333 | 1 |
| Obesity | LEC | *PCGF3* | 2.083525819 | 0.000858171 | 0.001432259 | 1 |
| Obesity | LEC | *PSMD13* | 2.06073944 | 0.000296797 | 0.000575203 | 1 |
| Obesity | LEC | *PBX1* | 1.327573636 | 0.000352558 | 0.000663036 | 1 |
| Obesity | LEC | *UBE2L3* | 1.990759425 | 0.00231493 | 0.003519701 | 1 |
| Obesity | LEC | *PMEL* | 1.053192808 | 2.74E-06 | 7.03E-05 | 0.017198272 |
| Obesity | LEC | *SNAI3* | 2.101798931 | 0.000443455 | 0.000805781 | 1 |
| Obesity | LEC | *C6orf201* | 1.225242458 | 0.000275499 | 0.000543477 | 1 |
| Obesity | LEC | *MYBL1* | 0.405705461 | 0.002190682 | 0.003352329 | 1 |
| Obesity | LEC | *C16orf52* | 1.825207191 | 0.000566967 | 0.000996233 | 1 |
| Obesity | LEC | *ADARB2* | 0.676506955 | 6.24E-05 | 0.000175889 | 0.392150628 |
| Obesity | LEC | *SRL* | 0.815618857 | 2.03E-06 | 6.08E-05 | 0.012766795 |
| Obesity | LEC | *KCNIP4* | 0.737865465 | 0.002707409 | 0.004050804 | 1 |
| Obesity | LEC | *SLC52A2* | -1.476629217 | 0.00595975 | 0.008369208 | 1 |
| Obesity | LEC | *ARL17A* | 1.371819839 | 0.000183561 | 0.000389486 | 1 |

| | | | | | | |
|---|---|---|---|---|---|---|
| Obesity | LEC | *NPIPB4* | 0.906575323 | 1.52E-05 | 0.000105108 | 0.095526237 |
| Obesity | LEC | *ZNF829* | 1.050457924 | 1.89E-05 | 0.000105211 | 0.118613357 |
| Obesity | LEC | *IFITM1* | 1.958185581 | 5.35E-05 | 0.00015899 | 0.336422259 |
| Obesity | LEC | *FFAR3* | 10.81334757 | 0.038521242 | 0.047673828 | 1 |
| Obesity | LEC | *POMK* | 1.120940135 | 1.25E-05 | 0.000105108 | 0.078432261 |
| Obesity | LEC | *LINC00839* | -0.237978124 | 9.76E-05 | 0.000240465 | 0.613908189 |
| Obesity | LEC | *PTCH1* | -0.969919826 | 0.019212119 | 0.024909588 | 1 |
| Obesity | LEC | *RTN4RL1* | 2.321645552 | 2.87E-05 | 0.000115038 | 0.180672655 |
| Obesity | LEC | *PLK5* | 2.042157489 | 0.000137501 | 0.00031029 | 0.864467298 |
| Obesity | LEC | *RASA3* | 0.964232789 | 1.18E-05 | 0.000105108 | 0.074065264 |
| Obesity | LEC | *ZNF566* | 1.195417632 | 1.21E-05 | 0.000105108 | 0.076029289 |
| Obesity | LEC | *ZNF529* | 1.733797743 | 0.000546101 | 0.000963145 | 1 |
| Obesity | LEC | *ZNF284* | 1.23993998 | 2.99E-05 | 0.000115831 | 0.188022297 |
| Obesity | LEC | *GSAP* | 1.313204857 | 0.000148687 | 0.000330667 | 0.934794681 |
| Obesity | LEC | *C2orf76* | 1.843934409 | 3.77E-06 | 8.03E-05 | 0.02368143 |
| Obesity | LEC | *WWOX* | 1.237358474 | 0.000116471 | 0.000273534 | 0.732251538 |
| Obesity | LEC | *CYP4Z1* | 1.420013496 | 2.09E-05 | 0.000106951 | 0.131229129 |
| Obesity | LEC | *CCDC84* | 1.737005749 | 3.47E-05 | 0.000121951 | 0.21796839 |
| Obesity | LEC | *FFAR4* | 0.528925635 | 0.007047601 | 0.009765983 | 1 |
| Obesity | LEC | *BPIFB4* | 47.28094122 | 5.70E-06 | 9.65E-05 | 0.035845234 |
| Obesity | LEC | *CYP4F12* | 1.258528028 | 6.88E-05 | 0.000187286 | 0.432790046 |
| Obesity | LEC | *ZNF749* | 0.769438389 | 0.030470423 | 0.038290536 | 1 |
| Obesity | LEC | *KLHL32* | 0.715620467 | 5.55E-05 | 0.000163627 | 0.349017344 |
| Obesity | LEC | *NBPF12* | 1.614628569 | 1.39E-05 | 0.000105108 | 0.087556165 |
| Obesity | LEC | *GPAT2* | -0.496470764 | 4.60E-05 | 0.000144505 | 0.289153545 |
| Obesity | LEC | *TOR3A* | 1.755641423 | 1.47E-05 | 0.000105108 | 0.092641933 |
| Obesity | LEC | *CYP4X1* | -0.64571824 | 0.000557156 | 0.000980363 | 1 |
| Obesity | LEC | *GOLGA8R* | -1.82795205 | 0.001404514 | 0.002236054 | 1 |
| Obesity | LEC | *CCDC30* | 1.571095351 | 2.65E-05 | 0.000112661 | 0.166439947 |
| Obesity | LEC | *GLDN* | 1.245607479 | 3.42E-06 | 7.73E-05 | 0.021527793 |
| Obesity | LEC | *FCAR* | 1.198620667 | 5.53E-05 | 0.000162968 | 0.347447129 |
| Obesity | LEC | *KPNA4* | 1.828427706 | 0.000159366 | 0.000348308 | 1 |
| Obesity | LEC | *ZNF501* | 2.345204942 | 0.000108647 | 0.000258788 | 0.683065137 |
| Obesity | LEC | *BTN3A2* | -1.050923489 | 5.07E-08 | 6.26E-06 | 0.000319037 |
| Obesity | LEC | *PCLO* | -0.380433739 | 0.000622636 | 0.001078675 | 1 |
| Obesity | LEC | *RGS7BP* | 1.239588676 | 0.00035591 | 0.000668602 | 1 |
| Obesity | LEC | *ZNF396* | 1.177829441 | 0.017379176 | 0.022668647 | 1 |
| Obesity | LEC | *TMEM222* | -1.204048095 | 0.035086573 | 0.043672398 | 1 |
| Obesity | LEC | *CLCNKA* | 2.605062186 | 0.003350446 | 0.004931926 | 1 |
| Obesity | LEC | *SMYD4* | 1.848360941 | 2.03E-05 | 0.000106065 | 0.127842368 |
| Obesity | LEC | *NF2* | 1.405922697 | 0.022651271 | 0.02899787 | 1 |
| Obesity | LEC | *MIR22HG* | 1.347674521 | 1.08E-05 | 0.000105108 | 0.06764941 |
| Obesity | LEC | *PDE2A* | -0.577486143 | 0.000197122 | 0.00041214 | 1 |
| Obesity | LEC | *C17orf58* | 1.038335258 | 0.000924022 | 0.001532803 | 1 |
| Obesity | LEC | *CYP27C1* | 0.743371322 | 0.000135893 | 0.000307546 | 0.854361769 |
| Obesity | LEC | *BCR* | -0.613258255 | 0.005228137 | 0.007413012 | 1 |
| Obesity | LEC | *HYAL3* | -1.686606858 | 7.65E-06 | 0.000105108 | 0.048102337 |
| Obesity | LEC | *LILRB4* | 0.644655754 | 1.15E-05 | 0.000105108 | 0.072355652 |
| Obesity | LEC | *KRT16* | 0.971248074 | 0.00745136 | 0.010277907 | 1 |
| Obesity | LEC | *TRABD2A* | 0.603475883 | 8.10E-07 | 3.79E-05 | 0.005093176 |

| | | | | | | |
|---|---|---|---|---|---|---|
| Obesity | LEC | *TMEM17* | -1.279881302 | 0.000822583 | 0.001377663 | 1 |
| Obesity | LEC | *TNFRSF18* | 1.896716684 | 2.72E-05 | 0.000113713 | 0.170983731 |
| Obesity | LEC | *RTN4RL2* | -1.099948576 | 0.000102156 | 0.000248262 | 0.6422546 |
| Obesity | LEC | *TMEM232* | 1.509088083 | 0.000589015 | 0.001028958 | 1 |
| Obesity | LEC | *EFCAB6* | 1.334626249 | 1.97E-05 | 0.00010532 | 0.123576679 |
| Obesity | LEC | *KANK3* | -0.301674811 | 0.000139057 | 0.000313096 | 0.87425268 |
| Obesity | LEC | *EMID1* | -0.399074158 | 0.000155697 | 0.000341783 | 0.978866603 |
| Obesity | LEC | *CYP4A11* | 1.667577882 | 4.48E-05 | 0.000141991 | 0.281567674 |
| Obesity | LEC | *TMEM216* | 1.983777132 | 0.00070514 | 0.001205661 | 1 |
| Obesity | LEC | *RPS19BP1* | 1.332085134 | 0.000257704 | 0.000513693 | 1 |
| Obesity | LEC | *C3orf70* | -0.600945619 | 0.000422979 | 0.000774845 | 1 |
| Obesity | LEC | *PLCD1* | 1.897221008 | 0.000302549 | 0.000584481 | 1 |
| Obesity | LEC | *ENTPD5* | 2.154902153 | 4.08E-05 | 0.000133256 | 0.256785071 |
| Obesity | LEC | *HEATR4* | 1.069429281 | 9.74E-06 | 0.000105108 | 0.061210018 |
| Obesity | LEC | *MRPS21* | 1.892591441 | 1.43E-06 | 5.18E-05 | 0.009016421 |
| Obesity | LEC | *ANGPTL5* | 0.604631826 | 3.14E-05 | 0.000116847 | 0.197588432 |
| Obesity | LEC | *TSPYL4* | -1.190976158 | 0.008729133 | 0.011899405 | 1 |
| Obesity | LEC | *MT1X* | 0.579055956 | 3.73E-05 | 0.000126137 | 0.23421229 |
| Obesity | LEC | *FNBP1* | 1.407134186 | 0.00064672 | 0.001113953 | 1 |
| Obesity | LEC | *WDR86* | 0.833151836 | 9.55E-05 | 0.000236504 | 0.600483677 |
| Obesity | LEC | *EPOR* | 1.027964032 | 5.60E-06 | 9.64E-05 | 0.03518564 |
| Obesity | LEC | *FPR3* | 0.422243799 | 4.68E-05 | 0.000145967 | 0.294266705 |
| Obesity | LEC | *GJA4* | -0.62126968 | 0.000254474 | 0.000508989 | 1 |
| Obesity | LEC | *IFT140* | 1.884450845 | 0.01197386 | 0.015989732 | 1 |
| Obesity | LEC | *TLR5* | 0.809981001 | 2.28E-05 | 0.000109111 | 0.143371944 |
| Obesity | LEC | *PLEKHN1* | -1.042818295 | 0.022115703 | 0.028352656 | 1 |
| Obesity | LEC | *ZNF385C* | 1.170299391 | 1.78E-05 | 0.000105108 | 0.111763694 |
| Obesity | LEC | *TET3* | 1.563478363 | 1.79E-05 | 0.000105108 | 0.112844624 |
| Obesity | LEC | *C17orf97* | 0.464480517 | 0.005796818 | 0.008162283 | 1 |
| Obesity | LEC | *DHRS4L2* | 0.812406946 | 0.021457882 | 0.0275768 | 1 |
| Obesity | LEC | *TRPV2* | 1.277623146 | 1.58E-05 | 0.000105108 | 0.099379747 |
| Obesity | LEC | *C2orf88* | 1.647041824 | 8.96E-05 | 0.000225541 | 0.563176532 |
| Obesity | LEC | *FANCA* | 0.809678731 | 0.011446031 | 0.015343539 | 1 |
| Obesity | LEC | *ADH1A* | 0.684087361 | 0.029916828 | 0.037647537 | 1 |
| Obesity | LEC | *FAM69C* | 1.491707897 | 0.000337945 | 0.00064131 | 1 |
| Obesity | LEC | *DNAH17* | -1.710666881 | 0.008292186 | 0.011345587 | 1 |
| Obesity | LEC | *ZFP69* | 1.833678633 | 0.001853466 | 0.002872256 | 1 |
| Obesity | LEC | *TMEM220* | 1.711453534 | 6.10E-05 | 0.000173892 | 0.38377894 |
| Obesity | LEC | *GFRAL* | 1.1998448 | 3.12E-05 | 0.000116624 | 0.195927481 |
| Obesity | LEC | *CPSF4L* | -0.704541146 | 0.000245216 | 0.00049381 | 1 |
| Obesity | LEC | *KLHL17* | -1.267990495 | 0.001573446 | 0.00247616 | 1 |
| Obesity | LEC | *ZSCAN23* | 0.732280012 | 5.61E-07 | 3.04E-05 | 0.003524184 |
| Obesity | LEC | *OR7D2* | -1.90559203 | 0.02043603 | 0.02638762 | 1 |
| Obesity | LEC | *MORN2* | 0.965815075 | 8.72E-05 | 0.000221061 | 0.548011291 |
| Obesity | LEC | *S100A3* | 1.288941496 | 0.006699249 | 0.009328911 | 1 |
| Obesity | LEC | *TMEM221* | 0.835167405 | 0.006012046 | 0.008436994 | 1 |
| Obesity | LEC | *C6orf132* | 0.830108771 | 6.24E-05 | 0.000175889 | 0.392035732 |
| Obesity | LEC | *MAPK12* | -0.772598877 | 0.000207378 | 0.000429583 | 1 |
| Obesity | LEC | *AGRN* | -0.470674567 | 0.000106563 | 0.000255389 | 0.66996118 |
| Obesity | LEC | *FAM166A* | 3.146349827 | 3.04E-05 | 0.000115831 | 0.191260679 |

| | | | | | | |
|---|---|---|---|---|---|---|
| Obesity | LEC | *SMTNL2* | 4.504864653 | 3.14E-05 | 0.000116847 | 0.19740165 |
| Obesity | LEC | *ZC3H6* | 1.781427633 | 5.10E-05 | 0.000153757 | 0.320430512 |
| Obesity | LEC | *LINC00265* | 1.518986742 | 3.72E-05 | 0.000126137 | 0.233681575 |
| Obesity | LEC | *LAMTOR4* | 1.589091371 | 0.00904963 | 0.012312275 | 1 |
| Obesity | LEC | *PRKAR1B* | -0.321331513 | 0.000150918 | 0.000334446 | 0.948823366 |
| Obesity | LEC | *NUTM2B* | -9.041616042 | 9.27E-05 | 0.000231516 | 0.582956644 |
| Obesity | LEC | *NCR3LG1* | 3.441217113 | 1.41E-05 | 0.000105108 | 0.08893467 |
| Obesity | LEC | *HYKK* | 1.579723606 | 6.60E-07 | 3.35E-05 | 0.004150165 |
| Obesity | LEC | *RUFY4* | 1.617330967 | 3.11E-05 | 0.000116567 | 0.195483176 |
| Obesity | LEC | *CENPP* | 1.191758519 | 7.13E-10 | 1.66E-07 | 4.48E-06 |
| Obesity | LEC | *PLSCR1* | 2.037937171 | 9.66E-05 | 0.000238637 | 0.607420617 |
| Obesity | LEC | *ZNF559* | 0.584888264 | 0.021420424 | 0.027539919 | 1 |
| Obesity | LEC | *JAKMIP3* | 0.531544954 | 0.012125947 | 0.016168786 | 1 |
| Obesity | LEC | *PDCD1* | 4.128168493 | 0.001421691 | 0.002261111 | 1 |
| Obesity | LEC | *C19orf54* | 0.762234786 | 0.000399075 | 0.00073707 | 1 |
| Obesity | LEC | *COL25A1* | 1.652890396 | 0.000208141 | 0.000430314 | 1 |
| Obesity | LEC | *SRSF10* | 1.845799185 | 8.54E-05 | 0.000217963 | 0.537060259 |
| Obesity | LEC | *NBR1* | 2.02595035 | 5.84E-05 | 0.000168851 | 0.366914048 |
| Obesity | LEC | *NDOR1* | 1.532694457 | 1.52E-05 | 0.000105108 | 0.095364059 |
| Obesity | LEC | *FAM72B* | 5.26044509 | 1.52E-05 | 0.000105108 | 0.09531419 |
| Obesity | LEC | *SUMO2* | 2.331659957 | 0.000162527 | 0.000353688 | 1 |
| Obesity | LEC | *GOLGA8M* | 1.940115156 | 0.027522361 | 0.034808507 | 1 |
| Obesity | LEC | *DPYD* | 1.076458632 | 2.06E-05 | 0.00010668 | 0.129722875 |
| Obesity | LEC | *S100A16* | -1.079520749 | 0.031486802 | 0.039473085 | 1 |
| Obesity | LEC | *RHCE* | 0.931729881 | 4.33E-05 | 0.000138781 | 0.272011646 |
| Obesity | LEC | *PARVB* | 1.132307835 | 0.000777315 | 0.001311237 | 1 |
| Obesity | LEC | *SLC4A5* | -0.885222238 | 0.000168076 | 0.000362876 | 1 |
| Obesity | LEC | *UROS* | 2.16353277 | 0.000143098 | 0.000320276 | 0.899656625 |
| Obesity | LEC | *FAM221A* | 1.881852659 | 3.62E-05 | 0.000124125 | 0.227644474 |
| Obesity | LEC | *NOXA1* | 2.089145532 | 1.55E-05 | 0.000105108 | 0.097182716 |
| Obesity | LEC | *SKOR1* | 3.100777802 | 5.66E-05 | 0.000165507 | 0.3558221 |
| Obesity | LEC | *TMEM201* | 2.378194197 | 1.55E-05 | 0.000105108 | 0.097685964 |
| Obesity | LEC | *ZDHHC11* | 2.036002184 | 9.15E-06 | 0.000105108 | 0.057542267 |
| Obesity | LEC | *FAM26F* | 0.868694554 | 2.70E-05 | 0.000113347 | 0.170020245 |
| Obesity | LEC | *BEND4* | 3.777268069 | 9.39E-06 | 0.000105108 | 0.059064918 |
| Obesity | LEC | *ZNF563* | 0.917338345 | 6.65E-05 | 0.000183524 | 0.418068153 |
| Obesity | LEC | *FBF1* | 1.017245807 | 3.62E-08 | 4.77E-06 | 0.000227645 |
| Obesity | LEC | *LRRK2* | 1.303719224 | 0.002400218 | 0.003637062 | 1 |
| Obesity | LEC | *FAM196A* | 2.113352852 | 2.48E-05 | 0.000110963 | 0.155681734 |
| Obesity | LEC | *UTS2B* | 0.68571863 | 0.001170685 | 0.0018984 | 1 |
| Obesity | LEC | *C9orf152* | 1.810759645 | 2.35E-05 | 0.000109118 | 0.147926058 |
| Obesity | LEC | *NELFB* | 1.659174248 | 1.39E-05 | 0.000105108 | 0.087200424 |
| Obesity | LEC | *SLC15A5* | 0.611897651 | 3.57E-05 | 0.000123048 | 0.224737073 |
| Obesity | LEC | *LIPI* | -0.610483041 | 0.017181631 | 0.022424935 | 1 |
| Obesity | LEC | *ADAT2* | 1.889655683 | 2.96E-06 | 7.23E-05 | 0.018587662 |
| Obesity | LEC | *ZNF567* | 2.3776282 | 8.33E-05 | 0.000214516 | 0.523848673 |
| Obesity | LEC | *ANKDD1B* | -0.469716912 | 0.000102258 | 0.000248318 | 0.642895795 |
| Obesity | LEC | *LITAF* | 1.252002034 | 7.16E-06 | 0.000105108 | 0.044989213 |
| Obesity | LEC | *VSTM1* | 2.03769681 | 0.002040454 | 0.00313728 | 1 |
| Obesity | LEC | *SF3B3* | 1.770568146 | 2.18E-05 | 0.000108281 | 0.137299751 |

| | | | | | | |
|---|---|---|---|---|---|---|
| Obesity | LEC | *SP6* | -3.0790763 | 4.06E-05 | 0.000132756 | 0.255157348 |
| Obesity | LEC | *PLAC9* | 1.45327161 | 2.41E-06 | 6.68E-05 | 0.015130104 |
| Obesity | LEC | *CLDN4* | 1.253648043 | 0.00857142 | 0.01170253 | 1 |
| Obesity | LEC | *ZNF573* | 2.146929112 | 6.07E-05 | 0.000173281 | 0.381737206 |
| Obesity | LEC | *S100A13* | 1.768325403 | 5.46E-06 | 9.50E-05 | 0.034302326 |
| Obesity | LEC | *LINC00994* | 2.418012734 | 0.000433288 | 0.000790736 | 1 |
| Obesity | LEC | *C15orf61* | 2.810446908 | 3.49E-05 | 0.000122199 | 0.219103371 |
| Obesity | LEC | *TSPYL1* | 1.565973678 | 0.005887036 | 0.008281896 | 1 |
| Obesity | LEC | *PNRC2* | 1.727635739 | 3.27E-05 | 0.000118624 | 0.205574574 |
| Obesity | LEC | *RRP7A* | 1.770692595 | 0.006166964 | 0.008640897 | 1 |
| Obesity | LEC | *SLC35E2B* | 1.40892966 | 1.60E-06 | 5.41E-05 | 0.010064333 |
| Obesity | LEC | *BLOC1S2* | 0.451437174 | 0.011154516 | 0.014987912 | 1 |
| Obesity | LEC | *HLA-DRB1* | 0.808913119 | 0.000569197 | 0.000999592 | 1 |
| Obesity | LEC | *WDSUB1* | 1.489380116 | 0.001452742 | 0.002305992 | 1 |
| Obesity | LEC | *ZNF79* | 1.384938159 | 0.000138626 | 0.000312492 | 0.87154371 |
| Obesity | LEC | *S100A4* | 0.933968978 | 0.000701573 | 0.001199888 | 1 |
| Obesity | LEC | *COLCA1* | 1.41644394 | 3.17E-05 | 0.000117011 | 0.199269921 |
| Obesity | LEC | *ACADSB* | 1.160239981 | 4.63E-05 | 0.000145187 | 0.29109924 |
| Obesity | LEC | *SEMA4A* | 0.752253178 | 7.29E-06 | 0.000105108 | 0.045835893 |
| Obesity | LEC | *MPHOSPH8* | 1.582264649 | 8.56E-05 | 0.000218223 | 0.538098006 |
| Obesity | LEC | *SIRPB2* | 0.626485486 | 3.07E-05 | 0.000115897 | 0.192799246 |
| Obesity | LEC | *SRGAP3* | -0.36453623 | 0.000962834 | 0.00158922 | 1 |
| Obesity | LEC | *SUPT5H* | 2.19197981 | 0.000181813 | 0.000386821 | 1 |
| Obesity | LEC | *ZNF107* | 0.888049829 | 0.023377414 | 0.029860586 | 1 |
| Obesity | LEC | *PPIA* | 1.887170274 | 7.13E-06 | 0.000105108 | 0.044806359 |
| Obesity | LEC | *GTF2IRD2* | 1.072472118 | 0.02444753 | 0.031145212 | 1 |
| Obesity | LEC | *GRM7* | 0.663194654 | 0.004045625 | 0.005860797 | 1 |
| Obesity | LEC | *SUPT3H* | 1.227611661 | 0.025748901 | 0.032730154 | 1 |
| Obesity | LEC | *POM121* | 1.42400856 | 0.000104853 | 0.000252502 | 0.659209539 |
| Obesity | LEC | *GIMAP5* | -0.466123745 | 0.000102839 | 0.000249503 | 0.64655118 |
| Obesity | LEC | *STK31* | 1.137418217 | 2.27E-05 | 0.000109111 | 0.142980992 |
| Obesity | LEC | *ZKSCAN7* | 0.707233926 | 2.72E-05 | 0.000113713 | 0.171249634 |
| Obesity | LEC | *ZNF565* | 1.875618667 | 2.16E-05 | 0.000107661 | 0.135867961 |
| Obesity | LEC | *NTNG2* | -1.957409939 | 0.030809345 | 0.038685511 | 1 |
| Obesity | LEC | *WDR5* | 2.404987006 | 8.90E-05 | 0.000224335 | 0.559634891 |
| Obesity | LEC | *LONP1* | 2.275023547 | 7.26E-06 | 0.000105108 | 0.045651659 |
| Obesity | LEC | *ASB13* | -1.25714656 | 0.032679701 | 0.040822031 | 1 |
| Obesity | LEC | *SLC35F1* | 1.868076234 | 4.26E-05 | 0.000137183 | 0.267782016 |
| Obesity | LEC | *INCA1* | 1.519168817 | 2.59E-06 | 6.85E-05 | 0.016292967 |
| Obesity | LEC | *ZNF774* | -1.982679814 | 0.000927991 | 0.00153817 | 1 |
| Obesity | LEC | *ZNF124* | 1.429541952 | 1.57E-05 | 0.000105108 | 0.098551214 |
| Obesity | LEC | *ZNF569* | 2.047047607 | 0.000197579 | 0.000412821 | 1 |
| Obesity | LEC | *PIK3R4* | -0.877838163 | 0.026173852 | 0.03320319 | 1 |
| Obesity | LEC | *RFX8* | 0.619234131 | 2.24E-05 | 0.000108832 | 0.14071927 |
| Obesity | LEC | *MYL6B* | -0.882272564 | 0.000506759 | 0.000902548 | 1 |
| Obesity | LEC | *ZNF799* | 1.011157783 | 0.000164565 | 0.000357012 | 1 |
| Obesity | LEC | *C20orf96* | 1.642551448 | 1.83E-05 | 0.000105108 | 0.114876997 |
| Obesity | LEC | *SULT1A1* | 0.329235691 | 0.018879654 | 0.024513917 | 1 |
| Obesity | LEC | *ANAPC7* | 1.871156454 | 0.000767486 | 0.001297442 | 1 |
| Obesity | LEC | *TPK1* | 1.768314536 | 5.17E-06 | 9.32E-05 | 0.032528624 |

| | | | | | | |
|---|---|---|---|---|---|---|
| Obesity | LEC | *C1orf186* | -1.716097269 | 0.007445189 | 0.010271648 | 1 |
| Obesity | LEC | *MYO18A* | 1.610888586 | 0.005821573 | 0.00819347 | 1 |
| Obesity | LEC | *SPTSSB* | 2.542210935 | 1.15E-05 | 0.000105108 | 0.072121201 |
| Obesity | LEC | *SULF2* | -0.466574389 | 0.000430124 | 0.000785416 | 1 |
| Obesity | LEC | *XRCC2* | -1.133571689 | 0.000279112 | 0.000548711 | 1 |
| Obesity | LEC | *HDAC2* | 1.639935139 | 0.022866288 | 0.029243359 | 1 |
| Obesity | LEC | *ADH1B* | 1.027552419 | 1.04E-05 | 0.000105108 | 0.065454909 |
| Obesity | LEC | *HRH1* | 0.616032579 | 1.86E-05 | 0.000105129 | 0.117008363 |
| Obesity | LEC | *ZNF502* | 2.104213156 | 7.31E-05 | 0.000195683 | 0.459463886 |
| Obesity | LEC | *TRAPPC4* | 1.217065166 | 0.000280325 | 0.000550751 | 1 |
| Obesity | LEC | *TECPR2* | 1.924183535 | 4.97E-05 | 0.000151436 | 0.312468413 |
| Obesity | LEC | *ERI2* | 1.625465377 | 1.27E-05 | 0.000105108 | 0.079950872 |
| Obesity | LEC | *TRPV1* | 0.895081182 | 5.15E-06 | 9.31E-05 | 0.032382314 |
| Obesity | LEC | *ZNF33B* | -0.784182792 | 0.005639141 | 0.00796524 | 1 |
| Obesity | LEC | *AMZ2* | 1.772492708 | 9.21E-06 | 0.000105108 | 0.057905339 |
| Obesity | LEC | *ZNF431* | 1.775678201 | 2.58E-05 | 0.000112124 | 0.162100461 |
| Obesity | LEC | *NF1* | 1.46450821 | 3.88E-05 | 0.000129427 | 0.243839762 |
| Obesity | LEC | *HLA-DQA1* | 1.022406521 | 0.000253175 | 0.000507076 | 1 |
| Obesity | LEC | *GM2A* | 1.812275911 | 3.36E-06 | 7.66E-05 | 0.021108546 |
| Obesity | LEC | *TLE1* | -0.766425016 | 0.010784329 | 0.014521541 | 1 |
| Obesity | LEC | *STRN3* | 1.501224419 | 0.000912576 | 0.001515817 | 1 |
| Obesity | LEC | *ZNF239* | 1.295750366 | 0.010042863 | 0.013572545 | 1 |
| Obesity | LEC | *C6orf106* | 2.178813685 | 1.29E-11 | 5.41E-09 | 8.11E-08 |
| Obesity | LEC | *PPTC7* | 0.831075445 | 3.92E-05 | 0.000130236 | 0.246407013 |
| Obesity | LEC | *NHLRC2* | 1.674092844 | 8.21E-05 | 0.000212304 | 0.515898261 |
| Obesity | LEC | *ZFP28* | 3.014009276 | 0.004199332 | 0.006062273 | 1 |
| Obesity | LEC | *LAMB3* | -1.233686797 | 0.000647633 | 0.001115058 | 1 |
| Obesity | LEC | *KPNA5* | 1.868943843 | 0.000103232 | 0.000250009 | 0.649022149 |
| Obesity | LEC | *HCAR1* | 1.142428631 | 1.81E-07 | 1.53E-05 | 0.001135196 |
| Obesity | LEC | *SRGAP1* | 1.234259405 | 8.04E-06 | 0.000105108 | 0.050546155 |
| Obesity | LEC | *SLC39A10* | 1.538722263 | 3.54E-05 | 0.000122996 | 0.222499456 |
| Obesity | LEC | *AP2A1* | 1.546264577 | 0.011114579 | 0.014937443 | 1 |
| Obesity | LEC | *HIST1H3E* | 1.290181152 | 0.00631014 | 0.008829702 | 1 |
| Obesity | LEC | *ZNF585A* | 1.138491206 | 2.29E-06 | 6.51E-05 | 0.01438307 |
| Obesity | LEC | *FUT11* | 3.162821756 | 4.39E-05 | 0.000140307 | 0.275843316 |
| Obesity | LEC | *ANXA4* | 1.240010928 | 0.020644573 | 0.026640482 | 1 |
| Obesity | LEC | *ZNF138* | 1.652531003 | 4.06E-06 | 8.46E-05 | 0.025537542 |
| Obesity | LEC | *ZNF429* | 1.521174003 | 0.000906179 | 0.001505589 | 1 |
| Obesity | LEC | *ZNF470* | 2.404314425 | 0.00105246 | 0.001726683 | 1 |
| Obesity | LEC | *ZNF100* | 0.814702335 | 0.021716859 | 0.02788113 | 1 |
| Obesity | LEC | *ZNF763* | 0.607090729 | 0.007869473 | 0.010797769 | 1 |
| Obesity | LEC | *ZMYM1* | 1.80254228 | 2.51E-05 | 0.000111345 | 0.157858714 |
| Obesity | LEC | *IGF2R* | 1.57771324 | 0.000221523 | 0.000453948 | 1 |
| Obesity | LEC | *GAL3ST4* | 1.232986078 | 1.31E-05 | 0.000105108 | 0.082617397 |
| Obesity | LEC | *SLC25A29* | 2.067848831 | 6.62E-07 | 3.35E-05 | 0.00416007 |
| Obesity | LEC | *ZNF772* | -1.004330152 | 0.001431257 | 0.002274024 | 1 |
| Obesity | LEC | *ZNF257* | -1.470255203 | 0.001877068 | 0.002906681 | 1 |
| Obesity | LEC | *ADAM32* | -1.130695038 | 0.003160413 | 0.00467518 | 1 |
| Obesity | LEC | *LRRC8B* | 1.083115796 | 7.02E-05 | 0.000189971 | 0.441467676 |
| Obesity | LEC | *ABCB8* | 1.418063889 | 7.73E-06 | 0.000105108 | 0.048610245 |

| | | | | | | |
|---|---|---|---|---|---|---|
| Obesity | LEC | *SND1* | 1.204981508 | 6.97E-06 | 0.000104897 | 0.043797434 |
| Obesity | LEC | *SULT1A2* | 1.223721295 | 2.71E-24 | 8.52E-21 | 1.70E-20 |
| Obesity | LEC | *NEK5* | 1.321011553 | 0.000271387 | 0.000536207 | 1 |
| Obesity | LEC | *PIWIL2* | 1.819638413 | 9.22E-05 | 0.000230354 | 0.579413696 |
| Obesity | LEC | *C1D* | 1.278786379 | 1.88E-05 | 0.000105211 | 0.117984323 |
| Obesity | LEC | *TBC1D9B* | 1.144705082 | 0.000368017 | 0.000688403 | 1 |
| Obesity | LEC | *FAM110D* | 1.132152211 | 2.04E-05 | 0.000106095 | 0.128219291 |
| Obesity | LEC | *KANK2* | 1.673192467 | 0.011678472 | 0.015635126 | 1 |
| Obesity | LEC | *C6orf141* | -0.099059442 | 5.58E-05 | 0.000164278 | 0.350732972 |
| Obesity | LEC | *CCL4L2* | 0.358438195 | 0.020983456 | 0.027022324 | 1 |
| Obesity | LEC | *GTF2E2* | 1.763651794 | 1.57E-06 | 5.41E-05 | 0.009893956 |
| Obesity | LEC | *ZNF165* | -2.870406301 | 3.59E-07 | 2.44E-05 | 0.002254252 |
| Obesity | LEC | *RAMP2-AS1* | -1.185443197 | 0.004422301 | 0.006362244 | 1 |
| Obesity | LEC | *DDI2* | 1.222864512 | 1.55E-05 | 0.000105108 | 0.097464391 |
| Obesity | LEC | *MRPL21* | -1.219638625 | 7.62E-07 | 3.73E-05 | 0.004788495 |
| Obesity | LEC | *ZNF786* | 1.450507302 | 5.65E-05 | 0.00016541 | 0.354913071 |
| Obesity | LEC | *SLC22A5* | 0.794483946 | 0.009783058 | 0.013244204 | 1 |
| Obesity | LEC | *C5AR1* | 1.010318664 | 4.58E-05 | 0.000144184 | 0.287865248 |
| Obesity | LEC | *VEPH1* | -1.07475345 | 0.001464088 | 0.002321634 | 1 |
| Obesity | LEC | *SHPK* | 0.754042033 | 3.45E-05 | 0.000121733 | 0.217136218 |
| Obesity | LEC | *MAP3K5* | 1.210058017 | 0.000154085 | 0.000339312 | 0.968734365 |
| Obesity | LEC | *STMN3* | 1.647740147 | 0.001385101 | 0.002207384 | 1 |
| Obesity | LEC | *GYPE* | 1.227933081 | 0.001431976 | 0.002274592 | 1 |
| Obesity | LEC | *PCDHB11* | -3.993483261 | 0.000548482 | 0.000966662 | 1 |
| Obesity | LEC | *SLC2A10* | 1.931762774 | 0.003932984 | 0.00570396 | 1 |
| Obesity | LEC | *ZNF665* | 1.108793974 | 0.000504541 | 0.000899107 | 1 |
| Obesity | LEC | *RPF2* | 1.406419315 | 3.30E-05 | 0.000119154 | 0.207695624 |
| Obesity | LEC | *MYO5A* | 0.959167001 | 1.19E-05 | 0.000105108 | 0.074559839 |
| Obesity | LEC | *ATG7* | 0.900590292 | 4.66E-05 | 0.000145628 | 0.292857956 |
| Obesity | LEC | *RAB40C* | 2.341326335 | 0.002505562 | 0.00378029 | 1 |
| Obesity | LEC | *PIGN* | 0.671220037 | 0.040457722 | 0.049874058 | 1 |
| Obesity | LEC | *ZNF624* | 2.860407604 | 0.000970413 | 0.001600889 | 1 |
| Obesity | LEC | *HHLA3* | 1.797953679 | 0.000398057 | 0.000735405 | 1 |
| Obesity | LEC | *HOXA4* | 1.166245875 | 3.11E-05 | 0.000116544 | 0.195223221 |
| Obesity | LEC | *BCO2* | -0.699217616 | 0.000299414 | 0.000579205 | 1 |
| Obesity | LEC | *ENTPD6* | 1.394777545 | 0.000253173 | 0.000507076 | 1 |
| Obesity | LEC | *CCDC154* | 1.585768972 | 0.000205688 | 0.000426504 | 1 |
| Obesity | LEC | *C5orf42* | 1.389532202 | 0.000470326 | 0.000846532 | 1 |
| Obesity | LEC | *MFAP5* | 0.960242518 | 1.44E-05 | 0.000105108 | 0.09042234 |
| Obesity | LEC | *CDC42SE1* | 2.181244381 | 0.000166346 | 0.000359816 | 1 |
| Obesity | LEC | *MPEG1* | 0.612386037 | 0.000436244 | 0.000794847 | 1 |
| Obesity | LEC | *PDCD1LG2* | 1.080912961 | 4.04E-05 | 0.00013241 | 0.253829186 |
| Obesity | LEC | *NMB* | 3.468735443 | 0.001205799 | 0.001948306 | 1 |
| Obesity | LEC | *KLHL14* | -0.1199698 | 6.95E-05 | 0.000188474 | 0.436693377 |
| Obesity | LEC | *FAM114A1* | 1.897374606 | 0.000135055 | 0.000305869 | 0.849092443 |
| Obesity | LEC | *RPS26* | -0.980478833 | 5.07E-05 | 0.0001531 | 0.318593132 |
| Obesity | LEC | *PSAP* | 1.146499775 | 1.41E-05 | 0.000105108 | 0.088463547 |
| Obesity | LEC | *S100A10* | 1.156302717 | 0.027458621 | 0.03473488 | 1 |
| Obesity | LEC | *MAP1LC3C* | 1.053681436 | 5.05E-05 | 0.000153026 | 0.317529686 |
| Obesity | LEC | *TAF13* | 1.716847268 | 0.000265751 | 0.000527146 | 1 |

| | | | | | | |
|---|---|---|---|---|---|---|
| Obesity | LEC | *ZNF780A* | -1.02845599 | 0.000308344 | 0.000593739 | 1 |
| Obesity | LEC | *AC122129.1* | 1.723456006 | 3.02E-05 | 0.000115831 | 0.189969385 |
| Obesity | LEC | *CCDC180* | 1.166894809 | 0.001468768 | 0.00232774 | 1 |
| Obesity | LEC | *OCLN* | -1.359573041 | 9.78E-05 | 0.000240649 | 0.614618269 |
| Obesity | LEC | *ZNF181* | 1.681055482 | 2.12E-05 | 0.00010722 | 0.133585048 |
| Obesity | LEC | *SGTB* | 1.712095429 | 7.69E-05 | 0.00020244 | 0.483170209 |
| Obesity | LEC | *ADH5* | 1.117831196 | 6.54E-05 | 0.000181705 | 0.411457733 |
| Obesity | LEC | *HRNR* | 0.490691627 | 0.000739378 | 0.001257363 | 1 |
| Obesity | LEC | *ZNF677* | 3.055229591 | 0.000296277 | 0.000574374 | 1 |
| Obesity | LEC | *ZNF311* | 1.11721579 | 1.72E-05 | 0.000105108 | 0.108044608 |
| Obesity | LEC | *FCHSD1* | 2.304629632 | 1.12E-05 | 0.000105108 | 0.070600456 |
| Obesity | LEC | *LEKR1* | 0.934641882 | 0.000784834 | 0.001322146 | 1 |
| Obesity | LEC | *NOL8* | 1.663349855 | 0.000400505 | 0.000739207 | 1 |
| Obesity | LEC | *ZNF335* | 1.135681454 | 9.69E-06 | 0.000105108 | 0.060926974 |
| Obesity | LEC | *AGAP9* | 2.391835988 | 4.53E-05 | 0.000143043 | 0.284656043 |
| Obesity | LEC | *MAK16* | 1.582926613 | 9.15E-05 | 0.000229207 | 0.575018669 |
| Obesity | LEC | *ZNF667* | 1.561525539 | 0.000486434 | 0.000871533 | 1 |
| Obesity | LEC | *PRIM1* | 1.923812373 | 0.000133204 | 0.000302876 | 0.837452595 |
| Obesity | LEC | *SULT1C4* | -0.57268646 | 0.000235278 | 0.000477005 | 1 |
| Obesity | LEC | *ZNF649* | 1.176661489 | 0.006540655 | 0.009121805 | 1 |
| Obesity | LEC | *HIBCH* | 1.146525458 | 0.002131809 | 0.003268948 | 1 |
| Obesity | LEC | *ZNF544* | 1.175484143 | 0.032804377 | 0.04096963 | 1 |
| Obesity | LEC | *SOWAHC* | -1.298169142 | 0.000392246 | 0.000726805 | 1 |
| Obesity | LEC | *AC135178.1* | 4.170201599 | 0.001498502 | 0.00237068 | 1 |
| Obesity | LEC | *MIER1* | 1.755679903 | 3.06E-05 | 0.000115831 | 0.192126294 |
| Obesity | LEC | *SVIP* | -0.849075503 | 0.001111918 | 0.001811043 | 1 |
| Obesity | LEC | *ZNF251* | 1.67840963 | 0.00011893 | 0.000278111 | 0.747711248 |
| Obesity | LEC | *DDRGK1* | 1.17146308 | 1.39E-05 | 0.000105108 | 0.087466404 |
| Obesity | LEC | *ZNF334* | 0.921112435 | 0.000279868 | 0.000550025 | 1 |
| Obesity | LEC | *HSD17B11* | 2.117339122 | 3.07E-05 | 0.000115897 | 0.192821286 |
| Obesity | LEC | *SULT1C2* | 0.754217519 | 0.000182708 | 0.000387938 | 1 |
| Obesity | LEC | *RPS6KL1* | -0.516182593 | 0.000260985 | 0.000519574 | 1 |
| Obesity | LEC | *QRICH1* | 1.821373596 | 7.17E-06 | 0.000105108 | 0.045061459 |
| Obesity | LEC | *DDX42* | 1.842946424 | 0.001670961 | 0.002614567 | 1 |
| Obesity | LEC | *SLC29A3* | 0.901901571 | 2.21E-05 | 0.000108445 | 0.139242959 |
| Obesity | LEC | *ANTXRL* | -13.27177744 | 0.000115332 | 0.000271366 | 0.725089884 |
| Obesity | LEC | *STYX* | 1.158910429 | 1.11E-05 | 0.000105108 | 0.069900922 |
| Obesity | LEC | *TMEM116* | 0.961583579 | 3.53E-05 | 0.000122857 | 0.221692357 |
| Obesity | LEC | *ZKSCAN8* | 3.293707978 | 3.86E-07 | 2.50E-05 | 0.002427537 |
| Obesity | LEC | *MYL4* | 1.909100187 | 0.011491287 | 0.015400921 | 1 |
| Obesity | LEC | *ZNF813* | 2.835816462 | 0.004731754 | 0.006759495 | 1 |
| Obesity | LEC | *GFPT1* | 1.434670076 | 1.90E-06 | 5.83E-05 | 0.011949496 |
| Obesity | LEC | *UVRAG* | 0.938886826 | 0.000117536 | 0.000275351 | 0.738949739 |
| Obesity | LEC | *MGEA5* | 1.121499119 | 0.000289915 | 0.000565868 | 1 |
| Obesity | LEC | *MT1F* | -1.246309674 | 6.14E-05 | 0.000174627 | 0.386100198 |
| Obesity | LEC | *ZNF587* | 2.317622233 | 7.03E-05 | 0.000189971 | 0.441682501 |
| Obesity | LEC | *SH3BGRL2* | -0.578340496 | 0.024091306 | 0.030703839 | 1 |
| Obesity | LEC | *ZNF808* | 2.090350808 | 0.001161971 | 0.001885242 | 1 |
| Obesity | LEC | *TMA16* | 2.263161464 | 3.95E-05 | 0.000130813 | 0.248545259 |
| Obesity | LEC | *C20orf203* | -2.232561035 | 0.010980894 | 0.014773567 | 1 |

| | | | | | | |
|---|---|---|---|---|---|---|
| Obesity | LEC | *ZNF627* | 1.800240421 | 4.48E-05 | 0.000142019 | 0.281907285 |
| Obesity | LEC | *ARC* | 13.67316827 | 0.000750499 | 0.001273519 | 1 |
| Obesity | LEC | *TLK1* | 1.347921357 | 0.010907159 | 0.014677506 | 1 |
| Obesity | LEC | *LRBA* | 1.458386474 | 5.05E-05 | 0.000152943 | 0.317204128 |
| Obesity | LEC | *MMP17* | 1.536537266 | 1.63E-05 | 0.000105108 | 0.102332013 |
| Obesity | LEC | *CCDC69* | 1.551817178 | 5.09E-05 | 0.000153738 | 0.320236017 |
| Obesity | LEC | *MDM4* | 1.477485797 | 3.19E-05 | 0.000117214 | 0.200361355 |
| Obesity | LEC | *ZNF534* | -1.830955066 | 0.000224368 | 0.000458137 | 1 |
| Obesity | LEC | *FAM19A2* | 1.307600598 | 3.02E-05 | 0.000115831 | 0.189685621 |
| Obesity | LEC | *TTC37* | 1.576086815 | 8.66E-05 | 0.00021996 | 0.544400955 |
| Obesity | LEC | *PAPSS2* | 0.620711757 | 1.62E-05 | 0.000105108 | 0.102003339 |
| Obesity | LEC | *FAN1* | 1.988748132 | 2.35E-05 | 0.000109118 | 0.147547287 |
| Obesity | LEC | *CEP290* | 1.851060417 | 0.000406216 | 0.00074892 | 1 |
| Obesity | LEC | *ANKRD13B* | 1.291901377 | 0.007764427 | 0.010665273 | 1 |
| Obesity | LEC | *ECI2* | 2.195433419 | 5.29E-05 | 0.000157771 | 0.332739458 |
| Obesity | LEC | *ZNF652* | 1.53269422 | 2.47E-05 | 0.000110668 | 0.155156373 |
| Obesity | LEC | *RPL10A* | 2.068710685 | 4.14E-05 | 0.00013454 | 0.260065912 |
| Obesity | LEC | *RASSF9* | -0.260964257 | 0.000130002 | 0.000297749 | 0.817320872 |
| Obesity | LEC | *FAM169A* | -0.467671038 | 0.000428274 | 0.000782494 | 1 |
| Obesity | LEC | *TMEM184B* | 1.723630651 | 0.011797578 | 0.015771077 | 1 |
| Obesity | LEC | *SCAMP5* | 0.744331219 | 0.010889542 | 0.014660075 | 1 |
| Obesity | LEC | *ALPK2* | 0.622097075 | 0.000386248 | 0.000716959 | 1 |
| Obesity | LEC | *SFT2D1* | 2.10864346 | 3.05E-05 | 0.000115831 | 0.191955702 |
| Obesity | LEC | *ARHGAP11A* | 1.061717907 | 3.30E-05 | 0.000119154 | 0.207496507 |
| Obesity | LEC | *HMGN2* | 1.430674079 | 0.010628994 | 0.014321578 | 1 |
| Obesity | LEC | *DENND4B* | 1.65493384 | 0.000577046 | 0.001012811 | 1 |
| Obesity | LEC | *CD3E* | 1.321262208 | 2.63E-05 | 0.000112582 | 0.165293706 |
| Obesity | LEC | *TSEN15* | 1.502114291 | 1.43E-05 | 0.000105108 | 0.090066423 |
| Obesity | LEC | *ITPRIPL1* | -1.042378795 | 0.000166842 | 0.000360584 | 1 |
| Obesity | LEC | *PRMT6* | 2.257101036 | 0.00045313 | 0.000820987 | 1 |
| Obesity | LEC | *SHISA4* | 2.632101132 | 0.000134768 | 0.00030533 | 0.8472895 |
| Obesity | LEC | *CAPZA2* | 1.317579246 | 1.61E-05 | 0.000105108 | 0.101339771 |
| Obesity | LEC | *C1orf174* | 2.28580631 | 0.001258777 | 0.002022988 | 1 |
| Obesity | LEC | *RASGEF1A* | 3.553881473 | 1.38E-05 | 0.000105108 | 0.086644226 |
| Obesity | LEC | *KIAA0753* | 1.491626458 | 0.025720966 | 0.032701257 | 1 |
| Obesity | LEC | *APRT* | 2.993064608 | 6.42E-05 | 0.000178992 | 0.403527348 |
| Obesity | LEC | *MFAP3L* | -1.292032758 | 0.001224568 | 0.00197356 | 1 |
| Obesity | LEC | *NAGA* | 2.615996904 | 1.03E-05 | 0.000105108 | 0.065026676 |
| Obesity | LEC | *SMG5* | 1.803378606 | 0.004504772 | 0.006472007 | 1 |
| Obesity | LEC | *PJA2* | 1.403026724 | 0.000579676 | 0.001016009 | 1 |
| Obesity | LEC | *AL590822.1* | 0.909320009 | 0.009648216 | 0.013070099 | 1 |
| Obesity | LEC | *AP001007.1* | -5.04445765 | 0.000497833 | 0.000889422 | 1 |
| Obesity | LEC | *ZNF525* | 1.275102702 | 1.43E-05 | 0.000105108 | 0.089669049 |
| Obesity | LEC | *AC105020.1* | 7.229859722 | 0.000411408 | 0.000756513 | 1 |
| Obesity | LEC | *EFCAB2* | 1.715963301 | 3.95E-05 | 0.000130742 | 0.248174512 |
| Obesity | LEC | *CHML* | 1.299796397 | 5.93E-06 | 9.90E-05 | 0.037307534 |
| Obesity | LEC | *CAPN8* | 2.086612605 | 0.000343532 | 0.00064995 | 1 |
| Obesity | LEC | *TATDN3* | 1.149107207 | 2.67E-05 | 0.000112975 | 0.167767991 |
| Obesity | LEC | *FAM229B* | 1.020542322 | 0.003261267 | 0.004813002 | 1 |
| Obesity | LEC | *DDO* | -1.277826064 | 0.002238624 | 0.003416055 | 1 |

| | | | | | | |
|---|---|---|---|---|---|---|
| Obesity | LEC | *HIST2H2BF* | 2.281005626 | 3.12E-05 | 0.000116624 | 0.195875671 |
| Obesity | LEC | *PCMTD2* | 2.0967579 | 0.000233372 | 0.0004736 | 1 |
| Obesity | LEC | *OOEP* | -0.763185459 | 0.000181759 | 0.000386821 | 1 |
| Obesity | LEC | *SYS1* | 3.735087167 | 0.000201963 | 0.000420167 | 1 |
| Obesity | LEC | *NEU4* | 5.6425603 | 0.002196715 | 0.003359461 | 1 |
| Obesity | LEC | *TRAF3IP1* | 1.886580197 | 0.000531559 | 0.000940588 | 1 |
| Obesity | LEC | *GIGYF2* | 1.617557342 | 0.000277163 | 0.000545903 | 1 |
| Obesity | LEC | *C2orf72* | -0.939183655 | 1.50E-05 | 0.000105108 | 0.094222381 |
| Obesity | LEC | *ASAH2B* | -0.811267768 | 0.007651152 | 0.010528079 | 1 |
| Obesity | LEC | *AGAP6* | 0.918989587 | 0.000121649 | 0.000282576 | 0.764805773 |
| Obesity | LEC | *C10orf128* | 2.031924333 | 2.33E-05 | 0.000109118 | 0.146478857 |
| Obesity | LEC | *SYT15* | -1.154079114 | 0.00570992 | 0.008059781 | 1 |
| Obesity | LEC | *RING1* | 1.573425438 | 0.002635022 | 0.00395002 | 1 |
| Obesity | LEC | *OXLD1* | 1.008558583 | 2.37E-05 | 0.000109118 | 0.149059812 |
| Obesity | LEC | *HLA-DOA* | 0.996949873 | 0.005618164 | 0.007939177 | 1 |
| Obesity | LEC | *HLA-DMA* | 1.499276601 | 1.12E-05 | 0.000105108 | 0.070348219 |
| Obesity | LEC | *HLA-DRA* | 0.722360889 | 0.003573185 | 0.005228123 | 1 |
| Obesity | LEC | *COL15A1* | 1.018547627 | 6.81E-05 | 0.00018633 | 0.428138242 |
| Obesity | LEC | *NOTCH4* | -0.378773884 | 2.16E-08 | 3.42E-06 | 0.000135485 |
| Obesity | LEC | *AGER* | 1.613776276 | 0.020447411 | 0.026396894 | 1 |
| Obesity | LEC | *PRRT1* | -0.768531909 | 0.0025238 | 0.003799601 | 1 |
| Obesity | LEC | *MRPL38* | -2.640352141 | 7.58E-06 | 0.000105108 | 0.047667534 |
| Obesity | LEC | *DXO* | 1.076746292 | 0.001692441 | 0.002645544 | 1 |
| Obesity | LEC | *SKIV2L* | -0.987692054 | 4.68E-06 | 9.00E-05 | 0.029412988 |
| Obesity | LEC | *NELFE* | 2.142268261 | 7.98E-09 | 1.62E-06 | 5.02E-05 |
| Obesity | LEC | *LAYN* | -0.161913255 | 0.000105643 | 0.000253987 | 0.664175286 |
| Obesity | LEC | *C6orf48* | 1.746576328 | 0.003193926 | 0.004721424 | 1 |
| Obesity | LEC | *CARD16* | -1.000027781 | 0.000637808 | 0.001100411 | 1 |
| Obesity | LEC | *MSH5* | 1.044702134 | 0.002379313 | 0.003608865 | 1 |
| Obesity | LEC | *BAG6* | 1.722999504 | 1.87E-05 | 0.000105211 | 0.117361258 |
| Obesity | LEC | *NFKBIL1* | 1.835551636 | 1.75E-05 | 0.000105108 | 0.110246157 |
| Obesity | LEC | *MICB* | 0.425017935 | 0.003509996 | 0.005145103 | 1 |
| Obesity | LEC | *HLA-C* | 1.129490835 | 5.63E-06 | 9.64E-05 | 0.035389144 |
| Obesity | LEC | *POU5F1* | 0.891524826 | 5.75E-07 | 3.09E-05 | 0.003617341 |
| Obesity | LEC | *CCHCR1* | 1.411443879 | 7.01E-10 | 1.66E-07 | 4.41E-06 |
| Obesity | LEC | *PSORS1C1* | -0.229489541 | 0.009102495 | 0.012378842 | 1 |
| Obesity | LEC | *ABCF1* | 2.190104289 | 2.37E-07 | 1.85E-05 | 0.001490621 |
| Obesity | LEC | *LILRB3* | 0.795637589 | 1.41E-05 | 0.000105108 | 0.088789397 |
| Obesity | LEC | *LINC01123* | 16.95467826 | 0.012926292 | 0.017145063 | 1 |
| Obesity | LEC | *HLA-E* | 2.679207957 | 6.77E-11 | 2.24E-08 | 4.26E-07 |
| Obesity | LEC | *ZNF468* | 1.17877928 | 2.53E-05 | 0.000111345 | 0.15883687 |
| Obesity | LEC | *HLA-G* | 0.643899862 | 0.033488586 | 0.041782643 | 1 |
| Obesity | LEC | *HLA-F* | 1.445976831 | 0.00022898 | 0.000465739 | 1 |
| Obesity | LEC | *ZFP57* | 1.227801439 | 1.26E-07 | 1.24E-05 | 0.000794169 |
| Obesity | LEC | *FAM153C* | -1.835971755 | 0.000288165 | 0.000563337 | 1 |
| Obesity | LEC | *TRIM27* | 1.209910577 | 0.00943206 | 0.012796582 | 1 |
| Obesity | LEC | *MALRD1* | 0.760082638 | 0.000179911 | 0.000383532 | 1 |
| Obesity | LEC | *ATXN2* | 1.68459335 | 1.34E-05 | 0.000105108 | 0.084230937 |
| Obesity | LEC | *FAM216A* | 1.634301165 | 0.000334127 | 0.000635408 | 1 |
| Obesity | LEC | *ZNF783* | 1.559813157 | 0.000519025 | 0.000921296 | 1 |

| | | | | | | |
|---|---|---|---|---|---|---|
| Obesity | LEC | *ZNF425* | 1.0509857 | 0.005498833 | 0.007779289 | 1 |
| Obesity | LEC | *LRRC10B* | 7.007944803 | 0.000807574 | 0.001357545 | 1 |
| Obesity | LEC | *C12orf73* | 1.405362873 | 0.00770516 | 0.01059312 | 1 |
| Obesity | LEC | *PCDHGA1* | -2.062898027 | 0.000617411 | 0.001071923 | 1 |
| Obesity | LEC | *PCDHA5* | -4.197335708 | 0.000817072 | 0.001369484 | 1 |
| Obesity | LEC | *PCDHA4* | 3.697980897 | 0.002141852 | 0.003282746 | 1 |
| Obesity | LEC | *TRIM13* | 1.354764965 | 8.39E-05 | 0.00021504 | 0.527649579 |
| Obesity | LEC | *SLFN12L* | 0.462941879 | 9.89E-05 | 0.000242506 | 0.621976028 |
| Obesity | LEC | *CLLU1OS* | 1.987950862 | 0.001181722 | 0.001913336 | 1 |
| Obesity | LEC | *SYCE1L* | 1.28002998 | 0.000489301 | 0.00087642 | 1 |
| Obesity | LEC | *FAM71F2* | 0.84340321 | 2.21E-05 | 0.000108445 | 0.139136127 |
| Obesity | LEC | *AC016586.1* | 10.07456342 | 1.68E-05 | 0.000105108 | 0.105830253 |
| Obesity | LEC | *TMEM170B* | 0.533417923 | 0.018140353 | 0.023612505 | 1 |
| Obesity | LEC | *PRR13* | 1.220838806 | 0.000437521 | 0.00079661 | 1 |
| Obesity | LEC | *MT1A* | 8.909429565 | 2.33E-05 | 0.000109118 | 0.146469563 |
| Obesity | LEC | *MT1M* | 1.573306252 | 1.85E-05 | 0.000105108 | 0.116016675 |
| Obesity | LEC | *CFI* | -0.456857676 | 0.000227011 | 0.000462781 | 1 |
| Obesity | LEC | *SAMD9* | 2.371468183 | 0.001836095 | 0.002849551 | 1 |
| Obesity | LEC | *EXOC3L4* | 1.419289628 | 8.79E-05 | 0.000222231 | 0.552465267 |
| Obesity | LEC | *TP53TG3D* | 3.689401087 | 0.004347657 | 0.006267764 | 1 |
| Obesity | LEC | *ATP6AP1L* | 1.045018243 | 0.000182274 | 0.000387409 | 1 |
| Obesity | LEC | *NAP1L4* | 1.137144049 | 0.011789277 | 0.015763331 | 1 |
| Obesity | LEC | *CPT1B* | 0.934472135 | 0.013569305 | 0.01794871 | 1 |
| Obesity | LEC | *SMN2* | 0.391757895 | 0.034475037 | 0.042962252 | 1 |
| Obesity | LEC | *SERF1B* | -1.039767018 | 1.08E-05 | 0.000105108 | 0.06809143 |
| Obesity | LEC | *DENND6B* | 2.228143655 | 1.67E-05 | 0.000105108 | 0.105003171 |
| Obesity | LEC | *EIF3CL* | 7.173974546 | 3.34E-16 | 3.50E-13 | 2.10E-12 |
| Obesity | LEC | *LCMT1* | 1.631198318 | 3.58E-05 | 0.000123158 | 0.225255641 |
| Obesity | LEC | *CDPF1* | 0.875382589 | 0.000503122 | 0.000897341 | 1 |
| Obesity | LEC | *LIN52* | 1.488182273 | 8.22E-06 | 0.000105108 | 0.051709898 |
| Obesity | LEC | *C17orf107* | 2.145149395 | 6.17E-06 | 9.96E-05 | 0.038770611 |
| Obesity | LEC | *RFPL3S* | -0.865564039 | 0.012582534 | 0.016710264 | 1 |
| Obesity | LEC | *C1QTNF9B* | 8.9734244 | 2.88E-05 | 0.000115038 | 0.180921625 |
| Obesity | LEC | *RNPS1* | 1.536447368 | 0.006313524 | 0.008832471 | 1 |
| Obesity | LEC | *NYNRIN* | 1.28841117 | 1.22E-05 | 0.000105108 | 0.076846025 |
| Obesity | LEC | *GOLGA8O* | 3.434226583 | 0.000439329 | 0.00079967 | 1 |
| Obesity | LEC | *HCG27* | 0.804373589 | 0.000100015 | 0.000244192 | 0.628793531 |
| Obesity | LEC | *LNP1* | -1.44539428 | 0.000946232 | 0.001565104 | 1 |
| Obesity | LEC | *VGLL3* | 0.41562637 | 0.00076921 | 0.001299722 | 1 |
| Obesity | LEC | *METTL6* | 2.199603491 | 6.08E-06 | 9.95E-05 | 0.038216061 |
| Obesity | LEC | *SACM1L* | 1.85891607 | 0.000321606 | 0.000614945 | 1 |
| Obesity | LEC | *SLC48A1* | 1.558700599 | 2.15E-05 | 0.000107557 | 0.135318939 |
| Obesity | LEC | *ZRSR1* | 3.680250155 | 3.28E-05 | 0.000118709 | 0.205944302 |
| Obesity | LEC | *C17orf51* | 1.891909128 | 9.70E-05 | 0.000239091 | 0.609681274 |
| Obesity | LEC | *OR2A42* | 0.763726197 | 0.010628412 | 0.014321578 | 1 |
| Obesity | LEC | *RNF208* | -4.370679314 | 0.039514027 | 0.048816013 | 1 |
| Obesity | LEC | *MAP10* | 1.134401 | 4.75E-05 | 0.000147095 | 0.298603773 |
| Obesity | LEC | *ZNF580* | 1.918301923 | 5.90E-05 | 0.000170005 | 0.371117623 |
| Obesity | LEC | *NUP62* | 1.563985163 | 2.77E-05 | 0.00011384 | 0.173912722 |
| Obesity | LEC | *FAM106A* | 0.849264044 | 0.02163171 | 0.027783159 | 1 |

| | | | | | | |
|---|---|---|---|---|---|---|
| Obesity | LEC | *ZNF254* | 1.598898303 | 0.005962457 | 0.00837114 | 1 |
| Obesity | LEC | *TCTEX1D2* | -0.665027023 | 0.000185821 | 0.000392823 | 1 |
| Obesity | LEC | *AC022498.1* | -1.284901469 | 1.13E-05 | 0.000105108 | 0.0713063 |
| Obesity | LEC | *ASIC3* | 2.525884823 | 9.13E-09 | 1.79E-06 | 5.74E-05 |
| Obesity | LEC | *GIMAP1* | -0.912804786 | 0.000594595 | 0.001037712 | 1 |
| Obesity | LEC | *CCDC183* | 2.394739884 | 2.55E-05 | 0.000111732 | 0.160220968 |
| Obesity | LEC | *ARHGEF35* | -1.540780508 | 6.99E-05 | 0.000189301 | 0.439308022 |
| Obesity | LEC | *NOTCH2NL* | 1.772536745 | 0.000310472 | 0.000597105 | 1 |
| Obesity | LEC | *MXD3* | 1.139244823 | 9.74E-06 | 0.000105108 | 0.061246741 |
| Obesity | LEC | *GSTM2* | 1.069453692 | 0.000837909 | 0.001401046 | 1 |
| Obesity | LEC | *SIPA1* | 3.233337495 | 8.48E-07 | 3.86E-05 | 0.005328441 |
| Obesity | LEC | *ERV3-1* | 0.616523418 | 9.99E-07 | 4.29E-05 | 0.006282466 |
| Obesity | LEC | *CCL14* | -0.341848176 | 0.007051859 | 0.009769731 | 1 |
| Obesity | LEC | *RBMXL1* | 2.613070786 | 2.36E-05 | 0.000109118 | 0.148133112 |
| Obesity | LEC | *SRA1* | 1.235855878 | 0.007589779 | 0.010448202 | 1 |
| Obesity | LEC | *TMEM110* | 2.63280953 | 9.21E-08 | 1.02E-05 | 0.000578978 |
| Obesity | LEC | *HEXA* | 2.263352993 | 1.96E-05 | 0.000105217 | 0.122982484 |
| Obesity | LEC | *LEPROT* | 1.585674296 | 0.000547153 | 0.000964652 | 1 |
| Obesity | LEC | *LBH* | 1.27358876 | 1.92E-05 | 0.000105211 | 0.121006731 |
| Obesity | LEC | *PPP1CB* | 1.316104798 | 0.001558717 | 0.002454823 | 1 |
| Obesity | LEC | *NCKIPSD* | 0.820190443 | 4.74E-05 | 0.000146993 | 0.298248951 |
| Obesity | LEC | *ATF6B* | 1.083301898 | 0.022295869 | 0.028566157 | 1 |
| Obesity | LEC | *SLC35F6* | 3.511044683 | 6.33E-05 | 0.000177484 | 0.397741529 |
| Obesity | LEC | *CLIC1* | 1.586339438 | 0.013404794 | 0.01774604 | 1 |
| Obesity | LEC | *KLRK1* | 1.558919934 | 0.034385191 | 0.042858782 | 1 |
| Obesity | LEC | *EMP2* | 1.614646448 | 0.004768231 | 0.006803874 | 1 |
| Obesity | LEC | *PPM1N* | 1.007512843 | 2.80E-05 | 0.000114038 | 0.176065117 |
| Obesity | LEC | *SLC23A3* | 1.038071965 | 8.62E-05 | 0.000219526 | 0.542228666 |
| Obesity | LEC | *LTB4R* | 1.092773466 | 0.00074115 | 0.001259402 | 1 |
| Obesity | LEC | *LTB4R2* | 1.386118882 | 0.001201219 | 0.001941904 | 1 |
| Obesity | LEC | *GALT* | 2.327027514 | 3.80E-07 | 2.50E-05 | 0.002389443 |
| Obesity | LEC | *ZNF726* | 3.067857646 | 4.51E-05 | 0.000142724 | 0.283735423 |
| Obesity | LEC | *TAX1BP3* | -0.750714409 | 0.000407424 | 0.000750725 | 1 |
| Obesity | LEC | *AP1G2* | -0.929133939 | 0.000604639 | 0.001054178 | 1 |
| Obesity | LEC | *ZNF90* | 2.481129784 | 0.000192529 | 0.000404684 | 1 |
| Obesity | LEC | *TM6SF2* | 10.60784253 | 2.22E-05 | 0.000108559 | 0.139781827 |
| Obesity | LEC | *SMIM7* | 1.17399063 | 9.13E-06 | 0.000105108 | 0.057393309 |
| Obesity | LEC | *AC079776.2* | -2.371090392 | 0.000876851 | 0.001461108 | 1 |
| Obesity | LEC | *WEE2* | 1.728526523 | 0.00524881 | 0.007440647 | 1 |
| Obesity | LEC | *PAXIP1-AS2* | 1.51489737 | 9.04E-06 | 0.000105108 | 0.056810991 |
| Obesity | LEC | *PRCD* | 0.732288182 | 5.24E-05 | 0.000156459 | 0.329190016 |
| Obesity | LEC | *C17orf67* | -1.237436732 | 0.000281157 | 0.000551868 | 1 |
| Obesity | LEC | *FIS1* | 1.367204373 | 0.00202472 | 0.003116899 | 1 |
| Obesity | LEC | *COLCA2* | 0.898444964 | 0.000116303 | 0.000273242 | 0.73119454 |
| Obesity | LEC | *NEURL1B* | 1.305798097 | 1.67E-05 | 0.000105108 | 0.105247812 |
| Obesity | LEC | *VSTM5* | 0.698420192 | 0.004764656 | 0.006800934 | 1 |
| Obesity | LEC | *PPME1* | 1.29761128 | 0.00358592 | 0.005242948 | 1 |
| Obesity | LEC | *NUTM2D* | 2.270850665 | 0.015934188 | 0.020883519 | 1 |
| Obesity | LEC | *CPEB1* | 0.777605183 | 3.05E-05 | 0.000115831 | 0.19168151 |
| Obesity | LEC | *C10orf105* | -1.710369137 | 0.000741148 | 0.001259402 | 1 |

| | | | | | | |
|---|---|---|---|---|---|---|
| Obesity | LEC | *ARHGEF33* | 0.904206325 | 0.006528707 | 0.009107162 | 1 |
| Obesity | LEC | *AC005082.1* | 3.081638905 | 2.41E-05 | 0.000109361 | 0.151700196 |
| Obesity | LEC | *SMTNL1* | 0.385698924 | 0.023800548 | 0.030364052 | 1 |
| Obesity | LEC | *TBC1D26* | 13.59347232 | 0.016585794 | 0.021701329 | 1 |
| Obesity | LEC | *ACSM4* | 0.606526086 | 0.027295387 | 0.034549245 | 1 |
| Obesity | LEC | *COL28A1* | 3.093055104 | 1.26E-06 | 4.90E-05 | 0.007903354 |
| Obesity | LEC | *PHB2* | 1.232457737 | 8.64E-05 | 0.000219858 | 0.543268143 |
| Obesity | LEC | *UBXN2B* | 1.46009695 | 0.005751794 | 0.008106149 | 1 |
| Obesity | LEC | *UBE2QL1* | -1.518954952 | 0.00052974 | 0.000937898 | 1 |
| Obesity | LEC | *KCNU1* | 1.84526161 | 3.04E-05 | 0.000115831 | 0.191111565 |
| Obesity | LEC | *MYL5* | 0.964988664 | 3.01E-05 | 0.000115831 | 0.189083184 |
| Obesity | LEC | *NPEPL1* | 0.866142291 | 2.33E-05 | 0.000109118 | 0.146416104 |
| Obesity | LEC | *LINC01139* | -0.611024306 | 0.000332343 | 0.000632398 | 1 |
| Obesity | LEC | *TSTD1* | 0.741121967 | 2.27E-05 | 0.000109111 | 0.142566911 |
| Obesity | LEC | *LINC01138* | 1.355558146 | 1.42E-05 | 0.000105108 | 0.089223443 |
| Obesity | LEC | *CYB5RL* | -0.552447429 | 0.000282137 | 0.000553446 | 1 |
| Obesity | LEC | *ATAD3C* | -0.91959192 | 0.002477974 | 0.003740461 | 1 |
| Obesity | LEC | *CCDC7* | 1.349168728 | 0.009018663 | 0.012275457 | 1 |
| Obesity | LEC | *AC016773.1* | 0.788507812 | 0.000251805 | 0.000504976 | 1 |
| Obesity | LEC | *AP000350.4* | 0.413423397 | 0.034658453 | 0.043165153 | 1 |
| Obesity | LEC | *RNASEK* | 2.362725246 | 4.17E-05 | 0.000135166 | 0.262087246 |
| Obesity | LEC | *NBPF1* | 1.019685643 | 1.73E-05 | 0.000105108 | 0.108610588 |
| Obesity | LEC | *FAM228B* | 0.512997818 | 0.039665755 | 0.048993831 | 1 |
| Obesity | LEC | *RNU6ATAC35I* | 10.67149444 | 1.49E-05 | 0.000105108 | 0.093673467 |
| Obesity | LEC | *C6orf226* | -1.360581214 | 0.001330078 | 0.002125623 | 1 |
| Obesity | LEC | *PPP3R1* | 1.457528232 | 5.56E-05 | 0.000163862 | 0.349681866 |
| Obesity | LEC | *FANCG* | -0.930739349 | 0.014909057 | 0.019609465 | 1 |
| Obesity | LEC | *C19orf73* | 5.021069775 | 8.78E-05 | 0.000222231 | 0.55180207 |
| Obesity | LEC | *OR2A25* | 27.91081419 | 0.00012994 | 0.000297717 | 0.816934396 |
| Obesity | LEC | *XKR9* | 1.435967465 | 8.96E-06 | 0.000105108 | 0.056361063 |
| Obesity | LEC | *C1orf229* | 1.866348064 | 0.006400427 | 0.008942107 | 1 |
| Obesity | LEC | *SLC12A8* | 0.770958823 | 0.020839029 | 0.026858338 | 1 |
| Obesity | LEC | *APOL6* | 1.060907579 | 1.41E-05 | 0.000105108 | 0.088926681 |
| Obesity | LEC | *TIAF1* | 1.343642554 | 0.000237762 | 0.000481111 | 1 |
| Obesity | LEC | *FAM185A* | 1.924520299 | 6.06E-06 | 9.95E-05 | 0.038115304 |
| Obesity | LEC | *AC062017.1* | 0.728039147 | 2.60E-05 | 0.000112197 | 0.163695243 |
| Obesity | LEC | *ADRA2B* | -6.056255239 | 0.004411033 | 0.006348938 | 1 |
| Obesity | LEC | *VPS52* | 2.303589695 | 0.00016602 | 0.000359423 | 1 |
| Obesity | LEC | *TINCR* | 2.575097111 | 0.027290043 | 0.034549245 | 1 |
| Obesity | LEC | *LINC00205* | 1.339087853 | 0.000518531 | 0.000920905 | 1 |
| Obesity | LEC | *AC147651.3* | 1.660839362 | 0.000318756 | 0.000609866 | 1 |
| Obesity | LEC | *HLA-DPB1* | 1.14848724 | 4.15E-05 | 0.000134809 | 0.260989279 |
| Obesity | LEC | *OSER1-AS1* | 1.869164404 | 1.85E-06 | 5.72E-05 | 0.011602787 |
| Obesity | LEC | *LINC01133* | 0.543645343 | 2.81E-05 | 0.000114038 | 0.176763257 |
| Obesity | LEC | *LINC01117* | -0.564783319 | 0.000389308 | 0.000722 | 1 |
| Obesity | LEC | *RASAL2-AS1* | -1.549138558 | 7.89E-05 | 0.00020643 | 0.49625694 |
| Obesity | LEC | *AC069213.1* | -0.809097072 | 3.12E-05 | 0.000116657 | 0.196217086 |
| Obesity | LEC | *LINC00240* | 1.316028837 | 1.05E-09 | 2.36E-07 | 6.62E-06 |
| Obesity | LEC | *LINC00863* | 1.207397816 | 2.47E-06 | 6.73E-05 | 0.01554233 |
| Obesity | LEC | *LINC00184* | 1.550956674 | 0.000232022 | 0.000471317 | 1 |

| | | | | | | |
|---|---|---|---|---|---|---|
| Obesity | LEC | *TMEM233* | 0.557600386 | 1.40E-05 | 0.000105108 | 0.088256905 |
| Obesity | LEC | *LINC00237* | -4.004097262 | 0.00028359 | 0.000555949 | 1 |
| Obesity | LEC | *PLEKHM1* | 1.80444342 | 8.61E-07 | 3.90E-05 | 0.005414071 |
| Obesity | LEC | *AP000350.6* | -0.587078729 | 0.012179322 | 0.016233071 | 1 |
| Obesity | LEC | *SFTA1P* | 2.381293738 | 0.000134168 | 0.000304408 | 0.843514094 |
| Obesity | LEC | *FAM225B* | 1.087838155 | 0.005010383 | 0.007120316 | 1 |
| Obesity | LEC | *SLC26A6* | -1.343780533 | 0.000953149 | 0.001575302 | 1 |
| Obesity | LEC | *MIAT* | 1.014972593 | 2.39E-05 | 0.000109361 | 0.15041233 |
| Obesity | LEC | *LINC00626* | -4.919601557 | 0.000153227 | 0.000337895 | 0.963337704 |
| Obesity | LEC | *ERCC6* | 2.139192103 | 7.43E-05 | 0.000197765 | 0.467318207 |
| Obesity | LEC | *NOL7* | 2.335156868 | 9.68E-05 | 0.000238821 | 0.608362441 |
| Obesity | LEC | *ELFN1* | 1.894285541 | 2.69E-05 | 0.000113184 | 0.16928081 |
| Obesity | LEC | *ZNF503-AS1* | 3.22550459 | 4.24E-05 | 0.000136977 | 0.266693904 |
| Obesity | LEC | *LINC00937* | 0.84794772 | 4.96E-05 | 0.000151155 | 0.311530439 |
| Obesity | LEC | *AC064836.3* | 1.488713168 | 2.93E-05 | 0.000115648 | 0.184433752 |
| Obesity | LEC | *TMEM185B* | -1.400144205 | 0.000577904 | 0.001013527 | 1 |
| Obesity | LEC | *HSBP1L1* | 2.858113273 | 3.65E-05 | 0.000124736 | 0.229228864 |
| Obesity | LEC | *TAS2R46* | -1.370853318 | 0.00046776 | 0.000842879 | 1 |
| Obesity | LEC | *AC093901.1* | 1.350894361 | 2.73E-05 | 0.000113803 | 0.171840399 |
| Obesity | LEC | *ZNF717* | 0.465116886 | 0.000240324 | 0.000485201 | 1 |
| Obesity | LEC | *AC009299.3* | 1.448432757 | 2.60E-05 | 0.000112153 | 0.163341674 |
| Obesity | LEC | *SLC2A1-AS1* | -1.877031903 | 0.000278538 | 0.000547753 | 1 |
| Obesity | LEC | *LINC01068* | 1.762515728 | 0.000115062 | 0.000271036 | 0.723395663 |
| Obesity | LEC | *AC108463.2* | -0.562002295 | 4.26E-05 | 0.000137183 | 0.267753558 |
| Obesity | LEC | *POLR2J2* | -0.515189178 | 0.007728441 | 0.010620483 | 1 |
| Obesity | LEC | *C19orf24* | 1.388109453 | 7.71E-05 | 0.000202909 | 0.484974695 |
| Obesity | LEC | *LINC01013* | -2.533404025 | 0.006393252 | 0.008934069 | 1 |
| Obesity | LEC | *AC073133.2* | 7.089483595 | 0.002913295 | 0.004336148 | 1 |
| Obesity | LEC | *PROB1* | 4.558313739 | 0.000244925 | 0.000493382 | 1 |
| Obesity | LEC | *ARL17B* | 1.543725044 | 0.000134032 | 0.000304249 | 0.842661267 |
| Obesity | LEC | *DHFR* | 0.458291376 | 0.013972929 | 0.018448699 | 1 |
| Obesity | LEC | *LINC00954* | 4.518404347 | 0.004601309 | 0.00659112 | 1 |
| Obesity | LEC | *AC067945.3* | 1.512886802 | 0.000170086 | 0.000365833 | 1 |
| Obesity | LEC | *RPL41* | 1.950180269 | 0.000111366 | 0.000263552 | 0.700158943 |
| Obesity | LEC | *PATL2* | 1.678105647 | 7.87E-06 | 0.000105108 | 0.049451784 |
| Obesity | LEC | *ZNF492* | 1.108847569 | 0.000348252 | 0.00065651 | 1 |
| Obesity | LEC | *U91324.1* | 4.627436259 | 1.33E-05 | 0.000105108 | 0.083671819 |
| Obesity | LEC | *ZNF688* | 1.891214106 | 7.11E-05 | 0.000191443 | 0.447019774 |
| Obesity | LEC | *CISD3* | -1.016088884 | 0.000491159 | 0.000879399 | 1 |
| Obesity | LEC | *KLHL7-AS1* | 1.763264262 | 2.96E-05 | 0.000115831 | 0.185827075 |
| Obesity | LEC | *AC092171.4* | 2.196976696 | 0.000220126 | 0.000451528 | 1 |
| Obesity | LEC | *HLA-DPA1* | 1.523097476 | 4.93E-06 | 9.12E-05 | 0.031007433 |
| Obesity | LEC | *RPS18* | 1.815824721 | 0.00055526 | 0.000977848 | 1 |
| Obesity | LEC | *DIRC3* | 0.947433195 | 4.72E-05 | 0.000146463 | 0.296440597 |
| Obesity | LEC | *LINC00865* | 1.017944897 | 8.49E-05 | 0.000216988 | 0.533573262 |
| Obesity | LEC | *AC009495.3* | 3.400856999 | 0.02132447 | 0.02742216 | 1 |
| Obesity | LEC | *GOLGA8N* | 1.312185066 | 7.73E-05 | 0.000203131 | 0.486091807 |
| Obesity | LEC | *LYRM9* | 0.945632023 | 3.35E-05 | 0.00011987 | 0.210491665 |
| Obesity | LEC | *LINC00342* | 2.622717094 | 0.000486359 | 0.000871533 | 1 |
| Obesity | LEC | *SNHG15* | 2.894948652 | 8.82E-07 | 3.91E-05 | 0.005546296 |

| | | | | | | |
|---|---|---|---|---|---|---|
| Obesity | LEC | *LINC00702* | 1.017879156 | 2.00E-05 | 0.000105913 | 0.125825019 |
| Obesity | LEC | *LINC00472* | 1.233257304 | 4.64E-05 | 0.000145502 | 0.292022783 |
| Obesity | LEC | *AC073133.1* | -1.089602173 | 0.008348396 | 0.01142001 | 1 |
| Obesity | LEC | *TRIM26* | 1.815376799 | 0.002663949 | 0.003990529 | 1 |
| Obesity | LEC | *LINC01114* | -4.480200493 | 0.036219244 | 0.045019847 | 1 |
| Obesity | LEC | *HLA-B* | 1.263973724 | 1.61E-06 | 5.41E-05 | 0.010124891 |
| Obesity | LEC | *C19orf81* | -2.074144637 | 0.023144801 | 0.029587508 | 1 |
| Obesity | LEC | *ANKRD65* | -0.890058482 | 0.000557127 | 0.000980363 | 1 |
| Obesity | LEC | *LINC00957* | 1.980607272 | 1.18E-05 | 0.000105108 | 0.074111047 |
| Obesity | LEC | *LINC00484* | -0.585368545 | 0.008261736 | 0.011306386 | 1 |
| Obesity | LEC | *KIAA0040* | 2.216498415 | 6.42E-06 | 0.000101236 | 0.040392996 |
| Obesity | LEC | *LINC00941* | -0.892831529 | 0.00011589 | 0.000272374 | 0.728601708 |
| Obesity | LEC | *ZBTB22* | 1.875700099 | 1.36E-05 | 0.000105108 | 0.085553024 |
| Obesity | LEC | *B3GNT9* | 1.972230891 | 1.19E-05 | 0.000105108 | 0.07508609 |
| Obesity | LEC | *CDKN2AIPNL* | 1.401572189 | 0.002508509 | 0.003782853 | 1 |
| Obesity | LEC | *GS1-124K5.4* | 0.543264851 | 0.001922792 | 0.002972361 | 1 |
| Obesity | LEC | *ZNF737* | 2.402144745 | 7.30E-05 | 0.000195658 | 0.459208515 |
| Obesity | LEC | *HLA-DQA2* | 0.519732638 | 0.023368315 | 0.029855028 | 1 |
| Obesity | LEC | *AC073257.2* | 7.364673612 | 7.59E-05 | 0.000200935 | 0.477421604 |
| Obesity | LEC | *LRRC37A2* | 1.211300283 | 2.56E-05 | 0.000111859 | 0.160760164 |
| Obesity | LEC | *TXNDC5* | -1.236549735 | 5.73E-05 | 0.000166529 | 0.360035151 |
| Obesity | LEC | *ASB14* | 1.873581802 | 0.001227275 | 0.001977416 | 1 |
| Obesity | LEC | *ADSL* | 0.882181083 | 4.69E-05 | 0.000146016 | 0.294951573 |
| Obesity | LEC | *LILRA4* | 21.6920998 | 0.005178866 | 0.007348123 | 1 |
| Obesity | LEC | *LILRA2* | 0.618556733 | 0.034923377 | 0.043477876 | 1 |
| Obesity | LEC | *PSMB9* | 1.611542062 | 1.57E-07 | 1.43E-05 | 0.000984143 |
| Obesity | LEC | *COX19* | 1.195270372 | 1.44E-05 | 0.000105108 | 0.090424287 |
| Obesity | LEC | *L1TD1* | 0.525200628 | 0.027669666 | 0.034973701 | 1 |
| Obesity | LEC | *AQP1* | 1.958883499 | 0.000883444 | 0.001470536 | 1 |
| Obesity | LEC | *C1QTNF9* | 2.943765088 | 0.000710509 | 0.00121418 | 1 |
| Obesity | LEC | *ARHGEF25* | 2.248816515 | 4.74E-05 | 0.000146992 | 0.29809965 |
| Obesity | LEC | *LINC00886* | 1.651265434 | 0.001293046 | 0.002073993 | 1 |
| Obesity | LEC | *MIF* | 0.578277615 | 0.000352375 | 0.000662891 | 1 |
| Obesity | LEC | *NSUN6* | 1.476562034 | 2.01E-05 | 0.000105931 | 0.12613786 |
| Obesity | LEC | *HLA-DOB* | -0.286010001 | 0.033241858 | 0.041499515 | 1 |
| Obesity | LEC | *TDGF1* | 1.1364072 | 0.028392569 | 0.035858594 | 1 |
| Obesity | LEC | *CD302* | 1.861346584 | 0.000121588 | 0.000282576 | 0.764423338 |
| Obesity | LEC | *ATP5J2* | 1.865575056 | 0.027457249 | 0.03473488 | 1 |
| Obesity | LEC | *ARHGAP8* | 3.488796591 | 2.64E-05 | 0.000112582 | 0.166130337 |
| Obesity | LEC | *PLEKHO2* | 2.704977555 | 2.99E-05 | 0.000115831 | 0.188168486 |
| Obesity | LEC | *PWP2* | 2.958652186 | 0.000257636 | 0.000513693 | 1 |
| Obesity | LEC | *PI4KA* | 2.01850562 | 8.31E-05 | 0.000214185 | 0.522484728 |
| Obesity | LEC | *AKAP2* | -0.618781912 | 9.36E-05 | 0.000233455 | 0.588772261 |
| Obesity | LEC | *LINC00969* | 0.989724706 | 0.004892288 | 0.006963508 | 1 |
| Obesity | LEC | *ARHGDIG* | 1.151452743 | 9.26E-05 | 0.000231285 | 0.582144706 |
| Obesity | LEC | *TCP10L* | 6.712553405 | 3.89E-05 | 0.000129587 | 0.244357015 |
| Obesity | LEC | *C22orf39* | 1.644570962 | 1.07E-05 | 0.000105108 | 0.067159668 |
| Obesity | LEC | *MRPL20* | 2.234616447 | 1.50E-05 | 0.000105108 | 0.094551745 |
| Obesity | LEC | *DECR2* | 0.870976055 | 2.96E-05 | 0.000115831 | 0.186067221 |
| Obesity | LEC | *CCDC169* | 1.855912143 | 4.65E-05 | 0.000145571 | 0.292598253 |

| | | | | | | |
|---|---|---|---|---|---|---|
| Obesity | LEC | *AP5Z1* | 1.63330333 | 4.80E-06 | 9.10E-05 | 0.030151664 |
| Obesity | LEC | *STRC* | -4.324188669 | 0.00121982 | 0.001966917 | 1 |
| Obesity | LEC | *PSG4* | 1.04944346 | 0.000521686 | 0.000924714 | 1 |
| Obesity | LEC | *VSIG8* | 1.151490047 | 4.98E-05 | 0.000151505 | 0.313312113 |
| Obesity | LEC | *C7orf73* | 1.461537611 | 1.61E-05 | 0.000105108 | 0.10123529 |
| Obesity | LEC | *FGF14-IT1* | 8.559719294 | 2.22E-05 | 0.000108559 | 0.139711766 |
| Obesity | LEC | *TICAM2* | 2.447859289 | 4.62E-05 | 0.000145061 | 0.290701801 |
| Obesity | LEC | *PALM2* | 0.927592528 | 0.011931441 | 0.015939857 | 1 |
| Obesity | LEC | *NAT6* | 1.926615508 | 2.41E-07 | 1.85E-05 | 0.001513147 |
| Obesity | LEC | *IL10RB* | 1.664572238 | 0.00015107 | 0.000334664 | 0.949776396 |
| Obesity | LEC | *WDR92* | 1.597063012 | 1.41E-05 | 0.000105108 | 0.088715523 |
| Obesity | LEC | *NME1-NME2* | -2.006734857 | 0.014290924 | 0.018834855 | 1 |
| Obesity | LEC | *LEFTY1* | -1.115523747 | 7.32E-05 | 0.000195742 | 0.460384678 |
| Obesity | LEC | *TTC4* | 0.612420662 | 0.004804056 | 0.006848776 | 1 |
| Obesity | LEC | *APOBEC3D* | 2.554426809 | 0.006040317 | 0.008472885 | 1 |
| Obesity | LEC | *RN7SL832P* | 1.332059652 | 4.76E-06 | 9.07E-05 | 0.029932241 |
| Obesity | LEC | *TMEM199* | 3.053901066 | 0.003052747 | 0.004527629 | 1 |
| Obesity | LEC | *P2RY11* | 1.467846863 | 0.001380384 | 0.002200982 | 1 |
| Obesity | LEC | *DBNDD2* | -1.799975597 | 0.002512172 | 0.003784814 | 1 |
| Obesity | LEC | *LILRA6* | 0.689359789 | 9.43E-05 | 0.000234161 | 0.592662456 |
| Obesity | LEC | *APOBEC3C* | 2.310999467 | 1.10E-05 | 0.000105108 | 0.069126443 |
| Obesity | LEC | *CCDC13* | 0.792963143 | 0.000830785 | 0.001390244 | 1 |
| Obesity | LEC | *ASPRV1* | -2.514409657 | 0.000219168 | 0.000449856 | 1 |
| Obesity | LEC | *UBE2V1* | -1.020327274 | 0.0005947 | 0.001037712 | 1 |
| Obesity | LEC | *CRYBB2* | 2.749492951 | 7.12E-05 | 0.000191506 | 0.44735821 |
| Obesity | LEC | *N4BP2L2* | 1.175119623 | 0.001464177 | 0.002321634 | 1 |
| Obesity | LEC | *LINC00847* | 0.818835092 | 2.35E-05 | 0.000109118 | 0.147820734 |
| Obesity | LEC | *NEAT1* | 0.888895303 | 4.39E-05 | 0.000140307 | 0.275772578 |
| Obesity | LEC | *ZNF585B* | 1.349793517 | 0.004078085 | 0.005901111 | 1 |
| Obesity | LEC | *RAB30-AS1* | 1.380379366 | 0.001260763 | 0.002025663 | 1 |
| Obesity | LEC | *LINC00920* | -0.719792471 | 6.24E-05 | 0.000175895 | 0.392598034 |
| Obesity | LEC | *PGAM5* | -1.095850201 | 0.000360869 | 0.000676845 | 1 |
| Obesity | LEC | *DNAJC3-AS1* | -1.596075487 | 0.004428672 | 0.006368495 | 1 |
| Obesity | LEC | *BCKDHA* | 3.011499393 | 0.001475786 | 0.002338273 | 1 |
| Obesity | LEC | *CDK11B* | 2.288402887 | 7.36E-05 | 0.000196337 | 0.462963557 |
| Obesity | LEC | *ABHD14A* | 1.896947206 | 4.70E-06 | 9.00E-05 | 0.029530426 |
| Obesity | LEC | *SRP14-AS1* | 0.709039858 | 2.32E-06 | 6.57E-05 | 0.014581119 |
| Obesity | LEC | *CCDC153* | 1.678459409 | 4.85E-06 | 9.10E-05 | 0.030463709 |
| Obesity | LEC | *C5orf17* | 2.138415026 | 0.010020337 | 0.013545013 | 1 |
| Obesity | LEC | *FMN1* | 0.525356644 | 8.56E-05 | 0.000218253 | 0.538429925 |
| Obesity | LEC | *NAIP* | 1.092956776 | 2.73E-05 | 0.000113713 | 0.171360391 |
| Obesity | LEC | *ZNF286B* | 1.089216547 | 2.07E-05 | 0.000106696 | 0.130368926 |
| Obesity | LEC | *LINC00942* | 2.457256792 | 0.003873461 | 0.005626722 | 1 |
| Obesity | LEC | *PDCD6* | 2.111848625 | 3.98E-05 | 0.000131395 | 0.250072119 |
| Obesity | LEC | *LINC00603* | 1.184576258 | 5.43E-05 | 0.000160788 | 0.341514567 |
| Obesity | LEC | *YJEFN3* | 1.049372031 | 7.49E-05 | 0.000199019 | 0.470680411 |
| Obesity | LEC | *DNAH10OS* | 6.308919749 | 2.88E-07 | 2.13E-05 | 0.001809866 |
| Obesity | LEC | *PCDHA10* | 1.68480199 | 1.43E-05 | 0.000105108 | 0.089952304 |
| Obesity | LEC | *LINC00989* | -1.420113074 | 0.003765494 | 0.005484267 | 1 |
| Obesity | LEC | *LINC01021* | 3.230831458 | 1.75E-05 | 0.000105108 | 0.11009389 |

| | | | | | | |
|---|---|---|---|---|---|---|
| Obesity | LEC | *FAM218A* | 1.409315976 | 3.03E-05 | 0.000115831 | 0.190468219 |
| Obesity | LEC | *ATP6V1E2* | 0.716382482 | 0.006518005 | 0.009094522 | 1 |
| Obesity | LEC | *GLI4* | 1.643516572 | 0.003857064 | 0.005605493 | 1 |
| Obesity | LEC | *SHANK3* | -0.336145029 | 0.000104816 | 0.000252502 | 0.658979948 |
| Obesity | LEC | *ZNF550* | 2.42912358 | 5.45E-05 | 0.000161076 | 0.342768743 |
| Obesity | LEC | *TMEM200B* | 1.494183341 | 0.0043585 | 0.006280516 | 1 |
| Obesity | LEC | *PCDHGB6* | 1.420016903 | 2.75E-05 | 0.00011384 | 0.172852357 |
| Obesity | LEC | *TRNP1* | 3.291675712 | 3.10E-05 | 0.000116544 | 0.195020478 |
| Obesity | LEC | *PCDHGA5* | 0.773510257 | 4.01E-05 | 0.000131815 | 0.252294226 |
| Obesity | LEC | *ALG11* | 0.979835619 | 0.001218395 | 0.001965122 | 1 |
| Obesity | LEC | *ATXN7L3B* | 2.521339212 | 0.000141346 | 0.000317372 | 0.888641577 |
| Obesity | LEC | *PRKDC* | 1.738237799 | 0.00036907 | 0.000689962 | 1 |
| Obesity | LEC | *PCDHGA11* | 1.184532034 | 0.0004459 | 0.000809521 | 1 |
| Obesity | LEC | *CLDN23* | 1.707196952 | 3.48E-06 | 7.79E-05 | 0.021870121 |
| Obesity | LEC | *ZNF260* | 2.232237024 | 0.004390008 | 0.006324468 | 1 |
| Obesity | LEC | *ARMS2* | 1.696283013 | 0.013882382 | 0.018335827 | 1 |
| Obesity | LEC | *FPGT* | 1.550780383 | 1.44E-05 | 0.000105108 | 0.090682473 |
| Obesity | LEC | *MPV17L2* | -0.557534593 | 0.005386503 | 0.007628958 | 1 |
| Obesity | LEC | *DPP3* | 1.404289229 | 1.64E-07 | 1.45E-05 | 0.001031179 |
| Obesity | LEC | *KRTAP5-9* | 4.892473562 | 0.019376521 | 0.025096866 | 1 |
| Obesity | LEC | *EID3* | 1.058654153 | 2.87E-05 | 0.000115038 | 0.180651732 |
| Obesity | LEC | *SNHG9* | 1.412568744 | 2.14E-05 | 0.000107354 | 0.134514707 |
| Obesity | LEC | *EBLN2* | 1.529285593 | 7.90E-06 | 0.000105108 | 0.049683858 |
| Obesity | LEC | *POLR2M* | 1.911759755 | 1.39E-05 | 0.000105108 | 0.087620934 |
| Obesity | LEC | *TAS2R20* | -0.440885033 | 0.000302606 | 0.000584481 | 1 |
| Obesity | LEC | *LINC00346* | 2.167811399 | 2.94E-05 | 0.000115699 | 0.185048533 |
| Obesity | LEC | *CTSO* | 1.843300913 | 1.72E-05 | 0.000105108 | 0.107911081 |
| Obesity | LEC | *ZNF432* | 1.944004901 | 3.08E-05 | 0.000116141 | 0.193607879 |
| Obesity | LEC | *LINC00944* | 1.08899507 | 5.89E-05 | 0.000169777 | 0.370112927 |
| Obesity | LEC | *ZNF10* | 1.046690744 | 0.009500672 | 0.012878552 | 1 |
| Obesity | LEC | *HMBS* | 2.466447979 | 8.31E-15 | 5.23E-12 | 5.23E-11 |
| Obesity | LEC | *TAS2R31* | -0.89462178 | 0.000396197 | 0.000732614 | 1 |
| Obesity | LEC | *AC093668.2* | -1.585227983 | 0.027678479 | 0.034977808 | 1 |
| Obesity | LEC | *PGA5* | 7.01535541 | 0.012189178 | 0.016242093 | 1 |
| Obesity | LEC | *ZNF253* | 1.904653599 | 0.006802603 | 0.009455663 | 1 |
| Obesity | LEC | *KIAA1147* | 2.195192412 | 0.000298676 | 0.000578133 | 1 |
| Obesity | LEC | *GATC* | 1.738871424 | 0.000434629 | 0.000792722 | 1 |
| Obesity | LEC | *ZBED6* | -3.686963784 | 0.009972969 | 0.013486784 | 1 |
| Obesity | LEC | *CUX1* | 1.345161775 | 0.003216247 | 0.004752184 | 1 |
| Obesity | LEC | *RTEL1* | 1.562389077 | 0.000423684 | 0.000775685 | 1 |
| Obesity | LEC | *ZNF578* | -1.084927438 | 0.005698218 | 0.00804507 | 1 |
| Obesity | LEC | *PDF* | -4.535416304 | 5.62E-05 | 0.000164918 | 0.353254658 |
| Obesity | LEC | *LINC00640* | 0.65885229 | 0.000491379 | 0.000879399 | 1 |
| Obesity | LEC | *LINC00638* | 2.776532605 | 7.02E-05 | 0.000189971 | 0.441636015 |
| Obesity | LEC | *DUXA* | -1.384327034 | 0.011014247 | 0.014812101 | 1 |
| Obesity | LEC | *CEP95* | 1.581739096 | 2.68E-05 | 0.000113184 | 0.16843433 |
| Obesity | LEC | *LINC00639* | 0.491840294 | 1.57E-05 | 0.000105108 | 0.098965689 |
| Obesity | LEC | *LINC00648* | 0.940678682 | 1.85E-05 | 0.000105108 | 0.116436562 |
| Obesity | LEC | *LINC00924* | -0.189972719 | 8.88E-05 | 0.000223816 | 0.557973823 |
| Obesity | LEC | *ITGB3* | 0.575147825 | 0.016662178 | 0.021792201 | 1 |

| | | | | | | |
|---|---|---|---|---|---|---|
| Obesity | LEC | *UBE2Q2L* | -1.912342779 | 3.10E-05 | 0.000116539 | 0.19484621 |
| Obesity | LEC | *IQCH-AS1* | 1.219263159 | 0.000493652 | 0.000883207 | 1 |
| Obesity | LEC | *AC083843.1* | -5.918218825 | 1.35E-05 | 0.000105108 | 0.084577144 |
| Obesity | LEC | *SH3RF3-AS1* | -2.561921578 | 8.42E-07 | 3.86E-05 | 0.005296544 |
| Obesity | LEC | *PWRN1* | 1.947947117 | 0.009411773 | 0.012771815 | 1 |
| Obesity | LEC | *HOXB7* | 2.822279613 | 1.11E-07 | 1.14E-05 | 0.000696438 |
| Obesity | LEC | *FRRS1L* | 1.815973969 | 1.54E-05 | 0.000105108 | 0.096996069 |
| Obesity | LEC | *AC009120.5* | -1.386509988 | 0.000523732 | 0.000927783 | 1 |
| Obesity | LEC | *LINC00565* | 7.993560191 | 0.001546138 | 0.002437587 | 1 |
| Obesity | LEC | *TMEM178B* | 1.185066674 | 1.33E-05 | 0.000105108 | 0.083655935 |
| Obesity | LEC | *LINC01003* | 1.028392171 | 3.98E-05 | 0.000131395 | 0.250176464 |
| Obesity | LEC | *HPR* | -0.965793323 | 0.000621026 | 0.001076776 | 1 |
| Obesity | LEC | *GOLGA8H* | 1.578880379 | 1.88E-06 | 5.79E-05 | 0.011808628 |
| Obesity | LEC | *LINC00662* | 1.928358876 | 0.000866115 | 0.001443984 | 1 |
| Obesity | LEC | *AC123768.4* | -3.714839727 | 0.000263767 | 0.000524117 | 1 |
| Obesity | LEC | *LINC00667* | 0.971819852 | 0.002530939 | 0.003809435 | 1 |
| Obesity | LEC | *LINC00672* | 1.612442714 | 3.55E-05 | 0.000123048 | 0.222995931 |
| Obesity | LEC | *LINC00909* | 1.334525016 | 0.028997374 | 0.036556345 | 1 |
| Obesity | LEC | *DYNLL2* | 1.882574969 | 0.000160947 | 0.000351102 | 1 |
| Obesity | LEC | *LINC00526* | 1.068608454 | 8.13E-05 | 0.000210521 | 0.511145054 |
| Obesity | LEC | *LINC00668* | 21.44975401 | 0.038080689 | 0.04716574 | 1 |
| Obesity | LEC | *STRADA* | 1.114629858 | 0.008772097 | 0.011955381 | 1 |
| Obesity | LEC | *MYO15B* | 1.72465054 | 0.007130877 | 0.009874851 | 1 |
| Obesity | LEC | *LINC00663* | 0.364655827 | 0.014998523 | 0.019723011 | 1 |
| Obesity | LEC | *ILF3-AS1* | 1.11870169 | 7.16E-05 | 0.000192609 | 0.450319899 |
| Obesity | LEC | *AC139100.2* | -1.328779473 | 0.000111746 | 0.000264116 | 0.702549009 |
| Obesity | LEC | *LINC01140* | 1.224053903 | 2.75E-06 | 7.03E-05 | 0.017291811 |
| Obesity | LEC | *LINC00906* | 5.580103518 | 3.10E-05 | 0.000116539 | 0.194853395 |
| Obesity | LEC | *ZNF582-AS1* | -1.389144687 | 0.000476261 | 0.000856234 | 1 |
| Obesity | LEC | *AC137932.1* | 2.560991772 | 0.001133895 | 0.001842067 | 1 |
| Obesity | LEC | *AC003005.2* | 1.705973976 | 0.00033138 | 0.000631148 | 1 |
| Obesity | LEC | *AL132989.1* | -0.848231762 | 0.000126426 | 0.000290753 | 0.79483882 |
| Obesity | LEC | *LINC00664* | -3.587951126 | 0.000330007 | 0.000629285 | 1 |
| Obesity | LEC | *AC009060.1* | 1.350450703 | 0.002571194 | 0.003860783 | 1 |
| Obesity | LEC | *SNHG8* | 1.764593479 | 0.000250762 | 0.000503044 | 1 |
| Obesity | LEC | *HCG17* | 1.661512329 | 0.000400265 | 0.000739051 | 1 |
| Obesity | LEC | *YTHDF3-AS1* | 1.522461522 | 9.54E-06 | 0.000105108 | 0.06000252 |
| Obesity | LEC | *LINC01050* | 6.382477737 | 2.79E-05 | 0.00011384 | 0.175183147 |
| Obesity | LEC | *MILR1* | 0.55589464 | 3.41E-05 | 0.000121059 | 0.214659905 |
| Obesity | LEC | *GTF2H5* | 2.744107533 | 3.05E-06 | 7.33E-05 | 0.019196794 |
| Obesity | LEC | *FGF14-AS2* | 2.885089341 | 8.57E-06 | 0.000105108 | 0.053875745 |
| Obesity | LEC | *CASC15* | -0.307568088 | 0.000235954 | 0.000478221 | 1 |
| Obesity | LEC | *LINC01023* | 1.249374118 | 2.75E-05 | 0.00011384 | 0.173027374 |
| Obesity | LEC | *AP000347.2* | 0.936110867 | 0.000485721 | 0.000870752 | 1 |
| Obesity | LEC | *DOC2B* | -0.349924325 | 0.000141735 | 0.000317678 | 0.891086341 |
| Obesity | LEC | *POLR2J4* | 1.108099699 | 1.91E-05 | 0.000105211 | 0.120277262 |
| Obesity | LEC | *AP000350.5* | 0.484770281 | 0.018516131 | 0.024076715 | 1 |
| Obesity | macrophage | *C1orf112* | 1.133715995 | 0.016622381 | 0.048225615 | 1 |
| Obesity | macrophage | *CFH* | 1.104531609 | 3.19E-05 | 0.000554992 | 0.200611098 |
| Obesity | macrophage | *NIPAL3* | 0.810489138 | 0.007548971 | 0.026981455 | 1 |

| | | | | | | |
|---|---|---|---|---|---|---|
| Obesity | macrophage | *SEMA3F* | -0.731494838 | 4.38E-20 | 2.75E-17 | 2.75E-16 |
| Obesity | macrophage | *RAD52* | 1.285842417 | 0.004195314 | 0.017619199 | 1 |
| Obesity | macrophage | *BAD* | 2.366240249 | 0.001275403 | 0.007536146 | 1 |
| Obesity | macrophage | *WNT16* | 0.87532612 | 4.72E-05 | 0.000735791 | 0.296523791 |
| Obesity | macrophage | *CYP26B1* | 1.436021059 | 4.76E-05 | 0.000738738 | 0.299188883 |
| Obesity | macrophage | *ALS2* | 1.088059623 | 0.006005733 | 0.022855957 | 1 |
| Obesity | macrophage | *POLDIP2* | 1.764706037 | 0.000174644 | 0.001794099 | 1 |
| Obesity | macrophage | *RBM6* | -0.583780441 | 3.75E-23 | 3.37E-20 | 2.36E-19 |
| Obesity | macrophage | *ARHGAP33* | 0.620893554 | 0.001384685 | 0.007986455 | 1 |
| Obesity | macrophage | *SLC22A16* | -0.526249504 | 2.73E-05 | 0.000519229 | 0.17134565 |
| Obesity | macrophage | *CDC27* | 0.688772336 | 0.000200452 | 0.001975299 | 1 |
| Obesity | macrophage | *CIAPIN1* | 1.557397938 | 0.00212405 | 0.010769278 | 1 |
| Obesity | macrophage | *COPZ2* | 0.463778793 | 6.25E-05 | 0.000878444 | 0.393115753 |
| Obesity | macrophage | *GCFC2* | 1.253422779 | 0.000429621 | 0.003494041 | 1 |
| Obesity | macrophage | *WDR54* | 1.263322928 | 0.002159844 | 0.01087936 | 1 |
| Obesity | macrophage | *ABCB4* | 1.836278414 | 8.41E-05 | 0.001070503 | 0.528828556 |
| Obesity | macrophage | *PDK2* | 0.758708017 | 0.011162376 | 0.036342755 | 1 |
| Obesity | macrophage | *GGNBP2* | -0.807442237 | 9.78E-06 | 0.000301542 | 0.061487911 |
| Obesity | macrophage | *TMEM98* | 0.444486895 | 0.000752587 | 0.00523977 | 1 |
| Obesity | macrophage | *ABCC8* | 1.900624305 | 0.003435123 | 0.01525185 | 1 |
| Obesity | macrophage | *TMEM132A* | 2.844256621 | 6.32E-05 | 0.000882572 | 0.397550875 |
| Obesity | macrophage | *ARHGAP44* | 1.235604122 | 2.78E-05 | 0.000522449 | 0.175020371 |
| Obesity | macrophage | *NOS2* | 7.075117658 | 0.000250411 | 0.002351705 | 1 |
| Obesity | macrophage | *TRAPPC6A* | 1.102155983 | 0.010364391 | 0.034331361 | 1 |
| Obesity | macrophage | *TSR3* | 1.707058718 | 0.004518534 | 0.018543096 | 1 |
| Obesity | macrophage | *MPND* | 0.717066103 | 0.016792082 | 0.048561095 | 1 |
| Obesity | macrophage | *MGST1* | 0.606279021 | 0.007580912 | 0.027049484 | 1 |
| Obesity | macrophage | *MMP25* | 0.960957387 | 0.000181556 | 0.001832175 | 1 |
| Obesity | macrophage | *PKD1* | 0.846113825 | 3.91E-05 | 0.000635507 | 0.2460604 |
| Obesity | macrophage | *MAPK8IP2* | 1.475721645 | 0.001760288 | 0.009491362 | 1 |
| Obesity | macrophage | *SEC62* | 1.009941197 | 0.008527956 | 0.029572676 | 1 |
| Obesity | macrophage | *MLXIPL* | 0.173146493 | 0.001776406 | 0.009539215 | 1 |
| Obesity | macrophage | *SPRTN* | 1.446983749 | 0.01103077 | 0.036007502 | 1 |
| Obesity | macrophage | *STARD3NL* | 1.249426875 | 0.002841905 | 0.013313753 | 1 |
| Obesity | macrophage | *CD4* | -0.471644658 | 0.004344917 | 0.018118457 | 1 |
| Obesity | macrophage | *HFE* | -0.848231582 | 3.47E-09 | 4.75E-07 | 2.18E-05 |
| Obesity | macrophage | *SCMH1* | 0.788692022 | 0.000599135 | 0.004416916 | 1 |
| Obesity | macrophage | *CLCN6* | 0.962523747 | 0.014878634 | 0.044617832 | 1 |
| Obesity | macrophage | *TSPAN9* | 0.490188983 | 0.001122782 | 0.006873351 | 1 |
| Obesity | macrophage | *MKS1* | 2.276679675 | 6.59E-05 | 0.000907009 | 0.414502899 |
| Obesity | macrophage | *TTC19* | 1.215478343 | 2.75E-07 | 2.16E-05 | 0.001730407 |
| Obesity | macrophage | *SYT7* | 0.896065158 | 0.002765965 | 0.013055269 | 1 |
| Obesity | macrophage | *PPP5C* | 0.935258408 | 0.002469564 | 0.012026452 | 1 |
| Obesity | macrophage | *ZBTB32* | 2.465038287 | 2.38E-05 | 0.000488143 | 0.149695861 |
| Obesity | macrophage | *LTF* | 0.648535995 | 0.006934855 | 0.025461273 | 1 |
| Obesity | macrophage | *ALOX5* | -0.594047109 | 0.001826284 | 0.009710845 | 1 |
| Obesity | macrophage | *MAP4K5* | 0.860865178 | 0.009290479 | 0.031453551 | 1 |
| Obesity | macrophage | *PSMC4* | 0.854574294 | 0.000137163 | 0.00150932 | 0.862345824 |
| Obesity | macrophage | *CLDN11* | 0.832359098 | 6.27E-05 | 0.000878444 | 0.394421543 |
| Obesity | macrophage | *MDH1* | 0.827183526 | 0.00041255 | 0.003421767 | 1 |

| | | | | | | |
|---|---|---|---|---|---|---|
| Obesity | macrophage | *SLC30A9* | 1.013891737 | 0.000775436 | 0.005351442 | 1 |
| Obesity | macrophage | *MTMR11* | 0.216773663 | 0.00227591 | 0.011266651 | 1 |
| Obesity | macrophage | *NUDCD3* | 0.486044858 | 0.000954402 | 0.006243833 | 1 |
| Obesity | macrophage | *CHDH* | 1.454489475 | 0.004002384 | 0.017013513 | 1 |
| Obesity | macrophage | *IL20RA* | 2.311687339 | 3.35E-05 | 0.000567846 | 0.210670737 |
| Obesity | macrophage | *WWTR1* | 0.985107289 | 0.000221829 | 0.002122734 | 1 |
| Obesity | macrophage | *MARCO* | -0.214487019 | 0.002006635 | 0.010374763 | 1 |
| Obesity | macrophage | *MNAT1* | 1.190914342 | 0.000484253 | 0.003819949 | 1 |
| Obesity | macrophage | *OSBPL5* | 1.565068473 | 0.000126038 | 0.001428394 | 0.792400017 |
| Obesity | macrophage | *CPS1* | 0.377511678 | 0.001731076 | 0.009374052 | 1 |
| Obesity | macrophage | *ZDHHC6* | 0.927173142 | 0.008975039 | 0.030616425 | 1 |
| Obesity | macrophage | *NDUFS1* | 1.044759156 | 0.000373729 | 0.003180829 | 1 |
| Obesity | macrophage | *AKAP11* | 1.058058526 | 0.011610518 | 0.037299606 | 1 |
| Obesity | macrophage | *GCLM* | 0.466938157 | 0.006968122 | 0.025544364 | 1 |
| Obesity | macrophage | *HSF2* | 1.391988384 | 0.006690541 | 0.024860184 | 1 |
| Obesity | macrophage | *TOMM34* | 2.072870099 | 0.001501467 | 0.008443402 | 1 |
| Obesity | macrophage | *KPNA6* | 0.864004571 | 0.005015565 | 0.020097422 | 1 |
| Obesity | macrophage | *VIM* | 0.859572482 | 0.003337225 | 0.014954477 | 1 |
| Obesity | macrophage | *BTN3A1* | -0.280933403 | 0.016588034 | 0.048170424 | 1 |
| Obesity | macrophage | *ARNTL2* | 1.72503487 | 4.26E-05 | 0.000681503 | 0.268058443 |
| Obesity | macrophage | *ANK1* | 1.367299388 | 0.005931871 | 0.022698522 | 1 |
| Obesity | macrophage | *RABEP1* | 0.954425702 | 8.16E-06 | 0.000274371 | 0.05130743 |
| Obesity | macrophage | *ZCCHC8* | 2.032036789 | 7.38E-07 | 5.11E-05 | 0.004638286 |
| Obesity | macrophage | *FUT8* | 1.065549395 | 0.002048443 | 0.010513714 | 1 |
| Obesity | macrophage | *SH3YL1* | -0.486971536 | 0.002391669 | 0.01171061 | 1 |
| Obesity | macrophage | *ADSS* | 1.105068764 | 0.015280885 | 0.045423605 | 1 |
| Obesity | macrophage | *STAP1* | -0.743824384 | 0.014948822 | 0.044753924 | 1 |
| Obesity | macrophage | *TBC1D23* | 0.923627455 | 0.014072649 | 0.042907248 | 1 |
| Obesity | macrophage | *NSUN2* | 1.065639158 | 0.00468028 | 0.019045254 | 1 |
| Obesity | macrophage | *MAT2B* | 1.748135295 | 0.000115844 | 0.001351584 | 0.72831313 |
| Obesity | macrophage | *SKIV2L2* | 1.05349688 | 0.001074149 | 0.006702419 | 1 |
| Obesity | macrophage | *DNAH5* | 1.486470499 | 1.23E-05 | 0.000347425 | 0.07739008 |
| Obesity | macrophage | *C6* | 0.965526448 | 0.000391569 | 0.003299993 | 1 |
| Obesity | macrophage | *LSG1* | 0.395426464 | 0.000476036 | 0.003774069 | 1 |
| Obesity | macrophage | *THAP3* | 1.543459764 | 0.005168541 | 0.020462606 | 1 |
| Obesity | macrophage | *MED17* | 0.836358882 | 0.016475428 | 0.047954174 | 1 |
| Obesity | macrophage | *RETSAT* | 0.349314511 | 0.009921633 | 0.033153454 | 1 |
| Obesity | macrophage | *LCP2* | -0.498437824 | 0.004509679 | 0.01852808 | 1 |
| Obesity | macrophage | *CTNNA1* | 1.05135323 | 0.000282435 | 0.002603616 | 1 |
| Obesity | macrophage | *EPHA3* | 1.386381094 | 3.40E-05 | 0.000571658 | 0.213560701 |
| Obesity | macrophage | *DSG2* | 1.295204029 | 4.78E-05 | 0.000739028 | 0.300784288 |
| Obesity | macrophage | *POLR2B* | 1.046819339 | 0.010510484 | 0.034669157 | 1 |
| Obesity | macrophage | *CP* | 1.631330085 | 0.007576531 | 0.027049205 | 1 |
| Obesity | macrophage | *ANO2* | 0.354449857 | 0.002862326 | 0.013389468 | 1 |
| Obesity | macrophage | *GOPC* | 0.917180905 | 0.008706072 | 0.029974191 | 1 |
| Obesity | macrophage | *TSPAN17* | 0.531677493 | 0.001633609 | 0.008954228 | 1 |
| Obesity | macrophage | *CC2D2A* | 1.23556675 | 0.000253783 | 0.002377846 | 1 |
| Obesity | macrophage | *CELF2* | 0.799400305 | 0.003772714 | 0.016271439 | 1 |
| Obesity | macrophage | *JKAMP* | 0.670171811 | 0.000565422 | 0.004201901 | 1 |
| Obesity | macrophage | *DKK3* | 1.524063325 | 2.96E-05 | 0.000541585 | 0.186305313 |

| | | | | | | |
|---|---|---|---|---|---|---|
| Obesity | macrophage | *ARHGEF5* | 1.788262851 | 0.00136465 | 0.007926286 | 1 |
| Obesity | macrophage | *PTGER3* | 1.047021621 | 2.66E-05 | 0.000515008 | 0.167402836 |
| Obesity | macrophage | *ALX4* | 2.452399535 | 0.00015018 | 0.001600682 | 0.944180922 |
| Obesity | macrophage | *AKR7A2* | 0.992024082 | 0.004345894 | 0.018118457 | 1 |
| Obesity | macrophage | *ANAPC4* | 1.247732373 | 0.000547398 | 0.00412076 | 1 |
| Obesity | macrophage | *SDCCAG8* | 0.653119749 | 6.99E-05 | 0.000946839 | 0.439333475 |
| Obesity | macrophage | *ATP9A* | 0.882310495 | 0.003094479 | 0.014180021 | 1 |
| Obesity | macrophage | *GINM1* | 1.159400228 | 0.001712158 | 0.009303662 | 1 |
| Obesity | macrophage | *ITIH4* | 0.380384162 | 6.57E-11 | 1.53E-08 | 4.13E-07 |
| Obesity | macrophage | *IL17RB* | 2.456582958 | 3.50E-05 | 0.000582975 | 0.219781492 |
| Obesity | macrophage | *TRAF3IP2* | 1.553237756 | 6.10E-05 | 0.000865942 | 0.383612504 |
| Obesity | macrophage | *DCBLD2* | 1.273043755 | 0.000711311 | 0.0050325 | 1 |
| Obesity | macrophage | *ATP11B* | 0.584017821 | 6.55E-05 | 0.000903457 | 0.411976428 |
| Obesity | macrophage | *CDK14* | 1.014126035 | 0.002223649 | 0.011077717 | 1 |
| Obesity | macrophage | *CROCC* | 0.301949812 | 0.007041513 | 0.025738368 | 1 |
| Obesity | macrophage | *ZC3H11A* | 1.472325126 | 0.009593617 | 0.03223681 | 1 |
| Obesity | macrophage | *YBX3* | 1.088981039 | 0.004360027 | 0.018131931 | 1 |
| Obesity | macrophage | *NCKAP1* | 0.900670752 | 0.000239577 | 0.002268409 | 1 |
| Obesity | macrophage | *ISOC2* | 2.605625054 | 3.56E-05 | 0.00059194 | 0.223753391 |
| Obesity | macrophage | *TM7SF3* | 0.964866388 | 0.001864163 | 0.009873622 | 1 |
| Obesity | macrophage | *DMRT3* | 1.631109111 | 0.000125318 | 0.00142473 | 0.787875487 |
| Obesity | macrophage | *TAF2* | 0.652237262 | 0.015526609 | 0.045843681 | 1 |
| Obesity | macrophage | *TMEM161A* | 1.60229601 | 6.41E-05 | 0.000889867 | 0.403109588 |
| Obesity | macrophage | *PMS1* | 1.119223676 | 0.006252523 | 0.023468426 | 1 |
| Obesity | macrophage | *UHRF1BP1* | 0.870912977 | 1.23E-23 | 1.29E-20 | 7.73E-20 |
| Obesity | macrophage | *IPO5* | 1.303255064 | 0.001941227 | 0.010161944 | 1 |
| Obesity | macrophage | *NDUFB4* | 1.512126681 | 0.001853334 | 0.009832834 | 1 |
| Obesity | macrophage | *MAP2K4* | 0.923018915 | 0.01361543 | 0.041776579 | 1 |
| Obesity | macrophage | *SEC61A2* | 1.741161107 | 0.000789262 | 0.005426203 | 1 |
| Obesity | macrophage | *PPP2R5A* | 1.109167573 | 0.001135248 | 0.006923965 | 1 |
| Obesity | macrophage | *NGEF* | 3.667683769 | 1.12E-05 | 0.000332862 | 0.070244284 |
| Obesity | macrophage | *CD84* | -0.352481046 | 0.001076855 | 0.006703154 | 1 |
| Obesity | macrophage | *GOLGA5* | 1.247081762 | 0.005781475 | 0.022244881 | 1 |
| Obesity | macrophage | *MYO9A* | 0.869145886 | 0.000423973 | 0.003466217 | 1 |
| Obesity | macrophage | *STOML1* | 2.69727648 | 0.000141022 | 0.001541919 | 0.886603409 |
| Obesity | macrophage | *TP53BP1* | 1.123442982 | 0.000121738 | 0.001399213 | 0.76536963 |
| Obesity | macrophage | *HDAC4* | 1.125049146 | 0.002161083 | 0.01087936 | 1 |
| Obesity | macrophage | *PITX1* | 3.685437397 | 2.41E-05 | 0.000492083 | 0.15162686 |
| Obesity | macrophage | *SDK2* | 1.595185027 | 0.001038491 | 0.006568405 | 1 |
| Obesity | macrophage | *NUCKS1* | 1.045367134 | 0.002472805 | 0.012032912 | 1 |
| Obesity | macrophage | *DNAJA2* | 1.779774379 | 0.001002551 | 0.006444825 | 1 |
| Obesity | macrophage | *KCNAB2* | -0.493627115 | 0.001850648 | 0.009826878 | 1 |
| Obesity | macrophage | *GAL* | -1.582508509 | 0.004053783 | 0.017173946 | 1 |
| Obesity | macrophage | *FGF10* | 0.272933931 | 0.011559035 | 0.037210267 | 1 |
| Obesity | macrophage | *TMEM260* | 1.052636336 | 0.010521766 | 0.03468817 | 1 |
| Obesity | macrophage | *CLTCL1* | 0.584854421 | 0.003290917 | 0.014831538 | 1 |
| Obesity | macrophage | *DGCR2* | 1.113607463 | 0.006082644 | 0.023009375 | 1 |
| Obesity | macrophage | *ZXDC* | 1.060083199 | 0.002325175 | 0.011447435 | 1 |
| Obesity | macrophage | *WIPI1* | 1.527504234 | 0.000420597 | 0.003447576 | 1 |
| Obesity | macrophage | *NDST1* | 1.165140887 | 0.00029495 | 0.002699202 | 1 |

| | | | | | | |
|---|---|---|---|---|---|---|
| Obesity | macrophage | *ASNS* | 1.295480602 | 0.000995307 | 0.006411368 | 1 |
| Obesity | macrophage | *EIF2B3* | 1.067675431 | 0.003571131 | 0.015656693 | 1 |
| Obesity | macrophage | *CAMK2A* | 4.317711151 | 6.51E-05 | 0.000899633 | 0.409332834 |
| Obesity | macrophage | *TRIP13* | 6.116078657 | 0.000592206 | 0.004380237 | 1 |
| Obesity | macrophage | *TCF3* | 1.084247028 | 0.014591053 | 0.043933885 | 1 |
| Obesity | macrophage | *MBD3* | 0.801335787 | 0.001673598 | 0.00912568 | 1 |
| Obesity | macrophage | *CYBRD1* | 0.5129231 | 0.011585494 | 0.037257291 | 1 |
| Obesity | macrophage | *EPN2* | 1.125137324 | 0.001001474 | 0.006444487 | 1 |
| Obesity | macrophage | *LIMS2* | 0.757372033 | 0.000467419 | 0.003719825 | 1 |
| Obesity | macrophage | *TRHDE* | 1.049822919 | 5.81E-05 | 0.000839679 | 0.365260341 |
| Obesity | macrophage | *TRNT1* | 1.239414021 | 0.005742675 | 0.022149813 | 1 |
| Obesity | macrophage | *PVR* | 0.743297801 | 0.000974798 | 0.006337699 | 1 |
| Obesity | macrophage | *TP63* | 0.881638701 | 0.013232415 | 0.04094104 | 1 |
| Obesity | macrophage | *PDE8A* | 0.50160122 | 0.017269323 | 0.049735335 | 1 |
| Obesity | macrophage | *SDHA* | 0.729554319 | 0.00035638 | 0.003077692 | 1 |
| Obesity | macrophage | *SMARCE1* | 1.274340538 | 0.000169369 | 0.001748473 | 1 |
| Obesity | macrophage | *CLNS1A* | 0.383434906 | 0.008114146 | 0.028419853 | 1 |
| Obesity | macrophage | *ATP2A3* | 0.72083497 | 0.00203508 | 0.0104647 | 1 |
| Obesity | macrophage | *NTN4* | 0.973027618 | 0.001086439 | 0.006736135 | 1 |
| Obesity | macrophage | *LMAN1* | 1.085854816 | 0.00680629 | 0.025156463 | 1 |
| Obesity | macrophage | *TIPIN* | 0.298522928 | 4.55E-06 | 0.000182223 | 0.028609009 |
| Obesity | macrophage | *SRI* | 1.637265669 | 0.000599273 | 0.004416916 | 1 |
| Obesity | macrophage | *SLC25A3* | 0.979607995 | 0.004103131 | 0.017350668 | 1 |
| Obesity | macrophage | *FSCN1* | -0.943244505 | 0.003361476 | 0.014997871 | 1 |
| Obesity | macrophage | *MOCOS* | 0.281850037 | 0.012687569 | 0.039665214 | 1 |
| Obesity | macrophage | *BCAP29* | 0.931027718 | 0.005181533 | 0.020488239 | 1 |
| Obesity | macrophage | *TUBA3D* | 1.130999581 | 0.01567745 | 0.046165867 | 1 |
| Obesity | macrophage | *MKRN2* | 1.930683552 | 1.59E-05 | 0.00040914 | 0.099830105 |
| Obesity | macrophage | *RBMS2* | 1.144663184 | 0.000563067 | 0.004199288 | 1 |
| Obesity | macrophage | *FMO4* | 1.293133452 | 0.0121544 | 0.038554345 | 1 |
| Obesity | macrophage | *GPATCH1* | 0.814231905 | 0.005966853 | 0.02277693 | 1 |
| Obesity | macrophage | *NT5C2* | 0.795433339 | 0.006121106 | 0.023099275 | 1 |
| Obesity | macrophage | *CAMSAP3* | 15.12194815 | 0.000367347 | 0.003142149 | 1 |
| Obesity | macrophage | *STXBP2* | -0.536188001 | 0.002439005 | 0.011896063 | 1 |
| Obesity | macrophage | *TM9SF3* | 0.790063954 | 0.015365509 | 0.045560262 | 1 |
| Obesity | macrophage | *DNAJC10* | 1.283034725 | 0.003630006 | 0.015881594 | 1 |
| Obesity | macrophage | *DYNC1I2* | 1.517844774 | 7.06E-06 | 0.000249485 | 0.044408385 |
| Obesity | macrophage | *NAALAD2* | 0.443700369 | 0.006284029 | 0.023572606 | 1 |
| Obesity | macrophage | *FBLN1* | 0.381216607 | 0.010637636 | 0.034960178 | 1 |
| Obesity | macrophage | *MCCC1* | 0.578796742 | 0.000957194 | 0.006255592 | 1 |
| Obesity | macrophage | *ACER3* | 0.687887371 | 0.001317347 | 0.007725898 | 1 |
| Obesity | macrophage | *TULP3* | 1.151747599 | 0.00241279 | 0.011795652 | 1 |
| Obesity | macrophage | *SYNJ2* | 0.660562414 | 0.017408187 | 0.049997839 | 1 |
| Obesity | macrophage | *FGF20* | -2.154962149 | 0.000348676 | 0.003044616 | 1 |
| Obesity | macrophage | *REXO1* | 1.641886139 | 0.005403114 | 0.021125234 | 1 |
| Obesity | macrophage | *SAR1A* | 1.130256972 | 0.002165495 | 0.01088709 | 1 |
| Obesity | macrophage | *FDFT1* | 1.674002297 | 0.000145542 | 0.001574915 | 0.915025659 |
| Obesity | macrophage | *DDX1* | 1.256504296 | 0.003113809 | 0.01424783 | 1 |
| Obesity | macrophage | *RIMS1* | 0.940219321 | 0.007618909 | 0.027138857 | 1 |
| Obesity | macrophage | *DCT* | 2.875482698 | 3.50E-06 | 0.000157071 | 0.021989918 |

| | | | | | | |
|---|---|---|---|---|---|---|
| Obesity | macrophage | *SLC35C2* | 1.406334574 | 0.005005127 | 0.020094018 | 1 |
| Obesity | macrophage | *SCTR* | -0.650928278 | 0.006907098 | 0.025409554 | 1 |
| Obesity | macrophage | *KCNN2* | 1.859960722 | 0.012956364 | 0.040285192 | 1 |
| Obesity | macrophage | *MOK* | 1.600173982 | 4.80E-05 | 0.000739768 | 0.30182541 |
| Obesity | macrophage | *MAGI3* | 1.181429108 | 0.000351829 | 0.003055847 | 1 |
| Obesity | macrophage | *COL4A4* | 0.725806626 | 0.000313264 | 0.002825671 | 1 |
| Obesity | macrophage | *ZNF506* | 0.990706326 | 0.005730315 | 0.022121059 | 1 |
| Obesity | macrophage | *PGR* | 0.490664219 | 0.000951059 | 0.006239888 | 1 |
| Obesity | macrophage | *C5orf22* | 1.517712383 | 0.001857314 | 0.009845642 | 1 |
| Obesity | macrophage | *CCNT2* | -0.495031807 | 0.00101327 | 0.006487201 | 1 |
| Obesity | macrophage | *COBLL1* | 0.608285173 | 9.01E-07 | 5.74E-05 | 0.005665825 |
| Obesity | macrophage | *ERC1* | 0.798933341 | 0.010143789 | 0.033780404 | 1 |
| Obesity | macrophage | *DOPEY1* | 0.437206116 | 0.000364727 | 0.003128295 | 1 |
| Obesity | macrophage | *LYRM2* | 0.380736294 | 0.013808809 | 0.042246221 | 1 |
| Obesity | macrophage | *TDRD3* | 0.846000348 | 0.016344542 | 0.047639376 | 1 |
| Obesity | macrophage | *ZNF324* | 2.222542598 | 0.000847332 | 0.005703616 | 1 |
| Obesity | macrophage | *ZNF446* | 2.505219038 | 5.73E-05 | 0.000831774 | 0.360158331 |
| Obesity | macrophage | *RPS5* | 1.176096528 | 0.011465274 | 0.036984187 | 1 |
| Obesity | macrophage | *PPIE* | 0.835771489 | 0.00034539 | 0.003037338 | 1 |
| Obesity | macrophage | *GSTP1* | 1.843956915 | 5.03E-05 | 0.000759323 | 0.316174898 |
| Obesity | macrophage | *WBP11* | 1.494813281 | 0.004546243 | 0.018620347 | 1 |
| Obesity | macrophage | *COL16A1* | -0.404885771 | 0.015981023 | 0.046796781 | 1 |
| Obesity | macrophage | *CD82* | 0.577551702 | 0.006032802 | 0.022901078 | 1 |
| Obesity | macrophage | *TAF9* | 1.97795258 | 0.007227957 | 0.026116187 | 1 |
| Obesity | macrophage | *SCAMP1* | 1.484081258 | 1.16E-05 | 0.000339028 | 0.073230143 |
| Obesity | macrophage | *HACE1* | 1.098131627 | 0.000871466 | 0.005828626 | 1 |
| Obesity | macrophage | *CTTN* | 1.210317677 | 2.78E-05 | 0.000522449 | 0.174497992 |
| Obesity | macrophage | *USP40* | 0.798810208 | 0.005068559 | 0.020181146 | 1 |
| Obesity | macrophage | *CHMP5* | 1.118262475 | 0.005128108 | 0.020353798 | 1 |
| Obesity | macrophage | *ITPKC* | 1.293614556 | 0.000122915 | 0.001406264 | 0.772765399 |
| Obesity | macrophage | *TMED2* | 1.399868299 | 0.0057585 | 0.022197235 | 1 |
| Obesity | macrophage | *ACOX3* | 0.654360446 | 0.009571661 | 0.032186845 | 1 |
| Obesity | macrophage | *HSD17B14* | -0.509303778 | 0.010274038 | 0.034103946 | 1 |
| Obesity | macrophage | *SRRT* | 0.567481048 | 0.005148247 | 0.020407961 | 1 |
| Obesity | macrophage | *OGFOD1* | -0.716260457 | 0.004458396 | 0.018419581 | 1 |
| Obesity | macrophage | *GMCL1* | 1.199131381 | 0.000702104 | 0.00499141 | 1 |
| Obesity | macrophage | *KLHL42* | 0.982391318 | 0.001283167 | 0.007560703 | 1 |
| Obesity | macrophage | *PTHLH* | 1.264452272 | 4.18E-06 | 0.000177641 | 0.026290865 |
| Obesity | macrophage | *ERGIC2* | 1.106889611 | 0.005315018 | 0.020851717 | 1 |
| Obesity | macrophage | *ASAP3* | 1.241682232 | 2.89E-06 | 0.000135608 | 0.018171485 |
| Obesity | macrophage | *C3orf18* | 1.754861143 | 1.72E-13 | 6.74E-11 | 1.08E-09 |
| Obesity | macrophage | *NSFL1C* | 0.932399458 | 0.015415563 | 0.04565127 | 1 |
| Obesity | macrophage | *ZNF343* | 1.041288003 | 0.001136944 | 0.006925546 | 1 |
| Obesity | macrophage | *LZTS3* | 1.048968103 | 0.003331699 | 0.014942136 | 1 |
| Obesity | macrophage | *TESC* | 2.924593398 | 0.000223062 | 0.002131298 | 1 |
| Obesity | macrophage | *TASP1* | 0.792137515 | 0.001916324 | 0.010065104 | 1 |
| Obesity | macrophage | *OAS1* | -0.795461862 | 0.008715615 | 0.029974191 | 1 |
| Obesity | macrophage | *GANAB* | 1.138055812 | 0.004913959 | 0.019791199 | 1 |
| Obesity | macrophage | *GMIP* | -0.717941702 | 0.007600286 | 0.027103231 | 1 |
| Obesity | macrophage | *MLF2* | 1.937474862 | 9.08E-05 | 0.001132299 | 0.570678656 |

| | | | | | | |
|---|---|---|---|---|---|---|
| Obesity | macrophage | *SLC9A1* | 1.375212736 | 0.007613915 | 0.027136441 | 1 |
| Obesity | macrophage | *RGS1* | -0.499281349 | 0.000614815 | 0.004510943 | 1 |
| Obesity | macrophage | *YPEL3* | 0.632894639 | 5.06E-10 | 8.83E-08 | 3.18E-06 |
| Obesity | macrophage | *PABPC4* | -1.100954132 | 8.06E-10 | 1.33E-07 | 5.07E-06 |
| Obesity | macrophage | *LAMB1* | 0.57686608 | 6.19E-05 | 0.000876225 | 0.389043848 |
| Obesity | macrophage | *SLC26A4* | 2.610422872 | 0.000113751 | 0.001344267 | 0.715149804 |
| Obesity | macrophage | *DLD* | 1.140642375 | 0.005224206 | 0.020631021 | 1 |
| Obesity | macrophage | *TXNL1* | 1.243207678 | 0.00317246 | 0.014442621 | 1 |
| Obesity | macrophage | *IL5RA* | 4.045074487 | 8.91E-05 | 0.001115582 | 0.560022162 |
| Obesity | macrophage | *ITGA6* | 1.051090496 | 0.000190393 | 0.001899997 | 1 |
| Obesity | macrophage | *RAPGEF4* | 0.816206901 | 0.002338451 | 0.011494793 | 1 |
| Obesity | macrophage | *TF* | 0.855064852 | 0.014185213 | 0.043124967 | 1 |
| Obesity | macrophage | *ALKBH5* | 0.64080078 | 0.001928979 | 0.010106241 | 1 |
| Obesity | macrophage | *SPAG7* | 0.389472702 | 0.006054197 | 0.022943181 | 1 |
| Obesity | macrophage | *ZFHX4* | 0.891984573 | 0.000482179 | 0.003808366 | 1 |
| Obesity | macrophage | *PPP2R3C* | 0.888860977 | 0.010370407 | 0.0343332 | 1 |
| Obesity | macrophage | *TOX4* | 1.408364917 | 0.000192787 | 0.001911756 | 1 |
| Obesity | macrophage | *RFFL* | 2.065028423 | 0.003518424 | 0.01552304 | 1 |
| Obesity | macrophage | *VNN3* | 2.684522041 | 0.001602653 | 0.008823011 | 1 |
| Obesity | macrophage | *SEC22C* | 1.018637087 | 0.008538105 | 0.029591545 | 1 |
| Obesity | macrophage | *FMO2* | 0.500999118 | 0.000323482 | 0.002892936 | 1 |
| Obesity | macrophage | *MSH2* | 1.313514855 | 0.011421807 | 0.036900771 | 1 |
| Obesity | macrophage | *PDE6C* | 1.31168896 | 2.87E-06 | 0.000135538 | 0.018026531 |
| Obesity | macrophage | *IKZF5* | 1.020480271 | 0.010349431 | 0.034299881 | 1 |
| Obesity | macrophage | *BLNK* | -0.253590357 | 0.012019083 | 0.038221536 | 1 |
| Obesity | macrophage | *TPSD1* | 3.647533752 | 0.003083571 | 0.014140343 | 1 |
| Obesity | macrophage | *ITPR3* | 1.615340456 | 2.38E-05 | 0.000488143 | 0.149859761 |
| Obesity | macrophage | *IFT74* | 0.409274476 | 0.006783478 | 0.025101662 | 1 |
| Obesity | macrophage | *IL12RB1* | -0.544962608 | 0.013158945 | 0.040773921 | 1 |
| Obesity | macrophage | *PCSK5* | 1.053909808 | 5.33E-05 | 0.000788276 | 0.335017419 |
| Obesity | macrophage | *PRTFDC1* | 1.40404619 | 0.000206063 | 0.002008997 | 1 |
| Obesity | macrophage | *PALMD* | 0.825928557 | 0.000250353 | 0.002351705 | 1 |
| Obesity | macrophage | *TSPAN15* | 1.34273844 | 0.000653623 | 0.004734252 | 1 |
| Obesity | macrophage | *STX1B* | -0.812205308 | 0.000645607 | 0.004688839 | 1 |
| Obesity | macrophage | *ATP5D* | 1.307789239 | 0.015672019 | 0.046165867 | 1 |
| Obesity | macrophage | *CDC34* | 2.073716558 | 0.000152711 | 0.001621783 | 0.960095385 |
| Obesity | macrophage | *MADCAM1* | 1.714766956 | 0.000399564 | 0.003349409 | 1 |
| Obesity | macrophage | *LZTR1* | 1.620055011 | 0.001990602 | 0.010317325 | 1 |
| Obesity | macrophage | *SMARCB1* | 0.62954159 | 0.000207548 | 0.002016776 | 1 |
| Obesity | macrophage | *GGT5* | 1.307246482 | 0.001570914 | 0.008716979 | 1 |
| Obesity | macrophage | *YPEL1* | 1.37599249 | 0.005141963 | 0.020395914 | 1 |
| Obesity | macrophage | *CYTH4* | -0.398269762 | 0.001713834 | 0.009304728 | 1 |
| Obesity | macrophage | *SLC25A1* | 1.255826733 | 0.00204856 | 0.010513714 | 1 |
| Obesity | macrophage | *LGALS2* | 0.565672444 | 0.013497867 | 0.041557829 | 1 |
| Obesity | macrophage | *SH3BP1* | -0.724154304 | 0.003320038 | 0.014909344 | 1 |
| Obesity | macrophage | *TFIP11* | 0.566563296 | 0.000982371 | 0.006347553 | 1 |
| Obesity | macrophage | *GCAT* | 0.691765254 | 9.52E-05 | 0.001170837 | 0.598297815 |
| Obesity | macrophage | *ANKRD54* | -0.440091526 | 0.001389755 | 0.00800127 | 1 |
| Obesity | macrophage | *CCDC134* | 2.046468044 | 0.001444753 | 0.008228849 | 1 |
| Obesity | macrophage | *FBXO7* | 1.372622906 | 1.07E-05 | 0.00032181 | 0.06725827 |

| | | | | | | |
|---|---|---|---|---|---|---|
| Obesity | macrophage | *TIMP3* | 1.00099782 | 0.000335601 | 0.002976611 | 1 |
| Obesity | macrophage | *CYB5R3* | 1.192490231 | 0.000527696 | 0.004006794 | 1 |
| Obesity | macrophage | *RHBDD3* | 1.338020739 | 0.001465275 | 0.00831075 | 1 |
| Obesity | macrophage | *TTLL1* | 2.03631591 | 0.000263342 | 0.00245576 | 1 |
| Obesity | macrophage | *HMGXB4* | 1.115219888 | 0.010914877 | 0.035718068 | 1 |
| Obesity | macrophage | *BIK* | 4.093893804 | 0.006042717 | 0.022913489 | 1 |
| Obesity | macrophage | *TSPO* | 0.897764946 | 0.001716891 | 0.009307938 | 1 |
| Obesity | macrophage | *RBFOX2* | 1.092205298 | 8.19E-05 | 0.001053048 | 0.514940475 |
| Obesity | macrophage | *IFT27* | 2.040230535 | 0.000143448 | 0.001561551 | 0.901858037 |
| Obesity | macrophage | *NCF4* | -0.223603648 | 0.014464968 | 0.043706831 | 1 |
| Obesity | macrophage | *ACO2* | 1.15562292 | 1.58E-05 | 0.00040914 | 0.099607612 |
| Obesity | macrophage | *DESI1* | 0.579486448 | 8.56E-06 | 0.000286211 | 0.053807662 |
| Obesity | macrophage | *SLC10A1* | 0.992573618 | 0.003307132 | 0.014872631 | 1 |
| Obesity | macrophage | *DICER1* | 0.89101258 | 0.000459398 | 0.003669931 | 1 |
| Obesity | macrophage | *ZFYVE21* | 1.759213511 | 5.17E-07 | 3.78E-05 | 0.003251227 |
| Obesity | macrophage | *MTHFD1* | 1.45813627 | 0.000300175 | 0.002731111 | 1 |
| Obesity | macrophage | *GSKIP* | 1.247333431 | 0.000815546 | 0.005525151 | 1 |
| Obesity | macrophage | *PSMB5* | 0.689310115 | 0.000259821 | 0.002427188 | 1 |
| Obesity | macrophage | *CCNB1IP1* | 0.799335623 | 0.003734568 | 0.016159137 | 1 |
| Obesity | macrophage | *TRIP11* | 1.148307954 | 0.002197624 | 0.01100831 | 1 |
| Obesity | macrophage | *PYGB* | 0.468202223 | 0.00674243 | 0.0249939 | 1 |
| Obesity | macrophage | *NDRG3* | 1.498136053 | 0.000629749 | 0.004603752 | 1 |
| Obesity | macrophage | *DOK5* | 1.557493996 | 2.30E-05 | 0.000484985 | 0.144525449 |
| Obesity | macrophage | *BMP7* | 2.570929814 | 3.00E-05 | 0.000542608 | 0.188827488 |
| Obesity | macrophage | *DIDO1* | 0.819719449 | 0.002247673 | 0.011154638 | 1 |
| Obesity | macrophage | *SLC17A9* | -0.478062322 | 0.005998934 | 0.022855957 | 1 |
| Obesity | macrophage | *PTK6* | -2.417739538 | 0.006840438 | 0.025252984 | 1 |
| Obesity | macrophage | *RNF24* | 0.945505558 | 0.001137918 | 0.006925546 | 1 |
| Obesity | macrophage | *TRIB3* | 0.951635472 | 0.011371023 | 0.036831336 | 1 |
| Obesity | macrophage | *SLC52A3* | 0.7758421 | 0.001171363 | 0.007079995 | 1 |
| Obesity | macrophage | *CCM2L* | 1.919998801 | 7.40E-06 | 0.000255529 | 0.046506298 |
| Obesity | macrophage | *MYL9* | 0.937007565 | 1.64E-05 | 0.00041726 | 0.103063184 |
| Obesity | macrophage | *MANBAL* | 0.333621473 | 0.003150747 | 0.014374998 | 1 |
| Obesity | macrophage | *IDH3B* | 1.565915134 | 0.008087328 | 0.028357517 | 1 |
| Obesity | macrophage | *PXMP4* | 0.918537741 | 0.00063048 | 0.004603752 | 1 |
| Obesity | macrophage | *MAP1LC3A* | 0.38672336 | 0.008612098 | 0.029769423 | 1 |
| Obesity | macrophage | *PIGU* | 0.800404631 | 0.007861485 | 0.027766941 | 1 |
| Obesity | macrophage | *APMAP* | 1.309248327 | 0.000351906 | 0.003055847 | 1 |
| Obesity | macrophage | *CELF4* | -0.697477473 | 0.004126222 | 0.017398764 | 1 |
| Obesity | macrophage | *METTL4* | 1.589889906 | 0.001907364 | 0.010034809 | 1 |
| Obesity | macrophage | *CEP76* | 0.669847353 | 0.008200742 | 0.02865929 | 1 |
| Obesity | macrophage | *LIPG* | 1.135961247 | 0.009055629 | 0.030807759 | 1 |
| Obesity | macrophage | *NALCN* | 1.392515547 | 0.00014663 | 0.001575829 | 0.921860248 |
| Obesity | macrophage | *HTR2A* | 3.182004043 | 1.88E-05 | 0.000445142 | 0.117962656 |
| Obesity | macrophage | *ARHGEF7* | 0.973535281 | 0.001710736 | 0.009303662 | 1 |
| Obesity | macrophage | *DHRS12* | 1.545736964 | 0.000265144 | 0.002465922 | 1 |
| Obesity | macrophage | *MEDAG* | 1.446076951 | 2.83E-05 | 0.00052574 | 0.177664332 |
| Obesity | macrophage | *CORO1A* | -1.764906074 | 1.99E-13 | 7.37E-11 | 1.25E-09 |
| Obesity | macrophage | *ELMO3* | 0.285724814 | 0.002445426 | 0.011918136 | 1 |
| Obesity | macrophage | *NFAT5* | 0.858266717 | 0.002989375 | 0.013798974 | 1 |

| | | | | | | |
|---|---|---|---|---|---|---|
| Obesity | macrophage | *PLLP* | 0.849233188 | 0.011969048 | 0.038139587 | 1 |
| Obesity | macrophage | *USB1* | 1.664756917 | 0.002102223 | 0.010680458 | 1 |
| Obesity | macrophage | *SETD6* | 1.334484197 | 0.000673659 | 0.004835717 | 1 |
| Obesity | macrophage | *COG4* | -0.875048085 | 9.66E-05 | 0.001183611 | 0.607192477 |
| Obesity | macrophage | *SLC7A6OS* | 0.255389561 | 0.007952185 | 0.028008622 | 1 |
| Obesity | macrophage | *HCFC1R1* | 2.31222796 | 0.000977362 | 0.006341253 | 1 |
| Obesity | macrophage | *NAGPA* | 0.889855379 | 0.006203613 | 0.023340582 | 1 |
| Obesity | macrophage | *LMF1* | 1.451487348 | 0.000168722 | 0.001744668 | 1 |
| Obesity | macrophage | *SLC7A5* | 5.280201583 | 7.87E-05 | 0.001026288 | 0.494670753 |
| Obesity | macrophage | *FBXO31* | 0.814206708 | 0.004239782 | 0.017758502 | 1 |
| Obesity | macrophage | *UBE2I* | 1.00167504 | 0.005960907 | 0.022768056 | 1 |
| Obesity | macrophage | *CLUAP1* | 0.541668533 | 6.90E-06 | 0.000246485 | 0.043381335 |
| Obesity | macrophage | *KAT8* | 2.121467578 | 1.97E-08 | 2.25E-06 | 0.000123849 |
| Obesity | macrophage | *KNOP1* | -0.647161028 | 0.013920152 | 0.042545454 | 1 |
| Obesity | macrophage | *IQCH* | 1.759251492 | 1.84E-05 | 0.000441618 | 0.115696025 |
| Obesity | macrophage | *TRIP4* | 0.569145775 | 0.001963983 | 0.010246937 | 1 |
| Obesity | macrophage | *RASL12* | 1.965446419 | 6.23E-05 | 0.000878061 | 0.391615164 |
| Obesity | macrophage | *FAH* | 1.625440917 | 0.001400509 | 0.008041095 | 1 |
| Obesity | macrophage | *HOMER2* | 0.611919941 | 0.01527679 | 0.045423605 | 1 |
| Obesity | macrophage | *ATP8B4* | -0.38201389 | 0.012008699 | 0.038207839 | 1 |
| Obesity | macrophage | *DTWD1* | 0.806957383 | 0.001601276 | 0.008823011 | 1 |
| Obesity | macrophage | *DNAJC17* | 1.343699048 | 0.005999244 | 0.022855957 | 1 |
| Obesity | macrophage | *PDGFRL* | 0.74588628 | 0.002619635 | 0.012495937 | 1 |
| Obesity | macrophage | *ZDHHC2* | -0.668683194 | 0.002711148 | 0.012844753 | 1 |
| Obesity | macrophage | *NBN* | 0.912721184 | 0.016718401 | 0.048437136 | 1 |
| Obesity | macrophage | *PLAT* | 1.559573813 | 4.06E-06 | 0.000173988 | 0.025554189 |
| Obesity | macrophage | *SPAG1* | 0.391768635 | 0.012893526 | 0.04018919 | 1 |
| Obesity | macrophage | *LEPROTL1* | 1.269117003 | 0.009865638 | 0.03300972 | 1 |
| Obesity | macrophage | *ERICH1* | 0.928844726 | 0.013595196 | 0.041758525 | 1 |
| Obesity | macrophage | *TUSC3* | 1.349753427 | 1.02E-05 | 0.000308603 | 0.064189482 |
| Obesity | macrophage | *ARHGEF10* | 0.490609882 | 0.001486064 | 0.008379267 | 1 |
| Obesity | macrophage | *MCM4* | 0.773295564 | 0.000781946 | 0.005390454 | 1 |
| Obesity | macrophage | *PPP1R37* | 0.976496974 | 0.000888764 | 0.005908605 | 1 |
| Obesity | macrophage | *ERCC2* | 1.756932561 | 0.01649181 | 0.047957451 | 1 |
| Obesity | macrophage | *FCER2* | -0.494738572 | 0.000525987 | 0.003998648 | 1 |
| Obesity | macrophage | *AES* | 1.23505072 | 0.007796813 | 0.027614456 | 1 |
| Obesity | macrophage | *SGTA* | 1.164394976 | 0.008039246 | 0.028236167 | 1 |
| Obesity | macrophage | *IL27RA* | 0.895352757 | 0.008898079 | 0.030418707 | 1 |
| Obesity | macrophage | *OLFM2* | 0.215729788 | 0.005819885 | 0.022351629 | 1 |
| Obesity | macrophage | *PDCD5* | 0.634654219 | 0.015370338 | 0.045560262 | 1 |
| Obesity | macrophage | *PRX* | 0.768633501 | 0.006026066 | 0.02289177 | 1 |
| Obesity | macrophage | *CACTIN* | 2.528679778 | 0.006776322 | 0.025089951 | 1 |
| Obesity | macrophage | *FZR1* | 1.70092017 | 0.011538652 | 0.03716368 | 1 |
| Obesity | macrophage | *BBC3* | 2.543699954 | 0.001322481 | 0.007748778 | 1 |
| Obesity | macrophage | *CEACAM4* | -1.463186757 | 0.000469375 | 0.003730668 | 1 |
| Obesity | macrophage | *MRPL4* | 1.325573263 | 0.0006149 | 0.004510943 | 1 |
| Obesity | macrophage | *ICAM5* | -2.925533021 | 0.001473472 | 0.00833068 | 1 |
| Obesity | macrophage | *CD33* | -0.307331614 | 0.016135007 | 0.047155364 | 1 |
| Obesity | macrophage | *BABAM1* | 2.415352605 | 0.000360416 | 0.003102809 | 1 |
| Obesity | macrophage | *PTPRS* | 0.62792748 | 0.000132407 | 0.00148121 | 0.83243981 |

| | | | | | | |
|---|---|---|---|---|---|---|
| Obesity | macrophage | *CYTH2* | 1.767830857 | 0.000182412 | 0.001834917 | 1 |
| Obesity | macrophage | *BCAT2* | 1.915734607 | 0.00093638 | 0.006157975 | 1 |
| Obesity | macrophage | *PPP2R1A* | 1.44207061 | 0.004026071 | 0.017076033 | 1 |
| Obesity | macrophage | *LILRB5* | -0.316112632 | 0.00108555 | 0.006736135 | 1 |
| Obesity | macrophage | *MAST1* | 4.271093681 | 2.22E-05 | 0.000483304 | 0.139884737 |
| Obesity | macrophage | *ELL* | -0.817742348 | 0.001928268 | 0.010106241 | 1 |
| Obesity | macrophage | *KXD1* | 1.487793127 | 0.016710417 | 0.048436327 | 1 |
| Obesity | macrophage | *HPN* | -0.547871864 | 0.001152546 | 0.006987517 | 1 |
| Obesity | macrophage | *ZNF14* | 1.817767072 | 0.00110945 | 0.00683165 | 1 |
| Obesity | macrophage | *ATP13A1* | -0.724010342 | 0.000166929 | 0.001733943 | 1 |
| Obesity | macrophage | *PON3* | 2.121363104 | 1.43E-05 | 0.00039059 | 0.089835724 |
| Obesity | macrophage | *PON2* | 1.286398593 | 0.002891178 | 0.013504334 | 1 |
| Obesity | macrophage | *WDR91* | -0.695788473 | 0.012142969 | 0.038537531 | 1 |
| Obesity | macrophage | *PTN* | 1.399391198 | 7.37E-05 | 0.000983662 | 0.463304882 |
| Obesity | macrophage | *MPP6* | 1.403379457 | 3.44E-05 | 0.000576402 | 0.216150931 |
| Obesity | macrophage | *ADAP1* | -1.554881705 | 0.007463837 | 0.026707538 | 1 |
| Obesity | macrophage | *CAV1* | 0.515598645 | 0.004713367 | 0.019130365 | 1 |
| Obesity | macrophage | *MET* | 1.018228889 | 0.000296952 | 0.002710134 | 1 |
| Obesity | macrophage | *HIBADH* | 0.995811259 | 0.003275909 | 0.014785097 | 1 |
| Obesity | macrophage | *CPVL* | -0.487727357 | 0.003512112 | 0.015506071 | 1 |
| Obesity | macrophage | *PLEKHA8* | 1.444696739 | 0.000517633 | 0.003973576 | 1 |
| Obesity | macrophage | *NOD1* | 1.118841616 | 0.003635944 | 0.015896509 | 1 |
| Obesity | macrophage | *EPHB6* | 0.610500082 | 0.009845961 | 0.032961424 | 1 |
| Obesity | macrophage | *NPTX2* | -4.411997115 | 0.005052615 | 0.020163061 | 1 |
| Obesity | macrophage | *CYP3A5* | 1.65157846 | 4.40E-05 | 0.000698582 | 0.276638281 |
| Obesity | macrophage | *LSM5* | 1.930405614 | 0.000162034 | 0.001693914 | 1 |
| Obesity | macrophage | *AP1S1* | 3.538800312 | 6.76E-05 | 0.000921647 | 0.424879426 |
| Obesity | macrophage | *RPA3* | 1.547972392 | 0.000176684 | 0.001808801 | 1 |
| Obesity | macrophage | *PHF14* | 0.873319108 | 0.009047071 | 0.030807759 | 1 |
| Obesity | macrophage | *ZNF862* | 0.978903965 | 0.000399555 | 0.003349409 | 1 |
| Obesity | macrophage | *POU6F2* | 1.634313898 | 2.42E-05 | 0.000492083 | 0.152219484 |
| Obesity | macrophage | *TSPAN13* | 2.268026919 | 2.81E-06 | 0.000134335 | 0.017653551 |
| Obesity | macrophage | *MRPL32* | 0.39630084 | 0.011406887 | 0.036871515 | 1 |
| Obesity | macrophage | *URGCP* | 1.147766314 | 0.002130533 | 0.01079344 | 1 |
| Obesity | macrophage | *TBL2* | 0.213431488 | 0.013106702 | 0.040632069 | 1 |
| Obesity | macrophage | *LIMK1* | -0.709950508 | 0.01411913 | 0.043007253 | 1 |
| Obesity | macrophage | *FKTN* | 1.258344068 | 0.001495321 | 0.008423911 | 1 |
| Obesity | macrophage | *OGN* | 1.30642609 | 3.07E-07 | 2.36E-05 | 0.001932754 |
| Obesity | macrophage | *ECM2* | 0.587457857 | 0.002674 | 0.012726295 | 1 |
| Obesity | macrophage | *TNFSF8* | -0.54543658 | 0.004303186 | 0.01796423 | 1 |
| Obesity | macrophage | *AK1* | 0.582856593 | 0.014370661 | 0.043555097 | 1 |
| Obesity | macrophage | *FUBP3* | 1.209495731 | 0.005553093 | 0.021590783 | 1 |
| Obesity | macrophage | *RGP1* | 1.880265784 | 0.000234947 | 0.002234659 | 1 |
| Obesity | macrophage | *MPDZ* | 1.121309797 | 3.29E-05 | 0.000560415 | 0.206793053 |
| Obesity | macrophage | *PTGDS* | 2.6914309 | 1.72E-05 | 0.000431251 | 0.108319809 |
| Obesity | macrophage | *PDLIM1* | 0.58044708 | 0.002227427 | 0.011087754 | 1 |
| Obesity | macrophage | *TRDMT1* | 1.570557323 | 7.59E-05 | 0.000996712 | 0.477424853 |
| Obesity | macrophage | *DDX50* | 1.315381177 | 0.001776749 | 0.009539215 | 1 |
| Obesity | macrophage | *MAPK8* | 0.88493369 | 0.008310236 | 0.028993594 | 1 |
| Obesity | macrophage | *ATE1* | 0.631161933 | 0.006521274 | 0.024376541 | 1 |

| | | | | | | |
|---|---|---|---|---|---|---|
| Obesity | macrophage | *NSMCE4A* | 1.501503149 | 0.000609662 | 0.004482975 | 1 |
| Obesity | macrophage | *MICU1* | 1.033814465 | 0.002538613 | 0.012272557 | 1 |
| Obesity | macrophage | *CCSER2* | 1.151548512 | 0.001135455 | 0.006923965 | 1 |
| Obesity | macrophage | *ACBD5* | 1.487433688 | 3.19E-05 | 0.000554992 | 0.200268317 |
| Obesity | macrophage | *MTPAP* | 1.32606672 | 0.005731701 | 0.022121059 | 1 |
| Obesity | macrophage | *PITRM1* | 1.562627717 | 0.001240192 | 0.00737662 | 1 |
| Obesity | macrophage | *SHOC2* | 0.849654643 | 0.013682459 | 0.041941303 | 1 |
| Obesity | macrophage | *RGS9* | 2.219535775 | 1.51E-05 | 0.000405213 | 0.094733525 |
| Obesity | macrophage | *WNT3* | 1.040541858 | 0.003720055 | 0.016136259 | 1 |
| Obesity | macrophage | *ASPA* | 1.252353454 | 2.77E-05 | 0.000522449 | 0.174037127 |
| Obesity | macrophage | *RAD51C* | -0.304107433 | 0.003985371 | 0.016987986 | 1 |
| Obesity | macrophage | *RPS6KB1* | 1.162195242 | 0.007203243 | 0.026071843 | 1 |
| Obesity | macrophage | *CBX1* | 0.856732742 | 0.000596983 | 0.004410376 | 1 |
| Obesity | macrophage | *PIGL* | 1.181648322 | 0.002083243 | 0.010636979 | 1 |
| Obesity | macrophage | *GALK1* | 2.972037353 | 0.00011442 | 0.001349643 | 0.719359486 |
| Obesity | macrophage | *HOXB6* | 1.247330329 | 3.91E-08 | 4.09E-06 | 0.000245685 |
| Obesity | macrophage | *RNF167* | 0.982076171 | 0.012432443 | 0.039075715 | 1 |
| Obesity | macrophage | *CHRNE* | 0.280243894 | 0.001125029 | 0.006880403 | 1 |
| Obesity | macrophage | *C1QBP* | 2.64212025 | 1.46E-06 | 8.04E-05 | 0.009194063 |
| Obesity | macrophage | *GOSR1* | 1.073068331 | 0.008783681 | 0.030148123 | 1 |
| Obesity | macrophage | *DRG2* | 2.097455513 | 0.000914205 | 0.006062876 | 1 |
| Obesity | macrophage | *UTP6* | 1.122518921 | 0.013558114 | 0.041687739 | 1 |
| Obesity | macrophage | *C17orf75* | 1.244327851 | 0.00100403 | 0.006447741 | 1 |
| Obesity | macrophage | *EZH1* | 0.883610549 | 0.001984108 | 0.010303093 | 1 |
| Obesity | macrophage | *DLX4* | -9.160907605 | 0.010437043 | 0.034481183 | 1 |
| Obesity | macrophage | *VAT1* | 0.785672935 | 0.005361619 | 0.021002181 | 1 |
| Obesity | macrophage | *LRRC59* | 2.296938964 | 0.001484582 | 0.008378428 | 1 |
| Obesity | macrophage | *MPP2* | 2.078074164 | 1.24E-05 | 0.000348861 | 0.078144806 |
| Obesity | macrophage | *EFNB3* | 4.712593089 | 0.000533963 | 0.004044368 | 1 |
| Obesity | macrophage | *FAM20A* | -0.437023342 | 0.016645569 | 0.048270614 | 1 |
| Obesity | macrophage | *DPH1* | 2.670410047 | 0.000340869 | 0.003009896 | 1 |
| Obesity | macrophage | *DHRS7B* | 1.079091533 | 0.009495024 | 0.031991006 | 1 |
| Obesity | macrophage | *SLAIN2* | 0.730612566 | 0.009166495 | 0.03113439 | 1 |
| Obesity | macrophage | *KIAA1211* | 0.215492696 | 0.005912496 | 0.022665771 | 1 |
| Obesity | macrophage | *CPE* | 0.861723693 | 0.005767327 | 0.022217638 | 1 |
| Obesity | macrophage | *WFS1* | 2.001389169 | 7.89E-06 | 0.000266545 | 0.049577298 |
| Obesity | macrophage | *GRPEL1* | 2.647601949 | 5.35E-05 | 0.000788949 | 0.336092063 |
| Obesity | macrophage | *AADAT* | 0.982310294 | 0.001775139 | 0.009539215 | 1 |
| Obesity | macrophage | *CPZ* | 14.83226101 | 5.31E-05 | 0.000788276 | 0.333616181 |
| Obesity | macrophage | *SLC2A9* | -0.381259535 | 0.003050739 | 0.014040994 | 1 |
| Obesity | macrophage | *SH3D19* | 0.426938673 | 0.004261466 | 0.017825574 | 1 |
| Obesity | macrophage | *LRP2BP* | 0.43782496 | 0.014154699 | 0.043073859 | 1 |
| Obesity | macrophage | *UFSP2* | 1.150528894 | 0.011741832 | 0.037606161 | 1 |
| Obesity | macrophage | *UGDH* | 1.178337812 | 0.004487791 | 0.018501472 | 1 |
| Obesity | macrophage | *CTSC* | -0.394024372 | 0.002397686 | 0.011730936 | 1 |
| Obesity | macrophage | *SC5D* | 2.174833876 | 7.96E-05 | 0.001034164 | 0.500535348 |
| Obesity | macrophage | *C11orf63* | 1.679394553 | 0.000139273 | 0.001528114 | 0.875609307 |
| Obesity | macrophage | *FOXRED1* | 0.58335665 | 0.000360769 | 0.003102809 | 1 |
| Obesity | macrophage | *NRXN2* | -0.908342253 | 0.001444989 | 0.008228849 | 1 |
| Obesity | macrophage | *MS4A4A* | -0.25075059 | 0.011192493 | 0.036406664 | 1 |

| | | | | | | |
|---|---|---|---|---|---|---|
| Obesity | macrophage | *CCDC86* | 4.803027869 | 0.014857457 | 0.044586556 | 1 |
| Obesity | macrophage | *CD5* | 0.442361307 | 0.012390971 | 0.039009532 | 1 |
| Obesity | macrophage | *NDUFS8* | 1.877749399 | 0.000831677 | 0.005616276 | 1 |
| Obesity | macrophage | *GTF2H1* | 0.904540478 | 0.001190662 | 0.007149657 | 1 |
| Obesity | macrophage | *CYP27B1* | -2.233452833 | 0.001556142 | 0.008680978 | 1 |
| Obesity | macrophage | *GLI1* | 3.473669152 | 2.12E-05 | 0.00046838 | 0.133020054 |
| Obesity | macrophage | *TRPV4* | -0.868207544 | 9.82E-07 | 6.11E-05 | 0.00617102 |
| Obesity | macrophage | *VPS29* | 1.016668534 | 0.011790696 | 0.037724227 | 1 |
| Obesity | macrophage | *NAA25* | 1.053312752 | 0.003060493 | 0.014075579 | 1 |
| Obesity | macrophage | *EIF2B1* | 1.452264769 | 0.004423727 | 0.018345628 | 1 |
| Obesity | macrophage | *DDX55* | 0.629938232 | 3.98E-06 | 0.000173912 | 0.025043335 |
| Obesity | macrophage | *NUP107* | 1.080767427 | 0.000827415 | 0.005593502 | 1 |
| Obesity | macrophage | *GNPTAB* | 0.881201198 | 0.01584688 | 0.046534019 | 1 |
| Obesity | macrophage | *SPSB2* | 1.013748652 | 0.004020188 | 0.017066118 | 1 |
| Obesity | macrophage | *PHC1* | 2.077559089 | 0.000418279 | 0.003437543 | 1 |
| Obesity | macrophage | *COL12A1* | 1.075157269 | 0.000119028 | 0.001375285 | 0.748327571 |
| Obesity | macrophage | *BTN3A3* | 0.282205682 | 0.002727495 | 0.012912472 | 1 |
| Obesity | macrophage | *SMIM8* | 1.018747747 | 0.000587441 | 0.004355237 | 1 |
| Obesity | macrophage | *MCM9* | 0.948697677 | 0.01693733 | 0.048913641 | 1 |
| Obesity | macrophage | *FAM184A* | 0.680056323 | 0.000297007 | 0.002710134 | 1 |
| Obesity | macrophage | *GABRR2* | 0.925866087 | 9.97E-05 | 0.001207859 | 0.626879067 |
| Obesity | macrophage | *MTRF1L* | 0.42626899 | 0.012347448 | 0.038930995 | 1 |
| Obesity | macrophage | *TULP1* | 10.4340008 | 8.74E-06 | 0.000289582 | 0.054959097 |
| Obesity | macrophage | *CD83* | -0.328189691 | 0.001469491 | 0.008323145 | 1 |
| Obesity | macrophage | *ALDH5A1* | 0.54604877 | 0.008789249 | 0.030148123 | 1 |
| Obesity | macrophage | *ACOT13* | 1.1707891 | 0.016761604 | 0.048495262 | 1 |
| Obesity | macrophage | *PACRG* | 1.925779077 | 0.000736985 | 0.005153973 | 1 |
| Obesity | macrophage | *BTN2A1* | -0.536912363 | 0.008191039 | 0.028641303 | 1 |
| Obesity | macrophage | *LAMA4* | 0.609752792 | 6.22E-06 | 0.000229467 | 0.039075732 |
| Obesity | macrophage | *LY86* | -0.535263585 | 0.007141238 | 0.025967011 | 1 |
| Obesity | macrophage | *PRSS16* | 1.636253931 | 6.75E-09 | 9.02E-07 | 4.24E-05 |
| Obesity | macrophage | *C7* | 1.018153202 | 0.001986217 | 0.010303093 | 1 |
| Obesity | macrophage | *NME5* | 0.774775227 | 1.43E-07 | 1.27E-05 | 0.000901944 |
| Obesity | macrophage | *PFDN1* | 0.652945769 | 0.001575791 | 0.008728633 | 1 |
| Obesity | macrophage | *LOX* | 1.642954238 | 5.54E-05 | 0.000810135 | 0.348357982 |
| Obesity | macrophage | *SPARC* | 0.753078557 | 0.003154742 | 0.014382787 | 1 |
| Obesity | macrophage | *PCDHB6* | 0.696914783 | 0.005591297 | 0.021699065 | 1 |
| Obesity | macrophage | *CLK4* | 1.17362094 | 0.003124672 | 0.014287135 | 1 |
| Obesity | macrophage | *CLINT1* | 0.848126517 | 0.010914333 | 0.035718068 | 1 |
| Obesity | macrophage | *RASGRF2* | 1.25363013 | 8.98E-06 | 0.000293144 | 0.056436713 |
| Obesity | macrophage | *DROSHA* | 0.568373143 | 0.014938127 | 0.044744487 | 1 |
| Obesity | macrophage | *SLC27A6* | 2.561869873 | 0.00017947 | 0.001819885 | 1 |
| Obesity | macrophage | *TARS* | 1.60843468 | 0.001465981 | 0.00831075 | 1 |
| Obesity | macrophage | *LNPEP* | 0.82537459 | 0.000833567 | 0.005623002 | 1 |
| Obesity | macrophage | *SKP1* | 1.549932317 | 0.000117286 | 0.001359647 | 0.737375139 |
| Obesity | macrophage | *C9* | -0.38150292 | 0.002623875 | 0.012506673 | 1 |
| Obesity | macrophage | *SMAD5* | 0.874929351 | 0.015716745 | 0.046238266 | 1 |
| Obesity | macrophage | *BNIP1* | -0.239511973 | 0.004593942 | 0.018742448 | 1 |
| Obesity | macrophage | *CNTN3* | 1.275162832 | 2.19E-05 | 0.00047915 | 0.137516108 |
| Obesity | macrophage | *NIT2* | 0.956490216 | 0.007783985 | 0.027586198 | 1 |

| | | | | | | |
|---|---|---|---|---|---|---|
| Obesity | macrophage | *PCCB* | 1.031891586 | 0.002911977 | 0.013582552 | 1 |
| Obesity | macrophage | *PRKAR2A* | 0.691735333 | 0.000129701 | 0.001453534 | 0.815432768 |
| Obesity | macrophage | *HYAL1* | -0.33604422 | 0.002602416 | 0.012457209 | 1 |
| Obesity | macrophage | *C3orf14* | -0.446649058 | 0.004582179 | 0.018718981 | 1 |
| Obesity | macrophage | *IFT57* | 1.675346393 | 0.000317532 | 0.002851892 | 1 |
| Obesity | macrophage | *GBE1* | 0.896940421 | 0.002580887 | 0.012386285 | 1 |
| Obesity | macrophage | *NCBP2* | 1.16844667 | 0.006875738 | 0.025353527 | 1 |
| Obesity | macrophage | *NEK11* | 1.051269589 | 0.009047548 | 0.030807759 | 1 |
| Obesity | macrophage | *PLSCR4* | 0.473953139 | 0.015155251 | 0.045156901 | 1 |
| Obesity | macrophage | *WDR48* | 0.644416494 | 0.009924416 | 0.033153454 | 1 |
| Obesity | macrophage | *COMMD2* | 1.128532483 | 0.00074946 | 0.005229581 | 1 |
| Obesity | macrophage | *ARHGEF26* | 0.971482962 | 1.95E-06 | 0.000102127 | 0.012255258 |
| Obesity | macrophage | *KLHL24* | 0.889374925 | 3.27E-05 | 0.000559429 | 0.205869845 |
| Obesity | macrophage | *PLCH1* | 2.167607409 | 8.21E-07 | 5.54E-05 | 0.005163562 |
| Obesity | macrophage | *ZBTB47* | 0.681378179 | 0.015804202 | 0.046442547 | 1 |
| Obesity | macrophage | *NKTR* | 0.896125379 | 0.001593526 | 0.008803601 | 1 |
| Obesity | macrophage | *CLCN2* | 2.568429426 | 7.38E-06 | 0.000255529 | 0.046411866 |
| Obesity | macrophage | *KCNIP3* | 1.434961534 | 0.000587186 | 0.004355237 | 1 |
| Obesity | macrophage | *FAHD2A* | 1.322974276 | 9.26E-06 | 0.000298528 | 0.058213039 |
| Obesity | macrophage | *ACTR1B* | 2.135593734 | 2.35E-07 | 1.94E-05 | 0.001476389 |
| Obesity | macrophage | *DNAJC27* | 1.161308527 | 4.54E-05 | 0.000713196 | 0.285278241 |
| Obesity | macrophage | *TANC1* | 0.479488749 | 0.008657866 | 0.029891271 | 1 |
| Obesity | macrophage | *GRB14* | 0.925463453 | 0.000958706 | 0.006255676 | 1 |
| Obesity | macrophage | *POLE4* | 2.113632441 | 0.000166726 | 0.001733943 | 1 |
| Obesity | macrophage | *LANCL1* | 1.560868778 | 0.000144264 | 0.00156451 | 0.906989648 |
| Obesity | macrophage | *WDR75* | 1.359739156 | 0.002963161 | 0.013718255 | 1 |
| Obesity | macrophage | *EFEMP1* | 0.548729119 | 0.004135555 | 0.017426431 | 1 |
| Obesity | macrophage | *GLS* | 0.827774698 | 0.010677032 | 0.035053002 | 1 |
| Obesity | macrophage | *GGCX* | 0.445598618 | 0.002270063 | 0.011246563 | 1 |
| Obesity | macrophage | *GNLY* | 0.915574638 | 0.013689129 | 0.041941303 | 1 |
| Obesity | macrophage | *PLCD4* | 1.266391724 | 0.003896874 | 0.016711903 | 1 |
| Obesity | macrophage | *CHMP3* | -0.949482924 | 9.17E-06 | 0.000297161 | 0.057649227 |
| Obesity | macrophage | *ZNF142* | 0.63868063 | 0.012968056 | 0.040301616 | 1 |
| Obesity | macrophage | *MLPH* | 2.37148614 | 9.81E-05 | 0.001194917 | 0.616577319 |
| Obesity | macrophage | *STK25* | 1.208599741 | 0.001564114 | 0.008709997 | 1 |
| Obesity | macrophage | *SOS1* | 0.500462134 | 0.015032188 | 0.044942875 | 1 |
| Obesity | macrophage | *COX7A2L* | 1.316615055 | 0.007693465 | 0.027352747 | 1 |
| Obesity | macrophage | *PNO1* | 1.822704501 | 0.005520478 | 0.021490552 | 1 |
| Obesity | macrophage | *ORC4* | 0.787067081 | 0.006535111 | 0.024398008 | 1 |
| Obesity | macrophage | *CD207* | 16.37763264 | 1.17E-07 | 1.05E-05 | 0.000732658 |
| Obesity | macrophage | *VAX2* | 2.359035351 | 6.24E-06 | 0.000229467 | 0.039238775 |
| Obesity | macrophage | *PLEKHA3* | 1.600535155 | 0.001570517 | 0.008716979 | 1 |
| Obesity | macrophage | *BCL9* | 0.714958575 | 0.008179929 | 0.02861837 | 1 |
| Obesity | macrophage | *SCP2* | 0.852353017 | 0.003332094 | 0.014942136 | 1 |
| Obesity | macrophage | *FAM20B* | 1.425342084 | 0.002263655 | 0.011223659 | 1 |
| Obesity | macrophage | *TMEM59* | 1.083425265 | 0.003358581 | 0.014997871 | 1 |
| Obesity | macrophage | *C1orf21* | 0.420787494 | 0.001006682 | 0.006457594 | 1 |
| Obesity | macrophage | *LEPR* | 0.867646813 | 0.001207412 | 0.007229522 | 1 |
| Obesity | macrophage | *NCF2* | -0.460783873 | 0.002141836 | 0.010824534 | 1 |
| Obesity | macrophage | *RGS2* | -0.960461912 | 0.004856547 | 0.019610217 | 1 |

| | | | | | | |
|---|---|---|---|---|---|---|
| Obesity | macrophage | *OLFML3* | 1.396379596 | 9.49E-05 | 0.001169956 | 0.596677492 |
| Obesity | macrophage | *WARS2* | 0.539317203 | 0.00085776 | 0.005761472 | 1 |
| Obesity | macrophage | *C1orf109* | 1.573651707 | 3.08E-05 | 0.000547478 | 0.193807195 |
| Obesity | macrophage | *ACADM* | 0.947290528 | 0.002387922 | 0.011702667 | 1 |
| Obesity | macrophage | *MFAP2* | -0.348457015 | 0.001925091 | 0.010102709 | 1 |
| Obesity | macrophage | *CD46* | 0.966058557 | 0.001901174 | 0.010019012 | 1 |
| Obesity | macrophage | *PRDX1* | 0.777899561 | 0.014466982 | 0.043706831 | 1 |
| Obesity | macrophage | *CNN3* | 1.332859875 | 3.16E-05 | 0.000554992 | 0.198465607 |
| Obesity | macrophage | *PRRC2C* | 1.007791494 | 0.001033267 | 0.006557047 | 1 |
| Obesity | macrophage | *ABCD3* | 1.196535642 | 0.004138616 | 0.01742765 | 1 |
| Obesity | macrophage | *VAMP4* | 0.695747162 | 0.011966212 | 0.038139587 | 1 |
| Obesity | macrophage | *PTBP2* | 0.652433731 | 0.000115083 | 0.001350604 | 0.723523845 |
| Obesity | macrophage | *DARS2* | -0.944008159 | 0.015520392 | 0.045843681 | 1 |
| Obesity | macrophage | *SERPINC1* | 0.79411174 | 1.69E-11 | 4.42E-09 | 1.06E-07 |
| Obesity | macrophage | *SLC5A9* | 2.164087395 | 1.74E-05 | 0.000431251 | 0.109106625 |
| Obesity | macrophage | *TXNDC12* | 1.29770369 | 0.003987048 | 0.016987986 | 1 |
| Obesity | macrophage | *IFT46* | 1.16566544 | 0.000415236 | 0.00342597 | 1 |
| Obesity | macrophage | *SPCS2* | 0.918669139 | 0.017068209 | 0.049246365 | 1 |
| Obesity | macrophage | *SGIP1* | 1.104339367 | 7.62E-05 | 0.00099826 | 0.479164878 |
| Obesity | macrophage | *SLC16A7* | 0.909215547 | 0.003820758 | 0.016430306 | 1 |
| Obesity | macrophage | *RAB3GAP2* | 1.019017296 | 0.006035802 | 0.022901078 | 1 |
| Obesity | macrophage | *WDR35* | 1.451356103 | 0.000495584 | 0.003870478 | 1 |
| Obesity | macrophage | *CPSF3* | 1.215509194 | 0.015183876 | 0.045220763 | 1 |
| Obesity | macrophage | *CCDC92* | 1.454826073 | 8.65E-15 | 3.63E-12 | 5.44E-11 |
| Obesity | macrophage | *CTNNAL1* | 0.207788459 | 0.009288731 | 0.031453551 | 1 |
| Obesity | macrophage | *BSPRY* | 4.505298709 | 0.000117128 | 0.001359647 | 0.736380665 |
| Obesity | macrophage | *PPP6C* | 1.135129669 | 0.00221129 | 0.011051175 | 1 |
| Obesity | macrophage | *INVS* | 1.326538952 | 4.06E-06 | 0.000173988 | 0.025528298 |
| Obesity | macrophage | *KDSR* | 1.419595723 | 0.001079244 | 0.006708303 | 1 |
| Obesity | macrophage | *DCAF4* | 0.977281224 | 1.10E-06 | 6.66E-05 | 0.006922562 |
| Obesity | macrophage | *ZC2HC1C* | 2.161789043 | 7.21E-05 | 0.000970403 | 0.453178048 |
| Obesity | macrophage | *KLHL29* | 0.951650179 | 0.000125138 | 0.00142473 | 0.786741903 |
| Obesity | macrophage | *ATAD2B* | 0.847456938 | 0.017311682 | 0.049788904 | 1 |
| Obesity | macrophage | *ATL2* | 0.936807934 | 0.006707385 | 0.024878661 | 1 |
| Obesity | macrophage | *CNRIP1* | -0.675941414 | 0.01562734 | 0.046061457 | 1 |
| Obesity | macrophage | *CUTC* | 0.777922334 | 0.011397865 | 0.036871515 | 1 |
| Obesity | macrophage | *PYROXD2* | 0.158222272 | 0.015336879 | 0.045525477 | 1 |
| Obesity | macrophage | *C10orf88* | 1.671591515 | 0.00228549 | 0.011269706 | 1 |
| Obesity | macrophage | *HOXB5* | 1.392613993 | 7.24E-08 | 6.80E-06 | 0.000455483 |
| Obesity | macrophage | *HOXB3* | 1.00676303 | 1.84E-09 | 2.69E-07 | 1.16E-05 |
| Obesity | macrophage | *CYSTM1* | 1.262039928 | 0.000354911 | 0.003073453 | 1 |
| Obesity | macrophage | *PCDHB14* | 1.392754149 | 2.48E-05 | 0.000492083 | 0.155990074 |
| Obesity | macrophage | *SNX19* | -0.575102534 | 0.0072351 | 0.026126983 | 1 |
| Obesity | macrophage | *MRC1* | -0.292036769 | 0.001043568 | 0.006593883 | 1 |
| Obesity | macrophage | *IQSEC3* | 2.310328449 | 0.000393395 | 0.003310939 | 1 |
| Obesity | macrophage | *ENOX1* | 1.174820459 | 9.11E-05 | 0.001134374 | 0.572859004 |
| Obesity | macrophage | *WBP4* | 0.75315199 | 0.015038781 | 0.044942875 | 1 |
| Obesity | macrophage | *SMAD9* | 1.597003706 | 2.00E-05 | 0.000456556 | 0.125428851 |
| Obesity | macrophage | *ADRA1A* | 0.133959926 | 1.81E-05 | 0.000441618 | 0.113587917 |
| Obesity | macrophage | *CRISPLD1* | 1.583975358 | 1.20E-05 | 0.000344993 | 0.075308666 |

| | | | | | | |
|---|---|---|---|---|---|---|
| Obesity | macrophage | *TEX14* | 0.68121126 | 0.007113887 | 0.025918616 | 1 |
| Obesity | macrophage | *TRIM6* | 0.42920554 | 0.013896013 | 0.042492331 | 1 |
| Obesity | macrophage | *PRB2* | 1.351186351 | 0.002112009 | 0.010716868 | 1 |
| Obesity | macrophage | *PYROXD1* | 0.603974845 | 0.00323767 | 0.014654596 | 1 |
| Obesity | macrophage | *ZSCAN18* | 1.036333638 | 0.003592185 | 0.015727067 | 1 |
| Obesity | macrophage | *PDZRN3* | 0.963486923 | 5.96E-05 | 0.000855087 | 0.374618471 |
| Obesity | macrophage | *B4GALT4* | 1.828236869 | 0.000810369 | 0.005513839 | 1 |
| Obesity | macrophage | *GJB6* | -6.534834685 | 0.002035681 | 0.0104647 | 1 |
| Obesity | macrophage | *RNF115* | 0.989268513 | 0.007893614 | 0.027864768 | 1 |
| Obesity | macrophage | *POLR3GL* | 1.543101223 | 0.002787774 | 0.013099205 | 1 |
| Obesity | macrophage | *TNFSF10* | 1.14557395 | 0.00042911 | 0.003494041 | 1 |
| Obesity | macrophage | *ZNF639* | 1.534379384 | 0.000185069 | 0.001858673 | 1 |
| Obesity | macrophage | *GPSM2* | 1.184967945 | 9.85E-06 | 0.000301542 | 0.06194014 |
| Obesity | macrophage | *FLT3* | 2.537660779 | 0.002741744 | 0.01296041 | 1 |
| Obesity | macrophage | *FMOD* | 0.984263238 | 0.004084854 | 0.017293924 | 1 |
| Obesity | macrophage | *FAM213A* | 0.49844598 | 0.002161336 | 0.01087936 | 1 |
| Obesity | macrophage | *ODF2L* | 0.724255279 | 0.000654414 | 0.004734526 | 1 |
| Obesity | macrophage | *RPAP2* | 1.164251576 | 0.001079816 | 0.006708303 | 1 |
| Obesity | macrophage | *PMS2* | 1.4586504 | 0.00010497 | 0.001259442 | 0.659947649 |
| Obesity | macrophage | *KLHL7* | 1.377350968 | 0.000981547 | 0.006347553 | 1 |
| Obesity | macrophage | *HERPUD2* | 1.043939667 | 0.017298189 | 0.049772866 | 1 |
| Obesity | macrophage | *FAM126A* | 0.738189688 | 0.008827407 | 0.03024409 | 1 |
| Obesity | macrophage | *FKBP9* | 0.694623828 | 0.005056729 | 0.020163061 | 1 |
| Obesity | macrophage | *CCZ1* | 0.74146893 | 0.000414984 | 0.00342597 | 1 |
| Obesity | macrophage | *SMU1* | 1.259549006 | 0.004103777 | 0.017350668 | 1 |
| Obesity | macrophage | *SLC25A51* | 0.194026697 | 0.003560598 | 0.015643243 | 1 |
| Obesity | macrophage | *DCAF10* | 1.078452258 | 0.002436943 | 0.011895232 | 1 |
| Obesity | macrophage | *BICC1* | 0.266398803 | 0.01608739 | 0.047064413 | 1 |
| Obesity | macrophage | *SLC25A16* | 0.339609289 | 0.00560198 | 0.021713716 | 1 |
| Obesity | macrophage | *ATP7B* | 0.127779567 | 0.006078945 | 0.023009228 | 1 |
| Obesity | macrophage | *ARHGAP9* | -0.754921734 | 2.22E-05 | 0.000483304 | 0.139701686 |
| Obesity | macrophage | *SMUG1* | 0.170081815 | 0.005044369 | 0.020161442 | 1 |
| Obesity | macrophage | *SARDH* | 4.637656863 | 1.78E-05 | 0.00044073 | 0.111950863 |
| Obesity | macrophage | *TTC21B* | 1.133763582 | 0.002586205 | 0.012402344 | 1 |
| Obesity | macrophage | *PLA2G12A* | 1.311439806 | 0.003988272 | 0.016987986 | 1 |
| Obesity | macrophage | *RAB38* | -0.635398747 | 0.000377748 | 0.003204992 | 1 |
| Obesity | macrophage | *DNPEP* | 1.833425887 | 0.000401219 | 0.003354341 | 1 |
| Obesity | macrophage | *FAM210B* | 0.536577802 | 0.012953375 | 0.040285192 | 1 |
| Obesity | macrophage | *SNX21* | 1.329387071 | 2.95E-05 | 0.000541496 | 0.185733157 |
| Obesity | macrophage | *ARFGEF2* | 0.921994725 | 0.01427096 | 0.043343731 | 1 |
| Obesity | macrophage | *CDH26* | 1.616475903 | 0.0033592 | 0.014997871 | 1 |
| Obesity | macrophage | *STX16* | 1.233712026 | 0.005099112 | 0.020277113 | 1 |
| Obesity | macrophage | *STAMBP* | 1.082602994 | 0.008611986 | 0.029769423 | 1 |
| Obesity | macrophage | *ATP8A1* | 0.414415757 | 0.012732668 | 0.039749528 | 1 |
| Obesity | macrophage | *HIF3A* | 0.592860906 | 5.04E-05 | 0.000759323 | 0.316979318 |
| Obesity | macrophage | *SNRPC* | -1.376329383 | 1.45E-21 | 1.01E-18 | 9.11E-18 |
| Obesity | macrophage | *RPS10* | 3.850506499 | 1.09E-05 | 0.00032774 | 0.068825449 |
| Obesity | macrophage | *HIST1H2BJ* | 1.138871304 | 4.22E-11 | 1.02E-08 | 2.66E-07 |
| Obesity | macrophage | *KLHL31* | 0.209784013 | 0.011269904 | 0.036579189 | 1 |
| Obesity | macrophage | *GLO1* | 1.505486134 | 4.95E-05 | 0.000749385 | 0.310994637 |

| | | | | | | |
|---|---|---|---|---|---|---|
| Obesity | macrophage | *NUP153* | 1.41738628 | 7.78E-05 | 0.001016521 | 0.488946407 |
| Obesity | macrophage | *EEF1E1* | 3.474958828 | 5.18E-06 | 0.000203227 | 0.032565585 |
| Obesity | macrophage | *BBS2* | 0.46276535 | 0.001531542 | 0.008574179 | 1 |
| Obesity | macrophage | *MT1G* | -2.564285174 | 0.001423886 | 0.008152981 | 1 |
| Obesity | macrophage | *TMTC4* | 1.671943447 | 0.002087805 | 0.010636979 | 1 |
| Obesity | macrophage | *GRK4* | 0.238946347 | 4.34E-06 | 0.000178092 | 0.027270303 |
| Obesity | macrophage | *MRPS7* | 1.255763719 | 0.002085636 | 0.010636979 | 1 |
| Obesity | macrophage | *OPRL1* | 0.483872991 | 0.002566752 | 0.0123416 | 1 |
| Obesity | macrophage | *SH2D3A* | 1.221572042 | 0.005355162 | 0.020989966 | 1 |
| Obesity | macrophage | *TRIP10* | 1.293378803 | 5.97E-05 | 0.000855087 | 0.375383275 |
| Obesity | macrophage | *SYMPK* | 1.015454911 | 0.007736225 | 0.02744788 | 1 |
| Obesity | macrophage | *BMP2* | 2.817157058 | 0.00397432 | 0.016987986 | 1 |
| Obesity | macrophage | *PCSK2* | 2.92339028 | 1.49E-10 | 3.02E-08 | 9.36E-07 |
| Obesity | macrophage | *DSTN* | 1.128829053 | 0.003577219 | 0.015672457 | 1 |
| Obesity | macrophage | *FAM110A* | -0.883196973 | 0.002218802 | 0.011071118 | 1 |
| Obesity | macrophage | *ZNF436* | 0.529710402 | 0.005943868 | 0.022716779 | 1 |
| Obesity | macrophage | *UROD* | 1.530851957 | 1.14E-05 | 0.000333865 | 0.071447209 |
| Obesity | macrophage | *KLC1* | 1.693307308 | 6.50E-07 | 4.64E-05 | 0.004084944 |
| Obesity | macrophage | *XRCC3* | 2.238944162 | 1.89E-08 | 2.20E-06 | 0.000118661 |
| Obesity | macrophage | *MCF2L* | 0.424655987 | 0.011099716 | 0.036176211 | 1 |
| Obesity | macrophage | *F10* | 1.740948677 | 6.41E-05 | 0.000889867 | 0.402844334 |
| Obesity | macrophage | *PCID2* | 0.778916702 | 0.004760435 | 0.019284057 | 1 |
| Obesity | macrophage | *PROZ* | 5.321247444 | 0.000158212 | 0.001666127 | 0.994677737 |
| Obesity | macrophage | *IGFLR1* | -2.418119558 | 0.00129105 | 0.007600031 | 1 |
| Obesity | macrophage | *COX6B1* | 1.563914845 | 0.004381172 | 0.018193148 | 1 |
| Obesity | macrophage | *PRDX5* | 1.332542186 | 0.00455851 | 0.018646295 | 1 |
| Obesity | macrophage | *SCAF1* | 2.055331104 | 0.002972809 | 0.013742684 | 1 |
| Obesity | macrophage | *TSKS* | 2.177740112 | 0.000324376 | 0.002896808 | 1 |
| Obesity | macrophage | *FLRT1* | 0.346932732 | 0.012812512 | 0.03995648 | 1 |
| Obesity | macrophage | *SBDS* | 1.150543345 | 0.00147232 | 0.00833068 | 1 |
| Obesity | macrophage | *BECN1* | 1.160668874 | 0.017356705 | 0.049895565 | 1 |
| Obesity | macrophage | *TRAP1* | 0.196298741 | 0.001066192 | 0.006674749 | 1 |
| Obesity | macrophage | *EMG1* | 0.899758705 | 0.016465028 | 0.047946101 | 1 |
| Obesity | macrophage | *ATG14* | 1.06322046 | 0.005525461 | 0.021496641 | 1 |
| Obesity | macrophage | *RHOJ* | 1.33012665 | 3.27E-05 | 0.000559429 | 0.205314279 |
| Obesity | macrophage | *HSPA2* | 1.033175194 | 0.001210686 | 0.007242229 | 1 |
| Obesity | macrophage | *PZP* | 1.216417343 | 0.000349787 | 0.003050081 | 1 |
| Obesity | macrophage | *RHOT1* | 0.550976708 | 0.008854508 | 0.030314414 | 1 |
| Obesity | macrophage | *AIF1L* | 1.670682924 | 8.99E-06 | 0.000293144 | 0.056528527 |
| Obesity | macrophage | *IPPK* | 1.370831853 | 0.00121673 | 0.007271467 | 1 |
| Obesity | macrophage | *FGD3* | -0.598367441 | 0.011083525 | 0.03614218 | 1 |
| Obesity | macrophage | *HRASLS* | 1.038684482 | 0.002203286 | 0.011019937 | 1 |
| Obesity | macrophage | *RAB3IP* | 0.831101853 | 0.006911614 | 0.025411298 | 1 |
| Obesity | macrophage | *YEATS4* | 0.451431232 | 0.001089842 | 0.006743934 | 1 |
| Obesity | macrophage | *AAMP* | 1.338744088 | 0.002285022 | 0.011269706 | 1 |
| Obesity | macrophage | *POR* | 1.266969704 | 0.002382355 | 0.011692323 | 1 |
| Obesity | macrophage | *GNAI1* | 0.73229192 | 0.002218247 | 0.011071118 | 1 |
| Obesity | macrophage | *SGCE* | 0.906462405 | 0.001827252 | 0.009710845 | 1 |
| Obesity | macrophage | *CASD1* | 1.195849643 | 0.002699931 | 0.012803881 | 1 |
| Obesity | macrophage | *MGAT3* | 1.666319682 | 1.41E-05 | 0.000389405 | 0.088873494 |

| | | | | | | |
|---|---|---|---|---|---|---|
| Obesity | macrophage | *BAIAP2L2* | 0.752167069 | 0.004273361 | 0.017851575 | 1 |
| Obesity | macrophage | *RIBC2* | 0.293464678 | 0.017126399 | 0.049391591 | 1 |
| Obesity | macrophage | *EMC4* | 1.107159466 | 0.000190714 | 0.001900193 | 1 |
| Obesity | macrophage | *SPECC1* | 0.955162557 | 0.010402061 | 0.034384884 | 1 |
| Obesity | macrophage | *CDHR3* | 0.507808241 | 0.004438249 | 0.018393717 | 1 |
| Obesity | macrophage | *HOXD1* | 1.925184207 | 0.009776576 | 0.032764037 | 1 |
| Obesity | macrophage | *HOXD3* | 0.786952233 | 0.001117372 | 0.006866976 | 1 |
| Obesity | macrophage | *WDFY4* | -0.453459875 | 0.000345427 | 0.003037338 | 1 |
| Obesity | macrophage | *CALML4* | -1.150045191 | 0.001021499 | 0.0065294 | 1 |
| Obesity | macrophage | *ACKR4* | 0.927066013 | 0.000306428 | 0.002779965 | 1 |
| Obesity | macrophage | *SUMF2* | 0.864357904 | 0.013216228 | 0.040911092 | 1 |
| Obesity | macrophage | *SERGEF* | 1.089170499 | 0.010204808 | 0.033927885 | 1 |
| Obesity | macrophage | *RPAIN* | -0.848466229 | 0.000455386 | 0.003647148 | 1 |
| Obesity | macrophage | *PHF20L1* | 0.92202491 | 0.010204601 | 0.033927885 | 1 |
| Obesity | macrophage | *SLC44A2* | 1.472684062 | 5.11E-05 | 0.000766665 | 0.321232697 |
| Obesity | macrophage | *CDKN2D* | 0.986737573 | 0.001432368 | 0.008183973 | 1 |
| Obesity | macrophage | *NGDN* | 1.600013673 | 0.002496395 | 0.012119562 | 1 |
| Obesity | macrophage | *RAB2B* | 1.06034806 | 0.015069312 | 0.044986118 | 1 |
| Obesity | macrophage | *BCL2L2* | 1.510053449 | 0.004944989 | 0.019890687 | 1 |
| Obesity | macrophage | *RNASE1* | -0.418296633 | 0.00717973 | 0.026046719 | 1 |
| Obesity | macrophage | *EPB41L4A* | 1.121795308 | 3.06E-05 | 0.000546074 | 0.192668523 |
| Obesity | macrophage | *CDO1* | 1.554398318 | 2.20E-06 | 0.000113382 | 0.013832645 |
| Obesity | macrophage | *REEP5* | 1.055897375 | 0.007221976 | 0.026109582 | 1 |
| Obesity | macrophage | *SHC2* | 0.635146024 | 0.000101026 | 0.001221438 | 0.635147816 |
| Obesity | macrophage | *PHF10* | 1.506273566 | 0.00130898 | 0.007683996 | 1 |
| Obesity | macrophage | *ELOF1* | 2.707221943 | 0.000420367 | 0.003447576 | 1 |
| Obesity | macrophage | *DPP6* | -0.412754175 | 0.002079776 | 0.010630532 | 1 |
| Obesity | macrophage | *RPL36* | 0.969882537 | 0.008857576 | 0.030314414 | 1 |
| Obesity | macrophage | *ATP8B3* | -0.528897415 | 1.11E-08 | 1.43E-06 | 7.00E-05 |
| Obesity | macrophage | *ACTN4* | 0.924394155 | 0.007316386 | 0.026254633 | 1 |
| Obesity | macrophage | *STK33* | 0.360616131 | 0.008492225 | 0.029481291 | 1 |
| Obesity | macrophage | *PXDN* | 1.126009675 | 3.20E-05 | 0.000554992 | 0.200963281 |
| Obesity | macrophage | *PGPEP1* | 0.558894549 | 0.003531138 | 0.015546405 | 1 |
| Obesity | macrophage | *LSM4* | 0.42988209 | 0.00277504 | 0.013067173 | 1 |
| Obesity | macrophage | *PAK4* | 1.544085789 | 0.007382685 | 0.026477432 | 1 |
| Obesity | macrophage | *TMEM160* | 2.485185997 | 0.00207594 | 0.010619555 | 1 |
| Obesity | macrophage | *SMPDL3B* | 4.818094697 | 3.92E-05 | 0.000635507 | 0.246576669 |
| Obesity | macrophage | *CLIP1* | 1.569588302 | 1.47E-06 | 8.04E-05 | 0.009242581 |
| Obesity | macrophage | *PPAN* | 3.564301495 | 4.93E-05 | 0.000749385 | 0.31014229 |
| Obesity | macrophage | *C12orf65* | 0.845417534 | 0.005013447 | 0.020097422 | 1 |
| Obesity | macrophage | *PKDREJ* | -0.492522686 | 0.001280128 | 0.007554732 | 1 |
| Obesity | macrophage | *HABP4* | 1.012363572 | 0.003305104 | 0.014872631 | 1 |
| Obesity | macrophage | *EPS8L1* | 3.71790795 | 0.013779237 | 0.042176273 | 1 |
| Obesity | macrophage | *LILRB2* | -0.572903433 | 0.001197466 | 0.007176804 | 1 |
| Obesity | macrophage | *COX4I2* | 1.55518145 | 0.002924072 | 0.013613165 | 1 |
| Obesity | macrophage | *RFTN1* | 0.977766607 | 8.13E-05 | 0.001048032 | 0.511439479 |
| Obesity | macrophage | *KCNC3* | -0.328564601 | 0.016579908 | 0.048169077 | 1 |
| Obesity | macrophage | *PDLIM4* | 0.643651803 | 0.004644506 | 0.018936453 | 1 |
| Obesity | macrophage | *KIF3A* | 0.921465471 | 6.33E-05 | 0.000882572 | 0.398040072 |
| Obesity | macrophage | *AOC2* | 2.093168647 | 4.18E-05 | 0.00067385 | 0.262801522 |

| | | | | | | |
|---|---|---|---|---|---|---|
| Obesity | macrophage | *EXOC4* | 0.961763818 | 0.000150964 | 0.001605939 | 0.949109968 |
| Obesity | macrophage | *ANO1* | 1.477089546 | 0.001101984 | 0.006798282 | 1 |
| Obesity | macrophage | *ZCCHC9* | 0.970578081 | 0.005081734 | 0.0202208 | 1 |
| Obesity | macrophage | *KHDRBS3* | 0.775465763 | 0.005452265 | 0.02127771 | 1 |
| Obesity | macrophage | *MATN3* | 3.934597674 | 0.000250101 | 0.002351705 | 1 |
| Obesity | macrophage | *EMILIN2* | 0.386386007 | 0.008444108 | 0.029362892 | 1 |
| Obesity | macrophage | *TRIM5* | 1.104798192 | 0.003134599 | 0.014322109 | 1 |
| Obesity | macrophage | *CNGA4* | 1.136327361 | 0.000519262 | 0.003978158 | 1 |
| Obesity | macrophage | *RPA1* | 0.685439181 | 0.008425861 | 0.029315655 | 1 |
| Obesity | macrophage | *SERPINF1* | 0.530243439 | 0.003013081 | 0.013888006 | 1 |
| Obesity | macrophage | *PTPRA* | 0.824033946 | 0.014748478 | 0.044322984 | 1 |
| Obesity | macrophage | *IGHMBP2* | 0.542469442 | 4.29E-06 | 0.000178092 | 0.026988927 |
| Obesity | macrophage | *MUTYH* | 2.199378356 | 2.48E-05 | 0.000492083 | 0.155859201 |
| Obesity | macrophage | *LPIN3* | 1.742739758 | 4.25E-05 | 0.000681503 | 0.26745363 |
| Obesity | macrophage | *DMGDH* | 0.908878727 | 0.001769456 | 0.009524458 | 1 |
| Obesity | macrophage | *KANK4* | 0.217566833 | 0.010699706 | 0.035109108 | 1 |
| Obesity | macrophage | *ATP8A2* | 2.878085381 | 0.000335679 | 0.002976611 | 1 |
| Obesity | macrophage | *WASF3* | 1.186820578 | 3.86E-05 | 0.000628283 | 0.242479143 |
| Obesity | macrophage | *TRPC4* | 1.803354923 | 2.82E-05 | 0.00052574 | 0.177443878 |
| Obesity | macrophage | *PRAM1* | -0.599849296 | 0.001356686 | 0.007904992 | 1 |
| Obesity | macrophage | *ZNF414* | 2.455003916 | 5.18E-05 | 0.000773626 | 0.325696747 |
| Obesity | macrophage | *RTN3* | 0.91720385 | 0.004679709 | 0.019045254 | 1 |
| Obesity | macrophage | *SLC2A11* | 2.084177794 | 0.000493858 | 0.003870478 | 1 |
| Obesity | macrophage | *C1QTNF6* | 2.098280215 | 1.92E-05 | 0.000450217 | 0.120658201 |
| Obesity | macrophage | *FAM83F* | 2.527438573 | 2.99E-05 | 0.000542547 | 0.187721367 |
| Obesity | macrophage | *ACTR3B* | 1.068174592 | 0.003196722 | 0.014511042 | 1 |
| Obesity | macrophage | *SFTPD* | 0.138911554 | 0.009431772 | 0.031846161 | 1 |
| Obesity | macrophage | *TMTC1* | 1.028338601 | 5.96E-05 | 0.000855087 | 0.374419751 |
| Obesity | macrophage | *LARS* | -0.280081751 | 0.003655911 | 0.015972699 | 1 |
| Obesity | macrophage | *RRAS2* | 0.27751714 | 0.003854863 | 0.01655432 | 1 |
| Obesity | macrophage | *DPF2* | 0.725494931 | 0.011597772 | 0.037277706 | 1 |
| Obesity | macrophage | *GSC* | 2.839498849 | 2.64E-05 | 0.000513064 | 0.165719825 |
| Obesity | macrophage | *MED6* | 0.471977398 | 0.008677849 | 0.029894594 | 1 |
| Obesity | macrophage | *ELP3* | 1.033293895 | 0.015154831 | 0.045156901 | 1 |
| Obesity | macrophage | *MRO* | -0.525549661 | 0.009402019 | 0.031779836 | 1 |
| Obesity | macrophage | *KATNBL1* | 1.072494963 | 0.002168319 | 0.010888355 | 1 |
| Obesity | macrophage | *GSTM5* | 0.428922269 | 0.000182148 | 0.001834917 | 1 |
| Obesity | macrophage | *SYT6* | 2.731026513 | 0.008253946 | 0.028813191 | 1 |
| Obesity | macrophage | *NAPG* | 0.936551823 | 0.009003795 | 0.030697862 | 1 |
| Obesity | macrophage | *PPHLN1* | 0.763907405 | 0.002997466 | 0.013826168 | 1 |
| Obesity | macrophage | *LDHA* | 1.567110441 | 8.29E-05 | 0.001061956 | 0.521420513 |
| Obesity | macrophage | *FST* | 1.074305845 | 0.000147541 | 0.001580225 | 0.927592164 |
| Obesity | macrophage | *FBXO18* | 1.031964481 | 0.014431315 | 0.043683041 | 1 |
| Obesity | macrophage | *IL15RA* | 1.709978969 | 0.000106595 | 0.001266847 | 0.670162173 |
| Obesity | macrophage | *KLRD1* | 1.419592754 | 3.65E-05 | 0.000604095 | 0.229556143 |
| Obesity | macrophage | *DDB2* | 0.971577036 | 0.00260359 | 0.012457209 | 1 |
| Obesity | macrophage | *ACP2* | 0.5682596 | 0.009889535 | 0.033072077 | 1 |
| Obesity | macrophage | *ZCCHC11* | 0.809980197 | 0.00523586 | 0.020651099 | 1 |
| Obesity | macrophage | *PRPF38A* | 1.206201743 | 0.002379913 | 0.011689461 | 1 |
| Obesity | macrophage | *TPGS2* | 0.816920149 | 0.006917053 | 0.02541643 | 1 |

| | | | | | | |
|---|---|---|---|---|---|---|
| Obesity | macrophage | *SLC43A3* | -0.616645001 | 0.011333864 | 0.03674884 | 1 |
| Obesity | macrophage | *DHX34* | -0.52237151 | 0.010394018 | 0.034384884 | 1 |
| Obesity | macrophage | *APLNR* | 1.354860614 | 0.000160065 | 0.001682826 | 1 |
| Obesity | macrophage | *FADS2* | 1.114826747 | 0.014371792 | 0.043555097 | 1 |
| Obesity | macrophage | *DZIP1* | 0.420253939 | 0.000981268 | 0.006347553 | 1 |
| Obesity | macrophage | *BIVM* | 2.006623067 | 2.48E-05 | 0.000492083 | 0.15599018 |
| Obesity | macrophage | *ERCC5* | 2.226883746 | 0.015042614 | 0.044942875 | 1 |
| Obesity | macrophage | *KLB* | 0.328482919 | 0.00105371 | 0.006631305 | 1 |
| Obesity | macrophage | *NAA35* | 1.153680403 | 0.003673937 | 0.015975945 | 1 |
| Obesity | macrophage | *TMEM60* | -0.923252369 | 0.00015506 | 0.001641177 | 0.974859166 |
| Obesity | macrophage | *POM121C* | -1.494478345 | 1.32E-08 | 1.65E-06 | 8.27E-05 |
| Obesity | macrophage | *SRPK2* | 0.801466673 | 0.013490602 | 0.041555813 | 1 |
| Obesity | macrophage | *MTO1* | 0.576571673 | 0.011193587 | 0.036406664 | 1 |
| Obesity | macrophage | *EPHA7* | 0.127032144 | 0.003934205 | 0.016837541 | 1 |
| Obesity | macrophage | *PPP1R1A* | 1.304835211 | 0.001361494 | 0.007925659 | 1 |
| Obesity | macrophage | *FAIM2* | 2.367588764 | 5.80E-05 | 0.000839679 | 0.364481636 |
| Obesity | macrophage | *HNRNPA1* | 1.276295345 | 0.00613654 | 0.023143626 | 1 |
| Obesity | macrophage | *SLC26A10* | 1.126021353 | 0.00743963 | 0.02663608 | 1 |
| Obesity | macrophage | *PRADC1* | 2.466626059 | 9.61E-06 | 0.000301542 | 0.060411724 |
| Obesity | macrophage | *SMYD5* | 0.804722482 | 0.014882073 | 0.044617832 | 1 |
| Obesity | macrophage | *KCNMB4* | -0.678215859 | 0.008393556 | 0.029235615 | 1 |
| Obesity | macrophage | *CCDC102A* | 0.581633429 | 0.010763403 | 0.035281292 | 1 |
| Obesity | macrophage | *NTPCR* | 1.051111678 | 0.000691468 | 0.00493446 | 1 |
| Obesity | macrophage | *PIGC* | 0.756059154 | 0.002545159 | 0.012280443 | 1 |
| Obesity | macrophage | *USP37* | 1.220857542 | 0.000273333 | 0.002530845 | 1 |
| Obesity | macrophage | *MRPS9* | 0.603720413 | 0.000177802 | 0.001808801 | 1 |
| Obesity | macrophage | *ANKRD36* | 1.052709944 | 0.002771026 | 0.013067173 | 1 |
| Obesity | macrophage | *VPS36* | 0.757761682 | 0.003101821 | 0.014203314 | 1 |
| Obesity | macrophage | *MED4* | 0.726349099 | 0.012617488 | 0.039524736 | 1 |
| Obesity | macrophage | *KDELR2* | 1.710425592 | 0.000523315 | 0.003992817 | 1 |
| Obesity | macrophage | *DDX56* | 1.920093773 | 0.001506337 | 0.008463218 | 1 |
| Obesity | macrophage | *RSAD1* | 1.477897926 | 0.002278774 | 0.011269706 | 1 |
| Obesity | macrophage | *NMT1* | 0.362816889 | 0.000925046 | 0.006115421 | 1 |
| Obesity | macrophage | *BLK* | -1.037375768 | 0.000220372 | 0.002115259 | 1 |
| Obesity | macrophage | *EPRS* | 1.294933694 | 0.004111355 | 0.017359363 | 1 |
| Obesity | macrophage | *IL1RN* | -0.58980741 | 0.003298086 | 0.014853201 | 1 |
| Obesity | macrophage | *ODF2* | 1.664646454 | 0.000444485 | 0.003573497 | 1 |
| Obesity | macrophage | *STX17* | 1.506854392 | 0.000315161 | 0.002838707 | 1 |
| Obesity | macrophage | *GABBR2* | 5.061036426 | 0.000173194 | 0.001782107 | 1 |
| Obesity | macrophage | *RABEPK* | 1.56774558 | 0.000842716 | 0.005678621 | 1 |
| Obesity | macrophage | *GOLGA1* | 1.099781176 | 0.007702598 | 0.027359456 | 1 |
| Obesity | macrophage | *DSCC1* | 0.143465681 | 0.001076554 | 0.006703154 | 1 |
| Obesity | macrophage | *NOV* | 0.777363641 | 0.003226476 | 0.014614449 | 1 |
| Obesity | macrophage | *SPAG8* | 0.654961825 | 0.015779979 | 0.046402585 | 1 |
| Obesity | macrophage | *TMEM8B* | 1.224916914 | 0.000502583 | 0.003900912 | 1 |
| Obesity | macrophage | *KIF13A* | 0.983501091 | 0.001426426 | 0.00816009 | 1 |
| Obesity | macrophage | *ZSCAN9* | 0.709775823 | 3.55E-06 | 0.000158104 | 0.022292646 |
| Obesity | macrophage | *FRS3* | 2.070536671 | 0.000299483 | 0.002728764 | 1 |
| Obesity | macrophage | *LRRC1* | 0.55942882 | 0.003773462 | 0.016271439 | 1 |
| Obesity | macrophage | *RIPK1* | 1.06033882 | 0.009974195 | 0.033302054 | 1 |

| | | | | | | |
|---|---|---|---|---|---|---|
| Obesity | macrophage | *TCF19* | 0.99016664 | 3.91E-06 | 0.000173246 | 0.024600976 |
| Obesity | macrophage | *FLOT1* | 1.045953703 | 0.000167409 | 0.001733943 | 1 |
| Obesity | macrophage | *NRM* | 0.58934958 | 0.000880581 | 0.005870854 | 1 |
| Obesity | macrophage | *FAM8A1* | 2.015676759 | 8.06E-05 | 0.001041118 | 0.5070245 |
| Obesity | macrophage | *FHDC1* | 2.232133592 | 2.33E-05 | 0.00486218 | 0.146351495 |
| Obesity | macrophage | *MYO7A* | -0.25914482 | 0.007805136 | 0.027614456 | 1 |
| Obesity | macrophage | *RAB30* | 0.849882367 | 0.001778665 | 0.009541357 | 1 |
| Obesity | macrophage | *NARS2* | 1.070647093 | 0.011920556 | 0.038062232 | 1 |
| Obesity | macrophage | *PI15* | 2.367417679 | 0.000883377 | 0.005883256 | 1 |
| Obesity | macrophage | *SULF1* | 0.731864793 | 0.000128847 | 0.001446542 | 0.810063296 |
| Obesity | macrophage | *NEK1* | 0.812877754 | 0.008038516 | 0.028236167 | 1 |
| Obesity | macrophage | *DDX60* | 1.248349821 | 0.012436855 | 0.039075715 | 1 |
| Obesity | macrophage | *DCUN1D5* | 0.60520832 | 0.000734403 | 0.005141641 | 1 |
| Obesity | macrophage | *PPP2R1B* | 0.995205431 | 0.000282321 | 0.002603616 | 1 |
| Obesity | macrophage | *TMPRSS13* | -0.762818396 | 0.011518212 | 0.037135897 | 1 |
| Obesity | macrophage | *MAP2K5* | 1.513192741 | 4.55E-06 | 0.000182223 | 0.028595185 |
| Obesity | macrophage | *ITGA11* | 0.67474824 | 0.007248162 | 0.026159126 | 1 |
| Obesity | macrophage | *PAQR5* | -0.693092421 | 0.009055328 | 0.030807759 | 1 |
| Obesity | macrophage | *RMDN3* | 0.895595061 | 0.003459147 | 0.015326043 | 1 |
| Obesity | macrophage | *UACA* | 1.056574279 | 0.001366645 | 0.007926286 | 1 |
| Obesity | macrophage | *SLC44A5* | 0.844985226 | 2.52E-05 | 0.000494987 | 0.158462145 |
| Obesity | macrophage | *DNASE2B* | -1.337659078 | 0.000135487 | 0.001502304 | 0.851806482 |
| Obesity | macrophage | *IFT172* | 1.954397753 | 9.15E-09 | 1.20E-06 | 5.75E-05 |
| Obesity | macrophage | *ADCY3* | 1.384704485 | 0.002821428 | 0.013237551 | 1 |
| Obesity | macrophage | *DYNC2LI1* | 0.375275349 | 0.012650346 | 0.039578981 | 1 |
| Obesity | macrophage | *LHCGR* | 0.704581461 | 0.014550163 | 0.043870231 | 1 |
| Obesity | macrophage | *ABCG5* | 0.614096114 | 0.004206889 | 0.017656015 | 1 |
| Obesity | macrophage | *CENPO* | 2.730627797 | 0.0007257 | 0.005108248 | 1 |
| Obesity | macrophage | *LRPPRC* | 0.922974322 | 0.016875331 | 0.048756987 | 1 |
| Obesity | macrophage | *STAMBPL1* | 0.720897684 | 0.004821732 | 0.019494682 | 1 |
| Obesity | macrophage | *BTBD16* | 0.580015847 | 0.005032536 | 0.020126943 | 1 |
| Obesity | macrophage | *TACC2* | 0.701412227 | 0.000176362 | 0.001808788 | 1 |
| Obesity | macrophage | *PLCE1* | 0.822990305 | 0.000875233 | 0.005847596 | 1 |
| Obesity | macrophage | *RBP4* | 1.329479075 | 0.000363879 | 0.00312528 | 1 |
| Obesity | macrophage | *CARF* | 0.876091955 | 4.56E-08 | 4.60E-06 | 0.000286456 |
| Obesity | macrophage | *METTL5* | 1.741149523 | 0.000405541 | 0.003372538 | 1 |
| Obesity | macrophage | *NAB1* | 1.275224912 | 0.003885978 | 0.016676548 | 1 |
| Obesity | macrophage | *MDH1B* | 1.340250983 | 0.000560095 | 0.004187058 | 1 |
| Obesity | macrophage | *FAM117B* | 1.055041739 | 0.014600509 | 0.043941312 | 1 |
| Obesity | macrophage | *COX17* | 1.498148749 | 0.004943096 | 0.019890687 | 1 |
| Obesity | macrophage | *UBL7* | 1.566356158 | 5.71E-06 | 0.000214064 | 0.035892425 |
| Obesity | macrophage | *PRKG2* | 1.184839557 | 0.000136515 | 0.001505734 | 0.858268557 |
| Obesity | macrophage | *BBS7* | 1.58859762 | 0.000930813 | 0.00614063 | 1 |
| Obesity | macrophage | *PRDM5* | 0.811861837 | 0.001176285 | 0.007097219 | 1 |
| Obesity | macrophage | *BMP2K* | -0.375958818 | 0.008606798 | 0.029769423 | 1 |
| Obesity | macrophage | *ANXA3* | 1.592398396 | 7.38E-06 | 0.000255529 | 0.046401009 |
| Obesity | macrophage | *ENPEP* | 0.917089871 | 0.000358601 | 0.003092627 | 1 |
| Obesity | macrophage | *PARVG* | -0.465418672 | 0.008110809 | 0.028419853 | 1 |
| Obesity | macrophage | *SLCO1C1* | 1.120042087 | 5.52E-05 | 0.000808285 | 0.346754369 |
| Obesity | macrophage | *PRICKLE1* | 0.727556174 | 0.002019053 | 0.01042183 | 1 |

| | | | | | | |
|---|---|---|---|---|---|---|
| Obesity | macrophage | *C1RL* | 0.548831884 | 0.003455067 | 0.015318763 | 1 |
| Obesity | macrophage | *CLSTN3* | 0.634939268 | 0.003977336 | 0.016987986 | 1 |
| Obesity | macrophage | *KLRG1* | 0.458404102 | 0.000179376 | 0.001819885 | 1 |
| Obesity | macrophage | *CD27* | 1.273665329 | 0.000219559 | 0.00211389 | 1 |
| Obesity | macrophage | *AMDHD1* | -0.427106848 | 0.005029709 | 0.020126943 | 1 |
| Obesity | macrophage | *GAS2L3* | -0.400406774 | 0.001402885 | 0.00804739 | 1 |
| Obesity | macrophage | *RITA1* | -0.285337026 | 0.011977903 | 0.038148469 | 1 |
| Obesity | macrophage | *GPR84* | -2.469656269 | 1.54E-05 | 0.000405213 | 0.096704495 |
| Obesity | macrophage | *CERS5* | 0.780047439 | 0.015501076 | 0.045839729 | 1 |
| Obesity | macrophage | *HNRNPA1L2* | 1.216835744 | 0.008212943 | 0.028685984 | 1 |
| Obesity | macrophage | *PTGR2* | -0.192395778 | 0.008457209 | 0.02939219 | 1 |
| Obesity | macrophage | *AK7* | 2.786241983 | 9.88E-06 | 0.000301542 | 0.062117718 |
| Obesity | macrophage | *FBLN5* | 1.426105226 | 5.72E-06 | 0.000214064 | 0.035962834 |
| Obesity | macrophage | *C14orf79* | 1.670585246 | 0.003779108 | 0.016284615 | 1 |
| Obesity | macrophage | *ZSCAN29* | 1.728917253 | 0.000225769 | 0.002150622 | 1 |
| Obesity | macrophage | *COMMD4* | 1.301157356 | 0.006907011 | 0.025409554 | 1 |
| Obesity | macrophage | *PSTPIP1* | -0.447852247 | 0.006025619 | 0.02289177 | 1 |
| Obesity | macrophage | *HMG20A* | 1.062236313 | 0.001378443 | 0.007972653 | 1 |
| Obesity | macrophage | *DNAJA4* | 2.181222327 | 0.001252859 | 0.007437889 | 1 |
| Obesity | macrophage | *IGF1R* | 0.846054702 | 0.015047678 | 0.044942875 | 1 |
| Obesity | macrophage | *CYP11A1* | 0.515766886 | 5.85E-05 | 0.000843961 | 0.367967214 |
| Obesity | macrophage | *BBS4* | 1.479706774 | 0.000135478 | 0.001502304 | 0.851748496 |
| Obesity | macrophage | *ADAMTS17* | 0.822971405 | 0.004027956 | 0.017076033 | 1 |
| Obesity | macrophage | *MFGE8* | 0.745193728 | 0.015257372 | 0.045396639 | 1 |
| Obesity | macrophage | *CRTC3* | 1.10685414 | 0.00224796 | 0.011154638 | 1 |
| Obesity | macrophage | *SH3GL3* | -0.418728621 | 0.001880357 | 0.009925948 | 1 |
| Obesity | macrophage | *TLDC1* | 1.72057956 | 0.000127602 | 0.001439194 | 0.802231794 |
| Obesity | macrophage | *NDUFB10* | 1.867926526 | 0.000156213 | 0.001647833 | 0.982108651 |
| Obesity | macrophage | *TIGD7* | 0.596073073 | 0.00071161 | 0.0050325 | 1 |
| Obesity | macrophage | *MYO19* | 2.240384553 | 2.10E-10 | 4.14E-08 | 1.32E-06 |
| Obesity | macrophage | *TOM1L1* | 1.616692506 | 7.30E-06 | 0.000255529 | 0.045899676 |
| Obesity | macrophage | *C17orf80* | 0.369712798 | 0.014550904 | 0.043870231 | 1 |
| Obesity | macrophage | *NPEPPS* | 1.063307112 | 0.003556741 | 0.015637225 | 1 |
| Obesity | macrophage | *ABCA8* | 0.419279061 | 0.00517401 | 0.020471367 | 1 |
| Obesity | macrophage | *IMPA2* | 1.33269626 | 0.016244897 | 0.047415902 | 1 |
| Obesity | macrophage | *GNAL* | 1.359870863 | 1.73E-05 | 0.000431251 | 0.108990086 |
| Obesity | macrophage | *C18orf21* | 1.089317924 | 0.006522594 | 0.024376541 | 1 |
| Obesity | macrophage | *ASXL3* | 1.139592685 | 0.000204814 | 0.002002802 | 1 |
| Obesity | macrophage | *C18orf8* | -1.38424375 | 1.08E-07 | 9.86E-06 | 0.0006804 |
| Obesity | macrophage | *NPC1* | -1.237688073 | 2.83E-05 | 0.00052574 | 0.177700136 |
| Obesity | macrophage | *SLC14A1* | 0.538825574 | 0.014445336 | 0.04370444 | 1 |
| Obesity | macrophage | *TRIM65* | 0.94521926 | 4.47E-06 | 0.000181209 | 0.028087345 |
| Obesity | macrophage | *CBX8* | 1.025989577 | 0.001738405 | 0.00940564 | 1 |
| Obesity | macrophage | *RNF157* | 0.171135646 | 0.013664181 | 0.041905712 | 1 |
| Obesity | macrophage | *RNF165* | 1.275484069 | 4.94E-05 | 0.000749385 | 0.310667941 |
| Obesity | macrophage | *PIP4K2B* | 1.015320985 | 0.007714389 | 0.027385863 | 1 |
| Obesity | macrophage | *MVB12A* | 0.555321451 | 0.012448283 | 0.039092085 | 1 |
| Obesity | macrophage | *SIRT3* | 0.779600244 | 0.000381942 | 0.003231858 | 1 |
| Obesity | macrophage | *IFITM3* | 1.598308227 | 0.000334897 | 0.002976611 | 1 |
| Obesity | macrophage | *COL6A1* | 0.38180493 | 0.009440234 | 0.031857624 | 1 |

| | | | | | | |
|---|---|---|---|---|---|---|
| Obesity | macrophage | *NTN5* | -0.549024552 | 0.017252294 | 0.049709062 | 1 |
| Obesity | macrophage | *WTIP* | 0.903174631 | 0.001303337 | 0.007658017 | 1 |
| Obesity | macrophage | *CACNG8* | 3.172562581 | 9.38E-06 | 0.000299477 | 0.058996995 |
| Obesity | macrophage | *EVI5L* | 1.261435138 | 0.007189661 | 0.02605268 | 1 |
| Obesity | macrophage | *RPS11* | 1.209682536 | 0.000133484 | 0.001490608 | 0.839212128 |
| Obesity | macrophage | *RCN3* | -0.518730422 | 0.007278706 | 0.026190797 | 1 |
| Obesity | macrophage | *SERBP1* | 1.042423728 | 0.015393974 | 0.04560882 | 1 |
| Obesity | macrophage | *RPS8* | 0.777720641 | 0.00224266 | 0.01114593 | 1 |
| Obesity | macrophage | *ATP1B1* | -0.481241209 | 0.000351581 | 0.003055847 | 1 |
| Obesity | macrophage | *TIPRL* | 1.388790534 | 0.002318167 | 0.011421877 | 1 |
| Obesity | macrophage | *RXRG* | 2.303743412 | 4.20E-05 | 0.000675506 | 0.264122694 |
| Obesity | macrophage | *TMCO1* | 1.412161275 | 0.000682717 | 0.004888659 | 1 |
| Obesity | macrophage | *MGST3* | 1.167032608 | 0.00094215 | 0.006189442 | 1 |
| Obesity | macrophage | *ADCY10* | 1.572436529 | 0.00046567 | 0.003715316 | 1 |
| Obesity | macrophage | *RGS5* | 0.759067301 | 0.001182803 | 0.007122879 | 1 |
| Obesity | macrophage | *SELENBP1* | 0.596704546 | 0.001657788 | 0.009055179 | 1 |
| Obesity | macrophage | *ENSA* | 1.007719738 | 0.012699733 | 0.039683511 | 1 |
| Obesity | macrophage | *SEMA6C* | 2.030347471 | 2.40E-07 | 1.96E-05 | 0.001507333 |
| Obesity | macrophage | *POGZ* | 0.700966185 | 0.002195461 | 0.01100707 | 1 |
| Obesity | macrophage | *HORMAD1* | -0.091554319 | 0.011681095 | 0.037488027 | 1 |
| Obesity | macrophage | *DYRK3* | 1.101754833 | 0.000466474 | 0.00371701 | 1 |
| Obesity | macrophage | *S100A8* | -0.946703447 | 0.009407792 | 0.031782261 | 1 |
| Obesity | macrophage | *HAX1* | 1.381229714 | 4.07E-06 | 0.000173988 | 0.02557619 |
| Obesity | macrophage | *KCNN3* | 1.534339178 | 1.52E-05 | 0.000405213 | 0.09585327 |
| Obesity | macrophage | *SCCPDH* | 1.373387421 | 0.000355916 | 0.003077692 | 1 |
| Obesity | macrophage | *CNIH4* | 1.372107643 | 0.003669824 | 0.015975945 | 1 |
| Obesity | macrophage | *GUK1* | 2.152418868 | 4.87E-06 | 0.000193818 | 0.030623182 |
| Obesity | macrophage | *CDC42BPA* | 1.150656744 | 2.50E-06 | 0.000125506 | 0.015688259 |
| Obesity | macrophage | *WNT9A* | 0.614134199 | 0.002950101 | 0.013677939 | 1 |
| Obesity | macrophage | *PLEKHA6* | 0.94370866 | 3.72E-05 | 0.000611689 | 0.233665268 |
| Obesity | macrophage | *CHAC2* | 1.456413511 | 0.000645862 | 0.004688839 | 1 |
| Obesity | macrophage | *WDPCP* | 0.222856699 | 0.012202926 | 0.038630309 | 1 |
| Obesity | macrophage | *TEX261* | 2.392865123 | 0.001870892 | 0.009900925 | 1 |
| Obesity | macrophage | *TMEM177* | 1.585373776 | 0.000564565 | 0.004201901 | 1 |
| Obesity | macrophage | *ZC3H8* | 0.866533921 | 0.00036791 | 0.003142149 | 1 |
| Obesity | macrophage | *AFF3* | -0.339347231 | 0.009573628 | 0.032186845 | 1 |
| Obesity | macrophage | *UBXN4* | 1.017898169 | 0.002779801 | 0.013071508 | 1 |
| Obesity | macrophage | *THSD7B* | 0.698246607 | 2.67E-06 | 0.000131017 | 0.016770217 |
| Obesity | macrophage | *ZNF385B* | 0.754460178 | 0.000160507 | 0.001684651 | 1 |
| Obesity | macrophage | *TMEFF2* | 1.004854055 | 0.000720962 | 0.005081491 | 1 |
| Obesity | macrophage | *GULP1* | 0.981645173 | 2.97E-05 | 0.000541797 | 0.186919853 |
| Obesity | macrophage | *FAM171B* | 0.575906116 | 0.001464935 | 0.00831075 | 1 |
| Obesity | macrophage | *HSPD1* | 1.505693468 | 0.000278091 | 0.002571115 | 1 |
| Obesity | macrophage | *NBEAL1* | 0.571380873 | 2.63E-07 | 2.10E-05 | 0.001655926 |
| Obesity | macrophage | *ESPNL* | 4.59826097 | 0.000619705 | 0.004535602 | 1 |
| Obesity | macrophage | *TAMM41* | 1.245754395 | 0.000494977 | 0.003870478 | 1 |
| Obesity | macrophage | *OSBPL10* | 0.781740642 | 0.001879702 | 0.009925948 | 1 |
| Obesity | macrophage | *STAC* | 1.529481054 | 2.25E-05 | 0.000483538 | 0.141193079 |
| Obesity | macrophage | *ARL6IP5* | 0.990463906 | 0.011905964 | 0.038034959 | 1 |
| Obesity | macrophage | *RABL3* | 1.803503458 | 0.000146629 | 0.001575829 | 0.921855322 |

| | | | | | | |
|---|---|---|---|---|---|---|
| Obesity | macrophage | *BOC* | 0.628514425 | 0.002539621 | 0.012272557 | 1 |
| Obesity | macrophage | *AMT* | -0.183253168 | 0.009540175 | 0.032125914 | 1 |
| Obesity | macrophage | *EAF2* | -0.785735527 | 0.000185708 | 0.001862115 | 1 |
| Obesity | macrophage | *LYAR* | 1.742574184 | 0.0092836 | 0.031453551 | 1 |
| Obesity | macrophage | *PYURF* | 1.12454741 | 0.006997709 | 0.025608031 | 1 |
| Obesity | macrophage | *USP53* | 1.136718829 | 2.75E-05 | 0.000521707 | 0.172965867 |
| Obesity | macrophage | *MARCH1* | -0.470282575 | 4.93E-05 | 0.000749385 | 0.309778373 |
| Obesity | macrophage | *CBR4* | 1.019278389 | 0.002851421 | 0.013348388 | 1 |
| Obesity | macrophage | *FAM105A* | -0.371579894 | 0.004271944 | 0.017851575 | 1 |
| Obesity | macrophage | *SSBP2* | 0.787052149 | 0.009806186 | 0.032845758 | 1 |
| Obesity | macrophage | *CRHBP* | -0.414378931 | 0.003442298 | 0.015272919 | 1 |
| Obesity | macrophage | *FEM1C* | 1.117413638 | 0.010248391 | 0.034036786 | 1 |
| Obesity | macrophage | *YIPF5* | 1.373309596 | 0.007921267 | 0.027915362 | 1 |
| Obesity | macrophage | *SLC25A48* | -0.309207787 | 0.003217548 | 0.014584518 | 1 |
| Obesity | macrophage | *GABRB2* | -0.265552112 | 0.001037337 | 0.006567709 | 1 |
| Obesity | macrophage | *PCYOX1L* | -0.599509269 | 0.000789721 | 0.005426203 | 1 |
| Obesity | macrophage | *TBC1D7* | 1.167065377 | 0.011953571 | 0.038139587 | 1 |
| Obesity | macrophage | *GFOD1* | 0.329306961 | 0.003840954 | 0.016505862 | 1 |
| Obesity | macrophage | *KLHL3* | 0.713774029 | 0.000561124 | 0.00418977 | 1 |
| Obesity | macrophage | *MUT* | 1.707429182 | 0.000653264 | 0.004734252 | 1 |
| Obesity | macrophage | *DOK3* | -0.320708741 | 0.001172304 | 0.007079995 | 1 |
| Obesity | macrophage | *ABT1* | 0.75162844 | 7.48E-05 | 0.000989215 | 0.47013601 |
| Obesity | macrophage | *FGD2* | -0.368648276 | 0.002543872 | 0.012280443 | 1 |
| Obesity | macrophage | *RPL7L1* | 0.699847959 | 0.000540043 | 0.004080826 | 1 |
| Obesity | macrophage | *TPBG* | 2.023152027 | 0.000136474 | 0.001505734 | 0.858009769 |
| Obesity | macrophage | *IRAK1BP1* | 1.60297221 | 2.55E-05 | 0.000497961 | 0.160343545 |
| Obesity | macrophage | *PM20D2* | 1.313372979 | 0.011001309 | 0.035929991 | 1 |
| Obesity | macrophage | *SLC18B1* | -0.17891114 | 0.002970555 | 0.013742367 | 1 |
| Obesity | macrophage | *AIG1* | 0.997491045 | 4.84E-05 | 0.000744502 | 0.304501409 |
| Obesity | macrophage | *DYNLT1* | 0.907978745 | 0.007177534 | 0.026046719 | 1 |
| Obesity | macrophage | *SLC22A3* | 1.264986166 | 2.37E-05 | 0.000488143 | 0.149191089 |
| Obesity | macrophage | *C7orf50* | 1.068281654 | 0.004230407 | 0.017731044 | 1 |
| Obesity | macrophage | *RBAK* | 1.982623686 | 0.001374965 | 0.007959856 | 1 |
| Obesity | macrophage | *EGFR* | 0.476741286 | 0.004352536 | 0.018123455 | 1 |
| Obesity | macrophage | *TMEM209* | 1.035116738 | 0.004463956 | 0.018427376 | 1 |
| Obesity | macrophage | *AGBL3* | 0.910074135 | 0.007262435 | 0.026171323 | 1 |
| Obesity | macrophage | *NOM1* | 1.267977501 | 0.01453126 | 0.043858871 | 1 |
| Obesity | macrophage | *FBXO25* | 1.295476147 | 0.002614754 | 0.012482126 | 1 |
| Obesity | macrophage | *SLC25A37* | 1.471885802 | 1.54E-05 | 0.000405213 | 0.096714468 |
| Obesity | macrophage | *ST18* | -0.34042971 | 0.00051201 | 0.00394003 | 1 |
| Obesity | macrophage | *TACC1* | 0.87547154 | 0.001986103 | 0.010303093 | 1 |
| Obesity | macrophage | *ADHFE1* | -0.13727974 | 0.016423778 | 0.047848142 | 1 |
| Obesity | macrophage | *ATP6V0D2* | -0.616429394 | 4.99E-08 | 4.90E-06 | 0.000313793 |
| Obesity | macrophage | *MAL2* | 0.538577522 | 0.000976709 | 0.006341253 | 1 |
| Obesity | macrophage | *HAUS6* | 1.461219773 | 0.00592607 | 0.022690134 | 1 |
| Obesity | macrophage | *CBWD5* | 1.876539272 | 0.001147549 | 0.006970668 | 1 |
| Obesity | macrophage | *ZNF462* | 1.374944693 | 0.000192521 | 0.001911756 | 1 |
| Obesity | macrophage | *WDR31* | 2.286796565 | 7.90E-07 | 5.40E-05 | 0.004966244 |
| Obesity | macrophage | *CIZ1* | 1.001748906 | 0.000798049 | 0.005447834 | 1 |
| Obesity | macrophage | *SH3GLB2* | 1.646795155 | 1.23E-05 | 0.000347425 | 0.077460468 |

| | | | | | | |
|---|---|---|---|---|---|---|
| Obesity | macrophage | *DPH7* | 1.870033657 | 2.34E-05 | 0.000486345 | 0.146876304 |
| Obesity | macrophage | *SLC39A12* | -0.472314833 | 0.005402829 | 0.021125234 | 1 |
| Obesity | macrophage | *PARD3* | 0.772849894 | 1.68E-05 | 0.000424589 | 0.105734114 |
| Obesity | macrophage | *NRBF2* | -0.371360356 | 0.005057603 | 0.020163061 | 1 |
| Obesity | macrophage | *ADIRF* | 1.190775004 | 0.00129713 | 0.007628679 | 1 |
| Obesity | macrophage | *HTR7* | -0.510940991 | 0.000755633 | 0.005249357 | 1 |
| Obesity | macrophage | *MTG1* | 0.41992137 | 0.000177647 | 0.001808801 | 1 |
| Obesity | macrophage | *RGS10* | -0.560446508 | 0.0027768 | 0.013067173 | 1 |
| Obesity | macrophage | *GAS2* | 2.067152468 | 0.000135976 | 0.001505074 | 0.85488188 |
| Obesity | macrophage | *LIN7C* | 1.76260448 | 0.002023825 | 0.0104219 | 1 |
| Obesity | macrophage | *HSD17B12* | -0.538605121 | 2.44E-10 | 4.64E-08 | 1.53E-06 |
| Obesity | macrophage | *GLYAT* | 0.386600713 | 0.002157764 | 0.01087936 | 1 |
| Obesity | macrophage | *ENDOD1* | 2.054745605 | 0.000334163 | 0.002976611 | 1 |
| Obesity | macrophage | *CAPN5* | 1.153811515 | 0.000206108 | 0.002008997 | 1 |
| Obesity | macrophage | *TTC12* | 0.361163625 | 9.57E-06 | 0.000301542 | 0.060135908 |
| Obesity | macrophage | *DRD2* | 3.094920928 | 0.000162089 | 0.001693914 | 1 |
| Obesity | macrophage | *C11orf52* | 7.270831308 | 2.31E-05 | 0.00048514 | 0.145056763 |
| Obesity | macrophage | *P4HA3* | 0.603462175 | 0.012040607 | 0.038264568 | 1 |
| Obesity | macrophage | *TMC2* | 3.227893324 | 2.27E-05 | 0.000484376 | 0.142905164 |
| Obesity | macrophage | *MS4A3* | -11.8264715 | 0.001033567 | 0.006557047 | 1 |
| Obesity | macrophage | *PLCH2* | 4.054658107 | 3.41E-06 | 0.00015608 | 0.021416428 |
| Obesity | macrophage | *CPSF7* | 1.114784186 | 0.012643656 | 0.039578981 | 1 |
| Obesity | macrophage | *CCDC15* | -0.596598969 | 2.27E-05 | 0.000484376 | 0.142919213 |
| Obesity | macrophage | *FEZ1* | 1.678322951 | 0.003922529 | 0.016799004 | 1 |
| Obesity | macrophage | *DUSP15* | -0.646520352 | 0.006510525 | 0.024364088 | 1 |
| Obesity | macrophage | *COMMD7* | 1.287583848 | 0.01660491 | 0.048197169 | 1 |
| Obesity | macrophage | *CNBD2* | 2.666904936 | 0.001263159 | 0.007491965 | 1 |
| Obesity | macrophage | *TBX6* | 0.559690773 | 8.50E-08 | 7.86E-06 | 0.000534285 |
| Obesity | macrophage | *DOC2A* | -1.875308814 | 6.25E-18 | 3.57E-15 | 3.93E-14 |
| Obesity | macrophage | *TAOK2* | 1.106288586 | 0.010838591 | 0.035509234 | 1 |
| Obesity | macrophage | *ARID5B* | 0.811593921 | 0.000748538 | 0.005228952 | 1 |
| Obesity | macrophage | *LYPD6B* | 1.402696643 | 0.00228067 | 0.011269706 | 1 |
| Obesity | macrophage | *CCDC102B* | 1.130066277 | 0.002698382 | 0.012803881 | 1 |
| Obesity | macrophage | *FSIP1* | 1.496634437 | 0.006694568 | 0.024860453 | 1 |
| Obesity | macrophage | *RGS18* | -0.597515349 | 0.006154058 | 0.023181881 | 1 |
| Obesity | macrophage | *PRSS23* | 0.559886579 | 0.001758335 | 0.009491362 | 1 |
| Obesity | macrophage | *C11orf53* | 3.756455282 | 0.00398651 | 0.016987986 | 1 |
| Obesity | macrophage | *CCT5* | 0.889680176 | 0.012653691 | 0.039578981 | 1 |
| Obesity | macrophage | *TEX12* | 15.22780716 | 4.93E-05 | 0.000749385 | 0.309712169 |
| Obesity | macrophage | *DHX37* | 1.674066352 | 0.014663787 | 0.044110635 | 1 |
| Obesity | macrophage | *PRSS53* | -0.990053709 | 9.32E-06 | 0.000298952 | 0.058594682 |
| Obesity | macrophage | *BTBD11* | 1.923520798 | 5.05E-05 | 0.000759323 | 0.317397139 |
| Obesity | macrophage | *UBE3B* | 0.578285268 | 0.000758214 | 0.00526147 | 1 |
| Obesity | macrophage | *FAM177A1* | 1.531828327 | 0.000123023 | 0.001406264 | 0.773445429 |
| Obesity | macrophage | *MIPOL1* | 0.943330235 | 3.12E-06 | 0.000145227 | 0.01960571 |
| Obesity | macrophage | *TMEM18* | 1.591812295 | 0.004449032 | 0.018414132 | 1 |
| Obesity | macrophage | *ME3* | 0.570307186 | 0.011227829 | 0.036480291 | 1 |
| Obesity | macrophage | *ADAMTS12* | 0.703209447 | 0.003309523 | 0.014872747 | 1 |
| Obesity | macrophage | *ANKRD50* | 1.376229436 | 0.003495571 | 0.015444234 | 1 |
| Obesity | macrophage | *VPS26B* | 1.305196058 | 0.002166335 | 0.01088709 | 1 |

| | | | | | | |
|---|---|---|---|---|---|---|
| Obesity | macrophage | *FAM160B1* | 1.396307423 | 0.000162198 | 0.001693914 | 1 |
| Obesity | macrophage | *ANO4* | 0.880417662 | 0.000557545 | 0.004172957 | 1 |
| Obesity | macrophage | *TEX9* | 1.126103749 | 0.001373496 | 0.007958683 | 1 |
| Obesity | macrophage | *AKR1C2* | 0.969122342 | 0.000729609 | 0.00511948 | 1 |
| Obesity | macrophage | *DPYSL4* | 0.262793041 | 0.000117431 | 0.001359647 | 0.738288287 |
| Obesity | macrophage | *MFSD6* | 1.251879591 | 0.002579325 | 0.012386285 | 1 |
| Obesity | macrophage | *RNF144A* | 0.942256574 | 0.000345074 | 0.003037338 | 1 |
| Obesity | macrophage | *ASAP2* | 1.027017427 | 0.001533828 | 0.008579339 | 1 |
| Obesity | macrophage | *RABGAP1L* | 0.810924207 | 0.001904613 | 0.010028728 | 1 |
| Obesity | macrophage | *PTPN14* | 0.773024453 | 6.08E-05 | 0.000865111 | 0.382379189 |
| Obesity | macrophage | *ARL14EP* | -0.830178629 | 0.002475627 | 0.01203733 | 1 |
| Obesity | macrophage | *G6PC2* | 35.70478483 | 2.69E-05 | 0.000516343 | 0.168931253 |
| Obesity | macrophage | *POC5* | -0.821250991 | 1.97E-05 | 0.000454265 | 0.124014247 |
| Obesity | macrophage | *TADA1* | 1.983082609 | 0.00052107 | 0.003985358 | 1 |
| Obesity | macrophage | *CWF19L2* | 1.249672038 | 0.000122888 | 0.001406264 | 0.772594732 |
| Obesity | macrophage | *ZFP36L2* | 0.864731394 | 0.013602904 | 0.041758525 | 1 |
| Obesity | macrophage | *SPARCL1* | 0.410150546 | 0.007489297 | 0.026783398 | 1 |
| Obesity | macrophage | *CLEC4F* | 6.084911516 | 7.19E-05 | 0.000970253 | 0.452138051 |
| Obesity | macrophage | *PELO* | 2.075316101 | 1.54E-05 | 0.000405213 | 0.096968481 |
| Obesity | macrophage | *SAR1B* | 0.753883589 | 0.004723706 | 0.01915996 | 1 |
| Obesity | macrophage | *TCTEX1D1* | 0.284448958 | 0.006959912 | 0.025529153 | 1 |
| Obesity | macrophage | *ZNF117* | 0.122545623 | 0.009485842 | 0.031977206 | 1 |
| Obesity | macrophage | *RAB3C* | 0.558275121 | 0.003273294 | 0.014783909 | 1 |
| Obesity | macrophage | *MED21* | 1.5483593 | 0.001186809 | 0.007140163 | 1 |
| Obesity | macrophage | *CENPH* | 0.348427463 | 0.011113 | 0.036200741 | 1 |
| Obesity | macrophage | *CARHSP1* | 1.068109844 | 0.003568536 | 0.015656238 | 1 |
| Obesity | macrophage | *BANK1* | 1.434153903 | 0.000203495 | 0.001999493 | 1 |
| Obesity | macrophage | *CLGN* | 6.481119044 | 1.16E-05 | 0.000339028 | 0.07299734 |
| Obesity | macrophage | *AHCTF1* | 1.266921524 | 0.001050112 | 0.006625322 | 1 |
| Obesity | macrophage | *PTPRR* | 1.588496046 | 9.26E-05 | 0.001145864 | 0.582099102 |
| Obesity | macrophage | *PLA2R1* | 0.691992686 | 0.00122218 | 0.007290175 | 1 |
| Obesity | macrophage | *FRMD1* | 1.834118522 | 1.79E-05 | 0.00044073 | 0.112386234 |
| Obesity | macrophage | *INO80C* | 1.294194503 | 0.003802986 | 0.016376284 | 1 |
| Obesity | macrophage | *TUBGCP5* | 1.422189379 | 0.001697056 | 0.00924557 | 1 |
| Obesity | macrophage | *LURAP1L* | 0.270398129 | 0.005106207 | 0.020292491 | 1 |
| Obesity | macrophage | *CFDP1* | 0.999929762 | 0.007218766 | 0.026109582 | 1 |
| Obesity | macrophage | *FAM92B* | 15.22977584 | 3.69E-05 | 0.000608669 | 0.231902982 |
| Obesity | macrophage | *JAZF1* | 1.234212488 | 5.37E-06 | 0.000207263 | 0.033783833 |
| Obesity | macrophage | *CACNA2D1* | 0.864070512 | 0.000127735 | 0.001439194 | 0.803070361 |
| Obesity | macrophage | *ROBO3* | 0.613964132 | 0.007409108 | 0.026557049 | 1 |
| Obesity | macrophage | *CC2D1B* | 0.439646426 | 0.002151537 | 0.010864829 | 1 |
| Obesity | macrophage | *CEP112* | 0.853667541 | 9.03E-07 | 5.74E-05 | 0.005678019 |
| Obesity | macrophage | *ENPP3* | 0.966661043 | 0.003567448 | 0.015656238 | 1 |
| Obesity | macrophage | *UCHL1* | 1.655074082 | 0.000434637 | 0.003516813 | 1 |
| Obesity | macrophage | *MIA3* | 0.904466875 | 0.009994789 | 0.033335405 | 1 |
| Obesity | macrophage | *FAM167A* | 1.102538125 | 0.003415455 | 0.015196719 | 1 |
| Obesity | macrophage | *NEIL2* | 0.597454734 | 9.65E-06 | 0.000301542 | 0.06070054 |
| Obesity | macrophage | *CCSAP* | -0.670090971 | 0.003708517 | 0.016101828 | 1 |
| Obesity | macrophage | *PDLIM3* | 0.662213795 | 0.000438044 | 0.003535281 | 1 |
| Obesity | macrophage | *NCAM2* | 0.347296154 | 0.01130455 | 0.036672706 | 1 |

| | | | | | | |
|---|---|---|---|---|---|---|
| Obesity | macrophage | *PDE1C* | 1.436991126 | 1.58E-06 | 8.48E-05 | 0.009920474 |
| Obesity | macrophage | *RABGEF1* | -0.219538627 | 0.0046482 | 0.018939229 | 1 |
| Obesity | macrophage | *FLCN* | 0.552348155 | 0.012289735 | 0.038826917 | 1 |
| Obesity | macrophage | *CYP2U1* | 1.829245731 | 0.00170256 | 0.009267527 | 1 |
| Obesity | macrophage | *FBXL18* | 1.682741908 | 0.0013505 | 0.00788266 | 1 |
| Obesity | macrophage | *AK9* | 0.91693072 | 0.000395215 | 0.003321814 | 1 |
| Obesity | macrophage | *MMS19* | 1.296038406 | 0.001023952 | 0.0065294 | 1 |
| Obesity | macrophage | *GOLGA7B* | -0.65131403 | 0.000934085 | 0.006149313 | 1 |
| Obesity | macrophage | *TRMT44* | 1.582641429 | 0.00031919 | 0.002862696 | 1 |
| Obesity | macrophage | *ZKSCAN2* | 0.356179294 | 0.001894686 | 0.009993197 | 1 |
| Obesity | macrophage | *FAM126B* | 0.945739589 | 0.009553431 | 0.032153329 | 1 |
| Obesity | macrophage | *ALS2CR12* | 0.62257108 | 0.005896144 | 0.022616875 | 1 |
| Obesity | macrophage | *SPAG17* | 0.890169104 | 0.000353882 | 0.00306877 | 1 |
| Obesity | macrophage | *UGT2B4* | 1.992869694 | 1.95E-05 | 0.000454076 | 0.122600588 |
| Obesity | macrophage | *ADAMTSL3* | 0.34356843 | 0.000868399 | 0.005820496 | 1 |
| Obesity | macrophage | *TDRD9* | -0.364502832 | 0.00780335 | 0.027614456 | 1 |
| Obesity | macrophage | *GDF6* | 1.220126123 | 0.007185914 | 0.02605268 | 1 |
| Obesity | macrophage | *NODAL* | 2.00356961 | 1.57E-05 | 0.00040914 | 0.098922064 |
| Obesity | macrophage | *ATAD2* | 1.342787719 | 0.007677779 | 0.027317599 | 1 |
| Obesity | macrophage | *RPUSD3* | 1.693377824 | 0.007429189 | 0.026613852 | 1 |
| Obesity | macrophage | *GHRL* | 2.06262023 | 0.00122185 | 0.007290175 | 1 |
| Obesity | macrophage | *CPT2* | 1.39358732 | 0.008078248 | 0.028341487 | 1 |
| Obesity | macrophage | *STEAP2* | -0.175171804 | 0.006618443 | 0.024635967 | 1 |
| Obesity | macrophage | *GATAD1* | 0.441750319 | 0.013174716 | 0.040802679 | 1 |
| Obesity | macrophage | *TMED6* | 6.634938644 | 2.44E-05 | 0.000492083 | 0.153209444 |
| Obesity | macrophage | *CLEC18A* | -1.500086305 | 7.13E-17 | 3.74E-14 | 4.49E-13 |
| Obesity | macrophage | *IL34* | 1.300207618 | 0.000915237 | 0.006063323 | 1 |
| Obesity | macrophage | *CREB3L1* | 0.589664387 | 0.004586891 | 0.018725834 | 1 |
| Obesity | macrophage | *WDR19* | 1.283866501 | 0.000798559 | 0.005447834 | 1 |
| Obesity | macrophage | *SLC37A3* | 1.356254605 | 0.000148152 | 0.001584064 | 0.931429755 |
| Obesity | macrophage | *C12orf43* | 0.530968668 | 0.010472992 | 0.034581774 | 1 |
| Obesity | macrophage | *PEX10* | 0.849354818 | 0.012918019 | 0.040205735 | 1 |
| Obesity | macrophage | *RADIL* | 1.139796664 | 0.000239448 | 0.002268409 | 1 |
| Obesity | macrophage | *LDLRAP1* | 1.948522149 | 0.000223431 | 0.002131576 | 1 |
| Obesity | macrophage | *HPD* | 0.702952313 | 1.25E-06 | 7.19E-05 | 0.007833135 |
| Obesity | macrophage | *RHPN1* | -0.567111829 | 0.000508425 | 0.003935678 | 1 |
| Obesity | macrophage | *LRRC43* | 6.032823374 | 3.18E-05 | 0.000554992 | 0.200153952 |
| Obesity | macrophage | *XKR8* | 0.789652527 | 0.004011413 | 0.017040375 | 1 |
| Obesity | macrophage | *CLSTN2* | 0.592836984 | 0.006525484 | 0.024376541 | 1 |
| Obesity | macrophage | *COLEC12* | -0.354478808 | 0.005420608 | 0.021167304 | 1 |
| Obesity | macrophage | *RNF207* | 2.20235821 | 2.28E-05 | 0.000484376 | 0.143375171 |
| Obesity | macrophage | *CD1D* | -0.840266183 | 0.000718162 | 0.005067436 | 1 |
| Obesity | macrophage | *FAM86C1* | 2.96329962 | 0.000337509 | 0.002988616 | 1 |
| Obesity | macrophage | *CD1E* | 1.256785233 | 0.000334701 | 0.002976611 | 1 |
| Obesity | macrophage | *NCF1* | -0.536336136 | 0.000641673 | 0.004674622 | 1 |
| Obesity | macrophage | *PPP1R9A* | 0.623287497 | 0.00726403 | 0.026171323 | 1 |
| Obesity | macrophage | *GDPD5* | 1.330890976 | 1.25E-05 | 0.000350095 | 0.078771472 |
| Obesity | macrophage | *DYNC1I1* | 1.534145974 | 0.000150215 | 0.001600682 | 0.9444023 |
| Obesity | macrophage | *TMED4* | 1.551890904 | 0.001993212 | 0.01032234 | 1 |
| Obesity | macrophage | *SPATA2L* | -1.149500742 | 0.007771844 | 0.027558706 | 1 |

| | | | | | | |
|---|---|---|---|---|---|---|
| Obesity | macrophage | *NIT1* | 0.396650332 | 0.008027993 | 0.028228183 | 1 |
| Obesity | macrophage | *DMTN* | 0.348065157 | 0.003366808 | 0.015001506 | 1 |
| Obesity | macrophage | *SLC5A11* | -0.303162377 | 4.64E-07 | 3.43E-05 | 0.002914057 |
| Obesity | macrophage | *EPB41* | 0.549272679 | 0.001061256 | 0.006658797 | 1 |
| Obesity | macrophage | *ALG8* | 1.05056409 | 0.01653566 | 0.048062733 | 1 |
| Obesity | macrophage | *SV2A* | 1.873849267 | 0.010935159 | 0.035751089 | 1 |
| Obesity | macrophage | *CLIC6* | 0.466575132 | 0.000511436 | 0.00394003 | 1 |
| Obesity | macrophage | *PADI4* | 1.975342471 | 0.006937357 | 0.025461273 | 1 |
| Obesity | macrophage | *PSMD4* | 1.393686269 | 0.005577847 | 0.021660237 | 1 |
| Obesity | macrophage | *STARD9* | 0.690646701 | 0.006624939 | 0.024645559 | 1 |
| Obesity | macrophage | *TCHH* | -1.012302442 | 6.03E-05 | 0.000861688 | 0.379142863 |
| Obesity | macrophage | *ATP6V0D1* | -0.666713011 | 0.003679647 | 0.015987518 | 1 |
| Obesity | macrophage | *ZFYVE28* | 0.856276495 | 0.001608898 | 0.008849645 | 1 |
| Obesity | macrophage | *NPR2* | 1.112168406 | 0.00066377 | 0.004791184 | 1 |
| Obesity | macrophage | *DFFA* | 1.088740919 | 9.75E-06 | 0.000301542 | 0.061322489 |
| Obesity | macrophage | *C21orf33* | -0.783701153 | 0.015220353 | 0.045307936 | 1 |
| Obesity | macrophage | *FAM207A* | 0.600439189 | 0.010605884 | 0.034876467 | 1 |
| Obesity | macrophage | *LSS* | 0.707323043 | 8.44E-05 | 0.00107172 | 0.530501549 |
| Obesity | macrophage | *PCNT* | 0.300473937 | 0.009989043 | 0.033333926 | 1 |
| Obesity | macrophage | *C19orf47* | 1.55178904 | 7.90E-05 | 0.001028385 | 0.49671017 |
| Obesity | macrophage | *RDH13* | 1.133515921 | 0.012176682 | 0.038593587 | 1 |
| Obesity | macrophage | *MPZL3* | 2.882149911 | 0.000505858 | 0.003921493 | 1 |
| Obesity | macrophage | *S100A1* | 0.908193145 | 0.012917662 | 0.040205735 | 1 |
| Obesity | macrophage | *ZBTB7B* | -1.37580645 | 0.004109437 | 0.017359363 | 1 |
| Obesity | macrophage | *PAQR6* | -0.763779986 | 0.005316574 | 0.020851717 | 1 |
| Obesity | macrophage | *PMF1* | 0.881018711 | 0.000347296 | 0.00303995 | 1 |
| Obesity | macrophage | *NBEAL2* | 0.833833079 | 0.013094457 | 0.040632069 | 1 |
| Obesity | macrophage | *ORAI2* | 1.559366705 | 0.000116748 | 0.001359254 | 0.733997075 |
| Obesity | macrophage | *RPL8* | 1.134793883 | 0.009751693 | 0.032698077 | 1 |
| Obesity | macrophage | *FBXL13* | 1.249378348 | 0.000347658 | 0.00303995 | 1 |
| Obesity | macrophage | *DVL3* | 1.127880673 | 0.001747206 | 0.009445127 | 1 |
| Obesity | macrophage | *AP2M1* | 1.025286885 | 0.006847777 | 0.025265244 | 1 |
| Obesity | macrophage | *FBXO27* | 0.262523119 | 0.002099108 | 0.010677257 | 1 |
| Obesity | macrophage | *BDH1* | 2.649084658 | 5.31E-05 | 0.000788276 | 0.333815475 |
| Obesity | macrophage | *PGAP3* | 2.417518893 | 0.00176313 | 0.009498543 | 1 |
| Obesity | macrophage | *IKZF3* | 1.155641746 | 0.000730874 | 0.00512264 | 1 |
| Obesity | macrophage | *ACOX1* | 1.004712655 | 0.003027984 | 0.013946471 | 1 |
| Obesity | macrophage | *TMEM143* | 0.566257116 | 0.003140256 | 0.014337537 | 1 |
| Obesity | macrophage | *SIGLEC11* | -0.503256086 | 0.000303558 | 0.002757903 | 1 |
| Obesity | macrophage | *MPP3* | 0.866208102 | 0.007062581 | 0.025789721 | 1 |
| Obesity | macrophage | *RACGAP1* | -0.963277602 | 0.005859187 | 0.022488834 | 1 |
| Obesity | macrophage | *SPC24* | 2.051054594 | 0.006234547 | 0.023428929 | 1 |
| Obesity | macrophage | *ASGR2* | -1.625279848 | 0.000959197 | 0.006255676 | 1 |
| Obesity | macrophage | *JMJD8* | 0.50736571 | 0.002557236 | 0.012310779 | 1 |
| Obesity | macrophage | *FLYWCH2* | 1.823692476 | 5.32E-05 | 0.000788276 | 0.334391041 |
| Obesity | macrophage | *ZG16B* | -3.55448273 | 0.000340283 | 0.003008943 | 1 |
| Obesity | macrophage | *TPCN2* | 0.737962786 | 0.003353716 | 0.014996312 | 1 |
| Obesity | macrophage | *NOL9* | 1.400054161 | 8.61E-05 | 0.001086501 | 0.541077394 |
| Obesity | macrophage | *ZSWIM5* | 0.72163392 | 0.001050651 | 0.006625322 | 1 |
| Obesity | macrophage | *RBP7* | 1.903000053 | 0.000804471 | 0.005479644 | 1 |

| | | | | | | |
|---|---|---|---|---|---|---|
| Obesity | macrophage | *SLC25A34* | -1.145740927 | 0.005241862 | 0.02066181 | 1 |
| Obesity | macrophage | *LRRC38* | 2.219330331 | 0.003079481 | 0.014140343 | 1 |
| Obesity | macrophage | *PEF1* | 2.982091233 | 6.45E-06 | 0.00023452 | 0.040571874 |
| Obesity | macrophage | *RBBP4* | 1.13464354 | 0.001243777 | 0.00739095 | 1 |
| Obesity | macrophage | *CAMK2N1* | 2.336987124 | 0.010078946 | 0.033598268 | 1 |
| Obesity | macrophage | *TTLL10* | 1.448745882 | 0.005668859 | 0.021945886 | 1 |
| Obesity | macrophage | *NFIA* | 0.934454719 | 0.000534575 | 0.004044368 | 1 |
| Obesity | macrophage | *TM2D1* | 1.086702712 | 0.005633298 | 0.021821653 | 1 |
| Obesity | macrophage | *NEXN* | 1.345994348 | 6.07E-05 | 0.000864708 | 0.381336352 |
| Obesity | macrophage | *B3GALT2* | 2.112457834 | 0.000142726 | 0.001557845 | 0.897318611 |
| Obesity | macrophage | *HFM1* | 1.337379673 | 3.26E-05 | 0.000559198 | 0.204666316 |
| Obesity | macrophage | *KCNT2* | 1.103919355 | 1.63E-05 | 0.000416657 | 0.102259976 |
| Obesity | macrophage | *IGSF8* | 2.308438432 | 7.47E-05 | 0.000989215 | 0.469821542 |
| Obesity | macrophage | *SNED1* | 1.059735989 | 0.000104274 | 0.001253481 | 0.655570369 |
| Obesity | macrophage | *C1orf115* | 0.522020937 | 0.001101575 | 0.006798282 | 1 |
| Obesity | macrophage | *PUS10* | 1.290196817 | 0.005470381 | 0.021335163 | 1 |
| Obesity | macrophage | *RFTN2* | 1.225210105 | 2.00E-05 | 0.000456556 | 0.126025358 |
| Obesity | macrophage | *FAM84A* | 2.371223111 | 2.51E-05 | 0.000494987 | 0.157968102 |
| Obesity | macrophage | *CLHC1* | 0.164018954 | 0.001541362 | 0.008613817 | 1 |
| Obesity | macrophage | *FBXO41* | -0.69517085 | 0.002087265 | 0.010636979 | 1 |
| Obesity | macrophage | *CCDC74A* | 5.023422152 | 2.90E-05 | 0.000534857 | 0.182386108 |
| Obesity | macrophage | *NOSTRIN* | 0.634533945 | 0.001807973 | 0.009649174 | 1 |
| Obesity | macrophage | *RPRD2* | 0.928961582 | 0.000418213 | 0.003437543 | 1 |
| Obesity | macrophage | *C1QTNF7* | 0.761354919 | 0.001365172 | 0.007926286 | 1 |
| Obesity | macrophage | *LYSMD1* | 1.418266308 | 0.000126095 | 0.001428394 | 0.792758736 |
| Obesity | macrophage | *DHX57* | 1.211025318 | 0.001036376 | 0.006567709 | 1 |
| Obesity | macrophage | *DCAF16* | 1.217056654 | 0.010581936 | 0.034831746 | 1 |
| Obesity | macrophage | *GNPDA2* | 0.289820582 | 0.011224343 | 0.036480291 | 1 |
| Obesity | macrophage | *ANTXR2* | 0.849749521 | 3.11E-05 | 0.000549896 | 0.195762859 |
| Obesity | macrophage | *PBXIP1* | 2.223263275 | 2.00E-05 | 0.000456556 | 0.125939607 |
| Obesity | macrophage | *DCST2* | 4.398935478 | 6.11E-06 | 0.000227466 | 0.038441819 |
| Obesity | macrophage | *LMOD3* | 2.769436326 | 3.65E-05 | 0.000604095 | 0.229291488 |
| Obesity | macrophage | *SLC15A2* | -0.612313688 | 6.71E-05 | 0.000919437 | 0.422021402 |
| Obesity | macrophage | *LMOD1* | -0.794427634 | 1.17E-09 | 1.76E-07 | 7.38E-06 |
| Obesity | macrophage | *SERPINI1* | 1.362690592 | 0.000665111 | 0.00479536 | 1 |
| Obesity | macrophage | *AIM2* | 2.394403329 | 1.19E-05 | 0.000343957 | 0.07463863 |
| Obesity | macrophage | *CD200R1* | -0.305269183 | 0.003528021 | 0.015543567 | 1 |
| Obesity | macrophage | *GTPBP8* | 1.383093247 | 0.006004079 | 0.022855957 | 1 |
| Obesity | macrophage | *CADPS* | 2.028404719 | 2.29E-05 | 0.00048472 | 0.143961697 |
| Obesity | macrophage | *DCLK3* | 0.931179377 | 0.012611481 | 0.039524736 | 1 |
| Obesity | macrophage | *DNASE1L3* | 1.134638577 | 0.000405523 | 0.003372538 | 1 |
| Obesity | macrophage | *CRELD1* | 0.496824319 | 0.002944773 | 0.013673405 | 1 |
| Obesity | macrophage | *CCDC158* | 0.921661353 | 0.00055283 | 0.004152498 | 1 |
| Obesity | macrophage | *GYG1* | 1.662885167 | 4.50E-05 | 0.000709767 | 0.283196968 |
| Obesity | macrophage | *RYK* | 1.120956958 | 0.000979741 | 0.006347553 | 1 |
| Obesity | macrophage | *PLB1* | -0.520779233 | 0.001265743 | 0.007493153 | 1 |
| Obesity | macrophage | *CDCP1* | -0.351766076 | 0.000556012 | 0.004171414 | 1 |
| Obesity | macrophage | *LZTFL1* | 1.995622135 | 0.000315765 | 0.00284008 | 1 |
| Obesity | macrophage | *FYCO1* | 1.351983552 | 0.000671829 | 0.004832714 | 1 |
| Obesity | macrophage | *NMNAT3* | 0.776893949 | 0.009334228 | 0.031584657 | 1 |

| | | | | | | |
|---|---|---|---|---|---|---|
| Obesity | macrophage | *SMIM12* | 1.338888101 | 0.006815686 | 0.025176392 | 1 |
| Obesity | macrophage | *TPRA1* | 1.180369603 | 0.015711134 | 0.046238266 | 1 |
| Obesity | macrophage | *RFT1* | -0.979882683 | 0.000105739 | 0.001261441 | 0.664779166 |
| Obesity | macrophage | *SLC51A* | -0.801012505 | 0.015131149 | 0.045149282 | 1 |
| Obesity | macrophage | *PIGX* | -0.596880801 | 0.00505519 | 0.020163061 | 1 |
| Obesity | macrophage | *ABLIM2* | 2.140276144 | 7.31E-05 | 0.000977963 | 0.459642782 |
| Obesity | macrophage | *DNAJB14* | 0.857396362 | 0.002925304 | 0.013613165 | 1 |
| Obesity | macrophage | *SLC9B2* | 1.034056174 | 2.17E-05 | 0.000477851 | 0.136665391 |
| Obesity | macrophage | *ATRIP* | 2.318587421 | 0.011984241 | 0.03814933 | 1 |
| Obesity | macrophage | *SHISA5* | 2.031579066 | 2.82E-06 | 0.000134335 | 0.017732182 |
| Obesity | macrophage | *APEH* | -1.047908591 | 7.30E-11 | 1.64E-08 | 4.59E-07 |
| Obesity | macrophage | *RNF123* | 1.064186174 | 8.41E-22 | 6.61E-19 | 5.29E-18 |
| Obesity | macrophage | *MST1R* | 0.782836638 | 3.31E-09 | 4.62E-07 | 2.08E-05 |
| Obesity | macrophage | *TEX264* | -0.538524745 | 0.001977874 | 0.010303093 | 1 |
| Obesity | macrophage | *GRM2* | -5.172198184 | 0.000263661 | 0.00245576 | 1 |
| Obesity | macrophage | *POC1A* | 1.193592656 | 6.79E-05 | 0.000924353 | 0.427051232 |
| Obesity | macrophage | *PPM1M* | -0.647285803 | 1.96E-05 | 0.000454265 | 0.123144197 |
| Obesity | macrophage | *C4orf3* | 1.240992383 | 0.00154291 | 0.00861481 | 1 |
| Obesity | macrophage | *MAP9* | 1.268088903 | 0.000313241 | 0.002825671 | 1 |
| Obesity | macrophage | *CEP44* | 1.410297833 | 0.00196058 | 0.010237679 | 1 |
| Obesity | macrophage | *C4orf45* | 0.908973786 | 0.010245211 | 0.034036786 | 1 |
| Obesity | macrophage | *NPY1R* | 1.222275366 | 5.13E-05 | 0.000767345 | 0.322284949 |
| Obesity | macrophage | *ARFIP1* | 0.682609093 | 0.014299121 | 0.043408292 | 1 |
| Obesity | macrophage | *OTUD4* | 1.21982291 | 0.006558204 | 0.024469691 | 1 |
| Obesity | macrophage | *ITGA2* | 1.842863593 | 1.55E-06 | 8.40E-05 | 0.009740629 |
| Obesity | macrophage | *CMBL* | 1.901888703 | 0.000114902 | 0.001350604 | 0.722388515 |
| Obesity | macrophage | *C5orf63* | 1.083148531 | 0.008621167 | 0.029780921 | 1 |
| Obesity | macrophage | *PRRC1* | 1.173384844 | 0.005383706 | 0.021075568 | 1 |
| Obesity | macrophage | *TIGD6* | 2.568117434 | 0.000524213 | 0.003994823 | 1 |
| Obesity | macrophage | *ENPP6* | 1.215710295 | 0.005597256 | 0.021708789 | 1 |
| Obesity | macrophage | *ERAP1* | -0.402218119 | 0.006103787 | 0.023061603 | 1 |
| Obesity | macrophage | *RICTOR* | 0.885165489 | 0.012434696 | 0.039075715 | 1 |
| Obesity | macrophage | *ANKRA2* | 1.538016088 | 0.000643607 | 0.004683283 | 1 |
| Obesity | macrophage | *TLR3* | 1.824543934 | 8.24E-05 | 0.001057659 | 0.518252825 |
| Obesity | macrophage | *CCDC127* | 1.080114229 | 0.003186028 | 0.014474798 | 1 |
| Obesity | macrophage | *TXLNB* | -0.503016954 | 0.002877632 | 0.013451057 | 1 |
| Obesity | macrophage | *TMEM200A* | 1.479425044 | 9.18E-05 | 0.001139978 | 0.577280946 |
| Obesity | macrophage | *STXBP5* | 0.86502645 | 0.011707519 | 0.037553658 | 1 |
| Obesity | macrophage | *PI16* | 1.271717556 | 0.000403677 | 0.003365937 | 1 |
| Obesity | macrophage | *STK17A* | 0.913202169 | 0.002735248 | 0.012939431 | 1 |
| Obesity | macrophage | *RPS14* | 0.821153418 | 0.014558247 | 0.043870231 | 1 |
| Obesity | macrophage | *SLU7* | 0.928960568 | 0.004961505 | 0.01994436 | 1 |
| Obesity | macrophage | *CAMLG* | 0.722654076 | 0.00148253 | 0.008374361 | 1 |
| Obesity | macrophage | *KCNK5* | 2.263104072 | 9.43E-05 | 0.001165236 | 0.593105248 |
| Obesity | macrophage | *ZNF704* | 0.837012139 | 0.002516729 | 0.012199439 | 1 |
| Obesity | macrophage | *FNDC1* | 0.423571795 | 0.007026211 | 0.025697376 | 1 |
| Obesity | macrophage | *LMTK2* | 1.337582985 | 5.71E-05 | 0.00083065 | 0.358840595 |
| Obesity | macrophage | *CTSB* | -0.265404307 | 0.003487169 | 0.015439316 | 1 |
| Obesity | macrophage | *TNFRSF11B* | 1.52815055 | 2.52E-06 | 0.000125539 | 0.01581792 |
| Obesity | macrophage | *SUN1* | 0.690690592 | 0.002015707 | 0.010413105 | 1 |

| | | | | | | |
|---|---|---|---|---|---|---|
| Obesity | macrophage | *GPER1* | 0.295032288 | 0.015875484 | 0.046596249 | 1 |
| Obesity | macrophage | *INTS1* | 1.458634378 | 0.015565298 | 0.045900108 | 1 |
| Obesity | macrophage | *COX6C* | 1.292345363 | 0.007281933 | 0.026190797 | 1 |
| Obesity | macrophage | *YWHAZ* | -0.408092686 | 0.002534244 | 0.012265427 | 1 |
| Obesity | macrophage | *DCAF13* | 1.764714362 | 0.001196541 | 0.007176804 | 1 |
| Obesity | macrophage | *FREM1* | 0.834220621 | 0.001119078 | 0.006867967 | 1 |
| Obesity | macrophage | *FAM219A* | 1.248136543 | 0.004498805 | 0.018522585 | 1 |
| Obesity | macrophage | *SYK* | -0.343645962 | 0.002550527 | 0.012296906 | 1 |
| Obesity | macrophage | *TMC1* | 1.562861522 | 1.85E-05 | 0.000441618 | 0.116077507 |
| Obesity | macrophage | *HNRNPK* | 1.021680456 | 0.010555572 | 0.034781385 | 1 |
| Obesity | macrophage | *SVEP1* | 1.071327919 | 1.63E-05 | 0.000416657 | 0.102497607 |
| Obesity | macrophage | *TRMT10B* | 1.204286395 | 0.005010475 | 0.020097422 | 1 |
| Obesity | macrophage | *LRFN5* | 0.458066265 | 9.46E-06 | 0.000300263 | 0.059452137 |
| Obesity | macrophage | *WRN* | 1.067582974 | 0.001007619 | 0.006457594 | 1 |
| Obesity | macrophage | *CFL2* | 0.91552944 | 0.006100412 | 0.023061603 | 1 |
| Obesity | macrophage | *ZCCHC24* | 1.254583477 | 0.001514521 | 0.008501604 | 1 |
| Obesity | macrophage | *TAF3* | 0.846493411 | 0.014375191 | 0.043555097 | 1 |
| Obesity | macrophage | *SLC18A2* | 0.879031284 | 0.015990345 | 0.046802279 | 1 |
| Obesity | macrophage | *QSOX2* | 2.099910915 | 0.006108195 | 0.023064398 | 1 |
| Obesity | macrophage | *AK8* | 1.082631657 | 0.001787287 | 0.009571271 | 1 |
| Obesity | macrophage | *TSC1* | 1.135678102 | 0.000666765 | 0.004801777 | 1 |
| Obesity | macrophage | *NSMF* | 2.3619276 | 0.000655372 | 0.004736002 | 1 |
| Obesity | macrophage | *ARHGAP42* | 1.184039801 | 7.51E-05 | 0.000989215 | 0.47185552 |
| Obesity | macrophage | *SLC39A13* | 3.487331807 | 2.07E-07 | 1.73E-05 | 0.001298307 |
| Obesity | macrophage | *PSMC3* | 0.803651898 | 6.66E-05 | 0.000913665 | 0.418458463 |
| Obesity | macrophage | *ABTB2* | 1.631367672 | 0.000123524 | 0.001409434 | 0.776597973 |
| Obesity | macrophage | *TCP11L2* | 1.458140971 | 0.000601497 | 0.004428117 | 1 |
| Obesity | macrophage | *JAM3* | 0.713689631 | 0.008902564 | 0.030418707 | 1 |
| Obesity | macrophage | *NDUFB8* | 1.59748189 | 0.003244208 | 0.014673621 | 1 |
| Obesity | macrophage | *TRMT61A* | -0.653763307 | 0.000930753 | 0.00614063 | 1 |
| Obesity | macrophage | *LARP6* | 0.810172461 | 0.000246165 | 0.002327276 | 1 |
| Obesity | macrophage | *ASPG* | 3.863648956 | 3.97E-06 | 0.000173912 | 0.024931411 |
| Obesity | macrophage | *TBATA* | 1.204331064 | 0.01514527 | 0.045156901 | 1 |
| Obesity | macrophage | *ZNF202* | 1.818926645 | 0.001366013 | 0.007926286 | 1 |
| Obesity | macrophage | *FAM227B* | 0.60066482 | 0.002933629 | 0.013641809 | 1 |
| Obesity | macrophage | *APBB1* | 1.174500009 | 0.00049891 | 0.003889873 | 1 |
| Obesity | macrophage | *ILK* | 1.180559901 | 0.002199211 | 0.01100831 | 1 |
| Obesity | macrophage | *C11orf74* | 1.353775126 | 0.000481927 | 0.003808366 | 1 |
| Obesity | macrophage | *WDR88* | 1.605132062 | 0.000525175 | 0.00399731 | 1 |
| Obesity | macrophage | *TUB* | 0.90109162 | 0.001060801 | 0.006658797 | 1 |
| Obesity | macrophage | *LMO1* | -0.567914808 | 0.001188676 | 0.007144555 | 1 |
| Obesity | macrophage | *PRTG* | 1.374124632 | 1.39E-05 | 0.000385193 | 0.087438738 |
| Obesity | macrophage | *C16orf46* | 8.743734255 | 8.75E-06 | 0.000289582 | 0.055020597 |
| Obesity | macrophage | *TMEM41B* | 1.165840305 | 0.009145273 | 0.031079097 | 1 |
| Obesity | macrophage | *ZSCAN21* | -0.242267892 | 0.014798597 | 0.044447445 | 1 |
| Obesity | macrophage | *BEAN1* | 1.525740053 | 0.001598959 | 0.008823011 | 1 |
| Obesity | macrophage | *SPDYE6* | 0.682519465 | 0.013559953 | 0.041687739 | 1 |
| Obesity | macrophage | *C16orf45* | 1.221303825 | 0.000208923 | 0.002027001 | 1 |
| Obesity | macrophage | *SAAL1* | 2.597170201 | 6.63E-07 | 4.68E-05 | 0.004166456 |
| Obesity | macrophage | *YPEL4* | 0.899962755 | 0.000702624 | 0.00499141 | 1 |

| | | | | | | |
|---|---|---|---|---|---|---|
| Obesity | macrophage | *MS4A6E* | -0.363570877 | 5.43E-05 | 0.000799526 | 0.341397468 |
| Obesity | macrophage | *MS4A7* | -0.366825105 | 0.000426582 | 0.0034785 | 1 |
| Obesity | macrophage | *MS4A14* | -0.303336707 | 8.90E-05 | 0.001115582 | 0.559773157 |
| Obesity | macrophage | *CCNDBP1* | 1.658962951 | 0.00123318 | 0.007341856 | 1 |
| Obesity | macrophage | *AKTIP* | 1.784136399 | 0.000134728 | 0.001501834 | 0.847034507 |
| Obesity | macrophage | *NKX3-1* | 2.189143701 | 0.01310194 | 0.040632069 | 1 |
| Obesity | macrophage | *TEF* | 0.324140303 | 0.006498768 | 0.024334577 | 1 |
| Obesity | macrophage | *SAMD14* | 0.873089032 | 0.003492267 | 0.015444234 | 1 |
| Obesity | macrophage | *ENDOG* | 0.411899392 | 0.010723318 | 0.035168233 | 1 |
| Obesity | macrophage | *ISLR2* | 0.692290851 | 0.000235914 | 0.002240474 | 1 |
| Obesity | macrophage | *SP2* | 1.902901736 | 7.64E-06 | 0.00025971 | 0.048046349 |
| Obesity | macrophage | *COQ7* | 1.974802354 | 0.000374394 | 0.003180829 | 1 |
| Obesity | macrophage | *GPRC5B* | 0.778770807 | 0.001878629 | 0.009925948 | 1 |
| Obesity | macrophage | *RBFOX3* | 4.277759441 | 1.26E-06 | 7.20E-05 | 0.007924967 |
| Obesity | macrophage | *ZNF646* | 1.838384072 | 1.17E-09 | 1.76E-07 | 7.36E-06 |
| Obesity | macrophage | *VKORC1* | -0.85395867 | 7.39E-07 | 5.11E-05 | 0.004647901 |
| Obesity | macrophage | *SMG8* | 6.595658063 | 0.000813611 | 0.005523945 | 1 |
| Obesity | macrophage | *ZNF528* | 1.172925124 | 0.005272227 | 0.020742485 | 1 |
| Obesity | macrophage | *SERTAD3* | 2.504647503 | 0.00785689 | 0.027766312 | 1 |
| Obesity | macrophage | *TTYH1* | 1.185869506 | 0.00964074 | 0.032377848 | 1 |
| Obesity | macrophage | *LENG8* | 0.781380922 | 0.004480693 | 0.018484331 | 1 |
| Obesity | macrophage | *CDC42EP5* | 0.851337539 | 0.003527785 | 0.015543567 | 1 |
| Obesity | macrophage | *PPP1R14A* | 2.024346713 | 0.000499304 | 0.003889873 | 1 |
| Obesity | macrophage | *PSCA* | 0.788996472 | 0.000415071 | 0.00342597 | 1 |
| Obesity | macrophage | *UBXN6* | -0.680147722 | 0.012320283 | 0.038884349 | 1 |
| Obesity | macrophage | *ZNF444* | 2.103141076 | 0.000164859 | 0.001718855 | 1 |
| Obesity | macrophage | *FAM57A* | 0.34012623 | 0.013420979 | 0.041361617 | 1 |
| Obesity | macrophage | *SRR* | 0.356284101 | 4.32E-06 | 0.000178092 | 0.027167065 |
| Obesity | macrophage | *CYB5D2* | 1.523188482 | 0.000374149 | 0.003180829 | 1 |
| Obesity | macrophage | *CD320* | 1.954518602 | 9.86E-06 | 0.000301542 | 0.061995693 |
| Obesity | macrophage | *SPRYD3* | 2.257087095 | 8.40E-05 | 0.001070503 | 0.528169795 |
| Obesity | macrophage | *NDUFV1* | 0.93691723 | 0.007555591 | 0.026989775 | 1 |
| Obesity | macrophage | *PRDX2* | 1.873052451 | 2.49E-06 | 0.000125506 | 0.015639073 |
| Obesity | macrophage | *GSDMA* | -0.610752783 | 0.002337111 | 0.011494793 | 1 |
| Obesity | macrophage | *VWCE* | 0.889069375 | 0.006885141 | 0.025373319 | 1 |
| Obesity | macrophage | *TRANK1* | 0.812963027 | 0.015808343 | 0.046442547 | 1 |
| Obesity | macrophage | *LTBP3* | 0.525512785 | 0.002949433 | 0.013677939 | 1 |
| Obesity | macrophage | *NAALADL1* | 1.051759388 | 0.003186436 | 0.014474798 | 1 |
| Obesity | macrophage | *SCARA5* | 0.753886478 | 0.012224148 | 0.038658559 | 1 |
| Obesity | macrophage | *NUDT16L1* | 1.340562279 | 0.007194497 | 0.026055186 | 1 |
| Obesity | macrophage | *ZCCHC4* | 1.00092466 | 0.007999331 | 0.028143142 | 1 |
| Obesity | macrophage | *GLYCTK* | 0.80187572 | 0.011431075 | 0.036911747 | 1 |
| Obesity | macrophage | *UBTD2* | 0.989876555 | 0.000753464 | 0.005240073 | 1 |
| Obesity | macrophage | *NT5DC2* | -0.721684263 | 0.0008134 | 0.005523945 | 1 |
| Obesity | macrophage | *KIF5C* | 1.142837283 | 0.002937151 | 0.013648092 | 1 |
| Obesity | macrophage | *FAM107A* | 0.421191431 | 0.006599901 | 0.024596076 | 1 |
| Obesity | macrophage | *TAP1* | 0.908068575 | 0.002139607 | 0.010824534 | 1 |
| Obesity | macrophage | *ING5* | 1.348077095 | 0.002606086 | 0.012459669 | 1 |
| Obesity | macrophage | *RFWD3* | 0.531817807 | 0.009726386 | 0.032630624 | 1 |
| Obesity | macrophage | *REEP4* | -1.147858296 | 0.002633079 | 0.012541038 | 1 |

| | | | | | | |
|---|---|---|---|---|---|---|
| Obesity | macrophage | *BMP1* | 0.548124341 | 0.011582512 | 0.037257291 | 1 |
| Obesity | macrophage | *FNTA* | 1.13857234 | 0.004695679 | 0.019095559 | 1 |
| Obesity | macrophage | *SLC20A2* | 1.236056133 | 0.000421247 | 0.003448415 | 1 |
| Obesity | macrophage | *ZSWIM1* | 1.312644348 | 0.000266738 | 0.002477077 | 1 |
| Obesity | macrophage | *NBPF9* | 2.019419945 | 0.002697464 | 0.012803881 | 1 |
| Obesity | macrophage | *WFDC13* | 2.375313051 | 0.010664659 | 0.035030676 | 1 |
| Obesity | macrophage | *NDUFS5* | 0.882426149 | 0.003185004 | 0.014474798 | 1 |
| Obesity | macrophage | *FAM84B* | 0.291367639 | 0.012370848 | 0.038985223 | 1 |
| Obesity | macrophage | *DNAJC21* | 1.351053676 | 0.003168066 | 0.014433066 | 1 |
| Obesity | macrophage | *NPNT* | 1.086000827 | 0.003995377 | 0.017000566 | 1 |
| Obesity | macrophage | *CNNM3* | 1.545401637 | 0.00044962 | 0.003605563 | 1 |
| Obesity | macrophage | *SNTB2* | 1.246965175 | 0.000876714 | 0.005851273 | 1 |
| Obesity | macrophage | *ZBTB49* | 1.615988943 | 0.011678418 | 0.037488027 | 1 |
| Obesity | macrophage | *BTNL3* | 0.134064095 | 0.006978643 | 0.025568023 | 1 |
| Obesity | macrophage | *ZNF608* | 1.100126114 | 2.15E-05 | 0.000473225 | 0.134869211 |
| Obesity | macrophage | *LETM1* | 1.851014267 | 0.000815315 | 0.005525151 | 1 |
| Obesity | macrophage | *STXBP6* | 1.018365354 | 0.000541192 | 0.004081114 | 1 |
| Obesity | macrophage | *CPLX1* | 0.362340639 | 0.004363546 | 0.018131931 | 1 |
| Obesity | macrophage | *SIGLEC7* | -0.445255978 | 0.004893 | 0.019719419 | 1 |
| Obesity | macrophage | *COL4A3* | 1.084051141 | 5.53E-06 | 0.000210874 | 0.034794152 |
| Obesity | macrophage | *C8orf46* | 0.09242494 | 0.000401094 | 0.003354341 | 1 |
| Obesity | macrophage | *FAM110B* | 1.386788215 | 0.000368342 | 0.003142149 | 1 |
| Obesity | macrophage | *RAB24* | -0.92809476 | 0.00017736 | 0.001808801 | 1 |
| Obesity | macrophage | *SNUPN* | 0.818236886 | 0.010574839 | 0.034826617 | 1 |
| Obesity | macrophage | *PTK2* | 0.771833218 | 0.008672675 | 0.029894594 | 1 |
| Obesity | macrophage | *PTAFR* | -0.383942802 | 0.010402425 | 0.034384884 | 1 |
| Obesity | macrophage | *RNASE6* | -0.819760006 | 0.00114601 | 0.00696805 | 1 |
| Obesity | macrophage | *NPR1* | 1.384032459 | 1.49E-05 | 0.000405213 | 0.093753004 |
| Obesity | macrophage | *CLIC4* | 0.851932123 | 0.008720025 | 0.029974191 | 1 |
| Obesity | macrophage | *SLC38A11* | 1.280888675 | 0.002064437 | 0.010569311 | 1 |
| Obesity | macrophage | *CCDC8* | 1.782746537 | 0.001416656 | 0.008118976 | 1 |
| Obesity | macrophage | *METTL15* | 0.679149005 | 0.000418145 | 0.003437543 | 1 |
| Obesity | macrophage | *INO80E* | -1.414206913 | 1.40E-10 | 2.93E-08 | 8.78E-07 |
| Obesity | macrophage | *FAM103A1* | -0.415741747 | 0.000143562 | 0.001561551 | 0.902576694 |
| Obesity | macrophage | *FASN* | 0.405447561 | 0.000923988 | 0.006114856 | 1 |
| Obesity | macrophage | *DCXR* | 1.692352418 | 0.000401937 | 0.00335588 | 1 |
| Obesity | macrophage | *UGP2* | 0.620370478 | 0.001382905 | 0.00798377 | 1 |
| Obesity | macrophage | *AVEN* | 1.459468566 | 0.00078713 | 0.005420247 | 1 |
| Obesity | macrophage | *BRD3* | 1.320633703 | 0.005808992 | 0.022323431 | 1 |
| Obesity | macrophage | *ZFPM2* | 0.920664282 | 7.49E-05 | 0.000989215 | 0.470867858 |
| Obesity | macrophage | *MYO7B* | 0.773660643 | 0.000458817 | 0.003669931 | 1 |
| Obesity | macrophage | *NIPA1* | 1.322718001 | 0.014397131 | 0.043600561 | 1 |
| Obesity | macrophage | *CRTAP* | 1.358953115 | 0.003663167 | 0.015975945 | 1 |
| Obesity | macrophage | *SEMA3E* | 1.288234061 | 9.00E-06 | 0.000293144 | 0.056576884 |
| Obesity | macrophage | *ADORA2B* | -0.484394093 | 1.50E-07 | 1.29E-05 | 0.000944716 |
| Obesity | macrophage | *METTL7B* | 4.465554477 | 9.65E-05 | 0.001183611 | 0.606495395 |
| Obesity | macrophage | *NFXL1* | 1.507362741 | 0.000492605 | 0.003870478 | 1 |
| Obesity | macrophage | *CD14* | -0.490643168 | 0.000119219 | 0.001375285 | 0.749530264 |
| Obesity | macrophage | *DLGAP1* | 1.11030417 | 0.001022119 | 0.0065294 | 1 |
| Obesity | macrophage | *NUDCD2* | 1.123393805 | 0.015966211 | 0.046775194 | 1 |

| | | | | | | |
|---|---|---|---|---|---|---|
| Obesity | macrophage | *SLC26A5* | 1.809841944 | 1.80E-05 | 0.000441618 | 0.113212042 |
| Obesity | macrophage | *ACYP2* | 0.735921847 | 0.014804032 | 0.044447445 | 1 |
| Obesity | macrophage | *TMEM133* | 3.902810165 | 1.93E-05 | 0.000450328 | 0.121138223 |
| Obesity | macrophage | *NDUFA3* | 1.060331944 | 0.013289977 | 0.041038351 | 1 |
| Obesity | macrophage | *PAQR8* | -0.863407753 | 0.00599098 | 0.022855151 | 1 |
| Obesity | macrophage | *NUDT6* | 0.813049575 | 0.004739181 | 0.019210336 | 1 |
| Obesity | macrophage | *ZNF160* | 0.820840612 | 0.003808512 | 0.016388852 | 1 |
| Obesity | macrophage | *TSNARE1* | 0.453962893 | 0.001600922 | 0.008823011 | 1 |
| Obesity | macrophage | *SOX7* | 2.184449645 | 1.63E-08 | 1.93E-06 | 0.000102486 |
| Obesity | macrophage | *C11orf24* | 1.456795909 | 0.01531862 | 0.045514255 | 1 |
| Obesity | macrophage | *TRMT61B* | 0.408444625 | 2.71E-06 | 0.000132196 | 0.017053332 |
| Obesity | macrophage | *MFN1* | 0.77196373 | 0.004545969 | 0.018620347 | 1 |
| Obesity | macrophage | *KCNMB3* | 0.443584645 | 0.008546479 | 0.02960425 | 1 |
| Obesity | macrophage | *KCND3* | 1.121181056 | 5.70E-06 | 0.000214064 | 0.035857128 |
| Obesity | macrophage | *MRPL36* | 0.728873072 | 0.003725514 | 0.016136259 | 1 |
| Obesity | macrophage | *ZNF581* | 2.35894245 | 0.001623567 | 0.008906951 | 1 |
| Obesity | macrophage | *MCC* | 1.134474137 | 0.000105294 | 0.001260164 | 0.661981055 |
| Obesity | macrophage | *HOPX* | 1.452273773 | 0.004815595 | 0.019482398 | 1 |
| Obesity | macrophage | *LRRC8C* | 1.771287178 | 0.003348251 | 0.014987476 | 1 |
| Obesity | macrophage | *RSL1D1* | 1.385322066 | 0.003661259 | 0.015975945 | 1 |
| Obesity | macrophage | *PLRG1* | 1.669107405 | 0.000988062 | 0.006371225 | 1 |
| Obesity | macrophage | *NMUR1* | 1.728203714 | 2.67E-05 | 0.000515008 | 0.167636402 |
| Obesity | macrophage | *P2RY6* | -0.336290365 | 0.00044202 | 0.003562795 | 1 |
| Obesity | macrophage | *TCEA2* | 2.180596856 | 2.94E-05 | 0.000541287 | 0.185120071 |
| Obesity | macrophage | *ANO5* | -0.746086449 | 0.01258152 | 0.03945138 | 1 |
| Obesity | macrophage | *VAT1L* | 0.659568666 | 0.007063766 | 0.025789721 | 1 |
| Obesity | macrophage | *CAMTA1* | 1.112731371 | 0.003279931 | 0.014792629 | 1 |
| Obesity | macrophage | *ZNF570* | 0.86911385 | 0.016485374 | 0.047957451 | 1 |
| Obesity | macrophage | *NINJ2* | 1.343564443 | 4.70E-05 | 0.000735154 | 0.295532014 |
| Obesity | macrophage | *FAM90A1* | 3.279398535 | 2.24E-05 | 0.000483304 | 0.140641443 |
| Obesity | macrophage | *TRAPPC12* | 0.87303225 | 0.016960831 | 0.048959021 | 1 |
| Obesity | macrophage | *C3AR1* | -0.426177882 | 0.000856813 | 0.005761263 | 1 |
| Obesity | macrophage | *AQP4* | 2.161027797 | 2.10E-05 | 0.000467599 | 0.132330643 |
| Obesity | macrophage | *ANKRD20A4* | 4.047322919 | 3.07E-05 | 0.000546074 | 0.192764023 |
| Obesity | macrophage | *SERF1A* | 5.644553968 | 3.22E-05 | 0.000554992 | 0.202153865 |
| Obesity | macrophage | *MTBP* | 1.182539992 | 0.011052298 | 0.036059054 | 1 |
| Obesity | macrophage | *CXCR6* | 0.533408984 | 0.000509701 | 0.003935678 | 1 |
| Obesity | macrophage | *C1QTNF4* | -0.325893153 | 2.42E-15 | 1.17E-12 | 1.52E-11 |
| Obesity | macrophage | *HINFP* | 1.850107521 | 0.001806744 | 0.009649174 | 1 |
| Obesity | macrophage | *COPRS* | 1.648111741 | 0.000435235 | 0.003517125 | 1 |
| Obesity | macrophage | *CSDC2* | 0.997281146 | 0.001057712 | 0.006649833 | 1 |
| Obesity | macrophage | *RCAN2* | 1.289098118 | 0.000105431 | 0.001260164 | 0.662846108 |
| Obesity | macrophage | *ZNF24* | 1.28029308 | 0.007280347 | 0.026190797 | 1 |
| Obesity | macrophage | *FIBP* | 2.626636495 | 1.35E-06 | 7.56E-05 | 0.008470878 |
| Obesity | macrophage | *CARNS1* | 2.276379549 | 2.88E-05 | 0.000534043 | 0.181040586 |
| Obesity | macrophage | *CTSW* | -0.170213514 | 3.36E-07 | 2.54E-05 | 0.002110576 |
| Obesity | macrophage | *EFEMP2* | 1.169307243 | 4.30E-10 | 7.72E-08 | 2.70E-06 |
| Obesity | macrophage | *AGAP5* | -16.27871642 | 0.001167159 | 0.007062494 | 1 |
| Obesity | macrophage | *ZMAT3* | 1.545218971 | 0.005681692 | 0.021982029 | 1 |
| Obesity | macrophage | *CCL19* | 4.785508401 | 0.013251661 | 0.040960272 | 1 |

| | | | | | | |
|---|---|---|---|---|---|---|
| Obesity | macrophage | *FUT10* | -0.367597148 | 0.001074604 | 0.006702419 | 1 |
| Obesity | macrophage | *RAB37* | 1.247004849 | 0.007696359 | 0.027352747 | 1 |
| Obesity | macrophage | *SNX32* | 0.905058297 | 4.60E-08 | 4.60E-06 | 0.000289501 |
| Obesity | macrophage | *METAP1D* | 1.730375209 | 1.67E-05 | 0.000423143 | 0.104939539 |
| Obesity | macrophage | *NADSYN1* | 0.898350476 | 0.004883956 | 0.019695594 | 1 |
| Obesity | macrophage | *MRGPRF* | 0.975318633 | 0.003495649 | 0.015444234 | 1 |
| Obesity | macrophage | *LCLAT1* | 0.903805508 | 0.015437651 | 0.045695156 | 1 |
| Obesity | macrophage | *GXYLT2* | 0.606453052 | 0.015549699 | 0.045875626 | 1 |
| Obesity | macrophage | *HPSE2* | 1.026336113 | 9.00E-13 | 3.14E-10 | 5.66E-09 |
| Obesity | macrophage | *BNC2* | 0.7539971 | 0.000430838 | 0.003495071 | 1 |
| Obesity | macrophage | *NOC3L* | 1.236743259 | 0.001385911 | 0.007986455 | 1 |
| Obesity | macrophage | *VANGL1* | 1.179720384 | 0.000702208 | 0.00499141 | 1 |
| Obesity | macrophage | *GLRX* | 1.284473674 | 0.000197248 | 0.001949844 | 1 |
| Obesity | macrophage | *GOLGB1* | 1.065067099 | 0.001957607 | 0.010230652 | 1 |
| Obesity | macrophage | *C1QA* | -0.465739445 | 0.005118248 | 0.020327497 | 1 |
| Obesity | macrophage | *NDNF* | 1.719356078 | 9.16E-07 | 5.76E-05 | 0.005761778 |
| Obesity | macrophage | *OLR1* | -0.963137876 | 0.005031004 | 0.020126943 | 1 |
| Obesity | macrophage | *GLIPR1L1* | 0.755623705 | 0.000726384 | 0.005108248 | 1 |
| Obesity | macrophage | *ARV1* | 1.14371894 | 0.000673785 | 0.004835717 | 1 |
| Obesity | macrophage | *CCDC36* | 1.202640725 | 2.92E-07 | 2.27E-05 | 0.001835255 |
| Obesity | macrophage | *THAP2* | 0.680312672 | 0.007296001 | 0.026208672 | 1 |
| Obesity | macrophage | *MST1* | -0.388962712 | 1.23E-05 | 0.000347425 | 0.077475679 |
| Obesity | macrophage | *GMPPB* | 1.442073164 | 7.54E-10 | 1.28E-07 | 4.74E-06 |
| Obesity | macrophage | *NUDT18* | 1.77702435 | 0.003199796 | 0.014514514 | 1 |
| Obesity | macrophage | *SCAI* | 1.186783486 | 0.001664123 | 0.009081895 | 1 |
| Obesity | macrophage | *C1orf100* | 3.446860908 | 9.86E-05 | 0.001199086 | 0.619927277 |
| Obesity | macrophage | *HOXB2* | 0.793656587 | 1.58E-05 | 0.00040914 | 0.099106444 |
| Obesity | macrophage | *CCS* | 1.331816639 | 0.000212224 | 0.002049538 | 1 |
| Obesity | macrophage | *SOCS7* | 1.520135039 | 0.001583969 | 0.008766211 | 1 |
| Obesity | macrophage | *FAM53A* | 1.502086158 | 0.00182396 | 0.009709767 | 1 |
| Obesity | macrophage | *CTU2* | 0.739969702 | 0.003252098 | 0.014698737 | 1 |
| Obesity | macrophage | *DDX23* | 1.24380994 | 0.00580797 | 0.022323431 | 1 |
| Obesity | macrophage | *LINGO2* | 1.844292817 | 1.48E-07 | 1.29E-05 | 0.000930204 |
| Obesity | macrophage | *BBS1* | 1.700841173 | 8.37E-07 | 5.54E-05 | 0.005260405 |
| Obesity | macrophage | *ZNF497* | 1.435277394 | 0.012429788 | 0.039075715 | 1 |
| Obesity | macrophage | *KLF14* | 7.067170707 | 5.38E-08 | 5.20E-06 | 0.000338199 |
| Obesity | macrophage | *ANGEL2* | 1.315563904 | 0.002000467 | 0.010351388 | 1 |
| Obesity | macrophage | *SH3PXD2B* | 0.342454345 | 0.004510846 | 0.01852808 | 1 |
| Obesity | macrophage | *FGFBP3* | 1.751661169 | 0.000902421 | 0.005991044 | 1 |
| Obesity | macrophage | *C4orf32* | 1.209021847 | 0.000293175 | 0.002686871 | 1 |
| Obesity | macrophage | *HRAS* | 1.178638665 | 0.00039133 | 0.003299993 | 1 |
| Obesity | macrophage | *SRP72* | 0.526306668 | 0.001150378 | 0.006981106 | 1 |
| Obesity | macrophage | *BTC* | 0.961573612 | 0.001329011 | 0.007772548 | 1 |
| Obesity | macrophage | *NLRP6* | -2.004142087 | 0.00425315 | 0.017802633 | 1 |
| Obesity | macrophage | *NDUFA11* | 1.379486087 | 0.006143191 | 0.023154822 | 1 |
| Obesity | macrophage | *FUT1* | 0.791372305 | 0.009280674 | 0.031453551 | 1 |
| Obesity | macrophage | *FBXW8* | 1.390452795 | 0.000798935 | 0.005447834 | 1 |
| Obesity | macrophage | *CHST2* | 3.558994713 | 0.000203543 | 0.001999493 | 1 |
| Obesity | macrophage | *ATR* | 0.933655248 | 0.005065256 | 0.020180774 | 1 |
| Obesity | macrophage | *CADM2* | 0.612851539 | 1.15E-06 | 6.77E-05 | 0.007249062 |

| | | | | | | |
|---|---|---|---|---|---|---|
| Obesity | macrophage | *PSMD2* | 0.550014562 | 0.00068971 | 0.004927507 | 1 |
| Obesity | macrophage | *GOLGA8A* | 0.924640171 | 5.31E-06 | 0.000206192 | 0.033403159 |
| Obesity | macrophage | *PHYHD1* | 0.663310104 | 0.003669709 | 0.015975945 | 1 |
| Obesity | macrophage | *ZNF519* | 1.225004808 | 0.000494259 | 0.003870478 | 1 |
| Obesity | macrophage | *TMEM9B* | 2.207246969 | 8.35E-07 | 5.54E-05 | 0.005248462 |
| Obesity | macrophage | *NRIP3* | -0.349606473 | 0.003745524 | 0.016195399 | 1 |
| Obesity | macrophage | *SCUBE2* | 0.65772002 | 0.006365205 | 0.023862876 | 1 |
| Obesity | macrophage | *PPP2R2D* | 0.511543907 | 0.014353417 | 0.043552092 | 1 |
| Obesity | macrophage | *LRRC25* | -0.174588989 | 0.016260328 | 0.047415902 | 1 |
| Obesity | macrophage | *UCP2* | -0.590221237 | 0.009110809 | 0.030978721 | 1 |
| Obesity | macrophage | *RPS6KB2* | 1.141863648 | 0.012044788 | 0.038264568 | 1 |
| Obesity | macrophage | *TOM1L2* | 1.139659132 | 0.002836916 | 0.013300291 | 1 |
| Obesity | macrophage | *TEX26* | 0.887134352 | 0.001567168 | 0.008716979 | 1 |
| Obesity | macrophage | *CTDNEP1* | 1.508734529 | 0.005711794 | 0.022071328 | 1 |
| Obesity | macrophage | *ETV4* | 0.643090119 | 0.001379752 | 0.007972889 | 1 |
| Obesity | macrophage | *LRRN1* | 2.302434093 | 0.000167238 | 0.001733943 | 1 |
| Obesity | macrophage | *IP6K1* | 0.880274682 | 0.003983247 | 0.016987986 | 1 |
| Obesity | macrophage | *SSNA1* | 2.235620808 | 0.001280952 | 0.007554732 | 1 |
| Obesity | macrophage | *GPX2* | 1.687747328 | 0.001031499 | 0.006557047 | 1 |
| Obesity | macrophage | *SMIM19* | 0.422039921 | 0.00112081 | 0.006867967 | 1 |
| Obesity | macrophage | *COX8A* | 2.098742776 | 0.001520985 | 0.008522665 | 1 |
| Obesity | macrophage | *B3GNT4* | 10.01722104 | 2.53E-05 | 0.000494987 | 0.158890842 |
| Obesity | macrophage | *PLA2G16* | 0.154961643 | 0.012733506 | 0.039749528 | 1 |
| Obesity | macrophage | *RNF152* | 0.975798112 | 0.008479243 | 0.029452485 | 1 |
| Obesity | macrophage | *ZNF843* | 2.324376352 | 0.000960438 | 0.006257277 | 1 |
| Obesity | macrophage | *C8orf59* | 1.357237439 | 9.80E-05 | 0.001194917 | 0.615907921 |
| Obesity | macrophage | *RUFY1* | 0.861767459 | 1.53E-05 | 0.000405213 | 0.09600315 |
| Obesity | macrophage | *IRX5* | 0.704386494 | 0.000752276 | 0.00523977 | 1 |
| Obesity | macrophage | *ANKLE2* | 1.398020114 | 0.001647052 | 0.009005362 | 1 |
| Obesity | macrophage | *GCNT4* | 3.596231588 | 2.90E-05 | 0.000534857 | 0.182101184 |
| Obesity | macrophage | *POLE* | 1.282602417 | 0.014522392 | 0.043853159 | 1 |
| Obesity | macrophage | *AP006621.1* | 0.728582846 | 1.23E-06 | 7.16E-05 | 0.007737365 |
| Obesity | macrophage | *MAN1B1* | 1.742542131 | 0.001647235 | 0.009005362 | 1 |
| Obesity | macrophage | *ZBTB38* | 0.85284743 | 0.00520579 | 0.020571212 | 1 |
| Obesity | macrophage | *C8orf31* | 2.172121897 | 1.29E-05 | 0.000359558 | 0.081260079 |
| Obesity | macrophage | *PPFIA3* | -0.459918846 | 0.008789806 | 0.030148123 | 1 |
| Obesity | macrophage | *MAGEF1* | 2.261262207 | 3.02E-05 | 0.000543887 | 0.189816705 |
| Obesity | macrophage | *PAWR* | 1.418288556 | 3.79E-05 | 0.000620889 | 0.238221517 |
| Obesity | macrophage | *NAP1L5* | 1.521408236 | 9.19E-05 | 0.001139978 | 0.577968914 |
| Obesity | macrophage | *RPLP2* | 1.115418197 | 0.003349357 | 0.014987476 | 1 |
| Obesity | macrophage | *C12orf54* | 1.158056365 | 0.015963568 | 0.046775194 | 1 |
| Obesity | macrophage | *CD163L1* | -0.209506459 | 0.010961699 | 0.035819233 | 1 |
| Obesity | macrophage | *CD151* | 1.354051172 | 3.45E-06 | 0.00015608 | 0.021695121 |
| Obesity | macrophage | *ZNF619* | 1.250844844 | 0.011445248 | 0.036938538 | 1 |
| Obesity | macrophage | *ZNF354C* | 1.916872151 | 8.05E-05 | 0.001041118 | 0.506315348 |
| Obesity | macrophage | *IMP3* | 1.624106203 | 0.001433204 | 0.008183973 | 1 |
| Obesity | macrophage | *ODF3B* | 0.696443553 | 0.014059783 | 0.042907248 | 1 |
| Obesity | macrophage | *C2orf73* | 1.968040955 | 1.97E-05 | 0.000454265 | 0.123858611 |
| Obesity | macrophage | *FAM26E* | 0.599467808 | 0.008895513 | 0.030418707 | 1 |
| Obesity | macrophage | *C2orf69* | 1.405626912 | 0.008710694 | 0.029974191 | 1 |

| | | | | | | |
|---|---|---|---|---|---|---|
| Obesity | macrophage | *GRAMD1C* | 1.756258877 | 0.001400291 | 0.008041095 | 1 |
| Obesity | macrophage | *ZNF454* | 0.433033322 | 0.005476269 | 0.021344886 | 1 |
| Obesity | macrophage | *SH2B1* | -1.127048769 | 3.73E-27 | 5.86E-24 | 2.35E-23 |
| Obesity | macrophage | *ZC3H12D* | -0.683999272 | 0.002388186 | 0.011702667 | 1 |
| Obesity | macrophage | *SH2D4B* | -1.884679723 | 0.006588397 | 0.024567764 | 1 |
| Obesity | macrophage | *PRSS36* | -0.45264671 | 0.000488718 | 0.003850341 | 1 |
| Obesity | macrophage | *AQP11* | 1.405499785 | 0.000378585 | 0.003207769 | 1 |
| Obesity | macrophage | *TMEM11* | 2.148106524 | 0.000953198 | 0.006242455 | 1 |
| Obesity | macrophage | *BEND3* | 2.684104306 | 0.001053554 | 0.006631305 | 1 |
| Obesity | macrophage | *LINC00843* | 2.536239267 | 0.001120118 | 0.006867967 | 1 |
| Obesity | macrophage | *COX14* | 0.865844619 | 0.000500416 | 0.003893704 | 1 |
| Obesity | macrophage | *ERBB4* | 1.019257205 | 9.95E-05 | 0.001207859 | 0.625676623 |
| Obesity | macrophage | *CSRNP3* | 0.839406317 | 0.01422787 | 0.043233745 | 1 |
| Obesity | macrophage | *RPP25* | -0.514003304 | 4.74E-05 | 0.000737654 | 0.298012273 |
| Obesity | macrophage | *STX19* | 1.719620099 | 1.13E-09 | 1.76E-07 | 7.11E-06 |
| Obesity | macrophage | *FAM219B* | 1.723021486 | 1.20E-05 | 0.000344993 | 0.075553554 |
| Obesity | macrophage | *GDPD4* | -0.975649446 | 0.000564802 | 0.004201901 | 1 |
| Obesity | macrophage | *MPI* | 1.219188582 | 2.32E-05 | 0.000485418 | 0.14562545 |
| Obesity | macrophage | *TRIM73* | -1.841001438 | 1.96E-12 | 6.15E-10 | 1.23E-08 |
| Obesity | macrophage | *TUFM* | 1.577454153 | 5.01E-25 | 6.30E-22 | 3.15E-21 |
| Obesity | macrophage | *CTC1* | 0.564159696 | 0.002611545 | 0.012476278 | 1 |
| Obesity | macrophage | *TRIML2* | -9.068548477 | 6.84E-05 | 0.000929432 | 0.430327033 |
| Obesity | macrophage | *SPTY2D1* | 1.384425087 | 0.00570503 | 0.022058749 | 1 |
| Obesity | macrophage | *ZNF664* | 1.353114416 | 0.008671073 | 0.029894594 | 1 |
| Obesity | macrophage | *NSUN7* | -0.184277727 | 0.002432521 | 0.011882874 | 1 |
| Obesity | macrophage | *EGR3* | -1.020914929 | 0.006438863 | 0.024124633 | 1 |
| Obesity | macrophage | *FJX1* | 0.468693936 | 0.015927445 | 0.046705152 | 1 |
| Obesity | macrophage | *CIITA* | -0.582328612 | 0.003670443 | 0.015975945 | 1 |
| Obesity | macrophage | *ZBTB42* | 2.255568316 | 1.89E-05 | 0.000447197 | 0.118954332 |
| Obesity | macrophage | *MAF1* | 1.992113839 | 0.005303901 | 0.020827997 | 1 |
| Obesity | macrophage | *PCED1B* | 0.670914162 | 0.005943528 | 0.022716779 | 1 |
| Obesity | macrophage | *CDH5* | 0.398882471 | 0.000793288 | 0.005438824 | 1 |
| Obesity | macrophage | *CITED4* | 0.646952679 | 0.015000133 | 0.044886166 | 1 |
| Obesity | macrophage | *B3GNT3* | 1.248233775 | 0.002022356 | 0.0104219 | 1 |
| Obesity | macrophage | *SEPT1* | 2.200143921 | 0.000179918 | 0.001821488 | 1 |
| Obesity | macrophage | *TDRP* | 1.123283115 | 0.000501444 | 0.003896887 | 1 |
| Obesity | macrophage | *HCLS1* | -0.489780528 | 0.012345173 | 0.038930995 | 1 |
| Obesity | macrophage | *SSTR2* | 3.173762044 | 1.83E-05 | 0.000441618 | 0.115041537 |
| Obesity | macrophage | *SHISA2* | 17.43904392 | 5.48E-05 | 0.000804841 | 0.344472105 |
| Obesity | macrophage | *SRSF8* | 1.822256609 | 0.002700484 | 0.012803881 | 1 |
| Obesity | macrophage | *CAPS2* | 1.230853143 | 0.000413217 | 0.003422789 | 1 |
| Obesity | macrophage | *KCTD2* | 1.838158044 | 0.001161794 | 0.007036799 | 1 |
| Obesity | macrophage | *ZFP82* | 1.248785379 | 0.002283504 | 0.011269706 | 1 |
| Obesity | macrophage | *FKRP* | 0.282276773 | 0.003726714 | 0.016136259 | 1 |
| Obesity | macrophage | *SLC26A11* | -0.697842124 | 0.007151765 | 0.025990258 | 1 |
| Obesity | macrophage | *POLR2A* | 1.029789598 | 0.004787028 | 0.019379293 | 1 |
| Obesity | macrophage | *SLC25A41* | 3.274906855 | 0.002221497 | 0.011075776 | 1 |
| Obesity | macrophage | *ZNF322* | -0.898653207 | 1.64E-06 | 8.74E-05 | 0.010313342 |
| Obesity | macrophage | *CCL13* | -0.439096069 | 0.00418682 | 0.01759528 | 1 |
| Obesity | macrophage | *ACBD4* | 1.262011136 | 0.000932967 | 0.006148392 | 1 |

| | | | | | | |
|---|---|---|---|---|---|---|
| Obesity | macrophage | *MRPS23* | 1.607307392 | 0.012090657 | 0.03839089 | 1 |
| Obesity | macrophage | *GPR135* | 0.368728338 | 0.006793699 | 0.025124699 | 1 |
| Obesity | macrophage | *ZFP41* | 0.378946369 | 0.001980162 | 0.010303093 | 1 |
| Obesity | macrophage | *ATG9B* | 3.837989166 | 3.21E-05 | 0.000554992 | 0.20181799 |
| Obesity | macrophage | *ZBTB20* | 0.69462881 | 0.00471321 | 0.019130365 | 1 |
| Obesity | macrophage | *AMIGO1* | 0.264094168 | 0.003761518 | 0.016242213 | 1 |
| Obesity | macrophage | *CLDN7* | -0.479398304 | 1.53E-05 | 0.000405213 | 0.096494995 |
| Obesity | macrophage | *SNRPE* | 1.822406278 | 0.000346666 | 0.00303995 | 1 |
| Obesity | macrophage | *FAM89A* | 0.37512307 | 0.003410884 | 0.015187132 | 1 |
| Obesity | macrophage | *HHIPL1* | -0.933902215 | 0.002680033 | 0.012745362 | 1 |
| Obesity | macrophage | *SYNM* | 1.690008326 | 2.28E-05 | 0.000484376 | 0.143326822 |
| Obesity | macrophage | *KBTBD3* | 1.810486802 | 0.001983976 | 0.010303093 | 1 |
| Obesity | macrophage | *GLRX5* | 1.345936601 | 0.002063399 | 0.010569311 | 1 |
| Obesity | macrophage | *CAV3* | -0.21221689 | 0.011383908 | 0.036854084 | 1 |
| Obesity | macrophage | *MFSD5* | 0.668977159 | 0.015484173 | 0.045811292 | 1 |
| Obesity | macrophage | *ADI1* | 1.042523973 | 0.012379161 | 0.038991878 | 1 |
| Obesity | macrophage | *SPNS3* | 1.749051103 | 0.000255261 | 0.002388133 | 1 |
| Obesity | macrophage | *CLEC4G* | -0.542226242 | 0.000443619 | 0.003571103 | 1 |
| Obesity | macrophage | *SKA2* | 1.635282344 | 0.000685614 | 0.004903817 | 1 |
| Obesity | macrophage | *BRICD5* | -0.393028959 | 0.002141415 | 0.010824534 | 1 |
| Obesity | macrophage | *TSKU* | 1.573190707 | 7.00E-06 | 0.000248772 | 0.044032709 |
| Obesity | macrophage | *COL18A1* | 0.568626332 | 0.00079082 | 0.005427825 | 1 |
| Obesity | macrophage | *C12orf60* | 0.824540111 | 0.009288667 | 0.031453551 | 1 |
| Obesity | macrophage | *LYSMD4* | 0.406981074 | 0.000195567 | 0.001936268 | 1 |
| Obesity | macrophage | *BEGAIN* | 3.219381393 | 3.10E-05 | 0.000548638 | 0.194766379 |
| Obesity | macrophage | *CSMD1* | 0.708460248 | 0.00355537 | 0.015637225 | 1 |
| Obesity | macrophage | *FANCF* | 3.10432799 | 0.001465476 | 0.00831075 | 1 |
| Obesity | macrophage | *C2CD4C* | 0.852580077 | 0.000519495 | 0.003978158 | 1 |
| Obesity | macrophage | *CTNNA3* | 0.466297976 | 0.00964978 | 0.032390906 | 1 |
| Obesity | macrophage | *PLGLB1* | 0.436279926 | 0.011758469 | 0.037640271 | 1 |
| Obesity | macrophage | *CCBE1* | 0.463874619 | 4.90E-05 | 0.000749385 | 0.308112147 |
| Obesity | macrophage | *GPR132* | -0.61018584 | 0.001264691 | 0.007493153 | 1 |
| Obesity | macrophage | *PSMG1* | 0.321300988 | 6.83E-06 | 0.000245531 | 0.042968007 |
| Obesity | macrophage | *SERHL2* | 1.999528753 | 4.31E-06 | 0.000178092 | 0.027084504 |
| Obesity | macrophage | *SFXN4* | 0.979239067 | 0.008339923 | 0.029081029 | 1 |
| Obesity | macrophage | *RP1L1* | 1.417125274 | 2.13E-11 | 5.35E-09 | 1.34E-07 |
| Obesity | macrophage | *FAM19A1* | 1.417928268 | 3.00E-05 | 0.000542608 | 0.188340995 |
| Obesity | macrophage | *BMP8A* | 2.798224043 | 2.10E-12 | 6.30E-10 | 1.32E-08 |
| Obesity | macrophage | *CMTM4* | 0.738905099 | 0.006610858 | 0.024622314 | 1 |
| Obesity | macrophage | *CBX6* | 2.069587878 | 0.00027084 | 0.002511459 | 1 |
| Obesity | macrophage | *AIFM3* | -0.841155349 | 0.001178373 | 0.007103002 | 1 |
| Obesity | macrophage | *KCTD16* | 1.057183942 | 1.12E-05 | 0.000332862 | 0.070566835 |
| Obesity | macrophage | *ANKRD45* | 3.275963897 | 0.000221827 | 0.002122734 | 1 |
| Obesity | macrophage | *MAATS1* | 0.480883594 | 0.001498458 | 0.008434025 | 1 |
| Obesity | macrophage | *PP2D1* | 0.699203033 | 0.005298846 | 0.020821152 | 1 |
| Obesity | macrophage | *TMEM236* | -0.731296574 | 0.000547949 | 0.00412076 | 1 |
| Obesity | macrophage | *UQCR10* | 1.437239481 | 0.007287144 | 0.026194555 | 1 |
| Obesity | macrophage | *EIF3C* | 1.203398672 | 1.72E-27 | 3.60E-24 | 1.08E-23 |
| Obesity | macrophage | *CNTN2* | 3.41152517 | 0.000248989 | 0.002350444 | 1 |
| Obesity | macrophage | *CRELD2* | 1.332792207 | 0.003724005 | 0.016136259 | 1 |

| | | | | | | |
|---|---|---|---|---|---|---|
| Obesity | macrophage | *PLA2G6* | 0.690923619 | 2.19E-09 | 3.14E-07 | 1.38E-05 |
| Obesity | macrophage | *TOP1MT* | -0.264304292 | 0.017225409 | 0.049654355 | 1 |
| Obesity | macrophage | *THAP7* | 1.775391831 | 0.010606618 | 0.034876467 | 1 |
| Obesity | macrophage | *CCR10* | 1.389076841 | 0.00181681 | 0.009679903 | 1 |
| Obesity | macrophage | *WDR27* | 1.20032018 | 0.000204836 | 0.002002802 | 1 |
| Obesity | macrophage | *HDDC3* | 0.582866956 | 0.010284657 | 0.03412118 | 1 |
| Obesity | macrophage | *SEPT5* | 2.871221511 | 3.39E-05 | 0.000571658 | 0.213109483 |
| Obesity | macrophage | *ATL3* | 0.741050159 | 0.00295253 | 0.013679112 | 1 |
| Obesity | macrophage | *NDUFA12* | 1.725257435 | 0.001615902 | 0.008876779 | 1 |
| Obesity | macrophage | *H1FX* | 1.746407295 | 7.45E-06 | 0.000255935 | 0.04683607 |
| Obesity | macrophage | *ZFP90* | 1.492751628 | 0.000816793 | 0.005527642 | 1 |
| Obesity | macrophage | *FAM227A* | 0.414386684 | 0.002597193 | 0.012445544 | 1 |
| Obesity | macrophage | *SORCS2* | 1.032451035 | 0.000365787 | 0.003133111 | 1 |
| Obesity | macrophage | *MAFF* | -0.705772485 | 0.013317232 | 0.041102327 | 1 |
| Obesity | macrophage | *BRF1* | 1.560302768 | 0.000692944 | 0.004939386 | 1 |
| Obesity | macrophage | *ADSSL1* | 1.981851703 | 0.000103088 | 0.001243986 | 0.6481165 |
| Obesity | macrophage | *IFITM2* | 0.656520424 | 0.014067274 | 0.042907248 | 1 |
| Obesity | macrophage | *GP1BA* | 0.864011517 | 1.08E-06 | 6.59E-05 | 0.006786994 |
| Obesity | macrophage | *KLHL33* | 0.424179856 | 0.000797504 | 0.005447834 | 1 |
| Obesity | macrophage | *HS6ST3* | 1.388020253 | 1.53E-05 | 0.000405213 | 0.096133734 |
| Obesity | macrophage | *MAPK11* | 1.028437129 | 0.001717386 | 0.009307938 | 1 |
| Obesity | macrophage | *AHNAK2* | 1.711999379 | 1.00E-05 | 0.00030506 | 0.063147342 |
| Obesity | macrophage | *SP1* | 0.730082982 | 0.001559604 | 0.008692581 | 1 |
| Obesity | macrophage | *PCGF3* | 1.071016404 | 0.000616427 | 0.004516873 | 1 |
| Obesity | macrophage | *PBX1* | 0.806844816 | 0.000156 | 0.001647833 | 0.980771954 |
| Obesity | macrophage | *PMEL* | 0.387655166 | 0.005408082 | 0.021131518 | 1 |
| Obesity | macrophage | *SRL* | 0.281036607 | 0.00298635 | 0.013795137 | 1 |
| Obesity | macrophage | *SLC52A2* | 2.368353239 | 0.001791727 | 0.009586885 | 1 |
| Obesity | macrophage | *ZNF829* | 0.480328896 | 0.017283319 | 0.049752852 | 1 |
| Obesity | macrophage | *RTN4RL1* | 1.897849665 | 0.000210783 | 0.002038763 | 1 |
| Obesity | macrophage | *RASA3* | 0.624208969 | 0.011863781 | 0.037919468 | 1 |
| Obesity | macrophage | *ZNF566* | 0.757489488 | 0.015329446 | 0.045524905 | 1 |
| Obesity | macrophage | *ZNF284* | 3.91322687 | 0.006699911 | 0.024865608 | 1 |
| Obesity | macrophage | *GSAP* | -0.461538246 | 0.004861187 | 0.019616354 | 1 |
| Obesity | macrophage | *C2orf76* | 0.65759168 | 0.005566753 | 0.021630518 | 1 |
| Obesity | macrophage | *WWOX* | 0.465745389 | 0.013279872 | 0.041027299 | 1 |
| Obesity | macrophage | *CYP4Z1* | 0.996584642 | 0.000250619 | 0.002351705 | 1 |
| Obesity | macrophage | *CCDC84* | 1.080013268 | 0.008675887 | 0.029894594 | 1 |
| Obesity | macrophage | *BPIFB4* | 22.54903838 | 0.000210699 | 0.002038763 | 1 |
| Obesity | macrophage | *NBPF12* | 0.985460561 | 0.004712105 | 0.019130365 | 1 |
| Obesity | macrophage | *CYP4X1* | 0.641068591 | 0.004496254 | 0.018522585 | 1 |
| Obesity | macrophage | *GLDN* | 0.596572742 | 0.000285611 | 0.002629042 | 1 |
| Obesity | macrophage | *BTN3A2* | -0.31408972 | 8.45E-11 | 1.83E-08 | 5.32E-07 |
| Obesity | macrophage | *PCLO* | 0.349811159 | 0.01589781 | 0.046640006 | 1 |
| Obesity | macrophage | *MIR22HG* | 0.6385045 | 0.006245113 | 0.023454614 | 1 |
| Obesity | macrophage | *BCR* | 0.930238964 | 5.62E-05 | 0.000820451 | 0.353614233 |
| Obesity | macrophage | *HYAL3* | -0.390500268 | 6.20E-05 | 0.000876317 | 0.38996087 |
| Obesity | macrophage | *ZSCAN30* | 1.276060758 | 0.007077729 | 0.0258257 | 1 |
| Obesity | macrophage | *TRABD2A* | 0.285384429 | 0.000412385 | 0.003421767 | 1 |
| Obesity | macrophage | *TNFRSF18* | 7.525658568 | 1.83E-05 | 0.000441618 | 0.115279841 |

| | | | | | | |
|---|---|---|---|---|---|---|
| Obesity | macrophage | *EFCAB6* | 0.761174002 | 0.01153071 | 0.037157136 | 1 |
| Obesity | macrophage | *KANK3* | 1.449795077 | 0.000109601 | 0.00129767 | 0.689062955 |
| Obesity | macrophage | *CYP4A11* | 0.408346877 | 0.01614098 | 0.047155364 | 1 |
| Obesity | macrophage | *ENTPD5* | 1.539655167 | 0.000773672 | 0.005345139 | 1 |
| Obesity | macrophage | *MRPS21* | 0.918501687 | 0.001810978 | 0.009657011 | 1 |
| Obesity | macrophage | *ANGPTL5* | 2.048469531 | 9.03E-05 | 0.001129168 | 0.56797166 |
| Obesity | macrophage | *FPR3* | -0.306155494 | 0.00195651 | 0.010230652 | 1 |
| Obesity | macrophage | *IFT140* | 1.365223183 | 0.000706798 | 0.005015397 | 1 |
| Obesity | macrophage | *TLR5* | -0.609151175 | 0.013250341 | 0.040960272 | 1 |
| Obesity | macrophage | *PLEKHN1* | 0.484073237 | 0.011404811 | 0.036871515 | 1 |
| Obesity | macrophage | *ZNF385C* | 2.091332724 | 0.002912664 | 0.013582552 | 1 |
| Obesity | macrophage | *TET3* | -0.461933363 | 0.009366883 | 0.031678101 | 1 |
| Obesity | macrophage | *FANCA* | -0.391706648 | 0.011349584 | 0.036780844 | 1 |
| Obesity | macrophage | *TMEM220* | 0.898107906 | 0.016137982 | 0.047155364 | 1 |
| Obesity | macrophage | *GFRAL* | 0.449466972 | 0.010487434 | 0.034611286 | 1 |
| Obesity | macrophage | *ZSCAN23* | 0.179522188 | 0.000286604 | 0.002634328 | 1 |
| Obesity | macrophage | *FAM166A* | 5.890704188 | 2.37E-05 | 0.000488143 | 0.149087535 |
| Obesity | macrophage | *ZNF626* | 0.989254985 | 0.001024016 | 0.0065294 | 1 |
| Obesity | macrophage | *SMTNL2* | 7.295147805 | 4.14E-05 | 0.000669716 | 0.260519503 |
| Obesity | macrophage | *ZC3H6* | 1.108458227 | 0.001132988 | 0.006922349 | 1 |
| Obesity | macrophage | *HYKK* | 1.175088999 | 1.07E-06 | 6.57E-05 | 0.00670064 |
| Obesity | macrophage | *CENPP* | 0.455016877 | 8.87E-07 | 5.74E-05 | 0.005578992 |
| Obesity | macrophage | *COL25A1* | 0.526129013 | 0.01231739 | 0.038884349 | 1 |
| Obesity | macrophage | *SRSF10* | 1.080662749 | 0.001984941 | 0.010303093 | 1 |
| Obesity | macrophage | *FAM72B* | -1.394401624 | 0.000769784 | 0.00532413 | 1 |
| Obesity | macrophage | *GOLGA8M* | 0.691462648 | 0.014938552 | 0.044744487 | 1 |
| Obesity | macrophage | *S100A16* | 1.24517324 | 0.000103559 | 0.001247276 | 0.651077813 |
| Obesity | macrophage | *SKOR1* | 1.891185977 | 0.013712027 | 0.041990994 | 1 |
| Obesity | macrophage | *ZDHHC11* | 0.669362873 | 0.004511928 | 0.01852808 | 1 |
| Obesity | macrophage | *BEND4* | 3.069152289 | 4.44E-05 | 0.000702455 | 0.278874716 |
| Obesity | macrophage | *FBF1* | 0.29634775 | 3.79E-05 | 0.000620889 | 0.238421466 |
| Obesity | macrophage | *FAM196A* | 1.235576812 | 4.65E-05 | 0.000728699 | 0.292208153 |
| Obesity | macrophage | *C9orf152* | 3.225415526 | 2.06E-05 | 0.000463953 | 0.129357607 |
| Obesity | macrophage | *ADAT2* | 0.731218946 | 0.003071547 | 0.014116093 | 1 |
| Obesity | macrophage | *ALKBH2* | 0.623565032 | 0.005246563 | 0.020667381 | 1 |
| Obesity | macrophage | *PLAC9* | 0.636266081 | 0.000146141 | 0.001575829 | 0.918785495 |
| Obesity | macrophage | *FAM47E* | 0.433804474 | 1.85E-05 | 0.000441618 | 0.116145517 |
| Obesity | macrophage | *S100A13* | 1.493118051 | 8.57E-05 | 0.001083923 | 0.53870956 |
| Obesity | macrophage | *C15orf61* | 1.672618008 | 0.000541379 | 0.004081114 | 1 |
| Obesity | macrophage | *SLC35E2B* | 0.342172401 | 0.003083271 | 0.014140343 | 1 |
| Obesity | macrophage | *BLOC1S2* | -0.236749607 | 0.012179048 | 0.038593587 | 1 |
| Obesity | macrophage | *AKR1C3* | 0.849594996 | 0.000430155 | 0.003494041 | 1 |
| Obesity | macrophage | *HRCT1* | 0.713944048 | 0.000889063 | 0.005908605 | 1 |
| Obesity | macrophage | *MPHOSPH8* | 0.860256022 | 0.007909712 | 0.027890273 | 1 |
| Obesity | macrophage | *SIRPB2* | -0.459376867 | 0.001063793 | 0.006668061 | 1 |
| Obesity | macrophage | *PPIA* | 1.428737215 | 0.001616655 | 0.008876779 | 1 |
| Obesity | macrophage | *STK31* | 2.674035533 | 0.004448866 | 0.018414132 | 1 |
| Obesity | macrophage | *ZKSCAN7* | 1.239638106 | 0.01675712 | 0.048495262 | 1 |
| Obesity | macrophage | *ZNF565* | 0.843693181 | 0.000434083 | 0.003516813 | 1 |
| Obesity | macrophage | *NTNG2* | 2.205527167 | 0.001101107 | 0.006798282 | 1 |

| | | | | | | |
|---|---|---|---|---|---|---|
| Obesity | macrophage | *WDR5* | 1.209418695 | 0.014562935 | 0.043870231 | 1 |
| Obesity | macrophage | *LONP1* | 0.59168462 | 0.003363607 | 0.014997871 | 1 |
| Obesity | macrophage | *ASB13* | 2.02161958 | 0.000951814 | 0.006239888 | 1 |
| Obesity | macrophage | *SLC35F1* | 1.058351825 | 0.005233488 | 0.020651099 | 1 |
| Obesity | macrophage | *INCA1* | 2.468906694 | 1.40E-08 | 1.72E-06 | 8.77E-05 |
| Obesity | macrophage | *ZNF605* | 1.832207152 | 4.78E-05 | 0.000739028 | 0.300317782 |
| Obesity | macrophage | *RFX8* | 0.618559334 | 0.004849483 | 0.019594279 | 1 |
| Obesity | macrophage | *MYL6B* | 1.437511921 | 0.002795166 | 0.013124127 | 1 |
| Obesity | macrophage | *SULT1A1* | -0.563920276 | 2.53E-07 | 2.04E-05 | 0.001592596 |
| Obesity | macrophage | *C1orf186* | 0.986033116 | 0.013598444 | 0.041758525 | 1 |
| Obesity | macrophage | *SPTSSB* | 1.820325115 | 0.002759266 | 0.01303344 | 1 |
| Obesity | macrophage | *ADH1B* | 1.103829713 | 2.07E-05 | 0.000465185 | 0.130335159 |
| Obesity | macrophage | *HRH1* | -0.390469285 | 0.007119683 | 0.025918616 | 1 |
| Obesity | macrophage | *ZNF502* | 0.447572211 | 0.007118765 | 0.025918616 | 1 |
| Obesity | macrophage | *TECPR2* | 1.260245978 | 0.004121921 | 0.017392293 | 1 |
| Obesity | macrophage | *FAM180B* | -0.382499431 | 0.01180949 | 0.037765139 | 1 |
| Obesity | macrophage | *ERI2* | 0.867723748 | 0.006011251 | 0.022863117 | 1 |
| Obesity | macrophage | *TOMM7* | 1.480311336 | 0.002775948 | 0.013067173 | 1 |
| Obesity | macrophage | *TRPV1* | 0.752933956 | 0.000177703 | 0.001808801 | 1 |
| Obesity | macrophage | *ZNF431* | 0.927837502 | 0.004582235 | 0.018718981 | 1 |
| Obesity | macrophage | *NF1* | 0.65961135 | 0.013053974 | 0.04052856 | 1 |
| Obesity | macrophage | *ZNF239* | 0.401426979 | 0.0013516 | 0.00788266 | 1 |
| Obesity | macrophage | *C6orf106* | 0.70884949 | 1.04E-09 | 1.67E-07 | 6.52E-06 |
| Obesity | macrophage | *ZFP28* | 2.0884596 | 1.91E-05 | 0.000450217 | 0.120239817 |
| Obesity | macrophage | *LAMB3* | 1.012008005 | 0.007898915 | 0.027867834 | 1 |
| Obesity | macrophage | *KPNA5* | 1.154429312 | 0.004506776 | 0.01852808 | 1 |
| Obesity | macrophage | *HCAR1* | 0.751648425 | 3.36E-06 | 0.000155177 | 0.021104121 |
| Obesity | macrophage | *FAM3C* | 1.411668544 | 1.55E-05 | 0.000405213 | 0.097251217 |
| Obesity | macrophage | *ZNF585A* | 0.437742787 | 0.005296239 | 0.020821152 | 1 |
| Obesity | macrophage | *FUT11* | 1.552409817 | 6.36E-06 | 0.000232518 | 0.03999312 |
| Obesity | macrophage | *ZNF138* | 1.086895769 | 0.000146131 | 0.001575829 | 0.918724672 |
| Obesity | macrophage | *ZMYM1* | 1.159376496 | 0.01249279 | 0.039212267 | 1 |
| Obesity | macrophage | *GAL3ST4* | 1.331036879 | 0.000204407 | 0.002002802 | 1 |
| Obesity | macrophage | *SLC25A29* | 1.100857538 | 2.50E-05 | 0.000494787 | 0.157342182 |
| Obesity | macrophage | *ZNF682* | 1.293443796 | 0.001066983 | 0.006674749 | 1 |
| Obesity | macrophage | *SULT1A2* | 0.544348997 | 9.80E-31 | 3.08E-27 | 6.16E-27 |
| Obesity | macrophage | *ENTPD4* | 0.81104962 | 0.004158081 | 0.017497893 | 1 |
| Obesity | macrophage | *FAM110D* | 1.557432287 | 7.56E-05 | 0.000994076 | 0.475168185 |
| Obesity | macrophage | *KANK2* | 1.641794329 | 7.14E-05 | 0.000965911 | 0.449148727 |
| Obesity | macrophage | *CCL4L2* | -0.517160203 | 4.17E-08 | 4.30E-06 | 0.00026243 |
| Obesity | macrophage | *GTF2E2* | 1.492333862 | 6.77E-06 | 0.000244627 | 0.042565061 |
| Obesity | macrophage | *ZNF165* | -0.69360585 | 0.000115875 | 0.001351584 | 0.728503555 |
| Obesity | macrophage | *MRPL21* | -0.251686722 | 0.00058828 | 0.00435632 | 1 |
| Obesity | macrophage | *ZNF517* | 2.72341265 | 3.06E-05 | 0.000546074 | 0.192498771 |
| Obesity | macrophage | *C5AR1* | -0.537772895 | 0.001087616 | 0.006736787 | 1 |
| Obesity | macrophage | *SLC2A10* | 0.604136422 | 0.008952624 | 0.03055654 | 1 |
| Obesity | macrophage | *RAB40C* | 1.740284122 | 0.000510192 | 0.003935678 | 1 |
| Obesity | macrophage | *HOXA4* | 1.376588118 | 0.000479379 | 0.003795788 | 1 |
| Obesity | macrophage | *CCDC154* | -0.967682923 | 0.006889313 | 0.025373819 | 1 |
| Obesity | macrophage | *MFAP5* | 1.088056289 | 0.000509083 | 0.003935678 | 1 |

| | | | | | | |
|---|---|---|---|---|---|---|
| Obesity | macrophage | *FAM114A1* | 1.189212444 | 0.000173111 | 0.001782107 | 1 |
| Obesity | macrophage | *RPS26* | -0.230233917 | 0.00015456 | 0.001638644 | 0.971716086 |
| Obesity | macrophage | *ZNF780A* | -0.308825849 | 1.82E-05 | 0.000441618 | 0.114609993 |
| Obesity | macrophage | *AC122129.1* | 2.595224612 | 2.67E-05 | 0.000515008 | 0.167892478 |
| Obesity | macrophage | *ZNF677* | 2.078912766 | 4.49E-05 | 0.000709767 | 0.28259273 |
| Obesity | macrophage | *ADH4* | -0.210257435 | 0.000635004 | 0.004631406 | 1 |
| Obesity | macrophage | *SOWAHC* | 2.895558014 | 0.001574472 | 0.008728633 | 1 |
| Obesity | macrophage | *MIER1* | 0.92700701 | 0.011250242 | 0.036534232 | 1 |
| Obesity | macrophage | *SVIP* | 1.882973247 | 0.000139635 | 0.001529414 | 0.87788368 |
| Obesity | macrophage | *DDRGK1* | 1.825912225 | 0.000727466 | 0.005110143 | 1 |
| Obesity | macrophage | *QRICH1* | 0.436540228 | 0.001269816 | 0.007510193 | 1 |
| Obesity | macrophage | *DDX42* | 0.900331183 | 0.001392258 | 0.008008348 | 1 |
| Obesity | macrophage | *SLC29A3* | -0.426991538 | 0.012803786 | 0.039949085 | 1 |
| Obesity | macrophage | *ANTXRL* | 26.02637273 | 9.80E-05 | 0.001194917 | 0.615854092 |
| Obesity | macrophage | *STYX* | 1.4437432 | 0.006175704 | 0.02324949 | 1 |
| Obesity | macrophage | *ZKSCAN8* | 1.684095454 | 2.36E-08 | 2.60E-06 | 0.000148255 |
| Obesity | macrophage | *ZNF813* | 1.261129317 | 0.013408883 | 0.041357287 | 1 |
| Obesity | macrophage | *GFPT1* | 1.409452751 | 3.44E-06 | 0.00015608 | 0.021608517 |
| Obesity | macrophage | *ZNF568* | 0.742247982 | 0.013412996 | 0.041357287 | 1 |
| Obesity | macrophage | *SH3BGRL2* | 0.480046412 | 0.000714943 | 0.005050387 | 1 |
| Obesity | macrophage | *TMA16* | 1.455893808 | 0.000761635 | 0.00527938 | 1 |
| Obesity | macrophage | *MAFK* | 1.635846441 | 0.008499554 | 0.029490449 | 1 |
| Obesity | macrophage | *ARC* | -2.940544499 | 0.000207426 | 0.002016776 | 1 |
| Obesity | macrophage | *TLK1* | 0.901422739 | 0.011966666 | 0.038139587 | 1 |
| Obesity | macrophage | *LRBA* | 0.891233819 | 0.005155623 | 0.020424323 | 1 |
| Obesity | macrophage | *MMP17* | 1.995653216 | 2.75E-05 | 0.000521707 | 0.173206723 |
| Obesity | macrophage | *CCDC69* | 0.610988428 | 0.012194322 | 0.038622519 | 1 |
| Obesity | macrophage | *TTC37* | 0.779231991 | 0.014729315 | 0.044286562 | 1 |
| Obesity | macrophage | *TUSC1* | 2.632318412 | 2.38E-05 | 0.000488143 | 0.149421933 |
| Obesity | macrophage | *FAN1* | 1.635045533 | 0.000347338 | 0.00303995 | 1 |
| Obesity | macrophage | *ANKRD13B* | 0.462489352 | 0.000120049 | 0.001382323 | 0.754748265 |
| Obesity | macrophage | *ECI2* | 1.583673404 | 0.000220375 | 0.002115259 | 1 |
| Obesity | macrophage | *RPL10A* | 0.566759872 | 0.01014433 | 0.033780404 | 1 |
| Obesity | macrophage | *APCDD1L* | 2.137241112 | 2.08E-05 | 0.000465185 | 0.130717038 |
| Obesity | macrophage | *RASSF9* | 1.513926392 | 2.06E-05 | 0.000463953 | 0.129442823 |
| Obesity | macrophage | *TMEM184B* | 1.175615659 | 0.001759954 | 0.009491362 | 1 |
| Obesity | macrophage | *SCAMP5* | -0.331317117 | 0.005921229 | 0.022685417 | 1 |
| Obesity | macrophage | *SFT2D1* | 1.141024505 | 0.010919378 | 0.035718068 | 1 |
| Obesity | macrophage | *DENND4B* | -0.550076011 | 0.015351986 | 0.045548814 | 1 |
| Obesity | macrophage | *TSEN15* | 1.560998771 | 0.00123217 | 0.007341856 | 1 |
| Obesity | macrophage | *PRMT6* | -2.707421824 | 0.002567649 | 0.0123416 | 1 |
| Obesity | macrophage | *RASGEF1A* | 5.264153723 | 7.54E-06 | 0.000257628 | 0.047403529 |
| Obesity | macrophage | *APRT* | 2.168433423 | 0.000556757 | 0.004172026 | 1 |
| Obesity | macrophage | *ZFP2* | 0.814563681 | 0.013523494 | 0.041616351 | 1 |
| Obesity | macrophage | *MFAP3L* | 2.028301119 | 5.09E-06 | 0.000201204 | 0.031991488 |
| Obesity | macrophage | *AP001007.1* | 3.94963999 | 3.86E-05 | 0.000628283 | 0.242517164 |
| Obesity | macrophage | *AC105020.1* | 2.080853373 | 0.000292318 | 0.002682925 | 1 |
| Obesity | macrophage | *EFCAB2* | 0.990990455 | 0.002062667 | 0.010569311 | 1 |
| Obesity | macrophage | *ECT2L* | 0.676690424 | 0.00668914 | 0.024860184 | 1 |
| Obesity | macrophage | *CENPW* | 1.288454972 | 2.33E-06 | 0.000118969 | 0.01463317 |

| | | | | | | |
|---|---|---|---|---|---|---|
| Obesity | macrophage | *HIST2H2BF* | -2.897357139 | 0.00622591 | 0.023410466 | 1 |
| Obesity | macrophage | *RBM20* | 0.834512773 | 0.014054892 | 0.042907248 | 1 |
| Obesity | macrophage | *PCMTD2* | 1.204381885 | 0.001326864 | 0.007767219 | 1 |
| Obesity | macrophage | *SYS1* | 2.609516484 | 7.40E-05 | 0.00098578 | 0.465287989 |
| Obesity | macrophage | *SYT15* | 1.068612426 | 0.000115361 | 0.001350604 | 0.725274186 |
| Obesity | macrophage | *ZDBF2* | 0.967762891 | 0.001028754 | 0.006551234 | 1 |
| Obesity | macrophage | *HLA-DRA* | -0.348923145 | 0.002181038 | 0.010943484 | 1 |
| Obesity | macrophage | *NOTCH4* | -0.086142237 | 3.68E-08 | 3.92E-06 | 0.000231559 |
| Obesity | macrophage | *AGER* | 0.631173095 | 0.000522888 | 0.003992817 | 1 |
| Obesity | macrophage | *MRPL38* | -0.442139565 | 0.006748462 | 0.025001522 | 1 |
| Obesity | macrophage | *SKIV2L* | -0.257548526 | 1.27E-06 | 7.20E-05 | 0.007995752 |
| Obesity | macrophage | *NELFE* | 0.788156875 | 5.58E-07 | 4.03E-05 | 0.00350587 |
| Obesity | macrophage | *NFKBIL1* | 0.556152716 | 0.005772736 | 0.022224856 | 1 |
| Obesity | macrophage | *HLA-C* | 0.321792187 | 0.001784772 | 0.00956595 | 1 |
| Obesity | macrophage | *POU5F1* | 0.221283835 | 4.37E-07 | 3.27E-05 | 0.002744796 |
| Obesity | macrophage | *CCHCR1* | 0.408821112 | 2.25E-08 | 2.53E-06 | 0.000141757 |
| Obesity | macrophage | *ABCF1* | 1.022699512 | 1.12E-06 | 6.71E-05 | 0.007044955 |
| Obesity | macrophage | *HLA-E* | 1.196640834 | 4.07E-15 | 1.83E-12 | 2.56E-11 |
| Obesity | macrophage | *ZFP57* | 0.31760493 | 1.47E-08 | 1.77E-06 | 9.21E-05 |
| Obesity | macrophage | *ATXN2* | 0.957325275 | 0.00042628 | 0.0034785 | 1 |
| Obesity | macrophage | *FAM71F2* | 2.115695335 | 0.000106401 | 0.001266847 | 0.668945524 |
| Obesity | macrophage | *PRR13* | -0.739292003 | 0.01399128 | 0.042742067 | 1 |
| Obesity | macrophage | *MT1A* | 2.242624969 | 2.23E-05 | 0.000483304 | 0.140381838 |
| Obesity | macrophage | *MT1M* | 2.564164704 | 2.10E-05 | 0.000467599 | 0.131886495 |
| Obesity | macrophage | *SMN2* | -0.219042543 | 0.014080891 | 0.042911568 | 1 |
| Obesity | macrophage | *EIF3CL* | 6.759191695 | 9.91E-35 | 6.23E-31 | 6.23E-31 |
| Obesity | macrophage | *LIN52* | 0.511654926 | 0.007958596 | 0.028015507 | 1 |
| Obesity | macrophage | *C17orf107* | 0.389692991 | 0.00110295 | 0.006798282 | 1 |
| Obesity | macrophage | *C1QTNF9B* | 9.86417173 | 3.21E-05 | 0.000554992 | 0.201724678 |
| Obesity | macrophage | *ZDHHC11B* | 0.585025568 | 0.009188076 | 0.031190839 | 1 |
| Obesity | macrophage | *METTL6* | 0.970652941 | 0.00018123 | 0.001831817 | 1 |
| Obesity | macrophage | *SLC48A1* | 1.314426404 | 0.008366282 | 0.02915677 | 1 |
| Obesity | macrophage | *AC084018.1* | -0.292511309 | 0.000489844 | 0.003854381 | 1 |
| Obesity | macrophage | *RNF208* | 6.958145045 | 0.000188097 | 0.00188307 | 1 |
| Obesity | macrophage | *AC022498.1* | -0.486054002 | 0.012287874 | 0.038826917 | 1 |
| Obesity | macrophage | *ASIC3* | 0.52716027 | 7.30E-05 | 0.000977963 | 0.458809029 |
| Obesity | macrophage | *CCDC183* | 2.240494832 | 0.000197727 | 0.001951502 | 1 |
| Obesity | macrophage | *NOTCH2NL* | 0.313097383 | 0.008420552 | 0.029313407 | 1 |
| Obesity | macrophage | *SUPT4H1* | 0.909967103 | 0.006078096 | 0.023009228 | 1 |
| Obesity | macrophage | *ERV3-1* | 0.128221642 | 0.007299409 | 0.026208672 | 1 |
| Obesity | macrophage | *TMEM110* | 1.807239169 | 1.43E-06 | 7.97E-05 | 0.009000545 |
| Obesity | macrophage | *LBH* | 1.733358847 | 0.001070063 | 0.006687359 | 1 |
| Obesity | macrophage | *SLC35F6* | 1.352990907 | 0.016255952 | 0.047415902 | 1 |
| Obesity | macrophage | *CLIC1* | -0.897267189 | 0.007842362 | 0.027730557 | 1 |
| Obesity | macrophage | *EMP2* | 0.637104394 | 0.004363201 | 0.018131931 | 1 |
| Obesity | macrophage | *PPM1N* | -0.644332767 | 0.007263276 | 0.026171323 | 1 |
| Obesity | macrophage | *GALT* | 1.471908865 | 4.36E-06 | 0.000178092 | 0.027426173 |
| Obesity | macrophage | *ITGA1* | 0.701095151 | 0.002090435 | 0.010641754 | 1 |
| Obesity | macrophage | *TAX1BP3* | 1.720173748 | 0.00011518 | 0.001350604 | 0.724136371 |
| Obesity | macrophage | *TM6SF2* | 10.67156134 | 2.71E-05 | 0.000517549 | 0.170273655 |

| | | | | | | |
|---|---|---|---|---|---|---|
| Obesity | macrophage | *SMIM7* | 0.765804787 | 0.017394002 | 0.049979932 | 1 |
| Obesity | macrophage | *AC079776.2* | 1.668879542 | 0.004352858 | 0.018123455 | 1 |
| Obesity | macrophage | *WEE2* | 0.442396827 | 0.005502064 | 0.021432141 | 1 |
| Obesity | macrophage | *PAXIP1-AS2* | 1.706359626 | 0.001639328 | 0.008977747 | 1 |
| Obesity | macrophage | *LYRM4* | 1.070740275 | 0.004459138 | 0.018419581 | 1 |
| Obesity | macrophage | *NEURL1B* | 1.605813612 | 0.000128553 | 0.001445816 | 0.808211201 |
| Obesity | macrophage | *COL28A1* | 1.878135572 | 1.80E-07 | 1.53E-05 | 0.001129822 |
| Obesity | macrophage | *UBE2QL1* | 1.912393514 | 2.46E-05 | 0.000492083 | 0.154439262 |
| Obesity | macrophage | *GOLGA8B* | 1.510372609 | 6.26E-05 | 0.000878444 | 0.393864632 |
| Obesity | macrophage | *KCNU1* | 0.549220765 | 0.011740968 | 0.037606161 | 1 |
| Obesity | macrophage | *LINC01138* | 1.52785593 | 0.00021787 | 0.00210084 | 1 |
| Obesity | macrophage | *ATAD3C* | 1.234327447 | 2.47E-05 | 0.000492083 | 0.155114971 |
| Obesity | macrophage | *CCDC7* | 0.749434601 | 0.008804484 | 0.030182003 | 1 |
| Obesity | macrophage | *RNU6ATAC35I* | 3.901980594 | 4.27E-05 | 0.000681503 | 0.268512082 |
| Obesity | macrophage | *XKR9* | 0.318494128 | 0.001797382 | 0.009608963 | 1 |
| Obesity | macrophage | *APOL6* | 1.310658365 | 0.000445552 | 0.003577502 | 1 |
| Obesity | macrophage | *FAM185A* | 1.407570106 | 7.48E-05 | 0.000989215 | 0.47049204 |
| Obesity | macrophage | *ADRA2B* | -2.11733995 | 0.000870834 | 0.005828626 | 1 |
| Obesity | macrophage | *EXOSC6* | -1.606905921 | 4.08E-10 | 7.54E-08 | 2.56E-06 |
| Obesity | macrophage | *AC147651.3* | -0.966401361 | 0.000475331 | 0.003773244 | 1 |
| Obesity | macrophage | *OSER1-AS1* | 1.850557144 | 2.02E-06 | 0.000104855 | 0.012687476 |
| Obesity | macrophage | *LINC01133* | 1.749826525 | 3.47E-05 | 0.000580869 | 0.218406847 |
| Obesity | macrophage | *LINC01117* | 1.030735047 | 3.40E-05 | 0.000571658 | 0.213800222 |
| Obesity | macrophage | *LINC00240* | 0.794035493 | 2.59E-12 | 7.40E-10 | 1.63E-08 |
| Obesity | macrophage | *LINC00863* | 0.662967309 | 0.000762537 | 0.005279812 | 1 |
| Obesity | macrophage | *TMEM233* | 0.360781695 | 0.008165373 | 0.028583352 | 1 |
| Obesity | macrophage | *AL049840.1* | -0.37932687 | 0.003674442 | 0.015975945 | 1 |
| Obesity | macrophage | *LINC00237* | -2.648593765 | 0.000797327 | 0.005447834 | 1 |
| Obesity | macrophage | *PLEKHM1* | 1.218704252 | 0.000126707 | 0.001432741 | 0.796603782 |
| Obesity | macrophage | *ERCC6* | 1.346601458 | 0.001112013 | 0.006840729 | 1 |
| Obesity | macrophage | *NOL7* | 1.38724956 | 0.002914405 | 0.013582552 | 1 |
| Obesity | macrophage | *ELFN1* | 5.062958859 | 4.32E-06 | 0.000178092 | 0.027163656 |
| Obesity | macrophage | *ZNF503-AS1* | 1.865484902 | 1.69E-05 | 0.000424589 | 0.106147205 |
| Obesity | macrophage | *AC064836.3* | 3.466749489 | 1.22E-05 | 0.000347425 | 0.076490871 |
| Obesity | macrophage | *C14orf132* | 0.928425417 | 0.000320316 | 0.002868697 | 1 |
| Obesity | macrophage | *LINC00954* | 3.48061908 | 8.83E-05 | 0.00111229 | 0.555032569 |
| Obesity | macrophage | *U91324.1* | 3.825674202 | 1.42E-05 | 0.000389405 | 0.089173733 |
| Obesity | macrophage | *CISD3* | 2.578119484 | 0.00049498 | 0.003870478 | 1 |
| Obesity | macrophage | *PSMG3-AS1* | 1.00675661 | 0.002557321 | 0.012310779 | 1 |
| Obesity | macrophage | *KLHL7-AS1* | 2.584719786 | 2.43E-05 | 0.000492083 | 0.152791875 |
| Obesity | macrophage | *RPS18* | 0.811769552 | 6.75E-05 | 0.000921647 | 0.424126467 |
| Obesity | macrophage | *TAPBP* | 0.569894318 | 0.009472585 | 0.031949649 | 1 |
| Obesity | macrophage | *LINC00865* | -0.376065652 | 0.003424761 | 0.015227348 | 1 |
| Obesity | macrophage | *AC009495.3* | 1.249718576 | 0.006991382 | 0.025599779 | 1 |
| Obesity | macrophage | *TNF* | -0.795445431 | 5.20E-06 | 0.000203227 | 0.032719513 |
| Obesity | macrophage | *SNHG15* | 1.478843469 | 7.25E-05 | 0.00097391 | 0.455789683 |
| Obesity | macrophage | *LINC00702* | 0.810648912 | 0.003905652 | 0.016738128 | 1 |
| Obesity | macrophage | *RPS28* | 2.096039914 | 0.001452598 | 0.008264692 | 1 |
| Obesity | macrophage | *AC123768.3* | 1.162629044 | 0.01553158 | 0.045843681 | 1 |
| Obesity | macrophage | *HLA-B* | 0.366656353 | 0.002230823 | 0.011095873 | 1 |

| | | | | | | |
|---|---|---|---|---|---|---|
| Obesity | macrophage | *ANKRD65* | 0.640653919 | 0.010198745 | 0.033927885 | 1 |
| Obesity | macrophage | *ZSCAN31* | 0.448287597 | 2.78E-06 | 0.000134335 | 0.017475639 |
| Obesity | macrophage | *KIAA0040* | 1.331930724 | 0.00013732 | 0.00150932 | 0.863331123 |
| Obesity | macrophage | *LINC00941* | 3.230082819 | 0.000106831 | 0.001267252 | 0.671643385 |
| Obesity | macrophage | *LINC00545* | -2.259832303 | 0.015509881 | 0.045843681 | 1 |
| Obesity | macrophage | *B3GNT9* | 1.150895993 | 0.001337983 | 0.007817749 | 1 |
| Obesity | macrophage | *ZNF737* | 1.718246752 | 8.41E-05 | 0.001070503 | 0.528728696 |
| Obesity | macrophage | *LINC00959* | 0.519885804 | 0.016728541 | 0.04844419 | 1 |
| Obesity | macrophage | *PSMB9* | 0.636474381 | 5.52E-06 | 0.000210874 | 0.034725543 |
| Obesity | macrophage | *AQP1* | 0.861391636 | 0.007114572 | 0.025918616 | 1 |
| Obesity | macrophage | *NSUN6* | 1.173477395 | 0.002521621 | 0.01221374 | 1 |
| Obesity | macrophage | *ARHGAP8* | -0.636208005 | 0.000709411 | 0.005028258 | 1 |
| Obesity | macrophage | *ATP5O* | 0.795563644 | 0.01414119 | 0.043053589 | 1 |
| Obesity | macrophage | *PWP2* | -2.163661525 | 0.005263117 | 0.020719608 | 1 |
| Obesity | macrophage | *AKAP2* | 3.536643804 | 2.47E-05 | 0.000492083 | 0.155131517 |
| Obesity | macrophage | *ARFGAP3* | 1.172843061 | 0.008789417 | 0.030148123 | 1 |
| Obesity | macrophage | *C22orf39* | 2.134149816 | 0.001029524 | 0.006551234 | 1 |
| Obesity | macrophage | *MRPL20* | 1.155586819 | 0.004214768 | 0.017677283 | 1 |
| Obesity | macrophage | *DECR2* | 2.949985584 | 0.000191562 | 0.001905618 | 1 |
| Obesity | macrophage | *CCDC169* | 0.352636329 | 0.007126143 | 0.025927118 | 1 |
| Obesity | macrophage | *C7orf73* | 1.328393083 | 0.002103135 | 0.010680458 | 1 |
| Obesity | macrophage | *FGF14-IT1* | 3.100324826 | 2.69E-05 | 0.000516343 | 0.169360599 |
| Obesity | macrophage | *NAT6* | 1.361217182 | 1.92E-06 | 0.00010131 | 0.012055937 |
| Obesity | macrophage | *RN7SL832P* | 0.516166812 | 0.004528388 | 0.018571411 | 1 |
| Obesity | macrophage | *APOBEC3C* | 3.088054091 | 0.000513533 | 0.003946923 | 1 |
| Obesity | macrophage | *UBE2V1* | 2.599411286 | 0.000144332 | 0.00156451 | 0.907416001 |
| Obesity | macrophage | *ZNF585B* | 0.349748864 | 0.004454792 | 0.018419581 | 1 |
| Obesity | macrophage | *LINC00920* | -0.661857646 | 0.000385909 | 0.003261031 | 1 |
| Obesity | macrophage | *PGAM5* | 3.300056355 | 0.000310423 | 0.002808097 | 1 |
| Obesity | macrophage | *PCDHAC1* | -1.09903552 | 0.003754068 | 0.016221187 | 1 |
| Obesity | macrophage | *ABHD14A* | 1.597951989 | 6.49E-05 | 0.000899313 | 0.408288177 |
| Obesity | macrophage | *SRP14-AS1* | 0.148501262 | 0.004550883 | 0.018627213 | 1 |
| Obesity | macrophage | *FMN1* | -0.269410314 | 0.003996634 | 0.017000566 | 1 |
| Obesity | macrophage | *NAIP* | -0.453016567 | 0.011723854 | 0.037586881 | 1 |
| Obesity | macrophage | *PDCD6* | 1.12911572 | 0.004170179 | 0.017537066 | 1 |
| Obesity | macrophage | *DNAH10OS* | 8.561973175 | 6.03E-12 | 1.65E-09 | 3.79E-08 |
| Obesity | macrophage | *PCDHA10* | 1.556354431 | 0.000147457 | 0.001580225 | 0.92706099 |
| Obesity | macrophage | *ZNF718* | 1.051965647 | 0.012916039 | 0.040205735 | 1 |
| Obesity | macrophage | *LINC00989* | 1.706993427 | 0.00202404 | 0.0104219 | 1 |
| Obesity | macrophage | *LINC01021* | 1.98499051 | 3.22E-05 | 0.000554992 | 0.202572075 |
| Obesity | macrophage | *FAM218A* | 3.053782631 | 2.01E-05 | 0.000456556 | 0.126466099 |
| Obesity | macrophage | *ZNF345* | 0.805901908 | 0.014459816 | 0.043706831 | 1 |
| Obesity | macrophage | *ZNF550* | 0.91260601 | 0.008063489 | 0.028305502 | 1 |
| Obesity | macrophage | *CCDC71L* | 0.838722341 | 0.016869879 | 0.048756987 | 1 |
| Obesity | macrophage | *TMEM200B* | 0.369042693 | 0.00717001 | 0.02604151 | 1 |
| Obesity | macrophage | *TRNP1* | 9.866488568 | 7.98E-05 | 0.001034173 | 0.501573966 |
| Obesity | macrophage | *ATXN7L3B* | 1.547235709 | 0.004395695 | 0.018241407 | 1 |
| Obesity | macrophage | *PCDHGA6* | 0.470786954 | 0.000675841 | 0.004844939 | 1 |
| Obesity | macrophage | *CLDN23* | 1.484620679 | 0.002516664 | 0.012199439 | 1 |
| Obesity | macrophage | *ZNF260* | 1.291508356 | 0.001439644 | 0.008213287 | 1 |

| | | | | | | |
|---|---|---|---|---|---|---|
| Obesity | macrophage | *MPV17L2* | 1.287671221 | 5.58E-08 | 5.31E-06 | 0.000350756 |
| Obesity | macrophage | *DPP3* | 0.328449837 | 0.00342921 | 0.015236357 | 1 |
| Obesity | macrophage | *SNHG9* | 0.31318963 | 0.002483488 | 0.012066218 | 1 |
| Obesity | macrophage | *EBLN2* | 1.013876805 | 0.016252419 | 0.047415902 | 1 |
| Obesity | macrophage | *POLR2M* | 2.244321873 | 0.000307578 | 0.002786372 | 1 |
| Obesity | macrophage | *CTSO* | 1.609261555 | 0.001913139 | 0.010056775 | 1 |
| Obesity | macrophage | *HMBS* | 0.753093179 | 1.14E-12 | 3.77E-10 | 7.17E-09 |
| Obesity | macrophage | *PGA5* | -4.544221375 | 0.000967384 | 0.006296006 | 1 |
| Obesity | macrophage | *HP* | -0.688599747 | 0.008910284 | 0.030428548 | 1 |
| Obesity | macrophage | *KIAA1147* | 0.758139323 | 0.014173637 | 0.043110622 | 1 |
| Obesity | macrophage | *ZBED6* | 5.901749552 | 8.87E-05 | 0.00111476 | 0.557379961 |
| Obesity | macrophage | *FNTB* | 2.360292048 | 0.000135373 | 0.001502304 | 0.851090462 |
| Obesity | macrophage | *OVCH1-AS1* | 1.493458749 | 0.00159233 | 0.008803601 | 1 |
| Obesity | macrophage | *DUXA* | -0.352051399 | 0.008613103 | 0.029769423 | 1 |
| Obesity | macrophage | *CEP95* | 0.885531375 | 0.007631139 | 0.027167028 | 1 |
| Obesity | macrophage | *LINC00648* | 1.518617856 | 1.87E-05 | 0.000445142 | 0.117697296 |
| Obesity | macrophage | *LINC00924* | 0.865596216 | 0.010347556 | 0.034299881 | 1 |
| Obesity | macrophage | *SH3RF3-AS1* | -0.443368541 | 0.00157086 | 0.008716979 | 1 |
| Obesity | macrophage | *PWRN1* | 0.531019825 | 0.012565431 | 0.039420592 | 1 |
| Obesity | macrophage | *HOXB7* | 2.134082832 | 2.76E-08 | 2.99E-06 | 0.000173357 |
| Obesity | macrophage | *FRRS1L* | 4.274241559 | 1.14E-06 | 6.75E-05 | 0.007157748 |
| Obesity | macrophage | *AC009120.5* | 2.64917894 | 3.06E-05 | 0.000546074 | 0.192086296 |
| Obesity | macrophage | *TMEM178B* | 1.30710838 | 3.33E-05 | 0.000565547 | 0.209252499 |
| Obesity | macrophage | *HPR* | 1.726537679 | 1.13E-05 | 0.000333865 | 0.071346086 |
| Obesity | macrophage | *GOLGA8H* | 0.351333048 | 0.000985343 | 0.006360219 | 1 |
| Obesity | macrophage | *STRADA* | -0.634102864 | 0.013021815 | 0.040448691 | 1 |
| Obesity | macrophage | *AC139100.2* | 10.25310867 | 0.000531174 | 0.004028339 | 1 |
| Obesity | macrophage | *LINC01140* | 1.418669464 | 8.66E-07 | 5.67E-05 | 0.005442201 |
| Obesity | macrophage | *LINC00906* | 6.047441879 | 4.29E-05 | 0.000682173 | 0.269458362 |
| Obesity | macrophage | *FBXO17* | 1.11454547 | 0.0001895 | 0.001894094 | 1 |
| Obesity | macrophage | *AC009060.1* | -1.546746552 | 0.000865273 | 0.005805731 | 1 |
| Obesity | macrophage | *YTHDF3-AS1* | 1.036732839 | 0.012218691 | 0.038658559 | 1 |
| Obesity | macrophage | *GTF2H5* | 0.965566534 | 2.66E-06 | 0.000131017 | 0.016716042 |
| Obesity | macrophage | *FGF14-AS2* | 0.881067057 | 0.001520895 | 0.008522665 | 1 |
| Obesity | macrophage | *CASC15* | 0.548754701 | 0.004989209 | 0.020042911 | 1 |
| Obesity | macrophage | *POLR2J4* | 2.402942082 | 8.53E-05 | 0.001081589 | 0.536468032 |
| Obesity | mast_cell | *SCYL3* | 4.189347184 | 4.17E-07 | 1.33E-06 | 0.002619126 |
| Obesity | mast_cell | *C1orf112* | 5.377639608 | 1.43E-09 | 4.06E-08 | 9.01E-06 |
| Obesity | mast_cell | *CFH* | 2.779783627 | 1.96E-09 | 4.32E-08 | 1.23E-05 |
| Obesity | mast_cell | *NFYA* | 7.179597986 | 1.50E-06 | 4.06E-06 | 0.009454711 |
| Obesity | mast_cell | *STPG1* | 3.605311566 | 4.89E-07 | 1.51E-06 | 0.003071948 |
| Obesity | mast_cell | *ENPP4* | 3.238976441 | 7.48E-06 | 1.70E-05 | 0.047013057 |
| Obesity | mast_cell | *SEMA3F* | -0.971453121 | 0.007737596 | 0.010647027 | 1 |
| Obesity | mast_cell | *KRIT1* | 5.498878203 | 2.13E-08 | 1.20E-07 | 0.000134156 |
| Obesity | mast_cell | *RAD52* | 7.050690665 | 4.87E-08 | 2.28E-07 | 0.000306213 |
| Obesity | mast_cell | *HS3ST1* | 9.040171918 | 2.45E-09 | 4.32E-08 | 1.54E-05 |
| Obesity | mast_cell | *WNT16* | 6.435193974 | 2.33E-10 | 1.41E-08 | 1.47E-06 |
| Obesity | mast_cell | *HECW1* | 1.367815108 | 0.006397567 | 0.008894627 | 1 |
| Obesity | mast_cell | *MAD1L1* | 4.306209477 | 0.005913594 | 0.008258277 | 1 |
| Obesity | mast_cell | *LASP1* | 4.27028764 | 0.005727295 | 0.008014134 | 1 |

| | | | | | | |
|---|---|---|---|---|---|---|
| Obesity | mast_cell | *TMEM176A* | 2.154545561 | 0.004268753 | 0.006082876 | 1 |
| Obesity | mast_cell | *M6PR* | 4.752220417 | 0.000596844 | 0.000962143 | 1 |
| Obesity | mast_cell | *CYP26B1* | 5.275076371 | 1.78E-09 | 4.31E-08 | 1.12E-05 |
| Obesity | mast_cell | *DBNDD1* | 2.044428299 | 0.007146569 | 0.009863991 | 1 |
| Obesity | mast_cell | *ALS2* | -2.027948085 | 1.90E-05 | 3.98E-05 | 0.119271514 |
| Obesity | mast_cell | *POLDIP2* | 9.444961239 | 4.08E-09 | 4.79E-08 | 2.57E-05 |
| Obesity | mast_cell | *CD38* | 2.479912371 | 3.30E-07 | 1.09E-06 | 0.002076582 |
| Obesity | mast_cell | *RBM6* | -3.171710314 | 1.40E-17 | 8.79E-15 | 8.79E-14 |
| Obesity | mast_cell | *RECQL* | 7.665310228 | 1.76E-07 | 6.41E-07 | 0.001104324 |
| Obesity | mast_cell | *PDK4* | 4.585980263 | 0.000337006 | 0.000565605 | 1 |
| Obesity | mast_cell | *SLC22A16* | -3.347471407 | 3.00E-08 | 1.55E-07 | 0.000188381 |
| Obesity | mast_cell | *CDC27* | 4.448370559 | 3.68E-06 | 8.92E-06 | 0.023134157 |
| Obesity | mast_cell | *MEOX1* | 1.917845534 | 0.000630774 | 0.001012944 | 1 |
| Obesity | mast_cell | *LIG3* | 2.027705261 | 0.006326227 | 0.008801281 | 1 |
| Obesity | mast_cell | *CIAPIN1* | 5.773802785 | 4.10E-09 | 4.79E-08 | 2.58E-05 |
| Obesity | mast_cell | *FAM214B* | 7.223291555 | 2.46E-10 | 1.46E-08 | 1.55E-06 |
| Obesity | mast_cell | *GCFC2* | 6.112434998 | 2.69E-08 | 1.42E-07 | 0.000169026 |
| Obesity | mast_cell | *WDR54* | 10.43568755 | 9.52E-06 | 2.12E-05 | 0.05984527 |
| Obesity | mast_cell | *ABCB4* | 3.944085066 | 5.63E-09 | 5.57E-08 | 3.54E-05 |
| Obesity | mast_cell | *ZNF195* | 4.049483872 | 1.76E-05 | 3.72E-05 | 0.110770869 |
| Obesity | mast_cell | *ASB4* | 2.058564322 | 0.033198221 | 0.041885855 | 1 |
| Obesity | mast_cell | *TMEM98* | 3.652015715 | 4.33E-09 | 4.90E-08 | 2.72E-05 |
| Obesity | mast_cell | *ABCC8* | 10.09950302 | 1.19E-08 | 8.50E-08 | 7.45E-05 |
| Obesity | mast_cell | *SYNRG* | 4.969218178 | 0.000281533 | 0.000477217 | 1 |
| Obesity | mast_cell | *ZNF263* | 7.696347917 | 1.44E-08 | 9.57E-08 | 9.07E-05 |
| Obesity | mast_cell | *SPATA20* | 6.254882906 | 7.26E-05 | 0.000135687 | 0.456451741 |
| Obesity | mast_cell | *MAP3K9* | 4.908770197 | 1.29E-05 | 2.80E-05 | 0.080960789 |
| Obesity | mast_cell | *RALA* | 6.529790264 | 1.28E-07 | 4.96E-07 | 0.000802322 |
| Obesity | mast_cell | *BAIAP2L1* | 3.69302868 | 6.98E-05 | 0.000130968 | 0.43913525 |
| Obesity | mast_cell | *FARP2* | 3.444813213 | 0.004225348 | 0.006027856 | 1 |
| Obesity | mast_cell | *IFRD1* | 3.146408594 | 0.000408016 | 0.000677011 | 1 |
| Obesity | mast_cell | *GTF2IRD1* | 3.404203472 | 6.68E-06 | 1.53E-05 | 0.042005867 |
| Obesity | mast_cell | *ARHGAP44* | 5.274548747 | 1.25E-09 | 3.78E-08 | 7.89E-06 |
| Obesity | mast_cell | *CCDC124* | 2.841670382 | 4.43E-08 | 2.10E-07 | 0.000278732 |
| Obesity | mast_cell | *CEACAM21* | 4.424829412 | 9.52E-05 | 0.000173653 | 0.598235923 |
| Obesity | mast_cell | *NOS2* | 15.23378888 | 9.14E-09 | 7.26E-08 | 5.74E-05 |
| Obesity | mast_cell | *TRAPPC6A* | 8.124786743 | 4.98E-08 | 2.32E-07 | 0.000313134 |
| Obesity | mast_cell | *ST7L* | 1.181415063 | 0.031657601 | 0.040054606 | 1 |
| Obesity | mast_cell | *PAX6* | 2.806076966 | 0.001546365 | 0.002342649 | 1 |
| Obesity | mast_cell | *RPUSD1* | 4.407334278 | 1.48E-08 | 9.68E-08 | 9.29E-05 |
| Obesity | mast_cell | *PIGQ* | 4.993668706 | 1.70E-07 | 6.25E-07 | 0.001068481 |
| Obesity | mast_cell | *PSMB1* | 4.836603149 | 0.003825532 | 0.005494887 | 1 |
| Obesity | mast_cell | *DLEC1* | 5.282021739 | 9.42E-08 | 3.88E-07 | 0.000592494 |
| Obesity | mast_cell | *MGST1* | 4.066041429 | 2.72E-06 | 6.84E-06 | 0.017129568 |
| Obesity | mast_cell | *NFIX* | 2.817827333 | 0.000419515 | 0.000694441 | 1 |
| Obesity | mast_cell | *MMP25* | 4.783054178 | 6.99E-10 | 2.77E-08 | 4.40E-06 |
| Obesity | mast_cell | *IL32* | 3.252540942 | 5.36E-06 | 1.25E-05 | 0.033708022 |
| Obesity | mast_cell | *PKD1* | 5.79514768 | 1.21E-09 | 3.76E-08 | 7.63E-06 |
| Obesity | mast_cell | *MAPK8IP2* | 12.89939463 | 2.94E-08 | 1.53E-07 | 0.00018463 |
| Obesity | mast_cell | *MED24* | 4.447825716 | 0.000461911 | 0.000758828 | 1 |

| | | | | | | |
|---|---|---|---|---|---|---|
| Obesity | mast_cell | *SEC62* | 5.827032038 | 1.40E-05 | 3.02E-05 | 0.08781702 |
| Obesity | mast_cell | *RPS20* | 4.40598527 | 0.005064431 | 0.007150254 | 1 |
| Obesity | mast_cell | *MLXIPL* | 1.608449289 | 4.67E-09 | 5.10E-08 | 2.94E-05 |
| Obesity | mast_cell | *SPRTN* | 6.418220351 | 1.87E-09 | 4.32E-08 | 1.17E-05 |
| Obesity | mast_cell | *METTL13* | 7.326266969 | 2.50E-05 | 5.10E-05 | 0.157303806 |
| Obesity | mast_cell | *DYRK4* | 3.589177762 | 4.18E-06 | 1.00E-05 | 0.026309629 |
| Obesity | mast_cell | *STARD3NL* | 8.576829865 | 1.61E-08 | 1.02E-07 | 0.000101041 |
| Obesity | mast_cell | *NCAPD2* | 1.618845826 | 0.006902465 | 0.009550132 | 1 |
| Obesity | mast_cell | *ZNF200* | 6.143905572 | 5.40E-08 | 2.46E-07 | 0.000339749 |
| Obesity | mast_cell | *CD4* | 8.802194662 | 9.17E-08 | 3.79E-07 | 0.000576643 |
| Obesity | mast_cell | *LRRC23* | 3.807200134 | 0.000876772 | 0.001377033 | 1 |
| Obesity | mast_cell | *SCMH1* | 4.653918078 | 1.35E-05 | 2.93E-05 | 0.085045598 |
| Obesity | mast_cell | *CLCN6* | 7.691409456 | 4.40E-06 | 1.05E-05 | 0.02764831 |
| Obesity | mast_cell | *MRC2* | 2.667181281 | 1.53E-06 | 4.11E-06 | 0.009591964 |
| Obesity | mast_cell | *TSPAN9* | 4.035938065 | 6.27E-07 | 1.87E-06 | 0.0039402 |
| Obesity | mast_cell | *MKS1* | 6.711599596 | 8.83E-10 | 3.12E-08 | 5.55E-06 |
| Obesity | mast_cell | *UTP18* | 5.985853355 | 2.36E-09 | 4.32E-08 | 1.49E-05 |
| Obesity | mast_cell | *TTC19* | 5.878137044 | 1.33E-07 | 5.15E-07 | 0.000839247 |
| Obesity | mast_cell | *SYT7* | 7.686518239 | 5.26E-07 | 1.60E-06 | 0.003305033 |
| Obesity | mast_cell | *LARS2* | 6.323192854 | 1.61E-06 | 4.29E-06 | 0.010090846 |
| Obesity | mast_cell | *PPP5C* | 4.717520838 | 0.00267116 | 0.003916189 | 1 |
| Obesity | mast_cell | *ZBTB32* | -5.034010756 | 0.000227268 | 0.000390819 | 1 |
| Obesity | mast_cell | *TMEM159* | 3.625651971 | 6.41E-08 | 2.81E-07 | 0.00040271 |
| Obesity | mast_cell | *LTF* | 5.998452334 | 7.76E-07 | 2.26E-06 | 0.004880853 |
| Obesity | mast_cell | *ALOX5* | -0.967670929 | 3.35E-07 | 1.10E-06 | 0.002106301 |
| Obesity | mast_cell | *MAP4K5* | 5.553500263 | 1.47E-06 | 3.98E-06 | 0.009243192 |
| Obesity | mast_cell | *PSMC4* | 4.686304812 | 1.54E-11 | 2.10E-09 | 9.65E-08 |
| Obesity | mast_cell | *MAN2B2* | -2.913292132 | 0.001451367 | 0.002205643 | 1 |
| Obesity | mast_cell | *CLDN11* | 4.221972326 | 8.95E-10 | 3.14E-08 | 5.62E-06 |
| Obesity | mast_cell | *ANGEL1* | 8.116114832 | 1.43E-05 | 3.08E-05 | 0.089964554 |
| Obesity | mast_cell | *RNF14* | 4.896983605 | 0.00267106 | 0.003916189 | 1 |
| Obesity | mast_cell | *HEBP1* | 3.88553314 | 1.17E-08 | 8.43E-08 | 7.33E-05 |
| Obesity | mast_cell | *TACC3* | 5.179459148 | 5.58E-05 | 0.000106921 | 0.350913747 |
| Obesity | mast_cell | *ZC3H3* | 2.666971716 | 0.026438951 | 0.033833088 | 1 |
| Obesity | mast_cell | *MDH1* | 4.796103664 | 1.90E-10 | 1.27E-08 | 1.19E-06 |
| Obesity | mast_cell | *SLC30A9* | 6.122762836 | 2.35E-07 | 8.20E-07 | 0.001480029 |
| Obesity | mast_cell | *MTMR11* | 1.975388287 | 4.22E-06 | 1.01E-05 | 0.026507301 |
| Obesity | mast_cell | *COX15* | 3.521841226 | 8.47E-08 | 3.54E-07 | 0.000532529 |
| Obesity | mast_cell | *CCDC88C* | -2.240087227 | 0.021562075 | 0.027944912 | 1 |
| Obesity | mast_cell | *NPC1L1* | 5.965557512 | 3.48E-07 | 1.14E-06 | 0.002187398 |
| Obesity | mast_cell | *NUDCD3* | 4.366242773 | 4.44E-09 | 4.98E-08 | 2.79E-05 |
| Obesity | mast_cell | *CHDH* | 6.574098504 | 6.36E-08 | 2.80E-07 | 0.000399598 |
| Obesity | mast_cell | *IL20RA* | 6.050590732 | 1.71E-08 | 1.05E-07 | 0.000107807 |
| Obesity | mast_cell | *GLT8D1* | 3.299597564 | 0.001996742 | 0.002974062 | 1 |
| Obesity | mast_cell | *RUFY3* | 4.122176152 | 3.43E-05 | 6.81E-05 | 0.215715918 |
| Obesity | mast_cell | *WWTR1* | 5.045266497 | 1.06E-08 | 8.02E-08 | 6.68E-05 |
| Obesity | mast_cell | *TTC27* | 4.2820072 | 7.74E-05 | 0.000143844 | 0.486624007 |
| Obesity | mast_cell | *VSIG2* | 5.779640719 | 5.03E-08 | 2.33E-07 | 0.000316309 |
| Obesity | mast_cell | *PHLDB1* | 1.723484154 | 0.000180415 | 0.000314988 | 1 |
| Obesity | mast_cell | *MARCO* | 1.417937592 | 0.001100761 | 0.001702037 | 1 |

| | | | | | | |
|---|---|---|---|---|---|---|
| Obesity | mast_cell | *MNAT1* | 5.924466961 | 1.49E-07 | 5.62E-07 | 0.000937115 |
| Obesity | mast_cell | *OSBPL5* | 7.592096067 | 1.04E-06 | 2.92E-06 | 0.006516294 |
| Obesity | mast_cell | *AQR* | 5.737400669 | 9.97E-08 | 4.08E-07 | 0.000626856 |
| Obesity | mast_cell | *CPS1* | 3.343601083 | 2.84E-07 | 9.60E-07 | 0.001787614 |
| Obesity | mast_cell | *NLRP2* | 3.127255519 | 2.50E-05 | 5.10E-05 | 0.157173089 |
| Obesity | mast_cell | *ZDHHC6* | 6.973166863 | 4.08E-07 | 1.31E-06 | 0.002563821 |
| Obesity | mast_cell | *RNH1* | 3.940563551 | 0.000231707 | 0.000398126 | 1 |
| Obesity | mast_cell | *NDUFS1* | 5.179996779 | 8.72E-05 | 0.000160098 | 0.548016461 |
| Obesity | mast_cell | *BIRC3* | -1.728607734 | 8.52E-07 | 2.46E-06 | 0.005358579 |
| Obesity | mast_cell | *AKAP11* | 6.704554946 | 1.41E-07 | 5.36E-07 | 0.000886201 |
| Obesity | mast_cell | *SNAPC1* | 5.580579925 | 1.30E-06 | 3.57E-06 | 0.008165351 |
| Obesity | mast_cell | *GCLM* | 3.898597194 | 4.30E-09 | 4.88E-08 | 2.71E-05 |
| Obesity | mast_cell | *UBR2* | 4.204107346 | 0.000675279 | 0.00107808 | 1 |
| Obesity | mast_cell | *HSF2* | 6.84418778 | 2.50E-09 | 4.32E-08 | 1.57E-05 |
| Obesity | mast_cell | *NR1H3* | 6.453109476 | 1.50E-11 | 2.10E-09 | 9.44E-08 |
| Obesity | mast_cell | *TYMP* | 3.897257688 | 0.0016038 | 0.002422073 | 1 |
| Obesity | mast_cell | *TOMM34* | 10.70002803 | 7.91E-05 | 0.000146662 | 0.497037392 |
| Obesity | mast_cell | *KPNA6* | 3.69794464 | 0.006899571 | 0.009550132 | 1 |
| Obesity | mast_cell | *VIM* | 2.471214323 | 0.024586859 | 0.031617424 | 1 |
| Obesity | mast_cell | *FAS* | 3.19972808 | 0.000161505 | 0.000284661 | 1 |
| Obesity | mast_cell | *CD44* | -1.982774234 | 0.000814763 | 0.001286716 | 1 |
| Obesity | mast_cell | *BTN3A1* | -1.523807617 | 0.036799209 | 0.046160541 | 1 |
| Obesity | mast_cell | *MIPEP* | 6.591362367 | 6.73E-09 | 6.06E-08 | 4.23E-05 |
| Obesity | mast_cell | *INSRR* | 4.118213364 | 0.000635641 | 0.001019979 | 1 |
| Obesity | mast_cell | *B4GALT7* | 3.363502044 | 7.83E-09 | 6.64E-08 | 4.92E-05 |
| Obesity | mast_cell | *VRK2* | 2.914038058 | 0.024615294 | 0.031641045 | 1 |
| Obesity | mast_cell | *VEZT* | 4.876667812 | 2.35E-05 | 4.84E-05 | 0.148029709 |
| Obesity | mast_cell | *SNX1* | 4.263668168 | 8.98E-05 | 0.000164459 | 0.564587388 |
| Obesity | mast_cell | *ARNTL2* | 8.579552303 | 1.65E-08 | 1.03E-07 | 0.000103742 |
| Obesity | mast_cell | *SLC39A9* | 4.129146499 | 5.29E-05 | 0.00010187 | 0.332300785 |
| Obesity | mast_cell | *ANK1* | -5.219393255 | 0.005177075 | 0.007289647 | 1 |
| Obesity | mast_cell | *RABEP1* | 6.117846581 | 2.61E-09 | 4.32E-08 | 1.64E-05 |
| Obesity | mast_cell | *GRN* | 6.435559013 | 1.00E-05 | 2.23E-05 | 0.062934097 |
| Obesity | mast_cell | *ARID4A* | 4.622366495 | 0.001027724 | 0.00159696 | 1 |
| Obesity | mast_cell | *IFT88* | 4.646980365 | 0.00016657 | 0.000292767 | 1 |
| Obesity | mast_cell | *ZCCHC8* | 8.587199985 | 5.95E-10 | 2.60E-08 | 3.74E-06 |
| Obesity | mast_cell | *ABCF2* | 4.539447789 | 8.12E-07 | 2.35E-06 | 0.005105062 |
| Obesity | mast_cell | *FUT8* | 5.358137291 | 1.55E-05 | 3.32E-05 | 0.097687221 |
| Obesity | mast_cell | *UBA6* | 2.866346998 | 0.012238049 | 0.016384288 | 1 |
| Obesity | mast_cell | *PIAS1* | 3.194539219 | 0.007985747 | 0.010968517 | 1 |
| Obesity | mast_cell | *SLC4A7* | 5.60967108 | 7.37E-08 | 3.14E-07 | 0.000463241 |
| Obesity | mast_cell | *ASTE1* | 2.204294825 | 0.002237071 | 0.003314745 | 1 |
| Obesity | mast_cell | *MYOC* | 2.397903388 | 1.57E-05 | 3.36E-05 | 0.098800888 |
| Obesity | mast_cell | *VCL* | 2.887284348 | 0.004503908 | 0.006402005 | 1 |
| Obesity | mast_cell | *ADSS* | 7.362305828 | 8.40E-08 | 3.52E-07 | 0.000528114 |
| Obesity | mast_cell | *STAP1* | -0.544676157 | 5.29E-08 | 2.42E-07 | 0.00033228 |
| Obesity | mast_cell | *TBC1D23* | 4.768514805 | 0.000849147 | 0.001336987 | 1 |
| Obesity | mast_cell | *CUL3* | 4.503620383 | 0.000392198 | 0.000652487 | 1 |
| Obesity | mast_cell | *MYOM2* | 2.841678251 | 3.81E-05 | 7.50E-05 | 0.239251187 |
| Obesity | mast_cell | *NSUN2* | 7.842197186 | 8.77E-08 | 3.65E-07 | 0.000551188 |

| | | | | | | |
|---|---|---|---|---|---|---|
| Obesity | mast_cell | *MFAP3* | 3.139986831 | 1.99E-05 | 4.15E-05 | 0.125210959 |
| Obesity | mast_cell | *MRI1* | 3.354476757 | 0.0002405 | 0.00041222 | 1 |
| Obesity | mast_cell | *AGA* | 7.114669733 | 2.99E-09 | 4.44E-08 | 1.88E-05 |
| Obesity | mast_cell | *BOD1L1* | 4.488848054 | 0.000161186 | 0.000284258 | 1 |
| Obesity | mast_cell | *CLEC16A* | 6.116090145 | 4.26E-07 | 1.35E-06 | 0.002679489 |
| Obesity | mast_cell | *SKIV2L2* | 5.109962578 | 7.17E-05 | 0.00013414 | 0.450711455 |
| Obesity | mast_cell | *DNAH5* | 5.269718494 | 7.68E-10 | 2.93E-08 | 4.83E-06 |
| Obesity | mast_cell | *C6* | 6.407961455 | 1.14E-06 | 3.18E-06 | 0.007147117 |
| Obesity | mast_cell | *RAI14* | 3.994605026 | 7.40E-05 | 0.000138099 | 0.465531086 |
| Obesity | mast_cell | *PNKP* | 2.413434354 | 0.005760773 | 0.008057392 | 1 |
| Obesity | mast_cell | *PHLPP2* | 5.048500096 | 0.000116931 | 0.000210824 | 0.73514374 |
| Obesity | mast_cell | *SPDL1* | 3.059072132 | 0.000137528 | 0.000245079 | 0.864637895 |
| Obesity | mast_cell | *PQLC2* | -4.570982004 | 0.035400039 | 0.044529821 | 1 |
| Obesity | mast_cell | *INPP4A* | 3.97104156 | 7.00E-06 | 1.60E-05 | 0.043978591 |
| Obesity | mast_cell | *RAB27B* | -0.211228926 | 1.09E-07 | 4.35E-07 | 0.000683115 |
| Obesity | mast_cell | *PSMA4* | 5.462759912 | 3.12E-08 | 1.59E-07 | 0.000195942 |
| Obesity | mast_cell | *LSG1* | 2.50775272 | 0.000359056 | 0.000600049 | 1 |
| Obesity | mast_cell | *THAP3* | 4.567096636 | 7.88E-09 | 6.64E-08 | 4.95E-05 |
| Obesity | mast_cell | *TDP1* | 7.625410611 | 0.000181702 | 0.000317147 | 1 |
| Obesity | mast_cell | *SPATA7* | 6.190429037 | 3.50E-05 | 6.93E-05 | 0.219858061 |
| Obesity | mast_cell | *MED17* | 5.039214072 | 2.90E-09 | 4.35E-08 | 1.82E-05 |
| Obesity | mast_cell | *RETSAT* | 3.171085384 | 5.61E-08 | 2.53E-07 | 0.000352989 |
| Obesity | mast_cell | *CAPG* | -2.576544378 | 8.77E-05 | 0.000160996 | 0.551250053 |
| Obesity | mast_cell | *AP2S1* | 7.140602193 | 0.000487904 | 0.00079757 | 1 |
| Obesity | mast_cell | *LCP2* | -2.355594603 | 1.59E-05 | 3.40E-05 | 0.100267849 |
| Obesity | mast_cell | *TRIT1* | 2.846718485 | 0.002713658 | 0.003973165 | 1 |
| Obesity | mast_cell | *CTNNA1* | 5.774417751 | 8.16E-08 | 3.43E-07 | 0.000513175 |
| Obesity | mast_cell | *CNTLN* | 4.685836934 | 0.000341146 | 0.000572248 | 1 |
| Obesity | mast_cell | *EPHA3* | 3.956602142 | 2.51E-09 | 4.32E-08 | 1.58E-05 |
| Obesity | mast_cell | *WDR37* | 3.410853102 | 0.014747224 | 0.019506796 | 1 |
| Obesity | mast_cell | *YTHDC2* | 5.364444696 | 6.08E-05 | 0.000115337 | 0.381996952 |
| Obesity | mast_cell | *ATP6V1H* | 6.234188892 | 1.08E-05 | 2.39E-05 | 0.06803544 |
| Obesity | mast_cell | *POLR2B* | 6.091003561 | 2.55E-05 | 5.19E-05 | 0.160394753 |
| Obesity | mast_cell | *CP* | 6.85343656 | 2.15E-09 | 4.32E-08 | 1.35E-05 |
| Obesity | mast_cell | *DTNBP1* | 3.794910673 | 0.014575599 | 0.019287896 | 1 |
| Obesity | mast_cell | *ANO2* | 2.691392472 | 0.000164394 | 0.000289185 | 1 |
| Obesity | mast_cell | *GOPC* | 4.660475518 | 0.001212184 | 0.001863781 | 1 |
| Obesity | mast_cell | *TSPAN17* | 4.412051228 | 7.27E-10 | 2.84E-08 | 4.57E-06 |
| Obesity | mast_cell | *CC2D2A* | 3.011351649 | 0.030150825 | 0.038225093 | 1 |
| Obesity | mast_cell | *LMO3* | 1.661870407 | 0.016030217 | 0.021128297 | 1 |
| Obesity | mast_cell | *CELF2* | 2.878614054 | 0.005216264 | 0.007338252 | 1 |
| Obesity | mast_cell | *ERCC8* | 5.324501629 | 1.82E-07 | 6.61E-07 | 0.0011427 |
| Obesity | mast_cell | *ADAMTS6* | 3.617726768 | 1.11E-07 | 4.43E-07 | 0.000697964 |
| Obesity | mast_cell | *H6PD* | 3.588128058 | 1.99E-06 | 5.16E-06 | 0.012488038 |
| Obesity | mast_cell | *UTS2* | 2.161496608 | 8.47E-05 | 0.000155882 | 0.532336711 |
| Obesity | mast_cell | *EPN3* | 8.823870667 | 0.005422175 | 0.007605804 | 1 |
| Obesity | mast_cell | *RFC2* | 8.554972511 | 1.32E-05 | 2.86E-05 | 0.08309851 |
| Obesity | mast_cell | *JKAMP* | 5.592114708 | 2.93E-08 | 1.52E-07 | 0.000184153 |
| Obesity | mast_cell | *DKK3* | 2.734871352 | 2.73E-09 | 4.32E-08 | 1.72E-05 |
| Obesity | mast_cell | *ARHGEF5* | 3.47900828 | 2.84E-08 | 1.48E-07 | 0.000178266 |

| | | | | | | |
|---|---|---|---|---|---|---|
| Obesity | mast_cell | *NFE2L3* | -3.395090949 | 0.000207006 | 0.000358525 | 1 |
| Obesity | mast_cell | *MCUR1* | 4.874241314 | 0.000873978 | 0.001373331 | 1 |
| Obesity | mast_cell | *LIMA1* | 1.55336541 | 0.000125529 | 0.000225035 | 0.789199329 |
| Obesity | mast_cell | *MAPK9* | 4.631163873 | 4.99E-06 | 1.17E-05 | 0.031392015 |
| Obesity | mast_cell | *FAM160A2* | 1.894486904 | 0.031698253 | 0.040097972 | 1 |
| Obesity | mast_cell | *HERPUD1* | 6.062842872 | 0.000111846 | 0.000202411 | 0.703177188 |
| Obesity | mast_cell | *PIK3CB* | 2.585818264 | 0.026925482 | 0.034403551 | 1 |
| Obesity | mast_cell | *CYBA* | 6.363179752 | 8.83E-07 | 2.54E-06 | 0.005553399 |
| Obesity | mast_cell | *THOC3* | 8.303093302 | 5.04E-08 | 2.33E-07 | 0.000316614 |
| Obesity | mast_cell | *RRP12* | 4.833668408 | 2.25E-07 | 7.90E-07 | 0.001414951 |
| Obesity | mast_cell | *ALX4* | 4.033813462 | 2.68E-08 | 1.42E-07 | 0.000168365 |
| Obesity | mast_cell | *FOXN3* | 5.007137529 | 1.37E-06 | 3.74E-06 | 0.008614144 |
| Obesity | mast_cell | *AKR7A2* | 5.508236341 | 5.52E-09 | 5.54E-08 | 3.47E-05 |
| Obesity | mast_cell | *MRTO4* | 4.989266103 | 1.43E-08 | 9.52E-08 | 8.97E-05 |
| Obesity | mast_cell | *MCF2L2* | -2.109090101 | 4.14E-06 | 9.93E-06 | 0.026035788 |
| Obesity | mast_cell | *ANAPC4* | 3.257404149 | 0.026438995 | 0.033833088 | 1 |
| Obesity | mast_cell | *LY75* | 5.193890579 | 0.000173446 | 0.000304257 | 1 |
| Obesity | mast_cell | *OPN3* | 1.876718319 | 0.016286777 | 0.021443972 | 1 |
| Obesity | mast_cell | *SDCCAG8* | 4.493147514 | 1.07E-07 | 4.30E-07 | 0.000672991 |
| Obesity | mast_cell | *TBC1D22A* | 3.9705889 | 0.000871029 | 0.001369039 | 1 |
| Obesity | mast_cell | *PLEKHH1* | 2.705565849 | 1.62E-06 | 4.33E-06 | 0.010199401 |
| Obesity | mast_cell | *ATP9A* | 4.82526794 | 5.37E-08 | 2.44E-07 | 0.000337332 |
| Obesity | mast_cell | *CHRDL2* | -3.447543476 | 3.58E-06 | 8.71E-06 | 0.02249659 |
| Obesity | mast_cell | *FAM114A2* | 3.946735887 | 0.008713081 | 0.011908509 | 1 |
| Obesity | mast_cell | *GINM1* | 6.924861352 | 1.17E-06 | 3.24E-06 | 0.007326559 |
| Obesity | mast_cell | *MRPL43* | 3.640822248 | 0.000164203 | 0.00028901 | 1 |
| Obesity | mast_cell | *ITIH4* | 1.556546712 | 1.55E-05 | 3.32E-05 | 0.097432567 |
| Obesity | mast_cell | *TRAF1* | 3.424555691 | 1.28E-08 | 8.92E-08 | 8.06E-05 |
| Obesity | mast_cell | *PCGF2* | 1.153726936 | 0.009597377 | 0.013034935 | 1 |
| Obesity | mast_cell | *IL17RB* | -6.444897878 | 0.000109768 | 0.000198823 | 0.690114543 |
| Obesity | mast_cell | *GDI2* | 3.27715582 | 0.009467957 | 0.012867981 | 1 |
| Obesity | mast_cell | *PITHD1* | 5.942166546 | 0.00106089 | 0.001644027 | 1 |
| Obesity | mast_cell | *ATP11B* | 4.463121038 | 2.15E-09 | 4.32E-08 | 1.35E-05 |
| Obesity | mast_cell | *CDK14* | 4.878215624 | 7.54E-09 | 6.47E-08 | 4.74E-05 |
| Obesity | mast_cell | *CROCC* | 3.096170681 | 1.10E-06 | 3.09E-06 | 0.006927101 |
| Obesity | mast_cell | *ZC3H11A* | 14.83853782 | 4.47E-08 | 2.11E-07 | 0.000280786 |
| Obesity | mast_cell | *FLYWCH1* | 1.783692561 | 0.003065559 | 0.004461381 | 1 |
| Obesity | mast_cell | *DNAJC25* | 6.801967392 | 3.96E-06 | 9.53E-06 | 0.024904754 |
| Obesity | mast_cell | *YBX3* | 6.639988372 | 2.79E-07 | 9.45E-07 | 0.001752403 |
| Obesity | mast_cell | *OGFR* | 5.578298677 | 1.64E-06 | 4.38E-06 | 0.010323533 |
| Obesity | mast_cell | *PIGV* | 4.192330676 | 0.000176511 | 0.000308772 | 1 |
| Obesity | mast_cell | *QSER1* | 6.575945516 | 2.37E-08 | 1.30E-07 | 0.000148974 |
| Obesity | mast_cell | *NCKAP1* | 5.255111582 | 8.39E-08 | 3.52E-07 | 0.000527445 |
| Obesity | mast_cell | *SFSWAP* | 3.649896716 | 0.009319551 | 0.012687747 | 1 |
| Obesity | mast_cell | *TNK2* | 3.376878914 | 0.000232896 | 0.00040006 | 1 |
| Obesity | mast_cell | *EIF4B* | 2.594047179 | 0.008536699 | 0.011680136 | 1 |
| Obesity | mast_cell | *SLC6A16* | 4.064345068 | 0.000186415 | 0.000324651 | 1 |
| Obesity | mast_cell | *SPHK2* | 5.235912756 | 1.85E-07 | 6.70E-07 | 0.001161331 |
| Obesity | mast_cell | *ISOC2* | 10.61712701 | 3.34E-09 | 4.52E-08 | 2.10E-05 |
| Obesity | mast_cell | *EPN1* | 9.598089854 | 6.59E-08 | 2.87E-07 | 0.000414597 |

| | | | | | | |
|---|---|---|---|---|---|---|
| Obesity | mast_cell | *ADCK1* | 5.638182829 | 0.001006149 | 0.001566144 | 1 |
| Obesity | mast_cell | *HAGH* | 3.051716368 | 9.07E-05 | 0.000165869 | 0.570167671 |
| Obesity | mast_cell | *CASP8* | 4.818354265 | 0.000233741 | 0.000401401 | 1 |
| Obesity | mast_cell | *TM7SF3* | 6.621921094 | 9.06E-09 | 7.25E-08 | 5.70E-05 |
| Obesity | mast_cell | *DMRT3* | 8.911784568 | 0.007862863 | 0.010807568 | 1 |
| Obesity | mast_cell | *ATP2C2* | -5.662951061 | 6.90E-05 | 0.000129593 | 0.433746588 |
| Obesity | mast_cell | *CDON* | 2.636753092 | 0.00012903 | 0.000230458 | 0.811213405 |
| Obesity | mast_cell | *TAF2* | 5.724352595 | 6.50E-08 | 2.84E-07 | 0.000408471 |
| Obesity | mast_cell | *TMEM161A* | 9.712378481 | 3.09E-08 | 1.58E-07 | 0.000194503 |
| Obesity | mast_cell | *LPAR2* | 4.296105519 | 0.01335005 | 0.017785922 | 1 |
| Obesity | mast_cell | *SUGP2* | 2.845688457 | 0.013027502 | 0.017393058 | 1 |
| Obesity | mast_cell | *DDX20* | 11.75897281 | 5.62E-08 | 2.54E-07 | 0.000353607 |
| Obesity | mast_cell | *BTBD1* | 7.400401722 | 1.56E-07 | 5.83E-07 | 0.000981995 |
| Obesity | mast_cell | *CHI3L2* | 2.037113005 | 0.0388038 | 0.048509374 | 1 |
| Obesity | mast_cell | *PMS1* | 5.987406062 | 4.32E-05 | 8.44E-05 | 0.27153765 |
| Obesity | mast_cell | *ANKS1A* | 5.782728705 | 2.37E-08 | 1.30E-07 | 0.000149241 |
| Obesity | mast_cell | *NTHL1* | 3.824553095 | 5.17E-06 | 1.21E-05 | 0.032530485 |
| Obesity | mast_cell | *UHRF1BP1* | 5.910843909 | 6.50E-29 | 1.02E-25 | 4.08E-25 |
| Obesity | mast_cell | *IPO5* | 7.510185466 | 1.50E-06 | 4.06E-06 | 0.009447743 |
| Obesity | mast_cell | *ADAT1* | 2.125056861 | 0.000356153 | 0.00059599 | 1 |
| Obesity | mast_cell | *NDUFB4* | 8.421650308 | 2.20E-05 | 4.54E-05 | 0.138193706 |
| Obesity | mast_cell | *ZC3H15* | 5.772941961 | 1.95E-05 | 4.06E-05 | 0.122313478 |
| Obesity | mast_cell | *MAP2K4* | 5.415086487 | 6.50E-05 | 0.000122666 | 0.408600579 |
| Obesity | mast_cell | *SEC61A2* | 8.859681709 | 3.13E-07 | 1.04E-06 | 0.001967118 |
| Obesity | mast_cell | *ASB1* | -4.298600751 | 0.004916747 | 0.006951111 | 1 |
| Obesity | mast_cell | *PPP2R5A* | 5.723445601 | 9.34E-07 | 2.66E-06 | 0.005869227 |
| Obesity | mast_cell | *TIE1* | 1.90948198 | 0.003513256 | 0.005074166 | 1 |
| Obesity | mast_cell | *NFYC* | 3.858602659 | 0.008253906 | 0.011307978 | 1 |
| Obesity | mast_cell | *ZMYND12* | 1.706249585 | 0.003945438 | 0.005658067 | 1 |
| Obesity | mast_cell | *NGEF* | -8.773282374 | 0.001756731 | 0.002635306 | 1 |
| Obesity | mast_cell | *CD84* | -1.590441196 | 9.49E-07 | 2.70E-06 | 0.005968983 |
| Obesity | mast_cell | *ZNRD1* | 2.54969346 | 0.005776129 | 0.008075278 | 1 |
| Obesity | mast_cell | *ATXN3* | 3.517945718 | 5.62E-06 | 1.31E-05 | 0.035341595 |
| Obesity | mast_cell | *GOLGA5* | 4.411558194 | 0.010852238 | 0.014645508 | 1 |
| Obesity | mast_cell | *EML1* | 3.17025563 | 6.85E-05 | 0.000128807 | 0.430729358 |
| Obesity | mast_cell | *ZFAT* | 5.268789801 | 5.59E-05 | 0.000107111 | 0.351752192 |
| Obesity | mast_cell | *MTFR1* | 4.178257065 | 0.001028867 | 0.001598341 | 1 |
| Obesity | mast_cell | *STAG3* | 4.187466803 | 3.22E-07 | 1.07E-06 | 0.002026136 |
| Obesity | mast_cell | *MYO9A* | 5.0090666 | 7.35E-07 | 2.16E-06 | 0.004622841 |
| Obesity | mast_cell | *SP100* | 4.631592321 | 0.000213297 | 0.000368203 | 1 |
| Obesity | mast_cell | *STOML1* | 2.883180585 | 2.00E-08 | 1.15E-07 | 0.000125936 |
| Obesity | mast_cell | *METTL22* | 4.855296596 | 0.020708639 | 0.026899837 | 1 |
| Obesity | mast_cell | *RRP15* | 1.406232555 | 0.010798647 | 0.014578289 | 1 |
| Obesity | mast_cell | *DHX8* | 5.4350749 | 2.51E-05 | 5.12E-05 | 0.157814276 |
| Obesity | mast_cell | *SYT1* | 3.161074582 | 4.50E-08 | 2.13E-07 | 0.000283088 |
| Obesity | mast_cell | *ROGDI* | 3.325696075 | 1.64E-08 | 1.03E-07 | 0.000103349 |
| Obesity | mast_cell | *HDAC4* | 6.071896419 | 4.15E-07 | 1.33E-06 | 0.002609622 |
| Obesity | mast_cell | *IFI35* | 2.647885914 | 5.82E-09 | 5.65E-08 | 3.66E-05 |
| Obesity | mast_cell | *HEATR6* | 9.202126403 | 1.54E-07 | 5.78E-07 | 0.000969621 |
| Obesity | mast_cell | *POLR1A* | 1.315335316 | 0.004051607 | 0.005798419 | 1 |

| | | | | | | |
|---|---|---|---|---|---|---|
| Obesity | mast_cell | *STON1-GTF2A* | 0.469948453 | 0.01576079 | 0.020781898 | 1 |
| Obesity | mast_cell | *SRBD1* | 5.263005883 | 0.000122516 | 0.000220011 | 0.770258791 |
| Obesity | mast_cell | *CYFIP1* | 5.449193871 | 0.000101713 | 0.000184711 | 0.639469708 |
| Obesity | mast_cell | *PSME4* | 4.882180575 | 2.58E-05 | 5.25E-05 | 0.162476686 |
| Obesity | mast_cell | *PITX1* | 26.79441008 | 0.000449451 | 0.000740875 | 1 |
| Obesity | mast_cell | *SDK2* | -0.751539518 | 2.50E-07 | 8.62E-07 | 0.001569226 |
| Obesity | mast_cell | *NUCKS1* | 4.252784879 | 0.001970559 | 0.002938545 | 1 |
| Obesity | mast_cell | *DNAJA2* | 8.82344071 | 1.61E-08 | 1.02E-07 | 0.000100916 |
| Obesity | mast_cell | *KCNAB2* | 2.887327011 | 0.029123304 | 0.037026939 | 1 |
| Obesity | mast_cell | *GAL* | 3.886993698 | 2.59E-08 | 1.38E-07 | 0.000162533 |
| Obesity | mast_cell | *NEDD4* | 4.261663428 | 1.90E-05 | 3.99E-05 | 0.119745228 |
| Obesity | mast_cell | *PIGB* | 4.837461151 | 0.000769895 | 0.001219226 | 1 |
| Obesity | mast_cell | *GNB5* | 3.489552407 | 0.002504082 | 0.003688651 | 1 |
| Obesity | mast_cell | *PFN2* | 5.250726027 | 3.50E-08 | 1.73E-07 | 0.000219873 |
| Obesity | mast_cell | *DAPP1* | 3.811636541 | 8.28E-05 | 0.00015264 | 0.520655613 |
| Obesity | mast_cell | *FGF10* | 2.354290307 | 1.02E-07 | 4.13E-07 | 0.000638535 |
| Obesity | mast_cell | *TMEM260* | 6.365116599 | 5.96E-06 | 1.38E-05 | 0.037448835 |
| Obesity | mast_cell | *CLTCL1* | 4.600145774 | 5.43E-08 | 2.47E-07 | 0.000341186 |
| Obesity | mast_cell | *DGCR2* | 6.991823344 | 3.03E-07 | 1.01E-06 | 0.001904915 |
| Obesity | mast_cell | *ZXDC* | 5.063461799 | 0.0002766 | 0.000469742 | 1 |
| Obesity | mast_cell | *POLB* | 4.651866079 | 5.63E-05 | 0.000107706 | 0.353922063 |
| Obesity | mast_cell | *NDST1* | 5.931188558 | 7.55E-09 | 6.47E-08 | 4.75E-05 |
| Obesity | mast_cell | *AP3M2* | 6.924310464 | 2.84E-07 | 9.59E-07 | 0.001785294 |
| Obesity | mast_cell | *ST6GALNAC2* | -2.476124832 | 0.000836449 | 0.001318315 | 1 |
| Obesity | mast_cell | *TESK2* | -2.079608702 | 1.07E-05 | 2.36E-05 | 0.067098476 |
| Obesity | mast_cell | *EIF2B3* | 4.815443805 | 3.66E-09 | 4.64E-08 | 2.30E-05 |
| Obesity | mast_cell | *CAMK2A* | 11.48366788 | 1.76E-08 | 1.05E-07 | 0.000110371 |
| Obesity | mast_cell | *CDC42* | 6.609141204 | 8.85E-08 | 3.68E-07 | 0.000556664 |
| Obesity | mast_cell | *OSBPL3* | 5.743582305 | 7.78E-09 | 6.64E-08 | 4.89E-05 |
| Obesity | mast_cell | *SNX13* | 5.240670719 | 7.29E-05 | 0.000136201 | 0.458451078 |
| Obesity | mast_cell | *VASH1* | 6.107655627 | 1.58E-08 | 1.01E-07 | 9.95E-05 |
| Obesity | mast_cell | *TRIP13* | -4.049138456 | 7.00E-07 | 2.06E-06 | 0.004402395 |
| Obesity | mast_cell | *TCF3* | 6.722645047 | 3.35E-09 | 4.52E-08 | 2.11E-05 |
| Obesity | mast_cell | *MBD3* | 7.455023576 | 1.73E-09 | 4.28E-08 | 1.09E-05 |
| Obesity | mast_cell | *CPSF1* | -4.705807369 | 0.000903251 | 0.001416144 | 1 |
| Obesity | mast_cell | *CYBRD1* | 4.394192816 | 3.51E-06 | 8.57E-06 | 0.022064118 |
| Obesity | mast_cell | *EPN2* | 3.914325136 | 0.004626863 | 0.006556027 | 1 |
| Obesity | mast_cell | *LIMS2* | 4.388947223 | 7.76E-09 | 6.63E-08 | 4.88E-05 |
| Obesity | mast_cell | *AFF4* | 3.235597005 | 0.005285847 | 0.007432815 | 1 |
| Obesity | mast_cell | *TRHDE* | 3.240102136 | 2.67E-09 | 4.32E-08 | 1.68E-05 |
| Obesity | mast_cell | *TRNT1* | 5.760556038 | 0.00049739 | 0.000811862 | 1 |
| Obesity | mast_cell | *ACADVL* | 6.028298008 | 1.03E-05 | 2.29E-05 | 0.064849355 |
| Obesity | mast_cell | *CRMP1* | 4.406750941 | 4.42E-07 | 1.40E-06 | 0.00277831 |
| Obesity | mast_cell | *DERL2* | 7.463324087 | 5.11E-07 | 1.56E-06 | 0.003210509 |
| Obesity | mast_cell | *NDE1* | 6.221756785 | 5.11E-09 | 5.35E-08 | 3.21E-05 |
| Obesity | mast_cell | *AP1M1* | 3.21162952 | 3.11E-05 | 6.23E-05 | 0.195476683 |
| Obesity | mast_cell | *PVR* | 6.421229098 | 5.85E-07 | 1.76E-06 | 0.003674857 |
| Obesity | mast_cell | *XRCC1* | 6.297395659 | 0.004165843 | 0.005951069 | 1 |
| Obesity | mast_cell | *MOV10L1* | 2.051972539 | 1.65E-06 | 4.38E-06 | 0.0103432 |
| Obesity | mast_cell | *TP63* | 2.862537027 | 1.38E-08 | 9.32E-08 | 8.71E-05 |

| | | | | | | |
|---|---|---|---|---|---|---|
| Obesity | mast_cell | *ALPK1* | 6.02424213 | 3.68E-06 | 8.92E-06 | 0.023142533 |
| Obesity | mast_cell | *LLGL2* | 3.636874472 | 4.73E-07 | 1.47E-06 | 0.002975061 |
| Obesity | mast_cell | *PDE8A* | 4.264421794 | 1.49E-05 | 3.21E-05 | 0.093988911 |
| Obesity | mast_cell | *SDHA* | 5.643180517 | 4.03E-08 | 1.95E-07 | 0.000253435 |
| Obesity | mast_cell | *SMARCE1* | 7.860011792 | 5.18E-08 | 2.38E-07 | 0.000325491 |
| Obesity | mast_cell | *GSDMB* | 2.800295922 | 7.67E-06 | 1.74E-05 | 0.04824728 |
| Obesity | mast_cell | *ADAM11* | 1.237995786 | 0.038171684 | 0.047758284 | 1 |
| Obesity | mast_cell | *PICALM* | 4.764946837 | 1.97E-06 | 5.11E-06 | 0.012368109 |
| Obesity | mast_cell | *GLI2* | 1.58332922 | 0.000179106 | 0.000312875 | 1 |
| Obesity | mast_cell | *CLNS1A* | 3.471051222 | 2.15E-05 | 4.46E-05 | 0.135346164 |
| Obesity | mast_cell | *ATP2A3* | -2.47262592 | 0.001111782 | 0.001716967 | 1 |
| Obesity | mast_cell | *CA12* | 2.863905714 | 2.83E-07 | 9.57E-07 | 0.001779739 |
| Obesity | mast_cell | *NTN4* | 3.431820491 | 8.48E-09 | 6.93E-08 | 5.33E-05 |
| Obesity | mast_cell | *LMAN1* | 5.874957166 | 0.000149744 | 0.000265195 | 0.941442278 |
| Obesity | mast_cell | *ENO1* | 6.951893649 | 2.58E-08 | 1.38E-07 | 0.00016198 |
| Obesity | mast_cell | *TIPIN* | 2.394754996 | 3.22E-09 | 4.49E-08 | 2.02E-05 |
| Obesity | mast_cell | *SRI* | 8.376612527 | 2.21E-07 | 7.77E-07 | 0.001388802 |
| Obesity | mast_cell | *EIF4G3* | 4.151323101 | 0.000247094 | 0.000422716 | 1 |
| Obesity | mast_cell | *TTC38* | 5.274771419 | 6.63E-08 | 2.88E-07 | 0.000416629 |
| Obesity | mast_cell | *CELSR1* | 11.6173835 | 4.34E-08 | 2.06E-07 | 0.000272792 |
| Obesity | mast_cell | *ADD2* | 3.994278408 | 1.62E-08 | 1.02E-07 | 0.000102147 |
| Obesity | mast_cell | *VPS9D1* | 6.020951003 | 9.76E-09 | 7.58E-08 | 6.14E-05 |
| Obesity | mast_cell | *MARK3* | 3.170111533 | 0.003794139 | 0.00545478 | 1 |
| Obesity | mast_cell | *SLC25A3* | 7.169386378 | 1.77E-08 | 1.06E-07 | 0.000111374 |
| Obesity | mast_cell | *MOCOS* | 2.076534227 | 4.16E-08 | 2.00E-07 | 0.000261483 |
| Obesity | mast_cell | *BCAP29* | 6.225025485 | 3.80E-07 | 1.23E-06 | 0.00238782 |
| Obesity | mast_cell | *SEC31B* | 1.262360243 | 0.000358202 | 0.000598781 | 1 |
| Obesity | mast_cell | *TUBA3D* | 7.834064013 | 0.013460324 | 0.017921443 | 1 |
| Obesity | mast_cell | *MKRN2* | 6.53619548 | 1.08E-10 | 9.17E-09 | 6.79E-07 |
| Obesity | mast_cell | *MCM6* | 4.068306311 | 7.80E-09 | 6.64E-08 | 4.90E-05 |
| Obesity | mast_cell | *RBMS2* | 6.073330834 | 4.98E-08 | 2.32E-07 | 0.000313326 |
| Obesity | mast_cell | *MLH1* | 3.370532956 | 0.004526652 | 0.006431426 | 1 |
| Obesity | mast_cell | *RGS11* | 1.747395879 | 0.00018531 | 0.000322816 | 1 |
| Obesity | mast_cell | *GPATCH1* | 5.321991877 | 5.49E-10 | 2.45E-08 | 3.45E-06 |
| Obesity | mast_cell | *ICAM3* | 3.672390354 | 8.79E-07 | 2.53E-06 | 0.005525463 |
| Obesity | mast_cell | *NT5C2* | 5.166344042 | 2.41E-05 | 4.94E-05 | 0.15154989 |
| Obesity | mast_cell | *CAMSAP3* | 15.32692623 | 1.70E-08 | 1.04E-07 | 0.000106662 |
| Obesity | mast_cell | *XAB2* | 3.517402339 | 1.44E-05 | 3.09E-05 | 0.090270852 |
| Obesity | mast_cell | *STXBP2* | -3.988238804 | 4.21E-05 | 8.24E-05 | 0.264624061 |
| Obesity | mast_cell | *MAP2K7* | 8.789375597 | 3.88E-08 | 1.89E-07 | 0.000243741 |
| Obesity | mast_cell | *NMRK2* | -3.771746693 | 7.57E-06 | 1.72E-05 | 0.047618181 |
| Obesity | mast_cell | *CTTNBP2* | -2.60740763 | 0.000598395 | 0.000964396 | 1 |
| Obesity | mast_cell | *TM9SF3* | 6.045345604 | 5.42E-07 | 1.64E-06 | 0.003407034 |
| Obesity | mast_cell | *DNAJC10* | 7.227080308 | 1.57E-07 | 5.85E-07 | 0.000988563 |
| Obesity | mast_cell | *DYNC1I2* | 7.275137879 | 1.54E-10 | 1.13E-08 | 9.68E-07 |
| Obesity | mast_cell | *APBB1IP* | 3.926582851 | 2.94E-08 | 1.53E-07 | 0.000185131 |
| Obesity | mast_cell | *SIRT6* | 13.54603301 | 1.02E-07 | 4.13E-07 | 0.00063923 |
| Obesity | mast_cell | *NAALAD2* | 2.454258495 | 0.013216768 | 0.017615819 | 1 |
| Obesity | mast_cell | *FGFR1* | 3.338824337 | 0.005409749 | 0.007593457 | 1 |
| Obesity | mast_cell | *FBLN1* | 2.942022988 | 9.19E-09 | 7.26E-08 | 5.78E-05 |

| | | | | | | |
|---|---|---|---|---|---|---|
| Obesity | mast_cell | *PIAS2* | 5.010742197 | 7.29E-06 | 1.66E-05 | 0.045820043 |
| Obesity | mast_cell | *AMPH* | 2.102301051 | 4.68E-06 | 1.11E-05 | 0.029446043 |
| Obesity | mast_cell | *MCCC1* | 4.557503628 | 2.10E-09 | 4.32E-08 | 1.32E-05 |
| Obesity | mast_cell | *FAP* | 2.351346666 | 3.80E-06 | 9.16E-06 | 0.023874127 |
| Obesity | mast_cell | *PIK3C3* | 4.943950926 | 0.000200368 | 0.000347602 | 1 |
| Obesity | mast_cell | *N4BP2* | 5.048089669 | 3.42E-06 | 8.38E-06 | 0.021509421 |
| Obesity | mast_cell | *TULP3* | 6.422829827 | 2.04E-07 | 7.25E-07 | 0.001284593 |
| Obesity | mast_cell | *SYNJ2* | 5.765050624 | 3.43E-06 | 8.39E-06 | 0.021556239 |
| Obesity | mast_cell | *ZCWPW1* | 5.840042997 | 6.86E-08 | 2.96E-07 | 0.000431126 |
| Obesity | mast_cell | *FGF20* | -22.43209874 | 2.22E-06 | 5.69E-06 | 0.013934698 |
| Obesity | mast_cell | *PCM1* | 4.003850125 | 0.000357551 | 0.000597853 | 1 |
| Obesity | mast_cell | *OSBPL6* | 7.000324969 | 1.02E-08 | 7.86E-08 | 6.43E-05 |
| Obesity | mast_cell | *XRCC5* | 4.524956473 | 0.000750607 | 0.001190782 | 1 |
| Obesity | mast_cell | *LXN* | -2.826969341 | 0.01520329 | 0.020076262 | 1 |
| Obesity | mast_cell | *REXO1* | 5.700959635 | 2.37E-09 | 4.32E-08 | 1.49E-05 |
| Obesity | mast_cell | *SAR1A* | 5.694758866 | 3.94E-09 | 4.78E-08 | 2.48E-05 |
| Obesity | mast_cell | *CDC14A* | -2.178181048 | 0.009246395 | 0.012596335 | 1 |
| Obesity | mast_cell | *RAPGEF3* | 1.011399381 | 0.003416747 | 0.004942726 | 1 |
| Obesity | mast_cell | *FDFT1* | 7.912255615 | 6.64E-08 | 2.88E-07 | 0.000417379 |
| Obesity | mast_cell | *SCGN* | 12.3290664 | 3.90E-09 | 4.78E-08 | 2.45E-05 |
| Obesity | mast_cell | *DDX1* | 6.845041558 | 5.61E-08 | 2.53E-07 | 0.000352426 |
| Obesity | mast_cell | *DNM2* | 3.770270091 | 0.010855426 | 0.014645508 | 1 |
| Obesity | mast_cell | *STX7* | 3.10556799 | 0.003730208 | 0.005372077 | 1 |
| Obesity | mast_cell | *DCT* | 3.545245194 | 1.07E-08 | 8.05E-08 | 6.71E-05 |
| Obesity | mast_cell | *SLC35C2* | 4.554549016 | 0.014397321 | 0.019088139 | 1 |
| Obesity | mast_cell | *SCTR* | 7.477760132 | 2.36E-08 | 1.30E-07 | 0.000148615 |
| Obesity | mast_cell | *RIF1* | 3.514256547 | 0.001048554 | 0.001626112 | 1 |
| Obesity | mast_cell | *KCNN2* | 5.286512286 | 4.97E-08 | 2.31E-07 | 0.000312217 |
| Obesity | mast_cell | *PSEN1* | 4.476860377 | 0.000127994 | 0.000228974 | 0.804695762 |
| Obesity | mast_cell | *CPOX* | 6.810027061 | 1.21E-07 | 4.76E-07 | 0.00076219 |
| Obesity | mast_cell | *MOK* | 6.582763407 | 3.62E-09 | 4.63E-08 | 2.28E-05 |
| Obesity | mast_cell | *RSBN1* | 5.323036963 | 1.69E-05 | 3.58E-05 | 0.106356544 |
| Obesity | mast_cell | *MAGI3* | 5.614790657 | 6.83E-06 | 1.57E-05 | 0.042913599 |
| Obesity | mast_cell | *COL4A4* | 2.726213497 | 3.61E-09 | 4.63E-08 | 2.27E-05 |
| Obesity | mast_cell | *PKP1* | -2.402129251 | 0.004019543 | 0.005755151 | 1 |
| Obesity | mast_cell | *STK17B* | 6.200508055 | 8.50E-07 | 2.45E-06 | 0.005343628 |
| Obesity | mast_cell | *ZNF506* | 6.363356948 | 5.47E-08 | 2.48E-07 | 0.000343845 |
| Obesity | mast_cell | *KIAA0141* | 6.078711925 | 2.83E-08 | 1.48E-07 | 0.000177941 |
| Obesity | mast_cell | *PCDHA6* | -8.149056004 | 1.59E-08 | 1.02E-07 | 9.97E-05 |
| Obesity | mast_cell | *PCDHGA2* | 4.001100684 | 0.002250511 | 0.003333089 | 1 |
| Obesity | mast_cell | *WDR70* | 4.736609902 | 3.12E-05 | 6.25E-05 | 0.196422307 |
| Obesity | mast_cell | *PGR* | 3.313747069 | 1.35E-08 | 9.19E-08 | 8.51E-05 |
| Obesity | mast_cell | *C5orf22* | 7.502925213 | 3.80E-09 | 4.74E-08 | 2.39E-05 |
| Obesity | mast_cell | *COBLL1* | 3.974446429 | 2.79E-09 | 4.32E-08 | 1.75E-05 |
| Obesity | mast_cell | *GEMIN5* | 2.499470168 | 0.025491958 | 0.032694398 | 1 |
| Obesity | mast_cell | *GSK3B* | 5.158412927 | 7.50E-07 | 2.20E-06 | 0.004718181 |
| Obesity | mast_cell | *ERC1* | 4.450192499 | 5.55E-05 | 0.000106425 | 0.348861496 |
| Obesity | mast_cell | *DOPEY1* | 3.312444335 | 3.59E-06 | 8.73E-06 | 0.022565772 |
| Obesity | mast_cell | *LYRM2* | 3.806464682 | 6.87E-09 | 6.14E-08 | 4.32E-05 |
| Obesity | mast_cell | *ULK2* | 5.067585812 | 3.29E-07 | 1.09E-06 | 0.002068864 |

| | | | | | | |
|---|---|---|---|---|---|---|
| Obesity | mast_cell | *PLOD1* | 10.45323591 | 4.53E-08 | 2.14E-07 | 0.000284991 |
| Obesity | mast_cell | *PIBF1* | 4.605289568 | 2.43E-05 | 4.98E-05 | 0.1528334 |
| Obesity | mast_cell | *PDS5B* | 4.284821023 | 0.001070316 | 0.001657816 | 1 |
| Obesity | mast_cell | *OXCT1* | 2.450828384 | 3.23E-06 | 7.95E-06 | 0.020304443 |
| Obesity | mast_cell | *CYLD* | 4.110135242 | 0.008458646 | 0.011575861 | 1 |
| Obesity | mast_cell | *ZNF324* | -4.135197394 | 0.000842516 | 0.001326878 | 1 |
| Obesity | mast_cell | *ZNF586* | 5.101567839 | 6.75E-07 | 2.00E-06 | 0.004246118 |
| Obesity | mast_cell | *ZNF446* | 9.641031355 | 2.61E-09 | 4.32E-08 | 1.64E-05 |
| Obesity | mast_cell | *RPS5* | 7.452692607 | 7.02E-06 | 1.61E-05 | 0.044154871 |
| Obesity | mast_cell | *CHMP2B* | 4.563004612 | 0.000571414 | 0.00092328 | 1 |
| Obesity | mast_cell | *PPIE* | 6.624435637 | 2.17E-09 | 4.32E-08 | 1.37E-05 |
| Obesity | mast_cell | *ZMPSTE24* | 7.517449474 | 1.11E-05 | 2.46E-05 | 0.06998735 |
| Obesity | mast_cell | *STARD7* | 8.679856115 | 6.85E-08 | 2.96E-07 | 0.000430376 |
| Obesity | mast_cell | *NOA1* | 3.574622302 | 0.000170615 | 0.000299458 | 1 |
| Obesity | mast_cell | *GSTP1* | 8.944317759 | 2.01E-07 | 7.18E-07 | 0.001265525 |
| Obesity | mast_cell | *WBP11* | 7.357636187 | 0.000118765 | 0.000213702 | 0.746673686 |
| Obesity | mast_cell | *APOB* | 2.496985978 | 1.42E-07 | 5.39E-07 | 0.000891154 |
| Obesity | mast_cell | *NCOA1* | 3.462608918 | 0.00243142 | 0.003586659 | 1 |
| Obesity | mast_cell | *CD59* | 5.581807896 | 4.16E-08 | 2.00E-07 | 0.000261226 |
| Obesity | mast_cell | *CD82* | -0.779112951 | 1.16E-06 | 3.23E-06 | 0.007283247 |
| Obesity | mast_cell | *TAF9* | 6.456363368 | 2.58E-09 | 4.32E-08 | 1.62E-05 |
| Obesity | mast_cell | *FCN1* | 2.391084365 | 6.37E-07 | 1.90E-06 | 0.004006423 |
| Obesity | mast_cell | *SCAMP1* | 7.904590184 | 6.95E-09 | 6.19E-08 | 4.37E-05 |
| Obesity | mast_cell | *HACE1* | 5.891454064 | 4.08E-09 | 4.79E-08 | 2.57E-05 |
| Obesity | mast_cell | *SEH1L* | 6.009458964 | 1.73E-08 | 1.05E-07 | 0.000108562 |
| Obesity | mast_cell | *OVGP1* | 3.787711403 | 0.00046946 | 0.00076942 | 1 |
| Obesity | mast_cell | *SLC25A24* | 5.413209486 | 1.54E-07 | 5.78E-07 | 0.000966885 |
| Obesity | mast_cell | *PILRA* | 0.844363863 | 0.001151229 | 0.001775271 | 1 |
| Obesity | mast_cell | *ZNF213* | 2.447109535 | 0.011195652 | 0.015068949 | 1 |
| Obesity | mast_cell | *CPNE3* | 2.377574144 | 0.01914958 | 0.024977886 | 1 |
| Obesity | mast_cell | *RRN3* | 5.807225398 | 5.59E-05 | 0.000107028 | 0.351373749 |
| Obesity | mast_cell | *CTTN* | 5.967728407 | 8.75E-09 | 7.07E-08 | 5.50E-05 |
| Obesity | mast_cell | *MTIF2* | 4.77948586 | 9.84E-06 | 2.19E-05 | 0.061859524 |
| Obesity | mast_cell | *EPS15* | 2.206841478 | 0.007527944 | 0.010374437 | 1 |
| Obesity | mast_cell | *ATG16L1* | 7.496494101 | 2.13E-05 | 4.42E-05 | 0.134079437 |
| Obesity | mast_cell | *USP40* | 5.594314514 | 5.23E-08 | 2.40E-07 | 0.000328623 |
| Obesity | mast_cell | *MAST2* | 6.488694634 | 4.66E-07 | 1.46E-06 | 0.002930178 |
| Obesity | mast_cell | *B4GALT1* | 5.56738078 | 9.78E-05 | 0.000178235 | 0.614912288 |
| Obesity | mast_cell | *CHMP5* | 8.010426314 | 1.76E-07 | 6.41E-07 | 0.001104364 |
| Obesity | mast_cell | *FOLH1* | 1.218659304 | 0.003331827 | 0.004827655 | 1 |
| Obesity | mast_cell | *EIF2AK1* | 6.339953196 | 8.10E-07 | 2.35E-06 | 0.005089873 |
| Obesity | mast_cell | *SEPHS1* | -2.735913216 | 0.038108693 | 0.047698458 | 1 |
| Obesity | mast_cell | *ITPKC* | 7.370059877 | 3.70E-06 | 8.97E-06 | 0.023285023 |
| Obesity | mast_cell | *TMED2* | 6.947305699 | 2.59E-09 | 4.32E-08 | 1.63E-05 |
| Obesity | mast_cell | *ACOX3* | 5.024529779 | 0.000113091 | 0.000204487 | 0.711001607 |
| Obesity | mast_cell | *HSD17B14* | 5.67209288 | 1.93E-06 | 5.03E-06 | 0.012139167 |
| Obesity | mast_cell | *TRIP6* | 3.601991724 | 0.003728284 | 0.005371156 | 1 |
| Obesity | mast_cell | *SRRT* | 4.184143085 | 0.000288717 | 0.000488473 | 1 |
| Obesity | mast_cell | *PIGS* | 2.61525353 | 3.23E-05 | 6.44E-05 | 0.202996552 |
| Obesity | mast_cell | *ATXN7L3* | 5.056865046 | 1.26E-09 | 3.78E-08 | 7.94E-06 |

| | | | | | | |
|---|---|---|---|---|---|---|
| Obesity | mast_cell | *PGS1* | 6.936901034 | 1.83E-06 | 4.81E-06 | 0.01152454 |
| Obesity | mast_cell | *PSMC5* | 3.296815564 | 0.001827401 | 0.002732842 | 1 |
| Obesity | mast_cell | *LPCAT2* | 4.806755019 | 6.96E-05 | 0.000130554 | 0.437487096 |
| Obesity | mast_cell | *TXNDC16* | 4.645205371 | 8.87E-07 | 2.55E-06 | 0.005576958 |
| Obesity | mast_cell | *GMCL1* | 6.635167658 | 5.49E-09 | 5.54E-08 | 3.45E-05 |
| Obesity | mast_cell | *PTHLH* | 8.03773066 | 1.18E-05 | 2.60E-05 | 0.074484989 |
| Obesity | mast_cell | *PHACTR3* | 1.782230242 | 3.52E-06 | 8.58E-06 | 0.022131905 |
| Obesity | mast_cell | *ERGIC2* | 5.786276532 | 0.000230407 | 0.000396107 | 1 |
| Obesity | mast_cell | *METTL2A* | 3.588815507 | 0.003178195 | 0.004615687 | 1 |
| Obesity | mast_cell | *ALG6* | 6.241552735 | 7.00E-08 | 3.01E-07 | 0.000439801 |
| Obesity | mast_cell | *DDX18* | 6.047544975 | 4.96E-06 | 1.17E-05 | 0.031207074 |
| Obesity | mast_cell | *ASAP3* | 5.075732598 | 4.59E-10 | 2.22E-08 | 2.88E-06 |
| Obesity | mast_cell | *EDEM2* | -2.548296262 | 0.001636306 | 0.002465243 | 1 |
| Obesity | mast_cell | *DOCK3* | 3.387860381 | 5.62E-08 | 2.53E-07 | 0.000353119 |
| Obesity | mast_cell | *FKBP1A* | 7.961656905 | 0.000177304 | 0.000309986 | 1 |
| Obesity | mast_cell | *NSFL1C* | 8.252192039 | 2.76E-09 | 4.32E-08 | 1.74E-05 |
| Obesity | mast_cell | *ZNF343* | 5.752834958 | 3.63E-09 | 4.63E-08 | 2.28E-05 |
| Obesity | mast_cell | *MAVS* | 5.357021672 | 0.000125983 | 0.000225785 | 0.792053822 |
| Obesity | mast_cell | *LZTS3* | -4.113584799 | 2.12E-06 | 5.46E-06 | 0.013315453 |
| Obesity | mast_cell | *XRN2* | 4.945554394 | 0.000698422 | 0.001112767 | 1 |
| Obesity | mast_cell | *TESC* | -1.437115521 | 4.81E-07 | 1.49E-06 | 0.003024545 |
| Obesity | mast_cell | *ANAPC5* | 3.867643124 | 0.005052466 | 0.007136565 | 1 |
| Obesity | mast_cell | *SLC23A2* | 6.58622942 | 3.65E-06 | 8.86E-06 | 0.022945461 |
| Obesity | mast_cell | *TMEM230* | 4.820201652 | 1.49E-07 | 5.61E-07 | 0.000933963 |
| Obesity | mast_cell | *DZANK1* | 4.774279201 | 0.007939299 | 0.010910246 | 1 |
| Obesity | mast_cell | *TASP1* | 5.43401359 | 3.08E-08 | 1.57E-07 | 0.000193522 |
| Obesity | mast_cell | *OAS1* | 8.017419482 | 0.000297498 | 0.000502664 | 1 |
| Obesity | mast_cell | *KIF16B* | 6.451207609 | 8.53E-07 | 2.46E-06 | 0.005365627 |
| Obesity | mast_cell | *FUS* | 5.540684114 | 6.49E-06 | 1.49E-05 | 0.04077474 |
| Obesity | mast_cell | *FXYD5* | 8.231719078 | 3.48E-05 | 6.90E-05 | 0.2186018 |
| Obesity | mast_cell | *FXYD3* | 9.498279269 | 7.50E-08 | 3.18E-07 | 0.000471272 |
| Obesity | mast_cell | *KCNH4* | 1.942513693 | 0.002194046 | 0.003256367 | 1 |
| Obesity | mast_cell | *GANAB* | 6.974060949 | 2.31E-09 | 4.32E-08 | 1.45E-05 |
| Obesity | mast_cell | *GMIP* | 11.98562861 | 1.83E-08 | 1.08E-07 | 0.000115015 |
| Obesity | mast_cell | *NECAP1* | 4.930907111 | 9.41E-05 | 0.000171977 | 0.591772664 |
| Obesity | mast_cell | *ANKRD24* | 4.878852867 | 2.38E-08 | 1.30E-07 | 0.00014989 |
| Obesity | mast_cell | *GPATCH2L* | 3.924474642 | 0.000343675 | 0.000576336 | 1 |
| Obesity | mast_cell | *SLC9A1* | -2.912467279 | 4.04E-05 | 7.94E-05 | 0.254239235 |
| Obesity | mast_cell | *SPTLC1* | 5.139490582 | 1.20E-05 | 2.64E-05 | 0.075716102 |
| Obesity | mast_cell | *PAPOLA* | 6.022826067 | 6.24E-08 | 2.76E-07 | 0.000392068 |
| Obesity | mast_cell | *RGS1* | -5.14768778 | 6.82E-05 | 0.000128212 | 0.428484963 |
| Obesity | mast_cell | *YPEL3* | 4.534342048 | 6.83E-16 | 3.31E-13 | 4.30E-12 |
| Obesity | mast_cell | *MRPS33* | 5.866233739 | 4.42E-08 | 2.09E-07 | 0.00027795 |
| Obesity | mast_cell | *MAEA* | 2.560698994 | 0.012217236 | 0.016363392 | 1 |
| Obesity | mast_cell | *STRN4* | 6.696817355 | 0.000636876 | 0.0010217 | 1 |
| Obesity | mast_cell | *MUL1* | 3.303858496 | 4.89E-08 | 2.29E-07 | 0.000307666 |
| Obesity | mast_cell | *SPG21* | 4.013489752 | 3.29E-07 | 1.09E-06 | 0.002065601 |
| Obesity | mast_cell | *GNPTG* | 6.96786291 | 4.00E-09 | 4.78E-08 | 2.52E-05 |
| Obesity | mast_cell | *ZNF268* | 7.599552072 | 5.17E-05 | 9.98E-05 | 0.325081742 |
| Obesity | mast_cell | *GOLGA3* | 3.805194426 | 0.002535866 | 0.003731099 | 1 |

| | | | | | | |
|---|---|---|---|---|---|---|
| Obesity | mast_cell | *CERS4* | -2.917129826 | 3.32E-05 | 6.61E-05 | 0.208823725 |
| Obesity | mast_cell | *PDPR* | 6.801549757 | 5.01E-06 | 1.18E-05 | 0.031505366 |
| Obesity | mast_cell | *EXOC1* | 5.242399567 | 7.04E-05 | 0.000131942 | 0.442796908 |
| Obesity | mast_cell | *NLRC4* | 7.514462323 | 2.85E-09 | 4.32E-08 | 1.79E-05 |
| Obesity | mast_cell | *LAMB1* | 4.299874738 | 1.31E-08 | 9.03E-08 | 8.26E-05 |
| Obesity | mast_cell | *SLC26A4* | 13.65289686 | 3.52E-09 | 4.62E-08 | 2.21E-05 |
| Obesity | mast_cell | *DLD* | 6.113777997 | 0.000147533 | 0.000261715 | 0.927536964 |
| Obesity | mast_cell | *TXNL1* | 7.387072994 | 1.10E-06 | 3.08E-06 | 0.006894897 |
| Obesity | mast_cell | *IL5RA* | -0.475627651 | 1.40E-07 | 5.34E-07 | 0.000880176 |
| Obesity | mast_cell | *RAPGEF4* | 4.862076363 | 1.12E-07 | 4.47E-07 | 0.000706569 |
| Obesity | mast_cell | *SEL1L3* | -2.560985401 | 0.039633045 | 0.049488173 | 1 |
| Obesity | mast_cell | *TF* | 4.459537137 | 4.01E-08 | 1.94E-07 | 0.000252045 |
| Obesity | mast_cell | *MYO15A* | -4.888881273 | 2.93E-05 | 5.90E-05 | 0.184165953 |
| Obesity | mast_cell | *ALKBH5* | 5.809394153 | 9.99E-09 | 7.72E-08 | 6.28E-05 |
| Obesity | mast_cell | *NLRP1* | 5.181385588 | 2.66E-07 | 9.10E-07 | 0.001672848 |
| Obesity | mast_cell | *SPAG7* | 2.399394187 | 0.005859787 | 0.008188593 | 1 |
| Obesity | mast_cell | *ZFHX4* | 5.343364638 | 3.89E-06 | 9.37E-06 | 0.024449552 |
| Obesity | mast_cell | *RGS17* | -1.887086507 | 1.55E-07 | 5.80E-07 | 0.000975504 |
| Obesity | mast_cell | *TMEM101* | 3.661635213 | 5.32E-08 | 2.43E-07 | 0.000334419 |
| Obesity | mast_cell | *CMA1* | -0.294024741 | 1.05E-07 | 4.21E-07 | 0.000657642 |
| Obesity | mast_cell | *PPP2R3C* | 6.711584187 | 5.31E-09 | 5.48E-08 | 3.34E-05 |
| Obesity | mast_cell | *HAUS4* | 1.347152802 | 0.010854124 | 0.014645508 | 1 |
| Obesity | mast_cell | *SLC22A17* | 2.864229949 | 6.66E-07 | 1.98E-06 | 0.004188068 |
| Obesity | mast_cell | *SCFD1* | 3.655164885 | 0.00049889 | 0.000813835 | 1 |
| Obesity | mast_cell | *RPGRIP1* | 4.897460432 | 3.36E-05 | 6.68E-05 | 0.211078183 |
| Obesity | mast_cell | *TOX4* | 7.05924111 | 1.59E-09 | 4.20E-08 | 1.00E-05 |
| Obesity | mast_cell | *TGM1* | 2.49444464 | 0.000284882 | 0.000482504 | 1 |
| Obesity | mast_cell | *AGO1* | 6.515020856 | 7.30E-05 | 0.000136228 | 0.458678343 |
| Obesity | mast_cell | *RFFL* | 8.823430507 | 2.59E-09 | 4.32E-08 | 1.63E-05 |
| Obesity | mast_cell | *MFSD11* | 3.924892711 | 1.62E-05 | 3.44E-05 | 0.101563727 |
| Obesity | mast_cell | *DPYSL2* | 5.950655864 | 4.23E-07 | 1.35E-06 | 0.002658995 |
| Obesity | mast_cell | *NUP50* | -2.334351137 | 0.019005836 | 0.024805831 | 1 |
| Obesity | mast_cell | *COMT* | 6.271965308 | 5.27E-09 | 5.45E-08 | 3.31E-05 |
| Obesity | mast_cell | *FMO2* | 3.547978457 | 3.82E-09 | 4.75E-08 | 2.40E-05 |
| Obesity | mast_cell | *MSH2* | 6.100811289 | 0.001308985 | 0.002001847 | 1 |
| Obesity | mast_cell | *MAP3K1* | 3.291362966 | 0.011399785 | 0.015333857 | 1 |
| Obesity | mast_cell | *TMEM38B* | 7.541227567 | 0.001494958 | 0.002267503 | 1 |
| Obesity | mast_cell | *PDE6C* | 8.007271994 | 2.71E-11 | 3.16E-09 | 1.70E-07 |
| Obesity | mast_cell | *IKZF5* | 7.132785703 | 1.84E-05 | 3.87E-05 | 0.115899154 |
| Obesity | mast_cell | *BLNK* | 4.206773946 | 1.34E-08 | 9.14E-08 | 8.44E-05 |
| Obesity | mast_cell | *WAC* | 5.207952089 | 2.71E-06 | 6.81E-06 | 0.017042743 |
| Obesity | mast_cell | *CREM* | 4.506003392 | 0.003480026 | 0.005027326 | 1 |
| Obesity | mast_cell | *TPSD1* | -0.551647437 | 1.18E-07 | 4.65E-07 | 0.000741482 |
| Obesity | mast_cell | *ITPR3* | 5.950696585 | 2.34E-09 | 4.32E-08 | 1.47E-05 |
| Obesity | mast_cell | *IFT74* | 3.380239024 | 0.000113658 | 0.000205453 | 0.714567178 |
| Obesity | mast_cell | *IL12RB1* | -3.096156319 | 0.026532554 | 0.033932093 | 1 |
| Obesity | mast_cell | *CDC7* | 1.476107688 | 0.027258233 | 0.034803516 | 1 |
| Obesity | mast_cell | *PCSK5* | 5.263100285 | 2.47E-07 | 8.55E-07 | 0.001552101 |
| Obesity | mast_cell | *RAB18* | 2.205106202 | 0.032584645 | 0.041161274 | 1 |
| Obesity | mast_cell | *NRP1* | 4.758893756 | 2.15E-06 | 5.54E-06 | 0.01353165 |

| | | | | | | |
|---|---|---|---|---|---|---|
| Obesity | mast_cell | *PRTFDC1* | 5.281364597 | 3.07E-09 | 4.48E-08 | 1.93E-05 |
| Obesity | mast_cell | *PALMD* | 4.653267104 | 7.12E-08 | 3.06E-07 | 0.000447337 |
| Obesity | mast_cell | *TSPAN15* | 5.407776834 | 1.26E-09 | 3.78E-08 | 7.90E-06 |
| Obesity | mast_cell | *OCEL1* | -2.214759729 | 0.036047397 | 0.045271671 | 1 |
| Obesity | mast_cell | *STX1B* | 2.218214881 | 0.00054723 | 0.000887168 | 1 |
| Obesity | mast_cell | *HSD3B7* | 4.815070594 | 1.41E-08 | 9.47E-08 | 8.89E-05 |
| Obesity | mast_cell | *ATP5D* | 9.126128552 | 1.66E-06 | 4.43E-06 | 0.010466434 |
| Obesity | mast_cell | *MARCH2* | 6.932356523 | 2.74E-07 | 9.32E-07 | 0.001722549 |
| Obesity | mast_cell | *CDC34* | 10.73541741 | 4.54E-06 | 1.08E-05 | 0.028513561 |
| Obesity | mast_cell | *MTAP* | 7.868159153 | 6.48E-09 | 5.97E-08 | 4.08E-05 |
| Obesity | mast_cell | *CEP170B* | 2.438935638 | 6.02E-07 | 1.80E-06 | 0.003784851 |
| Obesity | mast_cell | *POLR2E* | -3.531784316 | 0.004573042 | 0.006490003 | 1 |
| Obesity | mast_cell | *MADCAM1* | 3.366937193 | 5.65E-09 | 5.59E-08 | 3.55E-05 |
| Obesity | mast_cell | *TRMT2A* | 5.067704247 | 1.59E-07 | 5.89E-07 | 0.000996608 |
| Obesity | mast_cell | *ZDHHC8* | 5.507470019 | 6.50E-07 | 1.93E-06 | 0.004084043 |
| Obesity | mast_cell | *MED15* | 5.809431641 | 1.31E-07 | 5.06E-07 | 0.000820488 |
| Obesity | mast_cell | *LZTR1* | 7.337155508 | 5.80E-09 | 5.65E-08 | 3.65E-05 |
| Obesity | mast_cell | *CECR2* | 2.646003968 | 7.41E-07 | 2.17E-06 | 0.004658515 |
| Obesity | mast_cell | *SMARCB1* | 4.880908838 | 1.06E-06 | 2.97E-06 | 0.006636309 |
| Obesity | mast_cell | *SLC7A4* | -3.884669698 | 0.000101102 | 0.000183708 | 0.63562901 |
| Obesity | mast_cell | *DDT* | 5.556309485 | 7.27E-09 | 6.31E-08 | 4.57E-05 |
| Obesity | mast_cell | *GGT5* | 5.799994118 | 1.26E-08 | 8.78E-08 | 7.91E-05 |
| Obesity | mast_cell | *SPECC1L* | 2.734629519 | 1.27E-05 | 2.76E-05 | 0.079693384 |
| Obesity | mast_cell | *YPEL1* | 4.192722591 | 2.28E-09 | 4.32E-08 | 1.44E-05 |
| Obesity | mast_cell | *PRODH* | -9.389347566 | 0.000772741 | 0.001222935 | 1 |
| Obesity | mast_cell | *PPM1F* | 6.910406678 | 2.56E-06 | 6.47E-06 | 0.016105087 |
| Obesity | mast_cell | *CRYBB3* | 4.995825802 | 2.33E-06 | 5.95E-06 | 0.014639144 |
| Obesity | mast_cell | *CYTH4* | 6.238207704 | 2.48E-09 | 4.32E-08 | 1.56E-05 |
| Obesity | mast_cell | *CARD10* | -3.630900759 | 0.008649298 | 0.011826477 | 1 |
| Obesity | mast_cell | *LRP5L* | 5.753495892 | 0.000926167 | 0.001449904 | 1 |
| Obesity | mast_cell | *SLC25A1* | 7.300595472 | 3.14E-09 | 4.48E-08 | 1.97E-05 |
| Obesity | mast_cell | *GGA1* | 3.856979055 | 0.000864639 | 0.001359676 | 1 |
| Obesity | mast_cell | *SH3BP1* | 5.112210749 | 4.77E-09 | 5.11E-08 | 3.00E-05 |
| Obesity | mast_cell | *HPS4* | 7.558372125 | 7.17E-08 | 3.07E-07 | 0.000450559 |
| Obesity | mast_cell | *TRIOBP* | 1.950918428 | 0.031400955 | 0.039753888 | 1 |
| Obesity | mast_cell | *GCAT* | 4.128937488 | 0.000466686 | 0.000765472 | 1 |
| Obesity | mast_cell | *EIF3L* | 5.937438805 | 1.90E-07 | 6.84E-07 | 0.00119144 |
| Obesity | mast_cell | *CCDC134* | -2.853272098 | 2.49E-05 | 5.08E-05 | 0.156286178 |
| Obesity | mast_cell | *PICK1* | 1.196784552 | 0.028423789 | 0.036225494 | 1 |
| Obesity | mast_cell | *KDELR3* | -2.954554147 | 0.000152124 | 0.000269182 | 0.956402633 |
| Obesity | mast_cell | *DDX17* | 3.171329604 | 0.001006979 | 0.001567049 | 1 |
| Obesity | mast_cell | *FBXO7* | 7.668039533 | 2.78E-10 | 1.57E-08 | 1.75E-06 |
| Obesity | mast_cell | *POLDIP3* | 8.851226908 | 3.39E-06 | 8.32E-06 | 0.021334802 |
| Obesity | mast_cell | *PPP6R2* | 5.058207497 | 1.43E-05 | 3.08E-05 | 0.090013397 |
| Obesity | mast_cell | *CYB5R3* | 5.493774162 | 0.000408801 | 0.000678136 | 1 |
| Obesity | mast_cell | *RHBDD3* | 8.29114724 | 2.37E-09 | 4.32E-08 | 1.49E-05 |
| Obesity | mast_cell | *TTLL1* | 10.93511813 | 4.72E-07 | 1.47E-06 | 0.002965352 |
| Obesity | mast_cell | *RASL10A* | 3.960026522 | 2.84E-07 | 9.59E-07 | 0.001785074 |
| Obesity | mast_cell | *AP1B1* | 6.005456053 | 2.71E-05 | 5.49E-05 | 0.17039878 |
| Obesity | mast_cell | *HMGXB4* | 6.895292505 | 1.81E-05 | 3.81E-05 | 0.113997023 |

| | | | | | | |
|---|---|---|---|---|---|---|
| Obesity | mast_cell | *TOM1* | 1.81244382 | 0.026838754 | 0.034302754 | 1 |
| Obesity | mast_cell | *NEFH* | 4.311632123 | 0.025860984 | 0.033147403 | 1 |
| Obesity | mast_cell | *CHKB* | -1.81702908 | 0.013048751 | 0.017417727 | 1 |
| Obesity | mast_cell | *BIK* | -7.937068938 | 1.34E-06 | 3.68E-06 | 0.008452432 |
| Obesity | mast_cell | *MCAT* | 6.213017123 | 2.91E-07 | 9.81E-07 | 0.0018304 |
| Obesity | mast_cell | *THOC5* | 3.984669406 | 0.002088703 | 0.00310515 | 1 |
| Obesity | mast_cell | *TSPO* | 3.421996456 | 0.015696093 | 0.02070093 | 1 |
| Obesity | mast_cell | *TTLL12* | 3.975332597 | 1.65E-08 | 1.03E-07 | 0.000103562 |
| Obesity | mast_cell | *RBFOX2* | 5.278361794 | 3.43E-08 | 1.71E-07 | 0.000215859 |
| Obesity | mast_cell | *MTMR3* | 2.293377128 | 0.007578712 | 0.010442113 | 1 |
| Obesity | mast_cell | *SAMM50* | 7.441764906 | 5.32E-05 | 0.000102361 | 0.334210198 |
| Obesity | mast_cell | *TXN2* | 3.971610566 | 1.81E-06 | 4.76E-06 | 0.011366715 |
| Obesity | mast_cell | *IFT27* | 9.924238505 | 8.61E-08 | 3.59E-07 | 0.000541521 |
| Obesity | mast_cell | *KIAA0930* | 4.820267204 | 3.15E-05 | 6.29E-05 | 0.197894824 |
| Obesity | mast_cell | *NCF4* | 3.60104468 | 4.51E-06 | 1.07E-05 | 0.028348769 |
| Obesity | mast_cell | *SLC25A17* | 6.622514712 | 6.46E-06 | 1.49E-05 | 0.040631998 |
| Obesity | mast_cell | *FAM118A* | 1.724859489 | 0.006490533 | 0.009011921 | 1 |
| Obesity | mast_cell | *KCTD17* | 22.95296275 | 2.68E-08 | 1.42E-07 | 0.000168702 |
| Obesity | mast_cell | *EP300* | 6.077617539 | 1.92E-07 | 6.90E-07 | 0.001204178 |
| Obesity | mast_cell | *ACO2* | 5.2337254 | 8.21E-12 | 1.29E-09 | 5.16E-08 |
| Obesity | mast_cell | *DESI1* | 3.496480104 | 1.57E-05 | 3.36E-05 | 0.098957272 |
| Obesity | mast_cell | *HDAC10* | 5.42029331 | 3.24E-05 | 6.46E-05 | 0.203747553 |
| Obesity | mast_cell | *SDR39U1* | 1.76203875 | 0.003598483 | 0.005193678 | 1 |
| Obesity | mast_cell | *RBM23* | 6.579605686 | 7.25E-06 | 1.66E-05 | 0.045603552 |
| Obesity | mast_cell | *VCPKMT* | 3.0196286 | 0.003860647 | 0.005544059 | 1 |
| Obesity | mast_cell | *SOS2* | 3.61423513 | 0.004818919 | 0.00681587 | 1 |
| Obesity | mast_cell | *PYGL* | 0.955731723 | 0.017534199 | 0.022985302 | 1 |
| Obesity | mast_cell | *TRIM9* | 1.873421452 | 0.014428088 | 0.019120866 | 1 |
| Obesity | mast_cell | *DDHD1* | 5.043714378 | 1.77E-06 | 4.67E-06 | 0.011135786 |
| Obesity | mast_cell | *CNIH1* | 5.567839783 | 4.74E-07 | 1.47E-06 | 0.0029784 |
| Obesity | mast_cell | *PIGH* | 5.832815479 | 1.28E-07 | 4.99E-07 | 0.000807847 |
| Obesity | mast_cell | *VTI1B* | 7.465714008 | 9.62E-09 | 7.52E-08 | 6.05E-05 |
| Obesity | mast_cell | *TMED8* | -2.649646487 | 0.002927098 | 0.004271743 | 1 |
| Obesity | mast_cell | *DAAM1* | 4.145484155 | 2.11E-07 | 7.47E-07 | 0.0013286 |
| Obesity | mast_cell | *ISM2* | -4.21962563 | 0.001224907 | 0.001881963 | 1 |
| Obesity | mast_cell | *PPM1A* | 4.095250098 | 0.000389152 | 0.000648105 | 1 |
| Obesity | mast_cell | *SLC10A1* | 5.36176191 | 0.001931295 | 0.002882728 | 1 |
| Obesity | mast_cell | *DICER1* | 5.005436559 | 1.58E-05 | 3.37E-05 | 0.099351213 |
| Obesity | mast_cell | *ZFYVE21* | 9.543001209 | 2.58E-10 | 1.49E-08 | 1.62E-06 |
| Obesity | mast_cell | *MTHFD1* | 6.358278899 | 1.79E-09 | 4.32E-08 | 1.13E-05 |
| Obesity | mast_cell | *TELO2* | 4.936826153 | 1.29E-08 | 8.98E-08 | 8.13E-05 |
| Obesity | mast_cell | *VRK1* | 8.025575827 | 8.75E-07 | 2.52E-06 | 0.005503819 |
| Obesity | mast_cell | *PAPLN* | 5.563304729 | 2.50E-07 | 8.63E-07 | 0.001572507 |
| Obesity | mast_cell | *PSMB5* | 5.416531885 | 3.20E-07 | 1.06E-06 | 0.002014044 |
| Obesity | mast_cell | *CCNB1IP1* | 5.976600118 | 2.32E-08 | 1.28E-07 | 0.000146062 |
| Obesity | mast_cell | *TRIP11* | 3.673563817 | 0.009616854 | 0.013058566 | 1 |
| Obesity | mast_cell | *PCK2* | 3.602733026 | 4.36E-08 | 2.07E-07 | 0.000274209 |
| Obesity | mast_cell | *KIAA0391* | 11.62549378 | 8.82E-08 | 3.67E-07 | 0.000554663 |
| Obesity | mast_cell | *EMC9* | 2.093884433 | 0.012049892 | 0.016157446 | 1 |
| Obesity | mast_cell | *PLTP* | 3.272673557 | 1.36E-07 | 5.24E-07 | 0.000857227 |

| | | | | | | |
|---|---|---|---|---|---|---|
| Obesity | mast_cell | *GSS* | 5.750914355 | 2.59E-08 | 1.38E-07 | 0.000162987 |
| Obesity | mast_cell | *PYGB* | 4.022785795 | 4.44E-09 | 4.98E-08 | 2.79E-05 |
| Obesity | mast_cell | *ABHD12* | 7.017068609 | 1.14E-10 | 9.30E-09 | 7.16E-07 |
| Obesity | mast_cell | *NINL* | 4.849440562 | 8.46E-08 | 3.54E-07 | 0.000531631 |
| Obesity | mast_cell | *CD40* | 4.966952434 | 3.26E-08 | 1.64E-07 | 0.000204973 |
| Obesity | mast_cell | *UQCC1* | 4.286181834 | 7.47E-06 | 1.70E-05 | 0.046984401 |
| Obesity | mast_cell | *IFT52* | 5.471136968 | 7.35E-06 | 1.68E-05 | 0.046215589 |
| Obesity | mast_cell | *NDRG3* | 6.645083016 | 1.89E-10 | 1.27E-08 | 1.19E-06 |
| Obesity | mast_cell | *DOK5* | 5.068218691 | 2.55E-09 | 4.32E-08 | 1.60E-05 |
| Obesity | mast_cell | *CSTF1* | 6.851771955 | 6.94E-06 | 1.59E-05 | 0.043610234 |
| Obesity | mast_cell | *BMP7* | 2.586528948 | 1.70E-08 | 1.04E-07 | 0.000106716 |
| Obesity | mast_cell | *TPD52L2* | 6.678697778 | 1.15E-06 | 3.22E-06 | 0.007259824 |
| Obesity | mast_cell | *NELFCD* | 7.692714196 | 2.76E-06 | 6.91E-06 | 0.017347614 |
| Obesity | mast_cell | *CTSZ* | 5.106859024 | 2.02E-05 | 4.19E-05 | 0.126699939 |
| Obesity | mast_cell | *PRPF6* | 4.43993426 | 0.004744431 | 0.006713535 | 1 |
| Obesity | mast_cell | *MTG2* | 3.907758476 | 0.000950037 | 0.001484314 | 1 |
| Obesity | mast_cell | *TCFL5* | 3.392502876 | 3.62E-05 | 7.15E-05 | 0.227429966 |
| Obesity | mast_cell | *DIDO1* | 5.564254471 | 4.35E-06 | 1.04E-05 | 0.027324084 |
| Obesity | mast_cell | *SLC17A9* | 4.85238473 | 5.13E-09 | 5.36E-08 | 3.23E-05 |
| Obesity | mast_cell | *BIRC7* | -1.619842351 | 3.44E-07 | 1.13E-06 | 0.002162406 |
| Obesity | mast_cell | *NKAIN4* | 4.69205862 | 1.02E-05 | 2.27E-05 | 0.064364012 |
| Obesity | mast_cell | *PTK6* | -6.150038223 | 1.87E-06 | 4.89E-06 | 0.011738 |
| Obesity | mast_cell | *GMEB2* | 6.6817832 | 8.66E-09 | 7.05E-08 | 5.45E-05 |
| Obesity | mast_cell | *CDC25B* | 3.215092375 | 1.21E-08 | 8.63E-08 | 7.62E-05 |
| Obesity | mast_cell | *RNF24* | 5.6641895 | 3.68E-09 | 4.65E-08 | 2.32E-05 |
| Obesity | mast_cell | *NDUFAF5* | 0.63822722 | 0.03252215 | 0.041090586 | 1 |
| Obesity | mast_cell | *SLC52A3* | 6.601546064 | 1.01E-07 | 4.10E-07 | 0.000633094 |
| Obesity | mast_cell | *ANGPT4* | 4.454240981 | 1.93E-06 | 5.02E-06 | 0.012118767 |
| Obesity | mast_cell | *RSPO4* | 3.978820267 | 6.45E-07 | 1.92E-06 | 0.004054729 |
| Obesity | mast_cell | *SIRPB1* | 1.430181782 | 0.010767731 | 0.014542797 | 1 |
| Obesity | mast_cell | *SEC23B* | 2.237930531 | 0.023311947 | 0.030082556 | 1 |
| Obesity | mast_cell | *CCM2L* | 2.675093101 | 1.22E-08 | 8.65E-08 | 7.70E-05 |
| Obesity | mast_cell | *MYL9* | 5.717986576 | 5.98E-10 | 2.60E-08 | 3.76E-06 |
| Obesity | mast_cell | *MROH8* | 2.547761245 | 4.70E-06 | 1.11E-05 | 0.0295296 |
| Obesity | mast_cell | *MANBAL* | 2.942465696 | 2.76E-05 | 5.59E-05 | 0.173641323 |
| Obesity | mast_cell | *IDH3B* | 8.728520543 | 0.000211605 | 0.000365785 | 1 |
| Obesity | mast_cell | *MAPRE1* | 7.126874434 | 0.000329768 | 0.0005539 | 1 |
| Obesity | mast_cell | *TTI1* | 4.510212172 | 0.019361863 | 0.02524954 | 1 |
| Obesity | mast_cell | *RPRD1B* | 6.537921381 | 1.01E-06 | 2.86E-06 | 0.006341212 |
| Obesity | mast_cell | *PXMP4* | 7.419632109 | 3.36E-09 | 4.52E-08 | 2.11E-05 |
| Obesity | mast_cell | *BPI* | -4.044675685 | 2.17E-05 | 4.48E-05 | 0.136183683 |
| Obesity | mast_cell | *ACTR5* | 7.145127901 | 5.59E-08 | 2.53E-07 | 0.000351245 |
| Obesity | mast_cell | *DHX35* | 7.439228435 | 5.78E-08 | 2.59E-07 | 0.000363296 |
| Obesity | mast_cell | *MAP1LC3A* | 2.604012303 | 1.05E-08 | 7.96E-08 | 6.59E-05 |
| Obesity | mast_cell | *PIGU* | 6.550539172 | 1.90E-08 | 1.10E-07 | 0.000119177 |
| Obesity | mast_cell | *APMAP* | 4.285525842 | 1.74E-10 | 1.23E-08 | 1.09E-06 |
| Obesity | mast_cell | *CELF4* | -4.398718477 | 0.001033048 | 0.001604044 | 1 |
| Obesity | mast_cell | *RBFA* | 5.619011248 | 1.80E-06 | 4.74E-06 | 0.011306128 |
| Obesity | mast_cell | *LPIN2* | 6.045668665 | 3.89E-08 | 1.89E-07 | 0.000244492 |
| Obesity | mast_cell | *SMCHD1* | 5.227153681 | 4.66E-06 | 1.11E-05 | 0.029288089 |

| | | | | | | |
|---|---|---|---|---|---|---|
| Obesity | mast_cell | *MYL12A* | 6.760141418 | 6.39E-08 | 2.81E-07 | 0.00040152 |
| Obesity | mast_cell | *LIPG* | 7.074327671 | 1.75E-07 | 6.39E-07 | 0.001098976 |
| Obesity | mast_cell | *POLI* | 0.893537717 | 0.002389731 | 0.003528473 | 1 |
| Obesity | mast_cell | *NALCN* | 3.511426695 | 4.60E-09 | 5.08E-08 | 2.89E-05 |
| Obesity | mast_cell | *HTR2A* | 5.388749712 | 1.84E-08 | 1.08E-07 | 0.000115709 |
| Obesity | mast_cell | *KLF5* | 3.917051204 | 7.43E-07 | 2.18E-06 | 0.00467 |
| Obesity | mast_cell | *STK24* | 5.338022442 | 1.34E-05 | 2.89E-05 | 0.083986754 |
| Obesity | mast_cell | *DNAJC3* | 5.633091104 | 4.09E-05 | 8.01E-05 | 0.257046577 |
| Obesity | mast_cell | *ARHGEF7* | 5.506976624 | 3.98E-08 | 1.93E-07 | 0.000250416 |
| Obesity | mast_cell | *SGCG* | 2.130942391 | 3.03E-08 | 1.56E-07 | 0.000190788 |
| Obesity | mast_cell | *VWA8* | 4.013012896 | 0.000164346 | 0.000289181 | 1 |
| Obesity | mast_cell | *KATNAL1* | 4.58560291 | 8.19E-06 | 1.85E-05 | 0.051520007 |
| Obesity | mast_cell | *DHRS12* | 8.075587278 | 2.04E-08 | 1.16E-07 | 0.000128551 |
| Obesity | mast_cell | *MEDAG* | 3.708573784 | 3.18E-09 | 4.49E-08 | 2.00E-05 |
| Obesity | mast_cell | *TSC22D1* | 5.659838805 | 2.23E-07 | 7.84E-07 | 0.001401619 |
| Obesity | mast_cell | *CLN5* | 3.004900238 | 0.002920743 | 0.004264447 | 1 |
| Obesity | mast_cell | *MGRN1* | 2.504470326 | 0.009188058 | 0.012530438 | 1 |
| Obesity | mast_cell | *CORO1A* | -9.591473171 | 1.75E-11 | 2.30E-09 | 1.10E-07 |
| Obesity | mast_cell | *GDPD3* | 4.522025494 | 0.002883348 | 0.004211805 | 1 |
| Obesity | mast_cell | *ELMO3* | 2.247203397 | 0.000258472 | 0.00044086 | 1 |
| Obesity | mast_cell | *NFAT5* | 4.773449505 | 1.88E-06 | 4.92E-06 | 0.01181165 |
| Obesity | mast_cell | *LONP2* | 3.633875976 | 0.003745506 | 0.005389791 | 1 |
| Obesity | mast_cell | *PLLP* | -3.612573529 | 0.023240357 | 0.02999633 | 1 |
| Obesity | mast_cell | *DHODH* | 1.737701492 | 0.002668078 | 0.00391372 | 1 |
| Obesity | mast_cell | *CTCF* | 6.430855445 | 6.09E-08 | 2.71E-07 | 0.00038288 |
| Obesity | mast_cell | *ACD* | 1.993818776 | 0.023347661 | 0.030122459 | 1 |
| Obesity | mast_cell | *USB1* | 7.805577602 | 6.01E-10 | 2.60E-08 | 3.78E-06 |
| Obesity | mast_cell | *SETD6* | 8.800811576 | 9.39E-09 | 7.38E-08 | 5.90E-05 |
| Obesity | mast_cell | *SLC7A6OS* | 2.563971406 | 1.66E-07 | 6.11E-07 | 0.001042611 |
| Obesity | mast_cell | *HCFC1R1* | 10.6990281 | 8.67E-09 | 7.05E-08 | 5.45E-05 |
| Obesity | mast_cell | *NPRL3* | 2.715725019 | 0.003750818 | 0.005396199 | 1 |
| Obesity | mast_cell | *NAGPA* | 9.162924587 | 5.38E-11 | 5.48E-09 | 3.38E-07 |
| Obesity | mast_cell | *COTL1* | 5.962368078 | 2.02E-08 | 1.15E-07 | 0.000126695 |
| Obesity | mast_cell | *CRISPLD2* | 3.124027374 | 7.55E-07 | 2.21E-06 | 0.00474471 |
| Obesity | mast_cell | *NME4* | 8.034659285 | 1.73E-06 | 4.58E-06 | 0.010881157 |
| Obesity | mast_cell | *LMF1* | 7.33589724 | 1.23E-08 | 8.68E-08 | 7.75E-05 |
| Obesity | mast_cell | *NARFL* | 6.005409633 | 7.01E-09 | 6.20E-08 | 4.41E-05 |
| Obesity | mast_cell | *HAGHL* | -4.66953218 | 0.000114498 | 0.000206674 | 0.719845918 |
| Obesity | mast_cell | *SLC7A5* | 12.55600669 | 2.89E-09 | 4.35E-08 | 1.82E-05 |
| Obesity | mast_cell | *FBXO31* | 3.036955708 | 0.037641925 | 0.047142387 | 1 |
| Obesity | mast_cell | *STUB1* | 4.190731226 | 9.04E-07 | 2.59E-06 | 0.005680559 |
| Obesity | mast_cell | *NUBP1* | 6.601277468 | 2.68E-09 | 4.32E-08 | 1.69E-05 |
| Obesity | mast_cell | *UBE2I* | 5.371973603 | 0.000738168 | 0.001171638 | 1 |
| Obesity | mast_cell | *CLUAP1* | 3.437173192 | 9.95E-12 | 1.53E-09 | 6.25E-08 |
| Obesity | mast_cell | *CPPED1* | -2.510657429 | 0.000581893 | 0.000939004 | 1 |
| Obesity | mast_cell | *HMOX2* | 3.726704446 | 0.000863559 | 0.001358318 | 1 |
| Obesity | mast_cell | *BFAR* | -2.574906023 | 0.038145294 | 0.047734766 | 1 |
| Obesity | mast_cell | *RRN3P2* | 5.177453062 | 1.65E-06 | 4.40E-06 | 0.010385358 |
| Obesity | mast_cell | *RBL2* | 3.124317354 | 0.000215391 | 0.000371614 | 1 |
| Obesity | mast_cell | *STX4* | 6.375119701 | 1.46E-08 | 9.64E-08 | 9.20E-05 |

| | | | | | | |
|---|---|---|---|---|---|---|
| Obesity | mast_cell | *KAT8* | 10.69112364 | 3.85E-16 | 2.02E-13 | 2.42E-12 |
| Obesity | mast_cell | *TMC5* | 4.425163486 | 1.63E-07 | 6.04E-07 | 0.001027249 |
| Obesity | mast_cell | *RNF40* | 5.114451229 | 2.67E-06 | 6.73E-06 | 0.016806073 |
| Obesity | mast_cell | *KNOP1* | 2.576188266 | 0.020016687 | 0.026044062 | 1 |
| Obesity | mast_cell | *IQCH* | 4.146478687 | 0.016976289 | 0.022272523 | 1 |
| Obesity | mast_cell | *LACTB* | 7.180857009 | 2.17E-07 | 7.66E-07 | 0.001365125 |
| Obesity | mast_cell | *HERC1* | 2.947109756 | 0.013171976 | 0.017563565 | 1 |
| Obesity | mast_cell | *TRIP4* | 2.589427327 | 0.009766925 | 0.013256618 | 1 |
| Obesity | mast_cell | *RASL12* | 3.023692838 | 1.73E-08 | 1.05E-07 | 0.00010887 |
| Obesity | mast_cell | *AP3B2* | 1.125879824 | 0.003939645 | 0.005651049 | 1 |
| Obesity | mast_cell | *CTSH* | 1.560148972 | 0.000107041 | 0.000194051 | 0.672968524 |
| Obesity | mast_cell | *TTC23* | 5.022024539 | 1.27E-05 | 2.77E-05 | 0.080045188 |
| Obesity | mast_cell | *FAH* | 2.990730388 | 5.44E-09 | 5.53E-08 | 3.42E-05 |
| Obesity | mast_cell | *RPAP1* | 5.720601675 | 0.000858527 | 0.001351078 | 1 |
| Obesity | mast_cell | *EHD4* | 6.451416144 | 1.65E-09 | 4.26E-08 | 1.04E-05 |
| Obesity | mast_cell | *ATP8B4* | 4.761629443 | 0.000649212 | 0.001040161 | 1 |
| Obesity | mast_cell | *DTWD1* | 4.748210062 | 7.27E-05 | 0.0001358 | 0.456967562 |
| Obesity | mast_cell | *DNAJC17* | 7.471233374 | 1.19E-05 | 2.61E-05 | 0.074841864 |
| Obesity | mast_cell | *PDGFRL* | 3.146014529 | 6.99E-09 | 6.20E-08 | 4.40E-05 |
| Obesity | mast_cell | *ZDHHC2* | -3.604491356 | 0.00138381 | 0.002111141 | 1 |
| Obesity | mast_cell | *ZFAND1* | 3.810919631 | 0.00045261 | 0.000744521 | 1 |
| Obesity | mast_cell | *RIPK2* | 6.529082792 | 1.03E-07 | 4.18E-07 | 0.000649801 |
| Obesity | mast_cell | *EYA1* | 2.011173024 | 2.07E-06 | 5.35E-06 | 0.012999412 |
| Obesity | mast_cell | *NBN* | 3.376926752 | 0.02518683 | 0.032316245 | 1 |
| Obesity | mast_cell | *CPQ* | 3.875963942 | 2.52E-07 | 8.68E-07 | 0.001582428 |
| Obesity | mast_cell | *NIPAL2* | 4.914539446 | 5.71E-08 | 2.57E-07 | 0.000358694 |
| Obesity | mast_cell | *IKBKB* | 6.480480164 | 6.28E-06 | 1.45E-05 | 0.039507368 |
| Obesity | mast_cell | *PLAT* | 7.305965408 | 1.71E-09 | 4.28E-08 | 1.08E-05 |
| Obesity | mast_cell | *GDAP1* | -2.314671159 | 6.02E-05 | 0.000114316 | 0.378476333 |
| Obesity | mast_cell | *ZC2HC1A* | 5.096464608 | 7.68E-05 | 0.000142858 | 0.483001873 |
| Obesity | mast_cell | *SPAG1* | 3.901096753 | 1.04E-06 | 2.92E-06 | 0.006515871 |
| Obesity | mast_cell | *CHRAC1* | 4.591738448 | 6.51E-08 | 2.84E-07 | 0.000409421 |
| Obesity | mast_cell | *SNX16* | 6.782089389 | 1.22E-06 | 3.39E-06 | 0.007700099 |
| Obesity | mast_cell | *GSDMD* | 2.650677842 | 0.006890396 | 0.009541833 | 1 |
| Obesity | mast_cell | *TSTA3* | 3.7112517 | 5.39E-07 | 1.64E-06 | 0.003387604 |
| Obesity | mast_cell | *ERI1* | 7.726077739 | 3.35E-06 | 8.22E-06 | 0.021068177 |
| Obesity | mast_cell | *MTMR9* | 7.299068351 | 2.24E-06 | 5.75E-06 | 0.014097675 |
| Obesity | mast_cell | *LEPROTL1* | -2.721127623 | 0.02047577 | 0.026613844 | 1 |
| Obesity | mast_cell | *TNFRSF10A* | 1.698422162 | 0.018502487 | 0.024163925 | 1 |
| Obesity | mast_cell | *ERICH1* | 3.747877847 | 0.00675777 | 0.009368489 | 1 |
| Obesity | mast_cell | *TUSC3* | 3.433249443 | 3.24E-09 | 4.49E-08 | 2.04E-05 |
| Obesity | mast_cell | *ARHGEF10* | 4.041431232 | 1.44E-07 | 5.45E-07 | 0.000903367 |
| Obesity | mast_cell | *KLHDC4* | 4.408983231 | 1.03E-05 | 2.29E-05 | 0.064783748 |
| Obesity | mast_cell | *MCM4* | -1.857690018 | 0.016931322 | 0.022227443 | 1 |
| Obesity | mast_cell | *MAN2B1* | 2.779083016 | 0.002084523 | 0.003099668 | 1 |
| Obesity | mast_cell | *TULP2* | -1.014338157 | 1.90E-07 | 6.86E-07 | 0.001195153 |
| Obesity | mast_cell | *NUCB1* | 3.161005601 | 0.021578483 | 0.027960413 | 1 |
| Obesity | mast_cell | *NFKBIB* | 10.51607152 | 1.76E-05 | 3.72E-05 | 0.11091961 |
| Obesity | mast_cell | *SARS2* | 2.194710814 | 0.00169446 | 0.002547362 | 1 |
| Obesity | mast_cell | *FCGRT* | 3.80609178 | 1.06E-05 | 2.36E-05 | 0.06693703 |

| | | | | | | |
|---|---|---|---|---|---|---|
| Obesity | mast_cell | *ERCC2* | 10.49046379 | 2.48E-09 | 4.32E-08 | 1.56E-05 |
| Obesity | mast_cell | *PLEKHJ1* | 8.642677111 | 1.83E-08 | 1.08E-07 | 0.000114992 |
| Obesity | mast_cell | *CD37* | -2.686289934 | 0.001361171 | 0.002078115 | 1 |
| Obesity | mast_cell | *AMH* | 4.39487438 | 0.003358355 | 0.004863851 | 1 |
| Obesity | mast_cell | *DKKL1* | -3.245419915 | 3.27E-05 | 6.51E-05 | 0.205478844 |
| Obesity | mast_cell | *STX10* | 6.040152584 | 0.003759558 | 0.005407537 | 1 |
| Obesity | mast_cell | *IL4I1* | 3.433906618 | 1.20E-06 | 3.32E-06 | 0.007517087 |
| Obesity | mast_cell | *TLE6* | -3.817593262 | 0.000103439 | 0.000187737 | 0.650322386 |
| Obesity | mast_cell | *AES* | 4.394099518 | 0.013068388 | 0.017440237 | 1 |
| Obesity | mast_cell | *SGTA* | 7.614070155 | 6.95E-10 | 2.77E-08 | 4.37E-06 |
| Obesity | mast_cell | *C19orf53* | -4.050712011 | 0.003646499 | 0.005259357 | 1 |
| Obesity | mast_cell | *CCDC61* | 4.097193967 | 0.000990192 | 0.001543217 | 1 |
| Obesity | mast_cell | *IL27RA* | 8.018999962 | 2.98E-10 | 1.66E-08 | 1.87E-06 |
| Obesity | mast_cell | *C19orf44* | 2.156950699 | 0.002427364 | 0.003581516 | 1 |
| Obesity | mast_cell | *OLFM2* | 1.975764183 | 5.05E-05 | 9.77E-05 | 0.317463124 |
| Obesity | mast_cell | *RASAL3* | -2.403851683 | 0.011048175 | 0.01489277 | 1 |
| Obesity | mast_cell | *ZNF419* | 1.586202171 | 0.0028328 | 0.004139892 | 1 |
| Obesity | mast_cell | *SYDE1* | 3.304889497 | 3.56E-07 | 1.16E-06 | 0.002235725 |
| Obesity | mast_cell | *AURKC* | 2.552834604 | 0.000237726 | 0.000407687 | 1 |
| Obesity | mast_cell | *CCNE1* | -10.94936852 | 0.002192966 | 0.003255532 | 1 |
| Obesity | mast_cell | *PDCD5* | 4.420625023 | 7.31E-09 | 6.32E-08 | 4.59E-05 |
| Obesity | mast_cell | *ANKRD27* | -1.978127703 | 1.64E-05 | 3.48E-05 | 0.10293436 |
| Obesity | mast_cell | *PRX* | 6.593636461 | 0.000114005 | 0.000205979 | 0.716748762 |
| Obesity | mast_cell | *TBCB* | 5.026329667 | 0.001271977 | 0.001949517 | 1 |
| Obesity | mast_cell | *POLR2I* | 5.49300881 | 0.000513481 | 0.000836335 | 1 |
| Obesity | mast_cell | *SLC1A5* | 4.282372436 | 1.36E-06 | 3.72E-06 | 0.008573251 |
| Obesity | mast_cell | *PRKD2* | 3.447644229 | 4.69E-06 | 1.11E-05 | 0.029512683 |
| Obesity | mast_cell | *CACTIN* | 12.75016142 | 0.000163445 | 0.000287917 | 1 |
| Obesity | mast_cell | *CCDC9* | 4.104918847 | 1.96E-08 | 1.13E-07 | 0.000123158 |
| Obesity | mast_cell | *FZR1* | 7.941309023 | 8.39E-10 | 3.03E-08 | 5.28E-06 |
| Obesity | mast_cell | *BBC3* | 7.751442615 | 0.024557114 | 0.031598562 | 1 |
| Obesity | mast_cell | *CEACAM4* | 9.194580092 | 1.58E-08 | 1.01E-07 | 9.93E-05 |
| Obesity | mast_cell | *PLIN3* | 5.901402652 | 1.75E-09 | 4.28E-08 | 1.10E-05 |
| Obesity | mast_cell | *MRPL4* | 9.159509141 | 3.11E-07 | 1.04E-06 | 0.001956088 |
| Obesity | mast_cell | *ICAM5* | -23.6575466 | 3.78E-08 | 1.85E-07 | 0.000237583 |
| Obesity | mast_cell | *CD33* | -2.154937373 | 2.30E-05 | 4.74E-05 | 0.144698787 |
| Obesity | mast_cell | *BABAM1* | 8.669844867 | 2.03E-09 | 4.32E-08 | 1.28E-05 |
| Obesity | mast_cell | *TYK2* | 5.730096246 | 1.53E-09 | 4.16E-08 | 9.61E-06 |
| Obesity | mast_cell | *RABAC1* | 5.015829011 | 5.97E-08 | 2.67E-07 | 0.000375103 |
| Obesity | mast_cell | *PTPRS* | 3.940037936 | 1.68E-05 | 3.57E-05 | 0.105724407 |
| Obesity | mast_cell | *CYTH2* | 9.269784951 | 1.71E-10 | 1.22E-08 | 1.08E-06 |
| Obesity | mast_cell | *GRWD1* | -1.977114744 | 0.025251197 | 0.032392221 | 1 |
| Obesity | mast_cell | *GRIN2D* | -3.827873032 | 5.92E-06 | 1.37E-05 | 0.037219351 |
| Obesity | mast_cell | *CCDC114* | 1.759533068 | 0.037014848 | 0.046394009 | 1 |
| Obesity | mast_cell | *CARD8* | 3.143795156 | 0.017570835 | 0.023028526 | 1 |
| Obesity | mast_cell | *LIG1* | 8.587563288 | 4.77E-07 | 1.48E-06 | 0.002998081 |
| Obesity | mast_cell | *ZNF175* | -3.460517245 | 7.09E-06 | 1.62E-05 | 0.044601529 |
| Obesity | mast_cell | *PLA2G4C* | 4.283490453 | 4.93E-08 | 2.30E-07 | 0.00031001 |
| Obesity | mast_cell | *CAPS* | 5.173286382 | 4.67E-05 | 9.07E-05 | 0.293292797 |
| Obesity | mast_cell | *MIER2* | 3.963082387 | 1.67E-08 | 1.04E-07 | 0.000105246 |

| | | | | | | |
|---|---|---|---|---|---|---|
| Obesity | mast_cell | *PPP2R1A* | 8.431794787 | 5.84E-07 | 1.76E-06 | 0.003674712 |
| Obesity | mast_cell | *TNPO2* | 5.212462708 | 0.000222303 | 0.000382594 | 1 |
| Obesity | mast_cell | *GCDH* | 4.142289044 | 0.000523729 | 0.000851346 | 1 |
| Obesity | mast_cell | *LILRB5* | 4.481104423 | 2.67E-09 | 4.32E-08 | 1.68E-05 |
| Obesity | mast_cell | *PRPF31* | 4.128044755 | 0.000369132 | 0.000616233 | 1 |
| Obesity | mast_cell | *PDE4C* | 3.590496475 | 4.44E-06 | 1.06E-05 | 0.027916099 |
| Obesity | mast_cell | *UPK1A* | 6.824571893 | 9.83E-05 | 0.000179106 | 0.61827259 |
| Obesity | mast_cell | *COPE* | 7.102170173 | 3.16E-08 | 1.60E-07 | 0.000198624 |
| Obesity | mast_cell | *LSR* | -2.40900931 | 3.58E-05 | 7.07E-05 | 0.224778423 |
| Obesity | mast_cell | *KXD1* | 9.357021743 | 1.56E-05 | 3.35E-05 | 0.098377636 |
| Obesity | mast_cell | *HPN* | 2.967189741 | 1.10E-07 | 4.41E-07 | 0.000693921 |
| Obesity | mast_cell | *ZNF14* | 9.483198644 | 2.54E-07 | 8.73E-07 | 0.001596782 |
| Obesity | mast_cell | *SIPA1L3* | 4.915371674 | 0.000218043 | 0.000375881 | 1 |
| Obesity | mast_cell | *ZNF85* | 5.758193939 | 1.69E-08 | 1.04E-07 | 0.000106561 |
| Obesity | mast_cell | *ETHE1* | 3.690010212 | 1.44E-08 | 9.57E-08 | 9.08E-05 |
| Obesity | mast_cell | *SMG9* | 5.059183509 | 1.18E-07 | 4.66E-07 | 0.000742703 |
| Obesity | mast_cell | *GTPBP10* | 2.632438994 | 0.000462424 | 0.000759472 | 1 |
| Obesity | mast_cell | *RASA4* | 2.398915028 | 0.002630295 | 0.003861902 | 1 |
| Obesity | mast_cell | *PON3* | 2.698656573 | 1.27E-08 | 8.87E-08 | 8.01E-05 |
| Obesity | mast_cell | *PON2* | 6.60271324 | 1.32E-08 | 9.06E-08 | 8.32E-05 |
| Obesity | mast_cell | *DLX5* | 35.2098916 | 0.02365061 | 0.030488289 | 1 |
| Obesity | mast_cell | *STEAP1B* | 3.688690566 | 5.65E-06 | 1.32E-05 | 0.035532908 |
| Obesity | mast_cell | *PTN* | 5.947165582 | 5.73E-09 | 5.62E-08 | 3.60E-05 |
| Obesity | mast_cell | *MPP6* | 3.229994527 | 3.44E-09 | 4.57E-08 | 2.16E-05 |
| Obesity | mast_cell | *DFNA5* | 4.183335023 | 2.28E-06 | 5.84E-06 | 0.014347416 |
| Obesity | mast_cell | *OGDH* | 5.767749279 | 2.69E-05 | 5.45E-05 | 0.168919151 |
| Obesity | mast_cell | *ADAP1* | 14.94296198 | 0.008813823 | 0.012038346 | 1 |
| Obesity | mast_cell | *CAV1* | 3.831204256 | 3.49E-07 | 1.14E-06 | 0.002192167 |
| Obesity | mast_cell | *MET* | 3.574631396 | 6.11E-09 | 5.80E-08 | 3.84E-05 |
| Obesity | mast_cell | *RNF32* | 1.880537189 | 0.037202406 | 0.046619797 | 1 |
| Obesity | mast_cell | *LMBR1* | 4.970866526 | 6.94E-05 | 0.000130318 | 0.43656564 |
| Obesity | mast_cell | *HOXA2* | 5.304172799 | 2.74E-06 | 6.87E-06 | 0.017224128 |
| Obesity | mast_cell | *HOXA5* | 3.715397341 | 3.58E-05 | 7.07E-05 | 0.224884832 |
| Obesity | mast_cell | *HOXA6* | 4.129746559 | 3.01E-05 | 6.04E-05 | 0.188993723 |
| Obesity | mast_cell | *BRAT1* | 2.723631116 | 0.001111726 | 0.001716967 | 1 |
| Obesity | mast_cell | *CPED1* | 4.874667511 | 1.92E-05 | 4.02E-05 | 0.12072608 |
| Obesity | mast_cell | *HIBADH* | 5.777232801 | 1.41E-07 | 5.36E-07 | 0.000883577 |
| Obesity | mast_cell | *CPVL* | 4.003739155 | 2.67E-07 | 9.13E-07 | 0.001680331 |
| Obesity | mast_cell | *PLEKHA8* | 5.922209541 | 3.30E-10 | 1.77E-08 | 2.08E-06 |
| Obesity | mast_cell | *NOD1* | 6.077723517 | 2.53E-05 | 5.15E-05 | 0.158894054 |
| Obesity | mast_cell | *CRHR2* | -4.704356921 | 1.16E-06 | 3.24E-06 | 0.007314127 |
| Obesity | mast_cell | *EPHB6* | 5.179663637 | 2.58E-09 | 4.32E-08 | 1.63E-05 |
| Obesity | mast_cell | *NPTX2* | 46.18573114 | 0.000863211 | 0.001358109 | 1 |
| Obesity | mast_cell | *CYP3A5* | 8.38436619 | 3.70E-06 | 8.97E-06 | 0.023282568 |
| Obesity | mast_cell | *ZKSCAN1* | 5.868866834 | 8.47E-07 | 2.45E-06 | 0.005323153 |
| Obesity | mast_cell | *EIF3B* | 4.774778569 | 0.001257013 | 0.001927522 | 1 |
| Obesity | mast_cell | *SNX8* | 5.83954024 | 0.001010699 | 0.00157206 | 1 |
| Obesity | mast_cell | *TAF6* | 3.35179841 | 0.000617679 | 0.000993184 | 1 |
| Obesity | mast_cell | *AIMP2* | 8.536491905 | 1.41E-06 | 3.83E-06 | 0.008838341 |
| Obesity | mast_cell | *TFR2* | 6.575711422 | 4.90E-05 | 9.49E-05 | 0.307817805 |

| | | | | | | |
|---|---|---|---|---|---|---|
| Obesity | mast_cell | *RBM28* | 2.190371299 | 0.029785149 | 0.037784348 | 1 |
| Obesity | mast_cell | *USP42* | 7.034184718 | 1.59E-08 | 1.02E-07 | 0.000100141 |
| Obesity | mast_cell | *IMPDH1* | 3.126461581 | 0.02034578 | 0.026455826 | 1 |
| Obesity | mast_cell | *LSM5* | 8.491103253 | 6.22E-05 | 0.000117694 | 0.390743822 |
| Obesity | mast_cell | *AP1S1* | 3.184865329 | 1.67E-08 | 1.04E-07 | 0.00010481 |
| Obesity | mast_cell | *C1GALT1* | 5.952423592 | 1.08E-08 | 8.14E-08 | 6.82E-05 |
| Obesity | mast_cell | *PLOD3* | 4.980456763 | 4.18E-08 | 2.01E-07 | 0.00026308 |
| Obesity | mast_cell | *PHF14* | 5.35840324 | 9.01E-07 | 2.58E-06 | 0.005664186 |
| Obesity | mast_cell | *CEP41* | 4.545228815 | 1.91E-05 | 4.00E-05 | 0.120199758 |
| Obesity | mast_cell | *ZNF862* | 6.427698977 | 4.57E-07 | 1.44E-06 | 0.002873223 |
| Obesity | mast_cell | *MEST* | 1.058438155 | 0.010456657 | 0.014143933 | 1 |
| Obesity | mast_cell | *ACTR3C* | 2.989782103 | 8.60E-08 | 3.59E-07 | 0.000540405 |
| Obesity | mast_cell | *POU6F2* | 4.165331709 | 3.59E-09 | 4.63E-08 | 2.26E-05 |
| Obesity | mast_cell | *TSPAN13* | 10.5883079 | 1.03E-09 | 3.50E-08 | 6.47E-06 |
| Obesity | mast_cell | *RARRES2* | 5.021739984 | 1.86E-05 | 3.90E-05 | 0.116974558 |
| Obesity | mast_cell | *TMEM176B* | 1.924326486 | 0.000263326 | 0.000448411 | 1 |
| Obesity | mast_cell | *MRPL32* | 3.765692959 | 1.44E-07 | 5.45E-07 | 0.000903092 |
| Obesity | mast_cell | *BLVRA* | 2.897553631 | 0.002378502 | 0.003513544 | 1 |
| Obesity | mast_cell | *URGCP* | 4.049677464 | 3.87E-09 | 4.77E-08 | 2.43E-05 |
| Obesity | mast_cell | *POLD2* | 5.130341566 | 0.004532833 | 0.006438754 | 1 |
| Obesity | mast_cell | *GCK* | 112.7119426 | 5.49E-09 | 5.54E-08 | 3.45E-05 |
| Obesity | mast_cell | *BCL7B* | 2.02412007 | 0.016352293 | 0.02152122 | 1 |
| Obesity | mast_cell | *TBL2* | 2.344999595 | 8.02E-07 | 2.33E-06 | 0.00504351 |
| Obesity | mast_cell | *CLIP2* | 8.320130508 | 6.23E-08 | 2.76E-07 | 0.000391563 |
| Obesity | mast_cell | *LIMK1* | 6.582783283 | 2.32E-09 | 4.32E-08 | 1.46E-05 |
| Obesity | mast_cell | *SLC1A1* | 1.942823982 | 0.000129392 | 0.000231038 | 0.813486322 |
| Obesity | mast_cell | *FKTN* | 6.792336399 | 2.49E-06 | 6.32E-06 | 0.015656756 |
| Obesity | mast_cell | *NMRK1* | 4.85633725 | 1.04E-07 | 4.20E-07 | 0.000655213 |
| Obesity | mast_cell | *TMEM245* | 3.207996392 | 0.008051589 | 0.011050063 | 1 |
| Obesity | mast_cell | *MEGF9* | 3.025155167 | 1.11E-06 | 3.12E-06 | 0.006998972 |
| Obesity | mast_cell | *TRIM14* | 5.983672415 | 6.37E-08 | 2.81E-07 | 0.00040051 |
| Obesity | mast_cell | *CORO2A* | 6.224881295 | 0.002462133 | 0.00362941 | 1 |
| Obesity | mast_cell | *C5* | 2.842223297 | 0.007276084 | 0.010036144 | 1 |
| Obesity | mast_cell | *OGN* | 7.826164414 | 1.99E-10 | 1.30E-08 | 1.25E-06 |
| Obesity | mast_cell | *ECM2* | 4.211559084 | 8.45E-06 | 1.91E-05 | 0.053150565 |
| Obesity | mast_cell | *PTGR1* | 2.481177996 | 0.00020951 | 0.000362362 | 1 |
| Obesity | mast_cell | *TNFSF8* | 6.317029151 | 2.69E-09 | 4.32E-08 | 1.69E-05 |
| Obesity | mast_cell | *PLGRKT* | -2.739486551 | 1.97E-05 | 4.10E-05 | 0.123676879 |
| Obesity | mast_cell | *KDM4C* | 4.749054395 | 0.000126936 | 0.000227299 | 0.798047601 |
| Obesity | mast_cell | *DOCK8* | 3.029558127 | 5.19E-05 | 0.000100139 | 0.326352116 |
| Obesity | mast_cell | *ELAVL2* | 2.868985898 | 6.69E-05 | 0.000125982 | 0.420652914 |
| Obesity | mast_cell | *RGP1* | 11.58837108 | 5.43E-07 | 1.64E-06 | 0.003412705 |
| Obesity | mast_cell | *MPDZ* | 5.565306328 | 1.17E-08 | 8.45E-08 | 7.37E-05 |
| Obesity | mast_cell | *PTGDS* | 4.531162227 | 6.60E-09 | 6.01E-08 | 4.15E-05 |
| Obesity | mast_cell | *SHB* | -4.489891634 | 2.25E-06 | 5.77E-06 | 0.014150688 |
| Obesity | mast_cell | *DVL1* | 3.413146228 | 2.06E-08 | 1.16E-07 | 0.000129354 |
| Obesity | mast_cell | *PDLIM1* | 4.805601297 | 9.32E-07 | 2.66E-06 | 0.005857092 |
| Obesity | mast_cell | *ATRNL1* | 2.855400554 | 0.000298201 | 0.00050357 | 1 |
| Obesity | mast_cell | *HPS1* | 7.022177715 | 1.66E-08 | 1.03E-07 | 0.000104206 |
| Obesity | mast_cell | *PHYH* | 1.34446814 | 0.014157635 | 0.018781869 | 1 |

| | | | | | | |
|---|---|---|---|---|---|---|
| Obesity | mast_cell | *DNMBP* | 6.205298296 | 1.26E-08 | 8.78E-08 | 7.91E-05 |
| Obesity | mast_cell | *TRDMT1* | 5.987113196 | 0.000770856 | 0.001220442 | 1 |
| Obesity | mast_cell | *DDX50* | 7.003980218 | 1.31E-06 | 3.60E-06 | 0.008253139 |
| Obesity | mast_cell | *MAPK8* | 5.242629321 | 1.31E-05 | 2.85E-05 | 0.08256589 |
| Obesity | mast_cell | *ATE1* | 4.103757617 | 0.000159738 | 0.000281783 | 1 |
| Obesity | mast_cell | *NSMCE4A* | 6.61552397 | 5.42E-09 | 5.52E-08 | 3.40E-05 |
| Obesity | mast_cell | *PLEKHA1* | 5.339787534 | 5.17E-08 | 2.38E-07 | 0.000324849 |
| Obesity | mast_cell | *SPOCK2* | 2.855853934 | 0.000929068 | 0.001454083 | 1 |
| Obesity | mast_cell | *MICU1* | 6.399127074 | 3.48E-08 | 1.72E-07 | 0.000218782 |
| Obesity | mast_cell | *CCSER2* | 5.973383147 | 4.73E-09 | 5.10E-08 | 2.98E-05 |
| Obesity | mast_cell | *LIPA* | 4.31048738 | 3.13E-05 | 6.27E-05 | 0.197003662 |
| Obesity | mast_cell | *SFXN3* | 8.206072879 | 1.57E-10 | 1.14E-08 | 9.90E-07 |
| Obesity | mast_cell | *KAZALD1* | 4.884604796 | 3.02E-07 | 1.01E-06 | 0.001897282 |
| Obesity | mast_cell | *TNKS2* | 5.751860642 | 6.81E-06 | 1.56E-05 | 0.042810118 |
| Obesity | mast_cell | *ANKRD26* | 3.80220867 | 5.49E-05 | 0.000105314 | 0.344902805 |
| Obesity | mast_cell | *ACBD5* | 8.631843161 | 5.81E-09 | 5.65E-08 | 3.65E-05 |
| Obesity | mast_cell | *MTPAP* | 5.528565935 | 4.28E-10 | 2.12E-08 | 2.69E-06 |
| Obesity | mast_cell | *MAP3K8* | 5.560673378 | 4.48E-05 | 8.73E-05 | 0.28151188 |
| Obesity | mast_cell | *SHOC2* | 5.741976807 | 5.23E-07 | 1.60E-06 | 0.00328899 |
| Obesity | mast_cell | *TFAM* | 8.540128566 | 3.93E-06 | 9.47E-06 | 0.024733433 |
| Obesity | mast_cell | *RPL28* | 2.886557001 | 0.00297169 | 0.004331791 | 1 |
| Obesity | mast_cell | *PBLD* | 1.064668439 | 0.006768762 | 0.009381659 | 1 |
| Obesity | mast_cell | *MLLT6* | 7.116969963 | 7.38E-06 | 1.68E-05 | 0.046395004 |
| Obesity | mast_cell | *PSMB3* | 5.679929577 | 9.13E-09 | 7.26E-08 | 5.74E-05 |
| Obesity | mast_cell | *CASC3* | 4.396485522 | 0.002531561 | 0.003725637 | 1 |
| Obesity | mast_cell | *RAPGEFL1* | 2.777711525 | 7.77E-05 | 0.000144307 | 0.488480834 |
| Obesity | mast_cell | *RGS9* | 12.37144759 | 6.51E-05 | 0.000122809 | 0.409200143 |
| Obesity | mast_cell | *WNT3* | 4.25206739 | 1.97E-08 | 1.13E-07 | 0.000123838 |
| Obesity | mast_cell | *ASPA* | 7.026885326 | 1.38E-07 | 5.28E-07 | 0.000865554 |
| Obesity | mast_cell | *SEPT4* | 2.793833805 | 0.001426474 | 0.002171487 | 1 |
| Obesity | mast_cell | *TRIM37* | 6.29173286 | 3.84E-05 | 7.57E-05 | 0.241673071 |
| Obesity | mast_cell | *PNPO* | 2.676198518 | 0.002952513 | 0.004306001 | 1 |
| Obesity | mast_cell | *RPS6KB1* | 6.840344899 | 2.84E-06 | 7.09E-06 | 0.017837104 |
| Obesity | mast_cell | *TRIM16L* | 1.817850573 | 1.45E-06 | 3.94E-06 | 0.009130664 |
| Obesity | mast_cell | *CDK5RAP3* | -2.704802838 | 0.000924087 | 0.001447368 | 1 |
| Obesity | mast_cell | *CBX1* | 6.358979835 | 2.02E-06 | 5.24E-06 | 0.012720638 |
| Obesity | mast_cell | *RECQL5* | 2.294418697 | 0.014856416 | 0.019642962 | 1 |
| Obesity | mast_cell | *PIGL* | 5.691356588 | 7.94E-09 | 6.67E-08 | 4.99E-05 |
| Obesity | mast_cell | *GALK1* | -4.681603392 | 5.88E-05 | 0.000111956 | 0.369467222 |
| Obesity | mast_cell | *MED13* | 2.830175525 | 0.008142068 | 0.011162055 | 1 |
| Obesity | mast_cell | *HOXB6* | 7.02974176 | 4.23E-12 | 7.59E-10 | 2.66E-08 |
| Obesity | mast_cell | *RNF167* | 7.878619379 | 1.41E-05 | 3.04E-05 | 0.088606364 |
| Obesity | mast_cell | *C1QBP* | 6.105731397 | 0.010110987 | 0.013702905 | 1 |
| Obesity | mast_cell | *GOSR1* | 6.781589462 | 1.28E-07 | 4.98E-07 | 0.000805964 |
| Obesity | mast_cell | *DRG2* | 10.39811463 | 2.25E-08 | 1.25E-07 | 0.000141694 |
| Obesity | mast_cell | *AKAP10* | 5.228167963 | 2.89E-06 | 7.20E-06 | 0.018144121 |
| Obesity | mast_cell | *SMARCD2* | 6.93885462 | 0.000108999 | 0.000197486 | 0.685275185 |
| Obesity | mast_cell | *ICAM2* | 5.106840232 | 5.38E-07 | 1.64E-06 | 0.0033854 |
| Obesity | mast_cell | *UTP6* | 7.776025837 | 1.46E-07 | 5.53E-07 | 0.000919615 |
| Obesity | mast_cell | *DDX5* | 2.087476555 | 0.005086441 | 0.007176494 | 1 |

| | | | | | | |
|---|---|---|---|---|---|---|
| Obesity | mast_cell | *C17orf75* | 6.131676559 | 4.03E-09 | 4.78E-08 | 2.54E-05 |
| Obesity | mast_cell | *CYTH1* | 4.44105396 | 3.01E-05 | 6.06E-05 | 0.189522563 |
| Obesity | mast_cell | *DHX58* | 3.476227754 | 0.00042726 | 0.000706243 | 1 |
| Obesity | mast_cell | *NAGLU* | 7.278867714 | 1.25E-08 | 8.74E-08 | 7.85E-05 |
| Obesity | mast_cell | *HSD17B1* | -0.929257467 | 0.016373293 | 0.021544347 | 1 |
| Obesity | mast_cell | *EZH1* | 5.260266141 | 6.20E-09 | 5.83E-08 | 3.90E-05 |
| Obesity | mast_cell | *DLX4* | 24.15913966 | 1.85E-08 | 1.08E-07 | 0.000116179 |
| Obesity | mast_cell | *VAT1* | 6.446013609 | 1.74E-05 | 3.67E-05 | 0.109144531 |
| Obesity | mast_cell | *ALOX12* | 4.512485048 | 1.89E-07 | 6.84E-07 | 0.001190182 |
| Obesity | mast_cell | *ABCC3* | 1.961164099 | 0.000734357 | 0.00116618 | 1 |
| Obesity | mast_cell | *MPP2* | 4.566088525 | 1.11E-08 | 8.21E-08 | 6.95E-05 |
| Obesity | mast_cell | *EFTUD2* | 7.185495571 | 1.21E-06 | 3.36E-06 | 0.007619972 |
| Obesity | mast_cell | *EFNB3* | 30.77642906 | 1.38E-08 | 9.32E-08 | 8.70E-05 |
| Obesity | mast_cell | *FAM20A* | 4.266375323 | 2.32E-09 | 4.32E-08 | 1.46E-05 |
| Obesity | mast_cell | *YWHAE* | 6.024986691 | 2.37E-06 | 6.03E-06 | 0.014892784 |
| Obesity | mast_cell | *DPH1* | 6.404144517 | 2.21E-09 | 4.32E-08 | 1.39E-05 |
| Obesity | mast_cell | *DHRS7B* | 4.909096218 | 0.001273445 | 0.001951292 | 1 |
| Obesity | mast_cell | *RCVRN* | -3.790666901 | 1.03E-07 | 4.16E-07 | 0.000645081 |
| Obesity | mast_cell | *TMEM97* | 3.557446859 | 1.21E-07 | 4.73E-07 | 0.00075768 |
| Obesity | mast_cell | *RAB34* | 3.467610647 | 4.15E-07 | 1.33E-06 | 0.00260926 |
| Obesity | mast_cell | *SLAIN2* | 4.871174394 | 0.000128936 | 0.000230355 | 0.810620275 |
| Obesity | mast_cell | *OCIAD1* | 5.394495037 | 3.18E-07 | 1.06E-06 | 0.002001079 |
| Obesity | mast_cell | *KIAA1211* | 2.087602671 | 6.61E-06 | 1.52E-05 | 0.041549204 |
| Obesity | mast_cell | *AREG* | -1.60563896 | 0.001790814 | 0.002683233 | 1 |
| Obesity | mast_cell | *UBE2D3* | 5.232112972 | 1.98E-05 | 4.12E-05 | 0.12433786 |
| Obesity | mast_cell | *NDUFC1* | 7.403646535 | 0.00041001 | 0.000679962 | 1 |
| Obesity | mast_cell | *TBC1D9* | 5.746093431 | 1.68E-08 | 1.04E-07 | 0.000105898 |
| Obesity | mast_cell | *CPE* | 5.806126037 | 1.35E-06 | 3.68E-06 | 0.008462893 |
| Obesity | mast_cell | *WFS1* | 5.036401452 | 8.10E-10 | 3.01E-08 | 5.09E-06 |
| Obesity | mast_cell | *GRPEL1* | 11.8777604 | 1.04E-05 | 2.31E-05 | 0.065328213 |
| Obesity | mast_cell | *AADAT* | 3.628025562 | 2.04E-08 | 1.16E-07 | 0.000128066 |
| Obesity | mast_cell | *CPZ* | 48.11145167 | 1.85E-08 | 1.08E-07 | 0.000116411 |
| Obesity | mast_cell | *TRIM2* | 2.633513213 | 0.001013653 | 0.001576264 | 1 |
| Obesity | mast_cell | *SLC2A9* | 3.450311639 | 8.39E-09 | 6.89E-08 | 5.27E-05 |
| Obesity | mast_cell | *SH3D19* | 3.161628009 | 1.27E-07 | 4.93E-07 | 0.000796016 |
| Obesity | mast_cell | *STIM2* | 6.4959755 | 1.20E-07 | 4.73E-07 | 0.000757483 |
| Obesity | mast_cell | *RAPGEF2* | -1.913509513 | 0.007683443 | 0.010579459 | 1 |
| Obesity | mast_cell | *LRP2BP* | 4.371441153 | 9.23E-07 | 2.64E-06 | 0.005800567 |
| Obesity | mast_cell | *UFSP2* | 7.447900514 | 5.34E-06 | 1.25E-05 | 0.033558063 |
| Obesity | mast_cell | *KLF3* | 6.142355462 | 1.76E-08 | 1.05E-07 | 0.000110454 |
| Obesity | mast_cell | *KLHL5* | 2.605662104 | 0.005703381 | 0.007982448 | 1 |
| Obesity | mast_cell | *FAM149A* | 1.411724426 | 0.006480439 | 0.009001883 | 1 |
| Obesity | mast_cell | *UGDH* | 4.638742032 | 6.53E-09 | 6.00E-08 | 4.11E-05 |
| Obesity | mast_cell | *DDX25* | 5.241201126 | 5.85E-05 | 0.000111529 | 0.367821726 |
| Obesity | mast_cell | *HTATIP2* | 2.168282723 | 0.010919578 | 0.014728896 | 1 |
| Obesity | mast_cell | *CTSC* | 3.556785993 | 0.000490754 | 0.000801603 | 1 |
| Obesity | mast_cell | *SC5D* | 9.168257496 | 3.15E-09 | 4.48E-08 | 1.98E-05 |
| Obesity | mast_cell | *C11orf63* | 4.970381396 | 1.36E-09 | 3.95E-08 | 8.57E-06 |
| Obesity | mast_cell | *DTX4* | 7.420704671 | 7.13E-08 | 3.06E-07 | 0.000448229 |
| Obesity | mast_cell | *PUS3* | -3.583491804 | 0.000879411 | 0.001380833 | 1 |

| | | | | | | |
|---|---|---|---|---|---|---|
| Obesity | mast_cell | *DCPS* | 5.880254253 | 4.03E-06 | 9.69E-06 | 0.025351271 |
| Obesity | mast_cell | *FOXRED1* | 4.827248198 | 3.96E-09 | 4.78E-08 | 2.49E-05 |
| Obesity | mast_cell | *NRXN2* | 17.9969958 | 2.31E-05 | 4.75E-05 | 0.145033769 |
| Obesity | mast_cell | *MS4A4A* | 2.616198123 | 4.89E-07 | 1.51E-06 | 0.003073842 |
| Obesity | mast_cell | *CCDC86* | 29.06559381 | 1.47E-08 | 9.65E-08 | 9.22E-05 |
| Obesity | mast_cell | *PRPF19* | 3.877690737 | 4.95E-08 | 2.31E-07 | 0.000311265 |
| Obesity | mast_cell | *HPX* | 3.629386247 | 1.07E-06 | 3.01E-06 | 0.006732243 |
| Obesity | mast_cell | *PANX1* | 4.656183171 | 1.91E-06 | 4.99E-06 | 0.01202439 |
| Obesity | mast_cell | *EIF4G2* | 6.638405479 | 1.55E-06 | 4.17E-06 | 0.009765087 |
| Obesity | mast_cell | *CBL* | 5.53633384 | 1.04E-07 | 4.20E-07 | 0.000655542 |
| Obesity | mast_cell | *CD5* | 2.673841808 | 0.018892323 | 0.024662798 | 1 |
| Obesity | mast_cell | *MADD* | 7.750535874 | 4.76E-06 | 1.12E-05 | 0.029937525 |
| Obesity | mast_cell | *NAA40* | 6.38063579 | 5.33E-08 | 2.43E-07 | 0.00033509 |
| Obesity | mast_cell | *CARS* | 3.831321764 | 0.000558653 | 0.000903589 | 1 |
| Obesity | mast_cell | *SLC35F2* | 2.285671309 | 1.65E-05 | 3.51E-05 | 0.103928845 |
| Obesity | mast_cell | *C11orf21* | -3.348103704 | 0.000174816 | 0.000306062 | 1 |
| Obesity | mast_cell | *ELMOD1* | 8.112919325 | 0.00019687 | 0.000341818 | 1 |
| Obesity | mast_cell | *AIP* | 6.859342398 | 9.45E-07 | 2.69E-06 | 0.005943138 |
| Obesity | mast_cell | *NDUFS8* | 8.794952289 | 1.02E-08 | 7.86E-08 | 6.42E-05 |
| Obesity | mast_cell | *TCIRG1* | 5.919383127 | 0.000610718 | 0.000983248 | 1 |
| Obesity | mast_cell | *GTF2H1* | 6.081258821 | 2.40E-06 | 6.10E-06 | 0.015074458 |
| Obesity | mast_cell | *PTPN5* | 1.8537557 | 0.000282267 | 0.000478333 | 1 |
| Obesity | mast_cell | *PPFIBP1* | 4.137726374 | 9.39E-05 | 0.000171563 | 0.590175787 |
| Obesity | mast_cell | *PRDM4* | 6.064846998 | 0.000325514 | 0.000547048 | 1 |
| Obesity | mast_cell | *COQ5* | 0.795109082 | 0.003288633 | 0.004766167 | 1 |
| Obesity | mast_cell | *ASIC1* | 2.148643098 | 0.006845306 | 0.009483569 | 1 |
| Obesity | mast_cell | *CAPRIN2* | 6.024647717 | 8.68E-05 | 0.000159437 | 0.545593787 |
| Obesity | mast_cell | *CYP27B1* | 19.45002621 | 1.74E-08 | 1.05E-07 | 0.00010957 |
| Obesity | mast_cell | *ACSS3* | 0.955335091 | 0.019593164 | 0.025524705 | 1 |
| Obesity | mast_cell | *GLI1* | 11.79485131 | 3.14E-09 | 4.48E-08 | 1.98E-05 |
| Obesity | mast_cell | *METAP2* | 3.30757897 | 0.01706927 | 0.022385169 | 1 |
| Obesity | mast_cell | *LTA4H* | 6.87873565 | 1.14E-08 | 8.37E-08 | 7.19E-05 |
| Obesity | mast_cell | *WNT5B* | 3.341791502 | 6.92E-09 | 6.17E-08 | 4.35E-05 |
| Obesity | mast_cell | *TRPV4* | 2.756685811 | 4.86E-05 | 9.43E-05 | 0.305543248 |
| Obesity | mast_cell | *ITFG2* | 5.67412959 | 3.83E-05 | 7.55E-05 | 0.240961454 |
| Obesity | mast_cell | *PRR4* | 2.637768162 | 0.002461254 | 0.003628964 | 1 |
| Obesity | mast_cell | *PARP11* | 6.717104421 | 6.04E-07 | 1.81E-06 | 0.003798414 |
| Obesity | mast_cell | *ARPC3* | 7.104414108 | 1.16E-07 | 4.61E-07 | 0.000731334 |
| Obesity | mast_cell | *GPN3* | 8.653090893 | 7.60E-06 | 1.72E-05 | 0.047758925 |
| Obesity | mast_cell | *VPS29* | 7.042382078 | 1.07E-06 | 3.00E-06 | 0.00669981 |
| Obesity | mast_cell | *AKAP3* | 2.633993843 | 1.18E-06 | 3.27E-06 | 0.00740002 |
| Obesity | mast_cell | *KCNA1* | 5.010530066 | 2.40E-06 | 6.10E-06 | 0.015079295 |
| Obesity | mast_cell | *CREBL2* | 5.098269444 | 9.98E-08 | 4.08E-07 | 0.000627227 |
| Obesity | mast_cell | *NAA25* | 6.646137735 | 7.54E-07 | 2.21E-06 | 0.004741257 |
| Obesity | mast_cell | *SCNN1A* | 5.182369591 | 1.86E-07 | 6.75E-07 | 0.00117157 |
| Obesity | mast_cell | *LTBR* | 5.426407849 | 4.71E-06 | 1.11E-05 | 0.02961841 |
| Obesity | mast_cell | *CDK2AP1* | 1.585762395 | 0.001587966 | 0.00239989 | 1 |
| Obesity | mast_cell | *MGP* | 2.966885257 | 3.95E-05 | 7.77E-05 | 0.248519811 |
| Obesity | mast_cell | *RASAL1* | -7.597138194 | 1.65E-07 | 6.10E-07 | 0.001039473 |
| Obesity | mast_cell | *ARHGDIB* | -2.594424182 | 0.000394655 | 0.000656054 | 1 |

| | | | | | | |
|---|---|---|---|---|---|---|
| Obesity | mast_cell | *EIF2B1* | 8.288046279 | 3.22E-07 | 1.07E-06 | 0.002022763 |
| Obesity | mast_cell | *DDX55* | 4.546576315 | 2.54E-08 | 1.37E-07 | 0.000159466 |
| Obesity | mast_cell | *ENDOU* | -3.07806489 | 0.000278811 | 0.000472858 | 1 |
| Obesity | mast_cell | *STX2* | 3.584793955 | 0.000126455 | 0.000226566 | 0.795021592 |
| Obesity | mast_cell | *CAND1* | 3.081717928 | 0.005592372 | 0.007830567 | 1 |
| Obesity | mast_cell | *MDM1* | 3.783639616 | 1.29E-05 | 2.80E-05 | 0.081149071 |
| Obesity | mast_cell | *NUP107* | 4.760417938 | 0.000565463 | 0.000913899 | 1 |
| Obesity | mast_cell | *CNOT2* | 5.451099302 | 9.00E-07 | 2.58E-06 | 0.005660854 |
| Obesity | mast_cell | *KRR1* | 3.880903961 | 0.01239311 | 0.016570711 | 1 |
| Obesity | mast_cell | *MRPL51* | 1.17597427 | 0.035629127 | 0.044782152 | 1 |
| Obesity | mast_cell | *COPS7A* | 3.954118288 | 0.004581069 | 0.006499929 | 1 |
| Obesity | mast_cell | *GNPTAB* | -2.026651785 | 0.000128017 | 0.000228974 | 0.804842202 |
| Obesity | mast_cell | *SPSB2* | 5.99848983 | 1.47E-09 | 4.13E-08 | 9.25E-06 |
| Obesity | mast_cell | *ATN1* | 5.414531471 | 2.80E-08 | 1.47E-07 | 0.000176117 |
| Obesity | mast_cell | *C12orf57* | 5.454287708 | 1.93E-05 | 4.04E-05 | 0.121588674 |
| Obesity | mast_cell | *NT5DC3* | 2.835655052 | 0.001619085 | 0.002441052 | 1 |
| Obesity | mast_cell | *ST8SIA1* | -0.473954057 | 2.38E-07 | 8.29E-07 | 0.001498576 |
| Obesity | mast_cell | *C2CD5* | 5.568121519 | 2.46E-05 | 5.02E-05 | 0.154383088 |
| Obesity | mast_cell | *PHC1* | 6.267565756 | 4.07E-09 | 4.79E-08 | 2.56E-05 |
| Obesity | mast_cell | *COL12A1* | 5.573353458 | 1.37E-06 | 3.74E-06 | 0.008622695 |
| Obesity | mast_cell | *BTN3A3* | 2.627380894 | 6.34E-06 | 1.46E-05 | 0.039856664 |
| Obesity | mast_cell | *TDP2* | 5.516808105 | 0.001789118 | 0.002681331 | 1 |
| Obesity | mast_cell | *RWDD1* | 4.249206706 | 6.46E-08 | 2.83E-07 | 0.000406325 |
| Obesity | mast_cell | *RSPH4A* | 1.661136996 | 0.001614513 | 0.002434744 | 1 |
| Obesity | mast_cell | *SMIM8* | 6.502810434 | 7.34E-08 | 3.13E-07 | 0.00046137 |
| Obesity | mast_cell | *MCM9* | 5.798913903 | 4.70E-05 | 9.13E-05 | 0.295414887 |
| Obesity | mast_cell | *FAM184A* | 4.39067886 | 2.95E-09 | 4.42E-08 | 1.85E-05 |
| Obesity | mast_cell | *RNGTT* | 6.094686699 | 2.85E-08 | 1.49E-07 | 0.000179367 |
| Obesity | mast_cell | *GABRR2* | 3.27013517 | 6.78E-10 | 2.77E-08 | 4.26E-06 |
| Obesity | mast_cell | *TPD52L1* | 3.255742 | 0.000261718 | 0.000445914 | 1 |
| Obesity | mast_cell | *UST* | 3.55080254 | 1.38E-08 | 9.30E-08 | 8.66E-05 |
| Obesity | mast_cell | *MTRF1L* | 3.470022805 | 0.00071532 | 0.001137384 | 1 |
| Obesity | mast_cell | *TULP1* | 14.2924997 | 1.44E-08 | 9.57E-08 | 9.07E-05 |
| Obesity | mast_cell | *RNF8* | 4.734210605 | 5.71E-05 | 0.000108969 | 0.35883619 |
| Obesity | mast_cell | *PHACTR1* | 5.273001417 | 5.07E-08 | 2.34E-07 | 0.000318675 |
| Obesity | mast_cell | *MDGA1* | 8.039598111 | 1.21E-05 | 2.64E-05 | 0.075852918 |
| Obesity | mast_cell | *CD83* | -2.138482022 | 2.19E-05 | 4.52E-05 | 0.137501925 |
| Obesity | mast_cell | *MDN1* | 3.537147477 | 0.003184982 | 0.004624477 | 1 |
| Obesity | mast_cell | *SAYSD1* | 6.83318499 | 8.29E-09 | 6.85E-08 | 5.21E-05 |
| Obesity | mast_cell | *CAP2* | 2.260160112 | 0.000254833 | 0.000435008 | 1 |
| Obesity | mast_cell | *GPR63* | -1.780076362 | 0.010519662 | 0.014223035 | 1 |
| Obesity | mast_cell | *ASCC3* | 5.490521196 | 4.40E-06 | 1.05E-05 | 0.027656092 |
| Obesity | mast_cell | *BVES* | 1.491544641 | 0.001556895 | 0.002357466 | 1 |
| Obesity | mast_cell | *WASF1* | 1.305331885 | 0.009734212 | 0.01321507 | 1 |
| Obesity | mast_cell | *GPLD1* | 1.750578907 | 6.60E-07 | 1.96E-06 | 0.004147317 |
| Obesity | mast_cell | *VNN2* | 5.398322396 | 1.11E-08 | 8.21E-08 | 6.95E-05 |
| Obesity | mast_cell | *ACOT13* | 7.606382173 | 3.95E-06 | 9.49E-06 | 0.024806694 |
| Obesity | mast_cell | *C6orf62* | 5.184397824 | 3.28E-08 | 1.65E-07 | 0.000206305 |
| Obesity | mast_cell | *B3GAT2* | -4.958399655 | 0.000260219 | 0.0004436 | 1 |
| Obesity | mast_cell | *HBS1L* | 2.45828276 | 0.014999165 | 0.019819199 | 1 |

| | | | | | | |
|---|---|---|---|---|---|---|
| Obesity | mast_cell | *PEX7* | 1.631060231 | 0.014876789 | 0.019665764 | 1 |
| Obesity | mast_cell | *SLC16A10* | -1.81712014 | 0.015063782 | 0.019900398 | 1 |
| Obesity | mast_cell | *PACRG* | 4.928091141 | 2.54E-09 | 4.32E-08 | 1.59E-05 |
| Obesity | mast_cell | *TFEB* | 7.748371033 | 2.84E-08 | 1.48E-07 | 0.00017841 |
| Obesity | mast_cell | *TBP* | 5.363656936 | 1.43E-08 | 9.52E-08 | 8.97E-05 |
| Obesity | mast_cell | *DNPH1* | 1.547652682 | 0.029625065 | 0.037604035 | 1 |
| Obesity | mast_cell | *DUSP22* | 7.332775005 | 1.53E-06 | 4.12E-06 | 0.009620831 |
| Obesity | mast_cell | *EXOC2* | 6.247354857 | 4.60E-07 | 1.44E-06 | 0.002894371 |
| Obesity | mast_cell | *TMEM30A* | 5.82110945 | 1.70E-06 | 4.50E-06 | 0.010690505 |
| Obesity | mast_cell | *SENP6* | 4.796004773 | 4.22E-05 | 8.25E-05 | 0.265062032 |
| Obesity | mast_cell | *TTK* | 3.93320776 | 0.001196739 | 0.001842287 | 1 |
| Obesity | mast_cell | *BTN2A1* | 3.019762547 | 0.001344978 | 0.002054391 | 1 |
| Obesity | mast_cell | *LAMA4* | 3.593997769 | 2.31E-10 | 1.41E-08 | 1.45E-06 |
| Obesity | mast_cell | *CLIC5* | 2.925447442 | 5.47E-07 | 1.66E-06 | 0.003441765 |
| Obesity | mast_cell | *FBRSL1* | 4.496552898 | 0.009866682 | 0.013386238 | 1 |
| Obesity | mast_cell | *LY86* | 6.862509995 | 1.38E-09 | 3.97E-08 | 8.70E-06 |
| Obesity | mast_cell | *PRSS16* | 11.24879227 | 1.49E-10 | 1.12E-08 | 9.39E-07 |
| Obesity | mast_cell | *TBX18* | 2.819699991 | 0.005376908 | 0.00755073 | 1 |
| Obesity | mast_cell | *PCDHB2* | -1.74385782 | 0.00512574 | 0.007230233 | 1 |
| Obesity | mast_cell | *NUDT12* | 4.377571496 | 0.028484472 | 0.036288121 | 1 |
| Obesity | mast_cell | *CEP72* | -2.627432696 | 0.004540781 | 0.006448586 | 1 |
| Obesity | mast_cell | *C7* | 6.837856393 | 6.17E-07 | 1.84E-06 | 0.003881969 |
| Obesity | mast_cell | *PAPD7* | 5.575653633 | 3.19E-05 | 6.36E-05 | 0.200448107 |
| Obesity | mast_cell | *GHR* | 1.821293618 | 2.26E-05 | 4.65E-05 | 0.141830831 |
| Obesity | mast_cell | *NME5* | 5.618835651 | 5.42E-11 | 5.48E-09 | 3.41E-07 |
| Obesity | mast_cell | *NNT* | 2.647480679 | 0.000265552 | 0.000451834 | 1 |
| Obesity | mast_cell | *PFDN1* | 4.260354459 | 9.48E-05 | 0.000173088 | 0.595943 |
| Obesity | mast_cell | *LOX* | 8.376337873 | 2.42E-07 | 8.40E-07 | 0.001522531 |
| Obesity | mast_cell | *APBB3* | 2.598746882 | 1.18E-05 | 2.59E-05 | 0.074294216 |
| Obesity | mast_cell | *TMCO6* | 4.008052458 | 3.25E-07 | 1.08E-06 | 0.002042393 |
| Obesity | mast_cell | *SPARC* | 3.81805279 | 0.000385459 | 0.000642466 | 1 |
| Obesity | mast_cell | *PCDHB6* | 6.830257151 | 2.56E-06 | 6.46E-06 | 0.016079504 |
| Obesity | mast_cell | *PDE8B* | 2.976659742 | 5.02E-06 | 1.18E-05 | 0.031587467 |
| Obesity | mast_cell | *CLK4* | 6.353227541 | 3.79E-06 | 9.16E-06 | 0.023831238 |
| Obesity | mast_cell | *PCDHB15* | 1.031738586 | 0.016601795 | 0.021822179 | 1 |
| Obesity | mast_cell | *HAVCR1* | 2.065389087 | 9.05E-05 | 0.000165683 | 0.56911969 |
| Obesity | mast_cell | *ARSB* | 2.929490689 | 0.001706657 | 0.002563248 | 1 |
| Obesity | mast_cell | *CLINT1* | 5.637079835 | 3.01E-08 | 1.55E-07 | 0.000189228 |
| Obesity | mast_cell | *TTC1* | 6.617822548 | 2.71E-06 | 6.81E-06 | 0.017027733 |
| Obesity | mast_cell | *MSH3* | 1.734168162 | 0.016217889 | 0.021357744 | 1 |
| Obesity | mast_cell | *RASGRF2* | 5.189479046 | 1.72E-09 | 4.28E-08 | 1.08E-05 |
| Obesity | mast_cell | *POLR3G* | 4.360000252 | 0.000249787 | 0.000426626 | 1 |
| Obesity | mast_cell | *DROSHA* | 5.02780453 | 5.07E-06 | 1.19E-05 | 0.031893206 |
| Obesity | mast_cell | *CDH6* | 0.958064749 | 0.001488805 | 0.002259261 | 1 |
| Obesity | mast_cell | *SLC27A6* | 6.920794538 | 1.14E-08 | 8.37E-08 | 7.19E-05 |
| Obesity | mast_cell | *TARS* | 6.18197623 | 2.69E-09 | 4.32E-08 | 1.69E-05 |
| Obesity | mast_cell | *LNPEP* | 5.156627929 | 1.93E-09 | 4.32E-08 | 1.21E-05 |
| Obesity | mast_cell | *SLC12A7* | 7.036045437 | 1.44E-06 | 3.91E-06 | 0.009067021 |
| Obesity | mast_cell | *GNPDA1* | 8.69894881 | 1.86E-08 | 1.09E-07 | 0.000116937 |
| Obesity | mast_cell | *SKP1* | 5.473464337 | 0.000736053 | 0.001168578 | 1 |

| | | | | | | |
|---|---|---|---|---|---|---|
| Obesity | mast_cell | *PPWD1* | 2.408880231 | 0.00077282 | 0.001222935 | 1 |
| Obesity | mast_cell | *TRIM23* | 6.171191188 | 4.34E-05 | 8.49E-05 | 0.273169987 |
| Obesity | mast_cell | *TRAPPC13* | 3.473951472 | 0.001948105 | 0.00290644 | 1 |
| Obesity | mast_cell | *C9* | 2.774048737 | 1.46E-05 | 3.13E-05 | 0.091577981 |
| Obesity | mast_cell | *SMAD5* | 4.165345833 | 0.004218445 | 0.006019374 | 1 |
| Obesity | mast_cell | *HMGXB3* | 6.229902927 | 2.03E-08 | 1.16E-07 | 0.00012761 |
| Obesity | mast_cell | *ATP6V0E1* | 5.58452084 | 3.69E-05 | 7.28E-05 | 0.232125533 |
| Obesity | mast_cell | *CNTN3* | 5.048696077 | 2.68E-09 | 4.32E-08 | 1.68E-05 |
| Obesity | mast_cell | *ACTR8* | 7.789957608 | 1.96E-08 | 1.13E-07 | 0.000123108 |
| Obesity | mast_cell | *HGD* | 10.20819596 | 2.07E-09 | 4.32E-08 | 1.30E-05 |
| Obesity | mast_cell | *NIT2* | 5.310853335 | 8.29E-09 | 6.85E-08 | 5.21E-05 |
| Obesity | mast_cell | *FAM162A* | 3.707103301 | 0.016303615 | 0.021461647 | 1 |
| Obesity | mast_cell | *PCCB* | 4.649426274 | 0.004425984 | 0.006295511 | 1 |
| Obesity | mast_cell | *RBP1* | 4.736143007 | 2.44E-08 | 1.33E-07 | 0.000153148 |
| Obesity | mast_cell | *WNT5A* | 4.403736256 | 0.013653866 | 0.018152222 | 1 |
| Obesity | mast_cell | *PRKAR2A* | 4.692640311 | 1.41E-06 | 3.83E-06 | 0.008845765 |
| Obesity | mast_cell | *CYB561D2* | 4.188878905 | 3.16E-05 | 6.31E-05 | 0.198386948 |
| Obesity | mast_cell | *C3orf14* | 1.904874985 | 0.00052378 | 0.000851346 | 1 |
| Obesity | mast_cell | *IFT57* | 7.985433181 | 2.31E-08 | 1.28E-07 | 0.000145166 |
| Obesity | mast_cell | *IQCG* | 7.48968512 | 1.28E-07 | 4.98E-07 | 0.000806152 |
| Obesity | mast_cell | *NCBP2* | 8.229239562 | 2.38E-08 | 1.30E-07 | 0.00014977 |
| Obesity | mast_cell | *PLXNA1* | 4.112607609 | 3.19E-08 | 1.61E-07 | 0.000200404 |
| Obesity | mast_cell | *ABTB1* | 6.826904691 | 2.57E-08 | 1.38E-07 | 0.000161533 |
| Obesity | mast_cell | *PLSCR4* | 3.939872708 | 3.13E-06 | 7.74E-06 | 0.019704525 |
| Obesity | mast_cell | *HEMK1* | 5.002808722 | 9.70E-06 | 2.16E-05 | 0.06095482 |
| Obesity | mast_cell | *WDR48* | 5.480817585 | 8.11E-08 | 3.41E-07 | 0.000510161 |
| Obesity | mast_cell | *COMMD2* | 7.0967086 | 2.55E-08 | 1.37E-07 | 0.000160211 |
| Obesity | mast_cell | *GORASP1* | 8.53494775 | 4.00E-07 | 1.29E-06 | 0.002517354 |
| Obesity | mast_cell | *ABCC5* | 4.992567361 | 4.91E-07 | 1.51E-06 | 0.003084727 |
| Obesity | mast_cell | *ARHGEF26* | 6.301958881 | 1.74E-07 | 6.37E-07 | 0.001094291 |
| Obesity | mast_cell | *KLHL24* | 4.703643609 | 1.30E-05 | 2.82E-05 | 0.081609466 |
| Obesity | mast_cell | *PLCH1* | 12.08817454 | 6.71E-12 | 1.08E-09 | 4.22E-08 |
| Obesity | mast_cell | *ZBTB47* | 4.883735244 | 6.36E-09 | 5.93E-08 | 4.00E-05 |
| Obesity | mast_cell | *NKTR* | 3.689532151 | 0.001231419 | 0.001891044 | 1 |
| Obesity | mast_cell | *CLCN2* | 3.385877334 | 9.49E-09 | 7.44E-08 | 5.96E-05 |
| Obesity | mast_cell | *NEK4* | 6.475089384 | 1.11E-07 | 4.42E-07 | 0.000695963 |
| Obesity | mast_cell | *MOB1A* | 7.901794732 | 4.57E-07 | 1.44E-06 | 0.002873056 |
| Obesity | mast_cell | *PIKFYVE* | 3.895454828 | 0.002637783 | 0.003871992 | 1 |
| Obesity | mast_cell | *KCNIP3* | 3.481233813 | 1.04E-08 | 7.89E-08 | 6.51E-05 |
| Obesity | mast_cell | *FAHD2A* | 6.825139807 | 4.63E-11 | 4.86E-09 | 2.91E-07 |
| Obesity | mast_cell | *NCL* | 6.034582404 | 1.66E-08 | 1.04E-07 | 0.000104561 |
| Obesity | mast_cell | *ACTR1B* | 8.814145337 | 2.93E-12 | 5.42E-10 | 1.84E-08 |
| Obesity | mast_cell | *SLC35F5* | 3.400697391 | 0.00521586 | 0.007338252 | 1 |
| Obesity | mast_cell | *ACTR3* | 6.179158819 | 6.50E-07 | 1.93E-06 | 0.00408662 |
| Obesity | mast_cell | *STEAP3* | 1.539026497 | 0.000539373 | 0.000875785 | 1 |
| Obesity | mast_cell | *TFCP2L1* | 5.045256981 | 2.00E-10 | 1.30E-08 | 1.26E-06 |
| Obesity | mast_cell | *TP53I3* | 6.419583528 | 3.39E-05 | 6.73E-05 | 0.2128456 |
| Obesity | mast_cell | *DNAJC27* | 7.333443568 | 2.85E-07 | 9.61E-07 | 0.00178993 |
| Obesity | mast_cell | *STAM2* | 6.526661161 | 9.84E-08 | 4.03E-07 | 0.000618858 |
| Obesity | mast_cell | *GPD2* | 4.344146976 | 0.001410107 | 0.002148653 | 1 |

| | | | | | | |
|---|---|---|---|---|---|---|
| Obesity | mast_cell | *CENPA* | 7.443554835 | 0.000256598 | 0.000437901 | 1 |
| Obesity | mast_cell | *TANC1* | 4.127049142 | 3.49E-06 | 8.52E-06 | 0.02194329 |
| Obesity | mast_cell | *ITGB6* | -3.256999796 | 1.52E-06 | 4.10E-06 | 0.009569708 |
| Obesity | mast_cell | *PPM1G* | 6.094435589 | 1.02E-05 | 2.27E-05 | 0.064086463 |
| Obesity | mast_cell | *PCSK4* | 3.847081868 | 0.000261578 | 0.000445796 | 1 |
| Obesity | mast_cell | *INO80B* | -3.156221532 | 0.005903926 | 0.008246608 | 1 |
| Obesity | mast_cell | *MOGS* | 1.796353688 | 0.007028616 | 0.009716118 | 1 |
| Obesity | mast_cell | *GRB14* | 4.871670958 | 0.006988817 | 0.009665353 | 1 |
| Obesity | mast_cell | *CLIP4* | 5.972111476 | 3.66E-06 | 8.88E-06 | 0.022990157 |
| Obesity | mast_cell | *SPTBN1* | 3.336775284 | 4.87E-05 | 9.45E-05 | 0.306161872 |
| Obesity | mast_cell | *HTRA2* | 8.432466184 | 1.02E-07 | 4.15E-07 | 0.000642137 |
| Obesity | mast_cell | *GALNT3* | -1.79476172 | 2.49E-06 | 6.32E-06 | 0.01566964 |
| Obesity | mast_cell | *POLE4* | 7.415086699 | 2.30E-09 | 4.32E-08 | 1.44E-05 |
| Obesity | mast_cell | *MRPL19* | 4.103918774 | 0.000706577 | 0.001124621 | 1 |
| Obesity | mast_cell | *LANCL1* | 7.481485157 | 3.12E-08 | 1.59E-07 | 0.000196118 |
| Obesity | mast_cell | *WDR75* | 7.271180497 | 8.61E-09 | 7.01E-08 | 5.41E-05 |
| Obesity | mast_cell | *EFEMP1* | 4.101493312 | 9.29E-08 | 3.83E-07 | 0.00058407 |
| Obesity | mast_cell | *GLS* | 4.935713164 | 1.07E-05 | 2.37E-05 | 0.067306547 |
| Obesity | mast_cell | *DNAH6* | 0.607114186 | 0.001062609 | 0.001646285 | 1 |
| Obesity | mast_cell | *GGCX* | 4.263366292 | 5.03E-09 | 5.29E-08 | 3.16E-05 |
| Obesity | mast_cell | *GNLY* | 2.701993912 | 9.16E-09 | 7.26E-08 | 5.76E-05 |
| Obesity | mast_cell | *SF3B1* | 5.141396922 | 1.28E-05 | 2.79E-05 | 0.080707042 |
| Obesity | mast_cell | *ST3GAL5* | 5.881066839 | 1.15E-07 | 4.55E-07 | 0.00071998 |
| Obesity | mast_cell | *PLCD4* | 7.411190179 | 9.89E-08 | 4.05E-07 | 0.000621898 |
| Obesity | mast_cell | *CHMP3* | -5.368443361 | 5.82E-07 | 1.76E-06 | 0.003660939 |
| Obesity | mast_cell | *ZNF142* | 5.480201823 | 8.28E-05 | 0.00015264 | 0.520517963 |
| Obesity | mast_cell | *WNT6* | -10.22831832 | 0.000521824 | 0.000848825 | 1 |
| Obesity | mast_cell | *MLPH* | -0.465574305 | 1.92E-07 | 6.91E-07 | 0.0012072 |
| Obesity | mast_cell | *HDLBP* | 4.159353086 | 8.89E-05 | 0.000163081 | 0.55907093 |
| Obesity | mast_cell | *PASK* | 6.090588856 | 2.59E-07 | 8.87E-07 | 0.001626554 |
| Obesity | mast_cell | *STK25* | 7.030789735 | 3.51E-09 | 4.62E-08 | 2.21E-05 |
| Obesity | mast_cell | *TPO* | 1.381594661 | 0.003950933 | 0.005664656 | 1 |
| Obesity | mast_cell | *TAF1B* | 4.62304946 | 3.12E-08 | 1.59E-07 | 0.00019623 |
| Obesity | mast_cell | *STRN* | 4.704904677 | 0.001171481 | 0.001805171 | 1 |
| Obesity | mast_cell | *PRKD3* | 4.402668194 | 0.000607912 | 0.000979232 | 1 |
| Obesity | mast_cell | *SLC25A12* | 3.618860806 | 0.004205882 | 0.006005537 | 1 |
| Obesity | mast_cell | *RMDN2* | 7.235489946 | 1.81E-08 | 1.08E-07 | 0.000114085 |
| Obesity | mast_cell | *SDC1* | -3.772064078 | 1.04E-06 | 2.93E-06 | 0.006549608 |
| Obesity | mast_cell | *SOS1* | 4.111604292 | 6.20E-07 | 1.85E-06 | 0.003897309 |
| Obesity | mast_cell | *COX7A2L* | 6.425779946 | 4.38E-09 | 4.94E-08 | 2.76E-05 |
| Obesity | mast_cell | *ORC4* | 4.493500779 | 0.000666645 | 0.001066192 | 1 |
| Obesity | mast_cell | *THADA* | 5.592009343 | 4.69E-07 | 1.46E-06 | 0.002946876 |
| Obesity | mast_cell | *ARID3A* | 6.005490611 | 0.000694047 | 0.001106077 | 1 |
| Obesity | mast_cell | *CD207* | -10.81829157 | 5.12E-07 | 1.57E-06 | 0.00321584 |
| Obesity | mast_cell | *GRIN3B* | 4.03522698 | 3.17E-05 | 6.33E-05 | 0.199206311 |
| Obesity | mast_cell | *VAX2* | -6.242517201 | 0.00782784 | 0.010764135 | 1 |
| Obesity | mast_cell | *MSH6* | 6.482483872 | 0.000164157 | 0.00028901 | 1 |
| Obesity | mast_cell | *PLEKHA3* | 6.671490632 | 2.74E-09 | 4.32E-08 | 1.72E-05 |
| Obesity | mast_cell | *BCL9* | 5.828172754 | 1.29E-05 | 2.80E-05 | 0.08111242 |
| Obesity | mast_cell | *DHCR24* | 3.885238497 | 4.94E-08 | 2.30E-07 | 0.000310722 |

| | | | | | | |
|---|---|---|---|---|---|---|
| Obesity | mast_cell | *SCP2* | 5.542973814 | 3.44E-06 | 8.41E-06 | 0.021608945 |
| Obesity | mast_cell | *TPSG1* | -0.789353857 | 1.18E-07 | 4.65E-07 | 0.000741227 |
| Obesity | mast_cell | *RALGPS2* | 4.467183099 | 0.001344887 | 0.002054391 | 1 |
| Obesity | mast_cell | *FAM20B* | 6.285246397 | 3.38E-09 | 4.52E-08 | 2.13E-05 |
| Obesity | mast_cell | *TMEM59* | 6.200181015 | 3.84E-06 | 9.27E-06 | 0.024173135 |
| Obesity | mast_cell | *WRAP73* | 6.87719503 | 0.004559095 | 0.006471671 | 1 |
| Obesity | mast_cell | *MRPL37* | 4.463593596 | 4.56E-07 | 1.44E-06 | 0.0028686 |
| Obesity | mast_cell | *ICMT* | 4.428220971 | 1.12E-08 | 8.25E-08 | 7.03E-05 |
| Obesity | mast_cell | *QSOX1* | 7.106297447 | 9.04E-08 | 3.74E-07 | 0.000568173 |
| Obesity | mast_cell | *PARK7* | 5.951235052 | 6.15E-08 | 2.73E-07 | 0.00038645 |
| Obesity | mast_cell | *KCNC4* | -2.164386027 | 0.021432795 | 0.027788819 | 1 |
| Obesity | mast_cell | *EDEM3* | 5.455818486 | 7.24E-08 | 3.10E-07 | 0.000455059 |
| Obesity | mast_cell | *RAP1A* | 3.458973808 | 0.007053343 | 0.009748158 | 1 |
| Obesity | mast_cell | *CAPZA1* | 4.913369282 | 3.68E-05 | 7.25E-05 | 0.231059046 |
| Obesity | mast_cell | *DOCK7* | 5.293876203 | 3.36E-05 | 6.68E-05 | 0.210963378 |
| Obesity | mast_cell | *FBXO6* | 2.889777523 | 4.91E-07 | 1.51E-06 | 0.003084836 |
| Obesity | mast_cell | *C1orf21* | 1.861310526 | 0.014492102 | 0.019193564 | 1 |
| Obesity | mast_cell | *DNAJC6* | 4.212551662 | 3.42E-07 | 1.12E-06 | 0.00215218 |
| Obesity | mast_cell | *LEPR* | 3.598587929 | 0.004619527 | 0.006547107 | 1 |
| Obesity | mast_cell | *MIIP* | 5.202334616 | 5.95E-08 | 2.66E-07 | 0.000373948 |
| Obesity | mast_cell | *SMG7* | 6.144557683 | 7.14E-07 | 2.10E-06 | 0.004488616 |
| Obesity | mast_cell | *NCF2* | 4.163438223 | 3.25E-09 | 4.49E-08 | 2.04E-05 |
| Obesity | mast_cell | *OLFML3* | 4.372599194 | 1.68E-09 | 4.27E-08 | 1.06E-05 |
| Obesity | mast_cell | *TNNI3K* | 3.88032627 | 2.01E-05 | 4.18E-05 | 0.126384054 |
| Obesity | mast_cell | *CFHR3* | 1.662790156 | 0.0169982 | 0.022296616 | 1 |
| Obesity | mast_cell | *CRYZ* | 2.485009429 | 0.005375751 | 0.00755073 | 1 |
| Obesity | mast_cell | *TTF2* | 7.006268945 | 0.000131778 | 0.000235032 | 0.82848716 |
| Obesity | mast_cell | *WARS2* | 4.56102101 | 1.19E-08 | 8.50E-08 | 7.45E-05 |
| Obesity | mast_cell | *OSCP1* | -3.38635711 | 0.004586607 | 0.006506318 | 1 |
| Obesity | mast_cell | *GNPAT* | 5.694729983 | 1.37E-05 | 2.96E-05 | 0.086069757 |
| Obesity | mast_cell | *C1orf109* | 9.669121569 | 1.31E-09 | 3.89E-08 | 8.26E-06 |
| Obesity | mast_cell | *TBCE* | 5.421743853 | 0.000234812 | 0.000403021 | 1 |
| Obesity | mast_cell | *MTR* | 5.495734835 | 7.62E-07 | 2.23E-06 | 0.004792265 |
| Obesity | mast_cell | *ACADM* | 5.999884141 | 1.03E-08 | 7.88E-08 | 6.47E-05 |
| Obesity | mast_cell | *PADI2* | -0.755197458 | 1.31E-07 | 5.08E-07 | 0.000824646 |
| Obesity | mast_cell | *MFAP2* | -1.785370331 | 0.01422608 | 0.018865084 | 1 |
| Obesity | mast_cell | *CTBS* | 4.740168059 | 1.21E-06 | 3.35E-06 | 0.007598399 |
| Obesity | mast_cell | *KLHL12* | 5.208019003 | 1.10E-06 | 3.09E-06 | 0.006923892 |
| Obesity | mast_cell | *ZNHIT6* | 3.890014322 | 7.11E-05 | 0.000133039 | 0.446744453 |
| Obesity | mast_cell | *RBBP5* | 3.813082652 | 5.31E-06 | 1.24E-05 | 0.033358244 |
| Obesity | mast_cell | *KIF17* | -4.684167117 | 4.92E-05 | 9.53E-05 | 0.309226969 |
| Obesity | mast_cell | *CD46* | 5.16730531 | 6.57E-06 | 1.51E-05 | 0.041331304 |
| Obesity | mast_cell | *PRDX1* | 5.787426897 | 0.000191231 | 0.000332671 | 1 |
| Obesity | mast_cell | *FAAH* | 0.937621956 | 0.003148004 | 0.004573955 | 1 |
| Obesity | mast_cell | *NSUN4* | 4.229550737 | 0.000450181 | 0.000741495 | 1 |
| Obesity | mast_cell | *TMED5* | 6.665326507 | 1.89E-07 | 6.84E-07 | 0.00118835 |
| Obesity | mast_cell | *DR1* | 7.746358264 | 1.46E-06 | 3.95E-06 | 0.009172662 |
| Obesity | mast_cell | *CNN3* | 6.823916136 | 1.72E-08 | 1.05E-07 | 0.000108344 |
| Obesity | mast_cell | *PRRC2C* | 4.819185739 | 6.40E-05 | 0.000121 | 0.402567969 |
| Obesity | mast_cell | *ABCD3* | 5.891721204 | 5.71E-09 | 5.61E-08 | 3.59E-05 |

| | | | | | | |
|---|---|---|---|---|---|---|
| Obesity | mast_cell | *VAMP4* | 6.015634418 | 9.58E-08 | 3.93E-07 | 0.000602289 |
| Obesity | mast_cell | *PTBP2* | 4.454196981 | 7.36E-08 | 3.14E-07 | 0.000463033 |
| Obesity | mast_cell | *TNFSF4* | 2.349914692 | 1.56E-05 | 3.33E-05 | 0.097810543 |
| Obesity | mast_cell | *DARS2* | 4.136801812 | 6.94E-05 | 0.000130318 | 0.436513978 |
| Obesity | mast_cell | *DIEXF* | 4.8461106 | 2.58E-06 | 6.50E-06 | 0.016189079 |
| Obesity | mast_cell | *SERPINC1* | 5.697278964 | 1.55E-16 | 8.87E-14 | 9.76E-13 |
| Obesity | mast_cell | *RCAN3* | -2.359313947 | 0.00315685 | 0.004585748 | 1 |
| Obesity | mast_cell | *RCOR3* | 6.004859416 | 5.37E-07 | 1.63E-06 | 0.003378616 |
| Obesity | mast_cell | *NSL1* | 3.154434738 | 0.023532013 | 0.030347849 | 1 |
| Obesity | mast_cell | *RPA2* | 8.326839475 | 7.17E-08 | 3.07E-07 | 0.000450517 |
| Obesity | mast_cell | *SLC5A9* | 11.50172436 | 5.52E-05 | 0.000105845 | 0.346746951 |
| Obesity | mast_cell | *OSBPL9* | 4.054002859 | 0.001634972 | 0.002463824 | 1 |
| Obesity | mast_cell | *TXNDC12* | 7.716214304 | 1.31E-07 | 5.07E-07 | 0.000822249 |
| Obesity | mast_cell | *ESYT2* | 2.447123277 | 0.029409751 | 0.03734581 | 1 |
| Obesity | mast_cell | *CD3EAP* | 2.676763488 | 0.000328177 | 0.000551376 | 1 |
| Obesity | mast_cell | *COLEC11* | -4.977315494 | 6.09E-08 | 2.71E-07 | 0.000383138 |
| Obesity | mast_cell | *A4GNT* | 1.332231904 | 0.028455697 | 0.03625881 | 1 |
| Obesity | mast_cell | *KMT2A* | 5.407497726 | 2.74E-05 | 5.55E-05 | 0.17211164 |
| Obesity | mast_cell | *IFT46* | 6.542611815 | 5.22E-10 | 2.40E-08 | 3.28E-06 |
| Obesity | mast_cell | *CAMSAP2* | 5.411602433 | 1.97E-07 | 7.04E-07 | 0.001235722 |
| Obesity | mast_cell | *ATF6* | 5.715266615 | 1.84E-06 | 4.83E-06 | 0.011571312 |
| Obesity | mast_cell | *ATP10B* | 5.114056805 | 7.00E-09 | 6.20E-08 | 4.40E-05 |
| Obesity | mast_cell | *SPCS2* | 5.493927831 | 0.000580536 | 0.000937054 | 1 |
| Obesity | mast_cell | *ELOVL4* | 2.203845673 | 0.000497422 | 0.000811862 | 1 |
| Obesity | mast_cell | *FILIP1* | 2.804159637 | 2.22E-08 | 1.24E-07 | 0.000139373 |
| Obesity | mast_cell | *AKAP7* | 1.860125999 | 0.009528135 | 0.012946485 | 1 |
| Obesity | mast_cell | *MED28* | 4.823183722 | 1.70E-05 | 3.59E-05 | 0.106597897 |
| Obesity | mast_cell | *SLC16A7* | 4.576927815 | 1.24E-08 | 8.72E-08 | 7.81E-05 |
| Obesity | mast_cell | *VAMP8* | 5.261796494 | 6.42E-05 | 0.000121284 | 0.403754024 |
| Obesity | mast_cell | *FOXO3* | 5.160833685 | 3.14E-06 | 7.74E-06 | 0.019711929 |
| Obesity | mast_cell | *TGIF2* | -2.589821328 | 2.93E-06 | 7.29E-06 | 0.018414988 |
| Obesity | mast_cell | *ABCG2* | -4.429479685 | 0.011549399 | 0.015525139 | 1 |
| Obesity | mast_cell | *SPP1* | 5.81651917 | 5.12E-08 | 2.36E-07 | 0.000321611 |
| Obesity | mast_cell | *FAM47E-STBL* | 1.881416977 | 9.74E-07 | 2.77E-06 | 0.00612494 |
| Obesity | mast_cell | *MFSD1* | 6.868033772 | 9.04E-08 | 3.74E-07 | 0.00056853 |
| Obesity | mast_cell | *RAB3GAP2* | 6.114414755 | 6.99E-08 | 3.01E-07 | 0.000439292 |
| Obesity | mast_cell | *UBN1* | 4.394353406 | 0.000127798 | 0.000228711 | 0.803463191 |
| Obesity | mast_cell | *UCHL3* | -5.180380896 | 0.000979193 | 0.001527211 | 1 |
| Obesity | mast_cell | *WDR35* | 7.210936307 | 1.56E-08 | 1.00E-07 | 9.82E-05 |
| Obesity | mast_cell | *NDUFB3* | 5.409683778 | 5.78E-05 | 0.000110341 | 0.363684922 |
| Obesity | mast_cell | *C2orf40* | 3.200962581 | 2.83E-05 | 5.71E-05 | 0.177874789 |
| Obesity | mast_cell | *CPSF3* | 7.633857976 | 8.48E-09 | 6.93E-08 | 5.33E-05 |
| Obesity | mast_cell | *PIGZ* | 1.557158326 | 0.000821843 | 0.001296593 | 1 |
| Obesity | mast_cell | *SENP5* | 5.678892295 | 1.58E-05 | 3.38E-05 | 0.099567905 |
| Obesity | mast_cell | *CCDC92* | 8.100054061 | 7.27E-18 | 5.08E-15 | 4.57E-14 |
| Obesity | mast_cell | *PTBP3* | 4.012211367 | 0.005482328 | 0.007688467 | 1 |
| Obesity | mast_cell | *FKBP15* | 6.858405416 | 3.37E-09 | 4.52E-08 | 2.12E-05 |
| Obesity | mast_cell | *CTNNAL1* | 1.771313077 | 0.000410928 | 0.000681124 | 1 |
| Obesity | mast_cell | *WDR34* | 2.377858605 | 2.72E-07 | 9.27E-07 | 0.001712573 |
| Obesity | mast_cell | *CNTRL* | 3.16402063 | 0.028874246 | 0.036747446 | 1 |

| | | | | | | |
|---|---|---|---|---|---|---|
| Obesity | mast_cell | *TRIM32* | 7.245573721 | 2.31E-10 | 1.41E-08 | 1.45E-06 |
| Obesity | mast_cell | *PHF19* | 6.411516511 | 1.04E-07 | 4.20E-07 | 0.000656264 |
| Obesity | mast_cell | *BSPRY* | -0.501275341 | 1.26E-07 | 4.90E-07 | 0.000790653 |
| Obesity | mast_cell | *PPP6C* | 5.962135561 | 6.06E-09 | 5.78E-08 | 3.81E-05 |
| Obesity | mast_cell | *RBM18* | 5.558304372 | 1.38E-08 | 9.31E-08 | 8.67E-05 |
| Obesity | mast_cell | *HSDL2* | 2.386081722 | 0.000184331 | 0.000321288 | 1 |
| Obesity | mast_cell | *INVS* | 6.492866144 | 2.80E-09 | 4.32E-08 | 1.76E-05 |
| Obesity | mast_cell | *KDSR* | 6.357708539 | 9.94E-05 | 0.000180827 | 0.624937334 |
| Obesity | mast_cell | *DCAF4* | 3.267606498 | 1.98E-11 | 2.54E-09 | 1.24E-07 |
| Obesity | mast_cell | *IFI27L2* | 3.465128414 | 1.69E-06 | 4.49E-06 | 0.010630318 |
| Obesity | mast_cell | *DNAL1* | 3.017309041 | 1.31E-08 | 9.03E-08 | 8.21E-05 |
| Obesity | mast_cell | *IRF2BPL* | 6.589118478 | 0.002619468 | 0.003847327 | 1 |
| Obesity | mast_cell | *TTLL5* | 5.800641625 | 3.28E-05 | 6.52E-05 | 0.20596575 |
| Obesity | mast_cell | *FLVCR2* | 6.269630194 | 0.001893683 | 0.002829274 | 1 |
| Obesity | mast_cell | *PPP4R4* | 2.8171863 | 5.01E-06 | 1.18E-05 | 0.031481856 |
| Obesity | mast_cell | *ALDH6A1* | 2.646953131 | 3.25E-06 | 7.99E-06 | 0.020418978 |
| Obesity | mast_cell | *EIF2B2* | -2.844980749 | 9.06E-05 | 0.000165748 | 0.569509678 |
| Obesity | mast_cell | *COQ6* | 8.314306119 | 1.91E-07 | 6.88E-07 | 0.001198529 |
| Obesity | mast_cell | *GPR75* | 12.77878379 | 4.48E-09 | 4.99E-08 | 2.82E-05 |
| Obesity | mast_cell | *KLHL29* | 4.949938422 | 7.86E-09 | 6.64E-08 | 4.94E-05 |
| Obesity | mast_cell | *ATAD2B* | 4.817546976 | 0.000159251 | 0.000281003 | 1 |
| Obesity | mast_cell | *FKBP1B* | 2.41452141 | 0.000901676 | 0.001414381 | 1 |
| Obesity | mast_cell | *ATL2* | 5.217531511 | 1.75E-08 | 1.05E-07 | 0.000110071 |
| Obesity | mast_cell | *CNRIP1* | -3.317980124 | 0.003873948 | 0.005561889 | 1 |
| Obesity | mast_cell | *EPCAM* | -3.217860788 | 0.001163704 | 0.001793627 | 1 |
| Obesity | mast_cell | *SLC17A5* | 2.435160507 | 0.022195496 | 0.028706662 | 1 |
| Obesity | mast_cell | *OGFRL1* | 4.212720267 | 0.000121433 | 0.000218253 | 0.763449719 |
| Obesity | mast_cell | *CUTC* | 5.600199434 | 1.29E-07 | 5.00E-07 | 0.00081129 |
| Obesity | mast_cell | *PYROXD2* | 1.524691171 | 0.000170858 | 0.000299799 | 1 |
| Obesity | mast_cell | *MXI1* | 6.283817225 | 7.23E-09 | 6.29E-08 | 4.55E-05 |
| Obesity | mast_cell | *HELLS* | 3.016094496 | 0.038810586 | 0.048509374 | 1 |
| Obesity | mast_cell | *TCTN3* | 6.638588049 | 1.47E-06 | 3.98E-06 | 0.009236403 |
| Obesity | mast_cell | *WDR11* | 5.345178208 | 2.91E-05 | 5.86E-05 | 0.183001376 |
| Obesity | mast_cell | *C10orf76* | 6.063586974 | 1.40E-06 | 3.82E-06 | 0.008813356 |
| Obesity | mast_cell | *KCNIP2* | 2.07056623 | 2.26E-06 | 5.79E-06 | 0.014200437 |
| Obesity | mast_cell | *C10orf95* | 3.270345276 | 8.91E-06 | 2.00E-05 | 0.055993477 |
| Obesity | mast_cell | *KANSL1* | 3.796992196 | 9.03E-05 | 0.000165354 | 0.567824201 |
| Obesity | mast_cell | *HOXB5* | 9.273080481 | 2.66E-12 | 5.07E-10 | 1.67E-08 |
| Obesity | mast_cell | *HOXB3* | 6.202583121 | 8.55E-11 | 7.79E-09 | 5.38E-07 |
| Obesity | mast_cell | *CAAP1* | 3.474432817 | 0.015389587 | 0.020300952 | 1 |
| Obesity | mast_cell | *CD274* | -3.833690984 | 0.005289681 | 0.007436544 | 1 |
| Obesity | mast_cell | *NUP43* | 3.660582353 | 0.000466363 | 0.000765367 | 1 |
| Obesity | mast_cell | *LRP11* | 3.778576011 | 2.36E-05 | 4.84E-05 | 0.148237269 |
| Obesity | mast_cell | *CCDC170* | 2.289419679 | 3.14E-06 | 7.75E-06 | 0.019737796 |
| Obesity | mast_cell | *CYSTM1* | 6.187544507 | 5.70E-07 | 1.72E-06 | 0.00358324 |
| Obesity | mast_cell | *PCDHB10* | 2.037121769 | 0.004666435 | 0.00660763 | 1 |
| Obesity | mast_cell | *PCDHB14* | 4.735493147 | 7.89E-09 | 6.64E-08 | 4.96E-05 |
| Obesity | mast_cell | *GORAB* | 7.035789771 | 1.61E-07 | 5.98E-07 | 0.001013825 |
| Obesity | mast_cell | *ACAT2* | 6.18801622 | 0.000487311 | 0.000796905 | 1 |
| Obesity | mast_cell | *SNX19* | 2.260351717 | 0.010783788 | 0.014561356 | 1 |

| | | | | | | |
|---|---|---|---|---|---|---|
| Obesity | mast_cell | *MRC1* | 2.818737778 | 3.65E-08 | 1.80E-07 | 0.000229778 |
| Obesity | mast_cell | *IQSEC3* | 13.28107078 | 2.29E-08 | 1.27E-07 | 0.000143909 |
| Obesity | mast_cell | *CCDC77* | 5.538395133 | 3.08E-08 | 1.57E-07 | 0.000193403 |
| Obesity | mast_cell | *ENOX1* | 5.971689756 | 7.90E-09 | 6.64E-08 | 4.96E-05 |
| Obesity | mast_cell | *DNAJC15* | 7.171322006 | 1.19E-06 | 3.29E-06 | 0.007450484 |
| Obesity | mast_cell | *WBP4* | 5.870522848 | 1.56E-07 | 5.83E-07 | 0.000982074 |
| Obesity | mast_cell | *SMAD9* | 4.119072387 | 3.14E-09 | 4.48E-08 | 1.97E-05 |
| Obesity | mast_cell | *SIL1* | 5.005973142 | 1.24E-05 | 2.70E-05 | 0.077755486 |
| Obesity | mast_cell | *ZFP30* | 2.608624044 | 0.005170896 | 0.007282577 | 1 |
| Obesity | mast_cell | *ARL1* | 4.75728155 | 5.23E-07 | 1.60E-06 | 0.003286455 |
| Obesity | mast_cell | *APAF1* | 5.663560519 | 2.07E-08 | 1.17E-07 | 0.000129946 |
| Obesity | mast_cell | *ADRA1A* | 1.105033163 | 6.74E-09 | 6.06E-08 | 4.24E-05 |
| Obesity | mast_cell | *EPHX2* | 2.522988071 | 1.25E-05 | 2.73E-05 | 0.078868535 |
| Obesity | mast_cell | *ZNF706* | 3.875518654 | 0.00340175 | 0.004924431 | 1 |
| Obesity | mast_cell | *CRISPLD1* | 4.764321484 | 5.53E-09 | 5.54E-08 | 3.48E-05 |
| Obesity | mast_cell | *RDH10* | 1.812655158 | 0.013715927 | 0.018223169 | 1 |
| Obesity | mast_cell | *SCPEP1* | 1.823928408 | 7.17E-05 | 0.00013414 | 0.450675902 |
| Obesity | mast_cell | *TEX14* | 3.714521813 | 0.011607909 | 0.015600454 | 1 |
| Obesity | mast_cell | *CEP89* | 1.840181562 | 0.002704601 | 0.003961749 | 1 |
| Obesity | mast_cell | *PRB2* | 6.971107509 | 0.00423731 | 0.00604355 | 1 |
| Obesity | mast_cell | *PYROXD1* | 4.704523874 | 3.12E-05 | 6.25E-05 | 0.196286442 |
| Obesity | mast_cell | *A1BG* | -2.771665501 | 6.12E-06 | 1.42E-05 | 0.038488111 |
| Obesity | mast_cell | *ZNF211* | 2.758927618 | 3.11E-05 | 6.23E-05 | 0.19550327 |
| Obesity | mast_cell | *PDZRN3* | 4.480000667 | 1.09E-08 | 8.16E-08 | 6.87E-05 |
| Obesity | mast_cell | *SEC22A* | 1.978630155 | 0.032214452 | 0.040718186 | 1 |
| Obesity | mast_cell | *CSTA* | 3.637261759 | 1.48E-07 | 5.57E-07 | 0.000927746 |
| Obesity | mast_cell | *B4GALT4* | 9.24805423 | 6.13E-06 | 1.42E-05 | 0.038532735 |
| Obesity | mast_cell | *CD80* | 4.292188967 | 1.05E-08 | 7.97E-08 | 6.63E-05 |
| Obesity | mast_cell | *CAT* | 1.296898907 | 0.008700922 | 0.011894477 | 1 |
| Obesity | mast_cell | *PILRB* | 0.781583404 | 0.00141265 | 0.002152006 | 1 |
| Obesity | mast_cell | *TMEM39B* | 4.934314981 | 4.77E-07 | 1.48E-06 | 0.002996439 |
| Obesity | mast_cell | *RNF115* | 5.407936936 | 4.58E-05 | 8.93E-05 | 0.288221445 |
| Obesity | mast_cell | *POLR3GL* | 7.611436738 | 6.82E-09 | 6.11E-08 | 4.29E-05 |
| Obesity | mast_cell | *TNFSF10* | 5.180289451 | 0.000520557 | 0.000847201 | 1 |
| Obesity | mast_cell | *ZNF639* | 7.473088404 | 0.00012424 | 0.000222916 | 0.781098981 |
| Obesity | mast_cell | *LIAS* | 7.080124879 | 3.73E-06 | 9.02E-06 | 0.023438591 |
| Obesity | mast_cell | *GPSM2* | 6.765303863 | 4.13E-09 | 4.80E-08 | 2.60E-05 |
| Obesity | mast_cell | *ACVR2A* | 5.359290135 | 7.20E-05 | 0.000134598 | 0.452519152 |
| Obesity | mast_cell | *POLK* | 5.172845555 | 2.06E-05 | 4.28E-05 | 0.129751915 |
| Obesity | mast_cell | *FLT3* | 2.935212051 | 1.53E-08 | 9.90E-08 | 9.61E-05 |
| Obesity | mast_cell | *MTIF3* | 5.237693878 | 3.06E-05 | 6.13E-05 | 0.19226539 |
| Obesity | mast_cell | *RASL11A* | 7.819893968 | 9.58E-05 | 0.000174771 | 0.602434811 |
| Obesity | mast_cell | *FMOD* | 3.195521462 | 1.22E-08 | 8.65E-08 | 7.69E-05 |
| Obesity | mast_cell | *SERAC1* | 7.512011678 | 0.000144077 | 0.000256023 | 0.905808957 |
| Obesity | mast_cell | *LDB3* | 2.706854723 | 0.000156908 | 0.000277335 | 0.986479393 |
| Obesity | mast_cell | *OPN4* | -3.418591285 | 3.99E-05 | 7.85E-05 | 0.250983844 |
| Obesity | mast_cell | *FAM35A* | 2.539437793 | 0.022386204 | 0.028929509 | 1 |
| Obesity | mast_cell | *FAM213A* | 4.214190805 | 3.15E-08 | 1.60E-07 | 0.00019775 |
| Obesity | mast_cell | *ODF2L* | 5.083057778 | 4.20E-08 | 2.01E-07 | 0.000264194 |
| Obesity | mast_cell | *SPATA1* | 6.85572863 | 5.01E-07 | 1.54E-06 | 0.00314782 |

| | | | | | | |
|---|---|---|---|---|---|---|
| Obesity | mast_cell | *LRRC39* | 2.063404443 | 0.000103151 | 0.000187269 | 0.64851235 |
| Obesity | mast_cell | *CCDC18* | 4.755529206 | 0.000348826 | 0.000584663 | 1 |
| Obesity | mast_cell | *RPAP2* | 5.597025972 | 1.93E-05 | 4.03E-05 | 0.121345418 |
| Obesity | mast_cell | *BBS9* | 4.183347981 | 0.000276867 | 0.000470068 | 1 |
| Obesity | mast_cell | *PMS2* | 9.36098833 | 2.58E-08 | 1.38E-07 | 0.000161996 |
| Obesity | mast_cell | *KIAA0087* | 5.541298013 | 2.97E-06 | 7.38E-06 | 0.018689241 |
| Obesity | mast_cell | *KLHL7* | 6.114415284 | 5.11E-09 | 5.35E-08 | 3.21E-05 |
| Obesity | mast_cell | *CBX3* | 3.996043171 | 0.006424479 | 0.008928095 | 1 |
| Obesity | mast_cell | *FAM126A* | 5.382179255 | 3.47E-07 | 1.14E-06 | 0.002181933 |
| Obesity | mast_cell | *FKBP9* | 5.185710956 | 2.11E-07 | 7.47E-07 | 0.001326126 |
| Obesity | mast_cell | *CCZ1* | 6.219658232 | 6.41E-09 | 5.94E-08 | 4.03E-05 |
| Obesity | mast_cell | *SMU1* | 7.670030366 | 2.67E-08 | 1.41E-07 | 0.000167941 |
| Obesity | mast_cell | *GLIPR2* | 5.294475938 | 2.49E-07 | 8.60E-07 | 0.001564367 |
| Obesity | mast_cell | *DCAF10* | 5.442919678 | 1.30E-08 | 9.00E-08 | 8.17E-05 |
| Obesity | mast_cell | *TRIM24* | -2.524224113 | 0.001015732 | 0.001579107 | 1 |
| Obesity | mast_cell | *BICC1* | 2.545274647 | 3.76E-08 | 1.84E-07 | 0.000236512 |
| Obesity | mast_cell | *CISD1* | 1.888829939 | 0.013484197 | 0.017949428 | 1 |
| Obesity | mast_cell | *SLC25A16* | 2.944868111 | 3.66E-09 | 4.64E-08 | 2.30E-05 |
| Obesity | mast_cell | *ZWINT* | 2.843978161 | 8.88E-05 | 0.000162911 | 0.55829442 |
| Obesity | mast_cell | *RBM19* | 6.615780506 | 8.78E-09 | 7.08E-08 | 5.52E-05 |
| Obesity | mast_cell | *ACADS* | 6.76764047 | 9.87E-06 | 2.20E-05 | 0.062061427 |
| Obesity | mast_cell | *CCDC91* | 5.209835885 | 1.58E-07 | 5.89E-07 | 0.00099583 |
| Obesity | mast_cell | *EBPL* | 2.690666514 | 0.001099452 | 0.001700432 | 1 |
| Obesity | mast_cell | *ATP7B* | 0.678723423 | 0.025745322 | 0.033005881 | 1 |
| Obesity | mast_cell | *NLN* | 2.766753499 | 0.000991665 | 0.001544747 | 1 |
| Obesity | mast_cell | *OPTN* | 3.900865622 | 6.82E-05 | 0.000128283 | 0.428850799 |
| Obesity | mast_cell | *ATF1* | 5.818956344 | 1.70E-06 | 4.50E-06 | 0.010692092 |
| Obesity | mast_cell | *TSFM* | 5.360591279 | 2.58E-05 | 5.24E-05 | 0.162029556 |
| Obesity | mast_cell | *ARHGAP9* | 5.229843086 | 2.48E-05 | 5.06E-05 | 0.155829316 |
| Obesity | mast_cell | *SPATS2* | 5.057996481 | 2.90E-05 | 5.84E-05 | 0.182352867 |
| Obesity | mast_cell | *ORMDL2* | 5.200506456 | 5.01E-08 | 2.33E-07 | 0.000315083 |
| Obesity | mast_cell | *PDE1B* | 6.248359486 | 1.04E-07 | 4.20E-07 | 0.000653541 |
| Obesity | mast_cell | *IKZF4* | 3.964877619 | 1.78E-06 | 4.71E-06 | 0.01122046 |
| Obesity | mast_cell | *SMUG1* | 0.9794007 | 0.012387468 | 0.01656669 | 1 |
| Obesity | mast_cell | *SARDH* | -6.486340492 | 3.84E-06 | 9.26E-06 | 0.024145125 |
| Obesity | mast_cell | *STIL* | 7.676301321 | 4.75E-07 | 1.48E-06 | 0.002987406 |
| Obesity | mast_cell | *TTC21B* | 5.834433872 | 2.03E-05 | 4.22E-05 | 0.127511319 |
| Obesity | mast_cell | *ACVR1C* | 3.058893162 | 4.04E-07 | 1.30E-06 | 0.002538631 |
| Obesity | mast_cell | *PLA2G12A* | 5.212191393 | 0.003136436 | 0.004560308 | 1 |
| Obesity | mast_cell | *C4BPB* | 2.951909581 | 4.06E-07 | 1.30E-06 | 0.002551311 |
| Obesity | mast_cell | *RAB38* | -1.426059838 | 2.69E-07 | 9.17E-07 | 0.001688762 |
| Obesity | mast_cell | *AGO2* | 6.656028928 | 3.96E-07 | 1.27E-06 | 0.002490526 |
| Obesity | mast_cell | *CKS2* | 5.856695785 | 1.59E-06 | 4.27E-06 | 0.010017713 |
| Obesity | mast_cell | *CHPF* | 1.846002023 | 0.011382464 | 0.015317112 | 1 |
| Obesity | mast_cell | *DNPEP* | 8.772896411 | 3.38E-09 | 4.52E-08 | 2.12E-05 |
| Obesity | mast_cell | *MOGAT1* | 5.172267134 | 2.69E-05 | 5.46E-05 | 0.169171746 |
| Obesity | mast_cell | *FAM210B* | 4.865120398 | 4.40E-05 | 8.58E-05 | 0.276597439 |
| Obesity | mast_cell | *SNX21* | 5.068441725 | 2.65E-09 | 4.32E-08 | 1.66E-05 |
| Obesity | mast_cell | *SDC4* | 3.114650728 | 2.81E-08 | 1.48E-07 | 0.000176942 |
| Obesity | mast_cell | *VAPB* | 2.43897567 | 7.92E-05 | 0.000146953 | 0.49816927 |

| | | | | | | |
|---|---|---|---|---|---|---|
| Obesity | mast_cell | *TOX2* | -0.970030867 | 0.001922187 | 0.002869814 | 1 |
| Obesity | mast_cell | *ARFGEF2* | 6.418261585 | 5.42E-07 | 1.64E-06 | 0.003407461 |
| Obesity | mast_cell | *CDH26* | 5.33417171 | 2.23E-09 | 4.32E-08 | 1.40E-05 |
| Obesity | mast_cell | *STX16* | 6.209598096 | 5.97E-09 | 5.73E-08 | 3.76E-05 |
| Obesity | mast_cell | *RNF114* | 5.731176725 | 2.01E-08 | 1.15E-07 | 0.00012633 |
| Obesity | mast_cell | *NEURL2* | 1.998489605 | 0.004329559 | 0.006162539 | 1 |
| Obesity | mast_cell | *FASTKD3* | 6.291947303 | 0.001343641 | 0.002053846 | 1 |
| Obesity | mast_cell | *PEPD* | 4.816861573 | 7.98E-05 | 0.000147755 | 0.50148078 |
| Obesity | mast_cell | *CHST8* | -1.697780679 | 1.71E-06 | 4.53E-06 | 0.010754131 |
| Obesity | mast_cell | *STAMBP* | 7.102334917 | 2.32E-06 | 5.92E-06 | 0.014554637 |
| Obesity | mast_cell | *ATP8A1* | 3.41564039 | 0.000269357 | 0.00045806 | 1 |
| Obesity | mast_cell | *HIF3A* | 4.403557575 | 1.62E-07 | 5.99E-07 | 0.001016493 |
| Obesity | mast_cell | *ZNF45* | 6.042501577 | 3.13E-09 | 4.48E-08 | 1.97E-05 |
| Obesity | mast_cell | *BTN2A2* | -1.100120519 | 0.000387174 | 0.000644982 | 1 |
| Obesity | mast_cell | *MRS2* | 4.240420695 | 5.40E-05 | 0.00010384 | 0.339350145 |
| Obesity | mast_cell | *SNRPC* | -5.820387205 | 2.11E-11 | 2.61E-09 | 1.33E-07 |
| Obesity | mast_cell | *HIST1H1D* | -1.599138142 | 4.30E-06 | 1.03E-05 | 0.027019441 |
| Obesity | mast_cell | *PEX6* | 1.285161962 | 0.011953314 | 0.016040658 | 1 |
| Obesity | mast_cell | *NQO2* | 6.921406661 | 3.58E-06 | 8.71E-06 | 0.02250195 |
| Obesity | mast_cell | *ZNF391* | 0.905528979 | 0.004716574 | 0.006675619 | 1 |
| Obesity | mast_cell | *RPS10* | 5.136990233 | 5.03E-09 | 5.29E-08 | 3.16E-05 |
| Obesity | mast_cell | *HIST1H2BJ* | 6.728386842 | 4.73E-14 | 1.47E-11 | 2.97E-10 |
| Obesity | mast_cell | *MED20* | 4.592030503 | 7.61E-06 | 1.73E-05 | 0.047870339 |
| Obesity | mast_cell | *TCP11* | -2.491724586 | 1.64E-05 | 3.49E-05 | 0.103113232 |
| Obesity | mast_cell | *KLHDC3* | 4.852857708 | 4.69E-09 | 5.10E-08 | 2.95E-05 |
| Obesity | mast_cell | *GNMT* | -1.907467986 | 0.001590359 | 0.002402929 | 1 |
| Obesity | mast_cell | *KLHL31* | 1.681305112 | 1.61E-08 | 1.02E-07 | 0.000101339 |
| Obesity | mast_cell | *COL21A1* | 1.42209628 | 3.91E-08 | 1.90E-07 | 0.000245529 |
| Obesity | mast_cell | *GLO1* | 4.15913439 | 0.012180243 | 0.016320799 | 1 |
| Obesity | mast_cell | *CPNE5* | -0.514617969 | 1.00E-07 | 4.09E-07 | 0.000630689 |
| Obesity | mast_cell | *RIOK1* | 4.87014049 | 2.89E-06 | 7.20E-06 | 0.018158724 |
| Obesity | mast_cell | *RPP40* | 3.915088707 | 0.000655878 | 0.001050306 | 1 |
| Obesity | mast_cell | *NUP153* | 6.577049149 | 2.55E-10 | 1.48E-08 | 1.60E-06 |
| Obesity | mast_cell | *EEF1E1* | 18.38599538 | 1.87E-08 | 1.09E-07 | 0.000117722 |
| Obesity | mast_cell | *EREG* | 9.089114011 | 1.68E-08 | 1.04E-07 | 0.000105619 |
| Obesity | mast_cell | *CNOT1* | 5.963705778 | 1.65E-08 | 1.03E-07 | 0.000103432 |
| Obesity | mast_cell | *BBS2* | 3.885583252 | 5.91E-06 | 1.37E-05 | 0.037152883 |
| Obesity | mast_cell | *MT1G* | 33.87321242 | 1.68E-05 | 3.57E-05 | 0.105918686 |
| Obesity | mast_cell | *GOT2* | 10.10855298 | 9.54E-08 | 3.92E-07 | 0.000600005 |
| Obesity | mast_cell | *CLYBL* | 6.884935357 | 7.64E-07 | 2.23E-06 | 0.004804193 |
| Obesity | mast_cell | *KIF25* | 8.578612395 | 4.80E-07 | 1.49E-06 | 0.003014934 |
| Obesity | mast_cell | *GRK4* | 1.714911494 | 3.57E-07 | 1.16E-06 | 0.0022459 |
| Obesity | mast_cell | *MRPS7* | 7.439156523 | 0.000169119 | 0.000296914 | 1 |
| Obesity | mast_cell | *GGA3* | 7.537381179 | 1.35E-05 | 2.93E-05 | 0.085031401 |
| Obesity | mast_cell | *NUP85* | 4.129500026 | 0.005690027 | 0.007965531 | 1 |
| Obesity | mast_cell | *MIF4GD* | 4.910697234 | 5.17E-09 | 5.38E-08 | 3.25E-05 |
| Obesity | mast_cell | *NT5C* | 8.15450994 | 1.14E-07 | 4.52E-07 | 0.000714501 |
| Obesity | mast_cell | *DDX31* | 2.810213916 | 0.003700982 | 0.005335491 | 1 |
| Obesity | mast_cell | *PPP1R12C* | 6.414820145 | 1.14E-08 | 8.37E-08 | 7.17E-05 |
| Obesity | mast_cell | *OPRL1* | 3.754589851 | 0.00033905 | 0.000568883 | 1 |

| | | | | | | |
|---|---|---|---|---|---|---|
| Obesity | mast_cell | *CHCHD5* | 4.075412635 | 0.005085435 | 0.007176494 | 1 |
| Obesity | mast_cell | *PAX8* | 2.534195096 | 7.75E-05 | 0.00014396 | 0.487159648 |
| Obesity | mast_cell | *INSIG2* | 4.596390345 | 2.22E-09 | 4.32E-08 | 1.40E-05 |
| Obesity | mast_cell | *PSD4* | 9.009530795 | 1.90E-05 | 3.98E-05 | 0.119491333 |
| Obesity | mast_cell | *SLC25A23* | 2.396381418 | 5.45E-05 | 0.000104605 | 0.342370744 |
| Obesity | mast_cell | *CLPP* | 4.667415306 | 1.05E-07 | 4.22E-07 | 0.000660318 |
| Obesity | mast_cell | *SH2D3A* | 9.675076975 | 1.24E-06 | 3.44E-06 | 0.007820256 |
| Obesity | mast_cell | *TRIP10* | 5.856522861 | 2.16E-09 | 4.32E-08 | 1.36E-05 |
| Obesity | mast_cell | *SNRPD2* | 8.336057865 | 1.16E-07 | 4.59E-07 | 0.000728462 |
| Obesity | mast_cell | *RTN2* | 3.324361049 | 1.22E-08 | 8.63E-08 | 7.64E-05 |
| Obesity | mast_cell | *SYMPK* | 4.597215217 | 0.003144675 | 0.004570174 | 1 |
| Obesity | mast_cell | *GPCPD1* | 5.849503492 | 1.90E-06 | 4.96E-06 | 0.011943513 |
| Obesity | mast_cell | *SDCBP2* | -4.686415485 | 0.009272845 | 0.012627526 | 1 |
| Obesity | mast_cell | *PANK2* | 7.092160288 | 5.46E-08 | 2.48E-07 | 0.000343015 |
| Obesity | mast_cell | *DTD1* | 5.97885575 | 3.62E-05 | 7.15E-05 | 0.227476391 |
| Obesity | mast_cell | *RBCK1* | 4.787718966 | 1.19E-08 | 8.54E-08 | 7.50E-05 |
| Obesity | mast_cell | *RRBP1* | 3.92090281 | 0.001720606 | 0.002582964 | 1 |
| Obesity | mast_cell | *BMP2* | 7.067827842 | 1.48E-09 | 4.14E-08 | 9.30E-06 |
| Obesity | mast_cell | *PCSK2* | 6.573799939 | 2.69E-11 | 3.16E-09 | 1.69E-07 |
| Obesity | mast_cell | *MKKS* | 5.902814731 | 0.000543385 | 0.000882071 | 1 |
| Obesity | mast_cell | *DSTN* | 5.566714271 | 7.58E-05 | 0.000141223 | 0.476768855 |
| Obesity | mast_cell | *ITPA* | 8.51599312 | 1.66E-07 | 6.11E-07 | 0.001043038 |
| Obesity | mast_cell | *TMEM74B* | 6.559792704 | 9.09E-06 | 2.04E-05 | 0.057144128 |
| Obesity | mast_cell | *FAM110A* | -4.063128557 | 9.57E-06 | 2.13E-05 | 0.060136331 |
| Obesity | mast_cell | *MRPS26* | 1.03609367 | 0.009781943 | 0.013274136 | 1 |
| Obesity | mast_cell | *ZNF436* | 2.754057346 | 1.04E-08 | 7.93E-08 | 6.55E-05 |
| Obesity | mast_cell | *GDF5* | 2.110952299 | 0.002196718 | 0.003259562 | 1 |
| Obesity | mast_cell | *MMP24* | 4.161807828 | 0.00076151 | 0.001207469 | 1 |
| Obesity | mast_cell | *UROD* | 8.306087315 | 1.66E-05 | 3.52E-05 | 0.104219784 |
| Obesity | mast_cell | *KLC1* | 8.027206862 | 3.96E-09 | 4.78E-08 | 2.49E-05 |
| Obesity | mast_cell | *XRCC3* | 7.186063866 | 6.59E-13 | 1.54E-10 | 4.15E-09 |
| Obesity | mast_cell | *MCF2L* | 3.996774136 | 6.73E-07 | 1.99E-06 | 0.004229196 |
| Obesity | mast_cell | *F10* | 7.675566875 | 8.07E-09 | 6.74E-08 | 5.07E-05 |
| Obesity | mast_cell | *PCID2* | 5.059254375 | 1.89E-08 | 1.10E-07 | 0.000118896 |
| Obesity | mast_cell | *PROZ* | 14.92998821 | 8.08E-09 | 6.74E-08 | 5.08E-05 |
| Obesity | mast_cell | *IGFLR1* | 7.771653009 | 2.46E-08 | 1.34E-07 | 0.000154847 |
| Obesity | mast_cell | *CAPNS1* | 5.856928387 | 3.17E-05 | 6.33E-05 | 0.199408073 |
| Obesity | mast_cell | *COX6B1* | 5.686713273 | 0.005319913 | 0.007477374 | 1 |
| Obesity | mast_cell | *PRDX5* | 6.758991638 | 0.001793223 | 0.002686202 | 1 |
| Obesity | mast_cell | *SCAF1* | 3.176530687 | 8.03E-09 | 6.72E-08 | 5.05E-05 |
| Obesity | mast_cell | *PRR12* | 3.866513171 | 0.000144265 | 0.000256285 | 0.906993865 |
| Obesity | mast_cell | *TSKS* | 8.810046122 | 2.81E-09 | 4.32E-08 | 1.76E-05 |
| Obesity | mast_cell | *SBDS* | 7.802107819 | 4.96E-06 | 1.17E-05 | 0.031166776 |
| Obesity | mast_cell | *BECN1* | 8.280960399 | 1.92E-08 | 1.11E-07 | 0.000120894 |
| Obesity | mast_cell | *TRAP1* | 1.763647101 | 1.12E-06 | 3.13E-06 | 0.00703817 |
| Obesity | mast_cell | *DNAJC8* | 4.96740653 | 9.51E-06 | 2.12E-05 | 0.059811732 |
| Obesity | mast_cell | *ZNF384* | 7.605879356 | 7.48E-05 | 0.000139469 | 0.470569999 |
| Obesity | mast_cell | *EMG1* | 7.469854921 | 3.56E-08 | 1.76E-07 | 0.000224073 |
| Obesity | mast_cell | *ATG14* | 6.343122216 | 1.19E-07 | 4.70E-07 | 0.000751068 |
| Obesity | mast_cell | *RHOJ* | 7.547807171 | 1.41E-07 | 5.38E-07 | 0.000888901 |

| | | | | | | |
|---|---|---|---|---|---|---|
| Obesity | mast_cell | *HSPA2* | 6.408724899 | 4.06E-09 | 4.79E-08 | 2.55E-05 |
| Obesity | mast_cell | *ZBTB1* | 5.507703357 | 8.58E-06 | 1.93E-05 | 0.053920409 |
| Obesity | mast_cell | *PZP* | -0.64382977 | 3.51E-07 | 1.15E-06 | 0.002205191 |
| Obesity | mast_cell | *RHOT1* | 5.230040575 | 1.95E-07 | 7.01E-07 | 0.001227097 |
| Obesity | mast_cell | *WDR60* | 2.662993069 | 0.004007434 | 0.005740428 | 1 |
| Obesity | mast_cell | *AIF1L* | 2.731862935 | 9.10E-09 | 7.26E-08 | 5.72E-05 |
| Obesity | mast_cell | *MAP2K2* | 4.953245314 | 2.28E-05 | 4.69E-05 | 0.143080549 |
| Obesity | mast_cell | *CANX* | 4.163243736 | 0.006094053 | 0.00849519 | 1 |
| Obesity | mast_cell | *IPPK* | 5.4153488 | 0.003766693 | 0.00541656 | 1 |
| Obesity | mast_cell | *MASP1* | 2.508082075 | 1.96E-07 | 7.03E-07 | 0.001233681 |
| Obesity | mast_cell | *ATP13A4* | -2.686846861 | 1.41E-06 | 3.84E-06 | 0.008876719 |
| Obesity | mast_cell | *HRASLS* | 4.355111603 | 1.06E-08 | 7.98E-08 | 6.64E-05 |
| Obesity | mast_cell | *HELB* | 5.902981906 | 3.40E-09 | 4.52E-08 | 2.13E-05 |
| Obesity | mast_cell | *RAP1B* | 5.216779504 | 3.60E-07 | 1.17E-06 | 0.002260547 |
| Obesity | mast_cell | *RAB3IP* | 6.464384572 | 2.49E-08 | 1.35E-07 | 0.000156435 |
| Obesity | mast_cell | *TAS2R4* | 3.431413392 | 1.20E-07 | 4.71E-07 | 0.000753839 |
| Obesity | mast_cell | *LRRC61* | 1.360666164 | 0.000557487 | 0.000901936 | 1 |
| Obesity | mast_cell | *IDUA* | 6.352721868 | 2.43E-09 | 4.32E-08 | 1.53E-05 |
| Obesity | mast_cell | *TMEM175* | 5.340939392 | 4.20E-07 | 1.34E-06 | 0.002641829 |
| Obesity | mast_cell | *EMC1* | 4.404597723 | 0.000501606 | 0.000817841 | 1 |
| Obesity | mast_cell | *PLA2G5* | 2.307534593 | 0.000674535 | 0.001077166 | 1 |
| Obesity | mast_cell | *SIN3B* | 6.598780852 | 8.03E-09 | 6.72E-08 | 5.05E-05 |
| Obesity | mast_cell | *WDR24* | 9.157924082 | 5.02E-06 | 1.18E-05 | 0.031534062 |
| Obesity | mast_cell | *CHTF18* | 5.287518122 | 2.17E-09 | 4.32E-08 | 1.37E-05 |
| Obesity | mast_cell | *SMARCA4* | 5.482645296 | 0.00012661 | 0.000226779 | 0.795994674 |
| Obesity | mast_cell | *TICAM1* | 5.947672905 | 9.51E-09 | 7.45E-08 | 5.98E-05 |
| Obesity | mast_cell | *AAMP* | 8.764891614 | 1.75E-06 | 4.62E-06 | 0.010984944 |
| Obesity | mast_cell | *ZNF835* | 3.119349984 | 5.63E-05 | 0.000107715 | 0.354058 |
| Obesity | mast_cell | *HIP1* | 4.867615351 | 1.83E-08 | 1.08E-07 | 0.000115252 |
| Obesity | mast_cell | *PTPN12* | 5.499553349 | 1.76E-07 | 6.43E-07 | 0.00110886 |
| Obesity | mast_cell | *POR* | 6.787108698 | 1.67E-06 | 4.44E-06 | 0.010514966 |
| Obesity | mast_cell | *FGL2* | 4.656375571 | 7.07E-06 | 1.62E-05 | 0.044464772 |
| Obesity | mast_cell | *STYXL1* | 2.638864956 | 3.86E-05 | 7.59E-05 | 0.242432065 |
| Obesity | mast_cell | *STEAP4* | 3.684696356 | 1.16E-07 | 4.58E-07 | 0.000726436 |
| Obesity | mast_cell | *SGCE* | 5.902545364 | 4.89E-06 | 1.15E-05 | 0.030728243 |
| Obesity | mast_cell | *RBM48* | 4.300557686 | 1.21E-08 | 8.63E-08 | 7.62E-05 |
| Obesity | mast_cell | *CASD1* | 6.635501711 | 7.89E-07 | 2.30E-06 | 0.004962931 |
| Obesity | mast_cell | *SRD5A3* | -2.70769059 | 0.008018676 | 0.011007296 | 1 |
| Obesity | mast_cell | *DGCR6L* | 4.687630623 | 4.27E-05 | 8.36E-05 | 0.268697718 |
| Obesity | mast_cell | *VPREB3* | 19.74570025 | 0.000316888 | 0.000534122 | 1 |
| Obesity | mast_cell | *MGAT3* | 4.193665173 | 5.41E-09 | 5.52E-08 | 3.40E-05 |
| Obesity | mast_cell | *ATF4* | 5.54488722 | 0.001040081 | 0.001614167 | 1 |
| Obesity | mast_cell | *A4GALT* | 1.692494188 | 0.014444673 | 0.019138811 | 1 |
| Obesity | mast_cell | *CDC42EP1* | 6.082214983 | 2.84E-06 | 7.10E-06 | 0.017874293 |
| Obesity | mast_cell | *TPST2* | 5.765004342 | 0.000282401 | 0.00047843 | 1 |
| Obesity | mast_cell | *BAIAP2L2* | 5.619715384 | 7.64E-08 | 3.24E-07 | 0.000480362 |
| Obesity | mast_cell | *TST* | 2.426101371 | 1.04E-07 | 4.20E-07 | 0.000655245 |
| Obesity | mast_cell | *APOL5* | 4.230180718 | 0.028982362 | 0.036870116 | 1 |
| Obesity | mast_cell | *RIBC2* | 4.347810147 | 1.62E-08 | 1.02E-07 | 0.00010214 |
| Obesity | mast_cell | *EMC4* | 8.056817314 | 2.51E-07 | 8.66E-07 | 0.00157799 |

| | | | | | | |
|---|---|---|---|---|---|---|
| Obesity | mast_cell | *SPECC1* | -2.293661851 | 0.00036545 | 0.000610248 | 1 |
| Obesity | mast_cell | *POT1* | 4.716162327 | 0.00026483 | 0.000450849 | 1 |
| Obesity | mast_cell | *CDHR3* | 3.769526601 | 5.82E-08 | 2.61E-07 | 0.000365781 |
| Obesity | mast_cell | *PRKRIP1* | 3.21381487 | 0.003144652 | 0.004570174 | 1 |
| Obesity | mast_cell | *FOXP2* | 3.273164899 | 5.42E-06 | 1.27E-05 | 0.034075729 |
| Obesity | mast_cell | *STRIP2* | 3.228341972 | 7.67E-08 | 3.25E-07 | 0.000482523 |
| Obesity | mast_cell | *FLNC* | 3.092217103 | 1.71E-05 | 3.61E-05 | 0.107215805 |
| Obesity | mast_cell | *IRF5* | 1.384937902 | 0.012088593 | 0.0162049 | 1 |
| Obesity | mast_cell | *KLHDC10* | 5.357697431 | 9.39E-07 | 2.68E-06 | 0.005906266 |
| Obesity | mast_cell | *HOXD1* | 14.36931975 | 3.69E-07 | 1.20E-06 | 0.002320706 |
| Obesity | mast_cell | *HOXD3* | 5.883423073 | 2.93E-07 | 9.85E-07 | 0.001840762 |
| Obesity | mast_cell | *CHN1* | 4.021751904 | 0.000183324 | 0.000319799 | 1 |
| Obesity | mast_cell | *GAD1* | 9.549533043 | 7.84E-09 | 6.64E-08 | 4.93E-05 |
| Obesity | mast_cell | *HOXD9* | 2.615387754 | 0.021071657 | 0.027343139 | 1 |
| Obesity | mast_cell | *PSMG2* | 2.310952457 | 0.016960748 | 0.022256778 | 1 |
| Obesity | mast_cell | *EIF2AK4* | 4.580028111 | 4.56E-07 | 1.44E-06 | 0.002863966 |
| Obesity | mast_cell | *TMOD2* | 3.73927834 | 0.00200818 | 0.002989682 | 1 |
| Obesity | mast_cell | *ARPP19* | 6.961224426 | 6.73E-08 | 2.92E-07 | 0.000423397 |
| Obesity | mast_cell | *VPS13C* | 3.837432134 | 0.000798449 | 0.001261586 | 1 |
| Obesity | mast_cell | *CALML4* | -4.363602881 | 6.60E-07 | 1.96E-06 | 0.004147317 |
| Obesity | mast_cell | *ACKR4* | 5.086039875 | 2.49E-09 | 4.32E-08 | 1.57E-05 |
| Obesity | mast_cell | *SUMF2* | 5.648915868 | 7.23E-09 | 6.29E-08 | 4.55E-05 |
| Obesity | mast_cell | *SPCS3* | 6.789096883 | 5.81E-06 | 1.35E-05 | 0.036547237 |
| Obesity | mast_cell | *SERGEF* | 4.938219339 | 0.002073594 | 0.003084876 | 1 |
| Obesity | mast_cell | *DCTD* | 2.557792759 | 0.004340424 | 0.006176606 | 1 |
| Obesity | mast_cell | *PLD2* | 4.755459279 | 3.65E-10 | 1.88E-08 | 2.30E-06 |
| Obesity | mast_cell | *ATP1B2* | 4.995031361 | 1.53E-05 | 3.27E-05 | 0.096050755 |
| Obesity | mast_cell | *CCL4* | 2.717136462 | 0.006894243 | 0.009545057 | 1 |
| Obesity | mast_cell | *PHF20L1* | 5.69426331 | 1.36E-05 | 2.94E-05 | 0.085334357 |
| Obesity | mast_cell | *PUS7L* | 5.119816409 | 2.82E-05 | 5.70E-05 | 0.177303568 |
| Obesity | mast_cell | *KRI1* | 2.29789713 | 0.000143277 | 0.000254746 | 0.900780264 |
| Obesity | mast_cell | *SLC44A2* | 7.137758011 | 8.92E-09 | 7.14E-08 | 5.61E-05 |
| Obesity | mast_cell | *CDKN2D* | 6.488573187 | 5.48E-10 | 2.45E-08 | 3.44E-06 |
| Obesity | mast_cell | *MTUS1* | 4.933807092 | 1.23E-07 | 4.82E-07 | 0.000774747 |
| Obesity | mast_cell | *NGDN* | 8.236332672 | 2.06E-05 | 4.28E-05 | 0.129783077 |
| Obesity | mast_cell | *RIPK3* | 5.960929592 | 6.91E-07 | 2.04E-06 | 0.004341302 |
| Obesity | mast_cell | *ADCY4* | 3.934248132 | 3.50E-06 | 8.55E-06 | 0.022028388 |
| Obesity | mast_cell | *BCL2L2* | 8.391184903 | 2.95E-08 | 1.53E-07 | 0.000185357 |
| Obesity | mast_cell | *DTD2* | 1.813512215 | 1.77E-05 | 3.74E-05 | 0.111445212 |
| Obesity | mast_cell | *PARP2* | 4.15424665 | 5.83E-07 | 1.76E-06 | 0.003665111 |
| Obesity | mast_cell | *MIS18BP1* | 3.373149482 | 2.11E-05 | 4.38E-05 | 0.132940261 |
| Obesity | mast_cell | *RNASE1* | 3.592607547 | 0.000174322 | 0.000305283 | 1 |
| Obesity | mast_cell | *EPB41L4A* | 3.858757647 | 0.003707333 | 0.005342197 | 1 |
| Obesity | mast_cell | *CDO1* | 3.117976324 | 1.65E-09 | 4.26E-08 | 1.03E-05 |
| Obesity | mast_cell | *REEP5* | 5.342097395 | 0.000173543 | 0.000304304 | 1 |
| Obesity | mast_cell | *QRICH2* | 4.295937395 | 9.50E-07 | 2.70E-06 | 0.005969961 |
| Obesity | mast_cell | *SEC14L1* | 2.664635527 | 0.007986936 | 0.010968517 | 1 |
| Obesity | mast_cell | *TMEM8A* | 6.461079969 | 2.97E-06 | 7.37E-06 | 0.018648291 |
| Obesity | mast_cell | *SHC2* | 3.198926425 | 7.01E-10 | 2.77E-08 | 4.41E-06 |
| Obesity | mast_cell | *ERMARD* | 5.872532735 | 1.06E-05 | 2.34E-05 | 0.066464122 |

| | | | | | | |
|---|---|---|---|---|---|---|
| Obesity | mast_cell | *PHF10* | 6.607415064 | 5.76E-09 | 5.62E-08 | 3.62E-05 |
| Obesity | mast_cell | *GALNT8* | 4.255363924 | 1.42E-06 | 3.87E-06 | 0.008937321 |
| Obesity | mast_cell | *DOCK6* | 3.583966872 | 3.72E-06 | 9.00E-06 | 0.023379721 |
| Obesity | mast_cell | *ECSIT* | 7.373872564 | 3.34E-07 | 1.10E-06 | 0.002101991 |
| Obesity | mast_cell | *ELOF1* | 5.625058044 | 2.82E-09 | 4.32E-08 | 1.77E-05 |
| Obesity | mast_cell | *TOMM40* | 5.604747587 | 0.001135681 | 0.001751724 | 1 |
| Obesity | mast_cell | *DPP6* | 4.353924716 | 0.020843507 | 0.027058255 | 1 |
| Obesity | mast_cell | *FAM98C* | 4.263880917 | 0.001119261 | 0.001727668 | 1 |
| Obesity | mast_cell | *RPL36* | 6.933814986 | 3.63E-07 | 1.18E-06 | 0.00228514 |
| Obesity | mast_cell | *GTPBP3* | -2.759264773 | 0.005137809 | 0.007242467 | 1 |
| Obesity | mast_cell | *PLVAP* | 0.967032701 | 0.000389982 | 0.000649316 | 1 |
| Obesity | mast_cell | *BST2* | 8.670276958 | 2.21E-06 | 5.67E-06 | 0.013883776 |
| Obesity | mast_cell | *DDA1* | 5.237662144 | 3.35E-08 | 1.68E-07 | 0.000210727 |
| Obesity | mast_cell | *MRPL34* | 1.62060218 | 0.002205194 | 0.003269054 | 1 |
| Obesity | mast_cell | *LSM7* | 4.661356188 | 1.66E-05 | 3.52E-05 | 0.104188193 |
| Obesity | mast_cell | *TULP4* | 5.262195031 | 3.46E-06 | 8.46E-06 | 0.021763916 |
| Obesity | mast_cell | *RTN4IP1* | 6.354499487 | 4.58E-06 | 1.09E-05 | 0.028792932 |
| Obesity | mast_cell | *RSPH3* | 2.232447685 | 0.000317793 | 0.000534929 | 1 |
| Obesity | mast_cell | *ACTN4* | 5.506645314 | 5.65E-05 | 0.000107966 | 0.355100652 |
| Obesity | mast_cell | *STK33* | 2.823144296 | 3.44E-08 | 1.71E-07 | 0.000216165 |
| Obesity | mast_cell | *CACNG6* | 5.844843864 | 0.000373121 | 0.000622396 | 1 |
| Obesity | mast_cell | *PXDN* | 4.485498827 | 2.78E-09 | 4.32E-08 | 1.75E-05 |
| Obesity | mast_cell | *KIAA1683* | -2.087518666 | 5.61E-05 | 0.000107288 | 0.352440301 |
| Obesity | mast_cell | *TRPM4* | 4.395493647 | 6.96E-07 | 2.05E-06 | 0.004375956 |
| Obesity | mast_cell | *UBAC1* | 5.586955982 | 7.75E-08 | 3.27E-07 | 0.000486946 |
| Obesity | mast_cell | *TNNT3* | 3.170072401 | 1.83E-07 | 6.67E-07 | 0.001153213 |
| Obesity | mast_cell | *ATXN10* | 5.708501164 | 2.93E-07 | 9.87E-07 | 0.001844798 |
| Obesity | mast_cell | *TUBGCP2* | 3.533676904 | 0.012682302 | 0.016942973 | 1 |
| Obesity | mast_cell | *CYP2E1* | 2.618814717 | 1.92E-05 | 4.02E-05 | 0.120980644 |
| Obesity | mast_cell | *PAK4* | 5.176023335 | 0.016488278 | 0.021682034 | 1 |
| Obesity | mast_cell | *ZNF337* | 3.288189091 | 1.31E-05 | 2.84E-05 | 0.082363499 |
| Obesity | mast_cell | *CEP85* | 2.941681098 | 0.000875811 | 0.001375867 | 1 |
| Obesity | mast_cell | *POMT1* | 4.860945497 | 0.00020146 | 0.000349304 | 1 |
| Obesity | mast_cell | *UCK1* | 2.691199851 | 2.18E-07 | 7.68E-07 | 0.001369611 |
| Obesity | mast_cell | *YIPF2* | 3.668563561 | 0.009273308 | 0.012627526 | 1 |
| Obesity | mast_cell | *TMEM160* | 9.548357077 | 0.001610888 | 0.00243161 | 1 |
| Obesity | mast_cell | *MAP3K10* | 6.252357013 | 5.22E-07 | 1.59E-06 | 0.003281834 |
| Obesity | mast_cell | *SMPDL3B* | -14.29082806 | 1.69E-06 | 4.50E-06 | 0.010650571 |
| Obesity | mast_cell | *ATPIF1* | -2.843851112 | 0.005260675 | 0.007399075 | 1 |
| Obesity | mast_cell | *THEMIS2* | 3.936947057 | 4.39E-08 | 2.08E-07 | 0.000275933 |
| Obesity | mast_cell | *CLIP1* | 7.147468516 | 1.25E-09 | 3.78E-08 | 7.84E-06 |
| Obesity | mast_cell | *PPAN* | 4.2191849 | 1.09E-08 | 8.16E-08 | 6.87E-05 |
| Obesity | mast_cell | *ZNF236* | 5.26051195 | 7.45E-06 | 1.70E-05 | 0.046812223 |
| Obesity | mast_cell | *C12orf65* | 5.958735678 | 5.59E-09 | 5.57E-08 | 3.51E-05 |
| Obesity | mast_cell | *PKDREJ* | -3.411199528 | 0.000660712 | 0.001057509 | 1 |
| Obesity | mast_cell | *HSD17B3* | 2.442567149 | 0.010631567 | 0.014368156 | 1 |
| Obesity | mast_cell | *HABP4* | 6.543693194 | 5.02E-07 | 1.54E-06 | 0.00315584 |
| Obesity | mast_cell | *EPS8L1* | -17.83663372 | 6.89E-05 | 0.000129429 | 0.433067952 |
| Obesity | mast_cell | *LILRB2* | 4.023658802 | 7.23E-09 | 6.29E-08 | 4.55E-05 |
| Obesity | mast_cell | *COX4I2* | 6.558105446 | 7.01E-08 | 3.02E-07 | 0.000440642 |

| | | | | | | |
|---|---|---|---|---|---|---|
| Obesity | mast_cell | *HAUS8* | 5.907667873 | 8.81E-09 | 7.10E-08 | 5.54E-05 |
| Obesity | mast_cell | *TBC1D5* | 4.710629568 | 2.82E-06 | 7.06E-06 | 0.01776017 |
| Obesity | mast_cell | *RFTN1* | 5.375375111 | 3.33E-08 | 1.67E-07 | 0.000209362 |
| Obesity | mast_cell | *KCNC3* | 2.2696739 | 0.000436345 | 0.000720404 | 1 |
| Obesity | mast_cell | *PDLIM4* | 5.045379436 | 1.52E-08 | 9.86E-08 | 9.56E-05 |
| Obesity | mast_cell | *KIF3A* | 5.865672454 | 1.15E-09 | 3.68E-08 | 7.22E-06 |
| Obesity | mast_cell | *AOC3* | 3.877452864 | 0.000220085 | 0.000379088 | 1 |
| Obesity | mast_cell | *ACLY* | 5.363892125 | 3.44E-05 | 6.84E-05 | 0.216583984 |
| Obesity | mast_cell | *AOC2* | 11.28278152 | 3.56E-09 | 4.62E-08 | 2.24E-05 |
| Obesity | mast_cell | *EXOC4* | 4.498279214 | 1.22E-05 | 2.67E-05 | 0.076868368 |
| Obesity | mast_cell | *ACAP3* | 5.800498764 | 0.000115849 | 0.000208934 | 0.728343792 |
| Obesity | mast_cell | *C1orf159* | 5.603433513 | 0.002698546 | 0.0039538 | 1 |
| Obesity | mast_cell | *ANO1* | 5.787470168 | 6.83E-10 | 2.77E-08 | 4.30E-06 |
| Obesity | mast_cell | *PPFIA1* | 4.159576823 | 0.000161425 | 0.000284599 | 1 |
| Obesity | mast_cell | *NINJ1* | 6.518685771 | 3.54E-10 | 1.84E-08 | 2.23E-06 |
| Obesity | mast_cell | *NPHP4* | 9.174246883 | 4.01E-06 | 9.64E-06 | 0.025216395 |
| Obesity | mast_cell | *MAP1B* | 1.036691158 | 0.023542595 | 0.03035527 | 1 |
| Obesity | mast_cell | *CKMT2* | 2.501302201 | 0.020763679 | 0.026965761 | 1 |
| Obesity | mast_cell | *ZCCHC9* | 6.691380332 | 2.74E-08 | 1.45E-07 | 0.000172402 |
| Obesity | mast_cell | *KHDRBS3* | 5.575401283 | 5.13E-07 | 1.57E-06 | 0.00322623 |
| Obesity | mast_cell | *CHD1L* | 3.955467834 | 1.34E-06 | 3.67E-06 | 0.008434734 |
| Obesity | mast_cell | *RBM8A* | 2.953080307 | 0.000176726 | 0.000309062 | 1 |
| Obesity | mast_cell | *ZNF304* | 2.560793381 | 1.91E-06 | 4.99E-06 | 0.012029146 |
| Obesity | mast_cell | *ZSCAN5A* | 6.833457843 | 2.30E-06 | 5.88E-06 | 0.014445883 |
| Obesity | mast_cell | *RHPN2* | -4.672903267 | 0.000677915 | 0.001081465 | 1 |
| Obesity | mast_cell | *C19orf12* | 1.767599186 | 0.00017375 | 0.000304535 | 1 |
| Obesity | mast_cell | *ACTR10* | 5.620441496 | 3.43E-05 | 6.81E-05 | 0.215715147 |
| Obesity | mast_cell | *GCH1* | 5.733290728 | 2.26E-07 | 7.93E-07 | 0.001423996 |
| Obesity | mast_cell | *LGALS3* | -1.880580185 | 0.024337263 | 0.031328495 | 1 |
| Obesity | mast_cell | *MATN3* | 11.2729093 | 1.47E-08 | 9.66E-08 | 9.23E-05 |
| Obesity | mast_cell | *RAF1* | 4.928151747 | 0.0004665 | 0.000765367 | 1 |
| Obesity | mast_cell | *ENOSF1* | 3.985132574 | 1.21E-05 | 2.65E-05 | 0.076323429 |
| Obesity | mast_cell | *EMILIN2* | 3.770707315 | 6.19E-05 | 0.000117235 | 0.388984267 |
| Obesity | mast_cell | *TRIM5* | 6.6793943 | 5.87E-07 | 1.76E-06 | 0.003687348 |
| Obesity | mast_cell | *CNGA4* | 7.144351168 | 2.55E-08 | 1.37E-07 | 0.000160055 |
| Obesity | mast_cell | *TRIM22* | 5.670480461 | 8.35E-05 | 0.000153858 | 0.525118894 |
| Obesity | mast_cell | *RRP8* | 6.000883406 | 0.000295053 | 0.000498923 | 1 |
| Obesity | mast_cell | *IMMT* | 3.346787187 | 0.005536002 | 0.007756818 | 1 |
| Obesity | mast_cell | *CARD6* | 7.236414895 | 3.42E-10 | 1.82E-08 | 2.15E-06 |
| Obesity | mast_cell | *INPP5K* | 3.265809717 | 3.04E-06 | 7.54E-06 | 0.019130162 |
| Obesity | mast_cell | *MYBBP1A* | 3.947380536 | 0.012050623 | 0.016157446 | 1 |
| Obesity | mast_cell | *RPA1* | 5.926768744 | 3.59E-06 | 8.73E-06 | 0.022577435 |
| Obesity | mast_cell | *SERPINF1* | 3.917218892 | 4.96E-05 | 9.59E-05 | 0.311542381 |
| Obesity | mast_cell | *FIGNL1* | 4.979902423 | 0.02110404 | 0.027379508 | 1 |
| Obesity | mast_cell | *WBP2* | 8.310485262 | 1.81E-08 | 1.07E-07 | 0.000113684 |
| Obesity | mast_cell | *KDM6B* | 5.735219518 | 0.004024667 | 0.005761175 | 1 |
| Obesity | mast_cell | *XAF1* | 3.070005312 | 0.000248573 | 0.000424783 | 1 |
| Obesity | mast_cell | *PRMT7* | 2.127577469 | 0.005398633 | 0.007579545 | 1 |
| Obesity | mast_cell | *TERF2* | 6.66730246 | 1.17E-08 | 8.45E-08 | 7.37E-05 |
| Obesity | mast_cell | *BTBD3* | 5.87101265 | 1.36E-08 | 9.22E-08 | 8.56E-05 |

| | | | | | | |
|---|---|---|---|---|---|---|
| Obesity | mast_cell | *PCNA* | 4.388609132 | 3.55E-09 | 4.62E-08 | 2.23E-05 |
| Obesity | mast_cell | *PTPRA* | 4.955240653 | 4.36E-05 | 8.52E-05 | 0.274043382 |
| Obesity | mast_cell | *IGHMBP2* | 4.575051292 | 1.24E-10 | 9.74E-09 | 7.79E-07 |
| Obesity | mast_cell | *MMACHC* | -3.731404793 | 3.59E-07 | 1.17E-06 | 0.002255192 |
| Obesity | mast_cell | *CTNNBL1* | -1.895445826 | 1.42E-05 | 3.06E-05 | 0.089181345 |
| Obesity | mast_cell | *LPIN3* | 6.434747851 | 2.83E-09 | 4.32E-08 | 1.78E-05 |
| Obesity | mast_cell | *SERINC3* | 2.148299914 | 0.012528262 | 0.0167443 | 1 |
| Obesity | mast_cell | *DMGDH* | 4.844618482 | 3.55E-08 | 1.75E-07 | 0.000222987 |
| Obesity | mast_cell | *BHMT2* | 0.575089583 | 0.013625569 | 0.018122266 | 1 |
| Obesity | mast_cell | *KANK4* | 1.843256934 | 1.34E-08 | 9.13E-08 | 8.41E-05 |
| Obesity | mast_cell | *RSG1* | -3.938060101 | 0.005061057 | 0.007147095 | 1 |
| Obesity | mast_cell | *ATP8A2* | -2.829650802 | 2.64E-06 | 6.66E-06 | 0.016618555 |
| Obesity | mast_cell | *USPL1* | 6.750747526 | 8.05E-08 | 3.39E-07 | 0.000506038 |
| Obesity | mast_cell | *POMP* | 6.062689817 | 0.000679066 | 0.001083026 | 1 |
| Obesity | mast_cell | *WASF3* | 6.26090279 | 1.95E-08 | 1.12E-07 | 0.000122563 |
| Obesity | mast_cell | *PEMT* | 3.068130839 | 5.41E-05 | 0.000103934 | 0.33986542 |
| Obesity | mast_cell | *SCO1* | 5.634210094 | 2.07E-08 | 1.17E-07 | 0.000130329 |
| Obesity | mast_cell | *CHI3L1* | 1.488910764 | 4.32E-07 | 1.37E-06 | 0.002713219 |
| Obesity | mast_cell | *DSTYK* | 5.527039768 | 1.47E-05 | 3.17E-05 | 0.092679092 |
| Obesity | mast_cell | *TMCC2* | -3.275446742 | 0.000174179 | 0.000305116 | 1 |
| Obesity | mast_cell | *COG6* | 3.799838296 | 0.000773974 | 0.001224452 | 1 |
| Obesity | mast_cell | *TRPC4* | 12.38256496 | 5.90E-09 | 5.70E-08 | 3.71E-05 |
| Obesity | mast_cell | *RFC3* | 5.074745601 | 6.41E-06 | 1.48E-05 | 0.040320435 |
| Obesity | mast_cell | *BTBD2* | 3.432745429 | 0.012226058 | 0.01637172 | 1 |
| Obesity | mast_cell | *PRAM1* | 3.571238029 | 4.51E-05 | 8.80E-05 | 0.283847283 |
| Obesity | mast_cell | *CNDP2* | 7.910251104 | 5.31E-09 | 5.48E-08 | 3.34E-05 |
| Obesity | mast_cell | *RTN3* | 5.55615673 | 4.47E-07 | 1.41E-06 | 0.002811745 |
| Obesity | mast_cell | *SLC2A11* | 4.627912516 | 5.57E-09 | 5.56E-08 | 3.50E-05 |
| Obesity | mast_cell | *C1QTNF6* | 5.061636735 | 3.30E-09 | 4.52E-08 | 2.08E-05 |
| Obesity | mast_cell | *FAM83F* | -1.587571241 | 2.53E-07 | 8.71E-07 | 0.001591086 |
| Obesity | mast_cell | *GIMAP4* | 1.870999266 | 0.010363483 | 0.014027013 | 1 |
| Obesity | mast_cell | *AGAP3* | 3.347233727 | 0.033865211 | 0.042693118 | 1 |
| Obesity | mast_cell | *ACTR3B* | 5.814584842 | 5.99E-09 | 5.74E-08 | 3.77E-05 |
| Obesity | mast_cell | *SFTPD* | 1.242721799 | 0.000237444 | 0.000407415 | 1 |
| Obesity | mast_cell | *DYDC2* | 15.0162796 | 3.75E-05 | 7.40E-05 | 0.235927028 |
| Obesity | mast_cell | *TMTC1* | 5.149445369 | 9.21E-08 | 3.80E-07 | 0.000579124 |
| Obesity | mast_cell | *IPO8* | 6.731123545 | 1.14E-05 | 2.52E-05 | 0.071860384 |
| Obesity | mast_cell | *SPINK5* | 4.584425889 | 2.26E-07 | 7.92E-07 | 0.001419705 |
| Obesity | mast_cell | *LRRCC1* | 4.088071238 | 8.52E-06 | 1.92E-05 | 0.053547606 |
| Obesity | mast_cell | *SWAP70* | 6.270031344 | 2.47E-08 | 1.34E-07 | 0.000155087 |
| Obesity | mast_cell | *AMPD3* | 2.407127168 | 7.45E-09 | 6.41E-08 | 4.68E-05 |
| Obesity | mast_cell | *SBF2* | 4.734605829 | 1.23E-06 | 3.41E-06 | 0.007743905 |
| Obesity | mast_cell | *MICAL2* | 3.343995384 | 0.000760738 | 0.001206549 | 1 |
| Obesity | mast_cell | *RRAS2* | 2.514906818 | 4.92E-06 | 1.16E-05 | 0.03096247 |
| Obesity | mast_cell | *DPF2* | 5.481575722 | 1.43E-08 | 9.54E-08 | 9.02E-05 |
| Obesity | mast_cell | *GSC* | 6.847555845 | 1.48E-08 | 9.69E-08 | 9.32E-05 |
| Obesity | mast_cell | *CATSPERB* | 1.928295543 | 0.001513072 | 0.002293318 | 1 |
| Obesity | mast_cell | *COX16* | -1.577892978 | 5.34E-07 | 1.63E-06 | 0.003357982 |
| Obesity | mast_cell | *MED6* | 2.215841705 | 0.031311221 | 0.039648267 | 1 |
| Obesity | mast_cell | *EIF2S1* | 6.64049902 | 5.58E-08 | 2.52E-07 | 0.000350607 |

| | | | | | | |
|---|---|---|---|---|---|---|
| Obesity | mast_cell | *ELP3* | 6.707940598 | 1.84E-08 | 1.08E-07 | 0.000115807 |
| Obesity | mast_cell | *ADAMDEC1* | -4.237416908 | 3.56E-05 | 7.05E-05 | 0.223591977 |
| Obesity | mast_cell | *MRO* | 2.820201614 | 6.26E-06 | 1.45E-05 | 0.039358886 |
| Obesity | mast_cell | *THUMPD3* | 3.016426153 | 0.013081761 | 0.017454378 | 1 |
| Obesity | mast_cell | *CNTN6* | 2.629661141 | 4.89E-07 | 1.51E-06 | 0.003075313 |
| Obesity | mast_cell | *KATNBL1* | 6.421277386 | 9.47E-08 | 3.90E-07 | 0.000595602 |
| Obesity | mast_cell | *GSTM5* | 3.55802427 | 2.40E-07 | 8.33E-07 | 0.00150681 |
| Obesity | mast_cell | *SYT6* | 6.555329158 | 3.23E-08 | 1.63E-07 | 0.000203024 |
| Obesity | mast_cell | *PSRC1* | 3.1359016 | 0.000421798 | 0.000697854 | 1 |
| Obesity | mast_cell | *WNT2B* | 4.931572942 | 6.17E-05 | 0.000116875 | 0.387675069 |
| Obesity | mast_cell | *CEPT1* | 5.070302192 | 1.41E-05 | 3.04E-05 | 0.088619552 |
| Obesity | mast_cell | *AP4B1* | 5.024255071 | 0.005194698 | 0.007312823 | 1 |
| Obesity | mast_cell | *NAPG* | 6.336100057 | 2.31E-09 | 4.32E-08 | 1.45E-05 |
| Obesity | mast_cell | *PPHLN1* | 5.042412335 | 1.18E-05 | 2.59E-05 | 0.074224928 |
| Obesity | mast_cell | *FKBP11* | 5.001859661 | 3.33E-07 | 1.10E-06 | 0.002096193 |
| Obesity | mast_cell | *YWHAQ* | 5.225545597 | 8.60E-06 | 1.93E-05 | 0.054053129 |
| Obesity | mast_cell | *LPIN1* | 2.521648126 | 0.003225328 | 0.004677655 | 1 |
| Obesity | mast_cell | *ANO3* | 1.725526523 | 7.53E-06 | 1.71E-05 | 0.047314432 |
| Obesity | mast_cell | *FST* | 2.548982742 | 4.94E-09 | 5.24E-08 | 3.10E-05 |
| Obesity | mast_cell | *TIMM17A* | 5.659729489 | 4.94E-09 | 5.24E-08 | 3.11E-05 |
| Obesity | mast_cell | *FBXO18* | 5.798317766 | 9.13E-09 | 7.26E-08 | 5.74E-05 |
| Obesity | mast_cell | *ECHDC3* | 3.57599018 | 3.06E-06 | 7.57E-06 | 0.019208605 |
| Obesity | mast_cell | *IL15RA* | 3.98658775 | 2.97E-09 | 4.42E-08 | 1.86E-05 |
| Obesity | mast_cell | *TMEM241* | 5.934309369 | 3.66E-06 | 8.88E-06 | 0.022996916 |
| Obesity | mast_cell | *EMP1* | 2.648346788 | 0.000793955 | 0.001254801 | 1 |
| Obesity | mast_cell | *KLRD1* | 5.197021258 | 1.21E-09 | 3.76E-08 | 7.59E-06 |
| Obesity | mast_cell | *PRH2* | 4.825211974 | 1.89E-06 | 4.94E-06 | 0.011874632 |
| Obesity | mast_cell | *DDB2* | 6.916581845 | 6.03E-10 | 2.60E-08 | 3.79E-06 |
| Obesity | mast_cell | *ACP2* | 7.717719428 | 1.16E-12 | 2.43E-10 | 7.26E-09 |
| Obesity | mast_cell | *HOOK1* | 7.884589736 | 5.38E-07 | 1.63E-06 | 0.003379858 |
| Obesity | mast_cell | *ZCCHC11* | 5.061358602 | 1.22E-05 | 2.67E-05 | 0.076925713 |
| Obesity | mast_cell | *FHOD3* | 2.228742939 | 5.89E-06 | 1.37E-05 | 0.037023037 |
| Obesity | mast_cell | *TPGS2* | 6.133937701 | 5.10E-06 | 1.20E-05 | 0.032092605 |
| Obesity | mast_cell | *SLC43A3* | 3.456585926 | 0.01235956 | 0.016536402 | 1 |
| Obesity | mast_cell | *TIMM10* | 5.116754772 | 3.67E-07 | 1.19E-06 | 0.002304536 |
| Obesity | mast_cell | *DHX34* | 2.766585511 | 0.009079096 | 0.012384525 | 1 |
| Obesity | mast_cell | *APLNR* | 4.934271651 | 6.15E-09 | 5.81E-08 | 3.87E-05 |
| Obesity | mast_cell | *FADS2* | 7.976084678 | 9.14E-09 | 7.26E-08 | 5.75E-05 |
| Obesity | mast_cell | *TMEM165* | 2.752601552 | 0.009468312 | 0.012867981 | 1 |
| Obesity | mast_cell | *CLOCK* | 5.905534013 | 3.03E-08 | 1.56E-07 | 0.000190415 |
| Obesity | mast_cell | *DZIP1* | 3.356592908 | 1.44E-08 | 9.57E-08 | 9.07E-05 |
| Obesity | mast_cell | *BIVM* | 6.483260384 | 1.99E-09 | 4.32E-08 | 1.25E-05 |
| Obesity | mast_cell | *ERCC5* | 6.94545325 | 5.49E-09 | 5.54E-08 | 3.45E-05 |
| Obesity | mast_cell | *CARS2* | 3.503755346 | 0.002358096 | 0.003485856 | 1 |
| Obesity | mast_cell | *KLB* | 2.875718824 | 1.11E-08 | 8.22E-08 | 6.99E-05 |
| Obesity | mast_cell | *WDR36* | 7.02855407 | 2.52E-06 | 6.38E-06 | 0.01584487 |
| Obesity | mast_cell | *OSTF1* | 6.944678792 | 2.67E-06 | 6.73E-06 | 0.016816811 |
| Obesity | mast_cell | *UBQLN1* | 6.313145998 | 1.52E-06 | 4.09E-06 | 0.009557411 |
| Obesity | mast_cell | *NAA35* | 6.074772121 | 1.46E-08 | 9.63E-08 | 9.16E-05 |
| Obesity | mast_cell | *CTSL* | 4.573033588 | 0.002297274 | 0.003399144 | 1 |

| | | | | | | |
|---|---|---|---|---|---|---|
| Obesity | mast_cell | *TMEM2* | 5.965986224 | 6.00E-08 | 2.68E-07 | 0.000376918 |
| Obesity | mast_cell | *GOLM1* | 3.808139079 | 0.001414309 | 0.00215349 | 1 |
| Obesity | mast_cell | *USP30* | 3.719439035 | 2.57E-06 | 6.49E-06 | 0.016173527 |
| Obesity | mast_cell | *HRK* | 6.516193706 | 4.09E-05 | 8.01E-05 | 0.257034685 |
| Obesity | mast_cell | *P2RX4* | 5.281746861 | 4.69E-07 | 1.47E-06 | 0.002951361 |
| Obesity | mast_cell | *DTX1* | 2.393394604 | 0.001035297 | 0.001607139 | 1 |
| Obesity | mast_cell | *DMTF1* | 4.243333914 | 1.88E-06 | 4.92E-06 | 0.011821107 |
| Obesity | mast_cell | *CCDC146* | 5.57804012 | 8.90E-07 | 2.56E-06 | 0.005596538 |
| Obesity | mast_cell | *TMEM60* | -5.659569369 | 1.97E-05 | 4.11E-05 | 0.124046734 |
| Obesity | mast_cell | *SRPK2* | 5.1018598 | 3.71E-06 | 8.99E-06 | 0.023349467 |
| Obesity | mast_cell | *TES* | 3.541489754 | 4.34E-06 | 1.04E-05 | 0.02728949 |
| Obesity | mast_cell | *MDFIC* | 3.880639195 | 2.38E-08 | 1.30E-07 | 0.000149911 |
| Obesity | mast_cell | *MTO1* | 4.969275164 | 1.53E-08 | 9.91E-08 | 9.63E-05 |
| Obesity | mast_cell | *ANKRD6* | 6.420562349 | 7.73E-08 | 3.27E-07 | 0.000485951 |
| Obesity | mast_cell | *SNX14* | 5.356462634 | 2.10E-06 | 5.41E-06 | 0.01317589 |
| Obesity | mast_cell | *NT5E* | 4.016762205 | 2.40E-05 | 4.91E-05 | 0.150672364 |
| Obesity | mast_cell | *EPHA7* | 1.100669219 | 8.44E-05 | 0.000155492 | 0.530848693 |
| Obesity | mast_cell | *PRR5L* | 4.767550527 | 1.10E-08 | 8.18E-08 | 6.91E-05 |
| Obesity | mast_cell | *NAT10* | 5.413315343 | 6.64E-08 | 2.88E-07 | 0.00041715 |
| Obesity | mast_cell | *ATP5G2* | 7.202646954 | 9.75E-09 | 7.58E-08 | 6.13E-05 |
| Obesity | mast_cell | *ITGA7* | 1.200769133 | 0.000631045 | 0.001013121 | 1 |
| Obesity | mast_cell | *RDH5* | 2.569937382 | 1.33E-07 | 5.14E-07 | 0.000835128 |
| Obesity | mast_cell | *PPP1R1A* | 3.169461607 | 5.51E-09 | 5.54E-08 | 3.46E-05 |
| Obesity | mast_cell | *B4GALNT1* | 6.250232825 | 0.000241195 | 0.000413299 | 1 |
| Obesity | mast_cell | *FAIM2* | -2.696292358 | 4.97E-07 | 1.53E-06 | 0.003127752 |
| Obesity | mast_cell | *HNRNPA1* | 8.588131172 | 3.73E-09 | 4.68E-08 | 2.35E-05 |
| Obesity | mast_cell | *SLC26A10* | -3.835513655 | 3.31E-06 | 8.13E-06 | 0.020817094 |
| Obesity | mast_cell | *PKIB* | -2.827446553 | 0.001133141 | 0.001748235 | 1 |
| Obesity | mast_cell | *SMPD2* | -2.641585984 | 0.003342319 | 0.004841742 | 1 |
| Obesity | mast_cell | *PRADC1* | -4.256414387 | 0.00085409 | 0.001344433 | 1 |
| Obesity | mast_cell | *SMYD5* | 4.275006852 | 4.76E-09 | 5.11E-08 | 2.99E-05 |
| Obesity | mast_cell | *KCNMB4* | 3.758636829 | 1.21E-06 | 3.35E-06 | 0.007585915 |
| Obesity | mast_cell | *USP15* | 4.043483342 | 0.000594988 | 0.000959644 | 1 |
| Obesity | mast_cell | *KLHL36* | 4.070353123 | 0.008760383 | 0.011970556 | 1 |
| Obesity | mast_cell | *MPHOSPH6* | 8.091846024 | 8.14E-07 | 2.36E-06 | 0.005114823 |
| Obesity | mast_cell | *FHOD1* | 4.933084345 | 0.005925248 | 0.008272715 | 1 |
| Obesity | mast_cell | *CCDC102A* | 4.824761589 | 9.57E-08 | 3.93E-07 | 0.000601961 |
| Obesity | mast_cell | *RNASEL* | 4.69621711 | 2.31E-06 | 5.91E-06 | 0.014520661 |
| Obesity | mast_cell | *PIGC* | 5.347623873 | 9.29E-06 | 2.08E-05 | 0.05840584 |
| Obesity | mast_cell | *SP110* | 6.320263502 | 1.22E-05 | 2.66E-05 | 0.076641157 |
| Obesity | mast_cell | *USP37* | 4.198536646 | 0.003409798 | 0.004934944 | 1 |
| Obesity | mast_cell | *SLC19A3* | 1.313296332 | 2.38E-05 | 4.88E-05 | 0.149697794 |
| Obesity | mast_cell | *ARMC9* | 5.167337882 | 4.74E-09 | 5.10E-08 | 2.98E-05 |
| Obesity | mast_cell | *TMEM127* | 6.136085969 | 3.15E-07 | 1.05E-06 | 0.001983518 |
| Obesity | mast_cell | *GCC2* | 4.071379939 | 4.06E-05 | 7.96E-05 | 0.25507891 |
| Obesity | mast_cell | *MRPS9* | 3.695197968 | 9.90E-05 | 0.000180215 | 0.622462563 |
| Obesity | mast_cell | *ANKRD36* | 5.925783008 | 3.78E-06 | 9.12E-06 | 0.023741766 |
| Obesity | mast_cell | *USP44* | 3.081495957 | 3.64E-07 | 1.18E-06 | 0.002289683 |
| Obesity | mast_cell | *DRAM1* | 3.843213682 | 0.000182588 | 0.000318604 | 1 |
| Obesity | mast_cell | *SLC41A2* | 6.219680848 | 1.54E-08 | 9.97E-08 | 9.71E-05 |

| | | | | | | |
|---|---|---|---|---|---|---|
| Obesity | mast_cell | *NEK3* | 3.156068036 | 3.62E-05 | 7.15E-05 | 0.227645397 |
| Obesity | mast_cell | *VPS36* | 3.79601695 | 0.00159798 | 0.002413864 | 1 |
| Obesity | mast_cell | *TBC1D4* | 5.547340247 | 5.20E-08 | 2.39E-07 | 0.000327162 |
| Obesity | mast_cell | *SUCLA2* | 2.26857677 | 0.004606248 | 0.006531231 | 1 |
| Obesity | mast_cell | *RCBTB1* | 3.006753026 | 0.000767024 | 0.001215293 | 1 |
| Obesity | mast_cell | *RCBTB2* | 5.977367543 | 2.35E-08 | 1.29E-07 | 0.000147716 |
| Obesity | mast_cell | *SETDB2* | 5.088374421 | 4.55E-06 | 1.08E-05 | 0.028629275 |
| Obesity | mast_cell | *GPNMB* | 4.47546282 | 1.44E-05 | 3.10E-05 | 0.090671141 |
| Obesity | mast_cell | *KDELR2* | 6.445112191 | 0.000966996 | 0.001508936 | 1 |
| Obesity | mast_cell | *NUPL2* | 4.26226207 | 0.00305903 | 0.004452911 | 1 |
| Obesity | mast_cell | *AOAH* | 3.283648469 | 6.31E-05 | 0.000119262 | 0.396427127 |
| Obesity | mast_cell | *TBRG4* | 3.122173501 | 9.78E-08 | 4.01E-07 | 0.000614806 |
| Obesity | mast_cell | *DDX56* | 4.632251894 | 2.78E-09 | 4.32E-08 | 1.75E-05 |
| Obesity | mast_cell | *HUS1* | 4.961994118 | 0.000537436 | 0.000872865 | 1 |
| Obesity | mast_cell | *NACAD* | 7.559829916 | 2.56E-06 | 6.46E-06 | 0.016069911 |
| Obesity | mast_cell | *DBNL* | 3.62766544 | 0.013890491 | 0.01843951 | 1 |
| Obesity | mast_cell | *TTC5* | 5.152244088 | 0.000320491 | 0.000539039 | 1 |
| Obesity | mast_cell | *ALPK3* | 5.540795176 | 5.83E-06 | 1.35E-05 | 0.036672616 |
| Obesity | mast_cell | *CIB2* | 1.895997406 | 0.010699168 | 0.014456408 | 1 |
| Obesity | mast_cell | *RSAD1* | -3.443761663 | 0.004264591 | 0.006078324 | 1 |
| Obesity | mast_cell | *NMT1* | 3.243391268 | 4.10E-07 | 1.31E-06 | 0.002577891 |
| Obesity | mast_cell | *MYCBPAP* | -1.659419596 | 0.001753704 | 0.002631993 | 1 |
| Obesity | mast_cell | *BRIP1* | 5.097414807 | 0.02380084 | 0.030669376 | 1 |
| Obesity | mast_cell | *RTP4* | 2.621372053 | 0.001786388 | 0.002677879 | 1 |
| Obesity | mast_cell | *MRPL47* | 8.362566017 | 3.83E-07 | 1.24E-06 | 0.002409519 |
| Obesity | mast_cell | *BLK* | -5.672350765 | 0.001441718 | 0.002192039 | 1 |
| Obesity | mast_cell | *EPRS* | 7.334343626 | 1.54E-07 | 5.78E-07 | 0.000968407 |
| Obesity | mast_cell | *RPS6KC1* | 4.857820935 | 7.70E-05 | 0.000143099 | 0.483962316 |
| Obesity | mast_cell | *CBWD2* | 6.795800623 | 1.46E-07 | 5.52E-07 | 0.000917197 |
| Obesity | mast_cell | *IL1RN* | 3.680743513 | 5.55E-09 | 5.56E-08 | 3.49E-05 |
| Obesity | mast_cell | *UGGT1* | 5.933714293 | 0.000157723 | 0.000278619 | 0.991604214 |
| Obesity | mast_cell | *GYPC* | 5.942298473 | 8.11E-08 | 3.41E-07 | 0.000510163 |
| Obesity | mast_cell | *NIPSNAP3A* | 3.213486322 | 3.00E-06 | 7.44E-06 | 0.018839277 |
| Obesity | mast_cell | *TXN* | -3.278153932 | 0.000710556 | 0.001130667 | 1 |
| Obesity | mast_cell | *ODF2* | 8.485645177 | 4.00E-09 | 4.78E-08 | 2.51E-05 |
| Obesity | mast_cell | *ZFP37* | 3.6617986 | 7.05E-05 | 0.000132096 | 0.443444755 |
| Obesity | mast_cell | *ZNF189* | 5.874034256 | 1.07E-06 | 3.01E-06 | 0.006743132 |
| Obesity | mast_cell | *ALDOB* | -3.481752664 | 0.009238456 | 0.012588248 | 1 |
| Obesity | mast_cell | *STX17* | 4.628143621 | 0.003637395 | 0.00524743 | 1 |
| Obesity | mast_cell | *PRPF4* | 4.280753309 | 2.04E-06 | 5.29E-06 | 0.012846969 |
| Obesity | mast_cell | *FPGS* | 5.71732732 | 0.000114726 | 0.000207026 | 0.721279424 |
| Obesity | mast_cell | *TEX10* | 7.44146774 | 2.09E-06 | 5.39E-06 | 0.013130833 |
| Obesity | mast_cell | *GARNL3* | 4.645354338 | 2.70E-09 | 4.32E-08 | 1.70E-05 |
| Obesity | mast_cell | *GABBR2* | 9.405653565 | 4.25E-09 | 4.85E-08 | 2.67E-05 |
| Obesity | mast_cell | *PSMB7* | 6.767127845 | 4.44E-07 | 1.40E-06 | 0.002789081 |
| Obesity | mast_cell | *RABEPK* | 7.205040054 | 5.97E-09 | 5.73E-08 | 3.75E-05 |
| Obesity | mast_cell | *GOLGA1* | 6.306401502 | 2.90E-05 | 5.84E-05 | 0.182038226 |
| Obesity | mast_cell | *LMX1B* | 10.28675286 | 6.98E-08 | 3.01E-07 | 0.000438684 |
| Obesity | mast_cell | *DSCC1* | 1.000683391 | 0.000475498 | 0.000778707 | 1 |
| Obesity | mast_cell | *DERL1* | 6.848589171 | 6.41E-08 | 2.81E-07 | 0.000403251 |

| | | | | | | |
|---|---|---|---|---|---|---|
| Obesity | mast_cell | *IL11RA* | -2.854081332 | 0.027191578 | 0.034725462 | 1 |
| Obesity | mast_cell | *APTX* | 4.919773368 | 2.16E-05 | 4.48E-05 | 0.136097592 |
| Obesity | mast_cell | *TLN1* | 5.926372564 | 1.18E-05 | 2.60E-05 | 0.074340554 |
| Obesity | mast_cell | *SPAG8* | -2.941070498 | 0.000139295 | 0.000247947 | 0.87575012 |
| Obesity | mast_cell | *CD72* | 7.963083964 | 5.17E-08 | 2.38E-07 | 0.000324961 |
| Obesity | mast_cell | *TMEM8B* | 3.513297478 | 1.12E-08 | 8.25E-08 | 7.02E-05 |
| Obesity | mast_cell | *GRHPR* | 3.764543187 | 2.49E-05 | 5.09E-05 | 0.156796456 |
| Obesity | mast_cell | *ARHGEF39* | 2.412690791 | 5.38E-05 | 0.000103591 | 0.338432503 |
| Obesity | mast_cell | *DENND4C* | 3.649381026 | 0.002110344 | 0.003136581 | 1 |
| Obesity | mast_cell | *PPIL1* | 10.4514594 | 1.64E-09 | 4.26E-08 | 1.03E-05 |
| Obesity | mast_cell | *KIF13A* | 5.077797147 | 2.21E-06 | 5.69E-06 | 0.013918543 |
| Obesity | mast_cell | *ZSCAN9* | 4.865962471 | 2.82E-07 | 9.55E-07 | 0.001773642 |
| Obesity | mast_cell | *TMEM63B* | -3.591546272 | 0.00219922 | 0.003261736 | 1 |
| Obesity | mast_cell | *FRS3* | 7.055431942 | 1.93E-09 | 4.32E-08 | 1.21E-05 |
| Obesity | mast_cell | *SLC22A23* | 5.905200341 | 2.81E-08 | 1.48E-07 | 0.000176713 |
| Obesity | mast_cell | *RIPK1* | 5.887046886 | 5.85E-05 | 0.000111533 | 0.367948174 |
| Obesity | mast_cell | *UQCC2* | 4.273219616 | 2.95E-08 | 1.53E-07 | 0.0001857 |
| Obesity | mast_cell | *TCF19* | 5.717730096 | 3.54E-10 | 1.84E-08 | 2.23E-06 |
| Obesity | mast_cell | *FLOT1* | 3.975010832 | 0.004160662 | 0.005945018 | 1 |
| Obesity | mast_cell | *TPMT* | 4.603293997 | 0.019612449 | 0.025544534 | 1 |
| Obesity | mast_cell | *MTCH1* | 7.79294992 | 1.85E-07 | 6.71E-07 | 0.001164021 |
| Obesity | mast_cell | *FAM8A1* | 8.852478716 | 7.01E-09 | 6.20E-08 | 4.40E-05 |
| Obesity | mast_cell | *FGFBP2* | 2.434325772 | 4.74E-06 | 1.12E-05 | 0.029793171 |
| Obesity | mast_cell | *FHDC1* | 9.931093421 | 3.24E-09 | 4.49E-08 | 2.03E-05 |
| Obesity | mast_cell | *MYO7A* | 2.099641605 | 0.000297859 | 0.000503128 | 1 |
| Obesity | mast_cell | *ARRB1* | 3.737196026 | 0.006401794 | 0.008898537 | 1 |
| Obesity | mast_cell | *CCDC90B* | 7.175072719 | 7.86E-07 | 2.29E-06 | 0.004942159 |
| Obesity | mast_cell | *SYTL2* | -0.877575626 | 3.06E-07 | 1.02E-06 | 0.00192528 |
| Obesity | mast_cell | *RAB30* | 5.804968682 | 6.41E-06 | 1.48E-05 | 0.040289392 |
| Obesity | mast_cell | *PRCP* | 6.10376534 | 1.18E-05 | 2.59E-05 | 0.074250692 |
| Obesity | mast_cell | *NARS2* | 6.620921319 | 4.09E-06 | 9.82E-06 | 0.025717374 |
| Obesity | mast_cell | *PI15* | 13.18000906 | 3.05E-08 | 1.57E-07 | 0.000191571 |
| Obesity | mast_cell | *SULF1* | 3.199855501 | 1.49E-09 | 4.15E-08 | 9.37E-06 |
| Obesity | mast_cell | *TGS1* | 2.997831082 | 0.004260682 | 0.006074129 | 1 |
| Obesity | mast_cell | *NEK1* | 5.295381288 | 7.02E-06 | 1.61E-05 | 0.044118994 |
| Obesity | mast_cell | *DDX60* | 8.018898787 | 5.15E-07 | 1.58E-06 | 0.003240821 |
| Obesity | mast_cell | *MMP27* | 1.379966738 | 3.39E-05 | 6.74E-05 | 0.21327911 |
| Obesity | mast_cell | *DCUN1D5* | 3.590475528 | 2.71E-09 | 4.32E-08 | 1.70E-05 |
| Obesity | mast_cell | *TRIM29* | 2.180928864 | 0.000222811 | 0.000383365 | 1 |
| Obesity | mast_cell | *SLC37A4* | 10.40909873 | 0.000739644 | 0.001173685 | 1 |
| Obesity | mast_cell | *PPP2R1B* | 3.86806794 | 3.34E-09 | 4.52E-08 | 2.10E-05 |
| Obesity | mast_cell | *FDX1* | 6.971393408 | 4.54E-06 | 1.08E-05 | 0.028517393 |
| Obesity | mast_cell | *FXYD6* | -2.974553764 | 0.031225849 | 0.039556098 | 1 |
| Obesity | mast_cell | *TMPRSS13* | 6.383601354 | 3.12E-05 | 6.25E-05 | 0.196108402 |
| Obesity | mast_cell | *MAP2K5* | 6.738777042 | 1.95E-09 | 4.32E-08 | 1.22E-05 |
| Obesity | mast_cell | *NDUFAF1* | 4.440370601 | 3.76E-05 | 7.42E-05 | 0.23655435 |
| Obesity | mast_cell | *ITGA11* | 3.762729564 | 1.69E-08 | 1.04E-07 | 0.00010631 |
| Obesity | mast_cell | *PAQR5* | -0.601640981 | 1.93E-07 | 6.94E-07 | 0.001213584 |
| Obesity | mast_cell | *RMDN3* | 6.335911365 | 2.46E-09 | 4.32E-08 | 1.54E-05 |
| Obesity | mast_cell | *ITPKA* | 9.433886806 | 0.004209932 | 0.006008591 | 1 |

| | | | | | | |
|---|---|---|---|---|---|---|
| Obesity | mast_cell | *UACA* | 5.549247492 | 2.78E-07 | 9.43E-07 | 0.001746796 |
| Obesity | mast_cell | *RSL24D1* | 2.694154081 | 0.01416034 | 0.018781869 | 1 |
| Obesity | mast_cell | *SPTBN5* | 1.695052828 | 5.22E-05 | 0.000100748 | 0.328439855 |
| Obesity | mast_cell | *RABGGTB* | 7.904064553 | 1.41E-07 | 5.36E-07 | 0.000885391 |
| Obesity | mast_cell | *SLC44A5* | 2.91777672 | 0.0211568 | 0.027442294 | 1 |
| Obesity | mast_cell | *DNASE2B* | 12.03144402 | 1.79E-07 | 6.51E-07 | 0.001123294 |
| Obesity | mast_cell | *DBT* | 4.777653646 | 1.63E-07 | 6.01E-07 | 0.001022181 |
| Obesity | mast_cell | *RTCA* | 3.658695883 | 0.00107221 | 0.001659932 | 1 |
| Obesity | mast_cell | *IFT172* | 9.042865456 | 1.19E-11 | 1.70E-09 | 7.49E-08 |
| Obesity | mast_cell | *ADCY3* | 5.956142008 | 1.12E-09 | 3.63E-08 | 7.05E-06 |
| Obesity | mast_cell | *DYNC2LI1* | 2.932468569 | 0.000766418 | 0.001214639 | 1 |
| Obesity | mast_cell | *LHCGR* | 2.504730101 | 6.21E-08 | 2.75E-07 | 0.000390722 |
| Obesity | mast_cell | *THUMPD2* | 6.173409181 | 1.73E-05 | 3.65E-05 | 0.108561839 |
| Obesity | mast_cell | *PREB* | 1.94424943 | 0.033162635 | 0.041849355 | 1 |
| Obesity | mast_cell | *ABCG5* | 4.714941811 | 1.50E-08 | 9.74E-08 | 9.41E-05 |
| Obesity | mast_cell | *ATRAID* | 6.456129405 | 7.82E-07 | 2.28E-06 | 0.004919104 |
| Obesity | mast_cell | *CENPO* | 13.46896545 | 1.43E-08 | 9.52E-08 | 8.99E-05 |
| Obesity | mast_cell | *LRPPRC* | 6.147605835 | 9.07E-07 | 2.60E-06 | 0.005702966 |
| Obesity | mast_cell | *DTNB* | 3.991637633 | 0.000247458 | 0.000423224 | 1 |
| Obesity | mast_cell | *CYP2C9* | 1.554052849 | 0.00202614 | 0.003015706 | 1 |
| Obesity | mast_cell | *CYP2C8* | 3.887818872 | 4.77E-06 | 1.13E-05 | 0.029996758 |
| Obesity | mast_cell | *MYOF* | 1.923781689 | 0.007764839 | 0.010682175 | 1 |
| Obesity | mast_cell | *STAMBPL1* | 5.23052151 | 3.10E-08 | 1.58E-07 | 0.000195149 |
| Obesity | mast_cell | *ATAD1* | 6.572337968 | 2.07E-06 | 5.35E-06 | 0.013010978 |
| Obesity | mast_cell | *BTBD16* | 3.666383568 | 4.93E-08 | 2.30E-07 | 0.000309665 |
| Obesity | mast_cell | *TACC2* | 4.913166706 | 4.75E-07 | 1.48E-06 | 0.002987507 |
| Obesity | mast_cell | *CALHM2* | 4.466518019 | 3.11E-07 | 1.04E-06 | 0.001956218 |
| Obesity | mast_cell | *PLCE1* | 5.539426019 | 2.42E-07 | 8.39E-07 | 0.001520464 |
| Obesity | mast_cell | *RBP4* | 4.624438042 | 6.98E-10 | 2.77E-08 | 4.39E-06 |
| Obesity | mast_cell | *DNAJC13* | 3.143464906 | 0.028802032 | 0.036670387 | 1 |
| Obesity | mast_cell | *FAM149B1* | 3.54532244 | 0.001800761 | 0.002696209 | 1 |
| Obesity | mast_cell | *ASCC1* | 2.661924105 | 0.021931235 | 0.028388238 | 1 |
| Obesity | mast_cell | *RPS24* | 4.152537379 | 0.006076944 | 0.00847322 | 1 |
| Obesity | mast_cell | *BARD1* | 8.278652399 | 1.55E-06 | 4.17E-06 | 0.009747985 |
| Obesity | mast_cell | *CARF* | 6.00867696 | 1.16E-12 | 2.43E-10 | 7.28E-09 |
| Obesity | mast_cell | *METTL5* | 6.314045538 | 2.51E-09 | 4.32E-08 | 1.58E-05 |
| Obesity | mast_cell | *NAB1* | 6.044416329 | 4.46E-09 | 4.98E-08 | 2.80E-05 |
| Obesity | mast_cell | *FASTKD1* | 5.01359976 | 0.000161578 | 0.000284708 | 1 |
| Obesity | mast_cell | *MDH1B* | 5.637180635 | 3.44E-08 | 1.71E-07 | 0.000216508 |
| Obesity | mast_cell | *FAM117B* | 5.891873336 | 1.15E-09 | 3.69E-08 | 7.26E-06 |
| Obesity | mast_cell | *ITGAV* | 4.710908636 | 1.36E-07 | 5.23E-07 | 0.000855152 |
| Obesity | mast_cell | *SLC35A5* | 1.466300339 | 0.01455824 | 0.019268981 | 1 |
| Obesity | mast_cell | *SENP7* | 4.240974242 | 0.001102263 | 0.00170394 | 1 |
| Obesity | mast_cell | *USP8* | 4.800381597 | 3.47E-05 | 6.89E-05 | 0.218292108 |
| Obesity | mast_cell | *SPPL2A* | 4.911195153 | 0.000196755 | 0.000341712 | 1 |
| Obesity | mast_cell | *UBL7* | 6.642262672 | 0.000833967 | 0.001314732 | 1 |
| Obesity | mast_cell | *ARHGAP24* | 3.515232066 | 0.000838646 | 0.001321115 | 1 |
| Obesity | mast_cell | *FAM13A* | 3.390119452 | 0.000942423 | 0.001473517 | 1 |
| Obesity | mast_cell | *HERC3* | 3.079021175 | 0.028576468 | 0.036390572 | 1 |
| Obesity | mast_cell | *AP1AR* | 6.145686972 | 1.17E-08 | 8.44E-08 | 7.35E-05 |

| | | | | | | |
|---|---|---|---|---|---|---|
| Obesity | mast_cell | *PRKG2* | -0.577366748 | 2.80E-07 | 9.50E-07 | 0.001762338 |
| Obesity | mast_cell | *BBS7* | 6.516549623 | 3.65E-09 | 4.64E-08 | 2.29E-05 |
| Obesity | mast_cell | *KIAA1109* | 4.789810667 | 4.47E-05 | 8.71E-05 | 0.280751701 |
| Obesity | mast_cell | *MMRN1* | -1.058236752 | 0.002126941 | 0.0031605 | 1 |
| Obesity | mast_cell | *PRDM5* | 5.647812008 | 2.85E-06 | 7.10E-06 | 0.01788908 |
| Obesity | mast_cell | *TRPC3* | 3.577176082 | 2.69E-07 | 9.17E-07 | 0.001689224 |
| Obesity | mast_cell | *NAAA* | 3.1117435 | 0.016733202 | 0.021985714 | 1 |
| Obesity | mast_cell | *NUP54* | 3.780469136 | 0.001381555 | 0.002108213 | 1 |
| Obesity | mast_cell | *BMP2K* | -1.333202461 | 6.24E-07 | 1.86E-06 | 0.003923251 |
| Obesity | mast_cell | *G3BP2* | 4.414321898 | 0.002752894 | 0.004029673 | 1 |
| Obesity | mast_cell | *SCARB2* | 6.092197656 | 4.85E-07 | 1.50E-06 | 0.003052224 |
| Obesity | mast_cell | *ANXA3* | 10.39971916 | 1.77E-08 | 1.06E-07 | 0.000111241 |
| Obesity | mast_cell | *GSTCD* | 6.053299827 | 2.09E-08 | 1.18E-07 | 0.000131355 |
| Obesity | mast_cell | *PAPSS1* | 3.150269102 | 0.001193682 | 0.001838481 | 1 |
| Obesity | mast_cell | *SLC39A8* | 3.828092859 | 8.34E-07 | 2.41E-06 | 0.005244573 |
| Obesity | mast_cell | *FBN2* | 0.750111637 | 0.002952628 | 0.004306001 | 1 |
| Obesity | mast_cell | *RNF185* | 5.807074103 | 3.45E-06 | 8.43E-06 | 0.02167925 |
| Obesity | mast_cell | *B4GALNT3* | 4.713250258 | 0.001203203 | 0.00185133 | 1 |
| Obesity | mast_cell | *ERP27* | -4.760231285 | 0.001255338 | 0.001925423 | 1 |
| Obesity | mast_cell | *KIF21A* | 2.601588609 | 1.28E-05 | 2.78E-05 | 0.080374769 |
| Obesity | mast_cell | *CPNE8* | 2.932673676 | 0.000466496 | 0.000765367 | 1 |
| Obesity | mast_cell | *ALG10* | 10.27278755 | 2.76E-09 | 4.32E-08 | 1.73E-05 |
| Obesity | mast_cell | *SLCO1C1* | 3.404940361 | 2.77E-09 | 4.32E-08 | 1.74E-05 |
| Obesity | mast_cell | *ZCRB1* | 4.5836566 | 1.27E-06 | 3.49E-06 | 0.007963631 |
| Obesity | mast_cell | *PRICKLE1* | 3.831439373 | 2.50E-08 | 1.35E-07 | 0.000157463 |
| Obesity | mast_cell | *C1RL* | 4.412951317 | 8.54E-08 | 3.57E-07 | 0.000537202 |
| Obesity | mast_cell | *CLSTN3* | 5.654227324 | 1.24E-06 | 3.43E-06 | 0.007806002 |
| Obesity | mast_cell | *KLRG1* | -1.017207358 | 0.006726475 | 0.009329219 | 1 |
| Obesity | mast_cell | *CD27* | 5.346256262 | 1.08E-09 | 3.57E-08 | 6.79E-06 |
| Obesity | mast_cell | *PEX5* | 4.790208935 | 3.88E-05 | 7.64E-05 | 0.244120552 |
| Obesity | mast_cell | *PIANP* | 5.852347122 | 0.024800386 | 0.031846411 | 1 |
| Obesity | mast_cell | *PPFIA2* | 2.417128246 | 0.00035704 | 0.000597156 | 1 |
| Obesity | mast_cell | *GLIPR1* | 5.338328623 | 5.72E-08 | 2.57E-07 | 0.000359311 |
| Obesity | mast_cell | *LGR5* | 2.738777088 | 1.39E-07 | 5.31E-07 | 0.000872521 |
| Obesity | mast_cell | *POC1B* | 7.525312015 | 5.83E-08 | 2.61E-07 | 0.000366283 |
| Obesity | mast_cell | *AMDHD1* | 2.260395562 | 4.88E-05 | 9.47E-05 | 0.306821312 |
| Obesity | mast_cell | *GAS2L3* | 8.962577002 | 7.79E-08 | 3.29E-07 | 0.000489955 |
| Obesity | mast_cell | *TMEM132B* | 3.854867595 | 4.31E-07 | 1.37E-06 | 0.002711439 |
| Obesity | mast_cell | *TDG* | 5.846098124 | 1.44E-05 | 3.10E-05 | 0.090749831 |
| Obesity | mast_cell | *GLTP* | 5.421488365 | 1.57E-07 | 5.84E-07 | 0.000984637 |
| Obesity | mast_cell | *MTMR6* | 5.483357029 | 1.68E-05 | 3.56E-05 | 0.105562163 |
| Obesity | mast_cell | *SLC46A3* | 2.561010596 | 0.007728858 | 0.010637331 | 1 |
| Obesity | mast_cell | *ACVRL1* | 3.264511906 | 0.000114046 | 0.000205979 | 0.717010207 |
| Obesity | mast_cell | *GPR84* | -19.13557074 | 5.26E-08 | 2.41E-07 | 0.000330783 |
| Obesity | mast_cell | *BRCA2* | -3.5027328 | 0.001610409 | 0.00243147 | 1 |
| Obesity | mast_cell | *CERS5* | 6.203283582 | 5.45E-07 | 1.65E-06 | 0.003425913 |
| Obesity | mast_cell | *LMBR1L* | 6.60828859 | 3.02E-07 | 1.01E-06 | 0.00189869 |
| Obesity | mast_cell | *ZNF740* | 4.694490525 | 0.002580717 | 0.003795315 | 1 |
| Obesity | mast_cell | *HNRNPA1L2* | 5.307018607 | 1.95E-08 | 1.12E-07 | 0.000122347 |
| Obesity | mast_cell | *LPAR6* | 6.22781504 | 1.45E-06 | 3.94E-06 | 0.009135701 |

| | | | | | | |
|---|---|---|---|---|---|---|
| Obesity | mast_cell | *VPS33A* | 9.814784384 | 0.000353146 | 0.000591273 | 1 |
| Obesity | mast_cell | *TSSK4* | 8.458157316 | 1.04E-05 | 2.31E-05 | 0.06559932 |
| Obesity | mast_cell | *TMX1* | 8.141068113 | 2.69E-07 | 9.17E-07 | 0.001691835 |
| Obesity | mast_cell | *RTN1* | 4.940356303 | 4.21E-09 | 4.83E-08 | 2.64E-05 |
| Obesity | mast_cell | *C14orf37* | 4.217885661 | 6.72E-05 | 0.000126568 | 0.422736989 |
| Obesity | mast_cell | *RAB15* | 1.751179793 | 1.67E-06 | 4.43E-06 | 0.010488092 |
| Obesity | mast_cell | *STON2* | 3.891813002 | 3.99E-09 | 4.78E-08 | 2.51E-05 |
| Obesity | mast_cell | *EFCAB11* | 4.823508513 | 3.00E-09 | 4.44E-08 | 1.89E-05 |
| Obesity | mast_cell | *PTGR2* | -1.827655321 | 1.78E-05 | 3.75E-05 | 0.111866059 |
| Obesity | mast_cell | *AK7* | 7.517497783 | 4.29E-09 | 4.88E-08 | 2.70E-05 |
| Obesity | mast_cell | *SLC24A4* | 1.926283752 | 2.98E-06 | 7.41E-06 | 0.018757377 |
| Obesity | mast_cell | *FBLN5* | 5.841209285 | 8.31E-10 | 3.03E-08 | 5.23E-06 |
| Obesity | mast_cell | *C14orf79* | 11.09366698 | 5.87E-07 | 1.76E-06 | 0.003690577 |
| Obesity | mast_cell | *NIPA2* | 7.075896524 | 4.60E-08 | 2.17E-07 | 0.000289194 |
| Obesity | mast_cell | *MFAP1* | 5.925703046 | 3.42E-08 | 1.70E-07 | 0.000215114 |
| Obesity | mast_cell | *TCF12* | 4.459647598 | 1.77E-05 | 3.74E-05 | 0.111515628 |
| Obesity | mast_cell | *SORD* | 8.482115089 | 1.47E-08 | 9.66E-08 | 9.25E-05 |
| Obesity | mast_cell | *ZSCAN29* | 3.79224853 | 1.58E-09 | 4.20E-08 | 9.92E-06 |
| Obesity | mast_cell | *LYSMD2* | 4.242154783 | 2.28E-06 | 5.84E-06 | 0.014334902 |
| Obesity | mast_cell | *BAHD1* | 7.196742367 | 1.63E-08 | 1.02E-07 | 0.00010226 |
| Obesity | mast_cell | *ANP32A* | 4.374099738 | 0.00653701 | 0.009074449 | 1 |
| Obesity | mast_cell | *PSTPIP1* | 4.828566803 | 4.52E-05 | 8.80E-05 | 0.283960708 |
| Obesity | mast_cell | *HMG20A* | 6.746288194 | 5.74E-07 | 1.73E-06 | 0.003610685 |
| Obesity | mast_cell | *SCAPER* | 3.413592658 | 0.000489946 | 0.000800491 | 1 |
| Obesity | mast_cell | *NEIL1* | 3.961879573 | 0.001680793 | 0.002528025 | 1 |
| Obesity | mast_cell | *DNAJA4* | -4.089295176 | 0.000306283 | 0.000516689 | 1 |
| Obesity | mast_cell | *TPM1* | 4.757946807 | 1.78E-08 | 1.06E-07 | 0.000111727 |
| Obesity | mast_cell | *IGF1R* | 5.053086545 | 7.84E-09 | 6.64E-08 | 4.93E-05 |
| Obesity | mast_cell | *CYP11A1* | 4.178323254 | 3.20E-09 | 4.49E-08 | 2.01E-05 |
| Obesity | mast_cell | *ADAMTS17* | 4.669974197 | 2.71E-08 | 1.43E-07 | 0.000170491 |
| Obesity | mast_cell | *SCAMP2* | 5.205901541 | 0.00012806 | 0.000228985 | 0.805111544 |
| Obesity | mast_cell | *HAPLN3* | 3.712784197 | 8.09E-08 | 3.40E-07 | 0.00050836 |
| Obesity | mast_cell | *POLG* | 7.721456317 | 2.15E-06 | 5.53E-06 | 0.013488818 |
| Obesity | mast_cell | *WDR93* | 0.762223389 | 0.012840054 | 0.017150078 | 1 |
| Obesity | mast_cell | *TICRR* | 5.38570764 | 1.35E-07 | 5.20E-07 | 0.000849426 |
| Obesity | mast_cell | *MFGE8* | 4.929104348 | 1.39E-07 | 5.31E-07 | 0.000872551 |
| Obesity | mast_cell | *UNC45A* | 3.488579358 | 0.000198666 | 0.00034484 | 1 |
| Obesity | mast_cell | *ST8SIA2* | 16.64974879 | 3.03E-07 | 1.01E-06 | 0.001906565 |
| Obesity | mast_cell | *IQGAP1* | 3.319208324 | 0.000699556 | 0.001114291 | 1 |
| Obesity | mast_cell | *CRTC3* | 5.698515155 | 9.44E-06 | 2.11E-05 | 0.05935365 |
| Obesity | mast_cell | *SEC11A* | 6.088678975 | 5.05E-08 | 2.33E-07 | 0.000317203 |
| Obesity | mast_cell | *TGFB1I1* | 2.934026354 | 2.00E-06 | 5.18E-06 | 0.01256351 |
| Obesity | mast_cell | *PARN* | 4.609691609 | 5.53E-05 | 0.00010611 | 0.34772101 |
| Obesity | mast_cell | *FTO* | 3.317834527 | 0.005147283 | 0.007254195 | 1 |
| Obesity | mast_cell | *TXNL4B* | 4.262430014 | 2.96E-08 | 1.53E-07 | 0.000186182 |
| Obesity | mast_cell | *MARVELD3* | 5.887876923 | 1.01E-05 | 2.26E-05 | 0.06377071 |
| Obesity | mast_cell | *ZFHX3* | 3.853877692 | 0.001715653 | 0.002576143 | 1 |
| Obesity | mast_cell | *KIFC3* | 7.250235493 | 6.15E-08 | 2.73E-07 | 0.000386388 |
| Obesity | mast_cell | *ADAMTS18* | 2.691492403 | 6.26E-08 | 2.77E-07 | 0.000393846 |
| Obesity | mast_cell | *NUDT7* | 3.252184313 | 0.000924457 | 0.001447587 | 1 |

| | | | | | | |
|---|---|---|---|---|---|---|
| Obesity | mast_cell | *GCSH* | 3.327341363 | 0.000272149 | 0.000462433 | 1 |
| Obesity | mast_cell | *CMTM3* | 9.05073421 | 9.05E-06 | 2.03E-05 | 0.056870972 |
| Obesity | mast_cell | *MAP1LC3B* | 5.606428488 | 2.85E-06 | 7.12E-06 | 0.017937566 |
| Obesity | mast_cell | *TLDC1* | 5.638257413 | 1.09E-09 | 3.58E-08 | 6.87E-06 |
| Obesity | mast_cell | *TIGD7* | 4.279422715 | 8.21E-10 | 3.02E-08 | 5.16E-06 |
| Obesity | mast_cell | *DEF8* | 3.200694103 | 6.60E-07 | 1.96E-06 | 0.004149566 |
| Obesity | mast_cell | *GAS8* | 1.169185125 | 0.016166939 | 0.021295107 | 1 |
| Obesity | mast_cell | *COPS3* | 8.94820554 | 7.35E-08 | 3.13E-07 | 0.000462039 |
| Obesity | mast_cell | *RANBP10* | 5.110773347 | 6.95E-07 | 2.05E-06 | 0.004368016 |
| Obesity | mast_cell | *PRPSAP2* | 2.963569734 | 0.028047613 | 0.035767818 | 1 |
| Obesity | mast_cell | *MYO19* | 9.603086046 | 4.33E-14 | 1.43E-11 | 2.72E-10 |
| Obesity | mast_cell | *DDX52* | 5.746610602 | 0.000110269 | 0.000199671 | 0.693258291 |
| Obesity | mast_cell | *RASL10B* | 3.788442069 | 0.001611887 | 0.002432534 | 1 |
| Obesity | mast_cell | *UNC45B* | -7.254877322 | 1.14E-05 | 2.52E-05 | 0.07180006 |
| Obesity | mast_cell | *TOM1L1* | 5.933497509 | 3.07E-10 | 1.68E-08 | 1.93E-06 |
| Obesity | mast_cell | *C17orf80* | 3.7942407 | 6.14E-06 | 1.42E-05 | 0.038627341 |
| Obesity | mast_cell | *VPS53* | 4.859467173 | 5.04E-06 | 1.18E-05 | 0.031671913 |
| Obesity | mast_cell | *SPATA22* | 2.589668886 | 8.63E-05 | 0.000158747 | 0.542755051 |
| Obesity | mast_cell | *SGSM2* | 5.393963226 | 9.71E-05 | 0.000176971 | 0.610372061 |
| Obesity | mast_cell | *NPEPPS* | 5.964794756 | 1.76E-06 | 4.64E-06 | 0.011044436 |
| Obesity | mast_cell | *LRRC46* | 3.297670107 | 0.007001453 | 0.009680698 | 1 |
| Obesity | mast_cell | *SCRN2* | 1.026391511 | 0.000350173 | 0.000586764 | 1 |
| Obesity | mast_cell | *RHBDL3* | -2.539729837 | 1.33E-05 | 2.87E-05 | 0.083424614 |
| Obesity | mast_cell | *ABCA8* | 3.711291578 | 1.23E-06 | 3.40E-06 | 0.007717361 |
| Obesity | mast_cell | *IMPA2* | 6.873870835 | 1.67E-09 | 4.27E-08 | 1.05E-05 |
| Obesity | mast_cell | *GNAL* | 7.255961186 | 6.64E-09 | 6.03E-08 | 4.18E-05 |
| Obesity | mast_cell | *RPRD1A* | 4.405632623 | 0.00021909 | 0.000377479 | 1 |
| Obesity | mast_cell | *C18orf21* | 7.086453971 | 7.07E-10 | 2.78E-08 | 4.44E-06 |
| Obesity | mast_cell | *GALNT1* | 5.963641155 | 5.24E-07 | 1.60E-06 | 0.003296728 |
| Obesity | mast_cell | *ASXL3* | 2.776297031 | 1.62E-08 | 1.02E-07 | 0.000101669 |
| Obesity | mast_cell | *ADCYAP1* | -1.237639323 | 6.14E-08 | 2.73E-07 | 0.000386324 |
| Obesity | mast_cell | *ESCO1* | 6.14986012 | 9.06E-06 | 2.03E-05 | 0.05698492 |
| Obesity | mast_cell | *GREB1L* | 2.842205081 | 2.39E-05 | 4.89E-05 | 0.150142403 |
| Obesity | mast_cell | *NPC1* | 2.938058395 | 0.000150724 | 0.000266856 | 0.947604721 |
| Obesity | mast_cell | *SLC14A1* | 5.255786814 | 9.07E-05 | 0.000165869 | 0.570257987 |
| Obesity | mast_cell | *SLC13A5* | 6.867181139 | 0.016419435 | 0.021600542 | 1 |
| Obesity | mast_cell | *CCDC40* | 2.613269905 | 0.000179966 | 0.00031429 | 1 |
| Obesity | mast_cell | *CARD14* | -3.644401787 | 5.41E-05 | 0.000103934 | 0.339821971 |
| Obesity | mast_cell | *EIF4A3* | 4.160571158 | 1.84E-06 | 4.83E-06 | 0.011556223 |
| Obesity | mast_cell | *TBCD* | 3.268656141 | 0.016062901 | 0.021166938 | 1 |
| Obesity | mast_cell | *FN3KRP* | 1.847788408 | 0.0001462 | 0.000259504 | 0.91916248 |
| Obesity | mast_cell | *NARF* | 5.498959208 | 4.69E-05 | 9.12E-05 | 0.294799692 |
| Obesity | mast_cell | *TRIM65* | 5.590579989 | 2.13E-13 | 5.58E-11 | 1.34E-09 |
| Obesity | mast_cell | *CBX8* | 6.247008361 | 6.62E-10 | 2.76E-08 | 4.16E-06 |
| Obesity | mast_cell | *SECTM1* | 4.083918507 | 3.34E-08 | 1.67E-07 | 0.000209947 |
| Obesity | mast_cell | *RNF157* | 1.580725033 | 3.85E-08 | 1.88E-07 | 0.000242103 |
| Obesity | mast_cell | *WDR45B* | 3.22404809 | 0.023409779 | 0.030196406 | 1 |
| Obesity | mast_cell | *RNF165* | 8.673028456 | 6.05E-07 | 1.81E-06 | 0.003806013 |
| Obesity | mast_cell | *SMAD4* | 4.670965482 | 0.002203568 | 0.003267413 | 1 |
| Obesity | mast_cell | *PIP4K2B* | 6.198650492 | 3.77E-09 | 4.71E-08 | 2.37E-05 |

| | | | | | | |
|---|---|---|---|---|---|---|
| Obesity | mast_cell | *GRB7* | 5.165896828 | 9.98E-08 | 4.08E-07 | 0.00062766 |
| Obesity | mast_cell | *TXNL4A* | 1.978372254 | 0.037640376 | 0.047142387 | 1 |
| Obesity | mast_cell | *NFIC* | 3.675534113 | 6.15E-05 | 0.000116535 | 0.386431559 |
| Obesity | mast_cell | *TPGS1* | 6.939210421 | 6.46E-05 | 0.000121883 | 0.405870226 |
| Obesity | mast_cell | *PFKL* | 3.720733624 | 1.17E-05 | 2.58E-05 | 0.073741315 |
| Obesity | mast_cell | *MVB12A* | 5.334455517 | 4.11E-09 | 4.80E-08 | 2.59E-05 |
| Obesity | mast_cell | *DPP9* | 4.966356349 | 1.23E-08 | 8.65E-08 | 7.72E-05 |
| Obesity | mast_cell | *CCDC97* | 8.251425005 | 4.72E-05 | 9.17E-05 | 0.296945547 |
| Obesity | mast_cell | *TMEM91* | 4.077118001 | 0.000668708 | 0.001069218 | 1 |
| Obesity | mast_cell | *SIRT3* | 5.464535101 | 9.08E-06 | 2.04E-05 | 0.05708114 |
| Obesity | mast_cell | *IFITM3* | 7.754010808 | 2.37E-06 | 6.02E-06 | 0.014869041 |
| Obesity | mast_cell | *COL6A1* | 3.106960398 | 7.57E-05 | 0.000141082 | 0.4761502 |
| Obesity | mast_cell | *IFNAR1* | 2.376909778 | 0.008140006 | 0.011161662 | 1 |
| Obesity | mast_cell | *TMEM50B* | 1.133749235 | 0.031861434 | 0.04028008 | 1 |
| Obesity | mast_cell | *DOPEY2* | 5.365987007 | 1.16E-05 | 2.55E-05 | 0.072697184 |
| Obesity | mast_cell | *URB1* | 2.456214969 | 0.001306989 | 0.00199928 | 1 |
| Obesity | mast_cell | *AKT1* | 6.519471996 | 2.23E-09 | 4.32E-08 | 1.40E-05 |
| Obesity | mast_cell | *SAE1* | 4.966802902 | 7.68E-07 | 2.24E-06 | 0.004830118 |
| Obesity | mast_cell | *NTN5* | -5.733275539 | 1.09E-07 | 4.36E-07 | 0.000684041 |
| Obesity | mast_cell | *GEMIN7* | 6.188143772 | 8.01E-07 | 2.33E-06 | 0.005036542 |
| Obesity | mast_cell | *WTIP* | 4.198267888 | 1.08E-08 | 8.14E-08 | 6.82E-05 |
| Obesity | mast_cell | *MYO1F* | 4.868932379 | 0.000247966 | 0.000423977 | 1 |
| Obesity | mast_cell | *CACNG8* | -1.788631651 | 2.56E-07 | 8.80E-07 | 0.001609659 |
| Obesity | mast_cell | *FBN3* | -2.116799196 | 0.011094663 | 0.014945821 | 1 |
| Obesity | mast_cell | *EVI5L* | 5.838043305 | 6.44E-09 | 5.96E-08 | 4.05E-05 |
| Obesity | mast_cell | *RPS11* | 4.532507586 | 0.001250157 | 0.001918413 | 1 |
| Obesity | mast_cell | *CTU1* | 6.771313279 | 0.000520245 | 0.000846914 | 1 |
| Obesity | mast_cell | *RCN3* | 6.033983873 | 2.07E-09 | 4.32E-08 | 1.30E-05 |
| Obesity | mast_cell | *RERE* | 3.303007545 | 0.000946116 | 0.001478556 | 1 |
| Obesity | mast_cell | *PEX14* | 5.794043308 | 1.73E-07 | 6.35E-07 | 0.001088622 |
| Obesity | mast_cell | *SH3BGRL3* | 5.009493737 | 0.005499401 | 0.00771069 | 1 |
| Obesity | mast_cell | *CNKSR1* | 0.83117587 | 0.019546175 | 0.025468767 | 1 |
| Obesity | mast_cell | *IL22RA1* | 3.316221101 | 0.000201108 | 0.000348791 | 1 |
| Obesity | mast_cell | *ZNF593* | 3.594483095 | 0.000373087 | 0.000622396 | 1 |
| Obesity | mast_cell | *C1orf216* | 6.312308002 | 1.50E-08 | 9.77E-08 | 9.46E-05 |
| Obesity | mast_cell | *GPN2* | 7.741650507 | 8.31E-05 | 0.00015316 | 0.52258241 |
| Obesity | mast_cell | *NBPF3* | 1.54123103 | 0.000889045 | 0.001395612 | 1 |
| Obesity | mast_cell | *HSPG2* | 1.199153457 | 0.01399989 | 0.018580812 | 1 |
| Obesity | mast_cell | *ITGB3BP* | 2.890792724 | 0.000128493 | 0.000229695 | 0.807838184 |
| Obesity | mast_cell | *SERBP1* | 7.029237891 | 1.11E-07 | 4.42E-07 | 0.000696223 |
| Obesity | mast_cell | *PIGK* | 6.341288674 | 0.000151679 | 0.00026847 | 0.953607105 |
| Obesity | mast_cell | *RPS8* | 3.726149249 | 0.002960231 | 0.004316089 | 1 |
| Obesity | mast_cell | *MOB3C* | 6.891855938 | 1.96E-09 | 4.32E-08 | 1.23E-05 |
| Obesity | mast_cell | *SYPL2* | 2.573555229 | 1.07E-06 | 3.00E-06 | 0.006718924 |
| Obesity | mast_cell | *ZNF697* | 6.454954359 | 2.50E-08 | 1.35E-07 | 0.000157127 |
| Obesity | mast_cell | *GPR161* | 7.108020707 | 4.77E-10 | 2.29E-08 | 3.00E-06 |
| Obesity | mast_cell | *TIPRL* | 8.100511809 | 2.13E-09 | 4.32E-08 | 1.34E-05 |
| Obesity | mast_cell | *NME7* | 5.277236636 | 2.73E-06 | 6.85E-06 | 0.017147157 |
| Obesity | mast_cell | *CREG1* | 4.313422604 | 0.002606995 | 0.003830376 | 1 |
| Obesity | mast_cell | *GPA33* | 4.984879962 | 2.87E-07 | 9.69E-07 | 0.001806092 |

| | | | | | | |
|---|---|---|---|---|---|---|
| Obesity | mast_cell | *RXRG* | 6.465486715 | 1.25E-08 | 8.74E-08 | 7.86E-05 |
| Obesity | mast_cell | *TMCO1* | 5.675604164 | 0.000401149 | 0.000666497 | 1 |
| Obesity | mast_cell | *XCL2* | 2.753915071 | 0.00711811 | 0.009826868 | 1 |
| Obesity | mast_cell | *MGST3* | 6.02872578 | 2.87E-08 | 1.49E-07 | 0.000180133 |
| Obesity | mast_cell | *ADCY10* | 7.224611633 | 4.14E-09 | 4.80E-08 | 2.60E-05 |
| Obesity | mast_cell | *RFWD2* | 3.208349685 | 0.005982148 | 0.008346597 | 1 |
| Obesity | mast_cell | *PPOX* | 3.264903174 | 0.017853658 | 0.023374833 | 1 |
| Obesity | mast_cell | *RGS5* | 5.646708752 | 1.29E-08 | 8.97E-08 | 8.12E-05 |
| Obesity | mast_cell | *RRNAD1* | 3.278223556 | 0.00297865 | 0.004340929 | 1 |
| Obesity | mast_cell | *PIGM* | 4.240256891 | 0.031408109 | 0.039754938 | 1 |
| Obesity | mast_cell | *RGL1* | 5.079285293 | 7.87E-09 | 6.64E-08 | 4.95E-05 |
| Obesity | mast_cell | *TARS2* | 5.295225029 | 0.001230778 | 0.00189052 | 1 |
| Obesity | mast_cell | *SNX27* | 6.664910365 | 5.63E-06 | 1.31E-05 | 0.035411737 |
| Obesity | mast_cell | *SELENBP1* | 3.377296722 | 0.003199315 | 0.004643143 | 1 |
| Obesity | mast_cell | *ENSA* | 5.372447138 | 0.000272641 | 0.000463143 | 1 |
| Obesity | mast_cell | *SEMA6C* | 3.767838744 | 5.38E-10 | 2.45E-08 | 3.38E-06 |
| Obesity | mast_cell | *ARNT* | 4.854641271 | 5.58E-05 | 0.000106862 | 0.350613232 |
| Obesity | mast_cell | *POGZ* | 4.750438831 | 6.65E-06 | 1.53E-05 | 0.041817249 |
| Obesity | mast_cell | *GABPB2* | 1.921539427 | 0.011385688 | 0.015318172 | 1 |
| Obesity | mast_cell | *IKBKE* | 5.345156697 | 8.73E-09 | 7.06E-08 | 5.49E-05 |
| Obesity | mast_cell | *DYRK3* | 7.344015411 | 4.34E-08 | 2.06E-07 | 0.000272749 |
| Obesity | mast_cell | *HHIPL2* | -4.678466041 | 1.36E-05 | 2.93E-05 | 0.085274744 |
| Obesity | mast_cell | *TP53BP2* | 5.165130137 | 0.005347475 | 0.007514433 | 1 |
| Obesity | mast_cell | *RAB13* | 7.845832746 | 6.47E-07 | 1.93E-06 | 0.00406898 |
| Obesity | mast_cell | *HAX1* | 7.536943579 | 2.17E-06 | 5.57E-06 | 0.013613039 |
| Obesity | mast_cell | *KCNN3* | -2.95804284 | 0.000520917 | 0.000847569 | 1 |
| Obesity | mast_cell | *GATAD2B* | 4.42805672 | 4.56E-06 | 1.08E-05 | 0.028688811 |
| Obesity | mast_cell | *RIT1* | 7.149055103 | 2.24E-09 | 4.32E-08 | 1.41E-05 |
| Obesity | mast_cell | *GALNT2* | 5.410783411 | 0.000185178 | 0.000322675 | 1 |
| Obesity | mast_cell | *SCCPDH* | 6.681402155 | 6.46E-09 | 5.96E-08 | 4.06E-05 |
| Obesity | mast_cell | *CEP170* | 6.607766642 | 6.41E-09 | 5.94E-08 | 4.03E-05 |
| Obesity | mast_cell | *SNAP47* | 3.491686596 | 0.000450357 | 0.000741591 | 1 |
| Obesity | mast_cell | *NVL* | 6.501416641 | 2.51E-08 | 1.35E-07 | 0.000157598 |
| Obesity | mast_cell | *SDE2* | 8.316871083 | 3.86E-09 | 4.77E-08 | 2.43E-05 |
| Obesity | mast_cell | *FBXO28* | 6.155104598 | 3.52E-05 | 6.98E-05 | 0.221488725 |
| Obesity | mast_cell | *CNIH4* | 7.525529386 | 1.68E-05 | 3.56E-05 | 0.105317097 |
| Obesity | mast_cell | *GUK1* | 9.060552293 | 4.07E-10 | 2.04E-08 | 2.56E-06 |
| Obesity | mast_cell | *CDC42BPA* | 5.535623385 | 4.64E-09 | 5.09E-08 | 2.92E-05 |
| Obesity | mast_cell | *CNIH3* | 3.396654298 | 0.01476646 | 0.019528131 | 1 |
| Obesity | mast_cell | *PSEN2* | 4.44646737 | 7.68E-10 | 2.93E-08 | 4.83E-06 |
| Obesity | mast_cell | *PYCR2* | 3.935991942 | 1.93E-07 | 6.94E-07 | 0.001214346 |
| Obesity | mast_cell | *WNT9A* | 4.316540095 | 3.90E-08 | 1.90E-07 | 0.000245472 |
| Obesity | mast_cell | *ABCG8* | 8.105905714 | 7.02E-07 | 2.07E-06 | 0.004411796 |
| Obesity | mast_cell | *EML4* | 5.779260972 | 3.12E-08 | 1.59E-07 | 0.000196142 |
| Obesity | mast_cell | *CHAC2* | 9.242803458 | 5.96E-09 | 5.73E-08 | 3.75E-05 |
| Obesity | mast_cell | *WDPCP* | 2.325309575 | 1.23E-07 | 4.82E-07 | 0.000775295 |
| Obesity | mast_cell | *ASXL2* | 5.421952471 | 6.12E-06 | 1.41E-05 | 0.038445442 |
| Obesity | mast_cell | *CIAO1* | 5.619238365 | 1.19E-05 | 2.61E-05 | 0.074904563 |
| Obesity | mast_cell | *ZNF514* | 0.968263094 | 0.023855173 | 0.030733088 | 1 |
| Obesity | mast_cell | *TEX261* | 8.014848005 | 2.04E-09 | 4.32E-08 | 1.28E-05 |

| | | | | | | |
|---|---|---|---|---|---|---|
| Obesity | mast_cell | *RALB* | -2.583921309 | 0.017430605 | 0.022854268 | 1 |
| Obesity | mast_cell | *TMEM177* | 11.64710487 | 7.29E-08 | 3.12E-07 | 0.000458398 |
| Obesity | mast_cell | *RABL2A* | 5.52530847 | 9.80E-07 | 2.78E-06 | 0.006159185 |
| Obesity | mast_cell | *SLC20A1* | 4.917322163 | 1.52E-08 | 9.85E-08 | 9.55E-05 |
| Obesity | mast_cell | *LIPT1* | 2.440006575 | 0.000121697 | 0.000218664 | 0.765106519 |
| Obesity | mast_cell | *FAHD2B* | 2.354744026 | 2.03E-05 | 4.22E-05 | 0.127661518 |
| Obesity | mast_cell | *LYG1* | 3.038618853 | 0.000210699 | 0.000364319 | 1 |
| Obesity | mast_cell | *AFF3* | 1.406022774 | 0.018052256 | 0.023595537 | 1 |
| Obesity | mast_cell | *UBXN4* | 6.234935713 | 2.13E-07 | 7.52E-07 | 0.001336937 |
| Obesity | mast_cell | *THSD7B* | 3.021786187 | 2.24E-11 | 2.71E-09 | 1.41E-07 |
| Obesity | mast_cell | *GPR17* | 0.968614613 | 0.005169333 | 0.007282007 | 1 |
| Obesity | mast_cell | *POLR2D* | 6.991263528 | 5.70E-06 | 1.33E-05 | 0.035857212 |
| Obesity | mast_cell | *ZNF385B* | 1.612249358 | 8.09E-09 | 6.74E-08 | 5.09E-05 |
| Obesity | mast_cell | *TMEFF2* | -0.701086465 | 3.39E-07 | 1.11E-06 | 0.002128516 |
| Obesity | mast_cell | *GULP1* | 4.020698681 | 5.49E-10 | 2.45E-08 | 3.45E-06 |
| Obesity | mast_cell | *FAM171B* | 5.101240797 | 8.05E-07 | 2.34E-06 | 0.005059813 |
| Obesity | mast_cell | *HSPD1* | 8.603042796 | 5.50E-09 | 5.54E-08 | 3.46E-05 |
| Obesity | mast_cell | *PTH2R* | -1.757162962 | 2.76E-05 | 5.60E-05 | 0.173784419 |
| Obesity | mast_cell | *NBEAL1* | 3.655770038 | 2.00E-08 | 1.14E-07 | 0.000125566 |
| Obesity | mast_cell | *SUMF1* | 5.100978805 | 4.50E-07 | 1.42E-06 | 0.002830911 |
| Obesity | mast_cell | *ESPNL* | 24.7644639 | 2.47E-08 | 1.34E-07 | 0.000155572 |
| Obesity | mast_cell | *TAMM41* | 6.740727926 | 7.66E-10 | 2.93E-08 | 4.82E-06 |
| Obesity | mast_cell | *OSBPL10* | 4.789088295 | 6.19E-09 | 5.83E-08 | 3.89E-05 |
| Obesity | mast_cell | *FAM198A* | 2.284382745 | 4.48E-06 | 1.07E-05 | 0.028193071 |
| Obesity | mast_cell | *CSRNP1* | 4.8835802 | 0.001471262 | 0.002234797 | 1 |
| Obesity | mast_cell | *SLC25A38* | 3.393757408 | 2.45E-08 | 1.33E-07 | 0.000153796 |
| Obesity | mast_cell | *CTDSPL* | 5.850534107 | 6.15E-07 | 1.84E-06 | 0.003868208 |
| Obesity | mast_cell | *STAC* | 4.682053121 | 3.14E-09 | 4.48E-08 | 1.98E-05 |
| Obesity | mast_cell | *ARL6IP5* | 4.105907782 | 0.005977637 | 0.008344006 | 1 |
| Obesity | mast_cell | *TMF1* | 6.174706419 | 1.99E-06 | 5.16E-06 | 0.012489485 |
| Obesity | mast_cell | *COL8A1* | 2.372722429 | 7.90E-05 | 0.000146642 | 0.496823518 |
| Obesity | mast_cell | *NXPE3* | -3.164884017 | 0.005769611 | 0.00806796 | 1 |
| Obesity | mast_cell | *PLA1A* | 1.490318331 | 0.036588295 | 0.045905131 | 1 |
| Obesity | mast_cell | *RABL3* | 7.845415451 | 1.13E-09 | 3.65E-08 | 7.12E-06 |
| Obesity | mast_cell | *ADPRH* | -1.877929858 | 0.004494771 | 0.006390463 | 1 |
| Obesity | mast_cell | *BOC* | 4.309440244 | 3.73E-08 | 1.83E-07 | 0.000234386 |
| Obesity | mast_cell | *AGTR1* | 3.094666198 | 3.98E-08 | 1.93E-07 | 0.000250124 |
| Obesity | mast_cell | *EAF2* | 3.583289195 | 0.000176413 | 0.000308765 | 1 |
| Obesity | mast_cell | *LYAR* | -3.444817914 | 0.035320406 | 0.044447437 | 1 |
| Obesity | mast_cell | *ATP10D* | 3.336153736 | 0.000673651 | 0.001076027 | 1 |
| Obesity | mast_cell | *SLC10A4* | 7.25694266 | 0.003432144 | 0.004962716 | 1 |
| Obesity | mast_cell | *PYURF* | 6.919131749 | 0.000952546 | 0.001487865 | 1 |
| Obesity | mast_cell | *CISD2* | 7.183481311 | 1.77E-06 | 4.67E-06 | 0.011112429 |
| Obesity | mast_cell | *TIFA* | 6.066783352 | 4.72E-08 | 2.22E-07 | 0.00029646 |
| Obesity | mast_cell | *USP53* | 5.002017927 | 3.83E-09 | 4.75E-08 | 2.41E-05 |
| Obesity | mast_cell | *PDGFC* | 5.543211187 | 2.49E-07 | 8.60E-07 | 0.001563139 |
| Obesity | mast_cell | *CBR4* | 5.171532792 | 2.58E-05 | 5.24E-05 | 0.161985684 |
| Obesity | mast_cell | *NDUFS6* | 7.753841467 | 6.50E-06 | 1.49E-05 | 0.040844205 |
| Obesity | mast_cell | *FAM105A* | 3.895495033 | 2.42E-06 | 6.16E-06 | 0.015242745 |
| Obesity | mast_cell | *LHFPL2* | 4.813406687 | 1.99E-09 | 4.32E-08 | 1.25E-05 |

| | | | | | | |
|---|---|---|---|---|---|---|
| Obesity | mast_cell | *SSBP2* | 4.208326369 | 0.000120502 | 0.000216703 | 0.757593553 |
| Obesity | mast_cell | *BHMT* | 0.816541137 | 0.007079124 | 0.009781638 | 1 |
| Obesity | mast_cell | *CRHBP* | 3.400256293 | 0.000817426 | 0.001290597 | 1 |
| Obesity | mast_cell | *RASA1* | 2.153759758 | 0.018028926 | 0.023574846 | 1 |
| Obesity | mast_cell | *PPIP5K2* | 2.734935498 | 0.026097355 | 0.033429925 | 1 |
| Obesity | mast_cell | *GTF2H2* | 3.333159077 | 1.25E-07 | 4.86E-07 | 0.000783212 |
| Obesity | mast_cell | *SPATA9* | 3.588246776 | 1.10E-08 | 8.18E-08 | 6.91E-05 |
| Obesity | mast_cell | *TSLP* | -2.621185223 | 1.16E-05 | 2.56E-05 | 0.073088383 |
| Obesity | mast_cell | *FEM1C* | 7.223788645 | 1.64E-09 | 4.26E-08 | 1.03E-05 |
| Obesity | mast_cell | *YIPF5* | 6.116183387 | 7.09E-09 | 6.24E-08 | 4.46E-05 |
| Obesity | mast_cell | *GABRB2* | 1.667600832 | 1.59E-06 | 4.25E-06 | 0.009967945 |
| Obesity | mast_cell | *MYLK4* | 5.503683052 | 1.21E-08 | 8.62E-08 | 7.59E-05 |
| Obesity | mast_cell | *GFOD1* | 3.028878306 | 2.62E-06 | 6.60E-06 | 0.016451004 |
| Obesity | mast_cell | *PSD2* | -11.55293572 | 0.000677637 | 0.001081295 | 1 |
| Obesity | mast_cell | *ZMAT2* | 2.571919187 | 0.031824779 | 0.040249926 | 1 |
| Obesity | mast_cell | *KLHL3* | 3.696221716 | 0.002267904 | 0.003357267 | 1 |
| Obesity | mast_cell | *TRIM7* | 4.370543974 | 6.32E-08 | 2.79E-07 | 0.000397059 |
| Obesity | mast_cell | *HIGD2A* | 5.448346068 | 1.48E-06 | 4.01E-06 | 0.009324299 |
| Obesity | mast_cell | *FAM193B* | 6.222746659 | 2.03E-05 | 4.22E-05 | 0.127732455 |
| Obesity | mast_cell | *MUT* | 5.09445423 | 2.30E-09 | 4.32E-08 | 1.45E-05 |
| Obesity | mast_cell | *DOK3* | -2.598960877 | 2.06E-05 | 4.28E-05 | 0.129432289 |
| Obesity | mast_cell | *ABT1* | 2.350029803 | 0.034170245 | 0.0430604 | 1 |
| Obesity | mast_cell | *MLIP* | 4.231014855 | 1.04E-06 | 2.93E-06 | 0.006525624 |
| Obesity | mast_cell | *FGD2* | 3.704699858 | 4.09E-09 | 4.79E-08 | 2.57E-05 |
| Obesity | mast_cell | *ANO7* | 1.32764129 | 0.02929807 | 0.037211508 | 1 |
| Obesity | mast_cell | *RPL7L1* | 2.497478823 | 0.029203445 | 0.037113818 | 1 |
| Obesity | mast_cell | *TPBG* | 9.236335122 | 1.45E-08 | 9.62E-08 | 9.14E-05 |
| Obesity | mast_cell | *IRAK1BP1* | 7.389311725 | 1.64E-08 | 1.03E-07 | 0.000103243 |
| Obesity | mast_cell | *MMS22L* | 3.463265918 | 0.022121797 | 0.028617231 | 1 |
| Obesity | mast_cell | *PM20D2* | 8.731618356 | 2.69E-06 | 6.76E-06 | 0.016888863 |
| Obesity | mast_cell | *RNF217* | 5.496680481 | 8.08E-09 | 6.74E-08 | 5.08E-05 |
| Obesity | mast_cell | *ABRACL* | 3.594308162 | 0.004101009 | 0.00586378 | 1 |
| Obesity | mast_cell | *AIG1* | 5.881621789 | 1.20E-09 | 3.76E-08 | 7.56E-06 |
| Obesity | mast_cell | *PNLDC1* | 4.98172563 | 1.44E-06 | 3.91E-06 | 0.009069706 |
| Obesity | mast_cell | *WTAP* | 4.123631554 | 1.69E-05 | 3.58E-05 | 0.106139064 |
| Obesity | mast_cell | *SLC22A3* | 4.443844107 | 2.45E-09 | 4.32E-08 | 1.54E-05 |
| Obesity | mast_cell | *GNA12* | 4.739634001 | 1.43E-07 | 5.44E-07 | 0.000901628 |
| Obesity | mast_cell | *C7orf50* | 7.192460106 | 1.42E-08 | 9.48E-08 | 8.92E-05 |
| Obesity | mast_cell | *EGFR* | 3.645124479 | 4.87E-07 | 1.51E-06 | 0.003063095 |
| Obesity | mast_cell | *POMZP3* | 2.599592459 | 0.010363524 | 0.014027013 | 1 |
| Obesity | mast_cell | *CCT6A* | 7.110997348 | 3.77E-07 | 1.22E-06 | 0.002368603 |
| Obesity | mast_cell | *PSPH* | 2.484205071 | 4.20E-06 | 1.01E-05 | 0.026417154 |
| Obesity | mast_cell | *GIGYF1* | 6.869985339 | 4.75E-07 | 1.48E-06 | 0.002988887 |
| Obesity | mast_cell | *TMEM209* | 6.201275126 | 1.38E-09 | 3.97E-08 | 8.70E-06 |
| Obesity | mast_cell | *EPHA1* | -2.754829488 | 1.93E-05 | 4.03E-05 | 0.121338293 |
| Obesity | mast_cell | *NOM1* | 7.737520917 | 8.87E-09 | 7.12E-08 | 5.58E-05 |
| Obesity | mast_cell | *NCAPG2* | 3.151788448 | 0.025738233 | 0.033003523 | 1 |
| Obesity | mast_cell | *FBXO25* | 5.095995589 | 0.00352234 | 0.005086117 | 1 |
| Obesity | mast_cell | *CSGALNACT1* | 3.901979093 | 0.000478724 | 0.000783582 | 1 |
| Obesity | mast_cell | *CCDC25* | 4.594733667 | 8.86E-09 | 7.12E-08 | 5.57E-05 |

| | | | | | | |
|---|---|---|---|---|---|---|
| Obesity | mast_cell | *BIN3* | 5.418509183 | 2.54E-05 | 5.18E-05 | 0.159941171 |
| Obesity | mast_cell | *SLC25A37* | 7.032121769 | 5.43E-07 | 1.64E-06 | 0.003413768 |
| Obesity | mast_cell | *CHMP7* | 5.309525988 | 1.55E-05 | 3.32E-05 | 0.097415352 |
| Obesity | mast_cell | *DOCK5* | -2.485899637 | 0.004245009 | 0.006053157 | 1 |
| Obesity | mast_cell | *ST18* | 2.570828627 | 9.58E-08 | 3.93E-07 | 0.000602016 |
| Obesity | mast_cell | *TACC1* | 4.883984859 | 2.97E-06 | 7.37E-06 | 0.018655946 |
| Obesity | mast_cell | *SLC26A7* | 1.389952662 | 0.021207752 | 0.027502709 | 1 |
| Obesity | mast_cell | *MAL2* | 3.039416197 | 4.62E-09 | 5.08E-08 | 2.90E-05 |
| Obesity | mast_cell | *UTP23* | 6.875057341 | 1.65E-07 | 6.10E-07 | 0.001039155 |
| Obesity | mast_cell | *FAM135B* | -6.807592863 | 0.000214133 | 0.000369545 | 1 |
| Obesity | mast_cell | *ZNF7* | 5.102179793 | 0.000362907 | 0.000606324 | 1 |
| Obesity | mast_cell | *SLC39A4* | 5.296166616 | 1.51E-06 | 4.07E-06 | 0.009499356 |
| Obesity | mast_cell | *AK3* | 3.093383676 | 0.026928562 | 0.034403551 | 1 |
| Obesity | mast_cell | *CER1* | 5.421773953 | 3.32E-06 | 8.14E-06 | 0.020845396 |
| Obesity | mast_cell | *PLIN2* | -2.502446082 | 0.006287978 | 0.008750004 | 1 |
| Obesity | mast_cell | *HAUS6* | 7.976734144 | 1.97E-05 | 4.11E-05 | 0.123877316 |
| Obesity | mast_cell | *C9orf72* | 4.757482849 | 1.76E-07 | 6.43E-07 | 0.001109293 |
| Obesity | mast_cell | *CBWD5* | 5.163993608 | 2.53E-09 | 4.32E-08 | 1.59E-05 |
| Obesity | mast_cell | *ZNF462* | 5.376489536 | 3.28E-09 | 4.50E-08 | 2.06E-05 |
| Obesity | mast_cell | *ALAD* | 1.979478472 | 2.90E-05 | 5.84E-05 | 0.18238679 |
| Obesity | mast_cell | *ASTN2* | 7.065042154 | 4.22E-07 | 1.34E-06 | 0.002652179 |
| Obesity | mast_cell | *WDR31* | 10.6923429 | 1.36E-09 | 3.95E-08 | 8.57E-06 |
| Obesity | mast_cell | *POLE3* | -3.848584827 | 6.53E-05 | 0.000123128 | 0.410385816 |
| Obesity | mast_cell | *SURF1* | 3.081504374 | 0.000216647 | 0.000373679 | 1 |
| Obesity | mast_cell | *SURF6* | 9.537270145 | 1.01E-06 | 2.87E-06 | 0.006375526 |
| Obesity | mast_cell | *ASB6* | 6.714170747 | 2.11E-07 | 7.47E-07 | 0.001326701 |
| Obesity | mast_cell | *NTMT1* | 3.872788734 | 0.002928017 | 0.004272092 | 1 |
| Obesity | mast_cell | *CIZ1* | 7.301118941 | 3.44E-08 | 1.71E-07 | 0.000216546 |
| Obesity | mast_cell | *SH3GLB2* | 9.068972407 | 1.81E-10 | 1.26E-08 | 1.14E-06 |
| Obesity | mast_cell | *INPP5E* | 3.887085369 | 7.03E-08 | 3.03E-07 | 0.000442125 |
| Obesity | mast_cell | *DPH7* | 9.415464122 | 1.80E-07 | 6.57E-07 | 0.001133607 |
| Obesity | mast_cell | *SLC39A12* | 5.195859967 | 0.001994535 | 0.002971479 | 1 |
| Obesity | mast_cell | *RSU1* | 4.825250028 | 0.000322357 | 0.000541888 | 1 |
| Obesity | mast_cell | *PARD3* | 4.662848952 | 1.16E-08 | 8.42E-08 | 7.31E-05 |
| Obesity | mast_cell | *NRBF2* | -1.815806201 | 0.026529305 | 0.033932093 | 1 |
| Obesity | mast_cell | *GLUD1* | 5.902351564 | 6.56E-09 | 6.01E-08 | 4.13E-05 |
| Obesity | mast_cell | *HTR7* | 5.063560817 | 5.47E-09 | 5.54E-08 | 3.44E-05 |
| Obesity | mast_cell | *RPP30* | 5.63129106 | 0.000451288 | 0.000742929 | 1 |
| Obesity | mast_cell | *FRA10AC1* | 3.593551717 | 1.00E-07 | 4.09E-07 | 0.000631348 |
| Obesity | mast_cell | *DNAJB12* | 4.73536835 | 0.009351903 | 0.012729035 | 1 |
| Obesity | mast_cell | *PLEKHS1* | 8.397081801 | 6.56E-05 | 0.000123634 | 0.412442321 |
| Obesity | mast_cell | *FUOM* | -4.763106429 | 1.09E-05 | 2.41E-05 | 0.068464917 |
| Obesity | mast_cell | *LRRC27* | 1.678777695 | 0.000118653 | 0.000213562 | 0.745972866 |
| Obesity | mast_cell | *MTG1* | 1.784834882 | 0.010329101 | 0.013986444 | 1 |
| Obesity | mast_cell | *PAOX* | 8.905794194 | 0.003395603 | 0.004916664 | 1 |
| Obesity | mast_cell | *GSTO1* | 7.019747977 | 9.41E-07 | 2.68E-06 | 0.005913826 |
| Obesity | mast_cell | *PDCD11* | 4.429039484 | 0.00048351 | 0.000791003 | 1 |
| Obesity | mast_cell | *LIN7C* | 4.433795054 | 3.11E-09 | 4.48E-08 | 1.96E-05 |
| Obesity | mast_cell | *ZNF215* | 1.209190685 | 0.003266368 | 0.004736082 | 1 |
| Obesity | mast_cell | *HSD17B12* | -2.373009942 | 6.92E-06 | 1.59E-05 | 0.043528045 |

| | | | | | | |
|---|---|---|---|---|---|---|
| Obesity | mast_cell | *APIP* | 0.633752815 | 0.018427875 | 0.024076485 | 1 |
| Obesity | mast_cell | *EIF3M* | 5.277440867 | 2.28E-08 | 1.26E-07 | 0.000143045 |
| Obesity | mast_cell | *GLYAT* | 2.010149964 | 2.20E-09 | 4.32E-08 | 1.38E-05 |
| Obesity | mast_cell | *PTPRJ* | 4.626077893 | 1.41E-07 | 5.36E-07 | 0.000883916 |
| Obesity | mast_cell | *ENDOD1* | 7.892561254 | 0.000664496 | 0.001063025 | 1 |
| Obesity | mast_cell | *CCDC82* | 1.528496933 | 0.019943857 | 0.025954674 | 1 |
| Obesity | mast_cell | *CAPN5* | 7.802367922 | 2.98E-05 | 5.98E-05 | 0.187073352 |
| Obesity | mast_cell | *INTS4* | 4.515120512 | 4.13E-05 | 8.10E-05 | 0.259938076 |
| Obesity | mast_cell | *ZC3H12C* | 4.748933824 | 1.39E-09 | 3.97E-08 | 8.73E-06 |
| Obesity | mast_cell | *TTC12* | 3.038360921 | 1.21E-10 | 9.63E-09 | 7.61E-07 |
| Obesity | mast_cell | *DRD2* | -0.881728099 | 1.67E-07 | 6.14E-07 | 0.00104873 |
| Obesity | mast_cell | *C11orf52* | 17.90254273 | 1.55E-08 | 1.00E-07 | 9.76E-05 |
| Obesity | mast_cell | *GLB1L2* | -5.205438757 | 0.000147571 | 0.000261715 | 0.927779864 |
| Obesity | mast_cell | *P4HA3* | 6.019847164 | 6.41E-08 | 2.81E-07 | 0.0004032 |
| Obesity | mast_cell | *GRIK4* | 1.459659968 | 0.025988272 | 0.033296977 | 1 |
| Obesity | mast_cell | *ADAM33* | 3.710213238 | 1.33E-07 | 5.15E-07 | 0.000838431 |
| Obesity | mast_cell | *MTA2* | 4.986492046 | 0.000407589 | 0.000676481 | 1 |
| Obesity | mast_cell | *TMC2* | 17.32539337 | 9.64E-09 | 7.53E-08 | 6.06E-05 |
| Obesity | mast_cell | *ROM1* | -1.089682829 | 0.022081639 | 0.028571159 | 1 |
| Obesity | mast_cell | *EML3* | 5.733638114 | 4.64E-07 | 1.45E-06 | 0.002915461 |
| Obesity | mast_cell | *INCENP* | 7.844470346 | 2.35E-09 | 4.32E-08 | 1.48E-05 |
| Obesity | mast_cell | *MS4A3* | -6.746920751 | 1.56E-07 | 5.83E-07 | 0.000981656 |
| Obesity | mast_cell | *PLCH2* | 8.224254554 | 8.30E-09 | 6.85E-08 | 5.22E-05 |
| Obesity | mast_cell | *CPSF7* | 5.170497077 | 0.001558796 | 0.002359208 | 1 |
| Obesity | mast_cell | *CHEK1* | 1.62968011 | 0.035599392 | 0.04475748 | 1 |
| Obesity | mast_cell | *FEZ1* | 3.232891651 | 1.16E-08 | 8.42E-08 | 7.30E-05 |
| Obesity | mast_cell | *DUSP15* | -5.224180894 | 1.31E-08 | 9.03E-08 | 8.27E-05 |
| Obesity | mast_cell | *COMMD7* | 7.53209033 | 8.05E-05 | 0.000148943 | 0.505959422 |
| Obesity | mast_cell | *KIAA1755* | 2.112091651 | 5.27E-08 | 2.41E-07 | 0.000331137 |
| Obesity | mast_cell | *CNBD2* | 7.192149025 | 1.33E-09 | 3.93E-08 | 8.36E-06 |
| Obesity | mast_cell | *CABLES2* | 2.288932123 | 6.08E-05 | 0.000115377 | 0.382358018 |
| Obesity | mast_cell | *TRPT1* | 2.91656428 | 0.036063365 | 0.04528268 | 1 |
| Obesity | mast_cell | *TBX6* | 4.585681397 | 3.14E-13 | 7.90E-11 | 1.97E-09 |
| Obesity | mast_cell | *DOC2A* | -4.123222177 | 0.002303225 | 0.003407148 | 1 |
| Obesity | mast_cell | *TAOK2* | 6.819453612 | 4.02E-09 | 4.78E-08 | 2.53E-05 |
| Obesity | mast_cell | *MPP7* | 1.764404866 | 0.001111451 | 0.001716967 | 1 |
| Obesity | mast_cell | *CWC15* | 4.57057185 | 0.000242953 | 0.000415857 | 1 |
| Obesity | mast_cell | *ARID5B* | 5.161399816 | 1.88E-07 | 6.82E-07 | 0.001184383 |
| Obesity | mast_cell | *TMCO3* | 7.387919664 | 5.99E-08 | 2.68E-07 | 0.00037688 |
| Obesity | mast_cell | *LYPD6B* | 3.761964309 | 3.76E-08 | 1.84E-07 | 0.000236331 |
| Obesity | mast_cell | *PDCD4* | 2.892426084 | 0.031846209 | 0.040268929 | 1 |
| Obesity | mast_cell | *WDR17* | -3.563755562 | 0.005069799 | 0.007156225 | 1 |
| Obesity | mast_cell | *CCDC102B* | 5.488220591 | 1.16E-08 | 8.42E-08 | 7.28E-05 |
| Obesity | mast_cell | *CNDP1* | 2.575098234 | 0.001657933 | 0.002496031 | 1 |
| Obesity | mast_cell | *FSIP1* | -0.881992774 | 2.82E-07 | 9.54E-07 | 0.001771333 |
| Obesity | mast_cell | *RGS18* | 4.219827772 | 2.76E-09 | 4.32E-08 | 1.74E-05 |
| Obesity | mast_cell | *PRSS23* | 3.950014375 | 2.67E-07 | 9.13E-07 | 0.001680081 |
| Obesity | mast_cell | *MTMR12* | 5.32315315 | 1.33E-08 | 9.08E-08 | 8.35E-05 |
| Obesity | mast_cell | *PPP1R1C* | 3.12744203 | 9.38E-06 | 2.10E-05 | 0.058954017 |
| Obesity | mast_cell | *C11orf53* | 16.99729559 | 8.68E-08 | 3.62E-07 | 0.000545591 |

| | | | | | | |
|---|---|---|---|---|---|---|
| Obesity | mast_cell | *CCT5* | 5.764086157 | 0.000317026 | 0.000534211 | 1 |
| Obesity | mast_cell | *DOCK1* | 4.371737216 | 4.60E-07 | 1.44E-06 | 0.002889876 |
| Obesity | mast_cell | *TEX12* | -5.657783433 | 2.64E-07 | 9.04E-07 | 0.001659778 |
| Obesity | mast_cell | *CRIM1* | 3.694482536 | 0.001044864 | 0.001620789 | 1 |
| Obesity | mast_cell | *ABCB9* | 4.878128796 | 1.81E-07 | 6.57E-07 | 0.00113548 |
| Obesity | mast_cell | *DHX37* | 10.05493731 | 1.14E-08 | 8.37E-08 | 7.20E-05 |
| Obesity | mast_cell | *GPR158* | 2.874720723 | 2.50E-06 | 6.33E-06 | 0.015692946 |
| Obesity | mast_cell | *THRB* | 4.312248184 | 7.42E-05 | 0.000138302 | 0.466354405 |
| Obesity | mast_cell | *BTBD11* | 5.958552577 | 2.38E-09 | 4.32E-08 | 1.50E-05 |
| Obesity | mast_cell | *UBE3B* | 4.865933454 | 6.89E-09 | 6.15E-08 | 4.33E-05 |
| Obesity | mast_cell | *ANK3* | 3.038240748 | 3.91E-09 | 4.78E-08 | 2.46E-05 |
| Obesity | mast_cell | *IPMK* | 7.064365425 | 5.04E-08 | 2.33E-07 | 0.000316557 |
| Obesity | mast_cell | *DLG5* | 2.178399883 | 0.013709531 | 0.018218521 | 1 |
| Obesity | mast_cell | *FAM177A1* | 5.196899735 | 0.000899964 | 0.001412047 | 1 |
| Obesity | mast_cell | *MIPOL1* | 5.178063501 | 3.07E-10 | 1.68E-08 | 1.93E-06 |
| Obesity | mast_cell | *TMEM18* | 5.165914792 | 1.53E-09 | 4.16E-08 | 9.63E-06 |
| Obesity | mast_cell | *THRSP* | 2.505284619 | 0.000218567 | 0.000376681 | 1 |
| Obesity | mast_cell | *NDUFC2* | 7.276022523 | 1.89E-05 | 3.96E-05 | 0.118888937 |
| Obesity | mast_cell | *ME3* | 4.862056161 | 3.11E-08 | 1.59E-07 | 0.00019571 |
| Obesity | mast_cell | *ADAMTS12* | 4.120130977 | 4.63E-08 | 2.18E-07 | 0.000290894 |
| Obesity | mast_cell | *VIPAS39* | 6.123366466 | 1.71E-07 | 6.28E-07 | 0.001075909 |
| Obesity | mast_cell | *ANKRD50* | 7.833160339 | 9.13E-07 | 2.61E-06 | 0.005741355 |
| Obesity | mast_cell | *C4orf33* | -2.717697778 | 0.016912833 | 0.022207807 | 1 |
| Obesity | mast_cell | *PTPRO* | 6.319459568 | 4.53E-09 | 5.03E-08 | 2.85E-05 |
| Obesity | mast_cell | *VPS26B* | 8.185508262 | 1.14E-06 | 3.19E-06 | 0.007178056 |
| Obesity | mast_cell | *NCAPD3* | 6.349332643 | 2.61E-08 | 1.39E-07 | 0.000163845 |
| Obesity | mast_cell | *VTI1A* | 3.489966279 | 0.001579782 | 0.002388095 | 1 |
| Obesity | mast_cell | *FAM160B1* | 5.12991703 | 2.72E-09 | 4.32E-08 | 1.71E-05 |
| Obesity | mast_cell | *ANO4* | 2.53912661 | 4.16E-09 | 4.81E-08 | 2.61E-05 |
| Obesity | mast_cell | *TEX9* | 7.399808149 | 4.53E-07 | 1.43E-06 | 0.002848819 |
| Obesity | mast_cell | *AKR1C2* | 3.89203497 | 9.43E-09 | 7.41E-08 | 5.93E-05 |
| Obesity | mast_cell | *DPYSL4* | 1.920245598 | 1.61E-05 | 3.43E-05 | 0.101315223 |
| Obesity | mast_cell | *ADAM8* | 3.403610585 | 2.35E-09 | 4.32E-08 | 1.48E-05 |
| Obesity | mast_cell | *ITIH2* | 4.161210101 | 2.23E-08 | 1.24E-07 | 0.000140341 |
| Obesity | mast_cell | *KIN* | 5.356547633 | 5.36E-07 | 1.63E-06 | 0.003368265 |
| Obesity | mast_cell | *PIGF* | 6.326109506 | 9.44E-07 | 2.69E-06 | 0.005932443 |
| Obesity | mast_cell | *INPP1* | 5.689339674 | 5.75E-09 | 5.62E-08 | 3.62E-05 |
| Obesity | mast_cell | *RNF144A* | 5.56348628 | 1.03E-07 | 4.17E-07 | 0.000647909 |
| Obesity | mast_cell | *ASAP2* | 5.62933336 | 6.56E-08 | 2.86E-07 | 0.000412669 |
| Obesity | mast_cell | *ADAM17* | 5.913541708 | 4.20E-06 | 1.01E-05 | 0.026428956 |
| Obesity | mast_cell | *TMEM45B* | 8.947616678 | 0.004700612 | 0.006654525 | 1 |
| Obesity | mast_cell | *BICD1* | 5.827698614 | 1.35E-06 | 3.68E-06 | 0.008459723 |
| Obesity | mast_cell | *GABRA2* | -1.967904231 | 7.38E-05 | 0.00013777 | 0.464284599 |
| Obesity | mast_cell | *CENPJ* | 3.720092568 | 0.007662356 | 0.010555047 | 1 |
| Obesity | mast_cell | *PARP8* | 2.982150765 | 5.89E-05 | 0.000112044 | 0.370082703 |
| Obesity | mast_cell | *CACUL1* | 5.215324346 | 0.00014234 | 0.000253224 | 0.894893174 |
| Obesity | mast_cell | *RABGAP1L* | 4.107017982 | 0.000124217 | 0.000222916 | 0.780949782 |
| Obesity | mast_cell | *PTPN14* | 4.661415053 | 2.74E-09 | 4.32E-08 | 1.72E-05 |
| Obesity | mast_cell | *RNF219* | 7.385255777 | 4.90E-07 | 1.51E-06 | 0.003079362 |
| Obesity | mast_cell | *ARL14EP* | 3.126607856 | 0.004383565 | 0.006236586 | 1 |

| | | | | | | |
|---|---|---|---|---|---|---|
| Obesity | mast_cell | *G6PC2* | -75.83676957 | 8.95E-07 | 2.57E-06 | 0.005623986 |
| Obesity | mast_cell | *TADA1* | 5.472574633 | 2.40E-09 | 4.32E-08 | 1.51E-05 |
| Obesity | mast_cell | *GUCY1A2* | 3.479488144 | 2.33E-06 | 5.96E-06 | 0.014674184 |
| Obesity | mast_cell | *CWF19L2* | 5.996763697 | 1.70E-06 | 4.50E-06 | 0.010662858 |
| Obesity | mast_cell | *ZNF837* | 2.050626157 | 0.002780626 | 0.00406648 | 1 |
| Obesity | mast_cell | *CCDC50* | 2.393651454 | 0.00237922 | 0.003513778 | 1 |
| Obesity | mast_cell | *ZFP36L2* | 5.043803446 | 0.000124469 | 0.000223263 | 0.782537164 |
| Obesity | mast_cell | *TMEM123* | 7.129133421 | 4.74E-08 | 2.23E-07 | 0.000298281 |
| Obesity | mast_cell | *IGSF10* | 2.65764357 | 0.035685686 | 0.044826729 | 1 |
| Obesity | mast_cell | *SPEF2* | 4.306380658 | 1.39E-07 | 5.31E-07 | 0.000871807 |
| Obesity | mast_cell | *SPARCL1* | 2.736564551 | 5.39E-09 | 5.52E-08 | 3.39E-05 |
| Obesity | mast_cell | *MBNL1* | 4.010654666 | 7.76E-06 | 1.76E-05 | 0.048818507 |
| Obesity | mast_cell | *CCNO* | 8.990012824 | 0.011133905 | 0.014995471 | 1 |
| Obesity | mast_cell | *CLEC4F* | -6.458107213 | 2.17E-06 | 5.57E-06 | 0.013613198 |
| Obesity | mast_cell | *PELO* | 5.718592915 | 3.52E-09 | 4.62E-08 | 2.21E-05 |
| Obesity | mast_cell | *RASGRP3* | 2.874787067 | 8.91E-06 | 2.00E-05 | 0.056008404 |
| Obesity | mast_cell | *SAR1B* | 5.756841886 | 7.84E-08 | 3.30E-07 | 0.0004927 |
| Obesity | mast_cell | *GPR180* | 2.993243411 | 0.000246005 | 0.000420968 | 1 |
| Obesity | mast_cell | *TCTEX1D1* | -1.030278983 | 0.000297505 | 0.000502664 | 1 |
| Obesity | mast_cell | *WDR78* | 3.245466204 | 0.012662364 | 0.016919932 | 1 |
| Obesity | mast_cell | *PRDM8* | 1.968623575 | 1.60E-05 | 3.41E-05 | 0.1004744 |
| Obesity | mast_cell | *HHEX* | 5.333682842 | 1.96E-05 | 4.09E-05 | 0.12306811 |
| Obesity | mast_cell | *ZNF117* | 1.208958303 | 3.65E-05 | 7.21E-05 | 0.229438941 |
| Obesity | mast_cell | *MED21* | 8.167622428 | 3.13E-07 | 1.04E-06 | 0.001964909 |
| Obesity | mast_cell | *SRP19* | 8.513778294 | 4.92E-07 | 1.52E-06 | 0.003096108 |
| Obesity | mast_cell | *TEKT5* | 5.797507568 | 1.31E-08 | 9.03E-08 | 8.27E-05 |
| Obesity | mast_cell | *BANK1* | 3.066096174 | 3.25E-09 | 4.49E-08 | 2.04E-05 |
| Obesity | mast_cell | *SCOC* | 4.84313386 | 2.40E-07 | 8.34E-07 | 0.00150878 |
| Obesity | mast_cell | *CLGN* | -2.450492468 | 2.02E-07 | 7.20E-07 | 0.001270691 |
| Obesity | mast_cell | *SYCP2L* | -0.991589737 | 3.39E-07 | 1.12E-06 | 0.002132301 |
| Obesity | mast_cell | *AHCTF1* | 5.863638869 | 3.63E-09 | 4.63E-08 | 2.28E-05 |
| Obesity | mast_cell | *MERTK* | 0.6634804 | 0.00347722 | 0.005024427 | 1 |
| Obesity | mast_cell | *PTPRR* | 5.809921532 | 3.43E-09 | 4.56E-08 | 2.16E-05 |
| Obesity | mast_cell | *PLA2R1* | 4.327033025 | 6.28E-08 | 2.77E-07 | 0.000394716 |
| Obesity | mast_cell | *SLC25A27* | 1.639095499 | 0.001547369 | 0.002343607 | 1 |
| Obesity | mast_cell | *FRMD1* | 2.632981668 | 1.72E-08 | 1.05E-07 | 0.000108303 |
| Obesity | mast_cell | *ASAP1* | 4.62897725 | 2.11E-06 | 5.43E-06 | 0.013247171 |
| Obesity | mast_cell | *INO80C* | -3.43556423 | 0.006193944 | 0.00862106 | 1 |
| Obesity | mast_cell | *LPCAT1* | 5.802214983 | 1.83E-09 | 4.32E-08 | 1.15E-05 |
| Obesity | mast_cell | *TEX29* | 5.604778995 | 1.37E-07 | 5.25E-07 | 0.000860219 |
| Obesity | mast_cell | *ADPRHL1* | 2.371648506 | 4.96E-05 | 9.60E-05 | 0.311677244 |
| Obesity | mast_cell | *TUBGCP5* | 6.536145852 | 5.61E-09 | 5.57E-08 | 3.53E-05 |
| Obesity | mast_cell | *LURAP1L* | 2.410913678 | 2.70E-05 | 5.49E-05 | 0.170042366 |
| Obesity | mast_cell | *CFDP1* | 5.921465398 | 6.40E-08 | 2.81E-07 | 0.000402104 |
| Obesity | mast_cell | *ZDHHC7* | 6.501278136 | 3.16E-08 | 1.60E-07 | 0.000198618 |
| Obesity | mast_cell | *FAM92B* | -3.742680391 | 1.51E-07 | 5.67E-07 | 0.00094696 |
| Obesity | mast_cell | *C7orf31* | 5.435469051 | 1.79E-08 | 1.07E-07 | 0.000112482 |
| Obesity | mast_cell | *JAZF1* | 4.454104898 | 0.000101155 | 0.000183751 | 0.635962527 |
| Obesity | mast_cell | *FBXO36* | 6.13560991 | 2.75E-07 | 9.36E-07 | 0.001731841 |
| Obesity | mast_cell | *CACNA2D1* | 4.629702381 | 2.49E-07 | 8.60E-07 | 0.001565286 |

| | | | | | | |
|---|---|---|---|---|---|---|
| Obesity | mast_cell | *ZUFSP* | 7.513793011 | 1.02E-06 | 2.89E-06 | 0.006415729 |
| Obesity | mast_cell | *HS3ST3A1* | 2.956355566 | 2.97E-07 | 9.98E-07 | 0.001868356 |
| Obesity | mast_cell | *PPP2R5E* | 4.945865904 | 1.91E-06 | 4.97E-06 | 0.011984681 |
| Obesity | mast_cell | *ANKRD29* | 2.561655622 | 2.59E-08 | 1.38E-07 | 0.000163055 |
| Obesity | mast_cell | *THY1* | 3.946135111 | 2.43E-08 | 1.32E-07 | 0.000152736 |
| Obesity | mast_cell | *UBASH3B* | 7.77694043 | 3.38E-07 | 1.11E-06 | 0.002126614 |
| Obesity | mast_cell | *ROBO4* | 4.425180883 | 1.15E-05 | 2.54E-05 | 0.072488956 |
| Obesity | mast_cell | *ROBO3* | 4.455612721 | 4.23E-09 | 4.85E-08 | 2.66E-05 |
| Obesity | mast_cell | *CC2D1B* | 4.079418375 | 1.21E-08 | 8.63E-08 | 7.64E-05 |
| Obesity | mast_cell | *PRKCA* | -1.531183721 | 1.78E-05 | 3.75E-05 | 0.111790172 |
| Obesity | mast_cell | *LRRK1* | 6.724267249 | 6.82E-08 | 2.95E-07 | 0.000428607 |
| Obesity | mast_cell | *CEP112* | 4.639471299 | 5.49E-11 | 5.48E-09 | 3.45E-07 |
| Obesity | mast_cell | *GAL3ST2* | 9.677424833 | 0.00843612 | 0.011547547 | 1 |
| Obesity | mast_cell | *ENPP3* | -0.419511104 | 1.83E-07 | 6.64E-07 | 0.001147715 |
| Obesity | mast_cell | *UCHL1* | 10.45210745 | 1.56E-07 | 5.84E-07 | 0.000983875 |
| Obesity | mast_cell | *MIA3* | 5.729405107 | 2.32E-05 | 4.76E-05 | 0.145625821 |
| Obesity | mast_cell | *DISP1* | 2.646600299 | 0.001482915 | 0.002250867 | 1 |
| Obesity | mast_cell | *FAM167A* | 10.5629832 | 4.19E-08 | 2.01E-07 | 0.000263711 |
| Obesity | mast_cell | *NEIL2* | 3.383805221 | 3.19E-11 | 3.52E-09 | 2.01E-07 |
| Obesity | mast_cell | *CCSAP* | 5.275459396 | 3.98E-07 | 1.28E-06 | 0.002500304 |
| Obesity | mast_cell | *FAM69A* | 3.281407103 | 2.94E-06 | 7.31E-06 | 0.018475783 |
| Obesity | mast_cell | *ATP5G3* | 3.688393241 | 0.000497671 | 0.000812057 | 1 |
| Obesity | mast_cell | *PDLIM3* | 2.94183346 | 2.12E-09 | 4.32E-08 | 1.33E-05 |
| Obesity | mast_cell | *NCAM2* | 3.298236186 | 1.51E-06 | 4.08E-06 | 0.009509249 |
| Obesity | mast_cell | *L3MBTL4* | 5.50033971 | 8.16E-09 | 6.78E-08 | 5.13E-05 |
| Obesity | mast_cell | *PDE1C* | 3.260414751 | 7.81E-10 | 2.96E-08 | 4.91E-06 |
| Obesity | mast_cell | *MRPL39* | 1.435955859 | 0.038770554 | 0.048478615 | 1 |
| Obesity | mast_cell | *FLCN* | 5.514659522 | 1.34E-06 | 3.66E-06 | 0.008406722 |
| Obesity | mast_cell | *DPH3* | 6.270453932 | 2.43E-09 | 4.32E-08 | 1.53E-05 |
| Obesity | mast_cell | *CXXC1* | 9.233353181 | 4.05E-07 | 1.30E-06 | 0.00254623 |
| Obesity | mast_cell | *PPP4R1* | 6.583104589 | 2.34E-06 | 5.97E-06 | 0.014708956 |
| Obesity | mast_cell | *APCDD1* | 1.893886221 | 1.21E-05 | 2.64E-05 | 0.075803971 |
| Obesity | mast_cell | *MPPE1* | 1.733310871 | 0.01062021 | 0.014355894 | 1 |
| Obesity | mast_cell | *EPHB1* | 6.260852732 | 0.000166482 | 0.000292694 | 1 |
| Obesity | mast_cell | *ANKRD40* | 5.99952508 | 3.29E-07 | 1.09E-06 | 0.002069079 |
| Obesity | mast_cell | *CYP2U1* | 8.057626819 | 3.22E-09 | 4.49E-08 | 2.02E-05 |
| Obesity | mast_cell | *FBXL18* | 8.096217361 | 2.69E-05 | 5.45E-05 | 0.168851609 |
| Obesity | mast_cell | *PROM2* | 8.384885936 | 3.44E-06 | 8.41E-06 | 0.021610456 |
| Obesity | mast_cell | *AK9* | 4.155925516 | 0.001612981 | 0.002433601 | 1 |
| Obesity | mast_cell | *CDK19* | 4.922976279 | 0.00017357 | 0.000304304 | 1 |
| Obesity | mast_cell | *GTF3C6* | 5.621726452 | 1.82E-08 | 1.08E-07 | 0.000114311 |
| Obesity | mast_cell | *TTC39B* | -2.398961055 | 5.68E-05 | 0.000108515 | 0.357122142 |
| Obesity | mast_cell | *AGPAT5* | -2.314565301 | 0.001947935 | 0.00290644 | 1 |
| Obesity | mast_cell | *GOLGA7B* | 4.454266509 | 1.73E-06 | 4.57E-06 | 0.010865941 |
| Obesity | mast_cell | *TRMT44* | 8.427750455 | 2.64E-09 | 4.32E-08 | 1.66E-05 |
| Obesity | mast_cell | *C16orf87* | 4.652428309 | 1.56E-08 | 1.00E-07 | 9.81E-05 |
| Obesity | mast_cell | *HEATR3* | 1.999871228 | 0.000516345 | 0.000840782 | 1 |
| Obesity | mast_cell | *CNOT8* | 5.426511449 | 9.09E-06 | 2.04E-05 | 0.057177855 |
| Obesity | mast_cell | *ZKSCAN2* | 2.679832385 | 0.00028018 | 0.000475051 | 1 |
| Obesity | mast_cell | *PDIA4* | 8.475297695 | 6.71E-09 | 6.06E-08 | 4.22E-05 |

| | | | | | | |
|---|---|---|---|---|---|---|
| Obesity | mast_cell | *FAM126B* | 5.860986842 | 3.15E-07 | 1.05E-06 | 0.001983498 |
| Obesity | mast_cell | *ALS2CR12* | 5.790201662 | 2.11E-08 | 1.19E-07 | 0.000132797 |
| Obesity | mast_cell | *SPAG17* | 5.175360091 | 4.11E-08 | 1.98E-07 | 0.000258532 |
| Obesity | mast_cell | *DEPTOR* | 7.30915127 | 1.81E-08 | 1.07E-07 | 0.000113662 |
| Obesity | mast_cell | *RRAGA* | 8.677217654 | 4.10E-07 | 1.31E-06 | 0.002574879 |
| Obesity | mast_cell | *FAM161B* | 3.841107902 | 2.50E-09 | 4.32E-08 | 1.57E-05 |
| Obesity | mast_cell | *WIF1* | 8.021193776 | 0.000196705 | 0.000341712 | 1 |
| Obesity | mast_cell | *UGT2B4* | 3.532473457 | 1.72E-08 | 1.05E-07 | 0.000107848 |
| Obesity | mast_cell | *MMP16* | 2.532355197 | 1.79E-05 | 3.76E-05 | 0.112434361 |
| Obesity | mast_cell | *KCNMA1* | -1.889260376 | 0.001845497 | 0.002759249 | 1 |
| Obesity | mast_cell | *BATF* | -0.95796754 | 2.57E-07 | 8.83E-07 | 0.001617179 |
| Obesity | mast_cell | *DCK* | 6.60254746 | 1.74E-06 | 4.59E-06 | 0.010926732 |
| Obesity | mast_cell | *DPY19L4* | 4.969714744 | 0.000386846 | 0.000644606 | 1 |
| Obesity | mast_cell | *NDUFAF6* | 4.304530213 | 0.000166949 | 0.000293351 | 1 |
| Obesity | mast_cell | *DRAM2* | 4.987403791 | 3.57E-06 | 8.69E-06 | 0.022432369 |
| Obesity | mast_cell | *C8orf37* | 3.483887477 | 6.36E-05 | 0.000120206 | 0.399803865 |
| Obesity | mast_cell | *ADAMTSL3* | 1.415279991 | 0.027713143 | 0.035369982 | 1 |
| Obesity | mast_cell | *CXCL13* | -6.044904172 | 7.24E-05 | 0.000135267 | 0.454901275 |
| Obesity | mast_cell | *N6AMT1* | 1.72117681 | 0.001290927 | 0.001975672 | 1 |
| Obesity | mast_cell | *RWDD2B* | -1.834934618 | 0.031120245 | 0.039430266 | 1 |
| Obesity | mast_cell | *ANKRD9* | 5.992580149 | 8.76E-07 | 2.52E-06 | 0.005509278 |
| Obesity | mast_cell | *SFXN2* | 4.85283155 | 3.18E-05 | 6.34E-05 | 0.199819005 |
| Obesity | mast_cell | *C14orf2* | 2.642209695 | 0.006185625 | 0.008611387 | 1 |
| Obesity | mast_cell | *GDF6* | 5.522542936 | 1.03E-07 | 4.16E-07 | 0.000645136 |
| Obesity | mast_cell | *NODAL* | 9.695991707 | 2.55E-09 | 4.32E-08 | 1.61E-05 |
| Obesity | mast_cell | *WDYHV1* | 3.834999248 | 4.83E-06 | 1.14E-05 | 0.030348773 |
| Obesity | mast_cell | *ATAD2* | 6.727115812 | 4.75E-09 | 5.10E-08 | 2.98E-05 |
| Obesity | mast_cell | *ITGAD* | -2.173136544 | 4.10E-05 | 8.02E-05 | 0.2574884 |
| Obesity | mast_cell | *VPS8* | 4.117858751 | 0.000147038 | 0.000260917 | 0.924428567 |
| Obesity | mast_cell | *RPUSD3* | 9.856755489 | 8.35E-06 | 1.88E-05 | 0.052519029 |
| Obesity | mast_cell | *GHRL* | 9.5660913 | 7.50E-09 | 6.45E-08 | 4.72E-05 |
| Obesity | mast_cell | *EXOG* | 6.135884245 | 2.69E-07 | 9.17E-07 | 0.001692301 |
| Obesity | mast_cell | *NTAN1* | 5.059865164 | 6.45E-09 | 5.96E-08 | 4.06E-05 |
| Obesity | mast_cell | *FCHO2* | 5.650265088 | 3.18E-07 | 1.06E-06 | 0.002000163 |
| Obesity | mast_cell | *TIMP4* | -3.735467368 | 0.013635819 | 0.018132063 | 1 |
| Obesity | mast_cell | *CPT2* | 7.35350391 | 0.001785987 | 0.002677879 | 1 |
| Obesity | mast_cell | *NECAP2* | 7.416724721 | 6.00E-09 | 5.74E-08 | 3.77E-05 |
| Obesity | mast_cell | *LRP8* | 7.41566241 | 3.79E-08 | 1.85E-07 | 0.000238074 |
| Obesity | mast_cell | *GATAD1* | 4.191849013 | 4.85E-06 | 1.14E-05 | 0.030499884 |
| Obesity | mast_cell | *TMED6* | 34.29628682 | 2.77E-09 | 4.32E-08 | 1.74E-05 |
| Obesity | mast_cell | *CLEC18A* | -7.031119673 | 9.88E-21 | 8.87E-18 | 6.21E-17 |
| Obesity | mast_cell | *IL34* | 3.509780906 | 2.14E-08 | 1.20E-07 | 0.00013428 |
| Obesity | mast_cell | *AASDH* | 3.660245568 | 0.004120315 | 0.005890046 | 1 |
| Obesity | mast_cell | *RNF111* | 5.142030824 | 6.14E-08 | 2.73E-07 | 0.000386081 |
| Obesity | mast_cell | *DYRK1A* | 4.731900657 | 5.88E-05 | 0.000111956 | 0.369568056 |
| Obesity | mast_cell | *CREB3L1* | 3.200044703 | 0.018580363 | 0.02426059 | 1 |
| Obesity | mast_cell | *C2CD2* | 1.926563669 | 9.54E-05 | 0.000174139 | 0.60008374 |
| Obesity | mast_cell | *SLC38A10* | 5.866387008 | 9.75E-09 | 7.58E-08 | 6.13E-05 |
| Obesity | mast_cell | *C9orf43* | 3.765790635 | 1.88E-05 | 3.95E-05 | 0.118353004 |
| Obesity | mast_cell | *WDR19* | 3.635622377 | 0.022298583 | 0.028828129 | 1 |

| | | | | | | |
|---|---|---|---|---|---|---|
| Obesity | mast_cell | *SLC37A3* | 6.432354744 | 1.60E-08 | 1.02E-07 | 0.00010048 |
| Obesity | mast_cell | *SPPL3* | 4.568395744 | 8.19E-05 | 0.000151298 | 0.515170536 |
| Obesity | mast_cell | *FAM213B* | -2.92733093 | 3.05E-05 | 6.11E-05 | 0.191503802 |
| Obesity | mast_cell | *C12orf43* | 5.386600785 | 4.22E-06 | 1.01E-05 | 0.026511232 |
| Obesity | mast_cell | *PEX10* | -3.242644111 | 0.001702437 | 0.002558131 | 1 |
| Obesity | mast_cell | *TRIM63* | -0.338429412 | 2.26E-07 | 7.92E-07 | 0.001420726 |
| Obesity | mast_cell | *WDR66* | -2.167287241 | 0.001209585 | 0.00186024 | 1 |
| Obesity | mast_cell | *GALNT14* | -1.519069079 | 3.00E-07 | 1.00E-06 | 0.001884355 |
| Obesity | mast_cell | *HPD* | 5.658833037 | 6.12E-10 | 2.62E-08 | 3.85E-06 |
| Obesity | mast_cell | *RHPN1* | 2.320298479 | 0.008310686 | 0.011378329 | 1 |
| Obesity | mast_cell | *TPRG1L* | 8.485822962 | 0.00027835 | 0.000472331 | 1 |
| Obesity | mast_cell | *LRRC43* | 6.103372418 | 1.64E-08 | 1.03E-07 | 0.00010305 |
| Obesity | mast_cell | *EYA3* | 5.089762499 | 3.07E-05 | 6.15E-05 | 0.192946285 |
| Obesity | mast_cell | *MRAS* | 2.449100538 | 1.62E-06 | 4.33E-06 | 0.010205972 |
| Obesity | mast_cell | *ABHD3* | 2.556910068 | 0.012398838 | 0.016574844 | 1 |
| Obesity | mast_cell | *CLSTN2* | 3.012214086 | 3.97E-08 | 1.93E-07 | 0.000249724 |
| Obesity | mast_cell | *COLEC12* | 3.357456484 | 2.48E-09 | 4.32E-08 | 1.56E-05 |
| Obesity | mast_cell | *RNF207* | 13.56925728 | 4.47E-07 | 1.41E-06 | 0.002811314 |
| Obesity | mast_cell | *AUTS2* | 4.263800527 | 3.73E-06 | 9.03E-06 | 0.023478571 |
| Obesity | mast_cell | *MITD1* | 4.386671253 | 0.000639173 | 0.001025123 | 1 |
| Obesity | mast_cell | *AHCYL2* | 5.216720773 | 7.13E-07 | 2.10E-06 | 0.004480525 |
| Obesity | mast_cell | *CD1D* | 3.205043105 | 7.80E-05 | 0.000144792 | 0.490266216 |
| Obesity | mast_cell | *FAM86C1* | 4.940300988 | 1.73E-08 | 1.05E-07 | 0.000109055 |
| Obesity | mast_cell | *CD1E* | 10.56233568 | 3.33E-09 | 4.52E-08 | 2.09E-05 |
| Obesity | mast_cell | *NCF1* | 4.761019852 | 8.73E-09 | 7.06E-08 | 5.49E-05 |
| Obesity | mast_cell | *PPP1R9A* | 3.31612906 | 2.00E-08 | 1.14E-07 | 0.000125715 |
| Obesity | mast_cell | *ZFAND2B* | 4.067815476 | 8.99E-07 | 2.58E-06 | 0.00565393 |
| Obesity | mast_cell | *GDPD5* | 5.799725302 | 1.24E-09 | 3.78E-08 | 7.77E-06 |
| Obesity | mast_cell | *DYNC1I1* | 7.571836677 | 3.57E-08 | 1.76E-07 | 0.000224529 |
| Obesity | mast_cell | *TMED4* | 6.14358681 | 0.001196053 | 0.001841682 | 1 |
| Obesity | mast_cell | *ELK4* | 6.615919465 | 0.000202256 | 0.000350589 | 1 |
| Obesity | mast_cell | *SPATA2L* | -4.949997426 | 3.40E-06 | 8.34E-06 | 0.021399657 |
| Obesity | mast_cell | *NIT1* | 3.316995618 | 0.000184234 | 0.000321208 | 1 |
| Obesity | mast_cell | *ZNF276* | 5.248510094 | 0.000672312 | 0.001074161 | 1 |
| Obesity | mast_cell | *FGF17* | 6.016787281 | 0.003934714 | 0.005645264 | 1 |
| Obesity | mast_cell | *CDA* | 2.758899736 | 0.002373492 | 0.003506966 | 1 |
| Obesity | mast_cell | *B4GALT3* | 4.990166868 | 0.000438309 | 0.000723078 | 1 |
| Obesity | mast_cell | *DMTN* | 2.247615747 | 4.35E-09 | 4.91E-08 | 2.73E-05 |
| Obesity | mast_cell | *FAM160B2* | 4.343401815 | 1.21E-08 | 8.63E-08 | 7.63E-05 |
| Obesity | mast_cell | *NDUFS2* | 4.206304325 | 4.03E-05 | 7.91E-05 | 0.253315604 |
| Obesity | mast_cell | *FCER1G* | 3.234151535 | 2.23E-05 | 4.60E-05 | 0.140196122 |
| Obesity | mast_cell | *CCAR2* | 7.699092814 | 1.83E-05 | 3.84E-05 | 0.114906327 |
| Obesity | mast_cell | *RAPGEF6* | -6.070573498 | 0.000467109 | 0.000765965 | 1 |
| Obesity | mast_cell | *EPB41* | 3.947358797 | 1.09E-09 | 3.58E-08 | 6.87E-06 |
| Obesity | mast_cell | *ALG8* | 4.990099845 | 5.05E-09 | 5.30E-08 | 3.18E-05 |
| Obesity | mast_cell | *IFNAR2* | 2.863592747 | 0.000317683 | 0.000534889 | 1 |
| Obesity | mast_cell | *MRPL10* | 2.957416336 | 0.002219814 | 0.00328995 | 1 |
| Obesity | mast_cell | *SV2A* | 10.23126989 | 5.04E-08 | 2.33E-07 | 0.000316833 |
| Obesity | mast_cell | *UBE2Z* | 5.896623148 | 5.24E-06 | 1.23E-05 | 0.032965184 |
| Obesity | mast_cell | *CBR3* | -1.98974797 | 0.024683171 | 0.031708847 | 1 |

| | | | | | | |
|---|---|---|---|---|---|---|
| Obesity | mast_cell | *ARHGAP27* | 6.477948974 | 2.40E-09 | 4.32E-08 | 1.51E-05 |
| Obesity | mast_cell | *PADI4* | 6.272503978 | 2.05E-08 | 1.16E-07 | 0.000128794 |
| Obesity | mast_cell | *PSMD4* | 7.842318962 | 3.26E-08 | 1.64E-07 | 0.000205018 |
| Obesity | mast_cell | *STARD9* | 4.428744626 | 3.47E-08 | 1.72E-07 | 0.000218212 |
| Obesity | mast_cell | *THEM4* | 4.727652944 | 1.11E-07 | 4.42E-07 | 0.00069704 |
| Obesity | mast_cell | *TCHH* | 4.666520207 | 4.71E-05 | 9.15E-05 | 0.296160131 |
| Obesity | mast_cell | *MED8* | 4.098893061 | 6.42E-06 | 1.48E-05 | 0.040378852 |
| Obesity | mast_cell | *RSPRY1* | 4.641007194 | 0.000800729 | 0.00126487 | 1 |
| Obesity | mast_cell | *CCDC17* | 4.292186092 | 2.29E-07 | 8.01E-07 | 0.001441871 |
| Obesity | mast_cell | *ACE* | 3.821190552 | 1.49E-08 | 9.72E-08 | 9.36E-05 |
| Obesity | mast_cell | *TEPP* | 13.41800064 | 0.000393871 | 0.000654924 | 1 |
| Obesity | mast_cell | *EFCAB14* | 5.693330002 | 0.000608845 | 0.000980484 | 1 |
| Obesity | mast_cell | *ATP6V0D1* | 3.06848411 | 0.017867878 | 0.02338858 | 1 |
| Obesity | mast_cell | *C16orf86* | -3.016419697 | 6.58E-05 | 0.000123945 | 0.413604661 |
| Obesity | mast_cell | *LYPD5* | -2.636444393 | 2.12E-06 | 5.46E-06 | 0.01330923 |
| Obesity | mast_cell | *CCDC117* | 9.151123202 | 3.43E-07 | 1.13E-06 | 0.00215899 |
| Obesity | mast_cell | *CCDC107* | 2.781669278 | 0.004083567 | 0.005840169 | 1 |
| Obesity | mast_cell | *NPR2* | 6.156633715 | 2.04E-08 | 1.16E-07 | 0.000128451 |
| Obesity | mast_cell | *CALM3* | 6.638837476 | 0.000107213 | 0.000194306 | 0.674047177 |
| Obesity | mast_cell | *DFFA* | 5.612264285 | 5.05E-05 | 9.77E-05 | 0.317599999 |
| Obesity | mast_cell | *ATAD3B* | 4.766566829 | 1.40E-08 | 9.36E-08 | 8.77E-05 |
| Obesity | mast_cell | *SSU72* | 7.954533947 | 2.19E-08 | 1.23E-07 | 0.000137644 |
| Obesity | mast_cell | *NR2F6* | 6.020658329 | 9.76E-07 | 2.77E-06 | 0.006133806 |
| Obesity | mast_cell | *CILP2* | 17.2256058 | 3.07E-07 | 1.02E-06 | 0.001927356 |
| Obesity | mast_cell | *ABCG1* | 3.187638813 | 4.64E-07 | 1.45E-06 | 0.002917914 |
| Obesity | mast_cell | *SLC37A1* | 3.974873174 | 2.33E-09 | 4.32E-08 | 1.47E-05 |
| Obesity | mast_cell | *U2AF1* | 2.993420875 | 1.43E-05 | 3.08E-05 | 0.089943453 |
| Obesity | mast_cell | *HSF2BP* | 3.619275884 | 6.70E-08 | 2.91E-07 | 0.000421296 |
| Obesity | mast_cell | *CSTB* | 3.16894676 | 0.002364063 | 0.003493856 | 1 |
| Obesity | mast_cell | *ICOSLG* | -4.158718056 | 0.0006704 | 0.001071652 | 1 |
| Obesity | mast_cell | *C21orf2* | 1.269952126 | 0.009465333 | 0.012867981 | 1 |
| Obesity | mast_cell | *ZNF66* | 6.393613583 | 1.81E-05 | 3.81E-05 | 0.113714445 |
| Obesity | mast_cell | *LRRC3* | 3.441682101 | 9.86E-06 | 2.20E-05 | 0.062005053 |
| Obesity | mast_cell | *ITGB2* | 1.932885878 | 0.005415262 | 0.007597803 | 1 |
| Obesity | mast_cell | *FAM207A* | 6.637221218 | 3.09E-08 | 1.58E-07 | 0.000194335 |
| Obesity | mast_cell | *LSS* | 5.167535023 | 4.65E-09 | 5.10E-08 | 2.92E-05 |
| Obesity | mast_cell | *VAV2* | 6.157753867 | 6.38E-08 | 2.81E-07 | 0.000400938 |
| Obesity | mast_cell | *PCNT* | 2.890953518 | 2.31E-05 | 4.75E-05 | 0.145067795 |
| Obesity | mast_cell | *DIP2A* | 3.43071797 | 0.002249087 | 0.003331765 | 1 |
| Obesity | mast_cell | *PRMT2* | 2.682462927 | 0.006707365 | 0.009304767 | 1 |
| Obesity | mast_cell | *ZNF208* | 3.316126849 | 0.000212166 | 0.000366452 | 1 |
| Obesity | mast_cell | *ADAMTS13* | 3.748018737 | 8.54E-05 | 0.000157162 | 0.536865107 |
| Obesity | mast_cell | *FCN2* | 1.460893681 | 0.024610791 | 0.031641045 | 1 |
| Obesity | mast_cell | *ZNF714* | 5.294033734 | 2.43E-05 | 4.97E-05 | 0.152737166 |
| Obesity | mast_cell | *GPSM1* | -2.281914 | 0.024645277 | 0.031673111 | 1 |
| Obesity | mast_cell | *C19orf47* | 5.186331461 | 8.23E-11 | 7.61E-09 | 5.18E-07 |
| Obesity | mast_cell | *HIPK4* | 4.321196374 | 1.61E-07 | 5.97E-07 | 0.001012012 |
| Obesity | mast_cell | *RDH13* | 5.091559655 | 5.97E-09 | 5.73E-08 | 3.75E-05 |
| Obesity | mast_cell | *MPZL3* | -4.663307675 | 4.38E-05 | 8.55E-05 | 0.275251291 |
| Obesity | mast_cell | *PCSK7* | 4.617692402 | 0.002313539 | 0.003421599 | 1 |

| | | | | | | |
|---|---|---|---|---|---|---|
| Obesity | mast_cell | *VPS11* | 4.676444157 | 1.40E-05 | 3.03E-05 | 0.088261246 |
| Obesity | mast_cell | *FDPS* | 5.296502456 | 2.19E-05 | 4.53E-05 | 0.137881483 |
| Obesity | mast_cell | *PAQR6* | -5.797423277 | 0.000980679 | 0.00152915 | 1 |
| Obesity | mast_cell | *PMF1* | 4.617392934 | 0.001390695 | 0.002120616 | 1 |
| Obesity | mast_cell | *SLC25A44* | 6.067165779 | 2.48E-06 | 6.30E-06 | 0.015606928 |
| Obesity | mast_cell | *FCRL3* | 2.429573626 | 5.77E-06 | 1.34E-05 | 0.036278961 |
| Obesity | mast_cell | *NACC1* | 7.924459563 | 3.92E-09 | 4.78E-08 | 2.47E-05 |
| Obesity | mast_cell | *LY6K* | -3.011196077 | 2.17E-07 | 7.65E-07 | 0.001361666 |
| Obesity | mast_cell | *LY6E* | -3.100891698 | 0.006127694 | 0.008536409 | 1 |
| Obesity | mast_cell | *VPS28* | 2.391462763 | 0.024436115 | 0.031449305 | 1 |
| Obesity | mast_cell | *TONSL* | -2.476457345 | 3.56E-07 | 1.16E-06 | 0.002239859 |
| Obesity | mast_cell | *ZNF333* | 7.428988321 | 1.57E-07 | 5.84E-07 | 0.00098658 |
| Obesity | mast_cell | *PPP1R16A* | 2.108706481 | 0.001754104 | 0.002631993 | 1 |
| Obesity | mast_cell | *ORAI2* | 8.109308978 | 8.81E-05 | 0.000161636 | 0.553602369 |
| Obesity | mast_cell | *LRWD1* | -2.424548757 | 1.68E-05 | 3.57E-05 | 0.105911868 |
| Obesity | mast_cell | *MFSD12* | 6.660549411 | 2.43E-09 | 4.32E-08 | 1.53E-05 |
| Obesity | mast_cell | *YDJC* | 18.75891285 | 1.56E-09 | 4.18E-08 | 9.79E-06 |
| Obesity | mast_cell | *CCDC116* | -3.959881066 | 0.003423141 | 0.004950836 | 1 |
| Obesity | mast_cell | *DVL3* | 5.431049737 | 2.33E-09 | 4.32E-08 | 1.46E-05 |
| Obesity | mast_cell | *AP2M1* | 7.501151749 | 3.49E-06 | 8.52E-06 | 0.021911563 |
| Obesity | mast_cell | *ABCF3* | 2.375327537 | 0.003414868 | 0.004941144 | 1 |
| Obesity | mast_cell | *FBXO27* | 2.314304696 | 1.73E-08 | 1.05E-07 | 0.000108697 |
| Obesity | mast_cell | *DMKN* | 2.103846184 | 0.001080531 | 0.001671991 | 1 |
| Obesity | mast_cell | *U2AF1L4* | 0.898612541 | 0.009021623 | 0.01231147 | 1 |
| Obesity | mast_cell | *BDH1* | 9.691400668 | 2.70E-09 | 4.32E-08 | 1.70E-05 |
| Obesity | mast_cell | *COX7A1* | 3.270353662 | 0.000119729 | 0.000215375 | 0.752735305 |
| Obesity | mast_cell | *LRRC56* | 2.777303482 | 0.000899566 | 0.001411775 | 1 |
| Obesity | mast_cell | *IKZF3* | 4.542463971 | 3.57E-09 | 4.63E-08 | 2.24E-05 |
| Obesity | mast_cell | *ACOX1* | 5.893855774 | 1.37E-08 | 9.27E-08 | 8.61E-05 |
| Obesity | mast_cell | *PRPSAP1* | 6.791532379 | 1.75E-07 | 6.41E-07 | 0.001103294 |
| Obesity | mast_cell | *ZNF577* | 3.894202259 | 5.79E-06 | 1.35E-05 | 0.036416678 |
| Obesity | mast_cell | *TMEM143* | 3.608245863 | 1.43E-09 | 4.06E-08 | 8.97E-06 |
| Obesity | mast_cell | *CCL16* | -1.188157732 | 0.006363601 | 0.008849361 | 1 |
| Obesity | mast_cell | *SIGLEC11* | 3.624954727 | 1.03E-05 | 2.28E-05 | 0.064668337 |
| Obesity | mast_cell | *MPP3* | 6.561551935 | 3.59E-07 | 1.17E-06 | 0.002256284 |
| Obesity | mast_cell | *NAGS* | 11.63745532 | 0.001174926 | 0.001810037 | 1 |
| Obesity | mast_cell | *PLCD3* | 2.335772197 | 0.000729996 | 0.001159839 | 1 |
| Obesity | mast_cell | *RACGAP1* | 6.598432211 | 2.29E-07 | 8.01E-07 | 0.001439879 |
| Obesity | mast_cell | *LARP4* | 3.934118558 | 3.82E-07 | 1.23E-06 | 0.00240028 |
| Obesity | mast_cell | *SPC24* | 4.527037029 | 8.87E-09 | 7.12E-08 | 5.58E-05 |
| Obesity | mast_cell | *IP6K3* | -4.806488708 | 0.002799288 | 0.004092819 | 1 |
| Obesity | mast_cell | *ASGR2* | 8.271025582 | 1.84E-08 | 1.08E-07 | 0.000115952 |
| Obesity | mast_cell | *TNFSF13* | -2.910012211 | 0.001403394 | 0.002138942 | 1 |
| Obesity | mast_cell | *SENP3* | 9.072465564 | 1.82E-05 | 3.83E-05 | 0.114368105 |
| Obesity | mast_cell | *SNRNP25* | -4.632318605 | 0.000425963 | 0.000704373 | 1 |
| Obesity | mast_cell | *WDR90* | 1.944116899 | 0.000449239 | 0.000740719 | 1 |
| Obesity | mast_cell | *JMJD8* | 4.67797582 | 1.99E-06 | 5.16E-06 | 0.012510037 |
| Obesity | mast_cell | *FLYWCH2* | 9.749690762 | 5.27E-09 | 5.45E-08 | 3.31E-05 |
| Obesity | mast_cell | *ZG16B* | 65.1801222 | 7.22E-09 | 6.29E-08 | 4.54E-05 |
| Obesity | mast_cell | *NXF1* | 4.696564395 | 6.39E-09 | 5.94E-08 | 4.02E-05 |

| | | | | | | |
|---|---|---|---|---|---|---|
| Obesity | mast_cell | *RPL29* | 4.465012358 | 0.001822605 | 0.002726967 | 1 |
| Obesity | mast_cell | *TPCN2* | 7.12082548 | 1.18E-08 | 8.50E-08 | 7.45E-05 |
| Obesity | mast_cell | *CYP4A22* | -10.40295939 | 0.001812769 | 0.002712897 | 1 |
| Obesity | mast_cell | *PDZK1IP1* | 44.23865722 | 4.02E-05 | 7.89E-05 | 0.252661319 |
| Obesity | mast_cell | *COA7* | 5.219566973 | 6.34E-09 | 5.93E-08 | 3.98E-05 |
| Obesity | mast_cell | *ACOT11* | -1.229869996 | 1.25E-06 | 3.44E-06 | 0.007840908 |
| Obesity | mast_cell | *USP24* | 3.445050738 | 5.70E-05 | 0.00010877 | 0.358070079 |
| Obesity | mast_cell | *NOL9* | 8.008955866 | 5.12E-10 | 2.40E-08 | 3.22E-06 |
| Obesity | mast_cell | *ZSWIM5* | 4.406467578 | 4.14E-09 | 4.80E-08 | 2.60E-05 |
| Obesity | mast_cell | *RBP7* | 3.938913871 | 3.37E-09 | 4.52E-08 | 2.12E-05 |
| Obesity | mast_cell | *FBLIM1* | 3.14192488 | 3.07E-08 | 1.57E-07 | 0.000192942 |
| Obesity | mast_cell | *SLC25A34* | 5.818588225 | 1.15E-08 | 8.39E-08 | 7.23E-05 |
| Obesity | mast_cell | *PDPN* | 2.291870091 | 1.25E-05 | 2.73E-05 | 0.07886081 |
| Obesity | mast_cell | *LRRC38* | 9.166308983 | 4.35E-08 | 2.07E-07 | 0.000273754 |
| Obesity | mast_cell | *LAPTM5* | -2.25761682 | 0.017992216 | 0.023536634 | 1 |
| Obesity | mast_cell | *PEF1* | 10.29873452 | 7.95E-05 | 0.00014735 | 0.499809522 |
| Obesity | mast_cell | *SYNC* | 2.443399006 | 0.036219419 | 0.045469546 | 1 |
| Obesity | mast_cell | *RBBP4* | 6.910332158 | 3.93E-09 | 4.78E-08 | 2.47E-05 |
| Obesity | mast_cell | *TSSK3* | 2.56821495 | 0.019440456 | 0.025336266 | 1 |
| Obesity | mast_cell | *CAMK2N1* | 6.186711934 | 1.51E-09 | 4.16E-08 | 9.52E-06 |
| Obesity | mast_cell | *TTLL10* | 6.116867249 | 3.33E-08 | 1.67E-07 | 0.000209173 |
| Obesity | mast_cell | *NFIA* | 4.519224204 | 4.66E-06 | 1.11E-05 | 0.029274396 |
| Obesity | mast_cell | *OMA1* | 2.355928593 | 0.0019689 | 0.002936768 | 1 |
| Obesity | mast_cell | *TM2D1* | 6.331563024 | 8.33E-07 | 2.41E-06 | 0.005238085 |
| Obesity | mast_cell | *NEXN* | 6.797345755 | 5.53E-08 | 2.50E-07 | 0.000347645 |
| Obesity | mast_cell | *B3GALT2* | 5.392623983 | 9.75E-09 | 7.58E-08 | 6.13E-05 |
| Obesity | mast_cell | *HFM1* | 4.626514752 | 0.020777342 | 0.026977933 | 1 |
| Obesity | mast_cell | *KCNT2* | 6.100882207 | 7.20E-09 | 6.29E-08 | 4.53E-05 |
| Obesity | mast_cell | *VCAM1* | 2.817982581 | 2.35E-08 | 1.29E-07 | 0.000147992 |
| Obesity | mast_cell | *ARPC5* | 2.68889115 | 0.008058573 | 0.011057235 | 1 |
| Obesity | mast_cell | *ZNF496* | 9.64640873 | 3.17E-07 | 1.05E-06 | 0.001994195 |
| Obesity | mast_cell | *IGSF8* | 11.44790989 | 1.23E-06 | 3.39E-06 | 0.007703834 |
| Obesity | mast_cell | *VANGL2* | 5.391960548 | 2.48E-07 | 8.59E-07 | 0.001560351 |
| Obesity | mast_cell | *LMX1A* | 1.807110575 | 7.27E-06 | 1.66E-05 | 0.04572282 |
| Obesity | mast_cell | *TDRD5* | 2.691741586 | 2.78E-05 | 5.63E-05 | 0.174894682 |
| Obesity | mast_cell | *SNED1* | 5.249839244 | 2.03E-07 | 7.22E-07 | 0.001274576 |
| Obesity | mast_cell | *C1orf115* | 3.56303308 | 2.70E-09 | 4.32E-08 | 1.70E-05 |
| Obesity | mast_cell | *BROX* | 1.780345879 | 0.029671019 | 0.037654762 | 1 |
| Obesity | mast_cell | *ACP6* | 2.593573707 | 0.000421455 | 0.000697469 | 1 |
| Obesity | mast_cell | *PM20D1* | 3.316719032 | 1.34E-08 | 9.15E-08 | 8.45E-05 |
| Obesity | mast_cell | *HAAO* | 6.64122565 | 1.19E-10 | 9.56E-09 | 7.46E-07 |
| Obesity | mast_cell | *B3GALNT2* | 4.400792073 | 4.61E-07 | 1.45E-06 | 0.002899441 |
| Obesity | mast_cell | *PUS10* | 7.478755295 | 2.81E-06 | 7.04E-06 | 0.017689143 |
| Obesity | mast_cell | *RFTN2* | 5.52323025 | 3.01E-09 | 4.44E-08 | 1.89E-05 |
| Obesity | mast_cell | *FAM84A* | 4.674989664 | 1.69E-08 | 1.04E-07 | 0.000106145 |
| Obesity | mast_cell | *CLHC1* | 1.511704553 | 2.90E-06 | 7.22E-06 | 0.018205103 |
| Obesity | mast_cell | *FRZB* | 5.495622145 | 2.92E-06 | 7.27E-06 | 0.018349487 |
| Obesity | mast_cell | *NUP35* | 5.882011127 | 1.90E-08 | 1.10E-07 | 0.000119368 |
| Obesity | mast_cell | *FBXO41* | -3.651484265 | 4.31E-06 | 1.03E-05 | 0.027080969 |
| Obesity | mast_cell | *CCDC74A* | 10.11746314 | 1.98E-08 | 1.14E-07 | 0.000124513 |

| | | | | | | |
|---|---|---|---|---|---|---|
| Obesity | mast_cell | *NOSTRIN* | 4.009325762 | 2.56E-08 | 1.38E-07 | 0.000161047 |
| Obesity | mast_cell | *BBS5* | 3.409215308 | 1.04E-08 | 7.94E-08 | 6.56E-05 |
| Obesity | mast_cell | *HPGDS* | -0.368017766 | 2.57E-07 | 8.82E-07 | 0.001614666 |
| Obesity | mast_cell | *PDLIM5* | 5.224722701 | 1.89E-06 | 4.94E-06 | 0.011867719 |
| Obesity | mast_cell | *RPRD2* | 5.721853639 | 8.84E-08 | 3.67E-07 | 0.000555886 |
| Obesity | mast_cell | *PACRGL* | 5.374410151 | 7.35E-08 | 3.13E-07 | 0.000462198 |
| Obesity | mast_cell | *C1QTNF7* | 5.100372435 | 5.35E-06 | 1.25E-05 | 0.033642083 |
| Obesity | mast_cell | *LYSMD1* | 10.63935388 | 9.45E-08 | 3.89E-07 | 0.000594275 |
| Obesity | mast_cell | *ERCC3* | 3.486425472 | 9.44E-06 | 2.11E-05 | 0.059356797 |
| Obesity | mast_cell | *RNF149* | 5.441431426 | 2.38E-05 | 4.87E-05 | 0.149385759 |
| Obesity | mast_cell | *DHX57* | 4.947453431 | 4.89E-10 | 2.33E-08 | 3.08E-06 |
| Obesity | mast_cell | *S100A9* | 5.473121827 | 8.60E-05 | 0.000158124 | 0.540469094 |
| Obesity | mast_cell | *S100A12* | 18.46044103 | 8.23E-07 | 2.38E-06 | 0.005176485 |
| Obesity | mast_cell | *TDRD10* | 3.556013771 | 3.62E-09 | 4.63E-08 | 2.28E-05 |
| Obesity | mast_cell | *DCAF16* | 6.326515433 | 1.05E-08 | 7.97E-08 | 6.62E-05 |
| Obesity | mast_cell | *GNPDA2* | 2.40479836 | 0.000671794 | 0.001073607 | 1 |
| Obesity | mast_cell | *NIPAL1* | 11.53031001 | 8.32E-09 | 6.87E-08 | 5.23E-05 |
| Obesity | mast_cell | *ANTXR2* | 5.028642536 | 5.80E-07 | 1.75E-06 | 0.003648624 |
| Obesity | mast_cell | *PBXIP1* | 4.745490013 | 0.024671131 | 0.031699856 | 1 |
| Obesity | mast_cell | *DCST2* | 6.248170287 | 1.66E-09 | 4.26E-08 | 1.04E-05 |
| Obesity | mast_cell | *LMOD3* | 6.504856576 | 3.72E-09 | 4.67E-08 | 2.34E-05 |
| Obesity | mast_cell | *NBPF10* | 3.249454123 | 3.83E-07 | 1.24E-06 | 0.002407301 |
| Obesity | mast_cell | *POGLUT1* | 1.990030628 | 0.009209542 | 0.012554291 | 1 |
| Obesity | mast_cell | *SLC22A15* | 4.199676484 | 3.89E-10 | 1.97E-08 | 2.45E-06 |
| Obesity | mast_cell | *SLC15A2* | -2.573710668 | 0.01804492 | 0.023590853 | 1 |
| Obesity | mast_cell | *LRRC58* | 8.137318649 | 1.64E-06 | 4.37E-06 | 0.010310056 |
| Obesity | mast_cell | *FSTL1* | 2.852155408 | 0.000418218 | 0.000692659 | 1 |
| Obesity | mast_cell | *LMOD1* | -2.510891049 | 0.000769516 | 0.001218934 | 1 |
| Obesity | mast_cell | *TMEM183A* | 5.7550236 | 0.026141166 | 0.033479224 | 1 |
| Obesity | mast_cell | *IGFBP7* | 3.100151705 | 0.000100449 | 0.000182626 | 0.631520104 |
| Obesity | mast_cell | *ARPC2* | 6.09781106 | 6.27E-07 | 1.87E-06 | 0.003939049 |
| Obesity | mast_cell | *CCT3* | 3.506603025 | 0.001901122 | 0.002839038 | 1 |
| Obesity | mast_cell | *GLB1L* | -3.419320018 | 0.000188754 | 0.000328452 | 1 |
| Obesity | mast_cell | *NUAK2* | 2.568924162 | 7.96E-05 | 0.000147434 | 0.500244164 |
| Obesity | mast_cell | *PYHIN1* | 2.762832979 | 3.19E-06 | 7.86E-06 | 0.020060647 |
| Obesity | mast_cell | *AIM2* | -4.874257571 | 0.000787286 | 0.001244573 | 1 |
| Obesity | mast_cell | *EFHB* | 0.748021813 | 0.001088169 | 0.001683395 | 1 |
| Obesity | mast_cell | *CD200R1* | -1.458399351 | 1.61E-06 | 4.30E-06 | 0.010121487 |
| Obesity | mast_cell | *SPICE1* | 3.575000337 | 0.008263441 | 0.011316108 | 1 |
| Obesity | mast_cell | *CADPS* | -0.720989912 | 2.42E-07 | 8.39E-07 | 0.001518643 |
| Obesity | mast_cell | *PPM1K* | 6.821469533 | 1.73E-08 | 1.05E-07 | 0.000108747 |
| Obesity | mast_cell | *TIPARP* | 7.110633188 | 1.54E-07 | 5.78E-07 | 0.000969987 |
| Obesity | mast_cell | *DCLK3* | -4.009705846 | 0.018013044 | 0.023558978 | 1 |
| Obesity | mast_cell | *RPP14* | 7.998252684 | 6.80E-07 | 2.01E-06 | 0.004275161 |
| Obesity | mast_cell | *DNASE1L3* | 3.316246308 | 8.82E-09 | 7.10E-08 | 5.55E-05 |
| Obesity | mast_cell | *CRELD1* | 4.389605281 | 9.95E-09 | 7.69E-08 | 6.25E-05 |
| Obesity | mast_cell | *TTC14* | 3.180401624 | 0.001020764 | 0.001586538 | 1 |
| Obesity | mast_cell | *RCHY1* | 4.45942114 | 0.000944146 | 0.001475844 | 1 |
| Obesity | mast_cell | *CCDC158* | 2.857029398 | 6.52E-09 | 5.99E-08 | 4.10E-05 |
| Obesity | mast_cell | *GYG1* | 4.395952173 | 7.73E-11 | 7.25E-09 | 4.86E-07 |

| | | | | | | |
|---|---|---|---|---|---|---|
| Obesity | mast_cell | *HPS3* | 6.03964593 | 1.21E-05 | 2.64E-05 | 0.075886148 |
| Obesity | mast_cell | *RYK* | 5.759458459 | 9.18E-06 | 2.06E-05 | 0.057714209 |
| Obesity | mast_cell | *PLB1* | 4.32000602 | 5.14E-09 | 5.36E-08 | 3.23E-05 |
| Obesity | mast_cell | *CDCP1* | 12.2175693 | 2.99E-07 | 1.00E-06 | 0.001878991 |
| Obesity | mast_cell | *LZTFL1* | 9.378607704 | 1.05E-05 | 2.32E-05 | 0.065713362 |
| Obesity | mast_cell | *FYCO1* | 7.439623026 | 3.67E-08 | 1.81E-07 | 0.000230952 |
| Obesity | mast_cell | *LRRC2* | 2.71095857 | 0.000251184 | 0.000428895 | 1 |
| Obesity | mast_cell | *DTX3L* | 4.951843352 | 0.000870274 | 0.001368195 | 1 |
| Obesity | mast_cell | *NMNAT3* | 5.021578229 | 3.12E-09 | 4.48E-08 | 1.96E-05 |
| Obesity | mast_cell | *SMIM12* | 7.93635526 | 1.92E-07 | 6.91E-07 | 0.001205737 |
| Obesity | mast_cell | *TPRA1* | 6.371998316 | 1.66E-09 | 4.26E-08 | 1.04E-05 |
| Obesity | mast_cell | *YEATS2* | 6.525408059 | 4.63E-08 | 2.18E-07 | 0.000291037 |
| Obesity | mast_cell | *MEAF6* | 6.287250446 | 0.000821537 | 0.001296436 | 1 |
| Obesity | mast_cell | *DNALI1* | 3.030547085 | 1.57E-05 | 3.36E-05 | 0.098918128 |
| Obesity | mast_cell | *RPN1* | 6.173414635 | 0.00064261 | 0.001030372 | 1 |
| Obesity | mast_cell | *SFMBT1* | 7.36270217 | 1.03E-08 | 7.88E-08 | 6.48E-05 |
| Obesity | mast_cell | *GNL3* | 4.596090055 | 0.001897492 | 0.002834291 | 1 |
| Obesity | mast_cell | *UVSSA* | 3.614201321 | 0.000127453 | 0.000228159 | 0.80129576 |
| Obesity | mast_cell | *FAM208A* | 4.229122456 | 0.00030057 | 0.000507298 | 1 |
| Obesity | mast_cell | *LRPAP1* | 5.894037084 | 1.39E-08 | 9.36E-08 | 8.77E-05 |
| Obesity | mast_cell | *PIGX* | 5.735904956 | 2.36E-06 | 6.00E-06 | 0.01481869 |
| Obesity | mast_cell | *ABLIM2* | 6.764416541 | 1.25E-09 | 3.78E-08 | 7.85E-06 |
| Obesity | mast_cell | *EXO5* | -3.043938928 | 1.48E-05 | 3.17E-05 | 0.092913454 |
| Obesity | mast_cell | *C1orf50* | -6.715374711 | 8.89E-05 | 0.000163081 | 0.559204685 |
| Obesity | mast_cell | *ERMAP* | 3.347268196 | 2.06E-05 | 4.28E-05 | 0.129614799 |
| Obesity | mast_cell | *METAP1* | 4.556469562 | 1.66E-07 | 6.12E-07 | 0.001044958 |
| Obesity | mast_cell | *DNAJB14* | 6.082256046 | 2.08E-08 | 1.17E-07 | 0.000130649 |
| Obesity | mast_cell | *SLC9B2* | 5.332596379 | 3.30E-10 | 1.77E-08 | 2.08E-06 |
| Obesity | mast_cell | *ATRIP* | 14.14774043 | 2.10E-07 | 7.44E-07 | 0.001320453 |
| Obesity | mast_cell | *SHISA5* | 8.652400019 | 5.17E-10 | 2.40E-08 | 3.25E-06 |
| Obesity | mast_cell | *APEH* | -6.554043666 | 5.81E-15 | 2.28E-12 | 3.65E-11 |
| Obesity | mast_cell | *RNF123* | 6.919822908 | 5.63E-28 | 7.08E-25 | 3.54E-24 |
| Obesity | mast_cell | *MST1R* | 5.592429516 | 1.17E-11 | 1.70E-09 | 7.34E-08 |
| Obesity | mast_cell | *RAD54L2* | 4.651945735 | 7.18E-05 | 0.000134252 | 0.451219862 |
| Obesity | mast_cell | *POC1A* | 5.214818935 | 3.32E-11 | 3.60E-09 | 2.09E-07 |
| Obesity | mast_cell | *PPM1M* | -3.754109236 | 8.84E-05 | 0.000162296 | 0.556025103 |
| Obesity | mast_cell | *C4orf3* | 7.181470109 | 4.96E-05 | 9.61E-05 | 0.312084618 |
| Obesity | mast_cell | *ANXA5* | 4.581285427 | 6.42E-05 | 0.000121239 | 0.403483118 |
| Obesity | mast_cell | *MAP9* | -4.003628456 | 0.000611267 | 0.000983877 | 1 |
| Obesity | mast_cell | *CEP44* | 6.986179014 | 1.25E-08 | 8.73E-08 | 7.83E-05 |
| Obesity | mast_cell | *TMEM144* | 6.683058987 | 1.17E-08 | 8.43E-08 | 7.34E-05 |
| Obesity | mast_cell | *NPY1R* | 3.107257435 | 3.02E-09 | 4.44E-08 | 1.90E-05 |
| Obesity | mast_cell | *IL15* | 3.604999522 | 2.84E-09 | 4.32E-08 | 1.79E-05 |
| Obesity | mast_cell | *ARFIP1* | 4.87305832 | 2.96E-05 | 5.96E-05 | 0.18623759 |
| Obesity | mast_cell | *OTUD4* | 7.166262372 | 1.61E-06 | 4.30E-06 | 0.010111004 |
| Obesity | mast_cell | *ITGA2* | 8.17102858 | 2.70E-10 | 1.54E-08 | 1.70E-06 |
| Obesity | mast_cell | *NDUFAF2* | 4.343232102 | 3.40E-05 | 6.76E-05 | 0.213797363 |
| Obesity | mast_cell | *LMBRD2* | 6.213611645 | 0.000454337 | 0.000747166 | 1 |
| Obesity | mast_cell | *PGGT1B* | 4.330323978 | 0.009973205 | 0.013522006 | 1 |
| Obesity | mast_cell | *C5orf63* | 8.253057364 | 2.83E-05 | 5.71E-05 | 0.177678739 |

| | | | | | | |
|---|---|---|---|---|---|---|
| Obesity | mast_cell | *PRRC1* | 6.596066158 | 3.27E-09 | 4.50E-08 | 2.06E-05 |
| Obesity | mast_cell | *WDR41* | 2.775974634 | 0.013751149 | 0.018262246 | 1 |
| Obesity | mast_cell | *NDUFS4* | 5.720891164 | 4.07E-05 | 7.97E-05 | 0.255582793 |
| Obesity | mast_cell | *SERINC5* | 5.657830207 | 1.15E-06 | 3.22E-06 | 0.007255207 |
| Obesity | mast_cell | *ENPP6* | 11.27098723 | 1.70E-06 | 4.50E-06 | 0.010666642 |
| Obesity | mast_cell | *CASP3* | 8.335355437 | 1.28E-06 | 3.52E-06 | 0.008036172 |
| Obesity | mast_cell | *ERAP1* | -2.090092958 | 0.010994321 | 0.014823353 | 1 |
| Obesity | mast_cell | *ERAP2* | 1.785072674 | 0.001296354 | 0.001983494 | 1 |
| Obesity | mast_cell | *RICTOR* | 4.687209657 | 0.000248137 | 0.000424154 | 1 |
| Obesity | mast_cell | *ANKRA2* | 7.434130493 | 5.66E-09 | 5.59E-08 | 3.56E-05 |
| Obesity | mast_cell | *UBLCP1* | 2.586109486 | 0.024717442 | 0.031746385 | 1 |
| Obesity | mast_cell | *TLR3* | 4.955541548 | 2.15E-09 | 4.32E-08 | 1.35E-05 |
| Obesity | mast_cell | *KLKB1* | 1.971132041 | 2.29E-05 | 4.72E-05 | 0.144048304 |
| Obesity | mast_cell | *NSA2* | 1.566104775 | 0.001570194 | 0.002374172 | 1 |
| Obesity | mast_cell | *CCDC127* | 3.458709621 | 1.05E-08 | 7.97E-08 | 6.62E-05 |
| Obesity | mast_cell | *GDF9* | 4.157049238 | 0.001628045 | 0.002453972 | 1 |
| Obesity | mast_cell | *GRIK2* | 2.40030066 | 6.27E-05 | 0.000118592 | 0.394080252 |
| Obesity | mast_cell | *MB21D1* | 8.540050251 | 1.71E-05 | 3.62E-05 | 0.1075405 |
| Obesity | mast_cell | *TXLNB* | -2.88848705 | 3.39E-06 | 8.32E-06 | 0.021341445 |
| Obesity | mast_cell | *DCBLD1* | 6.082154291 | 2.73E-05 | 5.53E-05 | 0.171606217 |
| Obesity | mast_cell | *PDSS2* | 5.32032009 | 2.00E-05 | 4.16E-05 | 0.125505925 |
| Obesity | mast_cell | *STXBP5* | -1.587762697 | 2.20E-05 | 4.55E-05 | 0.138517445 |
| Obesity | mast_cell | *PI16* | 4.241838609 | 2.78E-09 | 4.32E-08 | 1.75E-05 |
| Obesity | mast_cell | *DAGLB* | 2.739128118 | 0.004854654 | 0.00686487 | 1 |
| Obesity | mast_cell | *STK17A* | 5.999288521 | 1.03E-06 | 2.90E-06 | 0.00645402 |
| Obesity | mast_cell | *GALNT10* | 5.199128783 | 6.07E-06 | 1.41E-05 | 0.038185012 |
| Obesity | mast_cell | *SAP30L* | 7.040521266 | 3.33E-08 | 1.67E-07 | 0.000209452 |
| Obesity | mast_cell | *HCN1* | -5.40171519 | 0.002858833 | 0.004176966 | 1 |
| Obesity | mast_cell | *SLU7* | 6.502835982 | 3.70E-09 | 4.66E-08 | 2.33E-05 |
| Obesity | mast_cell | *RP9* | 5.572584343 | 0.000287241 | 0.000486106 | 1 |
| Obesity | mast_cell | *CAMLG* | 5.16823464 | 5.30E-05 | 0.000102148 | 0.333411266 |
| Obesity | mast_cell | *KCNK5* | 17.48878748 | 1.60E-07 | 5.94E-07 | 0.001005514 |
| Obesity | mast_cell | *KIAA1324L* | 2.507527675 | 0.000544537 | 0.000883713 | 1 |
| Obesity | mast_cell | *USP49* | 4.539253718 | 0.00537105 | 0.007545876 | 1 |
| Obesity | mast_cell | *ZNF704* | 2.530754577 | 0.032873952 | 0.041518388 | 1 |
| Obesity | mast_cell | *FNDC1* | 2.839232856 | 1.01E-08 | 7.80E-08 | 6.37E-05 |
| Obesity | mast_cell | *BRI3* | 5.66156831 | 1.59E-06 | 4.26E-06 | 0.009992229 |
| Obesity | mast_cell | *LMTK2* | 6.841116959 | 2.68E-09 | 4.32E-08 | 1.69E-05 |
| Obesity | mast_cell | *PEX2* | 6.529446045 | 2.21E-08 | 1.23E-07 | 0.000138633 |
| Obesity | mast_cell | *TNFRSF11B* | 5.193953924 | 0.024982037 | 0.032066571 | 1 |
| Obesity | mast_cell | *DEFB1* | 1.975593313 | 0.000826355 | 0.001303385 | 1 |
| Obesity | mast_cell | *SUN1* | 4.942502212 | 6.10E-07 | 1.82E-06 | 0.003837487 |
| Obesity | mast_cell | *OXR1* | 5.249572594 | 2.04E-07 | 7.25E-07 | 0.001283909 |
| Obesity | mast_cell | *TMEM184A* | 3.502797365 | 0.004613662 | 0.006540269 | 1 |
| Obesity | mast_cell | *INTS1* | 7.73075673 | 2.72E-09 | 4.32E-08 | 1.71E-05 |
| Obesity | mast_cell | *SLC4A2* | 8.91850458 | 4.70E-09 | 5.10E-08 | 2.96E-05 |
| Obesity | mast_cell | *FASTK* | 5.401141508 | 0.007973373 | 0.010954676 | 1 |
| Obesity | mast_cell | *ALDH7A1* | 2.833331938 | 0.002619756 | 0.003847327 | 1 |
| Obesity | mast_cell | *COX6C* | 7.714106743 | 7.47E-08 | 3.18E-07 | 0.000469597 |
| Obesity | mast_cell | *SLC25A32* | 4.752786441 | 1.66E-07 | 6.11E-07 | 0.001042218 |

| | | | | | | |
|---|---|---|---|---|---|---|
| Obesity | mast_cell | *DCAF13* | 9.525938443 | 7.29E-09 | 6.32E-08 | 4.58E-05 |
| Obesity | mast_cell | *FREM1* | -2.044119676 | 0.021608406 | 0.027993416 | 1 |
| Obesity | mast_cell | *RPP25L* | 6.755338811 | 4.71E-05 | 9.16E-05 | 0.296268424 |
| Obesity | mast_cell | *FAM219A* | 5.905707358 | 4.00E-09 | 4.78E-08 | 2.52E-05 |
| Obesity | mast_cell | *TMEM65* | 5.664421023 | 2.79E-08 | 1.47E-07 | 0.000175685 |
| Obesity | mast_cell | *CCDC171* | 5.283697195 | 1.20E-05 | 2.63E-05 | 0.075418067 |
| Obesity | mast_cell | *SYK* | 6.457641773 | 3.46E-07 | 1.14E-06 | 0.002177105 |
| Obesity | mast_cell | *METTL2B* | 1.291995045 | 7.62E-05 | 0.000141888 | 0.479298287 |
| Obesity | mast_cell | *HGSNAT* | 4.324038743 | 0.000352625 | 0.000590558 | 1 |
| Obesity | mast_cell | *C9orf64* | -2.34464121 | 0.026359134 | 0.033751502 | 1 |
| Obesity | mast_cell | *HNRNPK* | 5.661464949 | 0.000208507 | 0.000361025 | 1 |
| Obesity | mast_cell | *SVEP1* | 5.260816297 | 2.56E-06 | 6.47E-06 | 0.016103325 |
| Obesity | mast_cell | *ZNF367* | 4.748745528 | 2.69E-07 | 9.17E-07 | 0.001689067 |
| Obesity | mast_cell | *NOL6* | 6.852737225 | 0.001123187 | 0.001733302 | 1 |
| Obesity | mast_cell | *AQP3* | -3.890467618 | 0.000410914 | 0.000681124 | 1 |
| Obesity | mast_cell | *TRMT10B* | 6.002273325 | 0.000112746 | 0.000203923 | 0.708835513 |
| Obesity | mast_cell | *VCP* | 4.483059277 | 4.70E-06 | 1.11E-05 | 0.029524886 |
| Obesity | mast_cell | *PIGO* | 6.600946066 | 3.41E-08 | 1.70E-07 | 0.000214432 |
| Obesity | mast_cell | *LRFN5* | 1.628954713 | 2.27E-10 | 1.41E-08 | 1.43E-06 |
| Obesity | mast_cell | *WRN* | 5.954047173 | 9.43E-07 | 2.69E-06 | 0.0059262 |
| Obesity | mast_cell | *MARCH8* | -2.646426535 | 0.003741628 | 0.005386676 | 1 |
| Obesity | mast_cell | *CFL2* | 4.3100386 | 3.59E-09 | 4.63E-08 | 2.26E-05 |
| Obesity | mast_cell | *ZCCHC24* | 6.551255621 | 3.64E-06 | 8.84E-06 | 0.022882511 |
| Obesity | mast_cell | *PGM2L1* | 4.908751982 | 5.44E-08 | 2.47E-07 | 0.000341792 |
| Obesity | mast_cell | *CRYL1* | 4.422965729 | 7.62E-06 | 1.73E-05 | 0.047899129 |
| Obesity | mast_cell | *REEP3* | 3.355750773 | 0.000429462 | 0.000709413 | 1 |
| Obesity | mast_cell | *LRR1* | 7.265383519 | 2.55E-05 | 5.18E-05 | 0.160005667 |
| Obesity | mast_cell | *RPL36AL* | 3.843813259 | 1.15E-05 | 2.54E-05 | 0.072524353 |
| Obesity | mast_cell | *NOXRED1* | -3.609347559 | 0.000203186 | 0.000352103 | 1 |
| Obesity | mast_cell | *AKR1E2* | 3.769427346 | 0.000168519 | 0.000295944 | 1 |
| Obesity | mast_cell | *ATP5C1* | 5.435783065 | 1.01E-07 | 4.11E-07 | 0.000634455 |
| Obesity | mast_cell | *TAF3* | 4.980135279 | 0.000237819 | 0.000407736 | 1 |
| Obesity | mast_cell | *VDAC2* | 5.575538675 | 2.46E-08 | 1.34E-07 | 0.000154521 |
| Obesity | mast_cell | *SLC18A2* | -0.251866701 | 1.52E-07 | 5.74E-07 | 0.000958642 |
| Obesity | mast_cell | *PDZD8* | 3.240394274 | 0.002179663 | 0.003236547 | 1 |
| Obesity | mast_cell | *QSOX2* | 6.259798184 | 0.031250649 | 0.039579538 | 1 |
| Obesity | mast_cell | *CLEC1B* | -1.018533314 | 0.000100249 | 0.000182315 | 0.630264235 |
| Obesity | mast_cell | *SDCCAG3* | 8.62220838 | 8.25E-06 | 1.86E-05 | 0.051843952 |
| Obesity | mast_cell | *TSC1* | 7.270586021 | 1.13E-07 | 4.48E-07 | 0.000707632 |
| Obesity | mast_cell | *ZMYND19* | 3.413940676 | 1.68E-08 | 1.04E-07 | 0.000105825 |
| Obesity | mast_cell | *STOX1* | 1.232694031 | 4.00E-05 | 7.85E-05 | 0.251291398 |
| Obesity | mast_cell | *RET* | -2.86185597 | 3.35E-07 | 1.10E-06 | 0.002108211 |
| Obesity | mast_cell | *STK32C* | -3.945726219 | 0.000599347 | 0.000965683 | 1 |
| Obesity | mast_cell | *NSMF* | 7.958123939 | 2.23E-09 | 4.32E-08 | 1.40E-05 |
| Obesity | mast_cell | *VWA2* | 2.985206958 | 0.005747964 | 0.008041266 | 1 |
| Obesity | mast_cell | *METTL3* | 6.679142052 | 2.23E-09 | 4.32E-08 | 1.40E-05 |
| Obesity | mast_cell | *TRUB1* | 3.629931284 | 9.94E-05 | 0.000180806 | 0.624683347 |
| Obesity | mast_cell | *HSPA12A* | 1.076308105 | 0.034929983 | 0.043991347 | 1 |
| Obesity | mast_cell | *UBTD1* | 4.340746394 | 2.64E-09 | 4.32E-08 | 1.66E-05 |
| Obesity | mast_cell | *ARHGAP42* | 5.999234752 | 1.17E-07 | 4.63E-07 | 0.000736836 |

| | | | | | | |
|---|---|---|---|---|---|---|
| Obesity | mast_cell | *OTOGL* | 5.594769481 | 5.46E-06 | 1.27E-05 | 0.034308683 |
| Obesity | mast_cell | *PACSIN3* | 1.705973779 | 0.000733357 | 0.001164885 | 1 |
| Obesity | mast_cell | *TTC7B* | -2.146341499 | 0.006050671 | 0.008438458 | 1 |
| Obesity | mast_cell | *SLC39A13* | 3.886324984 | 9.18E-10 | 3.21E-08 | 5.77E-06 |
| Obesity | mast_cell | *PSMC3* | 6.038669787 | 2.07E-10 | 1.33E-08 | 1.30E-06 |
| Obesity | mast_cell | *TC2N* | 1.489228915 | 2.64E-05 | 5.37E-05 | 0.166272589 |
| Obesity | mast_cell | *CPSF2* | 1.442579373 | 0.007112828 | 0.009821732 | 1 |
| Obesity | mast_cell | *CLMN* | 3.858906282 | 1.61E-06 | 4.31E-06 | 0.010138343 |
| Obesity | mast_cell | *PTER* | 4.538595527 | 0.004660537 | 0.006602253 | 1 |
| Obesity | mast_cell | *ABTB2* | 4.693874004 | 0.02236244 | 0.028904741 | 1 |
| Obesity | mast_cell | *LIPC* | 3.839337029 | 4.55E-08 | 2.14E-07 | 0.000285914 |
| Obesity | mast_cell | *CEP57* | 4.223391272 | 0.000117073 | 0.000211007 | 0.73604 |
| Obesity | mast_cell | *TCP11L2* | 5.530782468 | 2.75E-09 | 4.32E-08 | 1.73E-05 |
| Obesity | mast_cell | *JAM3* | 4.919878454 | 1.03E-07 | 4.16E-07 | 0.000645787 |
| Obesity | mast_cell | *GPT2* | 2.051826826 | 2.98E-08 | 1.54E-07 | 0.000187452 |
| Obesity | mast_cell | *IKBIP* | 5.317934393 | 1.47E-07 | 5.54E-07 | 0.000922126 |
| Obesity | mast_cell | *HIF1AN* | 7.102236099 | 6.55E-09 | 6.00E-08 | 4.12E-05 |
| Obesity | mast_cell | *ZFYVE19* | 3.627939156 | 4.11E-08 | 1.98E-07 | 0.0002585 |
| Obesity | mast_cell | *SPINT1* | -3.796240922 | 3.07E-05 | 6.15E-05 | 0.192819881 |
| Obesity | mast_cell | *AVPR1A* | 2.051030555 | 0.00400966 | 0.005742308 | 1 |
| Obesity | mast_cell | *BTRC* | 4.932433249 | 6.13E-05 | 0.0001163 | 0.385532984 |
| Obesity | mast_cell | *LARP6* | 4.50663734 | 6.07E-09 | 5.78E-08 | 3.81E-05 |
| Obesity | mast_cell | *ASPG* | 8.650565797 | 4.55E-09 | 5.05E-08 | 2.86E-05 |
| Obesity | mast_cell | *GABRB3* | 3.12510819 | 0.000209348 | 0.000362182 | 1 |
| Obesity | mast_cell | *TBATA* | 12.57991812 | 2.16E-05 | 4.46E-05 | 0.135530458 |
| Obesity | mast_cell | *COX11* | 5.210282494 | 1.04E-06 | 2.93E-06 | 0.006537857 |
| Obesity | mast_cell | *ZNF202* | 5.259384256 | 1.07E-09 | 3.57E-08 | 6.70E-06 |
| Obesity | mast_cell | *FAM227B* | 3.568775284 | 0.001030816 | 0.001600973 | 1 |
| Obesity | mast_cell | *STXBP4* | 5.188782179 | 1.92E-06 | 4.99E-06 | 0.012049035 |
| Obesity | mast_cell | *C2* | 4.473739478 | 0.002595169 | 0.003814783 | 1 |
| Obesity | mast_cell | *APBB1* | 3.028130325 | 6.99E-09 | 6.20E-08 | 4.39E-05 |
| Obesity | mast_cell | *ILK* | 5.680274937 | 2.79E-09 | 4.32E-08 | 1.75E-05 |
| Obesity | mast_cell | *TAF10* | 8.432419469 | 5.52E-07 | 1.67E-06 | 0.003472104 |
| Obesity | mast_cell | *DCHS1* | 5.168846209 | 2.05E-09 | 4.32E-08 | 1.29E-05 |
| Obesity | mast_cell | *NETO1* | 5.018760971 | 2.73E-06 | 6.86E-06 | 0.017186772 |
| Obesity | mast_cell | *USP54* | 4.502713842 | 0.000511916 | 0.000834003 | 1 |
| Obesity | mast_cell | *C11orf74* | 6.705897746 | 2.45E-09 | 4.32E-08 | 1.54E-05 |
| Obesity | mast_cell | *WDR88* | 4.745105356 | 4.99E-09 | 5.28E-08 | 3.14E-05 |
| Obesity | mast_cell | *CYB5R2* | -2.377487135 | 0.018444627 | 0.024093365 | 1 |
| Obesity | mast_cell | *TUB* | -4.316829648 | 0.001413684 | 0.002153061 | 1 |
| Obesity | mast_cell | *RIC3* | -2.892729415 | 0.001558648 | 0.002359208 | 1 |
| Obesity | mast_cell | *LMO1* | 5.040702152 | 4.58E-08 | 2.16E-07 | 0.000287705 |
| Obesity | mast_cell | *TRIM66* | 0.750267059 | 0.002340878 | 0.003461218 | 1 |
| Obesity | mast_cell | *RPL27A* | 5.98252204 | 0.000148241 | 0.000262829 | 0.931990738 |
| Obesity | mast_cell | *CDYL2* | 1.926736581 | 0.000392935 | 0.00065354 | 1 |
| Obesity | mast_cell | *PRTG* | 4.119233576 | 3.84E-09 | 4.75E-08 | 2.42E-05 |
| Obesity | mast_cell | *AKIP1* | 9.842156642 | 6.39E-09 | 5.94E-08 | 4.02E-05 |
| Obesity | mast_cell | *C16orf46* | 30.96961099 | 2.98E-09 | 4.43E-08 | 1.87E-05 |
| Obesity | mast_cell | *TMEM41B* | 7.062279495 | 5.48E-08 | 2.48E-07 | 0.000344619 |
| Obesity | mast_cell | *ZNF143* | 6.471093875 | 4.29E-08 | 2.05E-07 | 0.000269816 |

| | | | | | | |
|---|---|---|---|---|---|---|
| Obesity | mast_cell | *TMX3* | 6.383353796 | 1.23E-07 | 4.82E-07 | 0.000775859 |
| Obesity | mast_cell | *CCDC68* | -1.205587454 | 3.00E-06 | 7.45E-06 | 0.018880122 |
| Obesity | mast_cell | *CLEC4E* | 2.229222782 | 1.06E-05 | 2.35E-05 | 0.0665768 |
| Obesity | mast_cell | *CLEC4D* | 8.893639698 | 2.57E-08 | 1.38E-07 | 0.000161647 |
| Obesity | mast_cell | *ZSCAN21* | -1.93582474 | 0.001826263 | 0.00273179 | 1 |
| Obesity | mast_cell | *BEAN1* | 8.765483153 | 4.87E-09 | 5.19E-08 | 3.06E-05 |
| Obesity | mast_cell | *TK2* | 3.390352099 | 0.000117099 | 0.000211007 | 0.736203018 |
| Obesity | mast_cell | *SEC11C* | 10.46356637 | 6.26E-05 | 0.000118461 | 0.393528431 |
| Obesity | mast_cell | *GALR1* | 2.638735195 | 1.91E-05 | 3.99E-05 | 0.119948734 |
| Obesity | mast_cell | *CENPV* | 5.178271673 | 3.27E-07 | 1.08E-06 | 0.002053968 |
| Obesity | mast_cell | *CHRFAM7A* | 2.36991926 | 0.008943681 | 0.012213058 | 1 |
| Obesity | mast_cell | *SPDYE6* | 4.207941211 | 4.83E-09 | 5.16E-08 | 3.04E-05 |
| Obesity | mast_cell | *ATF7IP2* | 4.585143586 | 1.52E-07 | 5.72E-07 | 0.000955301 |
| Obesity | mast_cell | *ACSM1* | -3.414457647 | 0.013132637 | 0.017518542 | 1 |
| Obesity | mast_cell | *AP1G1* | 5.512194882 | 1.72E-08 | 1.05E-07 | 0.000108374 |
| Obesity | mast_cell | *SLFN5* | 3.806660777 | 0.003814305 | 0.005480013 | 1 |
| Obesity | mast_cell | *CATSPER2* | 6.209532523 | 2.46E-05 | 5.04E-05 | 0.154956791 |
| Obesity | mast_cell | *C16orf45* | 5.921923286 | 1.51E-05 | 3.23E-05 | 0.094834124 |
| Obesity | mast_cell | *SAAL1* | 9.168395556 | 3.75E-05 | 7.40E-05 | 0.235900603 |
| Obesity | mast_cell | *LDHC* | 1.829037112 | 2.97E-05 | 5.97E-05 | 0.18672795 |
| Obesity | mast_cell | *FAM111A* | 4.02368219 | 2.53E-05 | 5.15E-05 | 0.158900725 |
| Obesity | mast_cell | *SCNN1G* | 9.431863175 | 4.64E-05 | 9.03E-05 | 0.291916569 |
| Obesity | mast_cell | *ANKDD1A* | 4.421991213 | 3.89E-07 | 1.25E-06 | 0.002447244 |
| Obesity | mast_cell | *DCTN5* | 2.93996506 | 4.21E-05 | 8.24E-05 | 0.264540418 |
| Obesity | mast_cell | *STX3* | -0.423031894 | 1.55E-07 | 5.80E-07 | 0.000974476 |
| Obesity | mast_cell | *MTMR10* | 5.801778785 | 5.07E-07 | 1.55E-06 | 0.003186728 |
| Obesity | mast_cell | *YWHAB* | 4.780889948 | 0.000370905 | 0.000619029 | 1 |
| Obesity | mast_cell | *TSC22D4* | -3.286908006 | 0.008169654 | 0.011197431 | 1 |
| Obesity | mast_cell | *MS4A6E* | 1.255940355 | 0.005015256 | 0.007088784 | 1 |
| Obesity | mast_cell | *MS4A7* | 2.255346822 | 8.90E-08 | 3.69E-07 | 0.000559602 |
| Obesity | mast_cell | *DIS3L* | 3.878819217 | 0.002197758 | 0.003260336 | 1 |
| Obesity | mast_cell | *CCNDBP1* | 6.573078134 | 1.76E-09 | 4.28E-08 | 1.10E-05 |
| Obesity | mast_cell | *SMAD3* | 3.892439499 | 1.92E-05 | 4.01E-05 | 0.120482829 |
| Obesity | mast_cell | *AKTIP* | 6.192903782 | 3.09E-09 | 4.48E-08 | 1.94E-05 |
| Obesity | mast_cell | *NKX3-1* | 17.18301271 | 1.09E-07 | 4.36E-07 | 0.000685692 |
| Obesity | mast_cell | *DUSP18* | 5.700856199 | 4.23E-06 | 1.01E-05 | 0.026575832 |
| Obesity | mast_cell | *TEF* | 2.908129662 | 5.09E-05 | 9.83E-05 | 0.319934261 |
| Obesity | mast_cell | *MEI1* | 2.106711922 | 0.003072125 | 0.004468869 | 1 |
| Obesity | mast_cell | *SNRPD1* | 8.872396467 | 2.37E-05 | 4.87E-05 | 0.149196862 |
| Obesity | mast_cell | *SAMD14* | -2.409965162 | 7.97E-06 | 1.80E-05 | 0.05010244 |
| Obesity | mast_cell | *PIP5KL1* | 1.706384349 | 0.000612501 | 0.000985109 | 1 |
| Obesity | mast_cell | *URM1* | 3.193053112 | 5.65E-05 | 0.000108031 | 0.355420929 |
| Obesity | mast_cell | *ENDOG* | 2.373574618 | 0.02418052 | 0.0311331 | 1 |
| Obesity | mast_cell | *ISLR2* | 5.624261911 | 6.76E-09 | 6.07E-08 | 4.25E-05 |
| Obesity | mast_cell | *SP2* | 5.793544783 | 3.10E-11 | 3.52E-09 | 1.95E-07 |
| Obesity | mast_cell | *COQ7* | 5.069800822 | 2.59E-09 | 4.32E-08 | 1.63E-05 |
| Obesity | mast_cell | *GPRC5B* | 6.048767932 | 4.30E-11 | 4.58E-09 | 2.70E-07 |
| Obesity | mast_cell | *KATNAL2* | 1.337004197 | 0.02854503 | 0.036357902 | 1 |
| Obesity | mast_cell | *HDHD2* | 1.821811565 | 0.001989181 | 0.002964205 | 1 |
| Obesity | mast_cell | *RNF214* | 6.308012693 | 6.74E-05 | 0.000126899 | 0.423970447 |

| | | | | | | |
|---|---|---|---|---|---|---|
| Obesity | mast_cell | *ENGASE* | 4.948985209 | 3.60E-08 | 1.78E-07 | 0.000226492 |
| Obesity | mast_cell | *RBFOX3* | 13.52802405 | 0.000902315 | 0.001415029 | 1 |
| Obesity | mast_cell | *RRM1* | 2.157745295 | 1.52E-05 | 3.25E-05 | 0.095297835 |
| Obesity | mast_cell | *TRIM68* | 2.733714271 | 6.82E-06 | 1.56E-05 | 0.042867109 |
| Obesity | mast_cell | *FN3K* | 5.246868453 | 8.72E-07 | 2.51E-06 | 0.005480171 |
| Obesity | mast_cell | *ZNF23* | 2.920686478 | 1.93E-06 | 5.03E-06 | 0.012161746 |
| Obesity | mast_cell | *ZNF229* | -4.16203239 | 0.01784272 | 0.023365378 | 1 |
| Obesity | mast_cell | *ZNF646* | 12.99476807 | 4.92E-14 | 1.47E-11 | 3.10E-10 |
| Obesity | mast_cell | *SMG8* | 6.376640517 | 1.70E-08 | 1.04E-07 | 0.000106743 |
| Obesity | mast_cell | *RAB8A* | 8.071255495 | 1.73E-08 | 1.05E-07 | 0.000108768 |
| Obesity | mast_cell | *GATAD2A* | 4.549431765 | 0.001563259 | 0.002365393 | 1 |
| Obesity | mast_cell | *PROCA1* | 3.847060182 | 0.004599745 | 0.006523482 | 1 |
| Obesity | mast_cell | *RPL13* | 4.795565807 | 0.00055149 | 0.000893383 | 1 |
| Obesity | mast_cell | *TP53I13* | 3.609197595 | 2.89E-05 | 5.82E-05 | 0.181498872 |
| Obesity | mast_cell | *ZNF528* | 5.505966666 | 0.002285618 | 0.003382694 | 1 |
| Obesity | mast_cell | *ZNF701* | 6.258716424 | 5.27E-07 | 1.61E-06 | 0.003316365 |
| Obesity | mast_cell | *SERTAD3* | 3.61290896 | 1.13E-08 | 8.34E-08 | 7.11E-05 |
| Obesity | mast_cell | *TMC4* | 6.445760368 | 0.015291639 | 0.02018445 | 1 |
| Obesity | mast_cell | *TTYH1* | 2.569004882 | 8.40E-09 | 6.89E-08 | 5.28E-05 |
| Obesity | mast_cell | *LENG8* | 4.404139371 | 0.000578054 | 0.000933528 | 1 |
| Obesity | mast_cell | *CDC42EP5* | -3.40488429 | 0.01446281 | 0.019158805 | 1 |
| Obesity | mast_cell | *KIR3DL1* | 2.912336544 | 0.019906616 | 0.025916938 | 1 |
| Obesity | mast_cell | *ZNF283* | 5.205923823 | 3.92E-07 | 1.26E-06 | 0.002462868 |
| Obesity | mast_cell | *SPINT2* | 2.423956569 | 0.000504581 | 0.000822265 | 1 |
| Obesity | mast_cell | *C19orf33* | 2.149696975 | 0.013211885 | 0.017613046 | 1 |
| Obesity | mast_cell | *DNAAF3* | 4.577872214 | 0.039291861 | 0.049071897 | 1 |
| Obesity | mast_cell | *PSCA* | 6.940054213 | 1.67E-06 | 4.43E-06 | 0.010477707 |
| Obesity | mast_cell | *UBXN6* | 3.131475402 | 0.00179424 | 0.002687086 | 1 |
| Obesity | mast_cell | *FAM57A* | 3.188668465 | 9.81E-09 | 7.60E-08 | 6.17E-05 |
| Obesity | mast_cell | *GLOD4* | 5.655450039 | 1.24E-08 | 8.68E-08 | 7.77E-05 |
| Obesity | mast_cell | *KIFC2* | 1.887664177 | 0.000258285 | 0.000440661 | 1 |
| Obesity | mast_cell | *SERPINF2* | 2.965016142 | 0.016442621 | 0.021626519 | 1 |
| Obesity | mast_cell | *SRR* | 2.931062422 | 2.04E-09 | 4.32E-08 | 1.28E-05 |
| Obesity | mast_cell | *CYB5D2* | 5.854743461 | 1.75E-09 | 4.28E-08 | 1.10E-05 |
| Obesity | mast_cell | *ZNF83* | 5.085140613 | 7.97E-07 | 2.32E-06 | 0.005009669 |
| Obesity | mast_cell | *CD320* | 7.410650384 | 1.74E-09 | 4.28E-08 | 1.09E-05 |
| Obesity | mast_cell | *SPRYD3* | 10.08477754 | 8.72E-09 | 7.06E-08 | 5.48E-05 |
| Obesity | mast_cell | *SOAT2* | 2.834381896 | 0.003025419 | 0.004407046 | 1 |
| Obesity | mast_cell | *NDUFV1* | 6.423493342 | 2.02E-05 | 4.19E-05 | 0.126746678 |
| Obesity | mast_cell | *CDK2AP2* | 9.292192557 | 1.40E-08 | 9.40E-08 | 8.82E-05 |
| Obesity | mast_cell | *PRDX2* | 9.370945866 | 3.45E-10 | 1.82E-08 | 2.17E-06 |
| Obesity | mast_cell | *ZNF232* | -1.620884711 | 0.001070857 | 0.001658247 | 1 |
| Obesity | mast_cell | *MIS12* | 3.857628942 | 1.18E-08 | 8.48E-08 | 7.41E-05 |
| Obesity | mast_cell | *HID1* | 3.722257243 | 1.29E-06 | 3.55E-06 | 0.008113933 |
| Obesity | mast_cell | *ATP5H* | 4.226620973 | 0.029746484 | 0.037742915 | 1 |
| Obesity | mast_cell | *SRP68* | 3.89116769 | 1.39E-07 | 5.32E-07 | 0.000874423 |
| Obesity | mast_cell | *TMC8* | -1.391884543 | 2.32E-05 | 4.76E-05 | 0.145621814 |
| Obesity | mast_cell | *TMEM68* | 5.716896051 | 2.91E-08 | 1.51E-07 | 0.00018274 |
| Obesity | mast_cell | *CYP7A1* | 6.433177653 | 3.15E-05 | 6.29E-05 | 0.197734733 |
| Obesity | mast_cell | *GSDMA* | 6.760977517 | 5.99E-08 | 2.68E-07 | 0.000376521 |

| | | | | | | |
|---|---|---|---|---|---|---|
| Obesity | mast_cell | *ZNF598* | 3.251215532 | 7.67E-09 | 6.56E-08 | 4.83E-05 |
| Obesity | mast_cell | *ECI1* | 4.40419282 | 1.25E-05 | 2.72E-05 | 0.078454936 |
| Obesity | mast_cell | *ABCA3* | 3.856054085 | 8.78E-07 | 2.53E-06 | 0.005519529 |
| Obesity | mast_cell | *KCTD5* | -4.741160858 | 0.0036983 | 0.005332847 | 1 |
| Obesity | mast_cell | *SDHAF2* | 6.501872928 | 3.15E-09 | 4.48E-08 | 1.98E-05 |
| Obesity | mast_cell | *DDB1* | 3.993741197 | 0.019932891 | 0.025945773 | 1 |
| Obesity | mast_cell | *VPS37C* | 4.847195009 | 3.90E-07 | 1.26E-06 | 0.002450093 |
| Obesity | mast_cell | *VWCE* | -2.363999873 | 0.035602379 | 0.04475748 | 1 |
| Obesity | mast_cell | *BSCL2* | -4.058867582 | 7.57E-08 | 3.21E-07 | 0.000475852 |
| Obesity | mast_cell | *HRASLS5* | 0.850585068 | 0.000521976 | 0.000848852 | 1 |
| Obesity | mast_cell | *TTC21A* | 7.501074846 | 2.23E-09 | 4.32E-08 | 1.40E-05 |
| Obesity | mast_cell | *ULK4* | 0.962380783 | 0.016948653 | 0.022245549 | 1 |
| Obesity | mast_cell | *LTBP3* | 4.183978841 | 6.38E-08 | 2.81E-07 | 0.000401085 |
| Obesity | mast_cell | *NAALADL1* | 4.613947917 | 2.25E-08 | 1.25E-07 | 0.000141205 |
| Obesity | mast_cell | *SCARA5* | 3.918921547 | 1.46E-08 | 9.64E-08 | 9.19E-05 |
| Obesity | mast_cell | *NUDT16L1* | 11.35605226 | 5.86E-07 | 1.76E-06 | 0.003681058 |
| Obesity | mast_cell | *KIAA1586* | 4.340371 | 0.00053166 | 0.000863706 | 1 |
| Obesity | mast_cell | *ZCCHC4* | 7.623810843 | 2.34E-06 | 5.98E-06 | 0.014729961 |
| Obesity | mast_cell | *GLYCTK* | 3.923809576 | 6.59E-10 | 2.76E-08 | 4.14E-06 |
| Obesity | mast_cell | *UBTD2* | 6.49691298 | 1.32E-08 | 9.04E-08 | 8.28E-05 |
| Obesity | mast_cell | *POLR2J3* | 2.290675307 | 0.001217723 | 0.00187184 | 1 |
| Obesity | mast_cell | *NT5DC2* | -4.402361463 | 5.40E-08 | 2.46E-07 | 0.000339527 |
| Obesity | mast_cell | *KIF5C* | 7.053018155 | 4.03E-08 | 1.95E-07 | 0.000253434 |
| Obesity | mast_cell | *PDHB* | 3.75149742 | 0.022034984 | 0.028516663 | 1 |
| Obesity | mast_cell | *ACOX2* | 1.478073413 | 0.00504989 | 0.007134529 | 1 |
| Obesity | mast_cell | *FAM107A* | 3.315679287 | 0.001285417 | 0.001967718 | 1 |
| Obesity | mast_cell | *CX3CR1* | 2.53295945 | 8.26E-06 | 1.87E-05 | 0.05193518 |
| Obesity | mast_cell | *SCN11A* | 0.891977582 | 0.000473346 | 0.000775385 | 1 |
| Obesity | mast_cell | *SEPT2* | 1.426690481 | 0.00512683 | 0.007230233 | 1 |
| Obesity | mast_cell | *DTYMK* | 7.505881965 | 4.56E-09 | 5.05E-08 | 2.87E-05 |
| Obesity | mast_cell | *TAP1* | 6.641729266 | 1.70E-05 | 3.60E-05 | 0.107056509 |
| Obesity | mast_cell | *ING5* | 7.305933049 | 1.26E-06 | 3.48E-06 | 0.007937347 |
| Obesity | mast_cell | *MLKL* | 1.745386856 | 0.000611422 | 0.000983877 | 1 |
| Obesity | mast_cell | *RFWD3* | 5.616135988 | 9.54E-10 | 3.30E-08 | 6.00E-06 |
| Obesity | mast_cell | *RHOH* | -0.777288389 | 3.49E-07 | 1.14E-06 | 0.002193562 |
| Obesity | mast_cell | *KLHL30* | 3.316933204 | 2.20E-07 | 7.73E-07 | 0.001380649 |
| Obesity | mast_cell | *REEP4* | 3.214088359 | 3.62E-07 | 1.18E-06 | 0.002276696 |
| Obesity | mast_cell | *BMP1* | 4.730757868 | 1.17E-06 | 3.25E-06 | 0.00734196 |
| Obesity | mast_cell | *ATXN2L* | 3.672755843 | 0.012252869 | 0.016400637 | 1 |
| Obesity | mast_cell | *FNTA* | 5.868420105 | 6.03E-10 | 2.60E-08 | 3.79E-06 |
| Obesity | mast_cell | *SERINC2* | 4.49150518 | 2.20E-07 | 7.74E-07 | 0.001382599 |
| Obesity | mast_cell | *TRAPPC11* | 5.420581092 | 2.78E-09 | 4.32E-08 | 1.75E-05 |
| Obesity | mast_cell | *SLC20A2* | 3.024796512 | 0.027347953 | 0.034910981 | 1 |
| Obesity | mast_cell | *ZSWIM1* | 11.15127314 | 4.11E-08 | 1.98E-07 | 0.00025839 |
| Obesity | mast_cell | *NBPF9* | 7.798028851 | 0.002418328 | 0.003569021 | 1 |
| Obesity | mast_cell | *WFDC13* | 18.40335297 | 2.64E-07 | 9.04E-07 | 0.001658565 |
| Obesity | mast_cell | *ZNF30* | 3.716325924 | 0.000320311 | 0.00053888 | 1 |
| Obesity | mast_cell | *SLC16A4* | 2.335991551 | 0.007841021 | 0.010779904 | 1 |
| Obesity | mast_cell | *LRP1B* | -1.319892725 | 0.001531818 | 0.002321171 | 1 |
| Obesity | mast_cell | *DNAJC21* | 7.39115627 | 7.42E-09 | 6.39E-08 | 4.67E-05 |

| | | | | | | |
|---|---|---|---|---|---|---|
| Obesity | mast_cell | *NPNT* | 2.161998785 | 1.89E-08 | 1.10E-07 | 0.000118867 |
| Obesity | mast_cell | *TCTN2* | -1.032412223 | 0.033918998 | 0.042752354 | 1 |
| Obesity | mast_cell | *ABHD15* | 3.363813453 | 1.73E-05 | 3.65E-05 | 0.108798564 |
| Obesity | mast_cell | *ADAL* | 1.213915306 | 4.36E-05 | 8.52E-05 | 0.274368053 |
| Obesity | mast_cell | *SNTB2* | 5.663681262 | 5.42E-09 | 5.52E-08 | 3.41E-05 |
| Obesity | mast_cell | *ZNF507* | 3.375590268 | 0.000346171 | 0.000580367 | 1 |
| Obesity | mast_cell | *STX18* | 4.944987002 | 0.001049005 | 0.001626411 | 1 |
| Obesity | mast_cell | *ZBTB49* | 7.443380246 | 2.84E-09 | 4.32E-08 | 1.79E-05 |
| Obesity | mast_cell | *GFM1* | 2.562168241 | 0.018079911 | 0.023626772 | 1 |
| Obesity | mast_cell | *TNIP2* | 3.379160745 | 0.000249234 | 0.000425797 | 1 |
| Obesity | mast_cell | *BTNL3* | 1.308233255 | 1.07E-05 | 2.36E-05 | 0.067152007 |
| Obesity | mast_cell | *ZNF608* | 3.439883388 | 1.89E-09 | 4.32E-08 | 1.19E-05 |
| Obesity | mast_cell | *INPP5D* | -2.503649298 | 0.000563175 | 0.000910669 | 1 |
| Obesity | mast_cell | *LETM1* | 4.682140115 | 2.79E-09 | 4.32E-08 | 1.75E-05 |
| Obesity | mast_cell | *TMEM129* | 2.238085367 | 0.01169091 | 0.015705289 | 1 |
| Obesity | mast_cell | *CEP120* | 6.343462899 | 1.51E-06 | 4.07E-06 | 0.009494907 |
| Obesity | mast_cell | *STXBP6* | -1.312302474 | 1.83E-06 | 4.81E-06 | 0.011524136 |
| Obesity | mast_cell | *MFF* | 6.114455982 | 5.55E-07 | 1.68E-06 | 0.003486466 |
| Obesity | mast_cell | *CPLX1* | 3.208157067 | 3.91E-08 | 1.90E-07 | 0.000245734 |
| Obesity | mast_cell | *SIGLEC7* | -3.618683766 | 0.000156466 | 0.000276709 | 0.983701003 |
| Obesity | mast_cell | *FEM1B* | 6.455925763 | 1.74E-09 | 4.28E-08 | 1.09E-05 |
| Obesity | mast_cell | *MFSD7* | 8.259998556 | 8.51E-08 | 3.56E-07 | 0.000535001 |
| Obesity | mast_cell | *COL4A3* | 4.059340089 | 1.90E-10 | 1.27E-08 | 1.19E-06 |
| Obesity | mast_cell | *UPF3A* | 2.108079437 | 0.01358373 | 0.018074267 | 1 |
| Obesity | mast_cell | *C8orf46* | 0.730710057 | 1.72E-05 | 3.64E-05 | 0.108413132 |
| Obesity | mast_cell | *HSPBAP1* | 1.854798546 | 0.037945862 | 0.047513569 | 1 |
| Obesity | mast_cell | *PARM1* | 2.391473595 | 0.006902131 | 0.009550132 | 1 |
| Obesity | mast_cell | *FAM110B* | 5.806425928 | 4.81E-09 | 5.15E-08 | 3.03E-05 |
| Obesity | mast_cell | *ATF5* | 8.65074706 | 7.04E-07 | 2.07E-06 | 0.004425941 |
| Obesity | mast_cell | *PCSK9* | 13.71880944 | 0.000318551 | 0.000536063 | 1 |
| Obesity | mast_cell | *XPO6* | 7.155287146 | 2.98E-07 | 1.00E-06 | 0.001875098 |
| Obesity | mast_cell | *RGS14* | 1.793651741 | 0.000465035 | 0.000763561 | 1 |
| Obesity | mast_cell | *RAB24* | 2.432710768 | 0.000931978 | 0.001457912 | 1 |
| Obesity | mast_cell | *CXCL10* | 1.542337228 | 0.017904917 | 0.023427308 | 1 |
| Obesity | mast_cell | *SH3TC2* | -3.060731322 | 3.79E-06 | 9.16E-06 | 0.0238432 |
| Obesity | mast_cell | *GPRIN1* | 12.03917822 | 0.000234499 | 0.000402594 | 1 |
| Obesity | mast_cell | *STK32A* | 1.240005751 | 0.02457239 | 0.031610494 | 1 |
| Obesity | mast_cell | *P2RY12* | 2.586054985 | 2.35E-08 | 1.29E-07 | 0.000147568 |
| Obesity | mast_cell | *SNUPN* | 3.911205879 | 0.015339113 | 0.020242864 | 1 |
| Obesity | mast_cell | *PTK2* | 4.87314616 | 1.44E-06 | 3.91E-06 | 0.009058026 |
| Obesity | mast_cell | *RNASE6* | 3.829373083 | 1.49E-08 | 9.73E-08 | 9.38E-05 |
| Obesity | mast_cell | *NPR1* | 5.605421926 | 2.31E-09 | 4.32E-08 | 1.45E-05 |
| Obesity | mast_cell | *CLIC4* | 5.542194462 | 1.05E-07 | 4.24E-07 | 0.000662893 |
| Obesity | mast_cell | *SLC38A11* | -0.699215276 | 2.04E-07 | 7.25E-07 | 0.001284255 |
| Obesity | mast_cell | *CCDC8* | 5.212989709 | 2.30E-08 | 1.27E-07 | 0.000144585 |
| Obesity | mast_cell | *METTL15* | 4.99669704 | 8.37E-10 | 3.03E-08 | 5.26E-06 |
| Obesity | mast_cell | *DTWD2* | 6.100525372 | 1.38E-05 | 2.98E-05 | 0.086596453 |
| Obesity | mast_cell | *INO80E* | -7.217833463 | 3.54E-09 | 4.62E-08 | 2.22E-05 |
| Obesity | mast_cell | *DFFB* | 7.776146174 | 1.15E-08 | 8.42E-08 | 7.26E-05 |
| Obesity | mast_cell | *FAM103A1* | -1.637906499 | 0.019079012 | 0.024891004 | 1 |

| | | | | | | |
|---|---|---|---|---|---|---|
| Obesity | mast_cell | *RGPD8* | -1.094733067 | 0.003703101 | 0.005337321 | 1 |
| Obesity | mast_cell | *LUZP1* | 5.981339616 | 1.19E-07 | 4.67E-07 | 0.00074636 |
| Obesity | mast_cell | *HEXDC* | 4.580169258 | 0.00415721 | 0.005941437 | 1 |
| Obesity | mast_cell | *LRRC45* | 1.643133093 | 0.014008365 | 0.018588137 | 1 |
| Obesity | mast_cell | *CHRNA5* | 3.105175615 | 1.12E-06 | 3.12E-06 | 0.00702064 |
| Obesity | mast_cell | *FASN* | 3.153613558 | 2.07E-09 | 4.32E-08 | 1.30E-05 |
| Obesity | mast_cell | *MT1E* | -4.301899143 | 0.020662054 | 0.026844872 | 1 |
| Obesity | mast_cell | *DCXR* | 6.705921099 | 1.34E-09 | 3.93E-08 | 8.40E-06 |
| Obesity | mast_cell | *LIMS1* | 3.65541249 | 1.85E-06 | 4.85E-06 | 0.011614438 |
| Obesity | mast_cell | *UGP2* | 4.303331329 | 1.38E-06 | 3.77E-06 | 0.008695052 |
| Obesity | mast_cell | *CSGALNACT2* | 6.147978203 | 5.37E-05 | 0.00010336 | 0.337574221 |
| Obesity | mast_cell | *AVEN* | 6.519637166 | 3.16E-09 | 4.48E-08 | 1.99E-05 |
| Obesity | mast_cell | *CTNND2* | 1.569834824 | 8.26E-05 | 0.000152465 | 0.519448481 |
| Obesity | mast_cell | *CALML6* | 3.195196756 | 0.015356088 | 0.020261012 | 1 |
| Obesity | mast_cell | *TPST1* | 1.845913122 | 0.002925948 | 0.004271055 | 1 |
| Obesity | mast_cell | *GUSB* | 7.665725244 | 2.52E-07 | 8.68E-07 | 0.001584729 |
| Obesity | mast_cell | *ZFPM2* | 4.438979388 | 2.54E-08 | 1.37E-07 | 0.000159921 |
| Obesity | mast_cell | *ZNF747* | -8.054684974 | 1.21E-05 | 2.64E-05 | 0.075867516 |
| Obesity | mast_cell | *MAP3K2* | 5.796681808 | 3.46E-08 | 1.72E-07 | 0.000217827 |
| Obesity | mast_cell | *PUSL1* | 5.230349647 | 1.33E-07 | 5.14E-07 | 0.000835653 |
| Obesity | mast_cell | *MYO7B* | 6.556390299 | 2.28E-07 | 7.99E-07 | 0.001435219 |
| Obesity | mast_cell | *CNTROB* | 8.660995972 | 0.000419156 | 0.000694031 | 1 |
| Obesity | mast_cell | *KCNAB3* | 2.158382144 | 0.009890795 | 0.013416058 | 1 |
| Obesity | mast_cell | *NIPA1* | 8.767734605 | 1.56E-07 | 5.81E-07 | 0.000977937 |
| Obesity | mast_cell | *RNF150* | 2.286109284 | 0.000727402 | 0.00115601 | 1 |
| Obesity | mast_cell | *HOXD4* | -4.058021485 | 0.000482525 | 0.000789598 | 1 |
| Obesity | mast_cell | *CHRNB1* | -2.328468923 | 0.000459355 | 0.000755023 | 1 |
| Obesity | mast_cell | *ANKK1* | -1.436444368 | 3.96E-07 | 1.27E-06 | 0.002489432 |
| Obesity | mast_cell | *PDCD6IP* | 3.840142597 | 0.000375066 | 0.000625474 | 1 |
| Obesity | mast_cell | *ZNF212* | 5.980742945 | 7.66E-08 | 3.24E-07 | 0.000481372 |
| Obesity | mast_cell | *FAM161A* | 4.61822642 | 9.28E-06 | 2.08E-05 | 0.058374281 |
| Obesity | mast_cell | *GLB1* | 6.261447447 | 3.44E-08 | 1.71E-07 | 0.000216541 |
| Obesity | mast_cell | *FAXDC2* | 4.576364838 | 5.95E-07 | 1.79E-06 | 0.003742461 |
| Obesity | mast_cell | *ELP5* | 4.628652405 | 0.000712908 | 0.001133836 | 1 |
| Obesity | mast_cell | *STX8* | 4.98337807 | 3.20E-05 | 6.38E-05 | 0.201301578 |
| Obesity | mast_cell | *NFRKB* | 4.519227198 | 3.57E-05 | 7.06E-05 | 0.224312343 |
| Obesity | mast_cell | *SP7* | 9.408529077 | 0.000436911 | 0.000720961 | 1 |
| Obesity | mast_cell | *SEMA3E* | 8.146326116 | 2.46E-07 | 8.51E-07 | 0.001545335 |
| Obesity | mast_cell | *DCLK2* | 3.789806561 | 0.000135995 | 0.000242484 | 0.854999 |
| Obesity | mast_cell | *MGMT* | 4.465764302 | 1.57E-06 | 4.23E-06 | 0.009900238 |
| Obesity | mast_cell | *METTL7B* | 7.236797896 | 2.02E-08 | 1.15E-07 | 0.000126821 |
| Obesity | mast_cell | *NFXL1* | 8.03373092 | 3.65E-07 | 1.18E-06 | 0.00229209 |
| Obesity | mast_cell | *CD14* | 2.477072771 | 0.005551893 | 0.007777351 | 1 |
| Obesity | mast_cell | *KRT72* | -5.321381833 | 0.012000468 | 0.0161005 | 1 |
| Obesity | mast_cell | *HSD17B13* | 1.384104896 | 0.003202873 | 0.004647234 | 1 |
| Obesity | mast_cell | *TMC7* | 2.546992624 | 1.21E-07 | 4.76E-07 | 0.000763374 |
| Obesity | mast_cell | *SERPINB9* | 5.766916891 | 3.90E-06 | 9.38E-06 | 0.024489381 |
| Obesity | mast_cell | *SMAGP* | -3.065016759 | 2.48E-06 | 6.30E-06 | 0.015619831 |
| Obesity | mast_cell | *IRX2* | 2.838639014 | 7.40E-06 | 1.69E-05 | 0.046548754 |
| Obesity | mast_cell | *EMB* | 1.724949343 | 0.00744295 | 0.010261805 | 1 |

| | | | | | | |
|---|---|---|---|---|---|---|
| Obesity | mast_cell | *STAT2* | 5.450207587 | 1.02E-06 | 2.88E-06 | 0.00641008 |
| Obesity | mast_cell | *NUDCD2* | 7.134410314 | 5.86E-08 | 2.62E-07 | 0.00036832 |
| Obesity | mast_cell | *HSPA4* | 5.162260401 | 0.00400302 | 0.005735412 | 1 |
| Obesity | mast_cell | *SLC26A5* | 8.070532578 | 1.91E-09 | 4.32E-08 | 1.20E-05 |
| Obesity | mast_cell | *COMMD5* | 7.056260214 | 2.88E-09 | 4.35E-08 | 1.81E-05 |
| Obesity | mast_cell | *GTSF1* | 35.10086181 | 1.80E-06 | 4.74E-06 | 0.011310647 |
| Obesity | mast_cell | *RNF34* | 5.439769216 | 8.59E-06 | 1.93E-05 | 0.054022094 |
| Obesity | mast_cell | *ACYP2* | 4.963794517 | 1.81E-06 | 4.77E-06 | 0.011393396 |
| Obesity | mast_cell | *TMEM133* | 5.571593775 | 1.72E-08 | 1.05E-07 | 0.000108107 |
| Obesity | mast_cell | *TTLL6* | -8.971795344 | 8.19E-07 | 2.37E-06 | 0.005147358 |
| Obesity | mast_cell | *KCNS3* | 1.489945112 | 0.002917922 | 0.004261319 | 1 |
| Obesity | mast_cell | *HTRA3* | 3.128621238 | 2.13E-05 | 4.42E-05 | 0.134098547 |
| Obesity | mast_cell | *FOXN2* | 4.33117762 | 0.001803642 | 0.00269988 | 1 |
| Obesity | mast_cell | *GPR27* | 6.510444136 | 0.000576094 | 0.000930602 | 1 |
| Obesity | mast_cell | *KBTBD2* | 5.481196641 | 3.71E-08 | 1.82E-07 | 0.000232997 |
| Obesity | mast_cell | *MTSS1* | 5.470677802 | 1.37E-07 | 5.27E-07 | 0.000862506 |
| Obesity | mast_cell | *RPS9* | 4.750426523 | 2.87E-05 | 5.78E-05 | 0.180300595 |
| Obesity | mast_cell | *TSEN34* | 5.598781822 | 3.24E-09 | 4.49E-08 | 2.04E-05 |
| Obesity | mast_cell | *NDUFA3* | 6.121957023 | 1.35E-08 | 9.19E-08 | 8.52E-05 |
| Obesity | mast_cell | *OSCAR* | 4.761141014 | 1.55E-07 | 5.79E-07 | 0.000971536 |
| Obesity | mast_cell | *NUDT6* | 3.259162288 | 1.33E-08 | 9.11E-08 | 8.39E-05 |
| Obesity | mast_cell | *ZNF160* | 5.862545563 | 2.04E-07 | 7.25E-07 | 0.001281478 |
| Obesity | mast_cell | *ZNF415* | -1.844540481 | 2.83E-05 | 5.71E-05 | 0.177706332 |
| Obesity | mast_cell | *TSNARE1* | 4.39746465 | 2.41E-08 | 1.32E-07 | 0.000151787 |
| Obesity | mast_cell | *FPR1* | 3.257162199 | 5.71E-06 | 1.33E-05 | 0.035895593 |
| Obesity | mast_cell | *FEZ2* | 4.885885094 | 0.000142271 | 0.000253172 | 0.894456468 |
| Obesity | mast_cell | *SOX7* | 16.06082628 | 5.70E-11 | 5.60E-09 | 3.58E-07 |
| Obesity | mast_cell | *ALK* | 3.861756224 | 0.004297193 | 0.006120629 | 1 |
| Obesity | mast_cell | *TRMT61B* | 3.262568686 | 6.83E-10 | 2.77E-08 | 4.29E-06 |
| Obesity | mast_cell | *MFN1* | 5.619599471 | 3.01E-08 | 1.55E-07 | 0.000189348 |
| Obesity | mast_cell | *GIMAP8* | 3.680713344 | 5.18E-05 | 9.99E-05 | 0.325388059 |
| Obesity | mast_cell | *ATP6V0E2* | 2.930089195 | 7.61E-08 | 3.23E-07 | 0.000478344 |
| Obesity | mast_cell | *CLDN20* | -2.414794319 | 5.82E-06 | 1.35E-05 | 0.036572558 |
| Obesity | mast_cell | *CANT1* | 5.25785661 | 2.51E-05 | 5.11E-05 | 0.157679354 |
| Obesity | mast_cell | *CHST11* | -1.628142216 | 0.005517741 | 0.007734679 | 1 |
| Obesity | mast_cell | *KCND3* | 3.824197817 | 6.80E-10 | 2.77E-08 | 4.27E-06 |
| Obesity | mast_cell | *ZNF581* | 4.895256373 | 1.14E-08 | 8.37E-08 | 7.19E-05 |
| Obesity | mast_cell | *MCC* | 5.431722669 | 4.20E-09 | 4.83E-08 | 2.64E-05 |
| Obesity | mast_cell | *DLK2* | -4.037047017 | 3.87E-08 | 1.89E-07 | 0.000243509 |
| Obesity | mast_cell | *ZNF561* | 3.632275789 | 5.11E-07 | 1.56E-06 | 0.003209968 |
| Obesity | mast_cell | *LRRC8C* | 8.341713136 | 4.34E-09 | 4.91E-08 | 2.73E-05 |
| Obesity | mast_cell | *PPID* | 6.457604788 | 1.87E-08 | 1.09E-07 | 0.000117716 |
| Obesity | mast_cell | *TBCA* | 4.499696558 | 3.77E-05 | 7.42E-05 | 0.236795858 |
| Obesity | mast_cell | *MAP6* | 2.858830383 | 8.90E-08 | 3.69E-07 | 0.000559485 |
| Obesity | mast_cell | *BCL2L1* | 5.661543183 | 5.92E-05 | 0.000112705 | 0.372489205 |
| Obesity | mast_cell | *PLRG1* | 7.509761816 | 1.99E-09 | 4.32E-08 | 1.25E-05 |
| Obesity | mast_cell | *NMUR1* | 2.775047279 | 1.46E-08 | 9.64E-08 | 9.20E-05 |
| Obesity | mast_cell | *P2RY6* | 2.31519238 | 2.22E-08 | 1.24E-07 | 0.000139441 |
| Obesity | mast_cell | *ZIK1* | -4.678474076 | 0.032503108 | 0.041074782 | 1 |
| Obesity | mast_cell | *RGS19* | 3.259262662 | 1.68E-05 | 3.56E-05 | 0.105572713 |

| | | | | | | |
|---|---|---|---|---|---|---|
| Obesity | mast_cell | *TCEA2* | 4.501656938 | 4.03E-09 | 4.78E-08 | 2.53E-05 |
| Obesity | mast_cell | *ANO5* | 3.574398772 | 6.15E-09 | 5.81E-08 | 3.86E-05 |
| Obesity | mast_cell | *GPHN* | 3.010622406 | 0.000149605 | 0.000265022 | 0.940564215 |
| Obesity | mast_cell | *VAT1L* | 5.490222243 | 0.000436891 | 0.000720961 | 1 |
| Obesity | mast_cell | *CAMTA1* | 5.641622865 | 5.46E-05 | 0.000104857 | 0.343301408 |
| Obesity | mast_cell | *LGALS4* | 3.02647835 | 0.002706408 | 0.003963472 | 1 |
| Obesity | mast_cell | *LRRC34* | 1.13656431 | 0.036921483 | 0.046286214 | 1 |
| Obesity | mast_cell | *SYCE1* | 2.610789254 | 0.000208589 | 0.000361068 | 1 |
| Obesity | mast_cell | *BCL2* | 1.300440333 | 0.000681636 | 0.001086849 | 1 |
| Obesity | mast_cell | *RHNO1* | 3.759230503 | 0.000223409 | 0.000384289 | 1 |
| Obesity | mast_cell | *KNDC1* | 1.578053432 | 0.032891041 | 0.041531628 | 1 |
| Obesity | mast_cell | *METTL18* | 4.055326887 | 6.60E-09 | 6.01E-08 | 4.15E-05 |
| Obesity | mast_cell | *ZNF570* | 7.820136315 | 1.95E-07 | 7.00E-07 | 0.001225817 |
| Obesity | mast_cell | *NINJ2* | 4.684318235 | 1.21E-09 | 3.76E-08 | 7.60E-06 |
| Obesity | mast_cell | *FAM90A1* | 8.108195102 | 1.31E-08 | 9.03E-08 | 8.26E-05 |
| Obesity | mast_cell | *TRAPPC12* | 4.052079807 | 0.005531767 | 0.007752613 | 1 |
| Obesity | mast_cell | *AQP4* | 2.851261511 | 1.63E-08 | 1.03E-07 | 0.000102593 |
| Obesity | mast_cell | *TLN2* | 1.213524892 | 0.021772072 | 0.028193825 | 1 |
| Obesity | mast_cell | *TVP23B* | 5.131672933 | 3.67E-08 | 1.81E-07 | 0.000231044 |
| Obesity | mast_cell | *SRGAP2C* | 4.524574195 | 1.83E-09 | 4.32E-08 | 1.15E-05 |
| Obesity | mast_cell | *ATPAF2* | 10.17175206 | 3.42E-07 | 1.12E-06 | 0.00214855 |
| Obesity | mast_cell | *CYP4F22* | -1.717593892 | 3.25E-08 | 1.64E-07 | 0.000204261 |
| Obesity | mast_cell | *C20orf196* | 6.930811614 | 2.25E-07 | 7.89E-07 | 0.001412012 |
| Obesity | mast_cell | *ZNF554* | 2.066676612 | 0.039020647 | 0.048762235 | 1 |
| Obesity | mast_cell | *ANKRD20A4* | -2.801089818 | 4.25E-07 | 1.35E-06 | 0.002670625 |
| Obesity | mast_cell | *LAMB2* | 3.584831979 | 0.00046905 | 0.000768948 | 1 |
| Obesity | mast_cell | *ORMDL3* | 1.364945178 | 0.017883758 | 0.023404493 | 1 |
| Obesity | mast_cell | *SERF1A* | 11.64647833 | 1.90E-08 | 1.10E-07 | 0.000119453 |
| Obesity | mast_cell | *LRRC15* | -5.041000888 | 0.000922857 | 0.001446238 | 1 |
| Obesity | mast_cell | *FRMD3* | 7.897047934 | 1.54E-09 | 4.16E-08 | 9.66E-06 |
| Obesity | mast_cell | *MTBP* | 7.996763161 | 4.35E-07 | 1.38E-06 | 0.00273178 |
| Obesity | mast_cell | *MALT1* | 6.848888106 | 4.96E-07 | 1.52E-06 | 0.003115394 |
| Obesity | mast_cell | *CXCR6* | 4.036350851 | 2.51E-09 | 4.32E-08 | 1.58E-05 |
| Obesity | mast_cell | *C5orf34* | 5.434593724 | 8.19E-09 | 6.79E-08 | 5.15E-05 |
| Obesity | mast_cell | *C1QTNF4* | -1.994204801 | 1.18E-13 | 3.37E-11 | 7.42E-10 |
| Obesity | mast_cell | *ZNF131* | 5.349655493 | 3.16E-05 | 6.31E-05 | 0.198680541 |
| Obesity | mast_cell | *HINFP* | 9.76816232 | 8.84E-08 | 3.67E-07 | 0.000555464 |
| Obesity | mast_cell | *SPTLC3* | 7.412193865 | 5.28E-07 | 1.61E-06 | 0.003322339 |
| Obesity | mast_cell | *COPRS* | -3.295364606 | 0.029149149 | 0.037052305 | 1 |
| Obesity | mast_cell | *SUCLG2* | 4.451260895 | 0.000117669 | 0.000211912 | 0.739786234 |
| Obesity | mast_cell | *RCAN2* | 2.598158672 | 4.39E-09 | 4.94E-08 | 2.76E-05 |
| Obesity | mast_cell | *IL16* | 3.880942381 | 1.49E-07 | 5.63E-07 | 0.000939842 |
| Obesity | mast_cell | *GNB2* | 5.884397014 | 1.53E-05 | 3.29E-05 | 0.096483134 |
| Obesity | mast_cell | *PRSS27* | 13.76357888 | 6.72E-08 | 2.91E-07 | 0.000422219 |
| Obesity | mast_cell | *ZNF24* | 7.770141924 | 1.28E-06 | 3.53E-06 | 0.008058053 |
| Obesity | mast_cell | *FIBP* | 4.084522869 | 2.42E-10 | 1.45E-08 | 1.52E-06 |
| Obesity | mast_cell | *CARNS1* | 6.948466631 | 2.15E-09 | 4.32E-08 | 1.35E-05 |
| Obesity | mast_cell | *CTSW* | -0.408489154 | 0.038408314 | 0.048044781 | 1 |
| Obesity | mast_cell | *SNTG2* | 2.886250027 | 0.000618436 | 0.000994146 | 1 |
| Obesity | mast_cell | *PDE3A* | -2.050501386 | 1.70E-05 | 3.60E-05 | 0.106861322 |

| | | | | | | |
|---|---|---|---|---|---|---|
| Obesity | mast_cell | *MRPL52* | 2.710253126 | 0.028313665 | 0.03609978 | 1 |
| Obesity | mast_cell | *SMPDL3A* | 4.59306389 | 2.55E-09 | 4.32E-08 | 1.60E-05 |
| Obesity | mast_cell | *RAD9A* | 1.824011842 | 0.0002632 | 0.000448316 | 1 |
| Obesity | mast_cell | *EFEMP2* | 7.332490318 | 4.38E-13 | 1.06E-10 | 2.75E-09 |
| Obesity | mast_cell | *AGAP5* | 69.37646295 | 1.68E-08 | 1.04E-07 | 0.000105311 |
| Obesity | mast_cell | *SLFN11* | 4.002693584 | 1.35E-07 | 5.21E-07 | 0.000851174 |
| Obesity | mast_cell | *CCL19* | 10.77229736 | 6.49E-08 | 2.83E-07 | 0.000407842 |
| Obesity | mast_cell | *FAM192A* | 3.139700024 | 3.29E-06 | 8.07E-06 | 0.020655424 |
| Obesity | mast_cell | *RAB43* | -14.02835083 | 4.12E-07 | 1.32E-06 | 0.002590614 |
| Obesity | mast_cell | *CBWD1* | 5.641969047 | 4.25E-09 | 4.85E-08 | 2.67E-05 |
| Obesity | mast_cell | *HOXC5* | 4.101261338 | 4.83E-05 | 9.38E-05 | 0.303590978 |
| Obesity | mast_cell | *RAB37* | -2.076177196 | 3.03E-06 | 7.52E-06 | 0.019063708 |
| Obesity | mast_cell | *DCP2* | 6.484996851 | 9.03E-07 | 2.59E-06 | 0.005679996 |
| Obesity | mast_cell | *SNX32* | 3.999211264 | 9.89E-05 | 0.000180055 | 0.621728864 |
| Obesity | mast_cell | *METAP1D* | 8.016733482 | 6.17E-08 | 2.74E-07 | 0.00038786 |
| Obesity | mast_cell | *NADSYN1* | 6.47337189 | 1.66E-09 | 4.26E-08 | 1.04E-05 |
| Obesity | mast_cell | *RNASEH2C* | 4.11877802 | 1.16E-06 | 3.22E-06 | 0.007271761 |
| Obesity | mast_cell | *MYEOV* | 0.969697203 | 7.86E-05 | 0.000145874 | 0.49407572 |
| Obesity | mast_cell | *MRGPRF* | 5.452209592 | 1.03E-08 | 7.88E-08 | 6.45E-05 |
| Obesity | mast_cell | *ADH6* | 0.989419371 | 0.004186377 | 0.005979045 | 1 |
| Obesity | mast_cell | *GXYLT2* | 4.464351644 | 2.41E-07 | 8.36E-07 | 0.001513076 |
| Obesity | mast_cell | *HPSE2* | 2.827775892 | 0.000352477 | 0.000590468 | 1 |
| Obesity | mast_cell | *HECTD4* | 4.510214175 | 0.000138163 | 0.000246001 | 0.86862974 |
| Obesity | mast_cell | *TRMT112* | 4.099957283 | 0.000396223 | 0.000658486 | 1 |
| Obesity | mast_cell | *NOC3L* | 6.153752289 | 1.30E-09 | 3.88E-08 | 8.18E-06 |
| Obesity | mast_cell | *PARP14* | 1.892834822 | 8.11E-05 | 0.000150068 | 0.510080246 |
| Obesity | mast_cell | *AHSA2* | 4.131060196 | 0.000128653 | 0.000229916 | 0.808844367 |
| Obesity | mast_cell | *ABLIM3* | 6.770147851 | 4.03E-07 | 1.29E-06 | 0.002534462 |
| Obesity | mast_cell | *VANGL1* | 6.733138971 | 0.000622479 | 0.00100039 | 1 |
| Obesity | mast_cell | *GOLGB1* | 5.875403231 | 3.53E-07 | 1.15E-06 | 0.002219549 |
| Obesity | mast_cell | *ZNF483* | 2.064207523 | 7.94E-05 | 0.00014723 | 0.499255779 |
| Obesity | mast_cell | *SLC2A14* | -1.651972393 | 0.030464028 | 0.038614384 | 1 |
| Obesity | mast_cell | *MZT2A* | 3.998474315 | 4.88E-07 | 1.51E-06 | 0.003070258 |
| Obesity | mast_cell | *MAP3K11* | 9.280168985 | 1.22E-07 | 4.78E-07 | 0.000767358 |
| Obesity | mast_cell | *C1QA* | 4.436000281 | 2.42E-09 | 4.32E-08 | 1.52E-05 |
| Obesity | mast_cell | *NDNF* | 3.76378498 | 1.91E-09 | 4.32E-08 | 1.20E-05 |
| Obesity | mast_cell | *OLR1* | 7.087416399 | 4.73E-09 | 5.10E-08 | 2.97E-05 |
| Obesity | mast_cell | *GLIPR1L1* | 3.996978228 | 1.73E-09 | 4.28E-08 | 1.09E-05 |
| Obesity | mast_cell | *ARV1* | 6.638212104 | 3.88E-10 | 1.97E-08 | 2.44E-06 |
| Obesity | mast_cell | *CCDC36* | 5.343675401 | 6.26E-11 | 6.05E-09 | 3.93E-07 |
| Obesity | mast_cell | *THAP2* | 4.028969317 | 1.61E-08 | 1.02E-07 | 0.000101226 |
| Obesity | mast_cell | *PPP1R14B* | 2.580340755 | 2.22E-08 | 1.24E-07 | 0.000139458 |
| Obesity | mast_cell | *SMARCC1* | 3.008798978 | 0.027944956 | 0.035651368 | 1 |
| Obesity | mast_cell | *ZNF417* | -4.457462723 | 0.023945531 | 0.03083686 | 1 |
| Obesity | mast_cell | *VEGFB* | 2.693796547 | 8.28E-05 | 0.00015264 | 0.520535508 |
| Obesity | mast_cell | *PEAK1* | 4.926427011 | 2.08E-06 | 5.38E-06 | 0.013098019 |
| Obesity | mast_cell | *CSPG4* | 3.318707647 | 8.58E-06 | 1.93E-05 | 0.053941964 |
| Obesity | mast_cell | *SNX33* | -2.73269126 | 0.00267163 | 0.003916189 | 1 |
| Obesity | mast_cell | *SULT1B1* | 3.983965361 | 0.001115768 | 0.001722699 | 1 |
| Obesity | mast_cell | *NUDT4* | 4.634668364 | 1.12E-05 | 2.46E-05 | 0.070102837 |

| | | | | | | |
|---|---|---|---|---|---|---|
| Obesity | mast_cell | *SCAI* | 6.219035463 | 1.49E-06 | 4.02E-06 | 0.009347443 |
| Obesity | mast_cell | *C1orf100* | 14.0482084 | 2.96E-09 | 4.42E-08 | 1.86E-05 |
| Obesity | mast_cell | *AGFG1* | 4.166488109 | 2.81E-06 | 7.03E-06 | 0.017670717 |
| Obesity | mast_cell | *RNF213* | 5.34590533 | 3.81E-07 | 1.23E-06 | 0.002395433 |
| Obesity | mast_cell | *TUBB8* | 8.427131504 | 0.001224359 | 0.00188158 | 1 |
| Obesity | mast_cell | *GPR160* | 6.507735009 | 1.10E-06 | 3.09E-06 | 0.006944113 |
| Obesity | mast_cell | *GOLIM4* | 4.9257528 | 5.78E-05 | 0.000110313 | 0.363480118 |
| Obesity | mast_cell | *USMG5* | 7.114018998 | 1.97E-07 | 7.05E-07 | 0.001238648 |
| Obesity | mast_cell | *HOXB2* | 4.763915568 | 9.37E-11 | 8.18E-09 | 5.89E-07 |
| Obesity | mast_cell | *XXYLT1* | 5.990280243 | 5.29E-06 | 1.24E-05 | 0.033263587 |
| Obesity | mast_cell | *UBXN2A* | 4.476211823 | 3.03E-05 | 6.08E-05 | 0.190406752 |
| Obesity | mast_cell | *CCS* | 7.571437844 | 1.31E-08 | 9.03E-08 | 8.26E-05 |
| Obesity | mast_cell | *NRROS* | 2.132052154 | 0.000317354 | 0.000534621 | 1 |
| Obesity | mast_cell | *LEMD3* | 6.326051607 | 1.50E-06 | 4.06E-06 | 0.009454993 |
| Obesity | mast_cell | *C16orf91* | 4.357824881 | 1.61E-08 | 1.02E-07 | 0.000101192 |
| Obesity | mast_cell | *SOCS7* | 6.608430478 | 4.06E-09 | 4.79E-08 | 2.55E-05 |
| Obesity | mast_cell | *TLR6* | 2.729244987 | 2.53E-09 | 4.32E-08 | 1.59E-05 |
| Obesity | mast_cell | *CTU2* | 7.134545771 | 7.27E-08 | 3.12E-07 | 0.000457344 |
| Obesity | mast_cell | *DDX23* | 6.780484654 | 0.000391608 | 0.000651774 | 1 |
| Obesity | mast_cell | *GLIS1* | 1.585814026 | 1.85E-06 | 4.86E-06 | 0.011645359 |
| Obesity | mast_cell | *RALGAPA1* | 3.069064034 | 0.003916067 | 0.005619793 | 1 |
| Obesity | mast_cell | *LIG4* | 4.298038312 | 3.56E-05 | 7.05E-05 | 0.223734523 |
| Obesity | mast_cell | *SNAPC5* | 3.574311321 | 0.022625596 | 0.029232865 | 1 |
| Obesity | mast_cell | *LINGO2* | -2.760852835 | 0.012212573 | 0.016360633 | 1 |
| Obesity | mast_cell | *BBS1* | 5.174638787 | 9.11E-11 | 8.17E-09 | 5.73E-07 |
| Obesity | mast_cell | *ANKRD36C* | 3.821764915 | 0.002763989 | 0.004043089 | 1 |
| Obesity | mast_cell | *TMEM81* | -6.069112495 | 6.04E-07 | 1.81E-06 | 0.003799652 |
| Obesity | mast_cell | *IL20RB* | 2.472087011 | 0.004764916 | 0.006741006 | 1 |
| Obesity | mast_cell | *ZNF497* | 12.79390546 | 8.26E-08 | 3.47E-07 | 0.000519442 |
| Obesity | mast_cell | *KLF14* | 22.87575768 | 2.08E-11 | 2.61E-09 | 1.31E-07 |
| Obesity | mast_cell | *ANGEL2* | 7.067808734 | 2.29E-07 | 8.01E-07 | 0.001441703 |
| Obesity | mast_cell | *ZNF266* | 2.102429795 | 0.001251824 | 0.001920503 | 1 |
| Obesity | mast_cell | *BRSK2* | 4.187358235 | 3.11E-06 | 7.69E-06 | 0.019562505 |
| Obesity | mast_cell | *SH3PXD2B* | 2.557767096 | 6.34E-09 | 5.93E-08 | 3.99E-05 |
| Obesity | mast_cell | *FGFBP3* | 4.164714138 | 1.24E-08 | 8.68E-08 | 7.77E-05 |
| Obesity | mast_cell | *C4orf32* | -2.092236875 | 0.029057335 | 0.036950538 | 1 |
| Obesity | mast_cell | *HRAS* | 4.440341791 | 1.12E-10 | 9.24E-09 | 7.03E-07 |
| Obesity | mast_cell | *SRP72* | 4.494979842 | 1.22E-06 | 3.37E-06 | 0.007640922 |
| Obesity | mast_cell | *C4orf26* | -6.135443191 | 0.000994808 | 0.001549259 | 1 |
| Obesity | mast_cell | *BTC* | 2.503504012 | 2.07E-08 | 1.17E-07 | 0.000130445 |
| Obesity | mast_cell | *NLRP6* | 9.882669487 | 0.009457928 | 0.012864991 | 1 |
| Obesity | mast_cell | *NDUFA11* | 5.183382661 | 0.017696098 | 0.023183032 | 1 |
| Obesity | mast_cell | *PTDSS2* | 4.54697411 | 3.91E-09 | 4.78E-08 | 2.46E-05 |
| Obesity | mast_cell | *FUT1* | -3.039686089 | 4.88E-05 | 9.47E-05 | 0.306892143 |
| Obesity | mast_cell | *DHX36* | 3.049003062 | 0.026720248 | 0.034158235 | 1 |
| Obesity | mast_cell | *CHST2* | -3.669755372 | 1.27E-06 | 3.49E-06 | 0.007955757 |
| Obesity | mast_cell | *ATR* | 5.438253143 | 6.15E-09 | 5.81E-08 | 3.87E-05 |
| Obesity | mast_cell | *TVP23C* | -2.745183184 | 6.34E-06 | 1.46E-05 | 0.039847927 |
| Obesity | mast_cell | *PACS1* | 4.587413942 | 6.65E-05 | 0.000125307 | 0.418273121 |
| Obesity | mast_cell | *PSMD2* | 4.58577179 | 2.25E-10 | 1.41E-08 | 1.42E-06 |

| | | | | | | |
|---|---|---|---|---|---|---|
| Obesity | mast_cell | *ARHGAP1* | 5.112298185 | 6.01E-07 | 1.80E-06 | 0.003779549 |
| Obesity | mast_cell | *GOLGA8A* | 5.329849416 | 1.48E-10 | 1.12E-08 | 9.32E-07 |
| Obesity | mast_cell | *TP53I11* | 7.115045356 | 7.07E-09 | 6.23E-08 | 4.44E-05 |
| Obesity | mast_cell | *PHYHD1* | 3.907350088 | 0.005981386 | 0.008346597 | 1 |
| Obesity | mast_cell | *ZNF519* | 5.272939596 | 0.000765747 | 0.001213881 | 1 |
| Obesity | mast_cell | *BANF1* | 2.139866269 | 8.18E-05 | 0.000151097 | 0.514334192 |
| Obesity | mast_cell | *CHRNA7* | -4.765976981 | 0.000148335 | 0.000262922 | 0.932582566 |
| Obesity | mast_cell | *TMEM9B* | 9.569056366 | 1.76E-12 | 3.58E-10 | 1.11E-08 |
| Obesity | mast_cell | *NRIP3* | 2.790657172 | 6.98E-05 | 0.000130857 | 0.438634301 |
| Obesity | mast_cell | *SCUBE2* | 4.861414148 | 1.28E-07 | 4.96E-07 | 0.000802361 |
| Obesity | mast_cell | *SMAD2* | 2.910959199 | 0.003571665 | 0.005156155 | 1 |
| Obesity | mast_cell | *EIF3F* | 6.205951717 | 9.16E-06 | 2.05E-05 | 0.057606163 |
| Obesity | mast_cell | *CLTB* | 3.39921392 | 0.007693273 | 0.010590674 | 1 |
| Obesity | mast_cell | *PCSK1* | -1.783823909 | 1.71E-07 | 6.28E-07 | 0.001074612 |
| Obesity | mast_cell | *LPL* | 0.966664256 | 0.019741281 | 0.025707008 | 1 |
| Obesity | mast_cell | *TBC1D10C* | 3.306771203 | 0.000363878 | 0.000607784 | 1 |
| Obesity | mast_cell | *PPP2R2D* | 4.179775266 | 0.000114047 | 0.000205979 | 0.717011758 |
| Obesity | mast_cell | *LRRC25* | 0.951087856 | 0.024979313 | 0.032066571 | 1 |
| Obesity | mast_cell | *UBQLNL* | 5.306925204 | 0.000241448 | 0.000413512 | 1 |
| Obesity | mast_cell | *LIPT2* | 5.599769198 | 7.67E-05 | 0.000142715 | 0.482375855 |
| Obesity | mast_cell | *KCNE3* | -4.113405248 | 0.006141522 | 0.008553776 | 1 |
| Obesity | mast_cell | *UCP3* | 3.654056646 | 4.56E-06 | 1.08E-05 | 0.028685753 |
| Obesity | mast_cell | *UCP2* | 4.791810874 | 0.006820554 | 0.009451361 | 1 |
| Obesity | mast_cell | *RAB6A* | 4.361398977 | 0.000433218 | 0.000715429 | 1 |
| Obesity | mast_cell | *RPS6KB2* | 9.565235679 | 4.96E-07 | 1.52E-06 | 0.003116038 |
| Obesity | mast_cell | *RMI2* | 2.848894122 | 0.000158255 | 0.000279401 | 0.994946984 |
| Obesity | mast_cell | *TOM1L2* | 6.229730501 | 2.67E-06 | 6.72E-06 | 0.016768611 |
| Obesity | mast_cell | *TEX26* | 6.053056537 | 9.36E-09 | 7.37E-08 | 5.89E-05 |
| Obesity | mast_cell | *B3GNTL1* | 7.598676182 | 8.01E-08 | 3.37E-07 | 0.000503325 |
| Obesity | mast_cell | *TTLL11* | 1.72167988 | 0.033040622 | 0.041703752 | 1 |
| Obesity | mast_cell | *LINC01106* | 17.41997838 | 0.000624705 | 0.001003455 | 1 |
| Obesity | mast_cell | *MSRA* | -2.21762449 | 5.08E-06 | 1.19E-05 | 0.031950028 |
| Obesity | mast_cell | *CTDNEP1* | 5.37263932 | 4.98E-10 | 2.35E-08 | 3.13E-06 |
| Obesity | mast_cell | *ETV4* | 5.38816999 | 6.77E-09 | 6.08E-08 | 4.26E-05 |
| Obesity | mast_cell | *SWI5* | 4.963348986 | 2.36E-06 | 6.00E-06 | 0.014812568 |
| Obesity | mast_cell | *ZDHHC21* | 5.204336923 | 4.47E-07 | 1.41E-06 | 0.002810305 |
| Obesity | mast_cell | *A2M* | -1.790479203 | 5.06E-05 | 9.78E-05 | 0.318081897 |
| Obesity | mast_cell | *ARL4D* | 3.015316456 | 2.61E-07 | 8.93E-07 | 0.001637789 |
| Obesity | mast_cell | *LRRN1* | 5.106258471 | 3.35E-09 | 4.52E-08 | 2.11E-05 |
| Obesity | mast_cell | *B3GALT6* | -4.00877518 | 3.03E-06 | 7.52E-06 | 0.01906729 |
| Obesity | mast_cell | *IP6K1* | 3.483500379 | 0.011185747 | 0.015058842 | 1 |
| Obesity | mast_cell | *CHMP6* | 5.288695032 | 5.34E-09 | 5.50E-08 | 3.36E-05 |
| Obesity | mast_cell | *TCP11L1* | 5.266118362 | 1.00E-06 | 2.83E-06 | 0.006284164 |
| Obesity | mast_cell | *GPX2* | -4.830011721 | 0.011946337 | 0.016034718 | 1 |
| Obesity | mast_cell | *CCDC57* | 2.223208523 | 0.019387591 | 0.025277848 | 1 |
| Obesity | mast_cell | *CIDEA* | 0.586491064 | 0.010253233 | 0.013889696 | 1 |
| Obesity | mast_cell | *SMIM19* | 3.666781194 | 1.32E-06 | 3.61E-06 | 0.008274949 |
| Obesity | mast_cell | *SLC35G1* | 2.41322942 | 6.03E-07 | 1.81E-06 | 0.003793344 |
| Obesity | mast_cell | *COX8A* | -4.01078853 | 0.004284122 | 0.006103393 | 1 |
| Obesity | mast_cell | *PRR18* | -5.506087021 | 7.03E-05 | 0.000131644 | 0.441665275 |

| | | | | | | |
|---|---|---|---|---|---|---|
| Obesity | mast_cell | *B3GNT4* | -10.64306861 | 4.88E-07 | 1.51E-06 | 0.003071002 |
| Obesity | mast_cell | *CRLF3* | 4.990036924 | 1.20E-06 | 3.34E-06 | 0.007552295 |
| Obesity | mast_cell | *RNPEP* | 4.109495372 | 2.17E-07 | 7.65E-07 | 0.001362852 |
| Obesity | mast_cell | *VPS37D* | 2.174899567 | 0.000138022 | 0.00024589 | 0.867746257 |
| Obesity | mast_cell | *SLCO3A1* | 3.073886172 | 0.02319931 | 0.02995565 | 1 |
| Obesity | mast_cell | *WDR25* | 6.726590231 | 2.38E-06 | 6.04E-06 | 0.014937329 |
| Obesity | mast_cell | *PLA2G16* | 1.616064775 | 9.45E-06 | 2.11E-05 | 0.059434372 |
| Obesity | mast_cell | *PHLDB3* | 5.180234953 | 4.29E-06 | 1.02E-05 | 0.026942137 |
| Obesity | mast_cell | *RMDN1* | 5.637916885 | 1.21E-08 | 8.63E-08 | 7.64E-05 |
| Obesity | mast_cell | *RNF152* | 4.416625978 | 2.58E-08 | 1.38E-07 | 0.000162449 |
| Obesity | mast_cell | *BOK* | 5.140558406 | 2.73E-06 | 6.86E-06 | 0.017192391 |
| Obesity | mast_cell | *ZNF843* | 16.16077928 | 4.26E-07 | 1.35E-06 | 0.002677425 |
| Obesity | mast_cell | *C8orf59* | 7.37391274 | 1.25E-07 | 4.88E-07 | 0.000787549 |
| Obesity | mast_cell | *PFN4* | 2.102289147 | 0.000212003 | 0.000366372 | 1 |
| Obesity | mast_cell | *RUFY1* | 6.198809263 | 1.18E-09 | 3.75E-08 | 7.42E-06 |
| Obesity | mast_cell | *LRRC37A3* | 5.630540493 | 3.59E-09 | 4.63E-08 | 2.26E-05 |
| Obesity | mast_cell | *VSIG10* | 1.585695037 | 0.009941206 | 0.013481527 | 1 |
| Obesity | mast_cell | *IRX5* | 5.917843771 | 2.06E-06 | 5.33E-06 | 0.012958131 |
| Obesity | mast_cell | *PNMA1* | 6.823503812 | 2.01E-07 | 7.18E-07 | 0.001263922 |
| Obesity | mast_cell | *ANKLE2* | 7.522950013 | 3.09E-06 | 7.64E-06 | 0.019403905 |
| Obesity | mast_cell | *GCNT4* | -1.490203167 | 1.73E-07 | 6.35E-07 | 0.001088815 |
| Obesity | mast_cell | *DPP7* | 7.781178045 | 2.65E-07 | 9.08E-07 | 0.001668865 |
| Obesity | mast_cell | *SEC24C* | 7.444925724 | 2.52E-06 | 6.38E-06 | 0.015837938 |
| Obesity | mast_cell | *MTHFR* | 7.689029611 | 1.17E-07 | 4.64E-07 | 0.000737973 |
| Obesity | mast_cell | *C19orf18* | -3.145533804 | 0.009392041 | 0.012780901 | 1 |
| Obesity | mast_cell | *WDR73* | 4.383742996 | 2.11E-07 | 7.47E-07 | 0.001327353 |
| Obesity | mast_cell | *DSCAML1* | 2.917751841 | 4.20E-09 | 4.83E-08 | 2.64E-05 |
| Obesity | mast_cell | *EPS8L2* | 5.186568417 | 2.97E-05 | 5.98E-05 | 0.186945913 |
| Obesity | mast_cell | *TALDO1* | 2.005529746 | 0.036885578 | 0.046251202 | 1 |
| Obesity | mast_cell | *RIMKLA* | 3.031136426 | 9.05E-06 | 2.03E-05 | 0.056917717 |
| Obesity | mast_cell | *B3GNT8* | 11.66760817 | 5.18E-09 | 5.38E-08 | 3.25E-05 |
| Obesity | mast_cell | *AP006621.1* | 2.253289846 | 1.10E-10 | 9.24E-09 | 6.93E-07 |
| Obesity | mast_cell | *TOP3A* | 5.700779834 | 2.12E-05 | 4.39E-05 | 0.133260153 |
| Obesity | mast_cell | *ZBTB38* | 5.93457806 | 2.31E-07 | 8.06E-07 | 0.001451549 |
| Obesity | mast_cell | *C8orf31* | 5.435021278 | 4.98E-09 | 5.27E-08 | 3.13E-05 |
| Obesity | mast_cell | *AC024940.1* | 5.809986521 | 9.15E-07 | 2.62E-06 | 0.00575239 |
| Obesity | mast_cell | *TIMM22* | -4.377050279 | 0.000549829 | 0.000890922 | 1 |
| Obesity | mast_cell | *PPFIA3* | 1.804304526 | 0.00135805 | 0.002073854 | 1 |
| Obesity | mast_cell | *MAGEF1* | -4.603064356 | 8.15E-05 | 0.000150626 | 0.512251512 |
| Obesity | mast_cell | *SAMD9L* | 2.504155013 | 0.006867012 | 0.009511545 | 1 |
| Obesity | mast_cell | *PAWR* | 4.252063018 | 2.26E-09 | 4.32E-08 | 1.42E-05 |
| Obesity | mast_cell | *NAP1L5* | -3.133346803 | 4.61E-05 | 8.98E-05 | 0.290108112 |
| Obesity | mast_cell | *NIM1K* | 3.110291721 | 4.04E-08 | 1.95E-07 | 0.000254019 |
| Obesity | mast_cell | *ACOT4* | 2.84310028 | 1.85E-06 | 4.86E-06 | 0.011649719 |
| Obesity | mast_cell | *ARIH2* | 2.846157253 | 0.000975272 | 0.001521472 | 1 |
| Obesity | mast_cell | *IRX3* | -3.075249136 | 0.003457546 | 0.004998297 | 1 |
| Obesity | mast_cell | *ATOX1* | 6.50311471 | 0.000271034 | 0.000460663 | 1 |
| Obesity | mast_cell | *TBL1XR1* | 4.140106506 | 8.90E-06 | 2.00E-05 | 0.055938679 |
| Obesity | mast_cell | *SAMD12* | 2.192027908 | 0.000682279 | 0.001087598 | 1 |
| Obesity | mast_cell | *C18orf32* | 6.863183784 | 2.77E-05 | 5.61E-05 | 0.174151372 |

| | | | | | | |
|---|---|---|---|---|---|---|
| Obesity | mast_cell | *RPLP2* | 3.804507178 | 0.007508198 | 0.010349493 | 1 |
| Obesity | mast_cell | *GBA* | 7.701089329 | 1.82E-08 | 1.08E-07 | 0.000114716 |
| Obesity | mast_cell | *CD163L1* | 1.754704256 | 1.30E-08 | 9.01E-08 | 8.18E-05 |
| Obesity | mast_cell | *THAP5* | 8.260969671 | 3.27E-07 | 1.08E-06 | 0.002058752 |
| Obesity | mast_cell | *CD151* | 4.995919117 | 0.000427318 | 0.000706243 | 1 |
| Obesity | mast_cell | *FAM87B* | 4.774878417 | 0.000183618 | 0.000320224 | 1 |
| Obesity | mast_cell | *ZCCHC3* | -5.015134891 | 0.010747496 | 0.014518588 | 1 |
| Obesity | mast_cell | *PCDHB9* | 2.55031111 | 2.52E-05 | 5.13E-05 | 0.15840522 |
| Obesity | mast_cell | *ZNF619* | 3.094395719 | 2.10E-08 | 1.18E-07 | 0.000132049 |
| Obesity | mast_cell | *ZNF354C* | 5.279856714 | 1.94E-09 | 4.32E-08 | 1.22E-05 |
| Obesity | mast_cell | *CENPBD1* | 3.536407484 | 0.006735027 | 0.00933902 | 1 |
| Obesity | mast_cell | *BET1L* | 5.091141782 | 4.01E-09 | 4.78E-08 | 2.52E-05 |
| Obesity | mast_cell | *RIC8A* | 5.225921925 | 1.69E-06 | 4.48E-06 | 0.01059605 |
| Obesity | mast_cell | *ODF3B* | 7.272772432 | 4.09E-07 | 1.31E-06 | 0.002568949 |
| Obesity | mast_cell | *DPY19L2* | -1.348212714 | 0.029912448 | 0.037930529 | 1 |
| Obesity | mast_cell | *C2orf73* | 8.004838765 | 3.28E-09 | 4.50E-08 | 2.06E-05 |
| Obesity | mast_cell | *FAM26E* | 5.318816957 | 6.73E-07 | 1.99E-06 | 0.004232993 |
| Obesity | mast_cell | *ALS2CL* | 3.276041711 | 9.82E-08 | 4.02E-07 | 0.000617429 |
| Obesity | mast_cell | *MLF1* | 6.475529819 | 1.44E-07 | 5.44E-07 | 0.000902187 |
| Obesity | mast_cell | *C2orf69* | 8.371845158 | 8.09E-06 | 1.83E-05 | 0.050841822 |
| Obesity | mast_cell | *GRAMD1C* | 7.098567466 | 9.17E-09 | 7.26E-08 | 5.76E-05 |
| Obesity | mast_cell | *STAP2* | 5.630967022 | 0.001041344 | 0.001615728 | 1 |
| Obesity | mast_cell | *TSSK6* | 4.11946588 | 0.00143667 | 0.002184892 | 1 |
| Obesity | mast_cell | *GOLGA8Q* | 47.1150298 | 0.001502042 | 0.00227715 | 1 |
| Obesity | mast_cell | *DALRD3* | 3.447145453 | 0.000115021 | 0.0002075 | 0.723136747 |
| Obesity | mast_cell | *ZNF114* | -7.643335264 | 2.49E-05 | 5.08E-05 | 0.156471547 |
| Obesity | mast_cell | *ZNF518B* | 5.0310751 | 1.44E-07 | 5.45E-07 | 0.000905877 |
| Obesity | mast_cell | *LCORL* | 2.86293049 | 0.033317751 | 0.0420198 | 1 |
| Obesity | mast_cell | *ZNF454* | 3.01384672 | 1.60E-08 | 1.02E-07 | 0.000100331 |
| Obesity | mast_cell | *SH2B1* | -3.147128972 | 1.96E-08 | 1.13E-07 | 0.000123279 |
| Obesity | mast_cell | *ZC3H12D* | 6.119715889 | 3.33E-09 | 4.52E-08 | 2.09E-05 |
| Obesity | mast_cell | *KDELC2* | 5.552952494 | 1.63E-07 | 6.01E-07 | 0.001022391 |
| Obesity | mast_cell | *PLEC* | 6.519821784 | 2.89E-07 | 9.75E-07 | 0.001819034 |
| Obesity | mast_cell | *SH2D4B* | -10.94204988 | 0.002754948 | 0.004031741 | 1 |
| Obesity | mast_cell | *RNF212* | 1.717720181 | 0.0001123 | 0.000203174 | 0.70602854 |
| Obesity | mast_cell | *PRSS36* | 3.625282122 | 1.58E-08 | 1.01E-07 | 9.92E-05 |
| Obesity | mast_cell | *GEN1* | 1.963166535 | 0.000487371 | 0.000796905 | 1 |
| Obesity | mast_cell | *TMPRSS9* | 3.814182997 | 0.00166378 | 0.002504233 | 1 |
| Obesity | mast_cell | *AQP11* | 5.428228891 | 0.004319919 | 0.006150211 | 1 |
| Obesity | mast_cell | *TMEM11* | 10.62188837 | 3.31E-08 | 1.66E-07 | 0.000208129 |
| Obesity | mast_cell | *KCNG2* | -1.537444758 | 0.001642516 | 0.002474005 | 1 |
| Obesity | mast_cell | *ZFAND2A* | 4.200003828 | 1.14E-08 | 8.37E-08 | 7.18E-05 |
| Obesity | mast_cell | *FAM220A* | 2.429833419 | 0.000114447 | 0.000206643 | 0.719530538 |
| Obesity | mast_cell | *BEND3* | -6.883006234 | 0.002507785 | 0.003693241 | 1 |
| Obesity | mast_cell | *NT5DC1* | 4.184648568 | 0.00123336 | 0.0018931 | 1 |
| Obesity | mast_cell | *AC006111.1* | 13.68738701 | 1.73E-08 | 1.05E-07 | 0.000108607 |
| Obesity | mast_cell | *LINC00843* | 6.979697917 | 0.020122191 | 0.026175919 | 1 |
| Obesity | mast_cell | *COX14* | 3.821923554 | 1.23E-09 | 3.78E-08 | 7.73E-06 |
| Obesity | mast_cell | *CTXN1* | -6.310112333 | 0.004467096 | 0.006352552 | 1 |
| Obesity | mast_cell | *CA8* | 2.28761903 | 0.000441145 | 0.000727565 | 1 |

| | | | | | | |
|---|---|---|---|---|---|---|
| Obesity | mast_cell | *ERBB4* | 3.890571923 | 3.16E-09 | 4.48E-08 | 1.99E-05 |
| Obesity | mast_cell | *CSRNP3* | 6.10386578 | 5.23E-08 | 2.40E-07 | 0.000329035 |
| Obesity | mast_cell | *PARP10* | 4.362765924 | 1.63E-05 | 3.47E-05 | 0.102422211 |
| Obesity | mast_cell | *STX19* | 7.925268929 | 4.32E-07 | 1.37E-06 | 0.002718912 |
| Obesity | mast_cell | *FAM219B* | 5.001476452 | 1.50E-10 | 1.12E-08 | 9.41E-07 |
| Obesity | mast_cell | *GDPD4* | -8.629494436 | 4.36E-05 | 8.52E-05 | 0.274381953 |
| Obesity | mast_cell | *MPI* | 7.481383548 | 8.57E-06 | 1.93E-05 | 0.053859901 |
| Obesity | mast_cell | *TRIM73* | -6.941954848 | 1.20E-05 | 2.63E-05 | 0.075558839 |
| Obesity | mast_cell | *C17orf62* | 8.467649142 | 1.96E-06 | 5.09E-06 | 0.012307165 |
| Obesity | mast_cell | *GAK* | 6.219100025 | 1.38E-07 | 5.28E-07 | 0.000866421 |
| Obesity | mast_cell | *TUFM* | 9.928120627 | 2.94E-29 | 9.25E-26 | 1.85E-25 |
| Obesity | mast_cell | *RMI1* | 3.276983771 | 4.77E-08 | 2.24E-07 | 0.000300105 |
| Obesity | mast_cell | *CTC1* | 4.532355848 | 5.96E-05 | 0.000113385 | 0.374963626 |
| Obesity | mast_cell | *SNX18* | 5.467594429 | 8.87E-08 | 3.68E-07 | 0.000557735 |
| Obesity | mast_cell | *C14orf39* | 1.587417572 | 0.001448577 | 0.002201935 | 1 |
| Obesity | mast_cell | *TRIML2* | 41.8878938 | 2.36E-06 | 6.01E-06 | 0.014843547 |
| Obesity | mast_cell | *FARSA* | 3.973040663 | 0.000565036 | 0.000913443 | 1 |
| Obesity | mast_cell | *SPTY2D1* | 8.308924916 | 7.09E-06 | 1.62E-05 | 0.044591423 |
| Obesity | mast_cell | *TCAIM* | 3.558610792 | 0.000122034 | 0.000219207 | 0.767225584 |
| Obesity | mast_cell | *FUCA1* | 4.992142458 | 2.26E-05 | 4.65E-05 | 0.141888207 |
| Obesity | mast_cell | *ZNF664* | 7.89247087 | 4.30E-08 | 2.05E-07 | 0.000270282 |
| Obesity | mast_cell | *CALR* | 6.976014664 | 7.43E-10 | 2.88E-08 | 4.67E-06 |
| Obesity | mast_cell | *CDH4* | -2.600173202 | 0.000555433 | 0.000899075 | 1 |
| Obesity | mast_cell | *EGR3* | -1.122715706 | 2.92E-07 | 9.84E-07 | 0.001837102 |
| Obesity | mast_cell | *GEMIN4* | 1.988771629 | 1.88E-05 | 3.95E-05 | 0.11832891 |
| Obesity | mast_cell | *FJX1* | 5.029273744 | 1.04E-06 | 2.92E-06 | 0.006512101 |
| Obesity | mast_cell | *CIITA* | 3.607709786 | 4.78E-07 | 1.48E-06 | 0.003004244 |
| Obesity | mast_cell | *PLD6* | -2.954094751 | 1.60E-05 | 3.41E-05 | 0.10055863 |
| Obesity | mast_cell | *ZBTB42* | 18.35389862 | 1.40E-07 | 5.34E-07 | 0.000878839 |
| Obesity | mast_cell | *LACC1* | 7.662964374 | 1.54E-09 | 4.16E-08 | 9.70E-06 |
| Obesity | mast_cell | *MAF1* | 8.689724452 | 5.69E-09 | 5.59E-08 | 3.57E-05 |
| Obesity | mast_cell | *FCER1A* | -1.051423093 | 1.85E-07 | 6.71E-07 | 0.001163167 |
| Obesity | mast_cell | *PCED1B* | 4.084070058 | 0.000118017 | 0.000212478 | 0.741972581 |
| Obesity | mast_cell | *PIPOX* | 3.810523015 | 5.92E-05 | 0.000112702 | 0.372367219 |
| Obesity | mast_cell | *FOXS1* | 2.436721891 | 0.033271296 | 0.041969631 | 1 |
| Obesity | mast_cell | *CDH5* | 3.391860064 | 5.25E-08 | 2.41E-07 | 0.0003298 |
| Obesity | mast_cell | *SERTAD2* | 6.546558893 | 3.43E-08 | 1.71E-07 | 0.000215578 |
| Obesity | mast_cell | *CITED4* | 3.295611756 | 6.44E-08 | 2.82E-07 | 0.000404816 |
| Obesity | mast_cell | *TIGD5* | -3.539790038 | 3.42E-06 | 8.37E-06 | 0.021483435 |
| Obesity | mast_cell | *PDXDC1* | 4.442672278 | 0.000449653 | 0.000741014 | 1 |
| Obesity | mast_cell | *ZNF154* | 3.050352514 | 3.69E-08 | 1.81E-07 | 0.000232244 |
| Obesity | mast_cell | *R3HDM2* | 4.456230043 | 0.000278735 | 0.000472857 | 1 |
| Obesity | mast_cell | *B3GNT3* | -3.183104016 | 0.039189135 | 0.048953326 | 1 |
| Obesity | mast_cell | *SSC5D* | 1.321725142 | 0.000622898 | 0.001000808 | 1 |
| Obesity | mast_cell | *ZADH2* | 5.983656457 | 0.000356822 | 0.000596951 | 1 |
| Obesity | mast_cell | *TMEM86B* | 2.731202496 | 1.97E-07 | 7.04E-07 | 0.00123704 |
| Obesity | mast_cell | *SEPT1* | 3.004719881 | 1.09E-08 | 8.15E-08 | 6.84E-05 |
| Obesity | mast_cell | *EXOC3* | 3.980319915 | 0.000530786 | 0.00086251 | 1 |
| Obesity | mast_cell | *TDRD6* | 1.507593882 | 0.00299637 | 0.004365742 | 1 |
| Obesity | mast_cell | *LYNX1* | 3.977211891 | 1.22E-05 | 2.66E-05 | 0.07647273 |

| | | | | | | |
|---|---|---|---|---|---|---|
| Obesity | mast_cell | *TDRP* | 7.612133982 | 1.24E-07 | 4.83E-07 | 0.000777554 |
| Obesity | mast_cell | *ZNF816* | 3.052570823 | 9.60E-06 | 2.14E-05 | 0.060324287 |
| Obesity | mast_cell | *FGD6* | 6.602855851 | 7.18E-09 | 6.29E-08 | 4.51E-05 |
| Obesity | mast_cell | *HCLS1* | 8.179610875 | 1.75E-07 | 6.39E-07 | 0.001098052 |
| Obesity | mast_cell | *ZNF609* | 4.252788925 | 7.32E-05 | 0.000136677 | 0.460326467 |
| Obesity | mast_cell | *CCDC66* | 3.451973692 | 0.000504137 | 0.000821755 | 1 |
| Obesity | mast_cell | *DEFB124* | -2.45918644 | 1.98E-07 | 7.07E-07 | 0.001243938 |
| Obesity | mast_cell | *MCFD2* | 6.881952928 | 4.64E-07 | 1.45E-06 | 0.002914928 |
| Obesity | mast_cell | *CYP8B1* | 3.759468876 | 1.29E-06 | 3.54E-06 | 0.008084345 |
| Obesity | mast_cell | *RNF182* | 2.081389741 | 0.000660615 | 0.001057509 | 1 |
| Obesity | mast_cell | *MB21D2* | 6.709140402 | 1.33E-08 | 9.09E-08 | 8.36E-05 |
| Obesity | mast_cell | *SSTR2* | 4.815207293 | 1.32E-08 | 9.05E-08 | 8.30E-05 |
| Obesity | mast_cell | *PCGF5* | 4.724246808 | 0.000676019 | 0.001078988 | 1 |
| Obesity | mast_cell | *SHISA2* | 57.77338262 | 1.58E-08 | 1.01E-07 | 9.94E-05 |
| Obesity | mast_cell | *CHST13* | -3.7061514 | 7.15E-07 | 2.10E-06 | 0.004492959 |
| Obesity | mast_cell | *SRSF8* | 9.564175286 | 0.000336209 | 0.000564418 | 1 |
| Obesity | mast_cell | *SLC36A4* | 5.772276339 | 4.69E-06 | 1.11E-05 | 0.029502923 |
| Obesity | mast_cell | *ZDHHC20* | 6.221106323 | 2.35E-06 | 5.98E-06 | 0.01475757 |
| Obesity | mast_cell | *OR51E1* | 1.561859968 | 0.005211882 | 0.007335371 | 1 |
| Obesity | mast_cell | *PSMG4* | 4.473057634 | 0.00071095 | 0.001131008 | 1 |
| Obesity | mast_cell | *CAPS2* | 5.592175009 | 5.56E-09 | 5.56E-08 | 3.50E-05 |
| Obesity | mast_cell | *ZNF792* | -3.154445563 | 1.31E-06 | 3.59E-06 | 0.008220802 |
| Obesity | mast_cell | *KCTD2* | 8.279628393 | 6.36E-09 | 5.93E-08 | 4.00E-05 |
| Obesity | mast_cell | *D2HGDH* | 4.563224785 | 2.44E-05 | 4.98E-05 | 0.153100051 |
| Obesity | mast_cell | *LRRC57* | 5.849823994 | 0.001073112 | 0.001660919 | 1 |
| Obesity | mast_cell | *MRPL14* | 6.338601558 | 5.98E-07 | 1.79E-06 | 0.003758297 |
| Obesity | mast_cell | *BBS12* | 3.467548378 | 9.22E-09 | 7.27E-08 | 5.80E-05 |
| Obesity | mast_cell | *ZFP82* | 4.788855731 | 1.78E-09 | 4.31E-08 | 1.12E-05 |
| Obesity | mast_cell | *LSMEM1* | 3.046355978 | 1.10E-07 | 4.40E-07 | 0.000691886 |
| Obesity | mast_cell | *AEN* | 3.590506889 | 3.37E-08 | 1.68E-07 | 0.000211745 |
| Obesity | mast_cell | *FKRP* | 2.060446618 | 0.000923135 | 0.001446238 | 1 |
| Obesity | mast_cell | *TRAPPC5* | 4.640163801 | 5.18E-06 | 1.21E-05 | 0.032576427 |
| Obesity | mast_cell | *RPH3AL* | 5.702034141 | 3.86E-07 | 1.25E-06 | 0.002428259 |
| Obesity | mast_cell | *ANKRD34A* | -5.443000477 | 0.004557147 | 0.006470367 | 1 |
| Obesity | mast_cell | *HIGD1A* | 3.319429278 | 2.85E-05 | 5.75E-05 | 0.17912267 |
| Obesity | mast_cell | *OR52N4* | 2.204292148 | 0.021620843 | 0.028003758 | 1 |
| Obesity | mast_cell | *ADIPOQ* | 0.870710543 | 0.004064402 | 0.005814084 | 1 |
| Obesity | mast_cell | *F2R* | 2.093406397 | 4.75E-06 | 1.12E-05 | 0.029874367 |
| Obesity | mast_cell | *HIST3H2A* | 2.62449545 | 0.003730633 | 0.005372077 | 1 |
| Obesity | mast_cell | *POLR2A* | 5.325723894 | 0.000489776 | 0.000800421 | 1 |
| Obesity | mast_cell | *TMEM132C* | 2.811049096 | 3.57E-06 | 8.69E-06 | 0.022441554 |
| Obesity | mast_cell | *TMEM136* | -3.013271274 | 6.36E-06 | 1.47E-05 | 0.039999478 |
| Obesity | mast_cell | *TMEM132E* | 9.053938488 | 0.001981553 | 0.002954239 | 1 |
| Obesity | mast_cell | *ZNF322* | -3.024062599 | 0.006095544 | 0.008495385 | 1 |
| Obesity | mast_cell | *DDX60L* | 5.907808484 | 4.20E-08 | 2.01E-07 | 0.000264092 |
| Obesity | mast_cell | *OGFOD3* | 2.288101235 | 2.18E-06 | 5.60E-06 | 0.013687673 |
| Obesity | mast_cell | *ACBD4* | 4.918358509 | 4.03E-09 | 4.78E-08 | 2.53E-05 |
| Obesity | mast_cell | *SGSH* | 10.97912256 | 1.53E-09 | 4.16E-08 | 9.62E-06 |
| Obesity | mast_cell | *SETD2* | 3.650818686 | 0.00098507 | 0.001535615 | 1 |
| Obesity | mast_cell | *MRPS23* | 6.74813042 | 8.46E-10 | 3.04E-08 | 5.32E-06 |

| | | | | | | |
|---|---|---|---|---|---|---|
| Obesity | mast_cell | *GPR135* | 3.509866089 | 1.23E-07 | 4.82E-07 | 0.000774715 |
| Obesity | mast_cell | *ZFP41* | 3.595699678 | 1.17E-07 | 4.63E-07 | 0.000736451 |
| Obesity | mast_cell | *ATG9B* | -8.589949308 | 0.0263708 | 0.033759564 | 1 |
| Obesity | mast_cell | *HKR1* | 6.181138117 | 6.61E-08 | 2.88E-07 | 0.000415854 |
| Obesity | mast_cell | *C3orf58* | 5.776740388 | 1.70E-06 | 4.50E-06 | 0.010693951 |
| Obesity | mast_cell | *AMIGO1* | 1.680469596 | 0.005156945 | 0.007266184 | 1 |
| Obesity | mast_cell | *ODF3L2* | 2.865410063 | 0.022656401 | 0.029266652 | 1 |
| Obesity | mast_cell | *RELL1* | 5.617510734 | 0.00013815 | 0.000246001 | 0.8685516 |
| Obesity | mast_cell | *RFX7* | 4.167562834 | 0.002079908 | 0.003093537 | 1 |
| Obesity | mast_cell | *CLDN7* | 1.747355541 | 0.000472315 | 0.000773898 | 1 |
| Obesity | mast_cell | *SNRPE* | 9.352253466 | 1.10E-09 | 3.59E-08 | 6.93E-06 |
| Obesity | mast_cell | *CHST15* | 5.884067506 | 1.83E-08 | 1.08E-07 | 0.000115005 |
| Obesity | mast_cell | *TNRC18* | 6.022329773 | 1.05E-06 | 2.94E-06 | 0.006574955 |
| Obesity | mast_cell | *FAM181B* | 6.202665241 | 0.000965145 | 0.001506422 | 1 |
| Obesity | mast_cell | *NOP10* | 3.69950308 | 0.000174086 | 0.000305039 | 1 |
| Obesity | mast_cell | *FAM89A* | 3.536942419 | 6.76E-08 | 2.93E-07 | 0.000424983 |
| Obesity | mast_cell | *ZNF708* | 4.614953042 | 2.55E-06 | 6.44E-06 | 0.016021246 |
| Obesity | mast_cell | *IST1* | 5.762759915 | 8.75E-07 | 2.52E-06 | 0.005500417 |
| Obesity | mast_cell | *ERCC6L2* | 4.807128286 | 1.35E-07 | 5.20E-07 | 0.000849613 |
| Obesity | mast_cell | *HHIPL1* | 2.82922018 | 5.75E-08 | 2.59E-07 | 0.000361787 |
| Obesity | mast_cell | *FAM153B* | 4.304593058 | 2.19E-06 | 5.62E-06 | 0.013753259 |
| Obesity | mast_cell | *BACE2* | -0.960247543 | 1.22E-06 | 3.37E-06 | 0.007659337 |
| Obesity | mast_cell | *SYNM* | 2.805255642 | 8.41E-09 | 6.89E-08 | 5.29E-05 |
| Obesity | mast_cell | *IZUMO1* | -1.074556351 | 0.034613551 | 0.0436103 | 1 |
| Obesity | mast_cell | *C8orf33* | 5.619601507 | 5.24E-05 | 0.00010097 | 0.329263035 |
| Obesity | mast_cell | *FBXL6* | 3.251073412 | 5.01E-07 | 1.54E-06 | 0.003149446 |
| Obesity | mast_cell | *KBTBD3* | 4.015993978 | 6.02E-09 | 5.76E-08 | 3.79E-05 |
| Obesity | mast_cell | *CLN8* | 6.174804676 | 5.18E-07 | 1.58E-06 | 0.003254566 |
| Obesity | mast_cell | *CACNB4* | 1.880628063 | 4.87E-07 | 1.51E-06 | 0.003060664 |
| Obesity | mast_cell | *TRAPPC6B* | 4.803262512 | 8.63E-07 | 2.49E-06 | 0.005423489 |
| Obesity | mast_cell | *TSHZ2* | 3.556721295 | 1.70E-05 | 3.59E-05 | 0.106744562 |
| Obesity | mast_cell | *FES* | 5.233846525 | 0.021518459 | 0.027894134 | 1 |
| Obesity | mast_cell | *GLRX5* | 4.992961557 | 4.72E-09 | 5.10E-08 | 2.97E-05 |
| Obesity | mast_cell | *MXRA7* | 2.222436544 | 3.22E-06 | 7.92E-06 | 0.020217346 |
| Obesity | mast_cell | *ADI1* | 7.142124205 | 3.57E-06 | 8.69E-06 | 0.02243635 |
| Obesity | mast_cell | *SPNS3* | 5.117785507 | 5.91E-09 | 5.71E-08 | 3.72E-05 |
| Obesity | mast_cell | *CLEC4G* | 6.579762569 | 2.05E-08 | 1.16E-07 | 0.000129153 |
| Obesity | mast_cell | *SATB1* | 6.105353226 | 1.01E-08 | 7.80E-08 | 6.37E-05 |
| Obesity | mast_cell | *ACTL10* | -4.996840663 | 3.05E-05 | 6.13E-05 | 0.191984413 |
| Obesity | mast_cell | *TRAK1* | 3.929633049 | 0.00070011 | 0.001114891 | 1 |
| Obesity | mast_cell | *SKA2* | 4.472027301 | 0.017791953 | 0.023303752 | 1 |
| Obesity | mast_cell | *TTC3* | 1.006115704 | 0.006678621 | 0.009266937 | 1 |
| Obesity | mast_cell | *KCNB2* | 1.760446752 | 1.46E-06 | 3.94E-06 | 0.009148428 |
| Obesity | mast_cell | *BRICD5* | -1.814489976 | 0.028842941 | 0.036715038 | 1 |
| Obesity | mast_cell | *TSKU* | 5.891395498 | 0.003462144 | 0.005003794 | 1 |
| Obesity | mast_cell | *GRID1* | -2.910492708 | 0.032965536 | 0.041617334 | 1 |
| Obesity | mast_cell | *PLCXD3* | 3.103492797 | 0.000208907 | 0.000361519 | 1 |
| Obesity | mast_cell | *GPIHBP1* | -1.192827284 | 0.012382067 | 0.016562991 | 1 |
| Obesity | mast_cell | *COL18A1* | 4.616589043 | 6.41E-08 | 2.81E-07 | 0.000402768 |
| Obesity | mast_cell | *SLC25A18* | 2.371452976 | 3.89E-05 | 7.65E-05 | 0.244729282 |

| | | | | | | |
|---|---|---|---|---|---|---|
| Obesity | mast_cell | *ZNF721* | 2.823089069 | 0.00708404 | 0.00978413 | 1 |
| Obesity | mast_cell | *LENG9* | 4.34643725 | 1.82E-06 | 4.78E-06 | 0.011438898 |
| Obesity | mast_cell | *HMGN4* | 5.778176477 | 7.48E-06 | 1.70E-05 | 0.047024555 |
| Obesity | mast_cell | *ZNF320* | 1.040189151 | 0.003956527 | 0.00567009 | 1 |
| Obesity | mast_cell | *C12orf60* | 4.812715499 | 4.40E-08 | 2.09E-07 | 0.000276918 |
| Obesity | mast_cell | *LYSMD4* | 3.080715723 | 1.06E-10 | 9.10E-09 | 6.64E-07 |
| Obesity | mast_cell | *WBP2NL* | 8.721204262 | 2.97E-07 | 9.98E-07 | 0.001867186 |
| Obesity | mast_cell | *GAS6* | 6.099398151 | 1.40E-07 | 5.34E-07 | 0.000879556 |
| Obesity | mast_cell | *BEGAIN* | -1.346410746 | 1.84E-07 | 6.69E-07 | 0.001157869 |
| Obesity | mast_cell | *ARHGEF37* | 8.923682795 | 6.00E-07 | 1.80E-06 | 0.003775238 |
| Obesity | mast_cell | *FAM43B* | 5.731840855 | 0.0085663 | 0.011715538 | 1 |
| Obesity | mast_cell | *CSMD1* | 5.207021219 | 1.21E-07 | 4.76E-07 | 0.000763635 |
| Obesity | mast_cell | *PTGDR2* | 52.91881406 | 2.20E-05 | 4.54E-05 | 0.138287688 |
| Obesity | mast_cell | *GPR19* | 8.034880363 | 7.40E-09 | 6.38E-08 | 4.65E-05 |
| Obesity | mast_cell | *FANCF* | 14.10637277 | 0.000222219 | 0.000382554 | 1 |
| Obesity | mast_cell | *SMDT1* | 1.37786356 | 0.00265564 | 0.003896385 | 1 |
| Obesity | mast_cell | *C2CD4C* | 5.883605146 | 4.64E-09 | 5.09E-08 | 2.91E-05 |
| Obesity | mast_cell | *RUVBL2* | -2.581282943 | 0.001430984 | 0.002177299 | 1 |
| Obesity | mast_cell | *GDPGP1* | 4.10687716 | 1.81E-08 | 1.07E-07 | 0.000113528 |
| Obesity | mast_cell | *CTNNA3* | 3.814270498 | 0.000157794 | 0.000278666 | 0.99205039 |
| Obesity | mast_cell | *CCBE1* | 2.273010831 | 6.53E-11 | 6.22E-09 | 4.11E-07 |
| Obesity | mast_cell | *ZNF623* | 6.830105682 | 2.81E-05 | 5.69E-05 | 0.176924581 |
| Obesity | mast_cell | *CCDC125* | 4.998079288 | 6.68E-09 | 6.04E-08 | 4.20E-05 |
| Obesity | mast_cell | *C10orf107* | 0.666931605 | 0.003952324 | 0.005665358 | 1 |
| Obesity | mast_cell | *FHL3* | -4.249442153 | 8.64E-05 | 0.000158842 | 0.543398537 |
| Obesity | mast_cell | *C19orf71* | 5.657910915 | 5.60E-09 | 5.57E-08 | 3.52E-05 |
| Obesity | mast_cell | *LRIT3* | 3.964699161 | 9.65E-05 | 0.00017594 | 0.606641256 |
| Obesity | mast_cell | *SF3A3* | 5.593244927 | 5.31E-08 | 2.43E-07 | 0.000333991 |
| Obesity | mast_cell | *GRIN2A* | 2.73991311 | 7.30E-07 | 2.14E-06 | 0.004591779 |
| Obesity | mast_cell | *URAD* | 9.53870078 | 9.69E-07 | 2.75E-06 | 0.006094923 |
| Obesity | mast_cell | *GPR132* | 5.973491763 | 4.93E-08 | 2.30E-07 | 0.000309704 |
| Obesity | mast_cell | *MX2* | 3.647306008 | 3.56E-05 | 7.05E-05 | 0.22361268 |
| Obesity | mast_cell | *PSMG1* | 2.026050545 | 1.31E-05 | 2.84E-05 | 0.082391574 |
| Obesity | mast_cell | *ACSM5* | 2.696369706 | 0.000546422 | 0.000886544 | 1 |
| Obesity | mast_cell | *AC131971.1* | -6.44425408 | 1.37E-06 | 3.73E-06 | 0.008592724 |
| Obesity | mast_cell | *SERHL2* | 2.931058245 | 3.93E-09 | 4.78E-08 | 2.47E-05 |
| Obesity | mast_cell | *TANGO2* | 7.472096614 | 2.74E-08 | 1.45E-07 | 0.00017245 |
| Obesity | mast_cell | *FAM167B* | 9.027758049 | 7.12E-06 | 1.63E-05 | 0.044791935 |
| Obesity | mast_cell | *RP1L1* | 9.055333182 | 3.00E-14 | 1.05E-11 | 1.89E-10 |
| Obesity | mast_cell | *NDUFB1* | 3.654721519 | 3.13E-08 | 1.59E-07 | 0.000196843 |
| Obesity | mast_cell | *FAM19A1* | 2.771616834 | 5.68E-09 | 5.59E-08 | 3.57E-05 |
| Obesity | mast_cell | *TRMT12* | 3.630749787 | 2.33E-07 | 8.13E-07 | 0.001466767 |
| Obesity | mast_cell | *CMTM4* | 3.227203801 | 4.70E-09 | 5.10E-08 | 2.96E-05 |
| Obesity | mast_cell | *FIGLA* | -10.3646892 | 0.000495415 | 0.000809006 | 1 |
| Obesity | mast_cell | *CBX6* | 8.829517279 | 4.39E-10 | 2.14E-08 | 2.76E-06 |
| Obesity | mast_cell | *KREMEN1* | -0.751080585 | 0.00047722 | 0.000781323 | 1 |
| Obesity | mast_cell | *KCTD16* | 4.546030684 | 2.22E-10 | 1.41E-08 | 1.39E-06 |
| Obesity | mast_cell | *B3GALT5* | 5.160163361 | 0.000415503 | 0.000688344 | 1 |
| Obesity | mast_cell | *ZNF703* | -2.444483768 | 3.20E-05 | 6.38E-05 | 0.201050027 |
| Obesity | mast_cell | *KCTD8* | 2.086955994 | 1.55E-07 | 5.80E-07 | 0.000973891 |

| | | | | | | |
|---|---|---|---|---|---|---|
| Obesity | mast_cell | *C9orf66* | 8.469359381 | 3.22E-08 | 1.63E-07 | 0.000202338 |
| Obesity | mast_cell | *TUBA8* | 3.243714093 | 0.02064571 | 0.026829182 | 1 |
| Obesity | mast_cell | *NPIPA5* | 0.866856046 | 0.009561613 | 0.012989165 | 1 |
| Obesity | mast_cell | *BTBD9* | 4.166326468 | 0.000143756 | 0.000255526 | 0.903795994 |
| Obesity | mast_cell | *ANKRD45* | 6.42860856 | 2.27E-08 | 1.26E-07 | 0.000142434 |
| Obesity | mast_cell | *MAATS1* | 2.371596551 | 0.013365224 | 0.017802366 | 1 |
| Obesity | mast_cell | *GPR39* | 2.873147589 | 5.31E-08 | 2.43E-07 | 0.000334036 |
| Obesity | mast_cell | *NPIPA7* | 8.15995645 | 6.27E-06 | 1.45E-05 | 0.039421066 |
| Obesity | mast_cell | *PP2D1* | 3.389028425 | 0.022269918 | 0.028796992 | 1 |
| Obesity | mast_cell | *DENND5A* | 5.821175437 | 2.86E-06 | 7.13E-06 | 0.017967598 |
| Obesity | mast_cell | *TMEM236* | 3.097511944 | 2.90E-08 | 1.51E-07 | 0.000182521 |
| Obesity | mast_cell | *VPS33B* | 6.667915673 | 8.18E-07 | 2.37E-06 | 0.005145234 |
| Obesity | mast_cell | *TBX1* | -3.745687025 | 5.99E-05 | 0.000113848 | 0.376610452 |
| Obesity | mast_cell | *UQCR10* | 6.541172926 | 6.23E-10 | 2.65E-08 | 3.91E-06 |
| Obesity | mast_cell | *EIF3C* | 6.710305719 | 2.81E-21 | 2.94E-18 | 1.76E-17 |
| Obesity | mast_cell | *CNTN2* | 5.314941559 | 8.44E-09 | 6.91E-08 | 5.31E-05 |
| Obesity | mast_cell | *CRELD2* | 5.63418003 | 1.19E-09 | 3.76E-08 | 7.50E-06 |
| Obesity | mast_cell | *UBE2F* | 4.506168094 | 0.001376289 | 0.002100687 | 1 |
| Obesity | mast_cell | *GOLGA6L4* | 5.444027089 | 6.03E-06 | 1.40E-05 | 0.037934 |
| Obesity | mast_cell | *PCDH9* | 2.147681474 | 1.20E-05 | 2.63E-05 | 0.075415048 |
| Obesity | mast_cell | *ACOT1* | 1.62920241 | 0.002516043 | 0.003704535 | 1 |
| Obesity | mast_cell | *OAF* | -2.677077466 | 0.000306299 | 0.000516689 | 1 |
| Obesity | mast_cell | *TM2D3* | 5.331582095 | 0.000830693 | 0.001309899 | 1 |
| Obesity | mast_cell | *TACSTD2* | -1.2813882 | 0.035649629 | 0.044798964 | 1 |
| Obesity | mast_cell | *CLECL1* | 1.808518988 | 0.000549388 | 0.000890436 | 1 |
| Obesity | mast_cell | *MROH7* | -1.091895823 | 2.24E-07 | 7.86E-07 | 0.001407001 |
| Obesity | mast_cell | *GDF3* | 5.198645769 | 0.0009637 | 0.001504539 | 1 |
| Obesity | mast_cell | *CSF1* | -0.886543562 | 1.58E-06 | 4.25E-06 | 0.009957168 |
| Obesity | mast_cell | *PLA2G6* | 4.658646766 | 2.79E-14 | 1.03E-11 | 1.75E-10 |
| Obesity | mast_cell | *A3GALT2* | 2.408628852 | 5.29E-05 | 0.000101967 | 0.332716736 |
| Obesity | mast_cell | *SS18L1* | -2.969184028 | 0.006147779 | 0.008560595 | 1 |
| Obesity | mast_cell | *THAP7* | 7.381915981 | 5.76E-09 | 5.62E-08 | 3.62E-05 |
| Obesity | mast_cell | *WDR27* | 3.514881414 | 0.012037933 | 0.01614732 | 1 |
| Obesity | mast_cell | *TXNRD2* | 7.96337063 | 4.10E-08 | 1.98E-07 | 0.000257871 |
| Obesity | mast_cell | *HDDC3* | 5.659589513 | 4.16E-06 | 9.96E-06 | 0.026124604 |
| Obesity | mast_cell | *LPAR5* | -1.351911441 | 4.68E-07 | 1.46E-06 | 0.002940185 |
| Obesity | mast_cell | *TMEM173* | -3.793114389 | 0.001386156 | 0.002114207 | 1 |
| Obesity | mast_cell | *SNN* | 13.26130834 | 7.54E-06 | 1.71E-05 | 0.047432605 |
| Obesity | mast_cell | *NELL2* | 2.402306894 | 2.38E-08 | 1.30E-07 | 0.000149559 |
| Obesity | mast_cell | *KRBA2* | 3.147354451 | 0.001337169 | 0.00204445 | 1 |
| Obesity | mast_cell | *ZNF93* | 10.16079264 | 6.27E-09 | 5.88E-08 | 3.94E-05 |
| Obesity | mast_cell | *SEPT5* | 20.26520411 | 4.26E-09 | 4.85E-08 | 2.68E-05 |
| Obesity | mast_cell | *ATL3* | 5.504890244 | 7.34E-08 | 3.13E-07 | 0.000461683 |
| Obesity | mast_cell | *NDUFA12* | 8.193772783 | 7.85E-09 | 6.64E-08 | 4.93E-05 |
| Obesity | mast_cell | *UBE2G2* | 5.084121224 | 3.12E-06 | 7.71E-06 | 0.019615452 |
| Obesity | mast_cell | *OSBP2* | 7.232434984 | 1.53E-09 | 4.16E-08 | 9.61E-06 |
| Obesity | mast_cell | *TUSC5* | 1.965743791 | 1.26E-06 | 3.47E-06 | 0.007915992 |
| Obesity | mast_cell | *RBM43* | 5.476729822 | 2.61E-08 | 1.39E-07 | 0.000163842 |
| Obesity | mast_cell | *ZFP90* | 7.584546735 | 6.44E-06 | 1.48E-05 | 0.040501638 |
| Obesity | mast_cell | *NDUFA6* | 1.385279979 | 0.000195078 | 0.000338987 | 1 |

| | | | | | | |
|---|---|---|---|---|---|---|
| Obesity | mast_cell | *SORCS2* | 3.549869574 | 9.52E-10 | 3.30E-08 | 5.98E-06 |
| Obesity | mast_cell | *TMEM106A* | 7.813135508 | 7.11E-06 | 1.63E-05 | 0.044711037 |
| Obesity | mast_cell | *SIVA1* | -2.602639224 | 0.002759789 | 0.004037886 | 1 |
| Obesity | mast_cell | *BRF1* | 6.379751937 | 3.54E-09 | 4.62E-08 | 2.22E-05 |
| Obesity | mast_cell | *CIB1* | 3.75315755 | 2.23E-06 | 5.73E-06 | 0.014041833 |
| Obesity | mast_cell | *FLRT2* | 2.68088694 | 8.35E-06 | 1.88E-05 | 0.052481848 |
| Obesity | mast_cell | *ADSSL1* | -3.950428004 | 9.04E-06 | 2.03E-05 | 0.05686413 |
| Obesity | mast_cell | *C6orf120* | 3.883432251 | 0.031476972 | 0.039834083 | 1 |
| Obesity | mast_cell | *IFITM2* | 5.374680347 | 6.53E-05 | 0.00012314 | 0.410548065 |
| Obesity | mast_cell | *GP1BA* | 6.837269693 | 4.86E-12 | 8.31E-10 | 3.06E-08 |
| Obesity | mast_cell | *KIAA0825* | 4.293840992 | 2.23E-08 | 1.24E-07 | 0.000140238 |
| Obesity | mast_cell | *CCDC137* | 3.940431385 | 0.000104436 | 0.000189437 | 0.656588078 |
| Obesity | mast_cell | *CDK10* | 3.061895327 | 1.17E-05 | 2.57E-05 | 0.073388494 |
| Obesity | mast_cell | *TMEM105* | -26.39884481 | 1.07E-06 | 3.00E-06 | 0.006719001 |
| Obesity | mast_cell | *ATP6V0A2* | -1.256098084 | 6.10E-07 | 1.82E-06 | 0.0038352 |
| Obesity | mast_cell | *TNFAIP8L1* | 8.549249623 | 2.52E-06 | 6.38E-06 | 0.015858857 |
| Obesity | mast_cell | *MAPK11* | 7.85149974 | 2.82E-07 | 9.54E-07 | 0.001771403 |
| Obesity | mast_cell | *SP140L* | 5.649779457 | 0.000154914 | 0.000274041 | 0.973942332 |
| Obesity | mast_cell | *PDE6G* | 5.587158428 | 0.000580428 | 0.000937054 | 1 |
| Obesity | mast_cell | *TLCD2* | 1.786708775 | 2.29E-07 | 8.01E-07 | 0.00144106 |
| Obesity | mast_cell | *LSAMP* | 4.574251414 | 3.27E-06 | 8.04E-06 | 0.020578422 |
| Obesity | mast_cell | *AHNAK2* | 9.28933448 | 9.83E-09 | 7.61E-08 | 6.18E-05 |
| Obesity | mast_cell | *SP1* | 5.928095876 | 8.55E-10 | 3.05E-08 | 5.37E-06 |
| Obesity | mast_cell | *PDIA2* | 3.383858463 | 0.000164854 | 0.000289913 | 1 |
| Obesity | mast_cell | *PCGF3* | 6.041562189 | 2.38E-05 | 4.88E-05 | 0.149797199 |
| Obesity | mast_cell | *PSMD13* | 4.0907375 | 3.02E-08 | 1.55E-07 | 0.000189757 |
| Obesity | mast_cell | *UBE2L3* | 5.688687756 | 1.48E-08 | 9.68E-08 | 9.30E-05 |
| Obesity | mast_cell | *PMEL* | 3.820454972 | 3.24E-06 | 7.96E-06 | 0.02035265 |
| Obesity | mast_cell | *SNAI3* | 3.736822766 | 0.014408979 | 0.019099567 | 1 |
| Obesity | mast_cell | *C6orf201* | 1.72991758 | 0.01176113 | 0.015796246 | 1 |
| Obesity | mast_cell | *MYBL1* | 1.214716521 | 0.001982697 | 0.002955243 | 1 |
| Obesity | mast_cell | *C16orf52* | 5.142177804 | 0.000407177 | 0.000675976 | 1 |
| Obesity | mast_cell | *ADARB2* | 6.454912547 | 3.09E-06 | 7.64E-06 | 0.019415344 |
| Obesity | mast_cell | *SRL* | 2.249621754 | 5.82E-09 | 5.65E-08 | 3.66E-05 |
| Obesity | mast_cell | *KCNIP4* | 1.295092842 | 0.028020254 | 0.035740178 | 1 |
| Obesity | mast_cell | *SLC52A2* | 7.565761874 | 2.19E-09 | 4.32E-08 | 1.38E-05 |
| Obesity | mast_cell | *ARL17A* | 3.49736207 | 1.41E-06 | 3.85E-06 | 0.008887977 |
| Obesity | mast_cell | *NPIPB4* | 4.249784178 | 8.37E-09 | 6.89E-08 | 5.27E-05 |
| Obesity | mast_cell | *IFITM1* | 4.435841866 | 0.02922906 | 0.037131359 | 1 |
| Obesity | mast_cell | *POMK* | 8.500334807 | 0.000104824 | 0.000190087 | 0.659030072 |
| Obesity | mast_cell | *LINC00839* | -2.99388145 | 0.002251686 | 0.003334043 | 1 |
| Obesity | mast_cell | *PTCH1* | 4.359783984 | 2.28E-05 | 4.70E-05 | 0.143549232 |
| Obesity | mast_cell | *RTN4RL1* | -3.01691981 | 0.035359821 | 0.044488132 | 1 |
| Obesity | mast_cell | *PLK5* | 3.261505665 | 0.013808834 | 0.018334982 | 1 |
| Obesity | mast_cell | *RASA3* | 4.953563606 | 4.74E-08 | 2.22E-07 | 0.000297867 |
| Obesity | mast_cell | *ZNF566* | 5.591419649 | 1.07E-08 | 8.08E-08 | 6.74E-05 |
| Obesity | mast_cell | *ZNF529* | 2.81726081 | 0.035276005 | 0.044416438 | 1 |
| Obesity | mast_cell | *ZNF284* | 12.16837165 | 2.90E-09 | 4.35E-08 | 1.82E-05 |
| Obesity | mast_cell | *GSAP* | 4.324405378 | 7.59E-05 | 0.000141394 | 0.477486327 |
| Obesity | mast_cell | *C2orf76* | 5.998949324 | 1.09E-08 | 8.15E-08 | 6.85E-05 |

| | | | | | | |
|---|---|---|---|---|---|---|
| Obesity | mast_cell | *WWOX* | 3.169110215 | 0.000212153 | 0.000366452 | 1 |
| Obesity | mast_cell | *CYP4Z1* | 2.855592738 | 1.75E-08 | 1.05E-07 | 0.000110088 |
| Obesity | mast_cell | *CCDC84* | 6.688363056 | 3.12E-07 | 1.04E-06 | 0.00196107 |
| Obesity | mast_cell | *POLR1D* | 5.444706 | 3.55E-09 | 4.62E-08 | 2.23E-05 |
| Obesity | mast_cell | *FFAR4* | 1.183753061 | 5.18E-05 | 9.99E-05 | 0.325444859 |
| Obesity | mast_cell | *BPIFB4* | 176.7807646 | 2.11E-05 | 4.37E-05 | 0.132539821 |
| Obesity | mast_cell | *ZNF749* | 1.907092402 | 0.000452262 | 0.000744338 | 1 |
| Obesity | mast_cell | *NBPF12* | 6.896499309 | 2.05E-07 | 7.29E-07 | 0.001291369 |
| Obesity | mast_cell | *GPAT2* | 3.895295655 | 0.001005034 | 0.001564797 | 1 |
| Obesity | mast_cell | *TOR3A* | 10.04905603 | 1.82E-08 | 1.08E-07 | 0.000114399 |
| Obesity | mast_cell | *CYP4X1* | 5.391915829 | 3.51E-06 | 8.57E-06 | 0.022073037 |
| Obesity | mast_cell | *CCDC30* | 6.025901394 | 3.14E-06 | 7.75E-06 | 0.019743212 |
| Obesity | mast_cell | *GLDN* | 3.580276989 | 1.35E-09 | 3.95E-08 | 8.50E-06 |
| Obesity | mast_cell | *FCAR* | 3.450940416 | 4.71E-07 | 1.47E-06 | 0.002961785 |
| Obesity | mast_cell | *KPNA4* | 3.735670694 | 0.011083909 | 0.014934534 | 1 |
| Obesity | mast_cell | *ZNF501* | 5.181924838 | 1.39E-07 | 5.31E-07 | 0.000871956 |
| Obesity | mast_cell | *ZNF197* | 2.80494197 | 0.019396128 | 0.025283735 | 1 |
| Obesity | mast_cell | *RPS23* | 4.808060933 | 1.43E-05 | 3.07E-05 | 0.089726108 |
| Obesity | mast_cell | *BTN3A2* | -1.970138963 | 4.89E-12 | 8.31E-10 | 3.07E-08 |
| Obesity | mast_cell | *PCLO* | 2.617571762 | 2.03E-08 | 1.16E-07 | 0.000127797 |
| Obesity | mast_cell | *RGS7BP* | 3.049842143 | 2.51E-06 | 6.37E-06 | 0.015803082 |
| Obesity | mast_cell | *ZNF396* | 2.455623137 | 3.68E-05 | 7.26E-05 | 0.231240615 |
| Obesity | mast_cell | *CLCNKA* | 5.602776636 | 0.000157455 | 0.000278223 | 0.989918466 |
| Obesity | mast_cell | *SEPT10* | 2.471529682 | 0.039060032 | 0.048801753 | 1 |
| Obesity | mast_cell | *SMYD4* | 4.998086721 | 7.78E-07 | 2.27E-06 | 0.004894399 |
| Obesity | mast_cell | *MIR22HG* | 4.857615337 | 2.65E-08 | 1.41E-07 | 0.000166665 |
| Obesity | mast_cell | *C17orf58* | 2.367807464 | 0.000145014 | 0.000257544 | 0.911704089 |
| Obesity | mast_cell | *CYP27C1* | 1.698815337 | 3.86E-06 | 9.30E-06 | 0.02425811 |
| Obesity | mast_cell | *BCR* | 5.939998248 | 3.91E-07 | 1.26E-06 | 0.00245593 |
| Obesity | mast_cell | *FOXI2* | -2.884586624 | 1.64E-05 | 3.48E-05 | 0.102960444 |
| Obesity | mast_cell | *HYAL3* | -2.673908629 | 1.60E-11 | 2.14E-09 | 1.00E-07 |
| Obesity | mast_cell | *ZSCAN30* | 5.476089663 | 3.24E-09 | 4.49E-08 | 2.03E-05 |
| Obesity | mast_cell | *LILRB4* | 2.704313199 | 4.90E-08 | 2.29E-07 | 0.00030793 |
| Obesity | mast_cell | *TRABD2A* | 2.510287049 | 1.75E-08 | 1.05E-07 | 0.000110033 |
| Obesity | mast_cell | *TMEM17* | 4.326217191 | 1.90E-06 | 4.96E-06 | 0.011932701 |
| Obesity | mast_cell | *TNFRSF18* | -4.7898098 | 9.26E-07 | 2.65E-06 | 0.005822814 |
| Obesity | mast_cell | *RTN4RL2* | -7.170841535 | 2.42E-06 | 6.16E-06 | 0.015243535 |
| Obesity | mast_cell | *TMEM232* | 3.620552369 | 1.32E-07 | 5.13E-07 | 0.000832883 |
| Obesity | mast_cell | *EFCAB6* | 4.782682979 | 0.001431939 | 0.002178225 | 1 |
| Obesity | mast_cell | *KANK3* | 4.962565235 | 4.16E-09 | 4.81E-08 | 2.62E-05 |
| Obesity | mast_cell | *EMID1* | -1.395480061 | 7.09E-07 | 2.09E-06 | 0.004456324 |
| Obesity | mast_cell | *RHD* | 2.410857283 | 0.004509178 | 0.006408047 | 1 |
| Obesity | mast_cell | *CYP4A11* | 3.49729279 | 0.001276328 | 0.001955232 | 1 |
| Obesity | mast_cell | *C3orf70* | 2.649427712 | 0.001259819 | 0.001931354 | 1 |
| Obesity | mast_cell | *PLCD1* | 2.652597477 | 0.025905645 | 0.033197878 | 1 |
| Obesity | mast_cell | *HEATR4* | 2.193272237 | 6.02E-05 | 0.000114316 | 0.378470625 |
| Obesity | mast_cell | *MRPS21* | 6.92452438 | 5.21E-10 | 2.40E-08 | 3.27E-06 |
| Obesity | mast_cell | *ANGPTL5* | 3.87822739 | 7.30E-09 | 6.32E-08 | 4.59E-05 |
| Obesity | mast_cell | *FNBP1* | 3.241767132 | 0.000123665 | 0.000222011 | 0.777482041 |
| Obesity | mast_cell | *WDR86* | 3.295236931 | 0.007996008 | 0.010978577 | 1 |

| | | | | | | |
|---|---|---|---|---|---|---|
| Obesity | mast_cell | *EPOR* | -1.88773553 | 1.26E-05 | 2.74E-05 | 0.079261858 |
| Obesity | mast_cell | *FPR3* | 3.379609954 | 1.98E-06 | 5.15E-06 | 0.012452524 |
| Obesity | mast_cell | *GJA4* | 3.14083801 | 1.85E-08 | 1.08E-07 | 0.000116338 |
| Obesity | mast_cell | *IFT140* | 5.225416201 | 3.99E-09 | 4.78E-08 | 2.51E-05 |
| Obesity | mast_cell | *TLR5* | 8.548594875 | 9.02E-07 | 2.58E-06 | 0.005669994 |
| Obesity | mast_cell | *PLEKHN1* | 4.556430989 | 8.59E-05 | 0.000158011 | 0.539923665 |
| Obesity | mast_cell | *ZNF385C* | 4.032848827 | 6.59E-09 | 6.01E-08 | 4.14E-05 |
| Obesity | mast_cell | *TET3* | 6.366130838 | 7.11E-09 | 6.25E-08 | 4.47E-05 |
| Obesity | mast_cell | *C17orf97* | 0.9438132 | 0.000313652 | 0.000528951 | 1 |
| Obesity | mast_cell | *DHRS4L2* | 1.65618331 | 0.012308547 | 0.016471656 | 1 |
| Obesity | mast_cell | *TRPV2* | -3.673812902 | 0.00022046 | 0.000379631 | 1 |
| Obesity | mast_cell | *C2orf88* | 4.076803902 | 0.000820692 | 0.001295428 | 1 |
| Obesity | mast_cell | *FANCA* | 2.00336146 | 0.002003335 | 0.002983176 | 1 |
| Obesity | mast_cell | *ADH1A* | 1.631427717 | 0.012173126 | 0.01631474 | 1 |
| Obesity | mast_cell | *SEMA4D* | -2.624888175 | 2.10E-05 | 4.36E-05 | 0.132249184 |
| Obesity | mast_cell | *FAM69C* | 4.918386831 | 0.000406125 | 0.000674407 | 1 |
| Obesity | mast_cell | *DNAH17* | -4.533212498 | 5.96E-05 | 0.000113366 | 0.374788791 |
| Obesity | mast_cell | *ZFP69* | 4.279245677 | 4.43E-06 | 1.06E-05 | 0.027859256 |
| Obesity | mast_cell | *TMEM220* | 5.840056353 | 5.88E-05 | 0.000112026 | 0.369908529 |
| Obesity | mast_cell | *CCDC157* | 9.822669763 | 1.16E-08 | 8.42E-08 | 7.26E-05 |
| Obesity | mast_cell | *GFRAL* | 3.475910785 | 2.35E-07 | 8.17E-07 | 0.001475455 |
| Obesity | mast_cell | *LDLRAD2* | -2.428107412 | 0.000129485 | 0.000231139 | 0.814073105 |
| Obesity | mast_cell | *C2orf66* | 9.811964951 | 0.000241451 | 0.000413512 | 1 |
| Obesity | mast_cell | *CPSF4L* | -2.925372534 | 0.035483274 | 0.044625594 | 1 |
| Obesity | mast_cell | *KLHL17* | 3.267876353 | 1.08E-08 | 8.14E-08 | 6.80E-05 |
| Obesity | mast_cell | *ZSCAN23* | 1.441316005 | 1.49E-06 | 4.02E-06 | 0.00935846 |
| Obesity | mast_cell | *OR7D2* | 5.928992795 | 0.000355362 | 0.000594825 | 1 |
| Obesity | mast_cell | *MORN2* | 2.213968904 | 0.001568256 | 0.002371813 | 1 |
| Obesity | mast_cell | *S100A3* | -1.743509602 | 0.035313005 | 0.04444702 | 1 |
| Obesity | mast_cell | *TMEM221* | 1.325935625 | 0.007668119 | 0.010560671 | 1 |
| Obesity | mast_cell | *MAPK12* | -1.563146914 | 4.58E-07 | 1.44E-06 | 0.00287821 |
| Obesity | mast_cell | *AGRN* | 6.128550664 | 1.33E-06 | 3.65E-06 | 0.0083662 |
| Obesity | mast_cell | *FAM166A* | -4.938310212 | 3.47E-07 | 1.14E-06 | 0.00217892 |
| Obesity | mast_cell | *ZNF626* | 7.428671171 | 1.61E-06 | 4.30E-06 | 0.010101325 |
| Obesity | mast_cell | *SMTNL2* | 15.73336118 | 1.64E-08 | 1.03E-07 | 0.000102806 |
| Obesity | mast_cell | *ZC3H6* | 6.039021885 | 1.11E-08 | 8.21E-08 | 6.97E-05 |
| Obesity | mast_cell | *LINC00265* | 3.467751378 | 2.45E-08 | 1.33E-07 | 0.00015423 |
| Obesity | mast_cell | *LAMTOR4* | 4.656289864 | 9.81E-06 | 2.19E-05 | 0.061652642 |
| Obesity | mast_cell | *NUTM2B* | 36.80933641 | 0.001688686 | 0.00253929 | 1 |
| Obesity | mast_cell | *NCR3LG1* | 19.44738314 | 2.06E-09 | 4.32E-08 | 1.30E-05 |
| Obesity | mast_cell | *HYKK* | 3.068203162 | 2.79E-10 | 1.57E-08 | 1.76E-06 |
| Obesity | mast_cell | *RUFY4* | 29.2863727 | 3.27E-08 | 1.65E-07 | 0.000205763 |
| Obesity | mast_cell | *CENPP* | 2.398469509 | 1.49E-05 | 3.19E-05 | 0.093599178 |
| Obesity | mast_cell | *PLSCR1* | 5.815281842 | 0.000187898 | 0.000327053 | 1 |
| Obesity | mast_cell | *ZP3* | 2.878068822 | 0.00120607 | 0.001855287 | 1 |
| Obesity | mast_cell | *JAKMIP3* | 1.350403513 | 0.001673193 | 0.002517197 | 1 |
| Obesity | mast_cell | *PDCD1* | 7.641368397 | 0.001492616 | 0.002264498 | 1 |
| Obesity | mast_cell | *CERKL* | 6.491959991 | 4.01E-09 | 4.78E-08 | 2.52E-05 |
| Obesity | mast_cell | *C22orf34* | -1.609292511 | 0.001695387 | 0.002548146 | 1 |
| Obesity | mast_cell | *COL25A1* | 4.550021122 | 6.58E-07 | 1.96E-06 | 0.004138475 |

| | | | | | | |
|---|---|---|---|---|---|---|
| Obesity | mast_cell | *SRSF10* | 3.052077719 | 0.026962906 | 0.034440429 | 1 |
| Obesity | mast_cell | *NBR1* | 6.910199938 | 1.16E-08 | 8.42E-08 | 7.30E-05 |
| Obesity | mast_cell | *NDOR1* | 7.845692623 | 2.45E-09 | 4.32E-08 | 1.54E-05 |
| Obesity | mast_cell | *FAM72B* | -4.599384679 | 5.36E-07 | 1.63E-06 | 0.003369149 |
| Obesity | mast_cell | *GOLGA8M* | 2.342036153 | 7.12E-08 | 3.06E-07 | 0.0004476 |
| Obesity | mast_cell | *DPYD* | 2.967339785 | 0.00541281 | 0.007596057 | 1 |
| Obesity | mast_cell | *S100A16* | 7.661228959 | 3.45E-09 | 4.57E-08 | 2.17E-05 |
| Obesity | mast_cell | *SLC4A5* | 4.21292145 | 5.72E-08 | 2.57E-07 | 0.000359352 |
| Obesity | mast_cell | *UROS* | 4.890863946 | 1.61E-08 | 1.02E-07 | 0.000100974 |
| Obesity | mast_cell | *FAM221A* | 4.739294527 | 1.18E-07 | 4.67E-07 | 0.000744632 |
| Obesity | mast_cell | *NOXA1* | 9.497465925 | 7.07E-09 | 6.23E-08 | 4.44E-05 |
| Obesity | mast_cell | *SKOR1* | 10.20795644 | 1.12E-07 | 4.45E-07 | 0.000702104 |
| Obesity | mast_cell | *TMEM201* | -3.654535918 | 8.15E-05 | 0.000150626 | 0.512278703 |
| Obesity | mast_cell | *ZDHHC11* | -2.264489706 | 0.034837086 | 0.043883141 | 1 |
| Obesity | mast_cell | *LINC00910* | -3.090029868 | 0.020905385 | 0.02713298 | 1 |
| Obesity | mast_cell | *BEND4* | 10.28707092 | 5.40E-09 | 5.52E-08 | 3.40E-05 |
| Obesity | mast_cell | *ZNF563* | 3.755974112 | 0.000411092 | 0.000681217 | 1 |
| Obesity | mast_cell | *FBF1* | 2.086224493 | 7.51E-06 | 1.71E-05 | 0.047244001 |
| Obesity | mast_cell | *MSL1* | 2.03490809 | 0.002139884 | 0.003178981 | 1 |
| Obesity | mast_cell | *LRRK2* | 3.55654103 | 0.000259909 | 0.000443191 | 1 |
| Obesity | mast_cell | *FAM196A* | 6.218453816 | 2.17E-09 | 4.32E-08 | 1.36E-05 |
| Obesity | mast_cell | *C9orf152* | -3.976049823 | 1.03E-06 | 2.91E-06 | 0.006477129 |
| Obesity | mast_cell | *NOC2L* | -2.009628008 | 0.000299367 | 0.000505403 | 1 |
| Obesity | mast_cell | *NELFB* | 4.368766372 | 3.05E-09 | 4.46E-08 | 1.92E-05 |
| Obesity | mast_cell | *SLC15A5* | 2.026973293 | 4.75E-06 | 1.12E-05 | 0.029855307 |
| Obesity | mast_cell | *LIPI* | 2.332313838 | 0.003204421 | 0.004648407 | 1 |
| Obesity | mast_cell | *ADAT2* | 6.356341422 | 1.67E-08 | 1.04E-07 | 0.000105037 |
| Obesity | mast_cell | *ZNF567* | 5.996612516 | 3.08E-09 | 4.48E-08 | 1.94E-05 |
| Obesity | mast_cell | *ANKDD1B* | -1.880120354 | 5.04E-05 | 9.75E-05 | 0.316812745 |
| Obesity | mast_cell | *ALKBH2* | 3.426837392 | 0.016494551 | 0.021685747 | 1 |
| Obesity | mast_cell | *LITAF* | -1.977199817 | 0.016688842 | 0.021932013 | 1 |
| Obesity | mast_cell | *VSTM1* | 4.145780511 | 2.86E-05 | 5.77E-05 | 0.179766656 |
| Obesity | mast_cell | *SF3B3* | 7.020612144 | 1.50E-08 | 9.76E-08 | 9.43E-05 |
| Obesity | mast_cell | *PLAC9* | 4.547323753 | 1.50E-08 | 9.74E-08 | 9.41E-05 |
| Obesity | mast_cell | *ZNF573* | 7.542123721 | 1.42E-07 | 5.39E-07 | 0.000891472 |
| Obesity | mast_cell | *FAM47E* | 3.383293003 | 9.23E-11 | 8.17E-09 | 5.80E-07 |
| Obesity | mast_cell | *ZNF527* | 8.05962463 | 5.74E-06 | 1.34E-05 | 0.036109924 |
| Obesity | mast_cell | *ZNF33A* | 2.831684378 | 0.028412695 | 0.036218697 | 1 |
| Obesity | mast_cell | *ZNF600* | 2.635395359 | 0.000124599 | 0.000223432 | 0.783354214 |
| Obesity | mast_cell | *LINC00994* | 2.86033865 | 0.037615954 | 0.047128638 | 1 |
| Obesity | mast_cell | *TSPYL1* | 2.479518629 | 0.016878233 | 0.022167005 | 1 |
| Obesity | mast_cell | *PNRC2* | 5.359643747 | 0.00810334 | 0.011116233 | 1 |
| Obesity | mast_cell | *ZKSCAN3* | 4.665015188 | 2.80E-06 | 7.02E-06 | 0.017625869 |
| Obesity | mast_cell | *RRP7A* | 4.141098562 | 1.99E-06 | 5.16E-06 | 0.012493829 |
| Obesity | mast_cell | *SLC35E2B* | 3.123528564 | 6.38E-06 | 1.47E-05 | 0.04014191 |
| Obesity | mast_cell | *AKR1C3* | 5.700485445 | 4.73E-07 | 1.47E-06 | 0.0029723 |
| Obesity | mast_cell | *ZNF79* | 2.472858995 | 0.00901771 | 0.012308803 | 1 |
| Obesity | mast_cell | *COLCA1* | 4.565811994 | 1.33E-07 | 5.14E-07 | 0.00083739 |
| Obesity | mast_cell | *ACADSB* | 4.140383674 | 2.37E-05 | 4.86E-05 | 0.148809227 |
| Obesity | mast_cell | *SEMA4A* | -1.974890774 | 0.009195424 | 0.012537765 | 1 |

| | | | | | | |
|---|---|---|---|---|---|---|
| Obesity | mast_cell | *HRCT1* | 4.450032738 | 4.60E-09 | 5.08E-08 | 2.89E-05 |
| Obesity | mast_cell | *MPHOSPH8* | 5.509696002 | 6.87E-07 | 2.03E-06 | 0.004318227 |
| Obesity | mast_cell | *SIRPB2* | 5.220666532 | 4.46E-09 | 4.98E-08 | 2.80E-05 |
| Obesity | mast_cell | *SRGAP3* | 1.571750271 | 0.024995171 | 0.032076881 | 1 |
| Obesity | mast_cell | *SUPT5H* | 6.998364589 | 2.20E-08 | 1.23E-07 | 0.000138211 |
| Obesity | mast_cell | *XPNPEP3* | 1.419119776 | 0.016105981 | 0.021219259 | 1 |
| Obesity | mast_cell | *PPIA* | 7.040255928 | 0.000168225 | 0.00029551 | 1 |
| Obesity | mast_cell | *ZNF471* | 3.644305378 | 9.22E-05 | 0.000168611 | 0.579852703 |
| Obesity | mast_cell | *GTF2IRD2* | 2.883176064 | 0.000176458 | 0.000308765 | 1 |
| Obesity | mast_cell | *GRM7* | 1.352181957 | 0.003805443 | 0.005469781 | 1 |
| Obesity | mast_cell | *SUPT3H* | 4.426248279 | 0.000931506 | 0.001457535 | 1 |
| Obesity | mast_cell | *STK31* | 10.58407174 | 2.34E-09 | 4.32E-08 | 1.47E-05 |
| Obesity | mast_cell | *ZKSCAN7* | 5.274434666 | 5.21E-09 | 5.41E-08 | 3.28E-05 |
| Obesity | mast_cell | *ZNF565* | 3.868513406 | 0.001285236 | 0.001967718 | 1 |
| Obesity | mast_cell | *NTNG2* | -1.56082565 | 4.80E-07 | 1.49E-06 | 0.00302035 |
| Obesity | mast_cell | *WDR5* | 8.158984477 | 1.21E-05 | 2.64E-05 | 0.075978521 |
| Obesity | mast_cell | *LONP1* | 4.430158994 | 0.000145968 | 0.000259165 | 0.917702873 |
| Obesity | mast_cell | *ASB13* | 3.362220948 | 6.12E-09 | 5.81E-08 | 3.85E-05 |
| Obesity | mast_cell | *SLC35F1* | 6.6769569 | 0.000586679 | 0.000946484 | 1 |
| Obesity | mast_cell | *ZNF34* | -3.883080676 | 3.39E-05 | 6.73E-05 | 0.212842492 |
| Obesity | mast_cell | *INCA1* | -3.34367535 | 0.021814814 | 0.028243355 | 1 |
| Obesity | mast_cell | *ZNF774* | 3.695868477 | 4.40E-08 | 2.09E-07 | 0.000276875 |
| Obesity | mast_cell | *EVL* | 2.735338666 | 0.007288276 | 0.010050755 | 1 |
| Obesity | mast_cell | *ZNF124* | 7.014351223 | 2.25E-05 | 4.65E-05 | 0.141723945 |
| Obesity | mast_cell | *ZNF569* | 5.458550501 | 8.59E-08 | 3.59E-07 | 0.000540255 |
| Obesity | mast_cell | *PIK3R4* | 3.116942787 | 2.23E-05 | 4.61E-05 | 0.140375104 |
| Obesity | mast_cell | *ZNF775* | 2.411721714 | 0.010208001 | 0.013831402 | 1 |
| Obesity | mast_cell | *ZNF605* | 7.944961508 | 4.70E-09 | 5.10E-08 | 2.95E-05 |
| Obesity | mast_cell | *RFX8* | 4.593549636 | 4.33E-08 | 2.06E-07 | 0.000272431 |
| Obesity | mast_cell | *MYL6B* | 9.027819122 | 8.97E-05 | 0.000164391 | 0.564188936 |
| Obesity | mast_cell | *C20orf96* | 4.717545986 | 6.67E-09 | 6.04E-08 | 4.19E-05 |
| Obesity | mast_cell | *SULT1A1* | -2.496741148 | 0.00028655 | 0.000485068 | 1 |
| Obesity | mast_cell | *ANAPC7* | 5.748933351 | 7.61E-06 | 1.73E-05 | 0.047836348 |
| Obesity | mast_cell | *TPK1* | 3.41085596 | 0.00311891 | 0.004535876 | 1 |
| Obesity | mast_cell | *C1orf186* | -0.231675165 | 1.34E-07 | 5.18E-07 | 0.000845522 |
| Obesity | mast_cell | *MYO18A* | 4.594116781 | 4.03E-05 | 7.91E-05 | 0.253128006 |
| Obesity | mast_cell | *SPTSSB* | 7.131710125 | 5.87E-09 | 5.69E-08 | 3.69E-05 |
| Obesity | mast_cell | *SULF2* | 4.711473789 | 9.82E-06 | 2.19E-05 | 0.06173365 |
| Obesity | mast_cell | *XRCC2* | 13.24907825 | 0.000149301 | 0.000264559 | 0.938655003 |
| Obesity | mast_cell | *HDAC2* | 7.07337 | 1.88E-08 | 1.09E-07 | 0.000118051 |
| Obesity | mast_cell | *ADH1B* | 4.796960876 | 3.68E-09 | 4.65E-08 | 2.32E-05 |
| Obesity | mast_cell | *HRH1* | 4.384940736 | 2.43E-09 | 4.32E-08 | 1.53E-05 |
| Obesity | mast_cell | *ZNF502* | 4.36499486 | 1.04E-07 | 4.19E-07 | 0.000652422 |
| Obesity | mast_cell | *TRAPPC4* | 1.957035065 | 0.011679808 | 0.015693728 | 1 |
| Obesity | mast_cell | *TECPR2* | 6.100232333 | 4.75E-09 | 5.10E-08 | 2.98E-05 |
| Obesity | mast_cell | *FAM180B* | 1.658913842 | 0.007167965 | 0.009891352 | 1 |
| Obesity | mast_cell | *ZFP62* | 10.43347843 | 6.33E-08 | 2.79E-07 | 0.000398261 |
| Obesity | mast_cell | *TOMM7* | 8.106188141 | 1.04E-06 | 2.93E-06 | 0.006540187 |
| Obesity | mast_cell | *TRPV1* | 3.024269599 | 0.016790718 | 0.022056674 | 1 |
| Obesity | mast_cell | *AMZ2* | 6.103382381 | 2.01E-07 | 7.18E-07 | 0.001265121 |

| | | | | | | |
|---|---|---|---|---|---|---|
| Obesity | mast_cell | *ZNF431* | 5.870786287 | 8.34E-06 | 1.88E-05 | 0.052452207 |
| Obesity | mast_cell | *NF1* | 4.759815074 | 7.34E-06 | 1.68E-05 | 0.046166605 |
| Obesity | mast_cell | *DAPK1* | 2.346461066 | 0.009070838 | 0.012375946 | 1 |
| Obesity | mast_cell | *TLE1* | 3.415207857 | 3.23E-06 | 7.94E-06 | 0.020278154 |
| Obesity | mast_cell | *STRN3* | 5.166807636 | 4.01E-06 | 9.64E-06 | 0.025222655 |
| Obesity | mast_cell | *ZNF239* | 3.398580575 | 1.68E-07 | 6.18E-07 | 0.001056828 |
| Obesity | mast_cell | *MVB12B* | 6.285327019 | 3.17E-06 | 7.82E-06 | 0.019937839 |
| Obesity | mast_cell | *C6orf106* | 4.605027351 | 3.14E-11 | 3.52E-09 | 1.98E-07 |
| Obesity | mast_cell | *PPTC7* | 4.624717593 | 0.001465323 | 0.002226314 | 1 |
| Obesity | mast_cell | *NHLRC2* | 5.823933368 | 3.14E-06 | 7.75E-06 | 0.019768822 |
| Obesity | mast_cell | *ZFP28* | 5.320366585 | 1.02E-09 | 3.48E-08 | 6.40E-06 |
| Obesity | mast_cell | *KIAA1211L* | 1.996809622 | 0.000163861 | 0.00028857 | 1 |
| Obesity | mast_cell | *LAMB3* | 4.042125913 | 2.95E-08 | 1.53E-07 | 0.000185402 |
| Obesity | mast_cell | *KPNA5* | 6.526142724 | 6.44E-08 | 2.82E-07 | 0.000404754 |
| Obesity | mast_cell | *HCAR1* | 3.540576593 | 1.08E-11 | 1.61E-09 | 6.77E-08 |
| Obesity | mast_cell | *SRGAP1* | 5.292026163 | 1.59E-09 | 4.20E-08 | 9.99E-06 |
| Obesity | mast_cell | *FAM3C* | 7.511831158 | 1.83E-10 | 1.26E-08 | 1.15E-06 |
| Obesity | mast_cell | *SLC39A10* | 4.621960587 | 0.002031963 | 0.003023657 | 1 |
| Obesity | mast_cell | *AP2A1* | 3.55484402 | 0.006352465 | 0.008835829 | 1 |
| Obesity | mast_cell | *PCDHB16* | 2.058734917 | 9.46E-06 | 2.11E-05 | 0.059489936 |
| Obesity | mast_cell | *ZNF585A* | 4.049215809 | 2.20E-05 | 4.54E-05 | 0.138035439 |
| Obesity | mast_cell | *FUT11* | 5.4977787 | 0.001199723 | 0.001846428 | 1 |
| Obesity | mast_cell | *ANXA4* | 4.32811313 | 0.000936796 | 0.001465084 | 1 |
| Obesity | mast_cell | *WDR5B* | -2.851570954 | 0.009442931 | 0.012847373 | 1 |
| Obesity | mast_cell | *ZNF138* | 4.725429985 | 0.000547027 | 0.000887067 | 1 |
| Obesity | mast_cell | *ZNF429* | 4.601629751 | 0.00012085 | 0.000217268 | 0.759784721 |
| Obesity | mast_cell | *ZNF470* | 4.507823138 | 1.84E-07 | 6.69E-07 | 0.0011587 |
| Obesity | mast_cell | *GMFB* | 6.635136053 | 0.000264921 | 0.000450882 | 1 |
| Obesity | mast_cell | *ZNF763* | 1.172866847 | 0.008237771 | 0.011288332 | 1 |
| Obesity | mast_cell | *ZMYM1* | 7.230552242 | 3.78E-06 | 9.12E-06 | 0.0237416 |
| Obesity | mast_cell | *ARRDC1* | 8.621535289 | 1.44E-07 | 5.45E-07 | 0.000905826 |
| Obesity | mast_cell | *IGF2R* | 4.921781824 | 5.78E-05 | 0.000110245 | 0.36314725 |
| Obesity | mast_cell | *SLC25A29* | 4.645192872 | 0.001009043 | 0.001569872 | 1 |
| Obesity | mast_cell | *ZNF682* | 5.164813272 | 4.48E-09 | 4.99E-08 | 2.81E-05 |
| Obesity | mast_cell | *ZNF772* | -2.018370001 | 1.14E-05 | 2.52E-05 | 0.071829443 |
| Obesity | mast_cell | *ZNF257* | -3.998463751 | 0.001887019 | 0.002819987 | 1 |
| Obesity | mast_cell | *ADAM32* | 4.596643565 | 8.17E-05 | 0.000150852 | 0.513348089 |
| Obesity | mast_cell | *LRRC8B* | 4.43166475 | 8.33E-06 | 1.88E-05 | 0.052339995 |
| Obesity | mast_cell | *ABCB8* | 3.22010022 | 1.13E-06 | 3.15E-06 | 0.007092816 |
| Obesity | mast_cell | *SND1* | 5.206891796 | 1.96E-06 | 5.10E-06 | 0.012331668 |
| Obesity | mast_cell | *SULT1A2* | 3.450449752 | 5.38E-29 | 1.02E-25 | 3.38E-25 |
| Obesity | mast_cell | *PIWIL2* | 4.490531629 | 4.68E-06 | 1.11E-05 | 0.029425785 |
| Obesity | mast_cell | *ENTPD4* | 6.332383055 | 4.77E-07 | 1.48E-06 | 0.002997083 |
| Obesity | mast_cell | *C1D* | 5.395907492 | 2.02E-07 | 7.21E-07 | 0.001271907 |
| Obesity | mast_cell | *TBC1D9B* | 2.696161128 | 0.002598632 | 0.003818981 | 1 |
| Obesity | mast_cell | *FAM110D* | 3.850052378 | 1.03E-08 | 7.88E-08 | 6.46E-05 |
| Obesity | mast_cell | *KANK2* | 7.724973784 | 3.07E-08 | 1.57E-07 | 0.000193159 |
| Obesity | mast_cell | *CCL4L2* | -2.982965493 | 1.51E-10 | 1.12E-08 | 9.50E-07 |
| Obesity | mast_cell | *GTF2E2* | 7.613690337 | 4.92E-09 | 5.23E-08 | 3.09E-05 |
| Obesity | mast_cell | *ZNF165* | -5.371919954 | 9.89E-08 | 4.05E-07 | 0.000621804 |

| | | | | | | |
|---|---|---|---|---|---|---|
| Obesity | mast_cell | *RAMP2-AS1* | 3.725458374 | 4.93E-07 | 1.52E-06 | 0.003096706 |
| Obesity | mast_cell | *ZNF720* | 4.118443005 | 0.000596605 | 0.000962004 | 1 |
| Obesity | mast_cell | *DDI2* | 5.580477936 | 2.03E-08 | 1.16E-07 | 0.000127777 |
| Obesity | mast_cell | *ZNF786* | 6.067740789 | 9.57E-07 | 2.72E-06 | 0.006018908 |
| Obesity | mast_cell | *ZNF517* | 9.271598044 | 0.006937215 | 0.009596099 | 1 |
| Obesity | mast_cell | *SLC22A5* | 1.669791148 | 0.001402858 | 0.002138643 | 1 |
| Obesity | mast_cell | *C5AR1* | 5.839244675 | 4.49E-05 | 8.75E-05 | 0.282238011 |
| Obesity | mast_cell | *DCHS2* | 1.321256117 | 1.57E-05 | 3.36E-05 | 0.098709002 |
| Obesity | mast_cell | *VEPH1* | 4.179115969 | 8.41E-07 | 2.43E-06 | 0.005286673 |
| Obesity | mast_cell | *SHPK* | 2.452322907 | 6.02E-05 | 0.000114316 | 0.378498711 |
| Obesity | mast_cell | *MAP3K5* | 3.302374135 | 0.001705725 | 0.002562459 | 1 |
| Obesity | mast_cell | *SLC2A10* | 3.490751968 | 0.013142825 | 0.017528413 | 1 |
| Obesity | mast_cell | *ZNF665* | 2.4993155 | 0.001344377 | 0.002054391 | 1 |
| Obesity | mast_cell | *RPF2* | 4.343694503 | 2.97E-08 | 1.54E-07 | 0.000186639 |
| Obesity | mast_cell | *MYO5A* | 2.819747902 | 0.003743706 | 0.005388433 | 1 |
| Obesity | mast_cell | *ATG7* | 3.734716155 | 6.31E-05 | 0.000119378 | 0.396930948 |
| Obesity | mast_cell | *RAB40C* | 5.954837096 | 2.39E-09 | 4.32E-08 | 1.50E-05 |
| Obesity | mast_cell | *ZNF624* | 9.402856939 | 5.12E-08 | 2.36E-07 | 0.000322061 |
| Obesity | mast_cell | *HHLA3* | 4.215493946 | 7.55E-07 | 2.21E-06 | 0.004744239 |
| Obesity | mast_cell | *HOXA4* | 10.92639776 | 1.16E-07 | 4.59E-07 | 0.000728726 |
| Obesity | mast_cell | *BCO2* | 6.047249001 | 4.93E-08 | 2.30E-07 | 0.000310241 |
| Obesity | mast_cell | *ENTPD6* | 4.104835818 | 2.22E-06 | 5.69E-06 | 0.013932848 |
| Obesity | mast_cell | *CCDC154* | 6.222358484 | 0.00031413 | 0.000529615 | 1 |
| Obesity | mast_cell | *C5orf42* | 3.80690835 | 1.68E-05 | 3.56E-05 | 0.105504412 |
| Obesity | mast_cell | *ZNF841* | 6.531723965 | 4.47E-07 | 1.41E-06 | 0.002809787 |
| Obesity | mast_cell | *MFAP5* | 5.371889558 | 1.60E-06 | 4.29E-06 | 0.010075503 |
| Obesity | mast_cell | *CDC42SE1* | 6.310863929 | 1.10E-08 | 8.18E-08 | 6.89E-05 |
| Obesity | mast_cell | *MPEG1* | 1.329252678 | 0.005866823 | 0.008196604 | 1 |
| Obesity | mast_cell | *NMB* | 3.007324767 | 1.85E-08 | 1.08E-07 | 0.000116458 |
| Obesity | mast_cell | *KLHL14* | 3.01882951 | 2.07E-06 | 5.36E-06 | 0.013026934 |
| Obesity | mast_cell | *FAM114A1* | 5.980233126 | 6.64E-09 | 6.03E-08 | 4.18E-05 |
| Obesity | mast_cell | *RPS26* | -1.139421109 | 0.003812126 | 0.005478134 | 1 |
| Obesity | mast_cell | *PSAP* | 4.704033351 | 0.000200003 | 0.000347066 | 1 |
| Obesity | mast_cell | *MAP1LC3C* | 3.05491573 | 6.04E-07 | 1.81E-06 | 0.003800106 |
| Obesity | mast_cell | *TAF13* | 4.164017969 | 5.95E-06 | 1.38E-05 | 0.037429166 |
| Obesity | mast_cell | *ZNF780A* | -1.14929639 | 0.011525062 | 0.015495737 | 1 |
| Obesity | mast_cell | *AC122129.1* | -4.169344592 | 6.93E-07 | 2.05E-06 | 0.004354007 |
| Obesity | mast_cell | *ZNF181* | 3.490996329 | 1.16E-08 | 8.42E-08 | 7.28E-05 |
| Obesity | mast_cell | *SGTB* | 4.146362264 | 0.029593203 | 0.037571177 | 1 |
| Obesity | mast_cell | *ADH5* | 4.305916462 | 5.99E-06 | 1.39E-05 | 0.037647517 |
| Obesity | mast_cell | *HRNR* | 0.588288664 | 0.023759704 | 0.030622644 | 1 |
| Obesity | mast_cell | *ZNF677* | 9.473820171 | 6.16E-09 | 5.81E-08 | 3.88E-05 |
| Obesity | mast_cell | *ZNF311* | 2.559419863 | 9.18E-08 | 3.79E-07 | 0.000577285 |
| Obesity | mast_cell | *FCHSD1* | 10.56214692 | 2.26E-07 | 7.92E-07 | 0.001420957 |
| Obesity | mast_cell | *VPS13A* | 2.111355614 | 0.011800245 | 0.015845395 | 1 |
| Obesity | mast_cell | *LEKR1* | -3.547666921 | 0.000317627 | 0.000534889 | 1 |
| Obesity | mast_cell | *NOL8* | 5.680756922 | 3.51E-06 | 8.57E-06 | 0.02209865 |
| Obesity | mast_cell | *MRPL42* | 2.52002459 | 0.000191287 | 0.000332675 | 1 |
| Obesity | mast_cell | *ZNF335* | 5.508476621 | 2.01E-06 | 5.20E-06 | 0.012625639 |
| Obesity | mast_cell | *ZNF273* | 5.965527217 | 1.39E-05 | 2.99E-05 | 0.087088026 |

| | | | | | | |
|---|---|---|---|---|---|---|
| Obesity | mast_cell | *ZNF84* | 3.722827157 | 0.013706175 | 0.018217912 | 1 |
| Obesity | mast_cell | *ZNF667* | 2.930276659 | 3.08E-07 | 1.03E-06 | 0.00193674 |
| Obesity | mast_cell | *PRIM1* | 5.301185727 | 0.000459684 | 0.000755367 | 1 |
| Obesity | mast_cell | *SULT1C4* | 5.695169233 | 4.24E-07 | 1.35E-06 | 0.002667622 |
| Obesity | mast_cell | *SFI1* | 7.7105348 | 1.92E-08 | 1.11E-07 | 0.000120497 |
| Obesity | mast_cell | *ZNF649* | 3.022991577 | 0.001764406 | 0.002646189 | 1 |
| Obesity | mast_cell | *ZNF248* | 2.603083731 | 0.002167421 | 0.00321913 | 1 |
| Obesity | mast_cell | *HIBCH* | 2.651504475 | 0.008539179 | 0.011680987 | 1 |
| Obesity | mast_cell | *ZNF544* | 2.546975153 | 0.009222075 | 0.012568651 | 1 |
| Obesity | mast_cell | *SOWAHC* | 8.641971144 | 5.03E-09 | 5.29E-08 | 3.16E-05 |
| Obesity | mast_cell | *AC135178.1* | 9.735808172 | 0.000427674 | 0.000706645 | 1 |
| Obesity | mast_cell | *MIER1* | 5.449007482 | 7.00E-05 | 0.000131152 | 0.439883585 |
| Obesity | mast_cell | *SVIP* | -3.184728957 | 0.013600372 | 0.018092581 | 1 |
| Obesity | mast_cell | *ZNF251* | 5.705885286 | 3.31E-07 | 1.09E-06 | 0.00207899 |
| Obesity | mast_cell | *DDRGK1* | 9.017337946 | 1.39E-08 | 9.32E-08 | 8.71E-05 |
| Obesity | mast_cell | *ZNF334* | 1.985705929 | 0.015185714 | 0.020057265 | 1 |
| Obesity | mast_cell | *HSD17B11* | 7.375190377 | 5.01E-06 | 1.18E-05 | 0.031479587 |
| Obesity | mast_cell | *SULT1C2* | 1.800855142 | 1.20E-05 | 2.63E-05 | 0.075507044 |
| Obesity | mast_cell | *QRICH1* | 2.92183892 | 0.000295163 | 0.000498975 | 1 |
| Obesity | mast_cell | *DDX42* | 6.529518224 | 3.08E-08 | 1.57E-07 | 0.000193481 |
| Obesity | mast_cell | *ANTXRL* | -97.99685154 | 1.89E-06 | 4.94E-06 | 0.011886959 |
| Obesity | mast_cell | *STYX* | 5.163317219 | 1.04E-09 | 3.52E-08 | 6.54E-06 |
| Obesity | mast_cell | *TMEM116* | 1.654339047 | 0.00282715 | 0.004132596 | 1 |
| Obesity | mast_cell | *ZKSCAN8* | 7.905151627 | 1.94E-13 | 5.31E-11 | 1.22E-09 |
| Obesity | mast_cell | *MYL4* | 2.995949064 | 0.015998073 | 0.021090351 | 1 |
| Obesity | mast_cell | *ZNF813* | -3.676655375 | 0.004055179 | 0.00580221 | 1 |
| Obesity | mast_cell | *GFPT1* | 6.810102973 | 8.14E-10 | 3.01E-08 | 5.12E-06 |
| Obesity | mast_cell | *UVRAG* | 2.860503856 | 4.05E-06 | 9.74E-06 | 0.025488643 |
| Obesity | mast_cell | *MGEA5* | 3.677597891 | 9.53E-07 | 2.71E-06 | 0.005992117 |
| Obesity | mast_cell | *MT1F* | -6.001707965 | 3.35E-07 | 1.10E-06 | 0.002106853 |
| Obesity | mast_cell | *ZNF568* | 5.266075878 | 1.26E-08 | 8.78E-08 | 7.92E-05 |
| Obesity | mast_cell | *ZNF587* | 6.515284387 | 3.01E-08 | 1.55E-07 | 0.00018954 |
| Obesity | mast_cell | *SH3BGRL2* | 3.382132729 | 0.000142945 | 0.000254227 | 0.898693444 |
| Obesity | mast_cell | *ZNF808* | 5.714145786 | 0.000111448 | 0.000201748 | 0.700670725 |
| Obesity | mast_cell | *TMA16* | 6.249627521 | 0.000277404 | 0.000470853 | 1 |
| Obesity | mast_cell | *HLA-DRB5* | -0.505302055 | 0.024576443 | 0.031610494 | 1 |
| Obesity | mast_cell | *CNGA1* | 2.142083827 | 0.007083812 | 0.00978413 | 1 |
| Obesity | mast_cell | *MAFK* | 5.794564244 | 2.34E-09 | 4.32E-08 | 1.47E-05 |
| Obesity | mast_cell | *C20orf203* | -4.74582504 | 1.79E-06 | 4.72E-06 | 0.011255796 |
| Obesity | mast_cell | *ZNF627* | 7.055783075 | 5.85E-05 | 0.000111474 | 0.36752894 |
| Obesity | mast_cell | *ARC* | 12.60895134 | 7.44E-08 | 3.16E-07 | 0.00046778 |
| Obesity | mast_cell | *TLK1* | 5.74901843 | 4.74E-07 | 1.47E-06 | 0.002978716 |
| Obesity | mast_cell | *LRBA* | 4.556215395 | 7.33E-05 | 0.000136853 | 0.461058504 |
| Obesity | mast_cell | *CCDC69* | 5.131414538 | 1.13E-05 | 2.50E-05 | 0.071112085 |
| Obesity | mast_cell | *MDM4* | 5.764244851 | 9.19E-08 | 3.79E-07 | 0.000577492 |
| Obesity | mast_cell | *ZNF534* | 4.309092657 | 1.02E-05 | 2.27E-05 | 0.06433761 |
| Obesity | mast_cell | *NCOA6* | 2.983581278 | 0.006102074 | 0.008502602 | 1 |
| Obesity | mast_cell | *FAM19A2* | 5.563394242 | 1.05E-07 | 4.24E-07 | 0.000662687 |
| Obesity | mast_cell | *TTC37* | 5.491033275 | 1.13E-06 | 3.17E-06 | 0.007123823 |
| Obesity | mast_cell | *TUSC1* | -6.191832572 | 0.000611917 | 0.000984421 | 1 |

| | | | | | | |
|---|---|---|---|---|---|---|
| Obesity | mast_cell | *PAPSS2* | 4.570810156 | 4.91E-05 | 9.52E-05 | 0.308826784 |
| Obesity | mast_cell | *FAN1* | 5.447460967 | 1.84E-09 | 4.32E-08 | 1.16E-05 |
| Obesity | mast_cell | *CEP290* | 6.16411819 | 2.54E-07 | 8.73E-07 | 0.001594318 |
| Obesity | mast_cell | *ANKRD13B* | 3.6902529 | 2.84E-09 | 4.32E-08 | 1.79E-05 |
| Obesity | mast_cell | *ZNF652* | 6.273345982 | 7.81E-08 | 3.30E-07 | 0.000490857 |
| Obesity | mast_cell | *RPL10A* | 4.712173972 | 6.94E-05 | 0.000130318 | 0.436395172 |
| Obesity | mast_cell | *APCDD1L* | 6.570327798 | 2.35E-09 | 4.32E-08 | 1.48E-05 |
| Obesity | mast_cell | *RCSD1* | -1.542688762 | 0.01521798 | 0.02009144 | 1 |
| Obesity | mast_cell | *RASSF9* | 7.673638294 | 3.55E-09 | 4.62E-08 | 2.23E-05 |
| Obesity | mast_cell | *FAM169A* | 4.574902562 | 5.82E-08 | 2.61E-07 | 0.000366009 |
| Obesity | mast_cell | *TMEM184B* | 6.283672452 | 3.32E-08 | 1.67E-07 | 0.000208895 |
| Obesity | mast_cell | *SCAMP5* | 1.759016805 | 0.000556173 | 0.000900042 | 1 |
| Obesity | mast_cell | *SFT2D1* | 5.55092402 | 0.001232062 | 0.001891569 | 1 |
| Obesity | mast_cell | *ARHGAP11A* | -2.521952459 | 1.90E-06 | 4.95E-06 | 0.011917504 |
| Obesity | mast_cell | *DENND4B* | 4.871743133 | 1.83E-06 | 4.81E-06 | 0.011506682 |
| Obesity | mast_cell | *CD3E* | 2.63044184 | 3.28E-08 | 1.65E-07 | 0.000206024 |
| Obesity | mast_cell | *TSEN15* | 8.127240411 | 4.66E-07 | 1.46E-06 | 0.002930026 |
| Obesity | mast_cell | *ITPRIPL1* | 3.416820779 | 6.95E-07 | 2.05E-06 | 0.004366412 |
| Obesity | mast_cell | *PRMT6* | 5.373412376 | 1.42E-06 | 3.87E-06 | 0.008947206 |
| Obesity | mast_cell | *SHISA4* | 5.019260726 | 1.89E-07 | 6.84E-07 | 0.001189452 |
| Obesity | mast_cell | *CAPZA2* | 6.170326922 | 1.60E-06 | 4.29E-06 | 0.010077175 |
| Obesity | mast_cell | *C1orf174* | 5.660369744 | 2.77E-07 | 9.42E-07 | 0.001744543 |
| Obesity | mast_cell | *RASGEF1A* | 5.687821857 | 1.35E-08 | 9.17E-08 | 8.48E-05 |
| Obesity | mast_cell | *KIAA0753* | 5.986676656 | 3.73E-07 | 1.21E-06 | 0.00234589 |
| Obesity | mast_cell | *DCLRE1A* | 5.062140219 | 0.000449871 | 0.000741178 | 1 |
| Obesity | mast_cell | *APRT* | 7.97150628 | 0.001279536 | 0.00195967 | 1 |
| Obesity | mast_cell | *TBKBP1* | 3.505525382 | 5.68E-07 | 1.72E-06 | 0.003571716 |
| Obesity | mast_cell | *ZFP2* | 4.328405192 | 2.65E-08 | 1.41E-07 | 0.00016662 |
| Obesity | mast_cell | *MFAP3L* | 15.44604065 | 1.23E-07 | 4.80E-07 | 0.000771084 |
| Obesity | mast_cell | *NAGA* | 7.514298529 | 2.70E-07 | 9.20E-07 | 0.001699538 |
| Obesity | mast_cell | *SMG5* | 5.548544047 | 8.03E-05 | 0.00014866 | 0.504848835 |
| Obesity | mast_cell | *TGM2* | 1.214122866 | 0.002529107 | 0.003722897 | 1 |
| Obesity | mast_cell | *PJA2* | 4.217104478 | 0.000266379 | 0.000453118 | 1 |
| Obesity | mast_cell | *AP001007.1* | 12.72148663 | 1.63E-08 | 1.02E-07 | 0.000102252 |
| Obesity | mast_cell | *ZNF525* | -3.470480139 | 0.000649143 | 0.001040161 | 1 |
| Obesity | mast_cell | *AC105020.1* | 6.661711818 | 8.49E-09 | 6.93E-08 | 5.34E-05 |
| Obesity | mast_cell | *EFCAB2* | 5.009080178 | 0.000117434 | 0.000211549 | 0.738307696 |
| Obesity | mast_cell | *CHML* | 3.62259541 | 8.21E-06 | 1.86E-05 | 0.051613145 |
| Obesity | mast_cell | *CAPN8* | -7.634687076 | 0.00095549 | 0.001492093 | 1 |
| Obesity | mast_cell | *TATDN3* | 2.50581695 | 8.15E-05 | 0.000150645 | 0.512495909 |
| Obesity | mast_cell | *ECT2L* | 6.133763448 | 2.65E-06 | 6.67E-06 | 0.016656648 |
| Obesity | mast_cell | *CENPW* | 6.32633582 | 0.000653066 | 0.00104607 | 1 |
| Obesity | mast_cell | *FAM229B* | 2.256860237 | 4.55E-06 | 1.08E-05 | 0.028593383 |
| Obesity | mast_cell | *DDO* | 11.67567437 | 2.64E-08 | 1.40E-07 | 0.000165836 |
| Obesity | mast_cell | *HIST2H2BF* | 7.936396859 | 1.71E-08 | 1.05E-07 | 0.000107747 |
| Obesity | mast_cell | *RBM20* | 5.047440638 | 7.30E-08 | 3.12E-07 | 0.000458915 |
| Obesity | mast_cell | *PCMTD2* | 7.006054524 | 3.64E-07 | 1.18E-06 | 0.002285913 |
| Obesity | mast_cell | *OOEP* | 2.446158928 | 6.87E-05 | 0.000129198 | 0.432166348 |
| Obesity | mast_cell | *SYS1* | 7.245909584 | 2.17E-09 | 4.32E-08 | 1.36E-05 |
| Obesity | mast_cell | *NEU4* | 10.05362355 | 0.002577847 | 0.003791981 | 1 |

| | | | | | | |
|---|---|---|---|---|---|---|
| Obesity | mast_cell | *GIGYF2* | 4.4774596 | 0.000452536 | 0.000744521 | 1 |
| Obesity | mast_cell | *RUFY2* | 3.953109071 | 0.003186938 | 0.004626248 | 1 |
| Obesity | mast_cell | *ASAH2B* | 2.41759326 | 3.41E-07 | 1.12E-06 | 0.002144093 |
| Obesity | mast_cell | *AGAP6* | 1.411667339 | 0.020267994 | 0.026360132 | 1 |
| Obesity | mast_cell | *SYT15* | 8.326678059 | 2.34E-08 | 1.29E-07 | 0.000147207 |
| Obesity | mast_cell | *RING1* | 3.765116458 | 3.66E-06 | 8.88E-06 | 0.023006294 |
| Obesity | mast_cell | *HSD17B8* | -2.901481192 | 5.27E-06 | 1.23E-05 | 0.033154647 |
| Obesity | mast_cell | *OXLD1* | 4.560502995 | 2.69E-07 | 9.17E-07 | 0.001692557 |
| Obesity | mast_cell | *HLA-DOA* | 1.737680075 | 0.011050834 | 0.014893161 | 1 |
| Obesity | mast_cell | *HLA-DRA* | -1.513155699 | 0.026697128 | 0.03413562 | 1 |
| Obesity | mast_cell | *COL15A1* | 3.518199312 | 1.15E-06 | 3.22E-06 | 0.007252035 |
| Obesity | mast_cell | *NOTCH4* | -0.535103111 | 3.33E-08 | 1.67E-07 | 0.000209471 |
| Obesity | mast_cell | *AGER* | 3.034492526 | 9.75E-09 | 7.58E-08 | 6.13E-05 |
| Obesity | mast_cell | *STK19* | 2.708319832 | 0.000787183 | 0.001244573 | 1 |
| Obesity | mast_cell | *SKIV2L* | -1.124953227 | 0.001429849 | 0.002176099 | 1 |
| Obesity | mast_cell | *NELFE* | 5.837610384 | 2.54E-10 | 1.48E-08 | 1.60E-06 |
| Obesity | mast_cell | *LAYN* | 4.894427541 | 4.84E-07 | 1.50E-06 | 0.003041239 |
| Obesity | mast_cell | *CARD16* | 4.977212799 | 8.13E-06 | 1.84E-05 | 0.051099368 |
| Obesity | mast_cell | *BAG6* | 4.414597263 | 2.15E-07 | 7.59E-07 | 0.001351733 |
| Obesity | mast_cell | *NFKBIL1* | 4.653741822 | 1.02E-06 | 2.87E-06 | 0.006384002 |
| Obesity | mast_cell | *ZNF814* | 6.273471763 | 4.35E-07 | 1.38E-06 | 0.002737015 |
| Obesity | mast_cell | *HLA-C* | 2.410807378 | 7.66E-05 | 0.000142545 | 0.481658417 |
| Obesity | mast_cell | *POU5F1* | 1.506187149 | 6.69E-09 | 6.04E-08 | 4.21E-05 |
| Obesity | mast_cell | *CCHCR1* | 2.729449664 | 4.26E-09 | 4.85E-08 | 2.68E-05 |
| Obesity | mast_cell | *ABCF1* | 6.422152338 | 7.98E-10 | 3.00E-08 | 5.02E-06 |
| Obesity | mast_cell | *LILRB3* | 3.497180268 | 1.54E-07 | 5.78E-07 | 0.000968922 |
| Obesity | mast_cell | *LINC01123* | 36.17553079 | 0.010393522 | 0.014063694 | 1 |
| Obesity | mast_cell | *HLA-E* | 6.507249024 | 5.68E-15 | 2.28E-12 | 3.57E-11 |
| Obesity | mast_cell | *ZNF468* | 7.263756201 | 2.32E-08 | 1.28E-07 | 0.000146121 |
| Obesity | mast_cell | *HCG9* | 1.183879662 | 0.010395098 | 0.014063694 | 1 |
| Obesity | mast_cell | *HLA-G* | -2.014003391 | 0.005988948 | 0.00835423 | 1 |
| Obesity | mast_cell | *TBC1D8* | 4.765515402 | 7.29E-08 | 3.12E-07 | 0.000458482 |
| Obesity | mast_cell | *ZFP57* | 2.060658664 | 1.14E-08 | 8.36E-08 | 7.14E-05 |
| Obesity | mast_cell | *FAM153C* | -6.330292217 | 1.82E-05 | 3.83E-05 | 0.114474894 |
| Obesity | mast_cell | *GABBR1* | 6.486077648 | 1.25E-06 | 3.46E-06 | 0.007880417 |
| Obesity | mast_cell | *MALRD1* | -1.123918202 | 0.001475213 | 0.002240257 | 1 |
| Obesity | mast_cell | *MROH6* | 2.629098724 | 2.32E-05 | 4.76E-05 | 0.145687936 |
| Obesity | mast_cell | *ATXN2* | 5.585216804 | 2.04E-07 | 7.25E-07 | 0.001283031 |
| Obesity | mast_cell | *FAM216A* | 4.857073925 | 1.11E-06 | 3.11E-06 | 0.006989753 |
| Obesity | mast_cell | *ZNF155* | 4.368638123 | 5.55E-06 | 1.29E-05 | 0.034878714 |
| Obesity | mast_cell | *ZNF783* | 4.193301926 | 2.76E-06 | 6.91E-06 | 0.017334632 |
| Obesity | mast_cell | *LRRC10B* | -6.239918203 | 4.58E-07 | 1.44E-06 | 0.002878714 |
| Obesity | mast_cell | *PCDHA5* | 21.24806577 | 1.32E-06 | 3.63E-06 | 0.008316912 |
| Obesity | mast_cell | *PCDHA4* | 8.870245091 | 2.68E-05 | 5.45E-05 | 0.168738846 |
| Obesity | mast_cell | *TRIM13* | 3.925249175 | 0.002437242 | 0.003594403 | 1 |
| Obesity | mast_cell | *SPIRE2* | 1.629574682 | 0.001056422 | 0.001637506 | 1 |
| Obesity | mast_cell | *SLFN12L* | 3.211777221 | 4.48E-05 | 8.74E-05 | 0.281932489 |
| Obesity | mast_cell | *TMEM231* | 3.084696486 | 7.21E-07 | 2.12E-06 | 0.004530233 |
| Obesity | mast_cell | *FAM71F2* | 3.752928963 | 5.51E-09 | 5.54E-08 | 3.46E-05 |
| Obesity | mast_cell | *AC016586.1* | 26.76980993 | 2.78E-07 | 9.43E-07 | 0.001745939 |

| | | | | | | |
|---|---|---|---|---|---|---|
| Obesity | mast_cell | *LINC00654* | 5.660705122 | 1.56E-06 | 4.20E-06 | 0.009822677 |
| Obesity | mast_cell | *TMEM170B* | 0.967530967 | 0.014597605 | 0.019312951 | 1 |
| Obesity | mast_cell | *NT5M* | -2.147454646 | 7.90E-06 | 1.79E-05 | 0.049681591 |
| Obesity | mast_cell | *PRR13* | 3.626642798 | 4.86E-06 | 1.15E-05 | 0.030555743 |
| Obesity | mast_cell | *MT1A* | -2.889909819 | 4.21E-07 | 1.34E-06 | 0.002643953 |
| Obesity | mast_cell | *MT1M* | -3.546134213 | 4.27E-06 | 1.02E-05 | 0.026818407 |
| Obesity | mast_cell | *CFI* | 3.828904496 | 1.37E-05 | 2.95E-05 | 0.08587867 |
| Obesity | mast_cell | *EXOC3L4* | 4.277735001 | 1.96E-07 | 7.03E-07 | 0.001234523 |
| Obesity | mast_cell | *TP53TG3D* | -5.394829986 | 1.83E-05 | 3.84E-05 | 0.114780795 |
| Obesity | mast_cell | *ATP6AP1L* | 2.957360706 | 2.96E-06 | 7.35E-06 | 0.018587416 |
| Obesity | mast_cell | *CPT1B* | -1.558993602 | 0.012895164 | 0.017220029 | 1 |
| Obesity | mast_cell | *DENND6B* | 3.471918727 | 1.04E-08 | 7.89E-08 | 6.51E-05 |
| Obesity | mast_cell | *EIF3CL* | 26.31383469 | 1.10E-30 | 6.89E-27 | 6.89E-27 |
| Obesity | mast_cell | *LCMT1* | 6.855105192 | 1.29E-06 | 3.56E-06 | 0.008137952 |
| Obesity | mast_cell | *CDPF1* | 2.37515512 | 7.48E-05 | 0.000139469 | 0.470525422 |
| Obesity | mast_cell | *LIN52* | 4.722021748 | 4.24E-07 | 1.35E-06 | 0.002667318 |
| Obesity | mast_cell | *C17orf107* | 3.450638375 | 9.10E-06 | 2.04E-05 | 0.057237695 |
| Obesity | mast_cell | *LINC00487* | -5.864945788 | 0.000499828 | 0.000815154 | 1 |
| Obesity | mast_cell | *RFPL3S* | 2.333079474 | 0.000286409 | 0.000484959 | 1 |
| Obesity | mast_cell | *C1QTNF9B* | -4.60308066 | 1.63E-07 | 6.01E-07 | 0.001022507 |
| Obesity | mast_cell | *RNPS1* | 4.624467333 | 4.81E-06 | 1.13E-05 | 0.030216855 |
| Obesity | mast_cell | *NYNRIN* | 5.234472381 | 2.34E-09 | 4.32E-08 | 1.47E-05 |
| Obesity | mast_cell | *ZDHHC11B* | -2.226703095 | 8.92E-05 | 0.000163441 | 0.560767663 |
| Obesity | mast_cell | *GOLGA8O* | -4.247788057 | 1.71E-07 | 6.28E-07 | 0.001075094 |
| Obesity | mast_cell | *HCG27* | 1.52024924 | 0.001482817 | 0.002250867 | 1 |
| Obesity | mast_cell | *LNP1* | 3.241572505 | 5.67E-08 | 2.55E-07 | 0.000356213 |
| Obesity | mast_cell | *VGLL3* | 1.173047213 | 0.013495568 | 0.017960761 | 1 |
| Obesity | mast_cell | *COLQ* | 4.589084372 | 1.00E-07 | 4.09E-07 | 0.000630039 |
| Obesity | mast_cell | *METTL6* | 6.293253604 | 1.08E-09 | 3.57E-08 | 6.76E-06 |
| Obesity | mast_cell | *SACM1L* | 6.053704344 | 4.12E-06 | 9.89E-06 | 0.025908391 |
| Obesity | mast_cell | *SLC48A1* | 4.720478146 | 7.16E-09 | 6.28E-08 | 4.50E-05 |
| Obesity | mast_cell | *TAS2R14* | 2.3189585 | 3.57E-05 | 7.07E-05 | 0.224462657 |
| Obesity | mast_cell | *ZRSR1* | 12.35542177 | 3.36E-08 | 1.68E-07 | 0.00021154 |
| Obesity | mast_cell | *AC084018.1* | -2.155652253 | 7.98E-05 | 0.000147859 | 0.501982204 |
| Obesity | mast_cell | *C17orf51* | 3.17948851 | 8.27E-08 | 3.47E-07 | 0.000519837 |
| Obesity | mast_cell | *OR2A42* | 1.640109455 | 0.008135279 | 0.011157613 | 1 |
| Obesity | mast_cell | *RNF208* | 6.617287184 | 1.82E-08 | 1.08E-07 | 0.000114574 |
| Obesity | mast_cell | *MAP10* | 4.653478386 | 0.000553898 | 0.000896917 | 1 |
| Obesity | mast_cell | *ZNF580* | 4.7791834 | 1.85E-05 | 3.89E-05 | 0.116512522 |
| Obesity | mast_cell | *FGFR1OP* | 5.703067051 | 0.000269591 | 0.000458334 | 1 |
| Obesity | mast_cell | *FAM106A* | 1.342853735 | 0.036340407 | 0.045612325 | 1 |
| Obesity | mast_cell | *ZNF254* | 6.036365609 | 7.42E-06 | 1.69E-05 | 0.046626129 |
| Obesity | mast_cell | *TCTEX1D2* | 3.055656226 | 5.49E-06 | 1.28E-05 | 0.034495906 |
| Obesity | mast_cell | *ASIC3* | 4.774920447 | 1.88E-09 | 4.32E-08 | 1.18E-05 |
| Obesity | mast_cell | *GIMAP1* | 4.087340332 | 0.001882134 | 0.002813356 | 1 |
| Obesity | mast_cell | *CCDC183* | -3.567748176 | 5.39E-07 | 1.64E-06 | 0.00338788 |
| Obesity | mast_cell | *ARHGEF35* | 3.789419143 | 0.005131722 | 0.007235509 | 1 |
| Obesity | mast_cell | *SUPT4H1* | 8.49095994 | 1.38E-08 | 9.30E-08 | 8.65E-05 |
| Obesity | mast_cell | *MXD3* | 3.735958307 | 3.81E-06 | 9.20E-06 | 0.023970708 |
| Obesity | mast_cell | *GSTM2* | -1.820439204 | 0.003284997 | 0.004761996 | 1 |

| | | | | | | |
|---|---|---|---|---|---|---|
| Obesity | mast_cell | *ARHGAP19* | 10.37288206 | 0.000194764 | 0.000338629 | 1 |
| Obesity | mast_cell | *LCAT* | 2.136111748 | 0.004665872 | 0.00660763 | 1 |
| Obesity | mast_cell | *SIPA1* | 7.187192836 | 4.36E-10 | 2.14E-08 | 2.74E-06 |
| Obesity | mast_cell | *ERV3-1* | 1.140259037 | 0.000187598 | 0.00032662 | 1 |
| Obesity | mast_cell | *SRA1* | 2.385889691 | 0.013323837 | 0.017754761 | 1 |
| Obesity | mast_cell | *HEXA* | 4.656908517 | 0.010493122 | 0.014190203 | 1 |
| Obesity | mast_cell | *LEPROT* | 5.060999805 | 0.000136387 | 0.000243115 | 0.857467443 |
| Obesity | mast_cell | *LBH* | 10.26832091 | 1.59E-06 | 4.26E-06 | 0.010003559 |
| Obesity | mast_cell | *NCKIPSD* | 5.853225206 | 1.79E-05 | 3.76E-05 | 0.11223591 |
| Obesity | mast_cell | *ATF6B* | 3.388462107 | 1.14E-05 | 2.50E-05 | 0.071363361 |
| Obesity | mast_cell | *SLC35F6* | 9.195656546 | 1.38E-10 | 1.07E-08 | 8.70E-07 |
| Obesity | mast_cell | *CLIC1* | -3.096137417 | 0.020403366 | 0.02652522 | 1 |
| Obesity | mast_cell | *ATP6V1G2* | -1.5863698 | 1.80E-06 | 4.75E-06 | 0.01133883 |
| Obesity | mast_cell | *EMP2* | 4.950771649 | 1.59E-08 | 1.02E-07 | 0.00010025 |
| Obesity | mast_cell | *PPM1N* | 8.156929608 | 2.84E-08 | 1.48E-07 | 0.000178587 |
| Obesity | mast_cell | *GALT* | 6.040057685 | 0.000391666 | 0.000651774 | 1 |
| Obesity | mast_cell | *ITGA1* | 4.21678606 | 1.70E-08 | 1.04E-07 | 0.000106768 |
| Obesity | mast_cell | *ZNF726* | -5.25723467 | 0.000333417 | 0.00055988 | 1 |
| Obesity | mast_cell | *TAX1BP3* | 8.932855129 | 0.000546702 | 0.000886769 | 1 |
| Obesity | mast_cell | *AP1G2* | -3.779729979 | 8.64E-05 | 0.000158816 | 0.543150461 |
| Obesity | mast_cell | *ZNF90* | 5.554698309 | 5.15E-08 | 2.38E-07 | 0.000324079 |
| Obesity | mast_cell | *TM6SF2* | 36.16932848 | 1.23E-08 | 8.65E-08 | 7.72E-05 |
| Obesity | mast_cell | *REPIN1* | 3.773245638 | 0.00149532 | 0.002267506 | 1 |
| Obesity | mast_cell | *SMIM7* | 4.913508283 | 1.94E-09 | 4.32E-08 | 1.22E-05 |
| Obesity | mast_cell | *AC079776.2* | 5.842964096 | 2.58E-08 | 1.38E-07 | 0.000162361 |
| Obesity | mast_cell | *WEE2* | 4.102623994 | 1.89E-08 | 1.10E-07 | 0.000119089 |
| Obesity | mast_cell | *PAXIP1-AS2* | 7.992623839 | 1.44E-09 | 4.06E-08 | 9.05E-06 |
| Obesity | mast_cell | *LYRM4* | 6.995885041 | 9.31E-09 | 7.33E-08 | 5.85E-05 |
| Obesity | mast_cell | *C17orf67* | -1.950409338 | 0.033648375 | 0.042428265 | 1 |
| Obesity | mast_cell | *FIS1* | 2.272728665 | 0.011852728 | 0.015912471 | 1 |
| Obesity | mast_cell | *NEURL1B* | 2.208374565 | 1.54E-08 | 9.93E-08 | 9.66E-05 |
| Obesity | mast_cell | *PPME1* | 3.484939528 | 3.09E-06 | 7.64E-06 | 0.01941686 |
| Obesity | mast_cell | *STARD10* | -1.102829494 | 0.010275514 | 0.013916879 | 1 |
| Obesity | mast_cell | *NUTM2D* | 5.122881303 | 0.00045486 | 0.000747832 | 1 |
| Obesity | mast_cell | *CPEB1* | -3.078497255 | 0.002655092 | 0.003896385 | 1 |
| Obesity | mast_cell | *C10orf105* | -4.43546747 | 0.000104204 | 0.000189072 | 0.655133668 |
| Obesity | mast_cell | *ARHGEF33* | 1.934071503 | 0.003612511 | 0.005212728 | 1 |
| Obesity | mast_cell | *AC005082.1* | -6.290625467 | 1.65E-06 | 4.39E-06 | 0.010364669 |
| Obesity | mast_cell | *TBC1D26* | 23.61798102 | 0.019015868 | 0.024813774 | 1 |
| Obesity | mast_cell | *ACSM4* | 1.341609688 | 0.001111561 | 0.001716967 | 1 |
| Obesity | mast_cell | *COL28A1* | 12.28717842 | 1.54E-07 | 5.78E-07 | 0.000967613 |
| Obesity | mast_cell | *PHB2* | 5.354037028 | 6.92E-07 | 2.05E-06 | 0.004353228 |
| Obesity | mast_cell | *GRID2IP* | 6.211305037 | 7.44E-07 | 2.18E-06 | 0.00468055 |
| Obesity | mast_cell | *UBXN2B* | 2.617723591 | 0.017614703 | 0.023081208 | 1 |
| Obesity | mast_cell | *UBE2QL1* | 3.619488681 | 1.78E-08 | 1.06E-07 | 0.000112105 |
| Obesity | mast_cell | *GOLGA8B* | 4.953763748 | 1.90E-09 | 4.32E-08 | 1.20E-05 |
| Obesity | mast_cell | *KCNU1* | 3.323470163 | 2.81E-08 | 1.48E-07 | 0.0001766 |
| Obesity | mast_cell | *MYL5* | 17.94721428 | 1.13E-07 | 4.48E-07 | 0.00070806 |
| Obesity | mast_cell | *NPEPL1* | 8.091681128 | 1.03E-07 | 4.18E-07 | 0.000650024 |
| Obesity | mast_cell | *LINC01139* | 3.447624038 | 0.001614313 | 0.002434744 | 1 |

| | | | | | | |
|---|---|---|---|---|---|---|
| Obesity | mast_cell | *TSTD1* | 11.80225293 | 1.87E-08 | 1.09E-07 | 0.000117292 |
| Obesity | mast_cell | *LINC01138* | 4.298990229 | 2.27E-09 | 4.32E-08 | 1.43E-05 |
| Obesity | mast_cell | *CYB5RL* | 5.188173133 | 7.34E-09 | 6.34E-08 | 4.62E-05 |
| Obesity | mast_cell | *ATAD3C* | -2.841626575 | 0.000383257 | 0.000638964 | 1 |
| Obesity | mast_cell | *CCDC7* | 4.291968233 | 0.00040504 | 0.000672784 | 1 |
| Obesity | mast_cell | *AP000350.4* | 1.077627204 | 0.00017893 | 0.000312655 | 1 |
| Obesity | mast_cell | *RNASEK* | 5.768465913 | 3.71E-09 | 4.67E-08 | 2.33E-05 |
| Obesity | mast_cell | *RNU6ATAC35I* | 12.55050604 | 1.62E-08 | 1.02E-07 | 0.000101875 |
| Obesity | mast_cell | *PPP3R1* | 4.868701453 | 1.21E-06 | 3.35E-06 | 0.007587913 |
| Obesity | mast_cell | *FANCG* | -2.053784076 | 6.21E-05 | 0.000117659 | 0.390511277 |
| Obesity | mast_cell | *AP4M1* | -2.092176464 | 5.55E-05 | 0.000106466 | 0.349101328 |
| Obesity | mast_cell | *C19orf73* | 15.63283789 | 5.84E-07 | 1.76E-06 | 0.003672994 |
| Obesity | mast_cell | *OR2A25* | -32.81884552 | 4.45E-07 | 1.41E-06 | 0.002795549 |
| Obesity | mast_cell | *XKR9* | 2.153035829 | 5.62E-09 | 5.57E-08 | 3.53E-05 |
| Obesity | mast_cell | *C1orf229* | 4.174085809 | 0.000990586 | 0.001543448 | 1 |
| Obesity | mast_cell | *SLC12A8* | 2.023640239 | 0.002479312 | 0.00365302 | 1 |
| Obesity | mast_cell | *APOL6* | 4.449179261 | 2.37E-09 | 4.32E-08 | 1.49E-05 |
| Obesity | mast_cell | *UBA52* | 2.849193107 | 0.030731222 | 0.038945211 | 1 |
| Obesity | mast_cell | *TIAF1* | 3.241829039 | 3.29E-05 | 6.56E-05 | 0.207021561 |
| Obesity | mast_cell | *BTBD19* | 5.60290819 | 8.34E-09 | 6.87E-08 | 5.25E-05 |
| Obesity | mast_cell | *FAM185A* | 6.84966676 | 8.14E-10 | 3.01E-08 | 5.12E-06 |
| Obesity | mast_cell | *AC062017.1* | 3.911434785 | 2.32E-07 | 8.08E-07 | 0.001456639 |
| Obesity | mast_cell | *ADRA2B* | 9.522273181 | 1.73E-08 | 1.05E-07 | 0.000108542 |
| Obesity | mast_cell | *VPS52* | 4.938459449 | 7.08E-09 | 6.24E-08 | 4.45E-05 |
| Obesity | mast_cell | *LINC00205* | 3.074462703 | 2.93E-06 | 7.29E-06 | 0.01841777 |
| Obesity | mast_cell | *AC147651.3* | -2.389868389 | 1.08E-07 | 4.34E-07 | 0.000680153 |
| Obesity | mast_cell | *HLA-DPB1* | 1.741231849 | 0.028911797 | 0.036787789 | 1 |
| Obesity | mast_cell | *OSER1-AS1* | 10.82627086 | 6.27E-10 | 2.65E-08 | 3.94E-06 |
| Obesity | mast_cell | *LINC01133* | 2.560153677 | 9.16E-09 | 7.26E-08 | 5.76E-05 |
| Obesity | mast_cell | *LINC01117* | 3.467436729 | 3.89E-09 | 4.78E-08 | 2.45E-05 |
| Obesity | mast_cell | *RASAL2-AS1* | 8.539461059 | 1.90E-06 | 4.96E-06 | 0.011959907 |
| Obesity | mast_cell | *AC069213.1* | -9.534431759 | 0.000100576 | 0.000182805 | 0.632323072 |
| Obesity | mast_cell | *LINC00240* | 4.180895795 | 3.44E-15 | 1.54E-12 | 2.16E-11 |
| Obesity | mast_cell | *LINC00863* | 5.576855998 | 1.68E-08 | 1.04E-07 | 0.000105465 |
| Obesity | mast_cell | *LINC00184* | 4.913562459 | 2.77E-06 | 6.93E-06 | 0.01738362 |
| Obesity | mast_cell | *TMEM233* | 2.605827359 | 1.48E-08 | 9.68E-08 | 9.31E-05 |
| Obesity | mast_cell | *LINC00237* | 12.01836581 | 2.02E-08 | 1.15E-07 | 0.000126714 |
| Obesity | mast_cell | *PLEKHM1* | 8.10974415 | 3.39E-09 | 4.52E-08 | 2.13E-05 |
| Obesity | mast_cell | *AP000350.6* | 2.292906897 | 2.44E-07 | 8.47E-07 | 0.001536392 |
| Obesity | mast_cell | *SFTA1P* | 7.701188664 | 7.73E-07 | 2.26E-06 | 0.004862777 |
| Obesity | mast_cell | *FAM225B* | 2.175791922 | 0.004308093 | 0.006134764 | 1 |
| Obesity | mast_cell | *LINC00626* | 50.46545781 | 4.00E-08 | 1.93E-07 | 0.000251343 |
| Obesity | mast_cell | *ERCC6* | 7.40163218 | 1.15E-07 | 4.55E-07 | 0.000720699 |
| Obesity | mast_cell | *ZNF503-AS1* | 2.258166094 | 1.70E-08 | 1.04E-07 | 0.000107148 |
| Obesity | mast_cell | *AC064836.3* | 14.29523656 | 2.75E-09 | 4.32E-08 | 1.73E-05 |
| Obesity | mast_cell | *TMEM185B* | -3.137776777 | 1.74E-05 | 3.67E-05 | 0.109186218 |
| Obesity | mast_cell | *AC093901.1* | 4.430443107 | 5.26E-08 | 2.41E-07 | 0.000330883 |
| Obesity | mast_cell | *C14orf132* | 3.085875455 | 6.62E-09 | 6.02E-08 | 4.16E-05 |
| Obesity | mast_cell | *ZNF717* | 0.628111362 | 0.006483858 | 0.009004643 | 1 |
| Obesity | mast_cell | *AC009299.3* | 7.827534695 | 3.00E-06 | 7.45E-06 | 0.018891638 |

| | | | | | | |
|---|---|---|---|---|---|---|
| Obesity | mast_cell | *SLC2A1-AS1* | 2.779985607 | 7.84E-07 | 2.28E-06 | 0.00492595 |
| Obesity | mast_cell | *LINC01068* | -2.499091307 | 4.42E-06 | 1.05E-05 | 0.027791333 |
| Obesity | mast_cell | *AC108463.2* | 4.604544535 | 9.51E-05 | 0.000173526 | 0.597623388 |
| Obesity | mast_cell | *HCG20* | -13.10688435 | 2.32E-08 | 1.28E-07 | 0.000145917 |
| Obesity | mast_cell | *C19orf24* | 4.899367107 | 0.000130069 | 0.000232115 | 0.817742377 |
| Obesity | mast_cell | *LINC01013* | 6.680303949 | 8.16E-09 | 6.78E-08 | 5.13E-05 |
| Obesity | mast_cell | *AC073133.2* | -9.839713106 | 6.05E-08 | 2.70E-07 | 0.000380515 |
| Obesity | mast_cell | *PROB1* | 10.49817956 | 9.81E-05 | 0.000178698 | 0.616685401 |
| Obesity | mast_cell | *ARL17B* | 2.833155593 | 0.0089931 | 0.012277877 | 1 |
| Obesity | mast_cell | *LINC00954* | -3.170202801 | 1.78E-06 | 4.71E-06 | 0.011220505 |
| Obesity | mast_cell | *AC067945.3* | 4.16316681 | 5.43E-05 | 0.000104387 | 0.341553647 |
| Obesity | mast_cell | *RPL41* | 4.587982773 | 7.82E-08 | 3.30E-07 | 0.000491841 |
| Obesity | mast_cell | *LINC00200* | -14.6330861 | 3.59E-05 | 7.10E-05 | 0.225767328 |
| Obesity | mast_cell | *PATL2* | 6.459767231 | 8.67E-10 | 3.08E-08 | 5.45E-06 |
| Obesity | mast_cell | *ZNF492* | 2.606182461 | 6.55E-05 | 0.000123437 | 0.411661506 |
| Obesity | mast_cell | *U91324.1* | 6.070768659 | 1.48E-08 | 9.68E-08 | 9.29E-05 |
| Obesity | mast_cell | *CISD3* | 7.990670479 | 2.16E-09 | 4.32E-08 | 1.36E-05 |
| Obesity | mast_cell | *PSMG3-AS1* | 7.713794349 | 9.00E-08 | 3.73E-07 | 0.000565977 |
| Obesity | mast_cell | *PPP5D1* | 6.622031995 | 4.11E-08 | 1.98E-07 | 0.000258661 |
| Obesity | mast_cell | *KLHL7-AS1* | 17.15822941 | 0.023217294 | 0.029972716 | 1 |
| Obesity | mast_cell | *AC092171.4* | 5.242861741 | 0.003068412 | 0.0044645 | 1 |
| Obesity | mast_cell | *HLA-DPA1* | 3.324048099 | 0.004207654 | 0.006006703 | 1 |
| Obesity | mast_cell | *RPS18* | 5.283471082 | 9.81E-07 | 2.78E-06 | 0.00616865 |
| Obesity | mast_cell | *DIRC3* | 3.804335362 | 6.88E-07 | 2.03E-06 | 0.004322764 |
| Obesity | mast_cell | *TAPBP* | 5.782585645 | 2.57E-08 | 1.38E-07 | 0.000161627 |
| Obesity | mast_cell | *AC107021.1* | 2.470861026 | 5.71E-06 | 1.33E-05 | 0.035882063 |
| Obesity | mast_cell | *LINC00865* | 3.158539052 | 4.59E-07 | 1.44E-06 | 0.002887302 |
| Obesity | mast_cell | *AC009495.3* | -6.101177824 | 0.000321223 | 0.000540125 | 1 |
| Obesity | mast_cell | *HLA-DQB2* | 0.705447312 | 0.000782171 | 0.001237109 | 1 |
| Obesity | mast_cell | *GOLGA8N* | 2.454318136 | 0.000158921 | 0.000280498 | 0.999134627 |
| Obesity | mast_cell | *LINC00665* | 2.522590337 | 0.000644618 | 0.001033328 | 1 |
| Obesity | mast_cell | *TNF* | -3.565186392 | 1.68E-08 | 1.04E-07 | 0.00010566 |
| Obesity | mast_cell | *LYRM9* | -3.399372969 | 0.037975664 | 0.047541418 | 1 |
| Obesity | mast_cell | *LINC00342* | 8.217668805 | 2.10E-06 | 5.42E-06 | 0.01320358 |
| Obesity | mast_cell | *SNHG15* | 4.981973365 | 1.94E-10 | 1.28E-08 | 1.22E-06 |
| Obesity | mast_cell | *LINC00702* | 2.434420014 | 1.74E-08 | 1.05E-07 | 0.000109157 |
| Obesity | mast_cell | *LINC00472* | 3.822398734 | 4.45E-07 | 1.41E-06 | 0.002798438 |
| Obesity | mast_cell | *RPS28* | 7.562745408 | 0.002684989 | 0.003934855 | 1 |
| Obesity | mast_cell | *AC123768.3* | 8.494309979 | 7.99E-09 | 6.71E-08 | 5.02E-05 |
| Obesity | mast_cell | *TRIM26* | 5.371712789 | 1.25E-06 | 3.46E-06 | 0.007872508 |
| Obesity | mast_cell | *LINC01114* | 11.28806333 | 0.009461711 | 0.012867353 | 1 |
| Obesity | mast_cell | *ZNF736* | 3.380187035 | 0.000203929 | 0.000353293 | 1 |
| Obesity | mast_cell | *HLA-B* | 3.017971574 | 1.40E-05 | 3.03E-05 | 0.088291417 |
| Obesity | mast_cell | *ANKRD65* | 5.706151549 | 3.61E-07 | 1.17E-06 | 0.002268816 |
| Obesity | mast_cell | *ZSCAN31* | 3.253955796 | 2.84E-08 | 1.48E-07 | 0.00017827 |
| Obesity | mast_cell | *LINC00957* | 9.374529311 | 4.58E-07 | 1.44E-06 | 0.002879979 |
| Obesity | mast_cell | *KIAA0040* | 6.298278784 | 0.000194827 | 0.000338645 | 1 |
| Obesity | mast_cell | *B3GALT4* | -3.93452927 | 8.91E-05 | 0.000163243 | 0.559924558 |
| Obesity | mast_cell | *LINC00941* | 9.655277438 | 1.22E-08 | 8.65E-08 | 7.69E-05 |
| Obesity | mast_cell | *LINC00545* | -23.6167121 | 7.73E-07 | 2.26E-06 | 0.004862268 |

| | | | | | | |
|---|---|---|---|---|---|---|
| Obesity | mast_cell | *CDKN2AIPNL* | 2.910667181 | 0.000230469 | 0.000396107 | 1 |
| Obesity | mast_cell | *ZNF737* | 6.487655749 | 1.71E-09 | 4.28E-08 | 1.07E-05 |
| Obesity | mast_cell | *LINC00959* | 5.791394468 | 1.62E-07 | 5.99E-07 | 0.001015791 |
| Obesity | mast_cell | *AC073257.2* | 20.64884325 | 1.54E-06 | 4.14E-06 | 0.009671489 |
| Obesity | mast_cell | *LRRC37A2* | 5.682535947 | 1.43E-06 | 3.88E-06 | 0.008985288 |
| Obesity | mast_cell | *TXNDC5* | 9.354044863 | 2.61E-06 | 6.58E-06 | 0.016416223 |
| Obesity | mast_cell | *ASB14* | 3.007409635 | 4.18E-08 | 2.00E-07 | 0.000262555 |
| Obesity | mast_cell | *CDRT4* | 5.129494877 | 5.02E-08 | 2.33E-07 | 0.00031588 |
| Obesity | mast_cell | *ADSL* | 3.528407099 | 2.97E-07 | 9.98E-07 | 0.001867323 |
| Obesity | mast_cell | *LILRA4* | -28.47425698 | 2.28E-06 | 5.85E-06 | 0.014364406 |
| Obesity | mast_cell | *LILRA2* | 1.076410291 | 0.029878549 | 0.037895187 | 1 |
| Obesity | mast_cell | *PSMB9* | 3.836758062 | 1.17E-05 | 2.57E-05 | 0.073527001 |
| Obesity | mast_cell | *COX19* | 8.18739128 | 2.01E-07 | 7.17E-07 | 0.001261445 |
| Obesity | mast_cell | *L1TD1* | 0.948504519 | 0.038484761 | 0.048130832 | 1 |
| Obesity | mast_cell | *AQP1* | 6.061816142 | 1.48E-05 | 3.18E-05 | 0.093073417 |
| Obesity | mast_cell | *C1QTNF9* | -6.678180174 | 0.001563705 | 0.002365499 | 1 |
| Obesity | mast_cell | *ARHGEF25* | 3.601351973 | 4.28E-08 | 2.04E-07 | 0.000268957 |
| Obesity | mast_cell | *LINC00886* | 3.806065816 | 0.006446482 | 0.008956693 | 1 |
| Obesity | mast_cell | *MIF* | 1.080277793 | 0.000725589 | 0.001153421 | 1 |
| Obesity | mast_cell | *NSUN6* | 3.001007689 | 0.039806158 | 0.049694462 | 1 |
| Obesity | mast_cell | *TDGF1* | -2.22210644 | 0.008261374 | 0.011315743 | 1 |
| Obesity | mast_cell | *ATP5J2* | 4.841275775 | 0.008766017 | 0.011975652 | 1 |
| Obesity | mast_cell | *ARHGAP8* | 4.437031401 | 2.42E-08 | 1.32E-07 | 0.000152428 |
| Obesity | mast_cell | *ATP5O* | 7.293195095 | 7.79E-06 | 1.76E-05 | 0.048966991 |
| Obesity | mast_cell | *PWP2* | 9.444742444 | 2.32E-06 | 5.92E-06 | 0.014577462 |
| Obesity | mast_cell | *PI4KA* | 6.161657315 | 3.09E-06 | 7.65E-06 | 0.019444903 |
| Obesity | mast_cell | *AKAP2* | 28.18953798 | 1.59E-09 | 4.20E-08 | 9.98E-06 |
| Obesity | mast_cell | *LINC00969* | 1.762610863 | 0.026479308 | 0.03387778 | 1 |
| Obesity | mast_cell | *ARHGDIG* | 3.462486561 | 0.003033422 | 0.004416657 | 1 |
| Obesity | mast_cell | *TCP10L* | 17.2486892 | 1.30E-06 | 3.57E-06 | 0.008166508 |
| Obesity | mast_cell | *ARFGAP3* | 6.547483937 | 9.18E-09 | 7.26E-08 | 5.77E-05 |
| Obesity | mast_cell | *C22orf39* | 12.28840325 | 1.22E-08 | 8.65E-08 | 7.69E-05 |
| Obesity | mast_cell | *EIF6* | -4.397481154 | 0.000425239 | 0.000703361 | 1 |
| Obesity | mast_cell | *MRPL20* | 7.711579944 | 6.10E-09 | 5.80E-08 | 3.84E-05 |
| Obesity | mast_cell | *DECR2* | 2.879032256 | 1.80E-08 | 1.07E-07 | 0.000112986 |
| Obesity | mast_cell | *AP5Z1* | 7.846062074 | 4.10E-10 | 2.05E-08 | 2.58E-06 |
| Obesity | mast_cell | *STRC* | -7.19039976 | 0.010936603 | 0.014748697 | 1 |
| Obesity | mast_cell | *PSG4* | 2.996335538 | 2.62E-05 | 5.33E-05 | 0.164818154 |
| Obesity | mast_cell | *STON1* | 3.535328889 | 1.34E-06 | 3.68E-06 | 0.008445762 |
| Obesity | mast_cell | *VSIG8* | -3.114742631 | 3.61E-06 | 8.76E-06 | 0.022667729 |
| Obesity | mast_cell | *C7orf73* | 6.902677132 | 5.14E-06 | 1.21E-05 | 0.032330915 |
| Obesity | mast_cell | *FGF14-IT1* | 6.999899309 | 1.88E-08 | 1.09E-07 | 0.000118082 |
| Obesity | mast_cell | *TICAM2* | -4.614485558 | 1.05E-05 | 2.33E-05 | 0.066210458 |
| Obesity | mast_cell | *PALM2* | 1.700311895 | 1.45E-06 | 3.93E-06 | 0.009113663 |
| Obesity | mast_cell | *NAT6* | -2.203110597 | 0.0066103 | 0.009174163 | 1 |
| Obesity | mast_cell | *UPK3B* | 2.317219573 | 0.002410355 | 0.003558089 | 1 |
| Obesity | mast_cell | *IL10RB* | 4.200223529 | 0.001665678 | 0.002506491 | 1 |
| Obesity | mast_cell | *WDR92* | 5.028811133 | 1.11E-08 | 8.21E-08 | 6.97E-05 |
| Obesity | mast_cell | *NME1-NME2* | 3.272095878 | 0.014499774 | 0.019199679 | 1 |
| Obesity | mast_cell | *TTC4* | 1.268757774 | 0.00024226 | 0.000414784 | 1 |

| | | | | | | |
|---|---|---|---|---|---|---|
| Obesity | mast_cell | *APOBEC3D* | 5.723353368 | 0.003902598 | 0.005601743 | 1 |
| Obesity | mast_cell | *TMEM199* | -2.885083538 | 0.028996872 | 0.036881112 | 1 |
| Obesity | mast_cell | *P2RY11* | -5.421948049 | 8.06E-05 | 0.000149129 | 0.50674055 |
| Obesity | mast_cell | *LILRA6* | 1.980575019 | 4.09E-05 | 8.02E-05 | 0.257333983 |
| Obesity | mast_cell | *APOBEC3C* | 4.273369426 | 6.41E-09 | 5.94E-08 | 4.03E-05 |
| Obesity | mast_cell | *ASPRV1* | 7.305251498 | 7.51E-07 | 2.20E-06 | 0.004724156 |
| Obesity | mast_cell | *OR2AE1* | -5.695681294 | 2.54E-07 | 8.73E-07 | 0.001596401 |
| Obesity | mast_cell | *UBE2V1* | 10.06437406 | 3.01E-09 | 4.44E-08 | 1.89E-05 |
| Obesity | mast_cell | *CRYBB2* | -2.807522029 | 8.27E-05 | 0.000152568 | 0.5199522 |
| Obesity | mast_cell | *N4BP2L2* | 3.62906309 | 0.000237502 | 0.000407415 | 1 |
| Obesity | mast_cell | *LINC00847* | 7.062451857 | 1.73E-09 | 4.28E-08 | 1.09E-05 |
| Obesity | mast_cell | *NEAT1* | 2.74749182 | 0.000837476 | 0.001319603 | 1 |
| Obesity | mast_cell | *ZNF585B* | 2.956597189 | 0.000130402 | 0.000232643 | 0.819835426 |
| Obesity | mast_cell | *RAB30-AS1* | 3.801988343 | 0.000700519 | 0.00111526 | 1 |
| Obesity | mast_cell | *LINC00920* | 4.251588718 | 5.64E-05 | 0.000107753 | 0.354292801 |
| Obesity | mast_cell | *UBAP1L* | 5.023887768 | 3.13E-05 | 6.27E-05 | 0.196877312 |
| Obesity | mast_cell | *PGAM5* | 3.085983981 | 1.70E-08 | 1.04E-07 | 0.000106615 |
| Obesity | mast_cell | *DNAJC3-AS1* | 6.766587996 | 2.10E-06 | 5.42E-06 | 0.013205711 |
| Obesity | mast_cell | *LINC00926* | 2.791538036 | 1.30E-08 | 9.01E-08 | 8.19E-05 |
| Obesity | mast_cell | *BCKDHA* | 5.773231644 | 1.65E-06 | 4.40E-06 | 0.010377942 |
| Obesity | mast_cell | *ABHD14A* | 8.294313054 | 2.90E-05 | 5.84E-05 | 0.182189922 |
| Obesity | mast_cell | *SRP14-AS1* | 1.326510427 | 6.08E-05 | 0.000115337 | 0.382110748 |
| Obesity | mast_cell | *C5orf17* | 8.782860303 | 4.28E-08 | 2.04E-07 | 0.000269065 |
| Obesity | mast_cell | *FMN1* | -1.533118055 | 4.75E-06 | 1.12E-05 | 0.029878653 |
| Obesity | mast_cell | *LINC01060* | 2.181817606 | 0.014889009 | 0.019677779 | 1 |
| Obesity | mast_cell | *NAIP* | 5.780673036 | 0.000635161 | 0.001019469 | 1 |
| Obesity | mast_cell | *LINC00942* | 4.971753733 | 0.003026323 | 0.004407341 | 1 |
| Obesity | mast_cell | *PDCD6* | 7.259225616 | 3.74E-08 | 1.83E-07 | 0.000235234 |
| Obesity | mast_cell | *LINC00603* | 3.515352558 | 2.06E-07 | 7.30E-07 | 0.001294326 |
| Obesity | mast_cell | *YJEFN3* | 3.405979325 | 1.96E-07 | 7.02E-07 | 0.001230639 |
| Obesity | mast_cell | *DNAH10OS* | 21.61366618 | 2.38E-12 | 4.68E-10 | 1.50E-08 |
| Obesity | mast_cell | *PCDHA10* | 7.391117762 | 5.40E-09 | 5.52E-08 | 3.39E-05 |
| Obesity | mast_cell | *ZNF718* | 6.893871743 | 6.22E-08 | 2.75E-07 | 0.000390825 |
| Obesity | mast_cell | *LINC00989* | 10.2595179 | 2.31E-07 | 8.06E-07 | 0.00145087 |
| Obesity | mast_cell | *LINC01021* | 18.28904976 | 0.005037495 | 0.007118617 | 1 |
| Obesity | mast_cell | *FAM218A* | -6.657058252 | 7.52E-06 | 1.71E-05 | 0.047282435 |
| Obesity | mast_cell | *GLI4* | 3.348200689 | 1.06E-06 | 2.97E-06 | 0.006642547 |
| Obesity | mast_cell | *SNHG18* | -4.826695593 | 2.22E-05 | 4.59E-05 | 0.139690337 |
| Obesity | mast_cell | *ZNF345* | 5.969035737 | 1.26E-05 | 2.74E-05 | 0.079076366 |
| Obesity | mast_cell | *ZNF550* | 6.968914815 | 5.42E-05 | 0.000104217 | 0.340895214 |
| Obesity | mast_cell | *CCDC71L* | 6.550838224 | 1.72E-07 | 6.30E-07 | 0.001080067 |
| Obesity | mast_cell | *TMEM200B* | 3.316794195 | 1.04E-07 | 4.19E-07 | 0.000651441 |
| Obesity | mast_cell | *TRNP1* | 24.23901332 | 3.81E-09 | 4.74E-08 | 2.40E-05 |
| Obesity | mast_cell | *PCDHGA5* | 2.42013864 | 0.036563522 | 0.045883207 | 1 |
| Obesity | mast_cell | *ALG11* | 2.366897244 | 0.000177832 | 0.000310823 | 1 |
| Obesity | mast_cell | *ATXN7L3B* | 8.801449852 | 1.18E-07 | 4.67E-07 | 0.000744145 |
| Obesity | mast_cell | *PRKDC* | 4.69592113 | 0.000662268 | 0.001059731 | 1 |
| Obesity | mast_cell | *PCDHGA6* | 4.097643741 | 1.59E-07 | 5.92E-07 | 0.001001094 |
| Obesity | mast_cell | *PCDHGA11* | 2.838715389 | 8.21E-05 | 0.000151491 | 0.515978761 |
| Obesity | mast_cell | *CLDN23* | -3.122717043 | 2.84E-05 | 5.72E-05 | 0.178242038 |

| | | | | | | |
|---|---|---|---|---|---|---|
| Obesity | mast_cell | *ZNF260* | 6.793743441 | 2.51E-09 | 4.32E-08 | 1.58E-05 |
| Obesity | mast_cell | *ARMS2* | -5.251963872 | 1.07E-07 | 4.31E-07 | 0.000674841 |
| Obesity | mast_cell | *FPGT* | 10.98030064 | 3.77E-07 | 1.22E-06 | 0.002368233 |
| Obesity | mast_cell | *DPP3* | 3.346733005 | 1.07E-09 | 3.57E-08 | 6.73E-06 |
| Obesity | mast_cell | *KRTAP5-9* | 7.663746212 | 0.035685983 | 0.044826729 | 1 |
| Obesity | mast_cell | *EID3* | 1.528999282 | 0.023887586 | 0.030768542 | 1 |
| Obesity | mast_cell | *EBLN2* | 8.602229963 | 5.56E-05 | 0.000106655 | 0.349829973 |
| Obesity | mast_cell | *POLR2M* | 10.23654987 | 1.19E-08 | 8.51E-08 | 7.47E-05 |
| Obesity | mast_cell | *LINC00346* | -2.95367644 | 1.54E-05 | 3.29E-05 | 0.096691504 |
| Obesity | mast_cell | *CTSO* | 6.373369603 | 0.00115612 | 0.001782375 | 1 |
| Obesity | mast_cell | *ZNF432* | 6.110407715 | 5.07E-05 | 9.80E-05 | 0.318765528 |
| Obesity | mast_cell | *LINC00944* | 3.206176204 | 4.66E-07 | 1.46E-06 | 0.002930791 |
| Obesity | mast_cell | *ZNF10* | 2.068642834 | 0.014522907 | 0.019226262 | 1 |
| Obesity | mast_cell | *HMBS* | 4.652990033 | 1.47E-18 | 1.16E-15 | 9.27E-15 |
| Obesity | mast_cell | *PGA5* | 17.07839767 | 0.005834457 | 0.008155009 | 1 |
| Obesity | mast_cell | *ZNF253* | 7.299354636 | 1.95E-07 | 7.01E-07 | 0.00122876 |
| Obesity | mast_cell | *KIAA1147* | 6.079754577 | 2.00E-05 | 4.16E-05 | 0.125628537 |
| Obesity | mast_cell | *GATC* | 4.631616787 | 1.43E-05 | 3.08E-05 | 0.090039616 |
| Obesity | mast_cell | *ZBED6* | -6.226666794 | 2.83E-06 | 7.08E-06 | 0.017808955 |
| Obesity | mast_cell | *OVCH1-AS1* | 6.371980017 | 3.11E-09 | 4.48E-08 | 1.95E-05 |
| Obesity | mast_cell | *CUX1* | 3.102026362 | 0.0055763 | 0.007809801 | 1 |
| Obesity | mast_cell | *CHURC1* | 3.195421879 | 2.47E-05 | 5.04E-05 | 0.155035514 |
| Obesity | mast_cell | *C17orf49* | 4.360275839 | 0.01146236 | 0.015414729 | 1 |
| Obesity | mast_cell | *LINC00640* | 1.113318724 | 0.035281643 | 0.044416438 | 1 |
| Obesity | mast_cell | *LINC00638* | 11.12604622 | 0.002466356 | 0.003634782 | 1 |
| Obesity | mast_cell | *CEP95* | 4.567735022 | 0.000923082 | 0.001446238 | 1 |
| Obesity | mast_cell | *LINC00639* | 3.425024789 | 4.35E-07 | 1.38E-06 | 0.002735564 |
| Obesity | mast_cell | *LINC00648* | 2.483798708 | 1.03E-08 | 7.88E-08 | 6.48E-05 |
| Obesity | mast_cell | *LINC00924* | 2.903473947 | 2.63E-08 | 1.40E-07 | 0.000165435 |
| Obesity | mast_cell | *ITGB3* | 1.351677279 | 0.011144398 | 0.015006389 | 1 |
| Obesity | mast_cell | *AC083843.1* | 21.65462607 | 0.007772044 | 0.010689748 | 1 |
| Obesity | mast_cell | *PWRN1* | 4.108934112 | 6.58E-08 | 2.86E-07 | 0.000413412 |
| Obesity | mast_cell | *HOXB7* | 9.954603527 | 8.03E-13 | 1.80E-10 | 5.05E-09 |
| Obesity | mast_cell | *FRRS1L* | 7.666076588 | 2.85E-09 | 4.32E-08 | 1.79E-05 |
| Obesity | mast_cell | *LINC00562* | 0.917562787 | 0.013373476 | 0.017809584 | 1 |
| Obesity | mast_cell | *AC009120.5* | 3.298732931 | 1.76E-08 | 1.05E-07 | 0.000110464 |
| Obesity | mast_cell | *LINC00565* | 12.92538014 | 0.002588096 | 0.003805276 | 1 |
| Obesity | mast_cell | *TMEM178B* | 4.345040371 | 0.029210848 | 0.037115724 | 1 |
| Obesity | mast_cell | *LINC01003* | 2.134132086 | 6.94E-08 | 2.99E-07 | 0.000436051 |
| Obesity | mast_cell | *HPR* | 6.5538276 | 9.71E-10 | 3.34E-08 | 6.11E-06 |
| Obesity | mast_cell | *GOLGA8H* | 2.622850642 | 0.000156817 | 0.000277251 | 0.985905813 |
| Obesity | mast_cell | *LINC00662* | 5.724264662 | 0.000213027 | 0.000367838 | 1 |
| Obesity | mast_cell | *LINC00667* | 2.129366832 | 0.000553957 | 0.000896917 | 1 |
| Obesity | mast_cell | *LINC00672* | 3.72873135 | 8.00E-07 | 2.33E-06 | 0.005030367 |
| Obesity | mast_cell | *LINC00909* | 4.462605687 | 5.34E-06 | 1.25E-05 | 0.03360276 |
| Obesity | mast_cell | *DYNLL2* | -5.106865517 | 0.001645006 | 0.002477163 | 1 |
| Obesity | mast_cell | *LINC00526* | 3.441724206 | 3.11E-07 | 1.04E-06 | 0.001955567 |
| Obesity | mast_cell | *LINC00668* | 34.97623441 | 0.036886198 | 0.046251202 | 1 |
| Obesity | mast_cell | *MYO15B* | 3.058024758 | 0.022879625 | 0.029548933 | 1 |
| Obesity | mast_cell | *LINC00663* | 0.709129483 | 0.007236977 | 0.009984393 | 1 |

| | | | | | | |
|---|---|---|---|---|---|---|
| Obesity | mast_cell | *ILF3-AS1* | 4.032754308 | 5.74E-06 | 1.33E-05 | 0.036070174 |
| Obesity | mast_cell | *AC139100.2* | 31.16436875 | 2.62E-09 | 4.32E-08 | 1.65E-05 |
| Obesity | mast_cell | *LINC01140* | -2.060405943 | 2.76E-05 | 5.58E-05 | 0.173353428 |
| Obesity | mast_cell | *LINC00906* | -28.64296364 | 1.14E-05 | 2.51E-05 | 0.071407373 |
| Obesity | mast_cell | *AC137932.1* | -10.02921006 | 1.30E-05 | 2.82E-05 | 0.081834383 |
| Obesity | mast_cell | *AC003005.2* | 4.162353912 | 4.92E-05 | 9.53E-05 | 0.309374022 |
| Obesity | mast_cell | *AL132989.1* | 5.634357725 | 1.68E-08 | 1.04E-07 | 0.000105616 |
| Obesity | mast_cell | *LINC00664* | 6.36861445 | 9.44E-05 | 0.00017247 | 0.593640674 |
| Obesity | mast_cell | *AC026740.1* | -7.611948332 | 0.000290412 | 0.000491208 | 1 |
| Obesity | mast_cell | *FBXO17* | 7.230077257 | 3.71E-07 | 1.20E-06 | 0.002329415 |
| Obesity | mast_cell | *AC009060.1* | -2.719013102 | 1.42E-08 | 9.47E-08 | 8.90E-05 |
| Obesity | mast_cell | *SNHG8* | 2.975579333 | 0.027766158 | 0.035430452 | 1 |
| Obesity | mast_cell | *YTHDF3-AS1* | 5.08424222 | 2.84E-09 | 4.32E-08 | 1.79E-05 |
| Obesity | mast_cell | *LINC01050* | 20.90954515 | 3.79E-08 | 1.85E-07 | 0.000238106 |
| Obesity | mast_cell | *MILR1* | -0.946252441 | 0.013739464 | 0.018250583 | 1 |
| Obesity | mast_cell | *GTF2H5* | 4.665930344 | 5.74E-12 | 9.50E-10 | 3.61E-08 |
| Obesity | mast_cell | *CASC15* | 4.778725091 | 1.93E-07 | 6.94E-07 | 0.001212475 |
| Obesity | mast_cell | *LINC01023* | 3.171015402 | 2.53E-08 | 1.36E-07 | 0.000158904 |
| Obesity | mast_cell | *AP000347.2* | 1.957112594 | 0.00021697 | 0.000374133 | 1 |
| Obesity | mast_cell | *POLR2J4* | 5.394606674 | 3.20E-09 | 4.49E-08 | 2.01E-05 |
| Obesity | mast_cell | *AC144652.1* | 1.946518123 | 6.24E-05 | 0.000118062 | 0.392083163 |
| Obesity | mast_cell | *AP000350.5* | 0.873134073 | 0.010051393 | 0.013625077 | 1 |
| Obesity | monocyte | *SEMA3F* | -1.569117601 | 8.77E-06 | 0.001125653 | 0.055156975 |
| Obesity | monocyte | *RBM6* | -3.375278879 | 3.29E-16 | 3.45E-13 | 2.07E-12 |
| Obesity | monocyte | *CDC27* | 3.864981433 | 0.000138125 | 0.009238229 | 0.868393535 |
| Obesity | monocyte | *TTC19* | 2.472437875 | 0.000649014 | 0.026953187 | 1 |
| Obesity | monocyte | *PSMC4* | 5.554783433 | 2.67E-05 | 0.002737082 | 0.167672434 |
| Obesity | monocyte | *NUDCD3* | 2.395185835 | 0.000708165 | 0.027653612 | 1 |
| Obesity | monocyte | *NDUFS1* | 3.663200378 | 0.001283666 | 0.038461027 | 1 |
| Obesity | monocyte | *RABEP1* | 4.779918248 | 6.44E-06 | 0.000880167 | 0.040487702 |
| Obesity | monocyte | *ZCCHC8* | 3.236203923 | 0.001444436 | 0.041007537 | 1 |
| Obesity | monocyte | *LSG1* | 1.999129158 | 0.000616712 | 0.026739789 | 1 |
| Obesity | monocyte | *JKAMP* | 2.238242202 | 0.001721425 | 0.045664964 | 1 |
| Obesity | monocyte | *ANAPC4* | 3.684188858 | 0.001100917 | 0.036816301 | 1 |
| Obesity | monocyte | *SDCCAG8* | 2.951672881 | 7.32E-05 | 0.005767034 | 0.460373887 |
| Obesity | monocyte | *ITIH4* | 2.807285954 | 4.12E-10 | 1.99E-07 | 2.59E-06 |
| Obesity | monocyte | *ATP11B* | 1.926452883 | 0.000853951 | 0.031033464 | 1 |
| Obesity | monocyte | *UHRF1BP1* | 6.168080544 | 1.68E-23 | 5.27E-20 | 1.05E-19 |
| Obesity | monocyte | *PVR* | 4.540223573 | 0.00175367 | 0.045938846 | 1 |
| Obesity | monocyte | *SDHA* | 4.032441094 | 0.000330371 | 0.017905531 | 1 |
| Obesity | monocyte | *SMARCE1* | 5.231102695 | 0.001166607 | 0.037806475 | 1 |
| Obesity | monocyte | *TIPIN* | 1.813881404 | 9.40E-06 | 0.001171369 | 0.059084028 |
| Obesity | monocyte | *SLC25A3* | 4.984785627 | 0.001841353 | 0.047445015 | 1 |
| Obesity | monocyte | *NT5C2* | 4.31827642 | 0.000630302 | 0.026953187 | 1 |
| Obesity | monocyte | *DYNC1I2* | 5.404261163 | 0.000114394 | 0.008172666 | 0.719194597 |
| Obesity | monocyte | *DOPEY1* | 2.938286334 | 0.000158621 | 0.01028091 | 0.997248233 |
| Obesity | monocyte | *SCAMP1* | 4.855005401 | 0.001161762 | 0.037806475 | 1 |
| Obesity | monocyte | *MAST2* | 4.051444231 | 0.001462259 | 0.041176982 | 1 |
| Obesity | monocyte | *C3orf18* | 12.15359636 | 1.73E-11 | 9.06E-09 | 1.09E-07 |
| Obesity | monocyte | *NSFL1C* | 4.210383511 | 0.001580769 | 0.043022929 | 1 |

| | | | | | | |
|---|---|---|---|---|---|---|
| Obesity | monocyte | *YPEL3* | 4.262973643 | 6.88E-10 | 3.09E-07 | 4.32E-06 |
| Obesity | monocyte | *PABPC4* | -4.686371287 | 7.11E-05 | 0.00573033 | 0.446965703 |
| Obesity | monocyte | *PPP2R3C* | 4.540706854 | 0.001470923 | 0.041176982 | 1 |
| Obesity | monocyte | *PDE6C* | 4.190227017 | 0.000635414 | 0.026953187 | 1 |
| Obesity | monocyte | *SMARCB1* | 4.200515354 | 3.93E-05 | 0.003533296 | 0.247330706 |
| Obesity | monocyte | *TFIP11* | 3.223569086 | 0.000656551 | 0.026953187 | 1 |
| Obesity | monocyte | *FBXO7* | 3.490666176 | 0.000806076 | 0.02998697 | 1 |
| Obesity | monocyte | *ACO2* | 3.082945939 | 0.000667116 | 0.026980352 | 1 |
| Obesity | monocyte | *DESI1* | 4.55155446 | 1.43E-06 | 0.000249041 | 0.008965491 |
| Obesity | monocyte | *DICER1* | 4.748697371 | 7.46E-05 | 0.005767034 | 0.468756762 |
| Obesity | monocyte | *ZFYVE21* | 3.059649577 | 0.001972697 | 0.049650616 | 1 |
| Obesity | monocyte | *ABHD12* | 4.829289493 | 8.13E-06 | 0.0010645 | 0.05109601 |
| Obesity | monocyte | *ELMO3* | 1.637183037 | 0.001278099 | 0.038461027 | 1 |
| Obesity | monocyte | *NAGPA* | 5.142070631 | 0.000265326 | 0.015303737 | 1 |
| Obesity | monocyte | *CLUAP1* | 2.673205064 | 6.83E-05 | 0.005728958 | 0.42967182 |
| Obesity | monocyte | *KAT8* | 10.84375072 | 1.31E-08 | 3.75E-06 | 8.25E-05 |
| Obesity | monocyte | *TRIP4* | 2.802348852 | 0.000842243 | 0.030965975 | 1 |
| Obesity | monocyte | *PPP1R37* | 6.49378313 | 0.000376338 | 0.020051164 | 1 |
| Obesity | monocyte | *OGN* | 2.317197597 | 0.001266255 | 0.038461027 | 1 |
| Obesity | monocyte | *MICU1* | 4.23253427 | 0.001776928 | 0.046163426 | 1 |
| Obesity | monocyte | *ACBD5* | 5.856458559 | 0.000120013 | 0.008412295 | 0.754519525 |
| Obesity | monocyte | *CBX1* | 4.287659528 | 0.000275637 | 0.01558828 | 1 |
| Obesity | monocyte | *HOXB6* | 3.641267387 | 0.000289187 | 0.01608955 | 1 |
| Obesity | monocyte | *CHRNE* | 2.031111202 | 0.00070048 | 0.027536006 | 1 |
| Obesity | monocyte | *C1QBP* | 9.493932275 | 0.000193067 | 0.012138106 | 1 |
| Obesity | monocyte | *EFTUD2* | 5.310913357 | 0.000774467 | 0.029156138 | 1 |
| Obesity | monocyte | *GTF2H1* | 3.161308828 | 0.001360101 | 0.039888699 | 1 |
| Obesity | monocyte | *DDX55* | 3.972026978 | 5.21E-06 | 0.000780024 | 0.03276102 |
| Obesity | monocyte | *BTN3A3* | 2.21458151 | 0.001679243 | 0.044925097 | 1 |
| Obesity | monocyte | *GABRR2* | 5.617847313 | 0.001394998 | 0.040047263 | 1 |
| Obesity | monocyte | *PRSS16* | 12.58391302 | 4.17E-09 | 1.46E-06 | 2.62E-05 |
| Obesity | monocyte | *PRKAR2A* | 3.129641494 | 0.00024849 | 0.014878638 | 1 |
| Obesity | monocyte | *KLHL24* | 2.027892262 | 0.00163081 | 0.044193553 | 1 |
| Obesity | monocyte | *ACTR1B* | 3.521581954 | 0.001404808 | 0.040145576 | 1 |
| Obesity | monocyte | *GGCX* | 3.392197346 | 0.000923348 | 0.032612851 | 1 |
| Obesity | monocyte | *WARS2* | 3.089375551 | 0.000922681 | 0.032612851 | 1 |
| Obesity | monocyte | *PRRC2C* | 3.314823236 | 0.001113598 | 0.037043348 | 1 |
| Obesity | monocyte | *PTBP2* | 2.505572308 | 0.000563451 | 0.025669695 | 1 |
| Obesity | monocyte | *SERPINC1* | 4.655768061 | 1.60E-09 | 6.71E-07 | 1.01E-05 |
| Obesity | monocyte | *RCAN3* | 5.512613579 | 0.001921189 | 0.048900869 | 1 |
| Obesity | monocyte | *VAMP8* | 4.342302416 | 0.000452862 | 0.023337245 | 1 |
| Obesity | monocyte | *CCDC92* | 2.111343688 | 7.02E-05 | 0.00572896 | 0.441129891 |
| Obesity | monocyte | *TRIM32* | 7.738688014 | 0.000866466 | 0.031307324 | 1 |
| Obesity | monocyte | *DCAF4* | 4.510990433 | 2.97E-05 | 0.002956625 | 0.186902361 |
| Obesity | monocyte | *HOXB3* | 1.602599122 | 0.001381534 | 0.040026294 | 1 |
| Obesity | monocyte | *ADRA1A* | 0.858000174 | 1.96E-05 | 0.002239593 | 0.123177611 |
| Obesity | monocyte | *TEX14* | 7.144445178 | 3.50E-05 | 0.003302815 | 0.220022971 |
| Obesity | monocyte | *PYROXD1* | 3.732300776 | 0.000589359 | 0.026278717 | 1 |
| Obesity | monocyte | *GJB6* | -46.36523844 | 0.000769931 | 0.029156138 | 1 |
| Obesity | monocyte | *ATP8A1* | 2.467019963 | 0.001746134 | 0.045932825 | 1 |

| | | | | | | |
|---|---|---|---|---|---|---|
| Obesity | monocyte | *SNRPC* | -8.461584423 | 5.24E-16 | 4.71E-13 | 3.30E-12 |
| Obesity | monocyte | *HIST1H2BJ* | 7.614668114 | 7.30E-09 | 2.29E-06 | 4.59E-05 |
| Obesity | monocyte | *GLO1* | 4.646916654 | 0.001219524 | 0.038409744 | 1 |
| Obesity | monocyte | *NUP153* | 3.461353666 | 0.001366787 | 0.039888699 | 1 |
| Obesity | monocyte | *BBS2* | 2.132110337 | 0.001099476 | 0.036816301 | 1 |
| Obesity | monocyte | *GRK4* | 1.189589371 | 3.51E-05 | 0.003302815 | 0.220599967 |
| Obesity | monocyte | *UROD* | 6.554944795 | 0.000256105 | 0.015189916 | 1 |
| Obesity | monocyte | *KLC1* | 2.904894114 | 0.001284442 | 0.038461027 | 1 |
| Obesity | monocyte | *XRCC3* | 3.594873388 | 0.00074299 | 0.0284828 | 1 |
| Obesity | monocyte | *TRAP1* | 1.468995798 | 0.000461236 | 0.023454078 | 1 |
| Obesity | monocyte | *RHOT1* | 3.024073934 | 0.000971297 | 0.03411476 | 1 |
| Obesity | monocyte | *CANX* | 4.868970642 | 0.001647481 | 0.044453716 | 1 |
| Obesity | monocyte | *IPPK* | 4.992314694 | 0.001898439 | 0.048518227 | 1 |
| Obesity | monocyte | *YEATS4* | 3.839123573 | 7.52E-05 | 0.005767034 | 0.472896815 |
| Obesity | monocyte | *EIF2AK4* | 2.82159279 | 0.001061332 | 0.036264099 | 1 |
| Obesity | monocyte | *ATP8B3* | -2.850913788 | 1.23E-05 | 0.00146109 | 0.07743775 |
| Obesity | monocyte | *PGPEP1* | 4.407092413 | 0.000513502 | 0.024101319 | 1 |
| Obesity | monocyte | *CLIP1* | 4.711473277 | 8.28E-05 | 0.006194044 | 0.520299696 |
| Obesity | monocyte | *EXOC4* | 2.528348765 | 0.000905889 | 0.032359795 | 1 |
| Obesity | monocyte | *RPA1* | 4.547915965 | 0.001166001 | 0.037806475 | 1 |
| Obesity | monocyte | *IGHMBP2* | 3.983536121 | 9.29E-07 | 0.000182505 | 0.005840152 |
| Obesity | monocyte | *MUTYH* | 6.434557747 | 0.001021531 | 0.035617998 | 1 |
| Obesity | monocyte | *DDB2* | 5.093430444 | 0.000568593 | 0.025717567 | 1 |
| Obesity | monocyte | *ACP2* | 7.571761046 | 2.61E-06 | 0.000409711 | 0.016388429 |
| Obesity | monocyte | *NTPCR* | 4.415075218 | 0.001142047 | 0.037591894 | 1 |
| Obesity | monocyte | *USP37* | 4.60260947 | 0.000202056 | 0.012577505 | 1 |
| Obesity | monocyte | *MRPS9* | 4.141488375 | 6.73E-05 | 0.005719678 | 0.423256172 |
| Obesity | monocyte | *NMT1* | 2.340453989 | 0.000337439 | 0.018132319 | 1 |
| Obesity | monocyte | *DSCC1* | 0.956857179 | 0.000880957 | 0.031649018 | 1 |
| Obesity | monocyte | *CD72* | 5.839942882 | 0.000452491 | 0.023337245 | 1 |
| Obesity | monocyte | *ZSCAN9* | 5.479975506 | 3.53E-07 | 8.54E-05 | 0.0022205 |
| Obesity | monocyte | *TCF19* | 5.541641223 | 9.69E-06 | 0.001171369 | 0.06091121 |
| Obesity | monocyte | *FLOT1* | 3.176111277 | 0.001390384 | 0.040047263 | 1 |
| Obesity | monocyte | *PRCP* | 4.386130183 | 0.001221878 | 0.038409744 | 1 |
| Obesity | monocyte | *MAP2K5* | 3.374285709 | 0.000537626 | 0.024671951 | 1 |
| Obesity | monocyte | *RTCA* | 3.203935943 | 0.001524061 | 0.041841793 | 1 |
| Obesity | monocyte | *CARF* | 4.425428682 | 8.03E-07 | 0.000166078 | 0.005046973 |
| Obesity | monocyte | *KLRG1* | 1.715108752 | 0.001477467 | 0.041176982 | 1 |
| Obesity | monocyte | *SCAMP2* | 4.460041336 | 0.001264245 | 0.038461027 | 1 |
| Obesity | monocyte | *MYO19* | 4.988061053 | 1.46E-05 | 0.001702121 | 0.091914556 |
| Obesity | monocyte | *C18orf21* | 6.798812504 | 0.001486746 | 0.041176982 | 1 |
| Obesity | monocyte | *SIRT3* | 4.436038209 | 0.001556254 | 0.042539861 | 1 |
| Obesity | monocyte | *RPS11* | 3.879885636 | 0.000687034 | 0.027337871 | 1 |
| Obesity | monocyte | *POGZ* | 3.088262054 | 0.001044557 | 0.035885968 | 1 |
| Obesity | monocyte | *HAX1* | 6.965402477 | 2.18E-05 | 0.002401636 | 0.136893272 |
| Obesity | monocyte | *CEP170* | 3.114633254 | 0.000784048 | 0.029341138 | 1 |
| Obesity | monocyte | *GUK1* | 5.301701708 | 0.000851707 | 0.031033464 | 1 |
| Obesity | monocyte | *NBEAL1* | 2.159711467 | 3.69E-05 | 0.003366738 | 0.232185079 |
| Obesity | monocyte | *ABT1* | 2.996432315 | 0.000639533 | 0.026953187 | 1 |
| Obesity | monocyte | *RPL7L1* | 3.119239592 | 0.001133733 | 0.037514634 | 1 |

| | | | | | | |
|---|---|---|---|---|---|---|
| Obesity | monocyte | *DPH7* | 6.402550131 | 0.001179635 | 0.03797484 | 1 |
| Obesity | monocyte | *MTG1* | 3.455647825 | 3.01E-05 | 0.002956625 | 0.189224009 |
| Obesity | monocyte | *HSD17B12* | -2.763049233 | 1.78E-06 | 0.000295174 | 0.011212287 |
| Obesity | monocyte | *TTC12* | 2.450764759 | 6.96E-06 | 0.000931027 | 0.043758251 |
| Obesity | monocyte | *TBX6* | 2.986435563 | 7.35E-07 | 0.000164159 | 0.004623133 |
| Obesity | monocyte | *PRSS53* | -5.120531395 | 0.000657947 | 0.026953187 | 1 |
| Obesity | monocyte | *DPYSL4* | 1.891826569 | 0.000277698 | 0.01558828 | 1 |
| Obesity | monocyte | *RABGAP1L* | 3.645443873 | 0.00049456 | 0.024101319 | 1 |
| Obesity | monocyte | *JAZF1* | 4.602908894 | 9.34E-05 | 0.006831498 | 0.58750885 |
| Obesity | monocyte | *FLCN* | 3.186979557 | 0.00197486 | 0.049650616 | 1 |
| Obesity | monocyte | *CLEC18A* | -9.300081443 | 3.37E-14 | 2.35E-11 | 2.12E-10 |
| Obesity | monocyte | *HPD* | 1.934584055 | 0.000447474 | 0.023337245 | 1 |
| Obesity | monocyte | *SLC5A11* | -1.390289723 | 0.000478697 | 0.023761694 | 1 |
| Obesity | monocyte | *DFFA* | 5.62477903 | 9.22E-05 | 0.006817709 | 0.57950523 |
| Obesity | monocyte | *C19orf47* | 9.229759012 | 2.70E-05 | 0.002737082 | 0.169695206 |
| Obesity | monocyte | *TPCN2* | 3.817941768 | 0.000574535 | 0.02580073 | 1 |
| Obesity | monocyte | *RBBP4* | 4.768174549 | 0.001982234 | 0.049650616 | 1 |
| Obesity | monocyte | *CLHC1* | 1.210214222 | 0.000521358 | 0.024101319 | 1 |
| Obesity | monocyte | *LYSMD1* | 8.978877602 | 5.79E-05 | 0.005056256 | 0.36405045 |
| Obesity | monocyte | *DHX57* | 4.370871503 | 0.001877066 | 0.048167815 | 1 |
| Obesity | monocyte | *ANTXR2* | 2.687761434 | 0.000248458 | 0.014878638 | 1 |
| Obesity | monocyte | *GYG1* | 8.267788988 | 2.52E-05 | 0.002689571 | 0.158684668 |
| Obesity | monocyte | *DNAJB14* | 3.844602099 | 0.001729966 | 0.045698731 | 1 |
| Obesity | monocyte | *SHISA5* | 6.463000044 | 0.001236427 | 0.038461027 | 1 |
| Obesity | monocyte | *APEH* | -3.024716222 | 0.000518519 | 0.024101319 | 1 |
| Obesity | monocyte | *RNF123* | 5.667042997 | 1.77E-17 | 2.22E-14 | 1.11E-13 |
| Obesity | monocyte | *MST1R* | 5.021583529 | 8.19E-07 | 0.000166078 | 0.005148421 |
| Obesity | monocyte | *LMTK2* | 3.982520146 | 0.001933364 | 0.049012333 | 1 |
| Obesity | monocyte | *PSMC3* | 5.145980958 | 6.08E-05 | 0.005238812 | 0.382433277 |
| Obesity | monocyte | *CATSPER2* | 4.401199906 | 0.000510041 | 0.024101319 | 1 |
| Obesity | monocyte | *MTMR10* | 3.240189678 | 0.001187141 | 0.03797484 | 1 |
| Obesity | monocyte | *SP2* | 6.575347702 | 0.000648559 | 0.026953187 | 1 |
| Obesity | monocyte | *SRR* | 2.523994815 | 1.17E-06 | 0.000221981 | 0.007325361 |
| Obesity | monocyte | *TRANK1* | 4.306297073 | 0.001353785 | 0.039888699 | 1 |
| Obesity | monocyte | *GLYCTK* | 6.21109482 | 0.000669466 | 0.026980352 | 1 |
| Obesity | monocyte | *UBTD2* | 4.099751678 | 0.001798882 | 0.046541436 | 1 |
| Obesity | monocyte | *RFWD3* | 3.237166055 | 0.001025427 | 0.035617998 | 1 |
| Obesity | monocyte | *FNTA* | 5.503044585 | 0.001702879 | 0.045364421 | 1 |
| Obesity | monocyte | *C8orf46* | 0.75121369 | 0.000134613 | 0.009155843 | 0.846310874 |
| Obesity | monocyte | *METTL15* | 2.350323433 | 0.001249921 | 0.038461027 | 1 |
| Obesity | monocyte | *ADORA2B* | -1.816580149 | 0.000120424 | 0.008412295 | 0.757106561 |
| Obesity | monocyte | *TSNARE1* | 3.164036721 | 0.000293521 | 0.016187446 | 1 |
| Obesity | monocyte | *TRMT61B* | 2.571622014 | 2.32E-06 | 0.000374046 | 0.01458779 |
| Obesity | monocyte | *BCL2L1* | 3.297716017 | 0.001508269 | 0.041589862 | 1 |
| Obesity | monocyte | *C1QTNF4* | -2.495662357 | 2.02E-15 | 1.59E-12 | 1.27E-11 |
| Obesity | monocyte | *FIBP* | 8.683179761 | 2.70E-05 | 0.002737082 | 0.169699091 |
| Obesity | monocyte | *CTSW* | -1.009699919 | 2.06E-05 | 0.002314494 | 0.12961168 |
| Obesity | monocyte | *SNX32* | 2.233742024 | 0.000263877 | 0.015303737 | 1 |
| Obesity | monocyte | *NADSYN1* | 5.028157501 | 0.001189923 | 0.03797484 | 1 |
| Obesity | monocyte | *GMPPB* | 2.513926072 | 0.000508009 | 0.024101319 | 1 |

| | | | | | | |
|---|---|---|---|---|---|---|
| Obesity | monocyte | *HOXB2* | 2.457557513 | 0.001770159 | 0.046163426 | 1 |
| Obesity | monocyte | *BBS1* | 4.239703434 | 0.001231856 | 0.038461027 | 1 |
| Obesity | monocyte | *SRP72* | 3.841349544 | 0.000170742 | 0.010842972 | 1 |
| Obesity | monocyte | *TMEM9B* | 6.373958349 | 6.99E-05 | 0.00572896 | 0.439466406 |
| Obesity | monocyte | *IP6K1* | 5.743963329 | 0.000107528 | 0.00777042 | 0.676026538 |
| Obesity | monocyte | *RUFY1* | 4.453648919 | 9.54E-06 | 0.001171369 | 0.059958864 |
| Obesity | monocyte | *SH2B1* | -2.319727307 | 7.57E-07 | 0.000164159 | 0.004760608 |
| Obesity | monocyte | *TRIM73* | -9.897705603 | 4.30E-08 | 1.18E-05 | 0.00027051 |
| Obesity | monocyte | *TUFM* | 10.6414488 | 8.61E-23 | 1.81E-19 | 5.42E-19 |
| Obesity | monocyte | *CTC1* | 4.522838949 | 0.000485795 | 0.023860873 | 1 |
| Obesity | monocyte | *CALR* | 5.040596351 | 0.000656766 | 0.026953187 | 1 |
| Obesity | monocyte | *FKRP* | 1.767977372 | 0.001279854 | 0.038461027 | 1 |
| Obesity | monocyte | *POLR2A* | 4.459300157 | 0.001294519 | 0.038571766 | 1 |
| Obesity | monocyte | *LYSMD4* | 2.923556635 | 0.000134698 | 0.009155843 | 0.846849371 |
| Obesity | monocyte | *PSMG1* | 2.290283079 | 2.40E-05 | 0.00259881 | 0.150730977 |
| Obesity | monocyte | *TANGO2* | 4.847543365 | 0.000825567 | 0.030531403 | 1 |
| Obesity | monocyte | *RP1L1* | 5.26438719 | 1.37E-06 | 0.000246413 | 0.008624445 |
| Obesity | monocyte | *BMP8A* | 11.23146773 | 6.22E-06 | 0.000868362 | 0.039076309 |
| Obesity | monocyte | *EIF3C* | 7.634461419 | 5.19E-24 | 3.26E-20 | 3.26E-20 |
| Obesity | monocyte | *PLA2G6* | 1.638578821 | 0.000150232 | 0.009838663 | 0.944511647 |
| Obesity | monocyte | *SNN* | 9.563511358 | 0.001673437 | 0.044925097 | 1 |
| Obesity | monocyte | *GP1BA* | 2.486430941 | 0.00046259 | 0.023454078 | 1 |
| Obesity | monocyte | *SP1* | 3.383520349 | 0.000728389 | 0.02809435 | 1 |
| Obesity | monocyte | *BTN3A2* | -1.871009851 | 3.16E-09 | 1.17E-06 | 1.99E-05 |
| Obesity | monocyte | *HYAL3* | -2.964265888 | 8.03E-05 | 0.006084523 | 0.505015436 |
| Obesity | monocyte | *TRABD2A* | 2.181785824 | 0.000479996 | 0.023761694 | 1 |
| Obesity | monocyte | *MRPS21* | 3.651736663 | 0.001218456 | 0.038409744 | 1 |
| Obesity | monocyte | *EPOR* | 5.124257231 | 0.00137044 | 0.039888699 | 1 |
| Obesity | monocyte | *ZSCAN23* | 1.113560024 | 0.000507873 | 0.024101319 | 1 |
| Obesity | monocyte | *CENPP* | 2.959420681 | 1.20E-07 | 3.14E-05 | 0.000753499 |
| Obesity | monocyte | *FBF1* | 2.261779814 | 3.52E-05 | 0.003302815 | 0.221288591 |
| Obesity | monocyte | *FAM47E* | 1.789784511 | 0.000430256 | 0.022731273 | 1 |
| Obesity | monocyte | *SLC35E2B* | 2.344554783 | 0.001328822 | 0.039407081 | 1 |
| Obesity | monocyte | *LONP1* | 3.811937388 | 0.000660218 | 0.026953187 | 1 |
| Obesity | monocyte | *C6orf106* | 4.214496438 | 2.80E-09 | 1.10E-06 | 1.76E-05 |
| Obesity | monocyte | *FAM3C* | 4.012748841 | 0.001087411 | 0.036816301 | 1 |
| Obesity | monocyte | *SLC25A29* | 3.493837954 | 0.000765606 | 0.029156138 | 1 |
| Obesity | monocyte | *SULT1A2* | 2.326254962 | 6.45E-19 | 1.01E-15 | 4.06E-15 |
| Obesity | monocyte | *GTF2E2* | 3.922622073 | 0.000595194 | 0.026351994 | 1 |
| Obesity | monocyte | *MRPL21* | -1.712880558 | 0.001486198 | 0.041176982 | 1 |
| Obesity | monocyte | *RPS26* | -1.704630912 | 0.000274812 | 0.01558828 | 1 |
| Obesity | monocyte | *ZNF780A* | -2.096812496 | 0.000135437 | 0.009155843 | 0.851493418 |
| Obesity | monocyte | *QRICH1* | 3.295824581 | 0.000324277 | 0.017728102 | 1 |
| Obesity | monocyte | *ZKSCAN8* | 7.490596064 | 1.78E-06 | 0.000295174 | 0.011216619 |
| Obesity | monocyte | *NOTCH4* | -0.641754607 | 1.41E-07 | 3.56E-05 | 0.000888843 |
| Obesity | monocyte | *SKIV2L* | -1.463535732 | 7.41E-05 | 0.005767034 | 0.465915273 |
| Obesity | monocyte | *NELFE* | 4.256778162 | 1.29E-06 | 0.000238667 | 0.008114679 |
| Obesity | monocyte | *HLA-C* | 2.467363551 | 0.00060099 | 0.026411709 | 1 |
| Obesity | monocyte | *POU5F1* | 1.593449698 | 6.66E-07 | 0.00015519 | 0.004190118 |
| Obesity | monocyte | *CCHCR1* | 2.935069209 | 8.41E-09 | 2.52E-06 | 5.29E-05 |

| | | | | | | |
|---|---|---|---|---|---|---|
| Obesity | monocyte | *ABCF1* | 2.494303358 | 0.000517666 | 0.024101319 | 1 |
| Obesity | monocyte | *HLA-E* | 5.610422107 | 1.61E-11 | 9.06E-09 | 1.01E-07 |
| Obesity | monocyte | *ZFP57* | 2.455645346 | 7.16E-09 | 2.29E-06 | 4.50E-05 |
| Obesity | monocyte | *EIF3CL* | 5.70732112 | 5.59E-06 | 0.000816655 | 0.035116182 |
| Obesity | monocyte | *C17orf107* | 3.379913534 | 0.000220819 | 0.013478539 | 1 |
| Obesity | monocyte | *ASIC3* | 3.391431615 | 0.000479896 | 0.023761694 | 1 |
| Obesity | monocyte | *SLC35F6* | 7.733411143 | 0.000604944 | 0.026411709 | 1 |
| Obesity | monocyte | *OSER1-AS1* | 7.728083789 | 0.000261592 | 0.015303737 | 1 |
| Obesity | monocyte | *LINC00240* | 1.011224569 | 0.001094153 | 0.036816301 | 1 |
| Obesity | monocyte | *LINC00863* | 4.341516047 | 0.001034397 | 0.035732155 | 1 |
| Obesity | monocyte | *PLEKHM1* | 6.002065668 | 0.000205337 | 0.01265639 | 1 |
| Obesity | monocyte | *RPS18* | 4.244395836 | 5.03E-05 | 0.00445217 | 0.31610404 |
| Obesity | monocyte | *HLA-B* | 2.688651301 | 0.000700773 | 0.027536006 | 1 |
| Obesity | monocyte | *ZSCAN31* | 1.889282473 | 0.000140536 | 0.00930051 | 0.883548463 |
| Obesity | monocyte | *PSMB9* | 4.518393274 | 5.84E-06 | 0.000834501 | 0.036718036 |
| Obesity | monocyte | *AP5Z1* | 5.36977236 | 0.001284685 | 0.038461027 | 1 |
| Obesity | monocyte | *PCDHAC1* | -8.441179973 | 0.000719182 | 0.027910479 | 1 |
| Obesity | monocyte | *MPV17L2* | 4.517209885 | 3.70E-05 | 0.003366738 | 0.232304937 |
| Obesity | monocyte | *DPP3* | 2.988000154 | 0.00016079 | 0.010315163 | 1 |
| Obesity | monocyte | *HMBS* | 5.440124138 | 3.57E-12 | 2.24E-09 | 2.24E-08 |
| Obesity | monocyte | *SH3RF3-AS1* | -3.414992143 | 0.001448015 | 0.041007537 | 1 |
| Obesity | monocyte | *HOXB7* | 9.680584952 | 4.60E-06 | 0.000705805 | 0.028938009 |
| Obesity | monocyte | *GTF2H5* | 2.742741824 | 0.000675582 | 0.027053417 | 1 |
| Obesity | neutrophil | *SCYL3* | 2.649855703 | 0.003821897 | 0.011289644 | 1 |
| Obesity | neutrophil | *C1orf112* | 1.722273016 | 0.004052305 | 0.011289644 | 1 |
| Obesity | neutrophil | *CFH* | 0.864383517 | 0.003960567 | 0.011289644 | 1 |
| Obesity | neutrophil | *NFYA* | 1.329563775 | 0.004055099 | 0.011289644 | 1 |
| Obesity | neutrophil | *STPG1* | 2.135631573 | 0.002476482 | 0.011289644 | 1 |
| Obesity | neutrophil | *NIPAL3* | 1.762419348 | 0.002910659 | 0.011289644 | 1 |
| Obesity | neutrophil | *ENPP4* | 1.739192987 | 0.003610155 | 0.011289644 | 1 |
| Obesity | neutrophil | *SEMA3F* | -3.609879932 | 3.43E-10 | 3.08E-07 | 2.16E-06 |
| Obesity | neutrophil | *KRIT1* | 2.016320123 | 0.005331206 | 0.011289644 | 1 |
| Obesity | neutrophil | *RAD52* | 2.125781922 | 0.004913596 | 0.011289644 | 1 |
| Obesity | neutrophil | *BAD* | 2.768677971 | 0.004286753 | 0.011289644 | 1 |
| Obesity | neutrophil | *HS3ST1* | 3.864804382 | 0.002487328 | 0.011289644 | 1 |
| Obesity | neutrophil | *WNT16* | 8.176281323 | 4.25E-05 | 0.00636747 | 0.267433745 |
| Obesity | neutrophil | *HECW1* | 1.191451319 | 0.024705843 | 0.031372578 | 1 |
| Obesity | neutrophil | *MAD1L1* | 1.902727457 | 0.006123497 | 0.011482393 | 1 |
| Obesity | neutrophil | *LASP1* | 3.503223648 | 0.024919477 | 0.031624698 | 1 |
| Obesity | neutrophil | *TMEM176A* | 2.175548914 | 0.002715567 | 0.011289644 | 1 |
| Obesity | neutrophil | *CYP26B1* | 1.881071027 | 0.003302703 | 0.011289644 | 1 |
| Obesity | neutrophil | *ICA1* | 3.200738736 | 0.00311403 | 0.011289644 | 1 |
| Obesity | neutrophil | *DBNDD1* | -1.466625756 | 0.013756187 | 0.018978526 | 1 |
| Obesity | neutrophil | *POLDIP2* | -1.875810544 | 0.011088161 | 0.015992491 | 1 |
| Obesity | neutrophil | *RBM6* | -1.355876907 | 0.011172942 | 0.016077886 | 1 |
| Obesity | neutrophil | *PDK4* | 1.428368135 | 0.00442157 | 0.011289644 | 1 |
| Obesity | neutrophil | *CDC27* | 1.762161027 | 0.001380972 | 0.011289644 | 1 |
| Obesity | neutrophil | *MEOX1* | 1.439457455 | 0.01690787 | 0.022516057 | 1 |
| Obesity | neutrophil | *LIG3* | 2.300995162 | 0.005543406 | 0.011295339 | 1 |
| Obesity | neutrophil | *CIAPIN1* | 1.888943217 | 0.00447314 | 0.011289644 | 1 |

| | | | | | | |
|---|---|---|---|---|---|---|
| Obesity | neutrophil | *COPZ2* | 1.694410175 | 0.000676893 | 0.011289644 | 1 |
| Obesity | neutrophil | *GCFC2* | 1.075610465 | 0.004124404 | 0.011289644 | 1 |
| Obesity | neutrophil | *ABCB4* | 2.038306276 | 0.003805133 | 0.011289644 | 1 |
| Obesity | neutrophil | *ZNF195* | 1.231135655 | 0.003973274 | 0.011289644 | 1 |
| Obesity | neutrophil | *PDK2* | 3.244134361 | 0.007100315 | 0.012084375 | 1 |
| Obesity | neutrophil | *TMEM98* | 2.163570126 | 0.000696728 | 0.011289644 | 1 |
| Obesity | neutrophil | *ABCC8* | 4.434414167 | 0.004065792 | 0.011289644 | 1 |
| Obesity | neutrophil | *CCL18* | 2.14519464 | 0.007183226 | 0.012155908 | 1 |
| Obesity | neutrophil | *SYNRG* | 1.997378522 | 0.004926837 | 0.011289644 | 1 |
| Obesity | neutrophil | *TMEM132A* | 11.13586196 | 0.002547394 | 0.011289644 | 1 |
| Obesity | neutrophil | *ZNF263* | 2.767383368 | 0.005229988 | 0.011289644 | 1 |
| Obesity | neutrophil | *SPATA20* | 2.301522712 | 0.002992814 | 0.011289644 | 1 |
| Obesity | neutrophil | *TNFRSF12A* | 2.584422271 | 0.011073993 | 0.015980311 | 1 |
| Obesity | neutrophil | *MAP3K9* | 3.295214115 | 0.003260222 | 0.011289644 | 1 |
| Obesity | neutrophil | *RALA* | 3.296107178 | 0.016621027 | 0.022223819 | 1 |
| Obesity | neutrophil | *BAIAP2L1* | 2.783272919 | 0.028106656 | 0.035263729 | 1 |
| Obesity | neutrophil | *FARP2* | 1.763837686 | 0.004895157 | 0.011289644 | 1 |
| Obesity | neutrophil | *IFRD1* | 1.270183758 | 0.006625582 | 0.011737119 | 1 |
| Obesity | neutrophil | *GTF2IRD1* | 3.814838493 | 0.002079451 | 0.011289644 | 1 |
| Obesity | neutrophil | *VPS41* | 1.36648114 | 0.004978037 | 0.011289644 | 1 |
| Obesity | neutrophil | *ARHGAP44* | 1.383920885 | 0.003585329 | 0.011289644 | 1 |
| Obesity | neutrophil | *ELAC2* | 2.56823078 | 0.022085117 | 0.028394505 | 1 |
| Obesity | neutrophil | *CCDC124* | 1.863831779 | 0.003854882 | 0.011289644 | 1 |
| Obesity | neutrophil | *CEACAM21* | 4.449657166 | 0.015846984 | 0.021343186 | 1 |
| Obesity | neutrophil | *NOS2* | 12.57575839 | 0.003410249 | 0.011289644 | 1 |
| Obesity | neutrophil | *TRAPPC6A* | 1.637649602 | 0.004298966 | 0.011289644 | 1 |
| Obesity | neutrophil | *ST7L* | 1.483747972 | 0.014817668 | 0.020151132 | 1 |
| Obesity | neutrophil | *PAX6* | -1.578827181 | 0.022262057 | 0.028592758 | 1 |
| Obesity | neutrophil | *RPUSD1* | 3.596123279 | 0.004084634 | 0.011289644 | 1 |
| Obesity | neutrophil | *BAIAP3* | 7.551745425 | 0.01341844 | 0.018575147 | 1 |
| Obesity | neutrophil | *TSR3* | 5.568341551 | 0.007285787 | 0.012230772 | 1 |
| Obesity | neutrophil | *PIGQ* | 2.90626178 | 0.003772876 | 0.011289644 | 1 |
| Obesity | neutrophil | *PSMB1* | 2.733386491 | 0.008776504 | 0.013623945 | 1 |
| Obesity | neutrophil | *CDK11A* | -1.440084868 | 0.006469135 | 0.01166174 | 1 |
| Obesity | neutrophil | *DLEC1* | 2.084168542 | 0.004426094 | 0.011289644 | 1 |
| Obesity | neutrophil | *MPND* | 2.460396487 | 0.002345451 | 0.011289644 | 1 |
| Obesity | neutrophil | *MGST1* | 1.063455027 | 0.004923641 | 0.011289644 | 1 |
| Obesity | neutrophil | *NFIX* | 1.183148326 | 0.008086976 | 0.012907544 | 1 |
| Obesity | neutrophil | *MMP25* | -0.392087724 | 0.006501987 | 0.011671674 | 1 |
| Obesity | neutrophil | *PKD1* | 2.619946501 | 0.001147371 | 0.011289644 | 1 |
| Obesity | neutrophil | *MAPK8IP2* | 26.61037568 | 0.000278905 | 0.011289644 | 1 |
| Obesity | neutrophil | *MED24* | 1.517831254 | 0.003259567 | 0.011289644 | 1 |
| Obesity | neutrophil | *SEC62* | 2.014908508 | 0.005175931 | 0.011289644 | 1 |
| Obesity | neutrophil | *RPS20* | 2.040170062 | 0.004919249 | 0.011289644 | 1 |
| Obesity | neutrophil | *POMT2* | 1.386174293 | 0.007636258 | 0.012499129 | 1 |
| Obesity | neutrophil | *MLXIPL* | 1.049306239 | 0.00054585 | 0.011289644 | 1 |
| Obesity | neutrophil | *METTL13* | 2.374619629 | 0.005666514 | 0.011300433 | 1 |
| Obesity | neutrophil | *DYRK4* | 2.473244018 | 0.003895293 | 0.011289644 | 1 |
| Obesity | neutrophil | *STARD3NL* | 3.993062512 | 0.003932798 | 0.011289644 | 1 |
| Obesity | neutrophil | *NCAPD2* | 1.464821912 | 0.003968447 | 0.011289644 | 1 |

| | | | | | | |
|---|---|---|---|---|---|---|
| Obesity | neutrophil | *ZNF200* | -1.816359828 | 0.011863627 | 0.016851926 | 1 |
| Obesity | neutrophil | *CD4* | 3.06450985 | 0.004955621 | 0.011289644 | 1 |
| Obesity | neutrophil | *LRRC23* | 1.506950564 | 0.002352745 | 0.011289644 | 1 |
| Obesity | neutrophil | *SCMH1* | 1.88747323 | 0.003319038 | 0.011289644 | 1 |
| Obesity | neutrophil | *CLCN6* | 1.850929909 | 0.002678652 | 0.011289644 | 1 |
| Obesity | neutrophil | *MRC2* | 1.043741454 | 0.006041918 | 0.011447874 | 1 |
| Obesity | neutrophil | *TSPAN9* | 2.702342374 | 0.002180934 | 0.011289644 | 1 |
| Obesity | neutrophil | *MKS1* | 1.94448868 | 0.003547696 | 0.011289644 | 1 |
| Obesity | neutrophil | *UTP18* | 1.307232448 | 0.004252673 | 0.011289644 | 1 |
| Obesity | neutrophil | *TTC19* | 1.827516668 | 0.001270551 | 0.011289644 | 1 |
| Obesity | neutrophil | *DPF1* | 5.783258486 | 0.032970846 | 0.040716501 | 1 |
| Obesity | neutrophil | *SYT7* | -2.431286278 | 0.01254983 | 0.0176354 | 1 |
| Obesity | neutrophil | *LARS2* | 3.242108434 | 0.004376145 | 0.011289644 | 1 |
| Obesity | neutrophil | *PPP5C* | 2.211956977 | 0.003271152 | 0.011289644 | 1 |
| Obesity | neutrophil | *CEP68* | 2.416227426 | 0.005852411 | 0.011378838 | 1 |
| Obesity | neutrophil | *ZBTB32* | -2.535834232 | 0.009356054 | 0.014181124 | 1 |
| Obesity | neutrophil | *TMEM159* | 1.473635489 | 0.00464615 | 0.011289644 | 1 |
| Obesity | neutrophil | *ERCC1* | 1.585653048 | 0.003777459 | 0.011289644 | 1 |
| Obesity | neutrophil | *LTF* | -1.065968372 | 0.007711762 | 0.012586669 | 1 |
| Obesity | neutrophil | *ALOX5* | -2.320584927 | 0.01019983 | 0.015049596 | 1 |
| Obesity | neutrophil | *MAP4K5* | 1.192012924 | 0.004283634 | 0.011289644 | 1 |
| Obesity | neutrophil | *MAN2B2* | 2.142427819 | 0.003993657 | 0.011289644 | 1 |
| Obesity | neutrophil | *CLDN11* | 1.184718586 | 0.002907828 | 0.011289644 | 1 |
| Obesity | neutrophil | *SLC25A39* | 4.801225387 | 0.033347292 | 0.041165212 | 1 |
| Obesity | neutrophil | *ANGEL1* | 3.500694078 | 0.005372071 | 0.011289644 | 1 |
| Obesity | neutrophil | *RNF14* | 1.433388705 | 0.003729569 | 0.011289644 | 1 |
| Obesity | neutrophil | *HEBP1* | 3.782090747 | 0.006913604 | 0.011943085 | 1 |
| Obesity | neutrophil | *TACC3* | -2.957165669 | 0.017178148 | 0.022823122 | 1 |
| Obesity | neutrophil | *ZC3H3* | 2.192745997 | 0.004449167 | 0.011289644 | 1 |
| Obesity | neutrophil | *MDH1* | 2.11670927 | 0.001984382 | 0.011289644 | 1 |
| Obesity | neutrophil | *SLC30A9* | 2.247456758 | 0.003304974 | 0.011289644 | 1 |
| Obesity | neutrophil | *MTMR11* | 2.526081211 | 0.000208438 | 0.011289644 | 1 |
| Obesity | neutrophil | *COX15* | 1.336878597 | 0.005861676 | 0.011378838 | 1 |
| Obesity | neutrophil | *NPC1L1* | -0.587154509 | 0.005488134 | 0.011289644 | 1 |
| Obesity | neutrophil | *NUDCD3* | 1.721188744 | 0.000941721 | 0.011289644 | 1 |
| Obesity | neutrophil | *CHDH* | 4.770701135 | 0.005475045 | 0.011289644 | 1 |
| Obesity | neutrophil | *IL20RA* | 4.91182602 | 0.004371579 | 0.011289644 | 1 |
| Obesity | neutrophil | *RUFY3* | 1.874279841 | 0.005248002 | 0.011289644 | 1 |
| Obesity | neutrophil | *WWTR1* | 0.905163972 | 0.00419845 | 0.011289644 | 1 |
| Obesity | neutrophil | *TTC27* | 1.388559907 | 0.004980568 | 0.011289644 | 1 |
| Obesity | neutrophil | *VSIG2* | -1.978156204 | 0.007889757 | 0.01274567 | 1 |
| Obesity | neutrophil | *PHLDB1* | 1.839096558 | 0.002925883 | 0.011289644 | 1 |
| Obesity | neutrophil | *MARCO* | 1.129841566 | 0.016359079 | 0.021952941 | 1 |
| Obesity | neutrophil | *MNAT1* | 1.666516411 | 0.00445374 | 0.011289644 | 1 |
| Obesity | neutrophil | *SLC7A9* | 3.105214047 | 0.001096069 | 0.011289644 | 1 |
| Obesity | neutrophil | *OSBPL5* | 2.761207177 | 0.005422249 | 0.011289644 | 1 |
| Obesity | neutrophil | *AQR* | 2.29481048 | 0.005769736 | 0.011357022 | 1 |
| Obesity | neutrophil | *CPS1* | 1.709145582 | 0.00143327 | 0.011289644 | 1 |
| Obesity | neutrophil | *NLRP2* | 2.963348917 | 0.008551348 | 0.01340038 | 1 |
| Obesity | neutrophil | *ZDHHC6* | 1.848834417 | 0.003531885 | 0.011289644 | 1 |

| | | | | | | |
|---|---|---|---|---|---|---|
| Obesity | neutrophil | *RNH1* | 3.274577772 | 0.00266719 | 0.011289644 | 1 |
| Obesity | neutrophil | *NDUFS1* | 1.649388229 | 0.00263144 | 0.011289644 | 1 |
| Obesity | neutrophil | *BIRC3* | -2.331510025 | 0.007776714 | 0.012646715 | 1 |
| Obesity | neutrophil | *AKAP11* | 2.751286909 | 0.006704139 | 0.011809729 | 1 |
| Obesity | neutrophil | *GLRX2* | 1.396406495 | 0.012473335 | 0.017531826 | 1 |
| Obesity | neutrophil | *SNAPC1* | 1.884160272 | 0.00715976 | 0.012141606 | 1 |
| Obesity | neutrophil | *GCLM* | 1.351040799 | 0.002262707 | 0.011289644 | 1 |
| Obesity | neutrophil | *UBR2* | 1.721144928 | 0.005348327 | 0.011289644 | 1 |
| Obesity | neutrophil | *HSF2* | 4.833456651 | 0.024825466 | 0.031518115 | 1 |
| Obesity | neutrophil | *NR1H3* | 2.615597391 | 0.001233259 | 0.011289644 | 1 |
| Obesity | neutrophil | *TOMM34* | 1.969819772 | 0.003477339 | 0.011289644 | 1 |
| Obesity | neutrophil | *KPNA6* | 2.233763486 | 0.002953295 | 0.011289644 | 1 |
| Obesity | neutrophil | *VIM* | 1.310029339 | 0.00379075 | 0.011289644 | 1 |
| Obesity | neutrophil | *FAS* | 1.467643739 | 0.002570123 | 0.011289644 | 1 |
| Obesity | neutrophil | *RNASET2* | 1.291147334 | 0.0052926 | 0.011289644 | 1 |
| Obesity | neutrophil | *CD44* | 1.856858962 | 0.005198614 | 0.011289644 | 1 |
| Obesity | neutrophil | *MIPEP* | 2.054527699 | 0.004550642 | 0.011289644 | 1 |
| Obesity | neutrophil | *INSRR* | 5.890365698 | 0.023991386 | 0.030514636 | 1 |
| Obesity | neutrophil | *B4GALT7* | 2.219181967 | 0.002994656 | 0.011289644 | 1 |
| Obesity | neutrophil | *VRK2* | 1.630435413 | 0.006557241 | 0.011698461 | 1 |
| Obesity | neutrophil | *VEZT* | 1.407604279 | 0.004800128 | 0.011289644 | 1 |
| Obesity | neutrophil | *SNX1* | 3.048358082 | 0.003693437 | 0.011289644 | 1 |
| Obesity | neutrophil | *SLC39A9* | 1.955294972 | 0.009099622 | 0.013900632 | 1 |
| Obesity | neutrophil | *ANK1* | 2.923574597 | 0.005617964 | 0.011295339 | 1 |
| Obesity | neutrophil | *RABEP1* | 2.561336565 | 0.000639952 | 0.011289644 | 1 |
| Obesity | neutrophil | *BAK1* | 4.026794541 | 0.000579517 | 0.011289644 | 1 |
| Obesity | neutrophil | *GRN* | 2.24669388 | 0.006207131 | 0.011535516 | 1 |
| Obesity | neutrophil | *CENPQ* | 1.194913928 | 0.031669373 | 0.039302279 | 1 |
| Obesity | neutrophil | *ARID4A* | 1.867042654 | 0.004690822 | 0.011289644 | 1 |
| Obesity | neutrophil | *IFT88* | 1.947634066 | 0.003728169 | 0.011289644 | 1 |
| Obesity | neutrophil | *ZCCHC8* | 1.739062002 | 0.002370935 | 0.011289644 | 1 |
| Obesity | neutrophil | *ABCF2* | 1.735785721 | 0.00483042 | 0.011289644 | 1 |
| Obesity | neutrophil | *FUT8* | 2.225958412 | 0.006059876 | 0.011450953 | 1 |
| Obesity | neutrophil | *UBA6* | 1.547945745 | 0.009986381 | 0.014846152 | 1 |
| Obesity | neutrophil | *PIAS1* | 1.570375577 | 0.004758602 | 0.011289644 | 1 |
| Obesity | neutrophil | *SLC4A7* | 1.887445876 | 0.005249447 | 0.011289644 | 1 |
| Obesity | neutrophil | *MYOC* | 1.380275954 | 0.003001447 | 0.011289644 | 1 |
| Obesity | neutrophil | *VCL* | 1.747477501 | 0.021833656 | 0.028099938 | 1 |
| Obesity | neutrophil | *ADSS* | 1.12549351 | 0.00418903 | 0.011289644 | 1 |
| Obesity | neutrophil | *STAP1* | -2.409389887 | 0.001245983 | 0.011289644 | 1 |
| Obesity | neutrophil | *TBC1D23* | 1.627575043 | 0.004651826 | 0.011289644 | 1 |
| Obesity | neutrophil | *CUL3* | 1.639747522 | 0.005237041 | 0.011289644 | 1 |
| Obesity | neutrophil | *MYOM2* | -2.121554403 | 0.004436981 | 0.011289644 | 1 |
| Obesity | neutrophil | *NSUN2* | 4.291809112 | 0.006713191 | 0.011817556 | 1 |
| Obesity | neutrophil | *MRI1* | 1.29588599 | 0.003713626 | 0.011289644 | 1 |
| Obesity | neutrophil | *AGA* | 3.125541927 | 0.004844879 | 0.011289644 | 1 |
| Obesity | neutrophil | *BOD1L1* | 1.554596325 | 0.003765492 | 0.011289644 | 1 |
| Obesity | neutrophil | *CLEC16A* | 1.661462883 | 0.004886932 | 0.011289644 | 1 |
| Obesity | neutrophil | *SKIV2L2* | 1.759304452 | 0.003768166 | 0.011289644 | 1 |
| Obesity | neutrophil | *DNAH5* | -0.796975616 | 0.007036371 | 0.012040736 | 1 |

| | | | | | | |
|---|---|---|---|---|---|---|
| Obesity | neutrophil | *C6* | 1.1574071 | 0.00463506 | 0.011289644 | 1 |
| Obesity | neutrophil | *RAI14* | 2.287571 | 0.003582163 | 0.011289644 | 1 |
| Obesity | neutrophil | *PNKP* | 2.370284653 | 0.003102477 | 0.011289644 | 1 |
| Obesity | neutrophil | *PHLPP2* | 1.167938483 | 0.004214903 | 0.011289644 | 1 |
| Obesity | neutrophil | *PQLC2* | 3.330507723 | 0.005264611 | 0.011289644 | 1 |
| Obesity | neutrophil | *CTNS* | 2.040420109 | 0.018450922 | 0.024278138 | 1 |
| Obesity | neutrophil | *INPP4A* | 3.29141174 | 0.01094865 | 0.015864061 | 1 |
| Obesity | neutrophil | *RAB27B* | -1.311111044 | 0.008975759 | 0.013797211 | 1 |
| Obesity | neutrophil | *LSG1* | 2.590011996 | 0.000280243 | 0.011289644 | 1 |
| Obesity | neutrophil | *THAP3* | 1.739828016 | 0.004573457 | 0.011289644 | 1 |
| Obesity | neutrophil | *TDP1* | 1.369248955 | 0.004018569 | 0.011289644 | 1 |
| Obesity | neutrophil | *SPATA7* | 3.360542068 | 0.013070962 | 0.018217055 | 1 |
| Obesity | neutrophil | *MED17* | 2.638619743 | 0.003217848 | 0.011289644 | 1 |
| Obesity | neutrophil | *RETSAT* | 1.007151812 | 0.002843965 | 0.011289644 | 1 |
| Obesity | neutrophil | *CAPG* | -1.163616384 | 0.00693285 | 0.011952575 | 1 |
| Obesity | neutrophil | *AP2S1* | 1.411562315 | 0.005308893 | 0.011289644 | 1 |
| Obesity | neutrophil | *LCP2* | -1.284306375 | 0.007083549 | 0.012072542 | 1 |
| Obesity | neutrophil | *TRIT1* | 1.376177261 | 0.004507705 | 0.011289644 | 1 |
| Obesity | neutrophil | *CUL7* | 1.512791291 | 0.010815931 | 0.015729761 | 1 |
| Obesity | neutrophil | *CTNNA1* | 1.834584799 | 0.003181279 | 0.011289644 | 1 |
| Obesity | neutrophil | *CNTLN* | 1.429180478 | 0.004373359 | 0.011289644 | 1 |
| Obesity | neutrophil | *EPHA3* | 1.101208501 | 0.004219303 | 0.011289644 | 1 |
| Obesity | neutrophil | *DSG2* | 1.795300246 | 0.003465116 | 0.011289644 | 1 |
| Obesity | neutrophil | *WDR37* | 2.412573589 | 0.009317234 | 0.014135486 | 1 |
| Obesity | neutrophil | *YTHDC2* | 2.317663656 | 0.004822827 | 0.011289644 | 1 |
| Obesity | neutrophil | *ATP6V1H* | 1.777888612 | 0.004318272 | 0.011289644 | 1 |
| Obesity | neutrophil | *POLR2B* | 2.419731824 | 0.005689811 | 0.011300433 | 1 |
| Obesity | neutrophil | *CP* | 2.640821667 | 0.004328468 | 0.011289644 | 1 |
| Obesity | neutrophil | *DTNBP1* | 1.699947159 | 0.004391155 | 0.011289644 | 1 |
| Obesity | neutrophil | *ANO2* | 1.062500345 | 0.002184441 | 0.011289644 | 1 |
| Obesity | neutrophil | *GOPC* | 1.355053549 | 0.004110797 | 0.011289644 | 1 |
| Obesity | neutrophil | *CC2D2A* | 1.477490385 | 0.004366521 | 0.011289644 | 1 |
| Obesity | neutrophil | *RRM2B* | 2.107685372 | 0.006105182 | 0.011478524 | 1 |
| Obesity | neutrophil | *SNX29* | 1.696998346 | 0.006464238 | 0.01166174 | 1 |
| Obesity | neutrophil | *LMO3* | 1.187340766 | 0.009206738 | 0.014011804 | 1 |
| Obesity | neutrophil | *CELF2* | 1.400691373 | 0.004868882 | 0.011289644 | 1 |
| Obesity | neutrophil | *ERCC8* | 1.487419526 | 0.004430895 | 0.011289644 | 1 |
| Obesity | neutrophil | *ADAMTS6* | 1.81435284 | 0.003484357 | 0.011289644 | 1 |
| Obesity | neutrophil | *H6PD* | 2.058186507 | 0.003484946 | 0.011289644 | 1 |
| Obesity | neutrophil | *UTS2* | -0.695091915 | 0.003294682 | 0.011289644 | 1 |
| Obesity | neutrophil | *EPN3* | -4.536890097 | 0.005200684 | 0.011289644 | 1 |
| Obesity | neutrophil | *RCN1* | 2.329678264 | 0.020378102 | 0.026459547 | 1 |
| Obesity | neutrophil | *RFC2* | 5.898616041 | 0.025660623 | 0.032460431 | 1 |
| Obesity | neutrophil | *JKAMP* | 1.87292341 | 0.001225852 | 0.011289644 | 1 |
| Obesity | neutrophil | *DKK3* | 1.748025458 | 0.00291919 | 0.011289644 | 1 |
| Obesity | neutrophil | *ARHGEF5* | 2.359774119 | 0.005228158 | 0.011289644 | 1 |
| Obesity | neutrophil | *MCUR1* | 3.103424632 | 0.006914718 | 0.011943085 | 1 |
| Obesity | neutrophil | *LIMA1* | 1.224824047 | 0.006716101 | 0.011817556 | 1 |
| Obesity | neutrophil | *PTGER3* | 0.826386241 | 0.003813219 | 0.011289644 | 1 |
| Obesity | neutrophil | *MAPK9* | 2.216294659 | 0.011166453 | 0.016072228 | 1 |

| | | | | | | |
|---|---|---|---|---|---|---|
| Obesity | neutrophil | *FAM160A2* | 1.855140192 | 0.00988437 | 0.014746804 | 1 |
| Obesity | neutrophil | *HERPUD1* | 3.443533738 | 0.010726553 | 0.015628699 | 1 |
| Obesity | neutrophil | *PIK3CB* | 1.740403189 | 0.008935357 | 0.013763799 | 1 |
| Obesity | neutrophil | *CYBA* | 2.012396595 | 0.005668128 | 0.011300433 | 1 |
| Obesity | neutrophil | *THOC3* | 6.49701713 | 0.011599851 | 0.01655567 | 1 |
| Obesity | neutrophil | *RRP12* | 2.205301452 | 0.006087478 | 0.011468977 | 1 |
| Obesity | neutrophil | *ALX4* | 3.292150657 | 0.004833293 | 0.011289644 | 1 |
| Obesity | neutrophil | *FOXN3* | 1.924020716 | 0.005244593 | 0.011289644 | 1 |
| Obesity | neutrophil | *AKR7A2* | 1.665478816 | 0.003796949 | 0.011289644 | 1 |
| Obesity | neutrophil | *MRTO4* | 5.112916559 | 0.026232218 | 0.03314348 | 1 |
| Obesity | neutrophil | *MCF2L2* | 2.02916903 | 0.005158618 | 0.011289644 | 1 |
| Obesity | neutrophil | *ANAPC4* | 2.285690822 | 0.002181086 | 0.011289644 | 1 |
| Obesity | neutrophil | *LY75* | 5.224417479 | 0.003921637 | 0.011289644 | 1 |
| Obesity | neutrophil | *OPN3* | 4.020648518 | 0.029891623 | 0.037305116 | 1 |
| Obesity | neutrophil | *SDCCAG8* | 1.865162163 | 0.000830936 | 0.011289644 | 1 |
| Obesity | neutrophil | *TBC1D22A* | 1.788550645 | 0.004740298 | 0.011289644 | 1 |
| Obesity | neutrophil | *PLEKHH1* | 2.395918337 | 0.009997399 | 0.014855507 | 1 |
| Obesity | neutrophil | *ATP9A* | 1.086274998 | 0.005129417 | 0.011289644 | 1 |
| Obesity | neutrophil | *GALC* | 2.409395659 | 0.009506207 | 0.014328823 | 1 |
| Obesity | neutrophil | *FAM114A2* | 2.805891029 | 0.00560584 | 0.011295339 | 1 |
| Obesity | neutrophil | *GINM1* | 1.439518521 | 0.004007878 | 0.011289644 | 1 |
| Obesity | neutrophil | *MRPL43* | 3.247801682 | 0.0300897 | 0.037527067 | 1 |
| Obesity | neutrophil | *TRAF1* | -2.017435808 | 0.011235083 | 0.01615252 | 1 |
| Obesity | neutrophil | *TRAF3IP2* | 2.235390602 | 0.003864704 | 0.011289644 | 1 |
| Obesity | neutrophil | *DCBLD2* | 1.392603843 | 0.004179295 | 0.011289644 | 1 |
| Obesity | neutrophil | *MSH4* | 2.339837257 | 0.001426112 | 0.011289644 | 1 |
| Obesity | neutrophil | *GDI2* | 1.752844419 | 0.007775186 | 0.012646715 | 1 |
| Obesity | neutrophil | *TMCC3* | 3.947672731 | 0.012308895 | 0.017362805 | 1 |
| Obesity | neutrophil | *PITHD1* | 5.049501676 | 0.013375984 | 0.018523086 | 1 |
| Obesity | neutrophil | *ATP11B* | 2.158255995 | 0.000669032 | 0.011289644 | 1 |
| Obesity | neutrophil | *LAMC2* | 1.564677636 | 0.025092291 | 0.03182474 | 1 |
| Obesity | neutrophil | *CDK14* | 2.663515208 | 0.012096021 | 0.017128656 | 1 |
| Obesity | neutrophil | *CROCC* | 1.334953202 | 0.001419119 | 0.011289644 | 1 |
| Obesity | neutrophil | *RIOK2* | 3.834633799 | 0.006843624 | 0.011894781 | 1 |
| Obesity | neutrophil | *FLYWCH1* | 1.81419475 | 0.002503121 | 0.011289644 | 1 |
| Obesity | neutrophil | *DNAJC25* | 1.956562762 | 0.00385488 | 0.011289644 | 1 |
| Obesity | neutrophil | *YBX3* | 1.503713189 | 0.003778981 | 0.011289644 | 1 |
| Obesity | neutrophil | *OGFR* | 1.360736099 | 0.003464891 | 0.011289644 | 1 |
| Obesity | neutrophil | *QSER1* | 1.032962218 | 0.004232585 | 0.011289644 | 1 |
| Obesity | neutrophil | *NCKAP1* | 2.327718 | 0.00628406 | 0.011565541 | 1 |
| Obesity | neutrophil | *SFSWAP* | 2.286797583 | 0.005901282 | 0.011378838 | 1 |
| Obesity | neutrophil | *TNK2* | 3.229859616 | 0.01902276 | 0.024926238 | 1 |
| Obesity | neutrophil | *VMP1* | 2.612136255 | 0.013553721 | 0.018740322 | 1 |
| Obesity | neutrophil | *EIF4B* | 2.045222144 | 0.001374936 | 0.011289644 | 1 |
| Obesity | neutrophil | *SLC6A16* | -2.296452208 | 0.018367685 | 0.024178734 | 1 |
| Obesity | neutrophil | *SPHK2* | 1.798982789 | 0.005902261 | 0.011378838 | 1 |
| Obesity | neutrophil | *ISOC2* | 2.130786895 | 0.00428724 | 0.011289644 | 1 |
| Obesity | neutrophil | *EPN1* | 3.507609365 | 0.003985587 | 0.011289644 | 1 |
| Obesity | neutrophil | *ADCK1* | 1.413189735 | 0.004063086 | 0.011289644 | 1 |
| Obesity | neutrophil | *HAGH* | 1.684381294 | 0.00222449 | 0.011289644 | 1 |

| | | | | | | |
|---|---|---|---|---|---|---|
| Obesity | neutrophil | *TM7SF3* | 1.743960256 | 0.002779675 | 0.011289644 | 1 |
| Obesity | neutrophil | *DMRT3* | -1.461344282 | 0.00637611 | 0.011639549 | 1 |
| Obesity | neutrophil | *ST3GAL6* | 1.394905382 | 0.013982823 | 0.019215303 | 1 |
| Obesity | neutrophil | *ATP2C2* | -2.712941637 | 0.006864384 | 0.011915069 | 1 |
| Obesity | neutrophil | *CDON* | 0.992890227 | 0.004389298 | 0.011289644 | 1 |
| Obesity | neutrophil | *TAF2* | 1.644511352 | 0.003222866 | 0.011289644 | 1 |
| Obesity | neutrophil | *TMEM161A* | 5.206079442 | 0.003840546 | 0.011289644 | 1 |
| Obesity | neutrophil | *SUGP2* | 1.362769831 | 0.006741587 | 0.011825993 | 1 |
| Obesity | neutrophil | *ABCA7* | 3.460316303 | 0.008158848 | 0.012979422 | 1 |
| Obesity | neutrophil | *DDX20* | -2.428950677 | 0.011132635 | 0.016038239 | 1 |
| Obesity | neutrophil | *BTBD1* | 1.436414089 | 0.003589515 | 0.011289644 | 1 |
| Obesity | neutrophil | *CHI3L2* | 3.136917203 | 0.003606002 | 0.011289644 | 1 |
| Obesity | neutrophil | *PMS1* | 2.574447406 | 0.005766326 | 0.011357022 | 1 |
| Obesity | neutrophil | *ANKS1A* | 2.566025149 | 0.006403137 | 0.011648199 | 1 |
| Obesity | neutrophil | *NTHL1* | -1.785059441 | 0.004782679 | 0.011289644 | 1 |
| Obesity | neutrophil | *UHRF1BP1* | 5.273712595 | 2.58E-12 | 3.24E-09 | 1.62E-08 |
| Obesity | neutrophil | *WDR18* | 2.539900389 | 0.019351182 | 0.025303844 | 1 |
| Obesity | neutrophil | *ANKRD44* | 2.861904486 | 0.006022744 | 0.011447874 | 1 |
| Obesity | neutrophil | *ZC3H15* | 1.75844119 | 0.00572299 | 0.01134195 | 1 |
| Obesity | neutrophil | *MAP2K4* | 1.502262618 | 0.004298053 | 0.011289644 | 1 |
| Obesity | neutrophil | *TMEM206* | 1.819003638 | 0.004084762 | 0.011289644 | 1 |
| Obesity | neutrophil | *SEC61A2* | 2.288535888 | 0.004448348 | 0.011289644 | 1 |
| Obesity | neutrophil | *ASB1* | 1.668136467 | 0.006768128 | 0.011851936 | 1 |
| Obesity | neutrophil | *PPP2R5A* | 2.190581607 | 0.005958364 | 0.011413844 | 1 |
| Obesity | neutrophil | *TIE1* | 1.245617768 | 0.013212486 | 0.0183817 | 1 |
| Obesity | neutrophil | *NFYC* | 2.106375224 | 0.009798253 | 0.014656582 | 1 |
| Obesity | neutrophil | *ZMYND12* | 2.045649282 | 0.030240947 | 0.037685794 | 1 |
| Obesity | neutrophil | *NGEF* | 3.603127176 | 0.00410118 | 0.011289644 | 1 |
| Obesity | neutrophil | *ZNRD1* | -5.266970951 | 0.004884044 | 0.011289644 | 1 |
| Obesity | neutrophil | *ATXN3* | 1.285652638 | 0.004111105 | 0.011289644 | 1 |
| Obesity | neutrophil | *EML1* | 1.643538782 | 0.007052995 | 0.01205278 | 1 |
| Obesity | neutrophil | *ZFAT* | 1.454538848 | 0.006084642 | 0.011467069 | 1 |
| Obesity | neutrophil | *MTFR1* | 2.296912686 | 0.016148602 | 0.021689011 | 1 |
| Obesity | neutrophil | *STAG3* | 4.250212479 | 0.022872269 | 0.029215351 | 1 |
| Obesity | neutrophil | *MYO9A* | 1.50527377 | 0.003096437 | 0.011289644 | 1 |
| Obesity | neutrophil | *SP100* | 1.745264825 | 0.004852065 | 0.011289644 | 1 |
| Obesity | neutrophil | *STOML1* | 6.191928521 | 0.007500485 | 0.012396306 | 1 |
| Obesity | neutrophil | *METTL22* | 4.364325156 | 0.018261782 | 0.024059476 | 1 |
| Obesity | neutrophil | *TP53BP1* | 1.864677611 | 0.002498172 | 0.011289644 | 1 |
| Obesity | neutrophil | *RRP15* | 1.070636729 | 0.007753445 | 0.012628474 | 1 |
| Obesity | neutrophil | *DHX8* | 2.928650877 | 0.00598383 | 0.011427806 | 1 |
| Obesity | neutrophil | *SYT1* | 1.176372187 | 0.003717434 | 0.011289644 | 1 |
| Obesity | neutrophil | *ROGDI* | 2.719809021 | 0.004159292 | 0.011289644 | 1 |
| Obesity | neutrophil | *HDAC4* | 1.615762461 | 0.004507794 | 0.011289644 | 1 |
| Obesity | neutrophil | *RASSF1* | 3.818988217 | 0.022798401 | 0.029156539 | 1 |
| Obesity | neutrophil | *FGFR3* | -1.96473664 | 0.016063139 | 0.021601915 | 1 |
| Obesity | neutrophil | *HEATR6* | 2.08164886 | 0.003962549 | 0.011289644 | 1 |
| Obesity | neutrophil | *STON1-GTF2A* | 0.908960876 | 0.011240898 | 0.016157184 | 1 |
| Obesity | neutrophil | *SRBD1* | 1.545457008 | 0.004375417 | 0.011289644 | 1 |
| Obesity | neutrophil | *CYFIP1* | 2.533737864 | 0.006150446 | 0.011508289 | 1 |

| | | | | | | |
|---|---|---|---|---|---|---|
| Obesity | neutrophil | *PSME4* | 1.293580833 | 0.004724325 | 0.011289644 | 1 |
| Obesity | neutrophil | *PITX1* | 5.056581625 | 0.004397361 | 0.011289644 | 1 |
| Obesity | neutrophil | *SDK2* | -1.666123803 | 0.007646246 | 0.012505709 | 1 |
| Obesity | neutrophil | *NUCKS1* | 1.671196674 | 0.003589271 | 0.011289644 | 1 |
| Obesity | neutrophil | *DNAJA2* | 4.70776881 | 0.038282031 | 0.046652283 | 1 |
| Obesity | neutrophil | *KCNAB2* | -1.80692489 | 0.006981335 | 0.01198898 | 1 |
| Obesity | neutrophil | *CLEC2D* | 2.320402939 | 0.011549728 | 0.016502986 | 1 |
| Obesity | neutrophil | *NEDD4* | 1.614350619 | 0.005287726 | 0.011289644 | 1 |
| Obesity | neutrophil | *PIGB* | 1.39896043 | 0.004543181 | 0.011289644 | 1 |
| Obesity | neutrophil | *DAPP1* | 4.043823049 | 0.036530439 | 0.04476937 | 1 |
| Obesity | neutrophil | *FGF10* | 1.310646299 | 0.002929922 | 0.011289644 | 1 |
| Obesity | neutrophil | *TMEM260* | 1.943964196 | 0.004754086 | 0.011289644 | 1 |
| Obesity | neutrophil | *CLTCL1* | 2.333668264 | 0.002998857 | 0.011289644 | 1 |
| Obesity | neutrophil | *DGCR2* | 3.660280285 | 0.018231683 | 0.024024856 | 1 |
| Obesity | neutrophil | *MNT* | 1.42448508 | 0.010180781 | 0.015028544 | 1 |
| Obesity | neutrophil | *ZXDC* | 1.818617911 | 0.00318417 | 0.011289644 | 1 |
| Obesity | neutrophil | *POLB* | 2.728143728 | 0.012663229 | 0.017743197 | 1 |
| Obesity | neutrophil | *NDST1* | 2.908271138 | 0.005679851 | 0.011300433 | 1 |
| Obesity | neutrophil | *ASNS* | -1.179965398 | 0.008076776 | 0.012901091 | 1 |
| Obesity | neutrophil | *AP3M2* | 1.60541818 | 0.005229796 | 0.011289644 | 1 |
| Obesity | neutrophil | *ST6GALNAC2* | 5.457398856 | 0.001441643 | 0.011289644 | 1 |
| Obesity | neutrophil | *TESK2* | 3.544516827 | 0.017266496 | 0.022906617 | 1 |
| Obesity | neutrophil | *EIF2B3* | 1.53980393 | 0.004465548 | 0.011289644 | 1 |
| Obesity | neutrophil | *CAMK2A* | 9.383922599 | 0.004226389 | 0.011289644 | 1 |
| Obesity | neutrophil | *CDC42* | 1.382021682 | 0.004406464 | 0.011289644 | 1 |
| Obesity | neutrophil | *OSBPL3* | 3.135720344 | 0.006779953 | 0.011856902 | 1 |
| Obesity | neutrophil | *RAD18* | 1.45687976 | 0.003980882 | 0.011289644 | 1 |
| Obesity | neutrophil | *SNX13* | 1.4376264 | 0.00438142 | 0.011289644 | 1 |
| Obesity | neutrophil | *VASH1* | 4.147660475 | 0.023581117 | 0.030017105 | 1 |
| Obesity | neutrophil | *TRIP13* | 4.673900052 | 0.004317106 | 0.011289644 | 1 |
| Obesity | neutrophil | *MBD3* | -1.690738405 | 0.027526473 | 0.034604866 | 1 |
| Obesity | neutrophil | *HLTF* | 1.442962185 | 0.0147053 | 0.020029988 | 1 |
| Obesity | neutrophil | *CPSF1* | 3.397470094 | 0.011048247 | 0.015961637 | 1 |
| Obesity | neutrophil | *CYBRD1* | 1.912562912 | 0.003475049 | 0.011289644 | 1 |
| Obesity | neutrophil | *EPN2* | 1.102713168 | 0.003778737 | 0.011289644 | 1 |
| Obesity | neutrophil | *LIMS2* | 2.078707582 | 0.003429625 | 0.011289644 | 1 |
| Obesity | neutrophil | *AFF4* | 1.585445952 | 0.002864349 | 0.011289644 | 1 |
| Obesity | neutrophil | *TRHDE* | 0.812628489 | 0.00428332 | 0.011289644 | 1 |
| Obesity | neutrophil | *NFATC3* | 1.481802968 | 0.00691704 | 0.011943814 | 1 |
| Obesity | neutrophil | *TRNT1* | 2.342002593 | 0.004700726 | 0.011289644 | 1 |
| Obesity | neutrophil | *ACADVL* | 1.403894976 | 0.005294569 | 0.011289644 | 1 |
| Obesity | neutrophil | *CRMP1* | 2.864995868 | 0.008687287 | 0.013539098 | 1 |
| Obesity | neutrophil | *NDE1* | 1.89443507 | 0.003240852 | 0.011289644 | 1 |
| Obesity | neutrophil | *AP1M1* | 1.867350276 | 0.008493114 | 0.013331367 | 1 |
| Obesity | neutrophil | *PVR* | 1.997318847 | 0.001414692 | 0.011289644 | 1 |
| Obesity | neutrophil | *XRCC1* | 4.162816922 | 0.008858546 | 0.013707527 | 1 |
| Obesity | neutrophil | *MOV10L1* | 2.188802013 | 0.004990365 | 0.011289644 | 1 |
| Obesity | neutrophil | *TP63* | 1.793423918 | 0.005099384 | 0.011289644 | 1 |
| Obesity | neutrophil | *ALPK1* | 2.026347634 | 0.004115885 | 0.011289644 | 1 |
| Obesity | neutrophil | *LLGL2* | 2.623952691 | 0.004067596 | 0.011289644 | 1 |

| | | | | | | |
|---|---|---|---|---|---|---|
| Obesity | neutrophil | *PDE8A* | 2.231062426 | 0.003045536 | 0.011289644 | 1 |
| Obesity | neutrophil | *NLE1* | 2.875241998 | 0.005091652 | 0.011289644 | 1 |
| Obesity | neutrophil | *SDHA* | 2.101634163 | 0.001542501 | 0.011289644 | 1 |
| Obesity | neutrophil | *SMARCE1* | 4.573207853 | 0.004929275 | 0.011289644 | 1 |
| Obesity | neutrophil | *GSDMB* | 1.321610997 | 0.008770247 | 0.013623871 | 1 |
| Obesity | neutrophil | *PICALM* | 1.49849418 | 0.004853374 | 0.011289644 | 1 |
| Obesity | neutrophil | *GLI2* | 1.188087653 | 0.013713014 | 0.018923117 | 1 |
| Obesity | neutrophil | *CLNS1A* | 1.797092064 | 0.001627196 | 0.011289644 | 1 |
| Obesity | neutrophil | *ATP2A3* | 1.820844171 | 0.003468372 | 0.011289644 | 1 |
| Obesity | neutrophil | *CA12* | 1.685606244 | 0.003647498 | 0.011289644 | 1 |
| Obesity | neutrophil | *NTN4* | 1.028742404 | 0.00471763 | 0.011289644 | 1 |
| Obesity | neutrophil | *LMAN1* | 1.387595335 | 0.004707458 | 0.011289644 | 1 |
| Obesity | neutrophil | *ENO1* | 1.796809055 | 0.004539595 | 0.011289644 | 1 |
| Obesity | neutrophil | *TIPIN* | 4.077092425 | 0.000162694 | 0.011289644 | 1 |
| Obesity | neutrophil | *EIF4G3* | 2.034748159 | 0.006003216 | 0.011438456 | 1 |
| Obesity | neutrophil | *TTC38* | 3.678694657 | 0.026922004 | 0.033919567 | 1 |
| Obesity | neutrophil | *GRAMD4* | 2.002139994 | 0.005083537 | 0.011289644 | 1 |
| Obesity | neutrophil | *CELSR1* | 1.764015168 | 0.004279419 | 0.011289644 | 1 |
| Obesity | neutrophil | *ADD2* | -2.678269401 | 0.008015672 | 0.012842642 | 1 |
| Obesity | neutrophil | *VPS9D1* | 1.430239692 | 0.004780605 | 0.011289644 | 1 |
| Obesity | neutrophil | *MARK3* | 1.328421262 | 0.009994581 | 0.01485483 | 1 |
| Obesity | neutrophil | *SLC25A3* | 1.983095138 | 0.003023531 | 0.011289644 | 1 |
| Obesity | neutrophil | *FSCN1* | 2.017915166 | 0.009016982 | 0.013836969 | 1 |
| Obesity | neutrophil | *MOCOS* | 2.6186441 | 0.001857547 | 0.011289644 | 1 |
| Obesity | neutrophil | *PLD1* | 2.703859184 | 0.007432 | 0.012335001 | 1 |
| Obesity | neutrophil | *BCAP29* | 1.591517678 | 0.003929315 | 0.011289644 | 1 |
| Obesity | neutrophil | *SEC31B* | 2.382059736 | 0.000310229 | 0.011289644 | 1 |
| Obesity | neutrophil | *TUBA3D* | -1.014061156 | 0.005884905 | 0.011378838 | 1 |
| Obesity | neutrophil | *MKRN2* | -2.177935588 | 0.028037711 | 0.035184249 | 1 |
| Obesity | neutrophil | *MCM6* | 2.099392402 | 0.003968624 | 0.011289644 | 1 |
| Obesity | neutrophil | *RBMS2* | 1.667792191 | 0.004268597 | 0.011289644 | 1 |
| Obesity | neutrophil | *MLH1* | 1.894598958 | 0.006580996 | 0.011713234 | 1 |
| Obesity | neutrophil | *FMO4* | 3.950078683 | 0.0114078 | 0.016351571 | 1 |
| Obesity | neutrophil | *RGS11* | 1.315199158 | 0.009443432 | 0.014259024 | 1 |
| Obesity | neutrophil | *GPATCH1* | -2.120873944 | 0.04118677 | 0.049844317 | 1 |
| Obesity | neutrophil | *ICAM3* | -2.356017189 | 0.0235153 | 0.02994545 | 1 |
| Obesity | neutrophil | *NT5C2* | 1.639626796 | 0.002293252 | 0.011289644 | 1 |
| Obesity | neutrophil | *CAMSAP3* | -1.367200204 | 0.00524566 | 0.011289644 | 1 |
| Obesity | neutrophil | *XAB2* | 1.890790712 | 0.013254864 | 0.018416905 | 1 |
| Obesity | neutrophil | *STXBP2* | -1.534419881 | 0.00407958 | 0.011289644 | 1 |
| Obesity | neutrophil | *MAP2K7* | 2.554505079 | 0.004581188 | 0.011289644 | 1 |
| Obesity | neutrophil | *NMRK2* | 12.73237721 | 0.004501281 | 0.011289644 | 1 |
| Obesity | neutrophil | *CTTNBP2* | 2.216053441 | 0.012421142 | 0.017468922 | 1 |
| Obesity | neutrophil | *TM9SF3* | 2.505454117 | 0.004706797 | 0.011289644 | 1 |
| Obesity | neutrophil | *DNAJC10* | 2.311032588 | 0.004826148 | 0.011289644 | 1 |
| Obesity | neutrophil | *DYNC1I2* | 1.759250528 | 0.002177283 | 0.011289644 | 1 |
| Obesity | neutrophil | *APBB1IP* | 1.161320821 | 0.005198887 | 0.011289644 | 1 |
| Obesity | neutrophil | *LRCH4* | -3.76769297 | 0.013322214 | 0.018473039 | 1 |
| Obesity | neutrophil | *SIRT6* | -2.814732246 | 0.010967911 | 0.015887886 | 1 |
| Obesity | neutrophil | *CAPZB* | 2.977206821 | 0.025330022 | 0.032100352 | 1 |

| | | | | | | |
|---|---|---|---|---|---|---|
| Obesity | neutrophil | *GPR137B* | 2.513019658 | 0.028834575 | 0.036112146 | 1 |
| Obesity | neutrophil | *NAALAD2* | 1.209350214 | 0.00298606 | 0.011289644 | 1 |
| Obesity | neutrophil | *FGFR1* | 2.130138063 | 0.004777397 | 0.011289644 | 1 |
| Obesity | neutrophil | *FBLN1* | 0.883080689 | 0.00318386 | 0.011289644 | 1 |
| Obesity | neutrophil | *AMPH* | 1.293694303 | 0.003552027 | 0.011289644 | 1 |
| Obesity | neutrophil | *MCCC1* | 1.794116149 | 0.001459841 | 0.011289644 | 1 |
| Obesity | neutrophil | *FAP* | 1.134778246 | 0.006401516 | 0.011648199 | 1 |
| Obesity | neutrophil | *ACER3* | 2.704630773 | 0.001985078 | 0.011289644 | 1 |
| Obesity | neutrophil | *PIK3C3* | 1.226119842 | 0.005460751 | 0.011289644 | 1 |
| Obesity | neutrophil | *N4BP2* | 2.584456594 | 0.012976824 | 0.018122011 | 1 |
| Obesity | neutrophil | *TULP3* | 1.658790715 | 0.005037407 | 0.011289644 | 1 |
| Obesity | neutrophil | *SYNJ2* | 1.6960946 | 0.005071538 | 0.011289644 | 1 |
| Obesity | neutrophil | *ZCWPW1* | 2.002669243 | 0.002292063 | 0.011289644 | 1 |
| Obesity | neutrophil | *FGF20* | 16.81982115 | 0.0044965 | 0.011289644 | 1 |
| Obesity | neutrophil | *PCM1* | 1.420826173 | 0.00566534 | 0.011300433 | 1 |
| Obesity | neutrophil | *MYH7B* | 1.768918728 | 0.0133499 | 0.018495112 | 1 |
| Obesity | neutrophil | *FKBP7* | 1.531300168 | 0.011681611 | 0.016646031 | 1 |
| Obesity | neutrophil | *OSBPL6* | 1.289723905 | 0.0045097 | 0.011289644 | 1 |
| Obesity | neutrophil | *XRCC5* | 1.621979837 | 0.004664878 | 0.011289644 | 1 |
| Obesity | neutrophil | *LXN* | 1.958646802 | 0.005251379 | 0.011289644 | 1 |
| Obesity | neutrophil | *REXO1* | 3.184695056 | 0.005156545 | 0.011289644 | 1 |
| Obesity | neutrophil | *SAR1A* | 1.431575025 | 0.003976191 | 0.011289644 | 1 |
| Obesity | neutrophil | *CDC14A* | 1.47252263 | 0.010061841 | 0.014923046 | 1 |
| Obesity | neutrophil | *RAPGEF3* | 1.463912704 | 0.002007629 | 0.011289644 | 1 |
| Obesity | neutrophil | *FDFT1* | 4.121490854 | 0.020268921 | 0.026334099 | 1 |
| Obesity | neutrophil | *SCGN* | -1.022906574 | 0.006206162 | 0.011535516 | 1 |
| Obesity | neutrophil | *DDX1* | 3.277184443 | 0.010325704 | 0.015181876 | 1 |
| Obesity | neutrophil | *DNM2* | 2.023294091 | 0.005516659 | 0.011290116 | 1 |
| Obesity | neutrophil | *RIMS1* | 1.984294578 | 0.004901679 | 0.011289644 | 1 |
| Obesity | neutrophil | *STX7* | 2.314128101 | 0.008771979 | 0.013623871 | 1 |
| Obesity | neutrophil | *RABL2B* | 1.411842768 | 0.005268852 | 0.011289644 | 1 |
| Obesity | neutrophil | *DDX43* | 2.076946509 | 0.005287274 | 0.011289644 | 1 |
| Obesity | neutrophil | *DCT* | 2.903673297 | 0.003646073 | 0.011289644 | 1 |
| Obesity | neutrophil | *SLC35C2* | 2.672291847 | 0.005096035 | 0.011289644 | 1 |
| Obesity | neutrophil | *SCTR* | 1.462522797 | 0.004622466 | 0.011289644 | 1 |
| Obesity | neutrophil | *RIF1* | 2.700761482 | 0.006005493 | 0.011438456 | 1 |
| Obesity | neutrophil | *KCNN2* | -0.741292942 | 0.005170315 | 0.011289644 | 1 |
| Obesity | neutrophil | *PSEN1* | 1.890151106 | 0.005057163 | 0.011289644 | 1 |
| Obesity | neutrophil | *MOK* | 2.159618249 | 0.004459058 | 0.011289644 | 1 |
| Obesity | neutrophil | *RSBN1* | 1.884768476 | 0.008445219 | 0.013293713 | 1 |
| Obesity | neutrophil | *MAGI3* | 1.347376083 | 0.004302011 | 0.011289644 | 1 |
| Obesity | neutrophil | *COL4A4* | 1.807060737 | 0.002180496 | 0.011289644 | 1 |
| Obesity | neutrophil | *ZNF506* | 1.988451441 | 0.004985556 | 0.011289644 | 1 |
| Obesity | neutrophil | *KIAA0141* | 2.137784203 | 0.005143547 | 0.011289644 | 1 |
| Obesity | neutrophil | *HSPB11* | 2.719810135 | 0.005888466 | 0.011378838 | 1 |
| Obesity | neutrophil | *WDR70* | 1.548190807 | 0.004015329 | 0.011289644 | 1 |
| Obesity | neutrophil | *PGR* | 1.304931857 | 0.002906603 | 0.011289644 | 1 |
| Obesity | neutrophil | *C5orf22* | 1.613320441 | 0.003978798 | 0.011289644 | 1 |
| Obesity | neutrophil | *COBLL1* | 2.324642567 | 0.000333368 | 0.011289644 | 1 |
| Obesity | neutrophil | *GEMIN5* | 1.15716042 | 0.006106675 | 0.011478524 | 1 |

| | | | | | | |
|---|---|---|---|---|---|---|
| Obesity | neutrophil | *GSK3B* | 1.926543204 | 0.006095925 | 0.011474575 | 1 |
| Obesity | neutrophil | *ERC1* | 1.544278867 | 0.004792851 | 0.011289644 | 1 |
| Obesity | neutrophil | *DOPEY1* | 2.545253212 | 0.000490951 | 0.011289644 | 1 |
| Obesity | neutrophil | *LYRM2* | 3.957226458 | 0.001523692 | 0.011289644 | 1 |
| Obesity | neutrophil | *ULK2* | 3.199837585 | 0.007839563 | 0.012699648 | 1 |
| Obesity | neutrophil | *PLOD1* | 1.528863127 | 0.003769002 | 0.011289644 | 1 |
| Obesity | neutrophil | *PIBF1* | 1.644147051 | 0.004950979 | 0.011289644 | 1 |
| Obesity | neutrophil | *TDRD3* | 2.851652495 | 0.017623298 | 0.023330738 | 1 |
| Obesity | neutrophil | *PDS5B* | 1.733227608 | 0.005055323 | 0.011289644 | 1 |
| Obesity | neutrophil | *OXCT1* | 0.963611465 | 0.006563145 | 0.011701779 | 1 |
| Obesity | neutrophil | *CYLD* | 1.71746977 | 0.005287413 | 0.011289644 | 1 |
| Obesity | neutrophil | *ZNF324* | 4.396403082 | 0.006050556 | 0.01145004 | 1 |
| Obesity | neutrophil | *ZNF586* | 1.518166152 | 0.003314427 | 0.011289644 | 1 |
| Obesity | neutrophil | *ZNF446* | -1.126073156 | 0.006325496 | 0.011605985 | 1 |
| Obesity | neutrophil | *ZNF264* | 1.221687059 | 0.004519667 | 0.011289644 | 1 |
| Obesity | neutrophil | *RPS5* | 2.873687788 | 0.005502004 | 0.011289644 | 1 |
| Obesity | neutrophil | *CHMP2B* | 3.223150953 | 0.005833368 | 0.011378838 | 1 |
| Obesity | neutrophil | *PPIE* | 5.885475445 | 0.013538234 | 0.018727145 | 1 |
| Obesity | neutrophil | *ZMPSTE24* | 2.099756871 | 0.004641118 | 0.011289644 | 1 |
| Obesity | neutrophil | *STARD7* | 1.277187551 | 0.003489974 | 0.011289644 | 1 |
| Obesity | neutrophil | *GSTP1* | 4.639423112 | 0.039669979 | 0.048184922 | 1 |
| Obesity | neutrophil | *WBP11* | 1.31639026 | 0.004219354 | 0.011289644 | 1 |
| Obesity | neutrophil | *COL16A1* | 0.940705037 | 0.028659436 | 0.035907109 | 1 |
| Obesity | neutrophil | *APOB* | -1.220298152 | 0.004776304 | 0.011289644 | 1 |
| Obesity | neutrophil | *NCOA1* | 1.626405901 | 0.005009787 | 0.011289644 | 1 |
| Obesity | neutrophil | *CD59* | 2.153557193 | 0.0048556 | 0.011289644 | 1 |
| Obesity | neutrophil | *TAF9* | 1.811826949 | 0.00427507 | 0.011289644 | 1 |
| Obesity | neutrophil | *FCN1* | -2.01498837 | 0.005843436 | 0.011378838 | 1 |
| Obesity | neutrophil | *SCAMP1* | 1.421732909 | 0.00235243 | 0.011289644 | 1 |
| Obesity | neutrophil | *HACE1* | 2.081184108 | 0.003797786 | 0.011289644 | 1 |
| Obesity | neutrophil | *SEH1L* | 3.012953031 | 0.00441034 | 0.011289644 | 1 |
| Obesity | neutrophil | *OVGP1* | 2.938497226 | 0.019928255 | 0.025961238 | 1 |
| Obesity | neutrophil | *SLC25A24* | 1.76078614 | 0.004196269 | 0.011289644 | 1 |
| Obesity | neutrophil | *ABCB1* | 1.392059917 | 0.003577914 | 0.011289644 | 1 |
| Obesity | neutrophil | *ZNF213* | 1.029360158 | 0.008778528 | 0.013623945 | 1 |
| Obesity | neutrophil | *CPNE3* | 1.869618424 | 0.012729636 | 0.017820357 | 1 |
| Obesity | neutrophil | *CTTN* | 1.796698988 | 0.004497603 | 0.011289644 | 1 |
| Obesity | neutrophil | *MTIF2* | 1.742659052 | 0.008028172 | 0.012852843 | 1 |
| Obesity | neutrophil | *EPS15* | 1.130297238 | 0.01007724 | 0.01492911 | 1 |
| Obesity | neutrophil | *ATG16L1* | 4.158205211 | 0.01197521 | 0.016991232 | 1 |
| Obesity | neutrophil | *USP40* | 1.668735517 | 0.003677775 | 0.011289644 | 1 |
| Obesity | neutrophil | *MAST2* | 2.596086229 | 0.00290865 | 0.011289644 | 1 |
| Obesity | neutrophil | *B4GALT1* | 1.321970939 | 0.004224344 | 0.011289644 | 1 |
| Obesity | neutrophil | *CHMP5* | 1.362507059 | 0.003792716 | 0.011289644 | 1 |
| Obesity | neutrophil | *FOLH1* | -0.604893946 | 0.017200566 | 0.022848079 | 1 |
| Obesity | neutrophil | *SEPHS1* | 2.220361697 | 0.006656875 | 0.011757198 | 1 |
| Obesity | neutrophil | *ITPKC* | 1.286262778 | 0.003700442 | 0.011289644 | 1 |
| Obesity | neutrophil | *CEACAM6* | -21.18912202 | 0.027209434 | 0.034233683 | 1 |
| Obesity | neutrophil | *TMED2* | 1.755930057 | 0.004045497 | 0.011289644 | 1 |
| Obesity | neutrophil | *HSD17B14* | 2.170316091 | 0.005085653 | 0.011289644 | 1 |

| | | | | | | |
|---|---|---|---|---|---|---|
| Obesity | neutrophil | *SRRT* | 4.380356694 | 0.001626912 | 0.011289644 | 1 |
| Obesity | neutrophil | *PIGS* | 1.556481815 | 0.005093794 | 0.011289644 | 1 |
| Obesity | neutrophil | *ATXN7L3* | 1.92877833 | 0.003303472 | 0.011289644 | 1 |
| Obesity | neutrophil | *PGS1* | 1.319544433 | 0.004765573 | 0.011289644 | 1 |
| Obesity | neutrophil | *PSMC5* | -1.231884753 | 0.005626032 | 0.011295339 | 1 |
| Obesity | neutrophil | *LPCAT2* | 2.952147826 | 0.020129155 | 0.026168735 | 1 |
| Obesity | neutrophil | *OGFOD1* | 1.49865223 | 0.030746744 | 0.038285755 | 1 |
| Obesity | neutrophil | *TXNDC16* | 3.286099612 | 0.013438018 | 0.018596702 | 1 |
| Obesity | neutrophil | *GMCL1* | 3.736213229 | 0.007263864 | 0.012218115 | 1 |
| Obesity | neutrophil | *KLHL42* | 1.445026678 | 0.003192733 | 0.011289644 | 1 |
| Obesity | neutrophil | *PTHLH* | 2.509344277 | 0.001467059 | 0.011289644 | 1 |
| Obesity | neutrophil | *PHACTR3* | 1.295623731 | 0.011753267 | 0.016717826 | 1 |
| Obesity | neutrophil | *ERGIC2* | 1.602420026 | 0.004057546 | 0.011289644 | 1 |
| Obesity | neutrophil | *AAMDC* | 1.008023821 | 0.018685267 | 0.024530021 | 1 |
| Obesity | neutrophil | *METTL2A* | -2.416267804 | 0.007607596 | 0.012461948 | 1 |
| Obesity | neutrophil | *ALG6* | 2.362738354 | 0.005244486 | 0.011289644 | 1 |
| Obesity | neutrophil | *GP6* | -1.36653654 | 0.006160364 | 0.011512519 | 1 |
| Obesity | neutrophil | *DDX18* | 3.174562027 | 0.005936796 | 0.011396756 | 1 |
| Obesity | neutrophil | *ASAP3* | 1.210539133 | 0.003439906 | 0.011289644 | 1 |
| Obesity | neutrophil | *EDEM2* | 2.237062368 | 0.010709172 | 0.015617853 | 1 |
| Obesity | neutrophil | *DOCK3* | 1.174863999 | 0.002036696 | 0.011289644 | 1 |
| Obesity | neutrophil | *C3orf18* | -0.825929741 | 0.02159309 | 0.027835915 | 1 |
| Obesity | neutrophil | *NSFL1C* | 3.804919197 | 0.003415964 | 0.011289644 | 1 |
| Obesity | neutrophil | *ZNF343* | 1.820575419 | 0.003458853 | 0.011289644 | 1 |
| Obesity | neutrophil | *MAVS* | 2.102593967 | 0.003338008 | 0.011289644 | 1 |
| Obesity | neutrophil | *LZTS3* | -1.825730067 | 0.005530169 | 0.011295339 | 1 |
| Obesity | neutrophil | *XRN2* | 2.193445693 | 0.004536726 | 0.011289644 | 1 |
| Obesity | neutrophil | *RBBP9* | -1.942655393 | 0.005920677 | 0.011385978 | 1 |
| Obesity | neutrophil | *ANAPC5* | 1.875145828 | 0.007531337 | 0.012412599 | 1 |
| Obesity | neutrophil | *SLC23A2* | 1.412637156 | 0.004107468 | 0.011289644 | 1 |
| Obesity | neutrophil | *TMEM230* | 2.172922166 | 0.004796151 | 0.011289644 | 1 |
| Obesity | neutrophil | *DZANK1* | 1.615138122 | 0.006804405 | 0.011873524 | 1 |
| Obesity | neutrophil | *TASP1* | 1.983188041 | 0.003479185 | 0.011289644 | 1 |
| Obesity | neutrophil | *OAS1* | 3.138687085 | 0.006252095 | 0.011555481 | 1 |
| Obesity | neutrophil | *KIF16B* | 2.772488766 | 0.006919904 | 0.011945479 | 1 |
| Obesity | neutrophil | *FUS* | 1.578808105 | 0.00419952 | 0.011289644 | 1 |
| Obesity | neutrophil | *FXYD5* | 4.405708657 | 0.005618657 | 0.011295339 | 1 |
| Obesity | neutrophil | *FXYD3* | 9.563741685 | 0.002370617 | 0.011289644 | 1 |
| Obesity | neutrophil | *GANAB* | 2.887489807 | 0.004112536 | 0.011289644 | 1 |
| Obesity | neutrophil | *MLF2* | 2.416229126 | 0.00414836 | 0.011289644 | 1 |
| Obesity | neutrophil | *NECAP1* | 1.388712879 | 0.006236043 | 0.01154827 | 1 |
| Obesity | neutrophil | *ANKRD24* | 2.444455784 | 0.002509466 | 0.011289644 | 1 |
| Obesity | neutrophil | *GPATCH2L* | 1.686997813 | 0.005635745 | 0.011295339 | 1 |
| Obesity | neutrophil | *SLC9A1* | 1.898464625 | 0.004318532 | 0.011289644 | 1 |
| Obesity | neutrophil | *SPTLC1* | 1.381987767 | 0.003380238 | 0.011289644 | 1 |
| Obesity | neutrophil | *PAPOLA* | 1.637304056 | 0.004115519 | 0.011289644 | 1 |
| Obesity | neutrophil | *RGS1* | 4.462755973 | 0.005092802 | 0.011289644 | 1 |
| Obesity | neutrophil | *MRPS33* | 1.621320295 | 0.004621921 | 0.011289644 | 1 |
| Obesity | neutrophil | *MAEA* | 2.099326297 | 0.006598666 | 0.01172215 | 1 |
| Obesity | neutrophil | *STRN4* | 1.441449842 | 0.005519453 | 0.011292159 | 1 |

| | | | | | | |
|---|---|---|---|---|---|---|
| Obesity | neutrophil | *LYZ* | 3.957012745 | 0.004391785 | 0.011289644 | 1 |
| Obesity | neutrophil | *MUL1* | 3.643265223 | 0.006980086 | 0.01198898 | 1 |
| Obesity | neutrophil | *SPG21* | 1.929580667 | 0.00793075 | 0.01278095 | 1 |
| Obesity | neutrophil | *GNPTG* | 3.685893096 | 0.008063546 | 0.01288711 | 1 |
| Obesity | neutrophil | *ZNF268* | -4.029444911 | 0.01135447 | 0.016301794 | 1 |
| Obesity | neutrophil | *GOLGA3* | 1.793070951 | 0.003644245 | 0.011289644 | 1 |
| Obesity | neutrophil | *CERS4* | 1.720471557 | 0.005898925 | 0.011378838 | 1 |
| Obesity | neutrophil | *USP48* | 1.665997516 | 0.006644126 | 0.011753146 | 1 |
| Obesity | neutrophil | *PDPR* | 1.54068468 | 0.003863923 | 0.011289644 | 1 |
| Obesity | neutrophil | *EXOC1* | 2.278371431 | 0.005963674 | 0.011420536 | 1 |
| Obesity | neutrophil | *NLRC4* | 4.339926053 | 0.0076592 | 0.012520383 | 1 |
| Obesity | neutrophil | *LAMB1* | 2.034967359 | 0.0009862 | 0.011289644 | 1 |
| Obesity | neutrophil | *SLC26A4* | -4.624559022 | 0.010029726 | 0.014892983 | 1 |
| Obesity | neutrophil | *DLD* | 4.026619895 | 0.019338255 | 0.025292201 | 1 |
| Obesity | neutrophil | *TXNL1* | 2.003348288 | 0.003717715 | 0.011289644 | 1 |
| Obesity | neutrophil | *IL5RA* | 4.065279388 | 0.004653631 | 0.011289644 | 1 |
| Obesity | neutrophil | *ITGA6* | 1.101335152 | 0.003378017 | 0.011289644 | 1 |
| Obesity | neutrophil | *RAPGEF4* | 0.95420975 | 0.005236658 | 0.011289644 | 1 |
| Obesity | neutrophil | *TF* | -2.029399756 | 0.007296871 | 0.012233922 | 1 |
| Obesity | neutrophil | *MYO15A* | -3.708322254 | 0.005726321 | 0.01134195 | 1 |
| Obesity | neutrophil | *NLRP1* | 2.415372114 | 0.010276393 | 0.015128491 | 1 |
| Obesity | neutrophil | *SPAG7* | 2.487399075 | 0.001009797 | 0.011289644 | 1 |
| Obesity | neutrophil | *ZFHX4* | 0.903805807 | 0.004537304 | 0.011289644 | 1 |
| Obesity | neutrophil | *RGS17* | 1.370162447 | 0.013022837 | 0.018158035 | 1 |
| Obesity | neutrophil | *TMEM101* | 4.863106351 | 0.012630534 | 0.017721036 | 1 |
| Obesity | neutrophil | *CMA1* | 5.100801655 | 0.031704002 | 0.039329981 | 1 |
| Obesity | neutrophil | *PPP2R3C* | 1.837643097 | 0.003058296 | 0.011289644 | 1 |
| Obesity | neutrophil | *HAUS4* | 1.999070748 | 0.026274134 | 0.033189769 | 1 |
| Obesity | neutrophil | *OSGEP* | 1.299182555 | 0.004331908 | 0.011289644 | 1 |
| Obesity | neutrophil | *SLC22A17* | 2.476669549 | 0.009816965 | 0.014678826 | 1 |
| Obesity | neutrophil | *SCFD1* | 1.564264259 | 0.005040773 | 0.011289644 | 1 |
| Obesity | neutrophil | *RPGRIP1* | -0.480729222 | 0.005399597 | 0.011289644 | 1 |
| Obesity | neutrophil | *TOX4* | 1.658483358 | 0.003230714 | 0.011289644 | 1 |
| Obesity | neutrophil | *TGM1* | 3.215585463 | 0.00895641 | 0.013787697 | 1 |
| Obesity | neutrophil | *AGO1* | 2.287682916 | 0.004470915 | 0.011289644 | 1 |
| Obesity | neutrophil | *RFFL* | -2.436432098 | 0.011593559 | 0.01655057 | 1 |
| Obesity | neutrophil | *MFSD11* | 1.628870184 | 0.005666704 | 0.011300433 | 1 |
| Obesity | neutrophil | *DPYSL2* | 1.62734354 | 0.004200378 | 0.011289644 | 1 |
| Obesity | neutrophil | *NUP50* | 1.334462799 | 0.004793888 | 0.011289644 | 1 |
| Obesity | neutrophil | *COMT* | 3.096619634 | 0.007186675 | 0.012155908 | 1 |
| Obesity | neutrophil | *VNN3* | -0.323452616 | 0.005264612 | 0.011289644 | 1 |
| Obesity | neutrophil | *SEC22C* | 2.521212159 | 0.00482376 | 0.011289644 | 1 |
| Obesity | neutrophil | *FMO2* | 2.774663657 | 0.001648207 | 0.011289644 | 1 |
| Obesity | neutrophil | *SUCO* | 1.314252708 | 0.004472495 | 0.011289644 | 1 |
| Obesity | neutrophil | *MSH2* | 3.828917031 | 0.008878432 | 0.013723778 | 1 |
| Obesity | neutrophil | *MAP3K1* | 2.329325887 | 0.01330245 | 0.018449703 | 1 |
| Obesity | neutrophil | *TMEM38B* | 2.13473276 | 0.004598746 | 0.011289644 | 1 |
| Obesity | neutrophil | *PDE6C* | 2.384180295 | 0.001262688 | 0.011289644 | 1 |
| Obesity | neutrophil | *CWF19L1* | 1.408125736 | 0.010300666 | 0.01515215 | 1 |
| Obesity | neutrophil | *IKZF5* | 2.372284674 | 0.00445195 | 0.011289644 | 1 |

| | | | | | | |
|---|---|---|---|---|---|---|
| Obesity | neutrophil | *BLNK* | 1.434700468 | 0.003037207 | 0.011289644 | 1 |
| Obesity | neutrophil | *WAC* | 1.731711752 | 0.003994576 | 0.011289644 | 1 |
| Obesity | neutrophil | *CREM* | 2.66126057 | 0.006548679 | 0.011694062 | 1 |
| Obesity | neutrophil | *TPSD1* | 13.19175843 | 0.006759126 | 0.011846705 | 1 |
| Obesity | neutrophil | *ITPR3* | 1.669610235 | 0.004126551 | 0.011289644 | 1 |
| Obesity | neutrophil | *DSP* | 2.85291852 | 0.006178343 | 0.011512519 | 1 |
| Obesity | neutrophil | *IFT74* | 2.256578992 | 0.001373581 | 0.011289644 | 1 |
| Obesity | neutrophil | *IL12RB1* | 2.236762807 | 0.01119029 | 0.016091801 | 1 |
| Obesity | neutrophil | *ACOT7* | 1.715130471 | 0.005166961 | 0.011289644 | 1 |
| Obesity | neutrophil | *PCSK5* | 2.34640022 | 0.007751334 | 0.012628308 | 1 |
| Obesity | neutrophil | *RAB18* | 1.865351181 | 0.007460044 | 0.01236443 | 1 |
| Obesity | neutrophil | *NRP1* | 1.6447894 | 0.005206413 | 0.011289644 | 1 |
| Obesity | neutrophil | *PRTFDC1* | -2.322803606 | 0.015894046 | 0.021392821 | 1 |
| Obesity | neutrophil | *PALMD* | 2.082849582 | 0.006407722 | 0.011648199 | 1 |
| Obesity | neutrophil | *HSD3B7* | 4.267203896 | 0.002920339 | 0.011289644 | 1 |
| Obesity | neutrophil | *MARCH2* | 1.448181672 | 0.003550785 | 0.011289644 | 1 |
| Obesity | neutrophil | *MTAP* | 1.371146955 | 0.004273223 | 0.011289644 | 1 |
| Obesity | neutrophil | *CEP170B* | -1.061412142 | 0.009585597 | 0.01441738 | 1 |
| Obesity | neutrophil | *POLR2E* | -1.235348774 | 0.006970194 | 0.011985556 | 1 |
| Obesity | neutrophil | *MADCAM1* | -2.910170269 | 0.021138237 | 0.027316772 | 1 |
| Obesity | neutrophil | *TRMT2A* | 1.67686492 | 0.005868223 | 0.011378838 | 1 |
| Obesity | neutrophil | *ZDHHC8* | 1.492508304 | 0.0034619 | 0.011289644 | 1 |
| Obesity | neutrophil | *MED15* | 2.660841975 | 0.007458992 | 0.01236443 | 1 |
| Obesity | neutrophil | *LZTR1* | 4.67255303 | 0.031454014 | 0.039058145 | 1 |
| Obesity | neutrophil | *CECR2* | 1.256299336 | 0.004477058 | 0.011289644 | 1 |
| Obesity | neutrophil | *P2RX6* | 2.183189479 | 0.007199725 | 0.012164652 | 1 |
| Obesity | neutrophil | *SLC7A4* | -4.722264417 | 0.032638842 | 0.040346127 | 1 |
| Obesity | neutrophil | *DDT* | 1.734060406 | 0.004414015 | 0.011289644 | 1 |
| Obesity | neutrophil | *GGT5* | 1.650473908 | 0.004888983 | 0.011289644 | 1 |
| Obesity | neutrophil | *PPIL2* | 1.409419366 | 0.007426712 | 0.012334552 | 1 |
| Obesity | neutrophil | *UPB1* | -0.739555345 | 0.003542223 | 0.011289644 | 1 |
| Obesity | neutrophil | *YPEL1* | 2.488589328 | 0.004115122 | 0.011289644 | 1 |
| Obesity | neutrophil | *PPM1F* | 2.123729354 | 0.005628253 | 0.011295339 | 1 |
| Obesity | neutrophil | *TOP3B* | 3.458573212 | 0.00269397 | 0.011289644 | 1 |
| Obesity | neutrophil | *CRYBB3* | 5.352293016 | 0.00593961 | 0.011396756 | 1 |
| Obesity | neutrophil | *CARD10* | -2.154188805 | 0.006909807 | 0.011943085 | 1 |
| Obesity | neutrophil | *SLC25A1* | -2.092913232 | 0.013617337 | 0.01880347 | 1 |
| Obesity | neutrophil | *LGALS2* | 2.525077942 | 0.001472808 | 0.011289644 | 1 |
| Obesity | neutrophil | *GGA1* | 3.284744835 | 0.006244576 | 0.011550353 | 1 |
| Obesity | neutrophil | *SH3BP1* | 2.277808278 | 0.004131207 | 0.011289644 | 1 |
| Obesity | neutrophil | *HPS4* | 2.501497504 | 0.004702419 | 0.011289644 | 1 |
| Obesity | neutrophil | *PIK3IP1* | 1.43747159 | 0.004791803 | 0.011289644 | 1 |
| Obesity | neutrophil | *TRIOBP* | 1.538953352 | 0.018966823 | 0.024863306 | 1 |
| Obesity | neutrophil | *TFIP11* | 4.307723822 | 0.000498178 | 0.011289644 | 1 |
| Obesity | neutrophil | *GCAT* | -0.739742436 | 0.011051536 | 0.015961637 | 1 |
| Obesity | neutrophil | *CCDC134* | 2.588736802 | 0.003116351 | 0.011289644 | 1 |
| Obesity | neutrophil | *PICK1* | 1.452375508 | 0.018184284 | 0.023972446 | 1 |
| Obesity | neutrophil | *SLC16A8* | 3.830053619 | 0.009397523 | 0.014216128 | 1 |
| Obesity | neutrophil | *DDX17* | 1.349601874 | 0.004866371 | 0.011289644 | 1 |
| Obesity | neutrophil | *FBXO7* | 4.705729465 | 0.003741786 | 0.011289644 | 1 |

| | | | | | | |
|---|---|---|---|---|---|---|
| Obesity | neutrophil | *POLDIP3* | 3.964950128 | 0.008109552 | 0.012939288 | 1 |
| Obesity | neutrophil | *TIMP3* | 2.035259282 | 0.006670645 | 0.01177382 | 1 |
| Obesity | neutrophil | *PPP6R2* | 1.674734434 | 0.005159287 | 0.011289644 | 1 |
| Obesity | neutrophil | *CYB5R3* | 1.895650856 | 0.00410606 | 0.011289644 | 1 |
| Obesity | neutrophil | *RHBDD3* | 5.720817736 | 0.014758256 | 0.020092065 | 1 |
| Obesity | neutrophil | *TTLL1* | 1.702509374 | 0.003982637 | 0.011289644 | 1 |
| Obesity | neutrophil | *RASL10A* | 3.150121071 | 0.004067601 | 0.011289644 | 1 |
| Obesity | neutrophil | *AP1B1* | 1.895984708 | 0.003701633 | 0.011289644 | 1 |
| Obesity | neutrophil | *HMGXB4* | 2.819673677 | 0.005886364 | 0.011378838 | 1 |
| Obesity | neutrophil | *TOM1* | 2.331345998 | 0.012821006 | 0.017920335 | 1 |
| Obesity | neutrophil | *CHKB* | 1.806805421 | 0.012791797 | 0.017887462 | 1 |
| Obesity | neutrophil | *BIK* | -0.630701424 | 0.005182838 | 0.011289644 | 1 |
| Obesity | neutrophil | *THOC5* | 1.459678243 | 0.005846085 | 0.011378838 | 1 |
| Obesity | neutrophil | *ARSA* | 2.305025558 | 0.016510611 | 0.022099683 | 1 |
| Obesity | neutrophil | *TSPO* | 5.037930131 | 0.008202545 | 0.013029156 | 1 |
| Obesity | neutrophil | *TTLL12* | -1.239996942 | 0.005889226 | 0.011378838 | 1 |
| Obesity | neutrophil | *RBFOX2* | 1.524297854 | 0.004540365 | 0.011289644 | 1 |
| Obesity | neutrophil | *MTMR3* | 1.489203272 | 0.007741348 | 0.012620869 | 1 |
| Obesity | neutrophil | *APOL4* | 2.097495855 | 0.001459115 | 0.011289644 | 1 |
| Obesity | neutrophil | *SAMM50* | 2.263710687 | 0.006063676 | 0.011451587 | 1 |
| Obesity | neutrophil | *FOXRED2* | 1.85351096 | 0.007022179 | 0.012029547 | 1 |
| Obesity | neutrophil | *IFT27* | 1.710754343 | 0.004223486 | 0.011289644 | 1 |
| Obesity | neutrophil | *KIAA0930* | 2.775916956 | 0.007896989 | 0.012749967 | 1 |
| Obesity | neutrophil | *NCF4* | 2.815329372 | 0.004034763 | 0.011289644 | 1 |
| Obesity | neutrophil | *SLC25A17* | 1.32126258 | 0.004304795 | 0.011289644 | 1 |
| Obesity | neutrophil | *FAM118A* | 1.944370868 | 0.012280182 | 0.017330079 | 1 |
| Obesity | neutrophil | *KCTD17* | 9.045224436 | 0.006483315 | 0.011671674 | 1 |
| Obesity | neutrophil | *EP300* | 1.440045585 | 0.003293001 | 0.011289644 | 1 |
| Obesity | neutrophil | *ACO2* | 3.102130611 | 0.000976721 | 0.011289644 | 1 |
| Obesity | neutrophil | *POLR3H* | 1.293033592 | 0.018634629 | 0.024483994 | 1 |
| Obesity | neutrophil | *DESI1* | 3.568876983 | 0.035097152 | 0.04313053 | 1 |
| Obesity | neutrophil | *ZBED4* | 2.521540606 | 0.004797955 | 0.011289644 | 1 |
| Obesity | neutrophil | *HDAC10* | -3.077106709 | 0.021316194 | 0.027518463 | 1 |
| Obesity | neutrophil | *KHNYN* | 2.315025419 | 0.00122306 | 0.011289644 | 1 |
| Obesity | neutrophil | *SDR39U1* | -1.986188419 | 0.015680302 | 0.021173122 | 1 |
| Obesity | neutrophil | *RBM23* | 3.700353148 | 0.036570959 | 0.044810294 | 1 |
| Obesity | neutrophil | *SOS2* | 1.745599078 | 0.004878916 | 0.011289644 | 1 |
| Obesity | neutrophil | *PYGL* | 0.940020498 | 0.016452222 | 0.022040298 | 1 |
| Obesity | neutrophil | *DDHD1* | 1.758511676 | 0.006326342 | 0.011605985 | 1 |
| Obesity | neutrophil | *CNIH1* | 1.351829897 | 0.005453259 | 0.011289644 | 1 |
| Obesity | neutrophil | *ATP6V1D* | 1.530990088 | 0.007088221 | 0.012073596 | 1 |
| Obesity | neutrophil | *PIGH* | 4.924292085 | 0.002003984 | 0.011289644 | 1 |
| Obesity | neutrophil | *GSTZ1* | 1.891566232 | 0.004010326 | 0.011289644 | 1 |
| Obesity | neutrophil | *TMED8* | 2.110405709 | 0.005767249 | 0.011357022 | 1 |
| Obesity | neutrophil | *DAAM1* | 2.269907958 | 0.007607028 | 0.012461948 | 1 |
| Obesity | neutrophil | *PPM1A* | 1.523806963 | 0.005942325 | 0.01139701 | 1 |
| Obesity | neutrophil | *DICER1* | 3.333937051 | 0.002098654 | 0.011289644 | 1 |
| Obesity | neutrophil | *ZFYVE21* | 2.080020869 | 0.001770735 | 0.011289644 | 1 |
| Obesity | neutrophil | *MTHFD1* | 3.101205367 | 0.005368052 | 0.011289644 | 1 |
| Obesity | neutrophil | *TELO2* | 1.901990657 | 0.004973392 | 0.011289644 | 1 |

| | | | | | | |
|---|---|---|---|---|---|---|
| Obesity | neutrophil | *GSKIP* | 5.682223746 | 0.003930252 | 0.011289644 | 1 |
| Obesity | neutrophil | *VRK1* | 3.506384376 | 0.007751198 | 0.012628308 | 1 |
| Obesity | neutrophil | *PSMB5* | 1.880390586 | 0.001228584 | 0.011289644 | 1 |
| Obesity | neutrophil | *CCNB1IP1* | 1.453738477 | 0.003509139 | 0.011289644 | 1 |
| Obesity | neutrophil | *TRIP11* | 2.23619597 | 0.005551673 | 0.011295339 | 1 |
| Obesity | neutrophil | *PCK2* | 1.798359745 | 0.005508798 | 0.011289644 | 1 |
| Obesity | neutrophil | *KIAA0391* | 2.668184462 | 0.005158945 | 0.011289644 | 1 |
| Obesity | neutrophil | *EMC9* | -0.97831944 | 0.007334766 | 0.012274068 | 1 |
| Obesity | neutrophil | *PLTP* | 3.579256295 | 0.000795505 | 0.011289644 | 1 |
| Obesity | neutrophil | *GSS* | 1.531524197 | 0.00331607 | 0.011289644 | 1 |
| Obesity | neutrophil | *PYGB* | 1.435457978 | 0.00213014 | 0.011289644 | 1 |
| Obesity | neutrophil | *ABHD12* | 1.549905687 | 0.0010921 | 0.011289644 | 1 |
| Obesity | neutrophil | *PROCR* | 2.617670949 | 0.018895948 | 0.024785901 | 1 |
| Obesity | neutrophil | *NINL* | 1.233575576 | 0.004511637 | 0.011289644 | 1 |
| Obesity | neutrophil | *UQCC1* | 1.552700797 | 0.004037918 | 0.011289644 | 1 |
| Obesity | neutrophil | *IFT52* | 1.649724103 | 0.004215132 | 0.011289644 | 1 |
| Obesity | neutrophil | *NDRG3* | 1.759056033 | 0.002998675 | 0.011289644 | 1 |
| Obesity | neutrophil | *DOK5* | 1.31700671 | 0.004223641 | 0.011289644 | 1 |
| Obesity | neutrophil | *CSTF1* | 1.886036259 | 0.004360557 | 0.011289644 | 1 |
| Obesity | neutrophil | *BMP7* | 2.056859623 | 0.004232376 | 0.011289644 | 1 |
| Obesity | neutrophil | *TPD52L2* | 4.269470599 | 0.020436348 | 0.026508814 | 1 |
| Obesity | neutrophil | *NELFCD* | 2.166565789 | 0.00490103 | 0.011289644 | 1 |
| Obesity | neutrophil | *CTSZ* | 1.637868718 | 0.003865808 | 0.011289644 | 1 |
| Obesity | neutrophil | *PRPF6* | 3.140026152 | 0.011437385 | 0.016381339 | 1 |
| Obesity | neutrophil | *MTG2* | 2.402471792 | 0.003106355 | 0.011289644 | 1 |
| Obesity | neutrophil | *DIDO1* | 2.443139203 | 0.003135569 | 0.011289644 | 1 |
| Obesity | neutrophil | *GID8* | 2.640405968 | 0.01088666 | 0.015803594 | 1 |
| Obesity | neutrophil | *SLC17A9* | 2.62269803 | 0.003181984 | 0.011289644 | 1 |
| Obesity | neutrophil | *NKAIN4* | 4.318866826 | 0.019090538 | 0.025004628 | 1 |
| Obesity | neutrophil | *PTK6* | -1.187198007 | 0.005428468 | 0.011289644 | 1 |
| Obesity | neutrophil | *GMEB2* | 1.691804072 | 0.004327106 | 0.011289644 | 1 |
| Obesity | neutrophil | *CDC25B* | 1.745333261 | 0.003947914 | 0.011289644 | 1 |
| Obesity | neutrophil | *TRIB3* | 2.787997867 | 0.004810427 | 0.011289644 | 1 |
| Obesity | neutrophil | *SLC52A3* | 9.317138686 | 0.000303088 | 0.011289644 | 1 |
| Obesity | neutrophil | *ANGPT4* | -0.535761314 | 0.004451074 | 0.011289644 | 1 |
| Obesity | neutrophil | *RSPO4* | 2.870421456 | 0.006551323 | 0.01169454 | 1 |
| Obesity | neutrophil | *SIRPB1* | 3.070171197 | 0.0327104 | 0.040418688 | 1 |
| Obesity | neutrophil | *SEC23B* | 3.082628043 | 0.039330374 | 0.047800128 | 1 |
| Obesity | neutrophil | *CCM2L* | 1.945380859 | 0.003920839 | 0.011289644 | 1 |
| Obesity | neutrophil | *MYL9* | 1.423892336 | 0.002306366 | 0.011289644 | 1 |
| Obesity | neutrophil | *MROH8* | 1.314429011 | 0.00901923 | 0.013836969 | 1 |
| Obesity | neutrophil | *MAPRE1* | 1.491028418 | 0.003770307 | 0.011289644 | 1 |
| Obesity | neutrophil | *TTI1* | 1.287683439 | 0.004116963 | 0.011289644 | 1 |
| Obesity | neutrophil | *RPRD1B* | 1.671126616 | 0.004617446 | 0.011289644 | 1 |
| Obesity | neutrophil | *PXMP4* | 6.662360208 | 0.001291933 | 0.011289644 | 1 |
| Obesity | neutrophil | *BPI* | 3.963158396 | 0.006113055 | 0.011478524 | 1 |
| Obesity | neutrophil | *WFDC2* | -6.827780791 | 0.013296344 | 0.018446412 | 1 |
| Obesity | neutrophil | *DHX35* | 2.677646978 | 0.006042103 | 0.011447874 | 1 |
| Obesity | neutrophil | *DNTTIP1* | 1.887855244 | 0.009840871 | 0.014706337 | 1 |
| Obesity | neutrophil | *MAP1LC3A* | 2.113344245 | 0.002601777 | 0.011289644 | 1 |

| | | | | | | |
|---|---|---|---|---|---|---|
| Obesity | neutrophil | *APMAP* | 2.465011214 | 0.002404435 | 0.011289644 | 1 |
| Obesity | neutrophil | *CELF4* | -2.651975189 | 0.010583655 | 0.01547069 | 1 |
| Obesity | neutrophil | *RBFA* | 3.865315923 | 0.012218226 | 0.01726202 | 1 |
| Obesity | neutrophil | *METTL4* | 2.572080191 | 0.004540854 | 0.011289644 | 1 |
| Obesity | neutrophil | *LPIN2* | 1.902535348 | 0.005361051 | 0.011289644 | 1 |
| Obesity | neutrophil | *SMCHD1* | 2.564870057 | 0.009969335 | 0.014825402 | 1 |
| Obesity | neutrophil | *CEP76* | 1.876945636 | 0.002358924 | 0.011289644 | 1 |
| Obesity | neutrophil | *CEP192* | 1.111616128 | 0.018527523 | 0.024368731 | 1 |
| Obesity | neutrophil | *LIPG* | -1.411444373 | 0.005457707 | 0.011289644 | 1 |
| Obesity | neutrophil | *POLI* | 1.940090035 | 0.001115738 | 0.011289644 | 1 |
| Obesity | neutrophil | *NALCN* | 1.771721038 | 0.004032167 | 0.011289644 | 1 |
| Obesity | neutrophil | *HTR2A* | 4.407853692 | 0.004298164 | 0.011289644 | 1 |
| Obesity | neutrophil | *KLF5* | 2.991199089 | 0.00366788 | 0.011289644 | 1 |
| Obesity | neutrophil | *STK24* | 1.908537303 | 0.007400141 | 0.012300617 | 1 |
| Obesity | neutrophil | *DNAJC3* | 2.226560114 | 0.007118831 | 0.012102783 | 1 |
| Obesity | neutrophil | *UGGT2* | 1.333696626 | 0.010196185 | 0.01504775 | 1 |
| Obesity | neutrophil | *ARHGEF7* | 1.393844702 | 0.003981367 | 0.011289644 | 1 |
| Obesity | neutrophil | *SGCG* | 1.074608767 | 0.005382545 | 0.011289644 | 1 |
| Obesity | neutrophil | *PARP4* | 1.408011382 | 0.004284841 | 0.011289644 | 1 |
| Obesity | neutrophil | *RGCC* | 1.008863346 | 0.011025356 | 0.015942137 | 1 |
| Obesity | neutrophil | *VWA8* | 1.501079285 | 0.005909071 | 0.011378838 | 1 |
| Obesity | neutrophil | *KATNAL1* | 1.231424715 | 0.005773868 | 0.011358045 | 1 |
| Obesity | neutrophil | *MEDAG* | 1.23208301 | 0.004135424 | 0.011289644 | 1 |
| Obesity | neutrophil | *TSC22D1* | 1.186838834 | 0.004310833 | 0.011289644 | 1 |
| Obesity | neutrophil | *CLN5* | 2.98466934 | 0.040068286 | 0.048631142 | 1 |
| Obesity | neutrophil | *MGRN1* | 1.632615595 | 0.00465395 | 0.011289644 | 1 |
| Obesity | neutrophil | *GDPD3* | -3.453815704 | 0.007000524 | 0.012002262 | 1 |
| Obesity | neutrophil | *ELMO3* | 3.969264692 | 5.39E-05 | 0.00737107 | 0.339069198 |
| Obesity | neutrophil | *NFAT5* | 1.42496806 | 0.004916049 | 0.011289644 | 1 |
| Obesity | neutrophil | *LONP2* | 1.413991373 | 0.005137687 | 0.011289644 | 1 |
| Obesity | neutrophil | *PLLP* | 2.055332962 | 0.004747635 | 0.011289644 | 1 |
| Obesity | neutrophil | *DHODH* | 1.714773766 | 0.002423084 | 0.011289644 | 1 |
| Obesity | neutrophil | *CTCF* | 1.236513849 | 0.003795619 | 0.011289644 | 1 |
| Obesity | neutrophil | *SETD6* | 3.266923198 | 0.003262002 | 0.011289644 | 1 |
| Obesity | neutrophil | *SLC7A6OS* | 1.70410532 | 0.00139352 | 0.011289644 | 1 |
| Obesity | neutrophil | *MON1B* | 1.394354087 | 0.00555019 | 0.011295339 | 1 |
| Obesity | neutrophil | *HCFC1R1* | -3.358096106 | 0.020261837 | 0.026330337 | 1 |
| Obesity | neutrophil | *NPRL3* | 3.115118623 | 0.007839546 | 0.012699648 | 1 |
| Obesity | neutrophil | *TAF1C* | 1.193442897 | 0.020400448 | 0.026477625 | 1 |
| Obesity | neutrophil | *NAGPA* | 2.78717949 | 0.001360094 | 0.011289644 | 1 |
| Obesity | neutrophil | *COTL1* | 1.948656332 | 0.004663557 | 0.011289644 | 1 |
| Obesity | neutrophil | *CRISPLD2* | 1.448226837 | 0.003113279 | 0.011289644 | 1 |
| Obesity | neutrophil | *ZNF500* | 2.577942469 | 0.004643254 | 0.011289644 | 1 |
| Obesity | neutrophil | *NME4* | 1.815837495 | 0.00591066 | 0.011378838 | 1 |
| Obesity | neutrophil | *NOMO3* | 2.504657583 | 0.003837095 | 0.011289644 | 1 |
| Obesity | neutrophil | *LMF1* | 3.361109763 | 0.00519958 | 0.011289644 | 1 |
| Obesity | neutrophil | *CLCN7* | 1.678359191 | 0.003245842 | 0.011289644 | 1 |
| Obesity | neutrophil | *HAGHL* | 2.975101104 | 0.003885777 | 0.011289644 | 1 |
| Obesity | neutrophil | *SLC7A5* | -1.349727344 | 0.005663704 | 0.011300433 | 1 |
| Obesity | neutrophil | *FBXO31* | 3.566026652 | 0.008971121 | 0.013797211 | 1 |

| | | | | | | |
|---|---|---|---|---|---|---|
| Obesity | neutrophil | *STUB1* | 2.773913374 | 0.016128623 | 0.021666806 | 1 |
| Obesity | neutrophil | *RHBDL1* | -7.045318851 | 0.004496828 | 0.011289644 | 1 |
| Obesity | neutrophil | *NUBP1* | 4.52783726 | 0.009619099 | 0.014453938 | 1 |
| Obesity | neutrophil | *CLUAP1* | 4.701549054 | 5.62E-05 | 0.007523175 | 0.353589218 |
| Obesity | neutrophil | *EARS2* | 1.347012743 | 0.007598806 | 0.012461707 | 1 |
| Obesity | neutrophil | *HMOX2* | 1.780892571 | 0.009661599 | 0.014501216 | 1 |
| Obesity | neutrophil | *BFAR* | 4.03398138 | 0.016374377 | 0.021964094 | 1 |
| Obesity | neutrophil | *RBL2* | 2.219194532 | 0.007308011 | 0.012245593 | 1 |
| Obesity | neutrophil | *STX4* | 1.921100556 | 0.003766722 | 0.011289644 | 1 |
| Obesity | neutrophil | *KAT8* | 3.983974089 | 0.000311975 | 0.011289644 | 1 |
| Obesity | neutrophil | *NOMO1* | 1.866047962 | 0.004337426 | 0.011289644 | 1 |
| Obesity | neutrophil | *TMC5* | -5.831222245 | 0.022322757 | 0.028653159 | 1 |
| Obesity | neutrophil | *RNF40* | 3.136947718 | 0.037934418 | 0.046282493 | 1 |
| Obesity | neutrophil | *KNOP1* | 1.868227031 | 0.036478395 | 0.044714305 | 1 |
| Obesity | neutrophil | *LACTB* | 1.202822002 | 0.004434848 | 0.011289644 | 1 |
| Obesity | neutrophil | *HERC1* | 1.915432634 | 0.005805531 | 0.011378838 | 1 |
| Obesity | neutrophil | *TRIP4* | 2.511876262 | 0.001184936 | 0.011289644 | 1 |
| Obesity | neutrophil | *RASL12* | 2.457190904 | 0.004330553 | 0.011289644 | 1 |
| Obesity | neutrophil | *TTC23* | 2.53961126 | 0.011016995 | 0.015933712 | 1 |
| Obesity | neutrophil | *FAH* | -2.736556883 | 0.031739139 | 0.039365549 | 1 |
| Obesity | neutrophil | *HOMER2* | 1.463654075 | 0.003086253 | 0.011289644 | 1 |
| Obesity | neutrophil | *EHD4* | 3.326266572 | 0.004293276 | 0.011289644 | 1 |
| Obesity | neutrophil | *ATP8B4* | 1.437393955 | 0.004221001 | 0.011289644 | 1 |
| Obesity | neutrophil | *DTWD1* | 1.57842965 | 0.002919052 | 0.011289644 | 1 |
| Obesity | neutrophil | *DNAJC17* | 1.549418461 | 0.004282153 | 0.011289644 | 1 |
| Obesity | neutrophil | *PDGFRL* | 1.263701972 | 0.003908019 | 0.011289644 | 1 |
| Obesity | neutrophil | *ZDHHC2* | 1.093925471 | 0.015687296 | 0.021178018 | 1 |
| Obesity | neutrophil | *ZFAND1* | 4.094132126 | 0.014573096 | 0.019883041 | 1 |
| Obesity | neutrophil | *EYA1* | 1.980834551 | 0.004849338 | 0.011289644 | 1 |
| Obesity | neutrophil | *NBN* | 1.539157215 | 0.004623705 | 0.011289644 | 1 |
| Obesity | neutrophil | *CPQ* | 2.014744573 | 0.0034013 | 0.011289644 | 1 |
| Obesity | neutrophil | *IKBKB* | 1.878136078 | 0.003930816 | 0.011289644 | 1 |
| Obesity | neutrophil | *PLAT* | 1.323052192 | 0.003809732 | 0.011289644 | 1 |
| Obesity | neutrophil | *GDAP1* | 4.711254762 | 0.00888922 | 0.013731333 | 1 |
| Obesity | neutrophil | *RAB2A* | 1.084703687 | 0.015014915 | 0.0203797 | 1 |
| Obesity | neutrophil | *ZC2HC1A* | 1.519627781 | 0.005248953 | 0.011289644 | 1 |
| Obesity | neutrophil | *ARMC1* | 2.363499877 | 0.007960769 | 0.012793301 | 1 |
| Obesity | neutrophil | *CHRAC1* | 2.094566043 | 0.00582419 | 0.011378838 | 1 |
| Obesity | neutrophil | *SNX16* | 3.182854442 | 0.005321882 | 0.011289644 | 1 |
| Obesity | neutrophil | *GSDMD* | 1.378768415 | 0.008575995 | 0.013428961 | 1 |
| Obesity | neutrophil | *TSTA3* | -3.347220973 | 0.01758102 | 0.023279669 | 1 |
| Obesity | neutrophil | *ERI1* | 4.063908768 | 0.030140602 | 0.037575643 | 1 |
| Obesity | neutrophil | *LEPROTL1* | 2.580350555 | 0.004844422 | 0.011289644 | 1 |
| Obesity | neutrophil | *TNFRSF10A* | 3.977279096 | 0.002963615 | 0.011289644 | 1 |
| Obesity | neutrophil | *ERICH1* | 2.26249865 | 0.005090257 | 0.011289644 | 1 |
| Obesity | neutrophil | *TUSC3* | 1.213406433 | 0.004017902 | 0.011289644 | 1 |
| Obesity | neutrophil | *ARHGEF10* | 4.205368954 | 0.027598928 | 0.034678048 | 1 |
| Obesity | neutrophil | *KLHDC4* | 2.774267812 | 0.002370131 | 0.011289644 | 1 |
| Obesity | neutrophil | *MCM4* | -0.97701958 | 0.013765386 | 0.018982887 | 1 |
| Obesity | neutrophil | *MAN2B1* | 1.839845081 | 0.012893125 | 0.018013129 | 1 |

| | | | | | | |
|---|---|---|---|---|---|---|
| Obesity | neutrophil | *NUCB1* | 2.866018367 | 0.006994124 | 0.012000111 | 1 |
| Obesity | neutrophil | *NFKBIB* | 5.831978034 | 0.008855068 | 0.013705518 | 1 |
| Obesity | neutrophil | *SARS2* | 2.618423461 | 0.019287383 | 0.025241419 | 1 |
| Obesity | neutrophil | *SNRNP70* | 1.154788251 | 0.010682858 | 0.015583092 | 1 |
| Obesity | neutrophil | *PPP1R37* | 3.530161007 | 0.000679089 | 0.011289644 | 1 |
| Obesity | neutrophil | *FCGRT* | 2.124768209 | 0.004960869 | 0.011289644 | 1 |
| Obesity | neutrophil | *PEX11G* | -1.83189662 | 0.006053979 | 0.01145004 | 1 |
| Obesity | neutrophil | *ERCC2* | 2.10513323 | 0.004000532 | 0.011289644 | 1 |
| Obesity | neutrophil | *AMH* | 8.18231676 | 0.008684745 | 0.013539098 | 1 |
| Obesity | neutrophil | *DKKL1* | 4.519523397 | 0.032182551 | 0.039821236 | 1 |
| Obesity | neutrophil | *STX10* | 2.24794127 | 0.004155193 | 0.011289644 | 1 |
| Obesity | neutrophil | *FCER2* | 2.810850573 | 0.004497698 | 0.011289644 | 1 |
| Obesity | neutrophil | *DMPK* | 4.030557402 | 0.015183702 | 0.020582134 | 1 |
| Obesity | neutrophil | *IL4I1* | 2.458787944 | 0.004218876 | 0.011289644 | 1 |
| Obesity | neutrophil | *TLE6* | -0.686405377 | 0.003861112 | 0.011289644 | 1 |
| Obesity | neutrophil | *AES* | 1.194445877 | 0.004043189 | 0.011289644 | 1 |
| Obesity | neutrophil | *SGTA* | 4.628649345 | 0.003475371 | 0.011289644 | 1 |
| Obesity | neutrophil | *C19orf53* | 5.981258641 | 0.022298261 | 0.028633408 | 1 |
| Obesity | neutrophil | *TIMM44* | 1.302909521 | 0.004096401 | 0.011289644 | 1 |
| Obesity | neutrophil | *CCDC61* | 3.623624792 | 0.021856078 | 0.028117283 | 1 |
| Obesity | neutrophil | *IL27RA* | 2.063476879 | 0.0021408 | 0.011289644 | 1 |
| Obesity | neutrophil | *C19orf44* | 1.575492096 | 0.00328108 | 0.011289644 | 1 |
| Obesity | neutrophil | *RASAL3* | -1.167074402 | 0.009901227 | 0.014768449 | 1 |
| Obesity | neutrophil | *ZNF419* | 2.176426419 | 0.004324648 | 0.011289644 | 1 |
| Obesity | neutrophil | *SYDE1* | 2.131317213 | 0.00868595 | 0.013539098 | 1 |
| Obesity | neutrophil | *AURKC* | 4.311469986 | 0.006271773 | 0.011565541 | 1 |
| Obesity | neutrophil | *CCNE1* | 3.554710233 | 0.007979099 | 0.012800355 | 1 |
| Obesity | neutrophil | *ANKRD27* | 3.120151873 | 0.008049477 | 0.012874994 | 1 |
| Obesity | neutrophil | *RPS16* | 1.36550076 | 0.005826283 | 0.011378838 | 1 |
| Obesity | neutrophil | *PRX* | -2.946872201 | 0.016799326 | 0.022395539 | 1 |
| Obesity | neutrophil | *CCDC94* | 1.864660524 | 0.003325306 | 0.011289644 | 1 |
| Obesity | neutrophil | *TBCB* | -1.327729208 | 0.007879501 | 0.012741363 | 1 |
| Obesity | neutrophil | *POLR2I* | 2.146565999 | 0.004649447 | 0.011289644 | 1 |
| Obesity | neutrophil | *CLIP3* | 2.094065917 | 0.001197869 | 0.011289644 | 1 |
| Obesity | neutrophil | *PRKD2* | 2.33600822 | 0.015393084 | 0.020821067 | 1 |
| Obesity | neutrophil | *CACTIN* | 3.089308189 | 0.00428243 | 0.011289644 | 1 |
| Obesity | neutrophil | *CCDC9* | 1.700185849 | 0.004950166 | 0.011289644 | 1 |
| Obesity | neutrophil | *FZR1* | 1.786326231 | 0.003841378 | 0.011289644 | 1 |
| Obesity | neutrophil | *BBC3* | -1.761870654 | 0.007246797 | 0.01221029 | 1 |
| Obesity | neutrophil | *TGFB1* | 1.891994683 | 0.005810081 | 0.011378838 | 1 |
| Obesity | neutrophil | *CEACAM4* | -1.55104212 | 0.005367812 | 0.011289644 | 1 |
| Obesity | neutrophil | *PLIN3* | 4.144026794 | 0.00483293 | 0.011289644 | 1 |
| Obesity | neutrophil | *CD33* | 3.012526 | 0.002795342 | 0.011289644 | 1 |
| Obesity | neutrophil | *BABAM1* | 1.848803126 | 0.004146814 | 0.011289644 | 1 |
| Obesity | neutrophil | *TYK2* | 3.200123967 | 0.003598085 | 0.011289644 | 1 |
| Obesity | neutrophil | *RABAC1* | 2.540309326 | 0.007669213 | 0.012523726 | 1 |
| Obesity | neutrophil | *PTPRS* | 1.130921491 | 0.002447719 | 0.011289644 | 1 |
| Obesity | neutrophil | *CYTH2* | 1.783579111 | 0.002856295 | 0.011289644 | 1 |
| Obesity | neutrophil | *GRWD1* | -5.618097275 | 0.002792408 | 0.011289644 | 1 |
| Obesity | neutrophil | *GRIN2D* | 2.161748552 | 0.004669195 | 0.011289644 | 1 |

| | | | | | | |
|---|---|---|---|---|---|---|
| Obesity | neutrophil | *CARD8* | 1.934141721 | 0.008759488 | 0.013611196 | 1 |
| Obesity | neutrophil | *LIG1* | 2.50087384 | 0.005593725 | 0.011295339 | 1 |
| Obesity | neutrophil | *ZNF175* | -4.694887178 | 0.005157727 | 0.011289644 | 1 |
| Obesity | neutrophil | *PLA2G4C* | 1.937114209 | 0.00185226 | 0.011289644 | 1 |
| Obesity | neutrophil | *BCAT2* | 1.915143338 | 0.004230352 | 0.011289644 | 1 |
| Obesity | neutrophil | *MIER2* | 3.442889409 | 0.008129663 | 0.012949841 | 1 |
| Obesity | neutrophil | *PPP2R1A* | 2.300235922 | 0.004274641 | 0.011289644 | 1 |
| Obesity | neutrophil | *TNPO2* | 1.363209942 | 0.005829832 | 0.011378838 | 1 |
| Obesity | neutrophil | *GCDH* | 1.329344835 | 0.008479681 | 0.013331272 | 1 |
| Obesity | neutrophil | *LILRB5* | 1.208979048 | 0.004274545 | 0.011289644 | 1 |
| Obesity | neutrophil | *MAST1* | 4.53640557 | 0.00430239 | 0.011289644 | 1 |
| Obesity | neutrophil | *PRPF31* | 2.995186811 | 0.012342002 | 0.017393895 | 1 |
| Obesity | neutrophil | *TFPT* | 3.556527764 | 0.005370629 | 0.011289644 | 1 |
| Obesity | neutrophil | *PDE4C* | 2.386745468 | 0.007976988 | 0.012800235 | 1 |
| Obesity | neutrophil | *ELL* | 2.566915963 | 0.028560549 | 0.035790347 | 1 |
| Obesity | neutrophil | *UPK1A* | -0.965533576 | 0.004278278 | 0.011289644 | 1 |
| Obesity | neutrophil | *COPE* | 1.48832089 | 0.004873326 | 0.011289644 | 1 |
| Obesity | neutrophil | *KXD1* | 3.417827311 | 0.005127612 | 0.011289644 | 1 |
| Obesity | neutrophil | *ZNF14* | 2.166498108 | 0.004361385 | 0.011289644 | 1 |
| Obesity | neutrophil | *ATP13A1* | -1.840314176 | 0.000911413 | 0.011289644 | 1 |
| Obesity | neutrophil | *ZNF85* | 1.529947717 | 0.003917573 | 0.011289644 | 1 |
| Obesity | neutrophil | *ETHE1* | 3.411344126 | 0.005109964 | 0.011289644 | 1 |
| Obesity | neutrophil | *GTPBP10* | 1.510792219 | 0.008547604 | 0.013397852 | 1 |
| Obesity | neutrophil | *PON3* | 2.158947901 | 0.003893898 | 0.011289644 | 1 |
| Obesity | neutrophil | *PON2* | 2.812446348 | 0.006358802 | 0.011618636 | 1 |
| Obesity | neutrophil | *HBP1* | 1.525534237 | 0.005005858 | 0.011289644 | 1 |
| Obesity | neutrophil | *DLX5* | -14.20170787 | 0.013567724 | 0.018755558 | 1 |
| Obesity | neutrophil | *STEAP1B* | 2.10414963 | 0.00400802 | 0.011289644 | 1 |
| Obesity | neutrophil | *PTN* | -1.814381546 | 0.007890273 | 0.01274567 | 1 |
| Obesity | neutrophil | *MPP6* | 1.099596972 | 0.004237865 | 0.011289644 | 1 |
| Obesity | neutrophil | *DFNA5* | 2.035155472 | 0.003305816 | 0.011289644 | 1 |
| Obesity | neutrophil | *OGDH* | 1.774654296 | 0.004641841 | 0.011289644 | 1 |
| Obesity | neutrophil | *ADAP1* | -1.59530968 | 0.005976011 | 0.011427806 | 1 |
| Obesity | neutrophil | *H2AFV* | 1.808316683 | 0.006074926 | 0.011459063 | 1 |
| Obesity | neutrophil | *MET* | 1.214027073 | 0.004154678 | 0.011289644 | 1 |
| Obesity | neutrophil | *LMBR1* | 2.256547844 | 0.006816253 | 0.011874479 | 1 |
| Obesity | neutrophil | *HOXA2* | 4.405253704 | 0.017696603 | 0.023417921 | 1 |
| Obesity | neutrophil | *HOXA5* | 1.33892843 | 0.006422556 | 0.011648199 | 1 |
| Obesity | neutrophil | *HOXA6* | 1.416055364 | 0.007517423 | 0.01240055 | 1 |
| Obesity | neutrophil | *CPED1* | 0.983892471 | 0.004361811 | 0.011289644 | 1 |
| Obesity | neutrophil | *HIBADH* | 1.377286784 | 0.004155243 | 0.011289644 | 1 |
| Obesity | neutrophil | *CPVL* | 1.583367994 | 0.006688497 | 0.011792087 | 1 |
| Obesity | neutrophil | *ABHD11* | -1.696053008 | 0.024985319 | 0.03169546 | 1 |
| Obesity | neutrophil | *PLEKHA8* | 1.46072846 | 0.003269568 | 0.011289644 | 1 |
| Obesity | neutrophil | *NOD1* | 2.784334703 | 0.00731831 | 0.012259479 | 1 |
| Obesity | neutrophil | *CRHR2* | 11.12872797 | 0.00386222 | 0.011289644 | 1 |
| Obesity | neutrophil | *EPHB6* | 5.066246396 | 0.001628085 | 0.011289644 | 1 |
| Obesity | neutrophil | *CHCHD2* | 2.015298819 | 0.005316758 | 0.011289644 | 1 |
| Obesity | neutrophil | *NPTX2* | 69.15389949 | 0.020983901 | 0.027128477 | 1 |
| Obesity | neutrophil | *CYP3A5* | 1.297987157 | 0.004256078 | 0.011289644 | 1 |

| | | | | | | |
|---|---|---|---|---|---|---|
| Obesity | neutrophil | *ZKSCAN1* | 2.723992803 | 0.011536151 | 0.016494833 | 1 |
| Obesity | neutrophil | *SNX8* | 1.653705749 | 0.004308854 | 0.011289644 | 1 |
| Obesity | neutrophil | *TAF6* | -7.356839773 | 0.013263652 | 0.018424344 | 1 |
| Obesity | neutrophil | *AIMP2* | 1.349214785 | 0.00455032 | 0.011289644 | 1 |
| Obesity | neutrophil | *TFR2* | 3.696463097 | 0.007373351 | 0.012294791 | 1 |
| Obesity | neutrophil | *RBM28* | 2.132679984 | 0.009796272 | 0.014656582 | 1 |
| Obesity | neutrophil | *USP42* | 3.848250312 | 0.01265718 | 0.017738674 | 1 |
| Obesity | neutrophil | *IMPDH1* | 1.650715712 | 0.004999849 | 0.011289644 | 1 |
| Obesity | neutrophil | *AP1S1* | 2.605554232 | 0.004170605 | 0.011289644 | 1 |
| Obesity | neutrophil | *C1GALT1* | 1.295678326 | 0.004072883 | 0.011289644 | 1 |
| Obesity | neutrophil | *PLOD3* | 1.605977201 | 0.005638362 | 0.011295339 | 1 |
| Obesity | neutrophil | *PHF14* | 1.837382151 | 0.00519872 | 0.011289644 | 1 |
| Obesity | neutrophil | *CEP41* | 2.878306549 | 0.003127368 | 0.011289644 | 1 |
| Obesity | neutrophil | *ZNF862* | 1.841339394 | 0.002440895 | 0.011289644 | 1 |
| Obesity | neutrophil | *ACTR3C* | 1.804103677 | 0.00435566 | 0.011289644 | 1 |
| Obesity | neutrophil | *POU6F2* | 1.433636598 | 0.004302606 | 0.011289644 | 1 |
| Obesity | neutrophil | *TSPAN13* | 7.050326553 | 0.039165307 | 0.047627135 | 1 |
| Obesity | neutrophil | *RARRES2* | 2.198115512 | 0.008792284 | 0.013641927 | 1 |
| Obesity | neutrophil | *TMEM176B* | 1.799804412 | 0.003413164 | 0.011289644 | 1 |
| Obesity | neutrophil | *MRPL32* | 2.520790971 | 0.002419788 | 0.011289644 | 1 |
| Obesity | neutrophil | *COA1* | 1.748380843 | 0.005047648 | 0.011289644 | 1 |
| Obesity | neutrophil | *BLVRA* | 2.561307818 | 0.001276333 | 0.011289644 | 1 |
| Obesity | neutrophil | *URGCP* | 3.797081537 | 0.038681812 | 0.047075601 | 1 |
| Obesity | neutrophil | *POLD2* | 4.092777202 | 0.011050965 | 0.015961637 | 1 |
| Obesity | neutrophil | *GCK* | -19.43498255 | 0.007255513 | 0.0122168 | 1 |
| Obesity | neutrophil | *BCL7B* | 1.473316538 | 0.009374293 | 0.014196251 | 1 |
| Obesity | neutrophil | *YKT6* | 2.27452676 | 0.004552512 | 0.011289644 | 1 |
| Obesity | neutrophil | *CLIP2* | 1.524487284 | 0.004272538 | 0.011289644 | 1 |
| Obesity | neutrophil | *LIMK1* | 1.592592361 | 0.004273818 | 0.011289644 | 1 |
| Obesity | neutrophil | *SPATA6L* | 2.5730608 | 0.027036688 | 0.034043593 | 1 |
| Obesity | neutrophil | *SLC1A1* | 1.77212545 | 0.002077489 | 0.011289644 | 1 |
| Obesity | neutrophil | *FKTN* | 2.244061395 | 0.0048058 | 0.011289644 | 1 |
| Obesity | neutrophil | *NMRK1* | 1.579770216 | 0.0031494 | 0.011289644 | 1 |
| Obesity | neutrophil | *TMEM245* | 2.294824832 | 0.00699608 | 0.012000111 | 1 |
| Obesity | neutrophil | *MEGF9* | 1.742832412 | 0.010644625 | 0.015545356 | 1 |
| Obesity | neutrophil | *TRIM14* | 1.513310652 | 0.003623439 | 0.011289644 | 1 |
| Obesity | neutrophil | *CORO2A* | 2.545612742 | 0.004693614 | 0.011289644 | 1 |
| Obesity | neutrophil | *C5* | 2.420743943 | 0.003759416 | 0.011289644 | 1 |
| Obesity | neutrophil | *OGN* | 1.694451814 | 0.001209715 | 0.011289644 | 1 |
| Obesity | neutrophil | *ECM2* | 1.158993572 | 0.003859898 | 0.011289644 | 1 |
| Obesity | neutrophil | *PTGR1* | 2.448476101 | 0.003211007 | 0.011289644 | 1 |
| Obesity | neutrophil | *TNFSF8* | -3.292617361 | 0.028004666 | 0.035149797 | 1 |
| Obesity | neutrophil | *AK1* | 2.507027706 | 0.002492875 | 0.011289644 | 1 |
| Obesity | neutrophil | *PLGRKT* | -3.666943377 | 0.034556438 | 0.042540367 | 1 |
| Obesity | neutrophil | *KDM4C* | 1.676404587 | 0.004841317 | 0.011289644 | 1 |
| Obesity | neutrophil | *ELAVL2* | 2.623464373 | 0.037117817 | 0.045391891 | 1 |
| Obesity | neutrophil | *FUBP3* | 3.128292069 | 0.00823883 | 0.013066984 | 1 |
| Obesity | neutrophil | *MPDZ* | 2.860292036 | 0.010547363 | 0.01543255 | 1 |
| Obesity | neutrophil | *PTGDS* | -1.28585098 | 0.006465386 | 0.01166174 | 1 |
| Obesity | neutrophil | *SHB* | 5.297798871 | 0.003069795 | 0.011289644 | 1 |

| | | | | | | |
|---|---|---|---|---|---|---|
| Obesity | neutrophil | *DVL1* | 1.72786752 | 0.004764531 | 0.011289644 | 1 |
| Obesity | neutrophil | *PDLIM1* | 3.181830804 | 0.003534814 | 0.011289644 | 1 |
| Obesity | neutrophil | *CCNJ* | 1.549624038 | 0.004869457 | 0.011289644 | 1 |
| Obesity | neutrophil | *HPS1* | 2.419663285 | 0.004373491 | 0.011289644 | 1 |
| Obesity | neutrophil | *PHYH* | 1.370627005 | 0.00186759 | 0.011289644 | 1 |
| Obesity | neutrophil | *DNMBP* | 1.58556429 | 0.004371015 | 0.011289644 | 1 |
| Obesity | neutrophil | *TRDMT1* | 2.704641647 | 0.003026964 | 0.011289644 | 1 |
| Obesity | neutrophil | *DDX50* | 1.946515326 | 0.004196555 | 0.011289644 | 1 |
| Obesity | neutrophil | *MAPK8* | 1.68599003 | 0.004582256 | 0.011289644 | 1 |
| Obesity | neutrophil | *ATE1* | 1.417131342 | 0.003100664 | 0.011289644 | 1 |
| Obesity | neutrophil | *PLEKHA1* | 1.188418043 | 0.005022699 | 0.011289644 | 1 |
| Obesity | neutrophil | *SPOCK2* | -0.81791299 | 0.007937162 | 0.01278095 | 1 |
| Obesity | neutrophil | *MICU1* | 1.740069417 | 0.003171516 | 0.011289644 | 1 |
| Obesity | neutrophil | *CCSER2* | 2.015726366 | 0.003847141 | 0.011289644 | 1 |
| Obesity | neutrophil | *BMPR1A* | 1.370527048 | 0.003586068 | 0.011289644 | 1 |
| Obesity | neutrophil | *LIPA* | 1.55770214 | 0.00305543 | 0.011289644 | 1 |
| Obesity | neutrophil | *SFXN3* | 3.364506051 | 0.002679237 | 0.011289644 | 1 |
| Obesity | neutrophil | *KAZALD1* | 4.220600695 | 0.007839381 | 0.012699648 | 1 |
| Obesity | neutrophil | *TNKS2* | 1.395764046 | 0.005144783 | 0.011289644 | 1 |
| Obesity | neutrophil | *ANKRD26* | 3.093565213 | 0.005514258 | 0.011290116 | 1 |
| Obesity | neutrophil | *ACBD5* | 2.212263696 | 0.001867628 | 0.011289644 | 1 |
| Obesity | neutrophil | *MTPAP* | 1.699721363 | 0.003291043 | 0.011289644 | 1 |
| Obesity | neutrophil | *MAP3K8* | 2.246238431 | 0.005269949 | 0.011289644 | 1 |
| Obesity | neutrophil | *SHOC2* | 1.877199935 | 0.004976633 | 0.011289644 | 1 |
| Obesity | neutrophil | *RPL28* | 1.80584627 | 0.00328516 | 0.011289644 | 1 |
| Obesity | neutrophil | *PBLD* | 1.359945908 | 0.003164456 | 0.011289644 | 1 |
| Obesity | neutrophil | *TADA2A* | 1.048319892 | 0.040648073 | 0.049249265 | 1 |
| Obesity | neutrophil | *DHRS11* | 1.376578721 | 0.0404169 | 0.048988057 | 1 |
| Obesity | neutrophil | *MLLT6* | 2.240957259 | 0.004670379 | 0.011289644 | 1 |
| Obesity | neutrophil | *PSMB3* | 1.810479375 | 0.004172011 | 0.011289644 | 1 |
| Obesity | neutrophil | *CASC3* | 1.140517068 | 0.005466729 | 0.011289644 | 1 |
| Obesity | neutrophil | *RAPGEFL1* | 2.447781592 | 0.024212556 | 0.030777262 | 1 |
| Obesity | neutrophil | *RGS9* | 4.220080184 | 0.00474419 | 0.011289644 | 1 |
| Obesity | neutrophil | *WNT3* | -1.046113547 | 0.006258927 | 0.011559892 | 1 |
| Obesity | neutrophil | *ASPA* | 1.068601665 | 0.004288542 | 0.011289644 | 1 |
| Obesity | neutrophil | *SEPT4* | 1.905658448 | 0.023110824 | 0.029490105 | 1 |
| Obesity | neutrophil | *TRIM37* | 1.088957044 | 0.004137061 | 0.011289644 | 1 |
| Obesity | neutrophil | *PNPO* | -1.751460345 | 0.006145009 | 0.011504964 | 1 |
| Obesity | neutrophil | *RPS6KB1* | 1.936261471 | 0.004677514 | 0.011289644 | 1 |
| Obesity | neutrophil | *CDK5RAP3* | 2.253600212 | 0.002605252 | 0.011289644 | 1 |
| Obesity | neutrophil | *CBX1* | 2.268570616 | 0.001221452 | 0.011289644 | 1 |
| Obesity | neutrophil | *PIGL* | 1.467778956 | 0.004321107 | 0.011289644 | 1 |
| Obesity | neutrophil | *GALK1* | 2.209788127 | 0.004225936 | 0.011289644 | 1 |
| Obesity | neutrophil | *CAMTA2* | 2.252674042 | 0.00391055 | 0.011289644 | 1 |
| Obesity | neutrophil | *MED13* | 1.287215515 | 0.007877507 | 0.012741363 | 1 |
| Obesity | neutrophil | *HOXB6* | 1.860821105 | 0.001111687 | 0.011289644 | 1 |
| Obesity | neutrophil | *RNF167* | 2.583456852 | 0.004085932 | 0.011289644 | 1 |
| Obesity | neutrophil | *CHRNE* | 5.558210415 | 1.85E-05 | 0.003363761 | 0.116275571 |
| Obesity | neutrophil | *NUP88* | 1.381475436 | 0.010985211 | 0.015902376 | 1 |
| Obesity | neutrophil | *GOSR1* | 2.764515484 | 0.006007805 | 0.011438846 | 1 |

| | | | | | | |
|---|---|---|---|---|---|---|
| Obesity | neutrophil | *DRG2* | -3.054125095 | 0.021864761 | 0.028122699 | 1 |
| Obesity | neutrophil | *AKAP10* | 1.565201155 | 0.005398458 | 0.011289644 | 1 |
| Obesity | neutrophil | *ALDH3A1* | -1.210177331 | 0.00585737 | 0.011378838 | 1 |
| Obesity | neutrophil | *SMARCD2* | 1.691491834 | 0.005237971 | 0.011289644 | 1 |
| Obesity | neutrophil | *ICAM2* | 4.482921151 | 0.030185489 | 0.037624142 | 1 |
| Obesity | neutrophil | *UTP6* | 2.333596667 | 0.004038918 | 0.011289644 | 1 |
| Obesity | neutrophil | *DDX5* | 1.619202868 | 0.007163399 | 0.012141606 | 1 |
| Obesity | neutrophil | *CYTH1* | 2.609960638 | 0.010264905 | 0.01511723 | 1 |
| Obesity | neutrophil | *PEX12* | 3.179961908 | 0.018832334 | 0.024707613 | 1 |
| Obesity | neutrophil | *DHX58* | 2.117370598 | 0.002207432 | 0.011289644 | 1 |
| Obesity | neutrophil | *RAB5C* | 2.649842478 | 0.012122196 | 0.017157191 | 1 |
| Obesity | neutrophil | *NAGLU* | 2.082043261 | 0.004489286 | 0.011289644 | 1 |
| Obesity | neutrophil | *EZH1* | 1.850650916 | 0.00380867 | 0.011289644 | 1 |
| Obesity | neutrophil | *DLX4* | 19.57329582 | 0.004503333 | 0.011289644 | 1 |
| Obesity | neutrophil | *VAT1* | -2.202608223 | 0.029620239 | 0.037007639 | 1 |
| Obesity | neutrophil | *LRRC59* | 3.617494411 | 0.004688329 | 0.011289644 | 1 |
| Obesity | neutrophil | *ALOX12* | -1.646576947 | 0.005223917 | 0.011289644 | 1 |
| Obesity | neutrophil | *ABCC3* | 1.423991947 | 0.001552491 | 0.011289644 | 1 |
| Obesity | neutrophil | *MPP2* | 2.990359794 | 0.004051861 | 0.011289644 | 1 |
| Obesity | neutrophil | *EFTUD2* | 1.20005322 | 0.003182695 | 0.011289644 | 1 |
| Obesity | neutrophil | *EFNB3* | 31.08799089 | 0.001163318 | 0.011289644 | 1 |
| Obesity | neutrophil | *FAM20A* | 1.882171583 | 0.004572245 | 0.011289644 | 1 |
| Obesity | neutrophil | *YWHAE* | 1.768346673 | 0.004657403 | 0.011289644 | 1 |
| Obesity | neutrophil | *DPH1* | 2.037940066 | 0.003997828 | 0.011289644 | 1 |
| Obesity | neutrophil | *DHRS7B* | 2.468345432 | 0.005613351 | 0.011295339 | 1 |
| Obesity | neutrophil | *MYH3* | -4.22234463 | 0.01702925 | 0.022644436 | 1 |
| Obesity | neutrophil | *TMEM104* | 1.516409911 | 0.004133034 | 0.011289644 | 1 |
| Obesity | neutrophil | *TMEM97* | 4.079924526 | 0.00174266 | 0.011289644 | 1 |
| Obesity | neutrophil | *RAB34* | 1.857518934 | 0.006033959 | 0.011447874 | 1 |
| Obesity | neutrophil | *SLAIN2* | 1.811901013 | 0.003118984 | 0.011289644 | 1 |
| Obesity | neutrophil | *OCIAD1* | 1.702243519 | 0.006175291 | 0.011512519 | 1 |
| Obesity | neutrophil | *CWH43* | 17.04600342 | 0.009958652 | 0.014818298 | 1 |
| Obesity | neutrophil | *KIAA1211* | 2.334376369 | 0.000468988 | 0.011289644 | 1 |
| Obesity | neutrophil | *AREG* | 2.759456988 | 0.031056647 | 0.038625745 | 1 |
| Obesity | neutrophil | *UBE2D3* | 1.393327712 | 0.003856538 | 0.011289644 | 1 |
| Obesity | neutrophil | *NDUFC1* | 1.4245729 | 0.004477478 | 0.011289644 | 1 |
| Obesity | neutrophil | *TBC1D9* | 2.077408242 | 0.005065241 | 0.011289644 | 1 |
| Obesity | neutrophil | *CPE* | 2.888354369 | 0.011127418 | 0.016036425 | 1 |
| Obesity | neutrophil | *WFS1* | 3.225664558 | 0.003285483 | 0.011289644 | 1 |
| Obesity | neutrophil | *GRPEL1* | 2.761075981 | 0.003634457 | 0.011289644 | 1 |
| Obesity | neutrophil | *AADAT* | 2.01780594 | 0.004472868 | 0.011289644 | 1 |
| Obesity | neutrophil | *CPZ* | -5.644989369 | 0.005297214 | 0.011289644 | 1 |
| Obesity | neutrophil | *TRIM2* | 1.613849894 | 0.009873036 | 0.01473538 | 1 |
| Obesity | neutrophil | *SLC2A9* | 2.398381657 | 0.004724509 | 0.011289644 | 1 |
| Obesity | neutrophil | *SH3D19* | 1.270258239 | 0.004841555 | 0.011289644 | 1 |
| Obesity | neutrophil | *STIM2* | 1.48881216 | 0.003305209 | 0.011289644 | 1 |
| Obesity | neutrophil | *RAPGEF2* | 1.89799336 | 0.005356148 | 0.011289644 | 1 |
| Obesity | neutrophil | *UFSP2* | 1.904750477 | 0.004063114 | 0.011289644 | 1 |
| Obesity | neutrophil | *KLF3* | 1.51086556 | 0.004308299 | 0.011289644 | 1 |
| Obesity | neutrophil | *DDX25* | 7.484822976 | 0.007364668 | 0.012289299 | 1 |

| | | | | | | |
|---|---|---|---|---|---|---|
| Obesity | neutrophil | *HTATIP2* | 1.764555143 | 0.004452248 | 0.011289644 | 1 |
| Obesity | neutrophil | *CTSC* | 1.855483424 | 0.009412542 | 0.014231998 | 1 |
| Obesity | neutrophil | *ELP4* | 2.237705108 | 0.00730128 | 0.012237576 | 1 |
| Obesity | neutrophil | *SC5D* | 1.521859832 | 0.003976518 | 0.011289644 | 1 |
| Obesity | neutrophil | *C11orf63* | 1.377196597 | 0.003963453 | 0.011289644 | 1 |
| Obesity | neutrophil | *DTX4* | 1.612645989 | 0.004830219 | 0.011289644 | 1 |
| Obesity | neutrophil | *DCPS* | 2.059145266 | 0.007332458 | 0.012273472 | 1 |
| Obesity | neutrophil | *FOXRED1* | 2.027548414 | 0.000979758 | 0.011289644 | 1 |
| Obesity | neutrophil | *NRXN2* | 2.936321796 | 0.004396315 | 0.011289644 | 1 |
| Obesity | neutrophil | *MS4A4A* | 1.307496902 | 0.002729235 | 0.011289644 | 1 |
| Obesity | neutrophil | *CCDC86* | 4.734629173 | 0.004349086 | 0.011289644 | 1 |
| Obesity | neutrophil | *PRPF19* | 1.731425082 | 0.005440479 | 0.011289644 | 1 |
| Obesity | neutrophil | *CHORDC1* | 1.322697137 | 0.004897082 | 0.011289644 | 1 |
| Obesity | neutrophil | *EIF4G2* | 2.391591738 | 0.005483109 | 0.011289644 | 1 |
| Obesity | neutrophil | *GALNT18* | 1.63918469 | 0.020627455 | 0.02674465 | 1 |
| Obesity | neutrophil | *KIAA1549L* | 3.021877967 | 0.004858752 | 0.011289644 | 1 |
| Obesity | neutrophil | *CD5* | 3.236554063 | 0.000993215 | 0.011289644 | 1 |
| Obesity | neutrophil | *ACCS* | 1.909659524 | 0.003688478 | 0.011289644 | 1 |
| Obesity | neutrophil | *MADD* | 1.830657555 | 0.002648537 | 0.011289644 | 1 |
| Obesity | neutrophil | *NAA40* | -1.337758981 | 0.007560783 | 0.012433859 | 1 |
| Obesity | neutrophil | *CARS* | 1.902131849 | 0.014507219 | 0.019815536 | 1 |
| Obesity | neutrophil | *SLC22A18* | 2.23499477 | 0.000630094 | 0.011289644 | 1 |
| Obesity | neutrophil | *SLC35F2* | -1.475003939 | 0.003581814 | 0.011289644 | 1 |
| Obesity | neutrophil | *C11orf21* | 2.975329435 | 0.039357046 | 0.047823299 | 1 |
| Obesity | neutrophil | *ELMOD1* | -0.990862114 | 0.005742355 | 0.01134195 | 1 |
| Obesity | neutrophil | *RPS13* | 1.429381067 | 0.006418228 | 0.011648199 | 1 |
| Obesity | neutrophil | *AIP* | 4.739733535 | 0.003839595 | 0.011289644 | 1 |
| Obesity | neutrophil | *NDUFS8* | 2.100698531 | 0.004294151 | 0.011289644 | 1 |
| Obesity | neutrophil | *TCIRG1* | 5.194566731 | 0.00822723 | 0.013055173 | 1 |
| Obesity | neutrophil | *GTF2H1* | 1.753962639 | 0.002244973 | 0.011289644 | 1 |
| Obesity | neutrophil | *PPFIBP1* | 1.083650285 | 0.005771644 | 0.011357223 | 1 |
| Obesity | neutrophil | *CAPRIN2* | 2.959723995 | 0.009035384 | 0.013848236 | 1 |
| Obesity | neutrophil | *CYP27B1* | -18.55902164 | 0.014008015 | 0.019237307 | 1 |
| Obesity | neutrophil | *ACSS3* | 0.860003208 | 0.003862489 | 0.011289644 | 1 |
| Obesity | neutrophil | *GLI1* | 3.432400646 | 0.004327983 | 0.011289644 | 1 |
| Obesity | neutrophil | *METAP2* | 1.279889147 | 0.004568981 | 0.011289644 | 1 |
| Obesity | neutrophil | *LTA4H* | 3.221174072 | 0.00699076 | 0.011998609 | 1 |
| Obesity | neutrophil | *WNT5B* | 1.376508594 | 0.004399263 | 0.011289644 | 1 |
| Obesity | neutrophil | *TRPV4* | 1.796715463 | 0.024292815 | 0.03087304 | 1 |
| Obesity | neutrophil | *ITFG2* | 2.340426031 | 0.006735179 | 0.011825602 | 1 |
| Obesity | neutrophil | *PRR4* | 2.366358795 | 0.003670544 | 0.011289644 | 1 |
| Obesity | neutrophil | *PARP11* | 2.127770444 | 0.005441805 | 0.011289644 | 1 |
| Obesity | neutrophil | *ARPC3* | 1.834067449 | 0.005166622 | 0.011289644 | 1 |
| Obesity | neutrophil | *GPN3* | 1.590293363 | 0.004678775 | 0.011289644 | 1 |
| Obesity | neutrophil | *VPS29* | 2.533409177 | 0.003836957 | 0.011289644 | 1 |
| Obesity | neutrophil | *AKAP3* | -1.664899434 | 0.012594519 | 0.017681329 | 1 |
| Obesity | neutrophil | *KCNA1* | 5.682166039 | 0.024207849 | 0.030777262 | 1 |
| Obesity | neutrophil | *NAA25* | 2.065662551 | 0.00327677 | 0.011289644 | 1 |
| Obesity | neutrophil | *SCNN1A* | 2.997870575 | 0.00388632 | 0.011289644 | 1 |
| Obesity | neutrophil | *LTBR* | 4.377044265 | 0.002614565 | 0.011289644 | 1 |

| | | | | | | |
|---|---|---|---|---|---|---|
| Obesity | neutrophil | *CDK2AP1* | 3.692615068 | 0.007758815 | 0.012633947 | 1 |
| Obesity | neutrophil | *MGP* | 0.798041094 | 0.00481925 | 0.011289644 | 1 |
| Obesity | neutrophil | *EIF2B1* | 1.304166288 | 0.004218968 | 0.011289644 | 1 |
| Obesity | neutrophil | *DDX55* | 5.069675583 | 6.67E-05 | 0.00802313 | 0.419053479 |
| Obesity | neutrophil | *ENDOU* | 3.101588021 | 0.028975875 | 0.036274657 | 1 |
| Obesity | neutrophil | *STX2* | 1.329377101 | 0.004069495 | 0.011289644 | 1 |
| Obesity | neutrophil | *CAND1* | 2.585385911 | 0.009798148 | 0.014656582 | 1 |
| Obesity | neutrophil | *MDM1* | 1.193404442 | 0.006892355 | 0.011932455 | 1 |
| Obesity | neutrophil | *NUP107* | 1.841249291 | 0.003082396 | 0.011289644 | 1 |
| Obesity | neutrophil | *CNOT2* | 2.066760426 | 0.006486326 | 0.011671674 | 1 |
| Obesity | neutrophil | *KRR1* | 1.831366506 | 0.006234362 | 0.01154827 | 1 |
| Obesity | neutrophil | *MRPL51* | 1.730219286 | 0.003052129 | 0.011289644 | 1 |
| Obesity | neutrophil | *COPS7A* | 3.263987358 | 0.010122484 | 0.014963568 | 1 |
| Obesity | neutrophil | *GNB3* | -0.500595447 | 0.008825345 | 0.01366961 | 1 |
| Obesity | neutrophil | *CHPT1* | 1.662204838 | 0.016811257 | 0.022401944 | 1 |
| Obesity | neutrophil | *GNPTAB* | 2.17081057 | 0.004453278 | 0.011289644 | 1 |
| Obesity | neutrophil | *SPSB2* | -2.495930177 | 0.02004267 | 0.02607787 | 1 |
| Obesity | neutrophil | *ATN1* | 2.966630966 | 0.005050883 | 0.011289644 | 1 |
| Obesity | neutrophil | *C12orf57* | -1.132952652 | 0.005687387 | 0.011300433 | 1 |
| Obesity | neutrophil | *ST8SIA1* | 3.572337004 | 0.032159489 | 0.039821236 | 1 |
| Obesity | neutrophil | *C2CD5* | 2.284719846 | 0.005816578 | 0.011378838 | 1 |
| Obesity | neutrophil | *PHC1* | 3.899839564 | 0.005198101 | 0.011289644 | 1 |
| Obesity | neutrophil | *COL12A1* | 1.604628242 | 0.004855439 | 0.011289644 | 1 |
| Obesity | neutrophil | *RWDD1* | 3.184904385 | 0.007339149 | 0.012278135 | 1 |
| Obesity | neutrophil | *RSPH4A* | 2.343898419 | 0.036843429 | 0.045091422 | 1 |
| Obesity | neutrophil | *SMIM8* | 1.268804382 | 0.00308696 | 0.011289644 | 1 |
| Obesity | neutrophil | *MCM9* | 2.401571356 | 0.005283795 | 0.011289644 | 1 |
| Obesity | neutrophil | *RNGTT* | 1.991487632 | 0.004692147 | 0.011289644 | 1 |
| Obesity | neutrophil | *GABRR2* | 3.665945349 | 0.000714493 | 0.011289644 | 1 |
| Obesity | neutrophil | *TPD52L1* | 1.379656961 | 0.003720594 | 0.011289644 | 1 |
| Obesity | neutrophil | *UST* | 1.160638242 | 0.004096513 | 0.011289644 | 1 |
| Obesity | neutrophil | *TULP1* | -3.708524723 | 0.005834153 | 0.011378838 | 1 |
| Obesity | neutrophil | *MAPK14* | 1.579836873 | 0.006420344 | 0.011648199 | 1 |
| Obesity | neutrophil | *MRPL18* | 2.054539332 | 0.003361957 | 0.011289644 | 1 |
| Obesity | neutrophil | *RNF8* | 1.856622876 | 0.003404454 | 0.011289644 | 1 |
| Obesity | neutrophil | *PHACTR1* | 2.153524181 | 0.006328504 | 0.011606566 | 1 |
| Obesity | neutrophil | *MDGA1* | 6.556385787 | 0.026943276 | 0.033939566 | 1 |
| Obesity | neutrophil | *CD83* | 1.079170871 | 0.006174304 | 0.011512519 | 1 |
| Obesity | neutrophil | *MDN1* | 1.774303589 | 0.006033413 | 0.011447874 | 1 |
| Obesity | neutrophil | *SAYSD1* | 4.477391313 | 0.005545433 | 0.011295339 | 1 |
| Obesity | neutrophil | *CAP2* | 0.894693944 | 0.013208191 | 0.018379791 | 1 |
| Obesity | neutrophil | *GPR63* | 3.746209747 | 0.000847433 | 0.011289644 | 1 |
| Obesity | neutrophil | *ASCC3* | 1.745915517 | 0.004671744 | 0.011289644 | 1 |
| Obesity | neutrophil | *WASF1* | 1.297414304 | 0.002435936 | 0.011289644 | 1 |
| Obesity | neutrophil | *GPLD1* | 1.847230156 | 0.004524632 | 0.011289644 | 1 |
| Obesity | neutrophil | *ALDH5A1* | 3.078107035 | 0.004303833 | 0.011289644 | 1 |
| Obesity | neutrophil | *VNN1* | 1.812228328 | 0.00864878 | 0.013505931 | 1 |
| Obesity | neutrophil | *VNN2* | -0.370027367 | 0.005865928 | 0.011378838 | 1 |
| Obesity | neutrophil | *C6orf62* | 1.342627874 | 0.003744988 | 0.011289644 | 1 |
| Obesity | neutrophil | *B3GAT2* | 1.80309518 | 0.009670595 | 0.014510509 | 1 |

| | | | | | | |
|---|---|---|---|---|---|---|
| Obesity | neutrophil | *HBS1L* | 1.877520104 | 0.004099104 | 0.011289644 | 1 |
| Obesity | neutrophil | *PERP* | 2.232033922 | 0.009444024 | 0.014259024 | 1 |
| Obesity | neutrophil | *SLC16A10* | 2.43703167 | 0.004206058 | 0.011289644 | 1 |
| Obesity | neutrophil | *PHACTR2* | 1.136358007 | 0.004370861 | 0.011289644 | 1 |
| Obesity | neutrophil | *PACRG* | 2.0777916 | 0.003868524 | 0.011289644 | 1 |
| Obesity | neutrophil | *TFEB* | 1.820377481 | 0.004215385 | 0.011289644 | 1 |
| Obesity | neutrophil | *FAM120B* | 2.880245529 | 0.018147476 | 0.023933959 | 1 |
| Obesity | neutrophil | *TBP* | 1.342758296 | 0.004730251 | 0.011289644 | 1 |
| Obesity | neutrophil | *GUCA1B* | 2.337776321 | 0.005872619 | 0.011378838 | 1 |
| Obesity | neutrophil | *PRPH2* | 2.102165678 | 0.019971937 | 0.026001982 | 1 |
| Obesity | neutrophil | *DUSP22* | 3.154125015 | 0.007544241 | 0.012419095 | 1 |
| Obesity | neutrophil | *EXOC2* | 2.632225742 | 0.007672317 | 0.01252554 | 1 |
| Obesity | neutrophil | *TMEM30A* | 1.548846961 | 0.00546741 | 0.011289644 | 1 |
| Obesity | neutrophil | *SENP6* | 1.304888543 | 0.004326724 | 0.011289644 | 1 |
| Obesity | neutrophil | *TTK* | 4.719143448 | 0.016740077 | 0.022326021 | 1 |
| Obesity | neutrophil | *LAMA4* | 0.879653385 | 0.001890124 | 0.011289644 | 1 |
| Obesity | neutrophil | *FBRSL1* | 2.184748495 | 0.00347949 | 0.011289644 | 1 |
| Obesity | neutrophil | *LY86* | 1.588691159 | 0.003952384 | 0.011289644 | 1 |
| Obesity | neutrophil | *PRSS16* | 28.14675939 | 3.18E-09 | 2.50E-06 | 2.00E-05 |
| Obesity | neutrophil | *TBX18* | 1.107802895 | 0.007476332 | 0.012375909 | 1 |
| Obesity | neutrophil | *CEP72* | 5.211332128 | 0.012201896 | 0.017242824 | 1 |
| Obesity | neutrophil | *C7* | 1.346887212 | 0.005037506 | 0.011289644 | 1 |
| Obesity | neutrophil | *PAPD7* | 1.375721892 | 0.005062999 | 0.011289644 | 1 |
| Obesity | neutrophil | *GHR* | 0.984423688 | 0.002760353 | 0.011289644 | 1 |
| Obesity | neutrophil | *DAP* | 2.849952892 | 0.003507365 | 0.011289644 | 1 |
| Obesity | neutrophil | *NME5* | 6.322877699 | 1.87E-05 | 0.003363761 | 0.11773162 |
| Obesity | neutrophil | *NNT* | 1.908201441 | 0.003068025 | 0.011289644 | 1 |
| Obesity | neutrophil | *PFDN1* | 1.750052716 | 0.001877373 | 0.011289644 | 1 |
| Obesity | neutrophil | *LOX* | 1.267157365 | 0.004265695 | 0.011289644 | 1 |
| Obesity | neutrophil | *APBB3* | -1.906258074 | 0.004372249 | 0.011289644 | 1 |
| Obesity | neutrophil | *TMCO6* | 2.838872318 | 0.009222981 | 0.014033018 | 1 |
| Obesity | neutrophil | *SPARC* | 1.893575235 | 0.007945336 | 0.01278207 | 1 |
| Obesity | neutrophil | *PDE8B* | 0.997435733 | 0.007346598 | 0.012284058 | 1 |
| Obesity | neutrophil | *CLK4* | 2.420936891 | 0.005812738 | 0.011378838 | 1 |
| Obesity | neutrophil | *HAVCR1* | 2.159653885 | 0.027165552 | 0.034192157 | 1 |
| Obesity | neutrophil | *RNF130* | 2.08092011 | 0.00595419 | 0.011409324 | 1 |
| Obesity | neutrophil | *ARSB* | 1.216321971 | 0.006797027 | 0.011870336 | 1 |
| Obesity | neutrophil | *CLINT1* | 3.151829143 | 0.025686514 | 0.032486645 | 1 |
| Obesity | neutrophil | *TTC1* | 1.893167436 | 0.004639514 | 0.011289644 | 1 |
| Obesity | neutrophil | *MSH3* | 1.299946765 | 0.014831871 | 0.020161723 | 1 |
| Obesity | neutrophil | *RASGRF2* | 1.165862761 | 0.003747946 | 0.011289644 | 1 |
| Obesity | neutrophil | *POLR3G* | -3.299533614 | 0.027102499 | 0.034119626 | 1 |
| Obesity | neutrophil | *DROSHA* | 1.629163641 | 0.003243713 | 0.011289644 | 1 |
| Obesity | neutrophil | *CDH6* | 1.146949785 | 0.008935874 | 0.013763799 | 1 |
| Obesity | neutrophil | *SLC27A6* | -1.810070728 | 0.006123597 | 0.011482393 | 1 |
| Obesity | neutrophil | *TARS* | 1.345541537 | 0.004222841 | 0.011289644 | 1 |
| Obesity | neutrophil | *LNPEP* | 1.146019433 | 0.003033667 | 0.011289644 | 1 |
| Obesity | neutrophil | *SLC12A7* | 1.387702729 | 0.005175115 | 0.011289644 | 1 |
| Obesity | neutrophil | *IL4* | -0.78365249 | 0.004479757 | 0.011289644 | 1 |
| Obesity | neutrophil | *GNPDA1* | 2.472774907 | 0.004317203 | 0.011289644 | 1 |

| | | | | | | |
|---|---|---|---|---|---|---|
| Obesity | neutrophil | *SKP1* | 1.340212084 | 0.00335967 | 0.011289644 | 1 |
| Obesity | neutrophil | *PPWD1* | 2.418689676 | 0.015195169 | 0.020593237 | 1 |
| Obesity | neutrophil | *TRIM23* | 2.356215937 | 0.005225526 | 0.011289644 | 1 |
| Obesity | neutrophil | *TRAPPC13* | 2.246999606 | 0.013102797 | 0.018254562 | 1 |
| Obesity | neutrophil | *C9* | 1.498355749 | 0.008953327 | 0.013786326 | 1 |
| Obesity | neutrophil | *TXNDC15* | 1.377869369 | 0.004520636 | 0.011289644 | 1 |
| Obesity | neutrophil | *SMAD5* | 2.855283788 | 0.009960574 | 0.014818298 | 1 |
| Obesity | neutrophil | *HMGXB3* | 2.94314133 | 0.007668405 | 0.012523726 | 1 |
| Obesity | neutrophil | *ATP6V0E1* | 3.111162049 | 0.022670948 | 0.029017152 | 1 |
| Obesity | neutrophil | *CNTN3* | 1.130975844 | 0.004269082 | 0.011289644 | 1 |
| Obesity | neutrophil | *ACTR8* | 2.310391801 | 0.005075254 | 0.011289644 | 1 |
| Obesity | neutrophil | *HGD* | 3.481215255 | 0.004047575 | 0.011289644 | 1 |
| Obesity | neutrophil | *NIT2* | 1.303065914 | 0.00420624 | 0.011289644 | 1 |
| Obesity | neutrophil | *FAM162A* | 3.489521106 | 0.008122142 | 0.012947238 | 1 |
| Obesity | neutrophil | *PCCB* | 3.624476766 | 0.003733106 | 0.011289644 | 1 |
| Obesity | neutrophil | *RBP1* | 4.26293641 | 0.002900062 | 0.011289644 | 1 |
| Obesity | neutrophil | *RNF7* | 3.598125066 | 0.010746451 | 0.015648165 | 1 |
| Obesity | neutrophil | *BCHE* | 1.873297153 | 0.038648147 | 0.047052847 | 1 |
| Obesity | neutrophil | *PDCD10* | 2.470169778 | 0.006399741 | 0.011648199 | 1 |
| Obesity | neutrophil | *WNT5A* | 2.640733112 | 0.008050199 | 0.012874994 | 1 |
| Obesity | neutrophil | *PRKAR2A* | 1.945790574 | 0.001089942 | 0.011289644 | 1 |
| Obesity | neutrophil | *CYB561D2* | 3.253219028 | 0.003008444 | 0.011289644 | 1 |
| Obesity | neutrophil | *IFT57* | 2.488911481 | 0.004890494 | 0.011289644 | 1 |
| Obesity | neutrophil | *IQCG* | 1.646048543 | 0.004292005 | 0.011289644 | 1 |
| Obesity | neutrophil | *GBE1* | 1.616024635 | 0.005097084 | 0.011289644 | 1 |
| Obesity | neutrophil | *UMPS* | -3.294985762 | 0.025458302 | 0.032256418 | 1 |
| Obesity | neutrophil | *NCBP2* | 1.379755315 | 0.003821294 | 0.011289644 | 1 |
| Obesity | neutrophil | *PLXNA1* | -1.254279044 | 0.020301899 | 0.026371496 | 1 |
| Obesity | neutrophil | *EFCC1* | 1.751810787 | 0.018994595 | 0.024894521 | 1 |
| Obesity | neutrophil | *NEK11* | 1.398813877 | 0.003994177 | 0.011289644 | 1 |
| Obesity | neutrophil | *PLSCR4* | 1.087185753 | 0.004972171 | 0.011289644 | 1 |
| Obesity | neutrophil | *HEMK1* | 1.736506469 | 0.002213767 | 0.011289644 | 1 |
| Obesity | neutrophil | *WDR48* | 2.575927934 | 0.003614708 | 0.011289644 | 1 |
| Obesity | neutrophil | *COMMD2* | 1.461924204 | 0.003366448 | 0.011289644 | 1 |
| Obesity | neutrophil | *GORASP1* | 1.559036261 | 0.004559724 | 0.011289644 | 1 |
| Obesity | neutrophil | *ABCC5* | 1.556473104 | 0.00581131 | 0.011378838 | 1 |
| Obesity | neutrophil | *ARHGEF26* | 1.215764638 | 0.002446921 | 0.011289644 | 1 |
| Obesity | neutrophil | *KLHL24* | 2.244499272 | 0.001108277 | 0.011289644 | 1 |
| Obesity | neutrophil | *PLCH1* | -1.373991605 | 0.01185267 | 0.016840166 | 1 |
| Obesity | neutrophil | *ZBTB47* | 2.563365769 | 0.002549786 | 0.011289644 | 1 |
| Obesity | neutrophil | *NKTR* | 1.585915358 | 0.003623196 | 0.011289644 | 1 |
| Obesity | neutrophil | *NEK4* | 1.72155768 | 0.005434721 | 0.011289644 | 1 |
| Obesity | neutrophil | *MOB1A* | 4.001369283 | 0.018756478 | 0.024613229 | 1 |
| Obesity | neutrophil | *LMAN2L* | 1.575141736 | 0.005679053 | 0.011300433 | 1 |
| Obesity | neutrophil | *PIKFYVE* | 2.62101796 | 0.009389398 | 0.014210675 | 1 |
| Obesity | neutrophil | *KCNIP3* | 2.910223382 | 0.003290067 | 0.011289644 | 1 |
| Obesity | neutrophil | *FAHD2A* | 3.803622479 | 0.00096065 | 0.011289644 | 1 |
| Obesity | neutrophil | *NCL* | 2.240231173 | 0.005163422 | 0.011289644 | 1 |
| Obesity | neutrophil | *ACTR1B* | 1.946845644 | 0.001942711 | 0.011289644 | 1 |
| Obesity | neutrophil | *SLC35F5* | 2.829631625 | 0.016090525 | 0.021630679 | 1 |

| | | | | | | |
|---|---|---|---|---|---|---|
| Obesity | neutrophil | *ACTR3* | 1.760482481 | 0.004479234 | 0.011289644 | 1 |
| Obesity | neutrophil | *TFCP2L1* | 3.200182753 | 0.001030226 | 0.011289644 | 1 |
| Obesity | neutrophil | *TP53I3* | 1.384108466 | 0.004398067 | 0.011289644 | 1 |
| Obesity | neutrophil | *DNAJC27* | 1.834888882 | 0.002295569 | 0.011289644 | 1 |
| Obesity | neutrophil | *STAM2* | 2.338279395 | 0.005419923 | 0.011289644 | 1 |
| Obesity | neutrophil | *GPD2* | 1.790296928 | 0.005690658 | 0.011300433 | 1 |
| Obesity | neutrophil | *CENPA* | 1.78528159 | 0.005980145 | 0.011427806 | 1 |
| Obesity | neutrophil | *TANC1* | 2.191899032 | 0.005245177 | 0.011289644 | 1 |
| Obesity | neutrophil | *NRBP1* | 1.678407027 | 0.005838761 | 0.011378838 | 1 |
| Obesity | neutrophil | *PCSK4* | 3.504654122 | 0.003960883 | 0.011289644 | 1 |
| Obesity | neutrophil | *INO80B* | 2.009272048 | 0.009156404 | 0.013958853 | 1 |
| Obesity | neutrophil | *MOGS* | 1.535018262 | 0.007775057 | 0.012646715 | 1 |
| Obesity | neutrophil | *GRB14* | 4.731948416 | 0.035994392 | 0.0441727 | 1 |
| Obesity | neutrophil | *CLIP4* | 2.417813502 | 0.005395233 | 0.011289644 | 1 |
| Obesity | neutrophil | *SPTBN1* | 1.179299047 | 0.00583333 | 0.011378838 | 1 |
| Obesity | neutrophil | *RTN4* | 1.327674205 | 0.014223486 | 0.01949064 | 1 |
| Obesity | neutrophil | *HTRA2* | 1.899642889 | 0.004745425 | 0.011289644 | 1 |
| Obesity | neutrophil | *GALNT3* | 1.623845093 | 0.006333549 | 0.011609044 | 1 |
| Obesity | neutrophil | *MRPL19* | 2.798321694 | 0.031540635 | 0.039157972 | 1 |
| Obesity | neutrophil | *LANCL1* | 1.456486691 | 0.003907847 | 0.011289644 | 1 |
| Obesity | neutrophil | *WDR75* | 1.50190298 | 0.003974193 | 0.011289644 | 1 |
| Obesity | neutrophil | *EFEMP1* | 1.045617799 | 0.003870562 | 0.011289644 | 1 |
| Obesity | neutrophil | *GLS* | 1.575643153 | 0.004733789 | 0.011289644 | 1 |
| Obesity | neutrophil | *DNAH6* | 1.066686637 | 0.002621573 | 0.011289644 | 1 |
| Obesity | neutrophil | *GGCX* | 3.372639423 | 0.000393414 | 0.011289644 | 1 |
| Obesity | neutrophil | *GNLY* | -1.368690368 | 0.008589882 | 0.013440663 | 1 |
| Obesity | neutrophil | *SF3B1* | 1.730328924 | 0.004982833 | 0.011289644 | 1 |
| Obesity | neutrophil | *ST3GAL5* | 2.925190953 | 0.010886809 | 0.015803594 | 1 |
| Obesity | neutrophil | *PLCD4* | 1.813387725 | 0.005788946 | 0.011372552 | 1 |
| Obesity | neutrophil | *ZNF142* | 1.77231116 | 0.002734139 | 0.011289644 | 1 |
| Obesity | neutrophil | *WNT6* | -1.64259942 | 0.005012937 | 0.011289644 | 1 |
| Obesity | neutrophil | *MLPH* | 1.827464874 | 0.004057875 | 0.011289644 | 1 |
| Obesity | neutrophil | *HDLBP* | 2.716571939 | 0.006833107 | 0.01188046 | 1 |
| Obesity | neutrophil | *PASK* | -2.557466311 | 0.019338081 | 0.025292201 | 1 |
| Obesity | neutrophil | *STK25* | 3.102999188 | 0.003985916 | 0.011289644 | 1 |
| Obesity | neutrophil | *TPO* | -2.656296048 | 0.006556935 | 0.011698461 | 1 |
| Obesity | neutrophil | *TAF1B* | 3.160371617 | 0.022522582 | 0.028850748 | 1 |
| Obesity | neutrophil | *STRN* | 2.241877207 | 0.005388056 | 0.011289644 | 1 |
| Obesity | neutrophil | *CEBPZ* | 3.062178743 | 0.013912347 | 0.019139371 | 1 |
| Obesity | neutrophil | *PRKD3* | 1.222932303 | 0.005402712 | 0.011289644 | 1 |
| Obesity | neutrophil | *SLC25A12* | 2.469055274 | 0.007821856 | 0.012690582 | 1 |
| Obesity | neutrophil | *RMDN2* | 1.760897527 | 0.004298284 | 0.011289644 | 1 |
| Obesity | neutrophil | *SDC1* | 10.72171243 | 0.016961796 | 0.022569061 | 1 |
| Obesity | neutrophil | *SOS1* | 1.323006673 | 0.004007615 | 0.011289644 | 1 |
| Obesity | neutrophil | *COX7A2L* | 1.825223779 | 0.004441836 | 0.011289644 | 1 |
| Obesity | neutrophil | *PNO1* | 2.800581034 | 0.00448499 | 0.011289644 | 1 |
| Obesity | neutrophil | *ORC4* | 1.534949055 | 0.003513475 | 0.011289644 | 1 |
| Obesity | neutrophil | *THADA* | 1.720703051 | 0.004797338 | 0.011289644 | 1 |
| Obesity | neutrophil | *ARID3A* | 2.22999837 | 0.004729046 | 0.011289644 | 1 |
| Obesity | neutrophil | *CD207* | -7.644412239 | 0.007170225 | 0.012147455 | 1 |

| | | | | | | |
|---|---|---|---|---|---|---|
| Obesity | neutrophil | *GRIN3B* | -0.955889837 | 0.010677163 | 0.0155784 | 1 |
| Obesity | neutrophil | *VAX2* | -1.347634539 | 0.006460148 | 0.01166174 | 1 |
| Obesity | neutrophil | *MSH6* | 2.227519178 | 0.004918951 | 0.011289644 | 1 |
| Obesity | neutrophil | *PLEKHA3* | 1.692317758 | 0.004164854 | 0.011289644 | 1 |
| Obesity | neutrophil | *ALMS1* | 1.35570389 | 0.004545614 | 0.011289644 | 1 |
| Obesity | neutrophil | *BCL9* | 1.518041013 | 0.005004169 | 0.011289644 | 1 |
| Obesity | neutrophil | *DHCR24* | 3.688273313 | 0.010208406 | 0.015055184 | 1 |
| Obesity | neutrophil | *GPX7* | 1.837642794 | 0.011718278 | 0.016679378 | 1 |
| Obesity | neutrophil | *SCP2* | 3.012592768 | 0.006505406 | 0.011671674 | 1 |
| Obesity | neutrophil | *TPSG1* | -2.184016665 | 0.00561031 | 0.011295339 | 1 |
| Obesity | neutrophil | *RALGPS2* | 1.06247178 | 0.005183526 | 0.011289644 | 1 |
| Obesity | neutrophil | *FAM20B* | 1.836565156 | 0.004279857 | 0.011289644 | 1 |
| Obesity | neutrophil | *TMEM59* | 1.911202364 | 0.004450802 | 0.011289644 | 1 |
| Obesity | neutrophil | *WRAP73* | 1.64787411 | 0.004934401 | 0.011289644 | 1 |
| Obesity | neutrophil | *MRPL37* | 1.576810102 | 0.005857502 | 0.011378838 | 1 |
| Obesity | neutrophil | *ICMT* | -2.912308857 | 0.014779656 | 0.020112489 | 1 |
| Obesity | neutrophil | *KCNC4* | 2.246731274 | 0.016809088 | 0.022401944 | 1 |
| Obesity | neutrophil | *EDEM3* | 1.973994683 | 0.006428395 | 0.011648199 | 1 |
| Obesity | neutrophil | *RAP1A* | 1.430948385 | 0.004654496 | 0.011289644 | 1 |
| Obesity | neutrophil | *CAPZA1* | 2.489298564 | 0.009052773 | 0.013861369 | 1 |
| Obesity | neutrophil | *DOCK7* | 2.006438708 | 0.005567541 | 0.011295339 | 1 |
| Obesity | neutrophil | *FBXO6* | -2.881916109 | 0.005349026 | 0.011289644 | 1 |
| Obesity | neutrophil | *C1orf21* | 1.19691285 | 0.001908754 | 0.011289644 | 1 |
| Obesity | neutrophil | *DNAJC6* | 1.724295564 | 0.006828202 | 0.011878501 | 1 |
| Obesity | neutrophil | *LEPR* | 1.070501806 | 0.003797938 | 0.011289644 | 1 |
| Obesity | neutrophil | *MFN2* | 1.450123049 | 0.004824791 | 0.011289644 | 1 |
| Obesity | neutrophil | *MIIP* | 1.838916883 | 0.004668786 | 0.011289644 | 1 |
| Obesity | neutrophil | *SMG7* | 2.225041203 | 0.005832088 | 0.011378838 | 1 |
| Obesity | neutrophil | *NCF2* | -2.644798618 | 0.021260013 | 0.027451572 | 1 |
| Obesity | neutrophil | *RGS2* | -0.580329529 | 0.00537002 | 0.011289644 | 1 |
| Obesity | neutrophil | *OLFML3* | 1.477560391 | 0.00356164 | 0.011289644 | 1 |
| Obesity | neutrophil | *TNNI3K* | -1.118067958 | 0.00442286 | 0.011289644 | 1 |
| Obesity | neutrophil | *CRYZ* | -1.520595802 | 0.021649275 | 0.027891187 | 1 |
| Obesity | neutrophil | *TTF2* | 2.609842278 | 0.00473279 | 0.011289644 | 1 |
| Obesity | neutrophil | *WARS2* | 1.174396915 | 0.001779847 | 0.011289644 | 1 |
| Obesity | neutrophil | *OSCP1* | 1.392579541 | 0.005630095 | 0.011295339 | 1 |
| Obesity | neutrophil | *GNPAT* | 2.450258398 | 0.002897598 | 0.011289644 | 1 |
| Obesity | neutrophil | *C1orf109* | 2.525604241 | 0.002132411 | 0.011289644 | 1 |
| Obesity | neutrophil | *TBCE* | 1.639967787 | 0.004713719 | 0.011289644 | 1 |
| Obesity | neutrophil | *LGALS8* | 2.132250744 | 0.011693869 | 0.016655838 | 1 |
| Obesity | neutrophil | *MTR* | 2.184496524 | 0.006313438 | 0.011593738 | 1 |
| Obesity | neutrophil | *KCNQ4* | 2.0093427 | 0.036163934 | 0.044346139 | 1 |
| Obesity | neutrophil | *ACADM* | 1.807208297 | 0.003561854 | 0.011289644 | 1 |
| Obesity | neutrophil | *PADI2* | -1.304325391 | 0.004781947 | 0.011289644 | 1 |
| Obesity | neutrophil | *MFAP2* | -7.030316027 | 1.66E-05 | 0.003153152 | 0.104054011 |
| Obesity | neutrophil | *CTBS* | 1.772982335 | 0.006522132 | 0.011673832 | 1 |
| Obesity | neutrophil | *KLHL12* | 1.266843334 | 0.005829498 | 0.011378838 | 1 |
| Obesity | neutrophil | *ZNHIT6* | 1.660649921 | 0.005360186 | 0.011289644 | 1 |
| Obesity | neutrophil | *RBBP5* | 1.322477093 | 0.005741939 | 0.01134195 | 1 |
| Obesity | neutrophil | *KIF17* | -2.159178652 | 0.003836127 | 0.011289644 | 1 |

| | | | | | | |
|---|---|---|---|---|---|---|
| Obesity | neutrophil | *CD46* | 1.623005574 | 0.004425215 | 0.011289644 | 1 |
| Obesity | neutrophil | *PRDX1* | 4.202124415 | 0.004251129 | 0.011289644 | 1 |
| Obesity | neutrophil | *FAAH* | 1.429779756 | 0.004327616 | 0.011289644 | 1 |
| Obesity | neutrophil | *NSUN4* | 1.24764722 | 0.007506547 | 0.012399504 | 1 |
| Obesity | neutrophil | *TMED5* | 1.681095747 | 0.004459109 | 0.011289644 | 1 |
| Obesity | neutrophil | *DR1* | 1.357146675 | 0.004635016 | 0.011289644 | 1 |
| Obesity | neutrophil | *CNN3* | 1.605435717 | 0.004304884 | 0.011289644 | 1 |
| Obesity | neutrophil | *PRRC2C* | 1.572037813 | 0.002934811 | 0.011289644 | 1 |
| Obesity | neutrophil | *ABCD3* | 1.502024418 | 0.004279802 | 0.011289644 | 1 |
| Obesity | neutrophil | *VAMP4* | 2.75044446 | 0.003711969 | 0.011289644 | 1 |
| Obesity | neutrophil | *PTBP2* | 1.319226845 | 0.001876785 | 0.011289644 | 1 |
| Obesity | neutrophil | *TNFSF4* | 1.971294857 | 0.02980231 | 0.037212934 | 1 |
| Obesity | neutrophil | *DARS2* | 1.787165522 | 0.016110819 | 0.021647514 | 1 |
| Obesity | neutrophil | *IRF6* | 3.766300591 | 0.003665135 | 0.011289644 | 1 |
| Obesity | neutrophil | *DIEXF* | 2.337143234 | 0.009970065 | 0.014825402 | 1 |
| Obesity | neutrophil | *SERPINC1* | 10.78849498 | 1.97E-11 | 2.06E-08 | 1.24E-07 |
| Obesity | neutrophil | *RCAN3* | -1.377783487 | 0.01392736 | 0.019155833 | 1 |
| Obesity | neutrophil | *RCOR3* | 2.145812705 | 0.005480551 | 0.011289644 | 1 |
| Obesity | neutrophil | *NSL1* | 1.750588138 | 0.005735056 | 0.01134195 | 1 |
| Obesity | neutrophil | *RPA2* | 4.198963462 | 0.012183804 | 0.017221128 | 1 |
| Obesity | neutrophil | *SLC5A9* | 2.119711185 | 0.003704126 | 0.011289644 | 1 |
| Obesity | neutrophil | *OSBPL9* | 1.269961307 | 0.004370559 | 0.011289644 | 1 |
| Obesity | neutrophil | *TXNDC12* | 1.81422652 | 0.004235067 | 0.011289644 | 1 |
| Obesity | neutrophil | *ESYT2* | 1.547336646 | 0.009677063 | 0.014513286 | 1 |
| Obesity | neutrophil | *CD3EAP* | 1.414669529 | 0.015604566 | 0.021079912 | 1 |
| Obesity | neutrophil | *KMT2A* | 2.122793277 | 0.005508096 | 0.011289644 | 1 |
| Obesity | neutrophil | *IFT46* | 3.384351318 | 0.002077091 | 0.011289644 | 1 |
| Obesity | neutrophil | *DDX59* | 1.236202675 | 0.004670791 | 0.011289644 | 1 |
| Obesity | neutrophil | *CAMSAP2* | 1.215482535 | 0.004258793 | 0.011289644 | 1 |
| Obesity | neutrophil | *ATF6* | 2.479438844 | 0.006896436 | 0.011932455 | 1 |
| Obesity | neutrophil | *LRMP* | 2.302222298 | 0.001219861 | 0.011289644 | 1 |
| Obesity | neutrophil | *ATP10B* | 3.506058456 | 0.003328302 | 0.011289644 | 1 |
| Obesity | neutrophil | *SPCS2* | 3.867487007 | 0.033633272 | 0.041493796 | 1 |
| Obesity | neutrophil | *FILIP1* | 2.405458429 | 0.005493934 | 0.011289644 | 1 |
| Obesity | neutrophil | *SGIP1* | 1.215931873 | 0.003376686 | 0.011289644 | 1 |
| Obesity | neutrophil | *AKAP7* | 1.636411822 | 0.005348705 | 0.011289644 | 1 |
| Obesity | neutrophil | *MED28* | 1.195648427 | 0.005143362 | 0.011289644 | 1 |
| Obesity | neutrophil | *SLC16A7* | 1.839459079 | 0.005762684 | 0.011357022 | 1 |
| Obesity | neutrophil | *FOXO3* | 1.850897343 | 0.005425364 | 0.011289644 | 1 |
| Obesity | neutrophil | *TGIF2* | -4.253325302 | 0.031704206 | 0.039329981 | 1 |
| Obesity | neutrophil | *ABCG2* | -1.12960169 | 0.006415066 | 0.011648199 | 1 |
| Obesity | neutrophil | *SPP1* | 2.008436846 | 0.005656392 | 0.011300433 | 1 |
| Obesity | neutrophil | *FAM47E-STBL* | 1.76364263 | 0.008142288 | 0.012962918 | 1 |
| Obesity | neutrophil | *RAB3GAP2* | 1.992148291 | 0.004594616 | 0.011289644 | 1 |
| Obesity | neutrophil | *UBN1* | 2.409642418 | 0.016911217 | 0.022516057 | 1 |
| Obesity | neutrophil | *UCHL3* | 2.270647759 | 0.004158526 | 0.011289644 | 1 |
| Obesity | neutrophil | *WDR35* | 1.463512069 | 0.004140925 | 0.011289644 | 1 |
| Obesity | neutrophil | *DNAH7* | 1.015411924 | 0.036156555 | 0.044345739 | 1 |
| Obesity | neutrophil | *NDUFB3* | -1.943475977 | 0.007818179 | 0.012687891 | 1 |
| Obesity | neutrophil | *GDA* | 5.63661225 | 0.005050031 | 0.011289644 | 1 |

| | | | | | | |
|---|---|---|---|---|---|---|
| Obesity | neutrophil | *C2orf40* | -2.317130666 | 0.007742754 | 0.012620869 | 1 |
| Obesity | neutrophil | *PIGZ* | 2.359173861 | 0.021648838 | 0.027891187 | 1 |
| Obesity | neutrophil | *SENP5* | 1.779788591 | 0.004335317 | 0.011289644 | 1 |
| Obesity | neutrophil | *CCDC92* | 2.735692176 | 7.20E-05 | 0.008226393 | 0.452451599 |
| Obesity | neutrophil | *PTBP3* | 2.550486926 | 0.006418759 | 0.011648199 | 1 |
| Obesity | neutrophil | *FKBP15* | 2.622960256 | 0.003146419 | 0.011289644 | 1 |
| Obesity | neutrophil | *CTNNAL1* | 1.265127366 | 0.001501144 | 0.011289644 | 1 |
| Obesity | neutrophil | *WDR34* | 1.970440727 | 0.006895169 | 0.011932455 | 1 |
| Obesity | neutrophil | *CNTRL* | 2.426677181 | 0.00586176 | 0.011378838 | 1 |
| Obesity | neutrophil | *TRIM32* | 3.370581432 | 0.002554688 | 0.011289644 | 1 |
| Obesity | neutrophil | *PHF19* | -4.508207728 | 0.017945233 | 0.023711997 | 1 |
| Obesity | neutrophil | *BSPRY* | -2.179334681 | 0.00536591 | 0.011289644 | 1 |
| Obesity | neutrophil | *PPP6C* | 2.311773856 | 0.004679186 | 0.011289644 | 1 |
| Obesity | neutrophil | *RBM18* | 3.139819988 | 0.006392498 | 0.011648199 | 1 |
| Obesity | neutrophil | *HSDL2* | 1.471850259 | 0.006222972 | 0.011544147 | 1 |
| Obesity | neutrophil | *INVS* | 1.7761559 | 0.002027506 | 0.011289644 | 1 |
| Obesity | neutrophil | *KDSR* | 1.314824782 | 0.004223707 | 0.011289644 | 1 |
| Obesity | neutrophil | *DCAF4* | 3.526604953 | 0.000212104 | 0.011289644 | 1 |
| Obesity | neutrophil | *IFI27L2* | 1.818844482 | 0.005413958 | 0.011289644 | 1 |
| Obesity | neutrophil | *IRF2BPL* | 4.114079998 | 0.012392461 | 0.017443326 | 1 |
| Obesity | neutrophil | *TTLL5* | 1.620851751 | 0.004409874 | 0.011289644 | 1 |
| Obesity | neutrophil | *PPP4R4* | 2.691042598 | 0.012267157 | 0.017319474 | 1 |
| Obesity | neutrophil | *ZC2HC1C* | 4.852283777 | 0.004220006 | 0.011289644 | 1 |
| Obesity | neutrophil | *COQ6* | 2.027435263 | 0.004983404 | 0.011289644 | 1 |
| Obesity | neutrophil | *GPR75* | 11.10606958 | 0.002141305 | 0.011289644 | 1 |
| Obesity | neutrophil | *KLHL29* | 1.687488864 | 0.004302359 | 0.011289644 | 1 |
| Obesity | neutrophil | *ATAD2B* | 2.272283571 | 0.006439533 | 0.011648199 | 1 |
| Obesity | neutrophil | *FKBP1B* | 3.934300657 | 0.017401783 | 0.023070361 | 1 |
| Obesity | neutrophil | *ATL2* | 1.215445039 | 0.004478312 | 0.011289644 | 1 |
| Obesity | neutrophil | *CNRIP1* | 2.377010374 | 0.026751314 | 0.033738317 | 1 |
| Obesity | neutrophil | *CRIPT* | 1.274500996 | 0.008751841 | 0.013602675 | 1 |
| Obesity | neutrophil | *EPCAM* | 4.439986553 | 0.002517426 | 0.011289644 | 1 |
| Obesity | neutrophil | *SLC17A5* | 1.542419635 | 0.003479597 | 0.011289644 | 1 |
| Obesity | neutrophil | *OGFRL1* | 3.070829507 | 0.029509866 | 0.036891733 | 1 |
| Obesity | neutrophil | *CUTC* | 2.885858526 | 0.00929208 | 0.014110944 | 1 |
| Obesity | neutrophil | *PYROXD2* | 1.857680653 | 0.000653128 | 0.011289644 | 1 |
| Obesity | neutrophil | *MXI1* | 1.116715098 | 0.004246189 | 0.011289644 | 1 |
| Obesity | neutrophil | *HELLS* | 1.525598761 | 0.004209864 | 0.011289644 | 1 |
| Obesity | neutrophil | *TCTN3* | 3.794778598 | 0.031813081 | 0.039446692 | 1 |
| Obesity | neutrophil | *AVPI1* | 2.557347447 | 0.003260523 | 0.011289644 | 1 |
| Obesity | neutrophil | *WDR11* | 1.206347952 | 0.004174534 | 0.011289644 | 1 |
| Obesity | neutrophil | *C10orf76* | 2.897491521 | 0.00500879 | 0.011289644 | 1 |
| Obesity | neutrophil | *KCNIP2* | 1.193163116 | 0.002961337 | 0.011289644 | 1 |
| Obesity | neutrophil | *C10orf95* | 3.94038712 | 0.001211253 | 0.011289644 | 1 |
| Obesity | neutrophil | *KANSL1* | 1.3807407 | 0.004410639 | 0.011289644 | 1 |
| Obesity | neutrophil | *HOXB3* | 1.92144238 | 0.000462636 | 0.011289644 | 1 |
| Obesity | neutrophil | *CAAP1* | 1.900518459 | 0.004561865 | 0.011289644 | 1 |
| Obesity | neutrophil | *CD274* | -2.621818316 | 0.007635334 | 0.012499129 | 1 |
| Obesity | neutrophil | *LRP11* | 1.883283194 | 0.01209771 | 0.017128656 | 1 |
| Obesity | neutrophil | *CCDC170* | 1.036265774 | 0.004642159 | 0.011289644 | 1 |

| | | | | | | |
|---|---|---|---|---|---|---|
| Obesity | neutrophil | *CYSTM1* | 2.190021603 | 0.004117946 | 0.011289644 | 1 |
| Obesity | neutrophil | *WDR55* | 1.455429985 | 0.01007685 | 0.01492911 | 1 |
| Obesity | neutrophil | *PCDHB14* | 2.382902884 | 0.00425945 | 0.011289644 | 1 |
| Obesity | neutrophil | *SEC16B* | -2.126694018 | 0.004148886 | 0.011289644 | 1 |
| Obesity | neutrophil | *GORAB* | 2.760537309 | 0.006495301 | 0.011671674 | 1 |
| Obesity | neutrophil | *ACAT2* | -2.651541416 | 0.014279589 | 0.019554732 | 1 |
| Obesity | neutrophil | *MRC1* | 0.884479163 | 0.005329685 | 0.011289644 | 1 |
| Obesity | neutrophil | *IQSEC3* | 3.921423175 | 0.003767036 | 0.011289644 | 1 |
| Obesity | neutrophil | *CCDC77* | 2.020449924 | 0.004400556 | 0.011289644 | 1 |
| Obesity | neutrophil | *ENOX1* | 1.356409566 | 0.004174992 | 0.011289644 | 1 |
| Obesity | neutrophil | *DNAJC15* | 2.168297711 | 0.004728239 | 0.011289644 | 1 |
| Obesity | neutrophil | *WBP4* | 2.923639738 | 0.0068189 | 0.011875318 | 1 |
| Obesity | neutrophil | *SMAD9* | 2.334421692 | 0.004322982 | 0.011289644 | 1 |
| Obesity | neutrophil | *SIL1* | 2.155967541 | 0.008167598 | 0.01298677 | 1 |
| Obesity | neutrophil | *ZFP30* | 1.74403856 | 0.002490951 | 0.011289644 | 1 |
| Obesity | neutrophil | *ARL1* | 1.377535536 | 0.004209648 | 0.011289644 | 1 |
| Obesity | neutrophil | *APAF1* | 2.526404342 | 0.004026171 | 0.011289644 | 1 |
| Obesity | neutrophil | *ADRA1A* | 1.509431901 | 6.36E-06 | 0.001734043 | 0.040005465 |
| Obesity | neutrophil | *EPHX2* | 1.226611788 | 0.007160522 | 0.012141606 | 1 |
| Obesity | neutrophil | *ZNF706* | 2.082977646 | 0.004253076 | 0.011289644 | 1 |
| Obesity | neutrophil | *CRISPLD1* | 2.12312464 | 0.004069595 | 0.011289644 | 1 |
| Obesity | neutrophil | *SCPEP1* | 1.476509483 | 0.001452607 | 0.011289644 | 1 |
| Obesity | neutrophil | *TBX4* | -0.797455176 | 0.004510202 | 0.011289644 | 1 |
| Obesity | neutrophil | *TEX14* | 2.073658473 | 0.001238121 | 0.011289644 | 1 |
| Obesity | neutrophil | *CEP89* | 1.454568458 | 0.003948029 | 0.011289644 | 1 |
| Obesity | neutrophil | *ECHDC2* | 0.947901668 | 0.028802692 | 0.036079403 | 1 |
| Obesity | neutrophil | *PRB2* | 2.071428959 | 0.003935995 | 0.011289644 | 1 |
| Obesity | neutrophil | *PYROXD1* | 3.840232797 | 0.001216229 | 0.011289644 | 1 |
| Obesity | neutrophil | *A1BG* | 5.356544524 | 0.001102585 | 0.011289644 | 1 |
| Obesity | neutrophil | *ZSCAN18* | 2.21891138 | 0.003328209 | 0.011289644 | 1 |
| Obesity | neutrophil | *ZNF211* | -0.676465944 | 0.010134783 | 0.014978228 | 1 |
| Obesity | neutrophil | *PDZRN3* | 1.038060929 | 0.004290095 | 0.011289644 | 1 |
| Obesity | neutrophil | *SEC22A* | 1.198137184 | 0.011834364 | 0.016817958 | 1 |
| Obesity | neutrophil | *CSTA* | 2.021006116 | 0.00199651 | 0.011289644 | 1 |
| Obesity | neutrophil | *POPDC2* | 2.41583625 | 0.004236925 | 0.011289644 | 1 |
| Obesity | neutrophil | *B4GALT4* | 1.98995646 | 0.004124504 | 0.011289644 | 1 |
| Obesity | neutrophil | *CD80* | 2.872879066 | 0.004135229 | 0.011289644 | 1 |
| Obesity | neutrophil | *CAT* | 1.316585985 | 0.004999966 | 0.011289644 | 1 |
| Obesity | neutrophil | *PILRB* | 1.140080764 | 0.004074536 | 0.011289644 | 1 |
| Obesity | neutrophil | *TMEM39B* | 1.960728992 | 0.008065974 | 0.01288711 | 1 |
| Obesity | neutrophil | *RNF115* | 1.801442418 | 0.004732969 | 0.011289644 | 1 |
| Obesity | neutrophil | *POLR3GL* | 1.966743309 | 0.004260431 | 0.011289644 | 1 |
| Obesity | neutrophil | *TNFSF10* | 1.667119304 | 0.003899579 | 0.011289644 | 1 |
| Obesity | neutrophil | *ZNF639* | 1.531153509 | 0.003344606 | 0.011289644 | 1 |
| Obesity | neutrophil | *LIAS* | 2.834986841 | 0.004771737 | 0.011289644 | 1 |
| Obesity | neutrophil | *CLCC1* | 4.361787223 | 0.031369849 | 0.038961327 | 1 |
| Obesity | neutrophil | *GPSM2* | 1.599734138 | 0.002163316 | 0.011289644 | 1 |
| Obesity | neutrophil | *CXCR4* | -3.830149086 | 0.004152001 | 0.011289644 | 1 |
| Obesity | neutrophil | *ACVR2A* | 0.952802697 | 0.004259515 | 0.011289644 | 1 |
| Obesity | neutrophil | *POLK* | 2.613498147 | 0.012325747 | 0.017378779 | 1 |

| | | | | | | |
|---|---|---|---|---|---|---|
| Obesity | neutrophil | *FLT3* | -4.987207549 | 0.025764319 | 0.032571943 | 1 |
| Obesity | neutrophil | *MTIF3* | 1.604461218 | 0.004449346 | 0.011289644 | 1 |
| Obesity | neutrophil | *RASL11A* | -1.301770384 | 0.00421347 | 0.011289644 | 1 |
| Obesity | neutrophil | *FMOD* | 4.581201527 | 0.017999971 | 0.023759356 | 1 |
| Obesity | neutrophil | *SERAC1* | 1.457602679 | 0.00423618 | 0.011289644 | 1 |
| Obesity | neutrophil | *LDB3* | 1.76109184 | 0.003188591 | 0.011289644 | 1 |
| Obesity | neutrophil | *FAM35A* | 1.583614274 | 0.004388365 | 0.011289644 | 1 |
| Obesity | neutrophil | *FAM213A* | 2.541691144 | 0.001880535 | 0.011289644 | 1 |
| Obesity | neutrophil | *NAA60* | 2.667715619 | 0.005683491 | 0.011300433 | 1 |
| Obesity | neutrophil | *ODF2L* | 1.84182559 | 0.002616975 | 0.011289644 | 1 |
| Obesity | neutrophil | *SPATA1* | 3.415704353 | 0.011447277 | 0.016387432 | 1 |
| Obesity | neutrophil | *RWDD3* | 2.485714666 | 0.006960361 | 0.011974691 | 1 |
| Obesity | neutrophil | *CCDC18* | 2.14727732 | 0.004948984 | 0.011289644 | 1 |
| Obesity | neutrophil | *RPAP2* | 2.178301043 | 0.005567544 | 0.011295339 | 1 |
| Obesity | neutrophil | *BBS9* | 1.525040491 | 0.005413339 | 0.011289644 | 1 |
| Obesity | neutrophil | *PMS2* | 1.980746097 | 0.002758193 | 0.011289644 | 1 |
| Obesity | neutrophil | *KIAA0087* | 4.782764783 | 0.016363955 | 0.021954799 | 1 |
| Obesity | neutrophil | *KLHL7* | 3.035537906 | 0.007380389 | 0.01229744 | 1 |
| Obesity | neutrophil | *HERPUD2* | 1.591130539 | 0.003968799 | 0.011289644 | 1 |
| Obesity | neutrophil | *CBX3* | 1.110151849 | 0.006024743 | 0.011447874 | 1 |
| Obesity | neutrophil | *FAM126A* | 1.193678486 | 0.003949365 | 0.011289644 | 1 |
| Obesity | neutrophil | *CCZ1* | 1.998634315 | 0.001258641 | 0.011289644 | 1 |
| Obesity | neutrophil | *SMU1* | 2.2251096 | 0.003848962 | 0.011289644 | 1 |
| Obesity | neutrophil | *GLIPR2* | 3.184136426 | 0.021254867 | 0.027450565 | 1 |
| Obesity | neutrophil | *SLC25A51* | 2.391578733 | 0.000211484 | 0.011289644 | 1 |
| Obesity | neutrophil | *DCAF10* | 0.91212582 | 0.00423402 | 0.011289644 | 1 |
| Obesity | neutrophil | *TRIM24* | 3.104312181 | 0.008973635 | 0.013797211 | 1 |
| Obesity | neutrophil | *BICC1* | 0.938590415 | 0.002565238 | 0.011289644 | 1 |
| Obesity | neutrophil | *SLC25A16* | 1.257235888 | 0.001905416 | 0.011289644 | 1 |
| Obesity | neutrophil | *ZWINT* | 3.792492049 | 0.00252812 | 0.011289644 | 1 |
| Obesity | neutrophil | *RBM19* | 1.74003785 | 0.004233631 | 0.011289644 | 1 |
| Obesity | neutrophil | *ACADS* | 2.698329487 | 0.006723127 | 0.011825602 | 1 |
| Obesity | neutrophil | *SSPN* | 1.218366873 | 0.003797345 | 0.011289644 | 1 |
| Obesity | neutrophil | *CCDC91* | 1.303704481 | 0.003646373 | 0.011289644 | 1 |
| Obesity | neutrophil | *PKN1* | 2.144007952 | 0.004647737 | 0.011289644 | 1 |
| Obesity | neutrophil | *ATP7B* | 2.118070899 | 0.000755285 | 0.011289644 | 1 |
| Obesity | neutrophil | *NLN* | 1.435613238 | 0.006683374 | 0.011792087 | 1 |
| Obesity | neutrophil | *CENPK* | 1.881592343 | 0.002664566 | 0.011289644 | 1 |
| Obesity | neutrophil | *OPTN* | 1.070982071 | 0.00705042 | 0.012051656 | 1 |
| Obesity | neutrophil | *ATF1* | 2.428038696 | 0.007269643 | 0.01222323 | 1 |
| Obesity | neutrophil | *TSFM* | 2.999715843 | 0.011696441 | 0.016655838 | 1 |
| Obesity | neutrophil | *ARHGAP9* | -2.145448703 | 0.00355371 | 0.011289644 | 1 |
| Obesity | neutrophil | *SPATS2* | 1.775776799 | 0.007886073 | 0.01274567 | 1 |
| Obesity | neutrophil | *ORMDL2* | 5.127714667 | 0.034132133 | 0.04207622 | 1 |
| Obesity | neutrophil | *PDE1B* | 1.636810974 | 0.004178402 | 0.011289644 | 1 |
| Obesity | neutrophil | *IKZF4* | 3.013688594 | 0.026801461 | 0.033788006 | 1 |
| Obesity | neutrophil | *SMUG1* | 2.055370187 | 0.000180151 | 0.011289644 | 1 |
| Obesity | neutrophil | *SARDH* | 4.418576534 | 0.004269301 | 0.011289644 | 1 |
| Obesity | neutrophil | *DBH* | -6.893438315 | 0.004430691 | 0.011289644 | 1 |
| Obesity | neutrophil | *STIL* | 2.01662276 | 0.00432665 | 0.011289644 | 1 |

| | | | | | | |
|---|---|---|---|---|---|---|
| Obesity | neutrophil | *TTC21B* | 1.479815483 | 0.004394576 | 0.011289644 | 1 |
| Obesity | neutrophil | *NMI* | -2.808519387 | 0.027949457 | 0.035094515 | 1 |
| Obesity | neutrophil | *ACVR1C* | 0.88915585 | 0.006254701 | 0.011555481 | 1 |
| Obesity | neutrophil | *SLC36A1* | 2.874313981 | 0.006925074 | 0.011946923 | 1 |
| Obesity | neutrophil | *PLA2G12A* | 1.193089559 | 0.004025691 | 0.011289644 | 1 |
| Obesity | neutrophil | *C4BPB* | 3.155952673 | 0.002965732 | 0.011289644 | 1 |
| Obesity | neutrophil | *RAB38* | 3.549464082 | 0.00644923 | 0.011649838 | 1 |
| Obesity | neutrophil | *AGO2* | 1.245570119 | 0.004366231 | 0.011289644 | 1 |
| Obesity | neutrophil | *CKS2* | -0.601064745 | 0.005054697 | 0.011289644 | 1 |
| Obesity | neutrophil | *CHPF* | 2.073463229 | 0.016197639 | 0.021750226 | 1 |
| Obesity | neutrophil | *DNPEP* | 3.150244976 | 0.003886027 | 0.011289644 | 1 |
| Obesity | neutrophil | *MOGAT1* | 1.420968347 | 0.005239441 | 0.011289644 | 1 |
| Obesity | neutrophil | *FAM124B* | -0.612155843 | 0.005609219 | 0.011295339 | 1 |
| Obesity | neutrophil | *SNX21* | 1.599035358 | 0.004317053 | 0.011289644 | 1 |
| Obesity | neutrophil | *SDC4* | -2.211170566 | 0.007587977 | 0.012456909 | 1 |
| Obesity | neutrophil | *VAPB* | 2.029435996 | 0.003301016 | 0.011289644 | 1 |
| Obesity | neutrophil | *ARFGEF2* | 1.631804675 | 0.003765095 | 0.011289644 | 1 |
| Obesity | neutrophil | *CSE1L* | 2.582871669 | 0.003836196 | 0.011289644 | 1 |
| Obesity | neutrophil | *CDH26* | 3.621992121 | 0.004057077 | 0.011289644 | 1 |
| Obesity | neutrophil | *STX16* | 1.482867784 | 0.004273218 | 0.011289644 | 1 |
| Obesity | neutrophil | *KCNK15* | 5.505694627 | 0.0072481 | 0.01221029 | 1 |
| Obesity | neutrophil | *NEURL2* | 2.677218733 | 0.000339011 | 0.011289644 | 1 |
| Obesity | neutrophil | *FASTKD3* | -3.211957092 | 0.012800321 | 0.017895401 | 1 |
| Obesity | neutrophil | *PEPD* | 2.095560065 | 0.002231484 | 0.011289644 | 1 |
| Obesity | neutrophil | *CHST8* | 3.307395959 | 0.001731066 | 0.011289644 | 1 |
| Obesity | neutrophil | *STAMBP* | 1.436247253 | 0.004074411 | 0.011289644 | 1 |
| Obesity | neutrophil | *ATP8A1* | 1.854545332 | 0.001753242 | 0.011289644 | 1 |
| Obesity | neutrophil | *USP22* | 1.789639332 | 0.005369479 | 0.011289644 | 1 |
| Obesity | neutrophil | *HIF3A* | 1.269770453 | 0.001957361 | 0.011289644 | 1 |
| Obesity | neutrophil | *ZNF45* | 1.483991585 | 0.004215716 | 0.011289644 | 1 |
| Obesity | neutrophil | *BTN2A2* | 1.928516809 | 0.001812655 | 0.011289644 | 1 |
| Obesity | neutrophil | *MRS2* | 3.382880612 | 0.011396571 | 0.016347306 | 1 |
| Obesity | neutrophil | *SERPINB6* | 1.989089015 | 0.004562602 | 0.011289644 | 1 |
| Obesity | neutrophil | *HIST1H1D* | 5.315244798 | 0.02088293 | 0.02701882 | 1 |
| Obesity | neutrophil | *PEX6* | 1.720191964 | 0.005459842 | 0.011289644 | 1 |
| Obesity | neutrophil | *NQO2* | 1.546998965 | 0.004195444 | 0.011289644 | 1 |
| Obesity | neutrophil | *RPS10* | -1.018199804 | 0.006351552 | 0.011618636 | 1 |
| Obesity | neutrophil | *HIST1H2BJ* | 12.05935084 | 3.91E-09 | 2.73E-06 | 2.46E-05 |
| Obesity | neutrophil | *MED20* | -0.955562506 | 0.00742891 | 0.012334552 | 1 |
| Obesity | neutrophil | *TBCC* | 2.243506987 | 0.008619748 | 0.013470632 | 1 |
| Obesity | neutrophil | *KLHDC3* | 1.705153785 | 0.004350602 | 0.011289644 | 1 |
| Obesity | neutrophil | *GNMT* | -0.819301885 | 0.002163915 | 0.011289644 | 1 |
| Obesity | neutrophil | *MEA1* | 4.626999704 | 0.014438321 | 0.019742002 | 1 |
| Obesity | neutrophil | *KLHL31* | 1.304076839 | 0.001646234 | 0.011289644 | 1 |
| Obesity | neutrophil | *COL21A1* | 1.158368131 | 0.004517287 | 0.011289644 | 1 |
| Obesity | neutrophil | *GLO1* | 2.204665888 | 0.002825821 | 0.011289644 | 1 |
| Obesity | neutrophil | *CPNE5* | 1.784915789 | 0.014222374 | 0.01949064 | 1 |
| Obesity | neutrophil | *RIOK1* | 2.133313517 | 0.004363577 | 0.011289644 | 1 |
| Obesity | neutrophil | *RPP40* | -0.956729901 | 0.00708007 | 0.012072542 | 1 |
| Obesity | neutrophil | *NUP153* | 1.459369336 | 0.002861338 | 0.011289644 | 1 |

| | | | | | | |
|---|---|---|---|---|---|---|
| Obesity | neutrophil | *EEF1E1* | -1.362838759 | 0.008896569 | 0.013733966 | 1 |
| Obesity | neutrophil | *EREG* | -1.924092235 | 0.005407189 | 0.011289644 | 1 |
| Obesity | neutrophil | *CNOT1* | 1.912632228 | 0.003754589 | 0.011289644 | 1 |
| Obesity | neutrophil | *BBS2* | 2.445896013 | 0.00069635 | 0.011289644 | 1 |
| Obesity | neutrophil | *MT1G* | -1.426974264 | 0.005171257 | 0.011289644 | 1 |
| Obesity | neutrophil | *GOT2* | 1.883188977 | 0.003604159 | 0.011289644 | 1 |
| Obesity | neutrophil | *TMTC4* | 1.571110171 | 0.004218086 | 0.011289644 | 1 |
| Obesity | neutrophil | *KIF25* | 4.596818146 | 0.004595637 | 0.011289644 | 1 |
| Obesity | neutrophil | *GRK4* | 2.260281022 | 0.006467962 | 0.01166174 | 1 |
| Obesity | neutrophil | *GGA3* | 2.085835684 | 0.004563326 | 0.011289644 | 1 |
| Obesity | neutrophil | *NUP85* | 1.450648655 | 0.004806642 | 0.011289644 | 1 |
| Obesity | neutrophil | *MIF4GD* | 2.113474311 | 0.004258386 | 0.011289644 | 1 |
| Obesity | neutrophil | *NT5C* | 4.162610077 | 0.014208565 | 0.019482933 | 1 |
| Obesity | neutrophil | *DDX31* | 1.885390973 | 0.019875969 | 0.02590923 | 1 |
| Obesity | neutrophil | *PPP1R12C* | 1.343205257 | 0.003897306 | 0.011289644 | 1 |
| Obesity | neutrophil | *OPRL1* | 5.794434353 | 7.85E-05 | 0.008815059 | 0.493643287 |
| Obesity | neutrophil | *PLGLB2* | -36.79503262 | 0.009565794 | 0.014397929 | 1 |
| Obesity | neutrophil | *CHCHD5* | -3.446277253 | 0.005197273 | 0.011289644 | 1 |
| Obesity | neutrophil | *PAX8* | 1.343992728 | 0.007064757 | 0.012063272 | 1 |
| Obesity | neutrophil | *INSIG2* | 3.379760788 | 0.005670418 | 0.011300433 | 1 |
| Obesity | neutrophil | *PSD4* | -1.286624997 | 0.0073929 | 0.012298069 | 1 |
| Obesity | neutrophil | *SLC25A23* | -1.560679576 | 0.007064891 | 0.012063272 | 1 |
| Obesity | neutrophil | *GTF2F1* | 5.222593064 | 0.01821557 | 0.024008656 | 1 |
| Obesity | neutrophil | *CLPP* | 2.183507124 | 0.004071408 | 0.011289644 | 1 |
| Obesity | neutrophil | *SH2D3A* | 6.081301071 | 0.020052712 | 0.026085537 | 1 |
| Obesity | neutrophil | *TRIP10* | 1.62971465 | 0.003731795 | 0.011289644 | 1 |
| Obesity | neutrophil | *SNRPD2* | 1.558711721 | 0.004816935 | 0.011289644 | 1 |
| Obesity | neutrophil | *RTN2* | 5.432718745 | 0.005415733 | 0.011289644 | 1 |
| Obesity | neutrophil | *SYMPK* | 1.330174007 | 0.003597278 | 0.011289644 | 1 |
| Obesity | neutrophil | *GPCPD1* | 1.656195555 | 0.004583402 | 0.011289644 | 1 |
| Obesity | neutrophil | *SDCBP2* | 3.416508305 | 0.004507086 | 0.011289644 | 1 |
| Obesity | neutrophil | *PANK2* | 3.015918452 | 0.006813279 | 0.011874479 | 1 |
| Obesity | neutrophil | *PSMF1* | 2.896809919 | 0.016404542 | 0.021990481 | 1 |
| Obesity | neutrophil | *DTD1* | 2.839048873 | 0.010235436 | 0.015084432 | 1 |
| Obesity | neutrophil | *RBCK1* | 3.688418723 | 0.004067942 | 0.011289644 | 1 |
| Obesity | neutrophil | *RRBP1* | 1.695241444 | 0.005115693 | 0.011289644 | 1 |
| Obesity | neutrophil | *BMP2* | 3.056953003 | 0.003599155 | 0.011289644 | 1 |
| Obesity | neutrophil | *PCSK2* | 24.39269885 | 0.000490437 | 0.011289644 | 1 |
| Obesity | neutrophil | *MKKS* | 1.157862602 | 0.004417209 | 0.011289644 | 1 |
| Obesity | neutrophil | *DSTN* | 1.078290862 | 0.004117276 | 0.011289644 | 1 |
| Obesity | neutrophil | *MGME1* | 2.109107363 | 0.004205995 | 0.011289644 | 1 |
| Obesity | neutrophil | *ITPA* | 1.402247048 | 0.004221072 | 0.011289644 | 1 |
| Obesity | neutrophil | *MCM8* | 1.527029928 | 0.003104066 | 0.011289644 | 1 |
| Obesity | neutrophil | *TMEM74B* | -0.910568566 | 0.007359206 | 0.012285536 | 1 |
| Obesity | neutrophil | *FAM110A* | 2.743199116 | 0.004907442 | 0.011289644 | 1 |
| Obesity | neutrophil | *ZNF436* | 3.172657974 | 0.00296808 | 0.011289644 | 1 |
| Obesity | neutrophil | *MMP24* | 3.019624789 | 0.010117073 | 0.014959086 | 1 |
| Obesity | neutrophil | *UROD* | 7.752222925 | 0.001913478 | 0.011289644 | 1 |
| Obesity | neutrophil | *ST3GAL3* | 1.696789549 | 0.007026497 | 0.012031939 | 1 |
| Obesity | neutrophil | *KLC1* | 2.265460312 | 0.001748964 | 0.011289644 | 1 |

| | | | | | | |
|---|---|---|---|---|---|---|
| Obesity | neutrophil | *XRCC3* | 2.19012559 | 0.001268885 | 0.011289644 | 1 |
| Obesity | neutrophil | *MCF2L* | 3.882052638 | 0.00568038 | 0.011300433 | 1 |
| Obesity | neutrophil | *F10* | 1.3190999 | 0.004238782 | 0.011289644 | 1 |
| Obesity | neutrophil | *PCID2* | 1.317433597 | 0.004097833 | 0.011289644 | 1 |
| Obesity | neutrophil | *PROZ* | 12.29247137 | 0.003212875 | 0.011289644 | 1 |
| Obesity | neutrophil | *IGFLR1* | -2.573097846 | 0.005560705 | 0.011295339 | 1 |
| Obesity | neutrophil | *CAPNS1* | 3.295427843 | 0.009613412 | 0.014448846 | 1 |
| Obesity | neutrophil | *PRDX5* | 2.330437334 | 0.004568684 | 0.011289644 | 1 |
| Obesity | neutrophil | *SCAF1* | 3.031963834 | 0.004478579 | 0.011289644 | 1 |
| Obesity | neutrophil | *PRR12* | -1.608106853 | 0.022864332 | 0.029211147 | 1 |
| Obesity | neutrophil | *BECN1* | 1.32581003 | 0.004028924 | 0.011289644 | 1 |
| Obesity | neutrophil | *TRAP1* | 2.440191093 | 0.000170366 | 0.011289644 | 1 |
| Obesity | neutrophil | *DNAJC8* | 1.533300092 | 0.006440068 | 0.011648199 | 1 |
| Obesity | neutrophil | *ZNF384* | 1.503386145 | 0.003537089 | 0.011289644 | 1 |
| Obesity | neutrophil | *EMG1* | 1.448749711 | 0.003747868 | 0.011289644 | 1 |
| Obesity | neutrophil | *ATG14* | 2.2937614 | 0.005535668 | 0.011295339 | 1 |
| Obesity | neutrophil | *RHOJ* | 1.133496505 | 0.004289239 | 0.011289644 | 1 |
| Obesity | neutrophil | *L3HYPDH* | 1.417539162 | 0.020815988 | 0.02695574 | 1 |
| Obesity | neutrophil | *HSPA2* | -2.724724166 | 0.022205373 | 0.028537444 | 1 |
| Obesity | neutrophil | *ZBTB1* | 1.346953497 | 0.003784767 | 0.011289644 | 1 |
| Obesity | neutrophil | *PZP* | 1.729118182 | 0.004059877 | 0.011289644 | 1 |
| Obesity | neutrophil | *RHOT1* | 1.921787826 | 0.00207073 | 0.011289644 | 1 |
| Obesity | neutrophil | *WDR60* | 2.169618747 | 0.019969888 | 0.026001982 | 1 |
| Obesity | neutrophil | *AIF1L* | 1.610371579 | 0.004006456 | 0.011289644 | 1 |
| Obesity | neutrophil | *MAP2K2* | 3.06126836 | 0.017069222 | 0.022683196 | 1 |
| Obesity | neutrophil | *CANX* | 1.974916685 | 0.003314664 | 0.011289644 | 1 |
| Obesity | neutrophil | *IPPK* | 4.340069897 | 0.00410883 | 0.011289644 | 1 |
| Obesity | neutrophil | *FGD3* | -1.175243881 | 0.007510935 | 0.012399504 | 1 |
| Obesity | neutrophil | *ABHD8* | 2.949448665 | 0.002886683 | 0.011289644 | 1 |
| Obesity | neutrophil | *MASP1* | 2.024229569 | 0.005598694 | 0.011295339 | 1 |
| Obesity | neutrophil | *ATP13A4* | -1.718975977 | 0.003625992 | 0.011289644 | 1 |
| Obesity | neutrophil | *HRASLS* | 3.806656076 | 0.002581311 | 0.011289644 | 1 |
| Obesity | neutrophil | *HELB* | 1.315608674 | 0.003851671 | 0.011289644 | 1 |
| Obesity | neutrophil | *RAP1B* | 1.274331349 | 0.004420706 | 0.011289644 | 1 |
| Obesity | neutrophil | *RAB3IP* | 1.746867453 | 0.003667279 | 0.011289644 | 1 |
| Obesity | neutrophil | *YEATS4* | 5.913143839 | 0.000145093 | 0.011289644 | 0.912197635 |
| Obesity | neutrophil | *TMEM175* | 1.320154936 | 0.004134332 | 0.011289644 | 1 |
| Obesity | neutrophil | *EMC1* | 3.23946775 | 0.025645484 | 0.032449697 | 1 |
| Obesity | neutrophil | *UBR4* | 1.328993475 | 0.009170477 | 0.013973531 | 1 |
| Obesity | neutrophil | *SIN3B* | 2.323984203 | 0.004899102 | 0.011289644 | 1 |
| Obesity | neutrophil | *WDR24* | -4.759931836 | 0.005366865 | 0.011289644 | 1 |
| Obesity | neutrophil | *CHTF18* | -3.461482326 | 0.035146505 | 0.043174302 | 1 |
| Obesity | neutrophil | *SMARCA4* | 3.276327804 | 0.013015818 | 0.018152273 | 1 |
| Obesity | neutrophil | *TICAM1* | 2.41835743 | 0.005172321 | 0.011289644 | 1 |
| Obesity | neutrophil | *PNKD* | 2.282879983 | 0.006177286 | 0.011512519 | 1 |
| Obesity | neutrophil | *ZNF835* | -1.88960823 | 0.007794973 | 0.012669854 | 1 |
| Obesity | neutrophil | *HIP1* | 1.770087469 | 0.005436145 | 0.011289644 | 1 |
| Obesity | neutrophil | *PTPN12* | 2.244923712 | 0.006568479 | 0.011705223 | 1 |
| Obesity | neutrophil | *POR* | 1.394933879 | 0.004261509 | 0.011289644 | 1 |
| Obesity | neutrophil | *FGL2* | 2.896099605 | 0.004313412 | 0.011289644 | 1 |

| | | | | | | |
|---|---|---|---|---|---|---|
| Obesity | neutrophil | *STYXL1* | 2.823318975 | 0.001695665 | 0.011289644 | 1 |
| Obesity | neutrophil | *STEAP4* | 1.291342079 | 0.0055733 | 0.011295339 | 1 |
| Obesity | neutrophil | *GNAI1* | 1.779675208 | 0.004320241 | 0.011289644 | 1 |
| Obesity | neutrophil | *SGCE* | 1.53525244 | 0.005321252 | 0.011289644 | 1 |
| Obesity | neutrophil | *RBM48* | 3.648241313 | 0.012041053 | 0.017069244 | 1 |
| Obesity | neutrophil | *CASD1* | 1.838965123 | 0.004356504 | 0.011289644 | 1 |
| Obesity | neutrophil | *DGCR6L* | -2.414043911 | 0.011125377 | 0.016036425 | 1 |
| Obesity | neutrophil | *VPREB3* | -3.085178107 | 0.004328576 | 0.011289644 | 1 |
| Obesity | neutrophil | *ATF4* | 3.02684044 | 0.008879963 | 0.013723778 | 1 |
| Obesity | neutrophil | *A4GALT* | 1.451899545 | 0.004273665 | 0.011289644 | 1 |
| Obesity | neutrophil | *CDC42EP1* | 5.397475836 | 0.025646977 | 0.032449697 | 1 |
| Obesity | neutrophil | *TPST2* | 1.896309551 | 0.003690956 | 0.011289644 | 1 |
| Obesity | neutrophil | *BAIAP2L2* | 1.385814094 | 0.004125942 | 0.011289644 | 1 |
| Obesity | neutrophil | *TST* | -1.784156083 | 0.01988887 | 0.025920144 | 1 |
| Obesity | neutrophil | *EMC4* | 3.277556674 | 0.001296134 | 0.011289644 | 1 |
| Obesity | neutrophil | *SPECC1* | 2.492621311 | 0.006856984 | 0.011908801 | 1 |
| Obesity | neutrophil | *POT1* | 1.093420826 | 0.005337913 | 0.011289644 | 1 |
| Obesity | neutrophil | *CDHR3* | 1.016970508 | 0.004002582 | 0.011289644 | 1 |
| Obesity | neutrophil | *PRKRIP1* | 2.19922126 | 0.015892105 | 0.021392821 | 1 |
| Obesity | neutrophil | *FOXP2* | 0.78513533 | 0.005821351 | 0.011378838 | 1 |
| Obesity | neutrophil | *STRIP2* | -0.396577384 | 0.006408985 | 0.011648199 | 1 |
| Obesity | neutrophil | *IRF5* | 1.721523161 | 0.012346503 | 0.017396338 | 1 |
| Obesity | neutrophil | *KLHDC10* | 1.642878675 | 0.004665084 | 0.011289644 | 1 |
| Obesity | neutrophil | *OPN1SW* | 1.515420042 | 0.034752839 | 0.042749188 | 1 |
| Obesity | neutrophil | *HOXD1* | 14.45376254 | 0.004035035 | 0.011289644 | 1 |
| Obesity | neutrophil | *HOXD3* | 3.721442255 | 0.008190566 | 0.013013417 | 1 |
| Obesity | neutrophil | *CHN1* | 1.679356484 | 0.005084782 | 0.011289644 | 1 |
| Obesity | neutrophil | *GAD1* | 5.035511086 | 0.004771968 | 0.011289644 | 1 |
| Obesity | neutrophil | *ORMDL1* | 1.171201451 | 0.007374528 | 0.012294791 | 1 |
| Obesity | neutrophil | *HOXD9* | 3.683169138 | 0.031647777 | 0.039283233 | 1 |
| Obesity | neutrophil | *TWSG1* | 1.237505199 | 0.00448341 | 0.011289644 | 1 |
| Obesity | neutrophil | *WDFY4* | 2.005910218 | 0.031975617 | 0.039619768 | 1 |
| Obesity | neutrophil | *EIF2AK4* | 2.898793101 | 0.001721187 | 0.011289644 | 1 |
| Obesity | neutrophil | *TMOD2* | 1.151907087 | 0.006359567 | 0.011618636 | 1 |
| Obesity | neutrophil | *IVD* | 2.679750084 | 0.003510083 | 0.011289644 | 1 |
| Obesity | neutrophil | *VPS13C* | 1.539027982 | 0.004399033 | 0.011289644 | 1 |
| Obesity | neutrophil | *ACKR4* | 2.822895212 | 0.002906301 | 0.011289644 | 1 |
| Obesity | neutrophil | *SUMF2* | 1.164105202 | 0.004037602 | 0.011289644 | 1 |
| Obesity | neutrophil | *SPCS3* | 1.718028693 | 0.005201546 | 0.011289644 | 1 |
| Obesity | neutrophil | *SERGEF* | 1.146613698 | 0.003823363 | 0.011289644 | 1 |
| Obesity | neutrophil | *DCTD* | 1.54852034 | 0.005198828 | 0.011289644 | 1 |
| Obesity | neutrophil | *RPAIN* | 1.251636562 | 0.02403571 | 0.030564828 | 1 |
| Obesity | neutrophil | *USP6* | 2.090948898 | 0.025544501 | 0.032352595 | 1 |
| Obesity | neutrophil | *PLD2* | 3.415861541 | 0.00163619 | 0.011289644 | 1 |
| Obesity | neutrophil | *ATP1B2* | -1.29123587 | 0.005270849 | 0.011289644 | 1 |
| Obesity | neutrophil | *CCL4* | 1.432607179 | 0.003278209 | 0.011289644 | 1 |
| Obesity | neutrophil | *PHF20L1* | 2.657319875 | 0.006230554 | 0.01154827 | 1 |
| Obesity | neutrophil | *PUS7L* | 1.472164311 | 0.004320807 | 0.011289644 | 1 |
| Obesity | neutrophil | *KRI1* | 1.459417143 | 0.004608762 | 0.011289644 | 1 |
| Obesity | neutrophil | *SLC44A2* | 3.998084187 | 0.036888439 | 0.045137722 | 1 |

| | | | | | | |
|---|---|---|---|---|---|---|
| Obesity | neutrophil | *CDKN2D* | 2.752801562 | 0.001684866 | 0.011289644 | 1 |
| Obesity | neutrophil | *MTUS1* | 1.704541392 | 0.00509567 | 0.011289644 | 1 |
| Obesity | neutrophil | *NGDN* | 3.752889106 | 0.00694379 | 0.01196044 | 1 |
| Obesity | neutrophil | *RIPK3* | 4.131175247 | 0.001232954 | 0.011289644 | 1 |
| Obesity | neutrophil | *ADCY4* | -2.197755174 | 0.01442745 | 0.019731429 | 1 |
| Obesity | neutrophil | *RAB2B* | 1.878029485 | 0.00427415 | 0.011289644 | 1 |
| Obesity | neutrophil | *BCL2L2* | 2.180614534 | 0.004433323 | 0.011289644 | 1 |
| Obesity | neutrophil | *PARP2* | 3.260801308 | 0.0092974 | 0.014112205 | 1 |
| Obesity | neutrophil | *MIS18BP1* | 1.873029675 | 0.002064866 | 0.011289644 | 1 |
| Obesity | neutrophil | *RNASE1* | 2.332905695 | 0.028845345 | 0.03611844 | 1 |
| Obesity | neutrophil | *DAD1* | 2.881770894 | 0.010418797 | 0.015304435 | 1 |
| Obesity | neutrophil | *TEP1* | 2.250802959 | 0.009262234 | 0.014079222 | 1 |
| Obesity | neutrophil | *EPB41L4A* | 2.242609124 | 0.00572611 | 0.01134195 | 1 |
| Obesity | neutrophil | *CDO1* | 1.397021856 | 0.003256388 | 0.011289644 | 1 |
| Obesity | neutrophil | *REEP5* | 1.874941004 | 0.004718018 | 0.011289644 | 1 |
| Obesity | neutrophil | *QRICH2* | 2.707942365 | 0.009722798 | 0.014561037 | 1 |
| Obesity | neutrophil | *RHBDF2* | -2.60895288 | 0.037219729 | 0.04550767 | 1 |
| Obesity | neutrophil | *TMEM8A* | 1.544439113 | 0.005678719 | 0.011300433 | 1 |
| Obesity | neutrophil | *SHC2* | 2.569116691 | 0.001060349 | 0.011289644 | 1 |
| Obesity | neutrophil | *ERMARD* | 1.611514894 | 0.005744038 | 0.01134195 | 1 |
| Obesity | neutrophil | *PHF10* | 1.202758144 | 0.004306846 | 0.011289644 | 1 |
| Obesity | neutrophil | *GALNT8* | 2.403794644 | 0.001788637 | 0.011289644 | 1 |
| Obesity | neutrophil | *DOCK6* | 1.549790882 | 0.006734885 | 0.011825602 | 1 |
| Obesity | neutrophil | *ECSIT* | -2.449659511 | 0.015910869 | 0.021410881 | 1 |
| Obesity | neutrophil | *ELOF1* | 2.233192078 | 0.003849029 | 0.011289644 | 1 |
| Obesity | neutrophil | *CDC16* | 1.698580983 | 0.014499134 | 0.019812227 | 1 |
| Obesity | neutrophil | *THEM6* | -1.197708376 | 0.004616649 | 0.011289644 | 1 |
| Obesity | neutrophil | *TOMM40* | 1.623843541 | 0.004158448 | 0.011289644 | 1 |
| Obesity | neutrophil | *DPP6* | 2.753709675 | 0.004599908 | 0.011289644 | 1 |
| Obesity | neutrophil | *FAM98C* | 1.901304186 | 0.004092027 | 0.011289644 | 1 |
| Obesity | neutrophil | *RPL36* | 1.356252514 | 0.003810228 | 0.011289644 | 1 |
| Obesity | neutrophil | *GTPBP3* | 1.844765835 | 0.003855549 | 0.011289644 | 1 |
| Obesity | neutrophil | *PLVAP* | 1.225269504 | 0.004726274 | 0.011289644 | 1 |
| Obesity | neutrophil | *BST2* | -2.484862496 | 0.018395412 | 0.024210164 | 1 |
| Obesity | neutrophil | *DDA1* | -1.302710966 | 0.00747973 | 0.012378274 | 1 |
| Obesity | neutrophil | *LSM7* | 2.158579544 | 0.003654698 | 0.011289644 | 1 |
| Obesity | neutrophil | *TULP4* | 3.017719401 | 0.004404007 | 0.011289644 | 1 |
| Obesity | neutrophil | *RTN4IP1* | 4.319865486 | 0.003157103 | 0.011289644 | 1 |
| Obesity | neutrophil | *RSPH3* | 2.063111628 | 0.00345294 | 0.011289644 | 1 |
| Obesity | neutrophil | *STK33* | 2.776946953 | 0.002078773 | 0.011289644 | 1 |
| Obesity | neutrophil | *NDUFA10* | 2.164239362 | 0.009849539 | 0.014715792 | 1 |
| Obesity | neutrophil | *CACNG6* | 7.430290475 | 0.024546015 | 0.031175918 | 1 |
| Obesity | neutrophil | *PXDN* | 1.371360364 | 0.004331783 | 0.011289644 | 1 |
| Obesity | neutrophil | *PGPEP1* | 6.449847205 | 0.000169479 | 0.011289644 | 1 |
| Obesity | neutrophil | *LSM4* | 1.810032253 | 0.000828754 | 0.011289644 | 1 |
| Obesity | neutrophil | *HRC* | -1.639794319 | 0.001227151 | 0.011289644 | 1 |
| Obesity | neutrophil | *TRPM4* | 1.44283118 | 0.004886953 | 0.011289644 | 1 |
| Obesity | neutrophil | *CAMSAP1* | 2.855486771 | 0.005376615 | 0.011289644 | 1 |
| Obesity | neutrophil | *UBAC1* | 1.809507195 | 0.004621333 | 0.011289644 | 1 |
| Obesity | neutrophil | *TNNT3* | 2.294231522 | 0.003571378 | 0.011289644 | 1 |

| | | | | | | |
|---|---|---|---|---|---|---|
| Obesity | neutrophil | *ATXN10* | 1.580585379 | 0.004614365 | 0.011289644 | 1 |
| Obesity | neutrophil | *TUBGCP2* | 2.674925819 | 0.003363707 | 0.011289644 | 1 |
| Obesity | neutrophil | *PAK4* | -2.892510753 | 0.026283441 | 0.033194856 | 1 |
| Obesity | neutrophil | *ZNF337* | 2.484294018 | 0.016494081 | 0.022086962 | 1 |
| Obesity | neutrophil | *CEP85* | 1.623888255 | 0.003944227 | 0.011289644 | 1 |
| Obesity | neutrophil | *POMT1* | 1.743561529 | 0.008269192 | 0.013095317 | 1 |
| Obesity | neutrophil | *UCK1* | 2.282120144 | 0.004498088 | 0.011289644 | 1 |
| Obesity | neutrophil | *YIPF2* | 1.548824458 | 0.007186816 | 0.012155908 | 1 |
| Obesity | neutrophil | *TMEM160* | -2.287677883 | 0.009462134 | 0.014278393 | 1 |
| Obesity | neutrophil | *SMPDL3B* | -0.915486544 | 0.005224105 | 0.011289644 | 1 |
| Obesity | neutrophil | *ATPIF1* | 2.400436219 | 0.016473794 | 0.022064495 | 1 |
| Obesity | neutrophil | *CLIP1* | 1.29122399 | 0.002281444 | 0.011289644 | 1 |
| Obesity | neutrophil | *PPAN* | -3.76266701 | 0.011250807 | 0.016167732 | 1 |
| Obesity | neutrophil | *ZNF236* | 1.316546922 | 0.004484849 | 0.011289644 | 1 |
| Obesity | neutrophil | *PKDREJ* | -1.407672552 | 0.001006542 | 0.011289644 | 1 |
| Obesity | neutrophil | *EPS8L1* | 12.17482112 | 0.004077138 | 0.011289644 | 1 |
| Obesity | neutrophil | *LILRB2* | -3.256934085 | 0.013776108 | 0.018989342 | 1 |
| Obesity | neutrophil | *COX4I2* | 3.84054833 | 0.00671336 | 0.011817556 | 1 |
| Obesity | neutrophil | *ATP6V1E1* | 1.496178699 | 0.005527961 | 0.011295339 | 1 |
| Obesity | neutrophil | *CHMP1A* | 1.66637466 | 0.004182416 | 0.011289644 | 1 |
| Obesity | neutrophil | *PPT1* | 2.582788155 | 0.00381012 | 0.011289644 | 1 |
| Obesity | neutrophil | *HAUS8* | 2.567454704 | 0.00344692 | 0.011289644 | 1 |
| Obesity | neutrophil | *TBC1D5* | 1.540802061 | 0.004531078 | 0.011289644 | 1 |
| Obesity | neutrophil | *RFTN1* | 2.433411009 | 0.004374425 | 0.011289644 | 1 |
| Obesity | neutrophil | *PDLIM4* | -1.80459573 | 0.016719684 | 0.022308289 | 1 |
| Obesity | neutrophil | *AOC3* | 2.318994375 | 0.011507912 | 0.01646569 | 1 |
| Obesity | neutrophil | *ACLY* | -1.73978752 | 0.009371164 | 0.014196251 | 1 |
| Obesity | neutrophil | *AOC2* | -1.606289129 | 0.006490914 | 0.011671674 | 1 |
| Obesity | neutrophil | *DIAPH1* | 2.483514378 | 0.005062393 | 0.011289644 | 1 |
| Obesity | neutrophil | *EXOC4* | 1.811585765 | 0.002193141 | 0.011289644 | 1 |
| Obesity | neutrophil | *C1orf159* | 2.090497466 | 0.005227518 | 0.011289644 | 1 |
| Obesity | neutrophil | *ANO1* | 3.325411378 | 0.003485812 | 0.011289644 | 1 |
| Obesity | neutrophil | *PPFIA1* | 2.197160025 | 0.005718513 | 0.01133784 | 1 |
| Obesity | neutrophil | *NPHP4* | 3.445488719 | 0.005632283 | 0.011295339 | 1 |
| Obesity | neutrophil | *CKMT2* | 2.435436552 | 0.023557012 | 0.029992494 | 1 |
| Obesity | neutrophil | *ZCCHC9* | 1.296892775 | 0.003998812 | 0.011289644 | 1 |
| Obesity | neutrophil | *KHDRBS3* | 1.355032972 | 0.005132245 | 0.011289644 | 1 |
| Obesity | neutrophil | *CHD1L* | 2.621435504 | 0.016907715 | 0.022516057 | 1 |
| Obesity | neutrophil | *RBM8A* | 1.311959414 | 0.009817793 | 0.014678826 | 1 |
| Obesity | neutrophil | *ZNF304* | -5.530884848 | 0.010661302 | 0.015558868 | 1 |
| Obesity | neutrophil | *ZSCAN5A* | 2.167377215 | 0.005015937 | 0.011289644 | 1 |
| Obesity | neutrophil | *ACTR10* | 2.452692316 | 0.007948526 | 0.01278393 | 1 |
| Obesity | neutrophil | *GCH1* | 3.224481669 | 0.004600101 | 0.011289644 | 1 |
| Obesity | neutrophil | *LGALS3* | 1.376813362 | 0.003525752 | 0.011289644 | 1 |
| Obesity | neutrophil | *FBXW9* | -1.977839027 | 0.007585285 | 0.012456909 | 1 |
| Obesity | neutrophil | *C19orf57* | 3.240278864 | 0.004185271 | 0.011289644 | 1 |
| Obesity | neutrophil | *MATN3* | 5.88647347 | 0.00484723 | 0.011289644 | 1 |
| Obesity | neutrophil | *CCT6B* | 2.232233654 | 0.004683474 | 0.011289644 | 1 |
| Obesity | neutrophil | *RAF1* | 1.881747023 | 0.004980522 | 0.011289644 | 1 |
| Obesity | neutrophil | *ENOSF1* | 1.743988678 | 0.005923896 | 0.011385978 | 1 |

| | | | | | | |
|---|---|---|---|---|---|---|
| Obesity | neutrophil | *EMILIN2* | 4.384418706 | 4.07E-05 | 0.006238739 | 0.255788302 |
| Obesity | neutrophil | *TRIM5* | 2.198525567 | 0.004372639 | 0.011289644 | 1 |
| Obesity | neutrophil | *CNGA4* | 3.454198667 | 0.003301451 | 0.011289644 | 1 |
| Obesity | neutrophil | *TRIM22* | 2.338014385 | 0.005890111 | 0.011378838 | 1 |
| Obesity | neutrophil | *RRP8* | 2.43364881 | 0.004428115 | 0.011289644 | 1 |
| Obesity | neutrophil | *IMMT* | 3.024421715 | 0.002908049 | 0.011289644 | 1 |
| Obesity | neutrophil | *MRPL35* | -2.115979359 | 0.012301732 | 0.017356595 | 1 |
| Obesity | neutrophil | *CARD6* | 1.432613851 | 0.003317564 | 0.011289644 | 1 |
| Obesity | neutrophil | *MYBBP1A* | 1.817785448 | 0.003876608 | 0.011289644 | 1 |
| Obesity | neutrophil | *SERPINF1* | 1.34518308 | 0.003090704 | 0.011289644 | 1 |
| Obesity | neutrophil | *TMEM128* | 3.773748356 | 0.010147866 | 0.014990516 | 1 |
| Obesity | neutrophil | *WBP2* | 3.018058407 | 0.005229353 | 0.011289644 | 1 |
| Obesity | neutrophil | *EIF5A* | -2.027344037 | 0.003724207 | 0.011289644 | 1 |
| Obesity | neutrophil | *XAF1* | 2.023140804 | 0.01912854 | 0.025049183 | 1 |
| Obesity | neutrophil | *REEP2* | -0.506870446 | 0.002916279 | 0.011289644 | 1 |
| Obesity | neutrophil | *PRMT7* | 1.608461788 | 0.008472677 | 0.013326276 | 1 |
| Obesity | neutrophil | *TERF2* | 2.309996567 | 0.004520419 | 0.011289644 | 1 |
| Obesity | neutrophil | *BTBD3* | 3.265586294 | 0.009644883 | 0.014489218 | 1 |
| Obesity | neutrophil | *PCNA* | 2.247943682 | 0.00268994 | 0.011289644 | 1 |
| Obesity | neutrophil | *PTPRA* | 1.434171917 | 0.004501 | 0.011289644 | 1 |
| Obesity | neutrophil | *IGHMBP2* | 2.710996103 | 4.37E-05 | 0.006388824 | 0.274719414 |
| Obesity | neutrophil | *MMACHC* | -1.603773331 | 0.002468909 | 0.011289644 | 1 |
| Obesity | neutrophil | *MUTYH* | 4.324643086 | 0.002380783 | 0.011289644 | 1 |
| Obesity | neutrophil | *CTNNBL1* | 1.689679296 | 0.004249395 | 0.011289644 | 1 |
| Obesity | neutrophil | *LPIN3* | 1.377556067 | 0.004264227 | 0.011289644 | 1 |
| Obesity | neutrophil | *SERINC3* | 1.770908352 | 0.008527929 | 0.013373682 | 1 |
| Obesity | neutrophil | *DMGDH* | 3.265231298 | 0.041171342 | 0.049835238 | 1 |
| Obesity | neutrophil | *BHMT2* | 0.672481623 | 0.036906469 | 0.045150997 | 1 |
| Obesity | neutrophil | *KANK4* | 1.384859695 | 0.001426957 | 0.011289644 | 1 |
| Obesity | neutrophil | *CASP9* | 1.297429245 | 0.019440597 | 0.025404912 | 1 |
| Obesity | neutrophil | *ATP8A2* | 2.130324666 | 0.004230816 | 0.011289644 | 1 |
| Obesity | neutrophil | *USPL1* | 1.941916388 | 0.004626162 | 0.011289644 | 1 |
| Obesity | neutrophil | *POMP* | 2.729795216 | 0.005263572 | 0.011289644 | 1 |
| Obesity | neutrophil | *WASF3* | 1.340079241 | 0.004367497 | 0.011289644 | 1 |
| Obesity | neutrophil | *PEMT* | 1.939670266 | 0.023193262 | 0.029566764 | 1 |
| Obesity | neutrophil | *SCO1* | 1.839292844 | 0.004676589 | 0.011289644 | 1 |
| Obesity | neutrophil | *DSTYK* | 1.492971882 | 0.004957866 | 0.011289644 | 1 |
| Obesity | neutrophil | *TMCC2* | 2.645951241 | 0.011136067 | 0.016039508 | 1 |
| Obesity | neutrophil | *COG6* | 1.368997641 | 0.00451931 | 0.011289644 | 1 |
| Obesity | neutrophil | *TRPC4* | 4.291864387 | 0.005173029 | 0.011289644 | 1 |
| Obesity | neutrophil | *RFC3* | 1.599914255 | 0.005011518 | 0.011289644 | 1 |
| Obesity | neutrophil | *PRAM1* | -1.568741776 | 0.003646684 | 0.011289644 | 1 |
| Obesity | neutrophil | *ZNF414* | 2.10582639 | 0.004242032 | 0.011289644 | 1 |
| Obesity | neutrophil | *CNDP2* | 1.422908837 | 0.004247134 | 0.011289644 | 1 |
| Obesity | neutrophil | *RTN3* | 1.07987778 | 0.004004384 | 0.011289644 | 1 |
| Obesity | neutrophil | *C1QTNF6* | -3.091285427 | 0.01665902 | 0.022264543 | 1 |
| Obesity | neutrophil | *FAM83F* | 3.926138757 | 0.004381323 | 0.011289644 | 1 |
| Obesity | neutrophil | *GIMAP4* | 4.124407762 | 0.003727054 | 0.011289644 | 1 |
| Obesity | neutrophil | *ACTR3B* | 2.796161817 | 0.004691216 | 0.011289644 | 1 |
| Obesity | neutrophil | *DYDC2* | 9.080762374 | 0.007465464 | 0.01236443 | 1 |

| | | | | | | |
|---|---|---|---|---|---|---|
| Obesity | neutrophil | *IPO8* | 2.136475142 | 0.004985876 | 0.011289644 | 1 |
| Obesity | neutrophil | *SPINK5* | -1.818676575 | 0.007488104 | 0.012385612 | 1 |
| Obesity | neutrophil | *LRRCC1* | 4.02165536 | 0.031018924 | 0.038586461 | 1 |
| Obesity | neutrophil | *SWAP70* | 1.793313268 | 0.004398262 | 0.011289644 | 1 |
| Obesity | neutrophil | *AMPD3* | 1.705088721 | 0.003329743 | 0.011289644 | 1 |
| Obesity | neutrophil | *SBF2* | 1.448213585 | 0.004716942 | 0.011289644 | 1 |
| Obesity | neutrophil | *MICAL2* | 3.182979085 | 0.01299346 | 0.018137186 | 1 |
| Obesity | neutrophil | *RRAS2* | 1.131244854 | 0.001692373 | 0.011289644 | 1 |
| Obesity | neutrophil | *HSD17B4* | 1.263895492 | 0.005318741 | 0.011289644 | 1 |
| Obesity | neutrophil | *DPF2* | 2.518128796 | 0.003872134 | 0.011289644 | 1 |
| Obesity | neutrophil | *GSC* | 4.953216294 | 0.004346347 | 0.011289644 | 1 |
| Obesity | neutrophil | *CATSPERB* | 1.209264952 | 0.005186408 | 0.011289644 | 1 |
| Obesity | neutrophil | *COX16* | 3.226737772 | 0.022775053 | 0.029132606 | 1 |
| Obesity | neutrophil | *MED6* | 1.560521501 | 0.002363676 | 0.011289644 | 1 |
| Obesity | neutrophil | *ELP3* | 2.533755237 | 0.005348129 | 0.011289644 | 1 |
| Obesity | neutrophil | *ADAMDEC1* | 8.732358616 | 0.01795683 | 0.023717351 | 1 |
| Obesity | neutrophil | *MRO* | 2.556931162 | 0.016662108 | 0.022264543 | 1 |
| Obesity | neutrophil | *CNTN6* | -3.621311913 | 0.008689895 | 0.013539098 | 1 |
| Obesity | neutrophil | *KATNBL1* | 1.388304855 | 0.003413413 | 0.011289644 | 1 |
| Obesity | neutrophil | *GSTM5* | 2.722269887 | 0.000301013 | 0.011289644 | 1 |
| Obesity | neutrophil | *GSTM3* | 1.665826565 | 0.009951062 | 0.014815909 | 1 |
| Obesity | neutrophil | *SYT6* | 5.54080793 | 0.004296631 | 0.011289644 | 1 |
| Obesity | neutrophil | *VAV3* | 2.907552957 | 0.012553583 | 0.017636732 | 1 |
| Obesity | neutrophil | *PSRC1* | -3.787369384 | 0.023907425 | 0.030413999 | 1 |
| Obesity | neutrophil | *CEPT1* | 1.586785592 | 0.005032488 | 0.011289644 | 1 |
| Obesity | neutrophil | *AP4B1* | -3.18421216 | 0.020437127 | 0.026508814 | 1 |
| Obesity | neutrophil | *NAPG* | 2.864436043 | 0.004026994 | 0.011289644 | 1 |
| Obesity | neutrophil | *SPIRE1* | 2.036093051 | 0.011448004 | 0.016387432 | 1 |
| Obesity | neutrophil | *PPHLN1* | 1.570668503 | 0.002876796 | 0.011289644 | 1 |
| Obesity | neutrophil | *FKBP11* | -1.291578693 | 0.007460198 | 0.01236443 | 1 |
| Obesity | neutrophil | *YWHAQ* | 1.061699442 | 0.005862144 | 0.011378838 | 1 |
| Obesity | neutrophil | *LPIN1* | 2.162305696 | 0.009205876 | 0.014011804 | 1 |
| Obesity | neutrophil | *LDHA* | 1.336669951 | 0.004090221 | 0.011289644 | 1 |
| Obesity | neutrophil | *ANO3* | 1.603111855 | 0.021853661 | 0.028117283 | 1 |
| Obesity | neutrophil | *FST* | 2.118080865 | 0.002496321 | 0.011289644 | 1 |
| Obesity | neutrophil | *TIMM17A* | 1.884615201 | 0.004106085 | 0.011289644 | 1 |
| Obesity | neutrophil | *NARS* | 3.257821117 | 0.006144377 | 0.011504964 | 1 |
| Obesity | neutrophil | *KIAA1468* | 1.656100191 | 0.005833796 | 0.011378838 | 1 |
| Obesity | neutrophil | *FBXO18* | 1.056514519 | 0.004380323 | 0.011289644 | 1 |
| Obesity | neutrophil | *ECHDC3* | 1.31730377 | 0.006737961 | 0.011825602 | 1 |
| Obesity | neutrophil | *IL15RA* | 1.259636181 | 0.004200024 | 0.011289644 | 1 |
| Obesity | neutrophil | *TMEM241* | 2.017605095 | 0.005230777 | 0.011289644 | 1 |
| Obesity | neutrophil | *EMP1* | 1.50079496 | 0.007952095 | 0.0127864 | 1 |
| Obesity | neutrophil | *KLRD1* | -1.31095387 | 0.01107663 | 0.015980311 | 1 |
| Obesity | neutrophil | *DDB2* | 1.151200143 | 0.00299188 | 0.011289644 | 1 |
| Obesity | neutrophil | *ACP2* | 9.359540479 | 0.000881969 | 0.011289644 | 1 |
| Obesity | neutrophil | *HOOK1* | 4.71955347 | 0.006545654 | 0.011694062 | 1 |
| Obesity | neutrophil | *ZCCHC11* | 1.788036279 | 0.003569506 | 0.011289644 | 1 |
| Obesity | neutrophil | *FHOD3* | 1.105213348 | 0.005602494 | 0.011295339 | 1 |
| Obesity | neutrophil | *TPGS2* | 3.766259293 | 0.010485918 | 0.015374292 | 1 |

| | | | | | | |
|---|---|---|---|---|---|---|
| Obesity | neutrophil | *SLC43A3* | 1.409820958 | 0.008064402 | 0.01288711 | 1 |
| Obesity | neutrophil | *TIMM10* | 6.055905076 | 0.004992509 | 0.011289644 | 1 |
| Obesity | neutrophil | *APLNR* | 2.387648143 | 0.003300729 | 0.011289644 | 1 |
| Obesity | neutrophil | *FADS2* | 3.346021362 | 0.004525366 | 0.011289644 | 1 |
| Obesity | neutrophil | *TMEM165* | 1.593518133 | 0.012028459 | 0.017055731 | 1 |
| Obesity | neutrophil | *CLOCK* | 1.125494497 | 0.004216611 | 0.011289644 | 1 |
| Obesity | neutrophil | *DZIP1* | 1.766365838 | 0.001259921 | 0.011289644 | 1 |
| Obesity | neutrophil | *BIVM* | 1.76364995 | 0.004146489 | 0.011289644 | 1 |
| Obesity | neutrophil | *STT3A* | 1.845244158 | 0.003427816 | 0.011289644 | 1 |
| Obesity | neutrophil | *KLB* | 2.527505428 | 0.000626585 | 0.011289644 | 1 |
| Obesity | neutrophil | *WDR36* | 1.090966958 | 0.004218902 | 0.011289644 | 1 |
| Obesity | neutrophil | *OSTF1* | 2.36650159 | 0.00489761 | 0.011289644 | 1 |
| Obesity | neutrophil | *UBQLN1* | 1.611199908 | 0.004568044 | 0.011289644 | 1 |
| Obesity | neutrophil | *NAA35* | 1.793621534 | 0.004552035 | 0.011289644 | 1 |
| Obesity | neutrophil | *CTSL* | 1.989554683 | 0.004266874 | 0.011289644 | 1 |
| Obesity | neutrophil | *TMEM2* | 1.275230825 | 0.004137475 | 0.011289644 | 1 |
| Obesity | neutrophil | *AGTPBP1* | 3.455375718 | 0.014769402 | 0.020102886 | 1 |
| Obesity | neutrophil | *GOLM1* | 1.670980394 | 0.009915888 | 0.014786809 | 1 |
| Obesity | neutrophil | *ADAM19* | -2.275802825 | 0.009268388 | 0.014085172 | 1 |
| Obesity | neutrophil | *USP30* | 2.074519046 | 0.003110671 | 0.011289644 | 1 |
| Obesity | neutrophil | *HRK* | -0.56503413 | 0.004738371 | 0.011289644 | 1 |
| Obesity | neutrophil | *P2RX4* | 6.049003439 | 0.002464925 | 0.011289644 | 1 |
| Obesity | neutrophil | *DTX1* | 1.349454502 | 0.004010756 | 0.011289644 | 1 |
| Obesity | neutrophil | *DMTF1* | 1.446568644 | 0.003476257 | 0.011289644 | 1 |
| Obesity | neutrophil | *CCDC146* | 2.066048947 | 0.006414735 | 0.011648199 | 1 |
| Obesity | neutrophil | *TMEM60* | -2.303724629 | 0.001178289 | 0.011289644 | 1 |
| Obesity | neutrophil | *SRPK2* | 2.042504408 | 0.005805445 | 0.011378838 | 1 |
| Obesity | neutrophil | *MDFIC* | 1.243910243 | 0.004603176 | 0.011289644 | 1 |
| Obesity | neutrophil | *MTO1* | 2.06050459 | 0.003049748 | 0.011289644 | 1 |
| Obesity | neutrophil | *ANKRD6* | 1.359771393 | 0.004499241 | 0.011289644 | 1 |
| Obesity | neutrophil | *KHDC1* | 1.509547828 | 0.005876014 | 0.011378838 | 1 |
| Obesity | neutrophil | *SNX14* | 1.865539798 | 0.00650883 | 0.011671674 | 1 |
| Obesity | neutrophil | *NT5E* | 1.293428526 | 0.005447434 | 0.011289644 | 1 |
| Obesity | neutrophil | *PRR5L* | 1.603747511 | 0.003845864 | 0.011289644 | 1 |
| Obesity | neutrophil | *ATP5G2* | 1.380923564 | 0.004114452 | 0.011289644 | 1 |
| Obesity | neutrophil | *ITGA7* | 1.132547233 | 0.009874369 | 0.01473538 | 1 |
| Obesity | neutrophil | *RDH5* | 0.948549242 | 0.006125352 | 0.011482393 | 1 |
| Obesity | neutrophil | *BLOC1S1* | 1.659355366 | 0.005141544 | 0.011289644 | 1 |
| Obesity | neutrophil | *PPP1R1A* | 2.052264989 | 0.004012789 | 0.011289644 | 1 |
| Obesity | neutrophil | *B4GALNT1* | 6.648473785 | 0.03807676 | 0.046429129 | 1 |
| Obesity | neutrophil | *FAIM2* | 3.849634089 | 0.004279552 | 0.011289644 | 1 |
| Obesity | neutrophil | *HNRNPA1* | 3.999014047 | 0.004585513 | 0.011289644 | 1 |
| Obesity | neutrophil | *SLC26A10* | 6.518170819 | 0.002513952 | 0.011289644 | 1 |
| Obesity | neutrophil | *MAP7* | 1.717300298 | 0.006685715 | 0.011792087 | 1 |
| Obesity | neutrophil | *AHI1* | 1.031906316 | 0.029896769 | 0.037305116 | 1 |
| Obesity | neutrophil | *PKIB* | 1.96813406 | 0.00329789 | 0.011289644 | 1 |
| Obesity | neutrophil | *MICAL1* | 4.996687434 | 0.01118195 | 0.016083486 | 1 |
| Obesity | neutrophil | *PRADC1* | 2.238674316 | 0.003390278 | 0.011289644 | 1 |
| Obesity | neutrophil | *SMYD5* | -1.266101871 | 0.008288446 | 0.013122504 | 1 |
| Obesity | neutrophil | *KCNMB4* | 1.397663926 | 0.007345834 | 0.012284058 | 1 |

| | | | | | | |
|---|---|---|---|---|---|---|
| Obesity | neutrophil | *USP15* | 2.498283085 | 0.014706287 | 0.020029988 | 1 |
| Obesity | neutrophil | *KLHL36* | 3.594140048 | 0.00406961 | 0.011289644 | 1 |
| Obesity | neutrophil | *MPHOSPH6* | 1.568361386 | 0.004159651 | 0.011289644 | 1 |
| Obesity | neutrophil | *FHOD1* | 1.810922331 | 0.003567862 | 0.011289644 | 1 |
| Obesity | neutrophil | *ZNF670* | 3.935216957 | 0.000892566 | 0.011289644 | 1 |
| Obesity | neutrophil | *NTPCR* | 3.514500092 | 0.002224302 | 0.011289644 | 1 |
| Obesity | neutrophil | *PIGC* | 2.274698508 | 0.003133194 | 0.011289644 | 1 |
| Obesity | neutrophil | *SP110* | 2.511440145 | 0.003362976 | 0.011289644 | 1 |
| Obesity | neutrophil | *USP37* | 2.29749175 | 0.001817969 | 0.011289644 | 1 |
| Obesity | neutrophil | *SLC19A3* | 0.849630408 | 0.005042867 | 0.011289644 | 1 |
| Obesity | neutrophil | *TMBIM1* | 1.751679204 | 0.006517042 | 0.011671674 | 1 |
| Obesity | neutrophil | *ARMC9* | 2.286613964 | 0.005054852 | 0.011289644 | 1 |
| Obesity | neutrophil | *GCC2* | 1.369047586 | 0.006053275 | 0.01145004 | 1 |
| Obesity | neutrophil | *MRPS9* | 2.905501985 | 0.000405918 | 0.011289644 | 1 |
| Obesity | neutrophil | *ANKRD36* | 1.293130781 | 0.00401972 | 0.011289644 | 1 |
| Obesity | neutrophil | *USP44* | 2.052419644 | 0.002129497 | 0.011289644 | 1 |
| Obesity | neutrophil | *APPL2* | 1.69416831 | 0.004720259 | 0.011289644 | 1 |
| Obesity | neutrophil | *DRAM1* | 1.148270194 | 0.004377072 | 0.011289644 | 1 |
| Obesity | neutrophil | *NEK3* | 3.619407923 | 0.007917037 | 0.012769218 | 1 |
| Obesity | neutrophil | *VPS36* | 2.364972425 | 0.002789476 | 0.011289644 | 1 |
| Obesity | neutrophil | *RNASEH2B* | 1.587966678 | 0.005532047 | 0.011295339 | 1 |
| Obesity | neutrophil | *CKAP2* | -2.561428981 | 0.007963989 | 0.012793301 | 1 |
| Obesity | neutrophil | *TBC1D4* | 1.791719208 | 0.003950543 | 0.011289644 | 1 |
| Obesity | neutrophil | *LRCH1* | 2.954986064 | 0.017239385 | 0.022885138 | 1 |
| Obesity | neutrophil | *SUCLA2* | 2.10004607 | 0.006687372 | 0.011792087 | 1 |
| Obesity | neutrophil | *RCBTB1* | 1.574207972 | 0.01025994 | 0.015113459 | 1 |
| Obesity | neutrophil | *MED4* | 2.562123201 | 0.003818473 | 0.011289644 | 1 |
| Obesity | neutrophil | *RCBTB2* | 1.226669187 | 0.004489041 | 0.011289644 | 1 |
| Obesity | neutrophil | *C7orf25* | -1.413502703 | 0.001814125 | 0.011289644 | 1 |
| Obesity | neutrophil | *CHST12* | 1.222949696 | 0.005044799 | 0.011289644 | 1 |
| Obesity | neutrophil | *GPNMB* | 1.5917189 | 0.006043601 | 0.011447874 | 1 |
| Obesity | neutrophil | *KDELR2* | 2.370685137 | 0.004549955 | 0.011289644 | 1 |
| Obesity | neutrophil | *NUPL2* | 2.104554046 | 0.007789208 | 0.012663758 | 1 |
| Obesity | neutrophil | *ZDHHC4* | 1.879942773 | 0.012773503 | 0.017865854 | 1 |
| Obesity | neutrophil | *TBRG4* | 1.675394505 | 0.006613281 | 0.011727259 | 1 |
| Obesity | neutrophil | *DDX56* | 3.005969117 | 0.004581763 | 0.011289644 | 1 |
| Obesity | neutrophil | *HUS1* | 2.126761004 | 0.004287224 | 0.011289644 | 1 |
| Obesity | neutrophil | *NACAD* | 6.777694612 | 0.013187622 | 0.018359296 | 1 |
| Obesity | neutrophil | *DBNL* | 1.970684726 | 0.003607957 | 0.011289644 | 1 |
| Obesity | neutrophil | *TTC5* | 4.011175674 | 0.040408044 | 0.048986767 | 1 |
| Obesity | neutrophil | *ALPK3* | 1.317569085 | 0.005112089 | 0.011289644 | 1 |
| Obesity | neutrophil | *NMT1* | 2.185765509 | 0.000448433 | 0.011289644 | 1 |
| Obesity | neutrophil | *MYCBPAP* | 1.896707549 | 0.037240668 | 0.045515567 | 1 |
| Obesity | neutrophil | *BRIP1* | 4.56917102 | 0.008374608 | 0.013232259 | 1 |
| Obesity | neutrophil | *MRPL47* | 2.835270866 | 0.004117824 | 0.011289644 | 1 |
| Obesity | neutrophil | *EPRS* | 1.831785171 | 0.004422193 | 0.011289644 | 1 |
| Obesity | neutrophil | *RPS6KC1* | 1.715347101 | 0.005750863 | 0.011349688 | 1 |
| Obesity | neutrophil | *IL1RN* | 2.971704028 | 0.004679892 | 0.011289644 | 1 |
| Obesity | neutrophil | *SAP130* | 2.227324892 | 0.006667025 | 0.011770735 | 1 |
| Obesity | neutrophil | *UGGT1* | 2.231822092 | 0.005027993 | 0.011289644 | 1 |

| | | | | | | |
|---|---|---|---|---|---|---|
| Obesity | neutrophil | *NIPSNAP3A* | 1.273339706 | 0.008901847 | 0.013733966 | 1 |
| Obesity | neutrophil | *TXN* | 1.440829858 | 0.00515866 | 0.011289644 | 1 |
| Obesity | neutrophil | *ODF2* | 2.737690307 | 0.00369004 | 0.011289644 | 1 |
| Obesity | neutrophil | *TOR1B* | 1.532660457 | 0.006732931 | 0.011825602 | 1 |
| Obesity | neutrophil | *TOR1A* | 3.055640861 | 0.020890906 | 0.027019364 | 1 |
| Obesity | neutrophil | *SLC2A8* | -2.182047422 | 0.03431652 | 0.042278652 | 1 |
| Obesity | neutrophil | *ZFP37* | 1.771459851 | 0.007974803 | 0.012799996 | 1 |
| Obesity | neutrophil | *ZNF189* | 1.386606527 | 0.004537527 | 0.011289644 | 1 |
| Obesity | neutrophil | *ALDOB* | 4.029938778 | 0.01974532 | 0.025754943 | 1 |
| Obesity | neutrophil | *STX17* | 2.272750043 | 0.003754664 | 0.011289644 | 1 |
| Obesity | neutrophil | *PRPF4* | 3.701404104 | 0.001148039 | 0.011289644 | 1 |
| Obesity | neutrophil | *FPGS* | 1.248040381 | 0.005160521 | 0.011289644 | 1 |
| Obesity | neutrophil | *TEX10* | 2.939200345 | 0.00651108 | 0.011671674 | 1 |
| Obesity | neutrophil | *GARNL3* | 1.379173564 | 0.004321989 | 0.011289644 | 1 |
| Obesity | neutrophil | *GABBR2* | 3.712630654 | 0.004202154 | 0.011289644 | 1 |
| Obesity | neutrophil | *PSMB7* | 2.098032663 | 0.005009859 | 0.011289644 | 1 |
| Obesity | neutrophil | *RABEPK* | 2.130333457 | 0.004295242 | 0.011289644 | 1 |
| Obesity | neutrophil | *GOLGA1* | 3.323430119 | 0.009545925 | 0.014371464 | 1 |
| Obesity | neutrophil | *DSCC1* | 2.767874507 | 8.74E-05 | 0.009468977 | 0.54920067 |
| Obesity | neutrophil | *DERL1* | 1.364056228 | 0.004690497 | 0.011289644 | 1 |
| Obesity | neutrophil | *NOV* | 5.509293625 | 0.005868089 | 0.011378838 | 1 |
| Obesity | neutrophil | *POLR1E* | -1.574944167 | 0.00845516 | 0.013302701 | 1 |
| Obesity | neutrophil | *IL11RA* | 1.422061154 | 0.008899475 | 0.013733966 | 1 |
| Obesity | neutrophil | *APTX* | 3.102992238 | 0.01702344 | 0.022641499 | 1 |
| Obesity | neutrophil | *TLN1* | 2.075886681 | 0.003762516 | 0.011289644 | 1 |
| Obesity | neutrophil | *SPAG8* | 3.130218162 | 0.002341565 | 0.011289644 | 1 |
| Obesity | neutrophil | *CD72* | 1.426493218 | 0.002697131 | 0.011289644 | 1 |
| Obesity | neutrophil | *TMEM8B* | 1.709836626 | 0.004497876 | 0.011289644 | 1 |
| Obesity | neutrophil | *GRHPR* | 1.762893594 | 0.004474198 | 0.011289644 | 1 |
| Obesity | neutrophil | *DENND4C* | 1.250282933 | 0.00403684 | 0.011289644 | 1 |
| Obesity | neutrophil | *KIF13A* | 1.441688012 | 0.004577752 | 0.011289644 | 1 |
| Obesity | neutrophil | *ZSCAN9* | 2.011752764 | 0.000242523 | 0.011289644 | 1 |
| Obesity | neutrophil | *TMEM63B* | 1.245184451 | 0.010437868 | 0.015321708 | 1 |
| Obesity | neutrophil | *FRS3* | -1.919010844 | 0.008863972 | 0.013712548 | 1 |
| Obesity | neutrophil | *SLC22A23* | 2.455062327 | 0.006613408 | 0.011727259 | 1 |
| Obesity | neutrophil | *LRRC1* | 3.919198575 | 0.00611628 | 0.011478524 | 1 |
| Obesity | neutrophil | *FOXF2* | 6.222562754 | 0.040334293 | 0.048925661 | 1 |
| Obesity | neutrophil | *BPHL* | 1.206224744 | 0.009485305 | 0.014307608 | 1 |
| Obesity | neutrophil | *RIPK1* | 2.349937095 | 0.00534443 | 0.011289644 | 1 |
| Obesity | neutrophil | *TCF19* | -0.786229334 | 0.013609691 | 0.018801171 | 1 |
| Obesity | neutrophil | *FLOT1* | 3.093478757 | 0.001836755 | 0.011289644 | 1 |
| Obesity | neutrophil | *NRM* | 4.797149366 | 0.000129652 | 0.011289644 | 0.815121796 |
| Obesity | neutrophil | *MTCH1* | 1.784147597 | 0.004134953 | 0.011289644 | 1 |
| Obesity | neutrophil | *FAM8A1* | 1.498039561 | 0.004223661 | 0.011289644 | 1 |
| Obesity | neutrophil | *FGFBP2* | -1.66660925 | 0.003256759 | 0.011289644 | 1 |
| Obesity | neutrophil | *MYO7A* | 1.61712702 | 0.011139239 | 0.016040402 | 1 |
| Obesity | neutrophil | *ARRB1* | 1.615740169 | 0.004467689 | 0.011289644 | 1 |
| Obesity | neutrophil | *CCDC90B* | 1.394805375 | 0.004561852 | 0.011289644 | 1 |
| Obesity | neutrophil | *SYTL2* | -1.84069527 | 0.007964527 | 0.012793301 | 1 |
| Obesity | neutrophil | *PRCP* | 2.081287641 | 0.002758587 | 0.011289644 | 1 |

| | | | | | | |
|---|---|---|---|---|---|---|
| Obesity | neutrophil | *NARS2* | 1.08154215 | 0.004228609 | 0.011289644 | 1 |
| Obesity | neutrophil | *PI15* | 12.31030928 | 0.002517046 | 0.011289644 | 1 |
| Obesity | neutrophil | *GGH* | -1.112758502 | 0.00680744 | 0.011874479 | 1 |
| Obesity | neutrophil | *SULF1* | 1.127711313 | 0.003281332 | 0.011289644 | 1 |
| Obesity | neutrophil | *NEK1* | 1.794192809 | 0.005118093 | 0.011289644 | 1 |
| Obesity | neutrophil | *DDX60* | 1.205785827 | 0.004176223 | 0.011289644 | 1 |
| Obesity | neutrophil | *MMP27* | 2.216210229 | 0.003404971 | 0.011289644 | 1 |
| Obesity | neutrophil | *DCUN1D5* | 1.764379423 | 0.002607548 | 0.011289644 | 1 |
| Obesity | neutrophil | *SLC37A4* | -1.162936548 | 0.006115777 | 0.011478524 | 1 |
| Obesity | neutrophil | *PPP2R1B* | 0.989186402 | 0.004271856 | 0.011289644 | 1 |
| Obesity | neutrophil | *FXYD6* | 2.553898842 | 0.003110205 | 0.011289644 | 1 |
| Obesity | neutrophil | *TMPRSS13* | 6.945671907 | 0.01478878 | 0.02012055 | 1 |
| Obesity | neutrophil | *MAP2K5* | 2.477828528 | 0.002398099 | 0.011289644 | 1 |
| Obesity | neutrophil | *NDUFAF1* | 1.615409648 | 0.002624968 | 0.011289644 | 1 |
| Obesity | neutrophil | *ITGA11* | 1.058621193 | 0.004461499 | 0.011289644 | 1 |
| Obesity | neutrophil | *UACA* | 1.496004893 | 0.004195055 | 0.011289644 | 1 |
| Obesity | neutrophil | *PLCB2* | -1.077506737 | 0.021762279 | 0.028025286 | 1 |
| Obesity | neutrophil | *RSL24D1* | 1.492471869 | 0.009071292 | 0.013872833 | 1 |
| Obesity | neutrophil | *SPTBN5* | 2.261994085 | 0.008020582 | 0.012843963 | 1 |
| Obesity | neutrophil | *RABGGTB* | 1.992638837 | 0.004357592 | 0.011289644 | 1 |
| Obesity | neutrophil | *SLC44A5* | 1.97053459 | 0.00142905 | 0.011289644 | 1 |
| Obesity | neutrophil | *DNASE2B* | -23.22133074 | 0.007534035 | 0.012412599 | 1 |
| Obesity | neutrophil | *DBT* | 1.74968401 | 0.004611224 | 0.011289644 | 1 |
| Obesity | neutrophil | *RTCA* | 2.809007889 | 0.00160423 | 0.011289644 | 1 |
| Obesity | neutrophil | *IFT172* | 1.454078885 | 0.001995742 | 0.011289644 | 1 |
| Obesity | neutrophil | *KHK* | -1.408563036 | 0.000762542 | 0.011289644 | 1 |
| Obesity | neutrophil | *ADCY3* | 1.284461066 | 0.003877218 | 0.011289644 | 1 |
| Obesity | neutrophil | *DYNC2LI1* | 1.940483675 | 0.003457637 | 0.011289644 | 1 |
| Obesity | neutrophil | *LHCGR* | 1.767232515 | 0.005913482 | 0.011379877 | 1 |
| Obesity | neutrophil | *THUMPD2* | 2.595151589 | 0.005637822 | 0.011295339 | 1 |
| Obesity | neutrophil | *ABCG5* | -2.351418135 | 0.032175304 | 0.039821236 | 1 |
| Obesity | neutrophil | *CENPO* | 2.043794712 | 0.004003897 | 0.011289644 | 1 |
| Obesity | neutrophil | *LRPPRC* | 1.458015496 | 0.004383062 | 0.011289644 | 1 |
| Obesity | neutrophil | *CYP2C9* | 2.733715401 | 0.020806929 | 0.02694956 | 1 |
| Obesity | neutrophil | *CYP2C8* | -2.818435524 | 0.015783586 | 0.021287893 | 1 |
| Obesity | neutrophil | *MYOF* | 1.446839203 | 0.012313394 | 0.017365256 | 1 |
| Obesity | neutrophil | *STAMBPL1* | 1.228077837 | 0.003869862 | 0.011289644 | 1 |
| Obesity | neutrophil | *ATAD1* | 3.270390268 | 0.014963618 | 0.020323237 | 1 |
| Obesity | neutrophil | *BTBD16* | 2.259983901 | 0.003480623 | 0.011289644 | 1 |
| Obesity | neutrophil | *TACC2* | 1.319540479 | 0.003128196 | 0.011289644 | 1 |
| Obesity | neutrophil | *CALHM2* | 1.622034111 | 0.003736554 | 0.011289644 | 1 |
| Obesity | neutrophil | *ARL3* | 3.93961247 | 0.006052222 | 0.01145004 | 1 |
| Obesity | neutrophil | *RBP4* | 1.588129742 | 0.003501163 | 0.011289644 | 1 |
| Obesity | neutrophil | *DNAJC13* | 2.290577383 | 0.00564203 | 0.011296638 | 1 |
| Obesity | neutrophil | *FAM149B1* | 1.510521566 | 0.004009184 | 0.011289644 | 1 |
| Obesity | neutrophil | *ASCC1* | 1.685232346 | 0.008813477 | 0.013657957 | 1 |
| Obesity | neutrophil | *RPS24* | 1.907005226 | 0.004755394 | 0.011289644 | 1 |
| Obesity | neutrophil | *DNA2* | 1.115791506 | 0.017264407 | 0.022906617 | 1 |
| Obesity | neutrophil | *SMARCAL1* | 2.458583849 | 0.005507544 | 0.011289644 | 1 |
| Obesity | neutrophil | *STAT4* | -1.180664986 | 0.010517069 | 0.015405595 | 1 |

| | | | | | | |
|---|---|---|---|---|---|---|
| Obesity | neutrophil | *CARF* | 4.902067386 | 3.55E-05 | 0.005728017 | 0.223392673 |
| Obesity | neutrophil | *ASNSD1* | -1.937944832 | 0.014034647 | 0.019269671 | 1 |
| Obesity | neutrophil | *NAB1* | 3.097965164 | 0.007668308 | 0.012523726 | 1 |
| Obesity | neutrophil | *FASTKD1* | 1.555611842 | 0.004055764 | 0.011289644 | 1 |
| Obesity | neutrophil | *MDH1B* | 4.805274072 | 0.004188914 | 0.011289644 | 1 |
| Obesity | neutrophil | *FAM117B* | 3.743641219 | 0.014385287 | 0.019678046 | 1 |
| Obesity | neutrophil | *ITGAV* | 2.91893696 | 0.016091408 | 0.021630679 | 1 |
| Obesity | neutrophil | *SENP7* | 1.275284377 | 0.004661177 | 0.011289644 | 1 |
| Obesity | neutrophil | *COX17* | 1.724120118 | 0.004044354 | 0.011289644 | 1 |
| Obesity | neutrophil | *USP8* | 2.725354693 | 0.00830001 | 0.013134197 | 1 |
| Obesity | neutrophil | *SPPL2A* | 1.835298357 | 0.005373125 | 0.011289644 | 1 |
| Obesity | neutrophil | *UBL7* | 3.271182873 | 0.000938774 | 0.011289644 | 1 |
| Obesity | neutrophil | *ARHGAP24* | 2.351772466 | 0.019856949 | 0.025889805 | 1 |
| Obesity | neutrophil | *FAM13A* | 1.415567297 | 0.006882632 | 0.011932455 | 1 |
| Obesity | neutrophil | *HERC3* | 2.480621181 | 0.006509472 | 0.011671674 | 1 |
| Obesity | neutrophil | *AP1AR* | 1.251994261 | 0.004437437 | 0.011289644 | 1 |
| Obesity | neutrophil | *PRKG2* | 1.42449512 | 0.003864152 | 0.011289644 | 1 |
| Obesity | neutrophil | *BBS7* | 1.344152796 | 0.004235968 | 0.011289644 | 1 |
| Obesity | neutrophil | *KIAA1109* | 1.439971924 | 0.004549626 | 0.011289644 | 1 |
| Obesity | neutrophil | *PRDM5* | 1.306855526 | 0.005032261 | 0.011289644 | 1 |
| Obesity | neutrophil | *TRPC3* | 3.055133053 | 0.007899385 | 0.01275056 | 1 |
| Obesity | neutrophil | *NAAA* | 4.008860458 | 0.025173613 | 0.031915004 | 1 |
| Obesity | neutrophil | *NUP54* | 1.648128488 | 0.005274372 | 0.011289644 | 1 |
| Obesity | neutrophil | *BMP2K* | 1.378206951 | 0.014190049 | 0.019461789 | 1 |
| Obesity | neutrophil | *G3BP2* | 3.480665201 | 0.004362264 | 0.011289644 | 1 |
| Obesity | neutrophil | *SCARB2* | 2.347855533 | 0.006518092 | 0.011671674 | 1 |
| Obesity | neutrophil | *CNOT6L* | 1.484041947 | 0.010835776 | 0.015751748 | 1 |
| Obesity | neutrophil | *ANXA3* | 1.658860299 | 0.003919522 | 0.011289644 | 1 |
| Obesity | neutrophil | *PPA2* | 1.801774728 | 0.006942776 | 0.01196044 | 1 |
| Obesity | neutrophil | *GSTCD* | 1.643343712 | 0.003701818 | 0.011289644 | 1 |
| Obesity | neutrophil | *ENPEP* | 1.626493157 | 0.003299467 | 0.011289644 | 1 |
| Obesity | neutrophil | *SLC39A8* | 2.187234506 | 0.004385026 | 0.011289644 | 1 |
| Obesity | neutrophil | *FBN2* | 1.089421035 | 0.036281981 | 0.044482218 | 1 |
| Obesity | neutrophil | *B4GALNT3* | -0.538634809 | 0.004463212 | 0.011289644 | 1 |
| Obesity | neutrophil | *ERP27* | -4.611886764 | 0.034655348 | 0.042637607 | 1 |
| Obesity | neutrophil | *ETV6* | 2.002163375 | 0.00567654 | 0.011300433 | 1 |
| Obesity | neutrophil | *KIF21A* | 1.490417334 | 0.007573455 | 0.012444932 | 1 |
| Obesity | neutrophil | *CPNE8* | 1.38785131 | 0.006827972 | 0.011878501 | 1 |
| Obesity | neutrophil | *ALG10* | 2.132801092 | 0.004297794 | 0.011289644 | 1 |
| Obesity | neutrophil | *SLCO1C1* | 2.501030175 | 0.00229232 | 0.011289644 | 1 |
| Obesity | neutrophil | *ZCRB1* | 1.369424033 | 0.003489451 | 0.011289644 | 1 |
| Obesity | neutrophil | *PRICKLE1* | 0.914580842 | 0.004794107 | 0.011289644 | 1 |
| Obesity | neutrophil | *C1RL* | 2.05722528 | 0.003831816 | 0.011289644 | 1 |
| Obesity | neutrophil | *CLSTN3* | 4.562443389 | 0.006487718 | 0.011671674 | 1 |
| Obesity | neutrophil | *KLRG1* | 1.437368591 | 0.00074674 | 0.011289644 | 1 |
| Obesity | neutrophil | *VAMP1* | 1.8581709 | 0.009060172 | 0.013865944 | 1 |
| Obesity | neutrophil | *TAPBPL* | 3.191728229 | 0.037979583 | 0.046319619 | 1 |
| Obesity | neutrophil | *CD27* | 2.743317843 | 0.002116267 | 0.011289644 | 1 |
| Obesity | neutrophil | *PEX5* | 1.760094462 | 0.004090217 | 0.011289644 | 1 |
| Obesity | neutrophil | *PPFIA2* | 0.982788241 | 0.004499189 | 0.011289644 | 1 |

| | | | | | | |
|---|---|---|---|---|---|---|
| Obesity | neutrophil | *GLIPR1* | 4.10932406 | 0.028524305 | 0.035752055 | 1 |
| Obesity | neutrophil | *LGR5* | -1.484256063 | 0.007805588 | 0.012674 | 1 |
| Obesity | neutrophil | *POC1B* | 2.015849114 | 0.004516518 | 0.011289644 | 1 |
| Obesity | neutrophil | *AMDHD1* | 2.116767773 | 0.03112458 | 0.038694925 | 1 |
| Obesity | neutrophil | *GAS2L3* | 1.369383518 | 0.004290069 | 0.011289644 | 1 |
| Obesity | neutrophil | *TMEM132B* | 2.555282138 | 0.00592836 | 0.011391075 | 1 |
| Obesity | neutrophil | *TDG* | 3.157398806 | 0.011602399 | 0.01655567 | 1 |
| Obesity | neutrophil | *GLTP* | 2.831853942 | 0.007036111 | 0.012040736 | 1 |
| Obesity | neutrophil | *MTMR6* | 2.784928187 | 0.004744232 | 0.011289644 | 1 |
| Obesity | neutrophil | *SLC46A3* | 2.003127459 | 0.005638464 | 0.011295339 | 1 |
| Obesity | neutrophil | *SUOX* | 1.955610104 | 0.014924033 | 0.020273854 | 1 |
| Obesity | neutrophil | *CCDC65* | 2.315823236 | 0.009939419 | 0.014804341 | 1 |
| Obesity | neutrophil | *ACVRL1* | 1.335045998 | 0.009857932 | 0.014724831 | 1 |
| Obesity | neutrophil | *GPR84* | -1.315038961 | 0.004067523 | 0.011289644 | 1 |
| Obesity | neutrophil | *CERS5* | 1.266105953 | 0.00387207 | 0.011289644 | 1 |
| Obesity | neutrophil | *LMBR1L* | 1.752581617 | 0.004741211 | 0.011289644 | 1 |
| Obesity | neutrophil | *ZNF740* | 1.475614898 | 0.003859496 | 0.011289644 | 1 |
| Obesity | neutrophil | *WDFY2* | 1.66427011 | 0.007797593 | 0.012670836 | 1 |
| Obesity | neutrophil | *HNRNPA1L2* | 4.605853898 | 0.003213382 | 0.011289644 | 1 |
| Obesity | neutrophil | *LPAR6* | 1.990872643 | 0.004927491 | 0.011289644 | 1 |
| Obesity | neutrophil | *VPS33A* | 6.005033087 | 0.020836674 | 0.026976339 | 1 |
| Obesity | neutrophil | *RHOF* | 1.6828316 | 0.02238061 | 0.028709834 | 1 |
| Obesity | neutrophil | *GRTP1* | 1.359422175 | 0.013163887 | 0.018334373 | 1 |
| Obesity | neutrophil | *TSSK4* | 7.96934252 | 0.016382714 | 0.021965903 | 1 |
| Obesity | neutrophil | *RTN1* | 1.32307019 | 0.003987815 | 0.011289644 | 1 |
| Obesity | neutrophil | *C14orf37* | 2.111041467 | 0.006596967 | 0.01172215 | 1 |
| Obesity | neutrophil | *RAB15* | 1.380748756 | 0.01224163 | 0.017287316 | 1 |
| Obesity | neutrophil | *STON2* | 2.291548915 | 0.005696415 | 0.011302651 | 1 |
| Obesity | neutrophil | *EFCAB11* | 4.489687439 | 0.002215577 | 0.011289644 | 1 |
| Obesity | neutrophil | *AK7* | 3.059631447 | 0.004024837 | 0.011289644 | 1 |
| Obesity | neutrophil | *SLC24A4* | -1.615946169 | 0.004624984 | 0.011289644 | 1 |
| Obesity | neutrophil | *FBLN5* | 1.967048748 | 0.003446796 | 0.011289644 | 1 |
| Obesity | neutrophil | *C14orf79* | -0.995251263 | 0.005908041 | 0.011378838 | 1 |
| Obesity | neutrophil | *NIPA2* | 2.74921518 | 0.004967729 | 0.011289644 | 1 |
| Obesity | neutrophil | *DUOXA1* | 2.628854627 | 0.033186092 | 0.040974266 | 1 |
| Obesity | neutrophil | *MFAP1* | 1.426579099 | 0.003574256 | 0.011289644 | 1 |
| Obesity | neutrophil | *TCF12* | 1.448892653 | 0.004762866 | 0.011289644 | 1 |
| Obesity | neutrophil | *SORD* | 2.437351543 | 0.003011024 | 0.011289644 | 1 |
| Obesity | neutrophil | *ZSCAN29* | 2.350268296 | 0.003056619 | 0.011289644 | 1 |
| Obesity | neutrophil | *LYSMD2* | 3.106338744 | 0.009931422 | 0.01479594 | 1 |
| Obesity | neutrophil | *BAHD1* | 4.749193323 | 0.00784382 | 0.01270327 | 1 |
| Obesity | neutrophil | *ANP32A* | 2.507139273 | 0.005459444 | 0.011289644 | 1 |
| Obesity | neutrophil | *COMMD4* | 4.327275097 | 0.011786073 | 0.016756907 | 1 |
| Obesity | neutrophil | *UBE2Q2* | 2.926501735 | 0.00965926 | 0.014501216 | 1 |
| Obesity | neutrophil | *PSTPIP1* | -0.806004015 | 0.005555983 | 0.011295339 | 1 |
| Obesity | neutrophil | *HMG20A* | 2.046162661 | 0.0029207 | 0.011289644 | 1 |
| Obesity | neutrophil | *SCAPER* | 1.673360064 | 0.007055719 | 0.012054159 | 1 |
| Obesity | neutrophil | *NEIL1* | -2.867764709 | 0.018126598 | 0.02391144 | 1 |
| Obesity | neutrophil | *MAN2C1* | 0.880162644 | 0.016516835 | 0.022103308 | 1 |
| Obesity | neutrophil | *DNAJA4* | 2.589945142 | 0.003866193 | 0.011289644 | 1 |

| | | | | | | |
|---|---|---|---|---|---|---|
| Obesity | neutrophil | *TPM1* | 1.373413919 | 0.005028154 | 0.011289644 | 1 |
| Obesity | neutrophil | *IGF1R* | 1.441619431 | 0.004325945 | 0.011289644 | 1 |
| Obesity | neutrophil | *CYP11A1* | 3.060523106 | 0.000209271 | 0.011289644 | 1 |
| Obesity | neutrophil | *BBS4* | 1.57223197 | 0.002764161 | 0.011289644 | 1 |
| Obesity | neutrophil | *ULK3* | 2.075959091 | 0.004810139 | 0.011289644 | 1 |
| Obesity | neutrophil | *SCAMP2* | 2.000874602 | 0.002494523 | 0.011289644 | 1 |
| Obesity | neutrophil | *HAPLN3* | 2.043065232 | 0.005398505 | 0.011289644 | 1 |
| Obesity | neutrophil | *POLG* | 2.402189227 | 0.003987522 | 0.011289644 | 1 |
| Obesity | neutrophil | *TICRR* | -4.433689458 | 0.007429767 | 0.012334552 | 1 |
| Obesity | neutrophil | *MFGE8* | 2.130232963 | 0.007812313 | 0.012681646 | 1 |
| Obesity | neutrophil | *UNC45A* | 4.108930253 | 0.010615417 | 0.015509906 | 1 |
| Obesity | neutrophil | *ST8SIA2* | 21.22649949 | 0.006384305 | 0.011647743 | 1 |
| Obesity | neutrophil | *IQGAP1* | 1.824964915 | 0.005896517 | 0.011378838 | 1 |
| Obesity | neutrophil | *CRTC3* | 1.49106647 | 0.00441988 | 0.011289644 | 1 |
| Obesity | neutrophil | *SEC11A* | 2.319786755 | 0.005967698 | 0.011423216 | 1 |
| Obesity | neutrophil | *TGFB1I1* | 1.913962149 | 0.009866129 | 0.014733576 | 1 |
| Obesity | neutrophil | *PARN* | 1.05457353 | 0.004352927 | 0.011289644 | 1 |
| Obesity | neutrophil | *FTO* | 1.286852514 | 0.005237028 | 0.011289644 | 1 |
| Obesity | neutrophil | *MARVELD3* | -8.429441478 | 0.009919905 | 0.014789292 | 1 |
| Obesity | neutrophil | *ZFHX3* | 1.122898538 | 0.004030283 | 0.011289644 | 1 |
| Obesity | neutrophil | *KIFC3* | 2.644951427 | 0.00534785 | 0.011289644 | 1 |
| Obesity | neutrophil | *ADAMTS18* | 1.336590448 | 0.004882393 | 0.011289644 | 1 |
| Obesity | neutrophil | *NUDT7* | -1.227130911 | 0.007396055 | 0.012298069 | 1 |
| Obesity | neutrophil | *GCSH* | 2.004754588 | 0.007830778 | 0.012699648 | 1 |
| Obesity | neutrophil | *CMTM3* | 5.008376554 | 0.011546904 | 0.016502986 | 1 |
| Obesity | neutrophil | *MAP1LC3B* | 1.530154345 | 0.005126575 | 0.011289644 | 1 |
| Obesity | neutrophil | *TLDC1* | 4.153690236 | 0.005232643 | 0.011289644 | 1 |
| Obesity | neutrophil | *RPS2* | 2.288853151 | 0.008260475 | 0.013088107 | 1 |
| Obesity | neutrophil | *NDUFB10* | 1.702208849 | 0.003718023 | 0.011289644 | 1 |
| Obesity | neutrophil | *DEF8* | 1.85187251 | 0.007463844 | 0.01236443 | 1 |
| Obesity | neutrophil | *GAS8* | 1.764838703 | 0.002549171 | 0.011289644 | 1 |
| Obesity | neutrophil | *COPS3* | 2.033514814 | 0.004313598 | 0.011289644 | 1 |
| Obesity | neutrophil | *RANBP10* | 1.961049902 | 0.007969767 | 0.01279518 | 1 |
| Obesity | neutrophil | *PRPSAP2* | 2.881760764 | 0.003315517 | 0.011289644 | 1 |
| Obesity | neutrophil | *MYO19* | 2.663866448 | 0.000733852 | 0.011289644 | 1 |
| Obesity | neutrophil | *DDX52* | 1.683670999 | 0.00459599 | 0.011289644 | 1 |
| Obesity | neutrophil | *UNC45B* | 8.763985086 | 0.020020967 | 0.026055024 | 1 |
| Obesity | neutrophil | *TOM1L1* | 4.048734066 | 0.011070814 | 0.015980311 | 1 |
| Obesity | neutrophil | *C17orf80* | 1.841092882 | 0.001918687 | 0.011289644 | 1 |
| Obesity | neutrophil | *VPS53* | 1.92957999 | 0.003480139 | 0.011289644 | 1 |
| Obesity | neutrophil | *SPATA22* | 1.199395607 | 0.004336449 | 0.011289644 | 1 |
| Obesity | neutrophil | *SGSM2* | 1.588883981 | 0.005249529 | 0.011289644 | 1 |
| Obesity | neutrophil | *NPEPPS* | 1.285891358 | 0.004015521 | 0.011289644 | 1 |
| Obesity | neutrophil | *LRRC46* | 5.015266997 | 0.007568843 | 0.012440606 | 1 |
| Obesity | neutrophil | *SCRN2* | 1.553192643 | 0.002401027 | 0.011289644 | 1 |
| Obesity | neutrophil | *RHBDL3* | 2.649690907 | 0.004377908 | 0.011289644 | 1 |
| Obesity | neutrophil | *ABCA8* | 1.742364214 | 0.002341386 | 0.011289644 | 1 |
| Obesity | neutrophil | *G6PC3* | -2.603397247 | 0.01310368 | 0.018254562 | 1 |
| Obesity | neutrophil | *IMPA2* | 1.368988259 | 0.004077617 | 0.011289644 | 1 |
| Obesity | neutrophil | *GNAL* | 4.03844948 | 0.008083828 | 0.012905797 | 1 |

| | | | | | | |
|---|---|---|---|---|---|---|
| Obesity | neutrophil | *RPRD1A* | 1.517425593 | 0.005007847 | 0.011289644 | 1 |
| Obesity | neutrophil | *C18orf21* | 7.394270587 | 0.016694715 | 0.022293898 | 1 |
| Obesity | neutrophil | *GALNT1* | 3.230530143 | 0.01140896 | 0.016351571 | 1 |
| Obesity | neutrophil | *ASXL3* | 1.57163728 | 0.004709915 | 0.011289644 | 1 |
| Obesity | neutrophil | *ADCYAP1* | -2.654849115 | 0.002662229 | 0.011289644 | 1 |
| Obesity | neutrophil | *ESCO1* | 3.209549312 | 0.011255438 | 0.01617069 | 1 |
| Obesity | neutrophil | *GREB1L* | 1.027714947 | 0.004925314 | 0.011289644 | 1 |
| Obesity | neutrophil | *C18orf8* | 1.251878492 | 0.038674497 | 0.047075601 | 1 |
| Obesity | neutrophil | *NPC1* | 1.413927016 | 0.022609712 | 0.028950562 | 1 |
| Obesity | neutrophil | *SLC14A1* | -1.333576806 | 0.010573967 | 0.01546372 | 1 |
| Obesity | neutrophil | *CCDC40* | 2.025582878 | 0.024926992 | 0.031627851 | 1 |
| Obesity | neutrophil | *TMC6* | 1.873934788 | 0.002159018 | 0.011289644 | 1 |
| Obesity | neutrophil | *CARD14* | 1.767619873 | 0.004215263 | 0.011289644 | 1 |
| Obesity | neutrophil | *TBCD* | 1.517626291 | 0.005104036 | 0.011289644 | 1 |
| Obesity | neutrophil | *FN3KRP* | 2.062797112 | 0.002688761 | 0.011289644 | 1 |
| Obesity | neutrophil | *NARF* | 1.302464422 | 0.003938454 | 0.011289644 | 1 |
| Obesity | neutrophil | *TRIM65* | 3.657300703 | 0.000136916 | 0.011289644 | 0.860788491 |
| Obesity | neutrophil | *SECTM1* | -2.648534157 | 0.008140076 | 0.012962679 | 1 |
| Obesity | neutrophil | *RNF157* | 1.764906669 | 0.001010142 | 0.011289644 | 1 |
| Obesity | neutrophil | *WDR45B* | 1.436789487 | 0.005273094 | 0.011289644 | 1 |
| Obesity | neutrophil | *RNF165* | -2.316446157 | 0.01853737 | 0.024376583 | 1 |
| Obesity | neutrophil | *MAPK4* | 2.492569896 | 0.013236259 | 0.018402557 | 1 |
| Obesity | neutrophil | *SMAD4* | 1.098494375 | 0.004861986 | 0.011289644 | 1 |
| Obesity | neutrophil | *NT5C3B* | 1.483927765 | 0.009928427 | 0.01479594 | 1 |
| Obesity | neutrophil | *PIP4K2B* | 1.27525238 | 0.003902179 | 0.011289644 | 1 |
| Obesity | neutrophil | *GRB7* | 5.668209566 | 0.001604137 | 0.011289644 | 1 |
| Obesity | neutrophil | *FKBP10* | 1.599319803 | 0.009776532 | 0.014634538 | 1 |
| Obesity | neutrophil | *TXNL4A* | 1.62857764 | 0.003632048 | 0.011289644 | 1 |
| Obesity | neutrophil | *NFIC* | 1.667406714 | 0.005226294 | 0.011289644 | 1 |
| Obesity | neutrophil | *TPGS1* | 3.924476173 | 0.002508845 | 0.011289644 | 1 |
| Obesity | neutrophil | *PRDM15* | 1.917767358 | 0.034864845 | 0.04287019 | 1 |
| Obesity | neutrophil | *PFKL* | 1.852916862 | 0.005478999 | 0.011289644 | 1 |
| Obesity | neutrophil | *MVB12A* | 4.781788848 | 0.002110533 | 0.011289644 | 1 |
| Obesity | neutrophil | *DUS3L* | 3.28062052 | 0.021618627 | 0.027863122 | 1 |
| Obesity | neutrophil | *DPP9* | 1.520513766 | 0.004640814 | 0.011289644 | 1 |
| Obesity | neutrophil | *CCDC97* | 4.095913217 | 0.00906567 | 0.013870775 | 1 |
| Obesity | neutrophil | *TMEM91* | -1.21173057 | 0.009079052 | 0.013881322 | 1 |
| Obesity | neutrophil | *COL6A1* | 1.848097857 | 0.004516245 | 0.011289644 | 1 |
| Obesity | neutrophil | *IFNAR1* | 1.985570386 | 0.027539915 | 0.034614843 | 1 |
| Obesity | neutrophil | *TRPM2* | 1.202068459 | 0.00735806 | 0.012285536 | 1 |
| Obesity | neutrophil | *TMEM50B* | 2.212185759 | 0.022239884 | 0.028570116 | 1 |
| Obesity | neutrophil | *DOPEY2* | 1.471343294 | 0.004699983 | 0.011289644 | 1 |
| Obesity | neutrophil | *URB1* | 1.661638399 | 0.006621819 | 0.011733759 | 1 |
| Obesity | neutrophil | *SAE1* | 1.534926174 | 0.004007122 | 0.011289644 | 1 |
| Obesity | neutrophil | *GEMIN7* | -2.055301296 | 0.005475901 | 0.011289644 | 1 |
| Obesity | neutrophil | *WTIP* | 2.296954074 | 0.005265057 | 0.011289644 | 1 |
| Obesity | neutrophil | *CAPN10* | 1.778521826 | 0.004628631 | 0.011289644 | 1 |
| Obesity | neutrophil | *MYO1F* | -1.167865676 | 0.010723326 | 0.015628699 | 1 |
| Obesity | neutrophil | *CACNG8* | 4.610812969 | 0.003793402 | 0.011289644 | 1 |
| Obesity | neutrophil | *FBN3* | 4.33200565 | 0.002297972 | 0.011289644 | 1 |

| | | | | | | |
|---|---|---|---|---|---|---|
| Obesity | neutrophil | *RPS11* | 2.315038023 | 0.002126106 | 0.011289644 | 1 |
| Obesity | neutrophil | *CTU1* | 2.248628986 | 0.007470403 | 0.01236935 | 1 |
| Obesity | neutrophil | *RCN3* | 2.501493609 | 0.004494154 | 0.011289644 | 1 |
| Obesity | neutrophil | *RERE* | 1.547910611 | 0.003592897 | 0.011289644 | 1 |
| Obesity | neutrophil | *PEX14* | 1.590521283 | 0.004556119 | 0.011289644 | 1 |
| Obesity | neutrophil | *SH3BGRL3* | 4.053176815 | 0.019514118 | 0.025490392 | 1 |
| Obesity | neutrophil | *IL22RA1* | 4.985066766 | 0.009710999 | 0.014553767 | 1 |
| Obesity | neutrophil | *ZNF593* | 2.720041666 | 0.004112128 | 0.011289644 | 1 |
| Obesity | neutrophil | *C1orf216* | 2.61616905 | 0.00276732 | 0.011289644 | 1 |
| Obesity | neutrophil | *MAP3K6* | 2.142016318 | 0.00479415 | 0.011289644 | 1 |
| Obesity | neutrophil | *GPN2* | 2.447418498 | 0.006715907 | 0.011817556 | 1 |
| Obesity | neutrophil | *NBPF3* | 2.515545132 | 0.002465183 | 0.011289644 | 1 |
| Obesity | neutrophil | *HSPG2* | 1.236480328 | 0.009463665 | 0.014278393 | 1 |
| Obesity | neutrophil | *ITGB3BP* | 2.681671382 | 0.015844375 | 0.021343186 | 1 |
| Obesity | neutrophil | *PIGK* | 1.672885446 | 0.004603408 | 0.011289644 | 1 |
| Obesity | neutrophil | *RPS8* | 2.046056475 | 0.002271631 | 0.011289644 | 1 |
| Obesity | neutrophil | *MOB3C* | 4.036447787 | 0.003847963 | 0.011289644 | 1 |
| Obesity | neutrophil | *CYP4B1* | 1.298825803 | 0.005626592 | 0.011295339 | 1 |
| Obesity | neutrophil | *SYPL2* | 2.267891484 | 0.001597643 | 0.011289644 | 1 |
| Obesity | neutrophil | *ZNF697* | 2.328611599 | 0.004981441 | 0.011289644 | 1 |
| Obesity | neutrophil | *CELSR2* | 5.483038886 | 0.009291077 | 0.014110944 | 1 |
| Obesity | neutrophil | *ATP1B1* | 1.352504744 | 0.005835723 | 0.011378838 | 1 |
| Obesity | neutrophil | *TIPRL* | 1.992233272 | 0.003232939 | 0.011289644 | 1 |
| Obesity | neutrophil | *NME7* | 1.194252073 | 0.005388018 | 0.011289644 | 1 |
| Obesity | neutrophil | *CREG1* | 2.362283529 | 0.004237247 | 0.011289644 | 1 |
| Obesity | neutrophil | *GPA33* | 4.302088524 | 0.007668186 | 0.012523726 | 1 |
| Obesity | neutrophil | *RXRG* | 5.37489492 | 0.003540916 | 0.011289644 | 1 |
| Obesity | neutrophil | *TMCO1* | 3.659335469 | 0.015878271 | 0.021380743 | 1 |
| Obesity | neutrophil | *MGST3* | 2.787127154 | 0.007445562 | 0.012350988 | 1 |
| Obesity | neutrophil | *ADCY10* | 1.955886825 | 0.003977465 | 0.011289644 | 1 |
| Obesity | neutrophil | *RFWD2* | 2.120305032 | 0.007242532 | 0.012207452 | 1 |
| Obesity | neutrophil | *PPOX* | 2.301927613 | 0.002930329 | 0.011289644 | 1 |
| Obesity | neutrophil | *RGS5* | 2.110202175 | 0.002504414 | 0.011289644 | 1 |
| Obesity | neutrophil | *RRNAD1* | 1.858282781 | 0.003211077 | 0.011289644 | 1 |
| Obesity | neutrophil | *PIGM* | 1.578780029 | 0.003847606 | 0.011289644 | 1 |
| Obesity | neutrophil | *RGL1* | 1.627834669 | 0.004492611 | 0.011289644 | 1 |
| Obesity | neutrophil | *TARS2* | 1.596674607 | 0.003922697 | 0.011289644 | 1 |
| Obesity | neutrophil | *SNX27* | 2.128852933 | 0.005505451 | 0.011289644 | 1 |
| Obesity | neutrophil | *ENSA* | 1.223567552 | 0.003987784 | 0.011289644 | 1 |
| Obesity | neutrophil | *SEMA6C* | 2.479274203 | 0.001775904 | 0.011289644 | 1 |
| Obesity | neutrophil | *ARNT* | 2.374650478 | 0.005380484 | 0.011289644 | 1 |
| Obesity | neutrophil | *POGZ* | 1.767550377 | 0.002462411 | 0.011289644 | 1 |
| Obesity | neutrophil | *GABPB2* | 1.636156488 | 0.013953212 | 0.019178803 | 1 |
| Obesity | neutrophil | *IKBKE* | -0.874338771 | 0.00594673 | 0.01140172 | 1 |
| Obesity | neutrophil | *DYRK3* | 5.427638132 | 0.01209931 | 0.017128656 | 1 |
| Obesity | neutrophil | *EIF2D* | 1.54194104 | 0.006564685 | 0.011701779 | 1 |
| Obesity | neutrophil | *HHIPL2* | 5.525688167 | 0.029899869 | 0.037305116 | 1 |
| Obesity | neutrophil | *TP53BP2* | 3.273745737 | 0.006793627 | 0.011870336 | 1 |
| Obesity | neutrophil | *S100A8* | -0.631049631 | 0.004027247 | 0.011289644 | 1 |
| Obesity | neutrophil | *HAX1* | 1.569780459 | 0.001900523 | 0.011289644 | 1 |

| | | | | | | |
|---|---|---|---|---|---|---|
| Obesity | neutrophil | *CREB3L4* | 2.237742508 | 0.016687489 | 0.02229372 | 1 |
| Obesity | neutrophil | *KCNN3* | 1.665343101 | 0.003214468 | 0.011289644 | 1 |
| Obesity | neutrophil | *GATAD2B* | 1.196207846 | 0.005390308 | 0.011289644 | 1 |
| Obesity | neutrophil | *RIT1* | 1.461369687 | 0.003864451 | 0.011289644 | 1 |
| Obesity | neutrophil | *GALNT2* | 2.310754082 | 0.005496768 | 0.011289644 | 1 |
| Obesity | neutrophil | *SCCPDH* | 2.525731891 | 0.004612476 | 0.011289644 | 1 |
| Obesity | neutrophil | *CEP170* | 2.030294523 | 0.001968296 | 0.011289644 | 1 |
| Obesity | neutrophil | *NVL* | 2.144381027 | 0.004919986 | 0.011289644 | 1 |
| Obesity | neutrophil | *FBXO28* | 2.052266764 | 0.004786274 | 0.011289644 | 1 |
| Obesity | neutrophil | *CNIH4* | 3.044249894 | 0.006267461 | 0.011565541 | 1 |
| Obesity | neutrophil | *GUK1* | 2.3213972 | 0.002744438 | 0.011289644 | 1 |
| Obesity | neutrophil | *CDC42BPA* | 1.73906349 | 0.002579475 | 0.011289644 | 1 |
| Obesity | neutrophil | *PSEN2* | 7.539502009 | 0.002487318 | 0.011289644 | 1 |
| Obesity | neutrophil | *PYCR2* | 1.870118191 | 0.005585779 | 0.011295339 | 1 |
| Obesity | neutrophil | *WNT9A* | 1.81430203 | 0.003604691 | 0.011289644 | 1 |
| Obesity | neutrophil | *PLEKHA6* | 1.483807191 | 0.002859558 | 0.011289644 | 1 |
| Obesity | neutrophil | *ABCG8* | -1.373925839 | 0.006125648 | 0.011482393 | 1 |
| Obesity | neutrophil | *EML4* | 2.811626985 | 0.010051589 | 0.0149184 | 1 |
| Obesity | neutrophil | *CHAC2* | -1.126042106 | 0.00738892 | 0.012298069 | 1 |
| Obesity | neutrophil | *WDPCP* | 1.107600157 | 0.001824073 | 0.011289644 | 1 |
| Obesity | neutrophil | *ASXL2* | 1.779375489 | 0.005713865 | 0.011332198 | 1 |
| Obesity | neutrophil | *CIAO1* | 1.230024839 | 0.003809043 | 0.011289644 | 1 |
| Obesity | neutrophil | *ZNF514* | 1.410740348 | 0.00699736 | 0.012000111 | 1 |
| Obesity | neutrophil | *EXOC6B* | 1.340669933 | 0.004401912 | 0.011289644 | 1 |
| Obesity | neutrophil | *TEX261* | 1.911051398 | 0.004178203 | 0.011289644 | 1 |
| Obesity | neutrophil | *THNSL2* | 1.183062974 | 0.002537464 | 0.011289644 | 1 |
| Obesity | neutrophil | *RALB* | 3.470408559 | 0.011377296 | 0.016327108 | 1 |
| Obesity | neutrophil | *TMEM177* | 2.716732882 | 0.003082003 | 0.011289644 | 1 |
| Obesity | neutrophil | *RABL2A* | 1.4695235 | 0.005025154 | 0.011289644 | 1 |
| Obesity | neutrophil | *SLC20A1* | 1.272588352 | 0.004889503 | 0.011289644 | 1 |
| Obesity | neutrophil | *FBLN7* | 1.305271088 | 0.029249279 | 0.036602011 | 1 |
| Obesity | neutrophil | *ZC3H8* | 2.786320211 | 0.001850449 | 0.011289644 | 1 |
| Obesity | neutrophil | *LIPT1* | -1.984645257 | 0.005346356 | 0.011289644 | 1 |
| Obesity | neutrophil | *FAHD2B* | -7.458499711 | 0.00179291 | 0.011289644 | 1 |
| Obesity | neutrophil | *AFF3* | 1.050390018 | 0.03422453 | 0.042173583 | 1 |
| Obesity | neutrophil | *UBXN4* | 1.924990741 | 0.003676921 | 0.011289644 | 1 |
| Obesity | neutrophil | *GPR17* | 1.603564455 | 0.008117969 | 0.012943867 | 1 |
| Obesity | neutrophil | *POLR2D* | 1.588358387 | 0.00646391 | 0.01166174 | 1 |
| Obesity | neutrophil | *ZNF385B* | 1.152785128 | 0.003371128 | 0.011289644 | 1 |
| Obesity | neutrophil | *TMEFF2* | 1.650992902 | 0.004406132 | 0.011289644 | 1 |
| Obesity | neutrophil | *GULP1* | 1.160019308 | 0.003097566 | 0.011289644 | 1 |
| Obesity | neutrophil | *FAM171B* | 2.026022143 | 0.001762085 | 0.011289644 | 1 |
| Obesity | neutrophil | *HSPD1* | 2.49706221 | 0.002429494 | 0.011289644 | 1 |
| Obesity | neutrophil | *METTL21A* | 1.32637311 | 0.003982911 | 0.011289644 | 1 |
| Obesity | neutrophil | *NBEAL1* | 2.121151378 | 0.000152894 | 0.011289644 | 0.961244451 |
| Obesity | neutrophil | *SUMF1* | 1.676874109 | 0.004035778 | 0.011289644 | 1 |
| Obesity | neutrophil | *ESPNL* | 22.3044949 | 0.002850057 | 0.011289644 | 1 |
| Obesity | neutrophil | *FANCD2* | 6.001528931 | 0.004518852 | 0.011289644 | 1 |
| Obesity | neutrophil | *TAMM41* | 1.26650715 | 0.003239548 | 0.011289644 | 1 |
| Obesity | neutrophil | *OSBPL10* | 2.191178676 | 0.003647138 | 0.011289644 | 1 |

| | | | | | | |
|---|---|---|---|---|---|---|
| Obesity | neutrophil | *FAM198A* | -4.021318054 | 0.010747843 | 0.015648165 | 1 |
| Obesity | neutrophil | *CSRNP1* | 1.662629381 | 0.007297154 | 0.012233922 | 1 |
| Obesity | neutrophil | *CTDSPL* | 1.150791106 | 0.00408843 | 0.011289644 | 1 |
| Obesity | neutrophil | *STAC* | 1.446736027 | 0.004311061 | 0.011289644 | 1 |
| Obesity | neutrophil | *ARL6IP5* | 2.907107078 | 0.011158857 | 0.016064972 | 1 |
| Obesity | neutrophil | *TMF1* | 1.675811074 | 0.005168443 | 0.011289644 | 1 |
| Obesity | neutrophil | *COL8A1* | 1.769526639 | 0.035614681 | 0.043732324 | 1 |
| Obesity | neutrophil | *NXPE3* | 1.458304065 | 0.004208192 | 0.011289644 | 1 |
| Obesity | neutrophil | *PLA1A* | -2.428954271 | 0.003458061 | 0.011289644 | 1 |
| Obesity | neutrophil | *ADPRH* | -0.887230794 | 0.004861819 | 0.011289644 | 1 |
| Obesity | neutrophil | *IGSF11* | 2.17846933 | 0.002794036 | 0.011289644 | 1 |
| Obesity | neutrophil | *BOC* | 1.233896273 | 0.004245145 | 0.011289644 | 1 |
| Obesity | neutrophil | *AGTR1* | 1.150997718 | 0.005128976 | 0.011289644 | 1 |
| Obesity | neutrophil | *EAF2* | 1.961212135 | 0.019742805 | 0.025754943 | 1 |
| Obesity | neutrophil | *MUC4* | 1.475648986 | 0.037508031 | 0.045806719 | 1 |
| Obesity | neutrophil | *DGKQ* | -2.098338225 | 0.004353032 | 0.011289644 | 1 |
| Obesity | neutrophil | *SLC26A1* | 4.576808852 | 0.007369418 | 0.012292791 | 1 |
| Obesity | neutrophil | *LYAR* | 3.945593294 | 0.005646101 | 0.011297933 | 1 |
| Obesity | neutrophil | *CENPC* | 1.313836604 | 0.030646875 | 0.038168958 | 1 |
| Obesity | neutrophil | *SLC10A4* | 15.64586544 | 0.012425802 | 0.017468922 | 1 |
| Obesity | neutrophil | *PYURF* | 1.712962556 | 0.004108578 | 0.011289644 | 1 |
| Obesity | neutrophil | *CISD2* | 4.589223231 | 0.013275886 | 0.018433192 | 1 |
| Obesity | neutrophil | *CCNA2* | 2.73403351 | 0.002723308 | 0.011289644 | 1 |
| Obesity | neutrophil | *USP53* | 1.180196649 | 0.003927439 | 0.011289644 | 1 |
| Obesity | neutrophil | *PDGFC* | 1.289189749 | 0.003637759 | 0.011289644 | 1 |
| Obesity | neutrophil | *CBR4* | 1.247270159 | 0.004271338 | 0.011289644 | 1 |
| Obesity | neutrophil | *CYP4V2* | 1.402159711 | 0.016691048 | 0.022293737 | 1 |
| Obesity | neutrophil | *FAM105A* | 2.138769685 | 0.007164841 | 0.012141606 | 1 |
| Obesity | neutrophil | *LHFPL2* | 3.222362518 | 0.007801052 | 0.012671131 | 1 |
| Obesity | neutrophil | *SSBP2* | 1.200719801 | 0.004325662 | 0.011289644 | 1 |
| Obesity | neutrophil | *CRHBP* | 1.736999194 | 0.007238767 | 0.012207452 | 1 |
| Obesity | neutrophil | *RASA1* | 1.753189157 | 0.01275498 | 0.017843917 | 1 |
| Obesity | neutrophil | *PPIP5K2* | 1.7211173 | 0.007906249 | 0.012758365 | 1 |
| Obesity | neutrophil | *SPATA9* | 2.648415664 | 0.005178452 | 0.011289644 | 1 |
| Obesity | neutrophil | *FEM1C* | 1.838434183 | 0.003398688 | 0.011289644 | 1 |
| Obesity | neutrophil | *COMMD10* | 1.663211327 | 0.010528197 | 0.015418304 | 1 |
| Obesity | neutrophil | *YIPF5* | 1.813145071 | 0.004760133 | 0.011289644 | 1 |
| Obesity | neutrophil | *GABRB2* | 1.422618893 | 0.00870338 | 0.013548613 | 1 |
| Obesity | neutrophil | *MYLK4* | 2.631049335 | 0.004019543 | 0.011289644 | 1 |
| Obesity | neutrophil | *PSD2* | 12.52966386 | 0.01609621 | 0.021632508 | 1 |
| Obesity | neutrophil | *ZMAT2* | 1.379544987 | 0.010648629 | 0.015546153 | 1 |
| Obesity | neutrophil | *KLHL3* | 1.270196841 | 0.002871426 | 0.011289644 | 1 |
| Obesity | neutrophil | *TRIM7* | -2.437416625 | 0.010215079 | 0.015061492 | 1 |
| Obesity | neutrophil | *HIGD2A* | -3.02892505 | 0.012381031 | 0.017441081 | 1 |
| Obesity | neutrophil | *FAM193B* | 2.079445143 | 0.00559396 | 0.011295339 | 1 |
| Obesity | neutrophil | *MUT* | 3.535971166 | 0.006760932 | 0.011846705 | 1 |
| Obesity | neutrophil | *DOK3* | -0.845360962 | 0.004536129 | 0.011289644 | 1 |
| Obesity | neutrophil | *MLIP* | -1.643487692 | 0.005600603 | 0.011295339 | 1 |
| Obesity | neutrophil | *FGD2* | 2.051538159 | 0.004091989 | 0.011289644 | 1 |
| Obesity | neutrophil | *ANO7* | 3.101863582 | 0.004041506 | 0.011289644 | 1 |

| | | | | | | |
|---|---|---|---|---|---|---|
| Obesity | neutrophil | *RPL7L1* | 4.127381553 | 0.000807144 | 0.011289644 | 1 |
| Obesity | neutrophil | *TPBG* | 1.522647354 | 0.004237888 | 0.011289644 | 1 |
| Obesity | neutrophil | *IRAK1BP1* | 1.623141999 | 0.003764752 | 0.011289644 | 1 |
| Obesity | neutrophil | *PHIP* | 1.688736524 | 0.004310523 | 0.011289644 | 1 |
| Obesity | neutrophil | *MMS22L* | 3.136820344 | 0.015124405 | 0.020510599 | 1 |
| Obesity | neutrophil | *PM20D2* | 2.468745582 | 0.005788922 | 0.011372552 | 1 |
| Obesity | neutrophil | *RNF217* | 2.58591393 | 0.007152525 | 0.012141606 | 1 |
| Obesity | neutrophil | *ABRACL* | 1.836399468 | 0.00759065 | 0.012456909 | 1 |
| Obesity | neutrophil | *AIG1* | 1.508385269 | 0.002175778 | 0.011289644 | 1 |
| Obesity | neutrophil | *DYNLT1* | 4.21463916 | 0.022641359 | 0.028985181 | 1 |
| Obesity | neutrophil | *PNLDC1* | 7.001215584 | 0.000982215 | 0.011289644 | 1 |
| Obesity | neutrophil | *WTAP* | 1.285804719 | 0.003747377 | 0.011289644 | 1 |
| Obesity | neutrophil | *SLC22A3* | 1.14816635 | 0.004306607 | 0.011289644 | 1 |
| Obesity | neutrophil | *GNA12* | 2.133934196 | 0.006732159 | 0.011825602 | 1 |
| Obesity | neutrophil | *C7orf50* | 1.998447786 | 0.003455296 | 0.011289644 | 1 |
| Obesity | neutrophil | *CCZ1B* | 1.100918496 | 0.012655071 | 0.017738674 | 1 |
| Obesity | neutrophil | *RBAK* | 1.833439512 | 0.003889887 | 0.011289644 | 1 |
| Obesity | neutrophil | *EGFR* | 1.064565071 | 0.004239311 | 0.011289644 | 1 |
| Obesity | neutrophil | *POMZP3* | 3.716857224 | 0.030389629 | 0.037863575 | 1 |
| Obesity | neutrophil | *CCT6A* | 2.716957209 | 0.005194256 | 0.011289644 | 1 |
| Obesity | neutrophil | *GIGYF1* | 4.272951563 | 0.00825926 | 0.013088107 | 1 |
| Obesity | neutrophil | *TRIM4* | 1.094065174 | 0.036827857 | 0.04508114 | 1 |
| Obesity | neutrophil | *TMEM209* | -1.582204615 | 0.013286618 | 0.01844003 | 1 |
| Obesity | neutrophil | *AGBL3* | 1.315978784 | 0.00436833 | 0.011289644 | 1 |
| Obesity | neutrophil | *EPHA1* | 4.441171584 | 0.002480277 | 0.011289644 | 1 |
| Obesity | neutrophil | *NOM1* | 1.308252672 | 0.004195322 | 0.011289644 | 1 |
| Obesity | neutrophil | *NCAPG2* | 1.445818376 | 0.004098216 | 0.011289644 | 1 |
| Obesity | neutrophil | *FBXO25* | 2.553144566 | 0.005117878 | 0.011289644 | 1 |
| Obesity | neutrophil | *CSGALNACT1* | 3.034284409 | 0.008453128 | 0.013302701 | 1 |
| Obesity | neutrophil | *BIN3* | 2.077184521 | 0.007645855 | 0.012505709 | 1 |
| Obesity | neutrophil | *CHMP7* | 4.029558385 | 0.006091173 | 0.011472499 | 1 |
| Obesity | neutrophil | *DOCK5* | -1.474830414 | 0.011392777 | 0.016345593 | 1 |
| Obesity | neutrophil | *ST18* | 1.498304484 | 0.006782628 | 0.011858283 | 1 |
| Obesity | neutrophil | *RGS20* | 1.488149564 | 0.008427641 | 0.01327269 | 1 |
| Obesity | neutrophil | *TACC1* | 1.720975953 | 0.004751359 | 0.011289644 | 1 |
| Obesity | neutrophil | *MTDH* | 1.906198871 | 0.005084961 | 0.011289644 | 1 |
| Obesity | neutrophil | *MAL2* | 1.811141804 | 0.001979526 | 0.011289644 | 1 |
| Obesity | neutrophil | *UTP23* | 2.497256262 | 0.006545353 | 0.011694062 | 1 |
| Obesity | neutrophil | *FAM135B* | -3.353440973 | 0.008038448 | 0.012866019 | 1 |
| Obesity | neutrophil | *ZNF7* | 2.911646064 | 0.007858463 | 0.01271463 | 1 |
| Obesity | neutrophil | *SLC39A4* | -3.889763462 | 0.008986857 | 0.013810895 | 1 |
| Obesity | neutrophil | *AK3* | 1.067140471 | 0.004568583 | 0.011289644 | 1 |
| Obesity | neutrophil | *CER1* | -0.992521176 | 0.007115905 | 0.01210108 | 1 |
| Obesity | neutrophil | *PLIN2* | 1.035324693 | 0.004269258 | 0.011289644 | 1 |
| Obesity | neutrophil | *HAUS6* | 1.776794248 | 0.00425497 | 0.011289644 | 1 |
| Obesity | neutrophil | *C9orf72* | 2.078727227 | 0.003793389 | 0.011289644 | 1 |
| Obesity | neutrophil | *SIGMAR1* | 2.05562997 | 0.007042663 | 0.012044945 | 1 |
| Obesity | neutrophil | *CBWD5* | -2.856885875 | 0.022835309 | 0.02918593 | 1 |
| Obesity | neutrophil | *ZNF462* | 2.240233116 | 0.005002951 | 0.011289644 | 1 |
| Obesity | neutrophil | *ASTN2* | 2.4345331 | 0.004151668 | 0.011289644 | 1 |

| | | | | | | |
|---|---|---|---|---|---|---|
| Obesity | neutrophil | *WDR31* | 2.283927168 | 0.002912487 | 0.011289644 | 1 |
| Obesity | neutrophil | *POLE3* | 1.97580166 | 0.006505304 | 0.011671674 | 1 |
| Obesity | neutrophil | *SURF1* | 1.632406452 | 0.002319731 | 0.011289644 | 1 |
| Obesity | neutrophil | *SURF6* | 1.793380228 | 0.004353317 | 0.011289644 | 1 |
| Obesity | neutrophil | *ASB6* | -1.663196814 | 0.010290662 | 0.01514452 | 1 |
| Obesity | neutrophil | *NTMT1* | 2.534113283 | 0.005684878 | 0.011300433 | 1 |
| Obesity | neutrophil | *CIZ1* | 1.515966354 | 0.002708353 | 0.011289644 | 1 |
| Obesity | neutrophil | *SH3GLB2* | 2.971234212 | 0.00186987 | 0.011289644 | 1 |
| Obesity | neutrophil | *LRSAM1* | 1.194112128 | 0.00429356 | 0.011289644 | 1 |
| Obesity | neutrophil | *SEC16A* | 1.148051969 | 0.008297624 | 0.013133727 | 1 |
| Obesity | neutrophil | *DPH7* | 1.414678036 | 0.003384946 | 0.011289644 | 1 |
| Obesity | neutrophil | *SLC39A12* | -0.906432321 | 0.005693417 | 0.011302341 | 1 |
| Obesity | neutrophil | *RSU1* | 1.397141038 | 0.004196659 | 0.011289644 | 1 |
| Obesity | neutrophil | *PARD3* | 1.245206302 | 0.002683623 | 0.011289644 | 1 |
| Obesity | neutrophil | *NRBF2* | -5.126245573 | 0.000651135 | 0.011289644 | 1 |
| Obesity | neutrophil | *ADIRF* | 1.513916597 | 0.004159404 | 0.011289644 | 1 |
| Obesity | neutrophil | *GLUD1* | 1.550729149 | 0.003847527 | 0.011289644 | 1 |
| Obesity | neutrophil | *HTR7* | 2.115148361 | 0.004426776 | 0.011289644 | 1 |
| Obesity | neutrophil | *RPP30* | 1.281171422 | 0.00473703 | 0.011289644 | 1 |
| Obesity | neutrophil | *FRA10AC1* | 1.700231031 | 0.005020769 | 0.011289644 | 1 |
| Obesity | neutrophil | *DNAJB12* | 4.205676547 | 0.027433564 | 0.034494963 | 1 |
| Obesity | neutrophil | *FUOM* | 2.440308864 | 0.00754983 | 0.012419095 | 1 |
| Obesity | neutrophil | *LRRC27* | 1.809714265 | 0.030791364 | 0.038333724 | 1 |
| Obesity | neutrophil | *PAOX* | -1.030186802 | 0.006336563 | 0.011611011 | 1 |
| Obesity | neutrophil | *GSTO1* | 3.20555167 | 0.008161806 | 0.012980843 | 1 |
| Obesity | neutrophil | *PDCD11* | 2.719465879 | 0.027756855 | 0.034859638 | 1 |
| Obesity | neutrophil | *GAS2* | 4.970706393 | 0.012992176 | 0.018137186 | 1 |
| Obesity | neutrophil | *LIN7C* | 2.340041158 | 0.004407098 | 0.011289644 | 1 |
| Obesity | neutrophil | *ZNF215* | 1.587463634 | 0.013268433 | 0.018426914 | 1 |
| Obesity | neutrophil | *APIP* | 1.326130445 | 0.007123603 | 0.012107622 | 1 |
| Obesity | neutrophil | *EIF3M* | 2.654044556 | 0.004923239 | 0.011289644 | 1 |
| Obesity | neutrophil | *GLYAT* | 1.163306541 | 0.001998437 | 0.011289644 | 1 |
| Obesity | neutrophil | *C11orf49* | 0.959055824 | 0.004304277 | 0.011289644 | 1 |
| Obesity | neutrophil | *CCDC82* | 1.253200947 | 0.013788407 | 0.01900213 | 1 |
| Obesity | neutrophil | *SERPINH1* | 1.859776159 | 0.013607944 | 0.018801171 | 1 |
| Obesity | neutrophil | *CAPN5* | 2.075825368 | 0.003078686 | 0.011289644 | 1 |
| Obesity | neutrophil | *INTS4* | 1.696020474 | 0.003478616 | 0.011289644 | 1 |
| Obesity | neutrophil | *ZC3H12C* | 3.186958437 | 0.007401501 | 0.012300617 | 1 |
| Obesity | neutrophil | *TTC12* | 2.812630535 | 1.61E-05 | 0.003153152 | 0.101532272 |
| Obesity | neutrophil | *DRD2* | 9.563026704 | 0.002303581 | 0.011289644 | 1 |
| Obesity | neutrophil | *C11orf52* | 14.52535864 | 0.004259078 | 0.011289644 | 1 |
| Obesity | neutrophil | *NPAT* | 1.352000168 | 0.004748967 | 0.011289644 | 1 |
| Obesity | neutrophil | *GLB1L2* | 8.459494272 | 0.031817111 | 0.039446692 | 1 |
| Obesity | neutrophil | *P4HA3* | 2.041851623 | 0.002219403 | 0.011289644 | 1 |
| Obesity | neutrophil | *GRIK4* | 1.705577033 | 0.0206971 | 0.026823924 | 1 |
| Obesity | neutrophil | *ADAM33* | 1.254925066 | 0.005588481 | 0.011295339 | 1 |
| Obesity | neutrophil | *MTA2* | -1.936061916 | 0.014562641 | 0.01987309 | 1 |
| Obesity | neutrophil | *TMC2* | 8.562351128 | 0.008795769 | 0.013643968 | 1 |
| Obesity | neutrophil | *ROM1* | 2.514611395 | 0.019955188 | 0.02599094 | 1 |
| Obesity | neutrophil | *EML3* | 1.432659414 | 0.005336567 | 0.011289644 | 1 |

| | | | | | | |
|---|---|---|---|---|---|---|
| Obesity | neutrophil | *INCENP* | 2.046613513 | 0.004233185 | 0.011289644 | 1 |
| Obesity | neutrophil | *MS4A3* | 37.01081969 | 0.004683398 | 0.011289644 | 1 |
| Obesity | neutrophil | *PLCH2* | -3.88025504 | 0.007242333 | 0.012207452 | 1 |
| Obesity | neutrophil | *CPSF7* | 2.743809401 | 0.006068125 | 0.011456547 | 1 |
| Obesity | neutrophil | *CHEK1* | 2.254667319 | 0.001792901 | 0.011289644 | 1 |
| Obesity | neutrophil | *FEZ1* | -1.37494648 | 0.006926441 | 0.011946923 | 1 |
| Obesity | neutrophil | *DUSP15* | -0.75508669 | 0.003219584 | 0.011289644 | 1 |
| Obesity | neutrophil | *COMMD7* | 1.721174122 | 0.004243718 | 0.011289644 | 1 |
| Obesity | neutrophil | *KIAA1755* | 2.026766833 | 0.002655696 | 0.011289644 | 1 |
| Obesity | neutrophil | *CNBD2* | 2.138528502 | 0.003865769 | 0.011289644 | 1 |
| Obesity | neutrophil | *TRPT1* | -6.3305945 | 0.001145604 | 0.011289644 | 1 |
| Obesity | neutrophil | *TAOK2* | 1.967165815 | 0.003800057 | 0.011289644 | 1 |
| Obesity | neutrophil | *MPP7* | 1.066033611 | 0.008810811 | 0.013657192 | 1 |
| Obesity | neutrophil | *CWC15* | 4.159574964 | 0.034450686 | 0.042435631 | 1 |
| Obesity | neutrophil | *ARID5B* | 1.549681985 | 0.002910875 | 0.011289644 | 1 |
| Obesity | neutrophil | *TMCO3* | 1.626273623 | 0.00426004 | 0.011289644 | 1 |
| Obesity | neutrophil | *LYPD6B* | 2.785369903 | 0.005335155 | 0.011289644 | 1 |
| Obesity | neutrophil | *PDCD4* | 1.509397056 | 0.004265943 | 0.011289644 | 1 |
| Obesity | neutrophil | *WDR17* | 4.335944218 | 0.011882768 | 0.016871491 | 1 |
| Obesity | neutrophil | *CCDC102B* | 1.796911221 | 0.005054666 | 0.011289644 | 1 |
| Obesity | neutrophil | *CNDP1* | -1.520134247 | 0.005370869 | 0.011289644 | 1 |
| Obesity | neutrophil | *FSIP1* | 6.25591607 | 0.005348278 | 0.011289644 | 1 |
| Obesity | neutrophil | *RGS18* | -0.806015603 | 0.006299735 | 0.011581641 | 1 |
| Obesity | neutrophil | *PRSS23* | 2.588609472 | 0.020886189 | 0.02701882 | 1 |
| Obesity | neutrophil | *MTMR12* | 1.77288096 | 0.004710077 | 0.011289644 | 1 |
| Obesity | neutrophil | *PPP1R1C* | -2.309620629 | 0.009007468 | 0.013829048 | 1 |
| Obesity | neutrophil | *CCT5* | 1.373083458 | 0.003834712 | 0.011289644 | 1 |
| Obesity | neutrophil | *FAM173B* | 2.798654369 | 0.006816446 | 0.011874479 | 1 |
| Obesity | neutrophil | *DOCK1* | 1.314944539 | 0.005895782 | 0.011378838 | 1 |
| Obesity | neutrophil | *TEX12* | 12.7916716 | 0.004373658 | 0.011289644 | 1 |
| Obesity | neutrophil | *PIP4K2A* | 1.756365242 | 0.007854931 | 0.01271463 | 1 |
| Obesity | neutrophil | *ABCB9* | 2.34175907 | 0.003658837 | 0.011289644 | 1 |
| Obesity | neutrophil | *RILPL2* | 4.633861343 | 0.001113991 | 0.011289644 | 1 |
| Obesity | neutrophil | *DHX37* | 1.649321344 | 0.004245823 | 0.011289644 | 1 |
| Obesity | neutrophil | *PRSS53* | -6.546188957 | 0.040164001 | 0.048737903 | 1 |
| Obesity | neutrophil | *GPR158* | -3.413761644 | 0.015381058 | 0.020813756 | 1 |
| Obesity | neutrophil | *THRB* | 1.496769336 | 0.004832481 | 0.011289644 | 1 |
| Obesity | neutrophil | *BTBD11* | -2.102889588 | 0.010379557 | 0.015253921 | 1 |
| Obesity | neutrophil | *UBE3B* | 1.227440517 | 0.001796211 | 0.011289644 | 1 |
| Obesity | neutrophil | *ANK3* | 1.950798066 | 0.002803102 | 0.011289644 | 1 |
| Obesity | neutrophil | *IPMK* | 1.875158704 | 0.004474907 | 0.011289644 | 1 |
| Obesity | neutrophil | *PLBD2* | 1.309044206 | 0.003005401 | 0.011289644 | 1 |
| Obesity | neutrophil | *DLG5* | 1.592425877 | 0.006529253 | 0.011679921 | 1 |
| Obesity | neutrophil | *GXYLT1* | 1.544685159 | 0.004232592 | 0.011289644 | 1 |
| Obesity | neutrophil | *FAM177A1* | 2.12484352 | 0.003023777 | 0.011289644 | 1 |
| Obesity | neutrophil | *MIPOL1* | 3.248039073 | 0.001998831 | 0.011289644 | 1 |
| Obesity | neutrophil | *EXT2* | 2.172475553 | 0.00366736 | 0.011289644 | 1 |
| Obesity | neutrophil | *KCTD14* | -5.37950182 | 0.004193617 | 0.011289644 | 1 |
| Obesity | neutrophil | *THRSP* | 1.544824779 | 0.002589289 | 0.011289644 | 1 |
| Obesity | neutrophil | *NDUFC2* | 1.583175447 | 0.005911132 | 0.011378838 | 1 |

| | | | | | | |
|---|---|---|---|---|---|---|
| Obesity | neutrophil | *ME3* | 2.283787164 | 0.003390037 | 0.011289644 | 1 |
| Obesity | neutrophil | *ADAMTS12* | 0.869600766 | 0.004717444 | 0.011289644 | 1 |
| Obesity | neutrophil | *VIPAS39* | 3.558118745 | 0.010998966 | 0.015918623 | 1 |
| Obesity | neutrophil | *ANKRD50* | 1.603029114 | 0.003968671 | 0.011289644 | 1 |
| Obesity | neutrophil | *C4orf33* | 1.569769968 | 0.003562937 | 0.011289644 | 1 |
| Obesity | neutrophil | *VPS26B* | 1.879804708 | 0.003585743 | 0.011289644 | 1 |
| Obesity | neutrophil | *NCAPD3* | 1.485533281 | 0.003752822 | 0.011289644 | 1 |
| Obesity | neutrophil | *VTI1A* | 1.309321778 | 0.003571788 | 0.011289644 | 1 |
| Obesity | neutrophil | *FAM160B1* | 1.941147105 | 0.004599293 | 0.011289644 | 1 |
| Obesity | neutrophil | *ANO4* | 1.491198863 | 0.003226501 | 0.011289644 | 1 |
| Obesity | neutrophil | *TEX9* | 1.631252246 | 0.003596648 | 0.011289644 | 1 |
| Obesity | neutrophil | *AKR1C2* | 1.555256248 | 0.005041268 | 0.011289644 | 1 |
| Obesity | neutrophil | *ADAM8* | -1.121511091 | 0.008414919 | 0.013262621 | 1 |
| Obesity | neutrophil | *ITIH2* | 2.665881484 | 0.004932106 | 0.011289644 | 1 |
| Obesity | neutrophil | *KIN* | 1.904179785 | 0.003984363 | 0.011289644 | 1 |
| Obesity | neutrophil | *PIGF* | 1.732152695 | 0.005317518 | 0.011289644 | 1 |
| Obesity | neutrophil | *INPP1* | 2.68451109 | 0.00429279 | 0.011289644 | 1 |
| Obesity | neutrophil | *MFSD6* | 1.604994552 | 0.003791884 | 0.011289644 | 1 |
| Obesity | neutrophil | *RNF144A* | 2.658184483 | 0.009113584 | 0.013903688 | 1 |
| Obesity | neutrophil | *ASAP2* | 2.266514576 | 0.006078114 | 0.011461638 | 1 |
| Obesity | neutrophil | *ADAM17* | 2.252319069 | 0.005735881 | 0.01134195 | 1 |
| Obesity | neutrophil | *TMEM45B* | -0.801704735 | 0.004993423 | 0.011289644 | 1 |
| Obesity | neutrophil | *CENPU* | 2.387783506 | 0.003292165 | 0.011289644 | 1 |
| Obesity | neutrophil | *ACSL1* | 1.254573569 | 0.022517361 | 0.028849938 | 1 |
| Obesity | neutrophil | *BICD1* | 1.236784129 | 0.004657679 | 0.011289644 | 1 |
| Obesity | neutrophil | *GUF1* | 2.035247826 | 0.00944275 | 0.014259024 | 1 |
| Obesity | neutrophil | *GABRA2* | 2.38075067 | 0.004010135 | 0.011289644 | 1 |
| Obesity | neutrophil | *CENPJ* | 2.961365068 | 0.008909883 | 0.013739621 | 1 |
| Obesity | neutrophil | *CACUL1* | 1.662458167 | 0.004630401 | 0.011289644 | 1 |
| Obesity | neutrophil | *RABGAP1L* | 2.057911691 | 0.002579676 | 0.011289644 | 1 |
| Obesity | neutrophil | *PTPN14* | 1.866814945 | 0.002602842 | 0.011289644 | 1 |
| Obesity | neutrophil | *RNF219* | 1.233598407 | 0.004657124 | 0.011289644 | 1 |
| Obesity | neutrophil | *ARL14EP* | 2.026633911 | 0.034476204 | 0.042458745 | 1 |
| Obesity | neutrophil | *EPG5* | 1.726211455 | 0.004618233 | 0.011289644 | 1 |
| Obesity | neutrophil | *HAUS1* | 1.419206535 | 0.005537625 | 0.011295339 | 1 |
| Obesity | neutrophil | *G6PC2* | 92.28834059 | 0.003983872 | 0.011289644 | 1 |
| Obesity | neutrophil | *TGOLN2* | 2.101817397 | 0.006106868 | 0.011478524 | 1 |
| Obesity | neutrophil | *POC5* | 1.103330904 | 0.037574203 | 0.04587862 | 1 |
| Obesity | neutrophil | *TADA1* | 5.107061697 | 0.023484891 | 0.029918847 | 1 |
| Obesity | neutrophil | *GUCY1A2* | 2.928802442 | 0.019210753 | 0.025146368 | 1 |
| Obesity | neutrophil | *CWF19L2* | 3.266737473 | 0.00627858 | 0.011565541 | 1 |
| Obesity | neutrophil | *ZNF547* | 3.653173248 | 0.002688289 | 0.011289644 | 1 |
| Obesity | neutrophil | *ZNF773* | 1.919694618 | 0.007083892 | 0.012072542 | 1 |
| Obesity | neutrophil | *NMT2* | 1.299156325 | 0.016540648 | 0.022129918 | 1 |
| Obesity | neutrophil | *ZNF837* | -1.839371364 | 0.003239398 | 0.011289644 | 1 |
| Obesity | neutrophil | *CCDC50* | 2.357661992 | 0.004034893 | 0.011289644 | 1 |
| Obesity | neutrophil | *ZFP36L2* | 1.596334698 | 0.003820627 | 0.011289644 | 1 |
| Obesity | neutrophil | *PLEKHH2* | 0.876884968 | 0.007356631 | 0.012285536 | 1 |
| Obesity | neutrophil | *IGSF10* | 1.459367732 | 0.004771535 | 0.011289644 | 1 |
| Obesity | neutrophil | *SPEF2* | 2.266448362 | 0.004869405 | 0.011289644 | 1 |

| | | | | | | |
|---|---|---|---|---|---|---|
| Obesity | neutrophil | *SPARCL1* | 2.151221739 | 0.003460633 | 0.011289644 | 1 |
| Obesity | neutrophil | *MBNL1* | 1.174530702 | 0.004556167 | 0.011289644 | 1 |
| Obesity | neutrophil | *CLEC4F* | -1.393462909 | 0.005605457 | 0.011295339 | 1 |
| Obesity | neutrophil | *PELO* | 1.999638697 | 0.004189126 | 0.011289644 | 1 |
| Obesity | neutrophil | *RASGRP3* | 1.98298262 | 0.001608288 | 0.011289644 | 1 |
| Obesity | neutrophil | *SAR1B* | 1.115128602 | 0.0037785 | 0.011289644 | 1 |
| Obesity | neutrophil | *GPR180* | 1.926433121 | 0.007320198 | 0.012259479 | 1 |
| Obesity | neutrophil | *TCTEX1D1* | -0.567477816 | 0.013879782 | 0.01909875 | 1 |
| Obesity | neutrophil | *WDR78* | 1.740620213 | 0.00614952 | 0.011508289 | 1 |
| Obesity | neutrophil | *IFIT5* | 2.093428671 | 0.006283188 | 0.011565541 | 1 |
| Obesity | neutrophil | *PRDM8* | 1.799632793 | 0.017449236 | 0.023124652 | 1 |
| Obesity | neutrophil | *HHEX* | 2.126005752 | 0.003509989 | 0.011289644 | 1 |
| Obesity | neutrophil | *ZNF117* | 1.337592267 | 0.000387107 | 0.011289644 | 1 |
| Obesity | neutrophil | *RAB3C* | -1.637772748 | 0.01543888 | 0.02087403 | 1 |
| Obesity | neutrophil | *MED21* | 1.966367727 | 0.004203452 | 0.011289644 | 1 |
| Obesity | neutrophil | *SRP19* | -1.385226483 | 0.007295265 | 0.012233922 | 1 |
| Obesity | neutrophil | *CENPH* | 2.781563001 | 0.00048127 | 0.011289644 | 1 |
| Obesity | neutrophil | *CARHSP1* | 3.666244769 | 0.032216306 | 0.039855159 | 1 |
| Obesity | neutrophil | *TEKT5* | 4.960049201 | 0.003504197 | 0.011289644 | 1 |
| Obesity | neutrophil | *BANK1* | 1.141713973 | 0.004204453 | 0.011289644 | 1 |
| Obesity | neutrophil | *CAST* | 1.473234002 | 0.011098992 | 0.01600444 | 1 |
| Obesity | neutrophil | *SCOC* | 1.458040074 | 0.004978926 | 0.011289644 | 1 |
| Obesity | neutrophil | *CLGN* | -0.767148617 | 0.005290317 | 0.011289644 | 1 |
| Obesity | neutrophil | *SYCP2L* | 1.659967504 | 0.004726586 | 0.011289644 | 1 |
| Obesity | neutrophil | *AHCTF1* | 3.064510955 | 0.00826731 | 0.013095317 | 1 |
| Obesity | neutrophil | *MERTK* | 1.558153608 | 0.003579381 | 0.011289644 | 1 |
| Obesity | neutrophil | *TMEM87B* | 2.863079994 | 0.037949035 | 0.046291344 | 1 |
| Obesity | neutrophil | *PTPRR* | 2.442032331 | 0.003233186 | 0.011289644 | 1 |
| Obesity | neutrophil | *PLA2R1* | 1.241132449 | 0.004950768 | 0.011289644 | 1 |
| Obesity | neutrophil | *FRMD1* | 7.223532146 | 0.03891528 | 0.047350565 | 1 |
| Obesity | neutrophil | *ASAP1* | 1.690930287 | 0.006084279 | 0.011467069 | 1 |
| Obesity | neutrophil | *INO80C* | 2.635249728 | 0.002383321 | 0.011289644 | 1 |
| Obesity | neutrophil | *LPCAT1* | 2.58594691 | 0.004174369 | 0.011289644 | 1 |
| Obesity | neutrophil | *C16orf89* | 2.242498307 | 0.001685914 | 0.011289644 | 1 |
| Obesity | neutrophil | *ADPRHL1* | 2.548292876 | 0.018520494 | 0.024364584 | 1 |
| Obesity | neutrophil | *TUBGCP5* | 1.910588404 | 0.004427934 | 0.011289644 | 1 |
| Obesity | neutrophil | *LURAP1L* | 1.070427331 | 0.002091175 | 0.011289644 | 1 |
| Obesity | neutrophil | *CFDP1* | 1.687711572 | 0.004587267 | 0.011289644 | 1 |
| Obesity | neutrophil | *ZDHHC7* | 3.069868949 | 0.00676332 | 0.011847587 | 1 |
| Obesity | neutrophil | *FAM92B* | 23.89613463 | 0.004478439 | 0.011289644 | 1 |
| Obesity | neutrophil | *JAZF1* | 1.682489548 | 0.0019631 | 0.011289644 | 1 |
| Obesity | neutrophil | *FBXO36* | 2.497951435 | 0.007942607 | 0.01278095 | 1 |
| Obesity | neutrophil | *MCOLN2* | -1.369198382 | 0.008246808 | 0.013073465 | 1 |
| Obesity | neutrophil | *CACNA2D1* | 1.243310046 | 0.004448865 | 0.011289644 | 1 |
| Obesity | neutrophil | *ZUFSP* | 4.655290807 | 0.036009137 | 0.044182171 | 1 |
| Obesity | neutrophil | *HS3ST3A1* | 3.030800288 | 0.003313938 | 0.011289644 | 1 |
| Obesity | neutrophil | *PPP2R5E* | 1.149569558 | 0.003212144 | 0.011289644 | 1 |
| Obesity | neutrophil | *ANKRD29* | 1.955717856 | 0.00524595 | 0.011289644 | 1 |
| Obesity | neutrophil | *THY1* | 1.312828327 | 0.005287626 | 0.011289644 | 1 |
| Obesity | neutrophil | *TBCEL* | 1.524633695 | 0.004359661 | 0.011289644 | 1 |

| | | | | | | |
|---|---|---|---|---|---|---|
| Obesity | neutrophil | *UBASH3B* | -2.496577077 | 0.029579158 | 0.036971008 | 1 |
| Obesity | neutrophil | *ROBO4* | 1.905452797 | 0.006867902 | 0.011917887 | 1 |
| Obesity | neutrophil | *ROBO3* | 1.414812984 | 0.002842781 | 0.011289644 | 1 |
| Obesity | neutrophil | *CC2D1B* | 3.479878667 | 0.000694561 | 0.011289644 | 1 |
| Obesity | neutrophil | *PRKCA* | 3.399584122 | 0.012084688 | 0.017119521 | 1 |
| Obesity | neutrophil | *LRRK1* | 3.298612389 | 0.012066199 | 0.017101035 | 1 |
| Obesity | neutrophil | *CEP112* | 2.642121876 | 0.00109706 | 0.011289644 | 1 |
| Obesity | neutrophil | *GAL3ST2* | -1.223449741 | 0.006018718 | 0.011447874 | 1 |
| Obesity | neutrophil | *ENPP3* | 4.695831679 | 0.021544177 | 0.027778557 | 1 |
| Obesity | neutrophil | *UCHL1* | 1.864118068 | 0.004429551 | 0.011289644 | 1 |
| Obesity | neutrophil | *MIA3* | 1.478378746 | 0.004639358 | 0.011289644 | 1 |
| Obesity | neutrophil | *DISP1* | 1.729203024 | 0.005514966 | 0.011290116 | 1 |
| Obesity | neutrophil | *FAM167A* | 15.90265917 | 9.90E-05 | 0.010546443 | 0.622240123 |
| Obesity | neutrophil | *NEIL2* | 8.234624567 | 2.11E-05 | 0.00368836 | 0.132780958 |
| Obesity | neutrophil | *LONRF1* | 1.577807025 | 0.004201819 | 0.011289644 | 1 |
| Obesity | neutrophil | *CCSAP* | -3.37420059 | 0.012578745 | 0.017664189 | 1 |
| Obesity | neutrophil | *GBP5* | 2.197947407 | 0.001936678 | 0.011289644 | 1 |
| Obesity | neutrophil | *FAM69A* | 1.690073256 | 0.013003181 | 0.018142699 | 1 |
| Obesity | neutrophil | *PDLIM3* | 1.663428681 | 0.002342108 | 0.011289644 | 1 |
| Obesity | neutrophil | *NCAM2* | 1.623660662 | 0.003096504 | 0.011289644 | 1 |
| Obesity | neutrophil | *L3MBTL4* | 1.561855277 | 0.004219479 | 0.011289644 | 1 |
| Obesity | neutrophil | *PDE1C* | 1.271898328 | 0.003232819 | 0.011289644 | 1 |
| Obesity | neutrophil | *TSEN2* | 1.489431585 | 0.013371045 | 0.018520326 | 1 |
| Obesity | neutrophil | *SLFN13* | -1.456862332 | 0.001543441 | 0.011289644 | 1 |
| Obesity | neutrophil | *DPH3* | 1.862815198 | 0.004199009 | 0.011289644 | 1 |
| Obesity | neutrophil | *CXXC1* | 2.30802113 | 0.005126242 | 0.011289644 | 1 |
| Obesity | neutrophil | *PPP4R1* | 2.39462293 | 0.005109541 | 0.011289644 | 1 |
| Obesity | neutrophil | *APCDD1* | 1.027341497 | 0.005200352 | 0.011289644 | 1 |
| Obesity | neutrophil | *EPHB1* | -0.519928551 | 0.005404843 | 0.011289644 | 1 |
| Obesity | neutrophil | *ANKRD40* | 1.730808421 | 0.004703403 | 0.011289644 | 1 |
| Obesity | neutrophil | *CYP2U1* | 1.481470079 | 0.004165785 | 0.011289644 | 1 |
| Obesity | neutrophil | *FBXL18* | 1.985534753 | 0.003961598 | 0.011289644 | 1 |
| Obesity | neutrophil | *PROM2* | 7.450206841 | 0.014176313 | 0.019447191 | 1 |
| Obesity | neutrophil | *AK9* | 2.384809233 | 0.00272386 | 0.011289644 | 1 |
| Obesity | neutrophil | *CDK19* | 2.628399554 | 0.008920323 | 0.01375229 | 1 |
| Obesity | neutrophil | *GTF3C6* | 1.820792028 | 0.003259393 | 0.011289644 | 1 |
| Obesity | neutrophil | *TTC39B* | 1.480181659 | 0.005468152 | 0.011289644 | 1 |
| Obesity | neutrophil | *AGPAT5* | 4.583953543 | 0.005220509 | 0.011289644 | 1 |
| Obesity | neutrophil | *MMS19* | 2.677914965 | 0.004387496 | 0.011289644 | 1 |
| Obesity | neutrophil | *GOLGA7B* | 5.531165366 | 0.028445866 | 0.035667962 | 1 |
| Obesity | neutrophil | *TRMT44* | 1.418208266 | 0.003432591 | 0.011289644 | 1 |
| Obesity | neutrophil | *C16orf87* | 1.440602968 | 0.004164488 | 0.011289644 | 1 |
| Obesity | neutrophil | *HEATR3* | 2.541615005 | 0.020398761 | 0.026477625 | 1 |
| Obesity | neutrophil | *CNOT8* | 2.766740619 | 0.010720776 | 0.015628699 | 1 |
| Obesity | neutrophil | *LRGUK* | 1.582471989 | 0.0101055 | 0.014952525 | 1 |
| Obesity | neutrophil | *PDIA4* | 4.738436656 | 0.013701778 | 0.018911762 | 1 |
| Obesity | neutrophil | *KDM8* | 2.096815805 | 0.004997824 | 0.011289644 | 1 |
| Obesity | neutrophil | *FAM126B* | 1.199276372 | 0.004441274 | 0.011289644 | 1 |
| Obesity | neutrophil | *SPAG17* | 0.955870815 | 0.004403769 | 0.011289644 | 1 |
| Obesity | neutrophil | *RRAGA* | 3.50022548 | 0.00659362 | 0.01172215 | 1 |

| | | | | | | |
|---|---|---|---|---|---|---|
| Obesity | neutrophil | *TMBIM4* | 3.168367685 | 0.002746473 | 0.011289644 | 1 |
| Obesity | neutrophil | *FAM161B* | -2.445890997 | 0.016943854 | 0.022554734 | 1 |
| Obesity | neutrophil | *WIF1* | 10.42842965 | 0.015785614 | 0.021287893 | 1 |
| Obesity | neutrophil | *UGT2B4* | 2.796171016 | 0.004306215 | 0.011289644 | 1 |
| Obesity | neutrophil | *MMP16* | 1.017602062 | 0.004597349 | 0.011289644 | 1 |
| Obesity | neutrophil | *KCNMA1* | 1.914435064 | 0.0046453 | 0.011289644 | 1 |
| Obesity | neutrophil | *BATF* | 2.292248946 | 0.004361683 | 0.011289644 | 1 |
| Obesity | neutrophil | *DPY19L4* | 1.384003041 | 0.006360181 | 0.011618636 | 1 |
| Obesity | neutrophil | *NDUFAF6* | 2.643620043 | 0.005751568 | 0.011349688 | 1 |
| Obesity | neutrophil | *DRAM2* | 1.354402625 | 0.004385835 | 0.011289644 | 1 |
| Obesity | neutrophil | *ADAMTSL3* | 1.053586976 | 0.00167745 | 0.011289644 | 1 |
| Obesity | neutrophil | *WHAMM* | 1.802939873 | 0.010545592 | 0.01543255 | 1 |
| Obesity | neutrophil | *CXCL13* | 3.179563965 | 0.004207849 | 0.011289644 | 1 |
| Obesity | neutrophil | *CCT8* | 1.251622335 | 0.038151942 | 0.046511783 | 1 |
| Obesity | neutrophil | *ANKRD9* | -4.39895636 | 0.011622736 | 0.016577164 | 1 |
| Obesity | neutrophil | *SFXN2* | 2.182336282 | 0.003653694 | 0.011289644 | 1 |
| Obesity | neutrophil | *C14orf2* | 1.661387016 | 0.006652206 | 0.011757198 | 1 |
| Obesity | neutrophil | *SH3RF2* | 1.051721715 | 0.016838246 | 0.022433154 | 1 |
| Obesity | neutrophil | *GDF6* | 4.659658212 | 0.006191781 | 0.011521947 | 1 |
| Obesity | neutrophil | *NODAL* | 2.450811278 | 0.004218801 | 0.011289644 | 1 |
| Obesity | neutrophil | *WDYHV1* | 1.461993827 | 0.004924669 | 0.011289644 | 1 |
| Obesity | neutrophil | *ATAD2* | 3.904187258 | 0.012072917 | 0.0171067 | 1 |
| Obesity | neutrophil | *SASS6* | -2.735846925 | 0.01322535 | 0.018395525 | 1 |
| Obesity | neutrophil | *ITGAD* | 1.996877185 | 0.016740014 | 0.022326021 | 1 |
| Obesity | neutrophil | *VPS8* | 2.27527711 | 0.004589686 | 0.011289644 | 1 |
| Obesity | neutrophil | *MPV17L* | 2.855546676 | 0.003892435 | 0.011289644 | 1 |
| Obesity | neutrophil | *RPUSD3* | 1.78154251 | 0.004226713 | 0.011289644 | 1 |
| Obesity | neutrophil | *GHRL* | 1.601345669 | 0.004203688 | 0.011289644 | 1 |
| Obesity | neutrophil | *EXOG* | 2.606950118 | 0.00908618 | 0.013885468 | 1 |
| Obesity | neutrophil | *NTAN1* | 4.234607482 | 0.032166282 | 0.039821236 | 1 |
| Obesity | neutrophil | *FCHO2* | 2.438351631 | 0.007461717 | 0.01236443 | 1 |
| Obesity | neutrophil | *TIMP4* | 1.457448088 | 0.008901116 | 0.013733966 | 1 |
| Obesity | neutrophil | *CPT2* | 2.121169558 | 0.004393441 | 0.011289644 | 1 |
| Obesity | neutrophil | *LRP8* | 2.095796156 | 0.005198034 | 0.011289644 | 1 |
| Obesity | neutrophil | *GATAD1* | 2.069309768 | 0.002488684 | 0.011289644 | 1 |
| Obesity | neutrophil | *SUSD3* | -0.542254098 | 0.011128825 | 0.016036425 | 1 |
| Obesity | neutrophil | *TMED6* | 8.091901121 | 0.00439343 | 0.011289644 | 1 |
| Obesity | neutrophil | *CLEC18A* | -28.06418937 | 2.76E-15 | 5.78E-12 | 1.74E-11 |
| Obesity | neutrophil | *IL34* | 1.744904984 | 0.00533796 | 0.011289644 | 1 |
| Obesity | neutrophil | *AASDH* | 2.53796833 | 0.00317348 | 0.011289644 | 1 |
| Obesity | neutrophil | *RNF111* | 1.54389894 | 0.004723938 | 0.011289644 | 1 |
| Obesity | neutrophil | *DYRK1A* | 2.162431631 | 0.005893028 | 0.011378838 | 1 |
| Obesity | neutrophil | *KCNJ15* | -0.146258209 | 0.004640136 | 0.011289644 | 1 |
| Obesity | neutrophil | *LCA5L* | 1.520976148 | 0.011918005 | 0.016913882 | 1 |
| Obesity | neutrophil | *CREB3L1* | 2.250336416 | 0.002429316 | 0.011289644 | 1 |
| Obesity | neutrophil | *C2CD2* | 1.298858561 | 0.003452819 | 0.011289644 | 1 |
| Obesity | neutrophil | *SLC38A10* | 1.617305146 | 0.004441068 | 0.011289644 | 1 |
| Obesity | neutrophil | *C9orf43* | 4.387186621 | 0.017657377 | 0.023370933 | 1 |
| Obesity | neutrophil | *WDR19* | 1.717666812 | 0.004622881 | 0.011289644 | 1 |
| Obesity | neutrophil | *SLC37A3* | 1.101639736 | 0.004320118 | 0.011289644 | 1 |

| | | | | | | |
|---|---|---|---|---|---|---|
| Obesity | neutrophil | *SPPL3* | 1.725153789 | 0.004082793 | 0.011289644 | 1 |
| Obesity | neutrophil | *FAM213B* | -0.696599558 | 0.004255395 | 0.011289644 | 1 |
| Obesity | neutrophil | *C12orf43* | 2.240310566 | 0.001949587 | 0.011289644 | 1 |
| Obesity | neutrophil | *PEX10* | 5.695352549 | 0.012388877 | 0.017443326 | 1 |
| Obesity | neutrophil | *RADIL* | 1.423991032 | 0.004433234 | 0.011289644 | 1 |
| Obesity | neutrophil | *LDLRAP1* | 1.794625665 | 0.004370257 | 0.011289644 | 1 |
| Obesity | neutrophil | *TRIM63* | 4.336685651 | 0.00244865 | 0.011289644 | 1 |
| Obesity | neutrophil | *GALNT14* | 2.844878992 | 0.004670473 | 0.011289644 | 1 |
| Obesity | neutrophil | *HPD* | 4.326266035 | 1.46E-05 | 0.002963751 | 0.091876284 |
| Obesity | neutrophil | *RHPN1* | 2.296484677 | 0.039596046 | 0.048104414 | 1 |
| Obesity | neutrophil | *LRRC43* | 4.943262295 | 0.004364064 | 0.011289644 | 1 |
| Obesity | neutrophil | *XKR8* | -0.615374146 | 0.006435029 | 0.011648199 | 1 |
| Obesity | neutrophil | *EYA3* | 1.642095017 | 0.003694322 | 0.011289644 | 1 |
| Obesity | neutrophil | *MRAS* | 1.304342706 | 0.004060887 | 0.011289644 | 1 |
| Obesity | neutrophil | *ABHD3* | 2.021653774 | 0.004721335 | 0.011289644 | 1 |
| Obesity | neutrophil | *CLSTN2* | 1.126685456 | 0.005688889 | 0.011300433 | 1 |
| Obesity | neutrophil | *COLEC12* | 1.107474791 | 0.004340494 | 0.011289644 | 1 |
| Obesity | neutrophil | *RNF207* | 2.053359504 | 0.004283613 | 0.011289644 | 1 |
| Obesity | neutrophil | *AUTS2* | 1.843094134 | 0.008417254 | 0.013262976 | 1 |
| Obesity | neutrophil | *MITD1* | 2.757744214 | 0.003317263 | 0.011289644 | 1 |
| Obesity | neutrophil | *TSPAN33* | 1.989182884 | 0.008656028 | 0.013510538 | 1 |
| Obesity | neutrophil | *AHCYL2* | 1.500949587 | 0.004763213 | 0.011289644 | 1 |
| Obesity | neutrophil | *B4GALT5* | 1.139663145 | 0.004252058 | 0.011289644 | 1 |
| Obesity | neutrophil | *CD1E* | 14.4795642 | 6.46E-05 | 0.00802313 | 0.405832979 |
| Obesity | neutrophil | *NCF1* | 2.64044437 | 0.004321248 | 0.011289644 | 1 |
| Obesity | neutrophil | *PPP1R9A* | 2.226672411 | 0.009523379 | 0.01434439 | 1 |
| Obesity | neutrophil | *GDPD5* | 1.178522132 | 0.003822821 | 0.011289644 | 1 |
| Obesity | neutrophil | *DYNC1I1* | 2.53786873 | 0.005271753 | 0.011289644 | 1 |
| Obesity | neutrophil | *TMED4* | 3.65435643 | 0.008005427 | 0.012829497 | 1 |
| Obesity | neutrophil | *ELK4* | 3.115463327 | 0.004644536 | 0.011289644 | 1 |
| Obesity | neutrophil | *DUSP23* | 2.579276357 | 0.00414627 | 0.011289644 | 1 |
| Obesity | neutrophil | *RNF166* | 1.954370427 | 0.004972272 | 0.011289644 | 1 |
| Obesity | neutrophil | *F11R* | 1.616461575 | 0.00531657 | 0.011289644 | 1 |
| Obesity | neutrophil | *SPATA2L* | -2.962241486 | 0.005629474 | 0.011295339 | 1 |
| Obesity | neutrophil | *NIT1* | 2.068737194 | 0.001141069 | 0.011289644 | 1 |
| Obesity | neutrophil | *ZNF276* | 1.281356771 | 0.005866256 | 0.011378838 | 1 |
| Obesity | neutrophil | *CDA* | -0.869244293 | 0.004164866 | 0.011289644 | 1 |
| Obesity | neutrophil | *B4GALT3* | 2.26624886 | 0.002076502 | 0.011289644 | 1 |
| Obesity | neutrophil | *DMTN* | 3.086820348 | 0.000857941 | 0.011289644 | 1 |
| Obesity | neutrophil | *FCER1G* | 5.377538069 | 0.001963746 | 0.011289644 | 1 |
| Obesity | neutrophil | *CCAR2* | 3.853439219 | 0.009958167 | 0.014818298 | 1 |
| Obesity | neutrophil | *CDC42SE2* | 2.12481366 | 0.005473953 | 0.011289644 | 1 |
| Obesity | neutrophil | *RAPGEF6* | 3.605942547 | 0.010082502 | 0.01492911 | 1 |
| Obesity | neutrophil | *EPB41* | 1.453341535 | 0.002108886 | 0.011289644 | 1 |
| Obesity | neutrophil | *ALG8* | 1.717008646 | 0.004677335 | 0.011289644 | 1 |
| Obesity | neutrophil | *IFNAR2* | 3.231280692 | 0.010512337 | 0.015402252 | 1 |
| Obesity | neutrophil | *MRPL10* | 1.936476097 | 0.007589766 | 0.012456909 | 1 |
| Obesity | neutrophil | *SV2A* | 8.88344992 | 0.004273146 | 0.011289644 | 1 |
| Obesity | neutrophil | *UBE2Z* | 1.460572842 | 0.006475129 | 0.011664509 | 1 |
| Obesity | neutrophil | *CLIC6* | 1.803181801 | 0.00091528 | 0.011289644 | 1 |

| | | | | | | |
|---|---|---|---|---|---|---|
| Obesity | neutrophil | *CBR3* | 1.577937947 | 0.005551918 | 0.011295339 | 1 |
| Obesity | neutrophil | *SIM2* | 2.459180269 | 0.034498272 | 0.042477603 | 1 |
| Obesity | neutrophil | *HLCS* | 1.512976776 | 0.008494466 | 0.013331367 | 1 |
| Obesity | neutrophil | *ARHGAP27* | -2.07682535 | 0.013419487 | 0.018575147 | 1 |
| Obesity | neutrophil | *PADI4* | -0.293471468 | 0.005200742 | 0.011289644 | 1 |
| Obesity | neutrophil | *STARD9* | 1.11808614 | 0.004324876 | 0.011289644 | 1 |
| Obesity | neutrophil | *THEM4* | 1.616293624 | 0.004146205 | 0.011289644 | 1 |
| Obesity | neutrophil | *TCHH* | 4.226467714 | 0.032848911 | 0.040573891 | 1 |
| Obesity | neutrophil | *RSPRY1* | 2.445884857 | 0.011680217 | 0.016646031 | 1 |
| Obesity | neutrophil | *CCDC17* | 5.019230642 | 0.001943033 | 0.011289644 | 1 |
| Obesity | neutrophil | *ACE* | 4.279184677 | 0.012155715 | 0.01719689 | 1 |
| Obesity | neutrophil | *EFCAB14* | 3.497995472 | 0.010913919 | 0.01582833 | 1 |
| Obesity | neutrophil | *ATP6V0D1* | 1.79837134 | 0.011797345 | 0.01676914 | 1 |
| Obesity | neutrophil | *NPR2* | 1.842303946 | 0.004631295 | 0.011289644 | 1 |
| Obesity | neutrophil | *ZNF235* | 1.333375418 | 0.004385232 | 0.011289644 | 1 |
| Obesity | neutrophil | *CALM3* | 1.549277743 | 0.005987609 | 0.011430582 | 1 |
| Obesity | neutrophil | *ATAD3B* | 1.630948263 | 0.004020744 | 0.011289644 | 1 |
| Obesity | neutrophil | *SSU72* | 1.942022212 | 0.004411188 | 0.011289644 | 1 |
| Obesity | neutrophil | *NR2F6* | 1.819153781 | 0.004234853 | 0.011289644 | 1 |
| Obesity | neutrophil | *ANKLE1* | -0.652242416 | 0.003531174 | 0.011289644 | 1 |
| Obesity | neutrophil | *CILP2* | 14.1878355 | 0.009573534 | 0.014406129 | 1 |
| Obesity | neutrophil | *ABCG1* | 2.969945416 | 0.006885236 | 0.011932455 | 1 |
| Obesity | neutrophil | *RSPH1* | -5.897162109 | 0.016990119 | 0.022601963 | 1 |
| Obesity | neutrophil | *SLC37A1* | 1.729717084 | 0.003648346 | 0.011289644 | 1 |
| Obesity | neutrophil | *NDUFV3* | 1.284607318 | 0.001871636 | 0.011289644 | 1 |
| Obesity | neutrophil | *U2AF1* | -0.879164666 | 0.004671141 | 0.011289644 | 1 |
| Obesity | neutrophil | *HSF2BP* | 3.551948438 | 0.002369371 | 0.011289644 | 1 |
| Obesity | neutrophil | *CSTB* | 2.504355552 | 0.00522509 | 0.011289644 | 1 |
| Obesity | neutrophil | *ICOSLG* | -0.91967228 | 0.004144573 | 0.011289644 | 1 |
| Obesity | neutrophil | *C21orf2* | 1.834372707 | 0.014624688 | 0.019936126 | 1 |
| Obesity | neutrophil | *ZNF66* | 1.3958773 | 0.00332134 | 0.011289644 | 1 |
| Obesity | neutrophil | *LRRC3* | -0.488179501 | 0.005033426 | 0.011289644 | 1 |
| Obesity | neutrophil | *FAM207A* | 2.856432952 | 0.001342557 | 0.011289644 | 1 |
| Obesity | neutrophil | *LSS* | 1.281940226 | 0.001887104 | 0.011289644 | 1 |
| Obesity | neutrophil | *VAV2* | 1.282929395 | 0.005014344 | 0.011289644 | 1 |
| Obesity | neutrophil | *PCNT* | 1.178779097 | 0.001800946 | 0.011289644 | 1 |
| Obesity | neutrophil | *DIP2A* | 2.031833893 | 0.003862088 | 0.011289644 | 1 |
| Obesity | neutrophil | *PRMT2* | 1.762129748 | 0.002532512 | 0.011289644 | 1 |
| Obesity | neutrophil | *ZNF208* | 1.036665434 | 0.007407619 | 0.012307532 | 1 |
| Obesity | neutrophil | *ADAMTS13* | 1.961146703 | 0.004547532 | 0.011289644 | 1 |
| Obesity | neutrophil | *ZNF714* | 1.240704723 | 0.004818351 | 0.011289644 | 1 |
| Obesity | neutrophil | *GPSM1* | 2.363336667 | 0.00273817 | 0.011289644 | 1 |
| Obesity | neutrophil | *C19orf47* | 2.64170232 | 0.001442519 | 0.011289644 | 1 |
| Obesity | neutrophil | *HIPK4* | -3.018892415 | 0.008587134 | 0.013439709 | 1 |
| Obesity | neutrophil | *RDH13* | 2.122018147 | 0.004204986 | 0.011289644 | 1 |
| Obesity | neutrophil | *MPZL3* | 2.225111994 | 0.00419927 | 0.011289644 | 1 |
| Obesity | neutrophil | *PCSK7* | 2.013010143 | 0.004841894 | 0.011289644 | 1 |
| Obesity | neutrophil | *S100A1* | 8.284884178 | 0.001596847 | 0.011289644 | 1 |
| Obesity | neutrophil | *ZBTB7B* | 2.187086045 | 0.013842234 | 0.019072129 | 1 |
| Obesity | neutrophil | *VPS11* | 1.567767851 | 0.007997986 | 0.012824111 | 1 |

| | | | | | | |
|---|---|---|---|---|---|---|
| Obesity | neutrophil | *FDPS* | 2.514251894 | 0.004565218 | 0.011289644 | 1 |
| Obesity | neutrophil | *PMF1* | 2.95202517 | 0.000948857 | 0.011289644 | 1 |
| Obesity | neutrophil | *SLC25A44* | 3.811707676 | 0.018269657 | 0.024064809 | 1 |
| Obesity | neutrophil | *FCRL3* | 3.117248848 | 0.002148021 | 0.011289644 | 1 |
| Obesity | neutrophil | *NACC1* | 2.148373729 | 0.003889408 | 0.011289644 | 1 |
| Obesity | neutrophil | *LY6E* | -1.399139011 | 0.004780179 | 0.011289644 | 1 |
| Obesity | neutrophil | *VPS28* | 1.289779842 | 0.003362151 | 0.011289644 | 1 |
| Obesity | neutrophil | *TONSL* | -0.626795384 | 0.005135197 | 0.011289644 | 1 |
| Obesity | neutrophil | *MUM1* | 1.414452866 | 0.007160621 | 0.012141606 | 1 |
| Obesity | neutrophil | *PPP1R16A* | 2.432979815 | 0.001641336 | 0.011289644 | 1 |
| Obesity | neutrophil | *RPL8* | 1.911490436 | 0.004449665 | 0.011289644 | 1 |
| Obesity | neutrophil | *LRWD1* | -2.420312396 | 0.002557049 | 0.011289644 | 1 |
| Obesity | neutrophil | *FBXL13* | 3.020288699 | 0.006899003 | 0.011932455 | 1 |
| Obesity | neutrophil | *MFSD12* | 3.980390209 | 0.00759952 | 0.012461707 | 1 |
| Obesity | neutrophil | *YDJC* | 4.238946876 | 0.004111377 | 0.011289644 | 1 |
| Obesity | neutrophil | *CCDC116* | -3.88979142 | 0.013875385 | 0.01909688 | 1 |
| Obesity | neutrophil | *DVL3* | 2.224022431 | 0.003620547 | 0.011289644 | 1 |
| Obesity | neutrophil | *AP2M1* | 2.604619242 | 0.003443617 | 0.011289644 | 1 |
| Obesity | neutrophil | *ABCF3* | 3.349583768 | 0.000957058 | 0.011289644 | 1 |
| Obesity | neutrophil | *DMKN* | -1.562330894 | 0.005801725 | 0.011378838 | 1 |
| Obesity | neutrophil | *BDH1* | 2.035368464 | 0.004254164 | 0.011289644 | 1 |
| Obesity | neutrophil | *COX7A1* | 1.476545421 | 0.00424344 | 0.011289644 | 1 |
| Obesity | neutrophil | *DUSP14* | 1.355391472 | 0.009458091 | 0.014276835 | 1 |
| Obesity | neutrophil | *LRRC56* | -0.976851348 | 0.01542179 | 0.020855408 | 1 |
| Obesity | neutrophil | *PGAP3* | -4.215814372 | 0.038629438 | 0.047039178 | 1 |
| Obesity | neutrophil | *ACOX1* | 1.97221464 | 0.004295005 | 0.011289644 | 1 |
| Obesity | neutrophil | *PRPSAP1* | 1.469623145 | 0.004085149 | 0.011289644 | 1 |
| Obesity | neutrophil | *CYGB* | 2.088837768 | 0.005063629 | 0.011289644 | 1 |
| Obesity | neutrophil | *ZNF577* | 1.436995463 | 0.002221576 | 0.011289644 | 1 |
| Obesity | neutrophil | *ALDH16A1* | 1.846894 | 0.003936902 | 0.011289644 | 1 |
| Obesity | neutrophil | *SIGLEC11* | 3.398320529 | 0.017723244 | 0.023447786 | 1 |
| Obesity | neutrophil | *MPP3* | 1.571862458 | 0.004935304 | 0.011289644 | 1 |
| Obesity | neutrophil | *NAGS* | -3.90797615 | 0.01533967 | 0.020762219 | 1 |
| Obesity | neutrophil | *ASB16* | 1.768038038 | 0.00888735 | 0.013731333 | 1 |
| Obesity | neutrophil | *PLCD3* | -1.601734445 | 0.003581531 | 0.011289644 | 1 |
| Obesity | neutrophil | *RACGAP1* | 3.322266383 | 0.009042012 | 0.013855015 | 1 |
| Obesity | neutrophil | *LARP4* | 1.763646911 | 0.004874813 | 0.011289644 | 1 |
| Obesity | neutrophil | *SPC24* | -2.162372822 | 0.008149774 | 0.012971552 | 1 |
| Obesity | neutrophil | *LEMD2* | 2.391572975 | 0.018109355 | 0.023893707 | 1 |
| Obesity | neutrophil | *ASGR2* | -1.603081452 | 0.005450311 | 0.011289644 | 1 |
| Obesity | neutrophil | *SENP3* | -3.846929997 | 0.011554569 | 0.016506153 | 1 |
| Obesity | neutrophil | *SNRNP25* | 2.743982712 | 0.005270451 | 0.011289644 | 1 |
| Obesity | neutrophil | *JMJD8* | 5.933475619 | 0.000647875 | 0.011289644 | 1 |
| Obesity | neutrophil | *MEIOB* | 3.143526301 | 0.00310143 | 0.011289644 | 1 |
| Obesity | neutrophil | *FLYWCH2* | 6.023123253 | 0.016276708 | 0.021851732 | 1 |
| Obesity | neutrophil | *ZG16B* | 27.77241284 | 0.004595624 | 0.011289644 | 1 |
| Obesity | neutrophil | *TTC9C* | -2.574431747 | 0.025808362 | 0.032621064 | 1 |
| Obesity | neutrophil | *RPL29* | 1.454494177 | 0.004446321 | 0.011289644 | 1 |
| Obesity | neutrophil | *TPCN2* | 4.66919683 | 0.000936068 | 0.011289644 | 1 |
| Obesity | neutrophil | *CYP4A22* | 12.33135801 | 0.007255851 | 0.0122168 | 1 |

| | | | | | | |
|---|---|---|---|---|---|---|
| Obesity | neutrophil | *PDZK1IP1* | 49.04619854 | 0.016301799 | 0.021880745 | 1 |
| Obesity | neutrophil | *COA7* | -2.590983973 | 0.009581804 | 0.014415124 | 1 |
| Obesity | neutrophil | *SLC1A7* | 1.476073306 | 0.01951946 | 0.025492074 | 1 |
| Obesity | neutrophil | *C1orf123* | 2.098107795 | 0.010153684 | 0.014992065 | 1 |
| Obesity | neutrophil | *ACOT11* | 1.508410837 | 0.004135144 | 0.011289644 | 1 |
| Obesity | neutrophil | *USP24* | 1.977684731 | 0.00719774 | 0.012164568 | 1 |
| Obesity | neutrophil | *NOL9* | 3.469581519 | 0.002252451 | 0.011289644 | 1 |
| Obesity | neutrophil | *ZSWIM5* | 1.161575164 | 0.003346183 | 0.011289644 | 1 |
| Obesity | neutrophil | *FBLIM1* | -1.70749316 | 0.009103831 | 0.013902304 | 1 |
| Obesity | neutrophil | *SLC25A34* | -2.722610125 | 0.007547828 | 0.012419095 | 1 |
| Obesity | neutrophil | *PDPN* | 1.58954493 | 0.014996476 | 0.020363465 | 1 |
| Obesity | neutrophil | *LRRC38* | -1.334932282 | 0.005335724 | 0.011289644 | 1 |
| Obesity | neutrophil | *PEF1* | 1.79345443 | 0.003312226 | 0.011289644 | 1 |
| Obesity | neutrophil | *SYNC* | 1.092784686 | 0.008113877 | 0.012940625 | 1 |
| Obesity | neutrophil | *RBBP4* | 3.614841034 | 0.004985819 | 0.011289644 | 1 |
| Obesity | neutrophil | *TSSK3* | 4.081352785 | 0.018908861 | 0.024797665 | 1 |
| Obesity | neutrophil | *CAMK2N1* | 2.481694176 | 0.003519278 | 0.011289644 | 1 |
| Obesity | neutrophil | *TTLL10* | 5.349011237 | 0.003689672 | 0.011289644 | 1 |
| Obesity | neutrophil | *NFIA* | 1.049365021 | 0.003718543 | 0.011289644 | 1 |
| Obesity | neutrophil | *OMA1* | 1.821419187 | 0.00397436 | 0.011289644 | 1 |
| Obesity | neutrophil | *TM2D1* | 1.610530062 | 0.00440831 | 0.011289644 | 1 |
| Obesity | neutrophil | *FUBP1* | 1.62727995 | 0.019623955 | 0.0256179 | 1 |
| Obesity | neutrophil | *NEXN* | 3.100341204 | 0.008734242 | 0.013585399 | 1 |
| Obesity | neutrophil | *B3GALT2* | -0.996368266 | 0.005681298 | 0.011300433 | 1 |
| Obesity | neutrophil | *NTNG1* | 1.345432634 | 0.018683246 | 0.024530021 | 1 |
| Obesity | neutrophil | *HFM1* | 1.343263791 | 0.004319348 | 0.011289644 | 1 |
| Obesity | neutrophil | *KCNT2* | 1.022428092 | 0.003885202 | 0.011289644 | 1 |
| Obesity | neutrophil | *VCAM1* | -0.795491014 | 0.006275309 | 0.011565541 | 1 |
| Obesity | neutrophil | *EXTL2* | 2.489674265 | 0.006421758 | 0.011648199 | 1 |
| Obesity | neutrophil | *ARPC5* | 2.013111475 | 0.02928302 | 0.036622707 | 1 |
| Obesity | neutrophil | *ZNF496* | 4.378569936 | 0.005108453 | 0.011289644 | 1 |
| Obesity | neutrophil | *TRIM58* | 2.518968667 | 0.04044302 | 0.049010268 | 1 |
| Obesity | neutrophil | *IGSF8* | 1.968838079 | 0.004245127 | 0.011289644 | 1 |
| Obesity | neutrophil | *VANGL2* | 4.680947342 | 0.004296902 | 0.011289644 | 1 |
| Obesity | neutrophil | *FCRLB* | 6.641372922 | 0.002113755 | 0.011289644 | 1 |
| Obesity | neutrophil | *KLHDC9* | -1.591436817 | 0.004553948 | 0.011289644 | 1 |
| Obesity | neutrophil | *LMX1A* | 1.370408093 | 0.009598615 | 0.014433507 | 1 |
| Obesity | neutrophil | *FLVCR1* | 2.713967126 | 0.010435663 | 0.015321708 | 1 |
| Obesity | neutrophil | *TDRD5* | -1.084745894 | 0.005630354 | 0.011295339 | 1 |
| Obesity | neutrophil | *SNED1* | 1.151694715 | 0.004290706 | 0.011289644 | 1 |
| Obesity | neutrophil | *C1orf115* | 5.837408606 | 0.000508346 | 0.011289644 | 1 |
| Obesity | neutrophil | *BROX* | 1.669327543 | 0.006517661 | 0.011671674 | 1 |
| Obesity | neutrophil | *ACP6* | 1.522156277 | 0.023121178 | 0.029491346 | 1 |
| Obesity | neutrophil | *CNST* | 1.495661061 | 0.004354736 | 0.011289644 | 1 |
| Obesity | neutrophil | *PM20D1* | -1.40890702 | 0.006549196 | 0.011694062 | 1 |
| Obesity | neutrophil | *HAAO* | 5.974427407 | 0.014507859 | 0.019815536 | 1 |
| Obesity | neutrophil | *B3GALNT2* | 2.673600485 | 0.004202735 | 0.011289644 | 1 |
| Obesity | neutrophil | *PUS10* | 2.561309196 | 0.005060606 | 0.011289644 | 1 |
| Obesity | neutrophil | *RFTN2* | 1.221557394 | 0.004224552 | 0.011289644 | 1 |
| Obesity | neutrophil | *DISC1* | 2.892807772 | 0.011673701 | 0.016642303 | 1 |

| | | | | | | |
|---|---|---|---|---|---|---|
| Obesity | neutrophil | *FAM84A* | 3.785544211 | 0.004407537 | 0.011289644 | 1 |
| Obesity | neutrophil | *CLHC1* | 1.792663458 | 0.000125055 | 0.011289644 | 0.786221015 |
| Obesity | neutrophil | *FRZB* | 2.326754855 | 0.006274921 | 0.011565541 | 1 |
| Obesity | neutrophil | *NUP35* | 3.245987182 | 0.008774587 | 0.013623945 | 1 |
| Obesity | neutrophil | *FBXO41* | 2.907216367 | 0.004295487 | 0.011289644 | 1 |
| Obesity | neutrophil | *SMC6* | 2.018201247 | 0.005866648 | 0.011378838 | 1 |
| Obesity | neutrophil | *CCDC74A* | 8.293448669 | 0.004348827 | 0.011289644 | 1 |
| Obesity | neutrophil | *H3F3A* | 1.646500612 | 0.002922825 | 0.011289644 | 1 |
| Obesity | neutrophil | *NOSTRIN* | 2.269591307 | 0.004474856 | 0.011289644 | 1 |
| Obesity | neutrophil | *SGPP2* | 3.524530594 | 0.004127603 | 0.011289644 | 1 |
| Obesity | neutrophil | *BBS5* | 2.398214003 | 0.002498481 | 0.011289644 | 1 |
| Obesity | neutrophil | *HPGDS* | 1.366411939 | 0.002941638 | 0.011289644 | 1 |
| Obesity | neutrophil | *PDLIM5* | 1.398822042 | 0.004389489 | 0.011289644 | 1 |
| Obesity | neutrophil | *RPRD2* | 2.2221072 | 0.002838444 | 0.011289644 | 1 |
| Obesity | neutrophil | *PACRGL* | 2.621005739 | 0.003064696 | 0.011289644 | 1 |
| Obesity | neutrophil | *C1QTNF7* | 0.939987653 | 0.004305304 | 0.011289644 | 1 |
| Obesity | neutrophil | *LYSMD1* | 2.412757134 | 0.001656305 | 0.011289644 | 1 |
| Obesity | neutrophil | *ERCC3* | 1.893211789 | 0.003492818 | 0.011289644 | 1 |
| Obesity | neutrophil | *RNF149* | 2.973565583 | 0.039937614 | 0.048481904 | 1 |
| Obesity | neutrophil | *DHX57* | 3.712749962 | 0.006039719 | 0.011447874 | 1 |
| Obesity | neutrophil | *S100A9* | -0.683386605 | 0.00466043 | 0.011289644 | 1 |
| Obesity | neutrophil | *S100A12* | -13.45794173 | 0.010108439 | 0.014953355 | 1 |
| Obesity | neutrophil | *TDRD10* | 1.31587363 | 0.004161671 | 0.011289644 | 1 |
| Obesity | neutrophil | *DCAF16* | 1.207762962 | 0.004548984 | 0.011289644 | 1 |
| Obesity | neutrophil | *GNPDA2* | 2.640895792 | 0.001219072 | 0.011289644 | 1 |
| Obesity | neutrophil | *NIPAL1* | -0.511905854 | 0.005563656 | 0.011295339 | 1 |
| Obesity | neutrophil | *ANTXR2* | 3.120030902 | 0.000724583 | 0.011289644 | 1 |
| Obesity | neutrophil | *CGGBP1* | 1.180646017 | 0.040342542 | 0.048926227 | 1 |
| Obesity | neutrophil | *PBXIP1* | 1.959224864 | 0.003502964 | 0.011289644 | 1 |
| Obesity | neutrophil | *DCST2* | -1.268531204 | 0.00683072 | 0.011879594 | 1 |
| Obesity | neutrophil | *LMOD3* | 2.263943096 | 0.004301439 | 0.011289644 | 1 |
| Obesity | neutrophil | *NBPF10* | 2.934216748 | 0.006972383 | 0.011985556 | 1 |
| Obesity | neutrophil | *POGLUT1* | 1.381288447 | 0.00478624 | 0.011289644 | 1 |
| Obesity | neutrophil | *SLC15A2* | -3.86115438 | 0.00083589 | 0.011289644 | 1 |
| Obesity | neutrophil | *LRRC58* | 2.602445196 | 0.004935372 | 0.011289644 | 1 |
| Obesity | neutrophil | *FSTL1* | 1.398434409 | 0.009017275 | 0.013836969 | 1 |
| Obesity | neutrophil | *TMEM183A* | 1.704445481 | 0.004267401 | 0.011289644 | 1 |
| Obesity | neutrophil | *IGFBP7* | 2.113225232 | 0.039735138 | 0.048254744 | 1 |
| Obesity | neutrophil | *ARPC2* | 2.886947769 | 0.010151406 | 0.014992065 | 1 |
| Obesity | neutrophil | *GLB1L* | 2.303420333 | 0.02926065 | 0.036602011 | 1 |
| Obesity | neutrophil | *SERPINI1* | 1.442262908 | 0.00427532 | 0.011289644 | 1 |
| Obesity | neutrophil | *NUAK2* | -1.162419813 | 0.00913166 | 0.013924509 | 1 |
| Obesity | neutrophil | *SPTA1* | 1.309988136 | 0.037805304 | 0.046142875 | 1 |
| Obesity | neutrophil | *PYHIN1* | -1.399730456 | 0.003551305 | 0.011289644 | 1 |
| Obesity | neutrophil | *AIM2* | -3.505010202 | 0.018739638 | 0.024596264 | 1 |
| Obesity | neutrophil | *EFHB* | 1.410554782 | 0.00546464 | 0.011289644 | 1 |
| Obesity | neutrophil | *CD200R1* | 1.36096372 | 0.005584649 | 0.011295339 | 1 |
| Obesity | neutrophil | *GTPBP8* | 2.213480256 | 0.004425038 | 0.011289644 | 1 |
| Obesity | neutrophil | *SPICE1* | 1.385874617 | 0.004074192 | 0.011289644 | 1 |
| Obesity | neutrophil | *CADPS* | 1.867902 | 0.004278314 | 0.011289644 | 1 |

| | | | | | | |
|---|---|---|---|---|---|---|
| Obesity | neutrophil | *GMPS* | 1.8440274 | 0.004470263 | 0.011289644 | 1 |
| Obesity | neutrophil | *TIPARP* | 1.470494296 | 0.002944796 | 0.011289644 | 1 |
| Obesity | neutrophil | *DCLK3* | -0.812939008 | 0.006584153 | 0.011713234 | 1 |
| Obesity | neutrophil | *RPP14* | 2.242855285 | 0.005273401 | 0.011289644 | 1 |
| Obesity | neutrophil | *DNASE1L3* | -1.399399841 | 0.008391799 | 0.013239459 | 1 |
| Obesity | neutrophil | *CRELD1* | -1.383609132 | 0.032050609 | 0.039697041 | 1 |
| Obesity | neutrophil | *PCOLCE2* | 1.324759591 | 0.00634296 | 0.011616135 | 1 |
| Obesity | neutrophil | *TTC14* | 1.681993341 | 0.004061446 | 0.011289644 | 1 |
| Obesity | neutrophil | *RCHY1* | 2.518938176 | 0.010538141 | 0.015429272 | 1 |
| Obesity | neutrophil | *CCDC158* | 2.110286776 | 0.002718971 | 0.011289644 | 1 |
| Obesity | neutrophil | *GYG1* | 1.607313924 | 0.002240828 | 0.011289644 | 1 |
| Obesity | neutrophil | *HPS3* | 1.603741032 | 0.004948728 | 0.011289644 | 1 |
| Obesity | neutrophil | *RYK* | 1.510768841 | 0.003736345 | 0.011289644 | 1 |
| Obesity | neutrophil | *PLB1* | -2.750426275 | 0.012410065 | 0.01745851 | 1 |
| Obesity | neutrophil | *KIAA1143* | 1.469386409 | 0.010277364 | 0.015128491 | 1 |
| Obesity | neutrophil | *CDCP1* | 1.842820903 | 0.004384064 | 0.011289644 | 1 |
| Obesity | neutrophil | *LZTFL1* | 4.994324275 | 0.040967851 | 0.049598475 | 1 |
| Obesity | neutrophil | *FYCO1* | 4.161320519 | 0.026471007 | 0.033411608 | 1 |
| Obesity | neutrophil | *LRRC2* | 1.313158394 | 0.006220051 | 0.011544147 | 1 |
| Obesity | neutrophil | *NMNAT3* | 4.078948204 | 0.023799974 | 0.030283432 | 1 |
| Obesity | neutrophil | *SMIM12* | 4.063632063 | 0.019647884 | 0.025643813 | 1 |
| Obesity | neutrophil | *TPRA1* | 4.179130009 | 0.004896857 | 0.011289644 | 1 |
| Obesity | neutrophil | *YEATS2* | 1.283623682 | 0.004365601 | 0.011289644 | 1 |
| Obesity | neutrophil | *MEAF6* | 2.751042087 | 0.006961587 | 0.011974691 | 1 |
| Obesity | neutrophil | *DNALI1* | -0.759758452 | 0.003398914 | 0.011289644 | 1 |
| Obesity | neutrophil | *RPN1* | 2.260472972 | 0.004675592 | 0.011289644 | 1 |
| Obesity | neutrophil | *SFMBT1* | 4.332028402 | 0.02557006 | 0.032378442 | 1 |
| Obesity | neutrophil | *GNL3* | 1.605882584 | 0.008663863 | 0.013519411 | 1 |
| Obesity | neutrophil | *UVSSA* | 2.325089556 | 0.002651215 | 0.011289644 | 1 |
| Obesity | neutrophil | *FAM208A* | 1.705024274 | 0.003645392 | 0.011289644 | 1 |
| Obesity | neutrophil | *LRPAP1* | 2.018241486 | 0.004501682 | 0.011289644 | 1 |
| Obesity | neutrophil | *ABLIM2* | 1.668478525 | 0.003930363 | 0.011289644 | 1 |
| Obesity | neutrophil | *C1orf50* | 2.893205271 | 0.026975807 | 0.033973738 | 1 |
| Obesity | neutrophil | *METAP1* | 1.399369292 | 0.006026329 | 0.011447874 | 1 |
| Obesity | neutrophil | *DNAJB14* | 2.023962716 | 0.003029283 | 0.011289644 | 1 |
| Obesity | neutrophil | *SLC9B2* | 2.092723856 | 0.002134599 | 0.011289644 | 1 |
| Obesity | neutrophil | *BDH2* | 1.238119557 | 0.040404962 | 0.048986767 | 1 |
| Obesity | neutrophil | *ZNF589* | 1.941248792 | 0.016446219 | 0.022037374 | 1 |
| Obesity | neutrophil | *ATRIP* | 4.728160321 | 0.006889231 | 0.011932455 | 1 |
| Obesity | neutrophil | *APEH* | -3.722240482 | 0.038432657 | 0.046826766 | 1 |
| Obesity | neutrophil | *RNF123* | 2.710900855 | 3.05E-07 | 0.000159897 | 0.001918762 |
| Obesity | neutrophil | *MST1R* | 5.634664198 | 1.95E-06 | 0.000719624 | 0.0122336 |
| Obesity | neutrophil | *RAD54L2* | 1.705351846 | 0.004787 | 0.011289644 | 1 |
| Obesity | neutrophil | *GRM2* | -2.092057333 | 0.005084515 | 0.011289644 | 1 |
| Obesity | neutrophil | *POC1A* | 3.384103982 | 0.000638216 | 0.011289644 | 1 |
| Obesity | neutrophil | *C4orf3* | 1.392807166 | 0.003644206 | 0.011289644 | 1 |
| Obesity | neutrophil | *ANXA5* | 1.685146914 | 0.004757629 | 0.011289644 | 1 |
| Obesity | neutrophil | *MAP9* | 1.571534731 | 0.004826873 | 0.011289644 | 1 |
| Obesity | neutrophil | *CEP44* | 1.902250943 | 0.004412941 | 0.011289644 | 1 |
| Obesity | neutrophil | *C4orf45* | 3.276608155 | 0.002518532 | 0.011289644 | 1 |

| | | | | | | |
|---|---|---|---|---|---|---|
| Obesity | neutrophil | *TMEM144* | 1.49764904 | 0.004286229 | 0.011289644 | 1 |
| Obesity | neutrophil | *NPY1R* | 0.957538823 | 0.004296341 | 0.011289644 | 1 |
| Obesity | neutrophil | *IL15* | 3.909342846 | 0.014127639 | 0.019388882 | 1 |
| Obesity | neutrophil | *ARFIP1* | 2.182450234 | 0.004654486 | 0.011289644 | 1 |
| Obesity | neutrophil | *OTUD4* | 1.944976066 | 0.004495964 | 0.011289644 | 1 |
| Obesity | neutrophil | *ITGA2* | 2.002329788 | 0.003084961 | 0.011289644 | 1 |
| Obesity | neutrophil | *NDUFAF2* | 1.105134042 | 0.004659171 | 0.011289644 | 1 |
| Obesity | neutrophil | *LMBRD2* | 1.662374995 | 0.004886981 | 0.011289644 | 1 |
| Obesity | neutrophil | *PGGT1B* | 2.539578718 | 0.005306517 | 0.011289644 | 1 |
| Obesity | neutrophil | *CMBL* | 1.588981322 | 0.004386444 | 0.011289644 | 1 |
| Obesity | neutrophil | *C5orf63* | 2.80237762 | 0.007935928 | 0.01278095 | 1 |
| Obesity | neutrophil | *PRRC1* | 1.228608195 | 0.003908868 | 0.011289644 | 1 |
| Obesity | neutrophil | *WDR41* | 1.640632495 | 0.003998352 | 0.011289644 | 1 |
| Obesity | neutrophil | *NDUFS4* | 1.867020735 | 0.004555728 | 0.011289644 | 1 |
| Obesity | neutrophil | *RHOBTB3* | 0.954882026 | 0.004420371 | 0.011289644 | 1 |
| Obesity | neutrophil | *TIGD6* | 2.717459831 | 0.004186378 | 0.011289644 | 1 |
| Obesity | neutrophil | *SERINC5* | 1.747572208 | 0.005351752 | 0.011289644 | 1 |
| Obesity | neutrophil | *ENPP6* | -2.078608091 | 0.00788666 | 0.01274567 | 1 |
| Obesity | neutrophil | *CASP3* | -2.273587641 | 0.015123919 | 0.020510599 | 1 |
| Obesity | neutrophil | *PRIMPOL* | 1.877240333 | 0.004615695 | 0.011289644 | 1 |
| Obesity | neutrophil | *ERAP2* | 1.620584917 | 0.001572535 | 0.011289644 | 1 |
| Obesity | neutrophil | *RICTOR* | 1.900743018 | 0.005141266 | 0.011289644 | 1 |
| Obesity | neutrophil | *ANKRA2* | 2.563358692 | 0.004441996 | 0.011289644 | 1 |
| Obesity | neutrophil | *UBLCP1* | 2.974353885 | 0.007285475 | 0.012230772 | 1 |
| Obesity | neutrophil | *UTP15* | 2.009988971 | 0.004081103 | 0.011289644 | 1 |
| Obesity | neutrophil | *TLR3* | 1.770597973 | 0.003767921 | 0.011289644 | 1 |
| Obesity | neutrophil | *KLKB1* | 1.999006277 | 0.005000904 | 0.011289644 | 1 |
| Obesity | neutrophil | *NSA2* | 1.425679998 | 0.001743436 | 0.011289644 | 1 |
| Obesity | neutrophil | *CCDC127* | 2.037868731 | 0.003921634 | 0.011289644 | 1 |
| Obesity | neutrophil | *GDF9* | -0.73647133 | 0.004823048 | 0.011289644 | 1 |
| Obesity | neutrophil | *GRIK2* | 1.179256037 | 0.009107953 | 0.013903688 | 1 |
| Obesity | neutrophil | *TXLNB* | 2.84096966 | 0.004051626 | 0.011289644 | 1 |
| Obesity | neutrophil | *DCBLD1* | 2.530983278 | 0.002905876 | 0.011289644 | 1 |
| Obesity | neutrophil | *TMEM200A* | 2.274030072 | 0.003478785 | 0.011289644 | 1 |
| Obesity | neutrophil | *PDSS2* | 2.079910686 | 0.005604154 | 0.011295339 | 1 |
| Obesity | neutrophil | *STXBP5* | 2.793879708 | 0.011437207 | 0.016381339 | 1 |
| Obesity | neutrophil | *PI16* | 1.003047225 | 0.004217977 | 0.011289644 | 1 |
| Obesity | neutrophil | *DAGLB* | 1.731911658 | 0.006825385 | 0.011878501 | 1 |
| Obesity | neutrophil | *STK17A* | 1.450946437 | 0.004140521 | 0.011289644 | 1 |
| Obesity | neutrophil | *GALNT10* | 2.122326483 | 0.008324652 | 0.013163252 | 1 |
| Obesity | neutrophil | *RPS14* | 1.774200242 | 0.004082573 | 0.011289644 | 1 |
| Obesity | neutrophil | *HCN1* | 2.308334862 | 0.007578027 | 0.012449192 | 1 |
| Obesity | neutrophil | *SLU7* | 3.395622084 | 0.003522015 | 0.011289644 | 1 |
| Obesity | neutrophil | *CAMLG* | 1.532358807 | 0.002624207 | 0.011289644 | 1 |
| Obesity | neutrophil | *KCNK5* | 2.954227223 | 0.004723692 | 0.011289644 | 1 |
| Obesity | neutrophil | *KIAA1324L* | 1.292039689 | 0.006110296 | 0.011478524 | 1 |
| Obesity | neutrophil | *USP49* | 3.214153956 | 0.013855864 | 0.019081987 | 1 |
| Obesity | neutrophil | *ZNF704* | 1.157123401 | 0.004368952 | 0.011289644 | 1 |
| Obesity | neutrophil | *FNDC1* | 1.480399509 | 0.002702661 | 0.011289644 | 1 |
| Obesity | neutrophil | *BRI3* | 1.951182907 | 0.003659222 | 0.011289644 | 1 |

| | | | | | | |
|---|---|---|---|---|---|---|
| Obesity | neutrophil | *LMTK2* | 1.585815389 | 0.003217018 | 0.011289644 | 1 |
| Obesity | neutrophil | *PEX2* | 1.248362532 | 0.004485086 | 0.011289644 | 1 |
| Obesity | neutrophil | *TNFRSF11B* | 7.962697142 | 0.007939112 | 0.01278095 | 1 |
| Obesity | neutrophil | *SUN1* | 2.691790289 | 0.0063005 | 0.011581641 | 1 |
| Obesity | neutrophil | *OXR1* | 1.267167583 | 0.003776016 | 0.011289644 | 1 |
| Obesity | neutrophil | *GPER1* | 2.262022829 | 0.000840265 | 0.011289644 | 1 |
| Obesity | neutrophil | *TMEM184A* | 2.151493766 | 0.020940726 | 0.027078228 | 1 |
| Obesity | neutrophil | *INTS1* | 3.210064563 | 0.004862624 | 0.011289644 | 1 |
| Obesity | neutrophil | *SLC4A2* | 1.589488536 | 0.00423558 | 0.011289644 | 1 |
| Obesity | neutrophil | *FASTK* | 3.079589149 | 0.005439785 | 0.011289644 | 1 |
| Obesity | neutrophil | *ALDH7A1* | 1.201507858 | 0.002824616 | 0.011289644 | 1 |
| Obesity | neutrophil | *COX6C* | -2.442294134 | 0.029767841 | 0.037177278 | 1 |
| Obesity | neutrophil | *YWHAZ* | 1.018241555 | 0.034619683 | 0.042602064 | 1 |
| Obesity | neutrophil | *SLC25A32* | 1.507336697 | 0.006223693 | 0.011544147 | 1 |
| Obesity | neutrophil | *DCAF13* | 1.665542008 | 0.003714743 | 0.011289644 | 1 |
| Obesity | neutrophil | *INTS8* | 2.06689574 | 0.010450833 | 0.015333579 | 1 |
| Obesity | neutrophil | *FREM1* | 0.871198296 | 0.004861961 | 0.011289644 | 1 |
| Obesity | neutrophil | *RPP25L* | 4.566397649 | 0.007381985 | 0.01229744 | 1 |
| Obesity | neutrophil | *NUDT2* | 1.066583178 | 0.014698985 | 0.020028722 | 1 |
| Obesity | neutrophil | *TMEM65* | 1.954771449 | 0.005448534 | 0.011289644 | 1 |
| Obesity | neutrophil | *CCDC171* | 1.640153877 | 0.007184906 | 0.012155908 | 1 |
| Obesity | neutrophil | *SYK* | 1.488779706 | 0.004280657 | 0.011289644 | 1 |
| Obesity | neutrophil | *NIPSNAP3B* | -3.296279142 | 0.031118688 | 0.038694925 | 1 |
| Obesity | neutrophil | *METTL2B* | 2.349558198 | 0.008840879 | 0.013690298 | 1 |
| Obesity | neutrophil | *TMC1* | -2.127315933 | 0.012596576 | 0.017681329 | 1 |
| Obesity | neutrophil | *HGSNAT* | 1.539745944 | 0.005733018 | 0.01134195 | 1 |
| Obesity | neutrophil | *HNRNPK* | 1.486285816 | 0.004288643 | 0.011289644 | 1 |
| Obesity | neutrophil | *SVEP1* | 0.814015439 | 0.003932174 | 0.011289644 | 1 |
| Obesity | neutrophil | *ZNF367* | 2.334876947 | 0.00208308 | 0.011289644 | 1 |
| Obesity | neutrophil | *NOL6* | 2.02236628 | 0.005890867 | 0.011378838 | 1 |
| Obesity | neutrophil | *AQP3* | 4.087186308 | 0.006169207 | 0.011512519 | 1 |
| Obesity | neutrophil | *TRMT10B* | 1.863935838 | 0.004501392 | 0.011289644 | 1 |
| Obesity | neutrophil | *VCP* | 1.297629281 | 0.005448268 | 0.011289644 | 1 |
| Obesity | neutrophil | *PIGO* | 3.780567658 | 0.006499393 | 0.011671674 | 1 |
| Obesity | neutrophil | *LRFN5* | 1.478030164 | 0.000614282 | 0.011289644 | 1 |
| Obesity | neutrophil | *WRN* | 1.604171886 | 0.003741565 | 0.011289644 | 1 |
| Obesity | neutrophil | *MARCH8* | 1.604186761 | 0.004326629 | 0.011289644 | 1 |
| Obesity | neutrophil | *CFL2* | 1.762327995 | 0.003676039 | 0.011289644 | 1 |
| Obesity | neutrophil | *ZCCHC24* | 2.114420443 | 0.004911026 | 0.011289644 | 1 |
| Obesity | neutrophil | *PGM2L1* | 1.470639669 | 0.004972878 | 0.011289644 | 1 |
| Obesity | neutrophil | *HEPACAM* | 1.050003781 | 0.01360646 | 0.018801171 | 1 |
| Obesity | neutrophil | *LRR1* | 1.753374145 | 0.004246694 | 0.011289644 | 1 |
| Obesity | neutrophil | *RPL36AL* | 3.237621861 | 0.014456195 | 0.019762143 | 1 |
| Obesity | neutrophil | *C10orf25* | -1.136390899 | 0.010295682 | 0.015148362 | 1 |
| Obesity | neutrophil | *KLHDC2* | 1.134239779 | 0.004716196 | 0.011289644 | 1 |
| Obesity | neutrophil | *NOXRED1* | 5.335934863 | 0.031876634 | 0.039504907 | 1 |
| Obesity | neutrophil | *AKR1E2* | 2.046378269 | 0.004862684 | 0.011289644 | 1 |
| Obesity | neutrophil | *ATP5C1* | 2.207040817 | 0.005126764 | 0.011289644 | 1 |
| Obesity | neutrophil | *TAF3* | 1.934963062 | 0.003599313 | 0.011289644 | 1 |
| Obesity | neutrophil | *VDAC2* | 2.863396154 | 0.007355221 | 0.012285536 | 1 |

| | | | | | | |
|---|---|---|---|---|---|---|
| Obesity | neutrophil | *SLC18A2* | 3.155006624 | 0.004803092 | 0.011289644 | 1 |
| Obesity | neutrophil | *PDZD8* | 1.259537666 | 0.003940536 | 0.011289644 | 1 |
| Obesity | neutrophil | *QSOX2* | 3.1726898 | 0.004594142 | 0.011289644 | 1 |
| Obesity | neutrophil | *SDCCAG3* | 1.368797204 | 0.004200141 | 0.011289644 | 1 |
| Obesity | neutrophil | *AK8* | 2.050047035 | 0.003735509 | 0.011289644 | 1 |
| Obesity | neutrophil | *TSC1* | 3.024057993 | 0.002991039 | 0.011289644 | 1 |
| Obesity | neutrophil | *ZMYND19* | 5.778191658 | 0.004842596 | 0.011289644 | 1 |
| Obesity | neutrophil | *STOX1* | 1.329334487 | 0.001416637 | 0.011289644 | 1 |
| Obesity | neutrophil | *RET* | 2.924551817 | 0.006628769 | 0.011739457 | 1 |
| Obesity | neutrophil | *STK32C* | 3.87203063 | 0.029140393 | 0.036473353 | 1 |
| Obesity | neutrophil | *CASP7* | 1.051258182 | 0.01426323 | 0.019537424 | 1 |
| Obesity | neutrophil | *VWA2* | -3.352614025 | 0.002356623 | 0.011289644 | 1 |
| Obesity | neutrophil | *METTL3* | 2.448010932 | 0.003938849 | 0.011289644 | 1 |
| Obesity | neutrophil | *TRUB1* | 1.406391749 | 0.00259024 | 0.011289644 | 1 |
| Obesity | neutrophil | *HSPA12A* | 0.940754885 | 0.011663049 | 0.016630889 | 1 |
| Obesity | neutrophil | *UBTD1* | 1.934710323 | 0.004415257 | 0.011289644 | 1 |
| Obesity | neutrophil | *ARHGAP42* | 1.582675533 | 0.004559906 | 0.011289644 | 1 |
| Obesity | neutrophil | *OTOGL* | 4.35207779 | 0.006770562 | 0.011851936 | 1 |
| Obesity | neutrophil | *TTC7B* | 1.946704111 | 0.008679123 | 0.013536504 | 1 |
| Obesity | neutrophil | *SLC39A13* | 4.056751294 | 0.002090678 | 0.011289644 | 1 |
| Obesity | neutrophil | *PSMC3* | 7.002554208 | 0.003487802 | 0.011289644 | 1 |
| Obesity | neutrophil | *TC2N* | 1.054558124 | 0.004542294 | 0.011289644 | 1 |
| Obesity | neutrophil | *IFI27L1* | -1.920078805 | 0.00820903 | 0.013036164 | 1 |
| Obesity | neutrophil | *CLMN* | 2.143054949 | 0.013858494 | 0.019081987 | 1 |
| Obesity | neutrophil | *PTER* | 2.130528455 | 0.00664106 | 0.011751293 | 1 |
| Obesity | neutrophil | *C1QL3* | 2.219052211 | 0.000691851 | 0.011289644 | 1 |
| Obesity | neutrophil | *ABTB2* | 1.350317455 | 0.004418305 | 0.011289644 | 1 |
| Obesity | neutrophil | *CEP57* | 1.770650949 | 0.006217965 | 0.011544147 | 1 |
| Obesity | neutrophil | *TCP11L2* | 1.78320697 | 0.004347784 | 0.011289644 | 1 |
| Obesity | neutrophil | *JAM3* | 1.437316129 | 0.005163708 | 0.011289644 | 1 |
| Obesity | neutrophil | *GPT2* | 1.310072009 | 0.00330752 | 0.011289644 | 1 |
| Obesity | neutrophil | *HIF1AN* | 2.234897053 | 0.00465947 | 0.011289644 | 1 |
| Obesity | neutrophil | *NDUFB8* | 2.330470052 | 0.003742172 | 0.011289644 | 1 |
| Obesity | neutrophil | *ZFYVE19* | 2.037558035 | 0.001126248 | 0.011289644 | 1 |
| Obesity | neutrophil | *SPINT1* | 4.414476558 | 0.004978145 | 0.011289644 | 1 |
| Obesity | neutrophil | *AVPR1A* | 2.550695571 | 0.002152902 | 0.011289644 | 1 |
| Obesity | neutrophil | *BTRC* | 1.784908173 | 0.00476949 | 0.011289644 | 1 |
| Obesity | neutrophil | *LARP6* | 1.524487921 | 0.003204742 | 0.011289644 | 1 |
| Obesity | neutrophil | *ASPG* | 7.217389619 | 0.00253885 | 0.011289644 | 1 |
| Obesity | neutrophil | *GABRB3* | 1.744975737 | 0.010555918 | 0.015440916 | 1 |
| Obesity | neutrophil | *TBATA* | 23.13648949 | 0.000849943 | 0.011289644 | 1 |
| Obesity | neutrophil | *PCBD1* | 1.635998865 | 0.00610791 | 0.011478524 | 1 |
| Obesity | neutrophil | *COX11* | 1.477412126 | 0.003442684 | 0.011289644 | 1 |
| Obesity | neutrophil | *FAM227B* | 3.13871269 | 0.004307126 | 0.011289644 | 1 |
| Obesity | neutrophil | *STXBP4* | 1.641243299 | 0.004729119 | 0.011289644 | 1 |
| Obesity | neutrophil | *C2* | 2.408650638 | 0.008706282 | 0.013548613 | 1 |
| Obesity | neutrophil | *APBB1* | 2.239997005 | 0.003418235 | 0.011289644 | 1 |
| Obesity | neutrophil | *ILK* | 2.292546774 | 0.004427168 | 0.011289644 | 1 |
| Obesity | neutrophil | *DCHS1* | 1.928392521 | 0.003436685 | 0.011289644 | 1 |
| Obesity | neutrophil | *USP54* | 1.649236645 | 0.003833308 | 0.011289644 | 1 |

| | | | | | | |
|---|---|---|---|---|---|---|
| Obesity | neutrophil | *C11orf74* | 1.388972846 | 0.003705125 | 0.011289644 | 1 |
| Obesity | neutrophil | *WDR88* | -2.302079401 | 0.010877003 | 0.015796655 | 1 |
| Obesity | neutrophil | *CYB5R2* | 3.67544304 | 0.007636161 | 0.012499129 | 1 |
| Obesity | neutrophil | *TUB* | 2.488115312 | 0.005910543 | 0.011378838 | 1 |
| Obesity | neutrophil | *RIC3* | 1.542068298 | 0.003459594 | 0.011289644 | 1 |
| Obesity | neutrophil | *LMO1* | 4.375678825 | 0.004063137 | 0.011289644 | 1 |
| Obesity | neutrophil | *XRRA1* | 1.641703759 | 0.017468373 | 0.023145134 | 1 |
| Obesity | neutrophil | *TRIM66* | 1.029911703 | 0.001285505 | 0.011289644 | 1 |
| Obesity | neutrophil | *RPL27A* | 3.371374379 | 0.008568965 | 0.013424641 | 1 |
| Obesity | neutrophil | *PRTG* | 5.433554546 | 0.033952141 | 0.041870756 | 1 |
| Obesity | neutrophil | *C16orf46* | 26.14667787 | 0.002154281 | 0.011289644 | 1 |
| Obesity | neutrophil | *TMEM41B* | 1.654978453 | 0.004331807 | 0.011289644 | 1 |
| Obesity | neutrophil | *ZNF143* | 1.441223059 | 0.004751442 | 0.011289644 | 1 |
| Obesity | neutrophil | *TMX3* | 2.354899003 | 0.006027209 | 0.011447874 | 1 |
| Obesity | neutrophil | *CCDC68* | 2.136138228 | 0.002845036 | 0.011289644 | 1 |
| Obesity | neutrophil | *CLEC4E* | -0.374789138 | 0.005038057 | 0.011289644 | 1 |
| Obesity | neutrophil | *CLEC4D* | -0.540797381 | 0.005558663 | 0.011295339 | 1 |
| Obesity | neutrophil | *BEAN1* | 2.12710869 | 0.003366344 | 0.011289644 | 1 |
| Obesity | neutrophil | *TK2* | 1.508221012 | 0.004677757 | 0.011289644 | 1 |
| Obesity | neutrophil | *SEC11C* | 2.056563727 | 0.00407961 | 0.011289644 | 1 |
| Obesity | neutrophil | *GALR1* | 2.504889874 | 0.010589976 | 0.015476331 | 1 |
| Obesity | neutrophil | *CENPV* | 1.583422602 | 0.005193446 | 0.011289644 | 1 |
| Obesity | neutrophil | *CHRFAM7A* | 6.608639191 | 0.008155444 | 0.012977291 | 1 |
| Obesity | neutrophil | *SPDYE6* | 8.037132132 | 0.001093449 | 0.011289644 | 1 |
| Obesity | neutrophil | *ATF7IP2* | 1.295366493 | 0.004804883 | 0.011289644 | 1 |
| Obesity | neutrophil | *ZNF592* | 2.121659128 | 0.010465942 | 0.015348583 | 1 |
| Obesity | neutrophil | *AP1G1* | 2.827949031 | 0.010060457 | 0.014923046 | 1 |
| Obesity | neutrophil | *SLFN5* | 1.711601944 | 0.006602524 | 0.01172215 | 1 |
| Obesity | neutrophil | *CATSPER2* | 2.215133318 | 0.001751417 | 0.011289644 | 1 |
| Obesity | neutrophil | *C16orf45* | 1.673447381 | 0.003845805 | 0.011289644 | 1 |
| Obesity | neutrophil | *SAAL1* | 1.648769957 | 0.002652345 | 0.011289644 | 1 |
| Obesity | neutrophil | *FAM111A* | 2.163449528 | 0.010140169 | 0.014982665 | 1 |
| Obesity | neutrophil | *SCNN1G* | 9.570998071 | 0.023221192 | 0.029588901 | 1 |
| Obesity | neutrophil | *ANKDD1A* | 2.478842319 | 0.007275386 | 0.01222653 | 1 |
| Obesity | neutrophil | *DCTN5* | 3.896415059 | 0.009117215 | 0.013905854 | 1 |
| Obesity | neutrophil | *MTMR10* | 4.24792624 | 0.007514253 | 0.012399504 | 1 |
| Obesity | neutrophil | *YWHAB* | 1.481795315 | 0.00693538 | 0.011952575 | 1 |
| Obesity | neutrophil | *TSC22D4* | 4.352339374 | 0.016709373 | 0.022303998 | 1 |
| Obesity | neutrophil | *MS4A6E* | -2.181527692 | 0.00093795 | 0.011289644 | 1 |
| Obesity | neutrophil | *DIS3L* | -1.865332305 | 0.005479847 | 0.011289644 | 1 |
| Obesity | neutrophil | *CCNDBP1* | 2.178762363 | 0.004181258 | 0.011289644 | 1 |
| Obesity | neutrophil | *SMAD3* | 1.465878994 | 0.008922473 | 0.01375229 | 1 |
| Obesity | neutrophil | *RCCD1* | 1.8246224 | 0.007048639 | 0.012051656 | 1 |
| Obesity | neutrophil | *AKTIP* | 3.475966513 | 0.005208098 | 0.011289644 | 1 |
| Obesity | neutrophil | *EVA1C* | 1.836909864 | 0.00438523 | 0.011289644 | 1 |
| Obesity | neutrophil | *CNPY4* | 1.675322583 | 0.010436787 | 0.015321708 | 1 |
| Obesity | neutrophil | *NUDT21* | 3.700668121 | 0.009109815 | 0.013903688 | 1 |
| Obesity | neutrophil | *NKX3-1* | -2.734597198 | 0.006353471 | 0.011618636 | 1 |
| Obesity | neutrophil | *DUSP18* | -2.567954179 | 0.01004411 | 0.014910819 | 1 |
| Obesity | neutrophil | *TEF* | 1.396836943 | 0.002248344 | 0.011289644 | 1 |

| | | | | | | |
|---|---|---|---|---|---|---|
| Obesity | neutrophil | *PBX3* | 1.805651707 | 0.015041158 | 0.020410913 | 1 |
| Obesity | neutrophil | *SNRPD1* | 1.473802939 | 0.004261698 | 0.011289644 | 1 |
| Obesity | neutrophil | *SAMD14* | 6.518938141 | 0.000853512 | 0.011289644 | 1 |
| Obesity | neutrophil | *PIP5KL1* | 3.905269986 | 0.002171635 | 0.011289644 | 1 |
| Obesity | neutrophil | *URM1* | 1.555277695 | 0.015808149 | 0.021309141 | 1 |
| Obesity | neutrophil | *CERCAM* | 1.670324005 | 0.002562859 | 0.011289644 | 1 |
| Obesity | neutrophil | *ENDOG* | 2.76274243 | 0.001485568 | 0.011289644 | 1 |
| Obesity | neutrophil | *ISLR2* | -1.024623498 | 0.014291788 | 0.019567175 | 1 |
| Obesity | neutrophil | *SP2* | 4.089080397 | 0.00165219 | 0.011289644 | 1 |
| Obesity | neutrophil | *COQ7* | 1.465006951 | 0.004210196 | 0.011289644 | 1 |
| Obesity | neutrophil | *KATNAL2* | 2.302481207 | 0.008613777 | 0.013467998 | 1 |
| Obesity | neutrophil | *RNF214* | 3.00239694 | 0.00748763 | 0.012385612 | 1 |
| Obesity | neutrophil | *ENGASE* | 2.794479162 | 0.001675469 | 0.011289644 | 1 |
| Obesity | neutrophil | *RBFOX3* | 2.884731546 | 0.003332104 | 0.011289644 | 1 |
| Obesity | neutrophil | *MYO5B* | 1.781610142 | 0.003796344 | 0.011289644 | 1 |
| Obesity | neutrophil | *RRM1* | 2.131583148 | 0.010113953 | 0.014957991 | 1 |
| Obesity | neutrophil | *TRIM68* | 2.974723145 | 0.026439033 | 0.033382535 | 1 |
| Obesity | neutrophil | *FN3K* | 1.520101567 | 0.006060731 | 0.011450953 | 1 |
| Obesity | neutrophil | *ZNF23* | 2.554662303 | 0.012394925 | 0.017443326 | 1 |
| Obesity | neutrophil | *ZNF229* | 10.25546269 | 0.000467986 | 0.011289644 | 1 |
| Obesity | neutrophil | *ZNF646* | 2.706528852 | 0.000331121 | 0.011289644 | 1 |
| Obesity | neutrophil | *SMG8* | 5.220223468 | 0.004179683 | 0.011289644 | 1 |
| Obesity | neutrophil | *RAB8A* | 5.385309937 | 0.005356884 | 0.011289644 | 1 |
| Obesity | neutrophil | *GATAD2A* | 1.852847978 | 0.005268383 | 0.011289644 | 1 |
| Obesity | neutrophil | *PROCA1* | -0.997356018 | 0.007180914 | 0.012155908 | 1 |
| Obesity | neutrophil | *RPL13* | 1.392830947 | 0.004966183 | 0.011289644 | 1 |
| Obesity | neutrophil | *ZNF641* | 2.558569732 | 0.02173999 | 0.028002318 | 1 |
| Obesity | neutrophil | *CACNB3* | 5.793874446 | 0.012395739 | 0.017443326 | 1 |
| Obesity | neutrophil | *TP53I13* | 2.411456697 | 0.009386913 | 0.014210335 | 1 |
| Obesity | neutrophil | *ZNF528* | -1.498168784 | 0.009871005 | 0.01473538 | 1 |
| Obesity | neutrophil | *ZNF701* | 2.784441783 | 0.005979557 | 0.011427806 | 1 |
| Obesity | neutrophil | *SERTAD3* | 5.622564452 | 0.005734797 | 0.01134195 | 1 |
| Obesity | neutrophil | *TMC4* | 2.699746466 | 0.005256275 | 0.011289644 | 1 |
| Obesity | neutrophil | *TTYH1* | 2.127656666 | 0.003192789 | 0.011289644 | 1 |
| Obesity | neutrophil | *LENG8* | 3.400588991 | 0.006523012 | 0.011673832 | 1 |
| Obesity | neutrophil | *CDC42EP5* | 2.042571474 | 0.004117831 | 0.011289644 | 1 |
| Obesity | neutrophil | *KIR3DL1* | -1.011527594 | 0.006958465 | 0.011974691 | 1 |
| Obesity | neutrophil | *ZNF283* | 2.248348021 | 0.004760001 | 0.011289644 | 1 |
| Obesity | neutrophil | *SPINT2* | 1.598863705 | 0.005092674 | 0.011289644 | 1 |
| Obesity | neutrophil | *PSCA* | 4.491386114 | 0.000132127 | 0.011289644 | 0.830680087 |
| Obesity | neutrophil | *UBXN6* | 1.368914419 | 0.014538262 | 0.019852594 | 1 |
| Obesity | neutrophil | *ZNF444* | -1.719899065 | 0.008743172 | 0.013595925 | 1 |
| Obesity | neutrophil | *FAM57A* | 4.718023755 | 0.000481478 | 0.011289644 | 1 |
| Obesity | neutrophil | *GLOD4* | 1.590230923 | 0.004388469 | 0.011289644 | 1 |
| Obesity | neutrophil | *KIFC2* | 1.5624587 | 0.012721488 | 0.017812916 | 1 |
| Obesity | neutrophil | *SERPINF2* | 2.999876172 | 0.001455439 | 0.011289644 | 1 |
| Obesity | neutrophil | *C19orf48* | 2.529167105 | 0.000650727 | 0.011289644 | 1 |
| Obesity | neutrophil | *ZNF83* | 1.483614188 | 0.004499176 | 0.011289644 | 1 |
| Obesity | neutrophil | *CD320* | 1.64518365 | 0.003998091 | 0.011289644 | 1 |
| Obesity | neutrophil | *SPRYD3* | 3.729526495 | 0.005094679 | 0.011289644 | 1 |

| | | | | | | |
|---|---|---|---|---|---|---|
| Obesity | neutrophil | *SOAT2* | 3.497472413 | 0.00292676 | 0.011289644 | 1 |
| Obesity | neutrophil | *ZNF558* | 1.49604824 | 0.005056184 | 0.011289644 | 1 |
| Obesity | neutrophil | *NDUFV1* | 1.419840248 | 0.003754117 | 0.011289644 | 1 |
| Obesity | neutrophil | *PRDX2* | 1.587677497 | 0.002884193 | 0.011289644 | 1 |
| Obesity | neutrophil | *ZNF232* | 1.602060674 | 0.022966696 | 0.029330006 | 1 |
| Obesity | neutrophil | *HID1* | 2.71183084 | 0.003907183 | 0.011289644 | 1 |
| Obesity | neutrophil | *ATP5H* | 3.61108199 | 0.007837527 | 0.012699648 | 1 |
| Obesity | neutrophil | *SRP68* | 1.536180889 | 0.006636989 | 0.011750704 | 1 |
| Obesity | neutrophil | *MGAT5B* | 4.84108209 | 0.00237675 | 0.011289644 | 1 |
| Obesity | neutrophil | *TMEM68* | 1.628153943 | 0.004532498 | 0.011289644 | 1 |
| Obesity | neutrophil | *CYP7A1* | 12.43148724 | 0.003878823 | 0.011289644 | 1 |
| Obesity | neutrophil | *GSDMA* | -1.426383665 | 0.006298522 | 0.011581641 | 1 |
| Obesity | neutrophil | *KRT24* | -1.715118472 | 0.001837788 | 0.011289644 | 1 |
| Obesity | neutrophil | *TMEM99* | -3.606682287 | 0.013011229 | 0.018149899 | 1 |
| Obesity | neutrophil | *ZNF598* | 6.002992707 | 0.004899099 | 0.011289644 | 1 |
| Obesity | neutrophil | *MLST8* | 2.366932063 | 0.007085681 | 0.012072542 | 1 |
| Obesity | neutrophil | *ECI1* | 1.481902521 | 0.003624964 | 0.011289644 | 1 |
| Obesity | neutrophil | *ABCA3* | 1.288085291 | 0.00658738 | 0.01171566 | 1 |
| Obesity | neutrophil | *KCTD5* | 1.966291486 | 0.004595459 | 0.011289644 | 1 |
| Obesity | neutrophil | *SDHAF2* | 2.810421736 | 0.004273469 | 0.011289644 | 1 |
| Obesity | neutrophil | *DDB1* | 1.715162529 | 0.004714548 | 0.011289644 | 1 |
| Obesity | neutrophil | *VPS37C* | 3.655528762 | 0.039151967 | 0.047620123 | 1 |
| Obesity | neutrophil | *VWCE* | 2.496135603 | 0.001424072 | 0.011289644 | 1 |
| Obesity | neutrophil | *HRASLS5* | 1.150031661 | 0.006038251 | 0.011447874 | 1 |
| Obesity | neutrophil | *TRANK1* | 1.797865215 | 0.002956471 | 0.011289644 | 1 |
| Obesity | neutrophil | *TTC21A* | 1.763969383 | 0.00386144 | 0.011289644 | 1 |
| Obesity | neutrophil | *RPSA* | 1.756723639 | 0.003401449 | 0.011289644 | 1 |
| Obesity | neutrophil | *ULK4* | 2.044787712 | 0.014082398 | 0.019331012 | 1 |
| Obesity | neutrophil | *LTBP3* | 2.389640518 | 0.002739013 | 0.011289644 | 1 |
| Obesity | neutrophil | *NAALADL1* | -2.052130265 | 0.007508589 | 0.012399504 | 1 |
| Obesity | neutrophil | *SCARA5* | 1.016778451 | 0.004664826 | 0.011289644 | 1 |
| Obesity | neutrophil | *COPS6* | 1.72055404 | 0.010905571 | 0.015819872 | 1 |
| Obesity | neutrophil | *NUDT16L1* | 2.336303314 | 0.003525632 | 0.011289644 | 1 |
| Obesity | neutrophil | *KIAA1586* | 1.68665072 | 0.007499656 | 0.012396306 | 1 |
| Obesity | neutrophil | *RNF187* | 1.9147296 | 0.011572583 | 0.01652813 | 1 |
| Obesity | neutrophil | *ZCCHC4* | 4.390033294 | 0.005456331 | 0.011289644 | 1 |
| Obesity | neutrophil | *PTGDR* | 1.443984514 | 0.006192566 | 0.011521947 | 1 |
| Obesity | neutrophil | *GLYCTK* | 7.513010008 | 0.002001507 | 0.011289644 | 1 |
| Obesity | neutrophil | *POLR2J3* | 1.605197552 | 0.006607226 | 0.011724423 | 1 |
| Obesity | neutrophil | *NT5DC2* | -0.916273643 | 0.002945699 | 0.011289644 | 1 |
| Obesity | neutrophil | *KIF5C* | 2.958494355 | 0.004897873 | 0.011289644 | 1 |
| Obesity | neutrophil | *PDHB* | 1.710944901 | 0.004515997 | 0.011289644 | 1 |
| Obesity | neutrophil | *ACOX2* | 1.538484176 | 0.013334757 | 0.018482279 | 1 |
| Obesity | neutrophil | *FAM107A* | 2.340496094 | 0.002305278 | 0.011289644 | 1 |
| Obesity | neutrophil | *CX3CR1* | 3.084619164 | 0.025114908 | 0.031847 | 1 |
| Obesity | neutrophil | *SCN11A* | 0.990403734 | 0.017951526 | 0.023715328 | 1 |
| Obesity | neutrophil | *SEPT2* | 1.617002318 | 0.001135059 | 0.011289644 | 1 |
| Obesity | neutrophil | *DTYMK* | 1.798850714 | 0.00360775 | 0.011289644 | 1 |
| Obesity | neutrophil | *TAP1* | 2.284450218 | 0.002747144 | 0.011289644 | 1 |
| Obesity | neutrophil | *ING5* | 1.370260496 | 0.004257142 | 0.011289644 | 1 |

| | | | | | | |
|---|---|---|---|---|---|---|
| Obesity | neutrophil | *MLKL* | 2.142554012 | 0.008691554 | 0.013539098 | 1 |
| Obesity | neutrophil | *RFWD3* | 2.618052951 | 0.001211938 | 0.011289644 | 1 |
| Obesity | neutrophil | *KLHL30* | 3.466878567 | 0.002515132 | 0.011289644 | 1 |
| Obesity | neutrophil | *REEP4* | -1.541780435 | 0.004171229 | 0.011289644 | 1 |
| Obesity | neutrophil | *BMP1* | 1.780244683 | 0.004855413 | 0.011289644 | 1 |
| Obesity | neutrophil | *SERINC2* | 3.285045436 | 0.00286471 | 0.011289644 | 1 |
| Obesity | neutrophil | *TRAPPC11* | 1.638208252 | 0.004197639 | 0.011289644 | 1 |
| Obesity | neutrophil | *TMEM223* | 2.708923654 | 0.007858924 | 0.01271463 | 1 |
| Obesity | neutrophil | *SLC20A2* | 1.363475749 | 0.003770138 | 0.011289644 | 1 |
| Obesity | neutrophil | *ZSWIM1* | 4.907045424 | 0.000839305 | 0.011289644 | 1 |
| Obesity | neutrophil | *NBPF9* | 5.018682975 | 0.008574872 | 0.013428961 | 1 |
| Obesity | neutrophil | *WFDC13* | 18.09826484 | 0.004032367 | 0.011289644 | 1 |
| Obesity | neutrophil | *NDUFS5* | 1.389237038 | 0.003180389 | 0.011289644 | 1 |
| Obesity | neutrophil | *ZNF30* | 1.736477117 | 0.003716076 | 0.011289644 | 1 |
| Obesity | neutrophil | *FAM84B* | 3.54432276 | 0.000609684 | 0.011289644 | 1 |
| Obesity | neutrophil | *SLC16A4* | 2.485148425 | 0.004347862 | 0.011289644 | 1 |
| Obesity | neutrophil | *TMEM208* | -3.089454353 | 0.027979238 | 0.035124895 | 1 |
| Obesity | neutrophil | *LRP1B* | 1.471566657 | 0.014162452 | 0.019432417 | 1 |
| Obesity | neutrophil | *DNAJC21* | 1.500675106 | 0.003940559 | 0.011289644 | 1 |
| Obesity | neutrophil | *NPNT* | 1.779735006 | 0.004260303 | 0.011289644 | 1 |
| Obesity | neutrophil | *CNNM3* | 5.225518744 | 0.003000275 | 0.011289644 | 1 |
| Obesity | neutrophil | *GSTM4* | 1.833833212 | 0.004188301 | 0.011289644 | 1 |
| Obesity | neutrophil | *ABHD15* | 1.694401482 | 0.006438214 | 0.011648199 | 1 |
| Obesity | neutrophil | *ADAL* | 1.361911566 | 0.001019835 | 0.011289644 | 1 |
| Obesity | neutrophil | *SNTB2* | 1.510672586 | 0.004216307 | 0.011289644 | 1 |
| Obesity | neutrophil | *ZNF507* | 1.068504822 | 0.008017836 | 0.012842837 | 1 |
| Obesity | neutrophil | *STX18* | 2.66655658 | 0.007389771 | 0.012298069 | 1 |
| Obesity | neutrophil | *GFM1* | 2.543666901 | 0.002294206 | 0.011289644 | 1 |
| Obesity | neutrophil | *ZNF608* | 3.402205049 | 0.006575787 | 0.011712453 | 1 |
| Obesity | neutrophil | *INPP5D* | 3.597834772 | 0.025643436 | 0.032449697 | 1 |
| Obesity | neutrophil | *TMEM129* | 1.976284493 | 0.003736783 | 0.011289644 | 1 |
| Obesity | neutrophil | *CEP120* | 2.960530536 | 0.006579219 | 0.011713234 | 1 |
| Obesity | neutrophil | *STXBP6* | 2.033200858 | 0.004911879 | 0.011289644 | 1 |
| Obesity | neutrophil | *MFF* | 1.91982464 | 0.004191869 | 0.011289644 | 1 |
| Obesity | neutrophil | *LGALS9* | 3.907215223 | 0.004898488 | 0.011289644 | 1 |
| Obesity | neutrophil | *CPLX1* | -0.961351202 | 0.017036397 | 0.022649149 | 1 |
| Obesity | neutrophil | *SIGLEC7* | -2.37227859 | 0.004976425 | 0.011289644 | 1 |
| Obesity | neutrophil | *FEM1B* | 4.331252858 | 0.003511406 | 0.011289644 | 1 |
| Obesity | neutrophil | *MFSD7* | 5.750137446 | 0.004046161 | 0.011289644 | 1 |
| Obesity | neutrophil | *COL4A3* | 1.686275454 | 0.001828551 | 0.011289644 | 1 |
| Obesity | neutrophil | *IRS1* | 1.225532059 | 0.000784442 | 0.011289644 | 1 |
| Obesity | neutrophil | *UPF3A* | 1.896740493 | 0.012998101 | 0.018139636 | 1 |
| Obesity | neutrophil | *C8orf46* | 1.765187851 | 1.35E-05 | 0.002828781 | 0.084863431 |
| Obesity | neutrophil | *HSPBAP1* | -1.976748562 | 0.011072943 | 0.015980311 | 1 |
| Obesity | neutrophil | *PARM1* | -1.854035062 | 0.037898428 | 0.046247558 | 1 |
| Obesity | neutrophil | *FAM110B* | 3.550650244 | 0.02738198 | 0.034436989 | 1 |
| Obesity | neutrophil | *ATF5* | 1.836256451 | 0.005564607 | 0.011295339 | 1 |
| Obesity | neutrophil | *CPT1C* | 3.298171407 | 0.004824235 | 0.011289644 | 1 |
| Obesity | neutrophil | *PCSK9* | 15.72784069 | 0.034562779 | 0.042540367 | 1 |
| Obesity | neutrophil | *XPO6* | 3.659871701 | 0.016790572 | 0.022388617 | 1 |

| | | | | | | |
|---|---|---|---|---|---|---|
| Obesity | neutrophil | *CXCL10* | -0.409309142 | 0.007525634 | 0.012408514 | 1 |
| Obesity | neutrophil | *GPRIN1* | 14.9220563 | 0.022367791 | 0.028699246 | 1 |
| Obesity | neutrophil | *P2RY12* | 2.0320382 | 0.002981471 | 0.011289644 | 1 |
| Obesity | neutrophil | *SNUPN* | 1.385438008 | 0.003463881 | 0.011289644 | 1 |
| Obesity | neutrophil | *PTK2* | 1.788783988 | 0.005426922 | 0.011289644 | 1 |
| Obesity | neutrophil | *PTAFR* | 1.75858568 | 0.005030573 | 0.011289644 | 1 |
| Obesity | neutrophil | *RNASE6* | 2.813695079 | 0.004415086 | 0.011289644 | 1 |
| Obesity | neutrophil | *NPR1* | 4.32549084 | 0.012654582 | 0.017738674 | 1 |
| Obesity | neutrophil | *CLIC4* | 1.115329854 | 0.004474264 | 0.011289644 | 1 |
| Obesity | neutrophil | *SLC38A11* | 3.832857844 | 0.007962848 | 0.012793301 | 1 |
| Obesity | neutrophil | *CCDC8* | 4.366337112 | 0.004116649 | 0.011289644 | 1 |
| Obesity | neutrophil | *METTL15* | 1.37797457 | 0.001838212 | 0.011289644 | 1 |
| Obesity | neutrophil | *HINT1* | 2.339486992 | 0.006498289 | 0.011671674 | 1 |
| Obesity | neutrophil | *DTWD2* | 1.380078503 | 0.003626029 | 0.011289644 | 1 |
| Obesity | neutrophil | *CLIC3* | 3.630132762 | 0.009404959 | 0.014223954 | 1 |
| Obesity | neutrophil | *DFFB* | 1.693987313 | 0.004271577 | 0.011289644 | 1 |
| Obesity | neutrophil | *C15orf40* | 1.79387531 | 0.003100508 | 0.011289644 | 1 |
| Obesity | neutrophil | *LUZP1* | 1.872887893 | 0.004421358 | 0.011289644 | 1 |
| Obesity | neutrophil | *HEXDC* | 2.135361149 | 0.005790287 | 0.011372552 | 1 |
| Obesity | neutrophil | *LRRC45* | -3.275678218 | 0.002423073 | 0.011289644 | 1 |
| Obesity | neutrophil | *FASN* | 1.386183381 | 0.001356251 | 0.011289644 | 1 |
| Obesity | neutrophil | *MT1E* | 1.78732817 | 0.00619608 | 0.011525075 | 1 |
| Obesity | neutrophil | *DCXR* | 6.258237814 | 0.013029969 | 0.01816395 | 1 |
| Obesity | neutrophil | *LIMS1* | 1.746947612 | 0.007084066 | 0.012072542 | 1 |
| Obesity | neutrophil | *UGP2* | 0.884695182 | 0.003436308 | 0.011289644 | 1 |
| Obesity | neutrophil | *CSGALNACT2* | 1.60841899 | 0.004306348 | 0.011289644 | 1 |
| Obesity | neutrophil | *AVEN* | 1.556193571 | 0.003995096 | 0.011289644 | 1 |
| Obesity | neutrophil | *CTNND2* | 1.197074907 | 0.022826289 | 0.029180333 | 1 |
| Obesity | neutrophil | *TPST1* | 1.163301321 | 0.008489963 | 0.013331367 | 1 |
| Obesity | neutrophil | *OTUD3* | 3.517234426 | 0.002491578 | 0.011289644 | 1 |
| Obesity | neutrophil | *GUSB* | 3.961585072 | 0.025253815 | 0.032010229 | 1 |
| Obesity | neutrophil | *BRD3* | 1.382833002 | 0.003967579 | 0.011289644 | 1 |
| Obesity | neutrophil | *ZFPM2* | 1.034812002 | 0.004285861 | 0.011289644 | 1 |
| Obesity | neutrophil | *ZNF747* | 4.024484078 | 0.009373299 | 0.014196251 | 1 |
| Obesity | neutrophil | *MAP3K2* | 1.682822196 | 0.004250862 | 0.011289644 | 1 |
| Obesity | neutrophil | *PUSL1* | 3.641530905 | 0.006935421 | 0.011952575 | 1 |
| Obesity | neutrophil | *MYO7B* | -0.804621995 | 0.009375351 | 0.014196251 | 1 |
| Obesity | neutrophil | *CNTROB* | -3.295900895 | 0.020756841 | 0.026895766 | 1 |
| Obesity | neutrophil | *NIPA1* | 2.186107889 | 0.004378292 | 0.011289644 | 1 |
| Obesity | neutrophil | *RNF150* | 1.172344487 | 0.012175543 | 0.017221067 | 1 |
| Obesity | neutrophil | *CHRNB1* | 2.060827802 | 0.016506458 | 0.022098829 | 1 |
| Obesity | neutrophil | *PDCD6IP* | 1.478845027 | 0.005097475 | 0.011289644 | 1 |
| Obesity | neutrophil | *FAM161A* | 2.453176041 | 0.007851145 | 0.012711859 | 1 |
| Obesity | neutrophil | *GLB1* | 1.82322228 | 0.003797777 | 0.011289644 | 1 |
| Obesity | neutrophil | *FAXDC2* | 2.414448904 | 0.010756824 | 0.015654665 | 1 |
| Obesity | neutrophil | *CRTAP* | 1.37600503 | 0.003909746 | 0.011289644 | 1 |
| Obesity | neutrophil | *ELP5* | 1.515301459 | 0.009740813 | 0.014584542 | 1 |
| Obesity | neutrophil | *STX8* | 1.967537605 | 0.00601646 | 0.011447874 | 1 |
| Obesity | neutrophil | *NFRKB* | 2.578162166 | 0.015325153 | 0.020747037 | 1 |
| Obesity | neutrophil | *SP7* | -1.59364277 | 0.005156597 | 0.011289644 | 1 |

| | | | | | | |
|---|---|---|---|---|---|---|
| Obesity | neutrophil | *SEMA3E* | 3.437888937 | 0.005668208 | 0.011300433 | 1 |
| Obesity | neutrophil | *DCLK2* | -1.036762813 | 0.006517921 | 0.011671674 | 1 |
| Obesity | neutrophil | *GPRC5C* | -2.048941264 | 0.017759061 | 0.023480802 | 1 |
| Obesity | neutrophil | *ADORA2B* | -5.315741544 | 3.59E-07 | 0.000173645 | 0.00225738 |
| Obesity | neutrophil | *MGMT* | 1.161076741 | 0.003457981 | 0.011289644 | 1 |
| Obesity | neutrophil | *METTL7B* | -1.887827564 | 0.005605803 | 0.011295339 | 1 |
| Obesity | neutrophil | *NFXL1* | 1.404968125 | 0.003584161 | 0.011289644 | 1 |
| Obesity | neutrophil | *CD14* | 1.053054012 | 0.010837866 | 0.015751748 | 1 |
| Obesity | neutrophil | *ARL6IP1* | 2.433193303 | 0.006279472 | 0.011565541 | 1 |
| Obesity | neutrophil | *SERPINB9* | 1.780459799 | 0.005817724 | 0.011378838 | 1 |
| Obesity | neutrophil | *SMAGP* | 1.928236671 | 0.015796729 | 0.021298314 | 1 |
| Obesity | neutrophil | *IRX2* | 2.011632067 | 0.004517022 | 0.011289644 | 1 |
| Obesity | neutrophil | *EMB* | 2.435215273 | 0.020337572 | 0.026412376 | 1 |
| Obesity | neutrophil | *DLGAP1* | 2.833880737 | 0.005616456 | 0.011295339 | 1 |
| Obesity | neutrophil | *STAT2* | 1.506375811 | 0.003992334 | 0.011289644 | 1 |
| Obesity | neutrophil | *NUDCD2* | 3.575348824 | 0.00851816 | 0.013361694 | 1 |
| Obesity | neutrophil | *SLC26A5* | 2.302583449 | 0.003426902 | 0.011289644 | 1 |
| Obesity | neutrophil | *GTSF1* | -3.346496259 | 0.007150895 | 0.012141606 | 1 |
| Obesity | neutrophil | *ARMC10* | 3.364725745 | 0.006854041 | 0.011906979 | 1 |
| Obesity | neutrophil | *RNF34* | 1.737629047 | 0.004262615 | 0.011289644 | 1 |
| Obesity | neutrophil | *ACYP2* | 1.67127221 | 0.00487022 | 0.011289644 | 1 |
| Obesity | neutrophil | *TRABD* | 2.403436669 | 0.006061519 | 0.011450953 | 1 |
| Obesity | neutrophil | *TMEM133* | -2.260013042 | 0.006158788 | 0.011512519 | 1 |
| Obesity | neutrophil | *TTLL6* | -1.179911259 | 0.005000996 | 0.011289644 | 1 |
| Obesity | neutrophil | *KCNS3* | 1.451531032 | 0.007652241 | 0.01251226 | 1 |
| Obesity | neutrophil | *AKAP13* | 1.953263844 | 0.004786652 | 0.011289644 | 1 |
| Obesity | neutrophil | *HTRA3* | 1.179664334 | 0.003977963 | 0.011289644 | 1 |
| Obesity | neutrophil | *FOXN2* | 2.180138089 | 0.005586649 | 0.011295339 | 1 |
| Obesity | neutrophil | *GPR27* | -2.052376883 | 0.012751537 | 0.017843069 | 1 |
| Obesity | neutrophil | *MTSS1* | 2.361742036 | 0.005257747 | 0.011289644 | 1 |
| Obesity | neutrophil | *RPS9* | 2.061854799 | 0.002742693 | 0.011289644 | 1 |
| Obesity | neutrophil | *TSEN34* | 2.086477298 | 0.003923611 | 0.011289644 | 1 |
| Obesity | neutrophil | *NDUFA3* | -2.863180581 | 0.01583808 | 0.021340336 | 1 |
| Obesity | neutrophil | *PAQR8* | 2.835223913 | 0.00881711 | 0.01366022 | 1 |
| Obesity | neutrophil | *NUDT6* | 1.232578255 | 0.004702221 | 0.011289644 | 1 |
| Obesity | neutrophil | *ZNF160* | 2.915663427 | 0.00532023 | 0.011289644 | 1 |
| Obesity | neutrophil | *TSNARE1* | 3.659573424 | 0.000335763 | 0.011289644 | 1 |
| Obesity | neutrophil | *FPR1* | 2.539501779 | 0.002584096 | 0.011289644 | 1 |
| Obesity | neutrophil | *FEZ2* | 2.69103523 | 0.010205852 | 0.015054948 | 1 |
| Obesity | neutrophil | *SOX7* | 3.167446151 | 0.001499923 | 0.011289644 | 1 |
| Obesity | neutrophil | *C11orf24* | 1.841375407 | 0.004292563 | 0.011289644 | 1 |
| Obesity | neutrophil | *ALK* | 1.37443194 | 0.00298902 | 0.011289644 | 1 |
| Obesity | neutrophil | *TRMT61B* | 4.424803406 | 4.42E-06 | 0.001388831 | 0.027776611 |
| Obesity | neutrophil | *MFN1* | 3.113683646 | 0.006241429 | 0.01154827 | 1 |
| Obesity | neutrophil | *KCNMB3* | 2.259792223 | 0.00181408 | 0.011289644 | 1 |
| Obesity | neutrophil | *ATP6V0E2* | 2.557422669 | 0.006739483 | 0.011825602 | 1 |
| Obesity | neutrophil | *GRIK1* | 2.486936139 | 0.011609888 | 0.016562598 | 1 |
| Obesity | neutrophil | *CLDN20* | 1.697289219 | 0.005878371 | 0.011378838 | 1 |
| Obesity | neutrophil | *GAA* | 1.812973008 | 0.016611132 | 0.022215313 | 1 |
| Obesity | neutrophil | *CANT1* | 1.784126758 | 0.006211007 | 0.011539184 | 1 |

| | | | | | | |
|---|---|---|---|---|---|---|
| Obesity | neutrophil | *KCND3* | 2.17461669 | 0.002401794 | 0.011289644 | 1 |
| Obesity | neutrophil | *MRPL36* | -1.969251078 | 0.019087325 | 0.025004628 | 1 |
| Obesity | neutrophil | *ZNF581* | 10.88632035 | 0.007540134 | 0.012419095 | 1 |
| Obesity | neutrophil | *MCC* | 1.71085122 | 0.003659385 | 0.011289644 | 1 |
| Obesity | neutrophil | *KRT27* | -4.011477412 | 0.009393062 | 0.014212799 | 1 |
| Obesity | neutrophil | *HOPX* | -3.811090843 | 0.010643881 | 0.015545356 | 1 |
| Obesity | neutrophil | *LRRC8C* | 1.483407456 | 0.004184928 | 0.011289644 | 1 |
| Obesity | neutrophil | *RSL1D1* | 3.073720927 | 0.003081666 | 0.011289644 | 1 |
| Obesity | neutrophil | *PPID* | 1.721540683 | 0.004547681 | 0.011289644 | 1 |
| Obesity | neutrophil | *COL24A1* | 0.990556432 | 0.010455407 | 0.015336711 | 1 |
| Obesity | neutrophil | *ETFDH* | 1.092364208 | 0.014908114 | 0.020256605 | 1 |
| Obesity | neutrophil | *TBCA* | 2.134060581 | 0.008606669 | 0.013460231 | 1 |
| Obesity | neutrophil | *MAP6* | -1.94955402 | 0.006559795 | 0.011699696 | 1 |
| Obesity | neutrophil | *BCL2L1* | 2.143538022 | 0.002277867 | 0.011289644 | 1 |
| Obesity | neutrophil | *PLRG1* | 4.751482081 | 0.022234864 | 0.028569506 | 1 |
| Obesity | neutrophil | *ZNF584* | 2.097430309 | 0.010945692 | 0.015863431 | 1 |
| Obesity | neutrophil | *NMUR1* | 2.306107536 | 0.004050434 | 0.011289644 | 1 |
| Obesity | neutrophil | *P2RY6* | 1.890293586 | 0.0045389 | 0.011289644 | 1 |
| Obesity | neutrophil | *ZIK1* | -0.616531847 | 0.005636492 | 0.011295339 | 1 |
| Obesity | neutrophil | *RGS19* | 2.896031473 | 0.000337787 | 0.011289644 | 1 |
| Obesity | neutrophil | *TCEA2* | 1.865281843 | 0.004154982 | 0.011289644 | 1 |
| Obesity | neutrophil | *ANO5* | -1.293869541 | 0.007095957 | 0.01208139 | 1 |
| Obesity | neutrophil | *GPHN* | 1.164557024 | 0.005385246 | 0.011289644 | 1 |
| Obesity | neutrophil | *VAT1L* | 5.92931252 | 0.009822359 | 0.014682161 | 1 |
| Obesity | neutrophil | *CAMTA1* | 1.433529956 | 0.00376544 | 0.011289644 | 1 |
| Obesity | neutrophil | *LRRC34* | 1.790474558 | 0.006406927 | 0.011648199 | 1 |
| Obesity | neutrophil | *SPATA5L1* | 5.080099238 | 0.014541307 | 0.019852594 | 1 |
| Obesity | neutrophil | *SYCE1* | 2.917522636 | 0.030088393 | 0.037527067 | 1 |
| Obesity | neutrophil | *BCL2* | 0.733256792 | 0.005918092 | 0.011385265 | 1 |
| Obesity | neutrophil | *RHNO1* | 3.736927453 | 0.021241123 | 0.027438451 | 1 |
| Obesity | neutrophil | *PWWP2B* | 1.937194125 | 0.004168479 | 0.011289644 | 1 |
| Obesity | neutrophil | *ZNF570* | 3.055674714 | 0.003830232 | 0.011289644 | 1 |
| Obesity | neutrophil | *NINJ2* | 2.365612141 | 0.003635606 | 0.011289644 | 1 |
| Obesity | neutrophil | *FAM90A1* | 5.709287557 | 0.004254579 | 0.011289644 | 1 |
| Obesity | neutrophil | *TRAPPC12* | 1.904947087 | 0.004647812 | 0.011289644 | 1 |
| Obesity | neutrophil | *C3AR1* | 1.812962103 | 0.008236444 | 0.013066495 | 1 |
| Obesity | neutrophil | *AQP4* | -1.252132193 | 0.006348852 | 0.011618636 | 1 |
| Obesity | neutrophil | *TLN2* | 0.93591686 | 0.017889747 | 0.02364365 | 1 |
| Obesity | neutrophil | *TVP23B* | 2.072124125 | 0.005879398 | 0.011378838 | 1 |
| Obesity | neutrophil | *SRGAP2C* | 3.443833795 | 0.00506389 | 0.011289644 | 1 |
| Obesity | neutrophil | *ATPAF2* | 1.956661855 | 0.003805341 | 0.011289644 | 1 |
| Obesity | neutrophil | *CYP4F22* | -0.775372109 | 0.003585505 | 0.011289644 | 1 |
| Obesity | neutrophil | *C20orf196* | 1.72999012 | 0.004410875 | 0.011289644 | 1 |
| Obesity | neutrophil | *ANKRD20A4* | 4.338739962 | 0.004403657 | 0.011289644 | 1 |
| Obesity | neutrophil | *LAMB2* | 1.055243693 | 0.004829261 | 0.011289644 | 1 |
| Obesity | neutrophil | *ORMDL3* | 1.636310832 | 0.001982605 | 0.011289644 | 1 |
| Obesity | neutrophil | *SERF1A* | 9.519618889 | 0.004348782 | 0.011289644 | 1 |
| Obesity | neutrophil | *LRRC15* | 7.082974919 | 0.033818719 | 0.041714398 | 1 |
| Obesity | neutrophil | *MOB3A* | 2.900583752 | 0.009084327 | 0.013885468 | 1 |
| Obesity | neutrophil | *NME6* | -1.762945479 | 0.01101489 | 0.015933712 | 1 |

| | | | | | | |
|---|---|---|---|---|---|---|
| Obesity | neutrophil | *FRMD3* | 3.835940819 | 0.003719875 | 0.011289644 | 1 |
| Obesity | neutrophil | *MTBP* | 1.276437094 | 0.004260348 | 0.011289644 | 1 |
| Obesity | neutrophil | *MALT1* | 3.696282826 | 0.022396864 | 0.028718964 | 1 |
| Obesity | neutrophil | *ISG20* | -2.25760272 | 0.005184594 | 0.011289644 | 1 |
| Obesity | neutrophil | *MBOAT1* | 1.998262251 | 0.004481935 | 0.011289644 | 1 |
| Obesity | neutrophil | *CXCR6* | 4.752820658 | 0.000238673 | 0.011289644 | 1 |
| Obesity | neutrophil | *C5orf34* | -1.845990526 | 0.014749323 | 0.020084253 | 1 |
| Obesity | neutrophil | *C1QTNF4* | -5.763730939 | 1.26E-14 | 1.98E-11 | 7.91E-11 |
| Obesity | neutrophil | *ZNF131* | 2.226737817 | 0.005612309 | 0.011295339 | 1 |
| Obesity | neutrophil | *HINFP* | 4.158145671 | 0.007180785 | 0.012155908 | 1 |
| Obesity | neutrophil | *SPTLC3* | 2.5498022 | 0.005125905 | 0.011289644 | 1 |
| Obesity | neutrophil | *COPRS* | 2.682237148 | 0.003503063 | 0.011289644 | 1 |
| Obesity | neutrophil | *CLEC12A* | -0.877022369 | 0.022486722 | 0.028822431 | 1 |
| Obesity | neutrophil | *SUCLG2* | 1.839601999 | 0.004934393 | 0.011289644 | 1 |
| Obesity | neutrophil | *CSDC2* | 5.491925487 | 0.000926649 | 0.011289644 | 1 |
| Obesity | neutrophil | *RCAN2* | 1.035344785 | 0.004257375 | 0.011289644 | 1 |
| Obesity | neutrophil | *IL16* | 1.793032323 | 0.003575199 | 0.011289644 | 1 |
| Obesity | neutrophil | *GNB2* | 1.74578141 | 0.007395574 | 0.012298069 | 1 |
| Obesity | neutrophil | *PRSS27* | 7.941726314 | 0.003175174 | 0.011289644 | 1 |
| Obesity | neutrophil | *FGGY* | 2.046264294 | 0.003757769 | 0.011289644 | 1 |
| Obesity | neutrophil | *ZNF24* | 3.178662018 | 0.005948408 | 0.01140172 | 1 |
| Obesity | neutrophil | *MANEA* | 1.829928977 | 0.00476763 | 0.011289644 | 1 |
| Obesity | neutrophil | *FIBP* | 10.63500602 | 0.000447986 | 0.011289644 | 1 |
| Obesity | neutrophil | *CARNS1* | 6.035844102 | 0.001612225 | 0.011289644 | 1 |
| Obesity | neutrophil | *CTSW* | -2.319977232 | 5.33E-05 | 0.00737107 | 0.335002595 |
| Obesity | neutrophil | *SNTG2* | 0.920443779 | 0.010057558 | 0.014923046 | 1 |
| Obesity | neutrophil | *PDE3A* | 0.877846873 | 0.006023138 | 0.011447874 | 1 |
| Obesity | neutrophil | *MRPL52* | 4.661573108 | 0.00164076 | 0.011289644 | 1 |
| Obesity | neutrophil | *SMPDL3A* | 4.259673621 | 0.00296749 | 0.011289644 | 1 |
| Obesity | neutrophil | *RAD9A* | 1.292317747 | 0.001961763 | 0.011289644 | 1 |
| Obesity | neutrophil | *EFEMP2* | 1.853575414 | 0.000448721 | 0.011289644 | 1 |
| Obesity | neutrophil | *AGAP5* | 55.9822379 | 0.004468132 | 0.011289644 | 1 |
| Obesity | neutrophil | *ZFAND4* | 2.975112812 | 0.015248982 | 0.020661713 | 1 |
| Obesity | neutrophil | *ZNF738* | 1.706331943 | 0.001937057 | 0.011289644 | 1 |
| Obesity | neutrophil | *SLFN11* | 3.338954948 | 0.007595248 | 0.012461202 | 1 |
| Obesity | neutrophil | *CCL19* | 8.708376527 | 0.006541334 | 0.011693308 | 1 |
| Obesity | neutrophil | *CFL1* | 3.425848894 | 0.009003168 | 0.013825823 | 1 |
| Obesity | neutrophil | *FAM192A* | 2.411678825 | 0.008600315 | 0.013453641 | 1 |
| Obesity | neutrophil | *RAB43* | -7.029954545 | 0.005922844 | 0.011385978 | 1 |
| Obesity | neutrophil | *CBWD1* | 2.267940138 | 0.004392913 | 0.011289644 | 1 |
| Obesity | neutrophil | *DCP2* | 2.549025412 | 0.006360958 | 0.011618636 | 1 |
| Obesity | neutrophil | *SNX32* | 7.095125321 | 2.40E-06 | 0.00083697 | 0.015065468 |
| Obesity | neutrophil | *CYP7B1* | 1.002083769 | 0.004741441 | 0.011289644 | 1 |
| Obesity | neutrophil | *METAP1D* | 1.715224421 | 0.003611509 | 0.011289644 | 1 |
| Obesity | neutrophil | *NADSYN1* | 2.528249934 | 0.002673624 | 0.011289644 | 1 |
| Obesity | neutrophil | *RNASEH2C* | 3.120803499 | 0.008879681 | 0.013723778 | 1 |
| Obesity | neutrophil | *ANKRD13D* | 2.124688606 | 0.006165912 | 0.011512519 | 1 |
| Obesity | neutrophil | *MRGPRF* | 1.888924454 | 0.003959581 | 0.011289644 | 1 |
| Obesity | neutrophil | *LCLAT1* | 1.770667339 | 0.004210935 | 0.011289644 | 1 |
| Obesity | neutrophil | *ADH6* | -1.638674469 | 0.005195448 | 0.011289644 | 1 |

| | | | | | | |
|---|---|---|---|---|---|---|
| Obesity | neutrophil | *GXYLT2* | 1.132747732 | 0.005385554 | 0.011289644 | 1 |
| Obesity | neutrophil | *HPSE2* | 1.225297338 | 0.000441116 | 0.011289644 | 1 |
| Obesity | neutrophil | *DCAKD* | 1.232875409 | 0.010725104 | 0.015628699 | 1 |
| Obesity | neutrophil | *HECTD4* | 1.59780279 | 0.006616286 | 0.011727259 | 1 |
| Obesity | neutrophil | *BNC2* | 1.057652163 | 0.003123789 | 0.011289644 | 1 |
| Obesity | neutrophil | *NOC3L* | 1.557419627 | 0.003489786 | 0.011289644 | 1 |
| Obesity | neutrophil | *PARP14* | 1.749633753 | 0.002707272 | 0.011289644 | 1 |
| Obesity | neutrophil | *PARP15* | 3.206802478 | 0.00869843 | 0.013546452 | 1 |
| Obesity | neutrophil | *AHSA2* | 2.610365765 | 0.003683744 | 0.011289644 | 1 |
| Obesity | neutrophil | *ABLIM3* | 1.495713218 | 0.004031239 | 0.011289644 | 1 |
| Obesity | neutrophil | *VANGL1* | 1.729904534 | 0.003576293 | 0.011289644 | 1 |
| Obesity | neutrophil | *GLRX* | 1.236186543 | 0.004078362 | 0.011289644 | 1 |
| Obesity | neutrophil | *IQCB1* | 1.209926059 | 0.026542072 | 0.03348786 | 1 |
| Obesity | neutrophil | *GOLGB1* | 1.488081734 | 0.003880598 | 0.011289644 | 1 |
| Obesity | neutrophil | *DMRT2* | 1.435721943 | 0.009722106 | 0.014561037 | 1 |
| Obesity | neutrophil | *ZNF483* | 1.126004728 | 0.004707937 | 0.011289644 | 1 |
| Obesity | neutrophil | *SLC2A14* | 2.738905062 | 0.010019392 | 0.014881153 | 1 |
| Obesity | neutrophil | *MZT2A* | 1.680993253 | 0.003967515 | 0.011289644 | 1 |
| Obesity | neutrophil | *MAP3K11* | 1.898334402 | 0.003997312 | 0.011289644 | 1 |
| Obesity | neutrophil | *C1QA* | 1.180104666 | 0.004252395 | 0.011289644 | 1 |
| Obesity | neutrophil | *NDNF* | 2.138891807 | 0.002689327 | 0.011289644 | 1 |
| Obesity | neutrophil | *OLR1* | -8.322681301 | 0.03612226 | 0.044312322 | 1 |
| Obesity | neutrophil | *GLIPR1L1* | 2.138890126 | 0.002824922 | 0.011289644 | 1 |
| Obesity | neutrophil | *ARV1* | 1.898420894 | 0.00264107 | 0.011289644 | 1 |
| Obesity | neutrophil | *CCDC36* | 1.178263346 | 0.002575145 | 0.011289644 | 1 |
| Obesity | neutrophil | *THAP2* | 1.674443238 | 0.003968399 | 0.011289644 | 1 |
| Obesity | neutrophil | *PPP1R14B* | 3.875988071 | 0.005878675 | 0.011378838 | 1 |
| Obesity | neutrophil | *SMARCC1* | 2.017606767 | 0.005381383 | 0.011289644 | 1 |
| Obesity | neutrophil | *ZNF417* | -2.469249699 | 0.002958172 | 0.011289644 | 1 |
| Obesity | neutrophil | *VEGFB* | 2.273223924 | 0.007195196 | 0.012163537 | 1 |
| Obesity | neutrophil | *PEAK1* | 1.315358512 | 0.004575626 | 0.011289644 | 1 |
| Obesity | neutrophil | *MST1* | -4.03063911 | 5.24E-06 | 0.001568228 | 0.032932781 |
| Obesity | neutrophil | *TNFRSF10C* | -0.460815979 | 0.003155102 | 0.011289644 | 1 |
| Obesity | neutrophil | *GMPPB* | 4.781331572 | 3.79E-05 | 0.005962109 | 0.238484364 |
| Obesity | neutrophil | *CSPG4* | 1.814722863 | 0.009716407 | 0.014558401 | 1 |
| Obesity | neutrophil | *SNX33* | 3.308845571 | 0.004461801 | 0.011289644 | 1 |
| Obesity | neutrophil | *NUDT18* | 4.475392198 | 0.001886957 | 0.011289644 | 1 |
| Obesity | neutrophil | *SULT1B1* | -2.609116019 | 0.011365002 | 0.01631319 | 1 |
| Obesity | neutrophil | *NUDT4* | 1.143573584 | 0.004762996 | 0.011289644 | 1 |
| Obesity | neutrophil | *SCAI* | 2.300940258 | 0.005509245 | 0.011289644 | 1 |
| Obesity | neutrophil | *C1orf100* | 5.418956552 | 0.004461177 | 0.011289644 | 1 |
| Obesity | neutrophil | *AGFG1* | 1.875987841 | 0.004820014 | 0.011289644 | 1 |
| Obesity | neutrophil | *HAP1* | -3.074534417 | 0.001357742 | 0.011289644 | 1 |
| Obesity | neutrophil | *RNF213* | 1.742526283 | 0.005410513 | 0.011289644 | 1 |
| Obesity | neutrophil | *DPY19L1* | 1.307523265 | 0.004942455 | 0.011289644 | 1 |
| Obesity | neutrophil | *TUBB8* | 4.980505328 | 0.012181546 | 0.017221128 | 1 |
| Obesity | neutrophil | *GPR160* | -1.995641648 | 0.005310797 | 0.011289644 | 1 |
| Obesity | neutrophil | *GOLIM4* | 1.213872686 | 0.00437418 | 0.011289644 | 1 |
| Obesity | neutrophil | *USMG5* | 1.354837309 | 0.004270212 | 0.011289644 | 1 |
| Obesity | neutrophil | *HOXB2* | 1.859199891 | 0.001048636 | 0.011289644 | 1 |

| | | | | | | |
|---|---|---|---|---|---|---|
| Obesity | neutrophil | *XXYLT1* | 1.099657177 | 0.004826318 | 0.011289644 | 1 |
| Obesity | neutrophil | *UBXN2A* | 1.383717137 | 0.005300433 | 0.011289644 | 1 |
| Obesity | neutrophil | *CCS* | 2.909208867 | 0.003082821 | 0.011289644 | 1 |
| Obesity | neutrophil | *CEP19* | -3.494993576 | 0.004438676 | 0.011289644 | 1 |
| Obesity | neutrophil | *SLC25A30* | 1.704688977 | 0.00374576 | 0.011289644 | 1 |
| Obesity | neutrophil | *CTSF* | 1.150373464 | 0.012396498 | 0.017443326 | 1 |
| Obesity | neutrophil | *LEMD3* | 1.800167912 | 0.004981926 | 0.011289644 | 1 |
| Obesity | neutrophil | *C16orf91* | 2.573647516 | 0.004536769 | 0.011289644 | 1 |
| Obesity | neutrophil | *SOCS7* | 1.526621529 | 0.004253953 | 0.011289644 | 1 |
| Obesity | neutrophil | *TLR6* | -1.441000603 | 0.014314193 | 0.019593584 | 1 |
| Obesity | neutrophil | *RGMB* | 1.581487578 | 0.005557444 | 0.011295339 | 1 |
| Obesity | neutrophil | *FAM53A* | 7.673286879 | 0.023676986 | 0.030133039 | 1 |
| Obesity | neutrophil | *MGA* | 1.597473847 | 0.007378266 | 0.01229744 | 1 |
| Obesity | neutrophil | *SNX31* | 3.099042536 | 0.008805616 | 0.013652505 | 1 |
| Obesity | neutrophil | *DDX23* | 2.11595044 | 0.004251475 | 0.011289644 | 1 |
| Obesity | neutrophil | *GLIS1* | 1.621154985 | 0.005938473 | 0.011396756 | 1 |
| Obesity | neutrophil | *RALGAPA1* | 1.566960283 | 0.00502541 | 0.011289644 | 1 |
| Obesity | neutrophil | *LIG4* | 1.700522354 | 0.004049657 | 0.011289644 | 1 |
| Obesity | neutrophil | *SNAPC5* | 1.648399713 | 0.008483227 | 0.013331367 | 1 |
| Obesity | neutrophil | *LINGO2* | 1.773484773 | 0.002521371 | 0.011289644 | 1 |
| Obesity | neutrophil | *BBS1* | 7.282400672 | 0.000414715 | 0.011289644 | 1 |
| Obesity | neutrophil | *ANKRD36C* | 0.988803481 | 0.005460554 | 0.011289644 | 1 |
| Obesity | neutrophil | *TMEM81* | 16.18116024 | 0.009662095 | 0.014501216 | 1 |
| Obesity | neutrophil | *ZNF497* | 14.16697721 | 0.001383927 | 0.011289644 | 1 |
| Obesity | neutrophil | *KLF14* | 20.41396632 | 0.000330391 | 0.011289644 | 1 |
| Obesity | neutrophil | *ZNF266* | 2.028254298 | 0.001478413 | 0.011289644 | 1 |
| Obesity | neutrophil | *BRSK2* | 3.959397926 | 0.010970147 | 0.015887886 | 1 |
| Obesity | neutrophil | *SH3PXD2B* | 1.034132533 | 0.002431343 | 0.011289644 | 1 |
| Obesity | neutrophil | *LARP7* | 2.635836324 | 0.006471339 | 0.01166174 | 1 |
| Obesity | neutrophil | *FGFBP3* | -2.430254375 | 0.007892856 | 0.012746567 | 1 |
| Obesity | neutrophil | *C4orf32* | 1.752479889 | 0.00393906 | 0.011289644 | 1 |
| Obesity | neutrophil | *HRAS* | 2.32639138 | 0.00139046 | 0.011289644 | 1 |
| Obesity | neutrophil | *SRP72* | 1.293074145 | 0.001539893 | 0.011289644 | 1 |
| Obesity | neutrophil | *C4orf26* | 12.07568074 | 0.00344529 | 0.011289644 | 1 |
| Obesity | neutrophil | *BTC* | -1.407714691 | 0.007518832 | 0.01240055 | 1 |
| Obesity | neutrophil | *NLRP6* | -1.479860483 | 0.004258671 | 0.011289644 | 1 |
| Obesity | neutrophil | *NDUFA11* | 1.525575784 | 0.004356967 | 0.011289644 | 1 |
| Obesity | neutrophil | *RSRC1* | 1.307050731 | 0.019534133 | 0.025505939 | 1 |
| Obesity | neutrophil | *PTDSS2* | -2.07986817 | 0.031213974 | 0.03879072 | 1 |
| Obesity | neutrophil | *FUT1* | -1.172902368 | 0.008799492 | 0.013646375 | 1 |
| Obesity | neutrophil | *DHX36* | 1.342221148 | 0.006736319 | 0.011825602 | 1 |
| Obesity | neutrophil | *FBXW8* | 2.546276151 | 0.005329755 | 0.011289644 | 1 |
| Obesity | neutrophil | *CHST2* | 3.797169144 | 0.004160289 | 0.011289644 | 1 |
| Obesity | neutrophil | *ATR* | 2.089407448 | 0.004572045 | 0.011289644 | 1 |
| Obesity | neutrophil | *TVP23C* | 2.713347206 | 0.003479168 | 0.011289644 | 1 |
| Obesity | neutrophil | *PACS1* | 1.343960573 | 0.005416571 | 0.011289644 | 1 |
| Obesity | neutrophil | *SH3BP5L* | 5.266634396 | 0.002930726 | 0.011289644 | 1 |
| Obesity | neutrophil | *CADM2* | 1.867026167 | 0.00040827 | 0.011289644 | 1 |
| Obesity | neutrophil | *PSMD2* | 2.501605102 | 0.000597877 | 0.011289644 | 1 |
| Obesity | neutrophil | *ARHGAP1* | 1.333358401 | 0.004626773 | 0.011289644 | 1 |

| | | | | | | |
|---|---|---|---|---|---|---|
| Obesity | neutrophil | *CHST1* | -1.215851282 | 0.003735126 | 0.011289644 | 1 |
| Obesity | neutrophil | *GOLGA8A* | 3.076579871 | 0.001132999 | 0.011289644 | 1 |
| Obesity | neutrophil | *PHYHD1* | 1.534257116 | 0.003701719 | 0.011289644 | 1 |
| Obesity | neutrophil | *ZNF519* | 1.1232657 | 0.003837973 | 0.011289644 | 1 |
| Obesity | neutrophil | *BANF1* | 2.474530995 | 0.017964548 | 0.023722561 | 1 |
| Obesity | neutrophil | *CHRNA7* | -3.677154157 | 0.001995476 | 0.011289644 | 1 |
| Obesity | neutrophil | *TMEM9B* | 5.938609105 | 0.001401311 | 0.011289644 | 1 |
| Obesity | neutrophil | *NRIP3* | 2.282753113 | 0.01261837 | 0.017707967 | 1 |
| Obesity | neutrophil | *SMAD2* | 2.189782372 | 0.006800891 | 0.011873524 | 1 |
| Obesity | neutrophil | *EIF3F* | -3.168555132 | 0.025605863 | 0.03241725 | 1 |
| Obesity | neutrophil | *CLTB* | 3.831068179 | 0.006584066 | 0.011713234 | 1 |
| Obesity | neutrophil | *LPL* | 1.122380424 | 0.012228493 | 0.017272643 | 1 |
| Obesity | neutrophil | *TBC1D10C* | -0.773960701 | 0.007989985 | 0.012814549 | 1 |
| Obesity | neutrophil | *PPP2R2D* | 2.006515592 | 0.002663875 | 0.011289644 | 1 |
| Obesity | neutrophil | *MCTP1* | 2.401299445 | 0.011295678 | 0.016224794 | 1 |
| Obesity | neutrophil | *UBQLNL* | 4.559601631 | 0.025500853 | 0.032303821 | 1 |
| Obesity | neutrophil | *KCNE3* | -1.351177549 | 0.00650475 | 0.011671674 | 1 |
| Obesity | neutrophil | *UCP3* | 3.34608142 | 0.014215893 | 0.019488731 | 1 |
| Obesity | neutrophil | *UCP2* | 2.381620968 | 0.006426235 | 0.011648199 | 1 |
| Obesity | neutrophil | *MRPL48* | 1.205934511 | 0.013286676 | 0.01844003 | 1 |
| Obesity | neutrophil | *RAB6A* | 2.444982745 | 0.008654059 | 0.013510538 | 1 |
| Obesity | neutrophil | *RPS6KB2* | 6.02267443 | 0.011896812 | 0.016887617 | 1 |
| Obesity | neutrophil | *TOM1L2* | 2.019147265 | 0.004598333 | 0.011289644 | 1 |
| Obesity | neutrophil | *TEX26* | 10.83052953 | 0.026442664 | 0.033382535 | 1 |
| Obesity | neutrophil | *GPR156* | -0.620615792 | 0.005653016 | 0.011300433 | 1 |
| Obesity | neutrophil | *B3GNTL1* | 4.199529992 | 0.004589875 | 0.011289644 | 1 |
| Obesity | neutrophil | *TTLL11* | 1.477124721 | 0.013677933 | 0.018882997 | 1 |
| Obesity | neutrophil | *LINC01106* | 28.95436816 | 0.008340317 | 0.013184706 | 1 |
| Obesity | neutrophil | *MSRA* | 1.440521055 | 0.008878311 | 0.013723778 | 1 |
| Obesity | neutrophil | *CTDNEP1* | 2.803726423 | 0.002791006 | 0.011289644 | 1 |
| Obesity | neutrophil | *ETV4* | -1.562782179 | 0.02135112 | 0.027546582 | 1 |
| Obesity | neutrophil | *SWI5* | 1.829592121 | 0.003351571 | 0.011289644 | 1 |
| Obesity | neutrophil | *ZDHHC21* | 1.169517936 | 0.004795797 | 0.011289644 | 1 |
| Obesity | neutrophil | *A2M* | 2.612851269 | 0.008402519 | 0.013249721 | 1 |
| Obesity | neutrophil | *ARL4D* | -1.221069893 | 0.005317482 | 0.011289644 | 1 |
| Obesity | neutrophil | *LRRN1* | 3.57565784 | 0.00494037 | 0.011289644 | 1 |
| Obesity | neutrophil | *ZNF613* | 2.134701069 | 0.002230013 | 0.011289644 | 1 |
| Obesity | neutrophil | *IP6K1* | 2.914127889 | 0.001666617 | 0.011289644 | 1 |
| Obesity | neutrophil | *SSNA1* | 1.746818015 | 0.004135073 | 0.011289644 | 1 |
| Obesity | neutrophil | *CHMP6* | 2.510182725 | 0.004017695 | 0.011289644 | 1 |
| Obesity | neutrophil | *UFSP1* | 5.180205514 | 0.004892739 | 0.011289644 | 1 |
| Obesity | neutrophil | *TMEM39A* | 2.214708553 | 0.005012384 | 0.011289644 | 1 |
| Obesity | neutrophil | *TCP11L1* | 2.543862212 | 0.002356673 | 0.011289644 | 1 |
| Obesity | neutrophil | *GPX2* | 2.109844766 | 0.004535034 | 0.011289644 | 1 |
| Obesity | neutrophil | *CCDC57* | 1.128312447 | 0.013934264 | 0.019161137 | 1 |
| Obesity | neutrophil | *CIDEA* | 0.760231793 | 0.02300783 | 0.029370605 | 1 |
| Obesity | neutrophil | *SMIM19* | 3.324534991 | 0.000479301 | 0.011289644 | 1 |
| Obesity | neutrophil | *ZNF404* | 1.823814419 | 0.008709993 | 0.013551034 | 1 |
| Obesity | neutrophil | *SLC35G1* | 1.30164573 | 0.006896666 | 0.011932455 | 1 |
| Obesity | neutrophil | *ZNF135* | 1.743002496 | 0.012567849 | 0.017652831 | 1 |

| | | | | | | |
|---|---|---|---|---|---|---|
| Obesity | neutrophil | *COX8A* | 1.656505753 | 0.004218708 | 0.011289644 | 1 |
| Obesity | neutrophil | *B3GNT4* | 15.08412392 | 0.004340014 | 0.011289644 | 1 |
| Obesity | neutrophil | *CRLF3* | 1.35816639 | 0.005047095 | 0.011289644 | 1 |
| Obesity | neutrophil | *SPRYD4* | 1.719522863 | 0.004631588 | 0.011289644 | 1 |
| Obesity | neutrophil | *VPS37D* | -0.880643859 | 0.02069713 | 0.026823924 | 1 |
| Obesity | neutrophil | *SLCO3A1* | 3.023900205 | 0.015392971 | 0.020821067 | 1 |
| Obesity | neutrophil | *WDR25* | 3.303222937 | 0.00906774 | 0.013870775 | 1 |
| Obesity | neutrophil | *PLA2G16* | 1.581918573 | 0.000821476 | 0.011289644 | 1 |
| Obesity | neutrophil | *PHLDB3* | 1.798834201 | 0.004171721 | 0.011289644 | 1 |
| Obesity | neutrophil | *PRR15* | 11.17214946 | 0.007189477 | 0.012157139 | 1 |
| Obesity | neutrophil | *RMDN1* | 2.252798769 | 0.003001523 | 0.011289644 | 1 |
| Obesity | neutrophil | *HORMAD2* | 2.691969332 | 0.007211152 | 0.012174144 | 1 |
| Obesity | neutrophil | *LRRC37A* | -4.013801592 | 0.031325487 | 0.038913917 | 1 |
| Obesity | neutrophil | *ACSF3* | 2.123880816 | 0.015261133 | 0.020673722 | 1 |
| Obesity | neutrophil | *BOK* | 3.5212176 | 0.022003809 | 0.028295756 | 1 |
| Obesity | neutrophil | *ZNF843* | -0.859142643 | 0.005616481 | 0.011295339 | 1 |
| Obesity | neutrophil | *C8orf59* | 1.907741213 | 0.003153923 | 0.011289644 | 1 |
| Obesity | neutrophil | *RUFY1* | 1.781622266 | 0.000863062 | 0.011289644 | 1 |
| Obesity | neutrophil | *VSIG10* | 3.021567141 | 0.006771458 | 0.011851936 | 1 |
| Obesity | neutrophil | *IRX5* | 2.418766951 | 0.001949437 | 0.011289644 | 1 |
| Obesity | neutrophil | *PNMA1* | -0.617885516 | 0.005044987 | 0.011289644 | 1 |
| Obesity | neutrophil | *ANKLE2* | 3.983938697 | 0.007162834 | 0.012141606 | 1 |
| Obesity | neutrophil | *GCNT4* | 6.936220878 | 0.004196711 | 0.011289644 | 1 |
| Obesity | neutrophil | *MUC20* | 2.599861493 | 0.007287494 | 0.012230772 | 1 |
| Obesity | neutrophil | *THAP4* | -3.242098239 | 0.019734278 | 0.025751225 | 1 |
| Obesity | neutrophil | *SHMT1* | 2.692849723 | 0.003985822 | 0.011289644 | 1 |
| Obesity | neutrophil | *DPP7* | 1.666603607 | 0.004296989 | 0.011289644 | 1 |
| Obesity | neutrophil | *SEC24C* | 3.882138585 | 0.014372917 | 0.019665404 | 1 |
| Obesity | neutrophil | *MTHFR* | 1.70289154 | 0.003660742 | 0.011289644 | 1 |
| Obesity | neutrophil | *C19orf18* | 1.60821965 | 0.010749869 | 0.015648165 | 1 |
| Obesity | neutrophil | *TMEM80* | 1.321687837 | 0.017829738 | 0.023569294 | 1 |
| Obesity | neutrophil | *WDR73* | 2.416915818 | 0.004869378 | 0.011289644 | 1 |
| Obesity | neutrophil | *POLE* | 2.080909822 | 0.004167259 | 0.011289644 | 1 |
| Obesity | neutrophil | *DSCAML1* | 1.752688116 | 0.004217377 | 0.011289644 | 1 |
| Obesity | neutrophil | *EPS8L2* | 2.313595376 | 0.003982003 | 0.011289644 | 1 |
| Obesity | neutrophil | *B3GNT8* | 4.999529454 | 0.004250118 | 0.011289644 | 1 |
| Obesity | neutrophil | *AP006621.1* | 6.956421788 | 0.001904078 | 0.011289644 | 1 |
| Obesity | neutrophil | *MAN1B1* | 1.943786935 | 0.004271218 | 0.011289644 | 1 |
| Obesity | neutrophil | *TOP3A* | 2.768686082 | 0.011875664 | 0.016865213 | 1 |
| Obesity | neutrophil | *ZBTB38* | 2.219775832 | 0.003997861 | 0.011289644 | 1 |
| Obesity | neutrophil | *C8orf31* | 2.264599305 | 0.00420845 | 0.011289644 | 1 |
| Obesity | neutrophil | *AC024940.1* | 7.440520567 | 0.001541991 | 0.011289644 | 1 |
| Obesity | neutrophil | *TIMM22* | -2.714967211 | 0.011406197 | 0.016351571 | 1 |
| Obesity | neutrophil | *MAGEF1* | -4.113477082 | 0.021350067 | 0.027546582 | 1 |
| Obesity | neutrophil | *SAMD9L* | 2.223478681 | 0.005140312 | 0.011289644 | 1 |
| Obesity | neutrophil | *PAWR* | 3.63869115 | 0.011746184 | 0.016711532 | 1 |
| Obesity | neutrophil | *NAP1L5* | 10.68450853 | 0.001546484 | 0.011289644 | 1 |
| Obesity | neutrophil | *NIM1K* | 2.608116771 | 0.004817961 | 0.011289644 | 1 |
| Obesity | neutrophil | *ACOT4* | 2.710186018 | 0.006187659 | 0.011519636 | 1 |
| Obesity | neutrophil | *ARIH2* | 1.36190457 | 0.007834088 | 0.012699648 | 1 |

| | | | | | | |
|---|---|---|---|---|---|---|
| Obesity | neutrophil | *RBM44* | 2.315157331 | 0.006433364 | 0.011648199 | 1 |
| Obesity | neutrophil | *IRX3* | 2.112093912 | 0.017752544 | 0.023477123 | 1 |
| Obesity | neutrophil | *ATOX1* | 3.676817483 | 0.004159152 | 0.011289644 | 1 |
| Obesity | neutrophil | *TBL1XR1* | 1.653288962 | 0.007915748 | 0.012769218 | 1 |
| Obesity | neutrophil | *SAMD12* | 1.572377434 | 0.022816877 | 0.029174233 | 1 |
| Obesity | neutrophil | *C18orf32* | 1.996481228 | 0.004793455 | 0.011289644 | 1 |
| Obesity | neutrophil | *RPLP2* | 2.022684682 | 0.004993588 | 0.011289644 | 1 |
| Obesity | neutrophil | *C12orf54* | 8.285848736 | 0.00515521 | 0.011289644 | 1 |
| Obesity | neutrophil | *ACAD9* | 1.44276428 | 0.015751297 | 0.021255292 | 1 |
| Obesity | neutrophil | *CD163L1* | 1.092805004 | 0.003302975 | 0.011289644 | 1 |
| Obesity | neutrophil | *THAP5* | 1.587528026 | 0.005100105 | 0.011289644 | 1 |
| Obesity | neutrophil | *CD151* | 4.827986227 | 0.003076839 | 0.011289644 | 1 |
| Obesity | neutrophil | *ZCCHC3* | -1.616626031 | 0.004818127 | 0.011289644 | 1 |
| Obesity | neutrophil | *PCDHB9* | 3.035094759 | 0.009441813 | 0.014259024 | 1 |
| Obesity | neutrophil | *ZNF619* | 2.514372555 | 0.004047888 | 0.011289644 | 1 |
| Obesity | neutrophil | *ZBTB41* | 3.264672434 | 0.008386236 | 0.013235441 | 1 |
| Obesity | neutrophil | *ZNF354C* | 1.412112252 | 0.004118943 | 0.011289644 | 1 |
| Obesity | neutrophil | *RIC8A* | 1.77829488 | 0.007512778 | 0.012399504 | 1 |
| Obesity | neutrophil | *IMP3* | 1.713575004 | 0.004048218 | 0.011289644 | 1 |
| Obesity | neutrophil | *ODF3B* | 1.9922645 | 0.002303885 | 0.011289644 | 1 |
| Obesity | neutrophil | *DPY19L2* | 1.507405808 | 0.010082552 | 0.01492911 | 1 |
| Obesity | neutrophil | *C2orf73* | 2.410249073 | 0.00431105 | 0.011289644 | 1 |
| Obesity | neutrophil | *FAM26E* | -0.792567385 | 0.006265399 | 0.011565541 | 1 |
| Obesity | neutrophil | *ALS2CL* | 2.11624377 | 0.002197353 | 0.011289644 | 1 |
| Obesity | neutrophil | *MLF1* | 2.419363628 | 0.005754244 | 0.011351407 | 1 |
| Obesity | neutrophil | *PRSS42* | -7.064895981 | 0.000331506 | 0.011289644 | 1 |
| Obesity | neutrophil | *C2orf69* | 1.350373001 | 0.004545732 | 0.011289644 | 1 |
| Obesity | neutrophil | *GRAMD1C* | -1.237828895 | 0.006442583 | 0.011648199 | 1 |
| Obesity | neutrophil | *TSSK6* | -6.179240103 | 0.001090655 | 0.011289644 | 1 |
| Obesity | neutrophil | *GOLGA8Q* | -21.14848568 | 0.023194414 | 0.029566764 | 1 |
| Obesity | neutrophil | *DALRD3* | 1.855462752 | 0.013344634 | 0.018491892 | 1 |
| Obesity | neutrophil | *ZNF114* | 3.587918351 | 0.007277187 | 0.01222653 | 1 |
| Obesity | neutrophil | *ZNF518B* | -2.523538952 | 0.008310394 | 0.013144161 | 1 |
| Obesity | neutrophil | *LCORL* | 2.084865592 | 0.005288371 | 0.011289644 | 1 |
| Obesity | neutrophil | *ZNF454* | 2.437184777 | 0.001866759 | 0.011289644 | 1 |
| Obesity | neutrophil | *ZC3H12D* | 2.834774711 | 0.004412425 | 0.011289644 | 1 |
| Obesity | neutrophil | *KDELC2* | 1.800489119 | 0.004108605 | 0.011289644 | 1 |
| Obesity | neutrophil | *PLEC* | 1.268553556 | 0.004651467 | 0.011289644 | 1 |
| Obesity | neutrophil | *SH2D4B* | -2.010432131 | 0.006029619 | 0.011447874 | 1 |
| Obesity | neutrophil | *RNF212* | 1.570418318 | 0.003616888 | 0.011289644 | 1 |
| Obesity | neutrophil | *PRSS36* | 2.716854774 | 0.004341588 | 0.011289644 | 1 |
| Obesity | neutrophil | *WDR6* | 2.019117119 | 0.003689855 | 0.011289644 | 1 |
| Obesity | neutrophil | *GEN1* | 1.211786569 | 0.005248896 | 0.011289644 | 1 |
| Obesity | neutrophil | *TMPRSS9* | -1.928075864 | 0.01271647 | 0.017809857 | 1 |
| Obesity | neutrophil | *AQP11* | 2.708988787 | 0.003530149 | 0.011289644 | 1 |
| Obesity | neutrophil | *TMEM11* | 1.655032092 | 0.004242509 | 0.011289644 | 1 |
| Obesity | neutrophil | *KCNG2* | -2.550328955 | 0.01835351 | 0.024165134 | 1 |
| Obesity | neutrophil | *ZFAND2A* | 2.197632315 | 0.003407901 | 0.011289644 | 1 |
| Obesity | neutrophil | *FAM220A* | -0.970628916 | 0.003715069 | 0.011289644 | 1 |
| Obesity | neutrophil | *BEND3* | -0.673271686 | 0.005557892 | 0.011295339 | 1 |

| | | | | | | |
|---|---|---|---|---|---|---|
| Obesity | neutrophil | *NT5DC1* | 1.173085278 | 0.005039092 | 0.011289644 | 1 |
| Obesity | neutrophil | *AC006111.1* | 9.509006858 | 0.003512499 | 0.011289644 | 1 |
| Obesity | neutrophil | *LINC00843* | 1.974678237 | 0.004263328 | 0.011289644 | 1 |
| Obesity | neutrophil | *COX14* | 1.579972915 | 0.003010343 | 0.011289644 | 1 |
| Obesity | neutrophil | *CTXN1* | 14.37091974 | 0.006845127 | 0.011894781 | 1 |
| Obesity | neutrophil | *CA8* | 2.47271153 | 0.004400745 | 0.011289644 | 1 |
| Obesity | neutrophil | *ERBB4* | 0.762455319 | 0.004260073 | 0.011289644 | 1 |
| Obesity | neutrophil | *GPR35* | -0.721427918 | 0.00681428 | 0.011874479 | 1 |
| Obesity | neutrophil | *ZNF713* | 2.863382116 | 0.014484944 | 0.01979714 | 1 |
| Obesity | neutrophil | *PARP10* | 2.477017817 | 0.006247986 | 0.011553261 | 1 |
| Obesity | neutrophil | *KCTD12* | 1.808593387 | 0.004954062 | 0.011289644 | 1 |
| Obesity | neutrophil | *RPP25* | -0.768716208 | 0.001840034 | 0.011289644 | 1 |
| Obesity | neutrophil | *STX19* | 3.049523468 | 0.000368079 | 0.011289644 | 1 |
| Obesity | neutrophil | *TRIM73* | -15.59570013 | 5.86E-05 | 0.007678595 | 0.368572579 |
| Obesity | neutrophil | *EFCAB13* | 1.695693812 | 0.018620963 | 0.024471153 | 1 |
| Obesity | neutrophil | *C17orf62* | 1.441734372 | 0.003200153 | 0.011289644 | 1 |
| Obesity | neutrophil | *GAK* | 2.713626155 | 0.007286086 | 0.012230772 | 1 |
| Obesity | neutrophil | *TUFM* | 2.276734067 | 6.62E-06 | 0.001734043 | 0.041617038 |
| Obesity | neutrophil | *RMI1* | 1.490164646 | 0.004329667 | 0.011289644 | 1 |
| Obesity | neutrophil | *CTC1* | 3.389349592 | 0.000589645 | 0.011289644 | 1 |
| Obesity | neutrophil | *SNX18* | 2.271735401 | 0.00607343 | 0.011459063 | 1 |
| Obesity | neutrophil | *C14orf39* | 1.853419109 | 0.026525158 | 0.033473237 | 1 |
| Obesity | neutrophil | *KLHDC7A* | -1.962872336 | 0.000612598 | 0.011289644 | 1 |
| Obesity | neutrophil | *TRIML2* | 35.70227906 | 0.01551634 | 0.020965233 | 1 |
| Obesity | neutrophil | *FARSA* | -2.961656819 | 0.002750159 | 0.011289644 | 1 |
| Obesity | neutrophil | *SPTY2D1* | 3.761462329 | 0.006614429 | 0.011727259 | 1 |
| Obesity | neutrophil | *TCAIM* | 1.737030858 | 0.004259934 | 0.011289644 | 1 |
| Obesity | neutrophil | *ZNF664* | -1.967761437 | 0.008995397 | 0.013817264 | 1 |
| Obesity | neutrophil | *CDH4* | 2.37805886 | 0.005431339 | 0.011289644 | 1 |
| Obesity | neutrophil | *ARID3B* | 1.328169346 | 0.004113765 | 0.011289644 | 1 |
| Obesity | neutrophil | *EGR3* | 2.334826376 | 0.004378153 | 0.011289644 | 1 |
| Obesity | neutrophil | *GEMIN4* | -2.80206666 | 0.004279657 | 0.011289644 | 1 |
| Obesity | neutrophil | *FJX1* | -0.923608794 | 0.009230756 | 0.014038163 | 1 |
| Obesity | neutrophil | *PLD6* | -2.290127242 | 0.00361049 | 0.011289644 | 1 |
| Obesity | neutrophil | *ZBTB42* | -2.370673003 | 0.009951898 | 0.014815909 | 1 |
| Obesity | neutrophil | *LACC1* | 1.97388542 | 0.003028059 | 0.011289644 | 1 |
| Obesity | neutrophil | *MAF1* | 2.244226427 | 0.004558746 | 0.011289644 | 1 |
| Obesity | neutrophil | *FCER1A* | -1.871386943 | 0.005137949 | 0.011289644 | 1 |
| Obesity | neutrophil | *PCED1B* | 1.49125436 | 0.004426064 | 0.011289644 | 1 |
| Obesity | neutrophil | *PIPOX* | -2.600001064 | 0.003506117 | 0.011289644 | 1 |
| Obesity | neutrophil | *CDH5* | 1.400468067 | 0.001441593 | 0.011289644 | 1 |
| Obesity | neutrophil | *SERTAD2* | 1.012496772 | 0.004215931 | 0.011289644 | 1 |
| Obesity | neutrophil | *GIPC3* | -5.103573663 | 0.000463654 | 0.011289644 | 1 |
| Obesity | neutrophil | *CITED4* | -2.618996955 | 0.008704766 | 0.013548613 | 1 |
| Obesity | neutrophil | *TIGD5* | -0.715456363 | 0.004519338 | 0.011289644 | 1 |
| Obesity | neutrophil | *PDXDC1* | 2.3337048 | 0.005970524 | 0.011423216 | 1 |
| Obesity | neutrophil | *ZNF154* | 3.014401073 | 0.004542767 | 0.011289644 | 1 |
| Obesity | neutrophil | *R3HDM2* | 1.909512962 | 0.005023784 | 0.011289644 | 1 |
| Obesity | neutrophil | *B3GNT3* | 23.16768499 | 3.82E-06 | 0.001264386 | 0.024023327 |
| Obesity | neutrophil | *SSC5D* | 1.557367378 | 0.005247005 | 0.011289644 | 1 |

| | | | | | | |
|---|---|---|---|---|---|---|
| Obesity | neutrophil | *ZADH2* | 2.255875438 | 0.006111131 | 0.011478524 | 1 |
| Obesity | neutrophil | *TMEM86B* | 2.845493262 | 0.002582039 | 0.011289644 | 1 |
| Obesity | neutrophil | *SEPT1* | 1.922319931 | 0.004389727 | 0.011289644 | 1 |
| Obesity | neutrophil | *EXOC3* | 3.412928608 | 0.007720912 | 0.012598333 | 1 |
| Obesity | neutrophil | *LYNX1* | -1.630014731 | 0.022410816 | 0.028730996 | 1 |
| Obesity | neutrophil | *TDRP* | 1.779270107 | 0.004358906 | 0.011289644 | 1 |
| Obesity | neutrophil | *ZNF816* | 2.604063132 | 0.014804822 | 0.020138017 | 1 |
| Obesity | neutrophil | *HCLS1* | -1.911282874 | 0.014836893 | 0.02016419 | 1 |
| Obesity | neutrophil | *ZNF609* | 1.359142218 | 0.00476491 | 0.011289644 | 1 |
| Obesity | neutrophil | *CCDC66* | 1.887229923 | 0.003866402 | 0.011289644 | 1 |
| Obesity | neutrophil | *DEFB124* | -1.050961207 | 0.005080229 | 0.011289644 | 1 |
| Obesity | neutrophil | *CYP8B1* | 2.129564471 | 0.006409331 | 0.011648199 | 1 |
| Obesity | neutrophil | *RNF182* | -5.393953397 | 0.013763409 | 0.018982887 | 1 |
| Obesity | neutrophil | *MB21D2* | 1.224937753 | 0.003383426 | 0.011289644 | 1 |
| Obesity | neutrophil | *SSTR2* | 5.175534766 | 0.004398846 | 0.011289644 | 1 |
| Obesity | neutrophil | *ZNF594* | 1.601504881 | 0.00625456 | 0.011555481 | 1 |
| Obesity | neutrophil | *PCGF5* | 1.383963754 | 0.004566494 | 0.011289644 | 1 |
| Obesity | neutrophil | *SHISA2* | 46.66321921 | 0.004384474 | 0.011289644 | 1 |
| Obesity | neutrophil | *CHST13* | 9.491375614 | 0.002641572 | 0.011289644 | 1 |
| Obesity | neutrophil | *SRSF8* | 4.766414427 | 0.009225141 | 0.014033018 | 1 |
| Obesity | neutrophil | *SLC36A4* | 2.195741048 | 0.006027274 | 0.011447874 | 1 |
| Obesity | neutrophil | *ZDHHC20* | 2.114710047 | 0.005060915 | 0.011289644 | 1 |
| Obesity | neutrophil | *ZNF443* | -1.369641214 | 0.010093984 | 0.014939001 | 1 |
| Obesity | neutrophil | *CAPS2* | 2.203924773 | 0.005010574 | 0.011289644 | 1 |
| Obesity | neutrophil | *ZNF792* | 2.908376518 | 0.018342825 | 0.024156125 | 1 |
| Obesity | neutrophil | *KCTD2* | 2.482926933 | 0.004536923 | 0.011289644 | 1 |
| Obesity | neutrophil | *D2HGDH* | 1.430695331 | 0.006074584 | 0.011459063 | 1 |
| Obesity | neutrophil | *CMTR2* | 3.103089225 | 0.005743237 | 0.01134195 | 1 |
| Obesity | neutrophil | *ST20* | -0.75517014 | 0.005093696 | 0.011289644 | 1 |
| Obesity | neutrophil | *LRRC57* | 4.042241753 | 0.014614843 | 0.019931349 | 1 |
| Obesity | neutrophil | *MRPL14* | -2.428616367 | 0.026584028 | 0.033534065 | 1 |
| Obesity | neutrophil | *BBS12* | -2.342037341 | 0.012179486 | 0.017221128 | 1 |
| Obesity | neutrophil | *ZFP82* | 3.157772401 | 0.004050993 | 0.011289644 | 1 |
| Obesity | neutrophil | *AEN* | 4.325222188 | 0.005574944 | 0.011295339 | 1 |
| Obesity | neutrophil | *FKRP* | 2.895687085 | 0.0003208 | 0.011289644 | 1 |
| Obesity | neutrophil | *TRAPPC5* | -1.106646095 | 0.005842951 | 0.011378838 | 1 |
| Obesity | neutrophil | *RPH3AL* | 3.941233077 | 0.003636219 | 0.011289644 | 1 |
| Obesity | neutrophil | *ANKRD34A* | -1.639335374 | 0.003473344 | 0.011289644 | 1 |
| Obesity | neutrophil | *SLC26A11* | 1.475453566 | 0.00523196 | 0.011289644 | 1 |
| Obesity | neutrophil | *HIGD1A* | -0.84359142 | 0.005176981 | 0.011289644 | 1 |
| Obesity | neutrophil | *ADIPOQ* | 0.76385815 | 0.007801805 | 0.012671131 | 1 |
| Obesity | neutrophil | *F2R* | 2.905621316 | 0.022748929 | 0.029105111 | 1 |
| Obesity | neutrophil | *POLR2A* | 2.022097354 | 0.003015516 | 0.011289644 | 1 |
| Obesity | neutrophil | *TMEM132C* | 0.788615911 | 0.005612667 | 0.011295339 | 1 |
| Obesity | neutrophil | *SLC25A41* | 9.667637067 | 0.006775356 | 0.011855459 | 1 |
| Obesity | neutrophil | *TMEM136* | -1.755369615 | 0.003634539 | 0.011289644 | 1 |
| Obesity | neutrophil | *TMEM132E* | -1.102498536 | 0.004824804 | 0.011289644 | 1 |
| Obesity | neutrophil | *CCL13* | 2.538556028 | 0.011438537 | 0.016381339 | 1 |
| Obesity | neutrophil | *DDX60L* | 3.298334743 | 0.032477377 | 0.040170228 | 1 |
| Obesity | neutrophil | *OGFOD3* | 2.090242016 | 0.002677521 | 0.011289644 | 1 |

| | | | | | | |
|---|---|---|---|---|---|---|
| Obesity | neutrophil | *ACBD4* | 1.451060036 | 0.00415103 | 0.011289644 | 1 |
| Obesity | neutrophil | *SGSH* | 3.885079384 | 0.003619521 | 0.011289644 | 1 |
| Obesity | neutrophil | *SETD2* | 1.448678079 | 0.006184942 | 0.011517988 | 1 |
| Obesity | neutrophil | *GPR135* | 1.233373772 | 0.002203166 | 0.011289644 | 1 |
| Obesity | neutrophil | *ZFP41* | 4.611852146 | 0.000147162 | 0.011289644 | 0.925206098 |
| Obesity | neutrophil | *ATG9B* | 4.456917396 | 0.003932492 | 0.011289644 | 1 |
| Obesity | neutrophil | *HKR1* | 1.296914758 | 0.004369731 | 0.011289644 | 1 |
| Obesity | neutrophil | *ZBTB20* | 1.088198344 | 0.004676124 | 0.011289644 | 1 |
| Obesity | neutrophil | *C3orf58* | 2.049356731 | 0.004781155 | 0.011289644 | 1 |
| Obesity | neutrophil | *RELL1* | 1.397287162 | 0.00408814 | 0.011289644 | 1 |
| Obesity | neutrophil | *RFX7* | 1.4026493 | 0.004464457 | 0.011289644 | 1 |
| Obesity | neutrophil | *CLDN7* | -2.710402381 | 0.000803653 | 0.011289644 | 1 |
| Obesity | neutrophil | *ADO* | -1.348703707 | 0.006860025 | 0.011910792 | 1 |
| Obesity | neutrophil | *SNRPE* | 3.268644142 | 0.003195037 | 0.011289644 | 1 |
| Obesity | neutrophil | *CHST15* | -2.339478725 | 0.028223211 | 0.035402899 | 1 |
| Obesity | neutrophil | *IDH2* | 2.793861162 | 0.007027403 | 0.012031939 | 1 |
| Obesity | neutrophil | *TNRC18* | 1.679510271 | 0.00453549 | 0.011289644 | 1 |
| Obesity | neutrophil | *FAM181B* | -1.052317125 | 0.004547701 | 0.011289644 | 1 |
| Obesity | neutrophil | *NOP10* | -5.185969019 | 0.016446534 | 0.022037374 | 1 |
| Obesity | neutrophil | *TDRKH* | 1.861586351 | 0.010865482 | 0.015783832 | 1 |
| Obesity | neutrophil | *ZNF708* | 1.801485996 | 0.006398906 | 0.011648199 | 1 |
| Obesity | neutrophil | *IST1* | 1.94663243 | 0.005600983 | 0.011295339 | 1 |
| Obesity | neutrophil | *ERCC6L2* | 3.314184674 | 0.013769726 | 0.018984708 | 1 |
| Obesity | neutrophil | *MRPL41* | 1.677111034 | 0.003998961 | 0.011289644 | 1 |
| Obesity | neutrophil | *HHIPL1* | 2.322409901 | 0.00594038 | 0.011396756 | 1 |
| Obesity | neutrophil | *FAM153B* | 3.480754024 | 0.006973622 | 0.011985556 | 1 |
| Obesity | neutrophil | *SYNM* | 1.545285135 | 0.004315885 | 0.011289644 | 1 |
| Obesity | neutrophil | *C8orf33* | 2.781381102 | 0.010764637 | 0.015662409 | 1 |
| Obesity | neutrophil | *FBXL6* | -1.026091832 | 0.00418382 | 0.011289644 | 1 |
| Obesity | neutrophil | *KBTBD3* | 1.61885715 | 0.004322783 | 0.011289644 | 1 |
| Obesity | neutrophil | *CLN8* | 1.440372597 | 0.003188255 | 0.011289644 | 1 |
| Obesity | neutrophil | *CACNB4* | 1.268951605 | 0.00489672 | 0.011289644 | 1 |
| Obesity | neutrophil | *TRAPPC6B* | 2.122386551 | 0.006804566 | 0.011873524 | 1 |
| Obesity | neutrophil | *NPLOC4* | 1.110152363 | 0.008474384 | 0.013326276 | 1 |
| Obesity | neutrophil | *TSHZ2* | 0.749235187 | 0.005344999 | 0.011289644 | 1 |
| Obesity | neutrophil | *CAPN12* | 2.102729366 | 0.006695236 | 0.011800659 | 1 |
| Obesity | neutrophil | *FES* | -3.016423937 | 0.027190948 | 0.034217272 | 1 |
| Obesity | neutrophil | *GLRX5* | 2.071404096 | 0.004261158 | 0.011289644 | 1 |
| Obesity | neutrophil | *ADI1* | 2.962509316 | 0.008310489 | 0.013144161 | 1 |
| Obesity | neutrophil | *SPNS3* | -0.805463579 | 0.006103088 | 0.011478524 | 1 |
| Obesity | neutrophil | *CLEC4G* | 5.502775168 | 0.003877098 | 0.011289644 | 1 |
| Obesity | neutrophil | *SATB1* | 1.355097003 | 0.00456241 | 0.011289644 | 1 |
| Obesity | neutrophil | *TRAK1* | 1.602694448 | 0.005782787 | 0.011370951 | 1 |
| Obesity | neutrophil | *SKA2* | 1.241421756 | 0.004239996 | 0.011289644 | 1 |
| Obesity | neutrophil | *TTC3* | 1.35016709 | 0.009035211 | 0.013848236 | 1 |
| Obesity | neutrophil | *KCNB2* | 1.189574715 | 0.003644396 | 0.011289644 | 1 |
| Obesity | neutrophil | *TSKU* | 2.614960756 | 0.004199545 | 0.011289644 | 1 |
| Obesity | neutrophil | *GRID1* | 1.487255278 | 0.020778241 | 0.026917947 | 1 |
| Obesity | neutrophil | *ACBD3* | 1.952766026 | 0.005430859 | 0.011289644 | 1 |
| Obesity | neutrophil | *GPIHBP1* | 2.095080274 | 0.002050111 | 0.011289644 | 1 |

| | | | | | | |
|---|---|---|---|---|---|---|
| Obesity | neutrophil | *VMO1* | 3.221342749 | 0.001003321 | 0.011289644 | 1 |
| Obesity | neutrophil | *COL18A1* | 2.644658549 | 0.001681731 | 0.011289644 | 1 |
| Obesity | neutrophil | *SLC25A18* | 2.047196127 | 0.005697169 | 0.011302651 | 1 |
| Obesity | neutrophil | *ZNF721* | 1.55731763 | 0.004450662 | 0.011289644 | 1 |
| Obesity | neutrophil | *LENG9* | -2.267519161 | 0.023208684 | 0.029578957 | 1 |
| Obesity | neutrophil | *C11orf54* | 1.902170438 | 0.012649334 | 0.017738674 | 1 |
| Obesity | neutrophil | *CEP63* | 1.658527502 | 0.020840477 | 0.026976339 | 1 |
| Obesity | neutrophil | *HMGN4* | 2.700906421 | 0.008973412 | 0.013797211 | 1 |
| Obesity | neutrophil | *ZNF320* | 1.572234332 | 0.004665513 | 0.011289644 | 1 |
| Obesity | neutrophil | *C12orf60* | 4.368122726 | 0.024384669 | 0.030983511 | 1 |
| Obesity | neutrophil | *LYSMD4* | 2.62212366 | 0.000196369 | 0.011289644 | 1 |
| Obesity | neutrophil | *GAS6* | 1.170866494 | 0.003677764 | 0.011289644 | 1 |
| Obesity | neutrophil | *BEGAIN* | 5.279653109 | 0.004420338 | 0.011289644 | 1 |
| Obesity | neutrophil | *ARHGEF37* | 1.374907346 | 0.00421196 | 0.011289644 | 1 |
| Obesity | neutrophil | *FAM43B* | 4.919728427 | 0.037234151 | 0.045515567 | 1 |
| Obesity | neutrophil | *CSMD1* | 1.873920979 | 0.004067767 | 0.011289644 | 1 |
| Obesity | neutrophil | *PTGDR2* | 68.85300003 | 0.006923688 | 0.011946923 | 1 |
| Obesity | neutrophil | *FANCF* | 2.601752218 | 0.004155083 | 0.011289644 | 1 |
| Obesity | neutrophil | *SMDT1* | 1.358516947 | 0.01670389 | 0.022301414 | 1 |
| Obesity | neutrophil | *C2CD4C* | 6.565451758 | 0.000433925 | 0.011289644 | 1 |
| Obesity | neutrophil | *RUVBL2* | 1.865716635 | 0.003216466 | 0.011289644 | 1 |
| Obesity | neutrophil | *GDPGP1* | 4.189146449 | 0.001329615 | 0.011289644 | 1 |
| Obesity | neutrophil | *CTNNA3* | 2.057379558 | 0.003382101 | 0.011289644 | 1 |
| Obesity | neutrophil | *PTTG1IP* | 2.229684635 | 0.012914853 | 0.018039475 | 1 |
| Obesity | neutrophil | *CCBE1* | 1.368882105 | 0.000779301 | 0.011289644 | 1 |
| Obesity | neutrophil | *ZNF623* | 1.508982458 | 0.003603592 | 0.011289644 | 1 |
| Obesity | neutrophil | *CCDC125* | 3.055908757 | 0.008492161 | 0.013331367 | 1 |
| Obesity | neutrophil | *FHL3* | 2.677921727 | 0.005038136 | 0.011289644 | 1 |
| Obesity | neutrophil | *C19orf71* | -4.093622744 | 0.021450446 | 0.027663375 | 1 |
| Obesity | neutrophil | *LRIT3* | -1.077190469 | 0.003530297 | 0.011289644 | 1 |
| Obesity | neutrophil | *SF3A3* | 1.332553371 | 0.003978207 | 0.011289644 | 1 |
| Obesity | neutrophil | *GRIN2A* | 1.988805021 | 0.007204895 | 0.012170118 | 1 |
| Obesity | neutrophil | *URAD* | 8.763262328 | 0.008749042 | 0.013601688 | 1 |
| Obesity | neutrophil | *GPR132* | 1.727512146 | 0.005237761 | 0.011289644 | 1 |
| Obesity | neutrophil | *MX2* | 2.456342217 | 0.033423538 | 0.041251234 | 1 |
| Obesity | neutrophil | *PSMG1* | 2.763530812 | 8.92E-06 | 0.002076086 | 0.056054327 |
| Obesity | neutrophil | *ACSM5* | 1.483659341 | 0.003914564 | 0.011289644 | 1 |
| Obesity | neutrophil | *AC131971.1* | 8.949230438 | 0.007606651 | 0.012461948 | 1 |
| Obesity | neutrophil | *PCBP3* | 0.585949319 | 0.03250948 | 0.040202026 | 1 |
| Obesity | neutrophil | *TANGO2* | 2.203443319 | 0.002351254 | 0.011289644 | 1 |
| Obesity | neutrophil | *SFXN4* | 1.376909002 | 0.003820752 | 0.011289644 | 1 |
| Obesity | neutrophil | *FAM167B* | 10.47016476 | 0.00600036 | 0.011438456 | 1 |
| Obesity | neutrophil | *MRPL54* | 1.558694261 | 0.010925352 | 0.015837604 | 1 |
| Obesity | neutrophil | *NDUFB1* | 1.490352406 | 0.00386558 | 0.011289644 | 1 |
| Obesity | neutrophil | *FAM19A1* | 1.34644921 | 0.004257428 | 0.011289644 | 1 |
| Obesity | neutrophil | *TRMT12* | 1.960094548 | 0.006093938 | 0.011474271 | 1 |
| Obesity | neutrophil | *BMP8A* | -0.719702329 | 0.010249747 | 0.015101983 | 1 |
| Obesity | neutrophil | *CMTM4* | 1.57481977 | 0.003352074 | 0.011289644 | 1 |
| Obesity | neutrophil | *CBX6* | 5.002808403 | 0.005682927 | 0.011300433 | 1 |
| Obesity | neutrophil | *KREMEN1* | 1.313711776 | 0.002068979 | 0.011289644 | 1 |

| | | | | | | |
|---|---|---|---|---|---|---|
| Obesity | neutrophil | *KCTD16* | 1.711646554 | 0.001991328 | 0.011289644 | 1 |
| Obesity | neutrophil | *B3GALT5* | 6.549366003 | 0.027726973 | 0.034829067 | 1 |
| Obesity | neutrophil | *ZNF703* | 4.838798564 | 0.007260464 | 0.012218025 | 1 |
| Obesity | neutrophil | *KCTD8* | 2.232265785 | 0.008893236 | 0.013733966 | 1 |
| Obesity | neutrophil | *C9orf66* | -0.609276242 | 0.005106531 | 0.011289644 | 1 |
| Obesity | neutrophil | *TUBA8* | 1.964363561 | 0.004111334 | 0.011289644 | 1 |
| Obesity | neutrophil | *NPIPA5* | 2.178728477 | 0.001738559 | 0.011289644 | 1 |
| Obesity | neutrophil | *FAM162B* | 2.462980494 | 0.003822404 | 0.011289644 | 1 |
| Obesity | neutrophil | *BTBD9* | 1.654250551 | 0.004879442 | 0.011289644 | 1 |
| Obesity | neutrophil | *NUDT14* | 2.221016786 | 0.012155147 | 0.01719689 | 1 |
| Obesity | neutrophil | *ANKRD45* | 5.312818743 | 0.004367976 | 0.011289644 | 1 |
| Obesity | neutrophil | *MAATS1* | 4.726509296 | 0.000450922 | 0.011289644 | 1 |
| Obesity | neutrophil | *GPR39* | -0.621837643 | 0.005206279 | 0.011289644 | 1 |
| Obesity | neutrophil | *NPIPA7* | -2.435288802 | 0.012424244 | 0.017468922 | 1 |
| Obesity | neutrophil | *TTC32* | 1.546767596 | 0.004604017 | 0.011289644 | 1 |
| Obesity | neutrophil | *PP2D1* | 5.07983075 | 0.003981948 | 0.011289644 | 1 |
| Obesity | neutrophil | *DENND5A* | 2.202945279 | 0.006274896 | 0.011565541 | 1 |
| Obesity | neutrophil | *TMEM236* | 2.524389017 | 0.004989256 | 0.011289644 | 1 |
| Obesity | neutrophil | *VPS33B* | 1.795556043 | 0.004693935 | 0.011289644 | 1 |
| Obesity | neutrophil | *TBX1* | 1.849989091 | 0.004306521 | 0.011289644 | 1 |
| Obesity | neutrophil | *UQCR10* | 2.061610422 | 0.003381246 | 0.011289644 | 1 |
| Obesity | neutrophil | *EIF3C* | 16.38948296 | 4.20E-24 | 2.64E-20 | 2.64E-20 |
| Obesity | neutrophil | *NIPSNAP1* | 4.985160479 | 0.022329753 | 0.028656289 | 1 |
| Obesity | neutrophil | *CNTN2* | -4.1629467 | 0.01232914 | 0.017379664 | 1 |
| Obesity | neutrophil | *CRELD2* | 1.77459828 | 0.003415191 | 0.011289644 | 1 |
| Obesity | neutrophil | *UBE2F* | -4.712932848 | 0.012025759 | 0.017055731 | 1 |
| Obesity | neutrophil | *PCDH9* | 1.609931309 | 0.014263843 | 0.019537424 | 1 |
| Obesity | neutrophil | *OAF* | 1.457182292 | 0.035112887 | 0.043141435 | 1 |
| Obesity | neutrophil | *TM2D3* | 2.537112643 | 0.005383766 | 0.011289644 | 1 |
| Obesity | neutrophil | *CLECL1* | 2.141893721 | 0.005232377 | 0.011289644 | 1 |
| Obesity | neutrophil | *MROH7* | 6.730923867 | 0.004446507 | 0.011289644 | 1 |
| Obesity | neutrophil | *GDF3* | 10.71661796 | 0.00754876 | 0.012419095 | 1 |
| Obesity | neutrophil | *CSF1* | 3.201465253 | 0.004946846 | 0.011289644 | 1 |
| Obesity | neutrophil | *PLA2G6* | 1.689579088 | 0.00014105 | 0.011289644 | 0.886780122 |
| Obesity | neutrophil | *A3GALT2* | 4.355752561 | 0.001568008 | 0.011289644 | 1 |
| Obesity | neutrophil | *THAP7* | 2.668345167 | 0.004734742 | 0.011289644 | 1 |
| Obesity | neutrophil | *CCR10* | 3.390080117 | 0.004634955 | 0.011289644 | 1 |
| Obesity | neutrophil | *WDR27* | 1.359481238 | 0.00361759 | 0.011289644 | 1 |
| Obesity | neutrophil | *TXNRD2* | 4.366001814 | 0.011737842 | 0.016703443 | 1 |
| Obesity | neutrophil | *PROS1* | 2.634556811 | 0.01573219 | 0.021234066 | 1 |
| Obesity | neutrophil | *HDDC3* | 3.799962087 | 0.00234358 | 0.011289644 | 1 |
| Obesity | neutrophil | *LPAR5* | 2.566240704 | 0.019892587 | 0.025920144 | 1 |
| Obesity | neutrophil | *TMEM173* | 1.42281299 | 0.005896242 | 0.011378838 | 1 |
| Obesity | neutrophil | *SNN* | 3.170907169 | 0.0018185 | 0.011289644 | 1 |
| Obesity | neutrophil | *NELL2* | 1.483281198 | 0.003019324 | 0.011289644 | 1 |
| Obesity | neutrophil | *KRBA2* | -2.368874412 | 0.002328503 | 0.011289644 | 1 |
| Obesity | neutrophil | *ZNF93* | 4.389293882 | 0.005805378 | 0.011378838 | 1 |
| Obesity | neutrophil | *SEPT5* | 6.370975358 | 0.004068593 | 0.011289644 | 1 |
| Obesity | neutrophil | *ATL3* | 3.286216721 | 0.00590643 | 0.011378838 | 1 |
| Obesity | neutrophil | *NDUFA12* | 1.38883243 | 0.004208473 | 0.011289644 | 1 |

| | | | | | | |
|---|---|---|---|---|---|---|
| Obesity | neutrophil | *OSBP2* | 3.479895045 | 0.004114103 | 0.011289644 | 1 |
| Obesity | neutrophil | *TUSC5* | 1.161668209 | 0.007271334 | 0.01222323 | 1 |
| Obesity | neutrophil | *TMED9* | 4.813965439 | 0.035045983 | 0.043084493 | 1 |
| Obesity | neutrophil | *BTBD6* | 1.356076549 | 0.015977694 | 0.021491605 | 1 |
| Obesity | neutrophil | *H1FX* | 1.545045089 | 0.002613529 | 0.011289644 | 1 |
| Obesity | neutrophil | *NUTM2A* | -5.574370241 | 0.001483553 | 0.011289644 | 1 |
| Obesity | neutrophil | *ZFP90* | 1.622209954 | 0.003748858 | 0.011289644 | 1 |
| Obesity | neutrophil | *FAM227A* | 2.21859425 | 0.001688672 | 0.011289644 | 1 |
| Obesity | neutrophil | *NOC4L* | -1.421827295 | 0.00194257 | 0.011289644 | 1 |
| Obesity | neutrophil | *TMEM106A* | 1.758901785 | 0.003004268 | 0.011289644 | 1 |
| Obesity | neutrophil | *SIVA1* | 2.040080108 | 0.003063717 | 0.011289644 | 1 |
| Obesity | neutrophil | *UBOX5* | 1.343456538 | 0.00644147 | 0.011648199 | 1 |
| Obesity | neutrophil | *MAFF* | -4.899418951 | 0.001157585 | 0.011289644 | 1 |
| Obesity | neutrophil | *BRF1* | 1.987159325 | 0.004384301 | 0.011289644 | 1 |
| Obesity | neutrophil | *CIB1* | 4.128499534 | 0.032660164 | 0.040364547 | 1 |
| Obesity | neutrophil | *FLRT2* | 0.875902949 | 0.00427134 | 0.011289644 | 1 |
| Obesity | neutrophil | *ADSSL1* | -2.560912953 | 0.009421721 | 0.014240375 | 1 |
| Obesity | neutrophil | *C6orf120* | 1.816511142 | 0.007225731 | 0.012192209 | 1 |
| Obesity | neutrophil | *DDX51* | 3.270484328 | 0.003256099 | 0.011289644 | 1 |
| Obesity | neutrophil | *IFITM2* | -2.292562402 | 0.019791879 | 0.025810319 | 1 |
| Obesity | neutrophil | *GP1BA* | 5.758648163 | 8.84E-06 | 0.002076086 | 0.055564188 |
| Obesity | neutrophil | *KIAA0825* | 1.438421344 | 0.005497338 | 0.011289644 | 1 |
| Obesity | neutrophil | *KLHL33* | 4.950011308 | 0.000233856 | 0.011289644 | 1 |
| Obesity | neutrophil | *RBM11* | -1.68423087 | 0.004203317 | 0.011289644 | 1 |
| Obesity | neutrophil | *CCDC137* | 3.613455234 | 0.006655487 | 0.011757198 | 1 |
| Obesity | neutrophil | *CDK10* | 3.195675002 | 0.03922431 | 0.04768966 | 1 |
| Obesity | neutrophil | *TMEM105* | 31.68141585 | 0.033511849 | 0.041352109 | 1 |
| Obesity | neutrophil | *TCN2* | 1.562360119 | 0.037780764 | 0.046121876 | 1 |
| Obesity | neutrophil | *GAS2L1* | 1.680232889 | 0.004540273 | 0.011289644 | 1 |
| Obesity | neutrophil | *TNFAIP8L1* | 5.050985995 | 0.007231386 | 0.012198477 | 1 |
| Obesity | neutrophil | *MAPK11* | 2.368196993 | 0.00434995 | 0.011289644 | 1 |
| Obesity | neutrophil | *SP140L* | 1.355661526 | 0.004494383 | 0.011289644 | 1 |
| Obesity | neutrophil | *FAM174B* | 2.506842811 | 0.004824266 | 0.011289644 | 1 |
| Obesity | neutrophil | *MUC1* | 3.306510493 | 0.01044363 | 0.015326588 | 1 |
| Obesity | neutrophil | *SV2B* | 1.952448891 | 0.004918016 | 0.011289644 | 1 |
| Obesity | neutrophil | *PDE6G* | 8.824269107 | 0.001056638 | 0.011289644 | 1 |
| Obesity | neutrophil | *TLCD2* | 2.034569681 | 0.012270054 | 0.017319675 | 1 |
| Obesity | neutrophil | *LSAMP* | 2.365930894 | 0.006430822 | 0.011648199 | 1 |
| Obesity | neutrophil | *AHNAK2* | 5.387878806 | 0.017331634 | 0.022988182 | 1 |
| Obesity | neutrophil | *SP1* | 3.946566857 | 0.001840272 | 0.011289644 | 1 |
| Obesity | neutrophil | *PCGF3* | 1.510924284 | 0.002509803 | 0.011289644 | 1 |
| Obesity | neutrophil | *PBX1* | 0.915684389 | 0.004008065 | 0.011289644 | 1 |
| Obesity | neutrophil | *UBE2L3* | 2.563120509 | 0.006004655 | 0.011438456 | 1 |
| Obesity | neutrophil | *PMEL* | 4.978937935 | 0.000377815 | 0.011289644 | 1 |
| Obesity | neutrophil | *SNAI3* | -1.490023575 | 0.006444645 | 0.011648199 | 1 |
| Obesity | neutrophil | *C6orf201* | -0.648620465 | 0.009323809 | 0.014142048 | 1 |
| Obesity | neutrophil | *MYBL1* | -3.507951342 | 0.013177034 | 0.018348619 | 1 |
| Obesity | neutrophil | *C16orf52* | 2.437913141 | 0.006166903 | 0.011512519 | 1 |
| Obesity | neutrophil | *ADARB2* | -3.198327578 | 0.007109643 | 0.012093703 | 1 |
| Obesity | neutrophil | *SRL* | 4.602884875 | 0.000481771 | 0.011289644 | 1 |

| | | | | | | |
|---|---|---|---|---|---|---|
| Obesity | neutrophil | *KCNIP4* | 0.831209599 | 0.007219613 | 0.012185156 | 1 |
| Obesity | neutrophil | *SLC52A2* | 1.889172178 | 0.004235305 | 0.011289644 | 1 |
| Obesity | neutrophil | *ARL17A* | 1.38596829 | 0.005869731 | 0.011378838 | 1 |
| Obesity | neutrophil | *NPIPB4* | 1.626212188 | 0.003661605 | 0.011289644 | 1 |
| Obesity | neutrophil | *ZNF829* | 4.238014134 | 0.004029156 | 0.011289644 | 1 |
| Obesity | neutrophil | *TRIM69* | 1.558973654 | 0.013549405 | 0.018738476 | 1 |
| Obesity | neutrophil | *IFITM1* | -2.458490049 | 0.009205641 | 0.014011804 | 1 |
| Obesity | neutrophil | *POMK* | 1.677028052 | 0.003711509 | 0.011289644 | 1 |
| Obesity | neutrophil | *LINC00839* | -3.349754028 | 0.015928584 | 0.021430132 | 1 |
| Obesity | neutrophil | *PTCH1* | 2.179154352 | 0.006517085 | 0.011671674 | 1 |
| Obesity | neutrophil | *RTN4RL1* | 1.402842519 | 0.004260851 | 0.011289644 | 1 |
| Obesity | neutrophil | *PLK5* | 6.332995254 | 0.005423171 | 0.011289644 | 1 |
| Obesity | neutrophil | *RASA3* | 2.409541657 | 0.004708464 | 0.011289644 | 1 |
| Obesity | neutrophil | *ZNF566* | 1.947832499 | 0.003895675 | 0.011289644 | 1 |
| Obesity | neutrophil | *ZNF529* | 1.912001488 | 0.006331847 | 0.011609044 | 1 |
| Obesity | neutrophil | *ZNF284* | 3.499934079 | 0.004265488 | 0.011289644 | 1 |
| Obesity | neutrophil | *GSAP* | 2.201910974 | 0.011008997 | 0.015929474 | 1 |
| Obesity | neutrophil | *C2orf76* | 1.473180968 | 0.002523378 | 0.011289644 | 1 |
| Obesity | neutrophil | *WWOX* | 1.453101817 | 0.004019863 | 0.011289644 | 1 |
| Obesity | neutrophil | *CYP4Z1* | 3.037965923 | 0.004706307 | 0.011289644 | 1 |
| Obesity | neutrophil | *CCDC84* | 1.443728877 | 0.0042332 | 0.011289644 | 1 |
| Obesity | neutrophil | *POLR1D* | 2.466697044 | 0.005224933 | 0.011289644 | 1 |
| Obesity | neutrophil | *FFAR4* | -1.454505469 | 0.002344472 | 0.011289644 | 1 |
| Obesity | neutrophil | *BPIFB4* | 144.5200186 | 0.000907592 | 0.011289644 | 1 |
| Obesity | neutrophil | *CYP4F12* | 1.995298039 | 0.002929709 | 0.011289644 | 1 |
| Obesity | neutrophil | *ZNF749* | -0.699741904 | 0.003948829 | 0.011289644 | 1 |
| Obesity | neutrophil | *KLHL32* | 2.023777371 | 0.005784047 | 0.011370951 | 1 |
| Obesity | neutrophil | *NBPF12* | 2.100071623 | 0.00401104 | 0.011289644 | 1 |
| Obesity | neutrophil | *TOR3A* | -3.155184325 | 0.021047167 | 0.027204674 | 1 |
| Obesity | neutrophil | *CYP4X1* | 4.117027635 | 0.022973072 | 0.029332191 | 1 |
| Obesity | neutrophil | *CCDC30* | 1.654741185 | 0.004342173 | 0.011289644 | 1 |
| Obesity | neutrophil | *FCAR* | 3.50352836 | 0.004705243 | 0.011289644 | 1 |
| Obesity | neutrophil | *KPNA4* | 2.027492514 | 0.004960988 | 0.011289644 | 1 |
| Obesity | neutrophil | *ZNF501* | 1.999543144 | 0.004532758 | 0.011289644 | 1 |
| Obesity | neutrophil | *FAM228A* | -1.016539736 | 0.017519686 | 0.023208232 | 1 |
| Obesity | neutrophil | *RPS23* | 1.598836115 | 0.003793401 | 0.011289644 | 1 |
| Obesity | neutrophil | *BTN3A2* | -1.641700034 | 0.010224513 | 0.015071867 | 1 |
| Obesity | neutrophil | *PCLO* | 0.951710334 | 0.003611261 | 0.011289644 | 1 |
| Obesity | neutrophil | *RGS7BP* | 3.095387202 | 0.022304721 | 0.028635855 | 1 |
| Obesity | neutrophil | *ZNF396* | 2.237345143 | 0.029259897 | 0.036602011 | 1 |
| Obesity | neutrophil | *CLCNKA* | 7.217070235 | 0.017208389 | 0.022853642 | 1 |
| Obesity | neutrophil | *SEPT10* | 1.953876612 | 0.008002019 | 0.012827306 | 1 |
| Obesity | neutrophil | *SMYD4* | 2.29571046 | 0.00272731 | 0.011289644 | 1 |
| Obesity | neutrophil | *CEACAM19* | 1.538486178 | 0.016543759 | 0.022129918 | 1 |
| Obesity | neutrophil | *MIR22HG* | 2.218872769 | 0.003265877 | 0.011289644 | 1 |
| Obesity | neutrophil | *PDE2A* | 1.45036249 | 0.006297165 | 0.011581641 | 1 |
| Obesity | neutrophil | *C17orf58* | 1.814675595 | 0.009034896 | 0.013848236 | 1 |
| Obesity | neutrophil | *CYP27C1* | 1.85238329 | 0.003763885 | 0.011289644 | 1 |
| Obesity | neutrophil | *BCR* | 1.598127583 | 0.002583893 | 0.011289644 | 1 |
| Obesity | neutrophil | *HYAL3* | -6.130446387 | 0.000489818 | 0.011289644 | 1 |

| | | | | | | |
|---|---|---|---|---|---|---|
| Obesity | neutrophil | *ZSCAN30* | 3.501433302 | 0.009673277 | 0.014511069 | 1 |
| Obesity | neutrophil | *TRABD2A* | 4.159866188 | 6.90E-05 | 0.008036154 | 0.433952306 |
| Obesity | neutrophil | *TMEM17* | -0.801104011 | 0.005367409 | 0.011289644 | 1 |
| Obesity | neutrophil | *TNFRSF18* | 5.351334205 | 0.004060105 | 0.011289644 | 1 |
| Obesity | neutrophil | *RTN4RL2* | 5.131961195 | 0.007039448 | 0.012042724 | 1 |
| Obesity | neutrophil | *TMEM232* | 1.616924717 | 0.00576583 | 0.011357022 | 1 |
| Obesity | neutrophil | *EFCAB6* | 1.219847466 | 0.004273629 | 0.011289644 | 1 |
| Obesity | neutrophil | *KANK3* | 1.711819956 | 0.004390714 | 0.011289644 | 1 |
| Obesity | neutrophil | *EMID1* | 2.66154403 | 0.002921045 | 0.011289644 | 1 |
| Obesity | neutrophil | *CYP4A11* | -1.115255715 | 0.008433177 | 0.013278082 | 1 |
| Obesity | neutrophil | *RPS19BP1* | 2.513973304 | 0.011760636 | 0.016724523 | 1 |
| Obesity | neutrophil | *C3orf70* | 1.9448221 | 0.005630449 | 0.011295339 | 1 |
| Obesity | neutrophil | *PLCD1* | 1.608321508 | 0.003693057 | 0.011289644 | 1 |
| Obesity | neutrophil | *ENTPD5* | 3.647069267 | 0.007925436 | 0.012779486 | 1 |
| Obesity | neutrophil | *HEATR4* | 2.386721527 | 0.000684488 | 0.011289644 | 1 |
| Obesity | neutrophil | *MRPS21* | 2.297449254 | 0.001961084 | 0.011289644 | 1 |
| Obesity | neutrophil | *ANGPTL5* | 3.367170251 | 0.00500466 | 0.011289644 | 1 |
| Obesity | neutrophil | *TSPYL4* | 4.188592624 | 0.018080953 | 0.023861241 | 1 |
| Obesity | neutrophil | *MT1X* | 1.564788124 | 0.004936095 | 0.011289644 | 1 |
| Obesity | neutrophil | *FNBP1* | 2.368130556 | 0.002477189 | 0.011289644 | 1 |
| Obesity | neutrophil | *WDR86* | 1.491525693 | 0.006961279 | 0.011974691 | 1 |
| Obesity | neutrophil | *EPOR* | 2.743342535 | 0.001782517 | 0.011289644 | 1 |
| Obesity | neutrophil | *FPR3* | 1.10000084 | 0.005548138 | 0.011295339 | 1 |
| Obesity | neutrophil | *GJA4* | 2.667232217 | 0.003277691 | 0.011289644 | 1 |
| Obesity | neutrophil | *IFT140* | 1.245702733 | 0.004318899 | 0.011289644 | 1 |
| Obesity | neutrophil | *TLR5* | 2.778002943 | 0.005109637 | 0.011289644 | 1 |
| Obesity | neutrophil | *PLEKHN1* | 5.486482495 | 0.001322102 | 0.011289644 | 1 |
| Obesity | neutrophil | *ZNF385C* | 3.3529012 | 0.002895926 | 0.011289644 | 1 |
| Obesity | neutrophil | *TET3* | 2.634453565 | 0.005768832 | 0.011357022 | 1 |
| Obesity | neutrophil | *C17orf97* | 1.643290108 | 0.002582436 | 0.011289644 | 1 |
| Obesity | neutrophil | *DHRS4L2* | 2.064817394 | 0.036822327 | 0.04508114 | 1 |
| Obesity | neutrophil | *C2orf88* | 1.064662642 | 0.004347832 | 0.011289644 | 1 |
| Obesity | neutrophil | *KBTBD12* | 3.883013802 | 0.00057281 | 0.011289644 | 1 |
| Obesity | neutrophil | *FANCA* | 1.350029663 | 0.019312657 | 0.025269235 | 1 |
| Obesity | neutrophil | *SEMA4D* | -2.201460622 | 0.013228333 | 0.018395604 | 1 |
| Obesity | neutrophil | *FAM69C* | -1.461334711 | 0.004245269 | 0.011289644 | 1 |
| Obesity | neutrophil | *DNAH17* | 1.954613052 | 0.005181263 | 0.011289644 | 1 |
| Obesity | neutrophil | *ZFP69* | 1.772126873 | 0.00617612 | 0.011512519 | 1 |
| Obesity | neutrophil | *TMEM220* | 1.876716154 | 0.005109425 | 0.011289644 | 1 |
| Obesity | neutrophil | *CCDC157* | -1.456448961 | 0.006530129 | 0.011679921 | 1 |
| Obesity | neutrophil | *GFRAL* | 3.56171491 | 0.003084878 | 0.011289644 | 1 |
| Obesity | neutrophil | *LDLRAD2* | 2.092665297 | 0.008413541 | 0.013262621 | 1 |
| Obesity | neutrophil | *C2orf66* | -5.445402455 | 0.006883805 | 0.011932455 | 1 |
| Obesity | neutrophil | *KLHL17* | 2.755594546 | 0.00306402 | 0.011289644 | 1 |
| Obesity | neutrophil | *ZSCAN23* | 1.869770792 | 6.32E-05 | 0.00802313 | 0.397431467 |
| Obesity | neutrophil | *OR7D2* | 9.833946773 | 0.007605592 | 0.012461948 | 1 |
| Obesity | neutrophil | *MORN2* | -1.851903897 | 0.032612041 | 0.040320925 | 1 |
| Obesity | neutrophil | *S100A3* | -2.187181719 | 0.011077174 | 0.015980311 | 1 |
| Obesity | neutrophil | *TMEM221* | -0.679920506 | 0.019452328 | 0.025414959 | 1 |
| Obesity | neutrophil | *TREML4* | -1.696954525 | 0.005354152 | 0.011289644 | 1 |

| | | | | | | |
|---|---|---|---|---|---|---|
| Obesity | neutrophil | *C6orf132* | 1.883959978 | 0.006486112 | 0.011671674 | 1 |
| Obesity | neutrophil | *MAPK12* | 3.9765869 | 0.003591723 | 0.011289644 | 1 |
| Obesity | neutrophil | *AGRN* | -2.066956214 | 0.00632402 | 0.011605985 | 1 |
| Obesity | neutrophil | *FAM166A* | 8.854501989 | 0.004385913 | 0.011289644 | 1 |
| Obesity | neutrophil | *ZNF626* | 1.622096563 | 0.003425397 | 0.011289644 | 1 |
| Obesity | neutrophil | *SMTNL2* | -1.025882667 | 0.005206511 | 0.011289644 | 1 |
| Obesity | neutrophil | *ZC3H6* | 2.038455495 | 0.003946564 | 0.011289644 | 1 |
| Obesity | neutrophil | *LINC00265* | 4.399389655 | 0.013938034 | 0.019162129 | 1 |
| Obesity | neutrophil | *LAMTOR4* | 1.87712671 | 0.00710233 | 0.012084533 | 1 |
| Obesity | neutrophil | *PRKAR1B* | 1.455754243 | 0.004314622 | 0.011289644 | 1 |
| Obesity | neutrophil | *NUTM2B* | -40.74974074 | 0.006055572 | 0.01145004 | 1 |
| Obesity | neutrophil | *NCR3LG1* | -5.117487887 | 0.010774995 | 0.015673853 | 1 |
| Obesity | neutrophil | *HYKK* | 1.872688447 | 0.001493639 | 0.011289644 | 1 |
| Obesity | neutrophil | *RUFY4* | -2.127874596 | 0.005970429 | 0.011423216 | 1 |
| Obesity | neutrophil | *ZNF669* | 3.046676084 | 0.003097077 | 0.011289644 | 1 |
| Obesity | neutrophil | *CENPP* | 2.850292675 | 1.00E-05 | 0.0022525 | 0.063069993 |
| Obesity | neutrophil | *PLSCR1* | 2.165785492 | 0.004107939 | 0.011289644 | 1 |
| Obesity | neutrophil | *ZP3* | 4.607969316 | 0.00112682 | 0.011289644 | 1 |
| Obesity | neutrophil | *PDCD1* | 13.26701084 | 0.017726631 | 0.023447786 | 1 |
| Obesity | neutrophil | *CERKL* | -1.979500398 | 0.009189754 | 0.013999511 | 1 |
| Obesity | neutrophil | *C19orf54* | -2.474886767 | 0.003072013 | 0.011289644 | 1 |
| Obesity | neutrophil | *C22orf34* | 2.536903576 | 0.002357895 | 0.011289644 | 1 |
| Obesity | neutrophil | *COL25A1* | -1.074889136 | 0.009530529 | 0.014351721 | 1 |
| Obesity | neutrophil | *SRSF10* | 1.744713364 | 0.003906599 | 0.011289644 | 1 |
| Obesity | neutrophil | *NBR1* | 2.033379939 | 0.004405098 | 0.011289644 | 1 |
| Obesity | neutrophil | *FAM72B* | -1.426990141 | 0.005526805 | 0.011295339 | 1 |
| Obesity | neutrophil | *SUMO2* | 4.196984726 | 0.01859275 | 0.024439184 | 1 |
| Obesity | neutrophil | *GOLGA8M* | 1.676397486 | 0.006360483 | 0.011618636 | 1 |
| Obesity | neutrophil | *DPYD* | 1.375099542 | 0.004702006 | 0.011289644 | 1 |
| Obesity | neutrophil | *S100A16* | -2.424160618 | 0.023180206 | 0.029560639 | 1 |
| Obesity | neutrophil | *RHCE* | -1.767889656 | 0.011050382 | 0.015961637 | 1 |
| Obesity | neutrophil | *SLC4A5* | -1.613301217 | 0.006207197 | 0.011535516 | 1 |
| Obesity | neutrophil | *UROS* | 1.741679573 | 0.004567822 | 0.011289644 | 1 |
| Obesity | neutrophil | *FAM221A* | 1.614992251 | 0.003373175 | 0.011289644 | 1 |
| Obesity | neutrophil | *NOXA1* | 3.07081641 | 0.004686608 | 0.011289644 | 1 |
| Obesity | neutrophil | *SKOR1* | 8.903441232 | 0.005215287 | 0.011289644 | 1 |
| Obesity | neutrophil | *TMEM201* | 2.729400574 | 0.003516751 | 0.011289644 | 1 |
| Obesity | neutrophil | *LINC00910* | -3.32561825 | 0.030906541 | 0.038461881 | 1 |
| Obesity | neutrophil | *BEND4* | 15.03229322 | 0.001343878 | 0.011289644 | 1 |
| Obesity | neutrophil | *FBF1* | 1.884745383 | 8.22E-05 | 0.009066777 | 0.516806286 |
| Obesity | neutrophil | *MSL1* | 2.079468973 | 0.011351395 | 0.016301101 | 1 |
| Obesity | neutrophil | *FAM196A* | 3.827062143 | 0.006161452 | 0.011512519 | 1 |
| Obesity | neutrophil | *UTS2B* | 1.721833176 | 0.024477806 | 0.031095569 | 1 |
| Obesity | neutrophil | *C9orf152* | 4.93896975 | 0.00358524 | 0.011289644 | 1 |
| Obesity | neutrophil | *NELFB* | 1.571920408 | 0.004208139 | 0.011289644 | 1 |
| Obesity | neutrophil | *SLC15A5* | -1.308256296 | 0.030117138 | 0.037553837 | 1 |
| Obesity | neutrophil | *LIPI* | 2.005416458 | 0.002162743 | 0.011289644 | 1 |
| Obesity | neutrophil | *ADAT2* | 1.392492922 | 0.002475866 | 0.011289644 | 1 |
| Obesity | neutrophil | *ANKDD1B* | 1.205535758 | 0.013254138 | 0.018416905 | 1 |
| Obesity | neutrophil | *ALKBH2* | 2.241333467 | 0.002664796 | 0.011289644 | 1 |

| | | | | | | |
|---|---|---|---|---|---|---|
| Obesity | neutrophil | *LITAF* | 3.441422761 | 0.006989636 | 0.011998609 | 1 |
| Obesity | neutrophil | *VSTM1* | 6.147573963 | 0.003534765 | 0.011289644 | 1 |
| Obesity | neutrophil | *SF3B3* | 1.648184256 | 0.004236358 | 0.011289644 | 1 |
| Obesity | neutrophil | *SP6* | -1.944552915 | 0.004678427 | 0.011289644 | 1 |
| Obesity | neutrophil | *PLAC9* | 2.208727154 | 0.001825544 | 0.011289644 | 1 |
| Obesity | neutrophil | *CLDN4* | 2.667275723 | 0.006302019 | 0.011581641 | 1 |
| Obesity | neutrophil | *ZNF573* | 3.25178226 | 0.006760599 | 0.011846705 | 1 |
| Obesity | neutrophil | *FAM47E* | 5.798331256 | 7.54E-06 | 0.001895264 | 0.047381607 |
| Obesity | neutrophil | *ZNF527* | 3.245061724 | 0.005817623 | 0.011378838 | 1 |
| Obesity | neutrophil | *S100A13* | 1.766929531 | 0.0031979 | 0.011289644 | 1 |
| Obesity | neutrophil | *ZNF33A* | 2.906939891 | 0.010865663 | 0.015783832 | 1 |
| Obesity | neutrophil | *LINC00994* | 8.278029485 | 0.005485671 | 0.011289644 | 1 |
| Obesity | neutrophil | *TSPYL1* | 1.924107291 | 0.002660633 | 0.011289644 | 1 |
| Obesity | neutrophil | *PNRC2* | 2.141191585 | 0.005417955 | 0.011289644 | 1 |
| Obesity | neutrophil | *ZKSCAN3* | 1.833812854 | 0.010983085 | 0.015902376 | 1 |
| Obesity | neutrophil | *RRP7A* | 1.540998198 | 0.006911666 | 0.011943085 | 1 |
| Obesity | neutrophil | *SLC35E2B* | 4.406469536 | 0.000871124 | 0.011289644 | 1 |
| Obesity | neutrophil | *HLA-DRB1* | 1.26923895 | 0.009692334 | 0.014532723 | 1 |
| Obesity | neutrophil | *AKR1C3* | 1.830369732 | 0.004473351 | 0.011289644 | 1 |
| Obesity | neutrophil | *WDSUB1* | 2.517090739 | 0.010902588 | 0.015819194 | 1 |
| Obesity | neutrophil | *ZNF79* | -1.149218767 | 0.008906817 | 0.013738263 | 1 |
| Obesity | neutrophil | *S100A4* | 1.555237018 | 0.004515706 | 0.011289644 | 1 |
| Obesity | neutrophil | *COLCA1* | -0.990610327 | 0.006314134 | 0.011593738 | 1 |
| Obesity | neutrophil | *ACADSB* | 1.746497087 | 0.005420422 | 0.011289644 | 1 |
| Obesity | neutrophil | *HRCT1* | 2.284433662 | 0.00202411 | 0.011289644 | 1 |
| Obesity | neutrophil | *MPHOSPH8* | 1.591563878 | 0.004635962 | 0.011289644 | 1 |
| Obesity | neutrophil | *SIRPB2* | -1.497716219 | 0.007324097 | 0.012262742 | 1 |
| Obesity | neutrophil | *SRGAP3* | 1.740442128 | 0.002284173 | 0.011289644 | 1 |
| Obesity | neutrophil | *SUPT5H* | 1.870811146 | 0.004075411 | 0.011289644 | 1 |
| Obesity | neutrophil | *XPNPEP3* | 1.116198314 | 0.008042807 | 0.01286972 | 1 |
| Obesity | neutrophil | *PPIA* | 4.02334679 | 0.004609734 | 0.011289644 | 1 |
| Obesity | neutrophil | *ZNF471* | 1.482025338 | 0.007018043 | 0.012025739 | 1 |
| Obesity | neutrophil | *GTF2IRD2* | 1.279903147 | 0.006005781 | 0.011438456 | 1 |
| Obesity | neutrophil | *GRM7* | 1.021722646 | 0.008110981 | 0.012939288 | 1 |
| Obesity | neutrophil | *SUPT3H* | 1.058322266 | 0.004374911 | 0.011289644 | 1 |
| Obesity | neutrophil | *POM121* | 1.088598402 | 0.004978668 | 0.011289644 | 1 |
| Obesity | neutrophil | *GIMAP5* | -1.556605925 | 0.005025444 | 0.011289644 | 1 |
| Obesity | neutrophil | *STK31* | -4.12081139 | 0.018944666 | 0.024839441 | 1 |
| Obesity | neutrophil | *ZKSCAN7* | 1.935512914 | 0.004168337 | 0.011289644 | 1 |
| Obesity | neutrophil | *ZNF565* | 2.895798622 | 0.002876578 | 0.011289644 | 1 |
| Obesity | neutrophil | *NTNG2* | -1.279355272 | 0.006596704 | 0.01172215 | 1 |
| Obesity | neutrophil | *LONP1* | 5.408827804 | 0.009356339 | 0.014181124 | 1 |
| Obesity | neutrophil | *ASB13* | 1.561326359 | 0.004258705 | 0.011289644 | 1 |
| Obesity | neutrophil | *SLC35F1* | -3.097108879 | 0.018676716 | 0.02452904 | 1 |
| Obesity | neutrophil | *ZNF34* | 2.282988972 | 0.004551178 | 0.011289644 | 1 |
| Obesity | neutrophil | *INCA1* | 3.992470661 | 0.000947756 | 0.011289644 | 1 |
| Obesity | neutrophil | *ZNF774* | 3.049455605 | 0.005298487 | 0.011289644 | 1 |
| Obesity | neutrophil | *EVL* | 2.633551536 | 0.008219134 | 0.013045619 | 1 |
| Obesity | neutrophil | *ZNF124* | 2.566511212 | 0.005340939 | 0.011289644 | 1 |
| Obesity | neutrophil | *ZNF775* | 4.956680891 | 0.000760828 | 0.011289644 | 1 |

| | | | | | | |
|---|---|---|---|---|---|---|
| Obesity | neutrophil | *ZNF605* | 2.28349461 | 0.004327747 | 0.011289644 | 1 |
| Obesity | neutrophil | *RFX8* | 1.757079745 | 0.003595754 | 0.011289644 | 1 |
| Obesity | neutrophil | *MYL6B* | 4.372875479 | 0.006224695 | 0.011544147 | 1 |
| Obesity | neutrophil | *C20orf96* | 2.851876748 | 0.004697708 | 0.011289644 | 1 |
| Obesity | neutrophil | *TPK1* | 3.690336732 | 0.002772336 | 0.011289644 | 1 |
| Obesity | neutrophil | *C1orf186* | -1.492376375 | 0.006657487 | 0.011757198 | 1 |
| Obesity | neutrophil | *MYO18A* | 3.342400005 | 0.035058528 | 0.043091489 | 1 |
| Obesity | neutrophil | *SPTSSB* | -0.912689826 | 0.006138154 | 0.011498979 | 1 |
| Obesity | neutrophil | *XRCC2* | 3.707893991 | 0.005347298 | 0.011289644 | 1 |
| Obesity | neutrophil | *MYO6* | 1.344318625 | 0.008962625 | 0.01379051 | 1 |
| Obesity | neutrophil | *HDAC2* | 1.97345393 | 0.00417748 | 0.011289644 | 1 |
| Obesity | neutrophil | *ADH1B* | 0.786351465 | 0.003962821 | 0.011289644 | 1 |
| Obesity | neutrophil | *HRH1* | 1.453487837 | 0.004270142 | 0.011289644 | 1 |
| Obesity | neutrophil | *TRAPPC4* | 1.607252363 | 0.004097357 | 0.011289644 | 1 |
| Obesity | neutrophil | *TECPR2* | -2.32921742 | 0.037429511 | 0.045728593 | 1 |
| Obesity | neutrophil | *ZFP62* | 5.09480539 | 0.006699247 | 0.011804419 | 1 |
| Obesity | neutrophil | *ERI2* | 1.999477708 | 0.003721726 | 0.011289644 | 1 |
| Obesity | neutrophil | *TOMM7* | 3.828740264 | 0.010093021 | 0.014939001 | 1 |
| Obesity | neutrophil | *TRPV1* | 1.67594603 | 0.00203063 | 0.011289644 | 1 |
| Obesity | neutrophil | *ZNF33B* | 2.723434253 | 0.040916271 | 0.04954557 | 1 |
| Obesity | neutrophil | *AMZ2* | -1.79481308 | 0.015295017 | 0.02071516 | 1 |
| Obesity | neutrophil | *ZNF431* | 2.444097128 | 0.004020031 | 0.011289644 | 1 |
| Obesity | neutrophil | *NF1* | 1.792233815 | 0.003918919 | 0.011289644 | 1 |
| Obesity | neutrophil | *DAPK1* | 1.706500773 | 0.006471737 | 0.01166174 | 1 |
| Obesity | neutrophil | *HLA-DQA1* | 1.369988672 | 0.003040397 | 0.011289644 | 1 |
| Obesity | neutrophil | *GM2A* | 2.5408362 | 0.001376061 | 0.011289644 | 1 |
| Obesity | neutrophil | *TLE1* | 1.365617688 | 0.006005527 | 0.011438456 | 1 |
| Obesity | neutrophil | *STRN3* | 1.694548996 | 0.005335072 | 0.011289644 | 1 |
| Obesity | neutrophil | *MVB12B* | 1.476502508 | 0.004623282 | 0.011289644 | 1 |
| Obesity | neutrophil | *C6orf106* | 3.363646061 | 6.54E-06 | 0.001734043 | 0.041137163 |
| Obesity | neutrophil | *PPTC7* | 2.332116676 | 0.008958871 | 0.013788109 | 1 |
| Obesity | neutrophil | *NHLRC2* | 2.074609294 | 0.005646271 | 0.011297933 | 1 |
| Obesity | neutrophil | *ZFP28* | 1.840757247 | 0.003328896 | 0.011289644 | 1 |
| Obesity | neutrophil | *KIAA1211L* | 2.946746494 | 0.000646715 | 0.011289644 | 1 |
| Obesity | neutrophil | *LAMB3* | 2.186081284 | 0.005016564 | 0.011289644 | 1 |
| Obesity | neutrophil | *KPNA5* | 1.508676723 | 0.004329478 | 0.011289644 | 1 |
| Obesity | neutrophil | *ANKRD36B* | 1.566763805 | 0.007294619 | 0.012233922 | 1 |
| Obesity | neutrophil | *HCAR1* | -0.983124738 | 0.03128802 | 0.038875056 | 1 |
| Obesity | neutrophil | *SRGAP1* | 2.494756877 | 0.005204946 | 0.011289644 | 1 |
| Obesity | neutrophil | *FAM3C* | 1.979253304 | 0.002221643 | 0.011289644 | 1 |
| Obesity | neutrophil | *SLC39A10* | 1.553108784 | 0.004696581 | 0.011289644 | 1 |
| Obesity | neutrophil | *AP2A1* | 2.094991948 | 0.005076331 | 0.011289644 | 1 |
| Obesity | neutrophil | *PCDHB16* | 2.285710124 | 0.006576262 | 0.011712453 | 1 |
| Obesity | neutrophil | *HIST1H3E* | 4.038411316 | 0.006349376 | 0.011618636 | 1 |
| Obesity | neutrophil | *ZNF585A* | 2.342789789 | 0.000859616 | 0.011289644 | 1 |
| Obesity | neutrophil | *FUT11* | 2.009824264 | 0.001810061 | 0.011289644 | 1 |
| Obesity | neutrophil | *ANXA4* | 1.728849044 | 0.004769584 | 0.011289644 | 1 |
| Obesity | neutrophil | *ZNF138* | 1.777601626 | 0.00267018 | 0.011289644 | 1 |
| Obesity | neutrophil | *ZNF429* | 1.704747882 | 0.00736537 | 0.012289299 | 1 |
| Obesity | neutrophil | *ZNF470* | 1.982169765 | 0.005515764 | 0.011290116 | 1 |

| | | | | | | |
|---|---|---|---|---|---|---|
| Obesity | neutrophil | *GMFB* | 2.241178076 | 0.004917672 | 0.011289644 | 1 |
| Obesity | neutrophil | *ZMYM1* | 1.718863601 | 0.004330838 | 0.011289644 | 1 |
| Obesity | neutrophil | *ARRDC1* | 3.072804671 | 0.004595969 | 0.011289644 | 1 |
| Obesity | neutrophil | *IGF2R* | 1.85667905 | 0.006240856 | 0.01154827 | 1 |
| Obesity | neutrophil | *GAL3ST4* | 3.468706439 | 0.002820619 | 0.011289644 | 1 |
| Obesity | neutrophil | *SLC25A29* | 1.67886815 | 0.001846971 | 0.011289644 | 1 |
| Obesity | neutrophil | *ZNF682* | 3.774598523 | 0.007942263 | 0.01278095 | 1 |
| Obesity | neutrophil | *ZNF257* | 3.051292312 | 0.018568769 | 0.024412766 | 1 |
| Obesity | neutrophil | *ADAM32* | 1.527744929 | 0.007390957 | 0.012298069 | 1 |
| Obesity | neutrophil | *LRRC8B* | -2.58412348 | 0.031209492 | 0.03879072 | 1 |
| Obesity | neutrophil | *ABCB8* | 2.088694205 | 0.001387353 | 0.011289644 | 1 |
| Obesity | neutrophil | *SND1* | 1.739625167 | 0.003096811 | 0.011289644 | 1 |
| Obesity | neutrophil | *SULT1A2* | 4.547586634 | 6.78E-19 | 2.13E-15 | 4.26E-15 |
| Obesity | neutrophil | *NEK5* | 1.477315917 | 0.006413462 | 0.011648199 | 1 |
| Obesity | neutrophil | *PIWIL2* | -1.42954601 | 0.009710465 | 0.014553767 | 1 |
| Obesity | neutrophil | *ENTPD4* | 1.808422898 | 0.003388865 | 0.011289644 | 1 |
| Obesity | neutrophil | *C1D* | 1.395525896 | 0.004026996 | 0.011289644 | 1 |
| Obesity | neutrophil | *TBC1D9B* | 1.452048283 | 0.007549762 | 0.012419095 | 1 |
| Obesity | neutrophil | *FAM110D* | 5.456302058 | 0.002541267 | 0.011289644 | 1 |
| Obesity | neutrophil | *KANK2* | 2.942274834 | 0.004138807 | 0.011289644 | 1 |
| Obesity | neutrophil | *C6orf141* | 1.777775407 | 0.036937539 | 0.045180215 | 1 |
| Obesity | neutrophil | *GTF2E2* | 1.806646066 | 0.002226277 | 0.011289644 | 1 |
| Obesity | neutrophil | *ZNF165* | -0.537558175 | 0.002713952 | 0.011289644 | 1 |
| Obesity | neutrophil | *RAMP2-AS1* | 1.362186767 | 0.006445695 | 0.011648199 | 1 |
| Obesity | neutrophil | *ZNF720* | 2.653061986 | 0.009243018 | 0.014053411 | 1 |
| Obesity | neutrophil | *DDI2* | 1.875563651 | 0.004639564 | 0.011289644 | 1 |
| Obesity | neutrophil | *MRPL21* | -1.494414093 | 0.000183209 | 0.011289644 | 1 |
| Obesity | neutrophil | *ZNF786* | 1.483089959 | 0.005503835 | 0.011289644 | 1 |
| Obesity | neutrophil | *ZNF517* | 2.252179397 | 0.004264868 | 0.011289644 | 1 |
| Obesity | neutrophil | *SLC22A5* | 1.453652525 | 0.004671327 | 0.011289644 | 1 |
| Obesity | neutrophil | *C5AR1* | -2.952465186 | 0.016948862 | 0.022556625 | 1 |
| Obesity | neutrophil | *DCHS2* | 1.779505452 | 0.034801238 | 0.042800348 | 1 |
| Obesity | neutrophil | *VEPH1* | 2.42407501 | 0.006308247 | 0.011589699 | 1 |
| Obesity | neutrophil | *SHPK* | 2.273836897 | 0.002801249 | 0.011289644 | 1 |
| Obesity | neutrophil | *MAP3K5* | 1.419042144 | 0.00640917 | 0.011648199 | 1 |
| Obesity | neutrophil | *STMN3* | 1.721228234 | 0.008397744 | 0.013245513 | 1 |
| Obesity | neutrophil | *PCDHB11* | 2.820581507 | 0.037319329 | 0.045602842 | 1 |
| Obesity | neutrophil | *SLC2A10* | 1.551331601 | 0.003813508 | 0.011289644 | 1 |
| Obesity | neutrophil | *ZNF665* | 2.277816466 | 0.013334244 | 0.018482279 | 1 |
| Obesity | neutrophil | *RPF2* | 2.738164194 | 0.006450308 | 0.011649838 | 1 |
| Obesity | neutrophil | *MYO5A* | 1.852695262 | 0.004329836 | 0.011289644 | 1 |
| Obesity | neutrophil | *ATG7* | 2.22785538 | 0.020214201 | 0.026273865 | 1 |
| Obesity | neutrophil | *RAB40C* | -2.895810668 | 0.038584431 | 0.046993475 | 1 |
| Obesity | neutrophil | *PIGN* | 1.277859162 | 0.020461553 | 0.026535021 | 1 |
| Obesity | neutrophil | *ZNF624* | 3.257158697 | 0.005480097 | 0.011289644 | 1 |
| Obesity | neutrophil | *HHLA3* | -2.475150627 | 0.006645846 | 0.011753146 | 1 |
| Obesity | neutrophil | *HOXA4* | -2.065374469 | 0.007077981 | 0.012072542 | 1 |
| Obesity | neutrophil | *BCO2* | 1.622959473 | 0.004861568 | 0.011289644 | 1 |
| Obesity | neutrophil | *ENTPD6* | 1.35881797 | 0.006731785 | 0.011825602 | 1 |
| Obesity | neutrophil | *CCDC154* | 2.562580522 | 0.009112698 | 0.013903688 | 1 |

| | | | | | | |
|---|---|---|---|---|---|---|
| Obesity | neutrophil | *C5orf42* | 1.539208437 | 0.004505178 | 0.011289644 | 1 |
| Obesity | neutrophil | *ZNF841* | 1.742932114 | 0.00487918 | 0.011289644 | 1 |
| Obesity | neutrophil | *MFAP5* | 2.080731031 | 0.005981584 | 0.011427806 | 1 |
| Obesity | neutrophil | *MPEG1* | 1.851260898 | 0.03856974 | 0.046984684 | 1 |
| Obesity | neutrophil | *NMB* | 2.149869617 | 0.004598768 | 0.011289644 | 1 |
| Obesity | neutrophil | *KLHL14* | -2.86137339 | 0.004690556 | 0.011289644 | 1 |
| Obesity | neutrophil | *FAM114A1* | 1.247157988 | 0.003813639 | 0.011289644 | 1 |
| Obesity | neutrophil | *PHF2* | 2.977714122 | 0.004549926 | 0.011289644 | 1 |
| Obesity | neutrophil | *PSAP* | 1.556028746 | 0.004366927 | 0.011289644 | 1 |
| Obesity | neutrophil | *S100A10* | 2.142564405 | 0.008517005 | 0.013361694 | 1 |
| Obesity | neutrophil | *MAP1LC3C* | -2.695559218 | 0.015452917 | 0.020884026 | 1 |
| Obesity | neutrophil | *AC122129.1* | 4.869279553 | 0.004271878 | 0.011289644 | 1 |
| Obesity | neutrophil | *CCDC180* | -1.340404865 | 0.004657956 | 0.011289644 | 1 |
| Obesity | neutrophil | *ZNF181* | 1.680757737 | 0.004136909 | 0.011289644 | 1 |
| Obesity | neutrophil | *SGTB* | 2.969231704 | 0.008494286 | 0.013331367 | 1 |
| Obesity | neutrophil | *ADH5* | 2.31656013 | 0.008635707 | 0.013492218 | 1 |
| Obesity | neutrophil | *HRNR* | 1.823007511 | 0.003177377 | 0.011289644 | 1 |
| Obesity | neutrophil | *ZNF677* | 1.589597486 | 0.004078563 | 0.011289644 | 1 |
| Obesity | neutrophil | *FCHSD1* | -1.661428562 | 0.008169865 | 0.01298709 | 1 |
| Obesity | neutrophil | *ZNF71* | 1.834198561 | 0.002878643 | 0.011289644 | 1 |
| Obesity | neutrophil | *RPL12* | 1.416990818 | 0.00628185 | 0.011565541 | 1 |
| Obesity | neutrophil | *VPS13A* | 1.263078479 | 0.009490517 | 0.014308604 | 1 |
| Obesity | neutrophil | *NOL8* | 1.418148385 | 0.006537113 | 0.011689086 | 1 |
| Obesity | neutrophil | *ZNF335* | 1.38252654 | 0.003483349 | 0.011289644 | 1 |
| Obesity | neutrophil | *AGAP9* | 2.260701051 | 0.00496331 | 0.011289644 | 1 |
| Obesity | neutrophil | *ZNF84* | 2.33862587 | 0.005730243 | 0.01134195 | 1 |
| Obesity | neutrophil | *MAK16* | 4.326126911 | 0.039250835 | 0.047712684 | 1 |
| Obesity | neutrophil | *ZNF667* | 3.658417469 | 0.013255367 | 0.018416905 | 1 |
| Obesity | neutrophil | *PRIM1* | 1.607641245 | 0.004897176 | 0.011289644 | 1 |
| Obesity | neutrophil | *SFI1* | 1.21348971 | 0.004205908 | 0.011289644 | 1 |
| Obesity | neutrophil | *ZNF649* | -5.241300139 | 0.002534523 | 0.011289644 | 1 |
| Obesity | neutrophil | *HIBCH* | 1.515178643 | 0.010003157 | 0.01486055 | 1 |
| Obesity | neutrophil | *ZNF544* | 3.202209131 | 0.020000669 | 0.026033997 | 1 |
| Obesity | neutrophil | *SOWAHC* | 2.780704726 | 0.004472933 | 0.011289644 | 1 |
| Obesity | neutrophil | *AC135178.1* | 12.75800419 | 0.022496346 | 0.028828888 | 1 |
| Obesity | neutrophil | *MIER1* | 1.228042879 | 0.003760199 | 0.011289644 | 1 |
| Obesity | neutrophil | *SVIP* | 2.841817059 | 0.004677775 | 0.011289644 | 1 |
| Obesity | neutrophil | *ZNF251* | 2.382421422 | 0.007068268 | 0.012065762 | 1 |
| Obesity | neutrophil | *ZNF334* | 1.553894259 | 0.008648482 | 0.013505931 | 1 |
| Obesity | neutrophil | *HSD17B11* | 1.240939725 | 0.003816428 | 0.011289644 | 1 |
| Obesity | neutrophil | *SULT1C2* | -1.625508932 | 0.021807768 | 0.028072366 | 1 |
| Obesity | neutrophil | *RPS6KL1* | 1.842283098 | 0.008382114 | 0.013235441 | 1 |
| Obesity | neutrophil | *QRICH1* | 1.503084081 | 0.001061114 | 0.011289644 | 1 |
| Obesity | neutrophil | *DDX42* | 2.112133155 | 0.002318829 | 0.011289644 | 1 |
| Obesity | neutrophil | *SLC29A3* | 1.849326969 | 0.003950569 | 0.011289644 | 1 |
| Obesity | neutrophil | *ANTXRL* | 68.93904797 | 0.005084712 | 0.011289644 | 1 |
| Obesity | neutrophil | *STYX* | 3.526819394 | 0.005341879 | 0.011289644 | 1 |
| Obesity | neutrophil | *TMEM116* | 1.362290229 | 0.001834577 | 0.011289644 | 1 |
| Obesity | neutrophil | *ZKSCAN8* | 1.784375765 | 0.001194436 | 0.011289644 | 1 |
| Obesity | neutrophil | *MYL4* | 7.062318274 | 0.034179743 | 0.042126651 | 1 |

| | | | | | | |
|---|---|---|---|---|---|---|
| Obesity | neutrophil | *ZNF813* | 1.785646984 | 0.003774514 | 0.011289644 | 1 |
| Obesity | neutrophil | *GFPT1* | 1.215147954 | 0.00267034 | 0.011289644 | 1 |
| Obesity | neutrophil | *UVRAG* | 1.251030015 | 0.006496124 | 0.011671674 | 1 |
| Obesity | neutrophil | *MGEA5* | 1.388105397 | 0.008056091 | 0.01288114 | 1 |
| Obesity | neutrophil | *MT1F* | 7.598097933 | 0.020406969 | 0.026480622 | 1 |
| Obesity | neutrophil | *ZNF568* | 1.222217329 | 0.004050908 | 0.011289644 | 1 |
| Obesity | neutrophil | *ZNF587* | 2.193114641 | 0.003738505 | 0.011289644 | 1 |
| Obesity | neutrophil | *FLVCR1-AS1* | 2.756453545 | 0.007491543 | 0.01238804 | 1 |
| Obesity | neutrophil | *SH3BGRL2* | 1.370807794 | 0.002210607 | 0.011289644 | 1 |
| Obesity | neutrophil | *ZNF808* | 1.737416422 | 0.00394561 | 0.011289644 | 1 |
| Obesity | neutrophil | *TMA16* | 1.229749221 | 0.003749224 | 0.011289644 | 1 |
| Obesity | neutrophil | *CNGA1* | 1.664958607 | 0.005988919 | 0.011430582 | 1 |
| Obesity | neutrophil | *HIST1H4E* | 6.314539157 | 0.004898923 | 0.011289644 | 1 |
| Obesity | neutrophil | *C20orf203* | 2.435168874 | 0.010581781 | 0.01547069 | 1 |
| Obesity | neutrophil | *ZNF627* | 3.004466065 | 0.007357006 | 0.012285536 | 1 |
| Obesity | neutrophil | *ARC* | 10.27222306 | 0.006604081 | 0.01172215 | 1 |
| Obesity | neutrophil | *TLK1* | 1.267189113 | 0.004290722 | 0.011289644 | 1 |
| Obesity | neutrophil | *LRBA* | 1.393808565 | 0.004084668 | 0.011289644 | 1 |
| Obesity | neutrophil | *CCDC69* | 1.893379038 | 0.004963612 | 0.011289644 | 1 |
| Obesity | neutrophil | *MDM4* | 1.422791021 | 0.004377889 | 0.011289644 | 1 |
| Obesity | neutrophil | *ZNF534* | 4.132592513 | 0.016713521 | 0.022304799 | 1 |
| Obesity | neutrophil | *NCOA6* | 1.593577926 | 0.011463615 | 0.016406044 | 1 |
| Obesity | neutrophil | *TTC37* | 1.44981259 | 0.004406459 | 0.011289644 | 1 |
| Obesity | neutrophil | *TUSC1* | -4.010062203 | 0.013297143 | 0.018446412 | 1 |
| Obesity | neutrophil | *PAPSS2* | 1.847568245 | 0.004734842 | 0.011289644 | 1 |
| Obesity | neutrophil | *FAN1* | 1.311110202 | 0.004130284 | 0.011289644 | 1 |
| Obesity | neutrophil | *CEP290* | 2.993632115 | 0.009059576 | 0.013865944 | 1 |
| Obesity | neutrophil | *ANKRD13B* | 4.159225336 | 0.000101748 | 0.010661544 | 0.63969264 |
| Obesity | neutrophil | *ECI2* | 1.313378813 | 0.004237471 | 0.011289644 | 1 |
| Obesity | neutrophil | *ZNF652* | 2.327229999 | 0.005674984 | 0.011300433 | 1 |
| Obesity | neutrophil | *RPL10A* | 4.535896795 | 0.004452719 | 0.011289644 | 1 |
| Obesity | neutrophil | *APCDD1L* | 2.915944974 | 0.004303686 | 0.011289644 | 1 |
| Obesity | neutrophil | *RASSF9* | -2.853231887 | 0.02850119 | 0.035730206 | 1 |
| Obesity | neutrophil | *FAM169A* | 2.456844944 | 0.002093825 | 0.011289644 | 1 |
| Obesity | neutrophil | *PNP* | 1.484264109 | 0.027270532 | 0.034303688 | 1 |
| Obesity | neutrophil | *SFT2D1* | 2.278410395 | 0.003956623 | 0.011289644 | 1 |
| Obesity | neutrophil | *ARHGAP11A* | 3.021589791 | 0.004252188 | 0.011289644 | 1 |
| Obesity | neutrophil | *HMGN2* | 1.740630346 | 0.008128032 | 0.012949841 | 1 |
| Obesity | neutrophil | *CD3E* | 3.184604166 | 0.002130644 | 0.011289644 | 1 |
| Obesity | neutrophil | *TYW1* | 1.196978448 | 0.023510062 | 0.029944846 | 1 |
| Obesity | neutrophil | *ITPRIPL1* | 3.038581278 | 0.009052644 | 0.013861369 | 1 |
| Obesity | neutrophil | *PRMT6* | -3.160693098 | 0.002965714 | 0.011289644 | 1 |
| Obesity | neutrophil | *SHISA4* | 4.276657896 | 0.007153203 | 0.012141606 | 1 |
| Obesity | neutrophil | *CAPZA2* | 1.84475782 | 0.004604821 | 0.011289644 | 1 |
| Obesity | neutrophil | *C1orf174* | 1.702624893 | 0.004044044 | 0.011289644 | 1 |
| Obesity | neutrophil | *RASGEF1A* | 4.608888558 | 0.004113488 | 0.011289644 | 1 |
| Obesity | neutrophil | *KIAA0753* | 2.814211024 | 0.006415388 | 0.011648199 | 1 |
| Obesity | neutrophil | *DCLRE1A* | -4.422430547 | 0.00934065 | 0.014164174 | 1 |
| Obesity | neutrophil | *APRT* | 2.300564069 | 0.004244441 | 0.011289644 | 1 |
| Obesity | neutrophil | *ZFP2* | 1.706596341 | 0.004866345 | 0.011289644 | 1 |

| | | | | | | |
|---|---|---|---|---|---|---|
| Obesity | neutrophil | *L3MBTL3* | 3.717783097 | 0.0039906 | 0.011289644 | 1 |
| Obesity | neutrophil | *MFAP3L* | 3.582133273 | 0.003765621 | 0.011289644 | 1 |
| Obesity | neutrophil | *NAGA* | 2.213122287 | 0.003097588 | 0.011289644 | 1 |
| Obesity | neutrophil | *SMG5* | 1.263626985 | 0.005334877 | 0.011289644 | 1 |
| Obesity | neutrophil | *PJA2* | 1.227081928 | 0.006375061 | 0.011639549 | 1 |
| Obesity | neutrophil | *AL590822.1* | -1.801095361 | 0.009304268 | 0.014119221 | 1 |
| Obesity | neutrophil | *AP001007.1* | 10.35790911 | 0.004241112 | 0.011289644 | 1 |
| Obesity | neutrophil | *AC105020.1* | 5.709951613 | 0.002650412 | 0.011289644 | 1 |
| Obesity | neutrophil | *EFCAB2* | 1.932691498 | 0.003911212 | 0.011289644 | 1 |
| Obesity | neutrophil | *CHML* | 2.101958125 | 0.001931179 | 0.011289644 | 1 |
| Obesity | neutrophil | *CAPN8* | 5.725287658 | 0.018173268 | 0.023962948 | 1 |
| Obesity | neutrophil | *TATDN3* | 2.422228342 | 0.002115542 | 0.011289644 | 1 |
| Obesity | neutrophil | *ECT2L* | 3.156553909 | 0.002994092 | 0.011289644 | 1 |
| Obesity | neutrophil | *CENPW* | -0.643647985 | 0.0085783 | 0.013429226 | 1 |
| Obesity | neutrophil | *DDO* | -1.633913393 | 0.009100525 | 0.013900632 | 1 |
| Obesity | neutrophil | *HIST2H2BF* | -0.763891661 | 0.005264321 | 0.011289644 | 1 |
| Obesity | neutrophil | *C6orf163* | 2.245620483 | 0.005689782 | 0.011300433 | 1 |
| Obesity | neutrophil | *PCMTD2* | 1.3294059 | 0.003760943 | 0.011289644 | 1 |
| Obesity | neutrophil | *OOEP* | 2.765912797 | 0.017225036 | 0.022870919 | 1 |
| Obesity | neutrophil | *FOXO6* | 2.055473598 | 0.015837813 | 0.021340336 | 1 |
| Obesity | neutrophil | *SYS1* | -3.589973723 | 0.019413031 | 0.02537944 | 1 |
| Obesity | neutrophil | *NEU4* | 18.61092641 | 0.02010756 | 0.026146067 | 1 |
| Obesity | neutrophil | *TRAF3IP1* | 2.900243678 | 0.009296055 | 0.014112205 | 1 |
| Obesity | neutrophil | *GIGYF2* | 1.515000625 | 0.004508041 | 0.011289644 | 1 |
| Obesity | neutrophil | *RUFY2* | 1.264942288 | 0.005492991 | 0.011289644 | 1 |
| Obesity | neutrophil | *PHACTR4* | 1.501188952 | 0.008494585 | 0.013331367 | 1 |
| Obesity | neutrophil | *ASAH2B* | 2.433435634 | 0.003877654 | 0.011289644 | 1 |
| Obesity | neutrophil | *AGAP6* | 2.365632241 | 0.005940321 | 0.011396756 | 1 |
| Obesity | neutrophil | *C10orf128* | 3.182407224 | 0.000717141 | 0.011289644 | 1 |
| Obesity | neutrophil | *ZDBF2* | 3.296095411 | 0.008181067 | 0.01300161 | 1 |
| Obesity | neutrophil | *RING1* | -2.503281067 | 0.013992488 | 0.019220182 | 1 |
| Obesity | neutrophil | *HSD17B8* | -4.762186249 | 0.036700278 | 0.044942471 | 1 |
| Obesity | neutrophil | *OXLD1* | 1.759306129 | 0.004168067 | 0.011289644 | 1 |
| Obesity | neutrophil | *HLA-DOA* | 2.54748875 | 0.011582058 | 0.016537906 | 1 |
| Obesity | neutrophil | *HLA-DMA* | 2.420849246 | 0.004358082 | 0.011289644 | 1 |
| Obesity | neutrophil | *HLA-DRA* | 0.795672561 | 0.013862243 | 0.019082969 | 1 |
| Obesity | neutrophil | *COL15A1* | 1.876146425 | 0.007643437 | 0.012505709 | 1 |
| Obesity | neutrophil | *NOTCH4* | -1.57536256 | 3.89E-07 | 0.000174874 | 0.00244824 |
| Obesity | neutrophil | *PRRT1* | -2.451148935 | 0.036690662 | 0.044939449 | 1 |
| Obesity | neutrophil | *MRPL38* | -3.15418512 | 0.00026795 | 0.011289644 | 1 |
| Obesity | neutrophil | *STK19* | 1.774205461 | 0.008852562 | 0.013705013 | 1 |
| Obesity | neutrophil | *DXO* | 2.046717331 | 0.016380703 | 0.021965903 | 1 |
| Obesity | neutrophil | *SKIV2L* | -4.122170051 | 1.20E-05 | 0.002612309 | 0.07575695 |
| Obesity | neutrophil | *NELFE* | 2.004079916 | 0.000310783 | 0.011289644 | 1 |
| Obesity | neutrophil | *LAYN* | 3.164189754 | 0.002361227 | 0.011289644 | 1 |
| Obesity | neutrophil | *C6orf48* | 1.632140833 | 0.005757722 | 0.011354705 | 1 |
| Obesity | neutrophil | *CARD16* | -1.556593639 | 0.007008504 | 0.012012668 | 1 |
| Obesity | neutrophil | *MSH5* | 2.48597518 | 0.01263332 | 0.017721036 | 1 |
| Obesity | neutrophil | *ABHD16A* | -0.788963936 | 0.009659595 | 0.014501216 | 1 |
| Obesity | neutrophil | *BAG6* | 1.814143869 | 0.002127787 | 0.011289644 | 1 |

| | | | | | | |
|---|---|---|---|---|---|---|
| Obesity | neutrophil | *NFKBIL1* | 3.202035508 | 0.00197535 | 0.011289644 | 1 |
| Obesity | neutrophil | *ZNF814* | 2.920841761 | 0.008129954 | 0.012949841 | 1 |
| Obesity | neutrophil | *MICB* | 1.528309574 | 0.013615565 | 0.01880347 | 1 |
| Obesity | neutrophil | *HLA-C* | 3.547539193 | 0.004638761 | 0.011289644 | 1 |
| Obesity | neutrophil | *POU5F1* | 1.476448215 | 0.029876021 | 0.037297566 | 1 |
| Obesity | neutrophil | *ABCF1* | 1.281377719 | 0.001373643 | 0.011289644 | 1 |
| Obesity | neutrophil | *LILRB3* | -1.010471219 | 0.010368521 | 0.015241266 | 1 |
| Obesity | neutrophil | *LINC01123* | -2.160054257 | 0.004823534 | 0.011289644 | 1 |
| Obesity | neutrophil | *HLA-E* | 6.084775517 | 8.17E-07 | 0.000321058 | 0.00513693 |
| Obesity | neutrophil | *ZNF468* | -2.41738692 | 0.008382051 | 0.013235441 | 1 |
| Obesity | neutrophil | *HLA-G* | -3.903164453 | 0.000659475 | 0.011289644 | 1 |
| Obesity | neutrophil | *HLA-F* | 2.24118177 | 0.004571004 | 0.011289644 | 1 |
| Obesity | neutrophil | *ZFP57* | 5.553444866 | 6.52E-09 | 4.10E-06 | 4.10E-05 |
| Obesity | neutrophil | *FAM153C* | 7.131946243 | 0.005118858 | 0.011289644 | 1 |
| Obesity | neutrophil | *GABBR1* | 1.170151033 | 0.005466915 | 0.011289644 | 1 |
| Obesity | neutrophil | *TRIM27* | 1.576826421 | 0.010078217 | 0.01492911 | 1 |
| Obesity | neutrophil | *MALRD1* | 1.168975651 | 0.004738842 | 0.011289644 | 1 |
| Obesity | neutrophil | *MROH6* | 3.356885119 | 0.006778208 | 0.011856902 | 1 |
| Obesity | neutrophil | *ATXN2* | 1.97860781 | 0.002596319 | 0.011289644 | 1 |
| Obesity | neutrophil | *FAM216A* | 1.79513305 | 0.00821215 | 0.013037825 | 1 |
| Obesity | neutrophil | *ZNF155* | 1.803661958 | 0.00519897 | 0.011289644 | 1 |
| Obesity | neutrophil | *ZNF783* | 1.262347609 | 0.006136757 | 0.011498979 | 1 |
| Obesity | neutrophil | *ZNF425* | 1.461530767 | 0.010547744 | 0.01543255 | 1 |
| Obesity | neutrophil | *LRRC10B* | -6.730876153 | 0.006655029 | 0.011757198 | 1 |
| Obesity | neutrophil | *C12orf73* | 2.497268686 | 0.014357225 | 0.019648209 | 1 |
| Obesity | neutrophil | *PCDHA5* | -1.976195815 | 0.005849374 | 0.011378838 | 1 |
| Obesity | neutrophil | *PCDHA4* | 7.838320253 | 0.029613456 | 0.037006519 | 1 |
| Obesity | neutrophil | *TRIM13* | 1.797043633 | 0.006896217 | 0.011932455 | 1 |
| Obesity | neutrophil | *SPIRE2* | 1.574837779 | 0.011517347 | 0.01647544 | 1 |
| Obesity | neutrophil | *SLFN12L* | 2.25334369 | 0.031841903 | 0.039469646 | 1 |
| Obesity | neutrophil | *CLLU1OS* | 6.38915703 | 0.014638586 | 0.019950746 | 1 |
| Obesity | neutrophil | *TMEM231* | 2.631864345 | 0.002647002 | 0.011289644 | 1 |
| Obesity | neutrophil | *FAM71F2* | 1.654063875 | 0.004214364 | 0.011289644 | 1 |
| Obesity | neutrophil | *AC016586.1* | 29.76196717 | 0.002429064 | 0.011289644 | 1 |
| Obesity | neutrophil | *LINC00654* | 4.141993845 | 0.006393432 | 0.011648199 | 1 |
| Obesity | neutrophil | *TMEM170B* | 1.060743894 | 0.006980154 | 0.01198898 | 1 |
| Obesity | neutrophil | *NT5M* | -2.534333019 | 0.005502354 | 0.011289644 | 1 |
| Obesity | neutrophil | *PRR13* | 2.641496864 | 0.020881966 | 0.02701882 | 1 |
| Obesity | neutrophil | *MT1A* | 5.093599349 | 0.004031383 | 0.011289644 | 1 |
| Obesity | neutrophil | *MT1M* | 2.38083636 | 0.004250609 | 0.011289644 | 1 |
| Obesity | neutrophil | *CFI* | 1.457207583 | 0.004766337 | 0.011289644 | 1 |
| Obesity | neutrophil | *SAMD9* | -2.61578294 | 0.023119829 | 0.029491346 | 1 |
| Obesity | neutrophil | *EXOC3L4* | 3.524013604 | 0.008369166 | 0.013226985 | 1 |
| Obesity | neutrophil | *TP53TG3D* | 12.05711152 | 0.032177086 | 0.039821236 | 1 |
| Obesity | neutrophil | *ATP6AP1L* | 2.601933642 | 0.022849894 | 0.029198634 | 1 |
| Obesity | neutrophil | *NAP1L4* | 1.12481285 | 0.007933224 | 0.01278095 | 1 |
| Obesity | neutrophil | *CPT1B* | -1.284503911 | 0.007969223 | 0.01279518 | 1 |
| Obesity | neutrophil | *EIF3CL* | -1.056827736 | 0.022150681 | 0.028472977 | 1 |
| Obesity | neutrophil | *LCMT1* | 3.652440955 | 0.040246513 | 0.048828604 | 1 |
| Obesity | neutrophil | *CDPF1* | 2.345653868 | 0.016628714 | 0.022229369 | 1 |

| | | | | | | |
|---|---|---|---|---|---|---|
| Obesity | neutrophil | *LIN52* | 2.497610541 | 0.002415189 | 0.011289644 | 1 |
| Obesity | neutrophil | *C17orf107* | 5.228616825 | 0.011409948 | 0.016351571 | 1 |
| Obesity | neutrophil | *RFPL3S* | 3.278324199 | 0.015447218 | 0.020880812 | 1 |
| Obesity | neutrophil | *C1QTNF9B* | 25.42908547 | 0.004092957 | 0.011289644 | 1 |
| Obesity | neutrophil | *RNPS1* | 3.095448816 | 0.029447356 | 0.036820909 | 1 |
| Obesity | neutrophil | *NYNRIN* | 2.307266153 | 0.003445871 | 0.011289644 | 1 |
| Obesity | neutrophil | *ZDHHC11B* | 2.244703113 | 0.002940008 | 0.011289644 | 1 |
| Obesity | neutrophil | *GOLGA8O* | 9.605248158 | 0.018642203 | 0.024488828 | 1 |
| Obesity | neutrophil | *HCG27* | 2.074446654 | 0.003272947 | 0.011289644 | 1 |
| Obesity | neutrophil | *LNP1* | 2.120852428 | 0.005639092 | 0.011295339 | 1 |
| Obesity | neutrophil | *VGLL3* | 0.794523875 | 0.015780454 | 0.021287893 | 1 |
| Obesity | neutrophil | *COLQ* | 1.232234569 | 0.005999446 | 0.011438456 | 1 |
| Obesity | neutrophil | *METTL6* | 1.550426472 | 0.002319088 | 0.011289644 | 1 |
| Obesity | neutrophil | *SACM1L* | 1.085203084 | 0.004397222 | 0.011289644 | 1 |
| Obesity | neutrophil | *SLC48A1* | -1.357015603 | 0.007394552 | 0.012298069 | 1 |
| Obesity | neutrophil | *TAS2R14* | 2.180313071 | 0.005619305 | 0.011295339 | 1 |
| Obesity | neutrophil | *ZRSR1* | 10.376473 | 0.004580044 | 0.011289644 | 1 |
| Obesity | neutrophil | *AC084018.1* | -5.650888156 | 0.000136447 | 0.011289644 | 0.857843877 |
| Obesity | neutrophil | *C17orf51* | 4.401630675 | 0.00951965 | 0.01434221 | 1 |
| Obesity | neutrophil | *RNF208* | 5.378718297 | 0.004420356 | 0.011289644 | 1 |
| Obesity | neutrophil | *MAP10* | 3.200923612 | 0.00473594 | 0.011289644 | 1 |
| Obesity | neutrophil | *ZNF580* | 1.768551272 | 0.003460991 | 0.011289644 | 1 |
| Obesity | neutrophil | *NUP62* | -3.580727579 | 0.025971524 | 0.032820698 | 1 |
| Obesity | neutrophil | *FAM106A* | -0.586657499 | 0.006037753 | 0.011447874 | 1 |
| Obesity | neutrophil | *ZNF254* | 1.446017812 | 0.004559175 | 0.011289644 | 1 |
| Obesity | neutrophil | *TCTEX1D2* | 3.245466134 | 0.008387147 | 0.013235441 | 1 |
| Obesity | neutrophil | *LINGO4* | -5.930056448 | 0.003089521 | 0.011289644 | 1 |
| Obesity | neutrophil | *ASIC3* | 9.526486751 | 5.94E-07 | 0.000248874 | 0.003733108 |
| Obesity | neutrophil | *GIMAP1* | 2.604258892 | 0.006602047 | 0.01172215 | 1 |
| Obesity | neutrophil | *CCDC183* | 6.793968317 | 0.003808215 | 0.011289644 | 1 |
| Obesity | neutrophil | *NOTCH2NL* | 8.432013724 | 3.05E-05 | 0.005039115 | 0.191486354 |
| Obesity | neutrophil | *SUPT4H1* | 1.912404501 | 0.002502829 | 0.011289644 | 1 |
| Obesity | neutrophil | *MXD3* | 4.287197183 | 0.001312585 | 0.011289644 | 1 |
| Obesity | neutrophil | *GSTM2* | 1.308281196 | 0.006816132 | 0.011874479 | 1 |
| Obesity | neutrophil | *ARHGAP19* | 2.987647586 | 0.00305742 | 0.011289644 | 1 |
| Obesity | neutrophil | *LCAT* | 2.458036336 | 0.036624162 | 0.04486674 | 1 |
| Obesity | neutrophil | *SIPA1* | 8.181436982 | 0.000618284 | 0.011289644 | 1 |
| Obesity | neutrophil | *ERV3-1* | 1.394567242 | 0.000270508 | 0.011289644 | 1 |
| Obesity | neutrophil | *GBP7* | -0.979685613 | 0.004126713 | 0.011289644 | 1 |
| Obesity | neutrophil | *RBMXL1* | 1.851949572 | 0.00423315 | 0.011289644 | 1 |
| Obesity | neutrophil | *SRA1* | -2.416848667 | 0.019144229 | 0.025064508 | 1 |
| Obesity | neutrophil | *TMEM110* | 8.163256805 | 6.71E-05 | 0.00802313 | 0.421617966 |
| Obesity | neutrophil | *HEXA* | 4.236875813 | 0.008936508 | 0.013763799 | 1 |
| Obesity | neutrophil | *LEPROT* | 2.343112955 | 0.010650116 | 0.015546153 | 1 |
| Obesity | neutrophil | *LBH* | 2.513455548 | 0.004668537 | 0.011289644 | 1 |
| Obesity | neutrophil | *PPP1CB* | 0.955207215 | 0.006820705 | 0.011875318 | 1 |
| Obesity | neutrophil | *NCKIPSD* | 3.867078886 | 0.014822986 | 0.020154003 | 1 |
| Obesity | neutrophil | *ATF6B* | 2.209215617 | 0.003547159 | 0.011289644 | 1 |
| Obesity | neutrophil | *SLC35F6* | 4.212744702 | 0.002914988 | 0.011289644 | 1 |
| Obesity | neutrophil | *KLRK1* | -1.056357972 | 0.00560254 | 0.011295339 | 1 |

| | | | | | | |
|---|---|---|---|---|---|---|
| Obesity | neutrophil | *EMP2* | 1.935222385 | 0.002953628 | 0.011289644 | 1 |
| Obesity | neutrophil | *PPM1N* | 2.85827599 | 0.003967387 | 0.011289644 | 1 |
| Obesity | neutrophil | *SLC23A3* | 3.129453023 | 0.003696776 | 0.011289644 | 1 |
| Obesity | neutrophil | *LTB4R* | 1.814824123 | 0.005870927 | 0.011378838 | 1 |
| Obesity | neutrophil | *LTB4R2* | 3.979115189 | 0.026908933 | 0.033909894 | 1 |
| Obesity | neutrophil | *GALT* | 1.896738022 | 0.001793571 | 0.011289644 | 1 |
| Obesity | neutrophil | *NUDT19* | 1.721390637 | 0.003163465 | 0.011289644 | 1 |
| Obesity | neutrophil | *ZNF726* | -3.304719387 | 0.010838565 | 0.015751748 | 1 |
| Obesity | neutrophil | *TAX1BP3* | 2.406644739 | 0.002995756 | 0.011289644 | 1 |
| Obesity | neutrophil | *AP1G2* | 4.495577848 | 0.006162859 | 0.011512519 | 1 |
| Obesity | neutrophil | *ZNF90* | 2.27054075 | 0.004383103 | 0.011289644 | 1 |
| Obesity | neutrophil | *TM6SF2* | 30.28170169 | 0.00337266 | 0.011289644 | 1 |
| Obesity | neutrophil | *REPIN1* | -2.91129893 | 0.004421084 | 0.011289644 | 1 |
| Obesity | neutrophil | *SMIM7* | 1.236905733 | 0.003643223 | 0.011289644 | 1 |
| Obesity | neutrophil | *TSPAN4* | 3.156060591 | 0.007155822 | 0.012141606 | 1 |
| Obesity | neutrophil | *ARL16* | -1.966883681 | 0.009931425 | 0.01479594 | 1 |
| Obesity | neutrophil | *AC079776.2* | -2.791764047 | 0.006951361 | 0.011970202 | 1 |
| Obesity | neutrophil | *WEE2* | 3.095848204 | 0.000874174 | 0.011289644 | 1 |
| Obesity | neutrophil | *PAXIP1-AS2* | 1.740840348 | 0.003721354 | 0.011289644 | 1 |
| Obesity | neutrophil | *LYRM4* | 1.644978197 | 0.00341731 | 0.011289644 | 1 |
| Obesity | neutrophil | *PRCD* | 1.995544496 | 0.006338315 | 0.011611011 | 1 |
| Obesity | neutrophil | *C17orf67* | 1.184601628 | 0.015679037 | 0.021173122 | 1 |
| Obesity | neutrophil | *FIS1* | 1.447245774 | 0.00427737 | 0.011289644 | 1 |
| Obesity | neutrophil | *COLCA2* | -1.373695464 | 0.005370092 | 0.011289644 | 1 |
| Obesity | neutrophil | *MBLAC1* | -2.735856245 | 0.010076646 | 0.01492911 | 1 |
| Obesity | neutrophil | *NEURL1B* | 3.153495586 | 0.004540937 | 0.011289644 | 1 |
| Obesity | neutrophil | *VSTM5* | -2.231209066 | 0.003269224 | 0.011289644 | 1 |
| Obesity | neutrophil | *PPME1* | 1.291760883 | 0.003131228 | 0.011289644 | 1 |
| Obesity | neutrophil | *NUTM2D* | 10.54795039 | 0.004556788 | 0.011289644 | 1 |
| Obesity | neutrophil | *CPEB1* | 2.217272089 | 0.005111423 | 0.011289644 | 1 |
| Obesity | neutrophil | *C10orf105* | 3.769251133 | 0.007726126 | 0.012603569 | 1 |
| Obesity | neutrophil | *ARHGEF33* | 1.749536326 | 0.0174046 | 0.023070361 | 1 |
| Obesity | neutrophil | *AC005082.1* | 8.838966641 | 0.003353391 | 0.011289644 | 1 |
| Obesity | neutrophil | *ACSM4* | 1.832021201 | 0.03273417 | 0.040440111 | 1 |
| Obesity | neutrophil | *COL28A1* | 7.347489563 | 0.002780114 | 0.011289644 | 1 |
| Obesity | neutrophil | *GRID2IP* | 3.944799887 | 0.003907773 | 0.011289644 | 1 |
| Obesity | neutrophil | *UBXN2B* | 1.98566523 | 0.006168826 | 0.011512519 | 1 |
| Obesity | neutrophil | *UBE2QL1* | 2.941680721 | 0.004396327 | 0.011289644 | 1 |
| Obesity | neutrophil | *KCNU1* | 2.600155471 | 0.003094198 | 0.011289644 | 1 |
| Obesity | neutrophil | *MYL5* | -1.754859718 | 0.006639784 | 0.011751293 | 1 |
| Obesity | neutrophil | *NPEPL1* | 1.780930528 | 0.004464612 | 0.011289644 | 1 |
| Obesity | neutrophil | *LINC01139* | 4.318231671 | 0.00395242 | 0.011289644 | 1 |
| Obesity | neutrophil | *TSTD1* | 1.766801909 | 0.003886254 | 0.011289644 | 1 |
| Obesity | neutrophil | *ATAD3C* | 7.189673523 | 0.000308716 | 0.011289644 | 1 |
| Obesity | neutrophil | *CCDC7* | 1.428274545 | 0.004322157 | 0.011289644 | 1 |
| Obesity | neutrophil | *PAM16* | 4.705471591 | 0.022396483 | 0.028718964 | 1 |
| Obesity | neutrophil | *AC016773.1* | 2.457174113 | 0.007769543 | 0.012646715 | 1 |
| Obesity | neutrophil | *RNASEK* | 2.430729103 | 0.004738857 | 0.011289644 | 1 |
| Obesity | neutrophil | *NBPF1* | 1.862133423 | 0.004199916 | 0.011289644 | 1 |
| Obesity | neutrophil | *RNU6ATAC35I* | -1.921726656 | 0.005408861 | 0.011289644 | 1 |

| | | | | | | |
|---|---|---|---|---|---|---|
| Obesity | neutrophil | *C6orf226* | 4.27090045 | 0.030945985 | 0.038503346 | 1 |
| Obesity | neutrophil | *PPP3R1* | 3.920127502 | 0.017258913 | 0.022906224 | 1 |
| Obesity | neutrophil | *C19orf73* | 14.03064636 | 0.008081272 | 0.012904993 | 1 |
| Obesity | neutrophil | *ZNF880* | 1.027213967 | 0.040801073 | 0.049415593 | 1 |
| Obesity | neutrophil | *OR2A25* | 79.24386096 | 0.009022984 | 0.013839351 | 1 |
| Obesity | neutrophil | *TIGD1* | 2.104258831 | 0.009050916 | 0.013861369 | 1 |
| Obesity | neutrophil | *XKR9* | 2.203622874 | 0.000840544 | 0.011289644 | 1 |
| Obesity | neutrophil | *C1orf229* | -3.37553685 | 0.005539244 | 0.011295339 | 1 |
| Obesity | neutrophil | *SLC12A8* | -6.394872472 | 0.00022111 | 0.011289644 | 1 |
| Obesity | neutrophil | *APOL6* | 1.932698506 | 0.004738379 | 0.011289644 | 1 |
| Obesity | neutrophil | *UBA52* | 2.973280458 | 0.004613107 | 0.011289644 | 1 |
| Obesity | neutrophil | *TIAF1* | -1.002216437 | 0.006215503 | 0.011544125 | 1 |
| Obesity | neutrophil | *BTBD19* | 1.771305857 | 0.00301956 | 0.011289644 | 1 |
| Obesity | neutrophil | *FAM185A* | 2.418584696 | 0.003045757 | 0.011289644 | 1 |
| Obesity | neutrophil | *AC062017.1* | 3.284681489 | 0.004657616 | 0.011289644 | 1 |
| Obesity | neutrophil | *ADRA2B* | 7.668238577 | 0.004554919 | 0.011289644 | 1 |
| Obesity | neutrophil | *EXOSC6* | -5.377239215 | 0.028424442 | 0.035648208 | 1 |
| Obesity | neutrophil | *VPS52* | 1.861532513 | 0.004642621 | 0.011289644 | 1 |
| Obesity | neutrophil | *HLA-DPB1* | 1.069696001 | 0.003173934 | 0.011289644 | 1 |
| Obesity | neutrophil | *OSER1-AS1* | 1.809693235 | 0.002566615 | 0.011289644 | 1 |
| Obesity | neutrophil | *LINC01133* | 4.757279002 | 0.035906772 | 0.044082381 | 1 |
| Obesity | neutrophil | *LINC01117* | 1.557848579 | 0.0036318 | 0.011289644 | 1 |
| Obesity | neutrophil | *RASAL2-AS1* | 7.451469894 | 0.013990075 | 0.019220182 | 1 |
| Obesity | neutrophil | *LINC00240* | 2.105281537 | 5.33E-05 | 0.00737107 | 0.335235557 |
| Obesity | neutrophil | *LINC00863* | 1.962304455 | 0.001683947 | 0.011289644 | 1 |
| Obesity | neutrophil | *LINC00184* | 4.23535784 | 0.015013855 | 0.0203797 | 1 |
| Obesity | neutrophil | *TMEM233* | 1.614951719 | 0.002413714 | 0.011289644 | 1 |
| Obesity | neutrophil | *LINC00237* | 21.51811266 | 0.006240485 | 0.01154827 | 1 |
| Obesity | neutrophil | *PLEKHM1* | 2.177390589 | 0.001738046 | 0.011289644 | 1 |
| Obesity | neutrophil | *AP000350.6* | 2.489620891 | 0.002679733 | 0.011289644 | 1 |
| Obesity | neutrophil | *SFTA1P* | 6.551598084 | 0.011050502 | 0.015961637 | 1 |
| Obesity | neutrophil | *SLC26A6* | 2.358599644 | 0.005085991 | 0.011289644 | 1 |
| Obesity | neutrophil | *MIAT* | 4.89200019 | 0.005743756 | 0.01134195 | 1 |
| Obesity | neutrophil | *LINC00626* | 46.22154718 | 0.003041793 | 0.011289644 | 1 |
| Obesity | neutrophil | *LINC00115* | 2.164373662 | 0.012028807 | 0.017055731 | 1 |
| Obesity | neutrophil | *NOL7* | 2.553849918 | 0.004332052 | 0.011289644 | 1 |
| Obesity | neutrophil | *ELFN1* | 5.431373695 | 0.003680228 | 0.011289644 | 1 |
| Obesity | neutrophil | *ZNF503-AS1* | 1.843368702 | 0.004244714 | 0.011289644 | 1 |
| Obesity | neutrophil | *LINC00937* | 3.801161453 | 0.009516607 | 0.014341061 | 1 |
| Obesity | neutrophil | *AC064836.3* | 5.758412819 | 0.004030062 | 0.011289644 | 1 |
| Obesity | neutrophil | *TMEM185B* | 3.328046639 | 0.01456063 | 0.01987309 | 1 |
| Obesity | neutrophil | *HSBP1L1* | 6.014624595 | 0.038223383 | 0.046589843 | 1 |
| Obesity | neutrophil | *TAS2R46* | -3.013100219 | 0.006438721 | 0.011648199 | 1 |
| Obesity | neutrophil | *AC093901.1* | 3.894177871 | 0.003523038 | 0.011289644 | 1 |
| Obesity | neutrophil | *ZNF717* | 1.118636395 | 0.000679551 | 0.011289644 | 1 |
| Obesity | neutrophil | *AC017074.2* | -3.599803234 | 0.005014952 | 0.011289644 | 1 |
| Obesity | neutrophil | *AC009299.3* | 3.323404238 | 0.006241613 | 0.01154827 | 1 |
| Obesity | neutrophil | *SLC2A1-AS1* | 2.337598774 | 0.011534641 | 0.016494833 | 1 |
| Obesity | neutrophil | *LINC01068* | -3.608683865 | 0.019423568 | 0.025387936 | 1 |
| Obesity | neutrophil | *AC108463.2* | 4.324278624 | 0.03537635 | 0.043448156 | 1 |

| | | | | | | |
|---|---|---|---|---|---|---|
| Obesity | neutrophil | *C19orf24* | -1.838053462 | 0.007354306 | 0.012285536 | 1 |
| Obesity | neutrophil | *LINC01013* | 3.244682406 | 0.004266784 | 0.011289644 | 1 |
| Obesity | neutrophil | *PROB1* | 13.88890194 | 0.008383771 | 0.013235441 | 1 |
| Obesity | neutrophil | *LINC00954* | -2.883100828 | 0.008670532 | 0.01352646 | 1 |
| Obesity | neutrophil | *AC067945.3* | 4.765544421 | 0.010076736 | 0.01492911 | 1 |
| Obesity | neutrophil | *RPL41* | 1.902746483 | 0.006381273 | 0.011645591 | 1 |
| Obesity | neutrophil | *PATL2* | 2.019649337 | 0.003524874 | 0.011289644 | 1 |
| Obesity | neutrophil | *ZNF492* | 3.367539512 | 0.010320787 | 0.015178195 | 1 |
| Obesity | neutrophil | *U91324.1* | -9.984592901 | 0.02684853 | 0.03384056 | 1 |
| Obesity | neutrophil | *ZNF688* | 1.998404111 | 0.005795955 | 0.011378838 | 1 |
| Obesity | neutrophil | *PSMG3-AS1* | 4.854739545 | 0.004750144 | 0.011289644 | 1 |
| Obesity | neutrophil | *PPP5D1* | 1.716798739 | 0.003445335 | 0.011289644 | 1 |
| Obesity | neutrophil | *AC112229.4* | -2.93122094 | 0.007503846 | 0.012398603 | 1 |
| Obesity | neutrophil | *KLHL7-AS1* | 4.983028793 | 0.004215218 | 0.011289644 | 1 |
| Obesity | neutrophil | *AC092171.4* | 2.271623558 | 0.005639585 | 0.011295339 | 1 |
| Obesity | neutrophil | *HLA-DPA1* | 1.534384206 | 0.002747228 | 0.011289644 | 1 |
| Obesity | neutrophil | *RPS18* | 2.768844127 | 0.000884902 | 0.011289644 | 1 |
| Obesity | neutrophil | *DIRC3* | 0.831875185 | 0.004986646 | 0.011289644 | 1 |
| Obesity | neutrophil | *TAPBP* | 3.731066051 | 0.002031529 | 0.011289644 | 1 |
| Obesity | neutrophil | *AC107021.1* | 1.504678595 | 0.008247075 | 0.013073465 | 1 |
| Obesity | neutrophil | *LINC00865* | 2.90570937 | 0.006796915 | 0.011870336 | 1 |
| Obesity | neutrophil | *AC009495.3* | -6.055959686 | 0.02136003 | 0.027552423 | 1 |
| Obesity | neutrophil | *HLA-DQB2* | 1.356615287 | 0.00552404 | 0.011295339 | 1 |
| Obesity | neutrophil | *GOLGA8N* | 2.447760471 | 0.001255104 | 0.011289644 | 1 |
| Obesity | neutrophil | *LINC00665* | 2.552894418 | 0.009168916 | 0.013973531 | 1 |
| Obesity | neutrophil | *LYRM9* | 4.927224356 | 0.01073443 | 0.015636552 | 1 |
| Obesity | neutrophil | *LINC00342* | -1.666989829 | 0.00720693 | 0.012170284 | 1 |
| Obesity | neutrophil | *SNHG15* | -1.602272376 | 0.015152773 | 0.020544637 | 1 |
| Obesity | neutrophil | *LINC00702* | 2.040436815 | 0.003692139 | 0.011289644 | 1 |
| Obesity | neutrophil | *AC073133.1* | -1.282005471 | 0.003939229 | 0.011289644 | 1 |
| Obesity | neutrophil | *RPS28* | 1.66399996 | 0.0041756 | 0.011289644 | 1 |
| Obesity | neutrophil | *ZNF736* | 3.204966447 | 0.002603134 | 0.011289644 | 1 |
| Obesity | neutrophil | *PINLYP* | 1.789326695 | 0.01169944 | 0.016656335 | 1 |
| Obesity | neutrophil | *HLA-B* | 3.46197584 | 0.003197087 | 0.011289644 | 1 |
| Obesity | neutrophil | *C19orf81* | 10.23299244 | 0.002879535 | 0.011289644 | 1 |
| Obesity | neutrophil | *ANKRD65* | 4.990171567 | 0.00227123 | 0.011289644 | 1 |
| Obesity | neutrophil | *LINC00957* | 5.820236365 | 0.001951305 | 0.011289644 | 1 |
| Obesity | neutrophil | *KIAA0040* | 1.688043291 | 0.003037974 | 0.011289644 | 1 |
| Obesity | neutrophil | *LINC00941* | 8.067724465 | 0.003321143 | 0.011289644 | 1 |
| Obesity | neutrophil | *ZBTB22* | 3.912009807 | 0.001030483 | 0.011289644 | 1 |
| Obesity | neutrophil | *B3GNT9* | 9.339573959 | 0.001769809 | 0.011289644 | 1 |
| Obesity | neutrophil | *GS1-124K5.4* | 2.233643418 | 0.000365542 | 0.011289644 | 1 |
| Obesity | neutrophil | *ZNF737* | 1.260416529 | 0.003977137 | 0.011289644 | 1 |
| Obesity | neutrophil | *LINC00959* | 1.997355324 | 0.001835777 | 0.011289644 | 1 |
| Obesity | neutrophil | *HLA-DQA2* | 1.369829982 | 0.039925311 | 0.048476328 | 1 |
| Obesity | neutrophil | *AC073257.2* | 21.46578897 | 0.005630524 | 0.011295339 | 1 |
| Obesity | neutrophil | *IRGM* | 2.985329377 | 0.011175788 | 0.016078302 | 1 |
| Obesity | neutrophil | *TXNDC5* | 5.52395959 | 0.011691927 | 0.016655838 | 1 |
| Obesity | neutrophil | *ASB14* | 3.051506138 | 0.00709664 | 0.01208139 | 1 |
| Obesity | neutrophil | *NME1* | 2.583573413 | 0.00138118 | 0.011289644 | 1 |

| | | | | | | |
|---|---|---|---|---|---|---|
| Obesity | neutrophil | *TNFSF12* | 2.070985518 | 0.003034741 | 0.011289644 | 1 |
| Obesity | neutrophil | *CDRT4* | 4.844870396 | 0.003016199 | 0.011289644 | 1 |
| Obesity | neutrophil | *APOBEC3G* | -1.380013809 | 0.035972869 | 0.044154906 | 1 |
| Obesity | neutrophil | *ADSL* | -1.992519601 | 0.00919291 | 0.014000927 | 1 |
| Obesity | neutrophil | *LILRA4* | 68.63799252 | 0.040791696 | 0.049413756 | 1 |
| Obesity | neutrophil | *LILRA2* | -1.313607589 | 0.011549216 | 0.016502986 | 1 |
| Obesity | neutrophil | *PSMB9* | 4.392393766 | 0.017984114 | 0.023743411 | 1 |
| Obesity | neutrophil | *COX19* | 1.572641789 | 0.004239559 | 0.011289644 | 1 |
| Obesity | neutrophil | *AQP1* | 1.761047824 | 0.005106477 | 0.011289644 | 1 |
| Obesity | neutrophil | *C1QTNF9* | 8.749644452 | 0.017574756 | 0.023276278 | 1 |
| Obesity | neutrophil | *ARHGEF25* | 3.153057492 | 0.004006558 | 0.011289644 | 1 |
| Obesity | neutrophil | *LINC00886* | 2.763633605 | 0.01460623 | 0.019923924 | 1 |
| Obesity | neutrophil | *NSUN6* | 1.929514299 | 0.004689422 | 0.011289644 | 1 |
| Obesity | neutrophil | *CD302* | 3.037622581 | 0.010493264 | 0.015381476 | 1 |
| Obesity | neutrophil | *ATP5J2* | 4.574066327 | 0.006797075 | 0.011870336 | 1 |
| Obesity | neutrophil | *ARHGAP8* | -0.665266917 | 0.005400545 | 0.011289644 | 1 |
| Obesity | neutrophil | *ATP5O* | 3.44333571 | 0.003098486 | 0.011289644 | 1 |
| Obesity | neutrophil | *PLEKHO2* | 1.954644078 | 0.003680387 | 0.011289644 | 1 |
| Obesity | neutrophil | *PWP2* | 8.179225521 | 0.014618151 | 0.019931537 | 1 |
| Obesity | neutrophil | *PI4KA* | 2.096755886 | 0.002892499 | 0.011289644 | 1 |
| Obesity | neutrophil | *LINC00969* | 1.881877398 | 0.015070791 | 0.020446712 | 1 |
| Obesity | neutrophil | *ARHGDIG* | 3.302602042 | 0.007072569 | 0.012069827 | 1 |
| Obesity | neutrophil | *TCP10L* | 20.56241842 | 0.002709162 | 0.011289644 | 1 |
| Obesity | neutrophil | *ARFGAP3* | 2.252964245 | 0.004878824 | 0.011289644 | 1 |
| Obesity | neutrophil | *C22orf39* | 2.333335078 | 0.003648618 | 0.011289644 | 1 |
| Obesity | neutrophil | *EIF6* | 2.098275959 | 0.013456199 | 0.018617765 | 1 |
| Obesity | neutrophil | *MRPL20* | 2.528920853 | 0.003338123 | 0.011289644 | 1 |
| Obesity | neutrophil | *DECR2* | 2.351477577 | 0.004287129 | 0.011289644 | 1 |
| Obesity | neutrophil | *GNG10* | -1.548918036 | 0.030540745 | 0.038044316 | 1 |
| Obesity | neutrophil | *AP5Z1* | 2.578347703 | 0.002875904 | 0.011289644 | 1 |
| Obesity | neutrophil | *ZNF709* | -3.266280621 | 0.003319905 | 0.011289644 | 1 |
| Obesity | neutrophil | *PSG4* | 2.807319334 | 0.026758603 | 0.033740741 | 1 |
| Obesity | neutrophil | *STON1* | -1.595816191 | 0.021321191 | 0.027519263 | 1 |
| Obesity | neutrophil | *VSIG8* | 3.261302932 | 0.005494879 | 0.011289644 | 1 |
| Obesity | neutrophil | *C7orf73* | 1.405369435 | 0.00425209 | 0.011289644 | 1 |
| Obesity | neutrophil | *FGF14-IT1* | 5.720612197 | 0.00434331 | 0.011289644 | 1 |
| Obesity | neutrophil | *TICAM2* | 7.240688369 | 0.003715599 | 0.011289644 | 1 |
| Obesity | neutrophil | *PALM2* | 1.210229368 | 0.009488936 | 0.014308604 | 1 |
| Obesity | neutrophil | *NAT6* | 3.422460946 | 0.000562 | 0.011289644 | 1 |
| Obesity | neutrophil | *WDR92* | 1.663630162 | 0.003767423 | 0.011289644 | 1 |
| Obesity | neutrophil | *PLA2G4B* | -2.011436283 | 0.000828817 | 0.011289644 | 1 |
| Obesity | neutrophil | *LEFTY1* | 17.43555585 | 0.03744042 | 0.045733033 | 1 |
| Obesity | neutrophil | *TTC4* | 1.874790521 | 0.004087786 | 0.011289644 | 1 |
| Obesity | neutrophil | *JMJD7* | -1.865095658 | 0.006181032 | 0.011514117 | 1 |
| Obesity | neutrophil | *RN7SL832P* | 4.066940087 | 0.000812024 | 0.011289644 | 1 |
| Obesity | neutrophil | *TMEM199* | 6.174639883 | 0.002438337 | 0.011289644 | 1 |
| Obesity | neutrophil | *DBNDD2* | -5.334628737 | 0.021803369 | 0.028072366 | 1 |
| Obesity | neutrophil | *APOBEC3C* | 2.415513195 | 0.003975262 | 0.011289644 | 1 |
| Obesity | neutrophil | *CCDC13* | 2.036905586 | 0.010898322 | 0.015816655 | 1 |
| Obesity | neutrophil | *ASPRV1* | 6.655853738 | 0.007931143 | 0.01278095 | 1 |

| | | | | | | |
|---|---|---|---|---|---|---|
| Obesity | neutrophil | *UBE2V1* | 1.94315579 | 0.004274993 | 0.011289644 | 1 |
| Obesity | neutrophil | *CRYBB2* | -1.143048942 | 0.008536803 | 0.013384259 | 1 |
| Obesity | neutrophil | *N4BP2L2* | 1.533777207 | 0.008425778 | 0.01327269 | 1 |
| Obesity | neutrophil | *LINC00847* | 1.95594156 | 0.003959445 | 0.011289644 | 1 |
| Obesity | neutrophil | *NEAT1* | 1.264167678 | 0.006045323 | 0.011447874 | 1 |
| Obesity | neutrophil | *ZNF585B* | 2.232152587 | 0.000678376 | 0.011289644 | 1 |
| Obesity | neutrophil | *LINC00920* | -1.818204538 | 0.003733978 | 0.011289644 | 1 |
| Obesity | neutrophil | *UBAP1L* | 1.744786072 | 0.007152614 | 0.012141606 | 1 |
| Obesity | neutrophil | *PGAM5* | 8.502359954 | 0.029691597 | 0.037089423 | 1 |
| Obesity | neutrophil | *DNAJC3-AS1* | -4.547051093 | 0.022692898 | 0.029039335 | 1 |
| Obesity | neutrophil | *LINC00926* | 2.185239218 | 0.003681734 | 0.011289644 | 1 |
| Obesity | neutrophil | *BCKDHA* | -2.624942278 | 0.004742902 | 0.011289644 | 1 |
| Obesity | neutrophil | *CDK11B* | 1.234949193 | 0.004223389 | 0.011289644 | 1 |
| Obesity | neutrophil | *PCDHAC1* | -26.12935042 | 2.92E-05 | 0.004961414 | 0.183572326 |
| Obesity | neutrophil | *ABHD14A* | 3.320037341 | 0.002734489 | 0.011289644 | 1 |
| Obesity | neutrophil | *SRP14-AS1* | 1.447142086 | 0.000349616 | 0.011289644 | 1 |
| Obesity | neutrophil | *C5orf17* | 1.355285249 | 0.004277582 | 0.011289644 | 1 |
| Obesity | neutrophil | *FMN1* | 0.974396182 | 0.007260188 | 0.012218025 | 1 |
| Obesity | neutrophil | *LINC01060* | 1.206509615 | 0.004619213 | 0.011289644 | 1 |
| Obesity | neutrophil | *ZNF286B* | 4.66042753 | 0.002857422 | 0.011289644 | 1 |
| Obesity | neutrophil | *LINC00942* | -1.207477993 | 0.00517028 | 0.011289644 | 1 |
| Obesity | neutrophil | *PDCD6* | 3.663914436 | 0.007729731 | 0.012606179 | 1 |
| Obesity | neutrophil | *LINC00603* | 2.632337614 | 0.005333056 | 0.011289644 | 1 |
| Obesity | neutrophil | *YJEFN3* | -2.094124141 | 0.005344936 | 0.011289644 | 1 |
| Obesity | neutrophil | *DNAH10OS* | -1.263549348 | 0.00674961 | 0.011836764 | 1 |
| Obesity | neutrophil | *PCDHA10* | 4.904175014 | 0.002263046 | 0.011289644 | 1 |
| Obesity | neutrophil | *ZNF718* | 2.013349159 | 0.004483831 | 0.011289644 | 1 |
| Obesity | neutrophil | *LINC00989* | 1.672100807 | 0.004981941 | 0.011289644 | 1 |
| Obesity | neutrophil | *LINC01021* | 4.265061512 | 0.004324908 | 0.011289644 | 1 |
| Obesity | neutrophil | *FAM218A* | 3.969048631 | 0.00435463 | 0.011289644 | 1 |
| Obesity | neutrophil | *ATP6V1E2* | 1.80074659 | 0.021163381 | 0.027343644 | 1 |
| Obesity | neutrophil | *SNHG18* | 1.760932651 | 0.023089875 | 0.029469355 | 1 |
| Obesity | neutrophil | *ZNF345* | 1.704717059 | 0.004502305 | 0.011289644 | 1 |
| Obesity | neutrophil | *SHANK3* | 1.058745839 | 0.008617889 | 0.013470632 | 1 |
| Obesity | neutrophil | *TMEM200B* | 2.239310673 | 0.001958958 | 0.011289644 | 1 |
| Obesity | neutrophil | *PCDHGB6* | 5.318159779 | 0.010506511 | 0.015397304 | 1 |
| Obesity | neutrophil | *TRNP1* | 23.00484769 | 0.006899074 | 0.011932455 | 1 |
| Obesity | neutrophil | *PCDHGA5* | 2.216159048 | 0.004514382 | 0.011289644 | 1 |
| Obesity | neutrophil | *ATXN7L3B* | 3.008449517 | 0.005210427 | 0.011289644 | 1 |
| Obesity | neutrophil | *PRKDC* | 2.342743915 | 0.00615286 | 0.011509382 | 1 |
| Obesity | neutrophil | *PCDHGA6* | 2.377095258 | 0.000725983 | 0.011289644 | 1 |
| Obesity | neutrophil | *ZNF260* | 1.367289945 | 0.003796173 | 0.011289644 | 1 |
| Obesity | neutrophil | *FPGT* | 3.383284984 | 0.004806863 | 0.011289644 | 1 |
| Obesity | neutrophil | *MPV17L2* | 4.88789325 | 6.76E-05 | 0.00802313 | 0.425225866 |
| Obesity | neutrophil | *EID3* | -1.243161486 | 0.020091707 | 0.026130857 | 1 |
| Obesity | neutrophil | *SNHG9* | 2.218847809 | 0.000754181 | 0.011289644 | 1 |
| Obesity | neutrophil | *EBLN2* | 2.017345967 | 0.00334925 | 0.011289644 | 1 |
| Obesity | neutrophil | *POLR2M* | 1.662461827 | 0.004225831 | 0.011289644 | 1 |
| Obesity | neutrophil | *TAS2R20* | -1.204175001 | 0.010398718 | 0.01527851 | 1 |
| Obesity | neutrophil | *LINC00346* | 6.606629563 | 0.002420019 | 0.011289644 | 1 |

| | | | | | | |
|---|---|---|---|---|---|---|
| Obesity | neutrophil | *CTSO* | 1.744584764 | 0.004234099 | 0.011289644 | 1 |
| Obesity | neutrophil | *ZNF432* | 1.611617263 | 0.004209887 | 0.011289644 | 1 |
| Obesity | neutrophil | *LINC00944* | -1.310315737 | 0.007565991 | 0.01243917 | 1 |
| Obesity | neutrophil | *ZNF10* | 1.609503426 | 0.013850797 | 0.019079746 | 1 |
| Obesity | neutrophil | *HMBS* | 4.024943618 | 1.45E-07 | 8.27E-05 | 0.000909284 |
| Obesity | neutrophil | *TAS2R31* | -2.268400191 | 0.007443219 | 0.012350362 | 1 |
| Obesity | neutrophil | *ZNF253* | 2.206341884 | 0.004854512 | 0.011289644 | 1 |
| Obesity | neutrophil | *KIAA1147* | 1.803843989 | 0.002594038 | 0.011289644 | 1 |
| Obesity | neutrophil | *ZBED6* | 4.518187017 | 0.004336426 | 0.011289644 | 1 |
| Obesity | neutrophil | *GALNT4* | -15.30241415 | 0.007775125 | 0.012646715 | 1 |
| Obesity | neutrophil | *OVCH1-AS1* | 2.777569573 | 0.004571191 | 0.011289644 | 1 |
| Obesity | neutrophil | *CUX1* | 1.949781719 | 0.006735971 | 0.011825602 | 1 |
| Obesity | neutrophil | *CHURC1* | 1.632256902 | 0.002413595 | 0.011289644 | 1 |
| Obesity | neutrophil | *C17orf49* | -1.452946403 | 0.006903527 | 0.011936874 | 1 |
| Obesity | neutrophil | *RTEL1* | 4.311221093 | 0.013196132 | 0.018367076 | 1 |
| Obesity | neutrophil | *ZNF578* | -2.767234965 | 0.010924463 | 0.015837604 | 1 |
| Obesity | neutrophil | *PDF* | -1.934308592 | 0.003904034 | 0.011289644 | 1 |
| Obesity | neutrophil | *LINC00640* | 1.988158116 | 0.012734968 | 0.017823852 | 1 |
| Obesity | neutrophil | *LINC00638* | -1.665688202 | 0.005154022 | 0.011289644 | 1 |
| Obesity | neutrophil | *CEP95* | 1.385951795 | 0.004074285 | 0.011289644 | 1 |
| Obesity | neutrophil | *LINC00639* | 1.168456276 | 0.003150199 | 0.011289644 | 1 |
| Obesity | neutrophil | *LINC00648* | 1.573324422 | 0.004048202 | 0.011289644 | 1 |
| Obesity | neutrophil | *LINC00924* | 2.361388209 | 0.005047888 | 0.011289644 | 1 |
| Obesity | neutrophil | *ITGB3* | -0.996300999 | 0.001838886 | 0.011289644 | 1 |
| Obesity | neutrophil | *UBE2Q2L* | 4.942266131 | 0.017046654 | 0.022657995 | 1 |
| Obesity | neutrophil | *IQCH-AS1* | 2.854540927 | 0.01266875 | 0.017746977 | 1 |
| Obesity | neutrophil | *AC083843.1* | -2.366776203 | 0.004134294 | 0.011289644 | 1 |
| Obesity | neutrophil | *PWRN1* | 2.134077677 | 0.003637111 | 0.011289644 | 1 |
| Obesity | neutrophil | *HOXB7* | 2.56664777 | 0.001225192 | 0.011289644 | 1 |
| Obesity | neutrophil | *FRRS1L* | 5.218580875 | 0.002627253 | 0.011289644 | 1 |
| Obesity | neutrophil | *AC009120.5* | -1.547787296 | 0.006283132 | 0.011565541 | 1 |
| Obesity | neutrophil | *LINC00565* | 29.925876 | 0.007264405 | 0.012218115 | 1 |
| Obesity | neutrophil | *TMEM178B* | 2.125877538 | 0.003805442 | 0.011289644 | 1 |
| Obesity | neutrophil | *LINC01003* | 1.864101935 | 0.004664895 | 0.011289644 | 1 |
| Obesity | neutrophil | *C15orf65* | -0.932106239 | 0.002969863 | 0.011289644 | 1 |
| Obesity | neutrophil | *HPR* | 4.815408018 | 0.00152634 | 0.011289644 | 1 |
| Obesity | neutrophil | *GOLGA8H* | -0.67168371 | 0.017352807 | 0.023011411 | 1 |
| Obesity | neutrophil | *LINC00662* | 1.161287337 | 0.00451994 | 0.011289644 | 1 |
| Obesity | neutrophil | *LINC00667* | 1.775960845 | 0.009422612 | 0.014240375 | 1 |
| Obesity | neutrophil | *LINC00672* | 2.370051777 | 0.00305205 | 0.011289644 | 1 |
| Obesity | neutrophil | *LINC00909* | 2.187343409 | 0.008992879 | 0.013816772 | 1 |
| Obesity | neutrophil | *DYNLL2* | 2.262073131 | 0.007533007 | 0.012412599 | 1 |
| Obesity | neutrophil | *LINC00526* | -6.779457282 | 0.034041183 | 0.041972331 | 1 |
| Obesity | neutrophil | *STRADA* | 1.49000988 | 0.015323229 | 0.020747037 | 1 |
| Obesity | neutrophil | *MYO15B* | 4.49025231 | 0.009608921 | 0.014445549 | 1 |
| Obesity | neutrophil | *LINC00663* | 1.164582672 | 0.024833084 | 0.031521421 | 1 |
| Obesity | neutrophil | *ILF3-AS1* | 1.827273754 | 0.006895354 | 0.011932455 | 1 |
| Obesity | neutrophil | *AC139100.2* | 8.503669839 | 0.004647882 | 0.011289644 | 1 |
| Obesity | neutrophil | *LINC01140* | -0.870327879 | 0.009278209 | 0.014096689 | 1 |
| Obesity | neutrophil | *LINC00906* | 52.62602903 | 0.025752181 | 0.032563147 | 1 |

| | | | | | | |
|---|---|---|---|---|---|---|
| Obesity | neutrophil | *ZNF582-AS1* | 4.220988513 | 0.005855609 | 0.011378838 | 1 |
| Obesity | neutrophil | *AC137932.1* | -2.264114749 | 0.003835319 | 0.011289644 | 1 |
| Obesity | neutrophil | *AC003005.2* | 4.971796552 | 0.012863075 | 0.01797514 | 1 |
| Obesity | neutrophil | *AL132989.1* | 4.566275101 | 0.004378131 | 0.011289644 | 1 |
| Obesity | neutrophil | *LINC00664* | 6.390136836 | 0.030902242 | 0.038461881 | 1 |
| Obesity | neutrophil | *FBXO17* | 1.194851047 | 0.004669939 | 0.011289644 | 1 |
| Obesity | neutrophil | *SNHG8* | 1.387229965 | 0.00511095 | 0.011289644 | 1 |
| Obesity | neutrophil | *YTHDF3-AS1* | 4.484043818 | 0.001617991 | 0.011289644 | 1 |
| Obesity | neutrophil | *LINC01050* | -5.62757861 | 0.005982699 | 0.011427806 | 1 |
| Obesity | neutrophil | *FGF14-AS2* | 3.15684336 | 0.00239442 | 0.011289644 | 1 |
| Obesity | neutrophil | *CASC15* | 1.259838648 | 0.003542228 | 0.011289644 | 1 |
| Obesity | neutrophil | *LINC01023* | -1.174383505 | 0.006233993 | 0.01154827 | 1 |
| Obesity | neutrophil | *AP000347.2* | 3.141261468 | 0.031987777 | 0.039627025 | 1 |
| Obesity | neutrophil | *DOC2B* | 1.556384226 | 0.022532121 | 0.028857088 | 1 |
| Obesity | neutrophil | *POLR2J4* | 3.701153698 | 0.004751683 | 0.011289644 | 1 |
| Obesity | neutrophil | *AC144652.1* | -2.266567056 | 0.027601233 | 0.034678048 | 1 |
| Obesity | neutrophil | *AP000350.5* | 1.824179719 | 0.03899647 | 0.047440171 | 1 |
| Obesity | neutrophil | *PAXIP1-AS1* | 1.779372893 | 0.003315497 | 0.011289644 | 1 |
| Obesity | nk_cell | *NIPAL3* | 2.807909691 | 0.000156195 | 0.00182528 | 0.982000834 |
| Obesity | nk_cell | *ENPP4* | 1.305551622 | 0.001216809 | 0.006675463 | 1 |
| Obesity | nk_cell | *SEMA3F* | -2.9150755 | 4.83E-24 | 6.08E-21 | 3.04E-20 |
| Obesity | nk_cell | *HECW1* | -0.590120845 | 0.000215911 | 0.00216071 | 1 |
| Obesity | nk_cell | *CYP26B1* | -1.051122615 | 0.007885987 | 0.024964352 | 1 |
| Obesity | nk_cell | *POLDIP2* | -1.161805479 | 0.016604206 | 0.043568715 | 1 |
| Obesity | nk_cell | *CD38* | 0.349356787 | 0.000293351 | 0.002579346 | 1 |
| Obesity | nk_cell | *RBM6* | -1.964269252 | 2.20E-21 | 1.98E-18 | 1.38E-17 |
| Obesity | nk_cell | *RECQL* | 2.694740924 | 0.000700476 | 0.004621087 | 1 |
| Obesity | nk_cell | *ARHGAP33* | 1.84938614 | 2.32E-06 | 0.000208434 | 0.014590365 |
| Obesity | nk_cell | *SLC22A16* | -1.826066273 | 5.22E-05 | 0.001143108 | 0.328072018 |
| Obesity | nk_cell | *MEOX1* | -1.016832632 | 0.008995288 | 0.02771453 | 1 |
| Obesity | nk_cell | *WDR54* | 1.247297759 | 7.82E-05 | 0.001307059 | 0.491454351 |
| Obesity | nk_cell | *PDK2* | -1.052438304 | 0.006294837 | 0.021220182 | 1 |
| Obesity | nk_cell | *GGNBP2* | -1.996188674 | 2.69E-05 | 0.000955271 | 0.169083018 |
| Obesity | nk_cell | *CCL18* | -0.63483663 | 0.005148597 | 0.018308386 | 1 |
| Obesity | nk_cell | *SYNRG* | 2.105076875 | 0.001130453 | 0.006311866 | 1 |
| Obesity | nk_cell | *TMEM132A* | -1.604085611 | 0.001911706 | 0.008955959 | 1 |
| Obesity | nk_cell | *ZNF263* | 2.644815035 | 0.004510045 | 0.016689025 | 1 |
| Obesity | nk_cell | *SPATA20* | 2.462846066 | 0.004885905 | 0.017607077 | 1 |
| Obesity | nk_cell | *DLX6* | 2.071283955 | 0.000288911 | 0.002572785 | 1 |
| Obesity | nk_cell | *BAIAP2L1* | -1.324838243 | 0.016022397 | 0.042395964 | 1 |
| Obesity | nk_cell | *ARHGAP44* | -0.855624736 | 0.009407034 | 0.028474733 | 1 |
| Obesity | nk_cell | *CEACAM21* | 0.242890304 | 0.000199751 | 0.002086106 | 1 |
| Obesity | nk_cell | *NOS2* | -4.251931389 | 0.001481893 | 0.007556092 | 1 |
| Obesity | nk_cell | *RPUSD1* | 3.693843509 | 3.67E-05 | 0.001056956 | 0.230622251 |
| Obesity | nk_cell | *BAIAP3* | 2.810341229 | 0.002758945 | 0.01154826 | 1 |
| Obesity | nk_cell | *TSR3* | 3.391834909 | 6.29E-05 | 0.001177776 | 0.395732821 |
| Obesity | nk_cell | *CDK11A* | -1.192528831 | 0.002712961 | 0.01143956 | 1 |
| Obesity | nk_cell | *NFIX* | -0.875672307 | 0.003043929 | 0.012483486 | 1 |
| Obesity | nk_cell | *MMP25* | 0.51354935 | 6.12E-05 | 0.001177776 | 0.385011658 |
| Obesity | nk_cell | *IL32* | 0.349427926 | 0.00036979 | 0.003025789 | 1 |

| | | | | | | |
|---|---|---|---|---|---|---|
| Obesity | nk_cell | *MAPK8IP2* | 2.502211326 | 1.84E-05 | 0.000788276 | 0.115955882 |
| Obesity | nk_cell | *RPS20* | 2.007789555 | 0.010311352 | 0.030564579 | 1 |
| Obesity | nk_cell | *STARD3NL* | 2.39919572 | 6.02E-06 | 0.00039412 | 0.03783555 |
| Obesity | nk_cell | *HFE* | -2.493861936 | 2.65E-09 | 7.93E-07 | 1.66E-05 |
| Obesity | nk_cell | *MRC2* | -0.849393057 | 0.01169037 | 0.033540239 | 1 |
| Obesity | nk_cell | *UTP18* | 2.651598045 | 0.00743774 | 0.023967746 | 1 |
| Obesity | nk_cell | *DPF1* | -2.22347263 | 0.00228771 | 0.010107402 | 1 |
| Obesity | nk_cell | *SYT7* | -1.235696138 | 0.006839341 | 0.022583476 | 1 |
| Obesity | nk_cell | *PPP5C* | 2.605523754 | 0.000150535 | 0.001809392 | 0.946413091 |
| Obesity | nk_cell | *TMEM159* | -0.943178012 | 0.009569651 | 0.028828173 | 1 |
| Obesity | nk_cell | *LTF* | 2.50319174 | 0.000144104 | 0.001755781 | 0.905982999 |
| Obesity | nk_cell | *PSMC4* | 2.897621936 | 1.62E-07 | 3.00E-05 | 0.001018795 |
| Obesity | nk_cell | *MAN2B2* | 2.413390998 | 0.009033853 | 0.027759449 | 1 |
| Obesity | nk_cell | *SLC25A39* | -1.344548125 | 0.005260845 | 0.018632362 | 1 |
| Obesity | nk_cell | *CCDC88C* | 0.382948446 | 0.00028043 | 0.002529499 | 1 |
| Obesity | nk_cell | *NPC1L1* | -1.72483908 | 0.003580677 | 0.014105087 | 1 |
| Obesity | nk_cell | *IL20RA* | -1.78621343 | 0.000722287 | 0.004678965 | 1 |
| Obesity | nk_cell | *GLT8D1* | -1.072831368 | 0.009207918 | 0.028115678 | 1 |
| Obesity | nk_cell | *OSBPL5* | 0.907445492 | 5.45E-05 | 0.00116249 | 0.34249401 |
| Obesity | nk_cell | *NLRP2* | 0.305998184 | 0.000225639 | 0.002220018 | 1 |
| Obesity | nk_cell | *GLRX2* | -1.330433721 | 0.007944274 | 0.025098316 | 1 |
| Obesity | nk_cell | *HSF2* | 2.50630308 | 0.015655705 | 0.041653583 | 1 |
| Obesity | nk_cell | *FAS* | 1.305868267 | 0.005517957 | 0.019261422 | 1 |
| Obesity | nk_cell | *BTN3A1* | 0.426686247 | 0.018061868 | 0.046455308 | 1 |
| Obesity | nk_cell | *ARNTL2* | -1.121497991 | 0.010708515 | 0.031452639 | 1 |
| Obesity | nk_cell | *ANK1* | -1.855503571 | 0.010535358 | 0.031038329 | 1 |
| Obesity | nk_cell | *RABEP1* | 2.347751197 | 2.61E-05 | 0.000948114 | 0.164201083 |
| Obesity | nk_cell | *BAK1* | 1.441966496 | 0.000181729 | 0.001987008 | 1 |
| Obesity | nk_cell | *SH3YL1* | -1.63319642 | 6.26E-05 | 0.001177776 | 0.393539978 |
| Obesity | nk_cell | *ADSS* | 2.530504948 | 0.001858765 | 0.008813011 | 1 |
| Obesity | nk_cell | *STAP1* | 0.812160381 | 0.009606239 | 0.028910686 | 1 |
| Obesity | nk_cell | *MYOM2* | 0.134807113 | 0.00029154 | 0.002579346 | 1 |
| Obesity | nk_cell | *TUBG2* | -0.647396835 | 0.017637495 | 0.045707721 | 1 |
| Obesity | nk_cell | *NSUN2* | 2.885706672 | 0.000316494 | 0.002696203 | 1 |
| Obesity | nk_cell | *MAT2B* | 3.398146802 | 3.47E-05 | 0.001039749 | 0.218347271 |
| Obesity | nk_cell | *DNAH5* | -1.009545796 | 0.012291671 | 0.034919897 | 1 |
| Obesity | nk_cell | *C6* | -0.75086545 | 0.002395227 | 0.010450237 | 1 |
| Obesity | nk_cell | *SPDL1* | 1.426323254 | 0.001245445 | 0.006767601 | 1 |
| Obesity | nk_cell | *PQLC2* | -1.238689379 | 0.001364281 | 0.007141964 | 1 |
| Obesity | nk_cell | *CTNS* | -0.635942393 | 0.018152267 | 0.046561934 | 1 |
| Obesity | nk_cell | *INPP4A* | 1.28689062 | 3.24E-05 | 0.001016497 | 0.203473829 |
| Obesity | nk_cell | *RAB27B* | 0.582632034 | 0.000177607 | 0.001972822 | 1 |
| Obesity | nk_cell | *PSMA4* | 2.264311776 | 0.000490905 | 0.003687363 | 1 |
| Obesity | nk_cell | *LSG1* | 0.901310986 | 0.01596425 | 0.042277692 | 1 |
| Obesity | nk_cell | *MED17* | 2.850639357 | 4.68E-05 | 0.001109982 | 0.294365503 |
| Obesity | nk_cell | *CAPG* | -0.944872167 | 0.006561973 | 0.021880442 | 1 |
| Obesity | nk_cell | *LCP2* | 0.873462593 | 0.00015287 | 0.001813379 | 0.961090703 |
| Obesity | nk_cell | *CUL7* | -1.165607289 | 0.00901416 | 0.027739611 | 1 |
| Obesity | nk_cell | *DSG2* | -1.036934606 | 0.009268878 | 0.028246938 | 1 |
| Obesity | nk_cell | *WDR37* | 1.893277499 | 0.000822759 | 0.005061335 | 1 |

| | | | | | | |
|---|---|---|---|---|---|---|
| Obesity | nk_cell | *CP* | 3.512182868 | 0.000801643 | 0.004960561 | 1 |
| Obesity | nk_cell | *DTNBP1* | 2.373415033 | 0.000204652 | 0.002119686 | 1 |
| Obesity | nk_cell | *TSPAN17* | 2.087457641 | 0.000304498 | 0.002622439 | 1 |
| Obesity | nk_cell | *LMO3* | -0.60873928 | 0.001713836 | 0.008346154 | 1 |
| Obesity | nk_cell | *CELF2* | 1.510641123 | 0.002128905 | 0.009629085 | 1 |
| Obesity | nk_cell | *UTS2* | -1.677892992 | 0.000168131 | 0.001904576 | 1 |
| Obesity | nk_cell | *EPN3* | 3.761584488 | 0.007965528 | 0.025140198 | 1 |
| Obesity | nk_cell | *RCN1* | -1.635709816 | 0.002131213 | 0.009632593 | 1 |
| Obesity | nk_cell | *ARHGEF5* | -1.799751604 | 0.002954341 | 0.012179634 | 1 |
| Obesity | nk_cell | *MCUR1* | 2.032787288 | 0.006941748 | 0.022803206 | 1 |
| Obesity | nk_cell | *PTGER3* | -0.563320763 | 0.006764826 | 0.022384453 | 1 |
| Obesity | nk_cell | *CYBA* | 0.589897679 | 0.000101597 | 0.001465003 | 0.638741437 |
| Obesity | nk_cell | *THOC3* | 3.803503512 | 0.000797789 | 0.004950654 | 1 |
| Obesity | nk_cell | *ALX4* | -1.398680042 | 0.000737575 | 0.004730741 | 1 |
| Obesity | nk_cell | *MRTO4* | -1.35648586 | 0.009340476 | 0.028400797 | 1 |
| Obesity | nk_cell | *ANAPC4* | 2.703104032 | 0.000591651 | 0.004179446 | 1 |
| Obesity | nk_cell | *LY75* | 0.623568185 | 0.000120764 | 0.001603969 | 0.759243436 |
| Obesity | nk_cell | *PLEKHH1* | -0.730563906 | 0.000825441 | 0.00507287 | 1 |
| Obesity | nk_cell | *ATP9A* | -0.849838179 | 0.004928713 | 0.017706755 | 1 |
| Obesity | nk_cell | *CHRDL2* | -1.600904561 | 0.004049261 | 0.015513531 | 1 |
| Obesity | nk_cell | *GALC* | 1.750336696 | 0.000577757 | 0.004109 | 1 |
| Obesity | nk_cell | *MRPL43* | -1.145269619 | 0.012710959 | 0.035867404 | 1 |
| Obesity | nk_cell | *ITIH4* | 1.34519822 | 3.71E-10 | 1.23E-07 | 2.33E-06 |
| Obesity | nk_cell | *TRAF1* | 1.456016412 | 9.50E-05 | 0.001425377 | 0.596979822 |
| Obesity | nk_cell | *IL17RB* | -2.175073042 | 0.007906053 | 0.025015278 | 1 |
| Obesity | nk_cell | *F7* | -8.047433774 | 0.018617876 | 0.047388901 | 1 |
| Obesity | nk_cell | *PITHD1* | 2.517299879 | 0.007342699 | 0.023795645 | 1 |
| Obesity | nk_cell | *ATP11B* | 1.851859187 | 1.71E-05 | 0.000750451 | 0.107314559 |
| Obesity | nk_cell | *ZC3H11A* | -1.579510223 | 0.015920829 | 0.042216049 | 1 |
| Obesity | nk_cell | *TNK2* | 1.323048032 | 0.005648215 | 0.019597312 | 1 |
| Obesity | nk_cell | *EIF4B* | 1.815980381 | 0.000639588 | 0.004418927 | 1 |
| Obesity | nk_cell | *ISOC2* | -1.161089688 | 0.001644198 | 0.008126629 | 1 |
| Obesity | nk_cell | *CASP8* | 0.764413191 | 0.000175601 | 0.00195745 | 1 |
| Obesity | nk_cell | *DMRT3* | -0.998088491 | 0.000902059 | 0.005385799 | 1 |
| Obesity | nk_cell | *ATP2C2* | -2.277796155 | 0.012688311 | 0.035820122 | 1 |
| Obesity | nk_cell | *CDON* | -0.549310203 | 0.008781981 | 0.027171414 | 1 |
| Obesity | nk_cell | *TAF2* | 1.54885643 | 0.018953586 | 0.048107064 | 1 |
| Obesity | nk_cell | *LPAR2* | 1.975268408 | 2.09E-06 | 0.000193269 | 0.013142258 |
| Obesity | nk_cell | *ABCA7* | 1.234005371 | 0.000160276 | 0.001852311 | 1 |
| Obesity | nk_cell | *BTBD1* | 2.171992451 | 0.007558554 | 0.024195839 | 1 |
| Obesity | nk_cell | *CHI3L2* | 1.211508134 | 0.000362283 | 0.002996942 | 1 |
| Obesity | nk_cell | *UHRF1BP1* | 2.518396337 | 5.48E-18 | 3.83E-15 | 3.45E-14 |
| Obesity | nk_cell | *ANKRD44* | 0.90065185 | 1.98E-05 | 0.000825847 | 0.124780725 |
| Obesity | nk_cell | *ASB1* | 2.115803153 | 0.002473238 | 0.010716228 | 1 |
| Obesity | nk_cell | *TIE1* | -0.975520999 | 0.016156915 | 0.042680053 | 1 |
| Obesity | nk_cell | *ZMYND12* | -0.835291336 | 0.001525609 | 0.007716413 | 1 |
| Obesity | nk_cell | *NGEF* | -2.696604615 | 0.009106774 | 0.027869914 | 1 |
| Obesity | nk_cell | *CD84* | -1.047901333 | 0.010621717 | 0.031248824 | 1 |
| Obesity | nk_cell | *EML1* | -0.715234324 | 0.001544921 | 0.007801543 | 1 |
| Obesity | nk_cell | *SP100* | 1.956187615 | 0.00025667 | 0.002380061 | 1 |

| | | | | | | |
|---|---|---|---|---|---|---|
| Obesity | nk_cell | *STOML1* | -1.879463514 | 0.005847596 | 0.020056648 | 1 |
| Obesity | nk_cell | *RASSF1* | 0.727070375 | 8.16E-05 | 0.001340853 | 0.512774498 |
| Obesity | nk_cell | *IFI35* | 1.77139293 | 0.003315379 | 0.013352843 | 1 |
| Obesity | nk_cell | *SRBD1* | 1.892765635 | 0.006498151 | 0.021753928 | 1 |
| Obesity | nk_cell | *PITX1* | -1.869579089 | 0.000733286 | 0.004709057 | 1 |
| Obesity | nk_cell | *SDK2* | -1.060635307 | 0.001927593 | 0.009006758 | 1 |
| Obesity | nk_cell | *KCNAB2* | 0.864595805 | 0.000326526 | 0.002755426 | 1 |
| Obesity | nk_cell | *GAL* | -2.244631951 | 0.001923274 | 0.008996743 | 1 |
| Obesity | nk_cell | *CLEC2D* | 0.799823309 | 0.000205186 | 0.002121722 | 1 |
| Obesity | nk_cell | *PIGB* | 1.900486851 | 0.009387504 | 0.02845359 | 1 |
| Obesity | nk_cell | *PFN2* | -1.396948953 | 0.011064395 | 0.032249352 | 1 |
| Obesity | nk_cell | *CLTCL1* | -0.662940129 | 0.012837274 | 0.036094786 | 1 |
| Obesity | nk_cell | *WIPI1* | 2.66297973 | 0.000292378 | 0.002579346 | 1 |
| Obesity | nk_cell | *NDST1* | -0.905567284 | 0.015675863 | 0.041689572 | 1 |
| Obesity | nk_cell | *ASNS* | -1.080692635 | 0.005443258 | 0.019033238 | 1 |
| Obesity | nk_cell | *AP3M2* | -1.157120664 | 0.011804028 | 0.033809532 | 1 |
| Obesity | nk_cell | *CAMK2A* | -3.367428628 | 0.000782194 | 0.004902943 | 1 |
| Obesity | nk_cell | *OSBPL3* | 1.29162878 | 3.01E-05 | 0.000991348 | 0.189347485 |
| Obesity | nk_cell | *TRIP13* | -2.14623711 | 0.00108504 | 0.006118067 | 1 |
| Obesity | nk_cell | *MBD3* | 2.589979187 | 0.001520186 | 0.007695177 | 1 |
| Obesity | nk_cell | *HLTF* | -1.25439111 | 0.009069577 | 0.027822276 | 1 |
| Obesity | nk_cell | *AFF4* | 1.56234733 | 0.009887823 | 0.029567433 | 1 |
| Obesity | nk_cell | *TRHDE* | -0.608680076 | 0.005417228 | 0.018984456 | 1 |
| Obesity | nk_cell | *NFATC3* | 1.263680323 | 0.000428444 | 0.003325466 | 1 |
| Obesity | nk_cell | *TRNT1* | 2.413186532 | 0.003137393 | 0.012758596 | 1 |
| Obesity | nk_cell | *CRMP1* | -0.907266191 | 0.002807034 | 0.011666744 | 1 |
| Obesity | nk_cell | *DERL2* | 2.631547032 | 0.002322212 | 0.010231075 | 1 |
| Obesity | nk_cell | *NDE1* | 2.43505357 | 0.001226055 | 0.006708198 | 1 |
| Obesity | nk_cell | *XRCC1* | 2.920799887 | 0.005763371 | 0.019865305 | 1 |
| Obesity | nk_cell | *TP63* | -1.140500391 | 0.012522696 | 0.035441515 | 1 |
| Obesity | nk_cell | *LLGL2* | 0.319929124 | 0.000138978 | 0.001735535 | 0.873753069 |
| Obesity | nk_cell | *GSDMB* | -1.070256166 | 0.011396728 | 0.03300379 | 1 |
| Obesity | nk_cell | *GLI2* | -0.690413816 | 0.005011377 | 0.017921801 | 1 |
| Obesity | nk_cell | *ATP2A3* | 0.664773257 | 6.29E-05 | 0.001177776 | 0.395395056 |
| Obesity | nk_cell | *CA12* | -0.542293326 | 0.002412163 | 0.010491891 | 1 |
| Obesity | nk_cell | *NTN4* | -0.806295296 | 0.004660248 | 0.017093919 | 1 |
| Obesity | nk_cell | *ENO1* | 2.342850598 | 0.007623307 | 0.024353521 | 1 |
| Obesity | nk_cell | *TIPIN* | 0.5472338 | 0.009777869 | 0.029328941 | 1 |
| Obesity | nk_cell | *EIF4G3* | 1.738447065 | 0.001345901 | 0.007104684 | 1 |
| Obesity | nk_cell | *TTC38* | 0.871661247 | 8.93E-05 | 0.001379496 | 0.56145506 |
| Obesity | nk_cell | *ADD2* | -1.348055484 | 0.001391806 | 0.007231203 | 1 |
| Obesity | nk_cell | *VPS9D1* | 2.942406203 | 0.002013942 | 0.009255595 | 1 |
| Obesity | nk_cell | *TUBA3D* | -2.00309059 | 0.004127381 | 0.015726573 | 1 |
| Obesity | nk_cell | *MCM6* | 2.740752313 | 0.002541256 | 0.010958077 | 1 |
| Obesity | nk_cell | *FMO4* | -1.03470136 | 0.013012336 | 0.036407902 | 1 |
| Obesity | nk_cell | *RGS11* | -0.620244455 | 0.005842524 | 0.020050191 | 1 |
| Obesity | nk_cell | *TPD52* | 0.747913959 | 0.000298035 | 0.002597629 | 1 |
| Obesity | nk_cell | *ICAM3* | 0.397838991 | 0.000121752 | 0.001608102 | 0.765456589 |
| Obesity | nk_cell | *CAMSAP3* | -4.544720889 | 0.00064203 | 0.004421078 | 1 |
| Obesity | nk_cell | *XAB2* | -2.222724879 | 0.00025653 | 0.002380061 | 1 |

| | | | | | | |
|---|---|---|---|---|---|---|
| Obesity | nk_cell | *MAP2K7* | 2.942863726 | 8.23E-05 | 0.001340853 | 0.517475876 |
| Obesity | nk_cell | *NMRK2* | 3.112667976 | 0.000210341 | 0.002150263 | 1 |
| Obesity | nk_cell | *APBB1IP* | 1.552406736 | 0.000397655 | 0.00314899 | 1 |
| Obesity | nk_cell | *CAPZB* | 1.642638051 | 0.011896643 | 0.033981915 | 1 |
| Obesity | nk_cell | *FAP* | -0.599529452 | 0.002887666 | 0.011951781 | 1 |
| Obesity | nk_cell | *N4BP2* | 1.687206926 | 0.008195232 | 0.025684658 | 1 |
| Obesity | nk_cell | *TULP3* | -0.946841834 | 0.015262596 | 0.040954307 | 1 |
| Obesity | nk_cell | *SYNJ2* | 1.920137404 | 0.015297639 | 0.041030827 | 1 |
| Obesity | nk_cell | *FGF20* | -6.180160247 | 0.000668516 | 0.004508971 | 1 |
| Obesity | nk_cell | *MYH7B* | -1.878321201 | 0.000161618 | 0.001860978 | 1 |
| Obesity | nk_cell | *FKBP7* | -0.685033028 | 0.015397615 | 0.041158506 | 1 |
| Obesity | nk_cell | *OSBPL6* | -1.154141956 | 0.016678256 | 0.043696927 | 1 |
| Obesity | nk_cell | *LXN* | -1.090265345 | 0.018301864 | 0.046773911 | 1 |
| Obesity | nk_cell | *REXO1* | 3.107024626 | 0.003562087 | 0.01405828 | 1 |
| Obesity | nk_cell | *CDC14A* | 1.0173533 | 0.017578532 | 0.045573703 | 1 |
| Obesity | nk_cell | *RIMS1* | -1.182046727 | 0.001707555 | 0.008334938 | 1 |
| Obesity | nk_cell | *DCT* | -1.138336456 | 0.00129147 | 0.006897913 | 1 |
| Obesity | nk_cell | *SCTR* | -1.157000905 | 0.006537821 | 0.021840213 | 1 |
| Obesity | nk_cell | *KCNN2* | -3.657751516 | 0.016143981 | 0.042663811 | 1 |
| Obesity | nk_cell | *MOK* | -1.039416226 | 0.016930309 | 0.044258151 | 1 |
| Obesity | nk_cell | *PKP1* | -1.05641667 | 0.004488295 | 0.016637918 | 1 |
| Obesity | nk_cell | *STK17B* | 2.102932383 | 0.000182831 | 0.001992126 | 1 |
| Obesity | nk_cell | *PCDHA6* | -9.911455383 | 0.000428395 | 0.003325466 | 1 |
| Obesity | nk_cell | *HSPB11* | 2.459591739 | 0.000871362 | 0.005271688 | 1 |
| Obesity | nk_cell | *PGR* | -0.516598792 | 0.007019994 | 0.023010795 | 1 |
| Obesity | nk_cell | *DOPEY1* | 0.945714899 | 0.013591971 | 0.037611233 | 1 |
| Obesity | nk_cell | *LYRM2* | 1.239817226 | 0.015315009 | 0.041059897 | 1 |
| Obesity | nk_cell | *PLOD1* | 3.778466551 | 0.000102715 | 0.001477728 | 0.645767223 |
| Obesity | nk_cell | *EPYC* | -7.243291073 | 0.000143974 | 0.001755781 | 0.905166589 |
| Obesity | nk_cell | *CYLD* | 2.037572755 | 0.00020692 | 0.002132632 | 1 |
| Obesity | nk_cell | *ZNF446* | -1.334974089 | 0.002653471 | 0.011279494 | 1 |
| Obesity | nk_cell | *RPS5* | 2.142995213 | 4.27E-05 | 0.001082521 | 0.268642684 |
| Obesity | nk_cell | *PPIE* | 2.879541433 | 3.61E-06 | 0.000272306 | 0.022674009 |
| Obesity | nk_cell | *NOA1* | -1.131028378 | 0.00073892 | 0.004730741 | 1 |
| Obesity | nk_cell | *GSTP1* | 2.667369215 | 0.004774493 | 0.017340982 | 1 |
| Obesity | nk_cell | *COL16A1* | -1.500961103 | 3.14E-06 | 0.000259505 | 0.01972237 |
| Obesity | nk_cell | *APOB* | -0.843324831 | 0.000330221 | 0.002779249 | 1 |
| Obesity | nk_cell | *NCOA1* | 1.512938467 | 0.00788175 | 0.024963506 | 1 |
| Obesity | nk_cell | *TAF9* | -1.36744967 | 0.006142706 | 0.020870709 | 1 |
| Obesity | nk_cell | *FCN1* | -0.617927483 | 0.000391326 | 0.003128541 | 1 |
| Obesity | nk_cell | *ABCB1* | 0.783678683 | 5.68E-05 | 0.001177776 | 0.35732723 |
| Obesity | nk_cell | *CPNE3* | -0.917461012 | 0.011843766 | 0.033907905 | 1 |
| Obesity | nk_cell | *CTTN* | -0.805815053 | 0.01050463 | 0.030996363 | 1 |
| Obesity | nk_cell | *ATG16L1* | 2.021799469 | 4.94E-05 | 0.001117657 | 0.310708549 |
| Obesity | nk_cell | *TMED2* | 2.957840292 | 8.83E-05 | 0.001377186 | 0.555005842 |
| Obesity | nk_cell | *HSD17B14* | -0.830743055 | 0.000818063 | 0.005042313 | 1 |
| Obesity | nk_cell | *TRIP6* | -1.490815111 | 0.002302284 | 0.010157517 | 1 |
| Obesity | nk_cell | *ACHE* | -1.04907116 | 0.000517839 | 0.003825682 | 1 |
| Obesity | nk_cell | *ATXN7L3* | 1.819208529 | 2.02E-05 | 0.000825847 | 0.127130101 |
| Obesity | nk_cell | *OGFOD1* | -2.309267116 | 3.99E-05 | 0.001082521 | 0.250650289 |

| | | | | | | |
|---|---|---|---|---|---|---|
| Obesity | nk_cell | *PTHLH* | -0.729805105 | 0.009302794 | 0.028322841 | 1 |
| Obesity | nk_cell | *PHACTR3* | -0.736345801 | 0.000333621 | 0.002792913 | 1 |
| Obesity | nk_cell | *GP6* | 1.13533286 | 0.000212892 | 0.00215532 | 1 |
| Obesity | nk_cell | *ASAP3* | -0.638112784 | 0.003990767 | 0.015354844 | 1 |
| Obesity | nk_cell | *CPXM1* | -0.933255165 | 3.37E-05 | 0.001029448 | 0.212066227 |
| Obesity | nk_cell | *LZTS3* | -1.039838597 | 0.000534397 | 0.003891554 | 1 |
| Obesity | nk_cell | *XRN2* | 2.100809765 | 0.003819027 | 0.014830279 | 1 |
| Obesity | nk_cell | *TESC* | 1.315283088 | 6.41E-05 | 0.001185978 | 0.403232418 |
| Obesity | nk_cell | *RBBP9* | -1.343526175 | 0.000942917 | 0.005545483 | 1 |
| Obesity | nk_cell | *SLC23A2* | 2.146092414 | 0.004784126 | 0.017355174 | 1 |
| Obesity | nk_cell | *DZANK1* | -1.612265499 | 0.007284006 | 0.023690917 | 1 |
| Obesity | nk_cell | *FUS* | 2.045137809 | 0.000741369 | 0.004741595 | 1 |
| Obesity | nk_cell | *FXYD5* | 1.092782133 | 4.09E-05 | 0.001082521 | 0.256971371 |
| Obesity | nk_cell | *FXYD3* | -2.197957744 | 0.014723622 | 0.039902773 | 1 |
| Obesity | nk_cell | *KCNH4* | -1.252399906 | 0.006669472 | 0.022127162 | 1 |
| Obesity | nk_cell | *GMIP* | 1.867919955 | 4.13E-05 | 0.001082521 | 0.259917224 |
| Obesity | nk_cell | *ANKRD24* | -0.716750862 | 0.004975215 | 0.017833053 | 1 |
| Obesity | nk_cell | *SLC9A1* | 2.896878329 | 0.005450193 | 0.019046893 | 1 |
| Obesity | nk_cell | *PAPOLA* | 1.691877609 | 0.014266886 | 0.038964341 | 1 |
| Obesity | nk_cell | *RGS1* | -1.187521775 | 0.001181124 | 0.006545408 | 1 |
| Obesity | nk_cell | *YPEL3* | 1.324207138 | 5.26E-05 | 0.001147343 | 0.330434914 |
| Obesity | nk_cell | *MRPS33* | -1.215061264 | 0.007687871 | 0.024472733 | 1 |
| Obesity | nk_cell | *ZNF268* | 2.554713986 | 0.012600006 | 0.035618811 | 1 |
| Obesity | nk_cell | *PABPC4* | -2.755486102 | 3.21E-06 | 0.000261852 | 0.020162638 |
| Obesity | nk_cell | *CERS4* | 1.850275184 | 0.000240745 | 0.002297638 | 1 |
| Obesity | nk_cell | *NLRC4* | -1.100747408 | 0.003993188 | 0.015354844 | 1 |
| Obesity | nk_cell | *SLC26A4* | -1.259076191 | 0.000599416 | 0.004224806 | 1 |
| Obesity | nk_cell | *IL5RA* | 5.911202606 | 0.000110207 | 0.001516917 | 0.692870761 |
| Obesity | nk_cell | *RAPGEF4* | -0.873059197 | 0.003971988 | 0.015310782 | 1 |
| Obesity | nk_cell | *SEL1L3* | 1.831340096 | 1.91E-05 | 0.000805343 | 0.11999606 |
| Obesity | nk_cell | *TF* | -1.243678104 | 0.001945898 | 0.00905541 | 1 |
| Obesity | nk_cell | *MYO15A* | -1.917630647 | 0.003274355 | 0.013209789 | 1 |
| Obesity | nk_cell | *ZFHX4* | -0.626659002 | 0.002740483 | 0.011505493 | 1 |
| Obesity | nk_cell | *CMA1* | -2.445562547 | 0.000271264 | 0.002457401 | 1 |
| Obesity | nk_cell | *PPP2R3C* | 2.538412053 | 7.79E-05 | 0.001306706 | 0.490014675 |
| Obesity | nk_cell | *SLC22A17* | -1.315864788 | 0.000621898 | 0.004339482 | 1 |
| Obesity | nk_cell | *RPGRIP1* | -1.374632718 | 0.006424882 | 0.021543058 | 1 |
| Obesity | nk_cell | *RFFL* | 3.733542822 | 0.004044439 | 0.015513531 | 1 |
| Obesity | nk_cell | *VNN3* | -2.651034298 | 0.002241466 | 0.009953339 | 1 |
| Obesity | nk_cell | *SUCO* | 2.217193912 | 0.003277759 | 0.013209789 | 1 |
| Obesity | nk_cell | *MSH2* | 2.654418047 | 5.91E-05 | 0.001177776 | 0.371599453 |
| Obesity | nk_cell | *WAC* | 1.512799884 | 0.012901188 | 0.036177418 | 1 |
| Obesity | nk_cell | *IL12RB1* | 0.358078574 | 0.000384779 | 0.003101414 | 1 |
| Obesity | nk_cell | *PCSK5* | -0.651662792 | 0.018056519 | 0.046455308 | 1 |
| Obesity | nk_cell | *PRTFDC1* | 3.127583303 | 0.000476953 | 0.003601102 | 1 |
| Obesity | nk_cell | *PALMD* | -0.501027991 | 0.003210852 | 0.012998472 | 1 |
| Obesity | nk_cell | *TSPAN15* | -0.795235157 | 0.009785188 | 0.029336899 | 1 |
| Obesity | nk_cell | *OCEL1* | -1.505571968 | 0.000251502 | 0.00235297 | 1 |
| Obesity | nk_cell | *CATSPERG* | 0.86043482 | 0.017910864 | 0.0462446 | 1 |
| Obesity | nk_cell | *STX1B* | -3.515996269 | 1.35E-06 | 0.00014943 | 0.008517491 |

| | | | | | | |
|---|---|---|---|---|---|---|
| Obesity | nk_cell | *HSD3B7* | -0.784369447 | 0.010973844 | 0.032000259 | 1 |
| Obesity | nk_cell | *MADCAM1* | -0.948525849 | 0.002650649 | 0.011279494 | 1 |
| Obesity | nk_cell | *MED15* | 1.756309627 | 0.012031027 | 0.034272347 | 1 |
| Obesity | nk_cell | *SERPIND1* | -1.509034087 | 0.001103896 | 0.006191074 | 1 |
| Obesity | nk_cell | *SMARCB1* | 1.595304759 | 0.003097708 | 0.012638085 | 1 |
| Obesity | nk_cell | *P2RX6* | -1.95213474 | 0.001313476 | 0.006974511 | 1 |
| Obesity | nk_cell | *DDT* | 3.013385764 | 0.007952263 | 0.025110937 | 1 |
| Obesity | nk_cell | *YPEL1* | 0.88138026 | 6.16E-05 | 0.001177776 | 0.38702572 |
| Obesity | nk_cell | *CYTH4* | 1.562943685 | 4.07E-05 | 0.001082521 | 0.255638377 |
| Obesity | nk_cell | *SH3BP1* | 0.512339997 | 0.000145771 | 0.001770291 | 0.916459381 |
| Obesity | nk_cell | *TRIOBP* | -1.223034666 | 0.006774856 | 0.02240585 | 1 |
| Obesity | nk_cell | *ANKRD54* | -1.825770257 | 3.41E-06 | 0.00026837 | 0.021416813 |
| Obesity | nk_cell | *EIF3L* | 1.968690463 | 0.01091231 | 0.031902117 | 1 |
| Obesity | nk_cell | *CCDC134* | 3.136801401 | 8.24E-06 | 0.000470803 | 0.05182308 |
| Obesity | nk_cell | *KDELR3* | -1.117155443 | 0.003861842 | 0.014950369 | 1 |
| Obesity | nk_cell | *CYP2D6* | -1.433861652 | 0.001050185 | 0.005980539 | 1 |
| Obesity | nk_cell | *RHBDD3* | -0.981859537 | 0.015560618 | 0.041523602 | 1 |
| Obesity | nk_cell | *RASL10A* | -1.077603775 | 0.008417666 | 0.026263955 | 1 |
| Obesity | nk_cell | *BIK* | 7.627548794 | 0.000240838 | 0.002297638 | 1 |
| Obesity | nk_cell | *MCAT* | -1.128536277 | 0.005795682 | 0.019935754 | 1 |
| Obesity | nk_cell | *TTLL12* | -1.64969937 | 0.002562673 | 0.011027434 | 1 |
| Obesity | nk_cell | *KCTD17* | -1.337468847 | 0.000705089 | 0.004623717 | 1 |
| Obesity | nk_cell | *POLR3H* | -0.938957872 | 0.000949242 | 0.005567056 | 1 |
| Obesity | nk_cell | *DESI1* | 1.246561638 | 0.002891711 | 0.011960649 | 1 |
| Obesity | nk_cell | *PYGL* | -0.485234762 | 0.017516529 | 0.045469207 | 1 |
| Obesity | nk_cell | *MTHFD1* | -0.844358015 | 0.014308567 | 0.039029559 | 1 |
| Obesity | nk_cell | *VRK1* | 2.84217715 | 0.00053038 | 0.003872819 | 1 |
| Obesity | nk_cell | *PCK2* | 2.248053904 | 0.01420826 | 0.03887177 | 1 |
| Obesity | nk_cell | *KIAA0391* | 2.83172341 | 0.000149604 | 0.001805299 | 0.940560947 |
| Obesity | nk_cell | *ABHD12* | 2.213372926 | 0.000303197 | 0.002622007 | 1 |
| Obesity | nk_cell | *CD40* | -1.09620326 | 0.002876095 | 0.011919586 | 1 |
| Obesity | nk_cell | *DOK5* | -0.791777182 | 0.002387039 | 0.010429592 | 1 |
| Obesity | nk_cell | *BMP7* | -1.494334025 | 0.005564683 | 0.019360907 | 1 |
| Obesity | nk_cell | *NELFCD* | 2.567153222 | 0.003549888 | 0.014018935 | 1 |
| Obesity | nk_cell | *TCFL5* | -0.984590483 | 0.008163826 | 0.025635297 | 1 |
| Obesity | nk_cell | *DIDO1* | 2.115184005 | 0.000963472 | 0.005629508 | 1 |
| Obesity | nk_cell | *NKAIN4* | -1.968785458 | 0.000250998 | 0.00235175 | 1 |
| Obesity | nk_cell | *PTK6* | -4.30668501 | 0.004443375 | 0.01652988 | 1 |
| Obesity | nk_cell | *GMEB2* | 2.446891405 | 6.28E-05 | 0.001177776 | 0.394779024 |
| Obesity | nk_cell | *CDC25B* | 1.805751294 | 8.26E-05 | 0.001340853 | 0.519532599 |
| Obesity | nk_cell | *ANGPT4* | -0.976468631 | 0.000142382 | 0.001748346 | 0.895152954 |
| Obesity | nk_cell | *RSPO4* | 1.880768764 | 0.002244446 | 0.009958244 | 1 |
| Obesity | nk_cell | *SEC23B* | 1.211277263 | 0.00623845 | 0.021109329 | 1 |
| Obesity | nk_cell | *CCM2L* | -1.446989169 | 0.014689731 | 0.039842252 | 1 |
| Obesity | nk_cell | *MROH8* | -1.125805669 | 0.005609678 | 0.019485108 | 1 |
| Obesity | nk_cell | *MAPRE1* | 3.298604701 | 0.000387585 | 0.003116046 | 1 |
| Obesity | nk_cell | *TTI1* | 3.019150919 | 0.000183227 | 0.001992987 | 1 |
| Obesity | nk_cell | *BPI* | -1.527032835 | 0.003833679 | 0.014877988 | 1 |
| Obesity | nk_cell | *PIGU* | 2.112892533 | 0.005392189 | 0.018907247 | 1 |
| Obesity | nk_cell | *APMAP* | 2.127983936 | 3.74E-06 | 0.00027661 | 0.023511867 |

| | | | | | | |
|---|---|---|---|---|---|---|
| Obesity | nk_cell | *CELF4* | -1.5571111 | 0.011654185 | 0.033502451 | 1 |
| Obesity | nk_cell | *RBFA* | -1.097784854 | 0.003673965 | 0.014337813 | 1 |
| Obesity | nk_cell | *SMCHD1* | 1.8824761 | 0.000933079 | 0.005492758 | 1 |
| Obesity | nk_cell | *MYL12A* | 1.271723113 | 6.19E-05 | 0.001177776 | 0.389350676 |
| Obesity | nk_cell | *HTR2A* | -1.58894684 | 0.000788056 | 0.004923569 | 1 |
| Obesity | nk_cell | *UGGT2* | -0.962927036 | 0.007535209 | 0.024145698 | 1 |
| Obesity | nk_cell | *SGCG* | -0.898593888 | 0.004173511 | 0.015844725 | 1 |
| Obesity | nk_cell | *DHRS12* | -0.839371009 | 0.005607482 | 0.019485108 | 1 |
| Obesity | nk_cell | *MEDAG* | -0.647798992 | 0.001759255 | 0.008514578 | 1 |
| Obesity | nk_cell | *CORO1A* | 0.229873756 | 0.001917227 | 0.008975136 | 1 |
| Obesity | nk_cell | *CTCF* | 2.396685873 | 0.000342971 | 0.002852192 | 1 |
| Obesity | nk_cell | *ACD* | -1.786991861 | 0.000501953 | 0.003743261 | 1 |
| Obesity | nk_cell | *USB1* | 2.248537063 | 1.46E-05 | 0.000718208 | 0.091822037 |
| Obesity | nk_cell | *COG4* | -1.854627877 | 0.004264032 | 0.016071923 | 1 |
| Obesity | nk_cell | *NME4* | -1.369599577 | 0.001415713 | 0.007337664 | 1 |
| Obesity | nk_cell | *CLCN7* | 2.597637388 | 0.006865102 | 0.022644183 | 1 |
| Obesity | nk_cell | *HAGHL* | 3.414957893 | 0.007493481 | 0.024073334 | 1 |
| Obesity | nk_cell | *SLC7A5* | 1.612864058 | 0.000121184 | 0.001603969 | 0.761885226 |
| Obesity | nk_cell | *RHBDL1* | -1.611676108 | 9.43E-05 | 0.001424919 | 0.592766309 |
| Obesity | nk_cell | *CLUAP1* | 0.945803503 | 0.016362433 | 0.043132335 | 1 |
| Obesity | nk_cell | *HMOX2* | 0.966023063 | 0.000683953 | 0.004555095 | 1 |
| Obesity | nk_cell | *RRN3P2* | 1.983414887 | 0.000471189 | 0.003577736 | 1 |
| Obesity | nk_cell | *RBL2* | 1.289877162 | 0.000873193 | 0.005271726 | 1 |
| Obesity | nk_cell | *SYT17* | -1.120455417 | 0.001883822 | 0.00888624 | 1 |
| Obesity | nk_cell | *TMC5* | -1.080701639 | 0.000506603 | 0.003755909 | 1 |
| Obesity | nk_cell | *RNF40* | -1.690000084 | 0.001765029 | 0.008522838 | 1 |
| Obesity | nk_cell | *KNOP1* | -1.518635886 | 0.000156886 | 0.001829953 | 0.986344423 |
| Obesity | nk_cell | *CSK* | 1.306829083 | 5.45E-05 | 0.00116249 | 0.342495802 |
| Obesity | nk_cell | *TRIP4* | 1.51601747 | 0.005564369 | 0.019360907 | 1 |
| Obesity | nk_cell | *RASL12* | -0.892934077 | 0.00070924 | 0.004630316 | 1 |
| Obesity | nk_cell | *FAH* | -0.944404961 | 0.004684233 | 0.017126618 | 1 |
| Obesity | nk_cell | *ATP8B4* | 1.307906504 | 4.02E-05 | 0.001082521 | 0.252626713 |
| Obesity | nk_cell | *PDGFRL* | -0.798099672 | 0.010732763 | 0.031501812 | 1 |
| Obesity | nk_cell | *IKBKB* | 2.474936472 | 0.000365638 | 0.003008859 | 1 |
| Obesity | nk_cell | *PLAT* | -0.875113566 | 0.00699311 | 0.0229466 | 1 |
| Obesity | nk_cell | *SPAG1* | 1.424622532 | 0.004214387 | 0.015942149 | 1 |
| Obesity | nk_cell | *ERI1* | 2.36605731 | 0.008771824 | 0.027153352 | 1 |
| Obesity | nk_cell | *LEPROTL1* | 1.957191383 | 3.27E-05 | 0.001016497 | 0.205332388 |
| Obesity | nk_cell | *KLHDC4* | 1.186905431 | 2.53E-05 | 0.000934443 | 0.158988296 |
| Obesity | nk_cell | *MCM4* | 2.394286352 | 0.002217274 | 0.009880646 | 1 |
| Obesity | nk_cell | *TULP2* | -1.250738835 | 0.003545508 | 0.014010438 | 1 |
| Obesity | nk_cell | *NFKBIB* | 3.895709894 | 0.002102751 | 0.009531359 | 1 |
| Obesity | nk_cell | *TUBB4A* | -7.697015693 | 0.000290509 | 0.002577201 | 1 |
| Obesity | nk_cell | *ERCC2* | 3.736729023 | 0.014902742 | 0.04026366 | 1 |
| Obesity | nk_cell | *CD37* | 1.072204516 | 4.22E-05 | 0.001082521 | 0.265380688 |
| Obesity | nk_cell | *AMH* | 1.674011732 | 0.000365292 | 0.003008859 | 1 |
| Obesity | nk_cell | *DKKL1* | -1.319277035 | 0.01386705 | 0.038104084 | 1 |
| Obesity | nk_cell | *FCER2* | -1.190078596 | 0.000923949 | 0.005460274 | 1 |
| Obesity | nk_cell | *DMPK* | -1.101072165 | 0.009650478 | 0.02902993 | 1 |
| Obesity | nk_cell | *TLE6* | -1.59437669 | 0.011427698 | 0.033063018 | 1 |

| | | | | | | |
|---|---|---|---|---|---|---|
| Obesity | nk_cell | *AES* | 2.520363483 | 4.61E-05 | 0.001109982 | 0.289782723 |
| Obesity | nk_cell | *C19orf53* | 2.147235581 | 4.89E-05 | 0.001116189 | 0.307681478 |
| Obesity | nk_cell | *IL27RA* | 0.760961525 | 3.44E-05 | 0.001033598 | 0.216022078 |
| Obesity | nk_cell | *RASAL3* | 0.238832145 | 0.000139682 | 0.001735535 | 0.87818054 |
| Obesity | nk_cell | *SYDE1* | -1.975089369 | 0.000593681 | 0.004189081 | 1 |
| Obesity | nk_cell | *AURKC* | 1.027657681 | 0.00068488 | 0.004556445 | 1 |
| Obesity | nk_cell | *CCNE1* | -3.432525995 | 0.000680063 | 0.004552412 | 1 |
| Obesity | nk_cell | *RPS16* | 1.466954207 | 0.000403465 | 0.003178675 | 1 |
| Obesity | nk_cell | *TBCB* | 2.324425703 | 0.001072988 | 0.006088333 | 1 |
| Obesity | nk_cell | *CLIP3* | 1.332930597 | 0.0001283 | 0.001670927 | 0.80662254 |
| Obesity | nk_cell | *PRKD2* | 1.122366819 | 0.00090201 | 0.005385799 | 1 |
| Obesity | nk_cell | *CACTIN* | 2.153271568 | 5.15E-05 | 0.001138987 | 0.323739955 |
| Obesity | nk_cell | *FZR1* | 2.57192514 | 1.80E-05 | 0.000781588 | 0.113330277 |
| Obesity | nk_cell | *BBC3* | -1.318183571 | 0.002113954 | 0.009568344 | 1 |
| Obesity | nk_cell | *TGFB1* | 1.741143976 | 0.000140694 | 0.001737559 | 0.8845434 |
| Obesity | nk_cell | *CEACAM4* | -2.728808415 | 0.000581999 | 0.004129831 | 1 |
| Obesity | nk_cell | *ICAM5* | -6.750563968 | 6.63E-06 | 0.000415115 | 0.041706547 |
| Obesity | nk_cell | *BABAM1* | 4.060756713 | 6.87E-05 | 0.001217457 | 0.432197363 |
| Obesity | nk_cell | *TYK2* | 2.794253914 | 0.000184394 | 0.001995324 | 1 |
| Obesity | nk_cell | *GRWD1* | -1.155739479 | 5.45E-05 | 0.00116249 | 0.34293446 |
| Obesity | nk_cell | *GRIN2D* | -1.609821391 | 0.003052734 | 0.012503282 | 1 |
| Obesity | nk_cell | *CARD8* | 1.564673154 | 0.011648933 | 0.033502451 | 1 |
| Obesity | nk_cell | *LIG1* | 1.819564927 | 0.000106539 | 0.001499525 | 0.669807689 |
| Obesity | nk_cell | *ZNF175* | -1.3575802 | 0.000194214 | 0.002052137 | 1 |
| Obesity | nk_cell | *PPP2R1A* | 2.713861664 | 5.97E-05 | 0.001177776 | 0.375279246 |
| Obesity | nk_cell | *TNPO2* | -1.166420668 | 0.011620115 | 0.033450396 | 1 |
| Obesity | nk_cell | *MAST1* | -1.639815718 | 0.000761958 | 0.004814503 | 1 |
| Obesity | nk_cell | *RPL18A* | 1.200277633 | 0.000518866 | 0.003826032 | 1 |
| Obesity | nk_cell | *PDE4C* | -1.261964858 | 0.001391975 | 0.007231203 | 1 |
| Obesity | nk_cell | *UPK1A* | -5.83056731 | 0.000706759 | 0.004623717 | 1 |
| Obesity | nk_cell | *COPE* | 1.942320645 | 9.98E-05 | 0.001447345 | 0.627192554 |
| Obesity | nk_cell | *LSR* | -1.017392876 | 0.009546422 | 0.028784049 | 1 |
| Obesity | nk_cell | *HPN* | 2.093949834 | 0.002204008 | 0.009841332 | 1 |
| Obesity | nk_cell | *ATP13A1* | -2.411949153 | 6.72E-05 | 0.001203893 | 0.422566308 |
| Obesity | nk_cell | *PON3* | -1.450170727 | 0.007531355 | 0.024145654 | 1 |
| Obesity | nk_cell | *PTN* | -0.762845543 | 0.000895028 | 0.005369192 | 1 |
| Obesity | nk_cell | *ADAP1* | 0.817259034 | 0.00015498 | 0.001823463 | 0.974357835 |
| Obesity | nk_cell | *MET* | -0.816745883 | 0.005003784 | 0.017904832 | 1 |
| Obesity | nk_cell | *HOXA2* | -2.225998976 | 7.47E-05 | 0.001276215 | 0.469516173 |
| Obesity | nk_cell | *HOXA5* | -0.832765173 | 0.002097718 | 0.009524895 | 1 |
| Obesity | nk_cell | *HOXA6* | -0.738443755 | 0.00056551 | 0.0040533 | 1 |
| Obesity | nk_cell | *BRAT1* | -1.314173707 | 0.003443435 | 0.013727886 | 1 |
| Obesity | nk_cell | *CRHR2* | 6.072366764 | 0.003146915 | 0.012789045 | 1 |
| Obesity | nk_cell | *CYP3A5* | -0.975586256 | 0.005863585 | 0.020078628 | 1 |
| Obesity | nk_cell | *TFR2* | 0.558448482 | 0.000232831 | 0.002269474 | 1 |
| Obesity | nk_cell | *C1GALT1* | 2.29937037 | 0.000370583 | 0.003025789 | 1 |
| Obesity | nk_cell | *MEST* | -0.821524582 | 1.52E-05 | 0.000718997 | 0.095667832 |
| Obesity | nk_cell | *ACTR3C* | 1.221257371 | 0.000235676 | 0.002286562 | 1 |
| Obesity | nk_cell | *POU6F2* | -0.878362035 | 0.002170679 | 0.009753811 | 1 |
| Obesity | nk_cell | *POLD2* | -1.119999351 | 0.005161033 | 0.018342236 | 1 |

| | | | | | | |
|---|---|---|---|---|---|---|
| Obesity | nk_cell | *GCK* | -7.739727358 | 0.000629239 | 0.004376136 | 1 |
| Obesity | nk_cell | *SPATA6L* | -1.559426261 | 0.003755542 | 0.01460179 | 1 |
| Obesity | nk_cell | *TRIM14* | 1.528191717 | 3.43E-05 | 0.001033598 | 0.215882402 |
| Obesity | nk_cell | *SUSD1* | 1.127816264 | 8.29E-05 | 0.001340853 | 0.52112632 |
| Obesity | nk_cell | *PLGRKT* | -1.062199573 | 0.014135292 | 0.038705827 | 1 |
| Obesity | nk_cell | *KDM4C* | 1.794695875 | 0.003632639 | 0.014226271 | 1 |
| Obesity | nk_cell | *DOCK8* | 1.127804465 | 0.005807072 | 0.019950307 | 1 |
| Obesity | nk_cell | *ELAVL2* | -1.411290894 | 0.00011437 | 0.001553214 | 0.71904699 |
| Obesity | nk_cell | *MPDZ* | -0.718704925 | 0.008829342 | 0.027291088 | 1 |
| Obesity | nk_cell | *PTGDS* | 1.298080729 | 7.51E-05 | 0.001276215 | 0.472199372 |
| Obesity | nk_cell | *SHB* | -1.772231329 | 0.001666005 | 0.008215037 | 1 |
| Obesity | nk_cell | *DVL1* | -1.393733039 | 0.007015648 | 0.023008545 | 1 |
| Obesity | nk_cell | *PLEKHA1* | 2.049514738 | 0.001291958 | 0.006897913 | 1 |
| Obesity | nk_cell | *SPOCK2* | 0.505714709 | 0.000115512 | 0.001558555 | 0.726221447 |
| Obesity | nk_cell | *CCSER2* | 2.016980943 | 0.002378245 | 0.010412275 | 1 |
| Obesity | nk_cell | *ACBD5* | 2.485714356 | 9.09E-07 | 0.000114881 | 0.005712033 |
| Obesity | nk_cell | *MAP3K8* | 1.780814099 | 0.013433561 | 0.037271315 | 1 |
| Obesity | nk_cell | *RPL28* | 1.01539585 | 3.23E-05 | 0.001016497 | 0.202834419 |
| Obesity | nk_cell | *DHRS11* | -0.859678271 | 0.000590795 | 0.004178096 | 1 |
| Obesity | nk_cell | *MLLT6* | 1.905483148 | 4.47E-05 | 0.001108799 | 0.280830483 |
| Obesity | nk_cell | *PSMB3* | 2.679952929 | 0.001003041 | 0.005796063 | 1 |
| Obesity | nk_cell | *RAPGEFL1* | -1.156921566 | 0.001129195 | 0.00631182 | 1 |
| Obesity | nk_cell | *RGS9* | 0.418181003 | 0.000157413 | 0.001832696 | 0.98965581 |
| Obesity | nk_cell | *WNT3* | -1.188690019 | 0.00157495 | 0.007908215 | 1 |
| Obesity | nk_cell | *ASPA* | -0.667208183 | 0.002428465 | 0.010539698 | 1 |
| Obesity | nk_cell | *RAD51C* | -1.30006903 | 5.21E-05 | 0.001143108 | 0.327785836 |
| Obesity | nk_cell | *SEPT4* | -1.001142117 | 6.18E-05 | 0.001177776 | 0.388256403 |
| Obesity | nk_cell | *TRIM37* | 2.089051807 | 0.009107895 | 0.027869914 | 1 |
| Obesity | nk_cell | *TRIM16L* | -1.049950507 | 0.001740387 | 0.008436248 | 1 |
| Obesity | nk_cell | *RNF167* | 2.712489046 | 8.53E-05 | 0.001360449 | 0.536115383 |
| Obesity | nk_cell | *DRG2* | 2.990957978 | 0.01513177 | 0.040672698 | 1 |
| Obesity | nk_cell | *FTSJ3* | -0.647201179 | 0.005266376 | 0.018632362 | 1 |
| Obesity | nk_cell | *ICAM2* | 1.317845794 | 6.04E-05 | 0.001177776 | 0.379973294 |
| Obesity | nk_cell | *C17orf75* | 3.293147101 | 0.000182536 | 0.001992126 | 1 |
| Obesity | nk_cell | *CYTH1* | 1.456682286 | 6.98E-05 | 0.001225599 | 0.438956957 |
| Obesity | nk_cell | *DLX4* | -7.190939432 | 0.000669976 | 0.004508971 | 1 |
| Obesity | nk_cell | *LRRC59* | -1.502013524 | 0.013149833 | 0.036645833 | 1 |
| Obesity | nk_cell | *ALOX12* | -1.779407977 | 0.002136348 | 0.009635023 | 1 |
| Obesity | nk_cell | *MPP2* | -2.295961947 | 0.013691201 | 0.037819234 | 1 |
| Obesity | nk_cell | *DPH1* | -1.596949357 | 0.014099697 | 0.038625183 | 1 |
| Obesity | nk_cell | *RCVRN* | -3.22199506 | 0.000683477 | 0.004555095 | 1 |
| Obesity | nk_cell | *SLC9A3R1* | 0.340595902 | 0.000163948 | 0.001873074 | 1 |
| Obesity | nk_cell | *MYH3* | -1.50835289 | 0.0085178 | 0.02649748 | 1 |
| Obesity | nk_cell | *TMEM104* | 2.353941063 | 0.000103523 | 0.001482565 | 0.650846008 |
| Obesity | nk_cell | *TMEM97* | 1.587547596 | 0.005387219 | 0.018900362 | 1 |
| Obesity | nk_cell | *SLAIN2* | 1.655219893 | 0.013742476 | 0.03784448 | 1 |
| Obesity | nk_cell | *CWH43* | -9.930780653 | 0.000761214 | 0.004814503 | 1 |
| Obesity | nk_cell | *AREG* | 0.709754836 | 0.01931317 | 0.048822638 | 1 |
| Obesity | nk_cell | *NDUFC1* | 2.797285685 | 0.007971188 | 0.02514544 | 1 |
| Obesity | nk_cell | *GRPEL1* | 4.644980315 | 0.000391627 | 0.003128541 | 1 |

| | | | | | | |
|---|---|---|---|---|---|---|
| Obesity | nk_cell | *AADAT* | -1.324467987 | 0.004745432 | 0.01728536 | 1 |
| Obesity | nk_cell | *CPZ* | -14.22441166 | 0.000700459 | 0.004621087 | 1 |
| Obesity | nk_cell | *TRIM2* | -0.880793949 | 0.007667027 | 0.024432855 | 1 |
| Obesity | nk_cell | *SH3D19* | -0.521613658 | 0.018008021 | 0.046411851 | 1 |
| Obesity | nk_cell | *STIM2* | 2.268928548 | 1.86E-05 | 0.000788276 | 0.116664792 |
| Obesity | nk_cell | *FAM149A* | -0.568424144 | 0.015333047 | 0.041073229 | 1 |
| Obesity | nk_cell | *UGDH* | -1.068735371 | 0.016997623 | 0.044378005 | 1 |
| Obesity | nk_cell | *CTSC* | 1.288465317 | 0.001567316 | 0.007889286 | 1 |
| Obesity | nk_cell | *SNX15* | -0.924067901 | 0.00766712 | 0.024432855 | 1 |
| Obesity | nk_cell | *DTX4* | -0.980508064 | 0.001948506 | 0.009060839 | 1 |
| Obesity | nk_cell | *PUS3* | -1.308974529 | 0.014949759 | 0.040338685 | 1 |
| Obesity | nk_cell | *DCPS* | 1.653796322 | 0.000663782 | 0.004504249 | 1 |
| Obesity | nk_cell | *NRXN2* | -1.958351677 | 0.002628997 | 0.01122098 | 1 |
| Obesity | nk_cell | *CCDC86* | -1.719658186 | 0.000730198 | 0.004698059 | 1 |
| Obesity | nk_cell | *KIAA1549L* | -0.96573933 | 0.002652823 | 0.011279494 | 1 |
| Obesity | nk_cell | *CD5* | 1.113653699 | 1.85E-05 | 0.000788276 | 0.116030126 |
| Obesity | nk_cell | *MADD* | 3.301728922 | 5.67E-05 | 0.001177776 | 0.356506116 |
| Obesity | nk_cell | *NAA40* | 2.731446733 | 0.002218424 | 0.009880646 | 1 |
| Obesity | nk_cell | *SLC22A18* | 0.900335746 | 0.000870929 | 0.005271688 | 1 |
| Obesity | nk_cell | *SLC35F2* | -1.166602365 | 0.006388608 | 0.021455757 | 1 |
| Obesity | nk_cell | *C11orf21* | 0.239423324 | 0.000408541 | 0.003214643 | 1 |
| Obesity | nk_cell | *ELMOD1* | 2.871891482 | 0.000370492 | 0.003025789 | 1 |
| Obesity | nk_cell | *RPS13* | 1.586954953 | 0.011930381 | 0.034062809 | 1 |
| Obesity | nk_cell | *AIP* | 1.72069458 | 5.11E-05 | 0.001135455 | 0.321333832 |
| Obesity | nk_cell | *TCIRG1* | 2.942526996 | 0.000975865 | 0.005680798 | 1 |
| Obesity | nk_cell | *PPFIBP1* | -0.857544058 | 0.016637779 | 0.043638598 | 1 |
| Obesity | nk_cell | *ASIC1* | -1.819760516 | 1.40E-05 | 0.000703079 | 0.08788493 |
| Obesity | nk_cell | *KCTD10* | -1.33197806 | 0.00271586 | 0.011444108 | 1 |
| Obesity | nk_cell | *CYP27B1* | -5.465995053 | 0.001386976 | 0.007224458 | 1 |
| Obesity | nk_cell | *GLI1* | -2.011604895 | 0.001963744 | 0.009094983 | 1 |
| Obesity | nk_cell | *TRPV4* | -1.73054299 | 1.55E-05 | 0.000721314 | 0.097605844 |
| Obesity | nk_cell | *ARPC3* | 2.576660603 | 0.001039535 | 0.005942712 | 1 |
| Obesity | nk_cell | *KCNA1* | -3.340695822 | 0.000116926 | 0.001569507 | 0.73511594 |
| Obesity | nk_cell | *SCNN1A* | 2.218941554 | 0.000263239 | 0.00241603 | 1 |
| Obesity | nk_cell | *RASAL1* | -3.602424014 | 0.000143826 | 0.001755781 | 0.904233776 |
| Obesity | nk_cell | *ARHGDIB* | 0.616101127 | 0.000135908 | 0.001719853 | 0.854453886 |
| Obesity | nk_cell | *GNPTAB* | 1.23879621 | 2.67E-05 | 0.000952182 | 0.167583987 |
| Obesity | nk_cell | *C12orf57* | 2.02663854 | 0.004862659 | 0.01753961 | 1 |
| Obesity | nk_cell | *NT5DC3* | -0.939039733 | 0.011148315 | 0.032388842 | 1 |
| Obesity | nk_cell | *ST8SIA1* | -0.892257748 | 0.005796494 | 0.019935754 | 1 |
| Obesity | nk_cell | *PHC1* | -1.507991675 | 0.011601818 | 0.033413024 | 1 |
| Obesity | nk_cell | *COL12A1* | -0.645454056 | 0.004672033 | 0.017126618 | 1 |
| Obesity | nk_cell | *BTN3A3* | 1.217573961 | 3.08E-05 | 0.000998128 | 0.193636842 |
| Obesity | nk_cell | *TDP2* | 2.348171996 | 0.006180265 | 0.020946269 | 1 |
| Obesity | nk_cell | *RNGTT* | 2.199130011 | 0.000663485 | 0.004504249 | 1 |
| Obesity | nk_cell | *UST* | 1.44794372 | 0.009920972 | 0.02963095 | 1 |
| Obesity | nk_cell | *MTRF1L* | 1.694620907 | 0.002571055 | 0.011041134 | 1 |
| Obesity | nk_cell | *TULP1* | 4.931804844 | 8.86E-05 | 0.001377463 | 0.556740578 |
| Obesity | nk_cell | *SLC26A8* | -1.328849049 | 2.56E-05 | 0.000935035 | 0.160826058 |
| Obesity | nk_cell | *SAYSD1* | -1.413352379 | 0.006247027 | 0.021117351 | 1 |

| | | | | | | |
|---|---|---|---|---|---|---|
| Obesity | nk_cell | *CAP2* | -0.884099513 | 0.000101496 | 0.001465003 | 0.638102416 |
| Obesity | nk_cell | *ASCC3* | 1.842164064 | 0.003606193 | 0.014161232 | 1 |
| Obesity | nk_cell | *VNN1* | -0.620696999 | 0.009889633 | 0.029567433 | 1 |
| Obesity | nk_cell | *VNN2* | 0.644017504 | 8.59E-05 | 0.001360449 | 0.540359353 |
| Obesity | nk_cell | *TFEB* | 2.429566385 | 0.010811951 | 0.031689853 | 1 |
| Obesity | nk_cell | *PRPH2* | -1.218403723 | 0.000539679 | 0.003902762 | 1 |
| Obesity | nk_cell | *TTK* | -1.444262088 | 0.006895038 | 0.02268399 | 1 |
| Obesity | nk_cell | *FBRSL1* | 2.249862328 | 0.01287531 | 0.036137085 | 1 |
| Obesity | nk_cell | *LY86* | -1.229397549 | 0.017718911 | 0.045824266 | 1 |
| Obesity | nk_cell | *PRSS16* | 5.096476619 | 4.75E-07 | 6.94E-05 | 0.00298337 |
| Obesity | nk_cell | *TBX18* | -0.894894662 | 0.001060734 | 0.006032101 | 1 |
| Obesity | nk_cell | *PCDHB2* | -0.924284758 | 0.000494815 | 0.003703453 | 1 |
| Obesity | nk_cell | *NUDT12* | -0.896136028 | 0.00095593 | 0.005590635 | 1 |
| Obesity | nk_cell | *DAP* | 2.148031236 | 0.012918377 | 0.036209466 | 1 |
| Obesity | nk_cell | *LOX* | -1.060543232 | 0.014382847 | 0.039161957 | 1 |
| Obesity | nk_cell | *APBB3* | -1.134800927 | 0.001195967 | 0.006595651 | 1 |
| Obesity | nk_cell | *PDE8B* | -1.11268465 | 0.003085317 | 0.012612085 | 1 |
| Obesity | nk_cell | *BTNL8* | -0.87345164 | 0.002564981 | 0.011027434 | 1 |
| Obesity | nk_cell | *RASGRF2* | -0.683719178 | 0.014881768 | 0.040224279 | 1 |
| Obesity | nk_cell | *POLR3G* | -0.99333952 | 0.002173404 | 0.009753811 | 1 |
| Obesity | nk_cell | *LNPEP* | 2.035242684 | 0.000239484 | 0.002295174 | 1 |
| Obesity | nk_cell | *SLC12A7* | 2.354288716 | 0.009422941 | 0.028509158 | 1 |
| Obesity | nk_cell | *IL4* | -2.157481932 | 0.012646979 | 0.035719478 | 1 |
| Obesity | nk_cell | *BNIP1* | -1.025123815 | 0.00063712 | 0.004416286 | 1 |
| Obesity | nk_cell | *CNTN3* | -0.665362468 | 0.002066479 | 0.009428125 | 1 |
| Obesity | nk_cell | *ACTR8* | 2.324146144 | 9.50E-05 | 0.001425377 | 0.597232854 |
| Obesity | nk_cell | *HGD* | -2.650917268 | 0.008359726 | 0.026122067 | 1 |
| Obesity | nk_cell | *RBP1* | 1.219380571 | 8.61E-05 | 0.001360449 | 0.541458743 |
| Obesity | nk_cell | *BCHE* | -1.024601253 | 8.60E-05 | 0.001360449 | 0.540457446 |
| Obesity | nk_cell | *PDCD10* | 1.963238634 | 0.014169403 | 0.03878234 | 1 |
| Obesity | nk_cell | *WNT5A* | -1.0686782 | 0.000377734 | 0.003052459 | 1 |
| Obesity | nk_cell | *PRKAR2A* | 2.098159022 | 5.03E-05 | 0.001126299 | 0.316489944 |
| Obesity | nk_cell | *HYAL1* | -1.520694478 | 2.74E-05 | 0.000956877 | 0.17219342 |
| Obesity | nk_cell | *CYB561D2* | 1.295741833 | 6.12E-05 | 0.001177776 | 0.384815448 |
| Obesity | nk_cell | *UMPS* | -1.246066936 | 0.007914238 | 0.02502858 | 1 |
| Obesity | nk_cell | *SLC41A3* | -1.388387449 | 0.010245308 | 0.030446473 | 1 |
| Obesity | nk_cell | *EFCC1* | -0.869629557 | 9.24E-05 | 0.001409361 | 0.580656634 |
| Obesity | nk_cell | *GORASP1* | -1.176559492 | 0.004987767 | 0.017857684 | 1 |
| Obesity | nk_cell | *MOB1A* | 2.530942959 | 0.004054341 | 0.01551395 | 1 |
| Obesity | nk_cell | *LMAN2L* | -0.909637122 | 0.014934657 | 0.040315237 | 1 |
| Obesity | nk_cell | *KCNIP3* | -1.025643392 | 0.001986019 | 0.00916075 | 1 |
| Obesity | nk_cell | *FAHD2A* | 2.883335629 | 0.002154825 | 0.009711387 | 1 |
| Obesity | nk_cell | *NCL* | 2.334881192 | 0.000857445 | 0.005217206 | 1 |
| Obesity | nk_cell | *ACTR1B* | 3.770648676 | 0.00017399 | 0.001949864 | 1 |
| Obesity | nk_cell | *ACTR3* | 2.046504944 | 8.32E-05 | 0.001340853 | 0.522922648 |
| Obesity | nk_cell | *TFCP2L1* | 3.853875048 | 0.000109098 | 0.001510797 | 0.685901836 |
| Obesity | nk_cell | *ITGB6* | -1.514391927 | 0.000292833 | 0.002579346 | 1 |
| Obesity | nk_cell | *ITGA4* | 0.558577152 | 7.10E-05 | 0.001229074 | 0.446153917 |
| Obesity | nk_cell | *PCSK4* | -0.883367846 | 0.010741591 | 0.031513012 | 1 |
| Obesity | nk_cell | *GRB14* | -0.670497324 | 0.001191195 | 0.006587533 | 1 |

| | | | | | | |
|---|---|---|---|---|---|---|
| Obesity | nk_cell | *SPTBN1* | -0.623707212 | 0.017955922 | 0.046341905 | 1 |
| Obesity | nk_cell | *GALNT3* | 0.497276613 | 0.000250411 | 0.002349751 | 1 |
| Obesity | nk_cell | *GNLY* | 0.092927002 | 0.000211833 | 0.00215532 | 1 |
| Obesity | nk_cell | *PLCD4* | -1.648884925 | 0.002056997 | 0.009405339 | 1 |
| Obesity | nk_cell | *CHMP3* | -2.246477293 | 2.58E-06 | 0.000225402 | 0.01622891 |
| Obesity | nk_cell | *WNT6* | -4.078814798 | 0.015553667 | 0.041522677 | 1 |
| Obesity | nk_cell | *MLPH* | -1.151548183 | 0.004088219 | 0.015605729 | 1 |
| Obesity | nk_cell | *TPO* | -0.770603491 | 0.00268301 | 0.011374296 | 1 |
| Obesity | nk_cell | *WIPF1* | 1.74484519 | 0.000152368 | 0.001810852 | 0.957940627 |
| Obesity | nk_cell | *COX7A2L* | 2.802356713 | 0.001352062 | 0.007121874 | 1 |
| Obesity | nk_cell | *GRIN3B* | 1.499922189 | 0.013976171 | 0.03835364 | 1 |
| Obesity | nk_cell | *VAX2* | -1.107179758 | 0.001173237 | 0.006516026 | 1 |
| Obesity | nk_cell | *DHCR24* | 2.332790996 | 0.000725623 | 0.004678965 | 1 |
| Obesity | nk_cell | *MORN1* | -1.472959668 | 0.003605394 | 0.014161232 | 1 |
| Obesity | nk_cell | *GPX7* | 0.58106982 | 0.00254373 | 0.010961225 | 1 |
| Obesity | nk_cell | *RALGPS2* | -1.031604924 | 0.018702887 | 0.047547533 | 1 |
| Obesity | nk_cell | *ICMT* | 2.686909991 | 0.000108379 | 0.001507478 | 0.68138004 |
| Obesity | nk_cell | *QSOX1* | 2.493196155 | 0.007426665 | 0.02394433 | 1 |
| Obesity | nk_cell | *PARK7* | 2.17154222 | 0.006696599 | 0.022205443 | 1 |
| Obesity | nk_cell | *RAP1A* | 1.85173109 | 0.001889417 | 0.008900845 | 1 |
| Obesity | nk_cell | *FBXO6* | 1.114862312 | 0.000618573 | 0.004321077 | 1 |
| Obesity | nk_cell | *C1orf21* | 1.367689365 | 9.23E-06 | 0.00050446 | 0.05801291 |
| Obesity | nk_cell | *DNAJC6* | -1.752096684 | 0.015776565 | 0.04192192 | 1 |
| Obesity | nk_cell | *MIIP* | 2.338724825 | 0.015635098 | 0.041618385 | 1 |
| Obesity | nk_cell | *NCF2* | -1.143746659 | 0.011850099 | 0.033910593 | 1 |
| Obesity | nk_cell | *RGS2* | -1.592686468 | 0.003847081 | 0.014911589 | 1 |
| Obesity | nk_cell | *OLFML3* | -0.668335928 | 0.002203802 | 0.009841332 | 1 |
| Obesity | nk_cell | *CFHR3* | -1.070817502 | 0.004273691 | 0.016080797 | 1 |
| Obesity | nk_cell | *TMEM9* | -0.948389525 | 0.003050251 | 0.012501258 | 1 |
| Obesity | nk_cell | *OSCP1* | -1.292912578 | 0.009849823 | 0.029488495 | 1 |
| Obesity | nk_cell | *C1orf109* | 3.928357457 | 7.05E-05 | 0.001228518 | 0.443533837 |
| Obesity | nk_cell | *MFAP2* | -1.531427767 | 0.000178782 | 0.001975395 | 1 |
| Obesity | nk_cell | *PLA2G2D* | -9.919661766 | 0.00147156 | 0.007525264 | 1 |
| Obesity | nk_cell | *KIF17* | 1.361155598 | 0.00374536 | 0.014571211 | 1 |
| Obesity | nk_cell | *NSUN4* | -1.382925089 | 0.018208297 | 0.046650516 | 1 |
| Obesity | nk_cell | *DR1* | 2.483201541 | 0.000174569 | 0.001952874 | 1 |
| Obesity | nk_cell | *CNN3* | -0.829975427 | 0.006304767 | 0.021230889 | 1 |
| Obesity | nk_cell | *PRRC2C* | 2.164399243 | 0.000281585 | 0.002536279 | 1 |
| Obesity | nk_cell | *TNFSF4* | -2.057524573 | 1.99E-05 | 0.000825847 | 0.125086332 |
| Obesity | nk_cell | *DARS2* | -2.177664731 | 0.014737433 | 0.039902773 | 1 |
| Obesity | nk_cell | *DIEXF* | -1.703307167 | 0.003481566 | 0.013836034 | 1 |
| Obesity | nk_cell | *SERPINC1* | 1.362396991 | 0.000298725 | 0.002597629 | 1 |
| Obesity | nk_cell | *MTFR1L* | -1.136767254 | 0.008495961 | 0.026468833 | 1 |
| Obesity | nk_cell | *RPA2* | 2.289093439 | 0.018454639 | 0.04708779 | 1 |
| Obesity | nk_cell | *SLC5A9* | -0.86157674 | 0.001349928 | 0.007119967 | 1 |
| Obesity | nk_cell | *CD3EAP* | -1.714356227 | 0.003471897 | 0.013806336 | 1 |
| Obesity | nk_cell | *COLEC11* | -1.557809549 | 1.56E-05 | 0.000721314 | 0.098098769 |
| Obesity | nk_cell | *KMT2A* | 1.764181377 | 0.008769097 | 0.027153352 | 1 |
| Obesity | nk_cell | *LRMP* | 1.181088519 | 9.77E-06 | 0.000524753 | 0.061396146 |
| Obesity | nk_cell | *ATP10B* | 6.236105984 | 0.000505112 | 0.003749277 | 1 |

| | | | | | | |
|---|---|---|---|---|---|---|
| Obesity | nk_cell | *TCF21* | -6.700848602 | 0.011097413 | 0.032288178 | 1 |
| Obesity | nk_cell | *VAMP8* | 2.532970446 | 3.42E-06 | 0.00026837 | 0.021503801 |
| Obesity | nk_cell | *TGIF2* | 3.339609784 | 0.006098772 | 0.020737146 | 1 |
| Obesity | nk_cell | *SPP1* | 1.82449526 | 0.01497145 | 0.040362567 | 1 |
| Obesity | nk_cell | *FAM47E-STBL* | -0.610438334 | 0.001698695 | 0.008317519 | 1 |
| Obesity | nk_cell | *UCHL3* | -1.797602982 | 0.011686562 | 0.033540239 | 1 |
| Obesity | nk_cell | *DNAH7* | -0.814928344 | 0.001037353 | 0.005939745 | 1 |
| Obesity | nk_cell | *TRPM6* | -0.863206278 | 0.000304457 | 0.002622439 | 1 |
| Obesity | nk_cell | *GDA* | -0.841004989 | 0.001677764 | 0.008249237 | 1 |
| Obesity | nk_cell | *C2orf40* | -1.090432023 | 0.016681291 | 0.043696927 | 1 |
| Obesity | nk_cell | *PTBP3* | 2.162037077 | 0.000117083 | 0.001569507 | 0.736098659 |
| Obesity | nk_cell | *CNTRL* | 2.21450634 | 0.000990216 | 0.005743071 | 1 |
| Obesity | nk_cell | *TRIM32* | 4.956748598 | 5.29E-05 | 0.001150301 | 0.332437027 |
| Obesity | nk_cell | *BSPRY* | 13.75469758 | 0.000424073 | 0.003303774 | 1 |
| Obesity | nk_cell | *PPP6C* | 1.879861103 | 0.014925248 | 0.040307146 | 1 |
| Obesity | nk_cell | *HDHD3* | -0.376530348 | 0.017696068 | 0.045805544 | 1 |
| Obesity | nk_cell | *DCAF4* | 2.691577918 | 8.13E-05 | 0.001340853 | 0.510963727 |
| Obesity | nk_cell | *VSX2* | -1.206649645 | 0.003569567 | 0.014078964 | 1 |
| Obesity | nk_cell | *IFI27L2* | 1.343018677 | 0.000895166 | 0.005369192 | 1 |
| Obesity | nk_cell | *IRF2BPL* | 2.432734074 | 0.000676846 | 0.004536603 | 1 |
| Obesity | nk_cell | *PPP4R4* | -1.249888669 | 0.001353689 | 0.007121874 | 1 |
| Obesity | nk_cell | *ZC2HC1C* | -1.292521679 | 0.015052006 | 0.040527606 | 1 |
| Obesity | nk_cell | *GPR75* | -3.100980325 | 0.004243315 | 0.016003431 | 1 |
| Obesity | nk_cell | *KLHL29* | -0.735521067 | 0.009501428 | 0.028682453 | 1 |
| Obesity | nk_cell | *CNRIP1* | -1.404728629 | 0.005880037 | 0.020124003 | 1 |
| Obesity | nk_cell | *CRIPT* | -1.77879548 | 0.007348619 | 0.023799152 | 1 |
| Obesity | nk_cell | *C10orf95* | 0.553760642 | 7.14E-05 | 0.001232439 | 0.448607816 |
| Obesity | nk_cell | *CD274* | -1.524305087 | 0.007038986 | 0.023037016 | 1 |
| Obesity | nk_cell | *LRP11* | -1.100626078 | 0.001890031 | 0.008900845 | 1 |
| Obesity | nk_cell | *WDR55* | -1.817801438 | 0.000283947 | 0.002550248 | 1 |
| Obesity | nk_cell | *PCDHB14* | -1.079602083 | 0.001158398 | 0.006444997 | 1 |
| Obesity | nk_cell | *PCDHB12* | -0.741040632 | 0.01045859 | 0.030921385 | 1 |
| Obesity | nk_cell | *SEC16B* | -1.644014581 | 0.000270874 | 0.002457401 | 1 |
| Obesity | nk_cell | *SNX19* | -1.578389684 | 0.005750301 | 0.019863814 | 1 |
| Obesity | nk_cell | *IQSEC3* | -1.037634592 | 0.001819198 | 0.008713534 | 1 |
| Obesity | nk_cell | *SMAD9* | -0.946470371 | 0.003634069 | 0.014226271 | 1 |
| Obesity | nk_cell | *EPHX2* | -0.74750124 | 0.001627048 | 0.008086367 | 1 |
| Obesity | nk_cell | *CRISPLD1* | -1.285676355 | 0.003230806 | 0.013070835 | 1 |
| Obesity | nk_cell | *ECHDC2* | -0.697738447 | 0.003371272 | 0.013508722 | 1 |
| Obesity | nk_cell | *PRB2* | -1.256810988 | 0.005314651 | 0.018721528 | 1 |
| Obesity | nk_cell | *PYROXD1* | 2.08801252 | 0.000588415 | 0.004165949 | 1 |
| Obesity | nk_cell | *A1BG* | 5.875384064 | 0.001875963 | 0.008867802 | 1 |
| Obesity | nk_cell | *PDZRN3* | -0.616472293 | 0.013760775 | 0.03787828 | 1 |
| Obesity | nk_cell | *B4GALT4* | 3.36487988 | 0.000224656 | 0.002219862 | 1 |
| Obesity | nk_cell | *GJB6* | -32.65934147 | 1.16E-05 | 0.000605066 | 0.073213023 |
| Obesity | nk_cell | *TMEM39B* | -1.638032784 | 0.006507253 | 0.021772804 | 1 |
| Obesity | nk_cell | *RNF115* | 1.855503491 | 0.006611071 | 0.022003073 | 1 |
| Obesity | nk_cell | *POLR3GL* | 2.531939215 | 5.90E-05 | 0.001177776 | 0.370809923 |
| Obesity | nk_cell | *ZNF639* | 3.419963057 | 0.000503989 | 0.003749277 | 1 |
| Obesity | nk_cell | *CXCR4* | 1.488406221 | 0.001083185 | 0.006118067 | 1 |

| | | | | | | |
|---|---|---|---|---|---|---|
| Obesity | nk_cell | *FLT3* | -1.026192192 | 0.000771046 | 0.004847568 | 1 |
| Obesity | nk_cell | *RASL11A* | -1.528988663 | 0.000208582 | 0.002140997 | 1 |
| Obesity | nk_cell | *LDB3* | -0.55338386 | 0.005030275 | 0.017979157 | 1 |
| Obesity | nk_cell | *NAA60* | 2.339373919 | 0.000133107 | 0.001695707 | 0.836842966 |
| Obesity | nk_cell | *RWDD3* | -2.508496296 | 0.004805997 | 0.017385099 | 1 |
| Obesity | nk_cell | *KIAA0087* | -2.229024895 | 0.000193841 | 0.002051645 | 1 |
| Obesity | nk_cell | *HERPUD2* | 2.38903961 | 0.002734077 | 0.011497754 | 1 |
| Obesity | nk_cell | *CCZ1* | 2.653404088 | 6.76E-06 | 0.000416575 | 0.042490673 |
| Obesity | nk_cell | *ACADS* | -1.164102288 | 0.007464581 | 0.024005023 | 1 |
| Obesity | nk_cell | *CCDC91* | 1.909358763 | 0.000300632 | 0.00260762 | 1 |
| Obesity | nk_cell | *PKN1* | 1.648521533 | 5.96E-05 | 0.001177776 | 0.374869088 |
| Obesity | nk_cell | *CENPK* | 0.621989159 | 0.000501887 | 0.003743261 | 1 |
| Obesity | nk_cell | *TSFM* | -1.319631751 | 0.014776863 | 0.03999231 | 1 |
| Obesity | nk_cell | *ARHGAP9* | 0.650925252 | 0.000502515 | 0.003743261 | 1 |
| Obesity | nk_cell | *ORMDL2* | 1.231852592 | 0.000136119 | 0.001719853 | 0.855778527 |
| Obesity | nk_cell | *IKZF4* | -1.764623453 | 0.000670572 | 0.004508971 | 1 |
| Obesity | nk_cell | *SARDH* | 4.1147764 | 4.86E-05 | 0.001116189 | 0.305467105 |
| Obesity | nk_cell | *STIL* | 1.961593329 | 9.76E-05 | 0.00144289 | 0.613600527 |
| Obesity | nk_cell | *NMI* | 2.420555775 | 0.000315735 | 0.002693389 | 1 |
| Obesity | nk_cell | *ACVR1C* | -0.668546285 | 0.001047884 | 0.005972841 | 1 |
| Obesity | nk_cell | *RAB38* | 2.701152361 | 0.000196518 | 0.002062614 | 1 |
| Obesity | nk_cell | *CKS2* | 1.938975516 | 0.00094726 | 0.005564354 | 1 |
| Obesity | nk_cell | *MOGAT1* | -0.771010522 | 0.001096584 | 0.006172088 | 1 |
| Obesity | nk_cell | *SNX21* | -0.805900517 | 0.005057858 | 0.018046966 | 1 |
| Obesity | nk_cell | *SDC4* | -0.865896653 | 0.000724774 | 0.004678965 | 1 |
| Obesity | nk_cell | *KCNK15* | -3.009414744 | 0.00433233 | 0.016241717 | 1 |
| Obesity | nk_cell | *FASTKD3* | -1.135802851 | 0.002219098 | 0.009880646 | 1 |
| Obesity | nk_cell | *ATP8A1* | 0.922677558 | 8.22E-06 | 0.000470803 | 0.051704609 |
| Obesity | nk_cell | *SNRPC* | -3.823623888 | 4.38E-20 | 3.44E-17 | 2.75E-16 |
| Obesity | nk_cell | *HIST1H1D* | 0.53976165 | 0.000193199 | 0.0020485 | 1 |
| Obesity | nk_cell | *RPS10* | 7.412923188 | 3.13E-06 | 0.000259505 | 0.019665716 |
| Obesity | nk_cell | *HIST1H2BJ* | 1.526887266 | 0.003021152 | 0.012414368 | 1 |
| Obesity | nk_cell | *GNMT* | -0.863417894 | 0.01214263 | 0.03454331 | 1 |
| Obesity | nk_cell | *CPNE5* | -1.399784792 | 0.00293162 | 0.012093894 | 1 |
| Obesity | nk_cell | *RPP40* | 1.674564498 | 0.006318028 | 0.021264155 | 1 |
| Obesity | nk_cell | *NUP153* | 1.853520283 | 0.010359757 | 0.030693589 | 1 |
| Obesity | nk_cell | *EEF1E1* | 6.576404785 | 2.83E-07 | 4.81E-05 | 0.001779205 |
| Obesity | nk_cell | *EREG* | -2.698244176 | 0.000556007 | 0.004002979 | 1 |
| Obesity | nk_cell | *MYRF* | 0.561300561 | 0.001390103 | 0.007231203 | 1 |
| Obesity | nk_cell | *CNOT1* | 1.79218216 | 0.008835821 | 0.027297693 | 1 |
| Obesity | nk_cell | *MT1G* | -5.391144992 | 0.000793572 | 0.00493026 | 1 |
| Obesity | nk_cell | *TMTC4* | 3.028924735 | 0.000114478 | 0.001553214 | 0.71972262 |
| Obesity | nk_cell | *GRK4* | 0.765171742 | 5.57E-05 | 0.001177776 | 0.350499487 |
| Obesity | nk_cell | *MIF4GD* | 4.13258662 | 6.49E-05 | 0.001192487 | 0.407918647 |
| Obesity | nk_cell | *NT5C* | 2.90527898 | 0.000521037 | 0.003826819 | 1 |
| Obesity | nk_cell | *PSD4* | 0.406747473 | 0.000136764 | 0.001723121 | 0.859837196 |
| Obesity | nk_cell | *SLC25A23* | -0.698396418 | 0.001470177 | 0.007525264 | 1 |
| Obesity | nk_cell | *CLPP* | 2.147067409 | 0.00209981 | 0.009524895 | 1 |
| Obesity | nk_cell | *SH2D3A* | 3.551247591 | 0.00185645 | 0.008808682 | 1 |
| Obesity | nk_cell | *SNRPD2* | 3.106931892 | 0.00129421 | 0.006901356 | 1 |

| | | | | | | |
|---|---|---|---|---|---|---|
| Obesity | nk_cell | *RTN2* | -1.644893354 | 0.01499156 | 0.040399458 | 1 |
| Obesity | nk_cell | *SDCBP2* | -1.300692354 | 0.001805758 | 0.008672881 | 1 |
| Obesity | nk_cell | *RBCK1* | 1.847282754 | 0.004226467 | 0.015959038 | 1 |
| Obesity | nk_cell | *BMP2* | -1.362486674 | 0.002445578 | 0.010603691 | 1 |
| Obesity | nk_cell | *MGME1* | 2.315810824 | 0.001668681 | 0.008220528 | 1 |
| Obesity | nk_cell | *TMEM74B* | 2.698993615 | 0.000286385 | 0.002561168 | 1 |
| Obesity | nk_cell | *FAM110A* | 4.298544722 | 0.004063003 | 0.015528327 | 1 |
| Obesity | nk_cell | *ZNF436* | -0.732714434 | 0.011498382 | 0.033221658 | 1 |
| Obesity | nk_cell | *XRCC3* | 3.754836134 | 0.000308289 | 0.002651453 | 1 |
| Obesity | nk_cell | *F10* | -0.923859372 | 0.004346923 | 0.016267324 | 1 |
| Obesity | nk_cell | *PROZ* | -4.10360052 | 0.001223988 | 0.006703144 | 1 |
| Obesity | nk_cell | *IGFLR1* | 1.107144138 | 0.000190055 | 0.002025211 | 1 |
| Obesity | nk_cell | *COX6B1* | 2.78383327 | 6.08E-05 | 0.001177776 | 0.382367969 |
| Obesity | nk_cell | *PRDX5* | 3.15717013 | 0.002391133 | 0.010439622 | 1 |
| Obesity | nk_cell | *TSKS* | -0.782030346 | 0.000875939 | 0.00527999 | 1 |
| Obesity | nk_cell | *RHOJ* | -0.915972441 | 0.008276061 | 0.025886365 | 1 |
| Obesity | nk_cell | *HSPA2* | -1.124064571 | 0.011981982 | 0.034179093 | 1 |
| Obesity | nk_cell | *PZP* | 0.570628766 | 8.57E-05 | 0.001360449 | 0.538735974 |
| Obesity | nk_cell | *RHOT1* | 1.323421314 | 0.019647327 | 0.049508115 | 1 |
| Obesity | nk_cell | *AIF1L* | -1.03784925 | 0.004694132 | 0.017137401 | 1 |
| Obesity | nk_cell | *MAP2K2* | 1.695876722 | 0.006799503 | 0.02246373 | 1 |
| Obesity | nk_cell | *CANX* | 2.706672189 | 0.000106615 | 0.001499525 | 0.670287786 |
| Obesity | nk_cell | *FGD3* | 0.754328045 | 8.36E-05 | 0.001341236 | 0.525520997 |
| Obesity | nk_cell | *MASP1* | -0.972955044 | 0.003172066 | 0.012866307 | 1 |
| Obesity | nk_cell | *HRASLS* | -1.267027128 | 0.009028286 | 0.027759449 | 1 |
| Obesity | nk_cell | *HELB* | 2.488072633 | 6.26E-05 | 0.001177776 | 0.393255976 |
| Obesity | nk_cell | *RAP1B* | 1.84043977 | 0.000571723 | 0.004089218 | 1 |
| Obesity | nk_cell | *RAB3IP* | 2.699231938 | 0.002376447 | 0.010411653 | 1 |
| Obesity | nk_cell | *PLA2G5* | -0.775951916 | 0.004436525 | 0.016514169 | 1 |
| Obesity | nk_cell | *WDR24* | -3.002077928 | 0.001340554 | 0.007082403 | 1 |
| Obesity | nk_cell | *CHTF18* | 4.048500013 | 9.41E-05 | 0.001424919 | 0.591345724 |
| Obesity | nk_cell | *TICAM1* | -1.498567159 | 0.011646919 | 0.033502451 | 1 |
| Obesity | nk_cell | *PTPN12* | 1.937758281 | 0.000253818 | 0.002370413 | 1 |
| Obesity | nk_cell | *SGCE* | -0.891149522 | 0.004759772 | 0.017317527 | 1 |
| Obesity | nk_cell | *RBM48* | 2.494577988 | 0.000583548 | 0.004136149 | 1 |
| Obesity | nk_cell | *DGCR6L* | 1.76401263 | 0.00096602 | 0.005633922 | 1 |
| Obesity | nk_cell | *VPREB3* | -11.20617528 | 0.000390972 | 0.003128541 | 1 |
| Obesity | nk_cell | *MGAT3* | -0.909825999 | 0.001362654 | 0.007141964 | 1 |
| Obesity | nk_cell | *CDC42EP1* | -1.055818466 | 0.005435937 | 0.019028807 | 1 |
| Obesity | nk_cell | *TPST2* | 1.314741041 | 2.25E-05 | 0.0008736 | 0.141523167 |
| Obesity | nk_cell | *BAIAP2L2* | -0.922684652 | 0.009918248 | 0.02963095 | 1 |
| Obesity | nk_cell | *TST* | -0.545424376 | 0.002429143 | 0.010539698 | 1 |
| Obesity | nk_cell | *APOL5* | -3.285835602 | 0.001573917 | 0.007908215 | 1 |
| Obesity | nk_cell | *FOXP2* | -0.701811544 | 0.004742645 | 0.017285221 | 1 |
| Obesity | nk_cell | *FLNC* | -1.04597283 | 0.003379374 | 0.013523949 | 1 |
| Obesity | nk_cell | *OPN1SW* | -0.867578551 | 0.004236737 | 0.015988215 | 1 |
| Obesity | nk_cell | *HOXD1* | -4.014635346 | 0.00568767 | 0.019701589 | 1 |
| Obesity | nk_cell | *HOXD3* | -0.874497256 | 0.005802971 | 0.019947118 | 1 |
| Obesity | nk_cell | *PDE11A* | -0.88990915 | 0.003900141 | 0.015070796 | 1 |
| Obesity | nk_cell | *GAD1* | -1.891586927 | 0.00053723 | 0.003891554 | 1 |

| | | | | | | |
|---|---|---|---|---|---|---|
| Obesity | nk_cell | *HOXD9* | -1.459565797 | 0.001679754 | 0.00825048 | 1 |
| Obesity | nk_cell | *WDFY4* | -1.017302852 | 0.000537223 | 0.003891554 | 1 |
| Obesity | nk_cell | *VPS13C* | 1.677585504 | 0.002776709 | 0.011561038 | 1 |
| Obesity | nk_cell | *CALML4* | -3.102402949 | 0.003243469 | 0.013113628 | 1 |
| Obesity | nk_cell | *ACKR4* | -0.734612612 | 0.010944735 | 0.031951962 | 1 |
| Obesity | nk_cell | *SPCS3* | 2.261195619 | 0.005312605 | 0.018721528 | 1 |
| Obesity | nk_cell | *SERGEF* | 2.668005819 | 0.000120458 | 0.001603969 | 0.757319574 |
| Obesity | nk_cell | *RPAIN* | -2.148547421 | 4.66E-05 | 0.001109982 | 0.293037362 |
| Obesity | nk_cell | *PLD2* | 2.903051074 | 0.00065765 | 0.004484432 | 1 |
| Obesity | nk_cell | *ATP1B2* | -1.809861643 | 0.002197334 | 0.009832484 | 1 |
| Obesity | nk_cell | *CCL4* | 0.322458808 | 9.69E-05 | 0.001442418 | 0.609251292 |
| Obesity | nk_cell | *KRI1* | 0.906650443 | 0.012178301 | 0.034629117 | 1 |
| Obesity | nk_cell | *SLC44A2* | 1.854759877 | 2.47E-05 | 0.000931493 | 0.155559305 |
| Obesity | nk_cell | *CDKN2D* | 1.068552418 | 2.14E-05 | 0.000858426 | 0.134701449 |
| Obesity | nk_cell | *NGDN* | 3.117517539 | 0.000784624 | 0.004909512 | 1 |
| Obesity | nk_cell | *RIPK3* | 2.511044566 | 0.000476413 | 0.003601102 | 1 |
| Obesity | nk_cell | *BCL2L2* | -1.120733352 | 0.019544985 | 0.049289739 | 1 |
| Obesity | nk_cell | *RHBDF2* | 0.718735901 | 0.00037167 | 0.003026798 | 1 |
| Obesity | nk_cell | *TMEM8A* | 2.259899877 | 0.000667546 | 0.004508311 | 1 |
| Obesity | nk_cell | *DOCK6* | -0.868444964 | 0.011595414 | 0.033409883 | 1 |
| Obesity | nk_cell | *THEM6* | -0.811627864 | 0.000537278 | 0.003891554 | 1 |
| Obesity | nk_cell | *DPP6* | -1.062367688 | 0.005734927 | 0.019821597 | 1 |
| Obesity | nk_cell | *FAM98C* | 1.910128278 | 0.005086364 | 0.018122781 | 1 |
| Obesity | nk_cell | *RPL36* | 2.041090285 | 2.65E-05 | 0.000951164 | 0.166453683 |
| Obesity | nk_cell | *ATP8B3* | -1.604210858 | 1.96E-06 | 0.000189978 | 0.012348578 |
| Obesity | nk_cell | *BST2* | 3.177854817 | 0.000294219 | 0.002579346 | 1 |
| Obesity | nk_cell | *LSM7* | 1.879593829 | 0.000725261 | 0.004678965 | 1 |
| Obesity | nk_cell | *ACTN4* | 1.857799382 | 0.009046217 | 0.027770295 | 1 |
| Obesity | nk_cell | *CACNG6* | -2.283685573 | 0.0084823 | 0.026439376 | 1 |
| Obesity | nk_cell | *PXDN* | -0.677768866 | 0.005071822 | 0.018086527 | 1 |
| Obesity | nk_cell | *PGPEP1* | 2.554822964 | 4.92E-05 | 0.001116189 | 0.309184477 |
| Obesity | nk_cell | *LSM4* | 1.726963322 | 1.27E-06 | 0.000142344 | 0.007971263 |
| Obesity | nk_cell | *HRC* | -1.70744829 | 1.53E-05 | 0.000718997 | 0.09634566 |
| Obesity | nk_cell | *TRPM4* | -1.005814675 | 0.014058639 | 0.038546298 | 1 |
| Obesity | nk_cell | *TUBGCP2* | 2.207185918 | 0.002711191 | 0.01143956 | 1 |
| Obesity | nk_cell | *CYP2E1* | -2.607169263 | 1.63E-05 | 0.000743507 | 0.10273322 |
| Obesity | nk_cell | *YIPF2* | -1.206007141 | 0.001296009 | 0.006905094 | 1 |
| Obesity | nk_cell | *TMEM160* | 4.578190624 | 0.00011577 | 0.001558555 | 0.727845214 |
| Obesity | nk_cell | *SMPDL3B* | -10.29203061 | 0.011073685 | 0.032259758 | 1 |
| Obesity | nk_cell | *THEMIS2* | 1.495008202 | 0.004862401 | 0.01753961 | 1 |
| Obesity | nk_cell | *ZNF317* | 1.769894685 | 0.01460367 | 0.039660162 | 1 |
| Obesity | nk_cell | *PPAN* | 4.920697735 | 4.25E-05 | 0.001082521 | 0.267162522 |
| Obesity | nk_cell | *ZNF236* | 1.624228853 | 0.016658692 | 0.043675229 | 1 |
| Obesity | nk_cell | *C12orf65* | 2.399422272 | 0.004649696 | 0.01708512 | 1 |
| Obesity | nk_cell | *PKDREJ* | -2.209315667 | 8.38E-05 | 0.001341236 | 0.527105717 |
| Obesity | nk_cell | *EPS8L1* | 2.067119466 | 0.000169455 | 0.001905836 | 1 |
| Obesity | nk_cell | *LILRB2* | -1.295026981 | 0.002504943 | 0.010823766 | 1 |
| Obesity | nk_cell | *CHMP1A* | 1.817025769 | 0.015635856 | 0.041618385 | 1 |
| Obesity | nk_cell | *KCNC3* | -1.250857219 | 0.007595864 | 0.024299842 | 1 |
| Obesity | nk_cell | *AOC3* | -1.07480659 | 0.017023252 | 0.044408791 | 1 |

| | | | | | | |
|---|---|---|---|---|---|---|
| Obesity | nk_cell | *AOC2* | -1.219744551 | 0.001017974 | 0.00586618 | 1 |
| Obesity | nk_cell | *DIAPH1* | 0.928474095 | 4.33E-05 | 0.001089332 | 0.27233297 |
| Obesity | nk_cell | *EXOC4* | 1.542164511 | 0.003621394 | 0.014203186 | 1 |
| Obesity | nk_cell | *NINJ1* | -0.98334901 | 0.006563313 | 0.021880442 | 1 |
| Obesity | nk_cell | *CKMT2* | -0.880516507 | 0.000714389 | 0.004649447 | 1 |
| Obesity | nk_cell | *KHDRBS3* | -0.965571393 | 0.007307474 | 0.023730419 | 1 |
| Obesity | nk_cell | *ZNF304* | 1.160257796 | 0.018819475 | 0.047805269 | 1 |
| Obesity | nk_cell | *RHPN2* | -1.471243311 | 0.000134541 | 0.001708809 | 0.845860645 |
| Obesity | nk_cell | *GCH1* | 1.193067326 | 7.06E-05 | 0.001228518 | 0.443884324 |
| Obesity | nk_cell | *DNAJB1* | -1.387553828 | 0.01641413 | 0.043196638 | 1 |
| Obesity | nk_cell | *C19orf57* | -1.801214783 | 0.002228641 | 0.009916113 | 1 |
| Obesity | nk_cell | *MATN3* | -4.778402566 | 0.004549608 | 0.016786025 | 1 |
| Obesity | nk_cell | *RAF1* | 1.750283364 | 0.014729304 | 0.039902773 | 1 |
| Obesity | nk_cell | *TRIM5* | 2.293541596 | 0.002000375 | 0.009206705 | 1 |
| Obesity | nk_cell | *TRIM22* | 1.958026215 | 0.000104878 | 0.001498568 | 0.659369918 |
| Obesity | nk_cell | *INPP5K* | -1.076917697 | 0.003054728 | 0.012503306 | 1 |
| Obesity | nk_cell | *RPA1* | 1.922628237 | 0.006459691 | 0.02164823 | 1 |
| Obesity | nk_cell | *TMEM128* | -1.495612966 | 0.012484704 | 0.035420277 | 1 |
| Obesity | nk_cell | *REEP2* | -1.023883162 | 0.000918327 | 0.005441587 | 1 |
| Obesity | nk_cell | *BTBD3* | -1.079060002 | 0.009356671 | 0.028400797 | 1 |
| Obesity | nk_cell | *PTPRA* | 1.787994912 | 0.00349096 | 0.013842201 | 1 |
| Obesity | nk_cell | *IGHMBP2* | 1.116867534 | 0.003036814 | 0.012462434 | 1 |
| Obesity | nk_cell | *LPIN3* | -0.784326194 | 0.001904679 | 0.008939412 | 1 |
| Obesity | nk_cell | *DMGDH* | -0.9856542 | 0.00923215 | 0.028175984 | 1 |
| Obesity | nk_cell | *BHMT2* | -0.380346153 | 0.001254555 | 0.006784987 | 1 |
| Obesity | nk_cell | *CASP9* | -0.949398651 | 0.010952199 | 0.031951962 | 1 |
| Obesity | nk_cell | *ATP8A2* | 2.864943446 | 3.69E-05 | 0.001056956 | 0.231787919 |
| Obesity | nk_cell | *WASF3* | -0.708724081 | 0.003400762 | 0.013592239 | 1 |
| Obesity | nk_cell | *RNF17* | -2.749102028 | 5.88E-05 | 0.001177776 | 0.369797868 |
| Obesity | nk_cell | *PEMT* | -1.083340051 | 0.000294571 | 0.002579346 | 1 |
| Obesity | nk_cell | *TRPC4* | -0.80889708 | 0.000693393 | 0.004590651 | 1 |
| Obesity | nk_cell | *PRAM1* | -0.873838431 | 0.000256228 | 0.002380061 | 1 |
| Obesity | nk_cell | *GSTT2B* | -0.709904302 | 0.00771349 | 0.024529445 | 1 |
| Obesity | nk_cell | *FAM83F* | -2.648467604 | 0.002754294 | 0.01154826 | 1 |
| Obesity | nk_cell | *GIMAP4* | 0.695012858 | 3.42E-05 | 0.001033598 | 0.214906546 |
| Obesity | nk_cell | *DYDC2* | 5.888959128 | 0.003004485 | 0.012353955 | 1 |
| Obesity | nk_cell | *IPO8* | 2.27742351 | 0.004450627 | 0.016533039 | 1 |
| Obesity | nk_cell | *LARS* | -1.017018849 | 0.001631761 | 0.00808663 | 1 |
| Obesity | nk_cell | *SPINK5* | -1.277533063 | 0.004338722 | 0.016246303 | 1 |
| Obesity | nk_cell | *COX16* | 0.998533145 | 0.011707047 | 0.033562338 | 1 |
| Obesity | nk_cell | *MRO* | -1.36367416 | 0.000340006 | 0.002838798 | 1 |
| Obesity | nk_cell | *THUMPD3* | -1.312354834 | 0.005537595 | 0.019309408 | 1 |
| Obesity | nk_cell | *CNTN6* | -0.858946903 | 0.00046049 | 0.003527575 | 1 |
| Obesity | nk_cell | *SYT6* | -3.637631114 | 0.014280902 | 0.038985684 | 1 |
| Obesity | nk_cell | *VAV3* | 0.701289954 | 0.000107721 | 0.001507391 | 0.677240032 |
| Obesity | nk_cell | *NAPG* | 1.912899033 | 0.016086599 | 0.042530046 | 1 |
| Obesity | nk_cell | *FKBP11* | 1.146951847 | 0.000140674 | 0.001737559 | 0.884420271 |
| Obesity | nk_cell | *ANO3* | -0.634013675 | 0.002239474 | 0.009953339 | 1 |
| Obesity | nk_cell | *TIMM17A* | 2.268444976 | 0.010092951 | 0.0301017 | 1 |
| Obesity | nk_cell | *ECHDC3* | -0.976208784 | 0.002670983 | 0.011338604 | 1 |

| | | | | | | |
|---|---|---|---|---|---|---|
| Obesity | nk_cell | *KLRD1* | 0.333619863 | 0.000110065 | 0.001516917 | 0.691978109 |
| Obesity | nk_cell | *PRH2* | -1.805959854 | 0.001809184 | 0.008676081 | 1 |
| Obesity | nk_cell | *DDB2* | 2.713541322 | 0.000242512 | 0.002299423 | 1 |
| Obesity | nk_cell | *ACP2* | 3.684282454 | 2.04E-06 | 0.000191198 | 0.012810249 |
| Obesity | nk_cell | *HOOK1* | 1.494037217 | 0.000237883 | 0.002290309 | 1 |
| Obesity | nk_cell | *ZCCHC11* | 1.813408501 | 0.002700427 | 0.011417341 | 1 |
| Obesity | nk_cell | *SLC43A3* | -1.553538621 | 0.008516236 | 0.02649748 | 1 |
| Obesity | nk_cell | *TIMM10* | 2.113828554 | 0.004304535 | 0.016166433 | 1 |
| Obesity | nk_cell | *APLNR* | -1.181644298 | 0.00862598 | 0.026781005 | 1 |
| Obesity | nk_cell | *CLDN10* | -2.796450658 | 0.012772402 | 0.035976743 | 1 |
| Obesity | nk_cell | *ERCC5* | 5.427367435 | 0.001383111 | 0.007216279 | 1 |
| Obesity | nk_cell | *OSTF1* | 1.735162161 | 3.18E-05 | 0.001016497 | 0.199949792 |
| Obesity | nk_cell | *TMEM2* | 2.082079085 | 0.001211029 | 0.006655368 | 1 |
| Obesity | nk_cell | *AGTPBP1* | 1.519183549 | 2.02E-05 | 0.000825847 | 0.127180434 |
| Obesity | nk_cell | *HRK* | -2.84683012 | 0.000433623 | 0.003349123 | 1 |
| Obesity | nk_cell | *P2RX4* | 1.445182113 | 0.010461235 | 0.030921385 | 1 |
| Obesity | nk_cell | *DTX1* | -0.649051342 | 0.017564826 | 0.045556955 | 1 |
| Obesity | nk_cell | *TMEM60* | -2.856588576 | 2.80E-05 | 0.000968751 | 0.175813465 |
| Obesity | nk_cell | *SRPK2* | 1.860301056 | 0.00049336 | 0.003696965 | 1 |
| Obesity | nk_cell | *TES* | 1.016682789 | 0.000650331 | 0.0044587 | 1 |
| Obesity | nk_cell | *NT5E* | -0.958296618 | 0.014331827 | 0.039056869 | 1 |
| Obesity | nk_cell | *PRR5L* | 0.725853633 | 6.05E-05 | 0.001177776 | 0.380129424 |
| Obesity | nk_cell | *ATP5G2* | 2.401389753 | 0.007126806 | 0.023293323 | 1 |
| Obesity | nk_cell | *ITGA7* | -0.452546932 | 0.01199086 | 0.034188905 | 1 |
| Obesity | nk_cell | *RDH5* | -0.617186454 | 0.000792031 | 0.00493026 | 1 |
| Obesity | nk_cell | *B4GALNT1* | -2.932008853 | 0.000870597 | 0.005271688 | 1 |
| Obesity | nk_cell | *FAIM2* | -1.380514658 | 0.00082153 | 0.005058726 | 1 |
| Obesity | nk_cell | *KRT7* | -2.23750958 | 0.003505241 | 0.013877487 | 1 |
| Obesity | nk_cell | *HNRNPA1* | 2.709185017 | 0.005961682 | 0.02036041 | 1 |
| Obesity | nk_cell | *SLC26A10* | -1.827876838 | 0.00629895 | 0.021222669 | 1 |
| Obesity | nk_cell | *SMPD2* | -1.26755626 | 0.006542349 | 0.021843732 | 1 |
| Obesity | nk_cell | *PRADC1* | -1.088743858 | 0.003118554 | 0.012698414 | 1 |
| Obesity | nk_cell | *KCNMB4* | -1.283071854 | 0.001230242 | 0.006719835 | 1 |
| Obesity | nk_cell | *CCDC102A* | 2.049481756 | 0.000258056 | 0.00238939 | 1 |
| Obesity | nk_cell | *RNASEL* | 1.573892908 | 0.017413335 | 0.045257395 | 1 |
| Obesity | nk_cell | *SP110* | 2.51310941 | 3.77E-05 | 0.001056956 | 0.236783451 |
| Obesity | nk_cell | *USP37* | 1.962622482 | 0.008997194 | 0.02771453 | 1 |
| Obesity | nk_cell | *MRPS9* | 1.400123636 | 0.006169584 | 0.020921346 | 1 |
| Obesity | nk_cell | *SLC41A2* | -0.809514363 | 0.009708111 | 0.029147513 | 1 |
| Obesity | nk_cell | *RNASEH2B* | 1.700403815 | 0.004139315 | 0.015752949 | 1 |
| Obesity | nk_cell | *CKAP2* | -0.58783956 | 0.017420672 | 0.045257755 | 1 |
| Obesity | nk_cell | *CHST12* | 0.880663786 | 0.000126705 | 0.00166304 | 0.796596093 |
| Obesity | nk_cell | *ZDHHC4* | 1.100397191 | 0.008613597 | 0.026755774 | 1 |
| Obesity | nk_cell | *AOAH* | 0.342319542 | 0.00015177 | 0.001809392 | 0.954176084 |
| Obesity | nk_cell | *NACAD* | -2.867523903 | 0.000392654 | 0.00312879 | 1 |
| Obesity | nk_cell | *ALPK3* | -0.784070914 | 0.001819766 | 0.008713534 | 1 |
| Obesity | nk_cell | *CIB2* | -1.020123204 | 0.013836613 | 0.038037072 | 1 |
| Obesity | nk_cell | *NMT1* | 1.12002647 | 0.002336412 | 0.010286432 | 1 |
| Obesity | nk_cell | *BRIP1* | 2.475339973 | 5.37E-05 | 0.00116249 | 0.337476981 |
| Obesity | nk_cell | *MRPL47* | 3.136604783 | 0.000540688 | 0.003902762 | 1 |

| | | | | | | |
|---|---|---|---|---|---|---|
| Obesity | nk_cell | *BLK* | -4.489514002 | 1.03E-06 | 0.000121705 | 0.006450384 |
| Obesity | nk_cell | *IL1RN* | -1.336571055 | 0.013204625 | 0.036717151 | 1 |
| Obesity | nk_cell | *GYPC* | 1.811761094 | 0.008655945 | 0.026847522 | 1 |
| Obesity | nk_cell | *ODF2* | 2.575835456 | 0.008876685 | 0.02741047 | 1 |
| Obesity | nk_cell | *TOR1B* | 1.253706557 | 0.007598774 | 0.024299842 | 1 |
| Obesity | nk_cell | *SLC2A8* | 2.355300915 | 0.000653247 | 0.004468952 | 1 |
| Obesity | nk_cell | *ALDOB* | -1.652416152 | 0.007730858 | 0.024572247 | 1 |
| Obesity | nk_cell | *PRPF4* | 1.566994576 | 0.004320447 | 0.016216509 | 1 |
| Obesity | nk_cell | *TEX10* | 2.329269228 | 0.006886413 | 0.022667476 | 1 |
| Obesity | nk_cell | *LMX1B* | 6.806898766 | 0.006942156 | 0.022803206 | 1 |
| Obesity | nk_cell | *DERL1* | 2.32647339 | 0.000188706 | 0.00201768 | 1 |
| Obesity | nk_cell | *NOV* | 2.700776278 | 0.001150747 | 0.006419476 | 1 |
| Obesity | nk_cell | *IL11RA* | -1.577009914 | 0.010507404 | 0.030996363 | 1 |
| Obesity | nk_cell | *TLN1* | 2.2771091 | 0.000951062 | 0.005567345 | 1 |
| Obesity | nk_cell | *CD72* | 2.899417646 | 3.51E-06 | 0.00026938 | 0.02208916 |
| Obesity | nk_cell | *TMEM8B* | -1.162533421 | 0.003979754 | 0.015330891 | 1 |
| Obesity | nk_cell | *PPIL1* | 1.241703476 | 9.34E-05 | 0.001420063 | 0.587163018 |
| Obesity | nk_cell | *ZSCAN9* | 2.579312495 | 7.52E-07 | 0.00010295 | 0.004727798 |
| Obesity | nk_cell | *BPHL* | -0.56930469 | 0.015328068 | 0.041073229 | 1 |
| Obesity | nk_cell | *TUBB2B* | -1.05053232 | 0.002313498 | 0.010199831 | 1 |
| Obesity | nk_cell | *UQCC2* | -1.351156434 | 0.003897401 | 0.015069471 | 1 |
| Obesity | nk_cell | *TCF19* | 2.283122054 | 0.001221898 | 0.006697536 | 1 |
| Obesity | nk_cell | *TPMT* | 2.437098734 | 0.00606215 | 0.020624344 | 1 |
| Obesity | nk_cell | *NRM* | 2.613756235 | 2.83E-05 | 0.000973139 | 0.178084519 |
| Obesity | nk_cell | *FAM8A1* | 3.182011594 | 6.34E-05 | 0.001179251 | 0.398333374 |
| Obesity | nk_cell | *FGFBP2* | 0.112568863 | 0.000298507 | 0.002597629 | 1 |
| Obesity | nk_cell | *FHDC1* | -0.872317822 | 0.001044328 | 0.005957978 | 1 |
| Obesity | nk_cell | *SYTL2* | 0.827825615 | 0.000215861 | 0.00216071 | 1 |
| Obesity | nk_cell | *RAB30* | -0.907327007 | 0.010212435 | 0.030390274 | 1 |
| Obesity | nk_cell | *TTPA* | -0.495590158 | 0.000169397 | 0.001905836 | 1 |
| Obesity | nk_cell | *GGH* | -0.467405879 | 0.016791158 | 0.043930925 | 1 |
| Obesity | nk_cell | *TRIM29* | -1.136572336 | 0.000165489 | 0.001884832 | 1 |
| Obesity | nk_cell | *PPP2R1B* | -0.643187701 | 0.007355138 | 0.023799152 | 1 |
| Obesity | nk_cell | *TMPRSS13* | 1.791613666 | 0.001227044 | 0.006708198 | 1 |
| Obesity | nk_cell | *CASP1* | 3.228999187 | 0.000168092 | 0.001904576 | 1 |
| Obesity | nk_cell | *ITGA11* | -0.749081836 | 0.005195761 | 0.018444115 | 1 |
| Obesity | nk_cell | *PAQR5* | -1.438813007 | 0.004207687 | 0.015935983 | 1 |
| Obesity | nk_cell | *RMDN3* | 2.365948231 | 0.004162989 | 0.015814327 | 1 |
| Obesity | nk_cell | *ITPKA* | -4.621478771 | 0.008886443 | 0.027427133 | 1 |
| Obesity | nk_cell | *DNASE2B* | -4.132332704 | 0.000293173 | 0.002579346 | 1 |
| Obesity | nk_cell | *LHCGR* | -0.719337516 | 0.000580677 | 0.004125102 | 1 |
| Obesity | nk_cell | *PREB* | -1.101291338 | 0.018250871 | 0.046672727 | 1 |
| Obesity | nk_cell | *DTNB* | 1.992454365 | 0.000366386 | 0.00301107 | 1 |
| Obesity | nk_cell | *BTBD16* | -0.761843229 | 0.002582546 | 0.011075354 | 1 |
| Obesity | nk_cell | *TACC2* | -0.60907843 | 0.019033392 | 0.048173483 | 1 |
| Obesity | nk_cell | *PLCE1* | -0.773820271 | 0.012866102 | 0.036127372 | 1 |
| Obesity | nk_cell | *RPS24* | 2.090919808 | 9.22E-05 | 0.001409361 | 0.579531285 |
| Obesity | nk_cell | *BARD1* | 2.89677028 | 0.001624432 | 0.008086149 | 1 |
| Obesity | nk_cell | *STAT4* | 0.192923802 | 0.000151445 | 0.001809392 | 0.952137405 |
| Obesity | nk_cell | *MDH1B* | -1.900122002 | 0.001764726 | 0.008522838 | 1 |

| | | | | | | |
|---|---|---|---|---|---|---|
| Obesity | nk_cell | *PRKG2* | -1.01114283 | 0.012531651 | 0.035441515 | 1 |
| Obesity | nk_cell | *BBS7* | -0.948197698 | 0.007167173 | 0.023383508 | 1 |
| Obesity | nk_cell | *MMRN1* | -0.618817975 | 4.98E-05 | 0.001118514 | 0.313183908 |
| Obesity | nk_cell | *PRDM5* | -0.92648565 | 0.012523847 | 0.035441515 | 1 |
| Obesity | nk_cell | *TRPC3* | 1.407031942 | 0.000375249 | 0.003040193 | 1 |
| Obesity | nk_cell | *G3BP2* | 2.570158336 | 0.000158969 | 0.001840581 | 0.999435614 |
| Obesity | nk_cell | *CNOT6L* | 1.025843366 | 0.004205922 | 0.015935983 | 1 |
| Obesity | nk_cell | *PARVG* | 2.047938645 | 0.00980313 | 0.02937668 | 1 |
| Obesity | nk_cell | *ERP27* | 0.435764694 | 0.000153566 | 0.001814794 | 0.965470629 |
| Obesity | nk_cell | *ALG10* | -1.823032856 | 0.015621525 | 0.041615479 | 1 |
| Obesity | nk_cell | *PRICKLE1* | -0.737158122 | 0.005340315 | 0.018786097 | 1 |
| Obesity | nk_cell | *CLSTN3* | 2.171101327 | 0.002855141 | 0.01184055 | 1 |
| Obesity | nk_cell | *KLRG1* | 0.923572583 | 1.63E-06 | 0.00017078 | 0.010246807 |
| Obesity | nk_cell | *VAMP1* | 1.027037011 | 0.006846078 | 0.022593854 | 1 |
| Obesity | nk_cell | *CD27* | 3.971072537 | 1.70E-05 | 0.000750451 | 0.106710506 |
| Obesity | nk_cell | *PIANP* | 3.255706055 | 0.010221822 | 0.030399524 | 1 |
| Obesity | nk_cell | *GLIPR1* | 2.072474964 | 0.000284933 | 0.002551819 | 1 |
| Obesity | nk_cell | *LGR5* | -0.805818481 | 0.002020326 | 0.009271381 | 1 |
| Obesity | nk_cell | *POC1B* | 2.431328714 | 7.76E-05 | 0.001306706 | 0.488058468 |
| Obesity | nk_cell | *TMEM132B* | -0.860075267 | 0.001194648 | 0.006595651 | 1 |
| Obesity | nk_cell | *RITA1* | -1.277790579 | 0.001669733 | 0.008220528 | 1 |
| Obesity | nk_cell | *GLTP* | 2.22542765 | 0.002188983 | 0.00981393 | 1 |
| Obesity | nk_cell | *SUOX* | -1.085722517 | 0.00462192 | 0.017012887 | 1 |
| Obesity | nk_cell | *ACVRL1* | -1.231146095 | 0.00047713 | 0.003601102 | 1 |
| Obesity | nk_cell | *GPR84* | -9.357606927 | 0.00012846 | 0.001670927 | 0.80762592 |
| Obesity | nk_cell | *CERS5* | 1.896968263 | 0.009358303 | 0.028400797 | 1 |
| Obesity | nk_cell | *METTL21C* | 2.17477783 | 0.002895396 | 0.011968016 | 1 |
| Obesity | nk_cell | *GRTP1* | -0.611423802 | 0.002717792 | 0.01144458 | 1 |
| Obesity | nk_cell | *CBLN3* | -0.317922631 | 0.00867423 | 0.02689097 | 1 |
| Obesity | nk_cell | *TSSK4* | -3.576314659 | 0.000237176 | 0.002288695 | 1 |
| Obesity | nk_cell | *TMX1* | 2.719215661 | 0.002516912 | 0.010868011 | 1 |
| Obesity | nk_cell | *PTGR2* | -0.852970918 | 0.000516097 | 0.003817297 | 1 |
| Obesity | nk_cell | *AK7* | -1.702923796 | 0.002482557 | 0.0107418 | 1 |
| Obesity | nk_cell | *SLC24A4* | -0.857427886 | 0.000669515 | 0.004508971 | 1 |
| Obesity | nk_cell | *C14orf79* | -1.460132026 | 0.004002996 | 0.015383153 | 1 |
| Obesity | nk_cell | *DUOXA1* | -1.806076278 | 0.000189913 | 0.002025211 | 1 |
| Obesity | nk_cell | *MFAP1* | 2.341198802 | 0.00195868 | 0.009094697 | 1 |
| Obesity | nk_cell | *LYSMD2* | 1.433879145 | 0.001117935 | 0.00626422 | 1 |
| Obesity | nk_cell | *SRP14* | 2.087421024 | 0.002407754 | 0.010490333 | 1 |
| Obesity | nk_cell | *PSTPIP1* | 0.845165011 | 0.000226411 | 0.002220662 | 1 |
| Obesity | nk_cell | *DNAJA4* | -1.363446016 | 0.008172072 | 0.025637633 | 1 |
| Obesity | nk_cell | *ULK3* | 3.327455352 | 0.002203444 | 0.009841332 | 1 |
| Obesity | nk_cell | *SCAMP2* | 1.763045548 | 0.015886655 | 0.042143208 | 1 |
| Obesity | nk_cell | *WDR93* | -0.680708243 | 6.38E-06 | 0.000413483 | 0.040107808 |
| Obesity | nk_cell | *TICRR* | -1.263799645 | 0.000645185 | 0.004437939 | 1 |
| Obesity | nk_cell | *MFGE8* | -1.146361599 | 0.008378903 | 0.026168984 | 1 |
| Obesity | nk_cell | *ST8SIA2* | 4.40876744 | 0.000161146 | 0.001858941 | 1 |
| Obesity | nk_cell | *SH3GL3* | -1.837787612 | 3.89E-07 | 6.27E-05 | 0.002444266 |
| Obesity | nk_cell | *TGFB1I1* | -1.354524811 | 0.001507727 | 0.007650588 | 1 |
| Obesity | nk_cell | *MARVELD3* | -1.640570822 | 0.000397693 | 0.00314899 | 1 |

| | | | | | | |
|---|---|---|---|---|---|---|
| Obesity | nk_cell | *KIFC3* | 2.622585684 | 0.00023662 | 0.002288661 | 1 |
| Obesity | nk_cell | *NUDT7* | -0.880139787 | 0.011880927 | 0.033952448 | 1 |
| Obesity | nk_cell | *GCSH* | -1.088381916 | 0.004748863 | 0.01728784 | 1 |
| Obesity | nk_cell | *CMTM3* | 3.184400294 | 2.96E-05 | 0.000988752 | 0.185946319 |
| Obesity | nk_cell | *RPS2* | 1.545754241 | 0.000702895 | 0.004623717 | 1 |
| Obesity | nk_cell | *NDUFB10* | 2.667809951 | 1.64E-05 | 0.000743507 | 0.103347457 |
| Obesity | nk_cell | *RASL10B* | -1.904129958 | 0.001902471 | 0.008939412 | 1 |
| Obesity | nk_cell | *UNC45B* | -3.170133681 | 0.0051985 | 0.018444115 | 1 |
| Obesity | nk_cell | *SPATA22* | -0.516805881 | 0.009002872 | 0.027718442 | 1 |
| Obesity | nk_cell | *RHBDL3* | 1.444314477 | 0.000321615 | 0.002726073 | 1 |
| Obesity | nk_cell | *ARSG* | 0.834683415 | 0.000169228 | 0.001905836 | 1 |
| Obesity | nk_cell | *G6PC3* | -1.338193973 | 0.011948222 | 0.034098261 | 1 |
| Obesity | nk_cell | *IMPA2* | -1.103224524 | 0.019399943 | 0.048982909 | 1 |
| Obesity | nk_cell | *GNAL* | 2.23711135 | 0.00940226 | 0.028473992 | 1 |
| Obesity | nk_cell | *C18orf21* | 3.045968904 | 0.007206162 | 0.023474165 | 1 |
| Obesity | nk_cell | *ASXL3* | -1.329313778 | 0.012402013 | 0.03520156 | 1 |
| Obesity | nk_cell | *ADCYAP1* | -3.416510985 | 0.002165797 | 0.009739891 | 1 |
| Obesity | nk_cell | *GREB1L* | -0.626016037 | 0.013008288 | 0.036407902 | 1 |
| Obesity | nk_cell | *SLC14A1* | 0.402962826 | 0.000121164 | 0.001603969 | 0.761756362 |
| Obesity | nk_cell | *SLC13A5* | -4.782599325 | 0.013022931 | 0.036421336 | 1 |
| Obesity | nk_cell | *CCDC40* | -1.146580823 | 0.001828349 | 0.008724464 | 1 |
| Obesity | nk_cell | *TMC6* | 0.870778858 | 3.22E-05 | 0.001016497 | 0.202543932 |
| Obesity | nk_cell | *CARD14* | 4.0884347 | 0.000325103 | 0.002747211 | 1 |
| Obesity | nk_cell | *TRIM65* | 3.457893528 | 2.36E-08 | 5.50E-06 | 0.000148513 |
| Obesity | nk_cell | *CBX8* | -1.011114101 | 0.019325923 | 0.048835241 | 1 |
| Obesity | nk_cell | *RNF157* | 0.778181918 | 0.000505051 | 0.003749277 | 1 |
| Obesity | nk_cell | *RNF165* | 0.480327948 | 9.71E-05 | 0.001442418 | 0.610740006 |
| Obesity | nk_cell | *MAPK4* | -0.917878371 | 0.001303079 | 0.006931014 | 1 |
| Obesity | nk_cell | *TXNL4A* | 1.188689882 | 0.018102565 | 0.046478962 | 1 |
| Obesity | nk_cell | *TPGS1* | 3.197715539 | 6.10E-05 | 0.001177776 | 0.383516062 |
| Obesity | nk_cell | *DPP9* | 2.260741359 | 0.000240261 | 0.002297638 | 1 |
| Obesity | nk_cell | *CCDC97* | 3.133838763 | 0.000398859 | 0.003150283 | 1 |
| Obesity | nk_cell | *TRPM2* | 0.752904415 | 0.003695557 | 0.014413132 | 1 |
| Obesity | nk_cell | *NTN5* | -1.801725044 | 1.49E-05 | 0.000718997 | 0.093491148 |
| Obesity | nk_cell | *WTIP* | -0.924627605 | 0.005932723 | 0.02029327 | 1 |
| Obesity | nk_cell | *CAPN10* | 1.255062218 | 0.017775032 | 0.045950504 | 1 |
| Obesity | nk_cell | *MYO1F* | 0.605287493 | 5.82E-05 | 0.001177776 | 0.365795061 |
| Obesity | nk_cell | *CACNG8* | -2.090075397 | 0.001834822 | 0.008745662 | 1 |
| Obesity | nk_cell | *FBN3* | 1.594847192 | 0.006394019 | 0.021462465 | 1 |
| Obesity | nk_cell | *EVI5L* | -1.171710706 | 0.003932264 | 0.01518559 | 1 |
| Obesity | nk_cell | *RPS11* | 2.405506849 | 2.36E-06 | 0.000208737 | 0.01482034 |
| Obesity | nk_cell | *PEX14* | 1.757002171 | 0.011563547 | 0.033359559 | 1 |
| Obesity | nk_cell | *SH3BGRL3* | 1.909350464 | 3.71E-05 | 0.001056956 | 0.233455012 |
| Obesity | nk_cell | *MAP3K6* | -1.59515098 | 0.0094363 | 0.028535843 | 1 |
| Obesity | nk_cell | *GPN2* | 2.472478163 | 0.016412637 | 0.043196638 | 1 |
| Obesity | nk_cell | *SERBP1* | 2.431800209 | 0.001745796 | 0.008455946 | 1 |
| Obesity | nk_cell | *RPS8* | 2.254783291 | 9.49E-06 | 0.000514073 | 0.059632433 |
| Obesity | nk_cell | *MOB3C* | -1.224369768 | 0.004787307 | 0.017355174 | 1 |
| Obesity | nk_cell | *CYP4B1* | -0.477983539 | 0.002737918 | 0.011505493 | 1 |
| Obesity | nk_cell | *ZNF697* | -1.393441706 | 0.011375149 | 0.032956481 | 1 |

| | | | | | | |
|---|---|---|---|---|---|---|
| Obesity | nk_cell | *CELSR2* | 2.211458017 | 0.00917042 | 0.028014786 | 1 |
| Obesity | nk_cell | *ATP1B1* | -1.355882186 | 0.005296254 | 0.018685492 | 1 |
| Obesity | nk_cell | *GPA33* | 1.237317301 | 0.000326952 | 0.002755426 | 1 |
| Obesity | nk_cell | *RXRG* | -1.813759158 | 0.001273168 | 0.006835529 | 1 |
| Obesity | nk_cell | *XCL2* | 0.373249954 | 0.00037253 | 0.003029878 | 1 |
| Obesity | nk_cell | *PPOX* | 2.038286642 | 0.004533844 | 0.016737686 | 1 |
| Obesity | nk_cell | *TUFT1* | -1.391884628 | 1.12E-05 | 0.000586358 | 0.070362908 |
| Obesity | nk_cell | *SELENBP1* | -0.628704309 | 0.019002673 | 0.048134855 | 1 |
| Obesity | nk_cell | *HORMAD1* | -0.356761367 | 0.005786915 | 0.019924607 | 1 |
| Obesity | nk_cell | *IKBKE* | 2.916243218 | 0.000155925 | 0.00182528 | 0.980298168 |
| Obesity | nk_cell | *DYRK3* | -1.120114788 | 0.004415715 | 0.016446446 | 1 |
| Obesity | nk_cell | *HHIPL2* | -2.003131021 | 0.002906661 | 0.012006688 | 1 |
| Obesity | nk_cell | *TP53BP2* | 2.237910397 | 0.016781847 | 0.043924842 | 1 |
| Obesity | nk_cell | *RAB13* | -1.26097397 | 0.001712714 | 0.008346154 | 1 |
| Obesity | nk_cell | *FLG* | -0.602335785 | 0.007925233 | 0.025050748 | 1 |
| Obesity | nk_cell | *FBXO28* | 1.892069691 | 0.015411581 | 0.041178331 | 1 |
| Obesity | nk_cell | *GUK1* | 3.505351957 | 0.000183724 | 0.001993342 | 1 |
| Obesity | nk_cell | *PYCR2* | 1.810192607 | 0.000855943 | 0.005214452 | 1 |
| Obesity | nk_cell | *WNT9A* | -0.918166957 | 0.014233042 | 0.038895317 | 1 |
| Obesity | nk_cell | *PLEKHA6* | -0.718054472 | 0.007329696 | 0.023765753 | 1 |
| Obesity | nk_cell | *EML4* | 1.882152184 | 0.003363863 | 0.013487633 | 1 |
| Obesity | nk_cell | *CHAC2* | -0.931214055 | 0.00270708 | 0.011437775 | 1 |
| Obesity | nk_cell | *TMEM177* | 3.287630068 | 2.62E-05 | 0.000948114 | 0.164971829 |
| Obesity | nk_cell | *RABL2A* | -1.15998014 | 0.013770115 | 0.037887402 | 1 |
| Obesity | nk_cell | *SLC20A1* | 1.29957108 | 8.31E-05 | 0.001340853 | 0.522561511 |
| Obesity | nk_cell | *FBLN7* | -1.324206822 | 7.49E-05 | 0.001276215 | 0.47098126 |
| Obesity | nk_cell | *LIPT1* | -0.899465037 | 0.001828987 | 0.008724464 | 1 |
| Obesity | nk_cell | *AFF3* | -1.034136345 | 0.002018643 | 0.009270422 | 1 |
| Obesity | nk_cell | *ZNF385B* | -0.590114286 | 0.006655615 | 0.022116201 | 1 |
| Obesity | nk_cell | *TMEFF2* | -1.037871172 | 0.013656554 | 0.037740113 | 1 |
| Obesity | nk_cell | *FAM198A* | -1.127757256 | 0.000900248 | 0.005385799 | 1 |
| Obesity | nk_cell | *SLC25A38* | 1.653983811 | 0.000136231 | 0.001719853 | 0.856486707 |
| Obesity | nk_cell | *STAC* | -0.942339275 | 0.002760975 | 0.011548764 | 1 |
| Obesity | nk_cell | *ZNF660* | -0.725306857 | 0.01902671 | 0.048173483 | 1 |
| Obesity | nk_cell | *COL8A1* | -0.63196425 | 0.000655909 | 0.004478712 | 1 |
| Obesity | nk_cell | *ADPRH* | -0.818314934 | 0.018017364 | 0.046411851 | 1 |
| Obesity | nk_cell | *IGSF11* | 1.343176182 | 4.90E-05 | 0.001116189 | 0.308177497 |
| Obesity | nk_cell | *BOC* | -0.750265989 | 0.01383042 | 0.03803668 | 1 |
| Obesity | nk_cell | *AMT* | -0.916284949 | 0.000266867 | 0.00243511 | 1 |
| Obesity | nk_cell | *EAF2* | -1.546088597 | 0.000534836 | 0.003891554 | 1 |
| Obesity | nk_cell | *DGKQ* | 0.747006831 | 0.017313399 | 0.045016268 | 1 |
| Obesity | nk_cell | *LYAR* | 1.605829341 | 4.68E-05 | 0.001109982 | 0.294421852 |
| Obesity | nk_cell | *CENPC* | -1.405189012 | 0.001848025 | 0.008795254 | 1 |
| Obesity | nk_cell | *TIFA* | 1.703508705 | 0.00015546 | 0.001823463 | 0.977375911 |
| Obesity | nk_cell | *CCNA2* | -0.846972945 | 0.0027698 | 0.011548764 | 1 |
| Obesity | nk_cell | *MARCH1* | -1.153643718 | 0.004886965 | 0.017607077 | 1 |
| Obesity | nk_cell | *BHMT* | -0.603203578 | 0.002771924 | 0.011548764 | 1 |
| Obesity | nk_cell | *CRHBP* | -1.036173396 | 0.001277709 | 0.006854059 | 1 |
| Obesity | nk_cell | *SPATA9* | -0.753073134 | 0.008087195 | 0.025434815 | 1 |
| Obesity | nk_cell | *TSLP* | -1.030774681 | 0.000309455 | 0.002656833 | 1 |

| | | | | | | |
|---|---|---|---|---|---|---|
| Obesity | nk_cell | *SLC25A48* | -1.513075259 | 4.66E-05 | 0.001109982 | 0.292754331 |
| Obesity | nk_cell | *GABRB2* | -0.890485044 | 0.012519065 | 0.035441515 | 1 |
| Obesity | nk_cell | *PCYOX1L* | -2.016768954 | 0.001236493 | 0.00673642 | 1 |
| Obesity | nk_cell | *GFOD1* | 1.419399958 | 8.90E-05 | 0.001378539 | 0.559686871 |
| Obesity | nk_cell | *PSD2* | 3.590238404 | 0.000991587 | 0.005745723 | 1 |
| Obesity | nk_cell | *TRIM7* | -1.006824618 | 0.002022265 | 0.00927351 | 1 |
| Obesity | nk_cell | *DOK3* | -1.074812654 | 0.005372995 | 0.01887152 | 1 |
| Obesity | nk_cell | *ABT1* | 2.872089174 | 1.29E-07 | 2.45E-05 | 0.000809488 |
| Obesity | nk_cell | *MLIP* | -1.20043336 | 0.001847025 | 0.008795254 | 1 |
| Obesity | nk_cell | *FGD2* | -0.981873013 | 0.008218791 | 0.025732839 | 1 |
| Obesity | nk_cell | *RPL7L1* | 2.310857231 | 0.000206623 | 0.002132632 | 1 |
| Obesity | nk_cell | *TPBG* | -1.165806707 | 0.007553625 | 0.024192381 | 1 |
| Obesity | nk_cell | *PM20D2* | 2.896474811 | 0.005043805 | 0.018007041 | 1 |
| Obesity | nk_cell | *ABRACL* | 1.370295962 | 0.001555999 | 0.007846195 | 1 |
| Obesity | nk_cell | *DYNLT1* | 2.180838649 | 3.75E-05 | 0.001056956 | 0.235988305 |
| Obesity | nk_cell | *PNLDC1* | 2.023304369 | 0.000260734 | 0.0024071 | 1 |
| Obesity | nk_cell | *SLC22A3* | -0.660419285 | 0.001799204 | 0.00864801 | 1 |
| Obesity | nk_cell | *SDK1* | -0.781011136 | 0.000429675 | 0.0033295 | 1 |
| Obesity | nk_cell | *EGFR* | -0.575313743 | 0.015825013 | 0.041997406 | 1 |
| Obesity | nk_cell | *GIGYF1* | 2.53516984 | 0.000989655 | 0.005743071 | 1 |
| Obesity | nk_cell | *ST18* | -1.056618713 | 0.002081978 | 0.009485068 | 1 |
| Obesity | nk_cell | *RGS20* | -0.802545638 | 0.001155101 | 0.006436666 | 1 |
| Obesity | nk_cell | *ADHFE1* | -0.6385331 | 0.000386432 | 0.003110756 | 1 |
| Obesity | nk_cell | *SLC26A7* | -0.739203729 | 0.000270375 | 0.00245643 | 1 |
| Obesity | nk_cell | *ATP6V0D2* | -2.725432801 | 1.53E-06 | 0.000163441 | 0.009642999 |
| Obesity | nk_cell | *MAL2* | -0.590532722 | 0.009170247 | 0.028014786 | 1 |
| Obesity | nk_cell | *FAM135B* | -1.020748568 | 0.00061484 | 0.004313769 | 1 |
| Obesity | nk_cell | *SLC39A4* | -1.658613898 | 0.003356134 | 0.013465229 | 1 |
| Obesity | nk_cell | *HAUS6* | 2.860278938 | 0.002480662 | 0.010740994 | 1 |
| Obesity | nk_cell | *CBWD5* | -1.232789158 | 0.016984723 | 0.044363505 | 1 |
| Obesity | nk_cell | *ZNF462* | -0.854018439 | 0.018465032 | 0.047095194 | 1 |
| Obesity | nk_cell | *WDR31* | -0.925350985 | 0.008539546 | 0.02655199 | 1 |
| Obesity | nk_cell | *POLE3* | -1.540506354 | 0.01841608 | 0.047008483 | 1 |
| Obesity | nk_cell | *SLC39A12* | -1.216176606 | 0.001504408 | 0.007639915 | 1 |
| Obesity | nk_cell | *NRBF2* | -1.52623614 | 5.74E-05 | 0.001177776 | 0.361088278 |
| Obesity | nk_cell | *HTR7* | -1.396957623 | 0.002338378 | 0.010287881 | 1 |
| Obesity | nk_cell | *FUOM* | -1.506055492 | 0.00068121 | 0.004552412 | 1 |
| Obesity | nk_cell | *MTG1* | 0.924533609 | 0.015791775 | 0.041926897 | 1 |
| Obesity | nk_cell | *PAOX* | -1.539853481 | 0.00740966 | 0.023914031 | 1 |
| Obesity | nk_cell | *RGS10* | 2.498730038 | 0.015390645 | 0.041158506 | 1 |
| Obesity | nk_cell | *ZNF215* | -0.445034441 | 0.01263953 | 0.035714484 | 1 |
| Obesity | nk_cell | *HSD17B12* | -1.929713271 | 1.78E-11 | 7.45E-09 | 1.12E-07 |
| Obesity | nk_cell | *PTPRJ* | 1.722330506 | 0.001254975 | 0.006784987 | 1 |
| Obesity | nk_cell | *ENDOD1* | -1.236675863 | 0.011308113 | 0.032777367 | 1 |
| Obesity | nk_cell | *SERPINH1* | -1.107565892 | 0.001216466 | 0.006675463 | 1 |
| Obesity | nk_cell | *CAPN5* | 2.970819531 | 1.27E-05 | 0.000650265 | 0.07993058 |
| Obesity | nk_cell | *RPS3* | 0.90317798 | 0.000242853 | 0.002299423 | 1 |
| Obesity | nk_cell | *TTC12* | 1.040063097 | 0.000283859 | 0.002550248 | 1 |
| Obesity | nk_cell | *DRD2* | -2.829402039 | 0.002758751 | 0.01154826 | 1 |
| Obesity | nk_cell | *C11orf52* | 3.837785106 | 0.000141351 | 0.001739089 | 0.888674726 |

| | | | | | | |
|---|---|---|---|---|---|---|
| Obesity | nk_cell | *NPAT* | 1.681452667 | 0.018998392 | 0.048134855 | 1 |
| Obesity | nk_cell | *GLB1L2* | 1.626193515 | 0.000520939 | 0.003826819 | 1 |
| Obesity | nk_cell | *GRIK4* | 0.388904626 | 0.001019026 | 0.005866867 | 1 |
| Obesity | nk_cell | *ADAM33* | -0.76604755 | 0.000903939 | 0.005391097 | 1 |
| Obesity | nk_cell | *MTA2* | 1.71737641 | 6.17E-05 | 0.001177776 | 0.387946154 |
| Obesity | nk_cell | *MS4A3* | -13.75900054 | 0.00060193 | 0.004237776 | 1 |
| Obesity | nk_cell | *PLCH2* | 0.300078363 | 0.000217461 | 0.002167168 | 1 |
| Obesity | nk_cell | *CCDC15* | -1.202843544 | 0.012121143 | 0.034497794 | 1 |
| Obesity | nk_cell | *FEZ1* | 3.960655099 | 0.000193218 | 0.0020485 | 1 |
| Obesity | nk_cell | *DUSP15* | -2.754551417 | 2.74E-05 | 0.000956877 | 0.1722378 |
| Obesity | nk_cell | *COMMD7* | 2.740914193 | 0.004462468 | 0.01656171 | 1 |
| Obesity | nk_cell | *CNBD2* | -1.618451613 | 0.013559029 | 0.037553134 | 1 |
| Obesity | nk_cell | *TRPT1* | -2.407837615 | 4.78E-05 | 0.001116189 | 0.30045499 |
| Obesity | nk_cell | *CWC15* | 1.661082129 | 0.015391509 | 0.041158506 | 1 |
| Obesity | nk_cell | *SAP18* | 2.746474556 | 0.004638928 | 0.017065499 | 1 |
| Obesity | nk_cell | *LYPD6B* | -1.066037134 | 0.000540224 | 0.003902762 | 1 |
| Obesity | nk_cell | *CNDP1* | 1.058348208 | 0.00816725 | 0.025635297 | 1 |
| Obesity | nk_cell | *FSIP1* | 4.217693031 | 7.00E-05 | 0.001225599 | 0.439989872 |
| Obesity | nk_cell | *RGS18* | -1.359364849 | 0.00687587 | 0.022656497 | 1 |
| Obesity | nk_cell | *PPP1R1C* | -0.993426704 | 0.00281328 | 0.011682357 | 1 |
| Obesity | nk_cell | *C11orf53* | -5.35677503 | 0.000661096 | 0.004503048 | 1 |
| Obesity | nk_cell | *TEX12* | -4.651990458 | 0.000727992 | 0.004689435 | 1 |
| Obesity | nk_cell | *PIP4K2A* | 0.83072081 | 0.000129432 | 0.001670927 | 0.81374122 |
| Obesity | nk_cell | *RILPL2* | 1.473552452 | 0.010127616 | 0.030176455 | 1 |
| Obesity | nk_cell | *DHX37* | -1.386574728 | 0.01641431 | 0.043196638 | 1 |
| Obesity | nk_cell | *PRSS53* | -2.42124056 | 0.001729901 | 0.008399259 | 1 |
| Obesity | nk_cell | *TMEM86A* | -1.067034765 | 0.000420158 | 0.003281407 | 1 |
| Obesity | nk_cell | *UBE3B* | 1.80234079 | 0.000834876 | 0.005115855 | 1 |
| Obesity | nk_cell | *ANK3* | 1.637136761 | 6.70E-05 | 0.001202623 | 0.420917893 |
| Obesity | nk_cell | *GXYLT1* | 2.515597405 | 0.002598381 | 0.011128079 | 1 |
| Obesity | nk_cell | *THRSP* | -0.472662718 | 0.018085895 | 0.046467521 | 1 |
| Obesity | nk_cell | *NDUFC2* | 2.148402298 | 0.000398597 | 0.003150283 | 1 |
| Obesity | nk_cell | *ADAMTS12* | -0.736689564 | 0.007509357 | 0.024099706 | 1 |
| Obesity | nk_cell | *PTPRO* | -1.090637492 | 0.016560524 | 0.043472239 | 1 |
| Obesity | nk_cell | *VPS26B* | 3.001243995 | 6.91E-05 | 0.001220269 | 0.434415632 |
| Obesity | nk_cell | *NCAPD3* | 2.055945623 | 0.010910875 | 0.031902117 | 1 |
| Obesity | nk_cell | *VTI1A* | 1.582260656 | 0.004696635 | 0.017137401 | 1 |
| Obesity | nk_cell | *ANO4* | -0.646610136 | 0.003583902 | 0.014108949 | 1 |
| Obesity | nk_cell | *AKR1C2* | -0.79911901 | 0.003610496 | 0.014169283 | 1 |
| Obesity | nk_cell | *DPYSL4* | 0.864203912 | 0.000557119 | 0.004002979 | 1 |
| Obesity | nk_cell | *ADAM8* | 0.40292395 | 0.000105628 | 0.001499525 | 0.664082663 |
| Obesity | nk_cell | *MFSD6* | 2.104028822 | 0.010767794 | 0.03157515 | 1 |
| Obesity | nk_cell | *TMEM45B* | -2.034310618 | 0.000556896 | 0.004002979 | 1 |
| Obesity | nk_cell | *ACSL1* | -0.751859124 | 0.018550632 | 0.047236865 | 1 |
| Obesity | nk_cell | *CCDC122* | -0.600072333 | 0.005543598 | 0.01931584 | 1 |
| Obesity | nk_cell | *GUF1* | -1.311600952 | 0.005229686 | 0.018533842 | 1 |
| Obesity | nk_cell | *GABRA2* | -0.657548957 | 0.006577603 | 0.021914887 | 1 |
| Obesity | nk_cell | *PARP8* | 0.80547468 | 0.000522422 | 0.003828635 | 1 |
| Obesity | nk_cell | *BEND6* | -0.887452673 | 7.81E-06 | 0.000463113 | 0.049089974 |
| Obesity | nk_cell | *RABGAP1L* | 1.318053631 | 4.42E-06 | 0.00031589 | 0.027798343 |

| | | | | | | |
|---|---|---|---|---|---|---|
| Obesity | nk_cell | *G6PC2* | 13.49763454 | 0.000169277 | 0.001905836 | 1 |
| Obesity | nk_cell | *TGOLN2* | 2.186949431 | 0.001307776 | 0.006950116 | 1 |
| Obesity | nk_cell | *POC5* | -2.330123169 | 1.32E-05 | 0.000668256 | 0.08286376 |
| Obesity | nk_cell | *NMT2* | -1.01106798 | 0.001785857 | 0.008605745 | 1 |
| Obesity | nk_cell | *ZFP36L2* | 1.867128658 | 0.005434731 | 0.019028807 | 1 |
| Obesity | nk_cell | *IGSF10* | -0.957463487 | 0.01559725 | 0.041586052 | 1 |
| Obesity | nk_cell | *MBNL1* | 1.472655882 | 0.000693672 | 0.004590651 | 1 |
| Obesity | nk_cell | *CLEC4F* | -1.885595721 | 0.000789517 | 0.004924302 | 1 |
| Obesity | nk_cell | *PELO* | -1.204808175 | 0.002491705 | 0.010773967 | 1 |
| Obesity | nk_cell | *CENPH* | 1.32079992 | 0.007351523 | 0.023799152 | 1 |
| Obesity | nk_cell | *CLGN* | -3.795230574 | 0.001083452 | 0.006118067 | 1 |
| Obesity | nk_cell | *SYCP2L* | -1.200076116 | 0.009390997 | 0.02845359 | 1 |
| Obesity | nk_cell | *PLA2R1* | -0.723321921 | 0.006330738 | 0.021295534 | 1 |
| Obesity | nk_cell | *SLC25A27* | -0.475639752 | 0.018696964 | 0.047547533 | 1 |
| Obesity | nk_cell | *FRMD1* | -0.777367912 | 0.000748559 | 0.004763352 | 1 |
| Obesity | nk_cell | *LPCAT1* | 0.475452073 | 0.000112488 | 0.001540771 | 0.707213934 |
| Obesity | nk_cell | *NMRAL1* | 0.794543784 | 0.009692555 | 0.029127124 | 1 |
| Obesity | nk_cell | *ADPRHL1* | -1.090894256 | 0.015096399 | 0.040594978 | 1 |
| Obesity | nk_cell | *FAM92B* | -8.769604365 | 0.000667606 | 0.004508311 | 1 |
| Obesity | nk_cell | *C7orf31* | 2.626272865 | 0.000214392 | 0.002158544 | 1 |
| Obesity | nk_cell | *JAZF1* | 1.431180057 | 1.68E-06 | 0.000172688 | 0.010533951 |
| Obesity | nk_cell | *FBXO36* | -1.321338736 | 0.003265584 | 0.013186079 | 1 |
| Obesity | nk_cell | *MCOLN2* | 0.920339029 | 0.000198164 | 0.002076426 | 1 |
| Obesity | nk_cell | *PPP2R5E* | 1.895870131 | 0.000685624 | 0.004556572 | 1 |
| Obesity | nk_cell | *ANKRD29* | -0.779828813 | 0.002577378 | 0.011060733 | 1 |
| Obesity | nk_cell | *UBASH3B* | 2.374813444 | 4.57E-05 | 0.001109982 | 0.28731075 |
| Obesity | nk_cell | *ROBO4* | -0.871115265 | 0.001327637 | 0.00703189 | 1 |
| Obesity | nk_cell | *CC2D1B* | 1.237735076 | 0.010125273 | 0.030176455 | 1 |
| Obesity | nk_cell | *UCHL1* | -0.984173541 | 0.001385123 | 0.007220786 | 1 |
| Obesity | nk_cell | *DISP1* | -0.774778018 | 0.010948885 | 0.031951962 | 1 |
| Obesity | nk_cell | *NEIL2* | 1.917764012 | 5.70E-05 | 0.001177776 | 0.358134558 |
| Obesity | nk_cell | *CCSAP* | -1.470533463 | 0.00560497 | 0.019485108 | 1 |
| Obesity | nk_cell | *GBP5* | 0.339847153 | 9.73E-05 | 0.001442418 | 0.611585227 |
| Obesity | nk_cell | *PDE1C* | -0.76068514 | 0.013070471 | 0.036492089 | 1 |
| Obesity | nk_cell | *RABGEF1* | -0.880841853 | 0.001040856 | 0.005943563 | 1 |
| Obesity | nk_cell | *TSEN2* | -1.097341356 | 0.008416299 | 0.026263955 | 1 |
| Obesity | nk_cell | *SLFN13* | 0.266396251 | 0.001807202 | 0.008673192 | 1 |
| Obesity | nk_cell | *DPH3* | 4.144564515 | 0.000204269 | 0.002119204 | 1 |
| Obesity | nk_cell | *APCDD1* | -0.501604434 | 0.017827469 | 0.046067118 | 1 |
| Obesity | nk_cell | *EPHB1* | -1.196178904 | 0.006466551 | 0.021659672 | 1 |
| Obesity | nk_cell | *FBXL18* | 2.479065333 | 0.014358551 | 0.039112743 | 1 |
| Obesity | nk_cell | *PROM2* | -4.517293826 | 0.000290637 | 0.002577201 | 1 |
| Obesity | nk_cell | *AK9* | 1.780551735 | 0.010407153 | 0.030804976 | 1 |
| Obesity | nk_cell | *CDK19* | 1.989256003 | 0.001509938 | 0.007655628 | 1 |
| Obesity | nk_cell | *GTF3C6* | 2.496538371 | 0.000575287 | 0.00410071 | 1 |
| Obesity | nk_cell | *TTC39B* | 1.54899888 | 0.000412313 | 0.003239798 | 1 |
| Obesity | nk_cell | *GOLGA7B* | 1.502267314 | 0.001236253 | 0.00673642 | 1 |
| Obesity | nk_cell | *C16orf87* | 2.080800016 | 0.009890299 | 0.029567433 | 1 |
| Obesity | nk_cell | *SETD9* | -0.864894813 | 5.79E-05 | 0.001177776 | 0.364311564 |
| Obesity | nk_cell | *PDIA4* | 3.059067312 | 0.004452113 | 0.016533039 | 1 |

| | | | | | | |
|---|---|---|---|---|---|---|
| Obesity | nk_cell | *KDM8* | 3.666784196 | 0.000416167 | 0.003260557 | 1 |
| Obesity | nk_cell | *SPAG17* | -0.658592704 | 0.003632024 | 0.014226271 | 1 |
| Obesity | nk_cell | *DEPTOR* | -0.883381445 | 0.0176971 | 0.045805544 | 1 |
| Obesity | nk_cell | *WIF1* | -2.886587849 | 0.014734511 | 0.039902773 | 1 |
| Obesity | nk_cell | *UGT2B4* | -1.012484249 | 0.00075635 | 0.004798355 | 1 |
| Obesity | nk_cell | *MMP16* | -0.593137843 | 0.013487886 | 0.037372561 | 1 |
| Obesity | nk_cell | *BATF* | 0.443148966 | 0.000163895 | 0.001873074 | 1 |
| Obesity | nk_cell | *C8orf37* | -1.043568636 | 0.010914817 | 0.031902117 | 1 |
| Obesity | nk_cell | *CXCL13* | -1.5156054 | 0.001474585 | 0.007531045 | 1 |
| Obesity | nk_cell | *RWDD2B* | -0.993249404 | 0.000913217 | 0.005416413 | 1 |
| Obesity | nk_cell | *ANKRD9* | -0.909269308 | 0.000473889 | 0.0035939 | 1 |
| Obesity | nk_cell | *TDRD9* | 0.533790172 | 0.011078228 | 0.032259758 | 1 |
| Obesity | nk_cell | *SH3RF2* | -0.669613648 | 0.000242085 | 0.002299077 | 1 |
| Obesity | nk_cell | *GDF6* | -1.768129814 | 0.000611305 | 0.004294164 | 1 |
| Obesity | nk_cell | *NODAL* | -0.880672665 | 0.000801561 | 0.004960561 | 1 |
| Obesity | nk_cell | *MED19* | -0.984380869 | 0.006144693 | 0.020870709 | 1 |
| Obesity | nk_cell | *ITGAD* | 0.306819699 | 0.000374173 | 0.003035387 | 1 |
| Obesity | nk_cell | *VPS8* | 1.394274883 | 0.015512656 | 0.041430785 | 1 |
| Obesity | nk_cell | *GHRL* | 2.464992219 | 3.31E-05 | 0.001021303 | 0.208345796 |
| Obesity | nk_cell | *SLC6A1* | -1.18087441 | 0.004766184 | 0.017320809 | 1 |
| Obesity | nk_cell | *TIMP4* | -1.316624031 | 0.000332794 | 0.002789704 | 1 |
| Obesity | nk_cell | *LRP8* | 1.886352293 | 0.00013324 | 0.001695707 | 0.837679034 |
| Obesity | nk_cell | *STEAP2* | -0.813516135 | 1.84E-06 | 0.000183668 | 0.011571058 |
| Obesity | nk_cell | *TMED6* | -2.946857059 | 0.000715369 | 0.004651008 | 1 |
| Obesity | nk_cell | *CLEC18A* | -5.989496381 | 6.42E-17 | 4.03E-14 | 4.03E-13 |
| Obesity | nk_cell | *IL34* | -1.681799382 | 0.00786037 | 0.024908338 | 1 |
| Obesity | nk_cell | *DYRK1A* | 1.475858563 | 0.017046849 | 0.044451904 | 1 |
| Obesity | nk_cell | *SLC38A10* | 2.117859858 | 0.008706982 | 0.0269792 | 1 |
| Obesity | nk_cell | *C9orf43* | -1.310833367 | 0.004714129 | 0.017191258 | 1 |
| Obesity | nk_cell | *WDR19* | -0.916786504 | 0.017205245 | 0.044809185 | 1 |
| Obesity | nk_cell | *FAM213B* | -1.1847173 | 0.00133081 | 0.007042765 | 1 |
| Obesity | nk_cell | *PEX10* | -1.01468859 | 0.005863144 | 0.020078628 | 1 |
| Obesity | nk_cell | *RADIL* | -0.838642134 | 0.002012624 | 0.009255595 | 1 |
| Obesity | nk_cell | *PAFAH2* | 1.357751456 | 0.000998011 | 0.005772304 | 1 |
| Obesity | nk_cell | *WDR66* | -1.020262919 | 0.007192851 | 0.023442953 | 1 |
| Obesity | nk_cell | *GALNT14* | 3.75097388 | 8.66E-05 | 0.001360888 | 0.544564075 |
| Obesity | nk_cell | *HPD* | 2.883856396 | 1.94E-08 | 4.80E-06 | 0.000121669 |
| Obesity | nk_cell | *RHPN1* | -1.281797397 | 6.51E-05 | 0.001192487 | 0.409023191 |
| Obesity | nk_cell | *LRRC43* | 2.000380701 | 0.000114632 | 0.001553214 | 0.720691428 |
| Obesity | nk_cell | *XKR8* | 2.908148363 | 0.001281777 | 0.006864168 | 1 |
| Obesity | nk_cell | *EYA3* | 1.570143686 | 0.017123099 | 0.044632224 | 1 |
| Obesity | nk_cell | *CLSTN2* | -0.615458411 | 0.001796216 | 0.008646868 | 1 |
| Obesity | nk_cell | *RNF207* | -0.832023311 | 0.000907104 | 0.005395421 | 1 |
| Obesity | nk_cell | *CD1D* | -2.656682852 | 0.002091404 | 0.009511079 | 1 |
| Obesity | nk_cell | *CD1E* | 5.119316555 | 3.06E-05 | 0.000996069 | 0.192241352 |
| Obesity | nk_cell | *NCF1* | 5.78061246 | 0.00012918 | 0.001670927 | 0.812153888 |
| Obesity | nk_cell | *PPP1R9A* | -0.716293073 | 0.005701592 | 0.019728072 | 1 |
| Obesity | nk_cell | *GDPD5* | -0.884754039 | 0.018067047 | 0.046455308 | 1 |
| Obesity | nk_cell | *DYNC1I1* | -0.898577478 | 0.005287922 | 0.018666573 | 1 |
| Obesity | nk_cell | *COPG2* | -0.635666691 | 2.74E-05 | 0.000956877 | 0.171978361 |

| | | | | | | |
|---|---|---|---|---|---|---|
| Obesity | nk_cell | *ELK4* | 2.636557372 | 0.003377092 | 0.013523423 | 1 |
| Obesity | nk_cell | *RNF166* | 0.78158278 | 0.000127621 | 0.001670927 | 0.80235397 |
| Obesity | nk_cell | *SPATA2L* | -2.227480973 | 0.001828274 | 0.008724464 | 1 |
| Obesity | nk_cell | *ZNF276* | 0.912491131 | 0.000212832 | 0.00215532 | 1 |
| Obesity | nk_cell | *CDA* | -1.394971149 | 0.01430939 | 0.039029559 | 1 |
| Obesity | nk_cell | *B4GALT3* | 1.967858582 | 3.04E-05 | 0.000995667 | 0.191168039 |
| Obesity | nk_cell | *FAM160B2* | -1.013686967 | 0.014671303 | 0.039809444 | 1 |
| Obesity | nk_cell | *SLC5A11* | -1.202092647 | 5.32E-09 | 1.45E-06 | 3.34E-05 |
| Obesity | nk_cell | *FCER1G* | 1.098382559 | 4.13E-06 | 0.000298533 | 0.025972412 |
| Obesity | nk_cell | *CDC42SE2* | 1.342920335 | 3.73E-05 | 0.001056956 | 0.23481439 |
| Obesity | nk_cell | *RAPGEF6* | 3.0252662 | 0.005038445 | 0.017998127 | 1 |
| Obesity | nk_cell | *EPB41* | 1.659695761 | 0.000338829 | 0.002832733 | 1 |
| Obesity | nk_cell | *IFNAR2* | 1.380844703 | 0.000151377 | 0.001809392 | 0.951705371 |
| Obesity | nk_cell | *SV2A* | -2.957763922 | 0.001479812 | 0.007551608 | 1 |
| Obesity | nk_cell | *SIM2* | -1.530895675 | 6.36E-05 | 0.001179251 | 0.399766125 |
| Obesity | nk_cell | *PADI4* | 4.826454197 | 0.000187184 | 0.002009553 | 1 |
| Obesity | nk_cell | *TCHH* | -2.844813952 | 0.000107988 | 0.001507391 | 0.678923149 |
| Obesity | nk_cell | *CCDC17* | 1.850782062 | 0.000295582 | 0.002584593 | 1 |
| Obesity | nk_cell | *ACE* | -1.120578876 | 0.004274059 | 0.016080797 | 1 |
| Obesity | nk_cell | *TEPP* | 3.300142708 | 0.002685075 | 0.01137538 | 1 |
| Obesity | nk_cell | *ZFYVE28* | 1.338130299 | 1.27E-05 | 0.000650265 | 0.079982643 |
| Obesity | nk_cell | *C16orf86* | -1.150607017 | 0.005093605 | 0.01813335 | 1 |
| Obesity | nk_cell | *LYPD5* | -1.216646726 | 0.008028616 | 0.025301207 | 1 |
| Obesity | nk_cell | *NPR2* | -1.179397922 | 0.002271298 | 0.010049015 | 1 |
| Obesity | nk_cell | *CALM3* | 2.21599646 | 0.001079047 | 0.006106182 | 1 |
| Obesity | nk_cell | *NR2F6* | -1.014229579 | 0.005777987 | 0.019904769 | 1 |
| Obesity | nk_cell | *ANKLE1* | -2.630763983 | 0.000369419 | 0.003025789 | 1 |
| Obesity | nk_cell | *CILP2* | -6.132278154 | 0.000235408 | 0.002286562 | 1 |
| Obesity | nk_cell | *RSPH1* | -1.348427816 | 0.01157864 | 0.033376849 | 1 |
| Obesity | nk_cell | *SLC37A1* | 3.56659784 | 0.00023019 | 0.002250704 | 1 |
| Obesity | nk_cell | *U2AF1* | -1.29825044 | 0.001094557 | 0.006166199 | 1 |
| Obesity | nk_cell | *C21orf33* | -3.759931054 | 0.000130289 | 0.001677518 | 0.819126345 |
| Obesity | nk_cell | *LRRC3* | -1.204450895 | 0.002527918 | 0.010908046 | 1 |
| Obesity | nk_cell | *ITGB2* | 0.515504431 | 0.000241212 | 0.002297638 | 1 |
| Obesity | nk_cell | *VAV2* | -1.199159286 | 0.010904278 | 0.031902117 | 1 |
| Obesity | nk_cell | *DIP2A* | 1.758761189 | 8.87E-05 | 0.001377463 | 0.557872534 |
| Obesity | nk_cell | *PRMT2* | 1.758290632 | 0.000396786 | 0.00314899 | 1 |
| Obesity | nk_cell | *ZNF208* | -0.941898401 | 0.001419036 | 0.007342779 | 1 |
| Obesity | nk_cell | *C19orf47* | 5.262708005 | 5.68E-06 | 0.000379812 | 0.035702296 |
| Obesity | nk_cell | *HIPK4* | -1.614146095 | 0.010287629 | 0.030523041 | 1 |
| Obesity | nk_cell | *RDH13* | 2.317507478 | 5.57E-05 | 0.001177776 | 0.350341771 |
| Obesity | nk_cell | *MPZL3* | 2.951504643 | 2.95E-05 | 0.000988752 | 0.185516639 |
| Obesity | nk_cell | *PCSK7* | 1.548706116 | 5.87E-05 | 0.001177776 | 0.369256091 |
| Obesity | nk_cell | *ZBTB7B* | -2.792248129 | 0.001009223 | 0.005825786 | 1 |
| Obesity | nk_cell | *PAQR6* | -2.962396707 | 1.07E-05 | 0.000566305 | 0.067390348 |
| Obesity | nk_cell | *SLC25A44* | 1.986302624 | 0.001374496 | 0.007177288 | 1 |
| Obesity | nk_cell | *NBEAL2* | 1.542357778 | 5.90E-06 | 0.000390715 | 0.037117901 |
| Obesity | nk_cell | *FCRL3* | 0.221813044 | 0.000115703 | 0.001558555 | 0.727425637 |
| Obesity | nk_cell | *LY6K* | -1.922696686 | 0.000412769 | 0.003239798 | 1 |
| Obesity | nk_cell | *LY6E* | 1.12231427 | 0.000906188 | 0.005395081 | 1 |

| | | | | | | |
|---|---|---|---|---|---|---|
| Obesity | nk_cell | *TONSL* | -1.918401874 | 0.000793609 | 0.00493026 | 1 |
| Obesity | nk_cell | *ORAI2* | 1.414781248 | 8.98E-06 | 0.000495064 | 0.056437299 |
| Obesity | nk_cell | *RPL8* | 2.294286884 | 7.26E-05 | 0.001247173 | 0.45646517 |
| Obesity | nk_cell | *YDJC* | 0.794779832 | 0.000155262 | 0.001823463 | 0.976129777 |
| Obesity | nk_cell | *DUSP14* | -0.96036215 | 0.000219848 | 0.002180098 | 1 |
| Obesity | nk_cell | *PGAP3* | 4.13094697 | 7.94E-05 | 0.00132492 | 0.49949485 |
| Obesity | nk_cell | *IKZF3* | 0.163330117 | 0.000185301 | 0.001998267 | 1 |
| Obesity | nk_cell | *CYGB* | -1.089147599 | 0.001364323 | 0.007141964 | 1 |
| Obesity | nk_cell | *ALDH16A1* | 3.294846173 | 0.000367583 | 0.003016966 | 1 |
| Obesity | nk_cell | *SIGLEC11* | -1.902391098 | 0.001023844 | 0.005889213 | 1 |
| Obesity | nk_cell | *MPP3* | -0.924649648 | 0.001486926 | 0.007569476 | 1 |
| Obesity | nk_cell | *NAGS* | 6.316039207 | 4.28E-05 | 0.001082521 | 0.268894489 |
| Obesity | nk_cell | *PLCD3* | -0.930934067 | 0.000321735 | 0.002726073 | 1 |
| Obesity | nk_cell | *RACGAP1* | -1.68727954 | 0.003399844 | 0.013592239 | 1 |
| Obesity | nk_cell | *SPC24* | 5.743681434 | 2.00E-05 | 0.000825847 | 0.125730218 |
| Obesity | nk_cell | *ASGR2* | -2.446201281 | 0.000681377 | 0.004552412 | 1 |
| Obesity | nk_cell | *SNRNP25* | -1.646647578 | 0.004298512 | 0.016153463 | 1 |
| Obesity | nk_cell | *WDR90* | -0.982674944 | 0.004826993 | 0.017440981 | 1 |
| Obesity | nk_cell | *MEIOB* | -1.003726419 | 0.001852347 | 0.008802498 | 1 |
| Obesity | nk_cell | *FLYWCH2* | -1.031330607 | 0.007399712 | 0.023906469 | 1 |
| Obesity | nk_cell | *ZG16B* | -10.24929919 | 0.00064898 | 0.004454301 | 1 |
| Obesity | nk_cell | *RPL29* | 1.472750805 | 6.12E-05 | 0.001177776 | 0.384784939 |
| Obesity | nk_cell | *CYP4A22* | -3.705809591 | 0.004482314 | 0.016625549 | 1 |
| Obesity | nk_cell | *PDZK1IP1* | -15.94826664 | 0.004408339 | 0.016428706 | 1 |
| Obesity | nk_cell | *COA7* | -1.634554904 | 0.006663019 | 0.022117425 | 1 |
| Obesity | nk_cell | *C1orf123* | -1.463803041 | 0.006062317 | 0.020624344 | 1 |
| Obesity | nk_cell | *RBP7* | 2.996518154 | 0.006401523 | 0.021476186 | 1 |
| Obesity | nk_cell | *PDPN* | -1.024745157 | 0.000255117 | 0.002376178 | 1 |
| Obesity | nk_cell | *LAPTM5* | 1.262480381 | 6.68E-05 | 0.001202521 | 0.41967968 |
| Obesity | nk_cell | *PEF1* | 4.114819027 | 0.001985249 | 0.00916075 | 1 |
| Obesity | nk_cell | *TTLL10* | -1.723544348 | 0.00192828 | 0.009006758 | 1 |
| Obesity | nk_cell | *B3GALT2* | -2.136440772 | 0.008082933 | 0.025434133 | 1 |
| Obesity | nk_cell | *NTNG1* | -0.688027032 | 0.002767905 | 0.011548764 | 1 |
| Obesity | nk_cell | *HFM1* | -0.834575936 | 0.002409452 | 0.010490459 | 1 |
| Obesity | nk_cell | *VCAM1* | -1.040359468 | 0.005834353 | 0.020033084 | 1 |
| Obesity | nk_cell | *TRIM58* | -1.36859922 | 0.00021538 | 0.00216071 | 1 |
| Obesity | nk_cell | *VANGL2* | 1.666364568 | 0.000180642 | 0.001987008 | 1 |
| Obesity | nk_cell | *FCRLB* | 1.954082868 | 8.03E-05 | 0.001335836 | 0.504945895 |
| Obesity | nk_cell | *LMX1A* | -0.670487416 | 0.001489264 | 0.007575247 | 1 |
| Obesity | nk_cell | *TDRD5* | -0.914823432 | 0.006024698 | 0.020551969 | 1 |
| Obesity | nk_cell | *ACP6* | -1.768380177 | 0.008177339 | 0.025641361 | 1 |
| Obesity | nk_cell | *PM20D1* | -1.619849276 | 0.002771589 | 0.011548764 | 1 |
| Obesity | nk_cell | *RFTN2* | -0.720160642 | 0.003115409 | 0.012695788 | 1 |
| Obesity | nk_cell | *FAM84A* | -3.293973236 | 0.009686064 | 0.029123045 | 1 |
| Obesity | nk_cell | *CLHC1* | 0.461352876 | 0.012720636 | 0.035867404 | 1 |
| Obesity | nk_cell | *FRZB* | -1.131000177 | 0.000924087 | 0.005460274 | 1 |
| Obesity | nk_cell | *FBXO41* | 4.252544082 | 0.00030976 | 0.002656833 | 1 |
| Obesity | nk_cell | *CCDC74A* | -2.988888677 | 0.000798467 | 0.004950654 | 1 |
| Obesity | nk_cell | *H3F3A* | 1.553455573 | 0.018208529 | 0.046650516 | 1 |
| Obesity | nk_cell | *NOSTRIN* | -0.725370854 | 0.009072 | 0.027822276 | 1 |

| | | | | | | |
|---|---|---|---|---|---|---|
| Obesity | nk_cell | *SGPP2* | -1.175995768 | 0.001427239 | 0.007348939 | 1 |
| Obesity | nk_cell | *BBS5* | -0.921783659 | 0.008807363 | 0.027236543 | 1 |
| Obesity | nk_cell | *C1QTNF7* | -0.754408539 | 0.010213878 | 0.030390274 | 1 |
| Obesity | nk_cell | *S100A12* | 2.899140191 | 0.000212233 | 0.00215532 | 1 |
| Obesity | nk_cell | *PBXIP1* | 3.367080486 | 0.001631726 | 0.00808663 | 1 |
| Obesity | nk_cell | *LMOD3* | -1.768302343 | 0.006614924 | 0.022004248 | 1 |
| Obesity | nk_cell | *SLC15A2* | -1.755447499 | 0.000304281 | 0.002622439 | 1 |
| Obesity | nk_cell | *FSTL1* | -0.770105935 | 0.01451674 | 0.039498211 | 1 |
| Obesity | nk_cell | *LMOD1* | -1.892899087 | 7.28E-12 | 3.27E-09 | 4.58E-08 |
| Obesity | nk_cell | *ARPC2* | 1.594160733 | 4.28E-05 | 0.001082521 | 0.268826219 |
| Obesity | nk_cell | *GLB1L* | -1.430060822 | 0.00903278 | 0.027759449 | 1 |
| Obesity | nk_cell | *NUAK2* | 0.963471113 | 0.001962512 | 0.009094983 | 1 |
| Obesity | nk_cell | *PYHIN1* | 0.140407738 | 0.000289634 | 0.002575574 | 1 |
| Obesity | nk_cell | *AIM2* | -1.157030903 | 0.002696942 | 0.01141028 | 1 |
| Obesity | nk_cell | *ICA1L* | -1.606015573 | 0.004221334 | 0.015958824 | 1 |
| Obesity | nk_cell | *GTPBP8* | 2.960218318 | 0.001964528 | 0.009094983 | 1 |
| Obesity | nk_cell | *PPM1K* | 2.519933493 | 0.001779363 | 0.008585459 | 1 |
| Obesity | nk_cell | *DCLK3* | 4.127953927 | 0.001181651 | 0.006545408 | 1 |
| Obesity | nk_cell | *DNASE1L3* | -1.009845286 | 0.004119592 | 0.015706412 | 1 |
| Obesity | nk_cell | *PCOLCE2* | -0.66839998 | 0.004859613 | 0.01753961 | 1 |
| Obesity | nk_cell | *CCDC158* | -0.67153792 | 0.001727474 | 0.008399259 | 1 |
| Obesity | nk_cell | *GYG1* | 2.798353596 | 4.05E-07 | 6.37E-05 | 0.00254653 |
| Obesity | nk_cell | *HPS3* | 2.163077665 | 0.00142103 | 0.00734332 | 1 |
| Obesity | nk_cell | *PLB1* | -1.357296111 | 0.004944565 | 0.017753559 | 1 |
| Obesity | nk_cell | *CDCP1* | -1.420092419 | 0.004784934 | 0.017355174 | 1 |
| Obesity | nk_cell | *LZTFL1* | 3.225133087 | 9.53E-05 | 0.001427161 | 0.599407742 |
| Obesity | nk_cell | *LRRC2* | -0.715840369 | 0.00189677 | 0.008925895 | 1 |
| Obesity | nk_cell | *DTX3L* | 2.045212485 | 0.000208072 | 0.002140992 | 1 |
| Obesity | nk_cell | *DNALI1* | -4.503663301 | 2.78E-06 | 0.000236232 | 0.017481188 |
| Obesity | nk_cell | *RFC4* | 2.266329705 | 6.66E-05 | 0.001202521 | 0.418623915 |
| Obesity | nk_cell | *RFT1* | -2.824155463 | 5.72E-05 | 0.001177776 | 0.359913252 |
| Obesity | nk_cell | *GNL3* | -1.375219604 | 0.004967062 | 0.017813987 | 1 |
| Obesity | nk_cell | *PIGX* | 1.977837962 | 0.002034076 | 0.009307304 | 1 |
| Obesity | nk_cell | *C1orf50* | -3.066267161 | 0.002024414 | 0.009273819 | 1 |
| Obesity | nk_cell | *DNAJB14* | 2.291367063 | 3.27E-05 | 0.001016497 | 0.205329325 |
| Obesity | nk_cell | *BDH2* | -1.041941415 | 0.000248041 | 0.002330994 | 1 |
| Obesity | nk_cell | *ATRIP* | -1.925319743 | 0.000330772 | 0.002780163 | 1 |
| Obesity | nk_cell | *APEH* | -1.549467151 | 0.001243192 | 0.006765451 | 1 |
| Obesity | nk_cell | *RNF123* | 3.834024388 | 1.97E-24 | 4.13E-21 | 1.24E-20 |
| Obesity | nk_cell | *MST1R* | 1.047297476 | 0.013110419 | 0.036552197 | 1 |
| Obesity | nk_cell | *POC1A* | 3.062056866 | 8.85E-08 | 1.74E-05 | 0.000556204 |
| Obesity | nk_cell | *PPM1M* | -1.310445847 | 0.015040459 | 0.040513868 | 1 |
| Obesity | nk_cell | *C4orf3* | 2.665818406 | 0.004212455 | 0.015942149 | 1 |
| Obesity | nk_cell | *MAP9* | -1.235476357 | 0.004027275 | 0.015467 | 1 |
| Obesity | nk_cell | *C4orf45* | -0.812719602 | 0.013191897 | 0.036711508 | 1 |
| Obesity | nk_cell | *NPY1R* | -0.689135363 | 0.004136089 | 0.015750206 | 1 |
| Obesity | nk_cell | *ITGA2* | -1.119471178 | 0.01894057 | 0.048093443 | 1 |
| Obesity | nk_cell | *LMBRD2* | 2.439024098 | 0.004502903 | 0.016672408 | 1 |
| Obesity | nk_cell | *CMBL* | -1.041739951 | 0.002628766 | 0.01122098 | 1 |
| Obesity | nk_cell | *F2RL1* | 1.428988473 | 0.014963989 | 0.040359758 | 1 |

| | | | | | | |
|---|---|---|---|---|---|---|
| Obesity | nk_cell | *TIGD6* | -1.204919348 | 0.001155877 | 0.006436666 | 1 |
| Obesity | nk_cell | *ENPP6* | -1.759753195 | 0.005545585 | 0.01931584 | 1 |
| Obesity | nk_cell | *CASP3* | 2.442640024 | 5.38E-05 | 0.00116249 | 0.3385098 |
| Obesity | nk_cell | *ERAP2* | 0.858948852 | 0.005087754 | 0.018122781 | 1 |
| Obesity | nk_cell | *RICTOR* | 1.853680473 | 0.00333191 | 0.013410831 | 1 |
| Obesity | nk_cell | *UBLCP1* | 1.486162494 | 0.012819404 | 0.036060669 | 1 |
| Obesity | nk_cell | *UTP15* | 3.044972273 | 0.002741407 | 0.011505493 | 1 |
| Obesity | nk_cell | *KLKB1* | 0.786578878 | 0.006192234 | 0.020975524 | 1 |
| Obesity | nk_cell | *GDF9* | -1.966507043 | 0.004918373 | 0.017689823 | 1 |
| Obesity | nk_cell | *GRIK2* | -0.716668419 | 0.000498566 | 0.003727092 | 1 |
| Obesity | nk_cell | *MB21D1* | 3.255894179 | 0.000332724 | 0.002789704 | 1 |
| Obesity | nk_cell | *TXLNB* | 2.740487948 | 0.00017344 | 0.001947173 | 1 |
| Obesity | nk_cell | *DCBLD1* | 2.585662825 | 0.000391483 | 0.003128541 | 1 |
| Obesity | nk_cell | *TMEM200A* | -1.235858036 | 0.006363423 | 0.021382598 | 1 |
| Obesity | nk_cell | *STK17A* | 0.941971518 | 4.90E-05 | 0.001116189 | 0.307990326 |
| Obesity | nk_cell | *GALNT10* | 1.720087308 | 0.000784803 | 0.004909512 | 1 |
| Obesity | nk_cell | *RPS14* | 1.665179392 | 2.95E-05 | 0.000988752 | 0.185392328 |
| Obesity | nk_cell | *HCN1* | -1.492025206 | 0.000566056 | 0.0040533 | 1 |
| Obesity | nk_cell | *SLU7* | 2.34985862 | 0.00526582 | 0.018632362 | 1 |
| Obesity | nk_cell | *RP9* | -1.169179616 | 0.00351931 | 0.013924419 | 1 |
| Obesity | nk_cell | *KCNK5* | -1.093471752 | 0.000648702 | 0.004454301 | 1 |
| Obesity | nk_cell | *LMTK2* | 2.469058927 | 0.00153765 | 0.007771066 | 1 |
| Obesity | nk_cell | *PEX2* | 2.308871212 | 9.87E-05 | 0.001446589 | 0.620586719 |
| Obesity | nk_cell | *TNFRSF11B* | -0.672994354 | 0.00168556 | 0.008272535 | 1 |
| Obesity | nk_cell | *DEFB1* | -1.225810986 | 0.001470261 | 0.007525264 | 1 |
| Obesity | nk_cell | *TMEM184A* | -1.421123263 | 4.98E-05 | 0.001118514 | 0.313114743 |
| Obesity | nk_cell | *INTS1* | 3.27047422 | 8.37E-05 | 0.001341236 | 0.526257739 |
| Obesity | nk_cell | *COX6C* | 2.484473382 | 0.004266898 | 0.016073092 | 1 |
| Obesity | nk_cell | *DCAF13* | 3.129540776 | 2.27E-05 | 0.000875176 | 0.142878939 |
| Obesity | nk_cell | *FREM1* | -0.742367453 | 0.006353681 | 0.021361281 | 1 |
| Obesity | nk_cell | *RPP25L* | 2.387785898 | 0.018073753 | 0.046455308 | 1 |
| Obesity | nk_cell | *SYK* | 2.2797456 | 0.000996935 | 0.005771389 | 1 |
| Obesity | nk_cell | *NIPSNAP3B* | -0.881432094 | 0.000904662 | 0.005391097 | 1 |
| Obesity | nk_cell | *TMC1* | -0.951466966 | 0.003353208 | 0.013462083 | 1 |
| Obesity | nk_cell | *HNRNPK* | 2.398248094 | 0.000438378 | 0.003381695 | 1 |
| Obesity | nk_cell | *SVEP1* | -0.540834005 | 0.004908964 | 0.017666087 | 1 |
| Obesity | nk_cell | *ZNF367* | 1.514761573 | 6.24E-05 | 0.001177776 | 0.392594526 |
| Obesity | nk_cell | *NOL6* | 2.616299476 | 0.001288579 | 0.006894719 | 1 |
| Obesity | nk_cell | *MARCH8* | 2.30075877 | 0.004337399 | 0.016246303 | 1 |
| Obesity | nk_cell | *GJB2* | -3.291966453 | 0.004963907 | 0.017812831 | 1 |
| Obesity | nk_cell | *LRR1* | -1.299012488 | 0.013108391 | 0.036552197 | 1 |
| Obesity | nk_cell | *RPL36AL* | 1.227869519 | 0.00053342 | 0.003890498 | 1 |
| Obesity | nk_cell | *NOXRED1* | -1.127994359 | 0.002597768 | 0.011128079 | 1 |
| Obesity | nk_cell | *AKR1E2* | -0.850735967 | 0.009347845 | 0.028400797 | 1 |
| Obesity | nk_cell | *TAF3* | 2.288855264 | 0.000485205 | 0.003657655 | 1 |
| Obesity | nk_cell | *QSOX2* | 1.433636829 | 6.66E-05 | 0.001202521 | 0.418647365 |
| Obesity | nk_cell | *ZMYND19* | 6.344161367 | 0.000118245 | 0.001581721 | 0.743408905 |
| Obesity | nk_cell | *RET* | -1.438762133 | 0.000430023 | 0.0033295 | 1 |
| Obesity | nk_cell | *NSMF* | 3.288847066 | 0.017458774 | 0.045338005 | 1 |
| Obesity | nk_cell | *CASP7* | -1.046762681 | 0.008040283 | 0.025325281 | 1 |

| | | | | | | |
|---|---|---|---|---|---|---|
| Obesity | nk_cell | *HSPA12A* | -0.65068817 | 0.001258279 | 0.006790385 | 1 |
| Obesity | nk_cell | *PACSIN3* | 0.726605705 | 0.012860766 | 0.036127372 | 1 |
| Obesity | nk_cell | *TC2N* | 0.573084153 | 0.001356774 | 0.007132138 | 1 |
| Obesity | nk_cell | *IFI27L1* | -1.306935396 | 0.006726256 | 0.022268546 | 1 |
| Obesity | nk_cell | *CLMN* | -0.892939301 | 0.004789443 | 0.017355174 | 1 |
| Obesity | nk_cell | *ABTB2* | -1.024389013 | 0.004680251 | 0.017126618 | 1 |
| Obesity | nk_cell | *LIPC* | 0.519655971 | 0.0001495 | 0.001805299 | 0.939903824 |
| Obesity | nk_cell | *GPT2* | -0.547503227 | 0.006779982 | 0.022411013 | 1 |
| Obesity | nk_cell | *HIF1AN* | 2.785612571 | 0.004365895 | 0.016310351 | 1 |
| Obesity | nk_cell | *ZFYVE19* | 1.491317538 | 0.001447266 | 0.007433791 | 1 |
| Obesity | nk_cell | *SPINT1* | 2.132955407 | 0.001208294 | 0.006646143 | 1 |
| Obesity | nk_cell | *TRMT61A* | -1.507554414 | 1.09E-06 | 0.000125031 | 0.006876688 |
| Obesity | nk_cell | *LARP6* | -0.637085721 | 0.003027026 | 0.01243038 | 1 |
| Obesity | nk_cell | *ASPG* | -2.245446652 | 0.002111958 | 0.009566196 | 1 |
| Obesity | nk_cell | *GABRB3* | -1.242895169 | 0.000656099 | 0.004478712 | 1 |
| Obesity | nk_cell | *TAF10* | 2.925391542 | 0.000235313 | 0.002286562 | 1 |
| Obesity | nk_cell | *DCHS1* | -1.02185255 | 0.004046929 | 0.015513531 | 1 |
| Obesity | nk_cell | *NETO1* | -1.150188223 | 0.000768055 | 0.004843289 | 1 |
| Obesity | nk_cell | *C11orf74* | -0.91886609 | 0.009604865 | 0.028910686 | 1 |
| Obesity | nk_cell | *WDR88* | 2.55224004 | 5.99E-05 | 0.001177776 | 0.37683322 |
| Obesity | nk_cell | *CYB5R2* | -1.08048264 | 0.006226815 | 0.021081306 | 1 |
| Obesity | nk_cell | *TUB* | -0.931402873 | 0.00087373 | 0.005271726 | 1 |
| Obesity | nk_cell | *RPL27A* | 2.454004419 | 0.000140324 | 0.001737559 | 0.8822193 |
| Obesity | nk_cell | *PRTG* | -0.906420908 | 0.002076817 | 0.009468418 | 1 |
| Obesity | nk_cell | *C16orf46* | -7.617058528 | 0.003195787 | 0.012945822 | 1 |
| Obesity | nk_cell | *CLEC4E* | -0.802091963 | 0.001937977 | 0.009038621 | 1 |
| Obesity | nk_cell | *CLEC4D* | -2.139391824 | 0.007281932 | 0.023690917 | 1 |
| Obesity | nk_cell | *ZSCAN21* | -1.13188202 | 0.000310837 | 0.00266244 | 1 |
| Obesity | nk_cell | *RIMKLB* | -1.002321658 | 0.00095093 | 0.005567345 | 1 |
| Obesity | nk_cell | *SEC11C* | 1.167081502 | 6.00E-05 | 0.001177776 | 0.377108679 |
| Obesity | nk_cell | *GALR1* | -0.907949199 | 0.001264918 | 0.00680868 | 1 |
| Obesity | nk_cell | *CENPV* | -1.28326376 | 0.010257828 | 0.030463375 | 1 |
| Obesity | nk_cell | *SPDYE6* | 3.04480802 | 2.54E-05 | 0.000934443 | 0.159789818 |
| Obesity | nk_cell | *ATF7IP2* | 1.518034508 | 0.014235426 | 0.038895317 | 1 |
| Obesity | nk_cell | *ACSM1* | -1.371573917 | 0.000244148 | 0.002308205 | 1 |
| Obesity | nk_cell | *SLFN5* | 1.464614954 | 0.0002479 | 0.002330994 | 1 |
| Obesity | nk_cell | *CATSPER2* | 2.465941635 | 0.001911033 | 0.008955959 | 1 |
| Obesity | nk_cell | *SCNN1G* | -4.057811606 | 0.000640312 | 0.004418927 | 1 |
| Obesity | nk_cell | *MS4A6E* | -0.915063019 | 0.015961879 | 0.042277692 | 1 |
| Obesity | nk_cell | *MS4A14* | -1.055278641 | 0.000674366 | 0.004524803 | 1 |
| Obesity | nk_cell | *DIS3L* | -1.374140368 | 0.015578806 | 0.0415545 | 1 |
| Obesity | nk_cell | *AKTIP* | 3.347667098 | 9.99E-05 | 0.001447345 | 0.628147765 |
| Obesity | nk_cell | *DUSP18* | -1.297719518 | 0.014545052 | 0.039535125 | 1 |
| Obesity | nk_cell | *MEI1* | 0.959777246 | 8.56E-06 | 0.000480375 | 0.053801967 |
| Obesity | nk_cell | *SNRPD1* | 2.368268342 | 3.88E-05 | 0.001073267 | 0.243631555 |
| Obesity | nk_cell | *SP2* | 3.610786539 | 0.000187626 | 0.002009553 | 1 |
| Obesity | nk_cell | *RBFOX3* | 0.694528692 | 0.000106193 | 0.001499525 | 0.667635982 |
| Obesity | nk_cell | *OR51E2* | -1.138686349 | 0.009347089 | 0.028400797 | 1 |
| Obesity | nk_cell | *ZNF646* | 1.950603079 | 0.012017221 | 0.034248536 | 1 |
| Obesity | nk_cell | *VKORC1* | -2.508819441 | 1.70E-05 | 0.000750451 | 0.107084531 |

| | | | | | | |
|---|---|---|---|---|---|---|
| Obesity | nk_cell | *RAB8A* | 1.883157719 | 2.14E-05 | 0.000858426 | 0.134578117 |
| Obesity | nk_cell | *GATAD2A* | 1.927812575 | 0.000151306 | 0.001809392 | 0.95125825 |
| Obesity | nk_cell | *RPL13* | 1.710223787 | 0.000118503 | 0.001581798 | 0.745027032 |
| Obesity | nk_cell | *SERTAD3* | -1.152834706 | 0.001422617 | 0.00734332 | 1 |
| Obesity | nk_cell | *CDC42EP5* | -0.874438051 | 0.001661011 | 0.008203278 | 1 |
| Obesity | nk_cell | *KIR3DL1* | 0.192843024 | 0.00020883 | 0.002140997 | 1 |
| Obesity | nk_cell | *UBXN6* | -2.397597521 | 0.000373895 | 0.003035387 | 1 |
| Obesity | nk_cell | *ZNF444* | -1.184657141 | 0.005367622 | 0.018863186 | 1 |
| Obesity | nk_cell | *FAM57A* | 1.375009765 | 0.005633597 | 0.019557385 | 1 |
| Obesity | nk_cell | *GLOD4* | 2.471733315 | 0.005119923 | 0.018216727 | 1 |
| Obesity | nk_cell | *SRR* | 1.409874423 | 4.70E-07 | 6.94E-05 | 0.00295194 |
| Obesity | nk_cell | *CD320* | 1.980845298 | 2.44E-05 | 0.000930247 | 0.153490807 |
| Obesity | nk_cell | *ZNF558* | -0.700134971 | 0.016494321 | 0.043358385 | 1 |
| Obesity | nk_cell | *CDK2AP2* | 2.390566919 | 2.49E-05 | 0.000933482 | 0.156825024 |
| Obesity | nk_cell | *NUDT8* | -1.500747225 | 0.000623828 | 0.004348124 | 1 |
| Obesity | nk_cell | *ATP5H* | 2.464313914 | 0.008980145 | 0.027689149 | 1 |
| Obesity | nk_cell | *MGAT5B* | -1.480435946 | 0.007423143 | 0.02394433 | 1 |
| Obesity | nk_cell | *TMC8* | 0.405330761 | 4.66E-05 | 0.001109982 | 0.29271696 |
| Obesity | nk_cell | *KRT24* | -1.922972874 | 0.003001153 | 0.012348331 | 1 |
| Obesity | nk_cell | *TMEM99* | -1.03262749 | 0.004657388 | 0.017093919 | 1 |
| Obesity | nk_cell | *ZNF598* | 3.074199614 | 0.005323842 | 0.01874076 | 1 |
| Obesity | nk_cell | *MLST8* | -1.985950241 | 0.000743557 | 0.004750753 | 1 |
| Obesity | nk_cell | *KCTD5* | 2.794066307 | 0.000112276 | 0.001540771 | 0.70587893 |
| Obesity | nk_cell | *TRANK1* | 2.635644102 | 4.87E-05 | 0.001116189 | 0.30612376 |
| Obesity | nk_cell | *RPSA* | 1.305723618 | 0.000237352 | 0.002288695 | 1 |
| Obesity | nk_cell | *NAALADL1* | 0.765592214 | 0.000151304 | 0.001809392 | 0.951247329 |
| Obesity | nk_cell | *SCARA5* | -0.900032834 | 0.014850993 | 0.040158362 | 1 |
| Obesity | nk_cell | *NUDT16L1* | 1.829036235 | 3.77E-05 | 0.001056956 | 0.236891925 |
| Obesity | nk_cell | *PTGDR* | 0.194349226 | 0.00021185 | 0.00215532 | 1 |
| Obesity | nk_cell | *GLYCTK* | 2.825598563 | 0.003491925 | 0.013842201 | 1 |
| Obesity | nk_cell | *COA6* | -1.378067948 | 0.004188567 | 0.015882702 | 1 |
| Obesity | nk_cell | *KIF5C* | 3.58592521 | 0.001061158 | 0.006032101 | 1 |
| Obesity | nk_cell | *ACOX2* | -0.629705616 | 0.00779766 | 0.024722083 | 1 |
| Obesity | nk_cell | *CX3CR1* | 0.135665612 | 0.000228685 | 0.002239478 | 1 |
| Obesity | nk_cell | *SCN11A* | -0.38271844 | 0.007629482 | 0.024360871 | 1 |
| Obesity | nk_cell | *DTYMK* | 3.508169822 | 0.000526332 | 0.00385221 | 1 |
| Obesity | nk_cell | *TAP1* | 1.010459245 | 2.15E-05 | 0.000858426 | 0.135149847 |
| Obesity | nk_cell | *RFWD3* | 2.520940043 | 1.02E-05 | 0.000543427 | 0.064124432 |
| Obesity | nk_cell | *RHOH* | 0.309141938 | 0.000162462 | 0.001867269 | 1 |
| Obesity | nk_cell | *ZSWIM1* | 3.05930957 | 1.93E-06 | 0.000189946 | 0.012156518 |
| Obesity | nk_cell | *NBPF9* | -1.331803259 | 0.007765268 | 0.024649868 | 1 |
| Obesity | nk_cell | *WFDC13* | -5.032202999 | 0.005511295 | 0.019249727 | 1 |
| Obesity | nk_cell | *TMEM208* | 2.028316199 | 0.01111117 | 0.032303597 | 1 |
| Obesity | nk_cell | *LRP1B* | -0.482379878 | 0.013737988 | 0.03784448 | 1 |
| Obesity | nk_cell | *NPNT* | -0.6726248 | 0.001030972 | 0.005924788 | 1 |
| Obesity | nk_cell | *CNNM3* | 4.072175417 | 0.00018151 | 0.001987008 | 1 |
| Obesity | nk_cell | *TCTN2* | -0.811712248 | 9.35E-05 | 0.001420063 | 0.587906267 |
| Obesity | nk_cell | *ZBTB49* | 3.001132442 | 0.011468308 | 0.033165248 | 1 |
| Obesity | nk_cell | *ZNF608* | -0.668113223 | 0.003844289 | 0.014909959 | 1 |
| Obesity | nk_cell | *INPP5D* | 0.82086591 | 5.66E-05 | 0.001177776 | 0.355703968 |

| | | | | | | |
|---|---|---|---|---|---|---|
| Obesity | nk_cell | *CEP120* | 2.297655962 | 0.001191351 | 0.006587533 | 1 |
| Obesity | nk_cell | *STXBP6* | -0.821329201 | 0.006045837 | 0.020612894 | 1 |
| Obesity | nk_cell | *LGALS9* | 1.653882915 | 0.000691592 | 0.004586537 | 1 |
| Obesity | nk_cell | *SIGLEC7* | 0.569784599 | 0.00042377 | 0.003303774 | 1 |
| Obesity | nk_cell | *C8orf46* | 0.300886173 | 0.00190533 | 0.008939412 | 1 |
| Obesity | nk_cell | *FAM110B* | -0.879137703 | 0.013334715 | 0.037062489 | 1 |
| Obesity | nk_cell | *CPT1C* | -1.174442352 | 0.003462661 | 0.013778322 | 1 |
| Obesity | nk_cell | *PCSK9* | -5.833898853 | 0.003423351 | 0.013673829 | 1 |
| Obesity | nk_cell | *XPO6* | 2.513101811 | 9.93E-05 | 0.001447345 | 0.6241785 |
| Obesity | nk_cell | *RGS14* | 0.616405008 | 0.002264747 | 0.010034089 | 1 |
| Obesity | nk_cell | *RAB24* | -1.535175116 | 4.92E-06 | 0.000332564 | 0.030928424 |
| Obesity | nk_cell | *SH3TC2* | -0.982232471 | 0.00021751 | 0.002167168 | 1 |
| Obesity | nk_cell | *GPRIN1* | -4.495760441 | 0.019604501 | 0.049420008 | 1 |
| Obesity | nk_cell | *PGM2* | 3.975930563 | 9.85E-05 | 0.001446589 | 0.619506071 |
| Obesity | nk_cell | *STK32A* | -0.825325746 | 0.000812942 | 0.005020594 | 1 |
| Obesity | nk_cell | *P2RY12* | 1.384348792 | 0.002972857 | 0.012239915 | 1 |
| Obesity | nk_cell | *RNASE6* | -2.017865103 | 0.003338152 | 0.013427358 | 1 |
| Obesity | nk_cell | *NPR1* | -0.801830337 | 0.002720203 | 0.011447067 | 1 |
| Obesity | nk_cell | *CD52* | 0.720183354 | 8.99E-05 | 0.001382099 | 0.56527859 |
| Obesity | nk_cell | *SLC38A11* | -1.024207591 | 0.00091932 | 0.00544234 | 1 |
| Obesity | nk_cell | *CCDC8* | -1.51254502 | 0.001100262 | 0.006181721 | 1 |
| Obesity | nk_cell | *CLIC3* | 0.180255858 | 0.000265306 | 0.002424392 | 1 |
| Obesity | nk_cell | *INO80E* | -2.862666862 | 2.22E-05 | 0.000868302 | 0.139796693 |
| Obesity | nk_cell | *DFFB* | 3.306669343 | 0.001427189 | 0.007348939 | 1 |
| Obesity | nk_cell | *FAM103A1* | -1.664533489 | 2.93E-05 | 0.000988752 | 0.184330077 |
| Obesity | nk_cell | *CHRNA5* | -1.73802167 | 0.00186922 | 0.008849465 | 1 |
| Obesity | nk_cell | *MT1E* | -1.575609451 | 0.002364995 | 0.010368706 | 1 |
| Obesity | nk_cell | *DCXR* | 3.894113536 | 0.000192637 | 0.0020485 | 1 |
| Obesity | nk_cell | *CSGALNACT2* | 1.941219475 | 0.01483499 | 0.040132351 | 1 |
| Obesity | nk_cell | *CTNND2* | -1.042878098 | 0.000153357 | 0.001814794 | 0.964154517 |
| Obesity | nk_cell | *TPST1* | -0.691443974 | 0.009106881 | 0.027869914 | 1 |
| Obesity | nk_cell | *ZNF747* | -2.009617492 | 0.000461396 | 0.003527575 | 1 |
| Obesity | nk_cell | *PUSL1* | -1.47869934 | 0.000572521 | 0.004090271 | 1 |
| Obesity | nk_cell | *MYO7B* | -0.624199231 | 0.0060592 | 0.020624344 | 1 |
| Obesity | nk_cell | *KCNAB3* | 0.888219063 | 0.008155467 | 0.025625613 | 1 |
| Obesity | nk_cell | *RNF150* | -0.839081451 | 0.006978614 | 0.022910992 | 1 |
| Obesity | nk_cell | *FAM161A* | -1.240331033 | 0.003351763 | 0.013462083 | 1 |
| Obesity | nk_cell | *STX8* | 1.811063174 | 0.002346345 | 0.010308507 | 1 |
| Obesity | nk_cell | *SP7* | -3.629690535 | 0.010490697 | 0.030979338 | 1 |
| Obesity | nk_cell | *SEMA3E* | -0.738148572 | 0.003341135 | 0.013430764 | 1 |
| Obesity | nk_cell | *DCLK2* | -0.862696218 | 0.008214679 | 0.025732777 | 1 |
| Obesity | nk_cell | *ADORA2B* | -1.520628331 | 7.95E-06 | 0.000467386 | 0.050010353 |
| Obesity | nk_cell | *CD14* | -1.264727308 | 0.012516153 | 0.035441515 | 1 |
| Obesity | nk_cell | *KRT72* | 0.536855224 | 0.000238915 | 0.002293215 | 1 |
| Obesity | nk_cell | *SERPINB9* | 1.773575005 | 0.000510016 | 0.003776765 | 1 |
| Obesity | nk_cell | *SMAGP* | 0.936349434 | 0.007025882 | 0.023016765 | 1 |
| Obesity | nk_cell | *EMB* | 0.723525084 | 0.000260474 | 0.0024071 | 1 |
| Obesity | nk_cell | *HSPA4* | 2.294836801 | 0.010633672 | 0.031269361 | 1 |
| Obesity | nk_cell | *GTSF1* | 4.684395114 | 3.30E-05 | 0.00102082 | 0.207226532 |
| Obesity | nk_cell | *TRABD* | 1.723879196 | 0.000703219 | 0.004623717 | 1 |

| | | | | | | |
|---|---|---|---|---|---|---|
| Obesity | nk_cell | *TMEM133* | -2.030820888 | 0.001034159 | 0.005930961 | 1 |
| Obesity | nk_cell | *TTLL6* | -4.597387777 | 0.000607864 | 0.004274768 | 1 |
| Obesity | nk_cell | *KCNS3* | -0.504469297 | 0.004036628 | 0.015493455 | 1 |
| Obesity | nk_cell | *MTSS1* | 1.664758868 | 0.005755857 | 0.01986502 | 1 |
| Obesity | nk_cell | *RPS9* | 2.104906779 | 9.86E-05 | 0.001446589 | 0.619885691 |
| Obesity | nk_cell | *NDUFA3* | 2.343882626 | 7.20E-05 | 0.001240599 | 0.452818667 |
| Obesity | nk_cell | *OSCAR* | -0.761421126 | 0.013723975 | 0.03784448 | 1 |
| Obesity | nk_cell | *PAQR8* | -1.407855097 | 0.000454627 | 0.003494183 | 1 |
| Obesity | nk_cell | *NUDT6* | -0.939949505 | 0.006281503 | 0.021197966 | 1 |
| Obesity | nk_cell | *ZNF415* | -0.765744028 | 0.000196387 | 0.002062614 | 1 |
| Obesity | nk_cell | *TSNARE1* | 1.484441254 | 0.002064455 | 0.009428125 | 1 |
| Obesity | nk_cell | *TRMT61B* | 1.106781646 | 0.000232656 | 0.002269474 | 1 |
| Obesity | nk_cell | *ATP6V0E2* | 2.609081619 | 0.000665832 | 0.004508311 | 1 |
| Obesity | nk_cell | *MORN4* | -0.563052097 | 0.001943827 | 0.009052475 | 1 |
| Obesity | nk_cell | *CHST11* | 0.98534562 | 0.000113504 | 0.001549327 | 0.713598744 |
| Obesity | nk_cell | *LURAP1* | -0.85093074 | 0.000263237 | 0.00241603 | 1 |
| Obesity | nk_cell | *KCND3* | -0.768860525 | 0.01668782 | 0.043696927 | 1 |
| Obesity | nk_cell | *MRPL36* | 2.902991094 | 8.81E-05 | 0.001377186 | 0.55377044 |
| Obesity | nk_cell | *ZNF581* | -1.259877494 | 0.001121817 | 0.006280376 | 1 |
| Obesity | nk_cell | *DLK2* | -2.898554664 | 5.07E-05 | 0.001131189 | 0.318995414 |
| Obesity | nk_cell | *HOPX* | 0.212128433 | 0.000214584 | 0.002158544 | 1 |
| Obesity | nk_cell | *RSL1D1* | 2.462121494 | 0.018223901 | 0.046650516 | 1 |
| Obesity | nk_cell | *MAP6* | -1.314388448 | 0.002915741 | 0.012036286 | 1 |
| Obesity | nk_cell | *BCL2L1* | 2.543464013 | 4.79E-06 | 0.000327735 | 0.030134389 |
| Obesity | nk_cell | *PLRG1* | 2.696071154 | 0.007279435 | 0.023690917 | 1 |
| Obesity | nk_cell | *NMUR1* | 0.292844636 | 0.000181177 | 0.001987008 | 1 |
| Obesity | nk_cell | *ZIK1* | -1.155900989 | 0.001323724 | 0.007017078 | 1 |
| Obesity | nk_cell | *RGS19* | 1.706279329 | 1.66E-05 | 0.000744197 | 0.104187593 |
| Obesity | nk_cell | *TCEA2* | -1.199513439 | 0.004580251 | 0.016889171 | 1 |
| Obesity | nk_cell | *ANO5* | -1.241067333 | 0.0045196 | 0.016714543 | 1 |
| Obesity | nk_cell | *LGALS4* | -0.663553459 | 0.012722175 | 0.035867404 | 1 |
| Obesity | nk_cell | *SPATA5L1* | 1.655398628 | 0.004397957 | 0.016399738 | 1 |
| Obesity | nk_cell | *SYCE1* | -1.174280056 | 0.001393649 | 0.007231203 | 1 |
| Obesity | nk_cell | *RHNO1* | -1.791510506 | 0.003872444 | 0.014982188 | 1 |
| Obesity | nk_cell | *METTL18* | 1.870279312 | 2.53E-05 | 0.000934443 | 0.1589924 |
| Obesity | nk_cell | *ZNF570* | 3.116531424 | 0.000634009 | 0.004399574 | 1 |
| Obesity | nk_cell | *FAM90A1* | 6.227727759 | 4.27E-05 | 0.001082521 | 0.268699611 |
| Obesity | nk_cell | *C3AR1* | -1.29336504 | 0.009451779 | 0.028555183 | 1 |
| Obesity | nk_cell | *AQP4* | -0.805384367 | 0.000748183 | 0.004763352 | 1 |
| Obesity | nk_cell | *TLN2* | -0.595415174 | 0.000661815 | 0.004503067 | 1 |
| Obesity | nk_cell | *CYP4F22* | -2.059499476 | 4.13E-05 | 0.001082521 | 0.259765099 |
| Obesity | nk_cell | *C20orf196* | 2.425082863 | 0.000176032 | 0.001958788 | 1 |
| Obesity | nk_cell | *ZNF554* | -1.656463011 | 0.002401576 | 0.010470671 | 1 |
| Obesity | nk_cell | *ANKRD20A4* | -1.585795156 | 0.00068953 | 0.004577692 | 1 |
| Obesity | nk_cell | *SERF1A* | -4.428933089 | 0.001169558 | 0.006501334 | 1 |
| Obesity | nk_cell | *MOB3A* | 1.976591445 | 0.000400339 | 0.003158005 | 1 |
| Obesity | nk_cell | *MTBP* | 2.33665442 | 0.013462947 | 0.037319906 | 1 |
| Obesity | nk_cell | *MALT1* | 1.936209264 | 0.011563677 | 0.033359559 | 1 |
| Obesity | nk_cell | *ISG20* | 0.529306665 | 0.000788617 | 0.004923569 | 1 |
| Obesity | nk_cell | *CXCR6* | 1.776166996 | 0.00169647 | 0.008313098 | 1 |

| | | | | | | |
|---|---|---|---|---|---|---|
| Obesity | nk_cell | *C1QTNF4* | -1.107284209 | 2.19E-13 | 1.06E-10 | 1.38E-09 |
| Obesity | nk_cell | *HINFP* | 3.132937879 | 0.004586798 | 0.0169034 | 1 |
| Obesity | nk_cell | *SPTLC3* | -0.935468563 | 0.018027317 | 0.046411851 | 1 |
| Obesity | nk_cell | *SUCLG2* | 1.757399171 | 0.002274384 | 0.01005559 | 1 |
| Obesity | nk_cell | *RCAN2* | -0.676322685 | 0.00311589 | 0.012695788 | 1 |
| Obesity | nk_cell | *GNB2* | 1.948210084 | 0.013171309 | 0.036677343 | 1 |
| Obesity | nk_cell | *PRSS27* | 6.735721594 | 0.000301407 | 0.002610116 | 1 |
| Obesity | nk_cell | *FIBP* | 2.508223229 | 6.49E-06 | 0.000413581 | 0.040827775 |
| Obesity | nk_cell | *CARNS1* | 1.352831145 | 4.86E-05 | 0.001116189 | 0.305360023 |
| Obesity | nk_cell | *CTSW* | 0.116976505 | 0.003577779 | 0.014102506 | 1 |
| Obesity | nk_cell | *SNTG2* | -0.799455416 | 0.000219352 | 0.002178618 | 1 |
| Obesity | nk_cell | *MRPL52* | 2.154506766 | 4.29E-05 | 0.001082521 | 0.2695447 |
| Obesity | nk_cell | *CCL19* | -3.5051749 | 0.000349919 | 0.002898476 | 1 |
| Obesity | nk_cell | *FUT10* | -1.310352639 | 0.000416451 | 0.003260557 | 1 |
| Obesity | nk_cell | *CFL1* | 1.468692236 | 4.10E-05 | 0.001082521 | 0.257979627 |
| Obesity | nk_cell | *EFCAB12* | -0.975606123 | 0.000638919 | 0.004418927 | 1 |
| Obesity | nk_cell | *RAB43* | 2.588708707 | 0.000224917 | 0.002219862 | 1 |
| Obesity | nk_cell | *RAB37* | 0.472462458 | 0.000113606 | 0.001549327 | 0.714239915 |
| Obesity | nk_cell | *SNX32* | 2.133674868 | 0.017300317 | 0.045002525 | 1 |
| Obesity | nk_cell | *NADSYN1* | 1.868432345 | 0.019435115 | 0.049052013 | 1 |
| Obesity | nk_cell | *MYEOV* | -0.341863519 | 0.008327694 | 0.026034913 | 1 |
| Obesity | nk_cell | *ANKRD13D* | 2.041202609 | 0.006536989 | 0.021840213 | 1 |
| Obesity | nk_cell | *MRGPRF* | -0.771908244 | 0.001483602 | 0.007558675 | 1 |
| Obesity | nk_cell | *LCLAT1* | 2.132940478 | 0.006911934 | 0.022727681 | 1 |
| Obesity | nk_cell | *ADH6* | -0.578486221 | 0.001632246 | 0.00808663 | 1 |
| Obesity | nk_cell | *GXYLT2* | -0.95087082 | 0.004390268 | 0.01638078 | 1 |
| Obesity | nk_cell | *RHOD* | -0.643314393 | 0.002135461 | 0.009635023 | 1 |
| Obesity | nk_cell | *PARP15* | 0.423142227 | 0.000128539 | 0.001670927 | 0.808122395 |
| Obesity | nk_cell | *VANGL1* | 3.117144496 | 0.001823708 | 0.008724464 | 1 |
| Obesity | nk_cell | *IQCB1* | -1.247300859 | 0.003245945 | 0.013115202 | 1 |
| Obesity | nk_cell | *DMRT2* | -0.62674444 | 0.000466209 | 0.003557104 | 1 |
| Obesity | nk_cell | *MZT2A* | 1.553113097 | 0.010545718 | 0.031054298 | 1 |
| Obesity | nk_cell | *NDNF* | -0.875084752 | 0.003661101 | 0.014305371 | 1 |
| Obesity | nk_cell | *OLR1* | 3.163337636 | 4.23E-05 | 0.001082521 | 0.265723917 |
| Obesity | nk_cell | *CCDC36* | -0.560186451 | 0.010640844 | 0.031275825 | 1 |
| Obesity | nk_cell | *SMARCC1* | 1.988536781 | 0.002387177 | 0.010429592 | 1 |
| Obesity | nk_cell | *ZNF417* | -1.167790238 | 0.000139415 | 0.001735535 | 0.876501969 |
| Obesity | nk_cell | *MST1* | -1.531120525 | 7.53E-07 | 0.00010295 | 0.0047357 |
| Obesity | nk_cell | *CSPG4* | -1.270998482 | 0.001709273 | 0.00833685 | 1 |
| Obesity | nk_cell | *SNX33* | -0.966200648 | 0.011775112 | 0.033742084 | 1 |
| Obesity | nk_cell | *NUDT18* | 3.506940034 | 3.94E-05 | 0.001079501 | 0.247486117 |
| Obesity | nk_cell | *SULT1B1* | 0.265118283 | 0.000199465 | 0.002086106 | 1 |
| Obesity | nk_cell | *C1orf100* | -2.140607489 | 0.004614681 | 0.016996191 | 1 |
| Obesity | nk_cell | *RNF213* | 1.422489067 | 5.79E-05 | 0.001177776 | 0.363971359 |
| Obesity | nk_cell | *TUBB8* | 1.3183752 | 0.000445827 | 0.003430738 | 1 |
| Obesity | nk_cell | *GPR160* | -2.108502535 | 0.0076677 | 0.024432855 | 1 |
| Obesity | nk_cell | *HOXB2* | 1.61972501 | 0.008597765 | 0.026719797 | 1 |
| Obesity | nk_cell | *NRROS* | 0.802568559 | 0.018780606 | 0.047725817 | 1 |
| Obesity | nk_cell | *SLC25A30* | 4.003545407 | 0.000314382 | 0.002685491 | 1 |
| Obesity | nk_cell | *CTSF* | -1.075029174 | 0.001596165 | 0.007977017 | 1 |

| | | | | | | |
|---|---|---|---|---|---|---|
| Obesity | nk_cell | *C16orf91* | -2.051712554 | 0.00950303 | 0.028682453 | 1 |
| Obesity | nk_cell | *RGMB* | -1.116379798 | 0.002092226 | 0.009511079 | 1 |
| Obesity | nk_cell | *FAM53A* | 4.21496352 | 3.65E-05 | 0.001056956 | 0.229252533 |
| Obesity | nk_cell | *DDX23* | 2.457774487 | 0.012845815 | 0.036102654 | 1 |
| Obesity | nk_cell | *GLIS1* | -0.513633673 | 0.013357678 | 0.037109909 | 1 |
| Obesity | nk_cell | *SNAPC5* | -1.812678457 | 0.016924756 | 0.044258151 | 1 |
| Obesity | nk_cell | *LINGO2* | 0.154624389 | 0.000158285 | 0.001836045 | 0.995136346 |
| Obesity | nk_cell | *TMEM81* | -3.863817386 | 0.001678189 | 0.008249237 | 1 |
| Obesity | nk_cell | *KLF14* | 20.98456718 | 3.97E-08 | 8.33E-06 | 0.000249847 |
| Obesity | nk_cell | *ZNF266* | 1.012771231 | 0.004180551 | 0.015861872 | 1 |
| Obesity | nk_cell | *BRSK2* | -1.530592546 | 0.000753116 | 0.004787501 | 1 |
| Obesity | nk_cell | *FGFBP3* | -1.309229162 | 0.0016336 | 0.008086957 | 1 |
| Obesity | nk_cell | *HRAS* | 2.330005581 | 0.018223786 | 0.046650516 | 1 |
| Obesity | nk_cell | *BTC* | -1.202933321 | 0.006884898 | 0.022667476 | 1 |
| Obesity | nk_cell | *NLRP6* | 3.238524636 | 0.004499352 | 0.01666908 | 1 |
| Obesity | nk_cell | *FUT1* | 2.742669038 | 0.014011755 | 0.038434513 | 1 |
| Obesity | nk_cell | *CHST2* | 0.254368882 | 0.000214409 | 0.002158544 | 1 |
| Obesity | nk_cell | *TVP23C* | -1.371542282 | 0.015786139 | 0.041926897 | 1 |
| Obesity | nk_cell | *PACS1* | 1.718090511 | 0.001084301 | 0.006118067 | 1 |
| Obesity | nk_cell | *SH3BP5L* | -1.145742432 | 0.01852721 | 0.047196341 | 1 |
| Obesity | nk_cell | *PSMD2* | 1.777528058 | 8.27E-07 | 0.000108297 | 0.005198243 |
| Obesity | nk_cell | *CHST1* | -2.516142322 | 0.002667526 | 0.011331578 | 1 |
| Obesity | nk_cell | *GOLGA8A* | 1.716023111 | 0.006661928 | 0.022117425 | 1 |
| Obesity | nk_cell | *PHYHD1* | -0.746276611 | 0.003982081 | 0.015330891 | 1 |
| Obesity | nk_cell | *ZNF519* | -0.839995033 | 0.017976162 | 0.046375106 | 1 |
| Obesity | nk_cell | *CHRNA7* | -2.082115821 | 0.001634885 | 0.008086957 | 1 |
| Obesity | nk_cell | *TMEM9B* | 4.410580469 | 4.50E-07 | 6.90E-05 | 0.00282747 |
| Obesity | nk_cell | *NRIP3* | -0.897737794 | 0.000730826 | 0.004698059 | 1 |
| Obesity | nk_cell | *SCUBE2* | -1.019126012 | 0.009343329 | 0.028400797 | 1 |
| Obesity | nk_cell | *ZNF25* | -1.206017102 | 0.002089397 | 0.009511079 | 1 |
| Obesity | nk_cell | *PCSK1* | -2.417129478 | 0.001292467 | 0.006897913 | 1 |
| Obesity | nk_cell | *TBC1D10C* | 0.263207116 | 0.000137789 | 0.001728977 | 0.866279742 |
| Obesity | nk_cell | *UBQLNL* | -2.710605959 | 0.000470097 | 0.00357376 | 1 |
| Obesity | nk_cell | *KCNE3* | -1.309589857 | 0.003715006 | 0.014471029 | 1 |
| Obesity | nk_cell | *UCP2* | 2.011572759 | 0.000342242 | 0.002852192 | 1 |
| Obesity | nk_cell | *RPS6KB2* | 2.970597589 | 6.13E-05 | 0.001177776 | 0.385545259 |
| Obesity | nk_cell | *TEX26* | 3.469073429 | 0.000226041 | 0.0022205 | 1 |
| Obesity | nk_cell | *GPR156* | 2.074827742 | 7.78E-05 | 0.001306706 | 0.489263717 |
| Obesity | nk_cell | *A2M* | 2.051570809 | 0.000214472 | 0.002158544 | 1 |
| Obesity | nk_cell | *ARL4D* | -0.897260485 | 0.000627525 | 0.004369046 | 1 |
| Obesity | nk_cell | *LRRN1* | -1.444940632 | 0.015258068 | 0.040954307 | 1 |
| Obesity | nk_cell | *B3GALT6* | -1.80526844 | 0.001967392 | 0.009100196 | 1 |
| Obesity | nk_cell | *IP6K1* | 2.9383022 | 4.42E-05 | 0.001101898 | 0.277678289 |
| Obesity | nk_cell | *SSNA1* | 4.047381389 | 0.000107673 | 0.001507391 | 0.676938869 |
| Obesity | nk_cell | *CCDC57* | -1.409936105 | 0.014525189 | 0.039498211 | 1 |
| Obesity | nk_cell | *SLC35G1* | -0.998686155 | 0.007128416 | 0.023293323 | 1 |
| Obesity | nk_cell | *ZNF135* | -1.396938022 | 0.001631738 | 0.00808663 | 1 |
| Obesity | nk_cell | *COX8A* | 2.895339408 | 3.74E-05 | 0.001056956 | 0.234994931 |
| Obesity | nk_cell | *B3GNT4* | -5.481775638 | 0.000725537 | 0.004678965 | 1 |
| Obesity | nk_cell | *CDC26* | -1.171348917 | 0.000806797 | 0.004987542 | 1 |

| | | | | | | |
|---|---|---|---|---|---|---|
| Obesity | nk_cell | *SPRYD4* | -0.653386005 | 0.005669093 | 0.019648064 | 1 |
| Obesity | nk_cell | *SLCO3A1* | 1.706366868 | 0.012976846 | 0.036340948 | 1 |
| Obesity | nk_cell | *RNF152* | -0.999449586 | 0.001591433 | 0.007959696 | 1 |
| Obesity | nk_cell | *BOK* | -0.85443999 | 0.001273134 | 0.006835529 | 1 |
| Obesity | nk_cell | *ZNF843* | -2.361219587 | 0.007443307 | 0.023973397 | 1 |
| Obesity | nk_cell | *C8orf59* | 2.536376723 | 0.001297852 | 0.006909056 | 1 |
| Obesity | nk_cell | *PFN4* | -0.818525474 | 0.001446173 | 0.007433791 | 1 |
| Obesity | nk_cell | *IRX5* | -0.672848186 | 0.00411922 | 0.015706412 | 1 |
| Obesity | nk_cell | *PNMA1* | -0.873314491 | 0.000419803 | 0.003281407 | 1 |
| Obesity | nk_cell | *GCNT4* | -4.867251484 | 0.004366203 | 0.016310351 | 1 |
| Obesity | nk_cell | *SEC24C* | 2.532693662 | 0.00037856 | 0.003055211 | 1 |
| Obesity | nk_cell | *C19orf18* | -1.465985274 | 0.002763815 | 0.011548764 | 1 |
| Obesity | nk_cell | *TMEM80* | -1.126099072 | 0.001435332 | 0.007384559 | 1 |
| Obesity | nk_cell | *EPS8L2* | -0.796571803 | 0.001869374 | 0.008849465 | 1 |
| Obesity | nk_cell | *TALDO1* | -1.415837558 | 0.017655477 | 0.045735468 | 1 |
| Obesity | nk_cell | *B3GNT8* | -2.158191027 | 0.001033595 | 0.005930961 | 1 |
| Obesity | nk_cell | *C8orf31* | 5.203377664 | 0.000106426 | 0.001499525 | 0.669097503 |
| Obesity | nk_cell | *PPFIA3* | -1.915052698 | 0.002692456 | 0.011398971 | 1 |
| Obesity | nk_cell | *MAGEF1* | -1.270399582 | 0.001968549 | 0.009100196 | 1 |
| Obesity | nk_cell | *SAMD9L* | 1.340022017 | 0.002757355 | 0.01154826 | 1 |
| Obesity | nk_cell | *NAP1L5* | -0.922634565 | 0.003428084 | 0.013684042 | 1 |
| Obesity | nk_cell | *NIM1K* | -0.931258492 | 0.000912776 | 0.005416413 | 1 |
| Obesity | nk_cell | *RBM44* | -2.109984587 | 0.0105228 | 0.031015866 | 1 |
| Obesity | nk_cell | *IRX3* | -0.981738934 | 0.000178384 | 0.001974475 | 1 |
| Obesity | nk_cell | *ATOX1* | 2.949208618 | 0.000858055 | 0.005217206 | 1 |
| Obesity | nk_cell | *RPLP2* | 2.181032705 | 0.00024692 | 0.002327411 | 1 |
| Obesity | nk_cell | *C12orf54* | -2.681660732 | 0.001972445 | 0.009111509 | 1 |
| Obesity | nk_cell | *GBA* | -0.89366233 | 0.010051847 | 0.029993336 | 1 |
| Obesity | nk_cell | *FAM87B* | -2.909688113 | 2.18E-05 | 0.000858426 | 0.137348193 |
| Obesity | nk_cell | *PCDHB9* | -0.817414833 | 0.01823922 | 0.046670727 | 1 |
| Obesity | nk_cell | *ZNF619* | -1.70771608 | 0.011567336 | 0.033359559 | 1 |
| Obesity | nk_cell | *CENPBD1* | -1.509351352 | 0.017842109 | 0.046086006 | 1 |
| Obesity | nk_cell | *RIC8A* | 1.793946371 | 0.001247632 | 0.006773631 | 1 |
| Obesity | nk_cell | *C2orf73* | -1.636304698 | 0.003162267 | 0.012843135 | 1 |
| Obesity | nk_cell | *FAM26E* | -0.901121863 | 0.003491283 | 0.013842201 | 1 |
| Obesity | nk_cell | *MLF1* | -1.111001892 | 0.019218864 | 0.04862334 | 1 |
| Obesity | nk_cell | *PRSS42* | -2.741115112 | 3.24E-05 | 0.001016497 | 0.203798179 |
| Obesity | nk_cell | *GRAMD1C* | 3.993471493 | 0.001034876 | 0.005930961 | 1 |
| Obesity | nk_cell | *STAP2* | -1.039057068 | 0.000872046 | 0.005271688 | 1 |
| Obesity | nk_cell | *NDUFV2* | -1.141145311 | 0.002066284 | 0.009428125 | 1 |
| Obesity | nk_cell | *DALRD3* | -1.303409995 | 0.001694429 | 0.008309574 | 1 |
| Obesity | nk_cell | *ZNF114* | -1.423602655 | 0.000460672 | 0.003527575 | 1 |
| Obesity | nk_cell | *ZNF518B* | -1.564624428 | 0.017004392 | 0.044378005 | 1 |
| Obesity | nk_cell | *LCORL* | 1.834553304 | 0.006270457 | 0.021172053 | 1 |
| Obesity | nk_cell | *SH2B1* | -3.417108671 | 1.48E-25 | 4.65E-22 | 9.30E-22 |
| Obesity | nk_cell | *SH2D4B* | -2.953350697 | 0.00280699 | 0.011666744 | 1 |
| Obesity | nk_cell | *PRSS36* | -1.724320088 | 0.002362574 | 0.010368706 | 1 |
| Obesity | nk_cell | *TMPRSS9* | -0.925512013 | 0.010606298 | 0.03121807 | 1 |
| Obesity | nk_cell | *TMEM11* | -1.375012725 | 0.013738659 | 0.03784448 | 1 |
| Obesity | nk_cell | *BEND3* | -1.851932013 | 0.00263587 | 0.011242681 | 1 |

| | | | | | | |
|---|---|---|---|---|---|---|
| Obesity | nk_cell | *LINC00843* | -1.594772768 | 0.00929025 | 0.028298354 | 1 |
| Obesity | nk_cell | *P4HTM* | -2.017951015 | 0.009757685 | 0.029282371 | 1 |
| Obesity | nk_cell | *ERBB4* | -0.563705103 | 0.005174509 | 0.018379739 | 1 |
| Obesity | nk_cell | *GPR35* | 0.560683215 | 0.007132858 | 0.023295729 | 1 |
| Obesity | nk_cell | *CSRNP3* | -0.90467649 | 0.00160969 | 0.008025473 | 1 |
| Obesity | nk_cell | *PARP10* | 1.589899267 | 0.009550426 | 0.028784049 | 1 |
| Obesity | nk_cell | *RPP25* | -1.839268291 | 2.28E-06 | 0.000207331 | 0.014305871 |
| Obesity | nk_cell | *STX19* | 2.399858524 | 0.001939882 | 0.009040797 | 1 |
| Obesity | nk_cell | *CPNE7* | -2.862890216 | 0.001855514 | 0.008808682 | 1 |
| Obesity | nk_cell | *TRIM73* | -6.854927424 | 5.08E-15 | 2.90E-12 | 3.19E-11 |
| Obesity | nk_cell | *EFCAB13* | -1.068593935 | 0.003454569 | 0.013754831 | 1 |
| Obesity | nk_cell | *HYI* | -0.762529654 | 0.004056773 | 0.01551395 | 1 |
| Obesity | nk_cell | *C17orf62* | 2.866583987 | 8.64E-06 | 0.000480879 | 0.054339289 |
| Obesity | nk_cell | *GAK* | 2.128703657 | 0.004292387 | 0.016140096 | 1 |
| Obesity | nk_cell | *TUFM* | 5.167453944 | 3.38E-24 | 5.31E-21 | 2.13E-20 |
| Obesity | nk_cell | *CTC1* | 2.54053928 | 4.76E-06 | 0.000327735 | 0.029929207 |
| Obesity | nk_cell | *C14orf39* | -0.553158914 | 0.002413118 | 0.010491891 | 1 |
| Obesity | nk_cell | *KLHDC7A* | -0.630300417 | 0.011533536 | 0.033307921 | 1 |
| Obesity | nk_cell | *TRIML2* | -16.92918935 | 0.000131157 | 0.001682831 | 0.824587015 |
| Obesity | nk_cell | *CALR* | 2.817351419 | 0.000106317 | 0.001499525 | 0.668413044 |
| Obesity | nk_cell | *CDH4* | -1.011059547 | 0.015227272 | 0.040894429 | 1 |
| Obesity | nk_cell | *ARID3B* | 1.943824767 | 6.78E-05 | 0.001207849 | 0.426370589 |
| Obesity | nk_cell | *EGR3* | -1.994084243 | 0.010276632 | 0.03050481 | 1 |
| Obesity | nk_cell | *LINC00174* | -0.812971495 | 0.001418319 | 0.007342779 | 1 |
| Obesity | nk_cell | *CIITA* | -1.270791785 | 0.013570326 | 0.037567873 | 1 |
| Obesity | nk_cell | *ZBTB42* | -1.885930077 | 0.011211524 | 0.03252739 | 1 |
| Obesity | nk_cell | *FCER1A* | -1.418712338 | 0.002652454 | 0.011279494 | 1 |
| Obesity | nk_cell | *PCED1B* | 1.596003045 | 0.002676219 | 0.011353165 | 1 |
| Obesity | nk_cell | *FOXS1* | -2.297925839 | 0.001177901 | 0.006536157 | 1 |
| Obesity | nk_cell | *CITED4* | -1.38662045 | 0.00552077 | 0.019261422 | 1 |
| Obesity | nk_cell | *TIGD5* | -1.03437029 | 0.000319255 | 0.00271237 | 1 |
| Obesity | nk_cell | *SEPT1* | 0.739736447 | 0.000110264 | 0.001516917 | 0.693231244 |
| Obesity | nk_cell | *TDRP* | -1.137287177 | 0.015086951 | 0.040594978 | 1 |
| Obesity | nk_cell | *ZNF816* | 1.270807974 | 0.013461588 | 0.037319906 | 1 |
| Obesity | nk_cell | *HCLS1* | 1.432471123 | 3.56E-05 | 0.001051568 | 0.223983977 |
| Obesity | nk_cell | *DEFB124* | -2.746286089 | 0.001588335 | 0.007950526 | 1 |
| Obesity | nk_cell | *CYP8B1* | -0.906427196 | 0.000972732 | 0.005667807 | 1 |
| Obesity | nk_cell | *RNF182* | -1.214376453 | 0.000376711 | 0.00304811 | 1 |
| Obesity | nk_cell | *SSTR2* | -1.203455543 | 0.000760046 | 0.004812092 | 1 |
| Obesity | nk_cell | *SHISA2* | -16.9984735 | 0.000701259 | 0.004621399 | 1 |
| Obesity | nk_cell | *CHST13* | -2.675795433 | 0.004355132 | 0.016288348 | 1 |
| Obesity | nk_cell | *ZDHHC20* | 2.179784267 | 0.00012878 | 0.001670927 | 0.809639705 |
| Obesity | nk_cell | *OR51E1* | -0.792577986 | 0.012186836 | 0.034637719 | 1 |
| Obesity | nk_cell | *ZNF792* | -1.427414238 | 0.00125511 | 0.006784987 | 1 |
| Obesity | nk_cell | *BBS12* | 3.899711476 | 0.000165874 | 0.001885808 | 1 |
| Obesity | nk_cell | *LSMEM1* | -0.772008247 | 0.006585194 | 0.021928557 | 1 |
| Obesity | nk_cell | *AEN* | 1.630023147 | 0.000130477 | 0.001677518 | 0.820306333 |
| Obesity | nk_cell | *TRAPPC5* | -1.424805999 | 0.016518164 | 0.043397283 | 1 |
| Obesity | nk_cell | *ANKRD34A* | -2.763035087 | 0.006148657 | 0.020872898 | 1 |
| Obesity | nk_cell | *OR52N4* | 1.322539383 | 0.002970316 | 0.012237467 | 1 |

| | | | | | | |
|---|---|---|---|---|---|---|
| Obesity | nk_cell | *F2R* | 0.536134442 | 0.000294453 | 0.002579346 | 1 |
| Obesity | nk_cell | *TMEM132C* | -0.556301637 | 0.001786305 | 0.008605745 | 1 |
| Obesity | nk_cell | *SLC25A41* | -5.808444354 | 0.000428389 | 0.003325466 | 1 |
| Obesity | nk_cell | *TMEM136* | -0.918743109 | 0.000225319 | 0.002220018 | 1 |
| Obesity | nk_cell | *TMEM132E* | -4.064986248 | 0.009245013 | 0.028201552 | 1 |
| Obesity | nk_cell | *ZNF322* | -2.647560525 | 4.59E-06 | 0.000324026 | 0.028838291 |
| Obesity | nk_cell | *OGFOD3* | 0.851384149 | 0.012754311 | 0.035941888 | 1 |
| Obesity | nk_cell | *ATG9B* | -2.06721027 | 0.002298262 | 0.010146889 | 1 |
| Obesity | nk_cell | *AMIGO1* | 0.794503166 | 0.019002817 | 0.048134855 | 1 |
| Obesity | nk_cell | *ODF3L2* | -1.995252092 | 0.006563785 | 0.021880442 | 1 |
| Obesity | nk_cell | *CLDN7* | -1.431894494 | 5.16E-05 | 0.001138987 | 0.324611397 |
| Obesity | nk_cell | *ADO* | 5.241022974 | 0.000831801 | 0.005101986 | 1 |
| Obesity | nk_cell | *IDH2* | 1.660869733 | 0.000181266 | 0.001987008 | 1 |
| Obesity | nk_cell | *FAM181B* | -3.148102398 | 0.001367851 | 0.007154476 | 1 |
| Obesity | nk_cell | *HHIPL1* | 3.14439679 | 0.006262514 | 0.021156597 | 1 |
| Obesity | nk_cell | *FAM153B* | -1.191440087 | 0.001663612 | 0.00820968 | 1 |
| Obesity | nk_cell | *TSHZ2* | -0.668603525 | 0.011858624 | 0.033914226 | 1 |
| Obesity | nk_cell | *CAPN12* | 0.65194152 | 0.000288385 | 0.002571741 | 1 |
| Obesity | nk_cell | *MFSD5* | 2.537632782 | 2.47E-05 | 0.000931493 | 0.155451537 |
| Obesity | nk_cell | *ADI1* | -1.196702955 | 0.012944293 | 0.036265941 | 1 |
| Obesity | nk_cell | *CLEC4G* | 3.195494215 | 9.47E-05 | 0.001425377 | 0.595094337 |
| Obesity | nk_cell | *SATB1* | 2.617077927 | 0.000183893 | 0.001993342 | 1 |
| Obesity | nk_cell | *BRICD5* | -1.530062692 | 0.000682624 | 0.004555095 | 1 |
| Obesity | nk_cell | *TSKU* | -1.040382328 | 0.013071447 | 0.036492089 | 1 |
| Obesity | nk_cell | *GRID1* | -2.232439393 | 0.000364718 | 0.003008859 | 1 |
| Obesity | nk_cell | *VMO1* | 0.858664671 | 0.013633385 | 0.037692652 | 1 |
| Obesity | nk_cell | *SLC25A18* | -0.684557146 | 0.015378416 | 0.041158506 | 1 |
| Obesity | nk_cell | *LENG9* | -0.832410742 | 0.015996505 | 0.042345276 | 1 |
| Obesity | nk_cell | *C12orf60* | -1.249725824 | 0.00705161 | 0.023066323 | 1 |
| Obesity | nk_cell | *BEGAIN* | -4.719245372 | 0.013196757 | 0.036711508 | 1 |
| Obesity | nk_cell | *FAM43B* | -2.803737677 | 5.43E-05 | 0.00116249 | 0.341591109 |
| Obesity | nk_cell | *GPR19* | 1.278185667 | 9.99E-05 | 0.001447345 | 0.628147116 |
| Obesity | nk_cell | *FANCF* | -1.600060847 | 0.002767266 | 0.011548764 | 1 |
| Obesity | nk_cell | *CTNNA3* | -0.590057763 | 0.010511253 | 0.030996363 | 1 |
| Obesity | nk_cell | *ZNF623* | 2.60917795 | 0.002837645 | 0.011775758 | 1 |
| Obesity | nk_cell | *FHL3* | 1.12062807 | 0.000158168 | 0.001836045 | 0.994399474 |
| Obesity | nk_cell | *C19orf71* | 2.993830217 | 3.80E-05 | 0.001056956 | 0.23862734 |
| Obesity | nk_cell | *LRIT3* | -2.118275168 | 3.80E-05 | 0.001056956 | 0.23887204 |
| Obesity | nk_cell | *NPIPA1* | -0.591740671 | 0.007484577 | 0.024057022 | 1 |
| Obesity | nk_cell | *GRIN2A* | -0.946340804 | 0.00155626 | 0.007846195 | 1 |
| Obesity | nk_cell | *URAD* | -3.205532233 | 0.001097599 | 0.006172278 | 1 |
| Obesity | nk_cell | *GPR132* | 1.044282538 | 0.00014095 | 0.001737559 | 0.886154906 |
| Obesity | nk_cell | *PSMG1* | 0.985779836 | 0.000143701 | 0.001755781 | 0.903446526 |
| Obesity | nk_cell | *KLRC4* | 0.30297211 | 0.000777476 | 0.004878238 | 1 |
| Obesity | nk_cell | *TANGO2* | 2.853851538 | 0.000154059 | 0.001817204 | 0.968569871 |
| Obesity | nk_cell | *FAM167B* | -2.641254746 | 0.011677705 | 0.033539392 | 1 |
| Obesity | nk_cell | *FAM19A1* | 0.874750359 | 6.17E-05 | 0.001177776 | 0.388111325 |
| Obesity | nk_cell | *BMP8A* | 8.018066009 | 3.33E-08 | 7.21E-06 | 0.000209163 |
| Obesity | nk_cell | *KCTD16* | -0.618188337 | 0.009843472 | 0.02948352 | 1 |
| Obesity | nk_cell | *KCTD8* | -0.503891391 | 0.001762309 | 0.008522797 | 1 |

| | | | | | | |
|---|---|---|---|---|---|---|
| Obesity | nk_cell | *ANKRD45* | -1.903345532 | 0.000851418 | 0.005191916 | 1 |
| Obesity | nk_cell | *GPR39* | -1.096848464 | 0.000889863 | 0.005348535 | 1 |
| Obesity | nk_cell | *FAM3B* | -0.506658159 | 0.001706776 | 0.008334938 | 1 |
| Obesity | nk_cell | *TMEM236* | -1.255327279 | 0.000815327 | 0.005030383 | 1 |
| Obesity | nk_cell | *TBX1* | -1.451764624 | 0.004693432 | 0.017137401 | 1 |
| Obesity | nk_cell | *UQCR10* | 3.82418614 | 7.48E-05 | 0.001276215 | 0.470163886 |
| Obesity | nk_cell | *EIF3C* | 4.512479783 | 1.16E-28 | 7.28E-25 | 7.28E-25 |
| Obesity | nk_cell | *CNTN2* | -1.851033989 | 0.002712255 | 0.01143956 | 1 |
| Obesity | nk_cell | *CRELD2* | 3.41065615 | 2.80E-05 | 0.000968751 | 0.176312734 |
| Obesity | nk_cell | *PCDH9* | -0.497047851 | 0.01142576 | 0.033063018 | 1 |
| Obesity | nk_cell | *OAF* | -1.140911924 | 0.006725143 | 0.022268546 | 1 |
| Obesity | nk_cell | *MROH7* | 2.940647312 | 0.005962067 | 0.02036041 | 1 |
| Obesity | nk_cell | *PROS1* | -1.005702931 | 0.012529611 | 0.035441515 | 1 |
| Obesity | nk_cell | *HDDC3* | -0.711994215 | 0.006287894 | 0.021208147 | 1 |
| Obesity | nk_cell | *LPAR5* | 0.570503954 | 0.000631121 | 0.004384372 | 1 |
| Obesity | nk_cell | *SNN* | 6.534975169 | 2.97E-05 | 0.000988752 | 0.187014927 |
| Obesity | nk_cell | *KRBA2* | -1.638768453 | 0.000131558 | 0.001684536 | 0.827107293 |
| Obesity | nk_cell | *SEPT5* | -3.277933639 | 0.002025283 | 0.009273819 | 1 |
| Obesity | nk_cell | *NDUFA12* | 2.929339682 | 6.47E-05 | 0.001192365 | 0.406596512 |
| Obesity | nk_cell | *TUSC5* | -0.676353339 | 0.00750524 | 0.024098796 | 1 |
| Obesity | nk_cell | *TMED9* | 2.480222085 | 0.007769506 | 0.024649868 | 1 |
| Obesity | nk_cell | *BTBD6* | -1.113808647 | 0.0145236 | 0.039498211 | 1 |
| Obesity | nk_cell | *H1FX* | 2.878901823 | 0.003499082 | 0.013861834 | 1 |
| Obesity | nk_cell | *NUTM2A* | -2.549524742 | 0.002550847 | 0.010984366 | 1 |
| Obesity | nk_cell | *NOC4L* | -1.554850484 | 0.000476172 | 0.003601102 | 1 |
| Obesity | nk_cell | *TMEM106A* | 2.221909284 | 0.018491079 | 0.047142504 | 1 |
| Obesity | nk_cell | *UBOX5* | -1.211781007 | 0.002266328 | 0.010034089 | 1 |
| Obesity | nk_cell | *MAFF* | -2.43014755 | 8.68E-05 | 0.001360888 | 0.545716009 |
| Obesity | nk_cell | *CIB1* | 1.348902187 | 0.000261744 | 0.002409349 | 1 |
| Obesity | nk_cell | *ADSSL1* | -1.037907098 | 0.001078656 | 0.006106182 | 1 |
| Obesity | nk_cell | *SIGIRR* | 0.578147945 | 0.000185943 | 0.002001757 | 1 |
| Obesity | nk_cell | *IFITM2* | 1.330930686 | 7.07E-05 | 0.001228518 | 0.444723447 |
| Obesity | nk_cell | *TMEM105* | -13.0484843 | 0.001201459 | 0.006620133 | 1 |
| Obesity | nk_cell | *TCN2* | -0.894718699 | 0.014076521 | 0.038578503 | 1 |
| Obesity | nk_cell | *GAS2L1* | -1.375214174 | 0.009524434 | 0.028733261 | 1 |
| Obesity | nk_cell | *HS6ST3* | -1.050352582 | 0.01044236 | 0.030894644 | 1 |
| Obesity | nk_cell | *MAPK11* | -1.135674294 | 0.00182506 | 0.008724464 | 1 |
| Obesity | nk_cell | *SP140L* | 2.285560655 | 0.000724788 | 0.004678965 | 1 |
| Obesity | nk_cell | *SV2B* | -1.306951008 | 0.007405755 | 0.023913705 | 1 |
| Obesity | nk_cell | *PDE6G* | 2.777018885 | 0.0002846 | 0.002551819 | 1 |
| Obesity | nk_cell | *TLCD2* | -1.07588584 | 0.000535947 | 0.003891554 | 1 |
| Obesity | nk_cell | *AHNAK2* | -0.923286257 | 0.005861704 | 0.020078628 | 1 |
| Obesity | nk_cell | *PDIA2* | -1.784916078 | 0.008510308 | 0.02649748 | 1 |
| Obesity | nk_cell | *PBX1* | -0.555977937 | 0.01464887 | 0.039765736 | 1 |
| Obesity | nk_cell | *SNAI3* | 1.914633948 | 0.001371282 | 0.00716646 | 1 |
| Obesity | nk_cell | *MYBL1* | 0.115261342 | 0.000397639 | 0.00314899 | 1 |
| Obesity | nk_cell | *KCNIP4* | -0.600249136 | 0.017229554 | 0.044853917 | 1 |
| Obesity | nk_cell | *NPIPB4* | 2.019029272 | 0.013172807 | 0.036677343 | 1 |
| Obesity | nk_cell | *TRIM69* | -0.844918458 | 0.001602791 | 0.00799742 | 1 |
| Obesity | nk_cell | *IFITM1* | 0.696633005 | 0.000151958 | 0.001809392 | 0.955359204 |

| | | | | | | |
|---|---|---|---|---|---|---|
| Obesity | nk_cell | *PTCH1* | 1.170375445 | 0.000254121 | 0.002370413 | 1 |
| Obesity | nk_cell | *RTN4RL1* | -1.159372563 | 0.011475254 | 0.033170078 | 1 |
| Obesity | nk_cell | *RASA3* | 0.874536422 | 4.71E-05 | 0.001109982 | 0.295842556 |
| Obesity | nk_cell | *ZNF284* | -2.494070173 | 0.004387495 | 0.016380154 | 1 |
| Obesity | nk_cell | *GSAP* | 0.882758576 | 0.000196136 | 0.002062614 | 1 |
| Obesity | nk_cell | *CYP4Z1* | -1.124177801 | 0.003646789 | 0.014258312 | 1 |
| Obesity | nk_cell | *FFAR4* | -0.668484185 | 0.000269041 | 0.002451391 | 1 |
| Obesity | nk_cell | *BPIFB4* | 6.524972226 | 0.000137691 | 0.001728977 | 0.865660527 |
| Obesity | nk_cell | *ZNF749* | -0.820176273 | 0.0012597 | 0.006792223 | 1 |
| Obesity | nk_cell | *KLHL32* | 1.003975834 | 0.000461777 | 0.003527575 | 1 |
| Obesity | nk_cell | *GPAT2* | -2.247538418 | 0.00023653 | 0.002288661 | 1 |
| Obesity | nk_cell | *TOR3A* | -1.401949729 | 0.018509372 | 0.047170013 | 1 |
| Obesity | nk_cell | *RPS23* | 1.55522709 | 6.18E-05 | 0.001177776 | 0.388777528 |
| Obesity | nk_cell | *PCLO* | -0.505977369 | 0.008072877 | 0.025415213 | 1 |
| Obesity | nk_cell | *RGS7BP* | -1.171850714 | 0.000459269 | 0.00352555 | 1 |
| Obesity | nk_cell | *ZNF396* | -1.222744154 | 0.000103393 | 0.001482565 | 0.65003268 |
| Obesity | nk_cell | *TMEM222* | -1.478399216 | 0.003642504 | 0.014250419 | 1 |
| Obesity | nk_cell | *C17orf58* | -0.706759562 | 0.017154852 | 0.044696458 | 1 |
| Obesity | nk_cell | *CYP27C1* | -0.425176409 | 0.018342711 | 0.046848924 | 1 |
| Obesity | nk_cell | *HYAL3* | -1.634438806 | 4.80E-06 | 0.000327735 | 0.030151625 |
| Obesity | nk_cell | *KRT16* | -1.21521261 | 0.001879067 | 0.008875803 | 1 |
| Obesity | nk_cell | *TMEM17* | -0.970008369 | 0.000774141 | 0.004862164 | 1 |
| Obesity | nk_cell | *TNFRSF18* | 0.437191396 | 0.000201338 | 0.002099189 | 1 |
| Obesity | nk_cell | *TMEM232* | -0.760241814 | 0.003446447 | 0.013731187 | 1 |
| Obesity | nk_cell | *EMID1* | -1.412758899 | 0.014593898 | 0.039650751 | 1 |
| Obesity | nk_cell | *PLCD1* | -0.833799891 | 0.015371599 | 0.041158506 | 1 |
| Obesity | nk_cell | *HEATR4* | 0.752616529 | 0.019291991 | 0.048788716 | 1 |
| Obesity | nk_cell | *MRPS21* | 2.189357579 | 0.004052433 | 0.01551395 | 1 |
| Obesity | nk_cell | *ANGPTL5* | -1.195395262 | 0.00375841 | 0.014603908 | 1 |
| Obesity | nk_cell | *FNBP1* | 1.489728365 | 4.11E-05 | 0.001082521 | 0.258213399 |
| Obesity | nk_cell | *WDR86* | -0.893063241 | 0.000841134 | 0.005149179 | 1 |
| Obesity | nk_cell | *GJA4* | 2.70274243 | 0.006719784 | 0.022268546 | 1 |
| Obesity | nk_cell | *IFT140* | -1.002730374 | 0.010859678 | 0.03180009 | 1 |
| Obesity | nk_cell | *PLEKHN1* | 1.812550251 | 8.21E-05 | 0.001340853 | 0.516052718 |
| Obesity | nk_cell | *TET3* | 2.501557305 | 0.000349161 | 0.00289601 | 1 |
| Obesity | nk_cell | *TRPV2* | 1.56122548 | 4.82E-05 | 0.001116189 | 0.302990501 |
| Obesity | nk_cell | *C2orf88* | -0.793588717 | 0.015954865 | 0.042277692 | 1 |
| Obesity | nk_cell | *ADH1A* | -1.2657038 | 6.51E-06 | 0.000413581 | 0.040944553 |
| Obesity | nk_cell | *SEMA4D* | 0.474043773 | 0.000125601 | 0.001651994 | 0.789653324 |
| Obesity | nk_cell | *FAM69C* | -1.801126317 | 0.000342589 | 0.002852192 | 1 |
| Obesity | nk_cell | *DNAH17* | 3.336548377 | 0.001570295 | 0.007897955 | 1 |
| Obesity | nk_cell | *CCDC157* | -1.374090151 | 0.001704288 | 0.008334938 | 1 |
| Obesity | nk_cell | *GFRAL* | -0.906808029 | 0.008475114 | 0.026430081 | 1 |
| Obesity | nk_cell | *LDLRAD2* | -0.709914951 | 0.001195728 | 0.006595651 | 1 |
| Obesity | nk_cell | *C2orf66* | -3.370136644 | 0.017711011 | 0.045822685 | 1 |
| Obesity | nk_cell | *C6orf132* | -0.803660201 | 0.000964713 | 0.005631526 | 1 |
| Obesity | nk_cell | *MAPK12* | -1.420279586 | 0.007447434 | 0.023974408 | 1 |
| Obesity | nk_cell | *AGRN* | -1.330786799 | 0.001454399 | 0.007464334 | 1 |
| Obesity | nk_cell | *FAM166A* | 19.19357804 | 0.000245214 | 0.002314803 | 1 |
| Obesity | nk_cell | *ZNF626* | 2.466169254 | 0.007361668 | 0.023808027 | 1 |

| | | | | | | |
|---|---|---|---|---|---|---|
| Obesity | nk_cell | *SMTNL2* | -4.64363628 | 0.000666504 | 0.004508311 | 1 |
| Obesity | nk_cell | *LAMTOR4* | 1.667949446 | 0.007637674 | 0.024374649 | 1 |
| Obesity | nk_cell | *NUTM2B* | -17.21445435 | 0.005661154 | 0.019631372 | 1 |
| Obesity | nk_cell | *NCR3LG1* | 2.772196407 | 6.35E-05 | 0.001179251 | 0.39948504 |
| Obesity | nk_cell | *RUFY4* | -2.414917556 | 0.001256199 | 0.006784987 | 1 |
| Obesity | nk_cell | *CENPP* | 1.234074744 | 4.70E-05 | 0.001109982 | 0.295551844 |
| Obesity | nk_cell | *JAKMIP3* | -0.867421402 | 1.47E-05 | 0.000718208 | 0.092518355 |
| Obesity | nk_cell | *PDCD1* | 2.58081394 | 0.000263886 | 0.002418442 | 1 |
| Obesity | nk_cell | *CERKL* | 1.451228883 | 4.65E-05 | 0.001109982 | 0.292229737 |
| Obesity | nk_cell | *C19orf54* | -1.47374723 | 0.001394022 | 0.007231203 | 1 |
| Obesity | nk_cell | *C22orf34* | 1.534602507 | 3.94E-05 | 0.001079501 | 0.247856648 |
| Obesity | nk_cell | *FAM72B* | -2.863922395 | 0.001075783 | 0.006098692 | 1 |
| Obesity | nk_cell | *SUMO2* | 2.26956295 | 0.01802335 | 0.046411851 | 1 |
| Obesity | nk_cell | *GOLGA8M* | -1.095418076 | 0.000849849 | 0.005190886 | 1 |
| Obesity | nk_cell | *DPYD* | 1.348963337 | 0.011862157 | 0.033914226 | 1 |
| Obesity | nk_cell | *S100A16* | -0.951671431 | 0.011197268 | 0.032501026 | 1 |
| Obesity | nk_cell | *RHCE* | -0.789625121 | 0.010024877 | 0.029927067 | 1 |
| Obesity | nk_cell | *SLC4A5* | -1.928872933 | 0.003091788 | 0.012630328 | 1 |
| Obesity | nk_cell | *UROS* | 1.980679707 | 0.018217284 | 0.046650516 | 1 |
| Obesity | nk_cell | *FSIP2* | -0.742463369 | 0.001280293 | 0.006862063 | 1 |
| Obesity | nk_cell | *NOXA1* | -1.236277424 | 0.005691887 | 0.019705337 | 1 |
| Obesity | nk_cell | *SKOR1* | 3.109431827 | 0.000209094 | 0.002140997 | 1 |
| Obesity | nk_cell | *TMEM201* | -1.328233913 | 0.011098259 | 0.032288178 | 1 |
| Obesity | nk_cell | *ZNF563* | -1.058023285 | 0.01570383 | 0.041746292 | 1 |
| Obesity | nk_cell | *UTS2B* | -0.887367226 | 0.001266937 | 0.006813714 | 1 |
| Obesity | nk_cell | *NELFB* | 3.27227367 | 0.001472256 | 0.007525264 | 1 |
| Obesity | nk_cell | *SLC15A5* | -0.464051797 | 0.008156013 | 0.025625613 | 1 |
| Obesity | nk_cell | *ANKDD1B* | -0.727653837 | 0.000928215 | 0.005475922 | 1 |
| Obesity | nk_cell | *ALKBH2* | 1.926822017 | 0.013946434 | 0.038288748 | 1 |
| Obesity | nk_cell | *LITAF* | 1.281061548 | 1.77E-05 | 0.00077431 | 0.111500673 |
| Obesity | nk_cell | *ZNF527* | -1.499194535 | 0.009131643 | 0.027923462 | 1 |
| Obesity | nk_cell | *ZNF600* | 0.735267249 | 0.001576108 | 0.007908215 | 1 |
| Obesity | nk_cell | *C15orf61* | -1.352771022 | 0.004657855 | 0.017093919 | 1 |
| Obesity | nk_cell | *ZKSCAN3* | -2.334504518 | 0.009268783 | 0.028246938 | 1 |
| Obesity | nk_cell | *S100A4* | 0.777660254 | 0.012492965 | 0.035427726 | 1 |
| Obesity | nk_cell | *COLCA1* | 2.505162681 | 0.003597047 | 0.014151835 | 1 |
| Obesity | nk_cell | *HRCT1* | -0.783349681 | 0.012315191 | 0.034970915 | 1 |
| Obesity | nk_cell | *SIRPB2* | -1.356627096 | 0.006517344 | 0.021794969 | 1 |
| Obesity | nk_cell | *PPIA* | 2.862527228 | 1.53E-05 | 0.000718997 | 0.09632045 |
| Obesity | nk_cell | *ZNF471* | -0.994970647 | 0.019543483 | 0.049289739 | 1 |
| Obesity | nk_cell | *GRM7* | -0.524842809 | 0.010943404 | 0.031951962 | 1 |
| Obesity | nk_cell | *STK31* | -1.844374975 | 0.013738886 | 0.03784448 | 1 |
| Obesity | nk_cell | *ZKSCAN7* | 2.459563951 | 4.51E-05 | 0.001109982 | 0.283842801 |
| Obesity | nk_cell | *NTNG2* | 1.072403127 | 8.97E-05 | 0.001382099 | 0.564178037 |
| Obesity | nk_cell | *ZNF781* | -0.773560049 | 0.010406583 | 0.030804976 | 1 |
| Obesity | nk_cell | *INCA1* | 6.781430383 | 1.41E-08 | 3.69E-06 | 8.85E-05 |
| Obesity | nk_cell | *ZNF774* | -1.180970699 | 0.000616333 | 0.004313769 | 1 |
| Obesity | nk_cell | *EVL* | 0.938664552 | 8.15E-05 | 0.001340853 | 0.512424924 |
| Obesity | nk_cell | *ZNF605* | -1.150949445 | 0.016456497 | 0.043289539 | 1 |
| Obesity | nk_cell | *SULT1A1* | -1.903091125 | 1.99E-08 | 4.80E-06 | 0.000124924 |

| | | | | | | |
|---|---|---|---|---|---|---|
| Obesity | nk_cell | *TPK1* | 2.143924671 | 0.00026518 | 0.002424392 | 1 |
| Obesity | nk_cell | *C1orf186* | -1.307581053 | 0.001935799 | 0.009035167 | 1 |
| Obesity | nk_cell | *SPTSSB* | 6.430704605 | 6.03E-05 | 0.001177776 | 0.379049629 |
| Obesity | nk_cell | *XRCC2* | -1.516069775 | 0.001243974 | 0.006765451 | 1 |
| Obesity | nk_cell | *HDAC2* | 2.697014924 | 0.000468849 | 0.003568782 | 1 |
| Obesity | nk_cell | *ADH1B* | -0.585463918 | 0.009078875 | 0.027829784 | 1 |
| Obesity | nk_cell | *FAM180B* | -1.73796934 | 4.29E-05 | 0.001082521 | 0.269547763 |
| Obesity | nk_cell | *TOMM7* | 2.768320098 | 0.00012297 | 0.001620774 | 0.773109429 |
| Obesity | nk_cell | *C6orf106* | 1.811235015 | 1.08E-06 | 0.000125031 | 0.00679385 |
| Obesity | nk_cell | *PPTC7* | 1.848163461 | 0.00544035 | 0.019033238 | 1 |
| Obesity | nk_cell | *HCAR1* | 2.438213673 | 3.37E-05 | 0.001029448 | 0.211846397 |
| Obesity | nk_cell | *SLC39A10* | 2.174582329 | 0.001583315 | 0.007938038 | 1 |
| Obesity | nk_cell | *FUT11* | 2.151270933 | 7.80E-07 | 0.000104325 | 0.004903266 |
| Obesity | nk_cell | *WDR5B* | -1.185564614 | 0.016294728 | 0.042971876 | 1 |
| Obesity | nk_cell | *GMFB* | 2.479099552 | 0.007029138 | 0.023016765 | 1 |
| Obesity | nk_cell | *ARRDC1* | 2.737165036 | 0.013735395 | 0.03784448 | 1 |
| Obesity | nk_cell | *IGF2R* | 1.853510326 | 0.00071765 | 0.004661015 | 1 |
| Obesity | nk_cell | *GAL3ST4* | -1.064948018 | 0.002627147 | 0.01122098 | 1 |
| Obesity | nk_cell | *ZNF772* | -0.756195959 | 0.016205283 | 0.042789843 | 1 |
| Obesity | nk_cell | *ZNF257* | -1.627709158 | 0.007325654 | 0.023765753 | 1 |
| Obesity | nk_cell | *ADAM32* | -1.124295576 | 0.002343149 | 0.010301663 | 1 |
| Obesity | nk_cell | *SULT1A2* | 1.619611177 | 1.75E-22 | 1.83E-19 | 1.10E-18 |
| Obesity | nk_cell | *PIWIL2* | -0.834186326 | 0.007770962 | 0.024649868 | 1 |
| Obesity | nk_cell | *C6orf141* | -1.212932513 | 0.00110333 | 0.006191074 | 1 |
| Obesity | nk_cell | *ZNF165* | -3.152226761 | 2.68E-07 | 4.67E-05 | 0.001682016 |
| Obesity | nk_cell | *RAMP2-AS1* | -0.719899495 | 0.000671347 | 0.004509359 | 1 |
| Obesity | nk_cell | *MRPL21* | -1.115736183 | 2.03E-06 | 0.000191198 | 0.012774043 |
| Obesity | nk_cell | *ZNF517* | -1.362977216 | 0.002252003 | 0.009984725 | 1 |
| Obesity | nk_cell | *C5AR1* | -1.235074335 | 0.006061758 | 0.020624344 | 1 |
| Obesity | nk_cell | *PCDHB11* | -1.981227268 | 1.47E-05 | 0.000718208 | 0.092648869 |
| Obesity | nk_cell | *TTC30A* | -0.928247911 | 0.014230166 | 0.038895317 | 1 |
| Obesity | nk_cell | *HHLA3* | -1.384550525 | 0.007453331 | 0.023981113 | 1 |
| Obesity | nk_cell | *HOXA4* | -1.523852511 | 0.002098624 | 0.009524895 | 1 |
| Obesity | nk_cell | *BCO2* | -1.362241093 | 0.01015415 | 0.030241185 | 1 |
| Obesity | nk_cell | *CCDC154* | -2.389160651 | 0.001255573 | 0.006784987 | 1 |
| Obesity | nk_cell | *CDC42SE1* | 1.429252555 | 4.69E-05 | 0.001109982 | 0.295119923 |
| Obesity | nk_cell | *NMB* | 2.197684081 | 8.24E-05 | 0.001340853 | 0.517871217 |
| Obesity | nk_cell | *KLHL14* | -1.253678456 | 0.000522502 | 0.003828635 | 1 |
| Obesity | nk_cell | *RPS26* | -0.51731722 | 0.015140565 | 0.040678944 | 1 |
| Obesity | nk_cell | *C14orf178* | -1.001432492 | 8.32E-05 | 0.001340853 | 0.522932686 |
| Obesity | nk_cell | *MAP1LC3C* | -0.909916314 | 0.003489266 | 0.013842201 | 1 |
| Obesity | nk_cell | *ZNF780A* | -1.141093909 | 1.45E-05 | 0.000718208 | 0.091166323 |
| Obesity | nk_cell | *AC122129.1* | -1.75856062 | 0.000767595 | 0.004843289 | 1 |
| Obesity | nk_cell | *OCLN* | -1.147170223 | 6.62E-05 | 0.001202521 | 0.415970478 |
| Obesity | nk_cell | *FCHSD1* | 3.443474447 | 4.73E-05 | 0.001109982 | 0.297475092 |
| Obesity | nk_cell | *LEKR1* | -1.390141276 | 0.000488172 | 0.003673835 | 1 |
| Obesity | nk_cell | *MAK16* | -1.810874519 | 0.005342698 | 0.018786097 | 1 |
| Obesity | nk_cell | *ZNF667* | -0.710689197 | 0.003060407 | 0.012518398 | 1 |
| Obesity | nk_cell | *SFI1* | 2.169994222 | 3.71E-05 | 0.001056956 | 0.233021414 |
| Obesity | nk_cell | *ADH4* | -0.984045842 | 1.44E-06 | 0.00015604 | 0.009050344 |

| | | | | | | |
|---|---|---|---|---|---|---|
| Obesity | nk_cell | *ZNF248* | -0.989091019 | 0.011113835 | 0.032303597 | 1 |
| Obesity | nk_cell | *SOWAHC* | -1.737846814 | 0.001999305 | 0.009206705 | 1 |
| Obesity | nk_cell | *AC135178.1* | -3.774285855 | 0.010247294 | 0.030446473 | 1 |
| Obesity | nk_cell | *MIER1* | 2.271455296 | 0.00057595 | 0.004100787 | 1 |
| Obesity | nk_cell | *DDRGK1* | 3.140346932 | 0.002879715 | 0.011926726 | 1 |
| Obesity | nk_cell | *ZNF334* | -0.782274966 | 0.002173547 | 0.009753811 | 1 |
| Obesity | nk_cell | *HSD17B11* | 2.827137251 | 0.000318034 | 0.00270566 | 1 |
| Obesity | nk_cell | *RPS6KL1* | -0.639779794 | 0.001626969 | 0.008086367 | 1 |
| Obesity | nk_cell | *QRICH1* | 1.276275328 | 0.003720424 | 0.014483162 | 1 |
| Obesity | nk_cell | *ANTXRL* | -25.87233381 | 0.00056021 | 0.004020591 | 1 |
| Obesity | nk_cell | *STYX* | 2.806451929 | 2.99E-05 | 0.000988752 | 0.187862785 |
| Obesity | nk_cell | *MT1F* | 1.466294221 | 0.000748543 | 0.004763352 | 1 |
| Obesity | nk_cell | *HIST1H4E* | 2.222794557 | 0.007301092 | 0.023721946 | 1 |
| Obesity | nk_cell | *C20orf203* | -2.105288745 | 0.000932964 | 0.005492758 | 1 |
| Obesity | nk_cell | *ARC* | -4.101088771 | 0.000392512 | 0.00312879 | 1 |
| Obesity | nk_cell | *LRBA* | 1.884821475 | 0.000184925 | 0.00199764 | 1 |
| Obesity | nk_cell | *MMP17* | -0.915545825 | 0.001358658 | 0.007136073 | 1 |
| Obesity | nk_cell | *CCDC69* | 1.896824514 | 0.001063034 | 0.006037304 | 1 |
| Obesity | nk_cell | *ZNF534* | -1.653091423 | 0.000880587 | 0.005302922 | 1 |
| Obesity | nk_cell | *TUSC1* | -0.918711717 | 0.000759891 | 0.004812092 | 1 |
| Obesity | nk_cell | *ANKRD13B* | 1.728489477 | 2.64E-06 | 0.000227383 | 0.016598952 |
| Obesity | nk_cell | *ZNF652* | 1.733800714 | 0.018120576 | 0.046499616 | 1 |
| Obesity | nk_cell | *RPL10A* | 1.928868916 | 7.66E-06 | 0.000458713 | 0.048164878 |
| Obesity | nk_cell | *RCSD1* | 0.917129833 | 0.000363861 | 0.003006038 | 1 |
| Obesity | nk_cell | *RASSF9* | -1.04355665 | 0.005756969 | 0.01986502 | 1 |
| Obesity | nk_cell | *FAM169A* | 0.246921694 | 0.000138054 | 0.001728977 | 0.867946505 |
| Obesity | nk_cell | *ALPK2* | -1.373917132 | 0.003943358 | 0.01520975 | 1 |
| Obesity | nk_cell | *SFT2D1* | 2.663383727 | 0.001255866 | 0.006784987 | 1 |
| Obesity | nk_cell | *ARHGAP11A* | -1.551980633 | 0.016943943 | 0.044275381 | 1 |
| Obesity | nk_cell | *CD3E* | 0.236274975 | 0.00013957 | 0.001735535 | 0.8774736 |
| Obesity | nk_cell | *TSEN15* | 2.776979193 | 0.001461394 | 0.007494115 | 1 |
| Obesity | nk_cell | *TYW1* | -0.906838834 | 0.015094118 | 0.040594978 | 1 |
| Obesity | nk_cell | *ITPRIPL1* | 0.875474615 | 0.000432079 | 0.003341304 | 1 |
| Obesity | nk_cell | *PRMT6* | -6.703067635 | 4.24E-05 | 0.001082521 | 0.266446217 |
| Obesity | nk_cell | *SHISA4* | 2.150175952 | 0.000443491 | 0.003416949 | 1 |
| Obesity | nk_cell | *RASGEF1A* | 0.383198284 | 0.000203168 | 0.002111264 | 1 |
| Obesity | nk_cell | *APRT* | 2.41995368 | 3.51E-05 | 0.001047134 | 0.220945319 |
| Obesity | nk_cell | *TBKBP1* | 0.986469438 | 4.37E-05 | 0.001095134 | 0.274878613 |
| Obesity | nk_cell | *ZFP2* | -1.19988679 | 0.009042413 | 0.027770295 | 1 |
| Obesity | nk_cell | *L3MBTL3* | 2.413725574 | 2.18E-05 | 0.000858426 | 0.13722761 |
| Obesity | nk_cell | *MFAP3L* | -1.075880001 | 0.001618324 | 0.008062125 | 1 |
| Obesity | nk_cell | *SMG5* | 2.03799285 | 0.004796308 | 0.017360041 | 1 |
| Obesity | nk_cell | *AP001007.1* | -3.736179642 | 0.000770226 | 0.004847261 | 1 |
| Obesity | nk_cell | *ZNF525* | -1.220983043 | 0.016236092 | 0.042835212 | 1 |
| Obesity | nk_cell | *AC105020.1* | -1.731863249 | 0.002623031 | 0.011218364 | 1 |
| Obesity | nk_cell | *CAPN8* | 2.161309243 | 0.002195272 | 0.009830254 | 1 |
| Obesity | nk_cell | *FAM229B* | -0.420545389 | 0.006161071 | 0.020903754 | 1 |
| Obesity | nk_cell | *HIST2H2BF* | -3.001644601 | 0.000928477 | 0.005475922 | 1 |
| Obesity | nk_cell | *RBM20* | -1.610842325 | 0.004819983 | 0.017425665 | 1 |
| Obesity | nk_cell | *C6orf163* | 2.403415439 | 0.000396086 | 0.00314899 | 1 |

| | | | | | | |
|---|---|---|---|---|---|---|
| Obesity | nk_cell | *OOEP* | 0.907992605 | 0.003708614 | 0.014455086 | 1 |
| Obesity | nk_cell | *FOXO6* | -1.01287924 | 0.000218055 | 0.002169167 | 1 |
| Obesity | nk_cell | *C2orf72* | -2.448475998 | 0.001352854 | 0.007121874 | 1 |
| Obesity | nk_cell | *C10orf128* | 1.578190676 | 0.003941817 | 0.01520975 | 1 |
| Obesity | nk_cell | *ZDBF2* | -0.915670285 | 0.002214546 | 0.009880646 | 1 |
| Obesity | nk_cell | *RING1* | -1.011538183 | 0.014318089 | 0.039036352 | 1 |
| Obesity | nk_cell | *TAP2* | 0.591587666 | 0.012885124 | 0.036148493 | 1 |
| Obesity | nk_cell | *COL15A1* | -0.706923048 | 0.007694242 | 0.024480618 | 1 |
| Obesity | nk_cell | *NOTCH4* | -0.311595285 | 5.51E-08 | 1.12E-05 | 0.000346111 |
| Obesity | nk_cell | *AGER* | 2.747800054 | 4.43E-09 | 1.27E-06 | 2.78E-05 |
| Obesity | nk_cell | *PRRT1* | -0.711640398 | 0.004325347 | 0.016225214 | 1 |
| Obesity | nk_cell | *SKIV2L* | -1.064235382 | 3.16E-08 | 7.09E-06 | 0.000198586 |
| Obesity | nk_cell | *NELFE* | 1.889291209 | 0.000220915 | 0.002187237 | 1 |
| Obesity | nk_cell | *CARD16* | 1.30578441 | 0.000241568 | 0.002297638 | 1 |
| Obesity | nk_cell | *MSH5* | -0.845480343 | 0.007388683 | 0.02388311 | 1 |
| Obesity | nk_cell | *GPANK1* | -1.750523899 | 0.003276898 | 0.013209789 | 1 |
| Obesity | nk_cell | *MICB* | 0.451186652 | 0.004905024 | 0.017662018 | 1 |
| Obesity | nk_cell | *HLA-C* | 0.818118808 | 9.83E-07 | 0.00011884 | 0.006179685 |
| Obesity | nk_cell | *POU5F1* | 0.972846837 | 1.06E-09 | 3.32E-07 | 6.64E-06 |
| Obesity | nk_cell | *CCHCR1* | 1.25600834 | 9.28E-07 | 0.000114881 | 0.005835225 |
| Obesity | nk_cell | *PSORS1C1* | -0.22431621 | 0.004532367 | 0.016737686 | 1 |
| Obesity | nk_cell | *ABCF1* | 1.837130323 | 0.002161862 | 0.009729153 | 1 |
| Obesity | nk_cell | *LINC01123* | -24.92867143 | 0.000850424 | 0.005190886 | 1 |
| Obesity | nk_cell | *HLA-E* | 2.570768224 | 9.44E-15 | 4.94E-12 | 5.93E-11 |
| Obesity | nk_cell | *HLA-F* | 0.598392126 | 0.000164158 | 0.001873074 | 1 |
| Obesity | nk_cell | *ZFP57* | 1.349007513 | 5.58E-11 | 1.95E-08 | 3.51E-07 |
| Obesity | nk_cell | *FAM153C* | -2.195726849 | 0.003171281 | 0.012866307 | 1 |
| Obesity | nk_cell | *FAM216A* | -1.651670148 | 0.016077343 | 0.042523455 | 1 |
| Obesity | nk_cell | *ZNF783* | -1.229731653 | 0.013422574 | 0.037257274 | 1 |
| Obesity | nk_cell | *PCDHGA1* | -1.757098106 | 6.99E-05 | 0.001225599 | 0.439197048 |
| Obesity | nk_cell | *SLFN12L* | 0.190991753 | 0.000296561 | 0.002589555 | 1 |
| Obesity | nk_cell | *SYCE1L* | -1.060407508 | 0.013047677 | 0.03645811 | 1 |
| Obesity | nk_cell | *TMEM231* | -0.641005866 | 0.010711007 | 0.031452639 | 1 |
| Obesity | nk_cell | *FAM71F2* | -1.02402838 | 0.00256611 | 0.011027434 | 1 |
| Obesity | nk_cell | *LINC00654* | -1.454511493 | 0.001232985 | 0.006728972 | 1 |
| Obesity | nk_cell | *NT5M* | -0.866554849 | 0.00179851 | 0.00864801 | 1 |
| Obesity | nk_cell | *MT1A* | 1.895733687 | 0.00010631 | 0.001499525 | 0.668368084 |
| Obesity | nk_cell | *MT1M* | -1.206561745 | 0.001426458 | 0.007348939 | 1 |
| Obesity | nk_cell | *CFI* | -0.818097059 | 0.004794719 | 0.017360041 | 1 |
| Obesity | nk_cell | *SAMD9* | 1.598850882 | 4.72E-05 | 0.001109982 | 0.296615986 |
| Obesity | nk_cell | *EXOC3L4* | -1.510457664 | 0.000300704 | 0.00260762 | 1 |
| Obesity | nk_cell | *SMN2* | -1.094575499 | 0.000527343 | 0.00385512 | 1 |
| Obesity | nk_cell | *SERF1B* | -3.006646454 | 8.31E-06 | 0.000470803 | 0.052259144 |
| Obesity | nk_cell | *C17orf107* | 1.334255861 | 0.002241757 | 0.009953339 | 1 |
| Obesity | nk_cell | *LINC00487* | -2.588940339 | 0.004446787 | 0.016532792 | 1 |
| Obesity | nk_cell | *C1QTNF9B* | 10.11626523 | 9.78E-05 | 0.00144289 | 0.614671253 |
| Obesity | nk_cell | *NYNRIN* | -0.999280843 | 0.003093799 | 0.012630332 | 1 |
| Obesity | nk_cell | *ZDHHC11B* | -0.80267986 | 0.011667113 | 0.033524287 | 1 |
| Obesity | nk_cell | *GOLGA8O* | 2.442166927 | 0.000985911 | 0.005728674 | 1 |
| Obesity | nk_cell | *HLA-A* | 0.572834273 | 0.000947896 | 0.005564354 | 1 |

| | | | | | | |
|---|---|---|---|---|---|---|
| Obesity | nk_cell | *LNP1* | -1.37537039 | 0.001332003 | 0.007043146 | 1 |
| Obesity | nk_cell | *VGLL3* | -0.676433095 | 0.000163389 | 0.001873074 | 1 |
| Obesity | nk_cell | *COLQ* | 1.903309657 | 0.000985871 | 0.005728674 | 1 |
| Obesity | nk_cell | *METTL6* | 2.234199877 | 0.001851966 | 0.008802498 | 1 |
| Obesity | nk_cell | *SACM1L* | 1.871720238 | 7.74E-05 | 0.001306706 | 0.486501754 |
| Obesity | nk_cell | *SLC48A1* | -0.947835096 | 0.002382093 | 0.010421865 | 1 |
| Obesity | nk_cell | *TAS2R14* | 0.888828639 | 0.016218802 | 0.042807559 | 1 |
| Obesity | nk_cell | *ZRSR1* | 8.233622129 | 0.000147849 | 0.00179099 | 0.929523667 |
| Obesity | nk_cell | *AC084018.1* | -1.227042275 | 3.30E-07 | 5.46E-05 | 0.002073276 |
| Obesity | nk_cell | *OR2A42* | -1.131884641 | 0.000574701 | 0.00410071 | 1 |
| Obesity | nk_cell | *RNF208* | -1.96303186 | 0.000706553 | 0.004623717 | 1 |
| Obesity | nk_cell | *AC022498.1* | -1.941685479 | 0.006016942 | 0.020536653 | 1 |
| Obesity | nk_cell | *GIMAP1* | 0.86100645 | 0.000187509 | 0.002009553 | 1 |
| Obesity | nk_cell | *CCDC183* | -3.34605009 | 0.004984709 | 0.017856902 | 1 |
| Obesity | nk_cell | *ARHGEF35* | -2.250593622 | 0.001736441 | 0.008423616 | 1 |
| Obesity | nk_cell | *NOTCH2NL* | 1.010018971 | 0.018963767 | 0.048113479 | 1 |
| Obesity | nk_cell | *SUPT4H1* | 2.200125866 | 8.11E-06 | 0.000470803 | 0.050992444 |
| Obesity | nk_cell | *LCAT* | -0.804860055 | 0.001989285 | 0.009169089 | 1 |
| Obesity | nk_cell | *SIPA1* | 0.983594262 | 7.40E-06 | 0.000447593 | 0.046549716 |
| Obesity | nk_cell | *LBH* | 0.993848256 | 9.04E-05 | 0.001386249 | 0.568361885 |
| Obesity | nk_cell | *NCKIPSD* | -1.272304543 | 0.005274304 | 0.018649577 | 1 |
| Obesity | nk_cell | *KLRK1* | 0.479783305 | 0.000291823 | 0.002579346 | 1 |
| Obesity | nk_cell | *PPM1N* | -1.623639223 | 0.007296134 | 0.023718093 | 1 |
| Obesity | nk_cell | *ITGA1* | -0.678738232 | 0.013623691 | 0.037682423 | 1 |
| Obesity | nk_cell | *NUDT19* | 1.694968364 | 0.003600466 | 0.014156428 | 1 |
| Obesity | nk_cell | *ZNF726* | -1.309435313 | 0.0017061 | 0.008334938 | 1 |
| Obesity | nk_cell | *AP1G2* | 2.903411988 | 6.63E-05 | 0.001202521 | 0.416669621 |
| Obesity | nk_cell | *TM6SF2* | -10.10059674 | 0.001361318 | 0.007141964 | 1 |
| Obesity | nk_cell | *AC079776.2* | -1.706097251 | 0.001010038 | 0.005825786 | 1 |
| Obesity | nk_cell | *PRCD* | -1.112789627 | 0.001565122 | 0.007884553 | 1 |
| Obesity | nk_cell | *FIS1* | 1.148551447 | 0.012805117 | 0.036036601 | 1 |
| Obesity | nk_cell | *COLCA2* | -0.922112886 | 0.001129441 | 0.00631182 | 1 |
| Obesity | nk_cell | *NEURL1B* | -0.900325348 | 0.001960663 | 0.009094983 | 1 |
| Obesity | nk_cell | *VSTM5* | -0.988594914 | 0.00048852 | 0.003673835 | 1 |
| Obesity | nk_cell | *CPEB1* | -1.009279066 | 0.004676813 | 0.017126618 | 1 |
| Obesity | nk_cell | *C10orf105* | -1.328152494 | 0.001422644 | 0.00734332 | 1 |
| Obesity | nk_cell | *AC005082.1* | -2.92474543 | 0.00131646 | 0.006984461 | 1 |
| Obesity | nk_cell | *TBC1D26* | 6.846083061 | 0.000371077 | 0.003025894 | 1 |
| Obesity | nk_cell | *GRID2IP* | 2.603649368 | 0.000864755 | 0.005252866 | 1 |
| Obesity | nk_cell | *UBE2QL1* | -1.07317767 | 0.000707972 | 0.004626839 | 1 |
| Obesity | nk_cell | *GOLGA8B* | 2.657948465 | 0.001716917 | 0.008354687 | 1 |
| Obesity | nk_cell | *KCNU1* | -0.970883196 | 0.016496462 | 0.043358385 | 1 |
| Obesity | nk_cell | *NPEPL1* | -1.246594008 | 0.018346119 | 0.046848924 | 1 |
| Obesity | nk_cell | *TSTD1* | 2.512450205 | 2.28E-05 | 0.000875176 | 0.143528943 |
| Obesity | nk_cell | *LINC01138* | 1.457029869 | 3.95E-05 | 0.001079501 | 0.248285135 |
| Obesity | nk_cell | *CYB5RL* | -0.904337913 | 0.005280141 | 0.018649577 | 1 |
| Obesity | nk_cell | *PAM16* | -0.939999794 | 0.007520318 | 0.02412257 | 1 |
| Obesity | nk_cell | *RNU6ATAC35l* | -8.426105011 | 0.006868531 | 0.022644183 | 1 |
| Obesity | nk_cell | *FANCG* | -0.76467489 | 0.003131142 | 0.012741418 | 1 |
| Obesity | nk_cell | *AP4M1* | -0.883892407 | 0.004923775 | 0.017699125 | 1 |

| | | | | | | |
|---|---|---|---|---|---|---|
| Obesity | nk_cell | *C19orf73* | -5.260677745 | 0.000792094 | 0.00493026 | 1 |
| Obesity | nk_cell | *ZNF880* | -0.595951275 | 0.015616982 | 0.041615479 | 1 |
| Obesity | nk_cell | *OR2A25* | -25.65695018 | 0.003851103 | 0.014917981 | 1 |
| Obesity | nk_cell | *C1orf229* | -1.53075657 | 0.000700304 | 0.004621087 | 1 |
| Obesity | nk_cell | *SLC12A8* | -1.452491135 | 7.27E-06 | 0.000443626 | 0.045693469 |
| Obesity | nk_cell | *UBA52* | 1.527581565 | 3.79E-05 | 0.001056956 | 0.238362049 |
| Obesity | nk_cell | *AC062017.1* | -0.815448901 | 0.007178585 | 0.023408591 | 1 |
| Obesity | nk_cell | *ADRA2B* | -2.828100875 | 0.00064023 | 0.004418927 | 1 |
| Obesity | nk_cell | *VPS52* | 2.536997888 | 0.003538669 | 0.013992208 | 1 |
| Obesity | nk_cell | *LINC00205* | -1.506042612 | 0.009376668 | 0.028437583 | 1 |
| Obesity | nk_cell | *HLA-DPB1* | 1.095904989 | 0.011191844 | 0.032500289 | 1 |
| Obesity | nk_cell | *LINC01133* | -0.541716717 | 0.000910737 | 0.005411912 | 1 |
| Obesity | nk_cell | *LINC01117* | -0.863346914 | 0.005762774 | 0.019865305 | 1 |
| Obesity | nk_cell | *RASAL2-AS1* | -3.210377342 | 0.000347609 | 0.002886945 | 1 |
| Obesity | nk_cell | *AC069213.1* | -4.373520124 | 0.000108133 | 0.001507391 | 0.67983346 |
| Obesity | nk_cell | *AL049840.1* | -1.265566476 | 1.71E-06 | 0.000173084 | 0.010731193 |
| Obesity | nk_cell | *LINC00237* | -3.589151445 | 0.000651713 | 0.004463313 | 1 |
| Obesity | nk_cell | *PLEKHM1* | 2.712855492 | 0.000615952 | 0.004313769 | 1 |
| Obesity | nk_cell | *SFTA1P* | -5.086526045 | 0.001601673 | 0.00799742 | 1 |
| Obesity | nk_cell | *MIAT* | 0.401200749 | 0.000132731 | 0.001695707 | 0.834477275 |
| Obesity | nk_cell | *LINC00626* | -13.37689238 | 0.00366607 | 0.01431589 | 1 |
| Obesity | nk_cell | *ZNF503-AS1* | -0.952772586 | 0.001490647 | 0.007576148 | 1 |
| Obesity | nk_cell | *LINC00937* | -1.605773167 | 0.002363639 | 0.010368706 | 1 |
| Obesity | nk_cell | *AC064836.3* | -1.501472938 | 0.000883958 | 0.005318127 | 1 |
| Obesity | nk_cell | *TMEM185B* | -1.202353944 | 0.001586899 | 0.00794967 | 1 |
| Obesity | nk_cell | *TAS2R46* | -1.380530447 | 0.018020776 | 0.046411851 | 1 |
| Obesity | nk_cell | *AC093901.1* | -1.297876202 | 0.002807664 | 0.011666744 | 1 |
| Obesity | nk_cell | *C14orf132* | -0.864491723 | 0.005731871 | 0.019821597 | 1 |
| Obesity | nk_cell | *AC108463.2* | -2.367395111 | 6.77E-05 | 0.001207849 | 0.425794532 |
| Obesity | nk_cell | *HCG20* | -10.71811687 | 3.64E-06 | 0.000272306 | 0.022873686 |
| Obesity | nk_cell | *C19orf24* | 1.247469396 | 0.000314147 | 0.002685491 | 1 |
| Obesity | nk_cell | *LINC01013* | -1.833023923 | 0.002158144 | 0.009719376 | 1 |
| Obesity | nk_cell | *AC073133.2* | -10.15378829 | 0.00026135 | 0.002409252 | 1 |
| Obesity | nk_cell | *LINC00954* | 1.446699702 | 8.66E-05 | 0.001360888 | 0.544368891 |
| Obesity | nk_cell | *AC067945.3* | 0.271706879 | 0.000212282 | 0.00215532 | 1 |
| Obesity | nk_cell | *RPL41* | -2.155980193 | 0.004154832 | 0.015802436 | 1 |
| Obesity | nk_cell | *LINC00200* | -5.965036527 | 0.007164413 | 0.023383508 | 1 |
| Obesity | nk_cell | *PATL2* | 0.824424036 | 5.95E-05 | 0.001177776 | 0.374138961 |
| Obesity | nk_cell | *U91324.1* | 0.535121329 | 0.000195779 | 0.002062614 | 1 |
| Obesity | nk_cell | *PSMG3-AS1* | -0.960540436 | 0.013943377 | 0.038288748 | 1 |
| Obesity | nk_cell | *AC092171.4* | -1.583190667 | 0.018105134 | 0.046478962 | 1 |
| Obesity | nk_cell | *HLA-DPA1* | 1.7486619 | 0.004158168 | 0.015805562 | 1 |
| Obesity | nk_cell | *RPS18* | 1.78078962 | 5.64E-07 | 8.06E-05 | 0.003546798 |
| Obesity | nk_cell | *TAPBP* | 1.665407632 | 3.95E-06 | 0.000288662 | 0.02482496 |
| Obesity | nk_cell | *AC107021.1* | -0.527060555 | 0.000845581 | 0.005171368 | 1 |
| Obesity | nk_cell | *AC009495.3* | 0.443900897 | 0.000202124 | 0.0021039 | 1 |
| Obesity | nk_cell | *GOLGA8N* | 1.104841526 | 0.000129129 | 0.001670927 | 0.811834345 |
| Obesity | nk_cell | *TNF* | 0.414321546 | 0.005277536 | 0.018649577 | 1 |
| Obesity | nk_cell | *LYRM9* | -1.1551198 | 0.002133171 | 0.009634514 | 1 |
| Obesity | nk_cell | *LINC00702* | -0.871980576 | 0.001870676 | 0.008849465 | 1 |

| | | | | | | |
|---|---|---|---|---|---|---|
| Obesity | nk_cell | *LINC00472* | -0.654432828 | 0.002643798 | 0.011268854 | 1 |
| Obesity | nk_cell | *RPS28* | 3.680787719 | 0.00260472 | 0.011147636 | 1 |
| Obesity | nk_cell | *AC123768.3* | -1.413056252 | 0.00732689 | 0.023765753 | 1 |
| Obesity | nk_cell | *HLA-B* | 0.824657133 | 3.46E-06 | 0.00026837 | 0.021737995 |
| Obesity | nk_cell | *ANKRD65* | -1.157112348 | 0.013703396 | 0.037836298 | 1 |
| Obesity | nk_cell | *KIAA0040* | 3.042933287 | 2.97E-05 | 0.000988752 | 0.187018091 |
| Obesity | nk_cell | *B3GALT4* | 1.428523873 | 0.000181468 | 0.001987008 | 1 |
| Obesity | nk_cell | *LINC00941* | -2.71052017 | 0.001208236 | 0.006646143 | 1 |
| Obesity | nk_cell | *LINC00545* | -10.98534434 | 0.000549755 | 0.003963659 | 1 |
| Obesity | nk_cell | *B3GNT9* | -1.36891619 | 0.007751722 | 0.024626112 | 1 |
| Obesity | nk_cell | *LINC00959* | 2.057475091 | 0.004225132 | 0.015959038 | 1 |
| Obesity | nk_cell | *AC073257.2* | -6.118695513 | 0.005383612 | 0.018898253 | 1 |
| Obesity | nk_cell | *TXNDC5* | 3.101201539 | 0.001955823 | 0.009088146 | 1 |
| Obesity | nk_cell | *CDRT4* | -1.294454274 | 0.006623414 | 0.022020838 | 1 |
| Obesity | nk_cell | *APOBEC3G* | 0.273990664 | 0.000177961 | 0.001973263 | 1 |
| Obesity | nk_cell | *PSMB9* | 1.047247044 | 1.93E-07 | 3.46E-05 | 0.001212236 |
| Obesity | nk_cell | *L1TD1* | -0.682830657 | 0.008023901 | 0.025299028 | 1 |
| Obesity | nk_cell | *C1QTNF9* | -2.704366326 | 0.00595127 | 0.020345642 | 1 |
| Obesity | nk_cell | *LINC00886* | -1.04929381 | 0.017300955 | 0.045002525 | 1 |
| Obesity | nk_cell | *CD302* | -1.442701304 | 0.010481638 | 0.030967131 | 1 |
| Obesity | nk_cell | *ARHGAP8* | -1.325658743 | 0.000748006 | 0.004763352 | 1 |
| Obesity | nk_cell | *PLEKHO2* | 2.641630271 | 7.63E-05 | 0.001292271 | 0.479432539 |
| Obesity | nk_cell | *PWP2* | 1.567686606 | 0.000492703 | 0.003696452 | 1 |
| Obesity | nk_cell | *PI4KA* | 2.457494482 | 0.000216174 | 0.00216071 | 1 |
| Obesity | nk_cell | *AKAP2* | -5.399742281 | 0.00864115 | 0.026814862 | 1 |
| Obesity | nk_cell | *AMACR* | -0.373153378 | 0.018664233 | 0.047487669 | 1 |
| Obesity | nk_cell | *C22orf39* | 1.882728 | 3.14E-05 | 0.001011141 | 0.197172554 |
| Obesity | nk_cell | *LINC00960* | -0.597061522 | 0.009450614 | 0.028555183 | 1 |
| Obesity | nk_cell | *DECR2* | -1.707435364 | 0.004685507 | 0.017126618 | 1 |
| Obesity | nk_cell | *PSG4* | -1.371300268 | 0.000390289 | 0.003128541 | 1 |
| Obesity | nk_cell | *VSIG8* | -1.273482579 | 0.001904167 | 0.008939412 | 1 |
| Obesity | nk_cell | *FGF14-IT1* | -2.068256448 | 0.000770167 | 0.004847261 | 1 |
| Obesity | nk_cell | *PALM2* | -0.510142259 | 0.000279975 | 0.002529499 | 1 |
| Obesity | nk_cell | *UPK3B* | -1.044712024 | 0.01031126 | 0.030564579 | 1 |
| Obesity | nk_cell | *NME1-NME2* | -1.81122125 | 0.012097443 | 0.034445935 | 1 |
| Obesity | nk_cell | *APOBEC3D* | 1.122865547 | 0.00086977 | 0.005271688 | 1 |
| Obesity | nk_cell | *RN7SL832P* | 2.08219238 | 6.81E-05 | 0.0012086 | 0.427844299 |
| Obesity | nk_cell | *P2RY11* | -1.749662971 | 0.000280166 | 0.002529499 | 1 |
| Obesity | nk_cell | *DBNDD2* | 1.630684444 | 0.019362284 | 0.048907464 | 1 |
| Obesity | nk_cell | *APOBEC3C* | 0.484226684 | 0.000145858 | 0.001770291 | 0.917010934 |
| Obesity | nk_cell | *CCDC13* | 0.740998086 | 0.007671458 | 0.024432855 | 1 |
| Obesity | nk_cell | *ASPRV1* | -2.465130246 | 0.000895862 | 0.005369192 | 1 |
| Obesity | nk_cell | *LINC00920* | -1.928952157 | 0.000324707 | 0.002747211 | 1 |
| Obesity | nk_cell | *UBAP1L* | -1.285232115 | 0.005315401 | 0.018721528 | 1 |
| Obesity | nk_cell | *PGAM5* | -1.130758419 | 0.001039762 | 0.005942712 | 1 |
| Obesity | nk_cell | *DNAJC3-AS1* | -1.769078251 | 0.004763855 | 0.017320809 | 1 |
| Obesity | nk_cell | *BCKDHA* | -2.69366126 | 0.005215587 | 0.018494302 | 1 |
| Obesity | nk_cell | *CDK11B* | 1.923399152 | 3.55E-05 | 0.001051568 | 0.223057458 |
| Obesity | nk_cell | *PCDHAC1* | -5.623802387 | 1.61E-05 | 0.000736656 | 0.10092189 |
| Obesity | nk_cell | *LINC01060* | -0.597502712 | 0.002190066 | 0.00981393 | 1 |

| | | | | | | |
|---|---|---|---|---|---|---|
| Obesity | nk_cell | *NAIP* | -1.274848485 | 0.013038427 | 0.036448462 | 1 |
| Obesity | nk_cell | *ZNF286B* | 1.969344428 | 4.48E-05 | 0.001108799 | 0.281634909 |
| Obesity | nk_cell | *LINC00942* | 1.915000535 | 0.007617197 | 0.024346374 | 1 |
| Obesity | nk_cell | *LINC00603* | -0.91225426 | 0.001264336 | 0.00680868 | 1 |
| Obesity | nk_cell | *DNAH10OS* | 26.22853722 | 4.55E-11 | 1.68E-08 | 2.86E-07 |
| Obesity | nk_cell | *ZNF718* | 2.460214078 | 0.000238324 | 0.002291043 | 1 |
| Obesity | nk_cell | *LINC00989* | -0.716613127 | 0.000641315 | 0.004420995 | 1 |
| Obesity | nk_cell | *LINC01021* | -1.547946815 | 0.000738676 | 0.004730741 | 1 |
| Obesity | nk_cell | *FAM218A* | 8.989447511 | 0.000829718 | 0.005094177 | 1 |
| Obesity | nk_cell | *GLI4* | -1.432668241 | 0.000713533 | 0.004649447 | 1 |
| Obesity | nk_cell | *SNHG18* | -1.830396392 | 6.60E-05 | 0.001202521 | 0.415109279 |
| Obesity | nk_cell | *SHANK3* | -0.737114268 | 0.00464327 | 0.017071484 | 1 |
| Obesity | nk_cell | *CCDC71L* | -1.229891436 | 0.01755892 | 0.045556955 | 1 |
| Obesity | nk_cell | *TMEM200B* | -0.554299752 | 0.008928636 | 0.027543834 | 1 |
| Obesity | nk_cell | *TRNP1* | -3.383618901 | 0.000664139 | 0.004504249 | 1 |
| Obesity | nk_cell | *ALG11* | -0.841333861 | 0.011301523 | 0.032773375 | 1 |
| Obesity | nk_cell | *PRKDC* | 1.684336707 | 0.016391306 | 0.043190336 | 1 |
| Obesity | nk_cell | *MPV17L2* | 2.164931468 | 0.004533792 | 0.016737686 | 1 |
| Obesity | nk_cell | *DPP3* | 1.452291461 | 0.000287654 | 0.002568868 | 1 |
| Obesity | nk_cell | *POLR2M* | 3.240465476 | 0.010832208 | 0.031734433 | 1 |
| Obesity | nk_cell | *LINC00944* | 0.278312398 | 0.00017556 | 0.00195745 | 1 |
| Obesity | nk_cell | *ZNF10* | -1.155155512 | 0.01338051 | 0.03715692 | 1 |
| Obesity | nk_cell | *HMBS* | 2.544515909 | 3.77E-11 | 1.48E-08 | 2.37E-07 |
| Obesity | nk_cell | *PGA5* | -13.20927592 | 4.06E-05 | 0.001082521 | 0.255115381 |
| Obesity | nk_cell | *ZBED6* | -3.408141641 | 0.004677478 | 0.017126618 | 1 |
| Obesity | nk_cell | *FNTB* | -1.144060019 | 0.000900611 | 0.005385799 | 1 |
| Obesity | nk_cell | *TEN1* | -2.969959487 | 0.001515025 | 0.007675232 | 1 |
| Obesity | nk_cell | *CHURC1* | 1.062955876 | 0.018254849 | 0.046672727 | 1 |
| Obesity | nk_cell | *RTEL1* | -1.306342246 | 0.006247538 | 0.021117351 | 1 |
| Obesity | nk_cell | *LINC00638* | -4.056114647 | 0.009696687 | 0.029127124 | 1 |
| Obesity | nk_cell | *DUXA* | -1.666859847 | 0.000468874 | 0.003568782 | 1 |
| Obesity | nk_cell | *LINC00639* | -0.492330626 | 0.009109698 | 0.027869914 | 1 |
| Obesity | nk_cell | *LINC00648* | -1.019862761 | 0.004066045 | 0.015530512 | 1 |
| Obesity | nk_cell | *LINC00924* | -0.866633752 | 0.000706423 | 0.004623717 | 1 |
| Obesity | nk_cell | *ITGB3* | -1.521788634 | 9.98E-05 | 0.001447345 | 0.627589754 |
| Obesity | nk_cell | *UBE2Q2L* | -2.455581384 | 8.06E-05 | 0.00133653 | 0.506544852 |
| Obesity | nk_cell | *AC083843.1* | -19.26348242 | 3.27E-06 | 0.000263534 | 0.020555686 |
| Obesity | nk_cell | *SH3RF3-AS1* | -2.223237013 | 9.32E-07 | 0.000114881 | 0.005858925 |
| Obesity | nk_cell | *HOXB7* | 5.422811056 | 6.67E-06 | 0.000415115 | 0.041926584 |
| Obesity | nk_cell | *LINC00562* | -1.263277247 | 1.52E-05 | 0.000718997 | 0.095875137 |
| Obesity | nk_cell | *AC009120.5* | -1.05366466 | 0.000755004 | 0.004794657 | 1 |
| Obesity | nk_cell | *TMEM178B* | -0.843599914 | 0.003194482 | 0.012945822 | 1 |
| Obesity | nk_cell | *C15orf65* | -1.036886765 | 0.000108702 | 0.001508632 | 0.683410293 |
| Obesity | nk_cell | *GOLGA8H* | 0.937699479 | 0.016528562 | 0.043406462 | 1 |
| Obesity | nk_cell | *LINC00672* | 1.547245662 | 0.000519712 | 0.003826032 | 1 |
| Obesity | nk_cell | *DYNLL2* | -2.045864132 | 0.013087124 | 0.03651964 | 1 |
| Obesity | nk_cell | *LINC00526* | -1.150754651 | 0.000616841 | 0.004313769 | 1 |
| Obesity | nk_cell | *LINC00668* | 18.31603671 | 9.69E-05 | 0.001442418 | 0.609042898 |
| Obesity | nk_cell | *AC139100.2* | -7.059033706 | 0.003437566 | 0.013713184 | 1 |
| Obesity | nk_cell | *LINC01140* | -0.790589492 | 0.012788163 | 0.036005008 | 1 |

| | | | | | | |
|---|---|---|---|---|---|---|
| Obesity | nk_cell | *LINC00906* | -5.724991602 | 0.000718777 | 0.004663518 | 1 |
| Obesity | nk_cell | *ZNF582-AS1* | -1.038726882 | 0.000714049 | 0.004649447 | 1 |
| Obesity | nk_cell | *AC137932.1* | -3.387431966 | 0.000269868 | 0.002455374 | 1 |
| Obesity | nk_cell | *AC003005.2* | -1.354444513 | 0.011694004 | 0.033540239 | 1 |
| Obesity | nk_cell | *AL132989.1* | -1.665133919 | 0.000704012 | 0.004623717 | 1 |
| Obesity | nk_cell | *LINC00664* | -2.901996682 | 0.000519422 | 0.003826032 | 1 |
| Obesity | nk_cell | *FBXO17* | -0.786562363 | 0.001884103 | 0.00888624 | 1 |
| Obesity | nk_cell | *AC009060.1* | -1.489194914 | 2.18E-05 | 0.000858426 | 0.137089111 |
| Obesity | nk_cell | *LINC01050* | -5.930008483 | 0.001730084 | 0.008399259 | 1 |
| Obesity | nk_cell | *SRXN1* | 3.855561093 | 0.009360021 | 0.028400797 | 1 |
| Obesity | nk_cell | *LINC01023* | -1.35243451 | 0.002415384 | 0.010494484 | 1 |
| Obesity | nk_cell | *DOC2B* | -0.515758252 | 0.018973021 | 0.048117539 | 1 |
| Obesity | nk_cell | *POLR2J4* | -1.37980724 | 0.00824495 | 0.025801891 | 1 |
| Obesity | pericyte | *SCYL3* | 2.210696013 | 0.00673285 | 0.009798479 | 1 |
| Obesity | pericyte | *C1orf112* | 8.119776843 | 4.40E-07 | 1.86E-06 | 0.002764918 |
| Obesity | pericyte | *CFH* | 5.735779799 | 1.61E-08 | 1.91E-07 | 0.000101092 |
| Obesity | pericyte | *STPG1* | 2.243802761 | 0.00067969 | 0.001161766 | 1 |
| Obesity | pericyte | *NIPAL3* | 4.136111486 | 1.88E-08 | 2.07E-07 | 0.000118508 |
| Obesity | pericyte | *ENPP4* | 2.090715228 | 4.53E-05 | 9.77E-05 | 0.284866015 |
| Obesity | pericyte | *SEMA3F* | -3.401945078 | 2.93E-25 | 3.68E-22 | 1.84E-21 |
| Obesity | pericyte | *KRIT1* | 4.763309063 | 0.000255372 | 0.00047069 | 1 |
| Obesity | pericyte | *RAD52* | 7.561766117 | 1.46E-07 | 7.76E-07 | 0.000915429 |
| Obesity | pericyte | *BAD* | 10.94576163 | 9.33E-09 | 1.70E-07 | 5.86E-05 |
| Obesity | pericyte | *HS3ST1* | 7.846474318 | 1.32E-07 | 7.28E-07 | 0.00082901 |
| Obesity | pericyte | *WNT16* | 4.550431679 | 5.54E-08 | 3.96E-07 | 0.000348593 |
| Obesity | pericyte | *HECW1* | 0.797547532 | 0.016344909 | 0.022553256 | 1 |
| Obesity | pericyte | *MAD1L1* | 5.077864387 | 0.000690271 | 0.001176896 | 1 |
| Obesity | pericyte | *LASP1* | 6.078749852 | 3.44E-08 | 2.89E-07 | 0.000216472 |
| Obesity | pericyte | *M6PR* | 4.401088916 | 2.09E-05 | 4.90E-05 | 0.131658196 |
| Obesity | pericyte | *CYP26B1* | -1.718663887 | 0.001795174 | 0.00285757 | 1 |
| Obesity | pericyte | *ICA1* | 5.682953604 | 1.07E-09 | 5.30E-08 | 6.73E-06 |
| Obesity | pericyte | *ALS2* | 6.675969648 | 4.28E-07 | 1.83E-06 | 0.00268974 |
| Obesity | pericyte | *POLDIP2* | 3.60205972 | 7.15E-08 | 4.70E-07 | 0.000449219 |
| Obesity | pericyte | *CD38* | 2.070444203 | 5.74E-05 | 0.000120289 | 0.361107028 |
| Obesity | pericyte | *RBM6* | -2.393355135 | 2.74E-19 | 1.91E-16 | 1.72E-15 |
| Obesity | pericyte | *RECQL* | 7.101439911 | 8.72E-09 | 1.70E-07 | 5.48E-05 |
| Obesity | pericyte | *ARHGAP33* | 2.050915272 | 0.01904613 | 0.026008475 | 1 |
| Obesity | pericyte | *PDK4* | 5.191735018 | 1.04E-08 | 1.71E-07 | 6.53E-05 |
| Obesity | pericyte | *SLC22A16* | -2.340662916 | 0.000629425 | 0.00108268 | 1 |
| Obesity | pericyte | *CDC27* | 4.184273968 | 2.29E-09 | 8.46E-08 | 1.44E-05 |
| Obesity | pericyte | *MEOX1* | -1.535693489 | 0.000584962 | 0.001011456 | 1 |
| Obesity | pericyte | *CIAPIN1* | 3.977948549 | 1.32E-07 | 7.28E-07 | 0.000831474 |
| Obesity | pericyte | *FAM214B* | 4.883278979 | 5.13E-09 | 1.30E-07 | 3.23E-05 |
| Obesity | pericyte | *COPZ2* | 2.444845074 | 3.73E-08 | 3.02E-07 | 0.000234237 |
| Obesity | pericyte | *GCFC2* | 6.186406514 | 4.51E-06 | 1.26E-05 | 0.028375348 |
| Obesity | pericyte | *WDR54* | 4.705753815 | 7.13E-07 | 2.70E-06 | 0.004484782 |
| Obesity | pericyte | *ABCB4* | 5.476231414 | 6.15E-08 | 4.24E-07 | 0.000386833 |
| Obesity | pericyte | *ZNF195* | 2.59824245 | 0.000923036 | 0.001534002 | 1 |
| Obesity | pericyte | *PDK2* | 3.928573983 | 1.51E-06 | 4.89E-06 | 0.009467038 |
| Obesity | pericyte | *ABCC8* | -2.255827873 | 8.46E-05 | 0.000171611 | 0.532164391 |

| | | | | | | |
|---|---|---|---|---|---|---|
| Obesity | pericyte | *SYNRG* | 5.123695885 | 0.000139974 | 0.000271694 | 0.880017473 |
| Obesity | pericyte | *TMEM132A* | -3.40800516 | 0.000622076 | 0.001072681 | 1 |
| Obesity | pericyte | *ZNF263* | 5.793330494 | 1.27E-08 | 1.73E-07 | 8.00E-05 |
| Obesity | pericyte | *SPATA20* | 4.425189158 | 5.71E-07 | 2.29E-06 | 0.003589251 |
| Obesity | pericyte | *TNFRSF12A* | 2.314317335 | 8.70E-05 | 0.000176043 | 0.547141814 |
| Obesity | pericyte | *DLX6* | -2.057305787 | 2.02E-05 | 4.74E-05 | 0.127082278 |
| Obesity | pericyte | *MAP3K9* | 2.702384519 | 1.03E-05 | 2.60E-05 | 0.064675273 |
| Obesity | pericyte | *RALA* | 5.176287438 | 9.30E-09 | 1.70E-07 | 5.85E-05 |
| Obesity | pericyte | *FARP2* | 3.438851805 | 0.000687194 | 0.001172744 | 1 |
| Obesity | pericyte | *IFRD1* | 2.351885158 | 8.18E-05 | 0.000166254 | 0.514388757 |
| Obesity | pericyte | *GTF2IRD1* | 2.211128566 | 0.00018641 | 0.000353213 | 1 |
| Obesity | pericyte | *VPS41* | 3.261395142 | 0.000471847 | 0.000828631 | 1 |
| Obesity | pericyte | *ARHGAP44* | -1.573146586 | 0.03213675 | 0.042472933 | 1 |
| Obesity | pericyte | *CCDC124* | 2.370444931 | 0.005159787 | 0.007650845 | 1 |
| Obesity | pericyte | *CEACAM21* | 2.007453698 | 1.04E-05 | 2.63E-05 | 0.065591521 |
| Obesity | pericyte | *NOS2* | 13.77810763 | 5.57E-07 | 2.24E-06 | 0.003503181 |
| Obesity | pericyte | *TRAPPC6A* | 4.173315262 | 0.02130295 | 0.028864579 | 1 |
| Obesity | pericyte | *PAX6* | -0.92464959 | 0.027774779 | 0.03710583 | 1 |
| Obesity | pericyte | *RPUSD1* | -3.238598853 | 0.000260365 | 0.000478771 | 1 |
| Obesity | pericyte | *BAIAP3* | -2.428008275 | 2.75E-05 | 6.24E-05 | 0.172964073 |
| Obesity | pericyte | *TSR3* | 7.331229917 | 6.76E-08 | 4.50E-07 | 0.000424962 |
| Obesity | pericyte | *PIGQ* | 3.429908889 | 7.48E-06 | 1.96E-05 | 0.047011801 |
| Obesity | pericyte | *CDK11A* | 2.694603809 | 1.57E-05 | 3.79E-05 | 0.098767651 |
| Obesity | pericyte | *DLEC1* | 3.266576707 | 2.09E-06 | 6.45E-06 | 0.013131072 |
| Obesity | pericyte | *SS18L2* | 4.303072534 | 0.001659215 | 0.002653646 | 1 |
| Obesity | pericyte | *MPND* | 5.306648606 | 2.91E-06 | 8.58E-06 | 0.01827523 |
| Obesity | pericyte | *MGST1* | 3.490078996 | 3.75E-07 | 1.64E-06 | 0.002360749 |
| Obesity | pericyte | *MMP25* | 3.61158775 | 6.78E-08 | 4.51E-07 | 0.000426304 |
| Obesity | pericyte | *PKD1* | 4.397462792 | 7.74E-09 | 1.65E-07 | 4.86E-05 |
| Obesity | pericyte | *MED24* | 2.990053981 | 0.002989975 | 0.004596632 | 1 |
| Obesity | pericyte | *RHOBTB2* | 3.48675194 | 0.000475973 | 0.000835411 | 1 |
| Obesity | pericyte | *SEC62* | 5.340632957 | 0.000111124 | 0.000219767 | 0.69863829 |
| Obesity | pericyte | *RPS20* | 6.289335315 | 2.34E-07 | 1.12E-06 | 0.001474271 |
| Obesity | pericyte | *MLXIPL* | 1.075559106 | 1.20E-05 | 2.97E-05 | 0.075300227 |
| Obesity | pericyte | *SPRTN* | 5.285237651 | 1.60E-08 | 1.91E-07 | 0.000100749 |
| Obesity | pericyte | *METTL13* | 4.538945225 | 0.007744887 | 0.011167914 | 1 |
| Obesity | pericyte | *STARD3NL* | 6.964987412 | 2.76E-10 | 2.12E-08 | 1.74E-06 |
| Obesity | pericyte | *ZNF200* | 4.692379378 | 0.000115752 | 0.000228058 | 0.727733037 |
| Obesity | pericyte | *CD4* | 3.777635343 | 5.42E-08 | 3.90E-07 | 0.000340674 |
| Obesity | pericyte | *LRRC23* | 2.419824179 | 0.004995866 | 0.007421789 | 1 |
| Obesity | pericyte | *SCMH1* | 4.510162424 | 8.32E-07 | 3.04E-06 | 0.005228414 |
| Obesity | pericyte | *CLCN6* | 8.247615919 | 2.48E-10 | 2.00E-08 | 1.56E-06 |
| Obesity | pericyte | *MRC2* | 1.642416131 | 0.014963161 | 0.020771338 | 1 |
| Obesity | pericyte | *TSPAN9* | 2.294411829 | 0.001061103 | 0.001748205 | 1 |
| Obesity | pericyte | *MKS1* | 4.907417634 | 2.63E-08 | 2.41E-07 | 0.000165434 |
| Obesity | pericyte | *UTP18* | 8.435955427 | 1.50E-08 | 1.88E-07 | 9.41E-05 |
| Obesity | pericyte | *TTC19* | 5.114102608 | 1.32E-07 | 7.28E-07 | 0.00083093 |
| Obesity | pericyte | *DPF1* | 2.407909405 | 0.034152135 | 0.04499465 | 1 |
| Obesity | pericyte | *SYT7* | 4.021894921 | 3.57E-06 | 1.03E-05 | 0.022471594 |
| Obesity | pericyte | *LARS2* | 4.709319611 | 1.96E-08 | 2.11E-07 | 0.000123228 |

| | | | | | | |
|---|---|---|---|---|---|---|
| Obesity | pericyte | *PPP5C* | 5.622940173 | 1.38E-07 | 7.49E-07 | 0.000869945 |
| Obesity | pericyte | *CEP68* | 2.185808126 | 0.001367204 | 0.002220514 | 1 |
| Obesity | pericyte | *ZBTB32* | 7.394907354 | 0.0159334 | 0.022040326 | 1 |
| Obesity | pericyte | *TMEM159* | 2.416032844 | 0.023886708 | 0.032164432 | 1 |
| Obesity | pericyte | *ERCC1* | 2.747185459 | 0.030810481 | 0.040823076 | 1 |
| Obesity | pericyte | *ALOX5* | 2.496434268 | 1.75E-06 | 5.54E-06 | 0.01097905 |
| Obesity | pericyte | *MAP4K5* | 5.639921409 | 5.63E-07 | 2.26E-06 | 0.003537027 |
| Obesity | pericyte | *PSMC4* | 4.973023408 | 2.68E-10 | 2.11E-08 | 1.69E-06 |
| Obesity | pericyte | *MAN2B2* | 6.92156205 | 6.59E-08 | 4.44E-07 | 0.000414488 |
| Obesity | pericyte | *CLDN11* | 3.950766189 | 0.000109284 | 0.000216599 | 0.687068914 |
| Obesity | pericyte | *ANGEL1* | 6.264284902 | 1.09E-08 | 1.71E-07 | 6.88E-05 |
| Obesity | pericyte | *RNF14* | 5.839636131 | 2.12E-08 | 2.17E-07 | 0.000133479 |
| Obesity | pericyte | *DDX11* | 0.930958026 | 0.033351836 | 0.044004826 | 1 |
| Obesity | pericyte | *HEBP1* | 4.548961535 | 0.000659542 | 0.00112954 | 1 |
| Obesity | pericyte | *TACC3* | 3.611994394 | 6.08E-05 | 0.000126856 | 0.382470441 |
| Obesity | pericyte | *ZC3H3* | 2.930520022 | 4.63E-06 | 1.29E-05 | 0.029120367 |
| Obesity | pericyte | *MDH1* | 4.744532862 | 9.64E-07 | 3.42E-06 | 0.006062668 |
| Obesity | pericyte | *SLC30A9* | 5.783211446 | 3.21E-09 | 1.02E-07 | 2.02E-05 |
| Obesity | pericyte | *MTMR11* | 1.312341153 | 9.22E-06 | 2.36E-05 | 0.057962372 |
| Obesity | pericyte | *COX15* | 4.067325519 | 0.000187187 | 0.000354472 | 1 |
| Obesity | pericyte | *CCDC88C* | 2.248500435 | 0.000779812 | 0.001318278 | 1 |
| Obesity | pericyte | *NPC1L1* | 4.200976654 | 9.21E-05 | 0.000185545 | 0.578900827 |
| Obesity | pericyte | *NUDCD3* | 3.001866642 | 2.44E-06 | 7.35E-06 | 0.015316381 |
| Obesity | pericyte | *CHDH* | -3.060967498 | 8.11E-05 | 0.000164994 | 0.509997466 |
| Obesity | pericyte | *IL20RA* | 20.69127129 | 2.17E-08 | 2.19E-07 | 0.000136305 |
| Obesity | pericyte | *GLT8D1* | 2.294952928 | 0.000254646 | 0.00046949 | 1 |
| Obesity | pericyte | *RUFY3* | 3.550179389 | 4.26E-05 | 9.23E-05 | 0.267597924 |
| Obesity | pericyte | *WWTR1* | 5.537554563 | 6.99E-07 | 2.66E-06 | 0.004392913 |
| Obesity | pericyte | *TTC27* | 3.616866992 | 3.55E-05 | 7.84E-05 | 0.223359832 |
| Obesity | pericyte | *VSIG2* | 5.578425673 | 0.000159512 | 0.000305746 | 1 |
| Obesity | pericyte | *MNAT1* | 5.732378409 | 9.65E-09 | 1.71E-07 | 6.07E-05 |
| Obesity | pericyte | *SLC7A9* | 1.3077672 | 0.000552593 | 0.000960773 | 1 |
| Obesity | pericyte | *OSBPL5* | 7.076604976 | 2.47E-05 | 5.68E-05 | 0.155267813 |
| Obesity | pericyte | *AQR* | 5.578518405 | 4.79E-06 | 1.33E-05 | 0.030124767 |
| Obesity | pericyte | *CPS1* | 2.069735126 | 0.000151692 | 0.000292543 | 0.953689509 |
| Obesity | pericyte | *NLRP2* | 1.897958993 | 0.000827561 | 0.001391516 | 1 |
| Obesity | pericyte | *ZDHHC6* | 6.402158067 | 6.64E-08 | 4.46E-07 | 0.00041745 |
| Obesity | pericyte | *RNH1* | 3.339830054 | 5.30E-05 | 0.000112195 | 0.333107829 |
| Obesity | pericyte | *NDUFS1* | 5.961115157 | 3.16E-09 | 1.02E-07 | 1.99E-05 |
| Obesity | pericyte | *BIRC3* | 1.891653725 | 4.68E-06 | 1.30E-05 | 0.029417679 |
| Obesity | pericyte | *AKAP11* | 6.125001863 | 9.48E-09 | 1.70E-07 | 5.96E-05 |
| Obesity | pericyte | *GLRX2* | 1.498203553 | 0.00484744 | 0.007218345 | 1 |
| Obesity | pericyte | *SNAPC1* | 3.167999298 | 3.66E-06 | 1.05E-05 | 0.022989875 |
| Obesity | pericyte | *GCLM* | 3.12898564 | 6.97E-07 | 2.66E-06 | 0.004379411 |
| Obesity | pericyte | *UBR2* | 5.47141952 | 1.79E-06 | 5.66E-06 | 0.01122374 |
| Obesity | pericyte | *HSF2* | 6.134413916 | 1.15E-08 | 1.72E-07 | 7.24E-05 |
| Obesity | pericyte | *TYMP* | 3.412268653 | 0.000658944 | 0.001128823 | 1 |
| Obesity | pericyte | *TOMM34* | 10.83771275 | 1.20E-09 | 5.53E-08 | 7.53E-06 |
| Obesity | pericyte | *KPNA6* | 4.929280993 | 2.34E-09 | 8.55E-08 | 1.47E-05 |
| Obesity | pericyte | *VIM* | 5.35515034 | 7.82E-08 | 4.96E-07 | 0.000491367 |

| | | | | | | |
|---|---|---|---|---|---|---|
| Obesity | pericyte | *RNASET2* | 2.596462868 | 8.03E-05 | 0.000163518 | 0.504616756 |
| Obesity | pericyte | *CD44* | 4.968555642 | 5.24E-06 | 1.44E-05 | 0.032932181 |
| Obesity | pericyte | *MIPEP* | 7.853490167 | 3.38E-07 | 1.51E-06 | 0.002126429 |
| Obesity | pericyte | *INSRR* | 2.048549001 | 0.028199186 | 0.037577 | 1 |
| Obesity | pericyte | *B4GALT7* | 4.863981612 | 7.44E-05 | 0.000152367 | 0.467765832 |
| Obesity | pericyte | *VRK2* | 4.164160145 | 0.000264183 | 0.000484939 | 1 |
| Obesity | pericyte | *VEZT* | 4.628918032 | 3.55E-06 | 1.02E-05 | 0.02229449 |
| Obesity | pericyte | *SNX1* | 4.260902842 | 7.88E-07 | 2.93E-06 | 0.004954498 |
| Obesity | pericyte | *ARNTL2* | 6.126802447 | 1.30E-08 | 1.74E-07 | 8.19E-05 |
| Obesity | pericyte | *SLC39A9* | 2.897286539 | 4.54E-05 | 9.78E-05 | 0.285367221 |
| Obesity | pericyte | *ANK1* | 2.183659808 | 2.53E-06 | 7.59E-06 | 0.015901004 |
| Obesity | pericyte | *RABEP1* | 5.639486906 | 3.10E-12 | 5.21E-10 | 1.95E-08 |
| Obesity | pericyte | *BAK1* | 1.864554379 | 4.13E-05 | 8.97E-05 | 0.25960181 |
| Obesity | pericyte | *GRN* | 6.005768717 | 1.70E-07 | 8.76E-07 | 0.001070963 |
| Obesity | pericyte | *ARID4A* | 5.96767313 | 1.09E-05 | 2.72E-05 | 0.06827227 |
| Obesity | pericyte | *IFT88* | 4.661470589 | 7.64E-06 | 2.00E-05 | 0.048047576 |
| Obesity | pericyte | *ZCCHC8* | 9.05143423 | 1.56E-12 | 2.88E-10 | 9.78E-09 |
| Obesity | pericyte | *FUT8* | 5.897822396 | 1.37E-06 | 4.52E-06 | 0.008626255 |
| Obesity | pericyte | *UBA6* | 2.379960104 | 0.013850126 | 0.019285878 | 1 |
| Obesity | pericyte | *PIAS1* | 5.543796047 | 1.57E-07 | 8.18E-07 | 0.000984955 |
| Obesity | pericyte | *SLC4A7* | 5.913402086 | 2.41E-07 | 1.14E-06 | 0.001515545 |
| Obesity | pericyte | *MYOC* | 1.329602839 | 0.004634778 | 0.006924631 | 1 |
| Obesity | pericyte | *ADSS* | 7.337254915 | 4.48E-06 | 1.25E-05 | 0.028144859 |
| Obesity | pericyte | *STAP1* | -6.114988245 | 4.43E-07 | 1.87E-06 | 0.002785486 |
| Obesity | pericyte | *TBC1D23* | 6.011715866 | 1.03E-07 | 6.12E-07 | 0.000647907 |
| Obesity | pericyte | *CUL3* | 3.716132195 | 0.002724394 | 0.004215669 | 1 |
| Obesity | pericyte | *NSUN2* | 4.350837705 | 3.90E-09 | 1.13E-07 | 2.45E-05 |
| Obesity | pericyte | *MFAP3* | 1.846496101 | 0.004206715 | 0.006330209 | 1 |
| Obesity | pericyte | *AGA* | 11.3428212 | 5.41E-07 | 2.19E-06 | 0.003399987 |
| Obesity | pericyte | *BOD1L1* | 4.099043734 | 6.75E-05 | 0.000139442 | 0.424322624 |
| Obesity | pericyte | *MAT2B* | 8.470376011 | 1.57E-09 | 6.58E-08 | 9.87E-06 |
| Obesity | pericyte | *CLEC16A* | 5.979754442 | 6.69E-08 | 4.48E-07 | 0.000420609 |
| Obesity | pericyte | *SKIV2L2* | 5.617717142 | 3.14E-06 | 9.19E-06 | 0.019744283 |
| Obesity | pericyte | *DNAH5* | 3.255867468 | 7.61E-08 | 4.88E-07 | 0.000478494 |
| Obesity | pericyte | *C6* | 1.935779633 | 6.34E-07 | 2.47E-06 | 0.003985004 |
| Obesity | pericyte | *RAI14* | 3.076273034 | 6.71E-05 | 0.000138762 | 0.421559352 |
| Obesity | pericyte | *PNKP* | 2.332812198 | 3.29E-05 | 7.30E-05 | 0.206886028 |
| Obesity | pericyte | *PHLPP2* | 4.40845243 | 4.68E-06 | 1.30E-05 | 0.029431615 |
| Obesity | pericyte | *PQLC2* | -2.199580551 | 0.000235275 | 0.000436463 | 1 |
| Obesity | pericyte | *INPP4A* | 2.658649269 | 0.000411544 | 0.000731725 | 1 |
| Obesity | pericyte | *RAB27B* | 1.634957236 | 0.001052802 | 0.001734984 | 1 |
| Obesity | pericyte | *PSMA4* | 4.730647246 | 0.003104801 | 0.004760948 | 1 |
| Obesity | pericyte | *LSG1* | 2.431196636 | 7.96E-07 | 2.95E-06 | 0.005002862 |
| Obesity | pericyte | *THAP3* | 2.774031346 | 7.15E-07 | 2.71E-06 | 0.004496072 |
| Obesity | pericyte | *TDP1* | 9.549734449 | 7.06E-07 | 2.68E-06 | 0.004435803 |
| Obesity | pericyte | *SPATA7* | 5.788674119 | 1.67E-08 | 1.93E-07 | 0.000104829 |
| Obesity | pericyte | *MED17* | 5.408256098 | 1.14E-07 | 6.58E-07 | 0.000716986 |
| Obesity | pericyte | *RETSAT* | 2.316348879 | 5.68E-06 | 1.54E-05 | 0.035735487 |
| Obesity | pericyte | *CAPG* | 2.601588561 | 0.002882144 | 0.004444454 | 1 |
| Obesity | pericyte | *AP2S1* | 7.707476254 | 1.94E-05 | 4.58E-05 | 0.122179769 |

| | | | | | | |
|---|---|---|---|---|---|---|
| Obesity | pericyte | *LCP2* | 2.733062241 | 2.14E-06 | 6.59E-06 | 0.013477985 |
| Obesity | pericyte | *TRIT1* | 1.802594124 | 0.013517563 | 0.018868765 | 1 |
| Obesity | pericyte | *CTNNA1* | 5.459931023 | 5.28E-10 | 3.35E-08 | 3.32E-06 |
| Obesity | pericyte | *CNTLN* | 6.038635613 | 7.54E-07 | 2.83E-06 | 0.004738378 |
| Obesity | pericyte | *EPHA3* | -0.828689548 | 0.000123682 | 0.000242618 | 0.777590978 |
| Obesity | pericyte | *DSG2* | -1.757057684 | 0.002027747 | 0.003200714 | 1 |
| Obesity | pericyte | *WDR37* | 4.529606456 | 3.40E-07 | 1.52E-06 | 0.002138239 |
| Obesity | pericyte | *YTHDC2* | 6.314308035 | 9.46E-09 | 1.70E-07 | 5.95E-05 |
| Obesity | pericyte | *ATP6V1H* | 6.182678284 | 6.44E-09 | 1.51E-07 | 4.05E-05 |
| Obesity | pericyte | *POLR2B* | 4.898309011 | 0.000824822 | 0.001387653 | 1 |
| Obesity | pericyte | *CP* | 5.695835117 | 0.007265097 | 0.010529199 | 1 |
| Obesity | pericyte | *DTNBP1* | 6.970625241 | 2.69E-08 | 2.43E-07 | 0.000168838 |
| Obesity | pericyte | *GOPC* | 4.080251767 | 0.002266708 | 0.003550273 | 1 |
| Obesity | pericyte | *TSPAN17* | 2.678247663 | 0.000448437 | 0.000791501 | 1 |
| Obesity | pericyte | *CC2D2A* | 5.932756731 | 1.46E-08 | 1.86E-07 | 9.15E-05 |
| Obesity | pericyte | *RRM2B* | 4.448703554 | 1.84E-05 | 4.35E-05 | 0.115434015 |
| Obesity | pericyte | *SNX29* | 4.542428749 | 9.88E-06 | 2.50E-05 | 0.062094252 |
| Obesity | pericyte | *LMO3* | -1.115256405 | 0.001894812 | 0.003003709 | 1 |
| Obesity | pericyte | *CELF2* | 4.440625092 | 1.78E-08 | 2.01E-07 | 0.000111902 |
| Obesity | pericyte | *ERCC8* | 4.576535952 | 1.71E-05 | 4.08E-05 | 0.10763413 |
| Obesity | pericyte | *ADAMTS6* | 2.511593631 | 0.00051237 | 0.000895294 | 1 |
| Obesity | pericyte | *H6PD* | 2.362529867 | 0.000313265 | 0.000567578 | 1 |
| Obesity | pericyte | *RFC2* | 7.435018007 | 7.21E-06 | 1.90E-05 | 0.045327904 |
| Obesity | pericyte | *JKAMP* | 4.408485609 | 7.90E-09 | 1.66E-07 | 4.97E-05 |
| Obesity | pericyte | *DKK3* | 4.630567535 | 1.07E-08 | 1.71E-07 | 6.71E-05 |
| Obesity | pericyte | *ARHGEF5* | 2.60408386 | 2.23E-06 | 6.82E-06 | 0.014030687 |
| Obesity | pericyte | *NFE2L3* | 7.870273724 | 0.000190322 | 0.000359867 | 1 |
| Obesity | pericyte | *MCUR1* | 4.310345014 | 7.79E-06 | 2.03E-05 | 0.048970468 |
| Obesity | pericyte | *LIMA1* | 0.998945002 | 0.001596384 | 0.002563658 | 1 |
| Obesity | pericyte | *PTGER3* | 5.149721762 | 8.47E-09 | 1.70E-07 | 5.33E-05 |
| Obesity | pericyte | *MAPK9* | 3.548009243 | 0.000728168 | 0.001235959 | 1 |
| Obesity | pericyte | *FAM160A2* | 1.640903161 | 0.005125499 | 0.007601795 | 1 |
| Obesity | pericyte | *HERPUD1* | 6.380360776 | 1.27E-08 | 1.73E-07 | 7.99E-05 |
| Obesity | pericyte | *PIK3CB* | 3.247621541 | 4.91E-05 | 0.000105117 | 0.308519591 |
| Obesity | pericyte | *CYBA* | 6.030913657 | 3.15E-07 | 1.43E-06 | 0.00198253 |
| Obesity | pericyte | *THOC3* | 2.53649143 | 2.14E-06 | 6.58E-06 | 0.013443358 |
| Obesity | pericyte | *RRP12* | 3.404463249 | 2.73E-06 | 8.12E-06 | 0.017190627 |
| Obesity | pericyte | *ALX4* | -2.845330725 | 0.000107098 | 0.000212808 | 0.673323515 |
| Obesity | pericyte | *FOXN3* | 5.115861615 | 7.09E-07 | 2.69E-06 | 0.004455122 |
| Obesity | pericyte | *MCF2L2* | 5.412102356 | 5.45E-06 | 1.49E-05 | 0.034263853 |
| Obesity | pericyte | *ANAPC4* | 6.981739962 | 5.31E-07 | 2.16E-06 | 0.003338742 |
| Obesity | pericyte | *OPN3* | 1.944635837 | 0.000272617 | 0.000498965 | 1 |
| Obesity | pericyte | *SDCCAG8* | 4.144052305 | 3.87E-10 | 2.73E-08 | 2.43E-06 |
| Obesity | pericyte | *TBC1D22A* | 5.506928827 | 1.68E-07 | 8.68E-07 | 0.001055143 |
| Obesity | pericyte | *PLEKHH1* | 1.715104335 | 8.84E-05 | 0.000178663 | 0.556000673 |
| Obesity | pericyte | *ATP9A* | 3.320837059 | 3.14E-07 | 1.42E-06 | 0.001975293 |
| Obesity | pericyte | *GALC* | 4.410497963 | 0.000198816 | 0.000374014 | 1 |
| Obesity | pericyte | *FAM114A2* | 4.157279086 | 0.000228493 | 0.000425262 | 1 |
| Obesity | pericyte | *GINM1* | 3.275019366 | 0.021244918 | 0.028793671 | 1 |
| Obesity | pericyte | *ITIH4* | 1.756132733 | 4.86E-11 | 5.18E-09 | 3.06E-07 |

| | | | | | | |
|---|---|---|---|---|---|---|
| Obesity | pericyte | *TRAF1* | 4.942796439 | 0.000108855 | 0.000215891 | 0.68437451 |
| Obesity | pericyte | *IL17RB* | 4.362694434 | 5.41E-07 | 2.19E-06 | 0.003404279 |
| Obesity | pericyte | *TRAF3IP2* | 5.768206303 | 3.90E-09 | 1.13E-07 | 2.45E-05 |
| Obesity | pericyte | *MSH4* | 2.115357951 | 1.08E-06 | 3.73E-06 | 0.00678633 |
| Obesity | pericyte | *TMCC3* | 5.992249226 | 2.09E-05 | 4.89E-05 | 0.131226666 |
| Obesity | pericyte | *PITHD1* | 5.084164276 | 7.62E-08 | 4.88E-07 | 0.000478985 |
| Obesity | pericyte | *ATP11B* | 3.384775333 | 3.92E-08 | 3.13E-07 | 0.000246283 |
| Obesity | pericyte | *CROCC* | 1.896158027 | 0.000115897 | 0.000228272 | 0.728642752 |
| Obesity | pericyte | *ZC3H11A* | 4.050947605 | 1.98E-07 | 9.80E-07 | 0.001243924 |
| Obesity | pericyte | *RIOK2* | 8.071619615 | 7.31E-07 | 2.76E-06 | 0.004598317 |
| Obesity | pericyte | *FLYWCH1* | 0.993382903 | 0.025706869 | 0.034526615 | 1 |
| Obesity | pericyte | *DNAJC25* | -1.690002477 | 0.002280597 | 0.003569359 | 1 |
| Obesity | pericyte | *YBX3* | 6.739136981 | 5.92E-07 | 2.36E-06 | 0.003721598 |
| Obesity | pericyte | *OGFR* | 4.12842557 | 1.60E-05 | 3.84E-05 | 0.10037702 |
| Obesity | pericyte | *PIGV* | 2.644862242 | 0.00028532 | 0.000520698 | 1 |
| Obesity | pericyte | *QSER1* | 7.109959342 | 1.15E-06 | 3.92E-06 | 0.00725601 |
| Obesity | pericyte | *NCKAP1* | 3.352682298 | 0.003073927 | 0.004715905 | 1 |
| Obesity | pericyte | *SFSWAP* | 5.517707665 | 5.99E-08 | 4.16E-07 | 0.000376287 |
| Obesity | pericyte | *VMP1* | 4.648860902 | 0.000147837 | 0.000285809 | 0.929451635 |
| Obesity | pericyte | *EIF4B* | 2.848154229 | 0.000159016 | 0.000304982 | 0.999732418 |
| Obesity | pericyte | *SPHK2* | 4.40278549 | 1.00E-05 | 2.53E-05 | 0.062840148 |
| Obesity | pericyte | *ISOC2* | 11.67862912 | 1.15E-08 | 1.72E-07 | 7.24E-05 |
| Obesity | pericyte | *EPN1* | 9.874665688 | 1.83E-08 | 2.04E-07 | 0.000115085 |
| Obesity | pericyte | *ADCK1* | 4.55615646 | 0.001877877 | 0.002979862 | 1 |
| Obesity | pericyte | *HAGH* | 1.778000359 | 0.003467259 | 0.005273018 | 1 |
| Obesity | pericyte | *CASP8* | 3.633697169 | 5.01E-06 | 1.38E-05 | 0.031481894 |
| Obesity | pericyte | *TM7SF3* | 6.169657657 | 4.58E-09 | 1.26E-07 | 2.88E-05 |
| Obesity | pericyte | *DMRT3* | -2.42194506 | 0.000325141 | 0.000587234 | 1 |
| Obesity | pericyte | *ATP2C2* | -3.461444751 | 0.001894816 | 0.003003709 | 1 |
| Obesity | pericyte | *TAF2* | 4.332523836 | 6.10E-05 | 0.000127215 | 0.383807978 |
| Obesity | pericyte | *TMEM161A* | 4.392967868 | 1.27E-08 | 1.73E-07 | 7.99E-05 |
| Obesity | pericyte | *LPAR2* | 6.253147195 | 2.97E-06 | 8.74E-06 | 0.018642742 |
| Obesity | pericyte | *SUGP2* | 2.724046103 | 0.002097963 | 0.003302427 | 1 |
| Obesity | pericyte | *ABCA7* | 4.691580135 | 0.000275637 | 0.000504345 | 1 |
| Obesity | pericyte | *DDX20* | 7.91610943 | 1.18E-08 | 1.73E-07 | 7.42E-05 |
| Obesity | pericyte | *BTBD1* | 7.130811732 | 2.84E-09 | 9.54E-08 | 1.78E-05 |
| Obesity | pericyte | *CHI3L2* | 1.68561812 | 0.01196528 | 0.016844092 | 1 |
| Obesity | pericyte | *PMS1* | 6.676419867 | 1.23E-08 | 1.73E-07 | 7.74E-05 |
| Obesity | pericyte | *ANKS1A* | 6.139107267 | 1.27E-08 | 1.73E-07 | 8.01E-05 |
| Obesity | pericyte | *NTHL1* | 2.582694662 | 0.00372111 | 0.005633186 | 1 |
| Obesity | pericyte | *UHRF1BP1* | 4.143422678 | 4.59E-30 | 1.44E-26 | 2.89E-26 |
| Obesity | pericyte | *IPO5* | 6.444929487 | 8.05E-05 | 0.000163958 | 0.506137719 |
| Obesity | pericyte | *WDR18* | 1.208668323 | 0.017169207 | 0.02361987 | 1 |
| Obesity | pericyte | *MCM10* | -0.986427862 | 0.000579504 | 0.001004229 | 1 |
| Obesity | pericyte | *ANKRD44* | 5.785683449 | 9.87E-09 | 1.71E-07 | 6.20E-05 |
| Obesity | pericyte | *NDUFB4* | 8.441061227 | 2.58E-09 | 9.22E-08 | 1.62E-05 |
| Obesity | pericyte | *ZC3H15* | 5.546438699 | 5.24E-06 | 1.44E-05 | 0.032949791 |
| Obesity | pericyte | *MAP2K4* | 6.30865969 | 8.59E-07 | 3.12E-06 | 0.005402838 |
| Obesity | pericyte | *TMEM206* | 3.45711262 | 1.07E-07 | 6.31E-07 | 0.000675737 |
| Obesity | pericyte | *SEC61A2* | 6.008385562 | 1.25E-08 | 1.73E-07 | 7.84E-05 |

| | | | | | | |
|---|---|---|---|---|---|---|
| Obesity | pericyte | *PPP2R5A* | 4.071520108 | 1.08E-08 | 1.71E-07 | 6.79E-05 |
| Obesity | pericyte | *NFYC* | 4.703857522 | 5.59E-05 | 0.000117538 | 0.35132046 |
| Obesity | pericyte | *ZMYND12* | 0.961553796 | 0.02237669 | 0.030234741 | 1 |
| Obesity | pericyte | *NGEF* | 4.038477871 | 8.57E-07 | 3.11E-06 | 0.005390337 |
| Obesity | pericyte | *CD84* | 1.97776768 | 0.000261673 | 0.000481035 | 1 |
| Obesity | pericyte | *ZNRD1* | 1.423997727 | 0.008734654 | 0.012500517 | 1 |
| Obesity | pericyte | *ATXN3* | 2.209895124 | 0.001824186 | 0.002899053 | 1 |
| Obesity | pericyte | *GOLGA5* | 6.941954955 | 2.47E-05 | 5.67E-05 | 0.155127124 |
| Obesity | pericyte | *MTFR1* | 3.87629262 | 0.000187164 | 0.000354472 | 1 |
| Obesity | pericyte | *STAG3* | -2.161301125 | 0.019632395 | 0.026768352 | 1 |
| Obesity | pericyte | *MYO9A* | 4.935983602 | 4.68E-08 | 3.54E-07 | 0.000294471 |
| Obesity | pericyte | *SP100* | 4.664807268 | 0.00015558 | 0.000299123 | 0.97813281 |
| Obesity | pericyte | *STOML1* | -3.419076805 | 0.00257632 | 0.004005273 | 1 |
| Obesity | pericyte | *METTL22* | 3.860119612 | 1.97E-07 | 9.76E-07 | 0.001236641 |
| Obesity | pericyte | *TP53BP1* | 4.982664806 | 2.33E-05 | 5.38E-05 | 0.146175246 |
| Obesity | pericyte | *DHX8* | 3.752927698 | 0.001019579 | 0.001682882 | 1 |
| Obesity | pericyte | *SYT1* | 1.944918068 | 0.004974211 | 0.007393113 | 1 |
| Obesity | pericyte | *ROGDI* | 6.323465725 | 1.11E-07 | 6.46E-07 | 0.000697444 |
| Obesity | pericyte | *HDAC4* | 6.203850777 | 3.24E-08 | 2.79E-07 | 0.000203705 |
| Obesity | pericyte | *RASSF1* | 4.37490551 | 0.000130424 | 0.000254809 | 0.81997532 |
| Obesity | pericyte | *HEATR6* | 9.014436859 | 5.15E-08 | 3.78E-07 | 0.000323807 |
| Obesity | pericyte | *POLR1A* | 0.741288096 | 0.027995059 | 0.03733116 | 1 |
| Obesity | pericyte | *SRBD1* | 5.466695966 | 8.29E-09 | 1.70E-07 | 5.21E-05 |
| Obesity | pericyte | *CYFIP1* | 6.342385765 | 3.02E-06 | 8.88E-06 | 0.018971701 |
| Obesity | pericyte | *PSME4* | 4.684444248 | 2.09E-06 | 6.45E-06 | 0.013139778 |
| Obesity | pericyte | *PITX1* | -7.796372595 | 0.009484041 | 0.013529877 | 1 |
| Obesity | pericyte | *SDK2* | 8.37204261 | 1.38E-07 | 7.48E-07 | 0.000866611 |
| Obesity | pericyte | *NUCKS1* | 6.428015248 | 1.59E-08 | 1.91E-07 | 9.98E-05 |
| Obesity | pericyte | *KCNAB2* | 2.597524456 | 3.02E-05 | 6.79E-05 | 0.190133117 |
| Obesity | pericyte | *GAL* | 3.511735275 | 1.78E-06 | 5.63E-06 | 0.011167977 |
| Obesity | pericyte | *CLEC2D* | 2.641324402 | 0.001119609 | 0.001837854 | 1 |
| Obesity | pericyte | *PIGB* | 4.783557996 | 7.85E-07 | 2.92E-06 | 0.004934243 |
| Obesity | pericyte | *PFN2* | 5.555537766 | 4.79E-06 | 1.33E-05 | 0.030106241 |
| Obesity | pericyte | *DAPP1* | 1.588957949 | 4.92E-06 | 1.36E-05 | 0.030927058 |
| Obesity | pericyte | *TMEM260* | 6.925378856 | 2.89E-07 | 1.33E-06 | 0.001815111 |
| Obesity | pericyte | *DGCR2* | 6.915823585 | 1.82E-06 | 5.77E-06 | 0.011456846 |
| Obesity | pericyte | *ZXDC* | 6.361346917 | 8.63E-10 | 4.60E-08 | 5.43E-06 |
| Obesity | pericyte | *POLB* | 3.46123705 | 0.000299012 | 0.000543635 | 1 |
| Obesity | pericyte | *WIPI1* | 8.024953411 | 2.06E-07 | 1.01E-06 | 0.001292585 |
| Obesity | pericyte | *NDST1* | 5.605556871 | 4.08E-08 | 3.21E-07 | 0.000256542 |
| Obesity | pericyte | *ASNS* | 4.117784357 | 1.58E-07 | 8.24E-07 | 0.000993747 |
| Obesity | pericyte | *AP3M2* | 6.486795675 | 7.15E-05 | 0.000146965 | 0.449270545 |
| Obesity | pericyte | *ST6GALNAC2* | 4.964246741 | 1.93E-07 | 9.63E-07 | 0.00121394 |
| Obesity | pericyte | *EIF2B3* | 5.770780222 | 1.52E-08 | 1.90E-07 | 9.57E-05 |
| Obesity | pericyte | *CAMK2A* | -1.249839249 | 1.75E-05 | 4.15E-05 | 0.109713676 |
| Obesity | pericyte | *CDC42* | 6.251990281 | 1.93E-08 | 2.08E-07 | 0.000121251 |
| Obesity | pericyte | *OSBPL3* | 4.889581258 | 2.54E-08 | 2.37E-07 | 0.000159437 |
| Obesity | pericyte | *RAD18* | 6.351279828 | 1.55E-08 | 1.90E-07 | 9.77E-05 |
| Obesity | pericyte | *SNX13* | 5.611107588 | 1.20E-08 | 1.73E-07 | 7.56E-05 |
| Obesity | pericyte | *TRIP13* | 5.241227293 | 1.03E-06 | 3.60E-06 | 0.006460468 |

| | | | | | | |
|---|---|---|---|---|---|---|
| Obesity | pericyte | *TCF3* | 7.361725822 | 5.42E-08 | 3.90E-07 | 0.000340898 |
| Obesity | pericyte | *MBD3* | 5.431651715 | 1.16E-08 | 1.72E-07 | 7.31E-05 |
| Obesity | pericyte | *CPSF1* | 5.386702932 | 1.65E-05 | 3.95E-05 | 0.103738496 |
| Obesity | pericyte | *CYBRD1* | 3.669967829 | 8.72E-07 | 3.16E-06 | 0.005480405 |
| Obesity | pericyte | *EPN2* | 6.40098775 | 9.66E-08 | 5.86E-07 | 0.000607049 |
| Obesity | pericyte | *AFF4* | 5.264600155 | 3.82E-08 | 3.07E-07 | 0.000240036 |
| Obesity | pericyte | *TRHDE* | 3.297518169 | 2.07E-08 | 2.16E-07 | 0.0001302 |
| Obesity | pericyte | *NFATC3* | 2.676409264 | 0.004910988 | 0.007302597 | 1 |
| Obesity | pericyte | *TRNT1* | 7.791058965 | 1.75E-07 | 8.95E-07 | 0.001102354 |
| Obesity | pericyte | *ACADVL* | 5.743320564 | 2.62E-05 | 5.98E-05 | 0.164859807 |
| Obesity | pericyte | *CRMP1* | 3.100508875 | 0.00314046 | 0.004810934 | 1 |
| Obesity | pericyte | *DERL2* | 5.457845651 | 4.48E-07 | 1.89E-06 | 0.002818932 |
| Obesity | pericyte | *NDE1* | 6.009488014 | 4.16E-07 | 1.79E-06 | 0.002614176 |
| Obesity | pericyte | *AP1M1* | 1.932240441 | 0.00571209 | 0.008418169 | 1 |
| Obesity | pericyte | *PVR* | 4.670308488 | 3.23E-08 | 2.79E-07 | 0.000202841 |
| Obesity | pericyte | *XRCC1* | 6.737595213 | 6.02E-06 | 1.62E-05 | 0.037856097 |
| Obesity | pericyte | *MOV10L1* | 1.284064461 | 0.000442284 | 0.000782176 | 1 |
| Obesity | pericyte | *TP63* | 2.312224894 | 7.87E-07 | 2.93E-06 | 0.004946156 |
| Obesity | pericyte | *ALPK1* | 5.06737257 | 2.66E-09 | 9.26E-08 | 1.67E-05 |
| Obesity | pericyte | *LLGL2* | 2.487657909 | 0.000351374 | 0.000631169 | 1 |
| Obesity | pericyte | *PDE8A* | 3.644874387 | 1.98E-05 | 4.67E-05 | 0.124772655 |
| Obesity | pericyte | *NLE1* | 2.888914519 | 5.31E-06 | 1.45E-05 | 0.033363751 |
| Obesity | pericyte | *SDHA* | 4.17140924 | 1.84E-06 | 5.81E-06 | 0.011559478 |
| Obesity | pericyte | *SMARCE1* | 4.944179318 | 3.17E-09 | 1.02E-07 | 2.00E-05 |
| Obesity | pericyte | *GSDMB* | 1.943304572 | 0.001375494 | 0.002232825 | 1 |
| Obesity | pericyte | *PICALM* | 5.080107638 | 3.67E-07 | 1.61E-06 | 0.0023104 |
| Obesity | pericyte | *GLI2* | 0.849723057 | 0.020287972 | 0.027590413 | 1 |
| Obesity | pericyte | *CLNS1A* | 2.788454053 | 2.62E-06 | 7.82E-06 | 0.016446144 |
| Obesity | pericyte | *ATP2A3* | 1.972825489 | 6.22E-07 | 2.43E-06 | 0.003912855 |
| Obesity | pericyte | *CA12* | 1.960985683 | 2.35E-05 | 5.42E-05 | 0.147715212 |
| Obesity | pericyte | *NTN4* | 3.360173084 | 1.73E-07 | 8.86E-07 | 0.001086189 |
| Obesity | pericyte | *LMAN1* | 4.97912718 | 0.001276873 | 0.002080793 | 1 |
| Obesity | pericyte | *ENO1* | 7.34799275 | 2.24E-07 | 1.08E-06 | 0.001407247 |
| Obesity | pericyte | *TIPIN* | 1.600238668 | 3.95E-08 | 3.15E-07 | 0.00024849 |
| Obesity | pericyte | *SRI* | 3.725036902 | 0.029889711 | 0.039695103 | 1 |
| Obesity | pericyte | *EIF4G3* | 4.599367923 | 2.19E-05 | 5.10E-05 | 0.137846642 |
| Obesity | pericyte | *TTC38* | 2.942136105 | 4.98E-07 | 2.05E-06 | 0.00313113 |
| Obesity | pericyte | *GRAMD4* | 6.916840617 | 5.02E-07 | 2.07E-06 | 0.003156228 |
| Obesity | pericyte | *CELSR1* | 8.15758875 | 1.21E-08 | 1.73E-07 | 7.64E-05 |
| Obesity | pericyte | *ADD2* | 14.03364747 | 2.76E-06 | 8.18E-06 | 0.017333754 |
| Obesity | pericyte | *VPS9D1* | 7.204664788 | 0.000304609 | 0.000553071 | 1 |
| Obesity | pericyte | *MARK3* | 2.56530837 | 0.014635327 | 0.020338705 | 1 |
| Obesity | pericyte | *SLC25A3* | 3.749661729 | 0.004272824 | 0.006425076 | 1 |
| Obesity | pericyte | *FSCN1* | 3.181940959 | 0.013844374 | 0.01928214 | 1 |
| Obesity | pericyte | *PLD1* | 4.588588481 | 0.000442545 | 0.000782418 | 1 |
| Obesity | pericyte | *BCAP29* | 5.932911838 | 6.29E-07 | 2.45E-06 | 0.00395215 |
| Obesity | pericyte | *SEC31B* | 0.792951138 | 0.001616242 | 0.002591511 | 1 |
| Obesity | pericyte | *TUBA3D* | -1.892118576 | 7.77E-05 | 0.000158459 | 0.488371075 |
| Obesity | pericyte | *MKRN2* | 3.546361692 | 9.34E-09 | 1.70E-07 | 5.87E-05 |
| Obesity | pericyte | *MCM6* | 5.011828286 | 6.92E-07 | 2.64E-06 | 0.004348746 |

| | | | | | | |
|---|---|---|---|---|---|---|
| Obesity | pericyte | *RBMS2* | 4.158292494 | 0.001921483 | 0.003042913 | 1 |
| Obesity | pericyte | *MLH1* | 3.096858504 | 2.48E-05 | 5.70E-05 | 0.155993262 |
| Obesity | pericyte | *FMO4* | 5.452647816 | 1.45E-08 | 1.85E-07 | 9.10E-05 |
| Obesity | pericyte | *RGS11* | 0.877520558 | 0.028440667 | 0.037861772 | 1 |
| Obesity | pericyte | *TPD52* | 1.723637666 | 1.52E-05 | 3.68E-05 | 0.095657007 |
| Obesity | pericyte | *GPATCH1* | 5.476913175 | 6.67E-06 | 1.78E-05 | 0.041936768 |
| Obesity | pericyte | *ICAM3* | 2.168819283 | 1.06E-05 | 2.67E-05 | 0.066579955 |
| Obesity | pericyte | *NT5C2* | 6.803681399 | 2.12E-11 | 2.67E-09 | 1.33E-07 |
| Obesity | pericyte | *CAMSAP3* | 13.89505232 | 1.18E-06 | 3.97E-06 | 0.007395423 |
| Obesity | pericyte | *MAP2K7* | 5.08210124 | 2.13E-08 | 2.17E-07 | 0.000133622 |
| Obesity | pericyte | *CTTNBP2* | 2.546530344 | 1.09E-05 | 2.74E-05 | 0.068753574 |
| Obesity | pericyte | *TM9SF3* | 5.880663555 | 2.34E-08 | 2.26E-07 | 0.000146963 |
| Obesity | pericyte | *DNAJC10* | 7.646703543 | 6.77E-08 | 4.50E-07 | 0.000425529 |
| Obesity | pericyte | *DYNC1I2* | 7.361153526 | 6.58E-09 | 1.52E-07 | 4.14E-05 |
| Obesity | pericyte | *APBB1IP* | 2.672212015 | 1.82E-07 | 9.18E-07 | 0.001142591 |
| Obesity | pericyte | *LRCH4* | 1.694129617 | 0.008508703 | 0.012202148 | 1 |
| Obesity | pericyte | *SIRT6* | 8.130234763 | 0.006197899 | 0.009080911 | 1 |
| Obesity | pericyte | *CAPZB* | 5.456486897 | 3.70E-07 | 1.62E-06 | 0.002326644 |
| Obesity | pericyte | *NAALAD2* | 2.802885688 | 4.46E-06 | 1.25E-05 | 0.028031239 |
| Obesity | pericyte | *FGFR1* | 4.267884749 | 5.32E-08 | 3.86E-07 | 0.000334592 |
| Obesity | pericyte | *FBLN1* | 2.279588251 | 2.11E-07 | 1.03E-06 | 0.001326303 |
| Obesity | pericyte | *PIAS2* | 4.480770843 | 5.50E-06 | 1.50E-05 | 0.034556708 |
| Obesity | pericyte | *AMPH* | 1.320739785 | 0.000439346 | 0.000777638 | 1 |
| Obesity | pericyte | *MCCC1* | 3.579842411 | 2.38E-08 | 2.29E-07 | 0.000149913 |
| Obesity | pericyte | *ACER3* | 3.9684232 | 1.35E-09 | 5.95E-08 | 8.51E-06 |
| Obesity | pericyte | *PIK3C3* | 4.651676775 | 0.000455767 | 0.000803761 | 1 |
| Obesity | pericyte | *N4BP2* | 4.034775364 | 1.84E-05 | 4.35E-05 | 0.115533681 |
| Obesity | pericyte | *TULP3* | 6.584064691 | 1.34E-06 | 4.42E-06 | 0.008399012 |
| Obesity | pericyte | *ZCWPW1* | 3.33105352 | 1.75E-07 | 8.93E-07 | 0.001099894 |
| Obesity | pericyte | *FGF20* | 43.2649304 | 1.32E-07 | 7.28E-07 | 0.000830948 |
| Obesity | pericyte | *PCM1* | 3.629171663 | 0.00082203 | 0.001383324 | 1 |
| Obesity | pericyte | *OSBPL6* | 7.028078734 | 0.000176894 | 0.000336399 | 1 |
| Obesity | pericyte | *XRCC5* | 5.948886829 | 2.42E-06 | 7.32E-06 | 0.015241539 |
| Obesity | pericyte | *LXN* | 4.855338228 | 6.15E-07 | 2.42E-06 | 0.003869379 |
| Obesity | pericyte | *REXO1* | 4.663722735 | 0.022029074 | 0.029803483 | 1 |
| Obesity | pericyte | *SAR1A* | 5.548131926 | 1.15E-08 | 1.72E-07 | 7.22E-05 |
| Obesity | pericyte | *CDC14A* | 2.018443902 | 0.000638452 | 0.001097006 | 1 |
| Obesity | pericyte | *FDFT1* | 6.881384361 | 6.44E-10 | 3.68E-08 | 4.05E-06 |
| Obesity | pericyte | *SCGN* | -1.104032436 | 3.15E-05 | 7.04E-05 | 0.19816876 |
| Obesity | pericyte | *DDX1* | 6.649062382 | 1.47E-05 | 3.57E-05 | 0.092327852 |
| Obesity | pericyte | *DNM2* | 6.30139873 | 1.28E-06 | 4.28E-06 | 0.008076287 |
| Obesity | pericyte | *STX7* | 3.334028656 | 0.000111988 | 0.000221336 | 0.70407121 |
| Obesity | pericyte | *RABL2B* | 3.620309467 | 8.28E-07 | 3.04E-06 | 0.005205197 |
| Obesity | pericyte | *DDX43* | 2.894277261 | 0.00353539 | 0.005366248 | 1 |
| Obesity | pericyte | *DCT* | 3.264588842 | 5.39E-07 | 2.19E-06 | 0.003391035 |
| Obesity | pericyte | *SLC35C2* | 7.302272055 | 1.03E-08 | 1.71E-07 | 6.50E-05 |
| Obesity | pericyte | *SCTR* | 3.150192676 | 2.75E-07 | 1.27E-06 | 0.001730917 |
| Obesity | pericyte | *RIF1* | 3.600679845 | 4.31E-06 | 1.21E-05 | 0.027093619 |
| Obesity | pericyte | *KCNN2* | 5.614888718 | 3.01E-06 | 8.86E-06 | 0.018913144 |
| Obesity | pericyte | *PSEN1* | 3.973627073 | 0.000150337 | 0.000290367 | 0.945167359 |

| | | | | | | |
|---|---|---|---|---|---|---|
| Obesity | pericyte | *CPOX* | 5.032784936 | 4.37E-07 | 1.85E-06 | 0.002746079 |
| Obesity | pericyte | *MOK* | 3.5397513 | 5.95E-08 | 4.15E-07 | 0.000374309 |
| Obesity | pericyte | *RSBN1* | 4.456433337 | 2.67E-05 | 6.07E-05 | 0.167562949 |
| Obesity | pericyte | *MAGI3* | 5.835214815 | 1.79E-08 | 2.02E-07 | 0.00011284 |
| Obesity | pericyte | *STK17B* | 4.507056258 | 1.69E-08 | 1.95E-07 | 0.000106254 |
| Obesity | pericyte | *KIAA0141* | 5.375404792 | 1.47E-07 | 7.81E-07 | 0.000923776 |
| Obesity | pericyte | *HSPB11* | 6.035151603 | 3.40E-08 | 2.86E-07 | 0.000213902 |
| Obesity | pericyte | *WDR70* | 5.307699505 | 1.62E-07 | 8.41E-07 | 0.001015603 |
| Obesity | pericyte | *C5orf22* | 3.366622882 | 7.92E-08 | 5.03E-07 | 0.000498131 |
| Obesity | pericyte | *COBLL1* | 3.220437161 | 6.40E-10 | 3.68E-08 | 4.03E-06 |
| Obesity | pericyte | *GEMIN5* | 2.42303243 | 0.000258959 | 0.000476603 | 1 |
| Obesity | pericyte | *GSK3B* | 4.611101469 | 6.75E-05 | 0.000139442 | 0.424302462 |
| Obesity | pericyte | *ERC1* | 5.039328725 | 2.98E-06 | 8.78E-06 | 0.018737432 |
| Obesity | pericyte | *DOPEY1* | 2.361736299 | 1.34E-05 | 3.29E-05 | 0.08424829 |
| Obesity | pericyte | *LYRM2* | 1.970840693 | 0.004343566 | 0.006522093 | 1 |
| Obesity | pericyte | *ULK2* | 3.292266444 | 0.000756374 | 0.001280377 | 1 |
| Obesity | pericyte | *PIBF1* | 4.617245488 | 1.72E-06 | 5.49E-06 | 0.010822851 |
| Obesity | pericyte | *TDRD3* | 5.036286526 | 9.33E-09 | 1.70E-07 | 5.87E-05 |
| Obesity | pericyte | *PDS5B* | 3.373669689 | 0.00714545 | 0.010360573 | 1 |
| Obesity | pericyte | *OXCT1* | 1.457917476 | 0.012869325 | 0.018048058 | 1 |
| Obesity | pericyte | *CYLD* | 6.011510412 | 5.39E-07 | 2.19E-06 | 0.003391163 |
| Obesity | pericyte | *ZNF324* | -2.523214609 | 0.010611144 | 0.01504901 | 1 |
| Obesity | pericyte | *ZNF586* | 4.047434928 | 4.12E-07 | 1.78E-06 | 0.002593175 |
| Obesity | pericyte | *ZNF446* | 2.825844499 | 6.22E-07 | 2.43E-06 | 0.003911382 |
| Obesity | pericyte | *ZNF264* | 6.004095005 | 3.76E-07 | 1.64E-06 | 0.002365849 |
| Obesity | pericyte | *RPS5* | 7.801991246 | 9.80E-08 | 5.91E-07 | 0.00061584 |
| Obesity | pericyte | *CHMP2B* | 3.950227405 | 5.04E-05 | 0.00010765 | 0.317089456 |
| Obesity | pericyte | *PPIE* | 4.034174214 | 9.12E-05 | 0.000184136 | 0.57358395 |
| Obesity | pericyte | *ZMPSTE24* | 4.937152779 | 2.25E-08 | 2.24E-07 | 0.000141518 |
| Obesity | pericyte | *STARD7* | 7.68080896 | 8.57E-10 | 4.60E-08 | 5.39E-06 |
| Obesity | pericyte | *GSTP1* | 8.163993604 | 2.42E-06 | 7.32E-06 | 0.015239696 |
| Obesity | pericyte | *WBP11* | 8.29724159 | 9.76E-06 | 2.48E-05 | 0.061347678 |
| Obesity | pericyte | *APOB* | 2.113981026 | 1.66E-05 | 3.96E-05 | 0.104063095 |
| Obesity | pericyte | *NCOA1* | 4.97073642 | 4.71E-06 | 1.31E-05 | 0.029620972 |
| Obesity | pericyte | *CD59* | 2.632321676 | 0.032590842 | 0.043045929 | 1 |
| Obesity | pericyte | *CD82* | 3.882675226 | 1.13E-07 | 6.53E-07 | 0.000710107 |
| Obesity | pericyte | *TAF9* | 7.413215835 | 0.004366868 | 0.006553951 | 1 |
| Obesity | pericyte | *SCAMP1* | 7.668901228 | 2.96E-11 | 3.51E-09 | 1.86E-07 |
| Obesity | pericyte | *HACE1* | 5.916483024 | 2.31E-08 | 2.25E-07 | 0.00014503 |
| Obesity | pericyte | *SEH1L* | 5.174750708 | 5.90E-05 | 0.000123456 | 0.371109226 |
| Obesity | pericyte | *OVGP1* | -2.897866239 | 0.004200324 | 0.006322106 | 1 |
| Obesity | pericyte | *SLC25A24* | 4.447075839 | 5.42E-05 | 0.000114558 | 0.340695593 |
| Obesity | pericyte | *ABCB1* | 2.206814195 | 0.000330657 | 0.000595997 | 1 |
| Obesity | pericyte | *CPNE3* | 1.841394016 | 0.020230093 | 0.027528987 | 1 |
| Obesity | pericyte | *RRN3* | 4.327288407 | 0.000560242 | 0.000973532 | 1 |
| Obesity | pericyte | *CTTN* | 6.998213894 | 1.46E-07 | 7.76E-07 | 0.000916267 |
| Obesity | pericyte | *MTIF2* | 3.628109831 | 0.000444957 | 0.00078624 | 1 |
| Obesity | pericyte | *ATG16L1* | 6.17107762 | 3.30E-08 | 2.82E-07 | 0.000207226 |
| Obesity | pericyte | *USP40* | 5.228390579 | 4.28E-07 | 1.83E-06 | 0.002690631 |
| Obesity | pericyte | *MAST2* | 4.901367085 | 1.13E-09 | 5.30E-08 | 7.09E-06 |

| | | | | | | |
|---|---|---|---|---|---|---|
| Obesity | pericyte | *B4GALT1* | 5.912837972 | 1.97E-08 | 2.11E-07 | 0.000124125 |
| Obesity | pericyte | *FOLH1* | 0.791799453 | 0.006515606 | 0.009495507 | 1 |
| Obesity | pericyte | *EIF2AK1* | 4.605215076 | 9.59E-08 | 5.84E-07 | 0.0006029 |
| Obesity | pericyte | *SEPHS1* | 3.55833617 | 5.26E-06 | 1.44E-05 | 0.033084162 |
| Obesity | pericyte | *ITPKC* | 2.937176654 | 7.48E-08 | 4.83E-07 | 0.000470277 |
| Obesity | pericyte | *TMED2* | 8.482591969 | 1.61E-08 | 1.91E-07 | 0.000101236 |
| Obesity | pericyte | *ACOX3* | 3.377738345 | 1.81E-07 | 9.15E-07 | 0.001137716 |
| Obesity | pericyte | *HSD17B14* | 2.727365741 | 6.75E-06 | 1.80E-05 | 0.042461339 |
| Obesity | pericyte | *ACHE* | -1.472516482 | 3.09E-05 | 6.92E-05 | 0.194198996 |
| Obesity | pericyte | *SRRT* | 3.058158303 | 0.000576725 | 0.001000241 | 1 |
| Obesity | pericyte | *ATXN7L3* | 4.357383034 | 3.93E-08 | 3.13E-07 | 0.000247053 |
| Obesity | pericyte | *PGS1* | 5.474620306 | 1.48E-07 | 7.86E-07 | 0.000931778 |
| Obesity | pericyte | *LPCAT2* | 3.922365948 | 0.000276344 | 0.000505492 | 1 |
| Obesity | pericyte | *TXNDC16* | 3.520142438 | 9.82E-06 | 2.49E-05 | 0.061719035 |
| Obesity | pericyte | *GMCL1* | 4.997994982 | 0.00046365 | 0.000816289 | 1 |
| Obesity | pericyte | *PTHLH* | -1.359223131 | 0.01430976 | 0.019899461 | 1 |
| Obesity | pericyte | *ERGIC2* | 5.06678574 | 0.000680174 | 0.001162026 | 1 |
| Obesity | pericyte | *ALG6* | 5.862303824 | 2.28E-06 | 6.95E-06 | 0.014357658 |
| Obesity | pericyte | *DDX18* | 5.889585992 | 2.41E-07 | 1.14E-06 | 0.001512366 |
| Obesity | pericyte | *ASAP3* | 6.954292961 | 2.45E-06 | 7.38E-06 | 0.015398772 |
| Obesity | pericyte | *EDEM2* | 2.093591527 | 0.007083265 | 0.01027989 | 1 |
| Obesity | pericyte | *TPX2* | -1.228900385 | 0.000143887 | 0.00027903 | 0.904614775 |
| Obesity | pericyte | *DOCK3* | 2.229468027 | 3.51E-07 | 1.55E-06 | 0.002209397 |
| Obesity | pericyte | *C3orf18* | 3.839432293 | 1.08E-12 | 2.10E-10 | 6.82E-09 |
| Obesity | pericyte | *FKBP1A* | 8.622793853 | 1.53E-08 | 1.90E-07 | 9.64E-05 |
| Obesity | pericyte | *NSFL1C* | 7.992472001 | 8.25E-10 | 4.51E-08 | 5.19E-06 |
| Obesity | pericyte | *ZNF343* | 4.325198009 | 1.16E-07 | 6.66E-07 | 0.000729624 |
| Obesity | pericyte | *MAVS* | 5.253985766 | 5.96E-08 | 4.15E-07 | 0.00037481 |
| Obesity | pericyte | *LZTS3* | 3.42570542 | 5.20E-06 | 1.43E-05 | 0.03271858 |
| Obesity | pericyte | *XRN2* | 6.940986438 | 2.11E-08 | 2.17E-07 | 0.000132752 |
| Obesity | pericyte | *TESC* | -0.914260769 | 4.38E-05 | 9.46E-05 | 0.275112705 |
| Obesity | pericyte | *RBBP9* | 4.506031314 | 0.004178509 | 0.006292284 | 1 |
| Obesity | pericyte | *SLC23A2* | 4.634683168 | 0.001605224 | 0.002576473 | 1 |
| Obesity | pericyte | *TMEM230* | 4.018161217 | 5.89E-06 | 1.59E-05 | 0.037034129 |
| Obesity | pericyte | *DZANK1* | 4.375825021 | 9.30E-05 | 0.000187152 | 0.584662931 |
| Obesity | pericyte | *TASP1* | 4.540338609 | 1.91E-08 | 2.08E-07 | 0.000119939 |
| Obesity | pericyte | *OAS1* | 8.713218418 | 3.61E-05 | 7.96E-05 | 0.227162421 |
| Obesity | pericyte | *KIF16B* | 6.729648943 | 6.15E-07 | 2.42E-06 | 0.003863503 |
| Obesity | pericyte | *FUS* | 5.239230052 | 3.23E-05 | 7.18E-05 | 0.203048072 |
| Obesity | pericyte | *FXYD5* | 6.235950367 | 0.000705538 | 0.001201115 | 1 |
| Obesity | pericyte | *FXYD3* | 7.283914653 | 4.20E-05 | 9.10E-05 | 0.263748782 |
| Obesity | pericyte | *GANAB* | 6.891523351 | 4.25E-09 | 1.20E-07 | 2.67E-05 |
| Obesity | pericyte | *GMIP* | 4.622099766 | 7.62E-08 | 4.88E-07 | 0.000478814 |
| Obesity | pericyte | *MLF2* | 6.951238918 | 4.71E-09 | 1.27E-07 | 2.96E-05 |
| Obesity | pericyte | *NECAP1* | 3.39240295 | 0.000700288 | 0.001193147 | 1 |
| Obesity | pericyte | *ANKRD24* | 3.24790359 | 1.07E-06 | 3.70E-06 | 0.006716466 |
| Obesity | pericyte | *GPATCH2L* | 3.51728308 | 0.000103241 | 0.000205926 | 0.649077512 |
| Obesity | pericyte | *SLC9A1* | 6.242896403 | 2.81E-08 | 2.52E-07 | 0.000176641 |
| Obesity | pericyte | *SPTLC1* | 4.165126937 | 2.49E-05 | 5.73E-05 | 0.156819669 |
| Obesity | pericyte | *PAPOLA* | 5.672110707 | 1.06E-05 | 2.68E-05 | 0.066899443 |

| | | | | | | |
|---|---|---|---|---|---|---|
| Obesity | pericyte | *RGS1* | 2.607139658 | 1.12E-06 | 3.84E-06 | 0.007064812 |
| Obesity | pericyte | *YPEL3* | 3.176705449 | 4.36E-12 | 6.38E-10 | 2.74E-08 |
| Obesity | pericyte | *MRPS33* | 4.93001576 | 2.04E-06 | 6.34E-06 | 0.01285546 |
| Obesity | pericyte | *LYZ* | 3.927276415 | 0.000128021 | 0.000250426 | 0.804869973 |
| Obesity | pericyte | *SPG21* | 3.331579398 | 0.001609676 | 0.0025823 | 1 |
| Obesity | pericyte | *GNPTG* | 8.145637309 | 1.25E-08 | 1.73E-07 | 7.88E-05 |
| Obesity | pericyte | *ZNF268* | 3.006266453 | 5.92E-06 | 1.60E-05 | 0.037214117 |
| Obesity | pericyte | *GOLGA3* | 3.926348328 | 4.81E-07 | 2.00E-06 | 0.003023406 |
| Obesity | pericyte | *CERS4* | 6.187150649 | 2.81E-05 | 6.37E-05 | 0.176963964 |
| Obesity | pericyte | *USP48* | 3.094860786 | 0.000325887 | 0.000588412 | 1 |
| Obesity | pericyte | *PDPR* | 5.826912963 | 2.00E-08 | 2.13E-07 | 0.000125877 |
| Obesity | pericyte | *EXOC1* | 6.275562306 | 1.02E-07 | 6.08E-07 | 0.000641331 |
| Obesity | pericyte | *NLRC4* | 3.787594005 | 1.10E-07 | 6.42E-07 | 0.000692516 |
| Obesity | pericyte | *SLC26A4* | 3.775302397 | 1.22E-06 | 4.08E-06 | 0.007648634 |
| Obesity | pericyte | *DLD* | 6.27169845 | 5.52E-09 | 1.35E-07 | 3.47E-05 |
| Obesity | pericyte | *TXNL1* | 7.736223087 | 1.35E-07 | 7.35E-07 | 0.000848512 |
| Obesity | pericyte | *IL5RA* | 4.574605653 | 1.30E-06 | 4.33E-06 | 0.008192556 |
| Obesity | pericyte | *ITGA6* | 3.595574199 | 0.002669686 | 0.004136467 | 1 |
| Obesity | pericyte | *RAPGEF4* | 2.503216445 | 6.69E-07 | 2.57E-06 | 0.004205902 |
| Obesity | pericyte | *SEL1L3* | 2.952626136 | 6.41E-08 | 4.37E-07 | 0.000403183 |
| Obesity | pericyte | *TF* | 3.054122598 | 2.26E-06 | 6.89E-06 | 0.014206115 |
| Obesity | pericyte | *MYO15A* | 2.072821505 | 1.62E-05 | 3.88E-05 | 0.101688763 |
| Obesity | pericyte | *ALKBH5* | 4.252931262 | 7.67E-07 | 2.87E-06 | 0.004821534 |
| Obesity | pericyte | *NLRP1* | 5.173687843 | 5.07E-06 | 1.40E-05 | 0.031865918 |
| Obesity | pericyte | *SPAG7* | 2.740777538 | 1.31E-06 | 4.36E-06 | 0.008261159 |
| Obesity | pericyte | *ZFHX4* | 3.703666599 | 0.002651961 | 0.004112699 | 1 |
| Obesity | pericyte | *RGS17* | 1.659972466 | 0.000260164 | 0.000478638 | 1 |
| Obesity | pericyte | *TMEM101* | -3.263808482 | 0.004559377 | 0.006824953 | 1 |
| Obesity | pericyte | *PPP2R3C* | 6.174979385 | 2.46E-06 | 7.41E-06 | 0.015462672 |
| Obesity | pericyte | *OSGEP* | 5.453816308 | 2.18E-08 | 2.20E-07 | 0.00013721 |
| Obesity | pericyte | *SLC22A17* | -2.663040755 | 0.000992321 | 0.001640042 | 1 |
| Obesity | pericyte | *SCFD1* | 3.111841393 | 0.000767617 | 0.001298711 | 1 |
| Obesity | pericyte | *RPGRIP1* | 3.017556946 | 0.001190078 | 0.001946415 | 1 |
| Obesity | pericyte | *TOX4* | 4.8914398 | 1.66E-09 | 6.90E-08 | 1.04E-05 |
| Obesity | pericyte | *AGO1* | 5.161547199 | 5.19E-08 | 3.79E-07 | 0.000326364 |
| Obesity | pericyte | *RFFL* | 3.329355042 | 2.23E-07 | 1.08E-06 | 0.00140327 |
| Obesity | pericyte | *MFSD11* | 2.921913141 | 5.11E-05 | 0.000108744 | 0.321230791 |
| Obesity | pericyte | *NUP50* | 3.347267694 | 5.83E-05 | 0.00012205 | 0.366760909 |
| Obesity | pericyte | *COMT* | 6.321179582 | 1.01E-08 | 1.71E-07 | 6.33E-05 |
| Obesity | pericyte | *VNN3* | 8.972793858 | 5.10E-07 | 2.09E-06 | 0.00320603 |
| Obesity | pericyte | *SEC22C* | 6.41767162 | 1.32E-05 | 3.25E-05 | 0.082981474 |
| Obesity | pericyte | *FMO2* | 2.776641907 | 4.81E-06 | 1.33E-05 | 0.030246811 |
| Obesity | pericyte | *SUCO* | 6.956361273 | 7.35E-07 | 2.77E-06 | 0.004620213 |
| Obesity | pericyte | *MSH2* | 4.754281793 | 1.85E-08 | 2.06E-07 | 0.000116439 |
| Obesity | pericyte | *MAP3K1* | 3.245251215 | 0.006722529 | 0.009785723 | 1 |
| Obesity | pericyte | *TMEM38B* | 5.124345563 | 7.12E-08 | 4.69E-07 | 0.000447414 |
| Obesity | pericyte | *PDE6C* | 5.932813021 | 4.28E-10 | 2.93E-08 | 2.69E-06 |
| Obesity | pericyte | *IKZF5* | 6.585657996 | 2.26E-07 | 1.08E-06 | 0.001419164 |
| Obesity | pericyte | *BLNK* | 2.783867321 | 6.23E-07 | 2.44E-06 | 0.003916689 |
| Obesity | pericyte | *WAC* | 5.433890068 | 1.85E-07 | 9.33E-07 | 0.001165406 |

| | | | | | | |
|---|---|---|---|---|---|---|
| Obesity | pericyte | *CREM* | 5.123969076 | 1.25E-08 | 1.73E-07 | 7.83E-05 |
| Obesity | pericyte | *TPSD1* | -3.997527144 | 1.81E-05 | 4.30E-05 | 0.113895954 |
| Obesity | pericyte | *ITPR3* | 6.173396693 | 7.92E-09 | 1.66E-07 | 4.98E-05 |
| Obesity | pericyte | *DSP* | 1.056127333 | 0.026725041 | 0.03580233 | 1 |
| Obesity | pericyte | *IFT74* | 2.97418297 | 1.08E-06 | 3.74E-06 | 0.006820344 |
| Obesity | pericyte | *ACOT7* | 3.602013958 | 6.39E-05 | 0.00013249 | 0.401444814 |
| Obesity | pericyte | *PCSK5* | 2.738835862 | 0.014640232 | 0.020341025 | 1 |
| Obesity | pericyte | *RAB18* | 2.155999998 | 0.005344889 | 0.007912248 | 1 |
| Obesity | pericyte | *NRP1* | 3.001162202 | 0.004570012 | 0.006837617 | 1 |
| Obesity | pericyte | *PRTFDC1* | -1.622687207 | 0.001234457 | 0.002016376 | 1 |
| Obesity | pericyte | *PALMD* | 3.015976888 | 3.87E-08 | 3.10E-07 | 0.000243046 |
| Obesity | pericyte | *TSPAN15* | -1.362634456 | 0.003135707 | 0.004805714 | 1 |
| Obesity | pericyte | *OCEL1* | -2.295706197 | 0.000123319 | 0.000242056 | 0.775304665 |
| Obesity | pericyte | *CATSPERG* | 1.520456714 | 0.005530427 | 0.008169595 | 1 |
| Obesity | pericyte | *STX1B* | -4.281637226 | 6.09E-05 | 0.000126915 | 0.382776105 |
| Obesity | pericyte | *HSD3B7* | 3.910286024 | 1.40E-06 | 4.60E-06 | 0.008777133 |
| Obesity | pericyte | *ATP5D* | 9.030432716 | 7.60E-07 | 2.85E-06 | 0.004777601 |
| Obesity | pericyte | *MARCH2* | 5.448436662 | 4.48E-08 | 3.42E-07 | 0.000281582 |
| Obesity | pericyte | *MTAP* | 7.169931483 | 0.000386821 | 0.000690109 | 1 |
| Obesity | pericyte | *CEP170B* | 1.630920311 | 0.000511614 | 0.000894222 | 1 |
| Obesity | pericyte | *POLR2E* | 3.955776472 | 2.01E-07 | 9.92E-07 | 0.001262249 |
| Obesity | pericyte | *MADCAM1* | 4.905581225 | 5.27E-08 | 3.83E-07 | 0.000331048 |
| Obesity | pericyte | *TRMT2A* | 3.804630103 | 0.015277974 | 0.021194312 | 1 |
| Obesity | pericyte | *ZDHHC8* | 3.354337078 | 7.72E-07 | 2.88E-06 | 0.004854709 |
| Obesity | pericyte | *MED15* | 4.954778901 | 6.18E-08 | 4.25E-07 | 0.000388815 |
| Obesity | pericyte | *SNAP29* | 0.947518019 | 0.010593712 | 0.01503107 | 1 |
| Obesity | pericyte | *CECR2* | 1.7575306 | 0.001677941 | 0.002682231 | 1 |
| Obesity | pericyte | *SMARCB1* | 3.989999107 | 1.28E-08 | 1.73E-07 | 8.02E-05 |
| Obesity | pericyte | *P2RX6* | 5.443397888 | 0.000236098 | 0.000437602 | 1 |
| Obesity | pericyte | *DDTL* | -1.078342426 | 0.00173485 | 0.002765468 | 1 |
| Obesity | pericyte | *DDT* | 7.929232733 | 1.56E-05 | 3.77E-05 | 0.098261012 |
| Obesity | pericyte | *SUSD2* | -0.59186273 | 0.009665684 | 0.013770259 | 1 |
| Obesity | pericyte | *GGT5* | -1.163850765 | 7.03E-05 | 0.000144946 | 0.442230638 |
| Obesity | pericyte | *UPB1* | -1.357235087 | 0.00094296 | 0.001565459 | 1 |
| Obesity | pericyte | *YPEL1* | 4.577820062 | 2.60E-08 | 2.39E-07 | 0.000163308 |
| Obesity | pericyte | *PRODH* | -5.41101973 | 0.002803419 | 0.004329426 | 1 |
| Obesity | pericyte | *PPM1F* | 6.179567811 | 5.92E-06 | 1.60E-05 | 0.037188865 |
| Obesity | pericyte | *CRYBB3* | 3.224194809 | 0.000491898 | 0.000861918 | 1 |
| Obesity | pericyte | *CYTH4* | 2.638765945 | 1.90E-07 | 9.50E-07 | 0.001196571 |
| Obesity | pericyte | *CARD10* | 2.190233178 | 1.10E-05 | 2.75E-05 | 0.06927677 |
| Obesity | pericyte | *SLC25A1* | 4.184146589 | 1.06E-07 | 6.23E-07 | 0.000663958 |
| Obesity | pericyte | *LGALS2* | 4.273624097 | 4.82E-06 | 1.33E-05 | 0.030284678 |
| Obesity | pericyte | *GGA1* | 3.438826512 | 3.75E-06 | 1.07E-05 | 0.023548893 |
| Obesity | pericyte | *SH3BP1* | 12.35339494 | 9.69E-06 | 2.47E-05 | 0.060926503 |
| Obesity | pericyte | *HPS4* | 7.580501679 | 2.42E-08 | 2.30E-07 | 0.000152279 |
| Obesity | pericyte | *PIK3IP1* | 2.794092946 | 0.002441597 | 0.003805235 | 1 |
| Obesity | pericyte | *TFIP11* | 3.100325566 | 8.48E-09 | 1.70E-07 | 5.33E-05 |
| Obesity | pericyte | *GCAT* | 3.527591617 | 1.15E-07 | 6.62E-07 | 0.000725224 |
| Obesity | pericyte | *ANKRD54* | -2.464614105 | 2.15E-05 | 5.01E-05 | 0.135276819 |
| Obesity | pericyte | *EIF3L* | 4.702153762 | 1.22E-06 | 4.11E-06 | 0.007694717 |

| | | | | | | |
|---|---|---|---|---|---|---|
| Obesity | pericyte | *CCDC134* | 6.950754944 | 2.04E-09 | 7.85E-08 | 1.28E-05 |
| Obesity | pericyte | *SLC16A8* | 3.217817441 | 0.000105398 | 0.000209695 | 0.662634898 |
| Obesity | pericyte | *DDX17* | 2.950157126 | 0.00068314 | 0.001166776 | 1 |
| Obesity | pericyte | *FBXO7* | 6.194712755 | 1.97E-11 | 2.53E-09 | 1.24E-07 |
| Obesity | pericyte | *TIMP3* | 5.00956213 | 2.62E-06 | 7.82E-06 | 0.016476113 |
| Obesity | pericyte | *PPP6R2* | 4.502985206 | 1.18E-07 | 6.74E-07 | 0.000742158 |
| Obesity | pericyte | *CYB5R3* | 5.994253689 | 2.44E-08 | 2.32E-07 | 0.000153531 |
| Obesity | pericyte | *RHBDD3* | 8.00640797 | 1.60E-08 | 1.91E-07 | 0.000100638 |
| Obesity | pericyte | *TTLL1* | 5.38032844 | 0.013403332 | 0.018725945 | 1 |
| Obesity | pericyte | *AP1B1* | 5.139284129 | 5.82E-05 | 0.000121878 | 0.366019877 |
| Obesity | pericyte | *HMGXB4* | 7.20740052 | 3.69E-06 | 1.06E-05 | 0.023183676 |
| Obesity | pericyte | *TOM1* | 1.395439458 | 0.018990173 | 0.025943333 | 1 |
| Obesity | pericyte | *CHKB* | 1.278903453 | 0.000905677 | 0.001508073 | 1 |
| Obesity | pericyte | *BIK* | 9.441287886 | 1.89E-06 | 5.93E-06 | 0.011879298 |
| Obesity | pericyte | *MCAT* | 3.986022647 | 9.63E-06 | 2.46E-05 | 0.060569495 |
| Obesity | pericyte | *ARSA* | 0.679550992 | 0.02819053 | 0.037573428 | 1 |
| Obesity | pericyte | *TSPO* | 5.606124046 | 2.00E-06 | 6.24E-06 | 0.012577624 |
| Obesity | pericyte | *RASD2* | -2.981109578 | 1.44E-06 | 4.71E-06 | 0.009052198 |
| Obesity | pericyte | *TTLL12* | 3.417036879 | 1.00E-06 | 3.54E-06 | 0.006306109 |
| Obesity | pericyte | *RBFOX2* | 4.584759198 | 0.000150379 | 0.000290367 | 0.945435903 |
| Obesity | pericyte | *MTMR3* | 2.097417341 | 0.000972087 | 0.00161126 | 1 |
| Obesity | pericyte | *APOL4* | 1.561076722 | 0.006387823 | 0.009322247 | 1 |
| Obesity | pericyte | *SAMM50* | 5.13400775 | 3.07E-07 | 1.40E-06 | 0.001930344 |
| Obesity | pericyte | *FOXRED2* | 1.431989904 | 0.000119364 | 0.000234586 | 0.750441135 |
| Obesity | pericyte | *IFT27* | 3.300750606 | 1.52E-07 | 8.02E-07 | 0.000957186 |
| Obesity | pericyte | *KIAA0930* | 3.487739757 | 1.36E-05 | 3.33E-05 | 0.085510074 |
| Obesity | pericyte | *NCF4* | 2.059812158 | 0.002813792 | 0.004344379 | 1 |
| Obesity | pericyte | *SLC25A17* | 6.799275322 | 1.02E-06 | 3.58E-06 | 0.006405364 |
| Obesity | pericyte | *KCTD17* | -3.903316121 | 0.00035156 | 0.000631321 | 1 |
| Obesity | pericyte | *EP300* | 6.016696051 | 2.17E-07 | 1.06E-06 | 0.001367004 |
| Obesity | pericyte | *ACO2* | 5.483042934 | 7.95E-07 | 2.95E-06 | 0.004996787 |
| Obesity | pericyte | *DESI1* | 3.426012972 | 1.12E-10 | 1.05E-08 | 7.02E-07 |
| Obesity | pericyte | *ZBED4* | 8.81817277 | 1.16E-07 | 6.68E-07 | 0.000732401 |
| Obesity | pericyte | *HDAC10* | 3.232290335 | 8.50E-06 | 2.20E-05 | 0.053451129 |
| Obesity | pericyte | *SDR39U1* | 1.091950487 | 0.011842347 | 0.016689718 | 1 |
| Obesity | pericyte | *RBM23* | 6.241755359 | 7.21E-06 | 1.90E-05 | 0.045325486 |
| Obesity | pericyte | *SOS2* | 5.675535166 | 2.50E-06 | 7.52E-06 | 0.015742946 |
| Obesity | pericyte | *TRIM9* | 1.565682492 | 0.000230457 | 0.000428409 | 1 |
| Obesity | pericyte | *DDHD1* | 4.467417709 | 5.53E-05 | 0.000116434 | 0.347712407 |
| Obesity | pericyte | *ATP6V1D* | 1.937653884 | 0.000209299 | 0.000392912 | 1 |
| Obesity | pericyte | *PIGH* | 2.487154116 | 0.013402491 | 0.018725945 | 1 |
| Obesity | pericyte | *VTI1B* | 3.549519203 | 1.02E-07 | 6.08E-07 | 0.000641661 |
| Obesity | pericyte | *GSTZ1* | 2.506875731 | 0.000111652 | 0.000220742 | 0.701959226 |
| Obesity | pericyte | *TMED8* | 2.67942779 | 0.025003087 | 0.033631666 | 1 |
| Obesity | pericyte | *DAAM1* | 3.550735754 | 2.37E-06 | 7.18E-06 | 0.014882855 |
| Obesity | pericyte | *ISM2* | 3.446030287 | 0.006268882 | 0.009174223 | 1 |
| Obesity | pericyte | *PPM1A* | 3.281068084 | 0.000888311 | 0.00148335 | 1 |
| Obesity | pericyte | *DICER1* | 5.649024693 | 2.47E-08 | 2.32E-07 | 0.000155278 |
| Obesity | pericyte | *ZFYVE21* | 6.969837208 | 9.54E-12 | 1.28E-09 | 6.00E-08 |
| Obesity | pericyte | *MTHFD1* | 3.85769287 | 1.39E-08 | 1.80E-07 | 8.72E-05 |

| | | | | | | |
|---|---|---|---|---|---|---|
| Obesity | pericyte | *TELO2* | 8.588736211 | 2.49E-05 | 5.72E-05 | 0.156547039 |
| Obesity | pericyte | *GSKIP* | 6.229747848 | 1.66E-08 | 1.93E-07 | 0.000104256 |
| Obesity | pericyte | *VRK1* | 7.290517188 | 1.07E-08 | 1.71E-07 | 6.72E-05 |
| Obesity | pericyte | *PAPLN* | 3.608147604 | 1.96E-06 | 6.13E-06 | 0.012334155 |
| Obesity | pericyte | *PSMB5* | 3.86021785 | 1.64E-08 | 1.93E-07 | 0.000103202 |
| Obesity | pericyte | *CCNB1IP1* | 5.012521921 | 3.25E-06 | 9.44E-06 | 0.020409125 |
| Obesity | pericyte | *TRIP11* | 6.538434 | 1.55E-07 | 8.15E-07 | 0.000975821 |
| Obesity | pericyte | *KIAA0391* | -4.135237614 | 0.002361245 | 0.003691904 | 1 |
| Obesity | pericyte | *PLTP* | 2.26195018 | 4.57E-07 | 1.92E-06 | 0.002874533 |
| Obesity | pericyte | *GSS* | 4.061128902 | 4.38E-07 | 1.86E-06 | 0.002755257 |
| Obesity | pericyte | *PYGB* | 2.8837442 | 2.62E-07 | 1.22E-06 | 0.001647184 |
| Obesity | pericyte | *ABHD12* | 5.492238906 | 1.71E-12 | 3.06E-10 | 1.07E-08 |
| Obesity | pericyte | *PROCR* | 1.613135075 | 0.002437768 | 0.00380257 | 1 |
| Obesity | pericyte | *NINL* | 3.672966468 | 1.19E-06 | 4.01E-06 | 0.007497683 |
| Obesity | pericyte | *UQCC1* | 2.968729559 | 0.000477036 | 0.00083681 | 1 |
| Obesity | pericyte | *NDRG3* | 8.961224535 | 1.97E-08 | 2.11E-07 | 0.000124067 |
| Obesity | pericyte | *DOK5* | 6.398831291 | 1.03E-08 | 1.71E-07 | 6.48E-05 |
| Obesity | pericyte | *CSTF1* | 3.592011162 | 1.32E-06 | 4.37E-06 | 0.00829371 |
| Obesity | pericyte | *TPD52L2* | 4.524624136 | 9.82E-08 | 5.92E-07 | 0.000617669 |
| Obesity | pericyte | *NELFCD* | 7.949197469 | 8.23E-07 | 3.02E-06 | 0.005175011 |
| Obesity | pericyte | *CTSZ* | 4.226804015 | 3.14E-06 | 9.19E-06 | 0.01975463 |
| Obesity | pericyte | *PRPF6* | 5.580912683 | 2.78E-06 | 8.22E-06 | 0.017446923 |
| Obesity | pericyte | *MTG2* | 3.092994019 | 2.91E-07 | 1.33E-06 | 0.001827136 |
| Obesity | pericyte | *DIDO1* | 4.865057945 | 5.16E-09 | 1.30E-07 | 3.25E-05 |
| Obesity | pericyte | *GID8* | 3.01567794 | 1.25E-05 | 3.08E-05 | 0.078560096 |
| Obesity | pericyte | *SLC17A9* | 7.566844814 | 1.37E-05 | 3.34E-05 | 0.085835368 |
| Obesity | pericyte | *BIRC7* | -1.833115861 | 0.008672521 | 0.012414422 | 1 |
| Obesity | pericyte | *NKAIN4* | -2.372053483 | 1.58E-05 | 3.82E-05 | 0.099601737 |
| Obesity | pericyte | *PTK6* | 15.62010408 | 7.54E-08 | 4.87E-07 | 0.000474288 |
| Obesity | pericyte | *GMEB2* | 6.630103786 | 1.34E-07 | 7.34E-07 | 0.000845459 |
| Obesity | pericyte | *TRIB3* | 3.010575478 | 3.34E-06 | 9.66E-06 | 0.020986543 |
| Obesity | pericyte | *CCM2L* | -1.716828242 | 0.0002812 | 0.000513626 | 1 |
| Obesity | pericyte | *MYL9* | 2.913715571 | 0.002847515 | 0.004393209 | 1 |
| Obesity | pericyte | *MANBAL* | 2.272640141 | 2.19E-07 | 1.06E-06 | 0.00137524 |
| Obesity | pericyte | *IDH3B* | 3.781788342 | 9.68E-08 | 5.87E-07 | 0.000608806 |
| Obesity | pericyte | *MAPRE1* | 6.586602589 | 1.35E-08 | 1.77E-07 | 8.48E-05 |
| Obesity | pericyte | *TTI1* | 5.866301425 | 9.46E-09 | 1.70E-07 | 5.95E-05 |
| Obesity | pericyte | *RPRD1B* | 6.155017998 | 9.22E-06 | 2.36E-05 | 0.057972901 |
| Obesity | pericyte | *PXMP4* | 5.351361006 | 3.52E-08 | 2.92E-07 | 0.000221301 |
| Obesity | pericyte | *ACTR5* | 4.684171852 | 8.28E-07 | 3.04E-06 | 0.005206508 |
| Obesity | pericyte | *DHX35* | 6.930205852 | 5.66E-08 | 4.01E-07 | 0.000355808 |
| Obesity | pericyte | *MAP1LC3A* | 2.453712443 | 5.27E-05 | 0.000111638 | 0.331230095 |
| Obesity | pericyte | *PIGU* | 4.339706356 | 1.54E-08 | 1.90E-07 | 9.71E-05 |
| Obesity | pericyte | *APMAP* | 7.250274465 | 1.48E-10 | 1.29E-08 | 9.30E-07 |
| Obesity | pericyte | *CELF4* | 2.261434181 | 1.26E-06 | 4.21E-06 | 0.007936938 |
| Obesity | pericyte | *METTL4* | 8.289203814 | 9.30E-09 | 1.70E-07 | 5.84E-05 |
| Obesity | pericyte | *LPIN2* | 6.144057508 | 1.29E-07 | 7.16E-07 | 0.000807935 |
| Obesity | pericyte | *SMCHD1* | 5.708488728 | 2.03E-07 | 1.00E-06 | 0.001275684 |
| Obesity | pericyte | *MYL12A* | 5.458891528 | 0.000798231 | 0.001346158 | 1 |
| Obesity | pericyte | *CEP76* | 4.810879257 | 1.58E-07 | 8.24E-07 | 0.000993571 |

| | | | | | | |
|---|---|---|---|---|---|---|
| Obesity | pericyte | *LIPG* | 6.325753724 | 0.000113417 | 0.000223948 | 0.713050404 |
| Obesity | pericyte | *POLI* | 0.571449512 | 0.006133656 | 0.008995171 | 1 |
| Obesity | pericyte | *NALCN* | 2.247529725 | 6.30E-07 | 2.46E-06 | 0.003961155 |
| Obesity | pericyte | *HTR2A* | 5.123256712 | 9.58E-07 | 3.41E-06 | 0.00602414 |
| Obesity | pericyte | *KLF5* | 2.57687053 | 5.53E-05 | 0.000116434 | 0.34778977 |
| Obesity | pericyte | *DNAJC3* | 5.947457541 | 1.92E-06 | 6.01E-06 | 0.012062277 |
| Obesity | pericyte | *ARHGEF7* | 5.351523528 | 3.27E-06 | 9.48E-06 | 0.020555598 |
| Obesity | pericyte | *SGCG* | 3.293492304 | 3.02E-06 | 8.88E-06 | 0.019001399 |
| Obesity | pericyte | *PARP4* | 6.485599972 | 8.05E-08 | 5.08E-07 | 0.000506204 |
| Obesity | pericyte | *RGCC* | 0.507413905 | 0.034749938 | 0.0457094 | 1 |
| Obesity | pericyte | *VWA8* | 3.683733949 | 0.000101877 | 0.000203851 | 0.640501246 |
| Obesity | pericyte | *KATNAL1* | 3.129918725 | 0.003976952 | 0.006006029 | 1 |
| Obesity | pericyte | *DHRS12* | 8.248112408 | 5.33E-08 | 3.86E-07 | 0.000335021 |
| Obesity | pericyte | *MEDAG* | 4.401430214 | 2.54E-08 | 2.37E-07 | 0.000159746 |
| Obesity | pericyte | *TSC22D1* | 5.16799961 | 2.22E-05 | 5.15E-05 | 0.139471702 |
| Obesity | pericyte | *MGRN1* | 1.584294789 | 0.034752812 | 0.0457094 | 1 |
| Obesity | pericyte | *CORO1A* | -3.639835297 | 0.000213729 | 0.000400511 | 1 |
| Obesity | pericyte | *GDPD3* | 2.434841436 | 2.81E-05 | 6.36E-05 | 0.176660016 |
| Obesity | pericyte | *ELMO3* | 1.899342712 | 5.83E-07 | 2.33E-06 | 0.003664569 |
| Obesity | pericyte | *NFAT5* | 4.925947927 | 1.39E-06 | 4.58E-06 | 0.008750509 |
| Obesity | pericyte | *LONP2* | 4.918695889 | 1.46E-05 | 3.55E-05 | 0.091659274 |
| Obesity | pericyte | *PLLP* | 3.751798938 | 1.90E-06 | 5.97E-06 | 0.011967461 |
| Obesity | pericyte | *DHODH* | 1.225030936 | 0.001958777 | 0.003095734 | 1 |
| Obesity | pericyte | *CTCF* | 6.112747089 | 2.45E-07 | 1.16E-06 | 0.001539537 |
| Obesity | pericyte | *ACD* | -2.051936546 | 8.74E-06 | 2.25E-05 | 0.054958831 |
| Obesity | pericyte | *USB1* | 9.632655687 | 7.32E-09 | 1.60E-07 | 4.61E-05 |
| Obesity | pericyte | *SETD6* | 7.419609781 | 7.01E-06 | 1.86E-05 | 0.044096804 |
| Obesity | pericyte | *SLC7A6OS* | 1.704224468 | 1.83E-06 | 5.80E-06 | 0.011528551 |
| Obesity | pericyte | *HCFC1R1* | 2.793563339 | 4.17E-07 | 1.79E-06 | 0.002623556 |
| Obesity | pericyte | *NPRL3* | 2.209582363 | 0.001692725 | 0.002703802 | 1 |
| Obesity | pericyte | *NAGPA* | 6.243087508 | 1.14E-10 | 1.05E-08 | 7.16E-07 |
| Obesity | pericyte | *COTL1* | 4.417681365 | 1.06E-07 | 6.24E-07 | 0.000665407 |
| Obesity | pericyte | *CRISPLD2* | 2.039876452 | 2.24E-06 | 6.84E-06 | 0.014095537 |
| Obesity | pericyte | *ZNF500* | 1.934406604 | 0.000709609 | 0.001207066 | 1 |
| Obesity | pericyte | *NOMO3* | 2.450599819 | 2.82E-06 | 8.35E-06 | 0.017758161 |
| Obesity | pericyte | *LMF1* | 7.697974681 | 1.87E-08 | 2.07E-07 | 0.000117732 |
| Obesity | pericyte | *NARFL* | 4.305129991 | 1.81E-07 | 9.14E-07 | 0.001134959 |
| Obesity | pericyte | *MTHFSD* | 0.920309297 | 0.023406624 | 0.031558534 | 1 |
| Obesity | pericyte | *CLCN7* | 4.094115836 | 1.03E-08 | 1.71E-07 | 6.47E-05 |
| Obesity | pericyte | *SLC7A5* | 15.13893445 | 0.012520438 | 0.017590167 | 1 |
| Obesity | pericyte | *FBXO31* | 4.703062749 | 4.09E-07 | 1.77E-06 | 0.002572232 |
| Obesity | pericyte | *RHBDL1* | -1.451341574 | 0.037110171 | 0.048593129 | 1 |
| Obesity | pericyte | *NUBP1* | 5.181038962 | 3.38E-08 | 2.85E-07 | 0.000212366 |
| Obesity | pericyte | *UBE2I* | 6.91121459 | 2.60E-08 | 2.39E-07 | 0.000163357 |
| Obesity | pericyte | *CLUAP1* | 2.911676432 | 9.09E-09 | 1.70E-07 | 5.71E-05 |
| Obesity | pericyte | *CPPED1* | 2.860368789 | 9.40E-08 | 5.76E-07 | 0.000591182 |
| Obesity | pericyte | *BFAR* | 6.297812476 | 2.12E-08 | 2.17E-07 | 0.000133167 |
| Obesity | pericyte | *RBL2* | 2.114281856 | 0.001979914 | 0.003127567 | 1 |
| Obesity | pericyte | *STX4* | 5.552349627 | 7.09E-05 | 0.00014586 | 0.445455408 |
| Obesity | pericyte | *KAT8* | 13.12345151 | 3.27E-18 | 1.71E-15 | 2.05E-14 |

| | | | | | | |
|---|---|---|---|---|---|---|
| Obesity | pericyte | *NOMO1* | 7.128455058 | 4.97E-07 | 2.05E-06 | 0.003122715 |
| Obesity | pericyte | *SYT17* | 1.592518661 | 4.74E-05 | 0.000101993 | 0.298227071 |
| Obesity | pericyte | *TMC5* | 2.265030763 | 8.95E-06 | 2.30E-05 | 0.056252166 |
| Obesity | pericyte | *RNF40* | -3.300468942 | 0.003161415 | 0.004841855 | 1 |
| Obesity | pericyte | *KNOP1* | -2.749099623 | 0.008246687 | 0.011842604 | 1 |
| Obesity | pericyte | *IQCH* | 8.158567094 | 6.15E-10 | 3.62E-08 | 3.87E-06 |
| Obesity | pericyte | *LACTB* | 5.402322446 | 6.51E-08 | 4.42E-07 | 0.000409323 |
| Obesity | pericyte | *CSK* | 10.17498079 | 3.47E-06 | 9.99E-06 | 0.021788833 |
| Obesity | pericyte | *HERC1* | 3.518681314 | 0.002044225 | 0.003223487 | 1 |
| Obesity | pericyte | *TRIP4* | 4.011236736 | 3.53E-08 | 2.92E-07 | 0.000222009 |
| Obesity | pericyte | *RASL12* | -1.4384305 | 6.42E-05 | 0.000133184 | 0.403681485 |
| Obesity | pericyte | *AP3B2* | -0.760339986 | 0.001906492 | 0.003020694 | 1 |
| Obesity | pericyte | *TTC23* | 4.093146198 | 3.83E-05 | 8.38E-05 | 0.240770488 |
| Obesity | pericyte | *FAH* | 3.054452904 | 1.27E-07 | 7.12E-07 | 0.000801242 |
| Obesity | pericyte | *RPAP1* | 4.337483922 | 3.16E-05 | 7.05E-05 | 0.198567188 |
| Obesity | pericyte | *HOMER2* | 2.693767198 | 0.01338728 | 0.018711833 | 1 |
| Obesity | pericyte | *EHD4* | 6.957064111 | 2.50E-08 | 2.35E-07 | 0.000157256 |
| Obesity | pericyte | *ATP8B4* | 5.742083406 | 5.96E-08 | 4.15E-07 | 0.000374593 |
| Obesity | pericyte | *DTWD1* | 4.747687846 | 3.47E-06 | 9.99E-06 | 0.021799236 |
| Obesity | pericyte | *PDGFRL* | 2.370807519 | 3.47E-07 | 1.54E-06 | 0.002178468 |
| Obesity | pericyte | *ZFAND1* | 3.142823419 | 2.01E-05 | 4.73E-05 | 0.126490379 |
| Obesity | pericyte | *RIPK2* | 5.86475727 | 7.59E-06 | 1.99E-05 | 0.047749427 |
| Obesity | pericyte | *EYA1* | 1.15408932 | 4.62E-05 | 9.96E-05 | 0.290588342 |
| Obesity | pericyte | *NBN* | 5.126994944 | 4.07E-08 | 3.21E-07 | 0.00025604 |
| Obesity | pericyte | *CPQ* | 3.195722971 | 9.48E-07 | 3.38E-06 | 0.005957029 |
| Obesity | pericyte | *NIPAL2* | 4.222004781 | 2.21E-05 | 5.12E-05 | 0.138667866 |
| Obesity | pericyte | *IKBKB* | 7.051606502 | 5.53E-07 | 2.23E-06 | 0.003479477 |
| Obesity | pericyte | *PLAT* | 9.674701543 | 1.04E-08 | 1.71E-07 | 6.51E-05 |
| Obesity | pericyte | *GDAP1* | 5.675949313 | 0.000141504 | 0.000274494 | 0.889635192 |
| Obesity | pericyte | *ZC2HC1A* | 4.861432138 | 1.72E-06 | 5.48E-06 | 0.010788546 |
| Obesity | pericyte | *ARMC1* | 1.214718818 | 0.018098512 | 0.024811458 | 1 |
| Obesity | pericyte | *SPAG1* | 2.66840771 | 5.63E-05 | 0.000118237 | 0.353765821 |
| Obesity | pericyte | *CHRAC1* | -3.036787189 | 0.000891165 | 0.001486931 | 1 |
| Obesity | pericyte | *SNX16* | 4.528719496 | 7.47E-08 | 4.83E-07 | 0.000469866 |
| Obesity | pericyte | *GSDMD* | 1.70279213 | 0.017496205 | 0.024043419 | 1 |
| Obesity | pericyte | *ERI1* | 7.202121191 | 1.06E-08 | 1.71E-07 | 6.66E-05 |
| Obesity | pericyte | *MTMR9* | 7.245124048 | 9.03E-09 | 1.70E-07 | 5.67E-05 |
| Obesity | pericyte | *LEPROTL1* | 5.459213506 | 1.22E-08 | 1.73E-07 | 7.67E-05 |
| Obesity | pericyte | *TNFRSF10A* | 1.696666173 | 0.000582845 | 0.001009184 | 1 |
| Obesity | pericyte | *ERICH1* | 6.347341894 | 1.09E-06 | 3.74E-06 | 0.006821619 |
| Obesity | pericyte | *TUSC3* | -1.328317659 | 0.000687927 | 0.001173677 | 1 |
| Obesity | pericyte | *ARHGEF10* | 2.868186958 | 9.25E-06 | 2.37E-05 | 0.058164615 |
| Obesity | pericyte | *KLHDC4* | 3.265389083 | 3.28E-06 | 9.50E-06 | 0.020611674 |
| Obesity | pericyte | *TULP2* | 2.343123875 | 0.000563094 | 0.000977948 | 1 |
| Obesity | pericyte | *NUCB1* | 3.720033181 | 6.49E-07 | 2.51E-06 | 0.004079993 |
| Obesity | pericyte | *NFKBIB* | 2.932825954 | 7.11E-07 | 2.70E-06 | 0.0044695 |
| Obesity | pericyte | *SARS2* | -1.324039736 | 0.007881539 | 0.011344147 | 1 |
| Obesity | pericyte | *PPP1R37* | 4.364854481 | 4.34E-10 | 2.93E-08 | 2.73E-06 |
| Obesity | pericyte | *FCGRT* | 1.986463114 | 0.010656174 | 0.015106058 | 1 |
| Obesity | pericyte | *ERCC2* | 5.467497141 | 6.50E-08 | 4.41E-07 | 0.000408808 |

| | | | | | | |
|---|---|---|---|---|---|---|
| Obesity | pericyte | *PLEKHJ1* | 4.009323661 | 1.49E-07 | 7.90E-07 | 0.000937855 |
| Obesity | pericyte | *CD37* | 4.816522951 | 4.25E-07 | 1.82E-06 | 0.002674028 |
| Obesity | pericyte | *STX10* | 3.142987025 | 2.86E-07 | 1.32E-06 | 0.00179816 |
| Obesity | pericyte | *FCER2* | 3.627691685 | 9.50E-07 | 3.38E-06 | 0.005973886 |
| Obesity | pericyte | *DMPK* | 4.503120574 | 3.02E-05 | 6.78E-05 | 0.18963324 |
| Obesity | pericyte | *IL4I1* | 1.810362447 | 0.018608443 | 0.025460561 | 1 |
| Obesity | pericyte | *TLE6* | -2.342956857 | 0.000452466 | 0.000798387 | 1 |
| Obesity | pericyte | *AES* | 7.509763817 | 6.97E-06 | 1.85E-05 | 0.043789535 |
| Obesity | pericyte | *SGTA* | 4.336457296 | 1.75E-08 | 1.99E-07 | 0.000109829 |
| Obesity | pericyte | *C19orf53* | -2.468030789 | 0.011931153 | 0.016799812 | 1 |
| Obesity | pericyte | *TIMM44* | 4.744364529 | 5.44E-05 | 0.000114843 | 0.341773185 |
| Obesity | pericyte | *CCDC61* | 2.646287392 | 0.000856934 | 0.001436296 | 1 |
| Obesity | pericyte | *IL27RA* | 4.382181484 | 9.66E-09 | 1.71E-07 | 6.07E-05 |
| Obesity | pericyte | *RASAL3* | 2.243775421 | 5.81E-07 | 2.32E-06 | 0.003652123 |
| Obesity | pericyte | *ZNF419* | 1.148338522 | 0.00047237 | 0.000829318 | 1 |
| Obesity | pericyte | *SYDE1* | -3.715816425 | 0.015358409 | 0.021296497 | 1 |
| Obesity | pericyte | *AURKC* | 1.387565421 | 0.008215991 | 0.011801219 | 1 |
| Obesity | pericyte | *CCNE1* | 5.159355357 | 2.26E-05 | 5.24E-05 | 0.142137611 |
| Obesity | pericyte | *ANKRD27* | 4.974012963 | 6.71E-08 | 4.48E-07 | 0.000421919 |
| Obesity | pericyte | *PRX* | 3.614408301 | 2.60E-06 | 7.79E-06 | 0.016365111 |
| Obesity | pericyte | *CCDC94* | 3.50787411 | 1.11E-07 | 6.44E-07 | 0.000695268 |
| Obesity | pericyte | *TBCB* | 3.735364495 | 0.003226794 | 0.004935974 | 1 |
| Obesity | pericyte | *POLR2I* | 3.316195879 | 0.007530177 | 0.010885773 | 1 |
| Obesity | pericyte | *SLC1A5* | 3.071416694 | 0.002944907 | 0.004533455 | 1 |
| Obesity | pericyte | *CACTIN* | 3.589473567 | 4.07E-07 | 1.76E-06 | 0.002556686 |
| Obesity | pericyte | *CCDC9* | 5.76860911 | 1.51E-06 | 4.89E-06 | 0.009471686 |
| Obesity | pericyte | *FZR1* | 12.45566527 | 4.12E-08 | 3.22E-07 | 0.000259064 |
| Obesity | pericyte | *BBC3* | -3.314321831 | 0.018452027 | 0.025279559 | 1 |
| Obesity | pericyte | *TGFB1* | 4.209966225 | 2.65E-07 | 1.23E-06 | 0.001665082 |
| Obesity | pericyte | *CEACAM4* | 8.350659722 | 1.12E-06 | 3.84E-06 | 0.007063022 |
| Obesity | pericyte | *PLIN3* | 5.596991371 | 1.86E-08 | 2.06E-07 | 0.00011682 |
| Obesity | pericyte | *MRPL4* | 6.460694405 | 5.62E-09 | 1.36E-07 | 3.53E-05 |
| Obesity | pericyte | *CD33* | 3.377189877 | 2.60E-06 | 7.79E-06 | 0.016360551 |
| Obesity | pericyte | *BABAM1* | 5.082699755 | 5.88E-08 | 4.13E-07 | 0.000369601 |
| Obesity | pericyte | *TYK2* | 5.871084485 | 1.60E-05 | 3.86E-05 | 0.100867596 |
| Obesity | pericyte | *RABAC1* | 5.469197426 | 7.12E-06 | 1.88E-05 | 0.044742786 |
| Obesity | pericyte | *PTPRS* | 3.286897537 | 2.33E-08 | 2.26E-07 | 0.000146216 |
| Obesity | pericyte | *GRWD1* | -1.744806322 | 4.47E-06 | 1.25E-05 | 0.028118715 |
| Obesity | pericyte | *GRIN2D* | 4.283011757 | 4.35E-07 | 1.85E-06 | 0.002734788 |
| Obesity | pericyte | *CARD8* | 4.026705933 | 0.000135437 | 0.000263212 | 0.851490262 |
| Obesity | pericyte | *LIG1* | -2.610617219 | 0.035980607 | 0.047235347 | 1 |
| Obesity | pericyte | *PLA2G4C* | 2.726696035 | 3.05E-07 | 1.39E-06 | 0.001915781 |
| Obesity | pericyte | *CAPS* | 3.754110042 | 6.70E-06 | 1.78E-05 | 0.042118524 |
| Obesity | pericyte | *BCAT2* | 4.621137542 | 4.22E-08 | 3.27E-07 | 0.000265237 |
| Obesity | pericyte | *PPP2R1A* | 8.806591479 | 1.18E-07 | 6.73E-07 | 0.000739737 |
| Obesity | pericyte | *TNPO2* | 4.097691415 | 6.68E-06 | 1.78E-05 | 0.042026336 |
| Obesity | pericyte | *GCDH* | 2.078735502 | 7.28E-05 | 0.000149393 | 0.457440773 |
| Obesity | pericyte | *LILRB5* | 2.954641605 | 1.12E-07 | 6.48E-07 | 0.000702285 |
| Obesity | pericyte | *MAST1* | -1.980651081 | 4.11E-05 | 8.95E-05 | 0.258708829 |
| Obesity | pericyte | *PRPF31* | 2.818411213 | 0.001515893 | 0.002446835 | 1 |

| | | | | | | |
|---|---|---|---|---|---|---|
| Obesity | pericyte | *RPL18A* | 1.575724704 | 0.031757289 | 0.04198908 | 1 |
| Obesity | pericyte | *PDE4C* | 2.215752558 | 0.012275564 | 0.017261568 | 1 |
| Obesity | pericyte | *ELL* | 2.955439946 | 0.000171682 | 0.000326981 | 1 |
| Obesity | pericyte | *UPK1A* | 3.920765848 | 0.015943085 | 0.02204616 | 1 |
| Obesity | pericyte | *COPE* | 7.668269543 | 2.07E-06 | 6.41E-06 | 0.013032783 |
| Obesity | pericyte | *KXD1* | 4.391444876 | 5.07E-08 | 3.74E-07 | 0.000318806 |
| Obesity | pericyte | *HPN* | 2.189759598 | 1.08E-05 | 2.72E-05 | 0.068119819 |
| Obesity | pericyte | *ATP13A1* | -2.845316845 | 0.000895667 | 0.001493253 | 1 |
| Obesity | pericyte | *SIPA1L3* | 5.273137823 | 7.38E-06 | 1.94E-05 | 0.046412165 |
| Obesity | pericyte | *SMG9* | 1.99304157 | 2.72E-06 | 8.10E-06 | 0.017130852 |
| Obesity | pericyte | *GTPBP10* | 1.782911137 | 0.001462024 | 0.002367786 | 1 |
| Obesity | pericyte | *PON3* | -1.203628436 | 8.15E-05 | 0.000165779 | 0.512589837 |
| Obesity | pericyte | *PON2* | 7.478901702 | 2.61E-07 | 1.22E-06 | 0.001637895 |
| Obesity | pericyte | *HBP1* | 5.893278145 | 1.49E-06 | 4.85E-06 | 0.009376082 |
| Obesity | pericyte | *DLX5* | -4.29532855 | 3.64E-05 | 8.01E-05 | 0.228683543 |
| Obesity | pericyte | *STEAP1B* | 2.309155421 | 0.000164077 | 0.000313829 | 1 |
| Obesity | pericyte | *PTN* | 2.623625252 | 7.99E-07 | 2.96E-06 | 0.005026215 |
| Obesity | pericyte | *MPP6* | 2.599242354 | 1.20E-07 | 6.79E-07 | 0.000752555 |
| Obesity | pericyte | *DFNA5* | 2.154640839 | 1.49E-06 | 4.85E-06 | 0.009369433 |
| Obesity | pericyte | *OGDH* | 6.53020638 | 5.73E-08 | 4.04E-07 | 0.000360107 |
| Obesity | pericyte | *ADAP1* | 4.462240064 | 8.26E-07 | 3.04E-06 | 0.005196124 |
| Obesity | pericyte | *H2AFV* | 5.089013229 | 2.38E-06 | 7.19E-06 | 0.01493544 |
| Obesity | pericyte | *CAV1* | 3.180149697 | 4.51E-06 | 1.26E-05 | 0.028338309 |
| Obesity | pericyte | *RNF32* | 1.593294022 | 0.003559967 | 0.005400945 | 1 |
| Obesity | pericyte | *LMBR1* | 4.668622851 | 5.51E-05 | 0.000116213 | 0.34666244 |
| Obesity | pericyte | *HOXA5* | 2.303526251 | 0.004977668 | 0.007396502 | 1 |
| Obesity | pericyte | *HOXA6* | 2.718015612 | 0.00021685 | 0.000405151 | 1 |
| Obesity | pericyte | *CPED1* | 4.106712287 | 1.49E-08 | 1.88E-07 | 9.37E-05 |
| Obesity | pericyte | *HIBADH* | 5.518791824 | 1.22E-08 | 1.73E-07 | 7.65E-05 |
| Obesity | pericyte | *CPVL* | 3.70504543 | 0.000226215 | 0.000421646 | 1 |
| Obesity | pericyte | *PLEKHA8* | 8.321932691 | 2.28E-08 | 2.24E-07 | 0.000143542 |
| Obesity | pericyte | *NOD1* | 6.391310315 | 5.84E-06 | 1.58E-05 | 0.036713118 |
| Obesity | pericyte | *EPHB6* | 3.987168538 | 0.00010276 | 0.000205095 | 0.646049736 |
| Obesity | pericyte | *NPTX2* | -20.86785312 | 0.000266879 | 0.00048946 | 1 |
| Obesity | pericyte | *CYP3A5* | 6.898190063 | 9.87E-09 | 1.71E-07 | 6.20E-05 |
| Obesity | pericyte | *ZKSCAN1* | 4.804686683 | 1.38E-07 | 7.49E-07 | 0.000869899 |
| Obesity | pericyte | *SNX8* | 5.292741434 | 1.31E-08 | 1.74E-07 | 8.22E-05 |
| Obesity | pericyte | *TAF6* | 1.92518628 | 0.003323475 | 0.005067836 | 1 |
| Obesity | pericyte | *AIMP2* | 7.529103743 | 5.71E-08 | 4.04E-07 | 0.000359003 |
| Obesity | pericyte | *TFR2* | 3.155574461 | 0.035374003 | 0.046458399 | 1 |
| Obesity | pericyte | *RBM28* | 2.28844459 | 0.00164086 | 0.002626295 | 1 |
| Obesity | pericyte | *USP42* | 7.361509382 | 2.18E-06 | 6.69E-06 | 0.013720184 |
| Obesity | pericyte | *LSM5* | 7.230939908 | 1.01E-08 | 1.71E-07 | 6.38E-05 |
| Obesity | pericyte | *C1GALT1* | 6.44722327 | 5.59E-08 | 3.98E-07 | 0.000351681 |
| Obesity | pericyte | *PLOD3* | 4.285807676 | 0.012932216 | 0.018128169 | 1 |
| Obesity | pericyte | *RPA3* | 7.55498393 | 6.57E-07 | 2.53E-06 | 0.004133094 |
| Obesity | pericyte | *PHF14* | 5.629885577 | 6.00E-07 | 2.38E-06 | 0.003775039 |
| Obesity | pericyte | *CEP41* | 2.470910166 | 0.005486171 | 0.00810803 | 1 |
| Obesity | pericyte | *ZNF862* | 4.584227507 | 6.45E-05 | 0.000133828 | 0.405767483 |
| Obesity | pericyte | *MEST* | -1.394548229 | 0.001696487 | 0.002709122 | 1 |

| | | | | | | |
|---|---|---|---|---|---|---|
| Obesity | pericyte | *ACTR3C* | 2.583318033 | 3.50E-05 | 7.73E-05 | 0.220211312 |
| Obesity | pericyte | *POU6F2* | 1.666589834 | 9.40E-07 | 3.36E-06 | 0.005911952 |
| Obesity | pericyte | *TSPAN13* | 2.241420119 | 5.29E-07 | 2.15E-06 | 0.003326808 |
| Obesity | pericyte | *RARRES2* | 4.021087756 | 1.68E-05 | 4.01E-05 | 0.105442724 |
| Obesity | pericyte | *TMEM176B* | 1.088742557 | 0.006247955 | 0.009152119 | 1 |
| Obesity | pericyte | *MRPL32* | 2.626441894 | 1.10E-05 | 2.74E-05 | 0.068869842 |
| Obesity | pericyte | *COA1* | 1.748001719 | 0.009677132 | 0.013783446 | 1 |
| Obesity | pericyte | *BLVRA* | 2.969921098 | 3.25E-07 | 1.46E-06 | 0.002045494 |
| Obesity | pericyte | *URGCP* | 6.138508006 | 8.03E-08 | 5.08E-07 | 0.000504917 |
| Obesity | pericyte | *GCK* | -5.904492197 | 2.44E-05 | 5.62E-05 | 0.153526504 |
| Obesity | pericyte | *YKT6* | 2.450102958 | 1.24E-06 | 4.14E-06 | 0.00776876 |
| Obesity | pericyte | *CLIP2* | -1.764492418 | 0.002852521 | 0.004399853 | 1 |
| Obesity | pericyte | *SPATA6L* | 2.078162912 | 0.000162356 | 0.000310727 | 1 |
| Obesity | pericyte | *SLC1A1* | 0.849510473 | 0.030292657 | 0.040204758 | 1 |
| Obesity | pericyte | *FKTN* | 4.910610585 | 1.13E-08 | 1.71E-07 | 7.08E-05 |
| Obesity | pericyte | *NMRK1* | 3.810820683 | 2.37E-06 | 7.18E-06 | 0.01488236 |
| Obesity | pericyte | *TMEM245* | 4.270309521 | 3.20E-06 | 9.34E-06 | 0.020149438 |
| Obesity | pericyte | *MEGF9* | 2.296166623 | 0.000300656 | 0.000546466 | 1 |
| Obesity | pericyte | *TRIM14* | 4.971436987 | 1.55E-06 | 5.02E-06 | 0.009761343 |
| Obesity | pericyte | *CORO2A* | 3.725854008 | 6.09E-07 | 2.40E-06 | 0.003829112 |
| Obesity | pericyte | *C5* | 2.293897972 | 0.003613274 | 0.005477852 | 1 |
| Obesity | pericyte | *OGN* | 5.017807651 | 3.15E-12 | 5.21E-10 | 1.98E-08 |
| Obesity | pericyte | *ECM2* | 2.847874112 | 0.000991206 | 0.001640042 | 1 |
| Obesity | pericyte | *PTGR1* | 1.718581277 | 0.000565757 | 0.000982301 | 1 |
| Obesity | pericyte | *SUSD1* | 2.181502833 | 6.81E-05 | 0.000140562 | 0.428011436 |
| Obesity | pericyte | *TNFSF8* | 2.381674407 | 5.10E-07 | 2.09E-06 | 0.003207966 |
| Obesity | pericyte | *AK1* | 3.670871635 | 4.84E-06 | 1.34E-05 | 0.030428306 |
| Obesity | pericyte | *KDM4C* | 5.759337212 | 7.79E-08 | 4.96E-07 | 0.000489844 |
| Obesity | pericyte | *DOCK8* | 2.217646705 | 0.000294175 | 0.000535615 | 1 |
| Obesity | pericyte | *FUBP3* | 5.551833126 | 9.13E-09 | 1.70E-07 | 5.74E-05 |
| Obesity | pericyte | *RGP1* | 12.51621603 | 4.20E-10 | 2.93E-08 | 2.64E-06 |
| Obesity | pericyte | *MPDZ* | 5.747225638 | 1.19E-08 | 1.73E-07 | 7.47E-05 |
| Obesity | pericyte | *PTGDS* | 2.551097674 | 9.24E-07 | 3.32E-06 | 0.005808109 |
| Obesity | pericyte | *SHB* | 6.701825407 | 3.44E-07 | 1.53E-06 | 0.002164712 |
| Obesity | pericyte | *DVL1* | 4.92238452 | 1.01E-06 | 3.55E-06 | 0.00633131 |
| Obesity | pericyte | *CCNJ* | 3.713438298 | 0.00786381 | 0.011321221 | 1 |
| Obesity | pericyte | *ATRNL1* | 1.710188897 | 0.000501803 | 0.000878296 | 1 |
| Obesity | pericyte | *HPS1* | 6.933937683 | 5.32E-06 | 1.45E-05 | 0.033421576 |
| Obesity | pericyte | *PHYH* | 1.258242528 | 0.000726827 | 0.001234683 | 1 |
| Obesity | pericyte | *DNMBP* | 6.936756426 | 1.06E-06 | 3.68E-06 | 0.00667256 |
| Obesity | pericyte | *TRDMT1* | 7.017201138 | 1.23E-09 | 5.64E-08 | 7.72E-06 |
| Obesity | pericyte | *DDX50* | 6.609823649 | 1.51E-05 | 3.66E-05 | 0.09483625 |
| Obesity | pericyte | *MAPK8* | 5.778021758 | 4.41E-07 | 1.87E-06 | 0.002775689 |
| Obesity | pericyte | *ATE1* | 4.34120687 | 1.56E-06 | 5.03E-06 | 0.009794451 |
| Obesity | pericyte | *NSMCE4A* | 5.097450367 | 1.32E-08 | 1.75E-07 | 8.31E-05 |
| Obesity | pericyte | *PLEKHA1* | 4.914267185 | 1.88E-07 | 9.41E-07 | 0.001182517 |
| Obesity | pericyte | *SPOCK2* | 1.704005257 | 1.30E-05 | 3.20E-05 | 0.081614172 |
| Obesity | pericyte | *MICU1* | 6.855642159 | 3.91E-09 | 1.13E-07 | 2.46E-05 |
| Obesity | pericyte | *CCSER2* | 6.439197021 | 2.27E-07 | 1.09E-06 | 0.001427221 |
| Obesity | pericyte | *LIPA* | 3.566017286 | 1.01E-05 | 2.55E-05 | 0.063415229 |

| | | | | | | |
|---|---|---|---|---|---|---|
| Obesity | pericyte | *SFXN3* | 4.035145124 | 5.22E-09 | 1.30E-07 | 3.28E-05 |
| Obesity | pericyte | *TNKS2* | 5.444406675 | 2.93E-07 | 1.35E-06 | 0.001843901 |
| Obesity | pericyte | *ANKRD26* | 3.039841136 | 4.86E-05 | 0.000104259 | 0.305792184 |
| Obesity | pericyte | *ACBD5* | 6.525881237 | 1.82E-11 | 2.39E-09 | 1.15E-07 |
| Obesity | pericyte | *MTPAP* | 5.95058062 | 1.73E-09 | 7.10E-08 | 1.09E-05 |
| Obesity | pericyte | *PITRM1* | 6.101955563 | 1.04E-08 | 1.71E-07 | 6.51E-05 |
| Obesity | pericyte | *MAP3K8* | 5.888040521 | 8.57E-08 | 5.35E-07 | 0.000539007 |
| Obesity | pericyte | *SHOC2* | 5.731748024 | 6.39E-08 | 4.36E-07 | 0.000401519 |
| Obesity | pericyte | *TFAM* | 7.798223207 | 1.24E-05 | 3.06E-05 | 0.07771304 |
| Obesity | pericyte | *MLLT6* | 3.538139125 | 0.025423719 | 0.034168218 | 1 |
| Obesity | pericyte | *PSMB3* | 4.241128762 | 1.88E-07 | 9.41E-07 | 0.001181386 |
| Obesity | pericyte | *CASC3* | 4.663729206 | 1.27E-05 | 3.13E-05 | 0.079877591 |
| Obesity | pericyte | *RAPGEFL1* | 1.614665861 | 0.000915442 | 0.001522588 | 1 |
| Obesity | pericyte | *RGS9* | 7.820835464 | 2.28E-08 | 2.24E-07 | 0.000143354 |
| Obesity | pericyte | *WNT3* | 3.623665181 | 2.18E-06 | 6.67E-06 | 0.013689448 |
| Obesity | pericyte | *ASPA* | 3.697594816 | 3.46E-08 | 2.89E-07 | 0.000217313 |
| Obesity | pericyte | *SEPT4* | -0.528634615 | 6.03E-06 | 1.63E-05 | 0.037931685 |
| Obesity | pericyte | *TRIM37* | 6.900138996 | 8.83E-08 | 5.48E-07 | 0.000555153 |
| Obesity | pericyte | *RPS6KB1* | 4.507489223 | 0.00341559 | 0.005199471 | 1 |
| Obesity | pericyte | *TRIM16L* | 1.345256319 | 0.000280533 | 0.000512558 | 1 |
| Obesity | pericyte | *CDK5RAP3* | 4.166387551 | 1.23E-05 | 3.04E-05 | 0.077125348 |
| Obesity | pericyte | *CBX1* | 5.397258369 | 1.18E-09 | 5.48E-08 | 7.39E-06 |
| Obesity | pericyte | *PIGL* | 6.661872225 | 7.39E-08 | 4.80E-07 | 0.000464368 |
| Obesity | pericyte | *GALK1* | 7.30982251 | 3.32E-08 | 2.83E-07 | 0.000208519 |
| Obesity | pericyte | *CAMTA2* | 2.898563365 | 0.013348159 | 0.018661302 | 1 |
| Obesity | pericyte | *MED13* | 2.562125531 | 0.006141724 | 0.009004901 | 1 |
| Obesity | pericyte | *HOXB6* | 5.960969648 | 3.10E-11 | 3.61E-09 | 1.95E-07 |
| Obesity | pericyte | *RNF167* | 6.585026406 | 4.29E-07 | 1.84E-06 | 0.002700203 |
| Obesity | pericyte | *CHRNE* | 1.64011573 | 1.72E-05 | 4.10E-05 | 0.108202745 |
| Obesity | pericyte | *C1QBP* | 7.85184234 | 0.000227254 | 0.000423332 | 1 |
| Obesity | pericyte | *GOSR1* | 7.228041872 | 6.61E-08 | 4.45E-07 | 0.000415676 |
| Obesity | pericyte | *DRG2* | 10.70424865 | 1.43E-08 | 1.83E-07 | 8.97E-05 |
| Obesity | pericyte | *AKAP10* | 4.919150657 | 6.66E-06 | 1.78E-05 | 0.041895652 |
| Obesity | pericyte | *SMARCD2* | 5.703905159 | 5.74E-07 | 2.30E-06 | 0.003610365 |
| Obesity | pericyte | *ICAM2* | 3.45768448 | 0.00066842 | 0.00114381 | 1 |
| Obesity | pericyte | *SYNGR2* | 1.496633113 | 0.000213108 | 0.000399467 | 1 |
| Obesity | pericyte | *UTP6* | 8.286201176 | 6.05E-08 | 4.19E-07 | 0.000380083 |
| Obesity | pericyte | *DDX5* | 1.788131758 | 0.002730235 | 0.004223668 | 1 |
| Obesity | pericyte | *C17orf75* | 7.045003551 | 1.83E-05 | 4.34E-05 | 0.115150117 |
| Obesity | pericyte | *CYTH1* | 4.075341951 | 1.74E-06 | 5.53E-06 | 0.010931718 |
| Obesity | pericyte | *PEX12* | 1.614530451 | 0.010638505 | 0.015084412 | 1 |
| Obesity | pericyte | *DHX58* | 2.372323608 | 0.000785838 | 0.001327038 | 1 |
| Obesity | pericyte | *RAB5C* | 3.292606982 | 0.00131914 | 0.00214689 | 1 |
| Obesity | pericyte | *NAGLU* | 7.821434387 | 4.32E-07 | 1.84E-06 | 0.002715488 |
| Obesity | pericyte | *EZH1* | 4.427705263 | 0.000107509 | 0.000213489 | 0.675907281 |
| Obesity | pericyte | *DLX4* | 21.97917498 | 1.18E-06 | 3.98E-06 | 0.007418134 |
| Obesity | pericyte | *SGCA* | -0.958032237 | 0.001572305 | 0.002530095 | 1 |
| Obesity | pericyte | *VAT1* | 3.51502403 | 5.62E-08 | 3.99E-07 | 0.000353452 |
| Obesity | pericyte | *LRRC59* | 12.70777483 | 4.57E-07 | 1.92E-06 | 0.002872974 |
| Obesity | pericyte | *ALOX12* | 3.872814366 | 0.033658877 | 0.044363387 | 1 |

| | | | | | | |
|---|---|---|---|---|---|---|
| Obesity | pericyte | *ABCC3* | 1.500004201 | 1.45E-05 | 3.54E-05 | 0.091269655 |
| Obesity | pericyte | *MPP2* | 6.147993643 | 1.43E-07 | 7.65E-07 | 0.000896761 |
| Obesity | pericyte | *EFTUD2* | 6.24154935 | 1.09E-09 | 5.30E-08 | 6.88E-06 |
| Obesity | pericyte | *EFNB3* | -0.862784743 | 1.62E-05 | 3.89E-05 | 0.102054614 |
| Obesity | pericyte | *FAM20A* | 6.702866543 | 2.33E-08 | 2.26E-07 | 0.000146773 |
| Obesity | pericyte | *YWHAE* | 6.455499943 | 1.09E-07 | 6.36E-07 | 0.00068359 |
| Obesity | pericyte | *DPH1* | 8.566846356 | 9.33E-09 | 1.70E-07 | 5.87E-05 |
| Obesity | pericyte | *DHRS7B* | 6.190555273 | 9.34E-09 | 1.70E-07 | 5.87E-05 |
| Obesity | pericyte | *SLC9A3R1* | 2.898555025 | 2.13E-06 | 6.55E-06 | 0.013368724 |
| Obesity | pericyte | *TMEM104* | 3.391503363 | 3.53E-07 | 1.56E-06 | 0.002220548 |
| Obesity | pericyte | *TMEM97* | 1.957054365 | 0.00710133 | 0.01030373 | 1 |
| Obesity | pericyte | *RAB34* | 2.159286046 | 0.036025604 | 0.047284545 | 1 |
| Obesity | pericyte | *SLAIN2* | 5.268473266 | 4.68E-07 | 1.96E-06 | 0.002944215 |
| Obesity | pericyte | *OCIAD1* | 5.285063766 | 2.73E-05 | 6.20E-05 | 0.1716498 |
| Obesity | pericyte | *CWH43* | 14.00862669 | 0.000262217 | 0.000481752 | 1 |
| Obesity | pericyte | *KIAA1211* | 1.263831147 | 0.000296565 | 0.000539498 | 1 |
| Obesity | pericyte | *AREG* | -0.890684408 | 0.000429689 | 0.000762261 | 1 |
| Obesity | pericyte | *UBE2D3* | 5.60919785 | 2.21E-09 | 8.32E-08 | 1.39E-05 |
| Obesity | pericyte | *TBC1D9* | 6.28524412 | 4.53E-08 | 3.45E-07 | 0.000284781 |
| Obesity | pericyte | *WFS1* | 7.384355269 | 0.000314573 | 0.000569784 | 1 |
| Obesity | pericyte | *AADAT* | 5.445532084 | 4.34E-06 | 1.22E-05 | 0.027279428 |
| Obesity | pericyte | *CPZ* | -25.43367676 | 7.46E-05 | 0.000152682 | 0.468885726 |
| Obesity | pericyte | *SLC2A9* | 5.184288237 | 0.000128009 | 0.000250426 | 0.804794867 |
| Obesity | pericyte | *SH3D19* | 2.443890688 | 0.000307098 | 0.000557207 | 1 |
| Obesity | pericyte | *STIM2* | 6.288939884 | 4.15E-07 | 1.79E-06 | 0.002611766 |
| Obesity | pericyte | *LRP2BP* | 1.710860282 | 0.036493968 | 0.047869305 | 1 |
| Obesity | pericyte | *UFSP2* | 3.63169089 | 0.021708569 | 0.029401502 | 1 |
| Obesity | pericyte | *KLF3* | 6.768828937 | 8.18E-07 | 3.01E-06 | 0.005141852 |
| Obesity | pericyte | *KLHL5* | 1.641853632 | 0.032314044 | 0.042698276 | 1 |
| Obesity | pericyte | *UGDH* | 5.5120313 | 8.50E-08 | 5.31E-07 | 0.000534444 |
| Obesity | pericyte | *DDX25* | -1.868585339 | 0.001481344 | 0.002395989 | 1 |
| Obesity | pericyte | *HTATIP2* | 1.658362691 | 0.005859229 | 0.008618851 | 1 |
| Obesity | pericyte | *ELP4* | 5.044355947 | 2.52E-06 | 7.56E-06 | 0.015826348 |
| Obesity | pericyte | *SC5D* | 9.972368968 | 4.37E-09 | 1.22E-07 | 2.75E-05 |
| Obesity | pericyte | *C11orf63* | 4.969582105 | 8.15E-09 | 1.69E-07 | 5.12E-05 |
| Obesity | pericyte | *PUS3* | 3.143312449 | 2.61E-06 | 7.80E-06 | 0.01640089 |
| Obesity | pericyte | *DCPS* | 3.334904236 | 1.31E-05 | 3.22E-05 | 0.082355245 |
| Obesity | pericyte | *FOXRED1* | 3.522291775 | 1.70E-07 | 8.74E-07 | 0.001066779 |
| Obesity | pericyte | *NRXN2* | 9.667623734 | 5.05E-08 | 3.73E-07 | 0.000317301 |
| Obesity | pericyte | *MS4A4A* | 1.539293016 | 3.25E-05 | 7.23E-05 | 0.204540121 |
| Obesity | pericyte | *CHORDC1* | 4.953966871 | 3.68E-07 | 1.61E-06 | 0.002313307 |
| Obesity | pericyte | *EIF4G2* | 7.072540885 | 1.15E-07 | 6.61E-07 | 0.000723306 |
| Obesity | pericyte | *CBL* | 4.555618148 | 1.73E-06 | 5.51E-06 | 0.010886044 |
| Obesity | pericyte | *KIAA1549L* | 2.528112738 | 0.000317517 | 0.000574454 | 1 |
| Obesity | pericyte | *CD5* | 2.998892463 | 5.39E-06 | 1.47E-05 | 0.033883642 |
| Obesity | pericyte | *ACCS* | 0.941287674 | 0.019024492 | 0.025984571 | 1 |
| Obesity | pericyte | *MADD* | 7.088563242 | 6.57E-06 | 1.76E-05 | 0.041298776 |
| Obesity | pericyte | *NAA40* | 4.63971993 | 1.36E-06 | 4.49E-06 | 0.008548673 |
| Obesity | pericyte | *CARS* | 2.594871745 | 0.010071081 | 0.014315371 | 1 |
| Obesity | pericyte | *SLC22A18* | 1.317102992 | 6.25E-05 | 0.000129806 | 0.392662281 |

| | | | | | | |
|---|---|---|---|---|---|---|
| Obesity | pericyte | *RPS13* | 2.580200998 | 0.020489921 | 0.027846981 | 1 |
| Obesity | pericyte | *NDUFS8* | 9.017665753 | 1.98E-05 | 4.66E-05 | 0.124413905 |
| Obesity | pericyte | *TCIRG1* | 6.465556491 | 6.33E-10 | 3.68E-08 | 3.98E-06 |
| Obesity | pericyte | *GTF2H1* | 5.827101566 | 1.24E-09 | 5.65E-08 | 7.80E-06 |
| Obesity | pericyte | *PTPN5* | 1.003048946 | 0.010077522 | 0.014321288 | 1 |
| Obesity | pericyte | *PRDM4* | 7.592588527 | 4.97E-09 | 1.29E-07 | 3.12E-05 |
| Obesity | pericyte | *COQ5* | 0.465905646 | 0.015960952 | 0.022059025 | 1 |
| Obesity | pericyte | *CAPRIN2* | 5.476051288 | 4.90E-05 | 0.000104906 | 0.307794783 |
| Obesity | pericyte | *KCTD10* | -1.51473711 | 0.012242815 | 0.017219369 | 1 |
| Obesity | pericyte | *CYP27B1* | 16.97826021 | 1.53E-06 | 4.96E-06 | 0.009616059 |
| Obesity | pericyte | *GLI1* | 3.849778948 | 1.03E-06 | 3.60E-06 | 0.006474301 |
| Obesity | pericyte | *METAP2* | 4.743057276 | 2.34E-05 | 5.41E-05 | 0.147188813 |
| Obesity | pericyte | *LTA4H* | 7.438293586 | 7.67E-06 | 2.00E-05 | 0.048236187 |
| Obesity | pericyte | *WNT5B* | -1.916554209 | 0.004093076 | 0.006166587 | 1 |
| Obesity | pericyte | *TRPV4* | -4.257141817 | 2.41E-06 | 7.28E-06 | 0.01514865 |
| Obesity | pericyte | *ITFG2* | 5.577592309 | 1.41E-05 | 3.45E-05 | 0.088761616 |
| Obesity | pericyte | *PRR4* | 2.217868665 | 0.000216478 | 0.000404577 | 1 |
| Obesity | pericyte | *PARP11* | 5.713380578 | 5.43E-08 | 3.90E-07 | 0.000341542 |
| Obesity | pericyte | *ARPC3* | 6.391690378 | 2.17E-07 | 1.06E-06 | 0.00136318 |
| Obesity | pericyte | *GPN3* | 7.110079419 | 1.13E-06 | 3.86E-06 | 0.007117438 |
| Obesity | pericyte | *VPS29* | 6.980273501 | 1.88E-06 | 5.92E-06 | 0.01184398 |
| Obesity | pericyte | *KCNA1* | 3.745968411 | 0.000462062 | 0.000814177 | 1 |
| Obesity | pericyte | *CREBL2* | 4.605632506 | 6.65E-06 | 1.78E-05 | 0.041789043 |
| Obesity | pericyte | *NAA25* | 6.783400757 | 3.48E-09 | 1.07E-07 | 2.19E-05 |
| Obesity | pericyte | *SCNN1A* | 3.352449938 | 0.005602496 | 0.008270227 | 1 |
| Obesity | pericyte | *LTBR* | 3.651508769 | 3.48E-08 | 2.89E-07 | 0.000218768 |
| Obesity | pericyte | *CDK2AP1* | 0.978314726 | 0.006114408 | 0.008973222 | 1 |
| Obesity | pericyte | *MGP* | 2.170187799 | 0.000219644 | 0.000409863 | 1 |
| Obesity | pericyte | *RASAL1* | -5.788564242 | 0.000101428 | 0.000203083 | 0.637680826 |
| Obesity | pericyte | *EIF2B1* | 6.965185259 | 8.28E-09 | 1.70E-07 | 5.20E-05 |
| Obesity | pericyte | *DDX55* | 3.540465113 | 3.29E-10 | 2.46E-08 | 2.07E-06 |
| Obesity | pericyte | *ENDOU* | -1.833481135 | 0.002212524 | 0.003470593 | 1 |
| Obesity | pericyte | *STX2* | 1.896283386 | 0.015429734 | 0.021390681 | 1 |
| Obesity | pericyte | *MDM1* | 2.102153308 | 0.021821072 | 0.029541146 | 1 |
| Obesity | pericyte | *NUP107* | 6.003749019 | 2.89E-09 | 9.63E-08 | 1.82E-05 |
| Obesity | pericyte | *CNOT2* | 5.376547521 | 8.40E-06 | 2.17E-05 | 0.052807398 |
| Obesity | pericyte | *KRR1* | 3.734044144 | 0.007688207 | 0.011106562 | 1 |
| Obesity | pericyte | *COPS7A* | 2.530354863 | 7.55E-06 | 1.98E-05 | 0.047491817 |
| Obesity | pericyte | *GNPTAB* | 6.176251264 | 2.24E-06 | 6.84E-06 | 0.014082827 |
| Obesity | pericyte | *SPSB2* | 6.644467846 | 1.07E-06 | 3.70E-06 | 0.006711903 |
| Obesity | pericyte | *ATN1* | 4.750798025 | 1.85E-06 | 5.84E-06 | 0.011645106 |
| Obesity | pericyte | *C12orf57* | 2.723816968 | 1.29E-05 | 3.19E-05 | 0.081411624 |
| Obesity | pericyte | *NT5DC3* | 1.913270521 | 0.002511223 | 0.003908903 | 1 |
| Obesity | pericyte | *PRKAB1* | 2.389475966 | 0.001702964 | 0.002718775 | 1 |
| Obesity | pericyte | *ST8SIA1* | 3.072207151 | 3.14E-07 | 1.42E-06 | 0.001972756 |
| Obesity | pericyte | *C2CD5* | 6.155834725 | 2.18E-08 | 2.20E-07 | 0.000136884 |
| Obesity | pericyte | *PHC1* | 10.01006437 | 9.41E-05 | 0.00018899 | 0.591349592 |
| Obesity | pericyte | *BTN3A3* | 1.794488447 | 9.86E-06 | 2.50E-05 | 0.061958732 |
| Obesity | pericyte | *TDP2* | 6.587927984 | 5.18E-06 | 1.42E-05 | 0.032577285 |
| Obesity | pericyte | *RWDD1* | 2.647918247 | 0.009281806 | 0.013251125 | 1 |

| | | | | | | |
|---|---|---|---|---|---|---|
| Obesity | pericyte | *SMIM8* | 6.035505617 | 1.90E-08 | 2.08E-07 | 0.000119487 |
| Obesity | pericyte | *MCM9* | 6.891981021 | 1.35E-07 | 7.34E-07 | 0.000846468 |
| Obesity | pericyte | *FAM184A* | 3.776026598 | 2.52E-07 | 1.18E-06 | 0.001582236 |
| Obesity | pericyte | *RNGTT* | 6.103738391 | 9.81E-09 | 1.71E-07 | 6.17E-05 |
| Obesity | pericyte | *GABRR2* | 2.912704013 | 3.02E-08 | 2.65E-07 | 0.000189572 |
| Obesity | pericyte | *HDDC2* | 2.692839613 | 2.57E-06 | 7.71E-06 | 0.016166155 |
| Obesity | pericyte | *TPD52L1* | 2.055532236 | 0.002824182 | 0.00435935 | 1 |
| Obesity | pericyte | *UST* | 2.636805253 | 1.96E-07 | 9.73E-07 | 0.001230954 |
| Obesity | pericyte | *MTRF1L* | 2.74562557 | 2.76E-07 | 1.27E-06 | 0.001734497 |
| Obesity | pericyte | *TULP1* | 13.13181376 | 7.70E-07 | 2.88E-06 | 0.004840619 |
| Obesity | pericyte | *MAPK14* | 3.857631172 | 0.000904884 | 0.001507421 | 1 |
| Obesity | pericyte | *RNF8* | 3.730977336 | 7.94E-07 | 2.95E-06 | 0.004990813 |
| Obesity | pericyte | *PHACTR1* | 3.273460392 | 0.011871997 | 0.016727756 | 1 |
| Obesity | pericyte | *MDGA1* | 2.763561187 | 9.87E-07 | 3.49E-06 | 0.006203708 |
| Obesity | pericyte | *CD83* | 1.776218393 | 1.77E-05 | 4.22E-05 | 0.111508808 |
| Obesity | pericyte | *SAYSD1* | 8.273080122 | 9.30E-07 | 3.33E-06 | 0.005844751 |
| Obesity | pericyte | *CAP2* | 1.352587404 | 0.001894568 | 0.003003709 | 1 |
| Obesity | pericyte | *GPR63* | -0.992479644 | 0.013768878 | 0.019181241 | 1 |
| Obesity | pericyte | *ASCC3* | 5.72953328 | 1.11E-06 | 3.82E-06 | 0.007005871 |
| Obesity | pericyte | *BVES* | -1.372015331 | 0.000334042 | 0.000601582 | 1 |
| Obesity | pericyte | *GPLD1* | 1.191455617 | 0.000446983 | 0.0007896 | 1 |
| Obesity | pericyte | *ALDH5A1* | 2.895334699 | 8.57E-07 | 3.11E-06 | 0.005384892 |
| Obesity | pericyte | *VNN1* | 1.106725385 | 0.001033139 | 0.001703026 | 1 |
| Obesity | pericyte | *VNN2* | 4.319402381 | 0.000189804 | 0.000358995 | 1 |
| Obesity | pericyte | *ACOT13* | 4.768428575 | 1.69E-08 | 1.95E-07 | 0.000106241 |
| Obesity | pericyte | *C6orf62* | 4.83272487 | 4.10E-07 | 1.77E-06 | 0.002574594 |
| Obesity | pericyte | *HBS1L* | 1.843864983 | 0.019787704 | 0.026950887 | 1 |
| Obesity | pericyte | *PERP* | 1.591986128 | 0.000596309 | 0.001029661 | 1 |
| Obesity | pericyte | *SLC16A10* | 2.166356413 | 1.63E-05 | 3.90E-05 | 0.10219207 |
| Obesity | pericyte | *PHACTR2* | 5.571019264 | 4.45E-08 | 3.41E-07 | 0.000279774 |
| Obesity | pericyte | *TFEB* | 6.555179678 | 0.000172632 | 0.000328691 | 1 |
| Obesity | pericyte | *FAM120B* | 3.389359985 | 1.58E-05 | 3.81E-05 | 0.099316048 |
| Obesity | pericyte | *TBP* | 6.32222567 | 4.34E-06 | 1.22E-05 | 0.027278475 |
| Obesity | pericyte | *DUSP22* | 6.817615068 | 1.40E-08 | 1.81E-07 | 8.79E-05 |
| Obesity | pericyte | *EXOC2* | 6.437043599 | 1.41E-06 | 4.64E-06 | 0.008868477 |
| Obesity | pericyte | *TMEM30A* | 3.587792574 | 5.03E-07 | 2.07E-06 | 0.003163672 |
| Obesity | pericyte | *SENP6* | 4.084146648 | 0.000515721 | 0.0009009 | 1 |
| Obesity | pericyte | *TTK* | 2.367876354 | 0.005988754 | 0.008799088 | 1 |
| Obesity | pericyte | *LAMA4* | 3.108189433 | 1.25E-07 | 6.99E-07 | 0.000784718 |
| Obesity | pericyte | *CLIC5* | 2.158582456 | 0.001533613 | 0.002474165 | 1 |
| Obesity | pericyte | *FBRSL1* | 5.671384231 | 1.87E-08 | 2.07E-07 | 0.00011753 |
| Obesity | pericyte | *LY86* | 9.593682878 | 1.88E-08 | 2.07E-07 | 0.0001182 |
| Obesity | pericyte | *PRSS16* | 7.822217136 | 8.92E-10 | 4.71E-08 | 5.61E-06 |
| Obesity | pericyte | *NUDT12* | 3.593814663 | 1.47E-06 | 4.79E-06 | 0.009219116 |
| Obesity | pericyte | *CEP72* | 2.515450645 | 5.52E-06 | 1.50E-05 | 0.034681005 |
| Obesity | pericyte | *C7* | 4.524689736 | 6.50E-07 | 2.51E-06 | 0.004085698 |
| Obesity | pericyte | *PAPD7* | 5.212213175 | 6.97E-06 | 1.85E-05 | 0.043795766 |
| Obesity | pericyte | *GHR* | 1.181689399 | 0.000248517 | 0.000459132 | 1 |
| Obesity | pericyte | *DAP* | 5.963138106 | 7.10E-05 | 0.000146105 | 0.446349733 |
| Obesity | pericyte | *NME5* | 2.057221521 | 1.36E-09 | 5.95E-08 | 8.55E-06 |

| | | | | | | |
|---|---|---|---|---|---|---|
| Obesity | pericyte | *NNT* | 2.069574936 | 7.27E-05 | 0.00014922 | 0.456761512 |
| Obesity | pericyte | *PFDN1* | 3.857362506 | 1.09E-05 | 2.72E-05 | 0.068221447 |
| Obesity | pericyte | *LOX* | 7.053266195 | 1.04E-08 | 1.71E-07 | 6.54E-05 |
| Obesity | pericyte | *TMCO6* | -2.967471048 | 0.000657212 | 0.001126162 | 1 |
| Obesity | pericyte | *SPARC* | 4.429826632 | 1.91E-06 | 5.99E-06 | 0.012010318 |
| Obesity | pericyte | *PCDHB6* | 3.401331042 | 0.003113721 | 0.004773461 | 1 |
| Obesity | pericyte | *CLK4* | 4.685408726 | 1.18E-08 | 1.73E-07 | 7.40E-05 |
| Obesity | pericyte | *RNF130* | 5.768029208 | 7.81E-08 | 4.96E-07 | 0.000491245 |
| Obesity | pericyte | *ARSB* | 2.114779979 | 0.001641662 | 0.002626911 | 1 |
| Obesity | pericyte | *CLINT1* | 5.609989274 | 4.94E-08 | 3.67E-07 | 0.000310285 |
| Obesity | pericyte | *BTNL8* | -0.812370825 | 3.83E-06 | 1.09E-05 | 0.024058654 |
| Obesity | pericyte | *TTC1* | 7.011837265 | 1.28E-07 | 7.15E-07 | 0.000804618 |
| Obesity | pericyte | *RASGRF2* | 3.747978659 | 5.13E-09 | 1.30E-07 | 3.22E-05 |
| Obesity | pericyte | *POLR3G* | 2.39919766 | 1.56E-05 | 3.78E-05 | 0.098379312 |
| Obesity | pericyte | *DROSHA* | 3.402422591 | 0.000542504 | 0.000944275 | 1 |
| Obesity | pericyte | *CDH6* | -0.541333111 | 0.000157359 | 0.000301989 | 0.989314625 |
| Obesity | pericyte | *SLC27A6* | -1.93289835 | 5.94E-05 | 0.000124074 | 0.373463605 |
| Obesity | pericyte | *TARS* | 4.694105477 | 2.60E-08 | 2.39E-07 | 0.000163326 |
| Obesity | pericyte | *LNPEP* | 5.017900376 | 3.89E-08 | 3.11E-07 | 0.000244559 |
| Obesity | pericyte | *GNPDA1* | 5.082979249 | 1.36E-08 | 1.78E-07 | 8.57E-05 |
| Obesity | pericyte | *SKP1* | 8.076249017 | 2.12E-09 | 8.12E-08 | 1.33E-05 |
| Obesity | pericyte | *PPWD1* | 1.322774962 | 0.018773649 | 0.025675425 | 1 |
| Obesity | pericyte | *TRIM23* | 6.013030442 | 5.85E-06 | 1.58E-05 | 0.036748834 |
| Obesity | pericyte | *C9* | 1.520354571 | 0.023966305 | 0.032264702 | 1 |
| Obesity | pericyte | *TXNDC15* | 4.380142109 | 1.22E-07 | 6.87E-07 | 0.000764266 |
| Obesity | pericyte | *SMAD5* | 5.696103105 | 1.54E-05 | 3.72E-05 | 0.096850551 |
| Obesity | pericyte | *HMGXB3* | 5.148976437 | 0.000706261 | 0.001201951 | 1 |
| Obesity | pericyte | *ATP6V0E1* | 5.543765604 | 9.67E-06 | 2.47E-05 | 0.060816005 |
| Obesity | pericyte | *BNIP1* | -0.99566187 | 0.009839521 | 0.014002053 | 1 |
| Obesity | pericyte | *CNTN3* | 4.946154771 | 1.58E-08 | 1.91E-07 | 9.96E-05 |
| Obesity | pericyte | *ACTR8* | 4.537104244 | 2.85E-07 | 1.31E-06 | 0.001788656 |
| Obesity | pericyte | *HGD* | 6.211692642 | 2.12E-07 | 1.04E-06 | 0.001334163 |
| Obesity | pericyte | *NIT2* | 5.57608657 | 0.000164129 | 0.000313831 | 1 |
| Obesity | pericyte | *FAM162A* | 4.249718682 | 0.000242116 | 0.000448097 | 1 |
| Obesity | pericyte | *PCCB* | 4.906769628 | 1.08E-08 | 1.71E-07 | 6.79E-05 |
| Obesity | pericyte | *RBP1* | 3.852094113 | 3.18E-06 | 9.27E-06 | 0.01996825 |
| Obesity | pericyte | *RNF7* | 2.182164661 | 0.015172728 | 0.021052955 | 1 |
| Obesity | pericyte | *PDCD10* | 6.679656429 | 2.39E-07 | 1.14E-06 | 0.001502694 |
| Obesity | pericyte | *WNT5A* | 2.459566505 | 5.07E-05 | 0.00010814 | 0.318797217 |
| Obesity | pericyte | *PRKAR2A* | 4.062612677 | 1.06E-09 | 5.30E-08 | 6.67E-06 |
| Obesity | pericyte | *HYAL1* | -1.224939477 | 0.015693034 | 0.021726955 | 1 |
| Obesity | pericyte | *CYB561D2* | 2.821059859 | 8.02E-06 | 2.09E-05 | 0.05044874 |
| Obesity | pericyte | *IFT57* | 5.692387419 | 2.07E-08 | 2.16E-07 | 0.000130145 |
| Obesity | pericyte | *IQCG* | 7.930895545 | 1.85E-07 | 9.31E-07 | 0.00116248 |
| Obesity | pericyte | *GBE1* | 3.568821236 | 1.09E-08 | 1.71E-07 | 6.86E-05 |
| Obesity | pericyte | *UMPS* | 3.76053428 | 3.36E-05 | 7.44E-05 | 0.211229734 |
| Obesity | pericyte | *NCBP2* | 7.151285878 | 2.12E-08 | 2.17E-07 | 0.000133357 |
| Obesity | pericyte | *SLC41A3* | 3.584728213 | 3.02E-05 | 6.78E-05 | 0.189833652 |
| Obesity | pericyte | *PLXNA1* | 2.476817116 | 0.000605911 | 0.001045666 | 1 |
| Obesity | pericyte | *ABTB1* | 6.036127696 | 0.000170053 | 0.000323978 | 1 |

| | | | | | | |
|---|---|---|---|---|---|---|
| Obesity | pericyte | *NEK11* | 6.331375301 | 5.30E-09 | 1.32E-07 | 3.33E-05 |
| Obesity | pericyte | *PLSCR4* | 2.388911672 | 0.007445309 | 0.010772994 | 1 |
| Obesity | pericyte | *HEMK1* | 2.802434315 | 0.001862194 | 0.002956468 | 1 |
| Obesity | pericyte | *WDR48* | 4.013796841 | 0.000110825 | 0.000219313 | 0.696755883 |
| Obesity | pericyte | *COMMD2* | 6.574286778 | 5.19E-08 | 3.79E-07 | 0.000326199 |
| Obesity | pericyte | *GORASP1* | 7.522811938 | 9.39E-07 | 3.36E-06 | 0.005903453 |
| Obesity | pericyte | *ABCC5* | 3.021493286 | 0.012708571 | 0.01783632 | 1 |
| Obesity | pericyte | *ARHGEF26* | 2.306453651 | 1.54E-08 | 1.90E-07 | 9.68E-05 |
| Obesity | pericyte | *KLHL24* | 5.021656128 | 9.88E-09 | 1.71E-07 | 6.21E-05 |
| Obesity | pericyte | *ZBTB47* | 4.678306693 | 2.14E-06 | 6.58E-06 | 0.013458244 |
| Obesity | pericyte | *NKTR* | 5.049314953 | 1.57E-06 | 5.07E-06 | 0.009878874 |
| Obesity | pericyte | *CLCN2* | 4.654340009 | 1.33E-07 | 7.31E-07 | 0.000835437 |
| Obesity | pericyte | *NEK4* | 5.931232933 | 7.38E-07 | 2.78E-06 | 0.0046398 |
| Obesity | pericyte | *MOB1A* | 6.973315492 | 6.75E-05 | 0.00013946 | 0.424514762 |
| Obesity | pericyte | *PIKFYVE* | 3.544732315 | 0.002689766 | 0.004165163 | 1 |
| Obesity | pericyte | *FAHD2A* | 3.272998557 | 8.78E-09 | 1.70E-07 | 5.52E-05 |
| Obesity | pericyte | *SLC35F5* | 3.558115082 | 0.000590237 | 0.001019737 | 1 |
| Obesity | pericyte | *ACTR3* | 6.533520019 | 9.80E-08 | 5.91E-07 | 0.000616136 |
| Obesity | pericyte | *TFCP2L1* | 3.300925955 | 2.15E-08 | 2.18E-07 | 0.000134994 |
| Obesity | pericyte | *DNAJC27* | 5.736907768 | 1.36E-09 | 5.95E-08 | 8.56E-06 |
| Obesity | pericyte | *STAM2* | 4.962079209 | 0.000617531 | 0.001065135 | 1 |
| Obesity | pericyte | *GPD2* | 4.039943066 | 6.18E-08 | 4.25E-07 | 0.000388605 |
| Obesity | pericyte | *CENPA* | 5.532895657 | 3.69E-05 | 8.11E-05 | 0.232064645 |
| Obesity | pericyte | *ITGA4* | 1.871783239 | 2.43E-07 | 1.15E-06 | 0.001528243 |
| Obesity | pericyte | *PCSK4* | 2.401217041 | 8.10E-05 | 0.000164713 | 0.50896234 |
| Obesity | pericyte | *INO80B* | -2.064340771 | 0.008496266 | 0.012187092 | 1 |
| Obesity | pericyte | *GRB14* | 1.749906145 | 1.35E-06 | 4.46E-06 | 0.008478911 |
| Obesity | pericyte | *CLIP4* | 4.800458723 | 5.83E-09 | 1.40E-07 | 3.66E-05 |
| Obesity | pericyte | *SPTBN1* | 2.70839582 | 0.000440316 | 0.000778915 | 1 |
| Obesity | pericyte | *HTRA2* | 2.661323433 | 1.23E-06 | 4.11E-06 | 0.007704381 |
| Obesity | pericyte | *GALNT3* | 1.983435929 | 0.004769277 | 0.007113748 | 1 |
| Obesity | pericyte | *POLE4* | 9.82708987 | 8.14E-09 | 1.69E-07 | 5.12E-05 |
| Obesity | pericyte | *LANCL1* | 5.283940153 | 4.85E-09 | 1.29E-07 | 3.05E-05 |
| Obesity | pericyte | *WDR75* | 8.104666817 | 3.18E-08 | 2.76E-07 | 0.000200139 |
| Obesity | pericyte | *EFEMP1* | 2.574529442 | 1.91E-07 | 9.55E-07 | 0.001203456 |
| Obesity | pericyte | *GLS* | 5.505064035 | 2.11E-07 | 1.03E-06 | 0.00132447 |
| Obesity | pericyte | *DNAH6* | 0.372620861 | 0.005118187 | 0.007592743 | 1 |
| Obesity | pericyte | *GGCX* | 2.524251355 | 0.000109329 | 0.000216599 | 0.687352638 |
| Obesity | pericyte | *GNLY* | 5.710389412 | 7.42E-05 | 0.000152078 | 0.466576495 |
| Obesity | pericyte | *SF3B1* | 5.32576923 | 3.99E-06 | 1.13E-05 | 0.025108029 |
| Obesity | pericyte | *ST3GAL5* | 3.2638267 | 5.86E-08 | 4.12E-07 | 0.000368465 |
| Obesity | pericyte | *PLCD4* | 4.251797289 | 1.97E-06 | 6.13E-06 | 0.012354335 |
| Obesity | pericyte | *CHMP3* | -2.243493251 | 0.016539888 | 0.022799008 | 1 |
| Obesity | pericyte | *ZNF142* | 2.746178124 | 7.75E-08 | 4.93E-07 | 0.000487062 |
| Obesity | pericyte | *MLPH* | 1.975003624 | 8.14E-07 | 3.00E-06 | 0.00511498 |
| Obesity | pericyte | *HDLBP* | 3.773633935 | 6.13E-06 | 1.65E-05 | 0.038548589 |
| Obesity | pericyte | *PASK* | 2.53033299 | 1.23E-06 | 4.12E-06 | 0.007736106 |
| Obesity | pericyte | *STK25* | 7.275335163 | 1.10E-07 | 6.40E-07 | 0.000689592 |
| Obesity | pericyte | *STRN* | 6.166482829 | 9.70E-06 | 2.47E-05 | 0.060969828 |
| Obesity | pericyte | *CEBPZ* | 3.943201363 | 5.49E-05 | 0.000115845 | 0.345335018 |

| | | | | | | |
|---|---|---|---|---|---|---|
| Obesity | pericyte | *PRKD3* | 3.779674807 | 0.002171841 | 0.00341018 | 1 |
| Obesity | pericyte | *SLC25A12* | 4.400844832 | 6.15E-07 | 2.42E-06 | 0.003863577 |
| Obesity | pericyte | *RMDN2* | 7.0416562 | 2.77E-05 | 6.28E-05 | 0.174009147 |
| Obesity | pericyte | *SOS1* | 3.630470451 | 2.29E-07 | 1.10E-06 | 0.001441949 |
| Obesity | pericyte | *WIPF1* | 4.909964417 | 8.68E-08 | 5.40E-07 | 0.000545641 |
| Obesity | pericyte | *COX7A2L* | 8.361101595 | 2.22E-07 | 1.07E-06 | 0.00139608 |
| Obesity | pericyte | *PNO1* | 7.034547612 | 1.33E-08 | 1.76E-07 | 8.38E-05 |
| Obesity | pericyte | *ORC4* | 4.984159776 | 1.16E-05 | 2.89E-05 | 0.072936188 |
| Obesity | pericyte | *THADA* | 5.875960805 | 7.29E-08 | 4.75E-07 | 0.000458039 |
| Obesity | pericyte | *ARID3A* | 4.506171447 | 2.27E-08 | 2.24E-07 | 0.000142923 |
| Obesity | pericyte | *CD207* | 49.47122523 | 8.52E-09 | 1.70E-07 | 5.35E-05 |
| Obesity | pericyte | *GRIN3B* | 2.054814245 | 0.006275436 | 0.009181677 | 1 |
| Obesity | pericyte | *VAX2* | 8.329689249 | 2.29E-08 | 2.25E-07 | 0.00014422 |
| Obesity | pericyte | *MSH6* | 5.499939311 | 3.80E-08 | 3.06E-07 | 0.000238959 |
| Obesity | pericyte | *PLEKHA3* | 8.337102611 | 1.11E-08 | 1.71E-07 | 7.00E-05 |
| Obesity | pericyte | *ALMS1* | 4.189386423 | 0.000230164 | 0.000428119 | 1 |
| Obesity | pericyte | *DHCR24* | 4.245880837 | 5.59E-06 | 1.52E-05 | 0.035113838 |
| Obesity | pericyte | *DNAJC16* | 2.169424565 | 0.006346513 | 0.009272722 | 1 |
| Obesity | pericyte | *MORN1* | 2.005266858 | 0.009819669 | 0.013976966 | 1 |
| Obesity | pericyte | *SCP2* | 5.093793185 | 3.13E-08 | 2.73E-07 | 0.000196849 |
| Obesity | pericyte | *TPSG1* | -2.238266423 | 0.000509581 | 0.000890916 | 1 |
| Obesity | pericyte | *CEP104* | 2.406780954 | 0.002551571 | 0.003969741 | 1 |
| Obesity | pericyte | *FAM20B* | 7.725208024 | 4.33E-06 | 1.22E-05 | 0.027244937 |
| Obesity | pericyte | *TMEM59* | 6.511898553 | 8.22E-08 | 5.17E-07 | 0.000516579 |
| Obesity | pericyte | *WRAP73* | 8.731893618 | 1.00E-06 | 3.54E-06 | 0.00629866 |
| Obesity | pericyte | *MRPL37* | 3.581560957 | 0.001608693 | 0.002581382 | 1 |
| Obesity | pericyte | *QSOX1* | 6.431331202 | 3.11E-05 | 6.95E-05 | 0.195426754 |
| Obesity | pericyte | *PARK7* | 5.394646665 | 0.00015466 | 0.000297536 | 0.972348209 |
| Obesity | pericyte | *EDEM3* | 5.596687281 | 4.97E-07 | 2.05E-06 | 0.003127212 |
| Obesity | pericyte | *RAP1A* | 5.134687549 | 2.20E-05 | 5.11E-05 | 0.138175336 |
| Obesity | pericyte | *CAPZA1* | 4.198857521 | 0.000238627 | 0.00044203 | 1 |
| Obesity | pericyte | *DOCK7* | 4.601047121 | 0.000391039 | 0.000697238 | 1 |
| Obesity | pericyte | *FBXO6* | 2.248213655 | 0.000113551 | 0.000224142 | 0.713893063 |
| Obesity | pericyte | *C1orf21* | 2.51929521 | 1.31E-06 | 4.36E-06 | 0.008248989 |
| Obesity | pericyte | *DNAJC6* | 4.067147089 | 0.000319969 | 0.000578724 | 1 |
| Obesity | pericyte | *LEPR* | 3.894413737 | 2.06E-08 | 2.16E-07 | 0.000129601 |
| Obesity | pericyte | *MFN2* | 6.291949938 | 2.62E-07 | 1.22E-06 | 0.001646407 |
| Obesity | pericyte | *MIIP* | 4.102966879 | 0.005944399 | 0.00873596 | 1 |
| Obesity | pericyte | *SMG7* | 6.323298997 | 4.95E-07 | 2.05E-06 | 0.003114431 |
| Obesity | pericyte | *NCF2* | 4.963305342 | 2.74E-08 | 2.47E-07 | 0.00017195 |
| Obesity | pericyte | *OLFML3* | 3.771582066 | 4.11E-08 | 3.22E-07 | 0.000258563 |
| Obesity | pericyte | *TNNI3K* | 2.386173562 | 0.003283012 | 0.005012214 | 1 |
| Obesity | pericyte | *CRYZ* | 2.234326316 | 8.55E-05 | 0.000173158 | 0.537309773 |
| Obesity | pericyte | *TTF2* | 6.22695096 | 2.09E-08 | 2.17E-07 | 0.000131608 |
| Obesity | pericyte | *TMEM9* | 1.379516104 | 0.001553513 | 0.002502418 | 1 |
| Obesity | pericyte | *WARS2* | 3.311518287 | 1.18E-06 | 3.98E-06 | 0.007423307 |
| Obesity | pericyte | *OSCP1* | 3.871550127 | 7.89E-06 | 2.05E-05 | 0.049590311 |
| Obesity | pericyte | *GNPAT* | 5.029855881 | 2.83E-07 | 1.31E-06 | 0.001779448 |
| Obesity | pericyte | *C1orf109* | 7.996380423 | 1.56E-09 | 6.58E-08 | 9.83E-06 |
| Obesity | pericyte | *TBCE* | 4.983054149 | 1.24E-06 | 4.14E-06 | 0.007783148 |

| | | | | | | |
|---|---|---|---|---|---|---|
| Obesity | pericyte | *LGALS8* | 1.502708168 | 0.011051592 | 0.015634869 | 1 |
| Obesity | pericyte | *MTR* | 5.217078589 | 5.96E-07 | 2.37E-06 | 0.003745053 |
| Obesity | pericyte | *KCNQ4* | -1.104591095 | 0.035221044 | 0.046276846 | 1 |
| Obesity | pericyte | *ACADM* | 4.52565892 | 7.88E-09 | 1.66E-07 | 4.95E-05 |
| Obesity | pericyte | *PADI2* | -2.275314009 | 2.47E-05 | 5.67E-05 | 0.154998928 |
| Obesity | pericyte | *MFAP2* | -1.431463018 | 0.005264552 | 0.007798831 | 1 |
| Obesity | pericyte | *CTBS* | 3.758366779 | 0.000380931 | 0.000680373 | 1 |
| Obesity | pericyte | *KLHL12* | 4.237511404 | 0.000461353 | 0.000813157 | 1 |
| Obesity | pericyte | *ZNHIT6* | 1.901275468 | 0.029954162 | 0.039772294 | 1 |
| Obesity | pericyte | *RBBP5* | 2.475495933 | 0.002243326 | 0.003515402 | 1 |
| Obesity | pericyte | *KIF17* | -2.783338457 | 0.00139802 | 0.002268806 | 1 |
| Obesity | pericyte | *CD46* | 5.238424379 | 1.36E-08 | 1.78E-07 | 8.55E-05 |
| Obesity | pericyte | *PRDX1* | 6.235964826 | 1.51E-07 | 7.97E-07 | 0.000950469 |
| Obesity | pericyte | *FAAH* | 0.613387211 | 0.005019187 | 0.007451152 | 1 |
| Obesity | pericyte | *TMED5* | 6.680932593 | 3.81E-06 | 1.09E-05 | 0.023973196 |
| Obesity | pericyte | *DR1* | 7.489625868 | 1.76E-06 | 5.59E-06 | 0.01107787 |
| Obesity | pericyte | *CNN3* | 7.372078779 | 2.82E-08 | 2.52E-07 | 0.000177 |
| Obesity | pericyte | *PRRC2C* | 6.124043856 | 4.26E-08 | 3.30E-07 | 0.000267588 |
| Obesity | pericyte | *ABCD3* | 7.152200464 | 3.37E-07 | 1.50E-06 | 0.002117524 |
| Obesity | pericyte | *VAMP4* | 4.128143859 | 6.16E-08 | 4.24E-07 | 0.00038713 |
| Obesity | pericyte | *PTBP2* | 3.122517721 | 1.62E-05 | 3.89E-05 | 0.102089336 |
| Obesity | pericyte | *TNFSF4* | -2.776923856 | 1.74E-06 | 5.53E-06 | 0.010913412 |
| Obesity | pericyte | *IRF6* | 1.092961812 | 0.004887833 | 0.007271606 | 1 |
| Obesity | pericyte | *DIEXF* | 3.636082992 | 0.003282298 | 0.005012214 | 1 |
| Obesity | pericyte | *SERPINC1* | 3.864222011 | 7.61E-14 | 2.28E-11 | 4.78E-10 |
| Obesity | pericyte | *RCAN3* | 5.439773963 | 9.09E-09 | 1.70E-07 | 5.71E-05 |
| Obesity | pericyte | *RCOR3* | 4.843133924 | 0.000286744 | 0.000522993 | 1 |
| Obesity | pericyte | *MTFR1L* | 5.700380351 | 0.0004434 | 0.000783711 | 1 |
| Obesity | pericyte | *NSL1* | 4.263069978 | 2.49E-06 | 7.50E-06 | 0.01568516 |
| Obesity | pericyte | *RPA2* | 6.272649209 | 1.11E-08 | 1.71E-07 | 7.01E-05 |
| Obesity | pericyte | *SLC5A9* | 12.64934266 | 9.47E-08 | 5.79E-07 | 0.000595346 |
| Obesity | pericyte | *OSBPL9* | 5.757886378 | 4.67E-08 | 3.53E-07 | 0.000293518 |
| Obesity | pericyte | *TXNDC12* | 5.328919254 | 1.23E-08 | 1.73E-07 | 7.72E-05 |
| Obesity | pericyte | *COLEC11* | -1.382483316 | 1.41E-06 | 4.64E-06 | 0.00887857 |
| Obesity | pericyte | *KMT2A* | 5.965358489 | 2.64E-06 | 7.86E-06 | 0.016574667 |
| Obesity | pericyte | *IFT46* | 6.811273481 | 6.96E-07 | 2.66E-06 | 0.004376915 |
| Obesity | pericyte | *DDX59* | 3.537105863 | 8.32E-05 | 0.000168813 | 0.522971845 |
| Obesity | pericyte | *CAMSAP2* | 5.089370113 | 2.03E-06 | 6.31E-06 | 0.012770542 |
| Obesity | pericyte | *ATF6* | 5.733987999 | 5.19E-08 | 3.79E-07 | 0.000326296 |
| Obesity | pericyte | *CASC1* | 1.844563439 | 0.014837546 | 0.020610616 | 1 |
| Obesity | pericyte | *LRMP* | 2.323060594 | 0.000332097 | 0.000598422 | 1 |
| Obesity | pericyte | *ATP10B* | 4.696576937 | 3.49E-07 | 1.55E-06 | 0.002192005 |
| Obesity | pericyte | *SPCS2* | 5.514622566 | 2.87E-08 | 2.55E-07 | 0.000180582 |
| Obesity | pericyte | *ELOVL4* | 1.415915454 | 0.001485882 | 0.002402712 | 1 |
| Obesity | pericyte | *SGIP1* | -1.185592996 | 0.001829961 | 0.002906763 | 1 |
| Obesity | pericyte | *AKAP7* | 1.546160182 | 0.001475252 | 0.002386749 | 1 |
| Obesity | pericyte | *MED28* | 3.404912633 | 0.001029348 | 0.001697669 | 1 |
| Obesity | pericyte | *SLC16A7* | 3.49452694 | 3.46E-08 | 2.89E-07 | 0.000217652 |
| Obesity | pericyte | *FOXO3* | 5.473509149 | 4.90E-08 | 3.66E-07 | 0.000308314 |
| Obesity | pericyte | *TGIF2* | 3.940376143 | 8.94E-07 | 3.23E-06 | 0.005618417 |

| | | | | | | |
|---|---|---|---|---|---|---|
| Obesity | pericyte | *ABCG2* | 6.368586757 | 5.86E-08 | 4.12E-07 | 0.000368487 |
| Obesity | pericyte | *SPP1* | 6.069887566 | 3.25E-06 | 9.46E-06 | 0.020451536 |
| Obesity | pericyte | *FAM47E-STBD* | 1.342178682 | 0.000212278 | 0.000398029 | 1 |
| Obesity | pericyte | *MFSD1* | 4.57449747 | 8.81E-09 | 1.70E-07 | 5.54E-05 |
| Obesity | pericyte | *RAB3GAP2* | 5.361546794 | 3.52E-09 | 1.08E-07 | 2.21E-05 |
| Obesity | pericyte | *UBN1* | 3.475153448 | 4.68E-05 | 0.000100858 | 0.294504122 |
| Obesity | pericyte | *UCHL3* | -2.785887625 | 0.001638314 | 0.002622888 | 1 |
| Obesity | pericyte | *WDR35* | 5.539573626 | 1.20E-08 | 1.73E-07 | 7.52E-05 |
| Obesity | pericyte | *NDUFB3* | -1.924849822 | 0.002998712 | 0.004607259 | 1 |
| Obesity | pericyte | *TRPM6* | 1.537077315 | 0.002667487 | 0.004134737 | 1 |
| Obesity | pericyte | *GDA* | 2.525698646 | 2.81E-06 | 8.32E-06 | 0.017676596 |
| Obesity | pericyte | *C2orf40* | 1.880949785 | 0.005640219 | 0.008323957 | 1 |
| Obesity | pericyte | *CPSF3* | 7.608079004 | 1.08E-08 | 1.71E-07 | 6.81E-05 |
| Obesity | pericyte | *PIGZ* | 1.060303938 | 0.000462917 | 0.000815456 | 1 |
| Obesity | pericyte | *SENP5* | 5.755616998 | 9.17E-06 | 2.35E-05 | 0.057660582 |
| Obesity | pericyte | *CCDC92* | 4.84187925 | 9.59E-10 | 4.98E-08 | 6.03E-06 |
| Obesity | pericyte | *PTBP3* | 6.4816108 | 1.69E-06 | 5.41E-06 | 0.010630717 |
| Obesity | pericyte | *FKBP15* | 6.729443005 | 1.67E-09 | 6.91E-08 | 1.05E-05 |
| Obesity | pericyte | *CTNNAL1* | 1.337159441 | 9.80E-05 | 0.000196442 | 0.616238597 |
| Obesity | pericyte | *WDR34* | 2.174164017 | 0.000156401 | 0.000300518 | 0.983295137 |
| Obesity | pericyte | *CNTRL* | 6.136828551 | 7.81E-06 | 2.04E-05 | 0.049083925 |
| Obesity | pericyte | *PHF19* | 2.661912372 | 3.12E-06 | 9.15E-06 | 0.019643023 |
| Obesity | pericyte | *BSPRY* | -3.047680925 | 2.81E-05 | 6.35E-05 | 0.176383413 |
| Obesity | pericyte | *PPP6C* | 6.433782982 | 3.00E-08 | 2.64E-07 | 0.000188368 |
| Obesity | pericyte | *RBM18* | 6.781506247 | 0.000102647 | 0.000205066 | 0.645343803 |
| Obesity | pericyte | *HSDL2* | 1.66462561 | 0.000214233 | 0.000401217 | 1 |
| Obesity | pericyte | *INVS* | 6.719868767 | 1.23E-10 | 1.12E-08 | 7.75E-07 |
| Obesity | pericyte | *KDSR* | 7.329215515 | 2.03E-08 | 2.16E-07 | 0.000127899 |
| Obesity | pericyte | *DCAF4* | 4.343500998 | 5.27E-11 | 5.52E-09 | 3.31E-07 |
| Obesity | pericyte | *DNAL1* | 3.769368628 | 3.27E-06 | 9.48E-06 | 0.020572542 |
| Obesity | pericyte | *IRF2BPL* | 2.22276902 | 4.12E-06 | 1.16E-05 | 0.025919534 |
| Obesity | pericyte | *TTLL5* | 5.629517882 | 8.75E-09 | 1.70E-07 | 5.50E-05 |
| Obesity | pericyte | *FLVCR2* | 4.20329187 | 1.81E-07 | 9.14E-07 | 0.001135698 |
| Obesity | pericyte | *PPP4R4* | -1.089868948 | 2.17E-05 | 5.04E-05 | 0.136342141 |
| Obesity | pericyte | *ZC2HC1C* | -2.150200495 | 0.004006273 | 0.006047404 | 1 |
| Obesity | pericyte | *ALDH6A1* | 1.412180644 | 0.020435721 | 0.027779325 | 1 |
| Obesity | pericyte | *ESRRB* | 1.331350046 | 0.000268647 | 0.000492416 | 1 |
| Obesity | pericyte | *COQ6* | 4.254920526 | 1.71E-07 | 8.79E-07 | 0.001075424 |
| Obesity | pericyte | *GPR75* | 17.27637634 | 3.89E-06 | 1.11E-05 | 0.024478905 |
| Obesity | pericyte | *KLHL29* | 5.485139128 | 3.88E-07 | 1.68E-06 | 0.002439524 |
| Obesity | pericyte | *ATAD2B* | 5.97104343 | 7.26E-08 | 4.75E-07 | 0.00045619 |
| Obesity | pericyte | *FKBP1B* | 1.292605889 | 0.018515493 | 0.025344417 | 1 |
| Obesity | pericyte | *ATL2* | 5.349120364 | 8.98E-08 | 5.56E-07 | 0.000564706 |
| Obesity | pericyte | *EPCAM* | 3.273055904 | 2.06E-05 | 4.84E-05 | 0.129773354 |
| Obesity | pericyte | *SLC17A5* | 2.930299701 | 0.000144893 | 0.000280722 | 0.910943488 |
| Obesity | pericyte | *OGFRL1* | 2.612600688 | 0.008120229 | 0.011674338 | 1 |
| Obesity | pericyte | *CUTC* | 5.011499818 | 2.95E-05 | 6.64E-05 | 0.185273585 |
| Obesity | pericyte | *PYROXD2* | 0.871172683 | 0.003277467 | 0.005007396 | 1 |
| Obesity | pericyte | *MXI1* | 7.047024136 | 5.28E-06 | 1.45E-05 | 0.033220291 |
| Obesity | pericyte | *C10orf88* | 8.425330243 | 1.21E-08 | 1.73E-07 | 7.63E-05 |

| | | | | | | |
|---|---|---|---|---|---|---|
| Obesity | pericyte | *HELLS* | 3.89942507 | 1.60E-05 | 3.84E-05 | 0.100347403 |
| Obesity | pericyte | *AVPI1* | 2.038997554 | 1.26E-06 | 4.20E-06 | 0.007914584 |
| Obesity | pericyte | *WDR11* | 5.424508598 | 5.73E-08 | 4.04E-07 | 0.000360319 |
| Obesity | pericyte | *C10orf76* | 6.542011741 | 1.12E-09 | 5.30E-08 | 7.03E-06 |
| Obesity | pericyte | *KCNIP2* | 1.103625437 | 0.005453512 | 0.008061658 | 1 |
| Obesity | pericyte | *C10orf95* | 2.029608724 | 8.67E-05 | 0.000175411 | 0.544884894 |
| Obesity | pericyte | *KANSL1* | 3.543684968 | 4.97E-05 | 0.000106196 | 0.312214933 |
| Obesity | pericyte | *HOXB5* | 3.186342539 | 6.09E-10 | 3.62E-08 | 3.83E-06 |
| Obesity | pericyte | *HOXB3* | 4.793978142 | 7.45E-13 | 1.56E-10 | 4.68E-09 |
| Obesity | pericyte | *CAAP1* | 4.220679397 | 0.000728591 | 0.001236343 | 1 |
| Obesity | pericyte | *CD274* | 4.088863753 | 0.000127008 | 0.000248676 | 0.798498691 |
| Obesity | pericyte | *LRP11* | 2.279269548 | 0.01366672 | 0.019051589 | 1 |
| Obesity | pericyte | *CYSTM1* | 6.256226244 | 1.01E-06 | 3.55E-06 | 0.006337752 |
| Obesity | pericyte | *WDR55* | 2.228963705 | 0.000207467 | 0.000389707 | 1 |
| Obesity | pericyte | *PCDHB14* | -2.926276521 | 0.00094454 | 0.001567666 | 1 |
| Obesity | pericyte | *SEC16B* | 2.989285799 | 0.000560015 | 0.000973408 | 1 |
| Obesity | pericyte | *GORAB* | 6.554525154 | 2.34E-06 | 7.11E-06 | 0.014721131 |
| Obesity | pericyte | *ACAT2* | 3.058053682 | 7.04E-07 | 2.68E-06 | 0.004423023 |
| Obesity | pericyte | *MRC1* | 3.050338992 | 2.72E-06 | 8.09E-06 | 0.017109606 |
| Obesity | pericyte | *IQSEC3* | -2.348258341 | 0.001160892 | 0.001901154 | 1 |
| Obesity | pericyte | *CCDC77* | 4.378867641 | 9.87E-07 | 3.49E-06 | 0.006203268 |
| Obesity | pericyte | *ENOX1* | -0.513692657 | 5.06E-05 | 0.000107954 | 0.318140379 |
| Obesity | pericyte | *DNAJC15* | 4.673294477 | 0.003041891 | 0.004671317 | 1 |
| Obesity | pericyte | *WBP4* | 5.2198327 | 4.60E-06 | 1.28E-05 | 0.028940948 |
| Obesity | pericyte | *SIL1* | 4.772244501 | 7.92E-06 | 2.06E-05 | 0.049813803 |
| Obesity | pericyte | *ZFP30* | 2.628186002 | 5.96E-05 | 0.000124465 | 0.374765565 |
| Obesity | pericyte | *ARL1* | 3.292091476 | 7.58E-06 | 1.99E-05 | 0.047679785 |
| Obesity | pericyte | *APAF1* | 5.048097793 | 3.89E-07 | 1.69E-06 | 0.002448502 |
| Obesity | pericyte | *ADRA1A* | 0.685163674 | 1.26E-06 | 4.20E-06 | 0.007905671 |
| Obesity | pericyte | *EPHX2* | 1.51881679 | 0.009621692 | 0.013716911 | 1 |
| Obesity | pericyte | *ZNF706* | 5.088774389 | 2.62E-06 | 7.82E-06 | 0.016475801 |
| Obesity | pericyte | *CRISPLD1* | 2.377071072 | 8.43E-07 | 3.08E-06 | 0.005297646 |
| Obesity | pericyte | *RDH10* | 1.602130091 | 3.74E-05 | 8.19E-05 | 0.234820549 |
| Obesity | pericyte | *SCPEP1* | 1.251742035 | 9.70E-06 | 2.47E-05 | 0.060966707 |
| Obesity | pericyte | *TRIM6* | 2.32184865 | 1.95E-06 | 6.10E-06 | 0.012271839 |
| Obesity | pericyte | *CEP89* | 1.018892539 | 0.026529979 | 0.035556167 | 1 |
| Obesity | pericyte | *PRB2* | 7.447253543 | 4.91E-07 | 2.03E-06 | 0.003087988 |
| Obesity | pericyte | *PYROXD1* | 3.901136427 | 6.79E-09 | 1.54E-07 | 4.27E-05 |
| Obesity | pericyte | *A1BG* | 12.81807678 | 3.65E-08 | 2.99E-07 | 0.000229705 |
| Obesity | pericyte | *ZSCAN18* | 6.744138184 | 2.39E-07 | 1.14E-06 | 0.001501857 |
| Obesity | pericyte | *ZNF211* | 1.766524276 | 9.08E-06 | 2.33E-05 | 0.0571043 |
| Obesity | pericyte | *PDZRN3* | 5.00060501 | 3.37E-08 | 2.85E-07 | 0.000211988 |
| Obesity | pericyte | *LHX4* | -2.656886388 | 0.027222091 | 0.036421639 | 1 |
| Obesity | pericyte | *CSTA* | 2.163575784 | 0.001509684 | 0.002438064 | 1 |
| Obesity | pericyte | *POPDC2* | 9.496807002 | 2.47E-06 | 7.43E-06 | 0.01552086 |
| Obesity | pericyte | *B4GALT4* | 5.923558309 | 1.57E-08 | 1.91E-07 | 9.88E-05 |
| Obesity | pericyte | *CD80* | 3.225769154 | 8.56E-07 | 3.11E-06 | 0.005383623 |
| Obesity | pericyte | *CAT* | 1.062505148 | 0.001954052 | 0.003089043 | 1 |
| Obesity | pericyte | *GJB6* | -23.64726327 | 0.01355311 | 0.018909987 | 1 |
| Obesity | pericyte | *TMEM39B* | 3.838156323 | 1.62E-05 | 3.88E-05 | 0.101593231 |

| | | | | | | |
|---|---|---|---|---|---|---|
| Obesity | pericyte | *RNF115* | 6.339934329 | 2.29E-07 | 1.10E-06 | 0.001436997 |
| Obesity | pericyte | *POLR3GL* | 7.210598708 | 1.49E-07 | 7.90E-07 | 0.000939622 |
| Obesity | pericyte | *TNFSF10* | 6.375900827 | 2.37E-07 | 1.13E-06 | 0.001488023 |
| Obesity | pericyte | *ZNF639* | 7.235173268 | 3.22E-09 | 1.02E-07 | 2.02E-05 |
| Obesity | pericyte | *LIAS* | 6.148223025 | 2.15E-05 | 5.00E-05 | 0.134948892 |
| Obesity | pericyte | *CLCC1* | 4.234573345 | 0.006460678 | 0.009419825 | 1 |
| Obesity | pericyte | *GPSM2* | 6.091800581 | 3.84E-10 | 2.73E-08 | 2.41E-06 |
| Obesity | pericyte | *CXCR4* | 2.476226245 | 5.25E-05 | 0.000111326 | 0.330080328 |
| Obesity | pericyte | *ACVR2A* | 6.254660161 | 8.84E-07 | 3.20E-06 | 0.005560263 |
| Obesity | pericyte | *POLK* | 5.17094812 | 1.68E-05 | 4.01E-05 | 0.105443879 |
| Obesity | pericyte | *FLT3* | -1.738135357 | 0.000107619 | 0.000213641 | 0.676601304 |
| Obesity | pericyte | *MTIF3* | 4.740483647 | 6.35E-07 | 2.47E-06 | 0.003991598 |
| Obesity | pericyte | *RASL11A* | -1.187473894 | 1.20E-05 | 2.97E-05 | 0.075242911 |
| Obesity | pericyte | *FMOD* | 5.052902177 | 6.93E-07 | 2.65E-06 | 0.004354955 |
| Obesity | pericyte | *OPN4* | -0.328198081 | 2.08E-05 | 4.86E-05 | 0.130491505 |
| Obesity | pericyte | *FAM35A* | 3.49725809 | 4.28E-05 | 9.27E-05 | 0.268926194 |
| Obesity | pericyte | *NAA60* | 8.438029943 | 1.18E-06 | 3.99E-06 | 0.007441207 |
| Obesity | pericyte | *ODF2L* | 3.927431682 | 1.61E-08 | 1.91E-07 | 0.000101116 |
| Obesity | pericyte | *SPATA1* | 6.683800899 | 3.74E-06 | 1.07E-05 | 0.023489862 |
| Obesity | pericyte | *LRRC39* | 1.21689266 | 0.002385751 | 0.003727439 | 1 |
| Obesity | pericyte | *RWDD3* | 4.447108466 | 0.000131023 | 0.000255583 | 0.823744684 |
| Obesity | pericyte | *CCDC18* | 5.398135719 | 1.12E-05 | 2.79E-05 | 0.070301271 |
| Obesity | pericyte | *RPAP2* | 5.7504604 | 1.28E-08 | 1.73E-07 | 8.04E-05 |
| Obesity | pericyte | *BBS9* | 3.94695974 | 0.00023159 | 0.000430135 | 1 |
| Obesity | pericyte | *PMS2* | 6.528233438 | 7.12E-05 | 0.000146417 | 0.447451778 |
| Obesity | pericyte | *KIAA0087* | -5.404894718 | 7.19E-05 | 0.000147793 | 0.452245546 |
| Obesity | pericyte | *KLHL7* | 3.880835561 | 0.010883188 | 0.015413141 | 1 |
| Obesity | pericyte | *HERPUD2* | 7.394617859 | 6.73E-09 | 1.54E-07 | 4.23E-05 |
| Obesity | pericyte | *CBX3* | 3.51863326 | 0.005808629 | 0.008550421 | 1 |
| Obesity | pericyte | *FAM126A* | 4.543063011 | 3.12E-05 | 6.97E-05 | 0.196161471 |
| Obesity | pericyte | *FKBP9* | 4.335897674 | 7.23E-06 | 1.91E-05 | 0.045458742 |
| Obesity | pericyte | *CCZ1* | 3.609462167 | 0.000104257 | 0.000207688 | 0.655464672 |
| Obesity | pericyte | *SMU1* | 6.010993814 | 2.99E-09 | 9.87E-08 | 1.88E-05 |
| Obesity | pericyte | *GLIPR2* | 4.833914211 | 7.50E-05 | 0.000153513 | 0.471744423 |
| Obesity | pericyte | *SLC25A51* | 1.226036491 | 6.86E-06 | 1.82E-05 | 0.04315058 |
| Obesity | pericyte | *DCAF10* | 6.114014499 | 6.72E-07 | 2.57E-06 | 0.004226796 |
| Obesity | pericyte | *TRIM24* | 5.288536915 | 9.85E-08 | 5.93E-07 | 0.000619287 |
| Obesity | pericyte | *BICC1* | 1.87060839 | 1.47E-06 | 4.81E-06 | 0.009267605 |
| Obesity | pericyte | *CISD1* | 1.477472558 | 0.001726259 | 0.002753168 | 1 |
| Obesity | pericyte | *SLC25A16* | 2.205617211 | 2.64E-07 | 1.23E-06 | 0.00166233 |
| Obesity | pericyte | *ZWINT* | 1.758536454 | 0.000534554 | 0.000931211 | 1 |
| Obesity | pericyte | *RBM19* | 6.310198161 | 2.46E-08 | 2.32E-07 | 0.00015455 |
| Obesity | pericyte | *ACADS* | 3.609948758 | 9.41E-07 | 3.36E-06 | 0.005916377 |
| Obesity | pericyte | *SSPN* | 2.338434599 | 0.000113877 | 0.000224716 | 0.715944252 |
| Obesity | pericyte | *CCDC91* | 4.980878535 | 1.33E-08 | 1.76E-07 | 8.37E-05 |
| Obesity | pericyte | *PKN1* | 5.573040659 | 0.000238982 | 0.000442427 | 1 |
| Obesity | pericyte | *ATP7B* | 0.712930627 | 0.000639178 | 0.001097654 | 1 |
| Obesity | pericyte | *NLN* | 1.929797937 | 0.000286576 | 0.00052284 | 1 |
| Obesity | pericyte | *CENPK* | 0.900191619 | 0.00113946 | 0.001867514 | 1 |
| Obesity | pericyte | *OPTN* | 2.85613763 | 0.002233333 | 0.003502361 | 1 |

| | | | | | | |
|---|---|---|---|---|---|---|
| Obesity | pericyte | *ATF1* | 5.083636943 | 8.44E-06 | 2.18E-05 | 0.05306764 |
| Obesity | pericyte | *TSFM* | 3.459321253 | 3.70E-05 | 8.12E-05 | 0.232366869 |
| Obesity | pericyte | *ARHGAP9* | 1.697128349 | 2.90E-05 | 6.54E-05 | 0.182153362 |
| Obesity | pericyte | *SPATS2* | 3.170953712 | 0.01812789 | 0.024846315 | 1 |
| Obesity | pericyte | *ORMDL2* | 5.569016008 | 0.000130077 | 0.000254211 | 0.817797216 |
| Obesity | pericyte | *PDE1B* | -1.251601341 | 0.000300894 | 0.00054674 | 1 |
| Obesity | pericyte | *IKZF4* | 3.155152535 | 0.000291485 | 0.000531024 | 1 |
| Obesity | pericyte | *SMUG1* | 0.880775779 | 0.001468056 | 0.00237633 | 1 |
| Obesity | pericyte | *SARDH* | -4.396861248 | 0.000302351 | 0.000549229 | 1 |
| Obesity | pericyte | *STIL* | 3.709387583 | 1.16E-06 | 3.93E-06 | 0.007299329 |
| Obesity | pericyte | *TTC21B* | 6.473978945 | 7.58E-07 | 2.84E-06 | 0.00476524 |
| Obesity | pericyte | *NMI* | 4.857164516 | 2.51E-07 | 1.18E-06 | 0.001577485 |
| Obesity | pericyte | *ACVR1C* | 1.90987142 | 8.12E-06 | 2.11E-05 | 0.051033998 |
| Obesity | pericyte | *SLC36A1* | 0.948301203 | 0.023218726 | 0.031311911 | 1 |
| Obesity | pericyte | *PLA2G12A* | 7.752313844 | 2.06E-06 | 6.38E-06 | 0.01293977 |
| Obesity | pericyte | *C4BPB* | 1.963140739 | 0.000104731 | 0.000208434 | 0.658442913 |
| Obesity | pericyte | *RAB38* | 5.32313122 | 1.06E-06 | 3.69E-06 | 0.006695632 |
| Obesity | pericyte | *AGO2* | 4.825237889 | 0.001268905 | 0.002068345 | 1 |
| Obesity | pericyte | *DNPEP* | 6.689268983 | 2.88E-09 | 9.62E-08 | 1.81E-05 |
| Obesity | pericyte | *MOGAT1* | 1.774601211 | 4.02E-06 | 1.14E-05 | 0.025291013 |
| Obesity | pericyte | *FAM124B* | 1.668988071 | 0.002403681 | 0.003754519 | 1 |
| Obesity | pericyte | *FAM210B* | 3.255370655 | 0.000893005 | 0.00148921 | 1 |
| Obesity | pericyte | *SNX21* | 7.003402256 | 1.66E-08 | 1.93E-07 | 0.000104614 |
| Obesity | pericyte | *SDC4* | -2.826388443 | 0.023161298 | 0.031247872 | 1 |
| Obesity | pericyte | *VAPB* | 1.481892107 | 0.002096383 | 0.003300766 | 1 |
| Obesity | pericyte | *ARFGEF2* | 6.260250385 | 1.89E-09 | 7.58E-08 | 1.19E-05 |
| Obesity | pericyte | *CSE1L* | 5.559117972 | 1.93E-06 | 6.03E-06 | 0.012111411 |
| Obesity | pericyte | *STX16* | 7.459315341 | 6.53E-08 | 4.42E-07 | 0.000410825 |
| Obesity | pericyte | *KCNK15* | 5.634375849 | 0.000321115 | 0.000580629 | 1 |
| Obesity | pericyte | *NEURL2* | 1.918676585 | 1.32E-07 | 7.28E-07 | 0.000830348 |
| Obesity | pericyte | *FASTKD3* | 3.019013329 | 3.06E-06 | 8.98E-06 | 0.019263682 |
| Obesity | pericyte | *PEPD* | 4.172916161 | 4.65E-09 | 1.27E-07 | 2.93E-05 |
| Obesity | pericyte | *CHST8* | 10.44505924 | 6.77E-11 | 6.87E-09 | 4.26E-07 |
| Obesity | pericyte | *STAMBP* | 5.67333168 | 1.69E-08 | 1.95E-07 | 0.000106444 |
| Obesity | pericyte | *ATP8A1* | 2.805619547 | 0.000102733 | 0.000205095 | 0.645883486 |
| Obesity | pericyte | *USP22* | 5.442639861 | 6.58E-06 | 1.76E-05 | 0.041359565 |
| Obesity | pericyte | *HIF3A* | 2.616128217 | 0.000235068 | 0.000436336 | 1 |
| Obesity | pericyte | *ZNF45* | 7.543534485 | 3.30E-08 | 2.82E-07 | 0.00020724 |
| Obesity | pericyte | *BTN2A2* | 1.395090436 | 1.61E-05 | 3.88E-05 | 0.101480548 |
| Obesity | pericyte | *MRS2* | 3.06641298 | 0.00019038 | 0.000359867 | 1 |
| Obesity | pericyte | *SNRPC* | -4.705439059 | 1.19E-13 | 3.26E-11 | 7.50E-10 |
| Obesity | pericyte | *SERPINB6* | 7.455042178 | 3.39E-07 | 1.51E-06 | 0.002130121 |
| Obesity | pericyte | *HIST1H1D* | 1.999739171 | 0.002438072 | 0.00380257 | 1 |
| Obesity | pericyte | *PEX6* | 0.844399349 | 0.01910439 | 0.026076704 | 1 |
| Obesity | pericyte | *NQO2* | 5.152776511 | 1.91E-08 | 2.08E-07 | 0.000120317 |
| Obesity | pericyte | *ZNF391* | 0.552590235 | 0.017728182 | 0.024331475 | 1 |
| Obesity | pericyte | *RPS10* | -2.497666882 | 0.000282748 | 0.000516155 | 1 |
| Obesity | pericyte | *HIST1H2BJ* | 5.299866231 | 2.79E-13 | 6.75E-11 | 1.76E-09 |
| Obesity | pericyte | *MED20* | 2.003873578 | 2.28E-06 | 6.95E-06 | 0.014345847 |
| Obesity | pericyte | *TBCC* | -1.683295706 | 0.027314948 | 0.036538101 | 1 |

| | | | | | | |
|---|---|---|---|---|---|---|
| Obesity | pericyte | *KLHDC3* | 6.195484524 | 0.000195314 | 0.000368309 | 1 |
| Obesity | pericyte | *KLHL31* | 1.327088721 | 4.18E-06 | 1.18E-05 | 0.026276329 |
| Obesity | pericyte | *GLO1* | 7.636439166 | 1.30E-09 | 5.85E-08 | 8.20E-06 |
| Obesity | pericyte | *CPNE5* | 1.56677955 | 0.005222664 | 0.007738602 | 1 |
| Obesity | pericyte | *RIOK1* | 3.343042102 | 3.34E-05 | 7.40E-05 | 0.209976469 |
| Obesity | pericyte | *NUP153* | 6.487030544 | 1.05E-06 | 3.65E-06 | 0.006580024 |
| Obesity | pericyte | *EREG* | -2.520192592 | 2.94E-05 | 6.62E-05 | 0.184581426 |
| Obesity | pericyte | *MYRF* | -1.848702468 | 0.000181508 | 0.000344652 | 1 |
| Obesity | pericyte | *CNOT1* | 5.716415515 | 1.20E-06 | 4.05E-06 | 0.007566498 |
| Obesity | pericyte | *BBS2* | 2.304176988 | 0.000375384 | 0.000671419 | 1 |
| Obesity | pericyte | *MT1G* | -6.550216777 | 5.74E-05 | 0.000120228 | 0.360803703 |
| Obesity | pericyte | *GOT2* | 6.024362709 | 2.17E-08 | 2.20E-07 | 0.000136674 |
| Obesity | pericyte | *CLYBL* | 7.045222329 | 3.54E-08 | 2.92E-07 | 0.000222691 |
| Obesity | pericyte | *TMTC4* | 4.104382152 | 4.62E-08 | 3.50E-07 | 0.000290327 |
| Obesity | pericyte | *KIF25* | 4.382851443 | 6.63E-06 | 1.77E-05 | 0.041675517 |
| Obesity | pericyte | *GRK4* | 1.273440361 | 1.95E-08 | 2.10E-07 | 0.000122875 |
| Obesity | pericyte | *TEKT3* | 1.769179888 | 6.41E-06 | 1.72E-05 | 0.040328367 |
| Obesity | pericyte | *MRPS7* | 5.933773241 | 1.67E-08 | 1.93E-07 | 0.00010478 |
| Obesity | pericyte | *GGA3* | 5.263690709 | 1.65E-08 | 1.93E-07 | 0.000103745 |
| Obesity | pericyte | *NT5C* | 7.978056028 | 5.33E-07 | 2.17E-06 | 0.003352906 |
| Obesity | pericyte | *PPDPF* | 6.551071014 | 8.57E-07 | 3.11E-06 | 0.005386512 |
| Obesity | pericyte | *PAX8* | 1.489855059 | 0.000314836 | 0.000570096 | 1 |
| Obesity | pericyte | *INSIG2* | 2.903489719 | 1.34E-07 | 7.34E-07 | 0.000844943 |
| Obesity | pericyte | *POLR1B* | 1.883580068 | 3.59E-05 | 7.91E-05 | 0.225527883 |
| Obesity | pericyte | *PSD4* | 2.862850116 | 5.06E-07 | 2.08E-06 | 0.003182283 |
| Obesity | pericyte | *SLC25A23* | 1.395800903 | 0.000230383 | 0.000428399 | 1 |
| Obesity | pericyte | *GTF2F1* | 4.460306519 | 0.000403189 | 0.000718088 | 1 |
| Obesity | pericyte | *CLPP* | 3.301425589 | 0.002737656 | 0.004233069 | 1 |
| Obesity | pericyte | *TRIP10* | 6.243421146 | 9.41E-09 | 1.70E-07 | 5.92E-05 |
| Obesity | pericyte | *GPR108* | 1.933055054 | 0.031610829 | 0.041813019 | 1 |
| Obesity | pericyte | *RTN2* | -2.766208352 | 0.003457847 | 0.005259977 | 1 |
| Obesity | pericyte | *SYMPK* | 6.774534284 | 1.68E-06 | 5.39E-06 | 0.010567855 |
| Obesity | pericyte | *GPCPD1* | 6.225711252 | 4.39E-07 | 1.86E-06 | 0.002758776 |
| Obesity | pericyte | *SDCBP2* | 5.333450903 | 0.00118502 | 0.001938648 | 1 |
| Obesity | pericyte | *PANK2* | 5.417971383 | 0.000523281 | 0.000913091 | 1 |
| Obesity | pericyte | *PSMF1* | 3.29676811 | 0.003039718 | 0.00466912 | 1 |
| Obesity | pericyte | *DTD1* | 5.714227374 | 7.58E-06 | 1.99E-05 | 0.047664592 |
| Obesity | pericyte | *RBCK1* | 3.897399679 | 5.62E-06 | 1.53E-05 | 0.03534852 |
| Obesity | pericyte | *RRBP1* | 4.389025673 | 7.50E-06 | 1.97E-05 | 0.04716155 |
| Obesity | pericyte | *BMP2* | -1.459195296 | 9.15E-05 | 0.00018442 | 0.574991798 |
| Obesity | pericyte | *MKKS* | 6.789366013 | 5.41E-08 | 3.90E-07 | 0.000339852 |
| Obesity | pericyte | *DSTN* | 6.635640077 | 7.27E-07 | 2.75E-06 | 0.004572534 |
| Obesity | pericyte | *MGME1* | 4.96635865 | 9.50E-07 | 3.38E-06 | 0.005973439 |
| Obesity | pericyte | *ITPA* | 8.925095952 | 1.46E-07 | 7.76E-07 | 0.0009167 |
| Obesity | pericyte | *MCM8* | 0.764987901 | 0.038056423 | 0.049711351 | 1 |
| Obesity | pericyte | *TMEM74B* | -0.404833305 | 2.59E-05 | 5.92E-05 | 0.162873356 |
| Obesity | pericyte | *FAM110A* | -3.433836708 | 0.031110589 | 0.041185992 | 1 |
| Obesity | pericyte | *NCLN* | 5.486083519 | 3.09E-07 | 1.40E-06 | 0.001939554 |
| Obesity | pericyte | *GDF5* | -1.241015418 | 0.035061348 | 0.046095922 | 1 |
| Obesity | pericyte | *MMP24* | 3.070274856 | 0.003162495 | 0.00484233 | 1 |

| | | | | | | |
|---|---|---|---|---|---|---|
| Obesity | pericyte | *UROD* | 6.672423741 | 1.73E-05 | 4.12E-05 | 0.108889704 |
| Obesity | pericyte | *ST3GAL3* | 3.509582194 | 0.000215372 | 0.000402995 | 1 |
| Obesity | pericyte | *KLC1* | 8.056548837 | 6.15E-10 | 3.62E-08 | 3.87E-06 |
| Obesity | pericyte | *XRCC3* | 7.316970871 | 5.50E-13 | 1.19E-10 | 3.46E-09 |
| Obesity | pericyte | *MCF2L* | 2.672317264 | 0.000265319 | 0.000486741 | 1 |
| Obesity | pericyte | *F10* | 4.311568157 | 3.80E-08 | 3.06E-07 | 0.000238923 |
| Obesity | pericyte | *PCID2* | 4.773178582 | 9.43E-07 | 3.37E-06 | 0.005929489 |
| Obesity | pericyte | *PROZ* | -4.012138449 | 5.04E-05 | 0.00010765 | 0.317136139 |
| Obesity | pericyte | *CAPNS1* | 4.383951535 | 1.24E-05 | 3.07E-05 | 0.078254096 |
| Obesity | pericyte | *SCAF1* | 11.60269873 | 2.93E-08 | 2.59E-07 | 0.000184065 |
| Obesity | pericyte | *PRR12* | 2.672166277 | 0.000584594 | 0.001011099 | 1 |
| Obesity | pericyte | *TSKS* | 3.274990965 | 4.42E-07 | 1.87E-06 | 0.002779973 |
| Obesity | pericyte | *FLRT1* | 1.80978803 | 0.004772213 | 0.007115954 | 1 |
| Obesity | pericyte | *SBDS* | 4.467762062 | 0.003867935 | 0.005845602 | 1 |
| Obesity | pericyte | *TRAP1* | 1.120701525 | 2.62E-05 | 5.98E-05 | 0.16466637 |
| Obesity | pericyte | *DNAJC8* | 3.403428747 | 0.003427205 | 0.005215888 | 1 |
| Obesity | pericyte | *ZNF384* | 4.622577073 | 2.01E-08 | 2.14E-07 | 0.000126482 |
| Obesity | pericyte | *ATG14* | 4.994779213 | 1.59E-08 | 1.91E-07 | 0.00010011 |
| Obesity | pericyte | *RHOJ* | -1.49174444 | 0.001637873 | 0.002622849 | 1 |
| Obesity | pericyte | *HSPA2* | 3.101968414 | 2.66E-07 | 1.24E-06 | 0.001674462 |
| Obesity | pericyte | *ZBTB1* | 5.440437737 | 2.67E-08 | 2.43E-07 | 0.000168161 |
| Obesity | pericyte | *PZP* | 3.024929438 | 1.01E-07 | 6.04E-07 | 0.000635693 |
| Obesity | pericyte | *RHOT1* | 4.256694469 | 4.91E-09 | 1.29E-07 | 3.09E-05 |
| Obesity | pericyte | *WDR60* | 1.916081519 | 0.011777039 | 0.016605124 | 1 |
| Obesity | pericyte | *AIF1L* | -2.228588618 | 0.006540896 | 0.009530153 | 1 |
| Obesity | pericyte | *MAP2K2* | 3.571324063 | 0.000588895 | 0.001017697 | 1 |
| Obesity | pericyte | *CANX* | 6.842187559 | 5.33E-10 | 3.35E-08 | 3.35E-06 |
| Obesity | pericyte | *IPPK* | 5.019227492 | 3.87E-09 | 1.13E-07 | 2.43E-05 |
| Obesity | pericyte | *FGD3* | 2.9342978 | 1.65E-07 | 8.54E-07 | 0.001034515 |
| Obesity | pericyte | *ABHD8* | 2.999087835 | 6.93E-06 | 1.84E-05 | 0.043547469 |
| Obesity | pericyte | *MASP1* | 1.837561214 | 2.09E-05 | 4.89E-05 | 0.131466279 |
| Obesity | pericyte | *HRASLS* | 3.750999739 | 7.30E-07 | 2.76E-06 | 0.00459026 |
| Obesity | pericyte | *HELB* | 5.059702589 | 2.55E-08 | 2.37E-07 | 0.000160019 |
| Obesity | pericyte | *RAP1B* | 5.093448528 | 7.55E-06 | 1.98E-05 | 0.047456298 |
| Obesity | pericyte | *RAB3IP* | 5.184003486 | 1.12E-06 | 3.82E-06 | 0.00702151 |
| Obesity | pericyte | *YEATS4* | 3.004771626 | 2.12E-08 | 2.17E-07 | 0.000133159 |
| Obesity | pericyte | *TAS2R4* | 1.989381851 | 3.35E-06 | 9.69E-06 | 0.021075084 |
| Obesity | pericyte | *IDUA* | 3.44717794 | 2.06E-07 | 1.01E-06 | 0.001292808 |
| Obesity | pericyte | *TMEM175* | 4.058322515 | 9.23E-05 | 0.000186028 | 0.580594247 |
| Obesity | pericyte | *EMC1* | 3.401656344 | 0.000387066 | 0.000690349 | 1 |
| Obesity | pericyte | *PLA2G5* | 1.397307616 | 0.000583436 | 0.001009652 | 1 |
| Obesity | pericyte | *SIN3B* | 7.153094381 | 1.43E-08 | 1.83E-07 | 8.97E-05 |
| Obesity | pericyte | *WFIKKN1* | -7.954857844 | 0.000109694 | 0.000217143 | 0.689647149 |
| Obesity | pericyte | *WDR24* | 2.929490806 | 1.36E-05 | 3.34E-05 | 0.085799508 |
| Obesity | pericyte | *CHTF18* | -2.073603437 | 0.00769928 | 0.011117448 | 1 |
| Obesity | pericyte | *SMARCA4* | 4.816194897 | 1.43E-06 | 4.68E-06 | 0.008978408 |
| Obesity | pericyte | *TICAM1* | 8.149352921 | 0.00029716 | 0.000540423 | 1 |
| Obesity | pericyte | *AAMP* | 4.681472816 | 2.85E-08 | 2.55E-07 | 0.000179446 |
| Obesity | pericyte | *PNKD* | 5.605002495 | 3.31E-06 | 9.59E-06 | 0.020828181 |
| Obesity | pericyte | *ZNF835* | 1.696063068 | 0.008635412 | 0.012364117 | 1 |

| | | | | | | |
|---|---|---|---|---|---|---|
| Obesity | pericyte | *HIP1* | 2.798627275 | 0.01650507 | 0.022760994 | 1 |
| Obesity | pericyte | *PTPN12* | 5.44943352 | 4.58E-06 | 1.28E-05 | 0.028784555 |
| Obesity | pericyte | *POR* | 6.676317745 | 1.54E-08 | 1.90E-07 | 9.67E-05 |
| Obesity | pericyte | *FGL2* | 3.275674087 | 8.80E-07 | 3.18E-06 | 0.005530575 |
| Obesity | pericyte | *STYXL1* | 1.802756239 | 4.97E-05 | 0.000106289 | 0.312702045 |
| Obesity | pericyte | *STEAP4* | -0.2799681 | 1.61E-05 | 3.87E-05 | 0.101269683 |
| Obesity | pericyte | *SGCE* | 5.185620108 | 0.000109013 | 0.000216135 | 0.685362565 |
| Obesity | pericyte | *RBM48* | 2.843496362 | 6.26E-07 | 2.44E-06 | 0.003933837 |
| Obesity | pericyte | *CASD1* | 6.796987279 | 2.15E-08 | 2.18E-07 | 0.000135176 |
| Obesity | pericyte | *SRD5A3* | 5.316328511 | 1.33E-07 | 7.32E-07 | 0.000837189 |
| Obesity | pericyte | *DGCR6L* | 2.598980828 | 4.12E-05 | 8.96E-05 | 0.259198145 |
| Obesity | pericyte | *VPREB3* | 9.970920611 | 0.036892921 | 0.048352261 | 1 |
| Obesity | pericyte | *MGAT3* | -1.185898988 | 8.32E-05 | 0.000168813 | 0.522982158 |
| Obesity | pericyte | *ATF4* | 5.755430464 | 3.17E-06 | 9.25E-06 | 0.019913045 |
| Obesity | pericyte | *TPST2* | 5.234360887 | 3.30E-09 | 1.04E-07 | 2.08E-05 |
| Obesity | pericyte | *BAIAP2L2* | 4.51365704 | 0.000101999 | 0.00020403 | 0.641265579 |
| Obesity | pericyte | *TST* | -1.039736566 | 0.013234241 | 0.018506155 | 1 |
| Obesity | pericyte | *RIBC2* | 2.779393435 | 9.39E-07 | 3.36E-06 | 0.005902059 |
| Obesity | pericyte | *EMC4* | 3.369987121 | 7.44E-09 | 1.61E-07 | 4.68E-05 |
| Obesity | pericyte | *SPECC1* | -1.412840447 | 0.037161304 | 0.048643164 | 1 |
| Obesity | pericyte | *PRKRIP1* | 2.614107485 | 0.003476787 | 0.005282398 | 1 |
| Obesity | pericyte | *FOXP2* | 2.373452097 | 2.14E-05 | 4.98E-05 | 0.134380368 |
| Obesity | pericyte | *KLHDC10* | 5.073305815 | 3.83E-07 | 1.67E-06 | 0.002406923 |
| Obesity | pericyte | *HOXD1* | 10.59525248 | 5.13E-05 | 0.000109088 | 0.322572225 |
| Obesity | pericyte | *CHN1* | 2.215163324 | 0.019094164 | 0.026068405 | 1 |
| Obesity | pericyte | *ORMDL1* | 1.119368147 | 0.027874687 | 0.037203246 | 1 |
| Obesity | pericyte | *HOXD9* | 1.645755111 | 0.028311565 | 0.037718757 | 1 |
| Obesity | pericyte | *TWSG1* | 6.173109256 | 1.29E-07 | 7.18E-07 | 0.000813018 |
| Obesity | pericyte | *EIF2AK4* | 2.5949276 | 0.000684125 | 0.00116814 | 1 |
| Obesity | pericyte | *TMOD2* | 2.923414609 | 0.001307075 | 0.002127804 | 1 |
| Obesity | pericyte | *IVD* | 2.236436243 | 0.002512034 | 0.003909197 | 1 |
| Obesity | pericyte | *ARPP19* | 5.822544179 | 4.73E-08 | 3.56E-07 | 0.000297242 |
| Obesity | pericyte | *VPS13C* | 4.987469285 | 1.01E-06 | 3.56E-06 | 0.006364775 |
| Obesity | pericyte | *CALML4* | 4.489401831 | 5.94E-06 | 1.60E-05 | 0.037333751 |
| Obesity | pericyte | *ACKR4* | 4.584869843 | 0.000105861 | 0.00021055 | 0.665547386 |
| Obesity | pericyte | *SUMF2* | 4.530663295 | 0.001077712 | 0.00177371 | 1 |
| Obesity | pericyte | *SPCS3* | 5.574558066 | 0.000537086 | 0.000935362 | 1 |
| Obesity | pericyte | *SERGEF* | 7.671868366 | 1.87E-07 | 9.41E-07 | 0.001178643 |
| Obesity | pericyte | *PLD2* | 4.901238802 | 6.60E-05 | 0.000136712 | 0.415194152 |
| Obesity | pericyte | *ATP1B2* | -1.437688464 | 2.56E-05 | 5.87E-05 | 0.161175579 |
| Obesity | pericyte | *CCL4* | 2.017020395 | 0.006797636 | 0.009885897 | 1 |
| Obesity | pericyte | *PHF20L1* | 6.40558641 | 9.96E-08 | 5.97E-07 | 0.000626499 |
| Obesity | pericyte | *PUS7L* | 2.505295373 | 0.026310227 | 0.035269168 | 1 |
| Obesity | pericyte | *KRI1* | 1.277531032 | 0.007285291 | 0.010556033 | 1 |
| Obesity | pericyte | *SLC44A2* | 7.204300969 | 7.41E-06 | 1.95E-05 | 0.0466006 |
| Obesity | pericyte | *CDKN2D* | 6.117236029 | 1.34E-07 | 7.34E-07 | 0.000843639 |
| Obesity | pericyte | *MTUS1* | 2.490002749 | 0.030668553 | 0.040660732 | 1 |
| Obesity | pericyte | *NGDN* | 7.469134224 | 8.69E-09 | 1.70E-07 | 5.46E-05 |
| Obesity | pericyte | *RIPK3* | 2.466312225 | 0.017589827 | 0.024156234 | 1 |
| Obesity | pericyte | *RAB2B* | 5.113001222 | 2.27E-08 | 2.24E-07 | 0.000142911 |

| | | | | | | |
|---|---|---|---|---|---|---|
| Obesity | pericyte | *BCL2L2* | 7.608573398 | 9.81E-09 | 1.71E-07 | 6.17E-05 |
| Obesity | pericyte | *DTD2* | 0.906650289 | 0.036554832 | 0.04792914 | 1 |
| Obesity | pericyte | *PARP2* | 2.934891509 | 0.001249775 | 0.002038748 | 1 |
| Obesity | pericyte | *MIS18BP1* | 2.676647668 | 3.70E-06 | 1.06E-05 | 0.023271504 |
| Obesity | pericyte | *RNASE1* | 2.526729203 | 0.002609875 | 0.00405434 | 1 |
| Obesity | pericyte | *DAD1* | 4.083737454 | 4.95E-06 | 1.37E-05 | 0.031092137 |
| Obesity | pericyte | *TEP1* | 2.836025916 | 4.35E-06 | 1.22E-05 | 0.027372127 |
| Obesity | pericyte | *EPB41L4A* | 5.935604236 | 1.54E-08 | 1.90E-07 | 9.66E-05 |
| Obesity | pericyte | *CDO1* | 2.933984502 | 3.64E-08 | 2.98E-07 | 0.000228601 |
| Obesity | pericyte | *REEP5* | 6.679054067 | 2.45E-07 | 1.16E-06 | 0.001539481 |
| Obesity | pericyte | *QRICH2* | 2.488728549 | 0.037114033 | 0.048593129 | 1 |
| Obesity | pericyte | *SEC14L1* | 1.903343609 | 0.018048908 | 0.024748851 | 1 |
| Obesity | pericyte | *TMEM8A* | 4.724377914 | 0.001291239 | 0.002103113 | 1 |
| Obesity | pericyte | *SHC2* | 2.262996528 | 0.004050443 | 0.006107889 | 1 |
| Obesity | pericyte | *ERMARD* | 4.63339965 | 6.88E-06 | 1.83E-05 | 0.043265404 |
| Obesity | pericyte | *PHF10* | 4.834528668 | 2.31E-08 | 2.25E-07 | 0.00014528 |
| Obesity | pericyte | *GALNT8* | 2.467773417 | 2.19E-06 | 6.70E-06 | 0.013764897 |
| Obesity | pericyte | *ECSIT* | 5.560903966 | 0.000271985 | 0.000498097 | 1 |
| Obesity | pericyte | *ELOF1* | 11.10205622 | 5.40E-09 | 1.33E-07 | 3.39E-05 |
| Obesity | pericyte | *THEM6* | 1.611624808 | 0.001006753 | 0.001662467 | 1 |
| Obesity | pericyte | *TOMM40* | 5.340009182 | 1.47E-05 | 3.57E-05 | 0.092158596 |
| Obesity | pericyte | *DPP6* | 2.664282352 | 6.17E-07 | 2.42E-06 | 0.003877586 |
| Obesity | pericyte | *FAM98C* | 2.828649987 | 1.10E-05 | 2.75E-05 | 0.069147511 |
| Obesity | pericyte | *RPL36* | 5.500939624 | 0.000104006 | 0.00020732 | 0.653886356 |
| Obesity | pericyte | *ATP8B3* | -2.457093916 | 6.04E-09 | 1.44E-07 | 3.80E-05 |
| Obesity | pericyte | *GTPBP3* | 3.414363427 | 4.57E-06 | 1.28E-05 | 0.028714236 |
| Obesity | pericyte | *BST2* | 5.317441672 | 0.006292654 | 0.009202586 | 1 |
| Obesity | pericyte | *DDA1* | 1.990382433 | 1.74E-06 | 5.53E-06 | 0.01092938 |
| Obesity | pericyte | *MRPL34* | 0.951221548 | 0.012709832 | 0.01783632 | 1 |
| Obesity | pericyte | *LSM7* | 3.292200134 | 3.16E-05 | 7.05E-05 | 0.198653884 |
| Obesity | pericyte | *TULP4* | 4.455630025 | 1.49E-07 | 7.90E-07 | 0.000939093 |
| Obesity | pericyte | *RTN4IP1* | 3.795827349 | 0.001210668 | 0.001979062 | 1 |
| Obesity | pericyte | *RSPH3* | 1.539282796 | 0.000195017 | 0.000367873 | 1 |
| Obesity | pericyte | *ACTN4* | 3.342096565 | 0.012294377 | 0.017284156 | 1 |
| Obesity | pericyte | *NDUFA10* | 0.84448158 | 0.034306151 | 0.045178629 | 1 |
| Obesity | pericyte | *CACNG6* | 3.021559199 | 0.023682443 | 0.031914064 | 1 |
| Obesity | pericyte | *PXDN* | 4.247991867 | 1.27E-08 | 1.73E-07 | 8.01E-05 |
| Obesity | pericyte | *PGPEP1* | 4.046639711 | 2.58E-06 | 7.74E-06 | 0.016250032 |
| Obesity | pericyte | *LSM4* | 2.390375748 | 0.000224597 | 0.000418878 | 1 |
| Obesity | pericyte | *HRC* | -0.829843302 | 1.20E-06 | 4.04E-06 | 0.007558656 |
| Obesity | pericyte | *TRPM4* | 3.116544513 | 0.001324841 | 0.002155052 | 1 |
| Obesity | pericyte | *CAMSAP1* | 5.992262974 | 1.03E-08 | 1.71E-07 | 6.45E-05 |
| Obesity | pericyte | *UBAC1* | 4.09573651 | 6.11E-07 | 2.41E-06 | 0.003844244 |
| Obesity | pericyte | *TNNT3* | 2.178797064 | 1.52E-05 | 3.67E-05 | 0.09527129 |
| Obesity | pericyte | *ATXN10* | 5.846166696 | 5.92E-07 | 2.36E-06 | 0.00372305 |
| Obesity | pericyte | *TUBGCP2* | 5.382608321 | 8.36E-06 | 2.16E-05 | 0.052548691 |
| Obesity | pericyte | *CYP2E1* | 1.785510966 | 0.016124123 | 0.022274745 | 1 |
| Obesity | pericyte | *PAK4* | 8.823432071 | 2.00E-05 | 4.71E-05 | 0.12593355 |
| Obesity | pericyte | *ZNF337* | 1.789944218 | 0.019138381 | 0.026117431 | 1 |
| Obesity | pericyte | *TMEM160* | 2.55869489 | 6.44E-07 | 2.49E-06 | 0.00404665 |

| | | | | | | |
|---|---|---|---|---|---|---|
| Obesity | pericyte | *MAP3K10* | 3.131515793 | 0.016273381 | 0.022461196 | 1 |
| Obesity | pericyte | *SMPDL3B* | 14.4500005 | 8.05E-07 | 2.98E-06 | 0.005062098 |
| Obesity | pericyte | *ATPIF1* | 2.185804232 | 0.010796288 | 0.015301232 | 1 |
| Obesity | pericyte | *THEMIS2* | 2.126659766 | 5.48E-07 | 2.22E-06 | 0.003443009 |
| Obesity | pericyte | *CLIP1* | 7.381846329 | 8.55E-13 | 1.73E-10 | 5.37E-09 |
| Obesity | pericyte | *ZNF317* | 3.362209983 | 0.000108062 | 0.000214385 | 0.67938606 |
| Obesity | pericyte | *PPAN* | 11.64421864 | 1.41E-08 | 1.82E-07 | 8.86E-05 |
| Obesity | pericyte | *ZNF236* | 4.327307808 | 3.82E-05 | 8.37E-05 | 0.240245778 |
| Obesity | pericyte | *C12orf65* | 4.262262344 | 6.37E-08 | 4.35E-07 | 0.000400196 |
| Obesity | pericyte | *PKDREJ* | -1.913675135 | 0.006792196 | 0.009880272 | 1 |
| Obesity | pericyte | *HSD17B3* | 1.712966404 | 0.001076884 | 0.001772811 | 1 |
| Obesity | pericyte | *HABP4* | 6.142802272 | 3.54E-06 | 1.02E-05 | 0.022280065 |
| Obesity | pericyte | *EPS8L1* | -2.943698304 | 2.64E-05 | 6.02E-05 | 0.165883006 |
| Obesity | pericyte | *LILRB2* | 2.642625999 | 6.83E-07 | 2.61E-06 | 0.004297113 |
| Obesity | pericyte | *COX4I2* | -0.3285434 | 1.32E-05 | 3.25E-05 | 0.083253807 |
| Obesity | pericyte | *ATP6V1E1* | 3.587966791 | 0.005874997 | 0.008638004 | 1 |
| Obesity | pericyte | *CHMP1A* | 3.843080713 | 4.15E-05 | 9.02E-05 | 0.261186834 |
| Obesity | pericyte | *PPT1* | 5.758261707 | 2.39E-08 | 2.29E-07 | 0.000150452 |
| Obesity | pericyte | *HAUS8* | 3.252654862 | 0.030732159 | 0.040736472 | 1 |
| Obesity | pericyte | *TBC1D5* | 4.021892731 | 0.000124143 | 0.000243445 | 0.780485692 |
| Obesity | pericyte | *RFTN1* | 3.604322591 | 0.000455521 | 0.000803552 | 1 |
| Obesity | pericyte | *KCNC3* | 1.151866015 | 0.035203195 | 0.046263061 | 1 |
| Obesity | pericyte | *PDLIM4* | 3.442665993 | 8.45E-07 | 3.08E-06 | 0.005314702 |
| Obesity | pericyte | *KIF3A* | 5.069724411 | 1.41E-07 | 7.59E-07 | 0.000886503 |
| Obesity | pericyte | *AOC3* | 2.691225348 | 0.000135122 | 0.000262763 | 0.84951313 |
| Obesity | pericyte | *ACLY* | 3.618220842 | 3.48E-06 | 1.00E-05 | 0.021868346 |
| Obesity | pericyte | *DIAPH1* | 5.359160901 | 1.75E-07 | 8.93E-07 | 0.001097276 |
| Obesity | pericyte | *EXOC4* | 5.730733803 | 4.80E-11 | 5.18E-09 | 3.02E-07 |
| Obesity | pericyte | *C1orf159* | 3.879886095 | 1.43E-07 | 7.67E-07 | 0.000899895 |
| Obesity | pericyte | *ANO1* | -1.198864522 | 0.000385079 | 0.000687392 | 1 |
| Obesity | pericyte | *PPFIA1* | 3.609921986 | 0.000153138 | 0.000294946 | 0.962777488 |
| Obesity | pericyte | *NINJ1* | 9.914367684 | 3.64E-07 | 1.60E-06 | 0.002291381 |
| Obesity | pericyte | *NPHP4* | 5.526452667 | 0.007006096 | 0.010172593 | 1 |
| Obesity | pericyte | *CKMT2* | 2.007356589 | 0.005069464 | 0.007522238 | 1 |
| Obesity | pericyte | *ZCCHC9* | 5.498585479 | 1.20E-07 | 6.80E-07 | 0.000754284 |
| Obesity | pericyte | *KHDRBS3* | -0.887303569 | 9.36E-05 | 0.000188277 | 0.588742141 |
| Obesity | pericyte | *CHD1L* | 2.784583481 | 0.006381744 | 0.009315539 | 1 |
| Obesity | pericyte | *ZSCAN5A* | 6.055238367 | 9.82E-08 | 5.92E-07 | 0.000617452 |
| Obesity | pericyte | *ZNF132* | 1.863685984 | 0.010092341 | 0.014339106 | 1 |
| Obesity | pericyte | *RHPN2* | -2.804369337 | 5.66E-05 | 0.000118858 | 0.356097192 |
| Obesity | pericyte | *C19orf12* | 0.973979946 | 0.020884484 | 0.02834033 | 1 |
| Obesity | pericyte | *ACTR10* | 5.617635391 | 1.94E-06 | 6.08E-06 | 0.012215813 |
| Obesity | pericyte | *GCH1* | 3.880142501 | 1.73E-06 | 5.51E-06 | 0.010869178 |
| Obesity | pericyte | *LGALS3* | 4.616477224 | 0.000251831 | 0.000464845 | 1 |
| Obesity | pericyte | *DNAJB1* | 1.73160549 | 0.00010208 | 0.000204064 | 0.641779701 |
| Obesity | pericyte | *FBXW9* | 3.065782158 | 0.000577357 | 0.00100106 | 1 |
| Obesity | pericyte | *C19orf57* | 4.983870621 | 7.87E-07 | 2.93E-06 | 0.004950077 |
| Obesity | pericyte | *MATN3* | 21.47173536 | 4.95E-07 | 2.05E-06 | 0.00311507 |
| Obesity | pericyte | *RAF1* | 6.269507598 | 9.23E-08 | 5.67E-07 | 0.000580532 |
| Obesity | pericyte | *ENOSF1* | 3.146203968 | 2.96E-05 | 6.67E-05 | 0.186133622 |

| | | | | | | |
|---|---|---|---|---|---|---|
| Obesity | pericyte | *EMILIN2* | 4.50210505 | 4.52E-14 | 1.50E-11 | 2.84E-10 |
| Obesity | pericyte | *TRIM5* | 6.692250156 | 1.69E-07 | 8.70E-07 | 0.001059771 |
| Obesity | pericyte | *CNGA4* | 5.344475582 | 2.02E-06 | 6.30E-06 | 0.012725011 |
| Obesity | pericyte | *TRIM22* | 5.73317126 | 9.90E-06 | 2.51E-05 | 0.062228827 |
| Obesity | pericyte | *RRP8* | 5.13173539 | 1.06E-05 | 2.68E-05 | 0.066859678 |
| Obesity | pericyte | *IMMT* | 3.90689938 | 7.16E-05 | 0.000147104 | 0.449842564 |
| Obesity | pericyte | *CARD6* | 6.153679475 | 1.04E-09 | 5.30E-08 | 6.54E-06 |
| Obesity | pericyte | *RPA1* | 4.714983268 | 1.31E-08 | 1.74E-07 | 8.22E-05 |
| Obesity | pericyte | *SERPINF1* | 2.080842711 | 0.008540853 | 0.012239878 | 1 |
| Obesity | pericyte | *TMEM128* | 4.239664238 | 1.05E-06 | 3.66E-06 | 0.006591327 |
| Obesity | pericyte | *FIGNL1* | 2.87406849 | 1.43E-06 | 4.68E-06 | 0.008967941 |
| Obesity | pericyte | *WBP2* | 5.479385194 | 1.80E-08 | 2.02E-07 | 0.000113143 |
| Obesity | pericyte | *KDM6B* | 6.057778757 | 6.14E-07 | 2.42E-06 | 0.003861793 |
| Obesity | pericyte | *PRMT7* | 1.556710154 | 0.002946085 | 0.004534158 | 1 |
| Obesity | pericyte | *BTBD3* | 5.886221045 | 1.01E-07 | 6.05E-07 | 0.000637524 |
| Obesity | pericyte | *PTPRA* | 4.580823801 | 0.000165916 | 0.000317119 | 1 |
| Obesity | pericyte | *IGHMBP2* | 3.1054658 | 3.05E-09 | 9.89E-08 | 1.92E-05 |
| Obesity | pericyte | *MMACHC* | 1.410206431 | 0.004803595 | 0.007156446 | 1 |
| Obesity | pericyte | *CTNNBL1* | 6.468492869 | 1.61E-05 | 3.88E-05 | 0.101375835 |
| Obesity | pericyte | *LPIN3* | 9.579076662 | 1.47E-07 | 7.81E-07 | 0.000922813 |
| Obesity | pericyte | *DMGDH* | 4.184721298 | 3.58E-07 | 1.58E-06 | 0.002253501 |
| Obesity | pericyte | *KANK4* | 1.163519772 | 1.02E-06 | 3.58E-06 | 0.006419536 |
| Obesity | pericyte | *ATP8A2* | 6.591898702 | 4.35E-08 | 3.34E-07 | 0.000273257 |
| Obesity | pericyte | *USPL1* | 7.083007426 | 3.64E-07 | 1.60E-06 | 0.002288812 |
| Obesity | pericyte | *POMP* | 3.384217315 | 0.037410102 | 0.048958641 | 1 |
| Obesity | pericyte | *WASF3* | 3.494485368 | 4.11E-08 | 3.22E-07 | 0.000258531 |
| Obesity | pericyte | *PEMT* | 1.977537947 | 0.002415821 | 0.003772546 | 1 |
| Obesity | pericyte | *SCO1* | 5.593698394 | 5.25E-07 | 2.14E-06 | 0.003301892 |
| Obesity | pericyte | *CHI3L1* | 0.888166697 | 0.000144542 | 0.000280215 | 0.908738452 |
| Obesity | pericyte | *DSTYK* | 3.116580464 | 0.01407915 | 0.019595909 | 1 |
| Obesity | pericyte | *TMCC2* | 1.822700075 | 0.007854871 | 0.011310943 | 1 |
| Obesity | pericyte | *COG6* | 3.597308736 | 0.000106971 | 0.000212624 | 0.672529225 |
| Obesity | pericyte | *TRPC4* | -1.442308649 | 7.76E-05 | 0.000158435 | 0.488136776 |
| Obesity | pericyte | *RFC3* | 4.118646719 | 1.34E-05 | 3.28E-05 | 0.083942291 |
| Obesity | pericyte | *BTBD2* | 2.822377664 | 0.006826917 | 0.009926186 | 1 |
| Obesity | pericyte | *PRAM1* | 1.97405879 | 0.017093859 | 0.023526508 | 1 |
| Obesity | pericyte | *RTN3* | 5.33455247 | 1.04E-08 | 1.71E-07 | 6.55E-05 |
| Obesity | pericyte | *SLC2A11* | 3.44218433 | 3.26E-07 | 1.46E-06 | 0.002048108 |
| Obesity | pericyte | *C1QTNF6* | 12.42096719 | 1.56E-07 | 8.16E-07 | 0.00097882 |
| Obesity | pericyte | *FAM83F* | 23.4789589 | 1.79E-05 | 4.25E-05 | 0.112540169 |
| Obesity | pericyte | *GIMAP4* | 1.258518596 | 0.016218817 | 0.022390799 | 1 |
| Obesity | pericyte | *AGAP3* | 4.348651461 | 1.71E-05 | 4.08E-05 | 0.107607116 |
| Obesity | pericyte | *ACTR3B* | 3.894788094 | 9.56E-08 | 5.83E-07 | 0.000600727 |
| Obesity | pericyte | *SFTPD* | 0.834248338 | 0.000363174 | 0.000650506 | 1 |
| Obesity | pericyte | *DYDC2* | 7.872336235 | 4.75E-05 | 0.000102177 | 0.298866364 |
| Obesity | pericyte | *TMTC1* | 5.364181258 | 2.79E-08 | 2.51E-07 | 0.000175357 |
| Obesity | pericyte | *IPO8* | 7.262540394 | 3.04E-07 | 1.39E-06 | 0.001914171 |
| Obesity | pericyte | *SPINK5* | -2.054250151 | 0.035122783 | 0.046167037 | 1 |
| Obesity | pericyte | *LRRCC1* | 2.707464821 | 0.000159101 | 0.000305052 | 1 |
| Obesity | pericyte | *SWAP70* | 4.738136984 | 0.000851082 | 0.001426867 | 1 |

| | | | | | | |
|---|---|---|---|---|---|---|
| Obesity | pericyte | *AMPD3* | 2.861528318 | 6.40E-07 | 2.48E-06 | 0.004021687 |
| Obesity | pericyte | *SBF2* | 4.915709378 | 1.28E-06 | 4.27E-06 | 0.008061673 |
| Obesity | pericyte | *MICAL2* | 2.081028819 | 0.007899688 | 0.011367668 | 1 |
| Obesity | pericyte | *RRAS2* | 1.738317508 | 2.75E-05 | 6.23E-05 | 0.172698844 |
| Obesity | pericyte | *HSD17B4* | 3.659893024 | 1.11E-06 | 3.81E-06 | 0.006966232 |
| Obesity | pericyte | *DPF2* | 4.141934122 | 0.000640919 | 0.001100343 | 1 |
| Obesity | pericyte | *GSC* | -1.545339368 | 2.80E-05 | 6.33E-05 | 0.175745184 |
| Obesity | pericyte | *CATSPERB* | 1.352943662 | 0.000667224 | 0.001142074 | 1 |
| Obesity | pericyte | *MED6* | 2.932591396 | 0.000183408 | 0.000347944 | 1 |
| Obesity | pericyte | *EIF2S1* | 6.428070358 | 3.17E-07 | 1.43E-06 | 0.001992619 |
| Obesity | pericyte | *ELP3* | 7.210234219 | 5.19E-08 | 3.79E-07 | 0.00032608 |
| Obesity | pericyte | *ADAMDEC1* | 3.005998931 | 0.017870019 | 0.024508903 | 1 |
| Obesity | pericyte | *MRO* | 1.880502546 | 0.001556931 | 0.002506914 | 1 |
| Obesity | pericyte | *EDEM1* | 8.6641973 | 1.70E-07 | 8.74E-07 | 0.001066917 |
| Obesity | pericyte | *CNTN6* | -1.33120223 | 3.84E-05 | 8.39E-05 | 0.24112103 |
| Obesity | pericyte | *KATNBL1* | 6.558995786 | 2.01E-09 | 7.85E-08 | 1.27E-05 |
| Obesity | pericyte | *GSTM5* | 2.105702808 | 4.91E-08 | 3.66E-07 | 0.00030891 |
| Obesity | pericyte | *GSTM3* | 0.75681982 | 0.003247921 | 0.004967084 | 1 |
| Obesity | pericyte | *SYT6* | 5.957268801 | 1.58E-06 | 5.09E-06 | 0.009930517 |
| Obesity | pericyte | *VAV3* | 3.295329572 | 4.61E-07 | 1.93E-06 | 0.002898348 |
| Obesity | pericyte | *PSRC1* | -1.377293131 | 0.000214358 | 0.000401331 | 1 |
| Obesity | pericyte | *WNT2B* | 2.895439045 | 0.004881512 | 0.007264904 | 1 |
| Obesity | pericyte | *CEPT1* | 4.789075977 | 8.50E-07 | 3.10E-06 | 0.005347081 |
| Obesity | pericyte | *NAPG* | 6.742914572 | 1.18E-07 | 6.74E-07 | 0.000744556 |
| Obesity | pericyte | *SPIRE1* | 2.149854456 | 0.018887562 | 0.025819984 | 1 |
| Obesity | pericyte | *PPHLN1* | 4.551922533 | 3.67E-09 | 1.12E-07 | 2.31E-05 |
| Obesity | pericyte | *FKBP11* | 3.66716647 | 0.000130712 | 0.000255212 | 0.82178421 |
| Obesity | pericyte | *YWHAQ* | 4.179650337 | 0.00167945 | 0.002683961 | 1 |
| Obesity | pericyte | *LPIN1* | 1.833620294 | 0.004279139 | 0.006433034 | 1 |
| Obesity | pericyte | *LDHA* | 5.482836055 | 7.27E-09 | 1.60E-07 | 4.57E-05 |
| Obesity | pericyte | *ANO3* | -0.620021762 | 3.69E-05 | 8.10E-05 | 0.231709588 |
| Obesity | pericyte | *FST* | -1.580930939 | 0.022092114 | 0.029876428 | 1 |
| Obesity | pericyte | *TIMM17A* | 4.597628337 | 7.62E-08 | 4.88E-07 | 0.00047921 |
| Obesity | pericyte | *NARS* | 7.167471043 | 3.10E-05 | 6.94E-05 | 0.195119722 |
| Obesity | pericyte | *KIAA1468* | 4.908494656 | 5.52E-07 | 2.23E-06 | 0.003467674 |
| Obesity | pericyte | *FBXO18* | 5.876424657 | 0.000126614 | 0.000247982 | 0.796021908 |
| Obesity | pericyte | *ECHDC3* | 2.376280362 | 0.003054299 | 0.004689225 | 1 |
| Obesity | pericyte | *IL15RA* | 7.807448398 | 9.25E-09 | 1.70E-07 | 5.82E-05 |
| Obesity | pericyte | *TMEM241* | 4.950004932 | 1.14E-08 | 1.72E-07 | 7.20E-05 |
| Obesity | pericyte | *EMP1* | 2.227958323 | 9.51E-05 | 0.000191029 | 0.59791964 |
| Obesity | pericyte | *KLRD1* | 3.581786483 | 1.70E-08 | 1.95E-07 | 0.000106984 |
| Obesity | pericyte | *PRH2* | -2.913594024 | 0.005348718 | 0.007916052 | 1 |
| Obesity | pericyte | *DDB2* | 5.700682304 | 5.68E-10 | 3.48E-08 | 3.57E-06 |
| Obesity | pericyte | *ACP2* | 3.9143249 | 8.22E-11 | 8.07E-09 | 5.17E-07 |
| Obesity | pericyte | *HOOK1* | 5.530641514 | 5.43E-05 | 0.000114775 | 0.341456416 |
| Obesity | pericyte | *ZCCHC11* | 4.721258022 | 1.73E-05 | 4.12E-05 | 0.108731441 |
| Obesity | pericyte | *PRPF38A* | 7.529548533 | 3.66E-07 | 1.61E-06 | 0.002300058 |
| Obesity | pericyte | *FHOD3* | 1.40605706 | 0.001087933 | 0.001789595 | 1 |
| Obesity | pericyte | *TPGS2* | 5.007587632 | 4.80E-07 | 2.00E-06 | 0.003017905 |
| Obesity | pericyte | *TIMM10* | -1.420483598 | 0.036288033 | 0.047609111 | 1 |

| | | | | | | |
|---|---|---|---|---|---|---|
| Obesity | pericyte | *APLNR* | 4.310057331 | 1.71E-07 | 8.79E-07 | 0.00107461 |
| Obesity | pericyte | *TMEM165* | 2.649237558 | 0.002898572 | 0.004467596 | 1 |
| Obesity | pericyte | *CLOCK* | 6.362577026 | 4.47E-08 | 3.42E-07 | 0.00028077 |
| Obesity | pericyte | *CLDN10* | 2.825735015 | 0.000914944 | 0.001522565 | 1 |
| Obesity | pericyte | *BIVM* | 3.493985438 | 1.29E-07 | 7.18E-07 | 0.00081168 |
| Obesity | pericyte | *ERCC5* | 4.867961409 | 3.36E-07 | 1.50E-06 | 0.002112724 |
| Obesity | pericyte | *CARS2* | 3.404730612 | 8.80E-05 | 0.000177899 | 0.553264892 |
| Obesity | pericyte | *STT3A* | 5.587426182 | 1.32E-07 | 7.28E-07 | 0.00083064 |
| Obesity | pericyte | *KLB* | 1.7617164 | 1.09E-07 | 6.39E-07 | 0.000686448 |
| Obesity | pericyte | *WDR36* | 5.761125884 | 1.15E-08 | 1.72E-07 | 7.24E-05 |
| Obesity | pericyte | *OSTF1* | 7.118776967 | 1.08E-08 | 1.71E-07 | 6.79E-05 |
| Obesity | pericyte | *UBQLN1* | 6.286150117 | 7.56E-08 | 4.87E-07 | 0.000475157 |
| Obesity | pericyte | *NAA35* | 6.843687696 | 2.30E-07 | 1.10E-06 | 0.001447485 |
| Obesity | pericyte | *CTSL* | 5.907943765 | 1.23E-08 | 1.73E-07 | 7.75E-05 |
| Obesity | pericyte | *TMEM2* | 4.619505589 | 6.27E-09 | 1.47E-07 | 3.94E-05 |
| Obesity | pericyte | *AGTPBP1* | 5.37918073 | 3.66E-07 | 1.61E-06 | 0.00229802 |
| Obesity | pericyte | *GOLM1* | -2.330671539 | 0.00269371 | 0.004170243 | 1 |
| Obesity | pericyte | *USP30* | 2.380060619 | 5.30E-06 | 1.45E-05 | 0.033322168 |
| Obesity | pericyte | *P2RX4* | 3.298081432 | 7.14E-10 | 4.01E-08 | 4.49E-06 |
| Obesity | pericyte | *DTX1* | 1.574274929 | 0.002420696 | 0.003778281 | 1 |
| Obesity | pericyte | *DMTF1* | 3.160608961 | 8.64E-05 | 0.000174848 | 0.542901935 |
| Obesity | pericyte | *CCDC146* | 5.314424466 | 6.35E-07 | 2.47E-06 | 0.003989904 |
| Obesity | pericyte | *POM121C* | -3.643683605 | 0.000794447 | 0.001340137 | 1 |
| Obesity | pericyte | *SRPK2* | 5.580680804 | 1.44E-07 | 7.70E-07 | 0.000905242 |
| Obesity | pericyte | *TES* | 1.929652777 | 0.029271747 | 0.038899403 | 1 |
| Obesity | pericyte | *MDFIC* | 3.935139665 | 7.04E-07 | 2.68E-06 | 0.004423199 |
| Obesity | pericyte | *MTO1* | 4.042993672 | 4.86E-07 | 2.02E-06 | 0.003053976 |
| Obesity | pericyte | *ANKRD6* | 6.513986213 | 3.46E-07 | 1.54E-06 | 0.002176308 |
| Obesity | pericyte | *SNX14* | 5.07188221 | 4.69E-06 | 1.30E-05 | 0.029475133 |
| Obesity | pericyte | *NT5E* | 2.389335203 | 1.90E-05 | 4.48E-05 | 0.119434169 |
| Obesity | pericyte | *EPHA7* | 0.513442731 | 0.010903241 | 0.015431939 | 1 |
| Obesity | pericyte | *PRR5L* | 3.956438874 | 4.64E-07 | 1.94E-06 | 0.002915389 |
| Obesity | pericyte | *NAT10* | 4.729165381 | 3.29E-05 | 7.30E-05 | 0.206703732 |
| Obesity | pericyte | *ATP5G2* | 6.515313378 | 8.75E-09 | 1.70E-07 | 5.50E-05 |
| Obesity | pericyte | *RDH5* | 1.699039764 | 5.84E-06 | 1.58E-05 | 0.036747198 |
| Obesity | pericyte | *PPP1R1A* | 4.269521839 | 8.64E-08 | 5.38E-07 | 0.000542981 |
| Obesity | pericyte | *FAIM2* | -3.950407032 | 0.000485039 | 0.000850136 | 1 |
| Obesity | pericyte | *HNRNPA1* | 4.198251284 | 1.05E-08 | 1.71E-07 | 6.62E-05 |
| Obesity | pericyte | *SLC26A10* | -3.143685897 | 0.003781721 | 0.005717576 | 1 |
| Obesity | pericyte | *MAP7* | 2.585828412 | 0.007371005 | 0.010670391 | 1 |
| Obesity | pericyte | *SMPD2* | 1.294860969 | 0.002102322 | 0.00330846 | 1 |
| Obesity | pericyte | *MICAL1* | 5.888955741 | 2.04E-06 | 6.34E-06 | 0.012854149 |
| Obesity | pericyte | *PRADC1* | 5.484370265 | 2.07E-08 | 2.16E-07 | 0.00013034 |
| Obesity | pericyte | *SMYD5* | 3.139438407 | 3.47E-07 | 1.54E-06 | 0.002183757 |
| Obesity | pericyte | *DYSF* | -1.275046107 | 0.001398465 | 0.002268942 | 1 |
| Obesity | pericyte | *KCNMB4* | -2.011054169 | 0.000150567 | 0.00029064 | 0.946615753 |
| Obesity | pericyte | *USP15* | 4.814743288 | 9.06E-06 | 2.33E-05 | 0.056952102 |
| Obesity | pericyte | *KLHL36* | 5.973086288 | 1.18E-06 | 3.97E-06 | 0.00739123 |
| Obesity | pericyte | *MPHOSPH6* | 7.418617083 | 8.32E-07 | 3.04E-06 | 0.005231125 |
| Obesity | pericyte | *NTPCR* | 4.399488291 | 3.82E-09 | 1.13E-07 | 2.40E-05 |

| | | | | | | |
|---|---|---|---|---|---|---|
| Obesity | pericyte | *RNASEL* | 3.016059174 | 0.000115207 | 0.000227127 | 0.724309489 |
| Obesity | pericyte | *PIGC* | 4.655263075 | 1.56E-07 | 8.17E-07 | 0.000982821 |
| Obesity | pericyte | *SP110* | 6.631937524 | 2.20E-07 | 1.06E-06 | 0.001384367 |
| Obesity | pericyte | *USP37* | 7.938139105 | 3.87E-10 | 2.73E-08 | 2.43E-06 |
| Obesity | pericyte | *SLC19A3* | 0.830371967 | 0.001621249 | 0.002598213 | 1 |
| Obesity | pericyte | *TMBIM1* | 2.347345772 | 0.000380903 | 0.000680373 | 1 |
| Obesity | pericyte | *ARMC9* | 6.982579847 | 5.39E-08 | 3.90E-07 | 0.000338566 |
| Obesity | pericyte | *TMEM127* | 4.148294497 | 0.011184105 | 0.015808109 | 1 |
| Obesity | pericyte | *MRPS9* | 3.610117464 | 6.49E-09 | 1.51E-07 | 4.08E-05 |
| Obesity | pericyte | *ANKRD36* | 6.197501432 | 6.37E-08 | 4.35E-07 | 0.000400254 |
| Obesity | pericyte | *USP44* | 2.045353599 | 0.000179291 | 0.00034075 | 1 |
| Obesity | pericyte | *APPL2* | 6.032729037 | 6.33E-08 | 4.34E-07 | 0.000398184 |
| Obesity | pericyte | *DRAM1* | 2.819934656 | 0.000322369 | 0.00058273 | 1 |
| Obesity | pericyte | *SLC41A2* | 2.910885384 | 0.036664896 | 0.048063428 | 1 |
| Obesity | pericyte | *NEK3* | 1.987944881 | 0.000857998 | 0.001437695 | 1 |
| Obesity | pericyte | *VPS36* | 4.983523531 | 8.98E-07 | 3.24E-06 | 0.005643779 |
| Obesity | pericyte | *RNASEH2B* | 3.952393833 | 7.32E-05 | 0.000150265 | 0.460260435 |
| Obesity | pericyte | *TBC1D4* | 5.481707149 | 4.15E-09 | 1.19E-07 | 2.61E-05 |
| Obesity | pericyte | *LRCH1* | 3.440519457 | 0.008027436 | 0.011543571 | 1 |
| Obesity | pericyte | *SUCLA2* | 1.63018546 | 0.005718109 | 0.008425065 | 1 |
| Obesity | pericyte | *MED4* | 5.369186988 | 1.68E-07 | 8.69E-07 | 0.001057674 |
| Obesity | pericyte | *RCBTB2* | 6.444629117 | 5.23E-07 | 2.13E-06 | 0.00328513 |
| Obesity | pericyte | *SETDB2* | 3.127975297 | 0.003072877 | 0.004715446 | 1 |
| Obesity | pericyte | *C7orf25* | -1.469797423 | 0.000197571 | 0.000371894 | 1 |
| Obesity | pericyte | *SPDYE1* | 3.243949917 | 0.000685955 | 0.001170947 | 1 |
| Obesity | pericyte | *CHST12* | 4.49925005 | 7.64E-06 | 2.00E-05 | 0.048047326 |
| Obesity | pericyte | *GPNMB* | 3.223027389 | 1.93E-06 | 6.04E-06 | 0.012133103 |
| Obesity | pericyte | *KDELR2* | 4.27002856 | 3.44E-08 | 2.89E-07 | 0.000216154 |
| Obesity | pericyte | *AOAH* | 2.193568384 | 7.81E-06 | 2.04E-05 | 0.049095223 |
| Obesity | pericyte | *HUS1* | 5.279780072 | 9.93E-07 | 3.51E-06 | 0.006243343 |
| Obesity | pericyte | *NACAD* | -6.251996078 | 0.004534446 | 0.006789251 | 1 |
| Obesity | pericyte | *DBNL* | 5.241480324 | 1.46E-06 | 4.76E-06 | 0.009153871 |
| Obesity | pericyte | *TTC5* | 4.340688145 | 4.98E-06 | 1.37E-05 | 0.031316431 |
| Obesity | pericyte | *ALPK3* | 1.585725907 | 2.15E-06 | 6.60E-06 | 0.013512911 |
| Obesity | pericyte | *TM6SF1* | 5.809702047 | 1.11E-09 | 5.30E-08 | 6.99E-06 |
| Obesity | pericyte | *RSAD1* | 2.758306749 | 2.47E-07 | 1.16E-06 | 0.001553585 |
| Obesity | pericyte | *NMT1* | 2.092126885 | 1.82E-05 | 4.31E-05 | 0.114243264 |
| Obesity | pericyte | *BRIP1* | 2.563053206 | 4.83E-07 | 2.01E-06 | 0.003035904 |
| Obesity | pericyte | *MRPL47* | 6.76860498 | 3.11E-07 | 1.41E-06 | 0.001953478 |
| Obesity | pericyte | *EPRS* | 5.661277447 | 9.98E-09 | 1.71E-07 | 6.27E-05 |
| Obesity | pericyte | *RPS6KC1* | 4.917464555 | 5.55E-05 | 0.000116863 | 0.349185678 |
| Obesity | pericyte | *CBWD2* | 6.218440192 | 4.53E-06 | 1.27E-05 | 0.0284564 |
| Obesity | pericyte | *IL1RN* | 9.51764247 | 1.16E-08 | 1.72E-07 | 7.30E-05 |
| Obesity | pericyte | *SAP130* | 4.5809305 | 1.52E-05 | 3.69E-05 | 0.095844732 |
| Obesity | pericyte | *UGGT1* | 5.005368047 | 0.001239086 | 0.002022886 | 1 |
| Obesity | pericyte | *GYPC* | 4.908244926 | 1.88E-06 | 5.92E-06 | 0.011842013 |
| Obesity | pericyte | *NIPSNAP3A* | 2.47083006 | 0.001463459 | 0.002369499 | 1 |
| Obesity | pericyte | *LRRC8A* | 2.893217248 | 9.78E-07 | 3.46E-06 | 0.006147764 |
| Obesity | pericyte | *TXN* | 4.498066719 | 7.47E-07 | 2.80E-06 | 0.004696866 |
| Obesity | pericyte | *ODF2* | 8.655961791 | 1.08E-06 | 3.73E-06 | 0.006772275 |

| | | | | | | |
|---|---|---|---|---|---|---|
| Obesity | pericyte | *TOR1B* | 1.684693816 | 0.036539564 | 0.047919116 | 1 |
| Obesity | pericyte | *TOR1A* | 1.992821735 | 0.003173088 | 0.004856184 | 1 |
| Obesity | pericyte | *SLC2A8* | 3.428644667 | 0.000262754 | 0.000482599 | 1 |
| Obesity | pericyte | *ZFP37* | 1.9381796 | 0.023685545 | 0.031914064 | 1 |
| Obesity | pericyte | *ZNF189* | 4.292249807 | 1.46E-06 | 4.76E-06 | 0.009155998 |
| Obesity | pericyte | *ALDOB* | -2.272755165 | 0.000629411 | 0.00108268 | 1 |
| Obesity | pericyte | *STX17* | 6.09291406 | 6.98E-05 | 0.000143856 | 0.438761334 |
| Obesity | pericyte | *PRPF4* | 1.977478303 | 0.006903031 | 0.010034533 | 1 |
| Obesity | pericyte | *TEX10* | 5.207733629 | 1.80E-08 | 2.02E-07 | 0.000112941 |
| Obesity | pericyte | *GARNL3* | 4.08781375 | 0.00772237 | 0.01114567 | 1 |
| Obesity | pericyte | *GABBR2* | 4.165947507 | 9.23E-07 | 3.32E-06 | 0.005805657 |
| Obesity | pericyte | *PSMB7* | 5.078918011 | 1.63E-08 | 1.92E-07 | 0.000102704 |
| Obesity | pericyte | *RABEPK* | 7.366936988 | 8.86E-09 | 1.70E-07 | 5.57E-05 |
| Obesity | pericyte | *GOLGA1* | 7.069539529 | 5.55E-08 | 3.96E-07 | 0.000349179 |
| Obesity | pericyte | *DSCC1* | 0.831475447 | 1.59E-05 | 3.82E-05 | 0.099802876 |
| Obesity | pericyte | *DERL1* | 6.896234685 | 1.00E-05 | 2.54E-05 | 0.063010249 |
| Obesity | pericyte | *NOV* | 4.532066523 | 0.000164036 | 0.000313829 | 1 |
| Obesity | pericyte | *APTX* | 2.849238908 | 0.012299894 | 0.017288047 | 1 |
| Obesity | pericyte | *CD72* | 6.330167027 | 2.72E-09 | 9.38E-08 | 1.71E-05 |
| Obesity | pericyte | *GRHPR* | 2.166722687 | 0.004300215 | 0.006463172 | 1 |
| Obesity | pericyte | *ARHGEF39* | 1.367554365 | 0.006390547 | 0.009324058 | 1 |
| Obesity | pericyte | *DENND4C* | 5.523985122 | 2.81E-08 | 2.52E-07 | 0.000176463 |
| Obesity | pericyte | *PPIL1* | 7.320017268 | 6.75E-08 | 4.50E-07 | 0.000424429 |
| Obesity | pericyte | *KIF13A* | 4.518458686 | 8.17E-05 | 0.000165969 | 0.513341693 |
| Obesity | pericyte | *ZSCAN9* | 3.779864096 | 3.96E-08 | 3.15E-07 | 0.000249254 |
| Obesity | pericyte | *TMEM63B* | -2.539019672 | 0.004782252 | 0.007128027 | 1 |
| Obesity | pericyte | *FRS3* | 4.11808738 | 1.22E-07 | 6.87E-07 | 0.000765305 |
| Obesity | pericyte | *SLC22A23* | 6.244189324 | 2.36E-07 | 1.12E-06 | 0.001483692 |
| Obesity | pericyte | *LRRC1* | 3.272653603 | 1.55E-06 | 5.01E-06 | 0.009716542 |
| Obesity | pericyte | *RIPK1* | 7.007757908 | 1.53E-07 | 8.03E-07 | 0.00095938 |
| Obesity | pericyte | *UQCC2* | 5.156687172 | 1.87E-06 | 5.90E-06 | 0.011777542 |
| Obesity | pericyte | *TCF19* | 4.448550402 | 9.86E-10 | 5.08E-08 | 6.20E-06 |
| Obesity | pericyte | *FLOT1* | 5.952703468 | 2.40E-08 | 2.29E-07 | 0.000150861 |
| Obesity | pericyte | *NRM* | 3.362494867 | 2.09E-08 | 2.17E-07 | 0.000131475 |
| Obesity | pericyte | *MTCH1* | 8.067891271 | 3.53E-08 | 2.92E-07 | 0.000222059 |
| Obesity | pericyte | *FHDC1* | 3.967004367 | 2.20E-07 | 1.06E-06 | 0.001382392 |
| Obesity | pericyte | *MYO7A* | 1.116335401 | 0.019729121 | 0.02687692 | 1 |
| Obesity | pericyte | *ARRB1* | 3.954525356 | 0.002034432 | 0.003209655 | 1 |
| Obesity | pericyte | *SYTL2* | -1.873309232 | 0.000776812 | 0.00131356 | 1 |
| Obesity | pericyte | *RAB30* | 3.573494512 | 0.006286352 | 0.009195508 | 1 |
| Obesity | pericyte | *PRCP* | 4.887336222 | 3.71E-09 | 1.12E-07 | 2.33E-05 |
| Obesity | pericyte | *NARS2* | 5.550038211 | 0.000250422 | 0.000462516 | 1 |
| Obesity | pericyte | *PI15* | 10.62566481 | 3.21E-06 | 9.34E-06 | 0.020154312 |
| Obesity | pericyte | *TGS1* | 2.039236083 | 0.014567135 | 0.020248414 | 1 |
| Obesity | pericyte | *NEK1* | 5.280331256 | 1.00E-06 | 3.54E-06 | 0.006310062 |
| Obesity | pericyte | *DDX60* | 7.218983345 | 4.85E-05 | 0.000103951 | 0.304680558 |
| Obesity | pericyte | *MMP27* | 0.78839285 | 0.00172071 | 0.002745712 | 1 |
| Obesity | pericyte | *DCUN1D5* | 2.497219167 | 8.34E-08 | 5.24E-07 | 0.000524128 |
| Obesity | pericyte | *TRIM29* | 1.143308175 | 0.029794892 | 0.039577537 | 1 |
| Obesity | pericyte | *SLC37A4* | 3.920964418 | 4.91E-07 | 2.03E-06 | 0.003086371 |

| | | | | | | |
|---|---|---|---|---|---|---|
| Obesity | pericyte | *PPP2R1B* | 2.716701634 | 4.82E-08 | 3.61E-07 | 0.000302757 |
| Obesity | pericyte | *FDX1* | 6.17189206 | 5.24E-06 | 1.44E-05 | 0.032912842 |
| Obesity | pericyte | *FXYD6* | -1.714345193 | 0.001587381 | 0.002552394 | 1 |
| Obesity | pericyte | *TMPRSS13* | 3.628963121 | 0.006165518 | 0.00903768 | 1 |
| Obesity | pericyte | *CASP1* | 6.895587771 | 1.32E-07 | 7.28E-07 | 0.000831645 |
| Obesity | pericyte | *MAP2K5* | 6.787688614 | 2.62E-12 | 4.58E-10 | 1.65E-08 |
| Obesity | pericyte | *NDUFAF1* | 3.062182878 | 0.000160794 | 0.000308018 | 1 |
| Obesity | pericyte | *ITGA11* | 1.813504313 | 6.12E-07 | 2.41E-06 | 0.003845593 |
| Obesity | pericyte | *PAQR5* | 5.211907418 | 4.06E-06 | 1.15E-05 | 0.02554741 |
| Obesity | pericyte | *RMDN3* | 5.554875829 | 1.02E-05 | 2.56E-05 | 0.063844556 |
| Obesity | pericyte | *UACA* | 5.865438709 | 2.68E-07 | 1.25E-06 | 0.001687301 |
| Obesity | pericyte | *PLCB2* | 0.837764446 | 0.011685282 | 0.016483142 | 1 |
| Obesity | pericyte | *RSL24D1* | 2.04507254 | 0.004319672 | 0.006490865 | 1 |
| Obesity | pericyte | *SPTBN5* | 0.956685664 | 0.004610155 | 0.006892757 | 1 |
| Obesity | pericyte | *RABGGTB* | 6.223044833 | 0.000350613 | 0.000629982 | 1 |
| Obesity | pericyte | *SLC44A5* | 2.761827058 | 1.74E-08 | 1.99E-07 | 0.000109584 |
| Obesity | pericyte | *DNASE2B* | 10.12820443 | 2.05E-05 | 4.80E-05 | 0.128644553 |
| Obesity | pericyte | *DBT* | 2.906739863 | 8.10E-07 | 2.99E-06 | 0.005094504 |
| Obesity | pericyte | *RTCA* | 3.797874888 | 5.02E-06 | 1.38E-05 | 0.031535529 |
| Obesity | pericyte | *IFT172* | 9.257588254 | 3.49E-12 | 5.63E-10 | 2.20E-08 |
| Obesity | pericyte | *KHK* | -0.582457865 | 0.017729042 | 0.024331475 | 1 |
| Obesity | pericyte | *DYNC2LI1* | 1.828523983 | 0.007172474 | 0.010397358 | 1 |
| Obesity | pericyte | *LHCGR* | 1.869494207 | 5.51E-06 | 1.50E-05 | 0.034633631 |
| Obesity | pericyte | *THUMPD2* | 5.927364523 | 2.66E-08 | 2.42E-07 | 0.000167115 |
| Obesity | pericyte | *ATRAID* | 4.948138754 | 0.000134774 | 0.000262166 | 0.847321882 |
| Obesity | pericyte | *CENPO* | 9.967296156 | 4.28E-09 | 1.20E-07 | 2.69E-05 |
| Obesity | pericyte | *LRPPRC* | 6.445604028 | 1.07E-07 | 6.30E-07 | 0.000672689 |
| Obesity | pericyte | *DTNB* | 3.933440759 | 1.32E-06 | 4.37E-06 | 0.008292256 |
| Obesity | pericyte | *CYP2C9* | 0.81775818 | 0.029067345 | 0.038643773 | 1 |
| Obesity | pericyte | *CYP2C8* | 2.449246269 | 0.000631951 | 0.00108643 | 1 |
| Obesity | pericyte | *CH25H* | 1.02481113 | 0.032863759 | 0.043397281 | 1 |
| Obesity | pericyte | *ATAD1* | 5.835616494 | 0.000117202 | 0.000230626 | 0.736850002 |
| Obesity | pericyte | *BTBD16* | 3.40913801 | 4.95E-05 | 0.00010592 | 0.311297836 |
| Obesity | pericyte | *TACC2* | 3.047424478 | 4.34E-08 | 3.34E-07 | 0.000272934 |
| Obesity | pericyte | *CALHM2* | 1.669153782 | 2.09E-06 | 6.46E-06 | 0.013153247 |
| Obesity | pericyte | *ARL3* | 2.924144271 | 0.015290142 | 0.021206512 | 1 |
| Obesity | pericyte | *PLCE1* | 4.171846605 | 1.63E-07 | 8.48E-07 | 0.001025933 |
| Obesity | pericyte | *RBP4* | 7.300549121 | 5.60E-08 | 3.99E-07 | 0.000352332 |
| Obesity | pericyte | *DNAJC13* | 5.976026519 | 1.72E-05 | 4.10E-05 | 0.108273959 |
| Obesity | pericyte | *FAM149B1* | 3.281991909 | 0.000270104 | 0.000494797 | 1 |
| Obesity | pericyte | *ASCC1* | 1.975255358 | 0.037915445 | 0.049547787 | 1 |
| Obesity | pericyte | *RPS24* | 5.037881579 | 0.000521729 | 0.000910636 | 1 |
| Obesity | pericyte | *SMARCAL1* | 5.860846416 | 2.15E-08 | 2.18E-07 | 0.000135029 |
| Obesity | pericyte | *BARD1* | 7.580723232 | 1.80E-07 | 9.12E-07 | 0.00113054 |
| Obesity | pericyte | *STAT4* | -1.180843585 | 0.004681775 | 0.006988206 | 1 |
| Obesity | pericyte | *CARF* | 3.991471806 | 1.67E-08 | 1.93E-07 | 0.00010505 |
| Obesity | pericyte | *ASNSD1* | 2.469243252 | 5.08E-06 | 1.40E-05 | 0.031933718 |
| Obesity | pericyte | *METTL5* | 6.291861925 | 1.12E-08 | 1.71E-07 | 7.03E-05 |
| Obesity | pericyte | *NAB1* | 4.601795164 | 0.003588958 | 0.0054423 | 1 |
| Obesity | pericyte | *FASTKD1* | 4.296653441 | 3.21E-07 | 1.44E-06 | 0.002016432 |

| | | | | | | |
|---|---|---|---|---|---|---|
| Obesity | pericyte | *MDH1B* | 4.864821477 | 3.26E-06 | 9.47E-06 | 0.020521952 |
| Obesity | pericyte | *FAM117B* | 7.174618862 | 2.28E-09 | 8.46E-08 | 1.44E-05 |
| Obesity | pericyte | *ITGAV* | 4.05125722 | 2.35E-05 | 5.42E-05 | 0.147554177 |
| Obesity | pericyte | *SLC35A5* | 0.940343226 | 0.028443001 | 0.037861772 | 1 |
| Obesity | pericyte | *SENP7* | 4.170321587 | 0.000888604 | 0.001483445 | 1 |
| Obesity | pericyte | *USP8* | 3.857655372 | 0.000340776 | 0.000612831 | 1 |
| Obesity | pericyte | *SPPL2A* | 4.834017815 | 1.24E-07 | 6.95E-07 | 0.000778665 |
| Obesity | pericyte | *UBL7* | 3.791009121 | 0.007815916 | 0.011257425 | 1 |
| Obesity | pericyte | *ARHGAP24* | 3.703986723 | 0.000316679 | 0.000573268 | 1 |
| Obesity | pericyte | *FAM13A* | 3.754494146 | 5.78E-06 | 1.57E-05 | 0.036356882 |
| Obesity | pericyte | *HERC3* | 5.992642702 | 9.58E-06 | 2.45E-05 | 0.060227431 |
| Obesity | pericyte | *AP1AR* | 7.093901405 | 1.05E-06 | 3.66E-06 | 0.00661146 |
| Obesity | pericyte | *PRKG2* | 2.35109615 | 3.20E-07 | 1.44E-06 | 0.002011399 |
| Obesity | pericyte | *BBS7* | 7.49523988 | 3.34E-05 | 7.41E-05 | 0.210233849 |
| Obesity | pericyte | *KIAA1109* | 4.988722075 | 1.82E-05 | 4.31E-05 | 0.114387445 |
| Obesity | pericyte | *MMRN1* | -0.702743029 | 0.003775067 | 0.005709368 | 1 |
| Obesity | pericyte | *PRDM5* | 4.414159392 | 8.36E-07 | 3.06E-06 | 0.00525507 |
| Obesity | pericyte | *TRPC3* | -1.447670246 | 2.87E-05 | 6.49E-05 | 0.180545135 |
| Obesity | pericyte | *NAAA* | 4.20947372 | 7.69E-06 | 2.01E-05 | 0.048339446 |
| Obesity | pericyte | *NUP54* | 3.13462544 | 0.001581393 | 0.002543417 | 1 |
| Obesity | pericyte | *BMP2K* | 1.479964601 | 0.019391914 | 0.026451934 | 1 |
| Obesity | pericyte | *G3BP2* | 5.172819247 | 1.28E-08 | 1.73E-07 | 8.06E-05 |
| Obesity | pericyte | *SCARB2* | 6.18250957 | 2.77E-06 | 8.21E-06 | 0.017404392 |
| Obesity | pericyte | *ANXA3* | -2.173025354 | 0.004673178 | 0.006978687 | 1 |
| Obesity | pericyte | *PPA2* | 3.899735046 | 2.77E-05 | 6.27E-05 | 0.173876976 |
| Obesity | pericyte | *GSTCD* | 5.471960763 | 9.11E-08 | 5.61E-07 | 0.000573012 |
| Obesity | pericyte | *ENPEP* | -0.391942004 | 3.38E-05 | 7.47E-05 | 0.212188019 |
| Obesity | pericyte | *PAPSS1* | 2.393457293 | 0.00114644 | 0.001878464 | 1 |
| Obesity | pericyte | *SLC39A8* | 2.676933105 | 3.60E-05 | 7.94E-05 | 0.226426939 |
| Obesity | pericyte | *RNF185* | 4.084312352 | 2.49E-07 | 1.17E-06 | 0.001567602 |
| Obesity | pericyte | *PARVG* | 3.539223108 | 4.50E-06 | 1.26E-05 | 0.028295237 |
| Obesity | pericyte | *B4GALNT3* | 2.449046151 | 0.000502223 | 0.000878786 | 1 |
| Obesity | pericyte | *ERP27* | 5.576623021 | 3.09E-07 | 1.41E-06 | 0.001943751 |
| Obesity | pericyte | *ETV6* | 5.667303125 | 4.18E-08 | 3.26E-07 | 0.000263027 |
| Obesity | pericyte | *ALG10* | -3.092037384 | 0.004440515 | 0.006654951 | 1 |
| Obesity | pericyte | *SLCO1C1* | -0.422375443 | 3.46E-05 | 7.65E-05 | 0.21759659 |
| Obesity | pericyte | *ZCRB1* | 2.368785929 | 0.00754698 | 0.010907555 | 1 |
| Obesity | pericyte | *PRICKLE1* | 3.986129113 | 9.92E-07 | 3.51E-06 | 0.006237823 |
| Obesity | pericyte | *C1RL* | 3.417779255 | 1.51E-05 | 3.67E-05 | 0.095240257 |
| Obesity | pericyte | *CLSTN3* | 2.662052722 | 0.016745494 | 0.023062195 | 1 |
| Obesity | pericyte | *KLRG1* | 2.674012114 | 2.42E-08 | 2.30E-07 | 0.000151946 |
| Obesity | pericyte | *VAMP1* | 1.538428484 | 0.003526844 | 0.005354569 | 1 |
| Obesity | pericyte | *TAPBPL* | 1.560786306 | 9.70E-05 | 0.000194607 | 0.610053776 |
| Obesity | pericyte | *CD27* | 4.698244767 | 4.99E-08 | 3.70E-07 | 0.000313438 |
| Obesity | pericyte | *GLIPR1* | 3.509629646 | 2.47E-07 | 1.16E-06 | 0.001550232 |
| Obesity | pericyte | *LGR5* | 2.140301792 | 0.01306367 | 0.018283903 | 1 |
| Obesity | pericyte | *POC1B* | 7.206183427 | 1.34E-08 | 1.76E-07 | 8.42E-05 |
| Obesity | pericyte | *GAS2L3* | 7.740815613 | 0.000137658 | 0.000267364 | 0.865457743 |
| Obesity | pericyte | *TMEM132B* | -1.93951388 | 0.018857958 | 0.02578512 | 1 |
| Obesity | pericyte | *MMAB* | 2.487716091 | 0.001526546 | 0.002463398 | 1 |

| | | | | | | |
|---|---|---|---|---|---|---|
| Obesity | pericyte | *MTMR6* | 3.721828918 | 0.001028358 | 0.00169648 | 1 |
| Obesity | pericyte | *SLC46A3* | 2.399031286 | 3.87E-05 | 8.46E-05 | 0.2433503 |
| Obesity | pericyte | *SUOX* | 1.431455428 | 0.011252603 | 0.015897779 | 1 |
| Obesity | pericyte | *ACVRL1* | -2.219478834 | 0.000212172 | 0.00039795 | 1 |
| Obesity | pericyte | *BRCA2* | 3.427055537 | 1.20E-07 | 6.81E-07 | 0.000756394 |
| Obesity | pericyte | *CERS5* | 4.909126743 | 0.000102128 | 0.000204094 | 0.642080781 |
| Obesity | pericyte | *LMBR1L* | 5.641456196 | 9.27E-05 | 0.000186591 | 0.582537804 |
| Obesity | pericyte | *ZNF740* | 4.723262125 | 2.60E-05 | 5.95E-05 | 0.163679671 |
| Obesity | pericyte | *WDFY2* | 3.109181438 | 0.006179457 | 0.009056002 | 1 |
| Obesity | pericyte | *HNRNPA1L2* | 6.084167315 | 0.005294183 | 0.007840879 | 1 |
| Obesity | pericyte | *VPS33A* | 2.781786968 | 1.46E-06 | 4.77E-06 | 0.009184289 |
| Obesity | pericyte | *RHOF* | 1.535103749 | 0.001149299 | 0.001882659 | 1 |
| Obesity | pericyte | *METTL21C* | -3.937037893 | 2.34E-05 | 5.41E-05 | 0.147284671 |
| Obesity | pericyte | *TSSK4* | 5.400652606 | 0.002779747 | 0.004294979 | 1 |
| Obesity | pericyte | *RTN1* | 5.45138816 | 7.23E-08 | 4.73E-07 | 0.000454454 |
| Obesity | pericyte | *C14orf37* | 3.493662011 | 0.000156716 | 0.000301031 | 0.985274103 |
| Obesity | pericyte | *STON2* | 5.872754823 | 1.88E-07 | 9.42E-07 | 0.001183473 |
| Obesity | pericyte | *EFCAB11* | 2.266588221 | 4.32E-07 | 1.84E-06 | 0.002717805 |
| Obesity | pericyte | *AK7* | -3.4250929 | 0.000788377 | 0.001330611 | 1 |
| Obesity | pericyte | *FBLN5* | 6.859008662 | 1.32E-09 | 5.90E-08 | 8.31E-06 |
| Obesity | pericyte | *NIPA2* | 6.901362424 | 5.53E-07 | 2.23E-06 | 0.00347785 |
| Obesity | pericyte | *DUOXA1* | 1.734257493 | 0.00841629 | 0.012080643 | 1 |
| Obesity | pericyte | *MFAP1* | 3.662349072 | 2.61E-07 | 1.22E-06 | 0.001638347 |
| Obesity | pericyte | *TCF12* | 4.477605252 | 2.20E-05 | 5.12E-05 | 0.138484764 |
| Obesity | pericyte | *SORD* | -2.016250248 | 0.014525376 | 0.020194834 | 1 |
| Obesity | pericyte | *LYSMD2* | -1.948216538 | 0.028716235 | 0.038217394 | 1 |
| Obesity | pericyte | *SRP14* | 3.594848733 | 0.002573169 | 0.004001364 | 1 |
| Obesity | pericyte | *BAHD1* | 5.303049805 | 1.86E-07 | 9.33E-07 | 0.001166613 |
| Obesity | pericyte | *ANP32A* | 6.846930102 | 5.53E-08 | 3.96E-07 | 0.000347361 |
| Obesity | pericyte | *COMMD4* | 6.40480877 | 3.00E-07 | 1.37E-06 | 0.001884696 |
| Obesity | pericyte | *UBE2Q2* | 4.460146242 | 0.00269525 | 0.0041716 | 1 |
| Obesity | pericyte | *PSTPIP1* | 2.1832795 | 1.08E-05 | 2.72E-05 | 0.068177537 |
| Obesity | pericyte | *HMG20A* | 6.605020615 | 5.01E-09 | 1.29E-07 | 3.15E-05 |
| Obesity | pericyte | *SCAPER* | 3.049055373 | 0.000190979 | 0.00036089 | 1 |
| Obesity | pericyte | *NEIL1* | 3.148564899 | 0.000217639 | 0.000406384 | 1 |
| Obesity | pericyte | *DNAJA4* | 11.40910093 | 6.81E-09 | 1.54E-07 | 4.28E-05 |
| Obesity | pericyte | *TPM1* | -1.183091378 | 0.000714148 | 0.001214129 | 1 |
| Obesity | pericyte | *IGF1R* | 4.25457632 | 0.00062758 | 0.001080393 | 1 |
| Obesity | pericyte | *CYP11A1* | 2.555456235 | 2.34E-08 | 2.26E-07 | 0.000147164 |
| Obesity | pericyte | *BBS4* | 5.850611783 | 2.02E-10 | 1.67E-08 | 1.27E-06 |
| Obesity | pericyte | *ADAMTS17* | 5.093350887 | 2.69E-05 | 6.13E-05 | 0.169236704 |
| Obesity | pericyte | *ULK3* | 2.917813994 | 4.69E-07 | 1.96E-06 | 0.002945463 |
| Obesity | pericyte | *SCAMP2* | 4.436359933 | 5.64E-05 | 0.000118342 | 0.35431582 |
| Obesity | pericyte | *HAPLN3* | 3.913305682 | 0.001131538 | 0.001856467 | 1 |
| Obesity | pericyte | *POLG* | 4.604890376 | 2.05E-08 | 2.16E-07 | 0.0001289 |
| Obesity | pericyte | *TICRR* | 3.338737567 | 1.12E-05 | 2.79E-05 | 0.070182463 |
| Obesity | pericyte | *UNC45A* | 2.521691426 | 0.000228991 | 0.000426062 | 1 |
| Obesity | pericyte | *ST8SIA2* | 10.59953875 | 2.10E-05 | 4.90E-05 | 0.131770829 |
| Obesity | pericyte | *IQGAP1* | 3.052154463 | 0.000288997 | 0.00052695 | 1 |
| Obesity | pericyte | *CRTC3* | 4.822283834 | 0.000291698 | 0.000531258 | 1 |

| | | | | | | |
|---|---|---|---|---|---|---|
| Obesity | pericyte | *SEC11A* | 6.421077665 | 2.49E-07 | 1.17E-06 | 0.00156543 |
| Obesity | pericyte | *TGFB1I1* | -2.161526166 | 0.000200798 | 0.000377518 | 1 |
| Obesity | pericyte | *PARN* | 4.226844561 | 5.59E-07 | 2.25E-06 | 0.003517304 |
| Obesity | pericyte | *FTO* | 4.862616217 | 5.89E-07 | 2.35E-06 | 0.003703542 |
| Obesity | pericyte | *TXNL4B* | 2.717504515 | 1.51E-06 | 4.91E-06 | 0.009497096 |
| Obesity | pericyte | *MARVELD3* | 3.315016203 | 0.000238857 | 0.000442325 | 1 |
| Obesity | pericyte | *ZFHX3* | 5.761740095 | 2.88E-07 | 1.32E-06 | 0.001811067 |
| Obesity | pericyte | *KIFC3* | 4.670507906 | 0.005729979 | 0.008440575 | 1 |
| Obesity | pericyte | *NUDT7* | 1.840795833 | 0.016347254 | 0.022553256 | 1 |
| Obesity | pericyte | *GCSH* | 2.045414045 | 0.000839162 | 0.001409515 | 1 |
| Obesity | pericyte | *CMTM3* | 10.09911201 | 2.23E-07 | 1.08E-06 | 0.001402538 |
| Obesity | pericyte | *MAP1LC3B* | 3.899243201 | 1.69E-06 | 5.40E-06 | 0.010608923 |
| Obesity | pericyte | *TLDC1* | 8.750717154 | 1.11E-08 | 1.71E-07 | 6.97E-05 |
| Obesity | pericyte | *NDUFB10* | -1.97555728 | 0.02884534 | 0.03837297 | 1 |
| Obesity | pericyte | *TIGD7* | 2.699151138 | 0.000706411 | 0.001201951 | 1 |
| Obesity | pericyte | *DEF8* | 2.47279806 | 0.00083197 | 0.001398181 | 1 |
| Obesity | pericyte | *COPS3* | 5.013612206 | 2.40E-08 | 2.29E-07 | 0.000150865 |
| Obesity | pericyte | *RANBP10* | 4.558145833 | 2.83E-05 | 6.41E-05 | 0.17819549 |
| Obesity | pericyte | *PRPSAP2* | 4.251510948 | 1.34E-07 | 7.34E-07 | 0.00084452 |
| Obesity | pericyte | *MYO19* | 10.35009743 | 3.49E-16 | 1.69E-13 | 2.19E-12 |
| Obesity | pericyte | *DDX52* | 5.309898153 | 2.39E-07 | 1.14E-06 | 0.001502799 |
| Obesity | pericyte | *UNC45B* | 4.573910101 | 0.006637561 | 0.009666516 | 1 |
| Obesity | pericyte | *TOM1L1* | 6.480786316 | 3.36E-10 | 2.49E-08 | 2.11E-06 |
| Obesity | pericyte | *C17orf80* | 2.44985314 | 2.31E-06 | 7.02E-06 | 0.014531272 |
| Obesity | pericyte | *VPS53* | 4.287754787 | 9.57E-07 | 3.41E-06 | 0.006016934 |
| Obesity | pericyte | *SGSM2* | 4.042694604 | 0.000784066 | 0.001324758 | 1 |
| Obesity | pericyte | *NPEPPS* | 6.394444172 | 2.28E-08 | 2.24E-07 | 0.000143094 |
| Obesity | pericyte | *LRRC46* | 2.342916515 | 0.002737659 | 0.004233069 | 1 |
| Obesity | pericyte | *RHBDL3* | 2.443571966 | 3.68E-05 | 8.08E-05 | 0.231137692 |
| Obesity | pericyte | *ARSG* | 3.748287344 | 0.000520201 | 0.000908473 | 1 |
| Obesity | pericyte | *G6PC3* | 4.059332744 | 2.49E-07 | 1.17E-06 | 0.001566704 |
| Obesity | pericyte | *IMPA2* | 5.326687804 | 1.17E-08 | 1.72E-07 | 7.34E-05 |
| Obesity | pericyte | *GNAL* | 4.155889102 | 1.10E-08 | 1.71E-07 | 6.89E-05 |
| Obesity | pericyte | *RPRD1A* | 3.41546336 | 0.001897261 | 0.003006826 | 1 |
| Obesity | pericyte | *C18orf21* | 2.259957762 | 2.65E-07 | 1.23E-06 | 0.001665883 |
| Obesity | pericyte | *GALNT1* | 5.699990163 | 5.45E-08 | 3.91E-07 | 0.000342782 |
| Obesity | pericyte | *ESCO1* | 6.569338757 | 6.17E-08 | 4.25E-07 | 0.000387972 |
| Obesity | pericyte | *GREB1L* | 1.912264477 | 3.90E-05 | 8.51E-05 | 0.245248741 |
| Obesity | pericyte | *SLC14A1* | 3.441739524 | 1.66E-05 | 3.97E-05 | 0.104172684 |
| Obesity | pericyte | *CCDC40* | 1.537684877 | 0.004621418 | 0.006906312 | 1 |
| Obesity | pericyte | *TMC6* | 1.876821254 | 0.000295918 | 0.000538477 | 1 |
| Obesity | pericyte | *CARD14* | 11.82946872 | 8.41E-08 | 5.28E-07 | 0.000529046 |
| Obesity | pericyte | *EIF4A3* | -2.118613843 | 0.005592822 | 0.008257884 | 1 |
| Obesity | pericyte | *TBCD* | 4.552988747 | 6.91E-06 | 1.83E-05 | 0.043432387 |
| Obesity | pericyte | *NARF* | 4.624054831 | 5.46E-05 | 0.00011527 | 0.343275318 |
| Obesity | pericyte | *TRIM65* | 4.265673709 | 3.99E-12 | 6.27E-10 | 2.51E-08 |
| Obesity | pericyte | *CBX8* | 4.49871988 | 5.94E-08 | 4.15E-07 | 0.000373517 |
| Obesity | pericyte | *SECTM1* | 3.509131028 | 3.26E-06 | 9.46E-06 | 0.020478683 |
| Obesity | pericyte | *RNF157* | 1.131413579 | 3.18E-05 | 7.09E-05 | 0.19986479 |
| Obesity | pericyte | *WDR45B* | 4.044556774 | 0.000294823 | 0.00053664 | 1 |

| | | | | | | |
|---|---|---|---|---|---|---|
| Obesity | pericyte | *RNF165* | 7.213634989 | 8.01E-08 | 5.08E-07 | 0.000503619 |
| Obesity | pericyte | *MAPK4* | 1.510933588 | 0.002476794 | 0.003858178 | 1 |
| Obesity | pericyte | *SMAD4* | 5.624515165 | 2.15E-05 | 5.00E-05 | 0.134974803 |
| Obesity | pericyte | *PIP4K2B* | 3.906461576 | 3.86E-08 | 3.10E-07 | 0.000242526 |
| Obesity | pericyte | *GRB7* | 3.599561121 | 2.22E-05 | 5.15E-05 | 0.139595558 |
| Obesity | pericyte | *TXNL4A* | 1.781142059 | 0.01040501 | 0.014766658 | 1 |
| Obesity | pericyte | *NFIC* | 2.935680127 | 5.14E-06 | 1.42E-05 | 0.03234485 |
| Obesity | pericyte | *TPGS1* | 4.053292705 | 7.36E-08 | 4.79E-07 | 0.000462968 |
| Obesity | pericyte | *PFKL* | 2.054006005 | 0.013709883 | 0.019107522 | 1 |
| Obesity | pericyte | *MVB12A* | 2.89800317 | 0.002997996 | 0.004607259 | 1 |
| Obesity | pericyte | *DUS3L* | 1.366160598 | 0.027368603 | 0.036602086 | 1 |
| Obesity | pericyte | *DPP9* | 5.173515034 | 0.000403851 | 0.000719063 | 1 |
| Obesity | pericyte | *CCDC97* | 6.121861703 | 4.16E-08 | 3.24E-07 | 0.000261419 |
| Obesity | pericyte | *TMEM91* | 2.817941037 | 9.35E-06 | 2.39E-05 | 0.058802355 |
| Obesity | pericyte | *SIRT3* | 4.586454893 | 2.26E-07 | 1.08E-06 | 0.001418873 |
| Obesity | pericyte | *IFITM3* | 3.980901349 | 0.017773305 | 0.024386898 | 1 |
| Obesity | pericyte | *COL6A1* | 2.349242482 | 0.000212127 | 0.00039795 | 1 |
| Obesity | pericyte | *DOPEY2* | 4.451834231 | 1.83E-06 | 5.79E-06 | 0.011503424 |
| Obesity | pericyte | *URB1* | 1.417572444 | 0.014973256 | 0.020780764 | 1 |
| Obesity | pericyte | *AKT1* | 7.400974127 | 1.16E-08 | 1.72E-07 | 7.31E-05 |
| Obesity | pericyte | *SAE1* | 3.757187295 | 1.74E-06 | 5.54E-06 | 0.010954442 |
| Obesity | pericyte | *NTN5* | -1.975198696 | 7.40E-07 | 2.78E-06 | 0.00465464 |
| Obesity | pericyte | *GEMIN7* | 4.695812828 | 0.000584079 | 0.001010485 | 1 |
| Obesity | pericyte | *CAPN10* | 2.058259104 | 9.15E-05 | 0.00018442 | 0.575022733 |
| Obesity | pericyte | *MYO1F* | 4.054387668 | 3.38E-08 | 2.86E-07 | 0.000212753 |
| Obesity | pericyte | *CACNG8* | 12.01838022 | 3.61E-08 | 2.96E-07 | 0.000226714 |
| Obesity | pericyte | *EVI5L* | -1.91838718 | 0.000872908 | 0.001459568 | 1 |
| Obesity | pericyte | *RPS11* | 5.365132804 | 1.55E-05 | 3.74E-05 | 0.097386941 |
| Obesity | pericyte | *CTU1* | 2.319178139 | 1.61E-05 | 3.87E-05 | 0.101256046 |
| Obesity | pericyte | *RCN3* | 9.162754356 | 1.69E-07 | 8.72E-07 | 0.001062693 |
| Obesity | pericyte | *RERE* | 4.24658062 | 8.32E-07 | 3.04E-06 | 0.005231576 |
| Obesity | pericyte | *PEX14* | 4.158189002 | 4.52E-08 | 3.45E-07 | 0.00028423 |
| Obesity | pericyte | *SH3BGRL3* | 7.593972247 | 1.53E-08 | 1.90E-07 | 9.62E-05 |
| Obesity | pericyte | *IL22RA1* | 1.820265325 | 0.008538043 | 0.01223864 | 1 |
| Obesity | pericyte | *C1orf216* | 4.338194571 | 5.68E-07 | 2.28E-06 | 0.003573086 |
| Obesity | pericyte | *MAP3K6* | -2.627568033 | 0.003990837 | 0.00602555 | 1 |
| Obesity | pericyte | *ITGB3BP* | 1.441736917 | 0.021951852 | 0.029705401 | 1 |
| Obesity | pericyte | *SERBP1* | 4.981449766 | 0.001474466 | 0.002386092 | 1 |
| Obesity | pericyte | *PIGK* | 7.381386461 | 8.49E-07 | 3.09E-06 | 0.005335965 |
| Obesity | pericyte | *SYPL2* | -0.685139345 | 0.009571679 | 0.013648706 | 1 |
| Obesity | pericyte | *ZNF697* | -0.63075766 | 3.14E-05 | 7.01E-05 | 0.197447679 |
| Obesity | pericyte | *CELSR2* | -2.061140069 | 9.53E-05 | 0.000191411 | 0.599365383 |
| Obesity | pericyte | *ATP1B1* | 3.171045928 | 5.65E-06 | 1.53E-05 | 0.035548373 |
| Obesity | pericyte | *TIPRL* | 8.007971667 | 1.78E-06 | 5.63E-06 | 0.011161432 |
| Obesity | pericyte | *NME7* | 4.351068929 | 0.000340015 | 0.000611814 | 1 |
| Obesity | pericyte | *CREG1* | 4.448973088 | 1.28E-07 | 7.16E-07 | 0.00080748 |
| Obesity | pericyte | *GPA33* | 3.858990128 | 6.18E-05 | 0.000128614 | 0.388542143 |
| Obesity | pericyte | *RXRG* | -5.463051344 | 0.005863618 | 0.008623291 | 1 |
| Obesity | pericyte | *TMCO1* | 7.439715415 | 6.56E-07 | 2.53E-06 | 0.004123771 |
| Obesity | pericyte | *MGST3* | 5.846171736 | 7.88E-06 | 2.05E-05 | 0.049559253 |

| | | | | | | |
|---|---|---|---|---|---|---|
| Obesity | pericyte | *ADCY10* | 3.5173977 | 2.72E-07 | 1.26E-06 | 0.001710614 |
| Obesity | pericyte | *RFWD2* | 4.742629388 | 1.01E-05 | 2.54E-05 | 0.063229013 |
| Obesity | pericyte | *PPOX* | 3.45310383 | 9.22E-07 | 3.31E-06 | 0.005796038 |
| Obesity | pericyte | *RGS5* | -0.418857404 | 5.23E-05 | 0.000110871 | 0.328622458 |
| Obesity | pericyte | *RRNAD1* | 2.907042737 | 3.10E-05 | 6.93E-05 | 0.194870006 |
| Obesity | pericyte | *PIGM* | 5.200342916 | 6.92E-08 | 4.57E-07 | 0.000434793 |
| Obesity | pericyte | *RGL1* | 5.220158295 | 1.53E-08 | 1.90E-07 | 9.63E-05 |
| Obesity | pericyte | *TARS2* | 3.739914416 | 5.56E-07 | 2.24E-06 | 0.003497338 |
| Obesity | pericyte | *SNX27* | 6.021370169 | 6.71E-08 | 4.48E-07 | 0.000421888 |
| Obesity | pericyte | *CERS2* | 3.103501605 | 0.030411121 | 0.04034495 | 1 |
| Obesity | pericyte | *ENSA* | 6.804133607 | 8.07E-09 | 1.68E-07 | 5.08E-05 |
| Obesity | pericyte | *ARNT* | 3.374473662 | 0.0021226 | 0.003335362 | 1 |
| Obesity | pericyte | *POGZ* | 4.768045543 | 5.65E-08 | 4.01E-07 | 0.000355234 |
| Obesity | pericyte | *IKBKE* | 2.181923266 | 1.41E-06 | 4.64E-06 | 0.008866545 |
| Obesity | pericyte | *HHIPL2* | 2.26633751 | 0.030754897 | 0.040758019 | 1 |
| Obesity | pericyte | *RAB13* | 4.855099506 | 0.017351687 | 0.023855249 | 1 |
| Obesity | pericyte | *HAX1* | 7.295315949 | 8.18E-11 | 8.07E-09 | 5.15E-07 |
| Obesity | pericyte | *KCNN3* | 4.157030833 | 1.82E-08 | 2.04E-07 | 0.00011439 |
| Obesity | pericyte | *GATAD2B* | 3.33253795 | 0.000982251 | 0.001626821 | 1 |
| Obesity | pericyte | *RIT1* | 8.465556506 | 1.05E-07 | 6.23E-07 | 0.000662885 |
| Obesity | pericyte | *GALNT2* | 4.986996466 | 9.11E-09 | 1.70E-07 | 5.72E-05 |
| Obesity | pericyte | *SCCPDH* | 7.130897125 | 3.67E-08 | 2.99E-07 | 0.000230625 |
| Obesity | pericyte | *CEP170* | 6.426295637 | 2.64E-09 | 9.26E-08 | 1.66E-05 |
| Obesity | pericyte | *NVL* | 6.831083293 | 2.85E-06 | 8.42E-06 | 0.017920653 |
| Obesity | pericyte | *SDE2* | 5.039970904 | 4.36E-08 | 3.35E-07 | 0.000274219 |
| Obesity | pericyte | *FBXO28* | 6.136263135 | 9.74E-09 | 1.71E-07 | 6.12E-05 |
| Obesity | pericyte | *CNIH4* | 7.49231006 | 4.23E-08 | 3.28E-07 | 0.000265893 |
| Obesity | pericyte | *GUK1* | 5.402258224 | 0.001511945 | 0.002441088 | 1 |
| Obesity | pericyte | *CNIH3* | 3.840671013 | 2.77E-08 | 2.50E-07 | 0.000174104 |
| Obesity | pericyte | *PSEN2* | 5.087077344 | 4.06E-08 | 3.21E-07 | 0.000255103 |
| Obesity | pericyte | *PYCR2* | -2.063176174 | 0.009661491 | 0.013767406 | 1 |
| Obesity | pericyte | *WNT9A* | 3.593628559 | 2.41E-05 | 5.56E-05 | 0.151783568 |
| Obesity | pericyte | *PLEKHA6* | 5.2639784 | 1.56E-07 | 8.17E-07 | 0.000982675 |
| Obesity | pericyte | *ABCG8* | 5.448995746 | 0.00015731 | 0.000301988 | 0.98901074 |
| Obesity | pericyte | *EML4* | 6.029340927 | 1.08E-08 | 1.71E-07 | 6.82E-05 |
| Obesity | pericyte | *WDPCP* | 1.604826938 | 1.13E-06 | 3.86E-06 | 0.007126723 |
| Obesity | pericyte | *ASXL2* | 5.249993417 | 1.15E-05 | 2.87E-05 | 0.072498278 |
| Obesity | pericyte | *EXOC6B* | 5.122288012 | 2.02E-07 | 9.99E-07 | 0.001272985 |
| Obesity | pericyte | *TEX261* | 5.648417757 | 4.58E-08 | 3.48E-07 | 0.000287714 |
| Obesity | pericyte | *THNSL2* | 0.558027763 | 0.005735912 | 0.008447337 | 1 |
| Obesity | pericyte | *RALB* | 3.55074071 | 0.006024787 | 0.008847895 | 1 |
| Obesity | pericyte | *TMEM177* | -2.496938735 | 0.021246038 | 0.028793671 | 1 |
| Obesity | pericyte | *SLC20A1* | 5.742549734 | 4.78E-07 | 1.99E-06 | 0.003007271 |
| Obesity | pericyte | *ZC3H8* | 5.016315894 | 1.29E-08 | 1.73E-07 | 8.10E-05 |
| Obesity | pericyte | *FAHD2B* | -2.815941947 | 2.04E-05 | 4.79E-05 | 0.128501614 |
| Obesity | pericyte | *LYG1* | 2.16783298 | 4.69E-05 | 0.000100888 | 0.294693237 |
| Obesity | pericyte | *UBXN4* | 6.334453619 | 1.01E-08 | 1.71E-07 | 6.37E-05 |
| Obesity | pericyte | *THSD7B* | -0.374059081 | 0.000170004 | 0.000323978 | 1 |
| Obesity | pericyte | *GPR17* | 0.639660252 | 0.006378829 | 0.00931512 | 1 |
| Obesity | pericyte | *POLR2D* | 5.391815243 | 0.002669919 | 0.004136467 | 1 |

| | | | | | | |
|---|---|---|---|---|---|---|
| Obesity | pericyte | *ZNF385B* | 2.192148266 | 1.24E-07 | 6.95E-07 | 0.000778351 |
| Obesity | pericyte | *TMEFF2* | 4.100261161 | 1.63E-07 | 8.47E-07 | 0.001024532 |
| Obesity | pericyte | *GULP1* | 5.222412409 | 2.87E-08 | 2.55E-07 | 0.000180525 |
| Obesity | pericyte | *FAM171B* | 2.726757251 | 1.93E-07 | 9.63E-07 | 0.001215618 |
| Obesity | pericyte | *HSPD1* | 6.564663373 | 4.31E-11 | 4.75E-09 | 2.71E-07 |
| Obesity | pericyte | *METTL21A* | 7.082601471 | 1.21E-05 | 2.99E-05 | 0.075817861 |
| Obesity | pericyte | *PTH2R* | 1.568389973 | 0.000197187 | 0.000371605 | 1 |
| Obesity | pericyte | *NBEAL1* | 1.875759518 | 8.72E-05 | 0.000176339 | 0.548238674 |
| Obesity | pericyte | *SUMF1* | 5.057203076 | 2.06E-08 | 2.16E-07 | 0.000129295 |
| Obesity | pericyte | *ESPNL* | 19.88086827 | 3.44E-06 | 9.93E-06 | 0.021634929 |
| Obesity | pericyte | *FANCD2* | 5.978064292 | 0.009181027 | 0.013115455 | 1 |
| Obesity | pericyte | *TAMM41* | 4.896508163 | 0.000327834 | 0.000591757 | 1 |
| Obesity | pericyte | *OSBPL10* | -1.086340922 | 0.002179706 | 0.003421675 | 1 |
| Obesity | pericyte | *CSRNP1* | 3.542225214 | 0.000845544 | 0.001418718 | 1 |
| Obesity | pericyte | *CTDSPL* | 5.01823677 | 2.96E-05 | 6.67E-05 | 0.186298816 |
| Obesity | pericyte | *STAC* | -1.401633799 | 0.000261921 | 0.00048135 | 1 |
| Obesity | pericyte | *ARL6IP5* | 6.230711879 | 1.77E-07 | 9.01E-07 | 0.001113196 |
| Obesity | pericyte | *TMF1* | 5.627117113 | 6.50E-05 | 0.000134704 | 0.408693234 |
| Obesity | pericyte | *ZNF660* | -1.018712301 | 0.011674857 | 0.016472133 | 1 |
| Obesity | pericyte | *COL8A1* | 1.353420722 | 0.022092494 | 0.029876428 | 1 |
| Obesity | pericyte | *NXPE3* | 6.775551181 | 1.09E-06 | 3.77E-06 | 0.006870894 |
| Obesity | pericyte | *RABL3* | 8.324170941 | 9.13E-10 | 4.78E-08 | 5.74E-06 |
| Obesity | pericyte | *ADPRH* | 1.233397117 | 0.002590934 | 0.004026997 | 1 |
| Obesity | pericyte | *BOC* | -1.249705223 | 0.002883172 | 0.004444949 | 1 |
| Obesity | pericyte | *AGTR1* | -0.387466886 | 2.23E-05 | 5.17E-05 | 0.140159249 |
| Obesity | pericyte | *AMT* | -0.783842555 | 0.015589654 | 0.021598095 | 1 |
| Obesity | pericyte | *TCTA* | 2.837412421 | 0.005544377 | 0.008188277 | 1 |
| Obesity | pericyte | *EAF2* | 2.261495959 | 0.00772797 | 0.011151193 | 1 |
| Obesity | pericyte | *ILDR1* | -1.357050071 | 0.03620069 | 0.047504433 | 1 |
| Obesity | pericyte | *DGKQ* | 1.236697009 | 0.013914247 | 0.019370875 | 1 |
| Obesity | pericyte | *LYAR* | 5.845627577 | 2.46E-08 | 2.32E-07 | 0.000154836 |
| Obesity | pericyte | *CISD2* | 5.876364022 | 4.06E-06 | 1.15E-05 | 0.025513805 |
| Obesity | pericyte | *TIFA* | 5.661821355 | 3.19E-06 | 9.31E-06 | 0.020071214 |
| Obesity | pericyte | *CCNA2* | 3.467310318 | 3.61E-07 | 1.59E-06 | 0.002272222 |
| Obesity | pericyte | *USP53* | 5.888411738 | 3.50E-08 | 2.91E-07 | 0.000220186 |
| Obesity | pericyte | *PDGFC* | 4.576573731 | 3.00E-09 | 9.87E-08 | 1.88E-05 |
| Obesity | pericyte | *CBR4* | 5.910765256 | 3.45E-07 | 1.54E-06 | 0.002170626 |
| Obesity | pericyte | *NDUFS6* | 5.965984114 | 0.000791323 | 0.001335226 | 1 |
| Obesity | pericyte | *FAM105A* | 2.718681565 | 0.000758945 | 0.001284385 | 1 |
| Obesity | pericyte | *LHFPL2* | 5.688980247 | 0.000151378 | 0.000292025 | 0.951710722 |
| Obesity | pericyte | *SSBP2* | 3.336819772 | 0.001924012 | 0.00304615 | 1 |
| Obesity | pericyte | *RASA1* | 2.402721133 | 0.000858927 | 0.001438486 | 1 |
| Obesity | pericyte | *PPIP5K2* | 3.302052795 | 0.001575735 | 0.002534966 | 1 |
| Obesity | pericyte | *GTF2H2* | 2.923049267 | 0.00093828 | 0.001558099 | 1 |
| Obesity | pericyte | *SPATA9* | 3.853476112 | 2.71E-06 | 8.07E-06 | 0.017056896 |
| Obesity | pericyte | *FEM1C* | 7.240879051 | 3.28E-09 | 1.04E-07 | 2.06E-05 |
| Obesity | pericyte | *COMMD10* | 3.10812581 | 0.000115523 | 0.000227678 | 0.726293785 |
| Obesity | pericyte | *YIPF5* | 4.186193299 | 1.14E-07 | 6.60E-07 | 0.000719645 |
| Obesity | pericyte | *GABRB2* | 1.26162889 | 0.000885276 | 0.001479067 | 1 |
| Obesity | pericyte | *TBC1D7* | 5.191236931 | 1.26E-08 | 1.73E-07 | 7.90E-05 |

| | | | | | | |
|---|---|---|---|---|---|---|
| Obesity | pericyte | *GFOD1* | 1.826252547 | 0.000358337 | 0.00064239 | 1 |
| Obesity | pericyte | *PSD2* | 7.054112988 | 0.003705509 | 0.005610919 | 1 |
| Obesity | pericyte | *ZMAT2* | 1.997750323 | 0.001603344 | 0.002574113 | 1 |
| Obesity | pericyte | *KLHL3* | 3.984007282 | 1.42E-07 | 7.62E-07 | 0.000892008 |
| Obesity | pericyte | *TRIM7* | 2.98611465 | 4.14E-06 | 1.17E-05 | 0.026048268 |
| Obesity | pericyte | *HIGD2A* | 2.918704312 | 0.023039521 | 0.031096924 | 1 |
| Obesity | pericyte | *FAM193B* | 6.292339048 | 1.67E-05 | 4.01E-05 | 0.105290041 |
| Obesity | pericyte | *MUT* | 7.622610206 | 9.31E-09 | 1.70E-07 | 5.85E-05 |
| Obesity | pericyte | *ABT1* | 2.163218971 | 0.007692298 | 0.011109918 | 1 |
| Obesity | pericyte | *MLIP* | -1.209687987 | 5.10E-05 | 0.000108675 | 0.320916977 |
| Obesity | pericyte | *FGD2* | 2.06527547 | 4.53E-07 | 1.91E-06 | 0.002850448 |
| Obesity | pericyte | *ANO7* | 1.240381006 | 0.001494737 | 0.002415167 | 1 |
| Obesity | pericyte | *RPL7L1* | 4.302489488 | 2.64E-08 | 2.41E-07 | 0.000165891 |
| Obesity | pericyte | *TPBG* | 8.64722921 | 9.71E-09 | 1.71E-07 | 6.10E-05 |
| Obesity | pericyte | *IRAK1BP1* | 8.006831286 | 2.61E-08 | 2.40E-07 | 0.000164339 |
| Obesity | pericyte | *PHIP* | 5.864971927 | 2.07E-08 | 2.16E-07 | 0.000130086 |
| Obesity | pericyte | *MMS22L* | 5.185145715 | 4.85E-05 | 0.000104072 | 0.305139566 |
| Obesity | pericyte | *PM20D2* | 5.577594087 | 2.12E-07 | 1.04E-06 | 0.00133423 |
| Obesity | pericyte | *RNF217* | 5.671045147 | 1.05E-08 | 1.71E-07 | 6.63E-05 |
| Obesity | pericyte | *AIG1* | 5.175616708 | 2.65E-10 | 2.11E-08 | 1.66E-06 |
| Obesity | pericyte | *DYNLT1* | 5.661266709 | 2.50E-05 | 5.74E-05 | 0.157454129 |
| Obesity | pericyte | *PNLDC1* | 2.969195984 | 0.000406909 | 0.000724293 | 1 |
| Obesity | pericyte | *WTAP* | 3.390890415 | 3.22E-06 | 9.39E-06 | 0.0202697 |
| Obesity | pericyte | *SLC22A3* | 2.908880755 | 1.04E-07 | 6.17E-07 | 0.000654468 |
| Obesity | pericyte | *GNA12* | 4.246778291 | 7.51E-05 | 0.000153574 | 0.472087144 |
| Obesity | pericyte | *C7orf50* | 6.197003793 | 2.15E-05 | 5.02E-05 | 0.135466311 |
| Obesity | pericyte | *RBAK* | 6.685071459 | 1.40E-08 | 1.81E-07 | 8.79E-05 |
| Obesity | pericyte | *EGFR* | 2.506620924 | 6.57E-07 | 2.53E-06 | 0.004131023 |
| Obesity | pericyte | *POMZP3* | 1.982969109 | 0.007341517 | 0.010630151 | 1 |
| Obesity | pericyte | *CCT6A* | 5.818378551 | 1.75E-07 | 8.93E-07 | 0.001097593 |
| Obesity | pericyte | *GIGYF1* | 5.49515291 | 1.25E-07 | 6.99E-07 | 0.000784654 |
| Obesity | pericyte | *TMEM209* | 3.947971354 | 2.31E-08 | 2.25E-07 | 0.000145313 |
| Obesity | pericyte | *AGBL3* | 5.006356681 | 7.15E-08 | 4.70E-07 | 0.00044945 |
| Obesity | pericyte | *EPHA1* | 3.360915607 | 1.49E-05 | 3.61E-05 | 0.093582585 |
| Obesity | pericyte | *NOM1* | 5.895253306 | 1.31E-08 | 1.74E-07 | 8.22E-05 |
| Obesity | pericyte | *NCAPG2* | 3.329038015 | 2.33E-06 | 7.06E-06 | 0.014623758 |
| Obesity | pericyte | *FBXO25* | 7.402124605 | 6.04E-08 | 4.19E-07 | 0.000379621 |
| Obesity | pericyte | *CSGALNACT1* | 3.89061661 | 1.32E-05 | 3.25E-05 | 0.083171172 |
| Obesity | pericyte | *CCDC25* | 5.221496206 | 1.04E-06 | 3.63E-06 | 0.006522216 |
| Obesity | pericyte | *BIN3* | 3.800415074 | 0.003771054 | 0.005704671 | 1 |
| Obesity | pericyte | *SLC25A37* | 6.496394337 | 1.42E-09 | 6.17E-08 | 8.95E-06 |
| Obesity | pericyte | *CHMP7* | 3.429806891 | 0.000817682 | 0.001376746 | 1 |
| Obesity | pericyte | *DOCK5* | 6.068039527 | 1.26E-08 | 1.73E-07 | 7.93E-05 |
| Obesity | pericyte | *ST18* | 3.039368657 | 0.002144462 | 0.003368874 | 1 |
| Obesity | pericyte | *TACC1* | 3.540207055 | 0.000956955 | 0.001587015 | 1 |
| Obesity | pericyte | *SLC26A7* | -0.783590318 | 1.07E-05 | 2.68E-05 | 0.06708992 |
| Obesity | pericyte | *ATP6V0D2* | -3.746209543 | 3.55E-07 | 1.57E-06 | 0.002229844 |
| Obesity | pericyte | *MTDH* | 6.009486154 | 4.85E-08 | 3.63E-07 | 0.000304844 |
| Obesity | pericyte | *MAL2* | 2.663562725 | 7.10E-07 | 2.69E-06 | 0.004465554 |
| Obesity | pericyte | *UTP23* | 6.357997136 | 8.01E-07 | 2.96E-06 | 0.005035707 |

| | | | | | | |
|---|---|---|---|---|---|---|
| Obesity | pericyte | *FAM135B* | 3.027839614 | 1.87E-06 | 5.90E-06 | 0.011767403 |
| Obesity | pericyte | *ZNF7* | 4.842884301 | 1.21E-05 | 3.00E-05 | 0.076139494 |
| Obesity | pericyte | *SLC39A4* | 3.519146431 | 0.000369673 | 0.000661392 | 1 |
| Obesity | pericyte | *AK3* | 3.169531515 | 0.008850653 | 0.012663645 | 1 |
| Obesity | pericyte | *CER1* | 3.33010296 | 0.000500709 | 0.000876869 | 1 |
| Obesity | pericyte | *PLIN2* | 7.104152445 | 5.23E-08 | 3.82E-07 | 0.000329005 |
| Obesity | pericyte | *HAUS6* | 8.787641288 | 2.93E-08 | 2.59E-07 | 0.000184034 |
| Obesity | pericyte | *C9orf72* | 3.566611307 | 1.69E-06 | 5.42E-06 | 0.010654629 |
| Obesity | pericyte | *SIGMAR1* | 2.131046103 | 0.000809438 | 0.001363594 | 1 |
| Obesity | pericyte | *CBWD5* | 5.560849348 | 2.10E-08 | 2.17E-07 | 0.000132132 |
| Obesity | pericyte | *ZNF462* | 6.557942197 | 9.79E-06 | 2.49E-05 | 0.061555844 |
| Obesity | pericyte | *ALAD* | 1.17433998 | 0.001084625 | 0.00178462 | 1 |
| Obesity | pericyte | *ASTN2* | 7.161974266 | 1.02E-08 | 1.71E-07 | 6.42E-05 |
| Obesity | pericyte | *POLE3* | 2.336759335 | 1.88E-05 | 4.44E-05 | 0.11809325 |
| Obesity | pericyte | *SURF6* | 7.975482791 | 3.25E-08 | 2.79E-07 | 0.000204446 |
| Obesity | pericyte | *ASB6* | 4.68014017 | 7.04E-05 | 0.0001451 | 0.442844751 |
| Obesity | pericyte | *NTMT1* | 2.82087356 | 0.001891091 | 0.003000074 | 1 |
| Obesity | pericyte | *CIZ1* | 5.108736352 | 1.56E-09 | 6.58E-08 | 9.79E-06 |
| Obesity | pericyte | *SH3GLB2* | 7.818265488 | 4.87E-10 | 3.22E-08 | 3.06E-06 |
| Obesity | pericyte | *LRSAM1* | 3.395712653 | 1.00E-07 | 6.00E-07 | 0.000629547 |
| Obesity | pericyte | *SEC16A* | 2.179903025 | 0.002040332 | 0.003218156 | 1 |
| Obesity | pericyte | *DPH7* | 8.424272554 | 3.17E-06 | 9.25E-06 | 0.019924025 |
| Obesity | pericyte | *SLC39A12* | -1.760057263 | 0.000153173 | 0.000294946 | 0.962998019 |
| Obesity | pericyte | *RSU1* | 4.557429144 | 0.000420666 | 0.000747309 | 1 |
| Obesity | pericyte | *ADIRF* | 2.929344805 | 0.028817013 | 0.038343399 | 1 |
| Obesity | pericyte | *GLUD1* | 6.160069034 | 1.12E-06 | 3.82E-06 | 0.007012548 |
| Obesity | pericyte | *HTR7* | 2.805357756 | 7.38E-07 | 2.78E-06 | 0.004638797 |
| Obesity | pericyte | *RPP30* | 4.578831751 | 0.002051272 | 0.003233789 | 1 |
| Obesity | pericyte | *FRA10AC1* | 3.02181681 | 7.98E-05 | 0.000162565 | 0.50151386 |
| Obesity | pericyte | *DNAJB12* | 6.785059223 | 8.59E-06 | 2.22E-05 | 0.053995858 |
| Obesity | pericyte | *PLEKHS1* | -2.69595323 | 0.009164283 | 0.013097487 | 1 |
| Obesity | pericyte | *FUOM* | -3.001708693 | 0.000433356 | 0.000767899 | 1 |
| Obesity | pericyte | *LRRC27* | 0.908230713 | 0.012827613 | 0.017993575 | 1 |
| Obesity | pericyte | *MTG1* | 1.720226998 | 0.000679837 | 0.001161766 | 1 |
| Obesity | pericyte | *GSTO1* | 5.478709104 | 4.07E-07 | 1.76E-06 | 0.002559253 |
| Obesity | pericyte | *PDCD11* | 2.559951848 | 0.034359111 | 0.045238897 | 1 |
| Obesity | pericyte | *RGS10* | 6.453167398 | 5.90E-07 | 2.35E-06 | 0.003712072 |
| Obesity | pericyte | *GAS2* | 7.717655419 | 6.19E-09 | 1.46E-07 | 3.89E-05 |
| Obesity | pericyte | *LIN7C* | 4.232808768 | 5.31E-08 | 3.86E-07 | 0.000333782 |
| Obesity | pericyte | *ZNF215* | 0.692781284 | 0.017862411 | 0.024503814 | 1 |
| Obesity | pericyte | *HSD17B12* | -2.359585127 | 1.10E-09 | 5.30E-08 | 6.92E-06 |
| Obesity | pericyte | *APIP* | 0.396720734 | 0.037106594 | 0.048593129 | 1 |
| Obesity | pericyte | *GLYAT* | 1.787561906 | 1.22E-07 | 6.89E-07 | 0.000769273 |
| Obesity | pericyte | *PTPRJ* | 3.593996625 | 4.31E-07 | 1.84E-06 | 0.002709385 |
| Obesity | pericyte | *C11orf49* | 4.437244589 | 0.000184844 | 0.000350351 | 1 |
| Obesity | pericyte | *ENDOD1* | -2.092273385 | 0.002743469 | 0.004241011 | 1 |
| Obesity | pericyte | *SERPINH1* | 2.092285328 | 0.021728191 | 0.029421739 | 1 |
| Obesity | pericyte | *CAPN5* | 2.263924558 | 3.78E-07 | 1.65E-06 | 0.002376241 |
| Obesity | pericyte | *INTS4* | 3.650531303 | 5.62E-05 | 0.00011819 | 0.353505181 |
| Obesity | pericyte | *RPS3* | 1.837662294 | 0.031323259 | 0.04145008 | 1 |

| | | | | | | |
|---|---|---|---|---|---|---|
| Obesity | pericyte | *ZC3H12C* | 7.381033783 | 6.42E-08 | 4.37E-07 | 0.000403499 |
| Obesity | pericyte | *TTC12* | 1.813193852 | 1.03E-06 | 3.60E-06 | 0.006476473 |
| Obesity | pericyte | *DRD2* | 9.783069599 | 4.00E-07 | 1.73E-06 | 0.002512892 |
| Obesity | pericyte | *C11orf52* | 22.15865946 | 4.73E-07 | 1.98E-06 | 0.002975209 |
| Obesity | pericyte | *NPAT* | 4.258225562 | 2.08E-06 | 6.43E-06 | 0.01306746 |
| Obesity | pericyte | *GRIK4* | 1.033292262 | 0.00574194 | 0.008454233 | 1 |
| Obesity | pericyte | *ADAM33* | 2.956215394 | 6.15E-06 | 1.65E-05 | 0.038660336 |
| Obesity | pericyte | *TMC2* | 3.30923867 | 9.73E-07 | 3.45E-06 | 0.006115609 |
| Obesity | pericyte | *EML3* | 3.619571292 | 0.012661277 | 0.017780087 | 1 |
| Obesity | pericyte | *INCENP* | -2.8346529 | 0.00305665 | 0.004691689 | 1 |
| Obesity | pericyte | *MS4A3* | 41.6450484 | 1.33E-06 | 4.40E-06 | 0.008361558 |
| Obesity | pericyte | *CPSF7* | 7.534327198 | 2.34E-07 | 1.12E-06 | 0.001469483 |
| Obesity | pericyte | *CHEK1* | 1.301489505 | 0.017550969 | 0.024108137 | 1 |
| Obesity | pericyte | *FEZ1* | 3.132021616 | 5.47E-07 | 2.21E-06 | 0.003436878 |
| Obesity | pericyte | *COMMD7* | 7.911384393 | 3.11E-05 | 6.94E-05 | 0.19522134 |
| Obesity | pericyte | *KIAA1755* | 1.680343094 | 1.84E-05 | 4.35E-05 | 0.115449878 |
| Obesity | pericyte | *CNBD2* | 2.831669051 | 4.35E-07 | 1.85E-06 | 0.002732316 |
| Obesity | pericyte | *TRPT1* | -4.472275262 | 6.54E-05 | 0.000135469 | 0.41114866 |
| Obesity | pericyte | *TBX6* | 1.718016086 | 0.000133467 | 0.000259786 | 0.839109768 |
| Obesity | pericyte | *DOC2A* | -2.594450332 | 0.003476656 | 0.005282398 | 1 |
| Obesity | pericyte | *CWC15* | 2.165452106 | 0.030082162 | 0.039933816 | 1 |
| Obesity | pericyte | *ARID5B* | 4.801936414 | 4.35E-08 | 3.34E-07 | 0.000273245 |
| Obesity | pericyte | *TMCO3* | 7.02079717 | 1.00E-08 | 1.71E-07 | 6.32E-05 |
| Obesity | pericyte | *LYPD6B* | 3.00949385 | 3.35E-06 | 9.69E-06 | 0.021074579 |
| Obesity | pericyte | *PDCD4* | 6.229492002 | 2.97E-08 | 2.62E-07 | 0.000186872 |
| Obesity | pericyte | *WDR17* | 4.637961199 | 9.33E-07 | 3.34E-06 | 0.005866562 |
| Obesity | pericyte | *FSIP1* | 2.623759562 | 7.24E-07 | 2.74E-06 | 0.004552991 |
| Obesity | pericyte | *RGS18* | 2.329742227 | 4.91E-07 | 2.03E-06 | 0.003087685 |
| Obesity | pericyte | *PRSS23* | 3.323597866 | 3.66E-06 | 1.05E-05 | 0.022998542 |
| Obesity | pericyte | *MTMR12* | 4.928041816 | 7.21E-08 | 4.73E-07 | 0.000453132 |
| Obesity | pericyte | *PPP1R1C* | 1.841977196 | 0.001436545 | 0.002328321 | 1 |
| Obesity | pericyte | *C11orf53* | -8.397329007 | 6.26E-05 | 0.000130111 | 0.393714497 |
| Obesity | pericyte | *CCT5* | 5.701871298 | 6.08E-08 | 4.21E-07 | 0.000382274 |
| Obesity | pericyte | *FAM173B* | 2.960648191 | 7.70E-07 | 2.88E-06 | 0.004838714 |
| Obesity | pericyte | *TEX12* | 14.31780612 | 1.10E-06 | 3.79E-06 | 0.006923781 |
| Obesity | pericyte | *PIP4K2A* | 4.167196726 | 0.000103805 | 0.000206984 | 0.65262195 |
| Obesity | pericyte | *CRIM1* | 4.2573443 | 1.21E-06 | 4.08E-06 | 0.007630226 |
| Obesity | pericyte | *RILPL2* | 3.770657464 | 6.82E-08 | 4.52E-07 | 0.00042858 |
| Obesity | pericyte | *DHX37* | -2.463019673 | 0.015960025 | 0.022059025 | 1 |
| Obesity | pericyte | *PRSS53* | -3.185887378 | 0.001165904 | 0.001908865 | 1 |
| Obesity | pericyte | *GPR158* | 1.717330734 | 9.11E-05 | 0.000184077 | 0.573031745 |
| Obesity | pericyte | *THRB* | 5.103456303 | 6.42E-07 | 2.49E-06 | 0.004036185 |
| Obesity | pericyte | *UBE3B* | 3.601712747 | 3.55E-08 | 2.92E-07 | 0.000223027 |
| Obesity | pericyte | *ANK3* | 2.046143303 | 2.29E-07 | 1.10E-06 | 0.001438743 |
| Obesity | pericyte | *IPMK* | 4.6027095 | 1.49E-08 | 1.88E-07 | 9.35E-05 |
| Obesity | pericyte | *PLBD2* | 3.426207652 | 0.000545522 | 0.000949266 | 1 |
| Obesity | pericyte | *GXYLT1* | 6.964922016 | 2.83E-08 | 2.53E-07 | 0.000177818 |
| Obesity | pericyte | *FAM177A1* | 7.448236069 | 4.36E-07 | 1.85E-06 | 0.002742699 |
| Obesity | pericyte | *MIPOL1* | 5.042294336 | 8.76E-09 | 1.70E-07 | 5.51E-05 |
| Obesity | pericyte | *EXT2* | 2.228133431 | 6.13E-05 | 0.000127673 | 0.385316516 |

| | | | | | | |
|---|---|---|---|---|---|---|
| Obesity | pericyte | *TMEM18* | 10.30040073 | 1.77E-08 | 2.01E-07 | 0.00011152 |
| Obesity | pericyte | *THRSP* | 1.721399646 | 1.48E-05 | 3.60E-05 | 0.093290693 |
| Obesity | pericyte | *NDUFC2* | 3.918157413 | 4.01E-06 | 1.14E-05 | 0.025193915 |
| Obesity | pericyte | *ME3* | 3.651062137 | 9.61E-08 | 5.84E-07 | 0.000604023 |
| Obesity | pericyte | *ADAMTS12* | -0.993611832 | 0.000411984 | 0.0007323 | 1 |
| Obesity | pericyte | *VIPAS39* | 5.236213962 | 0.000146623 | 0.000283637 | 0.921818655 |
| Obesity | pericyte | *ANKRD50* | 5.727686528 | 4.65E-09 | 1.27E-07 | 2.92E-05 |
| Obesity | pericyte | *C4orf33* | 5.647730649 | 3.23E-05 | 7.18E-05 | 0.202942446 |
| Obesity | pericyte | *PTPRO* | 2.625193703 | 3.00E-07 | 1.37E-06 | 0.001886321 |
| Obesity | pericyte | *VPS26B* | 4.299620714 | 0.008160425 | 0.011729446 | 1 |
| Obesity | pericyte | *NCAPD3* | 5.671704708 | 5.15E-06 | 1.42E-05 | 0.032349463 |
| Obesity | pericyte | *VTI1A* | 4.985131701 | 3.20E-07 | 1.44E-06 | 0.002013224 |
| Obesity | pericyte | *FAM160B1* | 7.159405343 | 5.02E-08 | 3.72E-07 | 0.000315758 |
| Obesity | pericyte | *ANO4* | -0.570225101 | 6.89E-05 | 0.000142047 | 0.432960075 |
| Obesity | pericyte | *TEX9* | 4.651678281 | 2.47E-08 | 2.32E-07 | 0.000155096 |
| Obesity | pericyte | *AKR1C2* | 2.091415856 | 4.73E-07 | 1.98E-06 | 0.002976337 |
| Obesity | pericyte | *DPYSL4* | 1.281561921 | 4.02E-05 | 8.77E-05 | 0.252890811 |
| Obesity | pericyte | *ADAM8* | 2.567719788 | 1.94E-07 | 9.64E-07 | 0.001217926 |
| Obesity | pericyte | *ITIH2* | 8.037759501 | 5.25E-07 | 2.14E-06 | 0.003302922 |
| Obesity | pericyte | *KIN* | 4.387792897 | 8.15E-06 | 2.12E-05 | 0.051213271 |
| Obesity | pericyte | *PIGF* | 5.884164581 | 7.30E-07 | 2.76E-06 | 0.004587765 |
| Obesity | pericyte | *MFSD6* | 7.493194707 | 1.98E-08 | 2.11E-07 | 0.00012422 |
| Obesity | pericyte | *RNF144A* | 3.715284955 | 9.41E-08 | 5.76E-07 | 0.000591352 |
| Obesity | pericyte | *ADAM17* | 6.136249201 | 2.27E-08 | 2.24E-07 | 0.000142664 |
| Obesity | pericyte | *TMEM45B* | -4.034438377 | 0.000166809 | 0.000318568 | 1 |
| Obesity | pericyte | *CENPU* | 1.734212308 | 0.001322855 | 0.002152378 | 1 |
| Obesity | pericyte | *BICD1* | 5.378963605 | 5.10E-05 | 0.000108662 | 0.320771556 |
| Obesity | pericyte | *GUF1* | 4.015838258 | 0.000899013 | 0.001498403 | 1 |
| Obesity | pericyte | *CENPJ* | 3.812020231 | 1.43E-05 | 3.48E-05 | 0.089820492 |
| Obesity | pericyte | *PARP8* | 1.878221148 | 0.005210648 | 0.007722617 | 1 |
| Obesity | pericyte | *CACUL1* | 4.577382269 | 1.32E-08 | 1.75E-07 | 8.31E-05 |
| Obesity | pericyte | *RABGAP1L* | 5.581852848 | 2.67E-09 | 9.26E-08 | 1.68E-05 |
| Obesity | pericyte | *PTPN14* | 3.848896271 | 2.02E-05 | 4.75E-05 | 0.12719369 |
| Obesity | pericyte | *RNF219* | 6.178692757 | 0.000341568 | 0.00061408 | 1 |
| Obesity | pericyte | *EPG5* | 3.457482697 | 2.60E-06 | 7.79E-06 | 0.01635837 |
| Obesity | pericyte | *HAUS1* | 1.628851681 | 0.005400961 | 0.00798961 | 1 |
| Obesity | pericyte | *G6PC2* | 105.4079007 | 6.17E-07 | 2.42E-06 | 0.003879139 |
| Obesity | pericyte | *TGOLN2* | 3.699897218 | 0.010601486 | 0.015038706 | 1 |
| Obesity | pericyte | *TADA1* | 9.903619875 | 1.89E-08 | 2.07E-07 | 0.000118812 |
| Obesity | pericyte | *GUCY1A2* | -0.435273951 | 3.19E-05 | 7.11E-05 | 0.200757736 |
| Obesity | pericyte | *CWF19L2* | 6.600037691 | 2.26E-08 | 2.24E-07 | 0.000141896 |
| Obesity | pericyte | *ZNF547* | 3.665088597 | 2.09E-06 | 6.45E-06 | 0.013129341 |
| Obesity | pericyte | *CCDC50* | 2.137352437 | 0.000126042 | 0.000247015 | 0.792425576 |
| Obesity | pericyte | *ZFP36L2* | 5.880276382 | 2.38E-09 | 8.62E-08 | 1.50E-05 |
| Obesity | pericyte | *TMEM123* | 7.329615692 | 8.48E-07 | 3.09E-06 | 0.0053303 |
| Obesity | pericyte | *IGSF10* | 2.022831404 | 0.034704651 | 0.045665161 | 1 |
| Obesity | pericyte | *SPEF2* | 3.351100085 | 5.17E-05 | 0.000109716 | 0.324760573 |
| Obesity | pericyte | *MBNL1* | 4.010491801 | 9.98E-06 | 2.53E-05 | 0.062725348 |
| Obesity | pericyte | *CLEC4F* | 5.988820077 | 7.82E-07 | 2.91E-06 | 0.004914294 |
| Obesity | pericyte | *RASGRP3* | 1.832351457 | 6.06E-05 | 0.000126467 | 0.381171167 |

| | | | | | | |
|---|---|---|---|---|---|---|
| Obesity | pericyte | *SAR1B* | 4.091678705 | 5.94E-08 | 4.15E-07 | 0.000373259 |
| Obesity | pericyte | *TCTEX1D1* | 1.435957095 | 0.002913796 | 0.004488859 | 1 |
| Obesity | pericyte | *IFIT5* | 2.347757681 | 0.000727172 | 0.001234936 | 1 |
| Obesity | pericyte | *ZNF117* | 0.791015992 | 0.000172924 | 0.000329148 | 1 |
| Obesity | pericyte | *RAB3C* | 3.238375384 | 5.43E-06 | 1.48E-05 | 0.034117523 |
| Obesity | pericyte | *RAD17* | 1.228254236 | 0.032987912 | 0.043552079 | 1 |
| Obesity | pericyte | *MED21* | 6.605164501 | 7.64E-09 | 1.63E-07 | 4.80E-05 |
| Obesity | pericyte | *CENPH* | 2.491027713 | 2.15E-06 | 6.59E-06 | 0.013486561 |
| Obesity | pericyte | *CARHSP1* | 4.306068149 | 7.58E-08 | 4.88E-07 | 0.000476331 |
| Obesity | pericyte | *TEKT5* | 5.285123901 | 6.32E-07 | 2.46E-06 | 0.003970443 |
| Obesity | pericyte | *BANK1* | 5.834255877 | 7.38E-09 | 1.60E-07 | 4.64E-05 |
| Obesity | pericyte | *CAST* | 1.677441604 | 0.014874519 | 0.02065285 | 1 |
| Obesity | pericyte | *SCOC* | 3.226769341 | 0.009227651 | 0.013179064 | 1 |
| Obesity | pericyte | *CLGN* | 12.49993008 | 4.68E-07 | 1.96E-06 | 0.002940912 |
| Obesity | pericyte | *SYCP2L* | 4.05569753 | 2.43E-05 | 5.60E-05 | 0.152807849 |
| Obesity | pericyte | *AHCTF1* | 6.852768644 | 1.12E-08 | 1.71E-07 | 7.04E-05 |
| Obesity | pericyte | *MERTK* | 0.340139038 | 0.037699976 | 0.049296952 | 1 |
| Obesity | pericyte | *PTPRR* | -2.375134743 | 0.011802078 | 0.016636696 | 1 |
| Obesity | pericyte | *PLA2R1* | 2.550619134 | 3.73E-07 | 1.63E-06 | 0.002346186 |
| Obesity | pericyte | *FRMD1* | 2.404027659 | 1.02E-06 | 3.58E-06 | 0.006413704 |
| Obesity | pericyte | *INO80C* | 8.214684762 | 1.11E-08 | 1.71E-07 | 6.96E-05 |
| Obesity | pericyte | *LPCAT1* | 7.591386207 | 9.22E-09 | 1.70E-07 | 5.80E-05 |
| Obesity | pericyte | *C16orf89* | 1.850791126 | 0.006557956 | 0.009552797 | 1 |
| Obesity | pericyte | *TEX29* | 3.065398367 | 9.00E-07 | 3.24E-06 | 0.005656684 |
| Obesity | pericyte | *ADPRHL1* | 1.245876955 | 0.026187659 | 0.035119841 | 1 |
| Obesity | pericyte | *LURAP1L* | 1.692922424 | 6.84E-06 | 1.82E-05 | 0.043013154 |
| Obesity | pericyte | *CFDP1* | 6.257232435 | 7.02E-07 | 2.67E-06 | 0.004412551 |
| Obesity | pericyte | *ZDHHC7* | 5.082501545 | 1.50E-08 | 1.88E-07 | 9.44E-05 |
| Obesity | pericyte | *FAM92B* | 26.81707631 | 1.17E-06 | 3.94E-06 | 0.007329485 |
| Obesity | pericyte | *JAZF1* | 6.409851638 | 1.77E-10 | 1.48E-08 | 1.11E-06 |
| Obesity | pericyte | *FBXO36* | 4.776409381 | 0.006076871 | 0.008920216 | 1 |
| Obesity | pericyte | *MCOLN2* | 2.07991956 | 1.12E-05 | 2.79E-05 | 0.070172583 |
| Obesity | pericyte | *CACNA2D1* | 4.353547001 | 3.24E-08 | 2.79E-07 | 0.000203955 |
| Obesity | pericyte | *ZUFSP* | 5.844049389 | 7.65E-08 | 4.90E-07 | 0.000480695 |
| Obesity | pericyte | *HS3ST3A1* | -1.238046535 | 0.002921136 | 0.004499065 | 1 |
| Obesity | pericyte | *PPP2R5E* | 3.808197685 | 4.94E-05 | 0.000105724 | 0.310404834 |
| Obesity | pericyte | *THY1* | -2.141400999 | 0.002438949 | 0.003802995 | 1 |
| Obesity | pericyte | *TBCEL* | 4.905069717 | 0.001937269 | 0.003065596 | 1 |
| Obesity | pericyte | *UBASH3B* | 2.767954833 | 1.25E-07 | 6.99E-07 | 0.000785822 |
| Obesity | pericyte | *ROBO3* | 2.837445653 | 1.36E-07 | 7.39E-07 | 0.000855486 |
| Obesity | pericyte | *CC2D1B* | 2.858457092 | 4.65E-07 | 1.95E-06 | 0.002925626 |
| Obesity | pericyte | *PRKCA* | 5.100327149 | 8.67E-05 | 0.000175411 | 0.545001646 |
| Obesity | pericyte | *LRRK1* | 6.354593006 | 1.11E-08 | 1.71E-07 | 6.97E-05 |
| Obesity | pericyte | *CEP112* | 4.185896689 | 6.88E-08 | 4.56E-07 | 0.000432672 |
| Obesity | pericyte | *GAL3ST2* | -4.306743338 | 0.02626245 | 0.03521263 | 1 |
| Obesity | pericyte | *ENPP3* | 1.689400273 | 1.44E-06 | 4.72E-06 | 0.00906726 |
| Obesity | pericyte | *UCHL1* | 9.338636487 | 4.88E-07 | 2.03E-06 | 0.003070131 |
| Obesity | pericyte | *MIA3* | 5.872029879 | 1.18E-06 | 3.97E-06 | 0.007405265 |
| Obesity | pericyte | *FAM167A* | 6.890648869 | 6.28E-06 | 1.69E-05 | 0.03946029 |
| Obesity | pericyte | *NEIL2* | 3.198287106 | 1.93E-08 | 2.08E-07 | 0.000121235 |

| | | | | | | |
|---|---|---|---|---|---|---|
| Obesity | pericyte | *CCSAP* | 3.937387865 | 0.007741965 | 0.011166261 | 1 |
| Obesity | pericyte | *GBP5* | 2.402502675 | 9.08E-08 | 5.60E-07 | 0.00057109 |
| Obesity | pericyte | *FAM69A* | 2.552500874 | 0.000258141 | 0.000475236 | 1 |
| Obesity | pericyte | *PDLIM3* | 3.532689344 | 4.35E-07 | 1.85E-06 | 0.002737865 |
| Obesity | pericyte | *NCAM2* | 2.056323532 | 3.27E-06 | 9.48E-06 | 0.020537843 |
| Obesity | pericyte | *L3MBTL4* | 4.967859677 | 4.56E-09 | 1.26E-07 | 2.87E-05 |
| Obesity | pericyte | *PDE1C* | -0.507396768 | 6.30E-05 | 0.0001308 | 0.395967651 |
| Obesity | pericyte | *SLFN13* | -1.212354061 | 6.78E-06 | 1.80E-05 | 0.042610876 |
| Obesity | pericyte | *FLCN* | 2.950698867 | 2.45E-08 | 2.32E-07 | 0.000154307 |
| Obesity | pericyte | *DPH3* | 4.203275201 | 1.07E-07 | 6.31E-07 | 0.000675845 |
| Obesity | pericyte | *PPP4R1* | 7.172374648 | 6.54E-08 | 4.42E-07 | 0.000411018 |
| Obesity | pericyte | *APCDD1* | 1.105170344 | 7.68E-05 | 0.000156734 | 0.482584455 |
| Obesity | pericyte | *MPPE1* | 1.336645145 | 0.00436502 | 0.006552742 | 1 |
| Obesity | pericyte | *EPHB1* | 3.659875813 | 7.00E-07 | 2.67E-06 | 0.004403069 |
| Obesity | pericyte | *ANKRD40* | 3.259820765 | 8.29E-07 | 3.04E-06 | 0.005209827 |
| Obesity | pericyte | *CYP2U1* | 9.681996544 | 1.63E-08 | 1.92E-07 | 0.000102433 |
| Obesity | pericyte | *FBXL18* | 8.902234241 | 6.81E-09 | 1.54E-07 | 4.28E-05 |
| Obesity | pericyte | *PROM2* | 5.97750797 | 0.000742657 | 0.001258174 | 1 |
| Obesity | pericyte | *AK9* | 5.056392334 | 3.24E-06 | 9.43E-06 | 0.020386728 |
| Obesity | pericyte | *CDK19* | 5.443408824 | 1.72E-05 | 4.11E-05 | 0.108427119 |
| Obesity | pericyte | *GTF3C6* | 3.270092928 | 0.007073028 | 0.010267404 | 1 |
| Obesity | pericyte | *AGPAT5* | 3.580843516 | 1.44E-07 | 7.70E-07 | 0.000905488 |
| Obesity | pericyte | *MMS19* | 7.349169349 | 2.35E-08 | 2.26E-07 | 0.00014751 |
| Obesity | pericyte | *GOLGA7B* | 3.326013756 | 0.000996494 | 0.001646506 | 1 |
| Obesity | pericyte | *TRMT44* | 8.656410858 | 4.92E-09 | 1.29E-07 | 3.09E-05 |
| Obesity | pericyte | *C16orf87* | 4.029204905 | 8.04E-07 | 2.97E-06 | 0.005056203 |
| Obesity | pericyte | *HEATR3* | 1.266033156 | 0.002619221 | 0.004065937 | 1 |
| Obesity | pericyte | *CNOT8* | 3.843256607 | 0.002068215 | 0.003258864 | 1 |
| Obesity | pericyte | *ZKSCAN2* | 1.958864289 | 1.15E-07 | 6.61E-07 | 0.000723277 |
| Obesity | pericyte | *PDIA4* | 10.04497642 | 5.01E-07 | 2.06E-06 | 0.003151907 |
| Obesity | pericyte | *KDM8* | 9.326353111 | 2.15E-06 | 6.60E-06 | 0.01352398 |
| Obesity | pericyte | *FAM126B* | 5.988323915 | 1.05E-07 | 6.18E-07 | 0.000657391 |
| Obesity | pericyte | *ALS2CR12* | 3.690522877 | 1.01E-07 | 6.05E-07 | 0.000636619 |
| Obesity | pericyte | *SPAG17* | 4.900773243 | 3.47E-05 | 7.67E-05 | 0.218263033 |
| Obesity | pericyte | *DEPTOR* | 6.045580875 | 0.000132037 | 0.00025732 | 0.830114238 |
| Obesity | pericyte | *RRAGA* | 6.537473608 | 1.58E-06 | 5.09E-06 | 0.009914771 |
| Obesity | pericyte | *TMBIM4* | 1.863486554 | 0.006487718 | 0.009457057 | 1 |
| Obesity | pericyte | *FAM161B* | 3.746356793 | 7.15E-08 | 4.70E-07 | 0.000449447 |
| Obesity | pericyte | *WIF1* | 4.259841622 | 0.014857327 | 0.020633535 | 1 |
| Obesity | pericyte | *UGT2B4* | 3.126073386 | 1.06E-06 | 3.68E-06 | 0.006644902 |
| Obesity | pericyte | *KCNMA1* | 3.255257905 | 5.71E-05 | 0.000119828 | 0.359243926 |
| Obesity | pericyte | *BATF* | 2.570721086 | 1.06E-06 | 3.68E-06 | 0.006654948 |
| Obesity | pericyte | *DCK* | 3.257627284 | 0.01276016 | 0.017902951 | 1 |
| Obesity | pericyte | *DPY19L4* | 4.502026221 | 0.000157076 | 0.00030163 | 0.987535171 |
| Obesity | pericyte | *DRAM2* | 3.687248285 | 0.000329095 | 0.000593522 | 1 |
| Obesity | pericyte | *C8orf37* | 2.109909005 | 0.001368594 | 0.002222198 | 1 |
| Obesity | pericyte | *ADAMTSL3* | 1.13486689 | 0.016809953 | 0.02314083 | 1 |
| Obesity | pericyte | *N6AMT1* | 0.888061099 | 0.027659906 | 0.036975936 | 1 |
| Obesity | pericyte | *ANKRD9* | 2.192079675 | 8.20E-06 | 2.13E-05 | 0.051576078 |
| Obesity | pericyte | *SFXN2* | 2.147585645 | 2.07E-06 | 6.40E-06 | 0.012991707 |

| | | | | | | |
|---|---|---|---|---|---|---|
| Obesity | pericyte | *GDF6* | 4.58007516 | 1.48E-05 | 3.60E-05 | 0.093129541 |
| Obesity | pericyte | *NODAL* | 4.776686507 | 2.18E-07 | 1.06E-06 | 0.001368205 |
| Obesity | pericyte | *WDYHV1* | 1.972035801 | 0.020567625 | 0.027946544 | 1 |
| Obesity | pericyte | *ATAD2* | 6.511111605 | 1.16E-08 | 1.72E-07 | 7.30E-05 |
| Obesity | pericyte | *SASS6* | 4.591811633 | 9.00E-06 | 2.31E-05 | 0.056609991 |
| Obesity | pericyte | *ITGAD* | 1.255735074 | 0.005010542 | 0.007440075 | 1 |
| Obesity | pericyte | *VPS8* | 4.250478448 | 1.72E-06 | 5.48E-06 | 0.010801586 |
| Obesity | pericyte | *RPUSD3* | 9.669894198 | 1.88E-05 | 4.45E-05 | 0.11836286 |
| Obesity | pericyte | *GHRL* | 6.446273037 | 1.60E-08 | 1.91E-07 | 0.000100799 |
| Obesity | pericyte | *EXOG* | 5.824624736 | 2.64E-06 | 7.86E-06 | 0.016584943 |
| Obesity | pericyte | *NTAN1* | 5.471488383 | 2.24E-07 | 1.08E-06 | 0.001409491 |
| Obesity | pericyte | *SLC6A1* | -1.241954873 | 9.33E-05 | 0.000187661 | 0.586627078 |
| Obesity | pericyte | *FCHO2* | 5.679456313 | 3.23E-06 | 9.40E-06 | 0.020287819 |
| Obesity | pericyte | *TIMP4* | -2.307220994 | 4.79E-05 | 0.000102777 | 0.300932471 |
| Obesity | pericyte | *CPT2* | 6.978708945 | 1.13E-07 | 6.53E-07 | 0.000710068 |
| Obesity | pericyte | *NECAP2* | 7.527270212 | 1.07E-05 | 2.69E-05 | 0.067335484 |
| Obesity | pericyte | *LRP8* | 6.349892001 | 1.57E-06 | 5.06E-06 | 0.009858318 |
| Obesity | pericyte | *GATAD1* | 3.274315131 | 5.53E-07 | 2.23E-06 | 0.00347883 |
| Obesity | pericyte | *SUSD3* | 1.805603755 | 1.65E-05 | 3.94E-05 | 0.10346255 |
| Obesity | pericyte | *TMED6* | -5.125935947 | 7.34E-05 | 0.000150567 | 0.461337394 |
| Obesity | pericyte | *CLEC18A* | -7.202784137 | 1.87E-19 | 1.47E-16 | 1.17E-15 |
| Obesity | pericyte | *AASDH* | 4.313697685 | 9.76E-06 | 2.48E-05 | 0.061336658 |
| Obesity | pericyte | *RNF111* | 4.704053241 | 9.96E-09 | 1.71E-07 | 6.26E-05 |
| Obesity | pericyte | *DYRK1A* | 4.539330134 | 8.09E-05 | 0.000164713 | 0.508902116 |
| Obesity | pericyte | *KCNJ15* | 1.757947493 | 0.000626254 | 0.001078997 | 1 |
| Obesity | pericyte | *CREB3L1* | 3.44201395 | 3.46E-06 | 9.98E-06 | 0.021753265 |
| Obesity | pericyte | *SLC38A10* | 4.871479949 | 3.72E-08 | 3.02E-07 | 0.000234004 |
| Obesity | pericyte | *C9orf43* | 2.300713024 | 0.000467227 | 0.000822357 | 1 |
| Obesity | pericyte | *WDR19* | 3.371208745 | 6.23E-08 | 4.28E-07 | 0.00039174 |
| Obesity | pericyte | *SLC37A3* | 7.245574626 | 3.22E-07 | 1.45E-06 | 0.002026368 |
| Obesity | pericyte | *SPPL3* | 4.955087172 | 1.14E-06 | 3.88E-06 | 0.007180491 |
| Obesity | pericyte | *FAM213B* | 1.238879437 | 0.027715134 | 0.037034016 | 1 |
| Obesity | pericyte | *PEX10* | 4.073059324 | 3.33E-07 | 1.49E-06 | 0.002094388 |
| Obesity | pericyte | *RADIL* | -1.921616036 | 0.03537066 | 0.046458399 | 1 |
| Obesity | pericyte | *LDLRAP1* | 7.792008943 | 2.24E-08 | 2.24E-07 | 0.000140921 |
| Obesity | pericyte | *PAFAH2* | 2.780832534 | 9.78E-05 | 0.000196119 | 0.615028102 |
| Obesity | pericyte | *TRIM63* | 1.772532095 | 0.000953888 | 0.001582346 | 1 |
| Obesity | pericyte | *GALNT14* | -0.585225925 | 1.97E-05 | 4.64E-05 | 0.123802874 |
| Obesity | pericyte | *HPD* | 3.241898592 | 2.30E-11 | 2.84E-09 | 1.45E-07 |
| Obesity | pericyte | *TPRG1L* | 5.972202414 | 3.46E-06 | 9.97E-06 | 0.021724013 |
| Obesity | pericyte | *LRRC43* | -7.469865337 | 0.033430103 | 0.044088554 | 1 |
| Obesity | pericyte | *XKR8* | 3.247958921 | 6.58E-07 | 2.53E-06 | 0.004134225 |
| Obesity | pericyte | *EYA3* | 4.954113612 | 1.56E-07 | 8.16E-07 | 0.00097955 |
| Obesity | pericyte | *MRAS* | 1.625624947 | 9.28E-05 | 0.000186874 | 0.58360777 |
| Obesity | pericyte | *ABHD3* | 2.814387689 | 3.94E-06 | 1.12E-05 | 0.024794428 |
| Obesity | pericyte | *CLSTN2* | 2.088276246 | 1.06E-06 | 3.68E-06 | 0.006663193 |
| Obesity | pericyte | *COLEC12* | 4.013183033 | 1.48E-08 | 1.87E-07 | 9.28E-05 |
| Obesity | pericyte | *RNF207* | 5.235536726 | 9.75E-08 | 5.90E-07 | 0.000613104 |
| Obesity | pericyte | *AUTS2* | 3.987280296 | 5.17E-05 | 0.000109716 | 0.324732744 |
| Obesity | pericyte | *MITD1* | 3.625453686 | 0.000603633 | 0.001042022 | 1 |

| | | | | | | |
|---|---|---|---|---|---|---|
| Obesity | pericyte | *TSPAN33* | 0.883872778 | 0.018597822 | 0.025451568 | 1 |
| Obesity | pericyte | *AHCYL2* | 4.869849534 | 7.04E-06 | 1.86E-05 | 0.044289209 |
| Obesity | pericyte | *B4GALT5* | 4.947618542 | 5.00E-08 | 3.71E-07 | 0.000314344 |
| Obesity | pericyte | *CD1D* | -3.524196974 | 1.06E-05 | 2.67E-05 | 0.066815902 |
| Obesity | pericyte | *FAM86C1* | 10.46976461 | 1.99E-07 | 9.84E-07 | 0.00124969 |
| Obesity | pericyte | *CD1E* | 7.072527774 | 6.50E-07 | 2.51E-06 | 0.004089616 |
| Obesity | pericyte | *NCF1* | -3.961955384 | 0.010857507 | 0.015381061 | 1 |
| Obesity | pericyte | *PPP1R9A* | 2.291635167 | 5.15E-07 | 2.10E-06 | 0.003234707 |
| Obesity | pericyte | *ZFAND2B* | 3.149487144 | 0.001953307 | 0.003088643 | 1 |
| Obesity | pericyte | *GDPD5* | 3.629181297 | 2.23E-08 | 2.23E-07 | 0.000139979 |
| Obesity | pericyte | *DYNC1I1* | 5.838738829 | 0.000740473 | 0.001255151 | 1 |
| Obesity | pericyte | *TMED4* | 6.940913226 | 8.76E-09 | 1.70E-07 | 5.51E-05 |
| Obesity | pericyte | *ELK4* | 4.072409867 | 0.011473767 | 0.016196129 | 1 |
| Obesity | pericyte | *DUSP23* | 10.62172346 | 1.93E-08 | 2.08E-07 | 0.000121311 |
| Obesity | pericyte | *RNF166* | -2.215111151 | 0.0249711 | 0.033595828 | 1 |
| Obesity | pericyte | *F11R* | 2.17760567 | 0.000627247 | 0.001080115 | 1 |
| Obesity | pericyte | *SPATA2L* | 4.997513747 | 5.36E-05 | 0.000113522 | 0.337273185 |
| Obesity | pericyte | *NIT1* | 2.239201322 | 0.000776734 | 0.00131356 | 1 |
| Obesity | pericyte | *ZNF276* | 4.240919748 | 2.31E-05 | 5.34E-05 | 0.145033133 |
| Obesity | pericyte | *B4GALT3* | 4.117118562 | 6.07E-06 | 1.63E-05 | 0.038142939 |
| Obesity | pericyte | *DMTN* | 2.09494183 | 8.40E-06 | 2.17E-05 | 0.052795903 |
| Obesity | pericyte | *FAM160B2* | 4.942940846 | 2.03E-06 | 6.30E-06 | 0.012741944 |
| Obesity | pericyte | *NDUFS2* | 2.850767379 | 0.000520382 | 0.000908536 | 1 |
| Obesity | pericyte | *SLC5A11* | -0.995263148 | 0.000197425 | 0.000371731 | 1 |
| Obesity | pericyte | *FCER1G* | 2.454100913 | 9.67E-07 | 3.43E-06 | 0.006080715 |
| Obesity | pericyte | *CCAR2* | 6.533505963 | 0.000279633 | 0.000511061 | 1 |
| Obesity | pericyte | *CDC42SE2* | 6.023718484 | 4.07E-08 | 3.21E-07 | 0.000255816 |
| Obesity | pericyte | *RAPGEF6* | 5.653440211 | 7.84E-06 | 2.04E-05 | 0.049318282 |
| Obesity | pericyte | *EPB41* | 2.356063808 | 1.33E-08 | 1.76E-07 | 8.38E-05 |
| Obesity | pericyte | *ALG8* | 6.851863967 | 1.40E-05 | 3.41E-05 | 0.087855897 |
| Obesity | pericyte | *IFNAR2* | 2.179088764 | 0.00019791 | 0.000372422 | 1 |
| Obesity | pericyte | *MRPL10* | 1.71513052 | 0.018931306 | 0.025874157 | 1 |
| Obesity | pericyte | *SV2A* | 8.251207992 | 8.61E-06 | 2.22E-05 | 0.054155596 |
| Obesity | pericyte | *UBE2Z* | 5.067463249 | 1.93E-05 | 4.56E-05 | 0.121431643 |
| Obesity | pericyte | *CLIC6* | 2.676513272 | 6.16E-07 | 2.42E-06 | 0.003872538 |
| Obesity | pericyte | *HLCS* | 0.874551661 | 0.033135439 | 0.043728486 | 1 |
| Obesity | pericyte | *ARHGAP27* | 5.190370396 | 1.87E-08 | 2.06E-07 | 0.000117291 |
| Obesity | pericyte | *PADI4* | 11.78564367 | 5.64E-06 | 1.53E-05 | 0.035430315 |
| Obesity | pericyte | *STARD9* | 3.322181569 | 0.002117925 | 0.003329754 | 1 |
| Obesity | pericyte | *THEM4* | 3.829948067 | 8.84E-06 | 2.28E-05 | 0.055565932 |
| Obesity | pericyte | *TCHH* | -5.294412358 | 0.000152223 | 0.000293475 | 0.957023253 |
| Obesity | pericyte | *MED8* | 2.801573213 | 9.59E-06 | 2.45E-05 | 0.060292994 |
| Obesity | pericyte | *RSPRY1* | 3.258120103 | 0.01306912 | 0.01828746 | 1 |
| Obesity | pericyte | *CCDC17* | 2.945560649 | 5.74E-05 | 0.000120228 | 0.360741225 |
| Obesity | pericyte | *ACE* | 7.29517387 | 4.97E-06 | 1.37E-05 | 0.031249299 |
| Obesity | pericyte | *TEPP* | -9.955161544 | 6.54E-06 | 1.75E-05 | 0.041132477 |
| Obesity | pericyte | *EFCAB14* | 6.980244956 | 1.12E-08 | 1.71E-07 | 7.06E-05 |
| Obesity | pericyte | *ZFYVE28* | 4.723132399 | 1.35E-08 | 1.77E-07 | 8.48E-05 |
| Obesity | pericyte | *C16orf86* | -1.786629293 | 0.002657267 | 0.00411991 | 1 |
| Obesity | pericyte | *CCDC117* | 6.183219035 | 7.57E-08 | 4.87E-07 | 0.000475772 |

| | | | | | | |
|---|---|---|---|---|---|---|
| Obesity | pericyte | *NPR2* | 7.156379623 | 9.78E-06 | 2.49E-05 | 0.061502054 |
| Obesity | pericyte | *ZNF235* | 4.293736856 | 6.44E-07 | 2.49E-06 | 0.004045702 |
| Obesity | pericyte | *CALM3* | 5.264656838 | 0.000436839 | 0.000773418 | 1 |
| Obesity | pericyte | *DFFA* | 3.706462724 | 1.07E-09 | 5.30E-08 | 6.75E-06 |
| Obesity | pericyte | *ATAD3B* | 4.932587408 | 3.04E-05 | 6.83E-05 | 0.191324967 |
| Obesity | pericyte | *SSU72* | 8.166224289 | 3.82E-08 | 3.07E-07 | 0.00024038 |
| Obesity | pericyte | *NR2F6* | 4.171200742 | 0.000364427 | 0.000652564 | 1 |
| Obesity | pericyte | *CILP2* | 14.04676276 | 4.99E-05 | 0.000106559 | 0.313603538 |
| Obesity | pericyte | *RSPH1* | -1.806033474 | 0.007444796 | 0.010772994 | 1 |
| Obesity | pericyte | *NDUFV3* | 0.553150582 | 0.01308751 | 0.018309118 | 1 |
| Obesity | pericyte | *HSF2BP* | -0.831706382 | 0.000130807 | 0.00025532 | 0.822386131 |
| Obesity | pericyte | *ZNF66* | 5.577504917 | 3.03E-06 | 8.89E-06 | 0.019049608 |
| Obesity | pericyte | *ITGB2* | 1.474013508 | 0.000430694 | 0.000763613 | 1 |
| Obesity | pericyte | *FAM207A* | 3.575489013 | 3.33E-08 | 2.84E-07 | 0.000209593 |
| Obesity | pericyte | *LSS* | 3.101965935 | 4.10E-09 | 1.18E-07 | 2.58E-05 |
| Obesity | pericyte | *VAV2* | 5.483702407 | 5.98E-07 | 2.37E-06 | 0.003762016 |
| Obesity | pericyte | *PCNT* | 1.915124883 | 0.000188125 | 0.000356033 | 1 |
| Obesity | pericyte | *DIP2A* | 3.525314093 | 8.45E-05 | 0.000171292 | 0.531005645 |
| Obesity | pericyte | *PRMT2* | 3.250529858 | 3.17E-05 | 7.06E-05 | 0.199003103 |
| Obesity | pericyte | *ADAMTS13* | 2.07341502 | 2.94E-05 | 6.62E-05 | 0.184526915 |
| Obesity | pericyte | *ZNF714* | 4.073694742 | 0.00012305 | 0.000241604 | 0.77361533 |
| Obesity | pericyte | *GPSM1* | 3.497322143 | 8.31E-06 | 2.15E-05 | 0.052213747 |
| Obesity | pericyte | *C19orf47* | 8.518978355 | 1.56E-10 | 1.34E-08 | 9.79E-07 |
| Obesity | pericyte | *RDH13* | 7.708458723 | 1.96E-06 | 6.13E-06 | 0.012332744 |
| Obesity | pericyte | *MPZL3* | 3.820801004 | 3.16E-07 | 1.43E-06 | 0.001985806 |
| Obesity | pericyte | *PCSK7* | 5.911944339 | 8.17E-07 | 3.01E-06 | 0.005136026 |
| Obesity | pericyte | *S100A1* | 5.593046352 | 1.12E-05 | 2.80E-05 | 0.070479849 |
| Obesity | pericyte | *ZBTB7B* | 3.292975834 | 0.000594365 | 0.001026585 | 1 |
| Obesity | pericyte | *VPS11* | 3.10609472 | 0.00019724 | 0.000371605 | 1 |
| Obesity | pericyte | *FDPS* | 4.304623515 | 5.29E-05 | 0.000111976 | 0.332344748 |
| Obesity | pericyte | *PMF1* | 5.451501078 | 3.73E-08 | 3.02E-07 | 0.00023455 |
| Obesity | pericyte | *NBEAL2* | 7.468578585 | 8.38E-09 | 1.70E-07 | 5.27E-05 |
| Obesity | pericyte | *FCRL3* | 1.493526631 | 0.000305547 | 0.000554554 | 1 |
| Obesity | pericyte | *NACC1* | 4.825494345 | 1.36E-07 | 7.38E-07 | 0.000853633 |
| Obesity | pericyte | *LY6K* | -3.143475235 | 3.09E-05 | 6.92E-05 | 0.194504036 |
| Obesity | pericyte | *LY6E* | 2.233791597 | 0.012604182 | 0.017703864 | 1 |
| Obesity | pericyte | *VPS28* | 2.589086336 | 0.000732054 | 0.00124155 | 1 |
| Obesity | pericyte | *TONSL* | 6.589181556 | 3.59E-06 | 1.03E-05 | 0.0225706 |
| Obesity | pericyte | *MUM1* | 2.749962794 | 0.000731233 | 0.001240492 | 1 |
| Obesity | pericyte | *PPP1R16A* | 1.634102119 | 8.09E-05 | 0.00016466 | 0.508471267 |
| Obesity | pericyte | *RPL8* | 7.450292562 | 4.99E-07 | 2.06E-06 | 0.003136581 |
| Obesity | pericyte | *LRWD1* | -1.277252914 | 3.64E-05 | 8.02E-05 | 0.229076342 |
| Obesity | pericyte | *FBXL13* | 6.875078621 | 1.88E-07 | 9.41E-07 | 0.001180244 |
| Obesity | pericyte | *MFSD12* | 4.797109306 | 2.53E-08 | 2.36E-07 | 0.00015878 |
| Obesity | pericyte | *YDJC* | 4.762461587 | 8.42E-07 | 3.08E-06 | 0.005292542 |
| Obesity | pericyte | *DVL3* | 5.967394669 | 6.13E-05 | 0.000127745 | 0.385661835 |
| Obesity | pericyte | *AP2M1* | 6.859537352 | 2.65E-08 | 2.41E-07 | 0.000166353 |
| Obesity | pericyte | *FBXO27* | 1.545781505 | 2.13E-05 | 4.96E-05 | 0.133612644 |
| Obesity | pericyte | *DMKN* | -0.970844381 | 8.54E-05 | 0.000173046 | 0.536787803 |
| Obesity | pericyte | *BDH1* | 6.671157834 | 3.89E-08 | 3.11E-07 | 0.000244522 |

| | | | | | | |
|---|---|---|---|---|---|---|
| Obesity | pericyte | *COX7A1* | 2.126160537 | 0.000279351 | 0.000510696 | 1 |
| Obesity | pericyte | *DUSP14* | 2.305886295 | 0.004650228 | 0.006946064 | 1 |
| Obesity | pericyte | *LRRC56* | 1.890530617 | 0.000976118 | 0.00161709 | 1 |
| Obesity | pericyte | *IKZF3* | 2.704243874 | 2.96E-07 | 1.35E-06 | 0.001858944 |
| Obesity | pericyte | *ACOX1* | 4.738852896 | 2.24E-08 | 2.24E-07 | 0.000140616 |
| Obesity | pericyte | *PRPSAP1* | 4.926835348 | 3.97E-08 | 3.16E-07 | 0.000249901 |
| Obesity | pericyte | *CYGB* | -1.084205145 | 4.54E-05 | 9.79E-05 | 0.28572527 |
| Obesity | pericyte | *ZNF577* | 2.680395181 | 6.72E-05 | 0.000139032 | 0.422519748 |
| Obesity | pericyte | *TMEM143* | 3.269650805 | 1.97E-07 | 9.79E-07 | 0.001241322 |
| Obesity | pericyte | *ALDH16A1* | 4.317885361 | 0.01584689 | 0.021925484 | 1 |
| Obesity | pericyte | *SIGLEC11* | 2.347137466 | 0.002111628 | 0.003322274 | 1 |
| Obesity | pericyte | *MPP3* | 3.719296956 | 1.53E-06 | 4.98E-06 | 0.009646743 |
| Obesity | pericyte | *NAGS* | 7.213814222 | 0.004416863 | 0.006625821 | 1 |
| Obesity | pericyte | *ASB16* | 2.348061065 | 0.009155186 | 0.013087461 | 1 |
| Obesity | pericyte | *PLCD3* | -1.461193069 | 0.021625262 | 0.029294984 | 1 |
| Obesity | pericyte | *RACGAP1* | 1.920424644 | 4.53E-06 | 1.27E-05 | 0.028475692 |
| Obesity | pericyte | *LARP4* | 3.097396323 | 2.02E-05 | 4.74E-05 | 0.127098901 |
| Obesity | pericyte | *SPC24* | 7.669027999 | 9.87E-08 | 5.93E-07 | 0.000620564 |
| Obesity | pericyte | *LEMD2* | 2.777799211 | 0.012218813 | 0.017189455 | 1 |
| Obesity | pericyte | *SCIMP* | 2.125537047 | 0.000328468 | 0.000592561 | 1 |
| Obesity | pericyte | *ASGR2* | 7.475012491 | 1.27E-06 | 4.25E-06 | 0.008009256 |
| Obesity | pericyte | *TNFSF13* | 3.346532926 | 0.00194442 | 0.003076137 | 1 |
| Obesity | pericyte | *SENP3* | 2.668897497 | 3.55E-06 | 1.02E-05 | 0.022335362 |
| Obesity | pericyte | *SNRNP25* | 2.78169096 | 5.44E-06 | 1.48E-05 | 0.034194296 |
| Obesity | pericyte | *WDR90* | 1.00393925 | 0.028873274 | 0.038402004 | 1 |
| Obesity | pericyte | *ZG16B* | -15.81697432 | 5.36E-05 | 0.000113487 | 0.337055517 |
| Obesity | pericyte | *TTC9C* | 4.956575506 | 0.00010208 | 0.000204064 | 0.641779991 |
| Obesity | pericyte | *NXF1* | 5.486744719 | 1.18E-06 | 3.97E-06 | 0.007388619 |
| Obesity | pericyte | *TPCN2* | 5.737020691 | 7.36E-09 | 1.60E-07 | 4.63E-05 |
| Obesity | pericyte | *CYP4A22* | 7.95831884 | 0.000469186 | 0.000824881 | 1 |
| Obesity | pericyte | *PDZK1IP1* | -10.6583171 | 2.37E-05 | 5.46E-05 | 0.148907289 |
| Obesity | pericyte | *COA7* | 4.245765084 | 3.80E-07 | 1.66E-06 | 0.002389342 |
| Obesity | pericyte | *SLC1A7* | 0.648780265 | 0.013729804 | 0.019131046 | 1 |
| Obesity | pericyte | *C1orf123* | -2.365277874 | 0.004190887 | 0.006309413 | 1 |
| Obesity | pericyte | *ACOT11* | 3.202596755 | 5.72E-05 | 0.00011995 | 0.359728812 |
| Obesity | pericyte | *USP24* | 2.602015393 | 0.000384563 | 0.000686666 | 1 |
| Obesity | pericyte | *NOL9* | 7.833691187 | 1.84E-09 | 7.42E-08 | 1.16E-05 |
| Obesity | pericyte | *ZSWIM5* | 4.22841377 | 5.60E-06 | 1.52E-05 | 0.03521186 |
| Obesity | pericyte | *RBP7* | 9.962743122 | 1.31E-06 | 4.35E-06 | 0.008236949 |
| Obesity | pericyte | *FBLIM1* | -1.361089334 | 0.000434238 | 0.000769247 | 1 |
| Obesity | pericyte | *SLC25A34* | 2.880193743 | 1.55E-06 | 5.02E-06 | 0.009753312 |
| Obesity | pericyte | *PDPN* | 1.361419604 | 0.000430841 | 0.000763659 | 1 |
| Obesity | pericyte | *LRRC38* | -7.027410491 | 0.006302737 | 0.009210903 | 1 |
| Obesity | pericyte | *LAPTM5* | 4.325720655 | 6.66E-07 | 2.56E-06 | 0.004187985 |
| Obesity | pericyte | *PEF1* | 3.740983697 | 3.48E-08 | 2.89E-07 | 0.000218616 |
| Obesity | pericyte | *RBBP4* | 6.935068332 | 5.06E-09 | 1.30E-07 | 3.18E-05 |
| Obesity | pericyte | *CAMK2N1* | -2.927606006 | 0.005677127 | 0.008373012 | 1 |
| Obesity | pericyte | *TTLL10* | 5.277907845 | 2.49E-06 | 7.48E-06 | 0.015638408 |
| Obesity | pericyte | *NFIA* | 4.978166898 | 1.95E-07 | 9.72E-07 | 0.001229028 |
| Obesity | pericyte | *TM2D1* | 6.660058864 | 5.11E-07 | 2.09E-06 | 0.003215478 |

| | | | | | | |
|---|---|---|---|---|---|---|
| Obesity | pericyte | *NEXN* | 7.202602929 | 6.42E-07 | 2.49E-06 | 0.00403722 |
| Obesity | pericyte | *B3GALT2* | -3.481081836 | 0.00134772 | 0.002190002 | 1 |
| Obesity | pericyte | *HFM1* | 2.714669154 | 2.35E-07 | 1.12E-06 | 0.001478813 |
| Obesity | pericyte | *KCNT2* | 6.107587306 | 1.25E-08 | 1.73E-07 | 7.83E-05 |
| Obesity | pericyte | *VCAM1* | 4.351067512 | 0.000468562 | 0.000824014 | 1 |
| Obesity | pericyte | *ZNF496* | 7.82416202 | 4.81E-05 | 0.000103201 | 0.30227477 |
| Obesity | pericyte | *TRIM58* | -1.874273481 | 1.29E-05 | 3.17E-05 | 0.08079327 |
| Obesity | pericyte | *IGSF8* | 5.247851542 | 5.11E-08 | 3.76E-07 | 0.000321138 |
| Obesity | pericyte | *VANGL2* | 4.367404367 | 0.000161646 | 0.000309556 | 1 |
| Obesity | pericyte | *FCRLB* | -2.501895435 | 0.023698291 | 0.031921993 | 1 |
| Obesity | pericyte | *LMX1A* | 1.100286607 | 0.007304499 | 0.010578988 | 1 |
| Obesity | pericyte | *FLVCR1* | 4.258878256 | 0.001134681 | 0.001861138 | 1 |
| Obesity | pericyte | *TDRD5* | 1.350387639 | 0.031103912 | 0.041185825 | 1 |
| Obesity | pericyte | *SNED1* | 5.18175213 | 1.71E-08 | 1.96E-07 | 0.000107645 |
| Obesity | pericyte | *C1orf115* | 2.479861927 | 0.000841408 | 0.001412532 | 1 |
| Obesity | pericyte | *BROX* | 1.263887021 | 0.034140607 | 0.044988891 | 1 |
| Obesity | pericyte | *CNST* | 6.080563013 | 1.91E-08 | 2.08E-07 | 0.000120051 |
| Obesity | pericyte | *PM20D1* | 3.62392287 | 4.74E-07 | 1.98E-06 | 0.002981305 |
| Obesity | pericyte | *HAAO* | 8.015293781 | 8.15E-08 | 5.13E-07 | 0.000512335 |
| Obesity | pericyte | *B3GALNT2* | 3.273969499 | 1.72E-05 | 4.10E-05 | 0.108275473 |
| Obesity | pericyte | *PUS10* | 6.339313963 | 9.62E-09 | 1.71E-07 | 6.05E-05 |
| Obesity | pericyte | *RFTN2* | 6.060280012 | 8.95E-09 | 1.70E-07 | 5.62E-05 |
| Obesity | pericyte | *DISC1* | 5.090844751 | 9.08E-08 | 5.60E-07 | 0.000570747 |
| Obesity | pericyte | *FAM84A* | 4.246387996 | 1.11E-06 | 3.81E-06 | 0.006959199 |
| Obesity | pericyte | *CLHC1* | 0.995033092 | 1.35E-05 | 3.30E-05 | 0.084639411 |
| Obesity | pericyte | *FRZB* | -2.364960616 | 0.001197291 | 0.001957703 | 1 |
| Obesity | pericyte | *NUP35* | 7.264322547 | 3.88E-06 | 1.10E-05 | 0.024395395 |
| Obesity | pericyte | *FBXO41* | 3.181876115 | 1.34E-06 | 4.44E-06 | 0.008444773 |
| Obesity | pericyte | *CCDC74A* | -10.28837459 | 0.000909684 | 0.001514213 | 1 |
| Obesity | pericyte | *H3F3A* | 3.13894996 | 0.000733892 | 0.00124433 | 1 |
| Obesity | pericyte | *NOSTRIN* | -1.26078436 | 0.005929801 | 0.008716544 | 1 |
| Obesity | pericyte | *SGPP2* | 3.361600541 | 5.42E-06 | 1.48E-05 | 0.034072303 |
| Obesity | pericyte | *BBS5* | -1.485345339 | 0.005647397 | 0.008332594 | 1 |
| Obesity | pericyte | *HPGDS* | 2.069181796 | 1.86E-07 | 9.33E-07 | 0.001167468 |
| Obesity | pericyte | *PDLIM5* | 4.446439046 | 0.000153404 | 0.000295301 | 0.964453946 |
| Obesity | pericyte | *RPRD2* | 5.451196624 | 1.57E-08 | 1.91E-07 | 9.87E-05 |
| Obesity | pericyte | *CTSS* | 0.692243865 | 0.03711535 | 0.048593129 | 1 |
| Obesity | pericyte | *PACRGL* | 4.114591151 | 1.04E-05 | 2.62E-05 | 0.065184117 |
| Obesity | pericyte | *C1QTNF7* | 3.681822099 | 2.49E-07 | 1.17E-06 | 0.001563744 |
| Obesity | pericyte | *ERCC3* | 1.888100482 | 0.004882172 | 0.007264904 | 1 |
| Obesity | pericyte | *RNF149* | 5.717554727 | 2.32E-08 | 2.25E-07 | 0.000145619 |
| Obesity | pericyte | *DHX57* | 6.827293453 | 2.32E-09 | 8.54E-08 | 1.46E-05 |
| Obesity | pericyte | *S100A9* | 1.956606872 | 2.68E-05 | 6.11E-05 | 0.168565636 |
| Obesity | pericyte | *S100A12* | -3.339503766 | 2.57E-05 | 5.87E-05 | 0.161413409 |
| Obesity | pericyte | *TDRD10* | 4.59653373 | 2.25E-08 | 2.24E-07 | 0.000141436 |
| Obesity | pericyte | *GNPDA2* | 1.744003116 | 0.000628945 | 0.001082446 | 1 |
| Obesity | pericyte | *NIPAL1* | -0.420635594 | 2.15E-05 | 5.01E-05 | 0.135391238 |
| Obesity | pericyte | *ANTXR2* | 4.612817109 | 4.72E-08 | 3.56E-07 | 0.000297004 |
| Obesity | pericyte | *DCST2* | 3.85542244 | 2.09E-07 | 1.03E-06 | 0.001315009 |
| Obesity | pericyte | *LMOD3* | 13.10989263 | 1.21E-08 | 1.73E-07 | 7.63E-05 |

| | | | | | | |
|---|---|---|---|---|---|---|
| Obesity | pericyte | *LRRC58* | 8.553444481 | 8.15E-07 | 3.00E-06 | 0.005122748 |
| Obesity | pericyte | *FSTL1* | 2.196874842 | 0.000946252 | 0.001570093 | 1 |
| Obesity | pericyte | *LMOD1* | -2.339652334 | 1.10E-12 | 2.10E-10 | 6.92E-09 |
| Obesity | pericyte | *TMEM183A* | 3.428431785 | 2.08E-07 | 1.02E-06 | 0.001309659 |
| Obesity | pericyte | *IGFBP7* | -1.101383502 | 0.000183202 | 0.000347658 | 1 |
| Obesity | pericyte | *ARPC2* | 6.33449089 | 3.86E-07 | 1.68E-06 | 0.00242489 |
| Obesity | pericyte | *CCT3* | 3.265179575 | 7.05E-05 | 0.000145235 | 0.443402665 |
| Obesity | pericyte | *GLB1L* | 1.406134426 | 0.001090851 | 0.001793926 | 1 |
| Obesity | pericyte | *SERPINI1* | 4.976639493 | 7.18E-08 | 4.71E-07 | 0.000451102 |
| Obesity | pericyte | *PYHIN1* | 1.942267926 | 0.000317479 | 0.000574454 | 1 |
| Obesity | pericyte | *AIM2* | 7.946563156 | 9.22E-09 | 1.70E-07 | 5.80E-05 |
| Obesity | pericyte | *CD200R1* | 1.392226699 | 7.04E-06 | 1.86E-05 | 0.044247598 |
| Obesity | pericyte | *GTPBP8* | 7.129404298 | 2.11E-08 | 2.17E-07 | 0.000132872 |
| Obesity | pericyte | *SPICE1* | 4.531234921 | 1.16E-06 | 3.92E-06 | 0.007272055 |
| Obesity | pericyte | *CADPS* | 5.098715782 | 7.66E-08 | 4.90E-07 | 0.00048135 |
| Obesity | pericyte | *PPM1K* | 6.085266673 | 4.07E-05 | 8.86E-05 | 0.255694055 |
| Obesity | pericyte | *GMPS* | 7.661251992 | 1.24E-06 | 4.14E-06 | 0.007772823 |
| Obesity | pericyte | *TIPARP* | 5.910304701 | 3.43E-06 | 9.90E-06 | 0.021552063 |
| Obesity | pericyte | *DCLK3* | -0.825599614 | 4.03E-05 | 8.78E-05 | 0.253274732 |
| Obesity | pericyte | *RPP14* | 5.710759203 | 0.001493555 | 0.002413876 | 1 |
| Obesity | pericyte | *CRELD1* | 3.168652127 | 3.96E-06 | 1.12E-05 | 0.024873318 |
| Obesity | pericyte | *PCOLCE2* | 2.875746733 | 0.000338227 | 0.000608944 | 1 |
| Obesity | pericyte | *TTC14* | 3.055467909 | 2.95E-05 | 6.65E-05 | 0.185634698 |
| Obesity | pericyte | *RCHY1* | 2.903490712 | 0.016418933 | 0.022647177 | 1 |
| Obesity | pericyte | *CCDC158* | 2.213601909 | 5.58E-07 | 2.25E-06 | 0.003509771 |
| Obesity | pericyte | *GYG1* | 8.456561254 | 4.31E-12 | 6.38E-10 | 2.71E-08 |
| Obesity | pericyte | *HPS3* | 5.316577539 | 2.32E-05 | 5.38E-05 | 0.146118997 |
| Obesity | pericyte | *RYK* | 6.357976028 | 9.57E-08 | 5.83E-07 | 0.000601712 |
| Obesity | pericyte | *PLB1* | 3.609317244 | 2.18E-07 | 1.06E-06 | 0.001372064 |
| Obesity | pericyte | *CDCP1* | 2.566143705 | 6.50E-07 | 2.51E-06 | 0.00408559 |
| Obesity | pericyte | *LZTFL1* | 6.993944734 | 1.15E-08 | 1.72E-07 | 7.21E-05 |
| Obesity | pericyte | *FYCO1* | 7.419593866 | 1.44E-07 | 7.70E-07 | 0.000906315 |
| Obesity | pericyte | *LRRC2* | 1.701187996 | 0.000623243 | 0.001074398 | 1 |
| Obesity | pericyte | *DTX3L* | 3.781715141 | 0.001828416 | 0.002905043 | 1 |
| Obesity | pericyte | *NMNAT3* | 5.290684871 | 6.86E-06 | 1.82E-05 | 0.043135152 |
| Obesity | pericyte | *SMIM12* | -1.917949322 | 0.016515837 | 0.022770848 | 1 |
| Obesity | pericyte | *YEATS2* | 6.351015956 | 2.22E-08 | 2.22E-07 | 0.000139311 |
| Obesity | pericyte | *MEAF6* | 6.493202307 | 0.000408123 | 0.000726053 | 1 |
| Obesity | pericyte | *DNALI1* | -6.150531288 | 0.000890171 | 0.001485667 | 1 |
| Obesity | pericyte | *RPN1* | 7.372849206 | 2.51E-08 | 2.35E-07 | 0.000157681 |
| Obesity | pericyte | *RFC4* | 3.410628202 | 6.21E-07 | 2.43E-06 | 0.003901342 |
| Obesity | pericyte | *SFMBT1* | 6.594428048 | 1.60E-08 | 1.91E-07 | 0.00010061 |
| Obesity | pericyte | *GNL3* | 3.910841082 | 3.23E-05 | 7.18E-05 | 0.202969624 |
| Obesity | pericyte | *UVSSA* | 2.789421913 | 0.000100452 | 0.000201191 | 0.631539562 |
| Obesity | pericyte | *FAM208A* | 3.644470738 | 0.000291442 | 0.000531024 | 1 |
| Obesity | pericyte | *LRPAP1* | 3.143138537 | 0.03499858 | 0.046023023 | 1 |
| Obesity | pericyte | *SLC51A* | 2.588548047 | 0.004614437 | 0.00689752 | 1 |
| Obesity | pericyte | *PIGX* | 4.53549668 | 6.02E-06 | 1.62E-05 | 0.037828015 |
| Obesity | pericyte | *C1orf50* | -3.789127955 | 1.43E-05 | 3.48E-05 | 0.089845257 |
| Obesity | pericyte | *ERMAP* | 2.267134062 | 0.000106428 | 0.000211611 | 0.669112569 |

| | | | | | | |
|---|---|---|---|---|---|---|
| Obesity | pericyte | *METAP1* | 4.565474252 | 0.000131179 | 0.000255808 | 0.824724997 |
| Obesity | pericyte | *DNAJB14* | 5.75869139 | 4.33E-08 | 3.34E-07 | 0.000272278 |
| Obesity | pericyte | *SLC9B2* | 4.287804979 | 0.000354279 | 0.00063566 | 1 |
| Obesity | pericyte | *ATRIP* | 5.116629523 | 7.26E-06 | 1.91E-05 | 0.045640315 |
| Obesity | pericyte | *SHISA5* | 7.381414817 | 2.07E-10 | 1.69E-08 | 1.30E-06 |
| Obesity | pericyte | *APEH* | -4.18563594 | 1.06E-13 | 3.02E-11 | 6.65E-10 |
| Obesity | pericyte | *RNF123* | 4.921319841 | 1.50E-22 | 1.58E-19 | 9.45E-19 |
| Obesity | pericyte | *MST1R* | 3.537408614 | 4.66E-12 | 6.66E-10 | 2.93E-08 |
| Obesity | pericyte | *RAD54L2* | 3.864941543 | 3.96E-05 | 8.64E-05 | 0.248975226 |
| Obesity | pericyte | *GRM2* | -3.371923382 | 1.24E-05 | 3.07E-05 | 0.07811079 |
| Obesity | pericyte | *POC1A* | 3.551159897 | 2.76E-09 | 9.48E-08 | 1.74E-05 |
| Obesity | pericyte | *C4orf3* | 7.39700837 | 1.35E-07 | 7.34E-07 | 0.000847336 |
| Obesity | pericyte | *ANXA5* | 3.979414669 | 0.000107772 | 0.000213878 | 0.677565201 |
| Obesity | pericyte | *CEP44* | 7.84657445 | 8.22E-07 | 3.02E-06 | 0.005167626 |
| Obesity | pericyte | *TMEM144* | 5.121113783 | 1.23E-08 | 1.73E-07 | 7.70E-05 |
| Obesity | pericyte | *NPY1R* | 4.652745901 | 1.28E-08 | 1.73E-07 | 8.05E-05 |
| Obesity | pericyte | *IL15* | 6.417069669 | 4.91E-08 | 3.66E-07 | 0.000308558 |
| Obesity | pericyte | *ARFIP1* | 4.48289878 | 2.86E-05 | 6.47E-05 | 0.179914523 |
| Obesity | pericyte | *OTUD4* | 7.264520122 | 1.93E-08 | 2.08E-07 | 0.000121408 |
| Obesity | pericyte | *ITGA2* | 3.979537862 | 2.60E-08 | 2.39E-07 | 0.000163336 |
| Obesity | pericyte | *NDUFAF2* | 2.199384025 | 0.02545396 | 0.034201549 | 1 |
| Obesity | pericyte | *LMBRD2* | 6.03968921 | 9.31E-08 | 5.71E-07 | 0.000585025 |
| Obesity | pericyte | *PGGT1B* | 7.359440936 | 2.86E-08 | 2.55E-07 | 0.000179848 |
| Obesity | pericyte | *CMBL* | 3.69535704 | 1.76E-07 | 8.97E-07 | 0.001105513 |
| Obesity | pericyte | *C5orf63* | 5.101295019 | 2.39E-06 | 7.22E-06 | 0.015011245 |
| Obesity | pericyte | *PRRC1* | 4.988708082 | 0.001742817 | 0.002777462 | 1 |
| Obesity | pericyte | *WDR41* | 3.387295403 | 0.00020498 | 0.000385265 | 1 |
| Obesity | pericyte | *NDUFS4* | 5.825557578 | 2.50E-05 | 5.74E-05 | 0.157177064 |
| Obesity | pericyte | *TIGD6* | -3.689428934 | 0.023004917 | 0.031063555 | 1 |
| Obesity | pericyte | *SERINC5* | 5.429619932 | 6.92E-08 | 4.57E-07 | 0.000434752 |
| Obesity | pericyte | *ENPP6* | 5.652714862 | 3.26E-06 | 9.46E-06 | 0.020479883 |
| Obesity | pericyte | *CASP3* | 6.833636875 | 3.35E-08 | 2.85E-07 | 0.000210532 |
| Obesity | pericyte | *PRIMPOL* | 3.886819751 | 7.83E-06 | 2.04E-05 | 0.049241823 |
| Obesity | pericyte | *ERAP2* | 1.13085402 | 0.004799773 | 0.007152446 | 1 |
| Obesity | pericyte | *RICTOR* | 5.944488366 | 4.92E-07 | 2.03E-06 | 0.003090265 |
| Obesity | pericyte | *ANKRA2* | 6.640787888 | 7.34E-09 | 1.60E-07 | 4.62E-05 |
| Obesity | pericyte | *UTP15* | 2.174304812 | 1.41E-06 | 4.64E-06 | 0.008893058 |
| Obesity | pericyte | *TLR3* | 9.537385337 | 2.76E-08 | 2.49E-07 | 0.00017345 |
| Obesity | pericyte | *CCDC127* | 3.508362095 | 4.57E-07 | 1.92E-06 | 0.002872472 |
| Obesity | pericyte | *GRIK2* | 1.43666724 | 0.005693942 | 0.008393391 | 1 |
| Obesity | pericyte | *MB21D1* | 7.39944816 | 7.46E-09 | 1.61E-07 | 4.69E-05 |
| Obesity | pericyte | *TXLNB* | 2.966393347 | 1.80E-06 | 5.69E-06 | 0.011289845 |
| Obesity | pericyte | *DCBLD1* | -0.565944089 | 7.43E-05 | 0.000152234 | 0.467205235 |
| Obesity | pericyte | *TMEM200A* | -1.980381467 | 0.000799 | 0.001347093 | 1 |
| Obesity | pericyte | *PDSS2* | 6.018665902 | 8.56E-08 | 5.35E-07 | 0.000538322 |
| Obesity | pericyte | *STXBP5* | 5.882071504 | 3.61E-08 | 2.96E-07 | 0.000227007 |
| Obesity | pericyte | *PI16* | 5.004593675 | 1.28E-08 | 1.73E-07 | 8.04E-05 |
| Obesity | pericyte | *DAGLB* | 1.613906823 | 0.03036925 | 0.040297905 | 1 |
| Obesity | pericyte | *STK17A* | 5.090273591 | 1.30E-07 | 7.23E-07 | 0.000819048 |
| Obesity | pericyte | *GALNT10* | 4.310114266 | 0.000401725 | 0.000715682 | 1 |

| | | | | | | |
|---|---|---|---|---|---|---|
| Obesity | pericyte | *RPS14* | 4.960407304 | 0.00010273 | 0.000205095 | 0.645865635 |
| Obesity | pericyte | *HCN1* | -3.552238311 | 0.003328875 | 0.00507484 | 1 |
| Obesity | pericyte | *SLU7* | 5.027662069 | 1.70E-08 | 1.95E-07 | 0.000107156 |
| Obesity | pericyte | *RP9* | 4.333723275 | 0.000703011 | 0.001197138 | 1 |
| Obesity | pericyte | *CAMLG* | 4.363448983 | 3.53E-07 | 1.56E-06 | 0.002216893 |
| Obesity | pericyte | *KCNK5* | -3.599442298 | 0.000422212 | 0.000749844 | 1 |
| Obesity | pericyte | *USP49* | 5.393770082 | 5.94E-07 | 2.36E-06 | 0.003732083 |
| Obesity | pericyte | *ZNF704* | 4.12538756 | 4.19E-08 | 3.26E-07 | 0.000263694 |
| Obesity | pericyte | *FNDC1* | 2.392286811 | 9.54E-07 | 3.40E-06 | 0.00599653 |
| Obesity | pericyte | *CHMP4C* | -0.879671324 | 0.005332087 | 0.007895156 | 1 |
| Obesity | pericyte | *BRI3* | 4.915928248 | 2.23E-06 | 6.83E-06 | 0.014048334 |
| Obesity | pericyte | *LMTK2* | 7.088470145 | 3.18E-08 | 2.76E-07 | 0.000200074 |
| Obesity | pericyte | *PEX2* | 6.060204652 | 0.000168741 | 0.000321867 | 1 |
| Obesity | pericyte | *TNFRSF11B* | 2.836494974 | 1.19E-07 | 6.74E-07 | 0.000745357 |
| Obesity | pericyte | *SUN1* | 4.145003726 | 8.51E-07 | 3.10E-06 | 0.005352835 |
| Obesity | pericyte | *OXR1* | 5.032350273 | 5.11E-07 | 2.09E-06 | 0.003215301 |
| Obesity | pericyte | *INTS1* | 7.840684369 | 6.78E-09 | 1.54E-07 | 4.26E-05 |
| Obesity | pericyte | *SLC4A2* | 9.696116936 | 3.44E-05 | 7.62E-05 | 0.216574049 |
| Obesity | pericyte | *FASTK* | 7.024755809 | 7.45E-06 | 1.96E-05 | 0.046813391 |
| Obesity | pericyte | *ALDH7A1* | 2.819476472 | 2.18E-06 | 6.69E-06 | 0.013736364 |
| Obesity | pericyte | *COX6C* | 7.730043544 | 1.91E-08 | 2.08E-07 | 0.000119768 |
| Obesity | pericyte | *INTS8* | 2.802548582 | 0.008016198 | 0.011530047 | 1 |
| Obesity | pericyte | *FREM1* | 3.306260648 | 1.72E-07 | 8.85E-07 | 0.001084494 |
| Obesity | pericyte | *RPP25L* | 4.260303549 | 0.001114602 | 0.001830113 | 1 |
| Obesity | pericyte | *FAM219A* | 4.909879548 | 0.001798015 | 0.002861078 | 1 |
| Obesity | pericyte | *TMEM65* | 6.219225937 | 1.31E-06 | 4.35E-06 | 0.008239136 |
| Obesity | pericyte | *CCDC171* | 3.430557943 | 0.013489273 | 0.018837641 | 1 |
| Obesity | pericyte | *SYK* | 3.423299013 | 5.47E-08 | 3.92E-07 | 0.000343977 |
| Obesity | pericyte | *NIPSNAP3B* | 1.350445452 | 0.002953443 | 0.004543259 | 1 |
| Obesity | pericyte | *METTL2B* | 0.84916796 | 0.000236005 | 0.000437559 | 1 |
| Obesity | pericyte | *TMC1* | 9.840228808 | 2.60E-07 | 1.22E-06 | 0.001633545 |
| Obesity | pericyte | *HGSNAT* | 4.051743495 | 0.000264907 | 0.000486127 | 1 |
| Obesity | pericyte | *C9orf64* | 1.925411024 | 7.38E-05 | 0.000151335 | 0.464143356 |
| Obesity | pericyte | *HNRNPK* | 4.95879154 | 0.000784389 | 0.001324947 | 1 |
| Obesity | pericyte | *SVEP1* | 3.682044592 | 5.37E-09 | 1.33E-07 | 3.38E-05 |
| Obesity | pericyte | *ZNF367* | 3.041281441 | 0.000252487 | 0.000465782 | 1 |
| Obesity | pericyte | *AQP3* | 2.861046542 | 0.000247258 | 0.000456941 | 1 |
| Obesity | pericyte | *TRMT10B* | 6.591195253 | 1.71E-05 | 4.07E-05 | 0.107230104 |
| Obesity | pericyte | *VCP* | 3.091507107 | 0.001833606 | 0.002911816 | 1 |
| Obesity | pericyte | *PIGO* | 7.633600795 | 3.87E-05 | 8.46E-05 | 0.243314137 |
| Obesity | pericyte | *LRFN5* | -0.477517807 | 0.005857873 | 0.008618851 | 1 |
| Obesity | pericyte | *WRN* | 5.615096452 | 6.84E-09 | 1.54E-07 | 4.30E-05 |
| Obesity | pericyte | *MARCH8* | 5.770628802 | 1.11E-08 | 1.71E-07 | 6.99E-05 |
| Obesity | pericyte | *CFL2* | 5.101956393 | 0.000340648 | 0.000612777 | 1 |
| Obesity | pericyte | *ZCCHC24* | 4.951376768 | 0.000915968 | 0.001523061 | 1 |
| Obesity | pericyte | *CRYL1* | 3.208446478 | 3.00E-05 | 6.74E-05 | 0.188645228 |
| Obesity | pericyte | *HEPACAM* | 0.356824035 | 0.022101448 | 0.029882109 | 1 |
| Obesity | pericyte | *LRR1* | 4.354054958 | 0.005037751 | 0.007476946 | 1 |
| Obesity | pericyte | *RPL36AL* | 1.887822646 | 0.025747637 | 0.034573984 | 1 |
| Obesity | pericyte | *KLHDC2* | 3.560543276 | 0.001642348 | 0.002627338 | 1 |

| | | | | | | |
|---|---|---|---|---|---|---|
| Obesity | pericyte | *NOXRED1* | 2.599470047 | 3.59E-06 | 1.03E-05 | 0.0225519 |
| Obesity | pericyte | *AKR1E2* | 1.837101213 | 0.030453761 | 0.040392994 | 1 |
| Obesity | pericyte | *ATP5C1* | 2.521523913 | 0.03095886 | 0.041011031 | 1 |
| Obesity | pericyte | *TAF3* | 4.795409176 | 0.000167496 | 0.000319784 | 1 |
| Obesity | pericyte | *VDAC2* | 6.421430974 | 2.70E-05 | 6.14E-05 | 0.169786076 |
| Obesity | pericyte | *SLC18A2* | 2.498275102 | 5.52E-07 | 2.23E-06 | 0.003471208 |
| Obesity | pericyte | *PDZD8* | 3.253191459 | 4.01E-06 | 1.14E-05 | 0.02520027 |
| Obesity | pericyte | *QSOX2* | 8.696405299 | 0.001929871 | 0.003054658 | 1 |
| Obesity | pericyte | *SDCCAG3* | 8.719365831 | 7.67E-06 | 2.00E-05 | 0.04823574 |
| Obesity | pericyte | *TSC1* | 3.716077925 | 0.003501105 | 0.005316775 | 1 |
| Obesity | pericyte | *ZMYND19* | 5.641220597 | 1.96E-07 | 9.74E-07 | 0.001232577 |
| Obesity | pericyte | *STOX1* | 0.562818671 | 0.016147841 | 0.02230261 | 1 |
| Obesity | pericyte | *RET* | -2.090661344 | 3.09E-05 | 6.91E-05 | 0.194036565 |
| Obesity | pericyte | *STK32C* | 2.775692198 | 2.64E-06 | 7.86E-06 | 0.016591613 |
| Obesity | pericyte | *NSMF* | 8.503432493 | 1.22E-08 | 1.73E-07 | 7.65E-05 |
| Obesity | pericyte | *VWA2* | -2.724242774 | 0.002354765 | 0.003682688 | 1 |
| Obesity | pericyte | *METTL3* | 7.925140653 | 9.03E-08 | 5.58E-07 | 0.000567713 |
| Obesity | pericyte | *TRUB1* | 2.357400991 | 0.000892188 | 0.001488242 | 1 |
| Obesity | pericyte | *UBTD1* | 6.433694057 | 1.09E-08 | 1.71E-07 | 6.88E-05 |
| Obesity | pericyte | *ARHGAP42* | -1.26780479 | 0.002643153 | 0.004101062 | 1 |
| Obesity | pericyte | *OTOGL* | -2.482552317 | 0.000419275 | 0.000745049 | 1 |
| Obesity | pericyte | *PACSIN3* | -0.653252309 | 0.002960869 | 0.004553567 | 1 |
| Obesity | pericyte | *SLC39A13* | 15.5903955 | 8.28E-12 | 1.13E-09 | 5.21E-08 |
| Obesity | pericyte | *TC2N* | 0.898379204 | 0.002839084 | 0.004381276 | 1 |
| Obesity | pericyte | *IFI27L1* | 3.78300535 | 1.06E-06 | 3.69E-06 | 0.006685688 |
| Obesity | pericyte | *CLMN* | -1.607428045 | 0.006379976 | 0.00931512 | 1 |
| Obesity | pericyte | *PTER* | 5.645022327 | 3.30E-06 | 9.57E-06 | 0.020771642 |
| Obesity | pericyte | *C1QL3* | 1.598477547 | 1.47E-05 | 3.58E-05 | 0.092605525 |
| Obesity | pericyte | *ABTB2* | 6.13244108 | 2.57E-08 | 2.38E-07 | 0.000161709 |
| Obesity | pericyte | *LIPC* | 3.345748155 | 1.17E-05 | 2.92E-05 | 0.073835823 |
| Obesity | pericyte | *CEP57* | 3.480014056 | 0.000112459 | 0.000222196 | 0.707026698 |
| Obesity | pericyte | *TCP11L2* | 4.959030969 | 1.19E-08 | 1.73E-07 | 7.47E-05 |
| Obesity | pericyte | *GPT2* | 1.545593021 | 4.67E-06 | 1.30E-05 | 0.029389942 |
| Obesity | pericyte | *IKBIP* | 4.257965077 | 3.50E-05 | 7.73E-05 | 0.21998699 |
| Obesity | pericyte | *HIF1AN* | 7.131433903 | 2.59E-08 | 2.39E-07 | 0.000163135 |
| Obesity | pericyte | *NDUFB8* | 9.2219392 | 1.34E-05 | 3.30E-05 | 0.084525888 |
| Obesity | pericyte | *ZFYVE19* | 2.111813952 | 0.00044786 | 0.000790734 | 1 |
| Obesity | pericyte | *SPINT1* | 3.43025175 | 0.000131767 | 0.000256874 | 0.828418197 |
| Obesity | pericyte | *AVPR1A* | -0.277657098 | 3.80E-05 | 8.33E-05 | 0.238858371 |
| Obesity | pericyte | *BTRC* | 5.20048628 | 5.38E-06 | 1.47E-05 | 0.033805834 |
| Obesity | pericyte | *LARP6* | 4.283647459 | 3.28E-07 | 1.47E-06 | 0.002065111 |
| Obesity | pericyte | *ASPG* | -4.870904124 | 0.001126507 | 0.001848695 | 1 |
| Obesity | pericyte | *TBATA* | 7.251338787 | 0.00096814 | 0.001605141 | 1 |
| Obesity | pericyte | *FAM227B* | 3.828601617 | 8.32E-07 | 3.04E-06 | 0.005232985 |
| Obesity | pericyte | *STXBP4* | 4.133655574 | 4.94E-05 | 0.000105743 | 0.310566305 |
| Obesity | pericyte | *C2* | 3.380424824 | 2.67E-05 | 6.07E-05 | 0.167579675 |
| Obesity | pericyte | *APBB1* | 6.637825769 | 1.85E-05 | 4.38E-05 | 0.11654542 |
| Obesity | pericyte | *ILK* | 6.882099454 | 7.46E-07 | 2.80E-06 | 0.004688448 |
| Obesity | pericyte | *TAF10* | 4.059717001 | 0.01849501 | 0.025327407 | 1 |
| Obesity | pericyte | *DCHS1* | 4.471645634 | 4.86E-08 | 3.63E-07 | 0.000305488 |

| | | | | | | |
|---|---|---|---|---|---|---|
| Obesity | pericyte | *NETO1* | 3.158186309 | 9.64E-05 | 0.000193401 | 0.605924899 |
| Obesity | pericyte | *USP54* | 4.279813538 | 4.31E-07 | 1.84E-06 | 0.002708973 |
| Obesity | pericyte | *C11orf74* | 4.811157846 | 2.09E-08 | 2.17E-07 | 0.000131694 |
| Obesity | pericyte | *CYB5R2* | -1.221877216 | 0.000144652 | 0.000280342 | 0.909428443 |
| Obesity | pericyte | *TUB* | 3.944937144 | 2.01E-06 | 6.26E-06 | 0.01262881 |
| Obesity | pericyte | *RIC3* | 4.808640609 | 3.47E-08 | 2.89E-07 | 0.00021796 |
| Obesity | pericyte | *LMO1* | -3.377433329 | 0.001553024 | 0.00250227 | 1 |
| Obesity | pericyte | *TRIM66* | 0.497981732 | 0.004211178 | 0.00633541 | 1 |
| Obesity | pericyte | *RPL27A* | 6.113158437 | 2.53E-05 | 5.80E-05 | 0.159125548 |
| Obesity | pericyte | *AKIP1* | 11.58952171 | 4.93E-08 | 3.66E-07 | 0.000309673 |
| Obesity | pericyte | *C16orf46* | 27.96965577 | 1.63E-07 | 8.46E-07 | 0.001021651 |
| Obesity | pericyte | *TMEM41B* | 5.23615561 | 0.000991244 | 0.001640042 | 1 |
| Obesity | pericyte | *TMX3* | 5.614710245 | 3.70E-08 | 3.01E-07 | 0.00023293 |
| Obesity | pericyte | *CLEC4E* | 1.34492277 | 0.003929347 | 0.005936987 | 1 |
| Obesity | pericyte | *ZNF3* | 3.21823217 | 2.37E-05 | 5.46E-05 | 0.148891938 |
| Obesity | pericyte | *CLEC4D* | -2.421267351 | 0.000184564 | 0.000349925 | 1 |
| Obesity | pericyte | *BEAN1* | 3.244812586 | 3.36E-08 | 2.85E-07 | 0.000211311 |
| Obesity | pericyte | *TK2* | 2.446951303 | 0.000139874 | 0.000271584 | 0.879388519 |
| Obesity | pericyte | *SEC11C* | 4.478474142 | 7.21E-08 | 4.73E-07 | 0.000452996 |
| Obesity | pericyte | *GALR1* | 1.636708608 | 0.001107993 | 0.001819736 | 1 |
| Obesity | pericyte | *SPDYE6* | 3.571254793 | 4.20E-07 | 1.80E-06 | 0.002642522 |
| Obesity | pericyte | *ATF7IP2* | 4.005252492 | 4.77E-06 | 1.32E-05 | 0.029989492 |
| Obesity | pericyte | *ZNF592* | 3.665678371 | 0.000133188 | 0.000259322 | 0.837351937 |
| Obesity | pericyte | *ACSM1* | -2.293142761 | 0.01594463 | 0.02204616 | 1 |
| Obesity | pericyte | *AP1G1* | 6.262429129 | 4.25E-07 | 1.82E-06 | 0.002670282 |
| Obesity | pericyte | *SLFN5* | 4.28132606 | 3.99E-06 | 1.13E-05 | 0.025100777 |
| Obesity | pericyte | *CATSPER2* | 3.208970507 | 0.00477302 | 0.007115954 | 1 |
| Obesity | pericyte | *C16orf45* | 6.506750277 | 7.29E-08 | 4.75E-07 | 0.000458134 |
| Obesity | pericyte | *SAAL1* | 9.627084217 | 5.17E-12 | 7.22E-10 | 3.25E-08 |
| Obesity | pericyte | *FAM111A* | 2.450721912 | 0.007466441 | 0.010801084 | 1 |
| Obesity | pericyte | *SCNN1G* | 5.601318717 | 0.006118674 | 0.008977387 | 1 |
| Obesity | pericyte | *ANKDD1A* | 3.167924032 | 7.48E-06 | 1.96E-05 | 0.047001694 |
| Obesity | pericyte | *DCTN5* | 1.813345298 | 0.000817045 | 0.001376041 | 1 |
| Obesity | pericyte | *STX3* | 3.893137283 | 6.30E-05 | 0.0001308 | 0.396063506 |
| Obesity | pericyte | *MTMR10* | 5.037623166 | 8.06E-09 | 1.68E-07 | 5.07E-05 |
| Obesity | pericyte | *TSC22D4* | 7.763120288 | 1.22E-07 | 6.87E-07 | 0.000765001 |
| Obesity | pericyte | *MS4A7* | 1.799613681 | 2.89E-05 | 6.52E-05 | 0.181757795 |
| Obesity | pericyte | *DIS3L* | 3.062005141 | 0.001067018 | 0.001757491 | 1 |
| Obesity | pericyte | *CCNDBP1* | 9.080504322 | 2.19E-08 | 2.20E-07 | 0.000137638 |
| Obesity | pericyte | *SMAD3* | 2.492407756 | 0.013661108 | 0.01904799 | 1 |
| Obesity | pericyte | *AKTIP* | 8.617917133 | 2.85E-08 | 2.55E-07 | 0.000179178 |
| Obesity | pericyte | *EVA1C* | 1.584466711 | 0.017124298 | 0.023563244 | 1 |
| Obesity | pericyte | *NUDT21* | 5.821065483 | 2.71E-07 | 1.26E-06 | 0.001705807 |
| Obesity | pericyte | *NKX3-1* | 12.45615526 | 2.45E-05 | 5.63E-05 | 0.153792643 |
| Obesity | pericyte | *DUSP18* | 2.92789953 | 0.035433061 | 0.046526243 | 1 |
| Obesity | pericyte | *MEI1* | 1.539769154 | 0.002147736 | 0.003373174 | 1 |
| Obesity | pericyte | *PBX3* | 2.555391042 | 0.010096449 | 0.014341702 | 1 |
| Obesity | pericyte | *SNRPD1* | 6.374183044 | 1.78E-08 | 2.01E-07 | 0.000111872 |
| Obesity | pericyte | *SAMD14* | 4.777842777 | 1.29E-06 | 4.31E-06 | 0.008130869 |
| Obesity | pericyte | *URM1* | 1.982576449 | 0.019214549 | 0.026215683 | 1 |

| | | | | | | |
|---|---|---|---|---|---|---|
| Obesity | pericyte | *CERCAM* | 1.165124727 | 0.0266862 | 0.035757915 | 1 |
| Obesity | pericyte | *ENDOG* | 1.669823259 | 0.025955406 | 0.034823226 | 1 |
| Obesity | pericyte | *ISLR2* | 3.041275763 | 6.12E-08 | 4.23E-07 | 0.000384731 |
| Obesity | pericyte | *SP2* | 9.932533045 | 1.06E-09 | 5.30E-08 | 6.69E-06 |
| Obesity | pericyte | *COQ7* | 6.653754405 | 1.11E-08 | 1.71E-07 | 6.98E-05 |
| Obesity | pericyte | *GPRC5B* | 5.509800229 | 3.17E-07 | 1.43E-06 | 0.001991705 |
| Obesity | pericyte | *RNF214* | 7.396441381 | 2.43E-07 | 1.15E-06 | 0.001529978 |
| Obesity | pericyte | *RBFOX3* | 3.959434458 | 1.80E-07 | 9.13E-07 | 0.001132662 |
| Obesity | pericyte | *MYO5B* | 1.018990047 | 0.008553278 | 0.01225489 | 1 |
| Obesity | pericyte | *RRM1* | 1.215157142 | 0.006410441 | 0.009350914 | 1 |
| Obesity | pericyte | *TRIM68* | -2.070183358 | 5.20E-05 | 0.000110388 | 0.327078189 |
| Obesity | pericyte | *FN3K* | 3.143358049 | 9.94E-06 | 2.52E-05 | 0.06246678 |
| Obesity | pericyte | *ZNF23* | 2.443487917 | 0.003087074 | 0.004734919 | 1 |
| Obesity | pericyte | *SMG8* | -2.097492165 | 3.89E-05 | 8.49E-05 | 0.244520507 |
| Obesity | pericyte | *RAB8A* | 5.676990464 | 6.72E-08 | 4.48E-07 | 0.000422341 |
| Obesity | pericyte | *GATAD2A* | 4.93964796 | 0.000227836 | 0.00042429 | 1 |
| Obesity | pericyte | *PROCA1* | 3.271912994 | 2.79E-05 | 6.33E-05 | 0.175539497 |
| Obesity | pericyte | *ZNF641* | 1.272260569 | 0.01607164 | 0.022207121 | 1 |
| Obesity | pericyte | *CACNB3* | 2.425097823 | 2.67E-06 | 7.96E-06 | 0.016810425 |
| Obesity | pericyte | *TP53I13* | 2.351249235 | 0.000858246 | 0.001437727 | 1 |
| Obesity | pericyte | *ZNF701* | 4.871103937 | 5.88E-07 | 2.35E-06 | 0.003698416 |
| Obesity | pericyte | *SERTAD3* | -2.210376681 | 0.000276787 | 0.000506154 | 1 |
| Obesity | pericyte | *TTYH1* | 4.35140666 | 0.020760377 | 0.028184084 | 1 |
| Obesity | pericyte | *LENG8* | 4.456135675 | 5.82E-05 | 0.000121878 | 0.366121206 |
| Obesity | pericyte | *CDC42EP5* | -2.115075987 | 0.011656377 | 0.016449751 | 1 |
| Obesity | pericyte | *ZNF283* | 3.721740346 | 0.000484514 | 0.000849453 | 1 |
| Obesity | pericyte | *PPP1R14A* | -2.80169705 | 0.004846307 | 0.007218345 | 1 |
| Obesity | pericyte | *SPINT2* | 1.367118189 | 0.009641691 | 0.013742306 | 1 |
| Obesity | pericyte | *PSCA* | 4.014175048 | 3.18E-10 | 2.41E-08 | 2.00E-06 |
| Obesity | pericyte | *ZNF444* | 5.053125045 | 4.29E-08 | 3.32E-07 | 0.000269735 |
| Obesity | pericyte | *FAM57A* | 2.215924114 | 1.01E-06 | 3.55E-06 | 0.006349241 |
| Obesity | pericyte | *GLOD4* | 5.423676683 | 4.42E-07 | 1.87E-06 | 0.002778086 |
| Obesity | pericyte | *SERPINF2* | 2.915369998 | 1.84E-06 | 5.80E-06 | 0.011541849 |
| Obesity | pericyte | *SRR* | 2.043902402 | 6.12E-10 | 3.62E-08 | 3.85E-06 |
| Obesity | pericyte | *CYB5D2* | 5.28865102 | 0.001557102 | 0.002506914 | 1 |
| Obesity | pericyte | *C19orf48* | 1.774536136 | 7.20E-06 | 1.90E-05 | 0.045235912 |
| Obesity | pericyte | *ZNF83* | 4.437265424 | 1.81E-05 | 4.30E-05 | 0.113910758 |
| Obesity | pericyte | *CD320* | 10.99793924 | 3.83E-06 | 1.09E-05 | 0.024109241 |
| Obesity | pericyte | *SPRYD3* | 6.006694023 | 0.013048928 | 0.018271406 | 1 |
| Obesity | pericyte | *SOAT2* | 2.141117974 | 9.54E-05 | 0.000191411 | 0.599500271 |
| Obesity | pericyte | *ZNF558* | 1.665307818 | 0.002345377 | 0.003668918 | 1 |
| Obesity | pericyte | *NDUFV1* | 5.170228801 | 0.000216206 | 0.000404188 | 1 |
| Obesity | pericyte | *CDK2AP2* | 6.033620653 | 3.14E-07 | 1.42E-06 | 0.001972974 |
| Obesity | pericyte | *PRDX2* | 4.444619461 | 0.007756405 | 0.011179395 | 1 |
| Obesity | pericyte | *MIS12* | 5.313172785 | 2.61E-07 | 1.22E-06 | 0.001642735 |
| Obesity | pericyte | *SRP68* | 4.218002672 | 0.000561599 | 0.000975621 | 1 |
| Obesity | pericyte | *MGAT5B* | -2.625169896 | 0.02795198 | 0.037295013 | 1 |
| Obesity | pericyte | *TMC8* | 3.707176744 | 8.41E-10 | 4.56E-08 | 5.29E-06 |
| Obesity | pericyte | *TMEM68* | 5.56327589 | 1.54E-06 | 4.99E-06 | 0.009679144 |
| Obesity | pericyte | *CYP7A1* | 3.861261637 | 0.000727486 | 0.001235134 | 1 |

| | | | | | | |
|---|---|---|---|---|---|---|
| Obesity | pericyte | *GSDMA* | 4.112968157 | 4.54E-06 | 1.27E-05 | 0.0285476 |
| Obesity | pericyte | *KRT24* | -1.509021391 | 1.23E-06 | 4.11E-06 | 0.007704865 |
| Obesity | pericyte | *ZNF598* | 3.84456944 | 0.019646733 | 0.026782093 | 1 |
| Obesity | pericyte | *MLST8* | 2.554096213 | 8.18E-06 | 2.12E-05 | 0.051396665 |
| Obesity | pericyte | *ABCA3* | 3.172240775 | 0.000570575 | 0.000990119 | 1 |
| Obesity | pericyte | *KCTD5* | 3.665095172 | 5.65E-07 | 2.27E-06 | 0.003552534 |
| Obesity | pericyte | *SDHAF2* | 12.3965179 | 1.02E-08 | 1.71E-07 | 6.43E-05 |
| Obesity | pericyte | *DDB1* | 6.621083286 | 5.09E-05 | 0.000108465 | 0.319863728 |
| Obesity | pericyte | *VPS37C* | 3.753493193 | 1.51E-05 | 3.67E-05 | 0.095196987 |
| Obesity | pericyte | *VWCE* | 2.995503845 | 2.41E-08 | 2.30E-07 | 0.000151306 |
| Obesity | pericyte | *TRANK1* | 6.817018015 | 1.64E-08 | 1.92E-07 | 0.000102978 |
| Obesity | pericyte | *TTC21A* | 6.568499134 | 1.18E-08 | 1.73E-07 | 7.45E-05 |
| Obesity | pericyte | *RPSA* | 1.832761767 | 0.006995766 | 0.010159941 | 1 |
| Obesity | pericyte | *ULK4* | 0.863876681 | 0.002117971 | 0.003329754 | 1 |
| Obesity | pericyte | *LTBP3* | 2.519787447 | 0.001594669 | 0.002562147 | 1 |
| Obesity | pericyte | *NAALADL1* | -2.723242244 | 0.002649114 | 0.004109296 | 1 |
| Obesity | pericyte | *SCARA5* | 2.718029253 | 3.30E-07 | 1.48E-06 | 0.002076079 |
| Obesity | pericyte | *NUDT16L1* | 5.580869532 | 1.01E-07 | 6.03E-07 | 0.000633447 |
| Obesity | pericyte | *ZCCHC4* | 5.777576603 | 4.20E-09 | 1.20E-07 | 2.64E-05 |
| Obesity | pericyte | *PTGDR* | -1.533748488 | 0.020234046 | 0.027528987 | 1 |
| Obesity | pericyte | *GLYCTK* | 4.940998741 | 0.000354751 | 0.000636144 | 1 |
| Obesity | pericyte | *UBTD2* | 6.036299819 | 1.52E-08 | 1.90E-07 | 9.58E-05 |
| Obesity | pericyte | *POLR2J3* | 1.500534042 | 0.000850296 | 0.00142593 | 1 |
| Obesity | pericyte | *COA6* | 12.07957507 | 1.10E-08 | 1.71E-07 | 6.91E-05 |
| Obesity | pericyte | *KIF5C* | 4.796486625 | 1.73E-06 | 5.51E-06 | 0.010883302 |
| Obesity | pericyte | *PDHB* | 4.518352009 | 3.66E-06 | 1.05E-05 | 0.023037339 |
| Obesity | pericyte | *PXK* | 2.402748598 | 0.004681442 | 0.006988206 | 1 |
| Obesity | pericyte | *CX3CR1* | 1.531726778 | 0.000700637 | 0.001193417 | 1 |
| Obesity | pericyte | *SEPT2* | 1.222110735 | 0.000899232 | 0.001498403 | 1 |
| Obesity | pericyte | *ING5* | 6.301557266 | 1.03E-08 | 1.71E-07 | 6.46E-05 |
| Obesity | pericyte | *RFWD3* | 4.204995705 | 2.01E-06 | 6.27E-06 | 0.012655538 |
| Obesity | pericyte | *RHOH* | 1.594636105 | 1.56E-06 | 5.03E-06 | 0.009794208 |
| Obesity | pericyte | *KLHL30* | 2.352542937 | 4.33E-05 | 9.37E-05 | 0.272334995 |
| Obesity | pericyte | *REEP4* | 3.923716198 | 0.002616486 | 0.004062695 | 1 |
| Obesity | pericyte | *BMP1* | 3.285709321 | 4.38E-06 | 1.23E-05 | 0.027566099 |
| Obesity | pericyte | *ATXN2L* | 3.280444798 | 0.011179967 | 0.015805813 | 1 |
| Obesity | pericyte | *FNTA* | 7.081665852 | 3.04E-09 | 9.89E-08 | 1.91E-05 |
| Obesity | pericyte | *SERINC2* | 2.966408053 | 0.00227806 | 0.003567023 | 1 |
| Obesity | pericyte | *TRAPPC11* | 7.032696518 | 4.19E-08 | 3.26E-07 | 0.000263152 |
| Obesity | pericyte | *TMEM223* | 4.402204911 | 0.000168516 | 0.000321536 | 1 |
| Obesity | pericyte | *SLC20A2* | 6.043249988 | 2.77E-09 | 9.48E-08 | 1.74E-05 |
| Obesity | pericyte | *ZSWIM1* | -1.682991002 | 0.00853009 | 0.012230028 | 1 |
| Obesity | pericyte | *NBPF9* | 6.190055685 | 3.05E-08 | 2.68E-07 | 0.000191983 |
| Obesity | pericyte | *WFDC13* | 13.23849157 | 5.51E-05 | 0.000116159 | 0.346384773 |
| Obesity | pericyte | *NDUFS5* | 5.739294676 | 2.13E-07 | 1.04E-06 | 0.001338911 |
| Obesity | pericyte | *ZNF30* | 2.094042065 | 0.009941043 | 0.014140122 | 1 |
| Obesity | pericyte | *FAM84B* | 1.92660502 | 1.32E-05 | 3.25E-05 | 0.083258314 |
| Obesity | pericyte | *SLC16A4* | 2.199515747 | 0.00019991 | 0.00037596 | 1 |
| Obesity | pericyte | *TMEM208* | 3.360966684 | 2.30E-06 | 6.99E-06 | 0.014450959 |
| Obesity | pericyte | *LRP1B* | 1.16549679 | 0.000332216 | 0.000598465 | 1 |

| | | | | | | |
|---|---|---|---|---|---|---|
| Obesity | pericyte | *DNAJC21* | 8.109987558 | 3.25E-08 | 2.79E-07 | 0.000204235 |
| Obesity | pericyte | *NPNT* | -2.150032631 | 0.0114378 | 0.016153187 | 1 |
| Obesity | pericyte | *GSTM4* | 1.509001318 | 0.003782312 | 0.005717576 | 1 |
| Obesity | pericyte | *ABHD15* | 1.556706863 | 3.07E-05 | 6.89E-05 | 0.193157569 |
| Obesity | pericyte | *ADAL* | 0.65068891 | 0.004069164 | 0.006133501 | 1 |
| Obesity | pericyte | *SNTB2* | 6.677761061 | 3.63E-08 | 2.98E-07 | 0.000228522 |
| Obesity | pericyte | *ZNF507* | 2.271200519 | 0.001538284 | 0.002480428 | 1 |
| Obesity | pericyte | *STX18* | 5.50118558 | 0.000178175 | 0.000338731 | 1 |
| Obesity | pericyte | *ZBTB49* | 3.626975266 | 1.34E-07 | 7.33E-07 | 0.000840808 |
| Obesity | pericyte | *GFM1* | 2.630273755 | 0.00176002 | 0.002804168 | 1 |
| Obesity | pericyte | *INPP5D* | 5.978376173 | 6.94E-09 | 1.56E-07 | 4.37E-05 |
| Obesity | pericyte | *LETM1* | 5.405268744 | 2.19E-08 | 2.20E-07 | 0.000137868 |
| Obesity | pericyte | *TMEM129* | 1.627550398 | 0.00936288 | 0.013363094 | 1 |
| Obesity | pericyte | *CEP120* | 5.812952174 | 7.49E-07 | 2.81E-06 | 0.004707872 |
| Obesity | pericyte | *STXBP6* | 5.800767297 | 4.94E-06 | 1.37E-05 | 0.031089128 |
| Obesity | pericyte | *MFF* | 5.626434163 | 9.62E-08 | 5.84E-07 | 0.000604848 |
| Obesity | pericyte | *LGALS9* | 2.752647184 | 0.000145405 | 0.00028154 | 0.914159743 |
| Obesity | pericyte | *SIGLEC7* | 2.066067448 | 0.004269746 | 0.006421983 | 1 |
| Obesity | pericyte | *FEM1B* | 5.905369963 | 9.22E-08 | 5.67E-07 | 0.000579956 |
| Obesity | pericyte | *COL4A3* | 4.061639902 | 0.000354611 | 0.000636074 | 1 |
| Obesity | pericyte | *IRS1* | 0.878940563 | 2.96E-05 | 6.67E-05 | 0.186184677 |
| Obesity | pericyte | *UPF3A* | 1.596256235 | 0.015751113 | 0.021802565 | 1 |
| Obesity | pericyte | *C8orf46* | 0.343917165 | 0.003747691 | 0.005670693 | 1 |
| Obesity | pericyte | *PARM1* | -0.767987299 | 0.00015227 | 0.000293478 | 0.957323966 |
| Obesity | pericyte | *ATF5* | 2.712514857 | 2.38E-06 | 7.21E-06 | 0.014965339 |
| Obesity | pericyte | *CPT1C* | 3.326828954 | 0.000260217 | 0.000478638 | 1 |
| Obesity | pericyte | *PCSK9* | -6.509417064 | 4.38E-05 | 9.47E-05 | 0.275442345 |
| Obesity | pericyte | *XPO6* | 7.417037623 | 7.13E-09 | 1.59E-07 | 4.48E-05 |
| Obesity | pericyte | *RAB24* | -3.5287398 | 1.04E-07 | 6.17E-07 | 0.000655012 |
| Obesity | pericyte | *CXCL10* | 0.956030311 | 0.037442476 | 0.048990811 | 1 |
| Obesity | pericyte | *SH3TC2* | -1.982454978 | 0.000625168 | 0.00107742 | 1 |
| Obesity | pericyte | *GPRIN1* | 6.619621093 | 0.012989691 | 0.018192513 | 1 |
| Obesity | pericyte | *PGM2* | 4.784386416 | 3.15E-08 | 2.74E-07 | 0.00019815 |
| Obesity | pericyte | *STK32A* | 0.765874096 | 0.03764885 | 0.049240342 | 1 |
| Obesity | pericyte | *P2RY12* | 1.927599198 | 2.72E-06 | 8.07E-06 | 0.017069741 |
| Obesity | pericyte | *SLC33A1* | 5.883974717 | 9.41E-09 | 1.70E-07 | 5.92E-05 |
| Obesity | pericyte | *SNUPN* | 5.811661043 | 1.56E-06 | 5.03E-06 | 0.009782619 |
| Obesity | pericyte | *PTAFR* | 3.31516886 | 0.000136973 | 0.000266115 | 0.861148565 |
| Obesity | pericyte | *RNASE6* | -3.625122507 | 0.001179246 | 0.001929703 | 1 |
| Obesity | pericyte | *NPR1* | 9.092064351 | 3.39E-08 | 2.86E-07 | 0.000213096 |
| Obesity | pericyte | *CLIC4* | 5.195545803 | 2.46E-05 | 5.66E-05 | 0.154762672 |
| Obesity | pericyte | *CCDC8* | 4.873938564 | 1.53E-06 | 4.98E-06 | 0.009650225 |
| Obesity | pericyte | *METTL15* | 4.147645291 | 1.40E-07 | 7.55E-07 | 0.000879164 |
| Obesity | pericyte | *HINT1* | 3.557659119 | 0.017543462 | 0.024103091 | 1 |
| Obesity | pericyte | *DTWD2* | 4.132666357 | 4.46E-08 | 3.41E-07 | 0.000280322 |
| Obesity | pericyte | *CLIC3* | 3.346396972 | 8.40E-05 | 0.000170466 | 0.528274932 |
| Obesity | pericyte | *INO80E* | -4.54276151 | 2.16E-06 | 6.61E-06 | 0.013553225 |
| Obesity | pericyte | *DFFB* | 6.173558305 | 6.56E-08 | 4.43E-07 | 0.000412375 |
| Obesity | pericyte | *FAM103A1* | -2.309100856 | 3.92E-07 | 1.70E-06 | 0.002463425 |
| Obesity | pericyte | *LUZP1* | 5.85624823 | 8.00E-07 | 2.96E-06 | 0.005030244 |

| | | | | | | |
|---|---|---|---|---|---|---|
| Obesity | pericyte | *HEXDC* | 5.120526542 | 3.95E-06 | 1.12E-05 | 0.024812002 |
| Obesity | pericyte | *CHRNA5* | 2.544548683 | 0.004333356 | 0.006509871 | 1 |
| Obesity | pericyte | *FASN* | 2.435499157 | 6.20E-07 | 2.43E-06 | 0.00389667 |
| Obesity | pericyte | *DCXR* | 3.109406247 | 1.65E-07 | 8.55E-07 | 0.001035773 |
| Obesity | pericyte | *LIMS1* | 2.907040478 | 4.77E-05 | 0.000102422 | 0.299756974 |
| Obesity | pericyte | *UGP2* | 3.138384699 | 2.06E-08 | 2.16E-07 | 0.000129819 |
| Obesity | pericyte | *CSGALNACT2* | 6.823103651 | 5.40E-08 | 3.90E-07 | 0.000339303 |
| Obesity | pericyte | *AVEN* | 5.932261078 | 3.62E-09 | 1.10E-07 | 2.27E-05 |
| Obesity | pericyte | *GUSB* | 7.498736523 | 5.93E-08 | 4.15E-07 | 0.000373025 |
| Obesity | pericyte | *BRD3* | 7.867849319 | 1.19E-05 | 2.96E-05 | 0.07494212 |
| Obesity | pericyte | *ZFPM2* | 4.757935147 | 3.36E-08 | 2.85E-07 | 0.00021147 |
| Obesity | pericyte | *ZNF747* | 3.168586532 | 1.19E-05 | 2.95E-05 | 0.074791783 |
| Obesity | pericyte | *MAP3K2* | 6.098527418 | 6.04E-07 | 2.39E-06 | 0.003796851 |
| Obesity | pericyte | *MYO7B* | 3.111930762 | 1.11E-07 | 6.47E-07 | 0.000700215 |
| Obesity | pericyte | *NIPA1* | 8.982095284 | 1.14E-07 | 6.56E-07 | 0.000714229 |
| Obesity | pericyte | *PDCD6IP* | 3.548535041 | 0.000112623 | 0.000222451 | 0.708060102 |
| Obesity | pericyte | *GLB1* | 4.301068106 | 1.16E-08 | 1.72E-07 | 7.28E-05 |
| Obesity | pericyte | *FAXDC2* | 3.691772734 | 4.73E-06 | 1.31E-05 | 0.029714775 |
| Obesity | pericyte | *CRTAP* | 6.308295224 | 3.05E-09 | 9.89E-08 | 1.92E-05 |
| Obesity | pericyte | *STX8* | 5.228799921 | 1.34E-07 | 7.33E-07 | 0.000839658 |
| Obesity | pericyte | *NFRKB* | 2.905808183 | 2.48E-05 | 5.70E-05 | 0.155842091 |
| Obesity | pericyte | *SP7* | -3.583438974 | 3.92E-05 | 8.55E-05 | 0.246478469 |
| Obesity | pericyte | *DCLK2* | 2.671795418 | 2.74E-05 | 6.22E-05 | 0.172072088 |
| Obesity | pericyte | *MGMT* | 3.174551467 | 0.000208388 | 0.00039132 | 1 |
| Obesity | pericyte | *METTL7B* | 8.141755473 | 9.31E-07 | 3.34E-06 | 0.005853734 |
| Obesity | pericyte | *NFXL1* | 6.337043906 | 9.14E-05 | 0.000184378 | 0.574520745 |
| Obesity | pericyte | *CD14* | 1.50400233 | 0.029718613 | 0.039484557 | 1 |
| Obesity | pericyte | *KRT4* | -4.68168443 | 0.000352973 | 0.000633498 | 1 |
| Obesity | pericyte | *HSD17B13* | -1.042655408 | 0.002252376 | 0.003528704 | 1 |
| Obesity | pericyte | *ARL6IP1* | 3.387367128 | 1.73E-05 | 4.13E-05 | 0.108988973 |
| Obesity | pericyte | *SERPINB9* | 4.410715162 | 2.13E-05 | 4.96E-05 | 0.133672171 |
| Obesity | pericyte | *IRX2* | 1.78075738 | 0.000848669 | 0.001423581 | 1 |
| Obesity | pericyte | *EMB* | 1.413763164 | 0.000230737 | 0.000428805 | 1 |
| Obesity | pericyte | *STAT2* | 4.758322413 | 1.65E-06 | 5.29E-06 | 0.010369467 |
| Obesity | pericyte | *NUDCD2* | 3.760308778 | 4.59E-07 | 1.93E-06 | 0.002888818 |
| Obesity | pericyte | *HSPA4* | 7.177230428 | 1.03E-08 | 1.71E-07 | 6.50E-05 |
| Obesity | pericyte | *SLC26A5* | 6.468247102 | 1.13E-09 | 5.30E-08 | 7.10E-06 |
| Obesity | pericyte | *COMMD5* | 5.930624415 | 0.000546364 | 0.000950469 | 1 |
| Obesity | pericyte | *GTSF1* | 7.677289821 | 5.15E-07 | 2.11E-06 | 0.003239272 |
| Obesity | pericyte | *ARMC10* | 3.282952507 | 0.000786405 | 0.001327638 | 1 |
| Obesity | pericyte | *RNF34* | 3.99902932 | 0.000427626 | 0.00075903 | 1 |
| Obesity | pericyte | *ACYP2* | 4.97070252 | 2.42E-06 | 7.32E-06 | 0.015232932 |
| Obesity | pericyte | *TRABD* | 3.288394076 | 0.000228354 | 0.00042513 | 1 |
| Obesity | pericyte | *TMEM133* | -6.597187736 | 0.004423923 | 0.006631666 | 1 |
| Obesity | pericyte | *TTLL6* | -11.63844613 | 0.000645599 | 0.001107773 | 1 |
| Obesity | pericyte | *KCNS3* | 0.961606558 | 0.000532677 | 0.000928456 | 1 |
| Obesity | pericyte | *AKAP13* | 4.677005303 | 7.68E-07 | 2.87E-06 | 0.004825324 |
| Obesity | pericyte | *HTRA3* | 1.998770854 | 0.000646862 | 0.001109637 | 1 |
| Obesity | pericyte | *FOXN2* | 5.08478692 | 9.50E-09 | 1.70E-07 | 5.97E-05 |
| Obesity | pericyte | *GPR27* | 3.853090969 | 0.004575924 | 0.006844834 | 1 |

| | | | | | | |
|---|---|---|---|---|---|---|
| Obesity | pericyte | *MTSS1* | 5.092058594 | 4.98E-09 | 1.29E-07 | 3.13E-05 |
| Obesity | pericyte | *RPS9* | 1.967838482 | 0.034278692 | 0.045151924 | 1 |
| Obesity | pericyte | *TSEN34* | 7.216072474 | 0.00249348 | 0.003883207 | 1 |
| Obesity | pericyte | *NDUFA3* | 6.989688785 | 5.40E-05 | 0.000114202 | 0.339522212 |
| Obesity | pericyte | *PAQR8* | 2.745718828 | 4.11E-05 | 8.95E-05 | 0.25848892 |
| Obesity | pericyte | *NUDT6* | 3.428356345 | 5.18E-07 | 2.12E-06 | 0.003257403 |
| Obesity | pericyte | *ZNF160* | 4.797696603 | 6.47E-08 | 4.40E-07 | 0.000406659 |
| Obesity | pericyte | *TSNARE1* | 2.89100947 | 4.67E-06 | 1.30E-05 | 0.029341485 |
| Obesity | pericyte | *FPR1* | 1.921242134 | 5.95E-06 | 1.61E-05 | 0.037431743 |
| Obesity | pericyte | *FEZ2* | 6.052132842 | 1.24E-07 | 6.95E-07 | 0.000777859 |
| Obesity | pericyte | *C11orf24* | 3.76594064 | 2.45E-07 | 1.16E-06 | 0.001538299 |
| Obesity | pericyte | *ALK* | 1.954412737 | 2.23E-07 | 1.08E-06 | 0.001400969 |
| Obesity | pericyte | *TRMT61B* | 2.240009062 | 6.50E-09 | 1.51E-07 | 4.08E-05 |
| Obesity | pericyte | *MFN1* | 4.383023098 | 7.65E-05 | 0.000156263 | 0.480820032 |
| Obesity | pericyte | *GIMAP8* | 2.634421822 | 7.35E-05 | 0.000150803 | 0.462362963 |
| Obesity | pericyte | *KCNMB3* | 2.795399375 | 0.000135365 | 0.000263154 | 0.851039109 |
| Obesity | pericyte | *GRIK1* | 2.804730129 | 1.54E-05 | 3.71E-05 | 0.096603967 |
| Obesity | pericyte | *CLDN20* | 2.475837783 | 6.84E-05 | 0.00014114 | 0.429913832 |
| Obesity | pericyte | *CHST11* | 2.420108967 | 1.31E-05 | 3.23E-05 | 0.082425662 |
| Obesity | pericyte | *KCND3* | 6.117624277 | 1.16E-08 | 1.72E-07 | 7.28E-05 |
| Obesity | pericyte | *ZNF581* | -1.786991472 | 7.67E-05 | 0.000156637 | 0.482127774 |
| Obesity | pericyte | *MCC* | 5.932346936 | 7.30E-08 | 4.76E-07 | 0.00045922 |
| Obesity | pericyte | *DLK2* | 2.622077661 | 0.038035935 | 0.049694913 | 1 |
| Obesity | pericyte | *HOPX* | -3.069484787 | 0.000829727 | 0.001394784 | 1 |
| Obesity | pericyte | *LRRC8C* | 10.24275616 | 1.16E-06 | 3.92E-06 | 0.007270399 |
| Obesity | pericyte | *RSL1D1* | 6.762884221 | 1.51E-09 | 6.47E-08 | 9.52E-06 |
| Obesity | pericyte | *PPID* | 6.240617288 | 9.21E-05 | 0.000185545 | 0.578786176 |
| Obesity | pericyte | *TBCA* | 3.631652048 | 0.000582714 | 0.001009184 | 1 |
| Obesity | pericyte | *MAP6* | 3.036734001 | 3.37E-05 | 7.46E-05 | 0.212076882 |
| Obesity | pericyte | *BCL2L1* | 5.468712574 | 8.73E-06 | 2.25E-05 | 0.05490261 |
| Obesity | pericyte | *PLRG1* | 6.453848012 | 4.86E-09 | 1.29E-07 | 3.06E-05 |
| Obesity | pericyte | *NMUR1* | 11.02292276 | 4.61E-08 | 3.50E-07 | 0.00028976 |
| Obesity | pericyte | *P2RY6* | 2.61016883 | 9.48E-07 | 3.38E-06 | 0.005957951 |
| Obesity | pericyte | *ZIK1* | 4.099931453 | 8.56E-07 | 3.11E-06 | 0.005384322 |
| Obesity | pericyte | *RGS19* | 1.936894486 | 3.52E-08 | 2.92E-07 | 0.000221552 |
| Obesity | pericyte | *TCEA2* | 5.050087716 | 4.68E-08 | 3.54E-07 | 0.000294393 |
| Obesity | pericyte | *ANO5* | 2.234036382 | 6.73E-07 | 2.57E-06 | 0.004228069 |
| Obesity | pericyte | *GPHN* | 1.962359571 | 0.004889667 | 0.007272613 | 1 |
| Obesity | pericyte | *VAT1L* | 3.2065449 | 4.32E-06 | 1.22E-05 | 0.027184151 |
| Obesity | pericyte | *CAMTA1* | 6.901975243 | 1.23E-08 | 1.73E-07 | 7.73E-05 |
| Obesity | pericyte | *LGALS4* | 2.437868335 | 5.18E-05 | 0.000110021 | 0.325795981 |
| Obesity | pericyte | *SPATA5L1* | 2.708305211 | 0.00053328 | 0.00092925 | 1 |
| Obesity | pericyte | *SYCE1* | 1.375527415 | 0.020723279 | 0.028151957 | 1 |
| Obesity | pericyte | *RHNO1* | 2.170211195 | 0.00605813 | 0.008894783 | 1 |
| Obesity | pericyte | *KNDC1* | -1.268658584 | 0.00018294 | 0.000347265 | 1 |
| Obesity | pericyte | *METTL18* | 3.502034264 | 5.37E-07 | 2.18E-06 | 0.00337459 |
| Obesity | pericyte | *PWWP2B* | 2.613686481 | 7.96E-07 | 2.95E-06 | 0.005003029 |
| Obesity | pericyte | *ZNF570* | 2.52535353 | 8.69E-07 | 3.15E-06 | 0.00546364 |
| Obesity | pericyte | *NINJ2* | 5.901311385 | 5.17E-09 | 1.30E-07 | 3.25E-05 |
| Obesity | pericyte | *FAM90A1* | -2.185067478 | 3.55E-05 | 7.83E-05 | 0.223013883 |

| | | | | | | |
|---|---|---|---|---|---|---|
| Obesity | pericyte | *TRAPPC12* | 6.004988312 | 4.20E-06 | 1.19E-05 | 0.02643552 |
| Obesity | pericyte | *C3AR1* | 2.262872075 | 8.29E-05 | 0.000168291 | 0.521027623 |
| Obesity | pericyte | *AQP4* | 4.856450946 | 1.80E-07 | 9.12E-07 | 0.00112878 |
| Obesity | pericyte | *TVP23B* | 5.592206549 | 4.21E-06 | 1.19E-05 | 0.026476026 |
| Obesity | pericyte | *SRGAP2C* | 5.858866407 | 2.79E-05 | 6.33E-05 | 0.175621463 |
| Obesity | pericyte | *ATPAF2* | 7.412406672 | 3.97E-09 | 1.14E-07 | 2.49E-05 |
| Obesity | pericyte | *CYP4F22* | -3.85888642 | 0.001343319 | 0.002183414 | 1 |
| Obesity | pericyte | *C20orf196* | 5.849032771 | 6.48E-08 | 4.41E-07 | 0.000407558 |
| Obesity | pericyte | *ZNF554* | 1.490228189 | 0.008180931 | 0.011753545 | 1 |
| Obesity | pericyte | *ANKRD20A4* | 4.884735972 | 1.05E-06 | 3.67E-06 | 0.006621635 |
| Obesity | pericyte | *LAMB2* | 2.820830342 | 0.000324636 | 0.000586491 | 1 |
| Obesity | pericyte | *ORMDL3* | 1.284795765 | 2.12E-05 | 4.96E-05 | 0.13350016 |
| Obesity | pericyte | *SERF1A* | 10.63131478 | 1.10E-06 | 3.80E-06 | 0.00693494 |
| Obesity | pericyte | *MOB3A* | 4.453192833 | 9.18E-07 | 3.30E-06 | 0.005769162 |
| Obesity | pericyte | *FRMD3* | -0.859050854 | 0.00065206 | 0.00111764 | 1 |
| Obesity | pericyte | *MTBP* | 5.324379122 | 0.002241031 | 0.003513556 | 1 |
| Obesity | pericyte | *MALT1* | 5.857330324 | 2.01E-09 | 7.85E-08 | 1.26E-05 |
| Obesity | pericyte | *ISG20* | 1.241893687 | 0.002610461 | 0.00405434 | 1 |
| Obesity | pericyte | *MBOAT1* | 6.656868069 | 9.48E-09 | 1.70E-07 | 5.96E-05 |
| Obesity | pericyte | *CXCR6* | 2.830593894 | 3.33E-05 | 7.38E-05 | 0.20919668 |
| Obesity | pericyte | *C5orf34* | 4.528043565 | 2.25E-07 | 1.08E-06 | 0.001415726 |
| Obesity | pericyte | *C1QTNF4* | -1.526621585 | 6.21E-19 | 3.55E-16 | 3.91E-15 |
| Obesity | pericyte | *ZNF131* | 5.578200333 | 5.10E-08 | 3.75E-07 | 0.00032043 |
| Obesity | pericyte | *HINFP* | 4.089824891 | 6.57E-08 | 4.43E-07 | 0.000412926 |
| Obesity | pericyte | *SPTLC3* | 6.666343875 | 5.18E-05 | 0.000110021 | 0.325881545 |
| Obesity | pericyte | *SUCLG2* | 3.906678043 | 0.000246234 | 0.000455182 | 1 |
| Obesity | pericyte | *CSDC2* | 5.150708235 | 9.90E-07 | 3.50E-06 | 0.006221572 |
| Obesity | pericyte | *RCAN2* | 6.701281916 | 3.02E-06 | 8.88E-06 | 0.018980002 |
| Obesity | pericyte | *IL16* | 2.273983045 | 0.00335826 | 0.005114675 | 1 |
| Obesity | pericyte | *GNB2* | 4.278523438 | 3.62E-05 | 7.96E-05 | 0.227281161 |
| Obesity | pericyte | *PRSS27* | -1.917844006 | 4.24E-05 | 9.19E-05 | 0.2664254 |
| Obesity | pericyte | *FGGY* | 5.003907979 | 1.08E-07 | 6.35E-07 | 0.000681085 |
| Obesity | pericyte | *ZNF24* | 6.019874658 | 4.25E-09 | 1.20E-07 | 2.67E-05 |
| Obesity | pericyte | *MANEA* | 4.768894268 | 4.33E-07 | 1.85E-06 | 0.002724996 |
| Obesity | pericyte | *CARNS1* | 6.309550807 | 7.44E-08 | 4.82E-07 | 0.000467552 |
| Obesity | pericyte | *CTSW* | -0.577977014 | 0.000180245 | 0.000342459 | 1 |
| Obesity | pericyte | *NIPAL4* | -2.767955188 | 0.001822693 | 0.002897414 | 1 |
| Obesity | pericyte | *SNTG2* | 1.888116086 | 0.000690376 | 0.001176896 | 1 |
| Obesity | pericyte | *MRPL52* | 3.301626968 | 0.000244216 | 0.000451717 | 1 |
| Obesity | pericyte | *SMPDL3A* | 3.974468447 | 1.05E-07 | 6.18E-07 | 0.000657055 |
| Obesity | pericyte | *RAD9A* | 1.090771378 | 0.003307748 | 0.005046302 | 1 |
| Obesity | pericyte | *EFEMP2* | 3.522878286 | 2.57E-05 | 5.87E-05 | 0.161287028 |
| Obesity | pericyte | *AGAP5* | 63.05176831 | 1.11E-06 | 3.81E-06 | 0.006976949 |
| Obesity | pericyte | *ZMAT3* | 9.244441181 | 1.72E-06 | 5.48E-06 | 0.010792196 |
| Obesity | pericyte | *ZFAND4* | 4.02791231 | 0.000103162 | 0.000205834 | 0.6485821 |
| Obesity | pericyte | *ZNF738* | 2.107297582 | 0.000309859 | 0.000561957 | 1 |
| Obesity | pericyte | *SLFN11* | 2.423808138 | 0.007519255 | 0.010872483 | 1 |
| Obesity | pericyte | *CCL19* | -3.147594806 | 1.78E-05 | 4.23E-05 | 0.112010285 |
| Obesity | pericyte | *FUT10* | -1.363944796 | 0.006919138 | 0.010055622 | 1 |
| Obesity | pericyte | *CFL1* | 6.625794266 | 2.54E-08 | 2.37E-07 | 0.000159715 |

| | | | | | | |
|---|---|---|---|---|---|---|
| Obesity | pericyte | *FAM192A* | 2.106702812 | 0.000688655 | 0.001174599 | 1 |
| Obesity | pericyte | *CBWD1* | 6.757219466 | 8.75E-08 | 5.44E-07 | 0.000550206 |
| Obesity | pericyte | *HOXC5* | 2.28951257 | 0.004007712 | 0.006048124 | 1 |
| Obesity | pericyte | *RAB37* | 3.4231204 | 1.40E-07 | 7.55E-07 | 0.000879866 |
| Obesity | pericyte | *DCP2* | 6.256801578 | 3.59E-08 | 2.95E-07 | 0.000225702 |
| Obesity | pericyte | *SNX32* | 4.204438594 | 4.34E-10 | 2.93E-08 | 2.73E-06 |
| Obesity | pericyte | *METAP1D* | 8.311399049 | 3.41E-09 | 1.06E-07 | 2.14E-05 |
| Obesity | pericyte | *NADSYN1* | 5.916832715 | 5.71E-09 | 1.38E-07 | 3.59E-05 |
| Obesity | pericyte | *RNASEH2C* | 2.898142088 | 3.21E-05 | 7.14E-05 | 0.201538814 |
| Obesity | pericyte | *MYEOV* | 0.533826926 | 0.008180297 | 0.011753545 | 1 |
| Obesity | pericyte | *ANKRD13D* | 4.892550788 | 1.47E-06 | 4.81E-06 | 0.009270272 |
| Obesity | pericyte | *MRGPRF* | 5.568923387 | 1.09E-06 | 3.74E-06 | 0.006822556 |
| Obesity | pericyte | *GXYLT2* | 2.997120234 | 3.00E-06 | 8.84E-06 | 0.018854209 |
| Obesity | pericyte | *HPSE2* | 4.008154535 | 5.11E-16 | 2.30E-13 | 3.22E-12 |
| Obesity | pericyte | *HECTD4* | 4.486615469 | 1.17E-05 | 2.91E-05 | 0.073681706 |
| Obesity | pericyte | *BNC2* | 3.641825928 | 5.13E-09 | 1.30E-07 | 3.23E-05 |
| Obesity | pericyte | *NOC3L* | 5.074183811 | 7.14E-09 | 1.59E-07 | 4.49E-05 |
| Obesity | pericyte | *PARP14* | 0.946950277 | 0.010960425 | 0.015509384 | 1 |
| Obesity | pericyte | *PARP15* | 2.038404891 | 0.000134093 | 0.000260924 | 0.843045049 |
| Obesity | pericyte | *AHSA2* | 3.740506609 | 1.10E-05 | 2.75E-05 | 0.069076302 |
| Obesity | pericyte | *VANGL1* | 2.665443862 | 3.22E-07 | 1.45E-06 | 0.002026756 |
| Obesity | pericyte | *GLRX* | 3.579726608 | 0.007292752 | 0.010564408 | 1 |
| Obesity | pericyte | *GOLGB1* | 6.168857797 | 7.62E-09 | 1.63E-07 | 4.79E-05 |
| Obesity | pericyte | *ZNF483* | 1.123366994 | 0.008624677 | 0.012354373 | 1 |
| Obesity | pericyte | *SLC2A14* | 1.527677923 | 0.001687842 | 0.002696687 | 1 |
| Obesity | pericyte | *MAP3K11* | 8.414545741 | 1.82E-05 | 4.31E-05 | 0.114163136 |
| Obesity | pericyte | *C1QA* | 4.835952421 | 1.67E-08 | 1.93E-07 | 0.00010507 |
| Obesity | pericyte | *NDNF* | 11.0167609 | 3.85E-09 | 1.13E-07 | 2.42E-05 |
| Obesity | pericyte | *OLR1* | 10.5894622 | 0.016550817 | 0.022809071 | 1 |
| Obesity | pericyte | *GLIPR1L1* | 4.348604782 | 2.55E-07 | 1.19E-06 | 0.001600127 |
| Obesity | pericyte | *ARV1* | 5.035359 | 5.00E-09 | 1.29E-07 | 3.15E-05 |
| Obesity | pericyte | *CCDC36* | 4.192297478 | 4.98E-10 | 3.23E-08 | 3.13E-06 |
| Obesity | pericyte | *THAP2* | 4.093471443 | 0.000312016 | 0.000565642 | 1 |
| Obesity | pericyte | *PPP1R14B* | 5.532543017 | 2.13E-07 | 1.04E-06 | 0.001338725 |
| Obesity | pericyte | *SMARCC1* | 6.135332441 | 9.43E-08 | 5.77E-07 | 0.000593111 |
| Obesity | pericyte | *ZNF417* | 2.096692294 | 0.000128908 | 0.000252083 | 0.810446639 |
| Obesity | pericyte | *VEGFB* | 1.565270987 | 0.004423765 | 0.006631666 | 1 |
| Obesity | pericyte | *PEAK1* | 5.215106638 | 5.75E-07 | 2.30E-06 | 0.003615379 |
| Obesity | pericyte | *MST1* | -0.859709683 | 0.024161406 | 0.032520394 | 1 |
| Obesity | pericyte | *TNFRSF10C* | 0.93914921 | 0.006370973 | 0.009306299 | 1 |
| Obesity | pericyte | *GMPPB* | 6.261902097 | 6.04E-14 | 1.90E-11 | 3.80E-10 |
| Obesity | pericyte | *CSPG4* | -1.809637718 | 0.000168393 | 0.000321399 | 1 |
| Obesity | pericyte | *SNX33* | 3.475065219 | 0.002779633 | 0.004294979 | 1 |
| Obesity | pericyte | *NUDT18* | 9.803887063 | 2.72E-07 | 1.26E-06 | 0.001710673 |
| Obesity | pericyte | *SULT1B1* | 2.457607379 | 0.005005755 | 0.007434723 | 1 |
| Obesity | pericyte | *NUDT4* | 3.318608525 | 0.000408311 | 0.000726181 | 1 |
| Obesity | pericyte | *SCAI* | 6.427627821 | 2.32E-08 | 2.25E-07 | 0.000145646 |
| Obesity | pericyte | *C1orf100* | -2.462998003 | 0.000156303 | 0.00030042 | 0.982675258 |
| Obesity | pericyte | *AGFG1* | 3.388405971 | 1.68E-06 | 5.39E-06 | 0.01058037 |
| Obesity | pericyte | *RNF213* | 5.283934495 | 7.58E-06 | 1.99E-05 | 0.04763536 |

| | | | | | | |
|---|---|---|---|---|---|---|
| Obesity | pericyte | *DPY19L1* | 5.267394675 | 0.001542254 | 0.002486193 | 1 |
| Obesity | pericyte | *TUBB8* | -3.488201699 | 2.31E-05 | 5.34E-05 | 0.145087623 |
| Obesity | pericyte | *GPR160* | 2.876248606 | 8.47E-06 | 2.19E-05 | 0.053266763 |
| Obesity | pericyte | *GOLIM4* | 3.63814181 | 0.000872741 | 0.001459568 | 1 |
| Obesity | pericyte | *HOXB2* | 4.146180354 | 7.85E-09 | 1.66E-07 | 4.93E-05 |
| Obesity | pericyte | *XXYLT1* | 4.291268147 | 6.54E-07 | 2.52E-06 | 0.004113543 |
| Obesity | pericyte | *UBXN2A* | 3.123804396 | 0.002094968 | 0.003299364 | 1 |
| Obesity | pericyte | *CCS* | 5.200361458 | 1.78E-09 | 7.22E-08 | 1.12E-05 |
| Obesity | pericyte | *NRROS* | 1.272809846 | 0.000673032 | 0.001151076 | 1 |
| Obesity | pericyte | *SLC25A30* | 6.936503012 | 7.60E-09 | 1.63E-07 | 4.78E-05 |
| Obesity | pericyte | *LEMD3* | 6.206101089 | 6.31E-07 | 2.46E-06 | 0.003965168 |
| Obesity | pericyte | *C16orf91* | 6.353837915 | 4.47E-07 | 1.89E-06 | 0.002811463 |
| Obesity | pericyte | *SOCS7* | 7.715015628 | 1.46E-08 | 1.86E-07 | 9.19E-05 |
| Obesity | pericyte | *TLR6* | 3.581791139 | 1.82E-07 | 9.18E-07 | 0.001143954 |
| Obesity | pericyte | *RGMB* | 4.434894708 | 3.66E-05 | 8.04E-05 | 0.2298323 |
| Obesity | pericyte | *FAM53A* | 2.726407241 | 2.95E-07 | 1.35E-06 | 0.001856242 |
| Obesity | pericyte | *CTU2* | 4.119623708 | 1.53E-07 | 8.04E-07 | 0.000961044 |
| Obesity | pericyte | *MGA* | 2.102489211 | 0.022404146 | 0.030265334 | 1 |
| Obesity | pericyte | *SNX31* | -0.761370439 | 3.08E-05 | 6.91E-05 | 0.19390285 |
| Obesity | pericyte | *DDX23* | 6.505509992 | 4.11E-08 | 3.22E-07 | 0.000258302 |
| Obesity | pericyte | *GLIS1* | 1.049428492 | 0.000377459 | 0.000674887 | 1 |
| Obesity | pericyte | *RALGAPA1* | 3.665575338 | 4.22E-05 | 9.16E-05 | 0.265526271 |
| Obesity | pericyte | *LIG4* | 2.851070783 | 3.84E-05 | 8.40E-05 | 0.241495099 |
| Obesity | pericyte | *SNAPC5* | 2.870025277 | 0.00671211 | 0.009772819 | 1 |
| Obesity | pericyte | *LINGO2* | 2.465253543 | 5.95E-08 | 4.15E-07 | 0.000373796 |
| Obesity | pericyte | *TMEM81* | 8.424666668 | 5.52E-06 | 1.50E-05 | 0.034681292 |
| Obesity | pericyte | *IL20RB* | 1.845637233 | 0.000130678 | 0.000255212 | 0.821574375 |
| Obesity | pericyte | *ZNF497* | 6.698228321 | 0.012006733 | 0.016898663 | 1 |
| Obesity | pericyte | *KLF14* | 20.64814728 | 6.67E-10 | 3.78E-08 | 4.19E-06 |
| Obesity | pericyte | *ANGEL2* | 7.169295613 | 2.45E-08 | 2.32E-07 | 0.000154005 |
| Obesity | pericyte | *ZNF266* | 1.725031509 | 5.45E-05 | 0.000115082 | 0.342600228 |
| Obesity | pericyte | *BRSK2* | -2.578312066 | 0.000145728 | 0.000281992 | 0.916193515 |
| Obesity | pericyte | *SH3PXD2B* | 1.788627017 | 0.001248028 | 0.002036587 | 1 |
| Obesity | pericyte | *LARP7* | 6.99111048 | 2.90E-06 | 8.56E-06 | 0.018223208 |
| Obesity | pericyte | *C4orf32* | -1.172132832 | 0.001335037 | 0.002171075 | 1 |
| Obesity | pericyte | *HRAS* | 6.286991836 | 2.77E-10 | 2.12E-08 | 1.74E-06 |
| Obesity | pericyte | *SRP72* | 2.325302438 | 0.001025073 | 0.001691505 | 1 |
| Obesity | pericyte | *C4orf26* | 6.276076302 | 0.000322912 | 0.000583543 | 1 |
| Obesity | pericyte | *BTC* | 2.090919831 | 1.65E-06 | 5.29E-06 | 0.01036249 |
| Obesity | pericyte | *NDUFA11* | 8.120720171 | 3.05E-05 | 6.85E-05 | 0.192063258 |
| Obesity | pericyte | *FUT1* | 4.996052119 | 3.83E-06 | 1.09E-05 | 0.02408081 |
| Obesity | pericyte | *DHX36* | 3.876437893 | 0.002416521 | 0.003772701 | 1 |
| Obesity | pericyte | *FBXW8* | 7.376735789 | 3.06E-08 | 2.68E-07 | 0.000192585 |
| Obesity | pericyte | *SLC22A1* | 2.8205252 | 0.0002312 | 0.000429538 | 1 |
| Obesity | pericyte | *CHST2* | -1.116939804 | 2.86E-05 | 6.47E-05 | 0.18011384 |
| Obesity | pericyte | *ATR* | 5.948522856 | 1.57E-08 | 1.91E-07 | 9.86E-05 |
| Obesity | pericyte | *TVP23C* | 3.201921609 | 1.12E-06 | 3.82E-06 | 0.00702204 |
| Obesity | pericyte | *PACS1* | 4.901545437 | 1.12E-06 | 3.83E-06 | 0.007034063 |
| Obesity | pericyte | *SH3BP5L* | 6.672994133 | 3.86E-07 | 1.68E-06 | 0.002423751 |
| Obesity | pericyte | *CADM2* | 1.4471022 | 0.006456723 | 0.009416241 | 1 |

| | | | | | | |
|---|---|---|---|---|---|---|
| Obesity | pericyte | *PSMD2* | 1.92674643 | 0.006256654 | 0.009160821 | 1 |
| Obesity | pericyte | *CHST1* | -2.428443541 | 1.35E-05 | 3.31E-05 | 0.085029376 |
| Obesity | pericyte | *GOLGA8A* | 4.120898952 | 1.36E-05 | 3.33E-05 | 0.08543349 |
| Obesity | pericyte | *TP53I11* | 6.824972671 | 5.15E-06 | 1.42E-05 | 0.032359681 |
| Obesity | pericyte | *PHYHD1* | 3.908425415 | 0.000149986 | 0.000289786 | 0.942962074 |
| Obesity | pericyte | *ZNF519* | 2.786377976 | 6.04E-08 | 4.19E-07 | 0.000379742 |
| Obesity | pericyte | *CHRNA7* | 1.836034125 | 0.002460515 | 0.003833769 | 1 |
| Obesity | pericyte | *TMEM9B* | 8.524734749 | 5.46E-13 | 1.19E-10 | 3.43E-09 |
| Obesity | pericyte | *NRIP3* | 1.638305408 | 0.001214115 | 0.001984181 | 1 |
| Obesity | pericyte | *SCUBE2* | 3.909790968 | 8.80E-06 | 2.27E-05 | 0.055353335 |
| Obesity | pericyte | *SMAD2* | 3.113038094 | 5.12E-05 | 0.000108871 | 0.321712633 |
| Obesity | pericyte | *EIF3F* | 4.572706617 | 8.34E-06 | 2.16E-05 | 0.052463832 |
| Obesity | pericyte | *CLTB* | 3.353258895 | 1.64E-06 | 5.27E-06 | 0.010322399 |
| Obesity | pericyte | *PCSK1* | 3.604364324 | 5.13E-05 | 0.000109051 | 0.322355312 |
| Obesity | pericyte | *TBC1D10C* | 1.763186598 | 0.012674474 | 0.017794644 | 1 |
| Obesity | pericyte | *PPP2R2D* | 3.496326013 | 6.75E-05 | 0.000139442 | 0.424080745 |
| Obesity | pericyte | *MCTP1* | 1.691141621 | 0.012953001 | 0.018149213 | 1 |
| Obesity | pericyte | *UBQLNL* | 2.803131533 | 0.027150699 | 0.036333853 | 1 |
| Obesity | pericyte | *LIPT2* | -5.509521422 | 1.49E-05 | 3.62E-05 | 0.093817174 |
| Obesity | pericyte | *KCNE3* | 2.499601265 | 1.07E-06 | 3.70E-06 | 0.006725063 |
| Obesity | pericyte | *UCP2* | 3.566539327 | 0.017495533 | 0.024043419 | 1 |
| Obesity | pericyte | *RAB6A* | 4.801567622 | 8.16E-06 | 2.12E-05 | 0.051304541 |
| Obesity | pericyte | *RPS6KB2* | -2.065596746 | 0.004450423 | 0.006666621 | 1 |
| Obesity | pericyte | *TOM1L2* | 6.340305663 | 1.22E-08 | 1.73E-07 | 7.69E-05 |
| Obesity | pericyte | *TEX26* | 4.489365974 | 2.46E-06 | 7.40E-06 | 0.015445981 |
| Obesity | pericyte | *GPR156* | 5.17423126 | 7.74E-07 | 2.88E-06 | 0.00486412 |
| Obesity | pericyte | *B3GNTL1* | 4.265599415 | 5.03E-08 | 3.72E-07 | 0.000315968 |
| Obesity | pericyte | *LINC01106* | 10.19468561 | 0.00625681 | 0.009160821 | 1 |
| Obesity | pericyte | *MSRA* | 3.455374355 | 0.004150705 | 0.006251913 | 1 |
| Obesity | pericyte | *CTDNEP1* | 10.09642347 | 3.83E-10 | 2.73E-08 | 2.41E-06 |
| Obesity | pericyte | *ETV4* | 3.710067853 | 2.70E-05 | 6.14E-05 | 0.169904055 |
| Obesity | pericyte | *SWI5* | 3.222542298 | 8.41E-06 | 2.18E-05 | 0.05289822 |
| Obesity | pericyte | *ZDHHC21* | 5.067961921 | 3.61E-06 | 1.04E-05 | 0.022704598 |
| Obesity | pericyte | *LRRN1* | 5.373668377 | 4.52E-08 | 3.45E-07 | 0.000283981 |
| Obesity | pericyte | *B3GALT6* | -3.257884345 | 0.000802408 | 0.001352115 | 1 |
| Obesity | pericyte | *ZNF613* | 2.318765052 | 0.007800492 | 0.011237785 | 1 |
| Obesity | pericyte | *IP6K1* | 5.789185841 | 3.65E-06 | 1.05E-05 | 0.022923421 |
| Obesity | pericyte | *SSNA1* | 11.96942387 | 1.59E-08 | 1.91E-07 | 0.000100185 |
| Obesity | pericyte | *CHMP6* | 5.951175959 | 1.48E-07 | 7.83E-07 | 0.000927655 |
| Obesity | pericyte | *UFSP1* | 1.374701048 | 0.007637014 | 0.011035143 | 1 |
| Obesity | pericyte | *TMEM39A* | 4.238955194 | 3.00E-05 | 6.74E-05 | 0.188449179 |
| Obesity | pericyte | *TCP11L1* | 3.695280163 | 6.72E-05 | 0.000139037 | 0.422671406 |
| Obesity | pericyte | *GPX2* | 5.498986113 | 3.08E-07 | 1.40E-06 | 0.001934559 |
| Obesity | pericyte | *SMIM19* | 2.063124673 | 0.000385532 | 0.000688005 | 1 |
| Obesity | pericyte | *ZNF404* | 0.842537122 | 0.003317693 | 0.005060246 | 1 |
| Obesity | pericyte | *SLC35G1* | 1.95097646 | 0.000264038 | 0.000484815 | 1 |
| Obesity | pericyte | *ZNF135* | 1.834932129 | 0.038198549 | 0.049886639 | 1 |
| Obesity | pericyte | *COX8A* | 8.351367998 | 8.48E-09 | 1.70E-07 | 5.33E-05 |
| Obesity | pericyte | *B3GNT4* | 16.94866225 | 1.02E-06 | 3.58E-06 | 0.006409352 |
| Obesity | pericyte | *CDC26* | 1.102319038 | 0.002052635 | 0.003235126 | 1 |

| | | | | | | |
|---|---|---|---|---|---|---|
| Obesity | pericyte | *CRLF3* | 3.271672333 | 0.003166648 | 0.004847508 | 1 |
| Obesity | pericyte | *RNPEP* | 3.94108746 | 0.000252455 | 0.000465782 | 1 |
| Obesity | pericyte | *SPRYD4* | 1.337530989 | 0.016610134 | 0.022880787 | 1 |
| Obesity | pericyte | *VPS37D* | 1.220975141 | 0.003490253 | 0.005301576 | 1 |
| Obesity | pericyte | *WDR25* | 5.766898395 | 1.61E-06 | 5.18E-06 | 0.010125398 |
| Obesity | pericyte | *PLA2G16* | 1.140916218 | 1.42E-05 | 3.46E-05 | 0.08914108 |
| Obesity | pericyte | *PRR15* | 4.11964727 | 0.007113105 | 0.010318434 | 1 |
| Obesity | pericyte | *RMDN1* | 5.073925447 | 4.03E-08 | 3.19E-07 | 0.00025325 |
| Obesity | pericyte | *HORMAD2* | 1.393474968 | 0.001296568 | 0.002111246 | 1 |
| Obesity | pericyte | *RNF152* | -1.715124297 | 0.000281368 | 0.000513784 | 1 |
| Obesity | pericyte | *BOK* | 2.607384882 | 4.87E-06 | 1.35E-05 | 0.03064785 |
| Obesity | pericyte | *ZNF843* | 9.753209673 | 0.005840885 | 0.00859589 | 1 |
| Obesity | pericyte | *C8orf59* | 7.143761336 | 1.25E-08 | 1.73E-07 | 7.89E-05 |
| Obesity | pericyte | *PFN4* | -1.138020909 | 0.019691648 | 0.026831685 | 1 |
| Obesity | pericyte | *RUFY1* | 4.995366572 | 3.49E-09 | 1.07E-07 | 2.19E-05 |
| Obesity | pericyte | *LRRC37A3* | 5.478520544 | 3.27E-08 | 2.80E-07 | 0.000205852 |
| Obesity | pericyte | *VSIG10* | 1.214816607 | 0.006005627 | 0.008821817 | 1 |
| Obesity | pericyte | *IRX5* | 2.364471648 | 4.90E-07 | 2.03E-06 | 0.003083143 |
| Obesity | pericyte | *PNMA1* | 4.04272156 | 6.68E-06 | 1.78E-05 | 0.041971623 |
| Obesity | pericyte | *ANKLE2* | 8.392227447 | 1.08E-07 | 6.34E-07 | 0.000679587 |
| Obesity | pericyte | *GCNT4* | 7.815759859 | 8.75E-07 | 3.17E-06 | 0.005498184 |
| Obesity | pericyte | *MUC20* | 3.387268503 | 2.21E-05 | 5.12E-05 | 0.138732681 |
| Obesity | pericyte | *THAP4* | 3.194954054 | 9.80E-06 | 2.49E-05 | 0.061637859 |
| Obesity | pericyte | *SHMT1* | 1.766969912 | 5.15E-05 | 0.000109521 | 0.323963363 |
| Obesity | pericyte | *DPP7* | 7.754903401 | 4.37E-06 | 1.23E-05 | 0.027496473 |
| Obesity | pericyte | *SEC24C* | 3.947596004 | 0.020080867 | 0.027338331 | 1 |
| Obesity | pericyte | *MTHFR* | 5.418060102 | 1.68E-08 | 1.94E-07 | 0.000105348 |
| Obesity | pericyte | *C19orf18* | 1.867977491 | 0.000120988 | 0.00023763 | 0.760652538 |
| Obesity | pericyte | *TMEM80* | -1.248283195 | 0.030556619 | 0.040520873 | 1 |
| Obesity | pericyte | *WDR73* | 3.10596946 | 0.000500899 | 0.000876957 | 1 |
| Obesity | pericyte | *POLE* | 8.248450722 | 8.73E-09 | 1.70E-07 | 5.49E-05 |
| Obesity | pericyte | *DSCAML1* | 3.578679445 | 0.012340574 | 0.017341348 | 1 |
| Obesity | pericyte | *RIMKLA* | 1.869593716 | 0.000874215 | 0.001461364 | 1 |
| Obesity | pericyte | *B3GNT8* | 11.81025316 | 1.13E-07 | 6.53E-07 | 0.000710281 |
| Obesity | pericyte | *AP006621.1* | 2.665285072 | 1.29E-09 | 5.85E-08 | 8.13E-06 |
| Obesity | pericyte | *MAN1B1* | 6.632952652 | 1.06E-08 | 1.71E-07 | 6.65E-05 |
| Obesity | pericyte | *FBXO39* | -0.908378217 | 0.000195025 | 0.000367873 | 1 |
| Obesity | pericyte | *TOP3A* | 4.568278283 | 1.23E-05 | 3.04E-05 | 0.077371607 |
| Obesity | pericyte | *ZBTB38* | 5.399089044 | 2.77E-06 | 8.21E-06 | 0.01743026 |
| Obesity | pericyte | *C8orf31* | 4.696689577 | 1.77E-07 | 9.01E-07 | 0.001111873 |
| Obesity | pericyte | *AC024940.1* | 3.729138959 | 0.00011661 | 0.000229533 | 0.733128277 |
| Obesity | pericyte | *MAGEF1* | 12.10250742 | 1.58E-08 | 1.91E-07 | 9.96E-05 |
| Obesity | pericyte | *SAMD9L* | 2.083446858 | 0.002440662 | 0.003804721 | 1 |
| Obesity | pericyte | *PAWR* | -1.275569495 | 0.000610798 | 0.001053811 | 1 |
| Obesity | pericyte | *NIM1K* | 2.795483891 | 4.10E-06 | 1.16E-05 | 0.025796003 |
| Obesity | pericyte | *ACOT4* | 1.827032015 | 0.000231896 | 0.000430576 | 1 |
| Obesity | pericyte | *ARIH2* | 1.59448749 | 0.033436333 | 0.044088554 | 1 |
| Obesity | pericyte | *RBM44* | -2.389279879 | 0.000110913 | 0.000219418 | 0.69731109 |
| Obesity | pericyte | *IRX3* | -1.744478212 | 0.02554001 | 0.034309839 | 1 |
| Obesity | pericyte | *ATOX1* | 5.676183357 | 5.20E-09 | 1.30E-07 | 3.27E-05 |

| | | | | | | |
|---|---|---|---|---|---|---|
| Obesity | pericyte | *TBL1XR1* | 3.314401067 | 0.000755575 | 0.00127937 | 1 |
| Obesity | pericyte | *C18orf32* | -2.113803861 | 0.001781654 | 0.002837198 | 1 |
| Obesity | pericyte | *RPLP2* | 6.410652169 | 1.32E-06 | 4.37E-06 | 0.00827993 |
| Obesity | pericyte | *C12orf54* | -3.884357731 | 0.000217442 | 0.000406137 | 1 |
| Obesity | pericyte | *GBA* | 4.893129339 | 1.08E-07 | 6.31E-07 | 0.000676305 |
| Obesity | pericyte | *CD163L1* | 1.166295406 | 0.02591643 | 0.034778356 | 1 |
| Obesity | pericyte | *THAP5* | 6.031239211 | 5.08E-07 | 2.09E-06 | 0.003194884 |
| Obesity | pericyte | *CD151* | 6.894507084 | 1.27E-10 | 1.14E-08 | 7.99E-07 |
| Obesity | pericyte | *FAM87B* | 2.622154959 | 0.029272002 | 0.038899403 | 1 |
| Obesity | pericyte | *ZCCHC3* | 5.290456254 | 0.001100522 | 0.001808411 | 1 |
| Obesity | pericyte | *ZNF619* | 2.715706881 | 1.37E-06 | 4.53E-06 | 0.00864441 |
| Obesity | pericyte | *ZBTB41* | 4.590887533 | 0.000742561 | 0.001258174 | 1 |
| Obesity | pericyte | *CENPBD1* | 2.334839025 | 0.000434528 | 0.000769543 | 1 |
| Obesity | pericyte | *BET1L* | 3.520900163 | 1.66E-07 | 8.62E-07 | 0.001045895 |
| Obesity | pericyte | *RIC8A* | 3.720972172 | 7.17E-05 | 0.000147282 | 0.450534336 |
| Obesity | pericyte | *ODF3B* | 3.055269559 | 8.45E-08 | 5.29E-07 | 0.000531471 |
| Obesity | pericyte | *DPY19L2* | -0.286388686 | 4.16E-06 | 1.17E-05 | 0.026164586 |
| Obesity | pericyte | *C2orf73* | 4.733586081 | 2.51E-07 | 1.18E-06 | 0.001578701 |
| Obesity | pericyte | *FAM26E* | 3.115614869 | 6.40E-06 | 1.72E-05 | 0.04022722 |
| Obesity | pericyte | *ALS2CL* | 2.06079083 | 0.001256168 | 0.002048114 | 1 |
| Obesity | pericyte | *C2orf69* | 8.505704574 | 7.82E-06 | 2.04E-05 | 0.049133378 |
| Obesity | pericyte | *GRAMD1C* | -2.229272082 | 0.000578201 | 0.001002247 | 1 |
| Obesity | pericyte | *GOLGA8Q* | 27.18853689 | 0.009804374 | 0.013961518 | 1 |
| Obesity | pericyte | *NDUFV2* | 6.184199049 | 1.83E-07 | 9.25E-07 | 0.001153011 |
| Obesity | pericyte | *ZNF114* | -1.952520123 | 2.61E-05 | 5.97E-05 | 0.164287724 |
| Obesity | pericyte | *LCORL* | 6.172514476 | 2.96E-07 | 1.36E-06 | 0.001861771 |
| Obesity | pericyte | *ZNF454* | 2.097197766 | 1.36E-06 | 4.49E-06 | 0.008557076 |
| Obesity | pericyte | *SH2B1* | -3.445136762 | 5.13E-15 | 1.90E-12 | 3.23E-11 |
| Obesity | pericyte | *ZC3H12D* | 2.189845965 | 1.02E-06 | 3.59E-06 | 0.006441564 |
| Obesity | pericyte | *KDELC2* | 4.251809381 | 6.51E-06 | 1.75E-05 | 0.04095331 |
| Obesity | pericyte | *PLEC* | 6.081699828 | 9.17E-06 | 2.35E-05 | 0.057648169 |
| Obesity | pericyte | *SH2D4B* | 7.327224236 | 3.78E-07 | 1.65E-06 | 0.002374111 |
| Obesity | pericyte | *RNF212* | 0.979057794 | 0.003553378 | 0.005392249 | 1 |
| Obesity | pericyte | *PRSS36* | 3.799165404 | 6.31E-07 | 2.46E-06 | 0.003969657 |
| Obesity | pericyte | *TMPRSS9* | 2.583084616 | 1.25E-05 | 3.09E-05 | 0.078622944 |
| Obesity | pericyte | *AQP11* | 3.674155846 | 4.75E-08 | 3.56E-07 | 0.00029835 |
| Obesity | pericyte | *TMEM11* | 4.433904318 | 7.68E-08 | 4.90E-07 | 0.000482552 |
| Obesity | pericyte | *ZFAND2A* | 4.962797572 | 0.000115175 | 0.000227127 | 0.724107228 |
| Obesity | pericyte | *BEND3* | 7.410241153 | 1.33E-07 | 7.32E-07 | 0.00083762 |
| Obesity | pericyte | *NT5DC1* | 4.887839878 | 3.76E-06 | 1.07E-05 | 0.023664866 |
| Obesity | pericyte | *AC006111.1* | 9.766605011 | 1.61E-06 | 5.18E-06 | 0.010113587 |
| Obesity | pericyte | *LINC00843* | 5.124673901 | 8.85E-08 | 5.48E-07 | 0.000556103 |
| Obesity | pericyte | *COX14* | 3.208928452 | 4.80E-08 | 3.60E-07 | 0.00030161 |
| Obesity | pericyte | *P4HTM* | 2.044275597 | 0.007751328 | 0.011174638 | 1 |
| Obesity | pericyte | *CA8* | 1.488286092 | 0.000529426 | 0.000923046 | 1 |
| Obesity | pericyte | *ERBB4* | 2.727842062 | 2.83E-08 | 2.53E-07 | 0.000177984 |
| Obesity | pericyte | *CSRNP3* | -0.753156103 | 4.12E-05 | 8.96E-05 | 0.259181996 |
| Obesity | pericyte | *PARP10* | 2.690590356 | 0.003280722 | 0.005011152 | 1 |
| Obesity | pericyte | *RPP25* | -2.397782505 | 2.53E-05 | 5.80E-05 | 0.159140167 |
| Obesity | pericyte | *STX19* | 7.653418262 | 1.10E-14 | 3.83E-12 | 6.90E-11 |

| | | | | | | |
|---|---|---|---|---|---|---|
| Obesity | pericyte | *FAM219B* | 4.705828758 | 8.24E-10 | 4.51E-08 | 5.18E-06 |
| Obesity | pericyte | *GDPD4* | 4.803713114 | 1.74E-06 | 5.53E-06 | 0.010945449 |
| Obesity | pericyte | *MPI* | 6.64352578 | 5.72E-08 | 4.04E-07 | 0.000359327 |
| Obesity | pericyte | *TRIM73* | -6.374058099 | 3.74E-08 | 3.03E-07 | 0.000235321 |
| Obesity | pericyte | *C17orf62* | 6.655155716 | 2.18E-09 | 8.29E-08 | 1.37E-05 |
| Obesity | pericyte | *GAK* | 6.096853609 | 7.35E-07 | 2.77E-06 | 0.0046196 |
| Obesity | pericyte | *TUFM* | 7.00827955 | 1.74E-29 | 3.64E-26 | 1.09E-25 |
| Obesity | pericyte | *CTC1* | 3.365763274 | 8.31E-09 | 1.70E-07 | 5.23E-05 |
| Obesity | pericyte | *SNX18* | 4.990004497 | 0.000272216 | 0.000498375 | 1 |
| Obesity | pericyte | *C14orf39* | 1.000609817 | 0.000626777 | 0.001079602 | 1 |
| Obesity | pericyte | *TRIML2* | -27.1913085 | 9.05E-06 | 2.32E-05 | 0.056893526 |
| Obesity | pericyte | *SPTY2D1* | 8.681663189 | 8.54E-07 | 3.11E-06 | 0.005366369 |
| Obesity | pericyte | *TCAIM* | 2.741439341 | 0.00015396 | 0.000296279 | 0.967944216 |
| Obesity | pericyte | *FUCA1* | 2.544065714 | 0.008975227 | 0.012836044 | 1 |
| Obesity | pericyte | *ZNF664* | 8.209246564 | 7.71E-07 | 2.88E-06 | 0.004849493 |
| Obesity | pericyte | *CALR* | 6.614458873 | 3.57E-07 | 1.58E-06 | 0.002245245 |
| Obesity | pericyte | *CDH4* | 2.201838264 | 0.001285336 | 0.002094042 | 1 |
| Obesity | pericyte | *ARID3B* | 5.201811257 | 4.28E-05 | 9.27E-05 | 0.269085058 |
| Obesity | pericyte | *EGR3* | -2.388614294 | 0.000309894 | 0.000561957 | 1 |
| Obesity | pericyte | *GEMIN4* | 1.222116941 | 0.002990333 | 0.004596632 | 1 |
| Obesity | pericyte | *FJX1* | -1.225639649 | 0.003620422 | 0.005487365 | 1 |
| Obesity | pericyte | *CIITA* | 2.248612656 | 1.07E-05 | 2.69E-05 | 0.067418978 |
| Obesity | pericyte | *PLD6* | 1.496279698 | 0.013495983 | 0.018842827 | 1 |
| Obesity | pericyte | *ZBTB42* | 7.77705878 | 9.86E-08 | 5.93E-07 | 0.000619837 |
| Obesity | pericyte | *LACC1* | 5.437404591 | 2.93E-08 | 2.59E-07 | 0.000184008 |
| Obesity | pericyte | *MAF1* | 4.857935643 | 1.39E-07 | 7.53E-07 | 0.000876272 |
| Obesity | pericyte | *FCER1A* | 2.290602595 | 0.000251795 | 0.000464845 | 1 |
| Obesity | pericyte | *PCED1B* | 3.527455538 | 5.18E-08 | 3.79E-07 | 0.000325385 |
| Obesity | pericyte | *PIPOX* | -2.893996673 | 0.001248126 | 0.002036587 | 1 |
| Obesity | pericyte | *FOXS1* | -1.252018794 | 3.95E-06 | 1.12E-05 | 0.024814355 |
| Obesity | pericyte | *SERTAD2* | 7.099590266 | 1.29E-07 | 7.18E-07 | 0.000812825 |
| Obesity | pericyte | *CITED4* | 3.777474323 | 3.83E-05 | 8.38E-05 | 0.240649197 |
| Obesity | pericyte | *PDXDC1* | 5.246631664 | 3.73E-06 | 1.07E-05 | 0.023479441 |
| Obesity | pericyte | *ZNF154* | 3.048411611 | 1.84E-05 | 4.35E-05 | 0.115474645 |
| Obesity | pericyte | *R3HDM2* | 5.797267703 | 4.11E-08 | 3.22E-07 | 0.000258634 |
| Obesity | pericyte | *B3GNT3* | 9.142522534 | 1.19E-07 | 6.74E-07 | 0.000745567 |
| Obesity | pericyte | *TMEM86B* | 2.019369904 | 7.48E-05 | 0.000153156 | 0.470493825 |
| Obesity | pericyte | *SEPT1* | 11.81071502 | 1.08E-06 | 3.73E-06 | 0.006779235 |
| Obesity | pericyte | *EXOC3* | 3.044159025 | 0.000447877 | 0.000790734 | 1 |
| Obesity | pericyte | *LYNX1* | 2.507288089 | 0.000104365 | 0.000207837 | 0.656142157 |
| Obesity | pericyte | *FGD6* | 3.385812749 | 4.35E-08 | 3.35E-07 | 0.000273693 |
| Obesity | pericyte | *HCLS1* | 6.312425207 | 4.93E-09 | 1.29E-07 | 3.10E-05 |
| Obesity | pericyte | *ZNF609* | 4.236772919 | 1.94E-05 | 4.58E-05 | 0.122280583 |
| Obesity | pericyte | *CCDC66* | 2.463789901 | 0.002607073 | 0.00405108 | 1 |
| Obesity | pericyte | *DEFB124* | -0.809874949 | 1.40E-05 | 3.43E-05 | 0.088304107 |
| Obesity | pericyte | *MCFD2* | 5.501017785 | 2.45E-07 | 1.16E-06 | 0.001540444 |
| Obesity | pericyte | *CYP8B1* | 2.381976118 | 7.73E-05 | 0.000157784 | 0.485973883 |
| Obesity | pericyte | *RNF182* | -1.208635547 | 9.81E-06 | 2.49E-05 | 0.061666181 |
| Obesity | pericyte | *MB21D2* | 4.950456262 | 0.000123579 | 0.00024249 | 0.77693835 |
| Obesity | pericyte | *SSTR2* | 3.72874464 | 1.01E-06 | 3.55E-06 | 0.006324666 |

| | | | | | | |
|---|---|---|---|---|---|---|
| Obesity | pericyte | *PCGF5* | 5.97862208 | 2.52E-06 | 7.58E-06 | 0.015869677 |
| Obesity | pericyte | *SHISA2* | 73.26297255 | 4.49E-07 | 1.89E-06 | 0.002821342 |
| Obesity | pericyte | *CHST13* | 8.520876394 | 2.77E-06 | 8.21E-06 | 0.017415849 |
| Obesity | pericyte | *SLC36A4* | 4.938609685 | 2.47E-06 | 7.42E-06 | 0.015506516 |
| Obesity | pericyte | *ZDHHC20* | 6.286563223 | 1.98E-08 | 2.11E-07 | 0.00012446 |
| Obesity | pericyte | *OR51E1* | -0.838992638 | 3.18E-05 | 7.10E-05 | 0.200237407 |
| Obesity | pericyte | *PSMG4* | 3.677475965 | 0.000841204 | 0.001412532 | 1 |
| Obesity | pericyte | *ZNF443* | 2.454694114 | 0.000209708 | 0.000393562 | 1 |
| Obesity | pericyte | *CAPS2* | 4.319460032 | 1.11E-08 | 1.71E-07 | 7.00E-05 |
| Obesity | pericyte | *ZNF792* | 1.828172649 | 0.003941148 | 0.005953387 | 1 |
| Obesity | pericyte | *KCTD2* | 10.07074853 | 1.65E-07 | 8.55E-07 | 0.001036828 |
| Obesity | pericyte | *D2HGDH* | 3.11353303 | 0.002378465 | 0.003717904 | 1 |
| Obesity | pericyte | *CMTR2* | 4.83516836 | 2.06E-08 | 2.16E-07 | 0.000129395 |
| Obesity | pericyte | *LRRC57* | 5.201714227 | 0.000215375 | 0.000402995 | 1 |
| Obesity | pericyte | *BBS12* | 3.060013433 | 7.05E-07 | 2.68E-06 | 0.004434008 |
| Obesity | pericyte | *ZFP82* | 3.641876561 | 0.023151105 | 0.031240824 | 1 |
| Obesity | pericyte | *LSMEM1* | 1.848323369 | 0.021069907 | 0.028579612 | 1 |
| Obesity | pericyte | *AEN* | 4.010377951 | 2.37E-06 | 7.19E-06 | 0.014924956 |
| Obesity | pericyte | *FKRP* | 1.635535222 | 0.000194062 | 0.000366277 | 1 |
| Obesity | pericyte | *TRAPPC5* | 2.90890995 | 0.000905755 | 0.001508073 | 1 |
| Obesity | pericyte | *RPH3AL* | 4.585078999 | 2.28E-07 | 1.10E-06 | 0.001435876 |
| Obesity | pericyte | *ANKRD34A* | 2.500937939 | 0.001254325 | 0.00204564 | 1 |
| Obesity | pericyte | *SLC26A11* | 6.529538887 | 3.70E-05 | 8.12E-05 | 0.23268271 |
| Obesity | pericyte | *HIGD1A* | -1.531890703 | 0.000235275 | 0.000436463 | 1 |
| Obesity | pericyte | *POLR2A* | 7.158191628 | 1.22E-07 | 6.89E-07 | 0.000768931 |
| Obesity | pericyte | *TMEM132C* | 1.54161371 | 8.35E-06 | 2.16E-05 | 0.052504847 |
| Obesity | pericyte | *SLC25A41* | 10.24637526 | 9.35E-06 | 2.39E-05 | 0.05875555 |
| Obesity | pericyte | *TMEM136* | 1.491295487 | 0.001724737 | 0.002751439 | 1 |
| Obesity | pericyte | *ZNF322* | -2.637642957 | 0.001434816 | 0.002326119 | 1 |
| Obesity | pericyte | *DDX60L* | 5.894090102 | 1.09E-07 | 6.39E-07 | 0.000687787 |
| Obesity | pericyte | *ACBD4* | 2.590175484 | 3.11E-07 | 1.41E-06 | 0.001955325 |
| Obesity | pericyte | *SGSH* | 12.2778679 | 3.86E-09 | 1.13E-07 | 2.43E-05 |
| Obesity | pericyte | *SETD2* | 3.957255495 | 1.48E-05 | 3.60E-05 | 0.093219885 |
| Obesity | pericyte | *MRPS23* | 5.254147311 | 1.48E-08 | 1.87E-07 | 9.29E-05 |
| Obesity | pericyte | *GPR135* | 2.300405595 | 5.98E-07 | 2.37E-06 | 0.003762083 |
| Obesity | pericyte | *ATG9B* | 4.600057331 | 7.41E-07 | 2.78E-06 | 0.004655545 |
| Obesity | pericyte | *HKR1* | 5.647755296 | 1.55E-07 | 8.15E-07 | 0.000976619 |
| Obesity | pericyte | *ZBTB20* | 3.906374639 | 8.96E-06 | 2.31E-05 | 0.056359062 |
| Obesity | pericyte | *C3orf58* | 4.441217278 | 5.10E-05 | 0.000108662 | 0.320723599 |
| Obesity | pericyte | *AMIGO1* | 1.498808156 | 0.00018794 | 0.00035579 | 1 |
| Obesity | pericyte | *ODF3L2* | -2.820259167 | 0.000715502 | 0.001216102 | 1 |
| Obesity | pericyte | *RELL1* | 6.691746698 | 1.62E-08 | 1.92E-07 | 0.000102072 |
| Obesity | pericyte | *RFX7* | 5.346804036 | 3.09E-08 | 2.70E-07 | 0.000194261 |
| Obesity | pericyte | *CLDN7* | -0.869799342 | 1.87E-06 | 5.90E-06 | 0.01177851 |
| Obesity | pericyte | *ADO* | 2.874401139 | 6.47E-07 | 2.50E-06 | 0.004067432 |
| Obesity | pericyte | *SNRPE* | 10.22557744 | 3.70E-08 | 3.01E-07 | 0.000232548 |
| Obesity | pericyte | *CHST15* | 5.859195847 | 1.60E-06 | 5.14E-06 | 0.01003752 |
| Obesity | pericyte | *IDH2* | 4.575447513 | 1.49E-06 | 4.85E-06 | 0.009355734 |
| Obesity | pericyte | *TNRC18* | 6.002881403 | 3.99E-06 | 1.13E-05 | 0.025084227 |
| Obesity | pericyte | *NOP10* | 2.114086229 | 0.020212767 | 0.027511943 | 1 |

| | | | | | | |
|---|---|---|---|---|---|---|
| Obesity | pericyte | *FAM89A* | 2.25754887 | 6.21E-05 | 0.000129164 | 0.390593129 |
| Obesity | pericyte | *TDRKH* | 1.723081022 | 0.00143099 | 0.002320514 | 1 |
| Obesity | pericyte | *ZNF708* | 3.722726411 | 2.51E-05 | 5.75E-05 | 0.157900087 |
| Obesity | pericyte | *IST1* | 5.165053667 | 1.62E-05 | 3.88E-05 | 0.101722606 |
| Obesity | pericyte | *ERCC6L2* | 3.584041183 | 8.84E-05 | 0.000178659 | 0.555808317 |
| Obesity | pericyte | *FAM153B* | 2.786232693 | 0.008452655 | 0.012130072 | 1 |
| Obesity | pericyte | *BACE2* | 1.502264977 | 0.008351454 | 0.011990315 | 1 |
| Obesity | pericyte | *SYNM* | 6.215554176 | 2.13E-08 | 2.17E-07 | 0.000134057 |
| Obesity | pericyte | *FBXL6* | -3.920055492 | 4.39E-05 | 9.49E-05 | 0.276175117 |
| Obesity | pericyte | *KBTBD3* | 9.938847511 | 7.73E-08 | 4.93E-07 | 0.000486275 |
| Obesity | pericyte | *CLN8* | 4.640705554 | 3.91E-07 | 1.70E-06 | 0.002457835 |
| Obesity | pericyte | *CACNB4* | 1.253474202 | 0.003136288 | 0.004805714 | 1 |
| Obesity | pericyte | *TRAPPC6B* | 3.803068323 | 1.05E-05 | 2.64E-05 | 0.065821883 |
| Obesity | pericyte | *NPLOC4* | 1.504238797 | 0.016786595 | 0.023113738 | 1 |
| Obesity | pericyte | *CAPN12* | 4.52884314 | 0.00093575 | 0.00155431 | 1 |
| Obesity | pericyte | *GLRX5* | 7.651742056 | 3.04E-05 | 6.83E-05 | 0.191259481 |
| Obesity | pericyte | *CAV3* | -1.140823948 | 0.002911307 | 0.004486125 | 1 |
| Obesity | pericyte | *MXRA7* | 1.279284881 | 0.001070847 | 0.001763335 | 1 |
| Obesity | pericyte | *MFSD5* | 4.55892945 | 1.10E-06 | 3.80E-06 | 0.006946958 |
| Obesity | pericyte | *ADI1* | 6.73016495 | 6.55E-06 | 1.75E-05 | 0.041203313 |
| Obesity | pericyte | *CLEC4G* | 5.75678138 | 2.00E-06 | 6.23E-06 | 0.012545413 |
| Obesity | pericyte | *SATB1* | 7.094976428 | 2.20E-07 | 1.06E-06 | 0.001381036 |
| Obesity | pericyte | *ACTL10* | -2.480692302 | 5.92E-05 | 0.000123804 | 0.372402044 |
| Obesity | pericyte | *TRAK1* | 2.944919889 | 0.006295296 | 0.009204308 | 1 |
| Obesity | pericyte | *SKA2* | 8.354634844 | 2.37E-08 | 2.28E-07 | 0.000148839 |
| Obesity | pericyte | *TTC3* | 0.568725172 | 0.037826747 | 0.049442154 | 1 |
| Obesity | pericyte | *KCNB2* | 1.118200414 | 5.63E-05 | 0.000118283 | 0.354022462 |
| Obesity | pericyte | *BRICD5* | -2.121722525 | 0.000126305 | 0.000247454 | 0.79408086 |
| Obesity | pericyte | *TSKU* | 6.152995319 | 5.46E-09 | 1.34E-07 | 3.43E-05 |
| Obesity | pericyte | *GRID1* | -0.604118337 | 4.86E-06 | 1.35E-05 | 0.030579895 |
| Obesity | pericyte | *ACBD3* | 5.546240328 | 7.91E-05 | 0.000161226 | 0.497188882 |
| Obesity | pericyte | *VMO1* | 1.444727742 | 0.000339833 | 0.00061166 | 1 |
| Obesity | pericyte | *COL18A1* | -0.723047251 | 0.00093307 | 0.001550268 | 1 |
| Obesity | pericyte | *ZNF721* | 3.253673872 | 9.38E-05 | 0.0001886 | 0.589941432 |
| Obesity | pericyte | *LENG9* | 2.125737808 | 0.018480834 | 0.025313508 | 1 |
| Obesity | pericyte | *CEP63* | 0.91578183 | 0.031067513 | 0.041146293 | 1 |
| Obesity | pericyte | *HMGN4* | 3.688891195 | 1.13E-05 | 2.81E-05 | 0.070733916 |
| Obesity | pericyte | *LYSMD4* | 1.767113593 | 0.000377537 | 0.000674887 | 1 |
| Obesity | pericyte | *WBP2NL* | 6.0911879 | 0.001564793 | 0.002518651 | 1 |
| Obesity | pericyte | *BEGAIN* | 7.401710947 | 6.62E-07 | 2.55E-06 | 0.004161423 |
| Obesity | pericyte | *ARHGEF37* | 4.536838307 | 3.87E-08 | 3.10E-07 | 0.000243151 |
| Obesity | pericyte | *FAM43B* | 3.336243303 | 0.011100949 | 0.015701163 | 1 |
| Obesity | pericyte | *CSMD1* | 2.811794219 | 1.78E-07 | 9.05E-07 | 0.00111941 |
| Obesity | pericyte | *PTGDR2* | -18.58347753 | 0.010174888 | 0.014449857 | 1 |
| Obesity | pericyte | *GPR19* | -3.895072538 | 0.001236538 | 0.00201925 | 1 |
| Obesity | pericyte | *FANCF* | 9.661299282 | 2.06E-08 | 2.16E-07 | 0.000129673 |
| Obesity | pericyte | *RUVBL2* | 3.178813868 | 1.36E-06 | 4.48E-06 | 0.008533765 |
| Obesity | pericyte | *GDPGP1* | 3.107958235 | 2.22E-06 | 6.80E-06 | 0.01397149 |
| Obesity | pericyte | *PTTG1IP* | 2.45574738 | 0.0040512 | 0.006107889 | 1 |
| Obesity | pericyte | *PLGLB1* | -1.033321377 | 0.005678785 | 0.008373012 | 1 |

| | | | | | | |
|---|---|---|---|---|---|---|
| Obesity | pericyte | *CCBE1* | 2.502272881 | 1.15E-06 | 3.92E-06 | 0.007249385 |
| Obesity | pericyte | *ZNF623* | 5.150492157 | 0.000219697 | 0.000409863 | 1 |
| Obesity | pericyte | *CCDC125* | 3.876673752 | 6.82E-08 | 4.52E-07 | 0.000428952 |
| Obesity | pericyte | *C10orf107* | 0.484734169 | 0.002385559 | 0.003727439 | 1 |
| Obesity | pericyte | *FHL3* | -2.326912593 | 0.000675416 | 0.001154839 | 1 |
| Obesity | pericyte | *LRIT3* | 2.314258044 | 0.015781803 | 0.021840236 | 1 |
| Obesity | pericyte | *GRIN2A* | -1.529971223 | 0.033586844 | 0.044277729 | 1 |
| Obesity | pericyte | *URAD* | 6.901042446 | 0.000188754 | 0.000357115 | 1 |
| Obesity | pericyte | *GPR132* | 2.538742494 | 2.13E-06 | 6.57E-06 | 0.0134097 |
| Obesity | pericyte | *PSMG1* | 1.766892263 | 2.83E-09 | 9.54E-08 | 1.78E-05 |
| Obesity | pericyte | *KLRC4* | -2.135575951 | 0.000140216 | 0.00027208 | 0.881538364 |
| Obesity | pericyte | *ACSM5* | 1.422310436 | 5.43E-06 | 1.48E-05 | 0.034158327 |
| Obesity | pericyte | *AC131971.1* | -3.376948768 | 5.09E-05 | 0.000108515 | 0.320118262 |
| Obesity | pericyte | *SERHL2* | 10.89151846 | 3.35E-08 | 2.85E-07 | 0.000210318 |
| Obesity | pericyte | *TANGO2* | 6.700602584 | 5.74E-08 | 4.05E-07 | 0.000361139 |
| Obesity | pericyte | *SFXN4* | 3.091206094 | 1.18E-07 | 6.74E-07 | 0.000743466 |
| Obesity | pericyte | *FAM167B* | 5.606997561 | 0.001168646 | 0.001912855 | 1 |
| Obesity | pericyte | *RP1L1* | 5.644038863 | 2.15E-13 | 5.40E-11 | 1.35E-09 |
| Obesity | pericyte | *NDUFB1* | 3.155408326 | 0.000118548 | 0.000233129 | 0.745311858 |
| Obesity | pericyte | *FAM19A1* | 4.17976582 | 3.44E-08 | 2.89E-07 | 0.000216385 |
| Obesity | pericyte | *TRMT12* | 2.842544576 | 0.027067263 | 0.036229909 | 1 |
| Obesity | pericyte | *CMTM4* | 3.684609056 | 0.002278538 | 0.003567023 | 1 |
| Obesity | pericyte | *CBX6* | 9.353253269 | 8.17E-06 | 2.12E-05 | 0.051358609 |
| Obesity | pericyte | *KREMEN1* | 0.750776547 | 0.002191687 | 0.003439625 | 1 |
| Obesity | pericyte | *KCTD16* | 5.380381348 | 3.57E-06 | 1.03E-05 | 0.022416836 |
| Obesity | pericyte | *ZNF703* | 2.119593317 | 0.026105527 | 0.035017164 | 1 |
| Obesity | pericyte | *KCTD8* | 1.762898114 | 4.27E-05 | 9.25E-05 | 0.268344958 |
| Obesity | pericyte | *C9orf66* | 15.43930327 | 5.16E-06 | 1.42E-05 | 0.032416801 |
| Obesity | pericyte | *TUBA8* | 3.176825009 | 2.68E-05 | 6.10E-05 | 0.168363873 |
| Obesity | pericyte | *NPIPA5* | 0.719662434 | 0.00153485 | 0.002475526 | 1 |
| Obesity | pericyte | *FAM162B* | -0.272162046 | 2.40E-05 | 5.53E-05 | 0.150670395 |
| Obesity | pericyte | *BTBD9* | 4.460923902 | 1.07E-05 | 2.69E-05 | 0.067316966 |
| Obesity | pericyte | *ANKRD45* | 5.832117764 | 1.30E-06 | 4.32E-06 | 0.008166508 |
| Obesity | pericyte | *MAATS1* | 2.606975501 | 8.43E-08 | 5.28E-07 | 0.000529785 |
| Obesity | pericyte | *GPR39* | 2.953898569 | 4.45E-06 | 1.25E-05 | 0.027950394 |
| Obesity | pericyte | *NPIPA7* | 5.034780928 | 0.000378516 | 0.000676444 | 1 |
| Obesity | pericyte | *TTC32* | 3.552248099 | 0.003740644 | 0.005661394 | 1 |
| Obesity | pericyte | *PP2D1* | 4.470781855 | 1.35E-06 | 4.46E-06 | 0.008486059 |
| Obesity | pericyte | *TMEM236* | 3.972108739 | 1.12E-06 | 3.82E-06 | 0.007019176 |
| Obesity | pericyte | *VPS33B* | 5.188836724 | 8.94E-06 | 2.30E-05 | 0.056198329 |
| Obesity | pericyte | *TBX1* | -2.710063762 | 0.022331636 | 0.03018684 | 1 |
| Obesity | pericyte | *EIF3C* | 4.446069748 | 4.99E-20 | 4.48E-17 | 3.14E-16 |
| Obesity | pericyte | *NIPSNAP1* | 3.098215267 | 4.31E-05 | 9.32E-05 | 0.270930982 |
| Obesity | pericyte | *CNTN2* | 3.947953896 | 6.75E-07 | 2.58E-06 | 0.004242156 |
| Obesity | pericyte | *CRELD2* | 8.650393531 | 1.22E-06 | 4.08E-06 | 0.007650153 |
| Obesity | pericyte | *PCDH9* | 1.454449336 | 0.000181007 | 0.000343805 | 1 |
| Obesity | pericyte | *ACOT1* | 0.893715688 | 0.023203632 | 0.03129827 | 1 |
| Obesity | pericyte | *OAF* | -1.554912325 | 0.009410662 | 0.013428242 | 1 |
| Obesity | pericyte | *TM2D3* | 4.994066665 | 0.000358662 | 0.00064279 | 1 |
| Obesity | pericyte | *CLECL1* | 1.152450765 | 0.000881516 | 0.001473177 | 1 |

| | | | | | | |
|---|---|---|---|---|---|---|
| Obesity | pericyte | *MROH7* | 4.289904577 | 0.000206811 | 0.00038859 | 1 |
| Obesity | pericyte | *GDF3* | 2.864365446 | 0.011438522 | 0.016153187 | 1 |
| Obesity | pericyte | *PKP3* | -2.042786956 | 0.001760501 | 0.002804224 | 1 |
| Obesity | pericyte | *CSF1* | 1.904418384 | 0.004442329 | 0.006656082 | 1 |
| Obesity | pericyte | *PLA2G6* | 3.290145045 | 4.98E-13 | 1.16E-10 | 3.13E-09 |
| Obesity | pericyte | *SS18L1* | 4.558802658 | 1.01E-05 | 2.54E-05 | 0.063237275 |
| Obesity | pericyte | *THAP7* | 7.810573329 | 6.64E-08 | 4.46E-07 | 0.000417458 |
| Obesity | pericyte | *CCR10* | -1.294450917 | 3.88E-05 | 8.48E-05 | 0.244172491 |
| Obesity | pericyte | *WDR27* | 4.877102779 | 0.000169619 | 0.000323445 | 1 |
| Obesity | pericyte | *TXNRD2* | 3.885895962 | 9.05E-08 | 5.59E-07 | 0.000569117 |
| Obesity | pericyte | *PROS1* | 3.343325837 | 6.68E-06 | 1.78E-05 | 0.04199417 |
| Obesity | pericyte | *HDDC3* | 3.874089074 | 7.20E-06 | 1.90E-05 | 0.045288301 |
| Obesity | pericyte | *ZFP1* | 4.105201757 | 0.009175833 | 0.013111014 | 1 |
| Obesity | pericyte | *LPAR5* | 1.163082996 | 0.032571123 | 0.043028924 | 1 |
| Obesity | pericyte | *TMEM173* | 3.371114629 | 1.16E-05 | 2.89E-05 | 0.07313601 |
| Obesity | pericyte | *NELL2* | 1.786100981 | 5.06E-07 | 2.08E-06 | 0.003181298 |
| Obesity | pericyte | *KRBA2* | 1.644416496 | 0.026778881 | 0.035859175 | 1 |
| Obesity | pericyte | *ZNF93* | 8.006915821 | 0.00219399 | 0.003442379 | 1 |
| Obesity | pericyte | *SEPT5* | 7.07295302 | 9.62E-07 | 3.42E-06 | 0.00604729 |
| Obesity | pericyte | *ATL3* | 4.688767819 | 7.29E-07 | 2.75E-06 | 0.004581976 |
| Obesity | pericyte | *UBE2G2* | 4.337283655 | 1.47E-05 | 3.57E-05 | 0.092177209 |
| Obesity | pericyte | *OSBP2* | 5.219166407 | 9.04E-09 | 1.70E-07 | 5.68E-05 |
| Obesity | pericyte | *TUSC5* | 1.357142299 | 0.000552239 | 0.000960423 | 1 |
| Obesity | pericyte | *TMED9* | 7.023853599 | 4.35E-07 | 1.85E-06 | 0.00273582 |
| Obesity | pericyte | *H1FX* | 3.941518706 | 2.80E-09 | 9.50E-08 | 1.76E-05 |
| Obesity | pericyte | *RBM43* | 4.972564119 | 0.003357382 | 0.005114577 | 1 |
| Obesity | pericyte | *ZFP90* | 7.307956041 | 1.12E-05 | 2.80E-05 | 0.070570769 |
| Obesity | pericyte | *SORCS2* | 6.020110584 | 3.23E-07 | 1.45E-06 | 0.002029155 |
| Obesity | pericyte | *TMEM106A* | 5.821957241 | 4.73E-09 | 1.27E-07 | 2.97E-05 |
| Obesity | pericyte | *SIVA1* | 3.693371181 | 2.68E-07 | 1.25E-06 | 0.001687271 |
| Obesity | pericyte | *MAFF* | 1.821293858 | 0.005382459 | 0.007964114 | 1 |
| Obesity | pericyte | *BRF1* | 7.476625327 | 1.12E-08 | 1.71E-07 | 7.04E-05 |
| Obesity | pericyte | *FLRT2* | 1.809425437 | 1.47E-05 | 3.58E-05 | 0.092570589 |
| Obesity | pericyte | *ADSSL1* | 11.41043944 | 4.86E-07 | 2.02E-06 | 0.003053206 |
| Obesity | pericyte | *C6orf120* | 3.204864645 | 0.000130919 | 0.000255458 | 0.823084862 |
| Obesity | pericyte | *DDX51* | 1.730393664 | 0.000537234 | 0.000935362 | 1 |
| Obesity | pericyte | *IFITM2* | 4.416538218 | 7.29E-06 | 1.92E-05 | 0.045816761 |
| Obesity | pericyte | *GP1BA* | 4.080152064 | 6.04E-11 | 6.23E-09 | 3.80E-07 |
| Obesity | pericyte | *KIAA0825* | 3.622571834 | 0.005174903 | 0.007671449 | 1 |
| Obesity | pericyte | *KLHL33* | 2.378286555 | 4.73E-06 | 1.31E-05 | 0.029708286 |
| Obesity | pericyte | *CDK10* | 1.500524971 | 0.023038769 | 0.031096924 | 1 |
| Obesity | pericyte | *TMEM105* | 14.65097419 | 0.023601605 | 0.031814599 | 1 |
| Obesity | pericyte | *TCN2* | 0.875495938 | 0.024858406 | 0.03345137 | 1 |
| Obesity | pericyte | *HS6ST3* | 8.396154475 | 3.74E-07 | 1.64E-06 | 0.002350483 |
| Obesity | pericyte | *MAPK11* | 6.235421648 | 0.000125245 | 0.000245531 | 0.787417833 |
| Obesity | pericyte | *SP140L* | 5.651967422 | 1.40E-06 | 4.62E-06 | 0.008825012 |
| Obesity | pericyte | *FAM174B* | 3.730270171 | 1.68E-07 | 8.68E-07 | 0.001055678 |
| Obesity | pericyte | *MUC1* | 1.591204152 | 0.004090614 | 0.006164356 | 1 |
| Obesity | pericyte | *SV2B* | 4.528665276 | 0.002242772 | 0.003515402 | 1 |
| Obesity | pericyte | *PDE6G* | 3.918141845 | 0.000183601 | 0.000348204 | 1 |

| | | | | | | |
|---|---|---|---|---|---|---|
| Obesity | pericyte | *TLCD2* | 2.452616321 | 0.003207389 | 0.004907485 | 1 |
| Obesity | pericyte | *AHNAK2* | 3.131424559 | 7.47E-08 | 4.83E-07 | 0.000469693 |
| Obesity | pericyte | *SP1* | 4.956945363 | 3.78E-09 | 1.13E-07 | 2.38E-05 |
| Obesity | pericyte | *PCGF3* | 6.442283982 | 5.29E-10 | 3.35E-08 | 3.32E-06 |
| Obesity | pericyte | *PSMD13* | 4.313168439 | 1.15E-06 | 3.91E-06 | 0.007228493 |
| Obesity | pericyte | *PBX1* | 4.232166481 | 4.10E-08 | 3.22E-07 | 0.000257875 |
| Obesity | pericyte | *UBE2L3* | 6.545603248 | 4.18E-06 | 1.18E-05 | 0.026282565 |
| Obesity | pericyte | *PMEL* | 2.226597781 | 0.000343958 | 0.000618201 | 1 |
| Obesity | pericyte | *C6orf201* | 1.076041182 | 0.027800631 | 0.037132477 | 1 |
| Obesity | pericyte | *MYBL1* | 0.676026582 | 0.01560116 | 0.021609274 | 1 |
| Obesity | pericyte | *C16orf52* | 5.277819668 | 0.000215819 | 0.000403705 | 1 |
| Obesity | pericyte | *ADARB2* | 5.489152225 | 0.000888266 | 0.00148335 | 1 |
| Obesity | pericyte | *SRL* | 1.629035011 | 1.17E-06 | 3.94E-06 | 0.007327587 |
| Obesity | pericyte | *KCNIP4* | 0.93131675 | 0.027996886 | 0.03733116 | 1 |
| Obesity | pericyte | *SLC52A2* | 4.431229986 | 1.19E-07 | 6.74E-07 | 0.000745472 |
| Obesity | pericyte | *ARL17A* | 2.440260874 | 0.000133172 | 0.000259322 | 0.83725145 |
| Obesity | pericyte | *NPIPB4* | 4.606596941 | 8.73E-06 | 2.25E-05 | 0.054858633 |
| Obesity | pericyte | *ZNF829* | 3.168605025 | 0.000213801 | 0.000400526 | 1 |
| Obesity | pericyte | *IFITM1* | -1.98630331 | 0.000329195 | 0.000593533 | 1 |
| Obesity | pericyte | *POMK* | 7.291722626 | 0.000225687 | 0.000420787 | 1 |
| Obesity | pericyte | *LINC00839* | 2.448427765 | 0.004765461 | 0.007109742 | 1 |
| Obesity | pericyte | *PTCH1* | 2.949338732 | 0.000752415 | 0.001274363 | 1 |
| Obesity | pericyte | *RTN4RL1* | 2.38093829 | 3.50E-07 | 1.55E-06 | 0.002201804 |
| Obesity | pericyte | *PLK5* | 1.9783476 | 0.034625717 | 0.045570836 | 1 |
| Obesity | pericyte | *RASA3* | 3.891815001 | 1.42E-07 | 7.62E-07 | 0.000891597 |
| Obesity | pericyte | *ZNF566* | 5.489296136 | 5.87E-07 | 2.34E-06 | 0.003687363 |
| Obesity | pericyte | *ZNF284* | 8.538791686 | 1.13E-07 | 6.53E-07 | 0.000709577 |
| Obesity | pericyte | *GSAP* | 3.414512064 | 6.34E-05 | 0.00013169 | 0.398889965 |
| Obesity | pericyte | *C2orf76* | 4.554561715 | 6.53E-08 | 4.42E-07 | 0.000410651 |
| Obesity | pericyte | *WWOX* | 3.426921698 | 2.15E-07 | 1.05E-06 | 0.001350215 |
| Obesity | pericyte | *CYP4Z1* | 2.936664868 | 8.19E-07 | 3.01E-06 | 0.005147061 |
| Obesity | pericyte | *CCDC84* | 7.031806323 | 1.23E-07 | 6.90E-07 | 0.000771338 |
| Obesity | pericyte | *POLR1D* | 4.273420519 | 3.01E-08 | 2.65E-07 | 0.000189137 |
| Obesity | pericyte | *BPIFB4* | -24.7618025 | 0.00015956 | 0.000305746 | 1 |
| Obesity | pericyte | *CYP4F12* | 1.672784338 | 0.000196464 | 0.000370365 | 1 |
| Obesity | pericyte | *ZNF749* | 0.95675675 | 0.034377501 | 0.045253632 | 1 |
| Obesity | pericyte | *KLHL32* | 2.011772589 | 5.91E-05 | 0.000123547 | 0.371506941 |
| Obesity | pericyte | *NBPF12* | 4.523893523 | 4.00E-08 | 3.17E-07 | 0.000251479 |
| Obesity | pericyte | *TOR3A* | 7.357093665 | 0.001946776 | 0.003079089 | 1 |
| Obesity | pericyte | *CCDC30* | 5.264711817 | 6.03E-07 | 2.38E-06 | 0.003791369 |
| Obesity | pericyte | *GLDN* | 2.208643983 | 5.80E-08 | 4.08E-07 | 0.000364609 |
| Obesity | pericyte | *KPNA4* | 6.721948172 | 1.03E-07 | 6.12E-07 | 0.000647845 |
| Obesity | pericyte | *ZNF501* | 3.99985466 | 1.86E-05 | 4.39E-05 | 0.116711824 |
| Obesity | pericyte | *ZNF197* | 3.282248367 | 1.07E-05 | 2.70E-05 | 0.067574688 |
| Obesity | pericyte | *RPS23* | 2.617190413 | 0.008633722 | 0.012364117 | 1 |
| Obesity | pericyte | *BTN3A2* | -1.25532595 | 1.46E-08 | 1.86E-07 | 9.18E-05 |
| Obesity | pericyte | *PCLO* | 1.336007427 | 9.76E-07 | 3.46E-06 | 0.006138817 |
| Obesity | pericyte | *RGS7BP* | 1.875953514 | 0.000129544 | 0.000253247 | 0.814443946 |
| Obesity | pericyte | *CLCNKA* | -2.50473409 | 0.01958298 | 0.026706767 | 1 |
| Obesity | pericyte | *SEPT10* | 2.982511774 | 0.001219224 | 0.001992012 | 1 |

| | | | | | | |
|---|---|---|---|---|---|---|
| Obesity | pericyte | *SMYD4* | 3.76201053 | 2.63E-06 | 7.85E-06 | 0.016541932 |
| Obesity | pericyte | *NF2* | 3.304623151 | 0.00150719 | 0.002434662 | 1 |
| Obesity | pericyte | *MIR22HG* | 2.90951267 | 1.40E-07 | 7.56E-07 | 0.00088242 |
| Obesity | pericyte | *C17orf58* | 1.40031123 | 0.000169682 | 0.000323467 | 1 |
| Obesity | pericyte | *CYP27C1* | 0.940729164 | 0.007706855 | 0.011125832 | 1 |
| Obesity | pericyte | *BCR* | 4.850764957 | 9.78E-09 | 1.71E-07 | 6.15E-05 |
| Obesity | pericyte | *FOXI2* | -1.741003317 | 0.001714774 | 0.002736934 | 1 |
| Obesity | pericyte | *HYAL3* | -1.695599334 | 0.000145145 | 0.000281124 | 0.912528381 |
| Obesity | pericyte | *ZSCAN30* | 7.871502684 | 9.49E-08 | 5.79E-07 | 0.000596737 |
| Obesity | pericyte | *LILRB4* | 1.777630421 | 2.22E-06 | 6.79E-06 | 0.013955316 |
| Obesity | pericyte | *TRABD2A* | 1.780882642 | 8.14E-08 | 5.13E-07 | 0.000512057 |
| Obesity | pericyte | *QRFPR* | -3.54678297 | 0.002031128 | 0.003205247 | 1 |
| Obesity | pericyte | *TMEM17* | 2.305860891 | 4.31E-05 | 9.32E-05 | 0.270748255 |
| Obesity | pericyte | *TNFRSF18* | 6.005406738 | 8.19E-07 | 3.01E-06 | 0.005150501 |
| Obesity | pericyte | *RTN4RL2* | -2.151893909 | 2.35E-05 | 5.42E-05 | 0.147628894 |
| Obesity | pericyte | *TMEM232* | 3.242550954 | 0.000152994 | 0.000294781 | 0.961870418 |
| Obesity | pericyte | *EFCAB6* | 4.645098697 | 0.000197416 | 0.000371731 | 1 |
| Obesity | pericyte | *KANK3* | 7.9205133 | 3.62E-05 | 7.98E-05 | 0.227823414 |
| Obesity | pericyte | *EMID1* | 6.77506705 | 5.41E-08 | 3.90E-07 | 0.000339861 |
| Obesity | pericyte | *CYP4A11* | 2.487149594 | 5.94E-05 | 0.000124052 | 0.37327272 |
| Obesity | pericyte | *TMEM216* | 2.334193807 | 8.97E-06 | 2.31E-05 | 0.056388685 |
| Obesity | pericyte | *RPS19BP1* | 2.303402905 | 3.28E-05 | 7.29E-05 | 0.206315183 |
| Obesity | pericyte | *C3orf70* | -1.113973072 | 0.016575347 | 0.022837872 | 1 |
| Obesity | pericyte | *PLCD1* | 2.942819598 | 0.000116231 | 0.000228857 | 0.730741831 |
| Obesity | pericyte | *ENTPD5* | 7.806282377 | 1.55E-08 | 1.90E-07 | 9.76E-05 |
| Obesity | pericyte | *HEATR4* | 1.278172999 | 0.001594425 | 0.002562147 | 1 |
| Obesity | pericyte | *MRPS21* | 6.176596355 | 1.76E-09 | 7.21E-08 | 1.11E-05 |
| Obesity | pericyte | *ANGPTL5* | 2.121629299 | 9.00E-07 | 3.24E-06 | 0.005655965 |
| Obesity | pericyte | *TSPYL4* | 3.001846362 | 0.000665539 | 0.001139499 | 1 |
| Obesity | pericyte | *MT1X* | 2.787487575 | 1.19E-05 | 2.94E-05 | 0.074515114 |
| Obesity | pericyte | *FNBP1* | 2.527478406 | 0.000132213 | 0.000257583 | 0.831221041 |
| Obesity | pericyte | *WDR86* | 2.413764694 | 0.000191577 | 0.000361833 | 1 |
| Obesity | pericyte | *EPOR* | -1.181482418 | 0.000869917 | 0.001455341 | 1 |
| Obesity | pericyte | *FPR3* | 2.233877281 | 0.003575825 | 0.005423694 | 1 |
| Obesity | pericyte | *GJA4* | -0.898959759 | 6.17E-05 | 0.000128468 | 0.387974004 |
| Obesity | pericyte | *TLR5* | 7.414570213 | 4.14E-05 | 9.00E-05 | 0.260327489 |
| Obesity | pericyte | *PLEKHN1* | 2.97062036 | 6.21E-05 | 0.000129164 | 0.390476439 |
| Obesity | pericyte | *ZNF385C* | 13.05244883 | 2.67E-08 | 2.43E-07 | 0.000167844 |
| Obesity | pericyte | *TET3* | 5.697686696 | 9.16E-09 | 1.70E-07 | 5.76E-05 |
| Obesity | pericyte | *TRPV2* | 5.444670753 | 3.11E-08 | 2.71E-07 | 0.000195249 |
| Obesity | pericyte | *C2orf88* | 3.736774324 | 5.53E-06 | 1.50E-05 | 0.03475764 |
| Obesity | pericyte | *KBTBD12* | 1.407341351 | 0.001461187 | 0.00236704 | 1 |
| Obesity | pericyte | *SEMA4D* | 3.094130921 | 2.18E-07 | 1.06E-06 | 0.00136987 |
| Obesity | pericyte | *MCRS1* | 4.112876789 | 6.71E-07 | 2.57E-06 | 0.004218379 |
| Obesity | pericyte | *TMEM220* | 5.227353697 | 0.000245892 | 0.000454684 | 1 |
| Obesity | pericyte | *CCDC157* | 5.620763326 | 3.96E-07 | 1.71E-06 | 0.002489822 |
| Obesity | pericyte | *GFRAL* | 2.513271838 | 3.83E-05 | 8.38E-05 | 0.240645211 |
| Obesity | pericyte | *C2orf66* | 5.09534805 | 0.017173742 | 0.02362094 | 1 |
| Obesity | pericyte | *CPSF4L* | -1.877140703 | 0.018502451 | 0.02533208 | 1 |
| Obesity | pericyte | *ZSCAN23* | 0.983830829 | 1.70E-06 | 5.44E-06 | 0.01069473 |

| | | | | | | |
|---|---|---|---|---|---|---|
| Obesity | pericyte | *OR7D2* | 3.178290681 | 0.012107962 | 0.017037322 | 1 |
| Obesity | pericyte | *MORN2* | 1.54339712 | 0.000694314 | 0.001183289 | 1 |
| Obesity | pericyte | *C6orf132* | 2.663869382 | 3.08E-05 | 6.91E-05 | 0.193702514 |
| Obesity | pericyte | *MAPK12* | -1.734370068 | 0.00025674 | 0.000472933 | 1 |
| Obesity | pericyte | *FAM166A* | 9.914714155 | 1.11E-06 | 3.82E-06 | 0.006998114 |
| Obesity | pericyte | *ZNF626* | 4.212916841 | 1.31E-07 | 7.28E-07 | 0.000825833 |
| Obesity | pericyte | *SMTNL2* | -2.447377114 | 1.99E-05 | 4.69E-05 | 0.125377674 |
| Obesity | pericyte | *ZC3H6* | 6.068664446 | 5.95E-09 | 1.42E-07 | 3.74E-05 |
| Obesity | pericyte | *PRKAR1B* | 3.229269246 | 1.42E-05 | 3.46E-05 | 0.08906744 |
| Obesity | pericyte | *NUTM2B* | -18.16879714 | 5.53E-05 | 0.000116434 | 0.347728182 |
| Obesity | pericyte | *NCR3LG1* | 4.350237535 | 9.01E-07 | 3.24E-06 | 0.005663011 |
| Obesity | pericyte | *AGAP4* | 3.151074448 | 1.19E-06 | 4.00E-06 | 0.007464437 |
| Obesity | pericyte | *HYKK* | 3.406933445 | 2.39E-09 | 8.62E-08 | 1.50E-05 |
| Obesity | pericyte | *RUFY4* | 5.139272513 | 1.05E-06 | 3.66E-06 | 0.006599285 |
| Obesity | pericyte | *ZNF669* | -2.220193654 | 0.027814882 | 0.03714362 | 1 |
| Obesity | pericyte | *CENPP* | 2.737965109 | 4.32E-12 | 6.38E-10 | 2.71E-08 |
| Obesity | pericyte | *PLSCR1* | 6.563720013 | 2.28E-09 | 8.46E-08 | 1.43E-05 |
| Obesity | pericyte | *ZP3* | 2.36084698 | 9.81E-05 | 0.000196489 | 0.616581435 |
| Obesity | pericyte | *PDCD1* | 4.121948359 | 0.021849379 | 0.029573099 | 1 |
| Obesity | pericyte | *CERKL* | 6.261137223 | 3.87E-07 | 1.68E-06 | 0.002435302 |
| Obesity | pericyte | *C19orf54* | 1.512196548 | 0.00613351 | 0.008995171 | 1 |
| Obesity | pericyte | *C22orf34* | 2.755063691 | 3.06E-07 | 1.39E-06 | 0.001923262 |
| Obesity | pericyte | *COL25A1* | -0.20101216 | 1.98E-05 | 4.66E-05 | 0.124467951 |
| Obesity | pericyte | *SRSF10* | 5.442057774 | 3.22E-05 | 7.16E-05 | 0.202363559 |
| Obesity | pericyte | *NBR1* | 7.796234851 | 1.87E-07 | 9.39E-07 | 0.001176013 |
| Obesity | pericyte | *FAM72B* | 7.197218194 | 8.95E-07 | 3.23E-06 | 0.005628947 |
| Obesity | pericyte | *SUMO2* | 5.846558933 | 2.62E-08 | 2.40E-07 | 0.000164791 |
| Obesity | pericyte | *GOLGA8M* | 4.595114348 | 0.000165949 | 0.000317119 | 1 |
| Obesity | pericyte | *DPYD* | 4.064027824 | 7.91E-05 | 0.000161226 | 0.497220173 |
| Obesity | pericyte | *S100A16* | 3.986410731 | 4.70E-08 | 3.55E-07 | 0.000295684 |
| Obesity | pericyte | *SLC4A5* | -2.999280633 | 0.00086433 | 0.001446378 | 1 |
| Obesity | pericyte | *UROS* | 5.086343414 | 1.04E-05 | 2.63E-05 | 0.065629654 |
| Obesity | pericyte | *FAM221A* | 3.487734875 | 4.18E-05 | 9.07E-05 | 0.262805086 |
| Obesity | pericyte | *NOXA1* | 4.287425114 | 8.64E-08 | 5.38E-07 | 0.000543124 |
| Obesity | pericyte | *SKOR1* | -2.005961471 | 2.17E-05 | 5.04E-05 | 0.136340103 |
| Obesity | pericyte | *TMEM201* | 5.831127601 | 3.13E-06 | 9.16E-06 | 0.01967286 |
| Obesity | pericyte | *ZDHHC11* | 3.653313876 | 1.12E-06 | 3.83E-06 | 0.007038607 |
| Obesity | pericyte | *FAM26F* | 1.122643695 | 0.000638934 | 0.001097535 | 1 |
| Obesity | pericyte | *LINC00910* | 3.788965128 | 4.00E-07 | 1.73E-06 | 0.002512321 |
| Obesity | pericyte | *BEND4* | 9.321638676 | 2.02E-07 | 9.99E-07 | 0.001271292 |
| Obesity | pericyte | *ZNF563* | 2.22475866 | 0.008878969 | 0.012701269 | 1 |
| Obesity | pericyte | *FBF1* | 1.457634626 | 9.55E-06 | 2.44E-05 | 0.060051349 |
| Obesity | pericyte | *MSL1* | 1.096174433 | 0.033065222 | 0.043644982 | 1 |
| Obesity | pericyte | *LRRK2* | 2.583365156 | 2.70E-05 | 6.15E-05 | 0.170009226 |
| Obesity | pericyte | *FAM196A* | 5.578140351 | 4.08E-05 | 8.87E-05 | 0.25624016 |
| Obesity | pericyte | *UTS2B* | 1.029266303 | 0.007791168 | 0.011226925 | 1 |
| Obesity | pericyte | *C9orf152* | 5.46892279 | 6.70E-07 | 2.57E-06 | 0.00421539 |
| Obesity | pericyte | *SLC15A5* | 1.203688886 | 0.001486361 | 0.002402867 | 1 |
| Obesity | pericyte | *ADAT2* | 3.898605493 | 0.000253723 | 0.000467926 | 1 |
| Obesity | pericyte | *ZNF567* | 5.98510613 | 1.83E-08 | 2.04E-07 | 0.000115113 |

| | | | | | | |
|---|---|---|---|---|---|---|
| Obesity | pericyte | *ALKBH2* | 2.753359896 | 6.65E-07 | 2.56E-06 | 0.004183498 |
| Obesity | pericyte | *LITAF* | 4.209447274 | 8.45E-09 | 1.70E-07 | 5.31E-05 |
| Obesity | pericyte | *VSTM1* | 2.324089372 | 0.002683505 | 0.00415649 | 1 |
| Obesity | pericyte | *SF3B3* | 7.723650401 | 3.24E-08 | 2.79E-07 | 0.00020361 |
| Obesity | pericyte | *SP6* | -18.18986617 | 1.92E-06 | 6.01E-06 | 0.012057058 |
| Obesity | pericyte | *PLAC9* | 3.615061369 | 2.07E-07 | 1.02E-06 | 0.001304038 |
| Obesity | pericyte | *CLDN4* | 1.684953277 | 0.000476821 | 0.000836665 | 1 |
| Obesity | pericyte | *ZNF573* | 7.29062407 | 3.03E-06 | 8.89E-06 | 0.019048318 |
| Obesity | pericyte | *FAM47E* | 1.938457292 | 2.60E-09 | 9.25E-08 | 1.64E-05 |
| Obesity | pericyte | *ZNF527* | 6.225699076 | 9.32E-05 | 0.000187551 | 0.586097931 |
| Obesity | pericyte | *S100A13* | 5.467151953 | 0.000398224 | 0.000709647 | 1 |
| Obesity | pericyte | *ZNF33A* | 4.939127984 | 3.17E-07 | 1.43E-06 | 0.00199106 |
| Obesity | pericyte | *ZNF600* | 1.593318054 | 0.001626971 | 0.00260672 | 1 |
| Obesity | pericyte | *C15orf61* | 8.824572869 | 1.42E-07 | 7.64E-07 | 0.000895293 |
| Obesity | pericyte | *TSPYL1* | 2.458850835 | 6.87E-05 | 0.000141846 | 0.432204691 |
| Obesity | pericyte | *PNRC2* | 7.565946007 | 1.00E-05 | 2.53E-05 | 0.062943675 |
| Obesity | pericyte | *RRP7A* | 2.76033341 | 0.01306249 | 0.018283903 | 1 |
| Obesity | pericyte | *SLC35E2B* | 2.035922646 | 7.34E-05 | 0.00015057 | 0.461497206 |
| Obesity | pericyte | *AKR1C3* | 3.76340498 | 1.18E-07 | 6.73E-07 | 0.000738857 |
| Obesity | pericyte | *WDSUB1* | 2.806029636 | 0.002121907 | 0.003335107 | 1 |
| Obesity | pericyte | *S100A4* | 0.983836176 | 0.023701585 | 0.031921993 | 1 |
| Obesity | pericyte | *COLCA1* | 3.488929163 | 2.22E-05 | 5.15E-05 | 0.139570131 |
| Obesity | pericyte | *SEMA4A* | 2.971465999 | 6.68E-08 | 4.48E-07 | 0.00041983 |
| Obesity | pericyte | *HRCT1* | 3.979027567 | 4.44E-05 | 9.58E-05 | 0.278985994 |
| Obesity | pericyte | *MPHOSPH8* | 4.401914847 | 0.000257289 | 0.000473807 | 1 |
| Obesity | pericyte | *SIRPB2* | 4.629829472 | 1.39E-07 | 7.52E-07 | 0.000874714 |
| Obesity | pericyte | *SRGAP3* | 1.803943565 | 3.81E-05 | 8.35E-05 | 0.23957437 |
| Obesity | pericyte | *SUPT5H* | 5.215295848 | 0.000726091 | 0.001233765 | 1 |
| Obesity | pericyte | *XPNPEP3* | 1.022010128 | 0.011916929 | 0.016783542 | 1 |
| Obesity | pericyte | *PPIA* | 8.586579606 | 7.92E-09 | 1.66E-07 | 4.98E-05 |
| Obesity | pericyte | *ZNF471* | 2.370397863 | 0.001402981 | 0.002275682 | 1 |
| Obesity | pericyte | *GTF2IRD2* | 1.710417491 | 0.005677829 | 0.008373012 | 1 |
| Obesity | pericyte | *GRM7* | 0.908641287 | 0.000783669 | 0.001324442 | 1 |
| Obesity | pericyte | *SUPT3H* | 5.470896364 | 2.11E-05 | 4.93E-05 | 0.132667154 |
| Obesity | pericyte | *POM121* | 3.645456679 | 2.69E-05 | 6.12E-05 | 0.168991674 |
| Obesity | pericyte | *GIMAP5* | 1.970348016 | 3.99E-05 | 8.71E-05 | 0.251095464 |
| Obesity | pericyte | *STK31* | 2.573120356 | 8.99E-07 | 3.24E-06 | 0.005649104 |
| Obesity | pericyte | *ZKSCAN7* | 6.488642311 | 0.002426022 | 0.003785655 | 1 |
| Obesity | pericyte | *ZNF565* | 4.869315386 | 5.56E-08 | 3.96E-07 | 0.000349538 |
| Obesity | pericyte | *NTNG2* | 5.739170442 | 1.37E-07 | 7.44E-07 | 0.000861898 |
| Obesity | pericyte | *WDR5* | 8.139307007 | 6.03E-07 | 2.38E-06 | 0.003788084 |
| Obesity | pericyte | *LONP1* | 4.142999308 | 1.51E-07 | 7.97E-07 | 0.000949424 |
| Obesity | pericyte | *SLC35F1* | -0.657845515 | 3.23E-05 | 7.18E-05 | 0.202899836 |
| Obesity | pericyte | *ZNF34* | 8.021029983 | 3.51E-07 | 1.55E-06 | 0.002207501 |
| Obesity | pericyte | *INCA1* | 7.652594713 | 1.66E-10 | 1.41E-08 | 1.04E-06 |
| Obesity | pericyte | *ZNF774* | 4.034260741 | 3.98E-06 | 1.13E-05 | 0.025025837 |
| Obesity | pericyte | *EVL* | 1.860368596 | 0.022362732 | 0.030222377 | 1 |
| Obesity | pericyte | *ZNF124* | 6.683154261 | 1.24E-08 | 1.73E-07 | 7.78E-05 |
| Obesity | pericyte | *ZNF569* | 4.267577323 | 2.35E-06 | 7.12E-06 | 0.014743869 |
| Obesity | pericyte | *PIK3R4* | 1.66112037 | 0.025186613 | 0.033864037 | 1 |

| | | | | | | |
|---|---|---|---|---|---|---|
| Obesity | pericyte | *ZNF775* | 2.226392536 | 0.000114936 | 0.000226735 | 0.722603995 |
| Obesity | pericyte | *ZNF605* | 5.322392231 | 1.77E-08 | 2.01E-07 | 0.000111403 |
| Obesity | pericyte | *RFX8* | 3.288704979 | 2.06E-06 | 6.38E-06 | 0.01294172 |
| Obesity | pericyte | *ZNF799* | -2.255137427 | 5.67E-05 | 0.00011902 | 0.35670182 |
| Obesity | pericyte | *C20orf96* | 3.798034071 | 2.95E-07 | 1.35E-06 | 0.001855379 |
| Obesity | pericyte | *SULT1A1* | -1.397556984 | 0.003339867 | 0.005089129 | 1 |
| Obesity | pericyte | *ANAPC7* | 3.746828227 | 5.90E-06 | 1.59E-05 | 0.037094651 |
| Obesity | pericyte | *TPK1* | 4.131424836 | 1.50E-05 | 3.63E-05 | 0.094069546 |
| Obesity | pericyte | *C1orf186* | 2.800469529 | 1.64E-06 | 5.26E-06 | 0.010295715 |
| Obesity | pericyte | *MYO18A* | 3.012311375 | 0.003469927 | 0.0052758 | 1 |
| Obesity | pericyte | *XRCC2* | 2.930107729 | 1.87E-06 | 5.88E-06 | 0.011730863 |
| Obesity | pericyte | *HDAC2* | 6.974004666 | 1.13E-05 | 2.82E-05 | 0.0712161 |
| Obesity | pericyte | *ADH1B* | 3.613556 | 4.90E-09 | 1.29E-07 | 3.08E-05 |
| Obesity | pericyte | *HRH1* | 6.667616885 | 5.81E-07 | 2.32E-06 | 0.00365189 |
| Obesity | pericyte | *ZNF502* | 2.908391071 | 6.22E-06 | 1.67E-05 | 0.039101134 |
| Obesity | pericyte | *TECPR2* | 7.212936559 | 2.06E-08 | 2.16E-07 | 0.000129251 |
| Obesity | pericyte | *FAM180B* | 0.88727165 | 0.037780489 | 0.049391959 | 1 |
| Obesity | pericyte | *ZFP62* | 10.19786006 | 2.57E-08 | 2.38E-07 | 0.000161598 |
| Obesity | pericyte | *ERI2* | 4.769210888 | 3.46E-08 | 2.89E-07 | 0.000217395 |
| Obesity | pericyte | *TRPV1* | 3.172900451 | 3.22E-08 | 2.79E-07 | 0.000202273 |
| Obesity | pericyte | *AMZ2* | 4.170714111 | 2.39E-06 | 7.24E-06 | 0.0150427 |
| Obesity | pericyte | *ZNF431* | 5.875924382 | 1.45E-06 | 4.76E-06 | 0.009140737 |
| Obesity | pericyte | *NF1* | 4.38774031 | 1.34E-05 | 3.30E-05 | 0.084525363 |
| Obesity | pericyte | *DAPK1* | 2.311722596 | 0.0002385 | 0.000441925 | 1 |
| Obesity | pericyte | *HLA-DQA1* | 1.321439061 | 0.000470045 | 0.000825928 | 1 |
| Obesity | pericyte | *GM2A* | 3.218107472 | 1.11E-07 | 6.47E-07 | 0.000699617 |
| Obesity | pericyte | *TLE1* | 2.325974138 | 0.000583229 | 0.00100957 | 1 |
| Obesity | pericyte | *STRN3* | 5.026425827 | 1.71E-05 | 4.08E-05 | 0.107578591 |
| Obesity | pericyte | *ZNF239* | 1.819183528 | 0.002076342 | 0.00327085 | 1 |
| Obesity | pericyte | *MVB12B* | 6.328250546 | 8.05E-08 | 5.08E-07 | 0.000506389 |
| Obesity | pericyte | *C6orf106* | 3.816596237 | 4.57E-15 | 1.79E-12 | 2.87E-11 |
| Obesity | pericyte | *PPTC7* | 3.835718176 | 4.46E-06 | 1.25E-05 | 0.028060128 |
| Obesity | pericyte | *NHLRC2* | 5.492479818 | 1.22E-05 | 3.01E-05 | 0.076550402 |
| Obesity | pericyte | *ZFP28* | 3.420643127 | 7.35E-08 | 4.79E-07 | 0.00046225 |
| Obesity | pericyte | *KIAA1211L* | 1.306982839 | 0.000580898 | 0.001006368 | 1 |
| Obesity | pericyte | *LAMB3* | 3.233247105 | 2.03E-06 | 6.32E-06 | 0.012782734 |
| Obesity | pericyte | *HCAR1* | 1.965861264 | 0.005448747 | 0.008056507 | 1 |
| Obesity | pericyte | *SRGAP1* | 5.967423514 | 6.14E-07 | 2.42E-06 | 0.003860345 |
| Obesity | pericyte | *FAM3C* | 6.814698476 | 4.14E-07 | 1.78E-06 | 0.002605097 |
| Obesity | pericyte | *SLC39A10* | 5.793700583 | 7.68E-08 | 4.90E-07 | 0.000482693 |
| Obesity | pericyte | *AP2A1* | 3.079370313 | 0.003273077 | 0.005001904 | 1 |
| Obesity | pericyte | *ZNF585A* | 2.655896931 | 2.00E-07 | 9.91E-07 | 0.001259974 |
| Obesity | pericyte | *FUT11* | 6.244304991 | 3.25E-11 | 3.72E-09 | 2.04E-07 |
| Obesity | pericyte | *ANXA4* | 5.575477905 | 1.47E-08 | 1.87E-07 | 9.26E-05 |
| Obesity | pericyte | *ZNF138* | 5.169794047 | 1.43E-09 | 6.18E-08 | 9.02E-06 |
| Obesity | pericyte | *ZNF429* | 3.695447813 | 1.25E-05 | 3.08E-05 | 0.07854378 |
| Obesity | pericyte | *ZNF470* | 3.287266298 | 1.78E-05 | 4.23E-05 | 0.112091976 |
| Obesity | pericyte | *ZNF100* | 1.205428028 | 0.015510634 | 0.021498094 | 1 |
| Obesity | pericyte | *GMFB* | 5.740820794 | 1.52E-07 | 8.02E-07 | 0.000957349 |
| Obesity | pericyte | *ZMYM1* | 6.978180308 | 2.08E-05 | 4.86E-05 | 0.130511009 |

| | | | | | | |
|---|---|---|---|---|---|---|
| Obesity | pericyte | *ARRDC1* | 3.088755026 | 4.18E-07 | 1.80E-06 | 0.002630459 |
| Obesity | pericyte | *GAL3ST4* | -2.352699452 | 0.002305052 | 0.003606736 | 1 |
| Obesity | pericyte | *SLC25A29* | 4.595837818 | 4.59E-10 | 3.07E-08 | 2.89E-06 |
| Obesity | pericyte | *ZNF682* | 4.365603807 | 0.010188258 | 0.014465578 | 1 |
| Obesity | pericyte | *ZNF257* | 3.108549356 | 0.000289104 | 0.000526992 | 1 |
| Obesity | pericyte | *ADAM32* | 2.200634195 | 1.05E-05 | 2.64E-05 | 0.065940814 |
| Obesity | pericyte | *LRRC8B* | 3.26786276 | 3.35E-05 | 7.42E-05 | 0.210614706 |
| Obesity | pericyte | *ABCB8* | 1.277536236 | 0.020750966 | 0.028177392 | 1 |
| Obesity | pericyte | *SND1* | 4.733087846 | 6.08E-09 | 1.44E-07 | 3.82E-05 |
| Obesity | pericyte | *SULT1A2* | 2.437672901 | 4.42E-34 | 2.78E-30 | 2.78E-30 |
| Obesity | pericyte | *PIWIL2* | 2.406298019 | 5.59E-06 | 1.52E-05 | 0.03517431 |
| Obesity | pericyte | *ENTPD4* | 5.198335342 | 1.75E-07 | 8.93E-07 | 0.001097391 |
| Obesity | pericyte | *C1D* | 4.137147797 | 1.16E-06 | 3.92E-06 | 0.00727807 |
| Obesity | pericyte | *TBC1D9B* | 1.627956434 | 0.020745028 | 0.028175414 | 1 |
| Obesity | pericyte | *FAM110D* | 9.729706683 | 3.85E-06 | 1.10E-05 | 0.024179777 |
| Obesity | pericyte | *KANK2* | 7.518908587 | 8.96E-07 | 3.23E-06 | 0.005635356 |
| Obesity | pericyte | *GTF2E2* | 6.880625794 | 1.05E-07 | 6.23E-07 | 0.000662985 |
| Obesity | pericyte | *ZNF165* | -2.057946436 | 0.011144282 | 0.015758907 | 1 |
| Obesity | pericyte | *RAMP2-AS1* | 2.406970312 | 0.027682184 | 0.036997851 | 1 |
| Obesity | pericyte | *ZNF720* | 3.483184781 | 3.20E-05 | 7.13E-05 | 0.201192188 |
| Obesity | pericyte | *DDI2* | 4.86429999 | 5.26E-08 | 3.83E-07 | 0.000330989 |
| Obesity | pericyte | *MRPL21* | -0.701117826 | 0.031742526 | 0.041978389 | 1 |
| Obesity | pericyte | *ZNF786* | 3.161333325 | 3.03E-06 | 8.89E-06 | 0.019040825 |
| Obesity | pericyte | *ZNF517* | 14.233917 | 1.58E-05 | 3.80E-05 | 0.099125792 |
| Obesity | pericyte | *C5AR1* | 4.740427802 | 0.000191593 | 0.000361833 | 1 |
| Obesity | pericyte | *DCHS2* | -0.751000567 | 0.001818008 | 0.002891428 | 1 |
| Obesity | pericyte | *VEPH1* | 1.763781435 | 6.43E-06 | 1.73E-05 | 0.040436339 |
| Obesity | pericyte | *MAP3K5* | 2.99013039 | 0.0004262 | 0.000756713 | 1 |
| Obesity | pericyte | *STMN3* | 2.529406534 | 0.000109415 | 0.00021666 | 0.687895108 |
| Obesity | pericyte | *GYPE* | 1.625682787 | 0.002793462 | 0.004315109 | 1 |
| Obesity | pericyte | *PCDHB11* | -4.36488335 | 6.07E-07 | 2.40E-06 | 0.003818702 |
| Obesity | pericyte | *SLC2A10* | 3.435494475 | 0.001102987 | 0.001811989 | 1 |
| Obesity | pericyte | *RPF2* | 4.334494133 | 2.50E-05 | 5.74E-05 | 0.157451259 |
| Obesity | pericyte | *MYO5A* | 3.396425709 | 2.87E-06 | 8.48E-06 | 0.018058963 |
| Obesity | pericyte | *ATG7* | 3.299205482 | 1.35E-05 | 3.30E-05 | 0.084709601 |
| Obesity | pericyte | *RAB40C* | 9.016509666 | 5.06E-08 | 3.73E-07 | 0.000318219 |
| Obesity | pericyte | *ZNF624* | 7.364881895 | 0.001006947 | 0.001662467 | 1 |
| Obesity | pericyte | *HHLA3* | 3.303607094 | 0.000166794 | 0.000318568 | 1 |
| Obesity | pericyte | *BCO2* | 2.708221983 | 1.25E-06 | 4.18E-06 | 0.007869443 |
| Obesity | pericyte | *C5orf42* | 3.055383765 | 2.55E-06 | 7.65E-06 | 0.016051476 |
| Obesity | pericyte | *ZNF841* | 3.20386757 | 2.04E-06 | 6.34E-06 | 0.012844779 |
| Obesity | pericyte | *MFAP5* | 4.73281194 | 9.85E-09 | 1.71E-07 | 6.20E-05 |
| Obesity | pericyte | *CDC42SE1* | 6.80044755 | 2.13E-07 | 1.04E-06 | 0.001340979 |
| Obesity | pericyte | *MPEG1* | 0.893393501 | 0.003677161 | 0.005569336 | 1 |
| Obesity | pericyte | *PDCD1LG2* | 1.809633383 | 0.001031072 | 0.001700066 | 1 |
| Obesity | pericyte | *NMB* | 9.163560675 | 1.03E-06 | 3.60E-06 | 0.006473127 |
| Obesity | pericyte | *KLHL14* | -2.384637393 | 0.00063094 | 0.001084989 | 1 |
| Obesity | pericyte | *FAM114A1* | 4.970955099 | 5.55E-09 | 1.35E-07 | 3.49E-05 |
| Obesity | pericyte | *PHF2* | 6.28312969 | 6.56E-09 | 1.52E-07 | 4.13E-05 |
| Obesity | pericyte | *RPS26* | -0.906479802 | 0.001092245 | 0.001795279 | 1 |

| | | | | | | |
|---|---|---|---|---|---|---|
| Obesity | pericyte | *PSAP* | 5.975634066 | 1.29E-07 | 7.18E-07 | 0.000813106 |
| Obesity | pericyte | *S100A10* | 3.454380785 | 0.000650491 | 0.001115559 | 1 |
| Obesity | pericyte | *MAP1LC3C* | 2.105126479 | 0.000175524 | 0.000333995 | 1 |
| Obesity | pericyte | *TAF13* | 2.765991106 | 4.49E-05 | 9.68E-05 | 0.282215811 |
| Obesity | pericyte | *ZNF780A* | -1.377848495 | 2.88E-05 | 6.50E-05 | 0.181110874 |
| Obesity | pericyte | *AC122129.1* | 11.71517028 | 1.20E-07 | 6.80E-07 | 0.000754205 |
| Obesity | pericyte | *CCDC180* | -1.769124623 | 0.001795355 | 0.00285757 | 1 |
| Obesity | pericyte | *ZNF181* | 5.278456322 | 2.31E-06 | 7.02E-06 | 0.014512555 |
| Obesity | pericyte | *SGTB* | 5.522930614 | 9.12E-05 | 0.000184083 | 0.573234825 |
| Obesity | pericyte | *ADH5* | 3.227360554 | 6.49E-05 | 0.000134569 | 0.408148371 |
| Obesity | pericyte | *ZNF677* | 8.495116639 | 0.00019204 | 0.000362569 | 1 |
| Obesity | pericyte | *ZNF311* | 1.57413402 | 0.004338009 | 0.006515304 | 1 |
| Obesity | pericyte | *FCHSD1* | 7.580160664 | 1.35E-07 | 7.34E-07 | 0.000845818 |
| Obesity | pericyte | *ZNF71* | 1.961510749 | 0.013537792 | 0.018892807 | 1 |
| Obesity | pericyte | *RPL12* | 1.641928088 | 0.015589241 | 0.021598095 | 1 |
| Obesity | pericyte | *VPS13A* | 1.750836771 | 0.002552494 | 0.003970195 | 1 |
| Obesity | pericyte | *LEKR1* | 1.741537118 | 0.001974623 | 0.003119994 | 1 |
| Obesity | pericyte | *ZNF335* | 3.832152496 | 4.96E-07 | 2.05E-06 | 0.003116521 |
| Obesity | pericyte | *AGAP9* | 2.45975857 | 2.24E-06 | 6.85E-06 | 0.014110662 |
| Obesity | pericyte | *ZNF273* | 3.738415681 | 1.14E-06 | 3.88E-06 | 0.007183165 |
| Obesity | pericyte | *ZNF84* | 5.640674899 | 3.05E-06 | 8.94E-06 | 0.019169248 |
| Obesity | pericyte | *MAK16* | -3.311227259 | 0.014082124 | 0.019595909 | 1 |
| Obesity | pericyte | *ZNF667* | 1.664223735 | 9.11E-06 | 2.33E-05 | 0.05726227 |
| Obesity | pericyte | *PRIM1* | 3.421709288 | 6.91E-06 | 1.83E-05 | 0.043457752 |
| Obesity | pericyte | *SFI1* | 6.718004807 | 7.36E-09 | 1.60E-07 | 4.63E-05 |
| Obesity | pericyte | *ZNF248* | 1.844695339 | 0.00440033 | 0.006602596 | 1 |
| Obesity | pericyte | *HIBCH* | 2.463530046 | 0.003623497 | 0.005490703 | 1 |
| Obesity | pericyte | *SOWAHC* | -1.820746247 | 7.58E-05 | 0.000154841 | 0.476290011 |
| Obesity | pericyte | *MIER1* | 6.527650807 | 3.84E-07 | 1.67E-06 | 0.002417228 |
| Obesity | pericyte | *SVIP* | 4.803656162 | 2.47E-08 | 2.32E-07 | 0.000155452 |
| Obesity | pericyte | *ZNF251* | 4.095020587 | 0.00198553 | 0.003135651 | 1 |
| Obesity | pericyte | *DDRGK1* | 8.452808445 | 3.83E-05 | 8.39E-05 | 0.24107132 |
| Obesity | pericyte | *HSD17B11* | 5.55192088 | 2.65E-09 | 9.26E-08 | 1.66E-05 |
| Obesity | pericyte | *SULT1C2* | 1.105283042 | 0.000313076 | 0.000567399 | 1 |
| Obesity | pericyte | *RPS6KL1* | 1.370466315 | 0.000863787 | 0.001445854 | 1 |
| Obesity | pericyte | *QRICH1* | 2.757748283 | 3.02E-06 | 8.88E-06 | 0.018997404 |
| Obesity | pericyte | *DDX42* | 5.113009889 | 6.92E-06 | 1.83E-05 | 0.043485197 |
| Obesity | pericyte | *SLC29A3* | 4.564971967 | 1.63E-06 | 5.25E-06 | 0.010259312 |
| Obesity | pericyte | *ANTXRL* | 76.99258803 | 1.95E-06 | 6.09E-06 | 0.012237174 |
| Obesity | pericyte | *STYX* | 6.664287029 | 2.21E-09 | 8.32E-08 | 1.39E-05 |
| Obesity | pericyte | *TMEM116* | 1.297446239 | 0.00050287 | 0.000879673 | 1 |
| Obesity | pericyte | *ZKSCAN8* | 7.238130162 | 1.22E-08 | 1.73E-07 | 7.69E-05 |
| Obesity | pericyte | *ZNF813* | 5.713257661 | 3.09E-08 | 2.70E-07 | 0.00019424 |
| Obesity | pericyte | *GFPT1* | 2.481809273 | 0.018651303 | 0.025513652 | 1 |
| Obesity | pericyte | *UVRAG* | 2.005926497 | 0.000802269 | 0.001352115 | 1 |
| Obesity | pericyte | *MGEA5* | 3.289132021 | 0.000175717 | 0.000334261 | 1 |
| Obesity | pericyte | *MT1F* | -4.655366759 | 5.59E-05 | 0.000117587 | 0.351584841 |
| Obesity | pericyte | *ZNF568* | 5.089118006 | 1.78E-05 | 4.23E-05 | 0.112027959 |
| Obesity | pericyte | *ZNF587* | 5.371207313 | 3.73E-07 | 1.63E-06 | 0.002347742 |
| Obesity | pericyte | *FLVCR1-AS1* | 0.918406434 | 0.00163529 | 0.00261938 | 1 |

| | | | | | | |
|---|---|---|---|---|---|---|
| Obesity | pericyte | *SH3BGRL2* | 1.766218148 | 0.00849192 | 0.012183638 | 1 |
| Obesity | pericyte | *TMA16* | 7.464542314 | 3.42E-09 | 1.06E-07 | 2.15E-05 |
| Obesity | pericyte | *CNGA1* | -0.501515596 | 3.25E-05 | 7.23E-05 | 0.204560821 |
| Obesity | pericyte | *MAFK* | 3.397770133 | 1.38E-07 | 7.49E-07 | 0.000870032 |
| Obesity | pericyte | *HIST1H4E* | -1.577676206 | 0.00010729 | 0.000213123 | 0.67453374 |
| Obesity | pericyte | *C20orf203* | 2.945846167 | 0.001818778 | 0.002891921 | 1 |
| Obesity | pericyte | *ZNF627* | 6.160247175 | 2.71E-07 | 1.26E-06 | 0.001702417 |
| Obesity | pericyte | *ARC* | -13.34791046 | 0.000119117 | 0.000234174 | 0.748886955 |
| Obesity | pericyte | *TLK1* | 5.727535964 | 1.86E-08 | 2.06E-07 | 0.000116624 |
| Obesity | pericyte | *LRBA* | 5.071977561 | 7.01E-06 | 1.86E-05 | 0.044100194 |
| Obesity | pericyte | *MMP17* | 11.45885106 | 2.57E-08 | 2.38E-07 | 0.000161325 |
| Obesity | pericyte | *CCDC69* | 2.722959506 | 6.26E-07 | 2.45E-06 | 0.003936767 |
| Obesity | pericyte | *MDM4* | 5.682626258 | 7.54E-06 | 1.98E-05 | 0.047419801 |
| Obesity | pericyte | *FAM19A2* | 5.363225066 | 1.09E-06 | 3.76E-06 | 0.006859651 |
| Obesity | pericyte | *TTC37* | 5.41240103 | 3.31E-07 | 1.48E-06 | 0.002078098 |
| Obesity | pericyte | *TUSC1* | 3.549436589 | 5.99E-07 | 2.37E-06 | 0.003768326 |
| Obesity | pericyte | *PAPSS2* | 4.488165923 | 6.69E-06 | 1.78E-05 | 0.042074075 |
| Obesity | pericyte | *FAN1* | 5.159761091 | 0.002949129 | 0.004537733 | 1 |
| Obesity | pericyte | *CEP290* | 5.804510176 | 7.05E-06 | 1.87E-05 | 0.044351017 |
| Obesity | pericyte | *ANKRD13B* | 2.512339968 | 4.59E-07 | 1.93E-06 | 0.002888046 |
| Obesity | pericyte | *ECI2* | 5.1759944 | 1.78E-08 | 2.01E-07 | 0.000111874 |
| Obesity | pericyte | *ZNF652* | 6.162343068 | 8.92E-06 | 2.30E-05 | 0.056072339 |
| Obesity | pericyte | *RPL10A* | 4.066311074 | 7.61E-06 | 1.99E-05 | 0.047842032 |
| Obesity | pericyte | *APCDD1L* | 7.187256119 | 1.61E-08 | 1.91E-07 | 0.000101032 |
| Obesity | pericyte | *RASSF9* | 6.260882907 | 1.12E-08 | 1.71E-07 | 7.02E-05 |
| Obesity | pericyte | *FAM169A* | 2.529606862 | 0.004960864 | 0.007375018 | 1 |
| Obesity | pericyte | *TMEM184B* | 6.470377952 | 2.54E-05 | 5.82E-05 | 0.159997673 |
| Obesity | pericyte | *SFT2D1* | 6.682564847 | 4.29E-05 | 9.30E-05 | 0.269967151 |
| Obesity | pericyte | *ARHGAP11A* | 2.917811489 | 5.72E-06 | 1.55E-05 | 0.035964797 |
| Obesity | pericyte | *HMGN2* | 2.88703938 | 0.000862177 | 0.001443543 | 1 |
| Obesity | pericyte | *DENND4B* | 3.518961795 | 1.93E-05 | 4.55E-05 | 0.121092187 |
| Obesity | pericyte | *CD3E* | 2.056206892 | 5.18E-06 | 1.42E-05 | 0.032578025 |
| Obesity | pericyte | *TSEN15* | 5.952886633 | 9.87E-09 | 1.71E-07 | 6.21E-05 |
| Obesity | pericyte | *ITPRIPL1* | -2.267717418 | 0.000368949 | 0.000660285 | 1 |
| Obesity | pericyte | *PRMT6* | 5.384902807 | 0.001618113 | 0.00259385 | 1 |
| Obesity | pericyte | *SHISA4* | 4.067253624 | 2.99E-05 | 6.73E-05 | 0.188021693 |
| Obesity | pericyte | *CAPZA2* | 5.392849434 | 7.31E-09 | 1.60E-07 | 4.60E-05 |
| Obesity | pericyte | *MAP3K3* | 3.693612493 | 0.006942187 | 0.010086788 | 1 |
| Obesity | pericyte | *C1orf174* | 3.59859378 | 0.001871487 | 0.002970471 | 1 |
| Obesity | pericyte | *RASGEF1A* | -3.131985065 | 0.000104413 | 0.000207867 | 0.656443817 |
| Obesity | pericyte | *KIAA0753* | 4.987311889 | 2.01E-05 | 4.73E-05 | 0.126459347 |
| Obesity | pericyte | *DCLRE1A* | 2.278685463 | 3.88E-05 | 8.48E-05 | 0.24405626 |
| Obesity | pericyte | *ZFP2* | 5.367765486 | 4.95E-05 | 0.000105885 | 0.311090873 |
| Obesity | pericyte | *L3MBTL3* | 5.601217619 | 2.09E-08 | 2.17E-07 | 0.000131561 |
| Obesity | pericyte | *MFAP3L* | 2.540924497 | 6.51E-07 | 2.51E-06 | 0.00409088 |
| Obesity | pericyte | *NAGA* | 4.511784434 | 2.44E-07 | 1.16E-06 | 0.001535335 |
| Obesity | pericyte | *TGM2* | 1.013545681 | 0.000104167 | 0.000207575 | 0.654899682 |
| Obesity | pericyte | *PJA2* | 3.192694991 | 0.004565716 | 0.006832815 | 1 |
| Obesity | pericyte | *AL590822.1* | 1.052477035 | 0.028338205 | 0.037738253 | 1 |
| Obesity | pericyte | *AP001007.1* | 11.6262668 | 9.50E-07 | 3.38E-06 | 0.005974386 |

| | | | | | | |
|---|---|---|---|---|---|---|
| Obesity | pericyte | *ZNF525* | 3.144857616 | 2.19E-07 | 1.06E-06 | 0.001376141 |
| Obesity | pericyte | *AC105020.1* | 11.60110091 | 2.14E-06 | 6.59E-06 | 0.013477165 |
| Obesity | pericyte | *EFCAB2* | 5.971199106 | 4.13E-08 | 3.23E-07 | 0.000259949 |
| Obesity | pericyte | *CHML* | 1.838529831 | 0.004865363 | 0.007243319 | 1 |
| Obesity | pericyte | *CAPN8* | -0.840249863 | 1.14E-05 | 2.84E-05 | 0.071759999 |
| Obesity | pericyte | *TATDN3* | 1.389151488 | 0.004485526 | 0.006717604 | 1 |
| Obesity | pericyte | *ECT2L* | 4.005396018 | 1.03E-06 | 3.60E-06 | 0.006456284 |
| Obesity | pericyte | *CENPW* | 4.324818408 | 4.42E-09 | 1.23E-07 | 2.78E-05 |
| Obesity | pericyte | *DDO* | 4.777471252 | 9.93E-08 | 5.96E-07 | 0.000624547 |
| Obesity | pericyte | *HIST2H2BF* | 7.21225629 | 1.13E-06 | 3.86E-06 | 0.007110368 |
| Obesity | pericyte | *RBM20* | -0.924179143 | 3.36E-05 | 7.43E-05 | 0.210977303 |
| Obesity | pericyte | *C6orf163* | 2.687469591 | 4.60E-06 | 1.28E-05 | 0.028918418 |
| Obesity | pericyte | *PCMTD2* | 6.054380288 | 5.68E-09 | 1.37E-07 | 3.57E-05 |
| Obesity | pericyte | *OOEP* | 1.322612373 | 0.011474083 | 0.016196129 | 1 |
| Obesity | pericyte | *FOXO6* | -2.237179908 | 0.000843273 | 0.001415284 | 1 |
| Obesity | pericyte | *NEU4* | 5.479513522 | 0.025760709 | 0.034584151 | 1 |
| Obesity | pericyte | *TRAF3IP1* | 4.687680498 | 2.03E-06 | 6.30E-06 | 0.012746557 |
| Obesity | pericyte | *GIGYF2* | 5.676923671 | 3.17E-06 | 9.25E-06 | 0.019920371 |
| Obesity | pericyte | *RUFY2* | 4.854347563 | 2.12E-06 | 6.52E-06 | 0.013302128 |
| Obesity | pericyte | *PHACTR4* | 2.134954973 | 0.012961141 | 0.018156571 | 1 |
| Obesity | pericyte | *AGAP6* | 1.074111664 | 0.010828863 | 0.01534394 | 1 |
| Obesity | pericyte | *C10orf128* | 3.291662185 | 1.63E-06 | 5.25E-06 | 0.010270571 |
| Obesity | pericyte | *SYT15* | 3.294515529 | 0.017187464 | 0.023634642 | 1 |
| Obesity | pericyte | *ZDBF2* | 4.329717762 | 5.28E-07 | 2.15E-06 | 0.003316523 |
| Obesity | pericyte | *RING1* | 2.33246137 | 0.005448196 | 0.008056507 | 1 |
| Obesity | pericyte | *OXLD1* | 3.016349474 | 6.52E-06 | 1.75E-05 | 0.040970216 |
| Obesity | pericyte | *HLA-DMA* | 4.994264406 | 1.41E-07 | 7.59E-07 | 0.000887581 |
| Obesity | pericyte | *NOTCH4* | -0.384117226 | 1.56E-08 | 1.91E-07 | 9.81E-05 |
| Obesity | pericyte | *AGER* | 3.194553827 | 7.51E-05 | 0.000153625 | 0.472395367 |
| Obesity | pericyte | *DXO* | -1.407239394 | 0.00173026 | 0.002758849 | 1 |
| Obesity | pericyte | *SKIV2L* | -1.124071498 | 3.57E-06 | 1.03E-05 | 0.02242731 |
| Obesity | pericyte | *NELFE* | 4.341328376 | 4.15E-11 | 4.66E-09 | 2.61E-07 |
| Obesity | pericyte | *C6orf48* | 2.241048117 | 0.017704 | 0.024307719 | 1 |
| Obesity | pericyte | *MSH5* | 1.140687207 | 0.022533687 | 0.03043379 | 1 |
| Obesity | pericyte | *ABHD16A* | 1.196234976 | 0.020286326 | 0.027590413 | 1 |
| Obesity | pericyte | *BAG6* | 2.537338982 | 0.00091532 | 0.001522588 | 1 |
| Obesity | pericyte | *NFKBIL1* | 3.185963984 | 5.86E-07 | 2.34E-06 | 0.003683193 |
| Obesity | pericyte | *ZNF814* | 4.851936498 | 1.22E-07 | 6.89E-07 | 0.000770156 |
| Obesity | pericyte | *HLA-C* | 1.64262822 | 0.000392085 | 0.000698905 | 1 |
| Obesity | pericyte | *POU5F1* | 0.437908175 | 0.020292442 | 0.027590524 | 1 |
| Obesity | pericyte | *CCHCR1* | 2.00451697 | 5.70E-10 | 3.48E-08 | 3.58E-06 |
| Obesity | pericyte | *ABCF1* | 5.459490053 | 4.91E-10 | 3.22E-08 | 3.09E-06 |
| Obesity | pericyte | *LILRB3* | 1.728415126 | 0.014237283 | 0.019807435 | 1 |
| Obesity | pericyte | *HLA-E* | 5.570760821 | 4.57E-19 | 2.88E-16 | 2.88E-15 |
| Obesity | pericyte | *ZNF468* | 4.562989257 | 1.17E-06 | 3.97E-06 | 0.007381332 |
| Obesity | pericyte | *TBC1D8* | 4.898390705 | 2.13E-06 | 6.57E-06 | 0.013403345 |
| Obesity | pericyte | *HLA-F* | 1.854316967 | 0.003336414 | 0.005085099 | 1 |
| Obesity | pericyte | *ZFP57* | 1.403022889 | 6.15E-08 | 4.24E-07 | 0.000386591 |
| Obesity | pericyte | *FAM153C* | 5.652323226 | 4.82E-05 | 0.000103328 | 0.302750498 |
| Obesity | pericyte | *GABBR1* | -2.115962424 | 0.006299934 | 0.009208947 | 1 |

| | | | | | | |
|---|---|---|---|---|---|---|
| Obesity | pericyte | *MALRD1* | 1.002073647 | 0.002011432 | 0.003175759 | 1 |
| Obesity | pericyte | *ATXN2* | 5.754273853 | 6.11E-09 | 1.44E-07 | 3.84E-05 |
| Obesity | pericyte | *FAM216A* | 3.585505359 | 3.45E-05 | 7.64E-05 | 0.217154306 |
| Obesity | pericyte | *ZNF155* | 2.921089183 | 0.00052726 | 0.000919607 | 1 |
| Obesity | pericyte | *ZNF425* | 1.336583799 | 0.015668902 | 0.021698323 | 1 |
| Obesity | pericyte | *LRRC10B* | -2.815716438 | 2.28E-05 | 5.28E-05 | 0.143276701 |
| Obesity | pericyte | *PCDHA4* | -1.860101123 | 6.26E-06 | 1.68E-05 | 0.039357853 |
| Obesity | pericyte | *CLLU1OS* | 2.167130786 | 0.019666254 | 0.026802892 | 1 |
| Obesity | pericyte | *SYCE1L* | -1.556377437 | 0.006790538 | 0.009880146 | 1 |
| Obesity | pericyte | *TMEM231* | 1.955297143 | 2.87E-05 | 6.47E-05 | 0.18013934 |
| Obesity | pericyte | *FAM71F2* | 10.69209027 | 3.17E-06 | 9.25E-06 | 0.019915245 |
| Obesity | pericyte | *AC016586.1* | -6.455732004 | 0.000304642 | 0.000553071 | 1 |
| Obesity | pericyte | *LINC00654* | 3.496079627 | 5.53E-05 | 0.000116434 | 0.347504988 |
| Obesity | pericyte | *SAP25* | -1.146056447 | 0.020051715 | 0.027304555 | 1 |
| Obesity | pericyte | *PRR13* | -3.020754174 | 0.011906777 | 0.016773002 | 1 |
| Obesity | pericyte | *MT1A* | -5.797990793 | 0.000991585 | 0.001640042 | 1 |
| Obesity | pericyte | *MT1M* | 10.89100135 | 1.19E-08 | 1.73E-07 | 7.49E-05 |
| Obesity | pericyte | *CFI* | -1.332647319 | 0.027972596 | 0.0373146 | 1 |
| Obesity | pericyte | *SAMD9* | 4.706859566 | 1.15E-07 | 6.61E-07 | 0.000723256 |
| Obesity | pericyte | *EXOC3L4* | 5.168923625 | 5.47E-05 | 0.000115515 | 0.344119791 |
| Obesity | pericyte | *ATP6AP1L* | 1.970676407 | 0.000155023 | 0.000298142 | 0.974627392 |
| Obesity | pericyte | *DENND6B* | 5.333098699 | 9.29E-08 | 5.71E-07 | 0.000584193 |
| Obesity | pericyte | *EIF3CL* | 17.05011779 | 3.33E-27 | 5.24E-24 | 2.10E-23 |
| Obesity | pericyte | *LCMT1* | 5.544414013 | 1.04E-07 | 6.17E-07 | 0.000654762 |
| Obesity | pericyte | *LIN52* | 3.68010358 | 1.48E-06 | 4.81E-06 | 0.009286178 |
| Obesity | pericyte | *C17orf107* | 2.220917586 | 5.03E-05 | 0.000107507 | 0.316501278 |
| Obesity | pericyte | *C1QTNF9B* | 116.8406759 | 0.001551045 | 0.002499723 | 1 |
| Obesity | pericyte | *NYNRIN* | 4.739552482 | 0.025046054 | 0.033682255 | 1 |
| Obesity | pericyte | *GOLGA8O* | -4.777768695 | 5.40E-05 | 0.000114196 | 0.339390241 |
| Obesity | pericyte | *HCG27* | 0.901899803 | 0.009541909 | 0.013609343 | 1 |
| Obesity | pericyte | *LNP1* | -2.998059678 | 0.011753216 | 0.016575251 | 1 |
| Obesity | pericyte | *COLQ* | 4.018668425 | 3.42E-06 | 9.88E-06 | 0.021499698 |
| Obesity | pericyte | *METTL6* | 4.426944614 | 2.52E-09 | 9.07E-08 | 1.59E-05 |
| Obesity | pericyte | *SACM1L* | 5.520487986 | 1.98E-05 | 4.65E-05 | 0.124201704 |
| Obesity | pericyte | *TAS2R14* | 1.159027061 | 0.018956511 | 0.025902974 | 1 |
| Obesity | pericyte | *ZRSR1* | 10.7429849 | 2.82E-06 | 8.35E-06 | 0.017742317 |
| Obesity | pericyte | *AC084018.1* | -1.656431304 | 7.83E-06 | 2.04E-05 | 0.049213154 |
| Obesity | pericyte | *C17orf51* | -2.037365827 | 0.013618967 | 0.018993444 | 1 |
| Obesity | pericyte | *RNF208* | -5.83004059 | 0.000365903 | 0.000655021 | 1 |
| Obesity | pericyte | *MAP10* | -1.852344454 | 0.013161072 | 0.018407933 | 1 |
| Obesity | pericyte | *ZNF580* | 3.117924256 | 0.000527307 | 0.000919607 | 1 |
| Obesity | pericyte | *NUP62* | 5.211641782 | 2.85E-07 | 1.31E-06 | 0.001794151 |
| Obesity | pericyte | *FGFR1OP* | 4.253652695 | 0.000226347 | 0.000421767 | 1 |
| Obesity | pericyte | *ZNF254* | 6.303287586 | 1.85E-06 | 5.84E-06 | 0.011623487 |
| Obesity | pericyte | *TCTEX1D2* | 1.60507795 | 0.028332309 | 0.037738253 | 1 |
| Obesity | pericyte | *LINGO4* | -14.00337201 | 0.003263429 | 0.004988374 | 1 |
| Obesity | pericyte | *ASIC3* | 3.114886125 | 9.68E-08 | 5.87E-07 | 0.000608415 |
| Obesity | pericyte | *GIMAP1* | 2.861414993 | 0.003305329 | 0.005043836 | 1 |
| Obesity | pericyte | *CCDC183* | 9.967786359 | 1.48E-06 | 4.82E-06 | 0.009304618 |
| Obesity | pericyte | *NOTCH2NL* | 2.261298196 | 3.71E-05 | 8.14E-05 | 0.233231269 |

| | | | | | | |
|---|---|---|---|---|---|---|
| Obesity | pericyte | *GSTM2* | 1.397706468 | 0.007730107 | 0.011151718 | 1 |
| Obesity | pericyte | *ARHGAP19* | 5.768976982 | 1.45E-07 | 7.73E-07 | 0.000910725 |
| Obesity | pericyte | *SIPA1* | 3.563293194 | 0.000439564 | 0.000777804 | 1 |
| Obesity | pericyte | *ERV3-1* | 0.812275905 | 0.000149239 | 0.000288431 | 0.938266049 |
| Obesity | pericyte | *RBMXL1* | 6.193039245 | 8.12E-08 | 5.12E-07 | 0.000510597 |
| Obesity | pericyte | *SRA1* | 1.81145844 | 0.009994171 | 0.014212476 | 1 |
| Obesity | pericyte | *TMEM110* | -1.362517338 | 0.007138079 | 0.010352273 | 1 |
| Obesity | pericyte | *HEXA* | 7.861030895 | 5.41E-06 | 1.48E-05 | 0.034019087 |
| Obesity | pericyte | *LEPROT* | 4.256706852 | 0.00092012 | 0.00152956 | 1 |
| Obesity | pericyte | *LBH* | -2.226045434 | 0.001342569 | 0.002182759 | 1 |
| Obesity | pericyte | *PPP1CB* | 3.411348595 | 0.000637327 | 0.001095373 | 1 |
| Obesity | pericyte | *NCKIPSD* | 4.24714185 | 0.000146785 | 0.000283862 | 0.922836839 |
| Obesity | pericyte | *ATF6B* | 2.362457743 | 2.74E-05 | 6.22E-05 | 0.172144438 |
| Obesity | pericyte | *SLC35F6* | 11.13757279 | 3.49E-07 | 1.55E-06 | 0.002192027 |
| Obesity | pericyte | *EMP2* | 2.760348722 | 0.004684156 | 0.0069901 | 1 |
| Obesity | pericyte | *PPM1N* | 4.49273467 | 1.13E-06 | 3.86E-06 | 0.00711249 |
| Obesity | pericyte | *LTB4R2* | 1.764447351 | 0.026751797 | 0.035830538 | 1 |
| Obesity | pericyte | *GALT* | 6.113701819 | 1.04E-10 | 1.01E-08 | 6.56E-07 |
| Obesity | pericyte | *ITGA1* | -1.166856047 | 0.028905639 | 0.038436918 | 1 |
| Obesity | pericyte | *TAX1BP3* | 8.058326999 | 0.000120289 | 0.000236331 | 0.75625766 |
| Obesity | pericyte | *AP1G2* | 6.628566235 | 6.63E-08 | 4.46E-07 | 0.000417085 |
| Obesity | pericyte | *ZNF90* | 3.566448107 | 0.012945495 | 0.018142739 | 1 |
| Obesity | pericyte | *TM6SF2* | 33.03707057 | 5.93E-07 | 2.36E-06 | 0.003730573 |
| Obesity | pericyte | *REPIN1* | 2.421093676 | 0.003476752 | 0.005282398 | 1 |
| Obesity | pericyte | *SMIM7* | 4.97263127 | 3.66E-08 | 2.99E-07 | 0.000230139 |
| Obesity | pericyte | *TSPAN4* | 7.660264404 | 1.49E-07 | 7.90E-07 | 0.000938501 |
| Obesity | pericyte | *AC079776.2* | 5.266833733 | 2.67E-06 | 7.96E-06 | 0.016795531 |
| Obesity | pericyte | *WEE2* | 2.844463768 | 6.71E-06 | 1.79E-05 | 0.04218936 |
| Obesity | pericyte | *PAXIP1-AS2* | 7.20961716 | 1.97E-09 | 7.78E-08 | 1.24E-05 |
| Obesity | pericyte | *LYRM4* | 4.060340632 | 0.003295484 | 0.005030033 | 1 |
| Obesity | pericyte | *PRCD* | -2.141523363 | 0.001348772 | 0.002191144 | 1 |
| Obesity | pericyte | *C17orf67* | -1.655628122 | 0.009809109 | 0.013965097 | 1 |
| Obesity | pericyte | *COLCA2* | 1.957847766 | 0.000216006 | 0.000403936 | 1 |
| Obesity | pericyte | *NEURL1B* | -0.785621262 | 4.97E-05 | 0.000106289 | 0.312609798 |
| Obesity | pericyte | *VSTM5* | -1.063374731 | 0.037541631 | 0.049110328 | 1 |
| Obesity | pericyte | *PPME1* | 2.146456007 | 0.000974768 | 0.001615278 | 1 |
| Obesity | pericyte | *NUTM2D* | 2.301908646 | 0.031791586 | 0.042025589 | 1 |
| Obesity | pericyte | *CPEB1* | 4.543485452 | 2.50E-05 | 5.74E-05 | 0.157248876 |
| Obesity | pericyte | *C10orf105* | -2.474519179 | 0.001610416 | 0.002582827 | 1 |
| Obesity | pericyte | *ARHGEF33* | -1.404983247 | 0.000992261 | 0.001640042 | 1 |
| Obesity | pericyte | *AC005082.1* | 8.86939551 | 1.82E-06 | 5.77E-06 | 0.01145221 |
| Obesity | pericyte | *TBC1D26* | -13.03416222 | 0.000162357 | 0.000310727 | 1 |
| Obesity | pericyte | *PHB2* | 4.60836753 | 0.000327965 | 0.000591823 | 1 |
| Obesity | pericyte | *GRID2IP* | -1.797601479 | 0.008002058 | 0.011512343 | 1 |
| Obesity | pericyte | *UBXN2B* | 2.438893809 | 0.001590609 | 0.002556931 | 1 |
| Obesity | pericyte | *UBE2QL1* | 3.293664568 | 1.12E-06 | 3.83E-06 | 0.007034194 |
| Obesity | pericyte | *GOLGA8B* | 7.662601018 | 2.02E-08 | 2.14E-07 | 0.00012696 |
| Obesity | pericyte | *KCNU1* | 2.430121389 | 2.74E-06 | 8.14E-06 | 0.017232852 |
| Obesity | pericyte | *MYL5* | 11.36245768 | 1.83E-08 | 2.04E-07 | 0.000115077 |
| Obesity | pericyte | *LINC01139* | 2.343599689 | 0.000470191 | 0.000825954 | 1 |

| | | | | | | |
|---|---|---|---|---|---|---|
| Obesity | pericyte | *TSTD1* | 3.609938739 | 1.11E-07 | 6.46E-07 | 0.000698491 |
| Obesity | pericyte | *LINC01138* | 7.841063249 | 6.92E-08 | 4.57E-07 | 0.000434981 |
| Obesity | pericyte | *ATAD3C* | 5.521173633 | 3.78E-08 | 3.06E-07 | 0.000237815 |
| Obesity | pericyte | *CCDC7* | 4.441596084 | 6.06E-05 | 0.000126366 | 0.380741384 |
| Obesity | pericyte | *PAM16* | 2.247701315 | 0.001815316 | 0.002887877 | 1 |
| Obesity | pericyte | *AC016773.1* | 0.957797392 | 0.003248895 | 0.004967365 | 1 |
| Obesity | pericyte | *RNASEK* | 4.818345435 | 5.55E-08 | 3.96E-07 | 0.000349091 |
| Obesity | pericyte | *NBPF1* | 3.495264988 | 0.00010935 | 0.000216599 | 0.687483984 |
| Obesity | pericyte | *RNU6ATAC35I* | 11.40234936 | 1.06E-06 | 3.68E-06 | 0.006671171 |
| Obesity | pericyte | *PPP3R1* | 2.994205421 | 1.45E-05 | 3.54E-05 | 0.091379218 |
| Obesity | pericyte | *FANCG* | 1.267271498 | 0.003451338 | 0.005251346 | 1 |
| Obesity | pericyte | *C19orf73* | -6.882307517 | 5.48E-05 | 0.000115654 | 0.344649339 |
| Obesity | pericyte | *OR2A25* | 51.15341316 | 0.000641974 | 0.001101853 | 1 |
| Obesity | pericyte | *TIGD1* | 1.312955108 | 0.001135129 | 0.001861387 | 1 |
| Obesity | pericyte | *C1orf229* | 2.536442229 | 0.00293398 | 0.00451774 | 1 |
| Obesity | pericyte | *APOL6* | 4.315046623 | 1.42E-08 | 1.83E-07 | 8.92E-05 |
| Obesity | pericyte | *UBA52* | 4.563597845 | 5.06E-06 | 1.39E-05 | 0.031799399 |
| Obesity | pericyte | *FAM185A* | 7.281485756 | 4.70E-09 | 1.27E-07 | 2.95E-05 |
| Obesity | pericyte | *AC062017.1* | 2.098240677 | 0.021040261 | 0.028545559 | 1 |
| Obesity | pericyte | *ADRA2B* | -3.672035703 | 4.32E-05 | 9.34E-05 | 0.271646066 |
| Obesity | pericyte | *EXOSC6* | -4.783160904 | 1.52E-05 | 3.69E-05 | 0.095838371 |
| Obesity | pericyte | *TINCR* | -2.8033432 | 0.025345407 | 0.034070254 | 1 |
| Obesity | pericyte | *LINC00205* | -1.864007663 | 0.031356726 | 0.041485634 | 1 |
| Obesity | pericyte | *AC147651.3* | -3.742602677 | 0.03131003 | 0.041441296 | 1 |
| Obesity | pericyte | *HLA-DPB1* | 1.935382504 | 0.000469482 | 0.000825171 | 1 |
| Obesity | pericyte | *OSER1-AS1* | 2.981258598 | 2.98E-08 | 2.63E-07 | 0.000187261 |
| Obesity | pericyte | *LINC01133* | 5.094447901 | 2.66E-08 | 2.42E-07 | 0.000167418 |
| Obesity | pericyte | *RASAL2-AS1* | 6.003638036 | 0.000505606 | 0.000884213 | 1 |
| Obesity | pericyte | *AC069213.1* | -2.767970788 | 2.09E-06 | 6.46E-06 | 0.013158837 |
| Obesity | pericyte | *LINC00240* | 4.190614723 | 1.69E-13 | 4.43E-11 | 1.06E-09 |
| Obesity | pericyte | *LINC00863* | 2.996220687 | 5.41E-08 | 3.90E-07 | 0.000340431 |
| Obesity | pericyte | *LINC00184* | 3.591083395 | 0.000992165 | 0.001640042 | 1 |
| Obesity | pericyte | *TMEM233* | 2.259093743 | 0.000117414 | 0.00023097 | 0.73817978 |
| Obesity | pericyte | *LINC00237* | -2.350464635 | 2.17E-05 | 5.05E-05 | 0.136610704 |
| Obesity | pericyte | *PLEKHM1* | 7.009812592 | 1.44E-10 | 1.27E-08 | 9.03E-07 |
| Obesity | pericyte | *AP000350.6* | 1.509719563 | 0.00065185 | 0.001117585 | 1 |
| Obesity | pericyte | *SFTA1P* | -0.490845836 | 1.19E-05 | 2.94E-05 | 0.074552472 |
| Obesity | pericyte | *SLC26A6* | -2.430015887 | 0.001596425 | 0.002563658 | 1 |
| Obesity | pericyte | *MIAT* | 2.006989955 | 2.44E-06 | 7.36E-06 | 0.015347539 |
| Obesity | pericyte | *LINC00626* | 39.94751114 | 5.70E-06 | 1.54E-05 | 0.035809429 |
| Obesity | pericyte | *ERCC6* | 6.466328482 | 4.53E-05 | 9.76E-05 | 0.284499016 |
| Obesity | pericyte | *LINC00115* | 1.67011276 | 0.000463643 | 0.000816289 | 1 |
| Obesity | pericyte | *NOL7* | 8.020256069 | 1.62E-06 | 5.21E-06 | 0.010172249 |
| Obesity | pericyte | *ZNF503-AS1* | 11.79168516 | 1.51E-08 | 1.90E-07 | 9.52E-05 |
| Obesity | pericyte | *LINC00937* | 2.195421441 | 5.51E-06 | 1.50E-05 | 0.034618591 |
| Obesity | pericyte | *TMEM185B* | 1.856148562 | 0.004039073 | 0.006092527 | 1 |
| Obesity | pericyte | *HSBP1L1* | -2.252365929 | 0.01003339 | 0.014265021 | 1 |
| Obesity | pericyte | *C14orf132* | 3.686621957 | 2.20E-07 | 1.06E-06 | 0.001381137 |
| Obesity | pericyte | *ZNF717* | 0.422352175 | 0.009282321 | 0.013251125 | 1 |
| Obesity | pericyte | *AC009299.3* | 5.612789286 | 2.27E-06 | 6.91E-06 | 0.014267515 |

| | | | | | | |
|---|---|---|---|---|---|---|
| Obesity | pericyte | *LINC01068* | 2.459917538 | 0.010307716 | 0.014631882 | 1 |
| Obesity | pericyte | *AC108463.2* | -1.898038022 | 4.53E-06 | 1.27E-05 | 0.028457671 |
| Obesity | pericyte | *HCG20* | -8.937451074 | 1.56E-07 | 8.16E-07 | 0.000979398 |
| Obesity | pericyte | *LINC01013* | 3.629576485 | 9.64E-07 | 3.42E-06 | 0.006060961 |
| Obesity | pericyte | *PROB1* | -3.783701667 | 0.000267245 | 0.000489987 | 1 |
| Obesity | pericyte | *ARL17B* | 2.927368707 | 9.70E-05 | 0.000194607 | 0.610092945 |
| Obesity | pericyte | *LINC00954* | 3.725311442 | 5.92E-07 | 2.36E-06 | 0.003720014 |
| Obesity | pericyte | *U91324.1* | 33.19422583 | 1.90E-07 | 9.50E-07 | 0.001195593 |
| Obesity | pericyte | *ZNF688* | 6.287341558 | 1.34E-05 | 3.29E-05 | 0.084311863 |
| Obesity | pericyte | *CISD3* | -3.047952202 | 0.026930985 | 0.036055175 | 1 |
| Obesity | pericyte | *PSMG3-AS1* | 2.20031614 | 3.19E-07 | 1.44E-06 | 0.002006091 |
| Obesity | pericyte | *PPP5D1* | -1.576893537 | 0.005499953 | 0.008126488 | 1 |
| Obesity | pericyte | *KLHL7-AS1* | 33.04646707 | 1.91E-08 | 2.08E-07 | 0.000120029 |
| Obesity | pericyte | *AC092171.4* | 3.758913404 | 5.99E-05 | 0.000125133 | 0.376900467 |
| Obesity | pericyte | *HLA-DPA1* | 3.843943996 | 1.09E-05 | 2.73E-05 | 0.068459776 |
| Obesity | pericyte | *RPS18* | 1.673852488 | 0.027877285 | 0.037203246 | 1 |
| Obesity | pericyte | *DIRC3* | 2.563292621 | 5.21E-07 | 2.13E-06 | 0.003272897 |
| Obesity | pericyte | *TAPBP* | 4.392141167 | 1.44E-06 | 4.71E-06 | 0.009029639 |
| Obesity | pericyte | *AC107021.1* | 1.426335064 | 5.26E-05 | 0.000111414 | 0.330452619 |
| Obesity | pericyte | *LINC00865* | -1.791684642 | 0.021107314 | 0.028624176 | 1 |
| Obesity | pericyte | *LINC00665* | 1.511282773 | 0.006265132 | 0.009170869 | 1 |
| Obesity | pericyte | *LYRM9* | 4.804027514 | 4.81E-06 | 1.33E-05 | 0.030228204 |
| Obesity | pericyte | *SNHG15* | 6.075467279 | 1.08E-09 | 5.30E-08 | 6.78E-06 |
| Obesity | pericyte | *LINC00702* | -1.537919836 | 0.000468215 | 0.000823865 | 1 |
| Obesity | pericyte | *LINC00472* | 2.690880637 | 5.65E-05 | 0.000118621 | 0.355270416 |
| Obesity | pericyte | *RPS28* | 3.180578901 | 2.18E-07 | 1.06E-06 | 0.001370022 |
| Obesity | pericyte | *TRIM26* | 3.880765595 | 1.31E-06 | 4.36E-06 | 0.008252244 |
| Obesity | pericyte | *ZNF736* | 2.652275766 | 3.67E-05 | 8.07E-05 | 0.23076046 |
| Obesity | pericyte | *HLA-B* | 2.349988231 | 6.17E-06 | 1.66E-05 | 0.038762467 |
| Obesity | pericyte | *C19orf81* | 5.026052389 | 0.000430581 | 0.000763613 | 1 |
| Obesity | pericyte | *ZSCAN31* | 2.306999859 | 9.73E-08 | 5.89E-07 | 0.000611611 |
| Obesity | pericyte | *LINC00957* | 4.978297157 | 1.70E-06 | 5.43E-06 | 0.010674044 |
| Obesity | pericyte | *B3GALT4* | 4.494794077 | 1.01E-05 | 2.54E-05 | 0.063234637 |
| Obesity | pericyte | *LINC00941* | 8.630450956 | 8.15E-07 | 3.00E-06 | 0.005121491 |
| Obesity | pericyte | *ZBTB22* | 2.346847023 | 1.88E-06 | 5.90E-06 | 0.011796858 |
| Obesity | pericyte | *ZNF737* | 7.337495752 | 3.77E-09 | 1.13E-07 | 2.37E-05 |
| Obesity | pericyte | *LINC00959* | 3.983179049 | 6.70E-06 | 1.78E-05 | 0.042117489 |
| Obesity | pericyte | *AC073257.2* | -5.183319406 | 4.71E-05 | 0.000101424 | 0.296361006 |
| Obesity | pericyte | *TXNDC5* | -3.938519435 | 9.10E-06 | 2.33E-05 | 0.057227672 |
| Obesity | pericyte | *NME1* | 1.279931036 | 0.00010183 | 0.000203823 | 0.64020834 |
| Obesity | pericyte | *CDRT4* | 3.866339264 | 1.16E-05 | 2.89E-05 | 0.07312874 |
| Obesity | pericyte | *LY6G5B* | -0.753610813 | 8.56E-05 | 0.000173372 | 0.538146439 |
| Obesity | pericyte | *PSMB9* | 3.03081536 | 8.12E-10 | 4.51E-08 | 5.10E-06 |
| Obesity | pericyte | *COX19* | 8.634350473 | 1.18E-07 | 6.74E-07 | 0.000744234 |
| Obesity | pericyte | *AQP1* | 5.053319156 | 7.70E-07 | 2.88E-06 | 0.004843227 |
| Obesity | pericyte | *C1QTNF9* | -3.118873955 | 8.25E-05 | 0.000167625 | 0.518798814 |
| Obesity | pericyte | *ARHGEF25* | -1.600165082 | 0.000269122 | 0.000493142 | 1 |
| Obesity | pericyte | *LINC00886* | 2.745994292 | 0.027821672 | 0.037144798 | 1 |
| Obesity | pericyte | *NSUN6* | 6.761790568 | 1.18E-07 | 6.74E-07 | 0.000742197 |
| Obesity | pericyte | *CD302* | 5.655996153 | 9.75E-06 | 2.48E-05 | 0.061291068 |

| | | | | | | |
|---|---|---|---|---|---|---|
| Obesity | pericyte | *ATP5J2* | 6.720605008 | 1.14E-06 | 3.87E-06 | 0.007147179 |
| Obesity | pericyte | *ARHGAP8* | 3.931148544 | 2.00E-06 | 6.24E-06 | 0.012570632 |
| Obesity | pericyte | *ATP5O* | 4.023586071 | 1.82E-07 | 9.18E-07 | 0.001143646 |
| Obesity | pericyte | *PLEKHO2* | 5.378681213 | 1.43E-06 | 4.69E-06 | 0.008995505 |
| Obesity | pericyte | *PWP2* | 6.895187258 | 0.000456031 | 0.000804001 | 1 |
| Obesity | pericyte | *PI4KA* | 6.623018692 | 1.10E-08 | 1.71E-07 | 6.92E-05 |
| Obesity | pericyte | *AKAP2* | 7.524382436 | 8.91E-07 | 3.22E-06 | 0.005600524 |
| Obesity | pericyte | *LINC00969* | 1.545498497 | 0.009085809 | 0.01299124 | 1 |
| Obesity | pericyte | *TCP10L* | 10.94680345 | 0.000255956 | 0.000471628 | 1 |
| Obesity | pericyte | *C22orf39* | 2.566282761 | 4.56E-07 | 1.92E-06 | 0.002865238 |
| Obesity | pericyte | *EIF6* | 2.353052972 | 0.011197915 | 0.01582407 | 1 |
| Obesity | pericyte | *MRPL20* | 5.029831167 | 4.73E-09 | 1.27E-07 | 2.98E-05 |
| Obesity | pericyte | *LINC00960* | -0.984134803 | 0.000145584 | 0.0002818 | 0.915285885 |
| Obesity | pericyte | *DECR2* | -3.334866498 | 0.003363365 | 0.00512121 | 1 |
| Obesity | pericyte | *CCDC169* | 2.016397866 | 0.000569992 | 0.000989382 | 1 |
| Obesity | pericyte | *AP5Z1* | 9.264212755 | 2.02E-09 | 7.85E-08 | 1.27E-05 |
| Obesity | pericyte | *PSG4* | -2.4787083 | 0.001913346 | 0.00303079 | 1 |
| Obesity | pericyte | *STON1* | 1.467858883 | 0.033380282 | 0.044033116 | 1 |
| Obesity | pericyte | *VSIG8* | 2.931448313 | 2.13E-05 | 4.98E-05 | 0.134118172 |
| Obesity | pericyte | *C7orf73* | 7.312582164 | 3.26E-08 | 2.80E-07 | 0.000204924 |
| Obesity | pericyte | *TICAM2* | -2.196207086 | 0.000127731 | 0.000250014 | 0.803046517 |
| Obesity | pericyte | *IL10RB* | 3.474318226 | 0.001135615 | 0.001861697 | 1 |
| Obesity | pericyte | *TTC4* | 0.761482156 | 0.002117843 | 0.003329754 | 1 |
| Obesity | pericyte | *RN7SL832P* | 2.960883012 | 3.24E-06 | 9.42E-06 | 0.020349184 |
| Obesity | pericyte | *LINC01011* | 3.868335695 | 4.05E-06 | 1.15E-05 | 0.025436515 |
| Obesity | pericyte | *TMEM199* | 4.865981181 | 8.86E-08 | 5.49E-07 | 0.000557029 |
| Obesity | pericyte | *P2RY11* | 2.286466044 | 0.020851764 | 0.028302038 | 1 |
| Obesity | pericyte | *LILRA6* | 1.170861889 | 0.001438783 | 0.002331348 | 1 |
| Obesity | pericyte | *APOBEC3C* | 13.86214646 | 4.43E-05 | 9.57E-05 | 0.278553506 |
| Obesity | pericyte | *CCDC13* | 1.007567686 | 0.012878576 | 0.018057004 | 1 |
| Obesity | pericyte | *ASPRV1* | -3.458494643 | 6.58E-05 | 0.000136195 | 0.413487386 |
| Obesity | pericyte | *N4BP2L2* | 3.424255395 | 0.000427899 | 0.0007593 | 1 |
| Obesity | pericyte | *LINC00847* | 2.412925507 | 5.99E-07 | 2.37E-06 | 0.00376765 |
| Obesity | pericyte | *NEAT1* | 3.351118665 | 2.69E-07 | 1.25E-06 | 0.001692015 |
| Obesity | pericyte | *ZNF585B* | 2.258124964 | 2.26E-06 | 6.89E-06 | 0.014202612 |
| Obesity | pericyte | *RAB30-AS1* | 2.957738489 | 0.002504578 | 0.003899524 | 1 |
| Obesity | pericyte | *LINC00920* | -1.918436479 | 1.33E-05 | 3.26E-05 | 0.0834158 |
| Obesity | pericyte | *UBAP1L* | 3.614174332 | 6.89E-05 | 0.000142056 | 0.433127342 |
| Obesity | pericyte | *PGAM5* | -3.73280619 | 0.006957275 | 0.010106375 | 1 |
| Obesity | pericyte | *DNAJC3-AS1* | 5.486789631 | 0.000991704 | 0.001640042 | 1 |
| Obesity | pericyte | *BCKDHA* | -3.673378691 | 0.00099176 | 0.001640042 | 1 |
| Obesity | pericyte | *CDK11B* | 6.365681346 | 8.42E-09 | 1.70E-07 | 5.30E-05 |
| Obesity | pericyte | *ABHD14A* | 2.794472607 | 8.04E-08 | 5.08E-07 | 0.00050549 |
| Obesity | pericyte | *SRP14-AS1* | 0.958031233 | 1.22E-05 | 3.02E-05 | 0.076591834 |
| Obesity | pericyte | *C5orf17* | -1.683287792 | 0.001001181 | 0.001653816 | 1 |
| Obesity | pericyte | *FMN1* | 1.446237279 | 0.000571128 | 0.000990807 | 1 |
| Obesity | pericyte | *NAIP* | 2.332133107 | 2.85E-06 | 8.41E-06 | 0.017887683 |
| Obesity | pericyte | *ZNF286B* | 3.318437848 | 7.19E-07 | 2.72E-06 | 0.004517612 |
| Obesity | pericyte | *PDCD6* | 5.203478542 | 9.13E-09 | 1.70E-07 | 5.74E-05 |
| Obesity | pericyte | *LINC00603* | 2.571070061 | 0.02111188 | 0.028624195 | 1 |

| | | | | | | |
|---|---|---|---|---|---|---|
| Obesity | pericyte | *YJEFN3* | -2.530356071 | 0.000497594 | 0.000871657 | 1 |
| Obesity | pericyte | *DNAH10OS* | 20.54772939 | 2.55E-11 | 3.08E-09 | 1.60E-07 |
| Obesity | pericyte | *PCDHA10* | -2.08645585 | 0.00035236 | 0.000632577 | 1 |
| Obesity | pericyte | *ZNF718* | 5.080997637 | 1.28E-08 | 1.73E-07 | 8.02E-05 |
| Obesity | pericyte | *LINC00989* | -0.265622386 | 1.69E-05 | 4.04E-05 | 0.106415248 |
| Obesity | pericyte | *LINC01021* | 4.773214084 | 1.06E-06 | 3.68E-06 | 0.006672079 |
| Obesity | pericyte | *GLI4* | 1.997391481 | 4.77E-05 | 0.000102422 | 0.29978816 |
| Obesity | pericyte | *ZNF345* | 4.686397325 | 0.000407019 | 0.000724293 | 1 |
| Obesity | pericyte | *ZNF550* | 4.239499331 | 2.78E-07 | 1.28E-06 | 0.001750093 |
| Obesity | pericyte | *CCDC71L* | 5.658641203 | 9.62E-06 | 2.45E-05 | 0.060475863 |
| Obesity | pericyte | *TRNP1* | -1.946852942 | 2.11E-05 | 4.93E-05 | 0.132661214 |
| Obesity | pericyte | *PCDHGA5* | 2.363153065 | 0.000825322 | 0.001388121 | 1 |
| Obesity | pericyte | *ATXN7L3B* | 5.053723869 | 2.33E-08 | 2.26E-07 | 0.000146352 |
| Obesity | pericyte | *PRKDC* | 6.259608397 | 6.28E-08 | 4.30E-07 | 0.00039458 |
| Obesity | pericyte | *CLDN23* | 5.223268612 | 1.56E-08 | 1.91E-07 | 9.83E-05 |
| Obesity | pericyte | *ZNF260* | 7.212551425 | 7.45E-06 | 1.96E-05 | 0.046809118 |
| Obesity | pericyte | *DPP3* | 1.832759723 | 0.000361003 | 0.000646801 | 1 |
| Obesity | pericyte | *SNHG9* | 1.583756927 | 0.000820507 | 0.001381132 | 1 |
| Obesity | pericyte | *LINC00346* | 3.89411247 | 0.000468448 | 0.000824014 | 1 |
| Obesity | pericyte | *ZNF432* | 5.215971054 | 3.01E-06 | 8.87E-06 | 0.018947656 |
| Obesity | pericyte | *HMBS* | 3.753160075 | 2.15E-15 | 9.00E-13 | 1.35E-11 |
| Obesity | pericyte | *TAS2R31* | -1.281527667 | 0.027423801 | 0.036668107 | 1 |
| Obesity | pericyte | *AC093668.2* | -2.927470848 | 1.16E-05 | 2.89E-05 | 0.072918327 |
| Obesity | pericyte | *ZNF253* | 6.941434686 | 9.07E-06 | 2.33E-05 | 0.057021416 |
| Obesity | pericyte | *HP* | -5.208655986 | 4.63E-08 | 3.51E-07 | 0.000291315 |
| Obesity | pericyte | *KIAA1147* | 6.489809016 | 1.38E-08 | 1.79E-07 | 8.66E-05 |
| Obesity | pericyte | *ZBED6* | 5.066795953 | 1.05E-06 | 3.65E-06 | 0.006576052 |
| Obesity | pericyte | *FNTB* | -3.827078006 | 0.00441916 | 0.006627686 | 1 |
| Obesity | pericyte | *OVCH1-AS1* | 6.016617515 | 2.28E-08 | 2.24E-07 | 0.00014356 |
| Obesity | pericyte | *CUX1* | 3.867181857 | 1.87E-06 | 5.90E-06 | 0.011785148 |
| Obesity | pericyte | *TEN1* | -4.229984429 | 6.20E-05 | 0.000128972 | 0.389753678 |
| Obesity | pericyte | *CHURC1* | 1.75050271 | 0.003659218 | 0.005543495 | 1 |
| Obesity | pericyte | *ZNF578* | 1.582067678 | 0.000218443 | 0.000407764 | 1 |
| Obesity | pericyte | *LINC00640* | 0.886457374 | 0.00747985 | 0.010817993 | 1 |
| Obesity | pericyte | *CEP95* | 5.150012799 | 4.72E-05 | 0.000101526 | 0.296760655 |
| Obesity | pericyte | *LINC00639* | 1.670806408 | 0.010885056 | 0.015413141 | 1 |
| Obesity | pericyte | *LINC00648* | 3.539106005 | 1.31E-07 | 7.24E-07 | 0.000821141 |
| Obesity | pericyte | *LINC00924* | -0.615012962 | 2.59E-05 | 5.91E-05 | 0.162677248 |
| Obesity | pericyte | *ITGB3* | -1.262144395 | 0.028061991 | 0.037410038 | 1 |
| Obesity | pericyte | *UBE2Q2L* | 4.078198295 | 0.001091775 | 0.001794977 | 1 |
| Obesity | pericyte | *IQCH-AS1* | 1.545352044 | 0.013572352 | 0.018932634 | 1 |
| Obesity | pericyte | *SH3RF3-AS1* | -1.8469617 | 0.004030952 | 0.006081737 | 1 |
| Obesity | pericyte | *PWRN1* | -1.476967098 | 0.002642263 | 0.004100694 | 1 |
| Obesity | pericyte | *HOXB7* | 4.38856083 | 5.55E-10 | 3.46E-08 | 3.49E-06 |
| Obesity | pericyte | *FRRS1L* | 5.851269542 | 1.77E-07 | 9.03E-07 | 0.001115862 |
| Obesity | pericyte | *LINC00562* | -1.077316383 | 0.010890535 | 0.015417428 | 1 |
| Obesity | pericyte | *AC009120.5* | -3.178918984 | 0.000587847 | 0.001016165 | 1 |
| Obesity | pericyte | *LINC00565* | 7.230901383 | 0.018320947 | 0.025105448 | 1 |
| Obesity | pericyte | *TMEM178B* | 7.082439877 | 2.33E-08 | 2.26E-07 | 0.000146372 |
| Obesity | pericyte | *C15orf65* | 1.063297916 | 0.025913895 | 0.034778356 | 1 |

| | | | | | | |
|---|---|---|---|---|---|---|
| Obesity | pericyte | *HPR* | 4.979648548 | 7.27E-08 | 4.75E-07 | 0.000456867 |
| Obesity | pericyte | *LINC00662* | 6.565457594 | 1.48E-07 | 7.87E-07 | 0.000933458 |
| Obesity | pericyte | *DYNLL2* | 2.975686368 | 7.21E-06 | 1.90E-05 | 0.045325274 |
| Obesity | pericyte | *LINC00526* | -2.884552278 | 0.001677929 | 0.002682231 | 1 |
| Obesity | pericyte | *STRADA* | 1.771037777 | 0.000834418 | 0.00140192 | 1 |
| Obesity | pericyte | *MYO15B* | 4.769275947 | 1.27E-05 | 3.14E-05 | 0.080143689 |
| Obesity | pericyte | *ILF3-AS1* | 2.777660448 | 0.000483605 | 0.000848095 | 1 |
| Obesity | pericyte | *AC139100.2* | -8.659797716 | 0.000235507 | 0.000436765 | 1 |
| Obesity | pericyte | *LINC01140* | 5.897460899 | 1.96E-09 | 7.78E-08 | 1.23E-05 |
| Obesity | pericyte | *LINC00906* | 17.58998985 | 1.13E-06 | 3.86E-06 | 0.00710677 |
| Obesity | pericyte | *ZNF582-AS1* | -3.043648658 | 0.004601983 | 0.006882175 | 1 |
| Obesity | pericyte | *ZNF285* | -0.700913951 | 0.009862853 | 0.014032079 | 1 |
| Obesity | pericyte | *AC137932.1* | -6.020296754 | 0.000243463 | 0.000450456 | 1 |
| Obesity | pericyte | *AL132989.1* | 30.83290435 | 8.47E-08 | 5.30E-07 | 0.000532507 |
| Obesity | pericyte | *LINC00664* | 3.5461413 | 0.014290699 | 0.019877351 | 1 |
| Obesity | pericyte | *FBXO17* | 6.258917527 | 1.75E-07 | 8.93E-07 | 0.001098181 |
| Obesity | pericyte | *SNHG8* | 2.572532484 | 0.013481579 | 0.018831079 | 1 |
| Obesity | pericyte | *HCG17* | 3.683865009 | 0.000710542 | 0.001208325 | 1 |
| Obesity | pericyte | *LINC01050* | 25.24738616 | 2.00E-05 | 4.71E-05 | 0.125978359 |
| Obesity | pericyte | *MILR1* | 1.014519889 | 0.000670086 | 0.001146349 | 1 |
| Obesity | pericyte | *GTF2H5* | 4.69414022 | 1.07E-10 | 1.02E-08 | 6.73E-07 |
| Obesity | pericyte | *FGF14-AS2* | 4.720723918 | 2.04E-06 | 6.34E-06 | 0.012834595 |
| Obesity | pericyte | *CASC15* | -0.745218402 | 0.000151 | 0.000291386 | 0.949335963 |
| Obesity | pericyte | *LINC01023* | -2.600499192 | 0.036968273 | 0.048440919 | 1 |
| Obesity | pericyte | *AP000347.2* | 1.009312284 | 0.016182117 | 0.02234504 | 1 |
| Obesity | pericyte | *POLR2J4* | 4.710914663 | 7.41E-08 | 4.81E-07 | 0.0004658 |
| Obesity | pericyte | *PAXIP1-AS1* | 2.999610571 | 0.000158582 | 0.000304243 | 0.99700351 |
| Obesity | SMC | *SEMA3F* | -2.471152986 | 9.06E-23 | 9.05E-20 | 5.69E-19 |
| Obesity | SMC | *WNT16* | 2.419404431 | 0.000378311 | 0.016403029 | 1 |
| Obesity | SMC | *RBM6* | -1.794765379 | 1.01E-22 | 9.05E-20 | 6.33E-19 |
| Obesity | SMC | *COPZ2* | 1.078037033 | 0.001155179 | 0.03508508 | 1 |
| Obesity | SMC | *GGNBP2* | -2.08894102 | 4.44E-05 | 0.002937559 | 0.279068082 |
| Obesity | SMC | *HFE* | -2.040962918 | 4.29E-06 | 0.000435115 | 0.026977158 |
| Obesity | SMC | *TTC19* | 2.222412124 | 0.000758787 | 0.026951928 | 1 |
| Obesity | SMC | *PSMC4* | 1.982571482 | 0.0002747 | 0.012698828 | 1 |
| Obesity | SMC | *NR1H3* | 2.076878364 | 0.000845055 | 0.028718162 | 1 |
| Obesity | SMC | *RABEP1* | 2.415580317 | 4.52E-05 | 0.002962752 | 0.284424199 |
| Obesity | SMC | *ZCCHC8* | 3.245754382 | 0.000985254 | 0.031127091 | 1 |
| Obesity | SMC | *LSG1* | 1.120624052 | 0.001301407 | 0.038064767 | 1 |
| Obesity | SMC | *TSPAN17* | 1.490919907 | 0.001631912 | 0.044760442 | 1 |
| Obesity | SMC | *JKAMP* | 1.786765366 | 0.001301722 | 0.038064767 | 1 |
| Obesity | SMC | *SDCCAG8* | 1.698626565 | 0.000171798 | 0.008710446 | 1 |
| Obesity | SMC | *ITIH4* | 0.692646585 | 5.50E-06 | 0.000548546 | 0.034558423 |
| Obesity | SMC | *ATP11B* | 1.459240058 | 0.00065794 | 0.024075912 | 1 |
| Obesity | SMC | *LPAR2* | 2.227199655 | 0.001528268 | 0.043042863 | 1 |
| Obesity | SMC | *UHRF1BP1* | 2.647932101 | 3.88E-23 | 4.88E-20 | 2.44E-19 |
| Obesity | SMC | *TIPIN* | 0.903765516 | 6.29E-06 | 0.000602125 | 0.039521784 |
| Obesity | SMC | *COBLL1* | 1.397116686 | 0.000198084 | 0.009883763 | 1 |
| Obesity | SMC | *COL16A1* | -2.257653606 | 2.26E-05 | 0.001728975 | 0.141775914 |
| Obesity | SMC | *SCAMP1* | 2.855974774 | 0.001417561 | 0.040548744 | 1 |

| | | | | | | |
|---|---|---|---|---|---|---|
| Obesity | SMC | *OGFOD1* | -2.800773738 | 6.85E-05 | 0.004099244 | 0.430420611 |
| Obesity | SMC | *C3orf18* | 4.691792595 | 2.77E-10 | 7.93E-08 | 1.74E-06 |
| Obesity | SMC | *YPEL3* | 1.869894147 | 2.21E-09 | 5.78E-07 | 1.39E-05 |
| Obesity | SMC | *PABPC4* | -3.068691071 | 6.52E-08 | 1.17E-05 | 0.000409859 |
| Obesity | SMC | *PDE6C* | 2.50368888 | 0.000436225 | 0.01812399 | 1 |
| Obesity | SMC | *SMARCB1* | 1.761401348 | 0.000636579 | 0.023822452 | 1 |
| Obesity | SMC | *TFIP11* | 1.597101358 | 0.001927026 | 0.04908793 | 1 |
| Obesity | SMC | *ANKRD54* | -1.336346725 | 0.001418916 | 0.040548744 | 1 |
| Obesity | SMC | *FBXO7* | 2.582833078 | 0.00095808 | 0.031008218 | 1 |
| Obesity | SMC | *ACO2* | 2.53226469 | 0.000827891 | 0.028443479 | 1 |
| Obesity | SMC | *DESI1* | 1.806501561 | 6.84E-06 | 0.000622953 | 0.042983775 |
| Obesity | SMC | *DICER1* | 2.203084808 | 0.001492587 | 0.042269788 | 1 |
| Obesity | SMC | *ZFYVE21* | 3.212332567 | 0.000428367 | 0.018101596 | 1 |
| Obesity | SMC | *ABHD12* | 2.366284156 | 2.18E-05 | 0.001695736 | 0.137354632 |
| Obesity | SMC | *CORO1A* | -3.213726066 | 2.94E-07 | 4.40E-05 | 0.001849885 |
| Obesity | SMC | *ELMO3* | 0.968014116 | 0.001047928 | 0.032777719 | 1 |
| Obesity | SMC | *NAGPA* | 2.933316573 | 0.000445341 | 0.018180897 | 1 |
| Obesity | SMC | *CLUAP1* | 1.446793698 | 9.17E-05 | 0.005240744 | 0.576481859 |
| Obesity | SMC | *KAT8* | 5.730772112 | 1.12E-08 | 2.42E-06 | 7.02E-05 |
| Obesity | SMC | *ZDHHC2* | -2.051396383 | 0.001793734 | 0.046793386 | 1 |
| Obesity | SMC | *MCM4* | 2.323371855 | 0.000553436 | 0.021746568 | 1 |
| Obesity | SMC | *PPP1R37* | 2.287069359 | 0.000365803 | 0.016082562 | 1 |
| Obesity | SMC | *OGN* | 2.841807502 | 0.000124014 | 0.006839246 | 0.779674037 |
| Obesity | SMC | *RGP1* | 3.142540593 | 0.000859869 | 0.028909087 | 1 |
| Obesity | SMC | *ACBD5* | 2.758979459 | 0.000868896 | 0.02899905 | 1 |
| Obesity | SMC | *HOXB6* | 2.125327862 | 0.000136991 | 0.007361215 | 0.86126221 |
| Obesity | SMC | *CHRNE* | 0.914295315 | 0.000673076 | 0.024460264 | 1 |
| Obesity | SMC | *FOXRED1* | 1.485771714 | 0.001726497 | 0.046547973 | 1 |
| Obesity | SMC | *DDX55* | 1.740475036 | 2.18E-05 | 0.001695736 | 0.136895032 |
| Obesity | SMC | *LAMA4* | 1.029811661 | 0.001960071 | 0.049288282 | 1 |
| Obesity | SMC | *PRSS16* | 5.081750709 | 8.47E-09 | 1.97E-06 | 5.32E-05 |
| Obesity | SMC | *NME5* | 1.716848436 | 4.85E-05 | 0.003143221 | 0.304892403 |
| Obesity | SMC | *PRKAR2A* | 1.684883485 | 0.001270443 | 0.037823018 | 1 |
| Obesity | SMC | *KLHL24* | 2.043676164 | 0.000618227 | 0.023274209 | 1 |
| Obesity | SMC | *PLCH1* | 3.183435317 | 0.000539082 | 0.021315761 | 1 |
| Obesity | SMC | *ACTR1B* | 3.935972791 | 0.000605593 | 0.023164859 | 1 |
| Obesity | SMC | *TFCP2L1* | 1.977738989 | 0.001912026 | 0.049064923 | 1 |
| Obesity | SMC | *CHMP3* | -2.066880748 | 0.000651301 | 0.024075912 | 1 |
| Obesity | SMC | *MFAP2* | -1.147367877 | 0.001136106 | 0.034842427 | 1 |
| Obesity | SMC | *DARS2* | -3.181615719 | 0.000990874 | 0.031148114 | 1 |
| Obesity | SMC | *SERPINC1* | 2.357569624 | 2.45E-10 | 7.34E-08 | 1.54E-06 |
| Obesity | SMC | *CCDC92* | 3.652301199 | 1.06E-10 | 3.33E-08 | 6.66E-07 |
| Obesity | SMC | *INVS* | 2.325679804 | 0.001854274 | 0.04817281 | 1 |
| Obesity | SMC | *DCAF4* | 2.581621398 | 3.47E-05 | 0.002395358 | 0.217977548 |
| Obesity | SMC | *HOXB5* | 3.241532478 | 8.61E-06 | 0.000761996 | 0.054101703 |
| Obesity | SMC | *HOXB3* | 2.244256287 | 3.80E-06 | 0.000397863 | 0.023871761 |
| Obesity | SMC | *ADRA1A* | 0.30303031 | 0.00105883 | 0.032954759 | 1 |
| Obesity | SMC | *TEX14* | 2.497053446 | 0.001533577 | 0.043042863 | 1 |
| Obesity | SMC | *GJB6* | -20.73836083 | 0.001611701 | 0.044637714 | 1 |
| Obesity | SMC | *SNRPC* | -3.954268213 | 2.16E-19 | 1.36E-16 | 1.36E-15 |

| | | | | | | |
|---|---|---|---|---|---|---|
| Obesity | SMC | *HIST1H2BJ* | 3.053893126 | 7.92E-10 | 2.17E-07 | 4.98E-06 |
| Obesity | SMC | *EEF1E1* | 6.629885827 | 0.000234334 | 0.011165214 | 1 |
| Obesity | SMC | *GRK4* | 0.684308012 | 2.81E-05 | 0.00203066 | 0.17666738 |
| Obesity | SMC | *OPRL1* | 1.699053218 | 0.000338796 | 0.015106475 | 1 |
| Obesity | SMC | *KLC1* | 3.034112903 | 0.000533276 | 0.021226733 | 1 |
| Obesity | SMC | *XRCC3* | 5.077507922 | 6.62E-06 | 0.000612041 | 0.041618818 |
| Obesity | SMC | *YEATS4* | 1.392977039 | 0.00117456 | 0.03550222 | 1 |
| Obesity | SMC | *RIBC2* | 1.43137847 | 0.000387995 | 0.01670772 | 1 |
| Obesity | SMC | *RPAIN* | -2.245051996 | 0.000977895 | 0.031050638 | 1 |
| Obesity | SMC | *ATP8B3* | -1.550015963 | 8.57E-08 | 1.46E-05 | 0.00053849 |
| Obesity | SMC | *PGPEP1* | 2.024189988 | 0.000429002 | 0.018101596 | 1 |
| Obesity | SMC | *CYP2E1* | -3.101783724 | 0.00035963 | 0.015922475 | 1 |
| Obesity | SMC | *CLIP1* | 2.632520014 | 0.000442168 | 0.01816933 | 1 |
| Obesity | SMC | *EMILIN2* | 2.307971253 | 5.43E-07 | 6.83E-05 | 0.003414422 |
| Obesity | SMC | *IGHMBP2* | 1.560625744 | 1.64E-05 | 0.001356185 | 0.103070094 |
| Obesity | SMC | *LARS* | -0.917516571 | 0.001949353 | 0.049288282 | 1 |
| Obesity | SMC | *ACP2* | 3.026747144 | 1.26E-05 | 0.001088183 | 0.079437385 |
| Obesity | SMC | *P2RX4* | 1.851006431 | 0.000533454 | 0.021226733 | 1 |
| Obesity | SMC | *TMEM60* | -2.723065372 | 0.000263421 | 0.012359158 | 1 |
| Obesity | SMC | *POM121C* | -4.309503266 | 7.68E-09 | 1.86E-06 | 4.83E-05 |
| Obesity | SMC | *MRPS9* | 1.546510312 | 0.001600122 | 0.044513122 | 1 |
| Obesity | SMC | *NMT1* | 1.042119344 | 0.00176064 | 0.046547973 | 1 |
| Obesity | SMC | *ZSCAN9* | 2.160883781 | 4.36E-07 | 5.96E-05 | 0.002741536 |
| Obesity | SMC | *TCF19* | 2.794179049 | 4.33E-05 | 0.002895259 | 0.272154355 |
| Obesity | SMC | *NRM* | 1.808140241 | 0.001106267 | 0.034093642 | 1 |
| Obesity | SMC | *MAP2K5* | 2.480495907 | 0.001911831 | 0.049064923 | 1 |
| Obesity | SMC | *IFT172* | 3.621145312 | 0.000157345 | 0.008175417 | 0.989225449 |
| Obesity | SMC | *CARF* | 2.265948281 | 2.30E-06 | 0.000253463 | 0.014447368 |
| Obesity | SMC | *UBL7* | 2.80493829 | 0.000371858 | 0.016235216 | 1 |
| Obesity | SMC | *KLRG1* | 1.240769224 | 0.000703996 | 0.025436907 | 1 |
| Obesity | SMC | *CYP11A1* | 1.144091773 | 0.00065867 | 0.024075912 | 1 |
| Obesity | SMC | *SH3GL3* | -1.582808159 | 8.76E-05 | 0.005052567 | 0.550729818 |
| Obesity | SMC | *MYO19* | 4.590948485 | 6.57E-07 | 7.94E-05 | 0.004128987 |
| Obesity | SMC | *C18orf8* | -3.718254138 | 7.27E-07 | 8.46E-05 | 0.00456933 |
| Obesity | SMC | *NPC1* | -2.588116351 | 0.00123523 | 0.037157386 | 1 |
| Obesity | SMC | *TRIM65* | 2.354767934 | 8.24E-06 | 0.000740026 | 0.051801785 |
| Obesity | SMC | *SEMA6C* | 4.235024814 | 0.000850516 | 0.028748356 | 1 |
| Obesity | SMC | *CEP170* | 2.137138836 | 0.001430462 | 0.040693727 | 1 |
| Obesity | SMC | *NBEAL1* | 1.109015025 | 0.000592918 | 0.023010359 | 1 |
| Obesity | SMC | *ABT1* | 2.121169846 | 0.000301821 | 0.013604545 | 1 |
| Obesity | SMC | *MTG1* | 1.232593168 | 0.000165218 | 0.00851413 | 1 |
| Obesity | SMC | *HSD17B12* | -1.751649611 | 4.02E-11 | 1.58E-08 | 2.53E-07 |
| Obesity | SMC | *TTC12* | 1.090063335 | 1.59E-05 | 0.001332445 | 0.099933376 |
| Obesity | SMC | *CCDC15* | -1.25827453 | 0.00176427 | 0.046547973 | 1 |
| Obesity | SMC | *TBX6* | 1.78530668 | 5.22E-08 | 9.64E-06 | 0.00032792 |
| Obesity | SMC | *DOC2A* | -4.871033521 | 2.65E-11 | 1.19E-08 | 1.67E-07 |
| Obesity | SMC | *PRSS53* | -2.936358409 | 1.36E-06 | 0.00015241 | 0.008534943 |
| Obesity | SMC | *DPYSL4* | 0.83048283 | 0.000103571 | 0.005762942 | 0.651150244 |
| Obesity | SMC | *POC5* | -2.025552058 | 0.000231518 | 0.011165214 | 1 |
| Obesity | SMC | *TCTEX1D1* | 0.943999824 | 0.00163953 | 0.044760442 | 1 |

| | | | | | | |
|---|---|---|---|---|---|---|
| Obesity | SMC | *JAZF1* | 2.229465975 | 0.001751274 | 0.046547973 | 1 |
| Obesity | SMC | *CEP112* | 1.713271168 | 0.001069404 | 0.033119923 | 1 |
| Obesity | SMC | *NEIL2* | 1.69015312 | 3.20E-05 | 0.002233972 | 0.201057453 |
| Obesity | SMC | *CLEC18A* | -4.615126714 | 3.23E-16 | 1.84E-13 | 2.03E-12 |
| Obesity | SMC | *HPD* | 2.161025337 | 8.26E-07 | 9.44E-05 | 0.005192576 |
| Obesity | SMC | *CD1E* | 3.512369929 | 0.000961763 | 0.031008218 | 1 |
| Obesity | SMC | *SLC5A11* | -0.991787473 | 1.39E-07 | 2.19E-05 | 0.000875217 |
| Obesity | SMC | *DFFA* | 1.93001616 | 0.001331709 | 0.038405754 | 1 |
| Obesity | SMC | *C19orf47* | 4.02332942 | 0.000611638 | 0.023164859 | 1 |
| Obesity | SMC | *PMF1* | 2.117093748 | 0.000933205 | 0.030399288 | 1 |
| Obesity | SMC | *ACP6* | -1.918611655 | 0.001769519 | 0.046547973 | 1 |
| Obesity | SMC | *CLHC1* | 0.506819524 | 0.00168256 | 0.045595935 | 1 |
| Obesity | SMC | *LYSMD1* | 3.228822643 | 0.0007514 | 0.02685041 | 1 |
| Obesity | SMC | *ANTXR2* | 1.906933652 | 0.000133039 | 0.00721047 | 0.836414542 |
| Obesity | SMC | *LMOD1* | -0.277779237 | 0.001928538 | 0.04908793 | 1 |
| Obesity | SMC | *RFT1* | -2.677310889 | 0.000302948 | 0.013604545 | 1 |
| Obesity | SMC | *APEH* | -2.19439942 | 4.13E-07 | 5.78E-05 | 0.002598795 |
| Obesity | SMC | *RNF123* | 3.206866007 | 1.43E-20 | 9.99E-18 | 8.99E-17 |
| Obesity | SMC | *MST1R* | 1.871868121 | 3.43E-07 | 5.02E-05 | 0.002158832 |
| Obesity | SMC | *POC1A* | 2.077019202 | 0.000437551 | 0.01812399 | 1 |
| Obesity | SMC | *ERAP1* | -1.406271503 | 0.001281422 | 0.037823018 | 1 |
| Obesity | SMC | *LRFN5* | 1.109757213 | 0.000895664 | 0.029481877 | 1 |
| Obesity | SMC | *SLC39A13* | 5.386412093 | 0.001791742 | 0.046793386 | 1 |
| Obesity | SMC | *TRMT61A* | -2.669099872 | 1.86E-05 | 0.001480538 | 0.116962505 |
| Obesity | SMC | *SP2* | 3.625117156 | 0.00131283 | 0.038211866 | 1 |
| Obesity | SMC | *ZNF646* | 4.104775516 | 3.41E-08 | 6.49E-06 | 0.000214186 |
| Obesity | SMC | *VKORC1* | -1.666781501 | 0.0002038 | 0.0100889 | 1 |
| Obesity | SMC | *PSCA* | 2.921393712 | 1.51E-05 | 0.001282276 | 0.094888451 |
| Obesity | SMC | *SRR* | 1.115366349 | 4.13E-06 | 0.000425864 | 0.025977715 |
| Obesity | SMC | *TMC8* | 1.864740447 | 0.000971838 | 0.031014948 | 1 |
| Obesity | SMC | *COL4A3* | 2.113417252 | 0.001745422 | 0.046547973 | 1 |
| Obesity | SMC | *C8orf46* | 0.311113579 | 0.000145266 | 0.007674703 | 0.913289619 |
| Obesity | SMC | *RAB24* | -2.851885728 | 0.000103581 | 0.005762942 | 0.651212438 |
| Obesity | SMC | *INO80E* | -4.388562829 | 2.86E-11 | 1.20E-08 | 1.80E-07 |
| Obesity | SMC | *FAM103A1* | -1.165860345 | 0.000609754 | 0.023164859 | 1 |
| Obesity | SMC | *ADORA2B* | -0.976810345 | 0.000177855 | 0.008945419 | 1 |
| Obesity | SMC | *TSNARE1* | 1.342781186 | 0.001968132 | 0.049288282 | 1 |
| Obesity | SMC | *TRMT61B* | 1.19230512 | 6.38E-06 | 0.000602125 | 0.040132721 |
| Obesity | SMC | *RGS19* | 1.16100655 | 0.001278752 | 0.037823018 | 1 |
| Obesity | SMC | *C1QTNF4* | -0.945144747 | 9.35E-14 | 4.90E-11 | 5.88E-10 |
| Obesity | SMC | *FIBP* | 6.7951979 | 5.82E-06 | 0.000571786 | 0.03659431 |
| Obesity | SMC | *CTSW* | -0.532297245 | 3.61E-07 | 5.16E-05 | 0.002270494 |
| Obesity | SMC | *EFEMP2* | 2.933828251 | 5.27E-07 | 6.76E-05 | 0.003313116 |
| Obesity | SMC | *FUT10* | -1.154435703 | 0.000778191 | 0.027332332 | 1 |
| Obesity | SMC | *SNX32* | 1.942612737 | 5.59E-05 | 0.003447171 | 0.351611463 |
| Obesity | SMC | *MST1* | -1.19676403 | 1.79E-05 | 0.00144068 | 0.112373009 |
| Obesity | SMC | *GMPPB* | 3.167399092 | 1.40E-07 | 2.19E-05 | 0.000877743 |
| Obesity | SMC | *HOXB2* | 1.891655171 | 0.000607746 | 0.023164859 | 1 |
| Obesity | SMC | *BBS1* | 3.73246136 | 0.000296986 | 0.013604545 | 1 |
| Obesity | SMC | *KLF14* | 7.877603989 | 0.000788388 | 0.027423133 | 1 |

| | | | | | | |
|---|---|---|---|---|---|---|
| Obesity | SMC | *HRAS* | 3.410810077 | 0.000574222 | 0.022423184 | 1 |
| Obesity | SMC | *CADM2* | 1.538611365 | 4.93E-05 | 0.003162531 | 0.309928067 |
| Obesity | SMC | *PSMD2* | 1.582122603 | 0.001320925 | 0.038270316 | 1 |
| Obesity | SMC | *TMEM9B* | 3.904615127 | 0.000883339 | 0.029229222 | 1 |
| Obesity | SMC | *RUFY1* | 2.327573212 | 5.36E-05 | 0.003336626 | 0.336999272 |
| Obesity | SMC | *AP006621.1* | 1.061476331 | 0.000751658 | 0.02685041 | 1 |
| Obesity | SMC | *SH2B1* | -3.428022193 | 1.21E-26 | 3.82E-23 | 7.64E-23 |
| Obesity | SMC | *STX19* | 2.735115438 | 0.000234422 | 0.011165214 | 1 |
| Obesity | SMC | *MPI* | 2.28044012 | 0.001251864 | 0.037478431 | 1 |
| Obesity | SMC | *TRIM73* | -5.134746419 | 1.05E-10 | 3.33E-08 | 6.62E-07 |
| Obesity | SMC | *TUFM* | 4.575122314 | 3.04E-22 | 2.39E-19 | 1.91E-18 |
| Obesity | SMC | *B3GNT3* | 4.728087414 | 6.92E-05 | 0.004106241 | 0.435261511 |
| Obesity | SMC | *ZNF322* | -2.63583251 | 3.44E-06 | 0.000366164 | 0.021603666 |
| Obesity | SMC | *BRICD5* | -1.237911473 | 0.001145578 | 0.034962384 | 1 |
| Obesity | SMC | *LYSMD4* | 1.144055363 | 0.000643808 | 0.023950423 | 1 |
| Obesity | SMC | *CCBE1* | 1.112867412 | 0.000969513 | 0.031014948 | 1 |
| Obesity | SMC | *PSMG1* | 0.947221722 | 2.37E-05 | 0.001792052 | 0.14874035 |
| Obesity | SMC | *RP1L1* | 3.459891108 | 1.16E-08 | 2.43E-06 | 7.28E-05 |
| Obesity | SMC | *BMP8A* | 4.963921314 | 8.33E-08 | 1.46E-05 | 0.000523807 |
| Obesity | SMC | *EIF3C* | 3.658792429 | 3.11E-26 | 6.52E-23 | 1.96E-22 |
| Obesity | SMC | *PLA2G6* | 1.823127907 | 1.38E-07 | 2.19E-05 | 0.000870649 |
| Obesity | SMC | *GP1BA* | 2.285692824 | 2.74E-05 | 0.002000282 | 0.172024276 |
| Obesity | SMC | *BTN3A2* | -0.987897293 | 8.15E-11 | 2.85E-08 | 5.12E-07 |
| Obesity | SMC | *HYAL3* | -1.215236321 | 7.10E-05 | 0.004170398 | 0.446232553 |
| Obesity | SMC | *TRABD2A* | 0.782679851 | 0.001738741 | 0.046547973 | 1 |
| Obesity | SMC | *ZSCAN23* | 0.512680048 | 0.00087177 | 0.02899905 | 1 |
| Obesity | SMC | *CENPP* | 1.438106337 | 4.47E-07 | 5.98E-05 | 0.002812715 |
| Obesity | SMC | *FBF1* | 0.835986789 | 0.000132307 | 0.00721047 | 0.831811672 |
| Obesity | SMC | *FAM47E* | 1.168543135 | 9.32E-05 | 0.005276853 | 0.585730662 |
| Obesity | SMC | *INCA1* | 5.812346467 | 0.000153881 | 0.008062071 | 0.967448565 |
| Obesity | SMC | *SULT1A1* | -1.495863564 | 6.42E-06 | 0.000602125 | 0.0403424 |
| Obesity | SMC | *C6orf106* | 2.049110084 | 3.67E-09 | 9.23E-07 | 2.31E-05 |
| Obesity | SMC | *HCAR1* | 1.591819593 | 7.23E-05 | 0.004206147 | 0.454263914 |
| Obesity | SMC | *FUT11* | 3.394886 | 0.000477258 | 0.01923411 | 1 |
| Obesity | SMC | *SLC25A29* | 2.175129138 | 0.000835132 | 0.028535194 | 1 |
| Obesity | SMC | *SULT1A2* | 1.628763333 | 1.84E-29 | 1.16E-25 | 1.16E-25 |
| Obesity | SMC | *CCL4L2* | -1.323805047 | 5.13E-05 | 0.00325731 | 0.322473659 |
| Obesity | SMC | *GTF2E2* | 2.503366747 | 0.00190457 | 0.049064923 | 1 |
| Obesity | SMC | *ZNF165* | -2.250904341 | 5.19E-05 | 0.003265389 | 0.32653886 |
| Obesity | SMC | *MRPL21* | -0.809315623 | 0.000412028 | 0.017621912 | 1 |
| Obesity | SMC | *PCDHB11* | -3.559299936 | 0.000931869 | 0.030399288 | 1 |
| Obesity | SMC | *RPS26* | -0.717705035 | 0.000166661 | 0.008518686 | 1 |
| Obesity | SMC | *ZNF780A* | -0.91880573 | 4.08E-05 | 0.002758129 | 0.256505984 |
| Obesity | SMC | *ADH4* | -0.777647642 | 5.79E-05 | 0.003536526 | 0.364262188 |
| Obesity | SMC | *ZKSCAN8* | 3.678559533 | 9.29E-06 | 0.000811068 | 0.058396863 |
| Obesity | SMC | *CENPW* | 1.628068512 | 0.00164461 | 0.044760442 | 1 |
| Obesity | SMC | *NOTCH4* | -0.267179901 | 2.83E-08 | 5.56E-06 | 0.000177954 |
| Obesity | SMC | *AGER* | 1.714942816 | 0.000255627 | 0.012083667 | 1 |
| Obesity | SMC | *SKIV2L* | -0.832482366 | 6.16E-07 | 7.60E-05 | 0.003873735 |
| Obesity | SMC | *NELFE* | 2.297937677 | 2.35E-06 | 0.000254885 | 0.014783316 |

| | | | | | | |
|---|---|---|---|---|---|---|
| Obesity | SMC | *POU5F1* | 0.698564517 | 5.26E-07 | 6.76E-05 | 0.003309649 |
| Obesity | SMC | *CCHCR1* | 1.225247023 | 1.95E-08 | 3.96E-06 | 0.000122863 |
| Obesity | SMC | *ABCF1* | 2.561798441 | 2.52E-05 | 0.001872538 | 0.158401423 |
| Obesity | SMC | *HLA-E* | 3.189257707 | 1.13E-12 | 5.45E-10 | 7.09E-09 |
| Obesity | SMC | *ZFP57* | 0.98721365 | 1.03E-08 | 2.30E-06 | 6.45E-05 |
| Obesity | SMC | *EIF3CL* | 15.52889868 | 2.46E-25 | 3.86E-22 | 1.54E-21 |
| Obesity | SMC | *C17orf107* | 1.429699763 | 0.000139983 | 0.007458234 | 0.880071577 |
| Obesity | SMC | *AC084018.1* | -0.867153647 | 0.000827924 | 0.028443479 | 1 |
| Obesity | SMC | *ASIC3* | 1.722348008 | 1.74E-05 | 0.00142137 | 0.109445458 |
| Obesity | SMC | *NOTCH2NL* | 1.366361982 | 0.000272259 | 0.012679191 | 1 |
| Obesity | SMC | *SIPA1* | 2.14294646 | 0.000461521 | 0.018719888 | 1 |
| Obesity | SMC | *TMEM110* | 3.518825004 | 0.000777851 | 0.027332332 | 1 |
| Obesity | SMC | *GALT* | 3.174840306 | 0.0007895 | 0.027423133 | 1 |
| Obesity | SMC | *COL28A1* | 4.178476808 | 0.001964432 | 0.049288282 | 1 |
| Obesity | SMC | *ATAD3C* | 2.204725884 | 0.001634817 | 0.044760442 | 1 |
| Obesity | SMC | *EXOSC6* | -3.642377758 | 1.94E-07 | 2.97E-05 | 0.001219145 |
| Obesity | SMC | *LINC00240* | 1.423191509 | 2.53E-05 | 0.001872538 | 0.159165754 |
| Obesity | SMC | *AL049840.1* | -1.508002921 | 0.000220756 | 0.010758882 | 1 |
| Obesity | SMC | *PLEKHM1* | 2.189668204 | 0.001574535 | 0.04399602 | 1 |
| Obesity | SMC | *ZSCAN31* | 1.245847459 | 2.90E-05 | 0.002074919 | 0.182592885 |
| Obesity | SMC | *PSMB9* | 1.716957716 | 6.45E-05 | 0.003897086 | 0.40529691 |
| Obesity | SMC | *NAT6* | 3.492338084 | 3.90E-05 | 0.002665523 | 0.245228124 |
| Obesity | SMC | *PCDHAC1* | -4.163599003 | 0.000438181 | 0.01812399 | 1 |
| Obesity | SMC | *MPV17L2* | 3.63658622 | 6.96E-07 | 8.25E-05 | 0.0043735 |
| Obesity | SMC | *HMBS* | 2.166749043 | 5.06E-11 | 1.87E-08 | 3.18E-07 |
| Obesity | SMC | *HP* | -2.521515404 | 0.001975608 | 0.049288282 | 1 |
| Obesity | SMC | *HOXB7* | 2.575090118 | 0.00029913 | 0.013604545 | 1 |
| Obesity | SMC | *AC009060.1* | -5.836520188 | 3.05E-05 | 0.002157459 | 0.192013855 |
| Obesity | SMC | *GTF2H5* | 1.87832548 | 0.000213256 | 0.010474512 | 1 |
| Obesity | t_cell | *CFH* | 3.998837615 | 1.49E-05 | 0.000149261 | 0.093775222 |
| Obesity | t_cell | *STPG1* | 2.636649358 | 0.001982643 | 0.004560876 | 1 |
| Obesity | t_cell | *NIPAL3* | -1.2636946 | 0.002682444 | 0.005921532 | 1 |
| Obesity | t_cell | *ENPP4* | 1.942185989 | 0.02438318 | 0.039973156 | 1 |
| Obesity | t_cell | *SEMA3F* | -2.408440896 | 1.07E-08 | 1.31E-06 | 6.70E-05 |
| Obesity | t_cell | *KRIT1* | 3.484481863 | 0.009800253 | 0.01828857 | 1 |
| Obesity | t_cell | *RAD52* | 5.347597826 | 0.000243152 | 0.000848332 | 1 |
| Obesity | t_cell | *BAD* | 8.492199156 | 2.53E-05 | 0.000184808 | 0.159340554 |
| Obesity | t_cell | *HS3ST1* | 6.831803746 | 3.99E-06 | 9.51E-05 | 0.025102609 |
| Obesity | t_cell | *WNT16* | 4.976647541 | 1.17E-07 | 8.28E-06 | 0.000735288 |
| Obesity | t_cell | *LASP1* | 3.842720701 | 0.011355323 | 0.020777333 | 1 |
| Obesity | t_cell | *TMEM176A* | 1.939742355 | 0.004300747 | 0.008847774 | 1 |
| Obesity | t_cell | *CYP26B1* | 4.39700593 | 5.66E-06 | 0.000109552 | 0.035554702 |
| Obesity | t_cell | *ICA1* | 5.277858627 | 4.59E-06 | 0.000100727 | 0.028836171 |
| Obesity | t_cell | *ALS2* | 4.523814806 | 5.13E-05 | 0.000272374 | 0.322763451 |
| Obesity | t_cell | *POLDIP2* | 6.898091642 | 8.99E-06 | 0.000137059 | 0.056513466 |
| Obesity | t_cell | *CD38* | 2.491227024 | 0.021824628 | 0.036366667 | 1 |
| Obesity | t_cell | *RBM6* | -3.30093725 | 2.44E-20 | 3.07E-17 | 1.53E-16 |
| Obesity | t_cell | *ARHGAP33* | 4.107964269 | 0.000478566 | 0.001408587 | 1 |
| Obesity | t_cell | *PDK4* | 3.777729027 | 2.25E-05 | 0.00017328 | 0.141223275 |
| Obesity | t_cell | *SLC22A16* | -2.144684207 | 0.028884906 | 0.046468629 | 1 |

| | | | | | | |
|---|---|---|---|---|---|---|
| Obesity | t_cell | *CDC27* | 3.527531719 | 0.000191029 | 0.00071065 | 1 |
| Obesity | t_cell | *CIAPIN1* | 5.749222692 | 2.86E-05 | 0.000196979 | 0.179920285 |
| Obesity | t_cell | *FAM214B* | 6.097308314 | 1.41E-06 | 5.20E-05 | 0.008837562 |
| Obesity | t_cell | *COPZ2* | 2.927783045 | 8.31E-07 | 3.58E-05 | 0.005226026 |
| Obesity | t_cell | *GCFC2* | 4.703444154 | 0.000117272 | 0.000487948 | 0.737289864 |
| Obesity | t_cell | *WDR54* | -2.402813877 | 0.001598949 | 0.003792001 | 1 |
| Obesity | t_cell | *ABCB4* | 7.97894407 | 1.64E-05 | 0.000155091 | 0.102907905 |
| Obesity | t_cell | *PDK2* | 4.168435635 | 6.58E-05 | 0.000320973 | 0.413431536 |
| Obesity | t_cell | *GGNBP2* | -3.201461055 | 0.000759986 | 0.002038409 | 1 |
| Obesity | t_cell | *TMEM98* | 2.873498951 | 5.42E-06 | 0.000108587 | 0.034081558 |
| Obesity | t_cell | *ABCC8* | 10.92834517 | 8.25E-05 | 0.000378401 | 0.518409965 |
| Obesity | t_cell | *CCL18* | 1.457282225 | 0.020031152 | 0.033781076 | 1 |
| Obesity | t_cell | *TMEM132A* | 17.63917118 | 7.42E-05 | 0.000350236 | 0.46660141 |
| Obesity | t_cell | *SPATA20* | 4.017894713 | 1.50E-05 | 0.000149261 | 0.094121114 |
| Obesity | t_cell | *TNFRSF12A* | 3.306637182 | 0.024766177 | 0.040537609 | 1 |
| Obesity | t_cell | *FARP2* | 3.194828389 | 9.57E-05 | 0.000418843 | 0.601772197 |
| Obesity | t_cell | *IFRD1* | 2.281728075 | 0.000857543 | 0.002252975 | 1 |
| Obesity | t_cell | *GTF2IRD1* | 2.591795745 | 1.21E-05 | 0.000145864 | 0.076260329 |
| Obesity | t_cell | *VPS41* | 2.493623 | 0.023455876 | 0.038664681 | 1 |
| Obesity | t_cell | *ARHGAP44* | 5.028560171 | 9.54E-06 | 0.000140299 | 0.05998642 |
| Obesity | t_cell | *CCDC124* | 3.128315508 | 0.001233547 | 0.003054593 | 1 |
| Obesity | t_cell | *CEACAM21* | -0.523292901 | 0.000181363 | 0.000683591 | 1 |
| Obesity | t_cell | *NOS2* | 17.29470222 | 1.50E-05 | 0.000149261 | 0.094061454 |
| Obesity | t_cell | *TRAPPC6A* | -2.494561034 | 0.009623456 | 0.017974649 | 1 |
| Obesity | t_cell | *CD79B* | -2.03259497 | 0.029130155 | 0.046791335 | 1 |
| Obesity | t_cell | *PAX6* | 2.605775816 | 0.000391756 | 0.001204975 | 1 |
| Obesity | t_cell | *RPUSD1* | -5.815010827 | 0.013550761 | 0.024093222 | 1 |
| Obesity | t_cell | *BAIAP3* | -4.509141641 | 0.000704729 | 0.00192135 | 1 |
| Obesity | t_cell | *PIGQ* | 3.553217001 | 8.74E-05 | 0.000392852 | 0.549599271 |
| Obesity | t_cell | *PSMB1* | 4.331346052 | 0.004446581 | 0.009100148 | 1 |
| Obesity | t_cell | *DLEC1* | 3.801343642 | 0.015300087 | 0.026846678 | 1 |
| Obesity | t_cell | *MPND* | 5.753551332 | 4.40E-05 | 0.00025064 | 0.276706833 |
| Obesity | t_cell | *MGST1* | 2.350297307 | 4.41E-05 | 0.00025117 | 0.277543045 |
| Obesity | t_cell | *NFIX* | 1.877294951 | 0.002915062 | 0.006352511 | 1 |
| Obesity | t_cell | *MMP25* | -1.14561844 | 0.001748765 | 0.00408565 | 1 |
| Obesity | t_cell | *IL32* | -0.890333317 | 7.86E-05 | 0.000366121 | 0.494263021 |
| Obesity | t_cell | *PKD1* | 4.230893844 | 8.13E-07 | 3.55E-05 | 0.005112683 |
| Obesity | t_cell | *MAPK8IP2* | 10.50637995 | 7.96E-06 | 0.000132019 | 0.050053781 |
| Obesity | t_cell | *MED24* | 4.146294657 | 0.00010927 | 0.000464491 | 0.686982287 |
| Obesity | t_cell | *RHOBTB2* | 3.633053203 | 0.004337164 | 0.008906169 | 1 |
| Obesity | t_cell | *SEC62* | 3.094743607 | 0.028839163 | 0.046406915 | 1 |
| Obesity | t_cell | *POMT2* | 2.408746407 | 0.001181313 | 0.002941351 | 1 |
| Obesity | t_cell | *MLXIPL* | 1.154795734 | 1.65E-06 | 5.77E-05 | 0.010391653 |
| Obesity | t_cell | *SPRTN* | 6.741935778 | 0.000426251 | 0.001285912 | 1 |
| Obesity | t_cell | *METTL13* | 5.098137508 | 0.001147128 | 0.002871017 | 1 |
| Obesity | t_cell | *DYRK4* | 2.36723993 | 0.016368081 | 0.028356606 | 1 |
| Obesity | t_cell | *STARD3NL* | 6.346722527 | 0.000132175 | 0.000533023 | 0.830982318 |
| Obesity | t_cell | *ZNF200* | 4.542119235 | 4.95E-05 | 0.000266759 | 0.310953101 |
| Obesity | t_cell | *CD4* | -1.622690917 | 0.001638242 | 0.003858983 | 1 |
| Obesity | t_cell | *LRRC23* | 3.861868879 | 0.000213484 | 0.000769155 | 1 |

| | | | | | | |
|---|---|---|---|---|---|---|
| Obesity | t_cell | *SCMH1* | 2.926862784 | 0.007096455 | 0.013765941 | 1 |
| Obesity | t_cell | *CLCN6* | 6.3799457 | 1.66E-06 | 5.77E-05 | 0.010451067 |
| Obesity | t_cell | *TSPAN9* | 2.991853634 | 1.83E-05 | 0.000158794 | 0.114966646 |
| Obesity | t_cell | *MKS1* | 8.799976419 | 8.83E-06 | 0.000135758 | 0.05552508 |
| Obesity | t_cell | *TTC19* | 4.898094302 | 8.22E-06 | 0.000132932 | 0.051677675 |
| Obesity | t_cell | *DPF1* | 3.226014652 | 0.013235193 | 0.023618978 | 1 |
| Obesity | t_cell | *SYT7* | 3.364074137 | 4.05E-05 | 0.000239096 | 0.254634563 |
| Obesity | t_cell | *LARS2* | 3.430787754 | 0.015569046 | 0.027242581 | 1 |
| Obesity | t_cell | *PPP5C* | 3.544060736 | 0.021383344 | 0.035707061 | 1 |
| Obesity | t_cell | *CEP68* | 2.05948038 | 0.018776096 | 0.0319906 | 1 |
| Obesity | t_cell | *ZBTB32* | 10.03484595 | 0.001297809 | 0.003182264 | 1 |
| Obesity | t_cell | *TMEM159* | 2.958420036 | 0.000153594 | 0.000598665 | 0.96564606 |
| Obesity | t_cell | *ERCC1* | 4.649611881 | 1.99E-05 | 0.000163419 | 0.124852195 |
| Obesity | t_cell | *LTF* | 3.649910079 | 2.57E-05 | 0.000186437 | 0.161344338 |
| Obesity | t_cell | *MAP4K5* | 3.590763211 | 0.004111859 | 0.008517713 | 1 |
| Obesity | t_cell | *PSMC4* | 5.741421224 | 2.28E-06 | 6.76E-05 | 0.014335835 |
| Obesity | t_cell | *CLDN11* | 3.99724637 | 5.26E-06 | 0.000107122 | 0.033100813 |
| Obesity | t_cell | *ANGEL1* | 4.662697033 | 0.016943554 | 0.029232746 | 1 |
| Obesity | t_cell | *RNF14* | 5.235807183 | 3.01E-05 | 0.000201672 | 0.189370263 |
| Obesity | t_cell | *HEBP1* | 4.624200309 | 0.000152128 | 0.000594055 | 0.956427956 |
| Obesity | t_cell | *TACC3* | -1.817267895 | 0.001724097 | 0.004037021 | 1 |
| Obesity | t_cell | *MDH1* | 3.901898898 | 1.30E-06 | 4.94E-05 | 0.00815918 |
| Obesity | t_cell | *SLC30A9* | 4.436387832 | 0.000345734 | 0.001102804 | 1 |
| Obesity | t_cell | *MTMR11* | 1.653013332 | 4.22E-05 | 0.000245741 | 0.265154033 |
| Obesity | t_cell | *COX15* | 3.47578281 | 0.003367511 | 0.0071841 | 1 |
| Obesity | t_cell | *CCDC88C* | -0.843540526 | 0.000196928 | 0.000725725 | 1 |
| Obesity | t_cell | *NPC1L1* | 4.673050721 | 0.000753338 | 0.002024898 | 1 |
| Obesity | t_cell | *NUDCD3* | 2.09248965 | 0.010417511 | 0.019280215 | 1 |
| Obesity | t_cell | *CHDH* | 9.388130113 | 0.003963417 | 0.008250994 | 1 |
| Obesity | t_cell | *IL20RA* | 3.915753388 | 4.53E-05 | 0.0002542 | 0.285044364 |
| Obesity | t_cell | *RUFY3* | 3.214792117 | 0.000361442 | 0.001137329 | 1 |
| Obesity | t_cell | *WWTR1* | 3.60116992 | 2.68E-05 | 0.000189777 | 0.168332184 |
| Obesity | t_cell | *PHLDB1* | 1.261746444 | 0.000442082 | 0.00132414 | 1 |
| Obesity | t_cell | *MNAT1* | 4.409371977 | 0.000102501 | 0.00044078 | 0.644420755 |
| Obesity | t_cell | *OSBPL5* | 5.384465024 | 0.000906816 | 0.002359749 | 1 |
| Obesity | t_cell | *CPS1* | 2.133403379 | 7.31E-06 | 0.000127691 | 0.045968851 |
| Obesity | t_cell | *NLRP2* | -1.418484335 | 0.000856192 | 0.002250368 | 1 |
| Obesity | t_cell | *ZDHHC6* | 5.080528119 | 0.000438382 | 0.001316629 | 1 |
| Obesity | t_cell | *RNH1* | 3.674240439 | 0.000187916 | 0.000701143 | 1 |
| Obesity | t_cell | *NDUFS1* | 4.515722262 | 0.000457488 | 0.001363786 | 1 |
| Obesity | t_cell | *BIRC3* | -0.985309511 | 0.00022057 | 0.000788405 | 1 |
| Obesity | t_cell | *GCLM* | 3.161537843 | 0.000892341 | 0.002327712 | 1 |
| Obesity | t_cell | *NR1H3* | 5.241456005 | 1.47E-07 | 9.91E-06 | 0.000922441 |
| Obesity | t_cell | *TYMP* | 3.967905397 | 0.000347295 | 0.001106102 | 1 |
| Obesity | t_cell | *TOMM34* | 9.527631723 | 0.000249704 | 0.000867503 | 1 |
| Obesity | t_cell | *VIM* | 3.578478896 | 0.000763493 | 0.002046071 | 1 |
| Obesity | t_cell | *CD44* | -1.334988462 | 0.007665735 | 0.014724863 | 1 |
| Obesity | t_cell | *BTN3A1* | -1.567646778 | 9.49E-06 | 0.000140299 | 0.059655197 |
| Obesity | t_cell | *MIPEP* | 5.324234548 | 0.00077348 | 0.002071068 | 1 |
| Obesity | t_cell | *INSRR* | 2.893257084 | 0.010513286 | 0.019434587 | 1 |

| | | | | | | |
|---|---|---|---|---|---|---|
| Obesity | t_cell | *B4GALT7* | 4.966655339 | 0.002462143 | 0.005493077 | 1 |
| Obesity | t_cell | *VEZT* | 3.503698398 | 0.002081743 | 0.004764441 | 1 |
| Obesity | t_cell | *SNX1* | 3.396585345 | 0.001496228 | 0.003579448 | 1 |
| Obesity | t_cell | *ARNTL2* | 5.398607253 | 1.41E-05 | 0.000148367 | 0.088810888 |
| Obesity | t_cell | *ANK1* | -3.493886228 | 0.002947569 | 0.006416679 | 1 |
| Obesity | t_cell | *RABEP1* | 3.657477005 | 0.000338527 | 0.001089212 | 1 |
| Obesity | t_cell | *GRN* | 4.712791448 | 0.000572292 | 0.001622182 | 1 |
| Obesity | t_cell | *IFT88* | 3.975598221 | 0.000310697 | 0.001018432 | 1 |
| Obesity | t_cell | *ZCCHC8* | 6.909044152 | 2.31E-06 | 6.81E-05 | 0.014495284 |
| Obesity | t_cell | *MYOC* | 1.768045883 | 0.00122921 | 0.003048844 | 1 |
| Obesity | t_cell | *STAP1* | -5.257100354 | 6.07E-07 | 2.93E-05 | 0.003814746 |
| Obesity | t_cell | *TBC1D23* | 4.461184436 | 0.000437189 | 0.001314957 | 1 |
| Obesity | t_cell | *MYOM2* | -2.334358296 | 0.002533633 | 0.005630594 | 1 |
| Obesity | t_cell | *MFAP3* | 2.009493902 | 0.012097134 | 0.021933027 | 1 |
| Obesity | t_cell | *MRI1* | 2.332355124 | 0.010926288 | 0.02010936 | 1 |
| Obesity | t_cell | *AGA* | -2.933166807 | 0.019501661 | 0.032994334 | 1 |
| Obesity | t_cell | *BOD1L1* | 2.931348276 | 0.01429166 | 0.025239232 | 1 |
| Obesity | t_cell | *SKIV2L2* | 4.21203533 | 0.000754054 | 0.002025955 | 1 |
| Obesity | t_cell | *DNAH5* | 5.508939018 | 1.72E-06 | 5.94E-05 | 0.010815392 |
| Obesity | t_cell | *C6* | 2.03388052 | 2.78E-05 | 0.000194081 | 0.174937704 |
| Obesity | t_cell | *RAI14* | 3.26141516 | 0.0001418 | 0.000566387 | 0.891493871 |
| Obesity | t_cell | *PHLPP2* | 3.831630389 | 0.002152193 | 0.004902477 | 1 |
| Obesity | t_cell | *PQLC2* | 2.866547891 | 3.85E-05 | 0.000232002 | 0.242210593 |
| Obesity | t_cell | *RAB27B* | -1.122819473 | 0.002177393 | 0.004941974 | 1 |
| Obesity | t_cell | *LSG1* | 2.53591757 | 0.000182149 | 0.00068614 | 1 |
| Obesity | t_cell | *THAP3* | 6.483580055 | 5.72E-05 | 0.000291834 | 0.359831768 |
| Obesity | t_cell | *SPATA7* | 4.543858616 | 2.85E-05 | 0.000196531 | 0.179236294 |
| Obesity | t_cell | *RETSAT* | 2.185143704 | 1.90E-05 | 0.000160838 | 0.119619959 |
| Obesity | t_cell | *AP2S1* | 4.247066178 | 8.19E-05 | 0.00037651 | 0.515065341 |
| Obesity | t_cell | *LCP2* | -1.022918416 | 0.000354858 | 0.001124112 | 1 |
| Obesity | t_cell | *CTNNA1* | 4.320982476 | 3.99E-06 | 9.51E-05 | 0.025081045 |
| Obesity | t_cell | *CNTLN* | 3.898959909 | 2.31E-05 | 0.000175493 | 0.145483772 |
| Obesity | t_cell | *EPHA3* | 5.380311343 | 0.000380893 | 0.001179556 | 1 |
| Obesity | t_cell | *DSG2* | 3.720199281 | 7.74E-06 | 0.000130812 | 0.048690225 |
| Obesity | t_cell | *YTHDC2* | 3.545055172 | 0.008711448 | 0.016466888 | 1 |
| Obesity | t_cell | *ATP6V1H* | 4.17580592 | 0.004264297 | 0.008781407 | 1 |
| Obesity | t_cell | *CP* | 7.211153446 | 0.00050554 | 0.001470767 | 1 |
| Obesity | t_cell | *ANO2* | 2.379689327 | 2.16E-05 | 0.000169311 | 0.13599281 |
| Obesity | t_cell | *GOPC* | 4.480746166 | 0.000730607 | 0.001973925 | 1 |
| Obesity | t_cell | *TSPAN17* | 2.851722421 | 0.004897246 | 0.009919132 | 1 |
| Obesity | t_cell | *CC2D2A* | 4.020200588 | 2.03E-05 | 0.000164924 | 0.127486506 |
| Obesity | t_cell | *RRM2B* | 3.775379868 | 0.010732489 | 0.019799048 | 1 |
| Obesity | t_cell | *SNX29* | 2.966043162 | 0.007657739 | 0.014713998 | 1 |
| Obesity | t_cell | *LMO3* | 1.306997881 | 0.002799442 | 0.006136713 | 1 |
| Obesity | t_cell | *ERCC8* | 3.865324799 | 0.000836913 | 0.002208003 | 1 |
| Obesity | t_cell | *ADAMTS6* | 2.670906474 | 0.000127808 | 0.000520758 | 0.803530199 |
| Obesity | t_cell | *H6PD* | 2.706827017 | 0.000462365 | 0.001372468 | 1 |
| Obesity | t_cell | *RCN1* | 2.513644768 | 0.023773343 | 0.039116202 | 1 |
| Obesity | t_cell | *RFC2* | 5.382990469 | 1.43E-05 | 0.000148367 | 0.089614884 |
| Obesity | t_cell | *JKAMP* | 4.505020726 | 8.02E-07 | 3.53E-05 | 0.005043642 |

| | | | | | | |
|---|---|---|---|---|---|---|
| Obesity | t_cell | *DKK3* | 7.31022348 | 8.47E-05 | 0.000384937 | 0.532367438 |
| Obesity | t_cell | *ARHGEF5* | 5.871226834 | 4.67E-05 | 0.000259221 | 0.293697349 |
| Obesity | t_cell | *NFE2L3* | -2.361935856 | 0.008364822 | 0.015892909 | 1 |
| Obesity | t_cell | *LIMA1* | 1.139994599 | 0.000706155 | 0.001924404 | 1 |
| Obesity | t_cell | *PTGER3* | 1.973745532 | 7.50E-06 | 0.000128894 | 0.047123971 |
| Obesity | t_cell | *HERPUD1* | 5.216610682 | 0.000183079 | 0.000688834 | 1 |
| Obesity | t_cell | *THOC3* | -0.819208741 | 0.000184932 | 0.000692706 | 1 |
| Obesity | t_cell | *RRP12* | 3.583121526 | 0.005204592 | 0.010483908 | 1 |
| Obesity | t_cell | *ALX4* | 7.59161253 | 3.18E-05 | 0.000209022 | 0.199937646 |
| Obesity | t_cell | *AKR7A2* | 5.064720176 | 2.09E-05 | 0.000167886 | 0.131701845 |
| Obesity | t_cell | *MRTO4* | 5.565151062 | 0.000127999 | 0.000520913 | 0.804726951 |
| Obesity | t_cell | *MCF2L2* | -1.664981077 | 0.001049922 | 0.002667014 | 1 |
| Obesity | t_cell | *ANAPC4* | 4.833776607 | 0.001379667 | 0.003349021 | 1 |
| Obesity | t_cell | *LY75* | -1.922369079 | 0.005028779 | 0.010172437 | 1 |
| Obesity | t_cell | *OPN3* | 2.483578488 | 5.30E-05 | 0.000277986 | 0.333209441 |
| Obesity | t_cell | *SDCCAG8* | 3.991143656 | 2.90E-07 | 1.64E-05 | 0.001822995 |
| Obesity | t_cell | *PLEKHH1* | 2.118055529 | 0.001267248 | 0.003126841 | 1 |
| Obesity | t_cell | *ATP9A* | 2.125177651 | 4.25E-05 | 0.000246282 | 0.267461834 |
| Obesity | t_cell | *FAM114A2* | 3.569172384 | 0.010570692 | 0.019529222 | 1 |
| Obesity | t_cell | *GINM1* | 4.615105329 | 0.002760004 | 0.006067184 | 1 |
| Obesity | t_cell | *MRPL43* | 2.557105674 | 0.005459303 | 0.01091687 | 1 |
| Obesity | t_cell | *ITIH4* | 2.049851177 | 1.35E-08 | 1.49E-06 | 8.51E-05 |
| Obesity | t_cell | *TRAF1* | -1.135531848 | 0.000501671 | 0.001462219 | 1 |
| Obesity | t_cell | *IL17RB* | 11.66215121 | 1.07E-05 | 0.000145864 | 0.067361143 |
| Obesity | t_cell | *TRAF3IP2* | 5.199042168 | 8.23E-06 | 0.000132932 | 0.051710642 |
| Obesity | t_cell | *DCBLD2* | 3.858441236 | 1.06E-05 | 0.000145864 | 0.06676657 |
| Obesity | t_cell | *MSH4* | 2.748324686 | 9.83E-05 | 0.000426666 | 0.618238845 |
| Obesity | t_cell | *TMCC3* | 5.005494701 | 0.00042341 | 0.001281028 | 1 |
| Obesity | t_cell | *PITHD1* | 4.893898319 | 0.005362142 | 0.01075336 | 1 |
| Obesity | t_cell | *ATP11B* | 1.784037971 | 0.018249189 | 0.031202788 | 1 |
| Obesity | t_cell | *CDK14* | 3.604845068 | 2.00E-05 | 0.000163758 | 0.125634392 |
| Obesity | t_cell | *CROCC* | 2.228171568 | 0.000732795 | 0.001978136 | 1 |
| Obesity | t_cell | *ZC3H11A* | 10.72429045 | 5.42E-06 | 0.000108587 | 0.034096243 |
| Obesity | t_cell | *RIOK2* | 6.206642091 | 5.83E-05 | 0.000295444 | 0.366350886 |
| Obesity | t_cell | *FLYWCH1* | 1.584884056 | 0.004778992 | 0.009695231 | 1 |
| Obesity | t_cell | *DNAJC25* | 3.446272491 | 2.47E-05 | 0.000181573 | 0.155426819 |
| Obesity | t_cell | *YBX3* | 4.855501629 | 2.09E-05 | 0.000167886 | 0.131376666 |
| Obesity | t_cell | *QSER1* | 5.077529928 | 9.71E-05 | 0.00042284 | 0.61045249 |
| Obesity | t_cell | *NCKAP1* | 3.32412276 | 1.88E-05 | 0.000160637 | 0.117991379 |
| Obesity | t_cell | *SFSWAP* | 3.032035863 | 0.029028549 | 0.046675828 | 1 |
| Obesity | t_cell | *SPHK2* | 4.264368592 | 0.003561265 | 0.007518358 | 1 |
| Obesity | t_cell | *ISOC2* | 8.645190065 | 1.55E-05 | 0.000151906 | 0.097675245 |
| Obesity | t_cell | *ADCK1* | 4.067830651 | 1.84E-05 | 0.000159202 | 0.115580944 |
| Obesity | t_cell | *CASP8* | -1.318599735 | 0.000517459 | 0.001495753 | 1 |
| Obesity | t_cell | *TM7SF3* | 4.809960292 | 0.00026931 | 0.000917201 | 1 |
| Obesity | t_cell | *DMRT3* | 3.797919724 | 2.72E-05 | 0.000191821 | 0.171044007 |
| Obesity | t_cell | *ST3GAL6* | 1.426109848 | 0.009948011 | 0.018514845 | 1 |
| Obesity | t_cell | *ATP2C2* | 5.469693472 | 0.000200341 | 0.000734159 | 1 |
| Obesity | t_cell | *CDON* | 1.829778718 | 0.000257156 | 0.000886857 | 1 |
| Obesity | t_cell | *TAF2* | 3.384435823 | 0.005937119 | 0.011741638 | 1 |

| | | | | | | |
|---|---|---|---|---|---|---|
| Obesity | t_cell | *TMEM161A* | 7.366726963 | 7.52E-06 | 0.000128894 | 0.047304042 |
| Obesity | t_cell | *ABCA7* | -1.622668207 | 0.00049651 | 0.001450537 | 1 |
| Obesity | t_cell | *DDX20* | 8.002104586 | 2.41E-05 | 0.00017926 | 0.151801082 |
| Obesity | t_cell | *ANKS1A* | 4.387154295 | 0.000163832 | 0.000625008 | 1 |
| Obesity | t_cell | *UHRF1BP1* | 5.330925166 | 5.64E-27 | 1.77E-23 | 3.55E-23 |
| Obesity | t_cell | *IPO5* | 5.5051284 | 8.41E-05 | 0.000383092 | 0.528667582 |
| Obesity | t_cell | *MCM10* | -1.230311486 | 0.004599104 | 0.009366558 | 1 |
| Obesity | t_cell | *ANKRD44* | -0.926701363 | 0.002219396 | 0.005026421 | 1 |
| Obesity | t_cell | *NDUFB4* | 7.274563097 | 7.29E-05 | 0.000345811 | 0.458200041 |
| Obesity | t_cell | *ZC3H15* | 3.79657429 | 0.007621405 | 0.014648663 | 1 |
| Obesity | t_cell | *MAP2K4* | 3.025093555 | 0.02911701 | 0.046782173 | 1 |
| Obesity | t_cell | *TMEM206* | 5.714385079 | 0.000735491 | 0.001984562 | 1 |
| Obesity | t_cell | *SEC61A2* | 6.545879065 | 0.000222865 | 0.000793855 | 1 |
| Obesity | t_cell | *PPP2R5A* | 4.082927302 | 0.00082144 | 0.002179069 | 1 |
| Obesity | t_cell | *CD84* | -1.719449984 | 0.000904029 | 0.002353469 | 1 |
| Obesity | t_cell | *ZNRD1* | -2.682685365 | 0.002173056 | 0.004934228 | 1 |
| Obesity | t_cell | *ATXN3* | 2.420771317 | 0.002030826 | 0.004659783 | 1 |
| Obesity | t_cell | *GOLGA5* | 5.156714089 | 5.40E-05 | 0.000281234 | 0.339730737 |
| Obesity | t_cell | *EML1* | 2.128961451 | 0.003559525 | 0.00751721 | 1 |
| Obesity | t_cell | *ZFAT* | 3.348090765 | 0.013006062 | 0.023291103 | 1 |
| Obesity | t_cell | *MTFR1* | -2.141707879 | 0.021189688 | 0.035435379 | 1 |
| Obesity | t_cell | *MYO9A* | 3.769124526 | 0.00023081 | 0.000813854 | 1 |
| Obesity | t_cell | *STOML1* | 9.123922899 | 0.008853069 | 0.016704455 | 1 |
| Obesity | t_cell | *METTL22* | 4.948388241 | 0.014819224 | 0.0260903 | 1 |
| Obesity | t_cell | *TP53BP1* | 5.201970142 | 3.58E-05 | 0.000222471 | 0.22491801 |
| Obesity | t_cell | *SYT1* | 2.648468527 | 0.000300095 | 0.00099457 | 1 |
| Obesity | t_cell | *ROGDI* | 7.875086195 | 1.03E-05 | 0.000145719 | 0.064502502 |
| Obesity | t_cell | *HDAC4* | 3.526888208 | 0.010033856 | 0.018652529 | 1 |
| Obesity | t_cell | *CYFIP1* | 4.345230466 | 1.98E-05 | 0.000163419 | 0.124716935 |
| Obesity | t_cell | *PITX1* | 8.182026161 | 2.35E-05 | 0.000176532 | 0.147934082 |
| Obesity | t_cell | *NUCKS1* | 3.162707098 | 0.020004901 | 0.033745858 | 1 |
| Obesity | t_cell | *DNAJA2* | 4.541367415 | 0.028335906 | 0.04565552 | 1 |
| Obesity | t_cell | *KCNAB2* | -1.752556573 | 0.000717849 | 0.001946148 | 1 |
| Obesity | t_cell | *GAL* | 12.78602252 | 0.00026996 | 0.000918918 | 1 |
| Obesity | t_cell | *CLEC2D* | -1.142270906 | 0.000740892 | 0.001996566 | 1 |
| Obesity | t_cell | *NEDD4* | 3.070666503 | 0.001841502 | 0.004273725 | 1 |
| Obesity | t_cell | *PFN2* | 4.651541124 | 0.000109826 | 0.000465907 | 0.690473923 |
| Obesity | t_cell | *DAPP1* | -1.617955789 | 0.023842925 | 0.039210168 | 1 |
| Obesity | t_cell | *FGF10* | 1.958541916 | 0.000203589 | 0.000741579 | 1 |
| Obesity | t_cell | *CLTCL1* | 3.014649467 | 1.36E-05 | 0.000147429 | 0.085803635 |
| Obesity | t_cell | *ZXDC* | 4.672802055 | 0.000566328 | 0.00161036 | 1 |
| Obesity | t_cell | *POLB* | 3.128648548 | 0.007585122 | 0.014587844 | 1 |
| Obesity | t_cell | *WIPI1* | 5.508053764 | 6.18E-05 | 0.000307471 | 0.388373827 |
| Obesity | t_cell | *NDST1* | 3.667099629 | 1.22E-05 | 0.000145864 | 0.076480841 |
| Obesity | t_cell | *AP3M2* | -2.39138363 | 0.01624561 | 0.028206614 | 1 |
| Obesity | t_cell | *ST6GALNAC2* | 5.461451118 | 0.000262102 | 0.000898004 | 1 |
| Obesity | t_cell | *TESK2* | 4.633382321 | 0.002471239 | 0.005511416 | 1 |
| Obesity | t_cell | *EIF2B3* | 4.130568848 | 3.03E-05 | 0.000202181 | 0.190475473 |
| Obesity | t_cell | *CAMK2A* | 18.28700578 | 2.49E-05 | 0.000182908 | 0.156752145 |
| Obesity | t_cell | *OSBPL3* | -1.303660556 | 0.002659249 | 0.005876519 | 1 |

| | | | | | | |
|---|---|---|---|---|---|---|
| Obesity | t_cell | *RAD18* | 5.663523774 | 3.95E-05 | 0.000235809 | 0.248306356 |
| Obesity | t_cell | *SNX13* | 3.119412841 | 0.02097388 | 0.035144664 | 1 |
| Obesity | t_cell | *VASH1* | 4.800794595 | 5.37E-05 | 0.000280585 | 0.337824784 |
| Obesity | t_cell | *TRIP13* | 5.024867552 | 3.38E-05 | 0.000214224 | 0.212451927 |
| Obesity | t_cell | *MBD3* | 3.747868447 | 0.008814833 | 0.016642299 | 1 |
| Obesity | t_cell | *CYBRD1* | 3.397708749 | 4.95E-05 | 0.000266759 | 0.311040614 |
| Obesity | t_cell | *EPN2* | 4.493745768 | 1.35E-05 | 0.000147409 | 0.08469517 |
| Obesity | t_cell | *LIMS2* | 3.36866343 | 1.37E-05 | 0.000147678 | 0.086391425 |
| Obesity | t_cell | *AFF4* | 3.812425047 | 0.000781401 | 0.002089373 | 1 |
| Obesity | t_cell | *TRHDE* | 1.923585762 | 1.38E-05 | 0.000148122 | 0.086947725 |
| Obesity | t_cell | *ACADVL* | 3.970938643 | 0.000112664 | 0.000474427 | 0.708319684 |
| Obesity | t_cell | *CRMP1* | 3.220821286 | 0.000291258 | 0.000974528 | 1 |
| Obesity | t_cell | *PVR* | 5.025698685 | 6.36E-07 | 3.02E-05 | 0.00399718 |
| Obesity | t_cell | *MOV10L1* | 1.485480961 | 0.001487737 | 0.003565917 | 1 |
| Obesity | t_cell | *TP63* | 2.137682395 | 3.72E-05 | 0.000227538 | 0.233827071 |
| Obesity | t_cell | *ALPK1* | 4.675823177 | 2.94E-05 | 0.000200331 | 0.184905416 |
| Obesity | t_cell | *LLGL2* | -1.178895672 | 0.00224063 | 0.005067209 | 1 |
| Obesity | t_cell | *PDE8A* | 3.456872252 | 0.000251732 | 0.00087294 | 1 |
| Obesity | t_cell | *NLE1* | 4.94512643 | 0.0003855 | 0.001189808 | 1 |
| Obesity | t_cell | *SDHA* | 3.977131907 | 0.000286778 | 0.000962101 | 1 |
| Obesity | t_cell | *GLI2* | 1.072654291 | 0.018751356 | 0.031957109 | 1 |
| Obesity | t_cell | *CLNS1A* | 2.682158373 | 0.000783508 | 0.002093462 | 1 |
| Obesity | t_cell | *ATP2A3* | -1.219796706 | 0.001401474 | 0.003390859 | 1 |
| Obesity | t_cell | *CA12* | 1.957447244 | 8.40E-05 | 0.000383091 | 0.528282799 |
| Obesity | t_cell | *NTN4* | 4.292269811 | 0.000220367 | 0.000788405 | 1 |
| Obesity | t_cell | *LMAN1* | 4.755577977 | 0.000224046 | 0.000796707 | 1 |
| Obesity | t_cell | *TIPIN* | 2.094790837 | 6.28E-08 | 5.26E-06 | 0.000394664 |
| Obesity | t_cell | *SRI* | 6.231053924 | 0.000194364 | 0.000720074 | 1 |
| Obesity | t_cell | *EIF4G3* | 2.797520865 | 0.01380069 | 0.024502948 | 1 |
| Obesity | t_cell | *TTC38* | 3.10846967 | 3.73E-05 | 0.000227746 | 0.234351083 |
| Obesity | t_cell | *GRAMD4* | 5.257960572 | 0.001221016 | 0.003034201 | 1 |
| Obesity | t_cell | *CELSR1* | 5.841392785 | 1.32E-05 | 0.000147409 | 0.082929584 |
| Obesity | t_cell | *ADD2* | 4.892096016 | 2.63E-05 | 0.000187791 | 0.165256369 |
| Obesity | t_cell | *SLC25A3* | 4.788720622 | 0.001346157 | 0.003277803 | 1 |
| Obesity | t_cell | *FSCN1* | 2.902294444 | 0.001424077 | 0.003434281 | 1 |
| Obesity | t_cell | *MOCOS* | 2.02135151 | 0.000204092 | 0.000742121 | 1 |
| Obesity | t_cell | *PLD1* | 4.40320428 | 3.31E-05 | 0.000212266 | 0.207797855 |
| Obesity | t_cell | *BCAP29* | 4.548911529 | 0.0001632 | 0.000623733 | 1 |
| Obesity | t_cell | *TUBA3D* | 5.787759267 | 5.49E-05 | 0.000284557 | 0.345167693 |
| Obesity | t_cell | *MKRN2* | 6.888330388 | 0.00033702 | 0.001084919 | 1 |
| Obesity | t_cell | *RBMS2* | 3.884753263 | 1.52E-05 | 0.000149721 | 0.095323515 |
| Obesity | t_cell | *FMO4* | 4.980979902 | 1.50E-05 | 0.000149261 | 0.094002081 |
| Obesity | t_cell | *RGS11* | 1.227044447 | 0.005677841 | 0.011285674 | 1 |
| Obesity | t_cell | *TPD52* | -1.051656273 | 0.000401268 | 0.001228605 | 1 |
| Obesity | t_cell | *GPATCH1* | 4.628408456 | 0.001724946 | 0.004037505 | 1 |
| Obesity | t_cell | *ICAM3* | -1.120944141 | 0.001155057 | 0.002883972 | 1 |
| Obesity | t_cell | *NT5C2* | 5.209950872 | 3.38E-06 | 8.72E-05 | 0.021279315 |
| Obesity | t_cell | *CAMSAP3* | -6.010085843 | 0.000237052 | 0.000832263 | 1 |
| Obesity | t_cell | *STXBP2* | -2.487889257 | 0.000341262 | 0.001093533 | 1 |
| Obesity | t_cell | *MAP2K7* | 6.110559669 | 2.42E-05 | 0.00017926 | 0.152347062 |

| | | | | | | |
|---|---|---|---|---|---|---|
| Obesity | t_cell | *NMRK2* | 6.367940327 | 0.000966003 | 0.002481384 | 1 |
| Obesity | t_cell | *CTTNBP2* | 2.792045844 | 0.003947565 | 0.008227929 | 1 |
| Obesity | t_cell | *TM9SF3* | 2.945882698 | 0.026403147 | 0.042982027 | 1 |
| Obesity | t_cell | *DNAJC10* | 5.358459843 | 0.000167588 | 0.000638174 | 1 |
| Obesity | t_cell | *DYNC1I2* | 6.051464268 | 1.97E-06 | 6.34E-05 | 0.012371454 |
| Obesity | t_cell | *LRCH4* | -2.106662908 | 0.006922134 | 0.01349859 | 1 |
| Obesity | t_cell | *SIRT6* | 6.342312054 | 1.26E-05 | 0.000146996 | 0.079277991 |
| Obesity | t_cell | *NAALAD2* | 2.372935428 | 1.91E-05 | 0.000160838 | 0.119985499 |
| Obesity | t_cell | *FGFR1* | 3.488056627 | 1.67E-05 | 0.00015608 | 0.104946206 |
| Obesity | t_cell | *FBLN1* | 2.593648568 | 0.000613837 | 0.00172055 | 1 |
| Obesity | t_cell | *PIAS2* | 3.073183921 | 0.012585883 | 0.022646666 | 1 |
| Obesity | t_cell | *MCCC1* | 3.576537578 | 3.46E-06 | 8.84E-05 | 0.021754157 |
| Obesity | t_cell | *FAP* | 1.76388913 | 0.003330442 | 0.00712921 | 1 |
| Obesity | t_cell | *ACER3* | 3.879686144 | 1.96E-06 | 6.34E-05 | 0.01229906 |
| Obesity | t_cell | *TULP3* | 4.147214992 | 4.47E-05 | 0.000252695 | 0.281249866 |
| Obesity | t_cell | *ZCWPW1* | 4.545331325 | 3.43E-06 | 8.80E-05 | 0.021549558 |
| Obesity | t_cell | *FGF20* | 13.38570184 | 4.88E-05 | 0.000265648 | 0.306746001 |
| Obesity | t_cell | *OSBPL6* | 4.277272299 | 2.21E-05 | 0.000171505 | 0.138747783 |
| Obesity | t_cell | *LXN* | 4.609035912 | 0.00030567 | 0.001008791 | 1 |
| Obesity | t_cell | *REXO1* | 6.735674576 | 0.000968242 | 0.002485641 | 1 |
| Obesity | t_cell | *SAR1A* | 4.809382419 | 3.34E-05 | 0.000213631 | 0.21029159 |
| Obesity | t_cell | *CDC14A* | -0.986877299 | 0.00020557 | 0.000745678 | 1 |
| Obesity | t_cell | *RAPGEF3* | 0.899420166 | 0.003624499 | 0.00764159 | 1 |
| Obesity | t_cell | *FDFT1* | 5.667731665 | 0.000469651 | 0.001390813 | 1 |
| Obesity | t_cell | *SCGN* | -4.7622063 | 0.011342171 | 0.020759309 | 1 |
| Obesity | t_cell | *DDX1* | 5.140847586 | 0.000185738 | 0.000695081 | 1 |
| Obesity | t_cell | *RIMS1* | 3.445586983 | 6.48E-05 | 0.000318079 | 0.407481978 |
| Obesity | t_cell | *STX7* | 2.437531963 | 0.017920759 | 0.030741558 | 1 |
| Obesity | t_cell | *RABL2B* | 5.112275888 | 0.000334841 | 0.001079561 | 1 |
| Obesity | t_cell | *DDX43* | 3.185400731 | 0.008192129 | 0.015616711 | 1 |
| Obesity | t_cell | *DCT* | 3.406219979 | 1.80E-05 | 0.000158468 | 0.112975741 |
| Obesity | t_cell | *SLC35C2* | 4.170610206 | 0.024182837 | 0.039675756 | 1 |
| Obesity | t_cell | *SCTR* | 3.4418749 | 2.29E-05 | 0.00017469 | 0.144262953 |
| Obesity | t_cell | *KCNN2* | 4.0307431 | 8.00E-05 | 0.000371034 | 0.503121872 |
| Obesity | t_cell | *MOK* | 3.696292413 | 1.10E-05 | 0.000145864 | 0.069307166 |
| Obesity | t_cell | *MAGI3* | 4.273618703 | 5.80E-05 | 0.000294793 | 0.364954283 |
| Obesity | t_cell | *COL4A4* | 3.330799198 | 5.03E-06 | 0.000105164 | 0.03165438 |
| Obesity | t_cell | *STK17B* | -1.189179506 | 0.001356528 | 0.003299223 | 1 |
| Obesity | t_cell | *ZNF506* | 3.611284934 | 0.027805381 | 0.044892766 | 1 |
| Obesity | t_cell | *KIAA0141* | 4.273786707 | 0.00871121 | 0.016466888 | 1 |
| Obesity | t_cell | *WDR70* | 2.763529426 | 0.017634064 | 0.030299361 | 1 |
| Obesity | t_cell | *PGR* | 2.188517079 | 1.71E-05 | 0.000156669 | 0.107532625 |
| Obesity | t_cell | *C5orf22* | 6.464151223 | 0.000446684 | 0.00133538 | 1 |
| Obesity | t_cell | *CCNT2* | -2.645853201 | 0.001091936 | 0.002753712 | 1 |
| Obesity | t_cell | *COBLL1* | 3.002428375 | 1.49E-08 | 1.62E-06 | 9.37E-05 |
| Obesity | t_cell | *GEMIN5* | 2.237647871 | 0.023956337 | 0.039355498 | 1 |
| Obesity | t_cell | *ERC1* | 2.967104985 | 0.0082769 | 0.015759197 | 1 |
| Obesity | t_cell | *DOPEY1* | 2.448873673 | 0.000484271 | 0.00142205 | 1 |
| Obesity | t_cell | *LYRM2* | 2.765017509 | 0.001796515 | 0.004178577 | 1 |
| Obesity | t_cell | *ULK2* | 3.767324668 | 6.72E-05 | 0.000325577 | 0.422598393 |

| | | | | | | |
|---|---|---|---|---|---|---|
| Obesity | t_cell | *PLOD1* | 4.934811092 | 4.23E-06 | 9.89E-05 | 0.026590683 |
| Obesity | t_cell | *TDRD3* | 2.992016313 | 0.019246128 | 0.032623459 | 1 |
| Obesity | t_cell | *CYLD* | -1.711220388 | 0.002868782 | 0.006261665 | 1 |
| Obesity | t_cell | *ZNF324* | 7.592476623 | 0.002303192 | 0.005188165 | 1 |
| Obesity | t_cell | *ZNF586* | 3.288392715 | 0.004124649 | 0.008535771 | 1 |
| Obesity | t_cell | *ZNF446* | 9.045239041 | 2.75E-05 | 0.000192994 | 0.172729346 |
| Obesity | t_cell | *ZNF264* | 4.724551048 | 0.002704914 | 0.005966944 | 1 |
| Obesity | t_cell | *RPS5* | -1.998066257 | 0.012045892 | 0.021869052 | 1 |
| Obesity | t_cell | *CHMP2B* | 3.524692716 | 0.005660629 | 0.011269275 | 1 |
| Obesity | t_cell | *PPIE* | 3.574765817 | 0.005757531 | 0.01143684 | 1 |
| Obesity | t_cell | *ZMPSTE24* | 5.569701926 | 4.88E-05 | 0.000265648 | 0.306823599 |
| Obesity | t_cell | *STARD7* | 6.605557363 | 5.54E-05 | 0.000285796 | 0.348380114 |
| Obesity | t_cell | *GSTP1* | 6.695637643 | 2.57E-06 | 7.34E-05 | 0.016150404 |
| Obesity | t_cell | *APOB* | 2.642643181 | 0.000330437 | 0.0010692 | 1 |
| Obesity | t_cell | *CD59* | 4.358058127 | 0.000221705 | 0.000791068 | 1 |
| Obesity | t_cell | *CD82* | -1.148910275 | 0.01334801 | 0.023793292 | 1 |
| Obesity | t_cell | *TAF9* | 8.742390089 | 0.000149646 | 0.000586915 | 0.940824634 |
| Obesity | t_cell | *SCAMP1* | 4.672624448 | 0.001241834 | 0.003071739 | 1 |
| Obesity | t_cell | *HACE1* | 4.962003444 | 4.90E-05 | 0.000265838 | 0.307903734 |
| Obesity | t_cell | *OVGP1* | -3.657073349 | 0.010277502 | 0.019054748 | 1 |
| Obesity | t_cell | *SLC25A24* | 3.720914395 | 0.002218523 | 0.005026254 | 1 |
| Obesity | t_cell | *CPNE3* | 2.172553631 | 0.008020209 | 0.015307545 | 1 |
| Obesity | t_cell | *RRN3* | 4.27051194 | 0.001960398 | 0.004517969 | 1 |
| Obesity | t_cell | *CTTN* | 2.840761564 | 1.13E-05 | 0.000145864 | 0.070738562 |
| Obesity | t_cell | *ATG16L1* | -2.192674184 | 0.022605733 | 0.037489382 | 1 |
| Obesity | t_cell | *USP40* | 3.881313467 | 1.94E-05 | 0.000161806 | 0.122163532 |
| Obesity | t_cell | *MAST2* | 5.212944733 | 1.47E-05 | 0.000149118 | 0.092520411 |
| Obesity | t_cell | *CHMP5* | 5.869869069 | 4.36E-05 | 0.000249187 | 0.273856907 |
| Obesity | t_cell | *FOLH1* | 1.131925723 | 0.002798993 | 0.006136713 | 1 |
| Obesity | t_cell | *EIF2AK1* | 3.575946868 | 0.015844601 | 0.027632457 | 1 |
| Obesity | t_cell | *SEPHS1* | 3.960179133 | 0.003111302 | 0.00671038 | 1 |
| Obesity | t_cell | *ITPKC* | 3.928190059 | 6.39E-06 | 0.000116604 | 0.040142764 |
| Obesity | t_cell | *TMED2* | 5.119483657 | 0.006973319 | 0.013583377 | 1 |
| Obesity | t_cell | *ACOX3* | 4.133377833 | 0.00044086 | 0.00132237 | 1 |
| Obesity | t_cell | *HSD17B14* | 3.923241185 | 0.014060634 | 0.02488527 | 1 |
| Obesity | t_cell | *TRIP6* | 2.510377146 | 0.015649294 | 0.027352547 | 1 |
| Obesity | t_cell | *SRRT* | 2.702734133 | 0.015616535 | 0.027310475 | 1 |
| Obesity | t_cell | *PIGS* | 1.62837058 | 0.020434555 | 0.034350815 | 1 |
| Obesity | t_cell | *PGS1* | 4.708823602 | 0.002835964 | 0.006203795 | 1 |
| Obesity | t_cell | *LPCAT2* | 3.243102646 | 0.000232349 | 0.000817903 | 1 |
| Obesity | t_cell | *GMCL1* | 4.641047273 | 0.00242163 | 0.005416148 | 1 |
| Obesity | t_cell | *KLHL42* | 3.828596951 | 0.012063749 | 0.021895148 | 1 |
| Obesity | t_cell | *PTHLH* | 2.777083295 | 2.22E-06 | 6.70E-05 | 0.013946193 |
| Obesity | t_cell | *GP6* | -1.851015356 | 0.000617354 | 0.001725026 | 1 |
| Obesity | t_cell | *ASAP3* | 2.931284578 | 3.88E-06 | 9.42E-05 | 0.024402834 |
| Obesity | t_cell | *DOCK3* | 2.402332814 | 0.000991179 | 0.002534177 | 1 |
| Obesity | t_cell | *C3orf18* | 10.42700527 | 6.62E-13 | 2.78E-10 | 4.16E-09 |
| Obesity | t_cell | *FKBP1A* | 6.615626587 | 2.53E-05 | 0.000184756 | 0.158990526 |
| Obesity | t_cell | *NSFL1C* | 6.622914935 | 7.16E-06 | 0.000126396 | 0.045021823 |
| Obesity | t_cell | *ZNF343* | 4.193196331 | 1.01E-05 | 0.000144166 | 0.063345748 |

| | | | | | | |
|---|---|---|---|---|---|---|
| Obesity | t_cell | *MAVS* | 4.734903064 | 9.28E-05 | 0.00040921 | 0.583534092 |
| Obesity | t_cell | *LZTS3* | 4.163778722 | 0.000117086 | 0.000487819 | 0.736118594 |
| Obesity | t_cell | *TESC* | 8.800395432 | 1.23E-05 | 0.000145864 | 0.077122323 |
| Obesity | t_cell | *RBBP9* | 4.814049393 | 0.004545523 | 0.009278475 | 1 |
| Obesity | t_cell | *SLC23A2* | 3.835313335 | 0.011598121 | 0.021141602 | 1 |
| Obesity | t_cell | *TMEM230* | 3.267892921 | 0.007222525 | 0.013975998 | 1 |
| Obesity | t_cell | *TASP1* | 2.899444566 | 0.014638469 | 0.025793737 | 1 |
| Obesity | t_cell | *OAS1* | 6.430495494 | 0.000205663 | 0.000745678 | 1 |
| Obesity | t_cell | *KIF16B* | 4.805735533 | 0.00020974 | 0.000758274 | 1 |
| Obesity | t_cell | *FXYD5* | -1.610893337 | 0.001813088 | 0.004212448 | 1 |
| Obesity | t_cell | *FXYD3* | 7.090018469 | 5.16E-05 | 0.000272935 | 0.324427975 |
| Obesity | t_cell | *GMIP* | -2.002364111 | 0.001067663 | 0.002702253 | 1 |
| Obesity | t_cell | *MLF2* | 5.674561558 | 9.23E-06 | 0.000138155 | 0.058025047 |
| Obesity | t_cell | *SLC9A1* | 6.511669501 | 0.000689412 | 0.001888598 | 1 |
| Obesity | t_cell | *RGS1* | -1.672193887 | 0.000371564 | 0.001158166 | 1 |
| Obesity | t_cell | *YPEL3* | 3.737668131 | 1.29E-09 | 1.93E-07 | 8.10E-06 |
| Obesity | t_cell | *STRN4* | 4.969481109 | 0.00067224 | 0.001848807 | 1 |
| Obesity | t_cell | *LYZ* | 4.098354644 | 0.000766642 | 0.002053635 | 1 |
| Obesity | t_cell | *SPG21* | 3.027592071 | 0.012069825 | 0.021899853 | 1 |
| Obesity | t_cell | *GNPTG* | 6.054278348 | 5.69E-05 | 0.00029093 | 0.357844047 |
| Obesity | t_cell | *GOLGA3* | 3.850117301 | 0.00071294 | 0.001936988 | 1 |
| Obesity | t_cell | *PABPC4* | -4.882577134 | 4.54E-06 | 0.000100727 | 0.028518862 |
| Obesity | t_cell | *PDPR* | 4.314415763 | 0.005778565 | 0.011471372 | 1 |
| Obesity | t_cell | *EXOC1* | 3.856254562 | 0.003381156 | 0.007210762 | 1 |
| Obesity | t_cell | *NLRC4* | 4.405226904 | 1.32E-05 | 0.000147409 | 0.083084456 |
| Obesity | t_cell | *LAMB1* | 3.126317482 | 8.86E-07 | 3.69E-05 | 0.005570892 |
| Obesity | t_cell | *SLC26A4* | 3.837620757 | 4.43E-05 | 0.000251727 | 0.278661575 |
| Obesity | t_cell | *DLD* | 5.559465034 | 5.42E-05 | 0.000282108 | 0.341068818 |
| Obesity | t_cell | *TXNL1* | 5.672756379 | 0.00010996 | 0.000466164 | 0.691321218 |
| Obesity | t_cell | *IL5RA* | -1.630695279 | 0.00019798 | 0.000728322 | 1 |
| Obesity | t_cell | *RAPGEF4* | 2.714925811 | 6.98E-05 | 0.000334531 | 0.438981272 |
| Obesity | t_cell | *SEL1L3* | -1.210255891 | 0.0009356 | 0.002417639 | 1 |
| Obesity | t_cell | *TF* | 3.332927788 | 8.02E-05 | 0.00037155 | 0.504193294 |
| Obesity | t_cell | *MYO15A* | 3.750066839 | 0.012177564 | 0.02205079 | 1 |
| Obesity | t_cell | *ALKBH5* | 3.230707024 | 0.004597715 | 0.009366558 | 1 |
| Obesity | t_cell | *SPAG7* | 2.879459857 | 0.000424169 | 0.001282092 | 1 |
| Obesity | t_cell | *ZFHX4* | 2.416674422 | 2.39E-05 | 0.000178067 | 0.150093525 |
| Obesity | t_cell | *RGS17* | 1.860831553 | 0.012256861 | 0.022175218 | 1 |
| Obesity | t_cell | *TMEM101* | 4.959564914 | 0.021093016 | 0.035316056 | 1 |
| Obesity | t_cell | *CMA1* | 3.372025544 | 0.006911192 | 0.013481435 | 1 |
| Obesity | t_cell | *PPP2R3C* | 3.485006982 | 0.013155526 | 0.023510173 | 1 |
| Obesity | t_cell | *OSGEP* | 4.362045075 | 0.017209761 | 0.02962699 | 1 |
| Obesity | t_cell | *SLC22A17* | 2.892239362 | 0.016269589 | 0.028232654 | 1 |
| Obesity | t_cell | *SCFD1* | 2.550068242 | 0.0149997 | 0.026371117 | 1 |
| Obesity | t_cell | *RPGRIP1* | 3.111771034 | 0.001022152 | 0.002605948 | 1 |
| Obesity | t_cell | *TOX4* | 5.476075412 | 0.000299717 | 0.000993841 | 1 |
| Obesity | t_cell | *AGO1* | 4.133265523 | 0.013543076 | 0.02408637 | 1 |
| Obesity | t_cell | *RFFL* | -2.970340804 | 0.018842207 | 0.032085849 | 1 |
| Obesity | t_cell | *DPYSL2* | 4.322106321 | 6.63E-05 | 0.00032236 | 0.41713391 |
| Obesity | t_cell | *COMT* | 5.391269325 | 0.000292424 | 0.00097706 | 1 |

| | | | | | | |
|---|---|---|---|---|---|---|
| Obesity | t_cell | *VNN3* | 7.23256401 | 2.17E-05 | 0.000169311 | 0.136295194 |
| Obesity | t_cell | *FMO2* | 2.484291765 | 3.36E-06 | 8.70E-05 | 0.021152785 |
| Obesity | t_cell | *TMEM38B* | -2.775379237 | 0.009514179 | 0.017805924 | 1 |
| Obesity | t_cell | *PDE6C* | 3.591045428 | 3.95E-07 | 2.14E-05 | 0.002485167 |
| Obesity | t_cell | *BLNK* | 2.951881222 | 1.53E-05 | 0.000150769 | 0.096341494 |
| Obesity | t_cell | *TPSD1* | 16.89272826 | 0.000352492 | 0.001120383 | 1 |
| Obesity | t_cell | *DSP* | 1.46945345 | 0.021059413 | 0.035269188 | 1 |
| Obesity | t_cell | *IFT74* | 3.353830311 | 1.41E-05 | 0.000148367 | 0.088730323 |
| Obesity | t_cell | *IL12RB1* | -1.404523116 | 0.000107363 | 0.000458556 | 0.674994312 |
| Obesity | t_cell | *CDC7* | -1.453447446 | 0.000932359 | 0.002412655 | 1 |
| Obesity | t_cell | *PCSK5* | 3.544782384 | 1.72E-05 | 0.000156669 | 0.1083828 |
| Obesity | t_cell | *NRP1* | 3.472524621 | 4.73E-05 | 0.000261365 | 0.297432855 |
| Obesity | t_cell | *PALMD* | 2.302951919 | 1.34E-05 | 0.000147409 | 0.084093846 |
| Obesity | t_cell | *TSPAN15* | 4.212952959 | 6.58E-06 | 0.000118963 | 0.04139922 |
| Obesity | t_cell | *OCEL1* | 1.314344649 | 0.02225033 | 0.036968241 | 1 |
| Obesity | t_cell | *HSD3B7* | 3.849061131 | 0.003394184 | 0.007231188 | 1 |
| Obesity | t_cell | *MARCH2* | 4.565972029 | 1.75E-05 | 0.000157583 | 0.109840041 |
| Obesity | t_cell | *CDC34* | 6.632734883 | 9.61E-06 | 0.000140555 | 0.060438675 |
| Obesity | t_cell | *MTAP* | 6.616286237 | 6.90E-05 | 0.000331465 | 0.433887536 |
| Obesity | t_cell | *CEP170B* | 1.791924626 | 0.009527176 | 0.017821291 | 1 |
| Obesity | t_cell | *POLR2E* | 5.501586395 | 0.003710541 | 0.007796848 | 1 |
| Obesity | t_cell | *MADCAM1* | 3.146410148 | 1.44E-05 | 0.000148509 | 0.090820592 |
| Obesity | t_cell | *ZDHHC8* | 4.095030179 | 0.000203965 | 0.000742086 | 1 |
| Obesity | t_cell | *LZTR1* | 5.952114047 | 0.002972407 | 0.006458173 | 1 |
| Obesity | t_cell | *CECR2* | 1.568429144 | 0.000148178 | 0.000583339 | 0.931592104 |
| Obesity | t_cell | *SMARCB1* | 4.473819594 | 6.95E-07 | 3.22E-05 | 0.004369645 |
| Obesity | t_cell | *P2RX6* | 2.010100534 | 0.000153831 | 0.000599217 | 0.967135895 |
| Obesity | t_cell | *SLC7A4* | 4.488088481 | 0.000535611 | 0.001539026 | 1 |
| Obesity | t_cell | *DDT* | 5.871441792 | 4.58E-05 | 0.000255414 | 0.287851984 |
| Obesity | t_cell | *GGT5* | 4.941630892 | 6.49E-05 | 0.000318079 | 0.407777205 |
| Obesity | t_cell | *SPECC1L* | 1.795310399 | 0.026880567 | 0.043612419 | 1 |
| Obesity | t_cell | *UPB1* | -1.625260656 | 0.000420833 | 0.001275074 | 1 |
| Obesity | t_cell | *PPM1F* | 4.241691735 | 9.19E-05 | 0.000407341 | 0.578009273 |
| Obesity | t_cell | *TOP3B* | 4.639194145 | 0.000254383 | 0.000880674 | 1 |
| Obesity | t_cell | *CRYBB3* | 3.779746259 | 0.000925965 | 0.002398657 | 1 |
| Obesity | t_cell | *CYTH4* | -2.162492086 | 0.009264588 | 0.017392196 | 1 |
| Obesity | t_cell | *CARD10* | 2.661466593 | 0.000201737 | 0.000737395 | 1 |
| Obesity | t_cell | *SLC25A1* | 4.206863096 | 8.02E-06 | 0.000132019 | 0.05043128 |
| Obesity | t_cell | *LGALS2* | 3.660510368 | 3.60E-06 | 9.08E-05 | 0.022649561 |
| Obesity | t_cell | *SH3BP1* | -1.513230772 | 0.000403606 | 0.001232979 | 1 |
| Obesity | t_cell | *HPS4* | 5.386402691 | 2.99E-05 | 0.000200768 | 0.187718314 |
| Obesity | t_cell | *PIK3IP1* | -1.571801891 | 0.006605717 | 0.012937738 | 1 |
| Obesity | t_cell | *TFIP11* | 4.336073902 | 5.13E-06 | 0.000105615 | 0.032268507 |
| Obesity | t_cell | *GCAT* | 4.467876563 | 5.09E-06 | 0.000105254 | 0.031997084 |
| Obesity | t_cell | *EIF3L* | 3.759853045 | 0.008418811 | 0.015980998 | 1 |
| Obesity | t_cell | *CCDC134* | -2.503183154 | 0.018819837 | 0.032056438 | 1 |
| Obesity | t_cell | *SLC16A8* | 4.074152555 | 0.004826073 | 0.009787588 | 1 |
| Obesity | t_cell | *CYP2D6* | -2.18411652 | 0.008404442 | 0.01595854 | 1 |
| Obesity | t_cell | *FBXO7* | 5.026388936 | 0.000379708 | 0.001176552 | 1 |
| Obesity | t_cell | *POLDIP3* | 6.662068507 | 0.000190558 | 0.000709737 | 1 |

| | | | | | | |
|---|---|---|---|---|---|---|
| Obesity | t_cell | *TIMP3* | 3.498647866 | 3.21E-05 | 0.000209022 | 0.201690042 |
| Obesity | t_cell | *CYB5R3* | 5.063368715 | 6.08E-05 | 0.000303619 | 0.38195272 |
| Obesity | t_cell | *RHBDD3* | 3.487558776 | 4.86E-06 | 0.000103821 | 0.030560834 |
| Obesity | t_cell | *RASL10A* | 3.445199853 | 0.002322132 | 0.005227082 | 1 |
| Obesity | t_cell | *AP1B1* | 4.752876028 | 0.000486215 | 0.001425761 | 1 |
| Obesity | t_cell | *HMGXB4* | 5.350022239 | 0.000423985 | 0.001282092 | 1 |
| Obesity | t_cell | *TOM1* | 1.801759737 | 0.006518636 | 0.012775145 | 1 |
| Obesity | t_cell | *BIK* | -11.20690181 | 0.002158996 | 0.004914412 | 1 |
| Obesity | t_cell | *MCAT* | 4.891940236 | 0.000415686 | 0.001263743 | 1 |
| Obesity | t_cell | *TSPO* | 5.483067107 | 1.52E-05 | 0.000149721 | 0.095349096 |
| Obesity | t_cell | *RASD2* | -6.817020949 | 2.92E-05 | 0.000199772 | 0.18368642 |
| Obesity | t_cell | *TTLL12* | 8.448622937 | 0.000213607 | 0.000769157 | 1 |
| Obesity | t_cell | *RBFOX2* | 3.467242468 | 1.61E-05 | 0.000154104 | 0.101151992 |
| Obesity | t_cell | *APOL4* | 2.543378521 | 9.32E-05 | 0.000410531 | 0.585828158 |
| Obesity | t_cell | *SAMM50* | 5.308295375 | 0.00333958 | 0.007136621 | 1 |
| Obesity | t_cell | *FOXRED2* | 1.734423734 | 0.011799008 | 0.021464226 | 1 |
| Obesity | t_cell | *IFT27* | 7.364537233 | 6.80E-05 | 0.00032817 | 0.427360127 |
| Obesity | t_cell | *KIAA0930* | 3.263901529 | 0.000257647 | 0.000887575 | 1 |
| Obesity | t_cell | *NCF4* | 2.482087387 | 0.005888405 | 0.01165997 | 1 |
| Obesity | t_cell | *SLC25A17* | 4.119883901 | 0.00834973 | 0.01587383 | 1 |
| Obesity | t_cell | *EP300* | 3.049265936 | 0.016409568 | 0.028420649 | 1 |
| Obesity | t_cell | *ACO2* | 5.889662301 | 1.62E-06 | 5.73E-05 | 0.010177479 |
| Obesity | t_cell | *DESI1* | 3.044514491 | 8.64E-05 | 0.000390568 | 0.543280196 |
| Obesity | t_cell | *ZBED4* | 5.626857953 | 0.002166979 | 0.004927231 | 1 |
| Obesity | t_cell | *SOS2* | 4.073872008 | 0.000420827 | 0.001275074 | 1 |
| Obesity | t_cell | *TRIM9* | 1.660685801 | 0.01927567 | 0.032664727 | 1 |
| Obesity | t_cell | *PIGH* | 4.184051086 | 0.000781646 | 0.002089373 | 1 |
| Obesity | t_cell | *VTI1B* | 5.789418072 | 0.001735319 | 0.004060271 | 1 |
| Obesity | t_cell | *GSTZ1* | 2.756512121 | 0.000917164 | 0.0023788 | 1 |
| Obesity | t_cell | *DAAM1* | 2.915838284 | 0.000164395 | 0.000626773 | 1 |
| Obesity | t_cell | *ISM2* | 4.701793795 | 0.002244804 | 0.005073201 | 1 |
| Obesity | t_cell | *SLC10A1* | 6.104621826 | 7.71E-06 | 0.000130736 | 0.048502893 |
| Obesity | t_cell | *DICER1* | 4.959029442 | 7.62E-06 | 0.000129898 | 0.047932361 |
| Obesity | t_cell | *ZFYVE21* | 5.21713134 | 9.25E-08 | 7.18E-06 | 0.000581712 |
| Obesity | t_cell | *MTHFD1* | 4.485627666 | 5.45E-06 | 0.000108648 | 0.034232879 |
| Obesity | t_cell | *PAPLN* | 3.792367833 | 0.000199945 | 0.000733405 | 1 |
| Obesity | t_cell | *PSMB5* | 4.522262574 | 4.68E-06 | 0.000100727 | 0.029412225 |
| Obesity | t_cell | *CCNB1IP1* | 4.405583162 | 1.81E-05 | 0.000158673 | 0.113649763 |
| Obesity | t_cell | *TRIP11* | 4.070438405 | 0.00292774 | 0.00637572 | 1 |
| Obesity | t_cell | *PCK2* | 4.242960396 | 0.000359627 | 0.001132754 | 1 |
| Obesity | t_cell | *PLTP* | 2.613013928 | 1.10E-05 | 0.000145864 | 0.068918213 |
| Obesity | t_cell | *GSS* | 4.392942428 | 1.63E-05 | 0.000155022 | 0.102624586 |
| Obesity | t_cell | *PYGB* | 3.345836336 | 0.000319142 | 0.00104015 | 1 |
| Obesity | t_cell | *ABHD12* | 5.700973299 | 3.05E-07 | 1.70E-05 | 0.001915744 |
| Obesity | t_cell | *PROCR* | 1.60108362 | 0.020183799 | 0.033983809 | 1 |
| Obesity | t_cell | *NINL* | 3.175247916 | 6.60E-05 | 0.000321156 | 0.414933963 |
| Obesity | t_cell | *CD40* | 3.218381676 | 6.06E-05 | 0.000303293 | 0.381238853 |
| Obesity | t_cell | *UQCC1* | 2.642477647 | 0.009298949 | 0.017446283 | 1 |
| Obesity | t_cell | *IFT52* | 3.995144047 | 0.000218162 | 0.000781977 | 1 |
| Obesity | t_cell | *DOK5* | 4.009982165 | 1.04E-05 | 0.000145864 | 0.0653861 |

| | | | | | | |
|---|---|---|---|---|---|---|
| Obesity | t_cell | *BMP7* | 3.461167009 | 1.87E-05 | 0.000160625 | 0.117577481 |
| Obesity | t_cell | *TPD52L2* | 4.965551154 | 0.000569027 | 0.001615184 | 1 |
| Obesity | t_cell | *NELFCD* | 4.173861509 | 0.022221497 | 0.036930096 | 1 |
| Obesity | t_cell | *CTSZ* | 3.495030642 | 0.004332721 | 0.0089019 | 1 |
| Obesity | t_cell | *MTG2* | 3.465280328 | 0.001779203 | 0.004144441 | 1 |
| Obesity | t_cell | *DIDO1* | 3.661329169 | 0.003061429 | 0.006616434 | 1 |
| Obesity | t_cell | *GID8* | 3.56859952 | 0.012980746 | 0.023263955 | 1 |
| Obesity | t_cell | *SLC17A9* | 4.644886403 | 1.39E-05 | 0.000148367 | 0.087245942 |
| Obesity | t_cell | *PTK6* | 17.26521803 | 3.21E-05 | 0.000209048 | 0.201940074 |
| Obesity | t_cell | *CDC25B* | -1.920809349 | 0.027162933 | 0.043991077 | 1 |
| Obesity | t_cell | *SLC52A3* | 5.201934942 | 7.20E-06 | 0.000126396 | 0.045249838 |
| Obesity | t_cell | *RSPO4* | 3.31183942 | 0.014404807 | 0.025417631 | 1 |
| Obesity | t_cell | *CCM2L* | 3.95125497 | 1.05E-05 | 0.000145864 | 0.065847018 |
| Obesity | t_cell | *MYL9* | 4.735318006 | 1.35E-06 | 5.11E-05 | 0.008496478 |
| Obesity | t_cell | *MROH8* | 1.829920824 | 0.012098575 | 0.021933027 | 1 |
| Obesity | t_cell | *MANBAL* | 2.695452568 | 4.14E-05 | 0.000242764 | 0.260216602 |
| Obesity | t_cell | *IDH3B* | 7.952251543 | 8.05E-05 | 0.000372155 | 0.506026565 |
| Obesity | t_cell | *MAPRE1* | 6.744836828 | 0.000100418 | 0.000433604 | 0.631327604 |
| Obesity | t_cell | *TTI1* | 4.378978803 | 0.022722562 | 0.037633494 | 1 |
| Obesity | t_cell | *RPRD1B* | 4.274993738 | 0.004029083 | 0.008371066 | 1 |
| Obesity | t_cell | *PXMP4* | 5.175986206 | 0.000638677 | 0.001772787 | 1 |
| Obesity | t_cell | *BPI* | 2.89419903 | 0.000162501 | 0.000622573 | 1 |
| Obesity | t_cell | *DHX35* | 4.695473631 | 0.010146891 | 0.018834811 | 1 |
| Obesity | t_cell | *MAP1LC3A* | 2.262950767 | 2.26E-05 | 0.00017343 | 0.142348654 |
| Obesity | t_cell | *APMAP* | 6.173953567 | 1.71E-05 | 0.000156669 | 0.107612916 |
| Obesity | t_cell | *CELF4* | 2.673893807 | 3.77E-05 | 0.000229622 | 0.236740044 |
| Obesity | t_cell | *RBFA* | 3.154818932 | 0.000144153 | 0.000572515 | 0.906291186 |
| Obesity | t_cell | *LPIN2* | -1.917553044 | 0.017392177 | 0.029916448 | 1 |
| Obesity | t_cell | *SMCHD1* | -1.368498897 | 0.023469072 | 0.038676292 | 1 |
| Obesity | t_cell | *CEP76* | 4.680974099 | 8.66E-06 | 0.000134958 | 0.054435393 |
| Obesity | t_cell | *LIPG* | -5.33734666 | 0.006098772 | 0.012038612 | 1 |
| Obesity | t_cell | *POLI* | 0.758003606 | 0.007386468 | 0.014262507 | 1 |
| Obesity | t_cell | *NALCN* | 4.048853764 | 1.69E-05 | 0.000156669 | 0.106350843 |
| Obesity | t_cell | *HTR2A* | 16.39379139 | 1.18E-05 | 0.000145864 | 0.073992163 |
| Obesity | t_cell | *DNAJC3* | 3.440828126 | 0.016683341 | 0.028839199 | 1 |
| Obesity | t_cell | *UGGT2* | 2.320615579 | 0.004310053 | 0.008858223 | 1 |
| Obesity | t_cell | *ARHGEF7* | 4.104554475 | 0.000241399 | 0.00084409 | 1 |
| Obesity | t_cell | *SGCG* | 2.259406412 | 7.03E-05 | 0.000335975 | 0.442143108 |
| Obesity | t_cell | *KATNAL1* | 3.103488906 | 0.003459419 | 0.007340319 | 1 |
| Obesity | t_cell | *DHRS12* | 6.199056218 | 0.000111538 | 0.000470947 | 0.701240061 |
| Obesity | t_cell | *MEDAG* | 5.376939194 | 1.61E-05 | 0.000154104 | 0.101285612 |
| Obesity | t_cell | *MGRN1* | 2.275828864 | 0.011464744 | 0.020947064 | 1 |
| Obesity | t_cell | *CORO1A* | -1.272226848 | 9.29E-07 | 3.84E-05 | 0.005842756 |
| Obesity | t_cell | *GDPD3* | 3.339161968 | 0.008919454 | 0.016814574 | 1 |
| Obesity | t_cell | *ELMO3* | 2.142589233 | 0.000281107 | 0.000946769 | 1 |
| Obesity | t_cell | *NFAT5* | 3.205005492 | 0.002735565 | 0.006020935 | 1 |
| Obesity | t_cell | *LONP2* | 3.270773848 | 0.006437846 | 0.012636509 | 1 |
| Obesity | t_cell | *PLLP* | 5.062174052 | 0.008215459 | 0.015651694 | 1 |
| Obesity | t_cell | *DHODH* | 1.295153516 | 0.019569821 | 0.033092006 | 1 |
| Obesity | t_cell | *ACD* | -2.636054115 | 0.015591749 | 0.027274715 | 1 |

| | | | | | | |
|---|---|---|---|---|---|---|
| Obesity | t_cell | *USB1* | 8.810526682 | 7.83E-05 | 0.000365265 | 0.492012544 |
| Obesity | t_cell | *SETD6* | 5.187457828 | 5.58E-06 | 0.000109386 | 0.03505169 |
| Obesity | t_cell | *COG4* | -3.568119146 | 0.00208774 | 0.00477469 | 1 |
| Obesity | t_cell | *SLC7A6OS* | 1.995277574 | 0.000104507 | 0.000448488 | 0.657034919 |
| Obesity | t_cell | *HCFC1R1* | 8.09870319 | 2.41E-05 | 0.000179129 | 0.151363945 |
| Obesity | t_cell | *NPRL3* | 2.288835669 | 0.008622336 | 0.016318069 | 1 |
| Obesity | t_cell | *NAGPA* | 7.082809151 | 4.99E-05 | 0.000267593 | 0.313931021 |
| Obesity | t_cell | *CRISPLD2* | 2.055436393 | 0.007793361 | 0.014933514 | 1 |
| Obesity | t_cell | *ZNF500* | 2.196797385 | 0.002988239 | 0.006487244 | 1 |
| Obesity | t_cell | *NME4* | 5.206332462 | 0.000568582 | 0.001615184 | 1 |
| Obesity | t_cell | *NOMO3* | 4.041872578 | 0.005429369 | 0.010871889 | 1 |
| Obesity | t_cell | *LMF1* | 5.167196041 | 0.000557261 | 0.001591647 | 1 |
| Obesity | t_cell | *NARFL* | 5.586355175 | 0.000151201 | 0.000591538 | 0.950602318 |
| Obesity | t_cell | *CLCN7* | 6.623845606 | 0.000116515 | 0.000486407 | 0.7325285 |
| Obesity | t_cell | *HAGHL* | -3.053332067 | 0.003261166 | 0.007004765 | 1 |
| Obesity | t_cell | *SLC7A5* | -1.524552278 | 0.000200384 | 0.000734159 | 1 |
| Obesity | t_cell | *FBXO31* | 4.337502426 | 0.000937387 | 0.002420267 | 1 |
| Obesity | t_cell | *NUBP1* | 6.85223112 | 0.000539596 | 0.001549058 | 1 |
| Obesity | t_cell | *CLUAP1* | 2.156356438 | 0.00272329 | 0.005999062 | 1 |
| Obesity | t_cell | *CPPED1* | 4.762202301 | 0.001019236 | 0.002599568 | 1 |
| Obesity | t_cell | *HMOX2* | -2.480736303 | 0.002158598 | 0.004914412 | 1 |
| Obesity | t_cell | *BFAR* | 5.422095283 | 0.001433907 | 0.003454013 | 1 |
| Obesity | t_cell | *STX4* | 4.900088193 | 2.71E-05 | 0.000191468 | 0.17029739 |
| Obesity | t_cell | *KAT8* | 11.10572367 | 1.35E-10 | 3.04E-08 | 8.50E-07 |
| Obesity | t_cell | *SYT17* | 2.065617562 | 0.002201116 | 0.004990414 | 1 |
| Obesity | t_cell | *TMC5* | 2.240252018 | 0.000139249 | 0.00055726 | 0.875455972 |
| Obesity | t_cell | *RNF40* | 3.612284613 | 0.005102279 | 0.010294617 | 1 |
| Obesity | t_cell | *IQCH* | 6.561304725 | 1.59E-05 | 0.000153107 | 0.099668338 |
| Obesity | t_cell | *LACTB* | 4.818695841 | 4.81E-05 | 0.000263447 | 0.302701076 |
| Obesity | t_cell | *CSK* | -2.489054054 | 0.007171522 | 0.013885852 | 1 |
| Obesity | t_cell | *TRIP4* | 3.837662349 | 8.17E-05 | 0.000376082 | 0.513728251 |
| Obesity | t_cell | *RASL12* | 10.59337932 | 0.000188412 | 0.00070258 | 1 |
| Obesity | t_cell | *TTC23* | 2.838133503 | 0.000123117 | 0.000506238 | 0.774037242 |
| Obesity | t_cell | *FAH* | 3.493027125 | 1.27E-05 | 0.000146996 | 0.079854626 |
| Obesity | t_cell | *HOMER2* | 3.878003084 | 1.25E-05 | 0.000146996 | 0.078683868 |
| Obesity | t_cell | *EHD4* | 5.578867624 | 2.07E-05 | 0.000167271 | 0.130304261 |
| Obesity | t_cell | *ATP8B4* | 4.172068496 | 3.23E-05 | 0.000209712 | 0.203201101 |
| Obesity | t_cell | *DTWD1* | 4.169805229 | 2.16E-05 | 0.000169226 | 0.135719557 |
| Obesity | t_cell | *DNAJC17* | 5.747014147 | 6.99E-05 | 0.000334531 | 0.439239366 |
| Obesity | t_cell | *PDGFRL* | 4.268898845 | 6.04E-05 | 0.000302484 | 0.379707959 |
| Obesity | t_cell | *ZDHHC2* | -3.454693321 | 0.001597187 | 0.003789488 | 1 |
| Obesity | t_cell | *ZFAND1* | 2.863155988 | 0.006106298 | 0.012045903 | 1 |
| Obesity | t_cell | *RIPK2* | 4.734227159 | 0.000835569 | 0.002207235 | 1 |
| Obesity | t_cell | *EYA1* | 1.4277774 | 0.001953108 | 0.004504139 | 1 |
| Obesity | t_cell | *NBN* | 4.636397619 | 0.000171721 | 0.000651938 | 1 |
| Obesity | t_cell | *CPQ* | 2.865257937 | 0.000320056 | 0.001042045 | 1 |
| Obesity | t_cell | *NIPAL2* | 3.819002719 | 0.000258896 | 0.000890417 | 1 |
| Obesity | t_cell | *PLAT* | 4.070338282 | 4.64E-06 | 0.000100727 | 0.02914725 |
| Obesity | t_cell | *ZC2HC1A* | 3.926839537 | 0.000716317 | 0.001942833 | 1 |
| Obesity | t_cell | *CHRAC1* | 5.423744811 | 0.00629875 | 0.012386689 | 1 |

| | | | | | | |
|---|---|---|---|---|---|---|
| Obesity | t_cell | *SNX16* | 4.732650888 | 4.18E-05 | 0.000244306 | 0.262961635 |
| Obesity | t_cell | *LEPROTL1* | -1.17368455 | 0.000736546 | 0.001986556 | 1 |
| Obesity | t_cell | *TNFRSF10A* | -0.797253092 | 0.019033213 | 0.032331858 | 1 |
| Obesity | t_cell | *ERICH1* | 3.311517343 | 0.015972302 | 0.027839719 | 1 |
| Obesity | t_cell | *TUSC3* | 4.007692763 | 6.84E-06 | 0.000122832 | 0.042995586 |
| Obesity | t_cell | *ARHGEF10* | 2.3104062 | 8.75E-06 | 0.000135109 | 0.055038966 |
| Obesity | t_cell | *MCM4* | 4.81192617 | 2.10E-07 | 1.29E-05 | 0.00131996 |
| Obesity | t_cell | *TULP2* | 2.618907172 | 0.014867333 | 0.026160347 | 1 |
| Obesity | t_cell | *NUCB1* | 3.884461498 | 0.001282339 | 0.003152939 | 1 |
| Obesity | t_cell | *NFKBIB* | -3.189367858 | 0.023046297 | 0.03806757 | 1 |
| Obesity | t_cell | *PPP1R37* | 6.416203962 | 0.000115764 | 0.000484382 | 0.72780556 |
| Obesity | t_cell | *FCGRT* | 2.743301503 | 0.0009624 | 0.002474686 | 1 |
| Obesity | t_cell | *PEX11G* | 2.698181332 | 0.000508931 | 0.001475171 | 1 |
| Obesity | t_cell | *PLEKHJ1* | 6.107929747 | 4.16E-05 | 0.000243781 | 0.26182052 |
| Obesity | t_cell | *CD37* | -1.515411929 | 0.003036174 | 0.006568625 | 1 |
| Obesity | t_cell | *DKKL1* | 2.192273919 | 0.019435082 | 0.032899397 | 1 |
| Obesity | t_cell | *FCER2* | 6.358506783 | 2.26E-05 | 0.00017343 | 0.142170489 |
| Obesity | t_cell | *DMPK* | 4.039064129 | 0.004642961 | 0.009443642 | 1 |
| Obesity | t_cell | *SGTA* | 6.016031472 | 0.003066131 | 0.006622042 | 1 |
| Obesity | t_cell | *TIMM44* | 4.520740332 | 0.000274104 | 0.000930505 | 1 |
| Obesity | t_cell | *IL27RA* | -0.851601261 | 0.00067178 | 0.001848352 | 1 |
| Obesity | t_cell | *C19orf44* | 1.918296211 | 0.000309394 | 0.001017151 | 1 |
| Obesity | t_cell | *OLFM2* | 1.708020021 | 5.65E-05 | 0.000289825 | 0.355325164 |
| Obesity | t_cell | *RASAL3* | -0.544980351 | 0.000309693 | 0.001017151 | 1 |
| Obesity | t_cell | *SYDE1* | 3.563192312 | 0.002330403 | 0.005240074 | 1 |
| Obesity | t_cell | *AURKC* | 1.951176978 | 0.002881245 | 0.006285353 | 1 |
| Obesity | t_cell | *CCNE1* | -5.044003806 | 0.000196272 | 0.000723743 | 1 |
| Obesity | t_cell | *PDCD5* | 4.352955427 | 0.000201066 | 0.000736228 | 1 |
| Obesity | t_cell | *ANKRD27* | 4.072584125 | 0.005798093 | 0.011499246 | 1 |
| Obesity | t_cell | *RPS16* | -2.086424457 | 0.008645886 | 0.016357714 | 1 |
| Obesity | t_cell | *PRX* | 5.222288781 | 0.001106201 | 0.002784101 | 1 |
| Obesity | t_cell | *TBCB* | 4.273940955 | 0.001910851 | 0.004418359 | 1 |
| Obesity | t_cell | *POLR2I* | 3.986057859 | 0.00094476 | 0.002434306 | 1 |
| Obesity | t_cell | *CLIP3* | 1.722116405 | 0.0156691 | 0.027379553 | 1 |
| Obesity | t_cell | *SLC1A5* | 3.062936628 | 0.001151401 | 0.002879418 | 1 |
| Obesity | t_cell | *PRKD2* | -1.930715473 | 0.01582621 | 0.027608042 | 1 |
| Obesity | t_cell | *CACTIN* | 8.413115044 | 1.33E-05 | 0.000147409 | 0.083671786 |
| Obesity | t_cell | *CCDC9* | 5.362188337 | 0.000179588 | 0.000677714 | 1 |
| Obesity | t_cell | *TGFB1* | -1.972883973 | 0.014052529 | 0.024879822 | 1 |
| Obesity | t_cell | *CEACAM4* | 7.439916232 | 5.00E-05 | 0.000267593 | 0.314415387 |
| Obesity | t_cell | *PLIN3* | 6.143679727 | 0.000144836 | 0.000574864 | 0.910586708 |
| Obesity | t_cell | *MRPL4* | 7.241858545 | 2.17E-05 | 0.000169311 | 0.13621643 |
| Obesity | t_cell | *ICAM5* | -19.39356806 | 0.00045216 | 0.001348542 | 1 |
| Obesity | t_cell | *CD33* | 3.414381437 | 4.05E-05 | 0.000239096 | 0.254637427 |
| Obesity | t_cell | *TYK2* | 5.030594601 | 0.001036575 | 0.002638439 | 1 |
| Obesity | t_cell | *PTPRS* | 2.412983708 | 1.87E-06 | 6.24E-05 | 0.01173192 |
| Obesity | t_cell | *CYTH2* | 5.262393807 | 1.04E-06 | 4.20E-05 | 0.00655882 |
| Obesity | t_cell | *GRIN2D* | 6.609661966 | 2.60E-05 | 0.000187334 | 0.163542697 |
| Obesity | t_cell | *PLA2G4C* | 3.294473371 | 4.15E-05 | 0.000243101 | 0.260846994 |
| Obesity | t_cell | *BCAT2* | 7.171894626 | 6.04E-05 | 0.000302484 | 0.379474668 |

| | | | | | | |
|---|---|---|---|---|---|---|
| Obesity | t_cell | *MIER2* | 4.399150117 | 8.51E-05 | 0.000386124 | 0.535167735 |
| Obesity | t_cell | *PPP2R1A* | 5.338794978 | 0.004081201 | 0.00846257 | 1 |
| Obesity | t_cell | *TNPO2* | 3.809335062 | 0.00286603 | 0.006258677 | 1 |
| Obesity | t_cell | *LILRB5* | 5.054352774 | 1.70E-05 | 0.000156669 | 0.107065227 |
| Obesity | t_cell | *MAST1* | 5.561176978 | 2.96E-05 | 0.000200768 | 0.18626949 |
| Obesity | t_cell | *RPL18A* | -1.630131586 | 0.011043168 | 0.020271066 | 1 |
| Obesity | t_cell | *UPK1A* | 4.325618414 | 0.006310957 | 0.012406813 | 1 |
| Obesity | t_cell | *KXD1* | 5.258914637 | 0.020093812 | 0.033868578 | 1 |
| Obesity | t_cell | *HPN* | 4.038728295 | 0.00175724 | 0.004097836 | 1 |
| Obesity | t_cell | *ATP13A1* | -4.361542082 | 2.12E-06 | 6.57E-05 | 0.013354232 |
| Obesity | t_cell | *ZNF85* | 4.934540001 | 0.001677111 | 0.003940207 | 1 |
| Obesity | t_cell | *ETHE1* | 4.249356658 | 0.000149741 | 0.000586923 | 0.941424332 |
| Obesity | t_cell | *RASA4* | 2.174643352 | 0.003830872 | 0.008019485 | 1 |
| Obesity | t_cell | *PON3* | 2.819666863 | 1.94E-05 | 0.000161606 | 0.121850644 |
| Obesity | t_cell | *PON2* | 4.487970872 | 1.71E-05 | 0.000156669 | 0.107397432 |
| Obesity | t_cell | *HBP1* | 4.209182889 | 0.00105636 | 0.002679039 | 1 |
| Obesity | t_cell | *WDR91* | 4.465863969 | 6.87E-05 | 0.000330754 | 0.43196517 |
| Obesity | t_cell | *PTN* | 5.032838118 | 1.52E-05 | 0.000149721 | 0.095372324 |
| Obesity | t_cell | *DFNA5* | 3.087943219 | 0.00051214 | 0.001481809 | 1 |
| Obesity | t_cell | *ADAP1* | -2.923302062 | 0.000441472 | 0.001323575 | 1 |
| Obesity | t_cell | *CAV1* | 2.68273742 | 7.99E-05 | 0.000370926 | 0.502164402 |
| Obesity | t_cell | *MET* | 3.107160204 | 1.78E-05 | 0.000157704 | 0.111654364 |
| Obesity | t_cell | *RNF32* | 1.674596755 | 0.027109272 | 0.043915484 | 1 |
| Obesity | t_cell | *HOXA2* | 4.623217823 | 0.007184468 | 0.013906634 | 1 |
| Obesity | t_cell | *HOXA5* | 2.109160492 | 0.000296684 | 0.000985738 | 1 |
| Obesity | t_cell | *HOXA6* | 1.973951824 | 0.000301204 | 0.000997277 | 1 |
| Obesity | t_cell | *CPED1* | 3.195762437 | 1.93E-05 | 0.000161378 | 0.12135651 |
| Obesity | t_cell | *HIBADH* | 4.326515926 | 0.000120549 | 0.000497306 | 0.757894557 |
| Obesity | t_cell | *CPVL* | 3.101732912 | 0.001108404 | 0.0027881 | 1 |
| Obesity | t_cell | *ABHD11* | 1.845133762 | 0.002511941 | 0.005594253 | 1 |
| Obesity | t_cell | *PLEKHA8* | 6.28960556 | 5.55E-05 | 0.00028622 | 0.349188286 |
| Obesity | t_cell | *NOD1* | 4.642949704 | 0.000915163 | 0.00237557 | 1 |
| Obesity | t_cell | *CRHR2* | 7.696991843 | 0.000109267 | 0.000464491 | 0.686963478 |
| Obesity | t_cell | *CHCHD2* | 2.53828935 | 0.019728349 | 0.033341971 | 1 |
| Obesity | t_cell | *NPTX2* | 33.77228346 | 0.009230089 | 0.017342968 | 1 |
| Obesity | t_cell | *CYP3A5* | 5.673177292 | 1.99E-05 | 0.000163528 | 0.125099083 |
| Obesity | t_cell | *ZKSCAN1* | 4.09081884 | 0.001581135 | 0.003757095 | 1 |
| Obesity | t_cell | *SNX8* | 5.733149881 | 0.000206115 | 0.000746453 | 1 |
| Obesity | t_cell | *TAF6* | -4.258091814 | 0.001549551 | 0.003695761 | 1 |
| Obesity | t_cell | *AIMP2* | 5.997762625 | 9.79E-05 | 0.000425311 | 0.615611875 |
| Obesity | t_cell | *TFR2* | -2.203686542 | 0.000386898 | 0.001192952 | 1 |
| Obesity | t_cell | *RBM28* | 2.384298044 | 0.003754601 | 0.007884161 | 1 |
| Obesity | t_cell | *USP42* | 3.812111845 | 0.02472269 | 0.04048751 | 1 |
| Obesity | t_cell | *IMPDH1* | 3.269417212 | 0.002967861 | 0.006454147 | 1 |
| Obesity | t_cell | *LSM5* | 6.9547936 | 0.000420011 | 0.00127381 | 1 |
| Obesity | t_cell | *PLOD3* | 4.623381055 | 0.000508628 | 0.001474976 | 1 |
| Obesity | t_cell | *PHF14* | 2.90494178 | 0.017285955 | 0.029750013 | 1 |
| Obesity | t_cell | *CEP41* | 3.45731412 | 0.000462216 | 0.001372468 | 1 |
| Obesity | t_cell | *ZNF862* | 4.885901291 | 9.23E-05 | 0.00040823 | 0.580502921 |
| Obesity | t_cell | *ACTR3C* | -1.544612925 | 0.003831801 | 0.008019485 | 1 |

| | | | | | | |
|---|---|---|---|---|---|---|
| Obesity | t_cell | *POU6F2* | 3.40524306 | 1.42E-05 | 0.000148367 | 0.08912539 |
| Obesity | t_cell | *TSPAN13* | 7.765220684 | 3.04E-06 | 8.16E-05 | 0.019086393 |
| Obesity | t_cell | *RARRES2* | 2.892004641 | 0.000125196 | 0.000512285 | 0.7871058 |
| Obesity | t_cell | *TMEM176B* | 1.370117492 | 0.008288006 | 0.015773185 | 1 |
| Obesity | t_cell | *MRPL32* | 2.935344438 | 0.000340329 | 0.001091101 | 1 |
| Obesity | t_cell | *BLVRA* | 3.296226161 | 8.92E-05 | 0.000397461 | 0.560817435 |
| Obesity | t_cell | *URGCP* | 4.509621519 | 0.00446953 | 0.009141162 | 1 |
| Obesity | t_cell | *POLD2* | 4.628179851 | 0.000308869 | 0.001016779 | 1 |
| Obesity | t_cell | *GCK* | 16.62481753 | 5.24E-05 | 0.000275475 | 0.329192221 |
| Obesity | t_cell | *YKT6* | 5.041803244 | 0.021885654 | 0.036420091 | 1 |
| Obesity | t_cell | *TBL2* | 1.798993601 | 0.000357149 | 0.001127773 | 1 |
| Obesity | t_cell | *CLIP2* | 5.643731694 | 2.67E-05 | 0.000189464 | 0.167654976 |
| Obesity | t_cell | *LIMK1* | 5.133170647 | 1.21E-05 | 0.000145864 | 0.076240389 |
| Obesity | t_cell | *SLC1A1* | 1.477588166 | 0.003339428 | 0.007136621 | 1 |
| Obesity | t_cell | *FKTN* | 5.086171567 | 0.000382496 | 0.001182276 | 1 |
| Obesity | t_cell | *TRIM14* | -1.672052978 | 0.011023921 | 0.020253474 | 1 |
| Obesity | t_cell | *CORO2A* | -1.762632273 | 0.000869172 | 0.002277819 | 1 |
| Obesity | t_cell | *C5* | 2.563126065 | 5.80E-05 | 0.000294556 | 0.364365804 |
| Obesity | t_cell | *OGN* | 6.15965776 | 1.15E-08 | 1.33E-06 | 7.21E-05 |
| Obesity | t_cell | *ECM2* | 2.761012946 | 1.98E-05 | 0.000163419 | 0.124766482 |
| Obesity | t_cell | *PTGR1* | 2.106274697 | 8.56E-05 | 0.000387733 | 0.53846161 |
| Obesity | t_cell | *TNFSF8* | -0.713992079 | 0.000246358 | 0.000856665 | 1 |
| Obesity | t_cell | *PLGRKT* | -1.889601859 | 0.01236461 | 0.022318778 | 1 |
| Obesity | t_cell | *ELAVL2* | -3.713314705 | 0.000410794 | 0.001250682 | 1 |
| Obesity | t_cell | *FUBP3* | 4.110674914 | 0.009873084 | 0.018409885 | 1 |
| Obesity | t_cell | *MPDZ* | 3.68410569 | 1.43E-05 | 0.000148367 | 0.089762303 |
| Obesity | t_cell | *DVL1* | 5.746291958 | 0.000339444 | 0.001091046 | 1 |
| Obesity | t_cell | *PDLIM1* | 3.489874096 | 2.81E-05 | 0.000194841 | 0.176720986 |
| Obesity | t_cell | *CCNJ* | 4.161412399 | 0.000242404 | 0.000846663 | 1 |
| Obesity | t_cell | *ATRNL1* | 1.835198086 | 0.0122844 | 0.02219311 | 1 |
| Obesity | t_cell | *HPS1* | 5.650672086 | 0.000506264 | 0.001471511 | 1 |
| Obesity | t_cell | *PHYH* | 1.555195962 | 0.001335643 | 0.00325725 | 1 |
| Obesity | t_cell | *DNMBP* | 3.996879085 | 1.45E-05 | 0.000148509 | 0.091056956 |
| Obesity | t_cell | *MAPK8* | 3.776217026 | 0.001876479 | 0.004350082 | 1 |
| Obesity | t_cell | *ATE1* | 3.647087048 | 0.000424663 | 0.001282967 | 1 |
| Obesity | t_cell | *NSMCE4A* | 4.58293985 | 1.40E-05 | 0.000148367 | 0.088266545 |
| Obesity | t_cell | *SPOCK2* | -0.405501943 | 0.000124639 | 0.000510827 | 0.783608128 |
| Obesity | t_cell | *MICU1* | 4.732445408 | 0.000158501 | 0.00061097 | 0.99649268 |
| Obesity | t_cell | *BMPR1A* | 1.560184452 | 0.007974384 | 0.01522933 | 1 |
| Obesity | t_cell | *LIPA* | 2.788794901 | 0.008060745 | 0.015380244 | 1 |
| Obesity | t_cell | *SFXN3* | 5.376190278 | 0.005225522 | 0.010516265 | 1 |
| Obesity | t_cell | *KAZALD1* | 4.258632762 | 0.0005059 | 0.001471134 | 1 |
| Obesity | t_cell | *ANKRD26* | 3.04876612 | 0.000864309 | 0.00226791 | 1 |
| Obesity | t_cell | *MTPAP* | 6.740481948 | 0.000340242 | 0.001091101 | 1 |
| Obesity | t_cell | *PITRM1* | 6.282914916 | 0.000324487 | 0.001053745 | 1 |
| Obesity | t_cell | *MAP3K8* | 4.403143705 | 4.56E-05 | 0.000254774 | 0.286620223 |
| Obesity | t_cell | *SHOC2* | 4.152707936 | 0.00049841 | 0.001455411 | 1 |
| Obesity | t_cell | *PBLD* | 0.975338254 | 0.00614164 | 0.012104229 | 1 |
| Obesity | t_cell | *PSMB3* | 4.384044438 | 0.010491997 | 0.019412356 | 1 |
| Obesity | t_cell | *RAPGEFL1* | 1.752606806 | 0.01843429 | 0.031502143 | 1 |

| | | | | | | |
|---|---|---|---|---|---|---|
| Obesity | t_cell | *RGS9* | -1.869015463 | 0.000594019 | 0.001674708 | 1 |
| Obesity | t_cell | *WNT3* | 3.34581073 | 4.46E-05 | 0.000252269 | 0.280125897 |
| Obesity | t_cell | *ASPA* | 2.751084937 | 1.29E-05 | 0.000147409 | 0.081373039 |
| Obesity | t_cell | *PNPO* | 1.858440048 | 0.001459612 | 0.003507867 | 1 |
| Obesity | t_cell | *RPS6KB1* | 3.732630341 | 0.019052109 | 0.032338178 | 1 |
| Obesity | t_cell | *TRIM16L* | 1.574919191 | 0.02236298 | 0.037135778 | 1 |
| Obesity | t_cell | *CDK5RAP3* | 3.262690495 | 0.028196154 | 0.045441995 | 1 |
| Obesity | t_cell | *CBX1* | 5.749567624 | 4.21E-06 | 9.89E-05 | 0.026455273 |
| Obesity | t_cell | *GALK1* | 6.41172258 | 1.11E-05 | 0.000145864 | 0.069994575 |
| Obesity | t_cell | *CAMTA2* | 4.301067043 | 0.000897554 | 0.002338551 | 1 |
| Obesity | t_cell | *HOXB6* | 3.641495488 | 4.41E-08 | 3.85E-06 | 0.000277465 |
| Obesity | t_cell | *CHRNE* | 2.283906279 | 5.14E-06 | 0.000105615 | 0.032318158 |
| Obesity | t_cell | *C1QBP* | 9.885994757 | 2.33E-05 | 0.000175508 | 0.146724917 |
| Obesity | t_cell | *GOSR1* | 4.794428758 | 0.000722906 | 0.001955845 | 1 |
| Obesity | t_cell | *DRG2* | 5.406248765 | 0.025557661 | 0.041724491 | 1 |
| Obesity | t_cell | *SMARCD2* | 4.989201752 | 0.004087003 | 0.008471805 | 1 |
| Obesity | t_cell | *UTP6* | 5.646515669 | 0.000365514 | 0.001147274 | 1 |
| Obesity | t_cell | *C17orf75* | 6.304594999 | 0.000154549 | 0.000601267 | 0.971647495 |
| Obesity | t_cell | *CYTH1* | -1.290874566 | 0.007899283 | 0.015095074 | 1 |
| Obesity | t_cell | *RAB5C* | 3.197985594 | 0.010392074 | 0.019238802 | 1 |
| Obesity | t_cell | *NAGLU* | 4.93645471 | 2.24E-05 | 0.000172813 | 0.140669428 |
| Obesity | t_cell | *EZH1* | 4.325531673 | 6.23E-05 | 0.000309181 | 0.391732184 |
| Obesity | t_cell | *DLX4* | 15.58154857 | 4.90E-05 | 0.000265838 | 0.30837033 |
| Obesity | t_cell | *VAT1* | 5.449009888 | 5.61E-05 | 0.000288055 | 0.352867423 |
| Obesity | t_cell | *LRRC59* | 6.381167159 | 0.020627423 | 0.034619489 | 1 |
| Obesity | t_cell | *ABCC3* | 1.709999655 | 0.001057741 | 0.002681458 | 1 |
| Obesity | t_cell | *MPP2* | 2.935518439 | 3.10E-05 | 0.000205911 | 0.194586281 |
| Obesity | t_cell | *EFTUD2* | 4.947888289 | 0.001104549 | 0.002781057 | 1 |
| Obesity | t_cell | *EFNB3* | -10.65309867 | 0.011247904 | 0.020598768 | 1 |
| Obesity | t_cell | *FAM20A* | 3.509030782 | 1.09E-05 | 0.000145864 | 0.068631807 |
| Obesity | t_cell | *YWHAE* | 3.910870469 | 0.004114179 | 0.008519712 | 1 |
| Obesity | t_cell | *DPH1* | 9.31901329 | 1.68E-05 | 0.000156468 | 0.105460717 |
| Obesity | t_cell | *DHRS7B* | 4.963302614 | 0.000137209 | 0.0005505 | 0.862634148 |
| Obesity | t_cell | *RCVRN* | 4.008852422 | 0.017033155 | 0.029371213 | 1 |
| Obesity | t_cell | *SLC9A3R1* | -0.907045302 | 0.000340002 | 0.001091101 | 1 |
| Obesity | t_cell | *TMEM97* | 2.981706934 | 0.000775272 | 0.002074982 | 1 |
| Obesity | t_cell | *RAB34* | 2.985466843 | 0.010842625 | 0.019972923 | 1 |
| Obesity | t_cell | *OCIAD1* | 3.29324538 | 0.01564286 | 0.027348905 | 1 |
| Obesity | t_cell | *CWH43* | 12.24667636 | 0.000548513 | 0.00157107 | 1 |
| Obesity | t_cell | *KIAA1211* | 1.621294536 | 0.000477383 | 0.001407742 | 1 |
| Obesity | t_cell | *TBC1D9* | 4.2728996 | 3.59E-05 | 0.000223048 | 0.225724644 |
| Obesity | t_cell | *CPE* | 4.076500745 | 0.000353883 | 0.001122566 | 1 |
| Obesity | t_cell | *WFS1* | 4.436586369 | 4.36E-06 | 9.97E-05 | 0.027421755 |
| Obesity | t_cell | *GRPEL1* | 7.410531633 | 5.08E-06 | 0.000105254 | 0.031956348 |
| Obesity | t_cell | *AADAT* | 4.30749358 | 5.33E-05 | 0.00027858 | 0.335131157 |
| Obesity | t_cell | *CPZ* | 30.93810987 | 5.04E-05 | 0.000268633 | 0.316986997 |
| Obesity | t_cell | *TRIM2* | 1.76500528 | 0.019234409 | 0.032612387 | 1 |
| Obesity | t_cell | *SLC2A9* | 4.819899817 | 0.00059142 | 0.001668129 | 1 |
| Obesity | t_cell | *SH3D19* | 2.015316943 | 4.55E-05 | 0.000254768 | 0.286359512 |
| Obesity | t_cell | *RAPGEF2* | 3.136231985 | 0.007012897 | 0.013652732 | 1 |

| | | | | | | |
|---|---|---|---|---|---|---|
| Obesity | t_cell | *LRP2BP* | 3.237430184 | 0.000211082 | 0.000761809 | 1 |
| Obesity | t_cell | *UFSP2* | 5.669974519 | 0.000413006 | 0.001256809 | 1 |
| Obesity | t_cell | *FAM149A* | 1.088233544 | 0.013942754 | 0.024727249 | 1 |
| Obesity | t_cell | *UGDH* | 5.191322883 | 0.000122477 | 0.000504266 | 0.770014696 |
| Obesity | t_cell | *SC5D* | 5.204384019 | 0.021851075 | 0.036372176 | 1 |
| Obesity | t_cell | *C11orf63* | 6.091921277 | 4.92E-05 | 0.000266107 | 0.309215915 |
| Obesity | t_cell | *DTX4* | 3.980319842 | 4.44E-05 | 0.000251953 | 0.2791874 |
| Obesity | t_cell | *PUS3* | 4.859252406 | 0.00128084 | 0.003150486 | 1 |
| Obesity | t_cell | *FOXRED1* | 3.632781652 | 4.75E-07 | 2.43E-05 | 0.002984763 |
| Obesity | t_cell | *NRXN2* | 5.864353013 | 1.95E-05 | 0.000161982 | 0.122620288 |
| Obesity | t_cell | *MS4A4A* | 1.817183152 | 0.001108677 | 0.0027881 | 1 |
| Obesity | t_cell | *CCDC86* | 5.472575278 | 3.24E-05 | 0.000209715 | 0.203589392 |
| Obesity | t_cell | *HPX* | 2.722012865 | 0.0001378 | 0.000552165 | 0.866346658 |
| Obesity | t_cell | *KIAA1549L* | 3.013368618 | 0.00127814 | 0.003148772 | 1 |
| Obesity | t_cell | *CD5* | -0.468190986 | 0.000383534 | 0.001184322 | 1 |
| Obesity | t_cell | *ACCS* | 1.355804825 | 0.00848386 | 0.016099617 | 1 |
| Obesity | t_cell | *MADD* | 6.050153359 | 0.000331073 | 0.001070159 | 1 |
| Obesity | t_cell | *SLC35F2* | -2.147708737 | 0.004401333 | 0.009025174 | 1 |
| Obesity | t_cell | *C11orf21* | -0.835328245 | 8.57E-05 | 0.000387733 | 0.538560802 |
| Obesity | t_cell | *ELMOD1* | 5.686197381 | 0.000450988 | 0.001346325 | 1 |
| Obesity | t_cell | *RPS13* | -2.024664312 | 0.013034174 | 0.023326459 | 1 |
| Obesity | t_cell | *NDUFS8* | 6.560963201 | 2.79E-05 | 0.000194081 | 0.175429941 |
| Obesity | t_cell | *TCIRG1* | 4.969281413 | 0.003431514 | 0.007295883 | 1 |
| Obesity | t_cell | *GTF2H1* | 4.232996194 | 0.001090291 | 0.002750667 | 1 |
| Obesity | t_cell | *PPFIBP1* | 2.815206161 | 0.005312692 | 0.010664398 | 1 |
| Obesity | t_cell | *PRDM4* | 5.992902918 | 0.000147619 | 0.000581505 | 0.928081444 |
| Obesity | t_cell | *CYP27B1* | 12.73974087 | 3.81E-05 | 0.000231281 | 0.239376108 |
| Obesity | t_cell | *ACSS3* | 1.047545083 | 0.001644204 | 0.003871339 | 1 |
| Obesity | t_cell | *GLI1* | 11.16526529 | 1.27E-05 | 0.000146996 | 0.079891055 |
| Obesity | t_cell | *LTA4H* | 5.602603387 | 0.000246066 | 0.000856598 | 1 |
| Obesity | t_cell | *WNT5B* | 5.141375022 | 4.93E-05 | 0.000266596 | 0.310050995 |
| Obesity | t_cell | *TRPV4* | 1.755648593 | 0.01290207 | 0.023136143 | 1 |
| Obesity | t_cell | *ARPC3* | 5.278947598 | 0.000892044 | 0.002327712 | 1 |
| Obesity | t_cell | *GPN3* | 5.16185 | 6.25E-05 | 0.000309439 | 0.392678018 |
| Obesity | t_cell | *VPS29* | 5.427808862 | 0.000158201 | 0.000610564 | 0.994609385 |
| Obesity | t_cell | *AKAP3* | 2.138303698 | 0.003021729 | 0.006544129 | 1 |
| Obesity | t_cell | *KCNA1* | 3.159720958 | 0.00101302 | 0.002584764 | 1 |
| Obesity | t_cell | *NAA25* | 4.365600592 | 0.001965444 | 0.00452628 | 1 |
| Obesity | t_cell | *SCNN1A* | 3.993433471 | 0.012106722 | 0.021935148 | 1 |
| Obesity | t_cell | *LTBR* | 4.146396098 | 4.22E-06 | 9.89E-05 | 0.026556797 |
| Obesity | t_cell | *CDK2AP1* | 1.565113718 | 0.000503232 | 0.001465411 | 1 |
| Obesity | t_cell | *MGP* | 2.009313135 | 0.00773031 | 0.014835305 | 1 |
| Obesity | t_cell | *ARHGDIB* | -1.032185955 | 0.000520576 | 0.001502692 | 1 |
| Obesity | t_cell | *EIF2B1* | 5.370128493 | 0.003494792 | 0.007402883 | 1 |
| Obesity | t_cell | *DDX55* | 3.834435548 | 1.38E-06 | 5.14E-05 | 0.008684073 |
| Obesity | t_cell | *ENDOU* | 1.80669509 | 0.012627659 | 0.022702342 | 1 |
| Obesity | t_cell | *STX2* | 2.082478138 | 0.030908022 | 0.049306962 | 1 |
| Obesity | t_cell | *NUP107* | 4.708111469 | 0.000369122 | 0.001153414 | 1 |
| Obesity | t_cell | *GNPTAB* | 3.635938201 | 0.009377947 | 0.017584001 | 1 |
| Obesity | t_cell | *SPSB2* | 6.683016816 | 0.000419712 | 0.001273519 | 1 |

| | | | | | | |
|---|---|---|---|---|---|---|
| Obesity | t_cell | *ATN1* | 3.934590944 | 3.19E-05 | 0.000209022 | 0.200489199 |
| Obesity | t_cell | *C12orf57* | -2.684685278 | 0.003612291 | 0.007618407 | 1 |
| Obesity | t_cell | *NT5DC3* | 1.882966933 | 0.029001094 | 0.046643612 | 1 |
| Obesity | t_cell | *PRKAB1* | 2.91751761 | 0.003459382 | 0.007340319 | 1 |
| Obesity | t_cell | *ST8SIA1* | -1.434625289 | 0.001228709 | 0.003048844 | 1 |
| Obesity | t_cell | *COL12A1* | 3.27312671 | 1.84E-05 | 0.000159202 | 0.11556788 |
| Obesity | t_cell | *SMIM8* | 4.804514344 | 4.55E-06 | 0.000100727 | 0.028626113 |
| Obesity | t_cell | *FAM184A* | 3.983110011 | 6.00E-05 | 0.000301181 | 0.377078664 |
| Obesity | t_cell | *GABRR2* | 5.891345301 | 3.20E-05 | 0.000209022 | 0.201253766 |
| Obesity | t_cell | *TPD52L1* | 2.551525695 | 0.000862942 | 0.002265267 | 1 |
| Obesity | t_cell | *MTRF1L* | 2.946651641 | 0.002965631 | 0.006451531 | 1 |
| Obesity | t_cell | *TULP1* | 13.3759144 | 2.64E-05 | 0.000188499 | 0.166075379 |
| Obesity | t_cell | *MRPL18* | 2.541133004 | 0.030075106 | 0.048161537 | 1 |
| Obesity | t_cell | *RNF8* | 3.043860672 | 0.010884111 | 0.02004347 | 1 |
| Obesity | t_cell | *PHACTR1* | 3.952406415 | 9.77E-05 | 0.000424749 | 0.614186621 |
| Obesity | t_cell | *MDGA1* | -2.475297036 | 0.005416693 | 0.010852375 | 1 |
| Obesity | t_cell | *CD83* | 2.249626017 | 0.007062431 | 0.013725349 | 1 |
| Obesity | t_cell | *SAYSD1* | 6.681647721 | 3.85E-05 | 0.00023194 | 0.241897283 |
| Obesity | t_cell | *CAP2* | 1.329907251 | 0.019900666 | 0.033579036 | 1 |
| Obesity | t_cell | *GPR63* | 2.744464152 | 0.00450973 | 0.009214388 | 1 |
| Obesity | t_cell | *WASF1* | 1.31389436 | 0.002044281 | 0.004685524 | 1 |
| Obesity | t_cell | *GPLD1* | 1.331000053 | 0.003487547 | 0.007390025 | 1 |
| Obesity | t_cell | *ALDH5A1* | 3.405202284 | 5.58E-05 | 0.000286975 | 0.350970996 |
| Obesity | t_cell | *VNN2* | -1.488571565 | 0.001977475 | 0.004550653 | 1 |
| Obesity | t_cell | *ACOT13* | 5.758248629 | 9.84E-05 | 0.000426795 | 0.618852081 |
| Obesity | t_cell | *C6orf62* | 3.877116653 | 0.000679533 | 0.001866415 | 1 |
| Obesity | t_cell | *PERP* | -1.568191149 | 0.017978961 | 0.030824577 | 1 |
| Obesity | t_cell | *SLC16A10* | -0.985838715 | 0.000935306 | 0.002417639 | 1 |
| Obesity | t_cell | *PHACTR2* | -1.476567234 | 0.03052687 | 0.048835224 | 1 |
| Obesity | t_cell | *PACRG* | 7.857291789 | 2.98E-05 | 0.000200768 | 0.187389292 |
| Obesity | t_cell | *TFEB* | 5.585540173 | 0.001247882 | 0.00308511 | 1 |
| Obesity | t_cell | *TBP* | 4.965357164 | 0.000206004 | 0.000746453 | 1 |
| Obesity | t_cell | *DUSP22* | 5.430714683 | 0.000118638 | 0.000491681 | 0.745880134 |
| Obesity | t_cell | *TMEM30A* | 3.224727685 | 0.029532553 | 0.047353013 | 1 |
| Obesity | t_cell | *TTK* | -2.716183718 | 0.005772071 | 0.0114621 | 1 |
| Obesity | t_cell | *LAMA4* | 2.542644423 | 2.11E-07 | 1.29E-05 | 0.001327827 |
| Obesity | t_cell | *LY86* | 4.413343713 | 8.50E-06 | 0.000134614 | 0.053438294 |
| Obesity | t_cell | *PRSS16* | 10.692281 | 1.47E-09 | 2.15E-07 | 9.23E-06 |
| Obesity | t_cell | *TBX18* | 2.0628375 | 0.016235234 | 0.028196385 | 1 |
| Obesity | t_cell | *NUDT12* | 3.408754639 | 0.000110893 | 0.000469168 | 0.69718296 |
| Obesity | t_cell | *C7* | 3.468917416 | 5.68E-05 | 0.000290507 | 0.357033077 |
| Obesity | t_cell | *GHR* | 1.328805432 | 0.000193891 | 0.000719168 | 1 |
| Obesity | t_cell | *DAP* | 5.479883398 | 0.000635027 | 0.001764213 | 1 |
| Obesity | t_cell | *NME5* | 2.935971513 | 3.14E-08 | 3.05E-06 | 0.000197468 |
| Obesity | t_cell | *NNT* | 1.745399663 | 0.016608882 | 0.028726284 | 1 |
| Obesity | t_cell | *PFDN1* | 4.246984313 | 1.08E-05 | 0.000145864 | 0.067838333 |
| Obesity | t_cell | *LOX* | 6.219439201 | 0.000255212 | 0.0008819 | 1 |
| Obesity | t_cell | *TMCO6* | 3.964674983 | 0.005781507 | 0.011473591 | 1 |
| Obesity | t_cell | *SPARC* | 3.08592122 | 0.000107791 | 0.000459758 | 0.677682598 |
| Obesity | t_cell | *PCDHB6* | 5.258540542 | 0.000296803 | 0.000985738 | 1 |

| | | | | | | |
|---|---|---|---|---|---|---|
| Obesity | t_cell | *PDE8B* | 1.607771125 | 0.000654343 | 0.001807494 | 1 |
| Obesity | t_cell | *CLK4* | 4.531721568 | 0.001311652 | 0.003208698 | 1 |
| Obesity | t_cell | *RNF130* | 3.993164692 | 6.90E-05 | 0.000331465 | 0.433791842 |
| Obesity | t_cell | *CLINT1* | 2.971703115 | 0.024632966 | 0.0403616 | 1 |
| Obesity | t_cell | *TTC1* | 4.768973255 | 0.000870064 | 0.002278406 | 1 |
| Obesity | t_cell | *POLR3G* | 3.064890332 | 0.005532807 | 0.011053307 | 1 |
| Obesity | t_cell | *SLC27A6* | 11.59888553 | 4.51E-05 | 0.000253545 | 0.283463278 |
| Obesity | t_cell | *TARS* | 4.853864671 | 0.012540177 | 0.022570883 | 1 |
| Obesity | t_cell | *SLC12A7* | 4.812314919 | 0.002963204 | 0.006448482 | 1 |
| Obesity | t_cell | *IL4* | -3.548179478 | 0.027930429 | 0.04507151 | 1 |
| Obesity | t_cell | *GNPDA1* | 5.254008849 | 9.45E-06 | 0.000140178 | 0.0594354 |
| Obesity | t_cell | *C9* | 1.892208918 | 0.007689404 | 0.014761308 | 1 |
| Obesity | t_cell | *SMAD5* | 4.351488299 | 0.00016152 | 0.000619947 | 1 |
| Obesity | t_cell | *HMGXB3* | 4.899980831 | 8.45E-05 | 0.00038486 | 0.531492111 |
| Obesity | t_cell | *BNIP1* | -1.206879024 | 0.020106862 | 0.033881491 | 1 |
| Obesity | t_cell | *CNTN3* | 2.048281103 | 1.81E-05 | 0.000158673 | 0.114056668 |
| Obesity | t_cell | *HGD* | 5.027259301 | 1.95E-05 | 0.00016194 | 0.122426772 |
| Obesity | t_cell | *NIT2* | 4.906306322 | 5.16E-05 | 0.000272935 | 0.324445793 |
| Obesity | t_cell | *FAM162A* | 3.287414756 | 0.026034017 | 0.042425056 | 1 |
| Obesity | t_cell | *PCCB* | 5.590886987 | 0.000145875 | 0.000577166 | 0.917116967 |
| Obesity | t_cell | *WNT5A* | 2.824116754 | 0.000628188 | 0.001749079 | 1 |
| Obesity | t_cell | *PRKAR2A* | 2.896621305 | 0.002329877 | 0.005240074 | 1 |
| Obesity | t_cell | *IQCG* | 4.967732494 | 0.002721476 | 0.005997168 | 1 |
| Obesity | t_cell | *GBE1* | 3.101853347 | 1.72E-05 | 0.000156669 | 0.108415243 |
| Obesity | t_cell | *NCBP2* | 5.413987595 | 0.003223988 | 0.006932015 | 1 |
| Obesity | t_cell | *PLXNA1* | 3.318454538 | 0.000160368 | 0.000615904 | 1 |
| Obesity | t_cell | *ABTB1* | 5.666429718 | 0.000567971 | 0.0016143 | 1 |
| Obesity | t_cell | *NEK11* | 4.015169563 | 8.54E-06 | 0.000134719 | 0.053711695 |
| Obesity | t_cell | *PLSCR4* | 2.746077335 | 0.000470586 | 0.001391248 | 1 |
| Obesity | t_cell | *COMMD2* | 5.51702497 | 3.70E-05 | 0.000227068 | 0.232745192 |
| Obesity | t_cell | *GORASP1* | 5.288673684 | 5.56E-05 | 0.000286272 | 0.349547664 |
| Obesity | t_cell | *ABCC5* | 3.627393186 | 0.001085745 | 0.002740298 | 1 |
| Obesity | t_cell | *ARHGEF26* | 4.411566957 | 4.24E-07 | 2.24E-05 | 0.002666782 |
| Obesity | t_cell | *KLHL24* | 4.817727362 | 1.64E-06 | 5.77E-05 | 0.010336039 |
| Obesity | t_cell | *PLCH1* | 8.556727041 | 0.000473374 | 0.001397887 | 1 |
| Obesity | t_cell | *ZBTB47* | 4.207117951 | 1.18E-05 | 0.000145864 | 0.074037133 |
| Obesity | t_cell | *CLCN2* | 8.273462069 | 3.61E-06 | 9.08E-05 | 0.022697044 |
| Obesity | t_cell | *NEK4* | 4.977535791 | 0.000512163 | 0.001481809 | 1 |
| Obesity | t_cell | *LMAN2L* | 2.675189745 | 0.000283806 | 0.000954164 | 1 |
| Obesity | t_cell | *KCNIP3* | 6.067111095 | 9.89E-06 | 0.000142631 | 0.062187029 |
| Obesity | t_cell | *FAHD2A* | 6.3506126 | 2.27E-07 | 1.32E-05 | 0.001426152 |
| Obesity | t_cell | *ACTR1B* | 7.033099102 | 0.000191289 | 0.000711197 | 1 |
| Obesity | t_cell | *SLC35F5* | 2.769018756 | 0.018427877 | 0.031499746 | 1 |
| Obesity | t_cell | *STEAP3* | 1.122513501 | 0.011765181 | 0.021415082 | 1 |
| Obesity | t_cell | *TFCP2L1* | 7.130890745 | 6.34E-07 | 3.02E-05 | 0.003982847 |
| Obesity | t_cell | *TP53I3* | 3.259969218 | 2.62E-05 | 0.000187653 | 0.164613333 |
| Obesity | t_cell | *DNAJC27* | 5.784289745 | 1.17E-06 | 4.67E-05 | 0.007374327 |
| Obesity | t_cell | *STAM2* | 4.764396397 | 0.000451669 | 0.001347718 | 1 |
| Obesity | t_cell | *GPD2* | 3.856117658 | 0.002407488 | 0.005386433 | 1 |
| Obesity | t_cell | *CENPA* | 4.423078759 | 0.000437343 | 0.001314957 | 1 |

| | | | | | | |
|---|---|---|---|---|---|---|
| Obesity | t_cell | *TANC1* | 3.032669928 | 0.000381651 | 0.001180246 | 1 |
| Obesity | t_cell | *ITGA4* | -0.52420886 | 0.000305149 | 0.001008129 | 1 |
| Obesity | t_cell | *PCSK4* | 3.026511929 | 0.000696545 | 0.00190399 | 1 |
| Obesity | t_cell | *GRB14* | 4.602459057 | 7.32E-05 | 0.00034686 | 0.460282632 |
| Obesity | t_cell | *CLIP4* | 3.134140055 | 0.022286514 | 0.037018576 | 1 |
| Obesity | t_cell | *SPTBN1* | 2.379733837 | 0.000394535 | 0.001211743 | 1 |
| Obesity | t_cell | *HTRA2* | 6.404922096 | 0.002591534 | 0.00574303 | 1 |
| Obesity | t_cell | *GALNT3* | -1.707772638 | 0.026625829 | 0.043277297 | 1 |
| Obesity | t_cell | *MRPL19* | 2.909589492 | 0.008947505 | 0.016852295 | 1 |
| Obesity | t_cell | *EFEMP1* | 2.871409586 | 3.48E-05 | 0.000218022 | 0.218621583 |
| Obesity | t_cell | *GGCX* | 3.272052591 | 7.53E-05 | 0.000354783 | 0.473635014 |
| Obesity | t_cell | *GNLY* | -1.257568918 | 0.000867938 | 0.002275532 | 1 |
| Obesity | t_cell | *ST3GAL5* | 4.14694856 | 0.001609173 | 0.003810497 | 1 |
| Obesity | t_cell | *PLCD4* | 2.059750298 | 8.29E-05 | 0.000379822 | 0.521496244 |
| Obesity | t_cell | *CHMP3* | -4.294670697 | 0.000305556 | 0.001008791 | 1 |
| Obesity | t_cell | *ZNF142* | 4.295735934 | 0.001585623 | 0.00376608 | 1 |
| Obesity | t_cell | *WNT6* | 7.344723344 | 0.012716949 | 0.022843274 | 1 |
| Obesity | t_cell | *MLPH* | 6.297613872 | 8.14E-06 | 0.000132809 | 0.051172677 |
| Obesity | t_cell | *HDLBP* | 3.212204444 | 0.001686136 | 0.003956976 | 1 |
| Obesity | t_cell | *PASK* | -1.485652957 | 0.00149381 | 0.003576451 | 1 |
| Obesity | t_cell | *STK25* | 4.678810861 | 6.42E-06 | 0.000116734 | 0.040389913 |
| Obesity | t_cell | *TPO* | 0.9589898 | 0.016319279 | 0.028303257 | 1 |
| Obesity | t_cell | *CEBPZ* | 2.825829222 | 0.023952748 | 0.039355498 | 1 |
| Obesity | t_cell | *PRKD3* | 3.422882426 | 0.004606405 | 0.009378389 | 1 |
| Obesity | t_cell | *SLC25A12* | 2.923500252 | 0.017304581 | 0.02977392 | 1 |
| Obesity | t_cell | *RMDN2* | 5.003411714 | 2.13E-05 | 0.00016914 | 0.133959076 |
| Obesity | t_cell | *WIPF1* | -1.529246086 | 0.023146007 | 0.03819395 | 1 |
| Obesity | t_cell | *COX7A2L* | 5.898716908 | 0.000417727 | 0.001268109 | 1 |
| Obesity | t_cell | *PNO1* | 7.215675671 | 2.62E-05 | 0.000187653 | 0.164759658 |
| Obesity | t_cell | *ORC4* | 3.682155958 | 0.004390733 | 0.009006375 | 1 |
| Obesity | t_cell | *ARID3A* | 5.096019107 | 2.33E-05 | 0.000175508 | 0.146383244 |
| Obesity | t_cell | *CD207* | 35.34840494 | 2.72E-06 | 7.61E-05 | 0.01711112 |
| Obesity | t_cell | *VAX2* | 12.14312839 | 6.86E-06 | 0.000122832 | 0.043113902 |
| Obesity | t_cell | *MSH6* | 5.466557534 | 0.000366382 | 0.001147429 | 1 |
| Obesity | t_cell | *BCL9* | 3.93050768 | 0.004689122 | 0.009525206 | 1 |
| Obesity | t_cell | *GPX7* | -0.986603599 | 0.013502748 | 0.024021442 | 1 |
| Obesity | t_cell | *SCP2* | 4.3239842 | 9.71E-05 | 0.00042284 | 0.610580533 |
| Obesity | t_cell | *RALGPS2* | 3.5824564 | 0.005227188 | 0.010516265 | 1 |
| Obesity | t_cell | *MRPL37* | 3.364584551 | 0.000435084 | 0.001310043 | 1 |
| Obesity | t_cell | *ICMT* | 5.183310709 | 4.24E-05 | 0.000246282 | 0.26655876 |
| Obesity | t_cell | *QSOX1* | 5.424791656 | 0.000699746 | 0.001910249 | 1 |
| Obesity | t_cell | *DOCK7* | 3.859011982 | 3.00E-05 | 0.000201262 | 0.188783292 |
| Obesity | t_cell | *C1orf21* | 2.222359154 | 0.002731333 | 0.006014673 | 1 |
| Obesity | t_cell | *LEPR* | 4.230562004 | 0.000183415 | 0.000688848 | 1 |
| Obesity | t_cell | *MFN2* | 5.841683415 | 0.000272654 | 0.000926582 | 1 |
| Obesity | t_cell | *MIIP* | 3.918530889 | 0.030895171 | 0.049306962 | 1 |
| Obesity | t_cell | *NCF2* | 5.732740162 | 2.32E-05 | 0.000175508 | 0.146092245 |
| Obesity | t_cell | *RGS2* | 6.861994359 | 0.000690841 | 0.001891688 | 1 |
| Obesity | t_cell | *OLFML3* | 6.478854683 | 0.000125945 | 0.000514166 | 0.791814983 |
| Obesity | t_cell | *TNNI3K* | 2.544198898 | 0.002168616 | 0.004927627 | 1 |

| | | | | | | |
|---|---|---|---|---|---|---|
| Obesity | t_cell | *CRYZ* | 2.402831469 | 0.002674564 | 0.005906211 | 1 |
| Obesity | t_cell | *WARS2* | 3.406965479 | 9.43E-05 | 0.000414398 | 0.59262101 |
| Obesity | t_cell | *OSCP1* | 2.432865889 | 0.00010647 | 0.000455356 | 0.669373961 |
| Obesity | t_cell | *GNPAT* | 4.129234 | 0.001625563 | 0.003836303 | 1 |
| Obesity | t_cell | *C1orf109* | 6.066026795 | 5.80E-07 | 2.83E-05 | 0.003648805 |
| Obesity | t_cell | *TBCE* | 3.455052903 | 0.021124798 | 0.035350441 | 1 |
| Obesity | t_cell | *MTR* | 3.11179723 | 0.015824514 | 0.027608042 | 1 |
| Obesity | t_cell | *KCNQ4* | 1.120272697 | 0.016854624 | 0.029095283 | 1 |
| Obesity | t_cell | *ACADM* | 4.355597122 | 0.000964088 | 0.002478012 | 1 |
| Obesity | t_cell | *PADI2* | 7.276025112 | 0.016359113 | 0.028348882 | 1 |
| Obesity | t_cell | *MFAP2* | -1.8889553 | 0.006726381 | 0.013140127 | 1 |
| Obesity | t_cell | *GBP3* | -0.751095334 | 0.017072108 | 0.029422243 | 1 |
| Obesity | t_cell | *CD46* | 3.952752089 | 5.21E-05 | 0.000274115 | 0.327293443 |
| Obesity | t_cell | *PRDX1* | 4.350928342 | 0.004369721 | 0.008969126 | 1 |
| Obesity | t_cell | *FAAH* | 0.82917616 | 0.003048612 | 0.006591 | 1 |
| Obesity | t_cell | *TMED5* | 4.357834129 | 0.003157572 | 0.006805295 | 1 |
| Obesity | t_cell | *CNN3* | 3.784342306 | 1.15E-05 | 0.000145864 | 0.072294702 |
| Obesity | t_cell | *ABCD3* | 4.910058443 | 0.000565363 | 0.001609623 | 1 |
| Obesity | t_cell | *VAMP4* | 3.741566909 | 0.005609363 | 0.011174292 | 1 |
| Obesity | t_cell | *PTBP2* | 3.459329623 | 4.49E-05 | 0.00025278 | 0.282102447 |
| Obesity | t_cell | *TNFSF4* | -4.151053577 | 0.005601988 | 0.011163138 | 1 |
| Obesity | t_cell | *IRF6* | 1.495272208 | 0.004227826 | 0.008714867 | 1 |
| Obesity | t_cell | *SERPINC1* | 4.834742174 | 1.15E-13 | 6.03E-11 | 7.24E-10 |
| Obesity | t_cell | *RCAN3* | -0.602904745 | 0.000417015 | 0.001266559 | 1 |
| Obesity | t_cell | *RCOR3* | 3.612005964 | 0.00746502 | 0.01438326 | 1 |
| Obesity | t_cell | *MTFR1L* | 5.747128941 | 0.000130022 | 0.000527045 | 0.817447105 |
| Obesity | t_cell | *SLC5A9* | 2.724123461 | 1.69E-05 | 0.000156669 | 0.106465976 |
| Obesity | t_cell | *OSBPL9* | 4.095228071 | 0.000378391 | 0.001174206 | 1 |
| Obesity | t_cell | *TXNDC12* | 5.66677669 | 5.31E-05 | 0.000278336 | 0.334024815 |
| Obesity | t_cell | *COLEC11* | -3.655314145 | 0.006677583 | 0.013054093 | 1 |
| Obesity | t_cell | *IFT46* | 4.625887089 | 6.13E-06 | 0.00011398 | 0.038525304 |
| Obesity | t_cell | *CAMSAP2* | 3.561310449 | 3.48E-05 | 0.000218022 | 0.218711383 |
| Obesity | t_cell | *CASC1* | 2.208541321 | 0.005872376 | 0.011639227 | 1 |
| Obesity | t_cell | *SPCS2* | 4.182092285 | 0.008201891 | 0.015630582 | 1 |
| Obesity | t_cell | *FILIP1* | 2.720381769 | 0.001290314 | 0.00316883 | 1 |
| Obesity | t_cell | *SGIP1* | 4.48401265 | 6.05E-06 | 0.000113242 | 0.038049202 |
| Obesity | t_cell | *TCF21* | -12.04563407 | 0.000560238 | 0.001597376 | 1 |
| Obesity | t_cell | *VAMP8* | 4.856130681 | 9.21E-05 | 0.000407599 | 0.579198086 |
| Obesity | t_cell | *FOXO3* | 3.431001795 | 0.003301567 | 0.007077038 | 1 |
| Obesity | t_cell | *ABCG2* | 7.854888145 | 0.002563293 | 0.005694496 | 1 |
| Obesity | t_cell | *FAM47E-STBD* | 1.422871311 | 0.000641559 | 0.001778432 | 1 |
| Obesity | t_cell | *MFSD1* | 4.470532327 | 0.002740768 | 0.006029114 | 1 |
| Obesity | t_cell | *RAB3GAP2* | 3.594726106 | 0.007807859 | 0.014956737 | 1 |
| Obesity | t_cell | *UBN1* | 2.731156738 | 0.019852255 | 0.033538163 | 1 |
| Obesity | t_cell | *UCHL3* | 6.524353329 | 1.61E-05 | 0.000154104 | 0.101086876 |
| Obesity | t_cell | *WDR35* | 4.561258996 | 0.008245845 | 0.015704825 | 1 |
| Obesity | t_cell | *GDA* | 4.483539171 | 0.001899988 | 0.004394859 | 1 |
| Obesity | t_cell | *CPSF3* | 4.597774215 | 0.013942602 | 0.024727249 | 1 |
| Obesity | t_cell | *PIGZ* | 1.247723617 | 0.002762653 | 0.006070885 | 1 |
| Obesity | t_cell | *SENP5* | 3.268974296 | 0.016915291 | 0.029191995 | 1 |

| | | | | | | |
|---|---|---|---|---|---|---|
| Obesity | t_cell | *CCDC92* | 6.900894161 | 2.58E-14 | 1.62E-11 | 1.62E-10 |
| Obesity | t_cell | *FKBP15* | 5.120285444 | 0.000251397 | 0.000872369 | 1 |
| Obesity | t_cell | *CTNNAL1* | 1.647391226 | 0.000156232 | 0.000604451 | 0.98223237 |
| Obesity | t_cell | *WDR34* | 2.46247382 | 0.005668473 | 0.011277749 | 1 |
| Obesity | t_cell | *TRIM32* | 9.494058987 | 2.15E-06 | 6.57E-05 | 0.013540923 |
| Obesity | t_cell | *PHF19* | -2.082050993 | 0.000366665 | 0.001147429 | 1 |
| Obesity | t_cell | *BSPRY* | -4.211132172 | 0.000202462 | 0.000738329 | 1 |
| Obesity | t_cell | *PPP6C* | 4.638053982 | 0.001266864 | 0.003126841 | 1 |
| Obesity | t_cell | *HSDL2* | 1.673666654 | 0.00216872 | 0.004927627 | 1 |
| Obesity | t_cell | *INVS* | 4.916971996 | 2.14E-05 | 0.000169226 | 0.1346626 |
| Obesity | t_cell | *KDSR* | 5.326501077 | 0.000508621 | 0.001474976 | 1 |
| Obesity | t_cell | *DCAF4* | 6.109675722 | 1.50E-07 | 9.91E-06 | 0.000941713 |
| Obesity | t_cell | *IFI27L2* | 2.460961403 | 0.012535792 | 0.022569452 | 1 |
| Obesity | t_cell | *DNAL1* | 3.384353147 | 3.66E-05 | 0.000225719 | 0.230233604 |
| Obesity | t_cell | *IRF2BPL* | 4.594464398 | 0.000425534 | 0.001284982 | 1 |
| Obesity | t_cell | *TTLL5* | 4.404716979 | 0.001385608 | 0.003359552 | 1 |
| Obesity | t_cell | *FLVCR2* | 5.599718325 | 9.55E-05 | 0.000418298 | 0.600257356 |
| Obesity | t_cell | *PPP4R4* | -2.258455869 | 0.000571539 | 0.001620999 | 1 |
| Obesity | t_cell | *ZC2HC1C* | 3.980900323 | 7.76E-06 | 0.000130812 | 0.048792822 |
| Obesity | t_cell | *ALDH6A1* | 1.947979187 | 0.001389606 | 0.003365351 | 1 |
| Obesity | t_cell | *COQ6* | 6.050244517 | 0.000192176 | 0.000714071 | 1 |
| Obesity | t_cell | *GPR75* | 20.23006326 | 0.000162196 | 0.000621783 | 1 |
| Obesity | t_cell | *KLHL29* | 3.076341845 | 1.62E-05 | 0.000154555 | 0.102160669 |
| Obesity | t_cell | *FKBP1B* | 1.844570161 | 0.007846824 | 0.01501765 | 1 |
| Obesity | t_cell | *ATL2* | 4.288717199 | 0.0003963 | 0.001216571 | 1 |
| Obesity | t_cell | *CNRIP1* | 2.519902436 | 0.014136494 | 0.02499329 | 1 |
| Obesity | t_cell | *SLC17A5* | 2.588042787 | 0.010270326 | 0.019047062 | 1 |
| Obesity | t_cell | *OGFRL1* | 3.089649448 | 0.001321138 | 0.003228136 | 1 |
| Obesity | t_cell | *CUTC* | 4.124106399 | 0.000443207 | 0.001326877 | 1 |
| Obesity | t_cell | *PYROXD2* | 1.319015894 | 0.000584655 | 0.001652754 | 1 |
| Obesity | t_cell | *C10orf88* | 6.38688326 | 4.07E-05 | 0.000239751 | 0.255574707 |
| Obesity | t_cell | *TCTN3* | 4.25013737 | 0.00585656 | 0.011611541 | 1 |
| Obesity | t_cell | *AVPI1* | 3.018620113 | 3.35E-05 | 0.000213631 | 0.210426634 |
| Obesity | t_cell | *WDR11* | 4.220056138 | 0.000481822 | 0.001416183 | 1 |
| Obesity | t_cell | *C10orf76* | 4.557851853 | 0.000307825 | 0.001014839 | 1 |
| Obesity | t_cell | *KCNIP2* | 1.423407316 | 0.000110739 | 0.000468834 | 0.696217763 |
| Obesity | t_cell | *HOXB5* | 3.021110139 | 1.15E-07 | 8.28E-06 | 0.000722006 |
| Obesity | t_cell | *HOXB3* | 3.734063621 | 1.20E-09 | 1.83E-07 | 7.52E-06 |
| Obesity | t_cell | *CAAP1* | 3.181910069 | 0.02305731 | 0.03806757 | 1 |
| Obesity | t_cell | *LRP11* | 2.437748708 | 0.009246322 | 0.017365722 | 1 |
| Obesity | t_cell | *CCDC170* | 1.5733421 | 0.000324816 | 0.001054267 | 1 |
| Obesity | t_cell | *CYSTM1* | 4.546209155 | 9.36E-06 | 0.000139159 | 0.058864234 |
| Obesity | t_cell | *PCDHB14* | 3.1541809 | 2.59E-05 | 0.000186717 | 0.162607625 |
| Obesity | t_cell | *GORAB* | 4.433819005 | 0.014841337 | 0.026121917 | 1 |
| Obesity | t_cell | *ACAT2* | 5.21760953 | 0.000354917 | 0.001124112 | 1 |
| Obesity | t_cell | *MRC1* | 2.888176825 | 0.000407129 | 0.001242535 | 1 |
| Obesity | t_cell | *IQSEC3* | 4.829717028 | 9.07E-06 | 0.000137059 | 0.057016569 |
| Obesity | t_cell | *ENOX1* | 3.493780397 | 1.17E-05 | 0.000145864 | 0.073493824 |
| Obesity | t_cell | *DNAJC15* | 5.205093478 | 0.000621268 | 0.00173442 | 1 |
| Obesity | t_cell | *WBP4* | 4.135884296 | 6.08E-05 | 0.000303699 | 0.382356577 |

| | | | | | | |
|---|---|---|---|---|---|---|
| Obesity | t_cell | *SMAD9* | 5.226085765 | 1.09E-05 | 0.000145864 | 0.068814927 |
| Obesity | t_cell | *SIL1* | 3.608970607 | 0.000629876 | 0.001753001 | 1 |
| Obesity | t_cell | *ZFP30* | 2.8481628 | 0.000710501 | 0.001932058 | 1 |
| Obesity | t_cell | *ARL1* | 3.373434778 | 0.004013561 | 0.008341573 | 1 |
| Obesity | t_cell | *APAF1* | 3.427738314 | 0.011700399 | 0.02131568 | 1 |
| Obesity | t_cell | *ADRA1A* | 0.943896645 | 7.52E-07 | 3.39E-05 | 0.004725448 |
| Obesity | t_cell | *EPHX2* | 1.642019481 | 0.029865962 | 0.047838803 | 1 |
| Obesity | t_cell | *ZNF706* | 3.77694599 | 0.002847692 | 0.006222139 | 1 |
| Obesity | t_cell | *CRISPLD1* | 9.617620104 | 0.002075426 | 0.004753444 | 1 |
| Obesity | t_cell | *RDH10* | 1.898436728 | 0.00025493 | 0.000881598 | 1 |
| Obesity | t_cell | *SCPEP1* | 1.487680943 | 0.000354072 | 0.001122566 | 1 |
| Obesity | t_cell | *TEX14* | 4.910936461 | 0.000810432 | 0.002154413 | 1 |
| Obesity | t_cell | *TRIM6* | 2.660986272 | 3.11E-05 | 0.000206427 | 0.195692358 |
| Obesity | t_cell | *CEP89* | 1.311807802 | 0.027717364 | 0.044796675 | 1 |
| Obesity | t_cell | *PRB2* | 4.470890969 | 1.88E-05 | 0.000160637 | 0.11798772 |
| Obesity | t_cell | *ZSCAN18* | 5.759729218 | 5.50E-05 | 0.000284705 | 0.345632222 |
| Obesity | t_cell | *PDZRN3* | 2.23691535 | 1.14E-05 | 0.000145864 | 0.071745263 |
| Obesity | t_cell | *LHX4* | 4.101871095 | 0.005733996 | 0.011393689 | 1 |
| Obesity | t_cell | *CSTA* | 1.960922053 | 1.91E-05 | 0.000160838 | 0.119868117 |
| Obesity | t_cell | *POPDC2* | 8.261076875 | 9.14E-05 | 0.000405946 | 0.574819463 |
| Obesity | t_cell | *B4GALT4* | 7.105493143 | 9.53E-05 | 0.000418071 | 0.599095921 |
| Obesity | t_cell | *CD80* | 11.10906317 | 2.06E-05 | 0.000166806 | 0.129677495 |
| Obesity | t_cell | *CAT* | 1.309081121 | 0.00175258 | 0.00409093 | 1 |
| Obesity | t_cell | *GJB6* | -28.0571814 | 0.027955086 | 0.045088154 | 1 |
| Obesity | t_cell | *RNF115* | 3.05179671 | 0.025593956 | 0.041772897 | 1 |
| Obesity | t_cell | *TNFSF10* | 4.745417014 | 0.000695568 | 0.001902147 | 1 |
| Obesity | t_cell | *LIAS* | 5.31795961 | 0.000214081 | 0.00077042 | 1 |
| Obesity | t_cell | *CLCC1* | 5.184329531 | 0.000523968 | 0.001509901 | 1 |
| Obesity | t_cell | *GPSM2* | 5.349016388 | 9.85E-07 | 4.05E-05 | 0.006195337 |
| Obesity | t_cell | *CXCR4* | -0.615178952 | 0.000108028 | 0.000460144 | 0.679172198 |
| Obesity | t_cell | *ACVR2A* | 3.138775323 | 0.023403829 | 0.038589005 | 1 |
| Obesity | t_cell | *POLK* | 3.196729056 | 0.012531954 | 0.022569004 | 1 |
| Obesity | t_cell | *FLT3* | 2.079881374 | 4.92E-05 | 0.000266107 | 0.309201485 |
| Obesity | t_cell | *MTIF3* | 3.758142803 | 0.002789845 | 0.006122079 | 1 |
| Obesity | t_cell | *RASL11A* | 4.776831332 | 0.005486813 | 0.010968393 | 1 |
| Obesity | t_cell | *FMOD* | 4.740484897 | 6.89E-05 | 0.000331264 | 0.432962605 |
| Obesity | t_cell | *SERAC1* | 6.124683967 | 0.000597391 | 0.001680447 | 1 |
| Obesity | t_cell | *OPN4* | 5.934781687 | 0.003765433 | 0.007904267 | 1 |
| Obesity | t_cell | *FAM213A* | 2.783230794 | 6.24E-06 | 0.000114728 | 0.039232268 |
| Obesity | t_cell | *SPATA1* | 4.817377967 | 2.91E-05 | 0.000199016 | 0.182696881 |
| Obesity | t_cell | *RWDD3* | 4.876856822 | 0.006688243 | 0.013070869 | 1 |
| Obesity | t_cell | *CCDC18* | 4.318049859 | 0.000225838 | 0.000801072 | 1 |
| Obesity | t_cell | *PMS2* | 7.10083096 | 4.71E-05 | 0.000260943 | 0.296067692 |
| Obesity | t_cell | *KIAA0087* | 4.425903772 | 0.003194477 | 0.006875616 | 1 |
| Obesity | t_cell | *KLHL7* | 4.71310773 | 0.002527305 | 0.005620504 | 1 |
| Obesity | t_cell | *FAM126A* | 3.937624289 | 5.70E-05 | 0.000291014 | 0.358238311 |
| Obesity | t_cell | *FKBP9* | 3.154436427 | 3.98E-05 | 0.000236184 | 0.249919819 |
| Obesity | t_cell | *CCZ1* | 2.680209827 | 0.020163625 | 0.033958937 | 1 |
| Obesity | t_cell | *SMU1* | 4.910063499 | 0.003216361 | 0.006917983 | 1 |
| Obesity | t_cell | *SLC25A51* | 1.362999229 | 0.000877931 | 0.002295986 | 1 |

| | | | | | | |
|---|---|---|---|---|---|---|
| Obesity | t_cell | *DCAF10* | 3.913529782 | 0.002368104 | 0.005311549 | 1 |
| Obesity | t_cell | *BICC1* | 2.149876256 | 2.93E-05 | 0.000200055 | 0.18425066 |
| Obesity | t_cell | *SLC25A16* | 2.465042754 | 5.18E-05 | 0.000273498 | 0.325848557 |
| Obesity | t_cell | *ZWINT* | 2.089228814 | 0.004106284 | 0.008508967 | 1 |
| Obesity | t_cell | *ACADS* | 4.637788348 | 0.000308584 | 0.001016779 | 1 |
| Obesity | t_cell | *SSPN* | 2.510063994 | 0.000342337 | 0.001095303 | 1 |
| Obesity | t_cell | *EBPL* | 2.122221024 | 0.006454136 | 0.012660577 | 1 |
| Obesity | t_cell | *ATP7B* | 0.995952649 | 0.000318391 | 0.00103824 | 1 |
| Obesity | t_cell | *NLN* | 2.055120062 | 0.00380922 | 0.007982855 | 1 |
| Obesity | t_cell | *CENPK* | -0.604423497 | 0.010648658 | 0.019655935 | 1 |
| Obesity | t_cell | *TSFM* | 3.507719941 | 0.00195113 | 0.004501561 | 1 |
| Obesity | t_cell | *ARHGAP9* | -1.535882788 | 7.69E-05 | 0.000360145 | 0.483314577 |
| Obesity | t_cell | *SPATS2* | 3.325780485 | 0.007592759 | 0.014598067 | 1 |
| Obesity | t_cell | *ORMDL2* | -2.922400828 | 0.011820741 | 0.021497542 | 1 |
| Obesity | t_cell | *PDE1B* | 4.410342753 | 8.19E-05 | 0.00037651 | 0.514954361 |
| Obesity | t_cell | *SMUG1* | 1.254713614 | 0.0008328 | 0.002202958 | 1 |
| Obesity | t_cell | *SARDH* | -5.444119738 | 0.001422396 | 0.003432224 | 1 |
| Obesity | t_cell | *STIL* | 4.18001225 | 5.01E-05 | 0.000267593 | 0.314689643 |
| Obesity | t_cell | *TTC21B* | 4.556103132 | 0.000403502 | 0.001232979 | 1 |
| Obesity | t_cell | *ACVR1C* | 1.753346165 | 0.00019836 | 0.000728524 | 1 |
| Obesity | t_cell | *SLC36A1* | 1.480225621 | 0.005251964 | 0.010559353 | 1 |
| Obesity | t_cell | *PLA2G12A* | 5.479695359 | 0.001153735 | 0.002881865 | 1 |
| Obesity | t_cell | *CHPF* | 1.391410642 | 0.015530438 | 0.02719016 | 1 |
| Obesity | t_cell | *DNPEP* | 6.924855403 | 0.000377438 | 0.001173323 | 1 |
| Obesity | t_cell | *MOGAT1* | 1.703032112 | 7.85E-05 | 0.000365978 | 0.493704385 |
| Obesity | t_cell | *FAM124B* | 1.879087142 | 0.019038101 | 0.032331858 | 1 |
| Obesity | t_cell | *FAM210B* | 2.930709887 | 6.52E-05 | 0.000318788 | 0.409642754 |
| Obesity | t_cell | *SNX21* | 3.058377386 | 1.18E-05 | 0.000145864 | 0.074282795 |
| Obesity | t_cell | *SDC4* | 5.97140909 | 0.00320724 | 0.006900724 | 1 |
| Obesity | t_cell | *ARFGEF2* | 4.109417036 | 0.003017931 | 0.006538157 | 1 |
| Obesity | t_cell | *CDH26* | 7.978681813 | 3.43E-05 | 0.000215812 | 0.215595829 |
| Obesity | t_cell | *STX16* | 4.053958405 | 0.012074779 | 0.02190252 | 1 |
| Obesity | t_cell | *RNF114* | 3.415708684 | 0.021298647 | 0.035575079 | 1 |
| Obesity | t_cell | *KCNK15* | 4.140619991 | 0.000170594 | 0.000648049 | 1 |
| Obesity | t_cell | *NEURL2* | 2.275324109 | 1.75E-06 | 6.00E-05 | 0.011031985 |
| Obesity | t_cell | *PEPD* | 4.138703848 | 3.23E-06 | 8.53E-05 | 0.020304058 |
| Obesity | t_cell | *CHST8* | 9.191541606 | 1.18E-07 | 8.28E-06 | 0.000744905 |
| Obesity | t_cell | *STAMBP* | 4.050755025 | 0.013137451 | 0.023484548 | 1 |
| Obesity | t_cell | *USP22* | 4.194412499 | 0.000486734 | 0.001426618 | 1 |
| Obesity | t_cell | *HIF3A* | 3.090852633 | 2.08E-06 | 6.49E-05 | 0.013101526 |
| Obesity | t_cell | *ZNF45* | 6.690165015 | 0.000227208 | 0.000804311 | 1 |
| Obesity | t_cell | *MRS2* | 3.117466935 | 0.00114042 | 0.00285878 | 1 |
| Obesity | t_cell | *SNRPC* | -7.514869346 | 4.38E-18 | 3.06E-15 | 2.75E-14 |
| Obesity | t_cell | *SERPINB6* | 5.159425661 | 6.32E-05 | 0.000312305 | 0.397251445 |
| Obesity | t_cell | *HIST1H1D* | -1.103942023 | 0.001174194 | 0.002928104 | 1 |
| Obesity | t_cell | *NQO2* | 4.059473065 | 1.37E-05 | 0.000147621 | 0.086210424 |
| Obesity | t_cell | *ZNF391* | 0.875114657 | 0.000743228 | 0.002002002 | 1 |
| Obesity | t_cell | *RPS10* | -3.680384413 | 0.002918797 | 0.006358447 | 1 |
| Obesity | t_cell | *HIST1H2BJ* | 6.615909094 | 4.15E-13 | 2.01E-10 | 2.61E-09 |
| Obesity | t_cell | *MED20* | 3.248709721 | 0.00229832 | 0.005179045 | 1 |

| | | | | | | |
|---|---|---|---|---|---|---|
| Obesity | t_cell | *KLHDC3* | 5.691703663 | 0.000626178 | 0.001744986 | 1 |
| Obesity | t_cell | *KLHL31* | 1.438311313 | 1.50E-05 | 0.000149261 | 0.094018796 |
| Obesity | t_cell | *COL21A1* | 2.165862861 | 0.00330567 | 0.007081003 | 1 |
| Obesity | t_cell | *GLO1* | 6.293203705 | 5.35E-06 | 0.000107882 | 0.033659136 |
| Obesity | t_cell | *RPP40* | 2.89724943 | 0.000366775 | 0.001147429 | 1 |
| Obesity | t_cell | *EEF1E1* | 14.96193216 | 4.34E-06 | 9.97E-05 | 0.027264643 |
| Obesity | t_cell | *EREG* | 9.207405305 | 5.05E-05 | 0.000268897 | 0.317567335 |
| Obesity | t_cell | *BBS2* | 3.206057691 | 0.000119401 | 0.00049419 | 0.750674612 |
| Obesity | t_cell | *MT1G* | 15.23165412 | 0.000124472 | 0.000510472 | 0.782553152 |
| Obesity | t_cell | *GRK4* | 1.658397155 | 1.12E-07 | 8.28E-06 | 0.000704686 |
| Obesity | t_cell | *TEKT3* | 2.189216514 | 0.000220769 | 0.000788623 | 1 |
| Obesity | t_cell | *MRPS7* | 6.198690918 | 9.33E-06 | 0.000138985 | 0.058651596 |
| Obesity | t_cell | *NUP85* | 4.017264924 | 0.002296317 | 0.005178244 | 1 |
| Obesity | t_cell | *MIF4GD* | 8.372508255 | 1.71E-05 | 0.000156669 | 0.107444973 |
| Obesity | t_cell | *OPRL1* | 3.756915352 | 1.67E-07 | 1.08E-05 | 0.001052755 |
| Obesity | t_cell | *CHCHD5* | -3.319882501 | 0.0187278 | 0.031925619 | 1 |
| Obesity | t_cell | *PSD4* | -1.063563027 | 0.000361982 | 0.001137892 | 1 |
| Obesity | t_cell | *SLC25A23* | 1.67897979 | 0.005445095 | 0.010898857 | 1 |
| Obesity | t_cell | *SH2D3A* | -1.212785601 | 0.000201413 | 0.000737068 | 1 |
| Obesity | t_cell | *TRIP10* | 3.776043189 | 6.24E-06 | 0.000114728 | 0.039236949 |
| Obesity | t_cell | *RTN2* | 4.372685423 | 1.76E-05 | 0.000157653 | 0.110501255 |
| Obesity | t_cell | *SYMPK* | 4.702019531 | 0.00200738 | 0.004612718 | 1 |
| Obesity | t_cell | *SDCBP2* | 3.177342344 | 6.50E-05 | 0.0003183 | 0.408697149 |
| Obesity | t_cell | *PANK2* | 3.82588277 | 0.017040443 | 0.029375724 | 1 |
| Obesity | t_cell | *RRBP1* | 3.616466164 | 0.00028379 | 0.000954164 | 1 |
| Obesity | t_cell | *BMP2* | 5.384424586 | 7.48E-06 | 0.000128894 | 0.047050547 |
| Obesity | t_cell | *PCSK2* | 18.80416073 | 1.02E-09 | 1.73E-07 | 6.40E-06 |
| Obesity | t_cell | *MKKS* | 5.365348493 | 0.00034904 | 0.001111036 | 1 |
| Obesity | t_cell | *DSTN* | 4.215675343 | 0.002518108 | 0.005603691 | 1 |
| Obesity | t_cell | *TMEM74B* | 4.1016525 | 0.011372707 | 0.020803087 | 1 |
| Obesity | t_cell | *FAM110A* | 8.630785374 | 8.82E-05 | 0.000394869 | 0.554790581 |
| Obesity | t_cell | *NCLN* | 4.445933961 | 0.003941762 | 0.008219522 | 1 |
| Obesity | t_cell | *ZNF436* | 3.042984342 | 3.39E-05 | 0.000214483 | 0.21341071 |
| Obesity | t_cell | *MMP24* | 2.773385679 | 0.023539087 | 0.03878151 | 1 |
| Obesity | t_cell | *UROD* | 6.683783296 | 3.05E-07 | 1.70E-05 | 0.001918962 |
| Obesity | t_cell | *KLC1* | 6.509927689 | 1.25E-06 | 4.81E-05 | 0.007845572 |
| Obesity | t_cell | *XRCC3* | 10.10143307 | 1.05E-09 | 1.73E-07 | 6.59E-06 |
| Obesity | t_cell | *MCF2L* | 2.806004419 | 0.000159623 | 0.000614168 | 1 |
| Obesity | t_cell | *F10* | 4.552997969 | 1.06E-05 | 0.000145864 | 0.066598072 |
| Obesity | t_cell | *PCID2* | 4.1010225 | 0.000945206 | 0.002434458 | 1 |
| Obesity | t_cell | *PROZ* | -12.09848504 | 0.005530721 | 0.011052653 | 1 |
| Obesity | t_cell | *IGFLR1* | -6.597396845 | 0.000470097 | 0.001390824 | 1 |
| Obesity | t_cell | *CAPNS1* | 3.669245148 | 0.012712692 | 0.022842153 | 1 |
| Obesity | t_cell | *COX6B1* | 5.953379119 | 0.002364678 | 0.005305757 | 1 |
| Obesity | t_cell | *PRDX5* | 4.87101952 | 2.15E-05 | 0.000169226 | 0.135218926 |
| Obesity | t_cell | *SCAF1* | 8.18374145 | 4.78E-05 | 0.000262855 | 0.300362871 |
| Obesity | t_cell | *PRR12* | 2.607232367 | 0.010387128 | 0.019235309 | 1 |
| Obesity | t_cell | *TSKS* | 5.201772128 | 1.27E-05 | 0.000146996 | 0.080056573 |
| Obesity | t_cell | *FLRT1* | 2.507755458 | 0.002972793 | 0.006458173 | 1 |
| Obesity | t_cell | *SBDS* | 6.068396555 | 7.40E-05 | 0.000349813 | 0.465251694 |

| | | | | | | |
|---|---|---|---|---|---|---|
| Obesity | t_cell | *BECN1* | 5.44265327 | 0.003806687 | 0.00798236 | 1 |
| Obesity | t_cell | *TRAP1* | 1.331823446 | 0.000269128 | 0.000917077 | 1 |
| Obesity | t_cell | *ZNF384* | 5.883949792 | 0.001933575 | 0.004466035 | 1 |
| Obesity | t_cell | *EMG1* | 4.001100201 | 0.024020603 | 0.039450766 | 1 |
| Obesity | t_cell | *ATG14* | 4.537052541 | 5.32E-05 | 0.000278336 | 0.334281286 |
| Obesity | t_cell | *RHOJ* | 5.079447126 | 2.96E-05 | 0.000200631 | 0.185783856 |
| Obesity | t_cell | *HSPA2* | 4.879879777 | 7.47E-06 | 0.000128894 | 0.046959988 |
| Obesity | t_cell | *PZP* | -0.670045495 | 0.000308899 | 0.001016779 | 1 |
| Obesity | t_cell | *RHOT1* | 3.988740806 | 9.54E-05 | 0.000418224 | 0.599733544 |
| Obesity | t_cell | *WDR60* | 2.159984435 | 0.007088472 | 0.013758945 | 1 |
| Obesity | t_cell | *AIF1L* | 3.617796796 | 9.86E-06 | 0.000142631 | 0.061965935 |
| Obesity | t_cell | *IPPK* | 6.737015778 | 8.76E-06 | 0.000135109 | 0.055096233 |
| Obesity | t_cell | *FGD3* | -0.90117075 | 0.000354051 | 0.001122566 | 1 |
| Obesity | t_cell | *ABHD8* | 3.431783316 | 0.001030313 | 0.002624627 | 1 |
| Obesity | t_cell | *MASP1* | 2.654216677 | 0.002736092 | 0.006020935 | 1 |
| Obesity | t_cell | *ATP13A4* | -2.333186142 | 0.000990341 | 0.002533066 | 1 |
| Obesity | t_cell | *HRASLS* | 3.338556918 | 1.93E-05 | 0.000161262 | 0.121108011 |
| Obesity | t_cell | *HELB* | -0.991952059 | 0.00070815 | 0.001927334 | 1 |
| Obesity | t_cell | *RAB3IP* | -1.696029732 | 0.002391236 | 0.005351976 | 1 |
| Obesity | t_cell | *TAS2R4* | 2.81421794 | 0.001120905 | 0.002816598 | 1 |
| Obesity | t_cell | *IDUA* | 7.990976211 | 3.03E-05 | 0.000202181 | 0.19072646 |
| Obesity | t_cell | *TMEM175* | 3.150405333 | 0.020244265 | 0.034067371 | 1 |
| Obesity | t_cell | *EMC1* | 3.163271336 | 0.006357296 | 0.012490101 | 1 |
| Obesity | t_cell | *PLA2G5* | 1.667052343 | 0.010310445 | 0.019105778 | 1 |
| Obesity | t_cell | *SIN3B* | 5.284605049 | 4.78E-05 | 0.000262855 | 0.300763193 |
| Obesity | t_cell | *WDR24* | 3.191210497 | 0.000231726 | 0.000816168 | 1 |
| Obesity | t_cell | *TICAM1* | 5.549140355 | 4.86E-05 | 0.000265084 | 0.305642369 |
| Obesity | t_cell | *AAMP* | 6.766676207 | 0.000130514 | 0.000528699 | 0.820540893 |
| Obesity | t_cell | *PNKD* | 4.347473349 | 0.000210518 | 0.000760646 | 1 |
| Obesity | t_cell | *HIP1* | 3.77112612 | 0.000847996 | 0.00223443 | 1 |
| Obesity | t_cell | *PTPN12* | 4.032713453 | 7.64E-05 | 0.000358637 | 0.480573604 |
| Obesity | t_cell | *POR* | 4.331466765 | 1.67E-05 | 0.00015608 | 0.105041814 |
| Obesity | t_cell | *FGL2* | 3.474466028 | 0.000468884 | 0.001389195 | 1 |
| Obesity | t_cell | *STYXL1* | 1.630938067 | 0.013847742 | 0.024579547 | 1 |
| Obesity | t_cell | *STEAP4* | 3.336328337 | 0.004481214 | 0.009162078 | 1 |
| Obesity | t_cell | *GNAI1* | 3.067441853 | 1.56E-05 | 0.000152297 | 0.098079384 |
| Obesity | t_cell | *SGCE* | 3.499636921 | 6.29E-05 | 0.000311063 | 0.395361312 |
| Obesity | t_cell | *RBM48* | 5.037080545 | 0.001518474 | 0.003627145 | 1 |
| Obesity | t_cell | *SRD5A3* | 4.950773418 | 2.46E-05 | 0.000181284 | 0.154691255 |
| Obesity | t_cell | *VPREB3* | 12.07709865 | 0.015808699 | 0.027592807 | 1 |
| Obesity | t_cell | *MGAT3* | 2.320240124 | 2.67E-05 | 0.000189777 | 0.16815055 |
| Obesity | t_cell | *ATF4* | 3.807228843 | 0.023821773 | 0.039185632 | 1 |
| Obesity | t_cell | *A4GALT* | 1.635381015 | 0.003998806 | 0.008316404 | 1 |
| Obesity | t_cell | *CDC42EP1* | 3.787078273 | 9.57E-05 | 0.000418843 | 0.601876845 |
| Obesity | t_cell | *BAIAP2L2* | 3.696346499 | 4.49E-05 | 0.00025278 | 0.282090138 |
| Obesity | t_cell | *TST* | 2.160314286 | 0.000235542 | 0.000827755 | 1 |
| Obesity | t_cell | *RIBC2* | 3.287832502 | 4.18E-05 | 0.000244306 | 0.262883298 |
| Obesity | t_cell | *EMC4* | 6.415347059 | 3.67E-05 | 0.000225805 | 0.230547238 |
| Obesity | t_cell | *SPECC1* | 4.441481337 | 0.000224871 | 0.000799188 | 1 |
| Obesity | t_cell | *POT1* | 3.084282293 | 0.018496771 | 0.031586303 | 1 |

| | | | | | | |
|---|---|---|---|---|---|---|
| Obesity | t_cell | *CDHR3* | 2.995636051 | 0.000313141 | 0.001024841 | 1 |
| Obesity | t_cell | *FOXP2* | 2.047217735 | 0.00035625 | 0.001126632 | 1 |
| Obesity | t_cell | *STRIP2* | 2.733712795 | 0.000118946 | 0.000492632 | 0.747814658 |
| Obesity | t_cell | *FLNC* | 2.112013139 | 0.005914222 | 0.011703718 | 1 |
| Obesity | t_cell | *KLHDC10* | 3.862602793 | 0.000608999 | 0.001709155 | 1 |
| Obesity | t_cell | *HOXD1* | 10.10754962 | 0.000102401 | 0.000440652 | 0.643791975 |
| Obesity | t_cell | *HOXD3* | 3.043163289 | 3.29E-05 | 0.000211764 | 0.206893575 |
| Obesity | t_cell | *CHN1* | 2.999260683 | 0.000972945 | 0.002495678 | 1 |
| Obesity | t_cell | *GAD1* | 4.640646132 | 5.17E-05 | 0.000273203 | 0.325111848 |
| Obesity | t_cell | *HOXD9* | 2.046712911 | 0.01247826 | 0.022478745 | 1 |
| Obesity | t_cell | *TWSG1* | 5.316846312 | 0.000633331 | 0.001761058 | 1 |
| Obesity | t_cell | *WDFY4* | 1.640347068 | 0.006967046 | 0.01357775 | 1 |
| Obesity | t_cell | *EIF2AK4* | 3.59790748 | 5.06E-06 | 0.000105254 | 0.031825122 |
| Obesity | t_cell | *IVD* | 2.456893001 | 0.009942729 | 0.018510494 | 1 |
| Obesity | t_cell | *CALML4* | 3.33859872 | 0.000101548 | 0.000438182 | 0.638431331 |
| Obesity | t_cell | *ACKR4* | 4.059308374 | 6.00E-06 | 0.000113017 | 0.037747639 |
| Obesity | t_cell | *RPAIN* | -2.905064799 | 0.020520451 | 0.034485987 | 1 |
| Obesity | t_cell | *PLD2* | 5.70724582 | 2.14E-06 | 6.57E-05 | 0.013467196 |
| Obesity | t_cell | *CCL4* | -0.90144163 | 0.001573516 | 0.003742979 | 1 |
| Obesity | t_cell | *PUS7L* | 2.951601773 | 0.023287029 | 0.038406493 | 1 |
| Obesity | t_cell | *MTUS1* | 3.733483349 | 0.000230543 | 0.000813368 | 1 |
| Obesity | t_cell | *NGDN* | 6.4902252 | 0.000349198 | 0.001111036 | 1 |
| Obesity | t_cell | *ADCY4* | 2.844682801 | 0.002826268 | 0.006186889 | 1 |
| Obesity | t_cell | *RAB2B* | 5.666998284 | 0.000148356 | 0.000583676 | 0.932714374 |
| Obesity | t_cell | *BCL2L2* | 3.98969967 | 1.16E-05 | 0.000145864 | 0.073071523 |
| Obesity | t_cell | *DTD2* | 1.216006497 | 0.020202787 | 0.034006673 | 1 |
| Obesity | t_cell | *PARP2* | 2.811517106 | 0.000202673 | 0.000738669 | 1 |
| Obesity | t_cell | *MIS18BP1* | 2.571402115 | 0.00104008 | 0.002646289 | 1 |
| Obesity | t_cell | *EPB41L4A* | 3.781371721 | 1.44E-05 | 0.000148509 | 0.090655002 |
| Obesity | t_cell | *CDO1* | 3.184500209 | 3.20E-06 | 8.50E-05 | 0.020139843 |
| Obesity | t_cell | *REEP5* | 4.673590759 | 0.000226521 | 0.000802329 | 1 |
| Obesity | t_cell | *QRICH2* | 3.147421684 | 0.016153216 | 0.028084974 | 1 |
| Obesity | t_cell | *SHC2* | 3.455630988 | 1.20E-06 | 4.67E-05 | 0.007523515 |
| Obesity | t_cell | *ERMARD* | 3.33416176 | 0.030247835 | 0.048413478 | 1 |
| Obesity | t_cell | *DOCK6* | 2.48669644 | 0.000900995 | 0.002346544 | 1 |
| Obesity | t_cell | *ECSIT* | 5.436548313 | 0.000729332 | 0.001971329 | 1 |
| Obesity | t_cell | *THEM6* | 1.769147537 | 0.024792914 | 0.040570809 | 1 |
| Obesity | t_cell | *TOMM40* | 4.818993817 | 0.00010764 | 0.000459424 | 0.676732198 |
| Obesity | t_cell | *DPP6* | 3.938864929 | 3.96E-05 | 0.00023583 | 0.248800261 |
| Obesity | t_cell | *RPL36* | -1.680887321 | 0.026481023 | 0.043086489 | 1 |
| Obesity | t_cell | *ATP8B3* | -2.611207481 | 4.98E-06 | 0.000104418 | 0.031286569 |
| Obesity | t_cell | *PLVAP* | 0.666573236 | 0.01409408 | 0.024932325 | 1 |
| Obesity | t_cell | *BST2* | 4.750103546 | 0.01937405 | 0.032804916 | 1 |
| Obesity | t_cell | *MRPL34* | 1.138180557 | 0.027531937 | 0.044534189 | 1 |
| Obesity | t_cell | *RTN4IP1* | 4.789907857 | 0.000485966 | 0.001425696 | 1 |
| Obesity | t_cell | *RSPH3* | 1.400533401 | 0.027170766 | 0.043992431 | 1 |
| Obesity | t_cell | *STK33* | 2.703828383 | 0.000124387 | 0.00051046 | 0.782024111 |
| Obesity | t_cell | *CACNG6* | -4.24915886 | 0.001970674 | 0.004536664 | 1 |
| Obesity | t_cell | *PXDN* | 2.220339905 | 1.33E-05 | 0.000147409 | 0.083753496 |
| Obesity | t_cell | *PGPEP1* | 5.161789722 | 5.70E-07 | 2.82E-05 | 0.00358626 |

| | | | | | | |
|---|---|---|---|---|---|---|
| Obesity | t_cell | *TRPM4* | 3.359684972 | 0.00131115 | 0.003208698 | 1 |
| Obesity | t_cell | *UBAC1* | 4.211424419 | 0.000202197 | 0.000738217 | 1 |
| Obesity | t_cell | *TNNT3* | 2.76498015 | 0.002350526 | 0.005281542 | 1 |
| Obesity | t_cell | *TUBGCP2* | 4.623365511 | 0.000626441 | 0.001744986 | 1 |
| Obesity | t_cell | *PAK4* | 6.934061146 | 0.000128012 | 0.000520913 | 0.8048106 |
| Obesity | t_cell | *MAP3K10* | 4.768399588 | 0.000479463 | 0.001410568 | 1 |
| Obesity | t_cell | *SMPDL3B* | 10.93299908 | 3.98E-05 | 0.000236184 | 0.250118575 |
| Obesity | t_cell | *CLIP1* | 5.505190786 | 1.06E-05 | 0.000145864 | 0.066551998 |
| Obesity | t_cell | *PKDREJ* | -2.384489826 | 0.012804095 | 0.02298818 | 1 |
| Obesity | t_cell | *EPS8L1* | -8.993647885 | 0.000699548 | 0.001910249 | 1 |
| Obesity | t_cell | *LILRB2* | 3.87647156 | 2.10E-05 | 0.000167886 | 0.131800469 |
| Obesity | t_cell | *COX4I2* | 8.803415971 | 0.000672824 | 0.001849603 | 1 |
| Obesity | t_cell | *CHMP1A* | 3.814617729 | 0.002518853 | 0.005603691 | 1 |
| Obesity | t_cell | *PPT1* | 5.001681534 | 0.003885131 | 0.008122986 | 1 |
| Obesity | t_cell | *HAUS8* | 4.787901231 | 0.008357388 | 0.015883585 | 1 |
| Obesity | t_cell | *TBC1D5* | 2.612548864 | 0.015004782 | 0.026372677 | 1 |
| Obesity | t_cell | *RFTN1* | 3.89411542 | 0.000289829 | 0.000971809 | 1 |
| Obesity | t_cell | *PDLIM4* | 4.730838113 | 8.04E-05 | 0.00037203 | 0.505216531 |
| Obesity | t_cell | *AOC3* | 2.615767923 | 0.002375278 | 0.00532384 | 1 |
| Obesity | t_cell | *AOC2* | 4.706206404 | 2.00E-05 | 0.000163758 | 0.125766133 |
| Obesity | t_cell | *DIAPH1* | -1.374105058 | 0.004439247 | 0.0090909 | 1 |
| Obesity | t_cell | *EXOC4* | 2.994023775 | 0.003158558 | 0.006805295 | 1 |
| Obesity | t_cell | *ACAP3* | 4.512264479 | 0.001798478 | 0.004181595 | 1 |
| Obesity | t_cell | *ANO1* | 5.979142283 | 0.002195842 | 0.004980253 | 1 |
| Obesity | t_cell | *PPFIA1* | 3.402549545 | 0.000840282 | 0.002215961 | 1 |
| Obesity | t_cell | *NINJ1* | 7.437051516 | 2.14E-06 | 6.57E-05 | 0.013477727 |
| Obesity | t_cell | *NPHP4* | 5.472330056 | 1.33E-05 | 0.000147409 | 0.083779862 |
| Obesity | t_cell | *CKMT2* | 2.093342987 | 0.01441336 | 0.025425587 | 1 |
| Obesity | t_cell | *ZCCHC9* | 5.229187412 | 0.000367017 | 0.001147429 | 1 |
| Obesity | t_cell | *KHDRBS3* | 2.532760524 | 4.94E-05 | 0.000266732 | 0.310475796 |
| Obesity | t_cell | *ZSCAN5A* | 4.967643449 | 0.000126325 | 0.000515383 | 0.79420592 |
| Obesity | t_cell | *ZNF132* | 2.226322672 | 0.001034781 | 0.002634941 | 1 |
| Obesity | t_cell | *ACTR10* | 3.903944932 | 0.004410426 | 0.009039138 | 1 |
| Obesity | t_cell | *GCH1* | -1.777637875 | 0.008928683 | 0.016826928 | 1 |
| Obesity | t_cell | *LGALS3* | 4.396910211 | 9.05E-06 | 0.000137059 | 0.056897762 |
| Obesity | t_cell | *FBXW9* | 3.347003716 | 0.005913884 | 0.011703718 | 1 |
| Obesity | t_cell | *C19orf57* | 8.290971349 | 4.31E-05 | 0.000247705 | 0.2709891 |
| Obesity | t_cell | *MATN3* | 4.757827662 | 5.88E-05 | 0.000297413 | 0.369684764 |
| Obesity | t_cell | *CCT6B* | 2.744178785 | 0.000153424 | 0.000598373 | 0.964577513 |
| Obesity | t_cell | *ENOSF1* | 2.614394325 | 0.007418323 | 0.014306441 | 1 |
| Obesity | t_cell | *EMILIN2* | 4.726485172 | 9.81E-11 | 2.57E-08 | 6.17E-07 |
| Obesity | t_cell | *CNGA4* | 3.641544926 | 2.72E-05 | 0.000191821 | 0.171296126 |
| Obesity | t_cell | *TRIM22* | -1.863464921 | 0.024964103 | 0.040819068 | 1 |
| Obesity | t_cell | *CARD6* | 5.278936036 | 1.44E-06 | 5.24E-05 | 0.009032371 |
| Obesity | t_cell | *INPP5K* | 2.218904937 | 0.022937807 | 0.037940013 | 1 |
| Obesity | t_cell | *SERPINF1* | 3.293780534 | 0.000157776 | 0.000609671 | 0.991935486 |
| Obesity | t_cell | *REEP2* | -1.499585265 | 0.007493426 | 0.014433569 | 1 |
| Obesity | t_cell | *BTBD3* | 4.004632553 | 3.56E-05 | 0.000221846 | 0.224064573 |
| Obesity | t_cell | *IGHMBP2* | 3.788293983 | 1.02E-07 | 7.73E-06 | 0.000641905 |
| Obesity | t_cell | *MUTYH* | 9.426056203 | 1.42E-05 | 0.000148367 | 0.089086164 |

| | | | | | | |
|---|---|---|---|---|---|---|
| Obesity | t_cell | *LPIN3* | 3.544034543 | 1.25E-05 | 0.000146996 | 0.078299391 |
| Obesity | t_cell | *DMGDH* | 3.665554366 | 8.79E-05 | 0.000394101 | 0.552430793 |
| Obesity | t_cell | *KANK4* | 1.309634688 | 1.11E-05 | 0.000145864 | 0.069697001 |
| Obesity | t_cell | *USPL1* | 4.235578528 | 0.005580163 | 0.011123586 | 1 |
| Obesity | t_cell | *POMP* | 4.439302127 | 0.012421653 | 0.022408876 | 1 |
| Obesity | t_cell | *WASF3* | 2.815265205 | 1.34E-05 | 0.000147409 | 0.084506086 |
| Obesity | t_cell | *PEMT* | 1.874720761 | 0.007076305 | 0.013748062 | 1 |
| Obesity | t_cell | *CHI3L1* | 1.118058617 | 0.000534199 | 0.001536076 | 1 |
| Obesity | t_cell | *DSTYK* | 3.463974293 | 0.010993139 | 0.020220558 | 1 |
| Obesity | t_cell | *TMCC2* | 2.099870167 | 0.02306871 | 0.038076393 | 1 |
| Obesity | t_cell | *TRPC4* | 3.164519477 | 2.82E-05 | 0.000195275 | 0.177309725 |
| Obesity | t_cell | *ZNF414* | 9.314392407 | 9.73E-05 | 0.000423296 | 0.611662394 |
| Obesity | t_cell | *RTN3* | 3.640059793 | 1.19E-05 | 0.000145864 | 0.075030153 |
| Obesity | t_cell | *SLC2A11* | 6.473741443 | 1.66E-05 | 0.000155845 | 0.104572179 |
| Obesity | t_cell | *FAM83F* | 3.71167888 | 4.03E-05 | 0.000238823 | 0.253643567 |
| Obesity | t_cell | *GIMAP4* | -0.831494818 | 0.023670228 | 0.03897714 | 1 |
| Obesity | t_cell | *AGAP3* | 3.549973846 | 0.001054617 | 0.002677449 | 1 |
| Obesity | t_cell | *ACTR3B* | 4.076435648 | 0.016970594 | 0.029271365 | 1 |
| Obesity | t_cell | *SFTPD* | 1.059037435 | 0.000625232 | 0.001743165 | 1 |
| Obesity | t_cell | *DYDC2* | 6.838157902 | 0.000163133 | 0.000623733 | 1 |
| Obesity | t_cell | *TMTC1* | 3.37433822 | 1.68E-05 | 0.000156468 | 0.105615779 |
| Obesity | t_cell | *IPO8* | 4.026069225 | 0.013125969 | 0.023473559 | 1 |
| Obesity | t_cell | *LARS* | -1.246287491 | 0.02934001 | 0.047104352 | 1 |
| Obesity | t_cell | *SPINK5* | 4.140673026 | 0.002764751 | 0.006073371 | 1 |
| Obesity | t_cell | *LRRCC1* | 2.678009294 | 0.010332417 | 0.019139631 | 1 |
| Obesity | t_cell | *SWAP70* | 4.47485347 | 2.13E-05 | 0.000169127 | 0.133756032 |
| Obesity | t_cell | *AMPD3* | 2.740351298 | 0.016303267 | 0.028283288 | 1 |
| Obesity | t_cell | *SBF2* | 3.492612813 | 0.000361774 | 0.001137805 | 1 |
| Obesity | t_cell | *MICAL2* | 2.242957786 | 0.024271595 | 0.039810988 | 1 |
| Obesity | t_cell | *RRAS2* | 1.769726153 | 0.001815822 | 0.004215685 | 1 |
| Obesity | t_cell | *HSD17B4* | 3.358044555 | 0.001618606 | 0.003824674 | 1 |
| Obesity | t_cell | *DPF2* | 4.13012116 | 0.003012714 | 0.006533372 | 1 |
| Obesity | t_cell | *GSC* | 8.146112096 | 2.25E-05 | 0.000173328 | 0.141436045 |
| Obesity | t_cell | *CATSPERB* | -1.331472981 | 0.000631422 | 0.001756527 | 1 |
| Obesity | t_cell | *MED6* | 3.16985285 | 0.001076944 | 0.002724648 | 1 |
| Obesity | t_cell | *EIF2S1* | 4.038652954 | 0.019855105 | 0.033538163 | 1 |
| Obesity | t_cell | *ELP3* | 4.770162161 | 0.001581243 | 0.003757095 | 1 |
| Obesity | t_cell | *MRO* | 2.019016402 | 0.003678936 | 0.007740787 | 1 |
| Obesity | t_cell | *CNTN6* | 1.687557851 | 0.000369688 | 0.001154035 | 1 |
| Obesity | t_cell | *KATNBL1* | 4.843838375 | 9.43E-05 | 0.000414398 | 0.5930034 |
| Obesity | t_cell | *GSTM5* | 2.823813686 | 3.01E-06 | 8.11E-05 | 0.018899016 |
| Obesity | t_cell | *SYT6* | 8.85284495 | 3.16E-05 | 0.000209021 | 0.198779186 |
| Obesity | t_cell | *VAV3* | -1.74716202 | 0.013010713 | 0.023291103 | 1 |
| Obesity | t_cell | *WNT2B* | 3.937155054 | 1.54E-05 | 0.00015118 | 0.096906615 |
| Obesity | t_cell | *CEPT1* | 3.542574609 | 0.002775463 | 0.006092644 | 1 |
| Obesity | t_cell | *SPIRE1* | 2.539367152 | 0.002085828 | 0.004772053 | 1 |
| Obesity | t_cell | *LDHA* | 4.978582186 | 1.45E-05 | 0.000148509 | 0.091050119 |
| Obesity | t_cell | *ANO3* | 1.224241993 | 0.003027526 | 0.006554427 | 1 |
| Obesity | t_cell | *FST* | 4.256309181 | 5.62E-06 | 0.000109386 | 0.035331566 |
| Obesity | t_cell | *TIMM17A* | 3.831226635 | 0.025016052 | 0.040882745 | 1 |

| | | | | | | |
|---|---|---|---|---|---|---|
| Obesity | t_cell | *NARS* | 5.117495606 | 0.003358175 | 0.007170309 | 1 |
| Obesity | t_cell | *FBXO18* | 4.933948869 | 0.000194234 | 0.000720016 | 1 |
| Obesity | t_cell | *ECHDC3* | 1.714324148 | 0.000211057 | 0.000761809 | 1 |
| Obesity | t_cell | *IL15RA* | 5.15984238 | 0.006094701 | 0.012036447 | 1 |
| Obesity | t_cell | *TMEM241* | 3.619389255 | 1.59E-05 | 0.000153107 | 0.099672566 |
| Obesity | t_cell | *EMP1* | 1.885571261 | 0.012595772 | 0.022657973 | 1 |
| Obesity | t_cell | *PRH2* | 3.953652396 | 0.004187064 | 0.008645015 | 1 |
| Obesity | t_cell | *ACP2* | 6.478576624 | 3.29E-09 | 4.40E-07 | 2.07E-05 |
| Obesity | t_cell | *HOOK1* | -1.309213471 | 0.000173352 | 0.000656545 | 1 |
| Obesity | t_cell | *FHOD3* | 1.587276578 | 0.003948411 | 0.008227929 | 1 |
| Obesity | t_cell | *TPGS2* | 4.560950171 | 0.000271681 | 0.000924274 | 1 |
| Obesity | t_cell | *SLC43A3* | 2.69641759 | 0.008303781 | 0.01579125 | 1 |
| Obesity | t_cell | *TIMM10* | 3.881292071 | 0.001179346 | 0.002937619 | 1 |
| Obesity | t_cell | *APLNR* | 3.347561391 | 1.19E-05 | 0.000145864 | 0.074679605 |
| Obesity | t_cell | *FADS2* | 5.103692036 | 1.15E-05 | 0.000145864 | 0.072102019 |
| Obesity | t_cell | *CLOCK* | 4.259461274 | 0.00119833 | 0.002979005 | 1 |
| Obesity | t_cell | *CLDN10* | 2.928282503 | 0.003268475 | 0.007017167 | 1 |
| Obesity | t_cell | *DZIP1* | 2.768207036 | 0.00037965 | 0.001176552 | 1 |
| Obesity | t_cell | *BIVM* | 7.23576981 | 1.89E-05 | 0.000160637 | 0.118782804 |
| Obesity | t_cell | *ERCC5* | 11.0180883 | 8.13E-05 | 0.000375456 | 0.510995998 |
| Obesity | t_cell | *CARS2* | 3.107139104 | 0.002896642 | 0.006314559 | 1 |
| Obesity | t_cell | *KLB* | 1.799323599 | 1.97E-06 | 6.34E-05 | 0.012356953 |
| Obesity | t_cell | *WDR36* | 5.156465796 | 0.000596879 | 0.001679758 | 1 |
| Obesity | t_cell | *UBQLN1* | 3.900798389 | 0.007131129 | 0.013828936 | 1 |
| Obesity | t_cell | *NAA35* | 3.81552136 | 0.00901124 | 0.016957099 | 1 |
| Obesity | t_cell | *CTSL* | 4.164426384 | 1.30E-05 | 0.000147409 | 0.081630717 |
| Obesity | t_cell | *GOLM1* | 2.584571052 | 0.007571682 | 0.014566452 | 1 |
| Obesity | t_cell | *ADAM19* | -0.91757112 | 0.000137535 | 0.000551456 | 0.864682785 |
| Obesity | t_cell | *USP30* | 2.726649695 | 0.000109512 | 0.000464891 | 0.688504198 |
| Obesity | t_cell | *HRK* | 4.181891623 | 0.003769682 | 0.007910544 | 1 |
| Obesity | t_cell | *P2RX4* | 2.899718033 | 0.004054881 | 0.008413544 | 1 |
| Obesity | t_cell | *DTX1* | 1.760513774 | 0.000404686 | 0.001235678 | 1 |
| Obesity | t_cell | *DMTF1* | 2.166966441 | 0.02778199 | 0.044866523 | 1 |
| Obesity | t_cell | *CCDC146* | 3.944542511 | 0.000223663 | 0.000795793 | 1 |
| Obesity | t_cell | *TMEM60* | -3.086005171 | 0.027533704 | 0.044534189 | 1 |
| Obesity | t_cell | *POM121C* | -5.770790649 | 1.42E-05 | 0.000148367 | 0.089434489 |
| Obesity | t_cell | *TES* | -2.029064996 | 0.013457213 | 0.023947211 | 1 |
| Obesity | t_cell | *ANKRD6* | 4.809443536 | 0.000531864 | 0.001530358 | 1 |
| Obesity | t_cell | *NT5E* | 2.841224818 | 0.002160484 | 0.00491602 | 1 |
| Obesity | t_cell | *EPHA7* | 0.991527765 | 0.00021608 | 0.000775841 | 1 |
| Obesity | t_cell | *ITGA7* | 0.838724184 | 0.01433667 | 0.02531161 | 1 |
| Obesity | t_cell | *RDH5* | 1.567432344 | 0.000146029 | 0.000577413 | 0.918087066 |
| Obesity | t_cell | *BLOC1S1* | 3.904260792 | 0.002714385 | 0.005983638 | 1 |
| Obesity | t_cell | *PPP1R1A* | 3.379089326 | 1.35E-05 | 0.000147409 | 0.084907212 |
| Obesity | t_cell | *FAIM2* | 3.064420704 | 4.39E-05 | 0.000250474 | 0.27579734 |
| Obesity | t_cell | *SLC26A10* | 7.76162101 | 0.0011427 | 0.002862214 | 1 |
| Obesity | t_cell | *MAP7* | 2.530299024 | 0.029457557 | 0.047268928 | 1 |
| Obesity | t_cell | *PKIB* | 4.92204414 | 0.001477258 | 0.003544856 | 1 |
| Obesity | t_cell | *MICAL1* | -2.217674997 | 0.016709479 | 0.028876441 | 1 |
| Obesity | t_cell | *PRADC1* | 6.047020142 | 2.47E-06 | 7.12E-05 | 0.015515968 |

| | | | | | | |
|---|---|---|---|---|---|---|
| Obesity | t_cell | *SMYD5* | 4.473048445 | 0.015979547 | 0.027841162 | 1 |
| Obesity | t_cell | *KCNMB4* | 2.988361423 | 0.009589428 | 0.017921741 | 1 |
| Obesity | t_cell | *KLHL36* | 4.752634016 | 0.001435264 | 0.003455958 | 1 |
| Obesity | t_cell | *FHOD1* | 5.751160691 | 0.000314051 | 0.001026805 | 1 |
| Obesity | t_cell | *ZNF670* | 1.960376403 | 0.00773579 | 0.014841291 | 1 |
| Obesity | t_cell | *NTPCR* | 5.657924992 | 6.63E-06 | 0.000119398 | 0.041670076 |
| Obesity | t_cell | *PIGC* | 4.089410577 | 1.22E-05 | 0.000145864 | 0.076794102 |
| Obesity | t_cell | *USP37* | 3.788134478 | 0.007320658 | 0.014139778 | 1 |
| Obesity | t_cell | *SLC19A3* | 0.952446922 | 0.001455947 | 0.003500897 | 1 |
| Obesity | t_cell | *TMBIM1* | 2.271802288 | 0.016175384 | 0.028115742 | 1 |
| Obesity | t_cell | *ARMC9* | 5.491115071 | 8.34E-05 | 0.000381479 | 0.524250975 |
| Obesity | t_cell | *TMEM127* | 4.564981497 | 0.00133418 | 0.003254944 | 1 |
| Obesity | t_cell | *MRPS9* | 3.766646824 | 3.35E-05 | 0.0002138 | 0.21080725 |
| Obesity | t_cell | *APPL2* | 4.223843793 | 0.000879462 | 0.002299033 | 1 |
| Obesity | t_cell | *DRAM1* | 2.986212869 | 0.001644719 | 0.003871339 | 1 |
| Obesity | t_cell | *SLC41A2* | 4.477015639 | 2.62E-05 | 0.000187708 | 0.164994999 |
| Obesity | t_cell | *NEK3* | 2.443096 | 0.000602801 | 0.001694149 | 1 |
| Obesity | t_cell | *VPS36* | 4.176736855 | 0.000281154 | 0.000946769 | 1 |
| Obesity | t_cell | *TBC1D4* | 4.224957986 | 5.73E-05 | 0.000292128 | 0.360486512 |
| Obesity | t_cell | *MED4* | 3.219571184 | 0.016152441 | 0.028084974 | 1 |
| Obesity | t_cell | *RCBTB2* | 4.716692549 | 0.000127196 | 0.000518602 | 0.799683845 |
| Obesity | t_cell | *SPDYE1* | 3.1299036 | 0.001141186 | 0.002859561 | 1 |
| Obesity | t_cell | *GPNMB* | 2.604405633 | 0.023719874 | 0.039048664 | 1 |
| Obesity | t_cell | *KDELR2* | 6.299974819 | 8.15E-05 | 0.000375666 | 0.512693995 |
| Obesity | t_cell | *AOAH* | -1.281922633 | 0.0055574 | 0.011091865 | 1 |
| Obesity | t_cell | *DDX56* | 7.212127369 | 3.21E-05 | 0.000209022 | 0.2017065 |
| Obesity | t_cell | *HUS1* | 3.68995253 | 0.009075755 | 0.017068283 | 1 |
| Obesity | t_cell | *NACAD* | 4.940057208 | 0.000701263 | 0.001913559 | 1 |
| Obesity | t_cell | *DBNL* | 4.205000104 | 0.00271315 | 0.005983015 | 1 |
| Obesity | t_cell | *ALPK3* | 1.916955769 | 5.51E-05 | 0.000285135 | 0.346466187 |
| Obesity | t_cell | *TM6SF1* | 4.769336657 | 1.19E-06 | 4.67E-05 | 0.007472242 |
| Obesity | t_cell | *RSAD1* | 6.739757824 | 4.63E-05 | 0.000257823 | 0.291082519 |
| Obesity | t_cell | *NMT1* | 2.502466472 | 8.46E-05 | 0.000384937 | 0.532087711 |
| Obesity | t_cell | *BRIP1* | -2.398291551 | 0.004914693 | 0.009951264 | 1 |
| Obesity | t_cell | *RTP4* | -2.64919869 | 3.72E-05 | 0.000227538 | 0.233572418 |
| Obesity | t_cell | *BLK* | -2.149339889 | 2.34E-06 | 6.88E-05 | 0.014732737 |
| Obesity | t_cell | *RPS6KC1* | 2.89215649 | 0.022819859 | 0.037774738 | 1 |
| Obesity | t_cell | *GYPC* | 4.044667031 | 0.001328124 | 0.003242686 | 1 |
| Obesity | t_cell | *TXN* | 4.614352707 | 0.000462302 | 0.001372468 | 1 |
| Obesity | t_cell | *ODF2* | 6.549249534 | 0.000277974 | 0.000938539 | 1 |
| Obesity | t_cell | *SLC2A8* | 4.334593309 | 3.46E-05 | 0.000217214 | 0.217431218 |
| Obesity | t_cell | *ZNF189* | 4.093851448 | 0.001752977 | 0.00409093 | 1 |
| Obesity | t_cell | *STX17* | 5.276117141 | 0.000564753 | 0.00160879 | 1 |
| Obesity | t_cell | *PRPF4* | 2.749331902 | 0.004133515 | 0.00854849 | 1 |
| Obesity | t_cell | *FPGS* | 3.596720651 | 0.018507792 | 0.031593399 | 1 |
| Obesity | t_cell | *TEX10* | 4.344665722 | 0.011560553 | 0.02109759 | 1 |
| Obesity | t_cell | *GARNL3* | 4.644735474 | 2.45E-05 | 0.000180484 | 0.153772733 |
| Obesity | t_cell | *GABBR2* | 9.999576017 | 1.12E-05 | 0.000145864 | 0.070674471 |
| Obesity | t_cell | *PSMB7* | 4.328882989 | 0.004506733 | 0.009211258 | 1 |
| Obesity | t_cell | *RABEPK* | 5.69547114 | 0.001084818 | 0.002739056 | 1 |

| | | | | | | |
|---|---|---|---|---|---|---|
| Obesity | t_cell | *GOLGA1* | 4.403790292 | 0.003989376 | 0.00829954 | 1 |
| Obesity | t_cell | *LMX1B* | 8.508922416 | 0.000174454 | 0.000660322 | 1 |
| Obesity | t_cell | *DSCC1* | 1.064974751 | 5.92E-05 | 0.000298485 | 0.37224369 |
| Obesity | t_cell | *NOV* | 4.448268857 | 7.85E-05 | 0.000365978 | 0.493378137 |
| Obesity | t_cell | *APTX* | 3.451929727 | 0.002273329 | 0.005133772 | 1 |
| Obesity | t_cell | *TLN1* | 3.182322941 | 0.022848334 | 0.037801968 | 1 |
| Obesity | t_cell | *SPAG8* | 4.219339463 | 2.23E-05 | 0.000172409 | 0.14016841 |
| Obesity | t_cell | *CD72* | 5.626343634 | 0.000264894 | 0.000905594 | 1 |
| Obesity | t_cell | *TMEM8B* | 5.386828472 | 4.53E-05 | 0.0002542 | 0.284759123 |
| Obesity | t_cell | *GRHPR* | 2.758659882 | 0.00070439 | 0.001921258 | 1 |
| Obesity | t_cell | *ARHGEF39* | 1.62681989 | 0.003843977 | 0.00804229 | 1 |
| Obesity | t_cell | *DENND4C* | 3.424089239 | 0.003093742 | 0.006677088 | 1 |
| Obesity | t_cell | *PPIL1* | -3.112100208 | 0.001058264 | 0.002681704 | 1 |
| Obesity | t_cell | *KIF13A* | 3.333119861 | 1.85E-05 | 0.000159754 | 0.116141408 |
| Obesity | t_cell | *ZSCAN9* | 4.638814146 | 1.27E-10 | 2.96E-08 | 7.99E-07 |
| Obesity | t_cell | *FRS3* | 5.534355707 | 1.04E-05 | 0.000145864 | 0.065353922 |
| Obesity | t_cell | *SLC22A23* | 4.48431036 | 0.000311514 | 0.001020577 | 1 |
| Obesity | t_cell | *LRRC1* | 3.632701417 | 0.000340102 | 0.001091101 | 1 |
| Obesity | t_cell | *BPHL* | 0.996054023 | 0.021292166 | 0.035573704 | 1 |
| Obesity | t_cell | *RIPK1* | 4.05842575 | 0.006170473 | 0.012149629 | 1 |
| Obesity | t_cell | *UQCC2* | 4.506739761 | 0.000186798 | 0.000698215 | 1 |
| Obesity | t_cell | *TCF19* | 6.592939513 | 4.29E-08 | 3.80E-06 | 0.00026985 |
| Obesity | t_cell | *FLOT1* | 5.556100752 | 1.36E-06 | 5.11E-05 | 0.008537705 |
| Obesity | t_cell | *FGFBP2* | -3.082073927 | 0.000226038 | 0.000801072 | 1 |
| Obesity | t_cell | *FHDC1* | 3.648022938 | 1.79E-05 | 0.000158305 | 0.112238315 |
| Obesity | t_cell | *MYO7A* | 1.422768643 | 0.008504687 | 0.016124537 | 1 |
| Obesity | t_cell | *ARRB1* | 4.219042937 | 6.56E-05 | 0.000320472 | 0.412127119 |
| Obesity | t_cell | *CCDC90B* | 4.928520406 | 0.002035487 | 0.004668774 | 1 |
| Obesity | t_cell | *SYTL2* | -1.538506441 | 0.000461481 | 0.001372468 | 1 |
| Obesity | t_cell | *RAB30* | 3.809710374 | 5.76E-05 | 0.000293011 | 0.361868536 |
| Obesity | t_cell | *PRCP* | 5.299605787 | 2.93E-05 | 0.00020007 | 0.184464803 |
| Obesity | t_cell | *NARS2* | 3.788683124 | 0.014214372 | 0.025116852 | 1 |
| Obesity | t_cell | *PI15* | 9.868709459 | 3.42E-05 | 0.000215393 | 0.214747195 |
| Obesity | t_cell | *SULF1* | 3.391909774 | 1.05E-05 | 0.000145864 | 0.066024916 |
| Obesity | t_cell | *NEK1* | 3.64872162 | 0.002641922 | 0.005844393 | 1 |
| Obesity | t_cell | *DCUN1D5* | 3.1309831 | 1.27E-05 | 0.000146996 | 0.079635883 |
| Obesity | t_cell | *SLC37A4* | 6.681286818 | 0.03104831 | 0.049505637 | 1 |
| Obesity | t_cell | *PPP2R1B* | 2.396347626 | 1.27E-05 | 0.000146996 | 0.080051305 |
| Obesity | t_cell | *FDX1* | 5.360595333 | 4.79E-05 | 0.000262855 | 0.30091402 |
| Obesity | t_cell | *FXYD6* | 4.530852955 | 2.31E-05 | 0.000175493 | 0.145177391 |
| Obesity | t_cell | *TMPRSS13* | -3.249391516 | 0.000246215 | 0.000856642 | 1 |
| Obesity | t_cell | *CASP1* | -3.061303582 | 0.006727862 | 0.013140127 | 1 |
| Obesity | t_cell | *MAP2K5* | 5.010096127 | 4.12E-05 | 0.000242079 | 0.258998867 |
| Obesity | t_cell | *NDUFAF1* | 2.465912382 | 0.027747869 | 0.044834452 | 1 |
| Obesity | t_cell | *ITGA11* | 3.237393271 | 6.36E-05 | 0.000313573 | 0.39980614 |
| Obesity | t_cell | *PAQR5* | 2.635260613 | 6.47E-05 | 0.000318079 | 0.406843537 |
| Obesity | t_cell | *RMDN3* | 4.767404496 | 5.16E-06 | 0.000105675 | 0.032442195 |
| Obesity | t_cell | *UACA* | 3.792856791 | 1.48E-05 | 0.000149118 | 0.092791996 |
| Obesity | t_cell | *SLC44A5* | 4.643994808 | 5.51E-07 | 2.75E-05 | 0.003463758 |
| Obesity | t_cell | *DNASE2B* | 7.89327783 | 0.000202009 | 0.000737962 | 1 |

| | | | | | | |
|---|---|---|---|---|---|---|
| Obesity | t_cell | *DBT* | 3.35915495 | 0.005580371 | 0.011123586 | 1 |
| Obesity | t_cell | *IFT172* | 5.445247057 | 1.77E-08 | 1.89E-06 | 0.000111379 |
| Obesity | t_cell | *KHK* | -0.790649818 | 0.01912396 | 0.032442616 | 1 |
| Obesity | t_cell | *DYNC2LI1* | 2.579494456 | 4.40E-05 | 0.000250553 | 0.276359515 |
| Obesity | t_cell | *LHCGR* | 1.416422637 | 9.24E-05 | 0.000408268 | 0.581160471 |
| Obesity | t_cell | *THUMPD2* | 4.689751294 | 3.61E-05 | 0.000223656 | 0.227010409 |
| Obesity | t_cell | *ABCG5* | 2.949029098 | 1.07E-05 | 0.000145864 | 0.067051922 |
| Obesity | t_cell | *ATRAID* | 4.546483599 | 0.001412167 | 0.003413416 | 1 |
| Obesity | t_cell | *CENPO* | 9.801833661 | 1.86E-05 | 0.000160344 | 0.116730409 |
| Obesity | t_cell | *CYP2C9* | 1.211662067 | 0.010797419 | 0.019901311 | 1 |
| Obesity | t_cell | *CYP2C8* | 2.93873633 | 0.000751194 | 0.002019998 | 1 |
| Obesity | t_cell | *CH25H* | 1.504400014 | 0.019658755 | 0.033233287 | 1 |
| Obesity | t_cell | *BTBD16* | 2.84729845 | 5.93E-05 | 0.000298485 | 0.372509604 |
| Obesity | t_cell | *TACC2* | 2.63162406 | 3.75E-06 | 9.34E-05 | 0.023565848 |
| Obesity | t_cell | *CALHM2* | 2.97832768 | 0.013169905 | 0.023522497 | 1 |
| Obesity | t_cell | *ARL3* | 3.959980914 | 0.005259717 | 0.010571561 | 1 |
| Obesity | t_cell | *PLCE1* | 3.397609298 | 2.30E-05 | 0.00017469 | 0.14429374 |
| Obesity | t_cell | *RBP4* | 4.379368257 | 4.17E-06 | 9.84E-05 | 0.026185495 |
| Obesity | t_cell | *FAM149B1* | 3.349338275 | 0.001026894 | 0.002616977 | 1 |
| Obesity | t_cell | *ATIC* | -1.832874867 | 0.022958137 | 0.037963653 | 1 |
| Obesity | t_cell | *SMARCAL1* | 5.350444115 | 0.000356487 | 0.001126815 | 1 |
| Obesity | t_cell | *STAT4* | -0.490722456 | 0.000298138 | 0.000989571 | 1 |
| Obesity | t_cell | *CARF* | 5.110192205 | 1.58E-09 | 2.26E-07 | 9.96E-06 |
| Obesity | t_cell | *METTL5* | 6.125603338 | 0.000988239 | 0.002529746 | 1 |
| Obesity | t_cell | *NAB1* | 5.29310641 | 0.000111265 | 0.000470426 | 0.699524081 |
| Obesity | t_cell | *FASTKD1* | 3.391094041 | 0.01126011 | 0.020615117 | 1 |
| Obesity | t_cell | *MDH1B* | 3.809306031 | 6.49E-05 | 0.000318122 | 0.408150238 |
| Obesity | t_cell | *ITGAV* | 3.550060687 | 0.000229705 | 0.000811325 | 1 |
| Obesity | t_cell | *COX17* | 5.156632561 | 0.01312757 | 0.023473559 | 1 |
| Obesity | t_cell | *UBL7* | 8.678202851 | 8.06E-06 | 0.000132316 | 0.05067717 |
| Obesity | t_cell | *ARHGAP24* | 3.309150902 | 5.06E-05 | 0.000269159 | 0.31841547 |
| Obesity | t_cell | *FAM13A* | 2.85094828 | 0.000724175 | 0.001958231 | 1 |
| Obesity | t_cell | *AP1AR* | 4.946482022 | 0.003954926 | 0.008236433 | 1 |
| Obesity | t_cell | *BBS7* | 5.520367175 | 2.38E-05 | 0.000178067 | 0.149764548 |
| Obesity | t_cell | *MMRN1* | -0.771414064 | 0.021836054 | 0.036372176 | 1 |
| Obesity | t_cell | *PRDM5* | 3.714873993 | 0.00014521 | 0.000575257 | 0.912933644 |
| Obesity | t_cell | *SCARB2* | 4.447559012 | 0.000184993 | 0.000692706 | 1 |
| Obesity | t_cell | *CNOT6L* | -1.629367871 | 0.000795965 | 0.002122237 | 1 |
| Obesity | t_cell | *ANXA3* | 3.226341422 | 1.10E-05 | 0.000145864 | 0.069194521 |
| Obesity | t_cell | *GSTCD* | 4.646916467 | 3.45E-05 | 0.000216978 | 0.216978055 |
| Obesity | t_cell | *ENPEP* | 4.558001709 | 1.48E-05 | 0.000149118 | 0.092900671 |
| Obesity | t_cell | *RNF185* | 2.974652046 | 0.030737321 | 0.049144918 | 1 |
| Obesity | t_cell | *PARVG* | -2.013593069 | 0.002323236 | 0.005227698 | 1 |
| Obesity | t_cell | *B4GALNT3* | -3.626795632 | 7.31E-05 | 0.000346629 | 0.459629654 |
| Obesity | t_cell | *ERP27* | -1.007930402 | 0.000260732 | 0.000894771 | 1 |
| Obesity | t_cell | *SLCO1C1* | 4.108564733 | 4.52E-06 | 0.000100727 | 0.028433769 |
| Obesity | t_cell | *ZCRB1* | 3.37187692 | 0.000358087 | 0.001129033 | 1 |
| Obesity | t_cell | *PRICKLE1* | 2.391990343 | 4.25E-05 | 0.000246282 | 0.267374169 |
| Obesity | t_cell | *C1RL* | 3.037149419 | 0.003829957 | 0.008019485 | 1 |
| Obesity | t_cell | *CLSTN3* | -1.780786092 | 0.029456566 | 0.047268928 | 1 |

| | | | | | | |
|---|---|---|---|---|---|---|
| Obesity | t_cell | *KLRG1* | -0.599490888 | 0.024381597 | 0.039973156 | 1 |
| Obesity | t_cell | *VAMP1* | -1.678020541 | 0.006466668 | 0.012681205 | 1 |
| Obesity | t_cell | *CD27* | -0.703454396 | 0.000351062 | 0.001116403 | 1 |
| Obesity | t_cell | *PEX5* | 3.45860726 | 0.002283363 | 0.00515113 | 1 |
| Obesity | t_cell | *PPFIA2* | 1.88320191 | 0.002373169 | 0.005321012 | 1 |
| Obesity | t_cell | *LGR5* | 2.94583916 | 0.001949798 | 0.004500141 | 1 |
| Obesity | t_cell | *AMDHD1* | -2.477600696 | 0.007314287 | 0.014131813 | 1 |
| Obesity | t_cell | *GAS2L3* | 5.106287634 | 1.37E-05 | 0.000147535 | 0.086012894 |
| Obesity | t_cell | *MMAB* | 2.544258222 | 0.008787439 | 0.016595562 | 1 |
| Obesity | t_cell | *GLTP* | 4.494077868 | 0.002578341 | 0.005721861 | 1 |
| Obesity | t_cell | *ACVRL1* | 1.989805669 | 0.005577257 | 0.011123586 | 1 |
| Obesity | t_cell | *LPAR6* | 4.044047985 | 0.004139617 | 0.008558295 | 1 |
| Obesity | t_cell | *VPS33A* | 7.563727143 | 0.000435475 | 0.001310596 | 1 |
| Obesity | t_cell | *RHOF* | -4.069384647 | 4.67E-05 | 0.000259121 | 0.29332511 |
| Obesity | t_cell | *TSSK4* | 5.341196772 | 0.002224246 | 0.005033014 | 1 |
| Obesity | t_cell | *RTN1* | 3.645379264 | 1.40E-05 | 0.000148367 | 0.088237563 |
| Obesity | t_cell | *C14orf37* | 2.684472656 | 0.014120499 | 0.024972034 | 1 |
| Obesity | t_cell | *STON2* | 4.458007728 | 0.000160065 | 0.000615257 | 1 |
| Obesity | t_cell | *EFCAB11* | 3.028560755 | 8.45E-06 | 0.000134614 | 0.053128834 |
| Obesity | t_cell | *PTGR2* | -1.05869066 | 0.020258532 | 0.034082256 | 1 |
| Obesity | t_cell | *AK7* | 8.961887067 | 7.55E-06 | 0.000129066 | 0.04749645 |
| Obesity | t_cell | *FBLN5* | 3.930763239 | 3.25E-06 | 8.53E-05 | 0.02043113 |
| Obesity | t_cell | *C14orf79* | 7.943138436 | 0.000302395 | 0.001000084 | 1 |
| Obesity | t_cell | *MFAP1* | 4.255578227 | 0.002585679 | 0.005733027 | 1 |
| Obesity | t_cell | *TCF12* | 2.347909374 | 0.030767361 | 0.049144918 | 1 |
| Obesity | t_cell | *SORD* | 6.789247834 | 0.000759259 | 0.002038198 | 1 |
| Obesity | t_cell | *ZSCAN29* | 7.258216658 | 6.51E-06 | 0.000117896 | 0.04090978 |
| Obesity | t_cell | *LYSMD2* | -2.514490016 | 0.004770715 | 0.009681564 | 1 |
| Obesity | t_cell | *SRP14* | 3.867187277 | 0.003321637 | 0.007112784 | 1 |
| Obesity | t_cell | *ANP32A* | 5.084475922 | 0.000203846 | 0.000742086 | 1 |
| Obesity | t_cell | *COMMD4* | 6.173932797 | 0.001174595 | 0.002928104 | 1 |
| Obesity | t_cell | *PSTPIP1* | -1.301102509 | 0.000388635 | 0.001196546 | 1 |
| Obesity | t_cell | *HMG20A* | 5.20887144 | 0.000104072 | 0.000447234 | 0.654303271 |
| Obesity | t_cell | *NEIL1* | 3.054216098 | 0.002244893 | 0.005073201 | 1 |
| Obesity | t_cell | *DNAJA4* | 9.163836277 | 4.19E-05 | 0.000244306 | 0.263117052 |
| Obesity | t_cell | *TPM1* | 3.910191622 | 0.000162819 | 0.000623412 | 1 |
| Obesity | t_cell | *IGF1R* | 3.802779115 | 0.002121576 | 0.004845024 | 1 |
| Obesity | t_cell | *CYP11A1* | 2.1496207 | 5.04E-07 | 2.53E-05 | 0.003167869 |
| Obesity | t_cell | *BBS4* | 6.427772601 | 6.33E-06 | 0.000115948 | 0.039770229 |
| Obesity | t_cell | *HAPLN3* | -1.856531731 | 0.000707873 | 0.001927334 | 1 |
| Obesity | t_cell | *TICRR* | 4.669840123 | 0.000357895 | 0.001128993 | 1 |
| Obesity | t_cell | *MFGE8* | 3.304948408 | 0.028374374 | 0.045705788 | 1 |
| Obesity | t_cell | *UNC45A* | 2.700609896 | 0.003191595 | 0.006871767 | 1 |
| Obesity | t_cell | *CRTC3* | 3.364249021 | 0.013007427 | 0.023291103 | 1 |
| Obesity | t_cell | *SH3GL3* | -2.45840655 | 0.003463327 | 0.007346132 | 1 |
| Obesity | t_cell | *TGFB1I1* | 2.363267689 | 0.005239138 | 0.010536935 | 1 |
| Obesity | t_cell | *PARN* | 2.605736882 | 0.028135034 | 0.045355117 | 1 |
| Obesity | t_cell | *MARVELD3* | 3.676007071 | 0.001339532 | 0.003265466 | 1 |
| Obesity | t_cell | *ZFHX3* | 3.74490588 | 1.75E-05 | 0.000157653 | 0.110202977 |
| Obesity | t_cell | *KIFC3* | 3.712581078 | 1.81E-05 | 0.000158673 | 0.113980494 |

| | | | | | | |
|---|---|---|---|---|---|---|
| Obesity | t_cell | *ADAMTS18* | 3.002347009 | 0.000295636 | 0.000983942 | 1 |
| Obesity | t_cell | *NUDT7* | 2.276005042 | 0.013092831 | 0.023424766 | 1 |
| Obesity | t_cell | *GCSH* | 1.610718656 | 0.000317816 | 0.001036901 | 1 |
| Obesity | t_cell | *MAP1LC3B* | 3.956119217 | 0.001485122 | 0.003561008 | 1 |
| Obesity | t_cell | *RPS2* | -1.940226283 | 0.001614285 | 0.003818287 | 1 |
| Obesity | t_cell | *NDUFB10* | 7.542992076 | 0.000218863 | 0.000783936 | 1 |
| Obesity | t_cell | *TIGD7* | 3.830170106 | 3.32E-06 | 8.62E-05 | 0.020851813 |
| Obesity | t_cell | *COPS3* | 5.06495453 | 0.013401068 | 0.023861699 | 1 |
| Obesity | t_cell | *MYO19* | 7.722467817 | 2.83E-07 | 1.62E-05 | 0.001777207 |
| Obesity | t_cell | *DDX52* | 4.481498409 | 0.001364943 | 0.003315392 | 1 |
| Obesity | t_cell | *RASL10B* | 2.56446549 | 0.017911554 | 0.030734153 | 1 |
| Obesity | t_cell | *TOM1L1* | 5.452534241 | 3.26E-06 | 8.53E-05 | 0.020465674 |
| Obesity | t_cell | *C17orf80* | 2.894227252 | 0.000145446 | 0.000575829 | 0.914417231 |
| Obesity | t_cell | *SPATA22* | 1.817148436 | 0.000507616 | 0.001474308 | 1 |
| Obesity | t_cell | *NPEPPS* | 4.498002935 | 0.000276018 | 0.000933674 | 1 |
| Obesity | t_cell | *SCRN2* | 0.702386292 | 0.015689452 | 0.027407498 | 1 |
| Obesity | t_cell | *RHBDL3* | 3.03577402 | 0.001784243 | 0.004153106 | 1 |
| Obesity | t_cell | *ARSG* | -2.031843337 | 0.004003409 | 0.008323225 | 1 |
| Obesity | t_cell | *ABCA8* | 2.75704884 | 5.48E-06 | 0.000108648 | 0.034441538 |
| Obesity | t_cell | *G6PC3* | 6.959812257 | 2.28E-05 | 0.000174055 | 0.143246911 |
| Obesity | t_cell | *IMPA2* | 4.68555857 | 7.19E-06 | 0.000126396 | 0.045213092 |
| Obesity | t_cell | *GNAL* | 4.546515669 | 4.62E-06 | 0.000100727 | 0.029024581 |
| Obesity | t_cell | *C18orf21* | 6.273009089 | 4.05E-06 | 9.60E-05 | 0.0254513 |
| Obesity | t_cell | *GALNT1* | 4.115762566 | 0.001309946 | 0.003207021 | 1 |
| Obesity | t_cell | *ASXL3* | 5.081529633 | 4.12E-05 | 0.000242079 | 0.259024303 |
| Obesity | t_cell | *GREB1L* | 1.98493971 | 0.000490892 | 0.001437465 | 1 |
| Obesity | t_cell | *C18orf8* | -5.111013154 | 0.000128846 | 0.000523631 | 0.810057561 |
| Obesity | t_cell | *NPC1* | -3.87065377 | 0.001136862 | 0.002852135 | 1 |
| Obesity | t_cell | *SLC14A1* | -1.108707686 | 0.000871056 | 0.002279903 | 1 |
| Obesity | t_cell | *SLC13A5* | -7.630810187 | 0.023919513 | 0.03931555 | 1 |
| Obesity | t_cell | *TMC6* | -0.89801474 | 0.015801701 | 0.027588252 | 1 |
| Obesity | t_cell | *NARF* | 3.954192402 | 0.003278756 | 0.007035338 | 1 |
| Obesity | t_cell | *TRIM65* | 6.302614731 | 3.20E-09 | 4.37E-07 | 2.01E-05 |
| Obesity | t_cell | *CBX8* | 6.299419177 | 1.20E-06 | 4.67E-05 | 0.007519368 |
| Obesity | t_cell | *SECTM1* | 6.049784233 | 0.000195028 | 0.000721543 | 1 |
| Obesity | t_cell | *RNF157* | -0.508022317 | 0.024077076 | 0.039522866 | 1 |
| Obesity | t_cell | *RNF165* | 4.748123812 | 5.87E-06 | 0.000112142 | 0.036894709 |
| Obesity | t_cell | *SMAD4* | 3.362261477 | 0.02583284 | 0.042119052 | 1 |
| Obesity | t_cell | *PIP4K2B* | 5.68063827 | 0.000131813 | 0.000532526 | 0.828710985 |
| Obesity | t_cell | *GRB7* | 3.976024797 | 6.13E-05 | 0.000305871 | 0.385396882 |
| Obesity | t_cell | *NFIC* | 2.734724365 | 0.000207398 | 0.000750667 | 1 |
| Obesity | t_cell | *SIRT3* | 4.689677113 | 9.89E-06 | 0.000142631 | 0.062171647 |
| Obesity | t_cell | *IFITM3* | 4.966732595 | 1.44E-05 | 0.000148509 | 0.090377943 |
| Obesity | t_cell | *COL6A1* | 2.397784635 | 0.000885127 | 0.002311922 | 1 |
| Obesity | t_cell | *TRPM2* | 1.305912522 | 0.005326562 | 0.010688827 | 1 |
| Obesity | t_cell | *DOPEY2* | 3.487468423 | 0.007017691 | 0.013655285 | 1 |
| Obesity | t_cell | *NTN5* | -5.275380544 | 5.92E-05 | 0.000298485 | 0.372483139 |
| Obesity | t_cell | *WTIP* | 3.962546772 | 6.80E-05 | 0.00032817 | 0.427214387 |
| Obesity | t_cell | *MYO1F* | -1.16941958 | 0.002283474 | 0.00515113 | 1 |
| Obesity | t_cell | *CACNG8* | 3.881680854 | 3.18E-05 | 0.000209022 | 0.1999599 |

| | | | | | | |
|---|---|---|---|---|---|---|
| Obesity | t_cell | *FBN3* | 2.895990865 | 0.001387904 | 0.003362525 | 1 |
| Obesity | t_cell | *EVI5L* | 6.596337801 | 0.000329484 | 0.001066744 | 1 |
| Obesity | t_cell | *RCN3* | 3.828984954 | 1.03E-05 | 0.000145864 | 0.065017024 |
| Obesity | t_cell | *RERE* | 2.925439288 | 0.002565813 | 0.00569808 | 1 |
| Obesity | t_cell | *IL22RA1* | 2.160093861 | 0.024156229 | 0.039642446 | 1 |
| Obesity | t_cell | *ZNF593* | 2.460109526 | 0.01510751 | 0.026523572 | 1 |
| Obesity | t_cell | *C1orf216* | 5.167238701 | 2.77E-05 | 0.000194081 | 0.174420171 |
| Obesity | t_cell | *MAP3K6* | 3.711928054 | 7.27E-05 | 0.000345512 | 0.457112361 |
| Obesity | t_cell | *GPN2* | 4.777957444 | 0.019880145 | 0.033562425 | 1 |
| Obesity | t_cell | *NBPF3* | 1.008944171 | 0.029277252 | 0.047015602 | 1 |
| Obesity | t_cell | *HSPG2* | 0.961338417 | 0.028541785 | 0.045963678 | 1 |
| Obesity | t_cell | *MOB3C* | 6.886878718 | 1.05E-05 | 0.000145864 | 0.06572193 |
| Obesity | t_cell | *CYP4B1* | 1.432685227 | 0.002579421 | 0.005722237 | 1 |
| Obesity | t_cell | *SYPL2* | 1.926233638 | 0.000155394 | 0.000603171 | 0.976964071 |
| Obesity | t_cell | *ZNF697* | 3.967178013 | 3.93E-05 | 0.000234849 | 0.247061037 |
| Obesity | t_cell | *GPR161* | 4.890618675 | 2.44E-06 | 7.07E-05 | 0.015344083 |
| Obesity | t_cell | *TIPRL* | 3.774176759 | 0.031354612 | 0.049956018 | 1 |
| Obesity | t_cell | *CREG1* | 4.177985327 | 2.54E-05 | 0.000184808 | 0.159489075 |
| Obesity | t_cell | *GPA33* | -1.823894788 | 0.000164573 | 0.000627072 | 1 |
| Obesity | t_cell | *RXRG* | 4.622265684 | 2.98E-05 | 0.000200768 | 0.187434214 |
| Obesity | t_cell | *TMCO1* | 4.041566364 | 0.011587357 | 0.021134236 | 1 |
| Obesity | t_cell | *XCL2* | -2.383692595 | 0.012922181 | 0.023165598 | 1 |
| Obesity | t_cell | *MGST3* | 4.384231712 | 2.47E-05 | 0.000181284 | 0.154997843 |
| Obesity | t_cell | *ADCY10* | 3.147163394 | 1.92E-05 | 0.000161262 | 0.120820066 |
| Obesity | t_cell | *RGS5* | 4.473448577 | 0.000148563 | 0.000584124 | 0.934014143 |
| Obesity | t_cell | *RRNAD1* | 3.244436791 | 0.000257541 | 0.000887575 | 1 |
| Obesity | t_cell | *PIGM* | 6.2175168 | 0.000665594 | 0.001832131 | 1 |
| Obesity | t_cell | *RGL1* | 3.978488177 | 2.99E-05 | 0.000200794 | 0.187943205 |
| Obesity | t_cell | *SNX27* | 3.869246732 | 0.015529905 | 0.02719016 | 1 |
| Obesity | t_cell | *SELENBP1* | 2.840898471 | 7.94E-06 | 0.000132019 | 0.049927905 |
| Obesity | t_cell | *SEMA6C* | 4.400845955 | 4.70E-07 | 2.42E-05 | 0.00295256 |
| Obesity | t_cell | *ARNT* | 3.797734482 | 0.00097177 | 0.00249368 | 1 |
| Obesity | t_cell | *POGZ* | 3.975361978 | 0.000114143 | 0.00047905 | 0.71761753 |
| Obesity | t_cell | *IKBKE* | -1.457663171 | 0.00041478 | 0.001261597 | 1 |
| Obesity | t_cell | *DYRK3* | 2.809717123 | 2.79E-05 | 0.000194081 | 0.17527591 |
| Obesity | t_cell | *HHIPL2* | 2.97054599 | 0.012348443 | 0.022302403 | 1 |
| Obesity | t_cell | *RAB13* | 3.914086886 | 8.88E-05 | 0.000396406 | 0.558140162 |
| Obesity | t_cell | *S100A8* | -6.545825555 | 0.00029081 | 0.000973548 | 1 |
| Obesity | t_cell | *HAX1* | 5.898198608 | 0.000147111 | 0.000580186 | 0.924886318 |
| Obesity | t_cell | *KCNN3* | 5.284017744 | 2.03E-06 | 6.45E-05 | 0.012779641 |
| Obesity | t_cell | *RIT1* | 6.793376352 | 8.78E-05 | 0.000394101 | 0.552052558 |
| Obesity | t_cell | *GALNT2* | 4.669498027 | 0.000504712 | 0.001469038 | 1 |
| Obesity | t_cell | *SCCPDH* | 3.713494147 | 9.02E-06 | 0.000137059 | 0.056707761 |
| Obesity | t_cell | *CEP170* | 5.148845661 | 6.50E-07 | 3.05E-05 | 0.004086151 |
| Obesity | t_cell | *SDE2* | 5.571368225 | 0.011420483 | 0.02087833 | 1 |
| Obesity | t_cell | *FBXO28* | 3.551798581 | 0.020746832 | 0.034801316 | 1 |
| Obesity | t_cell | *CNIH4* | 5.829569572 | 0.000325877 | 0.001057168 | 1 |
| Obesity | t_cell | *GUK1* | 6.661453355 | 0.000367024 | 0.001147429 | 1 |
| Obesity | t_cell | *CDC42BPA* | 3.846723933 | 8.27E-07 | 3.58E-05 | 0.005198358 |
| Obesity | t_cell | *CNIH3* | 4.27657319 | 2.53E-05 | 0.000184756 | 0.159074556 |

| | | | | | | |
|---|---|---|---|---|---|---|
| Obesity | t_cell | *PSEN2* | 5.532962335 | 3.29E-06 | 8.57E-05 | 0.020657094 |
| Obesity | t_cell | *PYCR2* | 3.284345363 | 0.022646074 | 0.037526585 | 1 |
| Obesity | t_cell | *WNT9A* | 3.53306153 | 5.94E-05 | 0.000298844 | 0.373256597 |
| Obesity | t_cell | *PLEKHA6* | 2.568723945 | 5.99E-06 | 0.000113017 | 0.037683427 |
| Obesity | t_cell | *ABCG8* | 5.504141608 | 0.000221641 | 0.000791068 | 1 |
| Obesity | t_cell | *EML4* | -1.322807522 | 0.007144966 | 0.013847226 | 1 |
| Obesity | t_cell | *CHAC2* | 7.354816177 | 2.75E-05 | 0.000193012 | 0.173132164 |
| Obesity | t_cell | *WDPCP* | 1.739106098 | 7.09E-05 | 0.000337886 | 0.445672197 |
| Obesity | t_cell | *CIAO1* | 4.181052962 | 0.000818043 | 0.002171891 | 1 |
| Obesity | t_cell | *EXOC6B* | 3.196351649 | 0.002570345 | 0.005706131 | 1 |
| Obesity | t_cell | *TEX261* | 9.240997509 | 0.000580096 | 0.001642083 | 1 |
| Obesity | t_cell | *THNSL2* | 0.806909035 | 0.000845101 | 0.002227735 | 1 |
| Obesity | t_cell | *RALB* | 4.075138446 | 0.001446335 | 0.003481283 | 1 |
| Obesity | t_cell | *TMEM177* | 6.37879831 | 8.60E-06 | 0.000134958 | 0.054071429 |
| Obesity | t_cell | *RABL2A* | 3.379136443 | 0.022138291 | 0.036811276 | 1 |
| Obesity | t_cell | *SLC20A1* | -2.214857905 | 0.014027695 | 0.024856854 | 1 |
| Obesity | t_cell | *LYG1* | 2.60290618 | 0.000184268 | 0.000691638 | 1 |
| Obesity | t_cell | *THSD7B* | 3.096268195 | 7.60E-08 | 6.13E-06 | 0.000477843 |
| Obesity | t_cell | *GPR17* | 0.837764509 | 0.003649852 | 0.00768731 | 1 |
| Obesity | t_cell | *ZNF385B* | 2.767514112 | 6.22E-06 | 0.000114728 | 0.039075622 |
| Obesity | t_cell | *TMEFF2* | 2.950318502 | 2.40E-05 | 0.000178908 | 0.150998286 |
| Obesity | t_cell | *GULP1* | 3.450570034 | 2.41E-06 | 7.02E-05 | 0.015152621 |
| Obesity | t_cell | *FAM171B* | 3.829599297 | 0.00026168 | 0.000897536 | 1 |
| Obesity | t_cell | *HSPD1* | 6.784957115 | 9.07E-06 | 0.000137059 | 0.056991699 |
| Obesity | t_cell | *METTL21A* | 4.671376913 | 0.004667064 | 0.009483463 | 1 |
| Obesity | t_cell | *PTH2R* | 1.81936198 | 0.001759158 | 0.004100789 | 1 |
| Obesity | t_cell | *NBEAL1* | 3.319333239 | 3.57E-09 | 4.67E-07 | 2.24E-05 |
| Obesity | t_cell | *SUMF1* | 3.354164394 | 0.002130957 | 0.004863674 | 1 |
| Obesity | t_cell | *ESPNL* | 16.40988444 | 3.92E-05 | 0.000234641 | 0.246607481 |
| Obesity | t_cell | *FANCD2* | -2.255075209 | 0.003360234 | 0.007171009 | 1 |
| Obesity | t_cell | *OSBPL10* | 3.633029743 | 1.40E-05 | 0.000148367 | 0.088053694 |
| Obesity | t_cell | *FAM198A* | 1.811774662 | 0.009197314 | 0.017286551 | 1 |
| Obesity | t_cell | *SLC25A38* | -3.439009163 | 0.016263136 | 0.028229247 | 1 |
| Obesity | t_cell | *CTDSPL* | 3.832334896 | 2.22E-05 | 0.000171828 | 0.139353104 |
| Obesity | t_cell | *STAC* | 6.201384677 | 2.14E-05 | 0.000169226 | 0.134646357 |
| Obesity | t_cell | *LIMD1* | 5.155054051 | 0.000110387 | 0.000467659 | 0.69400534 |
| Obesity | t_cell | *COL8A1* | 1.465310982 | 0.002742034 | 0.006029789 | 1 |
| Obesity | t_cell | *RABL3* | 4.334067781 | 0.020568884 | 0.03454891 | 1 |
| Obesity | t_cell | *BOC* | 2.49557668 | 2.79E-05 | 0.000194081 | 0.175451449 |
| Obesity | t_cell | *AGTR1* | 3.020396418 | 0.000143517 | 0.00057071 | 0.902292215 |
| Obesity | t_cell | *TCTA* | 2.757595153 | 0.018102762 | 0.031028371 | 1 |
| Obesity | t_cell | *SLC26A1* | 1.686560967 | 0.007827837 | 0.014990439 | 1 |
| Obesity | t_cell | *ATP10D* | 2.228474992 | 0.016530571 | 0.028598707 | 1 |
| Obesity | t_cell | *SLC10A4* | 5.804151101 | 0.01403888 | 0.024869665 | 1 |
| Obesity | t_cell | *PYURF* | 5.592809553 | 3.70E-05 | 0.000226954 | 0.232361608 |
| Obesity | t_cell | *CISD2* | 5.188056855 | 0.000576523 | 0.001633439 | 1 |
| Obesity | t_cell | *TIFA* | 7.206509525 | 0.013418105 | 0.023884379 | 1 |
| Obesity | t_cell | *CCNA2* | 5.882466778 | 4.03E-05 | 0.000238593 | 0.253147504 |
| Obesity | t_cell | *USP53* | 4.021647414 | 1.49E-05 | 0.000149261 | 0.093847569 |
| Obesity | t_cell | *PDGFC* | 3.715139173 | 8.34E-06 | 0.000134424 | 0.052425451 |

| | | | | | | |
|---|---|---|---|---|---|---|
| Obesity | t_cell | *CBR4* | 4.15681805 | 0.000211214 | 0.000761849 | 1 |
| Obesity | t_cell | *NDUFS6* | 5.556701893 | 0.001153754 | 0.002881865 | 1 |
| Obesity | t_cell | *LHFPL2* | 3.833401466 | 1.02E-05 | 0.000145643 | 0.064228719 |
| Obesity | t_cell | *SSBP2* | 3.431246478 | 0.001149678 | 0.002876254 | 1 |
| Obesity | t_cell | *CRHBP* | 2.330685265 | 0.019898583 | 0.033579036 | 1 |
| Obesity | t_cell | *GTF2H2* | 3.273347494 | 0.002361046 | 0.005301392 | 1 |
| Obesity | t_cell | *SPATA9* | 2.198568895 | 2.28E-05 | 0.000174055 | 0.143232289 |
| Obesity | t_cell | *FEM1C* | 6.376100527 | 4.50E-05 | 0.000253415 | 0.283064493 |
| Obesity | t_cell | *PCYOX1L* | -3.959079911 | 3.12E-06 | 8.33E-05 | 0.019641203 |
| Obesity | t_cell | *MYLK4* | 3.83810002 | 3.17E-05 | 0.000209022 | 0.199164914 |
| Obesity | t_cell | *TBC1D7* | 6.330728549 | 0.001299623 | 0.00318547 | 1 |
| Obesity | t_cell | *GFOD1* | 2.600427477 | 4.29E-05 | 0.000247159 | 0.269463044 |
| Obesity | t_cell | *PSD2* | -5.159792686 | 0.000262309 | 0.000898222 | 1 |
| Obesity | t_cell | *KLHL3* | 4.071378696 | 0.000183131 | 0.000688834 | 1 |
| Obesity | t_cell | *TRIM7* | 3.435974227 | 4.96E-05 | 0.000266905 | 0.311957159 |
| Obesity | t_cell | *FAM193B* | 4.5550662 | 0.00143355 | 0.003454013 | 1 |
| Obesity | t_cell | *MUT* | 5.543136895 | 1.79E-05 | 0.000158468 | 0.112811967 |
| Obesity | t_cell | *ABT1* | 4.659841322 | 3.33E-08 | 3.17E-06 | 0.000209126 |
| Obesity | t_cell | *MLIP* | 2.383497153 | 0.00025545 | 0.000881942 | 1 |
| Obesity | t_cell | *FGD2* | 3.807024952 | 1.69E-05 | 0.000156669 | 0.106375158 |
| Obesity | t_cell | *ANO7* | 1.511720271 | 0.006095761 | 0.012036447 | 1 |
| Obesity | t_cell | *RPL7L1* | 3.912445883 | 0.000601631 | 0.001691616 | 1 |
| Obesity | t_cell | *TPBG* | 5.112836067 | 1.08E-05 | 0.000145864 | 0.067970928 |
| Obesity | t_cell | *IRAK1BP1* | 5.016126539 | 7.24E-06 | 0.000126773 | 0.045511328 |
| Obesity | t_cell | *RNF217* | 3.98156877 | 1.81E-05 | 0.000158673 | 0.114041112 |
| Obesity | t_cell | *ABRACL* | -2.025622239 | 0.000587999 | 0.001659968 | 1 |
| Obesity | t_cell | *AIG1* | 3.902437293 | 4.64E-07 | 2.42E-05 | 0.002919235 |
| Obesity | t_cell | *DYNLT1* | -1.671194497 | 0.022827793 | 0.037777924 | 1 |
| Obesity | t_cell | *PNLDC1* | 2.904345533 | 0.021841025 | 0.036372176 | 1 |
| Obesity | t_cell | *SLC22A3* | 4.054272409 | 1.29E-05 | 0.000147332 | 0.080885174 |
| Obesity | t_cell | *GNA12* | 3.596888469 | 0.000477797 | 0.001408301 | 1 |
| Obesity | t_cell | *C7orf50* | 5.206989396 | 0.000809577 | 0.002153051 | 1 |
| Obesity | t_cell | *RBAK* | 8.715003244 | 0.000244408 | 0.000851297 | 1 |
| Obesity | t_cell | *EGFR* | 2.373205679 | 5.39E-05 | 0.000280871 | 0.33864134 |
| Obesity | t_cell | *POMZP3* | 2.568598863 | 0.001401262 | 0.003390859 | 1 |
| Obesity | t_cell | *AGBL3* | 4.504405998 | 0.000223127 | 0.000794338 | 1 |
| Obesity | t_cell | *EPHA1* | -2.068720066 | 0.004278588 | 0.008807951 | 1 |
| Obesity | t_cell | *NOM1* | 6.179917087 | 0.000657776 | 0.001815381 | 1 |
| Obesity | t_cell | *FBXO25* | 5.597169553 | 0.000156171 | 0.000604451 | 0.981847666 |
| Obesity | t_cell | *CCDC25* | 4.002180013 | 0.010807391 | 0.019913854 | 1 |
| Obesity | t_cell | *SLC25A37* | 4.979071535 | 8.55E-06 | 0.000134719 | 0.053752824 |
| Obesity | t_cell | *DOCK5* | 4.861551855 | 0.000377591 | 0.001173323 | 1 |
| Obesity | t_cell | *ST18* | 2.608103953 | 0.000387157 | 0.001193165 | 1 |
| Obesity | t_cell | *RGS20* | 1.692929831 | 0.012274829 | 0.022182194 | 1 |
| Obesity | t_cell | *TACC1* | 3.733180051 | 0.000122093 | 0.000503016 | 0.767601825 |
| Obesity | t_cell | *MAL2* | 1.939679786 | 1.00E-05 | 0.000143604 | 0.06289834 |
| Obesity | t_cell | *UTP23* | 4.03003976 | 0.023547526 | 0.038785249 | 1 |
| Obesity | t_cell | *FAM135B* | 2.672872583 | 5.32E-05 | 0.000278506 | 0.334764014 |
| Obesity | t_cell | *ZNF7* | 4.083410726 | 0.001588429 | 0.003771319 | 1 |
| Obesity | t_cell | *SLC39A4* | 4.018012979 | 0.0019978 | 0.004594063 | 1 |

| | | | | | | |
|---|---|---|---|---|---|---|
| Obesity | t_cell | *CER1* | 3.830062133 | 0.000381053 | 0.001179556 | 1 |
| Obesity | t_cell | *PLIN2* | 4.431317988 | 1.93E-05 | 0.00016149 | 0.121601913 |
| Obesity | t_cell | *C9orf72* | 3.406341459 | 0.000854734 | 0.002247475 | 1 |
| Obesity | t_cell | *ZNF462* | 3.930800946 | 1.21E-05 | 0.000145864 | 0.075998225 |
| Obesity | t_cell | *ALAD* | 1.514093016 | 0.00082118 | 0.002179069 | 1 |
| Obesity | t_cell | *ASTN2* | 5.354758884 | 0.000102204 | 0.000440108 | 0.642557485 |
| Obesity | t_cell | *WDR31* | 2.699216541 | 5.84E-06 | 0.000112142 | 0.03668533 |
| Obesity | t_cell | *SURF1* | 2.659719942 | 0.000352734 | 0.001120585 | 1 |
| Obesity | t_cell | *SURF6* | 6.863235654 | 8.68E-05 | 0.000391529 | 0.54579128 |
| Obesity | t_cell | *ASB6* | 5.001954263 | 0.00161881 | 0.003824674 | 1 |
| Obesity | t_cell | *CIZ1* | 5.777912181 | 5.95E-06 | 0.000112647 | 0.037398662 |
| Obesity | t_cell | *SH3GLB2* | 7.452226812 | 1.62E-06 | 5.73E-05 | 0.010191795 |
| Obesity | t_cell | *LRSAM1* | 4.928263805 | 7.56E-05 | 0.000355346 | 0.475086577 |
| Obesity | t_cell | *INPP5E* | 4.316710496 | 0.008289504 | 0.015773185 | 1 |
| Obesity | t_cell | *DPH7* | 6.976929447 | 4.57E-06 | 0.000100727 | 0.02873201 |
| Obesity | t_cell | *SLC39A12* | 3.899152276 | 0.000507929 | 0.001474308 | 1 |
| Obesity | t_cell | *PARD3* | 2.770263842 | 1.78E-06 | 6.03E-05 | 0.011221847 |
| Obesity | t_cell | *ADIRF* | 4.815821999 | 0.000116679 | 0.000486769 | 0.733560188 |
| Obesity | t_cell | *HTR7* | 6.470329345 | 1.76E-05 | 0.000157653 | 0.110774916 |
| Obesity | t_cell | *RPP30* | 4.698665668 | 0.000468828 | 0.001389195 | 1 |
| Obesity | t_cell | *FRA10AC1* | 2.238335574 | 0.026767194 | 0.043458555 | 1 |
| Obesity | t_cell | *DNAJB12* | 5.578868039 | 0.000117443 | 0.000488336 | 0.738363859 |
| Obesity | t_cell | *FUOM* | 3.349259598 | 0.000687501 | 0.001886596 | 1 |
| Obesity | t_cell | *MTG1* | 3.166612076 | 1.23E-06 | 4.77E-05 | 0.007720226 |
| Obesity | t_cell | *PAOX* | 9.748674847 | 4.95E-05 | 0.000266905 | 0.311509949 |
| Obesity | t_cell | *RGS10* | -2.495831007 | 0.005449121 | 0.010899975 | 1 |
| Obesity | t_cell | *GAS2* | 7.009520282 | 2.36E-05 | 0.000176965 | 0.148650954 |
| Obesity | t_cell | *LIN7C* | 5.55346299 | 0.00928099 | 0.017417786 | 1 |
| Obesity | t_cell | *HSD17B12* | -1.875936077 | 7.66E-05 | 0.000359016 | 0.481440187 |
| Obesity | t_cell | *GLYAT* | 1.861769534 | 5.33E-06 | 0.000107786 | 0.033521343 |
| Obesity | t_cell | *SERPINH1* | 2.255217369 | 0.01194571 | 0.021712252 | 1 |
| Obesity | t_cell | *CAPN5* | 5.052910331 | 4.93E-06 | 0.000104237 | 0.030964323 |
| Obesity | t_cell | *INTS4* | 2.439062766 | 0.030804746 | 0.049192135 | 1 |
| Obesity | t_cell | *RPS3* | -1.549226902 | 0.00195368 | 0.004504139 | 1 |
| Obesity | t_cell | *ZC3H12C* | 4.2446581 | 8.40E-06 | 0.000134614 | 0.052834541 |
| Obesity | t_cell | *TTC12* | 1.156483081 | 0.011559955 | 0.02109759 | 1 |
| Obesity | t_cell | *DRD2* | 15.769015 | 1.14E-05 | 0.000145864 | 0.071796163 |
| Obesity | t_cell | *C11orf52* | 11.54804511 | 4.39E-05 | 0.000250474 | 0.276022685 |
| Obesity | t_cell | *NPAT* | -1.875382558 | 0.028803898 | 0.046362034 | 1 |
| Obesity | t_cell | *GLB1L2* | -2.418668362 | 0.000464637 | 0.001378561 | 1 |
| Obesity | t_cell | *P4HA3* | 4.228792745 | 4.45E-06 | 0.000100727 | 0.027977775 |
| Obesity | t_cell | *ADAM33* | 3.153227372 | 0.000544595 | 0.001561271 | 1 |
| Obesity | t_cell | *ROM1* | 1.061691286 | 0.018721575 | 0.031923661 | 1 |
| Obesity | t_cell | *EML3* | 4.256629653 | 0.005206102 | 0.010483908 | 1 |
| Obesity | t_cell | *INCENP* | 6.308702007 | 1.12E-05 | 0.000145864 | 0.070535616 |
| Obesity | t_cell | *MS4A3* | 29.37364407 | 5.45E-05 | 0.000282992 | 0.342703129 |
| Obesity | t_cell | *PLCH2* | -1.021673348 | 0.000158957 | 0.000612355 | 0.999362716 |
| Obesity | t_cell | *CPSF7* | 5.259215743 | 0.000253977 | 0.000879753 | 1 |
| Obesity | t_cell | *CCDC15* | -2.566589111 | 0.002481647 | 0.005532386 | 1 |
| Obesity | t_cell | *CHEK1* | 1.826372051 | 0.012620447 | 0.022695868 | 1 |

| | | | | | | |
|---|---|---|---|---|---|---|
| Obesity | t_cell | *FEZ1* | 6.783024587 | 0.009559558 | 0.017876543 | 1 |
| Obesity | t_cell | *COMMD7* | 5.074111386 | 0.00867998 | 0.016417279 | 1 |
| Obesity | t_cell | *KIAA1755* | 1.985406624 | 0.006182009 | 0.012164724 | 1 |
| Obesity | t_cell | *CNBD2* | 4.87949573 | 7.91E-06 | 0.000132019 | 0.049713953 |
| Obesity | t_cell | *TBX6* | 4.073920477 | 1.47E-10 | 3.18E-08 | 9.22E-07 |
| Obesity | t_cell | *TAOK2* | 6.338622864 | 4.04E-05 | 0.000238823 | 0.253869164 |
| Obesity | t_cell | *MPP7* | -1.248154672 | 0.006792044 | 0.013261361 | 1 |
| Obesity | t_cell | *CWC15* | 3.441449778 | 0.005082681 | 0.01027075 | 1 |
| Obesity | t_cell | *TMCO3* | 5.557219005 | 0.000121829 | 0.000502254 | 0.765937349 |
| Obesity | t_cell | *SAP18* | 5.729990293 | 8.48E-05 | 0.000385355 | 0.533331422 |
| Obesity | t_cell | *LYPD6B* | 7.10366885 | 0.026835942 | 0.043551257 | 1 |
| Obesity | t_cell | *WDR17* | 3.894111883 | 5.54E-05 | 0.000285796 | 0.348385883 |
| Obesity | t_cell | *CCDC102B* | 4.484197882 | 9.99E-05 | 0.000431853 | 0.628346094 |
| Obesity | t_cell | *RGS18* | 9.475006871 | 0.00040307 | 0.00123254 | 1 |
| Obesity | t_cell | *PRSS23* | 2.440961901 | 1.72E-05 | 0.000156669 | 0.10794375 |
| Obesity | t_cell | *MTMR12* | 4.406554917 | 0.000524035 | 0.001509901 | 1 |
| Obesity | t_cell | *PPP1R1C* | 2.136318458 | 0.002222905 | 0.005032555 | 1 |
| Obesity | t_cell | *C11orf53* | 10.9704274 | 0.000115821 | 0.000484382 | 0.728166493 |
| Obesity | t_cell | *CCT5* | 5.143050665 | 8.05E-05 | 0.000372155 | 0.506130375 |
| Obesity | t_cell | *FAM173B* | 4.093379623 | 0.000208199 | 0.000753133 | 1 |
| Obesity | t_cell | *DOCK1* | 3.115096255 | 0.000410118 | 0.001249836 | 1 |
| Obesity | t_cell | *TEX12* | 10.20084785 | 4.53E-05 | 0.0002542 | 0.284962726 |
| Obesity | t_cell | *PIP4K2A* | -1.395094442 | 0.001542117 | 0.003679426 | 1 |
| Obesity | t_cell | *CRIM1* | 2.626900891 | 0.018234596 | 0.031186317 | 1 |
| Obesity | t_cell | *PRSS53* | -2.95037743 | 0.019225755 | 0.032606506 | 1 |
| Obesity | t_cell | *GPR158* | 2.201053754 | 0.003034337 | 0.006566911 | 1 |
| Obesity | t_cell | *THRB* | 3.118936932 | 1.80E-05 | 0.000158501 | 0.113169664 |
| Obesity | t_cell | *BTBD11* | -0.906496216 | 0.00029623 | 0.000984875 | 1 |
| Obesity | t_cell | *UBE3B* | 3.758288904 | 3.79E-05 | 0.000230345 | 0.238176634 |
| Obesity | t_cell | *ANK3* | -0.478412072 | 0.0003293 | 0.001066744 | 1 |
| Obesity | t_cell | *IPMK* | 5.209759393 | 0.00053434 | 0.001536076 | 1 |
| Obesity | t_cell | *PLBD2* | 4.19351349 | 1.14E-05 | 0.000145864 | 0.071412226 |
| Obesity | t_cell | *DLG5* | 2.013653737 | 0.005378547 | 0.01078282 | 1 |
| Obesity | t_cell | *FAM177A1* | 5.120980352 | 0.000892654 | 0.002327712 | 1 |
| Obesity | t_cell | *MIPOL1* | 3.424545833 | 7.12E-07 | 3.24E-05 | 0.004475504 |
| Obesity | t_cell | *EXT2* | 1.842259782 | 0.0226378 | 0.037522766 | 1 |
| Obesity | t_cell | *THRSP* | 2.050284358 | 0.000344035 | 0.001099058 | 1 |
| Obesity | t_cell | *NDUFC2* | 4.443214945 | 0.000343156 | 0.001097366 | 1 |
| Obesity | t_cell | *ME3* | 3.235381145 | 1.75E-05 | 0.000157583 | 0.109993258 |
| Obesity | t_cell | *ADAMTS12* | 2.044534015 | 3.87E-05 | 0.000232657 | 0.243591985 |
| Obesity | t_cell | *VIPAS39* | 3.905984651 | 0.015800318 | 0.027588252 | 1 |
| Obesity | t_cell | *ANKRD50* | 4.62435763 | 8.02E-06 | 0.000132019 | 0.050422136 |
| Obesity | t_cell | *C4orf33* | 5.045571413 | 0.002132286 | 0.00486418 | 1 |
| Obesity | t_cell | *PTPRO* | 5.896719691 | 7.06E-05 | 0.000337166 | 0.444047992 |
| Obesity | t_cell | *VPS26B* | 6.174483165 | 0.00024975 | 0.000867503 | 1 |
| Obesity | t_cell | *FAM160B1* | 4.963723065 | 4.98E-05 | 0.000267366 | 0.312817826 |
| Obesity | t_cell | *ANO4* | 4.077415501 | 8.67E-06 | 0.000134958 | 0.054522952 |
| Obesity | t_cell | *TEX9* | 5.239931369 | 0.000874802 | 0.002288755 | 1 |
| Obesity | t_cell | *AKR1C2* | 2.292204677 | 2.92E-05 | 0.000199772 | 0.183789958 |
| Obesity | t_cell | *DPYSL4* | 1.624766055 | 0.000149403 | 0.000586556 | 0.9392966 |

| | | | | | | |
|---|---|---|---|---|---|---|
| Obesity | t_cell | *ADAM8* | -1.134237643 | 0.000710021 | 0.00193159 | 1 |
| Obesity | t_cell | *ITIH2* | 5.646011939 | 4.86E-05 | 0.000265084 | 0.305564163 |
| Obesity | t_cell | *KIN* | 3.843079619 | 0.001129926 | 0.002836998 | 1 |
| Obesity | t_cell | *PIGF* | 3.834277178 | 0.011773045 | 0.021423193 | 1 |
| Obesity | t_cell | *INPP1* | 4.952196318 | 2.97E-05 | 0.000200768 | 0.186588484 |
| Obesity | t_cell | *RNF144A* | 3.590130703 | 0.008394555 | 0.015944582 | 1 |
| Obesity | t_cell | *ASAP2* | 3.722463764 | 1.92E-05 | 0.000161227 | 0.120597464 |
| Obesity | t_cell | *ADAM17* | 3.949002834 | 0.003383701 | 0.007211298 | 1 |
| Obesity | t_cell | *TMEM45B* | -0.987554561 | 0.000132126 | 0.000533023 | 0.830678179 |
| Obesity | t_cell | *ACSL1* | 1.110433345 | 0.022049665 | 0.03667361 | 1 |
| Obesity | t_cell | *GABRA2* | 2.421224906 | 0.000341879 | 0.001094952 | 1 |
| Obesity | t_cell | *CENPJ* | 3.657931657 | 0.000278262 | 0.000938539 | 1 |
| Obesity | t_cell | *PARP8* | -1.036210201 | 0.000485832 | 0.001425696 | 1 |
| Obesity | t_cell | *CACUL1* | 4.232319832 | 0.001301417 | 0.003188624 | 1 |
| Obesity | t_cell | *PTPN14* | 3.521817209 | 4.54E-06 | 0.000100727 | 0.028518952 |
| Obesity | t_cell | *G6PC2* | -30.379606 | 0.00020557 | 0.000745678 | 1 |
| Obesity | t_cell | *POC5* | -3.734396401 | 0.000476779 | 0.001406621 | 1 |
| Obesity | t_cell | *TADA1* | 7.334920545 | 0.000116482 | 0.000486407 | 0.732320442 |
| Obesity | t_cell | *GUCY1A2* | 2.291446406 | 0.000500255 | 0.001459445 | 1 |
| Obesity | t_cell | *CWF19L2* | 3.121556066 | 0.014063522 | 0.02488527 | 1 |
| Obesity | t_cell | *ZNF547* | 5.744388062 | 0.000314972 | 0.001029225 | 1 |
| Obesity | t_cell | *CCDC50* | 2.328807008 | 0.000799585 | 0.002130983 | 1 |
| Obesity | t_cell | *TMEM123* | -1.809132067 | 0.014775038 | 0.026019794 | 1 |
| Obesity | t_cell | *IGSF10* | 2.824367627 | 0.000276953 | 0.000935627 | 1 |
| Obesity | t_cell | *SPARCL1* | 2.729825841 | 0.000211926 | 0.000763977 | 1 |
| Obesity | t_cell | *CLEC4F* | 7.017371726 | 2.94E-05 | 0.000200353 | 0.185126245 |
| Obesity | t_cell | *PELO* | 5.469277749 | 1.07E-05 | 0.000145864 | 0.067316435 |
| Obesity | t_cell | *RASGRP3* | 1.849818556 | 0.004031508 | 0.008373337 | 1 |
| Obesity | t_cell | *SAR1B* | 4.172757084 | 0.000894983 | 0.002332818 | 1 |
| Obesity | t_cell | *GPR180* | 2.045528587 | 0.002173195 | 0.004934228 | 1 |
| Obesity | t_cell | *TCTEX1D1* | 2.435797071 | 9.19E-05 | 0.000407341 | 0.577970083 |
| Obesity | t_cell | *WDR78* | 2.820091166 | 0.000265482 | 0.000906604 | 1 |
| Obesity | t_cell | *IFIT5* | -2.127824564 | 0.018161807 | 0.031104135 | 1 |
| Obesity | t_cell | *PRDM8* | -1.60948625 | 0.00018325 | 0.000688834 | 1 |
| Obesity | t_cell | *HHEX* | 3.264297289 | 4.44E-05 | 0.000251953 | 0.279416274 |
| Obesity | t_cell | *ZNF117* | 1.045264772 | 0.000119747 | 0.000495295 | 0.752848006 |
| Obesity | t_cell | *RAB3C* | 2.926469585 | 2.79E-05 | 0.000194081 | 0.175413473 |
| Obesity | t_cell | *MED21* | 4.570179292 | 0.012892313 | 0.023125241 | 1 |
| Obesity | t_cell | *SRP19* | 4.600109266 | 0.026906448 | 0.043643146 | 1 |
| Obesity | t_cell | *CENPH* | 2.84315839 | 0.000623641 | 0.00174027 | 1 |
| Obesity | t_cell | *CARHSP1* | 4.755146451 | 0.000849344 | 0.002235172 | 1 |
| Obesity | t_cell | *TEKT5* | -4.740929708 | 0.003064019 | 0.006619755 | 1 |
| Obesity | t_cell | *BANK1* | 3.959385028 | 1.18E-05 | 0.000145864 | 0.074343613 |
| Obesity | t_cell | *SCOC* | 3.53540392 | 0.000335358 | 0.001080675 | 1 |
| Obesity | t_cell | *CLGN* | 8.747321359 | 3.19E-05 | 0.000209022 | 0.200430869 |
| Obesity | t_cell | *SYCP2L* | 3.18046004 | 0.000117068 | 0.000487819 | 0.736007719 |
| Obesity | t_cell | *AHCTF1* | 4.022948337 | 0.005140085 | 0.010357601 | 1 |
| Obesity | t_cell | *MERTK* | 0.537625089 | 0.013009785 | 0.023291103 | 1 |
| Obesity | t_cell | *PTPRR* | 2.888416605 | 1.31E-05 | 0.000147409 | 0.082584745 |
| Obesity | t_cell | *PLA2R1* | 2.528356394 | 3.77E-05 | 0.000229753 | 0.23710556 |

| | | | | | | |
|---|---|---|---|---|---|---|
| Obesity | t_cell | *SLC25A27* | 1.256879268 | 0.003332977 | 0.00712978 | 1 |
| Obesity | t_cell | *FRMD1* | 1.711098279 | 4.36E-05 | 0.000249288 | 0.27421711 |
| Obesity | t_cell | *ASAP1* | 2.77392792 | 0.01058638 | 0.01955246 | 1 |
| Obesity | t_cell | *INO80C* | 6.551444366 | 1.96E-06 | 6.34E-05 | 0.012300683 |
| Obesity | t_cell | *NMRAL1* | -2.159438707 | 0.000407527 | 0.001243145 | 1 |
| Obesity | t_cell | *C16orf89* | 2.559877023 | 0.004117497 | 0.008523774 | 1 |
| Obesity | t_cell | *TEX29* | 4.119129342 | 3.49E-05 | 0.000218123 | 0.219213741 |
| Obesity | t_cell | *TUBGCP5* | 5.487972957 | 6.58E-05 | 0.000320973 | 0.413680757 |
| Obesity | t_cell | *LURAP1L* | 1.834080471 | 3.24E-05 | 0.000209715 | 0.203633613 |
| Obesity | t_cell | *CFDP1* | 3.953295011 | 0.002821593 | 0.006180959 | 1 |
| Obesity | t_cell | *ZDHHC7* | 4.199694066 | 1.50E-05 | 0.000149282 | 0.094388531 |
| Obesity | t_cell | *FAM92B* | 18.98295594 | 4.90E-05 | 0.000265838 | 0.308371837 |
| Obesity | t_cell | *C7orf31* | 5.416869182 | 0.000649487 | 0.001795658 | 1 |
| Obesity | t_cell | *FBXO36* | 4.303334052 | 0.000339894 | 0.001091101 | 1 |
| Obesity | t_cell | *MCOLN2* | -0.787537159 | 0.000315151 | 0.001029276 | 1 |
| Obesity | t_cell | *CACNA2D1* | 2.340511058 | 1.31E-05 | 0.000147409 | 0.082049062 |
| Obesity | t_cell | *HS3ST3A1* | 2.486551167 | 0.000391339 | 0.001204282 | 1 |
| Obesity | t_cell | *ANKRD29* | 3.233037923 | 9.42E-05 | 0.000414398 | 0.59246319 |
| Obesity | t_cell | *THY1* | 4.521834937 | 0.006215488 | 0.012226774 | 1 |
| Obesity | t_cell | *TBCEL* | 4.967675078 | 4.27E-05 | 0.000246756 | 0.268470869 |
| Obesity | t_cell | *UBASH3B* | -1.336344075 | 0.001184732 | 0.002946365 | 1 |
| Obesity | t_cell | *ROBO4* | 2.953703973 | 0.000614902 | 0.001721824 | 1 |
| Obesity | t_cell | *ROBO3* | 4.049762383 | 2.78E-05 | 0.000194081 | 0.174703573 |
| Obesity | t_cell | *PRKCA* | -1.177362638 | 0.003536568 | 0.007475126 | 1 |
| Obesity | t_cell | *LRRK1* | 4.401626028 | 1.97E-05 | 0.000162709 | 0.12360788 |
| Obesity | t_cell | *CEP112* | 3.55628463 | 1.69E-07 | 1.08E-05 | 0.001059809 |
| Obesity | t_cell | *GAL3ST2* | 7.953178543 | 0.022628327 | 0.037516955 | 1 |
| Obesity | t_cell | *ENPP3* | 4.002964342 | 4.84E-05 | 0.000264476 | 0.30441147 |
| Obesity | t_cell | *UCHL1* | 2.291769786 | 3.41E-05 | 0.00021533 | 0.21446867 |
| Obesity | t_cell | *MIA3* | 3.367060524 | 0.018662768 | 0.03183202 | 1 |
| Obesity | t_cell | *DISP1* | 2.116956536 | 0.003807714 | 0.00798236 | 1 |
| Obesity | t_cell | *FAM167A* | 8.71172385 | 0.000112656 | 0.000474427 | 0.70827138 |
| Obesity | t_cell | *NEIL2* | 4.152693324 | 2.84E-07 | 1.62E-05 | 0.001783443 |
| Obesity | t_cell | *LONRF1* | 3.910741638 | 4.89E-05 | 0.000265802 | 0.30753329 |
| Obesity | t_cell | *CCSAP* | -2.809718974 | 0.003527659 | 0.007462303 | 1 |
| Obesity | t_cell | *GBP5* | -0.510689821 | 0.000333247 | 0.001076076 | 1 |
| Obesity | t_cell | *ATP5G3* | 3.384910425 | 7.61E-05 | 0.000357559 | 0.478413634 |
| Obesity | t_cell | *PDLIM3* | 4.027498742 | 7.51E-06 | 0.000128894 | 0.047224321 |
| Obesity | t_cell | *NCAM2* | 2.439186266 | 0.00011773 | 0.000488971 | 0.740167796 |
| Obesity | t_cell | *L3MBTL4* | 4.356908759 | 5.52E-05 | 0.000285135 | 0.347009716 |
| Obesity | t_cell | *PDE1C* | 5.778550659 | 2.27E-06 | 6.76E-05 | 0.014242682 |
| Obesity | t_cell | *SLFN13* | -1.472967073 | 2.86E-05 | 0.000196979 | 0.180121859 |
| Obesity | t_cell | *FLCN* | 4.368083371 | 7.67E-06 | 0.000130339 | 0.048225429 |
| Obesity | t_cell | *CXXC1* | 6.16198715 | 9.91E-05 | 0.000429287 | 0.622964834 |
| Obesity | t_cell | *PPP4R1* | 4.80815437 | 0.000693875 | 0.001898344 | 1 |
| Obesity | t_cell | *APCDD1* | 1.327064148 | 0.001077547 | 0.002725076 | 1 |
| Obesity | t_cell | *EPHB1* | 3.963795659 | 0.000159079 | 0.00061245 | 1 |
| Obesity | t_cell | *ANKRD40* | 4.317325635 | 0.001623354 | 0.003832968 | 1 |
| Obesity | t_cell | *CYP2U1* | 5.029332439 | 1.03E-05 | 0.000145719 | 0.064553536 |
| Obesity | t_cell | *FBXL18* | 6.033003948 | 1.19E-05 | 0.000145864 | 0.074891183 |

| | | | | | | |
|---|---|---|---|---|---|---|
| Obesity | t_cell | *PROM2* | -11.69317905 | 0.001227794 | 0.003048633 | 1 |
| Obesity | t_cell | *AK9* | 4.127321868 | 0.001254504 | 0.003100261 | 1 |
| Obesity | t_cell | *GTF3C6* | 4.643236712 | 0.000950035 | 0.002444892 | 1 |
| Obesity | t_cell | *TTC39B* | -1.860501986 | 0.007278513 | 0.014071344 | 1 |
| Obesity | t_cell | *MMS19* | 4.77564304 | 0.00181462 | 0.00421445 | 1 |
| Obesity | t_cell | *GOLGA7B* | -3.028923223 | 0.000459115 | 0.001367342 | 1 |
| Obesity | t_cell | *TRMT44* | 5.764892831 | 2.61E-06 | 7.41E-05 | 0.01638164 |
| Obesity | t_cell | *LRGUK* | 1.379236422 | 0.007166375 | 0.013880159 | 1 |
| Obesity | t_cell | *ZKSCAN2* | 2.574151724 | 0.000227409 | 0.000804569 | 1 |
| Obesity | t_cell | *KDM8* | -3.854121423 | 0.005658084 | 0.011267777 | 1 |
| Obesity | t_cell | *FAM126B* | 4.223597573 | 0.000689307 | 0.001888598 | 1 |
| Obesity | t_cell | *ALS2CR12* | 4.498968379 | 0.00011382 | 0.000478333 | 0.715585893 |
| Obesity | t_cell | *SPAG17* | 2.485884345 | 1.86E-05 | 0.000160456 | 0.117133193 |
| Obesity | t_cell | *DEPTOR* | 4.887550484 | 8.46E-06 | 0.000134614 | 0.053164323 |
| Obesity | t_cell | *RRAGA* | 6.564602146 | 0.001062979 | 0.002691483 | 1 |
| Obesity | t_cell | *TMBIM4* | 2.469402023 | 0.004764058 | 0.009671176 | 1 |
| Obesity | t_cell | *FAM161B* | 8.082121942 | 0.000162985 | 0.000623668 | 1 |
| Obesity | t_cell | *WIF1* | 5.30721525 | 0.00789329 | 0.015088207 | 1 |
| Obesity | t_cell | *UGT2B4* | 8.724317006 | 1.16E-05 | 0.000145864 | 0.073224019 |
| Obesity | t_cell | *MMP16* | 1.809672142 | 0.001456155 | 0.003500897 | 1 |
| Obesity | t_cell | *KCNMA1* | 2.86559987 | 0.000145155 | 0.000575257 | 0.91258642 |
| Obesity | t_cell | *BATF* | -0.764119194 | 0.000193189 | 0.000716989 | 1 |
| Obesity | t_cell | *NDUFAF6* | 3.050212473 | 0.007092571 | 0.013762653 | 1 |
| Obesity | t_cell | *DRAM2* | 3.609844464 | 0.000823013 | 0.002182322 | 1 |
| Obesity | t_cell | *C8orf37* | 2.3443443 | 0.009339102 | 0.017516389 | 1 |
| Obesity | t_cell | *ADAMTSL3* | 2.063332739 | 8.15E-06 | 0.000132809 | 0.051264114 |
| Obesity | t_cell | *CXCL13* | 7.460565931 | 2.33E-05 | 0.000175508 | 0.14622797 |
| Obesity | t_cell | *ANKRD9* | 3.355256221 | 0.000217073 | 0.00077896 | 1 |
| Obesity | t_cell | *SFXN2* | 2.998732297 | 0.017622177 | 0.030287213 | 1 |
| Obesity | t_cell | *GDF6* | 4.814293995 | 0.000158178 | 0.000610564 | 0.994466053 |
| Obesity | t_cell | *NODAL* | 4.318024731 | 1.77E-05 | 0.000157653 | 0.11130299 |
| Obesity | t_cell | *VPS8* | 2.473226661 | 0.022486444 | 0.037321086 | 1 |
| Obesity | t_cell | *MPV17L* | 1.625711228 | 0.003269172 | 0.007017167 | 1 |
| Obesity | t_cell | *RPUSD3* | 7.555147384 | 9.20E-05 | 0.000407341 | 0.578147711 |
| Obesity | t_cell | *NTAN1* | 5.076414738 | 0.000520011 | 0.001501751 | 1 |
| Obesity | t_cell | *FCHO2* | 4.133801151 | 6.45E-05 | 0.000317791 | 0.405819316 |
| Obesity | t_cell | *TIMP4* | 2.493904013 | 0.002533054 | 0.005630594 | 1 |
| Obesity | t_cell | *CPT2* | 6.928843994 | 0.000156232 | 0.000604451 | 0.982231646 |
| Obesity | t_cell | *LRP8* | -2.421340449 | 0.000571617 | 0.001620999 | 1 |
| Obesity | t_cell | *GATAD1* | 3.365446927 | 2.51E-05 | 0.000183564 | 0.157497835 |
| Obesity | t_cell | *SUSD3* | -0.66391613 | 0.002666115 | 0.005889622 | 1 |
| Obesity | t_cell | *TMED6* | 8.20823658 | 3.83E-05 | 0.00023194 | 0.240984272 |
| Obesity | t_cell | *CLEC18A* | -9.613773436 | 3.36E-18 | 2.64E-15 | 2.11E-14 |
| Obesity | t_cell | *IL34* | 2.959813433 | 5.78E-05 | 0.000293903 | 0.363264352 |
| Obesity | t_cell | *AASDH* | 3.811591579 | 0.001649853 | 0.003881972 | 1 |
| Obesity | t_cell | *RNF111* | 3.645650282 | 0.001140187 | 0.00285878 | 1 |
| Obesity | t_cell | *CREB3L1* | 4.063703695 | 7.16E-05 | 0.000340923 | 0.450018146 |
| Obesity | t_cell | *C2CD2* | 1.49255029 | 0.000653894 | 0.001807046 | 1 |
| Obesity | t_cell | *C9orf43* | 2.587495254 | 0.00122484 | 0.003042501 | 1 |
| Obesity | t_cell | *WDR19* | 3.711153763 | 1.65E-05 | 0.000155092 | 0.103482283 |

| | | | | | | |
|---|---|---|---|---|---|---|
| Obesity | t_cell | *SLC37A3* | 4.979406942 | 0.000923386 | 0.002393949 | 1 |
| Obesity | t_cell | *C12orf43* | 3.348544086 | 0.006831881 | 0.013335 | 1 |
| Obesity | t_cell | *PEX10* | 4.746779694 | 2.64E-05 | 0.000188499 | 0.166256192 |
| Obesity | t_cell | *RADIL* | 3.4267736 | 2.13E-05 | 0.000169213 | 0.134185868 |
| Obesity | t_cell | *LDLRAP1* | -1.990711286 | 0.001268758 | 0.003129339 | 1 |
| Obesity | t_cell | *TRIM63* | 1.962251137 | 0.012649548 | 0.022735194 | 1 |
| Obesity | t_cell | *GALNT14* | 11.73997001 | 0.000144927 | 0.000574864 | 0.911158887 |
| Obesity | t_cell | *HPD* | 4.619535251 | 9.63E-10 | 1.68E-07 | 6.05E-06 |
| Obesity | t_cell | *RHPN1* | -3.342317743 | 0.000377919 | 0.001173323 | 1 |
| Obesity | t_cell | *TPRG1L* | 6.187307319 | 0.001177648 | 0.002934552 | 1 |
| Obesity | t_cell | *XKR8* | 5.464387519 | 0.000664858 | 0.001832131 | 1 |
| Obesity | t_cell | *EYA3* | 2.807319094 | 0.024521767 | 0.040189872 | 1 |
| Obesity | t_cell | *MRAS* | 1.743114863 | 0.000197543 | 0.000727565 | 1 |
| Obesity | t_cell | *CLSTN2* | 1.658907277 | 5.99E-05 | 0.000301064 | 0.376631437 |
| Obesity | t_cell | *COLEC12* | 4.011524076 | 4.19E-05 | 0.000244655 | 0.263737857 |
| Obesity | t_cell | *RNF207* | 7.483893415 | 1.27E-05 | 0.000146996 | 0.079590253 |
| Obesity | t_cell | *AUTS2* | 3.107810809 | 0.000558541 | 0.001593983 | 1 |
| Obesity | t_cell | *TSPAN33* | 1.108211512 | 0.029659324 | 0.047544153 | 1 |
| Obesity | t_cell | *B4GALT5* | 3.375691754 | 0.01100719 | 0.020234562 | 1 |
| Obesity | t_cell | *FAM86C1* | 3.835296047 | 3.32E-05 | 0.000212553 | 0.208726562 |
| Obesity | t_cell | *CD1E* | 8.090525492 | 8.83E-07 | 3.69E-05 | 0.005553025 |
| Obesity | t_cell | *NCF1* | 4.908130872 | 2.01E-05 | 0.000163915 | 0.126083011 |
| Obesity | t_cell | *PPP1R9A* | 2.718850446 | 7.96E-05 | 0.000370276 | 0.500612784 |
| Obesity | t_cell | *GDPD5* | 4.896311814 | 7.38E-06 | 0.000128167 | 0.046396541 |
| Obesity | t_cell | *DYNC1I1* | 3.563141121 | 1.36E-05 | 0.000147409 | 0.085215376 |
| Obesity | t_cell | *TMED4* | 6.13748096 | 0.000113163 | 0.000475891 | 0.711457762 |
| Obesity | t_cell | *DUSP23* | 4.720341033 | 1.44E-05 | 0.000148509 | 0.090796728 |
| Obesity | t_cell | *RNF166* | -1.869202105 | 0.000747511 | 0.002011816 | 1 |
| Obesity | t_cell | *NIT1* | 3.190155438 | 0.000108028 | 0.000460144 | 0.679171357 |
| Obesity | t_cell | *ZNF276* | -2.638497131 | 0.003352734 | 0.007162297 | 1 |
| Obesity | t_cell | *FGF17* | -5.923663231 | 0.010151419 | 0.018837654 | 1 |
| Obesity | t_cell | *DMTN* | 2.500597186 | 0.000169091 | 0.000643117 | 1 |
| Obesity | t_cell | *FAM160B2* | 4.389509387 | 0.00146948 | 0.003528885 | 1 |
| Obesity | t_cell | *NDUFS2* | 3.143564488 | 0.002224709 | 0.005033014 | 1 |
| Obesity | t_cell | *SLC5A11* | -1.320728173 | 0.000241987 | 0.000845677 | 1 |
| Obesity | t_cell | *FCER1G* | 2.875494825 | 4.09E-05 | 0.000241098 | 0.257251478 |
| Obesity | t_cell | *CDC42SE2* | -1.084419375 | 0.001421915 | 0.003432224 | 1 |
| Obesity | t_cell | *RAPGEF6* | -4.091035452 | 0.001493838 | 0.003576451 | 1 |
| Obesity | t_cell | *ALG8* | 5.077941869 | 6.41E-05 | 0.00031572 | 0.402858686 |
| Obesity | t_cell | *SV2A* | -5.012472427 | 0.000615401 | 0.001721864 | 1 |
| Obesity | t_cell | *CLIC6* | 2.865656435 | 2.07E-06 | 6.49E-05 | 0.01304399 |
| Obesity | t_cell | *SIM2* | 1.665284465 | 0.006492856 | 0.012728588 | 1 |
| Obesity | t_cell | *PADI4* | 10.57033456 | 0.013677005 | 0.02429707 | 1 |
| Obesity | t_cell | *PSMD4* | 5.369621567 | 0.002827884 | 0.006188271 | 1 |
| Obesity | t_cell | *STARD9* | 3.62003982 | 0.000359107 | 0.001131683 | 1 |
| Obesity | t_cell | *TCHH* | 2.910663269 | 0.005095864 | 0.010291583 | 1 |
| Obesity | t_cell | *MED8* | 2.980667564 | 0.00052669 | 0.001516582 | 1 |
| Obesity | t_cell | *CCDC17* | 3.514683237 | 0.000344508 | 0.001100011 | 1 |
| Obesity | t_cell | *ACE* | 5.144215626 | 8.71E-05 | 0.000392351 | 0.547721375 |
| Obesity | t_cell | *TEPP* | -6.782608438 | 4.25E-05 | 0.000246282 | 0.267056159 |

| | | | | | | |
|---|---|---|---|---|---|---|
| Obesity | t_cell | *ZFYVE28* | -1.294401557 | 0.00884805 | 0.016699997 | 1 |
| Obesity | t_cell | *C16orf86* | 3.25476035 | 0.001508488 | 0.003606032 | 1 |
| Obesity | t_cell | *NPR2* | 4.837525456 | 4.96E-05 | 0.000266905 | 0.312011834 |
| Obesity | t_cell | *ZNF235* | 3.589214618 | 0.003773118 | 0.007915112 | 1 |
| Obesity | t_cell | *DFFA* | 5.209525277 | 1.40E-07 | 9.56E-06 | 0.000879357 |
| Obesity | t_cell | *NR2F6* | 3.808220171 | 8.21E-05 | 0.000377231 | 0.516429618 |
| Obesity | t_cell | *CILP2* | 11.55874161 | 0.000322553 | 0.001048175 | 1 |
| Obesity | t_cell | *NDUFV3* | 0.722353876 | 0.016124716 | 0.028050938 | 1 |
| Obesity | t_cell | *HSF2BP* | 3.208086546 | 0.003382942 | 0.007211298 | 1 |
| Obesity | t_cell | *CSTB* | 2.492291457 | 0.010566428 | 0.019527083 | 1 |
| Obesity | t_cell | *ITGB2* | -1.109943508 | 0.018868529 | 0.032105604 | 1 |
| Obesity | t_cell | *LSS* | 4.025337203 | 1.04E-06 | 4.20E-05 | 0.0065206 |
| Obesity | t_cell | *VAV2* | 4.970459425 | 0.000409 | 0.001247033 | 1 |
| Obesity | t_cell | *PCNT* | 2.202833663 | 0.000911171 | 0.00236912 | 1 |
| Obesity | t_cell | *ZNF208* | 2.089671897 | 0.019491898 | 0.032986692 | 1 |
| Obesity | t_cell | *ADAMTS13* | 2.600357165 | 0.007139017 | 0.013839962 | 1 |
| Obesity | t_cell | *ZNF714* | 3.744000793 | 0.001754373 | 0.004092668 | 1 |
| Obesity | t_cell | *GPSM1* | 3.365417394 | 2.15E-05 | 0.000169226 | 0.135017437 |
| Obesity | t_cell | *C19orf47* | 10.31514889 | 4.06E-07 | 2.18E-05 | 0.002552182 |
| Obesity | t_cell | *HIPK4* | 4.418248887 | 0.00807989 | 0.015412095 | 1 |
| Obesity | t_cell | *MPZL3* | -0.560787435 | 0.000176812 | 0.000668039 | 1 |
| Obesity | t_cell | *PCSK7* | -1.838561254 | 0.01795476 | 0.030791482 | 1 |
| Obesity | t_cell | *S100A1* | 6.857305167 | 0.000152034 | 0.000594055 | 0.955840795 |
| Obesity | t_cell | *FDPS* | 4.08142662 | 0.000719526 | 0.001949182 | 1 |
| Obesity | t_cell | *PAQR6* | -4.880776895 | 0.004197306 | 0.008660473 | 1 |
| Obesity | t_cell | *PMF1* | 3.200764877 | 0.018498611 | 0.031586303 | 1 |
| Obesity | t_cell | *SLC25A44* | -3.812938189 | 0.018888816 | 0.032130408 | 1 |
| Obesity | t_cell | *FCRL3* | -0.78226 | 0.00298577 | 0.006484124 | 1 |
| Obesity | t_cell | *NACC1* | 8.271457088 | 0.000157556 | 0.000609197 | 0.990554005 |
| Obesity | t_cell | *LY6K* | 5.655905982 | 0.00012524 | 0.000512285 | 0.787381817 |
| Obesity | t_cell | *TONSL* | 8.327526593 | 0.000642304 | 0.001779711 | 1 |
| Obesity | t_cell | *MUM1* | 2.570349357 | 0.009143693 | 0.017190909 | 1 |
| Obesity | t_cell | *ZNF333* | 5.289955297 | 1.45E-05 | 0.000148509 | 0.090887369 |
| Obesity | t_cell | *PPP1R16A* | 1.869231213 | 0.000373877 | 0.001164221 | 1 |
| Obesity | t_cell | *FBXL13* | 4.842591755 | 4.14E-05 | 0.000242764 | 0.260243231 |
| Obesity | t_cell | *SCGB3A1* | -2.824108188 | 0.001084653 | 0.002739056 | 1 |
| Obesity | t_cell | *YDJC* | -2.108115193 | 0.000278248 | 0.000938539 | 1 |
| Obesity | t_cell | *DVL3* | 4.509351018 | 9.55E-06 | 0.000140299 | 0.060045796 |
| Obesity | t_cell | *AP2M1* | 5.639688737 | 1.34E-05 | 0.000147409 | 0.08421361 |
| Obesity | t_cell | *ABCF3* | 2.660085029 | 5.85E-05 | 0.000296502 | 0.367958455 |
| Obesity | t_cell | *FBXO27* | 1.661582034 | 3.52E-06 | 8.91E-05 | 0.022108521 |
| Obesity | t_cell | *DMKN* | 1.444850461 | 0.026248383 | 0.042741151 | 1 |
| Obesity | t_cell | *BDH1* | -2.181242424 | 0.00093252 | 0.002412655 | 1 |
| Obesity | t_cell | *COX7A1* | 2.353604257 | 0.00356442 | 0.007522494 | 1 |
| Obesity | t_cell | *DUSP14* | 1.967628078 | 0.001326072 | 0.003238933 | 1 |
| Obesity | t_cell | *LRRC56* | 2.103791582 | 0.009857916 | 0.018390717 | 1 |
| Obesity | t_cell | *IKZF3* | -0.435006144 | 0.000199907 | 0.000733405 | 1 |
| Obesity | t_cell | *ACOX1* | 3.428187461 | 1.16E-05 | 0.000145864 | 0.073217955 |
| Obesity | t_cell | *PRPSAP1* | 5.020160747 | 0.00029406 | 0.000981293 | 1 |
| Obesity | t_cell | *CYGB* | 3.660724305 | 7.70E-05 | 0.000360281 | 0.483857363 |

| | | | | | | |
|---|---|---|---|---|---|---|
| Obesity | t_cell | *ZNF577* | 2.655143794 | 0.002586108 | 0.005733027 | 1 |
| Obesity | t_cell | *TMEM143* | 3.683734985 | 1.29E-05 | 0.000147409 | 0.08124937 |
| Obesity | t_cell | *ALDH16A1* | 6.441813527 | 0.000398336 | 0.001222224 | 1 |
| Obesity | t_cell | *SIGLEC11* | -3.194607956 | 0.02791927 | 0.045065071 | 1 |
| Obesity | t_cell | *MPP3* | 2.344314894 | 5.19E-05 | 0.000273636 | 0.326448031 |
| Obesity | t_cell | *NAGS* | 10.12458274 | 0.003137008 | 0.0067635 | 1 |
| Obesity | t_cell | *PLCD3* | 1.4793524 | 0.008936636 | 0.016836868 | 1 |
| Obesity | t_cell | *LARP4* | 2.475355792 | 0.012226476 | 0.022126613 | 1 |
| Obesity | t_cell | *SPC24* | 12.0689696 | 1.82E-05 | 0.000158673 | 0.114640494 |
| Obesity | t_cell | *ASGR2* | -10.25798367 | 0.00404873 | 0.008403555 | 1 |
| Obesity | t_cell | *SNRNP25* | 4.053992828 | 0.000132671 | 0.000534681 | 0.834101712 |
| Obesity | t_cell | *JMJD8* | 3.821720699 | 7.80E-05 | 0.000364521 | 0.490280885 |
| Obesity | t_cell | *MEIOB* | 3.715658246 | 1.72E-05 | 0.000156669 | 0.10824557 |
| Obesity | t_cell | *FLYWCH2* | 5.388212397 | 4.35E-06 | 9.97E-05 | 0.02736485 |
| Obesity | t_cell | *ZG16B* | 22.11630598 | 5.10E-05 | 0.000270777 | 0.320599521 |
| Obesity | t_cell | *TTC9C* | 3.797327868 | 0.011057435 | 0.02029133 | 1 |
| Obesity | t_cell | *NXF1* | 4.559870198 | 0.002363339 | 0.005304645 | 1 |
| Obesity | t_cell | *RPL29* | -1.753834121 | 0.018521816 | 0.031608756 | 1 |
| Obesity | t_cell | *TPCN2* | 5.401048622 | 4.69E-07 | 2.42E-05 | 0.002947442 |
| Obesity | t_cell | *CYP4A22* | 8.428735567 | 0.024027725 | 0.039452157 | 1 |
| Obesity | t_cell | *PDZK1IP1* | 30.10560584 | 0.0024485 | 0.005464578 | 1 |
| Obesity | t_cell | *COA7* | -2.336626031 | 0.001044342 | 0.002654987 | 1 |
| Obesity | t_cell | *ACOT11* | 3.36932989 | 0.001163007 | 0.002902669 | 1 |
| Obesity | t_cell | *NOL9* | 5.695273903 | 0.000382712 | 0.001182365 | 1 |
| Obesity | t_cell | *RBP7* | 6.803525829 | 1.64E-05 | 0.000155092 | 0.103190894 |
| Obesity | t_cell | *FBLIM1* | 3.109707824 | 7.56E-05 | 0.000355346 | 0.475097582 |
| Obesity | t_cell | *SLC25A34* | 7.246188975 | 3.52E-05 | 0.000219892 | 0.221431006 |
| Obesity | t_cell | *PDPN* | 1.67931549 | 0.014235154 | 0.025146506 | 1 |
| Obesity | t_cell | *LRRC38* | 13.90983479 | 0.000624253 | 0.001741205 | 1 |
| Obesity | t_cell | *LAPTM5* | -1.666795991 | 0.006117447 | 0.012064112 | 1 |
| Obesity | t_cell | *PEF1* | 10.21827004 | 8.40E-06 | 0.000134614 | 0.052811532 |
| Obesity | t_cell | *TSSK3* | 2.340892664 | 0.000177287 | 0.000669432 | 1 |
| Obesity | t_cell | *CAMK2N1* | 10.19958091 | 2.04E-06 | 6.45E-05 | 0.012844363 |
| Obesity | t_cell | *TTLL10* | -4.042326402 | 0.00238883 | 0.005348495 | 1 |
| Obesity | t_cell | *NFIA* | 3.446638808 | 1.51E-05 | 0.000149538 | 0.094807402 |
| Obesity | t_cell | *NEXN* | 4.429116926 | 1.92E-05 | 0.000161262 | 0.120971384 |
| Obesity | t_cell | *HFM1* | 3.499453291 | 1.42E-05 | 0.000148367 | 0.089080895 |
| Obesity | t_cell | *KCNT2* | 4.028916334 | 6.98E-06 | 0.000124621 | 0.043866445 |
| Obesity | t_cell | *VCAM1* | 2.593715785 | 4.79E-05 | 0.000262855 | 0.301403946 |
| Obesity | t_cell | *EXTL2* | 2.299748645 | 0.001132979 | 0.00284353 | 1 |
| Obesity | t_cell | *ZNF496* | 6.333989533 | 8.71E-06 | 0.000135109 | 0.054774142 |
| Obesity | t_cell | *IGSF8* | 7.713910958 | 2.26E-05 | 0.00017343 | 0.141914399 |
| Obesity | t_cell | *FCRLB* | 6.710372749 | 0.000228421 | 0.00080724 | 1 |
| Obesity | t_cell | *LMX1A* | 1.269792211 | 0.016417845 | 0.028421446 | 1 |
| Obesity | t_cell | *TDRD5* | 1.776989597 | 0.018170594 | 0.031110709 | 1 |
| Obesity | t_cell | *SNED1* | 3.434573291 | 1.90E-05 | 0.000160838 | 0.119242185 |
| Obesity | t_cell | *C1orf115* | 3.417580622 | 3.77E-06 | 9.34E-05 | 0.023725575 |
| Obesity | t_cell | *ACP6* | -3.494424144 | 0.002638713 | 0.005839349 | 1 |
| Obesity | t_cell | *PM20D1* | -4.204446755 | 0.0108933 | 0.020054517 | 1 |
| Obesity | t_cell | *HAAO* | 3.716317395 | 1.44E-06 | 5.24E-05 | 0.009062699 |

| | | | | | | |
|---|---|---|---|---|---|---|
| Obesity | t_cell | *B3GALNT2* | 3.269046215 | 0.000410369 | 0.001249996 | 1 |
| Obesity | t_cell | *PUS10* | 5.494066498 | 0.000633716 | 0.00176135 | 1 |
| Obesity | t_cell | *RFTN2* | 3.981706577 | 1.13E-05 | 0.000145864 | 0.071019276 |
| Obesity | t_cell | *FAM84A* | 5.793072234 | 2.55E-05 | 0.000185244 | 0.160050884 |
| Obesity | t_cell | *CLHC1* | 1.23250884 | 9.60E-05 | 0.000419385 | 0.603495496 |
| Obesity | t_cell | *FRZB* | 3.790319748 | 0.006610548 | 0.01294317 | 1 |
| Obesity | t_cell | *FBXO41* | -3.669479715 | 0.003419318 | 0.007282267 | 1 |
| Obesity | t_cell | *CCDC74A* | 20.61533029 | 1.49E-05 | 0.000149261 | 0.093868491 |
| Obesity | t_cell | *NOSTRIN* | 2.654062848 | 1.79E-05 | 0.000158468 | 0.112566678 |
| Obesity | t_cell | *SGPP2* | -0.964054553 | 0.000231193 | 0.000814748 | 1 |
| Obesity | t_cell | *BBS5* | 2.922651439 | 1.82E-05 | 0.000158673 | 0.114407797 |
| Obesity | t_cell | *HPGDS* | 3.945815385 | 2.31E-05 | 0.000175493 | 0.145478071 |
| Obesity | t_cell | *PDLIM5* | 3.94318338 | 0.00013842 | 0.000554297 | 0.870245738 |
| Obesity | t_cell | *PACRGL* | 3.929162568 | 0.000369656 | 0.001154035 | 1 |
| Obesity | t_cell | *C1QTNF7* | 2.189821189 | 2.62E-05 | 0.000187653 | 0.164745376 |
| Obesity | t_cell | *LYSMD1* | 7.413520197 | 1.04E-07 | 7.80E-06 | 0.000655197 |
| Obesity | t_cell | *DHX57* | 5.655997796 | 6.48E-05 | 0.000318079 | 0.407694497 |
| Obesity | t_cell | *S100A12* | 11.91403927 | 0.000437653 | 0.001315259 | 1 |
| Obesity | t_cell | *TDRD10* | 4.793675657 | 9.94E-06 | 0.000142984 | 0.062483913 |
| Obesity | t_cell | *DCAF16* | 5.539593474 | 0.000441778 | 0.001323861 | 1 |
| Obesity | t_cell | *GNPDA2* | 2.038822561 | 0.002628679 | 0.005819191 | 1 |
| Obesity | t_cell | *NIPAL1* | 7.392541131 | 2.11E-05 | 0.000168313 | 0.132462589 |
| Obesity | t_cell | *ANTXR2* | 2.404383634 | 0.009874047 | 0.018409885 | 1 |
| Obesity | t_cell | *PBXIP1* | -1.347737918 | 0.001082096 | 0.00273548 | 1 |
| Obesity | t_cell | *DCST2* | -2.857334501 | 0.002145751 | 0.004891348 | 1 |
| Obesity | t_cell | *NBPF10* | 2.999396875 | 0.008721221 | 0.016480408 | 1 |
| Obesity | t_cell | *SLC22A15* | 5.716824756 | 2.18E-06 | 6.63E-05 | 0.013732432 |
| Obesity | t_cell | *FSTL1* | 1.877299447 | 0.0218476 | 0.036372176 | 1 |
| Obesity | t_cell | *LMOD1* | -4.814637436 | 9.34E-10 | 1.68E-07 | 5.87E-06 |
| Obesity | t_cell | *CCT3* | 2.857589009 | 0.009041281 | 0.017008539 | 1 |
| Obesity | t_cell | *GLB1L* | 1.59335771 | 0.025873014 | 0.042173617 | 1 |
| Obesity | t_cell | *PYHIN1* | -0.57240337 | 8.89E-05 | 0.000396739 | 0.559004857 |
| Obesity | t_cell | *EFHB* | 0.586233718 | 0.007772561 | 0.01490063 | 1 |
| Obesity | t_cell | *CD200R1* | -1.421735472 | 0.001554348 | 0.003704392 | 1 |
| Obesity | t_cell | *SPICE1* | 4.119808736 | 0.000258654 | 0.000890417 | 1 |
| Obesity | t_cell | *CADPS* | 5.704447299 | 1.22E-05 | 0.000145864 | 0.076432645 |
| Obesity | t_cell | *GMPS* | 4.34846718 | 0.009998806 | 0.018592869 | 1 |
| Obesity | t_cell | *TIPARP* | 5.240608499 | 0.00019524 | 0.000721619 | 1 |
| Obesity | t_cell | *DCLK3* | 6.709928307 | 2.71E-05 | 0.000191468 | 0.170406819 |
| Obesity | t_cell | *RPL9* | -0.764324325 | 0.012454256 | 0.022454805 | 1 |
| Obesity | t_cell | *RPP14* | 5.551918835 | 0.001770397 | 0.004125458 | 1 |
| Obesity | t_cell | *DNASE1L3* | 2.880454716 | 1.70E-05 | 0.000156669 | 0.106942408 |
| Obesity | t_cell | *CRELD1* | 3.376775984 | 5.66E-06 | 0.000109552 | 0.035604546 |
| Obesity | t_cell | *PCOLCE2* | 2.50163793 | 0.000552233 | 0.001581006 | 1 |
| Obesity | t_cell | *CCDC158* | 2.106148572 | 1.57E-05 | 0.000152754 | 0.098526201 |
| Obesity | t_cell | *GYG1* | 5.281758001 | 0.003911907 | 0.008172639 | 1 |
| Obesity | t_cell | *RYK* | 4.341375959 | 0.000759655 | 0.002038392 | 1 |
| Obesity | t_cell | *PLB1* | 6.776517654 | 1.57E-05 | 0.000152971 | 0.09897205 |
| Obesity | t_cell | *CDCP1* | 3.233252714 | 2.20E-05 | 0.000171178 | 0.138312225 |
| Obesity | t_cell | *LZTFL1* | 7.277898984 | 0.000160102 | 0.000615257 | 1 |

| | | | | | | |
|---|---|---|---|---|---|---|
| Obesity | t_cell | *LRRC2* | 1.928593877 | 0.00149958 | 0.0035861 | 1 |
| Obesity | t_cell | *NMNAT3* | 4.077025371 | 1.17E-05 | 0.000145864 | 0.073750883 |
| Obesity | t_cell | *SMIM12* | 5.517633574 | 5.72E-05 | 0.000291742 | 0.35942572 |
| Obesity | t_cell | *TPRA1* | 5.678345415 | 9.75E-06 | 0.000141822 | 0.061266961 |
| Obesity | t_cell | *DNALI1* | 2.172500736 | 0.011018474 | 0.020249385 | 1 |
| Obesity | t_cell | *RPN1* | 4.497562598 | 0.012474187 | 0.022477848 | 1 |
| Obesity | t_cell | *RFC4* | 5.513252513 | 0.000685675 | 0.001882463 | 1 |
| Obesity | t_cell | *SFMBT1* | -2.082245107 | 0.010196939 | 0.01891654 | 1 |
| Obesity | t_cell | *GNL3* | 3.142003513 | 0.021720516 | 0.03621238 | 1 |
| Obesity | t_cell | *UVSSA* | 2.344845756 | 0.01333428 | 0.023775558 | 1 |
| Obesity | t_cell | *LRPAP1* | 5.073858231 | 0.000515362 | 0.001490378 | 1 |
| Obesity | t_cell | *ABLIM2* | 7.81835009 | 6.61E-05 | 0.000321248 | 0.415373185 |
| Obesity | t_cell | *ERMAP* | 2.346163789 | 0.004862272 | 0.009857822 | 1 |
| Obesity | t_cell | *ATRIP* | 9.28000093 | 0.000240519 | 0.000842421 | 1 |
| Obesity | t_cell | *SHISA5* | 5.465462479 | 0.004129807 | 0.008543631 | 1 |
| Obesity | t_cell | *APEH* | -6.37245936 | 1.97E-12 | 6.88E-10 | 1.24E-08 |
| Obesity | t_cell | *RNF123* | 5.910559705 | 1.30E-18 | 1.36E-15 | 8.16E-15 |
| Obesity | t_cell | *MST1R* | 4.383510753 | 1.10E-10 | 2.77E-08 | 6.93E-07 |
| Obesity | t_cell | *TEX264* | -3.619890385 | 6.93E-05 | 0.000332617 | 0.435728498 |
| Obesity | t_cell | *GRM2* | -4.561071892 | 0.000115295 | 0.00048324 | 0.724859831 |
| Obesity | t_cell | *POC1A* | 6.233074045 | 4.18E-08 | 3.76E-06 | 0.000262879 |
| Obesity | t_cell | *PPM1M* | -3.059061966 | 7.04E-07 | 3.23E-05 | 0.004423813 |
| Obesity | t_cell | *C4orf3* | 5.868196225 | 0.000644196 | 0.001782597 | 1 |
| Obesity | t_cell | *ANXA5* | 3.42010462 | 0.001878822 | 0.004352304 | 1 |
| Obesity | t_cell | *CEP44* | 4.762091062 | 0.004696696 | 0.009537508 | 1 |
| Obesity | t_cell | *C4orf45* | 4.312554162 | 1.93E-06 | 6.34E-05 | 0.012159361 |
| Obesity | t_cell | *TMEM144* | 3.084902645 | 1.20E-05 | 0.000145864 | 0.075435751 |
| Obesity | t_cell | *NPY1R* | 2.393155313 | 1.36E-05 | 0.000147409 | 0.085485907 |
| Obesity | t_cell | *ARFIP1* | 3.909830565 | 0.000184749 | 0.000692615 | 1 |
| Obesity | t_cell | *OTUD4* | 4.334582034 | 0.007858966 | 0.015031737 | 1 |
| Obesity | t_cell | *PGGT1B* | 5.049826835 | 0.001448056 | 0.003484091 | 1 |
| Obesity | t_cell | *CMBL* | 3.433737717 | 1.77E-05 | 0.000157653 | 0.111160573 |
| Obesity | t_cell | *PRRC1* | 5.61756014 | 2.36E-05 | 0.000176915 | 0.148431938 |
| Obesity | t_cell | *WDR41* | 3.471811701 | 0.000113116 | 0.000475891 | 0.711159714 |
| Obesity | t_cell | *NDUFS4* | 4.169967122 | 0.002793825 | 0.006128673 | 1 |
| Obesity | t_cell | *RHOBTB3* | 1.985082935 | 0.000101681 | 0.000438456 | 0.639268936 |
| Obesity | t_cell | *TIGD6* | 4.065405908 | 2.26E-05 | 0.00017343 | 0.141952994 |
| Obesity | t_cell | *SERINC5* | -1.602369112 | 0.015411074 | 0.027026338 | 1 |
| Obesity | t_cell | *ENPP6* | 5.426691275 | 3.72E-05 | 0.000227538 | 0.233908843 |
| Obesity | t_cell | *PRIMPOL* | 3.575173302 | 0.000360833 | 0.001135981 | 1 |
| Obesity | t_cell | *ERAP1* | -1.796353642 | 0.027040606 | 0.043826834 | 1 |
| Obesity | t_cell | *ANKRA2* | 5.739067035 | 0.000984889 | 0.002523226 | 1 |
| Obesity | t_cell | *TLR3* | 4.910247225 | 6.05E-06 | 0.000113242 | 0.038011071 |
| Obesity | t_cell | *GRIK2* | 1.563398713 | 0.005886007 | 0.011658892 | 1 |
| Obesity | t_cell | *DCBLD1* | 5.203043778 | 0.000136336 | 0.000547348 | 0.857147497 |
| Obesity | t_cell | *TMEM200A* | -1.082870675 | 0.000381394 | 0.00118003 | 1 |
| Obesity | t_cell | *PI16* | 4.892455565 | 0.000172359 | 0.000653379 | 1 |
| Obesity | t_cell | *STK17A* | -1.548066146 | 0.00741808 | 0.014306441 | 1 |
| Obesity | t_cell | *RPS14* | -1.542200912 | 0.019569919 | 0.033092006 | 1 |
| Obesity | t_cell | *SLU7* | 5.458200419 | 8.81E-05 | 0.000394839 | 0.553958558 |

| | | | | | | |
|---|---|---|---|---|---|---|
| Obesity | t_cell | *RP9* | 4.002384447 | 0.001321084 | 0.003228136 | 1 |
| Obesity | t_cell | *KCNK5* | 3.779118828 | 3.85E-05 | 0.00023194 | 0.241913128 |
| Obesity | t_cell | *USP49* | 3.997570056 | 9.69E-05 | 0.000422417 | 0.609125842 |
| Obesity | t_cell | *ZNF704* | 3.179980087 | 2.28E-05 | 0.000174138 | 0.143489405 |
| Obesity | t_cell | *FNDC1* | 3.084189427 | 0.000296046 | 0.000984784 | 1 |
| Obesity | t_cell | *BRI3* | 3.991093781 | 0.001365285 | 0.003315392 | 1 |
| Obesity | t_cell | *PEX2* | 5.116279729 | 0.001604752 | 0.003801461 | 1 |
| Obesity | t_cell | *TNFRSF11B* | 4.260031905 | 2.94E-06 | 8.04E-05 | 0.018514518 |
| Obesity | t_cell | *SUN1* | 3.580156351 | 5.56E-05 | 0.000286272 | 0.34982387 |
| Obesity | t_cell | *OXR1* | 3.943884431 | 0.000149461 | 0.000586556 | 0.939662354 |
| Obesity | t_cell | *GPER1* | 2.550889417 | 9.62E-05 | 0.000420184 | 0.605065357 |
| Obesity | t_cell | *SLC4A2* | 7.522807019 | 3.88E-05 | 0.000232738 | 0.243909243 |
| Obesity | t_cell | *ALDH7A1* | 2.547563359 | 1.92E-05 | 0.000161227 | 0.120540081 |
| Obesity | t_cell | *YWHAZ* | -2.084008558 | 0.004550393 | 0.009285401 | 1 |
| Obesity | t_cell | *SLC25A32* | 3.523000349 | 0.012864865 | 0.023082593 | 1 |
| Obesity | t_cell | *FREM1* | 2.394867389 | 3.31E-05 | 0.000212339 | 0.208304619 |
| Obesity | t_cell | *FAM219A* | 5.287466595 | 9.45E-05 | 0.000414733 | 0.593897304 |
| Obesity | t_cell | *CCDC171* | 3.501007293 | 0.000423072 | 0.001280622 | 1 |
| Obesity | t_cell | *SYK* | 3.592105167 | 2.06E-05 | 0.000166806 | 0.129775236 |
| Obesity | t_cell | *NIPSNAP3B* | 1.419591498 | 0.022978141 | 0.037986741 | 1 |
| Obesity | t_cell | *METTL2B* | 0.785349344 | 0.017098656 | 0.02945992 | 1 |
| Obesity | t_cell | *TMC1* | 2.926712653 | 1.28E-05 | 0.000146996 | 0.080259638 |
| Obesity | t_cell | *HGSNAT* | 3.448319326 | 0.001280158 | 0.00315004 | 1 |
| Obesity | t_cell | *C9orf64* | 2.357310316 | 0.002382443 | 0.005337997 | 1 |
| Obesity | t_cell | *SVEP1* | 2.286714124 | 6.40E-06 | 0.000116604 | 0.040228522 |
| Obesity | t_cell | *ZNF367* | -1.307941915 | 0.006424072 | 0.012613411 | 1 |
| Obesity | t_cell | *NOL6* | 4.722012157 | 0.020824962 | 0.034913744 | 1 |
| Obesity | t_cell | *AQP3* | -0.68276943 | 0.000161676 | 0.00062017 | 1 |
| Obesity | t_cell | *TRMT10B* | 4.914501826 | 0.000940444 | 0.002427163 | 1 |
| Obesity | t_cell | *VCP* | 3.115576864 | 0.001921187 | 0.004440626 | 1 |
| Obesity | t_cell | *LRFN5* | 2.417863054 | 1.50E-07 | 9.91E-06 | 0.000940806 |
| Obesity | t_cell | *WRN* | 4.127047035 | 0.001287843 | 0.003164824 | 1 |
| Obesity | t_cell | *CFL2* | 4.518873144 | 1.88E-05 | 0.000160637 | 0.118250154 |
| Obesity | t_cell | *ZCCHC24* | 3.627059276 | 1.36E-05 | 0.000147409 | 0.085480051 |
| Obesity | t_cell | *LRR1* | 5.379228165 | 0.000238532 | 0.000836393 | 1 |
| Obesity | t_cell | *C10orf25* | 1.268402044 | 0.017186299 | 0.029594703 | 1 |
| Obesity | t_cell | *KLHDC2* | 2.993546091 | 0.01821044 | 0.031161959 | 1 |
| Obesity | t_cell | *AKR1E2* | 2.464429372 | 0.000183302 | 0.000688834 | 1 |
| Obesity | t_cell | *ATP5C1* | 3.943605761 | 0.00162354 | 0.003832968 | 1 |
| Obesity | t_cell | *VDAC2* | 4.918998476 | 0.004561025 | 0.00930106 | 1 |
| Obesity | t_cell | *SLC18A2* | 4.833162035 | 4.26E-05 | 0.000246646 | 0.268104726 |
| Obesity | t_cell | *AK8* | 3.265963053 | 1.38E-05 | 0.000148122 | 0.086877634 |
| Obesity | t_cell | *TSC1* | 5.363904274 | 2.53E-06 | 7.25E-05 | 0.015879008 |
| Obesity | t_cell | *ZMYND19* | -4.414459501 | 0.005878351 | 0.011647398 | 1 |
| Obesity | t_cell | *STOX1* | 0.995321529 | 8.65E-05 | 0.000390748 | 0.543920859 |
| Obesity | t_cell | *RET* | 2.669406676 | 0.000131588 | 0.000532526 | 0.827293844 |
| Obesity | t_cell | *STK32C* | 4.354476288 | 0.009429674 | 0.017675718 | 1 |
| Obesity | t_cell | *NSMF* | 8.539582462 | 0.000688068 | 0.001886596 | 1 |
| Obesity | t_cell | *TRUB1* | 2.416855697 | 0.009516129 | 0.017805924 | 1 |
| Obesity | t_cell | *HSPA12A* | 0.896123383 | 0.022670263 | 0.037556771 | 1 |

| | | | | | | |
|---|---|---|---|---|---|---|
| Obesity | t_cell | *UBTD1* | 3.246845399 | 1.33E-05 | 0.000147409 | 0.083599055 |
| Obesity | t_cell | *ARHGAP42* | 2.702360484 | 1.25E-05 | 0.000146996 | 0.078368517 |
| Obesity | t_cell | *PACSIN3* | 1.232414367 | 0.013401568 | 0.023861699 | 1 |
| Obesity | t_cell | *TTC7B* | 2.545663554 | 0.000363719 | 0.001142208 | 1 |
| Obesity | t_cell | *SLC39A13* | 10.78772998 | 2.27E-07 | 1.32E-05 | 0.001425536 |
| Obesity | t_cell | *PSMC3* | 4.967190457 | 3.71E-07 | 2.05E-05 | 0.002332487 |
| Obesity | t_cell | *TC2N* | -0.569826959 | 0.000149225 | 0.000586362 | 0.938178772 |
| Obesity | t_cell | *IFI27L1* | 2.617006032 | 5.66E-05 | 0.000289932 | 0.355830771 |
| Obesity | t_cell | *CLMN* | 2.665703526 | 0.001629294 | 0.003842225 | 1 |
| Obesity | t_cell | *C1QL3* | 1.953304423 | 9.35E-05 | 0.000411545 | 0.587686849 |
| Obesity | t_cell | *ABTB2* | 4.482005891 | 1.62E-05 | 0.000154104 | 0.101708579 |
| Obesity | t_cell | *LIPC* | -1.41781579 | 0.00080013 | 0.002131532 | 1 |
| Obesity | t_cell | *JAM3* | 3.991551998 | 0.001597287 | 0.003789488 | 1 |
| Obesity | t_cell | *GPT2* | 1.896480657 | 6.04E-05 | 0.000302484 | 0.379919892 |
| Obesity | t_cell | *IKBIP* | 4.012572616 | 0.000731975 | 0.001976773 | 1 |
| Obesity | t_cell | *HIF1AN* | 6.128717308 | 0.000722981 | 0.001955845 | 1 |
| Obesity | t_cell | *ZFYVE19* | 1.810065959 | 0.027948458 | 0.045088154 | 1 |
| Obesity | t_cell | *SPINT1* | -2.898941599 | 0.003641015 | 0.007671268 | 1 |
| Obesity | t_cell | *AVPR1A* | 1.786819584 | 0.000197767 | 0.000727964 | 1 |
| Obesity | t_cell | *BTRC* | 3.14406554 | 0.012265139 | 0.022179687 | 1 |
| Obesity | t_cell | *LARP6* | 3.984546262 | 9.67E-06 | 0.000141127 | 0.060825787 |
| Obesity | t_cell | *ASPG* | 6.091152254 | 1.29E-05 | 0.000147409 | 0.08132356 |
| Obesity | t_cell | *TBATA* | 10.16591688 | 0.00028451 | 0.00095551 | 1 |
| Obesity | t_cell | *PCBD1* | 1.168276406 | 0.017730563 | 0.030456844 | 1 |
| Obesity | t_cell | *COX11* | 3.224946793 | 0.008504181 | 0.016124537 | 1 |
| Obesity | t_cell | *ZNF202* | 6.730288901 | 2.65E-06 | 7.48E-05 | 0.016685873 |
| Obesity | t_cell | *FAM227B* | 3.233330433 | 0.001783873 | 0.004153106 | 1 |
| Obesity | t_cell | *APBB1* | -2.08944805 | 0.007086316 | 0.013758945 | 1 |
| Obesity | t_cell | *ILK* | 5.119463193 | 6.16E-05 | 0.000306975 | 0.387402083 |
| Obesity | t_cell | *TAF10* | 6.181129426 | 0.000616284 | 0.001723566 | 1 |
| Obesity | t_cell | *DCHS1* | 5.990307436 | 5.29E-06 | 0.000107292 | 0.03326062 |
| Obesity | t_cell | *NETO1* | 4.115457415 | 0.001933601 | 0.004466035 | 1 |
| Obesity | t_cell | *USP54* | 4.340356865 | 0.000104855 | 0.000449676 | 0.659225198 |
| Obesity | t_cell | *C11orf74* | 3.724008795 | 7.16E-06 | 0.000126396 | 0.045032683 |
| Obesity | t_cell | *WDR88* | 6.350786425 | 0.005555391 | 0.011091376 | 1 |
| Obesity | t_cell | *CYB5R2* | 2.366874603 | 0.012457915 | 0.022454963 | 1 |
| Obesity | t_cell | *TUB* | 2.170588722 | 5.96E-05 | 0.000299804 | 0.374754568 |
| Obesity | t_cell | *LMO1* | 3.338486223 | 5.88E-05 | 0.000297403 | 0.369375056 |
| Obesity | t_cell | *TRIM66* | 0.628617639 | 0.008114097 | 0.01547265 | 1 |
| Obesity | t_cell | *RPL27A* | -1.834749293 | 0.014440487 | 0.025466295 | 1 |
| Obesity | t_cell | *PRTG* | 3.62662303 | 1.28E-05 | 0.000147259 | 0.08055065 |
| Obesity | t_cell | *AKIP1* | 5.033415486 | 1.20E-05 | 0.000145864 | 0.075293497 |
| Obesity | t_cell | *C16orf46* | 24.2815054 | 9.80E-06 | 0.000142239 | 0.061589601 |
| Obesity | t_cell | *CCDC68* | 2.336374943 | 5.16E-05 | 0.000272935 | 0.324453471 |
| Obesity | t_cell | *CLEC4E* | 1.553406642 | 0.007273536 | 0.014066047 | 1 |
| Obesity | t_cell | *CLEC4D* | 5.942282737 | 3.37E-05 | 0.000214224 | 0.211887707 |
| Obesity | t_cell | *BEAN1* | 3.574691823 | 4.33E-06 | 9.97E-05 | 0.027237528 |
| Obesity | t_cell | *TK2* | 2.390928351 | 0.000233935 | 0.000822566 | 1 |
| Obesity | t_cell | *SEC11C* | -2.143465206 | 0.001572944 | 0.003742979 | 1 |
| Obesity | t_cell | *GALR1* | 1.64354157 | 0.001113702 | 0.002799618 | 1 |

| | | | | | | |
|---|---|---|---|---|---|---|
| Obesity | t_cell | *CENPV* | 3.688717292 | 0.015981987 | 0.027841162 | 1 |
| Obesity | t_cell | *CHRFAM7A* | 2.246087437 | 0.009436059 | 0.017682415 | 1 |
| Obesity | t_cell | *SPDYE6* | 3.823870391 | 8.77E-06 | 0.000135109 | 0.055124478 |
| Obesity | t_cell | *ATF7IP2* | -1.543761603 | 0.021571224 | 0.035992114 | 1 |
| Obesity | t_cell | *ACSM1* | 2.144449788 | 0.014400829 | 0.025417631 | 1 |
| Obesity | t_cell | *SLFN5* | -1.875731161 | 0.012032487 | 0.021852208 | 1 |
| Obesity | t_cell | *CATSPER2* | 5.038143654 | 0.000470683 | 0.001391248 | 1 |
| Obesity | t_cell | *C16orf45* | 4.762246861 | 2.61E-05 | 0.000187492 | 0.164055362 |
| Obesity | t_cell | *SAAL1* | 9.110736721 | 2.00E-07 | 1.25E-05 | 0.001257495 |
| Obesity | t_cell | *YPEL4* | 5.557388108 | 1.92E-06 | 6.34E-05 | 0.012056831 |
| Obesity | t_cell | *ANKDD1A* | 3.23443134 | 0.001674454 | 0.003935437 | 1 |
| Obesity | t_cell | *DCTN5* | 2.093607569 | 0.003428454 | 0.007294766 | 1 |
| Obesity | t_cell | *MTMR10* | 4.756301602 | 2.06E-05 | 0.000166599 | 0.129281116 |
| Obesity | t_cell | *TSC22D4* | -2.435241422 | 0.01935723 | 0.032785265 | 1 |
| Obesity | t_cell | *MS4A7* | 1.961675191 | 0.013610836 | 0.024186355 | 1 |
| Obesity | t_cell | *CCNDBP1* | 5.224349187 | 0.006386165 | 0.012542898 | 1 |
| Obesity | t_cell | *RCCD1* | 3.675685798 | 0.003252641 | 0.006988844 | 1 |
| Obesity | t_cell | *AKTIP* | -2.516044394 | 0.00749928 | 0.01444042 | 1 |
| Obesity | t_cell | *NUDT21* | 5.008828043 | 0.002617232 | 0.005795892 | 1 |
| Obesity | t_cell | *NKX3-1* | 14.6167025 | 0.007516326 | 0.014468812 | 1 |
| Obesity | t_cell | *TEF* | 2.075299453 | 3.82E-05 | 0.00023194 | 0.240341638 |
| Obesity | t_cell | *SAMD14* | 4.581564735 | 7.17E-06 | 0.000126396 | 0.045097824 |
| Obesity | t_cell | *PIP5KL1* | 1.324159808 | 0.007014208 | 0.013652732 | 1 |
| Obesity | t_cell | *CERCAM* | 1.95233029 | 0.001331715 | 0.003250191 | 1 |
| Obesity | t_cell | *ENDOG* | 3.193237396 | 0.000448002 | 0.001338683 | 1 |
| Obesity | t_cell | *ISLR2* | 4.642097113 | 5.53E-06 | 0.000109386 | 0.034788655 |
| Obesity | t_cell | *SP2* | 5.302506922 | 0.003573717 | 0.007539584 | 1 |
| Obesity | t_cell | *COQ7* | 4.253906769 | 1.06E-05 | 0.000145864 | 0.066890335 |
| Obesity | t_cell | *GPRC5B* | 4.704970497 | 1.17E-07 | 8.28E-06 | 0.000738472 |
| Obesity | t_cell | *KATNAL2* | 1.480014412 | 0.005129003 | 0.010341899 | 1 |
| Obesity | t_cell | *RNF214* | 5.223403049 | 0.000131882 | 0.000532526 | 0.829140245 |
| Obesity | t_cell | *RBFOX3* | 6.060462916 | 4.33E-06 | 9.97E-05 | 0.02724964 |
| Obesity | t_cell | *TRIM68* | 1.819432946 | 0.002692195 | 0.005940971 | 1 |
| Obesity | t_cell | *FN3K* | 3.759610454 | 0.000850631 | 0.002237622 | 1 |
| Obesity | t_cell | *ZNF229* | 8.330687917 | 3.77E-06 | 9.34E-05 | 0.02369732 |
| Obesity | t_cell | *ZNF646* | 6.358879998 | 4.40E-11 | 1.32E-08 | 2.77E-07 |
| Obesity | t_cell | *SMG8* | 6.690321363 | 2.59E-05 | 0.000186717 | 0.162630676 |
| Obesity | t_cell | *RPL13* | -1.584946447 | 0.003745324 | 0.007867308 | 1 |
| Obesity | t_cell | *CACNB3* | 5.045410769 | 0.000565813 | 0.001609623 | 1 |
| Obesity | t_cell | *TP53I13* | 2.169199427 | 0.025832356 | 0.042119052 | 1 |
| Obesity | t_cell | *ZNF528* | 5.582751489 | 5.91E-05 | 0.000298221 | 0.371284872 |
| Obesity | t_cell | *ZNF701* | 4.430734502 | 0.001659674 | 0.003902159 | 1 |
| Obesity | t_cell | *SERTAD3* | 3.958776314 | 1.91E-05 | 0.000160838 | 0.119795849 |
| Obesity | t_cell | *TMC4* | 6.758719791 | 0.00074658 | 0.00201017 | 1 |
| Obesity | t_cell | *TTYH1* | 5.548014352 | 4.51E-06 | 0.000100727 | 0.028324536 |
| Obesity | t_cell | *LENG8* | 4.20007313 | 0.00061696 | 0.001724691 | 1 |
| Obesity | t_cell | *CDC42EP5* | 4.875386503 | 0.000135229 | 0.000543248 | 0.850182807 |
| Obesity | t_cell | *ZNF283* | 3.813420183 | 0.002331876 | 0.00524151 | 1 |
| Obesity | t_cell | *PPP1R14A* | 7.183114387 | 3.70E-05 | 0.000226954 | 0.232400813 |
| Obesity | t_cell | *PSCA* | 7.118109871 | 3.54E-08 | 3.32E-06 | 0.000222637 |

| | | | | | | |
|---|---|---|---|---|---|---|
| Obesity | t_cell | *FAM57A* | 2.851686573 | 0.000195629 | 0.000722631 | 1 |
| Obesity | t_cell | *SERPINF2* | 3.438070814 | 1.89E-05 | 0.000160637 | 0.118871638 |
| Obesity | t_cell | *SRR* | 2.297641958 | 1.78E-06 | 6.03E-05 | 0.01119836 |
| Obesity | t_cell | *CYB5D2* | 5.696658873 | 0.000565608 | 0.001609623 | 1 |
| Obesity | t_cell | *CD320* | 8.431589866 | 0.000128737 | 0.000523525 | 0.809370031 |
| Obesity | t_cell | *SPRYD3* | 8.319946759 | 0.000242968 | 0.000848161 | 1 |
| Obesity | t_cell | *ZNF558* | 1.908677189 | 0.004872705 | 0.009875789 | 1 |
| Obesity | t_cell | *NDUFV1* | 5.06338266 | 0.000462273 | 0.001372468 | 1 |
| Obesity | t_cell | *CDK2AP2* | -2.538373292 | 0.027381857 | 0.044322795 | 1 |
| Obesity | t_cell | *PRDX2* | 6.951015199 | 0.00047822 | 0.001408464 | 1 |
| Obesity | t_cell | *HID1* | 2.759327442 | 0.000507864 | 0.001474308 | 1 |
| Obesity | t_cell | *ATP5H* | 4.849423784 | 0.000125887 | 0.000514166 | 0.791450941 |
| Obesity | t_cell | *MGAT5B* | 4.452453636 | 1.50E-05 | 0.000149261 | 0.094183447 |
| Obesity | t_cell | *TMC8* | -0.533706691 | 0.000609228 | 0.001709155 | 1 |
| Obesity | t_cell | *TMEM68* | 4.385726556 | 0.00353722 | 0.007475126 | 1 |
| Obesity | t_cell | *CYP7A1* | 4.701593981 | 0.003428668 | 0.007294766 | 1 |
| Obesity | t_cell | *GSDMA* | 5.448317937 | 0.000147192 | 0.000580186 | 0.925396316 |
| Obesity | t_cell | *ZNF598* | -2.358455352 | 0.028567749 | 0.04599371 | 1 |
| Obesity | t_cell | *MLST8* | 5.992931816 | 0.000777417 | 0.002079839 | 1 |
| Obesity | t_cell | *ECI1* | 2.931587329 | 0.006974255 | 0.013583377 | 1 |
| Obesity | t_cell | *KCTD5* | 7.513682214 | 0.000205385 | 0.000745678 | 1 |
| Obesity | t_cell | *DDB1* | 5.224768841 | 0.000310389 | 0.001018034 | 1 |
| Obesity | t_cell | *VPS37C* | 3.903041186 | 0.003358765 | 0.007170309 | 1 |
| Obesity | t_cell | *VWCE* | 3.840899114 | 7.60E-07 | 3.39E-05 | 0.004780225 |
| Obesity | t_cell | *HRASLS5* | 0.58206103 | 0.016349761 | 0.028348304 | 1 |
| Obesity | t_cell | *TTC21A* | 5.425306355 | 9.26E-06 | 0.000138236 | 0.058197338 |
| Obesity | t_cell | *LTBP3* | 2.978919021 | 0.001947364 | 0.004496171 | 1 |
| Obesity | t_cell | *NAALADL1* | 2.674786576 | 4.66E-05 | 0.000259121 | 0.29328202 |
| Obesity | t_cell | *SCARA5* | 3.862802098 | 0.002388778 | 0.005348495 | 1 |
| Obesity | t_cell | *KIAA1586* | 3.092489403 | 0.003442836 | 0.007317482 | 1 |
| Obesity | t_cell | *RNF187* | 2.466156827 | 0.017764891 | 0.030503806 | 1 |
| Obesity | t_cell | *ZCCHC4* | 5.621223735 | 0.000559415 | 0.001595754 | 1 |
| Obesity | t_cell | *PTGDR* | -1.061991739 | 0.000355775 | 0.001125696 | 1 |
| Obesity | t_cell | *GLYCTK* | 5.021632714 | 0.003816421 | 0.00799528 | 1 |
| Obesity | t_cell | *UBTD2* | 4.734966138 | 5.47E-06 | 0.000108648 | 0.034412774 |
| Obesity | t_cell | *COA6* | 7.853367985 | 1.47E-05 | 0.000149118 | 0.092645135 |
| Obesity | t_cell | *KIF5C* | -0.929096993 | 0.000220583 | 0.000788405 | 1 |
| Obesity | t_cell | *PDHB* | 4.239550126 | 0.004196312 | 0.008660473 | 1 |
| Obesity | t_cell | *ACOX2* | 1.028526278 | 0.030961772 | 0.049380178 | 1 |
| Obesity | t_cell | *FAM107A* | 2.672705544 | 4.78E-05 | 0.000262855 | 0.300607159 |
| Obesity | t_cell | *CX3CR1* | -1.15564399 | 0.000605402 | 0.001700698 | 1 |
| Obesity | t_cell | *SCN11A* | 0.583596481 | 0.030446468 | 0.048718998 | 1 |
| Obesity | t_cell | *SEPT2* | 1.408937535 | 0.0038641 | 0.008081703 | 1 |
| Obesity | t_cell | *DTYMK* | 6.796428415 | 0.000594422 | 0.001675092 | 1 |
| Obesity | t_cell | *TAP1* | -1.721457286 | 0.022395786 | 0.037180436 | 1 |
| Obesity | t_cell | *ING5* | 5.208757577 | 0.000942934 | 0.002430596 | 1 |
| Obesity | t_cell | *RHOH* | -0.388158638 | 0.000174812 | 0.000661276 | 1 |
| Obesity | t_cell | *KLHL30* | 2.643631674 | 0.000142726 | 0.000568644 | 0.897320245 |
| Obesity | t_cell | *REEP4* | 3.796341024 | 0.006321275 | 0.012423212 | 1 |
| Obesity | t_cell | *BMP1* | 2.531611741 | 5.52E-05 | 0.000285135 | 0.346811949 |

| | | | | | | |
|---|---|---|---|---|---|---|
| Obesity | t_cell | *FNTA* | 6.032850025 | 0.00033908 | 0.001090434 | 1 |
| Obesity | t_cell | *SERINC2* | 3.509156587 | 0.000290771 | 0.000973548 | 1 |
| Obesity | t_cell | *TRAPPC11* | 4.732183428 | 0.001364367 | 0.003315392 | 1 |
| Obesity | t_cell | *TMEM223* | 3.791265256 | 0.011757836 | 0.021407911 | 1 |
| Obesity | t_cell | *SLC20A2* | 4.204551046 | 0.00189876 | 0.004394859 | 1 |
| Obesity | t_cell | *ZSWIM1* | 9.107157941 | 9.21E-06 | 0.000138155 | 0.057905899 |
| Obesity | t_cell | *NBPF9* | 5.246975725 | 1.21E-05 | 0.000145864 | 0.075830963 |
| Obesity | t_cell | *WFDC13* | 15.76113156 | 0.001429349 | 0.003445673 | 1 |
| Obesity | t_cell | *ZNF30* | 2.47824838 | 0.018914419 | 0.032165256 | 1 |
| Obesity | t_cell | *SLC16A4* | 1.945925485 | 0.018581181 | 0.031701462 | 1 |
| Obesity | t_cell | *TMEM208* | 4.165778821 | 0.001344882 | 0.003275967 | 1 |
| Obesity | t_cell | *LRP1B* | 1.258905778 | 0.000933727 | 0.002414786 | 1 |
| Obesity | t_cell | *NPNT* | 5.705707676 | 0.003423717 | 0.007289166 | 1 |
| Obesity | t_cell | *GSTM4* | 1.57827418 | 0.027481778 | 0.044473085 | 1 |
| Obesity | t_cell | *ADAL* | 0.825963124 | 0.007670649 | 0.014729802 | 1 |
| Obesity | t_cell | *SNTB2* | 4.705606026 | 7.29E-05 | 0.000345811 | 0.458042812 |
| Obesity | t_cell | *ZNF507* | -2.27000887 | 0.027777084 | 0.044866523 | 1 |
| Obesity | t_cell | *STX18* | 4.147485674 | 0.004543616 | 0.009277595 | 1 |
| Obesity | t_cell | *GFM1* | 3.464042274 | 0.000595259 | 0.0016767 | 1 |
| Obesity | t_cell | *BTNL3* | 1.070938023 | 0.000146217 | 0.000577793 | 0.919268235 |
| Obesity | t_cell | *ZNF608* | 2.303571939 | 9.61E-06 | 0.000140555 | 0.060399089 |
| Obesity | t_cell | *INPP5D* | -1.350818426 | 0.001748471 | 0.00408565 | 1 |
| Obesity | t_cell | *LETM1* | 5.685516841 | 1.46E-05 | 0.000148855 | 0.091545736 |
| Obesity | t_cell | *TMEM129* | 1.894618447 | 0.024938278 | 0.04078745 | 1 |
| Obesity | t_cell | *STXBP6* | 3.865448206 | 3.31E-05 | 0.000212266 | 0.207969668 |
| Obesity | t_cell | *MFF* | 4.312355271 | 0.000941041 | 0.002427707 | 1 |
| Obesity | t_cell | *CPLX1* | 2.616466692 | 0.0005814 | 0.001644291 | 1 |
| Obesity | t_cell | *SIGLEC7* | 2.229459358 | 0.02703732 | 0.043826834 | 1 |
| Obesity | t_cell | *FEM1B* | 5.050001232 | 0.0004783 | 0.001408464 | 1 |
| Obesity | t_cell | *MFSD7* | 3.191971827 | 1.77E-05 | 0.000157653 | 0.111114477 |
| Obesity | t_cell | *COL4A3* | 5.155005192 | 4.20E-07 | 2.24E-05 | 0.002643329 |
| Obesity | t_cell | *IRS1* | 1.017261608 | 0.000509182 | 0.001475219 | 1 |
| Obesity | t_cell | *UPF3A* | 1.744799965 | 0.02649502 | 0.043098109 | 1 |
| Obesity | t_cell | *C8orf46* | 0.70638784 | 1.65E-05 | 0.000155092 | 0.103467797 |
| Obesity | t_cell | *PARM1* | 2.260657856 | 0.004182085 | 0.008637572 | 1 |
| Obesity | t_cell | *FAM110B* | 3.972934588 | 1.42E-05 | 0.000148367 | 0.088992665 |
| Obesity | t_cell | *ATF5* | 6.287305106 | 0.000814895 | 0.002165361 | 1 |
| Obesity | t_cell | *CPT1C* | 3.824645034 | 0.000448478 | 0.001339469 | 1 |
| Obesity | t_cell | *PCSK9* | -9.000696014 | 0.000557469 | 0.001591647 | 1 |
| Obesity | t_cell | *RGS14* | -1.018717852 | 0.001182915 | 0.002944175 | 1 |
| Obesity | t_cell | *CXCL10* | 1.56892775 | 0.007456678 | 0.014371592 | 1 |
| Obesity | t_cell | *SH3TC2* | 1.46188087 | 0.013571766 | 0.024123747 | 1 |
| Obesity | t_cell | *PGM2* | 7.791559413 | 0.00044584 | 0.001333489 | 1 |
| Obesity | t_cell | *STK32A* | 0.932458423 | 0.023755348 | 0.039096825 | 1 |
| Obesity | t_cell | *P2RY12* | 2.588167983 | 0.000239996 | 0.000841056 | 1 |
| Obesity | t_cell | *SNUPN* | 4.913781638 | 2.16E-05 | 0.000169226 | 0.135623952 |
| Obesity | t_cell | *PTK2* | 3.555492517 | 6.85E-05 | 0.00032982 | 0.430414664 |
| Obesity | t_cell | *PTAFR* | 3.308430693 | 0.000494263 | 0.001445318 | 1 |
| Obesity | t_cell | *RNASE6* | 10.45245591 | 2.09E-05 | 0.000167886 | 0.131702217 |
| Obesity | t_cell | *NPR1* | 3.065331495 | 1.20E-05 | 0.000145864 | 0.07546964 |

| | | | | | | |
|---|---|---|---|---|---|---|
| Obesity | t_cell | *CD52* | -0.831305433 | 0.000614138 | 0.001720626 | 1 |
| Obesity | t_cell | *CLIC4* | 4.040996921 | 8.40E-05 | 0.000383091 | 0.528262336 |
| Obesity | t_cell | *CCDC8* | 3.427285803 | 4.57E-05 | 0.000255177 | 0.287329226 |
| Obesity | t_cell | *METTL15* | 3.859481474 | 3.63E-05 | 0.000224733 | 0.228497158 |
| Obesity | t_cell | *DTWD2* | 4.811427426 | 5.89E-05 | 0.000297613 | 0.37023037 |
| Obesity | t_cell | *INO80E* | -2.673170736 | 0.01117251 | 0.020484564 | 1 |
| Obesity | t_cell | *DFFB* | 6.706611834 | 0.000147038 | 0.000580186 | 0.924427354 |
| Obesity | t_cell | *C15orf40* | 2.652498604 | 0.003528778 | 0.007462303 | 1 |
| Obesity | t_cell | *FAM103A1* | -1.498216594 | 0.021272666 | 0.035550571 | 1 |
| Obesity | t_cell | *LUZP1* | 3.748040925 | 0.005783716 | 0.011474352 | 1 |
| Obesity | t_cell | *CHRNA5* | 2.328220835 | 0.000707382 | 0.001926912 | 1 |
| Obesity | t_cell | *FASN* | 2.46825282 | 2.02E-06 | 6.45E-05 | 0.012699535 |
| Obesity | t_cell | *MT1E* | 4.157130592 | 0.000227814 | 0.00080555 | 1 |
| Obesity | t_cell | *DCXR* | 6.014792085 | 5.85E-06 | 0.000112142 | 0.036807917 |
| Obesity | t_cell | *LIMS1* | 2.643934208 | 0.000715251 | 0.001940777 | 1 |
| Obesity | t_cell | *UGP2* | 3.173558008 | 1.76E-05 | 0.000157653 | 0.110597907 |
| Obesity | t_cell | *AVEN* | 5.613439208 | 7.88E-05 | 0.000366894 | 0.495673803 |
| Obesity | t_cell | *CALML6* | -2.830512925 | 0.004661247 | 0.009474704 | 1 |
| Obesity | t_cell | *TPST1* | 1.2638389 | 0.027057604 | 0.043843082 | 1 |
| Obesity | t_cell | *OTUD3* | 2.302003739 | 0.000989008 | 0.002530684 | 1 |
| Obesity | t_cell | *GUSB* | 5.416453889 | 7.37E-05 | 0.000348804 | 0.463560612 |
| Obesity | t_cell | *BRD3* | 5.695156407 | 0.00284613 | 0.006221703 | 1 |
| Obesity | t_cell | *ZFPM2* | 2.923041105 | 1.61E-05 | 0.000154104 | 0.101449949 |
| Obesity | t_cell | *ZNF747* | 5.619715386 | 0.000523614 | 0.001509901 | 1 |
| Obesity | t_cell | *MAP3K2* | 2.897413831 | 0.026197453 | 0.042680329 | 1 |
| Obesity | t_cell | *MYO7B* | 4.057302617 | 4.67E-06 | 0.000100727 | 0.029342385 |
| Obesity | t_cell | *CNTROB* | 6.826124866 | 2.73E-05 | 0.000192118 | 0.171753747 |
| Obesity | t_cell | *KCNAB3* | -2.061777254 | 0.002147619 | 0.004893831 | 1 |
| Obesity | t_cell | *RNF150* | 1.49891573 | 0.014898711 | 0.026208225 | 1 |
| Obesity | t_cell | *FAM161A* | 3.091921365 | 0.013716275 | 0.02435995 | 1 |
| Obesity | t_cell | *FAXDC2* | 3.04805854 | 0.000225238 | 0.000799844 | 1 |
| Obesity | t_cell | *CRTAP* | 5.897589385 | 3.86E-05 | 0.000232312 | 0.24288315 |
| Obesity | t_cell | *NFRKB* | 2.83832144 | 0.018862361 | 0.032105604 | 1 |
| Obesity | t_cell | *SEMA3E* | 3.286397092 | 7.90E-06 | 0.000132019 | 0.049687525 |
| Obesity | t_cell | *DCLK2* | 2.688478844 | 0.001751813 | 0.00409093 | 1 |
| Obesity | t_cell | *GPRC5C* | 3.340515429 | 0.004308999 | 0.008858223 | 1 |
| Obesity | t_cell | *ADORA2B* | -3.061808415 | 7.48E-08 | 6.10E-06 | 0.000470002 |
| Obesity | t_cell | *MGMT* | 3.08534324 | 0.001617957 | 0.003824674 | 1 |
| Obesity | t_cell | *METTL7B* | 5.520960909 | 4.83E-05 | 0.000263865 | 0.303445191 |
| Obesity | t_cell | *NFXL1* | 6.120777645 | 9.80E-05 | 0.000425311 | 0.615850153 |
| Obesity | t_cell | *KRT72* | -4.264446484 | 0.00992691 | 0.018486517 | 1 |
| Obesity | t_cell | *TMC7* | 2.272387361 | 0.010120861 | 0.018792041 | 1 |
| Obesity | t_cell | *SERPINB9* | -1.798270011 | 0.000993058 | 0.002537223 | 1 |
| Obesity | t_cell | *IRX2* | 2.052305841 | 0.003941062 | 0.008219522 | 1 |
| Obesity | t_cell | *EMB* | -0.774748363 | 0.001229333 | 0.003048844 | 1 |
| Obesity | t_cell | *STAT2* | 3.931369598 | 0.000639367 | 0.001773918 | 1 |
| Obesity | t_cell | *NUDCD2* | 5.74452934 | 0.002002036 | 0.004602121 | 1 |
| Obesity | t_cell | *SLC26A5* | 4.839573484 | 2.84E-06 | 7.84E-05 | 0.017875299 |
| Obesity | t_cell | *COMMD5* | 6.765567978 | 8.70E-05 | 0.000391899 | 0.546699257 |
| Obesity | t_cell | *GTSF1* | -2.335940269 | 0.000268493 | 0.000915906 | 1 |

| | | | | | | |
|---|---|---|---|---|---|---|
| Obesity | t_cell | *ACYP2* | 3.143326468 | 0.005099973 | 0.010294617 | 1 |
| Obesity | t_cell | *TMEM133* | 8.304495567 | 1.91E-05 | 0.000160838 | 0.119813035 |
| Obesity | t_cell | *TTLL6* | 7.921206322 | 0.000143115 | 0.00056947 | 0.899762066 |
| Obesity | t_cell | *HTRA3* | 1.998165532 | 0.012852142 | 0.023066348 | 1 |
| Obesity | t_cell | *GPR27* | 4.930626726 | 0.007876204 | 0.015060126 | 1 |
| Obesity | t_cell | *MTSS1* | 4.143156964 | 8.73E-05 | 0.000392666 | 0.548603421 |
| Obesity | t_cell | *TSEN34* | 8.532846158 | 0.000620122 | 0.001731988 | 1 |
| Obesity | t_cell | *OSCAR* | 2.37467232 | 1.68E-05 | 0.000156471 | 0.105774505 |
| Obesity | t_cell | *PAQR8* | -1.382346706 | 0.000129598 | 0.000526004 | 0.814780684 |
| Obesity | t_cell | *NUDT6* | 3.302583393 | 3.66E-05 | 0.000225644 | 0.229931299 |
| Obesity | t_cell | *ZNF160* | 4.344666999 | 0.000243723 | 0.000849854 | 1 |
| Obesity | t_cell | *TSNARE1* | 3.138218362 | 0.000180321 | 0.000680071 | 1 |
| Obesity | t_cell | *FPR1* | 2.33597088 | 0.000150395 | 0.000589118 | 0.945534777 |
| Obesity | t_cell | *FEZ2* | 4.135787071 | 0.000554306 | 0.001584774 | 1 |
| Obesity | t_cell | *SOX7* | 9.547171057 | 1.81E-08 | 1.90E-06 | 0.000114079 |
| Obesity | t_cell | *C11orf24* | 3.698218924 | 1.77E-05 | 0.000157704 | 0.111556176 |
| Obesity | t_cell | *ALK* | 3.967295716 | 1.71E-05 | 0.000156669 | 0.10722709 |
| Obesity | t_cell | *TRMT61B* | 2.789494068 | 4.68E-08 | 4.03E-06 | 0.000294251 |
| Obesity | t_cell | *MFN1* | 4.279020053 | 0.000275511 | 0.000933381 | 1 |
| Obesity | t_cell | *KCNMB3* | 3.191497713 | 4.80E-05 | 0.000262855 | 0.301494847 |
| Obesity | t_cell | *ATP6V0E2* | -3.579911092 | 0.000552491 | 0.001581025 | 1 |
| Obesity | t_cell | *CHST11* | -1.110654154 | 0.004584681 | 0.009343237 | 1 |
| Obesity | t_cell | *KCND3* | 2.769149503 | 3.79E-06 | 9.34E-05 | 0.023824545 |
| Obesity | t_cell | *MRPL36* | 5.115638735 | 0.000259781 | 0.000892484 | 1 |
| Obesity | t_cell | *ZNF581* | 7.308093515 | 1.13E-05 | 0.000145864 | 0.070896086 |
| Obesity | t_cell | *MCC* | 4.294549246 | 1.34E-05 | 0.000147409 | 0.084258955 |
| Obesity | t_cell | *HOPX* | -1.85150014 | 0.000327395 | 0.001061543 | 1 |
| Obesity | t_cell | *PPID* | 5.253797342 | 0.00168248 | 0.00395087 | 1 |
| Obesity | t_cell | *MAP6* | 2.522594272 | 0.000152128 | 0.000594055 | 0.956429225 |
| Obesity | t_cell | *NMUR1* | 8.469444454 | 2.15E-05 | 0.000169226 | 0.135181283 |
| Obesity | t_cell | *P2RY6* | 3.41817291 | 2.98E-05 | 0.000200768 | 0.18758499 |
| Obesity | t_cell | *TCEA2* | 8.511367976 | 0.000322605 | 0.001048175 | 1 |
| Obesity | t_cell | *ANO5* | 2.523962303 | 2.42E-05 | 0.00017926 | 0.151961408 |
| Obesity | t_cell | *GPHN* | 1.970396716 | 0.014044528 | 0.024872662 | 1 |
| Obesity | t_cell | *VAT1L* | 3.417901448 | 5.03E-05 | 0.000268036 | 0.316014904 |
| Obesity | t_cell | *CAMTA1* | 4.460305457 | 0.001271574 | 0.003135054 | 1 |
| Obesity | t_cell | *LGALS4* | 2.817255118 | 0.001682906 | 0.00395087 | 1 |
| Obesity | t_cell | *LRRC34* | 1.13011249 | 0.011034938 | 0.020261874 | 1 |
| Obesity | t_cell | *SPATA5L1* | 3.584303922 | 8.67E-05 | 0.000391094 | 0.544793663 |
| Obesity | t_cell | *SYCE1* | 1.757314047 | 0.012269915 | 0.022179687 | 1 |
| Obesity | t_cell | *PWWP2B* | 3.82324031 | 2.26E-05 | 0.00017343 | 0.142385677 |
| Obesity | t_cell | *ZNF570* | 4.521750934 | 0.021099704 | 0.035317849 | 1 |
| Obesity | t_cell | *NINJ2* | 5.300439615 | 0.000665592 | 0.001832131 | 1 |
| Obesity | t_cell | *TRAPPC12* | 3.995593044 | 0.004569083 | 0.00931447 | 1 |
| Obesity | t_cell | *AQP4* | 2.572669933 | 3.12E-05 | 0.000206483 | 0.195952431 |
| Obesity | t_cell | *TLN2* | 0.923987246 | 0.026221643 | 0.042708671 | 1 |
| Obesity | t_cell | *TVP23B* | 4.299283444 | 0.012267664 | 0.022179687 | 1 |
| Obesity | t_cell | *SRGAP2C* | 3.906935091 | 0.014460052 | 0.025493648 | 1 |
| Obesity | t_cell | *ATPAF2* | 7.605118531 | 3.64E-05 | 0.000224733 | 0.22855343 |
| Obesity | t_cell | *CYP4F22* | -3.724813755 | 1.58E-05 | 0.000152971 | 0.099216769 |

| | | | | | | |
|---|---|---|---|---|---|---|
| Obesity | t_cell | *C20orf196* | -1.982065005 | 0.031344432 | 0.049952457 | 1 |
| Obesity | t_cell | *ZNF554* | 1.541745628 | 0.027709378 | 0.044795284 | 1 |
| Obesity | t_cell | *ANKRD20A4* | 4.772887367 | 3.60E-05 | 0.000223494 | 0.226623347 |
| Obesity | t_cell | *LAMB2* | 2.856757758 | 0.000805522 | 0.002144339 | 1 |
| Obesity | t_cell | *ORMDL3* | 1.596784368 | 0.000805619 | 0.002144339 | 1 |
| Obesity | t_cell | *SERF1A* | -9.649534884 | 0.000786356 | 0.002100178 | 1 |
| Obesity | t_cell | *LRRC15* | 3.606752805 | 0.015476384 | 0.027118178 | 1 |
| Obesity | t_cell | *FRMD3* | 5.999180851 | 8.74E-07 | 3.69E-05 | 0.0054979 |
| Obesity | t_cell | *ISG20* | -0.770121206 | 0.000139967 | 0.000559781 | 0.879975293 |
| Obesity | t_cell | *CXCR6* | -0.891534502 | 0.021934047 | 0.036490964 | 1 |
| Obesity | t_cell | *C5orf34* | 4.890034653 | 3.11E-05 | 0.000206384 | 0.195445998 |
| Obesity | t_cell | *C1QTNF4* | -1.681326081 | 2.31E-11 | 7.64E-09 | 1.45E-07 |
| Obesity | t_cell | *HINFP* | 6.738511331 | 0.001629243 | 0.003842225 | 1 |
| Obesity | t_cell | *SPTLC3* | 4.353858493 | 1.19E-05 | 0.000145864 | 0.074590751 |
| Obesity | t_cell | *COPRS* | 4.130593282 | 8.49E-06 | 0.000134614 | 0.053374575 |
| Obesity | t_cell | *RCAN2* | 3.547367534 | 0.021253781 | 0.035528455 | 1 |
| Obesity | t_cell | *IL16* | 2.490200582 | 0.00754116 | 0.014512174 | 1 |
| Obesity | t_cell | *FGGY* | 4.210268334 | 0.00039425 | 0.001211743 | 1 |
| Obesity | t_cell | *FIBP* | 11.48724025 | 1.36E-05 | 0.000147409 | 0.085366134 |
| Obesity | t_cell | *CARNS1* | 10.72063382 | 0.003926072 | 0.008194959 | 1 |
| Obesity | t_cell | *CTSW* | -0.852630474 | 1.11E-08 | 1.33E-06 | 7.00E-05 |
| Obesity | t_cell | *SNTG2* | 1.678682531 | 0.026503263 | 0.043100367 | 1 |
| Obesity | t_cell | *PDE3A* | 2.490420305 | 0.004883459 | 0.009894395 | 1 |
| Obesity | t_cell | *SMPDL3A* | 4.640585711 | 1.27E-05 | 0.000146996 | 0.079741024 |
| Obesity | t_cell | *RAD9A* | 1.357363519 | 0.005565755 | 0.011105016 | 1 |
| Obesity | t_cell | *EFEMP2* | 4.244501264 | 4.21E-09 | 5.29E-07 | 2.65E-05 |
| Obesity | t_cell | *AGAP5* | 44.48123008 | 4.89E-05 | 0.000265802 | 0.307297378 |
| Obesity | t_cell | *ZMAT3* | 5.333874716 | 0.008882418 | 0.016749778 | 1 |
| Obesity | t_cell | *ZNF738* | 2.054106881 | 0.011409637 | 0.020864569 | 1 |
| Obesity | t_cell | *SLFN11* | 3.245050613 | 0.000322403 | 0.001048175 | 1 |
| Obesity | t_cell | *CCL19* | 6.868137001 | 0.000115103 | 0.000482756 | 0.723650737 |
| Obesity | t_cell | *CBWD1* | 6.051681086 | 0.000306881 | 0.001012256 | 1 |
| Obesity | t_cell | *HOXC5* | 2.924572472 | 0.00189929 | 0.004394859 | 1 |
| Obesity | t_cell | *RAB37* | -1.264329019 | 0.000635827 | 0.001765656 | 1 |
| Obesity | t_cell | *DCP2* | 3.653834922 | 0.013982989 | 0.024791612 | 1 |
| Obesity | t_cell | *SNX32* | 5.150118465 | 1.26E-07 | 8.74E-06 | 0.000795193 |
| Obesity | t_cell | *CYP7B1* | 2.953370364 | 0.000698842 | 0.00190944 | 1 |
| Obesity | t_cell | *METAP1D* | 4.859618074 | 3.98E-06 | 9.51E-05 | 0.025045847 |
| Obesity | t_cell | *NADSYN1* | 4.934233593 | 0.000827095 | 0.002190374 | 1 |
| Obesity | t_cell | *MRGPRF* | 4.063390693 | 2.20E-05 | 0.000171115 | 0.13808973 |
| Obesity | t_cell | *GXYLT2* | 2.778189156 | 0.000123688 | 0.000507921 | 0.777627237 |
| Obesity | t_cell | *HPSE2* | 4.78536618 | 4.58E-13 | 2.06E-10 | 2.88E-09 |
| Obesity | t_cell | *BNC2* | 2.730362568 | 3.13E-06 | 8.33E-05 | 0.019649168 |
| Obesity | t_cell | *PARP15* | -0.94130981 | 0.00174485 | 0.004079535 | 1 |
| Obesity | t_cell | *AHSA2* | 3.221964244 | 0.002202366 | 0.004991448 | 1 |
| Obesity | t_cell | *ABLIM3* | 3.743415316 | 7.95E-06 | 0.000132019 | 0.049977323 |
| Obesity | t_cell | *VANGL1* | 5.469200786 | 2.15E-05 | 0.000169226 | 0.135170932 |
| Obesity | t_cell | *GLRX* | 3.093675041 | 9.10E-06 | 0.000137234 | 0.057226424 |
| Obesity | t_cell | *GOLGB1* | 4.297104314 | 0.000363567 | 0.001142208 | 1 |
| Obesity | t_cell | *DMRT2* | 1.438568817 | 0.000997697 | 0.002547733 | 1 |

| | | | | | | |
|---|---|---|---|---|---|---|
| Obesity | t_cell | *ZNF483* | 1.381670473 | 0.010078652 | 0.018730265 | 1 |
| Obesity | t_cell | *MAP3K11* | 5.116014123 | 1.18E-05 | 0.000145864 | 0.074269486 |
| Obesity | t_cell | *C1QA* | 5.709174713 | 0.000458989 | 0.001367342 | 1 |
| Obesity | t_cell | *NDNF* | 4.982997468 | 1.38E-06 | 5.14E-05 | 0.008679488 |
| Obesity | t_cell | *OLR1* | 7.616060935 | 1.21E-05 | 0.000145864 | 0.075780355 |
| Obesity | t_cell | *GLIPR1L1* | 3.676884153 | 1.15E-05 | 0.000145864 | 0.072339335 |
| Obesity | t_cell | *CCDC36* | 2.582471185 | 1.49E-06 | 5.38E-05 | 0.009396862 |
| Obesity | t_cell | *THAP2* | 4.291090023 | 0.000986371 | 0.002525995 | 1 |
| Obesity | t_cell | *PPP1R14B* | 6.237265384 | 3.65E-05 | 0.000225279 | 0.229333993 |
| Obesity | t_cell | *ZNF417* | -5.024942171 | 0.003289067 | 0.007055054 | 1 |
| Obesity | t_cell | *VEGFB* | 1.853261631 | 0.00665116 | 0.013010531 | 1 |
| Obesity | t_cell | *PEAK1* | 3.642414053 | 0.000371355 | 0.001158089 | 1 |
| Obesity | t_cell | *MST1* | -1.301957861 | 0.012033139 | 0.021852208 | 1 |
| Obesity | t_cell | *GMPPB* | 3.119256537 | 0.008582109 | 0.016246829 | 1 |
| Obesity | t_cell | *CSPG4* | 2.201725478 | 0.011444989 | 0.020917048 | 1 |
| Obesity | t_cell | *SNX33* | 2.873864686 | 4.29E-05 | 0.000247159 | 0.26965078 |
| Obesity | t_cell | *NUDT18* | 6.007694126 | 1.47E-05 | 0.000149118 | 0.092517059 |
| Obesity | t_cell | *SULT1B1* | -1.303345062 | 0.000561985 | 0.001601632 | 1 |
| Obesity | t_cell | *NUDT4* | 3.166137535 | 0.003012213 | 0.006533372 | 1 |
| Obesity | t_cell | *SCAI* | 3.542857596 | 0.012358344 | 0.022313875 | 1 |
| Obesity | t_cell | *AGFG1* | 2.882519721 | 0.002277672 | 0.005141733 | 1 |
| Obesity | t_cell | *HAP1* | -2.73987155 | 0.010717667 | 0.019777509 | 1 |
| Obesity | t_cell | *DPY19L1* | 4.132106386 | 0.017162324 | 0.029561515 | 1 |
| Obesity | t_cell | *TUBB8* | 5.513071756 | 0.015912927 | 0.02774392 | 1 |
| Obesity | t_cell | *GPR160* | 4.640693616 | 0.021691617 | 0.036173792 | 1 |
| Obesity | t_cell | *GOLIM4* | 3.651746107 | 0.002162542 | 0.004918923 | 1 |
| Obesity | t_cell | *USMG5* | 5.148741778 | 7.99E-05 | 0.000370926 | 0.50223316 |
| Obesity | t_cell | *HOXB2* | 4.743106266 | 2.24E-07 | 1.32E-05 | 0.001410968 |
| Obesity | t_cell | *XXYLT1* | 3.993572194 | 0.000284045 | 0.000954459 | 1 |
| Obesity | t_cell | *UBXN2A* | 3.272799754 | 0.000471 | 0.001391531 | 1 |
| Obesity | t_cell | *CCS* | 5.455729506 | 4.65E-06 | 0.000100727 | 0.029234048 |
| Obesity | t_cell | *SLC25A30* | 6.540950441 | 0.005049573 | 0.010211214 | 1 |
| Obesity | t_cell | *C16orf91* | 7.756973823 | 6.34E-05 | 0.000312818 | 0.398422077 |
| Obesity | t_cell | *SOCS7* | 4.956558576 | 1.59E-05 | 0.000153197 | 0.100037641 |
| Obesity | t_cell | *TLR6* | 4.436377934 | 1.21E-05 | 0.000145864 | 0.076301082 |
| Obesity | t_cell | *RGMB* | 3.111349425 | 0.000117228 | 0.000487948 | 0.737014451 |
| Obesity | t_cell | *FAM53A* | 2.999172069 | 1.19E-05 | 0.000145864 | 0.074772791 |
| Obesity | t_cell | *CTU2* | 5.354363797 | 0.000198383 | 0.000728524 | 1 |
| Obesity | t_cell | *SNX31* | 2.40628451 | 0.003165787 | 0.006818535 | 1 |
| Obesity | t_cell | *GLIS1* | 1.191549021 | 0.001263359 | 0.003119692 | 1 |
| Obesity | t_cell | *LIG4* | 3.060828522 | 0.000294838 | 0.000982323 | 1 |
| Obesity | t_cell | *LINGO2* | 5.270673513 | 0.009925546 | 0.018486517 | 1 |
| Obesity | t_cell | *BBS1* | 7.785821893 | 7.99E-08 | 6.28E-06 | 0.000502189 |
| Obesity | t_cell | *TMEM81* | 7.184087722 | 9.68E-05 | 0.000422336 | 0.608585511 |
| Obesity | t_cell | *IL20RB* | 1.90462753 | 0.016042264 | 0.027930687 | 1 |
| Obesity | t_cell | *ZNF497* | 10.51618937 | 7.41E-05 | 0.000349972 | 0.465812158 |
| Obesity | t_cell | *KLF14* | 31.03019054 | 0.000274921 | 0.00093227 | 1 |
| Obesity | t_cell | *ANGEL2* | 4.163245486 | 0.008761063 | 0.016550723 | 1 |
| Obesity | t_cell | *BRSK2* | -3.642235853 | 0.015514928 | 0.027178143 | 1 |
| Obesity | t_cell | *SH3PXD2B* | 2.338585991 | 2.67E-05 | 0.000189464 | 0.1676755 |

| | | | | | | |
|---|---|---|---|---|---|---|
| Obesity | t_cell | *LARP7* | 4.713834369 | 0.001882155 | 0.004358418 | 1 |
| Obesity | t_cell | *C4orf32* | 4.883518851 | 3.39E-05 | 0.000214483 | 0.213197547 |
| Obesity | t_cell | *HRAS* | 7.195577731 | 9.26E-05 | 0.000408745 | 0.582461432 |
| Obesity | t_cell | *SRP72* | 3.0528277 | 0.000929894 | 0.002407844 | 1 |
| Obesity | t_cell | *BTC* | 2.617306081 | 3.54E-05 | 0.000220782 | 0.222769305 |
| Obesity | t_cell | *NLRP6* | -2.428586213 | 7.03E-05 | 0.000335867 | 0.44166556 |
| Obesity | t_cell | *NDUFA11* | 6.503135114 | 0.001055252 | 0.002677449 | 1 |
| Obesity | t_cell | *PTDSS2* | 4.390903032 | 0.000281037 | 0.000946769 | 1 |
| Obesity | t_cell | *FUT1* | 5.893983041 | 0.001233595 | 0.003054593 | 1 |
| Obesity | t_cell | *FBXW8* | 3.547258019 | 0.025768815 | 0.042045638 | 1 |
| Obesity | t_cell | *CHST2* | 5.606041669 | 3.00E-05 | 0.000201262 | 0.18863174 |
| Obesity | t_cell | *TVP23C* | 4.791766152 | 0.020893193 | 0.035018797 | 1 |
| Obesity | t_cell | *PACS1* | -1.530505958 | 0.013229017 | 0.02361466 | 1 |
| Obesity | t_cell | *SH3BP5L* | 6.617210255 | 8.93E-05 | 0.000397537 | 0.561322809 |
| Obesity | t_cell | *CADM2* | 3.577457017 | 1.13E-08 | 1.33E-06 | 7.11E-05 |
| Obesity | t_cell | *PSMD2* | 4.06042822 | 4.33E-06 | 9.97E-05 | 0.02720627 |
| Obesity | t_cell | *ARHGAP1* | 2.913941617 | 0.025519982 | 0.041673799 | 1 |
| Obesity | t_cell | *GOLGA8A* | 4.100106301 | 0.000388547 | 0.001196546 | 1 |
| Obesity | t_cell | *TP53I11* | 5.806553835 | 1.53E-05 | 0.000150391 | 0.095949322 |
| Obesity | t_cell | *PHYHD1* | 2.448007607 | 2.33E-05 | 0.000175508 | 0.146355374 |
| Obesity | t_cell | *ZNF519* | 5.140544531 | 2.81E-05 | 0.000194841 | 0.176704439 |
| Obesity | t_cell | *TMEM9B* | 8.72971129 | 2.53E-08 | 2.52E-06 | 0.000158762 |
| Obesity | t_cell | *SCUBE2* | 2.839150217 | 6.82E-05 | 0.000328606 | 0.428502348 |
| Obesity | t_cell | *EIF3F* | -2.433637256 | 0.026719212 | 0.043395423 | 1 |
| Obesity | t_cell | *CLTB* | 3.099642392 | 0.011029869 | 0.020258483 | 1 |
| Obesity | t_cell | *PCSK1* | 1.902123223 | 0.00022531 | 0.000799844 | 1 |
| Obesity | t_cell | *LPL* | 0.807962334 | 0.024804964 | 0.040579965 | 1 |
| Obesity | t_cell | *TBC1D10C* | -0.604167279 | 0.00044068 | 0.00132237 | 1 |
| Obesity | t_cell | *PPP2R2D* | 3.728949967 | 0.000244176 | 0.000850962 | 1 |
| Obesity | t_cell | *MCTP1* | 2.055059862 | 0.010111659 | 0.018786052 | 1 |
| Obesity | t_cell | *UBQLNL* | 3.52691322 | 0.015431168 | 0.02705403 | 1 |
| Obesity | t_cell | *KCNE3* | 3.313727634 | 3.97E-05 | 0.000236175 | 0.249400832 |
| Obesity | t_cell | *UCP2* | -2.107938717 | 0.001153157 | 0.002881865 | 1 |
| Obesity | t_cell | *TOM1L2* | 3.31100009 | 0.020618492 | 0.03461374 | 1 |
| Obesity | t_cell | *TEX26* | 4.528676748 | 1.80E-05 | 0.000158468 | 0.11298742 |
| Obesity | t_cell | *GPR156* | 3.410468617 | 2.66E-05 | 0.000189464 | 0.167494427 |
| Obesity | t_cell | *LINC01106* | 13.94853936 | 0.002514608 | 0.005598208 | 1 |
| Obesity | t_cell | *ETV4* | 4.397241893 | 1.65E-05 | 0.000155092 | 0.103601137 |
| Obesity | t_cell | *SWI5* | 3.708326095 | 0.000836591 | 0.002208003 | 1 |
| Obesity | t_cell | *ZDHHC21* | 3.774297197 | 0.000719588 | 0.001949182 | 1 |
| Obesity | t_cell | *ARL4D* | 3.137400817 | 0.002848309 | 0.006222139 | 1 |
| Obesity | t_cell | *LRRN1* | 3.985063458 | 1.39E-05 | 0.000148367 | 0.087607663 |
| Obesity | t_cell | *ZNF613* | 3.426873377 | 0.00203646 | 0.004669301 | 1 |
| Obesity | t_cell | *TMEM39A* | 3.713929975 | 0.000294675 | 0.000982301 | 1 |
| Obesity | t_cell | *TCP11L1* | 3.934814936 | 0.000310414 | 0.001018034 | 1 |
| Obesity | t_cell | *GPX2* | 6.086550358 | 3.84E-05 | 0.00023194 | 0.241544528 |
| Obesity | t_cell | *CCDC57* | -2.464231085 | 0.023155199 | 0.03819909 | 1 |
| Obesity | t_cell | *SMIM19* | 2.997459754 | 5.66E-05 | 0.000289932 | 0.356036051 |
| Obesity | t_cell | *SLC35G1* | 2.136508831 | 0.008291773 | 0.015773185 | 1 |
| Obesity | t_cell | *ZNF135* | 2.196569559 | 0.005662948 | 0.011270324 | 1 |

| | | | | | | |
|---|---|---|---|---|---|---|
| Obesity | t_cell | *COX8A* | 8.087137877 | 0.000356885 | 0.001127504 | 1 |
| Obesity | t_cell | *B3GNT4* | 11.9968966 | 4.54E-05 | 0.0002542 | 0.285212383 |
| Obesity | t_cell | *RNPEP* | 3.541209627 | 0.003431012 | 0.007295883 | 1 |
| Obesity | t_cell | *WDR25* | 4.849278979 | 0.000491391 | 0.001438256 | 1 |
| Obesity | t_cell | *PLA2G16* | 1.273851605 | 6.58E-05 | 0.000320973 | 0.413887744 |
| Obesity | t_cell | *PRR15* | 5.401701287 | 0.006441347 | 0.012639435 | 1 |
| Obesity | t_cell | *RMDN1* | 3.989659982 | 0.000849047 | 0.002235172 | 1 |
| Obesity | t_cell | *RNF152* | 4.669080453 | 0.002503872 | 0.005578258 | 1 |
| Obesity | t_cell | *BOK* | 1.829303679 | 8.82E-05 | 0.000394869 | 0.554725387 |
| Obesity | t_cell | *ZNF843* | -4.642375916 | 0.005083915 | 0.01027075 | 1 |
| Obesity | t_cell | *C8orf59* | 5.062783363 | 0.000859543 | 0.002257287 | 1 |
| Obesity | t_cell | *RUFY1* | 5.192198821 | 1.95E-07 | 1.22E-05 | 0.00122353 |
| Obesity | t_cell | *BASP1* | 2.561589482 | 0.003467394 | 0.007352279 | 1 |
| Obesity | t_cell | *LRRC37A3* | 5.925516062 | 5.44E-05 | 0.000282714 | 0.342083867 |
| Obesity | t_cell | *VSIG10* | 1.785160723 | 0.000255298 | 0.0008819 | 1 |
| Obesity | t_cell | *IRX5* | 2.542834205 | 1.06E-05 | 0.000145864 | 0.06687959 |
| Obesity | t_cell | *GCNT4* | -4.627277512 | 0.000374649 | 0.001165531 | 1 |
| Obesity | t_cell | *MUC20* | 4.044724233 | 0.000462198 | 0.001372468 | 1 |
| Obesity | t_cell | *THAP4* | 3.690815155 | 0.002648246 | 0.005856322 | 1 |
| Obesity | t_cell | *SHMT1* | 1.938566651 | 0.00286939 | 0.006261665 | 1 |
| Obesity | t_cell | *DPP7* | 3.922798173 | 0.0266481 | 0.043302302 | 1 |
| Obesity | t_cell | *SEC24C* | 4.287288775 | 0.01641907 | 0.028421446 | 1 |
| Obesity | t_cell | *MTHFR* | 5.679846542 | 0.000346165 | 0.001103062 | 1 |
| Obesity | t_cell | *C19orf18* | 2.193208581 | 0.003524083 | 0.00745739 | 1 |
| Obesity | t_cell | *POLE* | 6.629054678 | 0.000240757 | 0.000842784 | 1 |
| Obesity | t_cell | *DSCAML1* | 5.045524581 | 0.000301605 | 0.000997996 | 1 |
| Obesity | t_cell | *EPS8L2* | 3.662379627 | 0.001614144 | 0.003818287 | 1 |
| Obesity | t_cell | *AP006621.1* | 2.742008753 | 7.00E-08 | 5.79E-06 | 0.000440273 |
| Obesity | t_cell | *MAN1B1* | 6.820565164 | 0.000143052 | 0.00056947 | 0.899365946 |
| Obesity | t_cell | *ZBTB38* | 3.117262003 | 0.015165874 | 0.026618606 | 1 |
| Obesity | t_cell | *C8orf31* | 5.415751583 | 1.23E-05 | 0.000145864 | 0.077162196 |
| Obesity | t_cell | *AC024940.1* | 4.286579268 | 8.73E-05 | 0.000392666 | 0.548947279 |
| Obesity | t_cell | *TIMM22* | 6.033873565 | 5.06E-05 | 0.000269159 | 0.318193654 |
| Obesity | t_cell | *MAGEF1* | 6.97673454 | 1.45E-05 | 0.000148509 | 0.091184575 |
| Obesity | t_cell | *PAWR* | 4.972799346 | 2.05E-05 | 0.000166507 | 0.129043304 |
| Obesity | t_cell | *NAP1L5* | 8.676345993 | 0.000163826 | 0.000625008 | 1 |
| Obesity | t_cell | *ACOT4* | 2.227582955 | 0.00330355 | 0.007078875 | 1 |
| Obesity | t_cell | *RBM44* | 4.960276187 | 0.003047911 | 0.006591 | 1 |
| Obesity | t_cell | *IRX3* | 1.715429569 | 0.016819724 | 0.029043012 | 1 |
| Obesity | t_cell | *ATOX1* | 5.821199798 | 0.000675755 | 0.001856849 | 1 |
| Obesity | t_cell | *SAMD12* | -1.707715727 | 0.000643829 | 0.001782366 | 1 |
| Obesity | t_cell | *C12orf54* | 6.911795369 | 0.000399428 | 0.001224382 | 1 |
| Obesity | t_cell | *GBA* | 4.53635686 | 0.008536383 | 0.016174877 | 1 |
| Obesity | t_cell | *CD163L1* | 2.003168124 | 0.001103229 | 0.002778846 | 1 |
| Obesity | t_cell | *CD151* | 5.559101592 | 1.81E-07 | 1.15E-05 | 0.001136282 |
| Obesity | t_cell | *FAM87B* | 2.834821683 | 0.01453288 | 0.025614863 | 1 |
| Obesity | t_cell | *PCDHB9* | 1.755051855 | 0.004522393 | 0.00923726 | 1 |
| Obesity | t_cell | *ZNF354C* | 6.954182232 | 4.54E-05 | 0.000254238 | 0.285509827 |
| Obesity | t_cell | *CENPBD1* | 2.218427823 | 0.000581142 | 0.001644291 | 1 |
| Obesity | t_cell | *BET1L* | 5.106009116 | 4.72E-05 | 0.000261143 | 0.296919083 |

| | | | | | | |
|---|---|---|---|---|---|---|
| Obesity | t_cell | *IMP3* | 6.474901311 | 0.004611521 | 0.009385767 | 1 |
| Obesity | t_cell | *ODF3B* | 4.924003717 | 4.94E-06 | 0.000104237 | 0.03106264 |
| Obesity | t_cell | *C2orf73* | 2.58296616 | 3.38E-05 | 0.000214294 | 0.212793989 |
| Obesity | t_cell | *FAM26E* | 3.877550711 | 8.85E-05 | 0.000395711 | 0.55636971 |
| Obesity | t_cell | *ALS2CL* | 2.31631056 | 0.006548548 | 0.012829767 | 1 |
| Obesity | t_cell | *MLF1* | 4.551485249 | 0.0001088 | 0.000463119 | 0.68402676 |
| Obesity | t_cell | *GRAMD1C* | 6.607596473 | 3.87E-05 | 0.000232312 | 0.242998097 |
| Obesity | t_cell | *TSSK6* | -3.665157047 | 2.22E-05 | 0.000171828 | 0.139524702 |
| Obesity | t_cell | *GOLGA8Q* | 35.69520603 | 0.013180949 | 0.023535537 | 1 |
| Obesity | t_cell | *ZNF114* | 2.941263678 | 0.000155734 | 0.000604009 | 0.979099221 |
| Obesity | t_cell | *ZNF518B* | 4.262537478 | 0.010502895 | 0.019421088 | 1 |
| Obesity | t_cell | *LCORL* | 3.446993753 | 0.009705414 | 0.018122345 | 1 |
| Obesity | t_cell | *ZNF454* | 2.902079405 | 3.10E-05 | 0.000206015 | 0.194890578 |
| Obesity | t_cell | *SH2B1* | -5.284963402 | 2.72E-18 | 2.45E-15 | 1.71E-14 |
| Obesity | t_cell | *ZC3H12D* | -0.964345302 | 0.000258827 | 0.000890417 | 1 |
| Obesity | t_cell | *KDELC2* | 3.521743448 | 0.019083174 | 0.032382164 | 1 |
| Obesity | t_cell | *SH2D4B* | 16.97732009 | 0.011195013 | 0.020519839 | 1 |
| Obesity | t_cell | *RNF212* | 1.201214772 | 0.00777383 | 0.01490063 | 1 |
| Obesity | t_cell | *PRSS36* | 6.125950172 | 1.57E-05 | 0.000152914 | 0.098782668 |
| Obesity | t_cell | *TMPRSS9* | 2.617707394 | 0.00011391 | 0.000478391 | 0.716150757 |
| Obesity | t_cell | *AQP11* | 4.62340166 | 5.61E-06 | 0.000109386 | 0.035288163 |
| Obesity | t_cell | *TMEM11* | 7.979931691 | 5.00E-05 | 0.000267593 | 0.314518591 |
| Obesity | t_cell | *AC006111.1* | 8.393350749 | 3.38E-05 | 0.000214224 | 0.212509794 |
| Obesity | t_cell | *LINC00843* | 6.782811115 | 1.17E-05 | 0.000145864 | 0.073623151 |
| Obesity | t_cell | *COX14* | 2.849582429 | 6.23E-06 | 0.000114728 | 0.039150121 |
| Obesity | t_cell | *CA8* | 1.737024873 | 0.000713546 | 0.001936988 | 1 |
| Obesity | t_cell | *ERBB4* | 2.505188626 | 1.13E-05 | 0.000145864 | 0.071046765 |
| Obesity | t_cell | *CSRNP3* | 4.062208851 | 6.72E-05 | 0.000325577 | 0.422299364 |
| Obesity | t_cell | *ZNF713* | 1.873360129 | 0.001495473 | 0.003579002 | 1 |
| Obesity | t_cell | *PARP10* | 2.645173066 | 0.020146576 | 0.033939315 | 1 |
| Obesity | t_cell | *KCTD12* | 2.247014449 | 0.003296521 | 0.007068631 | 1 |
| Obesity | t_cell | *STX19* | 7.334446169 | 5.97E-11 | 1.63E-08 | 3.76E-07 |
| Obesity | t_cell | *FAM219B* | 4.756035438 | 6.96E-07 | 3.22E-05 | 0.004377116 |
| Obesity | t_cell | *GDPD4* | 13.29522738 | 0.000286601 | 0.000962018 | 1 |
| Obesity | t_cell | *MPI* | 6.185779984 | 1.63E-07 | 1.07E-05 | 0.001025743 |
| Obesity | t_cell | *TRIM73* | -10.06962483 | 5.37E-10 | 9.93E-08 | 3.38E-06 |
| Obesity | t_cell | *EFCAB13* | 1.885515656 | 0.01100538 | 0.020234562 | 1 |
| Obesity | t_cell | *TUFM* | 8.791571335 | 3.33E-22 | 5.23E-19 | 2.09E-18 |
| Obesity | t_cell | *RMI1* | 2.911350445 | 0.01218816 | 0.022063622 | 1 |
| Obesity | t_cell | *SNX18* | 4.065232523 | 0.006964566 | 0.013577125 | 1 |
| Obesity | t_cell | *C14orf39* | 1.144384186 | 0.002028831 | 0.004656905 | 1 |
| Obesity | t_cell | *TRIML2* | 26.44462624 | 0.000936252 | 0.002418331 | 1 |
| Obesity | t_cell | *SPTY2D1* | 6.468834381 | 8.79E-05 | 0.000394101 | 0.552529069 |
| Obesity | t_cell | *TCAIM* | 2.675607352 | 0.00333708 | 0.007136131 | 1 |
| Obesity | t_cell | *FUCA1* | 3.589330606 | 0.00201283 | 0.004621864 | 1 |
| Obesity | t_cell | *ZNF664* | 6.168011032 | 0.000293814 | 0.000980992 | 1 |
| Obesity | t_cell | *CALR* | 5.620210348 | 6.46E-05 | 0.000317904 | 0.406280912 |
| Obesity | t_cell | *CDH4* | 2.440842474 | 0.004461801 | 0.009128325 | 1 |
| Obesity | t_cell | *EGR3* | 11.07457598 | 0.000125779 | 0.000514155 | 0.790769816 |
| Obesity | t_cell | *FJX1* | 3.835897372 | 6.67E-05 | 0.000323332 | 0.419038202 |

| | | | | | | |
|---|---|---|---|---|---|---|
| Obesity | t_cell | *ZBTB42* | -3.604206608 | 0.012105142 | 0.021935148 | 1 |
| Obesity | t_cell | *LACC1* | 7.457239774 | 2.12E-05 | 0.000169127 | 0.133316186 |
| Obesity | t_cell | *MAF1* | 5.908540153 | 2.01E-05 | 0.00016409 | 0.126641868 |
| Obesity | t_cell | *FCER1A* | 2.660936028 | 0.004638327 | 0.009437269 | 1 |
| Obesity | t_cell | *CDH5* | 2.535098649 | 2.73E-06 | 7.61E-05 | 0.017133391 |
| Obesity | t_cell | *SERTAD2* | 4.993427263 | 0.000129325 | 0.000525237 | 0.813066917 |
| Obesity | t_cell | *CITED4* | 3.674798912 | 0.000129748 | 0.000526273 | 0.815723205 |
| Obesity | t_cell | *TIGD5* | 2.31320238 | 0.001575245 | 0.003745674 | 1 |
| Obesity | t_cell | *PDXDC1* | 3.918739328 | 0.000966189 | 0.002481384 | 1 |
| Obesity | t_cell | *ZNF154* | 3.455555165 | 0.000713309 | 0.001936988 | 1 |
| Obesity | t_cell | *ZADH2* | 3.880080169 | 0.022170942 | 0.03685582 | 1 |
| Obesity | t_cell | *SEPT1* | -1.389703123 | 0.000426015 | 0.001285816 | 1 |
| Obesity | t_cell | *TDRD6* | 1.152339206 | 0.017409097 | 0.029937361 | 1 |
| Obesity | t_cell | *LYNX1* | 3.025508364 | 0.000238239 | 0.000835832 | 1 |
| Obesity | t_cell | *TDRP* | 4.020882395 | 2.13E-05 | 0.000169127 | 0.133610528 |
| Obesity | t_cell | *FGD6* | 4.135371263 | 1.28E-05 | 0.000146996 | 0.080171855 |
| Obesity | t_cell | *HCLS1* | -1.647063917 | 0.002771528 | 0.006086132 | 1 |
| Obesity | t_cell | *CYP8B1* | 2.451738811 | 0.000693339 | 0.001897703 | 1 |
| Obesity | t_cell | *SSTR2* | 5.890923975 | 1.89E-05 | 0.000160637 | 0.11857142 |
| Obesity | t_cell | *ZNF594* | 4.19596513 | 0.005226181 | 0.010516265 | 1 |
| Obesity | t_cell | *SHISA2* | -6.226797239 | 0.00014207 | 0.000566748 | 0.893194871 |
| Obesity | t_cell | *CHST13* | 8.967328287 | 2.84E-05 | 0.000196345 | 0.178674341 |
| Obesity | t_cell | *SRSF8* | -3.194906605 | 0.003912779 | 0.008172639 | 1 |
| Obesity | t_cell | *PSMG4* | 3.271705529 | 0.009894179 | 0.018441953 | 1 |
| Obesity | t_cell | *ZNF443* | 2.954197339 | 0.000401727 | 0.001229032 | 1 |
| Obesity | t_cell | *CAPS2* | 4.345874184 | 2.32E-05 | 0.000175508 | 0.14613026 |
| Obesity | t_cell | *ZNF792* | -2.898380798 | 0.000836292 | 0.002208003 | 1 |
| Obesity | t_cell | *KCTD2* | 7.052045503 | 0.000155422 | 0.000603171 | 0.977136226 |
| Obesity | t_cell | *CMTR2* | 4.635368104 | 1.23E-05 | 0.00014611 | 0.077438496 |
| Obesity | t_cell | *MRPL14* | 4.624921152 | 0.000908719 | 0.002363723 | 1 |
| Obesity | t_cell | *BBS12* | 6.904701107 | 0.000303327 | 0.001002638 | 1 |
| Obesity | t_cell | *ZFP82* | 5.731444888 | 1.47E-05 | 0.000149118 | 0.092537879 |
| Obesity | t_cell | *LSMEM1* | 2.098786955 | 6.96E-05 | 0.000334008 | 0.43788429 |
| Obesity | t_cell | *AEN* | -3.252790031 | 0.001279815 | 0.00315004 | 1 |
| Obesity | t_cell | *FKRP* | 2.052148808 | 0.00054409 | 0.001560534 | 1 |
| Obesity | t_cell | *TRAPPC5* | 3.329772864 | 0.010311103 | 0.019105778 | 1 |
| Obesity | t_cell | *RPH3AL* | 4.454808552 | 3.17E-05 | 0.000209022 | 0.199508197 |
| Obesity | t_cell | *SLC26A11* | 5.039197222 | 0.00265287 | 0.005864484 | 1 |
| Obesity | t_cell | *F2R* | -1.083715416 | 0.03131558 | 0.049919131 | 1 |
| Obesity | t_cell | *HIST3H2A* | -3.105978968 | 0.006669677 | 0.013042694 | 1 |
| Obesity | t_cell | *POLR2A* | 3.801830545 | 0.011596353 | 0.021141602 | 1 |
| Obesity | t_cell | *TMEM132C* | 1.409009797 | 0.000124737 | 0.000510892 | 0.784218544 |
| Obesity | t_cell | *SLC25A41* | 8.086492247 | 0.000140494 | 0.000561529 | 0.883285128 |
| Obesity | t_cell | *TMEM132E* | 6.301136421 | 0.02058032 | 0.034558886 | 1 |
| Obesity | t_cell | *CCL13* | 2.185353829 | 0.025774438 | 0.042045638 | 1 |
| Obesity | t_cell | *ACBD4* | 6.079130075 | 0.000133464 | 0.000537531 | 0.839086478 |
| Obesity | t_cell | *GPR135* | 2.374621312 | 1.36E-05 | 0.000147409 | 0.085625703 |
| Obesity | t_cell | *ZFP41* | 2.920191241 | 3.84E-05 | 0.00023194 | 0.241698049 |
| Obesity | t_cell | *ATG9B* | 3.949581858 | 2.87E-05 | 0.000196979 | 0.180236177 |
| Obesity | t_cell | *HKR1* | 4.398465694 | 0.002317787 | 0.005219172 | 1 |

| | | | | | | |
|---|---|---|---|---|---|---|
| Obesity | t_cell | *ZBTB20* | 1.986746484 | 0.030878681 | 0.049297681 | 1 |
| Obesity | t_cell | *CHST15* | 3.848153424 | 2.61E-05 | 0.000187403 | 0.163789788 |
| Obesity | t_cell | *TNRC18* | 4.449741878 | 8.16E-05 | 0.000375666 | 0.512784171 |
| Obesity | t_cell | *FAM181B* | 4.061075591 | 0.019044823 | 0.03233454 | 1 |
| Obesity | t_cell | *FAM89A* | 2.447858626 | 5.62E-06 | 0.000109386 | 0.035316905 |
| Obesity | t_cell | *TDRKH* | -2.508957977 | 0.000915851 | 0.002376374 | 1 |
| Obesity | t_cell | *MRPL41* | 4.069222803 | 0.000587871 | 0.001659968 | 1 |
| Obesity | t_cell | *HHIPL1* | 3.184324258 | 0.000106958 | 0.000457135 | 0.67244504 |
| Obesity | t_cell | *FAM153B* | -0.998780115 | 0.000198139 | 0.000728481 | 1 |
| Obesity | t_cell | *BACE2* | 1.920355634 | 0.004254062 | 0.0087632 | 1 |
| Obesity | t_cell | *SYNM* | 4.478774039 | 1.31E-05 | 0.000147409 | 0.082137713 |
| Obesity | t_cell | *IZUMO1* | 1.999175645 | 0.002895514 | 0.00631429 | 1 |
| Obesity | t_cell | *C8orf33* | 3.811663925 | 0.001723662 | 0.004037021 | 1 |
| Obesity | t_cell | *FBXL6* | 3.757898969 | 0.01601965 | 0.027899042 | 1 |
| Obesity | t_cell | *CLN8* | 4.603018248 | 0.000386736 | 0.001192952 | 1 |
| Obesity | t_cell | *CACNB4* | 1.525390832 | 0.001241985 | 0.003071739 | 1 |
| Obesity | t_cell | *TRAPPC6B* | 3.133605371 | 0.010116303 | 0.018789127 | 1 |
| Obesity | t_cell | *TSHZ2* | 2.39352218 | 0.004286837 | 0.008822043 | 1 |
| Obesity | t_cell | *CAPN12* | -4.465878978 | 0.005265431 | 0.010579662 | 1 |
| Obesity | t_cell | *FES* | 7.366470818 | 2.08E-05 | 0.000167886 | 0.131022013 |
| Obesity | t_cell | *GLRX5* | 6.2935358 | 0.001098238 | 0.002768493 | 1 |
| Obesity | t_cell | *MXRA7* | 1.451344984 | 0.006840128 | 0.013346954 | 1 |
| Obesity | t_cell | *MFSD5* | 4.483702042 | 1.09E-05 | 0.000145864 | 0.068760619 |
| Obesity | t_cell | *ADI1* | 3.962083703 | 6.34E-05 | 0.000312818 | 0.398529671 |
| Obesity | t_cell | *SPNS3* | 9.228792071 | 0.002971337 | 0.006458173 | 1 |
| Obesity | t_cell | *CLEC4G* | 4.253232622 | 5.19E-05 | 0.000273498 | 0.326009335 |
| Obesity | t_cell | *SATB1* | -2.197446595 | 0.003661503 | 0.007706685 | 1 |
| Obesity | t_cell | *TRAK1* | 3.166528582 | 0.003687554 | 0.007756323 | 1 |
| Obesity | t_cell | *KCNB2* | 1.377847279 | 0.000342272 | 0.001095303 | 1 |
| Obesity | t_cell | *TSKU* | 2.719696119 | 1.13E-05 | 0.000145864 | 0.071062007 |
| Obesity | t_cell | *ACBD3* | 4.353779102 | 0.001006228 | 0.002568475 | 1 |
| Obesity | t_cell | *PLCXD3* | 2.634269716 | 0.000190835 | 0.000710348 | 1 |
| Obesity | t_cell | *GPIHBP1* | 1.783769777 | 0.000757032 | 0.002033087 | 1 |
| Obesity | t_cell | *VMO1* | 1.742663844 | 0.00201173 | 0.004621025 | 1 |
| Obesity | t_cell | *COL18A1* | 3.707546988 | 1.41E-05 | 0.000148367 | 0.08843401 |
| Obesity | t_cell | *SLC25A18* | 1.647643828 | 0.001383159 | 0.003356201 | 1 |
| Obesity | t_cell | *ZNF721* | 2.261691764 | 0.026420348 | 0.042998895 | 1 |
| Obesity | t_cell | *LENG9* | 3.178457902 | 0.000640386 | 0.001775963 | 1 |
| Obesity | t_cell | *C12orf60* | 3.861527615 | 9.91E-05 | 0.000429287 | 0.623324825 |
| Obesity | t_cell | *LYSMD4* | 2.783610338 | 2.00E-05 | 0.000163758 | 0.125556834 |
| Obesity | t_cell | *GAS6* | 4.217874344 | 1.82E-05 | 0.000158673 | 0.114153632 |
| Obesity | t_cell | *BEGAIN* | 6.66311511 | 3.25E-05 | 0.000210045 | 0.204522358 |
| Obesity | t_cell | *ARHGEF37* | 6.449877723 | 4.79E-05 | 0.000262855 | 0.301016591 |
| Obesity | t_cell | *FAM43B* | 3.42883089 | 0.006158419 | 0.012129694 | 1 |
| Obesity | t_cell | *CSMD1* | 3.113937271 | 1.62E-05 | 0.000154104 | 0.101568322 |
| Obesity | t_cell | *PTGDR2* | -19.29302042 | 0.002191509 | 0.004972218 | 1 |
| Obesity | t_cell | *GPR19* | -1.013782832 | 0.000195105 | 0.000721543 | 1 |
| Obesity | t_cell | *C2CD4C* | 4.406315294 | 2.04E-06 | 6.45E-05 | 0.012820867 |
| Obesity | t_cell | *GDPGP1* | 2.896873194 | 0.022603684 | 0.037489382 | 1 |
| Obesity | t_cell | *CTNNA3* | 2.512535785 | 0.012386936 | 0.022352659 | 1 |

| | | | | | | |
|---|---|---|---|---|---|---|
| Obesity | t_cell | *CCBE1* | 2.793829514 | 2.24E-07 | 1.32E-05 | 0.00141134 |
| Obesity | t_cell | *CCDC125* | 4.594788231 | 0.000175669 | 0.000664119 | 1 |
| Obesity | t_cell | *C10orf107* | 0.534702617 | 0.016213211 | 0.028165918 | 1 |
| Obesity | t_cell | *FHL3* | 4.714106602 | 0.001420098 | 0.003429948 | 1 |
| Obesity | t_cell | *LRIT3* | -4.635089905 | 1.73E-05 | 0.000157091 | 0.109021307 |
| Obesity | t_cell | *URAD* | 6.59370546 | 0.000416127 | 0.001264471 | 1 |
| Obesity | t_cell | *GPR132* | -1.823283083 | 0.000473707 | 0.001398215 | 1 |
| Obesity | t_cell | *PSMG1* | 2.12877737 | 1.59E-06 | 5.67E-05 | 0.009971905 |
| Obesity | t_cell | *KLRC4* | -1.469654641 | 2.42E-05 | 0.00017926 | 0.152370927 |
| Obesity | t_cell | *ACSM5* | 1.969982402 | 0.000292172 | 0.00097706 | 1 |
| Obesity | t_cell | *SERHL2* | 8.127123374 | 2.94E-06 | 8.04E-05 | 0.018466958 |
| Obesity | t_cell | *TANGO2* | 5.356282133 | 0.000275912 | 0.000933674 | 1 |
| Obesity | t_cell | *SFXN4* | 5.559327636 | 5.46E-05 | 0.000283204 | 0.343243789 |
| Obesity | t_cell | *RP1L1* | 8.929537212 | 1.05E-12 | 3.88E-10 | 6.60E-09 |
| Obesity | t_cell | *FAM19A1* | -0.671615382 | 0.000298272 | 0.000989571 | 1 |
| Obesity | t_cell | *TRMT12* | 3.327930131 | 0.026561417 | 0.043183767 | 1 |
| Obesity | t_cell | *BMP8A* | 7.314302504 | 2.02E-10 | 4.23E-08 | 1.27E-06 |
| Obesity | t_cell | *CMTM4* | 4.669057432 | 0.000154893 | 0.000602235 | 0.973813365 |
| Obesity | t_cell | *CBX6* | 8.061240423 | 8.28E-05 | 0.000379615 | 0.5208314 |
| Obesity | t_cell | *KCTD16* | 3.531470017 | 8.62E-07 | 3.69E-05 | 0.005419281 |
| Obesity | t_cell | *B3GALT5* | 3.563540636 | 0.008558557 | 0.016212006 | 1 |
| Obesity | t_cell | *ZNF703* | 3.192350622 | 0.000832898 | 0.002202958 | 1 |
| Obesity | t_cell | *KCTD8* | 1.564208141 | 0.000152531 | 0.00059526 | 0.958964228 |
| Obesity | t_cell | *C9orf66* | 4.252698588 | 8.50E-05 | 0.000385811 | 0.534347621 |
| Obesity | t_cell | *NPIPA5* | 0.734270412 | 0.022741009 | 0.037654129 | 1 |
| Obesity | t_cell | *FAM162B* | 2.416018467 | 0.000526836 | 0.001516582 | 1 |
| Obesity | t_cell | *ANKRD45* | 4.239604725 | 4.30E-05 | 0.000247251 | 0.270130249 |
| Obesity | t_cell | *MAATS1* | 3.689736055 | 2.39E-05 | 0.000178067 | 0.150110309 |
| Obesity | t_cell | *GPR39* | 2.31134235 | 0.000119905 | 0.000495625 | 0.753844924 |
| Obesity | t_cell | *NPIPA7* | 5.826016071 | 0.003966549 | 0.008254782 | 1 |
| Obesity | t_cell | *PP2D1* | 4.479754935 | 0.00129171 | 0.00317056 | 1 |
| Obesity | t_cell | *DENND5A* | 4.247901518 | 0.000511414 | 0.001481005 | 1 |
| Obesity | t_cell | *VPS33B* | 4.795457775 | 0.000925831 | 0.002398657 | 1 |
| Obesity | t_cell | *TBX1* | 5.806866879 | 1.42E-05 | 0.000148367 | 0.08899515 |
| Obesity | t_cell | *UQCR10* | 5.435782471 | 0.013257256 | 0.023651637 | 1 |
| Obesity | t_cell | *EIF3C* | 7.043127056 | 1.49E-24 | 3.12E-21 | 9.35E-21 |
| Obesity | t_cell | *CNTN2* | 4.512225317 | 1.89E-05 | 0.000160637 | 0.118656847 |
| Obesity | t_cell | *GOLGA6L4* | 2.752985433 | 0.000461264 | 0.001372468 | 1 |
| Obesity | t_cell | *PCDH9* | 1.492931862 | 0.001143806 | 0.002863843 | 1 |
| Obesity | t_cell | *MROH7* | 4.773685857 | 0.013901317 | 0.024667678 | 1 |
| Obesity | t_cell | *CSF1* | 2.63577267 | 0.000241228 | 0.000843961 | 1 |
| Obesity | t_cell | *PLA2G6* | 4.049084493 | 5.56E-11 | 1.59E-08 | 3.49E-07 |
| Obesity | t_cell | *A3GALT2* | 1.738470301 | 0.005279328 | 0.01060081 | 1 |
| Obesity | t_cell | *SS18L1* | 4.082606495 | 0.001385439 | 0.003359552 | 1 |
| Obesity | t_cell | *CCR10* | 7.299183306 | 0.000112273 | 0.000473732 | 0.705860799 |
| Obesity | t_cell | *WDR27* | 4.669708645 | 8.94E-06 | 0.000136739 | 0.056199623 |
| Obesity | t_cell | *TXNRD2* | 5.947149398 | 0.000659741 | 0.001820004 | 1 |
| Obesity | t_cell | *PROS1* | 2.887247072 | 0.000336593 | 0.001084099 | 1 |
| Obesity | t_cell | *HDDC3* | 4.250021201 | 0.00042977 | 0.001295285 | 1 |
| Obesity | t_cell | *ZFP1* | 5.720585766 | 5.39E-05 | 0.000280871 | 0.338730097 |

| | | | | | | |
|---|---|---|---|---|---|---|
| Obesity | t_cell | *LPAR5* | -1.368012486 | 0.000720117 | 0.001949775 | 1 |
| Obesity | t_cell | *TMEM173* | -2.884531707 | 0.009461539 | 0.017719599 | 1 |
| Obesity | t_cell | *SNN* | 10.42672625 | 0.000309821 | 0.001017151 | 1 |
| Obesity | t_cell | *NELL2* | -0.414078989 | 0.000169817 | 0.000645488 | 1 |
| Obesity | t_cell | *SEPT5* | 5.072944656 | 3.97E-05 | 0.000236184 | 0.249873815 |
| Obesity | t_cell | *ATL3* | 4.175911922 | 0.000215838 | 0.000775413 | 1 |
| Obesity | t_cell | *TUSC5* | 1.38954975 | 0.000738377 | 0.001990642 | 1 |
| Obesity | t_cell | *TMED9* | 5.495442242 | 0.001314273 | 0.00321386 | 1 |
| Obesity | t_cell | *NUTM2A* | -3.649990714 | 0.018869128 | 0.032105604 | 1 |
| Obesity | t_cell | *FAM227A* | 2.674021576 | 6.25E-05 | 0.000309456 | 0.393009308 |
| Obesity | t_cell | *NDUFA6* | 0.950582156 | 0.011108347 | 0.020378809 | 1 |
| Obesity | t_cell | *SORCS2* | 4.122518581 | 2.06E-06 | 6.47E-05 | 0.012932225 |
| Obesity | t_cell | *TMEM106A* | 6.099326795 | 4.94E-06 | 0.000104237 | 0.031046426 |
| Obesity | t_cell | *SIVA1* | 4.900839158 | 9.20E-05 | 0.000407341 | 0.578423853 |
| Obesity | t_cell | *BRF1* | 5.843774886 | 0.000172395 | 0.000653379 | 1 |
| Obesity | t_cell | *FLRT2* | 1.87353938 | 0.000688081 | 0.001886596 | 1 |
| Obesity | t_cell | *ADSSL1* | 4.789682138 | 1.48E-05 | 0.000149118 | 0.092841415 |
| Obesity | t_cell | *C6orf120* | -3.631467517 | 0.011962448 | 0.02173639 | 1 |
| Obesity | t_cell | *DDX51* | 2.074909165 | 0.002093177 | 0.004785383 | 1 |
| Obesity | t_cell | *SIGIRR* | -1.515708205 | 0.00149131 | 0.00357312 | 1 |
| Obesity | t_cell | *IFITM2* | 3.168278478 | 0.025323236 | 0.041374008 | 1 |
| Obesity | t_cell | *GP1BA* | 2.064084593 | 0.029516708 | 0.047339679 | 1 |
| Obesity | t_cell | *KLHL33* | 2.805370497 | 3.91E-06 | 9.47E-05 | 0.024612571 |
| Obesity | t_cell | *RBM11* | -4.317620137 | 0.001044343 | 0.002654987 | 1 |
| Obesity | t_cell | *TMEM105* | 18.74980974 | 0.008503681 | 0.016124537 | 1 |
| Obesity | t_cell | *GAS2L1* | 3.062297688 | 2.87E-05 | 0.00019705 | 0.180498209 |
| Obesity | t_cell | *HS6ST3* | 6.2314453 | 4.98E-06 | 0.000104418 | 0.031325421 |
| Obesity | t_cell | *TNFAIP8L1* | 5.308528244 | 9.09E-05 | 0.000404229 | 0.571628355 |
| Obesity | t_cell | *MAPK11* | 4.35280594 | 4.28E-05 | 0.000247159 | 0.269196287 |
| Obesity | t_cell | *SP140L* | -1.887116319 | 0.010500167 | 0.019421088 | 1 |
| Obesity | t_cell | *FAM174B* | 4.372552129 | 0.000120515 | 0.000497306 | 0.7576792 |
| Obesity | t_cell | *SV2B* | 4.282370465 | 0.000150681 | 0.00058987 | 0.947330578 |
| Obesity | t_cell | *TLCD2* | 2.120403404 | 0.001355812 | 0.003298759 | 1 |
| Obesity | t_cell | *LSAMP* | 2.71764968 | 6.16E-05 | 0.000306965 | 0.387083261 |
| Obesity | t_cell | *AHNAK2* | 6.172247264 | 4.42E-06 | 0.000100609 | 0.027768035 |
| Obesity | t_cell | *SP1* | 4.566526159 | 1.50E-06 | 5.38E-05 | 0.009410452 |
| Obesity | t_cell | *PCGF3* | 5.53844299 | 1.27E-06 | 4.88E-05 | 0.008000831 |
| Obesity | t_cell | *PBX1* | 2.410852805 | 1.05E-05 | 0.000145864 | 0.065948896 |
| Obesity | t_cell | *MYBL1* | -0.554777249 | 2.10E-05 | 0.000167886 | 0.131958444 |
| Obesity | t_cell | *ADARB2* | 3.795997368 | 0.000374668 | 0.001165531 | 1 |
| Obesity | t_cell | *SRL* | 2.219205147 | 5.00E-05 | 0.000267593 | 0.314321997 |
| Obesity | t_cell | *KCNIP4* | 1.149734491 | 0.026714721 | 0.043395423 | 1 |
| Obesity | t_cell | *SLC52A2* | 9.309073334 | 0.000109349 | 0.00046451 | 0.687474472 |
| Obesity | t_cell | *TRIM69* | -1.474775378 | 0.021742347 | 0.036239167 | 1 |
| Obesity | t_cell | *POMK* | 5.015435951 | 0.02251652 | 0.03736114 | 1 |
| Obesity | t_cell | *LINC00839* | 2.803792082 | 0.020798318 | 0.034878374 | 1 |
| Obesity | t_cell | *RTN4RL1* | 2.737192666 | 1.66E-05 | 0.000155785 | 0.10437626 |
| Obesity | t_cell | *RASA3* | -1.324323687 | 0.009460237 | 0.017719599 | 1 |
| Obesity | t_cell | *ZNF284* | 8.20009969 | 1.36E-05 | 0.000147409 | 0.08546478 |
| Obesity | t_cell | *C2orf76* | 4.679182457 | 2.43E-05 | 0.000179683 | 0.152909813 |

| | | | | | | |
|---|---|---|---|---|---|---|
| Obesity | t_cell | *WWOX* | 2.563883796 | 0.002143694 | 0.00488843 | 1 |
| Obesity | t_cell | *CYP4Z1* | 3.361480439 | 3.36E-05 | 0.000213956 | 0.211175016 |
| Obesity | t_cell | *POLR1D* | 3.854363853 | 0.020994099 | 0.035169172 | 1 |
| Obesity | t_cell | *BPIFB4* | 143.8946542 | 0.000232931 | 0.000819494 | 1 |
| Obesity | t_cell | *CYP4F12* | 2.032771533 | 0.000265622 | 0.000906604 | 1 |
| Obesity | t_cell | *KLHL32* | -1.619826118 | 0.002096807 | 0.00479194 | 1 |
| Obesity | t_cell | *NBPF12* | 4.642652095 | 0.002356364 | 0.005292769 | 1 |
| Obesity | t_cell | *TOR3A* | 7.805257752 | 6.59E-05 | 0.000321134 | 0.414583528 |
| Obesity | t_cell | *CYP4X1* | 3.027740993 | 7.34E-05 | 0.000347258 | 0.461158918 |
| Obesity | t_cell | *CCDC30* | 4.319512817 | 0.000146339 | 0.000577911 | 0.920034783 |
| Obesity | t_cell | *GLDN* | 3.527717503 | 3.84E-05 | 0.00023194 | 0.241458126 |
| Obesity | t_cell | *FCAR* | 2.538208271 | 0.018941677 | 0.032194193 | 1 |
| Obesity | t_cell | *ZNF501* | 3.32523248 | 7.17E-05 | 0.000341164 | 0.450678126 |
| Obesity | t_cell | *ZNF197* | 3.306725723 | 0.000828212 | 0.002192409 | 1 |
| Obesity | t_cell | *BTN3A2* | -1.610235933 | 8.70E-13 | 3.42E-10 | 5.47E-09 |
| Obesity | t_cell | *PCLO* | 1.63898456 | 3.37E-05 | 0.000214224 | 0.212130635 |
| Obesity | t_cell | *RGS7BP* | 2.427182439 | 0.023051339 | 0.03806757 | 1 |
| Obesity | t_cell | *TMEM222* | -2.287302863 | 0.029059421 | 0.046713521 | 1 |
| Obesity | t_cell | *SEPT10* | 2.908757754 | 0.000788135 | 0.002103143 | 1 |
| Obesity | t_cell | *SMYD4* | 2.992874573 | 0.007233685 | 0.013993286 | 1 |
| Obesity | t_cell | *MIR22HG* | 2.555134751 | 1.28E-05 | 0.000147332 | 0.080750675 |
| Obesity | t_cell | *PDE2A* | 1.920440157 | 0.02693425 | 0.043676975 | 1 |
| Obesity | t_cell | *CYP27C1* | 1.204176319 | 0.00035733 | 0.001127779 | 1 |
| Obesity | t_cell | *BCR* | 3.238827564 | 0.008533247 | 0.016173808 | 1 |
| Obesity | t_cell | *HYAL3* | -2.593330987 | 8.50E-06 | 0.000134614 | 0.053441685 |
| Obesity | t_cell | *ZSCAN30* | 3.910865861 | 1.42E-05 | 0.000148367 | 0.089565368 |
| Obesity | t_cell | *LILRB4* | 2.398473151 | 0.000495297 | 0.001447668 | 1 |
| Obesity | t_cell | *TRABD2A* | -0.622464522 | 0.011878852 | 0.021596977 | 1 |
| Obesity | t_cell | *TMEM17* | 2.607656263 | 0.000353676 | 0.001122566 | 1 |
| Obesity | t_cell | *TNFRSF18* | -0.622199339 | 0.000142046 | 0.000566748 | 0.893044976 |
| Obesity | t_cell | *RTN4RL2* | -5.352582566 | 0.000587224 | 0.00165927 | 1 |
| Obesity | t_cell | *TMEM232* | 2.172454973 | 8.37E-05 | 0.00038231 | 0.526440298 |
| Obesity | t_cell | *EFCAB6* | 3.221600583 | 2.16E-05 | 0.000169226 | 0.135551419 |
| Obesity | t_cell | *KANK3* | 6.360456665 | 0.000134327 | 0.000540661 | 0.84451274 |
| Obesity | t_cell | *EMID1* | 5.71380051 | 5.93E-06 | 0.000112637 | 0.037283011 |
| Obesity | t_cell | *CYP4A11* | 2.90715052 | 0.000168446 | 0.000641052 | 1 |
| Obesity | t_cell | *C3orf70* | 2.0375371 | 0.009505033 | 0.017795755 | 1 |
| Obesity | t_cell | *ENTPD5* | 5.344103627 | 1.87E-05 | 0.000160619 | 0.11741222 |
| Obesity | t_cell | *HEATR4* | 1.647061524 | 0.002431847 | 0.00543127 | 1 |
| Obesity | t_cell | *MRPS21* | 5.784108618 | 2.79E-06 | 7.74E-05 | 0.017565733 |
| Obesity | t_cell | *ANGPTL5* | 4.140504069 | 1.55E-05 | 0.00015141 | 0.097205499 |
| Obesity | t_cell | *TSPYL4* | 3.221464498 | 0.003498696 | 0.007408657 | 1 |
| Obesity | t_cell | *MT1X* | 2.707026053 | 0.00011787 | 0.000489143 | 0.741051402 |
| Obesity | t_cell | *WDR86* | 2.620681839 | 0.009619125 | 0.017971898 | 1 |
| Obesity | t_cell | *EPOR* | 5.423867189 | 7.60E-07 | 3.39E-05 | 0.004780631 |
| Obesity | t_cell | *FPR3* | 2.090613085 | 0.000260647 | 0.000894771 | 1 |
| Obesity | t_cell | *GJA4* | 4.859219108 | 7.78E-05 | 0.000363997 | 0.489211339 |
| Obesity | t_cell | *IFT140* | 5.509787072 | 0.00086491 | 0.002268541 | 1 |
| Obesity | t_cell | *TLR5* | 3.034595236 | 2.33E-05 | 0.000175508 | 0.146592037 |
| Obesity | t_cell | *PLEKHN1* | -1.06409685 | 0.00104757 | 0.002662115 | 1 |

| | | | | | | |
|---|---|---|---|---|---|---|
| Obesity | t_cell | *ZNF385C* | 6.233350137 | 4.53E-06 | 0.000100727 | 0.028495765 |
| Obesity | t_cell | *TRPV2* | -2.102865086 | 0.001877999 | 0.004352001 | 1 |
| Obesity | t_cell | *C2orf88* | 2.743114933 | 3.92E-05 | 0.000234516 | 0.246242143 |
| Obesity | t_cell | *KBTBD12* | 2.074711826 | 0.000215702 | 0.000775369 | 1 |
| Obesity | t_cell | *SEMA4D* | -0.760656545 | 0.000276078 | 0.000933674 | 1 |
| Obesity | t_cell | *FAM69C* | 2.886376458 | 0.002427768 | 0.005426013 | 1 |
| Obesity | t_cell | *MCRS1* | 4.896246973 | 0.000222052 | 0.000791854 | 1 |
| Obesity | t_cell | *TMEM220* | 3.759366809 | 3.25E-05 | 0.000210045 | 0.20439245 |
| Obesity | t_cell | *CCDC157* | -3.91743905 | 0.018192735 | 0.031140138 | 1 |
| Obesity | t_cell | *GFRAL* | 2.807890043 | 0.000105238 | 0.000450703 | 0.661631851 |
| Obesity | t_cell | *LDLRAD2* | 2.055693619 | 0.004230397 | 0.008717308 | 1 |
| Obesity | t_cell | *C2orf66* | 6.851314335 | 0.005927161 | 0.011725632 | 1 |
| Obesity | t_cell | *CPSF4L* | 4.587918033 | 0.001715769 | 0.004020513 | 1 |
| Obesity | t_cell | *ZSCAN23* | 1.254136986 | 8.13E-06 | 0.000132809 | 0.051108029 |
| Obesity | t_cell | *OR7D2* | 4.106952318 | 0.015536287 | 0.027192827 | 1 |
| Obesity | t_cell | *MORN2* | 1.782579878 | 0.005412194 | 0.010846816 | 1 |
| Obesity | t_cell | *C6orf132* | 2.207302763 | 0.000201601 | 0.000737326 | 1 |
| Obesity | t_cell | *MAPK12* | 4.778891245 | 0.003013644 | 0.006533372 | 1 |
| Obesity | t_cell | *FAM166A* | 10.23329304 | 3.36E-05 | 0.000214008 | 0.211440234 |
| Obesity | t_cell | *ZNF626* | 4.091033069 | 1.17E-05 | 0.000145864 | 0.073392599 |
| Obesity | t_cell | *SMTNL2* | 10.04196204 | 5.02E-05 | 0.000268036 | 0.315786135 |
| Obesity | t_cell | *ZC3H6* | 2.905658919 | 0.030168563 | 0.048298894 | 1 |
| Obesity | t_cell | *NUTM2B* | 25.69586613 | 0.018126004 | 0.031051277 | 1 |
| Obesity | t_cell | *NCR3LG1* | 18.87484914 | 0.000254803 | 0.000881598 | 1 |
| Obesity | t_cell | *AGAP4* | 6.145397271 | 0.000143816 | 0.000571536 | 0.904170699 |
| Obesity | t_cell | *HYKK* | 4.568591365 | 1.14E-07 | 8.28E-06 | 0.00071983 |
| Obesity | t_cell | *RUFY4* | 5.744111632 | 3.42E-05 | 0.000215756 | 0.215324234 |
| Obesity | t_cell | *ZNF669* | 5.502935807 | 2.04E-05 | 0.000166062 | 0.12853162 |
| Obesity | t_cell | *CENPP* | 3.1747016 | 1.92E-09 | 2.69E-07 | 1.21E-05 |
| Obesity | t_cell | *PLSCR1* | 5.035703225 | 1.34E-05 | 0.000147409 | 0.084255893 |
| Obesity | t_cell | *ZP3* | 2.658145482 | 1.82E-05 | 0.000158673 | 0.114720784 |
| Obesity | t_cell | *PDCD1* | -1.940862784 | 0.000317555 | 0.001036588 | 1 |
| Obesity | t_cell | *CERKL* | -1.202397843 | 0.00039453 | 0.001211743 | 1 |
| Obesity | t_cell | *COL25A1* | 3.30504014 | 0.000379587 | 0.001176552 | 1 |
| Obesity | t_cell | *NBR1* | 4.013634218 | 0.015093429 | 0.026506254 | 1 |
| Obesity | t_cell | *NDOR1* | 7.683127383 | 2.98E-05 | 0.000200768 | 0.187294271 |
| Obesity | t_cell | *FAM72B* | 5.078680099 | 4.32E-05 | 0.000247883 | 0.271432112 |
| Obesity | t_cell | *GOLGA8M* | 3.238455947 | 0.000292481 | 0.00097706 | 1 |
| Obesity | t_cell | *S100A16* | 5.806160141 | 5.91E-06 | 0.000112637 | 0.03718626 |
| Obesity | t_cell | *RHCE* | 2.289644412 | 0.001183929 | 0.002945533 | 1 |
| Obesity | t_cell | *SLC4A5* | 5.338800348 | 0.000501615 | 0.001462219 | 1 |
| Obesity | t_cell | *UROS* | 4.569680695 | 0.000253836 | 0.000879749 | 1 |
| Obesity | t_cell | *FAM221A* | 3.610109349 | 7.62E-05 | 0.000357593 | 0.478817471 |
| Obesity | t_cell | *NOXA1* | 7.910094608 | 8.87E-05 | 0.000396247 | 0.557519889 |
| Obesity | t_cell | *SKOR1* | 6.641285434 | 0.000118599 | 0.000491681 | 0.745634653 |
| Obesity | t_cell | *TMEM201* | 5.093633444 | 0.018111992 | 0.03103573 | 1 |
| Obesity | t_cell | *ZDHHC11* | 4.457482334 | 3.03E-05 | 0.000202181 | 0.190564296 |
| Obesity | t_cell | *LINC00910* | 5.438707831 | 2.72E-05 | 0.000191821 | 0.171230407 |
| Obesity | t_cell | *BEND4* | 16.76312218 | 4.87E-06 | 0.000103821 | 0.030627278 |
| Obesity | t_cell | *FBF1* | 1.913555608 | 1.74E-05 | 0.000157135 | 0.109208945 |

| | | | | | | |
|---|---|---|---|---|---|---|
| Obesity | t_cell | *LRRK2* | 2.515055271 | 0.003452593 | 0.007335738 | 1 |
| Obesity | t_cell | *FAM196A* | 3.833194497 | 3.81E-06 | 9.36E-05 | 0.023969665 |
| Obesity | t_cell | *UTS2B* | 1.210696481 | 0.007855948 | 0.015030537 | 1 |
| Obesity | t_cell | *C9orf152* | 3.90972227 | 3.31E-05 | 0.000212266 | 0.208021078 |
| Obesity | t_cell | *NOC2L* | -1.464600194 | 0.01211067 | 0.021935979 | 1 |
| Obesity | t_cell | *NELFB* | 6.784368179 | 0.000816542 | 0.00216882 | 1 |
| Obesity | t_cell | *SLC15A5* | 1.532412088 | 0.000912392 | 0.002370333 | 1 |
| Obesity | t_cell | *LIPI* | 2.102550657 | 0.002617032 | 0.005795892 | 1 |
| Obesity | t_cell | *ADAT2* | 4.382551053 | 0.000763302 | 0.002046071 | 1 |
| Obesity | t_cell | *ANKDD1B* | 1.065294751 | 0.021482361 | 0.03586288 | 1 |
| Obesity | t_cell | *ALKBH2* | 4.165497522 | 4.46E-05 | 0.000252269 | 0.280271245 |
| Obesity | t_cell | *PLAC9* | 3.501082914 | 2.62E-06 | 7.41E-05 | 0.016454734 |
| Obesity | t_cell | *CLDN4* | 1.84658947 | 0.018451708 | 0.03152334 | 1 |
| Obesity | t_cell | *ZNF573* | 4.899040196 | 0.004035501 | 0.008378863 | 1 |
| Obesity | t_cell | *FAM47E* | 2.589713612 | 6.17E-08 | 5.24E-06 | 0.000387685 |
| Obesity | t_cell | *S100A13* | 4.856703878 | 2.27E-06 | 6.76E-05 | 0.014256084 |
| Obesity | t_cell | *ZNF600* | -2.051926957 | 0.010975181 | 0.020193433 | 1 |
| Obesity | t_cell | *PNRC2* | 5.11910754 | 0.007050054 | 0.013705532 | 1 |
| Obesity | t_cell | *SLC35E2B* | 2.223900517 | 0.001355406 | 0.003298759 | 1 |
| Obesity | t_cell | *BLOC1S2* | -1.447057371 | 0.011742093 | 0.021385441 | 1 |
| Obesity | t_cell | *AKR1C3* | 2.853678324 | 1.75E-05 | 0.000157583 | 0.1099151 |
| Obesity | t_cell | *WDSUB1* | 2.82473666 | 0.004644654 | 0.00944403 | 1 |
| Obesity | t_cell | *COLCA1* | 2.895331322 | 8.34E-05 | 0.000381479 | 0.524534292 |
| Obesity | t_cell | *SEMA4A* | 4.585032064 | 4.74E-06 | 0.000101665 | 0.029787903 |
| Obesity | t_cell | *HRCT1* | 3.137606126 | 8.90E-06 | 0.000136451 | 0.055945049 |
| Obesity | t_cell | *SIRPB2* | 6.139309925 | 3.21E-05 | 0.000209022 | 0.20154047 |
| Obesity | t_cell | *SRGAP3* | 1.798266187 | 0.005670266 | 0.011277749 | 1 |
| Obesity | t_cell | *ZNF107* | -1.597776126 | 0.00739109 | 0.014267051 | 1 |
| Obesity | t_cell | *ZNF471* | 2.48028096 | 0.001632055 | 0.003846116 | 1 |
| Obesity | t_cell | *GRM7* | 1.052182412 | 0.008965282 | 0.016880721 | 1 |
| Obesity | t_cell | *GIMAP5* | -1.374386646 | 0.000187799 | 0.000701123 | 1 |
| Obesity | t_cell | *STK31* | -3.500551356 | 0.005273222 | 0.010591931 | 1 |
| Obesity | t_cell | *ZKSCAN7* | 6.543244139 | 3.26E-05 | 0.000210045 | 0.204734795 |
| Obesity | t_cell | *ZNF565* | 4.20630019 | 0.000187436 | 0.000700185 | 1 |
| Obesity | t_cell | *NTNG2* | -3.139713033 | 0.0022972 | 0.005178378 | 1 |
| Obesity | t_cell | *WDR5* | 6.158152494 | 0.000346149 | 0.001103062 | 1 |
| Obesity | t_cell | *LONP1* | 3.774797624 | 0.000808617 | 0.002151406 | 1 |
| Obesity | t_cell | *ASB13* | 5.147276155 | 1.15E-05 | 0.000145864 | 0.072353455 |
| Obesity | t_cell | *SLC35F1* | 5.275298751 | 0.00370402 | 0.007785748 | 1 |
| Obesity | t_cell | *ZNF34* | 4.247287822 | 2.57E-05 | 0.000186437 | 0.161647156 |
| Obesity | t_cell | *INCA1* | 3.81360722 | 1.01E-06 | 4.14E-05 | 0.006378326 |
| Obesity | t_cell | *ZNF774* | 3.924503139 | 7.80E-05 | 0.000364544 | 0.490675853 |
| Obesity | t_cell | *EVL* | -1.179253267 | 0.003458838 | 0.007340319 | 1 |
| Obesity | t_cell | *ZNF775* | 2.718436903 | 0.000958083 | 0.002464594 | 1 |
| Obesity | t_cell | *ZNF605* | 3.798100047 | 1.09E-05 | 0.000145864 | 0.068467663 |
| Obesity | t_cell | *RFX8* | 3.914898373 | 0.000134874 | 0.000542168 | 0.847950726 |
| Obesity | t_cell | *MYL6B* | 5.030099105 | 2.98E-05 | 0.000200768 | 0.187579982 |
| Obesity | t_cell | *C20orf96* | 6.213175689 | 1.86E-05 | 0.000160456 | 0.117123533 |
| Obesity | t_cell | *SULT1A1* | -2.914350131 | 1.22E-05 | 0.000145864 | 0.076989015 |
| Obesity | t_cell | *C1orf186* | 4.831088307 | 5.30E-05 | 0.000277986 | 0.33330542 |

| | | | | | | |
|---|---|---|---|---|---|---|
| Obesity | t_cell | *SULF2* | 3.459679781 | 0.000400291 | 0.001226427 | 1 |
| Obesity | t_cell | *XRCC2* | 10.11304901 | 0.001743127 | 0.004077022 | 1 |
| Obesity | t_cell | *MYO6* | 2.277800965 | 0.008562426 | 0.01621445 | 1 |
| Obesity | t_cell | *HDAC2* | 5.458946974 | 8.15E-05 | 0.000375666 | 0.512476325 |
| Obesity | t_cell | *ADH1B* | 1.902302976 | 8.65E-06 | 0.000134958 | 0.054412114 |
| Obesity | t_cell | *HRH1* | 3.47948503 | 1.09E-05 | 0.000145864 | 0.068672003 |
| Obesity | t_cell | *TECPR2* | 5.288028363 | 0.000115611 | 0.000484242 | 0.726847453 |
| Obesity | t_cell | *ZFP62* | 7.830218598 | 0.000122974 | 0.00050598 | 0.773138142 |
| Obesity | t_cell | *ERI2* | 2.854947225 | 8.63E-06 | 0.000134958 | 0.054267037 |
| Obesity | t_cell | *TRPV1* | 3.507045077 | 2.75E-06 | 7.66E-05 | 0.017317158 |
| Obesity | t_cell | *ZNF431* | 4.513029271 | 0.00057702 | 0.00163411 | 1 |
| Obesity | t_cell | *NF1* | 3.615681612 | 0.000519622 | 0.001501315 | 1 |
| Obesity | t_cell | *DAPK1* | 2.186980862 | 0.005972337 | 0.011807574 | 1 |
| Obesity | t_cell | *GM2A* | 2.710910409 | 0.009997042 | 0.018592869 | 1 |
| Obesity | t_cell | *TLE1* | 2.302887115 | 0.000643746 | 0.001782366 | 1 |
| Obesity | t_cell | *STRN3* | 3.34289399 | 0.005134471 | 0.010349605 | 1 |
| Obesity | t_cell | *ZNF239* | 2.172817015 | 0.007160222 | 0.013872516 | 1 |
| Obesity | t_cell | *MVB12B* | 3.214643605 | 0.031388733 | 0.049985046 | 1 |
| Obesity | t_cell | *C6orf106* | 4.308855393 | 1.23E-10 | 2.96E-08 | 7.76E-07 |
| Obesity | t_cell | *NHLRC2* | 4.16206154 | 0.000975925 | 0.002501281 | 1 |
| Obesity | t_cell | *ZFP28* | 4.139749764 | 4.61E-06 | 0.000100727 | 0.028987133 |
| Obesity | t_cell | *KIAA1211L* | 1.749462882 | 0.000615113 | 0.001721824 | 1 |
| Obesity | t_cell | *LAMB3* | 3.071170397 | 6.02E-05 | 0.000302106 | 0.378538361 |
| Obesity | t_cell | *HCAR1* | 3.948903755 | 1.19E-08 | 1.36E-06 | 7.45E-05 |
| Obesity | t_cell | *SRGAP1* | 3.6100184 | 3.85E-06 | 9.38E-05 | 0.024203553 |
| Obesity | t_cell | *FAM3C* | 6.109462283 | 4.33E-05 | 0.000248058 | 0.272119848 |
| Obesity | t_cell | *AP2A1* | 3.314938751 | 0.00512158 | 0.010330245 | 1 |
| Obesity | t_cell | *PCDHB16* | 1.338265321 | 0.020084241 | 0.033861524 | 1 |
| Obesity | t_cell | *ZNF585A* | 2.678204329 | 3.84E-06 | 9.38E-05 | 0.02412205 |
| Obesity | t_cell | *FUT11* | 3.648782449 | 0.021192465 | 0.035435379 | 1 |
| Obesity | t_cell | *ANXA4* | 4.234174843 | 0.000229868 | 0.000811442 | 1 |
| Obesity | t_cell | *ZNF138* | 5.120020181 | 1.82E-05 | 0.000158673 | 0.114667634 |
| Obesity | t_cell | *ZNF429* | 3.142220206 | 0.00389512 | 0.008141163 | 1 |
| Obesity | t_cell | *ZNF470* | 3.658800133 | 0.000536942 | 0.001542145 | 1 |
| Obesity | t_cell | *GMFB* | 5.243069444 | 6.21E-05 | 0.000308403 | 0.390200528 |
| Obesity | t_cell | *GAL3ST4* | -2.76556702 | 0.030764479 | 0.049144918 | 1 |
| Obesity | t_cell | *SLC25A29* | 5.479519254 | 9.90E-08 | 7.59E-06 | 0.000622263 |
| Obesity | t_cell | *ZNF682* | 5.711343391 | 0.000401392 | 0.001228605 | 1 |
| Obesity | t_cell | *ADAM32* | 2.73563141 | 0.000656507 | 0.001812675 | 1 |
| Obesity | t_cell | *ABCB8* | 2.340031247 | 0.000823863 | 0.002183654 | 1 |
| Obesity | t_cell | *SULT1A2* | 3.430394957 | 3.72E-32 | 2.34E-28 | 2.34E-28 |
| Obesity | t_cell | *NEK5* | 2.369266499 | 0.0131661 | 0.023522385 | 1 |
| Obesity | t_cell | *PIWIL2* | 2.704270921 | 0.000115876 | 0.000484382 | 0.728510316 |
| Obesity | t_cell | *FAM110D* | 6.03637446 | 8.20E-06 | 0.000132932 | 0.051562342 |
| Obesity | t_cell | *KANK2* | 4.326862868 | 2.37E-06 | 6.92E-05 | 0.014887096 |
| Obesity | t_cell | *CCL4L2* | -2.466737126 | 3.16E-08 | 3.05E-06 | 0.000198401 |
| Obesity | t_cell | *GTF2E2* | 5.092255011 | 0.000309543 | 0.001017151 | 1 |
| Obesity | t_cell | *RAMP2-AS1* | 2.03083309 | 0.000158399 | 0.000610952 | 0.995851841 |
| Obesity | t_cell | *DDI2* | 4.34815186 | 0.000542727 | 0.001557337 | 1 |
| Obesity | t_cell | *MRPL21* | -1.377534759 | 0.002431459 | 0.00543127 | 1 |

| | | | | | | |
|---|---|---|---|---|---|---|
| Obesity | t_cell | *ZNF517* | 5.604941846 | 1.31E-05 | 0.000147409 | 0.082084487 |
| Obesity | t_cell | *C5AR1* | 3.716538434 | 0.000522749 | 0.00150827 | 1 |
| Obesity | t_cell | *DCHS2* | 0.923991156 | 0.014694265 | 0.025884798 | 1 |
| Obesity | t_cell | *VEPH1* | 3.053813771 | 0.001293653 | 0.00317331 | 1 |
| Obesity | t_cell | *SHPK* | 1.696404424 | 0.000259523 | 0.000892083 | 1 |
| Obesity | t_cell | *STMN3* | -1.159162968 | 0.000112407 | 0.000473978 | 0.706701232 |
| Obesity | t_cell | *GYPE* | 1.939931619 | 0.004382387 | 0.008992188 | 1 |
| Obesity | t_cell | *SLC2A10* | 3.931803633 | 0.000120157 | 0.00049634 | 0.75542993 |
| Obesity | t_cell | *ZNF665* | 1.757563945 | 0.014934874 | 0.026264491 | 1 |
| Obesity | t_cell | *RPF2* | 3.627528198 | 0.00776865 | 0.014899788 | 1 |
| Obesity | t_cell | *TTC30A* | 3.379770906 | 0.000365766 | 0.001147419 | 1 |
| Obesity | t_cell | *RAB40C* | 6.393443937 | 0.000343763 | 0.001098749 | 1 |
| Obesity | t_cell | *ZNF624* | 6.859057162 | 0.001534942 | 0.00366509 | 1 |
| Obesity | t_cell | *HOXA4* | 3.475343915 | 3.67E-05 | 0.000226047 | 0.231020503 |
| Obesity | t_cell | *BCO2* | 4.219998787 | 8.36E-05 | 0.000381762 | 0.525304131 |
| Obesity | t_cell | *C5orf42* | 2.884070092 | 0.000294508 | 0.000982267 | 1 |
| Obesity | t_cell | *ZNF841* | 4.579523297 | 0.000251429 | 0.000872369 | 1 |
| Obesity | t_cell | *MFAP5* | 3.981308989 | 6.18E-05 | 0.000307471 | 0.388643847 |
| Obesity | t_cell | *CDC42SE1* | -1.775190048 | 0.003955112 | 0.008236433 | 1 |
| Obesity | t_cell | *PDCD1LG2* | 2.163415012 | 0.000750872 | 0.002019996 | 1 |
| Obesity | t_cell | *NMB* | 6.961367254 | 6.24E-05 | 0.000309323 | 0.392221897 |
| Obesity | t_cell | *FAM114A1* | 3.117678791 | 6.09E-06 | 0.000113565 | 0.038271569 |
| Obesity | t_cell | *PHF2* | 5.988292727 | 0.000163534 | 0.000624628 | 1 |
| Obesity | t_cell | *RPS26* | -1.5320638 | 5.28E-05 | 0.000277521 | 0.331918262 |
| Obesity | t_cell | *PSAP* | 4.166105074 | 0.000281795 | 0.000948419 | 1 |
| Obesity | t_cell | *MAP1LC3C* | 2.053285958 | 0.00023709 | 0.000832263 | 1 |
| Obesity | t_cell | *TAF13* | 2.951647914 | 0.00123271 | 0.003054593 | 1 |
| Obesity | t_cell | *ZNF780A* | -1.527278702 | 0.000611414 | 0.001714523 | 1 |
| Obesity | t_cell | *AC122129.1* | 3.881791642 | 4.33E-05 | 0.000248058 | 0.271965093 |
| Obesity | t_cell | *ZNF181* | 5.024026391 | 0.000105415 | 0.000451153 | 0.662743535 |
| Obesity | t_cell | *SGTB* | -2.114021894 | 0.001055308 | 0.002677449 | 1 |
| Obesity | t_cell | *ADH5* | 2.935604676 | 0.001480137 | 0.003550408 | 1 |
| Obesity | t_cell | *HRNR* | 0.616604352 | 0.011511168 | 0.021025773 | 1 |
| Obesity | t_cell | *ZNF677* | 7.482478077 | 2.42E-05 | 0.00017926 | 0.152246588 |
| Obesity | t_cell | *ZNF311* | 2.175479576 | 0.000786862 | 0.002100637 | 1 |
| Obesity | t_cell | *ZNF71* | 2.840636121 | 0.004217434 | 0.008699149 | 1 |
| Obesity | t_cell | *AGAP9* | 5.95502031 | 0.000117751 | 0.000488971 | 0.740301787 |
| Obesity | t_cell | *ZNF273* | 4.251616587 | 0.000445044 | 0.001331744 | 1 |
| Obesity | t_cell | *ZNF84* | 4.408334817 | 0.00082516 | 0.002186169 | 1 |
| Obesity | t_cell | *MAK16* | 4.730118278 | 0.016357773 | 0.028348882 | 1 |
| Obesity | t_cell | *ZNF667* | 2.428207967 | 0.000484147 | 0.00142205 | 1 |
| Obesity | t_cell | *SULT1C4* | 3.311547197 | 6.59E-05 | 0.000320973 | 0.414055691 |
| Obesity | t_cell | *SFI1* | -1.519596889 | 0.001632168 | 0.003846116 | 1 |
| Obesity | t_cell | *ZNF248* | 1.803676515 | 0.015446649 | 0.027073621 | 1 |
| Obesity | t_cell | *HIBCH* | 2.095010814 | 0.023052021 | 0.03806757 | 1 |
| Obesity | t_cell | *SOWAHC* | 4.030463119 | 2.95E-05 | 0.000200631 | 0.185630339 |
| Obesity | t_cell | *DDRGK1* | 7.010478773 | 0.000355326 | 0.001124841 | 1 |
| Obesity | t_cell | *ZNF334* | 1.674814856 | 0.003454794 | 0.007337935 | 1 |
| Obesity | t_cell | *HSD17B11* | 4.070669348 | 0.016062239 | 0.027957723 | 1 |
| Obesity | t_cell | *SULT1C2* | 1.175433784 | 0.023014783 | 0.038037313 | 1 |

| | | | | | | |
|---|---|---|---|---|---|---|
| Obesity | t_cell | *RPS6KL1* | 1.50692219 | 0.021149468 | 0.035382305 | 1 |
| Obesity | t_cell | *QRICH1* | 2.168912534 | 0.005077441 | 0.010264267 | 1 |
| Obesity | t_cell | *DDX42* | 5.084573757 | 2.13E-05 | 0.000169127 | 0.133779559 |
| Obesity | t_cell | *SLC29A3* | 5.091165399 | 0.000790878 | 0.002109567 | 1 |
| Obesity | t_cell | *ANTXRL* | -3.010074235 | 0.000123221 | 0.000506335 | 0.774692765 |
| Obesity | t_cell | *STYX* | -2.041145307 | 0.018990036 | 0.032267664 | 1 |
| Obesity | t_cell | *ZKSCAN8* | 7.519545329 | 3.83E-07 | 2.10E-05 | 0.002410776 |
| Obesity | t_cell | *ZNF813* | 6.914584601 | 0.00196536 | 0.00452628 | 1 |
| Obesity | t_cell | *GFPT1* | 5.217409172 | 7.85E-07 | 3.47E-05 | 0.004933437 |
| Obesity | t_cell | *UVRAG* | 2.006590254 | 0.001277517 | 0.003148471 | 1 |
| Obesity | t_cell | *MT1F* | -2.695743001 | 0.000104188 | 0.000447426 | 0.655032322 |
| Obesity | t_cell | *ZNF568* | 4.578359477 | 0.000265156 | 0.000905996 | 1 |
| Obesity | t_cell | *ZNF587* | 5.035393191 | 8.54E-05 | 0.000386921 | 0.536659786 |
| Obesity | t_cell | *FLVCR1-AS1* | 1.029896399 | 0.012804932 | 0.02298818 | 1 |
| Obesity | t_cell | *SH3BGRL2* | 2.983414054 | 2.84E-05 | 0.000196238 | 0.17838078 |
| Obesity | t_cell | *ZNF808* | 3.885944418 | 0.008998448 | 0.016938096 | 1 |
| Obesity | t_cell | *TMA16* | 5.70023727 | 1.08E-05 | 0.000145864 | 0.068138928 |
| Obesity | t_cell | *CNGA1* | 1.862885587 | 0.003484426 | 0.007385901 | 1 |
| Obesity | t_cell | *MAFK* | 6.769277559 | 2.53E-05 | 0.000184756 | 0.158947961 |
| Obesity | t_cell | *HIST1H4E* | 4.103456301 | 0.00702479 | 0.013664869 | 1 |
| Obesity | t_cell | *C20orf203* | 3.140151975 | 0.013376101 | 0.023829851 | 1 |
| Obesity | t_cell | *ZNF627* | 4.706257922 | 8.13E-05 | 0.000375475 | 0.511397343 |
| Obesity | t_cell | *ARC* | -19.61834897 | 0.000942692 | 0.002430596 | 1 |
| Obesity | t_cell | *MMP17* | 3.901583864 | 1.60E-05 | 0.000153978 | 0.100701612 |
| Obesity | t_cell | *FAM19A2* | 3.353710183 | 0.016181487 | 0.028118576 | 1 |
| Obesity | t_cell | *TUSC1* | 7.902426899 | 1.22E-05 | 0.000145864 | 0.076642144 |
| Obesity | t_cell | *PAPSS2* | 3.441053627 | 4.63E-05 | 0.00025778 | 0.290775492 |
| Obesity | t_cell | *FAN1* | 5.764594685 | 3.26E-05 | 0.000210045 | 0.205004116 |
| Obesity | t_cell | *ANKRD13B* | 2.256072616 | 6.39E-07 | 3.02E-05 | 0.004015003 |
| Obesity | t_cell | *APCDD1L* | 4.997773099 | 1.09E-05 | 0.000145864 | 0.068755239 |
| Obesity | t_cell | *RCSD1* | -1.066582084 | 0.001360335 | 0.003307202 | 1 |
| Obesity | t_cell | *RASSF9* | 3.872648104 | 1.18E-05 | 0.000145864 | 0.074024266 |
| Obesity | t_cell | *FAM169A* | -0.957462555 | 0.00097384 | 0.002496954 | 1 |
| Obesity | t_cell | *TMEM184B* | 4.244595827 | 4.35E-05 | 0.000249037 | 0.273442341 |
| Obesity | t_cell | *SFT2D1* | 5.588761117 | 1.88E-05 | 0.000160637 | 0.118212662 |
| Obesity | t_cell | *ARHGAP11A* | 4.435244978 | 0.014022768 | 0.024855128 | 1 |
| Obesity | t_cell | *HMGN2* | 2.744471263 | 0.00791923 | 0.015128592 | 1 |
| Obesity | t_cell | *CD3E* | -0.402338442 | 0.000131882 | 0.000532526 | 0.829143032 |
| Obesity | t_cell | *TSEN15* | 5.172521597 | 0.00414128 | 0.008558918 | 1 |
| Obesity | t_cell | *ITPRIPL1* | -2.619976002 | 0.00071165 | 0.001934347 | 1 |
| Obesity | t_cell | *PRMT6* | -12.25485731 | 0.011551352 | 0.021093043 | 1 |
| Obesity | t_cell | *SHISA4* | 5.98528231 | 0.002269956 | 0.005127996 | 1 |
| Obesity | t_cell | *CAPZA2* | 4.253884716 | 0.00178555 | 0.004154608 | 1 |
| Obesity | t_cell | *C1orf174* | 4.292268538 | 0.000344727 | 0.001100153 | 1 |
| Obesity | t_cell | *RASGEF1A* | -1.319614239 | 0.000186162 | 0.000696253 | 1 |
| Obesity | t_cell | *KIAA0753* | 4.252588378 | 0.001168792 | 0.002915951 | 1 |
| Obesity | t_cell | *TBKBP1* | 2.317656996 | 0.007988512 | 0.015251678 | 1 |
| Obesity | t_cell | *ZFP2* | 4.733653716 | 0.000314068 | 0.001026805 | 1 |
| Obesity | t_cell | *MFAP3L* | 5.108635966 | 7.94E-06 | 0.000132019 | 0.049897993 |
| Obesity | t_cell | *NAGA* | 5.764858504 | 9.10E-05 | 0.000404229 | 0.571984332 |

| | | | | | | |
|---|---|---|---|---|---|---|
| Obesity | t_cell | *SMG5* | 3.436248857 | 0.020302475 | 0.034147046 | 1 |
| Obesity | t_cell | *TGM2* | 1.206840668 | 0.001288179 | 0.003164824 | 1 |
| Obesity | t_cell | *AL590822.1* | 1.340290171 | 0.003546735 | 0.007492716 | 1 |
| Obesity | t_cell | *AP001007.1* | 8.907423325 | 4.00E-05 | 0.000237458 | 0.251705873 |
| Obesity | t_cell | *AC105020.1* | 5.1193051 | 1.98E-05 | 0.000163419 | 0.124774205 |
| Obesity | t_cell | *EFCAB2* | 3.87390927 | 7.18E-06 | 0.000126396 | 0.04511591 |
| Obesity | t_cell | *CHML* | 2.366030264 | 0.006103213 | 0.012043597 | 1 |
| Obesity | t_cell | *TATDN3* | 1.807666759 | 0.004556101 | 0.009294032 | 1 |
| Obesity | t_cell | *ECT2L* | 3.895200016 | 2.19E-05 | 0.000171115 | 0.137954422 |
| Obesity | t_cell | *FAM229B* | 1.658597447 | 0.000664993 | 0.001832131 | 1 |
| Obesity | t_cell | *DDO* | 5.656588247 | 5.26E-06 | 0.000107122 | 0.033046997 |
| Obesity | t_cell | *HIST2H2BF* | 29.90157521 | 0.000264731 | 0.000905528 | 1 |
| Obesity | t_cell | *RBM20* | 4.675262004 | 0.000312269 | 0.001022519 | 1 |
| Obesity | t_cell | *C6orf163* | 4.555135679 | 0.000172412 | 0.000653379 | 1 |
| Obesity | t_cell | *PCMTD2* | 4.388491863 | 0.004223612 | 0.008709036 | 1 |
| Obesity | t_cell | *FOXO6* | 1.733744707 | 0.030635676 | 0.04899682 | 1 |
| Obesity | t_cell | *SYS1* | 8.954914931 | 0.001473682 | 0.003537624 | 1 |
| Obesity | t_cell | *TRAF3IP1* | 4.040931049 | 0.000275545 | 0.000933381 | 1 |
| Obesity | t_cell | *GIGYF2* | 2.892587126 | 0.023912015 | 0.039313504 | 1 |
| Obesity | t_cell | *C2orf72* | -3.905466382 | 0.003920482 | 0.008186009 | 1 |
| Obesity | t_cell | *RUFY2* | 3.688026991 | 0.001292027 | 0.00317056 | 1 |
| Obesity | t_cell | *C10orf128* | 2.507956381 | 0.009247723 | 0.017365722 | 1 |
| Obesity | t_cell | *SYT15* | 5.313256791 | 3.49E-06 | 8.87E-05 | 0.021913563 |
| Obesity | t_cell | *ZDBF2* | 4.791207943 | 0.000134669 | 0.000541693 | 0.846666049 |
| Obesity | t_cell | *RING1* | 2.631754572 | 0.001422678 | 0.003432224 | 1 |
| Obesity | t_cell | *HSD17B8* | -2.22273476 | 0.003100754 | 0.006689925 | 1 |
| Obesity | t_cell | *HLA-DMA* | 4.088145987 | 0.012450894 | 0.022454805 | 1 |
| Obesity | t_cell | *COL15A1* | 2.204495 | 0.000301229 | 0.000997277 | 1 |
| Obesity | t_cell | *NOTCH4* | -0.540489786 | 7.70E-08 | 6.13E-06 | 0.000484397 |
| Obesity | t_cell | *PRRT1* | 1.745319487 | 0.020537116 | 0.034504771 | 1 |
| Obesity | t_cell | *MRPL38* | -2.882360558 | 0.004992262 | 0.010103914 | 1 |
| Obesity | t_cell | *SKIV2L* | -1.508749616 | 4.54E-06 | 0.000100727 | 0.028568606 |
| Obesity | t_cell | *NELFE* | 4.390226458 | 2.95E-06 | 8.04E-05 | 0.018574825 |
| Obesity | t_cell | *CARD16* | -1.410625043 | 0.000553158 | 0.001582212 | 1 |
| Obesity | t_cell | *BAG6* | 3.077178377 | 0.001340846 | 0.003267403 | 1 |
| Obesity | t_cell | *NFKBIL1* | 4.008013366 | 3.34E-05 | 0.000213631 | 0.210120612 |
| Obesity | t_cell | *ZNF814* | 4.313339922 | 0.001848479 | 0.004288336 | 1 |
| Obesity | t_cell | *POU5F1* | 1.501220639 | 1.31E-08 | 1.47E-06 | 8.22E-05 |
| Obesity | t_cell | *CCHCR1* | 2.671644465 | 3.76E-09 | 4.83E-07 | 2.37E-05 |
| Obesity | t_cell | *ABCF1* | 5.365773565 | 2.24E-06 | 6.72E-05 | 0.014053815 |
| Obesity | t_cell | *LILRB3* | 2.636826983 | 0.000155286 | 0.000603171 | 0.9762839 |
| Obesity | t_cell | *LINC01123* | -32.15083999 | 8.64E-05 | 0.000390568 | 0.543032111 |
| Obesity | t_cell | *HLA-E* | 3.252066133 | 1.24E-05 | 0.000146996 | 0.078142229 |
| Obesity | t_cell | *ZNF468* | 7.315250446 | 0.00207874 | 0.004759301 | 1 |
| Obesity | t_cell | *HLA-G* | -2.320207003 | 0.000438527 | 0.001316629 | 1 |
| Obesity | t_cell | *TBC1D8* | 3.575798791 | 0.00020495 | 0.000744808 | 1 |
| Obesity | t_cell | *HLA-F* | -1.140462607 | 0.016528745 | 0.028598707 | 1 |
| Obesity | t_cell | *ZFP57* | 2.100170263 | 1.18E-09 | 1.83E-07 | 7.42E-06 |
| Obesity | t_cell | *FAM153C* | -1.329351423 | 0.000194801 | 0.00072127 | 1 |
| Obesity | t_cell | *MALRD1* | 1.187697353 | 0.004411019 | 0.009039138 | 1 |

| | | | | | | |
|---|---|---|---|---|---|---|
| Obesity | t_cell | *MROH6* | 1.909923304 | 0.00544749 | 0.010899975 | 1 |
| Obesity | t_cell | *ATXN2* | 4.445358642 | 2.10E-05 | 0.000167886 | 0.131828874 |
| Obesity | t_cell | *ZNF155* | 3.079252223 | 0.000332944 | 0.001075653 | 1 |
| Obesity | t_cell | *ZNF425* | 1.612099869 | 0.01415898 | 0.025026007 | 1 |
| Obesity | t_cell | *PCDHA5* | 14.88065942 | 0.000222736 | 0.000793844 | 1 |
| Obesity | t_cell | *PCDHA4* | 5.500101638 | 0.004161078 | 0.00859701 | 1 |
| Obesity | t_cell | *SLFN12L* | -0.485480535 | 9.00E-05 | 0.000400571 | 0.566006692 |
| Obesity | t_cell | *CLLU1OS* | 3.05984079 | 0.006641112 | 0.012994919 | 1 |
| Obesity | t_cell | *FAM71F2* | 3.290928721 | 1.54E-05 | 0.000150967 | 0.09661875 |
| Obesity | t_cell | *AC016586.1* | 17.09966422 | 0.025001658 | 0.040869844 | 1 |
| Obesity | t_cell | *LINC00654* | 4.377113246 | 0.00126209 | 0.003117783 | 1 |
| Obesity | t_cell | *PRR13* | -4.101225926 | 0.00122994 | 0.003049145 | 1 |
| Obesity | t_cell | *MT1A* | 5.851112582 | 2.69E-05 | 0.000190422 | 0.169095125 |
| Obesity | t_cell | *MT1M* | 5.127485967 | 1.47E-05 | 0.000149118 | 0.092459274 |
| Obesity | t_cell | *CFI* | 2.332805176 | 0.000184405 | 0.000691739 | 1 |
| Obesity | t_cell | *SAMD9* | -1.858177785 | 0.003331816 | 0.007129724 | 1 |
| Obesity | t_cell | *EXOC3L4* | 3.620748437 | 0.000269035 | 0.000917077 | 1 |
| Obesity | t_cell | *ATP6AP1L* | 2.204137738 | 0.001855173 | 0.004302278 | 1 |
| Obesity | t_cell | *NAP1L4* | -2.018821378 | 0.025458358 | 0.041583969 | 1 |
| Obesity | t_cell | *SMN2* | -1.205643833 | 0.02984208 | 0.047812731 | 1 |
| Obesity | t_cell | *DENND6B* | 9.192235231 | 0.000399429 | 0.001224382 | 1 |
| Obesity | t_cell | *EIF3CL* | 13.44434227 | 4.10E-08 | 3.73E-06 | 0.000257656 |
| Obesity | t_cell | *LIN52* | 2.80172944 | 0.006076413 | 0.012005785 | 1 |
| Obesity | t_cell | *C17orf107* | 3.194848158 | 1.77E-05 | 0.000157653 | 0.111139146 |
| Obesity | t_cell | *C1QTNF9B* | 20.18652839 | 4.11E-05 | 0.00024182 | 0.258263998 |
| Obesity | t_cell | *NYNRIN* | 4.453513955 | 7.36E-06 | 0.000128167 | 0.04630256 |
| Obesity | t_cell | *ZDHHC11B* | 3.952389918 | 0.001416339 | 0.003422185 | 1 |
| Obesity | t_cell | *GOLGA8O* | 6.085340114 | 0.002836983 | 0.006203864 | 1 |
| Obesity | t_cell | *HCG27* | 1.038312771 | 0.027649414 | 0.044709841 | 1 |
| Obesity | t_cell | *HLA-A* | -1.409484101 | 0.00555234 | 0.011088806 | 1 |
| Obesity | t_cell | *LNP1* | 2.409911671 | 8.90E-05 | 0.000396981 | 0.559743078 |
| Obesity | t_cell | *METTL6* | 4.542035932 | 1.43E-06 | 5.24E-05 | 0.008977945 |
| Obesity | t_cell | *SLC48A1* | 2.972383246 | 2.35E-05 | 0.000176449 | 0.147687635 |
| Obesity | t_cell | *TAS2R14* | 1.438954078 | 0.026771907 | 0.043458555 | 1 |
| Obesity | t_cell | *ZRSR1* | 10.34338112 | 5.60E-05 | 0.000287814 | 0.352284201 |
| Obesity | t_cell | *AC084018.1* | -1.110288593 | 0.028119706 | 0.045342035 | 1 |
| Obesity | t_cell | *C17orf51* | 3.071503662 | 0.000154197 | 0.00060027 | 0.969435377 |
| Obesity | t_cell | *RNF208* | 7.511443071 | 2.79E-05 | 0.000194081 | 0.175643108 |
| Obesity | t_cell | *MAP10* | 2.895897723 | 0.000217997 | 0.000781828 | 1 |
| Obesity | t_cell | *ZNF580* | 3.584615383 | 0.000835148 | 0.00220705 | 1 |
| Obesity | t_cell | *NUP62* | 5.43132083 | 0.010793573 | 0.019900056 | 1 |
| Obesity | t_cell | *FGFR1OP* | 4.050197919 | 0.005677066 | 0.011285674 | 1 |
| Obesity | t_cell | *ZNF254* | 4.075043913 | 0.003634667 | 0.00766046 | 1 |
| Obesity | t_cell | *ASIC3* | 4.067650033 | 2.14E-07 | 1.29E-05 | 0.001345626 |
| Obesity | t_cell | *GIMAP1* | -1.872248545 | 0.001401756 | 0.003390859 | 1 |
| Obesity | t_cell | *CCDC183* | 7.926814865 | 3.19E-05 | 0.000209022 | 0.20060014 |
| Obesity | t_cell | *SUPT4H1* | 5.363523647 | 0.002437989 | 0.005443052 | 1 |
| Obesity | t_cell | *MXD3* | 2.102387555 | 0.02042809 | 0.034349132 | 1 |
| Obesity | t_cell | *GSTM2* | 1.642362212 | 0.007031876 | 0.013674422 | 1 |
| Obesity | t_cell | *LCAT* | 1.630100182 | 0.011566503 | 0.021102323 | 1 |

| | | | | | | |
|---|---|---|---|---|---|---|
| Obesity | t_cell | *ERV3-1* | 1.103746258 | 6.21E-05 | 0.000308403 | 0.390437882 |
| Obesity | t_cell | *SRA1* | 2.104977977 | 0.010778567 | 0.01987822 | 1 |
| Obesity | t_cell | *TMEM110* | 11.59064253 | 1.17E-09 | 1.83E-07 | 7.37E-06 |
| Obesity | t_cell | *HEXA* | 6.40538545 | 3.79E-05 | 0.000230345 | 0.238066125 |
| Obesity | t_cell | *LEPROT* | 3.194684204 | 0.018940383 | 0.032194193 | 1 |
| Obesity | t_cell | *LBH* | -1.132448497 | 0.000331052 | 0.001070159 | 1 |
| Obesity | t_cell | *NCKIPSD* | 3.541817011 | 0.000329508 | 0.001066744 | 1 |
| Obesity | t_cell | *SLC35F6* | 8.677702334 | 1.19E-06 | 4.67E-05 | 0.007461263 |
| Obesity | t_cell | *ATP6V1G2* | -1.309607262 | 1.15E-05 | 0.000145864 | 0.072367754 |
| Obesity | t_cell | *KLRK1* | -1.10426096 | 0.000142336 | 0.000567447 | 0.894863657 |
| Obesity | t_cell | *EMP2* | 3.606288594 | 2.01E-05 | 0.00016409 | 0.126677369 |
| Obesity | t_cell | *PPM1N* | -3.375754778 | 0.011118089 | 0.020390731 | 1 |
| Obesity | t_cell | *ITGA1* | 3.525802382 | 0.000111465 | 0.000470947 | 0.700780297 |
| Obesity | t_cell | *ZNF726* | 5.44728692 | 3.60E-05 | 0.000223213 | 0.226114446 |
| Obesity | t_cell | *TAX1BP3* | 8.785915651 | 1.06E-05 | 0.000145864 | 0.066760134 |
| Obesity | t_cell | *AP1G2* | -2.773093426 | 0.004993297 | 0.010103914 | 1 |
| Obesity | t_cell | *ZNF90* | 4.17312206 | 0.013302822 | 0.023726196 | 1 |
| Obesity | t_cell | *TM6SF2* | 24.11929185 | 2.60E-05 | 0.000187226 | 0.163260855 |
| Obesity | t_cell | *REPIN1* | 2.439079777 | 0.002990592 | 0.006490112 | 1 |
| Obesity | t_cell | *SMIM7* | 3.585853864 | 0.017532893 | 0.030142 | 1 |
| Obesity | t_cell | *TSPAN4* | 4.482760903 | 1.10E-05 | 0.000145864 | 0.069093518 |
| Obesity | t_cell | *AC079776.2* | 5.147921686 | 5.02E-05 | 0.000267973 | 0.315404119 |
| Obesity | t_cell | *WEE2* | 3.516305923 | 9.98E-05 | 0.000431423 | 0.627289096 |
| Obesity | t_cell | *LYRM4* | 4.888035467 | 5.79E-06 | 0.000111662 | 0.036401743 |
| Obesity | t_cell | *PRCD* | 3.195105634 | 0.000608652 | 0.001709063 | 1 |
| Obesity | t_cell | *C17orf67* | -2.168692092 | 0.010626804 | 0.019621357 | 1 |
| Obesity | t_cell | *COLCA2* | 2.07648258 | 0.000883823 | 0.002309475 | 1 |
| Obesity | t_cell | *NEURL1B* | 7.089807326 | 4.79E-05 | 0.000262855 | 0.301065818 |
| Obesity | t_cell | *PPME1* | 2.579400964 | 0.001061126 | 0.002687871 | 1 |
| Obesity | t_cell | *NUTM2D* | 4.28320212 | 0.002046144 | 0.004688087 | 1 |
| Obesity | t_cell | *CPEB1* | 4.449559916 | 0.000493329 | 0.001443257 | 1 |
| Obesity | t_cell | *C10orf105* | 3.292950907 | 0.0004999 | 0.001459087 | 1 |
| Obesity | t_cell | *ARHGEF33* | 1.360766705 | 0.008871097 | 0.016733449 | 1 |
| Obesity | t_cell | *AC005082.1* | 9.747377978 | 3.75E-05 | 0.000228647 | 0.235505997 |
| Obesity | t_cell | *ACSM4* | 0.90896638 | 0.019861202 | 0.033539451 | 1 |
| Obesity | t_cell | *COL28A1* | 4.942696318 | 8.79E-07 | 3.69E-05 | 0.005523988 |
| Obesity | t_cell | *GRID2IP* | 3.774894375 | 0.019286552 | 0.032674361 | 1 |
| Obesity | t_cell | *UBE2QL1* | 3.538930915 | 3.15E-05 | 0.000208616 | 0.198185007 |
| Obesity | t_cell | *GOLGA8B* | -1.85102047 | 0.007082524 | 0.013755893 | 1 |
| Obesity | t_cell | *KCNU1* | 2.481527232 | 4.25E-05 | 0.000246282 | 0.267246575 |
| Obesity | t_cell | *MYL5* | 10.36731956 | 1.43E-05 | 0.000148411 | 0.089936851 |
| Obesity | t_cell | *NPEPL1* | 5.958066522 | 9.58E-05 | 0.000418954 | 0.602456456 |
| Obesity | t_cell | *LINC01139* | 2.556503136 | 0.000431596 | 0.001300166 | 1 |
| Obesity | t_cell | *TSTD1* | 8.533919085 | 1.26E-05 | 0.000146996 | 0.079457301 |
| Obesity | t_cell | *LINC01138* | -1.738257715 | 0.004337632 | 0.008906169 | 1 |
| Obesity | t_cell | *CYB5RL* | 4.236690542 | 1.35E-05 | 0.000147409 | 0.084573611 |
| Obesity | t_cell | *PAM16* | 2.57564815 | 0.000365926 | 0.001147419 | 1 |
| Obesity | t_cell | *AP000350.4* | 0.85298085 | 0.001700686 | 0.003988144 | 1 |
| Obesity | t_cell | *RNASEK* | 6.320047067 | 0.000993178 | 0.002537223 | 1 |
| Obesity | t_cell | *NBPF1* | 3.265079314 | 8.26E-05 | 0.000378832 | 0.51937837 |

| | | | | | | |
|---|---|---|---|---|---|---|
| Obesity | t_cell | *RNU6ATAC35I* | 13.27792469 | 2.97E-05 | 0.000200768 | 0.186541262 |
| Obesity | t_cell | *C6orf226* | 2.968624306 | 0.004440616 | 0.0090909 | 1 |
| Obesity | t_cell | *PPP3R1* | 3.412850335 | 0.006639164 | 0.012994919 | 1 |
| Obesity | t_cell | *C19orf73* | -13.69814135 | 0.002818065 | 0.006175384 | 1 |
| Obesity | t_cell | *OR2A25* | 53.59484661 | 0.000467139 | 0.001385331 | 1 |
| Obesity | t_cell | *C1orf229* | 2.542544676 | 0.00155373 | 0.003704324 | 1 |
| Obesity | t_cell | *APOL6* | 4.403324844 | 2.46E-05 | 0.000181284 | 0.154901346 |
| Obesity | t_cell | *BTBD19* | 5.077232705 | 0.000334332 | 0.001079026 | 1 |
| Obesity | t_cell | *FAM185A* | 5.555052593 | 3.94E-06 | 9.50E-05 | 0.02480061 |
| Obesity | t_cell | *AC062017.1* | 2.75260799 | 9.95E-05 | 0.000430567 | 0.625614434 |
| Obesity | t_cell | *ADRA2B* | 6.647237214 | 4.77E-05 | 0.000262855 | 0.299967777 |
| Obesity | t_cell | *EXOSC6* | -8.245349274 | 3.84E-10 | 7.32E-08 | 2.42E-06 |
| Obesity | t_cell | *AC147651.3* | -2.349226346 | 9.25E-05 | 0.000408268 | 0.581373549 |
| Obesity | t_cell | *HLA-DPB1* | 2.238729728 | 0.001658649 | 0.003901206 | 1 |
| Obesity | t_cell | *OSER1-AS1* | 5.676477951 | 4.88E-07 | 2.47E-05 | 0.003066508 |
| Obesity | t_cell | *LINC01133* | 3.768216024 | 1.08E-05 | 0.000145864 | 0.068098435 |
| Obesity | t_cell | *LINC01117* | 2.323498321 | 1.42E-05 | 0.000148367 | 0.089340617 |
| Obesity | t_cell | *RASAL2-AS1* | 7.029417084 | 0.009899237 | 0.018445911 | 1 |
| Obesity | t_cell | *AC069213.1* | -6.810577145 | 1.59E-05 | 0.000153197 | 0.100025626 |
| Obesity | t_cell | *LINC00240* | 2.799357834 | 3.63E-11 | 1.14E-08 | 2.28E-07 |
| Obesity | t_cell | *LINC00863* | 4.538264616 | 3.90E-05 | 0.000233522 | 0.244964986 |
| Obesity | t_cell | *LINC00184* | 3.078765389 | 0.001125964 | 0.002828181 | 1 |
| Obesity | t_cell | *TMEM233* | 1.939592663 | 2.01E-05 | 0.000163915 | 0.126214634 |
| Obesity | t_cell | *LINC00237* | -4.572406109 | 0.00022596 | 0.000801072 | 1 |
| Obesity | t_cell | *PLEKHM1* | 6.499758067 | 2.98E-06 | 8.09E-05 | 0.01876235 |
| Obesity | t_cell | *AP000350.6* | 1.871623414 | 0.001709113 | 0.004006411 | 1 |
| Obesity | t_cell | *SFTA1P* | 6.240145189 | 0.001101136 | 0.002774685 | 1 |
| Obesity | t_cell | *SLC26A6* | 3.445564097 | 0.000131853 | 0.000532526 | 0.828962388 |
| Obesity | t_cell | *MIAT* | -0.605253425 | 0.000264113 | 0.000903908 | 1 |
| Obesity | t_cell | *LINC00626* | 33.85296844 | 4.48E-05 | 0.00025278 | 0.281652456 |
| Obesity | t_cell | *ERCC6* | 4.914677537 | 0.002482402 | 0.005532386 | 1 |
| Obesity | t_cell | *LINC00115* | -2.076921405 | 0.024665349 | 0.040404129 | 1 |
| Obesity | t_cell | *ZNF503-AS1* | 3.059857432 | 1.97E-05 | 0.000162709 | 0.123658547 |
| Obesity | t_cell | *AC010536.1* | -3.01122104 | 0.030761966 | 0.049144918 | 1 |
| Obesity | t_cell | *AC064836.3* | 6.140777813 | 1.96E-05 | 0.000162591 | 0.123244176 |
| Obesity | t_cell | *HSBP1L1* | 6.895853312 | 0.000273437 | 0.000928739 | 1 |
| Obesity | t_cell | *AC093901.1* | 3.475259772 | 7.01E-05 | 0.00033537 | 0.440676317 |
| Obesity | t_cell | *C14orf132* | 2.930591318 | 1.14E-05 | 0.000145864 | 0.071643946 |
| Obesity | t_cell | *ZNF717* | 0.671481459 | 0.00160257 | 0.003797722 | 1 |
| Obesity | t_cell | *AC108463.2* | 2.875470867 | 0.031368588 | 0.049965623 | 1 |
| Obesity | t_cell | *HCG20* | -17.27459702 | 3.76E-06 | 9.34E-05 | 0.023644929 |
| Obesity | t_cell | *POLR2J2* | -0.832072494 | 0.00393062 | 0.008201728 | 1 |
| Obesity | t_cell | *C19orf24* | 2.933864668 | 0.029103765 | 0.046772846 | 1 |
| Obesity | t_cell | *LINC01013* | 3.454562525 | 3.20E-05 | 0.000209022 | 0.201155829 |
| Obesity | t_cell | *AC073133.2* | 11.42688141 | 0.02064265 | 0.0346358 | 1 |
| Obesity | t_cell | *PROB1* | 7.408328555 | 0.00213129 | 0.004863674 | 1 |
| Obesity | t_cell | *ARL17B* | 2.795921854 | 0.000105083 | 0.000450346 | 0.660657455 |
| Obesity | t_cell | *LINC00954* | -1.817997381 | 0.000370449 | 0.001155839 | 1 |
| Obesity | t_cell | *AC067945.3* | -0.778446972 | 0.000196275 | 0.000723743 | 1 |
| Obesity | t_cell | *RPL41* | -4.131895721 | 0.001568138 | 0.003733011 | 1 |

| | | | | | | |
|---|---|---|---|---|---|---|
| Obesity | t_cell | *PATL2* | -1.542873339 | 0.001691355 | 0.003967741 | 1 |
| Obesity | t_cell | *ZNF492* | 1.871178575 | 0.003698255 | 0.007776231 | 1 |
| Obesity | t_cell | *U91324.1* | 8.045698614 | 2.21E-05 | 0.000171828 | 0.139238011 |
| Obesity | t_cell | *CISD3* | 4.949498091 | 1.33E-05 | 0.000147409 | 0.083418405 |
| Obesity | t_cell | *PSMG3-AS1* | 5.142200564 | 7.99E-06 | 0.000132019 | 0.050248921 |
| Obesity | t_cell | *PPP5D1* | 3.914799322 | 1.31E-05 | 0.000147409 | 0.082094415 |
| Obesity | t_cell | *KLHL7-AS1* | 5.357660615 | 3.20E-05 | 0.000209022 | 0.200966339 |
| Obesity | t_cell | *AC092171.4* | 4.1578694 | 0.000870121 | 0.002278406 | 1 |
| Obesity | t_cell | *HLA-DPA1* | 3.792082847 | 0.000334606 | 0.001079359 | 1 |
| Obesity | t_cell | *DIRC3* | 2.009899237 | 6.78E-05 | 0.000327936 | 0.426317205 |
| Obesity | t_cell | *AC107021.1* | 1.430382089 | 0.000547189 | 0.001567993 | 1 |
| Obesity | t_cell | *LINC00865* | 2.792222408 | 0.001539877 | 0.003675478 | 1 |
| Obesity | t_cell | *AC009495.3* | -1.908515063 | 0.000320753 | 0.001043774 | 1 |
| Obesity | t_cell | *HLA-DQB2* | 0.546127293 | 0.015033487 | 0.026415744 | 1 |
| Obesity | t_cell | *GOLGA8N* | 1.992933526 | 0.001614072 | 0.003818287 | 1 |
| Obesity | t_cell | *LINC00665* | 1.891873277 | 0.002824626 | 0.00618545 | 1 |
| Obesity | t_cell | *TNF* | -0.951515328 | 1.14E-05 | 0.000145864 | 0.071475067 |
| Obesity | t_cell | *LYRM9* | 5.013190841 | 0.000366413 | 0.001147429 | 1 |
| Obesity | t_cell | *LINC00342* | 5.623012818 | 0.003068488 | 0.006624858 | 1 |
| Obesity | t_cell | *SNHG15* | 4.840093096 | 0.011244403 | 0.020598357 | 1 |
| Obesity | t_cell | *LINC00702* | 3.461087843 | 3.95E-05 | 0.000235816 | 0.248549969 |
| Obesity | t_cell | *LINC00472* | 2.36757587 | 0.000102072 | 0.000439842 | 0.641728921 |
| Obesity | t_cell | *AC123768.3* | 3.41362094 | 1.72E-05 | 0.000156669 | 0.10796735 |
| Obesity | t_cell | *C19orf81* | 5.822162738 | 0.00029541 | 0.000983708 | 1 |
| Obesity | t_cell | *ANKRD65* | 3.43430248 | 4.24E-05 | 0.000246282 | 0.266466262 |
| Obesity | t_cell | *ZSCAN31* | 2.626267988 | 2.03E-08 | 2.09E-06 | 0.000127442 |
| Obesity | t_cell | *LINC00957* | 5.143127683 | 2.57E-05 | 0.000186437 | 0.161478081 |
| Obesity | t_cell | *LINC00484* | -1.005400808 | 0.003232119 | 0.006947122 | 1 |
| Obesity | t_cell | *KIAA0040* | 5.69669123 | 0.000470005 | 0.001390824 | 1 |
| Obesity | t_cell | *LINC00941* | 7.711297052 | 2.89E-05 | 0.000198227 | 0.181774597 |
| Obesity | t_cell | *LINC00545* | -8.297853812 | 1.73E-05 | 0.000157091 | 0.108942457 |
| Obesity | t_cell | *B3GNT9* | 6.650632386 | 0.003506416 | 0.007422505 | 1 |
| Obesity | t_cell | *GS1-124K5.4* | 0.778113439 | 0.004302548 | 0.008848583 | 1 |
| Obesity | t_cell | *ZNF737* | 6.109022633 | 0.000481182 | 0.001414963 | 1 |
| Obesity | t_cell | *AC073257.2* | -10.15502215 | 0.001600541 | 0.003794344 | 1 |
| Obesity | t_cell | *IRGM* | -1.298544492 | 0.016667894 | 0.028820421 | 1 |
| Obesity | t_cell | *TXNDC5* | 4.190668544 | 0.000502562 | 0.001464136 | 1 |
| Obesity | t_cell | *ASB14* | 3.242841042 | 0.003652557 | 0.007690432 | 1 |
| Obesity | t_cell | *CDRT4* | 5.075582835 | 0.000912083 | 0.002370333 | 1 |
| Obesity | t_cell | *APOBEC3G* | -0.688563565 | 0.015057837 | 0.026451138 | 1 |
| Obesity | t_cell | *ADSL* | -2.05702582 | 0.005429892 | 0.010871889 | 1 |
| Obesity | t_cell | *LILRA4* | -10.57295825 | 0.000195894 | 0.000723185 | 1 |
| Obesity | t_cell | *COX19* | 5.639890654 | 0.001517524 | 0.003626254 | 1 |
| Obesity | t_cell | *AQP1* | 3.558077093 | 4.71E-05 | 0.000260943 | 0.296431702 |
| Obesity | t_cell | *C1QTNF9* | 4.764841019 | 0.006157123 | 0.012129694 | 1 |
| Obesity | t_cell | *ARHGEF25* | 3.511333685 | 6.64E-05 | 0.000322471 | 0.417599566 |
| Obesity | t_cell | *MIF* | 0.760066813 | 0.017767606 | 0.030503806 | 1 |
| Obesity | t_cell | *CD302* | 4.727620944 | 0.00130821 | 0.003204019 | 1 |
| Obesity | t_cell | *ATP5J2* | 5.088266808 | 1.58E-05 | 0.000152971 | 0.099278269 |
| Obesity | t_cell | *ARHGAP8* | 6.221139932 | 5.16E-05 | 0.000272935 | 0.324520052 |

| | | | | | | |
|---|---|---|---|---|---|---|
| Obesity | t_cell | *ATP5O* | 5.024686414 | 0.002426729 | 0.00542562 | 1 |
| Obesity | t_cell | *PLEKHO2* | 3.861904264 | 1.64E-05 | 0.000155091 | 0.102980611 |
| Obesity | t_cell | *PWP2* | 6.644506617 | 0.001082756 | 0.002736048 | 1 |
| Obesity | t_cell | *AKAP2* | 5.308059529 | 4.30E-05 | 0.000247251 | 0.270245647 |
| Obesity | t_cell | *LINC00969* | 1.625278243 | 0.02153021 | 0.035933217 | 1 |
| Obesity | t_cell | *ARHGDIG* | 2.329060353 | 0.000255808 | 0.000882691 | 1 |
| Obesity | t_cell | *ARFGAP3* | 5.104302454 | 0.000849157 | 0.002235172 | 1 |
| Obesity | t_cell | *C22orf39* | 9.677499454 | 4.42E-05 | 0.000251331 | 0.277971842 |
| Obesity | t_cell | *LINC00960* | -0.900538345 | 0.024763759 | 0.040537609 | 1 |
| Obesity | t_cell | *DECR2* | 2.669782215 | 3.23E-05 | 0.000209712 | 0.203210939 |
| Obesity | t_cell | *CCDC169* | 2.787502343 | 0.000261927 | 0.000897893 | 1 |
| Obesity | t_cell | *ZNF709* | -2.387746409 | 0.00018954 | 0.000706366 | 1 |
| Obesity | t_cell | *PSG4* | 2.148892039 | 0.009965912 | 0.018542672 | 1 |
| Obesity | t_cell | *STON1* | 2.282668468 | 6.80E-05 | 0.00032817 | 0.427605653 |
| Obesity | t_cell | *VSIG8* | 3.290848981 | 0.000214219 | 0.000770478 | 1 |
| Obesity | t_cell | *C7orf73* | 5.21477657 | 6.76E-05 | 0.000327358 | 0.4252377 |
| Obesity | t_cell | *FGF14-IT1* | 6.291430429 | 3.26E-05 | 0.000210045 | 0.204968495 |
| Obesity | t_cell | *TICAM2* | 6.310089868 | 0.000488265 | 0.00143044 | 1 |
| Obesity | t_cell | *PALM2* | 1.279319604 | 0.001278744 | 0.003149027 | 1 |
| Obesity | t_cell | *UPK3B* | 1.550169285 | 0.029800211 | 0.047757819 | 1 |
| Obesity | t_cell | *WDR92* | 4.936542055 | 0.000596412 | 0.001679194 | 1 |
| Obesity | t_cell | *PLA2G4B* | -1.565515714 | 0.030764902 | 0.049144918 | 1 |
| Obesity | t_cell | *LEFTY1* | 8.847342588 | 0.016109916 | 0.028032947 | 1 |
| Obesity | t_cell | *APOBEC3D* | -2.954657993 | 4.81E-05 | 0.000263445 | 0.302435199 |
| Obesity | t_cell | *RN7SL832P* | 3.514643118 | 1.34E-05 | 0.000147409 | 0.08445668 |
| Obesity | t_cell | *LINC01011* | 4.616657932 | 3.53E-05 | 0.000220179 | 0.221940659 |
| Obesity | t_cell | *TMEM199* | 6.938102453 | 7.43E-05 | 0.000350236 | 0.467214262 |
| Obesity | t_cell | *P2RY11* | -3.922264533 | 0.000274809 | 0.00093227 | 1 |
| Obesity | t_cell | *LILRA6* | 1.295114616 | 0.015356185 | 0.026937594 | 1 |
| Obesity | t_cell | *APOBEC3C* | -3.885913425 | 0.018232644 | 0.031186317 | 1 |
| Obesity | t_cell | *ASPRV1* | 6.883284517 | 0.002101664 | 0.004801294 | 1 |
| Obesity | t_cell | *UBE2V1* | 9.16134215 | 0.000833769 | 0.002204335 | 1 |
| Obesity | t_cell | *CRYBB2* | 4.050839619 | 0.003016085 | 0.006536409 | 1 |
| Obesity | t_cell | *LINC00847* | 9.47420688 | 3.03E-05 | 0.000202181 | 0.190453217 |
| Obesity | t_cell | *NEAT1* | 2.471576223 | 0.000373637 | 0.001164052 | 1 |
| Obesity | t_cell | *ZNF585B* | 2.445195422 | 0.000946457 | 0.002436681 | 1 |
| Obesity | t_cell | *LINC00920* | -4.409739037 | 0.001561992 | 0.003721198 | 1 |
| Obesity | t_cell | *LINC00926* | 6.720325536 | 0.000888002 | 0.002318468 | 1 |
| Obesity | t_cell | *BCKDHA* | 3.866525898 | 0.00064936 | 0.001795658 | 1 |
| Obesity | t_cell | *CDK11B* | 4.067559141 | 0.029493481 | 0.047314497 | 1 |
| Obesity | t_cell | *ABHD14A* | 6.937414856 | 0.000320024 | 0.001042045 | 1 |
| Obesity | t_cell | *SRP14-AS1* | 1.06574047 | 0.000915126 | 0.00237557 | 1 |
| Obesity | t_cell | *CCDC153* | 3.253991111 | 7.08E-05 | 0.000337886 | 0.445398818 |
| Obesity | t_cell | *C5orf17* | 5.336469539 | 1.50E-05 | 0.000149282 | 0.094495779 |
| Obesity | t_cell | *LINC01060* | 2.015740018 | 0.000556592 | 0.001590587 | 1 |
| Obesity | t_cell | *NAIP* | 4.668879931 | 0.00066512 | 0.001832131 | 1 |
| Obesity | t_cell | *PDCD6* | 5.022020634 | 0.001467859 | 0.00352634 | 1 |
| Obesity | t_cell | *LINC00603* | 3.374199571 | 0.011236002 | 0.020588966 | 1 |
| Obesity | t_cell | *YJEFN3* | 3.661485817 | 0.0097235 | 0.018150726 | 1 |
| Obesity | t_cell | *DNAH10OS* | 31.22240486 | 2.56E-10 | 5.20E-08 | 1.61E-06 |

| | | | | | | |
|---|---|---|---|---|---|---|
| Obesity | t_cell | *PCDHA10* | 3.760196766 | 1.72E-05 | 0.000156669 | 0.108404216 |
| Obesity | t_cell | *ZNF718* | 4.212234119 | 0.007430409 | 0.014325354 | 1 |
| Obesity | t_cell | *LINC00989* | 5.895675863 | 7.26E-05 | 0.000345029 | 0.456128252 |
| Obesity | t_cell | *LINC01021* | 5.908958346 | 2.59E-05 | 0.000186717 | 0.162538976 |
| Obesity | t_cell | *FAM218A* | 4.31640455 | 3.48E-05 | 0.000218022 | 0.218894255 |
| Obesity | t_cell | *ZNF345* | 4.488248745 | 0.000202423 | 0.000738329 | 1 |
| Obesity | t_cell | *SHANK3* | 1.323311618 | 0.007311528 | 0.014130826 | 1 |
| Obesity | t_cell | *ZNF550* | 4.255988723 | 0.016744717 | 0.028929386 | 1 |
| Obesity | t_cell | *CCDC71L* | 4.43799701 | 7.43E-05 | 0.000350236 | 0.467025329 |
| Obesity | t_cell | *TMEM200B* | 2.089649184 | 3.04E-05 | 0.000202181 | 0.190858538 |
| Obesity | t_cell | *PCDHGB6* | 2.968596942 | 0.000155895 | 0.000604261 | 0.980111455 |
| Obesity | t_cell | *TRNP1* | 41.86500048 | 0.000192941 | 0.000716491 | 1 |
| Obesity | t_cell | *PCDHGA5* | 2.506597302 | 0.000289982 | 0.000971809 | 1 |
| Obesity | t_cell | *PCDHGA6* | 3.222250132 | 1.06E-05 | 0.000145864 | 0.066564781 |
| Obesity | t_cell | *PCDHGA11* | 1.955766663 | 0.005341807 | 0.010715999 | 1 |
| Obesity | t_cell | *CLDN23* | 12.96644003 | 5.74E-07 | 2.82E-05 | 0.003608432 |
| Obesity | t_cell | *ZNF260* | 5.913757878 | 0.000428121 | 0.001290933 | 1 |
| Obesity | t_cell | *FPGT* | 5.143971977 | 1.11E-05 | 0.000145864 | 0.06991893 |
| Obesity | t_cell | *MPV17L2* | 6.362240616 | 3.03E-10 | 5.96E-08 | 1.91E-06 |
| Obesity | t_cell | *DPP3* | 1.898613521 | 0.006131258 | 0.012087557 | 1 |
| Obesity | t_cell | *EID3* | 1.931368342 | 0.000591235 | 0.001668129 | 1 |
| Obesity | t_cell | *SNHG9* | 2.100263262 | 9.19E-06 | 0.000138155 | 0.057765354 |
| Obesity | t_cell | *EBLN2* | 5.144780081 | 9.55E-06 | 0.000140299 | 0.060048141 |
| Obesity | t_cell | *POLR2M* | 5.723606972 | 1.11E-05 | 0.000145864 | 0.069799577 |
| Obesity | t_cell | *LINC00346* | 4.213181493 | 8.00E-05 | 0.000371012 | 0.502721623 |
| Obesity | t_cell | *CTSO* | 6.312377725 | 0.000377758 | 0.001173323 | 1 |
| Obesity | t_cell | *LINC00944* | -0.57134848 | 0.000218959 | 0.000783936 | 1 |
| Obesity | t_cell | *HMBS* | 4.874916835 | 3.05E-14 | 1.75E-11 | 1.92E-10 |
| Obesity | t_cell | *AC093668.2* | -3.891795944 | 0.000309138 | 0.001017032 | 1 |
| Obesity | t_cell | *ZNF253* | 5.11588961 | 0.001386762 | 0.003361053 | 1 |
| Obesity | t_cell | *KIAA1147* | 5.066680564 | 0.000272057 | 0.000925052 | 1 |
| Obesity | t_cell | *ZBED6* | 4.320509526 | 3.85E-05 | 0.00023194 | 0.241891655 |
| Obesity | t_cell | *FNTB* | 5.460102025 | 2.57E-05 | 0.000186437 | 0.161827013 |
| Obesity | t_cell | *OVCH1-AS1* | 6.180925209 | 3.23E-05 | 0.000209682 | 0.202762792 |
| Obesity | t_cell | *C17orf49* | 3.75592538 | 0.011680545 | 0.021285677 | 1 |
| Obesity | t_cell | *CEP95* | 4.185612189 | 0.001235459 | 0.003058005 | 1 |
| Obesity | t_cell | *LINC00639* | 2.154651119 | 3.21E-05 | 0.000209022 | 0.201675295 |
| Obesity | t_cell | *LINC00648* | 3.963681351 | 1.01E-05 | 0.000144166 | 0.063432933 |
| Obesity | t_cell | *LINC00924* | 5.188324768 | 0.000276702 | 0.000935283 | 1 |
| Obesity | t_cell | *UBE2Q2L* | 4.275299115 | 0.004073359 | 0.008449096 | 1 |
| Obesity | t_cell | *PWRN1* | 2.755549945 | 5.28E-05 | 0.000277521 | 0.332192499 |
| Obesity | t_cell | *HOXB7* | 5.932570929 | 2.22E-08 | 2.25E-06 | 0.000139423 |
| Obesity | t_cell | *FRRS1L* | 4.152198403 | 1.65E-05 | 0.000155268 | 0.103874279 |
| Obesity | t_cell | *AC009120.5* | 3.794330183 | 2.85E-05 | 0.000196531 | 0.17920052 |
| Obesity | t_cell | *LINC00565* | 10.86812841 | 0.007836434 | 0.015002332 | 1 |
| Obesity | t_cell | *TMEM178B* | 6.549213523 | 4.66E-05 | 0.000259121 | 0.293203782 |
| Obesity | t_cell | *LINC01003* | 2.638191937 | 0.006173323 | 0.012151434 | 1 |
| Obesity | t_cell | *HPR* | 8.385286166 | 1.73E-06 | 5.95E-05 | 0.010884059 |
| Obesity | t_cell | *GOLGA8H* | 2.255540779 | 1.86E-06 | 6.24E-05 | 0.011681535 |
| Obesity | t_cell | *LINC00662* | 4.871565262 | 0.000421048 | 0.001275109 | 1 |

| | | | | | | |
|---|---|---|---|---|---|---|
| Obesity | t_cell | *LINC00667* | 1.406493682 | 0.021645596 | 0.036106623 | 1 |
| Obesity | t_cell | *LINC00672* | 2.405699896 | 0.017860967 | 0.03065572 | 1 |
| Obesity | t_cell | *LINC00909* | 2.945191273 | 0.009572042 | 0.017894566 | 1 |
| Obesity | t_cell | *DYNLL2* | -3.468460351 | 0.001402793 | 0.003392062 | 1 |
| Obesity | t_cell | *LINC00526* | 3.462024211 | 0.001810253 | 0.004207415 | 1 |
| Obesity | t_cell | *MYO15B* | 4.780226524 | 5.00E-05 | 0.000267593 | 0.314663881 |
| Obesity | t_cell | *ILF3-AS1* | 2.634888719 | 0.013360735 | 0.023809223 | 1 |
| Obesity | t_cell | *AC139100.2* | 6.735488947 | 5.40E-05 | 0.000281114 | 0.339304872 |
| Obesity | t_cell | *LINC01140* | 2.967734268 | 1.94E-06 | 6.34E-05 | 0.012225049 |
| Obesity | t_cell | *LINC00906* | 13.01309127 | 4.47E-05 | 0.000252695 | 0.280999585 |
| Obesity | t_cell | *ZNF582-AS1* | 5.293730667 | 0.000569052 | 0.001615184 | 1 |
| Obesity | t_cell | *AC137932.1* | 4.554721488 | 0.006059748 | 0.011976623 | 1 |
| Obesity | t_cell | *AL132989.1* | 4.998913936 | 3.50E-05 | 0.000218427 | 0.219738019 |
| Obesity | t_cell | *LINC00664* | 3.996691011 | 0.007393581 | 0.014267479 | 1 |
| Obesity | t_cell | *FBXO17* | 2.677101953 | 2.75E-05 | 0.000193012 | 0.173050236 |
| Obesity | t_cell | *HCG17* | 2.737919933 | 0.030902276 | 0.049306962 | 1 |
| Obesity | t_cell | *YTHDF3-AS1* | 4.719655593 | 5.56E-06 | 0.000109386 | 0.034963241 |
| Obesity | t_cell | *LINC01050* | 13.7690606 | 5.82E-05 | 0.000295444 | 0.366180326 |
| Obesity | t_cell | *MILR1* | 1.224021571 | 0.001565067 | 0.003727111 | 1 |
| Obesity | t_cell | *GTF2H5* | 4.022671543 | 3.83E-08 | 3.54E-06 | 0.000240671 |
| Obesity | t_cell | *FGF14-AS2* | 3.661202114 | 1.25E-05 | 0.000146996 | 0.078740343 |
| Obesity | t_cell | *CASC15* | 3.608286303 | 4.71E-05 | 0.000260943 | 0.296238188 |
| Obesity | t_cell | *LINC01023* | 3.959158695 | 4.24E-05 | 0.000246282 | 0.266398957 |
| Obesity | t_cell | *AP000347.2* | 1.341492492 | 0.016755205 | 0.028939553 | 1 |
| Obesity | t_cell | *POLR2J4* | 7.416283741 | 1.36E-05 | 0.000147409 | 0.085644416 |
| Obesity | t_cell | *AC144652.1* | 1.348744622 | 0.0051011 | 0.010294617 | 1 |
| Obesity | t_cell | *PAXIP1-AS1* | 3.087390999 | 0.004432566 | 0.009080333 | 1 |
| Stroke | ASPC | *C17orf75* | -0.489812137 | 7.91E-06 | 0.02487282 | 0.04974564 |
| Stroke | ASPC | *GSTO1* | -0.678977323 | 6.39E-08 | 0.000401944 | 0.000401944 |
| Stroke | SMC | *C17orf75* | -3.199429581 | 6.69E-07 | 0.002103898 | 0.004207796 |
| Stroke | SMC | *GSTO1* | -3.05393546 | 2.48E-07 | 0.00155801 | 0.00155801 |
| Stroke | SMC | *ICA1L* | -1.497682004 | 9.09E-06 | 0.014294659 | 0.057178635 |
| Stroke | SMC | *CLDN7* | -1.125629792 | 4.47E-06 | 0.009367074 | 0.028101222 |
| Heart_Failure | adipocyte | *SEMA3F* | -0.205540845 | 3.47E-06 | 0.001556654 | 0.021793161 |
| Heart_Failure | adipocyte | *RBM6* | -0.163269595 | 7.04E-07 | 0.000491742 | 0.00442568 |
| Heart_Failure | adipocyte | *FAM214B* | 0.980422872 | 0.000162428 | 0.034039441 | 1 |
| Heart_Failure | adipocyte | *COL16A1* | -0.406016022 | 5.62E-05 | 0.016831169 | 0.353454555 |
| Heart_Failure | adipocyte | *SLC17A9* | 0.78851771 | 0.000259232 | 0.046565509 | 1 |
| Heart_Failure | adipocyte | *TIMM44* | 0.691435622 | 9.85E-05 | 0.026557809 | 0.619515853 |
| Heart_Failure | adipocyte | *HEMK1* | 0.423575859 | 0.000186044 | 0.037730913 | 1 |
| Heart_Failure | adipocyte | *CHMP1A* | 0.593606719 | 0.00010983 | 0.026557809 | 0.690503043 |
| Heart_Failure | adipocyte | *PDLIM4* | 0.613878382 | 2.27E-06 | 0.001187042 | 0.014244498 |
| Heart_Failure | adipocyte | *REEP2* | -0.202662705 | 6.03E-05 | 0.017226927 | 0.378992405 |
| Heart_Failure | adipocyte | *POM121C* | -0.579542911 | 0.000236652 | 0.04375983 | 1 |
| Heart_Failure | adipocyte | *NMT1* | 0.224446527 | 0.000269765 | 0.047111483 | 1 |
| Heart_Failure | adipocyte | *CISD2* | 0.818869135 | 0.000106125 | 0.026557809 | 0.66720722 |
| Heart_Failure | adipocyte | *TSLP* | 0.271411573 | 2.38E-05 | 0.007880493 | 0.149729376 |
| Heart_Failure | adipocyte | *RNF123* | 0.289458484 | 2.90E-06 | 0.001401502 | 0.01821953 |
| Heart_Failure | adipocyte | *SAP30L* | 0.95798509 | 0.000126068 | 0.028306755 | 0.792589127 |
| Heart_Failure | adipocyte | *C2* | 0.763249782 | 8.00E-06 | 0.003143764 | 0.050300218 |

| | | | | | | |
|---|---|---|---|---|---|---|
| Heart_Failure | adipocyte | *ORMDL3* | -0.323480852 | 4.20E-11 | 5.28E-08 | 2.64E-07 |
| Heart_Failure | adipocyte | *DCAKD* | -0.184700838 | 0.000142993 | 0.030999934 | 0.898998081 |
| Heart_Failure | adipocyte | *GMPPB* | 0.482326029 | 0.000220655 | 0.043351833 | 1 |
| Heart_Failure | adipocyte | *GOLIM4* | 0.598565885 | 0.000109001 | 0.026557809 | 0.685287214 |
| Heart_Failure | adipocyte | *HLA-DQB1* | -0.35936211 | 1.89E-24 | 1.19E-20 | 1.19E-20 |
| Heart_Failure | adipocyte | *MUC1* | 0.340376892 | 2.99E-05 | 0.009403774 | 0.188075477 |
| Heart_Failure | adipocyte | *MCRS1* | 0.688752625 | 1.74E-05 | 0.006070187 | 0.109263364 |
| Heart_Failure | adipocyte | *HLA-DRB1* | -0.396948162 | 2.43E-10 | 2.54E-07 | 1.52E-06 |
| Heart_Failure | adipocyte | *HLA-DQA1* | -0.502598521 | 2.91E-19 | 6.09E-16 | 1.83E-15 |
| Heart_Failure | adipocyte | *SLC22A5* | 0.255049516 | 1.84E-07 | 0.000144905 | 0.001159244 |
| Heart_Failure | adipocyte | *RPS26* | 0.129885895 | 0.000117713 | 0.02740972 | 0.740062436 |
| Heart_Failure | adipocyte | *HLA-DRA* | 0.366657184 | 9.23E-08 | 8.29E-05 | 0.000580546 |
| Heart_Failure | adipocyte | *BAG6* | 0.475517685 | 9.57E-07 | 0.00060145 | 0.006014496 |
| Heart_Failure | adipocyte | *MICA* | 0.162260803 | 9.02E-06 | 0.003336846 | 0.056726389 |
| Heart_Failure | adipocyte | *FAM153C* | 0.970423293 | 0.000227827 | 0.043404439 | 1 |
| Heart_Failure | adipocyte | *ATF6B* | 0.408354004 | 1.55E-06 | 0.000885651 | 0.009742159 |
| Heart_Failure | adipocyte | *HLA-DQB2* | 0.155052412 | 4.39E-14 | 6.90E-11 | 2.76E-10 |
| Heart_Failure | adipocyte | *TNF* | 0.48708134 | 7.45E-06 | 0.003122286 | 0.046834285 |
| Heart_Failure | adipocyte | *HLA-DQA2* | 0.363493107 | 6.99E-23 | 2.20E-19 | 4.40E-19 |
| Heart_Failure | ASPC | *SEMA3F* | -0.091333352 | 1.34E-06 | 0.000645811 | 0.008395539 |
| Heart_Failure | ASPC | *RBM6* | -0.07373145 | 1.41E-07 | 9.82E-05 | 0.000884224 |
| Heart_Failure | ASPC | *MED24* | -0.221750026 | 0.000180301 | 0.035423481 | 1 |
| Heart_Failure | ASPC | *OXCT1* | -0.145289481 | 7.43E-05 | 0.017972566 | 0.467286725 |
| Heart_Failure | ASPC | *COL16A1* | -0.213808271 | 7.85E-07 | 0.000448775 | 0.004936527 |
| Heart_Failure | ASPC | *PABPC4* | -0.181591526 | 8.73E-05 | 0.019610147 | 0.549084109 |
| Heart_Failure | ASPC | *SLC17A9* | 0.420837092 | 0.00017634 | 0.035423481 | 1 |
| Heart_Failure | ASPC | *FLT3* | 3.156800023 | 2.24E-09 | 2.01E-06 | 1.41E-05 |
| Heart_Failure | ASPC | *ARHGAP9* | -0.507042243 | 4.93E-05 | 0.014081279 | 0.309788138 |
| Heart_Failure | ASPC | *PDLIM4* | 0.241924868 | 1.09E-05 | 0.00382452 | 0.068841362 |
| Heart_Failure | ASPC | *REEP2* | -0.086448219 | 5.37E-05 | 0.014667788 | 0.33735912 |
| Heart_Failure | ASPC | *POM121C* | -0.356155576 | 1.20E-06 | 0.000628001 | 0.007536014 |
| Heart_Failure | ASPC | *LGR5* | -0.174904237 | 0.000233885 | 0.043248123 | 1 |
| Heart_Failure | ASPC | *TSLP* | 0.126707981 | 6.70E-06 | 0.002476427 | 0.042099265 |
| Heart_Failure | ASPC | *FUBP1* | -0.512696128 | 0.000249832 | 0.044876902 | 1 |
| Heart_Failure | ASPC | *RNF123* | 0.118685398 | 6.43E-06 | 0.002476427 | 0.040412798 |
| Heart_Failure | ASPC | *C2* | 0.32845837 | 4.82E-05 | 0.014081279 | 0.303241197 |
| Heart_Failure | ASPC | *ORMDL3* | -0.146047787 | 9.12E-12 | 1.15E-08 | 5.74E-08 |
| Heart_Failure | ASPC | *DCAKD* | -0.083105352 | 5.93E-05 | 0.015527612 | 0.372662682 |
| Heart_Failure | ASPC | *ACSF3* | -0.207133405 | 0.000265647 | 0.04639232 | 1 |
| Heart_Failure | ASPC | *HLA-DQB1* | -0.145898436 | 5.07E-24 | 3.19E-20 | 3.19E-20 |
| Heart_Failure | ASPC | *MUC1* | 0.137369744 | 6.93E-05 | 0.017421959 | 0.435548975 |
| Heart_Failure | ASPC | *MCRS1* | 0.278473841 | 8.27E-05 | 0.019258461 | 0.519978456 |
| Heart_Failure | ASPC | *MSL1* | -0.111717645 | 0.000136828 | 0.029663302 | 0.860235745 |
| Heart_Failure | ASPC | *HLA-DRB1* | -0.17419927 | 2.35E-11 | 2.46E-08 | 1.47E-07 |
| Heart_Failure | ASPC | *HLA-DQA1* | -0.216331124 | 3.36E-20 | 7.05E-17 | 2.11E-16 |
| Heart_Failure | ASPC | *SLC22A5* | 0.112219606 | 1.19E-07 | 9.38E-05 | 0.000750036 |
| Heart_Failure | ASPC | *RPS26* | 0.054288117 | 0.000141559 | 0.029666149 | 0.889984475 |
| Heart_Failure | ASPC | *HLA-DRA* | 0.14800345 | 6.23E-07 | 0.000391909 | 0.003919093 |
| Heart_Failure | ASPC | *BAG6* | 0.196147569 | 3.19E-06 | 0.001431028 | 0.020034399 |
| Heart_Failure | ASPC | *MICA* | 0.066347107 | 1.75E-05 | 0.005779461 | 0.109809759 |

| | | | | | | |
|---|---|---|---|---|---|---|
| Heart_Failure | ASPC | *FAM153C* | 0.430959412 | 0.000203933 | 0.038852418 | 1 |
| Heart_Failure | ASPC | *ATF6B* | 0.169922814 | 4.66E-06 | 0.001953275 | 0.029299132 |
| Heart_Failure | ASPC | *HLA-DQB2* | 0.069052918 | 8.62E-15 | 1.35E-11 | 5.42E-11 |
| Heart_Failure | ASPC | *TNF* | 0.188437683 | 2.89E-05 | 0.009076613 | 0.181532265 |
| Heart_Failure | ASPC | *HLA-DQA2* | 0.153733048 | 9.02E-23 | 2.84E-19 | 5.67E-19 |
| Heart_Failure | b_cell | *C1orf112* | 4.635568178 | 0.00830303 | 0.018961551 | 1 |
| Heart_Failure | b_cell | *CFH* | -1.100599769 | 0.001564645 | 0.008654124 | 1 |
| Heart_Failure | b_cell | *NIPAL3* | -2.632791294 | 0.012932706 | 0.026093684 | 1 |
| Heart_Failure | b_cell | *SEMA3F* | -3.7094213 | 7.03E-08 | 7.36E-05 | 0.000441764 |
| Heart_Failure | b_cell | *RAD52* | -1.870646035 | 0.005184081 | 0.013940256 | 1 |
| Heart_Failure | b_cell | *BAD* | -3.088097979 | 0.004916903 | 0.01349375 | 1 |
| Heart_Failure | b_cell | *HS3ST1* | 3.034121734 | 7.10E-05 | 0.002889125 | 0.446225675 |
| Heart_Failure | b_cell | *SNX11* | -2.870194758 | 0.00081913 | 0.0080216 | 1 |
| Heart_Failure | b_cell | *TMEM176A* | -2.301222088 | 0.004152605 | 0.012205437 | 1 |
| Heart_Failure | b_cell | *CYP26B1* | -2.742405025 | 0.00289572 | 0.010115189 | 1 |
| Heart_Failure | b_cell | *ICA1* | -1.338202106 | 0.00189547 | 0.008705907 | 1 |
| Heart_Failure | b_cell | *DBNDD1* | 1.749982939 | 0.018202672 | 0.033492461 | 1 |
| Heart_Failure | b_cell | *ALS2* | -1.859979431 | 0.007956581 | 0.018397 | 1 |
| Heart_Failure | b_cell | *POLDIP2* | -2.128216591 | 0.002191418 | 0.00904032 | 1 |
| Heart_Failure | b_cell | *CD38* | 4.758132421 | 0.000396082 | 0.004942367 | 1 |
| Heart_Failure | b_cell | *RBM6* | -2.79537295 | 1.69E-06 | 0.000884152 | 0.010618865 |
| Heart_Failure | b_cell | *ARHGAP33* | -2.827612123 | 0.014854073 | 0.028957382 | 1 |
| Heart_Failure | b_cell | *PDK4* | -1.58909689 | 0.015327502 | 0.029614015 | 1 |
| Heart_Failure | b_cell | *MEOX1* | -2.059776361 | 0.002505324 | 0.009482826 | 1 |
| Heart_Failure | b_cell | *LIG3* | -3.115761203 | 0.000674074 | 0.0070164 | 1 |
| Heart_Failure | b_cell | *CIAPIN1* | -2.964079196 | 0.01257955 | 0.025561612 | 1 |
| Heart_Failure | b_cell | *COPZ2* | -1.421034231 | 0.011484248 | 0.023876146 | 1 |
| Heart_Failure | b_cell | *GCFC2* | -1.732177315 | 0.013148264 | 0.026443742 | 1 |
| Heart_Failure | b_cell | *WDR54* | -3.449186588 | 0.002007404 | 0.008788681 | 1 |
| Heart_Failure | b_cell | *ABCB4* | 2.164279211 | 8.53E-05 | 0.002889125 | 0.536532535 |
| Heart_Failure | b_cell | *ZNF195* | -2.112389552 | 0.010124586 | 0.021881638 | 1 |
| Heart_Failure | b_cell | *PDK2* | -1.868698902 | 0.002331246 | 0.009229563 | 1 |
| Heart_Failure | b_cell | *GGNBP2* | -1.952739497 | 0.009533898 | 0.020826829 | 1 |
| Heart_Failure | b_cell | *TMEM98* | -2.288094454 | 0.001322957 | 0.008654124 | 1 |
| Heart_Failure | b_cell | *ABCC8* | -5.131434242 | 0.003759213 | 0.011574032 | 1 |
| Heart_Failure | b_cell | *CCL18* | 3.99234197 | 0.0011953 | 0.008654124 | 1 |
| Heart_Failure | b_cell | *TMEM132A* | -6.241134707 | 0.001486563 | 0.008654124 | 1 |
| Heart_Failure | b_cell | *SPATA20* | -2.050726854 | 0.005179727 | 0.013936542 | 1 |
| Heart_Failure | b_cell | *TNFRSF12A* | 6.433710342 | 0.000365736 | 0.004752125 | 1 |
| Heart_Failure | b_cell | *MAP3K9* | 1.156812176 | 7.99E-05 | 0.002889125 | 0.502643777 |
| Heart_Failure | b_cell | *BAIAP2L1* | -1.943947294 | 0.001222933 | 0.008654124 | 1 |
| Heart_Failure | b_cell | *IFRD1* | -1.453777337 | 0.002398173 | 0.009333474 | 1 |
| Heart_Failure | b_cell | *GTF2IRD1* | -1.477183165 | 0.007919611 | 0.018343665 | 1 |
| Heart_Failure | b_cell | *ELAC2* | -2.007817471 | 0.007570417 | 0.017807857 | 1 |
| Heart_Failure | b_cell | *CEACAM21* | 4.001329565 | 0.029889294 | 0.049269081 | 1 |
| Heart_Failure | b_cell | *NOS2* | -15.08799879 | 0.002265819 | 0.009180605 | 1 |
| Heart_Failure | b_cell | *CD79B* | 0.241169344 | 0.000282254 | 0.004036731 | 1 |
| Heart_Failure | b_cell | *ST7L* | -2.341714317 | 0.001600955 | 0.008654124 | 1 |
| Heart_Failure | b_cell | *RPUSD1* | -4.537120851 | 0.00176572 | 0.008654124 | 1 |
| Heart_Failure | b_cell | *PIGQ* | 6.824903831 | 0.004373244 | 0.012620483 | 1 |

| | | | | | | |
|---|---|---|---|---|---|---|
| Heart_Failure | b_cell | *CDK11A* | -3.85871334 | 0.000898727 | 0.008433282 | 1 |
| Heart_Failure | b_cell | *DLEC1* | -2.179513119 | 0.001255738 | 0.008654124 | 1 |
| Heart_Failure | b_cell | *SS18L2* | -2.309052188 | 0.00191649 | 0.008705907 | 1 |
| Heart_Failure | b_cell | *MPND* | 3.292218248 | 4.68E-05 | 0.002889125 | 0.294178045 |
| Heart_Failure | b_cell | *MGST1* | -0.920468907 | 0.002527251 | 0.009525675 | 1 |
| Heart_Failure | b_cell | *NFIX* | -1.386599831 | 0.002652281 | 0.009688632 | 1 |
| Heart_Failure | b_cell | *MMP25* | -2.735104061 | 0.003317876 | 0.01083435 | 1 |
| Heart_Failure | b_cell | *IL32* | -2.893049364 | 0.001118092 | 0.008654124 | 1 |
| Heart_Failure | b_cell | *PKD1* | -1.819780038 | 0.008893559 | 0.019792498 | 1 |
| Heart_Failure | b_cell | *MAPK8IP2* | 1.642496143 | 0.000383206 | 0.00487696 | 1 |
| Heart_Failure | b_cell | *MED24* | -2.941760146 | 0.000544651 | 0.006125623 | 1 |
| Heart_Failure | b_cell | *RHOBTB2* | 5.862861846 | 8.06E-05 | 0.002889125 | 0.50694757 |
| Heart_Failure | b_cell | *POMT2* | -3.094963897 | 0.016358833 | 0.030978309 | 1 |
| Heart_Failure | b_cell | *MLXIPL* | -0.81797211 | 0.014227662 | 0.028040632 | 1 |
| Heart_Failure | b_cell | *METTL13* | -2.210595684 | 0.001430324 | 0.008654124 | 1 |
| Heart_Failure | b_cell | *DYRK4* | -2.97859525 | 0.001832677 | 0.008654124 | 1 |
| Heart_Failure | b_cell | *STARD3NL* | -1.971995152 | 0.00543808 | 0.014347469 | 1 |
| Heart_Failure | b_cell | *ZNF200* | -3.437503569 | 0.011993632 | 0.024641818 | 1 |
| Heart_Failure | b_cell | *CD4* | -1.723107922 | 0.001565134 | 0.008654124 | 1 |
| Heart_Failure | b_cell | *LRRC23* | 4.195062369 | 0.002861139 | 0.01001558 | 1 |
| Heart_Failure | b_cell | *HFE* | -2.303866608 | 0.019641727 | 0.03552402 | 1 |
| Heart_Failure | b_cell | *MRC2* | -1.401449501 | 0.00135947 | 0.008654124 | 1 |
| Heart_Failure | b_cell | *TSPAN9* | -1.331319017 | 0.00402775 | 0.012047229 | 1 |
| Heart_Failure | b_cell | *UTP18* | -2.072231093 | 0.003924803 | 0.011882923 | 1 |
| Heart_Failure | b_cell | *TTC19* | -1.493711076 | 0.021677768 | 0.038261687 | 1 |
| Heart_Failure | b_cell | *DPF1* | -8.174087131 | 0.02500471 | 0.042695441 | 1 |
| Heart_Failure | b_cell | *SYT7* | -4.904957339 | 0.005129539 | 0.013858793 | 1 |
| Heart_Failure | b_cell | *PPP5C* | -2.327084014 | 0.022253553 | 0.039080961 | 1 |
| Heart_Failure | b_cell | *CEP68* | -1.733730897 | 0.00725855 | 0.017331752 | 1 |
| Heart_Failure | b_cell | *ZBTB32* | -2.796938145 | 0.001703445 | 0.008654124 | 1 |
| Heart_Failure | b_cell | *TMEM159* | -1.92373387 | 0.00175342 | 0.008654124 | 1 |
| Heart_Failure | b_cell | *ERCC1* | -1.535300584 | 0.004270865 | 0.012408081 | 1 |
| Heart_Failure | b_cell | *LTF* | -4.836766963 | 0.007444812 | 0.017643448 | 1 |
| Heart_Failure | b_cell | *ALOX5* | 1.984950782 | 8.80E-05 | 0.002889125 | 0.552969383 |
| Heart_Failure | b_cell | *MAP4K5* | -1.682231284 | 0.02470727 | 0.042256423 | 1 |
| Heart_Failure | b_cell | *PSMC4* | -1.865549269 | 0.001873033 | 0.008701813 | 1 |
| Heart_Failure | b_cell | *CLDN11* | -1.485684393 | 0.001226772 | 0.008654124 | 1 |
| Heart_Failure | b_cell | *SLC25A39* | 5.48406025 | 0.018309478 | 0.033619068 | 1 |
| Heart_Failure | b_cell | *ANGEL1* | -2.499160775 | 0.019740539 | 0.035667565 | 1 |
| Heart_Failure | b_cell | *RNF14* | -1.78774881 | 0.007295887 | 0.017381435 | 1 |
| Heart_Failure | b_cell | *DDX11* | -2.158679539 | 0.017913362 | 0.033130632 | 1 |
| Heart_Failure | b_cell | *TACC3* | -2.020969775 | 0.002478991 | 0.009457169 | 1 |
| Heart_Failure | b_cell | *ZC3H3* | 4.720341063 | 0.007546331 | 0.017782527 | 1 |
| Heart_Failure | b_cell | *MDH1* | -1.297529334 | 0.007348943 | 0.017461378 | 1 |
| Heart_Failure | b_cell | *MTMR11* | -1.56106169 | 0.024581889 | 0.042145169 | 1 |
| Heart_Failure | b_cell | *NPC1L1* | 5.476303661 | 0.000132681 | 0.003196044 | 0.834167527 |
| Heart_Failure | b_cell | *CHDH* | -6.155057317 | 0.001786857 | 0.008654124 | 1 |
| Heart_Failure | b_cell | *IL20RA* | -6.236496286 | 0.001734087 | 0.008654124 | 1 |
| Heart_Failure | b_cell | *GLT8D1* | -1.93088132 | 0.000374659 | 0.004787561 | 1 |
| Heart_Failure | b_cell | *WWTR1* | -1.570085864 | 0.005510988 | 0.014478721 | 1 |

| | | | | | | |
|---|---|---|---|---|---|---|
| Heart_Failure | b_cell | *VSIG2* | -7.026340328 | 0.000627016 | 0.0066316 | 1 |
| Heart_Failure | b_cell | *PHLDB1* | -1.085585868 | 0.001570993 | 0.008654124 | 1 |
| Heart_Failure | b_cell | *MARCO* | -1.855918369 | 0.000387967 | 0.004897883 | 1 |
| Heart_Failure | b_cell | *MNAT1* | -1.533417323 | 0.007861158 | 0.01824361 | 1 |
| Heart_Failure | b_cell | *OSBPL5* | -1.987728273 | 0.007030284 | 0.016976281 | 1 |
| Heart_Failure | b_cell | *AQR* | -1.718838776 | 0.010311943 | 0.022142102 | 1 |
| Heart_Failure | b_cell | *CPS1* | -1.749825804 | 0.003724736 | 0.01154705 | 1 |
| Heart_Failure | b_cell | *NLRP2* | -2.932969304 | 0.015718697 | 0.0301291 | 1 |
| Heart_Failure | b_cell | *ZDHHC6* | -2.006520321 | 0.009202028 | 0.02024253 | 1 |
| Heart_Failure | b_cell | *BIRC3* | 1.154381858 | 0.000135041 | 0.003206095 | 0.849000794 |
| Heart_Failure | b_cell | *SNAPC1* | -2.231910865 | 0.00562094 | 0.014681699 | 1 |
| Heart_Failure | b_cell | *GCLM* | -1.56378972 | 0.00316468 | 0.010566301 | 1 |
| Heart_Failure | b_cell | *NR1H3* | -1.57708331 | 0.00213955 | 0.008997561 | 1 |
| Heart_Failure | b_cell | *TYMP* | -2.542506385 | 0.004182818 | 0.012265566 | 1 |
| Heart_Failure | b_cell | *TOMM34* | 4.509012026 | 2.66E-05 | 0.002889125 | 0.166979458 |
| Heart_Failure | b_cell | *FAS* | -1.518927035 | 0.016702871 | 0.031384026 | 1 |
| Heart_Failure | b_cell | *RNASET2* | 5.112560765 | 0.000251623 | 0.003766552 | 1 |
| Heart_Failure | b_cell | *CD44* | -1.508721906 | 0.008609366 | 0.019407344 | 1 |
| Heart_Failure | b_cell | *MIPEP* | -1.990442704 | 0.008261667 | 0.018897686 | 1 |
| Heart_Failure | b_cell | *SLC39A9* | -2.334982591 | 0.02349005 | 0.040685911 | 1 |
| Heart_Failure | b_cell | *ANK1* | -2.744165464 | 0.001506776 | 0.008654124 | 1 |
| Heart_Failure | b_cell | *BAK1* | -3.775317795 | 0.002007196 | 0.008788681 | 1 |
| Heart_Failure | b_cell | *CENPQ* | -2.26147567 | 0.006353102 | 0.015908678 | 1 |
| Heart_Failure | b_cell | *IFT88* | -1.829482152 | 0.027321488 | 0.045817604 | 1 |
| Heart_Failure | b_cell | *UBA6* | -1.956026471 | 0.021290868 | 0.037705827 | 1 |
| Heart_Failure | b_cell | *ASTE1* | -2.401603552 | 0.005952157 | 0.015286442 | 1 |
| Heart_Failure | b_cell | *MYOC* | -1.615390427 | 0.002059304 | 0.008831917 | 1 |
| Heart_Failure | b_cell | *SH3YL1* | -2.053085439 | 0.003763545 | 0.011581697 | 1 |
| Heart_Failure | b_cell | *VCL* | -1.856688972 | 0.019773288 | 0.035702086 | 1 |
| Heart_Failure | b_cell | *STAP1* | 0.261133931 | 0.000234404 | 0.003705045 | 1 |
| Heart_Failure | b_cell | *TBC1D23* | -1.867930733 | 0.020163124 | 0.036177385 | 1 |
| Heart_Failure | b_cell | *TUBG2* | -1.307169466 | 0.012271006 | 0.025105048 | 1 |
| Heart_Failure | b_cell | *MRI1* | -1.620749331 | 0.02002135 | 0.036009891 | 1 |
| Heart_Failure | b_cell | *AGA* | -2.565800578 | 0.003321419 | 0.01083435 | 1 |
| Heart_Failure | b_cell | *C6* | -1.493631158 | 0.001595184 | 0.008654124 | 1 |
| Heart_Failure | b_cell | *RAI14* | -1.532241707 | 0.011355136 | 0.023727222 | 1 |
| Heart_Failure | b_cell | *PNKP* | -1.717966189 | 0.004208981 | 0.012307842 | 1 |
| Heart_Failure | b_cell | *SPDL1* | -2.264387053 | 0.019581558 | 0.035447526 | 1 |
| Heart_Failure | b_cell | *PQLC2* | -2.808118332 | 0.001524545 | 0.008654124 | 1 |
| Heart_Failure | b_cell | *PSMA4* | -2.138883995 | 0.015549429 | 0.029868396 | 1 |
| Heart_Failure | b_cell | *THAP3* | -3.072952009 | 0.00374693 | 0.011556943 | 1 |
| Heart_Failure | b_cell | *TDP1* | 5.708046949 | 0.005107269 | 0.013822386 | 1 |
| Heart_Failure | b_cell | *SPATA7* | -1.984437584 | 0.019316265 | 0.035118958 | 1 |
| Heart_Failure | b_cell | *MED17* | -2.795774557 | 0.016675946 | 0.031361554 | 1 |
| Heart_Failure | b_cell | *RETSAT* | -1.191848986 | 0.002101251 | 0.00889896 | 1 |
| Heart_Failure | b_cell | *CAPG* | 4.486893292 | 0.000208882 | 0.003568596 | 1 |
| Heart_Failure | b_cell | *AP2S1* | 5.504406716 | 0.013529145 | 0.026983885 | 1 |
| Heart_Failure | b_cell | *LCP2* | -1.485919391 | 0.001984528 | 0.008775328 | 1 |
| Heart_Failure | b_cell | *CUL7* | -2.856539688 | 0.011463446 | 0.023848672 | 1 |
| Heart_Failure | b_cell | *CNTLN* | -1.654764945 | 0.029157959 | 0.048279191 | 1 |

| | | | | | | |
|---|---|---|---|---|---|---|
| Heart_Failure | b_cell | *EPHA3* | -1.402267775 | 0.001675193 | 0.008654124 | 1 |
| Heart_Failure | b_cell | *DSG2* | -2.24988874 | 0.001661083 | 0.008654124 | 1 |
| Heart_Failure | b_cell | *ATP6V1H* | -1.830390691 | 0.018139137 | 0.033417044 | 1 |
| Heart_Failure | b_cell | *ANO2* | -1.260255664 | 0.001509959 | 0.008654124 | 1 |
| Heart_Failure | b_cell | *TSPAN17* | -2.579348669 | 0.003135935 | 0.010520609 | 1 |
| Heart_Failure | b_cell | *CC2D2A* | -1.636806055 | 0.008594603 | 0.019399557 | 1 |
| Heart_Failure | b_cell | *LMO3* | -1.091708909 | 0.003335075 | 0.010858423 | 1 |
| Heart_Failure | b_cell | *ERCC8* | -1.897266348 | 0.016180721 | 0.030715034 | 1 |
| Heart_Failure | b_cell | *UTS2* | -3.034995085 | 0.004258747 | 0.012401455 | 1 |
| Heart_Failure | b_cell | *RFC2* | 6.292003287 | 7.36E-05 | 0.002889125 | 0.462743787 |
| Heart_Failure | b_cell | *JKAMP* | -1.682391787 | 0.013366693 | 0.026754664 | 1 |
| Heart_Failure | b_cell | *DKK3* | -2.385778591 | 0.009068673 | 0.020075614 | 1 |
| Heart_Failure | b_cell | *ARHGEF5* | 4.040564902 | 6.94E-05 | 0.002889125 | 0.43641284 |
| Heart_Failure | b_cell | *LIMA1* | -0.819195315 | 0.002744806 | 0.009854904 | 1 |
| Heart_Failure | b_cell | *PTGER3* | -1.039468994 | 0.00166121 | 0.008654124 | 1 |
| Heart_Failure | b_cell | *PIK3CB* | -1.768504049 | 0.00694432 | 0.01683723 | 1 |
| Heart_Failure | b_cell | *CYBA* | 4.525458197 | 0.000886477 | 0.008368291 | 1 |
| Heart_Failure | b_cell | *ALX4* | 4.887783461 | 8.05E-05 | 0.002889125 | 0.506149981 |
| Heart_Failure | b_cell | *AKR7A2* | -2.654330094 | 0.009054143 | 0.020057575 | 1 |
| Heart_Failure | b_cell | *MCF2L2* | -2.385348229 | 0.013952915 | 0.027646383 | 1 |
| Heart_Failure | b_cell | *LY75* | 6.979822773 | 0.006985489 | 0.016910961 | 1 |
| Heart_Failure | b_cell | *PLEKHH1* | -2.638418444 | 0.00053896 | 0.006083376 | 1 |
| Heart_Failure | b_cell | *ATP9A* | -1.245467562 | 0.001473509 | 0.008654124 | 1 |
| Heart_Failure | b_cell | *CHRDL2* | -5.61323746 | 0.008712368 | 0.019590364 | 1 |
| Heart_Failure | b_cell | *FAM114A2* | -2.198912132 | 0.018021761 | 0.033275421 | 1 |
| Heart_Failure | b_cell | *MRPL43* | -3.508961424 | 0.004767328 | 0.013262031 | 1 |
| Heart_Failure | b_cell | *IL17RB* | -4.676356944 | 0.00156714 | 0.008654124 | 1 |
| Heart_Failure | b_cell | *TRAF3IP2* | -1.937114298 | 0.007087609 | 0.01706618 | 1 |
| Heart_Failure | b_cell | *DCBLD2* | -1.577936713 | 0.004020499 | 0.012042342 | 1 |
| Heart_Failure | b_cell | *PITHD1* | -2.321263086 | 0.004673261 | 0.013085296 | 1 |
| Heart_Failure | b_cell | *LAMC2* | -2.880178341 | 2.55E-05 | 0.002889125 | 0.160533399 |
| Heart_Failure | b_cell | *ZC3H11A* | -4.709611953 | 0.002964861 | 0.010176497 | 1 |
| Heart_Failure | b_cell | *DNAJC25* | -2.24542924 | 0.003858331 | 0.011769687 | 1 |
| Heart_Failure | b_cell | *PIGV* | 3.840518555 | 0.000186642 | 0.00340015 | 1 |
| Heart_Failure | b_cell | *QSER1* | -1.941168625 | 0.0172304 | 0.032153797 | 1 |
| Heart_Failure | b_cell | *NCKAP1* | -1.421229067 | 0.004010801 | 0.012024754 | 1 |
| Heart_Failure | b_cell | *SFSWAP* | -1.806929717 | 0.030457663 | 0.04994453 | 1 |
| Heart_Failure | b_cell | *SLC6A16* | -2.150651757 | 0.006676355 | 0.016382674 | 1 |
| Heart_Failure | b_cell | *ADCK1* | -1.865444462 | 0.003969177 | 0.011945533 | 1 |
| Heart_Failure | b_cell | *HAGH* | -1.592844497 | 0.020296622 | 0.036333958 | 1 |
| Heart_Failure | b_cell | *DMRT3* | -3.585302648 | 0.001593586 | 0.008654124 | 1 |
| Heart_Failure | b_cell | *ST3GAL6* | -1.419805122 | 0.001717038 | 0.008654124 | 1 |
| Heart_Failure | b_cell | *ATP2C2* | -3.74057682 | 0.004404421 | 0.01267545 | 1 |
| Heart_Failure | b_cell | *CDON* | -1.121911751 | 0.002569684 | 0.009560634 | 1 |
| Heart_Failure | b_cell | *TMEM161A* | -2.70205495 | 0.00183128 | 0.008654124 | 1 |
| Heart_Failure | b_cell | *LPAR2* | -2.58825803 | 0.002589695 | 0.009588327 | 1 |
| Heart_Failure | b_cell | *ABCA7* | 4.977505599 | 4.36E-05 | 0.002889125 | 0.274186689 |
| Heart_Failure | b_cell | *DDX20* | -2.983027796 | 0.005576847 | 0.014607888 | 1 |
| Heart_Failure | b_cell | *CHI3L2* | 4.510123872 | 1.88E-05 | 0.002889125 | 0.118477351 |
| Heart_Failure | b_cell | *UHRF1BP1* | -1.659220646 | 0.002756077 | 0.009878822 | 1 |

| | | | | | | |
|---|---|---|---|---|---|---|
| Heart_Failure | b_cell | *WDR18* | -2.879967462 | 0.00041103 | 0.005033892 | 1 |
| Heart_Failure | b_cell | *MCM10* | 1.4698927 | 8.52E-05 | 0.002889125 | 0.535543966 |
| Heart_Failure | b_cell | *ANKRD44* | 2.969066591 | 0.028423 | 0.047304802 | 1 |
| Heart_Failure | b_cell | *ADAT1* | 3.148827142 | 0.017337139 | 0.032286313 | 1 |
| Heart_Failure | b_cell | *TMEM206* | -2.522849146 | 0.004480414 | 0.012757412 | 1 |
| Heart_Failure | b_cell | *SEC61A2* | -1.894775618 | 0.002443916 | 0.009414769 | 1 |
| Heart_Failure | b_cell | *ASB1* | -2.682158024 | 0.007435716 | 0.01763423 | 1 |
| Heart_Failure | b_cell | *PPP2R5A* | -1.608743447 | 0.014715666 | 0.028767846 | 1 |
| Heart_Failure | b_cell | *TIE1* | -2.149304578 | 0.000172901 | 0.003365162 | 1 |
| Heart_Failure | b_cell | *CD84* | -1.909298834 | 0.001018146 | 0.008654124 | 1 |
| Heart_Failure | b_cell | *ZNRD1* | -3.29302162 | 0.008424654 | 0.019121228 | 1 |
| Heart_Failure | b_cell | *ATXN3* | -1.886425559 | 0.012287296 | 0.025113859 | 1 |
| Heart_Failure | b_cell | *GOLGA5* | -1.849168033 | 0.003841549 | 0.011752708 | 1 |
| Heart_Failure | b_cell | *EML1* | -1.524593187 | 0.001153476 | 0.008654124 | 1 |
| Heart_Failure | b_cell | *ZFAT* | 2.517316861 | 0.000158668 | 0.003281394 | 0.99754383 |
| Heart_Failure | b_cell | *STOML1* | 3.944786403 | 7.25E-05 | 0.002889125 | 0.456108979 |
| Heart_Failure | b_cell | *RRP15* | -1.958555162 | 0.002564398 | 0.009560634 | 1 |
| Heart_Failure | b_cell | *DHX8* | -2.296365957 | 0.003514889 | 0.011161493 | 1 |
| Heart_Failure | b_cell | *SYT1* | -1.813015944 | 0.006360594 | 0.015909937 | 1 |
| Heart_Failure | b_cell | *ROGDI* | 4.410797327 | 6.32E-05 | 0.002889125 | 0.397127996 |
| Heart_Failure | b_cell | *HDAC4* | -1.731273543 | 0.026633208 | 0.044887581 | 1 |
| Heart_Failure | b_cell | *IFI35* | -2.278784862 | 0.009018023 | 0.020008264 | 1 |
| Heart_Failure | b_cell | *HEATR6* | -2.634972385 | 0.02328584 | 0.040429418 | 1 |
| Heart_Failure | b_cell | *POLR1A* | -2.209701992 | 0.000973166 | 0.008654124 | 1 |
| Heart_Failure | b_cell | *STON1-GTF2A* | -0.760317248 | 0.028607978 | 0.047543843 | 1 |
| Heart_Failure | b_cell | *CYFIP1* | -1.770354487 | 0.017799678 | 0.032971884 | 1 |
| Heart_Failure | b_cell | *PITX1* | -6.448325636 | 0.001680877 | 0.008654124 | 1 |
| Heart_Failure | b_cell | *SDK2* | 1.733879149 | 0.000153228 | 0.003239931 | 0.963347277 |
| Heart_Failure | b_cell | *DNAJA2* | -2.238783538 | 0.004299859 | 0.012474948 | 1 |
| Heart_Failure | b_cell | *KCNAB2* | -2.013162388 | 0.003746431 | 0.011556943 | 1 |
| Heart_Failure | b_cell | *GAL* | 2.350414607 | 0.000135138 | 0.003206095 | 0.84961528 |
| Heart_Failure | b_cell | *NEDD4* | -1.68584791 | 0.005530225 | 0.014516264 | 1 |
| Heart_Failure | b_cell | *PFN2* | -2.704356077 | 0.004901283 | 0.013473707 | 1 |
| Heart_Failure | b_cell | *DAPP1* | 5.27121129 | 2.11E-06 | 0.000884152 | 0.013262287 |
| Heart_Failure | b_cell | *FGF10* | -1.545979023 | 0.00125618 | 0.008654124 | 1 |
| Heart_Failure | b_cell | *CLTCL1* | -1.810768306 | 0.001902728 | 0.008705907 | 1 |
| Heart_Failure | b_cell | *DGCR2* | -2.131534216 | 0.025964423 | 0.043999549 | 1 |
| Heart_Failure | b_cell | *WIPI1* | -1.81884394 | 0.004118022 | 0.012172074 | 1 |
| Heart_Failure | b_cell | *NDST1* | -2.11684934 | 0.017944299 | 0.033161614 | 1 |
| Heart_Failure | b_cell | *ASNS* | -3.926662465 | 0.007569975 | 0.017807857 | 1 |
| Heart_Failure | b_cell | *ST6GALNAC2* | -2.344387897 | 0.003665963 | 0.011455224 | 1 |
| Heart_Failure | b_cell | *EIF2B3* | -1.470302335 | 0.004634854 | 0.013020253 | 1 |
| Heart_Failure | b_cell | *CAMK2A* | 5.068109863 | 0.000119566 | 0.003135985 | 0.751708474 |
| Heart_Failure | b_cell | *OSBPL3* | -1.884435773 | 0.010835826 | 0.022984088 | 1 |
| Heart_Failure | b_cell | *RAD18* | -2.238613707 | 0.019200355 | 0.034938533 | 1 |
| Heart_Failure | b_cell | *VASH1* | -2.541321393 | 0.019077989 | 0.034796436 | 1 |
| Heart_Failure | b_cell | *MBD3* | -2.890097426 | 0.027531634 | 0.046059442 | 1 |
| Heart_Failure | b_cell | *HLTF* | -2.288025183 | 0.01065125 | 0.022692107 | 1 |
| Heart_Failure | b_cell | *CPSF1* | -2.101882535 | 0.001871691 | 0.008701813 | 1 |
| Heart_Failure | b_cell | *CYBRD1* | -1.4309736 | 0.025663158 | 0.043618349 | 1 |

| | | | | | | |
|---|---|---|---|---|---|---|
| Heart_Failure | b_cell | *EPN2* | -1.727291756 | 0.023237772 | 0.040380285 | 1 |
| Heart_Failure | b_cell | *LIMS2* | -1.649424062 | 0.000490051 | 0.005622167 | 1 |
| Heart_Failure | b_cell | *TRHDE* | -1.036887812 | 0.001649328 | 0.008654124 | 1 |
| Heart_Failure | b_cell | *TRNT1* | -1.963339628 | 0.008737064 | 0.019617828 | 1 |
| Heart_Failure | b_cell | *CRMP1* | -1.843293637 | 0.000721097 | 0.007316224 | 1 |
| Heart_Failure | b_cell | *PVR* | -2.30239177 | 0.016201296 | 0.030741377 | 1 |
| Heart_Failure | b_cell | *MOV10L1* | -2.58769394 | 0.001787973 | 0.008654124 | 1 |
| Heart_Failure | b_cell | *TP63* | -1.588457769 | 0.002475645 | 0.009455883 | 1 |
| Heart_Failure | b_cell | *NLE1* | 3.237588894 | 6.20E-05 | 0.002889125 | 0.389546876 |
| Heart_Failure | b_cell | *SDHA* | -2.375864317 | 0.026443384 | 0.044654729 | 1 |
| Heart_Failure | b_cell | *SMARCE1* | -2.006059633 | 0.00557874 | 0.014607888 | 1 |
| Heart_Failure | b_cell | *GSDMB* | -1.647658491 | 0.000327627 | 0.004448791 | 1 |
| Heart_Failure | b_cell | *GLI2* | -1.719866625 | 0.001409765 | 0.008654124 | 1 |
| Heart_Failure | b_cell | *ATP2A3* | 4.864427354 | 9.25E-05 | 0.002932166 | 0.581817577 |
| Heart_Failure | b_cell | *CA12* | -1.696778723 | 0.009856761 | 0.021383525 | 1 |
| Heart_Failure | b_cell | *NTN4* | -1.332334002 | 0.001592504 | 0.008654124 | 1 |
| Heart_Failure | b_cell | *SRI* | -2.201331696 | 0.006865274 | 0.016690634 | 1 |
| Heart_Failure | b_cell | *TTC38* | -1.548871411 | 0.001310003 | 0.008654124 | 1 |
| Heart_Failure | b_cell | *ADD2* | 8.062218932 | 0.000195165 | 0.003449722 | 1 |
| Heart_Failure | b_cell | *FSCN1* | -2.553936199 | 0.004048723 | 0.012092315 | 1 |
| Heart_Failure | b_cell | *MOCOS* | 2.110790824 | 0.002493023 | 0.009470475 | 1 |
| Heart_Failure | b_cell | *PLD1* | -1.572133568 | 0.011939738 | 0.024539108 | 1 |
| Heart_Failure | b_cell | *SEC31B* | -1.693994851 | 0.009046945 | 0.020048693 | 1 |
| Heart_Failure | b_cell | *TUBA3D* | -5.796512316 | 0.028790673 | 0.047737586 | 1 |
| Heart_Failure | b_cell | *MKRN2* | -2.03773764 | 0.001674229 | 0.008654124 | 1 |
| Heart_Failure | b_cell | *MCM6* | 4.837089289 | 4.68E-05 | 0.002889125 | 0.293964461 |
| Heart_Failure | b_cell | *RBMS2* | -1.463132383 | 0.003869085 | 0.011796769 | 1 |
| Heart_Failure | b_cell | *MLH1* | -2.091106647 | 0.012668261 | 0.025683765 | 1 |
| Heart_Failure | b_cell | *FMO4* | -2.116798322 | 0.005975307 | 0.015314616 | 1 |
| Heart_Failure | b_cell | *RGS11* | -1.994674023 | 0.000390319 | 0.004909064 | 1 |
| Heart_Failure | b_cell | *TPD52* | 1.062265419 | 0.000196437 | 0.003449722 | 1 |
| Heart_Failure | b_cell | *GPATCH1* | -1.969802547 | 0.006963587 | 0.016877438 | 1 |
| Heart_Failure | b_cell | *ICAM3* | 4.7244607 | 1.01E-05 | 0.002447238 | 0.063628183 |
| Heart_Failure | b_cell | *CAMSAP3* | -15.64121086 | 0.001794816 | 0.008654124 | 1 |
| Heart_Failure | b_cell | *XAB2* | -2.895279702 | 0.000961474 | 0.008654124 | 1 |
| Heart_Failure | b_cell | *STXBP2* | -3.618827407 | 0.000790143 | 0.007798474 | 1 |
| Heart_Failure | b_cell | *MAP2K7* | -2.149213531 | 0.004008083 | 0.012024754 | 1 |
| Heart_Failure | b_cell | *NMRK2* | -12.3583343 | 0.003742174 | 0.011555523 | 1 |
| Heart_Failure | b_cell | *CTTNBP2* | -1.635522125 | 0.012141602 | 0.024897017 | 1 |
| Heart_Failure | b_cell | *APBB1IP* | -1.409433214 | 0.014752863 | 0.028816763 | 1 |
| Heart_Failure | b_cell | *NAALAD2* | -1.781066762 | 0.008833836 | 0.019729423 | 1 |
| Heart_Failure | b_cell | *FGFR1* | -1.445467094 | 0.009247731 | 0.020335952 | 1 |
| Heart_Failure | b_cell | *FBLN1* | -1.154764397 | 0.000811893 | 0.00796314 | 1 |
| Heart_Failure | b_cell | *PIAS2* | -1.773220138 | 0.012735724 | 0.025778976 | 1 |
| Heart_Failure | b_cell | *AMPH* | -1.628804628 | 0.001044366 | 0.008654124 | 1 |
| Heart_Failure | b_cell | *MCCC1* | -1.573551229 | 0.015320717 | 0.029614015 | 1 |
| Heart_Failure | b_cell | *FAP* | -1.53918081 | 0.001216216 | 0.008654124 | 1 |
| Heart_Failure | b_cell | *ACER3* | -1.384634677 | 0.002974099 | 0.010181586 | 1 |
| Heart_Failure | b_cell | *TULP3* | -1.873091885 | 0.008966627 | 0.019919853 | 1 |
| Heart_Failure | b_cell | *SYNJ2* | -1.616455635 | 0.002849332 | 0.010013275 | 1 |

| | | | | | | |
|---|---|---|---|---|---|---|
| Heart_Failure | b_cell | *FGF20* | -21.2853336 | 0.001839125 | 0.008661108 | 1 |
| Heart_Failure | b_cell | *MYH7B* | -4.567533184 | 0.00224061 | 0.009117613 | 1 |
| Heart_Failure | b_cell | *FKBP7* | -2.10187241 | 0.001611741 | 0.008654124 | 1 |
| Heart_Failure | b_cell | *OSBPL6* | -2.369217197 | 0.00812136 | 0.018655094 | 1 |
| Heart_Failure | b_cell | *LXN* | -2.056653022 | 0.006337527 | 0.015886776 | 1 |
| Heart_Failure | b_cell | *SAR1A* | -2.056015114 | 0.028284285 | 0.04713048 | 1 |
| Heart_Failure | b_cell | *FDFT1* | -2.09962505 | 0.017744633 | 0.032898999 | 1 |
| Heart_Failure | b_cell | *DDX1* | -1.997068027 | 0.023149892 | 0.040260961 | 1 |
| Heart_Failure | b_cell | *RABL2B* | -1.80215383 | 0.002082285 | 0.008855223 | 1 |
| Heart_Failure | b_cell | *DDX43* | -2.631673282 | 0.002023854 | 0.00882996 | 1 |
| Heart_Failure | b_cell | *DCT* | -3.559443104 | 0.002057533 | 0.008831917 | 1 |
| Heart_Failure | b_cell | *SLC35C2* | -2.18188966 | 0.008758832 | 0.019648535 | 1 |
| Heart_Failure | b_cell | *SCTR* | -1.76799391 | 0.001512542 | 0.008654124 | 1 |
| Heart_Failure | b_cell | *KCNN2* | -4.787752354 | 0.001797068 | 0.008654124 | 1 |
| Heart_Failure | b_cell | *CPOX* | -2.104575427 | 0.009408284 | 0.020609716 | 1 |
| Heart_Failure | b_cell | *MOK* | -2.003212407 | 0.004412008 | 0.012689063 | 1 |
| Heart_Failure | b_cell | *MAGI3* | -1.645418396 | 0.014787517 | 0.0288569 | 1 |
| Heart_Failure | b_cell | *COL4A4* | 0.900056123 | 0.000149523 | 0.003239931 | 0.940053306 |
| Heart_Failure | b_cell | *ZNF506* | -2.219651098 | 0.012995443 | 0.02618665 | 1 |
| Heart_Failure | b_cell | *KIAA0141* | -2.077058904 | 0.005096968 | 0.013812459 | 1 |
| Heart_Failure | b_cell | *PCDHGA2* | 5.670999375 | 0.00029742 | 0.004173837 | 1 |
| Heart_Failure | b_cell | *HSPB11* | -1.980654161 | 0.006903595 | 0.016764351 | 1 |
| Heart_Failure | b_cell | *WDR70* | -1.602819437 | 0.028541287 | 0.047458098 | 1 |
| Heart_Failure | b_cell | *PGR* | -1.504758881 | 0.002732503 | 0.009839202 | 1 |
| Heart_Failure | b_cell | *COBLL1* | -1.013535231 | 0.002546623 | 0.009545808 | 1 |
| Heart_Failure | b_cell | *DOPEY1* | -1.931555691 | 0.019948594 | 0.035925754 | 1 |
| Heart_Failure | b_cell | *LYRM2* | -1.993990709 | 0.003501848 | 0.011147503 | 1 |
| Heart_Failure | b_cell | *ULK2* | -1.72362754 | 0.006191785 | 0.015621087 | 1 |
| Heart_Failure | b_cell | *PLOD1* | -2.620464277 | 0.00525962 | 0.014066038 | 1 |
| Heart_Failure | b_cell | *OXCT1* | -2.466159579 | 0.000285597 | 0.004036731 | 1 |
| Heart_Failure | b_cell | *ZNF324* | -2.449525202 | 0.002484643 | 0.00946151 | 1 |
| Heart_Failure | b_cell | *ZNF586* | -2.304046711 | 0.0140836 | 0.027817654 | 1 |
| Heart_Failure | b_cell | *ZNF446* | 9.252015254 | 0.00036481 | 0.004752125 | 1 |
| Heart_Failure | b_cell | *RPS5* | 4.610071046 | 0.01151609 | 0.023934432 | 1 |
| Heart_Failure | b_cell | *CHMP2B* | -2.051578113 | 0.017252871 | 0.032158778 | 1 |
| Heart_Failure | b_cell | *ZMPSTE24* | -1.998331452 | 0.004921203 | 0.01349375 | 1 |
| Heart_Failure | b_cell | *STARD7* | 4.906740905 | 0.008823495 | 0.019720338 | 1 |
| Heart_Failure | b_cell | *NOA1* | 2.33972148 | 0.000390414 | 0.004909064 | 1 |
| Heart_Failure | b_cell | *GSTP1* | -1.933750113 | 0.006729807 | 0.016475972 | 1 |
| Heart_Failure | b_cell | *WBP11* | -2.4172833 | 0.017365449 | 0.032329457 | 1 |
| Heart_Failure | b_cell | *COL16A1* | -1.947479704 | 8.06E-05 | 0.002889125 | 0.506736364 |
| Heart_Failure | b_cell | *APOB* | -2.593802159 | 0.00343698 | 0.011064154 | 1 |
| Heart_Failure | b_cell | *FCN1* | -2.016178962 | 0.001909559 | 0.008705907 | 1 |
| Heart_Failure | b_cell | *SEH1L* | 4.784607161 | 0.016674713 | 0.031361554 | 1 |
| Heart_Failure | b_cell | *OVGP1* | 5.359348779 | 3.28E-05 | 0.002889125 | 0.206027949 |
| Heart_Failure | b_cell | *ABCB1* | -1.525173383 | 0.00542693 | 0.014347469 | 1 |
| Heart_Failure | b_cell | *ZNF213* | -1.714541591 | 0.004674556 | 0.013085296 | 1 |
| Heart_Failure | b_cell | *CPNE3* | -2.026439048 | 0.004720207 | 0.013183596 | 1 |
| Heart_Failure | b_cell | *CTTN* | -1.443610363 | 0.002591352 | 0.009588327 | 1 |
| Heart_Failure | b_cell | *ATG16L1* | 4.600490441 | 0.022028386 | 0.038728318 | 1 |

| | | | | | | |
|---|---|---|---|---|---|---|
| Heart_Failure | b_cell | *USP40* | -1.667479256 | 0.019489121 | 0.035341246 | 1 |
| Heart_Failure | b_cell | *EIF2AK1* | -2.058061565 | 0.02190142 | 0.03855901 | 1 |
| Heart_Failure | b_cell | *SEPHS1* | -2.065475316 | 0.004772286 | 0.013264085 | 1 |
| Heart_Failure | b_cell | *ITPKC* | -1.622624233 | 0.001589745 | 0.008654124 | 1 |
| Heart_Failure | b_cell | *CEACAM6* | 21.1686193 | 0.001062859 | 0.008654124 | 1 |
| Heart_Failure | b_cell | *HSD17B14* | -2.783086297 | 0.001755226 | 0.008654124 | 1 |
| Heart_Failure | b_cell | *TRIP6* | -2.544840956 | 0.018438901 | 0.033836943 | 1 |
| Heart_Failure | b_cell | *SRRT* | -2.410130653 | 0.016873929 | 0.031592136 | 1 |
| Heart_Failure | b_cell | *PIGS* | -1.864557448 | 0.01030982 | 0.022142102 | 1 |
| Heart_Failure | b_cell | *ATXN7L3* | 3.19015539 | 7.69E-05 | 0.002889125 | 0.483714876 |
| Heart_Failure | b_cell | *PSMC5* | -1.67408458 | 0.008331441 | 0.01899194 | 1 |
| Heart_Failure | b_cell | *LPCAT2* | -1.890561907 | 0.003047646 | 0.01033471 | 1 |
| Heart_Failure | b_cell | *PTHLH* | -2.882381162 | 0.001457847 | 0.008654124 | 1 |
| Heart_Failure | b_cell | *PHACTR3* | -1.920084513 | 0.000998726 | 0.008654124 | 1 |
| Heart_Failure | b_cell | *AAMDC* | -1.676487699 | 0.001131925 | 0.008654124 | 1 |
| Heart_Failure | b_cell | *GP6* | 7.73384409 | 0.006457887 | 0.01602239 | 1 |
| Heart_Failure | b_cell | *DDX18* | -1.816577238 | 0.012683122 | 0.025700288 | 1 |
| Heart_Failure | b_cell | *ASAP3* | -1.846198251 | 0.002293267 | 0.009185338 | 1 |
| Heart_Failure | b_cell | *DOCK3* | -1.584261848 | 0.000622378 | 0.006609617 | 1 |
| Heart_Failure | b_cell | *C3orf18* | 2.853862222 | 2.40E-05 | 0.002889125 | 0.150848958 |
| Heart_Failure | b_cell | *ZNF343* | -2.364878924 | 0.00525675 | 0.014066038 | 1 |
| Heart_Failure | b_cell | *MAVS* | -2.051891485 | 0.022516615 | 0.03940291 | 1 |
| Heart_Failure | b_cell | *TESC* | -2.736684786 | 0.001645671 | 0.008654124 | 1 |
| Heart_Failure | b_cell | *DZANK1* | -2.517289193 | 0.00394108 | 0.011883725 | 1 |
| Heart_Failure | b_cell | *OAS1* | 6.768761688 | 0.001994583 | 0.008775328 | 1 |
| Heart_Failure | b_cell | *KIF16B* | -1.763006825 | 0.013338227 | 0.026710527 | 1 |
| Heart_Failure | b_cell | *GANAB* | -2.221616941 | 0.02368309 | 0.040961647 | 1 |
| Heart_Failure | b_cell | *GMIP* | -3.394102416 | 0.010875187 | 0.02302693 | 1 |
| Heart_Failure | b_cell | *NECAP1* | 6.045925507 | 0.001610218 | 0.008654124 | 1 |
| Heart_Failure | b_cell | *ANKRD24* | -2.37362194 | 0.009878232 | 0.021422714 | 1 |
| Heart_Failure | b_cell | *SLC9A1* | -1.833183428 | 0.001786488 | 0.008654124 | 1 |
| Heart_Failure | b_cell | *RGS1* | -2.930942999 | 0.001903518 | 0.008705907 | 1 |
| Heart_Failure | b_cell | *STRN4* | 5.870159481 | 0.003534185 | 0.011199305 | 1 |
| Heart_Failure | b_cell | *MUL1* | -2.712948749 | 0.019563003 | 0.035424135 | 1 |
| Heart_Failure | b_cell | *GNPTG* | -2.365845202 | 0.023144326 | 0.040260961 | 1 |
| Heart_Failure | b_cell | *ZNF268* | -2.396466923 | 0.003467367 | 0.011112537 | 1 |
| Heart_Failure | b_cell | *LAMB1* | -1.460763438 | 0.001195894 | 0.008654124 | 1 |
| Heart_Failure | b_cell | *SLC26A4* | -3.810161426 | 0.002096169 | 0.008886458 | 1 |
| Heart_Failure | b_cell | *DLD* | -2.109279151 | 0.014034167 | 0.027754893 | 1 |
| Heart_Failure | b_cell | *IL5RA* | -5.171725229 | 0.001760819 | 0.008654124 | 1 |
| Heart_Failure | b_cell | *ITGA6* | -1.396101391 | 0.001367589 | 0.008654124 | 1 |
| Heart_Failure | b_cell | *RAPGEF4* | -1.233000014 | 0.001828392 | 0.008654124 | 1 |
| Heart_Failure | b_cell | *SEL1L3* | 0.792791077 | 0.000143796 | 0.003239931 | 0.904042785 |
| Heart_Failure | b_cell | *TF* | -2.247457315 | 0.002019619 | 0.008817602 | 1 |
| Heart_Failure | b_cell | *MYO15A* | -2.678546444 | 0.006390463 | 0.015942634 | 1 |
| Heart_Failure | b_cell | *ALKBH5* | -2.708402084 | 0.002543556 | 0.009545808 | 1 |
| Heart_Failure | b_cell | *ZFHX4* | -1.170857222 | 0.001489718 | 0.008654124 | 1 |
| Heart_Failure | b_cell | *RGS17* | -1.967790428 | 0.025341893 | 0.043177366 | 1 |
| Heart_Failure | b_cell | *TMEM101* | -2.795553052 | 0.001272974 | 0.008654124 | 1 |
| Heart_Failure | b_cell | *CMA1* | -7.31176437 | 0.004103697 | 0.012159073 | 1 |

| | | | | | | |
|---|---|---|---|---|---|---|
| Heart_Failure | b_cell | *HAUS4* | 3.942342931 | 0.000562794 | 0.006251391 | 1 |
| Heart_Failure | b_cell | *SLC22A17* | -2.894328555 | 0.008060996 | 0.018566285 | 1 |
| Heart_Failure | b_cell | *RPGRIP1* | -2.925505532 | 0.00589742 | 0.01518308 | 1 |
| Heart_Failure | b_cell | *TGM1* | -3.500558124 | 0.00477188 | 0.013264085 | 1 |
| Heart_Failure | b_cell | *RFFL* | -2.922734543 | 0.00525683 | 0.014066038 | 1 |
| Heart_Failure | b_cell | *DPYSL2* | -1.603549001 | 0.004923598 | 0.01349375 | 1 |
| Heart_Failure | b_cell | *COMT* | -2.096512823 | 0.023443419 | 0.040650211 | 1 |
| Heart_Failure | b_cell | *VNN3* | 2.252912572 | 0.0001499 | 0.003239931 | 0.942422326 |
| Heart_Failure | b_cell | *FMO2* | -1.240601528 | 0.003116936 | 0.010473639 | 1 |
| Heart_Failure | b_cell | *PDE6C* | 5.49355068 | 0.000129214 | 0.003185748 | 0.812365715 |
| Heart_Failure | b_cell | *IKZF5* | -2.338054504 | 0.024322941 | 0.041826677 | 1 |
| Heart_Failure | b_cell | *BLNK* | 3.699660885 | 0.00013814 | 0.003214256 | 0.868488233 |
| Heart_Failure | b_cell | *CREM* | -1.826556382 | 0.015520691 | 0.029840545 | 1 |
| Heart_Failure | b_cell | *TPSD1* | -17.24370671 | 0.001941015 | 0.008738197 | 1 |
| Heart_Failure | b_cell | *ITPR3* | -1.628918245 | 0.001645239 | 0.008654124 | 1 |
| Heart_Failure | b_cell | *DSP* | -3.271975779 | 0.005817624 | 0.015057803 | 1 |
| Heart_Failure | b_cell | *IFT74* | -2.145663044 | 0.004646267 | 0.013040864 | 1 |
| Heart_Failure | b_cell | *IL12RB1* | 4.020084739 | 0.001071211 | 0.008654124 | 1 |
| Heart_Failure | b_cell | *ACOT7* | -2.512305939 | 0.003146681 | 0.010534175 | 1 |
| Heart_Failure | b_cell | *PCSK5* | -1.146424012 | 0.002521369 | 0.009509206 | 1 |
| Heart_Failure | b_cell | *RAB18* | -2.055447869 | 0.006443716 | 0.016018516 | 1 |
| Heart_Failure | b_cell | *NRP1* | -1.384055549 | 0.023784463 | 0.041114358 | 1 |
| Heart_Failure | b_cell | *PRTFDC1* | -1.839836608 | 0.001467053 | 0.008654124 | 1 |
| Heart_Failure | b_cell | *PALMD* | -0.991669918 | 0.00182186 | 0.008654124 | 1 |
| Heart_Failure | b_cell | *TSPAN15* | -1.374736603 | 0.001667675 | 0.008654124 | 1 |
| Heart_Failure | b_cell | *STX1B* | -3.461592081 | 0.004753616 | 0.01322974 | 1 |
| Heart_Failure | b_cell | *HSD3B7* | -1.890190551 | 0.011533562 | 0.023954908 | 1 |
| Heart_Failure | b_cell | *ATP5D* | 5.379760473 | 0.018378174 | 0.033735352 | 1 |
| Heart_Failure | b_cell | *MARCH2* | -1.908925452 | 0.021709577 | 0.038307075 | 1 |
| Heart_Failure | b_cell | *CDC34* | -2.200288162 | 0.00166109 | 0.008654124 | 1 |
| Heart_Failure | b_cell | *MTAP* | -2.530069274 | 0.013426643 | 0.026825962 | 1 |
| Heart_Failure | b_cell | *POLR2E* | -1.756551956 | 0.001542729 | 0.008654124 | 1 |
| Heart_Failure | b_cell | *TRMT2A* | -2.763269749 | 0.001939909 | 0.008738197 | 1 |
| Heart_Failure | b_cell | *ZDHHC8* | -2.215885366 | 0.000903983 | 0.008457357 | 1 |
| Heart_Failure | b_cell | *MED15* | -1.782395831 | 0.026059031 | 0.04410238 | 1 |
| Heart_Failure | b_cell | *LZTR1* | -2.588498626 | 0.015698273 | 0.030117499 | 1 |
| Heart_Failure | b_cell | *CECR2* | -1.379934827 | 0.006219801 | 0.015685476 | 1 |
| Heart_Failure | b_cell | *P2RX6* | -2.816156406 | 0.002653702 | 0.009688632 | 1 |
| Heart_Failure | b_cell | *SLC7A4* | -7.428084472 | 0.008810304 | 0.019702926 | 1 |
| Heart_Failure | b_cell | *DDTL* | -2.848601964 | 0.009639108 | 0.021027437 | 1 |
| Heart_Failure | b_cell | *GGT5* | -2.150709176 | 0.001518925 | 0.008654124 | 1 |
| Heart_Failure | b_cell | *PRODH* | -25.10880455 | 0.002048392 | 0.008831917 | 1 |
| Heart_Failure | b_cell | *PPM1F* | 4.70450984 | 0.015708092 | 0.030119222 | 1 |
| Heart_Failure | b_cell | *TOP3B* | -2.150565011 | 0.00304352 | 0.010326287 | 1 |
| Heart_Failure | b_cell | *CRYBB3* | -6.117109804 | 0.003403487 | 0.010985835 | 1 |
| Heart_Failure | b_cell | *CYTH4* | -2.305614111 | 0.009355174 | 0.020514817 | 1 |
| Heart_Failure | b_cell | *CARD10* | -2.941192771 | 0.00147701 | 0.008654124 | 1 |
| Heart_Failure | b_cell | *SLC25A1* | -2.145763176 | 0.003281842 | 0.010813912 | 1 |
| Heart_Failure | b_cell | *LGALS2* | 4.286987396 | 9.29E-05 | 0.002932166 | 0.584102562 |
| Heart_Failure | b_cell | *GGA1* | -1.920797145 | 0.003154379 | 0.010554127 | 1 |

| | | | | | | |
|---|---|---|---|---|---|---|
| Heart_Failure | b_cell | *SH3BP1* | -2.730038003 | 0.001701342 | 0.008654124 | 1 |
| Heart_Failure | b_cell | *HPS4* | -2.150095684 | 0.016565224 | 0.031265555 | 1 |
| Heart_Failure | b_cell | *TFIP11* | 4.759349793 | 0.007862217 | 0.01824361 | 1 |
| Heart_Failure | b_cell | *GCAT* | -3.492330443 | 0.002617913 | 0.009647609 | 1 |
| Heart_Failure | b_cell | *CCDC134* | 7.5803514 | 4.55E-05 | 0.002889125 | 0.285974257 |
| Heart_Failure | b_cell | *PICK1* | -2.422522725 | 0.010878356 | 0.02302693 | 1 |
| Heart_Failure | b_cell | *SLC16A8* | -5.431955408 | 0.000722662 | 0.007316224 | 1 |
| Heart_Failure | b_cell | *KDELR3* | -2.45628249 | 0.00089843 | 0.008433282 | 1 |
| Heart_Failure | b_cell | *CYP2D6* | 2.746383992 | 0.018915213 | 0.034553614 | 1 |
| Heart_Failure | b_cell | *POLDIP3* | -2.045681586 | 0.003080681 | 0.010370986 | 1 |
| Heart_Failure | b_cell | *TIMP3* | -1.254096807 | 0.00588434 | 0.015168039 | 1 |
| Heart_Failure | b_cell | *CYB5R3* | -1.973940386 | 0.015824918 | 0.030235497 | 1 |
| Heart_Failure | b_cell | *RHBDD3* | -2.020089623 | 0.001554597 | 0.008654124 | 1 |
| Heart_Failure | b_cell | *RASL10A* | -3.459040818 | 0.002625773 | 0.009653089 | 1 |
| Heart_Failure | b_cell | *AP1B1* | 4.426890632 | 0.009102782 | 0.020122782 | 1 |
| Heart_Failure | b_cell | *TOM1* | -1.729451229 | 0.001062798 | 0.008654124 | 1 |
| Heart_Failure | b_cell | *NEFH* | 5.000992785 | 9.37E-05 | 0.002932183 | 0.589368869 |
| Heart_Failure | b_cell | *BIK* | -10.110464 | 0.003474187 | 0.011121289 | 1 |
| Heart_Failure | b_cell | *MCAT* | -2.959845486 | 0.003739856 | 0.011555523 | 1 |
| Heart_Failure | b_cell | *THOC5* | 5.327780624 | 0.009150534 | 0.020188366 | 1 |
| Heart_Failure | b_cell | *ARSA* | -1.650029356 | 0.003322511 | 0.01083435 | 1 |
| Heart_Failure | b_cell | *TTLL12* | -2.75129467 | 0.002185391 | 0.009035107 | 1 |
| Heart_Failure | b_cell | *RBFOX2* | -1.439279184 | 0.007487005 | 0.017675855 | 1 |
| Heart_Failure | b_cell | *SAMM50* | -1.757236264 | 0.001414547 | 0.008654124 | 1 |
| Heart_Failure | b_cell | *IFT27* | -2.274039734 | 0.003074267 | 0.01036364 | 1 |
| Heart_Failure | b_cell | *NCF4* | -2.291842998 | 0.000719975 | 0.007316224 | 1 |
| Heart_Failure | b_cell | *KCTD17* | -6.19871262 | 0.006032989 | 0.015368478 | 1 |
| Heart_Failure | b_cell | *ACO2* | -2.083190985 | 0.010526497 | 0.022472017 | 1 |
| Heart_Failure | b_cell | *ZBED4* | -2.390732693 | 0.027040919 | 0.045444067 | 1 |
| Heart_Failure | b_cell | *HDAC10* | -3.041839496 | 0.016099472 | 0.030579269 | 1 |
| Heart_Failure | b_cell | *RBM23* | -2.202275385 | 0.01580904 | 0.030228539 | 1 |
| Heart_Failure | b_cell | *VCPKMT* | -2.450519602 | 0.016514225 | 0.031210659 | 1 |
| Heart_Failure | b_cell | *PYGL* | -0.975811651 | 0.005416517 | 0.014338377 | 1 |
| Heart_Failure | b_cell | *CNIH1* | -2.538742227 | 0.023021525 | 0.040104275 | 1 |
| Heart_Failure | b_cell | *ATP6V1D* | -1.568229716 | 0.00263353 | 0.009653089 | 1 |
| Heart_Failure | b_cell | *PIGH* | 4.914879564 | 0.012405609 | 0.025289905 | 1 |
| Heart_Failure | b_cell | *GSTZ1* | -1.979353228 | 0.007962828 | 0.018398493 | 1 |
| Heart_Failure | b_cell | *DAAM1* | -1.295643606 | 0.007599848 | 0.017835103 | 1 |
| Heart_Failure | b_cell | *ISM2* | 8.007484278 | 1.87E-05 | 0.002889125 | 0.117519382 |
| Heart_Failure | b_cell | *SLC10A1* | -1.954909938 | 0.001152017 | 0.008654124 | 1 |
| Heart_Failure | b_cell | *DICER1* | -1.754874827 | 0.026779285 | 0.045088742 | 1 |
| Heart_Failure | b_cell | *MTHFD1* | -1.41555395 | 0.001725991 | 0.008654124 | 1 |
| Heart_Failure | b_cell | *CCNB1IP1* | -2.052851759 | 0.027737961 | 0.046355279 | 1 |
| Heart_Failure | b_cell | *TRIP11* | -1.756047772 | 0.018649428 | 0.034163447 | 1 |
| Heart_Failure | b_cell | *KIAA0391* | 10.76690093 | 0.000232291 | 0.003705045 | 1 |
| Heart_Failure | b_cell | *EMC9* | 4.356619345 | 0.005443601 | 0.014350989 | 1 |
| Heart_Failure | b_cell | *PLTP* | -1.388141612 | 0.010848036 | 0.023000028 | 1 |
| Heart_Failure | b_cell | *GSS* | 7.084118644 | 0.000121946 | 0.003135985 | 0.7666765 |
| Heart_Failure | b_cell | *PYGB* | -1.617946671 | 0.007312574 | 0.017394685 | 1 |
| Heart_Failure | b_cell | *PROCR* | -1.565612369 | 0.002614176 | 0.009639485 | 1 |

| | | | | | | |
|---|---|---|---|---|---|---|
| Heart_Failure | b_cell | *NINL* | -1.646958365 | 0.005402235 | 0.014312623 | 1 |
| Heart_Failure | b_cell | *CD40* | 6.358406013 | 0.000171453 | 0.003365162 | 1 |
| Heart_Failure | b_cell | *IFT52* | -2.408919166 | 0.016776878 | 0.031476046 | 1 |
| Heart_Failure | b_cell | *DOK5* | -1.984605407 | 0.002971465 | 0.01018071 | 1 |
| Heart_Failure | b_cell | *CSTF1* | -3.500610454 | 0.009022336 | 0.020008264 | 1 |
| Heart_Failure | b_cell | *BMP7* | -2.621415523 | 0.001660382 | 0.008654124 | 1 |
| Heart_Failure | b_cell | *TPD52L2* | -2.025264449 | 0.002346684 | 0.009261519 | 1 |
| Heart_Failure | b_cell | *PRPF6* | -2.29138302 | 0.005068623 | 0.013800966 | 1 |
| Heart_Failure | b_cell | *SLC17A9* | 13.45907143 | 9.59E-07 | 0.000602686 | 0.006026865 |
| Heart_Failure | b_cell | *BIRC7* | 1.97851461 | 0.006083205 | 0.015452569 | 1 |
| Heart_Failure | b_cell | *NKAIN4* | -5.982089831 | 0.003403914 | 0.010985835 | 1 |
| Heart_Failure | b_cell | *PTK6* | 1.523056688 | 0.000166984 | 0.00335409 | 1 |
| Heart_Failure | b_cell | *NDUFAF5* | -1.962742329 | 0.000191246 | 0.003445165 | 1 |
| Heart_Failure | b_cell | *TRIB3* | -3.59400746 | 0.001598362 | 0.008654124 | 1 |
| Heart_Failure | b_cell | *ANGPT4* | -2.865639755 | 0.003063994 | 0.01036364 | 1 |
| Heart_Failure | b_cell | *RSPO4* | 4.064086456 | 0.000149646 | 0.003239931 | 0.940825475 |
| Heart_Failure | b_cell | *SIRPB1* | 1.159783138 | 0.000415743 | 0.005049082 | 1 |
| Heart_Failure | b_cell | *SEC23B* | -1.741057598 | 0.016694448 | 0.031377576 | 1 |
| Heart_Failure | b_cell | *CCM2L* | -2.485606377 | 0.001583087 | 0.008654124 | 1 |
| Heart_Failure | b_cell | *IDH3B* | 5.81688201 | 0.024038807 | 0.041473509 | 1 |
| Heart_Failure | b_cell | *MAPRE1* | 5.537194893 | 0.010343066 | 0.02216321 | 1 |
| Heart_Failure | b_cell | *RPRD1B* | -1.947223963 | 0.007919202 | 0.018343665 | 1 |
| Heart_Failure | b_cell | *ACTR5* | -2.446448222 | 0.002741048 | 0.009847411 | 1 |
| Heart_Failure | b_cell | *WFDC2* | 6.287390306 | 0.000760413 | 0.007600503 | 1 |
| Heart_Failure | b_cell | *MAP1LC3A* | -2.19853197 | 0.015185905 | 0.029430883 | 1 |
| Heart_Failure | b_cell | *APMAP* | -1.590042199 | 0.003081438 | 0.010370986 | 1 |
| Heart_Failure | b_cell | *CELF4* | -2.319548729 | 0.001549944 | 0.008654124 | 1 |
| Heart_Failure | b_cell | *RBFA* | 5.886702507 | 0.008849627 | 0.019736647 | 1 |
| Heart_Failure | b_cell | *LIPG* | -6.914572423 | 0.007311434 | 0.017394685 | 1 |
| Heart_Failure | b_cell | *NALCN* | -2.139918444 | 0.002401061 | 0.009333474 | 1 |
| Heart_Failure | b_cell | *HTR2A* | -5.61767831 | 0.001673159 | 0.008654124 | 1 |
| Heart_Failure | b_cell | *KLF5* | -3.351974828 | 0.003599771 | 0.01134424 | 1 |
| Heart_Failure | b_cell | *UGGT2* | -1.551600645 | 0.013226411 | 0.026557652 | 1 |
| Heart_Failure | b_cell | *ARHGEF7* | -1.586527476 | 0.024379329 | 0.041889271 | 1 |
| Heart_Failure | b_cell | *SGCG* | -1.424082859 | 0.001423612 | 0.008654124 | 1 |
| Heart_Failure | b_cell | *RGCC* | -1.003882972 | 0.007685119 | 0.017987759 | 1 |
| Heart_Failure | b_cell | *VWA8* | -1.577713378 | 0.020272183 | 0.036321235 | 1 |
| Heart_Failure | b_cell | *KATNAL1* | -1.768428509 | 0.015979502 | 0.030392508 | 1 |
| Heart_Failure | b_cell | *DHRS12* | -1.654006297 | 0.001986637 | 0.008775328 | 1 |
| Heart_Failure | b_cell | *MEDAG* | -1.572393298 | 0.001586155 | 0.008654124 | 1 |
| Heart_Failure | b_cell | *TSC22D1* | -1.695444494 | 0.024249842 | 0.041712382 | 1 |
| Heart_Failure | b_cell | *MGRN1* | -1.581897719 | 0.017729631 | 0.032880882 | 1 |
| Heart_Failure | b_cell | *CORO1A* | 3.12242359 | 0.010670212 | 0.022717109 | 1 |
| Heart_Failure | b_cell | *GDPD3* | -2.572537048 | 0.001170836 | 0.008654124 | 1 |
| Heart_Failure | b_cell | *NFAT5* | -1.407020051 | 0.026295565 | 0.044458272 | 1 |
| Heart_Failure | b_cell | *PLLP* | -3.692581449 | 0.004319579 | 0.012520606 | 1 |
| Heart_Failure | b_cell | *ACD* | -3.106764412 | 0.006699912 | 0.016416518 | 1 |
| Heart_Failure | b_cell | *SETD6* | -2.077638889 | 0.002631271 | 0.009653089 | 1 |
| Heart_Failure | b_cell | *COG4* | -2.049786715 | 0.007959256 | 0.018397 | 1 |
| Heart_Failure | b_cell | *SLC7A6OS* | -1.088057732 | 0.002547007 | 0.009545808 | 1 |

| | | | | | | |
|---|---|---|---|---|---|---|
| Heart_Failure | b_cell | *HCFC1R1* | -2.27291473 | 0.001691381 | 0.008654124 | 1 |
| Heart_Failure | b_cell | *NPRL3* | -1.624175281 | 0.021849509 | 0.038510754 | 1 |
| Heart_Failure | b_cell | *TAF1C* | 2.918580675 | 0.005674318 | 0.014778142 | 1 |
| Heart_Failure | b_cell | *NAGPA* | -2.953657418 | 0.008801028 | 0.019698135 | 1 |
| Heart_Failure | b_cell | *CRISPLD2* | -1.675134446 | 0.002379521 | 0.009318117 | 1 |
| Heart_Failure | b_cell | *NME4* | -2.329623162 | 0.002242681 | 0.009120139 | 1 |
| Heart_Failure | b_cell | *NOMO3* | -2.871012874 | 0.003648295 | 0.011427162 | 1 |
| Heart_Failure | b_cell | *HAGHL* | 8.718004718 | 0.000194297 | 0.003449722 | 1 |
| Heart_Failure | b_cell | *SLC7A5* | -7.713310704 | 0.014247786 | 0.028053814 | 1 |
| Heart_Failure | b_cell | *FBXO31* | -1.812886281 | 0.008537119 | 0.019314107 | 1 |
| Heart_Failure | b_cell | *NUBP1* | 5.841971574 | 0.007196614 | 0.017242802 | 1 |
| Heart_Failure | b_cell | *UBE2I* | -1.994901797 | 0.009301098 | 0.020420316 | 1 |
| Heart_Failure | b_cell | *EARS2* | -1.768374673 | 0.004803251 | 0.013301535 | 1 |
| Heart_Failure | b_cell | *BFAR* | -2.232489893 | 0.011276085 | 0.023636218 | 1 |
| Heart_Failure | b_cell | *STX4* | 6.248862481 | 5.10E-05 | 0.002889125 | 0.320359278 |
| Heart_Failure | b_cell | *NOMO1* | -2.361750287 | 0.001799461 | 0.008654124 | 1 |
| Heart_Failure | b_cell | *SYT17* | -1.855071717 | 0.00245039 | 0.009416629 | 1 |
| Heart_Failure | b_cell | *TMC5* | -2.8891423 | 0.001636447 | 0.008654124 | 1 |
| Heart_Failure | b_cell | *RNF40* | 5.781212081 | 0.002516798 | 0.009509081 | 1 |
| Heart_Failure | b_cell | *IQCH* | -1.933439705 | 0.005847501 | 0.015110251 | 1 |
| Heart_Failure | b_cell | *LACTB* | -2.213225903 | 0.010324746 | 0.022154156 | 1 |
| Heart_Failure | b_cell | *CSK* | 6.329434038 | 0.00593191 | 0.01524067 | 1 |
| Heart_Failure | b_cell | *RASL12* | -3.065743718 | 0.002062232 | 0.008831917 | 1 |
| Heart_Failure | b_cell | *AP3B2* | 1.709474189 | 0.001351246 | 0.008654124 | 1 |
| Heart_Failure | b_cell | *CTSH* | -2.135990722 | 0.027072002 | 0.045484147 | 1 |
| Heart_Failure | b_cell | *TTC23* | -2.034466261 | 0.01269315 | 0.025709354 | 1 |
| Heart_Failure | b_cell | *FAH* | -1.629810367 | 0.001680886 | 0.008654124 | 1 |
| Heart_Failure | b_cell | *RPAP1* | -1.995650514 | 0.001442147 | 0.008654124 | 1 |
| Heart_Failure | b_cell | *EHD4* | -2.154532156 | 0.009957601 | 0.021572514 | 1 |
| Heart_Failure | b_cell | *ATP8B4* | -1.229451573 | 0.00171858 | 0.008654124 | 1 |
| Heart_Failure | b_cell | *DTWD1* | -1.537997229 | 0.009818809 | 0.021309791 | 1 |
| Heart_Failure | b_cell | *DNAJC17* | -2.203532885 | 0.013429187 | 0.026825962 | 1 |
| Heart_Failure | b_cell | *PDGFRL* | -1.531943345 | 0.002475329 | 0.009455883 | 1 |
| Heart_Failure | b_cell | *ZFAND1* | -1.868829611 | 0.003567771 | 0.011288665 | 1 |
| Heart_Failure | b_cell | *EYA1* | -1.618037248 | 0.0048384 | 0.013365122 | 1 |
| Heart_Failure | b_cell | *NBN* | -2.000795424 | 0.026638603 | 0.044887581 | 1 |
| Heart_Failure | b_cell | *NIPAL2* | -1.62158912 | 0.007484066 | 0.017675553 | 1 |
| Heart_Failure | b_cell | *PLAT* | -1.684032799 | 0.00155096 | 0.008654124 | 1 |
| Heart_Failure | b_cell | *RAB2A* | -1.525618394 | 0.020159055 | 0.036177385 | 1 |
| Heart_Failure | b_cell | *ARMC1* | -1.498590423 | 0.009889626 | 0.021432636 | 1 |
| Heart_Failure | b_cell | *SPAG1* | -2.127597872 | 0.01323941 | 0.026559084 | 1 |
| Heart_Failure | b_cell | *CHRAC1* | -2.636400927 | 0.002669104 | 0.009716653 | 1 |
| Heart_Failure | b_cell | *SNX16* | -2.238324354 | 0.007742736 | 0.018055855 | 1 |
| Heart_Failure | b_cell | *TSTA3* | -4.065997751 | 0.013789828 | 0.027392307 | 1 |
| Heart_Failure | b_cell | *MTMR9* | -1.924906226 | 0.007065287 | 0.017040532 | 1 |
| Heart_Failure | b_cell | *LEPROTL1* | -2.34195132 | 0.028995523 | 0.048035535 | 1 |
| Heart_Failure | b_cell | *TNFRSF10A* | -1.606395157 | 0.00950175 | 0.020778262 | 1 |
| Heart_Failure | b_cell | *TUSC3* | -1.548757995 | 0.0016177 | 0.008654124 | 1 |
| Heart_Failure | b_cell | *ARHGEF10* | -1.814342329 | 0.000588164 | 0.006397557 | 1 |
| Heart_Failure | b_cell | *MCM4* | -3.121021327 | 0.014243624 | 0.028053814 | 1 |

| | | | | | | |
|---|---|---|---|---|---|---|
| Heart_Failure | b_cell | *MAN2B1* | -1.644007326 | 0.024668867 | 0.042247613 | 1 |
| Heart_Failure | b_cell | *TULP2* | 4.417683161 | 0.000137966 | 0.003214256 | 0.867391814 |
| Heart_Failure | b_cell | *NUCB1* | -2.24207699 | 0.014007196 | 0.027720418 | 1 |
| Heart_Failure | b_cell | *TUBB4A* | -19.92199209 | 0.016383828 | 0.031016299 | 1 |
| Heart_Failure | b_cell | *PPP1R37* | 6.828377534 | 0.00778556 | 0.018115668 | 1 |
| Heart_Failure | b_cell | *ERCC2* | -3.869820687 | 0.008429516 | 0.019125357 | 1 |
| Heart_Failure | b_cell | *PLEKHJ1* | -2.705355709 | 0.011035223 | 0.023257944 | 1 |
| Heart_Failure | b_cell | *CD37* | 0.755612116 | 0.000122137 | 0.003135985 | 0.767875387 |
| Heart_Failure | b_cell | *DKKL1* | -4.745718099 | 0.022078616 | 0.038805775 | 1 |
| Heart_Failure | b_cell | *STX10* | 6.40562116 | 0.000590897 | 0.006416186 | 1 |
| Heart_Failure | b_cell | *FCER2* | 0.808464915 | 0.000171343 | 0.003365162 | 1 |
| Heart_Failure | b_cell | *IL4I1* | -2.858049274 | 0.002910397 | 0.010125993 | 1 |
| Heart_Failure | b_cell | *CCDC61* | 4.799222222 | 0.011647355 | 0.024119539 | 1 |
| Heart_Failure | b_cell | *OLFM2* | -1.447702501 | 0.004288381 | 0.012447393 | 1 |
| Heart_Failure | b_cell | *RASAL3* | -2.682982674 | 0.001354094 | 0.008654124 | 1 |
| Heart_Failure | b_cell | *SYDE1* | -2.898757113 | 0.001972073 | 0.008775328 | 1 |
| Heart_Failure | b_cell | *AURKC* | 1.543064888 | 0.000467664 | 0.005414745 | 1 |
| Heart_Failure | b_cell | *CCNE1* | -4.986442322 | 0.001301196 | 0.008654124 | 1 |
| Heart_Failure | b_cell | *ANKRD27* | -1.688549907 | 0.004192289 | 0.012283789 | 1 |
| Heart_Failure | b_cell | *PRX* | 3.566431319 | 4.57E-05 | 0.002889125 | 0.287138741 |
| Heart_Failure | b_cell | *CCDC94* | -3.859892083 | 0.00835987 | 0.019042936 | 1 |
| Heart_Failure | b_cell | *POLR2I* | -3.293953606 | 0.019145629 | 0.034876985 | 1 |
| Heart_Failure | b_cell | *CLIP3* | -1.959097365 | 0.006030551 | 0.015368478 | 1 |
| Heart_Failure | b_cell | *SLC1A5* | -2.021128769 | 0.000907704 | 0.008476467 | 1 |
| Heart_Failure | b_cell | *CACTIN* | -2.859655376 | 0.001748557 | 0.008654124 | 1 |
| Heart_Failure | b_cell | *CCDC9* | 3.877218499 | 0.000138208 | 0.003214256 | 0.868915092 |
| Heart_Failure | b_cell | *FZR1* | 8.365535105 | 0.000121349 | 0.003135985 | 0.76292289 |
| Heart_Failure | b_cell | *TGFB1* | -2.162457123 | 0.030071028 | 0.049439475 | 1 |
| Heart_Failure | b_cell | *PLIN3* | -2.064642258 | 0.002306651 | 0.009188443 | 1 |
| Heart_Failure | b_cell | *MRPL4* | -2.602575364 | 0.015744328 | 0.03014386 | 1 |
| Heart_Failure | b_cell | *ICAM5* | -16.85487386 | 0.00203696 | 0.008831917 | 1 |
| Heart_Failure | b_cell | *CD33* | -2.178955724 | 0.015858909 | 0.030277851 | 1 |
| Heart_Failure | b_cell | *TYK2* | -2.621986885 | 0.026045203 | 0.04410238 | 1 |
| Heart_Failure | b_cell | *RABAC1* | -1.982364022 | 0.005156531 | 0.013901849 | 1 |
| Heart_Failure | b_cell | *PTPRS* | -1.217481113 | 0.003559518 | 0.011273899 | 1 |
| Heart_Failure | b_cell | *CYTH2* | 7.490319848 | 0.001295762 | 0.008654124 | 1 |
| Heart_Failure | b_cell | *GRIN2D* | -2.788176624 | 0.001621051 | 0.008654124 | 1 |
| Heart_Failure | b_cell | *LIG1* | 2.205362027 | 9.07E-05 | 0.002925789 | 0.570528819 |
| Heart_Failure | b_cell | *PLA2G4C* | -2.395229451 | 0.004820836 | 0.01333878 | 1 |
| Heart_Failure | b_cell | *GCDH* | -1.873627853 | 0.001297139 | 0.008654124 | 1 |
| Heart_Failure | b_cell | *LILRB5* | -1.536863403 | 0.001706097 | 0.008654124 | 1 |
| Heart_Failure | b_cell | *MAST1* | -5.778653218 | 0.001712104 | 0.008654124 | 1 |
| Heart_Failure | b_cell | *PRPF31* | -2.725759452 | 0.013236893 | 0.026559084 | 1 |
| Heart_Failure | b_cell | *TFPT* | 6.101271329 | 0.001068532 | 0.008654124 | 1 |
| Heart_Failure | b_cell | *ELL* | -1.876445056 | 0.005496458 | 0.014464726 | 1 |
| Heart_Failure | b_cell | *UPK1A* | -11.58313702 | 0.000563901 | 0.006252638 | 1 |
| Heart_Failure | b_cell | *COPE* | 5.476066602 | 0.000108603 | 0.003034612 | 0.682787699 |
| Heart_Failure | b_cell | *LSR* | -2.513123903 | 0.000262298 | 0.003832809 | 1 |
| Heart_Failure | b_cell | *KXD1* | -2.282115027 | 0.003507336 | 0.011157849 | 1 |
| Heart_Failure | b_cell | *HPN* | -2.946908069 | 0.001208881 | 0.008654124 | 1 |

| | | | | | | |
|---|---|---|---|---|---|---|
| Heart_Failure | b_cell | *ZNF14* | 6.479589139 | 0.001058149 | 0.008654124 | 1 |
| Heart_Failure | b_cell | *SIPA1L3* | 3.97676366 | 0.000237366 | 0.003709453 | 1 |
| Heart_Failure | b_cell | *ZNF85* | 5.35853975 | 0.013814377 | 0.027432403 | 1 |
| Heart_Failure | b_cell | *SMG9* | -2.266488307 | 0.004571034 | 0.012898605 | 1 |
| Heart_Failure | b_cell | *PON3* | -2.708162969 | 0.001803614 | 0.008654124 | 1 |
| Heart_Failure | b_cell | *PON2* | -2.007404653 | 0.019016814 | 0.034704995 | 1 |
| Heart_Failure | b_cell | *HBP1* | -1.891748068 | 0.027422086 | 0.045921648 | 1 |
| Heart_Failure | b_cell | *WDR91* | -2.449780791 | 0.015166842 | 0.029403002 | 1 |
| Heart_Failure | b_cell | *DLX5* | 30.86576523 | 0.000576407 | 0.006331367 | 1 |
| Heart_Failure | b_cell | *STEAP1B* | 4.1348543 | 0.013640782 | 0.027139113 | 1 |
| Heart_Failure | b_cell | *PTN* | -2.65414989 | 0.001638637 | 0.008654124 | 1 |
| Heart_Failure | b_cell | *MPP6* | -1.730664411 | 0.003689662 | 0.011477934 | 1 |
| Heart_Failure | b_cell | *DFNA5* | -1.926960319 | 0.003014146 | 0.010270968 | 1 |
| Heart_Failure | b_cell | *ADAP1* | -6.713831788 | 0.011289064 | 0.023636218 | 1 |
| Heart_Failure | b_cell | *CAV1* | -1.126468558 | 0.002666976 | 0.009714529 | 1 |
| Heart_Failure | b_cell | *MET* | -1.532670655 | 0.001882359 | 0.008705907 | 1 |
| Heart_Failure | b_cell | *RNF32* | -1.931935096 | 0.002204482 | 0.009058938 | 1 |
| Heart_Failure | b_cell | *HOXA2* | -7.096247963 | 0.002110048 | 0.008921231 | 1 |
| Heart_Failure | b_cell | *HOXA5* | -1.917412459 | 0.000688214 | 0.007116449 | 1 |
| Heart_Failure | b_cell | *HOXA6* | -2.135410661 | 0.000464146 | 0.005392272 | 1 |
| Heart_Failure | b_cell | *HIBADH* | -1.542952028 | 0.027284459 | 0.045767715 | 1 |
| Heart_Failure | b_cell | *CPVL* | -1.561664254 | 0.003711357 | 0.011522617 | 1 |
| Heart_Failure | b_cell | *PLEKHA8* | -1.843039304 | 0.002719291 | 0.009828026 | 1 |
| Heart_Failure | b_cell | *CRHR2* | -11.70396477 | 0.005795644 | 0.015007089 | 1 |
| Heart_Failure | b_cell | *NPTX2* | -102.7660356 | 0.000364836 | 0.004752125 | 1 |
| Heart_Failure | b_cell | *CYP3A5* | -2.310346946 | 0.008266047 | 0.018897686 | 1 |
| Heart_Failure | b_cell | *ZKSCAN1* | -1.874763488 | 0.022444515 | 0.039336491 | 1 |
| Heart_Failure | b_cell | *EIF3B* | -2.20181386 | 0.011305528 | 0.023653196 | 1 |
| Heart_Failure | b_cell | *SNX8* | -2.219893425 | 0.028586544 | 0.047520783 | 1 |
| Heart_Failure | b_cell | *TAF6* | -2.728269303 | 0.001791119 | 0.008654124 | 1 |
| Heart_Failure | b_cell | *AIMP2* | -2.134578795 | 0.003256681 | 0.010781862 | 1 |
| Heart_Failure | b_cell | *RBM28* | -1.356568333 | 0.00121677 | 0.008654124 | 1 |
| Heart_Failure | b_cell | *USP42* | -2.044916044 | 0.011045408 | 0.02327161 | 1 |
| Heart_Failure | b_cell | *IMPDH1* | -2.801094241 | 0.013712078 | 0.027255085 | 1 |
| Heart_Failure | b_cell | *LSM5* | -2.567391776 | 0.008827663 | 0.019722642 | 1 |
| Heart_Failure | b_cell | *AP1S1* | -4.797973824 | 0.011739946 | 0.024247385 | 1 |
| Heart_Failure | b_cell | *C1GALT1* | 4.433766094 | 0.001225887 | 0.008654124 | 1 |
| Heart_Failure | b_cell | *MEST* | -1.198863895 | 0.002287459 | 0.009185338 | 1 |
| Heart_Failure | b_cell | *POU6F2* | -1.823960145 | 0.001689705 | 0.008654124 | 1 |
| Heart_Failure | b_cell | *TSPAN13* | 2.645812258 | 7.99E-05 | 0.002889125 | 0.502339718 |
| Heart_Failure | b_cell | *RARRES2* | -1.553545507 | 0.002147736 | 0.008999799 | 1 |
| Heart_Failure | b_cell | *TMEM176B* | -2.214124916 | 0.010951099 | 0.023134933 | 1 |
| Heart_Failure | b_cell | *URGCP* | -1.984678921 | 0.005232215 | 0.014033677 | 1 |
| Heart_Failure | b_cell | *POLD2* | -3.402960247 | 0.019149889 | 0.034876985 | 1 |
| Heart_Failure | b_cell | *GCK* | -26.65922395 | 0.001746611 | 0.008654124 | 1 |
| Heart_Failure | b_cell | *YKT6* | -2.277958047 | 0.002899994 | 0.010117265 | 1 |
| Heart_Failure | b_cell | *TBL2* | -2.059830755 | 0.015625549 | 0.030005445 | 1 |
| Heart_Failure | b_cell | *SPATA6L* | -1.838391227 | 0.001754128 | 0.008654124 | 1 |
| Heart_Failure | b_cell | *SLC1A1* | -1.243393672 | 0.007863875 | 0.01824361 | 1 |
| Heart_Failure | b_cell | *NMRK1* | -1.852937281 | 0.002948283 | 0.010151069 | 1 |

| | | | | | | |
|---|---|---|---|---|---|---|
| Heart_Failure | b_cell | *TRIM14* | -1.632078241 | 0.001395955 | 0.008654124 | 1 |
| Heart_Failure | b_cell | *CORO2A* | -2.363599764 | 0.001093509 | 0.008654124 | 1 |
| Heart_Failure | b_cell | *C5* | -1.695350516 | 0.00460917 | 0.012965482 | 1 |
| Heart_Failure | b_cell | *OGN* | -1.966355067 | 0.015376711 | 0.029672617 | 1 |
| Heart_Failure | b_cell | *ECM2* | -1.192712504 | 0.00262986 | 0.009653089 | 1 |
| Heart_Failure | b_cell | *PTGR1* | -1.277433203 | 0.010689104 | 0.022749627 | 1 |
| Heart_Failure | b_cell | *TNFSF8* | -2.130712178 | 0.00169441 | 0.008654124 | 1 |
| Heart_Failure | b_cell | *PLGRKT* | -1.602669895 | 0.021958731 | 0.038616655 | 1 |
| Heart_Failure | b_cell | *ELAVL2* | -4.814452885 | 0.000573839 | 0.006318264 | 1 |
| Heart_Failure | b_cell | *FUBP3* | -2.001136071 | 0.019362101 | 0.03517178 | 1 |
| Heart_Failure | b_cell | *MPDZ* | -1.333292118 | 0.002789539 | 0.009915204 | 1 |
| Heart_Failure | b_cell | *PTGDS* | -4.125938777 | 0.010873718 | 0.02302693 | 1 |
| Heart_Failure | b_cell | *DVL1* | -2.588964457 | 0.003513772 | 0.011161493 | 1 |
| Heart_Failure | b_cell | *PDLIM1* | -1.500625571 | 0.023763266 | 0.041089013 | 1 |
| Heart_Failure | b_cell | *ATRNL1* | -1.662236407 | 0.007243801 | 0.0173261 | 1 |
| Heart_Failure | b_cell | *PHYH* | -1.639170168 | 0.00777067 | 0.018094148 | 1 |
| Heart_Failure | b_cell | *DNMBP* | -1.695946731 | 0.005560057 | 0.014577179 | 1 |
| Heart_Failure | b_cell | *NSMCE4A* | -1.9914029 | 0.005437437 | 0.014347469 | 1 |
| Heart_Failure | b_cell | *SPOCK2* | -1.795445961 | 0.022319821 | 0.039164029 | 1 |
| Heart_Failure | b_cell | *BMPR1A* | -1.130150105 | 0.005491233 | 0.014457029 | 1 |
| Heart_Failure | b_cell | *SFXN3* | -2.594253359 | 0.015735173 | 0.03014386 | 1 |
| Heart_Failure | b_cell | *KAZALD1* | -5.450693981 | 0.00263696 | 0.009653089 | 1 |
| Heart_Failure | b_cell | *MAP3K8* | -1.855637514 | 0.027648431 | 0.046217943 | 1 |
| Heart_Failure | b_cell | *TFAM* | -2.490381505 | 0.006922702 | 0.016804258 | 1 |
| Heart_Failure | b_cell | *TADA2A* | -2.097603212 | 0.021041895 | 0.037349067 | 1 |
| Heart_Failure | b_cell | *DHRS11* | -2.225149961 | 0.002230257 | 0.009104951 | 1 |
| Heart_Failure | b_cell | *RAPGEFL1* | -2.520617531 | 0.001538284 | 0.008654124 | 1 |
| Heart_Failure | b_cell | *RGS9* | -2.647755176 | 0.001769554 | 0.008654124 | 1 |
| Heart_Failure | b_cell | *WNT3* | -4.496664661 | 0.00156575 | 0.008654124 | 1 |
| Heart_Failure | b_cell | *ASPA* | -1.360067982 | 0.001690635 | 0.008654124 | 1 |
| Heart_Failure | b_cell | *SEPT4* | 4.446780034 | 0.003406609 | 0.010988893 | 1 |
| Heart_Failure | b_cell | *RPS6KB1* | -1.995219345 | 0.026384817 | 0.044579775 | 1 |
| Heart_Failure | b_cell | *TRIM16L* | -1.97149617 | 0.00588032 | 0.015163893 | 1 |
| Heart_Failure | b_cell | *CDK5RAP3* | -2.691689974 | 0.001656282 | 0.008654124 | 1 |
| Heart_Failure | b_cell | *CBX1* | -2.282954513 | 0.003311276 | 0.01083435 | 1 |
| Heart_Failure | b_cell | *RECQL5* | -3.152619301 | 0.004110877 | 0.012159073 | 1 |
| Heart_Failure | b_cell | *PIGL* | -1.807405362 | 0.02121072 | 0.037592215 | 1 |
| Heart_Failure | b_cell | *GALK1* | -4.054657057 | 0.011432469 | 0.02381575 | 1 |
| Heart_Failure | b_cell | *HOXB6* | -1.909255429 | 0.003667854 | 0.01145544 | 1 |
| Heart_Failure | b_cell | *RNF167* | 5.674129977 | 0.013267031 | 0.026606004 | 1 |
| Heart_Failure | b_cell | *C1QBP* | -2.457007846 | 0.00227091 | 0.009180605 | 1 |
| Heart_Failure | b_cell | *GOSR1* | -1.839067096 | 0.013385172 | 0.026776033 | 1 |
| Heart_Failure | b_cell | *DRG2* | -2.028587403 | 0.001748107 | 0.008654124 | 1 |
| Heart_Failure | b_cell | *ALDH3A1* | 4.066167566 | 0.007721333 | 0.018026001 | 1 |
| Heart_Failure | b_cell | *SMARCD2* | -1.972343819 | 0.004716567 | 0.013183596 | 1 |
| Heart_Failure | b_cell | *ICAM2* | -1.744234225 | 0.002934395 | 0.010146595 | 1 |
| Heart_Failure | b_cell | *SYNGR2* | -2.98358561 | 0.001332315 | 0.008654124 | 1 |
| Heart_Failure | b_cell | *PEX12* | -5.884961654 | 4.42E-05 | 0.002889125 | 0.278080871 |
| Heart_Failure | b_cell | *DHX58* | -3.590800357 | 0.005024507 | 0.013710535 | 1 |
| Heart_Failure | b_cell | *RAB5C* | -2.148607735 | 0.011461119 | 0.023848672 | 1 |

| | | | | | | |
|---|---|---|---|---|---|---|
| Heart_Failure | b_cell | *NAGLU* | -1.820003385 | 0.003066061 | 0.01036364 | 1 |
| Heart_Failure | b_cell | *EZH1* | -1.909247263 | 0.019400437 | 0.035221065 | 1 |
| Heart_Failure | b_cell | *DLX4* | -24.95934227 | 0.001682452 | 0.008654124 | 1 |
| Heart_Failure | b_cell | *VAT1* | -1.988254348 | 0.001777573 | 0.008654124 | 1 |
| Heart_Failure | b_cell | *ALOX12* | -3.45592633 | 0.028631533 | 0.047570414 | 1 |
| Heart_Failure | b_cell | *ABCC3* | -1.784798485 | 0.022909401 | 0.039964319 | 1 |
| Heart_Failure | b_cell | *MPP2* | -3.806544054 | 0.001658861 | 0.008654124 | 1 |
| Heart_Failure | b_cell | *EFNB3* | 12.9515931 | 0.000154438 | 0.003239931 | 0.970951772 |
| Heart_Failure | b_cell | *FAM20A* | -1.715674608 | 0.006771204 | 0.016525838 | 1 |
| Heart_Failure | b_cell | *DPH1* | -3.204527975 | 0.004443176 | 0.012718545 | 1 |
| Heart_Failure | b_cell | *DHRS7B* | -1.85859703 | 0.011408615 | 0.023789705 | 1 |
| Heart_Failure | b_cell | *RCVRN* | -8.641697101 | 0.017190122 | 0.032098098 | 1 |
| Heart_Failure | b_cell | *MYH3* | -2.745044679 | 0.002666573 | 0.009714529 | 1 |
| Heart_Failure | b_cell | *TMEM104* | -2.979449876 | 0.008314879 | 0.018974446 | 1 |
| Heart_Failure | b_cell | *TMEM97* | 2.99666111 | 0.000177797 | 0.003387297 | 1 |
| Heart_Failure | b_cell | *RAB34* | -2.256936609 | 0.003586804 | 0.011326084 | 1 |
| Heart_Failure | b_cell | *CWH43* | -21.77007834 | 0.003450824 | 0.011091682 | 1 |
| Heart_Failure | b_cell | *KIAA1211* | 2.58489223 | 0.006623263 | 0.016294798 | 1 |
| Heart_Failure | b_cell | *AREG* | 2.351969915 | 0.000161898 | 0.003294027 | 1 |
| Heart_Failure | b_cell | *NDUFC1* | 6.362905988 | 0.000883419 | 0.008355409 | 1 |
| Heart_Failure | b_cell | *CPE* | -1.646051659 | 0.008483649 | 0.019227361 | 1 |
| Heart_Failure | b_cell | *WFS1* | -1.969849308 | 0.001983563 | 0.008775328 | 1 |
| Heart_Failure | b_cell | *GRPEL1* | -3.003051546 | 0.003295465 | 0.01083435 | 1 |
| Heart_Failure | b_cell | *AADAT* | -2.561761794 | 0.001740126 | 0.008654124 | 1 |
| Heart_Failure | b_cell | *CPZ* | -49.66344992 | 0.001727794 | 0.008654124 | 1 |
| Heart_Failure | b_cell | *TRIM2* | -1.428774921 | 0.007113787 | 0.017116105 | 1 |
| Heart_Failure | b_cell | *SLC2A9* | -1.793803482 | 0.007587521 | 0.017826138 | 1 |
| Heart_Failure | b_cell | *SH3D19* | -0.961390145 | 0.001646394 | 0.008654124 | 1 |
| Heart_Failure | b_cell | *LRP2BP* | -3.227633661 | 0.012178639 | 0.024946687 | 1 |
| Heart_Failure | b_cell | *FAM149A* | -1.564231861 | 0.002166925 | 0.009019688 | 1 |
| Heart_Failure | b_cell | *UGDH* | -2.030337198 | 0.002854311 | 0.010013685 | 1 |
| Heart_Failure | b_cell | *DDX25* | -6.534121719 | 0.016147417 | 0.030661073 | 1 |
| Heart_Failure | b_cell | *HTATIP2* | -2.469117895 | 0.014645778 | 0.028666876 | 1 |
| Heart_Failure | b_cell | *CTSC* | -1.740798334 | 0.001062699 | 0.008654124 | 1 |
| Heart_Failure | b_cell | *ELP4* | -1.720676725 | 0.022672194 | 0.039616477 | 1 |
| Heart_Failure | b_cell | *C11orf63* | -1.783434997 | 0.002934582 | 0.010146595 | 1 |
| Heart_Failure | b_cell | *PUS3* | -2.675844413 | 0.002777583 | 0.009913839 | 1 |
| Heart_Failure | b_cell | *DCPS* | 6.45151263 | 0.000866045 | 0.008300042 | 1 |
| Heart_Failure | b_cell | *FOXRED1* | -1.889584966 | 0.011373262 | 0.023747492 | 1 |
| Heart_Failure | b_cell | *MS4A4A* | -1.685750936 | 0.000570804 | 0.006303585 | 1 |
| Heart_Failure | b_cell | *CCDC86* | -6.031565428 | 0.001686005 | 0.008654124 | 1 |
| Heart_Failure | b_cell | *PRPF19* | -2.643251313 | 0.003075959 | 0.01036364 | 1 |
| Heart_Failure | b_cell | *PANX1* | -2.334766661 | 0.004453909 | 0.012718855 | 1 |
| Heart_Failure | b_cell | *GALNT18* | -1.5219639 | 0.000949555 | 0.008639437 | 1 |
| Heart_Failure | b_cell | *CBL* | -1.504779308 | 0.024674055 | 0.042247613 | 1 |
| Heart_Failure | b_cell | *KIAA1549L* | -3.87702365 | 0.001197947 | 0.008654124 | 1 |
| Heart_Failure | b_cell | *CD5* | 6.409471062 | 0.001395156 | 0.008654124 | 1 |
| Heart_Failure | b_cell | *ACCS* | -1.675776938 | 0.026051888 | 0.04410238 | 1 |
| Heart_Failure | b_cell | *NAA40* | -2.17291748 | 0.001440349 | 0.008654124 | 1 |
| Heart_Failure | b_cell | *SLC22A18* | -1.656411881 | 0.015535062 | 0.029859045 | 1 |

| | | | | | | |
|---|---|---|---|---|---|---|
| Heart_Failure | b_cell | *SLC35F2* | 3.672149515 | 0.02005026 | 0.036036587 | 1 |
| Heart_Failure | b_cell | *ELMOD1* | -5.20978334 | 0.006861981 | 0.016689532 | 1 |
| Heart_Failure | b_cell | *RPS13* | 4.619252829 | 0.003006999 | 0.010257735 | 1 |
| Heart_Failure | b_cell | *AIP* | 9.650807874 | 0.000142324 | 0.003237933 | 0.894792627 |
| Heart_Failure | b_cell | *TCIRG1* | -2.47416183 | 0.01725331 | 0.032158778 | 1 |
| Heart_Failure | b_cell | *PPFIBP1* | -1.635490361 | 0.015025913 | 0.029189286 | 1 |
| Heart_Failure | b_cell | *COQ5* | -1.391850256 | 0.001112609 | 0.008654124 | 1 |
| Heart_Failure | b_cell | *KCTD10* | -2.538902989 | 0.000316323 | 0.00432146 | 1 |
| Heart_Failure | b_cell | *CYP27B1* | 2.986244254 | 0.000195985 | 0.003449722 | 1 |
| Heart_Failure | b_cell | *ACSS3* | -0.992419153 | 0.002967866 | 0.010176497 | 1 |
| Heart_Failure | b_cell | *GLI1* | -4.357974794 | 0.001745025 | 0.008654124 | 1 |
| Heart_Failure | b_cell | *TRPV4* | -5.63300289 | 0.015952771 | 0.030375928 | 1 |
| Heart_Failure | b_cell | *VPS29* | -1.727394517 | 0.006165895 | 0.015568266 | 1 |
| Heart_Failure | b_cell | *KCNA1* | -6.418373448 | 0.001416001 | 0.008654124 | 1 |
| Heart_Failure | b_cell | *CREBL2* | -1.851951739 | 0.006514945 | 0.016123569 | 1 |
| Heart_Failure | b_cell | *LTBR* | -1.930232452 | 0.028678767 | 0.04763453 | 1 |
| Heart_Failure | b_cell | *MGP* | -1.261534302 | 0.013550283 | 0.026992647 | 1 |
| Heart_Failure | b_cell | *RASAL1* | 3.004457612 | 0.000131252 | 0.003196044 | 0.825183428 |
| Heart_Failure | b_cell | *DDX55* | -2.400320495 | 0.001245916 | 0.008654124 | 1 |
| Heart_Failure | b_cell | *ENDOU* | -5.395928846 | 0.000270314 | 0.003897855 | 1 |
| Heart_Failure | b_cell | *CAND1* | -1.863051626 | 0.007704413 | 0.017999868 | 1 |
| Heart_Failure | b_cell | *NUP107* | -1.712473671 | 0.010249872 | 0.022076377 | 1 |
| Heart_Failure | b_cell | *COPS7A* | -2.748815016 | 0.007133706 | 0.017137796 | 1 |
| Heart_Failure | b_cell | *CHPT1* | -1.887645282 | 0.000548179 | 0.006154285 | 1 |
| Heart_Failure | b_cell | *GNPTAB* | -1.83877841 | 0.02468196 | 0.042247613 | 1 |
| Heart_Failure | b_cell | *SPSB2* | -2.838950467 | 0.001951375 | 0.008753931 | 1 |
| Heart_Failure | b_cell | *ATN1* | -1.898792091 | 0.00449882 | 0.012775103 | 1 |
| Heart_Failure | b_cell | *NT5DC3* | -1.644629961 | 0.005114992 | 0.01383733 | 1 |
| Heart_Failure | b_cell | *PRKAB1* | 3.632208162 | 0.000331158 | 0.004450951 | 1 |
| Heart_Failure | b_cell | *ST8SIA1* | -1.48170643 | 0.002354193 | 0.009268076 | 1 |
| Heart_Failure | b_cell | *COL12A1* | -1.098368754 | 0.001688135 | 0.008654124 | 1 |
| Heart_Failure | b_cell | *TDP2* | -2.27726844 | 0.025865305 | 0.043878891 | 1 |
| Heart_Failure | b_cell | *RWDD1* | -2.296368211 | 0.003962112 | 0.011935695 | 1 |
| Heart_Failure | b_cell | *SMIM8* | -1.532951541 | 0.005367489 | 0.014250594 | 1 |
| Heart_Failure | b_cell | *FAM184A* | -1.369822448 | 0.003038784 | 0.010315787 | 1 |
| Heart_Failure | b_cell | *GABRR2* | -3.066899532 | 0.004750638 | 0.013227309 | 1 |
| Heart_Failure | b_cell | *HDDC2* | -2.184459705 | 0.009331536 | 0.020470122 | 1 |
| Heart_Failure | b_cell | *TPD52L1* | -1.66537129 | 0.002302505 | 0.009188443 | 1 |
| Heart_Failure | b_cell | *UST* | -1.099329246 | 0.00208666 | 0.008864076 | 1 |
| Heart_Failure | b_cell | *TULP1* | -18.65335374 | 0.004431224 | 0.012718545 | 1 |
| Heart_Failure | b_cell | *MAPK14* | -1.678720655 | 0.02991853 | 0.049278963 | 1 |
| Heart_Failure | b_cell | *RNF8* | -1.693541809 | 0.001613563 | 0.008654124 | 1 |
| Heart_Failure | b_cell | *MDGA1* | 4.049132834 | 3.71E-05 | 0.002889125 | 0.233322406 |
| Heart_Failure | b_cell | *CAP2* | -1.316226833 | 0.001563428 | 0.008654124 | 1 |
| Heart_Failure | b_cell | *GPR63* | -3.699994903 | 0.002113216 | 0.008922624 | 1 |
| Heart_Failure | b_cell | *BVES* | -3.093741975 | 0.027329408 | 0.045818664 | 1 |
| Heart_Failure | b_cell | *WASF1* | -1.416824157 | 0.002806403 | 0.009957028 | 1 |
| Heart_Failure | b_cell | *ALDH5A1* | -2.654539132 | 0.005587544 | 0.014618764 | 1 |
| Heart_Failure | b_cell | *VNN1* | -1.908934424 | 0.010314172 | 0.022142102 | 1 |
| Heart_Failure | b_cell | *VNN2* | 3.914655735 | 4.22E-05 | 0.002889125 | 0.265259283 |

| | | | | | | |
|---|---|---|---|---|---|---|
| Heart_Failure | b_cell | *ACOT13* | -2.092490433 | 0.008602195 | 0.019399557 | 1 |
| Heart_Failure | b_cell | *C6orf62* | -1.703636324 | 0.029978156 | 0.049351314 | 1 |
| Heart_Failure | b_cell | *HBS1L* | -2.284058589 | 0.029068645 | 0.048143987 | 1 |
| Heart_Failure | b_cell | *SLC16A10* | -1.597493469 | 0.009184083 | 0.020217203 | 1 |
| Heart_Failure | b_cell | *PACRG* | -2.339910502 | 0.002927764 | 0.010146595 | 1 |
| Heart_Failure | b_cell | *PRPH2* | -3.521270976 | 0.000445934 | 0.005230572 | 1 |
| Heart_Failure | b_cell | *DNPH1* | -2.874907257 | 0.013554285 | 0.026992647 | 1 |
| Heart_Failure | b_cell | *DUSP22* | -2.07011702 | 0.014088772 | 0.027819129 | 1 |
| Heart_Failure | b_cell | *TTK* | 1.53196486 | 0.000419113 | 0.005055487 | 1 |
| Heart_Failure | b_cell | *BTN2A1* | 4.736990354 | 0.000437874 | 0.005184391 | 1 |
| Heart_Failure | b_cell | *LAMA4* | -0.866203283 | 0.001559222 | 0.008654124 | 1 |
| Heart_Failure | b_cell | *CLIC5* | -1.489872286 | 0.008742125 | 0.019622186 | 1 |
| Heart_Failure | b_cell | *LY86* | 2.192512614 | 7.02E-05 | 0.002889125 | 0.441197497 |
| Heart_Failure | b_cell | *TBX18* | -1.514359593 | 0.002956853 | 0.010163879 | 1 |
| Heart_Failure | b_cell | *PCDHB2* | -2.73682522 | 0.014870978 | 0.028972371 | 1 |
| Heart_Failure | b_cell | *NUDT12* | -2.662630651 | 0.019742823 | 0.035667565 | 1 |
| Heart_Failure | b_cell | *CEP72* | 4.170845393 | 0.022774339 | 0.039763515 | 1 |
| Heart_Failure | b_cell | *C7* | -1.74709613 | 0.001691084 | 0.008654124 | 1 |
| Heart_Failure | b_cell | *GHR* | -0.894933338 | 0.000862797 | 0.008294193 | 1 |
| Heart_Failure | b_cell | *NME5* | -1.938514444 | 0.002776728 | 0.009913839 | 1 |
| Heart_Failure | b_cell | *NNT* | -1.740743599 | 0.027094667 | 0.045497909 | 1 |
| Heart_Failure | b_cell | *PFDN1* | -1.788817292 | 0.012219923 | 0.02501682 | 1 |
| Heart_Failure | b_cell | *LOX* | -1.61293125 | 0.001683066 | 0.008654124 | 1 |
| Heart_Failure | b_cell | *APBB3* | 4.328975308 | 0.001427695 | 0.008654124 | 1 |
| Heart_Failure | b_cell | *TMCO6* | 2.147347561 | 0.000219944 | 0.003667878 | 1 |
| Heart_Failure | b_cell | *SPARC* | -1.281320796 | 0.006025388 | 0.015367794 | 1 |
| Heart_Failure | b_cell | *IK* | -2.890769186 | 0.014199218 | 0.028002033 | 1 |
| Heart_Failure | b_cell | *PDE8B* | -1.382653415 | 0.00126127 | 0.008654124 | 1 |
| Heart_Failure | b_cell | *CLK4* | -1.859986996 | 0.02707956 | 0.045484689 | 1 |
| Heart_Failure | b_cell | *HAVCR1* | 2.666148039 | 0.003308463 | 0.01083435 | 1 |
| Heart_Failure | b_cell | *RNF130* | -1.487398453 | 0.009387288 | 0.020578062 | 1 |
| Heart_Failure | b_cell | *ARSB* | -2.623911985 | 0.013505896 | 0.026947498 | 1 |
| Heart_Failure | b_cell | *CLINT1* | -1.620921688 | 0.027369688 | 0.045861735 | 1 |
| Heart_Failure | b_cell | *BTNL8* | -1.988889408 | 0.010927723 | 0.023101074 | 1 |
| Heart_Failure | b_cell | *RASGRF2* | -1.145145288 | 0.001764199 | 0.008654124 | 1 |
| Heart_Failure | b_cell | *POLR3G* | -1.684763207 | 0.004126942 | 0.012181261 | 1 |
| Heart_Failure | b_cell | *SLC27A6* | -4.240803915 | 0.001650042 | 0.008654124 | 1 |
| Heart_Failure | b_cell | *TARS* | -2.091636291 | 0.005500302 | 0.014467111 | 1 |
| Heart_Failure | b_cell | *SLC12A7* | -2.589850913 | 0.016014816 | 0.030436865 | 1 |
| Heart_Failure | b_cell | *IL4* | 3.367317614 | 0.005532204 | 0.014516264 | 1 |
| Heart_Failure | b_cell | *TXNDC15* | -2.142394233 | 0.007640064 | 0.017902752 | 1 |
| Heart_Failure | b_cell | *CNTN3* | -1.440891746 | 0.00167134 | 0.008654124 | 1 |
| Heart_Failure | b_cell | *ACTR8* | 7.357346036 | 0.000236542 | 0.003708573 | 1 |
| Heart_Failure | b_cell | *HGD* | 9.099743898 | 4.94E-05 | 0.002889125 | 0.3106597 |
| Heart_Failure | b_cell | *NIT2* | -2.145082816 | 0.003523773 | 0.01117758 | 1 |
| Heart_Failure | b_cell | *FAM162A* | -2.081898767 | 0.002415698 | 0.009356237 | 1 |
| Heart_Failure | b_cell | *PCCB* | -2.035692538 | 0.009750306 | 0.021211134 | 1 |
| Heart_Failure | b_cell | *RBP1* | -5.163710017 | 0.001393309 | 0.008654124 | 1 |
| Heart_Failure | b_cell | *BCHE* | -3.206750375 | 0.000242042 | 0.003729703 | 1 |
| Heart_Failure | b_cell | *HYAL1* | -1.772663337 | 0.011660006 | 0.024137787 | 1 |

| | | | | | | |
|---|---|---|---|---|---|---|
| Heart_Failure | b_cell | *CYB561D2* | -2.944796348 | 0.007463885 | 0.017644281 | 1 |
| Heart_Failure | b_cell | *IFT57* | 5.10711805 | 9.57E-05 | 0.002977985 | 0.601553019 |
| Heart_Failure | b_cell | *GBE1* | -1.074551411 | 0.002688026 | 0.009763417 | 1 |
| Heart_Failure | b_cell | *UMPS* | -2.174217667 | 0.011559507 | 0.023995328 | 1 |
| Heart_Failure | b_cell | *SLC41A3* | -2.007351123 | 0.001962437 | 0.008766903 | 1 |
| Heart_Failure | b_cell | *PLXNA1* | -1.834409344 | 0.023098546 | 0.0401939 | 1 |
| Heart_Failure | b_cell | *NEK11* | -1.896333524 | 0.019642635 | 0.03552402 | 1 |
| Heart_Failure | b_cell | *PLSCR4* | -1.47890539 | 0.012570645 | 0.025551777 | 1 |
| Heart_Failure | b_cell | *COMMD2* | -2.037958004 | 0.004859286 | 0.013391387 | 1 |
| Heart_Failure | b_cell | *ABCC5* | -1.711380796 | 0.007010442 | 0.01695831 | 1 |
| Heart_Failure | b_cell | *ARHGEF26* | -1.628197023 | 0.002782502 | 0.009915204 | 1 |
| Heart_Failure | b_cell | *KLHL24* | -1.462289702 | 0.030177666 | 0.049575904 | 1 |
| Heart_Failure | b_cell | *PLCH1* | 7.860691232 | 3.09E-05 | 0.002889125 | 0.194224561 |
| Heart_Failure | b_cell | *KCNIP3* | -3.390837131 | 0.00295918 | 0.01016632 | 1 |
| Heart_Failure | b_cell | *FAHD2A* | 6.340855245 | 7.42E-05 | 0.002889125 | 0.466320715 |
| Heart_Failure | b_cell | *STEAP3* | -1.759856717 | 0.020838451 | 0.037061199 | 1 |
| Heart_Failure | b_cell | *TFCP2L1* | -3.572279694 | 0.001286643 | 0.008654124 | 1 |
| Heart_Failure | b_cell | *TP53I3* | -1.830243137 | 0.003345949 | 0.010865693 | 1 |
| Heart_Failure | b_cell | *CENPA* | -2.652923747 | 0.00119177 | 0.008654124 | 1 |
| Heart_Failure | b_cell | *TANC1* | -1.142397732 | 0.002987356 | 0.01021289 | 1 |
| Heart_Failure | b_cell | *PCSK4* | -3.835666002 | 0.003701444 | 0.011508892 | 1 |
| Heart_Failure | b_cell | *INO80B* | -3.266400034 | 0.000192312 | 0.003449722 | 1 |
| Heart_Failure | b_cell | *MOGS* | -1.462086553 | 0.001345583 | 0.008654124 | 1 |
| Heart_Failure | b_cell | *GRB14* | -1.909928599 | 0.001755367 | 0.008654124 | 1 |
| Heart_Failure | b_cell | *CLIP4* | -1.510066672 | 0.003857971 | 0.011769687 | 1 |
| Heart_Failure | b_cell | *SPTBN1* | -1.072511978 | 0.001960423 | 0.008766131 | 1 |
| Heart_Failure | b_cell | *RTN4* | -1.340216813 | 0.024222581 | 0.041688301 | 1 |
| Heart_Failure | b_cell | *GALNT3* | 1.545572944 | 0.000234559 | 0.003705045 | 1 |
| Heart_Failure | b_cell | *POLE4* | -2.842210491 | 0.011356006 | 0.023727222 | 1 |
| Heart_Failure | b_cell | *MRPL19* | -1.767248364 | 0.005482969 | 0.014441319 | 1 |
| Heart_Failure | b_cell | *EFEMP1* | -1.102263773 | 0.001314691 | 0.008654124 | 1 |
| Heart_Failure | b_cell | *GNLY* | -1.588263345 | 0.001517455 | 0.008654124 | 1 |
| Heart_Failure | b_cell | *ST3GAL5* | -1.328316293 | 0.002418783 | 0.009356237 | 1 |
| Heart_Failure | b_cell | *PLCD4* | -2.449113129 | 0.001291115 | 0.008654124 | 1 |
| Heart_Failure | b_cell | *WNT6* | -19.47703428 | 0.002731466 | 0.009839202 | 1 |
| Heart_Failure | b_cell | *MLPH* | -2.245378453 | 0.0016232 | 0.008654124 | 1 |
| Heart_Failure | b_cell | *STK25* | -1.852287725 | 0.003909132 | 0.011873672 | 1 |
| Heart_Failure | b_cell | *TPO* | -1.349511057 | 0.005682411 | 0.014784061 | 1 |
| Heart_Failure | b_cell | *SLC25A12* | -1.695501049 | 0.013767836 | 0.027357265 | 1 |
| Heart_Failure | b_cell | *SOS1* | -1.474170437 | 0.015658726 | 0.030050797 | 1 |
| Heart_Failure | b_cell | *WIPF1* | -1.58387751 | 0.020734618 | 0.036886967 | 1 |
| Heart_Failure | b_cell | *PNO1* | -2.027305001 | 0.001723127 | 0.008654124 | 1 |
| Heart_Failure | b_cell | *ARID3A* | -2.20928668 | 0.020134133 | 0.036146 | 1 |
| Heart_Failure | b_cell | *CD207* | -43.0474413 | 0.007250671 | 0.0173261 | 1 |
| Heart_Failure | b_cell | *VAX2* | -4.129753645 | 0.00318161 | 0.010589085 | 1 |
| Heart_Failure | b_cell | *PLEKHA3* | 4.445721587 | 0.025167944 | 0.042927527 | 1 |
| Heart_Failure | b_cell | *ALMS1* | -1.795806664 | 0.009612107 | 0.020990385 | 1 |
| Heart_Failure | b_cell | *DHCR24* | 7.362828228 | 0.00157532 | 0.008654124 | 1 |
| Heart_Failure | b_cell | *MORN1* | -1.962943806 | 0.003025288 | 0.010282127 | 1 |
| Heart_Failure | b_cell | *GPX7* | 2.571856758 | 0.000432984 | 0.005151018 | 1 |

| | | | | | | |
|---|---|---|---|---|---|---|
| Heart_Failure | b_cell | *SCP2* | -1.613627 | 0.016596412 | 0.031305623 | 1 |
| Heart_Failure | b_cell | *RALGPS2* | 0.443791951 | 0.000139617 | 0.003214256 | 0.877771843 |
| Heart_Failure | b_cell | *CEP104* | -1.744828166 | 0.004338328 | 0.012551802 | 1 |
| Heart_Failure | b_cell | *TMEM59* | -1.919881189 | 0.024205194 | 0.041687125 | 1 |
| Heart_Failure | b_cell | *WRAP73* | -3.03594394 | 0.007690647 | 0.017987759 | 1 |
| Heart_Failure | b_cell | *MRPL37* | -1.962079044 | 0.002932945 | 0.010146595 | 1 |
| Heart_Failure | b_cell | *ICMT* | -3.327849343 | 0.007144975 | 0.017158312 | 1 |
| Heart_Failure | b_cell | *KCNC4* | -2.743447895 | 0.001915936 | 0.008705907 | 1 |
| Heart_Failure | b_cell | *DOCK7* | -1.66501214 | 0.022641848 | 0.039596466 | 1 |
| Heart_Failure | b_cell | *FBXO6* | -3.108129983 | 0.01683924 | 0.031545978 | 1 |
| Heart_Failure | b_cell | *C1orf21* | -1.188143869 | 0.002714657 | 0.009825588 | 1 |
| Heart_Failure | b_cell | *MFN2* | -2.229083509 | 0.004094919 | 0.012159073 | 1 |
| Heart_Failure | b_cell | *MIIP* | -3.494754301 | 0.008270217 | 0.018900347 | 1 |
| Heart_Failure | b_cell | *NCF2* | -1.743530788 | 0.001806947 | 0.008654124 | 1 |
| Heart_Failure | b_cell | *RGS2* | -3.638441769 | 0.023444833 | 0.040650211 | 1 |
| Heart_Failure | b_cell | *OLFML3* | -1.813286396 | 0.002004358 | 0.008787585 | 1 |
| Heart_Failure | b_cell | *CFHR3* | -6.645130993 | 4.74E-06 | 0.001567127 | 0.029775407 |
| Heart_Failure | b_cell | *TTF2* | -2.423311879 | 0.016719476 | 0.031405839 | 1 |
| Heart_Failure | b_cell | *TMEM9* | -2.028875384 | 0.001830971 | 0.008654124 | 1 |
| Heart_Failure | b_cell | *WARS2* | -2.039504094 | 0.00250271 | 0.009482826 | 1 |
| Heart_Failure | b_cell | *OSCP1* | -2.059607142 | 0.000800904 | 0.007867627 | 1 |
| Heart_Failure | b_cell | *C1orf109* | -2.537880469 | 0.001884028 | 0.008705907 | 1 |
| Heart_Failure | b_cell | *LGALS8* | -1.572192182 | 0.02441266 | 0.041900735 | 1 |
| Heart_Failure | b_cell | *PADI2* | 4.072849683 | 8.24E-05 | 0.002889125 | 0.517787411 |
| Heart_Failure | b_cell | *PLA2G2D* | 4.038442365 | 0.000444306 | 0.005221219 | 1 |
| Heart_Failure | b_cell | *KIF17* | -4.414533599 | 0.004860688 | 0.013391387 | 1 |
| Heart_Failure | b_cell | *CD46* | -1.433837281 | 0.021343029 | 0.037776921 | 1 |
| Heart_Failure | b_cell | *TMED5* | -1.873225224 | 0.015094584 | 0.029290016 | 1 |
| Heart_Failure | b_cell | *DR1* | -2.309213866 | 0.015248224 | 0.029497102 | 1 |
| Heart_Failure | b_cell | *CNN3* | -1.745361319 | 0.004099126 | 0.012159073 | 1 |
| Heart_Failure | b_cell | *VAMP4* | -2.371360526 | 0.009723436 | 0.021175716 | 1 |
| Heart_Failure | b_cell | *TNFSF4* | -3.695363274 | 0.000340741 | 0.004538892 | 1 |
| Heart_Failure | b_cell | *DARS2* | -2.483272041 | 0.026645531 | 0.044887581 | 1 |
| Heart_Failure | b_cell | *DIEXF* | -3.308899866 | 0.008975415 | 0.019932333 | 1 |
| Heart_Failure | b_cell | *SERPINC1* | -3.99292778 | 0.006639772 | 0.016325477 | 1 |
| Heart_Failure | b_cell | *RCAN3* | -2.288061327 | 0.007713081 | 0.018013424 | 1 |
| Heart_Failure | b_cell | *MTFR1L* | 6.871545612 | 0.001495893 | 0.008654124 | 1 |
| Heart_Failure | b_cell | *SLC5A9* | 6.759298467 | 9.13E-05 | 0.002929012 | 0.574086312 |
| Heart_Failure | b_cell | *ESYT2* | -1.605226298 | 0.023992644 | 0.041417284 | 1 |
| Heart_Failure | b_cell | *CD3EAP* | -2.068116915 | 0.002015318 | 0.00881106 | 1 |
| Heart_Failure | b_cell | *IFT46* | -2.686976836 | 0.023264435 | 0.040409587 | 1 |
| Heart_Failure | b_cell | *CASC1* | 4.590318616 | 0.002281826 | 0.009182983 | 1 |
| Heart_Failure | b_cell | *LRMP* | 1.52502646 | 0.000125036 | 0.003176202 | 0.786104127 |
| Heart_Failure | b_cell | *ATP10B* | -4.374830962 | 0.001478066 | 0.008654124 | 1 |
| Heart_Failure | b_cell | *FILIP1* | -1.341512221 | 0.004329684 | 0.012544113 | 1 |
| Heart_Failure | b_cell | *SGIP1* | -1.715048902 | 0.002215739 | 0.009081423 | 1 |
| Heart_Failure | b_cell | *AKAP7* | -1.830720789 | 0.006518948 | 0.016123569 | 1 |
| Heart_Failure | b_cell | *ALDH8A1* | -2.437542794 | 0.000874813 | 0.00834507 | 1 |
| Heart_Failure | b_cell | *SLC16A7* | -1.574638869 | 0.026806248 | 0.045097908 | 1 |
| Heart_Failure | b_cell | *VAMP8* | -2.041642412 | 0.004074857 | 0.012126691 | 1 |

| | | | | | | |
|---|---|---|---|---|---|---|
| Heart_Failure | b_cell | *ABCG2* | -2.354342247 | 0.001891251 | 0.008705907 | 1 |
| Heart_Failure | b_cell | *SPP1* | 4.541851302 | 0.000107009 | 0.003034612 | 0.672767497 |
| Heart_Failure | b_cell | *FAM47E-STBD* | -1.822186855 | 0.01361277 | 0.027091955 | 1 |
| Heart_Failure | b_cell | *MFSD1* | -1.736909766 | 0.004738491 | 0.013205183 | 1 |
| Heart_Failure | b_cell | *UBN1* | -1.895233222 | 0.026538248 | 0.044754819 | 1 |
| Heart_Failure | b_cell | *UCHL3* | -2.834695749 | 0.001972646 | 0.008775328 | 1 |
| Heart_Failure | b_cell | *DNAH7* | -1.971858519 | 0.000202029 | 0.003508723 | 1 |
| Heart_Failure | b_cell | *TRPM6* | -2.545827303 | 0.001506771 | 0.008654124 | 1 |
| Heart_Failure | b_cell | *GDA* | 5.695670913 | 0.004446973 | 0.012718545 | 1 |
| Heart_Failure | b_cell | *C2orf40* | -2.352114219 | 0.005238987 | 0.014045848 | 1 |
| Heart_Failure | b_cell | *CPSF3* | -2.110150579 | 0.006674319 | 0.016382674 | 1 |
| Heart_Failure | b_cell | *PIGZ* | -1.537109994 | 0.003307118 | 0.01083435 | 1 |
| Heart_Failure | b_cell | *CCDC92* | -2.075002264 | 0.003770422 | 0.011591512 | 1 |
| Heart_Failure | b_cell | *WDR34* | 4.005013572 | 0.000151188 | 0.003239931 | 0.950517857 |
| Heart_Failure | b_cell | *TRIM32* | -3.26845916 | 0.023568845 | 0.040797723 | 1 |
| Heart_Failure | b_cell | *PHF19* | -2.414110474 | 0.002183625 | 0.009035107 | 1 |
| Heart_Failure | b_cell | *BSPRY* | 3.878782747 | 0.000147836 | 0.003239931 | 0.929446284 |
| Heart_Failure | b_cell | *PPP6C* | -1.795377685 | 0.009045151 | 0.020048693 | 1 |
| Heart_Failure | b_cell | *HDHD3* | -0.899179731 | 0.030051281 | 0.049419932 | 1 |
| Heart_Failure | b_cell | *HSDL2* | -1.297365969 | 0.0012851 | 0.008654124 | 1 |
| Heart_Failure | b_cell | *KDSR* | -1.820884215 | 0.004856108 | 0.013391387 | 1 |
| Heart_Failure | b_cell | *IFI27L2* | 4.427873675 | 0.02307829 | 0.040180894 | 1 |
| Heart_Failure | b_cell | *DNAL1* | -2.344083224 | 0.001965218 | 0.008766903 | 1 |
| Heart_Failure | b_cell | *FLVCR2* | -1.861910982 | 0.001696633 | 0.008654124 | 1 |
| Heart_Failure | b_cell | *PPP4R4* | 3.264334907 | 2.27E-05 | 0.002889125 | 0.142795297 |
| Heart_Failure | b_cell | *ZC2HC1C* | -2.36815266 | 0.001483546 | 0.008654124 | 1 |
| Heart_Failure | b_cell | *ALDH6A1* | -2.369663772 | 0.001203698 | 0.008654124 | 1 |
| Heart_Failure | b_cell | *ESRRB* | -2.493269062 | 0.000399496 | 0.00494416 | 1 |
| Heart_Failure | b_cell | *COQ6* | 5.12728153 | 6.40E-05 | 0.002889125 | 0.402311213 |
| Heart_Failure | b_cell | *GPR75* | -14.28949951 | 0.00047099 | 0.005443227 | 1 |
| Heart_Failure | b_cell | *KLHL29* | -1.596826774 | 0.010393503 | 0.022248537 | 1 |
| Heart_Failure | b_cell | *FKBP1B* | 4.08913731 | 0.000942382 | 0.0086241 | 1 |
| Heart_Failure | b_cell | *EPCAM* | -4.325620658 | 0.007393888 | 0.017548273 | 1 |
| Heart_Failure | b_cell | *OGFRL1* | -1.975853013 | 0.023866417 | 0.041222024 | 1 |
| Heart_Failure | b_cell | *PYROXD2* | -1.350818826 | 0.005212153 | 0.014007998 | 1 |
| Heart_Failure | b_cell | *C10orf88* | -2.168087281 | 0.004027879 | 0.012047229 | 1 |
| Heart_Failure | b_cell | *HELLS* | 3.274527875 | 1.55E-05 | 0.002889125 | 0.097496923 |
| Heart_Failure | b_cell | *C10orf76* | -1.697470975 | 0.025415693 | 0.04326793 | 1 |
| Heart_Failure | b_cell | *KCNIP2* | -1.222468916 | 0.006733827 | 0.016479397 | 1 |
| Heart_Failure | b_cell | *C10orf95* | 4.323560542 | 0.001231481 | 0.008654124 | 1 |
| Heart_Failure | b_cell | *HOXB5* | -2.368241287 | 0.004781826 | 0.01327886 | 1 |
| Heart_Failure | b_cell | *HOXB3* | -1.65537132 | 0.026986487 | 0.045364718 | 1 |
| Heart_Failure | b_cell | *CD274* | -2.213440871 | 0.006776926 | 0.016532941 | 1 |
| Heart_Failure | b_cell | *LRP11* | -1.667572075 | 0.002067543 | 0.008836604 | 1 |
| Heart_Failure | b_cell | *CCDC170* | -1.306082309 | 0.002380292 | 0.009318117 | 1 |
| Heart_Failure | b_cell | *PCDHB10* | 1.637670616 | 0.00012529 | 0.003176202 | 0.787697974 |
| Heart_Failure | b_cell | *PCDHB14* | -4.611813828 | 0.029896858 | 0.049269081 | 1 |
| Heart_Failure | b_cell | *PCDHB12* | 2.508967639 | 0.023468399 | 0.04066864 | 1 |
| Heart_Failure | b_cell | *SEC16B* | -5.909232239 | 0.000554615 | 0.006204391 | 1 |
| Heart_Failure | b_cell | *MASTL* | -1.971478725 | 0.01140556 | 0.023789705 | 1 |

| | | | | | | |
|---|---|---|---|---|---|---|
| Heart_Failure | b_cell | *MRC1* | -1.145138249 | 0.001837942 | 0.008661108 | 1 |
| Heart_Failure | b_cell | *IQSEC3* | -4.227428556 | 0.021535365 | 0.038042382 | 1 |
| Heart_Failure | b_cell | *ENOX1* | -1.789351906 | 0.004356864 | 0.012588053 | 1 |
| Heart_Failure | b_cell | *WBP4* | -1.968333167 | 0.009427785 | 0.020638052 | 1 |
| Heart_Failure | b_cell | *SMAD9* | -1.786066115 | 0.001694425 | 0.008654124 | 1 |
| Heart_Failure | b_cell | *ARL1* | -1.994784331 | 0.00841841 | 0.019121228 | 1 |
| Heart_Failure | b_cell | *APAF1* | -1.947043787 | 0.030051085 | 0.049419932 | 1 |
| Heart_Failure | b_cell | *EPHX2* | -1.474188201 | 0.004705801 | 0.013160752 | 1 |
| Heart_Failure | b_cell | *CRISPLD1* | -2.707147819 | 0.001638917 | 0.008654124 | 1 |
| Heart_Failure | b_cell | *RDH10* | -1.617318681 | 0.004148436 | 0.012205437 | 1 |
| Heart_Failure | b_cell | *TRIM6* | -3.434504933 | 0.000488663 | 0.005622167 | 1 |
| Heart_Failure | b_cell | *ECHDC2* | -1.296099964 | 0.000967023 | 0.008654124 | 1 |
| Heart_Failure | b_cell | *PRB2* | -2.480369596 | 0.002819622 | 0.009979564 | 1 |
| Heart_Failure | b_cell | *A1BG* | 13.5311533 | 0.017130646 | 0.032015569 | 1 |
| Heart_Failure | b_cell | *ZNF211* | -2.380514844 | 0.021082774 | 0.037389958 | 1 |
| Heart_Failure | b_cell | *PDZRN3* | -0.938504378 | 0.001689621 | 0.008654124 | 1 |
| Heart_Failure | b_cell | *LHX4* | 7.429786263 | 0.000364946 | 0.004752125 | 1 |
| Heart_Failure | b_cell | *CSTA* | -2.229075682 | 0.00247761 | 0.009457169 | 1 |
| Heart_Failure | b_cell | *POPDC2* | 4.288785763 | 0.000131707 | 0.003196044 | 0.828043458 |
| Heart_Failure | b_cell | *B4GALT4* | 5.596146918 | 6.80E-05 | 0.002889125 | 0.427451744 |
| Heart_Failure | b_cell | *CD80* | 2.941126057 | 9.33E-05 | 0.002932166 | 0.586433149 |
| Heart_Failure | b_cell | *CAT* | -1.153965408 | 0.008537284 | 0.019314107 | 1 |
| Heart_Failure | b_cell | *GJB6* | -65.61387791 | 0.024811932 | 0.042412349 | 1 |
| Heart_Failure | b_cell | *TMEM39B* | -2.422621387 | 0.002601115 | 0.00960823 | 1 |
| Heart_Failure | b_cell | *POLR3GL* | 6.776994957 | 6.18E-05 | 0.002889125 | 0.388283558 |
| Heart_Failure | b_cell | *TNFSF10* | -2.01296236 | 0.013133904 | 0.026423314 | 1 |
| Heart_Failure | b_cell | *LIAS* | -1.529617908 | 0.00283951 | 0.010002746 | 1 |
| Heart_Failure | b_cell | *GPSM2* | -1.614094402 | 0.020948273 | 0.037203897 | 1 |
| Heart_Failure | b_cell | *CXCR4* | 1.130892685 | 0.000235232 | 0.003705045 | 1 |
| Heart_Failure | b_cell | *FLT3* | 3.10726298 | 4.86E-05 | 0.002889125 | 0.305415611 |
| Heart_Failure | b_cell | *MTIF3* | -1.833903688 | 0.017842104 | 0.033040739 | 1 |
| Heart_Failure | b_cell | *RASL11A* | -4.011268628 | 0.001824607 | 0.008654124 | 1 |
| Heart_Failure | b_cell | *FMOD* | -1.86281958 | 0.001762544 | 0.008654124 | 1 |
| Heart_Failure | b_cell | *SERAC1* | -1.909327818 | 0.001617035 | 0.008654124 | 1 |
| Heart_Failure | b_cell | *FAM213A* | -1.161553553 | 0.003266888 | 0.010797038 | 1 |
| Heart_Failure | b_cell | *NAA60* | -2.405726192 | 0.001802883 | 0.008654124 | 1 |
| Heart_Failure | b_cell | *LRRC39* | -1.602892029 | 0.018112231 | 0.033413027 | 1 |
| Heart_Failure | b_cell | *RWDD3* | 10.41799415 | 0.000372106 | 0.004779534 | 1 |
| Heart_Failure | b_cell | *CCDC18* | -1.721660681 | 0.028426522 | 0.047304802 | 1 |
| Heart_Failure | b_cell | *RPAP2* | -1.718680434 | 0.026972136 | 0.045352719 | 1 |
| Heart_Failure | b_cell | *KIAA0087* | -8.297245271 | 0.000263365 | 0.003832809 | 1 |
| Heart_Failure | b_cell | *FKBP9* | -1.62648621 | 0.002955879 | 0.010163879 | 1 |
| Heart_Failure | b_cell | *BICC1* | -1.020739951 | 0.004435214 | 0.012718545 | 1 |
| Heart_Failure | b_cell | *CISD1* | -2.145311929 | 0.001404479 | 0.008654124 | 1 |
| Heart_Failure | b_cell | *SLC25A16* | -1.133274375 | 0.010538986 | 0.022491041 | 1 |
| Heart_Failure | b_cell | *ACADS* | -1.797620108 | 0.001454366 | 0.008654124 | 1 |
| Heart_Failure | b_cell | *SSPN* | -1.795539677 | 0.009062704 | 0.020069469 | 1 |
| Heart_Failure | b_cell | *PKN1* | -2.289466711 | 0.022518634 | 0.03940291 | 1 |
| Heart_Failure | b_cell | *EBPL* | -2.055937412 | 0.018745809 | 0.034310014 | 1 |
| Heart_Failure | b_cell | *ATP7B* | -1.061112535 | 0.027920784 | 0.04661125 | 1 |

| | | | | | | |
|---|---|---|---|---|---|---|
| Heart_Failure | b_cell | *NLN* | -1.543174094 | 0.001174381 | 0.008654124 | 1 |
| Heart_Failure | b_cell | *CENPK* | 1.850140378 | 0.000511368 | 0.005824218 | 1 |
| Heart_Failure | b_cell | *OPTN* | -1.828402072 | 0.004797857 | 0.013299878 | 1 |
| Heart_Failure | b_cell | *TSFM* | -2.372914103 | 0.008224253 | 0.018829526 | 1 |
| Heart_Failure | b_cell | *ARHGAP9* | -4.396523726 | 0.002942446 | 0.010151069 | 1 |
| Heart_Failure | b_cell | *SMUG1* | -1.618595111 | 0.008079663 | 0.018579678 | 1 |
| Heart_Failure | b_cell | *SARDH* | -5.630970396 | 0.001660834 | 0.008654124 | 1 |
| Heart_Failure | b_cell | *STIL* | -3.728941298 | 0.003729508 | 0.011549198 | 1 |
| Heart_Failure | b_cell | *ACVR1C* | -1.220432775 | 0.001161274 | 0.008654124 | 1 |
| Heart_Failure | b_cell | *C4BPB* | -3.795688832 | 0.000929578 | 0.008594492 | 1 |
| Heart_Failure | b_cell | *RAB38* | -2.941418187 | 0.002038061 | 0.008831917 | 1 |
| Heart_Failure | b_cell | *CKS2* | 7.946284115 | 0.016756319 | 0.03145625 | 1 |
| Heart_Failure | b_cell | *CHPF* | -2.731628164 | 0.001725105 | 0.008654124 | 1 |
| Heart_Failure | b_cell | *DNPEP* | -2.365130489 | 0.007085389 | 0.01706618 | 1 |
| Heart_Failure | b_cell | *MOGAT1* | -1.949730698 | 0.000880035 | 0.00834507 | 1 |
| Heart_Failure | b_cell | *FAM124B* | -4.079191631 | 0.001629255 | 0.008654124 | 1 |
| Heart_Failure | b_cell | *FAM210B* | -2.605258331 | 0.001865828 | 0.008690419 | 1 |
| Heart_Failure | b_cell | *SNX21* | -1.396948977 | 0.001682676 | 0.008654124 | 1 |
| Heart_Failure | b_cell | *WFDC3* | -1.511520207 | 0.009754864 | 0.021211919 | 1 |
| Heart_Failure | b_cell | *SDC4* | -2.545851726 | 0.000991952 | 0.008654124 | 1 |
| Heart_Failure | b_cell | *CSE1L* | 4.707805257 | 0.004225579 | 0.012327711 | 1 |
| Heart_Failure | b_cell | *CDH26* | -2.372709927 | 0.001412983 | 0.008654124 | 1 |
| Heart_Failure | b_cell | *KCNK15* | 2.476676562 | 0.000250536 | 0.003766552 | 1 |
| Heart_Failure | b_cell | *PEPD* | -1.61745616 | 0.001460744 | 0.008654124 | 1 |
| Heart_Failure | b_cell | *HIF3A* | -1.447787187 | 0.002180042 | 0.009028936 | 1 |
| Heart_Failure | b_cell | *ZNF45* | 7.094370443 | 0.000416808 | 0.005049082 | 1 |
| Heart_Failure | b_cell | *BTN2A2* | 2.015032764 | 8.92E-05 | 0.002889125 | 0.560490291 |
| Heart_Failure | b_cell | *SNRPC* | 3.765809764 | 0.021748027 | 0.038353392 | 1 |
| Heart_Failure | b_cell | *SERPINB6* | -1.981937796 | 0.014702957 | 0.028751941 | 1 |
| Heart_Failure | b_cell | *PEX6* | -1.470717845 | 0.006944285 | 0.01683723 | 1 |
| Heart_Failure | b_cell | *NQO2* | -1.970956077 | 0.008188325 | 0.018787704 | 1 |
| Heart_Failure | b_cell | *RPS10* | 3.418976085 | 0.000132528 | 0.003196044 | 0.833206201 |
| Heart_Failure | b_cell | *HIST1H2BJ* | 3.218654125 | 0.000256606 | 0.003777026 | 1 |
| Heart_Failure | b_cell | *MED20* | -3.014909124 | 0.000193357 | 0.003449722 | 1 |
| Heart_Failure | b_cell | *TCP11* | 6.215157222 | 0.004143241 | 0.012205437 | 1 |
| Heart_Failure | b_cell | *KLHDC3* | -1.986692074 | 0.00240203 | 0.009333474 | 1 |
| Heart_Failure | b_cell | *MEA1* | -3.742168023 | 0.000429277 | 0.005135387 | 1 |
| Heart_Failure | b_cell | *KLHL31* | -1.305718734 | 0.004673275 | 0.013085296 | 1 |
| Heart_Failure | b_cell | *COL21A1* | -1.646024325 | 0.002532453 | 0.00953574 | 1 |
| Heart_Failure | b_cell | *RPP40* | -3.384210466 | 0.006447356 | 0.016018516 | 1 |
| Heart_Failure | b_cell | *EEF1E1* | -3.271932073 | 0.001399494 | 0.008654124 | 1 |
| Heart_Failure | b_cell | *EREG* | 3.275646031 | 0.00014622 | 0.003239931 | 0.919286196 |
| Heart_Failure | b_cell | *MYRF* | -5.111100789 | 0.005642465 | 0.014725687 | 1 |
| Heart_Failure | b_cell | *MT1G* | -16.62610721 | 0.001554061 | 0.008654124 | 1 |
| Heart_Failure | b_cell | *TMTC4* | -1.859781792 | 0.002217913 | 0.009081423 | 1 |
| Heart_Failure | b_cell | *KIF25* | -5.526973982 | 0.002892083 | 0.010112641 | 1 |
| Heart_Failure | b_cell | *GGA3* | -2.25068573 | 0.011783183 | 0.024301648 | 1 |
| Heart_Failure | b_cell | *NUP85* | -2.387832794 | 0.008026964 | 0.018519458 | 1 |
| Heart_Failure | b_cell | *MIF4GD* | 9.568535689 | 0.001161797 | 0.008654124 | 1 |
| Heart_Failure | b_cell | *OPRL1* | -4.513942969 | 0.002637286 | 0.009653089 | 1 |

| | | | | | | |
|---|---|---|---|---|---|---|
| Heart_Failure | b_cell | *INSIG2* | 5.327561268 | 0.008863031 | 0.019752526 | 1 |
| Heart_Failure | b_cell | *POLR1B* | -2.028131345 | 0.003920869 | 0.011879761 | 1 |
| Heart_Failure | b_cell | *SLC25A23* | -3.398015038 | 0.00105841 | 0.008654124 | 1 |
| Heart_Failure | b_cell | *GTF2F1* | -3.176229716 | 0.017916995 | 0.033130632 | 1 |
| Heart_Failure | b_cell | *CLPP* | -2.240782805 | 0.001803148 | 0.008654124 | 1 |
| Heart_Failure | b_cell | *SH2D3A* | -2.917928495 | 0.001899582 | 0.008705907 | 1 |
| Heart_Failure | b_cell | *TRIP10* | -1.624580453 | 0.002738027 | 0.009847411 | 1 |
| Heart_Failure | b_cell | *GPR108* | -2.998973445 | 0.004785604 | 0.013283485 | 1 |
| Heart_Failure | b_cell | *RTN2* | -2.846157917 | 0.001372629 | 0.008654124 | 1 |
| Heart_Failure | b_cell | *SDCBP2* | -6.32570192 | 0.011784698 | 0.024301648 | 1 |
| Heart_Failure | b_cell | *PANK2* | -2.022217843 | 0.026482202 | 0.044708272 | 1 |
| Heart_Failure | b_cell | *PSMF1* | -1.938751823 | 0.019053572 | 0.034761987 | 1 |
| Heart_Failure | b_cell | *RBCK1* | 4.292466773 | 0.006743513 | 0.016490263 | 1 |
| Heart_Failure | b_cell | *BMP2* | -3.289217688 | 0.001780676 | 0.008654124 | 1 |
| Heart_Failure | b_cell | *PCSK2* | 17.16392609 | 0.00035461 | 0.004664086 | 1 |
| Heart_Failure | b_cell | *DSTN* | -1.78880958 | 0.022410434 | 0.039301087 | 1 |
| Heart_Failure | b_cell | *MGME1* | -1.762300302 | 0.001176556 | 0.008654124 | 1 |
| Heart_Failure | b_cell | *MCM8* | -2.380989486 | 0.003185015 | 0.010594809 | 1 |
| Heart_Failure | b_cell | *FAM110A* | -2.763142637 | 0.001640531 | 0.008654124 | 1 |
| Heart_Failure | b_cell | *MRPS26* | -1.945352233 | 0.006852142 | 0.016678055 | 1 |
| Heart_Failure | b_cell | *NCLN* | -3.022226645 | 0.014940501 | 0.029080782 | 1 |
| Heart_Failure | b_cell | *ZNF436* | -2.124738628 | 0.001654836 | 0.008654124 | 1 |
| Heart_Failure | b_cell | *GDF5* | -3.878012792 | 0.006128918 | 0.015518529 | 1 |
| Heart_Failure | b_cell | *MMP24* | -2.240302151 | 0.006592108 | 0.016253852 | 1 |
| Heart_Failure | b_cell | *XRCC3* | -2.280420076 | 0.002248255 | 0.0091251 | 1 |
| Heart_Failure | b_cell | *MCF2L* | -1.740604207 | 0.001736054 | 0.008654124 | 1 |
| Heart_Failure | b_cell | *F10* | -1.67998504 | 0.001674314 | 0.008654124 | 1 |
| Heart_Failure | b_cell | *PCID2* | -1.949226946 | 0.015238615 | 0.029487588 | 1 |
| Heart_Failure | b_cell | *PROZ* | 4.062778857 | 0.000155117 | 0.003239931 | 0.975219368 |
| Heart_Failure | b_cell | *IGFLR1* | 10.10990217 | 7.29E-05 | 0.002889125 | 0.458049767 |
| Heart_Failure | b_cell | *COX6B1* | 4.637209144 | 0.028323389 | 0.047183133 | 1 |
| Heart_Failure | b_cell | *TSKS* | -2.82300039 | 0.001471735 | 0.008654124 | 1 |
| Heart_Failure | b_cell | *FLRT1* | -3.091783609 | 0.000282712 | 0.004036731 | 1 |
| Heart_Failure | b_cell | *DNAJC8* | -2.149810601 | 0.023202216 | 0.040329646 | 1 |
| Heart_Failure | b_cell | *ATG14* | -1.766201869 | 0.010733638 | 0.022828952 | 1 |
| Heart_Failure | b_cell | *RHOJ* | -1.438568622 | 0.001737864 | 0.008654124 | 1 |
| Heart_Failure | b_cell | *L3HYPDH* | -1.532661912 | 0.001272195 | 0.008654124 | 1 |
| Heart_Failure | b_cell | *ZBTB1* | -1.779175679 | 0.021396071 | 0.037860146 | 1 |
| Heart_Failure | b_cell | *PZP* | -1.561675689 | 0.001965978 | 0.008766903 | 1 |
| Heart_Failure | b_cell | *WDR60* | -1.730864968 | 0.008529598 | 0.019314107 | 1 |
| Heart_Failure | b_cell | *AIF1L* | -2.058527166 | 0.001593376 | 0.008654124 | 1 |
| Heart_Failure | b_cell | *MAP2K2* | -2.332200972 | 0.010644504 | 0.022685422 | 1 |
| Heart_Failure | b_cell | *IPPK* | -2.314615673 | 0.005690899 | 0.014796808 | 1 |
| Heart_Failure | b_cell | *FGD3* | -2.196275671 | 0.004450551 | 0.012718545 | 1 |
| Heart_Failure | b_cell | *MASP1* | -2.572762308 | 0.00194529 | 0.008738197 | 1 |
| Heart_Failure | b_cell | *ATP13A4* | -3.117856719 | 0.002376372 | 0.009318117 | 1 |
| Heart_Failure | b_cell | *HRASLS* | -4.48351563 | 0.001755562 | 0.008654124 | 1 |
| Heart_Failure | b_cell | *RAB3IP* | 7.043656207 | 0.000120318 | 0.003135985 | 0.756440659 |
| Heart_Failure | b_cell | *YEATS4* | -2.630791244 | 0.012527984 | 0.025482916 | 1 |
| Heart_Failure | b_cell | *LRRC61* | -2.172255825 | 0.005180037 | 0.013936542 | 1 |

| | | | | | | |
|---|---|---|---|---|---|---|
| Heart_Failure | b_cell | *IDUA* | 4.823005975 | 5.68E-05 | 0.002889125 | 0.357021448 |
| Heart_Failure | b_cell | *TMEM175* | 4.399624231 | 0.015381458 | 0.029672668 | 1 |
| Heart_Failure | b_cell | *PLA2G5* | -2.715549675 | 0.001743426 | 0.008654124 | 1 |
| Heart_Failure | b_cell | *WDR24* | -4.767731008 | 0.004146078 | 0.012205437 | 1 |
| Heart_Failure | b_cell | *CHTF18* | 9.148485195 | 0.000318167 | 0.004329688 | 1 |
| Heart_Failure | b_cell | *SMARCA4* | -2.269429177 | 0.018481558 | 0.033895435 | 1 |
| Heart_Failure | b_cell | *TICAM1* | -3.309635788 | 0.019882157 | 0.035847181 | 1 |
| Heart_Failure | b_cell | *AAMP* | -2.38154124 | 0.00336389 | 0.010895814 | 1 |
| Heart_Failure | b_cell | *PNKD* | -2.289612839 | 0.028792894 | 0.047737586 | 1 |
| Heart_Failure | b_cell | *ZNF835* | -3.490664003 | 0.002999078 | 0.010236268 | 1 |
| Heart_Failure | b_cell | *HIP1* | -1.524008028 | 0.020505964 | 0.036583711 | 1 |
| Heart_Failure | b_cell | *POR* | -1.612654126 | 0.003641717 | 0.011427162 | 1 |
| Heart_Failure | b_cell | *FGL2* | -1.9764395 | 0.015883185 | 0.030314993 | 1 |
| Heart_Failure | b_cell | *STEAP4* | -2.286567716 | 0.020017447 | 0.036009891 | 1 |
| Heart_Failure | b_cell | *GNAI1* | -1.242064172 | 0.000940612 | 0.00862045 | 1 |
| Heart_Failure | b_cell | *SGCE* | -1.679856946 | 0.002464779 | 0.009438469 | 1 |
| Heart_Failure | b_cell | *RBM48* | -1.726436655 | 0.001894823 | 0.008705907 | 1 |
| Heart_Failure | b_cell | *SRD5A3* | -2.449362887 | 0.005652132 | 0.014744794 | 1 |
| Heart_Failure | b_cell | *DGCR6L* | 3.877707553 | 0.000282207 | 0.004036731 | 1 |
| Heart_Failure | b_cell | *VPREB3* | -26.09477 | 0.014991692 | 0.029144331 | 1 |
| Heart_Failure | b_cell | *MGAT3* | -2.36985485 | 0.001676611 | 0.008654124 | 1 |
| Heart_Failure | b_cell | *A4GALT* | -1.590418059 | 0.012528661 | 0.025482916 | 1 |
| Heart_Failure | b_cell | *CDC42EP1* | -2.271623431 | 0.002062137 | 0.008831917 | 1 |
| Heart_Failure | b_cell | *TPST2* | -1.594772605 | 0.002748506 | 0.009857305 | 1 |
| Heart_Failure | b_cell | *TST* | -2.093991578 | 0.002167832 | 0.009019688 | 1 |
| Heart_Failure | b_cell | *APOL5* | -9.360773076 | 0.019960348 | 0.035936629 | 1 |
| Heart_Failure | b_cell | *RIBC2* | 3.044266811 | 3.16E-05 | 0.002889125 | 0.198490119 |
| Heart_Failure | b_cell | *EMC4* | -2.824094163 | 0.00444113 | 0.012718545 | 1 |
| Heart_Failure | b_cell | *SPECC1* | -1.492783332 | 0.003797388 | 0.011651623 | 1 |
| Heart_Failure | b_cell | *CDHR3* | -1.366411853 | 0.004314778 | 0.012512459 | 1 |
| Heart_Failure | b_cell | *FOXP2* | -1.037289337 | 0.001592135 | 0.008654124 | 1 |
| Heart_Failure | b_cell | *FLNC* | -2.296578738 | 0.005907956 | 0.015203978 | 1 |
| Heart_Failure | b_cell | *IRF5* | -2.039094182 | 0.005586632 | 0.014618764 | 1 |
| Heart_Failure | b_cell | *KLHDC10* | -1.742238346 | 0.011428224 | 0.023814797 | 1 |
| Heart_Failure | b_cell | *HOXD3* | -1.720727714 | 0.002065444 | 0.008833636 | 1 |
| Heart_Failure | b_cell | *PDE11A* | -2.917361373 | 0.005142648 | 0.01388228 | 1 |
| Heart_Failure | b_cell | *CHN1* | -1.386925724 | 0.001942476 | 0.008738197 | 1 |
| Heart_Failure | b_cell | *GAD1* | 3.354077086 | 0.000118294 | 0.003135985 | 0.74371141 |
| Heart_Failure | b_cell | *ORMDL1* | -1.409329557 | 0.022312696 | 0.039162457 | 1 |
| Heart_Failure | b_cell | *HOXD9* | -4.016090622 | 0.022969315 | 0.040035509 | 1 |
| Heart_Failure | b_cell | *PSMG2* | -1.949083648 | 0.011226781 | 0.023563662 | 1 |
| Heart_Failure | b_cell | *WDFY4* | 1.632104349 | 5.19E-05 | 0.002889125 | 0.326339652 |
| Heart_Failure | b_cell | *TMOD2* | -1.922173316 | 0.019899977 | 0.035858743 | 1 |
| Heart_Failure | b_cell | *KNSTRN* | -1.851326024 | 0.000902812 | 0.008457357 | 1 |
| Heart_Failure | b_cell | *CALML4* | -7.16372488 | 0.018120694 | 0.033414978 | 1 |
| Heart_Failure | b_cell | *ACKR4* | -2.586468623 | 0.01002029 | 0.021685909 | 1 |
| Heart_Failure | b_cell | *SUMF2* | -2.193963714 | 0.021942259 | 0.038609286 | 1 |
| Heart_Failure | b_cell | *RPAIN* | -1.983445924 | 0.013904068 | 0.027584373 | 1 |
| Heart_Failure | b_cell | *PLD2* | 4.01349384 | 8.53E-05 | 0.002889125 | 0.536277914 |
| Heart_Failure | b_cell | *ATP1B2* | -8.267791945 | 0.000254623 | 0.003777026 | 1 |

| | | | | | | |
|---|---|---|---|---|---|---|
| Heart_Failure | b_cell | *CCL4* | -1.990757493 | 0.001905497 | 0.008705907 | 1 |
| Heart_Failure | b_cell | *CDKN2D* | 2.190690457 | 0.000173959 | 0.003365162 | 1 |
| Heart_Failure | b_cell | *MTUS1* | -1.64950546 | 0.021603824 | 0.038141881 | 1 |
| Heart_Failure | b_cell | *NGDN* | -2.290390897 | 0.005438209 | 0.014347469 | 1 |
| Heart_Failure | b_cell | *RIPK3* | -3.30345044 | 0.001561357 | 0.008654124 | 1 |
| Heart_Failure | b_cell | *ADCY4* | -2.473722773 | 0.000639502 | 0.006700919 | 1 |
| Heart_Failure | b_cell | *RAB2B* | -2.080167241 | 0.005753856 | 0.014941962 | 1 |
| Heart_Failure | b_cell | *EPB41L4A* | -1.19094406 | 0.002053831 | 0.008831917 | 1 |
| Heart_Failure | b_cell | *CDO1* | -1.700061661 | 0.001966174 | 0.008766903 | 1 |
| Heart_Failure | b_cell | *QRICH2* | -2.549063026 | 0.000773857 | 0.007698164 | 1 |
| Heart_Failure | b_cell | *SEC14L1* | -1.611357701 | 0.013938531 | 0.027635302 | 1 |
| Heart_Failure | b_cell | *RHBDF2* | 2.570710949 | 0.00037251 | 0.004779534 | 1 |
| Heart_Failure | b_cell | *AANAT* | 3.713863848 | 0.001587796 | 0.008654124 | 1 |
| Heart_Failure | b_cell | *TMEM8A* | -2.609069404 | 0.023267489 | 0.040409587 | 1 |
| Heart_Failure | b_cell | *SHC2* | -1.93434186 | 0.003280446 | 0.010813912 | 1 |
| Heart_Failure | b_cell | *GALNT8* | -2.735398917 | 0.001532117 | 0.008654124 | 1 |
| Heart_Failure | b_cell | *DOCK6* | -1.359661204 | 0.004182 | 0.012265566 | 1 |
| Heart_Failure | b_cell | *ELOF1* | -3.245030092 | 0.002788313 | 0.009915204 | 1 |
| Heart_Failure | b_cell | *THEM6* | -2.450935869 | 0.003031243 | 0.010295744 | 1 |
| Heart_Failure | b_cell | *TOMM40* | -4.089975075 | 0.003516689 | 0.011161493 | 1 |
| Heart_Failure | b_cell | *DPP6* | -2.525229957 | 0.003443879 | 0.011080691 | 1 |
| Heart_Failure | b_cell | *FAM98C* | 5.546263522 | 4.93E-05 | 0.002889125 | 0.309916827 |
| Heart_Failure | b_cell | *RPL36* | 4.472216792 | 0.006383561 | 0.015933676 | 1 |
| Heart_Failure | b_cell | *ATP8B3* | -2.193299786 | 0.024702184 | 0.042256423 | 1 |
| Heart_Failure | b_cell | *BST2* | -2.306025898 | 0.005329338 | 0.014191254 | 1 |
| Heart_Failure | b_cell | *DDA1* | -2.331034169 | 0.001209092 | 0.008654124 | 1 |
| Heart_Failure | b_cell | *TULP4* | -1.856605407 | 0.027427356 | 0.045921648 | 1 |
| Heart_Failure | b_cell | *RTN4IP1* | -2.525821232 | 0.016542108 | 0.0312313 | 1 |
| Heart_Failure | b_cell | *QRSL1* | 3.49133895 | 0.00056004 | 0.006242861 | 1 |
| Heart_Failure | b_cell | *PXDN* | -1.16893183 | 0.001687586 | 0.008654124 | 1 |
| Heart_Failure | b_cell | *PGPEP1* | 3.766235929 | 0.009458027 | 0.020689846 | 1 |
| Heart_Failure | b_cell | *CAMSAP1* | -2.253143821 | 0.012041159 | 0.024715235 | 1 |
| Heart_Failure | b_cell | *TNNT3* | -2.866548856 | 0.001188825 | 0.008654124 | 1 |
| Heart_Failure | b_cell | *TUBGCP2* | -1.941284521 | 0.007849228 | 0.018236548 | 1 |
| Heart_Failure | b_cell | *CYP2E1* | 5.521994791 | 0.005394759 | 0.014310906 | 1 |
| Heart_Failure | b_cell | *PAK4* | -2.250446301 | 0.003983249 | 0.011982146 | 1 |
| Heart_Failure | b_cell | *ZNF337* | -1.741162163 | 0.020446777 | 0.036509197 | 1 |
| Heart_Failure | b_cell | *CEP85* | 4.251958523 | 0.010708953 | 0.022784157 | 1 |
| Heart_Failure | b_cell | *POMT1* | -2.610843915 | 0.012776087 | 0.025844035 | 1 |
| Heart_Failure | b_cell | *UCK1* | 4.698628352 | 2.77E-06 | 0.001060433 | 0.017391913 |
| Heart_Failure | b_cell | *MAP3K10* | -2.169487686 | 0.004540754 | 0.01284469 | 1 |
| Heart_Failure | b_cell | *SMPDL3B* | 5.242070606 | 0.000152201 | 0.003239931 | 0.956890222 |
| Heart_Failure | b_cell | *THEMIS2* | -1.816750807 | 0.00227447 | 0.009182983 | 1 |
| Heart_Failure | b_cell | *ZNF317* | -1.906763336 | 0.002490989 | 0.009470475 | 1 |
| Heart_Failure | b_cell | *PPAN* | 7.514584849 | 5.33E-05 | 0.002889125 | 0.335093585 |
| Heart_Failure | b_cell | *HSD17B3* | -2.096151813 | 0.008200465 | 0.018789711 | 1 |
| Heart_Failure | b_cell | *HABP4* | -1.708804466 | 0.002442045 | 0.009414769 | 1 |
| Heart_Failure | b_cell | *EPS8L1* | 4.381233631 | 0.0001724 | 0.003365162 | 1 |
| Heart_Failure | b_cell | *LILRB2* | -2.501074963 | 0.001814939 | 0.008654124 | 1 |
| Heart_Failure | b_cell | *COX4I2* | 2.650219005 | 9.94E-05 | 0.002990199 | 0.624951568 |

| | | | | | | |
|---|---|---|---|---|---|---|
| Heart_Failure | b_cell | *ATP6V1E1* | -2.113272588 | 0.005102473 | 0.013821305 | 1 |
| Heart_Failure | b_cell | *CHMP1A* | 8.220453592 | 6.29E-06 | 0.001978716 | 0.039574315 |
| Heart_Failure | b_cell | *PPT1* | -1.833625268 | 0.003525854 | 0.011178539 | 1 |
| Heart_Failure | b_cell | *HAUS8* | -2.679523383 | 0.002173099 | 0.009019688 | 1 |
| Heart_Failure | b_cell | *RFTN1* | -1.417024157 | 0.005573648 | 0.014606723 | 1 |
| Heart_Failure | b_cell | *PDLIM4* | 14.09724296 | 7.46E-07 | 0.000586228 | 0.004689826 |
| Heart_Failure | b_cell | *AOC3* | -1.536159041 | 0.001704162 | 0.008654124 | 1 |
| Heart_Failure | b_cell | *AOC2* | -4.251943393 | 0.002396308 | 0.009333474 | 1 |
| Heart_Failure | b_cell | *ACAP3* | -1.998523423 | 0.003015797 | 0.010271026 | 1 |
| Heart_Failure | b_cell | *C1orf159* | -2.331452613 | 0.018039283 | 0.033297995 | 1 |
| Heart_Failure | b_cell | *NINJ1* | -2.266496148 | 0.004123512 | 0.012176854 | 1 |
| Heart_Failure | b_cell | *NPHP4* | -2.091950758 | 0.00285582 | 0.010013685 | 1 |
| Heart_Failure | b_cell | *MAP1B* | -0.838190662 | 0.022718822 | 0.039686923 | 1 |
| Heart_Failure | b_cell | *CKMT2* | -1.838099509 | 0.001368279 | 0.008654124 | 1 |
| Heart_Failure | b_cell | *KHDRBS3* | -1.51062434 | 0.002834864 | 0.010000392 | 1 |
| Heart_Failure | b_cell | *RBM8A* | -1.898129557 | 0.000715212 | 0.00731144 | 1 |
| Heart_Failure | b_cell | *ZNF304* | 7.102037111 | 0.000203163 | 0.00351869 | 1 |
| Heart_Failure | b_cell | *ZNF132* | -2.933665953 | 0.008068189 | 0.018566876 | 1 |
| Heart_Failure | b_cell | *RHPN2* | -3.312500231 | 0.001143233 | 0.008654124 | 1 |
| Heart_Failure | b_cell | *C19orf12* | -1.197747759 | 0.003180653 | 0.010589085 | 1 |
| Heart_Failure | b_cell | *GCH1* | -2.147360926 | 0.002449764 | 0.009416629 | 1 |
| Heart_Failure | b_cell | *DNAJB1* | -2.388546315 | 0.000608418 | 0.006565891 | 1 |
| Heart_Failure | b_cell | *FBXW9* | -3.116618693 | 0.003068357 | 0.01036364 | 1 |
| Heart_Failure | b_cell | *C19orf57* | -4.564987842 | 0.004446018 | 0.012718545 | 1 |
| Heart_Failure | b_cell | *MATN3* | -9.519690567 | 0.003370468 | 0.010905883 | 1 |
| Heart_Failure | b_cell | *CCT6B* | -3.410350265 | 0.002282469 | 0.009182983 | 1 |
| Heart_Failure | b_cell | *ENOSF1* | -1.439412111 | 0.008051284 | 0.018566285 | 1 |
| Heart_Failure | b_cell | *EMILIN2* | -1.461286966 | 0.013532716 | 0.026983885 | 1 |
| Heart_Failure | b_cell | *CNGA4* | -3.785746848 | 0.005074384 | 0.013804695 | 1 |
| Heart_Failure | b_cell | *IMMT* | -1.736350555 | 0.02233669 | 0.039182693 | 1 |
| Heart_Failure | b_cell | *MRPL35* | 3.325436314 | 0.025403157 | 0.043258301 | 1 |
| Heart_Failure | b_cell | *INPP5K* | -1.865186157 | 0.003610647 | 0.011367119 | 1 |
| Heart_Failure | b_cell | *MYBBP1A* | -2.477426667 | 0.000621667 | 0.006609617 | 1 |
| Heart_Failure | b_cell | *RPA1* | -1.785179171 | 0.019975551 | 0.035953705 | 1 |
| Heart_Failure | b_cell | *SERPINF1* | -1.715089792 | 0.002836122 | 0.010000392 | 1 |
| Heart_Failure | b_cell | *SEC61G* | -1.972310392 | 0.012104381 | 0.024836893 | 1 |
| Heart_Failure | b_cell | *FIGNL1* | -3.201357666 | 0.018917376 | 0.034553614 | 1 |
| Heart_Failure | b_cell | *WBP2* | -2.400637567 | 0.011020328 | 0.023235699 | 1 |
| Heart_Failure | b_cell | *XAF1* | -1.248195989 | 0.010415522 | 0.022280499 | 1 |
| Heart_Failure | b_cell | *REEP2* | -5.978267339 | 7.42E-07 | 0.000586228 | 0.004666963 |
| Heart_Failure | b_cell | *BTBD3* | -1.703881948 | 0.002082581 | 0.008855223 | 1 |
| Heart_Failure | b_cell | *PCNA* | 4.875739279 | 0.029375567 | 0.048574676 | 1 |
| Heart_Failure | b_cell | *MMACHC* | -3.307087451 | 0.000544162 | 0.006125623 | 1 |
| Heart_Failure | b_cell | *LPIN3* | -2.077495965 | 0.00298975 | 0.010215522 | 1 |
| Heart_Failure | b_cell | *DMGDH* | -1.519035459 | 0.001177239 | 0.008654124 | 1 |
| Heart_Failure | b_cell | *BHMT2* | -1.230255057 | 0.000354422 | 0.004664086 | 1 |
| Heart_Failure | b_cell | *KANK4* | -1.147164503 | 0.017573814 | 0.032659346 | 1 |
| Heart_Failure | b_cell | *ATP8A2* | -2.711528834 | 0.0016695 | 0.008654124 | 1 |
| Heart_Failure | b_cell | *WASF3* | -1.304133662 | 0.001605714 | 0.008654124 | 1 |
| Heart_Failure | b_cell | *RNF17* | 5.036141188 | 0.000877375 | 0.00834507 | 1 |

| | | | | | | |
|---|---|---|---|---|---|---|
| Heart_Failure | b_cell | *PEMT* | -1.908279912 | 0.001481872 | 0.008654124 | 1 |
| Heart_Failure | b_cell | *SCO1* | -2.468174865 | 0.030196193 | 0.049580429 | 1 |
| Heart_Failure | b_cell | *TMCC2* | -4.75177446 | 0.0004583 | 0.005355635 | 1 |
| Heart_Failure | b_cell | *TRPC4* | -3.079165339 | 0.002470038 | 0.00945169 | 1 |
| Heart_Failure | b_cell | *RFC3* | -2.055007872 | 0.007295945 | 0.017381435 | 1 |
| Heart_Failure | b_cell | *PRAM1* | -2.522497049 | 0.001252241 | 0.008654124 | 1 |
| Heart_Failure | b_cell | *ZNF414* | -2.421074538 | 0.001693316 | 0.008654124 | 1 |
| Heart_Failure | b_cell | *CNDP2* | -2.499509469 | 0.005501966 | 0.014467111 | 1 |
| Heart_Failure | b_cell | *RTN3* | -1.510020074 | 0.008954186 | 0.019901071 | 1 |
| Heart_Failure | b_cell | *C1QTNF6* | -2.254989963 | 0.001871253 | 0.008701813 | 1 |
| Heart_Failure | b_cell | *FAM83F* | -4.915122818 | 0.002028168 | 0.008831917 | 1 |
| Heart_Failure | b_cell | *GIMAP4* | -1.60519239 | 0.008200886 | 0.018789711 | 1 |
| Heart_Failure | b_cell | *AGAP3* | -2.0342244 | 0.009390989 | 0.020578998 | 1 |
| Heart_Failure | b_cell | *DYDC2* | -9.808323155 | 0.010136674 | 0.021896175 | 1 |
| Heart_Failure | b_cell | *TMTC1* | -1.116419666 | 0.002504173 | 0.009482826 | 1 |
| Heart_Failure | b_cell | *SPINK5* | -4.437560788 | 0.009113867 | 0.02012838 | 1 |
| Heart_Failure | b_cell | *LRRCC1* | -2.577923142 | 0.004193865 | 0.012283789 | 1 |
| Heart_Failure | b_cell | *AMPD3* | -2.089255308 | 0.000887818 | 0.008368383 | 1 |
| Heart_Failure | b_cell | *DPF2* | -2.293800367 | 0.004151659 | 0.012205437 | 1 |
| Heart_Failure | b_cell | *GSC* | -6.309099592 | 0.001698857 | 0.008654124 | 1 |
| Heart_Failure | b_cell | *CATSPERB* | -1.549723665 | 0.001724916 | 0.008654124 | 1 |
| Heart_Failure | b_cell | *COX16* | -3.774376173 | 0.008699621 | 0.019568701 | 1 |
| Heart_Failure | b_cell | *MED6* | -1.608642988 | 0.001814908 | 0.008654124 | 1 |
| Heart_Failure | b_cell | *ELP3* | -1.961202039 | 0.02307205 | 0.040180894 | 1 |
| Heart_Failure | b_cell | *MRO* | -3.270151178 | 0.006099714 | 0.015471746 | 1 |
| Heart_Failure | b_cell | *THUMPD3* | -2.612154199 | 0.005257118 | 0.014066038 | 1 |
| Heart_Failure | b_cell | *CNTN6* | -1.922914139 | 0.002188718 | 0.009035107 | 1 |
| Heart_Failure | b_cell | *KATNBL1* | -1.714661819 | 0.020849536 | 0.037070428 | 1 |
| Heart_Failure | b_cell | *GSTM1* | 3.648821722 | 0.005217226 | 0.014011405 | 1 |
| Heart_Failure | b_cell | *GSTM5* | -1.934211496 | 0.018203235 | 0.033492461 | 1 |
| Heart_Failure | b_cell | *SYT6* | -7.404779668 | 0.001135928 | 0.008654124 | 1 |
| Heart_Failure | b_cell | *VAV3* | 3.965677086 | 9.20E-05 | 0.002932166 | 0.578461769 |
| Heart_Failure | b_cell | *PSRC1* | 2.939214305 | 0.001109916 | 0.008654124 | 1 |
| Heart_Failure | b_cell | *WNT2B* | -1.998502744 | 0.00660987 | 0.016277421 | 1 |
| Heart_Failure | b_cell | *CEPT1* | -1.664988166 | 0.016204378 | 0.030741377 | 1 |
| Heart_Failure | b_cell | *AP4B1* | -2.810938597 | 0.008192315 | 0.018787704 | 1 |
| Heart_Failure | b_cell | *NAPG* | -2.219450049 | 0.020420402 | 0.036472462 | 1 |
| Heart_Failure | b_cell | *SPIRE1* | -1.964119824 | 0.003354733 | 0.010877363 | 1 |
| Heart_Failure | b_cell | *LPIN1* | -1.588877294 | 0.011288777 | 0.023636218 | 1 |
| Heart_Failure | b_cell | *LDHA* | -2.044592616 | 0.00988271 | 0.021425034 | 1 |
| Heart_Failure | b_cell | *ANO3* | -1.310426776 | 0.001123711 | 0.008654124 | 1 |
| Heart_Failure | b_cell | *FST* | -2.660009171 | 0.000967336 | 0.008654124 | 1 |
| Heart_Failure | b_cell | *NARS* | -2.11312224 | 0.012962241 | 0.026143821 | 1 |
| Heart_Failure | b_cell | *ECHDC3* | -1.685865345 | 0.002327891 | 0.0092234 | 1 |
| Heart_Failure | b_cell | *IL15RA* | -1.848408926 | 0.002713567 | 0.009825588 | 1 |
| Heart_Failure | b_cell | *TMEM241* | -1.586592785 | 0.006557594 | 0.016185231 | 1 |
| Heart_Failure | b_cell | *EMP1* | -1.532267265 | 0.011710492 | 0.024218377 | 1 |
| Heart_Failure | b_cell | *KLRD1* | -1.602113046 | 0.003468224 | 0.011112537 | 1 |
| Heart_Failure | b_cell | *PRH2* | -6.764037438 | 0.000532673 | 0.006044971 | 1 |
| Heart_Failure | b_cell | *ACP2* | 7.924096508 | 0.000190967 | 0.003445165 | 1 |

| | | | | | | |
|---|---|---|---|---|---|---|
| Heart_Failure | b_cell | *HOOK1* | 2.694803308 | 0.000145021 | 0.003239931 | 0.91174403 |
| Heart_Failure | b_cell | *FHOD3* | -1.23294518 | 0.003635592 | 0.011422772 | 1 |
| Heart_Failure | b_cell | *TPGS2* | -1.853130587 | 0.012490297 | 0.025429566 | 1 |
| Heart_Failure | b_cell | *TIMM10* | -2.465475669 | 0.01211311 | 0.024846696 | 1 |
| Heart_Failure | b_cell | *DHX34* | -2.676791242 | 0.00443435 | 0.012718545 | 1 |
| Heart_Failure | b_cell | *APLNR* | -3.064924307 | 0.001213861 | 0.008654124 | 1 |
| Heart_Failure | b_cell | *FADS2* | -2.094833143 | 0.002454522 | 0.009420988 | 1 |
| Heart_Failure | b_cell | *TMEM165* | -1.618351519 | 0.016580552 | 0.031285094 | 1 |
| Heart_Failure | b_cell | *CLDN10* | 2.110454347 | 0.000197249 | 0.003454339 | 1 |
| Heart_Failure | b_cell | *DZIP1* | -1.853484431 | 0.002739997 | 0.009847411 | 1 |
| Heart_Failure | b_cell | *CARS2* | -1.796039758 | 0.008717734 | 0.019595421 | 1 |
| Heart_Failure | b_cell | *STT3A* | -2.161094385 | 0.00792451 | 0.018343665 | 1 |
| Heart_Failure | b_cell | *KLB* | -1.170769788 | 0.00384555 | 0.011759227 | 1 |
| Heart_Failure | b_cell | *WDR36* | -2.130002904 | 0.021078516 | 0.037389958 | 1 |
| Heart_Failure | b_cell | *TMEM2* | -1.535275847 | 0.0069278 | 0.016810143 | 1 |
| Heart_Failure | b_cell | *AGTPBP1* | -1.790124215 | 0.017580921 | 0.032662899 | 1 |
| Heart_Failure | b_cell | *GOLM1* | -2.931637609 | 0.002292084 | 0.009185338 | 1 |
| Heart_Failure | b_cell | *ADAM19* | 1.054000634 | 1.20E-05 | 0.002644519 | 0.075171745 |
| Heart_Failure | b_cell | *HRK* | 2.914538396 | 0.000224422 | 0.003674323 | 1 |
| Heart_Failure | b_cell | *P2RX4* | -2.258499521 | 0.001895313 | 0.008705907 | 1 |
| Heart_Failure | b_cell | *DTX1* | -1.508774846 | 0.003092671 | 0.010397658 | 1 |
| Heart_Failure | b_cell | *CCDC146* | -1.738391266 | 0.011714478 | 0.024218654 | 1 |
| Heart_Failure | b_cell | *TMEM60* | 5.44564051 | 0.004454735 | 0.012718855 | 1 |
| Heart_Failure | b_cell | *POM121C* | -2.844178321 | 0.000697404 | 0.007199636 | 1 |
| Heart_Failure | b_cell | *MDFIC* | -1.265333845 | 0.016603709 | 0.031309994 | 1 |
| Heart_Failure | b_cell | *KHDC1* | -2.262255558 | 0.001245825 | 0.008654124 | 1 |
| Heart_Failure | b_cell | *NT5E* | -1.65987874 | 0.001958611 | 0.008764262 | 1 |
| Heart_Failure | b_cell | *EPHA7* | -1.590504995 | 0.006709493 | 0.016432639 | 1 |
| Heart_Failure | b_cell | *PRR5L* | -1.644359457 | 0.004059443 | 0.012101338 | 1 |
| Heart_Failure | b_cell | *NAT10* | -2.378610172 | 0.009796212 | 0.021288898 | 1 |
| Heart_Failure | b_cell | *ITGA7* | -1.280037467 | 0.000333351 | 0.004459106 | 1 |
| Heart_Failure | b_cell | *RDH5* | -1.228848474 | 0.002116917 | 0.008932253 | 1 |
| Heart_Failure | b_cell | *BLOC1S1* | -3.385278482 | 0.005259942 | 0.014066038 | 1 |
| Heart_Failure | b_cell | *PPP1R1A* | -2.491244316 | 0.004077905 | 0.012126691 | 1 |
| Heart_Failure | b_cell | *B4GALNT1* | 4.507232657 | 0.000140595 | 0.003214256 | 0.88392043 |
| Heart_Failure | b_cell | *FAIM2* | -4.96822886 | 0.0014482 | 0.008654124 | 1 |
| Heart_Failure | b_cell | *KRT7* | 2.17927813 | 0.000179373 | 0.00338933 | 1 |
| Heart_Failure | b_cell | *SLC26A10* | -7.574578342 | 0.001711674 | 0.008654124 | 1 |
| Heart_Failure | b_cell | *MAP7* | -1.494090857 | 0.002083168 | 0.008855223 | 1 |
| Heart_Failure | b_cell | *AHI1* | -1.726937604 | 0.000785754 | 0.007767351 | 1 |
| Heart_Failure | b_cell | *PKIB* | -2.257802909 | 0.003727857 | 0.011549198 | 1 |
| Heart_Failure | b_cell | *SMPD2* | 4.849579595 | 2.19E-05 | 0.002889125 | 0.137502453 |
| Heart_Failure | b_cell | *MICAL1* | -2.986604668 | 0.013198113 | 0.026527025 | 1 |
| Heart_Failure | b_cell | *PRADC1* | 5.358563611 | 6.31E-05 | 0.002889125 | 0.396656391 |
| Heart_Failure | b_cell | *SMYD5* | -2.597976849 | 0.002376917 | 0.009318117 | 1 |
| Heart_Failure | b_cell | *DYSF* | -1.847137718 | 0.002785975 | 0.009915204 | 1 |
| Heart_Failure | b_cell | *KCNMB4* | 2.819465082 | 0.000108563 | 0.003034612 | 0.682536108 |
| Heart_Failure | b_cell | *KLHL36* | -2.101007565 | 0.006842448 | 0.01666091 | 1 |
| Heart_Failure | b_cell | *MPHOSPH6* | 6.184276899 | 0.000429833 | 0.005135387 | 1 |
| Heart_Failure | b_cell | *FHOD1* | -1.946274735 | 0.001108382 | 0.008654124 | 1 |

| | | | | | | |
|---|---|---|---|---|---|---|
| Heart_Failure | b_cell | *CCDC102A* | -2.631384991 | 0.013935139 | 0.027635302 | 1 |
| Heart_Failure | b_cell | *ZNF670* | 3.384432789 | 0.000172496 | 0.003365162 | 1 |
| Heart_Failure | b_cell | *NTPCR* | -2.181755292 | 0.012894372 | 0.026041412 | 1 |
| Heart_Failure | b_cell | *RNASEL* | -2.382651838 | 0.001865111 | 0.008690419 | 1 |
| Heart_Failure | b_cell | *PIGC* | -2.036530914 | 0.020076509 | 0.036063147 | 1 |
| Heart_Failure | b_cell | *SP110* | 4.437591375 | 0.000165779 | 0.00334055 | 1 |
| Heart_Failure | b_cell | *SLC19A3* | -1.040379056 | 0.002345553 | 0.009261519 | 1 |
| Heart_Failure | b_cell | *TMBIM1* | -1.955159195 | 0.001718368 | 0.008654124 | 1 |
| Heart_Failure | b_cell | *ARMC9* | -2.327556089 | 0.022253832 | 0.039080961 | 1 |
| Heart_Failure | b_cell | *TMEM127* | -2.659437673 | 0.006879978 | 0.016714065 | 1 |
| Heart_Failure | b_cell | *USP44* | -2.489920306 | 0.009806246 | 0.021303341 | 1 |
| Heart_Failure | b_cell | *DRAM1* | -1.814268308 | 0.022854023 | 0.03987878 | 1 |
| Heart_Failure | b_cell | *SLC41A2* | -1.97599349 | 0.015137141 | 0.029354474 | 1 |
| Heart_Failure | b_cell | *TBC1D4* | -1.514292642 | 0.027822594 | 0.04647201 | 1 |
| Heart_Failure | b_cell | *SUCLA2* | -1.849418116 | 0.007066115 | 0.017040532 | 1 |
| Heart_Failure | b_cell | *RCBTB1* | -2.463897032 | 0.007266765 | 0.017344781 | 1 |
| Heart_Failure | b_cell | *RCBTB2* | -1.818884765 | 0.006380787 | 0.015933676 | 1 |
| Heart_Failure | b_cell | *SPDYE1* | -2.314272375 | 0.001718756 | 0.008654124 | 1 |
| Heart_Failure | b_cell | *CHST12* | -2.392738385 | 0.012838475 | 0.025945192 | 1 |
| Heart_Failure | b_cell | *GPNMB* | -1.458549425 | 0.00272021 | 0.009828026 | 1 |
| Heart_Failure | b_cell | *NUPL2* | -2.145525609 | 0.015447355 | 0.029763262 | 1 |
| Heart_Failure | b_cell | *AOAH* | -1.374982344 | 0.001482518 | 0.008654124 | 1 |
| Heart_Failure | b_cell | *TBRG4* | 4.214813171 | 0.000238368 | 0.003709453 | 1 |
| Heart_Failure | b_cell | *HUS1* | -1.833526306 | 0.005614469 | 0.014670893 | 1 |
| Heart_Failure | b_cell | *NACAD* | -10.06625055 | 0.001036239 | 0.008654124 | 1 |
| Heart_Failure | b_cell | *DBNL* | 5.32469217 | 0.00018517 | 0.00340015 | 1 |
| Heart_Failure | b_cell | *ALPK3* | -1.760327386 | 0.001179634 | 0.008654124 | 1 |
| Heart_Failure | b_cell | *TM6SF1* | -1.805358471 | 0.002268435 | 0.009180605 | 1 |
| Heart_Failure | b_cell | *RSAD1* | -2.527648509 | 0.01085493 | 0.02300133 | 1 |
| Heart_Failure | b_cell | *MYCBPAP* | -2.173091354 | 0.018781766 | 0.034365821 | 1 |
| Heart_Failure | b_cell | *BRIP1* | 2.869917571 | 7.44E-05 | 0.002889125 | 0.467496777 |
| Heart_Failure | b_cell | *BLK* | 0.111129502 | 0.000232103 | 0.003705045 | 1 |
| Heart_Failure | b_cell | *IL1RN* | 3.167673975 | 6.05E-05 | 0.002889125 | 0.380059665 |
| Heart_Failure | b_cell | *GYPC* | -1.743312325 | 0.010452832 | 0.022352707 | 1 |
| Heart_Failure | b_cell | *LRRC8A* | -1.973695128 | 0.006052967 | 0.015400648 | 1 |
| Heart_Failure | b_cell | *TXN* | -2.142811399 | 0.010604189 | 0.022622511 | 1 |
| Heart_Failure | b_cell | *TOR1B* | -2.181745023 | 0.003566902 | 0.011288665 | 1 |
| Heart_Failure | b_cell | *TOR1A* | -2.473545483 | 0.002194062 | 0.009044448 | 1 |
| Heart_Failure | b_cell | *SLC2A8* | -2.808978965 | 0.007830831 | 0.018207261 | 1 |
| Heart_Failure | b_cell | *ZFP37* | -2.508362868 | 0.001013393 | 0.008654124 | 1 |
| Heart_Failure | b_cell | *ALDOB* | -3.609064203 | 0.010826538 | 0.022972137 | 1 |
| Heart_Failure | b_cell | *PRPF4* | 4.835397292 | 0.001595213 | 0.008654124 | 1 |
| Heart_Failure | b_cell | *FPGS* | -2.06895253 | 0.006100598 | 0.015471746 | 1 |
| Heart_Failure | b_cell | *TEX10* | -1.968347783 | 0.005979306 | 0.015318111 | 1 |
| Heart_Failure | b_cell | *GARNL3* | -2.036900385 | 0.024404188 | 0.041897633 | 1 |
| Heart_Failure | b_cell | *GABBR2* | -4.702863549 | 0.001765667 | 0.008654124 | 1 |
| Heart_Failure | b_cell | *RABEPK* | -1.93009009 | 0.00378377 | 0.011617979 | 1 |
| Heart_Failure | b_cell | *GOLGA1* | -1.64230419 | 0.003687166 | 0.011475847 | 1 |
| Heart_Failure | b_cell | *LMX1B* | -5.927517087 | 0.001665998 | 0.008654124 | 1 |
| Heart_Failure | b_cell | *DSCC1* | -1.43607517 | 0.002639562 | 0.009653826 | 1 |

| | | | | | | |
|---|---|---|---|---|---|---|
| Heart_Failure | b_cell | *NOV* | -3.389122841 | 0.004143182 | 0.012205437 | 1 |
| Heart_Failure | b_cell | *POLR1E* | -2.462348092 | 0.006646779 | 0.016328403 | 1 |
| Heart_Failure | b_cell | *IL11RA* | -2.660615779 | 0.001910058 | 0.008705907 | 1 |
| Heart_Failure | b_cell | *TLN1* | -1.586706755 | 0.007459247 | 0.017643448 | 1 |
| Heart_Failure | b_cell | *SPAG8* | 6.635209645 | 0.000793553 | 0.007819858 | 1 |
| Heart_Failure | b_cell | *TMEM8B* | 6.53347498 | 0.000842996 | 0.008166279 | 1 |
| Heart_Failure | b_cell | *ARHGEF39* | 4.00325951 | 5.21E-05 | 0.002889125 | 0.327655192 |
| Heart_Failure | b_cell | *PPIL1* | 5.438851218 | 5.73E-05 | 0.002889125 | 0.360341 |
| Heart_Failure | b_cell | *KIF13A* | -1.432079701 | 0.012457767 | 0.025379773 | 1 |
| Heart_Failure | b_cell | *TMEM63B* | -1.849635681 | 0.005876263 | 0.015159648 | 1 |
| Heart_Failure | b_cell | *FRS3* | 7.731003458 | 0.000127591 | 0.003183203 | 0.802167122 |
| Heart_Failure | b_cell | *SLC22A23* | -1.552162451 | 0.004166294 | 0.012232927 | 1 |
| Heart_Failure | b_cell | *LRRC1* | -1.493980403 | 0.004519892 | 0.012811795 | 1 |
| Heart_Failure | b_cell | *BPHL* | -1.162446778 | 0.007201276 | 0.017247398 | 1 |
| Heart_Failure | b_cell | *RIPK1* | -1.849398552 | 0.013554011 | 0.026992647 | 1 |
| Heart_Failure | b_cell | *TUBB2B* | -3.559966507 | 0.007571243 | 0.017807857 | 1 |
| Heart_Failure | b_cell | *TCF19* | -5.368740732 | 0.000130347 | 0.003188681 | 0.819490984 |
| Heart_Failure | b_cell | *FGFBP2* | 5.556612053 | 0.005326218 | 0.014188955 | 1 |
| Heart_Failure | b_cell | *FHDC1* | -2.547326799 | 0.001711048 | 0.008654124 | 1 |
| Heart_Failure | b_cell | *MYO7A* | -2.180721052 | 0.002101949 | 0.00889896 | 1 |
| Heart_Failure | b_cell | *ARRB1* | -1.811872565 | 0.016851297 | 0.031559161 | 1 |
| Heart_Failure | b_cell | *CCDC90B* | -2.165180442 | 0.005213729 | 0.014007998 | 1 |
| Heart_Failure | b_cell | *SYTL2* | -1.637001879 | 0.00105033 | 0.008654124 | 1 |
| Heart_Failure | b_cell | *PI15* | 4.141968846 | 0.000121273 | 0.003135985 | 0.762445753 |
| Heart_Failure | b_cell | *TTPA* | -1.21979591 | 0.005285169 | 0.014124665 | 1 |
| Heart_Failure | b_cell | *GGH* | -2.333251896 | 0.000733533 | 0.007402443 | 1 |
| Heart_Failure | b_cell | *SULF1* | -1.100967755 | 0.001821174 | 0.008654124 | 1 |
| Heart_Failure | b_cell | *DCUN1D5* | -1.337937698 | 0.005324562 | 0.014188955 | 1 |
| Heart_Failure | b_cell | *TRIM29* | -3.217893176 | 0.005180483 | 0.013936542 | 1 |
| Heart_Failure | b_cell | *POU2F3* | 2.706056201 | 0.000234949 | 0.003705045 | 1 |
| Heart_Failure | b_cell | *PPP2R1B* | -1.061087628 | 0.001684814 | 0.008654124 | 1 |
| Heart_Failure | b_cell | *FDX1* | -2.207564578 | 0.012398648 | 0.025289758 | 1 |
| Heart_Failure | b_cell | *TMPRSS13* | -6.611980024 | 0.028405721 | 0.047295224 | 1 |
| Heart_Failure | b_cell | *NDUFAF1* | -3.117949699 | 0.004967403 | 0.013584195 | 1 |
| Heart_Failure | b_cell | *ITGA11* | -1.35687935 | 0.001638712 | 0.008654124 | 1 |
| Heart_Failure | b_cell | *PAQR5* | -3.004321522 | 0.003588814 | 0.011326743 | 1 |
| Heart_Failure | b_cell | *ITPKA* | 4.210704661 | 0.000559606 | 0.006242861 | 1 |
| Heart_Failure | b_cell | *UACA* | -1.490252222 | 0.012837711 | 0.025945192 | 1 |
| Heart_Failure | b_cell | *PLCB2* | -1.752616097 | 0.001931829 | 0.008732736 | 1 |
| Heart_Failure | b_cell | *DNASE2B* | -15.55084034 | 0.005358629 | 0.014239096 | 1 |
| Heart_Failure | b_cell | *ADCY3* | -2.240952088 | 0.008328467 | 0.01899194 | 1 |
| Heart_Failure | b_cell | *LHCGR* | -2.36794832 | 0.001384591 | 0.008654124 | 1 |
| Heart_Failure | b_cell | *PREB* | -1.784274188 | 0.007081952 | 0.01706618 | 1 |
| Heart_Failure | b_cell | *ABCG5* | -2.422345849 | 0.021894994 | 0.038558494 | 1 |
| Heart_Failure | b_cell | *CENPO* | 9.212428578 | 0.000578051 | 0.006331367 | 1 |
| Heart_Failure | b_cell | *DTNB* | -1.776278111 | 0.010175372 | 0.021953521 | 1 |
| Heart_Failure | b_cell | *CYP2C9* | 3.958063381 | 2.05E-06 | 0.000884152 | 0.012857398 |
| Heart_Failure | b_cell | *CYP2C8* | 7.874221331 | 0.003322039 | 0.01083435 | 1 |
| Heart_Failure | b_cell | *MYOF* | -1.728213131 | 0.011311993 | 0.02365885 | 1 |
| Heart_Failure | b_cell | *STAMBPL1* | 4.772101524 | 0.001129314 | 0.008654124 | 1 |

| | | | | | | |
|---|---|---|---|---|---|---|
| Heart_Failure | b_cell | *TACC2* | -1.218288967 | 0.00250897 | 0.009490913 | 1 |
| Heart_Failure | b_cell | *CALHM2* | -1.829457916 | 0.004403863 | 0.01267545 | 1 |
| Heart_Failure | b_cell | *ARL3* | -2.017836218 | 0.013950198 | 0.027646383 | 1 |
| Heart_Failure | b_cell | *PLCE1* | -1.425858301 | 0.003784575 | 0.011617979 | 1 |
| Heart_Failure | b_cell | *RBP4* | -1.766063471 | 0.003660498 | 0.011449527 | 1 |
| Heart_Failure | b_cell | *BARD1* | -2.550125333 | 0.009816589 | 0.021309791 | 1 |
| Heart_Failure | b_cell | *STAT4* | -1.480366699 | 0.002687202 | 0.009763417 | 1 |
| Heart_Failure | b_cell | *ASNSD1* | -1.921588939 | 0.001983498 | 0.008775328 | 1 |
| Heart_Failure | b_cell | *MDH1B* | 2.416690501 | 0.00021182 | 0.003589525 | 1 |
| Heart_Failure | b_cell | *FAM117B* | 3.694393658 | 0.026525578 | 0.044745455 | 1 |
| Heart_Failure | b_cell | *COX17* | 6.11538139 | 0.000137462 | 0.003214256 | 0.86422048 |
| Heart_Failure | b_cell | *USP8* | -1.628722802 | 0.016471608 | 0.031144961 | 1 |
| Heart_Failure | b_cell | *UBL7* | -2.913562772 | 0.004748695 | 0.013227309 | 1 |
| Heart_Failure | b_cell | *FAM13A* | -1.288584635 | 0.009330107 | 0.020470122 | 1 |
| Heart_Failure | b_cell | *PRKG2* | -1.784123323 | 0.001769999 | 0.008654124 | 1 |
| Heart_Failure | b_cell | *BBS7* | -2.064691724 | 0.005444108 | 0.014350989 | 1 |
| Heart_Failure | b_cell | *PRDM5* | -1.535854129 | 0.002370489 | 0.009308722 | 1 |
| Heart_Failure | b_cell | *TRPC3* | -3.936631103 | 0.002726273 | 0.009828026 | 1 |
| Heart_Failure | b_cell | *NAAA* | -2.038968907 | 0.02029269 | 0.036333958 | 1 |
| Heart_Failure | b_cell | *NUP54* | -1.805464312 | 0.019558978 | 0.035424135 | 1 |
| Heart_Failure | b_cell | *ANXA3* | -2.091754556 | 0.001572172 | 0.008654124 | 1 |
| Heart_Failure | b_cell | *ENPEP* | -2.03449866 | 0.00141696 | 0.008654124 | 1 |
| Heart_Failure | b_cell | *SLC39A8* | -2.519081596 | 0.018143004 | 0.033417044 | 1 |
| Heart_Failure | b_cell | *FBN2* | -1.679739606 | 0.007257846 | 0.017331752 | 1 |
| Heart_Failure | b_cell | *ERP27* | 3.321126968 | 0.000253017 | 0.003769479 | 1 |
| Heart_Failure | b_cell | *KIF21A* | -1.32694876 | 0.001617747 | 0.008654124 | 1 |
| Heart_Failure | b_cell | *ALG10* | -3.375691036 | 0.003937505 | 0.011883725 | 1 |
| Heart_Failure | b_cell | *SLCO1C1* | -2.575310757 | 0.004732038 | 0.013198901 | 1 |
| Heart_Failure | b_cell | *PRICKLE1* | -1.183021461 | 0.001631396 | 0.008654124 | 1 |
| Heart_Failure | b_cell | *C1RL* | -1.67315911 | 0.007974062 | 0.01841768 | 1 |
| Heart_Failure | b_cell | *CLSTN3* | -3.250227807 | 0.027971488 | 0.046671111 | 1 |
| Heart_Failure | b_cell | *KLRG1* | -2.067067391 | 0.003395479 | 0.010972165 | 1 |
| Heart_Failure | b_cell | *VAMP1* | -2.645852567 | 0.006355646 | 0.015908678 | 1 |
| Heart_Failure | b_cell | *CD27* | -2.939156953 | 0.004123162 | 0.012176854 | 1 |
| Heart_Failure | b_cell | *PEX5* | 4.049302128 | 0.000106284 | 0.003034612 | 0.668206572 |
| Heart_Failure | b_cell | *PPFIA2* | -1.245989882 | 0.001834889 | 0.008654124 | 1 |
| Heart_Failure | b_cell | *LGR5* | -4.117191187 | 3.28E-05 | 0.002889125 | 0.206077656 |
| Heart_Failure | b_cell | *AMDHD1* | -2.468404935 | 0.022494296 | 0.039393214 | 1 |
| Heart_Failure | b_cell | *GAS2L3* | -1.739989162 | 0.001718242 | 0.008654124 | 1 |
| Heart_Failure | b_cell | *TMEM132B* | -3.924342034 | 0.002791258 | 0.009915204 | 1 |
| Heart_Failure | b_cell | *TDG* | -2.025922855 | 0.019426266 | 0.035257775 | 1 |
| Heart_Failure | b_cell | *MMAB* | -2.129189185 | 0.003820369 | 0.011710708 | 1 |
| Heart_Failure | b_cell | *GLTP* | -2.343276955 | 0.001891273 | 0.008705907 | 1 |
| Heart_Failure | b_cell | *MTMR6* | -1.924667458 | 0.002739627 | 0.009847411 | 1 |
| Heart_Failure | b_cell | *SLC46A3* | -1.933677331 | 0.003741702 | 0.011555523 | 1 |
| Heart_Failure | b_cell | *CCDC65* | -2.046310233 | 0.005891439 | 0.015176115 | 1 |
| Heart_Failure | b_cell | *ACVRL1* | -2.566961351 | 0.004834331 | 0.013359754 | 1 |
| Heart_Failure | b_cell | *KANSL2* | -1.528028491 | 0.002443166 | 0.009414769 | 1 |
| Heart_Failure | b_cell | *CERS5* | -2.157889919 | 0.029776711 | 0.049109702 | 1 |
| Heart_Failure | b_cell | *ZNF740* | -2.166889343 | 0.004906511 | 0.013476294 | 1 |

| | | | | | | |
|---|---|---|---|---|---|---|
| Heart_Failure | b_cell | *HNRNPA1L2* | 8.78624367 | 3.70E-05 | 0.002889125 | 0.232627008 |
| Heart_Failure | b_cell | *LPAR6* | -1.793196055 | 0.016409155 | 0.031054893 | 1 |
| Heart_Failure | b_cell | *VPS33A* | -3.181084668 | 0.004102712 | 0.012159073 | 1 |
| Heart_Failure | b_cell | *METTL21C* | 13.11647036 | 0.000240678 | 0.003717799 | 1 |
| Heart_Failure | b_cell | *GRTP1* | -1.746957849 | 0.003576852 | 0.011306016 | 1 |
| Heart_Failure | b_cell | *CBLN3* | 1.459977457 | 0.00162415 | 0.008654124 | 1 |
| Heart_Failure | b_cell | *TSSK4* | -12.08583218 | 0.001088895 | 0.008654124 | 1 |
| Heart_Failure | b_cell | *TMX1* | -2.353476342 | 0.015805744 | 0.030228539 | 1 |
| Heart_Failure | b_cell | *RTN1* | -1.216349606 | 0.001765464 | 0.008654124 | 1 |
| Heart_Failure | b_cell | *C14orf37* | -1.888648079 | 0.01871523 | 0.034264022 | 1 |
| Heart_Failure | b_cell | *RAB15* | -1.958633777 | 0.004053575 | 0.012094678 | 1 |
| Heart_Failure | b_cell | *STON2* | -1.511641008 | 0.003909535 | 0.011873672 | 1 |
| Heart_Failure | b_cell | *PTGR2* | -1.884121393 | 0.005591192 | 0.014622222 | 1 |
| Heart_Failure | b_cell | *AK7* | -3.914103722 | 0.001584303 | 0.008654124 | 1 |
| Heart_Failure | b_cell | *FBLN5* | -1.330776834 | 0.001573502 | 0.008654124 | 1 |
| Heart_Failure | b_cell | *C14orf79* | -2.406494318 | 0.002534471 | 0.00953574 | 1 |
| Heart_Failure | b_cell | *NIPA2* | 4.894026064 | 0.010475259 | 0.022393047 | 1 |
| Heart_Failure | b_cell | *DUOXA1* | -3.190452945 | 0.019610799 | 0.035490239 | 1 |
| Heart_Failure | b_cell | *MFAP1* | -2.327152149 | 0.024000755 | 0.041419914 | 1 |
| Heart_Failure | b_cell | *SORD* | 9.110497068 | 3.19E-05 | 0.002889125 | 0.200740991 |
| Heart_Failure | b_cell | *ZSCAN29* | -2.632666161 | 0.012477344 | 0.025411422 | 1 |
| Heart_Failure | b_cell | *BAHD1* | -2.542017416 | 0.005034743 | 0.01372689 | 1 |
| Heart_Failure | b_cell | *DISP2* | 1.934275119 | 0.014621981 | 0.028629211 | 1 |
| Heart_Failure | b_cell | *ANP32A* | -1.900988036 | 0.007455331 | 0.017643448 | 1 |
| Heart_Failure | b_cell | *UBE2Q2* | -1.999071713 | 0.02452751 | 0.042074885 | 1 |
| Heart_Failure | b_cell | *PSTPIP1* | -2.564788119 | 0.004665538 | 0.013083069 | 1 |
| Heart_Failure | b_cell | *HMG20A* | -1.803350456 | 0.019889713 | 0.035850523 | 1 |
| Heart_Failure | b_cell | *NEIL1* | -1.783293201 | 0.000978365 | 0.008654124 | 1 |
| Heart_Failure | b_cell | *MAN2C1* | -1.15035511 | 0.015823915 | 0.030235497 | 1 |
| Heart_Failure | b_cell | *DNAJA4* | -2.322247869 | 0.002277785 | 0.009182983 | 1 |
| Heart_Failure | b_cell | *TPM1* | -1.718049172 | 0.021747341 | 0.038353392 | 1 |
| Heart_Failure | b_cell | *IGF1R* | -1.503314503 | 0.007414895 | 0.017591489 | 1 |
| Heart_Failure | b_cell | *CYP11A1* | -2.025394913 | 0.024358682 | 0.041865235 | 1 |
| Heart_Failure | b_cell | *BBS4* | -2.198086839 | 0.024678515 | 0.042247613 | 1 |
| Heart_Failure | b_cell | *ADAMTS17* | -1.61247991 | 0.00096534 | 0.008654124 | 1 |
| Heart_Failure | b_cell | *HAPLN3* | -2.610288039 | 0.002050582 | 0.008831917 | 1 |
| Heart_Failure | b_cell | *POLG* | -2.440727458 | 0.024649533 | 0.042238107 | 1 |
| Heart_Failure | b_cell | *WDR93* | -1.22360934 | 0.013211086 | 0.026544614 | 1 |
| Heart_Failure | b_cell | *TICRR* | -4.101242258 | 0.003434486 | 0.01106179 | 1 |
| Heart_Failure | b_cell | *MFGE8* | -1.655047737 | 0.001228158 | 0.008654124 | 1 |
| Heart_Failure | b_cell | *ST8SIA2* | -16.28710676 | 0.001117328 | 0.008654124 | 1 |
| Heart_Failure | b_cell | *SH3GL3* | 2.881827484 | 0.000137253 | 0.003214256 | 0.862908558 |
| Heart_Failure | b_cell | *SEC11A* | -1.886339414 | 0.017269059 | 0.032178593 | 1 |
| Heart_Failure | b_cell | *TGFB1I1* | -2.549776662 | 0.002571502 | 0.009560634 | 1 |
| Heart_Failure | b_cell | *PARN* | -1.731756893 | 0.029189897 | 0.048319348 | 1 |
| Heart_Failure | b_cell | *TXNL4B* | -2.720997639 | 0.018215074 | 0.033504439 | 1 |
| Heart_Failure | b_cell | *MARVELD3* | 2.150527331 | 0.000187665 | 0.00340015 | 1 |
| Heart_Failure | b_cell | *ZFHX3* | -1.497161902 | 0.014079153 | 0.027817609 | 1 |
| Heart_Failure | b_cell | *KIFC3* | -2.309811498 | 0.02869197 | 0.04763453 | 1 |
| Heart_Failure | b_cell | *ADAMTS18* | -1.890015963 | 0.000582692 | 0.006360038 | 1 |

| | | | | | | |
|---|---|---|---|---|---|---|
| Heart_Failure | b_cell | *GCSH* | -2.541219748 | 0.002810174 | 0.009959169 | 1 |
| Heart_Failure | b_cell | *CMTM3* | -1.8646264 | 0.001856048 | 0.008687253 | 1 |
| Heart_Failure | b_cell | *MAP1LC3B* | -2.269867829 | 0.022625213 | 0.039578385 | 1 |
| Heart_Failure | b_cell | *TIGD7* | -2.134739313 | 0.002465081 | 0.009438469 | 1 |
| Heart_Failure | b_cell | *DEF8* | -1.675369032 | 0.005960659 | 0.015295782 | 1 |
| Heart_Failure | b_cell | *GALNS* | -2.764188081 | 0.007331597 | 0.017433339 | 1 |
| Heart_Failure | b_cell | *COPS3* | -2.128997787 | 0.003751824 | 0.011556943 | 1 |
| Heart_Failure | b_cell | *PRPSAP2* | 4.65426497 | 0.001206947 | 0.008654124 | 1 |
| Heart_Failure | b_cell | *RASL10B* | -8.613683016 | 0.000259573 | 0.003804045 | 1 |
| Heart_Failure | b_cell | *UNC45B* | -10.01804322 | 0.011402079 | 0.023789705 | 1 |
| Heart_Failure | b_cell | *TOM1L1* | -1.689799457 | 0.004787825 | 0.013283784 | 1 |
| Heart_Failure | b_cell | *C17orf80* | -2.138692446 | 0.016290889 | 0.03086824 | 1 |
| Heart_Failure | b_cell | *VPS53* | -1.857398679 | 0.019348743 | 0.035157673 | 1 |
| Heart_Failure | b_cell | *SPATA22* | -1.310087448 | 0.006104305 | 0.015474906 | 1 |
| Heart_Failure | b_cell | *LRRC46* | -4.441235309 | 0.020865802 | 0.037084534 | 1 |
| Heart_Failure | b_cell | *SCRN2* | -1.088510794 | 0.011912418 | 0.024506994 | 1 |
| Heart_Failure | b_cell | *RHBDL3* | -4.398256077 | 0.019785821 | 0.035714457 | 1 |
| Heart_Failure | b_cell | *ARSG* | -2.494090743 | 0.009177182 | 0.020209087 | 1 |
| Heart_Failure | b_cell | *ABCA8* | -1.052500884 | 0.001693088 | 0.008654124 | 1 |
| Heart_Failure | b_cell | *IMPA2* | -2.497204453 | 0.010890383 | 0.02303763 | 1 |
| Heart_Failure | b_cell | *GNAL* | -2.032567847 | 0.007524522 | 0.017751095 | 1 |
| Heart_Failure | b_cell | *C18orf21* | -3.058549824 | 0.003931166 | 0.011882923 | 1 |
| Heart_Failure | b_cell | *GALNT1* | -1.627907532 | 0.005286359 | 0.014124665 | 1 |
| Heart_Failure | b_cell | *ASXL3* | -2.389159395 | 0.002174834 | 0.009019688 | 1 |
| Heart_Failure | b_cell | *ADCYAP1* | 10.73990787 | 0.004503642 | 0.012782708 | 1 |
| Heart_Failure | b_cell | *GREB1L* | -1.196802239 | 0.001043223 | 0.008654124 | 1 |
| Heart_Failure | b_cell | *SLC14A1* | 5.835301229 | 0.000228085 | 0.003705045 | 1 |
| Heart_Failure | b_cell | *SLC13A5* | -13.75101769 | 0.020334909 | 0.036366807 | 1 |
| Heart_Failure | b_cell | *CCDC40* | -1.663640195 | 0.003253431 | 0.010776776 | 1 |
| Heart_Failure | b_cell | *TMC6* | -2.003458157 | 0.028199939 | 0.047014855 | 1 |
| Heart_Failure | b_cell | *CARD14* | -2.265595084 | 0.001531386 | 0.008654124 | 1 |
| Heart_Failure | b_cell | *EIF4A3* | 4.685446271 | 0.012147651 | 0.024901299 | 1 |
| Heart_Failure | b_cell | *TBCD* | -1.795964832 | 0.027448458 | 0.045944744 | 1 |
| Heart_Failure | b_cell | *FN3KRP* | -2.415133055 | 0.020065978 | 0.036054531 | 1 |
| Heart_Failure | b_cell | *CBX8* | 1.25205348 | 0.000115759 | 0.003135985 | 0.727777762 |
| Heart_Failure | b_cell | *SECTM1* | -4.324872364 | 0.001881135 | 0.008705907 | 1 |
| Heart_Failure | b_cell | *RNF157* | -1.453449925 | 0.003593868 | 0.011337004 | 1 |
| Heart_Failure | b_cell | *RNF165* | -2.01230972 | 0.001639781 | 0.008654124 | 1 |
| Heart_Failure | b_cell | *MAPK4* | -4.362914909 | 0.002410484 | 0.009356237 | 1 |
| Heart_Failure | b_cell | *PIP4K2B* | -2.090796636 | 0.006362119 | 0.015909937 | 1 |
| Heart_Failure | b_cell | *GRB7* | 3.389338483 | 2.70E-05 | 0.002889125 | 0.169597248 |
| Heart_Failure | b_cell | *FKBP10* | -1.682048792 | 0.012386665 | 0.025289758 | 1 |
| Heart_Failure | b_cell | *TXNL4A* | -2.286524297 | 0.005179789 | 0.013936542 | 1 |
| Heart_Failure | b_cell | *NFIC* | -1.4040274 | 0.003268135 | 0.010797038 | 1 |
| Heart_Failure | b_cell | *TPGS1* | -2.224374149 | 0.003486163 | 0.011132829 | 1 |
| Heart_Failure | b_cell | *PRDM15* | -2.357044292 | 0.007030675 | 0.016976281 | 1 |
| Heart_Failure | b_cell | *MVB12A* | -2.72913536 | 0.008002019 | 0.018475465 | 1 |
| Heart_Failure | b_cell | *DPP9* | 5.390758558 | 0.000211156 | 0.003587948 | 1 |
| Heart_Failure | b_cell | *SIRT3* | -1.884198424 | 0.014754439 | 0.028816763 | 1 |
| Heart_Failure | b_cell | *IFITM3* | -1.92398177 | 0.004334725 | 0.012548093 | 1 |

| | | | | | | |
|---|---|---|---|---|---|---|
| Heart_Failure | b_cell | *COL6A1* | -1.185106157 | 0.001228875 | 0.008654124 | 1 |
| Heart_Failure | b_cell | *TRPM2* | -1.804010844 | 0.015954812 | 0.030375928 | 1 |
| Heart_Failure | b_cell | *AKT1* | -2.438597021 | 0.003359959 | 0.010888691 | 1 |
| Heart_Failure | b_cell | *SAE1* | -2.032093291 | 0.011263527 | 0.023628226 | 1 |
| Heart_Failure | b_cell | *GEMIN7* | 4.208230538 | 0.00012169 | 0.003135985 | 0.765065391 |
| Heart_Failure | b_cell | *WTIP* | -1.6593648 | 0.001475977 | 0.008654124 | 1 |
| Heart_Failure | b_cell | *CAPN10* | -2.057988535 | 0.003913959 | 0.011873672 | 1 |
| Heart_Failure | b_cell | *MYO1F* | -1.359650138 | 0.002487997 | 0.009468546 | 1 |
| Heart_Failure | b_cell | *CACNG8* | -5.917132709 | 0.001524885 | 0.008654124 | 1 |
| Heart_Failure | b_cell | *FBN3* | -4.4240517 | 0.000832153 | 0.008098678 | 1 |
| Heart_Failure | b_cell | *EVI5L* | 6.713153586 | 0.000264034 | 0.003833675 | 1 |
| Heart_Failure | b_cell | *CTU1* | -3.136226432 | 0.001196321 | 0.008654124 | 1 |
| Heart_Failure | b_cell | *RCN3* | -1.735910321 | 0.002129459 | 0.00896712 | 1 |
| Heart_Failure | b_cell | *PEX14* | -1.541085211 | 0.004267658 | 0.012408081 | 1 |
| Heart_Failure | b_cell | *C1orf216* | -2.719800162 | 0.001604372 | 0.008654124 | 1 |
| Heart_Failure | b_cell | *MAP3K6* | 6.660989151 | 0.00543784 | 0.014347469 | 1 |
| Heart_Failure | b_cell | *GPN2* | -1.924923897 | 0.001491723 | 0.008654124 | 1 |
| Heart_Failure | b_cell | *HSPG2* | -1.558821669 | 0.001992171 | 0.008775328 | 1 |
| Heart_Failure | b_cell | *ITGB3BP* | -1.900176934 | 0.024202871 | 0.041687125 | 1 |
| Heart_Failure | b_cell | *MOB3C* | 2.99443369 | 0.000117064 | 0.003135985 | 0.735983301 |
| Heart_Failure | b_cell | *CYP4B1* | -1.575558539 | 0.002823556 | 0.009984081 | 1 |
| Heart_Failure | b_cell | *ZNF697* | -2.140648254 | 0.003684598 | 0.011475847 | 1 |
| Heart_Failure | b_cell | *CELSR2* | -6.905436076 | 0.002393898 | 0.009333474 | 1 |
| Heart_Failure | b_cell | *ATP1B1* | -1.823633125 | 0.001303354 | 0.008654124 | 1 |
| Heart_Failure | b_cell | *CREG1* | -2.266734744 | 0.028053977 | 0.046796326 | 1 |
| Heart_Failure | b_cell | *GPA33* | -7.683336787 | 0.000222263 | 0.003674323 | 1 |
| Heart_Failure | b_cell | *RXRG* | 17.73541054 | 0.000441333 | 0.005211133 | 1 |
| Heart_Failure | b_cell | *MGST3* | -1.604112862 | 0.009158663 | 0.020189521 | 1 |
| Heart_Failure | b_cell | *ADCY10* | -2.66265656 | 0.000709344 | 0.0072916 | 1 |
| Heart_Failure | b_cell | *PIGM* | -2.360122571 | 0.002980563 | 0.010195212 | 1 |
| Heart_Failure | b_cell | *RGL1* | -1.268817309 | 0.002746269 | 0.009854904 | 1 |
| Heart_Failure | b_cell | *TUFT1* | -2.252787464 | 0.002386239 | 0.00932704 | 1 |
| Heart_Failure | b_cell | *TARS2* | -2.283097884 | 0.003345627 | 0.010865693 | 1 |
| Heart_Failure | b_cell | *SELENBP1* | -1.621866646 | 0.001159527 | 0.008654124 | 1 |
| Heart_Failure | b_cell | *SEMA6C* | -3.579666729 | 0.029289824 | 0.048455071 | 1 |
| Heart_Failure | b_cell | *GABPB2* | -1.793888316 | 0.014796028 | 0.028862125 | 1 |
| Heart_Failure | b_cell | *DYRK3* | -2.993507251 | 0.004493781 | 0.012772334 | 1 |
| Heart_Failure | b_cell | *EIF2D* | -2.617303999 | 0.012684586 | 0.025700288 | 1 |
| Heart_Failure | b_cell | *TP53BP2* | -2.293087873 | 0.020042043 | 0.03603212 | 1 |
| Heart_Failure | b_cell | *RAB13* | -2.185774037 | 0.002293796 | 0.009185338 | 1 |
| Heart_Failure | b_cell | *S100A8* | 2.781593174 | 5.19E-05 | 0.002889125 | 0.326456344 |
| Heart_Failure | b_cell | *HAX1* | -3.038996732 | 0.029730393 | 0.04907193 | 1 |
| Heart_Failure | b_cell | *CREB3L4* | -2.622178261 | 0.010516174 | 0.022457604 | 1 |
| Heart_Failure | b_cell | *KCNN3* | -1.977604869 | 0.002521041 | 0.009509206 | 1 |
| Heart_Failure | b_cell | *RIT1* | -2.53492477 | 0.008059654 | 0.018566285 | 1 |
| Heart_Failure | b_cell | *FLG* | -3.623672014 | 4.73E-06 | 0.001567127 | 0.029768023 |
| Heart_Failure | b_cell | *GALNT2* | -1.83524951 | 0.022308255 | 0.039162457 | 1 |
| Heart_Failure | b_cell | *SCCPDH* | -2.163384128 | 0.024751998 | 0.042321406 | 1 |
| Heart_Failure | b_cell | *NVL* | -1.930092634 | 0.029709443 | 0.049050228 | 1 |
| Heart_Failure | b_cell | *GUK1* | -1.932646265 | 0.00237762 | 0.009318117 | 1 |

| | | | | | | |
|---|---|---|---|---|---|---|
| Heart_Failure | b_cell | *CDC42BPA* | -1.431926922 | 0.006410609 | 0.015968105 | 1 |
| Heart_Failure | b_cell | *CNIH3* | -1.959427671 | 0.014604189 | 0.028612196 | 1 |
| Heart_Failure | b_cell | *PSEN2* | 6.322702745 | 0.002897639 | 0.010115189 | 1 |
| Heart_Failure | b_cell | *PYCR2* | 6.345524279 | 7.91E-05 | 0.002889125 | 0.497159498 |
| Heart_Failure | b_cell | *WNT9A* | -2.183008789 | 0.002292825 | 0.009185338 | 1 |
| Heart_Failure | b_cell | *PLEKHA6* | -1.749740903 | 0.002324231 | 0.0092234 | 1 |
| Heart_Failure | b_cell | *CHAC2* | -4.57434788 | 0.008792924 | 0.019698135 | 1 |
| Heart_Failure | b_cell | *WDPCP* | -0.69613514 | 0.007163047 | 0.017176525 | 1 |
| Heart_Failure | b_cell | *ZNF514* | -1.633716853 | 0.005859991 | 0.015130088 | 1 |
| Heart_Failure | b_cell | *RALB* | 4.406530613 | 0.009658 | 0.021054038 | 1 |
| Heart_Failure | b_cell | *TMEM177* | -3.309360068 | 0.001772665 | 0.008654124 | 1 |
| Heart_Failure | b_cell | *RABL2A* | -3.048891595 | 0.017880063 | 0.033091538 | 1 |
| Heart_Failure | b_cell | *FBLN7* | -1.841900659 | 0.006356386 | 0.015908678 | 1 |
| Heart_Failure | b_cell | *FAHD2B* | -3.921593588 | 0.000615442 | 0.006609617 | 1 |
| Heart_Failure | b_cell | *LYG1* | -1.771833673 | 0.008111354 | 0.018638918 | 1 |
| Heart_Failure | b_cell | *AFF3* | 2.197184184 | 0.000969602 | 0.008654124 | 1 |
| Heart_Failure | b_cell | *THSD7B* | -1.151118099 | 0.004540467 | 0.01284469 | 1 |
| Heart_Failure | b_cell | *GPR17* | -2.213037346 | 0.000250478 | 0.003766552 | 1 |
| Heart_Failure | b_cell | *POLR2D* | -2.865504223 | 0.0124954 | 0.02543172 | 1 |
| Heart_Failure | b_cell | *ZNF385B* | -1.421141538 | 0.001793553 | 0.008654124 | 1 |
| Heart_Failure | b_cell | *TMEFF2* | -1.708840394 | 0.001608361 | 0.008654124 | 1 |
| Heart_Failure | b_cell | *GULP1* | -1.299498056 | 0.001987011 | 0.008775328 | 1 |
| Heart_Failure | b_cell | *FAM171B* | -2.173179905 | 0.00314465 | 0.010533998 | 1 |
| Heart_Failure | b_cell | *HSPD1* | -2.256476645 | 0.026162219 | 0.044263151 | 1 |
| Heart_Failure | b_cell | *PTH2R* | -1.928547078 | 0.000435463 | 0.005165577 | 1 |
| Heart_Failure | b_cell | *ESPNL* | -23.53634896 | 0.004776571 | 0.013270129 | 1 |
| Heart_Failure | b_cell | *FANCD2* | 1.199165683 | 0.00012697 | 0.003180328 | 0.798262426 |
| Heart_Failure | b_cell | *OSBPL10* | 2.615243525 | 0.00010679 | 0.003034612 | 0.671388708 |
| Heart_Failure | b_cell | *SLC25A38* | 2.450517551 | 0.000122396 | 0.003135985 | 0.769505159 |
| Heart_Failure | b_cell | *STAC* | -1.841507091 | 0.001696907 | 0.008654124 | 1 |
| Heart_Failure | b_cell | *ARL6IP5* | -1.888014355 | 0.028932999 | 0.047957228 | 1 |
| Heart_Failure | b_cell | *TMF1* | -1.990386441 | 0.026799161 | 0.045097908 | 1 |
| Heart_Failure | b_cell | *COL8A1* | -1.112746073 | 0.001455486 | 0.008654124 | 1 |
| Heart_Failure | b_cell | *ADPRH* | -2.738300293 | 0.002567612 | 0.009560634 | 1 |
| Heart_Failure | b_cell | *IGSF11* | -3.404392906 | 0.028786928 | 0.047737586 | 1 |
| Heart_Failure | b_cell | *BOC* | -1.388733779 | 0.001919596 | 0.008713718 | 1 |
| Heart_Failure | b_cell | *AGTR1* | -1.536776472 | 0.001202598 | 0.008654124 | 1 |
| Heart_Failure | b_cell | *AMT* | -2.502091347 | 0.003059515 | 0.010358195 | 1 |
| Heart_Failure | b_cell | *TCTA* | -2.464216583 | 0.002955242 | 0.010163879 | 1 |
| Heart_Failure | b_cell | *EAF2* | 1.655569662 | 0.000103309 | 0.00302839 | 0.649503311 |
| Heart_Failure | b_cell | *ILDR1* | 2.567134664 | 0.001818769 | 0.008654124 | 1 |
| Heart_Failure | b_cell | *SLC26A1* | 2.771628311 | 0.010124652 | 0.021881638 | 1 |
| Heart_Failure | b_cell | *LYAR* | -2.131547512 | 0.001926066 | 0.008732736 | 1 |
| Heart_Failure | b_cell | *CCNA2* | 4.327066927 | 0.00057775 | 0.006331367 | 1 |
| Heart_Failure | b_cell | *USP53* | -1.419775711 | 0.009723921 | 0.021175716 | 1 |
| Heart_Failure | b_cell | *MARCH1* | 3.143054357 | 0.000626802 | 0.0066316 | 1 |
| Heart_Failure | b_cell | *PDGFC* | -1.318505466 | 0.004596684 | 0.012936147 | 1 |
| Heart_Failure | b_cell | *FAM105A* | -2.218624885 | 0.001536197 | 0.008654124 | 1 |
| Heart_Failure | b_cell | *BHMT* | -2.88253814 | 2.87E-06 | 0.001060433 | 0.018027365 |
| Heart_Failure | b_cell | *CRHBP* | -2.639651291 | 0.001065402 | 0.008654124 | 1 |

| | | | | | | |
|---|---|---|---|---|---|---|
| Heart_Failure | b_cell | *SPATA9* | -1.474048371 | 0.001780459 | 0.008654124 | 1 |
| Heart_Failure | b_cell | *YIPF5* | -2.37650346 | 0.007083466 | 0.01706618 | 1 |
| Heart_Failure | b_cell | *GABRB2* | -1.761964808 | 0.003915975 | 0.011873672 | 1 |
| Heart_Failure | b_cell | *MYLK4* | -3.590780126 | 0.011865314 | 0.024434075 | 1 |
| Heart_Failure | b_cell | *GFOD1* | -1.159875632 | 0.002484005 | 0.00946151 | 1 |
| Heart_Failure | b_cell | *PSD2* | 5.323209549 | 7.32E-05 | 0.002889125 | 0.460502828 |
| Heart_Failure | b_cell | *ZMAT2* | -2.002399134 | 0.001258698 | 0.008654124 | 1 |
| Heart_Failure | b_cell | *KLHL3* | -1.522300997 | 0.001944094 | 0.008738197 | 1 |
| Heart_Failure | b_cell | *TRIM7* | 3.799335748 | 4.02E-05 | 0.002889125 | 0.252475031 |
| Heart_Failure | b_cell | *HIGD2A* | -3.137578712 | 0.009114919 | 0.02012838 | 1 |
| Heart_Failure | b_cell | *FAM193B* | -2.000495415 | 0.024693732 | 0.042256259 | 1 |
| Heart_Failure | b_cell | *MUT* | -2.384296048 | 0.011927892 | 0.024530801 | 1 |
| Heart_Failure | b_cell | *DOK3* | -2.800341988 | 0.001149926 | 0.008654124 | 1 |
| Heart_Failure | b_cell | *ABT1* | -2.377204286 | 0.004554049 | 0.012862221 | 1 |
| Heart_Failure | b_cell | *ANO7* | 2.757485229 | 0.029523141 | 0.048768258 | 1 |
| Heart_Failure | b_cell | *TPBG* | -1.941556068 | 0.001668948 | 0.008654124 | 1 |
| Heart_Failure | b_cell | *IRAK1BP1* | -1.953643611 | 0.015946995 | 0.030375928 | 1 |
| Heart_Failure | b_cell | *RNF217* | -1.663714893 | 0.010201992 | 0.021988318 | 1 |
| Heart_Failure | b_cell | *ABRACL* | 4.756151815 | 0.018235999 | 0.033533116 | 1 |
| Heart_Failure | b_cell | *SLC18B1* | -2.406583565 | 0.000433416 | 0.005151018 | 1 |
| Heart_Failure | b_cell | *AIG1* | -1.265251565 | 0.00912488 | 0.020143301 | 1 |
| Heart_Failure | b_cell | *DYNLT1* | -1.857446136 | 0.002695411 | 0.009784092 | 1 |
| Heart_Failure | b_cell | *PNLDC1* | -4.988375501 | 0.018248868 | 0.033539895 | 1 |
| Heart_Failure | b_cell | *SLC22A3* | -1.459552878 | 0.0017116 | 0.008654124 | 1 |
| Heart_Failure | b_cell | *C7orf50* | -1.894152791 | 0.007246996 | 0.0173261 | 1 |
| Heart_Failure | b_cell | *SDK1* | -1.11641937 | 0.001663959 | 0.008654124 | 1 |
| Heart_Failure | b_cell | *EGFR* | -1.007578496 | 0.002239705 | 0.009117613 | 1 |
| Heart_Failure | b_cell | *PSPH* | -1.884130621 | 0.00859238 | 0.019399557 | 1 |
| Heart_Failure | b_cell | *TMEM209* | -1.779594262 | 0.002219565 | 0.009081423 | 1 |
| Heart_Failure | b_cell | *AGBL3* | -1.59774178 | 0.003840072 | 0.011752708 | 1 |
| Heart_Failure | b_cell | *NCAPG2* | -2.218295042 | 0.022445421 | 0.039336491 | 1 |
| Heart_Failure | b_cell | *FBXO25* | -2.08411256 | 0.007554601 | 0.017795346 | 1 |
| Heart_Failure | b_cell | *SLC25A37* | -1.9340373 | 0.011020973 | 0.023235699 | 1 |
| Heart_Failure | b_cell | *CHMP7* | 6.209090201 | 0.001025046 | 0.008654124 | 1 |
| Heart_Failure | b_cell | *ST18* | -1.983953895 | 0.001826892 | 0.008654124 | 1 |
| Heart_Failure | b_cell | *RGS20* | -1.544388508 | 0.015767107 | 0.030175891 | 1 |
| Heart_Failure | b_cell | *TACC1* | -1.552293319 | 0.013898666 | 0.027582359 | 1 |
| Heart_Failure | b_cell | *ADHFE1* | -1.279343189 | 0.00045958 | 0.005360633 | 1 |
| Heart_Failure | b_cell | *SLC26A7* | -1.448030562 | 0.01075067 | 0.022849717 | 1 |
| Heart_Failure | b_cell | *ATP6V0D2* | -2.728612199 | 0.005227604 | 0.014027294 | 1 |
| Heart_Failure | b_cell | *MAL2* | -2.042434334 | 0.002170611 | 0.009019688 | 1 |
| Heart_Failure | b_cell | *FAM135B* | 3.710787181 | 7.88E-05 | 0.002889125 | 0.495142252 |
| Heart_Failure | b_cell | *AK3* | -1.855306419 | 0.015827107 | 0.030235497 | 1 |
| Heart_Failure | b_cell | *CER1* | -6.141759129 | 0.008899959 | 0.019799731 | 1 |
| Heart_Failure | b_cell | *SIGMAR1* | -2.564841003 | 0.007063899 | 0.017040532 | 1 |
| Heart_Failure | b_cell | *CBWD5* | -2.058696428 | 0.002340133 | 0.009248374 | 1 |
| Heart_Failure | b_cell | *ZNF462* | -1.745809093 | 0.00630876 | 0.015846254 | 1 |
| Heart_Failure | b_cell | *WDR31* | -2.64782919 | 0.024051601 | 0.041473509 | 1 |
| Heart_Failure | b_cell | *POLE3* | 6.034316576 | 5.78E-05 | 0.002889125 | 0.363252001 |
| Heart_Failure | b_cell | *SURF6* | 4.693848175 | 4.88E-05 | 0.002889125 | 0.306846075 |

| | | | | | | |
|---|---|---|---|---|---|---|
| Heart_Failure | b_cell | *ASB6* | -2.898754758 | 0.001988557 | 0.008775328 | 1 |
| Heart_Failure | b_cell | *NTMT1* | -3.157798039 | 0.019195989 | 0.034938533 | 1 |
| Heart_Failure | b_cell | *LRSAM1* | 4.392084352 | 0.016234329 | 0.030779623 | 1 |
| Heart_Failure | b_cell | *INPP5E* | -3.095773873 | 0.028539161 | 0.047458098 | 1 |
| Heart_Failure | b_cell | *DPH7* | -2.583571541 | 0.015253971 | 0.029499144 | 1 |
| Heart_Failure | b_cell | *SLC39A12* | -5.025247665 | 0.017405229 | 0.032384338 | 1 |
| Heart_Failure | b_cell | *PARD3* | -1.121600044 | 0.001437346 | 0.008654124 | 1 |
| Heart_Failure | b_cell | *NRBF2* | -1.874848008 | 0.004498437 | 0.012775103 | 1 |
| Heart_Failure | b_cell | *ADIRF* | -1.75971691 | 0.012281371 | 0.025109911 | 1 |
| Heart_Failure | b_cell | *HTR7* | -2.662872583 | 0.001953729 | 0.008754876 | 1 |
| Heart_Failure | b_cell | *DNAJB12* | -2.408179401 | 0.021028719 | 0.037336221 | 1 |
| Heart_Failure | b_cell | *PLEKHS1* | 5.894966974 | 0.000307699 | 0.004261028 | 1 |
| Heart_Failure | b_cell | *LRRC27* | -2.150973993 | 0.025926624 | 0.04394734 | 1 |
| Heart_Failure | b_cell | *PAOX* | 2.683710216 | 7.51E-05 | 0.002889125 | 0.471902148 |
| Heart_Failure | b_cell | *GSTO1* | -2.073976493 | 0.003285631 | 0.010820724 | 1 |
| Heart_Failure | b_cell | *PDCD11* | -2.047064728 | 0.00411053 | 0.012159073 | 1 |
| Heart_Failure | b_cell | *RGS10* | -1.881319578 | 0.002083152 | 0.008855223 | 1 |
| Heart_Failure | b_cell | *GAS2* | -2.823162233 | 0.017523003 | 0.032574548 | 1 |
| Heart_Failure | b_cell | *ZNF215* | 1.567628339 | 0.004724791 | 0.01319039 | 1 |
| Heart_Failure | b_cell | *HSD17B12* | -2.334599077 | 0.022988292 | 0.040057481 | 1 |
| Heart_Failure | b_cell | *GLYAT* | -1.29246452 | 0.002778461 | 0.009913839 | 1 |
| Heart_Failure | b_cell | *PTPRJ* | -1.717950033 | 0.0116681 | 0.024146591 | 1 |
| Heart_Failure | b_cell | *ENDOD1* | 6.447712113 | 0.00533266 | 0.014194088 | 1 |
| Heart_Failure | b_cell | *SERPINH1* | -1.768610099 | 0.00377467 | 0.011598902 | 1 |
| Heart_Failure | b_cell | *CAPN5* | -2.470898705 | 0.002295203 | 0.009185338 | 1 |
| Heart_Failure | b_cell | *DRD2* | -11.21282115 | 0.001703925 | 0.008654124 | 1 |
| Heart_Failure | b_cell | *C11orf52* | -18.38984563 | 0.001714828 | 0.008654124 | 1 |
| Heart_Failure | b_cell | *GRIK4* | -2.53196703 | 0.002387884 | 0.00932704 | 1 |
| Heart_Failure | b_cell | *ADAM33* | -1.759308131 | 0.000784117 | 0.007767351 | 1 |
| Heart_Failure | b_cell | *ROM1* | 2.887945691 | 0.017439343 | 0.032438211 | 1 |
| Heart_Failure | b_cell | *EML3* | -2.730173101 | 0.001583561 | 0.008654124 | 1 |
| Heart_Failure | b_cell | *INCENP* | -2.750754945 | 0.002049672 | 0.008831917 | 1 |
| Heart_Failure | b_cell | *MS4A3* | -47.30933014 | 0.001688519 | 0.008654124 | 1 |
| Heart_Failure | b_cell | *PLCH2* | -8.301763074 | 0.001653397 | 0.008654124 | 1 |
| Heart_Failure | b_cell | *CHEK1* | 3.923015921 | 0.004822498 | 0.01333878 | 1 |
| Heart_Failure | b_cell | *FEZ1* | -2.526152427 | 0.001503617 | 0.008654124 | 1 |
| Heart_Failure | b_cell | *DUSP15* | 2.993931295 | 0.027632335 | 0.046215613 | 1 |
| Heart_Failure | b_cell | *COMMD7* | -2.30343549 | 0.008724478 | 0.019603572 | 1 |
| Heart_Failure | b_cell | *KIAA1755* | -2.065651454 | 0.00598889 | 0.01532444 | 1 |
| Heart_Failure | b_cell | *CNBD2* | 9.967325688 | 0.000237787 | 0.003709453 | 1 |
| Heart_Failure | b_cell | *CABLES2* | 2.772550868 | 0.002127172 | 0.008963493 | 1 |
| Heart_Failure | b_cell | *TRPT1* | -2.834953525 | 0.000713709 | 0.007307958 | 1 |
| Heart_Failure | b_cell | *DOC2A* | -5.989196046 | 0.001480202 | 0.008654124 | 1 |
| Heart_Failure | b_cell | *MPP7* | -1.689852418 | 0.020484728 | 0.036556198 | 1 |
| Heart_Failure | b_cell | *LYPD6B* | -3.484048244 | 0.002540507 | 0.009545808 | 1 |
| Heart_Failure | b_cell | *CCDC102B* | -1.493425343 | 0.003176011 | 0.010587263 | 1 |
| Heart_Failure | b_cell | *FSIP1* | 9.332915091 | 0.001254347 | 0.008654124 | 1 |
| Heart_Failure | b_cell | *RGS18* | -2.457512578 | 0.002564235 | 0.009560634 | 1 |
| Heart_Failure | b_cell | *PRSS23* | -1.014052025 | 0.004674658 | 0.013085296 | 1 |
| Heart_Failure | b_cell | *MTMR12* | -1.787626974 | 0.018700236 | 0.034246544 | 1 |

| | | | | | | |
|---|---|---|---|---|---|---|
| Heart_Failure | b_cell | *PPP1R1C* | 4.680419856 | 0.030367554 | 0.049822759 | 1 |
| Heart_Failure | b_cell | *C11orf53* | 24.99683252 | 0.000126043 | 0.003179208 | 0.792430984 |
| Heart_Failure | b_cell | *CCT5* | -1.911957993 | 0.011118398 | 0.023394033 | 1 |
| Heart_Failure | b_cell | *FAM173B* | -1.691101964 | 0.001297046 | 0.008654124 | 1 |
| Heart_Failure | b_cell | *DOCK1* | -1.505767794 | 0.015198792 | 0.029437116 | 1 |
| Heart_Failure | b_cell | *TEX12* | -16.32757149 | 0.001663129 | 0.008654124 | 1 |
| Heart_Failure | b_cell | *CRIM1* | -1.003647763 | 0.001972531 | 0.008775328 | 1 |
| Heart_Failure | b_cell | *RILPL2* | -2.023289002 | 0.020217454 | 0.036233504 | 1 |
| Heart_Failure | b_cell | *DHX37* | 6.208818461 | 0.015903681 | 0.030332981 | 1 |
| Heart_Failure | b_cell | *PRSS53* | -6.588171491 | 0.020726105 | 0.036882259 | 1 |
| Heart_Failure | b_cell | *GPR158* | -2.63665622 | 0.002169548 | 0.009019688 | 1 |
| Heart_Failure | b_cell | *THRB* | -1.310276588 | 0.008409401 | 0.019114209 | 1 |
| Heart_Failure | b_cell | *TMEM86A* | -3.366647678 | 0.003931363 | 0.011882923 | 1 |
| Heart_Failure | b_cell | *BTBD11* | -2.000568107 | 0.001781185 | 0.008654124 | 1 |
| Heart_Failure | b_cell | *ANK3* | -1.646606681 | 0.011617579 | 0.024073738 | 1 |
| Heart_Failure | b_cell | *IPMK* | -2.017032345 | 0.01478876 | 0.0288569 | 1 |
| Heart_Failure | b_cell | *DLG5* | -1.548391563 | 0.004218287 | 0.012320004 | 1 |
| Heart_Failure | b_cell | *MIPOL1* | -1.330377129 | 0.004882431 | 0.01343363 | 1 |
| Heart_Failure | b_cell | *TMEM18* | -3.044150684 | 0.020356705 | 0.036388076 | 1 |
| Heart_Failure | b_cell | *KCTD14* | 9.150660778 | 0.001378922 | 0.008654124 | 1 |
| Heart_Failure | b_cell | *THRSP* | -1.746542213 | 0.001989999 | 0.008775328 | 1 |
| Heart_Failure | b_cell | *ME3* | -1.703578423 | 0.01546107 | 0.029767756 | 1 |
| Heart_Failure | b_cell | *ADAMTS12* | -1.04679067 | 0.001877095 | 0.008703021 | 1 |
| Heart_Failure | b_cell | *VIPAS39* | 4.642387022 | 0.015361859 | 0.029662165 | 1 |
| Heart_Failure | b_cell | *C4orf33* | 6.389087175 | 0.003196703 | 0.010628066 | 1 |
| Heart_Failure | b_cell | *PTPRO* | -2.667119813 | 0.011910337 | 0.024506994 | 1 |
| Heart_Failure | b_cell | *NCAPD3* | -2.110491454 | 0.012856427 | 0.025973122 | 1 |
| Heart_Failure | b_cell | *QDPR* | -1.4385154 | 0.007946515 | 0.018387832 | 1 |
| Heart_Failure | b_cell | *FAM160B1* | -1.763145552 | 0.005930083 | 0.01524067 | 1 |
| Heart_Failure | b_cell | *ANO4* | -1.714379939 | 0.003315119 | 0.01083435 | 1 |
| Heart_Failure | b_cell | *AKR1C2* | -1.334811598 | 0.002001572 | 0.008783721 | 1 |
| Heart_Failure | b_cell | *DPYSL4* | 3.266697487 | 0.000449411 | 0.005261543 | 1 |
| Heart_Failure | b_cell | *ADAM8* | -2.74828977 | 0.002026481 | 0.008831917 | 1 |
| Heart_Failure | b_cell | *ITIH2* | 1.40663285 | 0.000150281 | 0.003239931 | 0.944816432 |
| Heart_Failure | b_cell | *INPP1* | -2.101620062 | 0.003229199 | 0.010707792 | 1 |
| Heart_Failure | b_cell | *RNF144A* | -1.224283588 | 0.002156807 | 0.009019688 | 1 |
| Heart_Failure | b_cell | *ASAP2* | -1.526531952 | 0.014662815 | 0.028682365 | 1 |
| Heart_Failure | b_cell | *TMEM45B* | -6.758552244 | 0.002437183 | 0.0094119 | 1 |
| Heart_Failure | b_cell | *ACSL1* | -1.071157197 | 0.00126382 | 0.008654124 | 1 |
| Heart_Failure | b_cell | *BICD1* | 3.782483863 | 0.007155461 | 0.017176525 | 1 |
| Heart_Failure | b_cell | *GABRA2* | -3.116790707 | 0.000767041 | 0.007642448 | 1 |
| Heart_Failure | b_cell | *CENPJ* | -1.594203278 | 0.002474755 | 0.009455883 | 1 |
| Heart_Failure | b_cell | *PARP8* | -1.591378569 | 0.004804801 | 0.013301535 | 1 |
| Heart_Failure | b_cell | *PTPN14* | -1.167166894 | 0.002595214 | 0.00959207 | 1 |
| Heart_Failure | b_cell | *ARL14EP* | -2.536723035 | 0.011897587 | 0.024492511 | 1 |
| Heart_Failure | b_cell | *SPC25* | -2.028404102 | 0.000224119 | 0.003674323 | 1 |
| Heart_Failure | b_cell | *G6PC2* | -116.9529147 | 0.001651611 | 0.008654124 | 1 |
| Heart_Failure | b_cell | *POC5* | -2.80051396 | 0.002771775 | 0.009906851 | 1 |
| Heart_Failure | b_cell | *GUCY1A2* | -1.393086926 | 0.002328223 | 0.0092234 | 1 |
| Heart_Failure | b_cell | *ZNF547* | 2.617981065 | 0.000331249 | 0.004450951 | 1 |

| | | | | | | |
|---|---|---|---|---|---|---|
| Heart_Failure | b_cell | *NMT2* | -1.669080458 | 0.004109435 | 0.012159073 | 1 |
| Heart_Failure | b_cell | *ZNF837* | 2.294603104 | 0.000916928 | 0.008523384 | 1 |
| Heart_Failure | b_cell | *PLEKHH2* | -0.845468398 | 0.01699682 | 0.031793814 | 1 |
| Heart_Failure | b_cell | *IGSF10* | -1.699036537 | 0.004268032 | 0.012408081 | 1 |
| Heart_Failure | b_cell | *SPEF2* | -1.92034164 | 0.005249415 | 0.014066038 | 1 |
| Heart_Failure | b_cell | *SPARCL1* | -1.066155141 | 0.001802357 | 0.008654124 | 1 |
| Heart_Failure | b_cell | *CCNO* | 17.80107685 | 0.002166665 | 0.009019688 | 1 |
| Heart_Failure | b_cell | *CLEC4F* | -6.649965937 | 0.001847484 | 0.00866783 | 1 |
| Heart_Failure | b_cell | *PELO* | -2.547042444 | 0.001675051 | 0.008654124 | 1 |
| Heart_Failure | b_cell | *GPR180* | -1.949968496 | 0.001356595 | 0.008654124 | 1 |
| Heart_Failure | b_cell | *TCTEX1D1* | -3.334407947 | 0.00175655 | 0.008654124 | 1 |
| Heart_Failure | b_cell | *WDR78* | -2.189981992 | 0.003944973 | 0.011889762 | 1 |
| Heart_Failure | b_cell | *PRDM8* | 2.106947762 | 0.00047735 | 0.005506599 | 1 |
| Heart_Failure | b_cell | *HHEX* | -2.624060187 | 0.001644926 | 0.008654124 | 1 |
| Heart_Failure | b_cell | *RAB3C* | -1.899927266 | 0.001173307 | 0.008654124 | 1 |
| Heart_Failure | b_cell | *RAD17* | -2.09559748 | 0.019518507 | 0.03537413 | 1 |
| Heart_Failure | b_cell | *MED21* | -2.287383005 | 0.019372588 | 0.035180664 | 1 |
| Heart_Failure | b_cell | *SRP19* | -2.715270989 | 0.00927027 | 0.020371264 | 1 |
| Heart_Failure | b_cell | *CENPH* | -2.718815825 | 0.00190572 | 0.008705907 | 1 |
| Heart_Failure | b_cell | *CARHSP1* | -2.043108866 | 0.00600002 | 0.015339468 | 1 |
| Heart_Failure | b_cell | *TEKT5* | -6.085902717 | 0.001476121 | 0.008654124 | 1 |
| Heart_Failure | b_cell | *BANK1* | 0.301526069 | 0.000175618 | 0.003366185 | 1 |
| Heart_Failure | b_cell | *SCOC* | -2.176507018 | 0.001469162 | 0.008654124 | 1 |
| Heart_Failure | b_cell | *CLGN* | -15.45340616 | 0.004104921 | 0.012159073 | 1 |
| Heart_Failure | b_cell | *SYCP2L* | -2.054489427 | 0.002416961 | 0.009356237 | 1 |
| Heart_Failure | b_cell | *AHCTF1* | -1.802988031 | 0.023850508 | 0.041205866 | 1 |
| Heart_Failure | b_cell | *PTPRR* | -3.14931995 | 0.001139456 | 0.008654124 | 1 |
| Heart_Failure | b_cell | *PLA2R1* | -1.235950609 | 0.004009743 | 0.012024754 | 1 |
| Heart_Failure | b_cell | *SLC25A27* | -1.421570842 | 0.001517073 | 0.008654124 | 1 |
| Heart_Failure | b_cell | *FRMD1* | -2.728136709 | 0.001706925 | 0.008654124 | 1 |
| Heart_Failure | b_cell | *ASAP1* | -1.423745757 | 0.017235294 | 0.032153797 | 1 |
| Heart_Failure | b_cell | *INO80C* | -2.899187212 | 0.001354109 | 0.008654124 | 1 |
| Heart_Failure | b_cell | *LPCAT1* | -2.783143582 | 0.025704774 | 0.043677275 | 1 |
| Heart_Failure | b_cell | *C16orf89* | -2.051277241 | 0.010287495 | 0.022119521 | 1 |
| Heart_Failure | b_cell | *TEX29* | -2.72751002 | 0.00514997 | 0.013890116 | 1 |
| Heart_Failure | b_cell | *ADPRHL1* | -2.802497264 | 0.015326705 | 0.029614015 | 1 |
| Heart_Failure | b_cell | *LURAP1L* | -1.349454046 | 0.000641456 | 0.006710204 | 1 |
| Heart_Failure | b_cell | *ZDHHC7* | -1.902768052 | 0.008797945 | 0.019698135 | 1 |
| Heart_Failure | b_cell | *FAM92B* | 10.03672182 | 0.000155723 | 0.003241816 | 0.979028521 |
| Heart_Failure | b_cell | *C7orf31* | 6.201888766 | 0.015465156 | 0.029767756 | 1 |
| Heart_Failure | b_cell | *FBXO36* | 5.44049175 | 0.011735997 | 0.024247194 | 1 |
| Heart_Failure | b_cell | *MCOLN2* | 1.741003583 | 0.000250814 | 0.003766552 | 1 |
| Heart_Failure | b_cell | *CACNA2D1* | -0.998528778 | 0.002069849 | 0.008840448 | 1 |
| Heart_Failure | b_cell | *ZUFSP* | -2.520113808 | 0.005970658 | 0.015314358 | 1 |
| Heart_Failure | b_cell | *HS3ST3A1* | -2.877560969 | 0.010747207 | 0.022849717 | 1 |
| Heart_Failure | b_cell | *ANKRD29* | -1.605533781 | 0.002111939 | 0.008922624 | 1 |
| Heart_Failure | b_cell | *THY1* | -1.639866794 | 0.001601753 | 0.008654124 | 1 |
| Heart_Failure | b_cell | *TBCEL* | -1.762534468 | 0.003330335 | 0.010848611 | 1 |
| Heart_Failure | b_cell | *UBASH3B* | -2.054578868 | 0.003052707 | 0.010340715 | 1 |
| Heart_Failure | b_cell | *ROBO4* | -1.793175829 | 0.00148318 | 0.008654124 | 1 |

| | | | | | | |
|---|---|---|---|---|---|---|
| Heart_Failure | b_cell | *ROBO3* | -1.943351339 | 0.001207746 | 0.008654124 | 1 |
| Heart_Failure | b_cell | *CEP112* | -1.265624783 | 0.003487477 | 0.011132829 | 1 |
| Heart_Failure | b_cell | *GAL3ST2* | -31.45750503 | 8.21E-05 | 0.002889125 | 0.516450235 |
| Heart_Failure | b_cell | *ENPP3* | 6.757143851 | 0.000306403 | 0.004261028 | 1 |
| Heart_Failure | b_cell | *UCHL1* | -2.352866833 | 0.001911798 | 0.008705907 | 1 |
| Heart_Failure | b_cell | *DISP1* | 4.659745311 | 0.002235415 | 0.009114171 | 1 |
| Heart_Failure | b_cell | *FAM167A* | 2.970865249 | 0.000183909 | 0.00340015 | 1 |
| Heart_Failure | b_cell | *NEIL2* | 4.779449337 | 0.000418023 | 0.005054057 | 1 |
| Heart_Failure | b_cell | *LONRF1* | -1.796185816 | 0.003305114 | 0.01083435 | 1 |
| Heart_Failure | b_cell | *CCSAP* | -3.675708186 | 0.006521607 | 0.016123569 | 1 |
| Heart_Failure | b_cell | *GBP5* | -2.543580187 | 0.001797099 | 0.008654124 | 1 |
| Heart_Failure | b_cell | *FAM69A* | -1.458111594 | 0.01893263 | 0.034571434 | 1 |
| Heart_Failure | b_cell | *PDLIM3* | -2.058710526 | 0.001022238 | 0.008654124 | 1 |
| Heart_Failure | b_cell | *NCAM2* | -1.549634128 | 0.001123356 | 0.008654124 | 1 |
| Heart_Failure | b_cell | *L3MBTL4* | -1.672631166 | 0.028520331 | 0.047448351 | 1 |
| Heart_Failure | b_cell | *PDE1C* | -1.547354769 | 0.001945837 | 0.008738197 | 1 |
| Heart_Failure | b_cell | *RABGEF1* | -2.495396484 | 0.000995641 | 0.008654124 | 1 |
| Heart_Failure | b_cell | *MRPL39* | 3.180160278 | 0.011286232 | 0.023636218 | 1 |
| Heart_Failure | b_cell | *TSEN2* | -2.324320256 | 0.002567671 | 0.009560634 | 1 |
| Heart_Failure | b_cell | *CXXC1* | -2.844212863 | 0.008317731 | 0.018974446 | 1 |
| Heart_Failure | b_cell | *PPP4R1* | -1.868283739 | 0.02361425 | 0.040853822 | 1 |
| Heart_Failure | b_cell | *APCDD1* | -1.165726372 | 0.005095972 | 0.013812459 | 1 |
| Heart_Failure | b_cell | *ANKRD40* | -2.029704122 | 0.004687236 | 0.013114666 | 1 |
| Heart_Failure | b_cell | *CYP2U1* | -2.716615612 | 0.014247684 | 0.028053814 | 1 |
| Heart_Failure | b_cell | *FBXL18* | -2.381227241 | 0.009107189 | 0.020125447 | 1 |
| Heart_Failure | b_cell | *PROM2* | -10.37368011 | 0.002584022 | 0.009577319 | 1 |
| Heart_Failure | b_cell | *AK9* | -1.964293599 | 0.004025297 | 0.012047229 | 1 |
| Heart_Failure | b_cell | *GTF3C6* | 4.149283428 | 0.00025273 | 0.003769479 | 1 |
| Heart_Failure | b_cell | *AGPAT5* | 5.284108676 | 0.000269668 | 0.003897471 | 1 |
| Heart_Failure | b_cell | *MMS19* | -2.093981732 | 0.016773413 | 0.031476046 | 1 |
| Heart_Failure | b_cell | *GOLGA7B* | 2.929052965 | 0.000103756 | 0.00302839 | 0.652313556 |
| Heart_Failure | b_cell | *TRMT44* | -1.998104972 | 0.003582002 | 0.011316607 | 1 |
| Heart_Failure | b_cell | *C16orf87* | 4.921416855 | 3.06E-05 | 0.002889125 | 0.192574696 |
| Heart_Failure | b_cell | *SETD9* | -1.4822049 | 0.004634202 | 0.013020253 | 1 |
| Heart_Failure | b_cell | *FAM126B* | -1.784582708 | 0.02033811 | 0.036366807 | 1 |
| Heart_Failure | b_cell | *ALS2CR12* | -1.979183127 | 0.001136089 | 0.008654124 | 1 |
| Heart_Failure | b_cell | *SPAG17* | -1.221532762 | 0.001668676 | 0.008654124 | 1 |
| Heart_Failure | b_cell | *DEPTOR* | -2.05261121 | 0.008067269 | 0.018566876 | 1 |
| Heart_Failure | b_cell | *FAM161B* | -2.707153478 | 0.004880223 | 0.013433434 | 1 |
| Heart_Failure | b_cell | *WIF1* | 6.447151533 | 0.000232076 | 0.003705045 | 1 |
| Heart_Failure | b_cell | *UGT2B4* | -3.567438398 | 0.001652715 | 0.008654124 | 1 |
| Heart_Failure | b_cell | *MMP16* | -1.368487735 | 0.000883784 | 0.008355409 | 1 |
| Heart_Failure | b_cell | *KCNMA1* | -1.157903287 | 0.002501672 | 0.009482826 | 1 |
| Heart_Failure | b_cell | *BATF* | 9.776685067 | 0.000169316 | 0.003365162 | 1 |
| Heart_Failure | b_cell | *DCK* | 3.952542327 | 1.79E-05 | 0.002889125 | 0.1126766 |
| Heart_Failure | b_cell | *DPY19L4* | -2.200748862 | 0.00915424 | 0.020188366 | 1 |
| Heart_Failure | b_cell | *NDUFAF6* | -2.332365203 | 0.006025246 | 0.015367794 | 1 |
| Heart_Failure | b_cell | *ADAMTSL3* | -1.170839001 | 0.002195707 | 0.009044448 | 1 |
| Heart_Failure | b_cell | *WHAMM* | -2.968034084 | 0.00116791 | 0.008654124 | 1 |
| Heart_Failure | b_cell | *CXCL13* | -3.978714029 | 0.001944286 | 0.008738197 | 1 |

| | | | | | | |
|---|---|---|---|---|---|---|
| Heart_Failure | b_cell | *N6AMT1* | -2.117901077 | 0.00257133 | 0.009560634 | 1 |
| Heart_Failure | b_cell | *RWDD2B* | -1.813210957 | 0.006593807 | 0.016253852 | 1 |
| Heart_Failure | b_cell | *CCT8* | -1.917934905 | 0.023881824 | 0.041237305 | 1 |
| Heart_Failure | b_cell | *ANKRD9* | -2.769648046 | 0.001781738 | 0.008654124 | 1 |
| Heart_Failure | b_cell | *SFXN2* | -1.901031727 | 0.004490727 | 0.012772334 | 1 |
| Heart_Failure | b_cell | *TDRD9* | 5.349156482 | 0.00108026 | 0.008654124 | 1 |
| Heart_Failure | b_cell | *SH3RF2* | -1.415531391 | 0.004728758 | 0.013195605 | 1 |
| Heart_Failure | b_cell | *GDF6* | -5.758275421 | 0.00316922 | 0.010575841 | 1 |
| Heart_Failure | b_cell | *NODAL* | -3.130042852 | 0.001631314 | 0.008654124 | 1 |
| Heart_Failure | b_cell | *MED19* | -1.988491061 | 0.004927409 | 0.013494248 | 1 |
| Heart_Failure | b_cell | *WDYHV1* | -2.588457066 | 0.020194212 | 0.036220112 | 1 |
| Heart_Failure | b_cell | *SASS6* | -2.462961907 | 0.005930979 | 0.01524067 | 1 |
| Heart_Failure | b_cell | *ITGAD* | -2.118925702 | 0.019143452 | 0.034876985 | 1 |
| Heart_Failure | b_cell | *MPV17L* | -2.383510215 | 0.018881228 | 0.034524832 | 1 |
| Heart_Failure | b_cell | *RPUSD3* | -2.821260693 | 0.011229001 | 0.023563662 | 1 |
| Heart_Failure | b_cell | *GHRL* | -2.465070485 | 0.00336755 | 0.010902052 | 1 |
| Heart_Failure | b_cell | *SLC6A1* | -4.863298874 | 0.002388506 | 0.00932704 | 1 |
| Heart_Failure | b_cell | *FCHO2* | -1.608539956 | 0.029387982 | 0.048574676 | 1 |
| Heart_Failure | b_cell | *TIMP4* | -2.035153679 | 0.0015536 | 0.008654124 | 1 |
| Heart_Failure | b_cell | *CPT2* | 4.423271237 | 0.000147513 | 0.003239931 | 0.927413867 |
| Heart_Failure | b_cell | *LRP8* | 9.386097596 | 0.000412897 | 0.005033892 | 1 |
| Heart_Failure | b_cell | *STEAP2* | -1.364084809 | 0.0221748 | 0.038963938 | 1 |
| Heart_Failure | b_cell | *GATAD1* | -1.625235315 | 0.012384169 | 0.025289758 | 1 |
| Heart_Failure | b_cell | *TMED6* | -10.33241382 | 0.00166517 | 0.008654124 | 1 |
| Heart_Failure | b_cell | *DHRS1* | -2.902113183 | 0.000851259 | 0.008216886 | 1 |
| Heart_Failure | b_cell | *RNF111* | -1.448089312 | 0.012915882 | 0.026068106 | 1 |
| Heart_Failure | b_cell | *MX1* | -1.609665481 | 0.02036158 | 0.036388076 | 1 |
| Heart_Failure | b_cell | *CREB3L1* | -2.373474977 | 0.005034867 | 0.01372689 | 1 |
| Heart_Failure | b_cell | *C2CD2* | -1.100961489 | 0.017460658 | 0.032468251 | 1 |
| Heart_Failure | b_cell | *C9orf43* | -3.770976747 | 0.00248123 | 0.009459973 | 1 |
| Heart_Failure | b_cell | *WDR19* | -2.018604619 | 0.017655464 | 0.032774862 | 1 |
| Heart_Failure | b_cell | *SLC37A3* | -1.986041403 | 0.014458584 | 0.028379992 | 1 |
| Heart_Failure | b_cell | *AP3S2* | -2.00799522 | 0.001885413 | 0.008705907 | 1 |
| Heart_Failure | b_cell | *FAM213B* | -3.575182971 | 0.006780612 | 0.016532941 | 1 |
| Heart_Failure | b_cell | *PEX10* | 6.487916792 | 0.00083398 | 0.008103916 | 1 |
| Heart_Failure | b_cell | *RADIL* | -1.806132793 | 0.001751437 | 0.008654124 | 1 |
| Heart_Failure | b_cell | *TRIM63* | 3.254557489 | 0.000425007 | 0.005099275 | 1 |
| Heart_Failure | b_cell | *WDR66* | -1.749324312 | 0.011440579 | 0.023824749 | 1 |
| Heart_Failure | b_cell | *GALNT14* | 9.556028669 | 5.22E-05 | 0.002889125 | 0.32834487 |
| Heart_Failure | b_cell | *RHPN1* | -3.441411531 | 0.004997245 | 0.013653925 | 1 |
| Heart_Failure | b_cell | *TPRG1L* | -3.457853046 | 0.007024735 | 0.016976281 | 1 |
| Heart_Failure | b_cell | *LRRC43* | -6.265894712 | 0.001728942 | 0.008654124 | 1 |
| Heart_Failure | b_cell | *XKR8* | -3.848472093 | 0.000831474 | 0.008098678 | 1 |
| Heart_Failure | b_cell | *MRAS* | -1.227973027 | 0.002332958 | 0.009230526 | 1 |
| Heart_Failure | b_cell | *CLSTN2* | -1.119084774 | 0.001529802 | 0.008654124 | 1 |
| Heart_Failure | b_cell | *COLEC12* | -1.173119862 | 0.002093601 | 0.00888156 | 1 |
| Heart_Failure | b_cell | *RNF207* | -3.900708548 | 0.027425777 | 0.045921648 | 1 |
| Heart_Failure | b_cell | *AUTS2* | -1.261125396 | 0.017240841 | 0.032154604 | 1 |
| Heart_Failure | b_cell | *B4GALT5* | -1.596916509 | 0.005862872 | 0.015131313 | 1 |
| Heart_Failure | b_cell | *CD1D* | -5.532336674 | 0.025601287 | 0.043540184 | 1 |

| | | | | | | |
|---|---|---|---|---|---|---|
| Heart_Failure | b_cell | *FAM86C1* | 15.04955591 | 6.45E-05 | 0.002889125 | 0.405299646 |
| Heart_Failure | b_cell | *NCF1* | 2.221921841 | 0.000108 | 0.003034612 | 0.6789945 |
| Heart_Failure | b_cell | *PPP1R9A* | -1.157410904 | 0.001301435 | 0.008654124 | 1 |
| Heart_Failure | b_cell | *ZFAND2B* | -3.248297459 | 0.015907084 | 0.030332981 | 1 |
| Heart_Failure | b_cell | *GDPD5* | -1.485638455 | 0.001629116 | 0.008654124 | 1 |
| Heart_Failure | b_cell | *DYNC1I1* | -2.188390626 | 0.018802302 | 0.034393388 | 1 |
| Heart_Failure | b_cell | *TMED4* | -2.28461138 | 0.011227317 | 0.023563662 | 1 |
| Heart_Failure | b_cell | *COPG2* | -1.123758202 | 0.008170421 | 0.018754084 | 1 |
| Heart_Failure | b_cell | *DUSP23* | -2.94387161 | 0.002055829 | 0.008831917 | 1 |
| Heart_Failure | b_cell | *F11R* | -1.972775595 | 0.002855614 | 0.010013685 | 1 |
| Heart_Failure | b_cell | *ZNF276* | -2.258234887 | 0.009665777 | 0.021063689 | 1 |
| Heart_Failure | b_cell | *FGF17* | -9.991481316 | 0.019117466 | 0.034858325 | 1 |
| Heart_Failure | b_cell | *B4GALT3* | -2.707739149 | 0.006499663 | 0.01611332 | 1 |
| Heart_Failure | b_cell | *DMTN* | -1.659606054 | 0.010994 | 0.023209967 | 1 |
| Heart_Failure | b_cell | *FAM160B2* | -2.199351463 | 0.006126941 | 0.015518529 | 1 |
| Heart_Failure | b_cell | *SLC5A11* | -1.780844708 | 0.002922176 | 0.010133328 | 1 |
| Heart_Failure | b_cell | *CCAR2* | -2.598044261 | 0.021913754 | 0.038569925 | 1 |
| Heart_Failure | b_cell | *RAPGEF6* | 8.673283028 | 0.004376118 | 0.012620483 | 1 |
| Heart_Failure | b_cell | *MRPL10* | -2.267125707 | 0.005683645 | 0.014784061 | 1 |
| Heart_Failure | b_cell | *SV2A* | -9.696684524 | 0.005522881 | 0.014503906 | 1 |
| Heart_Failure | b_cell | *RCAN1* | -2.031907189 | 0.007621328 | 0.017871778 | 1 |
| Heart_Failure | b_cell | *SIM2* | 5.11008191 | 0.000845159 | 0.008174641 | 1 |
| Heart_Failure | b_cell | *HLCS* | -1.563612399 | 0.024931231 | 0.042593112 | 1 |
| Heart_Failure | b_cell | *ARHGAP27* | -2.124578562 | 0.004198708 | 0.012289235 | 1 |
| Heart_Failure | b_cell | *PADI4* | -6.20356777 | 0.002661019 | 0.009709707 | 1 |
| Heart_Failure | b_cell | *STARD9* | -1.24993809 | 0.003732048 | 0.011549198 | 1 |
| Heart_Failure | b_cell | *THEM4* | -1.969414595 | 0.004151556 | 0.012205437 | 1 |
| Heart_Failure | b_cell | *TCHH* | 3.075532878 | 0.00018395 | 0.00340015 | 1 |
| Heart_Failure | b_cell | *MED8* | -1.927960782 | 0.013293984 | 0.026643059 | 1 |
| Heart_Failure | b_cell | *ACE* | -1.973308906 | 0.00131234 | 0.008654124 | 1 |
| Heart_Failure | b_cell | *TEPP* | 16.08647335 | 0.021302919 | 0.037716544 | 1 |
| Heart_Failure | b_cell | *CCDC107* | -1.774142461 | 0.005825016 | 0.015070731 | 1 |
| Heart_Failure | b_cell | *NPR2* | -2.374601781 | 0.001609585 | 0.008654124 | 1 |
| Heart_Failure | b_cell | *ZNF235* | -1.866147303 | 0.024643049 | 0.042238107 | 1 |
| Heart_Failure | b_cell | *CALM3* | 6.396184064 | 0.00297496 | 0.010181586 | 1 |
| Heart_Failure | b_cell | *DFFA* | -2.253571972 | 0.016077495 | 0.030546755 | 1 |
| Heart_Failure | b_cell | *SSU72* | -2.071080643 | 0.003878936 | 0.011815344 | 1 |
| Heart_Failure | b_cell | *NR2F6* | -2.553619696 | 0.005092457 | 0.013812459 | 1 |
| Heart_Failure | b_cell | *ANKLE1* | -7.20042919 | 0.005118169 | 0.01383997 | 1 |
| Heart_Failure | b_cell | *CILP2* | -17.87949214 | 0.004470679 | 0.01274701 | 1 |
| Heart_Failure | b_cell | *HSF2BP* | -3.652729005 | 0.001529326 | 0.008654124 | 1 |
| Heart_Failure | b_cell | *CSTB* | -1.831446102 | 0.003469685 | 0.011112537 | 1 |
| Heart_Failure | b_cell | *C21orf33* | -11.10931759 | 3.53E-05 | 0.002889125 | 0.221701351 |
| Heart_Failure | b_cell | *ICOSLG* | 4.402248118 | 0.009101669 | 0.020122782 | 1 |
| Heart_Failure | b_cell | *LRRC3* | -4.377246259 | 0.004055286 | 0.012094678 | 1 |
| Heart_Failure | b_cell | *LSS* | -1.815166773 | 0.00199872 | 0.008781237 | 1 |
| Heart_Failure | b_cell | *VAV2* | 5.434706321 | 0.000222665 | 0.003674323 | 1 |
| Heart_Failure | b_cell | *PCNT* | -2.08621397 | 0.009655293 | 0.021054038 | 1 |
| Heart_Failure | b_cell | *PRMT2* | -1.870679725 | 0.015737752 | 0.03014386 | 1 |
| Heart_Failure | b_cell | *ZNF208* | -1.582814472 | 0.002045261 | 0.008831917 | 1 |

| | | | | | | |
|---|---|---|---|---|---|---|
| Heart_Failure | b_cell | *ADAMTS13* | -2.292244998 | 0.003452882 | 0.011092625 | 1 |
| Heart_Failure | b_cell | *GPSM1* | -2.693797227 | 0.004394717 | 0.012662504 | 1 |
| Heart_Failure | b_cell | *C19orf47* | -3.001824747 | 0.001671121 | 0.008654124 | 1 |
| Heart_Failure | b_cell | *HIPK4* | 7.942712261 | 4.72E-05 | 0.002889125 | 0.296548701 |
| Heart_Failure | b_cell | *MPZL3* | -2.83729074 | 0.001699051 | 0.008654124 | 1 |
| Heart_Failure | b_cell | *S100A1* | 2.094394651 | 0.00039815 | 0.00494416 | 1 |
| Heart_Failure | b_cell | *VPS11* | -2.26515817 | 0.030117502 | 0.049502937 | 1 |
| Heart_Failure | b_cell | *FDPS* | -1.774737437 | 0.020619965 | 0.036734974 | 1 |
| Heart_Failure | b_cell | *PAQR6* | 1.638046116 | 0.000224065 | 0.003674323 | 1 |
| Heart_Failure | b_cell | *PMF1* | 6.86093095 | 0.001913097 | 0.008705907 | 1 |
| Heart_Failure | b_cell | *SLC25A44* | -2.918382158 | 0.00159556 | 0.008654124 | 1 |
| Heart_Failure | b_cell | *FCRL3* | 0.250667881 | 0.000370152 | 0.004768742 | 1 |
| Heart_Failure | b_cell | *FGFR4* | 6.272156353 | 0.00044205 | 0.005211133 | 1 |
| Heart_Failure | b_cell | *NACC1* | 3.421893517 | 6.94E-05 | 0.002889125 | 0.43625762 |
| Heart_Failure | b_cell | *LY6K* | 11.00356185 | 0.000175447 | 0.003366185 | 1 |
| Heart_Failure | b_cell | *TONSL* | -3.668869349 | 0.002427182 | 0.009379036 | 1 |
| Heart_Failure | b_cell | *MUM1* | -1.557109372 | 0.009166678 | 0.02019303 | 1 |
| Heart_Failure | b_cell | *PPP1R16A* | -1.990386076 | 0.001373887 | 0.008654124 | 1 |
| Heart_Failure | b_cell | *ORAI2* | 2.244839101 | 5.66E-05 | 0.002889125 | 0.355914676 |
| Heart_Failure | b_cell | *RPL8* | 4.938557851 | 0.001070735 | 0.008654124 | 1 |
| Heart_Failure | b_cell | *LRWD1* | 2.621315074 | 0.016835927 | 0.031545978 | 1 |
| Heart_Failure | b_cell | *FBXL13* | -1.989342217 | 0.009519285 | 0.020802136 | 1 |
| Heart_Failure | b_cell | *YDJC* | 18.63141938 | 0.000256188 | 0.003777026 | 1 |
| Heart_Failure | b_cell | *CCDC116* | 4.144666039 | 0.000635282 | 0.006667813 | 1 |
| Heart_Failure | b_cell | *FBXO27* | -1.803985672 | 0.002145014 | 0.008999799 | 1 |
| Heart_Failure | b_cell | *U2AF1L4* | 1.595081944 | 0.005774125 | 0.014976042 | 1 |
| Heart_Failure | b_cell | *BDH1* | -2.588915873 | 0.001698365 | 0.008654124 | 1 |
| Heart_Failure | b_cell | *COX7A1* | -2.062393261 | 0.01202109 | 0.024690165 | 1 |
| Heart_Failure | b_cell | *DUSP14* | -1.941486475 | 0.001242844 | 0.008654124 | 1 |
| Heart_Failure | b_cell | *LRRC56* | 3.181976412 | 0.000384503 | 0.004883575 | 1 |
| Heart_Failure | b_cell | *PGAP3* | -3.543823654 | 0.011640835 | 0.024113981 | 1 |
| Heart_Failure | b_cell | *IKZF3* | 0.561423834 | 0.000143376 | 0.003239931 | 0.901405819 |
| Heart_Failure | b_cell | *ACOX1* | -1.720321885 | 0.004946979 | 0.01354012 | 1 |
| Heart_Failure | b_cell | *PRPSAP1* | -2.033695444 | 0.017728119 | 0.032880882 | 1 |
| Heart_Failure | b_cell | *CYGB* | -2.848230326 | 0.0009388 | 0.008620216 | 1 |
| Heart_Failure | b_cell | *ALDH16A1* | -3.035680331 | 0.01890009 | 0.034542112 | 1 |
| Heart_Failure | b_cell | *SIGLEC11* | -4.94074057 | 0.001764031 | 0.008654124 | 1 |
| Heart_Failure | b_cell | *MPP3* | -2.08812336 | 0.001256365 | 0.008654124 | 1 |
| Heart_Failure | b_cell | *ASB16* | 4.514185446 | 0.000461835 | 0.005376958 | 1 |
| Heart_Failure | b_cell | *PLCD3* | -2.210277804 | 0.001278892 | 0.008654124 | 1 |
| Heart_Failure | b_cell | *RACGAP1* | 8.487552621 | 0.000822946 | 0.008046441 | 1 |
| Heart_Failure | b_cell | *SCIMP* | 3.62027231 | 0.000100304 | 0.002997224 | 0.630610136 |
| Heart_Failure | b_cell | *ASGR2* | -11.45162765 | 0.010261047 | 0.022085313 | 1 |
| Heart_Failure | b_cell | *TNFSF13* | 4.501633857 | 0.012034413 | 0.024709456 | 1 |
| Heart_Failure | b_cell | *SENP3* | 3.453786363 | 3.37E-05 | 0.002889125 | 0.211810349 |
| Heart_Failure | b_cell | *SNRNP25* | -4.696159883 | 0.001485481 | 0.008654124 | 1 |
| Heart_Failure | b_cell | *WDR90* | -2.162041014 | 0.01358152 | 0.027038321 | 1 |
| Heart_Failure | b_cell | *JMJD8* | 6.390565965 | 1.31E-05 | 0.002704195 | 0.082213016 |
| Heart_Failure | b_cell | *MEIOB* | -4.035581217 | 0.009084271 | 0.020103066 | 1 |
| Heart_Failure | b_cell | *ZG16B* | -35.41028371 | 0.001714234 | 0.008654124 | 1 |

| | | | | | | |
|---|---|---|---|---|---|---|
| Heart_Failure | b_cell | *NXF1* | -1.8694753 | 0.018250355 | 0.033539895 | 1 |
| Heart_Failure | b_cell | *TPCN2* | -1.652619814 | 0.001706402 | 0.008654124 | 1 |
| Heart_Failure | b_cell | *CYP4A22* | 12.64348842 | 0.000373286 | 0.004779733 | 1 |
| Heart_Failure | b_cell | *PDZK1IP1* | 50.32880228 | 0.003673942 | 0.011460681 | 1 |
| Heart_Failure | b_cell | *COA7* | 3.805226728 | 5.94E-05 | 0.002889125 | 0.373225236 |
| Heart_Failure | b_cell | *SLC1A7* | -1.496108761 | 0.021054788 | 0.037361403 | 1 |
| Heart_Failure | b_cell | *C1orf123* | -2.522426556 | 0.011611534 | 0.024069143 | 1 |
| Heart_Failure | b_cell | *ACOT11* | -2.237876423 | 0.004652341 | 0.013051881 | 1 |
| Heart_Failure | b_cell | *RBP7* | -2.037606451 | 0.002288866 | 0.009185338 | 1 |
| Heart_Failure | b_cell | *FBLIM1* | -2.222268648 | 0.002665033 | 0.009714529 | 1 |
| Heart_Failure | b_cell | *SLC25A34* | 11.71581605 | 0.000100885 | 0.002997224 | 0.634262003 |
| Heart_Failure | b_cell | *PDPN* | -2.38910625 | 0.001191319 | 0.008654124 | 1 |
| Heart_Failure | b_cell | *LRRC38* | -11.89911427 | 0.008760115 | 0.019648535 | 1 |
| Heart_Failure | b_cell | *LAPTM5* | 4.679514056 | 0.001595231 | 0.008654124 | 1 |
| Heart_Failure | b_cell | *PEF1* | -2.181275252 | 0.002054543 | 0.008831917 | 1 |
| Heart_Failure | b_cell | *SYNC* | -2.294658615 | 0.004829934 | 0.013353473 | 1 |
| Heart_Failure | b_cell | *TSSK3* | -2.424781734 | 0.019488415 | 0.035341246 | 1 |
| Heart_Failure | b_cell | *CAMK2N1* | -3.073817107 | 0.001807843 | 0.008654124 | 1 |
| Heart_Failure | b_cell | *TTLL10* | -5.8480195 | 0.00599327 | 0.01532941 | 1 |
| Heart_Failure | b_cell | *NEXN* | -1.626069468 | 0.003715153 | 0.011528709 | 1 |
| Heart_Failure | b_cell | *B3GALT2* | -3.171272574 | 0.001816551 | 0.008654124 | 1 |
| Heart_Failure | b_cell | *NTNG1* | -1.714575455 | 0.006061985 | 0.015411121 | 1 |
| Heart_Failure | b_cell | *HFM1* | -1.720491449 | 0.001586918 | 0.008654124 | 1 |
| Heart_Failure | b_cell | *KCNT2* | -1.323706383 | 0.001258985 | 0.008654124 | 1 |
| Heart_Failure | b_cell | *VCAM1* | -2.096241424 | 0.00249835 | 0.009479257 | 1 |
| Heart_Failure | b_cell | *EXTL2* | -1.932485021 | 0.02590601 | 0.04392424 | 1 |
| Heart_Failure | b_cell | *TRIM58* | 1.620874446 | 0.000304785 | 0.004248737 | 1 |
| Heart_Failure | b_cell | *IGSF8* | -2.205876551 | 0.001687922 | 0.008654124 | 1 |
| Heart_Failure | b_cell | *VANGL2* | -5.310610457 | 0.004002925 | 0.012024754 | 1 |
| Heart_Failure | b_cell | *FCRLB* | -6.242723618 | 0.008506501 | 0.019272206 | 1 |
| Heart_Failure | b_cell | *KLHDC9* | 2.11041327 | 0.025767059 | 0.043735827 | 1 |
| Heart_Failure | b_cell | *LMX1A* | -1.729084761 | 0.00350868 | 0.011157849 | 1 |
| Heart_Failure | b_cell | *FLVCR1* | -2.018539001 | 0.001774695 | 0.008654124 | 1 |
| Heart_Failure | b_cell | *TDRD5* | -4.641158055 | 0.000267507 | 0.003875152 | 1 |
| Heart_Failure | b_cell | *SNED1* | -1.488797301 | 0.016516281 | 0.031210659 | 1 |
| Heart_Failure | b_cell | *C1orf115* | -2.386747491 | 0.004034572 | 0.012061508 | 1 |
| Heart_Failure | b_cell | *PM20D1* | -3.529587598 | 0.001957041 | 0.008763475 | 1 |
| Heart_Failure | b_cell | *PUS10* | -1.986830103 | 0.006060354 | 0.015411121 | 1 |
| Heart_Failure | b_cell | *RFTN2* | -1.80277912 | 0.004576936 | 0.012902652 | 1 |
| Heart_Failure | b_cell | *FAM84A* | -4.799527582 | 0.00174189 | 0.008654124 | 1 |
| Heart_Failure | b_cell | *FBXO41* | 9.175684135 | 0.000284523 | 0.004036731 | 1 |
| Heart_Failure | b_cell | *CCDC74A* | -10.39483924 | 0.002042482 | 0.008831917 | 1 |
| Heart_Failure | b_cell | *NOSTRIN* | -1.379300722 | 0.005125353 | 0.013853437 | 1 |
| Heart_Failure | b_cell | *SGPP2* | 9.198985426 | 0.000416111 | 0.005049082 | 1 |
| Heart_Failure | b_cell | *BBS5* | -2.425169591 | 0.00730508 | 0.017394685 | 1 |
| Heart_Failure | b_cell | *C1QTNF7* | -1.195657064 | 0.001698562 | 0.008654124 | 1 |
| Heart_Failure | b_cell | *LYSMD1* | -2.525524672 | 0.004253536 | 0.01239202 | 1 |
| Heart_Failure | b_cell | *DHX57* | -2.012547615 | 0.008602826 | 0.019399557 | 1 |
| Heart_Failure | b_cell | *S100A9* | 4.744117367 | 2.38E-05 | 0.002889125 | 0.149422131 |
| Heart_Failure | b_cell | *S100A12* | -21.62445385 | 0.002787211 | 0.009915204 | 1 |

| | | | | | | |
|---|---|---|---|---|---|---|
| Heart_Failure | b_cell | *TDRD10* | -2.14518918 | 0.005315381 | 0.014178109 | 1 |
| Heart_Failure | b_cell | *DCAF16* | -2.107134158 | 0.006392776 | 0.015942634 | 1 |
| Heart_Failure | b_cell | *NIPAL1* | -2.508118134 | 0.00154991 | 0.008654124 | 1 |
| Heart_Failure | b_cell | *DCST2* | -3.803927571 | 0.001438089 | 0.008654124 | 1 |
| Heart_Failure | b_cell | *LMOD3* | -2.880521235 | 0.001698084 | 0.008654124 | 1 |
| Heart_Failure | b_cell | *NBPF10* | 2.538348647 | 0.000128734 | 0.003185748 | 0.809348114 |
| Heart_Failure | b_cell | *SLC22A15* | -1.626709713 | 0.002394436 | 0.009333474 | 1 |
| Heart_Failure | b_cell | *SLC15A2* | 1.922426832 | 0.000411887 | 0.005033892 | 1 |
| Heart_Failure | b_cell | *LRRC58* | -2.346013402 | 0.025603318 | 0.043540184 | 1 |
| Heart_Failure | b_cell | *FSTL1* | -1.170697246 | 0.004354333 | 0.012586524 | 1 |
| Heart_Failure | b_cell | *LMOD1* | -1.594267023 | 0.000608862 | 0.006565891 | 1 |
| Heart_Failure | b_cell | *TMEM183A* | -2.843298226 | 0.005089028 | 0.013812459 | 1 |
| Heart_Failure | b_cell | *IGFBP7* | -1.514733657 | 0.007367964 | 0.017499958 | 1 |
| Heart_Failure | b_cell | *GLB1L* | 3.900660381 | 0.001082143 | 0.008654124 | 1 |
| Heart_Failure | b_cell | *NUAK2* | 1.012610507 | 0.000316875 | 0.00432146 | 1 |
| Heart_Failure | b_cell | *SPTA1* | -1.550558043 | 0.009154928 | 0.020188366 | 1 |
| Heart_Failure | b_cell | *PYHIN1* | -1.820851521 | 0.000919113 | 0.008523384 | 1 |
| Heart_Failure | b_cell | *AIM2* | 2.534136097 | 8.69E-05 | 0.002889125 | 0.546620677 |
| Heart_Failure | b_cell | *ICA1L* | -2.156576178 | 0.003645906 | 0.011427162 | 1 |
| Heart_Failure | b_cell | *CD200R1* | -1.778761595 | 0.001576427 | 0.008654124 | 1 |
| Heart_Failure | b_cell | *GTPBP8* | -2.146791095 | 0.004006848 | 0.012024754 | 1 |
| Heart_Failure | b_cell | *SPICE1* | -2.454537406 | 0.006559562 | 0.016185231 | 1 |
| Heart_Failure | b_cell | *CADPS* | 4.600184373 | 5.85E-05 | 0.002889125 | 0.367616535 |
| Heart_Failure | b_cell | *PPM1K* | 5.181761623 | 0.00034076 | 0.004538892 | 1 |
| Heart_Failure | b_cell | *TIPARP* | 4.553939486 | 0.007750286 | 0.018066759 | 1 |
| Heart_Failure | b_cell | *DCLK3* | 4.917793402 | 1.63E-05 | 0.002889125 | 0.102182138 |
| Heart_Failure | b_cell | *RPL9* | 1.578683249 | 0.028948379 | 0.047970074 | 1 |
| Heart_Failure | b_cell | *DNASE1L3* | -3.057100714 | 0.001202572 | 0.008654124 | 1 |
| Heart_Failure | b_cell | *CRELD1* | -1.839032802 | 0.01428262 | 0.028113598 | 1 |
| Heart_Failure | b_cell | *PCOLCE2* | -1.071060584 | 0.002052925 | 0.008831917 | 1 |
| Heart_Failure | b_cell | *CCDC158* | -2.102090476 | 0.009289728 | 0.02040689 | 1 |
| Heart_Failure | b_cell | *GYG1* | -2.733939632 | 0.01486154 | 0.028962958 | 1 |
| Heart_Failure | b_cell | *PLB1* | -2.288712539 | 0.001799139 | 0.008654124 | 1 |
| Heart_Failure | b_cell | *KIAA1143* | 3.19432038 | 0.007133358 | 0.017137796 | 1 |
| Heart_Failure | b_cell | *CDCP1* | -2.328419038 | 0.001846148 | 0.00866783 | 1 |
| Heart_Failure | b_cell | *LZTFL1* | -1.883841768 | 0.001707074 | 0.008654124 | 1 |
| Heart_Failure | b_cell | *LRRC2* | -1.910077571 | 0.000489417 | 0.005622167 | 1 |
| Heart_Failure | b_cell | *SMIM12* | -2.290399497 | 0.015708753 | 0.030119222 | 1 |
| Heart_Failure | b_cell | *MEAF6* | -2.485606465 | 0.018129233 | 0.033414978 | 1 |
| Heart_Failure | b_cell | *DNALI1* | -4.393590539 | 0.000256021 | 0.003777026 | 1 |
| Heart_Failure | b_cell | *RFC4* | 2.144507307 | 0.000134974 | 0.003206095 | 0.848579611 |
| Heart_Failure | b_cell | *RFT1* | -2.847608022 | 0.010911586 | 0.02307472 | 1 |
| Heart_Failure | b_cell | *SFMBT1* | 5.473981244 | 0.00187545 | 0.008701813 | 1 |
| Heart_Failure | b_cell | *GNL3* | -2.145144651 | 0.014985482 | 0.029141269 | 1 |
| Heart_Failure | b_cell | *SLC51A* | -2.024762507 | 0.001088179 | 0.008654124 | 1 |
| Heart_Failure | b_cell | *PIGX* | -2.300736959 | 0.015203168 | 0.029437116 | 1 |
| Heart_Failure | b_cell | *ABLIM2* | -1.951949796 | 0.001653993 | 0.008654124 | 1 |
| Heart_Failure | b_cell | *EXO5* | 4.66865992 | 0.000149578 | 0.003239931 | 0.940398268 |
| Heart_Failure | b_cell | *ERMAP* | -2.024127048 | 0.008551044 | 0.019338278 | 1 |
| Heart_Failure | b_cell | *SLC9B2* | -2.237845323 | 0.00365969 | 0.011449527 | 1 |

| | | | | | | |
|---|---|---|---|---|---|---|
| Heart_Failure | b_cell | *BDH2* | -1.442821313 | 0.003021757 | 0.010281376 | 1 |
| Heart_Failure | b_cell | *ATRIP* | -6.416448184 | 0.001572067 | 0.008654124 | 1 |
| Heart_Failure | b_cell | *SHISA5* | -2.356214736 | 0.009997652 | 0.021644366 | 1 |
| Heart_Failure | b_cell | *RNF123* | 3.425947057 | 0.006131893 | 0.015519811 | 1 |
| Heart_Failure | b_cell | *TEX264* | 3.332789887 | 0.015453289 | 0.029765573 | 1 |
| Heart_Failure | b_cell | *GRM2* | 4.258403619 | 0.000233044 | 0.003705045 | 1 |
| Heart_Failure | b_cell | *POC1A* | 5.36094586 | 0.000161867 | 0.003294027 | 1 |
| Heart_Failure | b_cell | *PPM1M* | -2.435815737 | 0.014119661 | 0.027862621 | 1 |
| Heart_Failure | b_cell | *C4orf3* | -2.755046061 | 0.010823197 | 0.022972137 | 1 |
| Heart_Failure | b_cell | *ANXA5* | -1.624754587 | 0.025032854 | 0.042731891 | 1 |
| Heart_Failure | b_cell | *MAP9* | -2.189585762 | 0.002120724 | 0.008942317 | 1 |
| Heart_Failure | b_cell | *CEP44* | -2.136190433 | 0.017926733 | 0.033138892 | 1 |
| Heart_Failure | b_cell | *C4orf45* | -1.717692832 | 0.001932119 | 0.008732736 | 1 |
| Heart_Failure | b_cell | *TMEM144* | -1.578573425 | 0.002580371 | 0.009572739 | 1 |
| Heart_Failure | b_cell | *NPY1R* | -1.218226101 | 0.001694053 | 0.008654124 | 1 |
| Heart_Failure | b_cell | *IL15* | -1.959777569 | 0.007874359 | 0.018261192 | 1 |
| Heart_Failure | b_cell | *ITGA2* | -2.278607035 | 0.002047725 | 0.008831917 | 1 |
| Heart_Failure | b_cell | *NDUFAF2* | -1.747985116 | 0.011140005 | 0.023408158 | 1 |
| Heart_Failure | b_cell | *LMBRD2* | -2.311299668 | 0.029750222 | 0.049091771 | 1 |
| Heart_Failure | b_cell | *CMBL* | -2.114837038 | 0.001728658 | 0.008654124 | 1 |
| Heart_Failure | b_cell | *C5orf63* | -2.144972439 | 0.001272218 | 0.008654124 | 1 |
| Heart_Failure | b_cell | *PRRC1* | -2.098312852 | 0.007237352 | 0.017320605 | 1 |
| Heart_Failure | b_cell | *WDR41* | -1.606095885 | 0.01106462 | 0.023304276 | 1 |
| Heart_Failure | b_cell | *RHOBTB3* | -1.029666385 | 0.00201828 | 0.008817602 | 1 |
| Heart_Failure | b_cell | *TIGD6* | -3.361408114 | 0.002172333 | 0.009019688 | 1 |
| Heart_Failure | b_cell | *ENPP6* | -7.653301464 | 0.001498856 | 0.008654124 | 1 |
| Heart_Failure | b_cell | *PRIMPOL* | -2.147095722 | 0.017620655 | 0.032727048 | 1 |
| Heart_Failure | b_cell | *ANKRA2* | -2.017594144 | 0.005853145 | 0.015118619 | 1 |
| Heart_Failure | b_cell | *UTP15* | -2.698641266 | 0.004206628 | 0.012306687 | 1 |
| Heart_Failure | b_cell | *TLR3* | -2.209014124 | 0.001823104 | 0.008654124 | 1 |
| Heart_Failure | b_cell | *CCDC127* | -2.980100224 | 0.002073253 | 0.008848976 | 1 |
| Heart_Failure | b_cell | *GRIK2* | -1.593479609 | 0.002908718 | 0.010125753 | 1 |
| Heart_Failure | b_cell | *MB21D1* | 5.551883343 | 0.000116482 | 0.003135985 | 0.732321803 |
| Heart_Failure | b_cell | *DCBLD1* | -1.609923105 | 0.004469898 | 0.01274701 | 1 |
| Heart_Failure | b_cell | *TMEM200A* | -2.584228847 | 0.004038921 | 0.012068773 | 1 |
| Heart_Failure | b_cell | *PI16* | -1.28909232 | 0.00151391 | 0.008654124 | 1 |
| Heart_Failure | b_cell | *HCN1* | -3.215725303 | 0.001226706 | 0.008654124 | 1 |
| Heart_Failure | b_cell | *SLU7* | 6.322159935 | 0.000283478 | 0.004036731 | 1 |
| Heart_Failure | b_cell | *KCNK5* | -3.861214519 | 0.00145821 | 0.008654124 | 1 |
| Heart_Failure | b_cell | *KIAA1324L* | -1.635670154 | 0.001277385 | 0.008654124 | 1 |
| Heart_Failure | b_cell | *USP49* | -1.924336778 | 0.015981809 | 0.030392508 | 1 |
| Heart_Failure | b_cell | *ZNF704* | -1.307821709 | 0.004843989 | 0.013374687 | 1 |
| Heart_Failure | b_cell | *FNDC1* | -1.867274697 | 0.000938949 | 0.008620216 | 1 |
| Heart_Failure | b_cell | *LMTK2* | -2.207898903 | 0.021951536 | 0.038614803 | 1 |
| Heart_Failure | b_cell | *CTSB* | -1.447920102 | 0.029596276 | 0.048876224 | 1 |
| Heart_Failure | b_cell | *PEX2* | -2.16275232 | 0.006429359 | 0.015989469 | 1 |
| Heart_Failure | b_cell | *TNFRSF11B* | -2.578785105 | 0.001451813 | 0.008654124 | 1 |
| Heart_Failure | b_cell | *DEFB1* | -3.051235406 | 0.00962981 | 0.021014446 | 1 |
| Heart_Failure | b_cell | *TMEM184A* | 1.857755866 | 0.000226426 | 0.003697509 | 1 |
| Heart_Failure | b_cell | *FASTK* | -1.959605486 | 0.002712167 | 0.009825588 | 1 |

| | | | | | | |
|---|---|---|---|---|---|---|
| Heart_Failure | b_cell | *ALDH7A1* | -2.162530808 | 0.009757415 | 0.021211919 | 1 |
| Heart_Failure | b_cell | *COX6C* | -2.160555011 | 0.014912303 | 0.029034887 | 1 |
| Heart_Failure | b_cell | *SLC25A32* | -2.157761818 | 0.014964132 | 0.029113027 | 1 |
| Heart_Failure | b_cell | *INTS8* | -1.932178766 | 0.017847997 | 0.033041919 | 1 |
| Heart_Failure | b_cell | *FREM1* | -1.145186762 | 0.001379873 | 0.008654124 | 1 |
| Heart_Failure | b_cell | *RPP25L* | 11.45791588 | 7.69E-06 | 0.002102517 | 0.048357897 |
| Heart_Failure | b_cell | *CCDC171* | -1.928890112 | 0.010191845 | 0.021973981 | 1 |
| Heart_Failure | b_cell | *SYK* | 4.244492982 | 0.005718452 | 0.014862301 | 1 |
| Heart_Failure | b_cell | *NIPSNAP3B* | -1.338551492 | 0.007699447 | 0.017999868 | 1 |
| Heart_Failure | b_cell | *TMC1* | -2.365078918 | 0.002770991 | 0.009906851 | 1 |
| Heart_Failure | b_cell | *C9orf64* | -2.31227224 | 0.001265007 | 0.008654124 | 1 |
| Heart_Failure | b_cell | *SVEP1* | -1.026100201 | 0.001692786 | 0.008654124 | 1 |
| Heart_Failure | b_cell | *ZNF367* | 4.87847038 | 7.30E-05 | 0.002889125 | 0.459004223 |
| Heart_Failure | b_cell | *NOL6* | -3.237341685 | 0.004076935 | 0.012126691 | 1 |
| Heart_Failure | b_cell | *AQP3* | -4.708540036 | 0.004069256 | 0.012124841 | 1 |
| Heart_Failure | b_cell | *PIGO* | -3.536497191 | 0.004167796 | 0.012232927 | 1 |
| Heart_Failure | b_cell | *LRFN5* | -1.255140788 | 0.010276605 | 0.022103666 | 1 |
| Heart_Failure | b_cell | *CFL2* | 6.482284146 | 6.16E-05 | 0.002889125 | 0.387417434 |
| Heart_Failure | b_cell | *ZCCHC24* | -1.666117224 | 0.005662095 | 0.014758538 | 1 |
| Heart_Failure | b_cell | *PGM2L1* | 4.748103293 | 4.49E-05 | 0.002889125 | 0.282557806 |
| Heart_Failure | b_cell | *GJB2* | 6.548143931 | 0.000949183 | 0.008639437 | 1 |
| Heart_Failure | b_cell | *REEP3* | -1.828684052 | 0.010138344 | 0.021896175 | 1 |
| Heart_Failure | b_cell | *LRR1* | -2.603152807 | 0.001510312 | 0.008654124 | 1 |
| Heart_Failure | b_cell | *C10orf25* | -2.800344679 | 0.01436565 | 0.028224012 | 1 |
| Heart_Failure | b_cell | *KLHDC2* | -1.990102417 | 0.015656685 | 0.030050797 | 1 |
| Heart_Failure | b_cell | *AKR1E2* | -2.136184091 | 0.000360864 | 0.004736438 | 1 |
| Heart_Failure | b_cell | *VDAC2* | -2.012235921 | 0.00296567 | 0.010176497 | 1 |
| Heart_Failure | b_cell | *SLC18A2* | -2.273612178 | 0.004737911 | 0.013205183 | 1 |
| Heart_Failure | b_cell | *QSOX2* | 2.503084693 | 8.68E-05 | 0.002889125 | 0.545619465 |
| Heart_Failure | b_cell | *SNAPC4* | -2.267160375 | 0.03033962 | 0.049789921 | 1 |
| Heart_Failure | b_cell | *SDCCAG3* | -2.596844394 | 0.026964735 | 0.045352404 | 1 |
| Heart_Failure | b_cell | *AK8* | -3.273309209 | 0.005844568 | 0.015108882 | 1 |
| Heart_Failure | b_cell | *ZMYND19* | -5.115924061 | 0.011577369 | 0.02401416 | 1 |
| Heart_Failure | b_cell | *RET* | 2.151719698 | 0.000154494 | 0.003239931 | 0.971302327 |
| Heart_Failure | b_cell | *STK32C* | -2.225633835 | 0.003739602 | 0.011555523 | 1 |
| Heart_Failure | b_cell | *NSMF* | -2.457340845 | 0.002174629 | 0.009019688 | 1 |
| Heart_Failure | b_cell | *METTL3* | -2.343585954 | 0.020116352 | 0.036124394 | 1 |
| Heart_Failure | b_cell | *HSPA12A* | -0.955401385 | 0.023291769 | 0.040429418 | 1 |
| Heart_Failure | b_cell | *UBTD1* | -2.179347263 | 0.007764538 | 0.018086569 | 1 |
| Heart_Failure | b_cell | *ARHGAP42* | -1.411300541 | 0.003516927 | 0.011161493 | 1 |
| Heart_Failure | b_cell | *TTC7B* | -1.651501072 | 0.011415597 | 0.023796373 | 1 |
| Heart_Failure | b_cell | *SLC39A13* | -2.972192046 | 0.004543756 | 0.01284469 | 1 |
| Heart_Failure | b_cell | *PSMC3* | -2.471250582 | 0.002303303 | 0.009188443 | 1 |
| Heart_Failure | b_cell | *TC2N* | -1.255907484 | 0.002853282 | 0.010013685 | 1 |
| Heart_Failure | b_cell | *CLMN* | -1.32595536 | 0.002355355 | 0.009268076 | 1 |
| Heart_Failure | b_cell | *PTER* | -1.99920897 | 0.013230247 | 0.026557652 | 1 |
| Heart_Failure | b_cell | *C1QL3* | -2.529235733 | 0.00169854 | 0.008654124 | 1 |
| Heart_Failure | b_cell | *ABTB2* | -2.107448377 | 0.00370738 | 0.011521649 | 1 |
| Heart_Failure | b_cell | *LIPC* | -2.213159625 | 0.002564988 | 0.009560634 | 1 |
| Heart_Failure | b_cell | *JAM3* | -2.41245158 | 0.011273594 | 0.023636218 | 1 |

| | | | | | | |
|---|---|---|---|---|---|---|
| Heart_Failure | b_cell | *GPT2* | -1.439589153 | 0.004382013 | 0.012631689 | 1 |
| Heart_Failure | b_cell | *HIF1AN* | -2.277464032 | 0.006250315 | 0.015737177 | 1 |
| Heart_Failure | b_cell | *NDUFB8* | -3.319544712 | 0.011001971 | 0.023218998 | 1 |
| Heart_Failure | b_cell | *TRMT61A* | -4.577511168 | 0.004791732 | 0.01328876 | 1 |
| Heart_Failure | b_cell | *BTRC* | -1.663378289 | 0.015230523 | 0.029481003 | 1 |
| Heart_Failure | b_cell | *LARP6* | -1.862117971 | 0.001795314 | 0.008654124 | 1 |
| Heart_Failure | b_cell | *ASPG* | -8.462221223 | 0.002138121 | 0.008997561 | 1 |
| Heart_Failure | b_cell | *GABRB3* | 5.513387693 | 0.000153389 | 0.003239931 | 0.964358149 |
| Heart_Failure | b_cell | *PCBD1* | -2.499333184 | 0.000658848 | 0.006880692 | 1 |
| Heart_Failure | b_cell | *COX11* | -1.823328686 | 0.003343306 | 0.010865693 | 1 |
| Heart_Failure | b_cell | *FAM227B* | -1.539961286 | 0.007578366 | 0.017817947 | 1 |
| Heart_Failure | b_cell | *NUDT13* | -1.191067377 | 0.022666344 | 0.039616477 | 1 |
| Heart_Failure | b_cell | *ILK* | -1.886555401 | 0.010505439 | 0.022449931 | 1 |
| Heart_Failure | b_cell | *TAF10* | -2.286092145 | 0.00510609 | 0.013822386 | 1 |
| Heart_Failure | b_cell | *DCHS1* | -2.344858104 | 0.002173527 | 0.009019688 | 1 |
| Heart_Failure | b_cell | *NETO1* | 0.666715511 | 0.000186923 | 0.00340015 | 1 |
| Heart_Failure | b_cell | *USP54* | -1.885902319 | 0.007014741 | 0.016962182 | 1 |
| Heart_Failure | b_cell | *C11orf74* | -1.690167218 | 0.0022684 | 0.009180605 | 1 |
| Heart_Failure | b_cell | *WDR88* | 6.237597827 | 0.010945999 | 0.023131932 | 1 |
| Heart_Failure | b_cell | *CYB5R2* | 2.862250175 | 0.000140519 | 0.003214256 | 0.883443763 |
| Heart_Failure | b_cell | *TUB* | -2.318309006 | 0.001334993 | 0.008654124 | 1 |
| Heart_Failure | b_cell | *RIC3* | -2.637086717 | 0.00884646 | 0.019736647 | 1 |
| Heart_Failure | b_cell | *LMO1* | -4.748020929 | 0.006395826 | 0.015943916 | 1 |
| Heart_Failure | b_cell | *XRRA1* | -1.769200198 | 0.005507587 | 0.014475836 | 1 |
| Heart_Failure | b_cell | *PRTG* | -2.741287691 | 0.006538409 | 0.016158404 | 1 |
| Heart_Failure | b_cell | *C16orf46* | -30.74783402 | 0.001644299 | 0.008654124 | 1 |
| Heart_Failure | b_cell | *TMEM41B* | -2.050613443 | 0.025790009 | 0.043762965 | 1 |
| Heart_Failure | b_cell | *CCDC68* | -2.049363628 | 0.011097543 | 0.023365792 | 1 |
| Heart_Failure | b_cell | *CLEC4E* | -3.294182954 | 0.021506219 | 0.038012258 | 1 |
| Heart_Failure | b_cell | *ZNF3* | -2.502222726 | 0.008861231 | 0.019752526 | 1 |
| Heart_Failure | b_cell | *CLEC4D* | -9.667874495 | 0.001403802 | 0.008654124 | 1 |
| Heart_Failure | b_cell | *ZSCAN21* | -1.898078406 | 0.014316768 | 0.02816577 | 1 |
| Heart_Failure | b_cell | *RIMKLB* | -1.901354514 | 0.002534391 | 0.00953574 | 1 |
| Heart_Failure | b_cell | *BEAN1* | -2.239866325 | 0.008535222 | 0.019314107 | 1 |
| Heart_Failure | b_cell | *TK2* | -1.667529978 | 0.005225397 | 0.014027294 | 1 |
| Heart_Failure | b_cell | *SEC11C* | 4.12287917 | 5.85E-05 | 0.002889125 | 0.367561922 |
| Heart_Failure | b_cell | *GALR1* | -3.1664248 | 0.00344578 | 0.011081136 | 1 |
| Heart_Failure | b_cell | *CENPV* | -2.030673959 | 0.001897751 | 0.008705907 | 1 |
| Heart_Failure | b_cell | *CHRFAM7A* | -4.779725375 | 0.01731547 | 0.032255515 | 1 |
| Heart_Failure | b_cell | *ZNF592* | -1.894031614 | 0.008279002 | 0.018913548 | 1 |
| Heart_Failure | b_cell | *ACSM1* | 1.464389996 | 0.000128628 | 0.003185748 | 0.808686881 |
| Heart_Failure | b_cell | *SLFN5* | -1.649226871 | 0.008241016 | 0.018861036 | 1 |
| Heart_Failure | b_cell | *CATSPER2* | -2.348262656 | 0.003163282 | 0.010566301 | 1 |
| Heart_Failure | b_cell | *YPEL4* | -6.533296007 | 0.008389503 | 0.019082781 | 1 |
| Heart_Failure | b_cell | *SCNN1G* | -11.31160275 | 0.01265653 | 0.025680788 | 1 |
| Heart_Failure | b_cell | *DCTN5* | -2.196733205 | 0.005733318 | 0.014894782 | 1 |
| Heart_Failure | b_cell | *STX3* | -2.022405258 | 0.006051498 | 0.015400648 | 1 |
| Heart_Failure | b_cell | *TSC22D4* | -1.835615624 | 0.002325561 | 0.0092234 | 1 |
| Heart_Failure | b_cell | *MS4A6E* | -2.082795072 | 0.018601156 | 0.034094889 | 1 |
| Heart_Failure | b_cell | *MS4A7* | -1.482360895 | 0.001168295 | 0.008654124 | 1 |

| | | | | | | |
|---|---|---|---|---|---|---|
| Heart_Failure | b_cell | *MS4A14* | -1.712921452 | 0.008881395 | 0.01977943 | 1 |
| Heart_Failure | b_cell | *DIS3L* | -1.744371981 | 0.001068037 | 0.008654124 | 1 |
| Heart_Failure | b_cell | *CCNDBP1* | -2.261748886 | 0.00881257 | 0.019702926 | 1 |
| Heart_Failure | b_cell | *SMAD3* | -1.778828291 | 0.006017208 | 0.015361627 | 1 |
| Heart_Failure | b_cell | *RCCD1* | -2.433108622 | 0.001795521 | 0.008654124 | 1 |
| Heart_Failure | b_cell | *AKTIP* | -2.74830732 | 0.014609272 | 0.028613239 | 1 |
| Heart_Failure | b_cell | *EVA1C* | -1.462194177 | 0.005084138 | 0.013812459 | 1 |
| Heart_Failure | b_cell | *CNPY4* | -2.054497013 | 0.019560748 | 0.035424135 | 1 |
| Heart_Failure | b_cell | *NKX3-1* | -23.47816157 | 0.029907975 | 0.049274487 | 1 |
| Heart_Failure | b_cell | *TEF* | -1.514927282 | 0.003490187 | 0.011132829 | 1 |
| Heart_Failure | b_cell | *MEI1* | 3.841804453 | 5.21E-05 | 0.002889125 | 0.327283417 |
| Heart_Failure | b_cell | *SNRPD1* | -2.118609033 | 0.002446906 | 0.009416629 | 1 |
| Heart_Failure | b_cell | *SAMD14* | 10.46917791 | 0.009302336 | 0.020420316 | 1 |
| Heart_Failure | b_cell | *URM1* | -2.680524865 | 0.025752856 | 0.043723523 | 1 |
| Heart_Failure | b_cell | *CERCAM* | -1.826114864 | 0.003209556 | 0.010665158 | 1 |
| Heart_Failure | b_cell | *ENDOG* | 3.175473409 | 2.65E-05 | 0.002889125 | 0.166554326 |
| Heart_Failure | b_cell | *ISLR2* | -2.682943287 | 0.020210012 | 0.036230495 | 1 |
| Heart_Failure | b_cell | *SP2* | -2.686768437 | 0.015777405 | 0.030186412 | 1 |
| Heart_Failure | b_cell | *COQ7* | 6.54756515 | 0.000378751 | 0.004830036 | 1 |
| Heart_Failure | b_cell | *GPRC5B* | -2.322238603 | 0.005360898 | 0.014239105 | 1 |
| Heart_Failure | b_cell | *KATNAL2* | -1.581658252 | 0.000419749 | 0.005055487 | 1 |
| Heart_Failure | b_cell | *HDHD2* | 3.912900605 | 0.00433505 | 0.012548093 | 1 |
| Heart_Failure | b_cell | *RNF214* | -1.938468783 | 0.008848865 | 0.019736647 | 1 |
| Heart_Failure | b_cell | *ENGASE* | -1.940426845 | 0.003396188 | 0.010972165 | 1 |
| Heart_Failure | b_cell | *RBFOX3* | -3.51804908 | 0.001992231 | 0.008775328 | 1 |
| Heart_Failure | b_cell | *MYO5B* | -1.739341336 | 0.002938913 | 0.010146595 | 1 |
| Heart_Failure | b_cell | *TRIM68* | -2.966618682 | 0.010850658 | 0.023000028 | 1 |
| Heart_Failure | b_cell | *FN3K* | -1.999220711 | 0.001794506 | 0.008654124 | 1 |
| Heart_Failure | b_cell | *ZNF229* | -10.03236337 | 0.001539572 | 0.008654124 | 1 |
| Heart_Failure | b_cell | *VKORC1* | -1.983632913 | 0.002620742 | 0.009652375 | 1 |
| Heart_Failure | b_cell | *SMG8* | -6.719810966 | 0.001493211 | 0.008654124 | 1 |
| Heart_Failure | b_cell | *CACNB3* | 2.012031293 | 8.28E-05 | 0.002889125 | 0.520349355 |
| Heart_Failure | b_cell | *TP53I13* | -1.811358304 | 0.005462949 | 0.014394618 | 1 |
| Heart_Failure | b_cell | *ZNF528* | -2.036183621 | 0.003120045 | 0.010478484 | 1 |
| Heart_Failure | b_cell | *SERTAD3* | -4.333363715 | 0.021513107 | 0.038013746 | 1 |
| Heart_Failure | b_cell | *TMC4* | -3.238724853 | 0.002592676 | 0.009588327 | 1 |
| Heart_Failure | b_cell | *TTYH1* | -2.475895381 | 0.002947936 | 0.010151069 | 1 |
| Heart_Failure | b_cell | *LENG8* | -1.879544535 | 0.012277119 | 0.025109384 | 1 |
| Heart_Failure | b_cell | *CDC42EP5* | -2.359947745 | 0.004131862 | 0.01219006 | 1 |
| Heart_Failure | b_cell | *ZNF283* | 5.067670862 | 0.000250643 | 0.003766552 | 1 |
| Heart_Failure | b_cell | *SPINT2* | -2.459918211 | 0.000329439 | 0.004450951 | 1 |
| Heart_Failure | b_cell | *DNAAF3* | -13.08308795 | 0.007648449 | 0.017910677 | 1 |
| Heart_Failure | b_cell | *PSCA* | -4.134785506 | 0.001465804 | 0.008654124 | 1 |
| Heart_Failure | b_cell | *UBXN6* | -2.833051798 | 0.001089268 | 0.008654124 | 1 |
| Heart_Failure | b_cell | *ZNF444* | -2.296648042 | 0.00264773 | 0.009678068 | 1 |
| Heart_Failure | b_cell | *SERPINF2* | 5.755266601 | 0.002272152 | 0.009180605 | 1 |
| Heart_Failure | b_cell | *CYB5D2* | -2.246992675 | 0.024400757 | 0.041897633 | 1 |
| Heart_Failure | b_cell | *ZNF83* | -1.66940212 | 0.013397048 | 0.026789834 | 1 |
| Heart_Failure | b_cell | *SPRYD3* | -2.444192362 | 0.002539364 | 0.009545808 | 1 |
| Heart_Failure | b_cell | *SOAT2* | -3.413210996 | 0.006419844 | 0.015978447 | 1 |

| | | | | | | |
|---|---|---|---|---|---|---|
| Heart_Failure | b_cell | *ZNF558* | -2.642800723 | 0.007455599 | 0.017643448 | 1 |
| Heart_Failure | b_cell | *NDUFV1* | -2.279311966 | 0.013048693 | 0.026285529 | 1 |
| Heart_Failure | b_cell | *CDK2AP2* | -3.590777422 | 0.002218833 | 0.009081423 | 1 |
| Heart_Failure | b_cell | *NUDT8* | -4.76461525 | 6.52E-05 | 0.002889125 | 0.4096585 |
| Heart_Failure | b_cell | *MIS12* | -2.446196298 | 0.010030833 | 0.021701255 | 1 |
| Heart_Failure | b_cell | *HID1* | -3.106462601 | 0.003240573 | 0.010739844 | 1 |
| Heart_Failure | b_cell | *SRP68* | 6.264283407 | 0.000394332 | 0.004938574 | 1 |
| Heart_Failure | b_cell | *MGAT5B* | -7.58634143 | 0.014167816 | 0.027948874 | 1 |
| Heart_Failure | b_cell | *TMC8* | -2.016655957 | 0.00423261 | 0.012342494 | 1 |
| Heart_Failure | b_cell | *GSDMA* | -5.92405401 | 0.000386475 | 0.004897883 | 1 |
| Heart_Failure | b_cell | *KRT24* | -6.83847315 | 0.00059569 | 0.006457072 | 1 |
| Heart_Failure | b_cell | *ZNF598* | -2.635848555 | 0.001857113 | 0.008687253 | 1 |
| Heart_Failure | b_cell | *MLST8* | -3.25872401 | 0.001351838 | 0.008654124 | 1 |
| Heart_Failure | b_cell | *ABCA3* | -2.350759925 | 0.006515851 | 0.016123569 | 1 |
| Heart_Failure | b_cell | *KCTD5* | -2.940251157 | 0.004505564 | 0.012782708 | 1 |
| Heart_Failure | b_cell | *SDHAF2* | -3.920078664 | 0.013966447 | 0.027655764 | 1 |
| Heart_Failure | b_cell | *VPS37C* | -2.669533354 | 0.010147489 | 0.021908401 | 1 |
| Heart_Failure | b_cell | *ULK4* | -1.04997542 | 0.024431924 | 0.041922355 | 1 |
| Heart_Failure | b_cell | *LTBP3* | -1.808288555 | 0.004492348 | 0.012772334 | 1 |
| Heart_Failure | b_cell | *SCARA5* | -1.331888741 | 0.001379761 | 0.008654124 | 1 |
| Heart_Failure | b_cell | *COPS6* | -2.024390112 | 0.006260158 | 0.01575565 | 1 |
| Heart_Failure | b_cell | *NUDT16L1* | 3.15307345 | 8.76E-05 | 0.002889125 | 0.550670808 |
| Heart_Failure | b_cell | *KIAA1586* | -1.84665611 | 0.003313151 | 0.01083435 | 1 |
| Heart_Failure | b_cell | *RNF187* | 5.794886526 | 9.75E-05 | 0.002978205 | 0.613026646 |
| Heart_Failure | b_cell | *PTGDR* | -1.608224176 | 0.008098294 | 0.018615713 | 1 |
| Heart_Failure | b_cell | *GLYCTK* | -2.171764862 | 0.004220978 | 0.012320004 | 1 |
| Heart_Failure | b_cell | *UBTD2* | -1.817058249 | 0.008363674 | 0.0190447 | 1 |
| Heart_Failure | b_cell | *NT5DC2* | -3.802912991 | 0.00149472 | 0.008654124 | 1 |
| Heart_Failure | b_cell | *COA6* | 7.435162329 | 8.31E-05 | 0.002889125 | 0.522397067 |
| Heart_Failure | b_cell | *KIF5C* | -2.912147296 | 0.002722351 | 0.009828026 | 1 |
| Heart_Failure | b_cell | *PXK* | -2.03089735 | 0.022449392 | 0.039336491 | 1 |
| Heart_Failure | b_cell | *ACOX2* | -1.646567754 | 0.003853369 | 0.011769074 | 1 |
| Heart_Failure | b_cell | *FAM107A* | -3.10159693 | 0.000346709 | 0.004598651 | 1 |
| Heart_Failure | b_cell | *CX3CR1* | -1.999689547 | 0.006331818 | 0.015885132 | 1 |
| Heart_Failure | b_cell | *SCN11A* | -1.095882959 | 0.013113164 | 0.026406939 | 1 |
| Heart_Failure | b_cell | *SEPT2* | -1.436927077 | 0.023455217 | 0.040657003 | 1 |
| Heart_Failure | b_cell | *DTYMK* | 6.882128411 | 0.000148933 | 0.003239931 | 0.936341345 |
| Heart_Failure | b_cell | *TAP1* | -3.15005754 | 0.006602003 | 0.016264418 | 1 |
| Heart_Failure | b_cell | *ING5* | -1.998581199 | 0.009965913 | 0.021583085 | 1 |
| Heart_Failure | b_cell | *MLKL* | -1.509955569 | 0.010236516 | 0.022055166 | 1 |
| Heart_Failure | b_cell | *RHOH* | 1.374375003 | 0.000101068 | 0.002997224 | 0.635411569 |
| Heart_Failure | b_cell | *KLHL30* | -3.543126244 | 0.004479421 | 0.012757412 | 1 |
| Heart_Failure | b_cell | *REEP4* | 2.814471477 | 7.28E-05 | 0.002889125 | 0.457434166 |
| Heart_Failure | b_cell | *BMP1* | -1.970519763 | 0.006177778 | 0.015592008 | 1 |
| Heart_Failure | b_cell | *ATXN2L* | -2.006808995 | 0.007532785 | 0.017759692 | 1 |
| Heart_Failure | b_cell | *SERINC2* | -3.100338987 | 0.011819951 | 0.024364601 | 1 |
| Heart_Failure | b_cell | *TMEM223* | -2.848943749 | 0.002992417 | 0.010219081 | 1 |
| Heart_Failure | b_cell | *SLC20A2* | -1.764574273 | 0.024343797 | 0.041851094 | 1 |
| Heart_Failure | b_cell | *ZSWIM1* | 6.344038584 | 5.75E-05 | 0.002889125 | 0.361323545 |
| Heart_Failure | b_cell | *NBPF9* | -2.924522526 | 0.005432197 | 0.014347469 | 1 |

| | | | | | | |
|---|---|---|---|---|---|---|
| Heart_Failure | b_cell | *ZNF30* | -1.942786651 | 0.02416631 | 0.041648462 | 1 |
| Heart_Failure | b_cell | *FAM84B* | -3.564855092 | 0.000914273 | 0.008515602 | 1 |
| Heart_Failure | b_cell | *SLC16A4* | -1.690692734 | 0.025317976 | 0.04314831 | 1 |
| Heart_Failure | b_cell | *TMEM208* | -1.988041048 | 0.001088932 | 0.008654124 | 1 |
| Heart_Failure | b_cell | *LRP1B* | -0.996392939 | 0.001866083 | 0.008690419 | 1 |
| Heart_Failure | b_cell | *NPNT* | -2.586914725 | 0.00674218 | 0.016490263 | 1 |
| Heart_Failure | b_cell | *TCTN2* | 3.066074512 | 0.000289956 | 0.004087334 | 1 |
| Heart_Failure | b_cell | *ABHD15* | -2.35795795 | 0.000858582 | 0.008266314 | 1 |
| Heart_Failure | b_cell | *SNTB2* | -1.763853923 | 0.0181463 | 0.033417044 | 1 |
| Heart_Failure | b_cell | *ZNF507* | -2.484858667 | 0.006141042 | 0.015536714 | 1 |
| Heart_Failure | b_cell | *ZBTB49* | -2.959992288 | 0.021839984 | 0.038504761 | 1 |
| Heart_Failure | b_cell | *TNIP2* | -2.700575861 | 0.015958596 | 0.030375928 | 1 |
| Heart_Failure | b_cell | *ZNF608* | -1.372239688 | 0.002056407 | 0.008831917 | 1 |
| Heart_Failure | b_cell | *LETM1* | -1.802612887 | 0.001701759 | 0.008654124 | 1 |
| Heart_Failure | b_cell | *CEP120* | -2.286148334 | 0.018446742 | 0.033841455 | 1 |
| Heart_Failure | b_cell | *STXBP6* | -1.383400901 | 0.001815574 | 0.008654124 | 1 |
| Heart_Failure | b_cell | *CPLX1* | -2.681905228 | 0.003916971 | 0.011873672 | 1 |
| Heart_Failure | b_cell | *MFSD7* | 4.325506182 | 0.000114168 | 0.003135985 | 0.717774626 |
| Heart_Failure | b_cell | *COL4A3* | 4.051883155 | 7.09E-05 | 0.002889125 | 0.446010115 |
| Heart_Failure | b_cell | *IRS1* | -1.191772378 | 0.002387074 | 0.00932704 | 1 |
| Heart_Failure | b_cell | *UPF3A* | -1.692096539 | 0.007343552 | 0.017455165 | 1 |
| Heart_Failure | b_cell | *HSPBAP1* | -1.736545087 | 0.007739339 | 0.018054629 | 1 |
| Heart_Failure | b_cell | *PARM1* | -1.749160172 | 0.000796917 | 0.007840712 | 1 |
| Heart_Failure | b_cell | *FAM110B* | -1.460722007 | 0.002315492 | 0.009201959 | 1 |
| Heart_Failure | b_cell | *ATF5* | -2.446058902 | 0.001421488 | 0.008654124 | 1 |
| Heart_Failure | b_cell | *CPT1C* | -4.387146235 | 0.000864745 | 0.008300042 | 1 |
| Heart_Failure | b_cell | *PCSK9* | 5.942774676 | 0.000181346 | 0.00338933 | 1 |
| Heart_Failure | b_cell | *RGS14* | -3.707329443 | 1.38E-05 | 0.002704195 | 0.086534225 |
| Heart_Failure | b_cell | *RAB24* | -4.080080027 | 0.000755392 | 0.007586497 | 1 |
| Heart_Failure | b_cell | *SH3TC2* | -3.143791587 | 0.001155394 | 0.008654124 | 1 |
| Heart_Failure | b_cell | *GPRIN1* | -14.36047782 | 0.029508281 | 0.048756521 | 1 |
| Heart_Failure | b_cell | *PGM2* | 7.565690597 | 0.000627613 | 0.0066316 | 1 |
| Heart_Failure | b_cell | *STK32A* | -1.654556303 | 0.00139082 | 0.008654124 | 1 |
| Heart_Failure | b_cell | *P2RY12* | -1.82304076 | 0.00121319 | 0.008654124 | 1 |
| Heart_Failure | b_cell | *SNUPN* | -2.660593269 | 0.007847638 | 0.018236548 | 1 |
| Heart_Failure | b_cell | *PTAFR* | -1.884652923 | 0.002519407 | 0.009509206 | 1 |
| Heart_Failure | b_cell | *RNASE6* | 1.588576504 | 0.000130128 | 0.003188681 | 0.818117375 |
| Heart_Failure | b_cell | *NPR1* | -1.6697995 | 0.001657671 | 0.008654124 | 1 |
| Heart_Failure | b_cell | *CLIC4* | -1.781324989 | 0.020279214 | 0.036323481 | 1 |
| Heart_Failure | b_cell | *SLC38A11* | 1.165343281 | 0.000142661 | 0.003237933 | 0.896907423 |
| Heart_Failure | b_cell | *CCDC8* | -5.666860955 | 0.001239022 | 0.008654124 | 1 |
| Heart_Failure | b_cell | *DTWD2* | -1.521973325 | 0.003155998 | 0.010554127 | 1 |
| Heart_Failure | b_cell | *CLIC3* | 6.469719334 | 0.00071114 | 0.007293538 | 1 |
| Heart_Failure | b_cell | *FAM103A1* | 1.647563304 | 0.00021802 | 0.00365518 | 1 |
| Heart_Failure | b_cell | *RGPD8* | -1.66991033 | 0.002238117 | 0.009117613 | 1 |
| Heart_Failure | b_cell | *HEXDC* | -2.397332144 | 0.008733529 | 0.019616899 | 1 |
| Heart_Failure | b_cell | *LRRC45* | -2.669221968 | 0.002314486 | 0.009201959 | 1 |
| Heart_Failure | b_cell | *CHRNA5* | 5.04994659 | 0.000316656 | 0.00432146 | 1 |
| Heart_Failure | b_cell | *FASN* | -1.28710054 | 0.006761279 | 0.016509252 | 1 |
| Heart_Failure | b_cell | *MT1E* | -2.879046923 | 0.007163488 | 0.017176525 | 1 |

| | | | | | | |
|---|---|---|---|---|---|---|
| Heart_Failure | b_cell | *DCXR* | -3.33076889 | 0.015745576 | 0.03014386 | 1 |
| Heart_Failure | b_cell | *LIMS1* | -1.322901793 | 0.007702197 | 0.017999868 | 1 |
| Heart_Failure | b_cell | *UGP2* | -1.306062961 | 0.006453226 | 0.01602239 | 1 |
| Heart_Failure | b_cell | *CSGALNACT2* | -1.987064163 | 0.01403128 | 0.027754893 | 1 |
| Heart_Failure | b_cell | *AVEN* | -2.033917989 | 0.013432182 | 0.026825962 | 1 |
| Heart_Failure | b_cell | *CALML6* | -10.62669645 | 0.000146905 | 0.003239931 | 0.923591572 |
| Heart_Failure | b_cell | *TPST1* | -1.314018303 | 0.001021737 | 0.008654124 | 1 |
| Heart_Failure | b_cell | *OTUD3* | -1.939941403 | 0.006380675 | 0.015933676 | 1 |
| Heart_Failure | b_cell | *ZFPM2* | -1.115175045 | 0.00427089 | 0.012408081 | 1 |
| Heart_Failure | b_cell | *ZNF747* | -3.870769873 | 0.002637825 | 0.009653089 | 1 |
| Heart_Failure | b_cell | *PUSL1* | -4.962937154 | 0.00135303 | 0.008654124 | 1 |
| Heart_Failure | b_cell | *MYO7B* | -2.453124907 | 0.004527893 | 0.01282869 | 1 |
| Heart_Failure | b_cell | *CNTROB* | -2.637935357 | 0.003663876 | 0.011454394 | 1 |
| Heart_Failure | b_cell | *KCNAB3* | -3.769511157 | 0.030411959 | 0.049882595 | 1 |
| Heart_Failure | b_cell | *RNF150* | -0.978055268 | 0.00301126 | 0.010266699 | 1 |
| Heart_Failure | b_cell | *HOXD4* | -6.410795142 | 0.002204577 | 0.009058938 | 1 |
| Heart_Failure | b_cell | *CHRNB1* | 3.307902664 | 2.01E-06 | 0.000884152 | 0.012631226 |
| Heart_Failure | b_cell | *FAXDC2* | -1.769344541 | 0.012991864 | 0.02618665 | 1 |
| Heart_Failure | b_cell | *ELP5* | -3.430993839 | 0.023192899 | 0.040324601 | 1 |
| Heart_Failure | b_cell | *SEMA3E* | -1.595100481 | 0.001500508 | 0.008654124 | 1 |
| Heart_Failure | b_cell | *DCLK2* | -1.710497692 | 0.007517001 | 0.01774001 | 1 |
| Heart_Failure | b_cell | *GPRC5C* | -3.076240788 | 0.006761782 | 0.016509252 | 1 |
| Heart_Failure | b_cell | *ADORA2B* | -3.603856153 | 0.000247286 | 0.003766552 | 1 |
| Heart_Failure | b_cell | *MGMT* | -1.859944287 | 0.023491308 | 0.040685911 | 1 |
| Heart_Failure | b_cell | *METTL7B* | -7.272902897 | 0.002355494 | 0.009268076 | 1 |
| Heart_Failure | b_cell | *NFXL1* | 4.781244543 | 0.007457745 | 0.017643448 | 1 |
| Heart_Failure | b_cell | *CD14* | -1.777385559 | 0.006164751 | 0.015568266 | 1 |
| Heart_Failure | b_cell | *KRT72* | 6.509513438 | 0.00109602 | 0.008654124 | 1 |
| Heart_Failure | b_cell | *TMC7* | -2.417002347 | 0.001022352 | 0.008654124 | 1 |
| Heart_Failure | b_cell | *SMAGP* | -3.72206894 | 0.014765006 | 0.028828445 | 1 |
| Heart_Failure | b_cell | *IRX2* | -2.422355071 | 0.002221605 | 0.009081423 | 1 |
| Heart_Failure | b_cell | *EMB* | -1.509078154 | 0.000630931 | 0.00664433 | 1 |
| Heart_Failure | b_cell | *DLGAP1* | -1.509783932 | 0.002188081 | 0.009035107 | 1 |
| Heart_Failure | b_cell | *HSPA4* | -2.201193995 | 0.016281048 | 0.030858894 | 1 |
| Heart_Failure | b_cell | *COMMD5* | -3.203848783 | 0.029295009 | 0.048455071 | 1 |
| Heart_Failure | b_cell | *GTSF1* | 7.137424691 | 6.61E-05 | 0.002889125 | 0.415824819 |
| Heart_Failure | b_cell | *TMEM133* | -5.740057099 | 0.00177763 | 0.008654124 | 1 |
| Heart_Failure | b_cell | *TTLL6* | 6.424740373 | 9.71E-05 | 0.002978205 | 0.610697445 |
| Heart_Failure | b_cell | *KCNS3* | -1.833893061 | 0.00206482 | 0.008833636 | 1 |
| Heart_Failure | b_cell | *HTRA3* | -1.431324364 | 0.002270199 | 0.009180605 | 1 |
| Heart_Failure | b_cell | *FOXN2* | -1.715705587 | 0.019646416 | 0.03552402 | 1 |
| Heart_Failure | b_cell | *KBTBD2* | -2.280105158 | 0.014589098 | 0.02859154 | 1 |
| Heart_Failure | b_cell | *MTSS1* | -1.480676979 | 0.013677512 | 0.027194978 | 1 |
| Heart_Failure | b_cell | *TSEN34* | 5.852936953 | 6.46E-05 | 0.002889125 | 0.406023313 |
| Heart_Failure | b_cell | *NDUFA3* | -2.251316871 | 0.002827075 | 0.009985294 | 1 |
| Heart_Failure | b_cell | *NUDT6* | -1.721746406 | 0.001667608 | 0.008654124 | 1 |
| Heart_Failure | b_cell | *FPR1* | -1.741776353 | 0.001929952 | 0.008732736 | 1 |
| Heart_Failure | b_cell | *SOX7* | -3.432558282 | 0.003180009 | 0.010589085 | 1 |
| Heart_Failure | b_cell | *C11orf24* | -2.487328695 | 0.00217494 | 0.009019688 | 1 |
| Heart_Failure | b_cell | *ALK* | -1.509777123 | 0.004634761 | 0.013020253 | 1 |

| | | | | | | |
|---|---|---|---|---|---|---|
| Heart_Failure | b_cell | *TRMT61B* | -1.882222026 | 0.017197714 | 0.032102739 | 1 |
| Heart_Failure | b_cell | *GIMAP8* | -1.626323581 | 0.001520005 | 0.008654124 | 1 |
| Heart_Failure | b_cell | *KCNMB3* | -1.585983014 | 0.002724207 | 0.009828026 | 1 |
| Heart_Failure | b_cell | *MORN4* | -1.741290543 | 0.006595129 | 0.016253852 | 1 |
| Heart_Failure | b_cell | *GRIK1* | -1.687077084 | 0.002014952 | 0.00881106 | 1 |
| Heart_Failure | b_cell | *CLDN20* | -2.326104562 | 0.006751879 | 0.016504302 | 1 |
| Heart_Failure | b_cell | *CANT1* | -2.644121033 | 0.007286333 | 0.017377612 | 1 |
| Heart_Failure | b_cell | *PGAM1* | -2.02397106 | 0.023549258 | 0.040775045 | 1 |
| Heart_Failure | b_cell | *LURAP1* | -1.76274465 | 0.007119221 | 0.017122624 | 1 |
| Heart_Failure | b_cell | *KCND3* | -2.391408982 | 0.001780066 | 0.008654124 | 1 |
| Heart_Failure | b_cell | *MRPL36* | -4.106132354 | 0.001329949 | 0.008654124 | 1 |
| Heart_Failure | b_cell | *ZNF581* | -6.101783772 | 0.014325545 | 0.02817163 | 1 |
| Heart_Failure | b_cell | *KRT27* | 3.735023536 | 0.000369052 | 0.004764327 | 1 |
| Heart_Failure | b_cell | *DLK2* | -6.402027041 | 0.018949874 | 0.034592874 | 1 |
| Heart_Failure | b_cell | *HOPX* | 10.86391132 | 0.002196735 | 0.009044448 | 1 |
| Heart_Failure | b_cell | *COL24A1* | -1.235677736 | 0.002002079 | 0.008783721 | 1 |
| Heart_Failure | b_cell | *ETFDH* | -1.326012925 | 0.003939755 | 0.011883725 | 1 |
| Heart_Failure | b_cell | *MAP6* | -2.749554824 | 0.006066895 | 0.015417368 | 1 |
| Heart_Failure | b_cell | *BCL2L1* | -1.8061109 | 0.021544449 | 0.03804774 | 1 |
| Heart_Failure | b_cell | *ZNF584* | -1.883793009 | 0.008313404 | 0.018974446 | 1 |
| Heart_Failure | b_cell | *NMUR1* | -2.959739202 | 0.001174135 | 0.008654124 | 1 |
| Heart_Failure | b_cell | *P2RY6* | -2.337757451 | 0.002412996 | 0.009356237 | 1 |
| Heart_Failure | b_cell | *ZIK1* | 11.9133575 | 0.000187402 | 0.00340015 | 1 |
| Heart_Failure | b_cell | *RGS19* | 1.876817273 | 0.001127621 | 0.008654124 | 1 |
| Heart_Failure | b_cell | *TCEA2* | -2.194247214 | 0.001683202 | 0.008654124 | 1 |
| Heart_Failure | b_cell | *ANO5* | -2.086381392 | 0.001758781 | 0.008654124 | 1 |
| Heart_Failure | b_cell | *GPHN* | -1.399504376 | 0.006087527 | 0.015457303 | 1 |
| Heart_Failure | b_cell | *VAT1L* | -2.6671334 | 0.00472026 | 0.013183596 | 1 |
| Heart_Failure | b_cell | *CAMTA1* | -1.835032015 | 0.021499055 | 0.038012258 | 1 |
| Heart_Failure | b_cell | *LGALS4* | 2.357001098 | 3.34E-05 | 0.002889125 | 0.209684673 |
| Heart_Failure | b_cell | *SPATA5L1* | -2.269757645 | 0.002813452 | 0.009965169 | 1 |
| Heart_Failure | b_cell | *SYCE1* | 2.408973059 | 0.000161857 | 0.003294027 | 1 |
| Heart_Failure | b_cell | *BCL2* | -0.966347882 | 0.008419359 | 0.019121228 | 1 |
| Heart_Failure | b_cell | *RHNO1* | 1.602847511 | 0.000219773 | 0.003667878 | 1 |
| Heart_Failure | b_cell | *KNDC1* | 2.943952891 | 0.008683948 | 0.019540438 | 1 |
| Heart_Failure | b_cell | *METTL18* | -3.508243136 | 0.016614739 | 0.031312009 | 1 |
| Heart_Failure | b_cell | *PWWP2B* | -3.137221808 | 0.003327137 | 0.01084381 | 1 |
| Heart_Failure | b_cell | *FAM90A1* | -7.26438513 | 0.001685055 | 0.008654124 | 1 |
| Heart_Failure | b_cell | *C3AR1* | -1.890478644 | 0.00258513 | 0.009577319 | 1 |
| Heart_Failure | b_cell | *AQP4* | -2.802121897 | 0.00168075 | 0.008654124 | 1 |
| Heart_Failure | b_cell | *TLN2* | -1.009436246 | 0.001649419 | 0.008654124 | 1 |
| Heart_Failure | b_cell | *TVP23B* | -2.265383324 | 0.008157358 | 0.018730938 | 1 |
| Heart_Failure | b_cell | *ATPAF2* | -2.157558181 | 0.003345845 | 0.010865693 | 1 |
| Heart_Failure | b_cell | *CYP4F22* | -6.475933739 | 0.00072183 | 0.007316224 | 1 |
| Heart_Failure | b_cell | *ZNF554* | -2.461676314 | 0.001048477 | 0.008654124 | 1 |
| Heart_Failure | b_cell | *ANKRD20A4* | 2.82983408 | 0.000122706 | 0.003135985 | 0.771452252 |
| Heart_Failure | b_cell | *LAMB2* | -1.617026162 | 0.00293861 | 0.010146595 | 1 |
| Heart_Failure | b_cell | *ORMDL3* | -3.094156872 | 1.49E-08 | 1.88E-05 | 9.38E-05 |
| Heart_Failure | b_cell | *SERF1A* | -12.20012149 | 0.001608682 | 0.008654124 | 1 |
| Heart_Failure | b_cell | *LRRC15* | 2.293826605 | 0.000154995 | 0.003239931 | 0.974451554 |

| | | | | | | |
|---|---|---|---|---|---|---|
| Heart_Failure | b_cell | *MOB3A* | -1.948449549 | 0.007450574 | 0.017643448 | 1 |
| Heart_Failure | b_cell | *NME6* | -2.042912025 | 0.013385887 | 0.026776033 | 1 |
| Heart_Failure | b_cell | *FRMD3* | -1.469756565 | 0.001952125 | 0.008753931 | 1 |
| Heart_Failure | b_cell | *MTBP* | -2.40073674 | 0.0191845 | 0.034929902 | 1 |
| Heart_Failure | b_cell | *ISG20* | 1.59563308 | 0.000430468 | 0.005135387 | 1 |
| Heart_Failure | b_cell | *MBOAT1* | -1.852819917 | 0.008991184 | 0.019960302 | 1 |
| Heart_Failure | b_cell | *CXCR6* | -3.068510806 | 0.022968494 | 0.040035509 | 1 |
| Heart_Failure | b_cell | *C5orf34* | -2.187682308 | 0.002675018 | 0.009732547 | 1 |
| Heart_Failure | b_cell | *C1QTNF4* | -1.251557526 | 0.026171462 | 0.044266877 | 1 |
| Heart_Failure | b_cell | *HINFP* | -2.524378687 | 0.00609893 | 0.015471746 | 1 |
| Heart_Failure | b_cell | *CLEC12A* | -1.20358609 | 0.001110967 | 0.008654124 | 1 |
| Heart_Failure | b_cell | *CSDC2* | -5.511788551 | 0.002629853 | 0.009653089 | 1 |
| Heart_Failure | b_cell | *RCAN2* | -1.316133913 | 0.001701367 | 0.008654124 | 1 |
| Heart_Failure | b_cell | *IL16* | -1.48226059 | 0.004368157 | 0.012614884 | 1 |
| Heart_Failure | b_cell | *PRSS27* | -10.54482022 | 0.008955007 | 0.019901071 | 1 |
| Heart_Failure | b_cell | *FGGY* | -1.761387373 | 0.021245535 | 0.037636145 | 1 |
| Heart_Failure | b_cell | *ZNF24* | -2.112425615 | 0.011932986 | 0.024533251 | 1 |
| Heart_Failure | b_cell | *MANEA* | 6.489334052 | 0.000620397 | 0.006609617 | 1 |
| Heart_Failure | b_cell | *CARNS1* | -6.453504892 | 0.003218719 | 0.010689955 | 1 |
| Heart_Failure | b_cell | *SNTG2* | -1.133149265 | 0.001979533 | 0.008775328 | 1 |
| Heart_Failure | b_cell | *PDE3A* | -1.282477837 | 0.004578625 | 0.012902652 | 1 |
| Heart_Failure | b_cell | *EFEMP2* | -1.907767805 | 0.003313484 | 0.01083435 | 1 |
| Heart_Failure | b_cell | *AGAP5* | -70.96270316 | 0.001747034 | 0.008654124 | 1 |
| Heart_Failure | b_cell | *ZFAND4* | -2.093982597 | 0.01347746 | 0.02690784 | 1 |
| Heart_Failure | b_cell | *SLFN11* | -2.512100011 | 0.00356988 | 0.011289657 | 1 |
| Heart_Failure | b_cell | *CCL19* | -11.91959103 | 0.001285336 | 0.008654124 | 1 |
| Heart_Failure | b_cell | *CBWD1* | -2.579303558 | 0.02439653 | 0.041897633 | 1 |
| Heart_Failure | b_cell | *HOXC5* | 4.581788421 | 0.002076603 | 0.008851254 | 1 |
| Heart_Failure | b_cell | *SNX32* | 3.606057976 | 0.004957544 | 0.013563132 | 1 |
| Heart_Failure | b_cell | *CYP7B1* | -1.253305983 | 0.003631466 | 0.011415514 | 1 |
| Heart_Failure | b_cell | *METAP1D* | -1.572139262 | 0.002176811 | 0.009021496 | 1 |
| Heart_Failure | b_cell | *NADSYN1* | -2.258706894 | 0.010881662 | 0.02302693 | 1 |
| Heart_Failure | b_cell | *RNASEH2C* | 6.267203467 | 0.000167535 | 0.003354429 | 1 |
| Heart_Failure | b_cell | *MYEOV* | -1.150893497 | 0.020515716 | 0.036590724 | 1 |
| Heart_Failure | b_cell | *LCLAT1* | 4.094256886 | 0.012700786 | 0.025716535 | 1 |
| Heart_Failure | b_cell | *ADH6* | -1.756911245 | 0.013131858 | 0.026423314 | 1 |
| Heart_Failure | b_cell | *GXYLT2* | -1.475533439 | 0.001720244 | 0.008654124 | 1 |
| Heart_Failure | b_cell | *HPSE2* | -1.301750564 | 0.027218933 | 0.045694374 | 1 |
| Heart_Failure | b_cell | *DCAKD* | -3.464877674 | 4.17E-05 | 0.002889125 | 0.26237367 |
| Heart_Failure | b_cell | *BNC2* | -1.212718391 | 0.006283032 | 0.015800568 | 1 |
| Heart_Failure | b_cell | *TRMT112* | -2.826947533 | 0.001480328 | 0.008654124 | 1 |
| Heart_Failure | b_cell | *PARP15* | 5.036587088 | 7.18E-06 | 0.002102517 | 0.045132804 |
| Heart_Failure | b_cell | *AHSA2* | -2.03521267 | 0.014881751 | 0.028984376 | 1 |
| Heart_Failure | b_cell | *ABLIM3* | -1.301792306 | 0.001846784 | 0.00866783 | 1 |
| Heart_Failure | b_cell | *VANGL1* | -2.161232401 | 0.001643259 | 0.008654124 | 1 |
| Heart_Failure | b_cell | *GLRX* | -1.210324313 | 0.001486432 | 0.008654124 | 1 |
| Heart_Failure | b_cell | *DMRT2* | -2.168629732 | 0.00058074 | 0.006349757 | 1 |
| Heart_Failure | b_cell | *ZNF483* | -1.566184875 | 0.012244965 | 0.025059927 | 1 |
| Heart_Failure | b_cell | *SLC2A14* | -3.744970719 | 0.000839364 | 0.008143642 | 1 |
| Heart_Failure | b_cell | *MZT2A* | 3.236909041 | 5.28E-05 | 0.002889125 | 0.332207411 |

| | | | | | | |
|---|---|---|---|---|---|---|
| Heart_Failure | b_cell | *MAP3K11* | -2.914153166 | 0.01924458 | 0.035004259 | 1 |
| Heart_Failure | b_cell | *C1QA* | -2.225490155 | 0.020325834 | 0.036365544 | 1 |
| Heart_Failure | b_cell | *NDNF* | -2.567329617 | 0.001915778 | 0.008705907 | 1 |
| Heart_Failure | b_cell | *OLR1* | 5.290264446 | 8.78E-05 | 0.002889125 | 0.552191735 |
| Heart_Failure | b_cell | *GLIPR1L1* | -1.574111624 | 0.003258552 | 0.010782378 | 1 |
| Heart_Failure | b_cell | *ARV1* | 5.588130844 | 8.79E-05 | 0.002889125 | 0.552558834 |
| Heart_Failure | b_cell | *CCDC36* | -1.29916347 | 0.004052155 | 0.012094678 | 1 |
| Heart_Failure | b_cell | *THAP2* | 4.546521284 | 3.49E-05 | 0.002889125 | 0.219653395 |
| Heart_Failure | b_cell | *ZNF417* | -3.155255994 | 0.001674392 | 0.008654124 | 1 |
| Heart_Failure | b_cell | *VEGFB* | -2.65259139 | 0.006017147 | 0.015361627 | 1 |
| Heart_Failure | b_cell | *TNFRSF10D* | -1.603491393 | 0.009099298 | 0.020122782 | 1 |
| Heart_Failure | b_cell | *MST1* | -2.535953049 | 0.004264859 | 0.012408081 | 1 |
| Heart_Failure | b_cell | *CSPG4* | -1.937378125 | 0.003649712 | 0.011427162 | 1 |
| Heart_Failure | b_cell | *SNX33* | -2.506702225 | 0.005432562 | 0.014347469 | 1 |
| Heart_Failure | b_cell | *NUDT18* | -4.517147501 | 0.00445058 | 0.012718545 | 1 |
| Heart_Failure | b_cell | *SULT1B1* | -2.749912807 | 0.015932591 | 0.030363201 | 1 |
| Heart_Failure | b_cell | *NUDT4* | -1.790792202 | 0.008561679 | 0.019354198 | 1 |
| Heart_Failure | b_cell | *C1orf100* | -4.759375486 | 0.00329693 | 0.01083435 | 1 |
| Heart_Failure | b_cell | *AGFG1* | -1.359010568 | 0.020304937 | 0.036338497 | 1 |
| Heart_Failure | b_cell | *PLK3* | -2.245931441 | 0.001765324 | 0.008654124 | 1 |
| Heart_Failure | b_cell | *TUBB8* | -6.868239272 | 0.00244854 | 0.009416629 | 1 |
| Heart_Failure | b_cell | *HOXB2* | -2.447779197 | 0.028193327 | 0.047014855 | 1 |
| Heart_Failure | b_cell | *XXYLT1* | -1.960564727 | 0.006152632 | 0.015553518 | 1 |
| Heart_Failure | b_cell | *NRROS* | -3.067354859 | 1.76E-05 | 0.002889125 | 0.110816963 |
| Heart_Failure | b_cell | *CEP19* | -3.278692538 | 0.000239836 | 0.003713907 | 1 |
| Heart_Failure | b_cell | *CTSF* | -1.632374017 | 0.006241707 | 0.015734407 | 1 |
| Heart_Failure | b_cell | *SOCS7* | -1.930195247 | 0.004140011 | 0.012205437 | 1 |
| Heart_Failure | b_cell | *TLR6* | -2.328308358 | 0.020688527 | 0.036836241 | 1 |
| Heart_Failure | b_cell | *FAM53A* | -2.606245884 | 0.002276729 | 0.009182983 | 1 |
| Heart_Failure | b_cell | *CTU2* | -3.535929255 | 0.001043418 | 0.008654124 | 1 |
| Heart_Failure | b_cell | *SNX31* | -3.558103292 | 0.005765198 | 0.014965236 | 1 |
| Heart_Failure | b_cell | *GLIS1* | -2.359530633 | 0.000224228 | 0.003674323 | 1 |
| Heart_Failure | b_cell | *LIG4* | -2.285675639 | 0.005096318 | 0.013812459 | 1 |
| Heart_Failure | b_cell | *SNAPC5* | 5.410817496 | 0.024542928 | 0.04208985 | 1 |
| Heart_Failure | b_cell | *LINGO2* | -2.722711048 | 0.004475873 | 0.012756035 | 1 |
| Heart_Failure | b_cell | *BBS1* | 5.338233042 | 1.92E-05 | 0.002889125 | 0.120630642 |
| Heart_Failure | b_cell | *ANKRD36C* | -1.766207634 | 0.027254586 | 0.045729806 | 1 |
| Heart_Failure | b_cell | *IL20RB* | 4.205724913 | 1.36E-05 | 0.002704195 | 0.085722394 |
| Heart_Failure | b_cell | *ZNF497* | 2.19096658 | 0.000154498 | 0.003239931 | 0.971327435 |
| Heart_Failure | b_cell | *KLF14* | 3.53856009 | 0.00023265 | 0.003705045 | 1 |
| Heart_Failure | b_cell | *ANGEL2* | -1.935803991 | 0.011334391 | 0.02369781 | 1 |
| Heart_Failure | b_cell | *BRSK2* | -4.821759312 | 0.005347156 | 0.01421623 | 1 |
| Heart_Failure | b_cell | *SH3PXD2B* | -1.204330383 | 0.00214686 | 0.008999799 | 1 |
| Heart_Failure | b_cell | *LARP7* | -1.894063936 | 0.008216931 | 0.018819615 | 1 |
| Heart_Failure | b_cell | *FGFBP3* | -3.395224172 | 0.00350188 | 0.011147503 | 1 |
| Heart_Failure | b_cell | *C4orf32* | -1.334534765 | 0.001395472 | 0.008654124 | 1 |
| Heart_Failure | b_cell | *HRAS* | -3.158177233 | 0.009163816 | 0.02019303 | 1 |
| Heart_Failure | b_cell | *C4orf26* | 5.783702324 | 0.002901452 | 0.010117265 | 1 |
| Heart_Failure | b_cell | *BTC* | -2.473355998 | 0.001735336 | 0.008654124 | 1 |
| Heart_Failure | b_cell | *NDUFA11* | 6.682888255 | 0.000931021 | 0.008595199 | 1 |

| | | | | | | |
|---|---|---|---|---|---|---|
| Heart_Failure | b_cell | *PTDSS2* | -2.478102267 | 0.011123918 | 0.023397816 | 1 |
| Heart_Failure | b_cell | *FUT1* | -5.431579683 | 0.02872215 | 0.047657998 | 1 |
| Heart_Failure | b_cell | *FBXW8* | -1.98605862 | 0.017915438 | 0.033130632 | 1 |
| Heart_Failure | b_cell | *CHST2* | -4.947157852 | 0.002049691 | 0.008831917 | 1 |
| Heart_Failure | b_cell | *ATR* | -1.54059427 | 0.014510648 | 0.028464414 | 1 |
| Heart_Failure | b_cell | *SH3BP5L* | -3.09431595 | 0.003455985 | 0.01109586 | 1 |
| Heart_Failure | b_cell | *CADM2* | -1.199915224 | 0.003837061 | 0.011751405 | 1 |
| Heart_Failure | b_cell | *PSMD2* | -1.729062048 | 0.002304776 | 0.009188443 | 1 |
| Heart_Failure | b_cell | *CHST1* | -13.86777604 | 2.10E-05 | 0.002889125 | 0.132329893 |
| Heart_Failure | b_cell | *PHYHD1* | -1.892193673 | 0.001893005 | 0.008705907 | 1 |
| Heart_Failure | b_cell | *BANF1* | 3.300436005 | 5.00E-05 | 0.002889125 | 0.314240645 |
| Heart_Failure | b_cell | *CHRNA7* | -4.025642526 | 0.022775317 | 0.039763515 | 1 |
| Heart_Failure | b_cell | *TMEM9B* | 4.710509075 | 0.030163999 | 0.049566403 | 1 |
| Heart_Failure | b_cell | *NRIP3* | -3.779222676 | 0.000186507 | 0.00340015 | 1 |
| Heart_Failure | b_cell | *SCUBE2* | -1.967904503 | 0.000621633 | 0.006609617 | 1 |
| Heart_Failure | b_cell | *ZNF25* | -2.134118124 | 0.00826546 | 0.018897686 | 1 |
| Heart_Failure | b_cell | *CLTB* | -2.087022683 | 0.02776201 | 0.046383141 | 1 |
| Heart_Failure | b_cell | *PCSK1* | -2.773586945 | 0.001834184 | 0.008654124 | 1 |
| Heart_Failure | b_cell | *LPL* | -0.990114956 | 0.000965059 | 0.008654124 | 1 |
| Heart_Failure | b_cell | *TBC1D10C* | 2.419538724 | 0.000348165 | 0.004608244 | 1 |
| Heart_Failure | b_cell | *MCTP1* | -0.933879318 | 0.003676824 | 0.011460681 | 1 |
| Heart_Failure | b_cell | *LRRC25* | -2.761698942 | 0.005145629 | 0.013884366 | 1 |
| Heart_Failure | b_cell | *UBQLNL* | 1.859483363 | 0.000164155 | 0.003325199 | 1 |
| Heart_Failure | b_cell | *LIPT2* | -7.42683663 | 0.006510897 | 0.016123569 | 1 |
| Heart_Failure | b_cell | *KCNE3* | -2.060678764 | 0.001848825 | 0.00866783 | 1 |
| Heart_Failure | b_cell | *UCP3* | 1.855762704 | 0.000405757 | 0.005001954 | 1 |
| Heart_Failure | b_cell | *PAAF1* | -2.086262676 | 7.12E-05 | 0.002889125 | 0.44791526 |
| Heart_Failure | b_cell | *RAB6A* | -1.828962303 | 0.010086792 | 0.021814813 | 1 |
| Heart_Failure | b_cell | *RPS6KB2* | -2.579406895 | 0.011138354 | 0.023408158 | 1 |
| Heart_Failure | b_cell | *RMI2* | -4.070827905 | 0.002795457 | 0.009923793 | 1 |
| Heart_Failure | b_cell | *TOM1L2* | -1.733746123 | 0.01422771 | 0.028040632 | 1 |
| Heart_Failure | b_cell | *TEX26* | -5.827736417 | 0.004426485 | 0.012718545 | 1 |
| Heart_Failure | b_cell | *GPR156* | -3.628863005 | 0.010509536 | 0.022451054 | 1 |
| Heart_Failure | b_cell | *TTLL11* | -2.275086101 | 0.002035767 | 0.008831917 | 1 |
| Heart_Failure | b_cell | *ETV4* | 5.424324684 | 0.000213292 | 0.003595078 | 1 |
| Heart_Failure | b_cell | *SWI5* | -2.493887787 | 0.001452437 | 0.008654124 | 1 |
| Heart_Failure | b_cell | *A2M* | -1.43697313 | 0.005000907 | 0.013657993 | 1 |
| Heart_Failure | b_cell | *LRRN1* | -2.368449873 | 0.002235214 | 0.009114171 | 1 |
| Heart_Failure | b_cell | *B3GALT6* | -4.288660355 | 0.001360329 | 0.008654124 | 1 |
| Heart_Failure | b_cell | *ZNF613* | -2.58625471 | 0.001102421 | 0.008654124 | 1 |
| Heart_Failure | b_cell | *SSNA1* | -2.205651197 | 0.00177104 | 0.008654124 | 1 |
| Heart_Failure | b_cell | *UFSP1* | 2.631206895 | 0.001049418 | 0.008654124 | 1 |
| Heart_Failure | b_cell | *TMEM39A* | -2.173688562 | 0.019771038 | 0.035702086 | 1 |
| Heart_Failure | b_cell | *GPX2* | -3.499055267 | 0.008078787 | 0.018579678 | 1 |
| Heart_Failure | b_cell | *CCDC57* | -1.94754123 | 0.003675614 | 0.011460681 | 1 |
| Heart_Failure | b_cell | *CIDEA* | -0.925333845 | 0.008456357 | 0.019179334 | 1 |
| Heart_Failure | b_cell | *SMIM19* | -1.416755411 | 0.003808111 | 0.011678827 | 1 |
| Heart_Failure | b_cell | *ZNF404* | -2.537496819 | 0.00034273 | 0.004555486 | 1 |
| Heart_Failure | b_cell | *SLC35G1* | -2.2819079 | 0.028278763 | 0.04713048 | 1 |
| Heart_Failure | b_cell | *ZNF135* | -2.194549568 | 0.008794361 | 0.019698135 | 1 |

| | | | | | | |
|---|---|---|---|---|---|---|
| Heart_Failure | b_cell | *B3GNT4* | 2.331306182 | 0.000199949 | 0.003491879 | 1 |
| Heart_Failure | b_cell | *CDC26* | -3.248866644 | 0.001703214 | 0.008654124 | 1 |
| Heart_Failure | b_cell | *RNPEP* | -1.910704649 | 0.007533816 | 0.017759692 | 1 |
| Heart_Failure | b_cell | *SPRYD4* | -2.074631678 | 0.019500857 | 0.035352333 | 1 |
| Heart_Failure | b_cell | *SLCO3A1* | -1.635738685 | 0.010784921 | 0.022914767 | 1 |
| Heart_Failure | b_cell | *PLA2G16* | -0.943872161 | 0.004214876 | 0.012319352 | 1 |
| Heart_Failure | b_cell | *HORMAD2* | -2.739468298 | 0.008422619 | 0.019121228 | 1 |
| Heart_Failure | b_cell | *RNF152* | -2.05656298 | 0.004251136 | 0.012390772 | 1 |
| Heart_Failure | b_cell | *LRRC37A* | 5.727945866 | 0.004483195 | 0.012759551 | 1 |
| Heart_Failure | b_cell | *ACSF3* | -3.482370907 | 0.004851711 | 0.013390125 | 1 |
| Heart_Failure | b_cell | *BOK* | -1.697157906 | 0.001610494 | 0.008654124 | 1 |
| Heart_Failure | b_cell | *ZNF843* | -4.054180067 | 0.001283144 | 0.008654124 | 1 |
| Heart_Failure | b_cell | *C8orf59* | -1.904864464 | 0.006796779 | 0.016562537 | 1 |
| Heart_Failure | b_cell | *PFN4* | -2.035297935 | 0.007249103 | 0.0173261 | 1 |
| Heart_Failure | b_cell | *BASP1* | -1.340702339 | 0.003894471 | 0.011845448 | 1 |
| Heart_Failure | b_cell | *VSIG10* | -1.653468047 | 0.000565159 | 0.006255554 | 1 |
| Heart_Failure | b_cell | *IRX5* | -2.417049551 | 0.006545259 | 0.016162624 | 1 |
| Heart_Failure | b_cell | *ANKLE2* | -2.111556431 | 0.016610226 | 0.031312009 | 1 |
| Heart_Failure | b_cell | *GCNT4* | 2.051243411 | 0.000173597 | 0.003365162 | 1 |
| Heart_Failure | b_cell | *MUC20* | 3.33528036 | 7.17E-05 | 0.002889125 | 0.450944809 |
| Heart_Failure | b_cell | *SHMT1* | -1.370214228 | 0.003965194 | 0.011939259 | 1 |
| Heart_Failure | b_cell | *MTHFR* | -2.442897319 | 0.02037242 | 0.036397102 | 1 |
| Heart_Failure | b_cell | *C19orf18* | -1.745598663 | 0.004405258 | 0.01267545 | 1 |
| Heart_Failure | b_cell | *TMEM80* | -1.797883691 | 0.003928669 | 0.011882923 | 1 |
| Heart_Failure | b_cell | *POLE* | -2.539892759 | 0.030193966 | 0.049580429 | 1 |
| Heart_Failure | b_cell | *DSCAML1* | -1.764602572 | 0.006364481 | 0.015909937 | 1 |
| Heart_Failure | b_cell | *EPS8L2* | -2.915002378 | 0.001434364 | 0.008654124 | 1 |
| Heart_Failure | b_cell | *TALDO1* | -1.732260892 | 0.014342095 | 0.028195358 | 1 |
| Heart_Failure | b_cell | *B3GNT8* | 1.254323067 | 0.000180462 | 0.00338933 | 1 |
| Heart_Failure | b_cell | *AP006621.1* | -1.944943956 | 0.003321617 | 0.01083435 | 1 |
| Heart_Failure | b_cell | *C8orf31* | -2.88208783 | 0.001667759 | 0.008654124 | 1 |
| Heart_Failure | b_cell | *AC024940.1* | 0.769559017 | 0.000139723 | 0.003214256 | 0.87843587 |
| Heart_Failure | b_cell | *TIMM22* | -2.643101558 | 0.002947221 | 0.010151069 | 1 |
| Heart_Failure | b_cell | *PPFIA3* | -3.352833251 | 0.002208391 | 0.009068682 | 1 |
| Heart_Failure | b_cell | *MAGEF1* | -2.645833646 | 0.001749376 | 0.008654124 | 1 |
| Heart_Failure | b_cell | *NAP1L5* | -3.43867647 | 0.004013918 | 0.012028362 | 1 |
| Heart_Failure | b_cell | *NIM1K* | -2.743105693 | 0.016657367 | 0.031361554 | 1 |
| Heart_Failure | b_cell | *ACOT4* | -2.868892087 | 0.007593406 | 0.017833299 | 1 |
| Heart_Failure | b_cell | *RBM44* | -3.085200045 | 0.001691355 | 0.008654124 | 1 |
| Heart_Failure | b_cell | *IRX3* | -3.235431544 | 0.001120708 | 0.008654124 | 1 |
| Heart_Failure | b_cell | *ATOX1* | 6.479054684 | 0.000160449 | 0.003294027 | 1 |
| Heart_Failure | b_cell | *C18orf32* | 4.136782616 | 0.000176176 | 0.00336662 | 1 |
| Heart_Failure | b_cell | *C12orf54* | -8.639196348 | 0.00856424 | 0.019354198 | 1 |
| Heart_Failure | b_cell | *GBA* | -2.936217229 | 0.000678308 | 0.007037165 | 1 |
| Heart_Failure | b_cell | *ACAD9* | -1.516343394 | 0.001416033 | 0.008654124 | 1 |
| Heart_Failure | b_cell | *IL17RA* | -1.931473291 | 0.014449123 | 0.02837028 | 1 |
| Heart_Failure | b_cell | *CD163L1* | -1.458463953 | 0.000629384 | 0.006639156 | 1 |
| Heart_Failure | b_cell | *POLR2L* | -2.471277504 | 0.00517803 | 0.013936542 | 1 |
| Heart_Failure | b_cell | *ZCCHC3* | 8.26935136 | 0.002577756 | 0.009572565 | 1 |
| Heart_Failure | b_cell | *ZBTB41* | -2.160978626 | 0.007857502 | 0.01824361 | 1 |

| | | | | | | |
|---|---|---|---|---|---|---|
| Heart_Failure | b_cell | *RIC8A* | -3.655894003 | 0.002905926 | 0.010125753 | 1 |
| Heart_Failure | b_cell | *IMP3* | -2.143009395 | 0.001894446 | 0.008705907 | 1 |
| Heart_Failure | b_cell | *ODF3B* | -3.788725126 | 0.001457985 | 0.008654124 | 1 |
| Heart_Failure | b_cell | *C2orf73* | -3.09037546 | 0.001569483 | 0.008654124 | 1 |
| Heart_Failure | b_cell | *FAM26E* | -4.565406555 | 0.014062973 | 0.027794376 | 1 |
| Heart_Failure | b_cell | *ALS2CL* | -1.995140803 | 0.001746634 | 0.008654124 | 1 |
| Heart_Failure | b_cell | *MLF1* | -1.585617099 | 0.002306018 | 0.009188443 | 1 |
| Heart_Failure | b_cell | *C2orf69* | -2.432784872 | 0.012390274 | 0.025289758 | 1 |
| Heart_Failure | b_cell | *GRAMD1C* | -2.362503529 | 0.001847962 | 0.00866783 | 1 |
| Heart_Failure | b_cell | *TSSK6* | -6.816288282 | 0.007388548 | 0.017542222 | 1 |
| Heart_Failure | b_cell | *GOLGA8Q* | 45.10449687 | 0.000966718 | 0.008654124 | 1 |
| Heart_Failure | b_cell | *NDUFV2* | 6.753163019 | 0.022813692 | 0.039819457 | 1 |
| Heart_Failure | b_cell | *ZNF114* | -4.534355681 | 0.003349955 | 0.010873087 | 1 |
| Heart_Failure | b_cell | *ZNF518B* | -2.928830071 | 0.006972598 | 0.016886256 | 1 |
| Heart_Failure | b_cell | *SH2B1* | -1.563609801 | 0.001001274 | 0.008654124 | 1 |
| Heart_Failure | b_cell | *KDELC2* | -2.04872044 | 0.001407915 | 0.008654124 | 1 |
| Heart_Failure | b_cell | *SH2D4B* | 6.002492885 | 8.04E-05 | 0.002889125 | 0.505413186 |
| Heart_Failure | b_cell | *RNF212* | -1.531264932 | 0.008798469 | 0.019698135 | 1 |
| Heart_Failure | b_cell | *PRSS36* | -3.442728659 | 0.00178278 | 0.008654124 | 1 |
| Heart_Failure | b_cell | *TMPRSS9* | -2.223056618 | 0.00078546 | 0.007767351 | 1 |
| Heart_Failure | b_cell | *AQP11* | -2.442019753 | 0.004008062 | 0.012024754 | 1 |
| Heart_Failure | b_cell | *FAM220A* | -3.02122787 | 0.017677781 | 0.032804076 | 1 |
| Heart_Failure | b_cell | *BEND3* | -3.935163673 | 0.001860957 | 0.008690419 | 1 |
| Heart_Failure | b_cell | *AC006111.1* | -10.43788794 | 0.00531507 | 0.014178109 | 1 |
| Heart_Failure | b_cell | *COX14* | -1.476187696 | 0.002580848 | 0.009572739 | 1 |
| Heart_Failure | b_cell | *CA8* | -2.05309861 | 0.004513284 | 0.012798834 | 1 |
| Heart_Failure | b_cell | *ERBB4* | -0.970481422 | 0.001683369 | 0.008654124 | 1 |
| Heart_Failure | b_cell | *CSRNP3* | -2.42112894 | 0.01979636 | 0.035723225 | 1 |
| Heart_Failure | b_cell | *ZNF713* | -3.966600083 | 0.00293746 | 0.010146595 | 1 |
| Heart_Failure | b_cell | *KCTD12* | -1.586640137 | 0.004374557 | 0.012620483 | 1 |
| Heart_Failure | b_cell | *RPP25* | -3.884875331 | 0.027517776 | 0.046048511 | 1 |
| Heart_Failure | b_cell | *FAM219B* | -1.500130801 | 0.00199419 | 0.008775328 | 1 |
| Heart_Failure | b_cell | *CPNE7* | 2.935617481 | 0.002934789 | 0.010146595 | 1 |
| Heart_Failure | b_cell | *GDPD4* | 3.119956201 | 0.000101597 | 0.002998781 | 0.638740301 |
| Heart_Failure | b_cell | *TRIM73* | -13.54810293 | 7.42E-05 | 0.002889125 | 0.466211652 |
| Heart_Failure | b_cell | *HYI* | -1.853723024 | 0.001413519 | 0.008654124 | 1 |
| Heart_Failure | b_cell | *C17orf62* | 6.34198955 | 0.00094708 | 0.008639437 | 1 |
| Heart_Failure | b_cell | *CTC1* | 5.269217178 | 7.38E-05 | 0.002889125 | 0.463850524 |
| Heart_Failure | b_cell | *SNX18* | -2.249292344 | 0.016801875 | 0.031504142 | 1 |
| Heart_Failure | b_cell | *C14orf39* | -1.633298629 | 0.00327431 | 0.010803355 | 1 |
| Heart_Failure | b_cell | *KLHDC7A* | -3.194315859 | 0.001129817 | 0.008654124 | 1 |
| Heart_Failure | b_cell | *TRIML2* | 7.375340168 | 0.000209792 | 0.003574421 | 1 |
| Heart_Failure | b_cell | *TCAIM* | -1.786542054 | 0.007792598 | 0.018125069 | 1 |
| Heart_Failure | b_cell | *FUCA1* | -2.24647979 | 0.00291429 | 0.010128327 | 1 |
| Heart_Failure | b_cell | *ZNF664* | -2.75377591 | 0.016670803 | 0.031361554 | 1 |
| Heart_Failure | b_cell | *HLA-DQB1* | -1.375803576 | 0.012827175 | 0.025939032 | 1 |
| Heart_Failure | b_cell | *EGR3* | -4.33532805 | 0.018278154 | 0.033571357 | 1 |
| Heart_Failure | b_cell | *LINC00174* | -2.240833608 | 0.005959021 | 0.015295782 | 1 |
| Heart_Failure | b_cell | *GEMIN4* | 3.634353255 | 0.002148672 | 0.008999799 | 1 |
| Heart_Failure | b_cell | *CIITA* | 2.994805514 | 8.14E-05 | 0.002889125 | 0.511794192 |

| | | | | | | |
|---|---|---|---|---|---|---|
| Heart_Failure | b_cell | *PLD6* | -3.406385772 | 0.015068622 | 0.029248665 | 1 |
| Heart_Failure | b_cell | *ZBTB42* | -8.607625797 | 0.002312221 | 0.009200589 | 1 |
| Heart_Failure | b_cell | *LACC1* | -3.722715382 | 0.026355676 | 0.04454251 | 1 |
| Heart_Failure | b_cell | *MAF1* | -2.836731387 | 0.003619736 | 0.011384333 | 1 |
| Heart_Failure | b_cell | *FCER1A* | 6.276822617 | 0.000255559 | 0.003777026 | 1 |
| Heart_Failure | b_cell | *PCED1B* | -1.483522554 | 0.014966328 | 0.029113027 | 1 |
| Heart_Failure | b_cell | *CDH5* | -1.628011232 | 0.001293778 | 0.008654124 | 1 |
| Heart_Failure | b_cell | *GIPC3* | 2.230492915 | 0.002845751 | 0.010011882 | 1 |
| Heart_Failure | b_cell | *CITED4* | 5.84411743 | 0.000118849 | 0.003135985 | 0.747200774 |
| Heart_Failure | b_cell | *TIGD5* | -2.937699819 | 0.011445961 | 0.023828065 | 1 |
| Heart_Failure | b_cell | *ZNF154* | -1.978199587 | 0.006322204 | 0.015867344 | 1 |
| Heart_Failure | b_cell | *SEPT1* | -2.353752496 | 0.00152528 | 0.008654124 | 1 |
| Heart_Failure | b_cell | *EXOC3* | -2.315902295 | 0.03032912 | 0.049785687 | 1 |
| Heart_Failure | b_cell | *LYNX1* | -2.955366417 | 0.009200811 | 0.02024253 | 1 |
| Heart_Failure | b_cell | *FAHD1* | -1.70253185 | 0.011289894 | 0.023636218 | 1 |
| Heart_Failure | b_cell | *TDRP* | -2.052110262 | 0.000995235 | 0.008654124 | 1 |
| Heart_Failure | b_cell | *ZNF816* | 3.372393605 | 0.000366595 | 0.004752125 | 1 |
| Heart_Failure | b_cell | *FGD6* | -1.925184371 | 0.008654326 | 0.019487733 | 1 |
| Heart_Failure | b_cell | *HCLS1* | -1.980626506 | 0.004928073 | 0.013494248 | 1 |
| Heart_Failure | b_cell | *MCFD2* | -1.813819236 | 0.002550053 | 0.009548648 | 1 |
| Heart_Failure | b_cell | *CYP8B1* | -2.648985442 | 0.002763544 | 0.009899943 | 1 |
| Heart_Failure | b_cell | *GLIPR1L2* | -2.047671856 | 0.001465613 | 0.008654124 | 1 |
| Heart_Failure | b_cell | *RNF182* | -3.369774203 | 0.005375621 | 0.014266158 | 1 |
| Heart_Failure | b_cell | *MB21D2* | -2.215919668 | 0.012658623 | 0.025680788 | 1 |
| Heart_Failure | b_cell | *SSTR2* | 3.297305568 | 9.85E-05 | 0.002986471 | 0.619509285 |
| Heart_Failure | b_cell | *ZNF594* | -1.935010024 | 0.002848748 | 0.010013275 | 1 |
| Heart_Failure | b_cell | *SHISA2* | -58.08011284 | 0.002224398 | 0.009086934 | 1 |
| Heart_Failure | b_cell | *CHST13* | -11.67878769 | 0.001042579 | 0.008654124 | 1 |
| Heart_Failure | b_cell | *SRSF8* | -3.180605777 | 0.00597277 | 0.015314358 | 1 |
| Heart_Failure | b_cell | *PSMG4* | -1.504630489 | 0.002158388 | 0.009019688 | 1 |
| Heart_Failure | b_cell | *ZNF443* | 4.931628391 | 0.013963168 | 0.027655764 | 1 |
| Heart_Failure | b_cell | *CAPS2* | -1.822690886 | 0.029809785 | 0.049151355 | 1 |
| Heart_Failure | b_cell | *ZNF792* | -4.869718986 | 0.000331166 | 0.004450951 | 1 |
| Heart_Failure | b_cell | *KCTD2* | -2.541708002 | 0.007223784 | 0.017294717 | 1 |
| Heart_Failure | b_cell | *D2HGDH* | -1.972090121 | 0.002921689 | 0.010133328 | 1 |
| Heart_Failure | b_cell | *LRRC57* | -3.327397293 | 0.027647913 | 0.046217943 | 1 |
| Heart_Failure | b_cell | *MRPL14* | -2.109508265 | 0.004161195 | 0.01222497 | 1 |
| Heart_Failure | b_cell | *BBS12* | -3.216308622 | 0.004581842 | 0.012905932 | 1 |
| Heart_Failure | b_cell | *AEN* | -4.034158666 | 0.000919175 | 0.008523384 | 1 |
| Heart_Failure | b_cell | *RPH3AL* | -1.749907069 | 0.001251963 | 0.008654124 | 1 |
| Heart_Failure | b_cell | *ANKRD34A* | -4.810573402 | 0.001385872 | 0.008654124 | 1 |
| Heart_Failure | b_cell | *SLC26A11* | -2.25164877 | 0.006165627 | 0.015568266 | 1 |
| Heart_Failure | b_cell | *HIGD1A* | -5.351038211 | 0.000205951 | 0.003537741 | 1 |
| Heart_Failure | b_cell | *ADIPOQ* | -1.09715326 | 0.00061795 | 0.006609617 | 1 |
| Heart_Failure | b_cell | *F2R* | -2.114888874 | 0.002089165 | 0.008868723 | 1 |
| Heart_Failure | b_cell | *HIST3H2A* | 6.406807669 | 0.002917767 | 0.010133328 | 1 |
| Heart_Failure | b_cell | *POLR2A* | -2.149384852 | 0.013305052 | 0.026656744 | 1 |
| Heart_Failure | b_cell | *TMEM132C* | -0.966427496 | 0.003272799 | 0.010803355 | 1 |
| Heart_Failure | b_cell | *SLC25A41* | -13.00766112 | 0.001501249 | 0.008654124 | 1 |
| Heart_Failure | b_cell | *TMEM136* | -2.0963601 | 0.011220383 | 0.023563662 | 1 |

| | | | | | | |
|---|---|---|---|---|---|---|
| Heart_Failure | b_cell | *TMEM132E* | -16.04881783 | 0.007924364 | 0.018343665 | 1 |
| Heart_Failure | b_cell | *ZNF322* | -2.015804931 | 0.004079522 | 0.012126691 | 1 |
| Heart_Failure | b_cell | *CCL13* | -4.123224945 | 0.000229877 | 0.003705045 | 1 |
| Heart_Failure | b_cell | *OGFOD3* | 4.249929921 | 2.94E-05 | 0.002889125 | 0.185135364 |
| Heart_Failure | b_cell | *MRPS23* | -2.420781585 | 0.002968617 | 0.010176497 | 1 |
| Heart_Failure | b_cell | *GPR135* | -1.438225946 | 0.006624702 | 0.016294798 | 1 |
| Heart_Failure | b_cell | *ATG9B* | -5.249749681 | 0.00170916 | 0.008654124 | 1 |
| Heart_Failure | b_cell | *C3orf58* | -2.331091453 | 0.014112744 | 0.027857714 | 1 |
| Heart_Failure | b_cell | *RELL1* | -1.786797679 | 0.0094407 | 0.020659131 | 1 |
| Heart_Failure | b_cell | *CLDN7* | -3.386183104 | 0.000295127 | 0.004150931 | 1 |
| Heart_Failure | b_cell | *ADO* | -3.191088159 | 0.00192796 | 0.008732736 | 1 |
| Heart_Failure | b_cell | *SNRPE* | 7.071896768 | 0.001500533 | 0.008654124 | 1 |
| Heart_Failure | b_cell | *TNRC18* | -1.673305797 | 0.02574898 | 0.043723523 | 1 |
| Heart_Failure | b_cell | *FAM181B* | 1.968094726 | 0.000119494 | 0.003135985 | 0.751259972 |
| Heart_Failure | b_cell | *NOP10* | -2.340031053 | 0.00347714 | 0.011125078 | 1 |
| Heart_Failure | b_cell | *FAM89A* | -1.363516014 | 0.006032056 | 0.015368478 | 1 |
| Heart_Failure | b_cell | *TDRKH* | -6.050972264 | 0.000308846 | 0.004267502 | 1 |
| Heart_Failure | b_cell | *MRPL41* | -2.4656855 | 0.005607447 | 0.014658637 | 1 |
| Heart_Failure | b_cell | *FAM153B* | -5.359195687 | 0.008387733 | 0.019082781 | 1 |
| Heart_Failure | b_cell | *BACE2* | -1.593925943 | 0.0117722 | 0.024301648 | 1 |
| Heart_Failure | b_cell | *SYNM* | -1.960631826 | 0.001735439 | 0.008654124 | 1 |
| Heart_Failure | b_cell | *IZUMO1* | 2.403655157 | 0.001405134 | 0.008654124 | 1 |
| Heart_Failure | b_cell | *C8orf33* | -2.212376457 | 0.005434405 | 0.014347469 | 1 |
| Heart_Failure | b_cell | *KBTBD3* | 5.144764272 | 4.87E-05 | 0.002889125 | 0.306071968 |
| Heart_Failure | b_cell | *CLN8* | 5.368244383 | 9.88E-05 | 0.002986471 | 0.621185874 |
| Heart_Failure | b_cell | *CACNB4* | -1.33020212 | 0.001694972 | 0.008654124 | 1 |
| Heart_Failure | b_cell | *TRAPPC6B* | -2.065404947 | 0.027234334 | 0.045708024 | 1 |
| Heart_Failure | b_cell | *NPLOC4* | -1.786857479 | 0.000939216 | 0.008620216 | 1 |
| Heart_Failure | b_cell | *TSHZ2* | -1.073235253 | 0.002340411 | 0.009248374 | 1 |
| Heart_Failure | b_cell | *CAPN12* | 3.604973849 | 7.04E-05 | 0.002889125 | 0.44281502 |
| Heart_Failure | b_cell | *CAV3* | 2.248529577 | 0.001299895 | 0.008654124 | 1 |
| Heart_Failure | b_cell | *MXRA7* | -2.444138808 | 0.004906161 | 0.013476294 | 1 |
| Heart_Failure | b_cell | *MFSD5* | -2.566258429 | 0.003732777 | 0.011549198 | 1 |
| Heart_Failure | b_cell | *ADI1* | -2.359878522 | 0.008582503 | 0.0193885 | 1 |
| Heart_Failure | b_cell | *SPNS3* | 1.275478415 | 0.000164488 | 0.003325199 | 1 |
| Heart_Failure | b_cell | *CLEC4G* | -7.354083875 | 0.000747412 | 0.007518371 | 1 |
| Heart_Failure | b_cell | *SATB1* | -2.648251132 | 0.008812136 | 0.019702926 | 1 |
| Heart_Failure | b_cell | *ACTL10* | -6.845458213 | 0.020023951 | 0.036009891 | 1 |
| Heart_Failure | b_cell | *KCNB2* | -1.347197782 | 0.003222385 | 0.010690838 | 1 |
| Heart_Failure | b_cell | *BRICD5* | -4.014984052 | 0.007649144 | 0.017910677 | 1 |
| Heart_Failure | b_cell | *TSKU* | -1.705622036 | 0.001679421 | 0.008654124 | 1 |
| Heart_Failure | b_cell | *GRID1* | -2.074768199 | 0.00061822 | 0.006609617 | 1 |
| Heart_Failure | b_cell | *ACBD3* | -1.855596194 | 0.015836808 | 0.03024484 | 1 |
| Heart_Failure | b_cell | *PLCXD3* | -1.633552439 | 0.010795416 | 0.022921575 | 1 |
| Heart_Failure | b_cell | *GPIHBP1* | -1.952302006 | 0.008387781 | 0.019082781 | 1 |
| Heart_Failure | b_cell | *SLC25A18* | -1.621705514 | 0.019912896 | 0.035871742 | 1 |
| Heart_Failure | b_cell | *LYSMD4* | -1.765631725 | 0.004446406 | 0.012718545 | 1 |
| Heart_Failure | b_cell | *BEGAIN* | -6.716040301 | 0.00172713 | 0.008654124 | 1 |
| Heart_Failure | b_cell | *ARHGEF37* | -2.320996907 | 0.006646309 | 0.016328403 | 1 |
| Heart_Failure | b_cell | *CSMD1* | -1.340906083 | 0.002048085 | 0.008831917 | 1 |

| | | | | | | |
|---|---|---|---|---|---|---|
| Heart_Failure | b_cell | *GPR19* | -6.264268033 | 0.003158392 | 0.01055652 | 1 |
| Heart_Failure | b_cell | *FANCF* | 4.200281275 | 6.72E-05 | 0.002889125 | 0.422736721 |
| Heart_Failure | b_cell | *C2CD4C* | -4.811761362 | 0.017189086 | 0.032098098 | 1 |
| Heart_Failure | b_cell | *RUVBL2* | -2.423131557 | 0.002492394 | 0.009470475 | 1 |
| Heart_Failure | b_cell | *GDPGP1* | 2.936411984 | 7.88E-05 | 0.002889125 | 0.495227119 |
| Heart_Failure | b_cell | *CTNNA3* | -1.523620393 | 0.011526868 | 0.023948916 | 1 |
| Heart_Failure | b_cell | *CCBE1* | -1.355947232 | 0.003354669 | 0.010877363 | 1 |
| Heart_Failure | b_cell | *ZNF623* | -3.275678245 | 0.003085797 | 0.010380099 | 1 |
| Heart_Failure | b_cell | *CCDC125* | -1.458564577 | 0.002247809 | 0.0091251 | 1 |
| Heart_Failure | b_cell | *FHL3* | -4.337599082 | 0.001311765 | 0.008654124 | 1 |
| Heart_Failure | b_cell | *C19orf71* | -4.093807935 | 0.004194893 | 0.012283789 | 1 |
| Heart_Failure | b_cell | *LRIT3* | -4.588737121 | 0.018636794 | 0.034150255 | 1 |
| Heart_Failure | b_cell | *SF3A3* | -2.555787667 | 0.012177584 | 0.024946687 | 1 |
| Heart_Failure | b_cell | *GRIN2A* | -2.109510871 | 0.011785546 | 0.024301648 | 1 |
| Heart_Failure | b_cell | *URAD* | -12.80152226 | 0.000584295 | 0.006366488 | 1 |
| Heart_Failure | b_cell | *GPR132* | 6.927339326 | 0.008841404 | 0.019736647 | 1 |
| Heart_Failure | b_cell | *PSMG1* | -2.689027447 | 2.99E-05 | 0.002889125 | 0.187856482 |
| Heart_Failure | b_cell | *ACSM5* | -1.471671481 | 0.005780385 | 0.014979918 | 1 |
| Heart_Failure | b_cell | *AC131971.1* | -10.46410751 | 0.004457671 | 0.01272146 | 1 |
| Heart_Failure | b_cell | *SERHL2* | 4.311287271 | 7.31E-05 | 0.002889125 | 0.459372162 |
| Heart_Failure | b_cell | *PCBP3* | -0.655392939 | 0.019821212 | 0.035757808 | 1 |
| Heart_Failure | b_cell | *SFXN4* | -2.199142768 | 0.016815689 | 0.031520643 | 1 |
| Heart_Failure | b_cell | *FAM167B* | 5.642616507 | 8.70E-05 | 0.002889125 | 0.547051231 |
| Heart_Failure | b_cell | *MRPL54* | 4.052916712 | 0.00160451 | 0.008654124 | 1 |
| Heart_Failure | b_cell | *FAM19A1* | -1.555151482 | 0.001554403 | 0.008654124 | 1 |
| Heart_Failure | b_cell | *TRMT12* | 3.764624611 | 4.30E-05 | 0.002889125 | 0.27060969 |
| Heart_Failure | b_cell | *CMTM4* | -2.716878028 | 0.015200132 | 0.029437116 | 1 |
| Heart_Failure | b_cell | *KREMEN1* | -1.379515214 | 0.002415193 | 0.009356237 | 1 |
| Heart_Failure | b_cell | *KCTD16* | -2.167945643 | 0.001478049 | 0.008654124 | 1 |
| Heart_Failure | b_cell | *B3GALT5* | 2.110124953 | 0.000250731 | 0.003766552 | 1 |
| Heart_Failure | b_cell | *ZNF703* | -4.592958088 | 0.000238961 | 0.003709499 | 1 |
| Heart_Failure | b_cell | *KCTD8* | -1.316580586 | 0.003854386 | 0.011769074 | 1 |
| Heart_Failure | b_cell | *C9orf66* | -6.842068687 | 0.002304766 | 0.009188443 | 1 |
| Heart_Failure | b_cell | *TUBA8* | -2.207881842 | 0.004867839 | 0.013405215 | 1 |
| Heart_Failure | b_cell | *FAM162B* | -2.786311134 | 0.002860362 | 0.01001558 | 1 |
| Heart_Failure | b_cell | *ANKRD45* | 19.88880326 | 0.000257129 | 0.003777026 | 1 |
| Heart_Failure | b_cell | *MAATS1* | -2.571392082 | 0.012422016 | 0.025315142 | 1 |
| Heart_Failure | b_cell | *GPR39* | -2.789950439 | 0.013542938 | 0.026992647 | 1 |
| Heart_Failure | b_cell | *FAM3B* | -1.732718813 | 0.003686269 | 0.011475847 | 1 |
| Heart_Failure | b_cell | *TTC32* | -2.180847711 | 0.002443723 | 0.009414769 | 1 |
| Heart_Failure | b_cell | *PP2D1* | -2.275487713 | 0.011680705 | 0.024164722 | 1 |
| Heart_Failure | b_cell | *TMEM236* | -3.279970496 | 0.001614721 | 0.008654124 | 1 |
| Heart_Failure | b_cell | *TBX1* | -2.762906505 | 0.003144952 | 0.010533998 | 1 |
| Heart_Failure | b_cell | *UQCR10* | 5.631333586 | 0.019137287 | 0.034876985 | 1 |
| Heart_Failure | b_cell | *NIPSNAP1* | 5.287830612 | 0.016961565 | 0.031746758 | 1 |
| Heart_Failure | b_cell | *CNTN2* | -4.441741752 | 0.001656081 | 0.008654124 | 1 |
| Heart_Failure | b_cell | *UBE2F* | -3.375471733 | 0.010378246 | 0.022223445 | 1 |
| Heart_Failure | b_cell | *GOLGA6L4* | 9.80783719 | 0.000234497 | 0.003705045 | 1 |
| Heart_Failure | b_cell | *PCDH9* | -0.865279082 | 0.001739866 | 0.008654124 | 1 |
| Heart_Failure | b_cell | *ACOT1* | -4.068546617 | 1.67E-05 | 0.002889125 | 0.104815385 |

| | | | | | | |
|---|---|---|---|---|---|---|
| Heart_Failure | b_cell | *OAF* | -1.618859589 | 0.000777539 | 0.007722573 | 1 |
| Heart_Failure | b_cell | *CLECL1* | 1.519530414 | 0.002496977 | 0.009479257 | 1 |
| Heart_Failure | b_cell | *MROH7* | -6.352048001 | 0.012621314 | 0.025629909 | 1 |
| Heart_Failure | b_cell | *CSF1* | -1.28654476 | 0.025239648 | 0.043038153 | 1 |
| Heart_Failure | b_cell | *PLA2G6* | -1.373985554 | 0.006299718 | 0.015836196 | 1 |
| Heart_Failure | b_cell | *A3GALT2* | -4.543618174 | 0.000828063 | 0.008083898 | 1 |
| Heart_Failure | b_cell | *THAP7* | -3.417538273 | 0.007689216 | 0.017987759 | 1 |
| Heart_Failure | b_cell | *CCR10* | -4.071989652 | 0.003173662 | 0.010585045 | 1 |
| Heart_Failure | b_cell | *TXNRD2* | -2.550839848 | 0.012635483 | 0.025650397 | 1 |
| Heart_Failure | b_cell | *PROS1* | -1.795162766 | 0.002897336 | 0.010115189 | 1 |
| Heart_Failure | b_cell | *HDDC3* | -2.45759349 | 0.00270851 | 0.009825588 | 1 |
| Heart_Failure | b_cell | *ZFP1* | -2.491064328 | 0.001915497 | 0.008705907 | 1 |
| Heart_Failure | b_cell | *LPAR5* | 2.80988058 | 0.001431806 | 0.008654124 | 1 |
| Heart_Failure | b_cell | *NELL2* | 2.913893381 | 0.001069008 | 0.008654124 | 1 |
| Heart_Failure | b_cell | *KRBA2* | -5.666345916 | 0.000398769 | 0.00494416 | 1 |
| Heart_Failure | b_cell | *ZNF93* | -2.626060539 | 0.003267203 | 0.010797038 | 1 |
| Heart_Failure | b_cell | *SEPT5* | -8.102824299 | 0.001632456 | 0.008654124 | 1 |
| Heart_Failure | b_cell | *ATL3* | -1.904209934 | 0.009508524 | 0.020785845 | 1 |
| Heart_Failure | b_cell | *NDUFA12* | -2.567401614 | 0.016433504 | 0.031091616 | 1 |
| Heart_Failure | b_cell | *UBE2G2* | -1.763929187 | 0.016008479 | 0.030434021 | 1 |
| Heart_Failure | b_cell | *OSBP2* | -2.229491395 | 0.003751194 | 0.011556943 | 1 |
| Heart_Failure | b_cell | *TUSC5* | -1.285046697 | 0.008890455 | 0.019792498 | 1 |
| Heart_Failure | b_cell | *TMED9* | -2.563586148 | 0.003928858 | 0.011882923 | 1 |
| Heart_Failure | b_cell | *BTBD6* | -1.978250485 | 0.006384115 | 0.015933676 | 1 |
| Heart_Failure | b_cell | *H1FX* | -2.527227108 | 0.024860409 | 0.042483662 | 1 |
| Heart_Failure | b_cell | *RBM43* | -3.050839883 | 0.005083317 | 0.013812459 | 1 |
| Heart_Failure | b_cell | *FAM227A* | -2.860088819 | 0.000285724 | 0.004036731 | 1 |
| Heart_Failure | b_cell | *NOC4L* | -2.724487009 | 0.005347763 | 0.01421623 | 1 |
| Heart_Failure | b_cell | *SORCS2* | -2.125564054 | 0.001482324 | 0.008654124 | 1 |
| Heart_Failure | b_cell | *SIVA1* | -3.008475962 | 0.002300369 | 0.009188443 | 1 |
| Heart_Failure | b_cell | *UBOX5* | -1.94105058 | 0.001737829 | 0.008654124 | 1 |
| Heart_Failure | b_cell | *MAFF* | -2.448498567 | 0.004108039 | 0.012159073 | 1 |
| Heart_Failure | b_cell | *FLRT2* | -1.031748929 | 0.001985899 | 0.008775328 | 1 |
| Heart_Failure | b_cell | *ADSSL1* | -3.930434937 | 0.001023703 | 0.008654124 | 1 |
| Heart_Failure | b_cell | *DDX51* | -1.823186574 | 0.003499928 | 0.011147503 | 1 |
| Heart_Failure | b_cell | *GP1BA* | -3.779272174 | 0.006245879 | 0.015735731 | 1 |
| Heart_Failure | b_cell | *KIAA0825* | -1.725016919 | 0.019736352 | 0.035667565 | 1 |
| Heart_Failure | b_cell | *CDK10* | -3.268221421 | 0.018168933 | 0.033448925 | 1 |
| Heart_Failure | b_cell | *TMEM105* | -49.48558432 | 0.001370523 | 0.008654124 | 1 |
| Heart_Failure | b_cell | *GAS2L1* | -2.211878054 | 0.002307707 | 0.009188443 | 1 |
| Heart_Failure | b_cell | *ATP6V0A2* | -2.017019414 | 0.014584251 | 0.02859154 | 1 |
| Heart_Failure | b_cell | *HS6ST3* | 2.180399804 | 7.45E-05 | 0.002889125 | 0.468395568 |
| Heart_Failure | b_cell | *TNFAIP8L1* | -2.655986302 | 0.001598275 | 0.008654124 | 1 |
| Heart_Failure | b_cell | *MAPK11* | -3.109059608 | 0.00123644 | 0.008654124 | 1 |
| Heart_Failure | b_cell | *FAM174B* | -2.227584353 | 0.029267228 | 0.048434605 | 1 |
| Heart_Failure | b_cell | *MUC1* | 10.2940623 | 1.20E-08 | 1.88E-05 | 7.53E-05 |
| Heart_Failure | b_cell | *SV2B* | -2.930216803 | 0.008054406 | 0.018566285 | 1 |
| Heart_Failure | b_cell | *PDE6G* | 2.190559358 | 0.00036657 | 0.004752125 | 1 |
| Heart_Failure | b_cell | *TLCD2* | -2.686062049 | 0.0036435 | 0.011427162 | 1 |
| Heart_Failure | b_cell | *LSAMP* | -1.298450458 | 0.001613367 | 0.008654124 | 1 |

| | | | | | | |
|---|---|---|---|---|---|---|
| Heart_Failure | b_cell | *AHNAK2* | -1.873501273 | 0.00174825 | 0.008654124 | 1 |
| Heart_Failure | b_cell | *PDIA2* | -4.732992 | 0.007558102 | 0.017796924 | 1 |
| Heart_Failure | b_cell | *PSMD13* | 4.528312289 | 0.028707873 | 0.047646885 | 1 |
| Heart_Failure | b_cell | *PBX1* | -1.059198044 | 0.003134906 | 0.010520609 | 1 |
| Heart_Failure | b_cell | *PMEL* | 2.644678923 | 0.001378201 | 0.008654124 | 1 |
| Heart_Failure | b_cell | *SNAI3* | -2.713111899 | 0.000721557 | 0.007316224 | 1 |
| Heart_Failure | b_cell | *MYBL1* | -2.25426943 | 4.05E-05 | 0.002889125 | 0.254730223 |
| Heart_Failure | b_cell | *C16orf52* | -1.74595872 | 0.010332426 | 0.022163071 | 1 |
| Heart_Failure | b_cell | *ADARB2* | -2.304042811 | 0.00157544 | 0.008654124 | 1 |
| Heart_Failure | b_cell | *SRL* | 3.636264808 | 0.000307539 | 0.004261028 | 1 |
| Heart_Failure | b_cell | *KCNIP4* | -0.973782248 | 0.005927783 | 0.01524067 | 1 |
| Heart_Failure | b_cell | *SLC52A2* | -2.399335098 | 0.001710899 | 0.008654124 | 1 |
| Heart_Failure | b_cell | *ARL17A* | 4.066903446 | 0.026513294 | 0.044736736 | 1 |
| Heart_Failure | b_cell | *NPIPB4* | -2.276555984 | 0.003770334 | 0.011591512 | 1 |
| Heart_Failure | b_cell | *TRIM69* | 3.375709779 | 0.006880229 | 0.016714065 | 1 |
| Heart_Failure | b_cell | *RTN4RL1* | -1.786312152 | 0.00166916 | 0.008654124 | 1 |
| Heart_Failure | b_cell | *PLK5* | -7.587531793 | 0.000669404 | 0.006979344 | 1 |
| Heart_Failure | b_cell | *RASA3* | -1.892051418 | 0.01924761 | 0.035004259 | 1 |
| Heart_Failure | b_cell | *ZNF284* | -4.451409327 | 0.001703679 | 0.008654124 | 1 |
| Heart_Failure | b_cell | *GSAP* | -1.599337852 | 0.015414131 | 0.029717461 | 1 |
| Heart_Failure | b_cell | *CYP4Z1* | -2.351443569 | 0.001549473 | 0.008654124 | 1 |
| Heart_Failure | b_cell | *FFAR4* | -1.866176298 | 0.000561256 | 0.00624534 | 1 |
| Heart_Failure | b_cell | *BPIFB4* | -151.0255102 | 0.001668157 | 0.008654124 | 1 |
| Heart_Failure | b_cell | *CYP4F12* | -1.514067237 | 0.017016737 | 0.031821601 | 1 |
| Heart_Failure | b_cell | *KLHL32* | 6.620227455 | 0.000399033 | 0.00494416 | 1 |
| Heart_Failure | b_cell | *NBPF12* | -2.03332729 | 0.013155273 | 0.026449377 | 1 |
| Heart_Failure | b_cell | *GPAT2* | 4.223170029 | 0.000133466 | 0.003202685 | 0.839103359 |
| Heart_Failure | b_cell | *TOR3A* | 6.646367768 | 8.86E-05 | 0.002889125 | 0.556814731 |
| Heart_Failure | b_cell | *CYP4X1* | -1.712392572 | 0.001874518 | 0.008701813 | 1 |
| Heart_Failure | b_cell | *CCDC30* | -2.308373551 | 0.02059587 | 0.036702448 | 1 |
| Heart_Failure | b_cell | *GLDN* | -1.485156097 | 0.001772414 | 0.008654124 | 1 |
| Heart_Failure | b_cell | *FCAR* | 2.830314098 | 0.000179033 | 0.00338933 | 1 |
| Heart_Failure | b_cell | *ZNF501* | -2.676150939 | 0.000962715 | 0.008654124 | 1 |
| Heart_Failure | b_cell | *ZNF197* | -2.118710787 | 0.007615166 | 0.017864384 | 1 |
| Heart_Failure | b_cell | *PCLO* | -1.084992386 | 0.003872798 | 0.011802366 | 1 |
| Heart_Failure | b_cell | *ZNF396* | -3.574324614 | 0.001728288 | 0.008654124 | 1 |
| Heart_Failure | b_cell | *TMEM222* | -2.142591838 | 0.001625924 | 0.008654124 | 1 |
| Heart_Failure | b_cell | *SEPT10* | -1.918376137 | 0.005559995 | 0.014577179 | 1 |
| Heart_Failure | b_cell | *SMYD4* | -2.136754692 | 0.002826573 | 0.009985294 | 1 |
| Heart_Failure | b_cell | *CEACAM19* | -2.264793271 | 0.001519814 | 0.008654124 | 1 |
| Heart_Failure | b_cell | *MIR22HG* | -1.387597386 | 0.002060748 | 0.008831917 | 1 |
| Heart_Failure | b_cell | *PDE2A* | -1.961921316 | 0.001249447 | 0.008654124 | 1 |
| Heart_Failure | b_cell | *C17orf58* | -1.415894295 | 0.010153365 | 0.021913563 | 1 |
| Heart_Failure | b_cell | *CYP27C1* | -1.800037748 | 0.00791497 | 0.018343665 | 1 |
| Heart_Failure | b_cell | *BCR* | -1.860420287 | 0.006521749 | 0.016123569 | 1 |
| Heart_Failure | b_cell | *FOXI2* | -5.057426139 | 0.015468627 | 0.029767756 | 1 |
| Heart_Failure | b_cell | *HYAL3* | 1.529418316 | 0.001092574 | 0.008654124 | 1 |
| Heart_Failure | b_cell | *LILRB4* | -1.626368625 | 0.016795447 | 0.031501484 | 1 |
| Heart_Failure | b_cell | *TMEM17* | -2.584554961 | 0.016525778 | 0.031217066 | 1 |
| Heart_Failure | b_cell | *TNFRSF18* | -6.782962209 | 0.001642809 | 0.008654124 | 1 |

| | | | | | | |
|---|---|---|---|---|---|---|
| Heart_Failure | b_cell | *RTN4RL2* | -6.518295865 | 0.002688162 | 0.009763417 | 1 |
| Heart_Failure | b_cell | *TMEM232* | -1.674752121 | 0.025291038 | 0.043114087 | 1 |
| Heart_Failure | b_cell | *EFCAB6* | -2.093983711 | 0.006002082 | 0.015339468 | 1 |
| Heart_Failure | b_cell | *KANK3* | -1.817324302 | 0.001334725 | 0.008654124 | 1 |
| Heart_Failure | b_cell | *EMID1* | -4.169861519 | 0.006455933 | 0.01602239 | 1 |
| Heart_Failure | b_cell | *TMEM216* | -2.620816472 | 0.001642186 | 0.008654124 | 1 |
| Heart_Failure | b_cell | *RPS19BP1* | -2.115791644 | 0.002605739 | 0.00961966 | 1 |
| Heart_Failure | b_cell | *ENTPD5* | -1.906220786 | 0.005291111 | 0.014126188 | 1 |
| Heart_Failure | b_cell | *MRPS21* | -2.801568872 | 0.006619191 | 0.01629399 | 1 |
| Heart_Failure | b_cell | *ANGPTL5* | -2.187550843 | 0.001632909 | 0.008654124 | 1 |
| Heart_Failure | b_cell | *TSPYL4* | -3.599112818 | 0.004003553 | 0.012024754 | 1 |
| Heart_Failure | b_cell | *MT1X* | -1.942637779 | 0.004546028 | 0.012845337 | 1 |
| Heart_Failure | b_cell | *WDR86* | -1.807683018 | 0.005038415 | 0.013730609 | 1 |
| Heart_Failure | b_cell | *EPOR* | -2.627997338 | 0.00127555 | 0.008654124 | 1 |
| Heart_Failure | b_cell | *FPR3* | -1.537100799 | 0.000762828 | 0.007612537 | 1 |
| Heart_Failure | b_cell | *GJA4* | -2.876891298 | 0.005833523 | 0.015086532 | 1 |
| Heart_Failure | b_cell | *TLR5* | -1.902307256 | 0.001457502 | 0.008654124 | 1 |
| Heart_Failure | b_cell | *PLEKHN1* | -4.267095042 | 0.015964634 | 0.030378224 | 1 |
| Heart_Failure | b_cell | *ZNF385C* | 5.177409328 | 0.000173234 | 0.003365162 | 1 |
| Heart_Failure | b_cell | *TET3* | -1.398339037 | 0.001865953 | 0.008690419 | 1 |
| Heart_Failure | b_cell | *C2orf88* | -1.565683233 | 0.002245644 | 0.0091251 | 1 |
| Heart_Failure | b_cell | *ADH1A* | -4.445512877 | 3.20E-05 | 0.002889125 | 0.201256089 |
| Heart_Failure | b_cell | *SEMA4D* | -2.084229158 | 0.00409263 | 0.012159073 | 1 |
| Heart_Failure | b_cell | *FAM69C* | -7.260174454 | 0.01716325 | 0.032066969 | 1 |
| Heart_Failure | b_cell | *DNAH17* | -2.583437797 | 0.001398966 | 0.008654124 | 1 |
| Heart_Failure | b_cell | *MCRS1* | 8.947794794 | 0.000106976 | 0.003034612 | 0.672558313 |
| Heart_Failure | b_cell | *ZFP69* | -2.612452079 | 0.003852876 | 0.011769074 | 1 |
| Heart_Failure | b_cell | *TMEM220* | -1.926576065 | 0.027954589 | 0.046655296 | 1 |
| Heart_Failure | b_cell | *CCDC157* | 1.674218068 | 0.000113965 | 0.003135985 | 0.71649656 |
| Heart_Failure | b_cell | *GFRAL* | 1.086264942 | 0.000196187 | 0.003449722 | 1 |
| Heart_Failure | b_cell | *C2orf66* | -13.79887543 | 0.011862007 | 0.024434075 | 1 |
| Heart_Failure | b_cell | *CPSF4L* | -6.608321658 | 0.00456832 | 0.012896736 | 1 |
| Heart_Failure | b_cell | *C6orf132* | -2.086518459 | 0.002147358 | 0.008999799 | 1 |
| Heart_Failure | b_cell | *MAPK12* | -4.17791973 | 0.006782003 | 0.016532941 | 1 |
| Heart_Failure | b_cell | *AGRN* | -4.077656928 | 0.001384778 | 0.008654124 | 1 |
| Heart_Failure | b_cell | *FAM166A* | -11.26575043 | 0.001687956 | 0.008654124 | 1 |
| Heart_Failure | b_cell | *ZNF626* | -2.506750773 | 0.000945118 | 0.008636566 | 1 |
| Heart_Failure | b_cell | *SMTNL2* | -16.13624862 | 0.001793969 | 0.008654124 | 1 |
| Heart_Failure | b_cell | *PRKAR1B* | -1.925775191 | 0.001035346 | 0.008654124 | 1 |
| Heart_Failure | b_cell | *NCR3LG1* | -4.871383079 | 0.001846543 | 0.00866783 | 1 |
| Heart_Failure | b_cell | *AGAP4* | -2.546558688 | 0.001525946 | 0.008654124 | 1 |
| Heart_Failure | b_cell | *HYKK* | -2.747870171 | 0.003069887 | 0.01036364 | 1 |
| Heart_Failure | b_cell | *RUFY4* | -5.779066519 | 0.001747026 | 0.008654124 | 1 |
| Heart_Failure | b_cell | *CENPP* | -1.523165008 | 0.003384833 | 0.01094673 | 1 |
| Heart_Failure | b_cell | *JAKMIP3* | -2.238196163 | 0.003289516 | 0.010827847 | 1 |
| Heart_Failure | b_cell | *PDCD1* | 9.607914603 | 0.000401498 | 0.004959169 | 1 |
| Heart_Failure | b_cell | *CERKL* | -5.73454754 | 0.0049199 | 0.01349375 | 1 |
| Heart_Failure | b_cell | *COL25A1* | -1.279763591 | 0.001620777 | 0.008654124 | 1 |
| Heart_Failure | b_cell | *NBR1* | -2.015589625 | 0.014007916 | 0.027720418 | 1 |
| Heart_Failure | b_cell | *SUMO2* | -2.378473733 | 0.018259898 | 0.033547627 | 1 |

| | | | | | | |
|---|---|---|---|---|---|---|
| Heart_Failure | b_cell | *GOLGA8M* | -2.260202216 | 0.001471963 | 0.008654124 | 1 |
| Heart_Failure | b_cell | *S100A16* | -2.538809535 | 0.001910284 | 0.008705907 | 1 |
| Heart_Failure | b_cell | *PARVB* | -1.70277503 | 0.005769681 | 0.014970691 | 1 |
| Heart_Failure | b_cell | *SLC4A5* | 1.440085406 | 0.000246515 | 0.003766552 | 1 |
| Heart_Failure | b_cell | *UROS* | -2.249696968 | 0.020634666 | 0.03675075 | 1 |
| Heart_Failure | b_cell | *FAM221A* | -2.062784734 | 0.001105697 | 0.008654124 | 1 |
| Heart_Failure | b_cell | *FSIP2* | -1.296830992 | 0.003457419 | 0.01109586 | 1 |
| Heart_Failure | b_cell | *NOXA1* | -2.260192195 | 0.002282965 | 0.009182983 | 1 |
| Heart_Failure | b_cell | *SKOR1* | 1.27987971 | 0.000227588 | 0.003705045 | 1 |
| Heart_Failure | b_cell | *TMEM201* | -2.951393723 | 0.006045655 | 0.015394505 | 1 |
| Heart_Failure | b_cell | *ZDHHC11* | -2.435805594 | 0.008952677 | 0.019901071 | 1 |
| Heart_Failure | b_cell | *FAM26F* | 0.971715736 | 0.001117122 | 0.008654124 | 1 |
| Heart_Failure | b_cell | *BEND4* | 1.274477899 | 0.000201355 | 0.003506707 | 1 |
| Heart_Failure | b_cell | *ZNF563* | -2.411504855 | 0.006966346 | 0.016877617 | 1 |
| Heart_Failure | b_cell | *FBF1* | -1.170311852 | 0.02404545 | 0.041473509 | 1 |
| Heart_Failure | b_cell | *MSL1* | -3.483265808 | 0.000878851 | 0.00834507 | 1 |
| Heart_Failure | b_cell | *LRRK2* | -1.906364574 | 0.016653109 | 0.031361554 | 1 |
| Heart_Failure | b_cell | *FAM196A* | -1.503952996 | 0.002417607 | 0.009356237 | 1 |
| Heart_Failure | b_cell | *UTS2B* | -3.262361616 | 8.87E-05 | 0.002889125 | 0.557857611 |
| Heart_Failure | b_cell | *C9orf152* | -6.078913528 | 0.001944111 | 0.008738197 | 1 |
| Heart_Failure | b_cell | *NOC2L* | 2.557475633 | 0.029390499 | 0.048574676 | 1 |
| Heart_Failure | b_cell | *LIPI* | -2.11454742 | 0.003609259 | 0.011367119 | 1 |
| Heart_Failure | b_cell | *ADAT2* | 5.964865655 | 0.00024451 | 0.00375852 | 1 |
| Heart_Failure | b_cell | *ZNF567* | -2.297701151 | 0.007273675 | 0.017354686 | 1 |
| Heart_Failure | b_cell | *ANKDD1B* | -2.158407808 | 0.015895895 | 0.030330043 | 1 |
| Heart_Failure | b_cell | *LITAF* | -1.661350102 | 0.006513809 | 0.016123569 | 1 |
| Heart_Failure | b_cell | *PLAC9* | -1.308604283 | 0.012965871 | 0.026143821 | 1 |
| Heart_Failure | b_cell | *CLDN4* | 4.589216271 | 0.001659808 | 0.008654124 | 1 |
| Heart_Failure | b_cell | *ZNF527* | -3.048416949 | 0.006448682 | 0.016018516 | 1 |
| Heart_Failure | b_cell | *S100A13* | -1.667362956 | 0.001695341 | 0.008654124 | 1 |
| Heart_Failure | b_cell | *C15orf61* | 7.186832421 | 0.011301195 | 0.023652002 | 1 |
| Heart_Failure | b_cell | *ZKSCAN3* | -2.391260143 | 0.010260227 | 0.022085313 | 1 |
| Heart_Failure | b_cell | *AKR1C3* | -1.463186122 | 0.002711445 | 0.009825588 | 1 |
| Heart_Failure | b_cell | *WDSUB1* | -2.127508032 | 0.017049669 | 0.031873705 | 1 |
| Heart_Failure | b_cell | *S100A4* | -1.524949672 | 0.012297703 | 0.025126961 | 1 |
| Heart_Failure | b_cell | *COLCA1* | -4.602594132 | 0.002360126 | 0.00927962 | 1 |
| Heart_Failure | b_cell | *ACADSB* | -1.911876184 | 0.007312295 | 0.017394685 | 1 |
| Heart_Failure | b_cell | *HRCT1* | -2.671320367 | 0.005072637 | 0.013804695 | 1 |
| Heart_Failure | b_cell | *SIRPB2* | -2.269705877 | 0.001524087 | 0.008654124 | 1 |
| Heart_Failure | b_cell | *SRGAP3* | -1.784405319 | 0.014318085 | 0.02816577 | 1 |
| Heart_Failure | b_cell | *XPNPEP3* | -1.677100412 | 0.00509701 | 0.013812459 | 1 |
| Heart_Failure | b_cell | *PPIA* | -2.214553099 | 0.007727774 | 0.018034341 | 1 |
| Heart_Failure | b_cell | *ZNF471* | -2.172999654 | 0.007481352 | 0.017675553 | 1 |
| Heart_Failure | b_cell | *GRM7* | -1.253831279 | 0.003709278 | 0.011521853 | 1 |
| Heart_Failure | b_cell | *GIMAP5* | -2.366168681 | 0.001606866 | 0.008654124 | 1 |
| Heart_Failure | b_cell | *STK31* | -2.928441375 | 0.001626793 | 0.008654124 | 1 |
| Heart_Failure | b_cell | *ZKSCAN7* | 5.405748555 | 7.10E-05 | 0.002889125 | 0.446656659 |
| Heart_Failure | b_cell | *ZNF565* | -1.453294544 | 0.003837384 | 0.011751405 | 1 |
| Heart_Failure | b_cell | *NTNG2* | 12.43196545 | 0.001297576 | 0.008654124 | 1 |
| Heart_Failure | b_cell | *WDR5* | -2.319877582 | 0.01136482 | 0.023737749 | 1 |

| | | | | | | |
|---|---|---|---|---|---|---|
| Heart_Failure | b_cell | *SLC35F1* | -2.370480816 | 0.003597367 | 0.01134235 | 1 |
| Heart_Failure | b_cell | *ZNF34* | -3.608903361 | 0.0067003 | 0.016416518 | 1 |
| Heart_Failure | b_cell | *ZNF781* | -2.249412354 | 0.015052065 | 0.02922555 | 1 |
| Heart_Failure | b_cell | *INCA1* | -5.850363401 | 0.010662495 | 0.022708369 | 1 |
| Heart_Failure | b_cell | *ZNF774* | -5.516274148 | 0.010292827 | 0.022123419 | 1 |
| Heart_Failure | b_cell | *ZNF124* | -2.170066613 | 0.023426329 | 0.040640544 | 1 |
| Heart_Failure | b_cell | *ZNF569* | -1.82018714 | 0.00141209 | 0.008654124 | 1 |
| Heart_Failure | b_cell | *ZNF605* | -2.013977419 | 0.002834889 | 0.010000392 | 1 |
| Heart_Failure | b_cell | *RFX8* | -2.233160808 | 0.001313047 | 0.008654124 | 1 |
| Heart_Failure | b_cell | *MYL6B* | -2.236028003 | 0.002634962 | 0.009653089 | 1 |
| Heart_Failure | b_cell | *ZNF799* | -3.972054467 | 0.001583204 | 0.008654124 | 1 |
| Heart_Failure | b_cell | *C20orf96* | -2.403821188 | 0.001506373 | 0.008654124 | 1 |
| Heart_Failure | b_cell | *SULT1A1* | -1.792942727 | 0.000890449 | 0.008380617 | 1 |
| Heart_Failure | b_cell | *ANAPC7* | -1.901968324 | 0.010957045 | 0.023139718 | 1 |
| Heart_Failure | b_cell | *C1orf186* | 0.337967127 | 0.000180832 | 0.00338933 | 1 |
| Heart_Failure | b_cell | *SPTSSB* | -7.131379587 | 0.002074716 | 0.008849214 | 1 |
| Heart_Failure | b_cell | *SULF2* | -1.953239362 | 0.005632383 | 0.014705479 | 1 |
| Heart_Failure | b_cell | *XRCC2* | -4.278589976 | 0.007192581 | 0.017239708 | 1 |
| Heart_Failure | b_cell | *MYO6* | -1.662214035 | 0.003342705 | 0.010865693 | 1 |
| Heart_Failure | b_cell | *HDAC2* | -2.024100485 | 0.014586853 | 0.02859154 | 1 |
| Heart_Failure | b_cell | *ADH1B* | -1.000403771 | 0.001548402 | 0.008654124 | 1 |
| Heart_Failure | b_cell | *ZNF502* | -2.278052688 | 0.001605172 | 0.008654124 | 1 |
| Heart_Failure | b_cell | *TECPR2* | -1.99609199 | 0.013897696 | 0.027582359 | 1 |
| Heart_Failure | b_cell | *FAM180B* | -3.440887509 | 8.60E-05 | 0.002889125 | 0.540905273 |
| Heart_Failure | b_cell | *ZFP62* | -3.049742414 | 0.01473327 | 0.028793307 | 1 |
| Heart_Failure | b_cell | *ERI2* | -1.4047281 | 0.001990319 | 0.008775328 | 1 |
| Heart_Failure | b_cell | *TRPV1* | -2.680305211 | 0.010271331 | 0.022099884 | 1 |
| Heart_Failure | b_cell | *ZNF33B* | -2.294578582 | 0.022507272 | 0.03940291 | 1 |
| Heart_Failure | b_cell | *AMZ2* | -2.02342375 | 0.006428695 | 0.015989469 | 1 |
| Heart_Failure | b_cell | *ZNF431* | -1.822371147 | 0.020539781 | 0.036623256 | 1 |
| Heart_Failure | b_cell | *DAPK1* | -1.38586779 | 0.005892301 | 0.015176115 | 1 |
| Heart_Failure | b_cell | *GM2A* | 4.147356704 | 0.015503517 | 0.029816644 | 1 |
| Heart_Failure | b_cell | *TLE1* | -1.240927361 | 0.002542213 | 0.009545808 | 1 |
| Heart_Failure | b_cell | *ZNF239* | -1.829860093 | 0.029760709 | 0.049096189 | 1 |
| Heart_Failure | b_cell | *ZFP28* | -2.434515223 | 0.002820691 | 0.009979564 | 1 |
| Heart_Failure | b_cell | *LAMB3* | -2.674674923 | 0.002864634 | 0.010022233 | 1 |
| Heart_Failure | b_cell | *HCAR1* | -2.796658065 | 0.005291423 | 0.014126188 | 1 |
| Heart_Failure | b_cell | *SRGAP1* | -1.515748325 | 0.01554729 | 0.029868396 | 1 |
| Heart_Failure | b_cell | *SLC39A10* | 4.456483671 | 0.001361284 | 0.008654124 | 1 |
| Heart_Failure | b_cell | *HIST1H3E* | 2.234784048 | 0.000108448 | 0.003034612 | 0.681810682 |
| Heart_Failure | b_cell | *ZNF585A* | -2.055519469 | 0.005656042 | 0.014748875 | 1 |
| Heart_Failure | b_cell | *FUT11* | -1.807729769 | 0.003025598 | 0.010282127 | 1 |
| Heart_Failure | b_cell | *WDR5B* | 5.189332661 | 0.000121776 | 0.003135985 | 0.765604115 |
| Heart_Failure | b_cell | *ZNF470* | -3.569384428 | 0.001702286 | 0.008654124 | 1 |
| Heart_Failure | b_cell | *GMFB* | -2.571054248 | 0.02294833 | 0.040021123 | 1 |
| Heart_Failure | b_cell | *ZNF763* | -2.447645043 | 0.008615437 | 0.019414068 | 1 |
| Heart_Failure | b_cell | *ARRDC1* | -2.252186582 | 0.00393675 | 0.011883725 | 1 |
| Heart_Failure | b_cell | *GAL3ST4* | -4.084318927 | 0.002279111 | 0.009182983 | 1 |
| Heart_Failure | b_cell | *SLC25A29* | -2.14528867 | 0.01147787 | 0.023870781 | 1 |
| Heart_Failure | b_cell | *ZNF682* | -1.650996844 | 0.002321828 | 0.00922131 | 1 |

| | | | | | | |
|---|---|---|---|---|---|---|
| Heart_Failure | b_cell | *ADAM32* | -1.770221277 | 0.001505966 | 0.008654124 | 1 |
| Heart_Failure | b_cell | *LRRC8B* | -1.453698029 | 0.001569684 | 0.008654124 | 1 |
| Heart_Failure | b_cell | *NEK5* | -2.566744396 | 0.000758138 | 0.007595773 | 1 |
| Heart_Failure | b_cell | *PIWIL2* | -2.104639491 | 0.001716656 | 0.008654124 | 1 |
| Heart_Failure | b_cell | *C1D* | -2.100221284 | 0.021492915 | 0.038012258 | 1 |
| Heart_Failure | b_cell | *TBC1D9B* | -2.084387755 | 0.017763196 | 0.032923706 | 1 |
| Heart_Failure | b_cell | *FAM110D* | -3.771232267 | 0.00371743 | 0.011530086 | 1 |
| Heart_Failure | b_cell | *KANK2* | -1.635144912 | 0.002808608 | 0.009959169 | 1 |
| Heart_Failure | b_cell | *C6orf141* | -2.74776215 | 0.002843174 | 0.010008417 | 1 |
| Heart_Failure | b_cell | *GTF2E2* | -1.899252686 | 0.013488108 | 0.026920551 | 1 |
| Heart_Failure | b_cell | *ZNF165* | 5.500032325 | 0.001024561 | 0.008654124 | 1 |
| Heart_Failure | b_cell | *RAMP2-AS1* | -1.751143362 | 0.002401525 | 0.009333474 | 1 |
| Heart_Failure | b_cell | *ZNF720* | -1.942462576 | 0.016683569 | 0.031366507 | 1 |
| Heart_Failure | b_cell | *MRPL21* | 2.431071559 | 0.010617809 | 0.022643883 | 1 |
| Heart_Failure | b_cell | *ZNF786* | -2.726796664 | 0.020587727 | 0.036698338 | 1 |
| Heart_Failure | b_cell | *ZNF517* | -2.871988757 | 0.001655127 | 0.008654124 | 1 |
| Heart_Failure | b_cell | *C5AR1* | -2.239941034 | 0.003066698 | 0.01036364 | 1 |
| Heart_Failure | b_cell | *DCHS2* | -1.440996467 | 0.01512837 | 0.029346518 | 1 |
| Heart_Failure | b_cell | *SHPK* | -2.464807148 | 0.002724996 | 0.009828026 | 1 |
| Heart_Failure | b_cell | *GYPE* | 3.182795736 | 0.005085152 | 0.013812459 | 1 |
| Heart_Failure | b_cell | *PCDHB11* | -5.613624426 | 0.000251081 | 0.003766552 | 1 |
| Heart_Failure | b_cell | *SLC2A10* | -1.792239042 | 0.003670219 | 0.011457134 | 1 |
| Heart_Failure | b_cell | *ZNF665* | -2.266001174 | 0.027864566 | 0.046529754 | 1 |
| Heart_Failure | b_cell | *RPF2* | -2.384781381 | 0.006148064 | 0.015548221 | 1 |
| Heart_Failure | b_cell | *MYO5A* | -1.496075714 | 0.012401111 | 0.025289758 | 1 |
| Heart_Failure | b_cell | *ATG7* | -1.455369373 | 0.011604292 | 0.024062066 | 1 |
| Heart_Failure | b_cell | *TTC30A* | -3.91222693 | 0.002419793 | 0.009356237 | 1 |
| Heart_Failure | b_cell | *RAB40C* | -2.190984896 | 0.005136818 | 0.013872497 | 1 |
| Heart_Failure | b_cell | *ZNF624* | 7.297935902 | 0.000495489 | 0.005674204 | 1 |
| Heart_Failure | b_cell | *HHLA3* | -4.086497989 | 0.022657236 | 0.039612359 | 1 |
| Heart_Failure | b_cell | *HOXA4* | -4.009974768 | 0.002573709 | 0.009563183 | 1 |
| Heart_Failure | b_cell | *ENTPD6* | -2.347572162 | 0.001365596 | 0.008654124 | 1 |
| Heart_Failure | b_cell | *MFAP5* | -1.043744946 | 0.001682645 | 0.008654124 | 1 |
| Heart_Failure | b_cell | *PDCD1LG2* | -1.355068231 | 0.010342821 | 0.02216321 | 1 |
| Heart_Failure | b_cell | *KLHL14* | 0.538745897 | 0.000331326 | 0.004450951 | 1 |
| Heart_Failure | b_cell | *FAM114A1* | -1.806541093 | 0.015912587 | 0.030334273 | 1 |
| Heart_Failure | b_cell | *RPS26* | 4.03680373 | 9.42E-07 | 0.000602686 | 0.005922573 |
| Heart_Failure | b_cell | *S100A10* | -1.87810816 | 0.018885144 | 0.034524832 | 1 |
| Heart_Failure | b_cell | *MAP1LC3C* | -2.658503342 | 0.025126987 | 0.042869299 | 1 |
| Heart_Failure | b_cell | *TAF13* | -1.696634412 | 0.004530734 | 0.012830956 | 1 |
| Heart_Failure | b_cell | *AC122129.1* | 10.39522088 | 6.15E-05 | 0.002889125 | 0.386621736 |
| Heart_Failure | b_cell | *OCLN* | -2.234229792 | 0.008661772 | 0.019497516 | 1 |
| Heart_Failure | b_cell | *ZNF181* | 5.069685183 | 3.61E-05 | 0.002889125 | 0.226696556 |
| Heart_Failure | b_cell | *SGTB* | -2.14776914 | 0.001811012 | 0.008654124 | 1 |
| Heart_Failure | b_cell | *ADH5* | -1.722195981 | 0.007031372 | 0.016976281 | 1 |
| Heart_Failure | b_cell | *ZNF677* | -2.003215887 | 0.001793587 | 0.008654124 | 1 |
| Heart_Failure | b_cell | *ZNF311* | -2.529676819 | 0.005413366 | 0.01433607 | 1 |
| Heart_Failure | b_cell | *ZNF71* | -1.991309002 | 0.004446834 | 0.012718545 | 1 |
| Heart_Failure | b_cell | *RPL12* | 3.681262128 | 0.006350198 | 0.015908678 | 1 |
| Heart_Failure | b_cell | *VPS13A* | -1.736382805 | 0.029997636 | 0.049370454 | 1 |

| | | | | | | |
|---|---|---|---|---|---|---|
| Heart_Failure | b_cell | *LEKR1* | -2.082208286 | 0.000596751 | 0.006457437 | 1 |
| Heart_Failure | b_cell | *ZNF335* | -2.65197737 | 0.014847561 | 0.028953665 | 1 |
| Heart_Failure | b_cell | *AGAP9* | 4.614724744 | 3.66E-05 | 0.002889125 | 0.230099227 |
| Heart_Failure | b_cell | *ZNF84* | -2.171175375 | 0.015028745 | 0.029189286 | 1 |
| Heart_Failure | b_cell | *MAK16* | -2.06373274 | 0.001981675 | 0.008775328 | 1 |
| Heart_Failure | b_cell | *ZNF667* | -2.119387654 | 0.014661092 | 0.028682365 | 1 |
| Heart_Failure | b_cell | *PRIM1* | -2.698521337 | 0.026298761 | 0.044458272 | 1 |
| Heart_Failure | b_cell | *SULT1C4* | -2.801322158 | 0.002045369 | 0.008831917 | 1 |
| Heart_Failure | b_cell | *ADH4* | -1.742454556 | 0.001640751 | 0.008654124 | 1 |
| Heart_Failure | b_cell | *ZNF248* | -1.784339682 | 0.00444742 | 0.012718545 | 1 |
| Heart_Failure | b_cell | *HIBCH* | -1.508514774 | 0.015361791 | 0.029662165 | 1 |
| Heart_Failure | b_cell | *ZNF544* | -2.535858616 | 0.003908457 | 0.011873672 | 1 |
| Heart_Failure | b_cell | *SOWAHC* | -3.551580959 | 0.001689052 | 0.008654124 | 1 |
| Heart_Failure | b_cell | *AC135178.1* | -13.22144845 | 0.021136034 | 0.037473843 | 1 |
| Heart_Failure | b_cell | *SVIP* | -2.217397578 | 0.004072167 | 0.012126691 | 1 |
| Heart_Failure | b_cell | *DDRGK1* | -2.668832773 | 0.020455553 | 0.036514498 | 1 |
| Heart_Failure | b_cell | *ZNF334* | -1.795069921 | 0.007123182 | 0.017125602 | 1 |
| Heart_Failure | b_cell | *RPS6KL1* | -1.801715118 | 0.019323273 | 0.035121542 | 1 |
| Heart_Failure | b_cell | *SLC29A3* | -2.291627527 | 0.001934544 | 0.008737414 | 1 |
| Heart_Failure | b_cell | *ANTXRL* | -88.76444474 | 0.001664598 | 0.008654124 | 1 |
| Heart_Failure | b_cell | *TMEM116* | -1.951076437 | 0.001151014 | 0.008654124 | 1 |
| Heart_Failure | b_cell | *UVRAG* | -1.415877984 | 0.026010453 | 0.044065674 | 1 |
| Heart_Failure | b_cell | *MGEA5* | -1.346407747 | 0.015373523 | 0.029672617 | 1 |
| Heart_Failure | b_cell | *MT1F* | -10.98876472 | 0.001805079 | 0.008654124 | 1 |
| Heart_Failure | b_cell | *SH3BGRL2* | -1.507234874 | 0.003308739 | 0.01083435 | 1 |
| Heart_Failure | b_cell | *CNGA1* | -2.217329015 | 0.001081872 | 0.008654124 | 1 |
| Heart_Failure | b_cell | *MAFK* | -2.854551812 | 0.026561848 | 0.044782606 | 1 |
| Heart_Failure | b_cell | *C20orf203* | 4.557675212 | 0.000303569 | 0.004241191 | 1 |
| Heart_Failure | b_cell | *ZNF627* | -2.036907921 | 0.005004103 | 0.013660787 | 1 |
| Heart_Failure | b_cell | *ARC* | -13.62308877 | 0.001796976 | 0.008654124 | 1 |
| Heart_Failure | b_cell | *MMP17* | -3.415828399 | 0.026503575 | 0.044732342 | 1 |
| Heart_Failure | b_cell | *CCDC69* | -1.707757597 | 0.005679547 | 0.014784061 | 1 |
| Heart_Failure | b_cell | *ZNF534* | -6.942048595 | 0.01259329 | 0.025581265 | 1 |
| Heart_Failure | b_cell | *NCOA6* | -1.757949612 | 0.021891364 | 0.038558494 | 1 |
| Heart_Failure | b_cell | *FAM19A2* | -1.669661351 | 0.006318972 | 0.015865567 | 1 |
| Heart_Failure | b_cell | *TUSC1* | 10.56748999 | 0.00626541 | 0.015762559 | 1 |
| Heart_Failure | b_cell | *PAPSS2* | -1.364783745 | 0.00577958 | 0.014979918 | 1 |
| Heart_Failure | b_cell | *FAN1* | -1.950643494 | 0.004575464 | 0.012902652 | 1 |
| Heart_Failure | b_cell | *ANKRD13B* | -2.658221631 | 0.006305163 | 0.015843549 | 1 |
| Heart_Failure | b_cell | *ECI2* | -2.045507879 | 0.004859077 | 0.013391387 | 1 |
| Heart_Failure | b_cell | *APCDD1L* | 6.5694735 | 7.88E-05 | 0.002889125 | 0.495533823 |
| Heart_Failure | b_cell | *RCSD1* | 3.543436304 | 0.000622102 | 0.006609617 | 1 |
| Heart_Failure | b_cell | *RASSF9* | -1.811694139 | 0.001892245 | 0.008705907 | 1 |
| Heart_Failure | b_cell | *FAM169A* | 4.486463667 | 0.02542835 | 0.043277757 | 1 |
| Heart_Failure | b_cell | *TMEM184B* | -2.156880615 | 0.015437266 | 0.02975294 | 1 |
| Heart_Failure | b_cell | *SCAMP5* | -2.686487649 | 0.01177956 | 0.024301648 | 1 |
| Heart_Failure | b_cell | *ALPK2* | -3.070014832 | 0.000682536 | 0.007069364 | 1 |
| Heart_Failure | b_cell | *SFT2D1* | -2.176881258 | 0.015406551 | 0.029711959 | 1 |
| Heart_Failure | b_cell | *ARHGAP11A* | -3.743072273 | 0.001927384 | 0.008732736 | 1 |
| Heart_Failure | b_cell | *HMGN2* | -1.814353209 | 0.002914251 | 0.010128327 | 1 |

| | | | | | | |
|---|---|---|---|---|---|---|
| Heart_Failure | b_cell | *CD3E* | -2.655846195 | 0.003615267 | 0.011375969 | 1 |
| Heart_Failure | b_cell | *TSEN15* | -2.091918827 | 0.006335354 | 0.015886776 | 1 |
| Heart_Failure | b_cell | *ITPRIPL1* | 7.268536831 | 4.00E-05 | 0.002889125 | 0.251546344 |
| Heart_Failure | b_cell | *PRMT6* | 4.296591307 | 0.000216873 | 0.003645673 | 1 |
| Heart_Failure | b_cell | *SHISA4* | -5.534604076 | 0.002369562 | 0.009308722 | 1 |
| Heart_Failure | b_cell | *MAP3K3* | -1.849634753 | 0.029952981 | 0.049322785 | 1 |
| Heart_Failure | b_cell | *C1orf174* | -2.689019828 | 0.004887503 | 0.013441703 | 1 |
| Heart_Failure | b_cell | *RASGEF1A* | -5.812493342 | 0.001738999 | 0.008654124 | 1 |
| Heart_Failure | b_cell | *DCLRE1A* | 7.104860948 | 0.008601222 | 0.019399557 | 1 |
| Heart_Failure | b_cell | *APRT* | 7.101505142 | 0.000204057 | 0.003524466 | 1 |
| Heart_Failure | b_cell | *TBKBP1* | -2.729499008 | 0.003469059 | 0.011112537 | 1 |
| Heart_Failure | b_cell | *ZFP2* | -2.293957267 | 0.016453081 | 0.03111929 | 1 |
| Heart_Failure | b_cell | *L3MBTL3* | 4.92793446 | 3.05E-05 | 0.002889125 | 0.191754006 |
| Heart_Failure | b_cell | *MFAP3L* | -3.616172599 | 0.006018083 | 0.015361627 | 1 |
| Heart_Failure | b_cell | *NAGA* | -2.148300067 | 0.004434974 | 0.012718545 | 1 |
| Heart_Failure | b_cell | *SMG5* | -2.322614967 | 0.023348391 | 0.040516515 | 1 |
| Heart_Failure | b_cell | *TGM2* | -1.306762579 | 0.026060179 | 0.04410238 | 1 |
| Heart_Failure | b_cell | *AL590822.1* | -2.079815235 | 0.007465212 | 0.017644281 | 1 |
| Heart_Failure | b_cell | *AP001007.1* | 2.434139976 | 0.000181665 | 0.00338933 | 1 |
| Heart_Failure | b_cell | *AC105020.1* | 5.42413492 | 0.00012642 | 0.003179208 | 0.794802121 |
| Heart_Failure | b_cell | *EFCAB2* | -1.712078413 | 0.02834609 | 0.047208442 | 1 |
| Heart_Failure | b_cell | *CHML* | 3.162518359 | 0.000313934 | 0.0043094 | 1 |
| Heart_Failure | b_cell | *CAPN8* | -6.335843723 | 0.018020493 | 0.033275421 | 1 |
| Heart_Failure | b_cell | *ECT2L* | 7.372098419 | 4.34E-05 | 0.002889125 | 0.272837793 |
| Heart_Failure | b_cell | *CENPW* | -3.48590339 | 0.016297592 | 0.030871637 | 1 |
| Heart_Failure | b_cell | *DDO* | -3.351568765 | 0.002355716 | 0.009268076 | 1 |
| Heart_Failure | b_cell | *HIST2H2BF* | -8.505817521 | 0.002200113 | 0.009052426 | 1 |
| Heart_Failure | b_cell | *RBM20* | -2.652761672 | 0.001398989 | 0.008654124 | 1 |
| Heart_Failure | b_cell | *C6orf163* | -3.069278806 | 0.001066613 | 0.008654124 | 1 |
| Heart_Failure | b_cell | *PCMTD2* | -1.768051675 | 0.006682171 | 0.016384872 | 1 |
| Heart_Failure | b_cell | *OOEP* | 2.28398653 | 6.68E-05 | 0.002889125 | 0.42015643 |
| Heart_Failure | b_cell | *FOXO6* | 5.777309214 | 0.0034335 | 0.01106179 | 1 |
| Heart_Failure | b_cell | *NEU4* | -26.79808607 | 0.000175563 | 0.003366185 | 1 |
| Heart_Failure | b_cell | *TRAF3IP1* | -1.885978131 | 0.008346836 | 0.019020136 | 1 |
| Heart_Failure | b_cell | *ASAH2B* | -2.363537851 | 0.010341286 | 0.02216321 | 1 |
| Heart_Failure | b_cell | *C10orf128* | -1.439721997 | 0.006990625 | 0.016916883 | 1 |
| Heart_Failure | b_cell | *SYT15* | -7.329820226 | 0.001823543 | 0.008654124 | 1 |
| Heart_Failure | b_cell | *ZDBF2* | -2.489382981 | 0.011568113 | 0.024002881 | 1 |
| Heart_Failure | b_cell | *RING1* | -2.262180254 | 0.007586018 | 0.017826138 | 1 |
| Heart_Failure | b_cell | *OXLD1* | -2.629029439 | 0.020894341 | 0.037118599 | 1 |
| Heart_Failure | b_cell | *HLA-DOA* | 0.774075338 | 0.000620772 | 0.006609617 | 1 |
| Heart_Failure | b_cell | *HLA-DMA* | 5.426836529 | 0.001916104 | 0.008705907 | 1 |
| Heart_Failure | b_cell | *HLA-DRA* | 5.273045813 | 4.54E-09 | 9.51E-06 | 2.85E-05 |
| Heart_Failure | b_cell | *COL15A1* | -1.468334312 | 0.006418035 | 0.015978447 | 1 |
| Heart_Failure | b_cell | *MRPL38* | 2.656130713 | 0.002458172 | 0.00942924 | 1 |
| Heart_Failure | b_cell | *SKIV2L* | -1.362820972 | 0.006108443 | 0.015479154 | 1 |
| Heart_Failure | b_cell | *LAYN* | -4.079711338 | 0.000413152 | 0.005033892 | 1 |
| Heart_Failure | b_cell | *C6orf48* | 5.03119493 | 0.009617969 | 0.020995893 | 1 |
| Heart_Failure | b_cell | *CARD16* | 5.394969649 | 0.000144983 | 0.003239931 | 0.911508064 |
| Heart_Failure | b_cell | *MSH5* | -3.10428555 | 0.028692853 | 0.04763453 | 1 |

| | | | | | | |
|---|---|---|---|---|---|---|
| Heart_Failure | b_cell | *ABHD16A* | 2.059006485 | 0.000161562 | 0.003294027 | 1 |
| Heart_Failure | b_cell | *BAG6* | 3.729113494 | 0.017950058 | 0.033162508 | 1 |
| Heart_Failure | b_cell | *ZNF814* | -1.783933124 | 0.008062026 | 0.018566285 | 1 |
| Heart_Failure | b_cell | *MICB* | 2.430844267 | 0.000554088 | 0.006204391 | 1 |
| Heart_Failure | b_cell | *HLA-C* | -2.690759472 | 0.002626231 | 0.009653089 | 1 |
| Heart_Failure | b_cell | *CCHCR1* | -2.848385478 | 9.41E-06 | 0.002447238 | 0.05914081 |
| Heart_Failure | b_cell | *LILRB3* | -2.140130975 | 0.004277774 | 0.012422339 | 1 |
| Heart_Failure | b_cell | *LINC01123* | -76.60770834 | 0.008475068 | 0.01921484 | 1 |
| Heart_Failure | b_cell | *HLA-E* | -2.032992309 | 0.005320063 | 0.014184578 | 1 |
| Heart_Failure | b_cell | *ZNF468* | -3.418477614 | 0.00185358 | 0.008683648 | 1 |
| Heart_Failure | b_cell | *FAM153C* | 2.476229345 | 5.21E-05 | 0.002889125 | 0.327829526 |
| Heart_Failure | b_cell | *TRIM27* | -1.91817554 | 0.010315606 | 0.022142102 | 1 |
| Heart_Failure | b_cell | *MALRD1* | -1.527588849 | 0.000924216 | 0.008557508 | 1 |
| Heart_Failure | b_cell | *MROH6* | -3.264629644 | 0.009742398 | 0.021208607 | 1 |
| Heart_Failure | b_cell | *ZNF155* | -2.350047354 | 0.018076653 | 0.033357181 | 1 |
| Heart_Failure | b_cell | *LRRC10B* | -22.41741716 | 0.009819383 | 0.021309791 | 1 |
| Heart_Failure | b_cell | *PCDHGA1* | -4.533087328 | 0.007754509 | 0.018069904 | 1 |
| Heart_Failure | b_cell | *PCDHA4* | -14.07293783 | 0.000534754 | 0.006057654 | 1 |
| Heart_Failure | b_cell | *SPIRE2* | -2.139275883 | 0.012771129 | 0.025842321 | 1 |
| Heart_Failure | b_cell | *SLFN12L* | -1.802209981 | 0.000408906 | 0.005022023 | 1 |
| Heart_Failure | b_cell | *SYCE1L* | 4.151610401 | 0.001199126 | 0.008654124 | 1 |
| Heart_Failure | b_cell | *TMEM231* | -2.443600198 | 0.01078884 | 0.02291535 | 1 |
| Heart_Failure | b_cell | *FAM71F2* | -2.32710196 | 0.002400673 | 0.009333474 | 1 |
| Heart_Failure | b_cell | *FAM205A* | -28.88536258 | 0.000758732 | 0.007595773 | 1 |
| Heart_Failure | b_cell | *LINC00654* | -4.840167144 | 0.004542654 | 0.01284469 | 1 |
| Heart_Failure | b_cell | *TMEM170B* | -1.214325001 | 0.006755207 | 0.016506018 | 1 |
| Heart_Failure | b_cell | *NT5M* | -2.026879776 | 0.018487146 | 0.033895797 | 1 |
| Heart_Failure | b_cell | *PRR13* | -3.217845347 | 0.030019784 | 0.049393976 | 1 |
| Heart_Failure | b_cell | *MT1A* | -6.399338009 | 0.001754943 | 0.008654124 | 1 |
| Heart_Failure | b_cell | *MT1M* | -3.028012588 | 0.001708289 | 0.008654124 | 1 |
| Heart_Failure | b_cell | *CFI* | -1.858824131 | 0.001765946 | 0.008654124 | 1 |
| Heart_Failure | b_cell | *SAMD9* | 5.398861517 | 0.00794977 | 0.018388596 | 1 |
| Heart_Failure | b_cell | *EXOC3L4* | -4.529237186 | 0.003298873 | 0.01083435 | 1 |
| Heart_Failure | b_cell | *TP53TG3D* | 4.018387398 | 0.000442619 | 0.005211133 | 1 |
| Heart_Failure | b_cell | *ATP6AP1L* | -2.045844306 | 0.005399841 | 0.014312311 | 1 |
| Heart_Failure | b_cell | *CPT1B* | -3.296009635 | 0.023821832 | 0.041167636 | 1 |
| Heart_Failure | b_cell | *SERF1B* | -9.889033069 | 0.000139122 | 0.003214256 | 0.874662182 |
| Heart_Failure | b_cell | *EIF3CL* | -6.79711889 | 0.00598491 | 0.015320492 | 1 |
| Heart_Failure | b_cell | *LCMT1* | -2.090151345 | 0.013292277 | 0.026643059 | 1 |
| Heart_Failure | b_cell | *LIN52* | -1.488907595 | 0.025385439 | 0.043239842 | 1 |
| Heart_Failure | b_cell | *C17orf107* | 2.035521996 | 0.006462111 | 0.016026545 | 1 |
| Heart_Failure | b_cell | *C1QTNF9B* | -32.4685681 | 0.001604104 | 0.008654124 | 1 |
| Heart_Failure | b_cell | *NYNRIN* | -3.619937146 | 0.009265391 | 0.020367663 | 1 |
| Heart_Failure | b_cell | *ZDHHC11B* | -2.293592502 | 0.006543925 | 0.016162624 | 1 |
| Heart_Failure | b_cell | *GOLGA8O* | 11.66795441 | 0.000963797 | 0.008654124 | 1 |
| Heart_Failure | b_cell | *LNP1* | -2.841411403 | 0.001278869 | 0.008654124 | 1 |
| Heart_Failure | b_cell | *VGLL3* | -1.01681765 | 0.006648753 | 0.016328403 | 1 |
| Heart_Failure | b_cell | *COLQ* | -2.102102799 | 0.003884967 | 0.011827985 | 1 |
| Heart_Failure | b_cell | *METTL6* | -2.143127263 | 0.025520464 | 0.043422776 | 1 |
| Heart_Failure | b_cell | *SLC48A1* | -1.99212548 | 0.001121354 | 0.008654124 | 1 |

| | | | | | | |
|---|---|---|---|---|---|---|
| Heart_Failure | b_cell | *TAS2R14* | 4.894921866 | 7.93E-05 | 0.002889125 | 0.49868921 |
| Heart_Failure | b_cell | *ZRSR1* | -13.36219583 | 0.001502082 | 0.008654124 | 1 |
| Heart_Failure | b_cell | *AC084018.1* | -3.072765147 | 0.000965214 | 0.008654124 | 1 |
| Heart_Failure | b_cell | *C17orf51* | -3.21052556 | 0.001321058 | 0.008654124 | 1 |
| Heart_Failure | b_cell | *OR2A42* | -3.877795889 | 0.00221367 | 0.009081423 | 1 |
| Heart_Failure | b_cell | *RNF208* | -6.84310555 | 0.001723608 | 0.008654124 | 1 |
| Heart_Failure | b_cell | *MAP10* | 7.795585734 | 0.003140361 | 0.010529839 | 1 |
| Heart_Failure | b_cell | *NUP62* | -2.908235731 | 0.008404697 | 0.019110428 | 1 |
| Heart_Failure | b_cell | *FGFR1OP* | -2.626475719 | 0.011721332 | 0.024224858 | 1 |
| Heart_Failure | b_cell | *ZNF254* | -1.901065292 | 0.023580724 | 0.040807049 | 1 |
| Heart_Failure | b_cell | *TCTEX1D2* | -3.14522663 | 0.016996446 | 0.031793814 | 1 |
| Heart_Failure | b_cell | *GIMAP1* | 5.030794618 | 0.021856773 | 0.038512762 | 1 |
| Heart_Failure | b_cell | *NOTCH2NL* | 4.442978994 | 0.01113883 | 0.023408158 | 1 |
| Heart_Failure | b_cell | *GSTM2* | -1.786720443 | 0.008623158 | 0.019424505 | 1 |
| Heart_Failure | b_cell | *ARHGAP19* | 8.931925847 | 0.00153965 | 0.008654124 | 1 |
| Heart_Failure | b_cell | *LCAT* | -1.905413436 | 0.003072148 | 0.01036364 | 1 |
| Heart_Failure | b_cell | *SIPA1* | 5.046984006 | 0.001084131 | 0.008654124 | 1 |
| Heart_Failure | b_cell | *RBMXL1* | -2.742418223 | 0.002188132 | 0.009035107 | 1 |
| Heart_Failure | b_cell | *SRA1* | -2.030270484 | 0.010377069 | 0.022223445 | 1 |
| Heart_Failure | b_cell | *TMEM110* | -7.517032567 | 0.000908723 | 0.008476467 | 1 |
| Heart_Failure | b_cell | *LEPROT* | -2.057778789 | 0.019450849 | 0.035292204 | 1 |
| Heart_Failure | b_cell | *LBH* | 2.959605316 | 7.27E-05 | 0.002889125 | 0.457072056 |
| Heart_Failure | b_cell | *NCKIPSD* | -2.440981843 | 0.006678669 | 0.016382674 | 1 |
| Heart_Failure | b_cell | *SLC35F6* | -3.082231743 | 0.015488253 | 0.029796403 | 1 |
| Heart_Failure | b_cell | *ATP6V1G2* | 2.624016393 | 7.48E-06 | 0.002102517 | 0.046995787 |
| Heart_Failure | b_cell | *EMP2* | -1.71930465 | 0.004923129 | 0.01349375 | 1 |
| Heart_Failure | b_cell | *LTB4R2* | -4.387929873 | 0.019857802 | 0.03581354 | 1 |
| Heart_Failure | b_cell | *GALT* | -2.25560124 | 0.012664177 | 0.025683765 | 1 |
| Heart_Failure | b_cell | *ITGA1* | -1.136873662 | 0.001997659 | 0.008781237 | 1 |
| Heart_Failure | b_cell | *NUDT19* | -2.368374193 | 0.002839972 | 0.010002746 | 1 |
| Heart_Failure | b_cell | *ZNF726* | -2.804890529 | 0.001396029 | 0.008654124 | 1 |
| Heart_Failure | b_cell | *TAX1BP3* | -3.114359781 | 0.000872606 | 0.008337498 | 1 |
| Heart_Failure | b_cell | *AP1G2* | 4.98026918 | 4.20E-05 | 0.002889125 | 0.264250366 |
| Heart_Failure | b_cell | *ZNF90* | 5.681754558 | 4.18E-05 | 0.002889125 | 0.263009821 |
| Heart_Failure | b_cell | *TM6SF2* | -36.70939114 | 0.002165674 | 0.009019688 | 1 |
| Heart_Failure | b_cell | *REPIN1* | -2.048783844 | 0.00279146 | 0.009915204 | 1 |
| Heart_Failure | b_cell | *SMIM7* | -2.038204366 | 0.027337897 | 0.045820678 | 1 |
| Heart_Failure | b_cell | *TSPAN4* | -1.884165414 | 0.005397782 | 0.014312311 | 1 |
| Heart_Failure | b_cell | *CPNE1* | -3.043626987 | 0.000869974 | 0.008325007 | 1 |
| Heart_Failure | b_cell | *ARL16* | -1.202245573 | 0.011560656 | 0.023995328 | 1 |
| Heart_Failure | b_cell | *WEE2* | -2.618086308 | 0.010405703 | 0.022267071 | 1 |
| Heart_Failure | b_cell | *PAXIP1-AS2* | -2.484554818 | 0.014476065 | 0.028405437 | 1 |
| Heart_Failure | b_cell | *PRCD* | -2.531144364 | 0.002625164 | 0.009653089 | 1 |
| Heart_Failure | b_cell | *C17orf67* | -1.809702071 | 0.002858128 | 0.01001558 | 1 |
| Heart_Failure | b_cell | *COLCA2* | -3.220148298 | 0.002768558 | 0.009906851 | 1 |
| Heart_Failure | b_cell | *NEURL1B* | -2.111134324 | 0.001396631 | 0.008654124 | 1 |
| Heart_Failure | b_cell | *VSTM5* | 2.451950179 | 0.002351917 | 0.009268076 | 1 |
| Heart_Failure | b_cell | *STARD10* | 3.049986 | 0.001387161 | 0.008654124 | 1 |
| Heart_Failure | b_cell | *NUTM2D* | -7.359386472 | 0.020705053 | 0.036855229 | 1 |
| Heart_Failure | b_cell | *CPEB1* | -1.92476813 | 0.001251143 | 0.008654124 | 1 |

| | | | | | | |
|---|---|---|---|---|---|---|
| Heart_Failure | b_cell | *C10orf105* | -4.698129209 | 0.003074865 | 0.01036364 | 1 |
| Heart_Failure | b_cell | *AC005082.1* | 9.118866313 | 0.000368092 | 0.004761711 | 1 |
| Heart_Failure | b_cell | *COL28A1* | -2.847345799 | 0.003648442 | 0.011427162 | 1 |
| Heart_Failure | b_cell | *PHB2* | 6.434030635 | 0.01018109 | 0.021958323 | 1 |
| Heart_Failure | b_cell | *GRID2IP* | -5.138192966 | 0.00709563 | 0.017078953 | 1 |
| Heart_Failure | b_cell | *UBE2QL1* | -3.741629561 | 0.001706341 | 0.008654124 | 1 |
| Heart_Failure | b_cell | *GOLGA8B* | -2.067107211 | 0.007452418 | 0.017643448 | 1 |
| Heart_Failure | b_cell | *KCNU1* | -3.763023111 | 0.00015692 | 0.003255968 | 0.986558202 |
| Heart_Failure | b_cell | *MYL5* | -3.706600641 | 0.002052384 | 0.008831917 | 1 |
| Heart_Failure | b_cell | *NPEPL1* | -2.673030613 | 0.014356792 | 0.028215427 | 1 |
| Heart_Failure | b_cell | *LINC01139* | -3.807799514 | 0.015022105 | 0.029189286 | 1 |
| Heart_Failure | b_cell | *TSTD1* | -2.501994682 | 0.003275202 | 0.010803355 | 1 |
| Heart_Failure | b_cell | *LINC01138* | -1.979940764 | 0.007624003 | 0.017871778 | 1 |
| Heart_Failure | b_cell | *CYB5RL* | 4.108699124 | 9.88E-06 | 0.002447238 | 0.062130668 |
| Heart_Failure | b_cell | *ATAD3C* | 1.313065162 | 0.000284776 | 0.004036731 | 1 |
| Heart_Failure | b_cell | *CCDC7* | -1.724820348 | 0.01342405 | 0.026825962 | 1 |
| Heart_Failure | b_cell | *PAM16* | 7.161912543 | 0.000391919 | 0.004918151 | 1 |
| Heart_Failure | b_cell | *AC016773.1* | 2.870953568 | 0.000263111 | 0.003832809 | 1 |
| Heart_Failure | b_cell | *RNASEK* | -2.653964065 | 0.018126529 | 0.033414978 | 1 |
| Heart_Failure | b_cell | *RNU6ATAC35I* | -12.8334112 | 0.00183003 | 0.008654124 | 1 |
| Heart_Failure | b_cell | *PPP3R1* | 4.32607202 | 5.81E-05 | 0.002889125 | 0.365167213 |
| Heart_Failure | b_cell | *FANCG* | 1.266825512 | 0.000333098 | 0.004459106 | 1 |
| Heart_Failure | b_cell | *C19orf73* | -17.1266779 | 0.003888598 | 0.011833309 | 1 |
| Heart_Failure | b_cell | *OR2A25* | -79.77556006 | 0.014387398 | 0.028257911 | 1 |
| Heart_Failure | b_cell | *XKR9* | -2.378703571 | 0.01184672 | 0.024411777 | 1 |
| Heart_Failure | b_cell | *C1orf229* | -5.772545115 | 0.003643634 | 0.011427162 | 1 |
| Heart_Failure | b_cell | *SLC12A8* | -2.530975341 | 0.000709791 | 0.0072916 | 1 |
| Heart_Failure | b_cell | *APOL6* | -1.246811096 | 0.001690046 | 0.008654124 | 1 |
| Heart_Failure | b_cell | *TIAF1* | -3.657657571 | 0.020869267 | 0.037084534 | 1 |
| Heart_Failure | b_cell | *BTBD19* | -2.036645273 | 0.003052558 | 0.010340715 | 1 |
| Heart_Failure | b_cell | *AC062017.1* | -2.151470924 | 0.001388361 | 0.008654124 | 1 |
| Heart_Failure | b_cell | *EXOSC6* | -2.292042196 | 0.004646339 | 0.013040864 | 1 |
| Heart_Failure | b_cell | *VPS52* | 5.748970924 | 0.003749011 | 0.011556943 | 1 |
| Heart_Failure | b_cell | *TINCR* | 2.956438868 | 0.000245706 | 0.003766552 | 1 |
| Heart_Failure | b_cell | *LINC00205* | -3.834594081 | 0.00050417 | 0.005763125 | 1 |
| Heart_Failure | b_cell | *AC147651.3* | -7.076304977 | 0.000933746 | 0.008607719 | 1 |
| Heart_Failure | b_cell | *HLA-DPB1* | 2.787324828 | 0.001725397 | 0.008654124 | 1 |
| Heart_Failure | b_cell | *OSER1-AS1* | -2.211376641 | 0.001439522 | 0.008654124 | 1 |
| Heart_Failure | b_cell | *LINC01133* | -1.786344254 | 0.001646854 | 0.008654124 | 1 |
| Heart_Failure | b_cell | *LINC01117* | -1.876464639 | 0.002512339 | 0.00949794 | 1 |
| Heart_Failure | b_cell | *RASAL2-AS1* | 1.781686959 | 0.000185048 | 0.00340015 | 1 |
| Heart_Failure | b_cell | *LINC00240* | -1.60484971 | 0.002047468 | 0.008831917 | 1 |
| Heart_Failure | b_cell | *LINC00184* | -6.469586248 | 0.001058523 | 0.008654124 | 1 |
| Heart_Failure | b_cell | *TMEM233* | -1.976805863 | 0.001106268 | 0.008654124 | 1 |
| Heart_Failure | b_cell | *AL049840.1* | -2.229854238 | 0.001203328 | 0.008654124 | 1 |
| Heart_Failure | b_cell | *LINC00237* | -12.41760657 | 0.001734617 | 0.008654124 | 1 |
| Heart_Failure | b_cell | *PLEKHM1* | -2.136148789 | 0.014051997 | 0.027781417 | 1 |
| Heart_Failure | b_cell | *AP000350.6* | 3.302746859 | 3.11E-05 | 0.002889125 | 0.195499097 |
| Heart_Failure | b_cell | *SFTA1P* | -9.555142619 | 0.001127798 | 0.008654124 | 1 |
| Heart_Failure | b_cell | *SLC26A6* | 6.406444958 | 0.000117232 | 0.003135985 | 0.737039244 |

| | | | | | | |
|---|---|---|---|---|---|---|
| Heart_Failure | b_cell | *LINC00626* | 73.77381623 | 0.007160758 | 0.017176525 | 1 |
| Heart_Failure | b_cell | *ERCC6* | -2.082067691 | 0.024208636 | 0.041687125 | 1 |
| Heart_Failure | b_cell | *ELFN1* | -8.749061003 | 0.003488986 | 0.011132829 | 1 |
| Heart_Failure | b_cell | *ZNF503-AS1* | 5.454136924 | 5.69E-05 | 0.002889125 | 0.357851576 |
| Heart_Failure | b_cell | *FTCDNL1* | -1.484331866 | 0.00144157 | 0.008654124 | 1 |
| Heart_Failure | b_cell | *AC064836.3* | 4.141626106 | 0.000104045 | 0.00302839 | 0.654132252 |
| Heart_Failure | b_cell | *TMEM185B* | -5.13405086 | 0.000424079 | 0.005097871 | 1 |
| Heart_Failure | b_cell | *HSBP1L1* | -3.5651482 | 0.025726566 | 0.043702491 | 1 |
| Heart_Failure | b_cell | *TAS2R46* | 2.662081589 | 0.002295239 | 0.009185338 | 1 |
| Heart_Failure | b_cell | *AC093901.1* | 1.145988189 | 0.000235727 | 0.003705045 | 1 |
| Heart_Failure | b_cell | *C14orf132* | -1.890080123 | 0.002547776 | 0.009545808 | 1 |
| Heart_Failure | b_cell | *AC009299.3* | -2.805143918 | 0.008868373 | 0.019757427 | 1 |
| Heart_Failure | b_cell | *AC108463.2* | -7.148737945 | 0.001502744 | 0.008654124 | 1 |
| Heart_Failure | b_cell | *HCG20* | -6.81274748 | 0.011010753 | 0.023229733 | 1 |
| Heart_Failure | b_cell | *C19orf24* | -3.590674138 | 0.000852141 | 0.008216886 | 1 |
| Heart_Failure | b_cell | *LINC01013* | 2.874635092 | 0.000121318 | 0.003135985 | 0.762724693 |
| Heart_Failure | b_cell | *AC073133.2* | -28.04032906 | 0.008797819 | 0.019698135 | 1 |
| Heart_Failure | b_cell | *ARL17B* | -1.779194933 | 0.003499885 | 0.011147503 | 1 |
| Heart_Failure | b_cell | *RPL41* | 2.824245007 | 6.08E-05 | 0.002889125 | 0.382471472 |
| Heart_Failure | b_cell | *PATL2* | 4.465089951 | 4.50E-05 | 0.002889125 | 0.282739717 |
| Heart_Failure | b_cell | *ZNF492* | -4.156547033 | 0.001793688 | 0.008654124 | 1 |
| Heart_Failure | b_cell | *U91324.1* | -6.230198556 | 0.00167957 | 0.008654124 | 1 |
| Heart_Failure | b_cell | *CISD3* | -2.913610893 | 0.002221289 | 0.009081423 | 1 |
| Heart_Failure | b_cell | *PSMG3-AS1* | -3.162845174 | 0.006247238 | 0.015735731 | 1 |
| Heart_Failure | b_cell | *PPP5D1* | 5.665712607 | 0.011386228 | 0.023766672 | 1 |
| Heart_Failure | b_cell | *AC112229.4* | 1.511637151 | 0.012401515 | 0.025289758 | 1 |
| Heart_Failure | b_cell | *KLHL7-AS1* | 5.017839214 | 9.76E-05 | 0.002978205 | 0.613510234 |
| Heart_Failure | b_cell | *AC092171.4* | -4.422056847 | 0.022457781 | 0.039340226 | 1 |
| Heart_Failure | b_cell | *HLA-DPA1* | 4.731901806 | 0.000119916 | 0.003135985 | 0.753910489 |
| Heart_Failure | b_cell | *RPS18* | 5.069972357 | 0.000204634 | 0.003524752 | 1 |
| Heart_Failure | b_cell | *DIRC3* | -0.942954484 | 0.001598334 | 0.008654124 | 1 |
| Heart_Failure | b_cell | *AC107021.1* | -1.266437524 | 0.002570145 | 0.009560634 | 1 |
| Heart_Failure | b_cell | *LINC00865* | -4.908239215 | 0.016529603 | 0.031217066 | 1 |
| Heart_Failure | b_cell | *AC009495.3* | 8.273893747 | 0.000193064 | 0.003449722 | 1 |
| Heart_Failure | b_cell | *HLA-DQB2* | 3.535003897 | 2.26E-15 | 1.42E-11 | 1.42E-11 |
| Heart_Failure | b_cell | *LINC00342* | -2.089293223 | 0.001811239 | 0.008654124 | 1 |
| Heart_Failure | b_cell | *SNHG15* | 8.632581638 | 0.000704114 | 0.007256988 | 1 |
| Heart_Failure | b_cell | *LINC00702* | -2.635677315 | 0.001113176 | 0.008654124 | 1 |
| Heart_Failure | b_cell | *LINC00472* | -1.85944413 | 0.001508469 | 0.008654124 | 1 |
| Heart_Failure | b_cell | *RPS28* | -2.114413973 | 0.001677759 | 0.008654124 | 1 |
| Heart_Failure | b_cell | *AC123768.3* | -2.799425141 | 0.001761781 | 0.008654124 | 1 |
| Heart_Failure | b_cell | *PINLYP* | -1.742315234 | 0.021501773 | 0.038012258 | 1 |
| Heart_Failure | b_cell | *C19orf81* | -7.839976056 | 0.025105844 | 0.042844854 | 1 |
| Heart_Failure | b_cell | *ANKRD65* | -4.023191435 | 0.024219722 | 0.041688301 | 1 |
| Heart_Failure | b_cell | *LINC00957* | -6.680204293 | 0.001528203 | 0.008654124 | 1 |
| Heart_Failure | b_cell | *LINC00484* | -1.361156763 | 0.025639838 | 0.043590497 | 1 |
| Heart_Failure | b_cell | *KIAA0040* | 5.233097287 | 3.90E-05 | 0.002889125 | 0.245407523 |
| Heart_Failure | b_cell | *B3GALT4* | 5.615082031 | 0.005667402 | 0.014766249 | 1 |
| Heart_Failure | b_cell | *LINC00941* | 1.860010722 | 0.000194958 | 0.003449722 | 1 |
| Heart_Failure | b_cell | *ZBTB22* | 1.761874206 | 1.22E-05 | 0.002644519 | 0.07669105 |

| | | | | | | |
|---|---|---|---|---|---|---|
| Heart_Failure | b_cell | *B3GNT9* | -5.942522172 | 0.000571504 | 0.006303585 | 1 |
| Heart_Failure | b_cell | *CDKN2AIPNL* | -2.44670034 | 0.001499485 | 0.008654124 | 1 |
| Heart_Failure | b_cell | *GS1-124K5.4* | 1.280163642 | 0.01366771 | 0.027184085 | 1 |
| Heart_Failure | b_cell | *HLA-DQA2* | 3.21116684 | 1.75E-14 | 5.51E-11 | 1.10E-10 |
| Heart_Failure | b_cell | *AC073257.2* | -21.25908701 | 0.011116277 | 0.023394033 | 1 |
| Heart_Failure | b_cell | *IRGM* | 1.299913344 | 9.70E-05 | 0.002978205 | 0.609700771 |
| Heart_Failure | b_cell | *TXNDC5* | -7.866757181 | 0.001783672 | 0.008654124 | 1 |
| Heart_Failure | b_cell | *CDRT4* | -4.537950427 | 0.012181697 | 0.024946687 | 1 |
| Heart_Failure | b_cell | *LILRA4* | 17.78006416 | 6.05E-05 | 0.002889125 | 0.380232554 |
| Heart_Failure | b_cell | *AQP1* | -2.082749535 | 0.002718686 | 0.009828026 | 1 |
| Heart_Failure | b_cell | *C1QTNF9* | -9.171842212 | 0.016674124 | 0.031361554 | 1 |
| Heart_Failure | b_cell | *ARHGEF25* | 6.252102954 | 0.002554248 | 0.009552978 | 1 |
| Heart_Failure | b_cell | *HLA-DOB* | 0.578250603 | 7.81E-05 | 0.002889125 | 0.490888438 |
| Heart_Failure | b_cell | *TDGF1* | -6.103099281 | 0.00215589 | 0.009019688 | 1 |
| Heart_Failure | b_cell | *ATP5J2* | -2.382456831 | 0.003912341 | 0.011873672 | 1 |
| Heart_Failure | b_cell | *ARHGAP8* | -4.627769477 | 0.001758085 | 0.008654124 | 1 |
| Heart_Failure | b_cell | *ATP5O* | 3.413475188 | 0.000276892 | 0.003983564 | 1 |
| Heart_Failure | b_cell | *PLEKHO2* | -3.357335549 | 0.020198459 | 0.036220112 | 1 |
| Heart_Failure | b_cell | *PWP2* | -13.44114167 | 0.000396207 | 0.004942367 | 1 |
| Heart_Failure | b_cell | *AKAP2* | -8.444450267 | 0.001777411 | 0.008654124 | 1 |
| Heart_Failure | b_cell | *ARHGDIG* | -4.637156567 | 0.000538319 | 0.006083376 | 1 |
| Heart_Failure | b_cell | *EIF6* | -2.716089468 | 0.000350713 | 0.004632212 | 1 |
| Heart_Failure | b_cell | *MRPL20* | -2.449775224 | 0.024977939 | 0.042661315 | 1 |
| Heart_Failure | b_cell | *AP5Z1* | -1.942141598 | 0.003221952 | 0.010690838 | 1 |
| Heart_Failure | b_cell | *PSG4* | -3.86793442 | 0.005343409 | 0.01421623 | 1 |
| Heart_Failure | b_cell | *STON1* | -1.794187709 | 0.00974829 | 0.021211134 | 1 |
| Heart_Failure | b_cell | *FGF14-IT1* | -7.287570476 | 0.001690938 | 0.008654124 | 1 |
| Heart_Failure | b_cell | *TICAM2* | -6.532352218 | 0.016660549 | 0.031361554 | 1 |
| Heart_Failure | b_cell | *PALM2* | -1.842865033 | 0.000520914 | 0.005922221 | 1 |
| Heart_Failure | b_cell | *NAT6* | -2.709409031 | 0.016215044 | 0.030752333 | 1 |
| Heart_Failure | b_cell | *UPK3B* | -6.363483497 | 0.002250855 | 0.00912976 | 1 |
| Heart_Failure | b_cell | *WDR92* | -2.825591874 | 0.026804962 | 0.045097908 | 1 |
| Heart_Failure | b_cell | *NME1-NME2* | -7.590187496 | 0.006862167 | 0.016689532 | 1 |
| Heart_Failure | b_cell | *TTC4* | -2.840003531 | 1.10E-05 | 0.002552674 | 0.068922188 |
| Heart_Failure | b_cell | *JMJD7* | 4.052672273 | 2.68E-05 | 0.002889125 | 0.168486709 |
| Heart_Failure | b_cell | *APOBEC3D* | 2.999558897 | 0.000311639 | 0.004287253 | 1 |
| Heart_Failure | b_cell | *LINC01011* | -3.74609353 | 0.00458674 | 0.012913944 | 1 |
| Heart_Failure | b_cell | *TMEM199* | -2.73203354 | 0.002461076 | 0.009434624 | 1 |
| Heart_Failure | b_cell | *P2RY11* | 3.939011078 | 0.000150663 | 0.003239931 | 0.947220205 |
| Heart_Failure | b_cell | *APOBEC3C* | 6.186766787 | 4.36E-05 | 0.002889125 | 0.274209871 |
| Heart_Failure | b_cell | *CCDC13* | -2.23901633 | 0.008194033 | 0.018787704 | 1 |
| Heart_Failure | b_cell | *ASPRV1* | -8.958026404 | 0.001498941 | 0.008654124 | 1 |
| Heart_Failure | b_cell | *OR2AE1* | 3.751655922 | 0.000212534 | 0.003591933 | 1 |
| Heart_Failure | b_cell | *LINC00847* | 3.409214536 | 7.34E-05 | 0.002889125 | 0.461602158 |
| Heart_Failure | b_cell | *NEAT1* | -1.082408405 | 0.012898695 | 0.026041778 | 1 |
| Heart_Failure | b_cell | *ZNF585B* | -2.227772846 | 0.006838982 | 0.016658924 | 1 |
| Heart_Failure | b_cell | *LINC00920* | 9.319458667 | 0.000387751 | 0.004897883 | 1 |
| Heart_Failure | b_cell | *PGAM5* | 9.253410882 | 7.12E-05 | 0.002889125 | 0.447808873 |
| Heart_Failure | b_cell | *DNAJC3-AS1* | -2.896351959 | 0.006407976 | 0.015967875 | 1 |
| Heart_Failure | b_cell | *LINC00926* | 0.324496621 | 0.000208295 | 0.003568265 | 1 |

| | | | | | | |
|---|---|---|---|---|---|---|
| Heart_Failure | b_cell | *BCKDHA* | -6.054264596 | 0.010632112 | 0.022666696 | 1 |
| Heart_Failure | b_cell | *PCDHAC1* | 3.377539279 | 0.000721529 | 0.007316224 | 1 |
| Heart_Failure | b_cell | *ABHD14A* | -2.104315065 | 0.00255284 | 0.009552978 | 1 |
| Heart_Failure | b_cell | *CCDC153* | -3.033936267 | 0.009424016 | 0.020636987 | 1 |
| Heart_Failure | b_cell | *C5orf17* | -1.953465598 | 0.002564376 | 0.009560634 | 1 |
| Heart_Failure | b_cell | *FMN1* | -1.258329716 | 0.001983678 | 0.008775328 | 1 |
| Heart_Failure | b_cell | *NAIP* | -3.047147628 | 0.000879586 | 0.00834507 | 1 |
| Heart_Failure | b_cell | *ZNF286B* | 8.240791615 | 0.000740734 | 0.007463129 | 1 |
| Heart_Failure | b_cell | *LINC00603* | -3.209103007 | 0.002770824 | 0.009906851 | 1 |
| Heart_Failure | b_cell | *YJEFN3* | -2.667617913 | 0.002780122 | 0.009914138 | 1 |
| Heart_Failure | b_cell | *DNAH10OS* | -20.13212476 | 0.000464866 | 0.005392272 | 1 |
| Heart_Failure | b_cell | *PCDHA10* | -5.381634229 | 0.003413045 | 0.011004007 | 1 |
| Heart_Failure | b_cell | *LINC01021* | -5.43350764 | 0.001680958 | 0.008654124 | 1 |
| Heart_Failure | b_cell | *FAM218A* | -5.048190865 | 0.001700256 | 0.008654124 | 1 |
| Heart_Failure | b_cell | *GLI4* | -2.220345001 | 0.00261285 | 0.009639485 | 1 |
| Heart_Failure | b_cell | *SNHG18* | -3.056370029 | 0.00051105 | 0.005824218 | 1 |
| Heart_Failure | b_cell | *ZNF345* | -2.12474724 | 0.02309209 | 0.04019379 | 1 |
| Heart_Failure | b_cell | *SHANK3* | -1.304002254 | 0.004219243 | 0.012320004 | 1 |
| Heart_Failure | b_cell | *CCDC71L* | -1.959060009 | 0.001676707 | 0.008654124 | 1 |
| Heart_Failure | b_cell | *TMEM200B* | 6.455123238 | 1.30E-06 | 0.000743789 | 0.008181676 |
| Heart_Failure | b_cell | *PCDHGB6* | -2.388031373 | 0.007596276 | 0.017833379 | 1 |
| Heart_Failure | b_cell | *TRNP1* | 2.259815266 | 0.000181677 | 0.00338933 | 1 |
| Heart_Failure | b_cell | *PCDHGA5* | -3.187918355 | 0.000302227 | 0.004231847 | 1 |
| Heart_Failure | b_cell | *ATXN7L3B* | -2.338422345 | 0.007288812 | 0.017377612 | 1 |
| Heart_Failure | b_cell | *PCDHGA6* | -2.078182639 | 0.016749154 | 0.03145219 | 1 |
| Heart_Failure | b_cell | *PCDHGA11* | -5.287507862 | 0.000725142 | 0.007329526 | 1 |
| Heart_Failure | b_cell | *CLDN23* | -4.963071075 | 0.004343391 | 0.012560671 | 1 |
| Heart_Failure | b_cell | *ARMS2* | -11.89541521 | 5.84E-05 | 0.002889125 | 0.366946688 |
| Heart_Failure | b_cell | *EID3* | -2.084537795 | 0.005793103 | 0.015006692 | 1 |
| Heart_Failure | b_cell | *EBLN2* | -3.52422406 | 0.003489993 | 0.011132829 | 1 |
| Heart_Failure | b_cell | *POLR2M* | -2.236909821 | 0.002029 | 0.008831917 | 1 |
| Heart_Failure | b_cell | *LINC00346* | 7.582474313 | 6.04E-05 | 0.002889125 | 0.379454638 |
| Heart_Failure | b_cell | *LINC00944* | -4.190945115 | 0.000676103 | 0.007025879 | 1 |
| Heart_Failure | b_cell | *HMBS* | -1.886229546 | 0.021214757 | 0.037592215 | 1 |
| Heart_Failure | b_cell | *AC093668.2* | 2.944909172 | 0.000248679 | 0.003766552 | 1 |
| Heart_Failure | b_cell | *CLEC12B* | -1.431038264 | 0.017783974 | 0.032952504 | 1 |
| Heart_Failure | b_cell | *PGA5* | -36.91621796 | 0.002907674 | 0.010125753 | 1 |
| Heart_Failure | b_cell | *HP* | -7.80595079 | 0.004978423 | 0.013608411 | 1 |
| Heart_Failure | b_cell | *KIAA1147* | -1.836096898 | 0.009019205 | 0.020008264 | 1 |
| Heart_Failure | b_cell | *ZBED6* | -5.752208644 | 0.001679553 | 0.008654124 | 1 |
| Heart_Failure | b_cell | *FNTB* | -3.097707839 | 0.001682584 | 0.008654124 | 1 |
| Heart_Failure | b_cell | *OVCH1-AS1* | -2.254567078 | 0.005062777 | 0.013791022 | 1 |
| Heart_Failure | b_cell | *TEN1* | 6.746139847 | 0.000108541 | 0.003034612 | 0.682397118 |
| Heart_Failure | b_cell | *CHURC1* | -1.821102181 | 0.017656825 | 0.032774862 | 1 |
| Heart_Failure | b_cell | *C17orf49* | -8.86538629 | 0.008020116 | 0.018510451 | 1 |
| Heart_Failure | b_cell | *ZNF578* | -2.633632537 | 0.01254211 | 0.02550202 | 1 |
| Heart_Failure | b_cell | *LINC00640* | -2.172348561 | 0.007785674 | 0.018115668 | 1 |
| Heart_Failure | b_cell | *LINC00638* | -12.08031156 | 0.000185191 | 0.00340015 | 1 |
| Heart_Failure | b_cell | *CEP95* | -1.812338187 | 0.026436556 | 0.044654729 | 1 |
| Heart_Failure | b_cell | *LINC00639* | -1.315230326 | 0.004111689 | 0.012159073 | 1 |

| | | | | | | |
|---|---|---|---|---|---|---|
| Heart_Failure | b_cell | *LINC00648* | -2.01934225 | 0.00144825 | 0.008654124 | 1 |
| Heart_Failure | b_cell | *LINC00924* | 2.14233759 | 0.000173404 | 0.003365162 | 1 |
| Heart_Failure | b_cell | *ITGB3* | -2.365215083 | 0.026693276 | 0.044955968 | 1 |
| Heart_Failure | b_cell | *UBE2Q2L* | 15.07084524 | 0.000310054 | 0.004274795 | 1 |
| Heart_Failure | b_cell | *AC083843.1* | -51.784784 | 0.001498268 | 0.008654124 | 1 |
| Heart_Failure | b_cell | *PWRN1* | -2.536515652 | 0.002452281 | 0.009418137 | 1 |
| Heart_Failure | b_cell | *HOXB7* | -2.736452747 | 0.003022106 | 0.010281376 | 1 |
| Heart_Failure | b_cell | *FRRS1L* | -5.978379914 | 0.00294505 | 0.010151069 | 1 |
| Heart_Failure | b_cell | *LINC00562* | -2.586229878 | 0.000408983 | 0.005022023 | 1 |
| Heart_Failure | b_cell | *AC009120.5* | -3.95347028 | 0.002921652 | 0.010133328 | 1 |
| Heart_Failure | b_cell | *TMEM178B* | -1.811348605 | 0.001744975 | 0.008654124 | 1 |
| Heart_Failure | b_cell | *LINC01003* | 6.509539183 | 7.51E-05 | 0.002889125 | 0.472183634 |
| Heart_Failure | b_cell | *C15orf65* | -3.846194142 | 0.000169044 | 0.003365162 | 1 |
| Heart_Failure | b_cell | *HPR* | -7.866609595 | 0.025882128 | 0.043895586 | 1 |
| Heart_Failure | b_cell | *AC123768.4* | 15.97104662 | 0.00063411 | 0.006666636 | 1 |
| Heart_Failure | b_cell | *LINC00667* | -2.156908785 | 0.002148492 | 0.008999799 | 1 |
| Heart_Failure | b_cell | *LINC00672* | -2.618663652 | 0.003484787 | 0.011132829 | 1 |
| Heart_Failure | b_cell | *LINC00909* | -2.197619317 | 0.013340393 | 0.026710527 | 1 |
| Heart_Failure | b_cell | *LINC00526* | -4.086596045 | 0.001327457 | 0.008654124 | 1 |
| Heart_Failure | b_cell | *LINC00668* | 26.85301266 | 0.000223014 | 0.003674323 | 1 |
| Heart_Failure | b_cell | *STRADA* | -2.182780503 | 0.001367111 | 0.008654124 | 1 |
| Heart_Failure | b_cell | *ILF3-AS1* | -2.315599191 | 0.02408058 | 0.041512094 | 1 |
| Heart_Failure | b_cell | *AC139100.2* | 8.346665559 | 8.61E-05 | 0.002889125 | 0.541052051 |
| Heart_Failure | b_cell | *LINC00906* | -20.0374357 | 0.001679573 | 0.008654124 | 1 |
| Heart_Failure | b_cell | *ZNF582-AS1* | -3.348821085 | 0.003933591 | 0.011883725 | 1 |
| Heart_Failure | b_cell | *AC137932.1* | -10.44179239 | 0.003303467 | 0.01083435 | 1 |
| Heart_Failure | b_cell | *AC003005.2* | -5.119966388 | 0.01321708 | 0.026548173 | 1 |
| Heart_Failure | b_cell | *AL132989.1* | 14.3665454 | 5.31E-05 | 0.002889125 | 0.333829449 |
| Heart_Failure | b_cell | *LINC00664* | -7.589676275 | 0.017388009 | 0.032361874 | 1 |
| Heart_Failure | b_cell | *FBXO17* | -1.520301073 | 0.001860752 | 0.008690419 | 1 |
| Heart_Failure | b_cell | *YTHDF3-AS1* | 4.67629304 | 7.70E-05 | 0.002889125 | 0.483935381 |
| Heart_Failure | b_cell | *LINC01050* | 3.424752713 | 0.000232896 | 0.003705045 | 1 |
| Heart_Failure | b_cell | *MILR1* | -1.582839232 | 0.024233369 | 0.041695454 | 1 |
| Heart_Failure | b_cell | *GTF2H5* | -1.552115774 | 0.013119701 | 0.026411643 | 1 |
| Heart_Failure | b_cell | *FGF14-AS2* | -3.75701138 | 0.001494285 | 0.008654124 | 1 |
| Heart_Failure | b_cell | *CASC15* | -1.415292876 | 0.005981543 | 0.015318111 | 1 |
| Heart_Failure | b_cell | *LINC01023* | -3.270223271 | 0.002105381 | 0.00890749 | 1 |
| Heart_Failure | b_cell | *AP000347.2* | -3.114156363 | 0.004803408 | 0.013301535 | 1 |
| Heart_Failure | b_cell | *POLR2J4* | -2.736971465 | 0.003307097 | 0.01083435 | 1 |
| Heart_Failure | dendritic_cell | *SCYL3* | -3.037917217 | 1.10E-09 | 1.82E-09 | 6.92E-06 |
| Heart_Failure | dendritic_cell | *C1orf112* | -5.35453187 | 3.43E-12 | 7.06E-12 | 2.16E-08 |
| Heart_Failure | dendritic_cell | *CFH* | -1.637137583 | 5.91E-20 | 9.53E-19 | 3.72E-16 |
| Heart_Failure | dendritic_cell | *NFYA* | -5.699799953 | 2.87E-10 | 4.95E-10 | 1.80E-06 |
| Heart_Failure | dendritic_cell | *STPG1* | -3.436742854 | 1.85E-13 | 4.39E-13 | 1.16E-09 |
| Heart_Failure | dendritic_cell | *NIPAL3* | -4.556233373 | 2.70E-06 | 3.65E-06 | 0.017003393 |
| Heart_Failure | dendritic_cell | *ENPP4* | -3.603435211 | 2.60E-14 | 7.00E-14 | 1.63E-10 |
| Heart_Failure | dendritic_cell | *SEMA3F* | -2.498679019 | 2.64E-24 | 7.47E-21 | 1.66E-20 |
| Heart_Failure | dendritic_cell | *KRIT1* | -4.094734716 | 6.06E-14 | 1.54E-13 | 3.81E-10 |
| Heart_Failure | dendritic_cell | *RAD52* | -4.747995368 | 1.66E-15 | 5.54E-15 | 1.04E-11 |
| Heart_Failure | dendritic_cell | *BAD* | -8.627615164 | 1.67E-13 | 4.00E-13 | 1.05E-09 |

| | | | | | | |
|---|---|---|---|---|---|---|
| Heart_Failure | dendritic_cell | *HS3ST1* | 5.983113073 | 7.44E-06 | 9.84E-06 | 0.046754248 |
| Heart_Failure | dendritic_cell | *WNT16* | -4.838992276 | 2.73E-11 | 5.13E-11 | 1.72E-07 |
| Heart_Failure | dendritic_cell | *HECW1* | -1.755234 | 9.88E-16 | 3.46E-15 | 6.21E-12 |
| Heart_Failure | dendritic_cell | *MAD1L1* | -4.900613468 | 1.60E-12 | 3.41E-12 | 1.01E-08 |
| Heart_Failure | dendritic_cell | *LASP1* | -4.895352733 | 2.21E-12 | 4.63E-12 | 1.39E-08 |
| Heart_Failure | dendritic_cell | *SNX11* | -2.954934594 | 1.62E-09 | 2.64E-09 | 1.02E-05 |
| Heart_Failure | dendritic_cell | *M6PR* | 3.394777712 | 0.000253777 | 0.000313149 | 1 |
| Heart_Failure | dendritic_cell | *CYP26B1* | -6.69383343 | 1.92E-14 | 5.28E-14 | 1.21E-10 |
| Heart_Failure | dendritic_cell | *ICA1* | -3.258078313 | 3.29E-17 | 1.60E-16 | 2.07E-13 |
| Heart_Failure | dendritic_cell | *DBNDD1* | -2.436508689 | 1.74E-05 | 2.27E-05 | 0.109444141 |
| Heart_Failure | dendritic_cell | *ALS2* | -3.89346864 | 4.42E-17 | 2.10E-16 | 2.78E-13 |
| Heart_Failure | dendritic_cell | *POLDIP2* | -4.309028701 | 1.95E-18 | 1.37E-17 | 1.23E-14 |
| Heart_Failure | dendritic_cell | *CD38* | 4.039497059 | 1.37E-15 | 4.67E-15 | 8.64E-12 |
| Heart_Failure | dendritic_cell | *RBM6* | -1.981463643 | 8.66E-16 | 3.07E-15 | 5.44E-12 |
| Heart_Failure | dendritic_cell | *RECQL* | -5.439149453 | 1.90E-09 | 3.08E-09 | 1.19E-05 |
| Heart_Failure | dendritic_cell | *PDK4* | -4.101850316 | 4.87E-10 | 8.26E-10 | 3.06E-06 |
| Heart_Failure | dendritic_cell | *SLC22A16* | 2.621837037 | 3.65E-19 | 3.42E-18 | 2.29E-15 |
| Heart_Failure | dendritic_cell | *CDC27* | -2.67302204 | 1.77E-06 | 2.41E-06 | 0.011143117 |
| Heart_Failure | dendritic_cell | *MEOX1* | -2.159243552 | 1.29E-17 | 7.08E-17 | 8.11E-14 |
| Heart_Failure | dendritic_cell | *CIAPIN1* | -3.528724207 | 0.020165603 | 0.022712495 | 1 |
| Heart_Failure | dendritic_cell | *FAM214B* | -5.239930716 | 3.46E-11 | 6.45E-11 | 2.18E-07 |
| Heart_Failure | dendritic_cell | *COPZ2* | -2.161002911 | 4.58E-11 | 8.43E-11 | 2.88E-07 |
| Heart_Failure | dendritic_cell | *GCFC2* | -4.199346129 | 2.28E-14 | 6.19E-14 | 1.43E-10 |
| Heart_Failure | dendritic_cell | *WDR54* | -7.472967954 | 3.24E-06 | 4.36E-06 | 0.020356139 |
| Heart_Failure | dendritic_cell | *ABCB4* | 2.899846933 | 8.21E-21 | 4.66E-19 | 5.16E-17 |
| Heart_Failure | dendritic_cell | *ZNF195* | -3.535504228 | 1.05E-13 | 2.57E-13 | 6.63E-10 |
| Heart_Failure | dendritic_cell | *PDK2* | -3.902621951 | 3.76E-17 | 1.80E-16 | 2.36E-13 |
| Heart_Failure | dendritic_cell | *GGNBP2* | -3.595800919 | 7.09E-12 | 1.41E-11 | 4.46E-08 |
| Heart_Failure | dendritic_cell | *ASB4* | -1.751924001 | 0.011506153 | 0.013107299 | 1 |
| Heart_Failure | dendritic_cell | *TMEM98* | -2.473718316 | 1.36E-17 | 7.38E-17 | 8.53E-14 |
| Heart_Failure | dendritic_cell | *ABCC8* | 8.859950539 | 2.69E-20 | 6.44E-19 | 1.69E-16 |
| Heart_Failure | dendritic_cell | *CCL18* | -2.276499468 | 3.13E-10 | 5.39E-10 | 1.97E-06 |
| Heart_Failure | dendritic_cell | *SYNRG* | -3.974284904 | 4.25E-15 | 1.31E-14 | 2.67E-11 |
| Heart_Failure | dendritic_cell | *TMEM132A* | -6.812481895 | 5.41E-20 | 9.02E-19 | 3.40E-16 |
| Heart_Failure | dendritic_cell | *TAC1* | -4.732285744 | 0.012535536 | 0.014261837 | 1 |
| Heart_Failure | dendritic_cell | *ZNF263* | -5.078965408 | 7.26E-16 | 2.61E-15 | 4.56E-12 |
| Heart_Failure | dendritic_cell | *SPATA20* | -2.783307656 | 0.008266961 | 0.009498243 | 1 |
| Heart_Failure | dendritic_cell | *TNFRSF12A* | 6.717044962 | 1.76E-08 | 2.68E-08 | 0.000110825 |
| Heart_Failure | dendritic_cell | *RALA* | -4.689914372 | 4.10E-09 | 6.49E-09 | 2.58E-05 |
| Heart_Failure | dendritic_cell | *BAIAP2L1* | -4.271207571 | 1.47E-15 | 4.95E-15 | 9.23E-12 |
| Heart_Failure | dendritic_cell | *FARP2* | -1.827884879 | 0.027269086 | 0.030489195 | 1 |
| Heart_Failure | dendritic_cell | *IFRD1* | -3.374580765 | 5.93E-13 | 1.32E-12 | 3.73E-09 |
| Heart_Failure | dendritic_cell | *GTF2IRD1* | -2.493852598 | 1.37E-11 | 2.65E-11 | 8.59E-08 |
| Heart_Failure | dendritic_cell | *VPS41* | -3.475670353 | 5.62E-08 | 8.33E-08 | 0.00035354 |
| Heart_Failure | dendritic_cell | *ARHGAP44* | -5.692040375 | 2.27E-12 | 4.76E-12 | 1.43E-08 |
| Heart_Failure | dendritic_cell | *ELAC2* | -3.510323486 | 5.88E-05 | 7.47E-05 | 0.36942881 |
| Heart_Failure | dendritic_cell | *CCDC124* | -4.062665374 | 9.00E-13 | 1.96E-12 | 5.66E-09 |
| Heart_Failure | dendritic_cell | *CEACAM21* | -4.272711833 | 2.35E-19 | 2.41E-18 | 1.48E-15 |
| Heart_Failure | dendritic_cell | *NOS2* | -17.07064889 | 1.49E-19 | 1.77E-18 | 9.35E-16 |
| Heart_Failure | dendritic_cell | *TRAPPC6A* | -6.423974113 | 5.43E-14 | 1.39E-13 | 3.41E-10 |

| | | | | | | |
|---|---|---|---|---|---|---|
| Heart_Failure | dendritic_cell | *CD79B* | -4.056865453 | 3.24E-13 | 7.48E-13 | 2.04E-09 |
| Heart_Failure | dendritic_cell | *ST7L* | -2.447173905 | 3.04E-12 | 6.28E-12 | 1.91E-08 |
| Heart_Failure | dendritic_cell | *PAX6* | -3.221813842 | 2.61E-10 | 4.53E-10 | 1.64E-06 |
| Heart_Failure | dendritic_cell | *RPUSD1* | 8.005333176 | 2.81E-20 | 6.44E-19 | 1.77E-16 |
| Heart_Failure | dendritic_cell | *BAIAP3* | 8.844916494 | 2.21E-16 | 8.86E-16 | 1.39E-12 |
| Heart_Failure | dendritic_cell | *TSR3* | -8.681712194 | 2.61E-15 | 8.38E-15 | 1.64E-11 |
| Heart_Failure | dendritic_cell | *PIGQ* | -4.030348571 | 2.14E-11 | 4.08E-11 | 1.35E-07 |
| Heart_Failure | dendritic_cell | *PSMB1* | -5.062094746 | 4.99E-13 | 1.12E-12 | 3.14E-09 |
| Heart_Failure | dendritic_cell | *CDK11A* | -4.704088055 | 4.20E-19 | 3.81E-18 | 2.64E-15 |
| Heart_Failure | dendritic_cell | *DLEC1* | -4.74372714 | 2.19E-17 | 1.11E-16 | 1.38E-13 |
| Heart_Failure | dendritic_cell | *SS18L2* | -5.099248212 | 1.28E-16 | 5.37E-16 | 8.03E-13 |
| Heart_Failure | dendritic_cell | *MPND* | -5.14539223 | 1.55E-07 | 2.24E-07 | 0.000975736 |
| Heart_Failure | dendritic_cell | *MGST1* | -1.842363302 | 3.47E-18 | 2.21E-17 | 2.18E-14 |
| Heart_Failure | dendritic_cell | *NFIX* | -2.63635663 | 5.95E-17 | 2.73E-16 | 3.74E-13 |
| Heart_Failure | dendritic_cell | *MMP25* | 5.963327289 | 5.73E-12 | 1.15E-11 | 3.61E-08 |
| Heart_Failure | dendritic_cell | *IL32* | -6.020289745 | 4.73E-15 | 1.45E-14 | 2.98E-11 |
| Heart_Failure | dendritic_cell | *PKD1* | -3.521386923 | 1.97E-13 | 4.66E-13 | 1.24E-09 |
| Heart_Failure | dendritic_cell | *MAPK8IP2* | 7.763449625 | 6.35E-06 | 8.43E-06 | 0.039932129 |
| Heart_Failure | dendritic_cell | *MED24* | -4.070275319 | 6.77E-06 | 8.97E-06 | 0.042546427 |
| Heart_Failure | dendritic_cell | *RHOBTB2* | -5.590298711 | 1.37E-10 | 2.43E-10 | 8.64E-07 |
| Heart_Failure | dendritic_cell | *SEC62* | -4.40317703 | 1.25E-13 | 3.04E-13 | 7.89E-10 |
| Heart_Failure | dendritic_cell | *RPS20* | -4.754430213 | 2.95E-07 | 4.21E-07 | 0.001856315 |
| Heart_Failure | dendritic_cell | *POMT2* | -5.038196874 | 8.94E-15 | 2.61E-14 | 5.62E-11 |
| Heart_Failure | dendritic_cell | *MLXIPL* | -1.078629119 | 2.83E-12 | 5.87E-12 | 1.78E-08 |
| Heart_Failure | dendritic_cell | *SPRTN* | -6.777992396 | 2.18E-11 | 4.15E-11 | 1.37E-07 |
| Heart_Failure | dendritic_cell | *METTL13* | 6.734056693 | 8.74E-10 | 1.46E-09 | 5.49E-06 |
| Heart_Failure | dendritic_cell | *DYRK4* | 3.439396574 | 8.82E-14 | 2.18E-13 | 5.54E-10 |
| Heart_Failure | dendritic_cell | *STARD3NL* | 5.389746397 | 3.71E-07 | 5.25E-07 | 0.002330737 |
| Heart_Failure | dendritic_cell | *ZNF200* | -4.640814121 | 7.96E-18 | 4.60E-17 | 5.01E-14 |
| Heart_Failure | dendritic_cell | *CD4* | 1.516057547 | 1.12E-20 | 4.99E-19 | 7.06E-17 |
| Heart_Failure | dendritic_cell | *LRRC23* | -1.646428342 | 0.041356814 | 0.045832944 | 1 |
| Heart_Failure | dendritic_cell | *HFE* | 2.553030454 | 6.56E-06 | 8.70E-06 | 0.041236646 |
| Heart_Failure | dendritic_cell | *SCMH1* | -3.69105895 | 1.00E-12 | 2.18E-12 | 6.30E-09 |
| Heart_Failure | dendritic_cell | *CLCN6* | -5.479326212 | 1.23E-12 | 2.65E-12 | 7.73E-09 |
| Heart_Failure | dendritic_cell | *MRC2* | -2.408473808 | 3.78E-18 | 2.39E-17 | 2.38E-14 |
| Heart_Failure | dendritic_cell | *TSPAN9* | -2.56113453 | 8.38E-15 | 2.46E-14 | 5.27E-11 |
| Heart_Failure | dendritic_cell | *MKS1* | -7.630279819 | 1.58E-14 | 4.41E-14 | 9.96E-11 |
| Heart_Failure | dendritic_cell | *UTP18* | -6.320231624 | 2.43E-10 | 4.22E-10 | 1.53E-06 |
| Heart_Failure | dendritic_cell | *TTC19* | -3.446340349 | 2.92E-11 | 5.46E-11 | 1.83E-07 |
| Heart_Failure | dendritic_cell | *DPF1* | 5.581346321 | 2.96E-12 | 6.13E-12 | 1.86E-08 |
| Heart_Failure | dendritic_cell | *SYT7* | 6.274836835 | 1.01E-17 | 5.69E-17 | 6.33E-14 |
| Heart_Failure | dendritic_cell | *LARS2* | -4.425292564 | 3.42E-09 | 5.45E-09 | 2.15E-05 |
| Heart_Failure | dendritic_cell | *PPP5C* | -4.420773909 | 3.16E-15 | 9.99E-15 | 1.99E-11 |
| Heart_Failure | dendritic_cell | *CEP68* | -3.02159916 | 1.96E-09 | 3.17E-09 | 1.23E-05 |
| Heart_Failure | dendritic_cell | *ZBTB32* | -4.56832052 | 1.08E-19 | 1.41E-18 | 6.82E-16 |
| Heart_Failure | dendritic_cell | *TMEM159* | -4.075040452 | 5.15E-16 | 1.90E-15 | 3.24E-12 |
| Heart_Failure | dendritic_cell | *ERCC1* | -4.200098738 | 3.70E-14 | 9.71E-14 | 2.33E-10 |
| Heart_Failure | dendritic_cell | *LTF* | -4.998318915 | 3.55E-14 | 9.33E-14 | 2.23E-10 |
| Heart_Failure | dendritic_cell | *ALOX5* | 2.246599708 | 3.49E-20 | 6.98E-19 | 2.19E-16 |
| Heart_Failure | dendritic_cell | *MAP4K5* | -4.129399255 | 2.29E-09 | 3.69E-09 | 1.44E-05 |

| | | | | | | |
|---|---|---|---|---|---|---|
| Heart_Failure | dendritic_cell | *PSMC4* | -4.174334325 | 1.21E-08 | 1.85E-08 | 7.60E-05 |
| Heart_Failure | dendritic_cell | *MAN2B2* | -5.434271491 | 6.24E-14 | 1.58E-13 | 3.92E-10 |
| Heart_Failure | dendritic_cell | *CLDN11* | -2.968236541 | 8.14E-19 | 6.55E-18 | 5.12E-15 |
| Heart_Failure | dendritic_cell | *SLC25A39* | 3.446252235 | 0.00474251 | 0.005520489 | 1 |
| Heart_Failure | dendritic_cell | *ANGEL1* | -4.09795103 | 4.87E-18 | 2.97E-17 | 3.06E-14 |
| Heart_Failure | dendritic_cell | *RNF14* | -4.119254755 | 1.07E-05 | 1.41E-05 | 0.067355275 |
| Heart_Failure | dendritic_cell | *DDX11* | 1.631218199 | 0.004662405 | 0.005429253 | 1 |
| Heart_Failure | dendritic_cell | *HEBP1* | -4.577991577 | 2.92E-17 | 1.44E-16 | 1.83E-13 |
| Heart_Failure | dendritic_cell | *TACC3* | -2.34675069 | 0.02124789 | 0.023897224 | 1 |
| Heart_Failure | dendritic_cell | *ZC3H3* | -3.349984339 | 2.26E-09 | 3.64E-09 | 1.42E-05 |
| Heart_Failure | dendritic_cell | *MDH1* | -2.807108735 | 2.43E-14 | 6.58E-14 | 1.53E-10 |
| Heart_Failure | dendritic_cell | *SLC30A9* | -3.826559302 | 4.59E-15 | 1.41E-14 | 2.89E-11 |
| Heart_Failure | dendritic_cell | *MTMR11* | -1.522649405 | 4.04E-10 | 6.87E-10 | 2.54E-06 |
| Heart_Failure | dendritic_cell | *COX15* | -4.348091936 | 1.35E-10 | 2.39E-10 | 8.47E-07 |
| Heart_Failure | dendritic_cell | *CCDC88C* | -3.410446309 | 1.10E-09 | 1.81E-09 | 6.90E-06 |
| Heart_Failure | dendritic_cell | *NPC1L1* | 7.225370279 | 2.15E-16 | 8.63E-16 | 1.35E-12 |
| Heart_Failure | dendritic_cell | *NUDCD3* | -2.608336433 | 1.47E-09 | 2.41E-09 | 9.26E-06 |
| Heart_Failure | dendritic_cell | *CHDH* | -6.774745713 | 1.53E-19 | 1.81E-18 | 9.60E-16 |
| Heart_Failure | dendritic_cell | *IL20RA* | -7.045816601 | 2.60E-20 | 6.44E-19 | 1.63E-16 |
| Heart_Failure | dendritic_cell | *GLT8D1* | -4.499610344 | 2.51E-18 | 1.69E-17 | 1.58E-14 |
| Heart_Failure | dendritic_cell | *RUFY3* | -2.575422741 | 0.00010756 | 0.000135219 | 0.676230612 |
| Heart_Failure | dendritic_cell | *WWTR1* | -4.115603135 | 1.14E-13 | 2.78E-13 | 7.20E-10 |
| Heart_Failure | dendritic_cell | *TTC27* | -3.841755834 | 7.53E-14 | 1.88E-13 | 4.73E-10 |
| Heart_Failure | dendritic_cell | *VSIG2* | -6.836838245 | 2.41E-19 | 2.46E-18 | 1.51E-15 |
| Heart_Failure | dendritic_cell | *PHLDB1* | -1.710036446 | 5.93E-14 | 1.51E-13 | 3.73E-10 |
| Heart_Failure | dendritic_cell | *MNAT1* | -3.28522522 | 8.87E-17 | 3.90E-16 | 5.58E-13 |
| Heart_Failure | dendritic_cell | *OSBPL5* | -3.533946459 | 2.48E-18 | 1.68E-17 | 1.56E-14 |
| Heart_Failure | dendritic_cell | *AQR* | -4.415234787 | 1.27E-10 | 2.27E-10 | 8.01E-07 |
| Heart_Failure | dendritic_cell | *CPS1* | -2.451371192 | 9.76E-13 | 2.12E-12 | 6.13E-09 |
| Heart_Failure | dendritic_cell | *NLRP2* | 3.40227972 | 9.36E-07 | 1.29E-06 | 0.005883429 |
| Heart_Failure | dendritic_cell | *ZDHHC6* | -4.352634665 | 3.79E-06 | 5.09E-06 | 0.023841458 |
| Heart_Failure | dendritic_cell | *RNH1* | -2.651526865 | 0.000129649 | 0.000162339 | 0.815105339 |
| Heart_Failure | dendritic_cell | *NDUFS1* | -4.103592151 | 9.77E-11 | 1.75E-10 | 6.14E-07 |
| Heart_Failure | dendritic_cell | *BIRC3* | 1.018453747 | 1.48E-20 | 5.69E-19 | 9.33E-17 |
| Heart_Failure | dendritic_cell | *AKAP11* | -4.612490148 | 1.06E-07 | 1.55E-07 | 0.000667639 |
| Heart_Failure | dendritic_cell | *GLRX2* | -3.409926233 | 5.83E-10 | 9.84E-10 | 3.66E-06 |
| Heart_Failure | dendritic_cell | *SNAPC1* | -5.602726833 | 5.40E-13 | 1.21E-12 | 3.40E-09 |
| Heart_Failure | dendritic_cell | *GCLM* | -2.779798009 | 1.02E-15 | 3.55E-15 | 6.38E-12 |
| Heart_Failure | dendritic_cell | *UBR2* | -3.430955931 | 1.83E-15 | 6.05E-15 | 1.15E-11 |
| Heart_Failure | dendritic_cell | *HSF2* | -4.791659346 | 1.07E-17 | 6.00E-17 | 6.73E-14 |
| Heart_Failure | dendritic_cell | *NR1H3* | -4.034143957 | 1.14E-06 | 1.56E-06 | 0.007152821 |
| Heart_Failure | dendritic_cell | *TYMP* | 2.842470664 | 0.000114044 | 0.000143198 | 0.716994889 |
| Heart_Failure | dendritic_cell | *KPNA6* | -4.471049349 | 4.44E-11 | 8.17E-11 | 2.79E-07 |
| Heart_Failure | dendritic_cell | *VIM* | -3.339847165 | 7.00E-08 | 1.03E-07 | 0.000440251 |
| Heart_Failure | dendritic_cell | *FAS* | -2.418463693 | 3.91E-12 | 7.98E-12 | 2.46E-08 |
| Heart_Failure | dendritic_cell | *RNASET2* | 2.224351431 | 3.52E-20 | 6.98E-19 | 2.21E-16 |
| Heart_Failure | dendritic_cell | *BTN3A1* | -1.816022129 | 7.61E-05 | 9.63E-05 | 0.478635676 |
| Heart_Failure | dendritic_cell | *MIPEP* | -5.258781058 | 1.93E-08 | 2.92E-08 | 0.000121236 |
| Heart_Failure | dendritic_cell | *INSRR* | -4.233662137 | 9.21E-07 | 1.27E-06 | 0.005789402 |
| Heart_Failure | dendritic_cell | *B4GALT7* | -4.734028606 | 7.90E-06 | 1.04E-05 | 0.049636392 |

| | | | | | | |
|---|---|---|---|---|---|---|
| Heart_Failure | dendritic_cell | *VRK2* | -3.715681201 | 8.16E-07 | 1.13E-06 | 0.005127969 |
| Heart_Failure | dendritic_cell | *VEZT* | -3.840454369 | 7.78E-15 | 2.30E-14 | 4.89E-11 |
| Heart_Failure | dendritic_cell | *SNX1* | -3.48806308 | 5.88E-09 | 9.21E-09 | 3.70E-05 |
| Heart_Failure | dendritic_cell | *ARNTL2* | -4.077812795 | 0.003047836 | 0.003581635 | 1 |
| Heart_Failure | dendritic_cell | *ANK1* | -6.375656273 | 4.47E-18 | 2.76E-17 | 2.81E-14 |
| Heart_Failure | dendritic_cell | *RABEP1* | -3.664127592 | 2.11E-13 | 4.96E-13 | 1.32E-09 |
| Heart_Failure | dendritic_cell | *GRN* | 5.302734478 | 7.39E-14 | 1.85E-13 | 4.65E-10 |
| Heart_Failure | dendritic_cell | *CENPQ* | -2.867509711 | 2.61E-12 | 5.44E-12 | 1.64E-08 |
| Heart_Failure | dendritic_cell | *ARID4A* | -4.433340648 | 4.01E-09 | 6.35E-09 | 2.52E-05 |
| Heart_Failure | dendritic_cell | *IFT88* | -3.453519982 | 3.57E-16 | 1.37E-15 | 2.24E-12 |
| Heart_Failure | dendritic_cell | *ZCCHC8* | -4.503926906 | 1.95E-15 | 6.43E-15 | 1.23E-11 |
| Heart_Failure | dendritic_cell | *ABCF2* | -4.019769139 | 3.44E-06 | 4.62E-06 | 0.021621353 |
| Heart_Failure | dendritic_cell | *FUT8* | -3.837247066 | 1.65E-14 | 4.58E-14 | 1.04E-10 |
| Heart_Failure | dendritic_cell | *UBA6* | -3.365940938 | 1.14E-05 | 1.50E-05 | 0.071909462 |
| Heart_Failure | dendritic_cell | *PIAS1* | -3.548104684 | 2.92E-07 | 4.17E-07 | 0.001838203 |
| Heart_Failure | dendritic_cell | *EFCAB1* | 5.365294413 | 9.30E-12 | 1.82E-11 | 5.85E-08 |
| Heart_Failure | dendritic_cell | *ASTE1* | -3.296804206 | 9.01E-11 | 1.62E-10 | 5.66E-07 |
| Heart_Failure | dendritic_cell | *MYOC* | -2.269332336 | 2.26E-08 | 3.41E-08 | 0.000142153 |
| Heart_Failure | dendritic_cell | *SH3YL1* | -3.259946437 | 3.50E-15 | 1.10E-14 | 2.20E-11 |
| Heart_Failure | dendritic_cell | *VCL* | -3.916750751 | 3.82E-14 | 1.00E-13 | 2.40E-10 |
| Heart_Failure | dendritic_cell | *ADSS* | -5.32248077 | 8.03E-13 | 1.76E-12 | 5.05E-09 |
| Heart_Failure | dendritic_cell | *STAP1* | -6.015571348 | 3.79E-18 | 2.39E-17 | 2.38E-14 |
| Heart_Failure | dendritic_cell | *TBC1D23* | -4.302127887 | 6.16E-15 | 1.85E-14 | 3.87E-11 |
| Heart_Failure | dendritic_cell | *CUL3* | -4.126084352 | 1.77E-12 | 3.76E-12 | 1.12E-08 |
| Heart_Failure | dendritic_cell | *MYOM2* | -2.719985117 | 1.29E-14 | 3.65E-14 | 8.10E-11 |
| Heart_Failure | dendritic_cell | *TUBG2* | -1.390217713 | 3.51E-06 | 4.72E-06 | 0.022083231 |
| Heart_Failure | dendritic_cell | *NSUN2* | -4.925596656 | 2.21E-07 | 3.17E-07 | 0.001388189 |
| Heart_Failure | dendritic_cell | *MRI1* | -2.847041164 | 3.63E-10 | 6.21E-10 | 2.28E-06 |
| Heart_Failure | dendritic_cell | *AGA* | -8.048357011 | 3.97E-10 | 6.77E-10 | 2.50E-06 |
| Heart_Failure | dendritic_cell | *BOD1L1* | -2.184223573 | 0.006991923 | 0.008073135 | 1 |
| Heart_Failure | dendritic_cell | *CLEC16A* | -4.670603844 | 1.77E-10 | 3.10E-10 | 1.11E-06 |
| Heart_Failure | dendritic_cell | *SKIV2L2* | -3.568624692 | 4.50E-16 | 1.68E-15 | 2.83E-12 |
| Heart_Failure | dendritic_cell | *DNAH5* | -2.846070093 | 2.26E-20 | 6.44E-19 | 1.42E-16 |
| Heart_Failure | dendritic_cell | *C6* | -2.619257103 | 1.82E-19 | 2.05E-18 | 1.14E-15 |
| Heart_Failure | dendritic_cell | *RAI14* | -3.074099299 | 5.12E-08 | 7.59E-08 | 0.000321641 |
| Heart_Failure | dendritic_cell | *PHLPP2* | -3.0440853 | 8.36E-05 | 0.000105555 | 0.525877068 |
| Heart_Failure | dendritic_cell | *SPDL1* | -1.694376124 | 0.003401391 | 0.003988166 | 1 |
| Heart_Failure | dendritic_cell | *PQLC2* | -4.383739505 | 0.032857146 | 0.03660319 | 1 |
| Heart_Failure | dendritic_cell | *CTNS* | -2.216096187 | 4.42E-11 | 8.15E-11 | 2.78E-07 |
| Heart_Failure | dendritic_cell | *RAB27B* | -2.733077338 | 2.64E-10 | 4.58E-10 | 1.66E-06 |
| Heart_Failure | dendritic_cell | *LSG1* | -1.669989182 | 0.000399388 | 0.000488798 | 1 |
| Heart_Failure | dendritic_cell | *THAP3* | 7.622585703 | 1.44E-13 | 3.46E-13 | 9.05E-10 |
| Heart_Failure | dendritic_cell | *SPATA7* | -4.923546365 | 7.40E-13 | 1.63E-12 | 4.65E-09 |
| Heart_Failure | dendritic_cell | *RETSAT* | -2.041806559 | 7.47E-17 | 3.36E-16 | 4.70E-13 |
| Heart_Failure | dendritic_cell | *CAPG* | -4.009609314 | 4.69E-14 | 1.21E-13 | 2.95E-10 |
| Heart_Failure | dendritic_cell | *AP2S1* | 7.07093624 | 3.98E-14 | 1.04E-13 | 2.50E-10 |
| Heart_Failure | dendritic_cell | *LCP2* | 3.724425925 | 1.02E-17 | 5.78E-17 | 6.44E-14 |
| Heart_Failure | dendritic_cell | *CUL7* | 2.728136219 | 3.55E-05 | 4.55E-05 | 0.223272443 |
| Heart_Failure | dendritic_cell | *CTNNA1* | -3.780293406 | 1.34E-11 | 2.60E-11 | 8.42E-08 |
| Heart_Failure | dendritic_cell | *CNTLN* | -3.052649217 | 0.000315521 | 0.000387892 | 1 |

| | | | | | | |
|---|---|---|---|---|---|---|
| Heart_Failure | dendritic_cell | *EPHA3* | -2.309056029 | 1.13E-19 | 1.44E-18 | 7.07E-16 |
| Heart_Failure | dendritic_cell | *DSG2* | -3.276595482 | 2.32E-19 | 2.41E-18 | 1.46E-15 |
| Heart_Failure | dendritic_cell | *WDR37* | -4.012809738 | 7.08E-07 | 9.87E-07 | 0.004453883 |
| Heart_Failure | dendritic_cell | *YTHDC2* | -4.027428518 | 2.36E-14 | 6.39E-14 | 1.48E-10 |
| Heart_Failure | dendritic_cell | *ATP6V1H* | -4.40041044 | 2.19E-07 | 3.15E-07 | 0.001379639 |
| Heart_Failure | dendritic_cell | *POLR2B* | -4.812724995 | 1.15E-11 | 2.25E-11 | 7.26E-08 |
| Heart_Failure | dendritic_cell | *ANO2* | -1.614069072 | 8.27E-19 | 6.63E-18 | 5.20E-15 |
| Heart_Failure | dendritic_cell | *GOPC* | -4.580201185 | 5.22E-12 | 1.05E-11 | 3.28E-08 |
| Heart_Failure | dendritic_cell | *TSPAN17* | -3.096550684 | 1.10E-16 | 4.71E-16 | 6.92E-13 |
| Heart_Failure | dendritic_cell | *CC2D2A* | -3.623654411 | 2.07E-16 | 8.34E-16 | 1.30E-12 |
| Heart_Failure | dendritic_cell | *RRM2B* | 3.915840792 | 0.000408978 | 0.000500339 | 1 |
| Heart_Failure | dendritic_cell | *SNX29* | -2.839844898 | 0.000201863 | 0.000250467 | 1 |
| Heart_Failure | dendritic_cell | *LMO3* | -1.651835481 | 2.69E-17 | 1.34E-16 | 1.69E-13 |
| Heart_Failure | dendritic_cell | *MRPS10* | -1.26195296 | 1.83E-05 | 2.37E-05 | 0.11484142 |
| Heart_Failure | dendritic_cell | *CELF2* | -3.064164646 | 3.19E-14 | 8.49E-14 | 2.00E-10 |
| Heart_Failure | dendritic_cell | *ERCC8* | -4.077040711 | 5.91E-08 | 8.74E-08 | 0.000371282 |
| Heart_Failure | dendritic_cell | *ADAMTS6* | -2.748625433 | 2.39E-16 | 9.47E-16 | 1.50E-12 |
| Heart_Failure | dendritic_cell | *UTS2* | -3.58840963 | 3.33E-12 | 6.86E-12 | 2.09E-08 |
| Heart_Failure | dendritic_cell | *EPN3* | -11.1091151 | 1.02E-06 | 1.40E-06 | 0.006404636 |
| Heart_Failure | dendritic_cell | *RCN1* | -5.594263162 | 2.90E-12 | 6.01E-12 | 1.83E-08 |
| Heart_Failure | dendritic_cell | *RFC2* | 7.363003304 | 7.47E-10 | 1.25E-09 | 4.70E-06 |
| Heart_Failure | dendritic_cell | *JKAMP* | -3.365994478 | 1.56E-10 | 2.75E-10 | 9.83E-07 |
| Heart_Failure | dendritic_cell | *DKK3* | -2.521574524 | 4.65E-19 | 4.14E-18 | 2.92E-15 |
| Heart_Failure | dendritic_cell | *ARHGEF5* | -3.716682533 | 4.75E-20 | 8.27E-19 | 2.99E-16 |
| Heart_Failure | dendritic_cell | *NFE2L3* | 4.871769837 | 1.45E-18 | 1.07E-17 | 9.12E-15 |
| Heart_Failure | dendritic_cell | *LIMA1* | -1.495660882 | 9.51E-14 | 2.34E-13 | 5.98E-10 |
| Heart_Failure | dendritic_cell | *PTGER3* | -1.838848219 | 1.95E-19 | 2.15E-18 | 1.23E-15 |
| Heart_Failure | dendritic_cell | *MAPK9* | -2.673952695 | 0.006598988 | 0.007625039 | 1 |
| Heart_Failure | dendritic_cell | *FAM160A2* | -3.037439143 | 7.20E-09 | 1.12E-08 | 4.53E-05 |
| Heart_Failure | dendritic_cell | *CYBA* | 4.6253534 | 2.36E-15 | 7.63E-15 | 1.48E-11 |
| Heart_Failure | dendritic_cell | *THOC3* | 5.762119319 | 3.02E-18 | 1.97E-17 | 1.90E-14 |
| Heart_Failure | dendritic_cell | *RRP12* | 4.934995157 | 4.03E-20 | 7.61E-19 | 2.53E-16 |
| Heart_Failure | dendritic_cell | *ALX4* | -4.768965141 | 2.73E-20 | 6.44E-19 | 1.71E-16 |
| Heart_Failure | dendritic_cell | *FOXN3* | -3.536587494 | 2.06E-08 | 3.11E-08 | 0.000129351 |
| Heart_Failure | dendritic_cell | *AKR7A2* | -5.6330245 | 2.16E-13 | 5.08E-13 | 1.36E-09 |
| Heart_Failure | dendritic_cell | *MRTO4* | -3.741182345 | 6.99E-19 | 5.75E-18 | 4.39E-15 |
| Heart_Failure | dendritic_cell | *MCF2L2* | 4.665101806 | 2.57E-18 | 1.72E-17 | 1.62E-14 |
| Heart_Failure | dendritic_cell | *ANAPC4* | -4.628563173 | 1.34E-13 | 3.23E-13 | 8.39E-10 |
| Heart_Failure | dendritic_cell | *LY75* | 1.485503176 | 1.02E-20 | 4.95E-19 | 6.43E-17 |
| Heart_Failure | dendritic_cell | *OPN3* | 1.901012772 | 4.05E-16 | 1.53E-15 | 2.55E-12 |
| Heart_Failure | dendritic_cell | *SDCCAG8* | -1.814043038 | 0.000223821 | 0.000277109 | 1 |
| Heart_Failure | dendritic_cell | *TBC1D22A* | 1.76070092 | 0.034236533 | 0.038123465 | 1 |
| Heart_Failure | dendritic_cell | *PLEKHH1* | -4.252815569 | 1.72E-15 | 5.72E-15 | 1.08E-11 |
| Heart_Failure | dendritic_cell | *ATP9A* | -3.012894571 | 1.28E-18 | 9.50E-18 | 8.06E-15 |
| Heart_Failure | dendritic_cell | *GALC* | -4.534328926 | 2.47E-11 | 4.66E-11 | 1.55E-07 |
| Heart_Failure | dendritic_cell | *FAM114A2* | -4.432267255 | 1.99E-13 | 4.69E-13 | 1.25E-09 |
| Heart_Failure | dendritic_cell | *GINM1* | -4.547436286 | 1.18E-14 | 3.36E-14 | 7.41E-11 |
| Heart_Failure | dendritic_cell | *MRPL43* | -4.370814456 | 1.21E-16 | 5.11E-16 | 7.58E-13 |
| Heart_Failure | dendritic_cell | *TRAF1* | 1.978478231 | 9.27E-20 | 1.27E-18 | 5.83E-16 |
| Heart_Failure | dendritic_cell | *PCGF2* | -0.826630537 | 0.012531249 | 0.01425954 | 1 |

| | | | | | | |
|---|---|---|---|---|---|---|
| Heart_Failure | dendritic_cell | *IL17RB* | -5.211773419 | 2.68E-20 | 6.44E-19 | 1.68E-16 |
| Heart_Failure | dendritic_cell | *TRAF3IP2* | -5.284852407 | 9.01E-12 | 1.77E-11 | 5.66E-08 |
| Heart_Failure | dendritic_cell | *DCBLD2* | -3.578899594 | 2.67E-17 | 1.33E-16 | 1.68E-13 |
| Heart_Failure | dendritic_cell | *MSH4* | -2.877756275 | 3.19E-15 | 1.01E-14 | 2.01E-11 |
| Heart_Failure | dendritic_cell | *F7* | 19.49708499 | 1.51E-14 | 4.22E-14 | 9.48E-11 |
| Heart_Failure | dendritic_cell | *GDI2* | -2.977867225 | 0.000427124 | 0.000522234 | 1 |
| Heart_Failure | dendritic_cell | *TMCC3* | 4.216484515 | 6.88E-18 | 4.03E-17 | 4.33E-14 |
| Heart_Failure | dendritic_cell | *PITHD1* | -4.374997714 | 0.000162305 | 0.000202422 | 1 |
| Heart_Failure | dendritic_cell | *ATP11B* | -2.303186339 | 4.00E-07 | 5.66E-07 | 0.002516693 |
| Heart_Failure | dendritic_cell | *LAMC2* | -1.87014084 | 2.34E-15 | 7.57E-15 | 1.47E-11 |
| Heart_Failure | dendritic_cell | *CDK14* | -3.540747233 | 3.08E-15 | 9.77E-15 | 1.94E-11 |
| Heart_Failure | dendritic_cell | *ZC3H11A* | 6.554582967 | 2.33E-19 | 2.41E-18 | 1.47E-15 |
| Heart_Failure | dendritic_cell | *RIOK2* | -4.380889708 | 5.47E-17 | 2.53E-16 | 3.44E-13 |
| Heart_Failure | dendritic_cell | *FLYWCH1* | -1.397308292 | 0.000528398 | 0.000643059 | 1 |
| Heart_Failure | dendritic_cell | *DNAJC25* | -4.555274697 | 4.50E-06 | 6.01E-06 | 0.028318978 |
| Heart_Failure | dendritic_cell | *YBX3* | -3.599737317 | 0.000112519 | 0.00014134 | 0.707404635 |
| Heart_Failure | dendritic_cell | *PIGV* | -4.310682579 | 2.06E-15 | 6.75E-15 | 1.29E-11 |
| Heart_Failure | dendritic_cell | *NCKAP1* | -3.478693629 | 9.54E-16 | 3.35E-15 | 6.00E-12 |
| Heart_Failure | dendritic_cell | *SFSWAP* | -4.445874915 | 1.22E-09 | 2.01E-09 | 7.69E-06 |
| Heart_Failure | dendritic_cell | *TNK2* | -2.548387806 | 0.000357703 | 0.000438635 | 1 |
| Heart_Failure | dendritic_cell | *VMP1* | -3.626876576 | 9.53E-07 | 1.32E-06 | 0.005991689 |
| Heart_Failure | dendritic_cell | *EIF4B* | -1.563658575 | 0.018924315 | 0.021360354 | 1 |
| Heart_Failure | dendritic_cell | *SLC6A16* | -3.632432102 | 1.88E-14 | 5.18E-14 | 1.18E-10 |
| Heart_Failure | dendritic_cell | *ISOC2* | -3.329766019 | 3.60E-20 | 6.99E-19 | 2.26E-16 |
| Heart_Failure | dendritic_cell | *ADCK1* | -5.127185407 | 1.10E-09 | 1.82E-09 | 6.95E-06 |
| Heart_Failure | dendritic_cell | *TM7SF3* | -4.582187051 | 1.37E-10 | 2.43E-10 | 8.62E-07 |
| Heart_Failure | dendritic_cell | *DMRT3* | -3.988077737 | 3.50E-20 | 6.98E-19 | 2.20E-16 |
| Heart_Failure | dendritic_cell | *ST3GAL6* | -1.617594247 | 0.003839554 | 0.004491863 | 1 |
| Heart_Failure | dendritic_cell | *ATP2C2* | -4.252028548 | 1.00E-18 | 7.77E-18 | 6.31E-15 |
| Heart_Failure | dendritic_cell | *CDON* | -1.894133561 | 6.25E-17 | 2.85E-16 | 3.93E-13 |
| Heart_Failure | dendritic_cell | *TAF2* | -4.821745984 | 1.63E-11 | 3.14E-11 | 1.03E-07 |
| Heart_Failure | dendritic_cell | *TMEM161A* | -4.894430001 | 1.24E-19 | 1.55E-18 | 7.82E-16 |
| Heart_Failure | dendritic_cell | *LPAR2* | 4.592543046 | 0.000105547 | 0.000132795 | 0.663575828 |
| Heart_Failure | dendritic_cell | *SUGP2* | -2.289877224 | 0.004612268 | 0.005372861 | 1 |
| Heart_Failure | dendritic_cell | *ABCA7* | 6.874900393 | 2.08E-10 | 3.63E-10 | 1.31E-06 |
| Heart_Failure | dendritic_cell | *CHI3L2* | -2.760754999 | 5.63E-11 | 1.03E-10 | 3.54E-07 |
| Heart_Failure | dendritic_cell | *PMS1* | -4.525539839 | 7.93E-14 | 1.98E-13 | 4.98E-10 |
| Heart_Failure | dendritic_cell | *TAF11* | -1.625301744 | 0.014412798 | 0.016356184 | 1 |
| Heart_Failure | dendritic_cell | *ANKS1A* | -2.439657872 | 0.004102668 | 0.004796109 | 1 |
| Heart_Failure | dendritic_cell | *NTHL1* | -5.637446516 | 2.20E-09 | 3.56E-09 | 1.39E-05 |
| Heart_Failure | dendritic_cell | *UHRF1BP1* | -1.720170414 | 9.29E-07 | 1.28E-06 | 0.005840743 |
| Heart_Failure | dendritic_cell | *IPO5* | -4.377168532 | 8.17E-17 | 3.65E-16 | 5.13E-13 |
| Heart_Failure | dendritic_cell | *WDR18* | -2.677365706 | 1.11E-06 | 1.53E-06 | 0.006991231 |
| Heart_Failure | dendritic_cell | *MCM10* | 2.017569896 | 5.88E-09 | 9.21E-09 | 3.69E-05 |
| Heart_Failure | dendritic_cell | *ANKRD44* | 2.487845494 | 4.20E-20 | 7.75E-19 | 2.64E-16 |
| Heart_Failure | dendritic_cell | *NDUFB4* | -5.82300547 | 6.48E-15 | 1.95E-14 | 4.07E-11 |
| Heart_Failure | dendritic_cell | *ZC3H15* | -4.979997458 | 3.62E-14 | 9.52E-14 | 2.28E-10 |
| Heart_Failure | dendritic_cell | *MAP2K4* | -4.471065006 | 6.57E-11 | 1.19E-10 | 4.13E-07 |
| Heart_Failure | dendritic_cell | *TMEM206* | -4.739892923 | 1.84E-18 | 1.30E-17 | 1.16E-14 |
| Heart_Failure | dendritic_cell | *SEC61A2* | -5.592963266 | 1.25E-15 | 4.28E-15 | 7.86E-12 |

| | | | | | | |
|---|---|---|---|---|---|---|
| Heart_Failure | dendritic_cell | *ASB1* | -5.844314866 | 6.50E-08 | 9.59E-08 | 0.000408346 |
| Heart_Failure | dendritic_cell | *TBC1D1* | -3.463187712 | 0.000528911 | 0.000643558 | 1 |
| Heart_Failure | dendritic_cell | *PPP2R5A* | -3.65221313 | 1.54E-15 | 5.18E-15 | 9.70E-12 |
| Heart_Failure | dendritic_cell | *TIE1* | -2.651567346 | 1.75E-20 | 6.17E-19 | 1.10E-16 |
| Heart_Failure | dendritic_cell | *ZMYND12* | 2.014491488 | 0.000118502 | 0.000148648 | 0.745022525 |
| Heart_Failure | dendritic_cell | *SLC9A3* | -1.468641436 | 0.001701385 | 0.002025105 | 1 |
| Heart_Failure | dendritic_cell | *NGEF* | -5.117252014 | 3.18E-20 | 6.76E-19 | 2.00E-16 |
| Heart_Failure | dendritic_cell | *CD84* | 2.309591939 | 2.30E-17 | 1.16E-16 | 1.45E-13 |
| Heart_Failure | dendritic_cell | *ZNRD1* | 2.88162362 | 6.82E-07 | 9.51E-07 | 0.004287967 |
| Heart_Failure | dendritic_cell | *ATXN3* | -3.360435058 | 9.98E-12 | 1.95E-11 | 6.28E-08 |
| Heart_Failure | dendritic_cell | *GOLGA5* | -5.507719546 | 1.08E-12 | 2.33E-12 | 6.76E-09 |
| Heart_Failure | dendritic_cell | *EML1* | -3.464796386 | 1.52E-14 | 4.24E-14 | 9.54E-11 |
| Heart_Failure | dendritic_cell | *ZFAT* | -4.031212239 | 9.37E-19 | 7.37E-18 | 5.89E-15 |
| Heart_Failure | dendritic_cell | *MTFR1* | -4.488227864 | 4.54E-14 | 1.17E-13 | 2.86E-10 |
| Heart_Failure | dendritic_cell | *STAG3* | -5.160113229 | 5.39E-12 | 1.09E-11 | 3.39E-08 |
| Heart_Failure | dendritic_cell | *MYO9A* | -3.464369168 | 1.05E-13 | 2.56E-13 | 6.58E-10 |
| Heart_Failure | dendritic_cell | *SP100* | -4.041435034 | 3.91E-10 | 6.67E-10 | 2.46E-06 |
| Heart_Failure | dendritic_cell | *STOML1* | -11.22224446 | 4.58E-13 | 1.03E-12 | 2.88E-09 |
| Heart_Failure | dendritic_cell | *METTL22* | -5.578185004 | 1.17E-15 | 4.02E-15 | 7.34E-12 |
| Heart_Failure | dendritic_cell | *TP53BP1* | -3.360191589 | 7.15E-18 | 4.17E-17 | 4.49E-14 |
| Heart_Failure | dendritic_cell | *RRP15* | -2.242001405 | 5.39E-10 | 9.11E-10 | 3.39E-06 |
| Heart_Failure | dendritic_cell | *DHX8* | -4.825014384 | 2.89E-15 | 9.22E-15 | 1.81E-11 |
| Heart_Failure | dendritic_cell | *SYT1* | -3.032842123 | 1.00E-14 | 2.90E-14 | 6.29E-11 |
| Heart_Failure | dendritic_cell | *ROGDI* | 6.23177163 | 1.92E-20 | 6.44E-19 | 1.21E-16 |
| Heart_Failure | dendritic_cell | *HDAC4* | -4.047542508 | 1.17E-14 | 3.35E-14 | 7.37E-11 |
| Heart_Failure | dendritic_cell | *RASSF1* | -3.42891864 | 5.60E-05 | 7.13E-05 | 0.352312925 |
| Heart_Failure | dendritic_cell | *FGFR3* | -0.684544988 | 0.003809207 | 0.00445802 | 1 |
| Heart_Failure | dendritic_cell | *IFI35* | -3.463428924 | 6.93E-12 | 1.38E-11 | 4.36E-08 |
| Heart_Failure | dendritic_cell | *HEATR6* | -4.869696491 | 4.63E-17 | 2.17E-16 | 2.91E-13 |
| Heart_Failure | dendritic_cell | *POLR1A* | -2.224882327 | 3.89E-13 | 8.86E-13 | 2.44E-09 |
| Heart_Failure | dendritic_cell | *STON1-GTF2A* | -0.581148461 | 8.28E-06 | 1.09E-05 | 0.052074788 |
| Heart_Failure | dendritic_cell | *SRBD1* | -4.443526275 | 1.78E-10 | 3.12E-10 | 1.12E-06 |
| Heart_Failure | dendritic_cell | *CYFIP1* | -4.486227706 | 2.52E-09 | 4.05E-09 | 1.58E-05 |
| Heart_Failure | dendritic_cell | *PSME4* | -3.976655144 | 1.12E-09 | 1.84E-09 | 7.02E-06 |
| Heart_Failure | dendritic_cell | *PITX1* | -7.247917622 | 2.71E-20 | 6.44E-19 | 1.71E-16 |
| Heart_Failure | dendritic_cell | *SDK2* | 6.370715748 | 4.50E-17 | 2.13E-16 | 2.83E-13 |
| Heart_Failure | dendritic_cell | *NUCKS1* | -4.191267647 | 2.48E-09 | 3.99E-09 | 1.56E-05 |
| Heart_Failure | dendritic_cell | *DNAJA2* | -5.270102653 | 8.40E-17 | 3.73E-16 | 5.28E-13 |
| Heart_Failure | dendritic_cell | *KCNAB2* | 4.044894341 | 8.92E-15 | 2.61E-14 | 5.61E-11 |
| Heart_Failure | dendritic_cell | *GAL* | 6.830752838 | 1.32E-19 | 1.61E-18 | 8.28E-16 |
| Heart_Failure | dendritic_cell | *NEDD4* | -3.664404524 | 3.20E-13 | 7.38E-13 | 2.01E-09 |
| Heart_Failure | dendritic_cell | *PIGB* | -4.810981551 | 6.72E-15 | 2.01E-14 | 4.22E-11 |
| Heart_Failure | dendritic_cell | *PFN2* | -3.246767351 | 3.10E-19 | 2.98E-18 | 1.95E-15 |
| Heart_Failure | dendritic_cell | *DAPP1* | 0.482803381 | 2.17E-21 | 4.66E-19 | 1.36E-17 |
| Heart_Failure | dendritic_cell | *FGF10* | -1.928110552 | 5.36E-18 | 3.24E-17 | 3.37E-14 |
| Heart_Failure | dendritic_cell | *TMEM260* | -4.789084649 | 6.66E-12 | 1.33E-11 | 4.19E-08 |
| Heart_Failure | dendritic_cell | *CLTCL1* | -4.087428048 | 1.35E-10 | 2.39E-10 | 8.49E-07 |
| Heart_Failure | dendritic_cell | *DGCR2* | -4.842981567 | 3.99E-15 | 1.24E-14 | 2.51E-11 |
| Heart_Failure | dendritic_cell | *MNT* | -4.239841537 | 1.70E-09 | 2.78E-09 | 1.07E-05 |
| Heart_Failure | dendritic_cell | *ZXDC* | -4.168486489 | 1.15E-08 | 1.76E-08 | 7.23E-05 |

| | | | | | | |
|---|---|---|---|---|---|---|
| Heart_Failure | dendritic_cell | *POLB* | -4.251032228 | 1.29E-16 | 5.44E-16 | 8.14E-13 |
| Heart_Failure | dendritic_cell | *WIPI1* | -4.938812587 | 3.51E-15 | 1.10E-14 | 2.21E-11 |
| Heart_Failure | dendritic_cell | *NDST1* | -4.147879271 | 4.50E-16 | 1.68E-15 | 2.83E-12 |
| Heart_Failure | dendritic_cell | *ASNS* | -4.654603294 | 5.06E-17 | 2.36E-16 | 3.18E-13 |
| Heart_Failure | dendritic_cell | *AP3M2* | -4.601723566 | 6.76E-17 | 3.07E-16 | 4.25E-13 |
| Heart_Failure | dendritic_cell | *ST6GALNAC2* | -3.724990155 | 1.14E-16 | 4.87E-16 | 7.18E-13 |
| Heart_Failure | dendritic_cell | *TESK2* | 2.877306919 | 0.010177923 | 0.011627949 | 1 |
| Heart_Failure | dendritic_cell | *EIF2B3* | -3.630646287 | 3.71E-16 | 1.42E-15 | 2.34E-12 |
| Heart_Failure | dendritic_cell | *CAMK2A* | -14.15770395 | 1.82E-19 | 2.05E-18 | 1.14E-15 |
| Heart_Failure | dendritic_cell | *OSBPL3* | 4.975911187 | 9.67E-13 | 2.10E-12 | 6.08E-09 |
| Heart_Failure | dendritic_cell | *RAD18* | -5.468833732 | 1.56E-11 | 3.00E-11 | 9.80E-08 |
| Heart_Failure | dendritic_cell | *SNX13* | -3.340186939 | 9.61E-05 | 0.000121068 | 0.604008065 |
| Heart_Failure | dendritic_cell | *VASH1* | 6.096434917 | 1.32E-07 | 1.91E-07 | 0.000827792 |
| Heart_Failure | dendritic_cell | *TRIP13* | -6.68490044 | 2.76E-20 | 6.44E-19 | 1.73E-16 |
| Heart_Failure | dendritic_cell | *TCF3* | -6.523175249 | 2.52E-15 | 8.11E-15 | 1.58E-11 |
| Heart_Failure | dendritic_cell | *MBD3* | -4.249447942 | 7.55E-15 | 2.23E-14 | 4.75E-11 |
| Heart_Failure | dendritic_cell | *HLTF* | -4.149583656 | 7.26E-13 | 1.60E-12 | 4.56E-09 |
| Heart_Failure | dendritic_cell | *CPSF1* | -6.94472469 | 1.24E-12 | 2.68E-12 | 7.82E-09 |
| Heart_Failure | dendritic_cell | *CYBRD1* | -2.596592933 | 2.65E-15 | 8.51E-15 | 1.67E-11 |
| Heart_Failure | dendritic_cell | *EPN2* | -4.046555374 | 1.52E-13 | 3.65E-13 | 9.57E-10 |
| Heart_Failure | dendritic_cell | *LIMS2* | -3.210651566 | 7.84E-21 | 4.66E-19 | 4.93E-17 |
| Heart_Failure | dendritic_cell | *AFF4* | -3.704682477 | 3.51E-09 | 5.59E-09 | 2.21E-05 |
| Heart_Failure | dendritic_cell | *TRHDE* | -1.723645748 | 1.09E-19 | 1.41E-18 | 6.86E-16 |
| Heart_Failure | dendritic_cell | *NFATC3* | -3.334733093 | 4.91E-13 | 1.10E-12 | 3.08E-09 |
| Heart_Failure | dendritic_cell | *TRNT1* | -5.172955452 | 8.90E-09 | 1.38E-08 | 5.59E-05 |
| Heart_Failure | dendritic_cell | *ACADVL* | -5.089477673 | 5.07E-13 | 1.14E-12 | 3.19E-09 |
| Heart_Failure | dendritic_cell | *CRMP1* | -4.563069117 | 3.02E-18 | 1.97E-17 | 1.90E-14 |
| Heart_Failure | dendritic_cell | *DERL2* | -5.863606699 | 1.79E-15 | 5.92E-15 | 1.12E-11 |
| Heart_Failure | dendritic_cell | *AP1M1* | -2.572655407 | 0.000171425 | 0.000213627 | 1 |
| Heart_Failure | dendritic_cell | *PVR* | 3.853648396 | 2.31E-05 | 2.98E-05 | 0.144954889 |
| Heart_Failure | dendritic_cell | *XRCC1* | 7.642562284 | 2.02E-08 | 3.05E-08 | 0.00012672 |
| Heart_Failure | dendritic_cell | *MOV10L1* | -2.368582677 | 3.14E-15 | 9.94E-15 | 1.97E-11 |
| Heart_Failure | dendritic_cell | *TP63* | -5.123862969 | 4.60E-12 | 9.34E-12 | 2.89E-08 |
| Heart_Failure | dendritic_cell | *PDE8A* | -3.021912522 | 2.18E-10 | 3.79E-10 | 1.37E-06 |
| Heart_Failure | dendritic_cell | *NLE1* | -5.751797102 | 1.96E-15 | 6.46E-15 | 1.23E-11 |
| Heart_Failure | dendritic_cell | *SDHA* | -4.170733609 | 4.85E-12 | 9.81E-12 | 3.05E-08 |
| Heart_Failure | dendritic_cell | *SMARCE1* | -2.973812614 | 0.006281284 | 0.007264612 | 1 |
| Heart_Failure | dendritic_cell | *GSDMB* | -2.855532449 | 1.06E-21 | 3.34E-19 | 6.68E-18 |
| Heart_Failure | dendritic_cell | *ADAM11* | -1.426317404 | 0.00232253 | 0.002745205 | 1 |
| Heart_Failure | dendritic_cell | *PICALM* | -3.260244911 | 2.88E-07 | 4.10E-07 | 0.001809216 |
| Heart_Failure | dendritic_cell | *GLI2* | -1.724693403 | 7.21E-18 | 4.20E-17 | 4.53E-14 |
| Heart_Failure | dendritic_cell | *ATP2A3* | -2.70495605 | 3.54E-18 | 2.26E-17 | 2.23E-14 |
| Heart_Failure | dendritic_cell | *CA12* | -2.01108339 | 3.27E-16 | 1.27E-15 | 2.06E-12 |
| Heart_Failure | dendritic_cell | *NTN4* | -2.520852819 | 2.66E-19 | 2.66E-18 | 1.67E-15 |
| Heart_Failure | dendritic_cell | *LMAN1* | -3.822410368 | 1.50E-17 | 8.04E-17 | 9.40E-14 |
| Heart_Failure | dendritic_cell | *ENO1* | -5.284609561 | 7.27E-12 | 1.44E-11 | 4.57E-08 |
| Heart_Failure | dendritic_cell | *TIPIN* | -0.78230819 | 0.002111568 | 0.002500552 | 1 |
| Heart_Failure | dendritic_cell | *SRI* | -5.935524637 | 1.01E-12 | 2.19E-12 | 6.33E-09 |
| Heart_Failure | dendritic_cell | *EIF4G3* | -3.518811955 | 3.00E-08 | 4.49E-08 | 0.000188471 |
| Heart_Failure | dendritic_cell | *TTC38* | -4.18309099 | 2.13E-17 | 1.09E-16 | 1.34E-13 |

| | | | | | | |
|---|---|---|---|---|---|---|
| Heart_Failure | dendritic_cell | *GRAMD4* | -3.231235128 | 0.009580798 | 0.010969673 | 1 |
| Heart_Failure | dendritic_cell | *CELSR1* | 6.678986445 | 0.00011617 | 0.000145752 | 0.730362393 |
| Heart_Failure | dendritic_cell | *ADD2* | 10.9494827 | 6.22E-14 | 1.58E-13 | 3.91E-10 |
| Heart_Failure | dendritic_cell | *VPS9D1* | 7.218656515 | 5.25E-15 | 1.60E-14 | 3.30E-11 |
| Heart_Failure | dendritic_cell | *MARK3* | -3.957469056 | 1.31E-12 | 2.82E-12 | 8.26E-09 |
| Heart_Failure | dendritic_cell | *SLC25A3* | -4.522319123 | 2.53E-17 | 1.27E-16 | 1.59E-13 |
| Heart_Failure | dendritic_cell | *FSCN1* | 3.234949509 | 4.57E-21 | 4.66E-19 | 2.87E-17 |
| Heart_Failure | dendritic_cell | *MOCOS* | -2.815457721 | 5.81E-15 | 1.76E-14 | 3.65E-11 |
| Heart_Failure | dendritic_cell | *PLD1* | -3.960218871 | 1.52E-13 | 3.65E-13 | 9.59E-10 |
| Heart_Failure | dendritic_cell | *BCAP29* | -4.292875326 | 1.48E-07 | 2.13E-07 | 0.000928531 |
| Heart_Failure | dendritic_cell | *SEC31B* | -1.130514533 | 3.25E-06 | 4.37E-06 | 0.020424129 |
| Heart_Failure | dendritic_cell | *TUBA3D* | -7.45359052 | 4.88E-14 | 1.25E-13 | 3.07E-10 |
| Heart_Failure | dendritic_cell | *MKRN2* | -5.124852615 | 4.20E-18 | 2.62E-17 | 2.64E-14 |
| Heart_Failure | dendritic_cell | *MCM6* | -5.530704691 | 2.93E-08 | 4.39E-08 | 0.000184294 |
| Heart_Failure | dendritic_cell | *RBMS2* | -4.221675476 | 2.76E-13 | 6.42E-13 | 1.73E-09 |
| Heart_Failure | dendritic_cell | *MLH1* | -4.027685622 | 1.36E-11 | 2.64E-11 | 8.56E-08 |
| Heart_Failure | dendritic_cell | *FMO4* | -4.099975813 | 6.20E-18 | 3.69E-17 | 3.90E-14 |
| Heart_Failure | dendritic_cell | *RGS11* | -2.381784838 | 5.42E-11 | 9.91E-11 | 3.41E-07 |
| Heart_Failure | dendritic_cell | *TPD52* | 2.803319892 | 3.46E-05 | 4.43E-05 | 0.217323474 |
| Heart_Failure | dendritic_cell | *GPATCH1* | -3.126031816 | 0.001296749 | 0.001548758 | 1 |
| Heart_Failure | dendritic_cell | *NT5C2* | -2.997617114 | 8.28E-05 | 0.000104529 | 0.520658258 |
| Heart_Failure | dendritic_cell | *CAMSAP3* | -17.78144439 | 2.41E-20 | 6.44E-19 | 1.52E-16 |
| Heart_Failure | dendritic_cell | *XAB2* | -4.986269969 | 1.53E-18 | 1.12E-17 | 9.64E-15 |
| Heart_Failure | dendritic_cell | *STXBP2* | 3.683694656 | 2.29E-15 | 7.45E-15 | 1.44E-11 |
| Heart_Failure | dendritic_cell | *MAP2K7* | -6.431718222 | 1.36E-12 | 2.91E-12 | 8.52E-09 |
| Heart_Failure | dendritic_cell | *NMRK2* | -9.02618489 | 6.37E-10 | 1.07E-09 | 4.00E-06 |
| Heart_Failure | dendritic_cell | *CTTNBP2* | -3.203230138 | 1.13E-05 | 1.49E-05 | 0.071105567 |
| Heart_Failure | dendritic_cell | *TM9SF3* | -4.541428087 | 2.65E-11 | 4.97E-11 | 1.66E-07 |
| Heart_Failure | dendritic_cell | *DYNC1I2* | -3.816429883 | 1.77E-15 | 5.88E-15 | 1.11E-11 |
| Heart_Failure | dendritic_cell | *APBB1IP* | -3.500436478 | 3.90E-12 | 7.95E-12 | 2.45E-08 |
| Heart_Failure | dendritic_cell | *LRCH4* | 3.179522011 | 8.13E-17 | 3.63E-16 | 5.11E-13 |
| Heart_Failure | dendritic_cell | *SIRT6* | -9.809548928 | 2.94E-12 | 6.08E-12 | 1.85E-08 |
| Heart_Failure | dendritic_cell | *CAPZB* | -4.066806206 | 5.22E-11 | 9.55E-11 | 3.28E-07 |
| Heart_Failure | dendritic_cell | *GPR137B* | 2.299720649 | 2.23E-05 | 2.89E-05 | 0.140460937 |
| Heart_Failure | dendritic_cell | *NAALAD2* | -2.635954091 | 8.58E-16 | 3.04E-15 | 5.40E-12 |
| Heart_Failure | dendritic_cell | *FGFR1* | -2.801323531 | 2.28E-16 | 9.11E-16 | 1.44E-12 |
| Heart_Failure | dendritic_cell | *FBLN1* | -1.876089575 | 5.18E-20 | 8.74E-19 | 3.26E-16 |
| Heart_Failure | dendritic_cell | *PIAS2* | -3.920385793 | 7.57E-07 | 1.05E-06 | 0.004759248 |
| Heart_Failure | dendritic_cell | *AMPH* | -1.645738793 | 7.28E-19 | 5.95E-18 | 4.57E-15 |
| Heart_Failure | dendritic_cell | *MCCC1* | -2.901877628 | 1.48E-12 | 3.17E-12 | 9.33E-09 |
| Heart_Failure | dendritic_cell | *FAP* | -1.624682118 | 1.30E-19 | 1.59E-18 | 8.16E-16 |
| Heart_Failure | dendritic_cell | *ACER3* | -3.311812996 | 1.26E-07 | 1.83E-07 | 0.00079279 |
| Heart_Failure | dendritic_cell | *PIK3C3* | -4.607719066 | 4.04E-13 | 9.18E-13 | 2.54E-09 |
| Heart_Failure | dendritic_cell | *N4BP2* | 4.770363357 | 1.91E-08 | 2.89E-08 | 0.000119775 |
| Heart_Failure | dendritic_cell | *TULP3* | -2.834121704 | 7.31E-19 | 5.97E-18 | 4.60E-15 |
| Heart_Failure | dendritic_cell | *SYNJ2* | -2.436817248 | 1.72E-18 | 1.24E-17 | 1.08E-14 |
| Heart_Failure | dendritic_cell | *ZCWPW1* | 3.614959913 | 5.87E-06 | 7.80E-06 | 0.036890583 |
| Heart_Failure | dendritic_cell | *FGF20* | 14.56081852 | 7.06E-21 | 4.66E-19 | 4.44E-17 |
| Heart_Failure | dendritic_cell | *PCM1* | -3.417891763 | 6.70E-14 | 1.69E-13 | 4.21E-10 |
| Heart_Failure | dendritic_cell | *MYH7B* | -6.852315547 | 2.70E-11 | 5.06E-11 | 1.69E-07 |

| | | | | | | |
|---|---|---|---|---|---|---|
| Heart_Failure | dendritic_cell | *FKBP7* | -2.282333035 | 1.81E-14 | 4.99E-14 | 1.14E-10 |
| Heart_Failure | dendritic_cell | *OSBPL6* | -5.489581575 | 4.69E-16 | 1.75E-15 | 2.95E-12 |
| Heart_Failure | dendritic_cell | *XRCC5* | -3.903361059 | 7.82E-07 | 1.09E-06 | 0.004916245 |
| Heart_Failure | dendritic_cell | *LXN* | -2.805770851 | 0.013255215 | 0.015066993 | 1 |
| Heart_Failure | dendritic_cell | *REXO1* | -4.566612853 | 3.60E-18 | 2.29E-17 | 2.26E-14 |
| Heart_Failure | dendritic_cell | *SAR1A* | -4.153123635 | 1.54E-16 | 6.39E-16 | 9.71E-13 |
| Heart_Failure | dendritic_cell | *RAPGEF3* | -1.004153735 | 5.23E-07 | 7.34E-07 | 0.003288869 |
| Heart_Failure | dendritic_cell | *FDFT1* | -4.482168744 | 6.57E-16 | 2.38E-15 | 4.13E-12 |
| Heart_Failure | dendritic_cell | *SCGN* | 10.94849316 | 4.68E-22 | 2.45E-19 | 2.94E-18 |
| Heart_Failure | dendritic_cell | *DDX1* | -3.742235425 | 2.03E-17 | 1.05E-16 | 1.28E-13 |
| Heart_Failure | dendritic_cell | *DNM2* | -4.601675736 | 3.88E-09 | 6.15E-09 | 2.44E-05 |
| Heart_Failure | dendritic_cell | *RIMS1* | -4.760749771 | 7.42E-18 | 4.31E-17 | 4.67E-14 |
| Heart_Failure | dendritic_cell | *STX7* | -2.571222205 | 0.000185152 | 0.000230323 | 1 |
| Heart_Failure | dendritic_cell | *RABL2B* | -5.346839164 | 1.63E-15 | 5.45E-15 | 1.02E-11 |
| Heart_Failure | dendritic_cell | *DDX43* | 3.926399307 | 8.53E-18 | 4.90E-17 | 5.37E-14 |
| Heart_Failure | dendritic_cell | *DCT* | -4.034953674 | 6.07E-20 | 9.70E-19 | 3.81E-16 |
| Heart_Failure | dendritic_cell | *SLC35C2* | -3.141221499 | 0.011974812 | 0.013631295 | 1 |
| Heart_Failure | dendritic_cell | *SCTR* | -4.059107424 | 7.05E-19 | 5.79E-18 | 4.43E-15 |
| Heart_Failure | dendritic_cell | *RIF1* | -3.000942063 | 2.35E-06 | 3.18E-06 | 0.014743849 |
| Heart_Failure | dendritic_cell | *KCNN2* | -5.320174299 | 6.16E-20 | 9.76E-19 | 3.87E-16 |
| Heart_Failure | dendritic_cell | *PSEN1* | -3.134090924 | 3.64E-05 | 4.66E-05 | 0.228786919 |
| Heart_Failure | dendritic_cell | *CPOX* | -3.60939367 | 5.75E-17 | 2.65E-16 | 3.62E-13 |
| Heart_Failure | dendritic_cell | *MOK* | -5.681277201 | 8.81E-14 | 2.18E-13 | 5.54E-10 |
| Heart_Failure | dendritic_cell | *RSBN1* | -4.467656583 | 6.99E-07 | 9.74E-07 | 0.004393959 |
| Heart_Failure | dendritic_cell | *MAGI3* | -3.087914443 | 1.70E-17 | 9.00E-17 | 1.07E-13 |
| Heart_Failure | dendritic_cell | *COL4A4* | -3.411797117 | 5.25E-17 | 2.44E-16 | 3.30E-13 |
| Heart_Failure | dendritic_cell | *STK17B* | 2.784774353 | 7.14E-20 | 1.07E-18 | 4.49E-16 |
| Heart_Failure | dendritic_cell | *ZNF506* | -5.379570143 | 2.80E-10 | 4.84E-10 | 1.76E-06 |
| Heart_Failure | dendritic_cell | *KIAA0141* | 5.343745034 | 7.56E-08 | 1.11E-07 | 0.000475509 |
| Heart_Failure | dendritic_cell | *PCDHA6* | -13.43460563 | 0.000255321 | 0.000314993 | 1 |
| Heart_Failure | dendritic_cell | *PCDHGA2* | -7.719076726 | 1.57E-16 | 6.49E-16 | 9.87E-13 |
| Heart_Failure | dendritic_cell | *HSPB11* | -4.783012879 | 1.00E-06 | 1.38E-06 | 0.0063164 |
| Heart_Failure | dendritic_cell | *WDR70* | -3.939253544 | 1.33E-11 | 2.57E-11 | 8.33E-08 |
| Heart_Failure | dendritic_cell | *PGR* | -2.336719515 | 3.16E-17 | 1.55E-16 | 1.99E-13 |
| Heart_Failure | dendritic_cell | *C5orf22* | -5.819294846 | 1.18E-06 | 1.62E-06 | 0.007409321 |
| Heart_Failure | dendritic_cell | *CCNT2* | -2.870968366 | 2.47E-09 | 3.98E-09 | 1.55E-05 |
| Heart_Failure | dendritic_cell | *COBLL1* | -2.008934573 | 1.03E-16 | 4.45E-16 | 6.49E-13 |
| Heart_Failure | dendritic_cell | *GEMIN5* | -3.253792668 | 4.43E-06 | 5.92E-06 | 0.027868658 |
| Heart_Failure | dendritic_cell | *GSK3B* | -4.005824232 | 2.63E-11 | 4.95E-11 | 1.66E-07 |
| Heart_Failure | dendritic_cell | *ERC1* | -3.613211475 | 1.41E-11 | 2.72E-11 | 8.85E-08 |
| Heart_Failure | dendritic_cell | *DOPEY1* | -2.334660017 | 5.90E-07 | 8.25E-07 | 0.003710892 |
| Heart_Failure | dendritic_cell | *LYRM2* | -3.273900735 | 4.87E-13 | 1.10E-12 | 3.06E-09 |
| Heart_Failure | dendritic_cell | *ULK2* | -3.499079829 | 3.41E-15 | 1.07E-14 | 2.14E-11 |
| Heart_Failure | dendritic_cell | *PLOD1* | -7.272080147 | 2.03E-12 | 4.27E-12 | 1.28E-08 |
| Heart_Failure | dendritic_cell | *PIBF1* | -3.528566418 | 9.93E-14 | 2.43E-13 | 6.24E-10 |
| Heart_Failure | dendritic_cell | *TDRD3* | -3.770368051 | 1.36E-14 | 3.83E-14 | 8.54E-11 |
| Heart_Failure | dendritic_cell | *PDS5B* | -4.189897957 | 9.25E-11 | 1.66E-10 | 5.82E-07 |
| Heart_Failure | dendritic_cell | *OXCT1* | -2.87758456 | 2.90E-22 | 1.82E-19 | 1.82E-18 |
| Heart_Failure | dendritic_cell | *EPYC* | 9.084330306 | 1.35E-18 | 9.94E-18 | 8.47E-15 |
| Heart_Failure | dendritic_cell | *CYLD* | 5.415041189 | 2.23E-10 | 3.88E-10 | 1.40E-06 |

| | | | | | | |
|---|---|---|---|---|---|---|
| Heart_Failure | dendritic_cell | *ZNF324* | -3.19794339 | 6.49E-20 | 1.01E-18 | 4.08E-16 |
| Heart_Failure | dendritic_cell | *ZNF586* | -4.194041952 | 9.35E-09 | 1.44E-08 | 5.88E-05 |
| Heart_Failure | dendritic_cell | *ZNF446* | -5.845400799 | 1.03E-18 | 7.98E-18 | 6.50E-15 |
| Heart_Failure | dendritic_cell | *ZNF264* | -5.198755536 | 1.17E-15 | 4.02E-15 | 7.34E-12 |
| Heart_Failure | dendritic_cell | *RPS5* | -5.642899528 | 3.58E-09 | 5.70E-09 | 2.25E-05 |
| Heart_Failure | dendritic_cell | *CHMP2B* | -3.67430581 | 1.25E-14 | 3.56E-14 | 7.87E-11 |
| Heart_Failure | dendritic_cell | *PPIE* | -3.512992919 | 1.65E-05 | 2.15E-05 | 0.103807809 |
| Heart_Failure | dendritic_cell | *ZMPSTE24* | -5.834536555 | 1.18E-10 | 2.11E-10 | 7.43E-07 |
| Heart_Failure | dendritic_cell | *NOA1* | -4.515127749 | 1.57E-17 | 8.36E-17 | 9.85E-14 |
| Heart_Failure | dendritic_cell | *WBP11* | 3.470724891 | 0.009624232 | 0.011015392 | 1 |
| Heart_Failure | dendritic_cell | *COL16A1* | -2.696163922 | 9.38E-24 | 1.18E-20 | 5.90E-20 |
| Heart_Failure | dendritic_cell | *APOB* | -2.999263722 | 6.70E-19 | 5.51E-18 | 4.21E-15 |
| Heart_Failure | dendritic_cell | *NCOA1* | -3.631373224 | 6.14E-11 | 1.12E-10 | 3.86E-07 |
| Heart_Failure | dendritic_cell | *CD59* | -3.82781222 | 2.36E-17 | 1.19E-16 | 1.48E-13 |
| Heart_Failure | dendritic_cell | *CD82* | -3.568768923 | 2.47E-12 | 5.15E-12 | 1.55E-08 |
| Heart_Failure | dendritic_cell | *TAF9* | -4.478000127 | 0.01444711 | 0.016392164 | 1 |
| Heart_Failure | dendritic_cell | *FCN1* | 0.667146174 | 1.03E-20 | 4.95E-19 | 6.50E-17 |
| Heart_Failure | dendritic_cell | *SCAMP1* | -4.520024305 | 5.44E-13 | 1.21E-12 | 3.42E-09 |
| Heart_Failure | dendritic_cell | *HACE1* | -3.920665291 | 1.43E-16 | 5.98E-16 | 9.02E-13 |
| Heart_Failure | dendritic_cell | *SEH1L* | -3.650238661 | 0.000441383 | 0.000539354 | 1 |
| Heart_Failure | dendritic_cell | *SLC25A24* | -3.850796981 | 1.24E-10 | 2.20E-10 | 7.79E-07 |
| Heart_Failure | dendritic_cell | *PILRA* | 0.764224801 | 8.42E-07 | 1.17E-06 | 0.005291802 |
| Heart_Failure | dendritic_cell | *ABCB1* | -2.116310995 | 3.54E-16 | 1.36E-15 | 2.22E-12 |
| Heart_Failure | dendritic_cell | *ZNF213* | -3.351239818 | 8.35E-08 | 1.22E-07 | 0.000525105 |
| Heart_Failure | dendritic_cell | *CPNE3* | -2.095934041 | 0.004395969 | 0.005126592 | 1 |
| Heart_Failure | dendritic_cell | *RRN3* | -4.32468344 | 5.92E-12 | 1.19E-11 | 3.72E-08 |
| Heart_Failure | dendritic_cell | *CTTN* | -3.761134542 | 6.85E-17 | 3.11E-16 | 4.30E-13 |
| Heart_Failure | dendritic_cell | *MTIF2* | -4.639147315 | 5.15E-14 | 1.32E-13 | 3.24E-10 |
| Heart_Failure | dendritic_cell | *EPS15* | -3.067344967 | 1.50E-10 | 2.64E-10 | 9.41E-07 |
| Heart_Failure | dendritic_cell | *ATG16L1* | -5.054357647 | 9.27E-16 | 3.27E-15 | 5.83E-12 |
| Heart_Failure | dendritic_cell | *USP40* | -3.19572758 | 2.44E-16 | 9.65E-16 | 1.53E-12 |
| Heart_Failure | dendritic_cell | *B4GALT1* | 4.522641415 | 8.34E-07 | 1.16E-06 | 0.005246064 |
| Heart_Failure | dendritic_cell | *CHMP5* | -3.927428604 | 0.000953443 | 0.001145261 | 1 |
| Heart_Failure | dendritic_cell | *FOLH1* | 1.084854768 | 0.000249311 | 0.00030788 | 1 |
| Heart_Failure | dendritic_cell | *EIF2AK1* | -4.393689491 | 2.35E-14 | 6.39E-14 | 1.48E-10 |
| Heart_Failure | dendritic_cell | *EPDR1* | -1.339840213 | 0.000681137 | 0.000823362 | 1 |
| Heart_Failure | dendritic_cell | *SEPHS1* | -4.830718085 | 3.58E-07 | 5.08E-07 | 0.002251844 |
| Heart_Failure | dendritic_cell | *ITPKC* | -4.099610818 | 0.000216515 | 0.00026817 | 1 |
| Heart_Failure | dendritic_cell | *CEACAM6* | -12.11449187 | 0.002082469 | 0.002467952 | 1 |
| Heart_Failure | dendritic_cell | *TMED2* | -6.254722777 | 3.73E-11 | 6.93E-11 | 2.34E-07 |
| Heart_Failure | dendritic_cell | *ACOX3* | -3.398161909 | 1.07E-12 | 2.32E-12 | 6.73E-09 |
| Heart_Failure | dendritic_cell | *HSD17B14* | 3.431558378 | 2.79E-19 | 2.77E-18 | 1.75E-15 |
| Heart_Failure | dendritic_cell | *TRIP6* | -3.655114709 | 4.73E-17 | 2.21E-16 | 2.97E-13 |
| Heart_Failure | dendritic_cell | *ACHE* | -1.363881287 | 0.000273229 | 0.000336575 | 1 |
| Heart_Failure | dendritic_cell | *SRRT* | -3.539310121 | 2.73E-07 | 3.90E-07 | 0.001718513 |
| Heart_Failure | dendritic_cell | *PIGS* | -2.345160141 | 1.42E-07 | 2.05E-07 | 0.000893974 |
| Heart_Failure | dendritic_cell | *PGS1* | -4.975902188 | 1.10E-06 | 1.51E-06 | 0.006889372 |
| Heart_Failure | dendritic_cell | *PSMC5* | -3.075654247 | 2.18E-12 | 4.58E-12 | 1.37E-08 |
| Heart_Failure | dendritic_cell | *LPCAT2* | 4.081905105 | 8.63E-14 | 2.14E-13 | 5.43E-10 |
| Heart_Failure | dendritic_cell | *TXNDC16* | 3.067121431 | 0.000193317 | 0.000240242 | 1 |

| | | | | | | |
|---|---|---|---|---|---|---|
| Heart_Failure | dendritic_cell | *GMCL1* | -4.286073638 | 2.06E-17 | 1.06E-16 | 1.30E-13 |
| Heart_Failure | dendritic_cell | *KLHL42* | -5.109837958 | 7.31E-11 | 1.32E-10 | 4.60E-07 |
| Heart_Failure | dendritic_cell | *PTHLH* | -3.122022443 | 1.03E-19 | 1.37E-18 | 6.49E-16 |
| Heart_Failure | dendritic_cell | *PHACTR3* | -2.769556507 | 6.03E-17 | 2.76E-16 | 3.79E-13 |
| Heart_Failure | dendritic_cell | *ERGIC2* | -4.591930767 | 5.69E-14 | 1.45E-13 | 3.57E-10 |
| Heart_Failure | dendritic_cell | *AAMDC* | -2.145387361 | 7.54E-12 | 1.49E-11 | 4.74E-08 |
| Heart_Failure | dendritic_cell | *METTL2A* | -4.101324611 | 1.33E-10 | 2.37E-10 | 8.38E-07 |
| Heart_Failure | dendritic_cell | *ALG6* | -4.177404075 | 7.05E-05 | 8.93E-05 | 0.443160159 |
| Heart_Failure | dendritic_cell | *GP6* | 4.152710035 | 4.79E-19 | 4.22E-18 | 3.01E-15 |
| Heart_Failure | dendritic_cell | *DDX18* | -3.950961609 | 2.02E-06 | 2.75E-06 | 0.012717097 |
| Heart_Failure | dendritic_cell | *ASAP3* | -3.201638759 | 2.45E-19 | 2.49E-18 | 1.54E-15 |
| Heart_Failure | dendritic_cell | *EDEM2* | -4.096447656 | 2.64E-11 | 4.96E-11 | 1.66E-07 |
| Heart_Failure | dendritic_cell | *TPX2* | -2.24363097 | 1.86E-11 | 3.57E-11 | 1.17E-07 |
| Heart_Failure | dendritic_cell | *DOCK3* | -2.205160206 | 3.51E-05 | 4.49E-05 | 0.220404807 |
| Heart_Failure | dendritic_cell | *C3orf18* | 5.656131904 | 6.11E-08 | 9.02E-08 | 0.000383862 |
| Heart_Failure | dendritic_cell | *FKBP1A* | 7.484848374 | 2.24E-11 | 4.24E-11 | 1.41E-07 |
| Heart_Failure | dendritic_cell | *NSFL1C* | -4.592462631 | 1.28E-13 | 3.10E-13 | 8.05E-10 |
| Heart_Failure | dendritic_cell | *ZNF343* | -2.708189893 | 1.09E-18 | 8.36E-18 | 6.88E-15 |
| Heart_Failure | dendritic_cell | *CPXM1* | -1.828274621 | 4.58E-07 | 6.45E-07 | 0.002878812 |
| Heart_Failure | dendritic_cell | *MAVS* | -3.825106851 | 1.31E-05 | 1.72E-05 | 0.082545231 |
| Heart_Failure | dendritic_cell | *LZTS3* | -4.29591743 | 2.74E-19 | 2.73E-18 | 1.72E-15 |
| Heart_Failure | dendritic_cell | *XRN2* | -3.466105708 | 0.00019743 | 0.000245159 | 1 |
| Heart_Failure | dendritic_cell | *TESC* | -9.50080342 | 1.44E-14 | 4.03E-14 | 9.04E-11 |
| Heart_Failure | dendritic_cell | *RBBP9* | -7.295946747 | 7.14E-12 | 1.42E-11 | 4.49E-08 |
| Heart_Failure | dendritic_cell | *ANAPC5* | -4.190850363 | 5.99E-12 | 1.20E-11 | 3.77E-08 |
| Heart_Failure | dendritic_cell | *TMEM230* | -4.60673999 | 1.13E-14 | 3.24E-14 | 7.12E-11 |
| Heart_Failure | dendritic_cell | *DZANK1* | -5.511540036 | 2.42E-06 | 3.28E-06 | 0.015224043 |
| Heart_Failure | dendritic_cell | *TASP1* | -3.571957858 | 3.29E-11 | 6.14E-11 | 2.07E-07 |
| Heart_Failure | dendritic_cell | *OAS1* | 5.367127788 | 1.27E-18 | 9.45E-18 | 7.97E-15 |
| Heart_Failure | dendritic_cell | *FUS* | -3.812989488 | 4.49E-07 | 6.34E-07 | 0.002825848 |
| Heart_Failure | dendritic_cell | *FXYD5* | 6.983093305 | 7.91E-08 | 1.16E-07 | 0.000497397 |
| Heart_Failure | dendritic_cell | *ZNF302* | -2.114807712 | 8.16E-09 | 1.27E-08 | 5.13E-05 |
| Heart_Failure | dendritic_cell | *FXYD3* | -8.507924798 | 1.42E-11 | 2.75E-11 | 8.95E-08 |
| Heart_Failure | dendritic_cell | *KCNH4* | 2.921369987 | 4.14E-22 | 2.37E-19 | 2.60E-18 |
| Heart_Failure | dendritic_cell | *GANAB* | -5.228179531 | 3.25E-13 | 7.50E-13 | 2.05E-09 |
| Heart_Failure | dendritic_cell | *GMIP* | 5.263652067 | 1.57E-19 | 1.85E-18 | 9.90E-16 |
| Heart_Failure | dendritic_cell | *MLF2* | -5.914021521 | 1.33E-06 | 1.82E-06 | 0.008338186 |
| Heart_Failure | dendritic_cell | *GPATCH2L* | -2.795623524 | 6.68E-05 | 8.47E-05 | 0.419997593 |
| Heart_Failure | dendritic_cell | *SLC9A1* | -6.64056731 | 1.59E-11 | 3.06E-11 | 1.00E-07 |
| Heart_Failure | dendritic_cell | *SPTLC1* | -4.034179187 | 4.98E-11 | 9.13E-11 | 3.13E-07 |
| Heart_Failure | dendritic_cell | *PAPOLA* | -3.81682952 | 4.27E-06 | 5.71E-06 | 0.026862715 |
| Heart_Failure | dendritic_cell | *RGS1* | 0.888421307 | 6.31E-21 | 4.66E-19 | 3.97E-17 |
| Heart_Failure | dendritic_cell | *YPEL3* | -1.756607621 | 7.66E-06 | 1.01E-05 | 0.048181688 |
| Heart_Failure | dendritic_cell | *MRPS33* | -3.711935915 | 0.004917747 | 0.005719178 | 1 |
| Heart_Failure | dendritic_cell | *MAEA* | -3.266745877 | 2.64E-08 | 3.96E-08 | 0.000165778 |
| Heart_Failure | dendritic_cell | *STRN4* | -5.500502 | 4.23E-18 | 2.63E-17 | 2.66E-14 |
| Heart_Failure | dendritic_cell | *LYZ* | 1.182947518 | 4.64E-20 | 8.15E-19 | 2.92E-16 |
| Heart_Failure | dendritic_cell | *GNPTG* | -3.705474905 | 0.002991269 | 0.003517791 | 1 |
| Heart_Failure | dendritic_cell | *ZNF268* | -4.857429871 | 1.05E-16 | 4.52E-16 | 6.60E-13 |
| Heart_Failure | dendritic_cell | *GOLGA3* | -4.108514411 | 5.01E-11 | 9.18E-11 | 3.15E-07 |

| | | | | | | |
|---|---|---|---|---|---|---|
| Heart_Failure | dendritic_cell | *PABPC4* | 1.493575946 | 0.026555436 | 0.029712409 | 1 |
| Heart_Failure | dendritic_cell | *CERS4* | -3.237870876 | 5.37E-19 | 4.66E-18 | 3.38E-15 |
| Heart_Failure | dendritic_cell | *USP48* | -2.931731902 | 7.04E-07 | 9.81E-07 | 0.004424213 |
| Heart_Failure | dendritic_cell | *PDPR* | -4.86237878 | 1.12E-11 | 2.19E-11 | 7.07E-08 |
| Heart_Failure | dendritic_cell | *EXOC1* | -4.199207761 | 1.43E-08 | 2.18E-08 | 8.98E-05 |
| Heart_Failure | dendritic_cell | *NLRC4* | 4.202056122 | 7.81E-20 | 1.13E-18 | 4.91E-16 |
| Heart_Failure | dendritic_cell | *LAMB1* | -3.151774092 | 5.65E-12 | 1.14E-11 | 3.55E-08 |
| Heart_Failure | dendritic_cell | *SLC26A4* | 11.8718804 | 1.08E-17 | 6.04E-17 | 6.79E-14 |
| Heart_Failure | dendritic_cell | *DLD* | -4.354635896 | 1.42E-16 | 5.95E-16 | 8.95E-13 |
| Heart_Failure | dendritic_cell | *TXNL1* | -5.125195308 | 2.96E-10 | 5.10E-10 | 1.86E-06 |
| Heart_Failure | dendritic_cell | *ITGA6* | -3.276709726 | 1.95E-18 | 1.37E-17 | 1.22E-14 |
| Heart_Failure | dendritic_cell | *RAPGEF4* | -3.081676345 | 7.16E-18 | 4.17E-17 | 4.50E-14 |
| Heart_Failure | dendritic_cell | *SEL1L3* | 3.753380231 | 5.67E-19 | 4.86E-18 | 3.56E-15 |
| Heart_Failure | dendritic_cell | *TF* | -2.537268332 | 6.80E-20 | 1.03E-18 | 4.28E-16 |
| Heart_Failure | dendritic_cell | *ALKBH5* | -4.577363642 | 1.93E-12 | 4.08E-12 | 1.22E-08 |
| Heart_Failure | dendritic_cell | *NLRP1* | -4.289654881 | 6.49E-12 | 1.30E-11 | 4.08E-08 |
| Heart_Failure | dendritic_cell | *SPAG7* | -1.383319449 | 0.019031966 | 0.021478006 | 1 |
| Heart_Failure | dendritic_cell | *ZFHX4* | -1.790916971 | 5.15E-20 | 8.72E-19 | 3.24E-16 |
| Heart_Failure | dendritic_cell | *RGS17* | -2.213537229 | 3.88E-18 | 2.43E-17 | 2.44E-14 |
| Heart_Failure | dendritic_cell | *PPP2R3C* | -4.427954312 | 1.23E-09 | 2.03E-09 | 7.73E-06 |
| Heart_Failure | dendritic_cell | *HAUS4* | -1.229375555 | 0.00088427 | 0.001064001 | 1 |
| Heart_Failure | dendritic_cell | *SLC22A17* | -4.877940825 | 1.35E-09 | 2.22E-09 | 8.51E-06 |
| Heart_Failure | dendritic_cell | *SCFD1* | -3.411378765 | 7.26E-10 | 1.21E-09 | 4.56E-06 |
| Heart_Failure | dendritic_cell | *RPGRIP1* | 2.486547409 | 1.13E-20 | 4.99E-19 | 7.09E-17 |
| Heart_Failure | dendritic_cell | *TOX4* | -4.873246623 | 3.42E-10 | 5.86E-10 | 2.15E-06 |
| Heart_Failure | dendritic_cell | *TGM1* | -2.981506524 | 2.62E-10 | 4.53E-10 | 1.65E-06 |
| Heart_Failure | dendritic_cell | *AGO1* | -5.276336079 | 2.92E-09 | 4.66E-09 | 1.83E-05 |
| Heart_Failure | dendritic_cell | *RFFL* | 8.913120521 | 9.65E-10 | 1.60E-09 | 6.07E-06 |
| Heart_Failure | dendritic_cell | *MFSD11* | -3.347141281 | 1.97E-08 | 2.98E-08 | 0.000123751 |
| Heart_Failure | dendritic_cell | *NUP50* | 3.3114037 | 0.000197484 | 0.000245178 | 1 |
| Heart_Failure | dendritic_cell | *VNN3* | 2.039990907 | 4.57E-21 | 4.66E-19 | 2.87E-17 |
| Heart_Failure | dendritic_cell | *SEC22C* | -4.794574347 | 8.13E-16 | 2.89E-15 | 5.11E-12 |
| Heart_Failure | dendritic_cell | *FMO2* | -2.692717509 | 6.99E-12 | 1.39E-11 | 4.40E-08 |
| Heart_Failure | dendritic_cell | *SUCO* | -4.913985372 | 5.30E-13 | 1.18E-12 | 3.33E-09 |
| Heart_Failure | dendritic_cell | *MSH2* | 6.314496281 | 6.24E-09 | 9.76E-09 | 3.92E-05 |
| Heart_Failure | dendritic_cell | *MAP3K1* | 3.64551945 | 1.88E-05 | 2.43E-05 | 0.11788532 |
| Heart_Failure | dendritic_cell | *TMEM38B* | 6.109023716 | 6.59E-18 | 3.89E-17 | 4.14E-14 |
| Heart_Failure | dendritic_cell | *PDE6C* | -3.497581892 | 6.70E-19 | 5.51E-18 | 4.21E-15 |
| Heart_Failure | dendritic_cell | *CWF19L1* | -2.08604964 | 5.06E-09 | 7.97E-09 | 3.18E-05 |
| Heart_Failure | dendritic_cell | *IKZF5* | -5.519738153 | 1.55E-10 | 2.72E-10 | 9.72E-07 |
| Heart_Failure | dendritic_cell | *BLNK* | 2.41646104 | 3.20E-19 | 3.06E-18 | 2.01E-15 |
| Heart_Failure | dendritic_cell | *WAC* | -3.561810011 | 1.11E-07 | 1.62E-07 | 0.00070047 |
| Heart_Failure | dendritic_cell | *TPSD1* | 40.83022374 | 1.01E-13 | 2.46E-13 | 6.33E-10 |
| Heart_Failure | dendritic_cell | *ITPR3* | -2.115980944 | 4.60E-20 | 8.12E-19 | 2.89E-16 |
| Heart_Failure | dendritic_cell | *DSP* | -2.518628365 | 2.11E-09 | 3.42E-09 | 1.33E-05 |
| Heart_Failure | dendritic_cell | *IFT74* | -3.228015864 | 8.71E-14 | 2.16E-13 | 5.48E-10 |
| Heart_Failure | dendritic_cell | *IL12RB1* | 4.351745161 | 1.33E-08 | 2.04E-08 | 8.36E-05 |
| Heart_Failure | dendritic_cell | *ACOT7* | -4.933040389 | 5.40E-14 | 1.38E-13 | 3.39E-10 |
| Heart_Failure | dendritic_cell | *PCSK5* | -2.430322283 | 2.69E-18 | 1.79E-17 | 1.69E-14 |
| Heart_Failure | dendritic_cell | *RAB18* | -3.345996021 | 2.81E-07 | 4.01E-07 | 0.001764393 |

| | | | | | | |
|---|---|---|---|---|---|---|
| Heart_Failure | dendritic_cell | *NRP1* | -3.487719184 | 4.01E-12 | 8.17E-12 | 2.52E-08 |
| Heart_Failure | dendritic_cell | *PRTFDC1* | -3.36111853 | 1.91E-19 | 2.12E-18 | 1.20E-15 |
| Heart_Failure | dendritic_cell | *PALMD* | -1.854744012 | 6.37E-19 | 5.32E-18 | 4.01E-15 |
| Heart_Failure | dendritic_cell | *TSPAN15* | -3.161009095 | 1.75E-18 | 1.25E-17 | 1.10E-14 |
| Heart_Failure | dendritic_cell | *OCEL1* | -2.478090038 | 3.69E-18 | 2.35E-17 | 2.32E-14 |
| Heart_Failure | dendritic_cell | *CATSPERG* | -2.890847334 | 8.95E-08 | 1.31E-07 | 0.000562758 |
| Heart_Failure | dendritic_cell | *STX1B* | -4.78067621 | 1.06E-12 | 2.30E-12 | 6.67E-09 |
| Heart_Failure | dendritic_cell | *HSD3B7* | -3.733288499 | 3.62E-10 | 6.19E-10 | 2.27E-06 |
| Heart_Failure | dendritic_cell | *ATP5D* | -4.874808211 | 6.75E-18 | 3.97E-17 | 4.25E-14 |
| Heart_Failure | dendritic_cell | *MARCH2* | -3.965681893 | 2.79E-06 | 3.77E-06 | 0.017551994 |
| Heart_Failure | dendritic_cell | *CDC34* | -7.233878049 | 4.20E-15 | 1.30E-14 | 2.64E-11 |
| Heart_Failure | dendritic_cell | *MTAP* | -6.224657441 | 6.03E-14 | 1.53E-13 | 3.79E-10 |
| Heart_Failure | dendritic_cell | *CEP170B* | -2.588741497 | 8.09E-17 | 3.62E-16 | 5.09E-13 |
| Heart_Failure | dendritic_cell | *POLR2E* | -4.123717495 | 8.70E-19 | 6.92E-18 | 5.47E-15 |
| Heart_Failure | dendritic_cell | *MADCAM1* | -6.417564392 | 1.24E-16 | 5.23E-16 | 7.78E-13 |
| Heart_Failure | dendritic_cell | *TRMT2A* | -6.983728507 | 3.62E-10 | 6.19E-10 | 2.28E-06 |
| Heart_Failure | dendritic_cell | *ZDHHC8* | -4.885526921 | 3.17E-16 | 1.23E-15 | 1.99E-12 |
| Heart_Failure | dendritic_cell | *MED15* | -4.198046135 | 7.63E-13 | 1.67E-12 | 4.80E-09 |
| Heart_Failure | dendritic_cell | *SERPIND1* | -3.589979485 | 4.60E-11 | 8.46E-11 | 2.89E-07 |
| Heart_Failure | dendritic_cell | *LZTR1* | -6.329781703 | 1.38E-13 | 3.34E-13 | 8.69E-10 |
| Heart_Failure | dendritic_cell | *CECR2* | -2.470669395 | 8.20E-09 | 1.27E-08 | 5.16E-05 |
| Heart_Failure | dendritic_cell | *SMARCB1* | -2.654505804 | 1.07E-07 | 1.56E-07 | 0.000674237 |
| Heart_Failure | dendritic_cell | *P2RX6* | -3.232802367 | 1.55E-19 | 1.83E-18 | 9.73E-16 |
| Heart_Failure | dendritic_cell | *SLC7A4* | 3.720256294 | 1.73E-18 | 1.24E-17 | 1.09E-14 |
| Heart_Failure | dendritic_cell | *DDT* | -5.432524321 | 2.20E-17 | 1.12E-16 | 1.38E-13 |
| Heart_Failure | dendritic_cell | *GGT5* | -4.461518398 | 4.52E-19 | 4.05E-18 | 2.84E-15 |
| Heart_Failure | dendritic_cell | *SPECC1L* | -2.814207906 | 5.30E-07 | 7.44E-07 | 0.003335229 |
| Heart_Failure | dendritic_cell | *PPIL2* | -1.069426603 | 0.037975789 | 0.042160301 | 1 |
| Heart_Failure | dendritic_cell | *UPB1* | 1.861865596 | 5.45E-05 | 6.94E-05 | 0.342642634 |
| Heart_Failure | dendritic_cell | *YPEL1* | 3.097004248 | 0.0266256 | 0.029785613 | 1 |
| Heart_Failure | dendritic_cell | *PPM1F* | 4.128593477 | 0.000569747 | 0.000691505 | 1 |
| Heart_Failure | dendritic_cell | *TOP3B* | -4.735573371 | 8.29E-12 | 1.63E-11 | 5.21E-08 |
| Heart_Failure | dendritic_cell | *CYTH4* | 2.279404033 | 1.40E-20 | 5.50E-19 | 8.81E-17 |
| Heart_Failure | dendritic_cell | *CARD10* | -3.169976469 | 1.16E-19 | 1.47E-18 | 7.31E-16 |
| Heart_Failure | dendritic_cell | *LRP5L* | -5.000960389 | 3.40E-14 | 8.98E-14 | 2.14E-10 |
| Heart_Failure | dendritic_cell | *SLC25A1* | -5.318588631 | 1.43E-14 | 4.01E-14 | 8.98E-11 |
| Heart_Failure | dendritic_cell | *LGALS2* | 0.891815328 | 7.60E-21 | 4.66E-19 | 4.78E-17 |
| Heart_Failure | dendritic_cell | *GGA1* | -2.242078138 | 0.005500356 | 0.00637668 | 1 |
| Heart_Failure | dendritic_cell | *SH3BP1* | 2.93308214 | 1.18E-20 | 5.00E-19 | 7.39E-17 |
| Heart_Failure | dendritic_cell | *HPS4* | -4.527284438 | 1.95E-05 | 2.52E-05 | 0.122321764 |
| Heart_Failure | dendritic_cell | *PIK3IP1* | -3.760725944 | 7.51E-19 | 6.11E-18 | 4.72E-15 |
| Heart_Failure | dendritic_cell | *TRIOBP* | -3.187180561 | 1.64E-11 | 3.14E-11 | 1.03E-07 |
| Heart_Failure | dendritic_cell | *TFIP11* | 3.939182423 | 4.72E-10 | 8.00E-10 | 2.97E-06 |
| Heart_Failure | dendritic_cell | *GCAT* | -3.954764579 | 5.44E-09 | 8.55E-09 | 3.42E-05 |
| Heart_Failure | dendritic_cell | *ANKRD54* | -2.966178017 | 2.07E-08 | 3.13E-08 | 0.000130143 |
| Heart_Failure | dendritic_cell | *EIF3L* | -4.692690082 | 5.32E-10 | 8.99E-10 | 3.34E-06 |
| Heart_Failure | dendritic_cell | *CCDC134* | -7.774151709 | 2.69E-10 | 4.65E-10 | 1.69E-06 |
| Heart_Failure | dendritic_cell | *SLC16A8* | 4.029392766 | 6.93E-18 | 4.05E-17 | 4.35E-14 |
| Heart_Failure | dendritic_cell | *SLC5A4* | -0.202702219 | 0.023388844 | 0.026220696 | 1 |
| Heart_Failure | dendritic_cell | *KDELR3* | -3.579536015 | 4.97E-17 | 2.32E-16 | 3.13E-13 |

| | | | | | | |
|---|---|---|---|---|---|---|
| Heart_Failure | dendritic_cell | *CYP2D6* | -3.448563295 | 2.39E-09 | 3.86E-09 | 1.51E-05 |
| Heart_Failure | dendritic_cell | *DDX17* | -3.339291014 | 1.61E-13 | 3.85E-13 | 1.01E-09 |
| Heart_Failure | dendritic_cell | *CBY1* | -0.873135198 | 0.011322177 | 0.012902397 | 1 |
| Heart_Failure | dendritic_cell | *FBXO7* | -3.687879078 | 1.16E-05 | 1.52E-05 | 0.072916139 |
| Heart_Failure | dendritic_cell | *POLDIP3* | -3.853812198 | 0.005239288 | 0.006079624 | 1 |
| Heart_Failure | dendritic_cell | *TIMP3* | -3.366876507 | 5.47E-14 | 1.40E-13 | 3.44E-10 |
| Heart_Failure | dendritic_cell | *PPP6R2* | -4.015765695 | 1.98E-11 | 3.77E-11 | 1.24E-07 |
| Heart_Failure | dendritic_cell | *CYB5R3* | -4.486693613 | 7.73E-07 | 1.07E-06 | 0.004861245 |
| Heart_Failure | dendritic_cell | *RHBDD3* | -3.722977193 | 2.47E-19 | 2.50E-18 | 1.55E-15 |
| Heart_Failure | dendritic_cell | *TTLL1* | -4.305859012 | 4.02E-19 | 3.70E-18 | 2.53E-15 |
| Heart_Failure | dendritic_cell | *RASL10A* | -3.794102819 | 3.40E-17 | 1.65E-16 | 2.14E-13 |
| Heart_Failure | dendritic_cell | *AP1B1* | -2.796261398 | 0.005237391 | 0.006078544 | 1 |
| Heart_Failure | dendritic_cell | *HMGXB4* | -3.871373317 | 8.04E-17 | 3.60E-16 | 5.05E-13 |
| Heart_Failure | dendritic_cell | *TOM1* | -3.153775666 | 8.07E-14 | 2.02E-13 | 5.08E-10 |
| Heart_Failure | dendritic_cell | *NEFH* | 3.887471961 | 1.21E-20 | 5.08E-19 | 7.62E-17 |
| Heart_Failure | dendritic_cell | *CHKB* | -1.773852355 | 0.002540471 | 0.002997174 | 1 |
| Heart_Failure | dendritic_cell | *BIK* | -12.12784349 | 1.60E-19 | 1.87E-18 | 1.01E-15 |
| Heart_Failure | dendritic_cell | *MCAT* | 3.050730067 | 0.019906587 | 0.022436843 | 1 |
| Heart_Failure | dendritic_cell | *THOC5* | -2.70738573 | 0.002364095 | 0.002793284 | 1 |
| Heart_Failure | dendritic_cell | *ARSA* | -1.580790534 | 7.81E-10 | 1.31E-09 | 4.91E-06 |
| Heart_Failure | dendritic_cell | *TSPO* | -4.643619105 | 1.29E-09 | 2.12E-09 | 8.12E-06 |
| Heart_Failure | dendritic_cell | *TTLL12* | 6.494798165 | 5.76E-20 | 9.45E-19 | 3.62E-16 |
| Heart_Failure | dendritic_cell | *RBFOX2* | -3.112449001 | 8.57E-17 | 3.79E-16 | 5.39E-13 |
| Heart_Failure | dendritic_cell | *MTMR3* | -2.60964819 | 2.88E-08 | 4.32E-08 | 0.000180885 |
| Heart_Failure | dendritic_cell | *SAMM50* | -5.463404672 | 5.76E-17 | 2.65E-16 | 3.62E-13 |
| Heart_Failure | dendritic_cell | *TXN2* | -4.368141341 | 1.11E-10 | 1.98E-10 | 6.96E-07 |
| Heart_Failure | dendritic_cell | *FOXRED2* | -2.808560638 | 1.79E-08 | 2.72E-08 | 0.000112719 |
| Heart_Failure | dendritic_cell | *IFT27* | -4.879622094 | 0.001099342 | 0.001314736 | 1 |
| Heart_Failure | dendritic_cell | *KIAA0930* | 2.907856149 | 0.001877874 | 0.002230108 | 1 |
| Heart_Failure | dendritic_cell | *NCF4* | 2.801623252 | 3.98E-14 | 1.04E-13 | 2.50E-10 |
| Heart_Failure | dendritic_cell | *SLC25A17* | -3.95582773 | 2.35E-16 | 9.33E-16 | 1.48E-12 |
| Heart_Failure | dendritic_cell | *KCTD17* | 12.6772355 | 6.67E-19 | 5.50E-18 | 4.19E-15 |
| Heart_Failure | dendritic_cell | *EP300* | -3.750330323 | 2.22E-07 | 3.18E-07 | 0.001393207 |
| Heart_Failure | dendritic_cell | *ACO2* | -4.160697441 | 1.11E-14 | 3.19E-14 | 7.01E-11 |
| Heart_Failure | dendritic_cell | *POLR3H* | -1.669805054 | 8.71E-05 | 0.000109851 | 0.547717012 |
| Heart_Failure | dendritic_cell | *DESI1* | -2.123001543 | 3.79E-08 | 5.65E-08 | 0.000238333 |
| Heart_Failure | dendritic_cell | *ZBED4* | -5.332514894 | 3.61E-06 | 4.84E-06 | 0.022677534 |
| Heart_Failure | dendritic_cell | *HDAC10* | 5.200679885 | 2.50E-14 | 6.77E-14 | 1.57E-10 |
| Heart_Failure | dendritic_cell | *KHNYN* | -1.861913354 | 6.02E-06 | 7.99E-06 | 0.037833678 |
| Heart_Failure | dendritic_cell | *SDR39U1* | -2.339630733 | 4.83E-12 | 9.77E-12 | 3.03E-08 |
| Heart_Failure | dendritic_cell | *RBM23* | -5.296989024 | 8.80E-10 | 1.47E-09 | 5.53E-06 |
| Heart_Failure | dendritic_cell | *VCPKMT* | -3.268817006 | 1.63E-05 | 2.13E-05 | 0.102688379 |
| Heart_Failure | dendritic_cell | *SOS2* | -3.530893407 | 1.22E-15 | 4.19E-15 | 7.68E-12 |
| Heart_Failure | dendritic_cell | *PYGL* | -1.435718947 | 2.62E-12 | 5.44E-12 | 1.64E-08 |
| Heart_Failure | dendritic_cell | *TRIM9* | -2.522295836 | 4.08E-13 | 9.25E-13 | 2.57E-09 |
| Heart_Failure | dendritic_cell | *DDHD1* | -4.171362883 | 5.21E-13 | 1.17E-12 | 3.28E-09 |
| Heart_Failure | dendritic_cell | *CNIH1* | -4.97614256 | 9.46E-15 | 2.75E-14 | 5.95E-11 |
| Heart_Failure | dendritic_cell | *ATP6V1D* | -2.631068851 | 1.59E-06 | 2.16E-06 | 0.009965432 |
| Heart_Failure | dendritic_cell | *PIGH* | -4.145786944 | 2.81E-13 | 6.53E-13 | 1.77E-09 |
| Heart_Failure | dendritic_cell | *VTI1B* | -5.021871804 | 3.63E-16 | 1.38E-15 | 2.28E-12 |

| | | | | | | |
|---|---|---|---|---|---|---|
| Heart_Failure | dendritic_cell | *TMED8* | -3.394753586 | 0.000306943 | 0.000377495 | 1 |
| Heart_Failure | dendritic_cell | *DAAM1* | -2.867898891 | 1.16E-14 | 3.31E-14 | 7.28E-11 |
| Heart_Failure | dendritic_cell | *ISM2* | -3.982214566 | 0.000684562 | 0.000827343 | 1 |
| Heart_Failure | dendritic_cell | *PPM1A* | -4.038687602 | 8.55E-11 | 1.54E-10 | 5.38E-07 |
| Heart_Failure | dendritic_cell | *SLC10A1* | -4.262584088 | 8.94E-19 | 7.09E-18 | 5.62E-15 |
| Heart_Failure | dendritic_cell | *DICER1* | -3.947483922 | 9.39E-13 | 2.05E-12 | 5.90E-09 |
| Heart_Failure | dendritic_cell | *ZFYVE21* | -4.538998441 | 1.70E-13 | 4.05E-13 | 1.07E-09 |
| Heart_Failure | dendritic_cell | *MTHFD1* | -3.6359132 | 8.91E-18 | 5.09E-17 | 5.60E-14 |
| Heart_Failure | dendritic_cell | *TELO2* | -5.982123051 | 3.10E-18 | 2.01E-17 | 1.95E-14 |
| Heart_Failure | dendritic_cell | *GSKIP* | -7.2461592 | 9.60E-14 | 2.36E-13 | 6.04E-10 |
| Heart_Failure | dendritic_cell | *VRK1* | -2.998706734 | 0.015965346 | 0.018062648 | 1 |
| Heart_Failure | dendritic_cell | *CCNB1IP1* | -4.267283844 | 1.07E-09 | 1.77E-09 | 6.73E-06 |
| Heart_Failure | dendritic_cell | *TRIP11* | -3.961031955 | 1.88E-15 | 6.20E-15 | 1.18E-11 |
| Heart_Failure | dendritic_cell | *KIAA0391* | -3.893395022 | 2.49E-20 | 6.44E-19 | 1.57E-16 |
| Heart_Failure | dendritic_cell | *EMC9* | -2.592562003 | 1.81E-14 | 4.99E-14 | 1.14E-10 |
| Heart_Failure | dendritic_cell | *PLTP* | -1.257786485 | 0.007126458 | 0.008225453 | 1 |
| Heart_Failure | dendritic_cell | *GSS* | -3.818744759 | 1.32E-06 | 1.81E-06 | 0.008314393 |
| Heart_Failure | dendritic_cell | *PYGB* | -3.078946969 | 1.96E-11 | 3.73E-11 | 1.23E-07 |
| Heart_Failure | dendritic_cell | *PROCR* | -2.643819959 | 4.05E-11 | 7.50E-11 | 2.55E-07 |
| Heart_Failure | dendritic_cell | *NINL* | -2.799509559 | 2.48E-17 | 1.24E-16 | 1.56E-13 |
| Heart_Failure | dendritic_cell | *CD40* | 6.584662598 | 2.42E-14 | 6.56E-14 | 1.52E-10 |
| Heart_Failure | dendritic_cell | *UQCC1* | -3.494654149 | 4.37E-10 | 7.41E-10 | 2.75E-06 |
| Heart_Failure | dendritic_cell | *IFT52* | -4.440176805 | 3.07E-15 | 9.75E-15 | 1.93E-11 |
| Heart_Failure | dendritic_cell | *NDRG3* | -5.686060271 | 1.43E-10 | 2.53E-10 | 9.00E-07 |
| Heart_Failure | dendritic_cell | *DOK5* | -3.109159331 | 2.41E-19 | 2.46E-18 | 1.52E-15 |
| Heart_Failure | dendritic_cell | *CSTF1* | 5.757064186 | 2.85E-09 | 4.57E-09 | 1.79E-05 |
| Heart_Failure | dendritic_cell | *BMP7* | -2.931801317 | 3.11E-20 | 6.68E-19 | 1.96E-16 |
| Heart_Failure | dendritic_cell | *TPD52L2* | -3.446117826 | 0.00127014 | 0.001517266 | 1 |
| Heart_Failure | dendritic_cell | *NELFCD* | -3.671135189 | 5.97E-18 | 3.57E-17 | 3.75E-14 |
| Heart_Failure | dendritic_cell | *CTSZ* | 3.658037652 | 8.65E-17 | 3.82E-16 | 5.44E-13 |
| Heart_Failure | dendritic_cell | *PRPF6* | -5.126215459 | 6.89E-17 | 3.12E-16 | 4.33E-13 |
| Heart_Failure | dendritic_cell | *MTG2* | -2.544911683 | 0.000909238 | 0.001093207 | 1 |
| Heart_Failure | dendritic_cell | *TCFL5* | -3.875145774 | 9.40E-13 | 2.05E-12 | 5.91E-09 |
| Heart_Failure | dendritic_cell | *DIDO1* | -3.690457899 | 1.83E-07 | 2.64E-07 | 0.001152662 |
| Heart_Failure | dendritic_cell | *GID8* | -5.751284618 | 3.27E-08 | 4.89E-08 | 0.000205392 |
| Heart_Failure | dendritic_cell | *SLC17A9* | 1.516961658 | 7.70E-22 | 3.01E-19 | 4.84E-18 |
| Heart_Failure | dendritic_cell | *BIRC7* | -5.046385858 | 3.26E-09 | 5.21E-09 | 2.05E-05 |
| Heart_Failure | dendritic_cell | *NKAIN4* | -6.317585046 | 8.28E-17 | 3.69E-16 | 5.20E-13 |
| Heart_Failure | dendritic_cell | *PTK6* | 3.217981464 | 7.12E-21 | 4.66E-19 | 4.47E-17 |
| Heart_Failure | dendritic_cell | *GMEB2* | 5.008351401 | 0.00029423 | 0.000362001 | 1 |
| Heart_Failure | dendritic_cell | *CDC25B* | -4.044651245 | 1.93E-17 | 1.00E-16 | 1.21E-13 |
| Heart_Failure | dendritic_cell | *RNF24* | -4.16584064 | 1.35E-10 | 2.39E-10 | 8.49E-07 |
| Heart_Failure | dendritic_cell | *NDUFAF5* | -1.290034471 | 1.20E-09 | 1.98E-09 | 7.57E-06 |
| Heart_Failure | dendritic_cell | *TRIB3* | -5.242180905 | 4.26E-19 | 3.86E-18 | 2.68E-15 |
| Heart_Failure | dendritic_cell | *SLC52A3* | 5.232910423 | 2.93E-16 | 1.15E-15 | 1.84E-12 |
| Heart_Failure | dendritic_cell | *ANGPT4* | -6.059254702 | 4.24E-16 | 1.59E-15 | 2.67E-12 |
| Heart_Failure | dendritic_cell | *RSPO4* | -5.515609891 | 1.76E-13 | 4.19E-13 | 1.11E-09 |
| Heart_Failure | dendritic_cell | *SIRPB1* | 1.869378491 | 8.04E-12 | 1.59E-11 | 5.05E-08 |
| Heart_Failure | dendritic_cell | *SEC23B* | -2.568348537 | 2.44E-05 | 3.16E-05 | 0.153610814 |
| Heart_Failure | dendritic_cell | *CCM2L* | -2.7522963 | 3.24E-20 | 6.76E-19 | 2.04E-16 |

| | | | | | | |
|---|---|---|---|---|---|---|
| Heart_Failure | dendritic_cell | *MYL9* | -2.888780888 | 4.45E-05 | 5.68E-05 | 0.27977858 |
| Heart_Failure | dendritic_cell | *MROH8* | -3.685867886 | 4.40E-09 | 6.95E-09 | 2.77E-05 |
| Heart_Failure | dendritic_cell | *MANBAL* | -2.116147217 | 8.30E-09 | 1.29E-08 | 5.22E-05 |
| Heart_Failure | dendritic_cell | *IDH3B* | -6.169801975 | 1.93E-05 | 2.50E-05 | 0.121357062 |
| Heart_Failure | dendritic_cell | *MAPRE1* | 6.436880563 | 4.98E-09 | 7.84E-09 | 3.13E-05 |
| Heart_Failure | dendritic_cell | *TTI1* | -5.636053926 | 5.51E-07 | 7.71E-07 | 0.003462437 |
| Heart_Failure | dendritic_cell | *RPRD1B* | -5.128900614 | 4.28E-11 | 7.90E-11 | 2.69E-07 |
| Heart_Failure | dendritic_cell | *BPI* | -4.973021699 | 1.80E-16 | 7.37E-16 | 1.13E-12 |
| Heart_Failure | dendritic_cell | *ACTR5* | 8.268587201 | 1.88E-11 | 3.60E-11 | 1.18E-07 |
| Heart_Failure | dendritic_cell | *WFDC2* | -6.82778149 | 0.000815217 | 0.000982416 | 1 |
| Heart_Failure | dendritic_cell | *DHX35* | -5.931692168 | 1.21E-11 | 2.35E-11 | 7.61E-08 |
| Heart_Failure | dendritic_cell | *DNTTIP1* | -2.849175404 | 4.22E-07 | 5.96E-07 | 0.002654387 |
| Heart_Failure | dendritic_cell | *MAP1LC3A* | -2.443143518 | 6.81E-05 | 8.63E-05 | 0.427846626 |
| Heart_Failure | dendritic_cell | *PIGU* | -3.934780997 | 1.71E-14 | 4.74E-14 | 1.08E-10 |
| Heart_Failure | dendritic_cell | *APMAP* | -4.877816549 | 7.31E-11 | 1.32E-10 | 4.60E-07 |
| Heart_Failure | dendritic_cell | *CELF4* | -3.28656559 | 5.48E-20 | 9.06E-19 | 3.44E-16 |
| Heart_Failure | dendritic_cell | *RBFA* | -4.572525407 | 2.95E-06 | 3.98E-06 | 0.018572183 |
| Heart_Failure | dendritic_cell | *METTL4* | -5.998219641 | 8.42E-09 | 1.31E-08 | 5.29E-05 |
| Heart_Failure | dendritic_cell | *LPIN2* | -4.991049049 | 1.08E-10 | 1.93E-10 | 6.79E-07 |
| Heart_Failure | dendritic_cell | *SMCHD1* | -3.14121146 | 3.17E-05 | 4.08E-05 | 0.199533891 |
| Heart_Failure | dendritic_cell | *MYL12A* | -4.676713069 | 5.19E-06 | 6.91E-06 | 0.032629513 |
| Heart_Failure | dendritic_cell | *CEP76* | -4.584150944 | 9.72E-12 | 1.90E-11 | 6.11E-08 |
| Heart_Failure | dendritic_cell | *CEP192* | -1.809084996 | 0.000561456 | 0.000681837 | 1 |
| Heart_Failure | dendritic_cell | *LIPG* | 12.5547367 | 3.52E-15 | 1.10E-14 | 2.21E-11 |
| Heart_Failure | dendritic_cell | *POLI* | -0.578294709 | 0.002168273 | 0.002566252 | 1 |
| Heart_Failure | dendritic_cell | *NALCN* | -3.905315045 | 5.95E-19 | 5.06E-18 | 3.74E-15 |
| Heart_Failure | dendritic_cell | *HTR2A* | -6.292701275 | 3.10E-20 | 6.68E-19 | 1.95E-16 |
| Heart_Failure | dendritic_cell | *STK24* | -4.470003803 | 7.31E-10 | 1.22E-09 | 4.59E-06 |
| Heart_Failure | dendritic_cell | *DNAJC3* | -4.327553977 | 7.11E-07 | 9.91E-07 | 0.004472705 |
| Heart_Failure | dendritic_cell | *UGGT2* | -3.231404894 | 1.05E-15 | 3.65E-15 | 6.59E-12 |
| Heart_Failure | dendritic_cell | *ARHGEF7* | -3.962037246 | 7.82E-11 | 1.41E-10 | 4.92E-07 |
| Heart_Failure | dendritic_cell | *SGCG* | -1.57900653 | 1.85E-20 | 6.38E-19 | 1.16E-16 |
| Heart_Failure | dendritic_cell | *PARP4* | -4.438864758 | 3.62E-10 | 6.19E-10 | 2.28E-06 |
| Heart_Failure | dendritic_cell | *VWA8* | -3.62946014 | 5.29E-12 | 1.07E-11 | 3.32E-08 |
| Heart_Failure | dendritic_cell | *KATNAL1* | -3.954803981 | 9.99E-14 | 2.45E-13 | 6.28E-10 |
| Heart_Failure | dendritic_cell | *DHRS12* | -3.939347937 | 6.26E-18 | 3.71E-17 | 3.93E-14 |
| Heart_Failure | dendritic_cell | *MEDAG* | -1.755888114 | 2.53E-20 | 6.44E-19 | 1.59E-16 |
| Heart_Failure | dendritic_cell | *TSC22D1* | -3.688917308 | 3.47E-15 | 1.09E-14 | 2.18E-11 |
| Heart_Failure | dendritic_cell | *CLN5* | -3.779952774 | 7.96E-13 | 1.75E-12 | 5.00E-09 |
| Heart_Failure | dendritic_cell | *MGRN1* | -2.140806394 | 0.00050897 | 0.000620015 | 1 |
| Heart_Failure | dendritic_cell | *CORO1A* | 2.572346878 | 1.81E-15 | 5.97E-15 | 1.14E-11 |
| Heart_Failure | dendritic_cell | *GDPD3* | -5.238653141 | 1.33E-06 | 1.82E-06 | 0.008383015 |
| Heart_Failure | dendritic_cell | *NFAT5* | -3.330739021 | 2.65E-14 | 7.11E-14 | 1.66E-10 |
| Heart_Failure | dendritic_cell | *LONP2* | -3.784155105 | 2.80E-13 | 6.52E-13 | 1.76E-09 |
| Heart_Failure | dendritic_cell | *PLLP* | -6.944839093 | 1.46E-13 | 3.50E-13 | 9.16E-10 |
| Heart_Failure | dendritic_cell | *CTCF* | -4.587497194 | 1.85E-09 | 3.00E-09 | 1.16E-05 |
| Heart_Failure | dendritic_cell | *ACD* | -3.499489299 | 8.64E-08 | 1.26E-07 | 0.000543271 |
| Heart_Failure | dendritic_cell | *USB1* | -7.664793934 | 5.38E-15 | 1.64E-14 | 3.39E-11 |
| Heart_Failure | dendritic_cell | *SETD6* | -6.118412187 | 2.48E-09 | 3.99E-09 | 1.56E-05 |
| Heart_Failure | dendritic_cell | *COG4* | -3.943187802 | 1.22E-13 | 2.97E-13 | 7.68E-10 |

| | | | | | | |
|---|---|---|---|---|---|---|
| Heart_Failure | dendritic_cell | *SLC7A6OS* | -1.95492548 | 7.15E-12 | 1.42E-11 | 4.50E-08 |
| Heart_Failure | dendritic_cell | *MON1B* | -6.036144614 | 2.61E-12 | 5.44E-12 | 1.64E-08 |
| Heart_Failure | dendritic_cell | *HCFC1R1* | -6.317052822 | 2.06E-17 | 1.06E-16 | 1.29E-13 |
| Heart_Failure | dendritic_cell | *NPRL3* | -2.862949708 | 2.27E-09 | 3.66E-09 | 1.43E-05 |
| Heart_Failure | dendritic_cell | *TAF1C* | -2.135616781 | 4.41E-14 | 1.14E-13 | 2.77E-10 |
| Heart_Failure | dendritic_cell | *NAGPA* | -5.913732753 | 8.76E-12 | 1.72E-11 | 5.51E-08 |
| Heart_Failure | dendritic_cell | *COTL1* | 3.160726374 | 2.93E-18 | 1.93E-17 | 1.85E-14 |
| Heart_Failure | dendritic_cell | *CRISPLD2* | -2.517648789 | 1.65E-15 | 5.52E-15 | 1.04E-11 |
| Heart_Failure | dendritic_cell | *NME4* | -5.190957741 | 1.96E-17 | 1.01E-16 | 1.23E-13 |
| Heart_Failure | dendritic_cell | *NOMO3* | -5.073024241 | 1.49E-09 | 2.44E-09 | 9.38E-06 |
| Heart_Failure | dendritic_cell | *LMF1* | -4.54117868 | 4.20E-13 | 9.50E-13 | 2.64E-09 |
| Heart_Failure | dendritic_cell | *NARFL* | -5.61387439 | 1.41E-10 | 2.49E-10 | 8.86E-07 |
| Heart_Failure | dendritic_cell | *MTHFSD* | -1.598005607 | 2.82E-07 | 4.02E-07 | 0.001772335 |
| Heart_Failure | dendritic_cell | *CLCN7* | -4.833570538 | 2.50E-05 | 3.23E-05 | 0.157436591 |
| Heart_Failure | dendritic_cell | *HAGHL* | -8.16002398 | 1.90E-08 | 2.88E-08 | 0.000119547 |
| Heart_Failure | dendritic_cell | *SLC7A5* | -6.019728728 | 2.34E-20 | 6.44E-19 | 1.47E-16 |
| Heart_Failure | dendritic_cell | *FBXO31* | -3.649989934 | 3.08E-15 | 9.77E-15 | 1.94E-11 |
| Heart_Failure | dendritic_cell | *STUB1* | -5.494628412 | 1.93E-13 | 4.57E-13 | 1.21E-09 |
| Heart_Failure | dendritic_cell | *NUBP1* | 6.535039833 | 3.75E-07 | 5.31E-07 | 0.002356999 |
| Heart_Failure | dendritic_cell | *UBE2I* | -4.473759804 | 6.65E-15 | 1.99E-14 | 4.18E-11 |
| Heart_Failure | dendritic_cell | *EARS2* | -2.17458409 | 1.32E-07 | 1.91E-07 | 0.000828694 |
| Heart_Failure | dendritic_cell | *CPPED1* | 5.171435229 | 2.18E-13 | 5.12E-13 | 1.37E-09 |
| Heart_Failure | dendritic_cell | *HMOX2* | -4.074805717 | 1.68E-14 | 4.66E-14 | 1.06E-10 |
| Heart_Failure | dendritic_cell | *BFAR* | -5.808102659 | 2.88E-10 | 4.97E-10 | 1.81E-06 |
| Heart_Failure | dendritic_cell | *RBL2* | -1.79471495 | 0.001775451 | 0.002110068 | 1 |
| Heart_Failure | dendritic_cell | *KAT8* | -4.911852193 | 3.69E-12 | 7.57E-12 | 2.32E-08 |
| Heart_Failure | dendritic_cell | *NOMO1* | 4.134380174 | 0.011216962 | 0.012787133 | 1 |
| Heart_Failure | dendritic_cell | *RNF40* | -6.224954349 | 1.48E-13 | 3.55E-13 | 9.31E-10 |
| Heart_Failure | dendritic_cell | *KNOP1* | -4.061015741 | 3.91E-13 | 8.91E-13 | 2.46E-09 |
| Heart_Failure | dendritic_cell | *IQCH* | -5.407438798 | 3.21E-11 | 6.01E-11 | 2.02E-07 |
| Heart_Failure | dendritic_cell | *LACTB* | 5.076319637 | 2.40E-06 | 3.25E-06 | 0.015063678 |
| Heart_Failure | dendritic_cell | *CSK* | 7.035111121 | 3.83E-16 | 1.46E-15 | 2.41E-12 |
| Heart_Failure | dendritic_cell | *HERC1* | -3.734228641 | 3.80E-09 | 6.04E-09 | 2.39E-05 |
| Heart_Failure | dendritic_cell | *TRIP4* | -1.858483432 | 0.007232901 | 0.008340654 | 1 |
| Heart_Failure | dendritic_cell | *RASL12* | -10.50404683 | 8.38E-14 | 2.09E-13 | 5.27E-10 |
| Heart_Failure | dendritic_cell | *AP3B2* | -1.41467391 | 3.05E-06 | 4.11E-06 | 0.019182968 |
| Heart_Failure | dendritic_cell | *IGDCC4* | 0.422612795 | 0.009910237 | 0.011334484 | 1 |
| Heart_Failure | dendritic_cell | *CTSH* | 1.197270741 | 7.04E-08 | 1.04E-07 | 0.000442327 |
| Heart_Failure | dendritic_cell | *TTC23* | -4.155564426 | 2.22E-15 | 7.25E-15 | 1.40E-11 |
| Heart_Failure | dendritic_cell | *FAH* | -2.157429868 | 4.39E-20 | 7.97E-19 | 2.76E-16 |
| Heart_Failure | dendritic_cell | *RPAP1* | -4.660147171 | 3.77E-18 | 2.38E-17 | 2.37E-14 |
| Heart_Failure | dendritic_cell | *HOMER2* | 3.359400769 | 2.28E-19 | 2.38E-18 | 1.43E-15 |
| Heart_Failure | dendritic_cell | *EHD4* | -2.445574471 | 0.021284044 | 0.023933604 | 1 |
| Heart_Failure | dendritic_cell | *ATP8B4* | 2.803556704 | 2.33E-19 | 2.41E-18 | 1.47E-15 |
| Heart_Failure | dendritic_cell | *DTWD1* | -3.26936133 | 7.55E-15 | 2.23E-14 | 4.75E-11 |
| Heart_Failure | dendritic_cell | *DNAJC17* | -5.454428138 | 8.26E-14 | 2.06E-13 | 5.19E-10 |
| Heart_Failure | dendritic_cell | *PDGFRL* | -2.006490566 | 4.55E-19 | 4.07E-18 | 2.86E-15 |
| Heart_Failure | dendritic_cell | *ZDHHC2* | -4.305003261 | 1.34E-15 | 4.55E-15 | 8.41E-12 |
| Heart_Failure | dendritic_cell | *ZFAND1* | -3.257114634 | 2.85E-06 | 3.84E-06 | 0.017899735 |
| Heart_Failure | dendritic_cell | *RIPK2* | 4.835795257 | 3.86E-15 | 1.20E-14 | 2.43E-11 |

| | | | | | | |
|---|---|---|---|---|---|---|
| Heart_Failure | dendritic_cell | *EYA1* | -1.75040945 | 3.25E-16 | 1.26E-15 | 2.04E-12 |
| Heart_Failure | dendritic_cell | *NBN* | -4.642731225 | 1.07E-07 | 1.56E-07 | 0.000673628 |
| Heart_Failure | dendritic_cell | *CPQ* | -2.486446023 | 1.12E-08 | 1.72E-08 | 7.02E-05 |
| Heart_Failure | dendritic_cell | *NIPAL2* | -3.535309337 | 1.90E-14 | 5.23E-14 | 1.20E-10 |
| Heart_Failure | dendritic_cell | *PLAT* | -2.776303154 | 1.22E-19 | 1.53E-18 | 7.66E-16 |
| Heart_Failure | dendritic_cell | *GDAP1* | 5.271282222 | 7.05E-07 | 9.82E-07 | 0.004431059 |
| Heart_Failure | dendritic_cell | *RAB2A* | -2.886389594 | 2.60E-13 | 6.06E-13 | 1.64E-09 |
| Heart_Failure | dendritic_cell | *ZC2HC1A* | -3.337390667 | 1.96E-16 | 7.96E-16 | 1.23E-12 |
| Heart_Failure | dendritic_cell | *ARMC1* | -1.923334697 | 1.72E-05 | 2.25E-05 | 0.1084403 |
| Heart_Failure | dendritic_cell | *SPAG1* | -3.120017161 | 1.41E-14 | 3.98E-14 | 8.89E-11 |
| Heart_Failure | dendritic_cell | *CHRAC1* | 8.903547063 | 2.79E-09 | 4.47E-09 | 1.76E-05 |
| Heart_Failure | dendritic_cell | *SNX16* | -4.851597324 | 3.76E-15 | 1.18E-14 | 2.37E-11 |
| Heart_Failure | dendritic_cell | *GSDMD* | -1.948953701 | 0.006312797 | 0.007299715 | 1 |
| Heart_Failure | dendritic_cell | *ERI1* | -5.230571172 | 1.01E-06 | 1.40E-06 | 0.006368061 |
| Heart_Failure | dendritic_cell | *LEPROTL1* | -5.811424113 | 1.52E-12 | 3.25E-12 | 9.57E-09 |
| Heart_Failure | dendritic_cell | *ERICH1* | -4.303066964 | 4.09E-12 | 8.33E-12 | 2.57E-08 |
| Heart_Failure | dendritic_cell | *TUSC3* | -3.665087017 | 2.94E-18 | 1.93E-17 | 1.85E-14 |
| Heart_Failure | dendritic_cell | *ARHGEF10* | -2.592851468 | 2.20E-19 | 2.32E-18 | 1.39E-15 |
| Heart_Failure | dendritic_cell | *KLHDC4* | -3.341679295 | 1.37E-07 | 1.98E-07 | 0.000862676 |
| Heart_Failure | dendritic_cell | *MAN2B1* | 3.080496851 | 1.73E-20 | 6.17E-19 | 1.09E-16 |
| Heart_Failure | dendritic_cell | *TULP2* | -4.354578107 | 5.24E-14 | 1.34E-13 | 3.29E-10 |
| Heart_Failure | dendritic_cell | *NUCB1* | -4.630885441 | 4.32E-12 | 8.78E-12 | 2.71E-08 |
| Heart_Failure | dendritic_cell | *NFKBIB* | 8.17428049 | 2.58E-17 | 1.29E-16 | 1.62E-13 |
| Heart_Failure | dendritic_cell | *TUBB4A* | 13.61443235 | 4.16E-11 | 7.70E-11 | 2.62E-07 |
| Heart_Failure | dendritic_cell | *SARS2* | -2.668483381 | 2.46E-11 | 4.65E-11 | 1.55E-07 |
| Heart_Failure | dendritic_cell | *SNRNP70* | -4.293685358 | 1.44E-13 | 3.46E-13 | 9.05E-10 |
| Heart_Failure | dendritic_cell | *PPP1R37* | 6.160901171 | 5.93E-09 | 9.29E-09 | 3.73E-05 |
| Heart_Failure | dendritic_cell | *PEX11G* | -4.39909829 | 2.64E-15 | 8.46E-15 | 1.66E-11 |
| Heart_Failure | dendritic_cell | *ERCC2* | -9.8371601 | 9.66E-13 | 2.10E-12 | 6.07E-09 |
| Heart_Failure | dendritic_cell | *PLEKHJ1* | 7.614772788 | 1.43E-14 | 4.01E-14 | 8.97E-11 |
| Heart_Failure | dendritic_cell | *CD37* | 3.791326408 | 7.03E-20 | 1.06E-18 | 4.42E-16 |
| Heart_Failure | dendritic_cell | *AMH* | 3.473964422 | 2.63E-20 | 6.44E-19 | 1.65E-16 |
| Heart_Failure | dendritic_cell | *DKKL1* | -4.191307732 | 5.50E-09 | 8.63E-09 | 3.45E-05 |
| Heart_Failure | dendritic_cell | *STX10* | -5.69028998 | 2.35E-05 | 3.04E-05 | 0.147852022 |
| Heart_Failure | dendritic_cell | *FCER2* | -10.25813556 | 1.52E-15 | 5.12E-15 | 9.57E-12 |
| Heart_Failure | dendritic_cell | *DMPK* | -3.687591664 | 5.00E-18 | 3.05E-17 | 3.14E-14 |
| Heart_Failure | dendritic_cell | *IL4I1* | 1.169115001 | 1.72E-20 | 6.17E-19 | 1.08E-16 |
| Heart_Failure | dendritic_cell | *TLE6* | -3.447208501 | 5.49E-08 | 8.13E-08 | 0.00034497 |
| Heart_Failure | dendritic_cell | *AES* | -5.195983883 | 3.64E-07 | 5.17E-07 | 0.002290343 |
| Heart_Failure | dendritic_cell | *SGTA* | 3.872300806 | 0.006882076 | 0.007947761 | 1 |
| Heart_Failure | dendritic_cell | *C19orf53* | -5.668028412 | 8.32E-18 | 4.78E-17 | 5.23E-14 |
| Heart_Failure | dendritic_cell | *TIMM44* | -3.657804032 | 6.55E-12 | 1.31E-11 | 4.12E-08 |
| Heart_Failure | dendritic_cell | *CCDC61* | -3.072177535 | 0.003427892 | 0.00401774 | 1 |
| Heart_Failure | dendritic_cell | *IL27RA* | 5.065535888 | 6.41E-17 | 2.92E-16 | 4.03E-13 |
| Heart_Failure | dendritic_cell | *C19orf44* | -2.246537378 | 1.43E-12 | 3.05E-12 | 8.97E-09 |
| Heart_Failure | dendritic_cell | *OLFM2* | -1.698940578 | 8.32E-11 | 1.50E-10 | 5.23E-07 |
| Heart_Failure | dendritic_cell | *RASAL3* | -4.834144971 | 5.13E-08 | 7.61E-08 | 0.000322292 |
| Heart_Failure | dendritic_cell | *ZNF419* | -1.807170284 | 1.54E-08 | 2.34E-08 | 9.66E-05 |
| Heart_Failure | dendritic_cell | *SYDE1* | -3.293240692 | 1.12E-19 | 1.43E-18 | 7.01E-16 |
| Heart_Failure | dendritic_cell | *CCNE1* | -7.270853021 | 4.57E-20 | 8.12E-19 | 2.87E-16 |

| | | | | | | |
|---|---|---|---|---|---|---|
| Heart_Failure | dendritic_cell | *PDCD5* | -4.29564129 | 8.02E-16 | 2.86E-15 | 5.04E-12 |
| Heart_Failure | dendritic_cell | *ANKRD27* | -2.937188683 | 0.003736173 | 0.004374175 | 1 |
| Heart_Failure | dendritic_cell | *RPS16* | -4.582026644 | 1.36E-11 | 2.64E-11 | 8.56E-08 |
| Heart_Failure | dendritic_cell | *PRX* | 7.018222669 | 8.35E-10 | 1.39E-09 | 5.25E-06 |
| Heart_Failure | dendritic_cell | *CCDC94* | 6.219567119 | 1.12E-07 | 1.62E-07 | 0.000702073 |
| Heart_Failure | dendritic_cell | *TBCB* | -4.667502711 | 3.29E-14 | 8.72E-14 | 2.07E-10 |
| Heart_Failure | dendritic_cell | *POLR2I* | 6.220850168 | 9.02E-12 | 1.77E-11 | 5.67E-08 |
| Heart_Failure | dendritic_cell | *CLIP3* | -2.286294341 | 9.48E-08 | 1.38E-07 | 0.000595938 |
| Heart_Failure | dendritic_cell | *SLC1A5* | -5.294526047 | 1.07E-09 | 1.77E-09 | 6.74E-06 |
| Heart_Failure | dendritic_cell | *PRKD2* | -3.200803322 | 1.40E-06 | 1.92E-06 | 0.008807768 |
| Heart_Failure | dendritic_cell | *CACTIN* | 8.551421933 | 3.37E-18 | 2.16E-17 | 2.12E-14 |
| Heart_Failure | dendritic_cell | *FZR1* | -6.991188991 | 2.29E-14 | 6.22E-14 | 1.44E-10 |
| Heart_Failure | dendritic_cell | *BBC3* | -11.615003 | 3.07E-10 | 5.29E-10 | 1.93E-06 |
| Heart_Failure | dendritic_cell | *TGFB1* | 4.210135332 | 5.19E-06 | 6.91E-06 | 0.032631173 |
| Heart_Failure | dendritic_cell | *CEACAM4* | 1.208948124 | 6.40E-21 | 4.66E-19 | 4.02E-17 |
| Heart_Failure | dendritic_cell | *PLIN3* | 5.842185998 | 6.86E-08 | 1.01E-07 | 0.000431375 |
| Heart_Failure | dendritic_cell | *MRPL4* | -4.353489235 | 0.00033742 | 0.000414085 | 1 |
| Heart_Failure | dendritic_cell | *ICAM5* | -19.55115847 | 1.81E-08 | 2.75E-08 | 0.000113743 |
| Heart_Failure | dendritic_cell | *CD33* | 1.786986593 | 9.09E-22 | 3.01E-19 | 5.72E-18 |
| Heart_Failure | dendritic_cell | *BABAM1* | 9.14062319 | 2.30E-14 | 6.24E-14 | 1.44E-10 |
| Heart_Failure | dendritic_cell | *TYK2* | 4.790881557 | 2.35E-07 | 3.37E-07 | 0.001478888 |
| Heart_Failure | dendritic_cell | *RABAC1* | -5.040800835 | 3.77E-14 | 9.88E-14 | 2.37E-10 |
| Heart_Failure | dendritic_cell | *PTPRS* | -1.69612615 | 1.23E-18 | 9.23E-18 | 7.76E-15 |
| Heart_Failure | dendritic_cell | *CYTH2* | 6.869876285 | 5.40E-07 | 7.56E-07 | 0.003391892 |
| Heart_Failure | dendritic_cell | *GRWD1* | -3.183990011 | 1.97E-19 | 2.15E-18 | 1.24E-15 |
| Heart_Failure | dendritic_cell | *GRIN2D* | -5.137145247 | 1.89E-19 | 2.11E-18 | 1.19E-15 |
| Heart_Failure | dendritic_cell | *CCDC114* | 2.583030749 | 2.07E-12 | 4.34E-12 | 1.30E-08 |
| Heart_Failure | dendritic_cell | *CARD8* | -2.833378831 | 0.001141465 | 0.001364592 | 1 |
| Heart_Failure | dendritic_cell | *ZNF175* | -4.173053144 | 5.39E-12 | 1.09E-11 | 3.39E-08 |
| Heart_Failure | dendritic_cell | *PLA2G4C* | 1.656761098 | 0.015372238 | 0.017407287 | 1 |
| Heart_Failure | dendritic_cell | *CAPS* | -2.857170422 | 3.13E-16 | 1.22E-15 | 1.97E-12 |
| Heart_Failure | dendritic_cell | *BCAT2* | -3.830307029 | 4.46E-19 | 4.01E-18 | 2.80E-15 |
| Heart_Failure | dendritic_cell | *MIER2* | 6.783777728 | 1.03E-11 | 2.00E-11 | 6.45E-08 |
| Heart_Failure | dendritic_cell | *PPP2R1A* | -5.356011989 | 1.29E-15 | 4.39E-15 | 8.09E-12 |
| Heart_Failure | dendritic_cell | *TNPO2* | -4.60317027 | 4.07E-14 | 1.06E-13 | 2.56E-10 |
| Heart_Failure | dendritic_cell | *GCDH* | -5.086687764 | 1.99E-14 | 5.45E-14 | 1.25E-10 |
| Heart_Failure | dendritic_cell | *MAST1* | 16.39244088 | 1.21E-18 | 9.07E-18 | 7.59E-15 |
| Heart_Failure | dendritic_cell | *PRPF31* | -4.665815933 | 3.10E-10 | 5.34E-10 | 1.95E-06 |
| Heart_Failure | dendritic_cell | *TFPT* | -5.328184754 | 4.61E-19 | 4.10E-18 | 2.90E-15 |
| Heart_Failure | dendritic_cell | *RPL18A* | -3.454961449 | 1.78E-07 | 2.56E-07 | 0.001120413 |
| Heart_Failure | dendritic_cell | *PDE4C* | -5.332247285 | 1.17E-13 | 2.85E-13 | 7.38E-10 |
| Heart_Failure | dendritic_cell | *UPK1A* | 10.01074339 | 1.15E-12 | 2.49E-12 | 7.24E-09 |
| Heart_Failure | dendritic_cell | *COPE* | 7.097371397 | 2.11E-09 | 3.41E-09 | 1.33E-05 |
| Heart_Failure | dendritic_cell | *LSR* | -3.398424066 | 7.21E-08 | 1.06E-07 | 0.000453441 |
| Heart_Failure | dendritic_cell | *KXD1* | -4.223523326 | 0.005115507 | 0.005944767 | 1 |
| Heart_Failure | dendritic_cell | *HPN* | -3.179076254 | 3.53E-20 | 6.98E-19 | 2.22E-16 |
| Heart_Failure | dendritic_cell | *ZNF14* | -4.7428636 | 5.96E-18 | 3.57E-17 | 3.75E-14 |
| Heart_Failure | dendritic_cell | *ATP13A1* | -2.539017664 | 0.001709493 | 0.002033986 | 1 |
| Heart_Failure | dendritic_cell | *SIPA1L3* | -1.956619477 | 0.039960802 | 0.044324905 | 1 |
| Heart_Failure | dendritic_cell | *ZNF85* | 6.112398318 | 4.06E-11 | 7.52E-11 | 2.55E-07 |

| | | | | | | |
|---|---|---|---|---|---|---|
| Heart_Failure | dendritic_cell | *ETHE1* | 4.074012443 | 2.27E-17 | 1.15E-16 | 1.43E-13 |
| Heart_Failure | dendritic_cell | *SMG9* | -2.737403515 | 0.023078869 | 0.025891658 | 1 |
| Heart_Failure | dendritic_cell | *GTPBP10* | -2.831392993 | 2.16E-08 | 3.26E-08 | 0.000135628 |
| Heart_Failure | dendritic_cell | *RASA4* | 2.819893315 | 1.25E-07 | 1.82E-07 | 0.000786486 |
| Heart_Failure | dendritic_cell | *PON3* | 8.647027972 | 1.57E-17 | 8.36E-17 | 9.85E-14 |
| Heart_Failure | dendritic_cell | *PON2* | 2.907864718 | 0.008860927 | 0.01016299 | 1 |
| Heart_Failure | dendritic_cell | *HBP1* | -4.598797362 | 2.20E-13 | 5.15E-13 | 1.38E-09 |
| Heart_Failure | dendritic_cell | *WDR91* | -5.570784571 | 1.57E-14 | 4.38E-14 | 9.88E-11 |
| Heart_Failure | dendritic_cell | *PTN* | -2.958820473 | 3.58E-20 | 6.99E-19 | 2.25E-16 |
| Heart_Failure | dendritic_cell | *MPP6* | -2.571240134 | 2.09E-19 | 2.24E-18 | 1.32E-15 |
| Heart_Failure | dendritic_cell | *DFNA5* | 2.218202132 | 0.00381616 | 0.004465326 | 1 |
| Heart_Failure | dendritic_cell | *OGDH* | -4.457126175 | 9.13E-09 | 1.41E-08 | 5.74E-05 |
| Heart_Failure | dendritic_cell | *ADAP1* | 8.727283199 | 4.06E-20 | 7.63E-19 | 2.55E-16 |
| Heart_Failure | dendritic_cell | *H2AFV* | -3.657610009 | 0.000553284 | 0.000672304 | 1 |
| Heart_Failure | dendritic_cell | *CAV1* | -2.509369325 | 9.45E-17 | 4.11E-16 | 5.94E-13 |
| Heart_Failure | dendritic_cell | *MET* | -2.33630529 | 2.59E-19 | 2.60E-18 | 1.63E-15 |
| Heart_Failure | dendritic_cell | *RNF32* | -1.829362261 | 0.005599697 | 0.006489455 | 1 |
| Heart_Failure | dendritic_cell | *LMBR1* | -3.858764203 | 2.14E-08 | 3.24E-08 | 0.000134713 |
| Heart_Failure | dendritic_cell | *HOXA2* | -7.26595702 | 1.88E-19 | 2.11E-18 | 1.18E-15 |
| Heart_Failure | dendritic_cell | *HOXA5* | -3.055524805 | 7.83E-20 | 1.13E-18 | 4.93E-16 |
| Heart_Failure | dendritic_cell | *HOXA6* | -2.904137028 | 2.08E-21 | 4.66E-19 | 1.31E-17 |
| Heart_Failure | dendritic_cell | *BRAT1* | -2.297537103 | 0.002267289 | 0.002682432 | 1 |
| Heart_Failure | dendritic_cell | *CPED1* | -3.084881905 | 1.20E-16 | 5.10E-16 | 7.56E-13 |
| Heart_Failure | dendritic_cell | *HIBADH* | -3.506466501 | 1.41E-14 | 3.96E-14 | 8.86E-11 |
| Heart_Failure | dendritic_cell | *CPVL* | 0.78072067 | 7.28E-21 | 4.66E-19 | 4.58E-17 |
| Heart_Failure | dendritic_cell | *ABHD11* | 1.719208672 | 0.001045248 | 0.001251947 | 1 |
| Heart_Failure | dendritic_cell | *PLEKHA8* | -3.851541809 | 0.000613814 | 0.000743842 | 1 |
| Heart_Failure | dendritic_cell | *NOD1* | -4.823281712 | 2.21E-10 | 3.84E-10 | 1.39E-06 |
| Heart_Failure | dendritic_cell | *CRHR2* | -12.41074337 | 2.01E-15 | 6.62E-15 | 1.26E-11 |
| Heart_Failure | dendritic_cell | *EPHB6* | 4.822472231 | 3.82E-13 | 8.72E-13 | 2.40E-09 |
| Heart_Failure | dendritic_cell | *CHCHD2* | -3.313047539 | 2.77E-06 | 3.74E-06 | 0.01740479 |
| Heart_Failure | dendritic_cell | *CCL24* | -1.947782407 | 0.009586689 | 0.010974419 | 1 |
| Heart_Failure | dendritic_cell | *NPTX2* | -68.8429041 | 3.46E-12 | 7.12E-12 | 2.18E-08 |
| Heart_Failure | dendritic_cell | *CYP3A5* | -5.004249227 | 1.15E-16 | 4.90E-16 | 7.24E-13 |
| Heart_Failure | dendritic_cell | *ZKSCAN1* | -4.606722576 | 1.57E-12 | 3.35E-12 | 9.87E-09 |
| Heart_Failure | dendritic_cell | *SNX8* | 4.992858063 | 1.04E-05 | 1.37E-05 | 0.065340655 |
| Heart_Failure | dendritic_cell | *TAF6* | -4.692615676 | 1.18E-14 | 3.36E-14 | 7.39E-11 |
| Heart_Failure | dendritic_cell | *AIMP2* | -5.095537679 | 2.94E-17 | 1.46E-16 | 1.85E-13 |
| Heart_Failure | dendritic_cell | *TFR2* | -4.586612261 | 2.98E-18 | 1.95E-17 | 1.87E-14 |
| Heart_Failure | dendritic_cell | *RBM28* | -3.04668495 | 3.15E-17 | 1.55E-16 | 1.98E-13 |
| Heart_Failure | dendritic_cell | *USP42* | -4.745257966 | 7.40E-15 | 2.19E-14 | 4.65E-11 |
| Heart_Failure | dendritic_cell | *IMPDH1* | 4.547731022 | 1.41E-09 | 2.31E-09 | 8.84E-06 |
| Heart_Failure | dendritic_cell | *LSM5* | -5.146067987 | 2.19E-17 | 1.11E-16 | 1.38E-13 |
| Heart_Failure | dendritic_cell | *AP1S1* | 12.65544386 | 8.40E-08 | 1.23E-07 | 0.00052797 |
| Heart_Failure | dendritic_cell | *C1GALT1* | -4.223186977 | 9.25E-15 | 2.70E-14 | 5.82E-11 |
| Heart_Failure | dendritic_cell | *RPA3* | -3.729763289 | 2.37E-17 | 1.19E-16 | 1.49E-13 |
| Heart_Failure | dendritic_cell | *ZNHIT1* | -3.013216373 | 1.26E-07 | 1.82E-07 | 0.000790555 |
| Heart_Failure | dendritic_cell | *PHF14* | -3.710586035 | 7.45E-14 | 1.87E-13 | 4.68E-10 |
| Heart_Failure | dendritic_cell | *CEP41* | -3.405401356 | 3.10E-18 | 2.01E-17 | 1.95E-14 |
| Heart_Failure | dendritic_cell | *ZNF862* | -3.774163366 | 1.43E-15 | 4.85E-15 | 9.02E-12 |

| | | | | | | |
|---|---|---|---|---|---|---|
| Heart_Failure | dendritic_cell | *MEST* | -1.690161996 | 7.59E-17 | 3.41E-16 | 4.77E-13 |
| Heart_Failure | dendritic_cell | *ACTR3C* | -3.752328847 | 2.25E-15 | 7.34E-15 | 1.42E-11 |
| Heart_Failure | dendritic_cell | *POU6F2* | -2.90179919 | 8.50E-20 | 1.20E-18 | 5.34E-16 |
| Heart_Failure | dendritic_cell | *TSPAN13* | -7.250075009 | 2.20E-16 | 8.81E-16 | 1.38E-12 |
| Heart_Failure | dendritic_cell | *RARRES2* | -3.996625322 | 3.80E-16 | 1.45E-15 | 2.39E-12 |
| Heart_Failure | dendritic_cell | *TMEM176B* | 1.731145504 | 2.04E-08 | 3.09E-08 | 0.000128246 |
| Heart_Failure | dendritic_cell | *MRPL32* | -2.896161318 | 1.75E-09 | 2.85E-09 | 1.10E-05 |
| Heart_Failure | dendritic_cell | *COA1* | -2.972741996 | 5.23E-10 | 8.84E-10 | 3.29E-06 |
| Heart_Failure | dendritic_cell | *BLVRA* | -2.363564308 | 6.40E-05 | 8.13E-05 | 0.402307547 |
| Heart_Failure | dendritic_cell | *URGCP* | -4.810911518 | 9.89E-16 | 3.46E-15 | 6.22E-12 |
| Heart_Failure | dendritic_cell | *POLD2* | -6.533508767 | 3.39E-13 | 7.79E-13 | 2.13E-09 |
| Heart_Failure | dendritic_cell | *GCK* | -30.18149035 | 2.49E-20 | 6.44E-19 | 1.57E-16 |
| Heart_Failure | dendritic_cell | *BCL7B* | -3.199785842 | 5.87E-10 | 9.90E-10 | 3.69E-06 |
| Heart_Failure | dendritic_cell | *YKT6* | 8.185882953 | 2.18E-15 | 7.13E-15 | 1.37E-11 |
| Heart_Failure | dendritic_cell | *TBL2* | -1.948477838 | 2.88E-10 | 4.98E-10 | 1.81E-06 |
| Heart_Failure | dendritic_cell | *CLIP2* | 6.283938578 | 2.30E-08 | 3.46E-08 | 0.0001445 |
| Heart_Failure | dendritic_cell | *LIMK1* | 2.832370896 | 1.48E-20 | 5.69E-19 | 9.27E-17 |
| Heart_Failure | dendritic_cell | *SPATA6L* | -3.494668586 | 1.95E-17 | 1.01E-16 | 1.22E-13 |
| Heart_Failure | dendritic_cell | *SLC1A1* | -1.022686362 | 0.004876396 | 0.005672138 | 1 |
| Heart_Failure | dendritic_cell | *FKTN* | -4.677786935 | 1.26E-14 | 3.56E-14 | 7.90E-11 |
| Heart_Failure | dendritic_cell | *NMRK1* | -3.917908948 | 7.32E-09 | 1.14E-08 | 4.60E-05 |
| Heart_Failure | dendritic_cell | *TMEM245* | -3.778365851 | 2.22E-11 | 4.22E-11 | 1.40E-07 |
| Heart_Failure | dendritic_cell | *CORO2A* | -4.021840449 | 1.06E-19 | 1.39E-18 | 6.68E-16 |
| Heart_Failure | dendritic_cell | *C5* | -3.045153826 | 1.34E-14 | 3.79E-14 | 8.45E-11 |
| Heart_Failure | dendritic_cell | *OGN* | -2.893452148 | 1.88E-17 | 9.81E-17 | 1.18E-13 |
| Heart_Failure | dendritic_cell | *ECM2* | -2.41316866 | 3.00E-17 | 1.48E-16 | 1.89E-13 |
| Heart_Failure | dendritic_cell | *PTGR1* | -2.070153375 | 1.74E-11 | 3.34E-11 | 1.10E-07 |
| Heart_Failure | dendritic_cell | *SUSD1* | 2.57000834 | 1.30E-06 | 1.78E-06 | 0.008178911 |
| Heart_Failure | dendritic_cell | *TNFSF8* | 2.292303535 | 1.22E-20 | 5.08E-19 | 7.69E-17 |
| Heart_Failure | dendritic_cell | *AK1* | -2.481598873 | 0.020264787 | 0.022820118 | 1 |
| Heart_Failure | dendritic_cell | *PLGRKT* | -1.581254048 | 0.002895423 | 0.003406986 | 1 |
| Heart_Failure | dendritic_cell | *KDM4C* | -3.812546097 | 1.25E-07 | 1.82E-07 | 0.000785914 |
| Heart_Failure | dendritic_cell | *DOCK8* | 2.798191878 | 9.36E-10 | 1.56E-09 | 5.88E-06 |
| Heart_Failure | dendritic_cell | *ELAVL2* | -4.519044897 | 1.73E-18 | 1.24E-17 | 1.08E-14 |
| Heart_Failure | dendritic_cell | *FUBP3* | -5.139841903 | 5.57E-11 | 1.02E-10 | 3.50E-07 |
| Heart_Failure | dendritic_cell | *RGP1* | -6.81911139 | 1.15E-18 | 8.73E-18 | 7.21E-15 |
| Heart_Failure | dendritic_cell | *MPDZ* | -3.724833473 | 6.29E-16 | 2.30E-15 | 3.96E-12 |
| Heart_Failure | dendritic_cell | *PTGDS* | -10.66945952 | 3.83E-13 | 8.74E-13 | 2.41E-09 |
| Heart_Failure | dendritic_cell | *SHB* | -7.167310444 | 6.78E-20 | 1.03E-18 | 4.26E-16 |
| Heart_Failure | dendritic_cell | *DVL1* | -5.940882357 | 8.83E-06 | 1.17E-05 | 0.055503281 |
| Heart_Failure | dendritic_cell | *PDLIM1* | -2.564954913 | 5.11E-05 | 6.52E-05 | 0.321328651 |
| Heart_Failure | dendritic_cell | *CCNJ* | 5.369158458 | 5.26E-07 | 7.37E-07 | 0.003304434 |
| Heart_Failure | dendritic_cell | *ATRNL1* | -2.228868053 | 0.000273243 | 0.000336575 | 1 |
| Heart_Failure | dendritic_cell | *HPS1* | -5.653234118 | 2.78E-09 | 4.45E-09 | 1.75E-05 |
| Heart_Failure | dendritic_cell | *PHYH* | -1.804845996 | 2.68E-12 | 5.56E-12 | 1.68E-08 |
| Heart_Failure | dendritic_cell | *DNMBP* | -4.650341796 | 6.31E-08 | 9.32E-08 | 0.000396897 |
| Heart_Failure | dendritic_cell | *TRDMT1* | -3.584139009 | 5.89E-18 | 3.54E-17 | 3.71E-14 |
| Heart_Failure | dendritic_cell | *DDX50* | -4.750888236 | 9.41E-14 | 2.32E-13 | 5.91E-10 |
| Heart_Failure | dendritic_cell | *MAPK8* | -3.823473163 | 6.61E-08 | 9.75E-08 | 0.000415467 |
| Heart_Failure | dendritic_cell | *ATE1* | -3.391667641 | 8.35E-12 | 1.64E-11 | 5.25E-08 |

| | | | | | | |
|---|---|---|---|---|---|---|
| Heart_Failure | dendritic_cell | *NSMCE4A* | -4.599038263 | 1.12E-16 | 4.78E-16 | 7.03E-13 |
| Heart_Failure | dendritic_cell | *PLEKHA1* | -4.775524019 | 4.41E-09 | 6.97E-09 | 2.77E-05 |
| Heart_Failure | dendritic_cell | *SPOCK2* | -2.801425282 | 8.75E-10 | 1.46E-09 | 5.50E-06 |
| Heart_Failure | dendritic_cell | *MICU1* | -3.966678656 | 5.89E-11 | 1.07E-10 | 3.70E-07 |
| Heart_Failure | dendritic_cell | *CCSER2* | -4.274283363 | 6.02E-12 | 1.21E-11 | 3.78E-08 |
| Heart_Failure | dendritic_cell | *BMPR1A* | -2.045425045 | 6.28E-12 | 1.25E-11 | 3.95E-08 |
| Heart_Failure | dendritic_cell | *KAZALD1* | -5.987884424 | 1.58E-18 | 1.15E-17 | 9.95E-15 |
| Heart_Failure | dendritic_cell | *TNKS2* | -4.587620902 | 8.22E-09 | 1.28E-08 | 5.17E-05 |
| Heart_Failure | dendritic_cell | *ANKRD26* | -3.249302665 | 8.50E-11 | 1.53E-10 | 5.34E-07 |
| Heart_Failure | dendritic_cell | *PITRM1* | -5.778452262 | 3.25E-14 | 8.63E-14 | 2.04E-10 |
| Heart_Failure | dendritic_cell | *MAP3K8* | -3.175692634 | 0.000543454 | 0.000660614 | 1 |
| Heart_Failure | dendritic_cell | *SHOC2* | -4.112251926 | 5.12E-11 | 9.37E-11 | 3.22E-07 |
| Heart_Failure | dendritic_cell | *TFAM* | -4.619663061 | 1.17E-17 | 6.51E-17 | 7.38E-14 |
| Heart_Failure | dendritic_cell | *RPL28* | 2.612459881 | 3.27E-05 | 4.19E-05 | 0.205454785 |
| Heart_Failure | dendritic_cell | *PBLD* | -0.730748796 | 0.004277773 | 0.004993382 | 1 |
| Heart_Failure | dendritic_cell | *TADA2A* | -1.493705527 | 2.84E-05 | 3.66E-05 | 0.17848491 |
| Heart_Failure | dendritic_cell | *DHRS11* | -2.13133901 | 1.07E-15 | 3.72E-15 | 6.73E-12 |
| Heart_Failure | dendritic_cell | *MLLT6* | -5.095147873 | 8.56E-14 | 2.12E-13 | 5.38E-10 |
| Heart_Failure | dendritic_cell | *PSMB3* | 5.254701688 | 2.54E-18 | 1.71E-17 | 1.60E-14 |
| Heart_Failure | dendritic_cell | *RAPGEFL1* | -3.537590498 | 3.54E-16 | 1.36E-15 | 2.23E-12 |
| Heart_Failure | dendritic_cell | *RGS9* | -9.24248436 | 7.26E-15 | 2.16E-14 | 4.56E-11 |
| Heart_Failure | dendritic_cell | *WNT3* | 5.657722334 | 6.66E-18 | 3.93E-17 | 4.19E-14 |
| Heart_Failure | dendritic_cell | *ASPA* | -2.814770093 | 4.86E-19 | 4.27E-18 | 3.05E-15 |
| Heart_Failure | dendritic_cell | *RAD51C* | -1.367225999 | 0.003140748 | 0.003687373 | 1 |
| Heart_Failure | dendritic_cell | *SEPT4* | -3.301090909 | 6.48E-20 | 1.01E-18 | 4.08E-16 |
| Heart_Failure | dendritic_cell | *TRIM37* | -5.027928681 | 1.64E-10 | 2.88E-10 | 1.03E-06 |
| Heart_Failure | dendritic_cell | *PNPO* | 3.120655338 | 1.99E-17 | 1.03E-16 | 1.25E-13 |
| Heart_Failure | dendritic_cell | *RPS6KB1* | -4.035854887 | 1.14E-16 | 4.87E-16 | 7.18E-13 |
| Heart_Failure | dendritic_cell | *CDK5RAP3* | -2.920124185 | 1.85E-19 | 2.08E-18 | 1.16E-15 |
| Heart_Failure | dendritic_cell | *CBX1* | -3.979138295 | 4.17E-06 | 5.58E-06 | 0.026192187 |
| Heart_Failure | dendritic_cell | *RECQL5* | -4.212592143 | 1.66E-10 | 2.91E-10 | 1.04E-06 |
| Heart_Failure | dendritic_cell | *PIGL* | -4.21912135 | 9.77E-15 | 2.83E-14 | 6.14E-11 |
| Heart_Failure | dendritic_cell | *GALK1* | -9.550464146 | 3.64E-15 | 1.14E-14 | 2.29E-11 |
| Heart_Failure | dendritic_cell | *CAMTA2* | -3.989005476 | 1.16E-16 | 4.92E-16 | 7.28E-13 |
| Heart_Failure | dendritic_cell | *MED13* | -3.491135662 | 4.57E-09 | 7.22E-09 | 2.87E-05 |
| Heart_Failure | dendritic_cell | *HOXB6* | -2.682872848 | 1.06E-17 | 5.95E-17 | 6.67E-14 |
| Heart_Failure | dendritic_cell | *RNF167* | 3.071542874 | 0.022642577 | 0.025420336 | 1 |
| Heart_Failure | dendritic_cell | *NUP88* | -4.332145579 | 5.17E-13 | 1.16E-12 | 3.25E-09 |
| Heart_Failure | dendritic_cell | *C1QBP* | 8.158615751 | 9.13E-14 | 2.25E-13 | 5.74E-10 |
| Heart_Failure | dendritic_cell | *GOSR1* | -4.721332208 | 3.79E-09 | 6.02E-09 | 2.38E-05 |
| Heart_Failure | dendritic_cell | *DRG2* | -4.21469935 | 0.0097229 | 0.011126297 | 1 |
| Heart_Failure | dendritic_cell | *AKAP10* | -2.671927457 | 0.000908459 | 0.001092479 | 1 |
| Heart_Failure | dendritic_cell | *ALDH3A1* | 4.101407651 | 8.63E-11 | 1.55E-10 | 5.43E-07 |
| Heart_Failure | dendritic_cell | *SMARCD2* | -4.865467886 | 1.90E-05 | 2.46E-05 | 0.119223934 |
| Heart_Failure | dendritic_cell | *ICAM2* | -4.158084516 | 3.44E-10 | 5.90E-10 | 2.16E-06 |
| Heart_Failure | dendritic_cell | *UTP6* | -5.129050322 | 7.74E-12 | 1.53E-11 | 4.87E-08 |
| Heart_Failure | dendritic_cell | *DDX5* | -2.399184265 | 3.97E-10 | 6.77E-10 | 2.50E-06 |
| Heart_Failure | dendritic_cell | *C17orf75* | -3.681940952 | 5.91E-20 | 9.53E-19 | 3.71E-16 |
| Heart_Failure | dendritic_cell | *PEX12* | 2.952677096 | 8.95E-11 | 1.61E-10 | 5.63E-07 |
| Heart_Failure | dendritic_cell | *RAB5C* | -2.379579701 | 0.028418251 | 0.031734555 | 1 |

| | | | | | | |
|---|---|---|---|---|---|---|
| Heart_Failure | dendritic_cell | *NAGLU* | -3.745263617 | 1.41E-17 | 7.61E-17 | 8.85E-14 |
| Heart_Failure | dendritic_cell | *EZH1* | -3.599792838 | 3.31E-17 | 1.61E-16 | 2.08E-13 |
| Heart_Failure | dendritic_cell | *DLX4* | -28.06057444 | 2.83E-20 | 6.44E-19 | 1.78E-16 |
| Heart_Failure | dendritic_cell | *VAT1* | -3.428468061 | 4.07E-18 | 2.54E-17 | 2.56E-14 |
| Heart_Failure | dendritic_cell | *LRRC59* | -3.53077663 | 6.97E-20 | 1.05E-18 | 4.38E-16 |
| Heart_Failure | dendritic_cell | *ALOX12* | -7.546436159 | 1.83E-10 | 3.19E-10 | 1.15E-06 |
| Heart_Failure | dendritic_cell | *MPP2* | -9.155313441 | 0.001930329 | 0.002291537 | 1 |
| Heart_Failure | dendritic_cell | *EFNB3* | -30.16278215 | 4.41E-15 | 1.36E-14 | 2.77E-11 |
| Heart_Failure | dendritic_cell | *FAM20A* | 2.54503146 | 0.014015052 | 0.015916299 | 1 |
| Heart_Failure | dendritic_cell | *YWHAE* | -4.419628495 | 1.46E-09 | 2.39E-09 | 9.18E-06 |
| Heart_Failure | dendritic_cell | *DPH1* | -3.776778815 | 1.02E-19 | 1.36E-18 | 6.41E-16 |
| Heart_Failure | dendritic_cell | *DHRS7B* | -4.341423647 | 2.27E-15 | 7.38E-15 | 1.42E-11 |
| Heart_Failure | dendritic_cell | *RCVRN* | -8.699660826 | 2.21E-11 | 4.20E-11 | 1.39E-07 |
| Heart_Failure | dendritic_cell | *MYH3* | -5.04086005 | 1.19E-15 | 4.09E-15 | 7.48E-12 |
| Heart_Failure | dendritic_cell | *TMEM104* | -6.329474279 | 1.10E-09 | 1.81E-09 | 6.90E-06 |
| Heart_Failure | dendritic_cell | *TMEM97* | 2.56448879 | 2.07E-18 | 1.44E-17 | 1.30E-14 |
| Heart_Failure | dendritic_cell | *RAB34* | -3.200570899 | 0.000554476 | 0.000673622 | 1 |
| Heart_Failure | dendritic_cell | *SLAIN2* | -3.863551092 | 2.92E-10 | 5.04E-10 | 1.84E-06 |
| Heart_Failure | dendritic_cell | *OCIAD1* | -4.613695961 | 6.62E-12 | 1.32E-11 | 4.16E-08 |
| Heart_Failure | dendritic_cell | *CWH43* | 5.220969434 | 8.04E-21 | 4.66E-19 | 5.06E-17 |
| Heart_Failure | dendritic_cell | *KIAA1211* | 1.817717378 | 2.61E-11 | 4.91E-11 | 1.64E-07 |
| Heart_Failure | dendritic_cell | *AREG* | 1.06669777 | 1.52E-20 | 5.74E-19 | 9.58E-17 |
| Heart_Failure | dendritic_cell | *UBE2D3* | -3.978217574 | 9.01E-10 | 1.50E-09 | 5.66E-06 |
| Heart_Failure | dendritic_cell | *NDUFC1* | -6.265982246 | 5.48E-15 | 1.66E-14 | 3.45E-11 |
| Heart_Failure | dendritic_cell | *TBC1D9* | 3.739333482 | 2.23E-05 | 2.89E-05 | 0.140148848 |
| Heart_Failure | dendritic_cell | *CPE* | -3.420031011 | 3.03E-16 | 1.18E-15 | 1.90E-12 |
| Heart_Failure | dendritic_cell | *WFS1* | -4.22057012 | 2.53E-18 | 1.70E-17 | 1.59E-14 |
| Heart_Failure | dendritic_cell | *GRPEL1* | -6.335946607 | 4.26E-18 | 2.64E-17 | 2.68E-14 |
| Heart_Failure | dendritic_cell | *AADAT* | -2.85314733 | 5.96E-20 | 9.56E-19 | 3.75E-16 |
| Heart_Failure | dendritic_cell | *CPZ* | -55.71410912 | 3.44E-20 | 6.93E-19 | 2.16E-16 |
| Heart_Failure | dendritic_cell | *TRIM2* | -2.666820917 | 2.28E-13 | 5.35E-13 | 1.43E-09 |
| Heart_Failure | dendritic_cell | *SLC2A9* | 1.826643743 | 1.68E-21 | 4.58E-19 | 1.05E-17 |
| Heart_Failure | dendritic_cell | *SH3D19* | -2.270033516 | 1.75E-17 | 9.25E-17 | 1.10E-13 |
| Heart_Failure | dendritic_cell | *RAPGEF2* | -3.808894256 | 4.78E-11 | 8.78E-11 | 3.00E-07 |
| Heart_Failure | dendritic_cell | *LRP2BP* | -4.078335264 | 5.21E-16 | 1.92E-15 | 3.27E-12 |
| Heart_Failure | dendritic_cell | *UFSP2* | -4.012758545 | 8.32E-17 | 3.70E-16 | 5.23E-13 |
| Heart_Failure | dendritic_cell | *KLF3* | -4.630722216 | 5.11E-09 | 8.04E-09 | 3.21E-05 |
| Heart_Failure | dendritic_cell | *KLHL5* | 2.242901066 | 0.001177742 | 0.001407426 | 1 |
| Heart_Failure | dendritic_cell | *FAM149A* | -2.024890873 | 6.43E-13 | 1.42E-12 | 4.04E-09 |
| Heart_Failure | dendritic_cell | *UGDH* | -3.573345533 | 3.00E-19 | 2.90E-18 | 1.88E-15 |
| Heart_Failure | dendritic_cell | *DDX25* | -5.393037249 | 8.19E-09 | 1.27E-08 | 5.15E-05 |
| Heart_Failure | dendritic_cell | *HTATIP2* | -2.383665895 | 1.12E-17 | 6.26E-17 | 7.07E-14 |
| Heart_Failure | dendritic_cell | *CTSC* | -4.332299334 | 6.45E-11 | 1.17E-10 | 4.05E-07 |
| Heart_Failure | dendritic_cell | *ELP4* | -4.142233477 | 5.22E-13 | 1.17E-12 | 3.28E-09 |
| Heart_Failure | dendritic_cell | *SC5D* | -4.84391633 | 5.16E-18 | 3.14E-17 | 3.25E-14 |
| Heart_Failure | dendritic_cell | *C11orf63* | -4.093125733 | 1.23E-17 | 6.78E-17 | 7.72E-14 |
| Heart_Failure | dendritic_cell | *DTX4* | -5.889360564 | 4.94E-15 | 1.51E-14 | 3.10E-11 |
| Heart_Failure | dendritic_cell | *PUS3* | -5.456802624 | 3.56E-07 | 5.05E-07 | 0.002237925 |
| Heart_Failure | dendritic_cell | *DCPS* | -5.155416341 | 1.38E-05 | 1.80E-05 | 0.086713567 |
| Heart_Failure | dendritic_cell | *FOXRED1* | -3.127295675 | 3.78E-10 | 6.45E-10 | 2.38E-06 |

| | | | | | | |
|---|---|---|---|---|---|---|
| Heart_Failure | dendritic_cell | *NRXN2* | 9.617116843 | 2.95E-19 | 2.88E-18 | 1.86E-15 |
| Heart_Failure | dendritic_cell | *MS4A4A* | 1.830845448 | 1.06E-07 | 1.54E-07 | 0.000665227 |
| Heart_Failure | dendritic_cell | *CCDC86* | -6.780857904 | 2.69E-20 | 6.44E-19 | 1.69E-16 |
| Heart_Failure | dendritic_cell | *PRPF19* | -6.473644784 | 6.48E-10 | 1.09E-09 | 4.08E-06 |
| Heart_Failure | dendritic_cell | *HPX* | -3.029933381 | 1.27E-08 | 1.95E-08 | 8.00E-05 |
| Heart_Failure | dendritic_cell | *FOLR3* | 1.783138163 | 3.61E-12 | 7.40E-12 | 2.27E-08 |
| Heart_Failure | dendritic_cell | *PANX1* | -4.662739029 | 3.91E-07 | 5.53E-07 | 0.002455913 |
| Heart_Failure | dendritic_cell | *EIF4G2* | -4.847115448 | 6.53E-12 | 1.30E-11 | 4.10E-08 |
| Heart_Failure | dendritic_cell | *GALNT18* | -2.152049747 | 5.31E-18 | 3.21E-17 | 3.34E-14 |
| Heart_Failure | dendritic_cell | *KIAA1549L* | -2.23598733 | 0.018558816 | 0.02095533 | 1 |
| Heart_Failure | dendritic_cell | *CD5* | 3.680160386 | 5.30E-07 | 7.43E-07 | 0.003332239 |
| Heart_Failure | dendritic_cell | *ACCS* | -1.78088781 | 4.19E-07 | 5.91E-07 | 0.00263241 |
| Heart_Failure | dendritic_cell | *MADD* | 5.900932196 | 8.59E-16 | 3.05E-15 | 5.40E-12 |
| Heart_Failure | dendritic_cell | *NAA40* | -4.278736305 | 2.86E-18 | 1.88E-17 | 1.80E-14 |
| Heart_Failure | dendritic_cell | *CARS* | -3.427935182 | 6.46E-15 | 1.94E-14 | 4.06E-11 |
| Heart_Failure | dendritic_cell | *SLC22A18* | 1.718757568 | 1.31E-08 | 2.00E-08 | 8.22E-05 |
| Heart_Failure | dendritic_cell | *C11orf21* | 3.444110818 | 5.60E-19 | 4.82E-18 | 3.52E-15 |
| Heart_Failure | dendritic_cell | *ELMOD1* | -5.973133327 | 1.49E-16 | 6.18E-16 | 9.36E-13 |
| Heart_Failure | dendritic_cell | *RPS13* | -3.675871233 | 5.38E-06 | 7.16E-06 | 0.033815163 |
| Heart_Failure | dendritic_cell | *AIP* | -3.778595485 | 4.43E-15 | 1.37E-14 | 2.79E-11 |
| Heart_Failure | dendritic_cell | *NDUFS8* | 6.829902964 | 2.43E-07 | 3.48E-07 | 0.001527532 |
| Heart_Failure | dendritic_cell | *TCIRG1* | 5.137479601 | 1.25E-06 | 1.71E-06 | 0.007841512 |
| Heart_Failure | dendritic_cell | *GTF2H1* | -4.358966987 | 7.79E-13 | 1.71E-12 | 4.90E-09 |
| Heart_Failure | dendritic_cell | *PTPN5* | -1.150460999 | 0.005184863 | 0.006020915 | 1 |
| Heart_Failure | dendritic_cell | *PPFIBP1* | -2.689358799 | 8.57E-18 | 4.91E-17 | 5.39E-14 |
| Heart_Failure | dendritic_cell | *PRDM4* | -4.840345069 | 4.94E-12 | 9.99E-12 | 3.11E-08 |
| Heart_Failure | dendritic_cell | *COQ5* | -0.660630743 | 0.001511802 | 0.001802522 | 1 |
| Heart_Failure | dendritic_cell | *ASIC1* | -3.328574606 | 4.65E-13 | 1.05E-12 | 2.92E-09 |
| Heart_Failure | dendritic_cell | *CAPRIN2* | -4.603975293 | 5.77E-15 | 1.74E-14 | 3.63E-11 |
| Heart_Failure | dendritic_cell | *KCTD10* | -3.37140017 | 1.90E-17 | 9.86E-17 | 1.19E-13 |
| Heart_Failure | dendritic_cell | *CYP27B1* | -22.24149842 | 1.10E-19 | 1.42E-18 | 6.91E-16 |
| Heart_Failure | dendritic_cell | *ACSS3* | -1.353940385 | 4.37E-14 | 1.13E-13 | 2.75E-10 |
| Heart_Failure | dendritic_cell | *GLI1* | 8.775663465 | 4.98E-20 | 8.51E-19 | 3.13E-16 |
| Heart_Failure | dendritic_cell | *METAP2* | -4.258626031 | 1.76E-08 | 2.68E-08 | 0.000110858 |
| Heart_Failure | dendritic_cell | *WNT5B* | 2.751593871 | 7.71E-20 | 1.12E-18 | 4.85E-16 |
| Heart_Failure | dendritic_cell | *TRPV4* | -6.953222884 | 1.37E-10 | 2.42E-10 | 8.60E-07 |
| Heart_Failure | dendritic_cell | *ITFG2* | -4.711217112 | 2.30E-13 | 5.38E-13 | 1.44E-09 |
| Heart_Failure | dendritic_cell | *PRR4* | -2.224682613 | 2.65E-05 | 3.42E-05 | 0.166535058 |
| Heart_Failure | dendritic_cell | *PARP11* | -3.012108425 | 0.004869936 | 0.005665672 | 1 |
| Heart_Failure | dendritic_cell | *ARPC3* | 5.858956711 | 3.95E-15 | 1.23E-14 | 2.48E-11 |
| Heart_Failure | dendritic_cell | *GPN3* | -5.958455641 | 2.45E-17 | 1.23E-16 | 1.54E-13 |
| Heart_Failure | dendritic_cell | *VPS29* | -2.50450306 | 0.014735972 | 0.016707855 | 1 |
| Heart_Failure | dendritic_cell | *AKAP3* | -3.187836063 | 1.74E-14 | 4.80E-14 | 1.09E-10 |
| Heart_Failure | dendritic_cell | *KCNA1* | -6.844946323 | 2.00E-19 | 2.16E-18 | 1.26E-15 |
| Heart_Failure | dendritic_cell | *CREBL2* | 5.025408571 | 4.69E-09 | 7.41E-09 | 2.95E-05 |
| Heart_Failure | dendritic_cell | *NAA25* | -4.448452504 | 7.31E-12 | 1.45E-11 | 4.60E-08 |
| Heart_Failure | dendritic_cell | *SCNN1A* | -4.039329077 | 2.56E-18 | 1.72E-17 | 1.61E-14 |
| Heart_Failure | dendritic_cell | *CDK2AP1* | 2.435520777 | 3.78E-15 | 1.18E-14 | 2.37E-11 |
| Heart_Failure | dendritic_cell | *MGP* | -2.284065003 | 1.07E-14 | 3.08E-14 | 6.73E-11 |
| Heart_Failure | dendritic_cell | *RASAL1* | 2.606462988 | 6.43E-21 | 4.66E-19 | 4.04E-17 |

| | | | | | | |
|---|---|---|---|---|---|---|
| Heart_Failure | dendritic_cell | *ARHGDIB* | 3.058588926 | 1.17E-17 | 6.48E-17 | 7.33E-14 |
| Heart_Failure | dendritic_cell | *EIF2B1* | -4.736094927 | 8.41E-17 | 3.73E-16 | 5.29E-13 |
| Heart_Failure | dendritic_cell | *DDX55* | -3.253671878 | 4.82E-11 | 8.86E-11 | 3.03E-07 |
| Heart_Failure | dendritic_cell | *ENDOU* | -4.380324716 | 2.18E-15 | 7.11E-15 | 1.37E-11 |
| Heart_Failure | dendritic_cell | *STX2* | -2.593611789 | 2.76E-05 | 3.56E-05 | 0.173417982 |
| Heart_Failure | dendritic_cell | *CAND1* | -3.884879656 | 1.10E-10 | 1.96E-10 | 6.91E-07 |
| Heart_Failure | dendritic_cell | *MDM1* | -3.405830849 | 2.71E-06 | 3.65E-06 | 0.017014229 |
| Heart_Failure | dendritic_cell | *NUP107* | -4.24674643 | 9.73E-12 | 1.90E-11 | 6.11E-08 |
| Heart_Failure | dendritic_cell | *CNOT2* | -4.095342196 | 1.71E-09 | 2.79E-09 | 1.08E-05 |
| Heart_Failure | dendritic_cell | *MRPL51* | 0.910169523 | 0.02112607 | 0.023764466 | 1 |
| Heart_Failure | dendritic_cell | *COPS7A* | -4.638232603 | 1.70E-07 | 2.44E-07 | 0.001067016 |
| Heart_Failure | dendritic_cell | *CDCA3* | -1.854564172 | 0.000495584 | 0.000604058 | 1 |
| Heart_Failure | dendritic_cell | *CHPT1* | -2.092456086 | 2.09E-13 | 4.93E-13 | 1.31E-09 |
| Heart_Failure | dendritic_cell | *GNPTAB* | -4.374175797 | 3.40E-08 | 5.08E-08 | 0.000213652 |
| Heart_Failure | dendritic_cell | *SPSB2* | 5.437832185 | 1.39E-17 | 7.55E-17 | 8.77E-14 |
| Heart_Failure | dendritic_cell | *ATN1* | -3.989976423 | 5.21E-16 | 1.92E-15 | 3.27E-12 |
| Heart_Failure | dendritic_cell | *C12orf57* | 3.466052433 | 0.003373641 | 0.003957843 | 1 |
| Heart_Failure | dendritic_cell | *NT5DC3* | -2.214694295 | 0.000882703 | 0.001062319 | 1 |
| Heart_Failure | dendritic_cell | *PRKAB1* | -3.609631463 | 4.51E-12 | 9.15E-12 | 2.83E-08 |
| Heart_Failure | dendritic_cell | *ST8SIA1* | -4.553184418 | 4.96E-13 | 1.11E-12 | 3.12E-09 |
| Heart_Failure | dendritic_cell | *C2CD5* | -4.373576991 | 2.47E-10 | 4.28E-10 | 1.55E-06 |
| Heart_Failure | dendritic_cell | *PHC1* | -7.162737001 | 1.27E-15 | 4.34E-15 | 7.97E-12 |
| Heart_Failure | dendritic_cell | *COL12A1* | -1.979812213 | 1.88E-19 | 2.11E-18 | 1.18E-15 |
| Heart_Failure | dendritic_cell | *TDP2* | -4.926168951 | 7.00E-15 | 2.08E-14 | 4.40E-11 |
| Heart_Failure | dendritic_cell | *RWDD1* | -4.381176519 | 1.89E-14 | 5.19E-14 | 1.19E-10 |
| Heart_Failure | dendritic_cell | *SMIM8* | -4.074422471 | 4.74E-11 | 8.72E-11 | 2.98E-07 |
| Heart_Failure | dendritic_cell | *MCM9* | -4.602405452 | 1.96E-09 | 3.18E-09 | 1.23E-05 |
| Heart_Failure | dendritic_cell | *FAM184A* | -3.080177776 | 2.56E-14 | 6.90E-14 | 1.61E-10 |
| Heart_Failure | dendritic_cell | *RNGTT* | -3.379909195 | 0.00013931 | 0.000174167 | 0.875838874 |
| Heart_Failure | dendritic_cell | *GABRR2* | -3.261712913 | 5.92E-18 | 3.55E-17 | 3.72E-14 |
| Heart_Failure | dendritic_cell | *HDDC2* | -3.755209403 | 7.32E-15 | 2.17E-14 | 4.60E-11 |
| Heart_Failure | dendritic_cell | *TPD52L1* | -2.409518354 | 4.54E-17 | 2.14E-16 | 2.86E-13 |
| Heart_Failure | dendritic_cell | *UST* | -3.088925509 | 5.04E-15 | 1.54E-14 | 3.17E-11 |
| Heart_Failure | dendritic_cell | *MTRF1L* | -4.042512834 | 3.76E-12 | 7.69E-12 | 2.36E-08 |
| Heart_Failure | dendritic_cell | *TULP1* | 11.72354929 | 7.23E-21 | 4.66E-19 | 4.55E-17 |
| Heart_Failure | dendritic_cell | *SLC26A8* | 1.307463675 | 0.006328359 | 0.007316365 | 1 |
| Heart_Failure | dendritic_cell | *MAPK14* | -4.089846525 | 1.16E-10 | 2.06E-10 | 7.27E-07 |
| Heart_Failure | dendritic_cell | *MRPL18* | -4.112576265 | 3.97E-09 | 6.30E-09 | 2.50E-05 |
| Heart_Failure | dendritic_cell | *RNF8* | -4.179693481 | 4.60E-10 | 7.79E-10 | 2.89E-06 |
| Heart_Failure | dendritic_cell | *PHACTR1* | -4.475961993 | 2.67E-09 | 4.28E-09 | 1.68E-05 |
| Heart_Failure | dendritic_cell | *MDGA1* | 7.750614201 | 2.72E-11 | 5.10E-11 | 1.71E-07 |
| Heart_Failure | dendritic_cell | *CD83* | 0.779096053 | 8.97E-21 | 4.70E-19 | 5.64E-17 |
| Heart_Failure | dendritic_cell | *MDN1* | -3.792225637 | 2.90E-12 | 6.00E-12 | 1.82E-08 |
| Heart_Failure | dendritic_cell | *SAYSD1* | -3.719953687 | 0.033434056 | 0.037236477 | 1 |
| Heart_Failure | dendritic_cell | *CAP2* | -3.173866352 | 5.14E-15 | 1.57E-14 | 3.23E-11 |
| Heart_Failure | dendritic_cell | *GPR63* | -3.54883995 | 6.39E-16 | 2.33E-15 | 4.02E-12 |
| Heart_Failure | dendritic_cell | *ASCC3* | -4.090314881 | 1.36E-12 | 2.91E-12 | 8.54E-09 |
| Heart_Failure | dendritic_cell | *BVES* | -1.16480615 | 0.019917218 | 0.022444802 | 1 |
| Heart_Failure | dendritic_cell | *WASF1* | -1.66897944 | 8.38E-15 | 2.46E-14 | 5.27E-11 |
| Heart_Failure | dendritic_cell | *GPLD1* | -1.387258904 | 5.92E-06 | 7.88E-06 | 0.037247587 |

| | | | | | | |
|---|---|---|---|---|---|---|
| Heart_Failure | dendritic_cell | *ALDH5A1* | -3.26728834 | 6.01E-19 | 5.10E-18 | 3.78E-15 |
| Heart_Failure | dendritic_cell | *VNN1* | 2.273561182 | 4.27E-11 | 7.89E-11 | 2.68E-07 |
| Heart_Failure | dendritic_cell | *VNN2* | 6.433651432 | 6.36E-12 | 1.27E-11 | 4.00E-08 |
| Heart_Failure | dendritic_cell | *ACOT13* | -5.612111856 | 2.17E-12 | 4.55E-12 | 1.36E-08 |
| Heart_Failure | dendritic_cell | *C6orf62* | -3.752830686 | 7.09E-09 | 1.11E-08 | 4.46E-05 |
| Heart_Failure | dendritic_cell | *B3GAT2* | -4.917414847 | 1.29E-16 | 5.43E-16 | 8.12E-13 |
| Heart_Failure | dendritic_cell | *HBS1L* | -3.668659553 | 5.86E-16 | 2.15E-15 | 3.68E-12 |
| Heart_Failure | dendritic_cell | *PEX7* | -1.337695661 | 0.003712007 | 0.004346692 | 1 |
| Heart_Failure | dendritic_cell | *PERP* | -2.575237351 | 1.38E-15 | 4.68E-15 | 8.67E-12 |
| Heart_Failure | dendritic_cell | *SLC16A10* | 2.306476071 | 6.94E-21 | 4.66E-19 | 4.36E-17 |
| Heart_Failure | dendritic_cell | *PACRG* | -4.436025501 | 2.62E-18 | 1.75E-17 | 1.65E-14 |
| Heart_Failure | dendritic_cell | *TFEB* | 6.398732467 | 1.41E-11 | 2.72E-11 | 8.86E-08 |
| Heart_Failure | dendritic_cell | *FAM120B* | -2.834045655 | 2.62E-09 | 4.20E-09 | 1.64E-05 |
| Heart_Failure | dendritic_cell | *GUCA1B* | -4.950344645 | 6.89E-18 | 4.04E-17 | 4.33E-14 |
| Heart_Failure | dendritic_cell | *PRPH2* | -4.378278442 | 5.79E-13 | 1.28E-12 | 3.64E-09 |
| Heart_Failure | dendritic_cell | *DNPH1* | -2.786621803 | 1.09E-09 | 1.80E-09 | 6.83E-06 |
| Heart_Failure | dendritic_cell | *DUSP22* | -3.77900087 | 0.000578502 | 0.000701996 | 1 |
| Heart_Failure | dendritic_cell | *EXOC2* | -4.633708268 | 9.45E-13 | 2.06E-12 | 5.94E-09 |
| Heart_Failure | dendritic_cell | *TMEM30A* | -4.069937373 | 2.05E-14 | 5.60E-14 | 1.29E-10 |
| Heart_Failure | dendritic_cell | *SENP6* | -3.801615675 | 2.95E-12 | 6.11E-12 | 1.86E-08 |
| Heart_Failure | dendritic_cell | *TTK* | -5.797752365 | 2.35E-15 | 7.61E-15 | 1.48E-11 |
| Heart_Failure | dendritic_cell | *BTN2A1* | 2.785536416 | 0.000578864 | 0.0007023 | 1 |
| Heart_Failure | dendritic_cell | *LAMA4* | -1.827481349 | 1.82E-17 | 9.54E-17 | 1.14E-13 |
| Heart_Failure | dendritic_cell | *CLIC5* | -1.778532433 | 9.13E-17 | 4.00E-16 | 5.74E-13 |
| Heart_Failure | dendritic_cell | *LY86* | 1.412376834 | 6.14E-21 | 4.66E-19 | 3.86E-17 |
| Heart_Failure | dendritic_cell | *PRSS16* | 6.663961794 | 2.22E-08 | 3.35E-08 | 0.000139733 |
| Heart_Failure | dendritic_cell | *TBX18* | -2.335265424 | 5.63E-18 | 3.39E-17 | 3.54E-14 |
| Heart_Failure | dendritic_cell | *PCDHB2* | -1.553339789 | 0.000500208 | 0.000609459 | 1 |
| Heart_Failure | dendritic_cell | *NUDT12* | -4.625067004 | 6.00E-18 | 3.58E-17 | 3.77E-14 |
| Heart_Failure | dendritic_cell | *CEP72* | 3.878425439 | 1.90E-09 | 3.09E-09 | 1.20E-05 |
| Heart_Failure | dendritic_cell | *C7* | -2.280227115 | 6.21E-20 | 9.79E-19 | 3.90E-16 |
| Heart_Failure | dendritic_cell | *PAPD7* | -3.965865923 | 5.76E-17 | 2.65E-16 | 3.62E-13 |
| Heart_Failure | dendritic_cell | *GHR* | -1.581408474 | 6.78E-17 | 3.08E-16 | 4.26E-13 |
| Heart_Failure | dendritic_cell | *NME5* | -2.925335923 | 5.56E-15 | 1.69E-14 | 3.50E-11 |
| Heart_Failure | dendritic_cell | *NNT* | -2.683297015 | 1.43E-13 | 3.44E-13 | 8.99E-10 |
| Heart_Failure | dendritic_cell | *PFDN1* | -3.57481409 | 3.64E-11 | 6.77E-11 | 2.29E-07 |
| Heart_Failure | dendritic_cell | *LOX* | -2.778592122 | 1.41E-19 | 1.69E-18 | 8.85E-16 |
| Heart_Failure | dendritic_cell | *APBB3* | -2.292099882 | 0.001586256 | 0.001889501 | 1 |
| Heart_Failure | dendritic_cell | *TMCO6* | -3.038531193 | 3.13E-21 | 4.66E-19 | 1.97E-17 |
| Heart_Failure | dendritic_cell | *SPARC* | -2.540246095 | 1.11E-17 | 6.20E-17 | 7.00E-14 |
| Heart_Failure | dendritic_cell | *IK* | -5.126126238 | 2.03E-14 | 5.56E-14 | 1.28E-10 |
| Heart_Failure | dendritic_cell | *PDE8B* | -2.135526161 | 1.11E-19 | 1.43E-18 | 6.99E-16 |
| Heart_Failure | dendritic_cell | *CLK4* | -3.923858348 | 3.49E-16 | 1.34E-15 | 2.20E-12 |
| Heart_Failure | dendritic_cell | *PCDHB15* | -1.464503746 | 2.07E-06 | 2.81E-06 | 0.013018156 |
| Heart_Failure | dendritic_cell | *HAVCR1* | 1.66213701 | 1.04E-17 | 5.83E-17 | 6.52E-14 |
| Heart_Failure | dendritic_cell | *RNF130* | -3.350464671 | 1.38E-05 | 1.81E-05 | 0.086966428 |
| Heart_Failure | dendritic_cell | *ARSB* | 2.549800509 | 1.50E-14 | 4.20E-14 | 9.44E-11 |
| Heart_Failure | dendritic_cell | *CLINT1* | -3.973810333 | 1.39E-11 | 2.68E-11 | 8.72E-08 |
| Heart_Failure | dendritic_cell | *TTC1* | -4.691062543 | 1.14E-07 | 1.66E-07 | 0.000716394 |
| Heart_Failure | dendritic_cell | *MSH3* | -2.51711999 | 4.26E-11 | 7.87E-11 | 2.68E-07 |

| | | | | | | |
|---|---|---|---|---|---|---|
| Heart_Failure | dendritic_cell | *RASGRF2* | -2.167007791 | 4.13E-19 | 3.75E-18 | 2.59E-15 |
| Heart_Failure | dendritic_cell | *POLR3G* | -3.929620674 | 3.12E-12 | 6.44E-12 | 1.96E-08 |
| Heart_Failure | dendritic_cell | *DROSHA* | -3.58101891 | 3.51E-09 | 5.59E-09 | 2.21E-05 |
| Heart_Failure | dendritic_cell | *CDH6* | -0.589312591 | 0.01454935 | 0.016505191 | 1 |
| Heart_Failure | dendritic_cell | *SLC27A6* | 15.71961142 | 2.35E-09 | 3.78E-09 | 1.47E-05 |
| Heart_Failure | dendritic_cell | *TARS* | -4.520539761 | 0.000259878 | 0.000320552 | 1 |
| Heart_Failure | dendritic_cell | *LNPEP* | -3.672157228 | 1.82E-10 | 3.19E-10 | 1.15E-06 |
| Heart_Failure | dendritic_cell | *SLC12A7* | 6.1209685 | 3.00E-15 | 9.56E-15 | 1.89E-11 |
| Heart_Failure | dendritic_cell | *IL4* | -6.168705496 | 7.92E-07 | 1.10E-06 | 0.004978701 |
| Heart_Failure | dendritic_cell | *GNPDA1* | -6.35164251 | 1.99E-10 | 3.47E-10 | 1.25E-06 |
| Heart_Failure | dendritic_cell | *SKP1* | -4.906862194 | 8.84E-14 | 2.18E-13 | 5.56E-10 |
| Heart_Failure | dendritic_cell | *PPWD1* | -1.754449322 | 0.00053659 | 0.000652649 | 1 |
| Heart_Failure | dendritic_cell | *TRIM23* | -4.549859279 | 9.67E-15 | 2.81E-14 | 6.08E-11 |
| Heart_Failure | dendritic_cell | *TRAPPC13* | -3.818809258 | 6.40E-15 | 1.92E-14 | 4.02E-11 |
| Heart_Failure | dendritic_cell | *C9* | -1.796730702 | 0.017673316 | 0.019966242 | 1 |
| Heart_Failure | dendritic_cell | *TXNDC15* | -5.279199174 | 3.31E-14 | 8.78E-14 | 2.08E-10 |
| Heart_Failure | dendritic_cell | *SMAD5* | -4.297709747 | 8.91E-15 | 2.61E-14 | 5.60E-11 |
| Heart_Failure | dendritic_cell | *HMGXB3* | -4.400882959 | 1.53E-15 | 5.12E-15 | 9.59E-12 |
| Heart_Failure | dendritic_cell | *ATP6V0E1* | -4.516646951 | 5.38E-08 | 7.98E-08 | 0.000338173 |
| Heart_Failure | dendritic_cell | *BNIP1* | -0.766955602 | 0.026675298 | 0.0298359 | 1 |
| Heart_Failure | dendritic_cell | *CNTN3* | -2.190212584 | 7.70E-20 | 1.12E-18 | 4.84E-16 |
| Heart_Failure | dendritic_cell | *ACTR8* | -7.580247661 | 3.39E-08 | 5.07E-08 | 0.000213332 |
| Heart_Failure | dendritic_cell | *HGD* | -14.15280668 | 7.95E-16 | 2.84E-15 | 5.00E-12 |
| Heart_Failure | dendritic_cell | *NPHP3* | -0.971296598 | 0.000910596 | 0.00109463 | 1 |
| Heart_Failure | dendritic_cell | *NIT2* | -5.511488574 | 3.90E-14 | 1.02E-13 | 2.45E-10 |
| Heart_Failure | dendritic_cell | *FAM162A* | -5.082914585 | 6.74E-09 | 1.05E-08 | 4.24E-05 |
| Heart_Failure | dendritic_cell | *PCCB* | -4.462160021 | 1.79E-14 | 4.95E-14 | 1.13E-10 |
| Heart_Failure | dendritic_cell | *RBP1* | -5.34847091 | 1.05E-18 | 8.06E-18 | 6.58E-15 |
| Heart_Failure | dendritic_cell | *RNF7* | 3.87528753 | 3.24E-07 | 4.61E-07 | 0.002037643 |
| Heart_Failure | dendritic_cell | *BCHE* | -2.985120908 | 3.28E-20 | 6.76E-19 | 2.06E-16 |
| Heart_Failure | dendritic_cell | *PDCD10* | -4.632656376 | 1.21E-14 | 3.46E-14 | 7.62E-11 |
| Heart_Failure | dendritic_cell | *PRKAR2A* | -2.830328767 | 5.70E-12 | 1.14E-11 | 3.58E-08 |
| Heart_Failure | dendritic_cell | *CYB561D2* | 1.70292202 | 0.023198681 | 0.026016787 | 1 |
| Heart_Failure | dendritic_cell | *C3orf14* | -3.025939683 | 8.64E-18 | 4.95E-17 | 5.43E-14 |
| Heart_Failure | dendritic_cell | *IQCG* | -5.460111362 | 3.06E-11 | 5.72E-11 | 1.92E-07 |
| Heart_Failure | dendritic_cell | *GBE1* | -2.601427693 | 1.79E-17 | 9.42E-17 | 1.12E-13 |
| Heart_Failure | dendritic_cell | *UMPS* | -4.176296683 | 1.38E-14 | 3.88E-14 | 8.66E-11 |
| Heart_Failure | dendritic_cell | *NCBP2* | -2.786628765 | 0.032243449 | 0.035942299 | 1 |
| Heart_Failure | dendritic_cell | *SLC41A3* | -5.499828624 | 5.61E-12 | 1.13E-11 | 3.52E-08 |
| Heart_Failure | dendritic_cell | *PLXNA1* | -2.469458606 | 2.69E-05 | 3.47E-05 | 0.169361823 |
| Heart_Failure | dendritic_cell | *ABTB1* | -5.761844798 | 4.12E-15 | 1.28E-14 | 2.59E-11 |
| Heart_Failure | dendritic_cell | *EFCC1* | -2.788988596 | 1.66E-16 | 6.81E-16 | 1.04E-12 |
| Heart_Failure | dendritic_cell | *NEK11* | -4.372158658 | 8.04E-07 | 1.11E-06 | 0.005055248 |
| Heart_Failure | dendritic_cell | *PLSCR4* | -2.741726921 | 4.15E-16 | 1.56E-15 | 2.61E-12 |
| Heart_Failure | dendritic_cell | *HEMK1* | -1.849446389 | 0.009428248 | 0.010796975 | 1 |
| Heart_Failure | dendritic_cell | *WDR48* | -4.061095243 | 2.45E-09 | 3.95E-09 | 1.54E-05 |
| Heart_Failure | dendritic_cell | *COMMD2* | -4.18623952 | 4.60E-17 | 2.16E-16 | 2.89E-13 |
| Heart_Failure | dendritic_cell | *GORASP1* | 6.636865321 | 2.47E-13 | 5.75E-13 | 1.55E-09 |
| Heart_Failure | dendritic_cell | *ABCC5* | -4.328827847 | 9.16E-11 | 1.64E-10 | 5.76E-07 |
| Heart_Failure | dendritic_cell | *AADAC* | -0.403177364 | 0.000894098 | 0.001075621 | 1 |

| | | | | | | |
|---|---|---|---|---|---|---|
| Heart_Failure | dendritic_cell | *ARHGEF26* | -2.664383735 | 2.35E-18 | 1.61E-17 | 1.48E-14 |
| Heart_Failure | dendritic_cell | *KLHL24* | -3.094471861 | 5.13E-11 | 9.39E-11 | 3.22E-07 |
| Heart_Failure | dendritic_cell | *PLCH1* | -6.597062894 | 1.08E-14 | 3.10E-14 | 6.79E-11 |
| Heart_Failure | dendritic_cell | *ZBTB47* | -4.694908754 | 5.60E-16 | 2.06E-15 | 3.52E-12 |
| Heart_Failure | dendritic_cell | *NKTR* | -3.655127661 | 6.59E-11 | 1.20E-10 | 4.14E-07 |
| Heart_Failure | dendritic_cell | *CLCN2* | -4.811584276 | 1.83E-19 | 2.06E-18 | 1.15E-15 |
| Heart_Failure | dendritic_cell | *NEK4* | -5.749540442 | 1.28E-12 | 2.75E-12 | 8.05E-09 |
| Heart_Failure | dendritic_cell | *MOB1A* | 6.184438724 | 5.23E-11 | 9.57E-11 | 3.29E-07 |
| Heart_Failure | dendritic_cell | *LMAN2L* | -3.531426086 | 3.79E-14 | 9.92E-14 | 2.38E-10 |
| Heart_Failure | dendritic_cell | *PIKFYVE* | -4.020667952 | 2.17E-09 | 3.51E-09 | 1.37E-05 |
| Heart_Failure | dendritic_cell | *KCNIP3* | -3.868430431 | 4.78E-19 | 4.22E-18 | 3.01E-15 |
| Heart_Failure | dendritic_cell | *FAHD2A* | 6.007159067 | 1.13E-08 | 1.73E-08 | 7.08E-05 |
| Heart_Failure | dendritic_cell | *NCL* | -4.680951963 | 1.49E-13 | 3.57E-13 | 9.35E-10 |
| Heart_Failure | dendritic_cell | *ACTR1B* | -4.489667043 | 0.000110963 | 0.000139413 | 0.697624594 |
| Heart_Failure | dendritic_cell | *SLC35F5* | -3.922535186 | 2.02E-12 | 4.25E-12 | 1.27E-08 |
| Heart_Failure | dendritic_cell | *ACTR3* | 4.82137747 | 3.47E-13 | 7.97E-13 | 2.18E-09 |
| Heart_Failure | dendritic_cell | *STEAP3* | 1.259438036 | 0.000137955 | 0.000172602 | 0.867325841 |
| Heart_Failure | dendritic_cell | *TFCP2L1* | -5.161531199 | 1.69E-17 | 8.96E-17 | 1.06E-13 |
| Heart_Failure | dendritic_cell | *TP53I3* | -5.118893857 | 8.42E-11 | 1.51E-10 | 5.29E-07 |
| Heart_Failure | dendritic_cell | *DNAJC27* | -3.51214485 | 9.88E-05 | 0.000124432 | 0.621290629 |
| Heart_Failure | dendritic_cell | *STAM2* | -4.616367298 | 5.09E-13 | 1.14E-12 | 3.20E-09 |
| Heart_Failure | dendritic_cell | *GPD2* | -4.282094993 | 9.45E-13 | 2.06E-12 | 5.94E-09 |
| Heart_Failure | dendritic_cell | *CENPA* | 6.957010523 | 9.91E-09 | 1.53E-08 | 6.23E-05 |
| Heart_Failure | dendritic_cell | *TANC1* | -2.680987334 | 1.82E-14 | 5.01E-14 | 1.14E-10 |
| Heart_Failure | dendritic_cell | *NRBP1* | -4.078494901 | 8.30E-13 | 1.82E-12 | 5.22E-09 |
| Heart_Failure | dendritic_cell | *ITGB6* | -2.813644271 | 1.15E-09 | 1.89E-09 | 7.21E-06 |
| Heart_Failure | dendritic_cell | *ITGA4* | 2.237843393 | 5.48E-21 | 4.66E-19 | 3.45E-17 |
| Heart_Failure | dendritic_cell | *PPM1G* | -4.290133987 | 6.18E-16 | 2.26E-15 | 3.88E-12 |
| Heart_Failure | dendritic_cell | *PCSK4* | -3.849551708 | 1.57E-15 | 5.28E-15 | 9.90E-12 |
| Heart_Failure | dendritic_cell | *INO80B* | -5.335132483 | 1.44E-18 | 1.06E-17 | 9.05E-15 |
| Heart_Failure | dendritic_cell | *MOGS* | -2.538523416 | 4.10E-10 | 6.97E-10 | 2.58E-06 |
| Heart_Failure | dendritic_cell | *GRB14* | -4.147203753 | 9.49E-18 | 5.39E-17 | 5.97E-14 |
| Heart_Failure | dendritic_cell | *CLIP4* | -3.410454159 | 2.05E-17 | 1.06E-16 | 1.29E-13 |
| Heart_Failure | dendritic_cell | *SPTBN1* | -2.530804305 | 8.01E-18 | 4.62E-17 | 5.04E-14 |
| Heart_Failure | dendritic_cell | *RTN4* | -2.820553198 | 2.12E-09 | 3.43E-09 | 1.33E-05 |
| Heart_Failure | dendritic_cell | *POLE4* | -5.11243961 | 7.50E-18 | 4.35E-17 | 4.72E-14 |
| Heart_Failure | dendritic_cell | *MRPL19* | -3.804701073 | 1.43E-15 | 4.83E-15 | 8.98E-12 |
| Heart_Failure | dendritic_cell | *LANCL1* | -4.937155986 | 3.77E-14 | 9.87E-14 | 2.37E-10 |
| Heart_Failure | dendritic_cell | *WDR75* | -5.297237593 | 1.18E-12 | 2.55E-12 | 7.43E-09 |
| Heart_Failure | dendritic_cell | *EFEMP1* | -2.147445989 | 6.10E-19 | 5.14E-18 | 3.83E-15 |
| Heart_Failure | dendritic_cell | *GLS* | -3.748056015 | 1.50E-09 | 2.45E-09 | 9.41E-06 |
| Heart_Failure | dendritic_cell | *GNLY* | -3.453005318 | 4.32E-19 | 3.91E-18 | 2.72E-15 |
| Heart_Failure | dendritic_cell | *SF3B1* | -3.860731049 | 8.27E-13 | 1.81E-12 | 5.20E-09 |
| Heart_Failure | dendritic_cell | *ST3GAL5* | -4.278710892 | 2.80E-13 | 6.51E-13 | 1.76E-09 |
| Heart_Failure | dendritic_cell | *PLCD4* | -9.393387861 | 1.23E-11 | 2.39E-11 | 7.74E-08 |
| Heart_Failure | dendritic_cell | *CHMP3* | -4.012609413 | 3.46E-07 | 4.91E-07 | 0.002173897 |
| Heart_Failure | dendritic_cell | *ZNF142* | -2.16759317 | 0.027680211 | 0.030932365 | 1 |
| Heart_Failure | dendritic_cell | *WNT6* | -15.0188957 | 3.09E-11 | 5.77E-11 | 1.94E-07 |
| Heart_Failure | dendritic_cell | *MLPH* | 4.808636764 | 0.008291111 | 0.009524249 | 1 |
| Heart_Failure | dendritic_cell | *HDLBP* | -3.285203194 | 1.44E-10 | 2.55E-10 | 9.08E-07 |

| | | | | | | |
|---|---|---|---|---|---|---|
| Heart_Failure | dendritic_cell | *PASK* | -2.393510918 | 0.034761958 | 0.038687985 | 1 |
| Heart_Failure | dendritic_cell | *STK25* | -4.559703726 | 1.03E-05 | 1.36E-05 | 0.065023802 |
| Heart_Failure | dendritic_cell | *TPO* | -1.475318106 | 2.41E-16 | 9.54E-16 | 1.51E-12 |
| Heart_Failure | dendritic_cell | *TAF1B* | -3.629839901 | 1.21E-15 | 4.16E-15 | 7.63E-12 |
| Heart_Failure | dendritic_cell | *STRN* | -4.125397078 | 3.42E-15 | 1.08E-14 | 2.15E-11 |
| Heart_Failure | dendritic_cell | *PRKD3* | -4.002833392 | 6.46E-09 | 1.01E-08 | 4.06E-05 |
| Heart_Failure | dendritic_cell | *SLC25A12* | -3.926551963 | 5.65E-13 | 1.26E-12 | 3.55E-09 |
| Heart_Failure | dendritic_cell | *RMDN2* | -5.420407511 | 6.16E-13 | 1.36E-12 | 3.87E-09 |
| Heart_Failure | dendritic_cell | *SDC1* | 5.83006518 | 4.79E-17 | 2.24E-16 | 3.01E-13 |
| Heart_Failure | dendritic_cell | *SOS1* | -2.986544107 | 6.83E-16 | 2.47E-15 | 4.29E-12 |
| Heart_Failure | dendritic_cell | *WIPF1* | 3.706372278 | 3.94E-06 | 5.28E-06 | 0.024778894 |
| Heart_Failure | dendritic_cell | *COX7A2L* | -5.358477851 | 5.84E-16 | 2.14E-15 | 3.67E-12 |
| Heart_Failure | dendritic_cell | *PNO1* | 7.538267593 | 8.90E-16 | 3.14E-15 | 5.60E-12 |
| Heart_Failure | dendritic_cell | *ORC4* | -2.980768398 | 5.20E-17 | 2.42E-16 | 3.27E-13 |
| Heart_Failure | dendritic_cell | *THADA* | -3.869385583 | 9.26E-14 | 2.28E-13 | 5.82E-10 |
| Heart_Failure | dendritic_cell | *ARID3A* | -4.515731348 | 1.10E-16 | 4.71E-16 | 6.91E-13 |
| Heart_Failure | dendritic_cell | *CD207* | -50.84157555 | 2.07E-17 | 1.06E-16 | 1.30E-13 |
| Heart_Failure | dendritic_cell | *GRIN3B* | 2.929020139 | 1.56E-21 | 4.44E-19 | 9.78E-18 |
| Heart_Failure | dendritic_cell | *VAX2* | -3.879469966 | 3.62E-20 | 7.00E-19 | 2.27E-16 |
| Heart_Failure | dendritic_cell | *MSH6* | -4.592826144 | 3.23E-05 | 4.15E-05 | 0.203371878 |
| Heart_Failure | dendritic_cell | *PLEKHA3* | -5.739648327 | 3.00E-14 | 8.01E-14 | 1.89E-10 |
| Heart_Failure | dendritic_cell | *ALMS1* | -4.11143472 | 5.80E-14 | 1.48E-13 | 3.65E-10 |
| Heart_Failure | dendritic_cell | *BCL9* | -3.051829597 | 6.11E-18 | 3.64E-17 | 3.84E-14 |
| Heart_Failure | dendritic_cell | *DHCR24* | -5.586936312 | 1.86E-17 | 9.73E-17 | 1.17E-13 |
| Heart_Failure | dendritic_cell | *DNAJC16* | -3.960807359 | 4.61E-08 | 6.86E-08 | 0.00029014 |
| Heart_Failure | dendritic_cell | *MORN1* | -4.045701867 | 9.28E-11 | 1.66E-10 | 5.83E-07 |
| Heart_Failure | dendritic_cell | *SCP2* | -3.58241106 | 2.92E-14 | 7.82E-14 | 1.84E-10 |
| Heart_Failure | dendritic_cell | *RALGPS2* | -3.047979213 | 3.45E-17 | 1.67E-16 | 2.17E-13 |
| Heart_Failure | dendritic_cell | *CEP104* | -3.875053094 | 5.99E-10 | 1.01E-09 | 3.76E-06 |
| Heart_Failure | dendritic_cell | *FAM20B* | -5.462064315 | 2.76E-08 | 4.15E-08 | 0.000173617 |
| Heart_Failure | dendritic_cell | *TMEM59* | -4.260366328 | 2.11E-15 | 6.92E-15 | 1.33E-11 |
| Heart_Failure | dendritic_cell | *WRAP73* | -4.262890359 | 0.01010585 | 0.011548365 | 1 |
| Heart_Failure | dendritic_cell | *MRPL37* | -4.741135191 | 2.42E-07 | 3.46E-07 | 0.001518772 |
| Heart_Failure | dendritic_cell | *ICMT* | -5.580765684 | 2.02E-17 | 1.04E-16 | 1.27E-13 |
| Heart_Failure | dendritic_cell | *PARK7* | -4.245213858 | 9.76E-05 | 0.000122961 | 0.613696876 |
| Heart_Failure | dendritic_cell | *KCNC4* | -2.75528447 | 2.04E-18 | 1.42E-17 | 1.28E-14 |
| Heart_Failure | dendritic_cell | *EDEM3* | -4.408721277 | 3.04E-14 | 8.11E-14 | 1.91E-10 |
| Heart_Failure | dendritic_cell | *CAPZA1* | -4.166546379 | 1.39E-07 | 2.01E-07 | 0.000875069 |
| Heart_Failure | dendritic_cell | *DOCK7* | -3.983986068 | 3.23E-14 | 8.59E-14 | 2.03E-10 |
| Heart_Failure | dendritic_cell | *FBXO6* | 5.750790173 | 1.70E-12 | 3.61E-12 | 1.07E-08 |
| Heart_Failure | dendritic_cell | *C1orf21* | -2.468303722 | 2.22E-12 | 4.65E-12 | 1.39E-08 |
| Heart_Failure | dendritic_cell | *DNAJC6* | 5.372914888 | 3.27E-18 | 2.10E-17 | 2.06E-14 |
| Heart_Failure | dendritic_cell | *LEPR* | -3.686346619 | 2.12E-15 | 6.96E-15 | 1.34E-11 |
| Heart_Failure | dendritic_cell | *MFN2* | 4.680051408 | 3.53E-19 | 3.32E-18 | 2.22E-15 |
| Heart_Failure | dendritic_cell | *MIIP* | -4.369039667 | 8.13E-19 | 6.55E-18 | 5.11E-15 |
| Heart_Failure | dendritic_cell | *SMG7* | -4.461259164 | 2.30E-08 | 3.47E-08 | 0.000144844 |
| Heart_Failure | dendritic_cell | *NCF2* | 3.009863813 | 2.70E-20 | 6.44E-19 | 1.70E-16 |
| Heart_Failure | dendritic_cell | *RGS2* | 1.019970331 | 8.34E-21 | 4.66E-19 | 5.24E-17 |
| Heart_Failure | dendritic_cell | *OLFML3* | -2.235377759 | 9.44E-20 | 1.28E-18 | 5.93E-16 |
| Heart_Failure | dendritic_cell | *TNNI3K* | -4.488637072 | 1.20E-10 | 2.14E-10 | 7.56E-07 |

| | | | | | | |
|---|---|---|---|---|---|---|
| Heart_Failure | dendritic_cell | *CFHR3* | -4.025027357 | 7.22E-16 | 2.60E-15 | 4.54E-12 |
| Heart_Failure | dendritic_cell | *CRYZ* | -2.474123257 | 4.50E-12 | 9.13E-12 | 2.83E-08 |
| Heart_Failure | dendritic_cell | *TMEM9* | -2.987884837 | 1.12E-13 | 2.73E-13 | 7.06E-10 |
| Heart_Failure | dendritic_cell | *WARS2* | -3.471330846 | 4.77E-13 | 1.08E-12 | 3.00E-09 |
| Heart_Failure | dendritic_cell | *OSCP1* | -4.721977474 | 1.15E-19 | 1.47E-18 | 7.24E-16 |
| Heart_Failure | dendritic_cell | *GNPAT* | -3.404415603 | 3.41E-05 | 4.37E-05 | 0.214371813 |
| Heart_Failure | dendritic_cell | *C1orf109* | -3.830875133 | 1.04E-19 | 1.38E-18 | 6.57E-16 |
| Heart_Failure | dendritic_cell | *TBCE* | -5.007995409 | 6.96E-15 | 2.07E-14 | 4.37E-11 |
| Heart_Failure | dendritic_cell | *LGALS8* | -2.568465299 | 6.85E-12 | 1.37E-11 | 4.31E-08 |
| Heart_Failure | dendritic_cell | *MTR* | -3.976497576 | 1.05E-14 | 3.02E-14 | 6.59E-11 |
| Heart_Failure | dendritic_cell | *ACADM* | -4.29117944 | 1.03E-09 | 1.70E-09 | 6.45E-06 |
| Heart_Failure | dendritic_cell | *PADI2* | 1.87586119 | 6.51E-21 | 4.66E-19 | 4.09E-17 |
| Heart_Failure | dendritic_cell | *MFAP2* | -1.379150078 | 0.008383236 | 0.009628316 | 1 |
| Heart_Failure | dendritic_cell | *KLHL12* | -4.642109845 | 9.29E-06 | 1.23E-05 | 0.058433995 |
| Heart_Failure | dendritic_cell | *ZNHIT6* | -3.834705248 | 1.87E-13 | 4.44E-13 | 1.18E-09 |
| Heart_Failure | dendritic_cell | *PLA2G2D* | 9.813502795 | 1.24E-17 | 6.81E-17 | 7.77E-14 |
| Heart_Failure | dendritic_cell | *RBBP5* | -3.288499137 | 1.93E-10 | 3.38E-10 | 1.22E-06 |
| Heart_Failure | dendritic_cell | *KIF17* | -5.294454199 | 1.31E-09 | 2.16E-09 | 8.25E-06 |
| Heart_Failure | dendritic_cell | *CD46* | -3.337868983 | 4.14E-14 | 1.08E-13 | 2.60E-10 |
| Heart_Failure | dendritic_cell | *PRDX1* | -2.476298636 | 0.016917712 | 0.019129794 | 1 |
| Heart_Failure | dendritic_cell | *FAAH* | -0.663810735 | 0.003107692 | 0.003649927 | 1 |
| Heart_Failure | dendritic_cell | *NSUN4* | -2.939736445 | 0.004188993 | 0.004893385 | 1 |
| Heart_Failure | dendritic_cell | *TMED5* | -4.160829449 | 7.63E-16 | 2.73E-15 | 4.80E-12 |
| Heart_Failure | dendritic_cell | *CNN3* | -4.534297475 | 8.85E-16 | 3.13E-15 | 5.56E-12 |
| Heart_Failure | dendritic_cell | *PRRC2C* | -3.756907461 | 5.86E-08 | 8.68E-08 | 0.000368432 |
| Heart_Failure | dendritic_cell | *ABCD3* | -4.008757695 | 1.83E-16 | 7.45E-16 | 1.15E-12 |
| Heart_Failure | dendritic_cell | *VAMP4* | -4.426127124 | 2.14E-07 | 3.07E-07 | 0.001344448 |
| Heart_Failure | dendritic_cell | *PTBP2* | -2.676724469 | 2.33E-11 | 4.41E-11 | 1.46E-07 |
| Heart_Failure | dendritic_cell | *TNFSF4* | -3.639320741 | 6.45E-21 | 4.66E-19 | 4.06E-17 |
| Heart_Failure | dendritic_cell | *DARS2* | -5.578737366 | 3.31E-11 | 6.18E-11 | 2.08E-07 |
| Heart_Failure | dendritic_cell | *IRF6* | 0.94433856 | 0.020533577 | 0.02311866 | 1 |
| Heart_Failure | dendritic_cell | *RCAN3* | 4.279125162 | 9.30E-19 | 7.34E-18 | 5.85E-15 |
| Heart_Failure | dendritic_cell | *RCOR3* | -4.110975325 | 4.41E-14 | 1.14E-13 | 2.77E-10 |
| Heart_Failure | dendritic_cell | *MTFR1L* | -6.191807967 | 6.59E-10 | 1.10E-09 | 4.14E-06 |
| Heart_Failure | dendritic_cell | *NSL1* | -3.92256704 | 1.96E-08 | 2.96E-08 | 0.000123071 |
| Heart_Failure | dendritic_cell | *RPA2* | -5.753878776 | 2.80E-08 | 4.21E-08 | 0.000176204 |
| Heart_Failure | dendritic_cell | *SLC5A9* | -9.51374806 | 1.26E-12 | 2.72E-12 | 7.94E-09 |
| Heart_Failure | dendritic_cell | *OSBPL9* | -3.759403347 | 8.14E-15 | 2.40E-14 | 5.12E-11 |
| Heart_Failure | dendritic_cell | *TXNDC12* | -5.3849993 | 1.86E-13 | 4.42E-13 | 1.17E-09 |
| Heart_Failure | dendritic_cell | *ESYT2* | -3.730164399 | 8.44E-11 | 1.52E-10 | 5.30E-07 |
| Heart_Failure | dendritic_cell | *CD3EAP* | 4.140954517 | 2.67E-12 | 5.54E-12 | 1.68E-08 |
| Heart_Failure | dendritic_cell | *COLEC11* | -5.150609577 | 7.32E-12 | 1.45E-11 | 4.60E-08 |
| Heart_Failure | dendritic_cell | *A4GNT* | 1.913389817 | 4.17E-06 | 5.58E-06 | 0.0262096 |
| Heart_Failure | dendritic_cell | *KMT2A* | -3.823307399 | 3.65E-15 | 1.14E-14 | 2.30E-11 |
| Heart_Failure | dendritic_cell | *IFT46* | -4.946235053 | 2.33E-15 | 7.56E-15 | 1.47E-11 |
| Heart_Failure | dendritic_cell | *DDX59* | -4.487770014 | 5.37E-11 | 9.82E-11 | 3.38E-07 |
| Heart_Failure | dendritic_cell | *CAMSAP2* | -4.048467801 | 4.23E-10 | 7.19E-10 | 2.66E-06 |
| Heart_Failure | dendritic_cell | *ATF6* | -3.151346232 | 0.000194969 | 0.000242247 | 1 |
| Heart_Failure | dendritic_cell | *CASC1* | 3.498209321 | 3.75E-06 | 5.03E-06 | 0.023562146 |
| Heart_Failure | dendritic_cell | *LRMP* | 3.001658311 | 1.73E-11 | 3.32E-11 | 1.09E-07 |

| | | | | | | |
|---|---|---|---|---|---|---|
| Heart_Failure | dendritic_cell | *ATP10B* | -12.32984324 | 4.05E-13 | 9.19E-13 | 2.54E-09 |
| Heart_Failure | dendritic_cell | *SPCS2* | -4.985658963 | 8.20E-15 | 2.42E-14 | 5.16E-11 |
| Heart_Failure | dendritic_cell | *ELOVL4* | -2.028711433 | 8.96E-07 | 1.24E-06 | 0.005636126 |
| Heart_Failure | dendritic_cell | *FILIP1* | -2.877328998 | 3.06E-12 | 6.32E-12 | 1.92E-08 |
| Heart_Failure | dendritic_cell | *SGIP1* | -3.854881695 | 1.03E-17 | 5.79E-17 | 6.45E-14 |
| Heart_Failure | dendritic_cell | *AKAP7* | -2.556866366 | 2.59E-14 | 6.97E-14 | 1.63E-10 |
| Heart_Failure | dendritic_cell | *ALDH8A1* | -0.784631352 | 0.002566376 | 0.003027168 | 1 |
| Heart_Failure | dendritic_cell | *TCF21* | 17.5574978 | 9.33E-16 | 3.29E-15 | 5.87E-12 |
| Heart_Failure | dendritic_cell | *SLC16A7* | -2.497071357 | 1.63E-18 | 1.18E-17 | 1.03E-14 |
| Heart_Failure | dendritic_cell | *VAMP8* | 4.596528728 | 1.19E-12 | 2.58E-12 | 7.51E-09 |
| Heart_Failure | dendritic_cell | *FOXO3* | -3.434777338 | 1.49E-06 | 2.03E-06 | 0.009368091 |
| Heart_Failure | dendritic_cell | *TGIF2* | -7.674704884 | 6.40E-08 | 9.45E-08 | 0.000402613 |
| Heart_Failure | dendritic_cell | *ABCG2* | -3.507068127 | 1.25E-19 | 1.55E-18 | 7.83E-16 |
| Heart_Failure | dendritic_cell | *SPP1* | -4.43658645 | 4.09E-08 | 6.08E-08 | 0.00025687 |
| Heart_Failure | dendritic_cell | *FAM47E-STBD* | -2.136130459 | 5.93E-15 | 1.79E-14 | 3.73E-11 |
| Heart_Failure | dendritic_cell | *MFSD1* | 4.482728264 | 3.01E-16 | 1.17E-15 | 1.89E-12 |
| Heart_Failure | dendritic_cell | *RAB3GAP2* | -4.248637392 | 8.66E-13 | 1.89E-12 | 5.44E-09 |
| Heart_Failure | dendritic_cell | *UBN1* | -4.295374279 | 2.79E-10 | 4.83E-10 | 1.76E-06 |
| Heart_Failure | dendritic_cell | *UCHL3* | 9.232991179 | 2.77E-17 | 1.38E-16 | 1.74E-13 |
| Heart_Failure | dendritic_cell | *WDR35* | -3.296003434 | 1.94E-18 | 1.36E-17 | 1.22E-14 |
| Heart_Failure | dendritic_cell | *DNAH7* | -2.119733147 | 2.43E-14 | 6.58E-14 | 1.53E-10 |
| Heart_Failure | dendritic_cell | *NDUFB3* | -4.266413682 | 1.31E-18 | 9.72E-18 | 8.26E-15 |
| Heart_Failure | dendritic_cell | *GDA* | -3.551591192 | 1.73E-20 | 6.17E-19 | 1.09E-16 |
| Heart_Failure | dendritic_cell | *C2orf40* | -3.286257137 | 3.67E-14 | 9.65E-14 | 2.31E-10 |
| Heart_Failure | dendritic_cell | *CPSF3* | -5.886700962 | 6.50E-11 | 1.18E-10 | 4.09E-07 |
| Heart_Failure | dendritic_cell | *PIGZ* | -1.592800804 | 2.28E-07 | 3.27E-07 | 0.001433574 |
| Heart_Failure | dendritic_cell | *SENP5* | -3.771156202 | 3.75E-15 | 1.17E-14 | 2.36E-11 |
| Heart_Failure | dendritic_cell | *CCDC92* | -3.624607526 | 1.70E-16 | 6.95E-16 | 1.07E-12 |
| Heart_Failure | dendritic_cell | *PTBP3* | 4.774681439 | 6.14E-14 | 1.56E-13 | 3.86E-10 |
| Heart_Failure | dendritic_cell | *CTNNAL1* | -0.670784143 | 0.038414456 | 0.042639775 | 1 |
| Heart_Failure | dendritic_cell | *WDR34* | -3.353918405 | 2.41E-05 | 3.12E-05 | 0.151587139 |
| Heart_Failure | dendritic_cell | *TRIM32* | -7.234482151 | 7.53E-13 | 1.65E-12 | 4.73E-09 |
| Heart_Failure | dendritic_cell | *PHF19* | 7.984630593 | 4.03E-16 | 1.53E-15 | 2.53E-12 |
| Heart_Failure | dendritic_cell | *BSPRY* | -12.17704198 | 1.43E-20 | 5.54E-19 | 8.98E-17 |
| Heart_Failure | dendritic_cell | *PPP6C* | -4.456366995 | 8.79E-14 | 2.17E-13 | 5.53E-10 |
| Heart_Failure | dendritic_cell | *HDHD3* | -0.748479701 | 3.46E-05 | 4.43E-05 | 0.21738337 |
| Heart_Failure | dendritic_cell | *HSDL2* | -2.247060802 | 1.09E-17 | 6.06E-17 | 6.82E-14 |
| Heart_Failure | dendritic_cell | *INVS* | -3.485400167 | 6.23E-13 | 1.38E-12 | 3.92E-09 |
| Heart_Failure | dendritic_cell | *KDSR* | -5.051283171 | 7.28E-14 | 1.83E-13 | 4.58E-10 |
| Heart_Failure | dendritic_cell | *DCAF4* | -3.670655894 | 3.64E-07 | 5.16E-07 | 0.002288058 |
| Heart_Failure | dendritic_cell | *IFI27L2* | 1.674413187 | 0.036400258 | 0.040439728 | 1 |
| Heart_Failure | dendritic_cell | *DNAL1* | -4.892310303 | 2.48E-14 | 6.71E-14 | 1.56E-10 |
| Heart_Failure | dendritic_cell | *IRF2BPL* | -3.869596418 | 2.69E-18 | 1.79E-17 | 1.69E-14 |
| Heart_Failure | dendritic_cell | *TTLL5* | -4.609267156 | 1.73E-11 | 3.33E-11 | 1.09E-07 |
| Heart_Failure | dendritic_cell | *FLVCR2* | 3.93207019 | 1.94E-19 | 2.14E-18 | 1.22E-15 |
| Heart_Failure | dendritic_cell | *PPP4R4* | 4.649665517 | 3.11E-08 | 4.66E-08 | 0.000195681 |
| Heart_Failure | dendritic_cell | *ZC2HC1C* | 8.499189322 | 6.02E-11 | 1.10E-10 | 3.78E-07 |
| Heart_Failure | dendritic_cell | *ALDH6A1* | -2.676204739 | 9.82E-19 | 7.64E-18 | 6.17E-15 |
| Heart_Failure | dendritic_cell | *ESRRB* | -2.49514031 | 7.88E-16 | 2.81E-15 | 4.96E-12 |
| Heart_Failure | dendritic_cell | *COQ6* | -4.132618006 | 0.003010846 | 0.003538828 | 1 |

| | | | | | | |
|---|---|---|---|---|---|---|
| Heart_Failure | dendritic_cell | *GPR75* | -14.44064434 | 9.88E-21 | 4.95E-19 | 6.21E-17 |
| Heart_Failure | dendritic_cell | *KLHL29* | -3.28385126 | 5.66E-16 | 2.08E-15 | 3.56E-12 |
| Heart_Failure | dendritic_cell | *ATAD2B* | -2.383386712 | 0.005913919 | 0.006847294 | 1 |
| Heart_Failure | dendritic_cell | *FKBP1B* | 2.901887622 | 6.74E-09 | 1.05E-08 | 4.24E-05 |
| Heart_Failure | dendritic_cell | *ATL2* | -3.622580312 | 2.29E-16 | 9.13E-16 | 1.44E-12 |
| Heart_Failure | dendritic_cell | *CNRIP1* | -4.288141042 | 2.89E-12 | 5.98E-12 | 1.82E-08 |
| Heart_Failure | dendritic_cell | *CRIPT* | -4.404228492 | 1.80E-14 | 4.98E-14 | 1.13E-10 |
| Heart_Failure | dendritic_cell | *EPCAM* | 4.244347211 | 4.31E-18 | 2.67E-17 | 2.71E-14 |
| Heart_Failure | dendritic_cell | *SLC17A5* | -3.081582321 | 2.38E-06 | 3.22E-06 | 0.014939076 |
| Heart_Failure | dendritic_cell | *OGFRL1* | 3.085314363 | 9.20E-18 | 5.24E-17 | 5.78E-14 |
| Heart_Failure | dendritic_cell | *CUTC* | -4.126445052 | 5.36E-13 | 1.20E-12 | 3.37E-09 |
| Heart_Failure | dendritic_cell | *PYROXD2* | -1.360473023 | 4.45E-08 | 6.62E-08 | 0.000279942 |
| Heart_Failure | dendritic_cell | *C10orf88* | -6.581983319 | 4.13E-10 | 7.02E-10 | 2.60E-06 |
| Heart_Failure | dendritic_cell | *TCTN3* | -5.359436003 | 1.03E-10 | 1.83E-10 | 6.45E-07 |
| Heart_Failure | dendritic_cell | *AVPI1* | -4.430380348 | 2.58E-12 | 5.38E-12 | 1.62E-08 |
| Heart_Failure | dendritic_cell | *WDR11* | -3.53466151 | 1.81E-05 | 2.35E-05 | 0.113492925 |
| Heart_Failure | dendritic_cell | *C10orf76* | -3.465301001 | 2.17E-05 | 2.81E-05 | 0.13665895 |
| Heart_Failure | dendritic_cell | *KCNIP2* | -1.379148111 | 2.73E-16 | 1.07E-15 | 1.71E-12 |
| Heart_Failure | dendritic_cell | *C10orf95* | -2.555293992 | 1.23E-06 | 1.68E-06 | 0.00771485 |
| Heart_Failure | dendritic_cell | *KANSL1* | -3.164307812 | 6.52E-10 | 1.09E-09 | 4.10E-06 |
| Heart_Failure | dendritic_cell | *HOXB5* | -3.527163912 | 1.63E-16 | 6.72E-16 | 1.03E-12 |
| Heart_Failure | dendritic_cell | *HOXB3* | -2.659968485 | 3.87E-15 | 1.20E-14 | 2.44E-11 |
| Heart_Failure | dendritic_cell | *CAAP1* | -4.279553082 | 1.43E-16 | 5.97E-16 | 9.00E-13 |
| Heart_Failure | dendritic_cell | *CD274* | 3.992866616 | 7.95E-22 | 3.01E-19 | 5.00E-18 |
| Heart_Failure | dendritic_cell | *NUP43* | -3.542450945 | 6.50E-14 | 1.64E-13 | 4.08E-10 |
| Heart_Failure | dendritic_cell | *LRP11* | -3.758657544 | 3.54E-16 | 1.36E-15 | 2.22E-12 |
| Heart_Failure | dendritic_cell | *CCDC170* | -2.050233817 | 1.08E-06 | 1.48E-06 | 0.006780076 |
| Heart_Failure | dendritic_cell | *CYSTM1* | -3.654726567 | 8.54E-16 | 3.03E-15 | 5.37E-12 |
| Heart_Failure | dendritic_cell | *WDR55* | -5.396372691 | 8.99E-19 | 7.12E-18 | 5.65E-15 |
| Heart_Failure | dendritic_cell | *PCDHB10* | 1.70346295 | 0.004181969 | 0.004886088 | 1 |
| Heart_Failure | dendritic_cell | *PCDHB14* | -3.406211811 | 2.77E-20 | 6.44E-19 | 1.74E-16 |
| Heart_Failure | dendritic_cell | *SEC16B* | -7.130911458 | 3.71E-18 | 2.36E-17 | 2.34E-14 |
| Heart_Failure | dendritic_cell | *GORAB* | -5.495234876 | 5.55E-07 | 7.77E-07 | 0.003487327 |
| Heart_Failure | dendritic_cell | *ACAT2* | -5.374090699 | 3.35E-14 | 8.86E-14 | 2.10E-10 |
| Heart_Failure | dendritic_cell | *SNX19* | -3.94050918 | 1.53E-09 | 2.50E-09 | 9.60E-06 |
| Heart_Failure | dendritic_cell | *MASTL* | -1.075730546 | 0.008589042 | 0.009857486 | 1 |
| Heart_Failure | dendritic_cell | *MRC1* | 1.286020998 | 1.09E-20 | 4.99E-19 | 6.88E-17 |
| Heart_Failure | dendritic_cell | *IQSEC3* | -5.178149429 | 1.98E-19 | 2.15E-18 | 1.24E-15 |
| Heart_Failure | dendritic_cell | *CCDC77* | -3.387276547 | 1.26E-17 | 6.91E-17 | 7.90E-14 |
| Heart_Failure | dendritic_cell | *ENOX1* | 2.160364063 | 0.041582287 | 0.04606658 | 1 |
| Heart_Failure | dendritic_cell | *DNAJC15* | -4.330286206 | 2.18E-05 | 2.82E-05 | 0.136845616 |
| Heart_Failure | dendritic_cell | *WBP4* | -3.725502422 | 4.83E-17 | 2.25E-16 | 3.04E-13 |
| Heart_Failure | dendritic_cell | *SMAD9* | -4.328617838 | 2.34E-18 | 1.61E-17 | 1.47E-14 |
| Heart_Failure | dendritic_cell | *SIL1* | -3.792168458 | 1.38E-15 | 4.69E-15 | 8.70E-12 |
| Heart_Failure | dendritic_cell | *ZFP30* | -2.750372039 | 3.49E-08 | 5.22E-08 | 0.000219719 |
| Heart_Failure | dendritic_cell | *ARL1* | -2.69973978 | 0.001386927 | 0.001655203 | 1 |
| Heart_Failure | dendritic_cell | *APAF1* | 4.636733786 | 4.28E-16 | 1.60E-15 | 2.69E-12 |
| Heart_Failure | dendritic_cell | *ZNF706* | -3.613949805 | 9.30E-15 | 2.71E-14 | 5.85E-11 |
| Heart_Failure | dendritic_cell | *CRISPLD1* | -9.643828457 | 1.66E-13 | 3.96E-13 | 1.04E-09 |
| Heart_Failure | dendritic_cell | *RDH10* | -2.36147122 | 9.72E-12 | 1.90E-11 | 6.11E-08 |

| | | | | | | |
|---|---|---|---|---|---|---|
| Heart_Failure | dendritic_cell | *SCPEP1* | -0.99674103 | 0.000929791 | 0.001117277 | 1 |
| Heart_Failure | dendritic_cell | *TBX4* | -3.785740946 | 0.000136336 | 0.00017061 | 0.857146056 |
| Heart_Failure | dendritic_cell | *TEX14* | -4.521268401 | 2.46E-09 | 3.96E-09 | 1.55E-05 |
| Heart_Failure | dendritic_cell | *TRIM6* | -3.668279549 | 9.26E-19 | 7.31E-18 | 5.82E-15 |
| Heart_Failure | dendritic_cell | *CEP89* | -1.495343442 | 0.000103375 | 0.000130088 | 0.649920844 |
| Heart_Failure | dendritic_cell | *ECHDC2* | -1.940453431 | 3.01E-16 | 1.17E-15 | 1.89E-12 |
| Heart_Failure | dendritic_cell | *PRB2* | -3.304754379 | 5.67E-19 | 4.86E-18 | 3.57E-15 |
| Heart_Failure | dendritic_cell | *PYROXD1* | -3.384118873 | 1.79E-11 | 3.43E-11 | 1.12E-07 |
| Heart_Failure | dendritic_cell | *A1BG* | 3.680346166 | 4.69E-21 | 4.66E-19 | 2.95E-17 |
| Heart_Failure | dendritic_cell | *ZSCAN18* | -5.123000536 | 7.79E-08 | 1.14E-07 | 0.00048973 |
| Heart_Failure | dendritic_cell | *PDZRN3* | -2.319705428 | 3.04E-18 | 1.98E-17 | 1.91E-14 |
| Heart_Failure | dendritic_cell | *LHX4* | -5.385465227 | 4.74E-16 | 1.76E-15 | 2.98E-12 |
| Heart_Failure | dendritic_cell | *SEC22A* | -3.35165715 | 5.02E-10 | 8.49E-10 | 3.15E-06 |
| Heart_Failure | dendritic_cell | *CSTA* | 3.002601081 | 1.35E-07 | 1.95E-07 | 0.000846092 |
| Heart_Failure | dendritic_cell | *POPDC2* | -5.00613903 | 4.36E-20 | 7.94E-19 | 2.74E-16 |
| Heart_Failure | dendritic_cell | *B4GALT4* | -6.671049186 | 1.75E-12 | 3.72E-12 | 1.10E-08 |
| Heart_Failure | dendritic_cell | *CD80* | 1.104488154 | 6.80E-21 | 4.66E-19 | 4.27E-17 |
| Heart_Failure | dendritic_cell | *CAT* | -1.60279833 | 3.30E-10 | 5.67E-10 | 2.07E-06 |
| Heart_Failure | dendritic_cell | *PILRB* | -0.616405178 | 0.001057954 | 0.001266443 | 1 |
| Heart_Failure | dendritic_cell | *GJB6* | -36.69688652 | 0.000135814 | 0.000169991 | 0.853863253 |
| Heart_Failure | dendritic_cell | *RNF115* | -4.088002385 | 1.86E-07 | 2.68E-07 | 0.001171904 |
| Heart_Failure | dendritic_cell | *POLR3GL* | -6.046589487 | 2.90E-17 | 1.44E-16 | 1.82E-13 |
| Heart_Failure | dendritic_cell | *TNFSF10* | -3.435446934 | 8.18E-18 | 4.71E-17 | 5.14E-14 |
| Heart_Failure | dendritic_cell | *ZNF639* | -3.587551645 | 2.84E-18 | 1.87E-17 | 1.78E-14 |
| Heart_Failure | dendritic_cell | *LIAS* | -3.862953066 | 7.75E-16 | 2.77E-15 | 4.87E-12 |
| Heart_Failure | dendritic_cell | *CLCC1* | -4.885622462 | 2.96E-17 | 1.46E-16 | 1.86E-13 |
| Heart_Failure | dendritic_cell | *GPSM2* | -3.572411922 | 1.43E-12 | 3.07E-12 | 9.02E-09 |
| Heart_Failure | dendritic_cell | *CXCR4* | 1.339982046 | 4.53E-20 | 8.07E-19 | 2.85E-16 |
| Heart_Failure | dendritic_cell | *ACVR2A* | -3.920202489 | 1.96E-06 | 2.67E-06 | 0.01232946 |
| Heart_Failure | dendritic_cell | *POLK* | -3.919616059 | 1.98E-12 | 4.18E-12 | 1.25E-08 |
| Heart_Failure | dendritic_cell | *FLT3* | 0.195129024 | 6.60E-21 | 4.66E-19 | 4.15E-17 |
| Heart_Failure | dendritic_cell | *MTIF3* | -3.073767658 | 2.44E-17 | 1.23E-16 | 1.53E-13 |
| Heart_Failure | dendritic_cell | *RASL11A* | -7.922642119 | 3.65E-17 | 1.76E-16 | 2.30E-13 |
| Heart_Failure | dendritic_cell | *FMOD* | -5.531255611 | 7.67E-15 | 2.26E-14 | 4.82E-11 |
| Heart_Failure | dendritic_cell | *SERAC1* | -6.181675931 | 1.08E-14 | 3.11E-14 | 6.81E-11 |
| Heart_Failure | dendritic_cell | *LDB3* | 3.199022221 | 8.09E-10 | 1.35E-09 | 5.08E-06 |
| Heart_Failure | dendritic_cell | *OPN4* | 4.565297311 | 2.15E-20 | 6.44E-19 | 1.35E-16 |
| Heart_Failure | dendritic_cell | *FAM35A* | -3.890304019 | 1.74E-10 | 3.05E-10 | 1.09E-06 |
| Heart_Failure | dendritic_cell | *FAM213A* | -2.477757577 | 9.64E-15 | 2.80E-14 | 6.06E-11 |
| Heart_Failure | dendritic_cell | *NAA60* | -7.415410598 | 1.15E-15 | 3.97E-15 | 7.24E-12 |
| Heart_Failure | dendritic_cell | *ODF2L* | -3.003411147 | 3.46E-14 | 9.12E-14 | 2.17E-10 |
| Heart_Failure | dendritic_cell | *SPATA1* | -5.004026213 | 4.51E-11 | 8.31E-11 | 2.84E-07 |
| Heart_Failure | dendritic_cell | *LRRC39* | -1.995615831 | 3.22E-10 | 5.53E-10 | 2.02E-06 |
| Heart_Failure | dendritic_cell | *RWDD3* | 7.80916595 | 4.77E-17 | 2.23E-16 | 3.00E-13 |
| Heart_Failure | dendritic_cell | *CCDC18* | -4.127507447 | 5.26E-13 | 1.18E-12 | 3.30E-09 |
| Heart_Failure | dendritic_cell | *RPAP2* | -3.707685475 | 5.64E-16 | 2.07E-15 | 3.54E-12 |
| Heart_Failure | dendritic_cell | *BBS9* | -3.736809467 | 3.99E-11 | 7.39E-11 | 2.51E-07 |
| Heart_Failure | dendritic_cell | *PMS2* | -4.92272576 | 1.36E-15 | 4.62E-15 | 8.54E-12 |
| Heart_Failure | dendritic_cell | *KIAA0087* | 5.008576127 | 1.03E-17 | 5.82E-17 | 6.49E-14 |
| Heart_Failure | dendritic_cell | *KLHL7* | -4.644983506 | 8.58E-15 | 2.52E-14 | 5.39E-11 |

| | | | | | | |
|---|---|---|---|---|---|---|
| Heart_Failure | dendritic_cell | *HERPUD2* | -5.096112502 | 1.89E-11 | 3.61E-11 | 1.19E-07 |
| Heart_Failure | dendritic_cell | *CBX3* | -4.866798683 | 2.96E-11 | 5.53E-11 | 1.86E-07 |
| Heart_Failure | dendritic_cell | *FAM126A* | -3.550341658 | 2.30E-15 | 7.48E-15 | 1.45E-11 |
| Heart_Failure | dendritic_cell | *FKBP9* | -4.052667728 | 1.23E-14 | 3.49E-14 | 7.72E-11 |
| Heart_Failure | dendritic_cell | *CCZ1* | -3.316788617 | 1.02E-12 | 2.22E-12 | 6.42E-09 |
| Heart_Failure | dendritic_cell | *POLM* | -0.931889052 | 0.000226388 | 0.000280122 | 1 |
| Heart_Failure | dendritic_cell | *GLIPR2* | 4.15209963 | 1.01E-05 | 1.34E-05 | 0.063742311 |
| Heart_Failure | dendritic_cell | *SLC25A51* | -1.292613231 | 1.88E-08 | 2.85E-08 | 0.00011795 |
| Heart_Failure | dendritic_cell | *DCAF10* | -3.951574647 | 1.97E-14 | 5.40E-14 | 1.24E-10 |
| Heart_Failure | dendritic_cell | *TRIM24* | -4.086769355 | 1.21E-05 | 1.59E-05 | 0.076337877 |
| Heart_Failure | dendritic_cell | *BICC1* | -1.508178394 | 3.00E-16 | 1.17E-15 | 1.89E-12 |
| Heart_Failure | dendritic_cell | *SLC25A16* | -2.075344557 | 1.31E-10 | 2.33E-10 | 8.25E-07 |
| Heart_Failure | dendritic_cell | *ZWINT* | -2.437548266 | 5.25E-08 | 7.79E-08 | 0.0003303 |
| Heart_Failure | dendritic_cell | *RBM19* | -5.011539579 | 3.45E-14 | 9.11E-14 | 2.17E-10 |
| Heart_Failure | dendritic_cell | *ACADS* | -3.681040667 | 5.30E-19 | 4.61E-18 | 3.33E-15 |
| Heart_Failure | dendritic_cell | *SSPN* | -2.593484224 | 4.77E-15 | 1.46E-14 | 3.00E-11 |
| Heart_Failure | dendritic_cell | *CCDC91* | -3.958836782 | 1.49E-10 | 2.63E-10 | 9.38E-07 |
| Heart_Failure | dendritic_cell | *PKN1* | -2.647677865 | 0.021532334 | 0.024198185 | 1 |
| Heart_Failure | dendritic_cell | *EBPL* | -2.283275316 | 2.62E-05 | 3.38E-05 | 0.164750339 |
| Heart_Failure | dendritic_cell | *ATP7B* | -0.804699525 | 5.02E-05 | 6.41E-05 | 0.315774245 |
| Heart_Failure | dendritic_cell | *NLN* | 2.047142084 | 0.000899957 | 0.001082463 | 1 |
| Heart_Failure | dendritic_cell | *CENPK* | 0.764261223 | 0.01396839 | 0.015866173 | 1 |
| Heart_Failure | dendritic_cell | *OPTN* | -2.751087848 | 2.35E-19 | 2.41E-18 | 1.48E-15 |
| Heart_Failure | dendritic_cell | *ATF1* | -4.243157499 | 8.86E-11 | 1.59E-10 | 5.57E-07 |
| Heart_Failure | dendritic_cell | *TSFM* | 5.758188929 | 9.84E-10 | 1.63E-09 | 6.18E-06 |
| Heart_Failure | dendritic_cell | *ARHGAP9* | 3.138075989 | 1.49E-17 | 8.02E-17 | 9.36E-14 |
| Heart_Failure | dendritic_cell | *SPATS2* | -3.600007434 | 1.92E-05 | 2.49E-05 | 0.12075984 |
| Heart_Failure | dendritic_cell | *ORMDL2* | 6.827930712 | 2.06E-07 | 2.95E-07 | 0.001292194 |
| Heart_Failure | dendritic_cell | *PDE1B* | 4.613881369 | 5.56E-05 | 7.08E-05 | 0.349762921 |
| Heart_Failure | dendritic_cell | *IKZF4* | -4.207840042 | 2.16E-15 | 7.08E-15 | 1.36E-11 |
| Heart_Failure | dendritic_cell | *SMUG1* | -1.108394793 | 2.12E-05 | 2.75E-05 | 0.133283125 |
| Heart_Failure | dendritic_cell | *SARDH* | -6.311837314 | 2.81E-20 | 6.44E-19 | 1.77E-16 |
| Heart_Failure | dendritic_cell | *DBH* | -5.914950284 | 0.001902474 | 0.002258896 | 1 |
| Heart_Failure | dendritic_cell | *STIL* | -7.559867465 | 6.59E-13 | 1.46E-12 | 4.14E-09 |
| Heart_Failure | dendritic_cell | *TTC21B* | -4.39533707 | 2.39E-13 | 5.59E-13 | 1.51E-09 |
| Heart_Failure | dendritic_cell | *NMI* | -5.363503217 | 1.98E-13 | 4.68E-13 | 1.25E-09 |
| Heart_Failure | dendritic_cell | *ACVR1C* | -1.747332754 | 2.80E-20 | 6.44E-19 | 1.76E-16 |
| Heart_Failure | dendritic_cell | *SLC36A1* | -1.421765874 | 0.000106458 | 0.000133914 | 0.669300433 |
| Heart_Failure | dendritic_cell | *PLA2G12A* | -5.32967464 | 4.29E-13 | 9.72E-13 | 2.70E-09 |
| Heart_Failure | dendritic_cell | *C4BPB* | -3.274465222 | 5.75E-08 | 8.52E-08 | 0.000361667 |
| Heart_Failure | dendritic_cell | *RAB38* | -9.397998055 | 1.11E-14 | 3.17E-14 | 6.96E-11 |
| Heart_Failure | dendritic_cell | *CKS2* | 4.35838174 | 4.91E-18 | 3.00E-17 | 3.09E-14 |
| Heart_Failure | dendritic_cell | *CHPF* | -2.583908249 | 8.19E-07 | 1.13E-06 | 0.005149289 |
| Heart_Failure | dendritic_cell | *DNPEP* | -4.804105752 | 0.000239097 | 0.000295499 | 1 |
| Heart_Failure | dendritic_cell | *MOGAT1* | -2.090897722 | 8.40E-21 | 4.66E-19 | 5.28E-17 |
| Heart_Failure | dendritic_cell | *FAM124B* | -3.526047737 | 2.22E-14 | 6.05E-14 | 1.40E-10 |
| Heart_Failure | dendritic_cell | *FAM210B* | -3.266903374 | 7.46E-20 | 1.09E-18 | 4.69E-16 |
| Heart_Failure | dendritic_cell | *SNX21* | -2.956844995 | 6.13E-19 | 5.15E-18 | 3.85E-15 |
| Heart_Failure | dendritic_cell | *WFDC3* | -0.696430062 | 0.00169559 | 0.00201859 | 1 |
| Heart_Failure | dendritic_cell | *SDC4* | 7.135818851 | 3.74E-09 | 5.94E-09 | 2.35E-05 |

| | | | | | | |
|---|---|---|---|---|---|---|
| Heart_Failure | dendritic_cell | *VAPB* | -1.555561775 | 0.000464174 | 0.000566653 | 1 |
| Heart_Failure | dendritic_cell | *TOX2* | -0.913930724 | 3.99E-05 | 5.10E-05 | 0.250579765 |
| Heart_Failure | dendritic_cell | *ARFGEF2* | -4.2072101 | 2.86E-07 | 4.08E-07 | 0.001797639 |
| Heart_Failure | dendritic_cell | *CSE1L* | -4.348617392 | 1.42E-06 | 1.95E-06 | 0.008954884 |
| Heart_Failure | dendritic_cell | *CDH26* | -5.039397259 | 4.52E-19 | 4.05E-18 | 2.84E-15 |
| Heart_Failure | dendritic_cell | *STX16* | -5.012933239 | 2.45E-13 | 5.72E-13 | 1.54E-09 |
| Heart_Failure | dendritic_cell | *NEURL2* | 2.602446621 | 1.00E-17 | 5.66E-17 | 6.29E-14 |
| Heart_Failure | dendritic_cell | *FASTKD3* | -4.60425532 | 9.48E-19 | 7.45E-18 | 5.96E-15 |
| Heart_Failure | dendritic_cell | *PEPD* | -3.967408457 | 1.66E-16 | 6.81E-16 | 1.04E-12 |
| Heart_Failure | dendritic_cell | *CHST8* | 11.14246893 | 1.33E-15 | 4.53E-15 | 8.36E-12 |
| Heart_Failure | dendritic_cell | *STAMBP* | -4.557705535 | 1.31E-15 | 4.48E-15 | 8.26E-12 |
| Heart_Failure | dendritic_cell | *ATP8A1* | -2.952843944 | 1.50E-09 | 2.46E-09 | 9.45E-06 |
| Heart_Failure | dendritic_cell | *USP22* | -4.825908218 | 2.49E-11 | 4.71E-11 | 1.57E-07 |
| Heart_Failure | dendritic_cell | *HIF3A* | -2.315030435 | 2.06E-17 | 1.06E-16 | 1.29E-13 |
| Heart_Failure | dendritic_cell | *BTN2A2* | 1.666636193 | 2.47E-18 | 1.68E-17 | 1.55E-14 |
| Heart_Failure | dendritic_cell | *MRS2* | -3.210408687 | 9.71E-08 | 1.42E-07 | 0.000610309 |
| Heart_Failure | dendritic_cell | *SERPINB6* | -4.966701798 | 2.77E-13 | 6.45E-13 | 1.74E-09 |
| Heart_Failure | dendritic_cell | *HIST1H1D* | 3.780452445 | 5.76E-10 | 9.72E-10 | 3.62E-06 |
| Heart_Failure | dendritic_cell | *PEX6* | -0.851126884 | 0.021077681 | 0.023714277 | 1 |
| Heart_Failure | dendritic_cell | *NQO2* | -4.606458337 | 1.84E-15 | 6.08E-15 | 1.16E-11 |
| Heart_Failure | dendritic_cell | *ZNF391* | -0.710379356 | 0.000837101 | 0.00100821 | 1 |
| Heart_Failure | dendritic_cell | *RPS10* | -5.795671662 | 2.27E-20 | 6.44E-19 | 1.43E-16 |
| Heart_Failure | dendritic_cell | *HIST1H2BJ* | 4.324026061 | 3.97E-09 | 6.30E-09 | 2.50E-05 |
| Heart_Failure | dendritic_cell | *MED20* | -5.137598374 | 1.89E-12 | 4.00E-12 | 1.19E-08 |
| Heart_Failure | dendritic_cell | *TBCC* | 2.80448203 | 2.01E-14 | 5.50E-14 | 1.26E-10 |
| Heart_Failure | dendritic_cell | *GNMT* | -1.260894126 | 0.004289638 | 0.005006303 | 1 |
| Heart_Failure | dendritic_cell | *MEA1* | -4.811261472 | 3.28E-13 | 7.56E-13 | 2.06E-09 |
| Heart_Failure | dendritic_cell | *KLHL31* | -1.494542039 | 1.27E-15 | 4.35E-15 | 7.99E-12 |
| Heart_Failure | dendritic_cell | *COL21A1* | -2.490255487 | 1.08E-16 | 4.64E-16 | 6.80E-13 |
| Heart_Failure | dendritic_cell | *GLO1* | -4.601299127 | 6.29E-08 | 9.29E-08 | 0.000395338 |
| Heart_Failure | dendritic_cell | *CPNE5* | -3.20870459 | 1.90E-05 | 2.47E-05 | 0.11957036 |
| Heart_Failure | dendritic_cell | *RIOK1* | 4.114096454 | 3.83E-09 | 6.08E-09 | 2.41E-05 |
| Heart_Failure | dendritic_cell | *RPP40* | -4.068664617 | 9.23E-13 | 2.01E-12 | 5.80E-09 |
| Heart_Failure | dendritic_cell | *NUP153* | -4.123348562 | 2.42E-13 | 5.64E-13 | 1.52E-09 |
| Heart_Failure | dendritic_cell | *EEF1E1* | -6.164660446 | 9.01E-20 | 1.25E-18 | 5.66E-16 |
| Heart_Failure | dendritic_cell | *EREG* | 8.006250692 | 7.31E-21 | 4.66E-19 | 4.60E-17 |
| Heart_Failure | dendritic_cell | *MYRF* | 2.215329089 | 0.000409657 | 0.000501073 | 1 |
| Heart_Failure | dendritic_cell | *CNOT1* | -4.30522668 | 1.81E-10 | 3.16E-10 | 1.14E-06 |
| Heart_Failure | dendritic_cell | *BBS2* | -3.091458511 | 9.65E-11 | 1.73E-10 | 6.07E-07 |
| Heart_Failure | dendritic_cell | *MT1G* | -18.12257222 | 1.02E-19 | 1.36E-18 | 6.41E-16 |
| Heart_Failure | dendritic_cell | *GOT2* | -6.896506751 | 7.24E-16 | 2.60E-15 | 4.55E-12 |
| Heart_Failure | dendritic_cell | *CLYBL* | -4.983352958 | 1.91E-12 | 4.03E-12 | 1.20E-08 |
| Heart_Failure | dendritic_cell | *TMTC4* | 4.819560547 | 0.00039504 | 0.000483664 | 1 |
| Heart_Failure | dendritic_cell | *KIF25* | -9.35574116 | 1.12E-11 | 2.18E-11 | 7.03E-08 |
| Heart_Failure | dendritic_cell | *GRK4* | -0.690437575 | 0.000564058 | 0.000684865 | 1 |
| Heart_Failure | dendritic_cell | *TEKT3* | 2.725589833 | 7.49E-14 | 1.88E-13 | 4.71E-10 |
| Heart_Failure | dendritic_cell | *MRPS7* | -5.200418014 | 3.45E-14 | 9.11E-14 | 2.17E-10 |
| Heart_Failure | dendritic_cell | *GGA3* | 3.011743568 | 0.016984128 | 0.019201441 | 1 |
| Heart_Failure | dendritic_cell | *NUP85* | -4.695801647 | 3.50E-16 | 1.34E-15 | 2.20E-12 |
| Heart_Failure | dendritic_cell | *MIF4GD* | 4.446583979 | 2.79E-20 | 6.44E-19 | 1.75E-16 |

| | | | | | | |
|---|---|---|---|---|---|---|
| Heart_Failure | dendritic_cell | *NT5C* | -6.971932956 | 3.47E-10 | 5.94E-10 | 2.18E-06 |
| Heart_Failure | dendritic_cell | *DDX31* | -4.31475156 | 3.91E-15 | 1.22E-14 | 2.46E-11 |
| Heart_Failure | dendritic_cell | *PPP1R12C* | 4.450099067 | 7.03E-05 | 8.92E-05 | 0.442273424 |
| Heart_Failure | dendritic_cell | *OPRL1* | 2.697062491 | 8.55E-16 | 3.04E-15 | 5.38E-12 |
| Heart_Failure | dendritic_cell | *PPDPF* | 6.245973925 | 3.94E-17 | 1.89E-16 | 2.48E-13 |
| Heart_Failure | dendritic_cell | *PLGLB2* | -16.19018337 | 0.000321842 | 0.000395431 | 1 |
| Heart_Failure | dendritic_cell | *CHCHD5* | -5.236511531 | 1.27E-13 | 3.07E-13 | 7.97E-10 |
| Heart_Failure | dendritic_cell | *INSIG2* | -5.204969524 | 7.99E-16 | 2.85E-15 | 5.02E-12 |
| Heart_Failure | dendritic_cell | *POLR1B* | -2.472876837 | 1.40E-07 | 2.03E-07 | 0.000882145 |
| Heart_Failure | dendritic_cell | *PSD4* | 5.610533336 | 3.25E-05 | 4.17E-05 | 0.204343406 |
| Heart_Failure | dendritic_cell | *SLC25A23* | -3.279139286 | 4.08E-14 | 1.06E-13 | 2.57E-10 |
| Heart_Failure | dendritic_cell | *GTF2F1* | -4.345468941 | 7.65E-18 | 4.42E-17 | 4.81E-14 |
| Heart_Failure | dendritic_cell | *CLPP* | -5.036494432 | 5.70E-12 | 1.14E-11 | 3.58E-08 |
| Heart_Failure | dendritic_cell | *SH2D3A* | -8.796819329 | 3.87E-08 | 5.77E-08 | 0.000243345 |
| Heart_Failure | dendritic_cell | *TRIP10* | -4.653240907 | 1.01E-14 | 2.93E-14 | 6.36E-11 |
| Heart_Failure | dendritic_cell | *GPR108* | -4.341766517 | 3.35E-14 | 8.88E-14 | 2.11E-10 |
| Heart_Failure | dendritic_cell | *RTN2* | -7.421969839 | 1.61E-17 | 8.56E-17 | 1.01E-13 |
| Heart_Failure | dendritic_cell | *SYMPK* | -4.554090638 | 4.31E-08 | 6.42E-08 | 0.000271236 |
| Heart_Failure | dendritic_cell | *SDCBP2* | -7.75820553 | 4.86E-16 | 1.81E-15 | 3.05E-12 |
| Heart_Failure | dendritic_cell | *PANK2* | -4.406746822 | 6.51E-06 | 8.63E-06 | 0.040911729 |
| Heart_Failure | dendritic_cell | *DTD1* | -4.84264143 | 5.08E-14 | 1.30E-13 | 3.19E-10 |
| Heart_Failure | dendritic_cell | *RBCK1* | -3.800362766 | 1.06E-07 | 1.55E-07 | 0.000668178 |
| Heart_Failure | dendritic_cell | *RRBP1* | -3.766578826 | 1.20E-09 | 1.98E-09 | 7.57E-06 |
| Heart_Failure | dendritic_cell | *BMP2* | -3.709943512 | 3.23E-20 | 6.76E-19 | 2.03E-16 |
| Heart_Failure | dendritic_cell | *PCSK2* | -7.360062127 | 8.07E-17 | 3.61E-16 | 5.07E-13 |
| Heart_Failure | dendritic_cell | *MKKS* | -4.781346217 | 3.51E-15 | 1.10E-14 | 2.21E-11 |
| Heart_Failure | dendritic_cell | *DSTN* | -4.196791396 | 1.55E-14 | 4.34E-14 | 9.77E-11 |
| Heart_Failure | dendritic_cell | *ITPA* | -6.105172604 | 1.33E-08 | 2.04E-08 | 8.39E-05 |
| Heart_Failure | dendritic_cell | *MCM8* | -1.726791362 | 7.24E-07 | 1.01E-06 | 0.004553111 |
| Heart_Failure | dendritic_cell | *TMEM74B* | 6.170124532 | 5.01E-14 | 1.29E-13 | 3.15E-10 |
| Heart_Failure | dendritic_cell | *FAM110A* | 7.758569453 | 2.56E-18 | 1.72E-17 | 1.61E-14 |
| Heart_Failure | dendritic_cell | *NCLN* | -5.474318599 | 7.94E-09 | 1.23E-08 | 4.99E-05 |
| Heart_Failure | dendritic_cell | *ZNF436* | -4.030501329 | 1.05E-13 | 2.56E-13 | 6.59E-10 |
| Heart_Failure | dendritic_cell | *GDF5* | 2.848723039 | 3.76E-12 | 7.70E-12 | 2.37E-08 |
| Heart_Failure | dendritic_cell | *MMP24* | -4.479987579 | 2.58E-09 | 4.15E-09 | 1.62E-05 |
| Heart_Failure | dendritic_cell | *UROD* | -3.550194689 | 9.80E-17 | 4.26E-16 | 6.16E-13 |
| Heart_Failure | dendritic_cell | *ST3GAL3* | -3.301370636 | 1.01E-10 | 1.80E-10 | 6.34E-07 |
| Heart_Failure | dendritic_cell | *KLC1* | -4.188169835 | 4.05E-13 | 9.19E-13 | 2.55E-09 |
| Heart_Failure | dendritic_cell | *XRCC3* | -6.183104297 | 3.09E-13 | 7.14E-13 | 1.94E-09 |
| Heart_Failure | dendritic_cell | *MCF2L* | -3.175892891 | 1.19E-15 | 4.10E-15 | 7.51E-12 |
| Heart_Failure | dendritic_cell | *F10* | -3.136582447 | 2.54E-19 | 2.56E-18 | 1.60E-15 |
| Heart_Failure | dendritic_cell | *PCID2* | -4.166231057 | 5.03E-14 | 1.29E-13 | 3.16E-10 |
| Heart_Failure | dendritic_cell | *PROZ* | 4.238797782 | 6.98E-21 | 4.66E-19 | 4.39E-17 |
| Heart_Failure | dendritic_cell | *IGFLR1* | 16.85518404 | 1.58E-19 | 1.85E-18 | 9.94E-16 |
| Heart_Failure | dendritic_cell | *CAPNS1* | -5.203931692 | 5.21E-15 | 1.59E-14 | 3.27E-11 |
| Heart_Failure | dendritic_cell | *COX6B1* | -4.907544225 | 3.52E-17 | 1.71E-16 | 2.21E-13 |
| Heart_Failure | dendritic_cell | *PRDX5* | -5.868670646 | 8.45E-17 | 3.75E-16 | 5.31E-13 |
| Heart_Failure | dendritic_cell | *SCAF1* | -5.751426164 | 9.58E-18 | 5.43E-17 | 6.02E-14 |
| Heart_Failure | dendritic_cell | *PRR12* | -3.430812356 | 2.59E-12 | 5.39E-12 | 1.63E-08 |
| Heart_Failure | dendritic_cell | *TSKS* | 7.868762962 | 2.12E-17 | 1.08E-16 | 1.33E-13 |

| | | | | | | |
|---|---|---|---|---|---|---|
| Heart_Failure | dendritic_cell | *FLRT1* | -3.706083869 | 1.41E-13 | 3.39E-13 | 8.85E-10 |
| Heart_Failure | dendritic_cell | *SBDS* | -4.308159039 | 1.98E-18 | 1.39E-17 | 1.24E-14 |
| Heart_Failure | dendritic_cell | *BECN1* | -5.322218224 | 2.05E-16 | 8.30E-16 | 1.29E-12 |
| Heart_Failure | dendritic_cell | *TRAP1* | -1.230782897 | 3.70E-07 | 5.25E-07 | 0.002329247 |
| Heart_Failure | dendritic_cell | *DNAJC8* | -4.088650893 | 3.63E-06 | 4.88E-06 | 0.022846186 |
| Heart_Failure | dendritic_cell | *ZNF384* | -5.657379944 | 1.05E-07 | 1.53E-07 | 0.00065825 |
| Heart_Failure | dendritic_cell | *EMG1* | -5.47275463 | 1.81E-14 | 4.99E-14 | 1.14E-10 |
| Heart_Failure | dendritic_cell | *ATG14* | -4.633185654 | 1.30E-11 | 2.52E-11 | 8.16E-08 |
| Heart_Failure | dendritic_cell | *RHOJ* | -3.865512816 | 8.47E-18 | 4.87E-17 | 5.33E-14 |
| Heart_Failure | dendritic_cell | *L3HYPDH* | -1.967154207 | 6.63E-15 | 1.99E-14 | 4.17E-11 |
| Heart_Failure | dendritic_cell | *ZBTB1* | -4.258921027 | 9.17E-11 | 1.64E-10 | 5.77E-07 |
| Heart_Failure | dendritic_cell | *PZP* | -3.869614355 | 2.95E-17 | 1.46E-16 | 1.86E-13 |
| Heart_Failure | dendritic_cell | *RHOT1* | -2.623399289 | 1.35E-05 | 1.76E-05 | 0.08464892 |
| Heart_Failure | dendritic_cell | *WDR60* | -3.042608751 | 5.07E-16 | 1.88E-15 | 3.19E-12 |
| Heart_Failure | dendritic_cell | *AIF1L* | -2.879529852 | 7.30E-20 | 1.08E-18 | 4.59E-16 |
| Heart_Failure | dendritic_cell | *MAP2K2* | 3.773008982 | 0.000210618 | 0.00026102 | 1 |
| Heart_Failure | dendritic_cell | *CANX* | -4.73297439 | 4.59E-09 | 7.24E-09 | 2.88E-05 |
| Heart_Failure | dendritic_cell | *IPPK* | -5.643807858 | 2.64E-14 | 7.11E-14 | 1.66E-10 |
| Heart_Failure | dendritic_cell | *FGD3* | 2.846523759 | 2.38E-20 | 6.44E-19 | 1.50E-16 |
| Heart_Failure | dendritic_cell | *ABHD8* | 4.070977873 | 5.49E-09 | 8.63E-09 | 3.45E-05 |
| Heart_Failure | dendritic_cell | *MASP1* | -2.753033713 | 1.04E-18 | 8.03E-18 | 6.55E-15 |
| Heart_Failure | dendritic_cell | *ATP13A4* | -1.83498272 | 0.015014545 | 0.017011433 | 1 |
| Heart_Failure | dendritic_cell | *HRASLS* | -4.825606894 | 5.08E-19 | 4.43E-18 | 3.19E-15 |
| Heart_Failure | dendritic_cell | *HELB* | -3.751226049 | 0.000446678 | 0.000545612 | 1 |
| Heart_Failure | dendritic_cell | *RAP1B* | -3.6137579 | 6.04E-08 | 8.93E-08 | 0.000380027 |
| Heart_Failure | dendritic_cell | *RAB3IP* | -5.051000373 | 4.99E-13 | 1.12E-12 | 3.14E-09 |
| Heart_Failure | dendritic_cell | *YEATS4* | -3.00045795 | 1.58E-13 | 3.77E-13 | 9.90E-10 |
| Heart_Failure | dendritic_cell | *TAS2R4* | 3.096271693 | 7.49E-06 | 9.91E-06 | 0.047105951 |
| Heart_Failure | dendritic_cell | *LRRC61* | -2.100922624 | 8.30E-14 | 2.07E-13 | 5.22E-10 |
| Heart_Failure | dendritic_cell | *IDUA* | -3.474820213 | 9.15E-20 | 1.26E-18 | 5.75E-16 |
| Heart_Failure | dendritic_cell | *TMEM175* | -4.742532813 | 2.85E-12 | 5.90E-12 | 1.79E-08 |
| Heart_Failure | dendritic_cell | *EMC1* | -4.470778929 | 3.12E-10 | 5.38E-10 | 1.96E-06 |
| Heart_Failure | dendritic_cell | *PLA2G5* | -2.624538888 | 5.85E-07 | 8.17E-07 | 0.003675766 |
| Heart_Failure | dendritic_cell | *UBR4* | -2.497482088 | 5.57E-07 | 7.80E-07 | 0.00350179 |
| Heart_Failure | dendritic_cell | *SIN3B* | -5.307134387 | 5.69E-12 | 1.14E-11 | 3.58E-08 |
| Heart_Failure | dendritic_cell | *WFIKKN1* | -9.464503772 | 0.000266523 | 0.000328555 | 1 |
| Heart_Failure | dendritic_cell | *WDR24* | 6.663184474 | 2.02E-19 | 2.17E-18 | 1.27E-15 |
| Heart_Failure | dendritic_cell | *CHTF18* | -6.325407171 | 4.31E-06 | 5.76E-06 | 0.027108062 |
| Heart_Failure | dendritic_cell | *SMARCA4* | -5.054785764 | 5.55E-09 | 8.72E-09 | 3.49E-05 |
| Heart_Failure | dendritic_cell | *TICAM1* | 7.580682627 | 1.40E-13 | 3.38E-13 | 8.82E-10 |
| Heart_Failure | dendritic_cell | *AAMP* | -4.767859466 | 6.71E-18 | 3.95E-17 | 4.22E-14 |
| Heart_Failure | dendritic_cell | *PNKD* | -4.44623059 | 1.08E-16 | 4.62E-16 | 6.76E-13 |
| Heart_Failure | dendritic_cell | *ZNF835* | 3.258125308 | 1.55E-07 | 2.24E-07 | 0.000976779 |
| Heart_Failure | dendritic_cell | *HIP1* | -3.474411467 | 1.97E-06 | 2.68E-06 | 0.012416157 |
| Heart_Failure | dendritic_cell | *PTPN12* | -3.69113706 | 4.81E-07 | 6.77E-07 | 0.00302674 |
| Heart_Failure | dendritic_cell | *POR* | -3.153524867 | 2.11E-18 | 1.47E-17 | 1.33E-14 |
| Heart_Failure | dendritic_cell | *FGL2* | 1.823096699 | 4.53E-20 | 8.07E-19 | 2.85E-16 |
| Heart_Failure | dendritic_cell | *STYXL1* | -1.218017008 | 0.011820339 | 0.013462767 | 1 |
| Heart_Failure | dendritic_cell | *STEAP4* | -3.435419811 | 1.80E-17 | 9.47E-17 | 1.13E-13 |
| Heart_Failure | dendritic_cell | *GNAI1* | -2.932034971 | 2.41E-19 | 2.46E-18 | 1.52E-15 |

| | | | | | | |
|---|---|---|---|---|---|---|
| Heart_Failure | dendritic_cell | *SGCE* | -3.399155368 | 1.16E-18 | 8.83E-18 | 7.31E-15 |
| Heart_Failure | dendritic_cell | *RBM48* | -5.778370979 | 2.13E-13 | 5.02E-13 | 1.34E-09 |
| Heart_Failure | dendritic_cell | *CASD1* | -4.597950473 | 1.10E-13 | 2.67E-13 | 6.89E-10 |
| Heart_Failure | dendritic_cell | *SRD5A3* | -4.881639036 | 3.13E-06 | 4.21E-06 | 0.019689463 |
| Heart_Failure | dendritic_cell | *DGCR6L* | -3.658174198 | 0.000158786 | 0.000198112 | 0.998285683 |
| Heart_Failure | dendritic_cell | *VPREB3* | -28.35774712 | 3.60E-13 | 8.24E-13 | 2.26E-09 |
| Heart_Failure | dendritic_cell | *MGAT3* | -2.655238669 | 2.91E-20 | 6.48E-19 | 1.83E-16 |
| Heart_Failure | dendritic_cell | *ATF4* | -5.926784742 | 1.09E-11 | 2.12E-11 | 6.84E-08 |
| Heart_Failure | dendritic_cell | *A4GALT* | -1.908143186 | 7.52E-06 | 9.95E-06 | 0.047271949 |
| Heart_Failure | dendritic_cell | *CDC42EP1* | -3.564826401 | 3.22E-18 | 2.07E-17 | 2.02E-14 |
| Heart_Failure | dendritic_cell | *TPST2* | -4.167559185 | 1.74E-16 | 7.11E-16 | 1.09E-12 |
| Heart_Failure | dendritic_cell | *BAIAP2L2* | -3.959670564 | 5.30E-18 | 3.21E-17 | 3.34E-14 |
| Heart_Failure | dendritic_cell | *TST* | -2.636152363 | 1.08E-16 | 4.63E-16 | 6.78E-13 |
| Heart_Failure | dendritic_cell | *APOL5* | 6.225908005 | 2.75E-16 | 1.08E-15 | 1.73E-12 |
| Heart_Failure | dendritic_cell | *RIBC2* | 3.115331311 | 1.34E-17 | 7.28E-17 | 8.41E-14 |
| Heart_Failure | dendritic_cell | *EMC4* | -5.585671592 | 1.66E-12 | 3.53E-12 | 1.04E-08 |
| Heart_Failure | dendritic_cell | *SPECC1* | 2.969814948 | 0.002163873 | 0.002561527 | 1 |
| Heart_Failure | dendritic_cell | *POT1* | -4.192034642 | 8.32E-14 | 2.07E-13 | 5.23E-10 |
| Heart_Failure | dendritic_cell | *CDHR3* | -2.436669016 | 4.51E-17 | 2.13E-16 | 2.83E-13 |
| Heart_Failure | dendritic_cell | *PRKRIP1* | -3.555375528 | 1.65E-13 | 3.94E-13 | 1.04E-09 |
| Heart_Failure | dendritic_cell | *FOXP2* | -1.833742415 | 3.91E-19 | 3.63E-18 | 2.46E-15 |
| Heart_Failure | dendritic_cell | *STRIP2* | -3.438779559 | 6.63E-16 | 2.40E-15 | 4.17E-12 |
| Heart_Failure | dendritic_cell | *FLNC* | -3.190246336 | 1.00E-13 | 2.45E-13 | 6.29E-10 |
| Heart_Failure | dendritic_cell | *IRF5* | 1.728215433 | 3.10E-16 | 1.21E-15 | 1.95E-12 |
| Heart_Failure | dendritic_cell | *KLHDC10* | -4.123146376 | 3.34E-14 | 8.85E-14 | 2.10E-10 |
| Heart_Failure | dendritic_cell | *OPN1SW* | -2.200813697 | 5.39E-15 | 1.64E-14 | 3.39E-11 |
| Heart_Failure | dendritic_cell | *HOXD1* | -14.30052216 | 2.40E-12 | 5.01E-12 | 1.51E-08 |
| Heart_Failure | dendritic_cell | *HOXD3* | -4.834997687 | 7.70E-12 | 1.52E-11 | 4.84E-08 |
| Heart_Failure | dendritic_cell | *PDE11A* | -2.46076147 | 1.92E-11 | 3.66E-11 | 1.21E-07 |
| Heart_Failure | dendritic_cell | *CHN1* | -3.313503029 | 2.69E-15 | 8.62E-15 | 1.69E-11 |
| Heart_Failure | dendritic_cell | *GAD1* | -7.872994074 | 2.00E-20 | 6.44E-19 | 1.26E-16 |
| Heart_Failure | dendritic_cell | *ORMDL1* | -2.132003186 | 1.99E-07 | 2.85E-07 | 0.001248657 |
| Heart_Failure | dendritic_cell | *PSMG2* | -3.372592892 | 7.53E-09 | 1.17E-08 | 4.73E-05 |
| Heart_Failure | dendritic_cell | *TWSG1* | 4.960561399 | 1.66E-05 | 2.16E-05 | 0.104380658 |
| Heart_Failure | dendritic_cell | *WDFY4* | 0.441141048 | 4.61E-21 | 4.66E-19 | 2.90E-17 |
| Heart_Failure | dendritic_cell | *EIF2AK4* | -2.395550368 | 0.000229822 | 0.000284204 | 1 |
| Heart_Failure | dendritic_cell | *TMOD2* | -3.737690853 | 6.60E-14 | 1.67E-13 | 4.15E-10 |
| Heart_Failure | dendritic_cell | *IVD* | -2.750254483 | 2.24E-05 | 2.90E-05 | 0.140913321 |
| Heart_Failure | dendritic_cell | *KNSTRN* | -1.370945819 | 3.62E-12 | 7.44E-12 | 2.28E-08 |
| Heart_Failure | dendritic_cell | *ARPP19* | -5.016298809 | 4.60E-13 | 1.04E-12 | 2.89E-09 |
| Heart_Failure | dendritic_cell | *VPS13C* | -3.621771423 | 1.25E-08 | 1.92E-08 | 7.85E-05 |
| Heart_Failure | dendritic_cell | *CALML4* | -6.119470838 | 2.61E-19 | 2.62E-18 | 1.64E-15 |
| Heart_Failure | dendritic_cell | *ACKR4* | -3.063118225 | 2.95E-19 | 2.88E-18 | 1.86E-15 |
| Heart_Failure | dendritic_cell | *ANAPC13* | 1.095368668 | 0.006063233 | 0.007017589 | 1 |
| Heart_Failure | dendritic_cell | *SUMF2* | -5.084008549 | 9.15E-10 | 1.52E-09 | 5.75E-06 |
| Heart_Failure | dendritic_cell | *SERGEF* | -3.236308946 | 0.001819729 | 0.002161874 | 1 |
| Heart_Failure | dendritic_cell | *DCTD* | -3.072014945 | 5.15E-07 | 7.23E-07 | 0.003235951 |
| Heart_Failure | dendritic_cell | *RPAIN* | -3.77479398 | 8.40E-14 | 2.09E-13 | 5.28E-10 |
| Heart_Failure | dendritic_cell | *USP6* | -1.739934588 | 1.45E-08 | 2.22E-08 | 9.13E-05 |
| Heart_Failure | dendritic_cell | *PLD2* | -4.18864625 | 2.84E-05 | 3.66E-05 | 0.178849115 |

| | | | | | | |
|---|---|---|---|---|---|---|
| Heart_Failure | dendritic_cell | *ATP1B2* | -7.010741279 | 1.36E-17 | 7.39E-17 | 8.55E-14 |
| Heart_Failure | dendritic_cell | *CCL4* | -3.51990612 | 6.11E-10 | 1.03E-09 | 3.84E-06 |
| Heart_Failure | dendritic_cell | *PHF20L1* | -4.478285493 | 5.70E-12 | 1.14E-11 | 3.58E-08 |
| Heart_Failure | dendritic_cell | *PUS7L* | -4.718378439 | 6.98E-11 | 1.27E-10 | 4.39E-07 |
| Heart_Failure | dendritic_cell | *KRI1* | -2.289192552 | 3.45E-06 | 4.63E-06 | 0.021666796 |
| Heart_Failure | dendritic_cell | *SLC44A2* | -4.948057344 | 1.13E-15 | 3.89E-15 | 7.08E-12 |
| Heart_Failure | dendritic_cell | *CDKN2D* | -5.772184015 | 1.72E-16 | 7.03E-16 | 1.08E-12 |
| Heart_Failure | dendritic_cell | *MTUS1* | -2.860354642 | 1.31E-16 | 5.52E-16 | 8.27E-13 |
| Heart_Failure | dendritic_cell | *NGDN* | -6.020064975 | 1.08E-14 | 3.10E-14 | 6.78E-11 |
| Heart_Failure | dendritic_cell | *ADCY4* | -2.714040424 | 3.21E-20 | 6.76E-19 | 2.02E-16 |
| Heart_Failure | dendritic_cell | *RAB2B* | -4.461326128 | 7.13E-17 | 3.22E-16 | 4.48E-13 |
| Heart_Failure | dendritic_cell | *BCL2L2* | -5.666789768 | 3.72E-15 | 1.16E-14 | 2.34E-11 |
| Heart_Failure | dendritic_cell | *DTD2* | -1.599886081 | 0.000816223 | 0.000983441 | 1 |
| Heart_Failure | dendritic_cell | *PARP2* | -3.766541866 | 2.85E-15 | 9.11E-15 | 1.79E-11 |
| Heart_Failure | dendritic_cell | *MIS18BP1* | 2.245603834 | 5.61E-05 | 7.14E-05 | 0.352674609 |
| Heart_Failure | dendritic_cell | *DAD1* | -4.540710774 | 2.15E-13 | 5.05E-13 | 1.35E-09 |
| Heart_Failure | dendritic_cell | *TEP1* | -3.945825714 | 0.000213003 | 0.000263924 | 1 |
| Heart_Failure | dendritic_cell | *EPB41L4A* | -3.688479687 | 1.72E-15 | 5.71E-15 | 1.08E-11 |
| Heart_Failure | dendritic_cell | *CDO1* | -1.922374741 | 4.96E-20 | 8.51E-19 | 3.12E-16 |
| Heart_Failure | dendritic_cell | *REEP5* | -4.596399696 | 4.00E-10 | 6.81E-10 | 2.51E-06 |
| Heart_Failure | dendritic_cell | *QRICH2* | -4.591350773 | 5.30E-19 | 4.61E-18 | 3.33E-15 |
| Heart_Failure | dendritic_cell | *SEC14L1* | -2.413452801 | 0.000197845 | 0.000245577 | 1 |
| Heart_Failure | dendritic_cell | *AANAT* | -1.841968096 | 0.0088617 | 0.01016299 | 1 |
| Heart_Failure | dendritic_cell | *ERMARD* | -4.693367702 | 1.99E-14 | 5.44E-14 | 1.25E-10 |
| Heart_Failure | dendritic_cell | *PHF10* | -5.826142957 | 6.89E-11 | 1.25E-10 | 4.33E-07 |
| Heart_Failure | dendritic_cell | *GALNT8* | -2.997811787 | 1.49E-05 | 1.94E-05 | 0.093576782 |
| Heart_Failure | dendritic_cell | *DOCK6* | -3.209122162 | 2.55E-11 | 4.81E-11 | 1.60E-07 |
| Heart_Failure | dendritic_cell | *ECSIT* | -5.80680132 | 1.87E-08 | 2.84E-08 | 0.000117648 |
| Heart_Failure | dendritic_cell | *ELOF1* | -5.791381075 | 5.55E-19 | 4.79E-18 | 3.49E-15 |
| Heart_Failure | dendritic_cell | *CDC16* | -3.373190546 | 2.18E-11 | 4.14E-11 | 1.37E-07 |
| Heart_Failure | dendritic_cell | *THEM6* | -3.22774129 | 6.60E-15 | 1.98E-14 | 4.15E-11 |
| Heart_Failure | dendritic_cell | *TOMM40* | -7.110851463 | 2.18E-16 | 8.73E-16 | 1.37E-12 |
| Heart_Failure | dendritic_cell | *DPP6* | -4.929027402 | 1.19E-17 | 6.58E-17 | 7.48E-14 |
| Heart_Failure | dendritic_cell | *ATP8B3* | -1.989793694 | 4.27E-09 | 6.75E-09 | 2.68E-05 |
| Heart_Failure | dendritic_cell | *GTPBP3* | -3.629915423 | 2.05E-19 | 2.21E-18 | 1.29E-15 |
| Heart_Failure | dendritic_cell | *PLVAP* | -0.927430869 | 1.49E-08 | 2.28E-08 | 9.39E-05 |
| Heart_Failure | dendritic_cell | *BST2* | -5.227402301 | 1.47E-16 | 6.10E-16 | 9.23E-13 |
| Heart_Failure | dendritic_cell | *DDA1* | -6.370451863 | 2.48E-11 | 4.68E-11 | 1.56E-07 |
| Heart_Failure | dendritic_cell | *TULP4* | -3.553820326 | 1.89E-15 | 6.25E-15 | 1.19E-11 |
| Heart_Failure | dendritic_cell | *RTN4IP1* | -3.43983544 | 0.000381691 | 0.000467502 | 1 |
| Heart_Failure | dendritic_cell | *QRSL1* | -0.952559645 | 0.022976411 | 0.025785915 | 1 |
| Heart_Failure | dendritic_cell | *ACTN4* | -4.19558239 | 7.39E-15 | 2.19E-14 | 4.64E-11 |
| Heart_Failure | dendritic_cell | *STK33* | -3.029102697 | 4.71E-18 | 2.90E-17 | 2.96E-14 |
| Heart_Failure | dendritic_cell | *NDUFA10* | -0.938257885 | 0.013625807 | 0.015485439 | 1 |
| Heart_Failure | dendritic_cell | *CACNG6* | 5.75024233 | 7.10E-05 | 8.99E-05 | 0.446508147 |
| Heart_Failure | dendritic_cell | *PXDN* | -2.508969742 | 6.59E-19 | 5.46E-18 | 4.14E-15 |
| Heart_Failure | dendritic_cell | *PGPEP1* | -4.156469416 | 1.51E-15 | 5.08E-15 | 9.50E-12 |
| Heart_Failure | dendritic_cell | *LSM4* | -1.374595247 | 0.023695818 | 0.026560101 | 1 |
| Heart_Failure | dendritic_cell | *HRC* | -2.819772454 | 1.08E-07 | 1.57E-07 | 0.00067867 |
| Heart_Failure | dendritic_cell | *TRPM4* | 3.846459585 | 1.18E-05 | 1.54E-05 | 0.073996239 |

| | | | | | | |
|---|---|---|---|---|---|---|
| Heart_Failure | dendritic_cell | *CAMSAP1* | -5.764060291 | 1.46E-08 | 2.22E-08 | 9.15E-05 |
| Heart_Failure | dendritic_cell | *UBAC1* | -5.121819863 | 2.63E-12 | 5.47E-12 | 1.65E-08 |
| Heart_Failure | dendritic_cell | *TNNT3* | -3.016299392 | 3.19E-18 | 2.06E-17 | 2.01E-14 |
| Heart_Failure | dendritic_cell | *ATXN10* | -4.430694923 | 8.59E-12 | 1.69E-11 | 5.40E-08 |
| Heart_Failure | dendritic_cell | *TUBGCP2* | -4.038791786 | 3.47E-06 | 4.67E-06 | 0.021841048 |
| Heart_Failure | dendritic_cell | *CYP2E1* | -6.369391982 | 1.18E-17 | 6.53E-17 | 7.42E-14 |
| Heart_Failure | dendritic_cell | *HBZ* | 13.62389101 | 1.10E-10 | 1.96E-10 | 6.90E-07 |
| Heart_Failure | dendritic_cell | *PAK4* | -6.257155508 | 1.08E-14 | 3.11E-14 | 6.80E-11 |
| Heart_Failure | dendritic_cell | *ZNF337* | -3.189427904 | 9.04E-11 | 1.62E-10 | 5.68E-07 |
| Heart_Failure | dendritic_cell | *CEP85* | -2.791533769 | 1.72E-13 | 4.10E-13 | 1.08E-09 |
| Heart_Failure | dendritic_cell | *POMT1* | -2.590309066 | 0.019944218 | 0.0224712 | 1 |
| Heart_Failure | dendritic_cell | *UCK1* | -2.604908048 | 3.45E-07 | 4.90E-07 | 0.00216683 |
| Heart_Failure | dendritic_cell | *YIPF2* | -4.940592547 | 1.17E-13 | 2.85E-13 | 7.38E-10 |
| Heart_Failure | dendritic_cell | *TMEM160* | -8.337685454 | 4.86E-06 | 6.48E-06 | 0.030567524 |
| Heart_Failure | dendritic_cell | *MAP3K10* | -4.726754177 | 3.93E-07 | 5.56E-07 | 0.002470666 |
| Heart_Failure | dendritic_cell | *ARHGEF16* | -1.312730107 | 0.002978033 | 0.00350288 | 1 |
| Heart_Failure | dendritic_cell | *SMPDL3B* | 8.2524907 | 6.41E-21 | 4.66E-19 | 4.03E-17 |
| Heart_Failure | dendritic_cell | *THEMIS2* | 1.369527754 | 2.81E-20 | 6.44E-19 | 1.77E-16 |
| Heart_Failure | dendritic_cell | *CLIP1* | -4.018171844 | 3.55E-13 | 8.14E-13 | 2.23E-09 |
| Heart_Failure | dendritic_cell | *ZNF317* | -5.12029469 | 6.78E-08 | 9.99E-08 | 0.000426103 |
| Heart_Failure | dendritic_cell | *PPAN* | 14.18932838 | 1.31E-12 | 2.81E-12 | 8.21E-09 |
| Heart_Failure | dendritic_cell | *ZNF236* | -4.26765101 | 3.82E-13 | 8.72E-13 | 2.40E-09 |
| Heart_Failure | dendritic_cell | *C12orf65* | -4.675591857 | 3.37E-11 | 6.29E-11 | 2.12E-07 |
| Heart_Failure | dendritic_cell | *HABP4* | -5.192751971 | 4.78E-14 | 1.23E-13 | 3.00E-10 |
| Heart_Failure | dendritic_cell | *EPS8L1* | -16.84545179 | 8.51E-20 | 1.20E-18 | 5.35E-16 |
| Heart_Failure | dendritic_cell | *LILRB2* | 1.212661535 | 6.02E-21 | 4.66E-19 | 3.78E-17 |
| Heart_Failure | dendritic_cell | *COX4I2* | -5.645302549 | 1.94E-19 | 2.14E-18 | 1.22E-15 |
| Heart_Failure | dendritic_cell | *ATP6V1E1* | -5.008547514 | 2.42E-13 | 5.64E-13 | 1.52E-09 |
| Heart_Failure | dendritic_cell | *PPT1* | 4.196007488 | 2.00E-19 | 2.16E-18 | 1.26E-15 |
| Heart_Failure | dendritic_cell | *HAUS8* | 4.761116367 | 3.18E-17 | 1.56E-16 | 2.00E-13 |
| Heart_Failure | dendritic_cell | *TBC1D5* | -3.263189036 | 4.33E-08 | 6.43E-08 | 0.000271938 |
| Heart_Failure | dendritic_cell | *RFTN1* | -3.671330678 | 2.95E-13 | 6.84E-13 | 1.85E-09 |
| Heart_Failure | dendritic_cell | *KCNC3* | 1.552178963 | 5.83E-21 | 4.66E-19 | 3.67E-17 |
| Heart_Failure | dendritic_cell | *KIF3A* | -2.644696155 | 1.33E-17 | 7.25E-17 | 8.36E-14 |
| Heart_Failure | dendritic_cell | *AOC3* | -3.210746191 | 2.30E-17 | 1.16E-16 | 1.45E-13 |
| Heart_Failure | dendritic_cell | *ACLY* | -4.632997647 | 4.90E-10 | 8.29E-10 | 3.08E-06 |
| Heart_Failure | dendritic_cell | *AOC2* | -4.850397245 | 1.22E-19 | 1.53E-18 | 7.68E-16 |
| Heart_Failure | dendritic_cell | *DIAPH1* | 4.574991328 | 5.02E-13 | 1.13E-12 | 3.16E-09 |
| Heart_Failure | dendritic_cell | *EXOC4* | -2.991789264 | 5.39E-10 | 9.11E-10 | 3.39E-06 |
| Heart_Failure | dendritic_cell | *ACAP3* | -2.190196186 | 1.23E-19 | 1.54E-18 | 7.75E-16 |
| Heart_Failure | dendritic_cell | *C1orf159* | -2.955793104 | 0.019547993 | 0.022052438 | 1 |
| Heart_Failure | dendritic_cell | *ANO1* | -6.810297113 | 1.96E-13 | 4.64E-13 | 1.23E-09 |
| Heart_Failure | dendritic_cell | *PPFIA1* | -3.450716364 | 9.18E-14 | 2.26E-13 | 5.77E-10 |
| Heart_Failure | dendritic_cell | *NINJ1* | -3.569426228 | 0.017578469 | 0.019862659 | 1 |
| Heart_Failure | dendritic_cell | *NPHP4* | 6.90558733 | 7.00E-08 | 1.03E-07 | 0.000440066 |
| Heart_Failure | dendritic_cell | *MAP1B* | -1.251048927 | 2.23E-07 | 3.20E-07 | 0.001403878 |
| Heart_Failure | dendritic_cell | *CKMT2* | -3.595985359 | 4.14E-16 | 1.56E-15 | 2.60E-12 |
| Heart_Failure | dendritic_cell | *ZCCHC9* | -4.292532674 | 2.31E-15 | 7.49E-15 | 1.45E-11 |
| Heart_Failure | dendritic_cell | *KHDRBS3* | -4.494154512 | 5.73E-13 | 1.27E-12 | 3.60E-09 |
| Heart_Failure | dendritic_cell | *CHD1L* | 3.858513072 | 1.07E-05 | 1.40E-05 | 0.067025085 |

| | | | | | | |
|---|---|---|---|---|---|---|
| Heart_Failure | dendritic_cell | *RBM8A* | -4.324634762 | 2.43E-11 | 4.60E-11 | 1.53E-07 |
| Heart_Failure | dendritic_cell | *ZNF304* | -3.704649116 | 1.66E-14 | 4.61E-14 | 1.04E-10 |
| Heart_Failure | dendritic_cell | *ZSCAN5A* | -5.123095504 | 1.76E-10 | 3.08E-10 | 1.11E-06 |
| Heart_Failure | dendritic_cell | *ZNF132* | -2.799275401 | 0.000138515 | 0.000173234 | 0.870845335 |
| Heart_Failure | dendritic_cell | *RHPN2* | -4.79044423 | 1.08E-17 | 6.05E-17 | 6.81E-14 |
| Heart_Failure | dendritic_cell | *C19orf12* | -2.096776518 | 2.51E-14 | 6.78E-14 | 1.58E-10 |
| Heart_Failure | dendritic_cell | *ACTR10* | -4.693667735 | 2.22E-11 | 4.22E-11 | 1.40E-07 |
| Heart_Failure | dendritic_cell | *GCH1* | 4.881954264 | 8.56E-09 | 1.33E-08 | 5.38E-05 |
| Heart_Failure | dendritic_cell | *LGALS3* | -4.148239288 | 7.05E-14 | 1.77E-13 | 4.43E-10 |
| Heart_Failure | dendritic_cell | *DNAJB1* | -4.817754157 | 1.11E-09 | 1.83E-09 | 6.96E-06 |
| Heart_Failure | dendritic_cell | *FBXW9* | 4.670398265 | 5.13E-16 | 1.89E-15 | 3.22E-12 |
| Heart_Failure | dendritic_cell | *C19orf57* | -10.85663078 | 3.33E-13 | 7.65E-13 | 2.09E-09 |
| Heart_Failure | dendritic_cell | *MATN3* | -8.562843249 | 1.86E-20 | 6.38E-19 | 1.17E-16 |
| Heart_Failure | dendritic_cell | *GAS2L2* | -1.125109433 | 6.05E-05 | 7.69E-05 | 0.380668186 |
| Heart_Failure | dendritic_cell | *CCT6B* | -4.046287464 | 7.89E-19 | 6.41E-18 | 4.96E-15 |
| Heart_Failure | dendritic_cell | *RAF1* | -4.515605449 | 2.34E-12 | 4.89E-12 | 1.47E-08 |
| Heart_Failure | dendritic_cell | *ENOSF1* | -3.008913425 | 1.92E-14 | 5.26E-14 | 1.21E-10 |
| Heart_Failure | dendritic_cell | *EMILIN2* | 2.862106719 | 3.98E-19 | 3.68E-18 | 2.50E-15 |
| Heart_Failure | dendritic_cell | *TRIM5* | -2.570676521 | 0.010292724 | 0.011754833 | 1 |
| Heart_Failure | dendritic_cell | *CNGA4* | 8.075049925 | 1.44E-18 | 1.06E-17 | 9.03E-15 |
| Heart_Failure | dendritic_cell | *TRIM22* | -4.81415948 | 1.11E-11 | 2.17E-11 | 6.99E-08 |
| Heart_Failure | dendritic_cell | *RRP8* | -3.167552632 | 0.006861629 | 0.007925604 | 1 |
| Heart_Failure | dendritic_cell | *IMMT* | -3.078605914 | 2.16E-05 | 2.80E-05 | 0.135975844 |
| Heart_Failure | dendritic_cell | *MRPL35* | -1.763197539 | 1.45E-05 | 1.89E-05 | 0.090874368 |
| Heart_Failure | dendritic_cell | *CARD6* | -5.589306626 | 3.51E-14 | 9.23E-14 | 2.20E-10 |
| Heart_Failure | dendritic_cell | *INPP5K* | -3.231069469 | 9.89E-17 | 4.29E-16 | 6.22E-13 |
| Heart_Failure | dendritic_cell | *RPA1* | -3.797208197 | 1.33E-11 | 2.58E-11 | 8.37E-08 |
| Heart_Failure | dendritic_cell | *SERPINF1* | -2.546752363 | 6.58E-19 | 5.46E-18 | 4.13E-15 |
| Heart_Failure | dendritic_cell | *TMEM128* | -4.669585479 | 1.88E-18 | 1.32E-17 | 1.18E-14 |
| Heart_Failure | dendritic_cell | *FIGNL1* | -5.906109076 | 6.95E-17 | 3.15E-16 | 4.37E-13 |
| Heart_Failure | dendritic_cell | *WBP2* | -6.349830645 | 4.75E-11 | 8.73E-11 | 2.99E-07 |
| Heart_Failure | dendritic_cell | *KDM6B* | 6.927415271 | 6.12E-10 | 1.03E-09 | 3.85E-06 |
| Heart_Failure | dendritic_cell | *XAF1* | -2.865143458 | 1.71E-08 | 2.60E-08 | 0.000107395 |
| Heart_Failure | dendritic_cell | *REEP2* | -2.829595344 | 1.70E-12 | 3.61E-12 | 1.07E-08 |
| Heart_Failure | dendritic_cell | *PRMT7* | -2.890562427 | 1.07E-09 | 1.77E-09 | 6.71E-06 |
| Heart_Failure | dendritic_cell | *TERF2* | -3.580270996 | 0.001366607 | 0.001631262 | 1 |
| Heart_Failure | dendritic_cell | *BTBD3* | -3.611710374 | 0.002750716 | 0.003240351 | 1 |
| Heart_Failure | dendritic_cell | *PCNA* | -5.854612707 | 4.50E-20 | 8.06E-19 | 2.83E-16 |
| Heart_Failure | dendritic_cell | *PTPRA* | -3.816295888 | 2.64E-14 | 7.11E-14 | 1.66E-10 |
| Heart_Failure | dendritic_cell | *MMACHC* | -3.252843486 | 9.29E-20 | 1.27E-18 | 5.84E-16 |
| Heart_Failure | dendritic_cell | *MUTYH* | -5.963643876 | 1.73E-15 | 5.73E-15 | 1.08E-11 |
| Heart_Failure | dendritic_cell | *LPIN3* | -2.732066215 | 8.76E-20 | 1.22E-18 | 5.51E-16 |
| Heart_Failure | dendritic_cell | *SERINC3* | -2.126532955 | 0.000196132 | 0.000243643 | 1 |
| Heart_Failure | dendritic_cell | *DMGDH* | -2.960648171 | 3.88E-20 | 7.39E-19 | 2.44E-16 |
| Heart_Failure | dendritic_cell | *BHMT2* | -1.15273983 | 8.81E-13 | 1.92E-12 | 5.54E-09 |
| Heart_Failure | dendritic_cell | *KANK4* | -1.38546015 | 1.29E-14 | 3.65E-14 | 8.10E-11 |
| Heart_Failure | dendritic_cell | *RSG1* | -4.933565283 | 1.01E-05 | 1.34E-05 | 0.063758118 |
| Heart_Failure | dendritic_cell | *CASP9* | -1.245582515 | 0.014682303 | 0.016650007 | 1 |
| Heart_Failure | dendritic_cell | *ATP8A2* | -4.64276109 | 1.29E-19 | 1.59E-18 | 8.11E-16 |
| Heart_Failure | dendritic_cell | *USPL1* | -4.946749706 | 1.88E-11 | 3.60E-11 | 1.18E-07 |

| | | | | | | |
|---|---|---|---|---|---|---|
| Heart_Failure | dendritic_cell | *POMP* | -5.620731279 | 1.59E-13 | 3.80E-13 | 1.00E-09 |
| Heart_Failure | dendritic_cell | *WASF3* | -2.328816463 | 1.29E-19 | 1.59E-18 | 8.14E-16 |
| Heart_Failure | dendritic_cell | *RNF17* | 2.842183169 | 2.92E-19 | 2.87E-18 | 1.84E-15 |
| Heart_Failure | dendritic_cell | *PEMT* | -4.095992093 | 5.50E-16 | 2.02E-15 | 3.46E-12 |
| Heart_Failure | dendritic_cell | *SCO1* | -5.347474504 | 4.39E-14 | 1.14E-13 | 2.76E-10 |
| Heart_Failure | dendritic_cell | *CHI3L1* | 0.577993924 | 0.024608902 | 0.027558989 | 1 |
| Heart_Failure | dendritic_cell | *DSTYK* | -4.912296835 | 6.14E-15 | 1.85E-14 | 3.86E-11 |
| Heart_Failure | dendritic_cell | *TMCC2* | 3.513679525 | 5.91E-11 | 1.08E-10 | 3.72E-07 |
| Heart_Failure | dendritic_cell | *COG6* | -3.346682858 | 2.36E-13 | 5.53E-13 | 1.49E-09 |
| Heart_Failure | dendritic_cell | *TRPC4* | -8.679916719 | 1.21E-16 | 5.13E-16 | 7.61E-13 |
| Heart_Failure | dendritic_cell | *RFC3* | -4.395843451 | 2.04E-14 | 5.57E-14 | 1.28E-10 |
| Heart_Failure | dendritic_cell | *BTBD2* | -5.845824696 | 6.31E-13 | 1.40E-12 | 3.97E-09 |
| Heart_Failure | dendritic_cell | *PRAM1* | 1.959619884 | 6.19E-19 | 5.19E-18 | 3.89E-15 |
| Heart_Failure | dendritic_cell | *ZNF414* | -8.997445802 | 8.61E-09 | 1.33E-08 | 5.41E-05 |
| Heart_Failure | dendritic_cell | *CNDP2* | 7.162945929 | 1.10E-10 | 1.96E-10 | 6.89E-07 |
| Heart_Failure | dendritic_cell | *RTN3* | -2.768171577 | 6.59E-18 | 3.89E-17 | 4.14E-14 |
| Heart_Failure | dendritic_cell | *GSTT2B* | -2.073959316 | 6.13E-09 | 9.60E-09 | 3.86E-05 |
| Heart_Failure | dendritic_cell | *SLC2A11* | -8.845636382 | 3.96E-10 | 6.75E-10 | 2.49E-06 |
| Heart_Failure | dendritic_cell | *C1QTNF6* | -3.421528154 | 1.15E-19 | 1.47E-18 | 7.23E-16 |
| Heart_Failure | dendritic_cell | *FAM83F* | -5.601834129 | 4.14E-20 | 7.75E-19 | 2.60E-16 |
| Heart_Failure | dendritic_cell | *GIMAP4* | 1.547417437 | 0.002445249 | 0.002886459 | 1 |
| Heart_Failure | dendritic_cell | *ACTR3B* | -4.410187289 | 1.26E-16 | 5.31E-16 | 7.92E-13 |
| Heart_Failure | dendritic_cell | *SFTPD* | -1.097571244 | 4.28E-07 | 6.05E-07 | 0.002693228 |
| Heart_Failure | dendritic_cell | *DYDC2* | -11.42711282 | 1.23E-15 | 4.21E-15 | 7.72E-12 |
| Heart_Failure | dendritic_cell | *TMTC1* | -3.442609649 | 1.09E-14 | 3.12E-14 | 6.83E-11 |
| Heart_Failure | dendritic_cell | *IPO8* | -4.343868092 | 3.15E-16 | 1.22E-15 | 1.98E-12 |
| Heart_Failure | dendritic_cell | *LARS* | -1.030423295 | 0.012178449 | 0.013860592 | 1 |
| Heart_Failure | dendritic_cell | *SPINK5* | -5.312761557 | 4.91E-07 | 6.91E-07 | 0.003089372 |
| Heart_Failure | dendritic_cell | *IMPA1* | -2.835029951 | 7.03E-09 | 1.10E-08 | 4.42E-05 |
| Heart_Failure | dendritic_cell | *LRRCC1* | 3.681702322 | 5.54E-09 | 8.70E-09 | 3.48E-05 |
| Heart_Failure | dendritic_cell | *SWAP70* | -4.556983238 | 3.54E-13 | 8.12E-13 | 2.23E-09 |
| Heart_Failure | dendritic_cell | *AMPD3* | 2.745734545 | 3.42E-15 | 1.08E-14 | 2.15E-11 |
| Heart_Failure | dendritic_cell | *SBF2* | -3.347127324 | 3.35E-13 | 7.71E-13 | 2.11E-09 |
| Heart_Failure | dendritic_cell | *MICAL2* | 2.675711672 | 2.20E-19 | 2.32E-18 | 1.38E-15 |
| Heart_Failure | dendritic_cell | *RRAS2* | -1.804175308 | 1.01E-11 | 1.97E-11 | 6.34E-08 |
| Heart_Failure | dendritic_cell | *HSD17B4* | -3.833083955 | 1.56E-11 | 2.99E-11 | 9.78E-08 |
| Heart_Failure | dendritic_cell | *DPF2* | -5.106701816 | 1.59E-09 | 2.61E-09 | 1.00E-05 |
| Heart_Failure | dendritic_cell | *GSC* | -11.10356768 | 1.61E-19 | 1.88E-18 | 1.02E-15 |
| Heart_Failure | dendritic_cell | *CATSPERB* | -2.185634479 | 3.16E-16 | 1.23E-15 | 1.99E-12 |
| Heart_Failure | dendritic_cell | *COX16* | 2.579834313 | 6.97E-16 | 2.51E-15 | 4.38E-12 |
| Heart_Failure | dendritic_cell | *MED6* | -3.604589469 | 3.78E-12 | 7.74E-12 | 2.38E-08 |
| Heart_Failure | dendritic_cell | *EIF2S1* | -3.625101077 | 0.002024644 | 0.002399875 | 1 |
| Heart_Failure | dendritic_cell | *ELP3* | -4.998494914 | 1.62E-10 | 2.84E-10 | 1.02E-06 |
| Heart_Failure | dendritic_cell | *ADAMDEC1* | 3.924971231 | 0.003413712 | 0.004001866 | 1 |
| Heart_Failure | dendritic_cell | *MRO* | -4.08896535 | 1.19E-14 | 3.39E-14 | 7.48E-11 |
| Heart_Failure | dendritic_cell | *THUMPD3* | -4.251770973 | 4.99E-07 | 7.01E-07 | 0.003136421 |
| Heart_Failure | dendritic_cell | *EDEM1* | 3.949188764 | 3.24E-19 | 3.08E-18 | 2.04E-15 |
| Heart_Failure | dendritic_cell | *CNTN6* | -2.668104005 | 1.25E-18 | 9.34E-18 | 7.87E-15 |
| Heart_Failure | dendritic_cell | *KATNBL1* | -4.251970125 | 7.86E-12 | 1.55E-11 | 4.94E-08 |
| Heart_Failure | dendritic_cell | *GSTM1* | 1.698485804 | 0.000151094 | 0.00018874 | 0.949927924 |

| | | | | | | |
|---|---|---|---|---|---|---|
| Heart_Failure | dendritic_cell | *GSTM5* | -2.172667586 | 3.75E-13 | 8.56E-13 | 2.36E-09 |
| Heart_Failure | dendritic_cell | *GSTM3* | -1.218538895 | 2.51E-07 | 3.59E-07 | 0.001576009 |
| Heart_Failure | dendritic_cell | *SYT6* | -7.841797926 | 4.43E-20 | 8.01E-19 | 2.79E-16 |
| Heart_Failure | dendritic_cell | *VAV3* | 2.303968001 | 5.03E-20 | 8.55E-19 | 3.16E-16 |
| Heart_Failure | dendritic_cell | *PSRC1* | -3.927647442 | 6.01E-10 | 1.01E-09 | 3.78E-06 |
| Heart_Failure | dendritic_cell | *CEPT1* | -3.843201086 | 8.45E-14 | 2.10E-13 | 5.31E-10 |
| Heart_Failure | dendritic_cell | *AP4B1* | -5.281217319 | 6.40E-16 | 2.33E-15 | 4.02E-12 |
| Heart_Failure | dendritic_cell | *NAPG* | -4.844589327 | 8.97E-15 | 2.62E-14 | 5.64E-11 |
| Heart_Failure | dendritic_cell | *SPIRE1* | -3.212087009 | 1.97E-19 | 2.15E-18 | 1.24E-15 |
| Heart_Failure | dendritic_cell | *PPHLN1* | -3.652262819 | 7.40E-09 | 1.15E-08 | 4.65E-05 |
| Heart_Failure | dendritic_cell | *FKBP11* | -3.128156434 | 0.002422322 | 0.002861006 | 1 |
| Heart_Failure | dendritic_cell | *YWHAQ* | -4.568633085 | 1.72E-07 | 2.48E-07 | 0.001083563 |
| Heart_Failure | dendritic_cell | *LPIN1* | -2.864536491 | 4.74E-12 | 9.60E-12 | 2.98E-08 |
| Heart_Failure | dendritic_cell | *LDHA* | -5.441427603 | 7.50E-10 | 1.25E-09 | 4.72E-06 |
| Heart_Failure | dendritic_cell | *ANO3* | -1.3959266 | 1.75E-19 | 2.01E-18 | 1.10E-15 |
| Heart_Failure | dendritic_cell | *FST* | -3.145580348 | 2.23E-20 | 6.44E-19 | 1.40E-16 |
| Heart_Failure | dendritic_cell | *TIMM17A* | -5.499895811 | 5.86E-11 | 1.07E-10 | 3.68E-07 |
| Heart_Failure | dendritic_cell | *NARS* | -5.482077896 | 3.66E-09 | 5.81E-09 | 2.30E-05 |
| Heart_Failure | dendritic_cell | *KIAA1468* | -3.781285062 | 9.05E-06 | 1.19E-05 | 0.056886034 |
| Heart_Failure | dendritic_cell | *FBXO18* | -4.260585871 | 6.58E-16 | 2.39E-15 | 4.14E-12 |
| Heart_Failure | dendritic_cell | *ECHDC3* | -2.092595189 | 4.03E-19 | 3.70E-18 | 2.53E-15 |
| Heart_Failure | dendritic_cell | *IL15RA* | -5.580813096 | 1.45E-14 | 4.07E-14 | 9.14E-11 |
| Heart_Failure | dendritic_cell | *TMEM241* | -3.982039174 | 3.93E-15 | 1.22E-14 | 2.47E-11 |
| Heart_Failure | dendritic_cell | *EMP1* | -2.767242475 | 4.66E-15 | 1.43E-14 | 2.93E-11 |
| Heart_Failure | dendritic_cell | *KLRD1* | -3.389797129 | 7.00E-18 | 4.09E-17 | 4.40E-14 |
| Heart_Failure | dendritic_cell | *PRH2* | 5.764917639 | 5.24E-08 | 7.78E-08 | 0.000329738 |
| Heart_Failure | dendritic_cell | *DDB2* | -4.278159435 | 6.16E-11 | 1.12E-10 | 3.88E-07 |
| Heart_Failure | dendritic_cell | *ACP2* | 4.499966739 | 3.67E-06 | 4.93E-06 | 0.023077593 |
| Heart_Failure | dendritic_cell | *HOOK1* | -6.578177435 | 9.99E-19 | 7.74E-18 | 6.28E-15 |
| Heart_Failure | dendritic_cell | *ZCCHC11* | -3.518184389 | 1.41E-15 | 4.76E-15 | 8.84E-12 |
| Heart_Failure | dendritic_cell | *PRPF38A* | -5.431597878 | 4.99E-16 | 1.85E-15 | 3.14E-12 |
| Heart_Failure | dendritic_cell | *FHOD3* | -1.781850547 | 1.26E-16 | 5.32E-16 | 7.93E-13 |
| Heart_Failure | dendritic_cell | *TPGS2* | -3.806389707 | 1.47E-05 | 1.92E-05 | 0.092185329 |
| Heart_Failure | dendritic_cell | *SLC43A3* | -4.656110762 | 4.60E-07 | 6.48E-07 | 0.002892375 |
| Heart_Failure | dendritic_cell | *TIMM10* | 5.244204908 | 2.63E-08 | 3.95E-08 | 0.00016513 |
| Heart_Failure | dendritic_cell | *DHX34* | 3.247874454 | 4.26E-06 | 5.70E-06 | 0.026800131 |
| Heart_Failure | dendritic_cell | *APLNR* | -4.50624391 | 1.78E-19 | 2.03E-18 | 1.12E-15 |
| Heart_Failure | dendritic_cell | *FADS2* | -5.683938402 | 1.03E-15 | 3.58E-15 | 6.46E-12 |
| Heart_Failure | dendritic_cell | *TMEM165* | -3.683456193 | 2.91E-10 | 5.03E-10 | 1.83E-06 |
| Heart_Failure | dendritic_cell | *CLOCK* | -4.269025391 | 2.03E-13 | 4.80E-13 | 1.28E-09 |
| Heart_Failure | dendritic_cell | *DZIP1* | -2.314304292 | 4.15E-16 | 1.56E-15 | 2.61E-12 |
| Heart_Failure | dendritic_cell | *BIVM* | -3.850784119 | 3.22E-19 | 3.06E-18 | 2.02E-15 |
| Heart_Failure | dendritic_cell | *ERCC5* | 10.35596919 | 1.22E-05 | 1.59E-05 | 0.076406154 |
| Heart_Failure | dendritic_cell | *CARS2* | -3.521731196 | 3.26E-15 | 1.03E-14 | 2.05E-11 |
| Heart_Failure | dendritic_cell | *STT3A* | -2.152231206 | 0.044354014 | 0.049093959 | 1 |
| Heart_Failure | dendritic_cell | *KLB* | -1.579207907 | 1.84E-16 | 7.47E-16 | 1.15E-12 |
| Heart_Failure | dendritic_cell | *WDR36* | -4.881799867 | 4.89E-07 | 6.88E-07 | 0.003073213 |
| Heart_Failure | dendritic_cell | *OSTF1* | -3.014387561 | 0.004769815 | 0.005551245 | 1 |
| Heart_Failure | dendritic_cell | *UBQLN1* | -4.527987052 | 9.67E-14 | 2.37E-13 | 6.08E-10 |
| Heart_Failure | dendritic_cell | *NAA35* | -4.592575063 | 5.38E-13 | 1.20E-12 | 3.38E-09 |

| | | | | | | |
|---|---|---|---|---|---|---|
| Heart_Failure | dendritic_cell | *CTSL* | -4.657758307 | 9.64E-14 | 2.36E-13 | 6.06E-10 |
| Heart_Failure | dendritic_cell | *TMEM2* | -3.292055371 | 7.13E-17 | 3.22E-16 | 4.49E-13 |
| Heart_Failure | dendritic_cell | *GOLM1* | -4.709951477 | 5.86E-19 | 5.00E-18 | 3.68E-15 |
| Heart_Failure | dendritic_cell | *ADAM19* | 1.779782649 | 4.19E-13 | 9.48E-13 | 2.63E-09 |
| Heart_Failure | dendritic_cell | *USP30* | -3.102043706 | 5.72E-12 | 1.15E-11 | 3.59E-08 |
| Heart_Failure | dendritic_cell | *HRK* | -9.731114089 | 1.88E-13 | 4.46E-13 | 1.18E-09 |
| Heart_Failure | dendritic_cell | *P2RX4* | 3.207471673 | 3.00E-11 | 5.61E-11 | 1.89E-07 |
| Heart_Failure | dendritic_cell | *DTX1* | -1.790697935 | 1.54E-17 | 8.28E-17 | 9.71E-14 |
| Heart_Failure | dendritic_cell | *DMTF1* | -3.249103214 | 6.78E-09 | 1.06E-08 | 4.26E-05 |
| Heart_Failure | dendritic_cell | *CCDC146* | -3.609876407 | 1.08E-16 | 4.63E-16 | 6.78E-13 |
| Heart_Failure | dendritic_cell | *TMEM60* | -5.118927941 | 1.99E-10 | 3.47E-10 | 1.25E-06 |
| Heart_Failure | dendritic_cell | *POM121C* | -6.007138695 | 2.78E-17 | 1.38E-16 | 1.74E-13 |
| Heart_Failure | dendritic_cell | *SRPK2* | -3.754859875 | 2.62E-11 | 4.93E-11 | 1.65E-07 |
| Heart_Failure | dendritic_cell | *TES* | -2.850170729 | 0.000243576 | 0.000300917 | 1 |
| Heart_Failure | dendritic_cell | *MDFIC* | -2.873391813 | 6.35E-14 | 1.61E-13 | 3.99E-10 |
| Heart_Failure | dendritic_cell | *MTO1* | -3.580457608 | 6.98E-15 | 2.08E-14 | 4.39E-11 |
| Heart_Failure | dendritic_cell | *ANKRD6* | -4.796855344 | 5.72E-14 | 1.46E-13 | 3.60E-10 |
| Heart_Failure | dendritic_cell | *KHDC1* | -2.27774613 | 5.56E-12 | 1.12E-11 | 3.49E-08 |
| Heart_Failure | dendritic_cell | *SNX14* | -4.015317411 | 3.54E-07 | 5.03E-07 | 0.002226742 |
| Heart_Failure | dendritic_cell | *NT5E* | -2.55433849 | 2.49E-18 | 1.69E-17 | 1.56E-14 |
| Heart_Failure | dendritic_cell | *EPHA7* | -0.895011268 | 3.90E-06 | 5.23E-06 | 0.024526476 |
| Heart_Failure | dendritic_cell | *PRR5L* | -3.677749919 | 2.48E-15 | 8.00E-15 | 1.56E-11 |
| Heart_Failure | dendritic_cell | *NAT10* | -4.567130208 | 1.42E-15 | 4.81E-15 | 8.94E-12 |
| Heart_Failure | dendritic_cell | *ATP5G2* | -5.704019142 | 1.45E-11 | 2.80E-11 | 9.13E-08 |
| Heart_Failure | dendritic_cell | *ITGA7* | -1.616627067 | 2.86E-17 | 1.42E-16 | 1.80E-13 |
| Heart_Failure | dendritic_cell | *RDH5* | -1.552528805 | 1.57E-19 | 1.85E-18 | 9.90E-16 |
| Heart_Failure | dendritic_cell | *BLOC1S1* | -5.425798196 | 4.06E-15 | 1.26E-14 | 2.55E-11 |
| Heart_Failure | dendritic_cell | *PPP1R1A* | -3.841604764 | 5.95E-19 | 5.06E-18 | 3.74E-15 |
| Heart_Failure | dendritic_cell | *B4GALNT1* | 6.521058869 | 2.42E-18 | 1.66E-17 | 1.52E-14 |
| Heart_Failure | dendritic_cell | *FAIM2* | -5.494052456 | 2.62E-20 | 6.44E-19 | 1.65E-16 |
| Heart_Failure | dendritic_cell | *KRT7* | -3.99401516 | 7.18E-05 | 9.09E-05 | 0.451295754 |
| Heart_Failure | dendritic_cell | *SLC26A10* | -9.2246273 | 5.85E-07 | 8.18E-07 | 0.003679142 |
| Heart_Failure | dendritic_cell | *MAP7* | -3.417815034 | 3.36E-16 | 1.30E-15 | 2.11E-12 |
| Heart_Failure | dendritic_cell | *AHI1* | -1.972089097 | 1.51E-12 | 3.22E-12 | 9.47E-09 |
| Heart_Failure | dendritic_cell | *PKIB* | 2.439967486 | 2.67E-20 | 6.44E-19 | 1.68E-16 |
| Heart_Failure | dendritic_cell | *SMPD2* | 3.872597048 | 1.12E-15 | 3.86E-15 | 7.01E-12 |
| Heart_Failure | dendritic_cell | *MICAL1* | 5.497132441 | 5.98E-16 | 2.19E-15 | 3.76E-12 |
| Heart_Failure | dendritic_cell | *PRADC1* | 10.05537949 | 1.74E-12 | 3.69E-12 | 1.09E-08 |
| Heart_Failure | dendritic_cell | *SMYD5* | -4.983736047 | 4.88E-16 | 1.81E-15 | 3.07E-12 |
| Heart_Failure | dendritic_cell | *KCNMB4* | 4.163194427 | 3.72E-05 | 4.76E-05 | 0.233872565 |
| Heart_Failure | dendritic_cell | *USP15* | -2.104967957 | 0.007717421 | 0.008886342 | 1 |
| Heart_Failure | dendritic_cell | *KLHL36* | -3.907675987 | 9.40E-17 | 4.09E-16 | 5.91E-13 |
| Heart_Failure | dendritic_cell | *MPHOSPH6* | 6.112611895 | 1.16E-15 | 3.99E-15 | 7.28E-12 |
| Heart_Failure | dendritic_cell | *FHOD1* | -5.399462701 | 2.10E-06 | 2.85E-06 | 0.01321328 |
| Heart_Failure | dendritic_cell | *CCDC102A* | -4.473977718 | 1.12E-09 | 1.84E-09 | 7.02E-06 |
| Heart_Failure | dendritic_cell | *ZNF670* | -2.338388524 | 2.13E-08 | 3.23E-08 | 0.000134208 |
| Heart_Failure | dendritic_cell | *KCNK1* | -0.458138175 | 0.008000508 | 0.009198829 | 1 |
| Heart_Failure | dendritic_cell | *NTPCR* | -4.745890506 | 2.87E-12 | 5.95E-12 | 1.80E-08 |
| Heart_Failure | dendritic_cell | *RNASEL* | 3.375800669 | 1.48E-05 | 1.93E-05 | 0.092765347 |
| Heart_Failure | dendritic_cell | *PIGC* | -3.576489684 | 1.15E-16 | 4.88E-16 | 7.21E-13 |

| | | | | | | |
|---|---|---|---|---|---|---|
| Heart_Failure | dendritic_cell | *USP37* | -3.732217875 | 9.55E-06 | 1.26E-05 | 0.060026341 |
| Heart_Failure | dendritic_cell | *SLC19A3* | -1.379345381 | 1.83E-14 | 5.03E-14 | 1.15E-10 |
| Heart_Failure | dendritic_cell | *TMBIM1* | -3.967748251 | 4.84E-13 | 1.09E-12 | 3.04E-09 |
| Heart_Failure | dendritic_cell | *ARMC9* | -5.61521208 | 1.96E-13 | 4.63E-13 | 1.23E-09 |
| Heart_Failure | dendritic_cell | *TMEM127* | 4.948965581 | 3.62E-05 | 4.63E-05 | 0.227512412 |
| Heart_Failure | dendritic_cell | *GCC2* | -3.560384108 | 6.38E-13 | 1.41E-12 | 4.01E-09 |
| Heart_Failure | dendritic_cell | *MRPS9* | -2.062673617 | 0.000642667 | 0.000778056 | 1 |
| Heart_Failure | dendritic_cell | *ANKRD36* | -4.490657894 | 8.70E-14 | 2.15E-13 | 5.47E-10 |
| Heart_Failure | dendritic_cell | *USP44* | -2.906493723 | 3.51E-10 | 6.01E-10 | 2.20E-06 |
| Heart_Failure | dendritic_cell | *APPL2* | -4.258015942 | 9.00E-12 | 1.77E-11 | 5.66E-08 |
| Heart_Failure | dendritic_cell | *DRAM1* | -2.638190207 | 0.000143499 | 0.000179288 | 0.902175682 |
| Heart_Failure | dendritic_cell | *SLC41A2* | 4.021669032 | 4.29E-17 | 2.05E-16 | 2.70E-13 |
| Heart_Failure | dendritic_cell | *NEK3* | -2.891781924 | 7.82E-12 | 1.54E-11 | 4.92E-08 |
| Heart_Failure | dendritic_cell | *VPS36* | -3.83952175 | 1.59E-14 | 4.43E-14 | 1.00E-10 |
| Heart_Failure | dendritic_cell | *RNASEH2B* | -2.715989649 | 0.002094204 | 0.002480923 | 1 |
| Heart_Failure | dendritic_cell | *CKAP2* | 1.43062293 | 6.92E-06 | 9.16E-06 | 0.043504666 |
| Heart_Failure | dendritic_cell | *TBC1D4* | -3.046252742 | 1.93E-16 | 7.86E-16 | 1.22E-12 |
| Heart_Failure | dendritic_cell | *LRCH1* | -4.376843979 | 9.10E-12 | 1.79E-11 | 5.72E-08 |
| Heart_Failure | dendritic_cell | *SUCLA2* | -2.85397304 | 1.52E-12 | 3.25E-12 | 9.58E-09 |
| Heart_Failure | dendritic_cell | *RCBTB1* | -2.883463776 | 6.62E-05 | 8.41E-05 | 0.416503465 |
| Heart_Failure | dendritic_cell | *MED4* | -4.017680612 | 1.63E-16 | 6.72E-16 | 1.03E-12 |
| Heart_Failure | dendritic_cell | *RCBTB2* | 3.875251116 | 0.00027352 | 0.00033685 | 1 |
| Heart_Failure | dendritic_cell | *C7orf25* | -1.236383714 | 0.008484004 | 0.009740491 | 1 |
| Heart_Failure | dendritic_cell | *SPDYE1* | -4.160586429 | 5.89E-18 | 3.53E-17 | 3.70E-14 |
| Heart_Failure | dendritic_cell | *CHST12* | -4.944682843 | 8.64E-15 | 2.53E-14 | 5.43E-11 |
| Heart_Failure | dendritic_cell | *GPNMB* | -3.795688478 | 9.32E-15 | 2.71E-14 | 5.86E-11 |
| Heart_Failure | dendritic_cell | *KDELR2* | 3.062571818 | 0.023843961 | 0.026721387 | 1 |
| Heart_Failure | dendritic_cell | *NUPL2* | -4.985436097 | 1.57E-11 | 3.02E-11 | 9.86E-08 |
| Heart_Failure | dendritic_cell | *ZDHHC4* | -3.60883283 | 7.40E-12 | 1.47E-11 | 4.65E-08 |
| Heart_Failure | dendritic_cell | *AOAH* | 1.366937828 | 2.21E-19 | 2.32E-18 | 1.39E-15 |
| Heart_Failure | dendritic_cell | *TBRG4* | -7.353783935 | 1.21E-10 | 2.15E-10 | 7.59E-07 |
| Heart_Failure | dendritic_cell | *DDX56* | -3.90424804 | 0.011426202 | 0.013018582 | 1 |
| Heart_Failure | dendritic_cell | *HUS1* | -3.762060642 | 3.18E-05 | 4.08E-05 | 0.199871338 |
| Heart_Failure | dendritic_cell | *NACAD* | -10.19778207 | 6.10E-19 | 5.14E-18 | 3.84E-15 |
| Heart_Failure | dendritic_cell | *DBNL* | -2.810702024 | 0.003396865 | 0.003984345 | 1 |
| Heart_Failure | dendritic_cell | *TTC5* | 5.004143772 | 3.66E-07 | 5.19E-07 | 0.002300703 |
| Heart_Failure | dendritic_cell | *MTHFS* | -1.369021307 | 3.03E-07 | 4.31E-07 | 0.001903679 |
| Heart_Failure | dendritic_cell | *TM6SF1* | 4.366540337 | 8.69E-17 | 3.83E-16 | 5.46E-13 |
| Heart_Failure | dendritic_cell | *CIB2* | 2.407526074 | 1.20E-05 | 1.58E-05 | 0.075748474 |
| Heart_Failure | dendritic_cell | *RSAD1* | -6.128401102 | 1.94E-13 | 4.59E-13 | 1.22E-09 |
| Heart_Failure | dendritic_cell | *NMT1* | -0.882897456 | 0.030339336 | 0.033843756 | 1 |
| Heart_Failure | dendritic_cell | *RTP4* | -4.96737204 | 1.37E-16 | 5.72E-16 | 8.60E-13 |
| Heart_Failure | dendritic_cell | *MRPL47* | -5.403270006 | 2.87E-05 | 3.69E-05 | 0.180232845 |
| Heart_Failure | dendritic_cell | *BLK* | -5.817407708 | 2.15E-22 | 1.50E-19 | 1.35E-18 |
| Heart_Failure | dendritic_cell | *EPRS* | -5.38456205 | 4.27E-10 | 7.25E-10 | 2.68E-06 |
| Heart_Failure | dendritic_cell | *RPS6KC1* | -2.971435454 | 0.000370909 | 0.000454474 | 1 |
| Heart_Failure | dendritic_cell | *CBWD2* | -5.753340937 | 1.96E-09 | 3.18E-09 | 1.23E-05 |
| Heart_Failure | dendritic_cell | *IL1RN* | -7.653558468 | 6.03E-10 | 1.02E-09 | 3.79E-06 |
| Heart_Failure | dendritic_cell | *SAP130* | -4.314716423 | 4.78E-15 | 1.47E-14 | 3.01E-11 |
| Heart_Failure | dendritic_cell | *UGGT1* | -4.551935222 | 2.38E-15 | 7.69E-15 | 1.49E-11 |

| | | | | | | |
|---|---|---|---|---|---|---|
| Heart_Failure | dendritic_cell | *GYPC* | -3.379615121 | 1.10E-15 | 3.81E-15 | 6.90E-12 |
| Heart_Failure | dendritic_cell | *NIPSNAP3A* | -3.715422275 | 5.65E-05 | 7.19E-05 | 0.355221182 |
| Heart_Failure | dendritic_cell | *LRRC8A* | -4.027594224 | 6.48E-13 | 1.43E-12 | 4.08E-09 |
| Heart_Failure | dendritic_cell | *TXN* | 6.152824389 | 3.77E-15 | 1.18E-14 | 2.37E-11 |
| Heart_Failure | dendritic_cell | *ODF2* | -6.228079392 | 7.28E-11 | 1.32E-10 | 4.58E-07 |
| Heart_Failure | dendritic_cell | *TOR1B* | -3.605314318 | 5.27E-14 | 1.35E-13 | 3.31E-10 |
| Heart_Failure | dendritic_cell | *TOR1A* | -3.851112998 | 2.75E-14 | 7.38E-14 | 1.73E-10 |
| Heart_Failure | dendritic_cell | *SLC2A8* | -4.283663996 | 4.77E-11 | 8.77E-11 | 3.00E-07 |
| Heart_Failure | dendritic_cell | *ZFP37* | -3.396317901 | 1.21E-18 | 9.07E-18 | 7.60E-15 |
| Heart_Failure | dendritic_cell | *ZNF189* | -4.307225664 | 2.10E-16 | 8.46E-16 | 1.32E-12 |
| Heart_Failure | dendritic_cell | *ALDOB* | -4.128939757 | 5.01E-15 | 1.53E-14 | 3.15E-11 |
| Heart_Failure | dendritic_cell | *STX17* | -4.337397552 | 1.29E-15 | 4.39E-15 | 8.10E-12 |
| Heart_Failure | dendritic_cell | *PRPF4* | -2.888606403 | 7.35E-07 | 1.02E-06 | 0.004620318 |
| Heart_Failure | dendritic_cell | *FPGS* | -4.018310034 | 7.79E-17 | 3.50E-16 | 4.90E-13 |
| Heart_Failure | dendritic_cell | *TEX10* | -5.099423183 | 8.15E-15 | 2.40E-14 | 5.12E-11 |
| Heart_Failure | dendritic_cell | *GARNL3* | -4.80162019 | 1.85E-14 | 5.09E-14 | 1.16E-10 |
| Heart_Failure | dendritic_cell | *GABBR2* | 3.15677918 | 6.80E-21 | 4.66E-19 | 4.27E-17 |
| Heart_Failure | dendritic_cell | *PSMB7* | -4.997727902 | 8.63E-13 | 1.88E-12 | 5.42E-09 |
| Heart_Failure | dendritic_cell | *RABEPK* | -5.38935064 | 1.07E-14 | 3.07E-14 | 6.70E-11 |
| Heart_Failure | dendritic_cell | *GOLGA1* | -4.267968324 | 5.72E-16 | 2.10E-15 | 3.60E-12 |
| Heart_Failure | dendritic_cell | *LMX1B* | -6.651088916 | 2.84E-20 | 6.44E-19 | 1.79E-16 |
| Heart_Failure | dendritic_cell | *DSCC1* | -0.465403164 | 0.022999605 | 0.025807339 | 1 |
| Heart_Failure | dendritic_cell | *DERL1* | -4.378241702 | 0.000205288 | 0.000254515 | 1 |
| Heart_Failure | dendritic_cell | *NOV* | -2.896396931 | 8.43E-20 | 1.19E-18 | 5.30E-16 |
| Heart_Failure | dendritic_cell | *POLR1E* | -2.674766905 | 2.23E-10 | 3.89E-10 | 1.41E-06 |
| Heart_Failure | dendritic_cell | *IL11RA* | -5.078229441 | 1.02E-14 | 2.93E-14 | 6.39E-11 |
| Heart_Failure | dendritic_cell | *APTX* | -3.805274724 | 5.19E-11 | 9.50E-11 | 3.26E-07 |
| Heart_Failure | dendritic_cell | *TLN1* | -4.122763312 | 1.74E-10 | 3.05E-10 | 1.09E-06 |
| Heart_Failure | dendritic_cell | *SPAG8* | -4.618301152 | 1.83E-17 | 9.59E-17 | 1.15E-13 |
| Heart_Failure | dendritic_cell | *CD72* | -5.858931634 | 4.31E-14 | 1.12E-13 | 2.71E-10 |
| Heart_Failure | dendritic_cell | *TMEM8B* | -5.70062996 | 9.74E-05 | 0.000122725 | 0.612397472 |
| Heart_Failure | dendritic_cell | *ARHGEF39* | -2.590377838 | 1.29E-09 | 2.12E-09 | 8.11E-06 |
| Heart_Failure | dendritic_cell | *DENND4C* | -3.691414311 | 2.01E-12 | 4.23E-12 | 1.26E-08 |
| Heart_Failure | dendritic_cell | *PPIL1* | 7.191542457 | 4.32E-20 | 7.89E-19 | 2.71E-16 |
| Heart_Failure | dendritic_cell | *KIF13A* | -3.165658432 | 5.00E-16 | 1.85E-15 | 3.14E-12 |
| Heart_Failure | dendritic_cell | *ZSCAN9* | 1.483048923 | 0.027740974 | 0.030994758 | 1 |
| Heart_Failure | dendritic_cell | *TMEM63B* | -3.357756612 | 2.92E-05 | 3.76E-05 | 0.183510522 |
| Heart_Failure | dendritic_cell | *FRS3* | 5.957145607 | 4.13E-19 | 3.75E-18 | 2.60E-15 |
| Heart_Failure | dendritic_cell | *SLC22A23* | -4.387768372 | 3.77E-14 | 9.88E-14 | 2.37E-10 |
| Heart_Failure | dendritic_cell | *LRRC1* | -3.211592323 | 6.74E-07 | 9.40E-07 | 0.004234478 |
| Heart_Failure | dendritic_cell | *FOXF2* | -5.22788926 | 1.74E-07 | 2.50E-07 | 0.001092784 |
| Heart_Failure | dendritic_cell | *BPHL* | -1.616956691 | 6.61E-14 | 1.67E-13 | 4.16E-10 |
| Heart_Failure | dendritic_cell | *RIPK1* | -4.050755951 | 4.28E-16 | 1.60E-15 | 2.69E-12 |
| Heart_Failure | dendritic_cell | *TUBB2B* | -2.970675512 | 1.60E-10 | 2.81E-10 | 1.00E-06 |
| Heart_Failure | dendritic_cell | *UQCC2* | -2.99178445 | 0.034642709 | 0.038562094 | 1 |
| Heart_Failure | dendritic_cell | *TCF19* | -5.191234575 | 7.70E-10 | 1.29E-09 | 4.84E-06 |
| Heart_Failure | dendritic_cell | *TPMT* | 3.003296526 | 2.44E-20 | 6.44E-19 | 1.53E-16 |
| Heart_Failure | dendritic_cell | *NRM* | 3.206226118 | 2.02E-16 | 8.17E-16 | 1.27E-12 |
| Heart_Failure | dendritic_cell | *VARS2* | -0.631652952 | 0.000967105 | 0.001160784 | 1 |
| Heart_Failure | dendritic_cell | *FAM8A1* | -3.381210897 | 1.67E-19 | 1.93E-18 | 1.05E-15 |

| | | | | | | |
|---|---|---|---|---|---|---|
| Heart_Failure | dendritic_cell | *FGFBP2* | -3.304426083 | 2.51E-18 | 1.69E-17 | 1.58E-14 |
| Heart_Failure | dendritic_cell | *FHDC1* | -8.448682349 | 3.59E-06 | 4.82E-06 | 0.022567874 |
| Heart_Failure | dendritic_cell | *CCDC90B* | -4.99707665 | 2.52E-16 | 9.95E-16 | 1.58E-12 |
| Heart_Failure | dendritic_cell | *SYTL2* | -3.69216325 | 9.05E-20 | 1.25E-18 | 5.69E-16 |
| Heart_Failure | dendritic_cell | *RAB30* | -3.694759257 | 4.32E-17 | 2.05E-16 | 2.72E-13 |
| Heart_Failure | dendritic_cell | *PRCP* | 4.859662756 | 1.32E-10 | 2.34E-10 | 8.29E-07 |
| Heart_Failure | dendritic_cell | *NARS2* | -3.894793019 | 8.28E-17 | 3.69E-16 | 5.20E-13 |
| Heart_Failure | dendritic_cell | *PI15* | -16.32712027 | 8.83E-10 | 1.47E-09 | 5.55E-06 |
| Heart_Failure | dendritic_cell | *TTPA* | -0.850769215 | 5.97E-06 | 7.93E-06 | 0.037531373 |
| Heart_Failure | dendritic_cell | *GGH* | -1.711120176 | 4.56E-12 | 9.25E-12 | 2.86E-08 |
| Heart_Failure | dendritic_cell | *SULF1* | -2.102292965 | 2.86E-18 | 1.88E-17 | 1.80E-14 |
| Heart_Failure | dendritic_cell | *TGS1* | -3.014108622 | 2.17E-11 | 4.12E-11 | 1.36E-07 |
| Heart_Failure | dendritic_cell | *NEK1* | -3.882798492 | 2.65E-12 | 5.52E-12 | 1.67E-08 |
| Heart_Failure | dendritic_cell | *DDX60* | -5.801287325 | 4.13E-11 | 7.65E-11 | 2.60E-07 |
| Heart_Failure | dendritic_cell | *MMP27* | -1.0854264 | 5.51E-06 | 7.33E-06 | 0.034636825 |
| Heart_Failure | dendritic_cell | *DCUN1D5* | -2.98521609 | 1.37E-12 | 2.93E-12 | 8.58E-09 |
| Heart_Failure | dendritic_cell | *TRIM29* | -3.315017477 | 3.47E-15 | 1.09E-14 | 2.18E-11 |
| Heart_Failure | dendritic_cell | *SLC37A4* | -6.385717444 | 1.67E-19 | 1.93E-18 | 1.05E-15 |
| Heart_Failure | dendritic_cell | *POU2F3* | -2.655829024 | 2.52E-07 | 3.61E-07 | 0.001587351 |
| Heart_Failure | dendritic_cell | *PPP2R1B* | -1.920255792 | 2.06E-19 | 2.21E-18 | 1.30E-15 |
| Heart_Failure | dendritic_cell | *FXYD6* | -5.537641234 | 3.52E-17 | 1.71E-16 | 2.22E-13 |
| Heart_Failure | dendritic_cell | *TMPRSS13* | 1.46391514 | 5.20E-21 | 4.66E-19 | 3.27E-17 |
| Heart_Failure | dendritic_cell | *CASP1* | 8.404964424 | 3.65E-16 | 1.39E-15 | 2.30E-12 |
| Heart_Failure | dendritic_cell | *MAP2K5* | -3.785117074 | 1.36E-12 | 2.91E-12 | 8.53E-09 |
| Heart_Failure | dendritic_cell | *NDUFAF1* | -4.367517555 | 1.87E-12 | 3.96E-12 | 1.18E-08 |
| Heart_Failure | dendritic_cell | *ITGA11* | -1.578815155 | 3.90E-20 | 7.41E-19 | 2.45E-16 |
| Heart_Failure | dendritic_cell | *PAQR5* | 3.862740728 | 0.008438899 | 0.009690476 | 1 |
| Heart_Failure | dendritic_cell | *RMDN3* | -3.162527894 | 5.33E-15 | 1.62E-14 | 3.35E-11 |
| Heart_Failure | dendritic_cell | *ITPKA* | -16.76033551 | 1.18E-12 | 2.55E-12 | 7.41E-09 |
| Heart_Failure | dendritic_cell | *UACA* | -3.40379595 | 1.92E-15 | 6.32E-15 | 1.21E-11 |
| Heart_Failure | dendritic_cell | *PLCB2* | 1.365612951 | 6.61E-15 | 1.98E-14 | 4.15E-11 |
| Heart_Failure | dendritic_cell | *RSL24D1* | -2.653383861 | 0.001754209 | 0.002086006 | 1 |
| Heart_Failure | dendritic_cell | *SPTBN5* | 1.124394935 | 0.00425416 | 0.004967664 | 1 |
| Heart_Failure | dendritic_cell | *RABGGTB* | 5.616510562 | 7.29E-07 | 1.01E-06 | 0.004585366 |
| Heart_Failure | dendritic_cell | *SLC44A5* | -3.851605057 | 1.03E-12 | 2.24E-12 | 6.50E-09 |
| Heart_Failure | dendritic_cell | *DNASE2B* | 7.284926858 | 6.70E-21 | 4.66E-19 | 4.21E-17 |
| Heart_Failure | dendritic_cell | *DBT* | -4.195760752 | 5.08E-14 | 1.30E-13 | 3.19E-10 |
| Heart_Failure | dendritic_cell | *RTCA* | -3.596754629 | 2.49E-09 | 4.00E-09 | 1.56E-05 |
| Heart_Failure | dendritic_cell | *IFT172* | -4.436546883 | 6.61E-10 | 1.11E-09 | 4.16E-06 |
| Heart_Failure | dendritic_cell | *ADCY3* | -4.616292016 | 9.05E-17 | 3.97E-16 | 5.69E-13 |
| Heart_Failure | dendritic_cell | *DYNC2LI1* | -2.572741882 | 2.14E-13 | 5.04E-13 | 1.35E-09 |
| Heart_Failure | dendritic_cell | *LHCGR* | -2.595527353 | 3.60E-20 | 6.99E-19 | 2.26E-16 |
| Heart_Failure | dendritic_cell | *THUMPD2* | -4.355776724 | 2.88E-13 | 6.68E-13 | 1.81E-09 |
| Heart_Failure | dendritic_cell | *PREB* | -2.7061381 | 4.23E-14 | 1.10E-13 | 2.66E-10 |
| Heart_Failure | dendritic_cell | *ABCG5* | -3.277054354 | 3.13E-05 | 4.02E-05 | 0.196824448 |
| Heart_Failure | dendritic_cell | *ATRAID* | 3.137624374 | 0.004845217 | 0.005637957 | 1 |
| Heart_Failure | dendritic_cell | *CENPO* | 10.31422266 | 9.59E-14 | 2.36E-13 | 6.03E-10 |
| Heart_Failure | dendritic_cell | *LRPPRC* | -4.432147133 | 5.52E-12 | 1.11E-11 | 3.47E-08 |
| Heart_Failure | dendritic_cell | *DTNB* | -3.19144407 | 1.70E-06 | 2.32E-06 | 0.010701655 |
| Heart_Failure | dendritic_cell | *CYP2C9* | 2.338403011 | 2.58E-10 | 4.46E-10 | 1.62E-06 |

| | | | | | | |
|---|---|---|---|---|---|---|
| Heart_Failure | dendritic_cell | *MYOF* | -2.80585331 | 6.42E-12 | 1.28E-11 | 4.04E-08 |
| Heart_Failure | dendritic_cell | *STAMBPL1* | -3.193484007 | 2.40E-15 | 7.77E-15 | 1.51E-11 |
| Heart_Failure | dendritic_cell | *CH25H* | -1.646311981 | 0.000169002 | 0.000210692 | 1 |
| Heart_Failure | dendritic_cell | *ATAD1* | -4.369968706 | 4.19E-16 | 1.58E-15 | 2.63E-12 |
| Heart_Failure | dendritic_cell | *BTBD16* | -2.73139336 | 2.23E-16 | 8.91E-16 | 1.40E-12 |
| Heart_Failure | dendritic_cell | *TACC2* | -3.000242939 | 8.22E-15 | 2.42E-14 | 5.17E-11 |
| Heart_Failure | dendritic_cell | *CALHM2* | -2.712765976 | 0.000879807 | 0.001059037 | 1 |
| Heart_Failure | dendritic_cell | *ARL3* | -4.352185379 | 6.45E-10 | 1.08E-09 | 4.06E-06 |
| Heart_Failure | dendritic_cell | *PLCE1* | -3.816491143 | 1.37E-13 | 3.32E-13 | 8.64E-10 |
| Heart_Failure | dendritic_cell | *RBP4* | -4.394527929 | 5.54E-16 | 2.04E-15 | 3.48E-12 |
| Heart_Failure | dendritic_cell | *DNAJC13* | -4.160622224 | 8.62E-07 | 1.19E-06 | 0.005417391 |
| Heart_Failure | dendritic_cell | *FAM149B1* | -3.711069996 | 2.32E-12 | 4.85E-12 | 1.46E-08 |
| Heart_Failure | dendritic_cell | *ASCC1* | -3.539973844 | 1.74E-15 | 5.76E-15 | 1.09E-11 |
| Heart_Failure | dendritic_cell | *DNA2* | -1.202428499 | 3.31E-09 | 5.29E-09 | 2.08E-05 |
| Heart_Failure | dendritic_cell | *ATIC* | -2.905639725 | 3.49E-12 | 7.18E-12 | 2.20E-08 |
| Heart_Failure | dendritic_cell | *SMARCAL1* | -3.764890388 | 4.75E-18 | 2.91E-17 | 2.99E-14 |
| Heart_Failure | dendritic_cell | *BARD1* | 6.595123413 | 3.56E-10 | 6.10E-10 | 2.24E-06 |
| Heart_Failure | dendritic_cell | *STAT4* | -1.99054947 | 1.23E-18 | 9.18E-18 | 7.70E-15 |
| Heart_Failure | dendritic_cell | *CARF* | -2.347656446 | 6.28E-06 | 8.33E-06 | 0.039460572 |
| Heart_Failure | dendritic_cell | *ASNSD1* | -4.157148663 | 1.35E-09 | 2.22E-09 | 8.51E-06 |
| Heart_Failure | dendritic_cell | *METTL5* | -5.957675979 | 1.49E-07 | 2.15E-07 | 0.000936827 |
| Heart_Failure | dendritic_cell | *NAB1* | -5.136267319 | 8.25E-13 | 1.81E-12 | 5.19E-09 |
| Heart_Failure | dendritic_cell | *FASTKD1* | -3.256123178 | 0.00036438 | 0.000446657 | 1 |
| Heart_Failure | dendritic_cell | *MDH1B* | -7.613497909 | 3.30E-20 | 6.76E-19 | 2.08E-16 |
| Heart_Failure | dendritic_cell | *FAM117B* | 4.980930666 | 7.59E-16 | 2.72E-15 | 4.77E-12 |
| Heart_Failure | dendritic_cell | *ITGAV* | -3.678845176 | 1.49E-08 | 2.28E-08 | 9.39E-05 |
| Heart_Failure | dendritic_cell | *SLC35A5* | -1.080787022 | 0.009253978 | 0.010603201 | 1 |
| Heart_Failure | dendritic_cell | *SENP7* | -3.991016588 | 4.58E-14 | 1.18E-13 | 2.88E-10 |
| Heart_Failure | dendritic_cell | *COX17* | 3.753831506 | 0.009400705 | 0.010769357 | 1 |
| Heart_Failure | dendritic_cell | *USP8* | -3.736674091 | 1.61E-08 | 2.45E-08 | 0.00010105 |
| Heart_Failure | dendritic_cell | *SPPL2A* | -4.308900552 | 3.81E-09 | 6.05E-09 | 2.40E-05 |
| Heart_Failure | dendritic_cell | *PPCDC* | -3.269280852 | 2.53E-12 | 5.27E-12 | 1.59E-08 |
| Heart_Failure | dendritic_cell | *UBL7* | 4.517721952 | 3.80E-20 | 7.26E-19 | 2.39E-16 |
| Heart_Failure | dendritic_cell | *ARHGAP24* | -3.056197935 | 7.50E-15 | 2.22E-14 | 4.71E-11 |
| Heart_Failure | dendritic_cell | *FAM13A* | -3.071181907 | 1.32E-14 | 3.73E-14 | 8.29E-11 |
| Heart_Failure | dendritic_cell | *HERC3* | -4.397060738 | 6.79E-09 | 1.06E-08 | 4.27E-05 |
| Heart_Failure | dendritic_cell | *AP1AR* | -5.225898201 | 1.34E-12 | 2.88E-12 | 8.43E-09 |
| Heart_Failure | dendritic_cell | *PRKG2* | -2.654316266 | 1.23E-19 | 1.54E-18 | 7.76E-16 |
| Heart_Failure | dendritic_cell | *BBS7* | -5.879345828 | 4.76E-12 | 9.63E-12 | 2.99E-08 |
| Heart_Failure | dendritic_cell | *KIAA1109* | -3.82862221 | 3.16E-12 | 6.52E-12 | 1.99E-08 |
| Heart_Failure | dendritic_cell | *MMRN1* | -1.06165136 | 2.05E-10 | 3.57E-10 | 1.29E-06 |
| Heart_Failure | dendritic_cell | *PRDM5* | -3.628216796 | 3.55E-17 | 1.71E-16 | 2.23E-13 |
| Heart_Failure | dendritic_cell | *TRPC3* | -6.829834857 | 2.09E-13 | 4.92E-13 | 1.31E-09 |
| Heart_Failure | dendritic_cell | *NAAA* | 4.567439124 | 1.59E-10 | 2.80E-10 | 1.00E-06 |
| Heart_Failure | dendritic_cell | *NUP54* | -3.169624977 | 4.10E-16 | 1.55E-15 | 2.57E-12 |
| Heart_Failure | dendritic_cell | *BMP2K* | 3.274009983 | 1.12E-14 | 3.20E-14 | 7.02E-11 |
| Heart_Failure | dendritic_cell | *G3BP2* | -3.455556628 | 7.30E-05 | 9.24E-05 | 0.459069231 |
| Heart_Failure | dendritic_cell | *SCARB2* | -4.227696484 | 2.53E-14 | 6.84E-14 | 1.59E-10 |
| Heart_Failure | dendritic_cell | *CNOT6L* | -3.278141074 | 2.21E-07 | 3.17E-07 | 0.00138684 |
| Heart_Failure | dendritic_cell | *ANXA3* | -2.630082698 | 4.87E-20 | 8.45E-19 | 3.06E-16 |

| | | | | | | |
|---|---|---|---|---|---|---|
| Heart_Failure | dendritic_cell | *PPA2* | -2.174043932 | 0.007991584 | 0.009190249 | 1 |
| Heart_Failure | dendritic_cell | *GSTCD* | -5.446105084 | 7.93E-11 | 1.43E-10 | 4.98E-07 |
| Heart_Failure | dendritic_cell | *ENPEP* | -2.686043017 | 6.68E-20 | 1.02E-18 | 4.20E-16 |
| Heart_Failure | dendritic_cell | *PAPSS1* | -3.197233916 | 2.32E-09 | 3.75E-09 | 1.46E-05 |
| Heart_Failure | dendritic_cell | *SLC39A8* | -2.009988462 | 0.00762161 | 0.008782453 | 1 |
| Heart_Failure | dendritic_cell | *FBN2* | 0.690961746 | 7.69E-05 | 9.71E-05 | 0.483192136 |
| Heart_Failure | dendritic_cell | *RNF185* | -3.975112525 | 1.48E-14 | 4.15E-14 | 9.31E-11 |
| Heart_Failure | dendritic_cell | *PARVG* | 1.562609094 | 6.55E-20 | 1.01E-18 | 4.12E-16 |
| Heart_Failure | dendritic_cell | *B4GALNT3* | -3.924825444 | 3.72E-17 | 1.79E-16 | 2.34E-13 |
| Heart_Failure | dendritic_cell | *ERP27* | 3.623931337 | 9.52E-19 | 7.47E-18 | 5.98E-15 |
| Heart_Failure | dendritic_cell | *ETV6* | 3.139401788 | 3.18E-18 | 2.05E-17 | 2.00E-14 |
| Heart_Failure | dendritic_cell | *KIF21A* | -2.160812344 | 1.33E-17 | 7.24E-17 | 8.33E-14 |
| Heart_Failure | dendritic_cell | *ALG10* | -8.133687203 | 6.38E-05 | 8.10E-05 | 0.400869399 |
| Heart_Failure | dendritic_cell | *SLCO1C1* | -5.100717932 | 2.85E-06 | 3.85E-06 | 0.017945455 |
| Heart_Failure | dendritic_cell | *ZCRB1* | -3.526991552 | 1.41E-07 | 2.03E-07 | 0.000885182 |
| Heart_Failure | dendritic_cell | *PRICKLE1* | -2.03485736 | 2.55E-19 | 2.57E-18 | 1.60E-15 |
| Heart_Failure | dendritic_cell | *C1RL* | -3.11816436 | 7.31E-15 | 2.17E-14 | 4.60E-11 |
| Heart_Failure | dendritic_cell | *CLSTN3* | -3.775093961 | 2.60E-18 | 1.74E-17 | 1.64E-14 |
| Heart_Failure | dendritic_cell | *KLRG1* | -2.604879104 | 1.82E-08 | 2.76E-08 | 0.000114181 |
| Heart_Failure | dendritic_cell | *TAPBPL* | -2.073921518 | 2.81E-06 | 3.79E-06 | 0.017652283 |
| Heart_Failure | dendritic_cell | *CD27* | -3.809038432 | 5.12E-18 | 3.11E-17 | 3.22E-14 |
| Heart_Failure | dendritic_cell | *PEX5* | -3.362057371 | 8.10E-17 | 3.62E-16 | 5.09E-13 |
| Heart_Failure | dendritic_cell | *PIANP* | 10.068383 | 1.36E-15 | 4.63E-15 | 8.56E-12 |
| Heart_Failure | dendritic_cell | *PPFIA2* | -2.454501631 | 3.26E-14 | 8.66E-14 | 2.05E-10 |
| Heart_Failure | dendritic_cell | *GLIPR1* | 2.345143421 | 1.82E-19 | 2.05E-18 | 1.14E-15 |
| Heart_Failure | dendritic_cell | *POC1B* | -5.528117458 | 6.92E-12 | 1.38E-11 | 4.35E-08 |
| Heart_Failure | dendritic_cell | *SNRPF* | -3.627645298 | 4.08E-07 | 5.77E-07 | 0.002565566 |
| Heart_Failure | dendritic_cell | *GAS2L3* | 7.181695831 | 1.59E-11 | 3.05E-11 | 9.98E-08 |
| Heart_Failure | dendritic_cell | *TMEM132B* | -3.421007167 | 1.75E-19 | 2.01E-18 | 1.10E-15 |
| Heart_Failure | dendritic_cell | *TDG* | -4.710922357 | 1.69E-09 | 2.76E-09 | 1.06E-05 |
| Heart_Failure | dendritic_cell | *RITA1* | -1.202084491 | 0.019334064 | 0.021815014 | 1 |
| Heart_Failure | dendritic_cell | *MMAB* | -4.474862061 | 7.17E-11 | 1.30E-10 | 4.50E-07 |
| Heart_Failure | dendritic_cell | *GLTP* | 5.890108558 | 4.58E-08 | 6.80E-08 | 0.000287668 |
| Heart_Failure | dendritic_cell | *MTMR6* | -3.072959604 | 0.001006135 | 0.00120602 | 1 |
| Heart_Failure | dendritic_cell | *SLC46A3* | 1.380396161 | 0.040717797 | 0.04514864 | 1 |
| Heart_Failure | dendritic_cell | *CCDC65* | -1.711752852 | 0.0362695 | 0.040301581 | 1 |
| Heart_Failure | dendritic_cell | *ACVRL1* | -2.584576021 | 2.18E-19 | 2.30E-18 | 1.37E-15 |
| Heart_Failure | dendritic_cell | *GPR84* | 7.166630257 | 8.85E-20 | 1.23E-18 | 5.56E-16 |
| Heart_Failure | dendritic_cell | *BRCA2* | 2.791782345 | 5.76E-20 | 9.45E-19 | 3.62E-16 |
| Heart_Failure | dendritic_cell | *KANSL2* | -3.071043812 | 4.14E-08 | 6.16E-08 | 0.000260294 |
| Heart_Failure | dendritic_cell | *CERS5* | -4.925331618 | 1.52E-10 | 2.68E-10 | 9.55E-07 |
| Heart_Failure | dendritic_cell | *LMBR1L* | -5.518367783 | 4.22E-09 | 6.67E-09 | 2.65E-05 |
| Heart_Failure | dendritic_cell | *ZNF740* | -4.566229089 | 1.04E-14 | 3.01E-14 | 6.55E-11 |
| Heart_Failure | dendritic_cell | *WDFY2* | 3.463259004 | 1.28E-05 | 1.68E-05 | 0.080405358 |
| Heart_Failure | dendritic_cell | *HNRNPA1L2* | 5.674473624 | 1.16E-20 | 5.00E-19 | 7.29E-17 |
| Heart_Failure | dendritic_cell | *LPAR6* | -2.309999781 | 0.018577638 | 0.020972816 | 1 |
| Heart_Failure | dendritic_cell | *VPS33A* | -4.647292512 | 0.017324394 | 0.019579087 | 1 |
| Heart_Failure | dendritic_cell | *RHOF* | -2.865385099 | 1.01E-20 | 4.95E-19 | 6.35E-17 |
| Heart_Failure | dendritic_cell | *METTL21C* | -6.173380364 | 0.001448617 | 0.001727842 | 1 |
| Heart_Failure | dendritic_cell | *GRTP1* | -2.003303355 | 2.21E-11 | 4.19E-11 | 1.39E-07 |

| | | | | | | |
|---|---|---|---|---|---|---|
| Heart_Failure | dendritic_cell | *TSSK4* | -11.87722261 | 4.37E-18 | 2.70E-17 | 2.75E-14 |
| Heart_Failure | dendritic_cell | *TMX1* | -5.088387277 | 3.83E-16 | 1.46E-15 | 2.41E-12 |
| Heart_Failure | dendritic_cell | *RTN1* | 0.94831449 | 1.02E-20 | 4.95E-19 | 6.41E-17 |
| Heart_Failure | dendritic_cell | *C14orf37* | -3.658720721 | 9.24E-16 | 3.26E-15 | 5.81E-12 |
| Heart_Failure | dendritic_cell | *RAB15* | -2.789038024 | 7.61E-07 | 1.06E-06 | 0.00478642 |
| Heart_Failure | dendritic_cell | *STON2* | -4.420062438 | 7.26E-08 | 1.07E-07 | 0.000456728 |
| Heart_Failure | dendritic_cell | *EFCAB11* | 7.41460929 | 2.38E-13 | 5.56E-13 | 1.50E-09 |
| Heart_Failure | dendritic_cell | *AK7* | -4.336665332 | 3.27E-20 | 6.76E-19 | 2.06E-16 |
| Heart_Failure | dendritic_cell | *SLC24A4* | 0.718628076 | 2.87E-20 | 6.44E-19 | 1.80E-16 |
| Heart_Failure | dendritic_cell | *FBLN5* | -2.640762344 | 3.13E-19 | 3.00E-18 | 1.97E-15 |
| Heart_Failure | dendritic_cell | *C14orf79* | 9.838872885 | 3.35E-16 | 1.30E-15 | 2.11E-12 |
| Heart_Failure | dendritic_cell | *NIPA2* | -5.135578401 | 2.71E-07 | 3.87E-07 | 0.001701609 |
| Heart_Failure | dendritic_cell | *DUOXA1* | -3.829616207 | 3.48E-14 | 9.16E-14 | 2.19E-10 |
| Heart_Failure | dendritic_cell | *MFAP1* | -4.661174903 | 4.54E-10 | 7.70E-10 | 2.85E-06 |
| Heart_Failure | dendritic_cell | *TCF12* | -3.470547301 | 2.08E-10 | 3.62E-10 | 1.31E-06 |
| Heart_Failure | dendritic_cell | *SORD* | 7.624471799 | 1.59E-17 | 8.50E-17 | 1.00E-13 |
| Heart_Failure | dendritic_cell | *LYSMD2* | 4.411519425 | 4.39E-06 | 5.86E-06 | 0.027575248 |
| Heart_Failure | dendritic_cell | *SRP14* | -3.590922297 | 4.66E-05 | 5.95E-05 | 0.293280029 |
| Heart_Failure | dendritic_cell | *BAHD1* | -6.284458624 | 1.20E-10 | 2.14E-10 | 7.56E-07 |
| Heart_Failure | dendritic_cell | *DISP2* | -1.165059551 | 0.000277137 | 0.000341238 | 1 |
| Heart_Failure | dendritic_cell | *ANP32A* | -4.961224633 | 1.83E-13 | 4.34E-13 | 1.15E-09 |
| Heart_Failure | dendritic_cell | *COMMD4* | -6.906323826 | 4.41E-09 | 6.96E-09 | 2.77E-05 |
| Heart_Failure | dendritic_cell | *UBE2Q2* | -4.970025541 | 1.52E-11 | 2.92E-11 | 9.53E-08 |
| Heart_Failure | dendritic_cell | *PSTPIP1* | 3.358806651 | 1.19E-18 | 9.00E-18 | 7.49E-15 |
| Heart_Failure | dendritic_cell | *HMG20A* | -4.335725902 | 1.86E-09 | 3.02E-09 | 1.17E-05 |
| Heart_Failure | dendritic_cell | *SCAPER* | -3.332096021 | 3.69E-12 | 7.57E-12 | 2.32E-08 |
| Heart_Failure | dendritic_cell | *NEIL1* | -3.901158921 | 5.33E-18 | 3.22E-17 | 3.35E-14 |
| Heart_Failure | dendritic_cell | *MAN2C1* | -0.566581052 | 0.010106426 | 0.011548365 | 1 |
| Heart_Failure | dendritic_cell | *TPM1* | -3.320348847 | 4.52E-17 | 2.13E-16 | 2.84E-13 |
| Heart_Failure | dendritic_cell | *IGF1R* | -3.539530456 | 1.00E-15 | 3.50E-15 | 6.30E-12 |
| Heart_Failure | dendritic_cell | *CYP11A1* | -2.385524847 | 4.85E-13 | 1.09E-12 | 3.05E-09 |
| Heart_Failure | dendritic_cell | *BBS4* | -4.54369481 | 6.45E-16 | 2.34E-15 | 4.05E-12 |
| Heart_Failure | dendritic_cell | *ADAMTS17* | -2.599778523 | 1.65E-20 | 6.06E-19 | 1.04E-16 |
| Heart_Failure | dendritic_cell | *ULK3* | -6.498741588 | 6.17E-16 | 2.26E-15 | 3.88E-12 |
| Heart_Failure | dendritic_cell | *HAPLN3* | -5.434603961 | 1.62E-14 | 4.51E-14 | 1.02E-10 |
| Heart_Failure | dendritic_cell | *POLG* | -5.814326269 | 1.88E-10 | 3.29E-10 | 1.18E-06 |
| Heart_Failure | dendritic_cell | *WDR93* | -1.460940999 | 7.91E-10 | 1.32E-09 | 4.98E-06 |
| Heart_Failure | dendritic_cell | *TICRR* | 10.14025115 | 2.11E-11 | 4.02E-11 | 1.33E-07 |
| Heart_Failure | dendritic_cell | *MFGE8* | -5.126338866 | 1.71E-16 | 6.99E-16 | 1.07E-12 |
| Heart_Failure | dendritic_cell | *UNC45A* | -2.008870091 | 0.001722784 | 0.002049412 | 1 |
| Heart_Failure | dendritic_cell | *ST8SIA2* | -16.37118454 | 1.62E-18 | 1.17E-17 | 1.02E-14 |
| Heart_Failure | dendritic_cell | *IQGAP1* | -2.856975011 | 1.49E-05 | 1.95E-05 | 0.0938271 |
| Heart_Failure | dendritic_cell | *CRTC3* | -4.299989672 | 2.88E-09 | 4.61E-09 | 1.81E-05 |
| Heart_Failure | dendritic_cell | *SH3GL3* | -2.678868948 | 2.91E-08 | 4.36E-08 | 0.000182754 |
| Heart_Failure | dendritic_cell | *SEC11A* | -4.713621716 | 1.17E-12 | 2.54E-12 | 7.38E-09 |
| Heart_Failure | dendritic_cell | *PARN* | -3.63992155 | 2.60E-14 | 7.00E-14 | 1.63E-10 |
| Heart_Failure | dendritic_cell | *FTO* | -3.429183773 | 7.34E-15 | 2.18E-14 | 4.62E-11 |
| Heart_Failure | dendritic_cell | *TXNL4B* | -5.883386107 | 1.32E-15 | 4.49E-15 | 8.28E-12 |
| Heart_Failure | dendritic_cell | *MARVELD3* | -5.850581783 | 1.98E-18 | 1.39E-17 | 1.25E-14 |
| Heart_Failure | dendritic_cell | *KIFC3* | -4.016598096 | 2.36E-17 | 1.19E-16 | 1.49E-13 |

| | | | | | | |
|---|---|---|---|---|---|---|
| Heart_Failure | dendritic_cell | *ADAMTS18* | -2.794134335 | 3.64E-20 | 7.01E-19 | 2.29E-16 |
| Heart_Failure | dendritic_cell | *NUDT7* | -2.785156334 | 1.32E-17 | 7.24E-17 | 8.32E-14 |
| Heart_Failure | dendritic_cell | *GCSH* | -2.969364808 | 1.33E-17 | 7.24E-17 | 8.34E-14 |
| Heart_Failure | dendritic_cell | *CMTM3* | 6.422136163 | 5.64E-07 | 7.90E-07 | 0.003548036 |
| Heart_Failure | dendritic_cell | *MAP1LC3B* | -4.38240108 | 1.61E-06 | 2.20E-06 | 0.010123184 |
| Heart_Failure | dendritic_cell | *TLDC1* | -6.26664347 | 1.38E-13 | 3.34E-13 | 8.71E-10 |
| Heart_Failure | dendritic_cell | *NDUFB10* | -5.547010952 | 3.36E-16 | 1.30E-15 | 2.11E-12 |
| Heart_Failure | dendritic_cell | *TIGD7* | -3.461367931 | 4.35E-15 | 1.34E-14 | 2.74E-11 |
| Heart_Failure | dendritic_cell | *DEF8* | 4.471462973 | 8.91E-18 | 5.09E-17 | 5.60E-14 |
| Heart_Failure | dendritic_cell | *GALNS* | -5.577690494 | 2.17E-13 | 5.10E-13 | 1.37E-09 |
| Heart_Failure | dendritic_cell | *COPS3* | 6.928937788 | 2.45E-09 | 3.94E-09 | 1.54E-05 |
| Heart_Failure | dendritic_cell | *RANBP10* | -4.944536758 | 4.59E-10 | 7.78E-10 | 2.88E-06 |
| Heart_Failure | dendritic_cell | *PRPSAP2* | -4.394775564 | 1.42E-13 | 3.42E-13 | 8.94E-10 |
| Heart_Failure | dendritic_cell | *MYO19* | -4.357898869 | 8.01E-08 | 1.17E-07 | 0.000503855 |
| Heart_Failure | dendritic_cell | *DDX52* | -5.010127161 | 2.30E-14 | 6.25E-14 | 1.45E-10 |
| Heart_Failure | dendritic_cell | *RASL10B* | -6.597850404 | 2.54E-15 | 8.18E-15 | 1.60E-11 |
| Heart_Failure | dendritic_cell | *UNC45B* | 8.890564466 | 6.42E-16 | 2.33E-15 | 4.04E-12 |
| Heart_Failure | dendritic_cell | *TOM1L1* | -3.725735471 | 7.08E-17 | 3.20E-16 | 4.45E-13 |
| Heart_Failure | dendritic_cell | *VPS53* | -3.862324435 | 7.17E-10 | 1.20E-09 | 4.51E-06 |
| Heart_Failure | dendritic_cell | *SPATA22* | -2.027263254 | 1.46E-14 | 4.08E-14 | 9.15E-11 |
| Heart_Failure | dendritic_cell | *SGSM2* | -5.158066683 | 7.37E-12 | 1.46E-11 | 4.63E-08 |
| Heart_Failure | dendritic_cell | *NPEPPS* | -4.32576547 | 2.30E-11 | 4.36E-11 | 1.45E-07 |
| Heart_Failure | dendritic_cell | *LRRC46* | -4.334821851 | 1.86E-10 | 3.26E-10 | 1.17E-06 |
| Heart_Failure | dendritic_cell | *SCRN2* | -0.688515575 | 0.001066532 | 0.001276225 | 1 |
| Heart_Failure | dendritic_cell | *RHBDL3* | -4.562193052 | 4.60E-15 | 1.42E-14 | 2.89E-11 |
| Heart_Failure | dendritic_cell | *ABCA8* | -1.708410553 | 6.13E-19 | 5.15E-18 | 3.85E-15 |
| Heart_Failure | dendritic_cell | *G6PC3* | 8.203726739 | 2.57E-09 | 4.13E-09 | 1.62E-05 |
| Heart_Failure | dendritic_cell | *IMPA2* | 6.712020455 | 1.19E-14 | 3.40E-14 | 7.51E-11 |
| Heart_Failure | dendritic_cell | *GNAL* | -3.883938291 | 2.52E-17 | 1.26E-16 | 1.59E-13 |
| Heart_Failure | dendritic_cell | *RPRD1A* | -3.990908726 | 6.22E-09 | 9.73E-09 | 3.91E-05 |
| Heart_Failure | dendritic_cell | *C18orf21* | 3.380720322 | 9.99E-20 | 1.34E-18 | 6.28E-16 |
| Heart_Failure | dendritic_cell | *GALNT1* | -4.321254835 | 4.12E-09 | 6.52E-09 | 2.59E-05 |
| Heart_Failure | dendritic_cell | *ASXL3* | -4.229176595 | 4.05E-19 | 3.71E-18 | 2.55E-15 |
| Heart_Failure | dendritic_cell | *ESCO1* | -4.190593208 | 1.07E-06 | 1.47E-06 | 0.006731082 |
| Heart_Failure | dendritic_cell | *GREB1L* | -1.746808159 | 1.36E-19 | 1.65E-18 | 8.52E-16 |
| Heart_Failure | dendritic_cell | *C18orf8* | -3.60948486 | 6.81E-05 | 8.63E-05 | 0.427971638 |
| Heart_Failure | dendritic_cell | *NPC1* | -4.327860561 | 2.09E-14 | 5.70E-14 | 1.31E-10 |
| Heart_Failure | dendritic_cell | *SLC13A5* | -12.40809175 | 1.55E-09 | 2.53E-09 | 9.71E-06 |
| Heart_Failure | dendritic_cell | *CCDC40* | -3.051905131 | 1.41E-13 | 3.40E-13 | 8.87E-10 |
| Heart_Failure | dendritic_cell | *EIF4A3* | 3.940317766 | 4.13E-15 | 1.28E-14 | 2.60E-11 |
| Heart_Failure | dendritic_cell | *TBCD* | -3.986467367 | 5.72E-13 | 1.27E-12 | 3.59E-09 |
| Heart_Failure | dendritic_cell | *FN3KRP* | -1.079909729 | 0.00570785 | 0.006612356 | 1 |
| Heart_Failure | dendritic_cell | *NARF* | -4.232074239 | 1.92E-11 | 3.67E-11 | 1.21E-07 |
| Heart_Failure | dendritic_cell | *TRIM65* | -1.668059224 | 0.034851857 | 0.038781172 | 1 |
| Heart_Failure | dendritic_cell | *CBX8* | -4.819226324 | 3.94E-15 | 1.22E-14 | 2.48E-11 |
| Heart_Failure | dendritic_cell | *SECTM1* | 4.656186112 | 1.26E-19 | 1.56E-18 | 7.95E-16 |
| Heart_Failure | dendritic_cell | *RNF157* | -1.527734574 | 6.31E-09 | 9.87E-09 | 3.97E-05 |
| Heart_Failure | dendritic_cell | *WDR45B* | -4.019175064 | 1.10E-05 | 1.45E-05 | 0.069399095 |
| Heart_Failure | dendritic_cell | *RNF165* | -4.170335002 | 1.80E-18 | 1.27E-17 | 1.13E-14 |
| Heart_Failure | dendritic_cell | *MAPK4* | 2.838960667 | 8.56E-11 | 1.54E-10 | 5.38E-07 |

| | | | | | | |
|---|---|---|---|---|---|---|
| Heart_Failure | dendritic_cell | *SMAD4* | -4.322986801 | 1.28E-15 | 4.37E-15 | 8.03E-12 |
| Heart_Failure | dendritic_cell | *GRB7* | -4.24847269 | 1.05E-09 | 1.73E-09 | 6.58E-06 |
| Heart_Failure | dendritic_cell | *FKBP10* | -1.867500381 | 3.22E-13 | 7.43E-13 | 2.02E-09 |
| Heart_Failure | dendritic_cell | *NFIC* | -3.306422412 | 8.45E-14 | 2.10E-13 | 5.31E-10 |
| Heart_Failure | dendritic_cell | *TPGS1* | 5.913994995 | 4.46E-17 | 2.11E-16 | 2.80E-13 |
| Heart_Failure | dendritic_cell | *PRDM15* | 1.279668655 | 0.011824709 | 0.013465305 | 1 |
| Heart_Failure | dendritic_cell | *PFKL* | -3.51254042 | 1.63E-09 | 2.66E-09 | 1.03E-05 |
| Heart_Failure | dendritic_cell | *MVB12A* | -4.433807585 | 8.07E-10 | 1.35E-09 | 5.08E-06 |
| Heart_Failure | dendritic_cell | *DUS3L* | -2.947954977 | 3.80E-06 | 5.09E-06 | 0.023878835 |
| Heart_Failure | dendritic_cell | *TMEM91* | -3.536485675 | 5.96E-06 | 7.92E-06 | 0.037445021 |
| Heart_Failure | dendritic_cell | *SIRT3* | -3.387874128 | 1.96E-13 | 4.64E-13 | 1.23E-09 |
| Heart_Failure | dendritic_cell | *IFITM3* | -5.376800811 | 4.20E-14 | 1.09E-13 | 2.64E-10 |
| Heart_Failure | dendritic_cell | *COL6A1* | -2.258147835 | 1.53E-18 | 1.12E-17 | 9.65E-15 |
| Heart_Failure | dendritic_cell | *IFNAR1* | -3.366045186 | 6.69E-11 | 1.21E-10 | 4.21E-07 |
| Heart_Failure | dendritic_cell | *TMEM50B* | -1.034109259 | 0.006064358 | 0.007017599 | 1 |
| Heart_Failure | dendritic_cell | *URB1* | -2.548628194 | 5.72E-08 | 8.47E-08 | 0.000359563 |
| Heart_Failure | dendritic_cell | *SAE1* | -3.849577803 | 8.66E-08 | 1.27E-07 | 0.000544281 |
| Heart_Failure | dendritic_cell | *NTN5* | -5.118942265 | 1.17E-18 | 8.86E-18 | 7.34E-15 |
| Heart_Failure | dendritic_cell | *GEMIN7* | -7.261713824 | 2.72E-07 | 3.89E-07 | 0.001710262 |
| Heart_Failure | dendritic_cell | *WTIP* | -5.457156314 | 5.93E-13 | 1.32E-12 | 3.73E-09 |
| Heart_Failure | dendritic_cell | *MYO1F* | 2.859979492 | 1.64E-19 | 1.91E-18 | 1.03E-15 |
| Heart_Failure | dendritic_cell | *CACNG8* | -19.70973305 | 1.64E-14 | 4.55E-14 | 1.03E-10 |
| Heart_Failure | dendritic_cell | *FBN3* | 2.793519046 | 7.53E-05 | 9.53E-05 | 0.473679968 |
| Heart_Failure | dendritic_cell | *EVI5L* | -2.715254509 | 4.98E-20 | 8.51E-19 | 3.13E-16 |
| Heart_Failure | dendritic_cell | *RPS11* | 2.519106369 | 0.007672307 | 0.008837632 | 1 |
| Heart_Failure | dendritic_cell | *CTU1* | 5.748208793 | 4.19E-17 | 2.00E-16 | 2.64E-13 |
| Heart_Failure | dendritic_cell | *RCN3* | 5.936427894 | 1.08E-16 | 4.64E-16 | 6.80E-13 |
| Heart_Failure | dendritic_cell | *RERE* | -2.836816244 | 3.30E-10 | 5.67E-10 | 2.07E-06 |
| Heart_Failure | dendritic_cell | *PEX14* | -4.275210224 | 5.63E-13 | 1.25E-12 | 3.54E-09 |
| Heart_Failure | dendritic_cell | *SH3BGRL3* | -3.892502732 | 0.000874395 | 0.001052723 | 1 |
| Heart_Failure | dendritic_cell | *CNKSR1* | 1.222353069 | 1.20E-07 | 1.74E-07 | 0.000752135 |
| Heart_Failure | dendritic_cell | *IL22RA1* | -2.9602933 | 3.61E-06 | 4.84E-06 | 0.022666022 |
| Heart_Failure | dendritic_cell | *ZNF593* | -3.392609993 | 7.52E-07 | 1.04E-06 | 0.004724917 |
| Heart_Failure | dendritic_cell | *C1orf216* | -3.589105526 | 0.003766706 | 0.004409101 | 1 |
| Heart_Failure | dendritic_cell | *MAP3K6* | -5.128634077 | 7.98E-20 | 1.14E-18 | 5.02E-16 |
| Heart_Failure | dendritic_cell | *GPN2* | -5.095807732 | 0.000340606 | 0.000417913 | 1 |
| Heart_Failure | dendritic_cell | *NBPF3* | -1.57449578 | 1.67E-09 | 2.72E-09 | 1.05E-05 |
| Heart_Failure | dendritic_cell | *HSPG2* | -2.073164777 | 1.72E-13 | 4.10E-13 | 1.08E-09 |
| Heart_Failure | dendritic_cell | *ITGB3BP* | -2.718825624 | 1.29E-07 | 1.87E-07 | 0.000812553 |
| Heart_Failure | dendritic_cell | *SERBP1* | -3.68775973 | 0.000496522 | 0.000605085 | 1 |
| Heart_Failure | dendritic_cell | *PIGK* | -5.550393839 | 2.17E-13 | 5.10E-13 | 1.37E-09 |
| Heart_Failure | dendritic_cell | *RPS8* | -1.880098402 | 0.027489038 | 0.030724193 | 1 |
| Heart_Failure | dendritic_cell | *MOB3C* | -9.030028016 | 2.85E-09 | 4.56E-09 | 1.79E-05 |
| Heart_Failure | dendritic_cell | *CYP4B1* | -2.238888778 | 9.82E-12 | 1.92E-11 | 6.18E-08 |
| Heart_Failure | dendritic_cell | *SYPL2* | -1.798860938 | 1.97E-10 | 3.44E-10 | 1.24E-06 |
| Heart_Failure | dendritic_cell | *ZNF697* | -5.187194162 | 4.28E-15 | 1.32E-14 | 2.69E-11 |
| Heart_Failure | dendritic_cell | *CELSR2* | 3.590357307 | 6.16E-19 | 5.17E-18 | 3.88E-15 |
| Heart_Failure | dendritic_cell | *GPR161* | -7.243253888 | 4.37E-11 | 8.07E-11 | 2.75E-07 |
| Heart_Failure | dendritic_cell | *ATP1B1* | 2.263084837 | 6.36E-20 | 9.97E-19 | 4.00E-16 |
| Heart_Failure | dendritic_cell | *TIPRL* | -5.797880761 | 1.49E-09 | 2.44E-09 | 9.38E-06 |

| | | | | | | |
|---|---|---|---|---|---|---|
| Heart_Failure | dendritic_cell | *NME7* | -4.834880255 | 3.03E-15 | 9.63E-15 | 1.91E-11 |
| Heart_Failure | dendritic_cell | *CREG1* | -2.134000352 | 0.03183718 | 0.035495718 | 1 |
| Heart_Failure | dendritic_cell | *GPA33* | 3.549176194 | 3.46E-18 | 2.21E-17 | 2.17E-14 |
| Heart_Failure | dendritic_cell | *RXRG* | -7.504828704 | 2.19E-20 | 6.44E-19 | 1.38E-16 |
| Heart_Failure | dendritic_cell | *TMCO1* | -3.998220986 | 7.25E-17 | 3.27E-16 | 4.56E-13 |
| Heart_Failure | dendritic_cell | *XCL2* | 3.290824595 | 2.16E-15 | 7.06E-15 | 1.36E-11 |
| Heart_Failure | dendritic_cell | *MGST3* | -3.827510423 | 4.04E-15 | 1.25E-14 | 2.54E-11 |
| Heart_Failure | dendritic_cell | *ADCY10* | -5.996343747 | 3.09E-20 | 6.68E-19 | 1.94E-16 |
| Heart_Failure | dendritic_cell | *RFWD2* | -3.194107138 | 5.23E-06 | 6.96E-06 | 0.032870676 |
| Heart_Failure | dendritic_cell | *PPOX* | -3.400916615 | 5.44E-10 | 9.19E-10 | 3.42E-06 |
| Heart_Failure | dendritic_cell | *RGS5* | -3.488265099 | 1.36E-05 | 1.78E-05 | 0.085266454 |
| Heart_Failure | dendritic_cell | *PIGM* | -4.621291428 | 1.15E-17 | 6.39E-17 | 7.22E-14 |
| Heart_Failure | dendritic_cell | *TUFT1* | 2.320339379 | 2.81E-05 | 3.63E-05 | 0.176941772 |
| Heart_Failure | dendritic_cell | *TARS2* | -5.4243833 | 9.95E-11 | 1.78E-10 | 6.26E-07 |
| Heart_Failure | dendritic_cell | *SELENBP1* | -2.946877646 | 1.71E-18 | 1.23E-17 | 1.08E-14 |
| Heart_Failure | dendritic_cell | *CERS2* | -3.64554696 | 1.46E-17 | 7.89E-17 | 9.19E-14 |
| Heart_Failure | dendritic_cell | *ENSA* | -4.514022532 | 1.02E-07 | 1.49E-07 | 0.000640287 |
| Heart_Failure | dendritic_cell | *SEMA6C* | -7.019528066 | 2.20E-09 | 3.55E-09 | 1.38E-05 |
| Heart_Failure | dendritic_cell | *ARNT* | -3.975392999 | 9.61E-17 | 4.18E-16 | 6.04E-13 |
| Heart_Failure | dendritic_cell | *POGZ* | -3.43105531 | 4.43E-11 | 8.16E-11 | 2.78E-07 |
| Heart_Failure | dendritic_cell | *GABPB2* | -2.799134098 | 4.28E-12 | 8.72E-12 | 2.69E-08 |
| Heart_Failure | dendritic_cell | *IKBKE* | -7.392531317 | 2.97E-12 | 6.13E-12 | 1.86E-08 |
| Heart_Failure | dendritic_cell | *DYRK3* | -7.074546726 | 7.04E-11 | 1.27E-10 | 4.43E-07 |
| Heart_Failure | dendritic_cell | *EIF2D* | -4.160644172 | 5.13E-13 | 1.15E-12 | 3.22E-09 |
| Heart_Failure | dendritic_cell | *HHIPL2* | -5.509231404 | 1.84E-09 | 2.99E-09 | 1.15E-05 |
| Heart_Failure | dendritic_cell | *TP53BP2* | 5.423844199 | 8.31E-17 | 3.70E-16 | 5.23E-13 |
| Heart_Failure | dendritic_cell | *RAB13* | -7.0099155 | 4.06E-10 | 6.92E-10 | 2.56E-06 |
| Heart_Failure | dendritic_cell | *S100A8* | 8.531116228 | 1.47E-13 | 3.54E-13 | 9.26E-10 |
| Heart_Failure | dendritic_cell | *CREB3L4* | -2.889299905 | 1.52E-14 | 4.25E-14 | 9.55E-11 |
| Heart_Failure | dendritic_cell | *KCNN3* | -2.259789505 | 1.39E-19 | 1.67E-18 | 8.72E-16 |
| Heart_Failure | dendritic_cell | *GATAD2B* | -3.243603024 | 1.48E-06 | 2.02E-06 | 0.009286608 |
| Heart_Failure | dendritic_cell | *RIT1* | 6.426180637 | 3.16E-15 | 9.99E-15 | 1.99E-11 |
| Heart_Failure | dendritic_cell | *FLG* | -1.047938701 | 0.001005851 | 0.001205908 | 1 |
| Heart_Failure | dendritic_cell | *GALNT2* | -4.411428433 | 4.51E-14 | 1.17E-13 | 2.84E-10 |
| Heart_Failure | dendritic_cell | *SCCPDH* | -2.956427627 | 8.81E-19 | 6.99E-18 | 5.54E-15 |
| Heart_Failure | dendritic_cell | *CEP170* | 4.129510038 | 4.05E-07 | 5.72E-07 | 0.002545185 |
| Heart_Failure | dendritic_cell | *SNAP47* | -3.236913751 | 1.27E-18 | 9.46E-18 | 8.00E-15 |
| Heart_Failure | dendritic_cell | *NVL* | -4.526018926 | 1.30E-14 | 3.68E-14 | 8.18E-11 |
| Heart_Failure | dendritic_cell | *SDE2* | -7.214048033 | 1.60E-15 | 5.36E-15 | 1.01E-11 |
| Heart_Failure | dendritic_cell | *FBXO28* | -4.87475488 | 1.65E-09 | 2.69E-09 | 1.04E-05 |
| Heart_Failure | dendritic_cell | *CNIH4* | -5.507536535 | 2.17E-13 | 5.10E-13 | 1.37E-09 |
| Heart_Failure | dendritic_cell | *GUK1* | -5.060093309 | 6.28E-17 | 2.87E-16 | 3.95E-13 |
| Heart_Failure | dendritic_cell | *CDC42BPA* | -3.187841495 | 2.65E-16 | 1.04E-15 | 1.66E-12 |
| Heart_Failure | dendritic_cell | *CNIH3* | -4.270722359 | 8.10E-15 | 2.39E-14 | 5.10E-11 |
| Heart_Failure | dendritic_cell | *PSEN2* | -5.801235544 | 1.48E-11 | 2.85E-11 | 9.30E-08 |
| Heart_Failure | dendritic_cell | *WNT9A* | -2.99107024 | 3.85E-18 | 2.42E-17 | 2.42E-14 |
| Heart_Failure | dendritic_cell | *PLEKHA6* | -2.585598694 | 1.07E-18 | 8.20E-18 | 6.72E-15 |
| Heart_Failure | dendritic_cell | *ABCG8* | 8.784781751 | 1.07E-18 | 8.23E-18 | 6.75E-15 |
| Heart_Failure | dendritic_cell | *EML4* | -4.118568541 | 3.69E-10 | 6.31E-10 | 2.32E-06 |
| Heart_Failure | dendritic_cell | *CHAC2* | -8.838807907 | 7.54E-15 | 2.23E-14 | 4.74E-11 |

| | | | | | | |
|---|---|---|---|---|---|---|
| Heart_Failure | dendritic_cell | *WDPCP* | -1.302745543 | 5.50E-14 | 1.41E-13 | 3.46E-10 |
| Heart_Failure | dendritic_cell | *ASXL2* | -4.375494192 | 3.83E-10 | 6.53E-10 | 2.41E-06 |
| Heart_Failure | dendritic_cell | *CIAO1* | -4.433202497 | 8.42E-09 | 1.31E-08 | 5.29E-05 |
| Heart_Failure | dendritic_cell | *ZNF514* | -1.101449333 | 0.000261993 | 0.000323097 | 1 |
| Heart_Failure | dendritic_cell | *EXOC6B* | -3.252105785 | 2.35E-14 | 6.38E-14 | 1.48E-10 |
| Heart_Failure | dendritic_cell | *TEX261* | -9.141045077 | 9.84E-10 | 1.63E-09 | 6.19E-06 |
| Heart_Failure | dendritic_cell | *THNSL2* | -0.941334642 | 1.70E-10 | 2.99E-10 | 1.07E-06 |
| Heart_Failure | dendritic_cell | *RALB* | -5.249643941 | 9.07E-14 | 2.24E-13 | 5.70E-10 |
| Heart_Failure | dendritic_cell | *TMEM177* | 3.839206663 | 3.49E-20 | 6.98E-19 | 2.19E-16 |
| Heart_Failure | dendritic_cell | *RABL2A* | -4.045471988 | 7.36E-19 | 6.01E-18 | 4.63E-15 |
| Heart_Failure | dendritic_cell | *FBLN7* | -2.822738146 | 4.25E-09 | 6.72E-09 | 2.67E-05 |
| Heart_Failure | dendritic_cell | *ZC3H8* | -3.679008631 | 1.57E-07 | 2.27E-07 | 0.000989081 |
| Heart_Failure | dendritic_cell | *LIPT1* | -3.330255336 | 6.50E-10 | 1.09E-09 | 4.08E-06 |
| Heart_Failure | dendritic_cell | *FAHD2B* | -4.852373941 | 3.40E-17 | 1.65E-16 | 2.14E-13 |
| Heart_Failure | dendritic_cell | *LYG1* | -2.083114097 | 2.85E-16 | 1.12E-15 | 1.79E-12 |
| Heart_Failure | dendritic_cell | *AFF3* | -2.093001917 | 1.82E-05 | 2.37E-05 | 0.114490312 |
| Heart_Failure | dendritic_cell | *UBXN4* | -4.179348389 | 7.56E-11 | 1.36E-10 | 4.75E-07 |
| Heart_Failure | dendritic_cell | *THSD7B* | -1.52714794 | 4.79E-18 | 2.94E-17 | 3.01E-14 |
| Heart_Failure | dendritic_cell | *GPR17* | -1.463547869 | 7.03E-10 | 1.18E-09 | 4.42E-06 |
| Heart_Failure | dendritic_cell | *ZNF385B* | -1.581241093 | 6.21E-20 | 9.79E-19 | 3.90E-16 |
| Heart_Failure | dendritic_cell | *TMEFF2* | -5.569238772 | 8.79E-09 | 1.36E-08 | 5.52E-05 |
| Heart_Failure | dendritic_cell | *PHOSPHO2* | -1.697632902 | 2.34E-07 | 3.35E-07 | 0.001469291 |
| Heart_Failure | dendritic_cell | *GULP1* | -3.380405566 | 2.31E-16 | 9.19E-16 | 1.45E-12 |
| Heart_Failure | dendritic_cell | *FAM171B* | -2.867580586 | 0.000128017 | 0.000160359 | 0.804843651 |
| Heart_Failure | dendritic_cell | *HSPD1* | -5.447070351 | 2.41E-12 | 5.03E-12 | 1.51E-08 |
| Heart_Failure | dendritic_cell | *PTH2R* | -3.306435886 | 2.80E-11 | 5.24E-11 | 1.76E-07 |
| Heart_Failure | dendritic_cell | *NBEAL1* | -1.836596302 | 1.60E-09 | 2.62E-09 | 1.01E-05 |
| Heart_Failure | dendritic_cell | *SUMF1* | -3.305998873 | 3.98E-08 | 5.93E-08 | 0.00025029 |
| Heart_Failure | dendritic_cell | *ESPNL* | -26.5986991 | 3.01E-17 | 1.48E-16 | 1.89E-13 |
| Heart_Failure | dendritic_cell | *FANCD2* | -7.671406348 | 5.67E-15 | 1.72E-14 | 3.57E-11 |
| Heart_Failure | dendritic_cell | *TAMM41* | -4.435647974 | 1.41E-14 | 3.96E-14 | 8.86E-11 |
| Heart_Failure | dendritic_cell | *OSBPL10* | -3.944451967 | 3.42E-15 | 1.08E-14 | 2.15E-11 |
| Heart_Failure | dendritic_cell | *FAM198A* | -3.069261148 | 1.23E-06 | 1.69E-06 | 0.007734632 |
| Heart_Failure | dendritic_cell | *CSRNP1* | -5.266494326 | 4.32E-15 | 1.33E-14 | 2.71E-11 |
| Heart_Failure | dendritic_cell | *SLC25A38* | 6.279673631 | 2.39E-05 | 3.09E-05 | 0.150401091 |
| Heart_Failure | dendritic_cell | *CTDSPL* | -4.179573869 | 1.58E-14 | 4.41E-14 | 9.96E-11 |
| Heart_Failure | dendritic_cell | *STAC* | -3.136711525 | 1.27E-19 | 1.57E-18 | 7.98E-16 |
| Heart_Failure | dendritic_cell | *ARL6IP5* | -4.018821952 | 3.56E-16 | 1.36E-15 | 2.24E-12 |
| Heart_Failure | dendritic_cell | *TMF1* | -4.790058358 | 1.24E-08 | 1.91E-08 | 7.82E-05 |
| Heart_Failure | dendritic_cell | *LIMD1* | -3.859375449 | 1.41E-05 | 1.84E-05 | 0.08865084 |
| Heart_Failure | dendritic_cell | *ZNF660* | -1.873210516 | 8.93E-06 | 1.18E-05 | 0.056125491 |
| Heart_Failure | dendritic_cell | *COL8A1* | -1.991466559 | 1.65E-18 | 1.19E-17 | 1.04E-14 |
| Heart_Failure | dendritic_cell | *NXPE3* | -6.386230163 | 6.81E-11 | 1.23E-10 | 4.28E-07 |
| Heart_Failure | dendritic_cell | *PLA1A* | 2.313906311 | 3.36E-16 | 1.30E-15 | 2.11E-12 |
| Heart_Failure | dendritic_cell | *RABL3* | -5.871374064 | 1.07E-11 | 2.09E-11 | 6.74E-08 |
| Heart_Failure | dendritic_cell | *ADPRH* | -1.874916238 | 0.000473611 | 0.000577612 | 1 |
| Heart_Failure | dendritic_cell | *IGSF11* | 4.84242174 | 3.92E-13 | 8.93E-13 | 2.46E-09 |
| Heart_Failure | dendritic_cell | *BOC* | -2.275624087 | 8.07E-19 | 6.52E-18 | 5.07E-15 |
| Heart_Failure | dendritic_cell | *AGTR1* | -1.96148136 | 2.00E-20 | 6.44E-19 | 1.26E-16 |
| Heart_Failure | dendritic_cell | *EIF2A* | -2.039835121 | 1.46E-08 | 2.22E-08 | 9.15E-05 |

| | | | | | | |
|---|---|---|---|---|---|---|
| Heart_Failure | dendritic_cell | *AMT* | -1.808630438 | 3.70E-16 | 1.41E-15 | 2.33E-12 |
| Heart_Failure | dendritic_cell | *TCTA* | -4.585799452 | 3.67E-17 | 1.77E-16 | 2.31E-13 |
| Heart_Failure | dendritic_cell | *ILDR1* | 3.149268701 | 8.32E-09 | 1.29E-08 | 5.23E-05 |
| Heart_Failure | dendritic_cell | *MUC4* | -1.372660285 | 9.14E-08 | 1.34E-07 | 0.000574912 |
| Heart_Failure | dendritic_cell | *SLC26A1* | 1.920031084 | 6.49E-06 | 8.61E-06 | 0.040802447 |
| Heart_Failure | dendritic_cell | *LYAR* | -7.650761783 | 9.16E-09 | 1.42E-08 | 5.76E-05 |
| Heart_Failure | dendritic_cell | *CENPC* | -3.164127188 | 1.33E-11 | 2.59E-11 | 8.38E-08 |
| Heart_Failure | dendritic_cell | *ATP10D* | 2.476737705 | 0.001098936 | 0.0013145 | 1 |
| Heart_Failure | dendritic_cell | *PYURF* | -6.165002965 | 1.69E-16 | 6.94E-16 | 1.06E-12 |
| Heart_Failure | dendritic_cell | *CISD2* | 7.192607989 | 3.56E-17 | 1.72E-16 | 2.24E-13 |
| Heart_Failure | dendritic_cell | *TIFA* | 3.880047324 | 7.61E-20 | 1.11E-18 | 4.79E-16 |
| Heart_Failure | dendritic_cell | *CCNA2* | -4.425628207 | 2.68E-20 | 6.44E-19 | 1.69E-16 |
| Heart_Failure | dendritic_cell | *USP53* | -2.870831769 | 9.14E-17 | 4.00E-16 | 5.75E-13 |
| Heart_Failure | dendritic_cell | *MARCH1* | 1.702676595 | 9.60E-19 | 7.49E-18 | 6.04E-15 |
| Heart_Failure | dendritic_cell | *PDGFC* | -3.084226606 | 2.04E-16 | 8.27E-16 | 1.29E-12 |
| Heart_Failure | dendritic_cell | *CBR4* | -4.014467454 | 5.43E-13 | 1.21E-12 | 3.41E-09 |
| Heart_Failure | dendritic_cell | *CYP4V2* | -1.898209315 | 1.35E-08 | 2.07E-08 | 8.51E-05 |
| Heart_Failure | dendritic_cell | *NDUFS6* | -5.8912337 | 5.43E-12 | 1.09E-11 | 3.42E-08 |
| Heart_Failure | dendritic_cell | *NKD2* | 11.81807977 | 2.16E-13 | 5.08E-13 | 1.36E-09 |
| Heart_Failure | dendritic_cell | *FAM105A* | 1.470056546 | 1.02E-19 | 1.36E-18 | 6.43E-16 |
| Heart_Failure | dendritic_cell | *LHFPL2* | -4.576791244 | 1.74E-06 | 2.37E-06 | 0.010918673 |
| Heart_Failure | dendritic_cell | *SSBP2* | -3.536441885 | 5.89E-11 | 1.07E-10 | 3.70E-07 |
| Heart_Failure | dendritic_cell | *BHMT* | -1.927906446 | 6.59E-19 | 5.46E-18 | 4.14E-15 |
| Heart_Failure | dendritic_cell | *CRHBP* | -4.549583886 | 1.77E-13 | 4.20E-13 | 1.11E-09 |
| Heart_Failure | dendritic_cell | *RASA1* | -3.126523921 | 3.94E-11 | 7.31E-11 | 2.48E-07 |
| Heart_Failure | dendritic_cell | *PPIP5K2* | -4.001691169 | 1.37E-09 | 2.26E-09 | 8.64E-06 |
| Heart_Failure | dendritic_cell | *GTF2H2* | -4.474005887 | 3.87E-13 | 8.83E-13 | 2.43E-09 |
| Heart_Failure | dendritic_cell | *SPATA9* | -1.829830231 | 9.23E-20 | 1.27E-18 | 5.80E-16 |
| Heart_Failure | dendritic_cell | *TSLP* | -1.448851586 | 0.000204377 | 0.000253436 | 1 |
| Heart_Failure | dendritic_cell | *COMMD10* | -3.340314455 | 9.58E-10 | 1.59E-09 | 6.02E-06 |
| Heart_Failure | dendritic_cell | *SLC25A48* | -1.618306564 | 0.00044008 | 0.000537971 | 1 |
| Heart_Failure | dendritic_cell | *GABRB2* | -2.162533798 | 3.24E-05 | 4.16E-05 | 0.203955501 |
| Heart_Failure | dendritic_cell | *PCYOX1L* | 2.880016782 | 1.32E-15 | 4.48E-15 | 8.27E-12 |
| Heart_Failure | dendritic_cell | *MYLK4* | -4.750179886 | 1.04E-17 | 5.83E-17 | 6.51E-14 |
| Heart_Failure | dendritic_cell | *TBC1D7* | -3.412365927 | 9.58E-19 | 7.49E-18 | 6.02E-15 |
| Heart_Failure | dendritic_cell | *GFOD1* | -2.213819119 | 2.39E-13 | 5.59E-13 | 1.50E-09 |
| Heart_Failure | dendritic_cell | *PSD2* | -12.55796519 | 3.67E-10 | 6.26E-10 | 2.30E-06 |
| Heart_Failure | dendritic_cell | *ZMAT2* | -3.746598845 | 2.45E-05 | 3.16E-05 | 0.153786321 |
| Heart_Failure | dendritic_cell | *KLHL3* | -3.51727364 | 5.37E-15 | 1.63E-14 | 3.37E-11 |
| Heart_Failure | dendritic_cell | *FAM193B* | -5.038896274 | 2.75E-10 | 4.76E-10 | 1.73E-06 |
| Heart_Failure | dendritic_cell | *MUT* | -5.66650524 | 5.89E-15 | 1.78E-14 | 3.71E-11 |
| Heart_Failure | dendritic_cell | *ABT1* | 3.861494211 | 7.48E-08 | 1.10E-07 | 0.000470506 |
| Heart_Failure | dendritic_cell | *MLIP* | 5.503361117 | 1.33E-10 | 2.35E-10 | 8.33E-07 |
| Heart_Failure | dendritic_cell | *FGD2* | 0.812720812 | 6.71E-21 | 4.66E-19 | 4.22E-17 |
| Heart_Failure | dendritic_cell | *ANO7* | -1.771455834 | 8.25E-07 | 1.14E-06 | 0.00518914 |
| Heart_Failure | dendritic_cell | *RPL7L1* | -3.851400354 | 1.12E-12 | 2.42E-12 | 7.03E-09 |
| Heart_Failure | dendritic_cell | *TPBG* | -4.063908889 | 6.23E-19 | 5.21E-18 | 3.91E-15 |
| Heart_Failure | dendritic_cell | *IRAK1BP1* | -4.004722595 | 1.96E-16 | 7.94E-16 | 1.23E-12 |
| Heart_Failure | dendritic_cell | *PHIP* | -3.676037746 | 9.00E-08 | 1.31E-07 | 0.000565538 |
| Heart_Failure | dendritic_cell | *MMS22L* | -3.809473573 | 1.19E-17 | 6.58E-17 | 7.48E-14 |

| | | | | | | |
|---|---|---|---|---|---|---|
| Heart_Failure | dendritic_cell | *PM20D2* | -7.099542189 | 2.07E-11 | 3.94E-11 | 1.30E-07 |
| Heart_Failure | dendritic_cell | *RNF217* | -4.436984724 | 1.35E-10 | 2.39E-10 | 8.49E-07 |
| Heart_Failure | dendritic_cell | *ABRACL* | 4.163512044 | 5.95E-08 | 8.80E-08 | 0.000374161 |
| Heart_Failure | dendritic_cell | *SLC18B1* | -0.916847338 | 0.001972563 | 0.002340348 | 1 |
| Heart_Failure | dendritic_cell | *AIG1* | -3.038333571 | 1.32E-12 | 2.84E-12 | 8.32E-09 |
| Heart_Failure | dendritic_cell | *DYNLT1* | -5.204888257 | 4.01E-12 | 8.17E-12 | 2.52E-08 |
| Heart_Failure | dendritic_cell | *PNLDC1* | -4.496023293 | 2.43E-09 | 3.92E-09 | 1.53E-05 |
| Heart_Failure | dendritic_cell | *VIP* | -1.739379072 | 0.00202243 | 0.002398155 | 1 |
| Heart_Failure | dendritic_cell | *SLC22A3* | -4.463487075 | 1.02E-16 | 4.40E-16 | 6.39E-13 |
| Heart_Failure | dendritic_cell | *C7orf50* | -4.852641916 | 7.10E-12 | 1.41E-11 | 4.47E-08 |
| Heart_Failure | dendritic_cell | *SDK1* | -1.631755317 | 2.63E-08 | 3.96E-08 | 0.000165496 |
| Heart_Failure | dendritic_cell | *CCZ1B* | -2.690584729 | 0.000182249 | 0.000226891 | 1 |
| Heart_Failure | dendritic_cell | *RBAK* | -6.842257352 | 1.01E-16 | 4.37E-16 | 6.35E-13 |
| Heart_Failure | dendritic_cell | *EGFR* | -2.146813996 | 4.68E-17 | 2.20E-16 | 2.94E-13 |
| Heart_Failure | dendritic_cell | *POMZP3* | -3.208807689 | 1.39E-07 | 2.01E-07 | 0.000876367 |
| Heart_Failure | dendritic_cell | *CCT6A* | -5.721434056 | 6.05E-10 | 1.02E-09 | 3.80E-06 |
| Heart_Failure | dendritic_cell | *PSPH* | -2.367551867 | 7.73E-05 | 9.77E-05 | 0.485968632 |
| Heart_Failure | dendritic_cell | *GIGYF1* | -4.523345026 | 3.55E-11 | 6.61E-11 | 2.23E-07 |
| Heart_Failure | dendritic_cell | *AGBL3* | -4.4706433 | 1.47E-13 | 3.52E-13 | 9.22E-10 |
| Heart_Failure | dendritic_cell | *EPHA1* | 4.856086297 | 3.47E-21 | 4.66E-19 | 2.18E-17 |
| Heart_Failure | dendritic_cell | *NCAPG2* | 2.667937734 | 0.007148943 | 0.008246863 | 1 |
| Heart_Failure | dendritic_cell | *FBXO25* | -5.586798909 | 8.41E-12 | 1.65E-11 | 5.29E-08 |
| Heart_Failure | dendritic_cell | *CSGALNACT1* | -3.547128879 | 3.71E-13 | 8.48E-13 | 2.33E-09 |
| Heart_Failure | dendritic_cell | *CCDC25* | -4.627537131 | 4.76E-13 | 1.07E-12 | 2.99E-09 |
| Heart_Failure | dendritic_cell | *BIN3* | -4.529894442 | 6.82E-16 | 2.46E-15 | 4.28E-12 |
| Heart_Failure | dendritic_cell | *SLC25A37* | -4.623138381 | 7.05E-15 | 2.10E-14 | 4.43E-11 |
| Heart_Failure | dendritic_cell | *CHMP7* | -3.228612994 | 3.61E-16 | 1.38E-15 | 2.27E-12 |
| Heart_Failure | dendritic_cell | *DOCK5* | 4.093309788 | 1.07E-17 | 6.00E-17 | 6.74E-14 |
| Heart_Failure | dendritic_cell | *ST18* | 0.574500065 | 8.31E-21 | 4.66E-19 | 5.23E-17 |
| Heart_Failure | dendritic_cell | *RGS20* | 1.38918656 | 0.038953112 | 0.043222417 | 1 |
| Heart_Failure | dendritic_cell | *TACC1* | -3.627132052 | 9.40E-15 | 2.73E-14 | 5.91E-11 |
| Heart_Failure | dendritic_cell | *ADHFE1* | -1.3314168 | 6.90E-13 | 1.52E-12 | 4.34E-09 |
| Heart_Failure | dendritic_cell | *SLC26A7* | -2.022410652 | 1.08E-07 | 1.57E-07 | 0.000678655 |
| Heart_Failure | dendritic_cell | *ATP6V0D2* | 3.524315297 | 5.59E-07 | 7.82E-07 | 0.003511817 |
| Heart_Failure | dendritic_cell | *MTDH* | -4.17004507 | 1.24E-09 | 2.04E-09 | 7.80E-06 |
| Heart_Failure | dendritic_cell | *MAL2* | -3.069886595 | 8.67E-17 | 3.83E-16 | 5.45E-13 |
| Heart_Failure | dendritic_cell | *UTP23* | -3.736052548 | 1.52E-18 | 1.11E-17 | 9.53E-15 |
| Heart_Failure | dendritic_cell | *FAM135B* | 13.29282181 | 2.45E-13 | 5.72E-13 | 1.54E-09 |
| Heart_Failure | dendritic_cell | *ZNF7* | -4.493431231 | 5.82E-14 | 1.49E-13 | 3.66E-10 |
| Heart_Failure | dendritic_cell | *SLC39A4* | -5.431994444 | 4.10E-12 | 8.35E-12 | 2.58E-08 |
| Heart_Failure | dendritic_cell | *AK3* | -4.206639216 | 1.45E-12 | 3.10E-12 | 9.13E-09 |
| Heart_Failure | dendritic_cell | *CER1* | -5.579665302 | 1.84E-11 | 3.53E-11 | 1.16E-07 |
| Heart_Failure | dendritic_cell | *PLIN2* | -4.356846689 | 1.00E-15 | 3.50E-15 | 6.30E-12 |
| Heart_Failure | dendritic_cell | *HAUS6* | -5.918697557 | 8.35E-08 | 1.22E-07 | 0.000525194 |
| Heart_Failure | dendritic_cell | *C9orf72* | 4.495521217 | 8.79E-17 | 3.87E-16 | 5.53E-13 |
| Heart_Failure | dendritic_cell | *SIGMAR1* | -3.291223223 | 2.98E-15 | 9.50E-15 | 1.87E-11 |
| Heart_Failure | dendritic_cell | *CBWD5* | -6.014765382 | 6.82E-16 | 2.46E-15 | 4.29E-12 |
| Heart_Failure | dendritic_cell | *ZNF462* | -3.613349781 | 0.000249302 | 0.00030788 | 1 |
| Heart_Failure | dendritic_cell | *ASTN2* | -3.709887799 | 1.54E-16 | 6.36E-16 | 9.65E-13 |
| Heart_Failure | dendritic_cell | *WDR31* | -4.460891033 | 1.79E-17 | 9.42E-17 | 1.12E-13 |

| | | | | | | |
|---|---|---|---|---|---|---|
| Heart_Failure | dendritic_cell | *SURF6* | -5.004100284 | 2.03E-17 | 1.05E-16 | 1.28E-13 |
| Heart_Failure | dendritic_cell | *ASB6* | 3.423418115 | 0.003603296 | 0.004221752 | 1 |
| Heart_Failure | dendritic_cell | *NTMT1* | 4.488883981 | 4.44E-09 | 7.00E-09 | 2.79E-05 |
| Heart_Failure | dendritic_cell | *CIZ1* | -4.186869085 | 5.40E-12 | 1.09E-11 | 3.40E-08 |
| Heart_Failure | dendritic_cell | *SH3GLB2* | -5.191666677 | 2.45E-15 | 7.92E-15 | 1.54E-11 |
| Heart_Failure | dendritic_cell | *LRSAM1* | -5.173633618 | 3.70E-11 | 6.88E-11 | 2.32E-07 |
| Heart_Failure | dendritic_cell | *INPP5E* | -5.434974571 | 6.78E-15 | 2.03E-14 | 4.26E-11 |
| Heart_Failure | dendritic_cell | *DPH7* | -5.359323223 | 3.47E-16 | 1.33E-15 | 2.18E-12 |
| Heart_Failure | dendritic_cell | *SLC39A12* | -4.868877298 | 2.52E-17 | 1.26E-16 | 1.59E-13 |
| Heart_Failure | dendritic_cell | *RSU1* | -4.094589064 | 5.43E-13 | 1.21E-12 | 3.41E-09 |
| Heart_Failure | dendritic_cell | *PARD3* | -2.611757645 | 3.91E-18 | 2.45E-17 | 2.46E-14 |
| Heart_Failure | dendritic_cell | *NRBF2* | -2.777580461 | 4.00E-10 | 6.81E-10 | 2.51E-06 |
| Heart_Failure | dendritic_cell | *ADIRF* | -3.769445072 | 3.91E-16 | 1.49E-15 | 2.46E-12 |
| Heart_Failure | dendritic_cell | *HTR7* | 0.626712439 | 6.22E-21 | 4.66E-19 | 3.91E-17 |
| Heart_Failure | dendritic_cell | *RPP30* | -3.360824304 | 0.001543732 | 0.001840243 | 1 |
| Heart_Failure | dendritic_cell | *FRA10AC1* | -3.189516661 | 1.25E-09 | 2.05E-09 | 7.83E-06 |
| Heart_Failure | dendritic_cell | *DNAJB12* | -5.654523407 | 5.17E-12 | 1.05E-11 | 3.25E-08 |
| Heart_Failure | dendritic_cell | *PLEKHS1* | -7.367015389 | 9.89E-07 | 1.36E-06 | 0.006216159 |
| Heart_Failure | dendritic_cell | *FUOM* | 6.782518542 | 3.45E-09 | 5.49E-09 | 2.17E-05 |
| Heart_Failure | dendritic_cell | *LRRC27* | -1.722502176 | 3.02E-05 | 3.88E-05 | 0.189565777 |
| Heart_Failure | dendritic_cell | *MTG1* | -1.997742264 | 1.35E-09 | 2.22E-09 | 8.47E-06 |
| Heart_Failure | dendritic_cell | *PAOX* | -9.284207086 | 1.08E-08 | 1.67E-08 | 6.80E-05 |
| Heart_Failure | dendritic_cell | *GSTO1* | 5.767607372 | 3.65E-09 | 5.81E-09 | 2.30E-05 |
| Heart_Failure | dendritic_cell | *PDCD11* | -4.940270416 | 1.66E-08 | 2.53E-08 | 0.000104351 |
| Heart_Failure | dendritic_cell | *RGS10* | 2.94887811 | 1.24E-20 | 5.10E-19 | 7.81E-17 |
| Heart_Failure | dendritic_cell | *GAS2* | -2.917595537 | 7.70E-20 | 1.12E-18 | 4.84E-16 |
| Heart_Failure | dendritic_cell | *LIN7C* | -5.798842116 | 4.97E-16 | 1.84E-15 | 3.12E-12 |
| Heart_Failure | dendritic_cell | *APIP* | -0.76709182 | 1.82E-05 | 2.36E-05 | 0.114186425 |
| Heart_Failure | dendritic_cell | *EIF3M* | -3.284301468 | 0.000459254 | 0.000560755 | 1 |
| Heart_Failure | dendritic_cell | *GLYAT* | -1.500932782 | 1.52E-18 | 1.11E-17 | 9.57E-15 |
| Heart_Failure | dendritic_cell | *PTPRJ* | 3.014982091 | 1.56E-17 | 8.35E-17 | 9.82E-14 |
| Heart_Failure | dendritic_cell | *C11orf49* | -3.611890595 | 1.07E-15 | 3.71E-15 | 6.71E-12 |
| Heart_Failure | dendritic_cell | *ENDOD1* | -7.415388202 | 7.04E-11 | 1.27E-10 | 4.43E-07 |
| Heart_Failure | dendritic_cell | *CCDC82* | -2.130603861 | 4.44E-07 | 6.27E-07 | 0.00279423 |
| Heart_Failure | dendritic_cell | *SERPINH1* | -4.004710716 | 1.92E-13 | 4.54E-13 | 1.20E-09 |
| Heart_Failure | dendritic_cell | *CAPN5* | -5.057342797 | 3.98E-16 | 1.51E-15 | 2.50E-12 |
| Heart_Failure | dendritic_cell | *INTS4* | -3.477326553 | 3.65E-11 | 6.80E-11 | 2.30E-07 |
| Heart_Failure | dendritic_cell | *RPS3* | -2.722021534 | 5.49E-05 | 6.99E-05 | 0.345337347 |
| Heart_Failure | dendritic_cell | *ZC3H12C* | -2.881159774 | 0.005941828 | 0.006878341 | 1 |
| Heart_Failure | dendritic_cell | *DRD2* | 3.666114529 | 1.31E-20 | 5.24E-19 | 8.22E-17 |
| Heart_Failure | dendritic_cell | *C11orf52* | -20.80440877 | 2.33E-20 | 6.44E-19 | 1.47E-16 |
| Heart_Failure | dendritic_cell | *GLB1L2* | -6.115987977 | 1.58E-06 | 2.15E-06 | 0.009905982 |
| Heart_Failure | dendritic_cell | *P4HA3* | -3.959989426 | 1.59E-12 | 3.39E-12 | 9.99E-09 |
| Heart_Failure | dendritic_cell | *GRIK4* | -2.452479418 | 2.24E-07 | 3.21E-07 | 0.001407178 |
| Heart_Failure | dendritic_cell | *ADAM33* | -2.068766312 | 3.92E-21 | 4.66E-19 | 2.47E-17 |
| Heart_Failure | dendritic_cell | *MTA2* | -4.45333906 | 9.08E-17 | 3.98E-16 | 5.71E-13 |
| Heart_Failure | dendritic_cell | *TMC2* | -4.504085276 | 3.32E-20 | 6.76E-19 | 2.09E-16 |
| Heart_Failure | dendritic_cell | *ROM1* | -0.855202479 | 0.012936444 | 0.014709969 | 1 |
| Heart_Failure | dendritic_cell | *EML3* | -5.443568935 | 6.53E-19 | 5.43E-18 | 4.10E-15 |
| Heart_Failure | dendritic_cell | *INCENP* | 5.607382869 | 8.10E-20 | 1.15E-18 | 5.09E-16 |

| | | | | | | |
|---|---|---|---|---|---|---|
| Heart_Failure | dendritic_cell | *MS4A3* | -53.39528389 | 2.40E-20 | 6.44E-19 | 1.51E-16 |
| Heart_Failure | dendritic_cell | *PLCH2* | -9.406736904 | 1.74E-20 | 6.17E-19 | 1.10E-16 |
| Heart_Failure | dendritic_cell | *CPSF7* | -5.201134718 | 2.11E-11 | 4.02E-11 | 1.33E-07 |
| Heart_Failure | dendritic_cell | *CCDC15* | -1.919046358 | 0.000984059 | 0.001180458 | 1 |
| Heart_Failure | dendritic_cell | *CHEK1* | -1.513932802 | 0.00306379 | 0.00359971 | 1 |
| Heart_Failure | dendritic_cell | *FEZ1* | -8.301618961 | 9.06E-15 | 2.64E-14 | 5.70E-11 |
| Heart_Failure | dendritic_cell | *DUSP15* | -7.369317032 | 6.09E-17 | 2.78E-16 | 3.83E-13 |
| Heart_Failure | dendritic_cell | *COMMD7* | -5.849262345 | 3.17E-14 | 8.45E-14 | 1.99E-10 |
| Heart_Failure | dendritic_cell | *KIAA1755* | -2.592956029 | 5.93E-12 | 1.19E-11 | 3.73E-08 |
| Heart_Failure | dendritic_cell | *CNBD2* | -9.579784577 | 2.32E-11 | 4.39E-11 | 1.46E-07 |
| Heart_Failure | dendritic_cell | *CABLES2* | -3.690954888 | 3.77E-07 | 5.33E-07 | 0.002367901 |
| Heart_Failure | dendritic_cell | *TRPT1* | -5.926299694 | 5.76E-17 | 2.65E-16 | 3.62E-13 |
| Heart_Failure | dendritic_cell | *TBX6* | 1.313708807 | 0.007143117 | 0.008243168 | 1 |
| Heart_Failure | dendritic_cell | *DOC2A* | 4.632805361 | 3.14E-14 | 8.36E-14 | 1.97E-10 |
| Heart_Failure | dendritic_cell | *TAOK2* | -4.358383381 | 2.36E-16 | 9.38E-16 | 1.48E-12 |
| Heart_Failure | dendritic_cell | *MPP7* | -1.518344856 | 0.000525113 | 0.000639308 | 1 |
| Heart_Failure | dendritic_cell | *CWC15* | -3.586019681 | 1.21E-05 | 1.59E-05 | 0.076058585 |
| Heart_Failure | dendritic_cell | *ARID5B* | -2.972342488 | 4.02E-14 | 1.05E-13 | 2.53E-10 |
| Heart_Failure | dendritic_cell | *TMCO3* | -5.394565158 | 4.02E-12 | 8.20E-12 | 2.53E-08 |
| Heart_Failure | dendritic_cell | *SAP18* | -5.685329402 | 7.96E-16 | 2.84E-15 | 5.00E-12 |
| Heart_Failure | dendritic_cell | *LYPD6B* | -7.994563697 | 0.000213429 | 0.000264399 | 1 |
| Heart_Failure | dendritic_cell | *PDCD4* | -4.158489804 | 3.72E-13 | 8.51E-13 | 2.34E-09 |
| Heart_Failure | dendritic_cell | *CCDC102B* | -3.006972561 | 1.78E-18 | 1.26E-17 | 1.12E-14 |
| Heart_Failure | dendritic_cell | *CNDP1* | -3.126451908 | 9.77E-08 | 1.43E-07 | 0.000614159 |
| Heart_Failure | dendritic_cell | *FSIP1* | 8.450997688 | 5.15E-07 | 7.23E-07 | 0.003238444 |
| Heart_Failure | dendritic_cell | *RGS18* | 0.644417857 | 5.05E-21 | 4.66E-19 | 3.18E-17 |
| Heart_Failure | dendritic_cell | *PRSS23* | -2.287925903 | 5.70E-15 | 1.73E-14 | 3.58E-11 |
| Heart_Failure | dendritic_cell | *MTMR12* | -4.276859183 | 2.67E-12 | 5.54E-12 | 1.68E-08 |
| Heart_Failure | dendritic_cell | *PPP1R1C* | -3.712150341 | 3.47E-12 | 7.14E-12 | 2.18E-08 |
| Heart_Failure | dendritic_cell | *C11orf53* | -20.49057834 | 1.16E-19 | 1.47E-18 | 7.31E-16 |
| Heart_Failure | dendritic_cell | *CCT5* | 4.231412424 | 0.000127057 | 0.000159188 | 0.798806138 |
| Heart_Failure | dendritic_cell | *FAM173B* | -5.224456837 | 6.23E-15 | 1.87E-14 | 3.92E-11 |
| Heart_Failure | dendritic_cell | *DOCK1* | -3.257217826 | 1.47E-15 | 4.94E-15 | 9.22E-12 |
| Heart_Failure | dendritic_cell | *TEX12* | -18.32878153 | 2.68E-20 | 6.44E-19 | 1.68E-16 |
| Heart_Failure | dendritic_cell | *PIP4K2A* | 3.893949413 | 2.88E-07 | 4.11E-07 | 0.001811421 |
| Heart_Failure | dendritic_cell | *CRIM1* | -3.322355535 | 6.29E-14 | 1.59E-13 | 3.96E-10 |
| Heart_Failure | dendritic_cell | *ABCB9* | -4.865449562 | 1.66E-08 | 2.52E-08 | 0.000104063 |
| Heart_Failure | dendritic_cell | *RILPL2* | 3.708870423 | 4.72E-12 | 9.57E-12 | 2.97E-08 |
| Heart_Failure | dendritic_cell | *PRSS53* | -4.552563321 | 1.89E-06 | 2.57E-06 | 0.011873633 |
| Heart_Failure | dendritic_cell | *GPR158* | -2.775263429 | 7.50E-18 | 4.35E-17 | 4.72E-14 |
| Heart_Failure | dendritic_cell | *THRB* | -2.77876054 | 6.69E-17 | 3.04E-16 | 4.21E-13 |
| Heart_Failure | dendritic_cell | *TMEM86A* | 2.279097451 | 5.24E-10 | 8.87E-10 | 3.30E-06 |
| Heart_Failure | dendritic_cell | *BTBD11* | 1.795294455 | 8.03E-21 | 4.66E-19 | 5.05E-17 |
| Heart_Failure | dendritic_cell | *UBE3B* | -2.782830876 | 6.20E-08 | 9.16E-08 | 0.000389957 |
| Heart_Failure | dendritic_cell | *IPMK* | 5.178487862 | 1.85E-07 | 2.66E-07 | 0.001161488 |
| Heart_Failure | dendritic_cell | *PLBD2* | -3.886608799 | 1.66E-15 | 5.53E-15 | 1.04E-11 |
| Heart_Failure | dendritic_cell | *DLG5* | -2.769908093 | 2.17E-14 | 5.93E-14 | 1.37E-10 |
| Heart_Failure | dendritic_cell | *GXYLT1* | 3.108528158 | 0.008229911 | 0.009457403 | 1 |
| Heart_Failure | dendritic_cell | *FAM177A1* | -4.758651406 | 6.73E-11 | 1.22E-10 | 4.23E-07 |
| Heart_Failure | dendritic_cell | *MIPOL1* | -3.343402992 | 3.89E-12 | 7.95E-12 | 2.45E-08 |

| | | | | | | |
|---|---|---|---|---|---|---|
| Heart_Failure | dendritic_cell | *EXT2* | -2.761144821 | 2.38E-08 | 3.58E-08 | 0.00014957 |
| Heart_Failure | dendritic_cell | *TMEM18* | -4.405871953 | 0.005169894 | 0.006005751 | 1 |
| Heart_Failure | dendritic_cell | *KCTD14* | -7.256953328 | 0.010464898 | 0.011949294 | 1 |
| Heart_Failure | dendritic_cell | *THRSP* | -1.835937263 | 3.77E-18 | 2.38E-17 | 2.37E-14 |
| Heart_Failure | dendritic_cell | *NDUFC2* | -5.806660522 | 6.42E-16 | 2.33E-15 | 4.03E-12 |
| Heart_Failure | dendritic_cell | *ME3* | -3.230952185 | 1.45E-15 | 4.90E-15 | 9.14E-12 |
| Heart_Failure | dendritic_cell | *ADAMTS12* | -2.100990142 | 8.19E-19 | 6.58E-18 | 5.15E-15 |
| Heart_Failure | dendritic_cell | *VIPAS39* | -5.731544336 | 2.70E-11 | 5.07E-11 | 1.70E-07 |
| Heart_Failure | dendritic_cell | *ANKRD50* | -4.124603132 | 0.000184368 | 0.000229439 | 1 |
| Heart_Failure | dendritic_cell | *C4orf33* | -4.491750293 | 3.08E-15 | 9.76E-15 | 1.93E-11 |
| Heart_Failure | dendritic_cell | *PTPRO* | 2.012274751 | 1.16E-20 | 5.00E-19 | 7.29E-17 |
| Heart_Failure | dendritic_cell | *VPS26B* | -5.485576218 | 1.69E-12 | 3.59E-12 | 1.06E-08 |
| Heart_Failure | dendritic_cell | *NCAPD3* | 5.104083614 | 7.32E-08 | 1.08E-07 | 0.000460283 |
| Heart_Failure | dendritic_cell | *VTI1A* | -3.673494075 | 2.23E-12 | 4.67E-12 | 1.40E-08 |
| Heart_Failure | dendritic_cell | *QDPR* | -0.961465779 | 0.000140767 | 0.000175909 | 0.884999463 |
| Heart_Failure | dendritic_cell | *FAM160B1* | -4.586590774 | 1.01E-14 | 2.93E-14 | 6.36E-11 |
| Heart_Failure | dendritic_cell | *ANO4* | -1.981739474 | 4.97E-19 | 4.34E-18 | 3.12E-15 |
| Heart_Failure | dendritic_cell | *TEX9* | -4.958797002 | 3.47E-07 | 4.92E-07 | 0.002179239 |
| Heart_Failure | dendritic_cell | *AKR1C2* | -2.057595531 | 2.34E-19 | 2.41E-18 | 1.47E-15 |
| Heart_Failure | dendritic_cell | *ADAM8* | 1.597575455 | 9.33E-21 | 4.77E-19 | 5.87E-17 |
| Heart_Failure | dendritic_cell | *ITIH2* | 11.46465915 | 1.32E-14 | 3.73E-14 | 8.29E-11 |
| Heart_Failure | dendritic_cell | *KIN* | -3.697364368 | 1.94E-08 | 2.95E-08 | 0.000122245 |
| Heart_Failure | dendritic_cell | *PIGF* | -5.05515017 | 3.52E-11 | 6.57E-11 | 2.22E-07 |
| Heart_Failure | dendritic_cell | *INPP1* | -3.508192812 | 0.003611379 | 0.004230434 | 1 |
| Heart_Failure | dendritic_cell | *MFSD6* | -3.91044098 | 0.000122564 | 0.000153651 | 0.770558496 |
| Heart_Failure | dendritic_cell | *RNF144A* | -3.413242728 | 3.53E-16 | 1.36E-15 | 2.22E-12 |
| Heart_Failure | dendritic_cell | *ASAP2* | -3.243530715 | 9.57E-16 | 3.36E-15 | 6.02E-12 |
| Heart_Failure | dendritic_cell | *ADAM17* | -3.826439371 | 5.03E-06 | 6.70E-06 | 0.031594306 |
| Heart_Failure | dendritic_cell | *TMEM45B* | -7.421144746 | 1.27E-18 | 9.45E-18 | 7.98E-15 |
| Heart_Failure | dendritic_cell | *CENPU* | 1.652371499 | 0.006570417 | 0.007593421 | 1 |
| Heart_Failure | dendritic_cell | *ACSL1* | -2.175223921 | 3.92E-16 | 1.49E-15 | 2.46E-12 |
| Heart_Failure | dendritic_cell | *BICD1* | -4.506281811 | 1.01E-09 | 1.68E-09 | 6.37E-06 |
| Heart_Failure | dendritic_cell | *CCDC122* | -1.368666149 | 5.05E-08 | 7.49E-08 | 0.000317226 |
| Heart_Failure | dendritic_cell | *GUF1* | -5.100668079 | 2.11E-08 | 3.19E-08 | 0.000132736 |
| Heart_Failure | dendritic_cell | *GABRA2* | -3.169588456 | 6.81E-18 | 4.00E-17 | 4.28E-14 |
| Heart_Failure | dendritic_cell | *CENPJ* | -4.287686687 | 1.03E-09 | 1.70E-09 | 6.47E-06 |
| Heart_Failure | dendritic_cell | *PARP8* | 2.923651862 | 2.93E-09 | 4.68E-09 | 1.84E-05 |
| Heart_Failure | dendritic_cell | *RABGAP1L* | -3.682148889 | 6.93E-10 | 1.16E-09 | 4.36E-06 |
| Heart_Failure | dendritic_cell | *PTPN14* | -3.277597307 | 8.97E-12 | 1.76E-11 | 5.64E-08 |
| Heart_Failure | dendritic_cell | *RNF219* | -4.334606292 | 0.000240107 | 0.000296689 | 1 |
| Heart_Failure | dendritic_cell | *EPG5* | -2.89105912 | 0.000173682 | 0.000216354 | 1 |
| Heart_Failure | dendritic_cell | *SPC25* | -1.364904317 | 1.27E-07 | 1.85E-07 | 0.00080086 |
| Heart_Failure | dendritic_cell | *G6PC2* | -131.4341871 | 2.17E-20 | 6.44E-19 | 1.36E-16 |
| Heart_Failure | dendritic_cell | *TGOLN2* | -4.770768222 | 2.24E-07 | 3.21E-07 | 0.00140689 |
| Heart_Failure | dendritic_cell | *ATG10* | -0.723928864 | 0.00140746 | 0.001679389 | 1 |
| Heart_Failure | dendritic_cell | *POC5* | -4.395905758 | 4.27E-14 | 1.11E-13 | 2.68E-10 |
| Heart_Failure | dendritic_cell | *TADA1* | 6.363005096 | 1.16E-05 | 1.53E-05 | 0.073206652 |
| Heart_Failure | dendritic_cell | *GUCY1A2* | -1.713186161 | 3.86E-19 | 3.59E-18 | 2.43E-15 |
| Heart_Failure | dendritic_cell | *CWF19L2* | -3.846437335 | 4.87E-14 | 1.25E-13 | 3.06E-10 |
| Heart_Failure | dendritic_cell | *ZNF547* | -6.427929912 | 4.80E-06 | 6.40E-06 | 0.030187344 |

| | | | | | | |
|---|---|---|---|---|---|---|
| Heart_Failure | dendritic_cell | *NMT2* | -3.035544185 | 2.18E-14 | 5.94E-14 | 1.37E-10 |
| Heart_Failure | dendritic_cell | *ZNF837* | -3.302603969 | 8.21E-12 | 1.62E-11 | 5.16E-08 |
| Heart_Failure | dendritic_cell | *CCDC50* | -2.879069695 | 3.92E-09 | 6.22E-09 | 2.47E-05 |
| Heart_Failure | dendritic_cell | *ZFP36L2* | 1.855883818 | 0.044148789 | 0.048884015 | 1 |
| Heart_Failure | dendritic_cell | *PLEKHH2* | -0.962986017 | 8.88E-09 | 1.38E-08 | 5.58E-05 |
| Heart_Failure | dendritic_cell | *TMEM123* | 4.84772312 | 2.81E-06 | 3.79E-06 | 0.017646496 |
| Heart_Failure | dendritic_cell | *IGSF10* | -2.05456144 | 0.013894387 | 0.015784968 | 1 |
| Heart_Failure | dendritic_cell | *SPEF2* | -3.982158412 | 1.43E-11 | 2.76E-11 | 8.98E-08 |
| Heart_Failure | dendritic_cell | *SPARCL1* | -2.167581248 | 7.30E-17 | 3.29E-16 | 4.59E-13 |
| Heart_Failure | dendritic_cell | *MBNL1* | -2.323739459 | 0.00011952 | 0.000149895 | 0.751425102 |
| Heart_Failure | dendritic_cell | *CCNO* | 11.00435459 | 3.45E-17 | 1.67E-16 | 2.17E-13 |
| Heart_Failure | dendritic_cell | *CLEC4F* | 1.39583598 | 5.94E-21 | 4.66E-19 | 3.74E-17 |
| Heart_Failure | dendritic_cell | *PELO* | 4.30576818 | 0.041392818 | 0.04586476 | 1 |
| Heart_Failure | dendritic_cell | *RASGRP3* | -2.62392345 | 1.43E-10 | 2.53E-10 | 8.98E-07 |
| Heart_Failure | dendritic_cell | *SAR1B* | -4.002652719 | 5.10E-15 | 1.56E-14 | 3.21E-11 |
| Heart_Failure | dendritic_cell | *GPR180* | -3.214749212 | 2.94E-17 | 1.45E-16 | 1.85E-13 |
| Heart_Failure | dendritic_cell | *WDR78* | -3.726110969 | 5.23E-18 | 3.17E-17 | 3.29E-14 |
| Heart_Failure | dendritic_cell | *IFIT5* | -3.808420576 | 1.73E-09 | 2.82E-09 | 1.09E-05 |
| Heart_Failure | dendritic_cell | *PRDM8* | -2.819839337 | 1.47E-16 | 6.10E-16 | 9.23E-13 |
| Heart_Failure | dendritic_cell | *HHEX* | 4.698859351 | 7.19E-12 | 1.43E-11 | 4.52E-08 |
| Heart_Failure | dendritic_cell | *ZNF117* | -0.739052145 | 1.61E-05 | 2.10E-05 | 0.101406952 |
| Heart_Failure | dendritic_cell | *RAB3C* | -3.064024075 | 2.01E-18 | 1.40E-17 | 1.26E-14 |
| Heart_Failure | dendritic_cell | *RAD17* | -3.118548914 | 3.87E-12 | 7.92E-12 | 2.43E-08 |
| Heart_Failure | dendritic_cell | *MED21* | -2.749336396 | 0.028443146 | 0.031756715 | 1 |
| Heart_Failure | dendritic_cell | *CENPH* | -3.089657184 | 3.54E-13 | 8.12E-13 | 2.23E-09 |
| Heart_Failure | dendritic_cell | *CARHSP1* | -3.927111229 | 6.68E-18 | 3.93E-17 | 4.20E-14 |
| Heart_Failure | dendritic_cell | *TEKT5* | -6.828309497 | 4.19E-20 | 7.75E-19 | 2.63E-16 |
| Heart_Failure | dendritic_cell | *BANK1* | -4.077067296 | 1.12E-16 | 4.80E-16 | 7.07E-13 |
| Heart_Failure | dendritic_cell | *CAST* | -2.772731497 | 2.63E-09 | 4.21E-09 | 1.65E-05 |
| Heart_Failure | dendritic_cell | *SCOC* | -5.620206094 | 2.17E-12 | 4.55E-12 | 1.36E-08 |
| Heart_Failure | dendritic_cell | *CLGN* | 5.388672395 | 5.17E-21 | 4.66E-19 | 3.25E-17 |
| Heart_Failure | dendritic_cell | *SYCP2L* | -3.586675284 | 0.001243351 | 0.001485547 | 1 |
| Heart_Failure | dendritic_cell | *AHCTF1* | -4.452460239 | 4.71E-13 | 1.06E-12 | 2.96E-09 |
| Heart_Failure | dendritic_cell | *PTPRR* | -3.56089765 | 2.78E-20 | 6.44E-19 | 1.75E-16 |
| Heart_Failure | dendritic_cell | *PLA2R1* | -2.74517195 | 5.47E-16 | 2.01E-15 | 3.44E-12 |
| Heart_Failure | dendritic_cell | *SLC25A27* | -2.014705094 | 6.78E-09 | 1.06E-08 | 4.26E-05 |
| Heart_Failure | dendritic_cell | *FRMD1* | -7.840145438 | 3.05E-17 | 1.50E-16 | 1.91E-13 |
| Heart_Failure | dendritic_cell | *ASAP1* | -1.993308236 | 0.007891235 | 0.009079831 | 1 |
| Heart_Failure | dendritic_cell | *INO80C* | -5.997240872 | 2.04E-18 | 1.43E-17 | 1.29E-14 |
| Heart_Failure | dendritic_cell | *LPCAT1* | -4.71385433 | 5.84E-18 | 3.51E-17 | 3.67E-14 |
| Heart_Failure | dendritic_cell | *NMRAL1* | -3.046102032 | 9.60E-08 | 1.40E-07 | 0.000603741 |
| Heart_Failure | dendritic_cell | *C16orf89* | -2.358489727 | 2.25E-15 | 7.34E-15 | 1.42E-11 |
| Heart_Failure | dendritic_cell | *TEX29* | 3.670405186 | 9.95E-05 | 0.000125315 | 0.625821894 |
| Heart_Failure | dendritic_cell | *ADPRHL1* | 2.920581248 | 9.42E-07 | 1.30E-06 | 0.005924093 |
| Heart_Failure | dendritic_cell | *TUBGCP5* | -4.974756819 | 6.57E-15 | 1.97E-14 | 4.13E-11 |
| Heart_Failure | dendritic_cell | *LURAP1L* | -2.381728481 | 3.29E-13 | 7.59E-13 | 2.07E-09 |
| Heart_Failure | dendritic_cell | *CFDP1* | -4.287063569 | 1.61E-12 | 3.42E-12 | 1.01E-08 |
| Heart_Failure | dendritic_cell | *FAM92B* | -34.41485302 | 2.29E-20 | 6.44E-19 | 1.44E-16 |
| Heart_Failure | dendritic_cell | *C7orf31* | -3.970085392 | 1.31E-17 | 7.19E-17 | 8.26E-14 |
| Heart_Failure | dendritic_cell | *JAZF1* | -3.9290766 | 1.81E-12 | 3.83E-12 | 1.14E-08 |

| | | | | | | |
|---|---|---|---|---|---|---|
| Heart_Failure | dendritic_cell | *FBXO36* | -4.940902889 | 1.95E-17 | 1.01E-16 | 1.22E-13 |
| Heart_Failure | dendritic_cell | *MCOLN2* | 0.681089287 | 2.10E-20 | 6.44E-19 | 1.32E-16 |
| Heart_Failure | dendritic_cell | *CACNA2D1* | -2.168776468 | 1.38E-18 | 1.02E-17 | 8.67E-15 |
| Heart_Failure | dendritic_cell | *ZUFSP* | -5.088113478 | 2.55E-06 | 3.45E-06 | 0.016056452 |
| Heart_Failure | dendritic_cell | *HS3ST3A1* | -3.133902498 | 3.22E-14 | 8.57E-14 | 2.02E-10 |
| Heart_Failure | dendritic_cell | *PPP2R5E* | -3.380939228 | 1.25E-06 | 1.71E-06 | 0.007847373 |
| Heart_Failure | dendritic_cell | *ANKRD29* | -2.509489388 | 6.38E-19 | 5.32E-18 | 4.01E-15 |
| Heart_Failure | dendritic_cell | *THY1* | -3.672996799 | 1.62E-18 | 1.17E-17 | 1.02E-14 |
| Heart_Failure | dendritic_cell | *TBCEL* | -5.24958163 | 8.83E-09 | 1.37E-08 | 5.55E-05 |
| Heart_Failure | dendritic_cell | *UBASH3B* | 3.324892046 | 6.05E-19 | 5.12E-18 | 3.80E-15 |
| Heart_Failure | dendritic_cell | *ROBO4* | -4.283901855 | 4.32E-15 | 1.33E-14 | 2.72E-11 |
| Heart_Failure | dendritic_cell | *ROBO3* | -4.646526675 | 1.37E-12 | 2.93E-12 | 8.59E-09 |
| Heart_Failure | dendritic_cell | *CC2D1B* | -3.171352157 | 3.85E-10 | 6.56E-10 | 2.42E-06 |
| Heart_Failure | dendritic_cell | *PRKCA* | 3.226074583 | 0.000405062 | 0.000495645 | 1 |
| Heart_Failure | dendritic_cell | *LRRK1* | 2.856687735 | 9.29E-20 | 1.27E-18 | 5.84E-16 |
| Heart_Failure | dendritic_cell | *CEP112* | -3.04934795 | 2.05E-13 | 4.83E-13 | 1.29E-09 |
| Heart_Failure | dendritic_cell | *ENPP3* | -2.535528414 | 1.52E-19 | 1.81E-18 | 9.57E-16 |
| Heart_Failure | dendritic_cell | *MIA3* | -4.508419127 | 1.77E-13 | 4.21E-13 | 1.11E-09 |
| Heart_Failure | dendritic_cell | *DISP1* | -1.749814518 | 0.004997967 | 0.005811396 | 1 |
| Heart_Failure | dendritic_cell | *FAM167A* | 3.778337232 | 1.32E-20 | 5.25E-19 | 8.30E-17 |
| Heart_Failure | dendritic_cell | *NEIL2* | -2.048589921 | 2.78E-05 | 3.58E-05 | 0.174603595 |
| Heart_Failure | dendritic_cell | *LONRF1* | 2.937215306 | 0.001340543 | 0.001600455 | 1 |
| Heart_Failure | dendritic_cell | *CCSAP* | 4.769476781 | 8.23E-15 | 2.42E-14 | 5.18E-11 |
| Heart_Failure | dendritic_cell | *GBP5* | 6.268272192 | 3.95E-13 | 8.99E-13 | 2.49E-09 |
| Heart_Failure | dendritic_cell | *FAM69A* | -3.014760729 | 1.68E-06 | 2.30E-06 | 0.010592754 |
| Heart_Failure | dendritic_cell | *ATP5G3* | -1.985561326 | 0.004482565 | 0.005225642 | 1 |
| Heart_Failure | dendritic_cell | *NCAM2* | -2.142091119 | 9.24E-19 | 7.30E-18 | 5.81E-15 |
| Heart_Failure | dendritic_cell | *L3MBTL4* | -4.04251266 | 8.54E-14 | 2.12E-13 | 5.37E-10 |
| Heart_Failure | dendritic_cell | *PDE1C* | -4.43319759 | 1.37E-05 | 1.79E-05 | 0.086068831 |
| Heart_Failure | dendritic_cell | *RABGEF1* | -1.271584732 | 0.000308565 | 0.000379415 | 1 |
| Heart_Failure | dendritic_cell | *MRPL39* | -2.057481802 | 3.46E-14 | 9.13E-14 | 2.18E-10 |
| Heart_Failure | dendritic_cell | *TSEN2* | -3.339635142 | 7.18E-19 | 5.88E-18 | 4.51E-15 |
| Heart_Failure | dendritic_cell | *SLFN13* | -1.580885175 | 0.000225436 | 0.000279054 | 1 |
| Heart_Failure | dendritic_cell | *FLCN* | -3.151052101 | 2.17E-14 | 5.92E-14 | 1.36E-10 |
| Heart_Failure | dendritic_cell | *DPH3* | 9.114972626 | 3.32E-13 | 7.64E-13 | 2.08E-09 |
| Heart_Failure | dendritic_cell | *CXXC1* | -6.889381191 | 1.59E-14 | 4.42E-14 | 9.99E-11 |
| Heart_Failure | dendritic_cell | *APCDD1* | -1.769382946 | 4.05E-13 | 9.20E-13 | 2.55E-09 |
| Heart_Failure | dendritic_cell | *MPPE1* | -1.109212528 | 0.024385262 | 0.02732314 | 1 |
| Heart_Failure | dendritic_cell | *EPHB1* | -5.994894421 | 1.42E-14 | 4.00E-14 | 8.94E-11 |
| Heart_Failure | dendritic_cell | *ANKRD40* | -4.982670869 | 1.87E-12 | 3.95E-12 | 1.17E-08 |
| Heart_Failure | dendritic_cell | *CYP2U1* | -7.057274961 | 4.29E-11 | 7.92E-11 | 2.70E-07 |
| Heart_Failure | dendritic_cell | *FBXL18* | -6.045964983 | 6.93E-14 | 1.74E-13 | 4.35E-10 |
| Heart_Failure | dendritic_cell | *PROM2* | -11.17685346 | 4.35E-18 | 2.69E-17 | 2.73E-14 |
| Heart_Failure | dendritic_cell | *AK9* | -4.644482224 | 2.33E-11 | 4.42E-11 | 1.47E-07 |
| Heart_Failure | dendritic_cell | *CDK19* | -2.842303396 | 0.001053368 | 0.001261434 | 1 |
| Heart_Failure | dendritic_cell | *GTF3C6* | -5.295738461 | 3.21E-09 | 5.12E-09 | 2.02E-05 |
| Heart_Failure | dendritic_cell | *TTC39B* | -3.285077807 | 3.36E-06 | 4.52E-06 | 0.0211526 |
| Heart_Failure | dendritic_cell | *AGPAT5* | 3.199941873 | 0.000667638 | 0.000807822 | 1 |
| Heart_Failure | dendritic_cell | *MMS19* | -3.959317027 | 1.67E-17 | 8.85E-17 | 1.05E-13 |
| Heart_Failure | dendritic_cell | *GOLGA7B* | 6.533755404 | 4.98E-16 | 1.85E-15 | 3.13E-12 |

| | | | | | | |
|---|---|---|---|---|---|---|
| Heart_Failure | dendritic_cell | *TRMT44* | -4.060358912 | 7.61E-18 | 4.41E-17 | 4.79E-14 |
| Heart_Failure | dendritic_cell | *C16orf87* | -4.487450035 | 3.03E-07 | 4.31E-07 | 0.001905852 |
| Heart_Failure | dendritic_cell | *CNOT8* | -4.710079359 | 2.63E-10 | 4.56E-10 | 1.66E-06 |
| Heart_Failure | dendritic_cell | *LRGUK* | -2.040388616 | 5.06E-07 | 7.10E-07 | 0.003179607 |
| Heart_Failure | dendritic_cell | *SETD9* | -1.8355544 | 2.61E-10 | 4.53E-10 | 1.64E-06 |
| Heart_Failure | dendritic_cell | *ZKSCAN2* | -1.955597259 | 3.76E-05 | 4.81E-05 | 0.236433675 |
| Heart_Failure | dendritic_cell | *PDIA4* | -6.946213888 | 3.59E-11 | 6.69E-11 | 2.26E-07 |
| Heart_Failure | dendritic_cell | *KDM8* | -7.985836496 | 2.99E-15 | 9.53E-15 | 1.88E-11 |
| Heart_Failure | dendritic_cell | *FAM126B* | -3.557665845 | 4.10E-17 | 1.96E-16 | 2.58E-13 |
| Heart_Failure | dendritic_cell | *ALS2CR12* | -3.91392991 | 3.73E-06 | 5.01E-06 | 0.023454919 |
| Heart_Failure | dendritic_cell | *SPAG17* | -1.707610573 | 6.44E-20 | 1.01E-18 | 4.05E-16 |
| Heart_Failure | dendritic_cell | *DEPTOR* | 3.510349475 | 0.001615525 | 0.001923637 | 1 |
| Heart_Failure | dendritic_cell | *RRAGA* | -6.112013642 | 8.90E-15 | 2.60E-14 | 5.59E-11 |
| Heart_Failure | dendritic_cell | *TMBIM4* | -1.470105255 | 0.012656646 | 0.01439702 | 1 |
| Heart_Failure | dendritic_cell | *FAM161B* | 7.801902285 | 2.10E-07 | 3.01E-07 | 0.001318901 |
| Heart_Failure | dendritic_cell | *WIF1* | 8.136374914 | 2.23E-16 | 8.93E-16 | 1.40E-12 |
| Heart_Failure | dendritic_cell | *UGT2B4* | -3.996827138 | 2.83E-20 | 6.44E-19 | 1.78E-16 |
| Heart_Failure | dendritic_cell | *MMP16* | -2.119156771 | 3.74E-18 | 2.37E-17 | 2.35E-14 |
| Heart_Failure | dendritic_cell | *KCNMA1* | -2.892389651 | 7.45E-15 | 2.21E-14 | 4.69E-11 |
| Heart_Failure | dendritic_cell | *BATF* | -11.22439399 | 1.04E-10 | 1.85E-10 | 6.52E-07 |
| Heart_Failure | dendritic_cell | *DCK* | 4.705604823 | 5.29E-20 | 8.88E-19 | 3.33E-16 |
| Heart_Failure | dendritic_cell | *DPY19L4* | -5.209096536 | 1.10E-13 | 2.69E-13 | 6.94E-10 |
| Heart_Failure | dendritic_cell | *NDUFAF6* | -4.57965243 | 2.40E-16 | 9.50E-16 | 1.51E-12 |
| Heart_Failure | dendritic_cell | *C8orf37* | -3.473701466 | 3.03E-10 | 5.21E-10 | 1.90E-06 |
| Heart_Failure | dendritic_cell | *ADAMTSL3* | -1.726064482 | 4.30E-17 | 2.05E-16 | 2.70E-13 |
| Heart_Failure | dendritic_cell | *WHAMM* | -3.332601787 | 1.37E-11 | 2.65E-11 | 8.62E-08 |
| Heart_Failure | dendritic_cell | *CXCL13* | -11.68680315 | 1.27E-13 | 3.08E-13 | 7.99E-10 |
| Heart_Failure | dendritic_cell | *N6AMT1* | -1.688947943 | 3.26E-05 | 4.19E-05 | 0.205033994 |
| Heart_Failure | dendritic_cell | *RWDD2B* | -2.287196818 | 3.46E-05 | 4.43E-05 | 0.217607063 |
| Heart_Failure | dendritic_cell | *CCT8* | -1.477044102 | 0.000318134 | 0.000391028 | 1 |
| Heart_Failure | dendritic_cell | *ANKRD9* | -6.562930011 | 1.22E-06 | 1.67E-06 | 0.007640473 |
| Heart_Failure | dendritic_cell | *SFXN2* | -3.508436377 | 4.76E-06 | 6.34E-06 | 0.029903105 |
| Heart_Failure | dendritic_cell | *C14orf2* | -2.669807407 | 2.34E-10 | 4.06E-10 | 1.47E-06 |
| Heart_Failure | dendritic_cell | *TDRD9* | 2.72763671 | 4.29E-06 | 5.74E-06 | 0.026990211 |
| Heart_Failure | dendritic_cell | *SH3RF2* | -2.16373931 | 6.01E-08 | 8.88E-08 | 0.000377679 |
| Heart_Failure | dendritic_cell | *GDF6* | -6.529525347 | 1.10E-18 | 8.38E-18 | 6.91E-15 |
| Heart_Failure | dendritic_cell | *NODAL* | -3.493446875 | 2.99E-20 | 6.61E-19 | 1.88E-16 |
| Heart_Failure | dendritic_cell | *MED19* | -2.374044854 | 1.78E-08 | 2.71E-08 | 0.00011205 |
| Heart_Failure | dendritic_cell | *WDYHV1* | -3.86599945 | 5.08E-09 | 7.99E-09 | 3.19E-05 |
| Heart_Failure | dendritic_cell | *ATAD2* | 3.530018836 | 0.005407886 | 0.006270635 | 1 |
| Heart_Failure | dendritic_cell | *SASS6* | -4.394605602 | 4.49E-17 | 2.12E-16 | 2.82E-13 |
| Heart_Failure | dendritic_cell | *ITGAD* | 2.870410856 | 7.48E-16 | 2.68E-15 | 4.70E-12 |
| Heart_Failure | dendritic_cell | *VPS8* | -3.058272168 | 2.38E-06 | 3.23E-06 | 0.014971426 |
| Heart_Failure | dendritic_cell | *MPV17L* | 1.571216127 | 0.000981051 | 0.001177074 | 1 |
| Heart_Failure | dendritic_cell | *RPUSD3* | -6.226417232 | 4.45E-16 | 1.67E-15 | 2.80E-12 |
| Heart_Failure | dendritic_cell | *GHRL* | -7.057419566 | 6.77E-07 | 9.45E-07 | 0.004259079 |
| Heart_Failure | dendritic_cell | *NTAN1* | -5.371090881 | 2.05E-12 | 4.31E-12 | 1.29E-08 |
| Heart_Failure | dendritic_cell | *SLC6A1* | 5.079333715 | 6.33E-09 | 9.90E-09 | 3.98E-05 |
| Heart_Failure | dendritic_cell | *FCHO2* | -1.793295551 | 0.040971711 | 0.045422174 | 1 |
| Heart_Failure | dendritic_cell | *TIMP4* | -5.627150264 | 4.81E-12 | 9.74E-12 | 3.02E-08 |

| | | | | | | |
|---|---|---|---|---|---|---|
| Heart_Failure | dendritic_cell | *CPT2* | -3.243446768 | 1.61E-20 | 5.94E-19 | 1.01E-16 |
| Heart_Failure | dendritic_cell | *NECAP2* | -6.061727571 | 1.29E-14 | 3.66E-14 | 8.13E-11 |
| Heart_Failure | dendritic_cell | *LRP8* | 9.502230383 | 8.30E-09 | 1.29E-08 | 5.22E-05 |
| Heart_Failure | dendritic_cell | *STEAP2* | -1.374361137 | 2.68E-14 | 7.21E-14 | 1.69E-10 |
| Heart_Failure | dendritic_cell | *GATAD1* | -3.036598804 | 1.87E-08 | 2.83E-08 | 0.000117402 |
| Heart_Failure | dendritic_cell | *SUSD3* | 1.840349194 | 7.69E-17 | 3.45E-16 | 4.83E-13 |
| Heart_Failure | dendritic_cell | *TMED6* | -11.58611512 | 2.80E-20 | 6.44E-19 | 1.76E-16 |
| Heart_Failure | dendritic_cell | *CLEC18A* | 2.792414369 | 3.56E-15 | 1.11E-14 | 2.24E-11 |
| Heart_Failure | dendritic_cell | *IL34* | -3.52871284 | 1.34E-19 | 1.64E-18 | 8.41E-16 |
| Heart_Failure | dendritic_cell | *DHRS1* | -2.875848918 | 4.22E-14 | 1.10E-13 | 2.65E-10 |
| Heart_Failure | dendritic_cell | *AASDH* | -4.50054061 | 7.47E-13 | 1.64E-12 | 4.70E-09 |
| Heart_Failure | dendritic_cell | *RNF111* | -3.616445243 | 9.27E-14 | 2.28E-13 | 5.83E-10 |
| Heart_Failure | dendritic_cell | *DYRK1A* | -3.870892105 | 1.23E-10 | 2.18E-10 | 7.71E-07 |
| Heart_Failure | dendritic_cell | *LCA5L* | -1.323018741 | 9.58E-06 | 1.26E-05 | 0.060209914 |
| Heart_Failure | dendritic_cell | *MX1* | -0.989471812 | 0.000107762 | 0.000135446 | 0.677498709 |
| Heart_Failure | dendritic_cell | *CREB3L1* | -2.707523671 | 1.97E-17 | 1.02E-16 | 1.24E-13 |
| Heart_Failure | dendritic_cell | *C2CD2* | -1.462423792 | 6.59E-06 | 8.74E-06 | 0.041457013 |
| Heart_Failure | dendritic_cell | *SLC38A10* | -4.716291046 | 9.37E-15 | 2.73E-14 | 5.89E-11 |
| Heart_Failure | dendritic_cell | *C9orf43* | 4.320787213 | 8.63E-11 | 1.55E-10 | 5.43E-07 |
| Heart_Failure | dendritic_cell | *WDR19* | -4.213835273 | 1.55E-16 | 6.42E-16 | 9.75E-13 |
| Heart_Failure | dendritic_cell | *SLC37A3* | -2.786872731 | 5.71E-19 | 4.89E-18 | 3.59E-15 |
| Heart_Failure | dendritic_cell | *AP3S2* | -2.2055767 | 8.98E-09 | 1.39E-08 | 5.65E-05 |
| Heart_Failure | dendritic_cell | *SPPL3* | 1.896917068 | 0.02317152 | 0.025990963 | 1 |
| Heart_Failure | dendritic_cell | *FAM213B* | 3.269209006 | 1.60E-09 | 2.62E-09 | 1.01E-05 |
| Heart_Failure | dendritic_cell | *C12orf43* | -3.072396486 | 1.23E-13 | 2.99E-13 | 7.75E-10 |
| Heart_Failure | dendritic_cell | *PEX10* | -4.429928438 | 3.37E-18 | 2.16E-17 | 2.12E-14 |
| Heart_Failure | dendritic_cell | *RADIL* | -3.004952551 | 1.42E-19 | 1.70E-18 | 8.90E-16 |
| Heart_Failure | dendritic_cell | *LDLRAP1* | -5.137402446 | 2.72E-18 | 1.81E-17 | 1.71E-14 |
| Heart_Failure | dendritic_cell | *TRIM63* | -2.362487974 | 9.76E-06 | 1.29E-05 | 0.061338573 |
| Heart_Failure | dendritic_cell | *WDR66* | -1.018214058 | 0.037402034 | 0.041545333 | 1 |
| Heart_Failure | dendritic_cell | *UBXN11* | 3.172841035 | 3.80E-18 | 2.39E-17 | 2.39E-14 |
| Heart_Failure | dendritic_cell | *GALNT14* | 3.620500987 | 1.05E-20 | 4.95E-19 | 6.57E-17 |
| Heart_Failure | dendritic_cell | *HPD* | -2.547980133 | 7.62E-11 | 1.37E-10 | 4.79E-07 |
| Heart_Failure | dendritic_cell | *RHPN1* | 3.782610613 | 1.22E-15 | 4.17E-15 | 7.65E-12 |
| Heart_Failure | dendritic_cell | *TPRG1L* | -8.300468712 | 2.12E-12 | 4.45E-12 | 1.33E-08 |
| Heart_Failure | dendritic_cell | *LRRC43* | -7.096511471 | 2.35E-20 | 6.44E-19 | 1.48E-16 |
| Heart_Failure | dendritic_cell | *XKR8* | -5.90262496 | 1.78E-07 | 2.56E-07 | 0.001117 |
| Heart_Failure | dendritic_cell | *EYA3* | -3.624416976 | 5.71E-06 | 7.60E-06 | 0.035908644 |
| Heart_Failure | dendritic_cell | *MRAS* | -2.068931128 | 9.76E-15 | 2.83E-14 | 6.14E-11 |
| Heart_Failure | dendritic_cell | *CLSTN2* | -2.02787466 | 1.90E-19 | 2.11E-18 | 1.19E-15 |
| Heart_Failure | dendritic_cell | *COLEC12* | -4.089137879 | 2.92E-13 | 6.77E-13 | 1.84E-09 |
| Heart_Failure | dendritic_cell | *RNF207* | 8.811075974 | 2.98E-17 | 1.47E-16 | 1.88E-13 |
| Heart_Failure | dendritic_cell | *AUTS2* | -3.145528098 | 3.13E-13 | 7.23E-13 | 1.97E-09 |
| Heart_Failure | dendritic_cell | *MITD1* | -3.572183812 | 5.38E-07 | 7.54E-07 | 0.003384128 |
| Heart_Failure | dendritic_cell | *TSPAN33* | 0.837014335 | 1.37E-19 | 1.66E-18 | 8.61E-16 |
| Heart_Failure | dendritic_cell | *B4GALT5* | -3.952094675 | 6.39E-15 | 1.92E-14 | 4.02E-11 |
| Heart_Failure | dendritic_cell | *CD1D* | 0.625216585 | 1.00E-20 | 4.95E-19 | 6.31E-17 |
| Heart_Failure | dendritic_cell | *FAM86C1* | -5.686753233 | 8.33E-20 | 1.18E-18 | 5.23E-16 |
| Heart_Failure | dendritic_cell | *CD1E* | 0.415027053 | 5.73E-21 | 4.66E-19 | 3.60E-17 |
| Heart_Failure | dendritic_cell | *NCF1* | 4.955093501 | 1.50E-20 | 5.70E-19 | 9.40E-17 |

| | | | | | | |
|---|---|---|---|---|---|---|
| Heart_Failure | dendritic_cell | *PPP1R9A* | -2.553067078 | 3.61E-19 | 3.39E-18 | 2.27E-15 |
| Heart_Failure | dendritic_cell | *ZFAND2B* | -5.587940419 | 3.11E-14 | 8.29E-14 | 1.95E-10 |
| Heart_Failure | dendritic_cell | *GDPD5* | -2.841455142 | 2.96E-19 | 2.88E-18 | 1.86E-15 |
| Heart_Failure | dendritic_cell | *DYNC1I1* | -5.694832241 | 2.23E-11 | 4.23E-11 | 1.40E-07 |
| Heart_Failure | dendritic_cell | *TMED4* | -5.721016087 | 5.80E-14 | 1.48E-13 | 3.64E-10 |
| Heart_Failure | dendritic_cell | *COPG2* | -1.153592463 | 1.90E-10 | 3.32E-10 | 1.19E-06 |
| Heart_Failure | dendritic_cell | *ELK4* | -5.069847293 | 2.51E-07 | 3.59E-07 | 0.001578781 |
| Heart_Failure | dendritic_cell | *DUSP23* | 4.413586893 | 2.19E-20 | 6.44E-19 | 1.38E-16 |
| Heart_Failure | dendritic_cell | *RNF166* | 5.763576478 | 9.50E-17 | 4.13E-16 | 5.97E-13 |
| Heart_Failure | dendritic_cell | *F11R* | 3.057464976 | 1.01E-05 | 1.33E-05 | 0.063624582 |
| Heart_Failure | dendritic_cell | *SPATA2L* | 5.118991964 | 0.006632313 | 0.007662138 | 1 |
| Heart_Failure | dendritic_cell | *NIT1* | -2.262477298 | 4.21E-08 | 6.27E-08 | 0.0002649 |
| Heart_Failure | dendritic_cell | *FGF17* | -9.098183047 | 1.05E-09 | 1.73E-09 | 6.58E-06 |
| Heart_Failure | dendritic_cell | *CDA* | -3.101741831 | 8.95E-17 | 3.93E-16 | 5.63E-13 |
| Heart_Failure | dendritic_cell | *DMTN* | -2.051358289 | 1.34E-14 | 3.77E-14 | 8.40E-11 |
| Heart_Failure | dendritic_cell | *FAM160B2* | 3.281611925 | 0.00230919 | 0.002730464 | 1 |
| Heart_Failure | dendritic_cell | *NDUFS2* | -3.514335867 | 2.46E-08 | 3.70E-08 | 0.000154495 |
| Heart_Failure | dendritic_cell | *SLC5A11* | -1.310721847 | 7.99E-08 | 1.17E-07 | 0.000502342 |
| Heart_Failure | dendritic_cell | *FCER1G* | 1.964694755 | 8.77E-23 | 7.88E-20 | 5.52E-19 |
| Heart_Failure | dendritic_cell | *CCAR2* | -6.055118424 | 7.48E-14 | 1.87E-13 | 4.70E-10 |
| Heart_Failure | dendritic_cell | *CDC42SE2* | 4.520779297 | 1.25E-12 | 2.68E-12 | 7.83E-09 |
| Heart_Failure | dendritic_cell | *RAPGEF6* | -5.04846702 | 1.87E-18 | 1.32E-17 | 1.18E-14 |
| Heart_Failure | dendritic_cell | *EPB41* | -2.350270946 | 1.86E-09 | 3.02E-09 | 1.17E-05 |
| Heart_Failure | dendritic_cell | *ALG8* | -5.278454539 | 2.67E-12 | 5.54E-12 | 1.68E-08 |
| Heart_Failure | dendritic_cell | *MRPL10* | -3.514465847 | 3.27E-11 | 6.11E-11 | 2.06E-07 |
| Heart_Failure | dendritic_cell | *SV2A* | -12.10887779 | 2.54E-17 | 1.27E-16 | 1.60E-13 |
| Heart_Failure | dendritic_cell | *CLIC6* | -2.185507425 | 2.70E-16 | 1.06E-15 | 1.70E-12 |
| Heart_Failure | dendritic_cell | *CBR3* | -3.504315748 | 7.18E-14 | 1.80E-13 | 4.51E-10 |
| Heart_Failure | dendritic_cell | *SIM2* | -4.465032288 | 2.60E-16 | 1.02E-15 | 1.63E-12 |
| Heart_Failure | dendritic_cell | *HLCS* | -2.029097783 | 1.69E-10 | 2.96E-10 | 1.06E-06 |
| Heart_Failure | dendritic_cell | *PADI4* | 13.81455172 | 3.29E-17 | 1.61E-16 | 2.07E-13 |
| Heart_Failure | dendritic_cell | *PSMD4* | -5.798615382 | 1.02E-08 | 1.57E-08 | 6.40E-05 |
| Heart_Failure | dendritic_cell | *STARD9* | -3.396504784 | 2.15E-13 | 5.06E-13 | 1.35E-09 |
| Heart_Failure | dendritic_cell | *THEM4* | -3.135030279 | 0.00025198 | 0.000311054 | 1 |
| Heart_Failure | dendritic_cell | *TCHH* | 1.764441352 | 1.29E-20 | 5.18E-19 | 8.09E-17 |
| Heart_Failure | dendritic_cell | *MED8* | -1.558879302 | 0.028895628 | 0.032256181 | 1 |
| Heart_Failure | dendritic_cell | *RSPRY1* | -4.205563277 | 1.50E-06 | 2.05E-06 | 0.009455583 |
| Heart_Failure | dendritic_cell | *CCDC17* | 3.2594431 | 5.62E-21 | 4.66E-19 | 3.53E-17 |
| Heart_Failure | dendritic_cell | *ACE* | -4.569098809 | 9.95E-19 | 7.74E-18 | 6.26E-15 |
| Heart_Failure | dendritic_cell | *TEPP* | -24.02693776 | 3.82E-19 | 3.56E-18 | 2.40E-15 |
| Heart_Failure | dendritic_cell | *EFCAB14* | -5.316088907 | 1.41E-10 | 2.50E-10 | 8.89E-07 |
| Heart_Failure | dendritic_cell | *ATP6V0D1* | -4.323606145 | 1.00E-07 | 1.46E-07 | 0.000630855 |
| Heart_Failure | dendritic_cell | *ZFYVE28* | -3.650262223 | 7.38E-16 | 2.65E-15 | 4.64E-12 |
| Heart_Failure | dendritic_cell | *C16orf86* | -4.901450074 | 2.54E-09 | 4.08E-09 | 1.60E-05 |
| Heart_Failure | dendritic_cell | *FAM131B* | 11.05468269 | 1.65E-15 | 5.50E-15 | 1.04E-11 |
| Heart_Failure | dendritic_cell | *LYPD5* | 2.959605791 | 3.26E-17 | 1.59E-16 | 2.05E-13 |
| Heart_Failure | dendritic_cell | *CCDC117* | -4.642043454 | 0.00157206 | 0.001873658 | 1 |
| Heart_Failure | dendritic_cell | *CCDC107* | -3.63275507 | 3.61E-14 | 9.48E-14 | 2.27E-10 |
| Heart_Failure | dendritic_cell | *NPR2* | -4.158033829 | 3.47E-19 | 3.28E-18 | 2.18E-15 |
| Heart_Failure | dendritic_cell | *ZNF235* | -4.156947276 | 8.40E-09 | 1.30E-08 | 5.28E-05 |

| | | | | | | |
|---|---|---|---|---|---|---|
| Heart_Failure | dendritic_cell | *CALM3* | 4.734308064 | 0.000335893 | 0.000412373 | 1 |
| Heart_Failure | dendritic_cell | *DFFA* | -4.182791073 | 1.83E-13 | 4.34E-13 | 1.15E-09 |
| Heart_Failure | dendritic_cell | *ATAD3B* | 5.835408926 | 9.24E-17 | 4.04E-16 | 5.81E-13 |
| Heart_Failure | dendritic_cell | *SSU72* | -3.904702273 | 0.002530735 | 0.002986249 | 1 |
| Heart_Failure | dendritic_cell | *NR2F6* | -4.47502781 | 2.05E-15 | 6.74E-15 | 1.29E-11 |
| Heart_Failure | dendritic_cell | *ANKLE1* | -5.231024899 | 2.94E-09 | 4.70E-09 | 1.85E-05 |
| Heart_Failure | dendritic_cell | *CILP2* | 9.476258032 | 2.44E-21 | 4.66E-19 | 1.54E-17 |
| Heart_Failure | dendritic_cell | *RSPH1* | -2.076483259 | 0.002820803 | 0.003321047 | 1 |
| Heart_Failure | dendritic_cell | *NDUFV3* | -0.776427688 | 0.000135773 | 0.000169973 | 0.853604762 |
| Heart_Failure | dendritic_cell | *U2AF1* | -3.166717366 | 0.000267035 | 0.000329122 | 1 |
| Heart_Failure | dendritic_cell | *HSF2BP* | -4.481978888 | 3.05E-11 | 5.71E-11 | 1.92E-07 |
| Heart_Failure | dendritic_cell | *C21orf33* | -8.522685906 | 1.12E-18 | 8.56E-18 | 7.06E-15 |
| Heart_Failure | dendritic_cell | *C21orf2* | -1.273325819 | 0.00056953 | 0.000691376 | 1 |
| Heart_Failure | dendritic_cell | *ZNF66* | -5.053746858 | 2.97E-12 | 6.13E-12 | 1.86E-08 |
| Heart_Failure | dendritic_cell | *LRRC3* | -4.314265002 | 7.49E-15 | 2.22E-14 | 4.71E-11 |
| Heart_Failure | dendritic_cell | *ITGB2* | 2.73549081 | 2.76E-16 | 1.08E-15 | 1.73E-12 |
| Heart_Failure | dendritic_cell | *FAM207A* | 3.584228101 | 4.53E-17 | 2.14E-16 | 2.85E-13 |
| Heart_Failure | dendritic_cell | *LSS* | -3.300036521 | 5.37E-17 | 2.49E-16 | 3.37E-13 |
| Heart_Failure | dendritic_cell | *VAV2* | 4.181010845 | 0.000679065 | 0.000821173 | 1 |
| Heart_Failure | dendritic_cell | *MCM3AP* | -1.63013371 | 7.33E-05 | 9.27E-05 | 0.460890391 |
| Heart_Failure | dendritic_cell | *PCNT* | -2.43015264 | 4.99E-08 | 7.41E-08 | 0.00031382 |
| Heart_Failure | dendritic_cell | *S100B* | -0.95253955 | 2.19E-06 | 2.97E-06 | 0.013750022 |
| Heart_Failure | dendritic_cell | *PRMT2* | -1.958575071 | 0.004125761 | 0.004822209 | 1 |
| Heart_Failure | dendritic_cell | *ZNF208* | -3.047941672 | 1.89E-17 | 9.82E-17 | 1.19E-13 |
| Heart_Failure | dendritic_cell | *ADAMTS13* | 3.708184679 | 1.83E-07 | 2.62E-07 | 0.001147382 |
| Heart_Failure | dendritic_cell | *GPSM1* | -3.742024539 | 1.19E-15 | 4.10E-15 | 7.51E-12 |
| Heart_Failure | dendritic_cell | *C19orf47* | 6.934089481 | 2.82E-14 | 7.56E-14 | 1.77E-10 |
| Heart_Failure | dendritic_cell | *HIPK4* | -4.38903466 | 8.34E-17 | 3.70E-16 | 5.24E-13 |
| Heart_Failure | dendritic_cell | *RDH13* | -6.289892953 | 1.50E-13 | 3.60E-13 | 9.43E-10 |
| Heart_Failure | dendritic_cell | *MPZL3* | 4.875376303 | 2.00E-20 | 6.44E-19 | 1.26E-16 |
| Heart_Failure | dendritic_cell | *PCSK7* | -5.20938402 | 1.05E-11 | 2.04E-11 | 6.58E-08 |
| Heart_Failure | dendritic_cell | *S100A1* | 5.658930802 | 6.52E-15 | 1.95E-14 | 4.10E-11 |
| Heart_Failure | dendritic_cell | *ZBTB7B* | -5.960942473 | 1.19E-15 | 4.10E-15 | 7.51E-12 |
| Heart_Failure | dendritic_cell | *VPS11* | 2.874173844 | 0.015679914 | 0.017752497 | 1 |
| Heart_Failure | dendritic_cell | *FDPS* | -3.854399742 | 2.17E-08 | 3.28E-08 | 0.000136438 |
| Heart_Failure | dendritic_cell | *PAQR6* | -6.129824345 | 2.16E-11 | 4.11E-11 | 1.36E-07 |
| Heart_Failure | dendritic_cell | *PMF1* | -4.12376086 | 8.26E-12 | 1.63E-11 | 5.19E-08 |
| Heart_Failure | dendritic_cell | *SLC25A44* | 6.356091509 | 2.05E-08 | 3.10E-08 | 0.000128757 |
| Heart_Failure | dendritic_cell | *NBEAL2* | -3.182234336 | 0.007834607 | 0.009016323 | 1 |
| Heart_Failure | dendritic_cell | *FCRL3* | -3.114620319 | 3.54E-14 | 9.32E-14 | 2.23E-10 |
| Heart_Failure | dendritic_cell | *FGFR4* | 13.23499205 | 1.23E-14 | 3.49E-14 | 7.71E-11 |
| Heart_Failure | dendritic_cell | *NACC1* | 5.429590475 | 2.16E-19 | 2.29E-18 | 1.36E-15 |
| Heart_Failure | dendritic_cell | *LY6K* | 6.966488714 | 1.15E-19 | 1.47E-18 | 7.25E-16 |
| Heart_Failure | dendritic_cell | *LY6E* | -4.743816211 | 7.13E-09 | 1.11E-08 | 4.48E-05 |
| Heart_Failure | dendritic_cell | *VPS28* | -4.06855497 | 6.48E-11 | 1.18E-10 | 4.08E-07 |
| Heart_Failure | dendritic_cell | *TONSL* | -4.272889938 | 9.27E-20 | 1.27E-18 | 5.83E-16 |
| Heart_Failure | dendritic_cell | *MUM1* | -3.070273254 | 2.81E-14 | 7.55E-14 | 1.77E-10 |
| Heart_Failure | dendritic_cell | *ZNF333* | -4.276305358 | 4.91E-06 | 6.55E-06 | 0.030884329 |
| Heart_Failure | dendritic_cell | *PPP1R16A* | -2.490848847 | 1.54E-14 | 4.30E-14 | 9.68E-11 |
| Heart_Failure | dendritic_cell | *ORAI2* | 6.424108125 | 2.97E-18 | 1.94E-17 | 1.86E-14 |

| | | | | | | |
|---|---|---|---|---|---|---|
| Heart_Failure | dendritic_cell | *RPL8* | -4.158693387 | 5.13E-05 | 6.54E-05 | 0.322727129 |
| Heart_Failure | dendritic_cell | *LRWD1* | -2.82278044 | 5.50E-10 | 9.30E-10 | 3.46E-06 |
| Heart_Failure | dendritic_cell | *FBXL13* | -4.941496849 | 1.21E-13 | 2.93E-13 | 7.58E-10 |
| Heart_Failure | dendritic_cell | *SCGB3A1* | -3.990636463 | 0.00175808 | 0.002090214 | 1 |
| Heart_Failure | dendritic_cell | *MFSD12* | -4.127947671 | 0.002417282 | 0.002855591 | 1 |
| Heart_Failure | dendritic_cell | *CCDC116* | 5.772795001 | 1.21E-10 | 2.16E-10 | 7.64E-07 |
| Heart_Failure | dendritic_cell | *DVL3* | -5.529267365 | 1.75E-10 | 3.07E-10 | 1.10E-06 |
| Heart_Failure | dendritic_cell | *AP2M1* | -5.294779915 | 2.78E-14 | 7.45E-14 | 1.75E-10 |
| Heart_Failure | dendritic_cell | *ABCF3* | -2.74482683 | 9.72E-10 | 1.61E-09 | 6.11E-06 |
| Heart_Failure | dendritic_cell | *FBXO27* | -1.452699612 | 7.98E-05 | 0.000100758 | 0.501575262 |
| Heart_Failure | dendritic_cell | *DMKN* | 2.115146781 | 1.79E-05 | 2.33E-05 | 0.112634119 |
| Heart_Failure | dendritic_cell | *U2AF1L4* | 0.610856612 | 0.035337535 | 0.039307694 | 1 |
| Heart_Failure | dendritic_cell | *BDH1* | 10.62391885 | 1.62E-12 | 3.45E-12 | 1.02E-08 |
| Heart_Failure | dendritic_cell | *COX7A1* | -3.110588268 | 2.83E-11 | 5.31E-11 | 1.78E-07 |
| Heart_Failure | dendritic_cell | *DUSP14* | -4.26748959 | 1.47E-16 | 6.10E-16 | 9.23E-13 |
| Heart_Failure | dendritic_cell | *LRRC56* | 2.491003108 | 1.07E-05 | 1.40E-05 | 0.067105198 |
| Heart_Failure | dendritic_cell | *PGAP3* | -6.212673336 | 4.00E-18 | 2.50E-17 | 2.52E-14 |
| Heart_Failure | dendritic_cell | *IKZF3* | -2.927756475 | 1.28E-18 | 9.50E-18 | 8.05E-15 |
| Heart_Failure | dendritic_cell | *ACOX1* | -3.387167978 | 8.75E-18 | 5.01E-17 | 5.50E-14 |
| Heart_Failure | dendritic_cell | *PRPSAP1* | -4.842426245 | 6.19E-11 | 1.13E-10 | 3.89E-07 |
| Heart_Failure | dendritic_cell | *CYGB* | -4.726681001 | 7.27E-20 | 1.08E-18 | 4.57E-16 |
| Heart_Failure | dendritic_cell | *ZNF577* | 3.192974247 | 6.72E-16 | 2.43E-15 | 4.23E-12 |
| Heart_Failure | dendritic_cell | *TMEM143* | -3.603407769 | 2.92E-13 | 6.77E-13 | 1.83E-09 |
| Heart_Failure | dendritic_cell | *ALDH16A1* | -6.314411282 | 1.47E-12 | 3.15E-12 | 9.26E-09 |
| Heart_Failure | dendritic_cell | *SIGLEC11* | 3.027919014 | 1.20E-18 | 9.02E-18 | 7.52E-15 |
| Heart_Failure | dendritic_cell | *MPP3* | -5.502792485 | 5.90E-06 | 7.84E-06 | 0.037068734 |
| Heart_Failure | dendritic_cell | *NAGS* | 11.03221989 | 1.89E-17 | 9.83E-17 | 1.19E-13 |
| Heart_Failure | dendritic_cell | *ASB16* | -4.688581959 | 7.81E-16 | 2.79E-15 | 4.91E-12 |
| Heart_Failure | dendritic_cell | *DBF4B* | -1.532260031 | 3.50E-07 | 4.96E-07 | 0.002197828 |
| Heart_Failure | dendritic_cell | *PLCD3* | -2.840376067 | 4.70E-17 | 2.20E-16 | 2.96E-13 |
| Heart_Failure | dendritic_cell | *RACGAP1* | 4.741374048 | 2.21E-20 | 6.44E-19 | 1.39E-16 |
| Heart_Failure | dendritic_cell | *LARP4* | -2.927433479 | 5.42E-12 | 1.09E-11 | 3.41E-08 |
| Heart_Failure | dendritic_cell | *SPC24* | 6.815204053 | 6.47E-21 | 4.66E-19 | 4.07E-17 |
| Heart_Failure | dendritic_cell | *IP6K3* | -3.054999939 | 0.021920345 | 0.024618293 | 1 |
| Heart_Failure | dendritic_cell | *LEMD2* | -4.536422268 | 4.35E-10 | 7.38E-10 | 2.73E-06 |
| Heart_Failure | dendritic_cell | *SCIMP* | 4.601561128 | 1.91E-12 | 4.03E-12 | 1.20E-08 |
| Heart_Failure | dendritic_cell | *ASGR2* | 1.816556497 | 6.35E-21 | 4.66E-19 | 3.99E-17 |
| Heart_Failure | dendritic_cell | *TNFSF13* | -5.126277056 | 1.36E-06 | 1.86E-06 | 0.008532925 |
| Heart_Failure | dendritic_cell | *SENP3* | -3.551339646 | 2.15E-18 | 1.49E-17 | 1.35E-14 |
| Heart_Failure | dendritic_cell | *SNRNP25* | 3.831420816 | 2.36E-18 | 1.62E-17 | 1.48E-14 |
| Heart_Failure | dendritic_cell | *WDR90* | -2.609725682 | 3.69E-11 | 6.87E-11 | 2.32E-07 |
| Heart_Failure | dendritic_cell | *JMJD8* | 1.603411119 | 0.035244076 | 0.039210672 | 1 |
| Heart_Failure | dendritic_cell | *MEIOB* | 4.510519985 | 1.05E-20 | 4.95E-19 | 6.58E-17 |
| Heart_Failure | dendritic_cell | *FLYWCH2* | -6.565645716 | 3.08E-15 | 9.77E-15 | 1.94E-11 |
| Heart_Failure | dendritic_cell | *ZG16B* | -39.89741266 | 2.90E-20 | 6.48E-19 | 1.82E-16 |
| Heart_Failure | dendritic_cell | *TTC9C* | -4.040455553 | 4.57E-19 | 4.08E-18 | 2.87E-15 |
| Heart_Failure | dendritic_cell | *NXF1* | -4.42823327 | 5.10E-10 | 8.63E-10 | 3.21E-06 |
| Heart_Failure | dendritic_cell | *RPL29* | -4.171262542 | 4.72E-13 | 1.06E-12 | 2.96E-09 |
| Heart_Failure | dendritic_cell | *TPCN2* | -3.71582231 | 1.54E-05 | 2.01E-05 | 0.096664634 |
| Heart_Failure | dendritic_cell | *CYP4A22* | 5.535107552 | 5.28E-20 | 8.88E-19 | 3.32E-16 |

| | | | | | | |
|---|---|---|---|---|---|---|
| Heart_Failure | dendritic_cell | *PDZK1IP1* | -61.12490861 | 2.24E-15 | 7.32E-15 | 1.41E-11 |
| Heart_Failure | dendritic_cell | *COA7* | 10.94042696 | 2.63E-12 | 5.48E-12 | 1.66E-08 |
| Heart_Failure | dendritic_cell | *SLC1A7* | -1.503835408 | 5.02E-11 | 9.21E-11 | 3.16E-07 |
| Heart_Failure | dendritic_cell | *ACOT11* | -2.510415816 | 0.003278847 | 0.003848069 | 1 |
| Heart_Failure | dendritic_cell | *USP24* | -2.643494716 | 2.21E-11 | 4.19E-11 | 1.39E-07 |
| Heart_Failure | dendritic_cell | *NOL9* | -4.646980168 | 7.00E-15 | 2.08E-14 | 4.40E-11 |
| Heart_Failure | dendritic_cell | *ZSWIM5* | -3.873810645 | 4.07E-13 | 9.23E-13 | 2.56E-09 |
| Heart_Failure | dendritic_cell | *RBP7* | -6.356683458 | 3.91E-07 | 5.54E-07 | 0.002460799 |
| Heart_Failure | dendritic_cell | *FBLIM1* | -4.166002629 | 2.51E-15 | 8.10E-15 | 1.58E-11 |
| Heart_Failure | dendritic_cell | *SLC25A34* | -6.56931692 | 2.30E-18 | 1.59E-17 | 1.44E-14 |
| Heart_Failure | dendritic_cell | *PDPN* | -2.494593079 | 3.46E-19 | 3.27E-18 | 2.18E-15 |
| Heart_Failure | dendritic_cell | *LRRC38* | 8.774720783 | 5.33E-20 | 8.91E-19 | 3.35E-16 |
| Heart_Failure | dendritic_cell | *LAPTM5* | 4.598335057 | 3.47E-09 | 5.53E-09 | 2.18E-05 |
| Heart_Failure | dendritic_cell | *PEF1* | 9.329318778 | 1.14E-10 | 2.04E-10 | 7.18E-07 |
| Heart_Failure | dendritic_cell | *SYNC* | -4.460392194 | 6.12E-12 | 1.22E-11 | 3.85E-08 |
| Heart_Failure | dendritic_cell | *RBBP4* | -4.113186796 | 1.86E-06 | 2.53E-06 | 0.011707253 |
| Heart_Failure | dendritic_cell | *TSSK3* | -3.468408537 | 9.92E-09 | 1.53E-08 | 6.24E-05 |
| Heart_Failure | dendritic_cell | *CAMK2N1* | -4.725840538 | 1.37E-19 | 1.66E-18 | 8.62E-16 |
| Heart_Failure | dendritic_cell | *TTLL10* | -6.691415682 | 4.07E-17 | 1.94E-16 | 2.56E-13 |
| Heart_Failure | dendritic_cell | *NFIA* | -2.815013864 | 1.13E-15 | 3.90E-15 | 7.10E-12 |
| Heart_Failure | dendritic_cell | *OMA1* | -2.861363778 | 7.44E-11 | 1.34E-10 | 4.68E-07 |
| Heart_Failure | dendritic_cell | *TM2D1* | -4.513135525 | 7.36E-13 | 1.62E-12 | 4.63E-09 |
| Heart_Failure | dendritic_cell | *FUBP1* | -4.588175868 | 8.44E-09 | 1.31E-08 | 5.31E-05 |
| Heart_Failure | dendritic_cell | *NEXN* | -4.079888012 | 1.60E-16 | 6.62E-16 | 1.01E-12 |
| Heart_Failure | dendritic_cell | *B3GALT2* | -7.464226755 | 2.97E-18 | 1.94E-17 | 1.87E-14 |
| Heart_Failure | dendritic_cell | *NTNG1* | -1.961996506 | 3.33E-11 | 6.22E-11 | 2.09E-07 |
| Heart_Failure | dendritic_cell | *HFM1* | -4.174764079 | 1.79E-18 | 1.27E-17 | 1.12E-14 |
| Heart_Failure | dendritic_cell | *KCNT2* | -1.742641979 | 1.66E-20 | 6.08E-19 | 1.05E-16 |
| Heart_Failure | dendritic_cell | *VCAM1* | -4.765541346 | 6.13E-14 | 1.56E-13 | 3.85E-10 |
| Heart_Failure | dendritic_cell | *EXTL2* | -2.909176495 | 3.23E-12 | 6.65E-12 | 2.03E-08 |
| Heart_Failure | dendritic_cell | *ZNF496* | -5.855660278 | 2.70E-15 | 8.67E-15 | 1.70E-11 |
| Heart_Failure | dendritic_cell | *TRIM58* | -4.220678781 | 4.33E-14 | 1.12E-13 | 2.72E-10 |
| Heart_Failure | dendritic_cell | *IGSF8* | -4.28996334 | 2.86E-19 | 2.82E-18 | 1.80E-15 |
| Heart_Failure | dendritic_cell | *VANGL2* | -7.31685304 | 1.98E-10 | 3.46E-10 | 1.25E-06 |
| Heart_Failure | dendritic_cell | *FCRLB* | 5.367884556 | 2.26E-19 | 2.36E-18 | 1.42E-15 |
| Heart_Failure | dendritic_cell | *KLHDC9* | -1.760858082 | 0.014849211 | 0.016833211 | 1 |
| Heart_Failure | dendritic_cell | *LMX1A* | -1.841186254 | 7.21E-17 | 3.25E-16 | 4.53E-13 |
| Heart_Failure | dendritic_cell | *TDRD5* | -3.985901919 | 5.14E-13 | 1.15E-12 | 3.23E-09 |
| Heart_Failure | dendritic_cell | *SNED1* | -2.25176837 | 1.03E-18 | 7.97E-18 | 6.49E-15 |
| Heart_Failure | dendritic_cell | *C1orf115* | -3.406399974 | 2.92E-11 | 5.47E-11 | 1.84E-07 |
| Heart_Failure | dendritic_cell | *BROX* | -1.846652957 | 0.001043268 | 0.001249815 | 1 |
| Heart_Failure | dendritic_cell | *ACP6* | -3.882243191 | 8.90E-17 | 3.91E-16 | 5.60E-13 |
| Heart_Failure | dendritic_cell | *CNST* | -4.385694155 | 1.33E-13 | 3.22E-13 | 8.36E-10 |
| Heart_Failure | dendritic_cell | *PM20D1* | -6.417090235 | 3.21E-19 | 3.06E-18 | 2.02E-15 |
| Heart_Failure | dendritic_cell | *B3GALNT2* | -3.318256138 | 8.04E-07 | 1.11E-06 | 0.005053863 |
| Heart_Failure | dendritic_cell | *PUS10* | -5.580490131 | 1.43E-11 | 2.77E-11 | 9.02E-08 |
| Heart_Failure | dendritic_cell | *RFTN2* | -3.393302334 | 2.57E-18 | 1.72E-17 | 1.61E-14 |
| Heart_Failure | dendritic_cell | *DISC1* | 3.2262619 | 4.34E-18 | 2.68E-17 | 2.73E-14 |
| Heart_Failure | dendritic_cell | *FAM84A* | -5.441653233 | 2.33E-20 | 6.44E-19 | 1.47E-16 |
| Heart_Failure | dendritic_cell | *FRZB* | -6.630769889 | 2.26E-09 | 3.65E-09 | 1.42E-05 |

| | | | | | | |
|---|---|---|---|---|---|---|
| Heart_Failure | dendritic_cell | *FBXO41* | 1.399351091 | 1.26E-20 | 5.13E-19 | 7.90E-17 |
| Heart_Failure | dendritic_cell | *SMC6* | -4.908425656 | 3.30E-11 | 6.16E-11 | 2.07E-07 |
| Heart_Failure | dendritic_cell | *CCDC74A* | -11.73641219 | 6.93E-20 | 1.05E-18 | 4.36E-16 |
| Heart_Failure | dendritic_cell | *H3F3A* | 4.123342414 | 2.17E-09 | 3.51E-09 | 1.36E-05 |
| Heart_Failure | dendritic_cell | *NOSTRIN* | -2.462170557 | 6.29E-16 | 2.29E-15 | 3.95E-12 |
| Heart_Failure | dendritic_cell | *SGPP2* | -6.454406247 | 1.50E-15 | 5.03E-15 | 9.40E-12 |
| Heart_Failure | dendritic_cell | *BBS5* | 5.044824884 | 4.81E-18 | 2.94E-17 | 3.02E-14 |
| Heart_Failure | dendritic_cell | *HPGDS* | -3.540037148 | 3.56E-13 | 8.16E-13 | 2.24E-09 |
| Heart_Failure | dendritic_cell | *PDLIM5* | -3.781449285 | 2.06E-12 | 4.34E-12 | 1.30E-08 |
| Heart_Failure | dendritic_cell | *RPRD2* | -3.992078604 | 4.64E-13 | 1.05E-12 | 2.92E-09 |
| Heart_Failure | dendritic_cell | *CTSS* | 1.806331531 | 2.09E-15 | 6.87E-15 | 1.32E-11 |
| Heart_Failure | dendritic_cell | *PACRGL* | -3.709842829 | 1.71E-17 | 9.03E-17 | 1.07E-13 |
| Heart_Failure | dendritic_cell | *C1QTNF7* | -2.424581423 | 9.55E-19 | 7.48E-18 | 6.00E-15 |
| Heart_Failure | dendritic_cell | *LYSMD1* | -2.949515062 | 1.58E-18 | 1.15E-17 | 9.93E-15 |
| Heart_Failure | dendritic_cell | *ERCC3* | -2.297571946 | 4.30E-06 | 5.75E-06 | 0.027028897 |
| Heart_Failure | dendritic_cell | *DHX57* | -4.491390506 | 8.09E-16 | 2.88E-15 | 5.09E-12 |
| Heart_Failure | dendritic_cell | *S100A9* | -5.352940009 | 1.37E-06 | 1.88E-06 | 0.00862423 |
| Heart_Failure | dendritic_cell | *S100A12* | -21.89990852 | 2.18E-16 | 8.73E-16 | 1.37E-12 |
| Heart_Failure | dendritic_cell | *TDRD10* | -5.154485393 | 4.94E-16 | 1.83E-15 | 3.10E-12 |
| Heart_Failure | dendritic_cell | *DCAF16* | -5.723929893 | 1.07E-08 | 1.65E-08 | 6.72E-05 |
| Heart_Failure | dendritic_cell | *GNPDA2* | -2.067595267 | 1.16E-06 | 1.59E-06 | 0.007289841 |
| Heart_Failure | dendritic_cell | *NIPAL1* | -4.093563191 | 5.16E-20 | 8.72E-19 | 3.24E-16 |
| Heart_Failure | dendritic_cell | *CGGBP1* | -0.81008115 | 0.005173296 | 0.006008592 | 1 |
| Heart_Failure | dendritic_cell | *DCST2* | -6.266329832 | 7.79E-20 | 1.13E-18 | 4.90E-16 |
| Heart_Failure | dendritic_cell | *LMOD3* | -10.01999967 | 8.99E-15 | 2.63E-14 | 5.65E-11 |
| Heart_Failure | dendritic_cell | *NBPF10* | 5.312427615 | 1.43E-10 | 2.54E-10 | 9.02E-07 |
| Heart_Failure | dendritic_cell | *POGLUT1* | 2.446222328 | 7.88E-08 | 1.16E-07 | 0.000495309 |
| Heart_Failure | dendritic_cell | *SLC22A15* | 5.570899358 | 1.31E-10 | 2.33E-10 | 8.26E-07 |
| Heart_Failure | dendritic_cell | *SLC15A2* | -3.235905546 | 1.57E-05 | 2.04E-05 | 0.098535664 |
| Heart_Failure | dendritic_cell | *LRRC58* | -4.044334148 | 0.000954175 | 0.001145921 | 1 |
| Heart_Failure | dendritic_cell | *FSTL1* | -2.030797063 | 1.51E-17 | 8.10E-17 | 9.48E-14 |
| Heart_Failure | dendritic_cell | *TMEM183A* | -7.916332216 | 1.06E-13 | 2.58E-13 | 6.65E-10 |
| Heart_Failure | dendritic_cell | *IGFBP7* | -2.703367226 | 9.89E-17 | 4.29E-16 | 6.22E-13 |
| Heart_Failure | dendritic_cell | *CCT3* | 2.105848174 | 0.007820796 | 0.009002077 | 1 |
| Heart_Failure | dendritic_cell | *GLB1L* | -2.813087706 | 4.07E-19 | 3.72E-18 | 2.56E-15 |
| Heart_Failure | dendritic_cell | *SERPINI1* | -5.209112288 | 8.83E-17 | 3.89E-16 | 5.55E-13 |
| Heart_Failure | dendritic_cell | *SPTA1* | -1.155700903 | 1.39E-08 | 2.12E-08 | 8.74E-05 |
| Heart_Failure | dendritic_cell | *PYHIN1* | -2.204772389 | 8.31E-21 | 4.66E-19 | 5.23E-17 |
| Heart_Failure | dendritic_cell | *AIM2* | 3.414244932 | 1.07E-20 | 4.95E-19 | 6.73E-17 |
| Heart_Failure | dendritic_cell | *EFHB* | -0.404489245 | 0.006103527 | 0.007060326 | 1 |
| Heart_Failure | dendritic_cell | *ICA1L* | -4.349702298 | 4.46E-13 | 1.01E-12 | 2.80E-09 |
| Heart_Failure | dendritic_cell | *CD200R1* | 1.502327114 | 9.38E-20 | 1.27E-18 | 5.90E-16 |
| Heart_Failure | dendritic_cell | *GTPBP8* | -6.205123691 | 1.87E-08 | 2.84E-08 | 0.000117531 |
| Heart_Failure | dendritic_cell | *SPICE1* | -4.94854954 | 5.60E-15 | 1.70E-14 | 3.52E-11 |
| Heart_Failure | dendritic_cell | *CADPS* | -2.671080811 | 2.69E-20 | 6.44E-19 | 1.69E-16 |
| Heart_Failure | dendritic_cell | *PPM1K* | -5.302726107 | 9.51E-09 | 1.47E-08 | 5.98E-05 |
| Heart_Failure | dendritic_cell | *GMPS* | -5.094648676 | 3.37E-14 | 8.92E-14 | 2.12E-10 |
| Heart_Failure | dendritic_cell | *TIPARP* | -4.564016806 | 7.99E-12 | 1.58E-11 | 5.02E-08 |
| Heart_Failure | dendritic_cell | *DCLK3* | 6.041186948 | 0.000599521 | 0.00072694 | 1 |
| Heart_Failure | dendritic_cell | *RPP14* | -6.141327085 | 1.00E-06 | 1.38E-06 | 0.006299976 |

| | | | | | | |
|---|---|---|---|---|---|---|
| Heart_Failure | dendritic_cell | *DNASE1L3* | 1.198241233 | 1.95E-20 | 6.44E-19 | 1.23E-16 |
| Heart_Failure | dendritic_cell | *CRELD1* | -2.948432638 | 3.98E-13 | 9.04E-13 | 2.50E-09 |
| Heart_Failure | dendritic_cell | *PCOLCE2* | -1.759058592 | 4.11E-19 | 3.75E-18 | 2.59E-15 |
| Heart_Failure | dendritic_cell | *TTC14* | -2.894975834 | 8.61E-14 | 2.14E-13 | 5.41E-10 |
| Heart_Failure | dendritic_cell | *RCHY1* | -5.188604858 | 1.28E-15 | 4.39E-15 | 8.08E-12 |
| Heart_Failure | dendritic_cell | *CCDC158* | -2.605815499 | 2.14E-17 | 1.09E-16 | 1.34E-13 |
| Heart_Failure | dendritic_cell | *GYG1* | -6.05854411 | 1.73E-12 | 3.68E-12 | 1.09E-08 |
| Heart_Failure | dendritic_cell | *RYK* | -4.372797756 | 7.96E-11 | 1.43E-10 | 5.01E-07 |
| Heart_Failure | dendritic_cell | *PLB1* | 1.330176451 | 6.42E-21 | 4.66E-19 | 4.04E-17 |
| Heart_Failure | dendritic_cell | *KIAA1143* | -1.661376618 | 0.00023139 | 0.000286086 | 1 |
| Heart_Failure | dendritic_cell | *CDCP1* | 1.07737017 | 6.40E-21 | 4.66E-19 | 4.02E-17 |
| Heart_Failure | dendritic_cell | *LZTFL1* | -5.743271998 | 1.07E-16 | 4.59E-16 | 6.71E-13 |
| Heart_Failure | dendritic_cell | *FYCO1* | -5.6001032 | 4.53E-11 | 8.34E-11 | 2.85E-07 |
| Heart_Failure | dendritic_cell | *LRRC2* | -1.933746775 | 8.85E-21 | 4.70E-19 | 5.56E-17 |
| Heart_Failure | dendritic_cell | *DTX3L* | 2.867707736 | 0.008099396 | 0.009310825 | 1 |
| Heart_Failure | dendritic_cell | *NMNAT3* | -3.901911945 | 6.07E-06 | 8.06E-06 | 0.038132404 |
| Heart_Failure | dendritic_cell | *SMIM12* | -5.235498645 | 2.82E-15 | 9.03E-15 | 1.77E-11 |
| Heart_Failure | dendritic_cell | *TPRA1* | -6.380816885 | 3.05E-16 | 1.19E-15 | 1.92E-12 |
| Heart_Failure | dendritic_cell | *YEATS2* | -2.201489821 | 0.040139031 | 0.044514744 | 1 |
| Heart_Failure | dendritic_cell | *MEAF6* | -5.761178007 | 5.46E-15 | 1.66E-14 | 3.43E-11 |
| Heart_Failure | dendritic_cell | *DNALI1* | -5.8414072 | 5.85E-22 | 2.63E-19 | 3.68E-18 |
| Heart_Failure | dendritic_cell | *RPN1* | -5.262859449 | 4.95E-07 | 6.96E-07 | 0.003111293 |
| Heart_Failure | dendritic_cell | *RFC4* | -5.373856359 | 1.61E-17 | 8.56E-17 | 1.01E-13 |
| Heart_Failure | dendritic_cell | *RFT1* | -2.910706621 | 0.002892353 | 0.003404011 | 1 |
| Heart_Failure | dendritic_cell | *SFMBT1* | -6.145071342 | 8.58E-13 | 1.88E-12 | 5.40E-09 |
| Heart_Failure | dendritic_cell | *UVSSA* | -2.72608405 | 1.81E-05 | 2.35E-05 | 0.113661969 |
| Heart_Failure | dendritic_cell | *FAM208A* | -3.46584701 | 1.75E-07 | 2.52E-07 | 0.001102004 |
| Heart_Failure | dendritic_cell | *LRPAP1* | -3.102609359 | 0.007629266 | 0.008789664 | 1 |
| Heart_Failure | dendritic_cell | *SLC51A* | -3.477632144 | 2.17E-20 | 6.44E-19 | 1.36E-16 |
| Heart_Failure | dendritic_cell | *PIGX* | -5.320329153 | 3.84E-11 | 7.13E-11 | 2.42E-07 |
| Heart_Failure | dendritic_cell | *ABLIM2* | -6.020277931 | 2.60E-16 | 1.02E-15 | 1.63E-12 |
| Heart_Failure | dendritic_cell | *EXO5* | -3.912915278 | 1.00E-12 | 2.18E-12 | 6.31E-09 |
| Heart_Failure | dendritic_cell | *C1orf50* | 5.448779374 | 2.01E-09 | 3.25E-09 | 1.26E-05 |
| Heart_Failure | dendritic_cell | *ERMAP* | -3.024270132 | 1.35E-07 | 1.96E-07 | 0.000851783 |
| Heart_Failure | dendritic_cell | *METAP1* | -3.838998984 | 0.000335963 | 0.000412377 | 1 |
| Heart_Failure | dendritic_cell | *DNAJB14* | -3.984214346 | 6.18E-12 | 1.24E-11 | 3.89E-08 |
| Heart_Failure | dendritic_cell | *SLC9B2* | -4.236526919 | 2.85E-16 | 1.12E-15 | 1.79E-12 |
| Heart_Failure | dendritic_cell | *BDH2* | -2.157097033 | 1.01E-14 | 2.93E-14 | 6.36E-11 |
| Heart_Failure | dendritic_cell | *CDC25A* | -2.531304349 | 2.29E-07 | 3.27E-07 | 0.001436999 |
| Heart_Failure | dendritic_cell | *ZNF589* | -2.247445986 | 3.67E-09 | 5.83E-09 | 2.30E-05 |
| Heart_Failure | dendritic_cell | *ATRIP* | 4.511236169 | 8.91E-21 | 4.70E-19 | 5.60E-17 |
| Heart_Failure | dendritic_cell | *SHISA5* | -6.037179067 | 6.05E-12 | 1.21E-11 | 3.80E-08 |
| Heart_Failure | dendritic_cell | *APEH* | -4.359320485 | 4.24E-15 | 1.31E-14 | 2.67E-11 |
| Heart_Failure | dendritic_cell | *MST1R* | 1.884651086 | 0.002905876 | 0.003418646 | 1 |
| Heart_Failure | dendritic_cell | *RAD54L2* | -3.524939835 | 3.14E-07 | 4.46E-07 | 0.001971368 |
| Heart_Failure | dendritic_cell | *TEX264* | -2.930262175 | 6.50E-05 | 8.25E-05 | 0.408422105 |
| Heart_Failure | dendritic_cell | *GRM2* | 3.735155194 | 7.43E-21 | 4.66E-19 | 4.67E-17 |
| Heart_Failure | dendritic_cell | *POC1A* | 3.368919996 | 1.69E-19 | 1.95E-18 | 1.07E-15 |
| Heart_Failure | dendritic_cell | *PPM1M* | 2.263100411 | 8.71E-19 | 6.92E-18 | 5.47E-15 |
| Heart_Failure | dendritic_cell | *C4orf3* | -6.303915312 | 2.41E-13 | 5.62E-13 | 1.51E-09 |

| | | | | | | |
|---|---|---|---|---|---|---|
| Heart_Failure | dendritic_cell | *ANXA5* | -3.659467172 | 8.48E-13 | 1.85E-12 | 5.33E-09 |
| Heart_Failure | dendritic_cell | *MAP9* | -4.384512948 | 1.19E-18 | 9.00E-18 | 7.48E-15 |
| Heart_Failure | dendritic_cell | *CEP44* | -5.456205158 | 6.46E-12 | 1.29E-11 | 4.06E-08 |
| Heart_Failure | dendritic_cell | *C4orf45* | -4.389747625 | 2.12E-16 | 8.53E-16 | 1.33E-12 |
| Heart_Failure | dendritic_cell | *TMEM144* | -5.16865307 | 5.46E-13 | 1.22E-12 | 3.43E-09 |
| Heart_Failure | dendritic_cell | *NPY1R* | -2.394964047 | 3.06E-19 | 2.95E-18 | 1.92E-15 |
| Heart_Failure | dendritic_cell | *IL15* | 2.705724281 | 2.85E-20 | 6.44E-19 | 1.79E-16 |
| Heart_Failure | dendritic_cell | *ARFIP1* | -3.576122339 | 1.23E-14 | 3.49E-14 | 7.72E-11 |
| Heart_Failure | dendritic_cell | *OTUD4* | -5.204952757 | 2.71E-12 | 5.62E-12 | 1.70E-08 |
| Heart_Failure | dendritic_cell | *ITGA2* | 8.609118672 | 3.44E-14 | 9.08E-14 | 2.16E-10 |
| Heart_Failure | dendritic_cell | *NDUFAF2* | -3.755021415 | 5.30E-09 | 8.34E-09 | 3.33E-05 |
| Heart_Failure | dendritic_cell | *LMBRD2* | -5.097267077 | 1.83E-15 | 6.05E-15 | 1.15E-11 |
| Heart_Failure | dendritic_cell | *PGGT1B* | -4.803437 | 9.54E-07 | 1.32E-06 | 0.006000183 |
| Heart_Failure | dendritic_cell | *CMBL* | -2.280234237 | 2.44E-20 | 6.44E-19 | 1.53E-16 |
| Heart_Failure | dendritic_cell | *C5orf63* | -4.804363783 | 2.39E-19 | 2.45E-18 | 1.50E-15 |
| Heart_Failure | dendritic_cell | *PRRC1* | -3.840249528 | 0.000669575 | 0.000810009 | 1 |
| Heart_Failure | dendritic_cell | *F2RL1* | 4.64761726 | 6.86E-12 | 1.37E-11 | 4.31E-08 |
| Heart_Failure | dendritic_cell | *WDR41* | -3.5020003 | 4.16E-11 | 7.69E-11 | 2.61E-07 |
| Heart_Failure | dendritic_cell | *NDUFS4* | -4.493901425 | 5.77E-13 | 1.28E-12 | 3.63E-09 |
| Heart_Failure | dendritic_cell | *RHOBTB3* | -2.357106643 | 1.78E-15 | 5.91E-15 | 1.12E-11 |
| Heart_Failure | dendritic_cell | *TIGD6* | -10.90610385 | 6.65E-13 | 1.47E-12 | 4.18E-09 |
| Heart_Failure | dendritic_cell | *SERINC5* | -3.323181166 | 8.27E-05 | 0.000104367 | 0.519747038 |
| Heart_Failure | dendritic_cell | *PRIMPOL* | -4.211037048 | 2.00E-14 | 5.48E-14 | 1.26E-10 |
| Heart_Failure | dendritic_cell | *ERAP1* | -1.541257785 | 0.00368994 | 0.004321657 | 1 |
| Heart_Failure | dendritic_cell | *RICTOR* | -4.032658988 | 6.58E-13 | 1.46E-12 | 4.14E-09 |
| Heart_Failure | dendritic_cell | *ANKRA2* | -5.20797016 | 1.80E-14 | 4.97E-14 | 1.13E-10 |
| Heart_Failure | dendritic_cell | *UBLCP1* | -3.312076996 | 1.62E-09 | 2.65E-09 | 1.02E-05 |
| Heart_Failure | dendritic_cell | *UTP15* | -5.189034206 | 6.08E-16 | 2.22E-15 | 3.82E-12 |
| Heart_Failure | dendritic_cell | *TLR3* | -6.620303554 | 2.90E-15 | 9.26E-15 | 1.82E-11 |
| Heart_Failure | dendritic_cell | *KLKB1* | -2.053418518 | 4.93E-11 | 9.05E-11 | 3.10E-07 |
| Heart_Failure | dendritic_cell | *NSA2* | -1.537498196 | 2.62E-06 | 3.54E-06 | 0.016469787 |
| Heart_Failure | dendritic_cell | *CCDC127* | 3.827187472 | 4.70E-19 | 4.16E-18 | 2.95E-15 |
| Heart_Failure | dendritic_cell | *GDF9* | -5.101788536 | 5.94E-08 | 8.78E-08 | 0.000373314 |
| Heart_Failure | dendritic_cell | *GRIK2* | -1.809804088 | 2.22E-18 | 1.54E-17 | 1.40E-14 |
| Heart_Failure | dendritic_cell | *MB21D1* | -5.405234108 | 2.13E-16 | 8.55E-16 | 1.34E-12 |
| Heart_Failure | dendritic_cell | *TXLNB* | 5.104579855 | 6.76E-20 | 1.03E-18 | 4.25E-16 |
| Heart_Failure | dendritic_cell | *DCBLD1* | -4.203915125 | 1.74E-07 | 2.50E-07 | 0.001092804 |
| Heart_Failure | dendritic_cell | *TMEM200A* | -3.476882664 | 2.37E-18 | 1.62E-17 | 1.49E-14 |
| Heart_Failure | dendritic_cell | *PDSS2* | -4.112428242 | 2.46E-12 | 5.14E-12 | 1.55E-08 |
| Heart_Failure | dendritic_cell | *STXBP5* | -3.886497447 | 3.98E-13 | 9.05E-13 | 2.51E-09 |
| Heart_Failure | dendritic_cell | *PI16* | -2.60310146 | 2.95E-19 | 2.88E-18 | 1.85E-15 |
| Heart_Failure | dendritic_cell | *DAGLB* | -2.19683076 | 0.002023894 | 0.002399438 | 1 |
| Heart_Failure | dendritic_cell | *STK17A* | -4.481878274 | 1.74E-15 | 5.78E-15 | 1.10E-11 |
| Heart_Failure | dendritic_cell | *GALNT10* | -4.361065046 | 1.38E-12 | 2.96E-12 | 8.68E-09 |
| Heart_Failure | dendritic_cell | *SAP30L* | -4.676833282 | 1.11E-12 | 2.40E-12 | 6.97E-09 |
| Heart_Failure | dendritic_cell | *RPS14* | -4.370456826 | 3.18E-09 | 5.08E-09 | 2.00E-05 |
| Heart_Failure | dendritic_cell | *HCN1* | -3.4915859 | 2.65E-20 | 6.44E-19 | 1.67E-16 |
| Heart_Failure | dendritic_cell | *SLU7* | -2.409954673 | 0.022968781 | 0.025781954 | 1 |
| Heart_Failure | dendritic_cell | *RP9* | -4.750576243 | 4.25E-16 | 1.60E-15 | 2.67E-12 |
| Heart_Failure | dendritic_cell | *CAMLG* | -3.889390402 | 2.31E-16 | 9.19E-16 | 1.45E-12 |

| | | | | | | |
|---|---|---|---|---|---|---|
| Heart_Failure | dendritic_cell | *KCNK5* | -12.21727854 | 1.46E-16 | 6.07E-16 | 9.16E-13 |
| Heart_Failure | dendritic_cell | *KIAA1324L* | -1.663505914 | 0.004208619 | 0.004915399 | 1 |
| Heart_Failure | dendritic_cell | *USP49* | -4.061250081 | 1.70E-15 | 5.65E-15 | 1.07E-11 |
| Heart_Failure | dendritic_cell | *ZNF704* | -3.500662991 | 6.26E-14 | 1.59E-13 | 3.93E-10 |
| Heart_Failure | dendritic_cell | *FNDC1* | -2.407500016 | 1.97E-19 | 2.15E-18 | 1.24E-15 |
| Heart_Failure | dendritic_cell | *CHMP4C* | -0.916744219 | 0.020142419 | 0.022690447 | 1 |
| Heart_Failure | dendritic_cell | *BRI3* | -3.52661104 | 2.67E-06 | 3.60E-06 | 0.016766259 |
| Heart_Failure | dendritic_cell | *LMTK2* | -4.614634758 | 1.72E-15 | 5.71E-15 | 1.08E-11 |
| Heart_Failure | dendritic_cell | *PEX2* | -5.196101831 | 5.70E-15 | 1.73E-14 | 3.59E-11 |
| Heart_Failure | dendritic_cell | *TNFRSF11B* | -4.380591308 | 2.01E-19 | 2.17E-18 | 1.26E-15 |
| Heart_Failure | dendritic_cell | *DEFB1* | -2.973077595 | 1.79E-12 | 3.80E-12 | 1.13E-08 |
| Heart_Failure | dendritic_cell | *SUN1* | -2.926283843 | 9.53E-14 | 2.34E-13 | 5.99E-10 |
| Heart_Failure | dendritic_cell | *OXR1* | -3.779836341 | 3.03E-13 | 7.02E-13 | 1.91E-09 |
| Heart_Failure | dendritic_cell | *GPER1* | -2.113176257 | 2.83E-13 | 6.58E-13 | 1.78E-09 |
| Heart_Failure | dendritic_cell | *TMEM184A* | 3.207765155 | 1.75E-18 | 1.25E-17 | 1.10E-14 |
| Heart_Failure | dendritic_cell | *INTS1* | -6.087302748 | 1.44E-15 | 4.86E-15 | 9.06E-12 |
| Heart_Failure | dendritic_cell | *SLC4A2* | 7.970936479 | 1.09E-08 | 1.68E-08 | 6.88E-05 |
| Heart_Failure | dendritic_cell | *FASTK* | -5.475070391 | 3.14E-14 | 8.38E-14 | 1.98E-10 |
| Heart_Failure | dendritic_cell | *ALDH7A1* | -3.412545295 | 9.81E-12 | 1.92E-11 | 6.17E-08 |
| Heart_Failure | dendritic_cell | *COX6C* | -5.622653236 | 2.11E-11 | 4.02E-11 | 1.33E-07 |
| Heart_Failure | dendritic_cell | *YWHAZ* | -2.78867247 | 3.26E-09 | 5.20E-09 | 2.05E-05 |
| Heart_Failure | dendritic_cell | *SLC25A32* | -5.119477858 | 3.92E-09 | 6.22E-09 | 2.47E-05 |
| Heart_Failure | dendritic_cell | *DCAF13* | -6.71479079 | 3.96E-11 | 7.34E-11 | 2.49E-07 |
| Heart_Failure | dendritic_cell | *INTS8* | -4.140551233 | 3.76E-12 | 7.69E-12 | 2.36E-08 |
| Heart_Failure | dendritic_cell | *FREM1* | -2.14658001 | 9.99E-20 | 1.34E-18 | 6.28E-16 |
| Heart_Failure | dendritic_cell | *RPP25L* | 8.009624813 | 3.37E-09 | 5.38E-09 | 2.12E-05 |
| Heart_Failure | dendritic_cell | *FAM219A* | -4.914502562 | 6.87E-14 | 1.73E-13 | 4.32E-10 |
| Heart_Failure | dendritic_cell | *NUDT2* | -0.617002159 | 0.041193664 | 0.045660184 | 1 |
| Heart_Failure | dendritic_cell | *TMEM65* | 3.16137516 | 0.001585358 | 0.00188879 | 1 |
| Heart_Failure | dendritic_cell | *CCDC171* | -4.428203866 | 9.88E-16 | 3.46E-15 | 6.21E-12 |
| Heart_Failure | dendritic_cell | *SYK* | 1.102826369 | 1.18E-20 | 5.00E-19 | 7.39E-17 |
| Heart_Failure | dendritic_cell | *NIPSNAP3B* | -2.434185454 | 1.62E-10 | 2.84E-10 | 1.02E-06 |
| Heart_Failure | dendritic_cell | *METTL2B* | -0.914724057 | 9.46E-05 | 0.000119261 | 0.59475476 |
| Heart_Failure | dendritic_cell | *TMC1* | -3.680148476 | 2.34E-19 | 2.41E-18 | 1.47E-15 |
| Heart_Failure | dendritic_cell | *HGSNAT* | -4.189225523 | 1.50E-10 | 2.65E-10 | 9.45E-07 |
| Heart_Failure | dendritic_cell | *C9orf64* | -3.852813732 | 1.16E-15 | 3.98E-15 | 7.27E-12 |
| Heart_Failure | dendritic_cell | *HNRNPK* | -4.730631634 | 6.01E-08 | 8.89E-08 | 0.000378041 |
| Heart_Failure | dendritic_cell | *SVEP1* | -1.8644102 | 2.24E-19 | 2.34E-18 | 1.41E-15 |
| Heart_Failure | dendritic_cell | *ZNF367* | -3.535054841 | 4.95E-07 | 6.96E-07 | 0.003111435 |
| Heart_Failure | dendritic_cell | *NOL6* | 7.785315667 | 1.92E-09 | 3.12E-09 | 1.21E-05 |
| Heart_Failure | dendritic_cell | *AQP3* | 3.521888337 | 9.56E-19 | 7.48E-18 | 6.01E-15 |
| Heart_Failure | dendritic_cell | *TRMT10B* | -4.021302332 | 1.03E-16 | 4.45E-16 | 6.48E-13 |
| Heart_Failure | dendritic_cell | *VCP* | -3.859362974 | 3.33E-14 | 8.82E-14 | 2.09E-10 |
| Heart_Failure | dendritic_cell | *PIGO* | 8.472918611 | 1.11E-14 | 3.19E-14 | 7.00E-11 |
| Heart_Failure | dendritic_cell | *LRFN5* | -1.788579682 | 8.46E-14 | 2.10E-13 | 5.32E-10 |
| Heart_Failure | dendritic_cell | *WRN* | -3.780828743 | 9.27E-13 | 2.02E-12 | 5.83E-09 |
| Heart_Failure | dendritic_cell | *CFL2* | -3.909207747 | 0.000226343 | 0.000280122 | 1 |
| Heart_Failure | dendritic_cell | *ZCCHC24* | -3.727502899 | 1.14E-16 | 4.86E-16 | 7.15E-13 |
| Heart_Failure | dendritic_cell | *PGM2L1* | -4.856408698 | 1.23E-14 | 3.49E-14 | 7.73E-11 |
| Heart_Failure | dendritic_cell | *GJB2* | 4.783352787 | 2.74E-18 | 1.82E-17 | 1.72E-14 |

| | | | | | | |
|---|---|---|---|---|---|---|
| Heart_Failure | dendritic_cell | *REEP3* | -4.075541462 | 1.19E-09 | 1.97E-09 | 7.51E-06 |
| Heart_Failure | dendritic_cell | *HEPACAM* | -0.626877266 | 8.52E-05 | 0.00010749 | 0.535730359 |
| Heart_Failure | dendritic_cell | *RPL36AL* | -2.194566065 | 0.00557639 | 0.006463636 | 1 |
| Heart_Failure | dendritic_cell | *C10orf25* | 1.943820946 | 9.61E-12 | 1.88E-11 | 6.04E-08 |
| Heart_Failure | dendritic_cell | *KLHDC2* | -4.18507985 | 1.56E-14 | 4.35E-14 | 9.81E-11 |
| Heart_Failure | dendritic_cell | *NOXRED1* | -3.102768091 | 0.010563695 | 0.012057725 | 1 |
| Heart_Failure | dendritic_cell | *OXGR1* | 4.586256853 | 3.12E-20 | 6.68E-19 | 1.96E-16 |
| Heart_Failure | dendritic_cell | *ATP5C1* | -3.049073297 | 0.000645225 | 0.000781002 | 1 |
| Heart_Failure | dendritic_cell | *TAF3* | -4.25483406 | 4.30E-14 | 1.12E-13 | 2.71E-10 |
| Heart_Failure | dendritic_cell | *VDAC2* | -5.811904543 | 3.41E-08 | 5.09E-08 | 0.00021413 |
| Heart_Failure | dendritic_cell | *SLC18A2* | 6.797004763 | 6.13E-11 | 1.12E-10 | 3.85E-07 |
| Heart_Failure | dendritic_cell | *PDZD8* | -2.122401494 | 0.002768169 | 0.003259689 | 1 |
| Heart_Failure | dendritic_cell | *QSOX2* | 9.540519018 | 7.63E-14 | 1.91E-13 | 4.80E-10 |
| Heart_Failure | dendritic_cell | *CLEC1B* | 0.707411348 | 0.00031981 | 0.000393011 | 1 |
| Heart_Failure | dendritic_cell | *SNAPC4* | -2.69322362 | 9.53E-08 | 1.39E-07 | 0.000599387 |
| Heart_Failure | dendritic_cell | *SDCCAG3* | -6.001639097 | 1.02E-14 | 2.93E-14 | 6.39E-11 |
| Heart_Failure | dendritic_cell | *AK8* | -3.526556585 | 0.013214054 | 0.015022922 | 1 |
| Heart_Failure | dendritic_cell | *TSC1* | -4.493867907 | 3.86E-10 | 6.58E-10 | 2.43E-06 |
| Heart_Failure | dendritic_cell | *ZMYND19* | -12.0998464 | 5.91E-15 | 1.78E-14 | 3.72E-11 |
| Heart_Failure | dendritic_cell | *STOX1* | -0.932972094 | 1.16E-06 | 1.59E-06 | 0.007292992 |
| Heart_Failure | dendritic_cell | *RET* | 8.485361547 | 0.000173672 | 0.000216354 | 1 |
| Heart_Failure | dendritic_cell | *STK32C* | 6.624869712 | 1.81E-12 | 3.83E-12 | 1.14E-08 |
| Heart_Failure | dendritic_cell | *CASP7* | -2.69008769 | 3.19E-08 | 4.77E-08 | 0.000200348 |
| Heart_Failure | dendritic_cell | *VWA2* | 5.379547193 | 4.98E-13 | 1.12E-12 | 3.13E-09 |
| Heart_Failure | dendritic_cell | *METTL3* | -5.05444859 | 5.67E-15 | 1.72E-14 | 3.57E-11 |
| Heart_Failure | dendritic_cell | *TRUB1* | -2.389434881 | 0.00019792 | 0.000245623 | 1 |
| Heart_Failure | dendritic_cell | *HSPA12A* | -1.490898333 | 2.45E-11 | 4.64E-11 | 1.54E-07 |
| Heart_Failure | dendritic_cell | *UBTD1* | -5.672468551 | 1.30E-07 | 1.88E-07 | 0.000814937 |
| Heart_Failure | dendritic_cell | *ARHGAP42* | -2.283750074 | 3.18E-19 | 3.05E-18 | 2.00E-15 |
| Heart_Failure | dendritic_cell | *OTOGL* | 6.267976181 | 2.19E-16 | 8.76E-16 | 1.37E-12 |
| Heart_Failure | dendritic_cell | *PACSIN3* | 1.012351719 | 0.010551723 | 0.012046247 | 1 |
| Heart_Failure | dendritic_cell | *TTC7B* | -3.038783037 | 2.87E-16 | 1.12E-15 | 1.80E-12 |
| Heart_Failure | dendritic_cell | *SLC39A13* | -5.742845306 | 4.72E-18 | 2.90E-17 | 2.97E-14 |
| Heart_Failure | dendritic_cell | *PSMC3* | -3.85266234 | 4.75E-07 | 6.69E-07 | 0.002986268 |
| Heart_Failure | dendritic_cell | *TC2N* | -1.633302571 | 6.20E-11 | 1.13E-10 | 3.90E-07 |
| Heart_Failure | dendritic_cell | *CPSF2* | -1.22555655 | 0.002316528 | 0.002738626 | 1 |
| Heart_Failure | dendritic_cell | *IFI27L1* | -5.83720352 | 3.48E-14 | 9.16E-14 | 2.19E-10 |
| Heart_Failure | dendritic_cell | *CLMN* | -3.4829136 | 1.79E-15 | 5.92E-15 | 1.13E-11 |
| Heart_Failure | dendritic_cell | *PTER* | -4.971069799 | 1.18E-09 | 1.94E-09 | 7.41E-06 |
| Heart_Failure | dendritic_cell | *C1QL3* | -2.041704464 | 3.94E-07 | 5.58E-07 | 0.002479892 |
| Heart_Failure | dendritic_cell | *ABTB2* | -6.093755114 | 3.39E-14 | 8.95E-14 | 2.13E-10 |
| Heart_Failure | dendritic_cell | *LIPC* | -2.490729055 | 4.70E-19 | 4.16E-18 | 2.96E-15 |
| Heart_Failure | dendritic_cell | *CEP57* | -4.088066896 | 1.38E-12 | 2.95E-12 | 8.65E-09 |
| Heart_Failure | dendritic_cell | *TCP11L2* | -5.201263876 | 5.93E-13 | 1.32E-12 | 3.73E-09 |
| Heart_Failure | dendritic_cell | *JAM3* | -3.916634811 | 6.69E-18 | 3.94E-17 | 4.21E-14 |
| Heart_Failure | dendritic_cell | *GPT2* | -1.950444155 | 4.72E-16 | 1.76E-15 | 2.97E-12 |
| Heart_Failure | dendritic_cell | *IKBIP* | -4.271076055 | 3.04E-08 | 4.55E-08 | 0.00019107 |
| Heart_Failure | dendritic_cell | *HIF1AN* | -5.815666977 | 5.42E-07 | 7.59E-07 | 0.003406546 |
| Heart_Failure | dendritic_cell | *NDUFB8* | -4.620088645 | 4.35E-19 | 3.93E-18 | 2.73E-15 |
| Heart_Failure | dendritic_cell | *ZFYVE19* | 1.24758564 | 0.02834509 | 0.031658479 | 1 |

| | | | | | | |
|---|---|---|---|---|---|---|
| Heart_Failure | dendritic_cell | *SPINT1* | 1.261736719 | 4.05E-20 | 7.61E-19 | 2.54E-16 |
| Heart_Failure | dendritic_cell | *BTRC* | -4.120366604 | 8.30E-12 | 1.63E-11 | 5.22E-08 |
| Heart_Failure | dendritic_cell | *LARP6* | -4.649507675 | 5.40E-13 | 1.21E-12 | 3.40E-09 |
| Heart_Failure | dendritic_cell | *ASPG* | 3.410821975 | 7.03E-21 | 4.66E-19 | 4.42E-17 |
| Heart_Failure | dendritic_cell | *GABRB3* | -2.025798679 | 0.016264514 | 0.018394495 | 1 |
| Heart_Failure | dendritic_cell | *TBATA* | -7.491273604 | 0.000398218 | 0.00048746 | 1 |
| Heart_Failure | dendritic_cell | *PCBD1* | -2.254640406 | 1.80E-12 | 3.82E-12 | 1.13E-08 |
| Heart_Failure | dendritic_cell | *COX11* | -3.943874455 | 8.65E-08 | 1.27E-07 | 0.000543631 |
| Heart_Failure | dendritic_cell | *ZNF202* | -7.266514992 | 1.81E-08 | 2.76E-08 | 0.000114079 |
| Heart_Failure | dendritic_cell | *FAM227B* | -3.274356483 | 2.00E-12 | 4.21E-12 | 1.26E-08 |
| Heart_Failure | dendritic_cell | *STXBP4* | -3.629073934 | 1.13E-15 | 3.91E-15 | 7.13E-12 |
| Heart_Failure | dendritic_cell | *C2* | -3.855505194 | 4.15E-16 | 1.56E-15 | 2.61E-12 |
| Heart_Failure | dendritic_cell | *APBB1* | -6.410076727 | 1.25E-12 | 2.69E-12 | 7.84E-09 |
| Heart_Failure | dendritic_cell | *ILK* | -4.732387793 | 3.56E-13 | 8.15E-13 | 2.24E-09 |
| Heart_Failure | dendritic_cell | *TAF10* | -5.88043026 | 1.73E-08 | 2.63E-08 | 0.000108562 |
| Heart_Failure | dendritic_cell | *DCHS1* | -5.623922567 | 1.00E-16 | 4.35E-16 | 6.31E-13 |
| Heart_Failure | dendritic_cell | *NETO1* | -4.040231512 | 6.87E-18 | 4.03E-17 | 4.32E-14 |
| Heart_Failure | dendritic_cell | *USP54* | -4.028549023 | 2.05E-13 | 4.84E-13 | 1.29E-09 |
| Heart_Failure | dendritic_cell | *C11orf74* | -3.873420271 | 2.65E-17 | 1.32E-16 | 1.66E-13 |
| Heart_Failure | dendritic_cell | *WDR88* | -4.002666774 | 2.53E-20 | 6.44E-19 | 1.59E-16 |
| Heart_Failure | dendritic_cell | *CYB5R2* | -2.701972956 | 0.000538941 | 0.000655382 | 1 |
| Heart_Failure | dendritic_cell | *TUB* | -2.990029327 | 3.40E-20 | 6.90E-19 | 2.14E-16 |
| Heart_Failure | dendritic_cell | *RIC3* | -5.185331424 | 1.10E-15 | 3.81E-15 | 6.91E-12 |
| Heart_Failure | dendritic_cell | *LMO1* | -8.690945727 | 3.39E-10 | 5.81E-10 | 2.13E-06 |
| Heart_Failure | dendritic_cell | *XRRA1* | -2.361814635 | 4.82E-12 | 9.75E-12 | 3.03E-08 |
| Heart_Failure | dendritic_cell | *TRIM66* | -0.56610701 | 0.001010092 | 0.001210531 | 1 |
| Heart_Failure | dendritic_cell | *RPL27A* | -4.383880641 | 8.98E-07 | 1.24E-06 | 0.005644482 |
| Heart_Failure | dendritic_cell | *CDYL2* | -2.018404569 | 1.85E-05 | 2.40E-05 | 0.116139374 |
| Heart_Failure | dendritic_cell | *PRTG* | 5.009395831 | 0.001942202 | 0.002305196 | 1 |
| Heart_Failure | dendritic_cell | *AKIP1* | -6.588034491 | 1.55E-16 | 6.41E-16 | 9.74E-13 |
| Heart_Failure | dendritic_cell | *C16orf46* | 16.09070831 | 1.57E-20 | 5.85E-19 | 9.88E-17 |
| Heart_Failure | dendritic_cell | *TMEM41B* | -4.812965004 | 1.36E-14 | 3.84E-14 | 8.58E-11 |
| Heart_Failure | dendritic_cell | *ZNF143* | -5.668800227 | 3.86E-12 | 7.90E-12 | 2.43E-08 |
| Heart_Failure | dendritic_cell | *TMX3* | -4.154878836 | 2.88E-16 | 1.13E-15 | 1.81E-12 |
| Heart_Failure | dendritic_cell | *CCDC68* | -2.370250712 | 2.04E-15 | 6.70E-15 | 1.28E-11 |
| Heart_Failure | dendritic_cell | *CLEC4E* | 1.090811504 | 1.80E-19 | 2.05E-18 | 1.13E-15 |
| Heart_Failure | dendritic_cell | *ZNF3* | -5.161839204 | 1.62E-11 | 3.10E-11 | 1.02E-07 |
| Heart_Failure | dendritic_cell | *CLEC4D* | -9.771140869 | 3.92E-18 | 2.46E-17 | 2.47E-14 |
| Heart_Failure | dendritic_cell | *ZSCAN21* | -1.712465635 | 1.05E-05 | 1.38E-05 | 0.065756846 |
| Heart_Failure | dendritic_cell | *RIMKLB* | -2.449117573 | 2.18E-14 | 5.95E-14 | 1.37E-10 |
| Heart_Failure | dendritic_cell | *BEAN1* | -5.433608594 | 1.53E-14 | 4.27E-14 | 9.61E-11 |
| Heart_Failure | dendritic_cell | *TK2* | -3.131620808 | 1.51E-12 | 3.21E-12 | 9.46E-09 |
| Heart_Failure | dendritic_cell | *SEC11C* | -6.965784384 | 2.11E-16 | 8.49E-16 | 1.32E-12 |
| Heart_Failure | dendritic_cell | *GALR1* | -3.160330443 | 1.11E-15 | 3.85E-15 | 7.00E-12 |
| Heart_Failure | dendritic_cell | *CENPV* | -2.80302801 | 0.015806688 | 0.017892806 | 1 |
| Heart_Failure | dendritic_cell | *CHRFAM7A* | -2.073874967 | 0.001444915 | 0.001723754 | 1 |
| Heart_Failure | dendritic_cell | *SPDYE6* | 7.181402117 | 7.51E-13 | 1.65E-12 | 4.72E-09 |
| Heart_Failure | dendritic_cell | *ATF7IP2* | -1.826513653 | 0.025769646 | 0.028843469 | 1 |
| Heart_Failure | dendritic_cell | *ZNF592* | -3.46662049 | 5.12E-05 | 6.53E-05 | 0.321915719 |
| Heart_Failure | dendritic_cell | *ACSM1* | 2.321877701 | 2.52E-20 | 6.44E-19 | 1.59E-16 |

| | | | | | | |
|---|---|---|---|---|---|---|
| Heart_Failure | dendritic_cell | *AP1G1* | -4.149368994 | 1.34E-15 | 4.55E-15 | 8.41E-12 |
| Heart_Failure | dendritic_cell | *SLFN5* | -3.425447986 | 2.11E-16 | 8.51E-16 | 1.33E-12 |
| Heart_Failure | dendritic_cell | *CATSPER2* | -5.053483093 | 3.25E-13 | 7.50E-13 | 2.05E-09 |
| Heart_Failure | dendritic_cell | *C16orf45* | -4.592183012 | 3.82E-11 | 7.09E-11 | 2.40E-07 |
| Heart_Failure | dendritic_cell | *SAAL1* | -5.096758316 | 0.000284187 | 0.000349781 | 1 |
| Heart_Failure | dendritic_cell | *YPEL4* | -4.861784322 | 1.66E-18 | 1.19E-17 | 1.04E-14 |
| Heart_Failure | dendritic_cell | *LDHC* | -1.348914184 | 0.000724748 | 0.000874566 | 1 |
| Heart_Failure | dendritic_cell | *SCNN1G* | 7.166204303 | 9.58E-20 | 1.29E-18 | 6.02E-16 |
| Heart_Failure | dendritic_cell | *ANKDD1A* | -3.62374665 | 2.38E-08 | 3.58E-08 | 0.000149449 |
| Heart_Failure | dendritic_cell | *MTMR10* | -3.257571933 | 1.21E-06 | 1.66E-06 | 0.007608542 |
| Heart_Failure | dendritic_cell | *YWHAB* | -5.283272595 | 3.44E-12 | 7.08E-12 | 2.17E-08 |
| Heart_Failure | dendritic_cell | *NYAP1* | -1.33084678 | 0.001574856 | 0.001876634 | 1 |
| Heart_Failure | dendritic_cell | *TSC22D4* | 3.798917521 | 0.006090608 | 0.007046679 | 1 |
| Heart_Failure | dendritic_cell | *MS4A6E* | -2.170762421 | 2.35E-10 | 4.08E-10 | 1.48E-06 |
| Heart_Failure | dendritic_cell | *MS4A7* | 2.035675188 | 3.70E-17 | 1.78E-16 | 2.32E-13 |
| Heart_Failure | dendritic_cell | *MS4A14* | 1.248991954 | 6.95E-20 | 1.05E-18 | 4.37E-16 |
| Heart_Failure | dendritic_cell | *DIS3L* | -5.25611388 | 2.35E-10 | 4.09E-10 | 1.48E-06 |
| Heart_Failure | dendritic_cell | *CCNDBP1* | -4.679913431 | 5.18E-17 | 2.41E-16 | 3.26E-13 |
| Heart_Failure | dendritic_cell | *SMAD3* | -3.555275847 | 1.65E-17 | 8.75E-17 | 1.04E-13 |
| Heart_Failure | dendritic_cell | *RCCD1* | -6.465437562 | 1.63E-09 | 2.65E-09 | 1.02E-05 |
| Heart_Failure | dendritic_cell | *AKTIP* | 6.554570177 | 1.25E-05 | 1.63E-05 | 0.078443505 |
| Heart_Failure | dendritic_cell | *EVA1C* | -2.699125044 | 7.34E-14 | 1.84E-13 | 4.61E-10 |
| Heart_Failure | dendritic_cell | *CNPY4* | -2.435781971 | 2.74E-09 | 4.39E-09 | 1.72E-05 |
| Heart_Failure | dendritic_cell | *NUDT21* | -6.108926986 | 2.59E-12 | 5.39E-12 | 1.63E-08 |
| Heart_Failure | dendritic_cell | *NKX3-1* | -19.65049484 | 1.77E-17 | 9.33E-17 | 1.11E-13 |
| Heart_Failure | dendritic_cell | *DUSP18* | 6.268413197 | 6.90E-10 | 1.16E-09 | 4.34E-06 |
| Heart_Failure | dendritic_cell | *TEF* | -2.234600064 | 2.70E-14 | 7.25E-14 | 1.70E-10 |
| Heart_Failure | dendritic_cell | *MEI1* | 2.613563243 | 5.55E-09 | 8.71E-09 | 3.49E-05 |
| Heart_Failure | dendritic_cell | *PBX3* | -2.858283324 | 6.86E-05 | 8.69E-05 | 0.431171203 |
| Heart_Failure | dendritic_cell | *SNRPD1* | -6.814957399 | 2.40E-08 | 3.61E-08 | 0.0001506 |
| Heart_Failure | dendritic_cell | *SAMD14* | 5.948266205 | 2.78E-19 | 2.77E-18 | 1.75E-15 |
| Heart_Failure | dendritic_cell | *PIP5KL1* | -1.021794912 | 0.004732067 | 0.005509352 | 1 |
| Heart_Failure | dendritic_cell | *URM1* | -3.793995328 | 8.10E-05 | 0.000102333 | 0.50951495 |
| Heart_Failure | dendritic_cell | *CERCAM* | -2.393626777 | 1.72E-13 | 4.10E-13 | 1.08E-09 |
| Heart_Failure | dendritic_cell | *ISLR2* | -3.291981391 | 9.61E-14 | 2.36E-13 | 6.04E-10 |
| Heart_Failure | dendritic_cell | *SP2* | -5.029683876 | 2.41E-05 | 3.12E-05 | 0.151588192 |
| Heart_Failure | dendritic_cell | *COQ7* | -3.438094524 | 2.22E-19 | 2.32E-18 | 1.39E-15 |
| Heart_Failure | dendritic_cell | *GPRC5B* | -3.224821932 | 3.06E-18 | 1.98E-17 | 1.92E-14 |
| Heart_Failure | dendritic_cell | *KATNAL2* | -2.082916129 | 1.61E-18 | 1.17E-17 | 1.01E-14 |
| Heart_Failure | dendritic_cell | *RNF214* | -5.176783864 | 2.15E-09 | 3.48E-09 | 1.35E-05 |
| Heart_Failure | dendritic_cell | *ENGASE* | -3.715848424 | 2.47E-11 | 4.66E-11 | 1.55E-07 |
| Heart_Failure | dendritic_cell | *RBFOX3* | -11.9554014 | 1.02E-13 | 2.49E-13 | 6.40E-10 |
| Heart_Failure | dendritic_cell | *MYO5B* | -1.765469731 | 2.60E-16 | 1.03E-15 | 1.64E-12 |
| Heart_Failure | dendritic_cell | *ART5* | 3.160027116 | 0.000203903 | 0.000252897 | 1 |
| Heart_Failure | dendritic_cell | *RRM1* | -2.238418482 | 1.38E-08 | 2.11E-08 | 8.66E-05 |
| Heart_Failure | dendritic_cell | *OR51E2* | -1.960009142 | 0.000693932 | 0.000838507 | 1 |
| Heart_Failure | dendritic_cell | *TRIM68* | 4.372725372 | 3.97E-17 | 1.90E-16 | 2.50E-13 |
| Heart_Failure | dendritic_cell | *FN3K* | -4.025095846 | 3.99E-18 | 2.49E-17 | 2.51E-14 |
| Heart_Failure | dendritic_cell | *ZNF23* | -3.87682859 | 8.30E-19 | 6.65E-18 | 5.22E-15 |
| Heart_Failure | dendritic_cell | *ZNF229* | -9.383136992 | 4.33E-16 | 1.62E-15 | 2.72E-12 |

| | | | | | | |
|---|---|---|---|---|---|---|
| Heart_Failure | dendritic_cell | *VKORC1* | -3.149989905 | 9.44E-07 | 1.30E-06 | 0.005933301 |
| Heart_Failure | dendritic_cell | *SMG8* | -7.451095688 | 2.39E-20 | 6.44E-19 | 1.50E-16 |
| Heart_Failure | dendritic_cell | *RAB8A* | 6.920186193 | 8.34E-13 | 1.82E-12 | 5.24E-09 |
| Heart_Failure | dendritic_cell | *GATAD2A* | -4.651992261 | 2.56E-09 | 4.12E-09 | 1.61E-05 |
| Heart_Failure | dendritic_cell | *PROCA1* | 5.009535033 | 6.75E-08 | 9.94E-08 | 0.000424149 |
| Heart_Failure | dendritic_cell | *RPL13* | -2.915268988 | 0.001707537 | 0.002032043 | 1 |
| Heart_Failure | dendritic_cell | *ZNF641* | 2.632177413 | 6.82E-14 | 1.72E-13 | 4.29E-10 |
| Heart_Failure | dendritic_cell | *TP53I13* | -3.449437874 | 1.45E-09 | 2.39E-09 | 9.15E-06 |
| Heart_Failure | dendritic_cell | *ZNF528* | -4.655392009 | 2.58E-17 | 1.29E-16 | 1.62E-13 |
| Heart_Failure | dendritic_cell | *ZNF701* | -4.635142229 | 2.61E-15 | 8.39E-15 | 1.64E-11 |
| Heart_Failure | dendritic_cell | *SERTAD3* | 10.67990633 | 5.38E-07 | 7.54E-07 | 0.003384487 |
| Heart_Failure | dendritic_cell | *TMC4* | -8.806765287 | 2.75E-10 | 4.76E-10 | 1.73E-06 |
| Heart_Failure | dendritic_cell | *TTYH1* | -2.856701963 | 2.40E-19 | 2.46E-18 | 1.51E-15 |
| Heart_Failure | dendritic_cell | *LENG8* | -4.262694806 | 2.56E-11 | 4.82E-11 | 1.61E-07 |
| Heart_Failure | dendritic_cell | *CDC42EP5* | -5.458744582 | 9.32E-12 | 1.83E-11 | 5.86E-08 |
| Heart_Failure | dendritic_cell | *PPP1R14A* | -6.369103249 | 0.000440472 | 0.000538345 | 1 |
| Heart_Failure | dendritic_cell | *SPINT2* | 2.158309535 | 2.38E-09 | 3.83E-09 | 1.49E-05 |
| Heart_Failure | dendritic_cell | *C19orf33* | -1.588916555 | 0.010757697 | 0.012274708 | 1 |
| Heart_Failure | dendritic_cell | *DNAAF3* | -5.878124043 | 0.00022292 | 0.000276048 | 1 |
| Heart_Failure | dendritic_cell | *PSCA* | 4.499740979 | 5.25E-09 | 8.26E-09 | 3.30E-05 |
| Heart_Failure | dendritic_cell | *UBXN6* | -3.986265911 | 4.47E-20 | 8.02E-19 | 2.81E-16 |
| Heart_Failure | dendritic_cell | *ZNF444* | -4.22991326 | 5.62E-19 | 4.83E-18 | 3.53E-15 |
| Heart_Failure | dendritic_cell | *FAM57A* | -2.194480767 | 1.52E-05 | 1.98E-05 | 0.095504531 |
| Heart_Failure | dendritic_cell | *GLOD4* | -5.341848296 | 3.26E-07 | 4.64E-07 | 0.002050415 |
| Heart_Failure | dendritic_cell | *KIFC2* | -1.836992951 | 4.82E-13 | 1.09E-12 | 3.03E-09 |
| Heart_Failure | dendritic_cell | *SERPINF2* | -4.083541851 | 1.97E-11 | 3.76E-11 | 1.24E-07 |
| Heart_Failure | dendritic_cell | *SRR* | -0.799334849 | 0.01777458 | 0.020077036 | 1 |
| Heart_Failure | dendritic_cell | *CYB5D2* | -4.494002796 | 1.63E-16 | 6.72E-16 | 1.03E-12 |
| Heart_Failure | dendritic_cell | *C19orf48* | 1.216170227 | 0.008512945 | 0.009771935 | 1 |
| Heart_Failure | dendritic_cell | *ZNF83* | -3.93373773 | 3.51E-10 | 6.02E-10 | 2.21E-06 |
| Heart_Failure | dendritic_cell | *CD320* | -4.241481174 | 5.33E-19 | 4.63E-18 | 3.35E-15 |
| Heart_Failure | dendritic_cell | *SPRYD3* | -3.013404048 | 5.46E-20 | 9.05E-19 | 3.43E-16 |
| Heart_Failure | dendritic_cell | *SOAT2* | -3.369030948 | 8.42E-14 | 2.09E-13 | 5.29E-10 |
| Heart_Failure | dendritic_cell | *ZNF558* | -3.10065057 | 2.52E-12 | 5.25E-12 | 1.58E-08 |
| Heart_Failure | dendritic_cell | *NDUFV1* | -5.255301112 | 3.96E-13 | 9.01E-13 | 2.49E-09 |
| Heart_Failure | dendritic_cell | *CDK2AP2* | 4.137761687 | 1.07E-19 | 1.39E-18 | 6.72E-16 |
| Heart_Failure | dendritic_cell | *NUDT8* | -2.623688361 | 1.69E-05 | 2.21E-05 | 0.10646178 |
| Heart_Failure | dendritic_cell | *PRDX2* | -4.213189941 | 2.19E-17 | 1.12E-16 | 1.38E-13 |
| Heart_Failure | dendritic_cell | *ZNF232* | -1.798213454 | 2.21E-06 | 3.00E-06 | 0.013911225 |
| Heart_Failure | dendritic_cell | *MIS12* | 5.408161886 | 1.99E-05 | 2.59E-05 | 0.125402556 |
| Heart_Failure | dendritic_cell | *CD300C* | -2.95865583 | 6.61E-05 | 8.39E-05 | 0.415810292 |
| Heart_Failure | dendritic_cell | *HID1* | -3.27540949 | 5.31E-17 | 2.47E-16 | 3.34E-13 |
| Heart_Failure | dendritic_cell | *ATP5H* | -5.49764804 | 3.11E-15 | 9.86E-15 | 1.96E-11 |
| Heart_Failure | dendritic_cell | *SRP68* | -2.533215258 | 0.032480212 | 0.036199804 | 1 |
| Heart_Failure | dendritic_cell | *MGAT5B* | -6.089067036 | 4.68E-19 | 4.15E-18 | 2.94E-15 |
| Heart_Failure | dendritic_cell | *TMEM68* | 4.122055085 | 0.000152998 | 0.00019108 | 0.96189589 |
| Heart_Failure | dendritic_cell | *CYP7A1* | 5.069958018 | 3.12E-14 | 8.33E-14 | 1.96E-10 |
| Heart_Failure | dendritic_cell | *GSDMA* | -5.998013073 | 3.03E-21 | 4.66E-19 | 1.91E-17 |
| Heart_Failure | dendritic_cell | *KRT24* | -4.756846784 | 4.85E-12 | 9.81E-12 | 3.05E-08 |
| Heart_Failure | dendritic_cell | *TMEM99* | -3.650576379 | 1.65E-14 | 4.57E-14 | 1.03E-10 |

| | | | | | | |
|---|---|---|---|---|---|---|
| Heart_Failure | dendritic_cell | *ZNF598* | -3.146927157 | 1.48E-19 | 1.77E-18 | 9.32E-16 |
| Heart_Failure | dendritic_cell | *MLST8* | -6.305424393 | 2.19E-19 | 2.31E-18 | 1.37E-15 |
| Heart_Failure | dendritic_cell | *ECI1* | -4.268085006 | 2.89E-12 | 5.98E-12 | 1.82E-08 |
| Heart_Failure | dendritic_cell | *ABCA3* | -4.710537625 | 1.34E-14 | 3.77E-14 | 8.41E-11 |
| Heart_Failure | dendritic_cell | *KCTD5* | -6.554779521 | 0.000106837 | 0.000134364 | 0.671685142 |
| Heart_Failure | dendritic_cell | *SDHAF2* | -6.509360481 | 2.43E-18 | 1.66E-17 | 1.53E-14 |
| Heart_Failure | dendritic_cell | *DDB1* | -4.332293471 | 6.07E-17 | 2.78E-16 | 3.81E-13 |
| Heart_Failure | dendritic_cell | *VPS37C* | 5.526158771 | 1.17E-07 | 1.70E-07 | 0.000735237 |
| Heart_Failure | dendritic_cell | *VWCE* | -3.793089775 | 2.00E-20 | 6.44E-19 | 1.26E-16 |
| Heart_Failure | dendritic_cell | *BSCL2* | 2.543701263 | 2.56E-20 | 6.44E-19 | 1.61E-16 |
| Heart_Failure | dendritic_cell | *HRASLS5* | -0.600762632 | 0.00013821 | 0.000172886 | 0.868925459 |
| Heart_Failure | dendritic_cell | *TRANK1* | -4.775153067 | 6.23E-10 | 1.05E-09 | 3.92E-06 |
| Heart_Failure | dendritic_cell | *TTC21A* | -7.01472047 | 2.98E-09 | 4.76E-09 | 1.87E-05 |
| Heart_Failure | dendritic_cell | *RPSA* | -3.203312522 | 1.12E-08 | 1.72E-08 | 7.02E-05 |
| Heart_Failure | dendritic_cell | *ULK4* | -1.174890202 | 1.00E-06 | 1.38E-06 | 0.006297037 |
| Heart_Failure | dendritic_cell | *LTBP3* | -3.543619533 | 7.35E-12 | 1.46E-11 | 4.62E-08 |
| Heart_Failure | dendritic_cell | *NAALADL1* | -4.607085165 | 2.46E-20 | 6.44E-19 | 1.55E-16 |
| Heart_Failure | dendritic_cell | *SCARA5* | -1.81718601 | 3.10E-20 | 6.68E-19 | 1.95E-16 |
| Heart_Failure | dendritic_cell | *COPS6* | -4.139621694 | 1.36E-06 | 1.86E-06 | 0.008531621 |
| Heart_Failure | dendritic_cell | *NUDT16L1* | 5.770380449 | 1.06E-18 | 8.17E-18 | 6.68E-15 |
| Heart_Failure | dendritic_cell | *KIAA1586* | -4.643389479 | 4.14E-10 | 7.04E-10 | 2.60E-06 |
| Heart_Failure | dendritic_cell | *RNF187* | -4.635740491 | 1.94E-11 | 3.70E-11 | 1.22E-07 |
| Heart_Failure | dendritic_cell | *ZCCHC4* | -3.626788139 | 3.59E-19 | 3.37E-18 | 2.26E-15 |
| Heart_Failure | dendritic_cell | *PTGDR* | -2.262565633 | 1.21E-16 | 5.13E-16 | 7.62E-13 |
| Heart_Failure | dendritic_cell | *GLYCTK* | -3.667358625 | 1.19E-16 | 5.06E-16 | 7.49E-13 |
| Heart_Failure | dendritic_cell | *UBTD2* | -4.250811859 | 2.40E-08 | 3.61E-08 | 0.00015063 |
| Heart_Failure | dendritic_cell | *POLR2J3* | 1.248860241 | 0.030784088 | 0.034333787 | 1 |
| Heart_Failure | dendritic_cell | *NT5DC2* | -3.720179386 | 1.34E-18 | 9.90E-18 | 8.43E-15 |
| Heart_Failure | dendritic_cell | *COA6* | 6.147998983 | 2.31E-19 | 2.40E-18 | 1.45E-15 |
| Heart_Failure | dendritic_cell | *KIF5C* | -5.353589539 | 2.15E-17 | 1.10E-16 | 1.35E-13 |
| Heart_Failure | dendritic_cell | *PDHB* | -4.745414095 | 1.14E-06 | 1.56E-06 | 0.007157834 |
| Heart_Failure | dendritic_cell | *PXK* | -4.008370855 | 7.28E-14 | 1.83E-13 | 4.58E-10 |
| Heart_Failure | dendritic_cell | *ACOX2* | -1.825101194 | 3.53E-15 | 1.11E-14 | 2.22E-11 |
| Heart_Failure | dendritic_cell | *FAM107A* | -2.942176006 | 6.10E-20 | 9.71E-19 | 3.84E-16 |
| Heart_Failure | dendritic_cell | *CX3CR1* | 1.962384834 | 3.25E-20 | 6.76E-19 | 2.04E-16 |
| Heart_Failure | dendritic_cell | *SCN11A* | -0.751407378 | 0.000964104 | 0.001157625 | 1 |
| Heart_Failure | dendritic_cell | *SEPT2* | -1.31902118 | 0.000102089 | 0.000128495 | 0.641833992 |
| Heart_Failure | dendritic_cell | *DTYMK* | 5.352951491 | 0.000190294 | 0.000236532 | 1 |
| Heart_Failure | dendritic_cell | *ING5* | -4.170198946 | 7.33E-17 | 3.30E-16 | 4.61E-13 |
| Heart_Failure | dendritic_cell | *RFWD3* | -3.419560509 | 7.00E-12 | 1.39E-11 | 4.40E-08 |
| Heart_Failure | dendritic_cell | *RHOH* | -5.176539347 | 1.87E-11 | 3.57E-11 | 1.17E-07 |
| Heart_Failure | dendritic_cell | *KLHL30* | 3.505368943 | 1.22E-14 | 3.46E-14 | 7.64E-11 |
| Heart_Failure | dendritic_cell | *REEP4* | 3.734474537 | 1.72E-20 | 6.17E-19 | 1.08E-16 |
| Heart_Failure | dendritic_cell | *BMP1* | -4.180052861 | 1.96E-11 | 3.73E-11 | 1.23E-07 |
| Heart_Failure | dendritic_cell | *ATXN2L* | -5.050168709 | 6.42E-14 | 1.63E-13 | 4.04E-10 |
| Heart_Failure | dendritic_cell | *SERINC2* | -4.008524305 | 4.99E-07 | 7.02E-07 | 0.003140078 |
| Heart_Failure | dendritic_cell | *TRAPPC11* | -4.643967192 | 8.41E-08 | 1.23E-07 | 0.000528799 |
| Heart_Failure | dendritic_cell | *TMEM223* | -5.370803515 | 2.48E-17 | 1.25E-16 | 1.56E-13 |
| Heart_Failure | dendritic_cell | *SLC20A2* | -4.409005659 | 7.46E-12 | 1.48E-11 | 4.69E-08 |
| Heart_Failure | dendritic_cell | *ZSWIM1* | -5.506738057 | 3.40E-16 | 1.31E-15 | 2.14E-12 |

| | | | | | | |
|---|---|---|---|---|---|---|
| Heart_Failure | dendritic_cell | *NBPF9* | -4.539414746 | 4.52E-19 | 4.05E-18 | 2.84E-15 |
| Heart_Failure | dendritic_cell | *WFDC13* | 23.26273408 | 5.88E-20 | 9.53E-19 | 3.70E-16 |
| Heart_Failure | dendritic_cell | *NDUFS5* | -5.209566893 | 3.11E-12 | 6.41E-12 | 1.95E-08 |
| Heart_Failure | dendritic_cell | *ZNF30* | -3.175161697 | 6.79E-12 | 1.35E-11 | 4.27E-08 |
| Heart_Failure | dendritic_cell | *FAM84B* | -2.897518912 | 2.36E-14 | 6.40E-14 | 1.48E-10 |
| Heart_Failure | dendritic_cell | *SLC16A4* | -2.791343615 | 6.69E-11 | 1.21E-10 | 4.20E-07 |
| Heart_Failure | dendritic_cell | *TMEM208* | -5.136658927 | 3.41E-07 | 4.85E-07 | 0.002145981 |
| Heart_Failure | dendritic_cell | *LRP1B* | -1.879431911 | 8.75E-12 | 1.72E-11 | 5.50E-08 |
| Heart_Failure | dendritic_cell | *DNAJC21* | -5.292180397 | 8.61E-15 | 2.53E-14 | 5.41E-11 |
| Heart_Failure | dendritic_cell | *NPNT* | -4.735135375 | 1.62E-16 | 6.68E-16 | 1.02E-12 |
| Heart_Failure | dendritic_cell | *CNNM3* | -7.762018775 | 4.32E-11 | 7.98E-11 | 2.72E-07 |
| Heart_Failure | dendritic_cell | *GSTM4* | -1.539828995 | 0.000973591 | 0.001168346 | 1 |
| Heart_Failure | dendritic_cell | *TCTN2* | -0.941964218 | 0.002450382 | 0.002891976 | 1 |
| Heart_Failure | dendritic_cell | *ADAL* | -0.546351605 | 0.015959379 | 0.018059146 | 1 |
| Heart_Failure | dendritic_cell | *SNTB2* | -4.52579003 | 1.40E-11 | 2.70E-11 | 8.77E-08 |
| Heart_Failure | dendritic_cell | *ZNF507* | -4.205205397 | 1.82E-16 | 7.42E-16 | 1.14E-12 |
| Heart_Failure | dendritic_cell | *STX18* | -4.67806149 | 3.01E-13 | 6.98E-13 | 1.90E-09 |
| Heart_Failure | dendritic_cell | *ZBTB49* | 5.206589569 | 0.000988582 | 0.00118543 | 1 |
| Heart_Failure | dendritic_cell | *GFM1* | -3.482827994 | 1.70E-08 | 2.59E-08 | 0.000107158 |
| Heart_Failure | dendritic_cell | *TNIP2* | 1.597094543 | 0.037461718 | 0.041604279 | 1 |
| Heart_Failure | dendritic_cell | *BTNL3* | -0.621534499 | 0.005358176 | 0.006215286 | 1 |
| Heart_Failure | dendritic_cell | *ZNF608* | -2.915939862 | 3.72E-18 | 2.36E-17 | 2.34E-14 |
| Heart_Failure | dendritic_cell | *INPP5D* | 3.471859634 | 4.39E-19 | 3.96E-18 | 2.76E-15 |
| Heart_Failure | dendritic_cell | *LETM1* | -6.343924183 | 1.58E-14 | 4.39E-14 | 9.90E-11 |
| Heart_Failure | dendritic_cell | *TMEM129* | -1.231038949 | 0.0377075 | 0.041869843 | 1 |
| Heart_Failure | dendritic_cell | *CEP120* | -4.948339098 | 2.27E-08 | 3.43E-08 | 0.000142833 |
| Heart_Failure | dendritic_cell | *STXBP6* | -2.428607275 | 3.21E-19 | 3.06E-18 | 2.02E-15 |
| Heart_Failure | dendritic_cell | *MFF* | -4.556991274 | 1.35E-11 | 2.63E-11 | 8.51E-08 |
| Heart_Failure | dendritic_cell | *LGALS9* | 3.533394825 | 2.88E-05 | 3.70E-05 | 0.180776474 |
| Heart_Failure | dendritic_cell | *CPLX1* | -2.67109436 | 2.53E-15 | 8.16E-15 | 1.59E-11 |
| Heart_Failure | dendritic_cell | *SIGLEC7* | 1.315640869 | 1.76E-20 | 6.17E-19 | 1.11E-16 |
| Heart_Failure | dendritic_cell | *FEM1B* | 6.086785607 | 2.04E-12 | 4.28E-12 | 1.28E-08 |
| Heart_Failure | dendritic_cell | *MFSD7* | -5.992882835 | 2.73E-06 | 3.69E-06 | 0.017192006 |
| Heart_Failure | dendritic_cell | *COL4A3* | -2.868523213 | 2.23E-18 | 1.54E-17 | 1.40E-14 |
| Heart_Failure | dendritic_cell | *IRS1* | -1.187176161 | 1.57E-10 | 2.77E-10 | 9.90E-07 |
| Heart_Failure | dendritic_cell | *UPF3A* | -2.98236641 | 4.39E-14 | 1.14E-13 | 2.76E-10 |
| Heart_Failure | dendritic_cell | *HSPBAP1* | 1.775794828 | 0.005381085 | 0.006240708 | 1 |
| Heart_Failure | dendritic_cell | *PARM1* | 2.019644586 | 0.000698291 | 0.00084345 | 1 |
| Heart_Failure | dendritic_cell | *FAM110B* | -3.755588055 | 2.40E-17 | 1.21E-16 | 1.51E-13 |
| Heart_Failure | dendritic_cell | *ATF5* | 6.996588083 | 1.43E-05 | 1.87E-05 | 0.089825895 |
| Heart_Failure | dendritic_cell | *CPT1C* | -5.254329273 | 2.79E-17 | 1.39E-16 | 1.76E-13 |
| Heart_Failure | dendritic_cell | *PCSK9* | -17.42771058 | 1.57E-10 | 2.77E-10 | 9.90E-07 |
| Heart_Failure | dendritic_cell | *XPO6* | -2.438461427 | 0.021813026 | 0.024502144 | 1 |
| Heart_Failure | dendritic_cell | *RGS14* | 1.597279742 | 7.62E-09 | 1.19E-08 | 4.79E-05 |
| Heart_Failure | dendritic_cell | *RAB24* | -5.138032477 | 1.80E-17 | 9.48E-17 | 1.13E-13 |
| Heart_Failure | dendritic_cell | *CXCL10* | -1.850924174 | 8.73E-07 | 1.21E-06 | 0.005487804 |
| Heart_Failure | dendritic_cell | *SH3TC2* | -3.381914672 | 2.77E-17 | 1.38E-16 | 1.74E-13 |
| Heart_Failure | dendritic_cell | *GPRIN1* | -14.4608158 | 6.27E-10 | 1.05E-09 | 3.94E-06 |
| Heart_Failure | dendritic_cell | *PGM2* | 5.542559841 | 0.000470452 | 0.000574094 | 1 |
| Heart_Failure | dendritic_cell | *STK32A* | -1.689769908 | 2.45E-18 | 1.67E-17 | 1.54E-14 |

| | | | | | | |
|---|---|---|---|---|---|---|
| Heart_Failure | dendritic_cell | *P2RY12* | 2.180844193 | 0.000189827 | 0.000235998 | 1 |
| Heart_Failure | dendritic_cell | *SLC33A1* | -4.851278381 | 1.51E-15 | 5.09E-15 | 9.52E-12 |
| Heart_Failure | dendritic_cell | *SNUPN* | -3.909472802 | 8.51E-19 | 6.79E-18 | 5.35E-15 |
| Heart_Failure | dendritic_cell | *PTK2* | -3.700915072 | 2.39E-11 | 4.53E-11 | 1.51E-07 |
| Heart_Failure | dendritic_cell | *PTAFR* | 2.371483645 | 3.21E-20 | 6.76E-19 | 2.02E-16 |
| Heart_Failure | dendritic_cell | *RNASE6* | 0.723723261 | 6.46E-21 | 4.66E-19 | 4.06E-17 |
| Heart_Failure | dendritic_cell | *NPR1* | -3.651510924 | 9.74E-19 | 7.59E-18 | 6.13E-15 |
| Heart_Failure | dendritic_cell | *CD52* | 3.73526544 | 1.95E-09 | 3.17E-09 | 1.23E-05 |
| Heart_Failure | dendritic_cell | *CLIC4* | -4.333267676 | 2.94E-10 | 5.06E-10 | 1.85E-06 |
| Heart_Failure | dendritic_cell | *SLC38A11* | -5.519179468 | 6.58E-19 | 5.46E-18 | 4.14E-15 |
| Heart_Failure | dendritic_cell | *CCDC8* | 6.178097609 | 2.14E-19 | 2.27E-18 | 1.34E-15 |
| Heart_Failure | dendritic_cell | *METTL15* | -3.249285044 | 3.82E-11 | 7.10E-11 | 2.40E-07 |
| Heart_Failure | dendritic_cell | *HINT1* | -2.42786034 | 0.034308696 | 0.038197055 | 1 |
| Heart_Failure | dendritic_cell | *DTWD2* | -4.014008529 | 2.91E-06 | 3.92E-06 | 0.018297954 |
| Heart_Failure | dendritic_cell | *CLIC3* | -8.031234884 | 1.99E-19 | 2.16E-18 | 1.25E-15 |
| Heart_Failure | dendritic_cell | *INO80E* | -4.117403 | 5.68E-10 | 9.59E-10 | 3.57E-06 |
| Heart_Failure | dendritic_cell | *DFFB* | -6.318552573 | 2.77E-07 | 3.95E-07 | 0.001741377 |
| Heart_Failure | dendritic_cell | *C15orf40* | -3.034006907 | 7.78E-08 | 1.14E-07 | 0.000489419 |
| Heart_Failure | dendritic_cell | *FAM103A1* | -1.765744548 | 0.000120323 | 0.000150872 | 0.756470345 |
| Heart_Failure | dendritic_cell | *LUZP1* | -4.35424565 | 5.49E-12 | 1.11E-11 | 3.45E-08 |
| Heart_Failure | dendritic_cell | *HEXDC* | -5.423399007 | 5.43E-13 | 1.21E-12 | 3.41E-09 |
| Heart_Failure | dendritic_cell | *LRRC45* | -2.888236584 | 7.95E-09 | 1.24E-08 | 5.00E-05 |
| Heart_Failure | dendritic_cell | *CHRNA5* | 5.255398256 | 2.43E-10 | 4.22E-10 | 1.53E-06 |
| Heart_Failure | dendritic_cell | *FASN* | -1.30604707 | 0.009250114 | 0.010600705 | 1 |
| Heart_Failure | dendritic_cell | *MT1E* | 6.602648811 | 1.57E-05 | 2.04E-05 | 0.098522072 |
| Heart_Failure | dendritic_cell | *DCXR* | -4.264162603 | 3.76E-19 | 3.51E-18 | 2.36E-15 |
| Heart_Failure | dendritic_cell | *LIMS1* | -3.045647687 | 1.02E-11 | 1.98E-11 | 6.38E-08 |
| Heart_Failure | dendritic_cell | *UGP2* | -2.451727288 | 9.17E-17 | 4.01E-16 | 5.76E-13 |
| Heart_Failure | dendritic_cell | *CSGALNACT2* | 5.082767111 | 1.68E-14 | 4.65E-14 | 1.06E-10 |
| Heart_Failure | dendritic_cell | *AVEN* | -5.147153423 | 1.18E-08 | 1.81E-08 | 7.42E-05 |
| Heart_Failure | dendritic_cell | *CTNND2* | -1.979910509 | 4.79E-19 | 4.22E-18 | 3.01E-15 |
| Heart_Failure | dendritic_cell | *CALML6* | -6.224202294 | 4.42E-11 | 8.15E-11 | 2.78E-07 |
| Heart_Failure | dendritic_cell | *TPST1* | -2.467228694 | 3.58E-16 | 1.37E-15 | 2.25E-12 |
| Heart_Failure | dendritic_cell | *OTUD3* | -2.348672268 | 2.65E-06 | 3.58E-06 | 0.016635504 |
| Heart_Failure | dendritic_cell | *BRD3* | -4.252222174 | 0.000841408 | 0.001013203 | 1 |
| Heart_Failure | dendritic_cell | *ZFPM2* | -2.517089338 | 3.27E-17 | 1.60E-16 | 2.06E-13 |
| Heart_Failure | dendritic_cell | *ZNF747* | -8.79319188 | 2.29E-16 | 9.13E-16 | 1.44E-12 |
| Heart_Failure | dendritic_cell | *MAP3K2* | -4.135345702 | 6.02E-12 | 1.21E-11 | 3.78E-08 |
| Heart_Failure | dendritic_cell | *PUSL1* | 5.837926763 | 5.39E-19 | 4.68E-18 | 3.39E-15 |
| Heart_Failure | dendritic_cell | *MYO7B* | -3.029914116 | 5.90E-17 | 2.71E-16 | 3.71E-13 |
| Heart_Failure | dendritic_cell | *CNTROB* | -8.024790097 | 7.04E-11 | 1.27E-10 | 4.42E-07 |
| Heart_Failure | dendritic_cell | *KCNAB3* | 1.847797123 | 5.75E-19 | 4.91E-18 | 3.62E-15 |
| Heart_Failure | dendritic_cell | *NIPA1* | 6.85345813 | 1.99E-09 | 3.23E-09 | 1.25E-05 |
| Heart_Failure | dendritic_cell | *RNF150* | -1.824842944 | 5.86E-17 | 2.70E-16 | 3.69E-13 |
| Heart_Failure | dendritic_cell | *HOXD4* | -4.556847021 | 1.54E-09 | 2.51E-09 | 9.66E-06 |
| Heart_Failure | dendritic_cell | *PDCD6IP* | -3.324183998 | 2.61E-14 | 7.02E-14 | 1.64E-10 |
| Heart_Failure | dendritic_cell | *ZNF212* | -4.594943477 | 1.05E-17 | 5.92E-17 | 6.62E-14 |
| Heart_Failure | dendritic_cell | *FAM161A* | 4.374687886 | 1.86E-17 | 9.73E-17 | 1.17E-13 |
| Heart_Failure | dendritic_cell | *GLB1* | -4.334903521 | 4.20E-08 | 6.25E-08 | 0.000264122 |
| Heart_Failure | dendritic_cell | *FAXDC2* | -3.520885689 | 6.27E-16 | 2.29E-15 | 3.94E-12 |

| | | | | | | |
|---|---|---|---|---|---|---|
| Heart_Failure | dendritic_cell | *CRTAP* | -3.914999362 | 0.000545154 | 0.000662553 | 1 |
| Heart_Failure | dendritic_cell | *ELP5* | -6.460129552 | 4.67E-11 | 8.59E-11 | 2.94E-07 |
| Heart_Failure | dendritic_cell | *STX8* | -4.034241739 | 3.71E-13 | 8.49E-13 | 2.33E-09 |
| Heart_Failure | dendritic_cell | *NFRKB* | -4.458961739 | 1.47E-07 | 2.13E-07 | 0.000926089 |
| Heart_Failure | dendritic_cell | *SP7* | -10.80618819 | 1.43E-08 | 2.18E-08 | 8.99E-05 |
| Heart_Failure | dendritic_cell | *SEMA3E* | -2.114531255 | 4.20E-20 | 7.75E-19 | 2.64E-16 |
| Heart_Failure | dendritic_cell | *GPRC5C* | -3.074001784 | 1.25E-17 | 6.87E-17 | 7.84E-14 |
| Heart_Failure | dendritic_cell | *ADORA2B* | 1.320794532 | 1.12E-15 | 3.86E-15 | 7.02E-12 |
| Heart_Failure | dendritic_cell | *MGMT* | -3.611526061 | 9.83E-14 | 2.41E-13 | 6.18E-10 |
| Heart_Failure | dendritic_cell | *METTL7B* | -8.361144075 | 7.21E-20 | 1.08E-18 | 4.53E-16 |
| Heart_Failure | dendritic_cell | *NFXL1* | -5.078824143 | 4.85E-14 | 1.25E-13 | 3.05E-10 |
| Heart_Failure | dendritic_cell | *CD14* | 3.334095446 | 2.06E-16 | 8.32E-16 | 1.29E-12 |
| Heart_Failure | dendritic_cell | *KRT72* | 4.949783913 | 6.19E-18 | 3.69E-17 | 3.89E-14 |
| Heart_Failure | dendritic_cell | *HSD17B13* | 1.01418625 | 0.019614953 | 0.022124006 | 1 |
| Heart_Failure | dendritic_cell | *TMC7* | -2.44430238 | 5.54E-19 | 4.79E-18 | 3.49E-15 |
| Heart_Failure | dendritic_cell | *ARL6IP1* | -4.501939395 | 4.27E-09 | 6.75E-09 | 2.68E-05 |
| Heart_Failure | dendritic_cell | *SERPINB9* | 4.911475509 | 7.27E-07 | 1.01E-06 | 0.004570403 |
| Heart_Failure | dendritic_cell | *SMAGP* | -3.949489988 | 2.94E-14 | 7.87E-14 | 1.85E-10 |
| Heart_Failure | dendritic_cell | *IRX2* | -2.515964089 | 1.42E-17 | 7.66E-17 | 8.92E-14 |
| Heart_Failure | dendritic_cell | *EMB* | 1.964779526 | 1.68E-09 | 2.75E-09 | 1.06E-05 |
| Heart_Failure | dendritic_cell | *DLGAP1* | -3.225770795 | 4.75E-18 | 2.91E-17 | 2.98E-14 |
| Heart_Failure | dendritic_cell | *STAT2* | -3.846655049 | 5.63E-13 | 1.25E-12 | 3.54E-09 |
| Heart_Failure | dendritic_cell | *NUDCD2* | 5.653931577 | 8.45E-05 | 0.000106559 | 0.530985783 |
| Heart_Failure | dendritic_cell | *HSPA4* | -3.293030212 | 0.005757233 | 0.006668336 | 1 |
| Heart_Failure | dendritic_cell | *SLC26A5* | -5.252878643 | 2.26E-13 | 5.29E-13 | 1.42E-09 |
| Heart_Failure | dendritic_cell | *COMMD5* | -6.792557339 | 7.97E-13 | 1.75E-12 | 5.01E-09 |
| Heart_Failure | dendritic_cell | *GTSF1* | -23.07111323 | 2.86E-10 | 4.93E-10 | 1.80E-06 |
| Heart_Failure | dendritic_cell | *ARMC10* | -4.31584106 | 5.58E-13 | 1.24E-12 | 3.51E-09 |
| Heart_Failure | dendritic_cell | *RNF34* | -4.16002919 | 1.37E-11 | 2.65E-11 | 8.62E-08 |
| Heart_Failure | dendritic_cell | *ACYP2* | -3.26480045 | 2.77E-15 | 8.87E-15 | 1.74E-11 |
| Heart_Failure | dendritic_cell | *TRABD* | 4.11348851 | 5.19E-07 | 7.29E-07 | 0.00326522 |
| Heart_Failure | dendritic_cell | *TMEM133* | -13.80399833 | 0.00216177 | 0.002559519 | 1 |
| Heart_Failure | dendritic_cell | *TTLL6* | -25.609885 | 6.59E-16 | 2.39E-15 | 4.15E-12 |
| Heart_Failure | dendritic_cell | *KCNS3* | -1.918672617 | 8.78E-15 | 2.57E-14 | 5.52E-11 |
| Heart_Failure | dendritic_cell | *GPR37* | -2.27355728 | 0.000185968 | 0.000231292 | 1 |
| Heart_Failure | dendritic_cell | *HTRA3* | -2.523925014 | 1.15E-15 | 3.96E-15 | 7.22E-12 |
| Heart_Failure | dendritic_cell | *KBTBD2* | -4.415121635 | 9.14E-06 | 1.21E-05 | 0.057435167 |
| Heart_Failure | dendritic_cell | *MTSS1* | -3.005834812 | 4.35E-05 | 5.56E-05 | 0.273662214 |
| Heart_Failure | dendritic_cell | *TSEN34* | 9.338599797 | 1.43E-10 | 2.53E-10 | 8.99E-07 |
| Heart_Failure | dendritic_cell | *NDUFA3* | 6.491624383 | 2.27E-06 | 3.08E-06 | 0.014285682 |
| Heart_Failure | dendritic_cell | *OSCAR* | 1.450841345 | 8.01E-21 | 4.66E-19 | 5.04E-17 |
| Heart_Failure | dendritic_cell | *PAQR8* | 3.783215521 | 4.99E-20 | 8.51E-19 | 3.13E-16 |
| Heart_Failure | dendritic_cell | *NUDT6* | -4.088177721 | 1.39E-17 | 7.52E-17 | 8.71E-14 |
| Heart_Failure | dendritic_cell | *ZNF160* | -4.147603608 | 1.25E-08 | 1.92E-08 | 7.88E-05 |
| Heart_Failure | dendritic_cell | *ZNF415* | -0.861357888 | 0.021924481 | 0.024618541 | 1 |
| Heart_Failure | dendritic_cell | *FPR1* | 1.122234248 | 5.44E-20 | 9.05E-19 | 3.42E-16 |
| Heart_Failure | dendritic_cell | *FEZ2* | -3.577032342 | 8.67E-16 | 3.07E-15 | 5.45E-12 |
| Heart_Failure | dendritic_cell | *SOX7* | -4.517274232 | 3.80E-18 | 2.39E-17 | 2.39E-14 |
| Heart_Failure | dendritic_cell | *C11orf24* | -7.336045033 | 3.03E-15 | 9.63E-15 | 1.91E-11 |
| Heart_Failure | dendritic_cell | *ALK* | -3.396558464 | 1.97E-13 | 4.65E-13 | 1.24E-09 |

| | | | | | | |
|---|---|---|---|---|---|---|
| Heart_Failure | dendritic_cell | *TRMT61B* | -2.005901543 | 3.70E-12 | 7.59E-12 | 2.33E-08 |
| Heart_Failure | dendritic_cell | *MFN1* | -3.778096283 | 1.92E-09 | 3.12E-09 | 1.21E-05 |
| Heart_Failure | dendritic_cell | *GIMAP8* | -3.101462249 | 7.38E-08 | 1.09E-07 | 0.000464004 |
| Heart_Failure | dendritic_cell | *KCNMB3* | -2.569217435 | 5.71E-05 | 7.26E-05 | 0.358876549 |
| Heart_Failure | dendritic_cell | *ATP6V0E2* | -6.381480805 | 1.83E-17 | 9.59E-17 | 1.15E-13 |
| Heart_Failure | dendritic_cell | *MORN4* | -1.258486669 | 2.40E-06 | 3.25E-06 | 0.015079802 |
| Heart_Failure | dendritic_cell | *GRIK1* | 5.059376443 | 7.24E-16 | 2.60E-15 | 4.55E-12 |
| Heart_Failure | dendritic_cell | *CLDN20* | -2.350173628 | 1.58E-18 | 1.15E-17 | 9.91E-15 |
| Heart_Failure | dendritic_cell | *CANT1* | -4.579351653 | 1.36E-06 | 1.86E-06 | 0.00855248 |
| Heart_Failure | dendritic_cell | *CHST11* | 2.720859575 | 3.86E-16 | 1.47E-15 | 2.42E-12 |
| Heart_Failure | dendritic_cell | *LURAP1* | -1.561978232 | 1.47E-05 | 1.92E-05 | 0.092441653 |
| Heart_Failure | dendritic_cell | *KCND3* | 4.176151711 | 4.49E-15 | 1.38E-14 | 2.82E-11 |
| Heart_Failure | dendritic_cell | *MRPL36* | -2.287974517 | 0.044219125 | 0.048953273 | 1 |
| Heart_Failure | dendritic_cell | *ZNF581* | -12.99552515 | 1.24E-11 | 2.42E-11 | 7.82E-08 |
| Heart_Failure | dendritic_cell | *MCC* | -3.099830286 | 4.69E-17 | 2.20E-16 | 2.95E-13 |
| Heart_Failure | dendritic_cell | *KRT27* | 5.484319954 | 6.89E-11 | 1.25E-10 | 4.33E-07 |
| Heart_Failure | dendritic_cell | *DLK2* | -7.661029138 | 7.81E-11 | 1.41E-10 | 4.91E-07 |
| Heart_Failure | dendritic_cell | *HOPX* | -4.798500567 | 2.09E-20 | 6.44E-19 | 1.31E-16 |
| Heart_Failure | dendritic_cell | *LRRC8C* | -6.634458151 | 2.95E-07 | 4.20E-07 | 0.001855214 |
| Heart_Failure | dendritic_cell | *RSL1D1* | -5.965653345 | 3.32E-13 | 7.65E-13 | 2.09E-09 |
| Heart_Failure | dendritic_cell | *PPID* | -5.936894651 | 2.54E-13 | 5.91E-13 | 1.60E-09 |
| Heart_Failure | dendritic_cell | *COL24A1* | -1.01221748 | 1.03E-08 | 1.59E-08 | 6.49E-05 |
| Heart_Failure | dendritic_cell | *ETFDH* | -1.346269432 | 1.34E-08 | 2.05E-08 | 8.43E-05 |
| Heart_Failure | dendritic_cell | *TBCA* | -3.42312124 | 1.31E-06 | 1.80E-06 | 0.00824575 |
| Heart_Failure | dendritic_cell | *MAP6* | -3.246679792 | 1.29E-17 | 7.10E-17 | 8.14E-14 |
| Heart_Failure | dendritic_cell | *BCL2L1* | -3.92650322 | 3.11E-07 | 4.42E-07 | 0.001954077 |
| Heart_Failure | dendritic_cell | *PLRG1* | -6.055729385 | 2.70E-14 | 7.25E-14 | 1.70E-10 |
| Heart_Failure | dendritic_cell | *ZNF584* | -1.253901193 | 0.007963795 | 0.009159967 | 1 |
| Heart_Failure | dendritic_cell | *NMUR1* | 7.904208526 | 4.07E-16 | 1.54E-15 | 2.56E-12 |
| Heart_Failure | dendritic_cell | *P2RY6* | 1.00488212 | 4.70E-21 | 4.66E-19 | 2.96E-17 |
| Heart_Failure | dendritic_cell | *ZIK1* | -6.925209825 | 1.54E-17 | 8.27E-17 | 9.69E-14 |
| Heart_Failure | dendritic_cell | *RGS19* | 2.282631858 | 4.46E-12 | 9.05E-12 | 2.80E-08 |
| Heart_Failure | dendritic_cell | *TCEA2* | 6.506739885 | 0.00015797 | 0.000197134 | 0.993160395 |
| Heart_Failure | dendritic_cell | *ANO5* | 6.319509598 | 3.68E-18 | 2.34E-17 | 2.31E-14 |
| Heart_Failure | dendritic_cell | *GPHN* | -2.884417434 | 2.23E-14 | 6.06E-14 | 1.40E-10 |
| Heart_Failure | dendritic_cell | *VAT1L* | 6.151099488 | 2.62E-09 | 4.21E-09 | 1.65E-05 |
| Heart_Failure | dendritic_cell | *CAMTA1* | -3.318822994 | 0.000304162 | 0.000374147 | 1 |
| Heart_Failure | dendritic_cell | *LGALS4* | -2.918042809 | 5.43E-09 | 8.54E-09 | 3.41E-05 |
| Heart_Failure | dendritic_cell | *LRRC34* | -1.276402466 | 1.59E-05 | 2.07E-05 | 0.099942031 |
| Heart_Failure | dendritic_cell | *SPATA5L1* | -3.753940433 | 1.88E-11 | 3.60E-11 | 1.18E-07 |
| Heart_Failure | dendritic_cell | *SYCE1* | -3.327849749 | 3.62E-11 | 6.74E-11 | 2.28E-07 |
| Heart_Failure | dendritic_cell | *NXNL1* | -3.874191127 | 1.61E-08 | 2.45E-08 | 0.00010118 |
| Heart_Failure | dendritic_cell | *BCL2* | -1.335434228 | 1.52E-07 | 2.20E-07 | 0.000957543 |
| Heart_Failure | dendritic_cell | *RHNO1* | -4.099340361 | 3.90E-05 | 4.98E-05 | 0.245027139 |
| Heart_Failure | dendritic_cell | *KNDC1* | -2.59766677 | 8.95E-07 | 1.24E-06 | 0.005624369 |
| Heart_Failure | dendritic_cell | *METTL18* | 7.673167024 | 3.55E-18 | 2.26E-17 | 2.23E-14 |
| Heart_Failure | dendritic_cell | *PWWP2B* | -3.381496924 | 5.84E-20 | 9.53E-19 | 3.67E-16 |
| Heart_Failure | dendritic_cell | *NINJ2* | -2.942979801 | 5.17E-18 | 3.14E-17 | 3.25E-14 |
| Heart_Failure | dendritic_cell | *FAM90A1* | -8.161892133 | 2.72E-20 | 6.44E-19 | 1.71E-16 |
| Heart_Failure | dendritic_cell | *TRAPPC12* | -4.207678623 | 2.08E-16 | 8.38E-16 | 1.31E-12 |

| | | | | | | |
|---|---|---|---|---|---|---|
| Heart_Failure | dendritic_cell | *C3AR1* | 2.249298781 | 3.09E-20 | 6.68E-19 | 1.94E-16 |
| Heart_Failure | dendritic_cell | *AQP4* | -3.146539535 | 2.80E-20 | 6.44E-19 | 1.76E-16 |
| Heart_Failure | dendritic_cell | *TLN2* | -1.897321373 | 4.66E-16 | 1.74E-15 | 2.93E-12 |
| Heart_Failure | dendritic_cell | *TVP23B* | -5.640111195 | 4.12E-10 | 7.01E-10 | 2.59E-06 |
| Heart_Failure | dendritic_cell | *SRGAP2C* | 3.29663318 | 0.002836044 | 0.003338366 | 1 |
| Heart_Failure | dendritic_cell | *ATPAF2* | 7.371803642 | 5.34E-08 | 7.92E-08 | 0.000335606 |
| Heart_Failure | dendritic_cell | *CYP4F22* | -6.955923051 | 2.99E-14 | 7.98E-14 | 1.88E-10 |
| Heart_Failure | dendritic_cell | *C20orf196* | -4.70389038 | 1.98E-15 | 6.51E-15 | 1.24E-11 |
| Heart_Failure | dendritic_cell | *ZNF554* | -3.241663618 | 4.71E-05 | 6.00E-05 | 0.2958604 |
| Heart_Failure | dendritic_cell | *ANKRD20A4* | 3.849797112 | 7.55E-21 | 4.66E-19 | 4.75E-17 |
| Heart_Failure | dendritic_cell | *LAMB2* | -3.099947082 | 9.52E-17 | 4.14E-16 | 5.98E-13 |
| Heart_Failure | dendritic_cell | *ORMDL3* | -3.627261659 | 7.25E-28 | 4.56E-24 | 4.56E-24 |
| Heart_Failure | dendritic_cell | *SERF1A* | -13.64437604 | 2.54E-20 | 6.44E-19 | 1.60E-16 |
| Heart_Failure | dendritic_cell | *LRRC15* | -6.109298065 | 7.50E-08 | 1.10E-07 | 0.00047154 |
| Heart_Failure | dendritic_cell | *MOB3A* | 4.895853167 | 1.90E-08 | 2.88E-08 | 0.000119247 |
| Heart_Failure | dendritic_cell | *SLFN12* | -2.162825696 | 2.82E-06 | 3.81E-06 | 0.017746491 |
| Heart_Failure | dendritic_cell | *FRMD3* | -3.605020483 | 6.93E-18 | 4.05E-17 | 4.35E-14 |
| Heart_Failure | dendritic_cell | *MTBP* | -5.759377931 | 1.75E-08 | 2.65E-08 | 0.00010975 |
| Heart_Failure | dendritic_cell | *MALT1* | 3.456484371 | 0.000636751 | 0.000771339 | 1 |
| Heart_Failure | dendritic_cell | *ISG20* | 1.435256464 | 0.010978043 | 0.01251931 | 1 |
| Heart_Failure | dendritic_cell | *C5orf34* | -4.449740873 | 2.95E-16 | 1.15E-15 | 1.85E-12 |
| Heart_Failure | dendritic_cell | *C1QTNF4* | 0.416392522 | 0.020992538 | 0.023624297 | 1 |
| Heart_Failure | dendritic_cell | *ZNF131* | -4.437590101 | 2.07E-10 | 3.61E-10 | 1.30E-06 |
| Heart_Failure | dendritic_cell | *HINFP* | -5.426678764 | 3.75E-17 | 1.80E-16 | 2.36E-13 |
| Heart_Failure | dendritic_cell | *SPTLC3* | -4.419398063 | 1.68E-16 | 6.89E-16 | 1.06E-12 |
| Heart_Failure | dendritic_cell | *COPRS* | -4.06943278 | 2.51E-18 | 1.69E-17 | 1.58E-14 |
| Heart_Failure | dendritic_cell | *CLEC12A* | 0.579448353 | 1.65E-08 | 2.51E-08 | 0.000103591 |
| Heart_Failure | dendritic_cell | *SUCLG2* | -3.742297629 | 5.98E-14 | 1.52E-13 | 3.76E-10 |
| Heart_Failure | dendritic_cell | *CSDC2* | 2.916948301 | 1.07E-19 | 1.39E-18 | 6.71E-16 |
| Heart_Failure | dendritic_cell | *RCAN2* | -4.451526878 | 3.63E-15 | 1.14E-14 | 2.28E-11 |
| Heart_Failure | dendritic_cell | *IL16* | -2.295907796 | 7.63E-18 | 4.42E-17 | 4.80E-14 |
| Heart_Failure | dendritic_cell | *GNB2* | -6.602936794 | 4.97E-13 | 1.12E-12 | 3.13E-09 |
| Heart_Failure | dendritic_cell | *PRSS27* | -13.55535684 | 1.39E-15 | 4.72E-15 | 8.75E-12 |
| Heart_Failure | dendritic_cell | *FGGY* | -3.924065391 | 5.59E-13 | 1.25E-12 | 3.52E-09 |
| Heart_Failure | dendritic_cell | *MANEA* | -5.583619959 | 6.43E-14 | 1.63E-13 | 4.04E-10 |
| Heart_Failure | dendritic_cell | *FIBP* | -5.950325534 | 4.19E-20 | 7.75E-19 | 2.63E-16 |
| Heart_Failure | dendritic_cell | *CARNS1* | -7.296359997 | 1.16E-18 | 8.81E-18 | 7.28E-15 |
| Heart_Failure | dendritic_cell | *CTSW* | -0.359990616 | 0.013653793 | 0.01551444 | 1 |
| Heart_Failure | dendritic_cell | *SNTG2* | -2.087662388 | 1.00E-18 | 7.77E-18 | 6.31E-15 |
| Heart_Failure | dendritic_cell | *PDE3A* | -3.411117664 | 9.64E-12 | 1.89E-11 | 6.06E-08 |
| Heart_Failure | dendritic_cell | *MRPL52* | -3.920875452 | 3.94E-07 | 5.57E-07 | 0.002475548 |
| Heart_Failure | dendritic_cell | *SMPDL3A* | -4.802598091 | 1.49E-10 | 2.63E-10 | 9.37E-07 |
| Heart_Failure | dendritic_cell | *RAD9A* | -0.949850256 | 0.005165146 | 0.006001344 | 1 |
| Heart_Failure | dendritic_cell | *EFEMP2* | -3.905818687 | 4.43E-14 | 1.14E-13 | 2.78E-10 |
| Heart_Failure | dendritic_cell | *AGAP5* | -80.46259097 | 2.41E-20 | 6.44E-19 | 1.51E-16 |
| Heart_Failure | dendritic_cell | *ZMAT3* | -4.135786621 | 0.002436706 | 0.002876915 | 1 |
| Heart_Failure | dendritic_cell | *ZFAND4* | -4.061719988 | 9.40E-16 | 3.31E-15 | 5.91E-12 |
| Heart_Failure | dendritic_cell | *ZNF738* | 2.420652318 | 4.24E-06 | 5.68E-06 | 0.026683718 |
| Heart_Failure | dendritic_cell | *SLFN11* | -4.081563786 | 7.40E-16 | 2.65E-15 | 4.65E-12 |
| Heart_Failure | dendritic_cell | *CCL19* | -13.19970298 | 1.12E-20 | 4.99E-19 | 7.05E-17 |

| | | | | | | |
|---|---|---|---|---|---|---|
| Heart_Failure | dendritic_cell | *FUT10* | -1.983543688 | 7.48E-08 | 1.10E-07 | 0.000470427 |
| Heart_Failure | dendritic_cell | *CFL1* | 2.596817444 | 0.041604504 | 0.046083072 | 1 |
| Heart_Failure | dendritic_cell | *EFCAB12* | -2.461265381 | 3.06E-10 | 5.28E-10 | 1.93E-06 |
| Heart_Failure | dendritic_cell | *FAM192A* | -2.194154136 | 0.000140357 | 0.000175432 | 0.882421348 |
| Heart_Failure | dendritic_cell | *RAB43* | 3.328828834 | 4.49E-21 | 4.66E-19 | 2.82E-17 |
| Heart_Failure | dendritic_cell | *CBWD1* | -3.994984018 | 0.001849753 | 0.002197128 | 1 |
| Heart_Failure | dendritic_cell | *HOXC5* | 1.982848233 | 0.024498005 | 0.027444575 | 1 |
| Heart_Failure | dendritic_cell | *RAB37* | 7.739169795 | 3.21E-15 | 1.01E-14 | 2.02E-11 |
| Heart_Failure | dendritic_cell | *SNX32* | -3.792731222 | 1.27E-07 | 1.85E-07 | 0.000801552 |
| Heart_Failure | dendritic_cell | *CYP7B1* | -2.61397009 | 1.41E-15 | 4.77E-15 | 8.86E-12 |
| Heart_Failure | dendritic_cell | *METAP1D* | -3.491132261 | 2.54E-18 | 1.71E-17 | 1.60E-14 |
| Heart_Failure | dendritic_cell | *MYEOV* | -1.150549104 | 5.87E-11 | 1.07E-10 | 3.69E-07 |
| Heart_Failure | dendritic_cell | *MRGPRF* | 7.238590863 | 2.58E-11 | 4.86E-11 | 1.62E-07 |
| Heart_Failure | dendritic_cell | *LCLAT1* | -5.420577396 | 8.01E-11 | 1.44E-10 | 5.04E-07 |
| Heart_Failure | dendritic_cell | *ADH6* | -1.533576385 | 5.69E-10 | 9.61E-10 | 3.58E-06 |
| Heart_Failure | dendritic_cell | *GXYLT2* | -2.082660624 | 1.38E-19 | 1.67E-18 | 8.70E-16 |
| Heart_Failure | dendritic_cell | *HPSE2* | -2.37194196 | 3.82E-11 | 7.10E-11 | 2.40E-07 |
| Heart_Failure | dendritic_cell | *DCAKD* | -2.571125122 | 1.58E-12 | 3.36E-12 | 9.91E-09 |
| Heart_Failure | dendritic_cell | *HECTD4* | -3.809578785 | 3.83E-13 | 8.74E-13 | 2.41E-09 |
| Heart_Failure | dendritic_cell | *BNC2* | -2.451575371 | 1.00E-16 | 4.33E-16 | 6.29E-13 |
| Heart_Failure | dendritic_cell | *TRMT112* | 3.79458639 | 8.40E-14 | 2.09E-13 | 5.28E-10 |
| Heart_Failure | dendritic_cell | *NOC3L* | -4.722894466 | 1.77E-06 | 2.41E-06 | 0.011118212 |
| Heart_Failure | dendritic_cell | *RHOD* | -1.327187674 | 1.49E-06 | 2.03E-06 | 0.009348551 |
| Heart_Failure | dendritic_cell | *AHSA2* | -3.733980092 | 7.30E-15 | 2.17E-14 | 4.59E-11 |
| Heart_Failure | dendritic_cell | *ABLIM3* | -2.077958864 | 2.07E-19 | 2.21E-18 | 1.30E-15 |
| Heart_Failure | dendritic_cell | *VANGL1* | -4.893943275 | 2.09E-17 | 1.07E-16 | 1.31E-13 |
| Heart_Failure | dendritic_cell | *GLRX* | -2.564727114 | 2.87E-19 | 2.83E-18 | 1.81E-15 |
| Heart_Failure | dendritic_cell | *IQCB1* | -2.675787377 | 6.92E-07 | 9.64E-07 | 0.004348246 |
| Heart_Failure | dendritic_cell | *GOLGB1* | -4.046049313 | 1.30E-10 | 2.31E-10 | 8.16E-07 |
| Heart_Failure | dendritic_cell | *DMRT2* | -2.179733141 | 3.43E-20 | 6.93E-19 | 2.16E-16 |
| Heart_Failure | dendritic_cell | *ZNF483* | -2.074207658 | 7.10E-11 | 1.28E-10 | 4.46E-07 |
| Heart_Failure | dendritic_cell | *SLC2A14* | 2.400943874 | 1.05E-06 | 1.45E-06 | 0.006631705 |
| Heart_Failure | dendritic_cell | *MZT2A* | 3.615828814 | 1.54E-06 | 2.10E-06 | 0.0096705 |
| Heart_Failure | dendritic_cell | *MAP3K11* | 6.291268525 | 3.24E-15 | 1.02E-14 | 2.03E-11 |
| Heart_Failure | dendritic_cell | *C1QA* | 5.681575477 | 1.58E-15 | 5.29E-15 | 9.93E-12 |
| Heart_Failure | dendritic_cell | *NDNF* | -3.884286758 | 3.34E-19 | 3.17E-18 | 2.10E-15 |
| Heart_Failure | dendritic_cell | *OLR1* | 0.839666076 | 6.49E-21 | 4.66E-19 | 4.08E-17 |
| Heart_Failure | dendritic_cell | *GLIPR1L1* | -3.984299958 | 3.75E-11 | 6.97E-11 | 2.36E-07 |
| Heart_Failure | dendritic_cell | *ARV1* | -4.529683438 | 9.67E-16 | 3.39E-15 | 6.08E-12 |
| Heart_Failure | dendritic_cell | *CCDC36* | -3.328820063 | 4.12E-14 | 1.07E-13 | 2.59E-10 |
| Heart_Failure | dendritic_cell | *THAP2* | -4.284896837 | 1.18E-13 | 2.88E-13 | 7.44E-10 |
| Heart_Failure | dendritic_cell | *PPP1R14B* | -5.373998201 | 0.005057196 | 0.00587809 | 1 |
| Heart_Failure | dendritic_cell | *SMARCC1* | -4.203452906 | 3.16E-08 | 4.72E-08 | 0.000198397 |
| Heart_Failure | dendritic_cell | *VEGFB* | -3.144467272 | 6.14E-13 | 1.36E-12 | 3.86E-09 |
| Heart_Failure | dendritic_cell | *PEAK1* | -3.635253581 | 1.33E-09 | 2.19E-09 | 8.38E-06 |
| Heart_Failure | dendritic_cell | *TNFRSF10D* | -2.087282045 | 5.77E-12 | 1.16E-11 | 3.63E-08 |
| Heart_Failure | dendritic_cell | *MST1* | -1.862892094 | 4.71E-07 | 6.64E-07 | 0.002961906 |
| Heart_Failure | dendritic_cell | *TNFRSF10C* | -1.306846614 | 0.004301661 | 0.005019403 | 1 |
| Heart_Failure | dendritic_cell | *GMPPB* | -2.85286743 | 6.45E-14 | 1.63E-13 | 4.06E-10 |
| Heart_Failure | dendritic_cell | *CSPG4* | -3.206951295 | 1.60E-16 | 6.62E-16 | 1.01E-12 |

| | | | | | | |
|---|---|---|---|---|---|---|
| Heart_Failure | dendritic_cell | *SNX33* | -2.885196227 | 4.02E-19 | 3.70E-18 | 2.53E-15 |
| Heart_Failure | dendritic_cell | *NUDT18* | -6.629463233 | 3.93E-18 | 2.46E-17 | 2.47E-14 |
| Heart_Failure | dendritic_cell | *SULT1B1* | 5.078575169 | 1.42E-09 | 2.32E-09 | 8.90E-06 |
| Heart_Failure | dendritic_cell | *NUDT4* | -3.211949714 | 4.34E-16 | 1.63E-15 | 2.73E-12 |
| Heart_Failure | dendritic_cell | *SCAI* | -3.751446627 | 3.16E-16 | 1.23E-15 | 1.99E-12 |
| Heart_Failure | dendritic_cell | *C1orf100* | -4.429130957 | 3.26E-20 | 6.76E-19 | 2.05E-16 |
| Heart_Failure | dendritic_cell | *AGFG1* | -2.978342797 | 8.40E-12 | 1.65E-11 | 5.28E-08 |
| Heart_Failure | dendritic_cell | *HAP1* | 3.463088458 | 4.67E-16 | 1.74E-15 | 2.94E-12 |
| Heart_Failure | dendritic_cell | *RNF213* | -3.788393851 | 5.86E-08 | 8.67E-08 | 0.000368158 |
| Heart_Failure | dendritic_cell | *PLK3* | -7.7363739 | 5.17E-12 | 1.04E-11 | 3.25E-08 |
| Heart_Failure | dendritic_cell | *DPY19L1* | -5.59212927 | 5.99E-14 | 1.52E-13 | 3.76E-10 |
| Heart_Failure | dendritic_cell | *TUBB8* | -8.988177868 | 5.25E-17 | 2.44E-16 | 3.30E-13 |
| Heart_Failure | dendritic_cell | *GPR160* | -8.163557509 | 7.12E-13 | 1.57E-12 | 4.47E-09 |
| Heart_Failure | dendritic_cell | *HOXB2* | 4.015056629 | 1.09E-11 | 2.12E-11 | 6.85E-08 |
| Heart_Failure | dendritic_cell | *XXYLT1* | -5.060442369 | 1.63E-12 | 3.48E-12 | 1.03E-08 |
| Heart_Failure | dendritic_cell | *UBXN2A* | -4.276527524 | 1.23E-13 | 2.98E-13 | 7.73E-10 |
| Heart_Failure | dendritic_cell | *CCS* | -3.971756057 | 3.66E-05 | 4.69E-05 | 0.230390648 |
| Heart_Failure | dendritic_cell | *NRROS* | 1.626199317 | 4.51E-06 | 6.01E-06 | 0.028325398 |
| Heart_Failure | dendritic_cell | *CEP19* | 2.68957259 | 8.04E-07 | 1.12E-06 | 0.005057438 |
| Heart_Failure | dendritic_cell | *SLC25A30* | -6.15864851 | 2.06E-05 | 2.67E-05 | 0.129405285 |
| Heart_Failure | dendritic_cell | *CTSF* | -2.547693343 | 2.92E-10 | 5.04E-10 | 1.84E-06 |
| Heart_Failure | dendritic_cell | *LEMD3* | -4.330404003 | 2.51E-14 | 6.78E-14 | 1.58E-10 |
| Heart_Failure | dendritic_cell | *C16orf91* | 8.262639522 | 1.02E-15 | 3.55E-15 | 6.38E-12 |
| Heart_Failure | dendritic_cell | *SOCS7* | -5.698063752 | 6.12E-13 | 1.36E-12 | 3.85E-09 |
| Heart_Failure | dendritic_cell | *TLR6* | 2.155633488 | 9.01E-22 | 3.01E-19 | 5.67E-18 |
| Heart_Failure | dendritic_cell | *RGMB* | -2.668173717 | 4.52E-19 | 4.05E-18 | 2.84E-15 |
| Heart_Failure | dendritic_cell | *FAM53A* | -6.434612356 | 1.70E-05 | 2.22E-05 | 0.107147274 |
| Heart_Failure | dendritic_cell | *CTU2* | 4.109081423 | 3.93E-16 | 1.49E-15 | 2.47E-12 |
| Heart_Failure | dendritic_cell | *MGA* | -3.502870146 | 1.78E-12 | 3.78E-12 | 1.12E-08 |
| Heart_Failure | dendritic_cell | *SNX31* | -2.075475493 | 0.014384448 | 0.016329907 | 1 |
| Heart_Failure | dendritic_cell | *DDX23* | -6.539804116 | 1.47E-12 | 3.15E-12 | 9.27E-09 |
| Heart_Failure | dendritic_cell | *GLIS1* | -2.417616581 | 7.31E-14 | 1.83E-13 | 4.59E-10 |
| Heart_Failure | dendritic_cell | *RALGAPA1* | -3.170716401 | 1.02E-13 | 2.49E-13 | 6.40E-10 |
| Heart_Failure | dendritic_cell | *LIG4* | -3.760914167 | 1.31E-14 | 3.69E-14 | 8.21E-11 |
| Heart_Failure | dendritic_cell | *SNAPC5* | -5.395979491 | 2.92E-14 | 7.82E-14 | 1.84E-10 |
| Heart_Failure | dendritic_cell | *LINGO2* | -6.858859083 | 5.61E-09 | 8.81E-09 | 3.53E-05 |
| Heart_Failure | dendritic_cell | *ANKRD36C* | -3.193948435 | 3.33E-17 | 1.62E-16 | 2.10E-13 |
| Heart_Failure | dendritic_cell | *TMEM81* | 8.763059303 | 2.85E-21 | 4.66E-19 | 1.79E-17 |
| Heart_Failure | dendritic_cell | *IL20RB* | -2.477180526 | 6.38E-07 | 8.91E-07 | 0.00401304 |
| Heart_Failure | dendritic_cell | *ZNF497* | -11.22404419 | 4.10E-11 | 7.58E-11 | 2.58E-07 |
| Heart_Failure | dendritic_cell | *KLF14* | -25.00333823 | 2.69E-19 | 2.70E-18 | 1.69E-15 |
| Heart_Failure | dendritic_cell | *ANGEL2* | -4.983800925 | 1.39E-08 | 2.12E-08 | 8.72E-05 |
| Heart_Failure | dendritic_cell | *ZNF266* | -1.959833403 | 2.63E-06 | 3.56E-06 | 0.016541268 |
| Heart_Failure | dendritic_cell | *BRSK2* | 6.489240015 | 1.16E-12 | 2.50E-12 | 7.28E-09 |
| Heart_Failure | dendritic_cell | *SH3PXD2B* | -1.977550198 | 1.37E-16 | 5.72E-16 | 8.61E-13 |
| Heart_Failure | dendritic_cell | *LARP7* | -5.008270995 | 3.78E-13 | 8.63E-13 | 2.38E-09 |
| Heart_Failure | dendritic_cell | *FGFBP3* | 12.03833056 | 2.91E-16 | 1.14E-15 | 1.83E-12 |
| Heart_Failure | dendritic_cell | *C4orf32* | -3.842617469 | 2.35E-17 | 1.19E-16 | 1.48E-13 |
| Heart_Failure | dendritic_cell | *HRAS* | 6.576387859 | 5.08E-15 | 1.55E-14 | 3.20E-11 |
| Heart_Failure | dendritic_cell | *SRP72* | -1.798411433 | 0.00399808 | 0.004676452 | 1 |

| | | | | | | |
|---|---|---|---|---|---|---|
| Heart_Failure | dendritic_cell | *BTC* | -2.762838255 | 7.98E-20 | 1.14E-18 | 5.02E-16 |
| Heart_Failure | dendritic_cell | *NLRP6* | 9.535522838 | 2.08E-17 | 1.07E-16 | 1.31E-13 |
| Heart_Failure | dendritic_cell | *NDUFA11* | -4.217113393 | 8.58E-19 | 6.83E-18 | 5.39E-15 |
| Heart_Failure | dendritic_cell | *RSRC1* | -3.444383935 | 9.29E-10 | 1.54E-09 | 5.84E-06 |
| Heart_Failure | dendritic_cell | *FUT1* | -6.05024914 | 1.26E-11 | 2.44E-11 | 7.90E-08 |
| Heart_Failure | dendritic_cell | *DHX36* | -4.069208377 | 9.77E-07 | 1.35E-06 | 0.006145027 |
| Heart_Failure | dendritic_cell | *FBXW8* | -4.58390323 | 4.86E-15 | 1.49E-14 | 3.05E-11 |
| Heart_Failure | dendritic_cell | *SLC22A1* | -3.856761903 | 5.86E-18 | 3.52E-17 | 3.69E-14 |
| Heart_Failure | dendritic_cell | *CHST2* | 4.310515476 | 1.16E-20 | 5.00E-19 | 7.32E-17 |
| Heart_Failure | dendritic_cell | *ATR* | -3.967922264 | 2.76E-11 | 5.17E-11 | 1.73E-07 |
| Heart_Failure | dendritic_cell | *TVP23C* | -6.216376553 | 2.98E-13 | 6.91E-13 | 1.87E-09 |
| Heart_Failure | dendritic_cell | *PACS1* | -3.99226724 | 4.57E-10 | 7.75E-10 | 2.87E-06 |
| Heart_Failure | dendritic_cell | *CADM2* | -1.987374305 | 2.01E-15 | 6.60E-15 | 1.26E-11 |
| Heart_Failure | dendritic_cell | *PSMD2* | -2.389861868 | 0.000209995 | 0.0002603 | 1 |
| Heart_Failure | dendritic_cell | *ARHGAP1* | -3.382649787 | 0.000116142 | 0.000145745 | 0.730181918 |
| Heart_Failure | dendritic_cell | *CHST1* | 4.141359049 | 2.07E-12 | 4.34E-12 | 1.30E-08 |
| Heart_Failure | dendritic_cell | *GOLGA8A* | -3.671633911 | 4.92E-15 | 1.51E-14 | 3.09E-11 |
| Heart_Failure | dendritic_cell | *TP53I11* | -5.342004185 | 4.69E-12 | 9.51E-12 | 2.95E-08 |
| Heart_Failure | dendritic_cell | *PHYHD1* | -4.433778502 | 8.28E-14 | 2.06E-13 | 5.20E-10 |
| Heart_Failure | dendritic_cell | *ZNF519* | -4.758326535 | 1.27E-14 | 3.60E-14 | 7.98E-11 |
| Heart_Failure | dendritic_cell | *BANF1* | 2.651566274 | 1.08E-18 | 8.28E-18 | 6.80E-15 |
| Heart_Failure | dendritic_cell | *CHRNA7* | -3.79078941 | 3.72E-17 | 1.79E-16 | 2.34E-13 |
| Heart_Failure | dendritic_cell | *TMEM9B* | -4.494706832 | 0.000253709 | 0.000313126 | 1 |
| Heart_Failure | dendritic_cell | *NRIP3* | -4.288525623 | 3.24E-14 | 8.61E-14 | 2.04E-10 |
| Heart_Failure | dendritic_cell | *SCUBE2* | -5.653720939 | 9.17E-16 | 3.23E-15 | 5.76E-12 |
| Heart_Failure | dendritic_cell | *SMAD2* | -2.991764277 | 3.83E-10 | 6.53E-10 | 2.41E-06 |
| Heart_Failure | dendritic_cell | *EIF3F* | -3.221266507 | 0.005207918 | 0.006046571 | 1 |
| Heart_Failure | dendritic_cell | *ZNF25* | -2.793005632 | 3.54E-10 | 6.07E-10 | 2.23E-06 |
| Heart_Failure | dendritic_cell | *CLTB* | -3.508595818 | 1.95E-06 | 2.65E-06 | 0.012247286 |
| Heart_Failure | dendritic_cell | *PCSK1* | -3.117607086 | 4.64E-20 | 8.15E-19 | 2.92E-16 |
| Heart_Failure | dendritic_cell | *LPL* | -1.609338339 | 1.00E-14 | 2.91E-14 | 6.31E-11 |
| Heart_Failure | dendritic_cell | *TBC1D10C* | -3.38140532 | 1.64E-08 | 2.50E-08 | 0.000103105 |
| Heart_Failure | dendritic_cell | *PPP2R2D* | -3.245964638 | 1.16E-12 | 2.50E-12 | 7.27E-09 |
| Heart_Failure | dendritic_cell | *MCTP1* | -1.796524136 | 0.000543181 | 0.00066041 | 1 |
| Heart_Failure | dendritic_cell | *LRRC25* | 1.305738296 | 6.69E-14 | 1.69E-13 | 4.20E-10 |
| Heart_Failure | dendritic_cell | *UBQLNL* | -6.563598649 | 2.84E-15 | 9.07E-15 | 1.78E-11 |
| Heart_Failure | dendritic_cell | *LIPT2* | 6.61919538 | 5.58E-19 | 4.81E-18 | 3.51E-15 |
| Heart_Failure | dendritic_cell | *KCNE3* | -7.201664666 | 5.56E-09 | 8.73E-09 | 3.50E-05 |
| Heart_Failure | dendritic_cell | *UCP3* | 4.98267905 | 2.14E-12 | 4.49E-12 | 1.34E-08 |
| Heart_Failure | dendritic_cell | *UCP2* | 4.445302676 | 8.61E-20 | 1.21E-18 | 5.41E-16 |
| Heart_Failure | dendritic_cell | *PAAF1* | -1.488862209 | 1.04E-11 | 2.02E-11 | 6.52E-08 |
| Heart_Failure | dendritic_cell | *RAB6A* | -4.450194801 | 2.28E-11 | 4.32E-11 | 1.43E-07 |
| Heart_Failure | dendritic_cell | *RPS6KB2* | -4.047857715 | 2.04E-17 | 1.05E-16 | 1.28E-13 |
| Heart_Failure | dendritic_cell | *RMI2* | -3.201779783 | 1.12E-16 | 4.77E-16 | 7.01E-13 |
| Heart_Failure | dendritic_cell | *TOM1L2* | -4.438848835 | 2.22E-12 | 4.65E-12 | 1.39E-08 |
| Heart_Failure | dendritic_cell | *TEX26* | -6.452974046 | 1.71E-09 | 2.79E-09 | 1.08E-05 |
| Heart_Failure | dendritic_cell | *GPR156* | -3.225977065 | 1.97E-19 | 2.15E-18 | 1.24E-15 |
| Heart_Failure | dendritic_cell | *B3GNTL1* | 5.273183309 | 1.81E-14 | 4.99E-14 | 1.14E-10 |
| Heart_Failure | dendritic_cell | *TTLL11* | -3.375898276 | 5.63E-15 | 1.71E-14 | 3.54E-11 |
| Heart_Failure | dendritic_cell | *LINC01106* | -14.32556098 | 4.34E-05 | 5.55E-05 | 0.272932366 |

| | | | | | | |
|---|---|---|---|---|---|---|
| Heart_Failure | dendritic_cell | *MSRA* | -4.81605159 | 8.03E-12 | 1.58E-11 | 5.05E-08 |
| Heart_Failure | dendritic_cell | *CTDNEP1* | -6.845900025 | 1.11E-08 | 1.71E-08 | 6.97E-05 |
| Heart_Failure | dendritic_cell | *ETV4* | -3.898598602 | 8.75E-14 | 2.17E-13 | 5.50E-10 |
| Heart_Failure | dendritic_cell | *SWI5* | -3.312817175 | 2.47E-18 | 1.68E-17 | 1.55E-14 |
| Heart_Failure | dendritic_cell | *ZDHHC21* | -4.04563526 | 3.52E-13 | 8.08E-13 | 2.21E-09 |
| Heart_Failure | dendritic_cell | *A2M* | -3.15509639 | 3.84E-17 | 1.84E-16 | 2.41E-13 |
| Heart_Failure | dendritic_cell | *ARL4D* | -4.52575925 | 9.07E-17 | 3.98E-16 | 5.70E-13 |
| Heart_Failure | dendritic_cell | *LRRN1* | -6.924843083 | 3.98E-16 | 1.51E-15 | 2.50E-12 |
| Heart_Failure | dendritic_cell | *B3GALT6* | -5.293503203 | 5.88E-20 | 9.53E-19 | 3.69E-16 |
| Heart_Failure | dendritic_cell | *ZNF613* | -4.185240687 | 6.95E-15 | 2.07E-14 | 4.37E-11 |
| Heart_Failure | dendritic_cell | *IP6K1* | -4.208710937 | 2.54E-08 | 3.82E-08 | 0.000159698 |
| Heart_Failure | dendritic_cell | *CHMP6* | 7.704778628 | 9.20E-09 | 1.42E-08 | 5.79E-05 |
| Heart_Failure | dendritic_cell | *UFSP1* | 1.979854033 | 6.85E-05 | 8.68E-05 | 0.430387928 |
| Heart_Failure | dendritic_cell | *TMEM39A* | -4.289823466 | 9.99E-15 | 2.90E-14 | 6.28E-11 |
| Heart_Failure | dendritic_cell | *TCP11L1* | -2.504085401 | 0.000602855 | 0.000730702 | 1 |
| Heart_Failure | dendritic_cell | *GPX2* | -5.269552407 | 1.74E-18 | 1.24E-17 | 1.09E-14 |
| Heart_Failure | dendritic_cell | *CCDC57* | -3.890990717 | 3.92E-14 | 1.02E-13 | 2.47E-10 |
| Heart_Failure | dendritic_cell | *CIDEA* | -0.902490965 | 1.13E-09 | 1.86E-09 | 7.09E-06 |
| Heart_Failure | dendritic_cell | *ZNF404* | -1.601924614 | 1.09E-06 | 1.50E-06 | 0.006876388 |
| Heart_Failure | dendritic_cell | *SLC35G1* | -2.846056454 | 4.32E-17 | 2.05E-16 | 2.71E-13 |
| Heart_Failure | dendritic_cell | *ZNF135* | -3.602227695 | 1.44E-15 | 4.86E-15 | 9.04E-12 |
| Heart_Failure | dendritic_cell | *COX8A* | -8.020210386 | 7.83E-11 | 1.41E-10 | 4.92E-07 |
| Heart_Failure | dendritic_cell | *B3GNT4* | -21.6286533 | 2.64E-20 | 6.44E-19 | 1.66E-16 |
| Heart_Failure | dendritic_cell | *CDC26* | -1.649126891 | 0.009077221 | 0.010404464 | 1 |
| Heart_Failure | dendritic_cell | *RNPEP* | -4.413834212 | 1.27E-07 | 1.84E-07 | 0.000796996 |
| Heart_Failure | dendritic_cell | *SPRYD4* | -2.524848132 | 7.51E-08 | 1.10E-07 | 0.000472128 |
| Heart_Failure | dendritic_cell | *SLCO3A1* | 4.387905002 | 3.77E-08 | 5.63E-08 | 0.000237288 |
| Heart_Failure | dendritic_cell | *WDR25* | -5.668689992 | 1.04E-13 | 2.54E-13 | 6.53E-10 |
| Heart_Failure | dendritic_cell | *PLA2G16* | -1.33631285 | 2.27E-13 | 5.32E-13 | 1.43E-09 |
| Heart_Failure | dendritic_cell | *PHLDB3* | 3.20104902 | 0.000334173 | 0.000410421 | 1 |
| Heart_Failure | dendritic_cell | *PRR15* | -6.137339426 | 1.16E-05 | 1.52E-05 | 0.072934356 |
| Heart_Failure | dendritic_cell | *RMDN1* | -3.551104161 | 7.07E-12 | 1.41E-11 | 4.44E-08 |
| Heart_Failure | dendritic_cell | *HORMAD2* | -2.59132765 | 3.40E-11 | 6.34E-11 | 2.14E-07 |
| Heart_Failure | dendritic_cell | *RNF152* | -3.725963955 | 0.003128814 | 0.003674048 | 1 |
| Heart_Failure | dendritic_cell | *LRRC37A* | -2.532414104 | 1.42E-05 | 1.86E-05 | 0.089559299 |
| Heart_Failure | dendritic_cell | *ACSF3* | -5.228006317 | 5.68E-17 | 2.63E-16 | 3.57E-13 |
| Heart_Failure | dendritic_cell | *BOK* | -3.032259529 | 7.65E-19 | 6.22E-18 | 4.81E-15 |
| Heart_Failure | dendritic_cell | *ZNF843* | 10.89575655 | 4.51E-16 | 1.68E-15 | 2.83E-12 |
| Heart_Failure | dendritic_cell | *C8orf59* | -4.41359002 | 3.29E-15 | 1.04E-14 | 2.07E-11 |
| Heart_Failure | dendritic_cell | *PFN4* | -2.249367687 | 3.10E-06 | 4.17E-06 | 0.019479285 |
| Heart_Failure | dendritic_cell | *RUFY1* | -2.948801166 | 4.19E-06 | 5.61E-06 | 0.02634305 |
| Heart_Failure | dendritic_cell | *BASP1* | 2.088537687 | 4.86E-21 | 4.66E-19 | 3.05E-17 |
| Heart_Failure | dendritic_cell | *LRRC37A3* | -4.887339545 | 4.77E-17 | 2.23E-16 | 3.00E-13 |
| Heart_Failure | dendritic_cell | *VSIG10* | -2.164211599 | 3.11E-17 | 1.53E-16 | 1.95E-13 |
| Heart_Failure | dendritic_cell | *IRX5* | -2.832124361 | 3.01E-17 | 1.48E-16 | 1.89E-13 |
| Heart_Failure | dendritic_cell | *PNMA1* | -3.612262219 | 1.80E-19 | 2.05E-18 | 1.13E-15 |
| Heart_Failure | dendritic_cell | *ANKLE2* | -4.684074507 | 9.13E-16 | 3.22E-15 | 5.74E-12 |
| Heart_Failure | dendritic_cell | *GCNT4* | 11.37491828 | 1.41E-20 | 5.50E-19 | 8.86E-17 |
| Heart_Failure | dendritic_cell | *THAP4* | -4.405881529 | 1.32E-12 | 2.84E-12 | 8.32E-09 |
| Heart_Failure | dendritic_cell | *SHMT1* | -2.249614056 | 3.04E-13 | 7.04E-13 | 1.91E-09 |

| | | | | | | |
|---|---|---|---|---|---|---|
| Heart_Failure | dendritic_cell | *DPP7* | -3.870966499 | 0.00079708 | 0.000961113 | 1 |
| Heart_Failure | dendritic_cell | *SEC24C* | -3.904667316 | 4.96E-19 | 4.34E-18 | 3.12E-15 |
| Heart_Failure | dendritic_cell | *MTHFR* | -3.306922642 | 0.004306733 | 0.005024389 | 1 |
| Heart_Failure | dendritic_cell | *C19orf18* | -3.435260022 | 1.23E-14 | 3.49E-14 | 7.70E-11 |
| Heart_Failure | dendritic_cell | *TMEM80* | -2.546092058 | 2.25E-07 | 3.23E-07 | 0.001417161 |
| Heart_Failure | dendritic_cell | *WDR73* | -3.565754508 | 7.38E-13 | 1.62E-12 | 4.64E-09 |
| Heart_Failure | dendritic_cell | *POLE* | -4.633408258 | 2.18E-17 | 1.11E-16 | 1.37E-13 |
| Heart_Failure | dendritic_cell | *DSCAML1* | -3.412840758 | 1.83E-17 | 9.59E-17 | 1.15E-13 |
| Heart_Failure | dendritic_cell | *EPS8L2* | -3.0973724 | 3.06E-19 | 2.95E-18 | 1.92E-15 |
| Heart_Failure | dendritic_cell | *TALDO1* | -3.299357797 | 2.34E-09 | 3.77E-09 | 1.47E-05 |
| Heart_Failure | dendritic_cell | *RIMKLA* | -2.506252814 | 3.01E-07 | 4.29E-07 | 0.001894678 |
| Heart_Failure | dendritic_cell | *B3GNT8* | 10.67735513 | 2.29E-20 | 6.44E-19 | 1.44E-16 |
| Heart_Failure | dendritic_cell | *AP006621.1* | -2.275135364 | 1.17E-17 | 6.50E-17 | 7.36E-14 |
| Heart_Failure | dendritic_cell | *MAN1B1* | -6.690102382 | 1.58E-11 | 3.04E-11 | 9.95E-08 |
| Heart_Failure | dendritic_cell | *FBXO39* | 2.55910526 | 1.04E-14 | 2.99E-14 | 6.51E-11 |
| Heart_Failure | dendritic_cell | *ZBTB38* | -3.287835278 | 4.68E-14 | 1.21E-13 | 2.94E-10 |
| Heart_Failure | dendritic_cell | *C8orf31* | -4.146920269 | 6.48E-20 | 1.01E-18 | 4.07E-16 |
| Heart_Failure | dendritic_cell | *AC024940.1* | -4.358343732 | 2.62E-07 | 3.75E-07 | 0.001648061 |
| Heart_Failure | dendritic_cell | *PPFIA3* | 2.779232572 | 1.33E-17 | 7.24E-17 | 8.33E-14 |
| Heart_Failure | dendritic_cell | *MAGEF1* | 10.70060422 | 2.32E-13 | 5.44E-13 | 1.46E-09 |
| Heart_Failure | dendritic_cell | *SAMD9L* | -3.222530849 | 9.31E-09 | 1.44E-08 | 5.85E-05 |
| Heart_Failure | dendritic_cell | *PAWR* | -5.026845679 | 1.32E-14 | 3.73E-14 | 8.30E-11 |
| Heart_Failure | dendritic_cell | *NAP1L5* | 9.313761952 | 1.04E-14 | 3.00E-14 | 6.54E-11 |
| Heart_Failure | dendritic_cell | *NIM1K* | 5.053477871 | 0.001314031 | 0.001569101 | 1 |
| Heart_Failure | dendritic_cell | *ACOT4* | -3.059431429 | 8.02E-15 | 2.36E-14 | 5.04E-11 |
| Heart_Failure | dendritic_cell | *ARIH2* | -2.830324617 | 4.76E-14 | 1.23E-13 | 2.99E-10 |
| Heart_Failure | dendritic_cell | *RBM44* | -7.785373833 | 9.09E-10 | 1.51E-09 | 5.71E-06 |
| Heart_Failure | dendritic_cell | *IRX3* | -3.234162351 | 2.51E-18 | 1.69E-17 | 1.58E-14 |
| Heart_Failure | dendritic_cell | *ATOX1* | -3.262299613 | 0.004388891 | 0.005119287 | 1 |
| Heart_Failure | dendritic_cell | *TBL1XR1* | -3.658683951 | 5.46E-12 | 1.10E-11 | 3.43E-08 |
| Heart_Failure | dendritic_cell | *C18orf32* | -6.043408309 | 1.43E-16 | 5.95E-16 | 8.97E-13 |
| Heart_Failure | dendritic_cell | *RPLP2* | -3.857541566 | 7.73E-06 | 1.02E-05 | 0.048608529 |
| Heart_Failure | dendritic_cell | *C12orf54* | -11.35226643 | 3.60E-12 | 7.40E-12 | 2.26E-08 |
| Heart_Failure | dendritic_cell | *GBA* | 4.581721574 | 4.08E-06 | 5.46E-06 | 0.025647489 |
| Heart_Failure | dendritic_cell | *ACAD9* | -1.383224647 | 5.85E-07 | 8.18E-07 | 0.003677263 |
| Heart_Failure | dendritic_cell | *CD163L1* | 1.639221798 | 0.000181318 | 0.000225776 | 1 |
| Heart_Failure | dendritic_cell | *THAP5* | -3.74357156 | 1.16E-19 | 1.47E-18 | 7.30E-16 |
| Heart_Failure | dendritic_cell | *CD151* | -4.251446242 | 1.80E-11 | 3.44E-11 | 1.13E-07 |
| Heart_Failure | dendritic_cell | *POLR2L* | -4.248875893 | 6.26E-18 | 3.71E-17 | 3.93E-14 |
| Heart_Failure | dendritic_cell | *FAM87B* | -7.241172597 | 1.74E-15 | 5.78E-15 | 1.10E-11 |
| Heart_Failure | dendritic_cell | *ZCCHC3* | 7.303858543 | 1.61E-16 | 6.64E-16 | 1.01E-12 |
| Heart_Failure | dendritic_cell | *PCDHB9* | 2.545878454 | 1.20E-05 | 1.57E-05 | 0.075328907 |
| Heart_Failure | dendritic_cell | *ZNF619* | -6.399748809 | 1.86E-17 | 9.73E-17 | 1.17E-13 |
| Heart_Failure | dendritic_cell | *ZBTB41* | -3.729446014 | 5.73E-18 | 3.45E-17 | 3.60E-14 |
| Heart_Failure | dendritic_cell | *ZNF354C* | -5.262118345 | 1.26E-16 | 5.31E-16 | 7.89E-13 |
| Heart_Failure | dendritic_cell | *CENPBD1* | -3.237815946 | 0.000251591 | 0.000310635 | 1 |
| Heart_Failure | dendritic_cell | *BET1L* | -4.494235618 | 2.07E-09 | 3.35E-09 | 1.30E-05 |
| Heart_Failure | dendritic_cell | *RIC8A* | 5.900830565 | 1.47E-07 | 2.12E-07 | 0.000921985 |
| Heart_Failure | dendritic_cell | *IMP3* | -6.930377735 | 5.91E-14 | 1.51E-13 | 3.72E-10 |
| Heart_Failure | dendritic_cell | *DPY19L2* | -1.923109156 | 1.05E-15 | 3.66E-15 | 6.61E-12 |

| | | | | | | |
|---|---|---|---|---|---|---|
| Heart_Failure | dendritic_cell | *C2orf73* | -3.455278313 | 2.30E-20 | 6.44E-19 | 1.44E-16 |
| Heart_Failure | dendritic_cell | *FAM26E* | -3.619883954 | 6.07E-19 | 5.13E-18 | 3.81E-15 |
| Heart_Failure | dendritic_cell | *ALS2CL* | -3.116843958 | 1.11E-11 | 2.17E-11 | 7.00E-08 |
| Heart_Failure | dendritic_cell | *MLF1* | -5.150838807 | 3.97E-09 | 6.29E-09 | 2.49E-05 |
| Heart_Failure | dendritic_cell | *PRSS42* | -5.228409801 | 2.59E-09 | 4.16E-09 | 1.63E-05 |
| Heart_Failure | dendritic_cell | *GRAMD1C* | -6.781189267 | 6.39E-16 | 2.33E-15 | 4.02E-12 |
| Heart_Failure | dendritic_cell | *STAP2* | 6.13794332 | 3.55E-14 | 9.34E-14 | 2.23E-10 |
| Heart_Failure | dendritic_cell | *TSSK6* | -4.407291911 | 0.002104334 | 0.002492454 | 1 |
| Heart_Failure | dendritic_cell | *GOLGA8Q* | -24.78441674 | 0.019830317 | 0.022358896 | 1 |
| Heart_Failure | dendritic_cell | *NDUFV2* | 8.328966513 | 1.71E-17 | 9.05E-17 | 1.08E-13 |
| Heart_Failure | dendritic_cell | *DALRD3* | -3.579384993 | 1.08E-15 | 3.74E-15 | 6.76E-12 |
| Heart_Failure | dendritic_cell | *ZNF114* | -5.143813211 | 1.28E-18 | 9.49E-18 | 8.03E-15 |
| Heart_Failure | dendritic_cell | *ZNF518B* | -6.236150212 | 1.49E-08 | 2.28E-08 | 9.39E-05 |
| Heart_Failure | dendritic_cell | *LCORL* | 2.653490547 | 0.004095981 | 0.004789183 | 1 |
| Heart_Failure | dendritic_cell | *SH2B1* | -2.412716522 | 2.47E-10 | 4.29E-10 | 1.56E-06 |
| Heart_Failure | dendritic_cell | *ZC3H12D* | 1.669377108 | 6.81E-21 | 4.66E-19 | 4.28E-17 |
| Heart_Failure | dendritic_cell | *KDELC2* | -5.404268818 | 1.52E-13 | 3.64E-13 | 9.54E-10 |
| Heart_Failure | dendritic_cell | *PLEC* | -5.243892191 | 1.39E-09 | 2.29E-09 | 8.76E-06 |
| Heart_Failure | dendritic_cell | *SH2D4B* | -6.368672476 | 2.86E-20 | 6.44E-19 | 1.80E-16 |
| Heart_Failure | dendritic_cell | *RNF212* | -1.70383177 | 1.19E-10 | 2.12E-10 | 7.49E-07 |
| Heart_Failure | dendritic_cell | *PRSS36* | 2.771902988 | 7.17E-21 | 4.66E-19 | 4.51E-17 |
| Heart_Failure | dendritic_cell | *TMPRSS9* | -3.084526372 | 1.05E-19 | 1.38E-18 | 6.60E-16 |
| Heart_Failure | dendritic_cell | *AQP11* | -5.276802418 | 8.50E-17 | 3.77E-16 | 5.35E-13 |
| Heart_Failure | dendritic_cell | *TMEM11* | -2.831690583 | 4.71E-20 | 8.25E-19 | 2.96E-16 |
| Heart_Failure | dendritic_cell | *KCNG2* | -1.407027858 | 6.55E-05 | 8.31E-05 | 0.411626655 |
| Heart_Failure | dendritic_cell | *ZFAND2A* | -5.135509206 | 8.44E-15 | 2.48E-14 | 5.31E-11 |
| Heart_Failure | dendritic_cell | *FAM220A* | -3.457727102 | 2.45E-11 | 4.63E-11 | 1.54E-07 |
| Heart_Failure | dendritic_cell | *BEND3* | 10.44880582 | 2.44E-18 | 1.66E-17 | 1.53E-14 |
| Heart_Failure | dendritic_cell | *NT5DC1* | -3.965044115 | 1.33E-12 | 2.85E-12 | 8.33E-09 |
| Heart_Failure | dendritic_cell | *AC006111.1* | -12.29555736 | 4.62E-18 | 2.84E-17 | 2.91E-14 |
| Heart_Failure | dendritic_cell | *LINC00843* | -7.6765455 | 1.27E-16 | 5.36E-16 | 8.00E-13 |
| Heart_Failure | dendritic_cell | *COX14* | -2.354362136 | 2.24E-18 | 1.55E-17 | 1.41E-14 |
| Heart_Failure | dendritic_cell | *P4HTM* | -4.924815209 | 1.49E-16 | 6.19E-16 | 9.38E-13 |
| Heart_Failure | dendritic_cell | *CTXN1* | 6.877566501 | 0.000184532 | 0.000229596 | 1 |
| Heart_Failure | dendritic_cell | *CA8* | -2.151875511 | 1.11E-15 | 3.85E-15 | 6.98E-12 |
| Heart_Failure | dendritic_cell | *ERBB4* | -2.100849078 | 7.18E-19 | 5.88E-18 | 4.51E-15 |
| Heart_Failure | dendritic_cell | *GPR35* | -1.453374287 | 2.05E-06 | 2.78E-06 | 0.012867077 |
| Heart_Failure | dendritic_cell | *CSRNP3* | -3.887308043 | 0.00109754 | 0.001313079 | 1 |
| Heart_Failure | dendritic_cell | *PARP10* | -2.611696859 | 0.001764307 | 0.002097221 | 1 |
| Heart_Failure | dendritic_cell | *KCTD12* | -1.316133434 | 0.027238632 | 0.030460561 | 1 |
| Heart_Failure | dendritic_cell | *RPP25* | 1.966634654 | 9.78E-05 | 0.000123145 | 0.614738297 |
| Heart_Failure | dendritic_cell | *STX19* | -2.488749113 | 0.017276085 | 0.019528002 | 1 |
| Heart_Failure | dendritic_cell | *FAM219B* | -4.270910132 | 4.27E-16 | 1.60E-15 | 2.68E-12 |
| Heart_Failure | dendritic_cell | *CPNE7* | -9.081358192 | 8.99E-14 | 2.22E-13 | 5.65E-10 |
| Heart_Failure | dendritic_cell | *GDPD4* | 15.99034411 | 4.31E-17 | 2.05E-16 | 2.71E-13 |
| Heart_Failure | dendritic_cell | *MPI* | -3.38374643 | 1.11E-17 | 6.18E-17 | 6.97E-14 |
| Heart_Failure | dendritic_cell | *TRIM73* | -7.097923222 | 4.35E-10 | 7.39E-10 | 2.74E-06 |
| Heart_Failure | dendritic_cell | *EFCAB13* | -3.071254935 | 5.08E-13 | 1.14E-12 | 3.19E-09 |
| Heart_Failure | dendritic_cell | *HYI* | -0.85218947 | 0.019860231 | 0.022388609 | 1 |
| Heart_Failure | dendritic_cell | *C17orf62* | 4.473158627 | 0.00036581 | 0.000448314 | 1 |

| | | | | | | |
|---|---|---|---|---|---|---|
| Heart_Failure | dendritic_cell | *TUFM* | 1.607840437 | 0.015260593 | 0.01728709 | 1 |
| Heart_Failure | dendritic_cell | *RMI1* | 2.383460187 | 0.007959218 | 0.009156377 | 1 |
| Heart_Failure | dendritic_cell | *CTC1* | 3.878325494 | 3.54E-08 | 5.29E-08 | 0.000222678 |
| Heart_Failure | dendritic_cell | *C14orf39* | -1.697919724 | 1.58E-16 | 6.51E-16 | 9.91E-13 |
| Heart_Failure | dendritic_cell | *KLHDC7A* | -1.398526929 | 1.21E-05 | 1.59E-05 | 0.076287326 |
| Heart_Failure | dendritic_cell | *TRIML2* | -56.49690692 | 3.88E-19 | 3.60E-18 | 2.44E-15 |
| Heart_Failure | dendritic_cell | *FARSA* | -5.295840109 | 3.12E-08 | 4.66E-08 | 0.000195944 |
| Heart_Failure | dendritic_cell | *SPTY2D1* | -5.957566309 | 1.22E-14 | 3.48E-14 | 7.68E-11 |
| Heart_Failure | dendritic_cell | *TCAIM* | -3.304076929 | 1.45E-14 | 4.07E-14 | 9.13E-11 |
| Heart_Failure | dendritic_cell | *FUCA1* | 2.070783747 | 0.018040558 | 0.020373808 | 1 |
| Heart_Failure | dendritic_cell | *ZNF664* | -6.525739628 | 2.50E-14 | 6.77E-14 | 1.57E-10 |
| Heart_Failure | dendritic_cell | *CALR* | -4.219304205 | 7.64E-07 | 1.06E-06 | 0.004805688 |
| Heart_Failure | dendritic_cell | *CDH4* | -4.366194304 | 8.53E-17 | 3.78E-16 | 5.36E-13 |
| Heart_Failure | dendritic_cell | *HLA-DQB1* | 0.485023573 | 8.29E-17 | 3.69E-16 | 5.21E-13 |
| Heart_Failure | dendritic_cell | *ARID3B* | -3.876967406 | 0.000122989 | 0.000154153 | 0.773233238 |
| Heart_Failure | dendritic_cell | *EGR3* | 2.194807537 | 7.58E-21 | 4.66E-19 | 4.77E-17 |
| Heart_Failure | dendritic_cell | *LINC00174* | -1.949233318 | 4.81E-09 | 7.59E-09 | 3.03E-05 |
| Heart_Failure | dendritic_cell | *GEMIN4* | -2.515232898 | 4.32E-17 | 2.05E-16 | 2.72E-13 |
| Heart_Failure | dendritic_cell | *CIITA* | 0.62480199 | 6.79E-21 | 4.66E-19 | 4.27E-17 |
| Heart_Failure | dendritic_cell | *PLD6* | -2.858022079 | 4.63E-05 | 5.91E-05 | 0.291192019 |
| Heart_Failure | dendritic_cell | *ZBTB42* | -6.917947771 | 0.012695407 | 0.014438499 | 1 |
| Heart_Failure | dendritic_cell | *LACC1* | 3.868109662 | 5.62E-19 | 4.83E-18 | 3.53E-15 |
| Heart_Failure | dendritic_cell | *MAF1* | 4.941009552 | 8.18E-20 | 1.16E-18 | 5.14E-16 |
| Heart_Failure | dendritic_cell | *FCER1A* | 0.250125665 | 7.03E-21 | 4.66E-19 | 4.42E-17 |
| Heart_Failure | dendritic_cell | *PCED1B* | -3.165344894 | 2.76E-15 | 8.84E-15 | 1.74E-11 |
| Heart_Failure | dendritic_cell | *PIPOX* | -4.036208531 | 1.06E-17 | 5.93E-17 | 6.64E-14 |
| Heart_Failure | dendritic_cell | *FOXS1* | 4.331698626 | 1.70E-19 | 1.95E-18 | 1.07E-15 |
| Heart_Failure | dendritic_cell | *ATOH7* | 15.07571763 | 2.13E-08 | 3.22E-08 | 0.000133864 |
| Heart_Failure | dendritic_cell | *CDH5* | -1.860990695 | 3.13E-18 | 2.02E-17 | 1.97E-14 |
| Heart_Failure | dendritic_cell | *SERTAD2* | 2.679098791 | 0.010621826 | 0.012121877 | 1 |
| Heart_Failure | dendritic_cell | *GIPC3* | -4.823606667 | 3.78E-12 | 7.73E-12 | 2.37E-08 |
| Heart_Failure | dendritic_cell | *CITED4* | -5.14780222 | 3.39E-16 | 1.31E-15 | 2.13E-12 |
| Heart_Failure | dendritic_cell | *TIGD5* | -3.388619551 | 1.27E-16 | 5.34E-16 | 7.96E-13 |
| Heart_Failure | dendritic_cell | *PDXDC1* | -3.655259943 | 4.90E-15 | 1.50E-14 | 3.08E-11 |
| Heart_Failure | dendritic_cell | *ZNF154* | -3.21056014 | 3.91E-05 | 5.01E-05 | 0.246097925 |
| Heart_Failure | dendritic_cell | *R3HDM2* | -3.908035275 | 1.30E-12 | 2.80E-12 | 8.19E-09 |
| Heart_Failure | dendritic_cell | *B3GNT3* | 6.103393854 | 2.60E-20 | 6.44E-19 | 1.64E-16 |
| Heart_Failure | dendritic_cell | *SSC5D* | -1.104441406 | 2.84E-06 | 3.83E-06 | 0.017853937 |
| Heart_Failure | dendritic_cell | *ZADH2* | -5.54372064 | 2.34E-12 | 4.89E-12 | 1.47E-08 |
| Heart_Failure | dendritic_cell | *SEPT1* | 7.632852513 | 2.13E-16 | 8.57E-16 | 1.34E-12 |
| Heart_Failure | dendritic_cell | *EXOC3* | -3.367908303 | 1.02E-05 | 1.34E-05 | 0.064115402 |
| Heart_Failure | dendritic_cell | *TDRD6* | -1.65652378 | 2.20E-10 | 3.83E-10 | 1.38E-06 |
| Heart_Failure | dendritic_cell | *LYNX1* | -3.294171715 | 2.80E-12 | 5.81E-12 | 1.76E-08 |
| Heart_Failure | dendritic_cell | *TDRP* | -3.509017351 | 3.75E-20 | 7.19E-19 | 2.36E-16 |
| Heart_Failure | dendritic_cell | *ZNF816* | 4.074936355 | 2.06E-06 | 2.79E-06 | 0.012922252 |
| Heart_Failure | dendritic_cell | *FGD6* | -5.218289885 | 4.20E-12 | 8.55E-12 | 2.64E-08 |
| Heart_Failure | dendritic_cell | *HCLS1* | 3.23047592 | 9.49E-20 | 1.28E-18 | 5.97E-16 |
| Heart_Failure | dendritic_cell | *ZNF609* | -3.314033219 | 4.10E-07 | 5.79E-07 | 0.002577825 |
| Heart_Failure | dendritic_cell | *CCDC66* | -3.47345526 | 1.27E-12 | 2.74E-12 | 8.01E-09 |
| Heart_Failure | dendritic_cell | *DEFB124* | -7.950455151 | 5.83E-11 | 1.06E-10 | 3.67E-07 |

| | | | | | | |
|---|---|---|---|---|---|---|
| Heart_Failure | dendritic_cell | *MCFD2* | -5.206214273 | 7.93E-08 | 1.16E-07 | 0.000498616 |
| Heart_Failure | dendritic_cell | *CYP8B1* | -2.943579413 | 1.80E-18 | 1.27E-17 | 1.13E-14 |
| Heart_Failure | dendritic_cell | *RNF182* | 3.097689573 | 3.92E-13 | 8.94E-13 | 2.47E-09 |
| Heart_Failure | dendritic_cell | *MB21D2* | -3.018635634 | 0.002596961 | 0.003061522 | 1 |
| Heart_Failure | dendritic_cell | *SSTR2* | -15.56184589 | 2.56E-08 | 3.85E-08 | 0.000160815 |
| Heart_Failure | dendritic_cell | *ZNF594* | -4.447158234 | 0.000129385 | 0.00016204 | 0.813440908 |
| Heart_Failure | dendritic_cell | *PCGF5* | -4.454728155 | 2.03E-10 | 3.54E-10 | 1.28E-06 |
| Heart_Failure | dendritic_cell | *SHISA2* | 7.180088158 | 6.59E-21 | 4.66E-19 | 4.14E-17 |
| Heart_Failure | dendritic_cell | *CHST13* | 10.90011134 | 1.08E-16 | 4.63E-16 | 6.77E-13 |
| Heart_Failure | dendritic_cell | *SRSF8* | 5.333606479 | 0.004548362 | 0.00530038 | 1 |
| Heart_Failure | dendritic_cell | *SLC36A4* | -4.052996474 | 8.61E-06 | 1.14E-05 | 0.054100992 |
| Heart_Failure | dendritic_cell | *ZDHHC20* | -2.510849229 | 0.008956102 | 0.010267508 | 1 |
| Heart_Failure | dendritic_cell | *OR51E1* | -1.036497928 | 0.029994322 | 0.03347671 | 1 |
| Heart_Failure | dendritic_cell | *PSMG4* | -4.048019249 | 7.47E-07 | 1.04E-06 | 0.004697867 |
| Heart_Failure | dendritic_cell | *ZNF443* | -2.958556724 | 0.000108495 | 0.00013634 | 0.682110028 |
| Heart_Failure | dendritic_cell | *CAPS2* | -3.807520909 | 3.46E-16 | 1.33E-15 | 2.18E-12 |
| Heart_Failure | dendritic_cell | *ZNF792* | -4.875426067 | 6.93E-16 | 2.50E-15 | 4.36E-12 |
| Heart_Failure | dendritic_cell | *KCTD2* | 3.214724528 | 0.035547029 | 0.03953373 | 1 |
| Heart_Failure | dendritic_cell | *D2HGDH* | -4.646859529 | 6.70E-10 | 1.12E-09 | 4.21E-06 |
| Heart_Failure | dendritic_cell | *CMTR2* | -6.094834078 | 1.89E-10 | 3.31E-10 | 1.19E-06 |
| Heart_Failure | dendritic_cell | *ST20* | -0.526588692 | 0.008188972 | 0.009412079 | 1 |
| Heart_Failure | dendritic_cell | *LRRC57* | -4.029214123 | 0.00208503 | 0.002470521 | 1 |
| Heart_Failure | dendritic_cell | *MRPL14* | -4.643359519 | 3.53E-10 | 6.05E-10 | 2.22E-06 |
| Heart_Failure | dendritic_cell | *BBS12* | 4.592454981 | 8.22E-21 | 4.66E-19 | 5.17E-17 |
| Heart_Failure | dendritic_cell | *ZFP82* | -4.902278615 | 1.37E-16 | 5.72E-16 | 8.59E-13 |
| Heart_Failure | dendritic_cell | *LSMEM1* | -3.162818145 | 2.12E-15 | 6.95E-15 | 1.33E-11 |
| Heart_Failure | dendritic_cell | *AEN* | 8.520196249 | 2.18E-14 | 5.95E-14 | 1.37E-10 |
| Heart_Failure | dendritic_cell | *FKRP* | -1.043565254 | 0.005227297 | 0.00606795 | 1 |
| Heart_Failure | dendritic_cell | *TRAPPC5* | 3.017255971 | 3.00E-19 | 2.90E-18 | 1.88E-15 |
| Heart_Failure | dendritic_cell | *RPH3AL* | -4.448569233 | 6.51E-15 | 1.95E-14 | 4.09E-11 |
| Heart_Failure | dendritic_cell | *ANKRD34A* | -5.012996109 | 8.19E-19 | 6.58E-18 | 5.15E-15 |
| Heart_Failure | dendritic_cell | *SLC26A11* | -6.001240988 | 1.54E-11 | 2.97E-11 | 9.70E-08 |
| Heart_Failure | dendritic_cell | *HIGD1A* | -4.689109564 | 8.36E-19 | 6.68E-18 | 5.26E-15 |
| Heart_Failure | dendritic_cell | *OR52N4* | -1.771712509 | 0.010017234 | 0.011454775 | 1 |
| Heart_Failure | dendritic_cell | *ADIPOQ* | -1.287771018 | 3.15E-13 | 7.27E-13 | 1.98E-09 |
| Heart_Failure | dendritic_cell | *F2R* | -2.472612974 | 8.04E-11 | 1.45E-10 | 5.05E-07 |
| Heart_Failure | dendritic_cell | *HIST3H2A* | -4.246833065 | 5.28E-11 | 9.65E-11 | 3.32E-07 |
| Heart_Failure | dendritic_cell | *POLR2A* | -5.153342001 | 5.70E-12 | 1.14E-11 | 3.58E-08 |
| Heart_Failure | dendritic_cell | *TMEM132C* | -1.743785515 | 3.21E-17 | 1.57E-16 | 2.02E-13 |
| Heart_Failure | dendritic_cell | *SLC25A41* | -14.52376643 | 2.52E-20 | 6.44E-19 | 1.58E-16 |
| Heart_Failure | dendritic_cell | *TMEM136* | 3.825163844 | 1.13E-08 | 1.73E-08 | 7.07E-05 |
| Heart_Failure | dendritic_cell | *TMEM132E* | 10.1145571 | 7.12E-15 | 2.12E-14 | 4.48E-11 |
| Heart_Failure | dendritic_cell | *ZNF322* | -3.650737366 | 5.07E-09 | 7.98E-09 | 3.19E-05 |
| Heart_Failure | dendritic_cell | *DDX60L* | -4.300521155 | 3.82E-10 | 6.51E-10 | 2.40E-06 |
| Heart_Failure | dendritic_cell | *OGFOD3* | 2.252872175 | 5.62E-08 | 8.33E-08 | 0.00035333 |
| Heart_Failure | dendritic_cell | *ACBD4* | -5.201064394 | 9.11E-14 | 2.25E-13 | 5.73E-10 |
| Heart_Failure | dendritic_cell | *SETD2* | -3.602722993 | 3.01E-13 | 6.96E-13 | 1.89E-09 |
| Heart_Failure | dendritic_cell | *MRPS23* | -6.895113579 | 1.83E-06 | 2.49E-06 | 0.011504798 |
| Heart_Failure | dendritic_cell | *GPR135* | -2.552907356 | 2.92E-11 | 5.47E-11 | 1.84E-07 |
| Heart_Failure | dendritic_cell | *ZFP41* | -1.964754577 | 4.41E-05 | 5.63E-05 | 0.277024263 |

| | | | | | | |
|---|---|---|---|---|---|---|
| Heart_Failure | dendritic_cell | *ATG9B* | -5.784165294 | 5.95E-20 | 9.56E-19 | 3.74E-16 |
| Heart_Failure | dendritic_cell | *HKR1* | -3.953147993 | 7.08E-17 | 3.20E-16 | 4.45E-13 |
| Heart_Failure | dendritic_cell | *ZBTB20* | -2.710424854 | 6.47E-15 | 1.94E-14 | 4.06E-11 |
| Heart_Failure | dendritic_cell | *C3orf58* | 4.96499032 | 3.46E-13 | 7.95E-13 | 2.18E-09 |
| Heart_Failure | dendritic_cell | *ODF3L2* | 2.78090533 | 0.008896922 | 0.010201522 | 1 |
| Heart_Failure | dendritic_cell | *RELL1* | -4.255560837 | 1.56E-06 | 2.13E-06 | 0.009794509 |
| Heart_Failure | dendritic_cell | *RFX7* | -3.224758254 | 0.000197268 | 0.000245007 | 1 |
| Heart_Failure | dendritic_cell | *ADO* | -9.242777032 | 6.54E-07 | 9.12E-07 | 0.004109203 |
| Heart_Failure | dendritic_cell | *SNRPE* | 5.452426678 | 5.20E-18 | 3.15E-17 | 3.27E-14 |
| Heart_Failure | dendritic_cell | *CHST15* | -4.851709175 | 2.91E-08 | 4.36E-08 | 0.0001827 |
| Heart_Failure | dendritic_cell | *IDH2* | -4.877800971 | 1.73E-17 | 9.14E-17 | 1.09E-13 |
| Heart_Failure | dendritic_cell | *PTCHD3* | 0.969310689 | 0.000163813 | 0.000204263 | 1 |
| Heart_Failure | dendritic_cell | *WRB* | 1.100792271 | 0.001723922 | 0.002050377 | 1 |
| Heart_Failure | dendritic_cell | *TNRC18* | -2.669821772 | 0.003084715 | 0.003623617 | 1 |
| Heart_Failure | dendritic_cell | *FAM181B* | -7.110907326 | 1.14E-07 | 1.66E-07 | 0.000716085 |
| Heart_Failure | dendritic_cell | *NOP10* | -4.519846902 | 4.32E-07 | 6.09E-07 | 0.002713586 |
| Heart_Failure | dendritic_cell | *FAM89A* | -2.090300216 | 1.15E-14 | 3.28E-14 | 7.21E-11 |
| Heart_Failure | dendritic_cell | *TDRKH* | -5.449712137 | 4.69E-17 | 2.20E-16 | 2.95E-13 |
| Heart_Failure | dendritic_cell | *ZNF708* | -3.904683684 | 7.56E-12 | 1.50E-11 | 4.76E-08 |
| Heart_Failure | dendritic_cell | *ERCC6L2* | -3.270097216 | 1.82E-10 | 3.19E-10 | 1.15E-06 |
| Heart_Failure | dendritic_cell | *MRPL41* | -4.443857286 | 2.28E-07 | 3.27E-07 | 0.001436362 |
| Heart_Failure | dendritic_cell | *HHIPL1* | -3.444680942 | 1.87E-20 | 6.38E-19 | 1.17E-16 |
| Heart_Failure | dendritic_cell | *FAM153B* | -4.592268652 | 9.66E-18 | 5.48E-17 | 6.08E-14 |
| Heart_Failure | dendritic_cell | *BACE2* | -2.450390332 | 1.52E-07 | 2.19E-07 | 0.000953021 |
| Heart_Failure | dendritic_cell | *SYNM* | -7.425838629 | 1.15E-12 | 2.48E-12 | 7.22E-09 |
| Heart_Failure | dendritic_cell | *IZUMO1* | -1.024551863 | 0.041968883 | 0.046478487 | 1 |
| Heart_Failure | dendritic_cell | *C8orf33* | -5.08253089 | 8.28E-14 | 2.06E-13 | 5.20E-10 |
| Heart_Failure | dendritic_cell | *FBXL6* | -8.503029847 | 1.13E-11 | 2.19E-11 | 7.08E-08 |
| Heart_Failure | dendritic_cell | *KBTBD3* | -5.978248406 | 2.31E-16 | 9.20E-16 | 1.45E-12 |
| Heart_Failure | dendritic_cell | *YBEY* | 0.783067117 | 0.002577317 | 0.003039504 | 1 |
| Heart_Failure | dendritic_cell | *CACNB4* | -2.04225419 | 3.79E-15 | 1.18E-14 | 2.38E-11 |
| Heart_Failure | dendritic_cell | *TRAPPC6B* | -4.203827021 | 2.90E-14 | 7.77E-14 | 1.82E-10 |
| Heart_Failure | dendritic_cell | *NPLOC4* | -3.589278881 | 2.16E-12 | 4.53E-12 | 1.36E-08 |
| Heart_Failure | dendritic_cell | *TSHZ2* | -2.961436677 | 1.07E-15 | 3.72E-15 | 6.73E-12 |
| Heart_Failure | dendritic_cell | *CAPN12* | -5.837992459 | 1.05E-16 | 4.52E-16 | 6.59E-13 |
| Heart_Failure | dendritic_cell | *GLRX5* | -7.821214747 | 4.48E-12 | 9.09E-12 | 2.81E-08 |
| Heart_Failure | dendritic_cell | *CAV3* | -1.876925393 | 1.13E-12 | 2.44E-12 | 7.08E-09 |
| Heart_Failure | dendritic_cell | *MXRA7* | -2.204225291 | 1.76E-13 | 4.19E-13 | 1.11E-09 |
| Heart_Failure | dendritic_cell | *MFSD5* | -3.730275458 | 3.99E-17 | 1.91E-16 | 2.51E-13 |
| Heart_Failure | dendritic_cell | *ADI1* | -3.44491903 | 4.51E-19 | 4.05E-18 | 2.83E-15 |
| Heart_Failure | dendritic_cell | *SPNS3* | 2.958755697 | 1.09E-20 | 4.99E-19 | 6.87E-17 |
| Heart_Failure | dendritic_cell | *CLEC4G* | -7.811847124 | 6.53E-21 | 4.66E-19 | 4.11E-17 |
| Heart_Failure | dendritic_cell | *SATB1* | 5.467479892 | 6.19E-06 | 8.21E-06 | 0.038901118 |
| Heart_Failure | dendritic_cell | *ACTL10* | -6.158733817 | 1.99E-09 | 3.22E-09 | 1.25E-05 |
| Heart_Failure | dendritic_cell | *TRAK1* | -3.391072287 | 4.67E-06 | 6.23E-06 | 0.029353801 |
| Heart_Failure | dendritic_cell | *SKA2* | -4.942975973 | 2.50E-16 | 9.89E-16 | 1.57E-12 |
| Heart_Failure | dendritic_cell | *TTC3* | -1.3689625 | 4.92E-09 | 7.76E-09 | 3.09E-05 |
| Heart_Failure | dendritic_cell | *KCNB2* | -1.411173729 | 9.07E-17 | 3.98E-16 | 5.70E-13 |
| Heart_Failure | dendritic_cell | *BRICD5* | 1.924478779 | 0.000112727 | 0.000141573 | 0.70871514 |
| Heart_Failure | dendritic_cell | *TSKU* | -3.541298505 | 4.68E-19 | 4.15E-18 | 2.94E-15 |

| | | | | | | |
|---|---|---|---|---|---|---|
| Heart_Failure | dendritic_cell | *GRID1* | -3.985392492 | 2.12E-19 | 2.26E-18 | 1.33E-15 |
| Heart_Failure | dendritic_cell | *ACBD3* | -4.184500024 | 2.28E-15 | 7.42E-15 | 1.43E-11 |
| Heart_Failure | dendritic_cell | *PLCXD3* | -2.107450153 | 1.70E-15 | 5.66E-15 | 1.07E-11 |
| Heart_Failure | dendritic_cell | *GPIHBP1* | -2.004024072 | 2.90E-07 | 4.14E-07 | 0.001826249 |
| Heart_Failure | dendritic_cell | *VMO1* | 1.137979071 | 1.37E-19 | 1.66E-18 | 8.63E-16 |
| Heart_Failure | dendritic_cell | *COL18A1* | -2.661757264 | 1.33E-06 | 1.82E-06 | 0.008332886 |
| Heart_Failure | dendritic_cell | *SLC25A18* | -1.990235861 | 1.33E-14 | 3.77E-14 | 8.39E-11 |
| Heart_Failure | dendritic_cell | *ZNF721* | -3.31301453 | 1.41E-11 | 2.72E-11 | 8.86E-08 |
| Heart_Failure | dendritic_cell | *LENG9* | -3.301616401 | 3.01E-13 | 6.97E-13 | 1.89E-09 |
| Heart_Failure | dendritic_cell | *C11orf54* | -1.320992987 | 1.83E-05 | 2.38E-05 | 0.115318694 |
| Heart_Failure | dendritic_cell | *CEP63* | -1.66861911 | 7.70E-07 | 1.07E-06 | 0.004843356 |
| Heart_Failure | dendritic_cell | *HMGN4* | 5.260610084 | 1.53E-14 | 4.28E-14 | 9.63E-11 |
| Heart_Failure | dendritic_cell | *C12orf60* | -4.758925363 | 7.81E-16 | 2.79E-15 | 4.91E-12 |
| Heart_Failure | dendritic_cell | *LYSMD4* | -1.589880968 | 0.000581648 | 0.000705541 | 1 |
| Heart_Failure | dendritic_cell | *WBP2NL* | 6.630208452 | 4.79E-09 | 7.55E-09 | 3.01E-05 |
| Heart_Failure | dendritic_cell | *BEGAIN* | 21.58410864 | 1.87E-17 | 9.76E-17 | 1.18E-13 |
| Heart_Failure | dendritic_cell | *ARHGEF37* | -4.59194486 | 2.20E-17 | 1.12E-16 | 1.38E-13 |
| Heart_Failure | dendritic_cell | *FAM43B* | 7.744820416 | 4.09E-21 | 4.66E-19 | 2.57E-17 |
| Heart_Failure | dendritic_cell | *CSMD1* | -1.991194285 | 3.47E-19 | 3.28E-18 | 2.18E-15 |
| Heart_Failure | dendritic_cell | *PTGDR2* | -50.89969163 | 7.65E-09 | 1.19E-08 | 4.81E-05 |
| Heart_Failure | dendritic_cell | *GPR19* | -7.134737716 | 8.14E-19 | 6.55E-18 | 5.11E-15 |
| Heart_Failure | dendritic_cell | *FANCF* | -12.18037172 | 8.32E-12 | 1.64E-11 | 5.23E-08 |
| Heart_Failure | dendritic_cell | *C2CD4C* | -5.116278446 | 1.61E-12 | 3.43E-12 | 1.01E-08 |
| Heart_Failure | dendritic_cell | *PLGLB1* | -2.33006155 | 0.002423012 | 0.002861284 | 1 |
| Heart_Failure | dendritic_cell | *CCBE1* | -1.545348884 | 1.55E-18 | 1.13E-17 | 9.78E-15 |
| Heart_Failure | dendritic_cell | *ZNF623* | -6.472074973 | 6.95E-16 | 2.51E-15 | 4.37E-12 |
| Heart_Failure | dendritic_cell | *CCDC125* | -4.744728724 | 4.37E-12 | 8.89E-12 | 2.75E-08 |
| Heart_Failure | dendritic_cell | *FHL3* | -3.788572418 | 0.01024847 | 0.011706419 | 1 |
| Heart_Failure | dendritic_cell | *C19orf71* | -5.035018293 | 1.76E-19 | 2.01E-18 | 1.10E-15 |
| Heart_Failure | dendritic_cell | *LRIT3* | -6.260409653 | 9.25E-09 | 1.43E-08 | 5.82E-05 |
| Heart_Failure | dendritic_cell | *NPIPA1* | -1.385692711 | 2.88E-08 | 4.32E-08 | 0.000181012 |
| Heart_Failure | dendritic_cell | *GRIN2A* | -3.206990782 | 5.13E-13 | 1.15E-12 | 3.23E-09 |
| Heart_Failure | dendritic_cell | *URAD* | -12.4172542 | 4.04E-19 | 3.70E-18 | 2.54E-15 |
| Heart_Failure | dendritic_cell | *GPR132* | 1.428375523 | 4.25E-21 | 4.66E-19 | 2.67E-17 |
| Heart_Failure | dendritic_cell | *MX2* | 3.914089881 | 8.75E-16 | 3.10E-15 | 5.50E-12 |
| Heart_Failure | dendritic_cell | *PSMG1* | -1.642752656 | 2.92E-07 | 4.16E-07 | 0.001834348 |
| Heart_Failure | dendritic_cell | *KLRC4* | 3.643843854 | 2.78E-11 | 5.22E-11 | 1.75E-07 |
| Heart_Failure | dendritic_cell | *ACSM5* | -2.179641508 | 6.63E-15 | 1.99E-14 | 4.17E-11 |
| Heart_Failure | dendritic_cell | *AC131971.1* | -11.08331343 | 5.31E-16 | 1.96E-15 | 3.34E-12 |
| Heart_Failure | dendritic_cell | *SERHL2* | -3.784844968 | 0.020993947 | 0.023624297 | 1 |
| Heart_Failure | dendritic_cell | *PCBP3* | -0.610784968 | 9.05E-08 | 1.32E-07 | 0.000568828 |
| Heart_Failure | dendritic_cell | *TANGO2* | -2.373933984 | 0.021450178 | 0.024111795 | 1 |
| Heart_Failure | dendritic_cell | *SFXN4* | -5.006314232 | 2.15E-08 | 3.25E-08 | 0.00013544 |
| Heart_Failure | dendritic_cell | *FAM167B* | -7.327632862 | 0.000157731 | 0.000196913 | 0.991653516 |
| Heart_Failure | dendritic_cell | *RP1L1* | -4.01662996 | 3.36E-10 | 5.77E-10 | 2.11E-06 |
| Heart_Failure | dendritic_cell | *NDUFB1* | -2.872493869 | 0.000115974 | 0.000145564 | 0.729130896 |
| Heart_Failure | dendritic_cell | *FAM19A1* | -4.745651268 | 6.03E-17 | 2.76E-16 | 3.79E-13 |
| Heart_Failure | dendritic_cell | *BMP8A* | 3.812079144 | 7.37E-23 | 7.72E-20 | 4.63E-19 |
| Heart_Failure | dendritic_cell | *FIGLA* | -9.686389559 | 2.43E-05 | 3.14E-05 | 0.152958258 |
| Heart_Failure | dendritic_cell | *CBX6* | 6.712410042 | 7.56E-07 | 1.05E-06 | 0.004751572 |

| | | | | | | |
|---|---|---|---|---|---|---|
| Heart_Failure | dendritic_cell | *KREMEN1* | -1.25349378 | 2.38E-08 | 3.58E-08 | 0.000149616 |
| Heart_Failure | dendritic_cell | *AIFM3* | 2.934433125 | 4.94E-19 | 4.33E-18 | 3.10E-15 |
| Heart_Failure | dendritic_cell | *KCTD16* | -2.182404233 | 2.31E-20 | 6.44E-19 | 1.45E-16 |
| Heart_Failure | dendritic_cell | *B3GALT5* | -4.685319289 | 0.00028864 | 0.000355193 | 1 |
| Heart_Failure | dendritic_cell | *ZNF703* | -3.46773455 | 8.66E-22 | 3.01E-19 | 5.44E-18 |
| Heart_Failure | dendritic_cell | *KCTD8* | -1.783156353 | 3.23E-17 | 1.58E-16 | 2.03E-13 |
| Heart_Failure | dendritic_cell | *C9orf66* | 17.70509235 | 1.50E-18 | 1.10E-17 | 9.42E-15 |
| Heart_Failure | dendritic_cell | *TUBA8* | -3.835657061 | 9.41E-14 | 2.32E-13 | 5.92E-10 |
| Heart_Failure | dendritic_cell | *NPIPA5* | -1.022925649 | 1.74E-06 | 2.37E-06 | 0.010917637 |
| Heart_Failure | dendritic_cell | *FAM162B* | -3.0968721 | 8.67E-08 | 1.27E-07 | 0.000545146 |
| Heart_Failure | dendritic_cell | *BTBD9* | -3.601845002 | 4.98E-11 | 9.13E-11 | 3.13E-07 |
| Heart_Failure | dendritic_cell | *NUDT14* | -1.757497318 | 0.030045563 | 0.033527947 | 1 |
| Heart_Failure | dendritic_cell | *ANKRD45* | -7.552568989 | 3.98E-20 | 7.54E-19 | 2.50E-16 |
| Heart_Failure | dendritic_cell | *MAATS1* | -3.121493017 | 2.18E-09 | 3.53E-09 | 1.37E-05 |
| Heart_Failure | dendritic_cell | *GPR39* | 3.587714185 | 4.46E-20 | 8.02E-19 | 2.80E-16 |
| Heart_Failure | dendritic_cell | *FAM3B* | -1.181035214 | 6.09E-09 | 9.53E-09 | 3.83E-05 |
| Heart_Failure | dendritic_cell | *NPIPA7* | -6.108707003 | 2.36E-06 | 3.20E-06 | 0.014844105 |
| Heart_Failure | dendritic_cell | *TTC32* | -4.495456738 | 3.86E-06 | 5.17E-06 | 0.024266819 |
| Heart_Failure | dendritic_cell | *PP2D1* | -4.276903433 | 4.45E-11 | 8.19E-11 | 2.80E-07 |
| Heart_Failure | dendritic_cell | *DENND5A* | -4.459997552 | 1.39E-09 | 2.28E-09 | 8.75E-06 |
| Heart_Failure | dendritic_cell | *TMEM236* | 10.13679346 | 1.81E-17 | 9.52E-17 | 1.14E-13 |
| Heart_Failure | dendritic_cell | *VPS33B* | -4.344760999 | 1.97E-05 | 2.55E-05 | 0.123677051 |
| Heart_Failure | dendritic_cell | *TBX1* | -3.190126609 | 2.58E-20 | 6.44E-19 | 1.62E-16 |
| Heart_Failure | dendritic_cell | *UQCR10* | -6.087665989 | 7.51E-16 | 2.69E-15 | 4.72E-12 |
| Heart_Failure | dendritic_cell | *NIPSNAP1* | -3.862719244 | 1.33E-17 | 7.24E-17 | 8.35E-14 |
| Heart_Failure | dendritic_cell | *CNTN2* | -4.970935194 | 3.02E-20 | 6.66E-19 | 1.90E-16 |
| Heart_Failure | dendritic_cell | *CRELD2* | -7.447948029 | 3.75E-14 | 9.83E-14 | 2.36E-10 |
| Heart_Failure | dendritic_cell | *UBE2F* | -5.563838985 | 8.27E-13 | 1.81E-12 | 5.20E-09 |
| Heart_Failure | dendritic_cell | *GOLGA6L4* | 6.594243121 | 9.22E-06 | 1.22E-05 | 0.057972154 |
| Heart_Failure | dendritic_cell | *PCDH9* | -1.402024788 | 1.20E-18 | 9.03E-18 | 7.54E-15 |
| Heart_Failure | dendritic_cell | *ACOT1* | -2.655496431 | 6.32E-16 | 2.30E-15 | 3.98E-12 |
| Heart_Failure | dendritic_cell | *OAF* | -3.058577164 | 1.07E-15 | 3.72E-15 | 6.73E-12 |
| Heart_Failure | dendritic_cell | *TACSTD2* | 2.135628019 | 3.16E-10 | 5.44E-10 | 1.99E-06 |
| Heart_Failure | dendritic_cell | *CLECL1* | 0.547334614 | 8.80E-20 | 1.22E-18 | 5.53E-16 |
| Heart_Failure | dendritic_cell | *GDF3* | 5.579024647 | 1.11E-17 | 6.18E-17 | 6.97E-14 |
| Heart_Failure | dendritic_cell | *PKP3* | -2.878048361 | 0.001055672 | 0.001263952 | 1 |
| Heart_Failure | dendritic_cell | *CSF1* | -2.094927176 | 3.12E-05 | 4.01E-05 | 0.195972038 |
| Heart_Failure | dendritic_cell | *PLA2G6* | -2.149617808 | 1.37E-11 | 2.65E-11 | 8.62E-08 |
| Heart_Failure | dendritic_cell | *SS18L1* | -4.531930921 | 1.04E-06 | 1.43E-06 | 0.006516953 |
| Heart_Failure | dendritic_cell | *TOP1MT* | -2.38040787 | 4.89E-09 | 7.72E-09 | 3.08E-05 |
| Heart_Failure | dendritic_cell | *THAP7* | -4.51199322 | 2.46E-19 | 2.49E-18 | 1.54E-15 |
| Heart_Failure | dendritic_cell | *CCR10* | -9.818630832 | 9.50E-09 | 1.47E-08 | 5.97E-05 |
| Heart_Failure | dendritic_cell | *WDR27* | -4.356279072 | 1.77E-09 | 2.88E-09 | 1.11E-05 |
| Heart_Failure | dendritic_cell | *TXNRD2* | -4.997335782 | 5.69E-17 | 2.63E-16 | 3.58E-13 |
| Heart_Failure | dendritic_cell | *PROS1* | -4.309959267 | 9.95E-16 | 3.48E-15 | 6.26E-12 |
| Heart_Failure | dendritic_cell | *HDDC3* | -4.107411897 | 1.26E-06 | 1.72E-06 | 0.007896226 |
| Heart_Failure | dendritic_cell | *ZFP1* | -6.107403235 | 6.70E-14 | 1.69E-13 | 4.21E-10 |
| Heart_Failure | dendritic_cell | *LPAR5* | 2.407467485 | 1.72E-15 | 5.73E-15 | 1.08E-11 |
| Heart_Failure | dendritic_cell | *TMEM173* | -4.71717668 | 5.21E-15 | 1.59E-14 | 3.28E-11 |
| Heart_Failure | dendritic_cell | *NELL2* | 2.553664602 | 3.60E-08 | 5.37E-08 | 0.000226122 |

| | | | | | | |
|---|---|---|---|---|---|---|
| Heart_Failure | dendritic_cell | *KRBA2* | -5.084333126 | 4.55E-18 | 2.80E-17 | 2.86E-14 |
| Heart_Failure | dendritic_cell | *ZNF93* | 6.974715619 | 1.19E-18 | 8.98E-18 | 7.45E-15 |
| Heart_Failure | dendritic_cell | *SEPT5* | -9.020653519 | 3.56E-20 | 6.99E-19 | 2.24E-16 |
| Heart_Failure | dendritic_cell | *ATL3* | -3.895751824 | 1.02E-14 | 2.96E-14 | 6.44E-11 |
| Heart_Failure | dendritic_cell | *NDUFA12* | -6.098268664 | 1.62E-14 | 4.50E-14 | 1.02E-10 |
| Heart_Failure | dendritic_cell | *UBE2G2* | -3.745222355 | 1.71E-14 | 4.74E-14 | 1.08E-10 |
| Heart_Failure | dendritic_cell | *OSBP2* | -5.965187994 | 6.19E-14 | 1.57E-13 | 3.89E-10 |
| Heart_Failure | dendritic_cell | *TUSC5* | -1.761725212 | 2.40E-15 | 7.74E-15 | 1.51E-11 |
| Heart_Failure | dendritic_cell | *TMED9* | -3.465279687 | 0.005255422 | 0.006097221 | 1 |
| Heart_Failure | dendritic_cell | *BTBD6* | -2.657628522 | 8.29E-15 | 2.44E-14 | 5.21E-11 |
| Heart_Failure | dendritic_cell | *H1FX* | 6.903982706 | 6.23E-12 | 1.25E-11 | 3.92E-08 |
| Heart_Failure | dendritic_cell | *RBM43* | 6.330070897 | 7.04E-11 | 1.27E-10 | 4.43E-07 |
| Heart_Failure | dendritic_cell | *NUTM2A* | -4.341441782 | 3.76E-05 | 4.81E-05 | 0.23614893 |
| Heart_Failure | dendritic_cell | *ZFP90* | -5.544474762 | 6.29E-10 | 1.06E-09 | 3.96E-06 |
| Heart_Failure | dendritic_cell | *FAM227A* | -3.76945938 | 3.21E-12 | 6.61E-12 | 2.02E-08 |
| Heart_Failure | dendritic_cell | *NOC4L* | 2.114814457 | 0.000383808 | 0.000470004 | 1 |
| Heart_Failure | dendritic_cell | *NDUFA6* | -0.561035 | 0.038831462 | 0.04309504 | 1 |
| Heart_Failure | dendritic_cell | *SORCS2* | -2.526509613 | 7.47E-20 | 1.09E-18 | 4.70E-16 |
| Heart_Failure | dendritic_cell | *TMEM106A* | 4.17925025 | 5.93E-19 | 5.05E-18 | 3.73E-15 |
| Heart_Failure | dendritic_cell | *SIVA1* | -4.885047614 | 2.47E-18 | 1.68E-17 | 1.55E-14 |
| Heart_Failure | dendritic_cell | *UBOX5* | -4.780267266 | 1.05E-14 | 3.03E-14 | 6.62E-11 |
| Heart_Failure | dendritic_cell | *MAFF* | -4.850553404 | 1.78E-14 | 4.92E-14 | 1.12E-10 |
| Heart_Failure | dendritic_cell | *BRF1* | -3.587983724 | 2.04E-18 | 1.42E-17 | 1.28E-14 |
| Heart_Failure | dendritic_cell | *CIB1* | -2.217720655 | 0.002205638 | 0.002609984 | 1 |
| Heart_Failure | dendritic_cell | *FLRT2* | -1.703330664 | 1.25E-17 | 6.87E-17 | 7.85E-14 |
| Heart_Failure | dendritic_cell | *ADSSL1* | -4.281696619 | 7.83E-21 | 4.66E-19 | 4.92E-17 |
| Heart_Failure | dendritic_cell | *C6orf120* | -5.976626995 | 2.33E-15 | 7.57E-15 | 1.47E-11 |
| Heart_Failure | dendritic_cell | *DDX51* | -2.440231354 | 3.66E-12 | 7.51E-12 | 2.30E-08 |
| Heart_Failure | dendritic_cell | *SIGIRR* | -3.994254587 | 3.55E-09 | 5.65E-09 | 2.23E-05 |
| Heart_Failure | dendritic_cell | *IFITM2* | -4.535884596 | 5.82E-12 | 1.17E-11 | 3.66E-08 |
| Heart_Failure | dendritic_cell | *GP1BA* | -3.783453858 | 4.35E-14 | 1.13E-13 | 2.74E-10 |
| Heart_Failure | dendritic_cell | *KIAA0825* | -4.176554901 | 1.70E-13 | 4.05E-13 | 1.07E-09 |
| Heart_Failure | dendritic_cell | *KLHL33* | -3.286110338 | 1.94E-12 | 4.08E-12 | 1.22E-08 |
| Heart_Failure | dendritic_cell | *RBM11* | -5.352234331 | 2.83E-05 | 3.65E-05 | 0.177971987 |
| Heart_Failure | dendritic_cell | *CCDC137* | -2.082154586 | 0.005857797 | 0.006783564 | 1 |
| Heart_Failure | dendritic_cell | *CDK10* | -3.985115205 | 1.38E-13 | 3.32E-13 | 8.65E-10 |
| Heart_Failure | dendritic_cell | *TMEM105* | -41.86434066 | 3.24E-14 | 8.62E-14 | 2.04E-10 |
| Heart_Failure | dendritic_cell | *TCN2* | -1.566331977 | 0.000726375 | 0.000876362 | 1 |
| Heart_Failure | dendritic_cell | *GAS2L1* | -5.468158279 | 2.54E-16 | 1.00E-15 | 1.60E-12 |
| Heart_Failure | dendritic_cell | *ATP6V0A2* | -3.942050957 | 7.10E-06 | 9.40E-06 | 0.044607969 |
| Heart_Failure | dendritic_cell | *HS6ST3* | 7.455499681 | 3.69E-09 | 5.87E-09 | 2.32E-05 |
| Heart_Failure | dendritic_cell | *TNFAIP8L1* | 3.485541925 | 1.05E-19 | 1.38E-18 | 6.59E-16 |
| Heart_Failure | dendritic_cell | *MAPK11* | -4.074490824 | 6.34E-20 | 9.96E-19 | 3.98E-16 |
| Heart_Failure | dendritic_cell | *SP140L* | 3.028439442 | 0.004613527 | 0.005373332 | 1 |
| Heart_Failure | dendritic_cell | *SH3BGR* | 0.846443141 | 8.69E-06 | 1.15E-05 | 0.054663029 |
| Heart_Failure | dendritic_cell | *FAM174B* | -4.04047821 | 1.95E-16 | 7.91E-16 | 1.23E-12 |
| Heart_Failure | dendritic_cell | *MUC1* | -1.875263338 | 7.83E-05 | 9.90E-05 | 0.492567603 |
| Heart_Failure | dendritic_cell | *SV2B* | -4.311210279 | 0.000602839 | 0.000730702 | 1 |
| Heart_Failure | dendritic_cell | *PDE6G* | -4.119792454 | 0.000443088 | 0.000541332 | 1 |
| Heart_Failure | dendritic_cell | *TLCD2* | -2.980891403 | 1.50E-19 | 1.79E-18 | 9.44E-16 |

| | | | | | | |
|---|---|---|---|---|---|---|
| Heart_Failure | dendritic_cell | *LSAMP* | -3.23638788 | 2.14E-16 | 8.60E-16 | 1.35E-12 |
| Heart_Failure | dendritic_cell | *AHNAK2* | -3.349605157 | 2.82E-19 | 2.79E-18 | 1.78E-15 |
| Heart_Failure | dendritic_cell | *SP1* | -3.713184291 | 2.72E-13 | 6.32E-13 | 1.71E-09 |
| Heart_Failure | dendritic_cell | *PDIA2* | -4.693889413 | 2.40E-13 | 5.60E-13 | 1.51E-09 |
| Heart_Failure | dendritic_cell | *PCGF3* | -3.312314547 | 0.000335386 | 0.00041183 | 1 |
| Heart_Failure | dendritic_cell | *PSMD13* | 3.687976859 | 0.004048543 | 0.004734596 | 1 |
| Heart_Failure | dendritic_cell | *PBX1* | -2.25383407 | 6.53E-18 | 3.87E-17 | 4.11E-14 |
| Heart_Failure | dendritic_cell | *UBE2L3* | -5.437463238 | 1.63E-09 | 2.66E-09 | 1.02E-05 |
| Heart_Failure | dendritic_cell | *PMEL* | -3.326866274 | 2.22E-14 | 6.05E-14 | 1.40E-10 |
| Heart_Failure | dendritic_cell | *SNAI3* | -4.206915629 | 1.58E-19 | 1.85E-18 | 9.91E-16 |
| Heart_Failure | dendritic_cell | *C6orf201* | -2.009597734 | 4.77E-07 | 6.72E-07 | 0.003001671 |
| Heart_Failure | dendritic_cell | *MYBL1* | -2.406810017 | 9.25E-17 | 4.04E-16 | 5.82E-13 |
| Heart_Failure | dendritic_cell | *C16orf52* | -4.60423954 | 2.05E-09 | 3.33E-09 | 1.29E-05 |
| Heart_Failure | dendritic_cell | *ADARB2* | -2.530535182 | 4.99E-20 | 8.51E-19 | 3.14E-16 |
| Heart_Failure | dendritic_cell | *KCNIP4* | -1.462632943 | 1.55E-15 | 5.20E-15 | 9.74E-12 |
| Heart_Failure | dendritic_cell | *ARL17A* | -2.665556947 | 0.000806919 | 0.000972789 | 1 |
| Heart_Failure | dendritic_cell | *NPIPB4* | -4.874317162 | 1.34E-08 | 2.06E-08 | 8.45E-05 |
| Heart_Failure | dendritic_cell | *ZNF829* | -3.267217094 | 2.17E-05 | 2.81E-05 | 0.136276338 |
| Heart_Failure | dendritic_cell | *TRIM69* | -2.506183549 | 1.14E-08 | 1.74E-08 | 7.14E-05 |
| Heart_Failure | dendritic_cell | *IFITM1* | -5.20397744 | 7.68E-05 | 9.70E-05 | 0.482616157 |
| Heart_Failure | dendritic_cell | *FFAR3* | -17.06003356 | 0.007357087 | 0.008480749 | 1 |
| Heart_Failure | dendritic_cell | *POMK* | 6.922790012 | 1.06E-17 | 5.94E-17 | 6.66E-14 |
| Heart_Failure | dendritic_cell | *LINC00839* | -4.684713368 | 2.53E-09 | 4.06E-09 | 1.59E-05 |
| Heart_Failure | dendritic_cell | *PTCH1* | -3.165055467 | 1.55E-17 | 8.29E-17 | 9.75E-14 |
| Heart_Failure | dendritic_cell | *RTN4RL1* | -2.223617815 | 3.55E-20 | 6.99E-19 | 2.23E-16 |
| Heart_Failure | dendritic_cell | *PLK5* | -5.195339711 | 8.62E-10 | 1.44E-09 | 5.42E-06 |
| Heart_Failure | dendritic_cell | *RASA3* | -3.332178842 | 2.99E-17 | 1.47E-16 | 1.88E-13 |
| Heart_Failure | dendritic_cell | *ZNF566* | -3.309719104 | 3.52E-17 | 1.70E-16 | 2.21E-13 |
| Heart_Failure | dendritic_cell | *ZNF529* | -4.255966074 | 1.26E-10 | 2.25E-10 | 7.95E-07 |
| Heart_Failure | dendritic_cell | *ZNF284* | 17.59447215 | 1.04E-08 | 1.60E-08 | 6.51E-05 |
| Heart_Failure | dendritic_cell | *GSAP* | 2.494321255 | 0.004256962 | 0.004970013 | 1 |
| Heart_Failure | dendritic_cell | *C2orf76* | -3.566517125 | 1.25E-07 | 1.81E-07 | 0.000784418 |
| Heart_Failure | dendritic_cell | *WWOX* | -2.418436432 | 3.95E-11 | 7.32E-11 | 2.48E-07 |
| Heart_Failure | dendritic_cell | *CYP4Z1* | -2.605266204 | 3.52E-20 | 6.98E-19 | 2.21E-16 |
| Heart_Failure | dendritic_cell | *CCDC84* | -4.281497108 | 2.91E-15 | 9.28E-15 | 1.83E-11 |
| Heart_Failure | dendritic_cell | *POLR1D* | -5.447643529 | 2.30E-11 | 4.36E-11 | 1.45E-07 |
| Heart_Failure | dendritic_cell | *FFAR4* | -1.956082558 | 8.64E-17 | 3.82E-16 | 5.43E-13 |
| Heart_Failure | dendritic_cell | *BPIFB4* | -144.0699603 | 1.86E-16 | 7.56E-16 | 1.17E-12 |
| Heart_Failure | dendritic_cell | *CYP4F12* | -2.030516506 | 2.52E-07 | 3.61E-07 | 0.001586884 |
| Heart_Failure | dendritic_cell | *KLHL32* | -3.097815415 | 4.60E-07 | 6.48E-07 | 0.002893285 |
| Heart_Failure | dendritic_cell | *NBPF12* | -4.867165209 | 2.46E-11 | 4.65E-11 | 1.55E-07 |
| Heart_Failure | dendritic_cell | *GPAT2* | 4.672452508 | 1.94E-18 | 1.37E-17 | 1.22E-14 |
| Heart_Failure | dendritic_cell | *TOR3A* | 7.819061916 | 4.75E-16 | 1.77E-15 | 2.99E-12 |
| Heart_Failure | dendritic_cell | *CYP4X1* | -1.921312531 | 7.23E-20 | 1.08E-18 | 4.55E-16 |
| Heart_Failure | dendritic_cell | *CCDC30* | -5.138494682 | 8.32E-13 | 1.82E-12 | 5.23E-09 |
| Heart_Failure | dendritic_cell | *GLDN* | -2.929686501 | 4.16E-06 | 5.58E-06 | 0.02618427 |
| Heart_Failure | dendritic_cell | *FCAR* | 4.04593025 | 1.82E-09 | 2.96E-09 | 1.14E-05 |
| Heart_Failure | dendritic_cell | *ZNF501* | -3.54930068 | 4.16E-20 | 7.75E-19 | 2.61E-16 |
| Heart_Failure | dendritic_cell | *ZNF197* | -3.721385886 | 1.08E-15 | 3.75E-15 | 6.80E-12 |
| Heart_Failure | dendritic_cell | *BTN3A2* | -0.874939511 | 7.10E-05 | 8.99E-05 | 0.446070734 |

| | | | | | | |
|---|---|---|---|---|---|---|
| Heart_Failure | dendritic_cell | *PCLO* | -1.668324224 | 1.96E-17 | 1.02E-16 | 1.23E-13 |
| Heart_Failure | dendritic_cell | *RGS7BP* | -2.499750161 | 0.014158009 | 0.016075745 | 1 |
| Heart_Failure | dendritic_cell | *ZNF396* | -3.649566016 | 7.42E-18 | 4.31E-17 | 4.67E-14 |
| Heart_Failure | dendritic_cell | *TMEM222* | -3.645385193 | 2.86E-16 | 1.12E-15 | 1.80E-12 |
| Heart_Failure | dendritic_cell | *CLCNKA* | -5.886987764 | 2.87E-08 | 4.31E-08 | 0.000180428 |
| Heart_Failure | dendritic_cell | *SEPT10* | -3.63844362 | 1.03E-16 | 4.45E-16 | 6.47E-13 |
| Heart_Failure | dendritic_cell | *SMYD4* | -4.152872998 | 4.06E-10 | 6.90E-10 | 2.55E-06 |
| Heart_Failure | dendritic_cell | *NF2* | -4.140521177 | 1.46E-13 | 3.51E-13 | 9.17E-10 |
| Heart_Failure | dendritic_cell | *MIR22HG* | -3.43877221 | 8.45E-16 | 3.00E-15 | 5.31E-12 |
| Heart_Failure | dendritic_cell | *PDE2A* | -2.197352193 | 2.02E-19 | 2.17E-18 | 1.27E-15 |
| Heart_Failure | dendritic_cell | *C17orf58* | -2.030624157 | 3.91E-14 | 1.02E-13 | 2.46E-10 |
| Heart_Failure | dendritic_cell | *CYP27C1* | -1.792443088 | 6.84E-10 | 1.15E-09 | 4.30E-06 |
| Heart_Failure | dendritic_cell | *BCR* | -3.804094219 | 9.71E-15 | 2.82E-14 | 6.11E-11 |
| Heart_Failure | dendritic_cell | *FOXI2* | -2.552613475 | 0.000179842 | 0.000223983 | 1 |
| Heart_Failure | dendritic_cell | *HYAL3* | 1.366656796 | 0.000527172 | 0.00064169 | 1 |
| Heart_Failure | dendritic_cell | *ZSCAN30* | -5.151826631 | 7.14E-15 | 2.12E-14 | 4.49E-11 |
| Heart_Failure | dendritic_cell | *LILRB4* | 2.867577379 | 6.81E-17 | 3.09E-16 | 4.28E-13 |
| Heart_Failure | dendritic_cell | *TRABD2A* | 1.370592482 | 0.000115283 | 0.000144725 | 0.724781228 |
| Heart_Failure | dendritic_cell | *QRFPR* | 4.458437268 | 2.96E-18 | 1.94E-17 | 1.86E-14 |
| Heart_Failure | dendritic_cell | *TMEM17* | -3.598009194 | 6.69E-15 | 2.00E-14 | 4.21E-11 |
| Heart_Failure | dendritic_cell | *TNFRSF18* | -7.625527074 | 2.31E-20 | 6.44E-19 | 1.45E-16 |
| Heart_Failure | dendritic_cell | *RTN4RL2* | -12.6740954 | 6.84E-13 | 1.51E-12 | 4.30E-09 |
| Heart_Failure | dendritic_cell | *TMEM232* | -2.77992005 | 4.94E-16 | 1.83E-15 | 3.10E-12 |
| Heart_Failure | dendritic_cell | *EFCAB6* | -3.050980852 | 8.33E-19 | 6.67E-18 | 5.24E-15 |
| Heart_Failure | dendritic_cell | *KANK3* | -2.650205933 | 2.76E-20 | 6.44E-19 | 1.74E-16 |
| Heart_Failure | dendritic_cell | *EMID1* | -7.619111603 | 5.29E-16 | 1.95E-15 | 3.32E-12 |
| Heart_Failure | dendritic_cell | *RHD* | -2.870712879 | 7.27E-08 | 1.07E-07 | 0.000456984 |
| Heart_Failure | dendritic_cell | *CYP4A11* | -3.772025387 | 1.33E-10 | 2.37E-10 | 8.39E-07 |
| Heart_Failure | dendritic_cell | *RPS19BP1* | 6.01177295 | 4.20E-08 | 6.25E-08 | 0.000264213 |
| Heart_Failure | dendritic_cell | *C3orf70* | -2.623217475 | 8.05E-16 | 2.87E-15 | 5.06E-12 |
| Heart_Failure | dendritic_cell | *PLCD1* | -3.804850527 | 9.95E-10 | 1.65E-09 | 6.26E-06 |
| Heart_Failure | dendritic_cell | *ENTPD5* | -5.548632372 | 1.57E-11 | 3.01E-11 | 9.84E-08 |
| Heart_Failure | dendritic_cell | *HEATR4* | -1.477248331 | 2.10E-06 | 2.85E-06 | 0.013179191 |
| Heart_Failure | dendritic_cell | *ANGPTL5* | -5.520425758 | 5.01E-18 | 3.05E-17 | 3.15E-14 |
| Heart_Failure | dendritic_cell | *FNBP1* | 3.029838373 | 4.49E-13 | 1.02E-12 | 2.82E-09 |
| Heart_Failure | dendritic_cell | *WDR86* | -2.920660076 | 2.73E-16 | 1.07E-15 | 1.71E-12 |
| Heart_Failure | dendritic_cell | *EPOR* | -4.967776173 | 2.87E-18 | 1.89E-17 | 1.80E-14 |
| Heart_Failure | dendritic_cell | *FPR3* | 0.899853106 | 6.65E-20 | 1.02E-18 | 4.18E-16 |
| Heart_Failure | dendritic_cell | *GJA4* | -5.082076618 | 1.57E-13 | 3.76E-13 | 9.88E-10 |
| Heart_Failure | dendritic_cell | *IFT140* | -5.50952727 | 5.07E-15 | 1.55E-14 | 3.19E-11 |
| Heart_Failure | dendritic_cell | *TLR5* | 4.033688869 | 6.49E-19 | 5.42E-18 | 4.08E-15 |
| Heart_Failure | dendritic_cell | *PLEKHN1* | 4.299427725 | 5.51E-09 | 8.65E-09 | 3.46E-05 |
| Heart_Failure | dendritic_cell | *ZNF385C* | -7.305304616 | 4.58E-20 | 8.12E-19 | 2.88E-16 |
| Heart_Failure | dendritic_cell | *TET3* | 3.102016712 | 1.23E-19 | 1.54E-18 | 7.73E-16 |
| Heart_Failure | dendritic_cell | *ISG15* | -3.866086304 | 2.28E-15 | 7.42E-15 | 1.43E-11 |
| Heart_Failure | dendritic_cell | *C17orf97* | 0.714262102 | 0.000423912 | 0.000518408 | 1 |
| Heart_Failure | dendritic_cell | *TRPV2* | 5.836950482 | 1.74E-18 | 1.24E-17 | 1.09E-14 |
| Heart_Failure | dendritic_cell | *C2orf88* | -2.660101613 | 9.60E-19 | 7.49E-18 | 6.03E-15 |
| Heart_Failure | dendritic_cell | *KBTBD12* | -1.472865124 | 0.000187543 | 0.000233204 | 1 |
| Heart_Failure | dendritic_cell | *FANCA* | 2.45382382 | 7.18E-05 | 9.09E-05 | 0.451455507 |

| | | | | | | |
|---|---|---|---|---|---|---|
| Heart_Failure | dendritic_cell | *ADH1A* | -3.660371458 | 1.00E-20 | 4.95E-19 | 6.29E-17 |
| Heart_Failure | dendritic_cell | *SEMA4D* | -4.958734183 | 5.18E-05 | 6.60E-05 | 0.325616206 |
| Heart_Failure | dendritic_cell | *FAM69C* | -7.186176567 | 5.80E-11 | 1.06E-10 | 3.65E-07 |
| Heart_Failure | dendritic_cell | *DNAH17* | -7.418728563 | 1.02E-16 | 4.42E-16 | 6.43E-13 |
| Heart_Failure | dendritic_cell | *ZFP69* | -2.52455851 | 2.54E-19 | 2.56E-18 | 1.59E-15 |
| Heart_Failure | dendritic_cell | *TMEM220* | -4.079627153 | 2.42E-15 | 7.82E-15 | 1.52E-11 |
| Heart_Failure | dendritic_cell | *CCDC157* | -4.935690005 | 8.02E-19 | 6.49E-18 | 5.04E-15 |
| Heart_Failure | dendritic_cell | *GFRAL* | -3.937457964 | 1.08E-14 | 3.10E-14 | 6.77E-11 |
| Heart_Failure | dendritic_cell | *LDLRAD2* | -2.74431829 | 8.48E-21 | 4.66E-19 | 5.33E-17 |
| Heart_Failure | dendritic_cell | *C2orf66* | -12.00108478 | 3.61E-10 | 6.18E-10 | 2.27E-06 |
| Heart_Failure | dendritic_cell | *CPSF4L* | -6.246496529 | 8.19E-14 | 2.04E-13 | 5.15E-10 |
| Heart_Failure | dendritic_cell | *KLHL17* | -7.063912979 | 3.26E-14 | 8.65E-14 | 2.05E-10 |
| Heart_Failure | dendritic_cell | *OR7D2* | -4.016321918 | 0.000717175 | 0.00086576 | 1 |
| Heart_Failure | dendritic_cell | *S100A3* | -2.207865232 | 0.000516413 | 0.000628837 | 1 |
| Heart_Failure | dendritic_cell | *TREML4* | 6.270129237 | 5.84E-20 | 9.53E-19 | 3.67E-16 |
| Heart_Failure | dendritic_cell | *C6orf132* | -2.588376207 | 3.09E-19 | 2.98E-18 | 1.94E-15 |
| Heart_Failure | dendritic_cell | *MAPK12* | -4.759046504 | 1.33E-16 | 5.56E-16 | 8.34E-13 |
| Heart_Failure | dendritic_cell | *AGRN* | -4.6247142 | 3.66E-19 | 3.42E-18 | 2.30E-15 |
| Heart_Failure | dendritic_cell | *FAM166A* | -12.72031627 | 2.29E-20 | 6.44E-19 | 1.44E-16 |
| Heart_Failure | dendritic_cell | *ZNF626* | -5.182463861 | 3.45E-19 | 3.26E-18 | 2.17E-15 |
| Heart_Failure | dendritic_cell | *SMTNL2* | -18.23528057 | 2.57E-20 | 6.44E-19 | 1.61E-16 |
| Heart_Failure | dendritic_cell | *ZC3H6* | -3.744671968 | 4.48E-17 | 2.12E-16 | 2.82E-13 |
| Heart_Failure | dendritic_cell | *LINC00265* | 5.387693846 | 3.16E-05 | 4.07E-05 | 0.198948571 |
| Heart_Failure | dendritic_cell | *LAMTOR4* | -3.208127306 | 0.000596436 | 0.00072334 | 1 |
| Heart_Failure | dendritic_cell | *PRKAR1B* | -4.786547132 | 1.00E-09 | 1.66E-09 | 6.29E-06 |
| Heart_Failure | dendritic_cell | *NUTM2B* | -48.02919897 | 9.21E-09 | 1.42E-08 | 5.79E-05 |
| Heart_Failure | dendritic_cell | *NCR3LG1* | 2.079309856 | 6.40E-21 | 4.66E-19 | 4.03E-17 |
| Heart_Failure | dendritic_cell | *ZNF793* | -1.875615437 | 7.91E-14 | 1.98E-13 | 4.97E-10 |
| Heart_Failure | dendritic_cell | *AGAP4* | 8.149816996 | 4.03E-09 | 6.39E-09 | 2.53E-05 |
| Heart_Failure | dendritic_cell | *HYKK* | -3.802948888 | 1.85E-18 | 1.31E-17 | 1.16E-14 |
| Heart_Failure | dendritic_cell | *RUFY4* | 7.767940856 | 1.22E-20 | 5.08E-19 | 7.69E-17 |
| Heart_Failure | dendritic_cell | *ZNF669* | -5.70164836 | 0.0002793 | 0.000343834 | 1 |
| Heart_Failure | dendritic_cell | *CENPP* | -1.845779584 | 3.94E-07 | 5.57E-07 | 0.002475619 |
| Heart_Failure | dendritic_cell | *PLSCR1* | 3.664425006 | 0.000494867 | 0.000603302 | 1 |
| Heart_Failure | dendritic_cell | *ZP3* | -2.176605155 | 0.000642363 | 0.000777838 | 1 |
| Heart_Failure | dendritic_cell | *JAKMIP3* | -2.393258714 | 1.26E-07 | 1.82E-07 | 0.000789092 |
| Heart_Failure | dendritic_cell | *PDCD1* | 9.319284545 | 9.03E-10 | 1.50E-09 | 5.67E-06 |
| Heart_Failure | dendritic_cell | *CERKL* | 3.709470475 | 1.06E-20 | 4.95E-19 | 6.66E-17 |
| Heart_Failure | dendritic_cell | *C19orf54* | 2.091185015 | 0.010822878 | 0.01234684 | 1 |
| Heart_Failure | dendritic_cell | *COL25A1* | -1.681790774 | 7.34E-20 | 1.08E-18 | 4.62E-16 |
| Heart_Failure | dendritic_cell | *SRSF10* | -4.368621275 | 3.02E-13 | 6.99E-13 | 1.90E-09 |
| Heart_Failure | dendritic_cell | *NBR1* | -5.135194922 | 8.95E-09 | 1.39E-08 | 5.63E-05 |
| Heart_Failure | dendritic_cell | *NDOR1* | 8.201581698 | 2.20E-15 | 7.20E-15 | 1.39E-11 |
| Heart_Failure | dendritic_cell | *FAM72B* | 5.47270124 | 6.62E-21 | 4.66E-19 | 4.16E-17 |
| Heart_Failure | dendritic_cell | *SUMO2* | -5.888966288 | 6.86E-13 | 1.51E-12 | 4.31E-09 |
| Heart_Failure | dendritic_cell | *GOLGA8M* | -3.340827151 | 1.53E-19 | 1.81E-18 | 9.63E-16 |
| Heart_Failure | dendritic_cell | *S100A16* | -5.660535534 | 2.48E-06 | 3.36E-06 | 0.01561093 |
| Heart_Failure | dendritic_cell | *RHCE* | -3.14374963 | 2.00E-16 | 8.10E-16 | 1.26E-12 |
| Heart_Failure | dendritic_cell | *SLC4A5* | 4.516487821 | 0.007145037 | 0.008243871 | 1 |
| Heart_Failure | dendritic_cell | *UROS* | -4.406080999 | 2.49E-06 | 3.36E-06 | 0.015628293 |

| | | | | | | |
|---|---|---|---|---|---|---|
| Heart_Failure | dendritic_cell | *FAM221A* | -4.239139167 | 2.35E-15 | 7.62E-15 | 1.48E-11 |
| Heart_Failure | dendritic_cell | *FSIP2* | -1.918287518 | 1.41E-13 | 3.40E-13 | 8.87E-10 |
| Heart_Failure | dendritic_cell | *NOXA1* | 7.671159843 | 6.97E-16 | 2.51E-15 | 4.38E-12 |
| Heart_Failure | dendritic_cell | *SKOR1* | 14.19431258 | 2.44E-16 | 9.65E-16 | 1.53E-12 |
| Heart_Failure | dendritic_cell | *TMEM201* | -5.274203809 | 1.29E-14 | 3.66E-14 | 8.14E-11 |
| Heart_Failure | dendritic_cell | *ZDHHC11* | -3.856745537 | 1.46E-13 | 3.51E-13 | 9.19E-10 |
| Heart_Failure | dendritic_cell | *FAM26F* | 1.33614738 | 5.32E-06 | 7.08E-06 | 0.033447549 |
| Heart_Failure | dendritic_cell | *LINC00910* | -4.332822093 | 3.53E-17 | 1.71E-16 | 2.22E-13 |
| Heart_Failure | dendritic_cell | *BEND4* | -14.38415551 | 6.89E-17 | 3.12E-16 | 4.33E-13 |
| Heart_Failure | dendritic_cell | *ZNF563* | -2.761710634 | 0.000712994 | 0.000860879 | 1 |
| Heart_Failure | dendritic_cell | *FBF1* | -0.911910457 | 0.002749408 | 0.003239417 | 1 |
| Heart_Failure | dendritic_cell | *MSL1* | -3.429715451 | 2.57E-14 | 6.93E-14 | 1.62E-10 |
| Heart_Failure | dendritic_cell | *LRRK2* | -1.654429609 | 0.032859616 | 0.03660319 | 1 |
| Heart_Failure | dendritic_cell | *FAM196A* | -4.50769149 | 1.77E-13 | 4.20E-13 | 1.11E-09 |
| Heart_Failure | dendritic_cell | *UTS2B* | -3.015684505 | 2.79E-18 | 1.85E-17 | 1.75E-14 |
| Heart_Failure | dendritic_cell | *C9orf152* | -11.55923652 | 1.30E-18 | 9.63E-18 | 8.18E-15 |
| Heart_Failure | dendritic_cell | *NOC2L* | -1.573925294 | 0.000203709 | 0.000252707 | 1 |
| Heart_Failure | dendritic_cell | *NELFB* | 6.874623653 | 1.37E-07 | 1.98E-07 | 0.000860976 |
| Heart_Failure | dendritic_cell | *SLC15A5* | -1.696286191 | 3.89E-12 | 7.94E-12 | 2.44E-08 |
| Heart_Failure | dendritic_cell | *LIPI* | -2.299321199 | 4.45E-16 | 1.67E-15 | 2.80E-12 |
| Heart_Failure | dendritic_cell | *ADAT2* | -3.435268449 | 1.07E-05 | 1.41E-05 | 0.0673142 |
| Heart_Failure | dendritic_cell | *ZNF567* | -6.194103898 | 6.47E-09 | 1.01E-08 | 4.07E-05 |
| Heart_Failure | dendritic_cell | *ANKDD1B* | -2.269464 | 4.18E-08 | 6.23E-08 | 0.000263093 |
| Heart_Failure | dendritic_cell | *ALKBH2* | -4.922437621 | 4.24E-11 | 7.85E-11 | 2.67E-07 |
| Heart_Failure | dendritic_cell | *LITAF* | -2.673099993 | 0.002595163 | 0.003059975 | 1 |
| Heart_Failure | dendritic_cell | *VSTM1* | 3.947040675 | 4.57E-07 | 6.44E-07 | 0.002870071 |
| Heart_Failure | dendritic_cell | *SF3B3* | -5.137831943 | 9.80E-14 | 2.40E-13 | 6.16E-10 |
| Heart_Failure | dendritic_cell | *PLAC9* | -2.551302694 | 1.31E-13 | 3.17E-13 | 8.24E-10 |
| Heart_Failure | dendritic_cell | *CLDN4* | 3.362848965 | 3.99E-11 | 7.39E-11 | 2.51E-07 |
| Heart_Failure | dendritic_cell | *ZNF573* | -5.442705553 | 6.58E-08 | 9.71E-08 | 0.000413794 |
| Heart_Failure | dendritic_cell | *FAM47E* | -1.695672267 | 3.19E-10 | 5.49E-10 | 2.01E-06 |
| Heart_Failure | dendritic_cell | *ZNF527* | -3.85412785 | 0.008703826 | 0.009987398 | 1 |
| Heart_Failure | dendritic_cell | *S100A13* | -2.980926226 | 1.87E-19 | 2.10E-18 | 1.18E-15 |
| Heart_Failure | dendritic_cell | *ZNF33A* | -3.483503036 | 7.87E-06 | 1.04E-05 | 0.049458812 |
| Heart_Failure | dendritic_cell | *LINC00994* | 4.374550022 | 5.35E-09 | 8.41E-09 | 3.36E-05 |
| Heart_Failure | dendritic_cell | *C15orf61* | -6.684744541 | 3.42E-05 | 4.38E-05 | 0.214808529 |
| Heart_Failure | dendritic_cell | *TSPYL1* | 1.689746091 | 0.023356001 | 0.026188546 | 1 |
| Heart_Failure | dendritic_cell | *PNRC2* | -6.455905857 | 1.59E-10 | 2.80E-10 | 1.00E-06 |
| Heart_Failure | dendritic_cell | *ZKSCAN3* | -5.776058721 | 7.25E-09 | 1.13E-08 | 4.56E-05 |
| Heart_Failure | dendritic_cell | *SLC35E2B* | -2.485856152 | 8.23E-10 | 1.37E-09 | 5.17E-06 |
| Heart_Failure | dendritic_cell | *ALG1L* | 6.179624786 | 6.59E-17 | 3.00E-16 | 4.14E-13 |
| Heart_Failure | dendritic_cell | *HLA-DRB1* | 0.738598526 | 2.08E-19 | 2.22E-18 | 1.31E-15 |
| Heart_Failure | dendritic_cell | *AKR1C3* | -2.773056194 | 4.40E-18 | 2.71E-17 | 2.77E-14 |
| Heart_Failure | dendritic_cell | *WDSUB1* | -3.232639191 | 9.13E-18 | 5.20E-17 | 5.74E-14 |
| Heart_Failure | dendritic_cell | *ZNF79* | -3.269972649 | 9.69E-10 | 1.61E-09 | 6.09E-06 |
| Heart_Failure | dendritic_cell | *S100A4* | -2.006177469 | 5.80E-09 | 9.09E-09 | 3.65E-05 |
| Heart_Failure | dendritic_cell | *COLCA1* | -4.82755809 | 1.22E-17 | 6.72E-17 | 7.65E-14 |
| Heart_Failure | dendritic_cell | *ACADSB* | -3.3990525 | 5.96E-07 | 8.33E-07 | 0.003749936 |
| Heart_Failure | dendritic_cell | *SEMA4A* | 1.919467579 | 1.12E-20 | 4.99E-19 | 7.05E-17 |
| Heart_Failure | dendritic_cell | *HRCT1* | -4.024850334 | 9.20E-15 | 2.68E-14 | 5.79E-11 |

| | | | | | | |
|---|---|---|---|---|---|---|
| Heart_Failure | dendritic_cell | *MPHOSPH8* | -3.715800789 | 3.93E-14 | 1.03E-13 | 2.47E-10 |
| Heart_Failure | dendritic_cell | *SIRPB2* | 1.382532285 | 9.29E-21 | 4.77E-19 | 5.84E-17 |
| Heart_Failure | dendritic_cell | *SUPT5H* | -5.038888647 | 2.64E-07 | 3.77E-07 | 0.001658854 |
| Heart_Failure | dendritic_cell | *XPNPEP3* | -2.113706626 | 2.27E-08 | 3.42E-08 | 0.000142438 |
| Heart_Failure | dendritic_cell | *ZNF107* | -2.415723424 | 8.38E-07 | 1.16E-06 | 0.005271615 |
| Heart_Failure | dendritic_cell | *PPIA* | -5.72630889 | 9.92E-09 | 1.53E-08 | 6.24E-05 |
| Heart_Failure | dendritic_cell | *ZNF471* | -3.64559332 | 5.96E-14 | 1.52E-13 | 3.74E-10 |
| Heart_Failure | dendritic_cell | *GTF2IRD2* | -2.674271302 | 1.26E-07 | 1.82E-07 | 0.000789827 |
| Heart_Failure | dendritic_cell | *GRM7* | -1.645880151 | 3.98E-13 | 9.04E-13 | 2.50E-09 |
| Heart_Failure | dendritic_cell | *SUPT3H* | -4.286701835 | 5.50E-11 | 1.01E-10 | 3.46E-07 |
| Heart_Failure | dendritic_cell | *POM121* | -3.752444341 | 6.59E-07 | 9.19E-07 | 0.004141643 |
| Heart_Failure | dendritic_cell | *GIMAP5* | -3.867996556 | 7.81E-18 | 4.52E-17 | 4.91E-14 |
| Heart_Failure | dendritic_cell | *STK31* | -3.266562776 | 2.99E-20 | 6.61E-19 | 1.88E-16 |
| Heart_Failure | dendritic_cell | *ZKSCAN7* | -4.953442932 | 1.20E-18 | 9.03E-18 | 7.55E-15 |
| Heart_Failure | dendritic_cell | *ZNF565* | -3.702730919 | 7.63E-13 | 1.67E-12 | 4.80E-09 |
| Heart_Failure | dendritic_cell | *NTNG2* | 9.284282291 | 3.45E-18 | 2.21E-17 | 2.17E-14 |
| Heart_Failure | dendritic_cell | *WDR5* | -4.330160613 | 4.25E-17 | 2.02E-16 | 2.67E-13 |
| Heart_Failure | dendritic_cell | *LONP1* | -2.542053124 | 0.000354101 | 0.000434302 | 1 |
| Heart_Failure | dendritic_cell | *ASB13* | -4.966676937 | 3.73E-18 | 2.36E-17 | 2.35E-14 |
| Heart_Failure | dendritic_cell | *SLC35F1* | -4.065220784 | 7.91E-19 | 6.42E-18 | 4.97E-15 |
| Heart_Failure | dendritic_cell | *ZNF34* | -7.801148248 | 1.14E-06 | 1.57E-06 | 0.007190753 |
| Heart_Failure | dendritic_cell | *ZNF781* | -1.92256097 | 2.87E-05 | 3.69E-05 | 0.180462199 |
| Heart_Failure | dendritic_cell | *INCA1* | 6.516307055 | 2.73E-18 | 1.81E-17 | 1.72E-14 |
| Heart_Failure | dendritic_cell | *ZNF774* | 2.88617516 | 1.41E-20 | 5.50E-19 | 8.86E-17 |
| Heart_Failure | dendritic_cell | *EVL* | -2.339896028 | 0.000326363 | 0.000400907 | 1 |
| Heart_Failure | dendritic_cell | *ZNF124* | 5.911973258 | 4.43E-13 | 1.00E-12 | 2.78E-09 |
| Heart_Failure | dendritic_cell | *ZNF569* | -5.073925733 | 3.35E-13 | 7.71E-13 | 2.11E-09 |
| Heart_Failure | dendritic_cell | *PIK3R4* | -3.079422432 | 2.30E-08 | 3.46E-08 | 0.000144367 |
| Heart_Failure | dendritic_cell | *ZNF775* | -1.68176964 | 0.007534518 | 0.008683688 | 1 |
| Heart_Failure | dendritic_cell | *ZNF605* | -6.148045235 | 3.24E-07 | 4.61E-07 | 0.002038424 |
| Heart_Failure | dendritic_cell | *RFX8* | -2.522272932 | 9.77E-20 | 1.31E-18 | 6.14E-16 |
| Heart_Failure | dendritic_cell | *MYL6B* | -6.757783898 | 5.80E-07 | 8.11E-07 | 0.003646319 |
| Heart_Failure | dendritic_cell | *ZNF799* | -6.008674364 | 1.46E-12 | 3.12E-12 | 9.18E-09 |
| Heart_Failure | dendritic_cell | *C20orf96* | -8.997268829 | 1.78E-12 | 3.78E-12 | 1.12E-08 |
| Heart_Failure | dendritic_cell | *SULT1A1* | -2.106195776 | 2.87E-06 | 3.87E-06 | 0.018067197 |
| Heart_Failure | dendritic_cell | *ANAPC7* | -4.609623352 | 4.73E-12 | 9.58E-12 | 2.97E-08 |
| Heart_Failure | dendritic_cell | *TPK1* | 3.327044948 | 1.93E-06 | 2.62E-06 | 0.01211558 |
| Heart_Failure | dendritic_cell | *C1orf186* | 1.176879336 | 3.08E-21 | 4.66E-19 | 1.94E-17 |
| Heart_Failure | dendritic_cell | *MYO18A* | 3.692577021 | 4.55E-05 | 5.81E-05 | 0.285965836 |
| Heart_Failure | dendritic_cell | *SPTSSB* | -13.12463153 | 2.45E-10 | 4.25E-10 | 1.54E-06 |
| Heart_Failure | dendritic_cell | *SULF2* | 3.196767584 | 2.68E-17 | 1.34E-16 | 1.68E-13 |
| Heart_Failure | dendritic_cell | *XRCC2* | -5.132332811 | 2.79E-19 | 2.77E-18 | 1.75E-15 |
| Heart_Failure | dendritic_cell | *MYO6* | -3.099154968 | 3.03E-18 | 1.97E-17 | 1.91E-14 |
| Heart_Failure | dendritic_cell | *HDAC2* | -5.058128229 | 2.76E-08 | 4.14E-08 | 0.000173367 |
| Heart_Failure | dendritic_cell | *ADH1B* | -1.7053705 | 9.31E-20 | 1.27E-18 | 5.85E-16 |
| Heart_Failure | dendritic_cell | *HRH1* | -3.962068571 | 1.06E-15 | 3.67E-15 | 6.64E-12 |
| Heart_Failure | dendritic_cell | *ZNF502* | -3.483028804 | 3.83E-08 | 5.70E-08 | 0.000240598 |
| Heart_Failure | dendritic_cell | *TRAPPC4* | -1.917587407 | 0.000182806 | 0.00022754 | 1 |
| Heart_Failure | dendritic_cell | *TECPR2* | -5.197996796 | 1.29E-10 | 2.29E-10 | 8.11E-07 |
| Heart_Failure | dendritic_cell | *FAM180B* | -3.510891035 | 5.58E-20 | 9.21E-19 | 3.51E-16 |

| | | | | | | |
|---|---|---|---|---|---|---|
| Heart_Failure | dendritic_cell | *ZFP62* | -6.889710961 | 4.48E-15 | 1.38E-14 | 2.82E-11 |
| Heart_Failure | dendritic_cell | *ERI2* | -3.193346823 | 1.41E-17 | 7.62E-17 | 8.86E-14 |
| Heart_Failure | dendritic_cell | *TOMM7* | -5.643857165 | 9.02E-14 | 2.23E-13 | 5.67E-10 |
| Heart_Failure | dendritic_cell | *TRPV1* | -3.350124428 | 1.49E-17 | 8.03E-17 | 9.38E-14 |
| Heart_Failure | dendritic_cell | *ZNF33B* | -2.732127286 | 7.11E-06 | 9.41E-06 | 0.044701053 |
| Heart_Failure | dendritic_cell | *AMZ2* | 5.710675184 | 3.00E-15 | 9.56E-15 | 1.89E-11 |
| Heart_Failure | dendritic_cell | *ZNF431* | -4.275899144 | 1.17E-12 | 2.54E-12 | 7.38E-09 |
| Heart_Failure | dendritic_cell | *NF1* | -3.390122371 | 6.93E-09 | 1.08E-08 | 4.36E-05 |
| Heart_Failure | dendritic_cell | *DAPK1* | 1.554632219 | 0.014924023 | 0.01691192 | 1 |
| Heart_Failure | dendritic_cell | *HLA-DQA1* | 0.382490462 | 8.35E-20 | 1.18E-18 | 5.25E-16 |
| Heart_Failure | dendritic_cell | *TLE1* | -2.837541843 | 1.07E-14 | 3.09E-14 | 6.76E-11 |
| Heart_Failure | dendritic_cell | *STRN3* | -3.683264289 | 1.08E-14 | 3.11E-14 | 6.80E-11 |
| Heart_Failure | dendritic_cell | *ZNF239* | -2.416042628 | 2.20E-09 | 3.55E-09 | 1.38E-05 |
| Heart_Failure | dendritic_cell | *MVB12B* | -4.718158523 | 9.69E-09 | 1.50E-08 | 6.09E-05 |
| Heart_Failure | dendritic_cell | *C6orf106* | -2.172349533 | 4.94E-10 | 8.37E-10 | 3.11E-06 |
| Heart_Failure | dendritic_cell | *PPTC7* | 5.225983975 | 3.51E-08 | 5.24E-08 | 0.00022086 |
| Heart_Failure | dendritic_cell | *NHLRC2* | -4.223019057 | 1.82E-09 | 2.96E-09 | 1.14E-05 |
| Heart_Failure | dendritic_cell | *ZFP28* | -6.614936177 | 7.00E-15 | 2.08E-14 | 4.40E-11 |
| Heart_Failure | dendritic_cell | *KIAA1211L* | -0.913949167 | 0.010883271 | 0.012413485 | 1 |
| Heart_Failure | dendritic_cell | *LAMB3* | -3.080887246 | 2.43E-19 | 2.47E-18 | 1.53E-15 |
| Heart_Failure | dendritic_cell | *KPNA5* | -4.763656194 | 1.04E-12 | 2.26E-12 | 6.55E-09 |
| Heart_Failure | dendritic_cell | *ANKRD36B* | -3.313919487 | 2.92E-15 | 9.30E-15 | 1.83E-11 |
| Heart_Failure | dendritic_cell | *HCAR1* | -2.977985384 | 1.02E-15 | 3.57E-15 | 6.43E-12 |
| Heart_Failure | dendritic_cell | *SRGAP1* | -3.874195017 | 8.24E-10 | 1.38E-09 | 5.18E-06 |
| Heart_Failure | dendritic_cell | *FAM3C* | -5.039062807 | 1.74E-10 | 3.05E-10 | 1.09E-06 |
| Heart_Failure | dendritic_cell | *AP2A1* | -3.363931815 | 5.95E-05 | 7.56E-05 | 0.374179214 |
| Heart_Failure | dendritic_cell | *PCDHB16* | 1.877842533 | 7.63E-05 | 9.65E-05 | 0.479904826 |
| Heart_Failure | dendritic_cell | *ZNF585A* | 3.558291919 | 2.85E-14 | 7.65E-14 | 1.79E-10 |
| Heart_Failure | dendritic_cell | *FUT11* | 6.442666005 | 6.20E-14 | 1.57E-13 | 3.90E-10 |
| Heart_Failure | dendritic_cell | *ANXA4* | -3.937098968 | 4.81E-14 | 1.24E-13 | 3.02E-10 |
| Heart_Failure | dendritic_cell | *WDR5B* | -4.099957116 | 2.98E-15 | 9.49E-15 | 1.87E-11 |
| Heart_Failure | dendritic_cell | *ZNF138* | -3.835978157 | 3.52E-12 | 7.24E-12 | 2.21E-08 |
| Heart_Failure | dendritic_cell | *ZNF429* | -3.277007061 | 4.25E-16 | 1.60E-15 | 2.67E-12 |
| Heart_Failure | dendritic_cell | *ZNF470* | 5.6557979 | 2.77E-10 | 4.79E-10 | 1.74E-06 |
| Heart_Failure | dendritic_cell | *ZNF100* | -2.618609549 | 2.11E-06 | 2.87E-06 | 0.013293753 |
| Heart_Failure | dendritic_cell | *GMFB* | -5.858003466 | 3.74E-14 | 9.82E-14 | 2.35E-10 |
| Heart_Failure | dendritic_cell | *ZMYM1* | -4.396238883 | 1.91E-16 | 7.78E-16 | 1.20E-12 |
| Heart_Failure | dendritic_cell | *ARRDC1* | 5.289685296 | 6.63E-20 | 1.02E-18 | 4.17E-16 |
| Heart_Failure | dendritic_cell | *IGF2R* | -4.046490605 | 6.04E-10 | 1.02E-09 | 3.80E-06 |
| Heart_Failure | dendritic_cell | *GAL3ST4* | -4.69469388 | 8.71E-20 | 1.22E-18 | 5.47E-16 |
| Heart_Failure | dendritic_cell | *ZNF682* | -4.546239204 | 6.49E-16 | 2.36E-15 | 4.08E-12 |
| Heart_Failure | dendritic_cell | *ZNF772* | -1.013115174 | 0.010072556 | 0.011515942 | 1 |
| Heart_Failure | dendritic_cell | *ADAM32* | -3.755071408 | 3.01E-18 | 1.96E-17 | 1.89E-14 |
| Heart_Failure | dendritic_cell | *LRRC8B* | -4.142709348 | 4.27E-14 | 1.11E-13 | 2.68E-10 |
| Heart_Failure | dendritic_cell | *ABCB8* | -1.916290645 | 2.06E-05 | 2.67E-05 | 0.129595825 |
| Heart_Failure | dendritic_cell | *SND1* | -2.678450643 | 0.000263746 | 0.000325195 | 1 |
| Heart_Failure | dendritic_cell | *SULT1A2* | -0.844637747 | 2.55E-06 | 3.45E-06 | 0.016048806 |
| Heart_Failure | dendritic_cell | *NEK5* | -4.653329849 | 8.90E-14 | 2.20E-13 | 5.60E-10 |
| Heart_Failure | dendritic_cell | *ENTPD4* | -4.76177207 | 1.09E-09 | 1.80E-09 | 6.85E-06 |
| Heart_Failure | dendritic_cell | *C1D* | -4.377716102 | 1.86E-12 | 3.94E-12 | 1.17E-08 |

| | | | | | | |
|---|---|---|---|---|---|---|
| Heart_Failure | dendritic_cell | *FAM110D* | -7.517865633 | 2.90E-14 | 7.76E-14 | 1.82E-10 |
| Heart_Failure | dendritic_cell | *KANK2* | -3.415472568 | 1.85E-18 | 1.31E-17 | 1.16E-14 |
| Heart_Failure | dendritic_cell | *C6orf141* | -2.668129972 | 2.53E-15 | 8.16E-15 | 1.59E-11 |
| Heart_Failure | dendritic_cell | *CCL4L2* | -2.164272144 | 8.04E-17 | 3.60E-16 | 5.06E-13 |
| Heart_Failure | dendritic_cell | *GTF2E2* | -3.98236166 | 9.57E-16 | 3.36E-15 | 6.02E-12 |
| Heart_Failure | dendritic_cell | *RAMP2-AS1* | -4.313876389 | 4.37E-13 | 9.90E-13 | 2.75E-09 |
| Heart_Failure | dendritic_cell | *ZNF720* | -3.673886546 | 1.74E-14 | 4.82E-14 | 1.10E-10 |
| Heart_Failure | dendritic_cell | *DDI2* | -3.675214812 | 3.12E-15 | 9.87E-15 | 1.96E-11 |
| Heart_Failure | dendritic_cell | *MRPL21* | -1.251524334 | 1.06E-05 | 1.39E-05 | 0.066544051 |
| Heart_Failure | dendritic_cell | *ZNF517* | -3.216677291 | 2.82E-20 | 6.44E-19 | 1.77E-16 |
| Heart_Failure | dendritic_cell | *C5AR1* | 4.287269889 | 1.50E-16 | 6.21E-16 | 9.41E-13 |
| Heart_Failure | dendritic_cell | *DCHS2* | -1.473734247 | 3.76E-07 | 5.33E-07 | 0.002363805 |
| Heart_Failure | dendritic_cell | *VEPH1* | -3.143137316 | 0.000710384 | 0.000857892 | 1 |
| Heart_Failure | dendritic_cell | *SHPK* | -2.420201375 | 1.10E-15 | 3.83E-15 | 6.94E-12 |
| Heart_Failure | dendritic_cell | *STMN3* | -4.03425049 | 1.06E-19 | 1.39E-18 | 6.69E-16 |
| Heart_Failure | dendritic_cell | *GYPE* | -3.257070837 | 1.36E-16 | 5.68E-16 | 8.52E-13 |
| Heart_Failure | dendritic_cell | *PCDHB11* | -8.760229072 | 2.55E-11 | 4.80E-11 | 1.60E-07 |
| Heart_Failure | dendritic_cell | *SLC2A10* | -2.433060837 | 2.15E-18 | 1.49E-17 | 1.35E-14 |
| Heart_Failure | dendritic_cell | *RPF2* | -5.172545971 | 1.13E-11 | 2.21E-11 | 7.12E-08 |
| Heart_Failure | dendritic_cell | *TTC30A* | -4.142022838 | 2.17E-15 | 7.10E-15 | 1.37E-11 |
| Heart_Failure | dendritic_cell | *RAB40C* | -4.139177695 | 2.90E-18 | 1.90E-17 | 1.82E-14 |
| Heart_Failure | dendritic_cell | *PIGN* | -1.533459586 | 0.000811445 | 0.000978059 | 1 |
| Heart_Failure | dendritic_cell | *HHLA3* | -4.985851991 | 1.52E-17 | 8.15E-17 | 9.55E-14 |
| Heart_Failure | dendritic_cell | *HOXA4* | -4.542368449 | 2.85E-19 | 2.81E-18 | 1.79E-15 |
| Heart_Failure | dendritic_cell | *BCO2* | -3.571568824 | 2.93E-19 | 2.87E-18 | 1.84E-15 |
| Heart_Failure | dendritic_cell | *ENTPD6* | -5.450152048 | 8.77E-10 | 1.46E-09 | 5.51E-06 |
| Heart_Failure | dendritic_cell | *CCDC154* | 3.997650344 | 1.44E-19 | 1.73E-18 | 9.06E-16 |
| Heart_Failure | dendritic_cell | *C5orf42* | -2.978176653 | 2.89E-09 | 4.63E-09 | 1.82E-05 |
| Heart_Failure | dendritic_cell | *ZNF841* | 6.828991093 | 2.38E-16 | 9.43E-16 | 1.49E-12 |
| Heart_Failure | dendritic_cell | *MFAP5* | -2.094531298 | 4.03E-19 | 3.70E-18 | 2.54E-15 |
| Heart_Failure | dendritic_cell | *CDC42SE1* | 3.921755945 | 0.000512238 | 0.000623875 | 1 |
| Heart_Failure | dendritic_cell | *MPEG1* | 1.507561744 | 1.35E-19 | 1.64E-18 | 8.47E-16 |
| Heart_Failure | dendritic_cell | *PDCD1LG2* | -1.863400654 | 0.000157128 | 0.0001962 | 0.98786486 |
| Heart_Failure | dendritic_cell | *KLHL14* | -4.405921945 | 1.52E-22 | 1.20E-19 | 9.57E-19 |
| Heart_Failure | dendritic_cell | *FAM114A1* | -3.194170442 | 1.49E-17 | 8.01E-17 | 9.34E-14 |
| Heart_Failure | dendritic_cell | *PHF2* | -4.408421808 | 5.31E-05 | 6.76E-05 | 0.333764499 |
| Heart_Failure | dendritic_cell | *RPS26* | 1.247147743 | 1.14E-05 | 1.49E-05 | 0.071371559 |
| Heart_Failure | dendritic_cell | *PSAP* | -2.858453089 | 0.000530702 | 0.000645612 | 1 |
| Heart_Failure | dendritic_cell | *S100A10* | -2.226623873 | 0.007682847 | 0.008848152 | 1 |
| Heart_Failure | dendritic_cell | *MAP1LC3C* | -3.218359852 | 1.76E-13 | 4.20E-13 | 1.11E-09 |
| Heart_Failure | dendritic_cell | *AC122129.1* | -6.96210036 | 2.73E-20 | 6.44E-19 | 1.72E-16 |
| Heart_Failure | dendritic_cell | *CCDC180* | -3.047215553 | 6.99E-14 | 1.76E-13 | 4.39E-10 |
| Heart_Failure | dendritic_cell | *OCLN* | -1.702893463 | 0.000228522 | 0.000282707 | 1 |
| Heart_Failure | dendritic_cell | *ZNF181* | 6.300449483 | 2.79E-08 | 4.18E-08 | 0.000175098 |
| Heart_Failure | dendritic_cell | *ADH5* | -3.151786871 | 3.48E-16 | 1.34E-15 | 2.19E-12 |
| Heart_Failure | dendritic_cell | *ZNF677* | -4.438799005 | 1.58E-18 | 1.15E-17 | 9.91E-15 |
| Heart_Failure | dendritic_cell | *ZNF311* | -1.68389962 | 0.002758326 | 0.003248707 | 1 |
| Heart_Failure | dendritic_cell | *FCHSD1* | -7.914391431 | 1.94E-09 | 3.14E-09 | 1.22E-05 |
| Heart_Failure | dendritic_cell | *ZNF71* | -3.131151545 | 7.13E-14 | 1.79E-13 | 4.48E-10 |
| Heart_Failure | dendritic_cell | *RPL12* | -2.873281079 | 1.09E-07 | 1.59E-07 | 0.000685099 |

| | | | | | | |
|---|---|---|---|---|---|---|
| Heart_Failure | dendritic_cell | *VPS13A* | -3.079480055 | 7.81E-12 | 1.54E-11 | 4.91E-08 |
| Heart_Failure | dendritic_cell | *LEKR1* | -4.214573242 | 1.65E-18 | 1.19E-17 | 1.03E-14 |
| Heart_Failure | dendritic_cell | *NOL8* | -4.266486853 | 2.25E-16 | 8.99E-16 | 1.42E-12 |
| Heart_Failure | dendritic_cell | *MRPL42* | -2.217590064 | 0.000641184 | 0.00077656 | 1 |
| Heart_Failure | dendritic_cell | *ZNF335* | -4.463442281 | 3.34E-08 | 4.99E-08 | 0.000209977 |
| Heart_Failure | dendritic_cell | *AGAP9* | 10.04401545 | 2.63E-11 | 4.95E-11 | 1.65E-07 |
| Heart_Failure | dendritic_cell | *ZNF273* | -4.467336912 | 8.90E-11 | 1.60E-10 | 5.60E-07 |
| Heart_Failure | dendritic_cell | *ZNF84* | -5.11697124 | 3.91E-12 | 7.99E-12 | 2.46E-08 |
| Heart_Failure | dendritic_cell | *MAK16* | -6.351452935 | 8.82E-15 | 2.58E-14 | 5.55E-11 |
| Heart_Failure | dendritic_cell | *ZNF667* | -2.706068485 | 9.41E-16 | 3.31E-15 | 5.92E-12 |
| Heart_Failure | dendritic_cell | *SULT1C4* | 3.466732521 | 0.002616717 | 0.003084233 | 1 |
| Heart_Failure | dendritic_cell | *SFI1* | -4.965564107 | 4.05E-16 | 1.53E-15 | 2.54E-12 |
| Heart_Failure | dendritic_cell | *ZNF649* | 4.830586373 | 7.75E-08 | 1.14E-07 | 0.000487077 |
| Heart_Failure | dendritic_cell | *ADH4* | -1.481892654 | 8.72E-14 | 2.16E-13 | 5.48E-10 |
| Heart_Failure | dendritic_cell | *ZNF248* | -3.113092659 | 1.23E-16 | 5.21E-16 | 7.73E-13 |
| Heart_Failure | dendritic_cell | *HIBCH* | -2.940578036 | 7.23E-16 | 2.60E-15 | 4.54E-12 |
| Heart_Failure | dendritic_cell | *ZNF544* | -3.506042085 | 5.57E-14 | 1.42E-13 | 3.50E-10 |
| Heart_Failure | dendritic_cell | *SOWAHC* | -4.620404025 | 4.72E-20 | 8.25E-19 | 2.97E-16 |
| Heart_Failure | dendritic_cell | *AC135178.1* | -12.69608076 | 1.96E-10 | 3.43E-10 | 1.24E-06 |
| Heart_Failure | dendritic_cell | *MIER1* | -3.952637946 | 7.57E-07 | 1.05E-06 | 0.004758112 |
| Heart_Failure | dendritic_cell | *SVIP* | -4.588062668 | 6.65E-18 | 3.92E-17 | 4.18E-14 |
| Heart_Failure | dendritic_cell | *ZNF251* | -4.393386721 | 8.67E-13 | 1.89E-12 | 5.45E-09 |
| Heart_Failure | dendritic_cell | *DDRGK1* | -3.921824516 | 1.09E-18 | 8.36E-18 | 6.87E-15 |
| Heart_Failure | dendritic_cell | *ZNF334* | -2.729336209 | 4.28E-11 | 7.90E-11 | 2.69E-07 |
| Heart_Failure | dendritic_cell | *HSD17B11* | -4.904242639 | 1.33E-06 | 1.82E-06 | 0.008382816 |
| Heart_Failure | dendritic_cell | *SULT1C2* | -1.915595637 | 7.25E-11 | 1.31E-10 | 4.56E-07 |
| Heart_Failure | dendritic_cell | *RPS6KL1* | -2.134407165 | 7.41E-14 | 1.86E-13 | 4.66E-10 |
| Heart_Failure | dendritic_cell | *QRICH1* | -1.478690639 | 0.004560999 | 0.005314122 | 1 |
| Heart_Failure | dendritic_cell | *DDX42* | -4.833137445 | 1.88E-13 | 4.46E-13 | 1.18E-09 |
| Heart_Failure | dendritic_cell | *SLC29A3* | 2.89040968 | 7.34E-20 | 1.08E-18 | 4.62E-16 |
| Heart_Failure | dendritic_cell | *ANTXRL* | -100.2196884 | 2.36E-20 | 6.44E-19 | 1.49E-16 |
| Heart_Failure | dendritic_cell | *STYX* | -4.364673117 | 0.000472397 | 0.000576356 | 1 |
| Heart_Failure | dendritic_cell | *TMEM116* | -2.057928224 | 2.63E-09 | 4.22E-09 | 1.65E-05 |
| Heart_Failure | dendritic_cell | *ZKSCAN8* | -4.822875607 | 1.40E-10 | 2.47E-10 | 8.78E-07 |
| Heart_Failure | dendritic_cell | *MYL4* | 3.439077933 | 1.25E-19 | 1.55E-18 | 7.87E-16 |
| Heart_Failure | dendritic_cell | *ZNF813* | -5.489479435 | 3.20E-18 | 2.06E-17 | 2.01E-14 |
| Heart_Failure | dendritic_cell | *GFPT1* | -3.126337186 | 5.75E-17 | 2.65E-16 | 3.62E-13 |
| Heart_Failure | dendritic_cell | *UVRAG* | -1.739843115 | 0.001104286 | 0.001320397 | 1 |
| Heart_Failure | dendritic_cell | *MGEA5* | -2.757466403 | 6.26E-16 | 2.29E-15 | 3.93E-12 |
| Heart_Failure | dendritic_cell | *MT1F* | -10.60762195 | 2.42E-16 | 9.57E-16 | 1.52E-12 |
| Heart_Failure | dendritic_cell | *ZNF568* | -4.416751596 | 1.17E-07 | 1.71E-07 | 0.000738482 |
| Heart_Failure | dendritic_cell | *SH3BGRL2* | -2.760712148 | 1.06E-12 | 2.31E-12 | 6.70E-09 |
| Heart_Failure | dendritic_cell | *ZNF808* | -5.221237671 | 1.24E-10 | 2.20E-10 | 7.78E-07 |
| Heart_Failure | dendritic_cell | *TMA16* | -3.779206841 | 0.000781308 | 0.000942276 | 1 |
| Heart_Failure | dendritic_cell | *HLA-DRB5* | 0.59419438 | 2.26E-16 | 9.04E-16 | 1.42E-12 |
| Heart_Failure | dendritic_cell | *MAFK* | -4.829819661 | 4.23E-18 | 2.63E-17 | 2.66E-14 |
| Heart_Failure | dendritic_cell | *HIST1H4E* | 3.782515081 | 7.33E-16 | 2.63E-15 | 4.61E-12 |
| Heart_Failure | dendritic_cell | *C20orf203* | -6.717938018 | 3.74E-08 | 5.58E-08 | 0.000235265 |
| Heart_Failure | dendritic_cell | *ARC* | 6.592189809 | 7.85E-21 | 4.66E-19 | 4.94E-17 |
| Heart_Failure | dendritic_cell | *TLK1* | -3.957120064 | 6.37E-14 | 1.61E-13 | 4.00E-10 |

| | | | | | | |
|---|---|---|---|---|---|---|
| Heart_Failure | dendritic_cell | *LRBA* | -3.676668887 | 1.08E-10 | 1.93E-10 | 6.77E-07 |
| Heart_Failure | dendritic_cell | *MMP17* | -2.555433038 | 2.87E-20 | 6.44E-19 | 1.80E-16 |
| Heart_Failure | dendritic_cell | *CCDC69* | -3.938507398 | 5.93E-08 | 8.78E-08 | 0.000372894 |
| Heart_Failure | dendritic_cell | *MDM4* | -4.065403793 | 2.87E-08 | 4.31E-08 | 0.000180745 |
| Heart_Failure | dendritic_cell | *ZNF534* | 3.27528277 | 0.009420549 | 0.010790124 | 1 |
| Heart_Failure | dendritic_cell | *NCOA6* | -3.745207395 | 5.07E-09 | 7.98E-09 | 3.18E-05 |
| Heart_Failure | dendritic_cell | *TTC37* | -3.634163353 | 3.98E-15 | 1.24E-14 | 2.50E-11 |
| Heart_Failure | dendritic_cell | *TUSC1* | 5.855775086 | 3.31E-20 | 6.76E-19 | 2.08E-16 |
| Heart_Failure | dendritic_cell | *PAPSS2* | 3.511891875 | 2.50E-06 | 3.38E-06 | 0.01570544 |
| Heart_Failure | dendritic_cell | *FAN1* | -5.632450536 | 7.51E-13 | 1.65E-12 | 4.72E-09 |
| Heart_Failure | dendritic_cell | *CEP290* | -4.534859788 | 1.14E-14 | 3.26E-14 | 7.16E-11 |
| Heart_Failure | dendritic_cell | *ANKRD13B* | -2.614655971 | 1.23E-13 | 2.98E-13 | 7.73E-10 |
| Heart_Failure | dendritic_cell | *ECI2* | -5.41509538 | 9.00E-15 | 2.63E-14 | 5.66E-11 |
| Heart_Failure | dendritic_cell | *ZNF652* | -4.594207721 | 1.52E-11 | 2.92E-11 | 9.53E-08 |
| Heart_Failure | dendritic_cell | *RPL10A* | -3.210885778 | 2.50E-05 | 3.22E-05 | 0.156997161 |
| Heart_Failure | dendritic_cell | *APCDD1L* | -4.078820505 | 1.76E-19 | 2.02E-18 | 1.11E-15 |
| Heart_Failure | dendritic_cell | *RCSD1* | 2.623596155 | 3.42E-13 | 7.85E-13 | 2.15E-09 |
| Heart_Failure | dendritic_cell | *RASSF9* | -6.613702734 | 1.25E-12 | 2.69E-12 | 7.85E-09 |
| Heart_Failure | dendritic_cell | *FAM169A* | -3.383430101 | 5.15E-05 | 6.56E-05 | 0.323715402 |
| Heart_Failure | dendritic_cell | *TMEM184B* | -4.582304017 | 4.17E-16 | 1.57E-15 | 2.62E-12 |
| Heart_Failure | dendritic_cell | *SCAMP5* | -1.502907045 | 0.007724187 | 0.008892504 | 1 |
| Heart_Failure | dendritic_cell | *ALPK2* | -2.736158828 | 2.14E-19 | 2.27E-18 | 1.34E-15 |
| Heart_Failure | dendritic_cell | *PNP* | 1.031770148 | 3.39E-05 | 4.35E-05 | 0.213069762 |
| Heart_Failure | dendritic_cell | *SFT2D1* | -5.399859999 | 3.67E-13 | 8.40E-13 | 2.31E-09 |
| Heart_Failure | dendritic_cell | *ARHGAP11A* | 5.598473877 | 2.33E-05 | 3.01E-05 | 0.146195193 |
| Heart_Failure | dendritic_cell | *HMGN2* | -4.286106271 | 3.23E-12 | 6.65E-12 | 2.03E-08 |
| Heart_Failure | dendritic_cell | *CD3E* | -3.314093389 | 1.09E-08 | 1.68E-08 | 6.85E-05 |
| Heart_Failure | dendritic_cell | *TSEN15* | -5.173588429 | 1.73E-15 | 5.74E-15 | 1.09E-11 |
| Heart_Failure | dendritic_cell | *TYW1* | -2.223376515 | 8.90E-11 | 1.60E-10 | 5.59E-07 |
| Heart_Failure | dendritic_cell | *ITPRIPL1* | -4.691941205 | 1.59E-13 | 3.81E-13 | 1.00E-09 |
| Heart_Failure | dendritic_cell | *PRMT6* | 4.345107806 | 6.87E-20 | 1.04E-18 | 4.32E-16 |
| Heart_Failure | dendritic_cell | *SHISA4* | -6.51980115 | 2.31E-18 | 1.59E-17 | 1.45E-14 |
| Heart_Failure | dendritic_cell | *CAPZA2* | -2.334475011 | 0.014663984 | 0.016632233 | 1 |
| Heart_Failure | dendritic_cell | *MAP3K3* | -4.046407904 | 2.77E-06 | 3.73E-06 | 0.01738686 |
| Heart_Failure | dendritic_cell | *C1orf174* | 4.209056378 | 1.26E-16 | 5.31E-16 | 7.91E-13 |
| Heart_Failure | dendritic_cell | *RASGEF1A* | 22.78689731 | 1.09E-08 | 1.68E-08 | 6.85E-05 |
| Heart_Failure | dendritic_cell | *KIAA0753* | -4.794311533 | 1.73E-14 | 4.77E-14 | 1.08E-10 |
| Heart_Failure | dendritic_cell | *DCLRE1A* | -4.924097975 | 8.73E-17 | 3.85E-16 | 5.49E-13 |
| Heart_Failure | dendritic_cell | *APRT* | -7.864017784 | 6.12E-10 | 1.03E-09 | 3.85E-06 |
| Heart_Failure | dendritic_cell | *TBKBP1* | -3.163089161 | 4.01E-14 | 1.05E-13 | 2.52E-10 |
| Heart_Failure | dendritic_cell | *ZFP2* | -4.620044868 | 4.38E-14 | 1.13E-13 | 2.75E-10 |
| Heart_Failure | dendritic_cell | *L3MBTL3* | 5.367025236 | 1.31E-07 | 1.90E-07 | 0.000825157 |
| Heart_Failure | dendritic_cell | *MFAP3L* | 11.62786081 | 2.07E-13 | 4.88E-13 | 1.30E-09 |
| Heart_Failure | dendritic_cell | *NAGA* | 6.705735289 | 1.89E-10 | 3.30E-10 | 1.19E-06 |
| Heart_Failure | dendritic_cell | *SMG5* | -2.780647894 | 0.009779712 | 0.011189272 | 1 |
| Heart_Failure | dendritic_cell | *TGM2* | -1.123141462 | 6.64E-06 | 8.80E-06 | 0.041726764 |
| Heart_Failure | dendritic_cell | *PJA2* | -4.192537714 | 5.98E-11 | 1.09E-10 | 3.76E-07 |
| Heart_Failure | dendritic_cell | *AL590822.1* | -2.37856519 | 4.40E-13 | 9.96E-13 | 2.77E-09 |
| Heart_Failure | dendritic_cell | *AP001007.1* | 28.10691951 | 6.80E-20 | 1.03E-18 | 4.28E-16 |
| Heart_Failure | dendritic_cell | *ZNF525* | -6.160059986 | 4.54E-16 | 1.69E-15 | 2.85E-12 |

| | | | | | | |
|---|---|---|---|---|---|---|
| Heart_Failure | dendritic_cell | *AC105020.1* | -7.440784792 | 1.93E-19 | 2.14E-18 | 1.22E-15 |
| Heart_Failure | dendritic_cell | *EFCAB2* | -4.184624603 | 1.93E-11 | 3.70E-11 | 1.22E-07 |
| Heart_Failure | dendritic_cell | *CHML* | 2.729038468 | 3.02E-15 | 9.60E-15 | 1.90E-11 |
| Heart_Failure | dendritic_cell | *TATDN3* | -1.407100671 | 0.003250813 | 0.003815882 | 1 |
| Heart_Failure | dendritic_cell | *ECT2L* | -3.907255617 | 8.10E-13 | 1.78E-12 | 5.09E-09 |
| Heart_Failure | dendritic_cell | *CENPW* | -4.245226568 | 5.89E-15 | 1.78E-14 | 3.70E-11 |
| Heart_Failure | dendritic_cell | *FAM229B* | -1.878819434 | 2.77E-10 | 4.79E-10 | 1.74E-06 |
| Heart_Failure | dendritic_cell | *DDO* | -7.971408098 | 1.11E-13 | 2.70E-13 | 6.98E-10 |
| Heart_Failure | dendritic_cell | *HIST2H2BF* | 4.999822099 | 6.79E-21 | 4.66E-19 | 4.27E-17 |
| Heart_Failure | dendritic_cell | *RBM20* | -3.317007188 | 3.75E-20 | 7.19E-19 | 2.36E-16 |
| Heart_Failure | dendritic_cell | *C6orf163* | -3.673822886 | 1.15E-20 | 5.00E-19 | 7.26E-17 |
| Heart_Failure | dendritic_cell | *PCMTD2* | -4.599864172 | 6.96E-08 | 1.02E-07 | 0.000437324 |
| Heart_Failure | dendritic_cell | *OOEP* | -2.756605189 | 3.16E-10 | 5.43E-10 | 1.98E-06 |
| Heart_Failure | dendritic_cell | *FOXO6* | -3.808522475 | 3.91E-09 | 6.20E-09 | 2.46E-05 |
| Heart_Failure | dendritic_cell | *SYS1* | 7.757103671 | 5.41E-05 | 6.89E-05 | 0.339931992 |
| Heart_Failure | dendritic_cell | *INPP5B* | -0.597459509 | 0.008808932 | 0.010106159 | 1 |
| Heart_Failure | dendritic_cell | *NEU4* | -14.83441337 | 5.27E-10 | 8.92E-10 | 3.32E-06 |
| Heart_Failure | dendritic_cell | *TRAF3IP1* | -4.493665934 | 7.36E-13 | 1.62E-12 | 4.63E-09 |
| Heart_Failure | dendritic_cell | *GIGYF2* | -4.026480155 | 1.29E-12 | 2.77E-12 | 8.09E-09 |
| Heart_Failure | dendritic_cell | *C2orf72* | -3.543691644 | 0.000345977 | 0.00042442 | 1 |
| Heart_Failure | dendritic_cell | *RUFY2* | -3.915794283 | 4.85E-15 | 1.49E-14 | 3.05E-11 |
| Heart_Failure | dendritic_cell | *PHACTR4* | -3.389483502 | 3.44E-09 | 5.48E-09 | 2.16E-05 |
| Heart_Failure | dendritic_cell | *ASAH2B* | -2.785345255 | 2.86E-09 | 4.57E-09 | 1.80E-05 |
| Heart_Failure | dendritic_cell | *C10orf128* | -2.747374094 | 1.10E-08 | 1.70E-08 | 6.94E-05 |
| Heart_Failure | dendritic_cell | *SYT15* | -6.262160321 | 1.82E-17 | 9.56E-17 | 1.15E-13 |
| Heart_Failure | dendritic_cell | *ZDBF2* | -5.9173903 | 1.85E-11 | 3.55E-11 | 1.16E-07 |
| Heart_Failure | dendritic_cell | *RING1* | -4.027235299 | 1.57E-08 | 2.40E-08 | 9.89E-05 |
| Heart_Failure | dendritic_cell | *HSD17B8* | -1.899179106 | 0.002327566 | 0.00275064 | 1 |
| Heart_Failure | dendritic_cell | *OXLD1* | 3.235293541 | 0.00045194 | 0.000551932 | 1 |
| Heart_Failure | dendritic_cell | *HLA-DOA* | 0.661595922 | 6.59E-20 | 1.02E-18 | 4.14E-16 |
| Heart_Failure | dendritic_cell | *HLA-DMA* | 2.100952347 | 1.85E-20 | 6.38E-19 | 1.17E-16 |
| Heart_Failure | dendritic_cell | *TAP2* | -1.550054791 | 4.32E-06 | 5.77E-06 | 0.027141706 |
| Heart_Failure | dendritic_cell | *HLA-DRA* | 0.731693806 | 1.75E-21 | 4.60E-19 | 1.10E-17 |
| Heart_Failure | dendritic_cell | *COL15A1* | -3.195116394 | 1.15E-12 | 2.49E-12 | 7.22E-09 |
| Heart_Failure | dendritic_cell | *NOTCH4* | -0.153395216 | 0.039288945 | 0.043587365 | 1 |
| Heart_Failure | dendritic_cell | *AGER* | 3.75080587 | 4.89E-07 | 6.88E-07 | 0.003073547 |
| Heart_Failure | dendritic_cell | *PRRT1* | 2.500749481 | 5.03E-05 | 6.42E-05 | 0.316489504 |
| Heart_Failure | dendritic_cell | *MRPL38* | -3.473183606 | 1.22E-06 | 1.67E-06 | 0.00765909 |
| Heart_Failure | dendritic_cell | *STK19* | -1.831394759 | 0.006498948 | 0.007512206 | 1 |
| Heart_Failure | dendritic_cell | *DXO* | -1.911642073 | 0.000694689 | 0.00083926 | 1 |
| Heart_Failure | dendritic_cell | *NELFE* | -2.310314019 | 1.42E-07 | 2.05E-07 | 0.000892532 |
| Heart_Failure | dendritic_cell | *LAYN* | -3.833037799 | 3.45E-05 | 4.42E-05 | 0.216694941 |
| Heart_Failure | dendritic_cell | *C6orf48* | -4.428520905 | 9.16E-08 | 1.34E-07 | 0.000575794 |
| Heart_Failure | dendritic_cell | *CARD16* | -4.919964013 | 2.71E-12 | 5.63E-12 | 1.71E-08 |
| Heart_Failure | dendritic_cell | *MSH5* | -3.664704896 | 2.68E-16 | 1.05E-15 | 1.68E-12 |
| Heart_Failure | dendritic_cell | *ABHD16A* | 2.490854994 | 3.79E-11 | 7.05E-11 | 2.39E-07 |
| Heart_Failure | dendritic_cell | *LY6G5C* | 1.155656453 | 0.000157912 | 0.0001971 | 0.992791226 |
| Heart_Failure | dendritic_cell | *GPANK1* | 1.653316081 | 0.01104024 | 0.012587956 | 1 |
| Heart_Failure | dendritic_cell | *BAG6* | 1.670311945 | 0.027848527 | 0.031109397 | 1 |
| Heart_Failure | dendritic_cell | *NFKBIL1* | -3.192177132 | 3.52E-08 | 5.25E-08 | 0.000221316 |

| | | | | | | |
|---|---|---|---|---|---|---|
| Heart_Failure | dendritic_cell | *ZNF814* | -4.607687914 | 3.68E-12 | 7.56E-12 | 2.32E-08 |
| Heart_Failure | dendritic_cell | *MICB* | 1.452609302 | 2.55E-11 | 4.81E-11 | 1.61E-07 |
| Heart_Failure | dendritic_cell | *HLA-C* | -2.003503409 | 6.33E-07 | 8.84E-07 | 0.003978573 |
| Heart_Failure | dendritic_cell | *CCHCR1* | -1.384075247 | 1.38E-05 | 1.80E-05 | 0.086446841 |
| Heart_Failure | dendritic_cell | *PSORS1C1* | -0.44665072 | 2.94E-05 | 3.79E-05 | 0.185107672 |
| Heart_Failure | dendritic_cell | *ABCF1* | -3.334057122 | 7.24E-07 | 1.01E-06 | 0.004549066 |
| Heart_Failure | dendritic_cell | *LILRB3* | 1.503320648 | 3.05E-20 | 6.68E-19 | 1.92E-16 |
| Heart_Failure | dendritic_cell | *LINC01123* | -63.3289191 | 3.60E-10 | 6.16E-10 | 2.26E-06 |
| Heart_Failure | dendritic_cell | *HLA-E* | -2.870230451 | 8.30E-12 | 1.63E-11 | 5.22E-08 |
| Heart_Failure | dendritic_cell | *ZNF468* | -3.798702336 | 9.64E-20 | 1.30E-18 | 6.06E-16 |
| Heart_Failure | dendritic_cell | *HCG9* | -1.408288952 | 5.17E-06 | 6.89E-06 | 0.032500309 |
| Heart_Failure | dendritic_cell | *TBC1D8* | 2.294090433 | 0.014856044 | 0.016837922 | 1 |
| Heart_Failure | dendritic_cell | *FAM153C* | 13.48125047 | 3.80E-21 | 4.66E-19 | 2.39E-17 |
| Heart_Failure | dendritic_cell | *GABBR1* | 3.837163374 | 0.000635157 | 0.000769557 | 1 |
| Heart_Failure | dendritic_cell | *TRIM27* | -3.098868134 | 1.19E-05 | 1.57E-05 | 0.075031827 |
| Heart_Failure | dendritic_cell | *MALRD1* | -1.889061905 | 5.73E-14 | 1.46E-13 | 3.60E-10 |
| Heart_Failure | dendritic_cell | *MROH6* | 1.786877624 | 2.43E-18 | 1.66E-17 | 1.53E-14 |
| Heart_Failure | dendritic_cell | *ATXN2* | -3.517490595 | 1.94E-12 | 4.08E-12 | 1.22E-08 |
| Heart_Failure | dendritic_cell | *FAM216A* | -4.72980253 | 6.44E-14 | 1.63E-13 | 4.05E-10 |
| Heart_Failure | dendritic_cell | *ZNF155* | -4.029552228 | 9.03E-10 | 1.50E-09 | 5.68E-06 |
| Heart_Failure | dendritic_cell | *ZNF783* | -3.592240598 | 6.46E-05 | 8.21E-05 | 0.406343187 |
| Heart_Failure | dendritic_cell | *ZNF425* | -2.475426871 | 3.08E-05 | 3.96E-05 | 0.193564674 |
| Heart_Failure | dendritic_cell | *LRRC10B* | -18.94004905 | 3.58E-10 | 6.12E-10 | 2.25E-06 |
| Heart_Failure | dendritic_cell | *C12orf73* | -4.282105103 | 1.94E-12 | 4.09E-12 | 1.22E-08 |
| Heart_Failure | dendritic_cell | *PCDHGA1* | 2.647501451 | 1.82E-16 | 7.42E-16 | 1.14E-12 |
| Heart_Failure | dendritic_cell | *PCDHA5* | 12.93442289 | 0.004465075 | 0.005206218 | 1 |
| Heart_Failure | dendritic_cell | *PCDHA4* | -13.69727903 | 1.39E-19 | 1.67E-18 | 8.74E-16 |
| Heart_Failure | dendritic_cell | *TRIM13* | -4.067152552 | 9.85E-14 | 2.41E-13 | 6.20E-10 |
| Heart_Failure | dendritic_cell | *SPIRE2* | -1.181313149 | 0.00725175 | 0.008360857 | 1 |
| Heart_Failure | dendritic_cell | *SLFN12L* | -3.129239305 | 2.48E-21 | 4.66E-19 | 1.56E-17 |
| Heart_Failure | dendritic_cell | *CLLU1OS* | -2.926087726 | 0.000681123 | 0.000823362 | 1 |
| Heart_Failure | dendritic_cell | *SYCE1L* | -3.505543238 | 9.77E-14 | 2.40E-13 | 6.15E-10 |
| Heart_Failure | dendritic_cell | *TMEM231* | -2.857711056 | 3.00E-10 | 5.17E-10 | 1.89E-06 |
| Heart_Failure | dendritic_cell | *FAM71F2* | -4.459324491 | 8.01E-19 | 6.49E-18 | 5.04E-15 |
| Heart_Failure | dendritic_cell | *FAM205A* | -11.1749396 | 5.75E-05 | 7.31E-05 | 0.361498395 |
| Heart_Failure | dendritic_cell | *AC016586.1* | 21.7132201 | 4.28E-20 | 7.85E-19 | 2.69E-16 |
| Heart_Failure | dendritic_cell | *LINC00654* | -5.349758951 | 5.59E-17 | 2.59E-16 | 3.52E-13 |
| Heart_Failure | dendritic_cell | *SAP25* | -1.372666192 | 0.015276413 | 0.017301894 | 1 |
| Heart_Failure | dendritic_cell | *NT5M* | -1.421165022 | 0.011854006 | 0.013496222 | 1 |
| Heart_Failure | dendritic_cell | *PRR13* | -3.915864554 | 0.000720789 | 0.000869956 | 1 |
| Heart_Failure | dendritic_cell | *MT1A* | 6.326168407 | 1.27E-20 | 5.17E-19 | 8.01E-17 |
| Heart_Failure | dendritic_cell | *MT1M* | 9.801236496 | 2.53E-05 | 3.26E-05 | 0.158757139 |
| Heart_Failure | dendritic_cell | *CFI* | -2.558814201 | 2.39E-18 | 1.64E-17 | 1.50E-14 |
| Heart_Failure | dendritic_cell | *SAMD9* | 4.936216051 | 1.69E-16 | 6.94E-16 | 1.06E-12 |
| Heart_Failure | dendritic_cell | *EXOC3L4* | -5.210054558 | 6.60E-19 | 5.46E-18 | 4.15E-15 |
| Heart_Failure | dendritic_cell | *TP53TG3D* | 6.71554637 | 2.59E-05 | 3.35E-05 | 0.163016356 |
| Heart_Failure | dendritic_cell | *ATP6AP1L* | -3.474714072 | 3.16E-11 | 5.91E-11 | 1.99E-07 |
| Heart_Failure | dendritic_cell | *NAP1L4* | -3.248886653 | 9.26E-07 | 1.28E-06 | 0.005822456 |
| Heart_Failure | dendritic_cell | *CPT1B* | 1.633315357 | 0.000364387 | 0.000446657 | 1 |
| Heart_Failure | dendritic_cell | *SMN2* | -1.401935894 | 0.000376689 | 0.000461466 | 1 |

| | | | | | | |
|---|---|---|---|---|---|---|
| Heart_Failure | dendritic_cell | *SERF1B* | -7.188371102 | 4.01E-15 | 1.24E-14 | 2.52E-11 |
| Heart_Failure | dendritic_cell | *DENND6B* | -9.309376915 | 1.75E-09 | 2.84E-09 | 1.10E-05 |
| Heart_Failure | dendritic_cell | *EIF3CL* | -7.728536961 | 9.34E-17 | 4.07E-16 | 5.87E-13 |
| Heart_Failure | dendritic_cell | *LCMT1* | -5.334704265 | 5.10E-11 | 9.34E-11 | 3.20E-07 |
| Heart_Failure | dendritic_cell | *LIN52* | -2.899381873 | 3.98E-12 | 8.12E-12 | 2.50E-08 |
| Heart_Failure | dendritic_cell | *C17orf107* | 2.252465013 | 1.39E-08 | 2.12E-08 | 8.74E-05 |
| Heart_Failure | dendritic_cell | *LINC00487* | -6.391835106 | 1.36E-07 | 1.97E-07 | 0.000856388 |
| Heart_Failure | dendritic_cell | *RFPL3S* | 2.791715361 | 1.56E-14 | 4.36E-14 | 9.83E-11 |
| Heart_Failure | dendritic_cell | *C1QTNF9B* | -36.26309313 | 2.46E-20 | 6.44E-19 | 1.55E-16 |
| Heart_Failure | dendritic_cell | *RNPS1* | -4.198889771 | 2.24E-12 | 4.69E-12 | 1.41E-08 |
| Heart_Failure | dendritic_cell | *NYNRIN* | 8.733244462 | 1.06E-13 | 2.59E-13 | 6.68E-10 |
| Heart_Failure | dendritic_cell | *ZDHHC11B* | -4.069199422 | 1.39E-13 | 3.34E-13 | 8.72E-10 |
| Heart_Failure | dendritic_cell | *GOLGA8O* | -9.398625304 | 0.000229124 | 0.000283396 | 1 |
| Heart_Failure | dendritic_cell | *HCG27* | -1.493490673 | 6.00E-06 | 7.97E-06 | 0.037714933 |
| Heart_Failure | dendritic_cell | *HLA-A* | -1.677443785 | 0.00106505 | 0.001274695 | 1 |
| Heart_Failure | dendritic_cell | *LNP1* | -3.559964265 | 3.58E-20 | 6.99E-19 | 2.25E-16 |
| Heart_Failure | dendritic_cell | *VGLL3* | -1.495268292 | 3.80E-15 | 1.18E-14 | 2.39E-11 |
| Heart_Failure | dendritic_cell | *COLQ* | -3.659599266 | 6.62E-19 | 5.47E-18 | 4.16E-15 |
| Heart_Failure | dendritic_cell | *METTL6* | -3.834025886 | 3.94E-06 | 5.28E-06 | 0.024751575 |
| Heart_Failure | dendritic_cell | *SACM1L* | -4.96692068 | 2.47E-14 | 6.69E-14 | 1.55E-10 |
| Heart_Failure | dendritic_cell | *SLC48A1* | -7.324803364 | 3.32E-14 | 8.80E-14 | 2.09E-10 |
| Heart_Failure | dendritic_cell | *TAS2R14* | -1.042279835 | 0.035654834 | 0.0396396 | 1 |
| Heart_Failure | dendritic_cell | *ZRSR1* | -14.56659757 | 7.45E-20 | 1.09E-18 | 4.68E-16 |
| Heart_Failure | dendritic_cell | *AC084018.1* | -2.189092027 | 8.66E-12 | 1.70E-11 | 5.45E-08 |
| Heart_Failure | dendritic_cell | *C17orf51* | -3.437139121 | 1.05E-19 | 1.38E-18 | 6.57E-16 |
| Heart_Failure | dendritic_cell | *OR2A42* | -3.107163417 | 2.50E-12 | 5.21E-12 | 1.57E-08 |
| Heart_Failure | dendritic_cell | *RNF208* | -14.55524956 | 5.51E-19 | 4.77E-18 | 3.47E-15 |
| Heart_Failure | dendritic_cell | *MAP10* | 5.345197401 | 1.56E-10 | 2.74E-10 | 9.79E-07 |
| Heart_Failure | dendritic_cell | *ZNF580* | -3.565247702 | 2.56E-11 | 4.82E-11 | 1.61E-07 |
| Heart_Failure | dendritic_cell | *NUP62* | 7.348061562 | 2.48E-09 | 3.99E-09 | 1.56E-05 |
| Heart_Failure | dendritic_cell | *FAM106A* | -1.602370022 | 0.000470357 | 0.00057409 | 1 |
| Heart_Failure | dendritic_cell | *ZNF254* | -4.680140861 | 4.10E-09 | 6.49E-09 | 2.58E-05 |
| Heart_Failure | dendritic_cell | *TCTEX1D2* | 3.886796879 | 5.63E-09 | 8.82E-09 | 3.54E-05 |
| Heart_Failure | dendritic_cell | *LINGO4* | -14.97821222 | 0.000750701 | 0.000905536 | 1 |
| Heart_Failure | dendritic_cell | *ASIC3* | -1.88760795 | 0.000914298 | 0.00109887 | 1 |
| Heart_Failure | dendritic_cell | *GIMAP1* | -2.095995908 | 0.035711004 | 0.039695029 | 1 |
| Heart_Failure | dendritic_cell | *CCDC183* | 10.60200893 | 4.06E-19 | 3.71E-18 | 2.55E-15 |
| Heart_Failure | dendritic_cell | *ARHGEF35* | -5.483629264 | 1.48E-08 | 2.26E-08 | 9.32E-05 |
| Heart_Failure | dendritic_cell | *SUPT4H1* | 5.356946094 | 2.85E-18 | 1.88E-17 | 1.79E-14 |
| Heart_Failure | dendritic_cell | *MXD3* | -3.020130879 | 3.76E-09 | 5.98E-09 | 2.37E-05 |
| Heart_Failure | dendritic_cell | *GSTM2* | -2.51656312 | 1.77E-12 | 3.74E-12 | 1.11E-08 |
| Heart_Failure | dendritic_cell | *ARHGAP19* | 7.927142385 | 1.46E-06 | 1.99E-06 | 0.009165058 |
| Heart_Failure | dendritic_cell | *LCAT* | -2.608792937 | 6.90E-07 | 9.63E-07 | 0.004340331 |
| Heart_Failure | dendritic_cell | *SIPA1* | 4.268359238 | 7.36E-11 | 1.33E-10 | 4.63E-07 |
| Heart_Failure | dendritic_cell | *ERV3-1* | -0.794369304 | 8.18E-06 | 1.08E-05 | 0.051400334 |
| Heart_Failure | dendritic_cell | *GBP7* | -1.16567697 | 0.000170532 | 0.000212557 | 1 |
| Heart_Failure | dendritic_cell | *RBMXL1* | 7.97717935 | 6.68E-08 | 9.84E-08 | 0.00041966 |
| Heart_Failure | dendritic_cell | *SRA1* | -3.324074014 | 5.91E-09 | 9.26E-09 | 3.72E-05 |
| Heart_Failure | dendritic_cell | *TMEM110* | 6.113508873 | 1.52E-15 | 5.12E-15 | 9.59E-12 |
| Heart_Failure | dendritic_cell | *HEXA* | -5.835641913 | 2.50E-11 | 4.72E-11 | 1.57E-07 |

| | | | | | | |
|---|---|---|---|---|---|---|
| Heart_Failure | dendritic_cell | *LEPROT* | -4.886351803 | 3.07E-12 | 6.34E-12 | 1.93E-08 |
| Heart_Failure | dendritic_cell | *LBH* | -2.855850998 | 3.32E-20 | 6.76E-19 | 2.08E-16 |
| Heart_Failure | dendritic_cell | *PPP1CB* | 4.286778511 | 9.33E-17 | 4.07E-16 | 5.87E-13 |
| Heart_Failure | dendritic_cell | *NCKIPSD* | -4.74262383 | 5.36E-15 | 1.63E-14 | 3.37E-11 |
| Heart_Failure | dendritic_cell | *SLC35F6* | -7.233155411 | 9.04E-14 | 2.23E-13 | 5.69E-10 |
| Heart_Failure | dendritic_cell | *CLIC1* | -6.472137223 | 5.85E-15 | 1.77E-14 | 3.68E-11 |
| Heart_Failure | dendritic_cell | *ATP6V1G2* | 0.722367527 | 0.021546364 | 0.024206932 | 1 |
| Heart_Failure | dendritic_cell | *EMP2* | -3.645736 | 7.51E-14 | 1.88E-13 | 4.72E-10 |
| Heart_Failure | dendritic_cell | *PPM1N* | 4.551671729 | 1.55E-19 | 1.83E-18 | 9.75E-16 |
| Heart_Failure | dendritic_cell | *LTB4R2* | -4.506659211 | 2.10E-11 | 4.00E-11 | 1.32E-07 |
| Heart_Failure | dendritic_cell | *GALT* | -4.279884717 | 1.37E-05 | 1.79E-05 | 0.086209096 |
| Heart_Failure | dendritic_cell | *ITGA1* | -2.997658188 | 4.91E-16 | 1.82E-15 | 3.09E-12 |
| Heart_Failure | dendritic_cell | *NUDT19* | -4.003581713 | 2.51E-10 | 4.35E-10 | 1.58E-06 |
| Heart_Failure | dendritic_cell | *ZNF726* | -3.750606485 | 2.77E-20 | 6.44E-19 | 1.74E-16 |
| Heart_Failure | dendritic_cell | *TAX1BP3* | -7.757534527 | 2.58E-18 | 1.72E-17 | 1.62E-14 |
| Heart_Failure | dendritic_cell | *AP1G2* | 7.62096309 | 6.06E-17 | 2.77E-16 | 3.81E-13 |
| Heart_Failure | dendritic_cell | *ZNF90* | -4.810508813 | 6.15E-15 | 1.85E-14 | 3.87E-11 |
| Heart_Failure | dendritic_cell | *TM6SF2* | -41.08453688 | 1.53E-19 | 1.81E-18 | 9.59E-16 |
| Heart_Failure | dendritic_cell | *REPIN1* | 2.243818311 | 0.031524907 | 0.035153794 | 1 |
| Heart_Failure | dendritic_cell | *SMIM7* | 3.116556293 | 0.003533542 | 0.004140798 | 1 |
| Heart_Failure | dendritic_cell | *TSPAN4* | -4.532808523 | 3.36E-16 | 1.30E-15 | 2.11E-12 |
| Heart_Failure | dendritic_cell | *CPNE1* | -2.115620462 | 2.37E-11 | 4.49E-11 | 1.49E-07 |
| Heart_Failure | dendritic_cell | *ARL16* | -0.670476456 | 0.001959837 | 0.002325688 | 1 |
| Heart_Failure | dendritic_cell | *AC079776.2* | -13.29564032 | 2.28E-08 | 3.44E-08 | 0.000143639 |
| Heart_Failure | dendritic_cell | *WEE2* | -3.003110492 | 4.81E-07 | 6.77E-07 | 0.003026546 |
| Heart_Failure | dendritic_cell | *PAXIP1-AS2* | -4.828118811 | 6.27E-17 | 2.86E-16 | 3.94E-13 |
| Heart_Failure | dendritic_cell | *LYRM4* | -4.343917312 | 4.65E-14 | 1.20E-13 | 2.93E-10 |
| Heart_Failure | dendritic_cell | *PRCD* | -2.873418692 | 1.19E-18 | 9.00E-18 | 7.50E-15 |
| Heart_Failure | dendritic_cell | *C17orf67* | -1.670201481 | 0.010075789 | 0.011517543 | 1 |
| Heart_Failure | dendritic_cell | *FIS1* | -2.792545857 | 4.27E-07 | 6.02E-07 | 0.002681817 |
| Heart_Failure | dendritic_cell | *COLCA2* | -3.893913651 | 3.78E-15 | 1.18E-14 | 2.38E-11 |
| Heart_Failure | dendritic_cell | *MBLAC1* | 1.844829929 | 7.48E-13 | 1.64E-12 | 4.70E-09 |
| Heart_Failure | dendritic_cell | *NEURL1B* | -4.295695998 | 2.98E-19 | 2.90E-18 | 1.87E-15 |
| Heart_Failure | dendritic_cell | *VSTM5* | -1.522390946 | 0.003400775 | 0.003988166 | 1 |
| Heart_Failure | dendritic_cell | *PPME1* | -3.01527285 | 1.44E-10 | 2.54E-10 | 9.04E-07 |
| Heart_Failure | dendritic_cell | *STARD10* | 0.798898711 | 0.01591113 | 0.01800779 | 1 |
| Heart_Failure | dendritic_cell | *NUTM2D* | 4.474898001 | 5.02E-09 | 7.91E-09 | 3.16E-05 |
| Heart_Failure | dendritic_cell | *CPEB1* | -2.107318355 | 1.95E-20 | 6.44E-19 | 1.23E-16 |
| Heart_Failure | dendritic_cell | *C10orf105* | 2.887339114 | 8.80E-20 | 1.22E-18 | 5.53E-16 |
| Heart_Failure | dendritic_cell | *ARHGEF33* | -2.482618191 | 9.93E-07 | 1.37E-06 | 0.006242517 |
| Heart_Failure | dendritic_cell | *AC005082.1* | -12.30414118 | 3.43E-21 | 4.66E-19 | 2.16E-17 |
| Heart_Failure | dendritic_cell | *SMTNL1* | -0.935068577 | 2.41E-05 | 3.11E-05 | 0.151275383 |
| Heart_Failure | dendritic_cell | *TBC1D26* | -39.89161258 | 3.33E-08 | 4.97E-08 | 0.000209193 |
| Heart_Failure | dendritic_cell | *ACSM4* | -1.527922132 | 3.29E-07 | 4.67E-07 | 0.002067278 |
| Heart_Failure | dendritic_cell | *COL28A1* | -2.749909715 | 7.60E-20 | 1.11E-18 | 4.78E-16 |
| Heart_Failure | dendritic_cell | *PHB2* | -5.824742869 | 1.31E-08 | 2.00E-08 | 8.21E-05 |
| Heart_Failure | dendritic_cell | *GRID2IP* | -3.605935941 | 3.13E-20 | 6.70E-19 | 1.97E-16 |
| Heart_Failure | dendritic_cell | *UBE2QL1* | -4.220710161 | 2.57E-20 | 6.44E-19 | 1.62E-16 |
| Heart_Failure | dendritic_cell | *GOLGA8B* | -4.035873552 | 1.39E-17 | 7.54E-17 | 8.73E-14 |
| Heart_Failure | dendritic_cell | *KCNU1* | -3.613490516 | 5.40E-22 | 2.61E-19 | 3.39E-18 |

| | | | | | | |
|---|---|---|---|---|---|---|
| Heart_Failure | dendritic_cell | *MYL5* | -4.692577838 | 4.92E-20 | 8.51E-19 | 3.09E-16 |
| Heart_Failure | dendritic_cell | *NPEPL1* | -5.563257031 | 1.49E-15 | 5.03E-15 | 9.39E-12 |
| Heart_Failure | dendritic_cell | *LINC01139* | 1.865173361 | 0.024679579 | 0.027633217 | 1 |
| Heart_Failure | dendritic_cell | *TSTD1* | 7.981129467 | 3.60E-17 | 1.74E-16 | 2.26E-13 |
| Heart_Failure | dendritic_cell | *LINC01138* | -5.442234975 | 9.54E-10 | 1.59E-09 | 6.00E-06 |
| Heart_Failure | dendritic_cell | *CYB5RL* | 6.530983778 | 7.66E-08 | 1.13E-07 | 0.000481811 |
| Heart_Failure | dendritic_cell | *ATAD3C* | 5.999983065 | 1.10E-08 | 1.69E-08 | 6.92E-05 |
| Heart_Failure | dendritic_cell | *CCDC7* | -3.771597337 | 2.04E-15 | 6.70E-15 | 1.28E-11 |
| Heart_Failure | dendritic_cell | *PAM16* | 4.434983915 | 6.53E-10 | 1.10E-09 | 4.11E-06 |
| Heart_Failure | dendritic_cell | *AC016773.1* | -1.21477305 | 5.91E-05 | 7.51E-05 | 0.371494973 |
| Heart_Failure | dendritic_cell | *AP000350.4* | -0.569672201 | 0.007191386 | 0.008294303 | 1 |
| Heart_Failure | dendritic_cell | *RNASEK* | 3.154575948 | 0.030232811 | 0.03373091 | 1 |
| Heart_Failure | dendritic_cell | *NBPF1* | -4.167782902 | 3.93E-13 | 8.95E-13 | 2.47E-09 |
| Heart_Failure | dendritic_cell | *FAM228B* | -0.84208606 | 0.005029209 | 0.005846641 | 1 |
| Heart_Failure | dendritic_cell | *RNU6ATAC35I* | 11.96450233 | 9.20E-21 | 4.77E-19 | 5.78E-17 |
| Heart_Failure | dendritic_cell | *C6orf226* | -5.182300675 | 2.50E-10 | 4.34E-10 | 1.57E-06 |
| Heart_Failure | dendritic_cell | *PPP3R1* | -4.693855258 | 4.20E-09 | 6.65E-09 | 2.64E-05 |
| Heart_Failure | dendritic_cell | *FANCG* | 2.259145194 | 1.88E-06 | 2.56E-06 | 0.011835626 |
| Heart_Failure | dendritic_cell | *C19orf73* | -18.6968845 | 4.56E-17 | 2.14E-16 | 2.86E-13 |
| Heart_Failure | dendritic_cell | *ZNF880* | -1.388741613 | 1.52E-05 | 1.98E-05 | 0.09524861 |
| Heart_Failure | dendritic_cell | *OR2A25* | -90.60714842 | 3.79E-14 | 9.92E-14 | 2.38E-10 |
| Heart_Failure | dendritic_cell | *TIGD1* | 2.203893009 | 1.26E-13 | 3.06E-13 | 7.94E-10 |
| Heart_Failure | dendritic_cell | *XKR9* | -2.333049048 | 1.33E-11 | 2.58E-11 | 8.35E-08 |
| Heart_Failure | dendritic_cell | *C1orf229* | -5.286623524 | 4.43E-17 | 2.10E-16 | 2.78E-13 |
| Heart_Failure | dendritic_cell | *SLC12A8* | -4.439549268 | 2.15E-11 | 4.09E-11 | 1.35E-07 |
| Heart_Failure | dendritic_cell | *APOL6* | -2.92217528 | 1.58E-18 | 1.15E-17 | 9.93E-15 |
| Heart_Failure | dendritic_cell | *UBA52* | 2.28075959 | 0.014396419 | 0.016340546 | 1 |
| Heart_Failure | dendritic_cell | *TIAF1* | -3.23913371 | 1.05E-06 | 1.45E-06 | 0.006605371 |
| Heart_Failure | dendritic_cell | *BTBD19* | -4.399369786 | 1.91E-06 | 2.60E-06 | 0.012008781 |
| Heart_Failure | dendritic_cell | *FAM185A* | -5.412985854 | 3.22E-13 | 7.43E-13 | 2.02E-09 |
| Heart_Failure | dendritic_cell | *AC062017.1* | -2.622656033 | 2.33E-19 | 2.41E-18 | 1.47E-15 |
| Heart_Failure | dendritic_cell | *ADRA2B* | 5.772673066 | 7.26E-21 | 4.66E-19 | 4.57E-17 |
| Heart_Failure | dendritic_cell | *EXOSC6* | -3.392579521 | 2.35E-17 | 1.19E-16 | 1.48E-13 |
| Heart_Failure | dendritic_cell | *VPS52* | -5.605406837 | 7.35E-13 | 1.62E-12 | 4.62E-09 |
| Heart_Failure | dendritic_cell | *TINCR* | 5.972453228 | 6.15E-17 | 2.81E-16 | 3.86E-13 |
| Heart_Failure | dendritic_cell | *LINC00205* | -4.27694729 | 1.25E-18 | 9.34E-18 | 7.86E-15 |
| Heart_Failure | dendritic_cell | *AC147651.3* | -7.359303002 | 2.63E-05 | 3.40E-05 | 0.165507972 |
| Heart_Failure | dendritic_cell | *HLA-DPB1* | 0.665600634 | 6.57E-21 | 4.66E-19 | 4.13E-17 |
| Heart_Failure | dendritic_cell | *OSER1-AS1* | -7.061101424 | 1.47E-10 | 2.59E-10 | 9.22E-07 |
| Heart_Failure | dendritic_cell | *LINC01133* | -1.994213577 | 3.18E-20 | 6.76E-19 | 2.00E-16 |
| Heart_Failure | dendritic_cell | *LINC01117* | -2.338359188 | 2.71E-19 | 2.71E-18 | 1.70E-15 |
| Heart_Failure | dendritic_cell | *RASAL2-AS1* | -10.72718753 | 3.69E-17 | 1.78E-16 | 2.32E-13 |
| Heart_Failure | dendritic_cell | *LINC00240* | -2.117131772 | 2.65E-15 | 8.50E-15 | 1.66E-11 |
| Heart_Failure | dendritic_cell | *LINC00863* | -2.742699293 | 0.000231469 | 0.000286127 | 1 |
| Heart_Failure | dendritic_cell | *LINC00184* | -6.804705529 | 7.94E-20 | 1.14E-18 | 4.99E-16 |
| Heart_Failure | dendritic_cell | *TMEM233* | -2.021831872 | 4.94E-19 | 4.33E-18 | 3.10E-15 |
| Heart_Failure | dendritic_cell | *LINC00237* | -14.02393177 | 2.73E-20 | 6.44E-19 | 1.72E-16 |
| Heart_Failure | dendritic_cell | *PLEKHM1* | -4.104224379 | 1.44E-15 | 4.86E-15 | 9.04E-12 |
| Heart_Failure | dendritic_cell | *LINC00092* | -1.325189917 | 0.024540198 | 0.027486945 | 1 |
| Heart_Failure | dendritic_cell | *SFTA1P* | -10.03904054 | 1.18E-19 | 1.48E-18 | 7.40E-16 |

| | | | | | | |
|---|---|---|---|---|---|---|
| Heart_Failure | dendritic_cell | *FAM225B* | 3.274270291 | 3.78E-08 | 5.63E-08 | 0.0002374 |
| Heart_Failure | dendritic_cell | *SLC26A6* | -4.325900552 | 2.74E-18 | 1.82E-17 | 1.72E-14 |
| Heart_Failure | dendritic_cell | *MIAT* | -4.474466723 | 7.23E-10 | 1.21E-09 | 4.54E-06 |
| Heart_Failure | dendritic_cell | *LINC00626* | -58.5386621 | 2.80E-19 | 2.77E-18 | 1.76E-15 |
| Heart_Failure | dendritic_cell | *ERCC6* | -4.797974903 | 2.36E-14 | 6.40E-14 | 1.49E-10 |
| Heart_Failure | dendritic_cell | *LINC00115* | -3.194764138 | 5.26E-14 | 1.35E-13 | 3.31E-10 |
| Heart_Failure | dendritic_cell | *NOL7* | -5.694324538 | 8.69E-11 | 1.56E-10 | 5.47E-07 |
| Heart_Failure | dendritic_cell | *ELFN1* | 26.17798032 | 8.82E-16 | 3.12E-15 | 5.54E-12 |
| Heart_Failure | dendritic_cell | *ZNF503-AS1* | -8.075123298 | 6.98E-15 | 2.08E-14 | 4.39E-11 |
| Heart_Failure | dendritic_cell | *LINC00937* | 5.484493235 | 2.73E-18 | 1.81E-17 | 1.72E-14 |
| Heart_Failure | dendritic_cell | *AC010536.1* | -3.390691921 | 0.000560613 | 0.000680945 | 1 |
| Heart_Failure | dendritic_cell | *AC064836.3* | -6.001368887 | 3.32E-20 | 6.76E-19 | 2.08E-16 |
| Heart_Failure | dendritic_cell | *HSBP1L1* | -6.496003253 | 2.04E-16 | 8.26E-16 | 1.28E-12 |
| Heart_Failure | dendritic_cell | *TAS2R46* | 3.506302417 | 3.94E-12 | 8.05E-12 | 2.48E-08 |
| Heart_Failure | dendritic_cell | *AC093901.1* | 4.157165882 | 1.22E-17 | 6.76E-17 | 7.70E-14 |
| Heart_Failure | dendritic_cell | *C14orf132* | -4.563088974 | 1.64E-14 | 4.54E-14 | 1.03E-10 |
| Heart_Failure | dendritic_cell | *ZNF717* | -0.406040066 | 0.011235766 | 0.012806247 | 1 |
| Heart_Failure | dendritic_cell | *AC009299.3* | -6.068283428 | 4.47E-07 | 6.31E-07 | 0.002811517 |
| Heart_Failure | dendritic_cell | *SLC2A1-AS1* | -3.921399021 | 1.54E-21 | 4.44E-19 | 9.70E-18 |
| Heart_Failure | dendritic_cell | *LINC01068* | -4.575712797 | 1.45E-10 | 2.56E-10 | 9.10E-07 |
| Heart_Failure | dendritic_cell | *AC108463.2* | 4.948624878 | 1.55E-17 | 8.29E-17 | 9.75E-14 |
| Heart_Failure | dendritic_cell | *HCG20* | -9.517419052 | 3.27E-18 | 2.10E-17 | 2.05E-14 |
| Heart_Failure | dendritic_cell | *C19orf24* | -3.815272687 | 1.06E-20 | 4.95E-19 | 6.69E-17 |
| Heart_Failure | dendritic_cell | *LINC01013* | 7.492257653 | 0.019739812 | 0.022260842 | 1 |
| Heart_Failure | dendritic_cell | *AC073133.2* | 23.12145667 | 2.25E-17 | 1.14E-16 | 1.42E-13 |
| Heart_Failure | dendritic_cell | *PROB1* | -8.527623688 | 4.46E-06 | 5.95E-06 | 0.028019744 |
| Heart_Failure | dendritic_cell | *ARL17B* | -3.546478834 | 7.99E-14 | 2.00E-13 | 5.02E-10 |
| Heart_Failure | dendritic_cell | *LINC00954* | 6.629440338 | 5.85E-20 | 9.53E-19 | 3.68E-16 |
| Heart_Failure | dendritic_cell | *AC067945.3* | -3.842978466 | 4.78E-07 | 6.73E-07 | 0.003006823 |
| Heart_Failure | dendritic_cell | *RPL41* | -5.165077367 | 0.00337232 | 0.003957031 | 1 |
| Heart_Failure | dendritic_cell | *PATL2* | -5.720795652 | 2.21E-06 | 3.00E-06 | 0.013886745 |
| Heart_Failure | dendritic_cell | *U91324.1* | -12.61321267 | 1.92E-19 | 2.13E-18 | 1.21E-15 |
| Heart_Failure | dendritic_cell | *ZNF688* | 7.920070263 | 6.79E-15 | 2.03E-14 | 4.27E-11 |
| Heart_Failure | dendritic_cell | *CISD3* | -6.791385028 | 0.000948254 | 0.001139246 | 1 |
| Heart_Failure | dendritic_cell | *PSMG3-AS1* | -2.582185898 | 1.53E-20 | 5.74E-19 | 9.59E-17 |
| Heart_Failure | dendritic_cell | *PPP5D1* | -4.303613381 | 5.36E-17 | 2.48E-16 | 3.37E-13 |
| Heart_Failure | dendritic_cell | *AC112229.4* | -2.001879583 | 9.03E-07 | 1.25E-06 | 0.00567659 |
| Heart_Failure | dendritic_cell | *KLHL7-AS1* | 14.7630787 | 0.004174902 | 0.004878738 | 1 |
| Heart_Failure | dendritic_cell | *AC092171.4* | -5.369317619 | 9.75E-18 | 5.52E-17 | 6.13E-14 |
| Heart_Failure | dendritic_cell | *HLA-DPA1* | 0.730226366 | 2.68E-21 | 4.66E-19 | 1.69E-17 |
| Heart_Failure | dendritic_cell | *RPS18* | -2.116024288 | 0.002302511 | 0.002723079 | 1 |
| Heart_Failure | dendritic_cell | *DIRC3* | -2.133379127 | 1.70E-18 | 1.22E-17 | 1.07E-14 |
| Heart_Failure | dendritic_cell | *AC107021.1* | -1.801862723 | 4.98E-18 | 3.04E-17 | 3.13E-14 |
| Heart_Failure | dendritic_cell | *LINC00865* | -5.263774529 | 5.91E-12 | 1.18E-11 | 3.71E-08 |
| Heart_Failure | dendritic_cell | *AC009495.3* | -4.363057168 | 2.37E-20 | 6.44E-19 | 1.49E-16 |
| Heart_Failure | dendritic_cell | *HLA-DQB2* | 0.506229521 | 3.56E-24 | 7.47E-21 | 2.24E-20 |
| Heart_Failure | dendritic_cell | *GOLGA8N* | 1.468917563 | 0.002022329 | 0.002398155 | 1 |
| Heart_Failure | dendritic_cell | *TNF* | 2.171958766 | 8.87E-21 | 4.70E-19 | 5.58E-17 |
| Heart_Failure | dendritic_cell | *LYRM9* | -4.611397747 | 6.03E-19 | 5.11E-18 | 3.79E-15 |
| Heart_Failure | dendritic_cell | *LINC00342* | -3.254157226 | 1.93E-19 | 2.13E-18 | 1.21E-15 |

| | | | | | | |
|---|---|---|---|---|---|---|
| Heart_Failure | dendritic_cell | *SNHG15* | 5.455820125 | 7.33E-05 | 9.27E-05 | 0.460943458 |
| Heart_Failure | dendritic_cell | *LINC00702* | -3.012525303 | 2.48E-20 | 6.44E-19 | 1.56E-16 |
| Heart_Failure | dendritic_cell | *LINC00472* | -3.71256506 | 4.63E-15 | 1.42E-14 | 2.91E-11 |
| Heart_Failure | dendritic_cell | *AC073133.1* | 2.103119891 | 1.39E-17 | 7.55E-17 | 8.76E-14 |
| Heart_Failure | dendritic_cell | *RPS28* | -6.45390598 | 1.35E-16 | 5.66E-16 | 8.49E-13 |
| Heart_Failure | dendritic_cell | *AC123768.3* | -3.623605974 | 1.09E-19 | 1.41E-18 | 6.85E-16 |
| Heart_Failure | dendritic_cell | *LINC01114* | -12.38111968 | 7.07E-05 | 8.95E-05 | 0.444356786 |
| Heart_Failure | dendritic_cell | *PINLYP* | 0.946253097 | 0.03563868 | 0.039628649 | 1 |
| Heart_Failure | dendritic_cell | *HLA-B* | -1.871985556 | 0.000161549 | 0.00020152 | 1 |
| Heart_Failure | dendritic_cell | *C19orf81* | 8.014242696 | 6.01E-14 | 1.53E-13 | 3.78E-10 |
| Heart_Failure | dendritic_cell | *ANKRD65* | -3.742055348 | 0.000475127 | 0.000579349 | 1 |
| Heart_Failure | dendritic_cell | *ZSCAN31* | -1.292652114 | 0.002623095 | 0.003091171 | 1 |
| Heart_Failure | dendritic_cell | *LINC00957* | 4.913235108 | 3.82E-18 | 2.40E-17 | 2.40E-14 |
| Heart_Failure | dendritic_cell | *LINC00484* | -1.048359671 | 7.79E-06 | 1.03E-05 | 0.04895084 |
| Heart_Failure | dendritic_cell | *KIAA0040* | -4.338909409 | 3.28E-15 | 1.03E-14 | 2.06E-11 |
| Heart_Failure | dendritic_cell | *B3GALT4* | -7.096801648 | 1.42E-09 | 2.33E-09 | 8.93E-06 |
| Heart_Failure | dendritic_cell | *LINC00941* | -16.4812355 | 0.00096609 | 0.001159788 | 1 |
| Heart_Failure | dendritic_cell | *LINC00545* | 12.14512445 | 3.00E-19 | 2.90E-18 | 1.88E-15 |
| Heart_Failure | dendritic_cell | *ZBTB22* | 4.255545442 | 1.64E-10 | 2.88E-10 | 1.03E-06 |
| Heart_Failure | dendritic_cell | *B3GNT9* | -7.220676509 | 9.97E-19 | 7.74E-18 | 6.27E-15 |
| Heart_Failure | dendritic_cell | *CDKN2AIPNL* | -2.513468324 | 8.30E-18 | 4.77E-17 | 5.22E-14 |
| Heart_Failure | dendritic_cell | *ZNF737* | -5.609889759 | 4.21E-14 | 1.10E-13 | 2.65E-10 |
| Heart_Failure | dendritic_cell | *HLA-DQA2* | 0.711620697 | 5.77E-24 | 9.07E-21 | 3.63E-20 |
| Heart_Failure | dendritic_cell | *AC073257.2* | -22.84637281 | 8.30E-14 | 2.07E-13 | 5.22E-10 |
| Heart_Failure | dendritic_cell | *IRGM* | -2.55113568 | 5.91E-10 | 9.96E-10 | 3.71E-06 |
| Heart_Failure | dendritic_cell | *LRRC37A2* | -4.02266521 | 2.40E-06 | 3.25E-06 | 0.015082787 |
| Heart_Failure | dendritic_cell | *TXNDC5* | -8.682056432 | 1.12E-19 | 1.43E-18 | 7.02E-16 |
| Heart_Failure | dendritic_cell | *ASB14* | -4.152166421 | 1.59E-09 | 2.61E-09 | 1.00E-05 |
| Heart_Failure | dendritic_cell | *NME1* | -1.515669746 | 8.44E-07 | 1.17E-06 | 0.005306127 |
| Heart_Failure | dendritic_cell | *CDRT4* | 7.279321161 | 3.28E-14 | 8.71E-14 | 2.06E-10 |
| Heart_Failure | dendritic_cell | *APOBEC3G* | -1.028556905 | 0.001032172 | 0.001236758 | 1 |
| Heart_Failure | dendritic_cell | *ADSL* | -3.267158269 | 4.23E-18 | 2.63E-17 | 2.66E-14 |
| Heart_Failure | dendritic_cell | *LILRA2* | 1.032457075 | 0.003003963 | 0.003532058 | 1 |
| Heart_Failure | dendritic_cell | *COX19* | -5.228009243 | 3.04E-06 | 4.09E-06 | 0.019116924 |
| Heart_Failure | dendritic_cell | *AQP1* | -2.634814228 | 2.82E-20 | 6.44E-19 | 1.77E-16 |
| Heart_Failure | dendritic_cell | *C1QTNF9* | -8.792743963 | 6.25E-11 | 1.14E-10 | 3.93E-07 |
| Heart_Failure | dendritic_cell | *ARHGEF25* | -6.850269727 | 5.60E-13 | 1.25E-12 | 3.52E-09 |
| Heart_Failure | dendritic_cell | *LINC00886* | -5.042586815 | 6.02E-12 | 1.21E-11 | 3.79E-08 |
| Heart_Failure | dendritic_cell | *NSUN6* | -3.856086604 | 3.54E-16 | 1.36E-15 | 2.22E-12 |
| Heart_Failure | dendritic_cell | *HLA-DOB* | 0.918424932 | 6.26E-18 | 3.71E-17 | 3.94E-14 |
| Heart_Failure | dendritic_cell | *TDGF1* | -3.288652635 | 5.77E-07 | 8.07E-07 | 0.003625911 |
| Heart_Failure | dendritic_cell | *ATP5J2* | 5.58194047 | 9.18E-07 | 1.27E-06 | 0.005769381 |
| Heart_Failure | dendritic_cell | *ARHGAP8* | -5.162150736 | 6.12E-20 | 9.72E-19 | 3.85E-16 |
| Heart_Failure | dendritic_cell | *ATP5O* | -4.780753135 | 3.01E-19 | 2.91E-18 | 1.89E-15 |
| Heart_Failure | dendritic_cell | *PLEKHO2* | 6.290921843 | 7.14E-09 | 1.11E-08 | 4.49E-05 |
| Heart_Failure | dendritic_cell | *PWP2* | 15.5302489 | 5.97E-09 | 9.35E-09 | 3.75E-05 |
| Heart_Failure | dendritic_cell | *PI4KA* | -4.175624868 | 1.01E-09 | 1.68E-09 | 6.36E-06 |
| Heart_Failure | dendritic_cell | *AKAP2* | 15.75398168 | 4.95E-20 | 8.51E-19 | 3.11E-16 |
| Heart_Failure | dendritic_cell | *LINC00969* | -2.240699709 | 5.25E-07 | 7.37E-07 | 0.003301826 |
| Heart_Failure | dendritic_cell | *AMACR* | -0.682987766 | 0.000988347 | 0.001185375 | 1 |

| | | | | | | |
|---|---|---|---|---|---|---|
| Heart_Failure | dendritic_cell | *ARHGDIG* | -4.232953979 | 8.97E-18 | 5.12E-17 | 5.64E-14 |
| Heart_Failure | dendritic_cell | *TCP10L* | 4.289254658 | 2.23E-21 | 4.66E-19 | 1.40E-17 |
| Heart_Failure | dendritic_cell | *ARFGAP3* | -4.975753655 | 7.84E-14 | 1.96E-13 | 4.93E-10 |
| Heart_Failure | dendritic_cell | *C22orf39* | -7.869875486 | 1.46E-07 | 2.10E-07 | 0.00091589 |
| Heart_Failure | dendritic_cell | *LINC00960* | -0.762052877 | 0.009798648 | 0.011208897 | 1 |
| Heart_Failure | dendritic_cell | *DECR2* | 11.32381338 | 3.77E-15 | 1.18E-14 | 2.37E-11 |
| Heart_Failure | dendritic_cell | *GNG10* | 1.047389134 | 0.000268226 | 0.000330525 | 1 |
| Heart_Failure | dendritic_cell | *CCDC169* | 2.237391378 | 1.52E-05 | 1.98E-05 | 0.095265141 |
| Heart_Failure | dendritic_cell | *AP5Z1* | -5.625774351 | 1.71E-13 | 4.08E-13 | 1.08E-09 |
| Heart_Failure | dendritic_cell | *ZNF709* | 2.269649643 | 0.021324557 | 0.023974873 | 1 |
| Heart_Failure | dendritic_cell | *STRC* | -4.653917698 | 0.032668902 | 0.036403649 | 1 |
| Heart_Failure | dendritic_cell | *PSG4* | -4.032663909 | 2.59E-15 | 8.32E-15 | 1.63E-11 |
| Heart_Failure | dendritic_cell | *STON1* | -2.47197906 | 2.98E-14 | 7.97E-14 | 1.88E-10 |
| Heart_Failure | dendritic_cell | *VSIG8* | 7.738195465 | 1.98E-14 | 5.44E-14 | 1.25E-10 |
| Heart_Failure | dendritic_cell | *C7orf73* | -4.54595814 | 1.69E-15 | 5.63E-15 | 1.06E-11 |
| Heart_Failure | dendritic_cell | *FGF14-IT1* | -8.17646862 | 3.09E-20 | 6.68E-19 | 1.94E-16 |
| Heart_Failure | dendritic_cell | *TICAM2* | 5.382865098 | 4.43E-20 | 8.01E-19 | 2.79E-16 |
| Heart_Failure | dendritic_cell | *PALM2* | -1.869163534 | 1.23E-20 | 5.08E-19 | 7.73E-17 |
| Heart_Failure | dendritic_cell | *NAT6* | 6.768763575 | 1.13E-08 | 1.74E-08 | 7.12E-05 |
| Heart_Failure | dendritic_cell | *UPK3B* | -2.705373499 | 8.54E-05 | 0.000107677 | 0.53677123 |
| Heart_Failure | dendritic_cell | *WDR92* | -3.658571398 | 8.23E-18 | 4.74E-17 | 5.18E-14 |
| Heart_Failure | dendritic_cell | *NME1-NME2* | -5.525919704 | 8.00E-09 | 1.24E-08 | 5.03E-05 |
| Heart_Failure | dendritic_cell | *PLA2G4B* | 2.802353463 | 7.49E-09 | 1.17E-08 | 4.71E-05 |
| Heart_Failure | dendritic_cell | *LEFTY1* | 18.34754173 | 1.89E-11 | 3.61E-11 | 1.19E-07 |
| Heart_Failure | dendritic_cell | *APOBEC3D* | -9.835860629 | 4.97E-11 | 9.12E-11 | 3.13E-07 |
| Heart_Failure | dendritic_cell | *RN7SL832P* | -3.142147711 | 5.63E-06 | 7.50E-06 | 0.035418123 |
| Heart_Failure | dendritic_cell | *LINC01011* | 4.844833877 | 2.60E-12 | 5.41E-12 | 1.63E-08 |
| Heart_Failure | dendritic_cell | *TMEM199* | 7.040899602 | 1.10E-13 | 2.68E-13 | 6.92E-10 |
| Heart_Failure | dendritic_cell | *P2RY11* | 5.232224157 | 6.64E-16 | 2.40E-15 | 4.18E-12 |
| Heart_Failure | dendritic_cell | *DBNDD2* | -3.168211812 | 0.001473523 | 0.001757216 | 1 |
| Heart_Failure | dendritic_cell | *LILRA6* | 2.201772716 | 1.04E-20 | 4.95E-19 | 6.55E-17 |
| Heart_Failure | dendritic_cell | *APOBEC3C* | 3.508032956 | 8.52E-21 | 4.66E-19 | 5.36E-17 |
| Heart_Failure | dendritic_cell | *CCDC13* | -2.212207497 | 4.10E-13 | 9.29E-13 | 2.58E-09 |
| Heart_Failure | dendritic_cell | *ASPRV1* | -9.197100195 | 2.52E-18 | 1.70E-17 | 1.59E-14 |
| Heart_Failure | dendritic_cell | *N4BP2L2* | -3.175944719 | 4.55E-14 | 1.17E-13 | 2.86E-10 |
| Heart_Failure | dendritic_cell | *LINC00847* | -8.934699733 | 1.74E-13 | 4.15E-13 | 1.10E-09 |
| Heart_Failure | dendritic_cell | *NEAT1* | -2.430632504 | 6.16E-16 | 2.25E-15 | 3.87E-12 |
| Heart_Failure | dendritic_cell | *ZNF585B* | -2.585154544 | 6.04E-10 | 1.02E-09 | 3.80E-06 |
| Heart_Failure | dendritic_cell | *RAB30-AS1* | -3.493517109 | 1.41E-11 | 2.72E-11 | 8.84E-08 |
| Heart_Failure | dendritic_cell | *UBAP1L* | -4.970307545 | 1.49E-10 | 2.62E-10 | 9.34E-07 |
| Heart_Failure | dendritic_cell | *PGAM5* | 9.719141022 | 3.85E-18 | 2.42E-17 | 2.42E-14 |
| Heart_Failure | dendritic_cell | *DNAJC3-AS1* | -4.110532304 | 3.92E-19 | 3.63E-18 | 2.46E-15 |
| Heart_Failure | dendritic_cell | *LINC00926* | -4.656245707 | 2.78E-19 | 2.77E-18 | 1.75E-15 |
| Heart_Failure | dendritic_cell | *BCKDHA* | -6.803683507 | 2.30E-15 | 7.46E-15 | 1.44E-11 |
| Heart_Failure | dendritic_cell | *CDK11B* | -3.68184692 | 0.003006104 | 0.003533914 | 1 |
| Heart_Failure | dendritic_cell | *PCDHAC1* | -5.877560521 | 0.000824875 | 0.000993675 | 1 |
| Heart_Failure | dendritic_cell | *ABHD14A* | -5.15311424 | 3.78E-16 | 1.44E-15 | 2.38E-12 |
| Heart_Failure | dendritic_cell | *SRP14-AS1* | -0.790707822 | 0.000677388 | 0.000819303 | 1 |
| Heart_Failure | dendritic_cell | *CCDC153* | 3.091918573 | 8.90E-16 | 3.14E-15 | 5.60E-12 |
| Heart_Failure | dendritic_cell | *C5orf17* | -4.492822114 | 5.96E-18 | 3.57E-17 | 3.75E-14 |

| | | | | | | |
|---|---|---|---|---|---|---|
| Heart_Failure | dendritic_cell | *FMN1* | 1.708390489 | 2.35E-18 | 1.62E-17 | 1.48E-14 |
| Heart_Failure | dendritic_cell | *LINC01060* | -2.738360157 | 2.06E-12 | 4.33E-12 | 1.29E-08 |
| Heart_Failure | dendritic_cell | *ZNF286B* | -4.296057245 | 1.54E-20 | 5.77E-19 | 9.69E-17 |
| Heart_Failure | dendritic_cell | *LINC00942* | -5.937691657 | 7.25E-07 | 1.01E-06 | 0.004560353 |
| Heart_Failure | dendritic_cell | *PDCD6* | -5.441178229 | 9.38E-11 | 1.68E-10 | 5.89E-07 |
| Heart_Failure | dendritic_cell | *LINC00603* | -3.47684768 | 6.50E-18 | 3.85E-17 | 4.09E-14 |
| Heart_Failure | dendritic_cell | *YJEFN3* | -5.18517227 | 6.10E-06 | 8.10E-06 | 0.038373223 |
| Heart_Failure | dendritic_cell | *DNAH10OS* | -20.62503633 | 3.35E-21 | 4.66E-19 | 2.11E-17 |
| Heart_Failure | dendritic_cell | *PCDHA10* | 10.34691839 | 2.27E-08 | 3.43E-08 | 0.000142784 |
| Heart_Failure | dendritic_cell | *LINC00989* | -4.448799696 | 3.66E-19 | 3.42E-18 | 2.30E-15 |
| Heart_Failure | dendritic_cell | *LINC01021* | -9.006669493 | 1.03E-19 | 1.36E-18 | 6.45E-16 |
| Heart_Failure | dendritic_cell | *FAM218A* | -8.706084421 | 1.17E-19 | 1.48E-18 | 7.37E-16 |
| Heart_Failure | dendritic_cell | *ATP6V1E2* | -1.70011364 | 5.65E-13 | 1.26E-12 | 3.55E-09 |
| Heart_Failure | dendritic_cell | *SNHG18* | -3.078248103 | 1.80E-20 | 6.29E-19 | 1.13E-16 |
| Heart_Failure | dendritic_cell | *ZNF345* | -3.632644849 | 1.29E-17 | 7.08E-17 | 8.11E-14 |
| Heart_Failure | dendritic_cell | *SHANK3* | -2.167431971 | 9.67E-15 | 2.81E-14 | 6.08E-11 |
| Heart_Failure | dendritic_cell | *ZNF550* | -3.785297085 | 1.31E-16 | 5.51E-16 | 8.25E-13 |
| Heart_Failure | dendritic_cell | *CCDC71L* | -4.931558705 | 8.19E-17 | 3.66E-16 | 5.15E-13 |
| Heart_Failure | dendritic_cell | *TMEM200B* | -2.126886828 | 1.76E-05 | 2.29E-05 | 0.110719635 |
| Heart_Failure | dendritic_cell | *PCDHGB6* | -3.348804288 | 1.54E-06 | 2.11E-06 | 0.009707363 |
| Heart_Failure | dendritic_cell | *TRNP1* | -13.3049521 | 2.44E-20 | 6.44E-19 | 1.53E-16 |
| Heart_Failure | dendritic_cell | *PCDHGA5* | -4.385676145 | 2.11E-17 | 1.08E-16 | 1.33E-13 |
| Heart_Failure | dendritic_cell | *ATXN7L3B* | -6.455679506 | 1.26E-11 | 2.45E-11 | 7.93E-08 |
| Heart_Failure | dendritic_cell | *PRKDC* | -4.279891595 | 2.58E-08 | 3.88E-08 | 0.000162139 |
| Heart_Failure | dendritic_cell | *PCDHGA6* | -2.630206584 | 2.62E-11 | 4.93E-11 | 1.65E-07 |
| Heart_Failure | dendritic_cell | *PCDHGA11* | -1.734631348 | 0.016220796 | 0.018348353 | 1 |
| Heart_Failure | dendritic_cell | *CLDN23* | 5.591119782 | 8.93E-21 | 4.70E-19 | 5.61E-17 |
| Heart_Failure | dendritic_cell | *ZNF260* | -4.722828867 | 4.30E-05 | 5.50E-05 | 0.270609961 |
| Heart_Failure | dendritic_cell | *SIGLEC14* | 0.904046051 | 0.021534729 | 0.024198185 | 1 |
| Heart_Failure | dendritic_cell | *ARMS2* | -8.658498113 | 1.24E-16 | 5.25E-16 | 7.81E-13 |
| Heart_Failure | dendritic_cell | *FPGT* | -7.582142732 | 6.66E-08 | 9.82E-08 | 0.00041847 |
| Heart_Failure | dendritic_cell | *DPP3* | 1.7418575 | 0.000654567 | 0.000792158 | 1 |
| Heart_Failure | dendritic_cell | *KRTAP5-9* | -5.774570521 | 0.027343965 | 0.03056748 | 1 |
| Heart_Failure | dendritic_cell | *EID3* | -2.264627067 | 8.78E-10 | 1.46E-09 | 5.52E-06 |
| Heart_Failure | dendritic_cell | *SNHG9* | -2.164901281 | 1.89E-12 | 3.99E-12 | 1.19E-08 |
| Heart_Failure | dendritic_cell | *TAS2R43* | 1.206896071 | 6.39E-06 | 8.48E-06 | 0.040166079 |
| Heart_Failure | dendritic_cell | *EBLN2* | -4.85325859 | 2.17E-19 | 2.30E-18 | 1.37E-15 |
| Heart_Failure | dendritic_cell | *POLR2M* | 8.705187108 | 7.50E-12 | 1.48E-11 | 4.71E-08 |
| Heart_Failure | dendritic_cell | *TAS2R20* | 2.657458877 | 3.38E-10 | 5.81E-10 | 2.13E-06 |
| Heart_Failure | dendritic_cell | *LINC00346* | -5.41027247 | 6.34E-11 | 1.15E-10 | 3.99E-07 |
| Heart_Failure | dendritic_cell | *CTSO* | -5.793388398 | 3.08E-07 | 4.38E-07 | 0.001936788 |
| Heart_Failure | dendritic_cell | *ZNF432* | -2.998240041 | 0.005638455 | 0.006533167 | 1 |
| Heart_Failure | dendritic_cell | *LINC00944* | -3.865816119 | 1.55E-17 | 8.29E-17 | 9.73E-14 |
| Heart_Failure | dendritic_cell | *ZNF10* | 3.21265029 | 5.48E-07 | 7.67E-07 | 0.003444062 |
| Heart_Failure | dendritic_cell | *HMBS* | -1.863051179 | 6.84E-06 | 9.06E-06 | 0.043014911 |
| Heart_Failure | dendritic_cell | *TAS2R31* | 2.58591575 | 9.59E-05 | 0.000120926 | 0.603178048 |
| Heart_Failure | dendritic_cell | *AC093668.2* | -4.127284129 | 2.19E-14 | 5.98E-14 | 1.38E-10 |
| Heart_Failure | dendritic_cell | *CLEC12B* | 0.746871073 | 8.77E-08 | 1.28E-07 | 0.000551489 |
| Heart_Failure | dendritic_cell | *PGA5* | -33.64842893 | 3.56E-14 | 9.37E-14 | 2.24E-10 |
| Heart_Failure | dendritic_cell | *ZNF253* | -5.483725794 | 6.51E-10 | 1.09E-09 | 4.09E-06 |

| | | | | | | |
|---|---|---|---|---|---|---|
| Heart_Failure | dendritic_cell | *HP* | -6.387755535 | 4.39E-11 | 8.09E-11 | 2.76E-07 |
| Heart_Failure | dendritic_cell | *KIAA1147* | -4.068456617 | 2.04E-14 | 5.58E-14 | 1.28E-10 |
| Heart_Failure | dendritic_cell | *GATC* | -3.930345955 | 0.000123838 | 0.000155187 | 0.778571552 |
| Heart_Failure | dendritic_cell | *ZBED6* | -6.469534867 | 2.62E-20 | 6.44E-19 | 1.65E-16 |
| Heart_Failure | dendritic_cell | *FNTB* | 9.247805059 | 0.000106979 | 0.000134515 | 0.672575732 |
| Heart_Failure | dendritic_cell | *GALNT4* | -10.99250768 | 0.001611873 | 0.001919652 | 1 |
| Heart_Failure | dendritic_cell | *OVCH1-AS1* | 5.451663229 | 5.99E-18 | 3.58E-17 | 3.77E-14 |
| Heart_Failure | dendritic_cell | *CUX1* | -2.548888966 | 0.000472763 | 0.00057669 | 1 |
| Heart_Failure | dendritic_cell | *TEN1* | -3.342748669 | 0.004543685 | 0.005295911 | 1 |
| Heart_Failure | dendritic_cell | *CHURC1* | -1.77748072 | 0.001147006 | 0.001370955 | 1 |
| Heart_Failure | dendritic_cell | *C17orf49* | -6.259771051 | 3.10E-08 | 4.64E-08 | 0.000194635 |
| Heart_Failure | dendritic_cell | *RTEL1* | -4.437932651 | 1.35E-11 | 2.63E-11 | 8.52E-08 |
| Heart_Failure | dendritic_cell | *ZNF578* | -3.011476825 | 1.32E-07 | 1.91E-07 | 0.000828477 |
| Heart_Failure | dendritic_cell | *LINC00640* | -1.976280868 | 7.23E-12 | 1.43E-11 | 4.55E-08 |
| Heart_Failure | dendritic_cell | *LINC00638* | -11.47972411 | 3.37E-21 | 4.66E-19 | 2.12E-17 |
| Heart_Failure | dendritic_cell | *DUXA* | -1.892141415 | 0.001990769 | 0.002361503 | 1 |
| Heart_Failure | dendritic_cell | *CEP95* | -4.015763169 | 1.26E-14 | 3.58E-14 | 7.93E-11 |
| Heart_Failure | dendritic_cell | *LINC00639* | -1.501715884 | 3.44E-18 | 2.21E-17 | 2.16E-14 |
| Heart_Failure | dendritic_cell | *LINC00648* | -2.242268605 | 1.95E-20 | 6.44E-19 | 1.23E-16 |
| Heart_Failure | dendritic_cell | *LINC00924* | -7.959935468 | 1.02E-15 | 3.56E-15 | 6.41E-12 |
| Heart_Failure | dendritic_cell | *ITGB3* | -2.546878798 | 2.38E-11 | 4.50E-11 | 1.50E-07 |
| Heart_Failure | dendritic_cell | *UBE2Q2L* | -7.869678341 | 9.35E-19 | 7.37E-18 | 5.88E-15 |
| Heart_Failure | dendritic_cell | *IQCH-AS1* | -2.303126985 | 4.34E-06 | 5.80E-06 | 0.027272012 |
| Heart_Failure | dendritic_cell | *AC083843.1* | 17.53568494 | 1.98E-19 | 2.15E-18 | 1.24E-15 |
| Heart_Failure | dendritic_cell | *SH3RF3-AS1* | 2.850490989 | 9.83E-09 | 1.52E-08 | 6.18E-05 |
| Heart_Failure | dendritic_cell | *PWRN1* | -3.803316439 | 1.81E-05 | 2.35E-05 | 0.11358527 |
| Heart_Failure | dendritic_cell | *HOXB7* | -6.360632523 | 3.95E-13 | 8.98E-13 | 2.48E-09 |
| Heart_Failure | dendritic_cell | *FRRS1L* | -6.849016279 | 3.39E-19 | 3.21E-18 | 2.13E-15 |
| Heart_Failure | dendritic_cell | *LINC00562* | -2.178332481 | 1.81E-16 | 7.37E-16 | 1.14E-12 |
| Heart_Failure | dendritic_cell | *AC009120.5* | -5.072154235 | 6.10E-20 | 9.71E-19 | 3.83E-16 |
| Heart_Failure | dendritic_cell | *TMEM178B* | -6.170037253 | 6.72E-13 | 1.49E-12 | 4.23E-09 |
| Heart_Failure | dendritic_cell | *LINC01003* | -3.335305812 | 1.02E-11 | 1.99E-11 | 6.41E-08 |
| Heart_Failure | dendritic_cell | *C15orf65* | -1.882808692 | 0.007066618 | 0.008157883 | 1 |
| Heart_Failure | dendritic_cell | *HPR* | 6.714114079 | 2.71E-17 | 1.35E-16 | 1.71E-13 |
| Heart_Failure | dendritic_cell | *LINC00662* | -5.142112482 | 4.47E-11 | 8.23E-11 | 2.81E-07 |
| Heart_Failure | dendritic_cell | *LINC00667* | -2.696984868 | 6.69E-14 | 1.69E-13 | 4.21E-10 |
| Heart_Failure | dendritic_cell | *LINC00672* | 3.53431895 | 3.94E-14 | 1.03E-13 | 2.48E-10 |
| Heart_Failure | dendritic_cell | *LINC00909* | -3.273236124 | 9.30E-18 | 5.29E-17 | 5.85E-14 |
| Heart_Failure | dendritic_cell | *DYNLL2* | -3.241043763 | 4.27E-20 | 7.85E-19 | 2.68E-16 |
| Heart_Failure | dendritic_cell | *LINC00526* | 5.962192368 | 1.23E-08 | 1.89E-08 | 7.73E-05 |
| Heart_Failure | dendritic_cell | *STRADA* | -5.004477141 | 8.77E-10 | 1.46E-09 | 5.51E-06 |
| Heart_Failure | dendritic_cell | *MYO15B* | -2.062429524 | 0.026497306 | 0.029652646 | 1 |
| Heart_Failure | dendritic_cell | *LINC00663* | -0.733870816 | 0.000139317 | 0.000174167 | 0.875884356 |
| Heart_Failure | dendritic_cell | *ILF3-AS1* | -4.156646394 | 7.19E-12 | 1.43E-11 | 4.52E-08 |
| Heart_Failure | dendritic_cell | *AC139100.2* | 2.947461043 | 6.78E-21 | 4.66E-19 | 4.26E-17 |
| Heart_Failure | dendritic_cell | *LINC01140* | -3.638306848 | 6.03E-17 | 2.76E-16 | 3.79E-13 |
| Heart_Failure | dendritic_cell | *LINC00906* | -22.51151742 | 2.80E-20 | 6.44E-19 | 1.76E-16 |
| Heart_Failure | dendritic_cell | *ZNF582-AS1* | -4.470943141 | 9.46E-18 | 5.38E-17 | 5.95E-14 |
| Heart_Failure | dendritic_cell | *AC137932.1* | -10.3479908 | 1.65E-15 | 5.50E-15 | 1.04E-11 |
| Heart_Failure | dendritic_cell | *AC003005.2* | -4.894470883 | 2.53E-11 | 4.78E-11 | 1.59E-07 |

| | | | | | | |
|---|---|---|---|---|---|---|
| Heart_Failure | dendritic_cell | *AL132989.1* | 13.4074 | 1.67E-19 | 1.93E-18 | 1.05E-15 |
| Heart_Failure | dendritic_cell | *LINC00664* | -8.531893025 | 2.34E-13 | 5.48E-13 | 1.47E-09 |
| Heart_Failure | dendritic_cell | *AC026740.1* | -5.605125355 | 0.020591661 | 0.023179906 | 1 |
| Heart_Failure | dendritic_cell | *FBXO17* | -2.543767634 | 1.90E-19 | 2.11E-18 | 1.19E-15 |
| Heart_Failure | dendritic_cell | *AC009060.1* | -6.629502598 | 1.80E-19 | 2.05E-18 | 1.13E-15 |
| Heart_Failure | dendritic_cell | *SNHG8* | -3.605179181 | 9.45E-16 | 3.32E-15 | 5.94E-12 |
| Heart_Failure | dendritic_cell | *HCG17* | -3.311054113 | 1.45E-16 | 6.05E-16 | 9.13E-13 |
| Heart_Failure | dendritic_cell | *YTHDF3-AS1* | -6.043344969 | 1.14E-16 | 4.87E-16 | 7.19E-13 |
| Heart_Failure | dendritic_cell | *LINC01050* | -24.98044396 | 2.75E-20 | 6.44E-19 | 1.73E-16 |
| Heart_Failure | dendritic_cell | *MILR1* | 1.442494075 | 5.15E-17 | 2.40E-16 | 3.24E-13 |
| Heart_Failure | dendritic_cell | *GTF2H5* | -2.226394347 | 0.002281994 | 0.002699322 | 1 |
| Heart_Failure | dendritic_cell | *FGF14-AS2* | -3.982226701 | 3.96E-19 | 3.66E-18 | 2.49E-15 |
| Heart_Failure | dendritic_cell | *CASC15* | 2.577544243 | 0.001779791 | 0.002114827 | 1 |
| Heart_Failure | dendritic_cell | *LINC01023* | -5.267753107 | 2.32E-18 | 1.60E-17 | 1.46E-14 |
| Heart_Failure | dendritic_cell | *DOC2B* | -1.484144355 | 3.04E-09 | 4.86E-09 | 1.91E-05 |
| Heart_Failure | dendritic_cell | *POLR2J4* | 4.202944815 | 0.024952383 | 0.027933695 | 1 |
| Heart_Failure | dendritic_cell | *AC144652.1* | 2.309014021 | 3.82E-12 | 7.80E-12 | 2.40E-08 |
| Heart_Failure | dendritic_cell | *AP000350.5* | 0.525462573 | 0.036182929 | 0.040212493 | 1 |
| Heart_Failure | dendritic_cell | *PAXIP1-AS1* | -3.844255814 | 4.18E-12 | 8.50E-12 | 2.62E-08 |
| Heart_Failure | endothelial | *RBM6* | -0.217488279 | 1.75E-05 | 0.01374068 | 0.109925439 |
| Heart_Failure | endothelial | *PDLIM4* | 0.858531588 | 2.13E-05 | 0.014911324 | 0.134201913 |
| Heart_Failure | endothelial | *REEP2* | -0.355182254 | 6.02E-05 | 0.029109193 | 0.378419515 |
| Heart_Failure | endothelial | *TSLP* | 0.486539864 | 1.65E-05 | 0.01374068 | 0.103549065 |
| Heart_Failure | endothelial | *RNF123* | 0.395591673 | 3.15E-05 | 0.019785527 | 0.197855267 |
| Heart_Failure | endothelial | *ORMDL3* | -0.30862498 | 3.82E-06 | 0.003999386 | 0.023996316 |
| Heart_Failure | endothelial | *HLA-DQB1* | -0.52573336 | 1.30E-21 | 8.17E-18 | 8.17E-18 |
| Heart_Failure | endothelial | *MUC1* | 0.537216462 | 5.16E-05 | 0.027047447 | 0.324569366 |
| Heart_Failure | endothelial | *HLA-DRB1* | -0.241096218 | 7.74E-05 | 0.03475452 | 0.486563283 |
| Heart_Failure | endothelial | *HLA-DQA1* | -0.622047082 | 1.86E-14 | 2.93E-11 | 1.17E-10 |
| Heart_Failure | endothelial | *SLC22A5* | 0.341330693 | 2.43E-06 | 0.003057721 | 0.015288603 |
| Heart_Failure | endothelial | *MICA* | 0.250568616 | 3.78E-05 | 0.021589793 | 0.237487722 |
| Heart_Failure | endothelial | *HLA-DQB2* | 0.297248918 | 6.56E-16 | 1.37E-12 | 4.12E-12 |
| Heart_Failure | endothelial | *HLA-DQA2* | 0.57032885 | 4.15E-21 | 1.31E-17 | 2.61E-17 |
| Heart_Failure | LEC | *SEMA3F* | -0.98248638 | 1.00E-06 | 0.001048136 | 0.006288814 |
| Heart_Failure | LEC | *RBM6* | -0.626143541 | 1.84E-06 | 0.001651182 | 0.011558275 |
| Heart_Failure | LEC | *ORMDL3* | -0.751822081 | 3.52E-07 | 0.000442952 | 0.002214759 |
| Heart_Failure | LEC | *HLA-DQB1* | -1.519276282 | 4.93E-23 | 3.10E-19 | 3.10E-19 |
| Heart_Failure | LEC | *HLA-DRB1* | -0.624936705 | 3.43E-05 | 0.026990517 | 0.215924139 |
| Heart_Failure | LEC | *HLA-DQA1* | -1.235109333 | 6.02E-12 | 9.47E-09 | 3.79E-08 |
| Heart_Failure | LEC | *MICA* | 0.684943889 | 3.87E-05 | 0.027034347 | 0.243309119 |
| Heart_Failure | LEC | *HLA-DQB2* | 0.708654176 | 6.77E-13 | 1.42E-09 | 4.26E-09 |
| Heart_Failure | LEC | *HLA-DQA2* | 1.093115043 | 1.16E-13 | 3.66E-10 | 7.32E-10 |
| Heart_Failure | macrophage | *SEMA3F* | -0.239714515 | 2.87E-06 | 0.002001609 | 0.018014478 |
| Heart_Failure | macrophage | *RBM6* | -0.190618105 | 4.77E-07 | 0.000374512 | 0.002996096 |
| Heart_Failure | macrophage | *PDLIM4* | 0.678020952 | 3.47E-06 | 0.002183893 | 0.021838932 |
| Heart_Failure | macrophage | *REEP2* | -0.231028118 | 8.35E-05 | 0.027625533 | 0.524885136 |
| Heart_Failure | macrophage | *TSLP* | 0.352330418 | 4.89E-06 | 0.00239386 | 0.030736774 |
| Heart_Failure | macrophage | *RNF123* | 0.325494442 | 4.95E-06 | 0.00239386 | 0.031120176 |
| Heart_Failure | macrophage | *ORMDL3* | -0.379603899 | 4.85E-11 | 6.10E-08 | 3.05E-07 |
| Heart_Failure | macrophage | *DCAKD* | -0.212705269 | 0.00014193 | 0.042491098 | 0.892313053 |

| | | | | | | |
|---|---|---|---|---|---|---|
| Heart_Failure | macrophage | *GMPPB* | 0.566191312 | 0.00016967 | 0.048487103 | 1 |
| Heart_Failure | macrophage | *HLA-DQB1* | -0.356883013 | 1.36E-21 | 4.27E-18 | 8.53E-18 |
| Heart_Failure | macrophage | *MUC1* | 0.390045423 | 3.16E-05 | 0.011690543 | 0.198739225 |
| Heart_Failure | macrophage | *MCRS1* | 0.714839072 | 6.95E-05 | 0.024258526 | 0.436653474 |
| Heart_Failure | macrophage | *HLA-DRB1* | -0.35506206 | 7.40E-09 | 7.76E-06 | 4.65E-05 |
| Heart_Failure | macrophage | *HLA-DQA1* | -0.49584453 | 2.59E-17 | 5.42E-14 | 1.63E-13 |
| Heart_Failure | macrophage | *SLC22A5* | 0.307522015 | 7.76E-08 | 6.97E-05 | 0.000487597 |
| Heart_Failure | macrophage | *RPS26* | 0.150364642 | 0.000120422 | 0.037854529 | 0.75709058 |
| Heart_Failure | macrophage | *HLA-DRA* | 0.321801168 | 1.09E-05 | 0.004289619 | 0.068633909 |
| Heart_Failure | macrophage | *BAG6* | 0.511312512 | 3.93E-06 | 0.002248112 | 0.024729232 |
| Heart_Failure | macrophage | *MICA* | 0.186039598 | 1.02E-05 | 0.004289619 | 0.064408055 |
| Heart_Failure | macrophage | *ATF6B* | 0.434680369 | 6.40E-06 | 0.002873394 | 0.040227517 |
| Heart_Failure | macrophage | *HLA-DQB2* | 0.189327724 | 1.40E-14 | 2.20E-11 | 8.80E-11 |
| Heart_Failure | macrophage | *HLA-DQA2* | 0.418716875 | 2.41E-22 | 1.51E-18 | 1.51E-18 |
| Heart_Failure | mast_cell | *RBM6* | -1.014671738 | 2.15E-05 | 0.01184547 | 0.134974081 |
| Heart_Failure | mast_cell | *TIMM44* | 3.438747024 | 0.000156492 | 0.042776618 | 0.983862205 |
| Heart_Failure | mast_cell | *HEMK1* | 2.804599611 | 0.000113766 | 0.035762478 | 0.715249561 |
| Heart_Failure | mast_cell | *CHMP1A* | 3.559053467 | 6.95E-05 | 0.025693544 | 0.436790244 |
| Heart_Failure | mast_cell | *NMT1* | 1.581050608 | 0.000122375 | 0.036636703 | 0.769370764 |
| Heart_Failure | mast_cell | *PPOX* | 3.313223288 | 0.000184063 | 0.046288093 | 1 |
| Heart_Failure | mast_cell | *TSLP* | 1.443745802 | 0.000171465 | 0.044916588 | 1 |
| Heart_Failure | mast_cell | *RNF123* | 1.758671494 | 1.45E-05 | 0.009110689 | 0.09110689 |
| Heart_Failure | mast_cell | *C2* | 3.912947681 | 4.15E-05 | 0.018628481 | 0.260798738 |
| Heart_Failure | mast_cell | *ORMDL3* | -2.284315534 | 6.98E-10 | 8.78E-07 | 4.39E-06 |
| Heart_Failure | mast_cell | *HLA-DQB1* | -2.079947014 | 5.61E-18 | 1.76E-14 | 3.53E-14 |
| Heart_Failure | mast_cell | *MUC1* | 2.492417557 | 2.26E-05 | 0.01184547 | 0.14214564 |
| Heart_Failure | mast_cell | *MCRS1* | 3.122832234 | 0.000135948 | 0.038850249 | 0.854705479 |
| Heart_Failure | mast_cell | *HLA-DRB1* | -1.774306354 | 6.42E-06 | 0.004481518 | 0.040333658 |
| Heart_Failure | mast_cell | *HLA-DQA1* | -2.23323339 | 4.56E-11 | 9.56E-08 | 2.87E-07 |
| Heart_Failure | mast_cell | *SLC22A5* | 1.697446324 | 4.37E-07 | 0.00039268 | 0.00274876 |
| Heart_Failure | mast_cell | *RPS26* | 1.026834001 | 4.96E-05 | 0.019667119 | 0.312073994 |
| Heart_Failure | mast_cell | *HLA-DRA* | 2.30393531 | 1.52E-07 | 0.000159384 | 0.000956305 |
| Heart_Failure | mast_cell | *PRRT1* | 2.264625743 | 9.36E-05 | 0.030977205 | 0.588566896 |
| Heart_Failure | mast_cell | *BAG6* | 2.21855221 | 5.01E-05 | 0.019667119 | 0.314673906 |
| Heart_Failure | mast_cell | *MICA* | 1.210823116 | 6.03E-06 | 0.004481518 | 0.037900259 |
| Heart_Failure | mast_cell | *ATF6B* | 2.069875256 | 3.01E-05 | 0.014567947 | 0.189383316 |
| Heart_Failure | mast_cell | *HLA-DQB2* | 0.843374801 | 4.13E-10 | 6.50E-07 | 2.60E-06 |
| Heart_Failure | mast_cell | *ZBTB22* | 2.541040427 | 7.72E-05 | 0.026979273 | 0.485626913 |
| Heart_Failure | mast_cell | *HLA-DQA2* | 2.81317297 | 2.10E-23 | 1.32E-19 | 1.32E-19 |
| Heart_Failure | monocyte | *MAD1L1* | 2.267709163 | 0.006180439 | 0.039608989 | 1 |
| Heart_Failure | monocyte | *LASP1* | 1.854986458 | 0.008809304 | 0.04936194 | 1 |
| Heart_Failure | monocyte | *RBM6* | -0.854950594 | 0.001294678 | 0.017046605 | 1 |
| Heart_Failure | monocyte | *CDC27* | 2.183204819 | 0.000808906 | 0.01444259 | 1 |
| Heart_Failure | monocyte | *ABCB4* | 6.667140705 | 0.00028202 | 0.011389066 | 1 |
| Heart_Failure | monocyte | *ZNF263* | 3.709771009 | 0.000749086 | 0.014314608 | 1 |
| Heart_Failure | monocyte | *VPS41* | 1.95860474 | 0.003468489 | 0.028730426 | 1 |
| Heart_Failure | monocyte | *PIGQ* | 2.519726917 | 0.002861649 | 0.025775336 | 1 |
| Heart_Failure | monocyte | *RHOBTB2* | 3.552086497 | 0.005025025 | 0.035417411 | 1 |
| Heart_Failure | monocyte | *SEC62* | 1.722464024 | 0.008160548 | 0.047530474 | 1 |
| Heart_Failure | monocyte | *RPS20* | 2.064302237 | 0.00833986 | 0.048191822 | 1 |

| | | | | | | |
|---|---|---|---|---|---|---|
| Heart_Failure | monocyte | *SPRTN* | 3.834834117 | 0.00146795 | 0.017885666 | 1 |
| Heart_Failure | monocyte | *UTP18* | 4.685542064 | 0.00027662 | 0.011389066 | 1 |
| Heart_Failure | monocyte | *LARS2* | 1.993086025 | 0.005603635 | 0.037565605 | 1 |
| Heart_Failure | monocyte | *MAN2B2* | 3.453209637 | 0.00136175 | 0.017365766 | 1 |
| Heart_Failure | monocyte | *ANGEL1* | 4.676053281 | 0.000354875 | 0.011389066 | 1 |
| Heart_Failure | monocyte | *RNF14* | 1.651683612 | 0.008588102 | 0.048922662 | 1 |
| Heart_Failure | monocyte | *ZC3H3* | 3.400649895 | 8.63E-05 | 0.011389066 | 0.542682961 |
| Heart_Failure | monocyte | *SLC30A9* | 1.886068167 | 0.006470911 | 0.040520538 | 1 |
| Heart_Failure | monocyte | *RUFY3* | 1.466482066 | 0.00652607 | 0.040809462 | 1 |
| Heart_Failure | monocyte | *SLC7A9* | 1.373622925 | 0.001418973 | 0.017630601 | 1 |
| Heart_Failure | monocyte | *OSBPL5* | 4.089908212 | 0.000274295 | 0.011389066 | 1 |
| Heart_Failure | monocyte | *AQR* | 2.507648782 | 0.001346811 | 0.01728185 | 1 |
| Heart_Failure | monocyte | *ZDHHC6* | 3.603059718 | 0.000538031 | 0.012528161 | 1 |
| Heart_Failure | monocyte | *NDUFS1* | 2.298126877 | 0.001674251 | 0.01917307 | 1 |
| Heart_Failure | monocyte | *AKAP11* | 1.878317972 | 0.006829981 | 0.041933684 | 1 |
| Heart_Failure | monocyte | *SNAPC1* | 3.743329035 | 0.001320409 | 0.017175592 | 1 |
| Heart_Failure | monocyte | *UBR2* | 2.373234292 | 0.000828084 | 0.014464609 | 1 |
| Heart_Failure | monocyte | *VIM* | 1.337593429 | 0.006850379 | 0.041937353 | 1 |
| Heart_Failure | monocyte | *FAS* | 2.198442115 | 0.001029689 | 0.015840489 | 1 |
| Heart_Failure | monocyte | *B4GALT7* | 3.343732077 | 0.008469432 | 0.04876128 | 1 |
| Heart_Failure | monocyte | *VRK2* | 2.830146942 | 0.000623432 | 0.013459015 | 1 |
| Heart_Failure | monocyte | *GRN* | 3.610202809 | 0.000700888 | 0.014081785 | 1 |
| Heart_Failure | monocyte | *ARID4A* | 3.274348437 | 0.000318703 | 0.011389066 | 1 |
| Heart_Failure | monocyte | *PIAS1* | 2.733077698 | 0.00043274 | 0.011479484 | 1 |
| Heart_Failure | monocyte | *SLC4A7* | 1.986081191 | 0.003204378 | 0.027825865 | 1 |
| Heart_Failure | monocyte | *ADSS* | 3.975361718 | 0.000397008 | 0.011389066 | 1 |
| Heart_Failure | monocyte | *TBC1D23* | 2.119049461 | 0.004542849 | 0.033430274 | 1 |
| Heart_Failure | monocyte | *CUL3* | 2.457378102 | 0.001300394 | 0.017068006 | 1 |
| Heart_Failure | monocyte | *BOD1L1* | 2.694944083 | 0.000649813 | 0.013585129 | 1 |
| Heart_Failure | monocyte | *CLEC16A* | 3.23964101 | 0.000609703 | 0.013402803 | 1 |
| Heart_Failure | monocyte | *PHLPP2* | 4.248322122 | 3.33E-06 | 0.002961882 | 0.020950971 |
| Heart_Failure | monocyte | *PSMA4* | 2.805425887 | 0.001537006 | 0.018371025 | 1 |
| Heart_Failure | monocyte | *TDP1* | 3.470512251 | 0.001920406 | 0.020611606 | 1 |
| Heart_Failure | monocyte | *TRIT1* | 1.765505584 | 0.008922025 | 0.049581438 | 1 |
| Heart_Failure | monocyte | *CNTLN* | 1.788464532 | 0.004646576 | 0.033929176 | 1 |
| Heart_Failure | monocyte | *WDR37* | 2.214232848 | 0.002506372 | 0.023895639 | 1 |
| Heart_Failure | monocyte | *YTHDC2* | 2.33560135 | 0.001655012 | 0.019022043 | 1 |
| Heart_Failure | monocyte | *POLR2B* | 2.015729035 | 0.00509031 | 0.035460966 | 1 |
| Heart_Failure | monocyte | *DTNBP1* | 2.37763371 | 0.004504908 | 0.033359668 | 1 |
| Heart_Failure | monocyte | *GOPC* | 2.882005385 | 0.000933159 | 0.015111629 | 1 |
| Heart_Failure | monocyte | *RRM2B* | 3.218865784 | 0.001626859 | 0.018937074 | 1 |
| Heart_Failure | monocyte | *SNX29* | 2.008410749 | 0.003283519 | 0.028091022 | 1 |
| Heart_Failure | monocyte | *CELF2* | 1.477663999 | 0.003628665 | 0.029323162 | 1 |
| Heart_Failure | monocyte | *ERCC8* | 1.779819928 | 0.008926241 | 0.049581438 | 1 |
| Heart_Failure | monocyte | *H6PD* | 2.030145223 | 0.000766689 | 0.014388582 | 1 |
| Heart_Failure | monocyte | *NFE2L3* | 4.289563916 | 0.002662672 | 0.024617972 | 1 |
| Heart_Failure | monocyte | *ALX4* | 9.840837598 | 0.001066804 | 0.015840489 | 1 |
| Heart_Failure | monocyte | *FOXN3* | 1.646244505 | 0.003873843 | 0.03075107 | 1 |
| Heart_Failure | monocyte | *ANAPC4* | 1.963864355 | 0.0067978 | 0.041858735 | 1 |
| Heart_Failure | monocyte | *SDCCAG8* | 1.955857037 | 4.34E-05 | 0.009491021 | 0.273093553 |

| | | | | | | |
|---|---|---|---|---|---|---|
| Heart_Failure | monocyte | *TBC1D22A* | 2.144316178 | 0.001755393 | 0.019662125 | 1 |
| Heart_Failure | monocyte | *GALC* | 4.064371738 | 9.81E-05 | 0.011389066 | 0.616605982 |
| Heart_Failure | monocyte | *FAM114A2* | 3.399096832 | 0.001072722 | 0.015840489 | 1 |
| Heart_Failure | monocyte | *CROCC* | 1.465805454 | 0.001239381 | 0.016649549 | 1 |
| Heart_Failure | monocyte | *YBX3* | 3.117540703 | 0.000797647 | 0.01444259 | 1 |
| Heart_Failure | monocyte | *QSER1* | 3.823715685 | 0.000229331 | 0.011389066 | 1 |
| Heart_Failure | monocyte | *SFSWAP* | 1.979995649 | 0.004610928 | 0.033747272 | 1 |
| Heart_Failure | monocyte | *EIF4B* | 1.852544657 | 0.007142138 | 0.043217151 | 1 |
| Heart_Failure | monocyte | *CASP8* | 1.905266714 | 0.005813977 | 0.03807549 | 1 |
| Heart_Failure | monocyte | *TM7SF3* | 2.71537834 | 0.003451486 | 0.028665114 | 1 |
| Heart_Failure | monocyte | *BTBD1* | 3.417458946 | 0.000341027 | 0.011389066 | 1 |
| Heart_Failure | monocyte | *PMS1* | 2.933169549 | 0.000783369 | 0.01444259 | 1 |
| Heart_Failure | monocyte | *ANKS1A* | 2.072185592 | 0.001966561 | 0.020744974 | 1 |
| Heart_Failure | monocyte | *IPO5* | 3.993113512 | 0.000448364 | 0.011600275 | 1 |
| Heart_Failure | monocyte | *ASB1* | 4.133785157 | 0.001565152 | 0.018607731 | 1 |
| Heart_Failure | monocyte | *TBC1D1* | 2.277542856 | 0.005378546 | 0.036675617 | 1 |
| Heart_Failure | monocyte | *NFYC* | 3.159140458 | 0.001590561 | 0.01864041 | 1 |
| Heart_Failure | monocyte | *NGEF* | 11.58914996 | 0.000830559 | 0.014464609 | 1 |
| Heart_Failure | monocyte | *SP100* | 2.955147684 | 0.000315687 | 0.011389066 | 1 |
| Heart_Failure | monocyte | *METTL22* | 4.950018662 | 0.000280402 | 0.011389066 | 1 |
| Heart_Failure | monocyte | *HDAC4* | 3.103081395 | 0.000363255 | 0.011389066 | 1 |
| Heart_Failure | monocyte | *RASSF1* | 3.979974604 | 4.44E-05 | 0.009491021 | 0.279063249 |
| Heart_Failure | monocyte | *HEATR6* | 4.648305127 | 0.000377656 | 0.011389066 | 1 |
| Heart_Failure | monocyte | *SRBD1* | 3.138255207 | 0.000476224 | 0.011604726 | 1 |
| Heart_Failure | monocyte | *PITX1* | 17.43204995 | 0.000391565 | 0.011389066 | 1 |
| Heart_Failure | monocyte | *DNAJA2* | 3.874293246 | 0.001154008 | 0.016135389 | 1 |
| Heart_Failure | monocyte | *PFN2* | 4.595248205 | 0.001737612 | 0.019607023 | 1 |
| Heart_Failure | monocyte | *TMEM260* | 2.402564983 | 0.00234406 | 0.022886147 | 1 |
| Heart_Failure | monocyte | *ZXDC* | 3.655851377 | 8.88E-05 | 0.011389066 | 0.558106989 |
| Heart_Failure | monocyte | *EIF2B3* | 1.581562349 | 0.008593795 | 0.048922662 | 1 |
| Heart_Failure | monocyte | *CDC42* | 3.369793409 | 0.00086127 | 0.014634599 | 1 |
| Heart_Failure | monocyte | *OSBPL3* | 3.26311228 | 0.000683774 | 0.013912252 | 1 |
| Heart_Failure | monocyte | *SNX13* | 2.112281796 | 0.002528543 | 0.023905185 | 1 |
| Heart_Failure | monocyte | *TRIP13* | 16.9404719 | 0.000332724 | 0.011389066 | 1 |
| Heart_Failure | monocyte | *NFATC3* | 2.598095174 | 0.000357364 | 0.011389066 | 1 |
| Heart_Failure | monocyte | *TRNT1* | 2.034536453 | 0.007089657 | 0.043107033 | 1 |
| Heart_Failure | monocyte | *NDE1* | 2.870089626 | 0.003916949 | 0.030937005 | 1 |
| Heart_Failure | monocyte | *PVR* | 2.5050174 | 0.007233747 | 0.043644967 | 1 |
| Heart_Failure | monocyte | *ALPK1* | 2.723816581 | 0.000842332 | 0.014523588 | 1 |
| Heart_Failure | monocyte | *PDE8A* | 1.420500072 | 0.004373021 | 0.032868331 | 1 |
| Heart_Failure | monocyte | *SMARCE1* | 3.453722348 | 0.000852426 | 0.014523588 | 1 |
| Heart_Failure | monocyte | *PICALM* | 2.689175815 | 0.000239004 | 0.011389066 | 1 |
| Heart_Failure | monocyte | *CLNS1A* | 1.268553422 | 0.006685219 | 0.041490592 | 1 |
| Heart_Failure | monocyte | *ENO1* | 4.261911015 | 0.000160958 | 0.011389066 | 1 |
| Heart_Failure | monocyte | *TIPIN* | 0.714198433 | 0.006642974 | 0.041426552 | 1 |
| Heart_Failure | monocyte | *EIF4G3* | 1.670775409 | 0.00371859 | 0.029934415 | 1 |
| Heart_Failure | monocyte | *GRAMD4* | 3.34846127 | 0.00303128 | 0.02671487 | 1 |
| Heart_Failure | monocyte | *VPS9D1* | 4.955823367 | 0.000202589 | 0.011389066 | 1 |
| Heart_Failure | monocyte | *NT5C2* | 2.973033969 | 0.000251486 | 0.011389066 | 1 |
| Heart_Failure | monocyte | *TM9SF3* | 2.171982126 | 0.005081246 | 0.035460966 | 1 |

| | | | | | | |
|---|---|---|---|---|---|---|
| Heart_Failure | monocyte | *DNAJC10* | 3.977731908 | 0.000218727 | 0.011389066 | 1 |
| Heart_Failure | monocyte | *DYNC1I2* | 3.187587991 | 0.000400259 | 0.011389066 | 1 |
| Heart_Failure | monocyte | *APBB1IP* | 2.739817989 | 0.000251964 | 0.011389066 | 1 |
| Heart_Failure | monocyte | *CAPZB* | 2.89812668 | 0.000379862 | 0.011389066 | 1 |
| Heart_Failure | monocyte | *GPR137B* | 1.713737714 | 0.002349526 | 0.022886147 | 1 |
| Heart_Failure | monocyte | *PIK3C3* | 1.95358539 | 0.006908779 | 0.042170383 | 1 |
| Heart_Failure | monocyte | *N4BP2* | 1.938207483 | 0.008963554 | 0.049628243 | 1 |
| Heart_Failure | monocyte | *PCM1* | 1.671117663 | 0.003566539 | 0.029170528 | 1 |
| Heart_Failure | monocyte | *XRCC5* | 2.872632847 | 0.000474182 | 0.011604726 | 1 |
| Heart_Failure | monocyte | *REXO1* | 3.21369412 | 0.005671905 | 0.037774645 | 1 |
| Heart_Failure | monocyte | *DDX1* | 3.325882908 | 0.00058124 | 0.013050905 | 1 |
| Heart_Failure | monocyte | *DNM2* | 3.656605091 | 0.000191099 | 0.011389066 | 1 |
| Heart_Failure | monocyte | *RIMS1* | 4.706335247 | 0.002678318 | 0.024726265 | 1 |
| Heart_Failure | monocyte | *STX7* | 1.931318279 | 0.002838504 | 0.025677234 | 1 |
| Heart_Failure | monocyte | *SLC35C2* | 4.298112465 | 0.000297627 | 0.011389066 | 1 |
| Heart_Failure | monocyte | *RIF1* | 1.802316714 | 0.003834362 | 0.030592173 | 1 |
| Heart_Failure | monocyte | *PSEN1* | 2.364621776 | 0.002049094 | 0.021295573 | 1 |
| Heart_Failure | monocyte | *CPOX* | 2.863051186 | 0.001510679 | 0.018237795 | 1 |
| Heart_Failure | monocyte | *KIAA0141* | 3.180561775 | 0.003344802 | 0.028241098 | 1 |
| Heart_Failure | monocyte | *C5orf22* | 3.336511498 | 0.003012479 | 0.026660729 | 1 |
| Heart_Failure | monocyte | *GSK3B* | 2.041019668 | 0.00307009 | 0.026958725 | 1 |
| Heart_Failure | monocyte | *PIBF1* | 1.631707088 | 0.005176246 | 0.035605096 | 1 |
| Heart_Failure | monocyte | *PDS5B* | 2.332844037 | 0.0013769 | 0.017452765 | 1 |
| Heart_Failure | monocyte | *CYLD* | 3.531976017 | 0.000391392 | 0.011389066 | 1 |
| Heart_Failure | monocyte | *ZNF446* | 5.911173858 | 0.000911569 | 0.015005461 | 1 |
| Heart_Failure | monocyte | *RPS5* | 2.776747426 | 0.003078792 | 0.026958725 | 1 |
| Heart_Failure | monocyte | *CHMP2B* | 2.660400786 | 0.0020214 | 0.021110533 | 1 |
| Heart_Failure | monocyte | *ZMPSTE24* | 3.673179482 | 0.000735568 | 0.014268876 | 1 |
| Heart_Failure | monocyte | *STARD7* | 3.913766489 | 0.001124907 | 0.016109997 | 1 |
| Heart_Failure | monocyte | *WBP11* | 3.423800272 | 0.001418249 | 0.017630601 | 1 |
| Heart_Failure | monocyte | *NCOA1* | 2.170225694 | 0.00092188 | 0.015067323 | 1 |
| Heart_Failure | monocyte | *SCAMP1* | 3.120445016 | 0.001163986 | 0.016152985 | 1 |
| Heart_Failure | monocyte | *ATG16L1* | 3.67325138 | 0.000830024 | 0.014464609 | 1 |
| Heart_Failure | monocyte | *MAST2* | 3.253248064 | 7.02E-05 | 0.011101372 | 0.441475169 |
| Heart_Failure | monocyte | *B4GALT1* | 3.328396738 | 0.000225634 | 0.011389066 | 1 |
| Heart_Failure | monocyte | *SEPHS1* | 2.702156013 | 0.003516368 | 0.028974316 | 1 |
| Heart_Failure | monocyte | *PGS1* | 3.921527947 | 0.000616051 | 0.013448301 | 1 |
| Heart_Failure | monocyte | *GMCL1* | 2.78108669 | 0.005110169 | 0.035460966 | 1 |
| Heart_Failure | monocyte | *ALG6* | 3.49448582 | 0.001158453 | 0.016135389 | 1 |
| Heart_Failure | monocyte | *DDX18* | 2.785904012 | 0.00096375 | 0.015363418 | 1 |
| Heart_Failure | monocyte | *C3orf18* | 4.061542747 | 0.000466863 | 0.011604726 | 1 |
| Heart_Failure | monocyte | *FKBP1A* | 2.579444579 | 0.008193221 | 0.047563045 | 1 |
| Heart_Failure | monocyte | *NSFL1C* | 2.780471728 | 0.001169384 | 0.016152985 | 1 |
| Heart_Failure | monocyte | *XRN2* | 3.198745514 | 0.000463808 | 0.011604726 | 1 |
| Heart_Failure | monocyte | *ANAPC5* | 1.463964552 | 0.008836651 | 0.049383136 | 1 |
| Heart_Failure | monocyte | *SLC23A2* | 3.349393913 | 0.000467682 | 0.011604726 | 1 |
| Heart_Failure | monocyte | *FUS* | 2.17021506 | 0.001921171 | 0.020611606 | 1 |
| Heart_Failure | monocyte | *NECAP1* | 3.437758864 | 0.001619866 | 0.018929553 | 1 |
| Heart_Failure | monocyte | *GPATCH2L* | 1.903472892 | 0.001342342 | 0.01728185 | 1 |
| Heart_Failure | monocyte | *SLC9A1* | 4.988169557 | 0.000358537 | 0.011389066 | 1 |

| | | | | | | |
|---|---|---|---|---|---|---|
| Heart_Failure | monocyte | *SPTLC1* | 2.132658567 | 0.005802477 | 0.038039808 | 1 |
| Heart_Failure | monocyte | *PAPOLA* | 2.250279151 | 0.002828218 | 0.025621046 | 1 |
| Heart_Failure | monocyte | *STRN4* | 3.94817846 | 0.005451759 | 0.036974334 | 1 |
| Heart_Failure | monocyte | *SPG21* | 2.495875078 | 0.007710964 | 0.045562812 | 1 |
| Heart_Failure | monocyte | *GNPTG* | 4.551725982 | 0.000261357 | 0.011389066 | 1 |
| Heart_Failure | monocyte | *USP48* | 1.796303804 | 0.000960608 | 0.015363418 | 1 |
| Heart_Failure | monocyte | *PDPR* | 4.013569613 | 0.000178661 | 0.011389066 | 1 |
| Heart_Failure | monocyte | *EXOC1* | 1.978139204 | 0.003424498 | 0.028571324 | 1 |
| Heart_Failure | monocyte | *PPP2R3C* | 3.386210832 | 0.000224585 | 0.011389066 | 1 |
| Heart_Failure | monocyte | *SCFD1* | 1.744331506 | 0.004056701 | 0.031448189 | 1 |
| Heart_Failure | monocyte | *TOX4* | 2.226739403 | 0.003395952 | 0.028543248 | 1 |
| Heart_Failure | monocyte | *AGO1* | 3.74999568 | 0.000743713 | 0.014268876 | 1 |
| Heart_Failure | monocyte | *RFFL* | 5.222061408 | 0.000821276 | 0.014464609 | 1 |
| Heart_Failure | monocyte | *COMT* | 2.341765001 | 0.004080621 | 0.031594658 | 1 |
| Heart_Failure | monocyte | *SUCO* | 3.255549478 | 0.000931813 | 0.015111629 | 1 |
| Heart_Failure | monocyte | *WAC* | 2.754338965 | 0.000358883 | 0.011389066 | 1 |
| Heart_Failure | monocyte | *CREM* | 1.735423319 | 0.007240608 | 0.043644967 | 1 |
| Heart_Failure | monocyte | *MARCH2* | 4.433409936 | 1.97E-05 | 0.006886504 | 0.123957072 |
| Heart_Failure | monocyte | *CDC34* | 5.496415618 | 0.000824777 | 0.014464609 | 1 |
| Heart_Failure | monocyte | *MED15* | 2.349132105 | 0.001526212 | 0.018285615 | 1 |
| Heart_Failure | monocyte | *SMARCB1* | 2.389629702 | 0.000273473 | 0.011389066 | 1 |
| Heart_Failure | monocyte | *CCDC134* | 3.868125465 | 0.003567923 | 0.029170528 | 1 |
| Heart_Failure | monocyte | *FBXO7* | 1.891535224 | 0.004725027 | 0.034184404 | 1 |
| Heart_Failure | monocyte | *PPP6R2* | 2.046067355 | 0.002589313 | 0.024224715 | 1 |
| Heart_Failure | monocyte | *HMGXB4* | 1.927618264 | 0.006641489 | 0.041426552 | 1 |
| Heart_Failure | monocyte | *THOC5* | 2.830028288 | 0.002027404 | 0.021138127 | 1 |
| Heart_Failure | monocyte | *TSPO* | 2.369397385 | 0.00862192 | 0.048964714 | 1 |
| Heart_Failure | monocyte | *MTMR3* | 1.701200858 | 0.00233177 | 0.022834639 | 1 |
| Heart_Failure | monocyte | *KIAA0930* | 3.009456462 | 0.000658258 | 0.013613382 | 1 |
| Heart_Failure | monocyte | *EP300* | 2.165158653 | 0.001725799 | 0.01958502 | 1 |
| Heart_Failure | monocyte | *DESI1* | 2.212502443 | 0.000265492 | 0.011389066 | 1 |
| Heart_Failure | monocyte | *ZBED4* | 2.602574636 | 0.004535035 | 0.033430274 | 1 |
| Heart_Failure | monocyte | *RBM23* | 2.479015942 | 0.005887589 | 0.038477411 | 1 |
| Heart_Failure | monocyte | *SOS2* | 1.984311706 | 0.002572063 | 0.024099196 | 1 |
| Heart_Failure | monocyte | *DDHD1* | 2.51734598 | 0.001739571 | 0.019607023 | 1 |
| Heart_Failure | monocyte | *VRK1* | 3.631419897 | 0.00078922 | 0.01444259 | 1 |
| Heart_Failure | monocyte | *CCNB1IP1* | 2.615411659 | 0.001638582 | 0.018937074 | 1 |
| Heart_Failure | monocyte | *PCK2* | 3.800079381 | 0.002612038 | 0.024364811 | 1 |
| Heart_Failure | monocyte | *ABHD12* | 2.278534254 | 0.001054497 | 0.015840489 | 1 |
| Heart_Failure | monocyte | *NDRG3* | 3.742171748 | 0.001056066 | 0.015840489 | 1 |
| Heart_Failure | monocyte | *NELFCD* | 2.100851299 | 0.007026098 | 0.042844887 | 1 |
| Heart_Failure | monocyte | *GID8* | 3.486943218 | 0.005055075 | 0.035460966 | 1 |
| Heart_Failure | monocyte | *BIRC7* | -2.745426371 | 0.003574275 | 0.029183725 | 1 |
| Heart_Failure | monocyte | *GMEB2* | 3.653460503 | 0.001996946 | 0.020959596 | 1 |
| Heart_Failure | monocyte | *RNF24* | 2.46783218 | 0.002285153 | 0.022589237 | 1 |
| Heart_Failure | monocyte | *MAPRE1* | 4.09588598 | 0.002445445 | 0.023508427 | 1 |
| Heart_Failure | monocyte | *PXMP4* | 2.963670507 | 0.002947172 | 0.026394399 | 1 |
| Heart_Failure | monocyte | *DHX35* | 3.25474196 | 0.001410976 | 0.017630601 | 1 |
| Heart_Failure | monocyte | *PIGU* | 2.977628317 | 0.000703305 | 0.014081785 | 1 |
| Heart_Failure | monocyte | *RBFA* | 3.842360469 | 0.000818532 | 0.014464609 | 1 |

| | | | | | | |
|---|---|---|---|---|---|---|
| Heart_Failure | monocyte | *SMCHD1* | 3.06516794 | 0.000156444 | 0.011389066 | 0.983562619 |
| Heart_Failure | monocyte | *MYL12A* | 4.252460304 | 0.000190552 | 0.011389066 | 1 |
| Heart_Failure | monocyte | *STK24* | 1.688719405 | 0.008070124 | 0.047065741 | 1 |
| Heart_Failure | monocyte | *DNAJC3* | 2.230837379 | 0.003587791 | 0.029256087 | 1 |
| Heart_Failure | monocyte | *PARP4* | 3.326861024 | 0.00024421 | 0.011389066 | 1 |
| Heart_Failure | monocyte | *MGRN1* | 1.910104651 | 0.003429094 | 0.028571324 | 1 |
| Heart_Failure | monocyte | *DHODH* | 1.47101317 | 0.001160044 | 0.016135389 | 1 |
| Heart_Failure | monocyte | *CTCF* | 3.485807243 | 0.000427897 | 0.011479484 | 1 |
| Heart_Failure | monocyte | *USB1* | 4.258393234 | 0.007545253 | 0.045092208 | 1 |
| Heart_Failure | monocyte | *HCFC1R1* | 4.628707165 | 0.002740448 | 0.025131074 | 1 |
| Heart_Failure | monocyte | *NPRL3* | 1.485788555 | 0.008761435 | 0.049269357 | 1 |
| Heart_Failure | monocyte | *CLCN7* | 4.438420692 | 0.000409405 | 0.011389066 | 1 |
| Heart_Failure | monocyte | *NUBP1* | 4.293648582 | 0.002245096 | 0.022298454 | 1 |
| Heart_Failure | monocyte | *RRN3P2* | 2.127937483 | 0.008867658 | 0.049424614 | 1 |
| Heart_Failure | monocyte | *RBL2* | 1.614987937 | 0.001545103 | 0.018432755 | 1 |
| Heart_Failure | monocyte | *KAT8* | 5.472807713 | 8.08E-06 | 0.004431396 | 0.050812633 |
| Heart_Failure | monocyte | *NOMO1* | 5.169918888 | 0.001466737 | 0.017885666 | 1 |
| Heart_Failure | monocyte | *LACTB* | 3.627837283 | 0.001258354 | 0.016796759 | 1 |
| Heart_Failure | monocyte | *HERC1* | 2.095487204 | 0.001464062 | 0.017885666 | 1 |
| Heart_Failure | monocyte | *ATP8B4* | 3.277133794 | 0.000396471 | 0.011389066 | 1 |
| Heart_Failure | monocyte | *DNAJC17* | 2.054477838 | 0.00892734 | 0.049581438 | 1 |
| Heart_Failure | monocyte | *NBN* | 3.157190057 | 0.001053897 | 0.015840489 | 1 |
| Heart_Failure | monocyte | *CPQ* | 1.608232501 | 0.00087108 | 0.014721728 | 1 |
| Heart_Failure | monocyte | *NIPAL2* | 2.029788461 | 0.002942357 | 0.026388872 | 1 |
| Heart_Failure | monocyte | *IKBKB* | 2.154143134 | 0.004283557 | 0.03244665 | 1 |
| Heart_Failure | monocyte | *CHRAC1* | 4.866389208 | 0.001902832 | 0.020519903 | 1 |
| Heart_Failure | monocyte | *MTMR9* | 2.205413951 | 0.005222758 | 0.035768499 | 1 |
| Heart_Failure | monocyte | *LEPROTL1* | 4.342245201 | 0.000370631 | 0.011389066 | 1 |
| Heart_Failure | monocyte | *ERICH1* | 3.441281572 | 0.000176071 | 0.011389066 | 1 |
| Heart_Failure | monocyte | *MAN2B1* | 1.65904177 | 0.005720193 | 0.037841897 | 1 |
| Heart_Failure | monocyte | *PPP1R37* | 3.449783638 | 0.003287587 | 0.028091022 | 1 |
| Heart_Failure | monocyte | *FCGRT* | 1.775857176 | 0.003656947 | 0.029513769 | 1 |
| Heart_Failure | monocyte | *C19orf53* | 4.563641621 | 0.003795274 | 0.030458888 | 1 |
| Heart_Failure | monocyte | *TIMM44* | 3.218202148 | 5.89E-05 | 0.011101372 | 0.370554786 |
| Heart_Failure | monocyte | *RPS16* | 2.329283571 | 0.004179087 | 0.03192457 | 1 |
| Heart_Failure | monocyte | *PRKD2* | 2.323126926 | 0.002052768 | 0.021295573 | 1 |
| Heart_Failure | monocyte | *FZR1* | 3.078449532 | 0.008858992 | 0.049424614 | 1 |
| Heart_Failure | monocyte | *MRPL4* | 2.325151383 | 0.007625658 | 0.045271496 | 1 |
| Heart_Failure | monocyte | *RABAC1* | 3.284159063 | 0.001071784 | 0.015840489 | 1 |
| Heart_Failure | monocyte | *CARD8* | 3.176515172 | 0.000462451 | 0.011604726 | 1 |
| Heart_Failure | monocyte | *LIG1* | 3.745109972 | 0.007562397 | 0.045116469 | 1 |
| Heart_Failure | monocyte | *PPP2R1A* | 4.49206061 | 0.000382681 | 0.011389066 | 1 |
| Heart_Failure | monocyte | *KXD1* | 3.016463824 | 0.004575754 | 0.033567985 | 1 |
| Heart_Failure | monocyte | *SIPA1L3* | 3.121390481 | 0.000762361 | 0.014350186 | 1 |
| Heart_Failure | monocyte | *ZNF85* | 2.910804786 | 0.004270542 | 0.032426208 | 1 |
| Heart_Failure | monocyte | *OGDH* | 2.275953709 | 0.00385412 | 0.030677814 | 1 |
| Heart_Failure | monocyte | *LMBR1* | 2.652858763 | 0.000404106 | 0.011389066 | 1 |
| Heart_Failure | monocyte | *ZKSCAN1* | 1.747415546 | 0.008736676 | 0.049218173 | 1 |
| Heart_Failure | monocyte | *USP42* | 3.057274067 | 0.001135728 | 0.016135389 | 1 |
| Heart_Failure | monocyte | *C1GALT1* | 2.91920403 | 0.000874774 | 0.014744519 | 1 |

| | | | | | | |
|---|---|---|---|---|---|---|
| Heart_Failure | monocyte | *PHF14* | 1.765175766 | 0.003617871 | 0.029323162 | 1 |
| Heart_Failure | monocyte | *URGCP* | 2.649767058 | 0.004460857 | 0.03311146 | 1 |
| Heart_Failure | monocyte | *YKT6* | 5.777723195 | 0.000173842 | 0.011389066 | 1 |
| Heart_Failure | monocyte | *CLIP2* | 3.578211884 | 0.00100585 | 0.015614266 | 1 |
| Heart_Failure | monocyte | *LIMK1* | 3.244009886 | 0.004549454 | 0.033430274 | 1 |
| Heart_Failure | monocyte | *KDM4C* | 3.03301756 | 0.000269106 | 0.011389066 | 1 |
| Heart_Failure | monocyte | *CCNJ* | 3.394683944 | 0.004385977 | 0.032905294 | 1 |
| Heart_Failure | monocyte | *HPS1* | 3.833273457 | 0.001324982 | 0.017175592 | 1 |
| Heart_Failure | monocyte | *DNMBP* | 2.130644935 | 0.004149981 | 0.031877388 | 1 |
| Heart_Failure | monocyte | *DDX50* | 2.053745032 | 0.005085567 | 0.035460966 | 1 |
| Heart_Failure | monocyte | *ATE1* | 1.342987819 | 0.008829999 | 0.049383136 | 1 |
| Heart_Failure | monocyte | *MICU1* | 3.464382591 | 6.92E-05 | 0.011101372 | 0.434798168 |
| Heart_Failure | monocyte | *TNKS2* | 2.419461917 | 0.002458762 | 0.023564385 | 1 |
| Heart_Failure | monocyte | *ACBD5* | 3.749245124 | 0.000127675 | 0.011389066 | 0.802694622 |
| Heart_Failure | monocyte | *MAP3K8* | 3.268223424 | 0.000415152 | 0.0114576 | 1 |
| Heart_Failure | monocyte | *SHOC2* | 3.192449348 | 0.000190795 | 0.011389066 | 1 |
| Heart_Failure | monocyte | *PBLD* | 0.778690576 | 0.007589013 | 0.04521281 | 1 |
| Heart_Failure | monocyte | *MLLT6* | 2.937859984 | 0.001432235 | 0.017690495 | 1 |
| Heart_Failure | monocyte | *PSMB3* | 3.896427408 | 0.000383573 | 0.011389066 | 1 |
| Heart_Failure | monocyte | *CASC3* | 1.948326938 | 0.003344612 | 0.028241098 | 1 |
| Heart_Failure | monocyte | *RPS6KB1* | 2.416501473 | 0.002812332 | 0.025513898 | 1 |
| Heart_Failure | monocyte | *GALK1* | 4.499470395 | 0.004894075 | 0.03492514 | 1 |
| Heart_Failure | monocyte | *MED13* | 1.560108101 | 0.006174033 | 0.039608989 | 1 |
| Heart_Failure | monocyte | *RNF167* | 3.492954117 | 0.008464403 | 0.04876128 | 1 |
| Heart_Failure | monocyte | *C1QBP* | 5.622562184 | 0.000593534 | 0.013277434 | 1 |
| Heart_Failure | monocyte | *GOSR1* | 2.022396918 | 0.004994832 | 0.035307337 | 1 |
| Heart_Failure | monocyte | *FTSJ3* | 0.75312229 | 0.006001274 | 0.038926702 | 1 |
| Heart_Failure | monocyte | *AKAP10* | 3.773561399 | 1.37E-05 | 0.00577283 | 0.086325598 |
| Heart_Failure | monocyte | *SMARCD2* | 3.560001353 | 0.000987854 | 0.015508077 | 1 |
| Heart_Failure | monocyte | *UTP6* | 3.533823192 | 0.000319952 | 0.011389066 | 1 |
| Heart_Failure | monocyte | *CYTH1* | 2.317284937 | 0.002130905 | 0.021667534 | 1 |
| Heart_Failure | monocyte | *RAB5C* | 3.280543421 | 0.000950421 | 0.015243105 | 1 |
| Heart_Failure | monocyte | *EFTUD2* | 3.805887379 | 0.000178232 | 0.011389066 | 1 |
| Heart_Failure | monocyte | *YWHAE* | 2.276220497 | 0.002053623 | 0.021295573 | 1 |
| Heart_Failure | monocyte | *DPH1* | 4.092459058 | 0.004433854 | 0.033015784 | 1 |
| Heart_Failure | monocyte | *DHRS7B* | 3.530947453 | 0.000427417 | 0.011479484 | 1 |
| Heart_Failure | monocyte | *SLAIN2* | 3.391103281 | 0.00011763 | 0.011389066 | 0.739540272 |
| Heart_Failure | monocyte | *UBE2D3* | 2.475945587 | 0.000751559 | 0.01431753 | 1 |
| Heart_Failure | monocyte | *NDUFC1* | 4.902987837 | 0.000506674 | 0.012204821 | 1 |
| Heart_Failure | monocyte | *TBC1D9* | 2.974184891 | 0.000441015 | 0.011517615 | 1 |
| Heart_Failure | monocyte | *GRPEL1* | 4.028670291 | 0.005498373 | 0.03716864 | 1 |
| Heart_Failure | monocyte | *STIM2* | 3.552670545 | 9.90E-05 | 0.011389066 | 0.622329654 |
| Heart_Failure | monocyte | *KLF3* | 2.796908043 | 0.001515866 | 0.018237795 | 1 |
| Heart_Failure | monocyte | *ELP4* | 2.498001615 | 0.001488047 | 0.018025719 | 1 |
| Heart_Failure | monocyte | *NRXN2* | 5.01361476 | 0.008335981 | 0.048191822 | 1 |
| Heart_Failure | monocyte | *CHORDC1* | 2.647702838 | 0.001115306 | 0.016083148 | 1 |
| Heart_Failure | monocyte | *PANX1* | 2.918441332 | 0.008012353 | 0.046815675 | 1 |
| Heart_Failure | monocyte | *EIF4G2* | 3.74291273 | 0.000233983 | 0.011389066 | 1 |
| Heart_Failure | monocyte | *CARS* | 2.155621297 | 0.002255957 | 0.022370982 | 1 |
| Heart_Failure | monocyte | *RPS13* | 3.027321984 | 0.001217733 | 0.016601601 | 1 |

| | | | | | | |
|---|---|---|---|---|---|---|
| Heart_Failure | monocyte | *NDUFS8* | 3.039033528 | 0.004086069 | 0.031597925 | 1 |
| Heart_Failure | monocyte | *PRDM4* | 2.74878188 | 0.001003649 | 0.015614266 | 1 |
| Heart_Failure | monocyte | *CAPRIN2* | 3.371184444 | 0.000809792 | 0.01444259 | 1 |
| Heart_Failure | monocyte | *ITFG2* | 2.4138806 | 0.003915266 | 0.030937005 | 1 |
| Heart_Failure | monocyte | *PRR4* | 1.682118404 | 0.005157697 | 0.035529945 | 1 |
| Heart_Failure | monocyte | *PARP11* | 1.90289921 | 0.007514864 | 0.044996142 | 1 |
| Heart_Failure | monocyte | *GPN3* | 4.031941242 | 0.00392815 | 0.030986549 | 1 |
| Heart_Failure | monocyte | *VPS29* | 3.655383605 | 0.000280618 | 0.011389066 | 1 |
| Heart_Failure | monocyte | *CREBL2* | 2.001824361 | 0.007259792 | 0.043718688 | 1 |
| Heart_Failure | monocyte | *NAA25* | 2.095485814 | 0.003349577 | 0.028241098 | 1 |
| Heart_Failure | monocyte | *LTBR* | 2.84623054 | 0.000887128 | 0.014874442 | 1 |
| Heart_Failure | monocyte | *CDK2AP1* | 1.129391172 | 0.001101615 | 0.016083148 | 1 |
| Heart_Failure | monocyte | *STX2* | 1.992408675 | 0.005590009 | 0.037565605 | 1 |
| Heart_Failure | monocyte | *MDM1* | 2.522977264 | 0.0026346 | 0.02448227 | 1 |
| Heart_Failure | monocyte | *CNOT2* | 2.540310578 | 0.000807103 | 0.01444259 | 1 |
| Heart_Failure | monocyte | *KRR1* | 3.299304941 | 0.000378679 | 0.011389066 | 1 |
| Heart_Failure | monocyte | *C2CD5* | 2.796629767 | 0.000639784 | 0.013540529 | 1 |
| Heart_Failure | monocyte | *MCM9* | 2.072954144 | 0.003798277 | 0.030458888 | 1 |
| Heart_Failure | monocyte | *RNGTT* | 3.001361998 | 0.000643163 | 0.013540529 | 1 |
| Heart_Failure | monocyte | *MAPK14* | 2.923527728 | 0.000409308 | 0.011389066 | 1 |
| Heart_Failure | monocyte | *MDN1* | 1.98581129 | 0.004313239 | 0.0326141 | 1 |
| Heart_Failure | monocyte | *ASCC3* | 3.047343988 | 0.000391465 | 0.011389066 | 1 |
| Heart_Failure | monocyte | *PHACTR2* | 2.9727036 | 0.000404576 | 0.011389066 | 1 |
| Heart_Failure | monocyte | *TFEB* | 2.772409438 | 0.007633281 | 0.045273997 | 1 |
| Heart_Failure | monocyte | *FAM120B* | 1.571998181 | 0.001450942 | 0.017792349 | 1 |
| Heart_Failure | monocyte | *EXOC2* | 2.030150735 | 0.004802926 | 0.034549195 | 1 |
| Heart_Failure | monocyte | *TMEM30A* | 1.833059536 | 0.005593103 | 0.037565605 | 1 |
| Heart_Failure | monocyte | *SENP6* | 2.731455293 | 0.000339337 | 0.011389066 | 1 |
| Heart_Failure | monocyte | *DAP* | 4.073234108 | 0.00029265 | 0.011389066 | 1 |
| Heart_Failure | monocyte | *RNF130* | 2.735331303 | 0.000603901 | 0.013396857 | 1 |
| Heart_Failure | monocyte | *CLINT1* | 1.675834979 | 0.004592657 | 0.033652724 | 1 |
| Heart_Failure | monocyte | *TTC1* | 2.072639577 | 0.005511288 | 0.03716864 | 1 |
| Heart_Failure | monocyte | *TARS* | 2.407261465 | 0.005515881 | 0.03716864 | 1 |
| Heart_Failure | monocyte | *SLC12A7* | 3.636660739 | 0.00231093 | 0.022701279 | 1 |
| Heart_Failure | monocyte | *ATP6V0E1* | 3.39713601 | 0.000557193 | 0.01265792 | 1 |
| Heart_Failure | monocyte | *PRKAR2A* | 1.624175995 | 0.003091487 | 0.027032234 | 1 |
| Heart_Failure | monocyte | *NCBP2* | 3.353824062 | 0.004634398 | 0.033879605 | 1 |
| Heart_Failure | monocyte | *ABTB1* | 3.544994954 | 0.004321226 | 0.0326141 | 1 |
| Heart_Failure | monocyte | *HEMK1* | 1.702016668 | 0.001233171 | 0.016601601 | 1 |
| Heart_Failure | monocyte | *WDR48* | 2.014427768 | 0.008507974 | 0.04884898 | 1 |
| Heart_Failure | monocyte | *NKTR* | 2.058021774 | 0.001451435 | 0.017792349 | 1 |
| Heart_Failure | monocyte | *NEK4* | 4.349238125 | 0.000388871 | 0.011389066 | 1 |
| Heart_Failure | monocyte | *PIKFYVE* | 3.131844925 | 0.000286441 | 0.011389066 | 1 |
| Heart_Failure | monocyte | *STAM2* | 3.399644297 | 0.000322882 | 0.011389066 | 1 |
| Heart_Failure | monocyte | *GPD2* | 2.177029359 | 0.002434792 | 0.023462924 | 1 |
| Heart_Failure | monocyte | *PPM1G* | 2.382661173 | 0.005895081 | 0.038486367 | 1 |
| Heart_Failure | monocyte | *GGCX* | 1.725251238 | 0.008752986 | 0.049265913 | 1 |
| Heart_Failure | monocyte | *SF3B1* | 1.665512768 | 0.004712314 | 0.034171068 | 1 |
| Heart_Failure | monocyte | *ST3GAL5* | 2.391332652 | 0.001752475 | 0.019662125 | 1 |
| Heart_Failure | monocyte | *HDLBP* | 2.583457724 | 0.000286198 | 0.011389066 | 1 |

| | | | | | | |
|---|---|---|---|---|---|---|
| Heart_Failure | monocyte | *TAF1B* | 3.121312263 | 0.000248854 | 0.011389066 | 1 |
| Heart_Failure | monocyte | *CEBPZ* | 1.998545218 | 0.001312566 | 0.01715614 | 1 |
| Heart_Failure | monocyte | *PRKD3* | 1.923842133 | 0.002568824 | 0.024099196 | 1 |
| Heart_Failure | monocyte | *SLC25A12* | 1.751962797 | 0.006845687 | 0.041937353 | 1 |
| Heart_Failure | monocyte | *RMDN2* | 2.002546359 | 0.008914973 | 0.049581438 | 1 |
| Heart_Failure | monocyte | *WIPF1* | 2.88797712 | 0.000400225 | 0.011389066 | 1 |
| Heart_Failure | monocyte | *ORC4* | 1.602847507 | 0.007609829 | 0.045263005 | 1 |
| Heart_Failure | monocyte | *THADA* | 1.692894255 | 0.005080698 | 0.035460966 | 1 |
| Heart_Failure | monocyte | *MSH6* | 3.049999493 | 0.002142104 | 0.021721622 | 1 |
| Heart_Failure | monocyte | *DNAJC16* | 2.217189972 | 0.004437464 | 0.033015784 | 1 |
| Heart_Failure | monocyte | *CEP104* | 2.31980732 | 0.006820153 | 0.041921553 | 1 |
| Heart_Failure | monocyte | *FAM20B* | 3.035492734 | 0.001758362 | 0.019662125 | 1 |
| Heart_Failure | monocyte | *TMEM59* | 2.213332393 | 0.004435631 | 0.033015784 | 1 |
| Heart_Failure | monocyte | *WRAP73* | 4.771147262 | 0.002094187 | 0.021513326 | 1 |
| Heart_Failure | monocyte | *MRPL37* | 3.927937364 | 0.000851539 | 0.014523588 | 1 |
| Heart_Failure | monocyte | *QSOX1* | 4.052534412 | 0.001092787 | 0.016014809 | 1 |
| Heart_Failure | monocyte | *EDEM3* | 3.054859214 | 0.000652796 | 0.013585129 | 1 |
| Heart_Failure | monocyte | *RAP1A* | 3.13677261 | 0.000213334 | 0.011389066 | 1 |
| Heart_Failure | monocyte | *SMG7* | 3.396627885 | 0.000380921 | 0.011389066 | 1 |
| Heart_Failure | monocyte | *TTF2* | 3.422543359 | 0.002412759 | 0.023372904 | 1 |
| Heart_Failure | monocyte | *KLHL12* | 2.963538672 | 0.007763972 | 0.045747038 | 1 |
| Heart_Failure | monocyte | *RBBP5* | 2.880188666 | 0.00030552 | 0.011389066 | 1 |
| Heart_Failure | monocyte | *DR1* | 4.851869303 | 0.000127419 | 0.011389066 | 0.801084822 |
| Heart_Failure | monocyte | *PRRC2C* | 1.836442737 | 0.004953043 | 0.035186193 | 1 |
| Heart_Failure | monocyte | *PTBP2* | 1.310087037 | 0.005020386 | 0.035417411 | 1 |
| Heart_Failure | monocyte | *RCAN3* | 3.570822908 | 0.001768862 | 0.019717796 | 1 |
| Heart_Failure | monocyte | *MTFR1L* | 4.683292473 | 0.000625105 | 0.013459015 | 1 |
| Heart_Failure | monocyte | *NSL1* | 2.806194388 | 0.000262192 | 0.011389066 | 1 |
| Heart_Failure | monocyte | *KMT2A* | 2.231174017 | 0.001878018 | 0.020319415 | 1 |
| Heart_Failure | monocyte | *ATF6* | 3.072559031 | 0.000329428 | 0.011389066 | 1 |
| Heart_Failure | monocyte | *MED28* | 3.953105688 | 2.63E-05 | 0.008253281 | 0.165065622 |
| Heart_Failure | monocyte | *SPP1* | 2.260592183 | 0.002093383 | 0.021513326 | 1 |
| Heart_Failure | monocyte | *MFSD1* | 2.283487443 | 0.004205025 | 0.032044837 | 1 |
| Heart_Failure | monocyte | *RAB3GAP2* | 2.51078607 | 0.001109124 | 0.016083148 | 1 |
| Heart_Failure | monocyte | *UBN1* | 2.021108529 | 0.005099871 | 0.035460966 | 1 |
| Heart_Failure | monocyte | *SENP5* | 2.58558351 | 0.000802968 | 0.01444259 | 1 |
| Heart_Failure | monocyte | *PTBP3* | 3.47044093 | 0.000285176 | 0.011389066 | 1 |
| Heart_Failure | monocyte | *FKBP15* | 3.246391525 | 0.000802288 | 0.01444259 | 1 |
| Heart_Failure | monocyte | *CTNNAL1* | 0.770033108 | 0.003976101 | 0.031151779 | 1 |
| Heart_Failure | monocyte | *CNTRL* | 2.047089809 | 0.005647386 | 0.037654428 | 1 |
| Heart_Failure | monocyte | *TRIM32* | 5.207793205 | 0.000487982 | 0.011845344 | 1 |
| Heart_Failure | monocyte | *RBM18* | 2.662359349 | 0.007411486 | 0.044461843 | 1 |
| Heart_Failure | monocyte | *ATAD2B* | 2.466522202 | 0.000922685 | 0.015067323 | 1 |
| Heart_Failure | monocyte | *OGFRL1* | 2.262500551 | 0.006445662 | 0.040520538 | 1 |
| Heart_Failure | monocyte | *MXI1* | 3.836082064 | 0.00045348 | 0.011604726 | 1 |
| Heart_Failure | monocyte | *HELLS* | 2.092170513 | 0.003216566 | 0.027893175 | 1 |
| Heart_Failure | monocyte | *TCTN3* | 3.34199098 | 0.002209188 | 0.022061708 | 1 |
| Heart_Failure | monocyte | *WDR11* | 3.227022184 | 0.000324669 | 0.011389066 | 1 |
| Heart_Failure | monocyte | *C10orf76* | 1.897807985 | 0.00424548 | 0.032274892 | 1 |
| Heart_Failure | monocyte | *KANSL1* | 1.438973544 | 0.005128143 | 0.035529945 | 1 |

| | | | | | | |
|---|---|---|---|---|---|---|
| Heart_Failure | monocyte | *GORAB* | 4.013548418 | 0.000678365 | 0.013847008 | 1 |
| Heart_Failure | monocyte | *DNAJC15* | 2.580605088 | 0.002633066 | 0.02448227 | 1 |
| Heart_Failure | monocyte | *ARL1* | 2.946533493 | 0.001591077 | 0.01864041 | 1 |
| Heart_Failure | monocyte | *SCPEP1* | 1.022900796 | 0.004812416 | 0.034577894 | 1 |
| Heart_Failure | monocyte | *TEX14* | 3.246790219 | 0.003435648 | 0.028571324 | 1 |
| Heart_Failure | monocyte | *CEP89* | 1.320444133 | 0.005229688 | 0.035776983 | 1 |
| Heart_Failure | monocyte | *PYROXD1* | 2.016782203 | 0.003859252 | 0.030677814 | 1 |
| Heart_Failure | monocyte | *B4GALT4* | 2.395569955 | 0.008967343 | 0.049628243 | 1 |
| Heart_Failure | monocyte | *TNFSF10* | 3.038414812 | 0.001881008 | 0.020319415 | 1 |
| Heart_Failure | monocyte | *ACVR2A* | 3.219325589 | 0.000280784 | 0.011389066 | 1 |
| Heart_Failure | monocyte | *POLK* | 2.445058456 | 0.000615701 | 0.013448301 | 1 |
| Heart_Failure | monocyte | *HERPUD2* | 3.663979488 | 0.000307962 | 0.011389066 | 1 |
| Heart_Failure | monocyte | *CBX3* | 3.463033384 | 0.000784413 | 0.01444259 | 1 |
| Heart_Failure | monocyte | *FAM126A* | 1.767721718 | 0.006470239 | 0.040520538 | 1 |
| Heart_Failure | monocyte | *SMU1* | 4.231990807 | 0.000181611 | 0.011389066 | 1 |
| Heart_Failure | monocyte | *TRIM24* | 3.281796011 | 0.001066837 | 0.015840489 | 1 |
| Heart_Failure | monocyte | *RBM19* | 3.270560361 | 0.001356626 | 0.017335584 | 1 |
| Heart_Failure | monocyte | *ATF1* | 1.875061826 | 0.003033946 | 0.02671487 | 1 |
| Heart_Failure | monocyte | *SPATS2* | 1.419955319 | 0.008642782 | 0.048996544 | 1 |
| Heart_Failure | monocyte | *PDE1B* | 2.959222125 | 0.002056653 | 0.021295573 | 1 |
| Heart_Failure | monocyte | *TTC21B* | 2.539101328 | 0.001348886 | 0.01728185 | 1 |
| Heart_Failure | monocyte | *NMI* | 4.131550116 | 0.000530162 | 0.012398944 | 1 |
| Heart_Failure | monocyte | *AGO2* | 3.676899502 | 0.000258498 | 0.011389066 | 1 |
| Heart_Failure | monocyte | *CSE1L* | 2.782606147 | 0.005401935 | 0.036755376 | 1 |
| Heart_Failure | monocyte | *STX16* | 2.801044861 | 0.001592158 | 0.01864041 | 1 |
| Heart_Failure | monocyte | *RNF114* | 1.841903425 | 0.007799194 | 0.045868657 | 1 |
| Heart_Failure | monocyte | *MRS2* | 1.435066642 | 0.006706595 | 0.041554619 | 1 |
| Heart_Failure | monocyte | *SNRPC* | 2.044759331 | 0.00230375 | 0.022666165 | 1 |
| Heart_Failure | monocyte | *GLO1* | 3.550199826 | 0.000120812 | 0.011389066 | 0.759542289 |
| Heart_Failure | monocyte | *NUP153* | 2.089350754 | 0.002639814 | 0.02448227 | 1 |
| Heart_Failure | monocyte | *CNOT1* | 2.059727246 | 0.004419591 | 0.032999965 | 1 |
| Heart_Failure | monocyte | *CLYBL* | 2.390918385 | 0.002978595 | 0.026522845 | 1 |
| Heart_Failure | monocyte | *GGA3* | 3.470948238 | 0.001750357 | 0.019662125 | 1 |
| Heart_Failure | monocyte | *PPP1R12C* | 3.965877033 | 0.000744424 | 0.014268876 | 1 |
| Heart_Failure | monocyte | *PPDPF* | 3.997438255 | 0.001590026 | 0.01864041 | 1 |
| Heart_Failure | monocyte | *INSIG2* | 4.980855483 | 0.000157355 | 0.011389066 | 0.989292523 |
| Heart_Failure | monocyte | *SNRPD2* | 3.38902796 | 0.003350454 | 0.028241098 | 1 |
| Heart_Failure | monocyte | *RTN2* | 4.935567567 | 0.006156765 | 0.039578303 | 1 |
| Heart_Failure | monocyte | *SYMPK* | 3.45137931 | 0.00037528 | 0.011389066 | 1 |
| Heart_Failure | monocyte | *PANK2* | 2.91524901 | 0.00197634 | 0.020812817 | 1 |
| Heart_Failure | monocyte | *PSMF1* | 3.029087859 | 0.000965254 | 0.015363418 | 1 |
| Heart_Failure | monocyte | *MKKS* | 2.328521986 | 0.004224507 | 0.032154331 | 1 |
| Heart_Failure | monocyte | *ITPA* | 3.713404907 | 0.00179317 | 0.019813108 | 1 |
| Heart_Failure | monocyte | *UROD* | 4.407901763 | 0.000130608 | 0.011389066 | 0.821133984 |
| Heart_Failure | monocyte | *ST3GAL3* | 1.226538548 | 0.007048547 | 0.04294013 | 1 |
| Heart_Failure | monocyte | *SCAF1* | 3.668880636 | 0.006668676 | 0.0414698 | 1 |
| Heart_Failure | monocyte | *SBDS* | 3.075962901 | 0.008727097 | 0.049218173 | 1 |
| Heart_Failure | monocyte | *BECN1* | 3.955577392 | 0.001389309 | 0.017539327 | 1 |
| Heart_Failure | monocyte | *ZNF384* | 5.029640002 | 0.00016104 | 0.011389066 | 1 |
| Heart_Failure | monocyte | *EMG1* | 3.131654258 | 0.005092369 | 0.035460966 | 1 |

| | | | | | | |
|---|---|---|---|---|---|---|
| Heart_Failure | monocyte | *RHOT1* | 2.125645721 | 0.000309231 | 0.011389066 | 1 |
| Heart_Failure | monocyte | *CANX* | 3.625971684 | 0.000266191 | 0.011389066 | 1 |
| Heart_Failure | monocyte | *IPPK* | 2.893042642 | 0.005106978 | 0.035460966 | 1 |
| Heart_Failure | monocyte | *RAP1B* | 2.912357777 | 0.000231295 | 0.011389066 | 1 |
| Heart_Failure | monocyte | *EMC1* | 2.696095612 | 0.003407771 | 0.02856621 | 1 |
| Heart_Failure | monocyte | *SIN3B* | 2.47633306 | 0.003235627 | 0.028019816 | 1 |
| Heart_Failure | monocyte | *TICAM1* | 4.410211186 | 0.002969603 | 0.026522845 | 1 |
| Heart_Failure | monocyte | *PTPN12* | 2.231194036 | 0.001423747 | 0.017655026 | 1 |
| Heart_Failure | monocyte | *SPECC1* | 3.470438345 | 0.000260401 | 0.011389066 | 1 |
| Heart_Failure | monocyte | *POT1* | 2.409133817 | 0.002004879 | 0.021007792 | 1 |
| Heart_Failure | monocyte | *TWSG1* | 4.183794871 | 0.000374621 | 0.011389066 | 1 |
| Heart_Failure | monocyte | *TMOD2* | 2.681742381 | 0.00212743 | 0.021667534 | 1 |
| Heart_Failure | monocyte | *ARPP19* | 2.653954247 | 0.002283403 | 0.022589237 | 1 |
| Heart_Failure | monocyte | *VPS13C* | 2.51805263 | 0.000702222 | 0.014081785 | 1 |
| Heart_Failure | monocyte | *SUMF2* | 2.459512796 | 0.006322935 | 0.040153831 | 1 |
| Heart_Failure | monocyte | *SPCS3* | 3.520108186 | 0.000661042 | 0.013626132 | 1 |
| Heart_Failure | monocyte | *SERGEF* | 3.903539274 | 0.000128651 | 0.011389066 | 0.808829029 |
| Heart_Failure | monocyte | *MIS18BP1* | 1.524241727 | 0.005410473 | 0.036773672 | 1 |
| Heart_Failure | monocyte | *REEP5* | 1.928964513 | 0.004950208 | 0.035186193 | 1 |
| Heart_Failure | monocyte | *SEC14L1* | 2.308604535 | 0.001565689 | 0.018607731 | 1 |
| Heart_Failure | monocyte | *TMEM8A* | 4.835858847 | 0.000249445 | 0.011389066 | 1 |
| Heart_Failure | monocyte | *ECSIT* | 3.085723276 | 0.006821337 | 0.041921553 | 1 |
| Heart_Failure | monocyte | *FAM98C* | 2.804934571 | 0.006114557 | 0.039427916 | 1 |
| Heart_Failure | monocyte | *RPL36* | 3.635471807 | 0.000425267 | 0.011479484 | 1 |
| Heart_Failure | monocyte | *BST2* | 3.727011666 | 0.001273392 | 0.016854347 | 1 |
| Heart_Failure | monocyte | *TUBGCP2* | 2.050597646 | 0.008229925 | 0.047732048 | 1 |
| Heart_Failure | monocyte | *UCK1* | 2.909551783 | 0.000255574 | 0.011389066 | 1 |
| Heart_Failure | monocyte | *ATPIF1* | 2.220722899 | 0.005710174 | 0.037841897 | 1 |
| Heart_Failure | monocyte | *CLIP1* | 2.445342886 | 0.001481334 | 0.01797905 | 1 |
| Heart_Failure | monocyte | *ZNF317* | 4.21532589 | 0.000556502 | 0.01265792 | 1 |
| Heart_Failure | monocyte | *ZNF236* | 2.15682904 | 0.006680978 | 0.041490592 | 1 |
| Heart_Failure | monocyte | *CHMP1A* | 4.314722734 | 3.77E-06 | 0.002961882 | 0.023695053 |
| Heart_Failure | monocyte | *TBC1D5* | 1.778360491 | 0.001681207 | 0.019217722 | 1 |
| Heart_Failure | monocyte | *EXOC4* | 1.596746695 | 0.001112323 | 0.016083148 | 1 |
| Heart_Failure | monocyte | *ACAP3* | 2.497901225 | 0.00853889 | 0.048892536 | 1 |
| Heart_Failure | monocyte | *C1orf159* | 3.920127394 | 0.000743892 | 0.014268876 | 1 |
| Heart_Failure | monocyte | *CHD1L* | 2.631736756 | 0.001947317 | 0.020744974 | 1 |
| Heart_Failure | monocyte | *ZSCAN5A* | 2.706644718 | 0.001821679 | 0.020006349 | 1 |
| Heart_Failure | monocyte | *RAF1* | 2.995443717 | 0.000630351 | 0.013525646 | 1 |
| Heart_Failure | monocyte | *TRIM5* | 2.607748986 | 0.001159664 | 0.016135389 | 1 |
| Heart_Failure | monocyte | *TRIM22* | 3.573966709 | 0.000408806 | 0.011389066 | 1 |
| Heart_Failure | monocyte | *IMMT* | 1.813608041 | 0.004802878 | 0.034549195 | 1 |
| Heart_Failure | monocyte | *MRPL35* | 1.806624697 | 0.00052574 | 0.012379507 | 1 |
| Heart_Failure | monocyte | *RPA1* | 3.617549432 | 5.84E-05 | 0.011101372 | 0.367448345 |
| Heart_Failure | monocyte | *SEC61G* | 2.632721343 | 0.002063382 | 0.021295573 | 1 |
| Heart_Failure | monocyte | *WBP2* | 2.279321454 | 0.008606404 | 0.048922662 | 1 |
| Heart_Failure | monocyte | *XAF1* | 1.219520772 | 0.005459692 | 0.036988234 | 1 |
| Heart_Failure | monocyte | *REEP2* | -1.405522097 | 0.000940574 | 0.015162532 | 1 |
| Heart_Failure | monocyte | *TERF2* | 2.960083073 | 0.001267572 | 0.016848255 | 1 |
| Heart_Failure | monocyte | *BTBD3* | 3.511479236 | 0.001255021 | 0.016787905 | 1 |

| | | | | | | |
|---|---|---|---|---|---|---|
| Heart_Failure | monocyte | *MUTYH* | 4.036655056 | 0.001345429 | 0.01728185 | 1 |
| Heart_Failure | monocyte | *CTNNBL1* | 3.276590739 | 0.000784119 | 0.01444259 | 1 |
| Heart_Failure | monocyte | *USPL1* | 2.337239643 | 0.003075535 | 0.026958725 | 1 |
| Heart_Failure | monocyte | *POMP* | 2.494968011 | 0.00575375 | 0.037938944 | 1 |
| Heart_Failure | monocyte | *COG6* | 1.488996504 | 0.007528279 | 0.045033576 | 1 |
| Heart_Failure | monocyte | *ZNF414* | 5.374901778 | 0.0010699 | 0.015840489 | 1 |
| Heart_Failure | monocyte | *CNDP2* | 3.543419905 | 0.00440667 | 0.03298183 | 1 |
| Heart_Failure | monocyte | *ACTR3B* | 3.412041975 | 0.002236252 | 0.02224575 | 1 |
| Heart_Failure | monocyte | *IPO8* | 2.7918328 | 0.001716829 | 0.019518456 | 1 |
| Heart_Failure | monocyte | *SWAP70* | 1.979767267 | 0.004986958 | 0.035307337 | 1 |
| Heart_Failure | monocyte | *SBF2* | 2.450861001 | 0.000343668 | 0.011389066 | 1 |
| Heart_Failure | monocyte | *GSC* | 10.23625759 | 0.004722137 | 0.034184404 | 1 |
| Heart_Failure | monocyte | *EDEM1* | 2.950848291 | 0.006649884 | 0.041426552 | 1 |
| Heart_Failure | monocyte | *PPHLN1* | 1.59122981 | 0.006379954 | 0.040408293 | 1 |
| Heart_Failure | monocyte | *NARS* | 3.962581933 | 0.000260566 | 0.011389066 | 1 |
| Heart_Failure | monocyte | *KIAA1468* | 2.902950165 | 0.001429131 | 0.017686906 | 1 |
| Heart_Failure | monocyte | *FBXO18* | 2.791530009 | 0.001439328 | 0.017743239 | 1 |
| Heart_Failure | monocyte | *DDB2* | 3.689562119 | 0.000102869 | 0.011389066 | 0.646740425 |
| Heart_Failure | monocyte | *ACP2* | 3.714094217 | 0.000334572 | 0.011389066 | 1 |
| Heart_Failure | monocyte | *TMEM165* | 1.74825884 | 0.008177194 | 0.047530474 | 1 |
| Heart_Failure | monocyte | *CLOCK* | 2.556160392 | 0.001133685 | 0.016135389 | 1 |
| Heart_Failure | monocyte | *OSTF1* | 2.663997479 | 0.00326458 | 0.028091022 | 1 |
| Heart_Failure | monocyte | *NAA35* | 1.898899981 | 0.005763148 | 0.037938944 | 1 |
| Heart_Failure | monocyte | *TMEM2* | 1.895197999 | 0.003705759 | 0.029869367 | 1 |
| Heart_Failure | monocyte | *ADAM19* | 0.765681811 | 0.003934737 | 0.030999611 | 1 |
| Heart_Failure | monocyte | *SRPK2* | 2.201025552 | 0.000991609 | 0.015508077 | 1 |
| Heart_Failure | monocyte | *ANKRD6* | 2.442694851 | 0.00316634 | 0.02753358 | 1 |
| Heart_Failure | monocyte | *SNX14* | 1.770878677 | 0.006206903 | 0.039738083 | 1 |
| Heart_Failure | monocyte | *PRR5L* | 2.539143771 | 0.002756946 | 0.025193195 | 1 |
| Heart_Failure | monocyte | *ATP5G2* | 2.480476576 | 0.004965209 | 0.035225419 | 1 |
| Heart_Failure | monocyte | *USP15* | 1.597692325 | 0.006422756 | 0.040520538 | 1 |
| Heart_Failure | monocyte | *NTPCR* | 2.499427968 | 0.004166845 | 0.031908589 | 1 |
| Heart_Failure | monocyte | *USP37* | 2.841042465 | 0.0003579 | 0.011389066 | 1 |
| Heart_Failure | monocyte | *GCC2* | 1.735319297 | 0.005687855 | 0.037800788 | 1 |
| Heart_Failure | monocyte | *MRPS9* | 1.919416199 | 0.004055209 | 0.031448189 | 1 |
| Heart_Failure | monocyte | *APPL2* | 2.520229032 | 0.000914123 | 0.015005461 | 1 |
| Heart_Failure | monocyte | *DRAM1* | 2.049723598 | 0.003326899 | 0.028241098 | 1 |
| Heart_Failure | monocyte | *SETDB2* | 2.157969947 | 0.00279616 | 0.025477477 | 1 |
| Heart_Failure | monocyte | *KDELR2* | 2.877578696 | 0.003242709 | 0.028042522 | 1 |
| Heart_Failure | monocyte | *ZDHHC4* | 3.610558713 | 0.000277323 | 0.011389066 | 1 |
| Heart_Failure | monocyte | *DDX56* | 4.472636525 | 0.000843631 | 0.014523588 | 1 |
| Heart_Failure | monocyte | *HUS1* | 2.148030954 | 0.005769099 | 0.037938944 | 1 |
| Heart_Failure | monocyte | *DBNL* | 3.001675814 | 0.000821578 | 0.014464609 | 1 |
| Heart_Failure | monocyte | *TTC5* | 3.841513749 | 0.000128964 | 0.011389066 | 0.810797849 |
| Heart_Failure | monocyte | *NMT1* | 1.772387622 | 2.40E-05 | 0.007936725 | 0.150797768 |
| Heart_Failure | monocyte | *RPS6KC1* | 2.08785079 | 0.002552514 | 0.024059456 | 1 |
| Heart_Failure | monocyte | *CBWD2* | 3.21556218 | 0.004038726 | 0.031425084 | 1 |
| Heart_Failure | monocyte | *IL1RN* | 3.975889925 | 0.001962279 | 0.020744974 | 1 |
| Heart_Failure | monocyte | *SAP130* | 2.982152748 | 0.002843641 | 0.025686744 | 1 |
| Heart_Failure | monocyte | *UGGT1* | 2.634852281 | 0.002126849 | 0.021667534 | 1 |

| | | | | | | |
|---|---|---|---|---|---|---|
| Heart_Failure | monocyte | *GABBR2* | 8.563327705 | 0.002523732 | 0.023895639 | 1 |
| Heart_Failure | monocyte | *APTX* | 1.393461966 | 0.008591125 | 0.048922662 | 1 |
| Heart_Failure | monocyte | *TLN1* | 3.077299694 | 0.000196865 | 0.011389066 | 1 |
| Heart_Failure | monocyte | *PPIL1* | 10.1007368 | 0.000310156 | 0.011389066 | 1 |
| Heart_Failure | monocyte | *TMEM63B* | 2.403029109 | 0.002976858 | 0.026522845 | 1 |
| Heart_Failure | monocyte | *UQCC2* | 2.973866012 | 0.006031629 | 0.039013224 | 1 |
| Heart_Failure | monocyte | *FLOT1* | 2.039445707 | 0.001403923 | 0.017630601 | 1 |
| Heart_Failure | monocyte | *PRCP* | 2.521453373 | 0.003809813 | 0.030495028 | 1 |
| Heart_Failure | monocyte | *TGS1* | 1.516121162 | 0.005515309 | 0.03716864 | 1 |
| Heart_Failure | monocyte | *NEK1* | 1.50478093 | 0.007101868 | 0.043108986 | 1 |
| Heart_Failure | monocyte | *DDX60* | 3.190773723 | 0.002400562 | 0.023290634 | 1 |
| Heart_Failure | monocyte | *MAP2K5* | 1.940282986 | 0.001957147 | 0.020744974 | 1 |
| Heart_Failure | monocyte | *CENPO* | 4.224840556 | 0.004412152 | 0.032983587 | 1 |
| Heart_Failure | monocyte | *LRPPRC* | 1.634161811 | 0.00798678 | 0.046815675 | 1 |
| Heart_Failure | monocyte | *CYP2C9* | 1.000992299 | 0.005724136 | 0.037841897 | 1 |
| Heart_Failure | monocyte | *ARL3* | 3.166373177 | 0.001018507 | 0.015771801 | 1 |
| Heart_Failure | monocyte | *DNAJC13* | 2.386238474 | 0.001870729 | 0.020319415 | 1 |
| Heart_Failure | monocyte | *RPS24* | 2.568275687 | 0.001629933 | 0.018937074 | 1 |
| Heart_Failure | monocyte | *CARF* | 1.888431326 | 0.001051358 | 0.015840489 | 1 |
| Heart_Failure | monocyte | *NAB1* | 2.385391124 | 0.003462738 | 0.028720623 | 1 |
| Heart_Failure | monocyte | *FASTKD1* | 2.622189602 | 0.005098445 | 0.035460966 | 1 |
| Heart_Failure | monocyte | *SLC35A5* | 1.756721027 | 0.000364828 | 0.011389066 | 1 |
| Heart_Failure | monocyte | *COX17* | 3.804938139 | 0.001068567 | 0.015840489 | 1 |
| Heart_Failure | monocyte | *SPPL2A* | 3.059161411 | 0.000515132 | 0.012314203 | 1 |
| Heart_Failure | monocyte | *HERC3* | 2.953212211 | 0.00077949 | 0.01444259 | 1 |
| Heart_Failure | monocyte | *BBS7* | 4.49789525 | 0.00024617 | 0.011389066 | 1 |
| Heart_Failure | monocyte | *KIAA1109* | 2.336368986 | 0.000928114 | 0.015111629 | 1 |
| Heart_Failure | monocyte | *BMP2K* | 2.192147867 | 0.000340381 | 0.011389066 | 1 |
| Heart_Failure | monocyte | *G3BP2* | 1.630397781 | 0.005977395 | 0.038926702 | 1 |
| Heart_Failure | monocyte | *PPA2* | 1.650281046 | 0.008998191 | 0.049751338 | 1 |
| Heart_Failure | monocyte | *PAPSS1* | 2.26661679 | 0.000810039 | 0.01444259 | 1 |
| Heart_Failure | monocyte | *CPNE8* | 2.622289462 | 6.28E-05 | 0.011101372 | 0.395079709 |
| Heart_Failure | monocyte | *ALG10* | 6.989335233 | 0.000641018 | 0.013540529 | 1 |
| Heart_Failure | monocyte | *CD27* | 3.1518563 | 0.008629379 | 0.048964714 | 1 |
| Heart_Failure | monocyte | *POC1B* | 3.544076121 | 0.000618415 | 0.0134532 | 1 |
| Heart_Failure | monocyte | *LMBR1L* | 3.432641417 | 0.002734128 | 0.025130796 | 1 |
| Heart_Failure | monocyte | *LPAR6* | 2.376630749 | 0.001705578 | 0.019460925 | 1 |
| Heart_Failure | monocyte | *TMX1* | 4.464823379 | 0.000347452 | 0.011389066 | 1 |
| Heart_Failure | monocyte | *NIPA2* | 4.350657414 | 0.000248786 | 0.011389066 | 1 |
| Heart_Failure | monocyte | *MFAP1* | 3.433552852 | 0.001223339 | 0.016601601 | 1 |
| Heart_Failure | monocyte | *TCF12* | 1.340461678 | 0.006778971 | 0.041824721 | 1 |
| Heart_Failure | monocyte | *BAHD1* | 3.824797385 | 0.003272544 | 0.028091022 | 1 |
| Heart_Failure | monocyte | *ANP32A* | 3.420131366 | 0.000812 | 0.01444259 | 1 |
| Heart_Failure | monocyte | *UBE2Q2* | 2.786466023 | 0.001472142 | 0.01790205 | 1 |
| Heart_Failure | monocyte | *SCAMP2* | 2.940158471 | 0.000943464 | 0.015170228 | 1 |
| Heart_Failure | monocyte | *POLG* | 3.874133648 | 0.001175841 | 0.016176179 | 1 |
| Heart_Failure | monocyte | *SEC11A* | 3.430511216 | 0.000424978 | 0.011479484 | 1 |
| Heart_Failure | monocyte | *ZFHX3* | 1.825807175 | 0.003029846 | 0.02671487 | 1 |
| Heart_Failure | monocyte | *RPS2* | 3.121891708 | 0.0030774 | 0.026958725 | 1 |
| Heart_Failure | monocyte | *RANBP10* | 2.27018099 | 0.00447702 | 0.033192249 | 1 |

| | | | | | | |
|---|---|---|---|---|---|---|
| Heart_Failure | monocyte | *MYO19* | 2.265511493 | 0.002191365 | 0.022058295 | 1 |
| Heart_Failure | monocyte | *DDX52* | 2.443302448 | 0.007653531 | 0.045351322 | 1 |
| Heart_Failure | monocyte | *NPEPPS* | 2.178979004 | 0.0023861 | 0.02318611 | 1 |
| Heart_Failure | monocyte | *G6PC3* | 5.2694647 | 0.00141652 | 0.017630601 | 1 |
| Heart_Failure | monocyte | *RPRD1A* | 2.497409269 | 0.001793114 | 0.019813108 | 1 |
| Heart_Failure | monocyte | *GALNT1* | 1.919440401 | 0.006463628 | 0.040520538 | 1 |
| Heart_Failure | monocyte | *ESCO1* | 1.769397954 | 0.007136716 | 0.043217151 | 1 |
| Heart_Failure | monocyte | *TBCD* | 3.075278056 | 0.000475725 | 0.011604726 | 1 |
| Heart_Failure | monocyte | *NARF* | 2.699884575 | 0.001039845 | 0.015840489 | 1 |
| Heart_Failure | monocyte | *SMAD4* | 2.806206685 | 0.001654805 | 0.019022043 | 1 |
| Heart_Failure | monocyte | *PIP4K2B* | 3.586355966 | 0.001035845 | 0.015840489 | 1 |
| Heart_Failure | monocyte | *PFKL* | 1.992148686 | 0.007411171 | 0.044461843 | 1 |
| Heart_Failure | monocyte | *DPP9* | 2.14275419 | 0.008012044 | 0.046815675 | 1 |
| Heart_Failure | monocyte | *CCDC97* | 4.130554329 | 0.00191094 | 0.020572053 | 1 |
| Heart_Failure | monocyte | *SIRT3* | 3.262190129 | 0.000294803 | 0.011389066 | 1 |
| Heart_Failure | monocyte | *DOPEY2* | 2.290359522 | 0.006786529 | 0.041830301 | 1 |
| Heart_Failure | monocyte | *AKT1* | 3.481551021 | 0.003568019 | 0.029170528 | 1 |
| Heart_Failure | monocyte | *RERE* | 1.433228939 | 0.001514036 | 0.018237795 | 1 |
| Heart_Failure | monocyte | *SH3BGRL3* | 4.372287353 | 0.000237441 | 0.011389066 | 1 |
| Heart_Failure | monocyte | *PIGK* | 3.091254408 | 0.002165915 | 0.021927707 | 1 |
| Heart_Failure | monocyte | *RFWD2* | 2.407239767 | 0.000893447 | 0.014889101 | 1 |
| Heart_Failure | monocyte | *PPOX* | 2.513926194 | 0.000403457 | 0.011389066 | 1 |
| Heart_Failure | monocyte | *RRNAD1* | 2.768638105 | 0.000431257 | 0.011479484 | 1 |
| Heart_Failure | monocyte | *PIGM* | 3.270049389 | 0.006354248 | 0.040311967 | 1 |
| Heart_Failure | monocyte | *CERS2* | 3.338366485 | 0.001814833 | 0.020006349 | 1 |
| Heart_Failure | monocyte | *POGZ* | 1.761938523 | 0.003614999 | 0.029323162 | 1 |
| Heart_Failure | monocyte | *TP53BP2* | 3.01524133 | 0.001948576 | 0.020744974 | 1 |
| Heart_Failure | monocyte | *GATAD2B* | 1.685809058 | 0.003431904 | 0.028571324 | 1 |
| Heart_Failure | monocyte | *GALNT2* | 1.851278303 | 0.006251827 | 0.039944348 | 1 |
| Heart_Failure | monocyte | *CEP170* | 2.096399732 | 0.000436383 | 0.011517615 | 1 |
| Heart_Failure | monocyte | *FBXO28* | 2.745789134 | 0.001312009 | 0.01715614 | 1 |
| Heart_Failure | monocyte | *CNIH4* | 4.731057936 | 0.000106122 | 0.011389066 | 0.667188061 |
| Heart_Failure | monocyte | *GUK1* | 2.872648023 | 0.004927414 | 0.03508341 | 1 |
| Heart_Failure | monocyte | *EML4* | 3.162496895 | 0.000240451 | 0.011389066 | 1 |
| Heart_Failure | monocyte | *ASXL2* | 2.374724522 | 0.002130522 | 0.021667534 | 1 |
| Heart_Failure | monocyte | *UBXN4* | 1.967025229 | 0.002908136 | 0.026119216 | 1 |
| Heart_Failure | monocyte | *POLR2D* | 4.715531637 | 0.000905642 | 0.014983607 | 1 |
| Heart_Failure | monocyte | *NBEAL1* | 0.918588509 | 0.006321869 | 0.040153831 | 1 |
| Heart_Failure | monocyte | *SUMF1* | 2.194171496 | 0.000502485 | 0.012150464 | 1 |
| Heart_Failure | monocyte | *FANCD2* | 6.170230136 | 0.000643967 | 0.013540529 | 1 |
| Heart_Failure | monocyte | *ARL6IP5* | 2.312771878 | 0.002851665 | 0.025722268 | 1 |
| Heart_Failure | monocyte | *TMF1* | 2.883519954 | 0.001417664 | 0.017630601 | 1 |
| Heart_Failure | monocyte | *NXPE3* | 3.757757426 | 0.003909848 | 0.030937005 | 1 |
| Heart_Failure | monocyte | *ATP10D* | 2.270415072 | 0.004341765 | 0.032729825 | 1 |
| Heart_Failure | monocyte | *CISD2* | 4.936331635 | 4.43E-06 | 0.003093116 | 0.027838042 |
| Heart_Failure | monocyte | *PPIP5K2* | 2.570148354 | 0.001760741 | 0.019662125 | 1 |
| Heart_Failure | monocyte | *TSLP* | 1.716140076 | 7.79E-05 | 0.011389066 | 0.489668051 |
| Heart_Failure | monocyte | *COMMD10* | 1.920860325 | 0.001879357 | 0.020319415 | 1 |
| Heart_Failure | monocyte | *TRIM7* | 3.600710278 | 0.00685059 | 0.041937353 | 1 |
| Heart_Failure | monocyte | *MUT* | 3.710631781 | 0.000991295 | 0.015508077 | 1 |

| | | | | | | |
|---|---|---|---|---|---|---|
| Heart_Failure | monocyte | *PHIP* | 3.055612068 | 0.000159106 | 0.011389066 | 1 |
| Heart_Failure | monocyte | *MMS22L* | 2.51511473 | 0.00538918 | 0.036708316 | 1 |
| Heart_Failure | monocyte | *DYNLT1* | 2.826699456 | 0.005093378 | 0.035460966 | 1 |
| Heart_Failure | monocyte | *WTAP* | 1.778620818 | 0.005687647 | 0.037800788 | 1 |
| Heart_Failure | monocyte | *NOM1* | 2.859358902 | 0.005849678 | 0.038269434 | 1 |
| Heart_Failure | monocyte | *FBXO25* | 3.915824713 | 0.000524734 | 0.012379507 | 1 |
| Heart_Failure | monocyte | *CCDC25* | 4.089543064 | 0.000166459 | 0.011389066 | 1 |
| Heart_Failure | monocyte | *BIN3* | 2.356221591 | 0.008348737 | 0.048198815 | 1 |
| Heart_Failure | monocyte | *MTDH* | 2.40386158 | 0.001159986 | 0.016135389 | 1 |
| Heart_Failure | monocyte | *UTP23* | 3.695043225 | 0.001245641 | 0.016697963 | 1 |
| Heart_Failure | monocyte | *FAM135B* | 6.188714125 | 0.004669049 | 0.034014266 | 1 |
| Heart_Failure | monocyte | *HAUS6* | 3.692545684 | 0.000980472 | 0.015508077 | 1 |
| Heart_Failure | monocyte | *CBWD5* | 4.29269685 | 0.000870614 | 0.014721728 | 1 |
| Heart_Failure | monocyte | *LRSAM1* | 3.426652076 | 0.001115357 | 0.016083148 | 1 |
| Heart_Failure | monocyte | *SEC16A* | 2.615213853 | 0.002172945 | 0.02196351 | 1 |
| Heart_Failure | monocyte | *DPH7* | 4.315431858 | 0.000670627 | 0.01373365 | 1 |
| Heart_Failure | monocyte | *RSU1* | 3.355964596 | 0.000143826 | 0.011389066 | 0.904231082 |
| Heart_Failure | monocyte | *RPP30* | 2.066207734 | 0.005075511 | 0.035460966 | 1 |
| Heart_Failure | monocyte | *DNAJB12* | 2.746882562 | 0.003859734 | 0.030677814 | 1 |
| Heart_Failure | monocyte | *MTG1* | 1.557612602 | 0.003421664 | 0.028571324 | 1 |
| Heart_Failure | monocyte | *PAOX* | 5.840558151 | 0.001145593 | 0.016135389 | 1 |
| Heart_Failure | monocyte | *PDCD11* | 2.378099368 | 0.006407807 | 0.040488323 | 1 |
| Heart_Failure | monocyte | *RGS10* | 2.817820107 | 0.007751013 | 0.045713524 | 1 |
| Heart_Failure | monocyte | *INTS4* | 2.020128115 | 0.002205686 | 0.022061708 | 1 |
| Heart_Failure | monocyte | *RPS3* | 1.828533986 | 0.004375822 | 0.032868331 | 1 |
| Heart_Failure | monocyte | *NPAT* | 2.365106367 | 0.003329593 | 0.028241098 | 1 |
| Heart_Failure | monocyte | *ROM1* | 0.942355987 | 0.005604666 | 0.037565605 | 1 |
| Heart_Failure | monocyte | *DUSP15* | -3.431066095 | 0.00356577 | 0.029170528 | 1 |
| Heart_Failure | monocyte | *TAOK2* | 2.15883107 | 0.007333792 | 0.04412206 | 1 |
| Heart_Failure | monocyte | *ARID5B* | 1.218234507 | 0.007848772 | 0.046073974 | 1 |
| Heart_Failure | monocyte | *PDCD4* | 1.972860448 | 0.003346413 | 0.028241098 | 1 |
| Heart_Failure | monocyte | *WDR17* | 3.115487865 | 0.008714988 | 0.049218173 | 1 |
| Heart_Failure | monocyte | *CCT5* | 2.865881798 | 0.000723276 | 0.014254661 | 1 |
| Heart_Failure | monocyte | *GXYLT1* | 3.864728934 | 0.000842804 | 0.014523588 | 1 |
| Heart_Failure | monocyte | *FAM177A1* | 2.014644664 | 0.003978826 | 0.031151779 | 1 |
| Heart_Failure | monocyte | *NDUFC2* | 4.207066179 | 0.002214552 | 0.022064796 | 1 |
| Heart_Failure | monocyte | *ANKRD50* | 4.470713862 | 0.000122534 | 0.011389066 | 0.770368287 |
| Heart_Failure | monocyte | *C4orf33* | 4.033071496 | 0.000913293 | 0.015005461 | 1 |
| Heart_Failure | monocyte | *NCAPD3* | 3.405728241 | 0.000813214 | 0.01444259 | 1 |
| Heart_Failure | monocyte | *VTI1A* | 2.219472954 | 0.001981355 | 0.020830739 | 1 |
| Heart_Failure | monocyte | *ITIH2* | 7.352405852 | 0.002436979 | 0.023462924 | 1 |
| Heart_Failure | monocyte | *KIN* | 2.639311149 | 0.000653971 | 0.013585129 | 1 |
| Heart_Failure | monocyte | *MFSD6* | 3.425036934 | 0.000475428 | 0.011604726 | 1 |
| Heart_Failure | monocyte | *ADAM17* | 2.078954968 | 0.003009494 | 0.026660729 | 1 |
| Heart_Failure | monocyte | *GUF1* | 2.647503225 | 0.005775023 | 0.037938944 | 1 |
| Heart_Failure | monocyte | *CENPJ* | 1.876277961 | 0.008686768 | 0.049157255 | 1 |
| Heart_Failure | monocyte | *CACUL1* | 1.843332286 | 0.005147015 | 0.035529945 | 1 |
| Heart_Failure | monocyte | *TGOLN2* | 3.078039296 | 0.002683901 | 0.024741478 | 1 |
| Heart_Failure | monocyte | *TADA1* | 5.354008003 | 0.000325598 | 0.011389066 | 1 |
| Heart_Failure | monocyte | *ZFP36L2* | 3.01014964 | 0.000472713 | 0.011604726 | 1 |

| | | | | | | |
|---|---|---|---|---|---|---|
| Heart_Failure | monocyte | *TMEM123* | 2.909105333 | 0.000998896 | 0.015583272 | 1 |
| Heart_Failure | monocyte | *MBNL1* | 2.295734596 | 0.00021644 | 0.011389066 | 1 |
| Heart_Failure | monocyte | *PELO* | 7.381131591 | 0.000243855 | 0.011389066 | 1 |
| Heart_Failure | monocyte | *SAR1B* | 2.330056676 | 0.005647865 | 0.037654428 | 1 |
| Heart_Failure | monocyte | *CAST* | 1.49428446 | 0.005152507 | 0.035529945 | 1 |
| Heart_Failure | monocyte | *AHCTF1* | 2.044646338 | 0.003616956 | 0.029323162 | 1 |
| Heart_Failure | monocyte | *ASAP1* | 2.096542712 | 0.001266856 | 0.016848255 | 1 |
| Heart_Failure | monocyte | *JAZF1* | 2.186065212 | 0.003895095 | 0.030880785 | 1 |
| Heart_Failure | monocyte | *PRKCA* | 2.826655192 | 0.001075853 | 0.015840489 | 1 |
| Heart_Failure | monocyte | *LRRK1* | 2.332922005 | 0.004851199 | 0.034777066 | 1 |
| Heart_Failure | monocyte | *GAL3ST2* | -7.37315327 | 0.006723717 | 0.041557112 | 1 |
| Heart_Failure | monocyte | *FLCN* | 1.956672178 | 0.00311909 | 0.027197947 | 1 |
| Heart_Failure | monocyte | *FBXL18* | 2.331553268 | 0.008723158 | 0.049218173 | 1 |
| Heart_Failure | monocyte | *CDK19* | 3.413105286 | 0.00013251 | 0.011389066 | 0.833091014 |
| Heart_Failure | monocyte | *TTC39B* | 2.01936516 | 0.00080205 | 0.01444259 | 1 |
| Heart_Failure | monocyte | *HEATR3* | 1.264927594 | 0.00622063 | 0.039785454 | 1 |
| Heart_Failure | monocyte | *PDIA4* | 3.59809057 | 0.001934732 | 0.020703807 | 1 |
| Heart_Failure | monocyte | *FAM126B* | 1.835310891 | 0.007103692 | 0.043108986 | 1 |
| Heart_Failure | monocyte | *RRAGA* | 3.750423405 | 0.002060688 | 0.021295573 | 1 |
| Heart_Failure | monocyte | *TMBIM4* | 1.828053744 | 0.001776355 | 0.01973135 | 1 |
| Heart_Failure | monocyte | *DCK* | 2.820768848 | 0.003999736 | 0.031237687 | 1 |
| Heart_Failure | monocyte | *DRAM2* | 1.851092609 | 0.003400548 | 0.02854372 | 1 |
| Heart_Failure | monocyte | *C14orf2* | 2.807952781 | 0.000124217 | 0.011389066 | 0.780955344 |
| Heart_Failure | monocyte | *ATAD2* | 2.525737287 | 0.004704411 | 0.034153156 | 1 |
| Heart_Failure | monocyte | *VPS8* | 2.670046682 | 0.000426368 | 0.011479484 | 1 |
| Heart_Failure | monocyte | *RPUSD3* | 4.973901335 | 0.000711278 | 0.0141962 | 1 |
| Heart_Failure | monocyte | *EXOG* | 4.035078253 | 0.000293842 | 0.011389066 | 1 |
| Heart_Failure | monocyte | *RNF111* | 1.489691907 | 0.006393684 | 0.04043973 | 1 |
| Heart_Failure | monocyte | *DYRK1A* | 2.552194027 | 0.000666706 | 0.013697975 | 1 |
| Heart_Failure | monocyte | *SLC38A10* | 3.780628783 | 0.000360549 | 0.011389066 | 1 |
| Heart_Failure | monocyte | *SPPL3* | 2.819653294 | 8.35E-05 | 0.011389066 | 0.52506794 |
| Heart_Failure | monocyte | *C12orf43* | 2.625384818 | 0.001403245 | 0.017630601 | 1 |
| Heart_Failure | monocyte | *LDLRAP1* | 5.551011016 | 0.000542683 | 0.012549055 | 1 |
| Heart_Failure | monocyte | *ABHD3* | 2.666035772 | 0.000124912 | 0.011389066 | 0.785321117 |
| Heart_Failure | monocyte | *RNF207* | 6.294922581 | 0.000989018 | 0.015508077 | 1 |
| Heart_Failure | monocyte | *MITD1* | 2.15210234 | 0.00220918 | 0.022061708 | 1 |
| Heart_Failure | monocyte | *AHCYL2* | 2.77926871 | 0.000809484 | 0.01444259 | 1 |
| Heart_Failure | monocyte | *FAM86C1* | 9.498883524 | 0.001740213 | 0.019607023 | 1 |
| Heart_Failure | monocyte | *ELK4* | 4.882793637 | 4.12E-05 | 0.009491021 | 0.259117154 |
| Heart_Failure | monocyte | *NIT1* | 1.402759793 | 0.001776301 | 0.01973135 | 1 |
| Heart_Failure | monocyte | *ZNF276* | 2.414446866 | 0.006005869 | 0.038926702 | 1 |
| Heart_Failure | monocyte | *CDC42SE2* | 2.927505854 | 0.000719537 | 0.014254661 | 1 |
| Heart_Failure | monocyte | *EPB41* | 0.970108595 | 0.004690921 | 0.034134048 | 1 |
| Heart_Failure | monocyte | *ALG8* | 2.326324418 | 0.004519394 | 0.033427566 | 1 |
| Heart_Failure | monocyte | *RCAN1* | 2.670268957 | 0.003474868 | 0.028745389 | 1 |
| Heart_Failure | monocyte | *UBE2Z* | 3.846780418 | 0.000731393 | 0.014268876 | 1 |
| Heart_Failure | monocyte | *PSMD4* | 2.354446156 | 0.00606884 | 0.039173303 | 1 |
| Heart_Failure | monocyte | *EFCAB14* | 2.444231952 | 0.003329873 | 0.028241098 | 1 |
| Heart_Failure | monocyte | *CALM3* | 2.649270147 | 0.008866411 | 0.049424614 | 1 |
| Heart_Failure | monocyte | *DFFA* | 2.660034632 | 0.004009481 | 0.031274945 | 1 |

| | | | | | | |
|---|---|---|---|---|---|---|
| Heart_Failure | monocyte | *SSU72* | 3.70828291 | 0.001120626 | 0.016109997 | 1 |
| Heart_Failure | monocyte | *ZNF714* | 2.138171771 | 0.006858761 | 0.041946527 | 1 |
| Heart_Failure | monocyte | *PCSK7* | 2.504880935 | 0.006268345 | 0.039968644 | 1 |
| Heart_Failure | monocyte | *ZNF333* | 4.07797616 | 6.96E-05 | 0.011101372 | 0.437560377 |
| Heart_Failure | monocyte | *RPL8* | 3.975181133 | 0.000241943 | 0.011389066 | 1 |
| Heart_Failure | monocyte | *MFSD12* | 2.657376138 | 0.006428389 | 0.040520538 | 1 |
| Heart_Failure | monocyte | *PGAP3* | 6.681439629 | 0.000336596 | 0.011389066 | 1 |
| Heart_Failure | monocyte | *PRPSAP1* | 3.725393954 | 0.000401604 | 0.011389066 | 1 |
| Heart_Failure | monocyte | *RACGAP1* | 4.854761392 | 0.00081177 | 0.01444259 | 1 |
| Heart_Failure | monocyte | *LARP4* | 1.472127587 | 0.003968073 | 0.031145159 | 1 |
| Heart_Failure | monocyte | *NXF1* | 2.601110816 | 0.000845539 | 0.014523588 | 1 |
| Heart_Failure | monocyte | *RPL29* | 2.327034858 | 0.002294541 | 0.022610939 | 1 |
| Heart_Failure | monocyte | *USP24* | 1.239178221 | 0.004546354 | 0.033430274 | 1 |
| Heart_Failure | monocyte | *SLC25A34* | 9.748402869 | 0.000105788 | 0.011389066 | 0.665086573 |
| Heart_Failure | monocyte | *RBBP4* | 3.735800537 | 0.000163248 | 0.011389066 | 1 |
| Heart_Failure | monocyte | *TM2D1* | 2.018474918 | 0.00415701 | 0.031877388 | 1 |
| Heart_Failure | monocyte | *B3GALT2* | 5.613353635 | 0.00272207 | 0.025056591 | 1 |
| Heart_Failure | monocyte | *ZNF496* | 3.866519367 | 0.001580222 | 0.01864041 | 1 |
| Heart_Failure | monocyte | *SNED1* | 1.627028252 | 0.004797264 | 0.034549195 | 1 |
| Heart_Failure | monocyte | *PDLIM5* | 2.225149732 | 0.001090438 | 0.016014809 | 1 |
| Heart_Failure | monocyte | *LYSMD1* | 5.266630462 | 0.000242404 | 0.011389066 | 1 |
| Heart_Failure | monocyte | *ERCC3* | 2.383508454 | 0.00026585 | 0.011389066 | 1 |
| Heart_Failure | monocyte | *DHX57* | 2.62865695 | 0.003625422 | 0.029323162 | 1 |
| Heart_Failure | monocyte | *ANTXR2* | 1.302755361 | 0.005722902 | 0.037841897 | 1 |
| Heart_Failure | monocyte | *CCT3* | 2.006528056 | 0.008520004 | 0.048873415 | 1 |
| Heart_Failure | monocyte | *EFHB* | 0.482251122 | 0.002569994 | 0.024099196 | 1 |
| Heart_Failure | monocyte | *GMPS* | 3.55926291 | 0.000555858 | 0.01265792 | 1 |
| Heart_Failure | monocyte | *RPP14* | 4.826049602 | 0.001124512 | 0.016109997 | 1 |
| Heart_Failure | monocyte | *GYG1* | 3.931354845 | 0.001831979 | 0.0200307 | 1 |
| Heart_Failure | monocyte | *HPS3* | 3.882972986 | 0.000257737 | 0.011389066 | 1 |
| Heart_Failure | monocyte | *RYK* | 1.654869562 | 0.008601103 | 0.048922662 | 1 |
| Heart_Failure | monocyte | *CDCP1* | 5.643431838 | 0.00053051 | 0.012398944 | 1 |
| Heart_Failure | monocyte | *DTX3L* | 3.457878979 | 0.000754649 | 0.01431753 | 1 |
| Heart_Failure | monocyte | *YEATS2* | 3.76779267 | 0.000274012 | 0.011389066 | 1 |
| Heart_Failure | monocyte | *FAM208A* | 1.9444347 | 0.002981196 | 0.026522845 | 1 |
| Heart_Failure | monocyte | *DNAJB14* | 2.586656946 | 0.001040548 | 0.015840489 | 1 |
| Heart_Failure | monocyte | *SHISA5* | 4.670962407 | 0.000281544 | 0.011389066 | 1 |
| Heart_Failure | monocyte | *RNF123* | 1.951826908 | 5.12E-06 | 0.003220133 | 0.032201331 |
| Heart_Failure | monocyte | *RAD54L2* | 2.105337554 | 0.001627467 | 0.018937074 | 1 |
| Heart_Failure | monocyte | *ANXA5* | 1.806462675 | 0.004100273 | 0.031637758 | 1 |
| Heart_Failure | monocyte | *OTUD4* | 3.375333894 | 0.000935013 | 0.015111629 | 1 |
| Heart_Failure | monocyte | *PGGT1B* | 2.67022265 | 0.003604367 | 0.029323162 | 1 |
| Heart_Failure | monocyte | *TIGD6* | 9.079590187 | 0.000213735 | 0.011389066 | 1 |
| Heart_Failure | monocyte | *SERINC5* | 1.659491589 | 0.006454258 | 0.040520538 | 1 |
| Heart_Failure | monocyte | *CASP3* | 3.349759893 | 0.001661445 | 0.019061143 | 1 |
| Heart_Failure | monocyte | *ERAP1* | 1.150062927 | 0.005997207 | 0.038926702 | 1 |
| Heart_Failure | monocyte | *RICTOR* | 2.026292895 | 0.00248813 | 0.023773364 | 1 |
| Heart_Failure | monocyte | *UBLCP1* | 2.728434732 | 0.00057613 | 0.012982535 | 1 |
| Heart_Failure | monocyte | *DCBLD1* | 2.718918447 | 0.000887214 | 0.014874442 | 1 |
| Heart_Failure | monocyte | *PDSS2* | 3.066676779 | 0.000278699 | 0.011389066 | 1 |

| | | | | | | |
|---|---|---|---|---|---|---|
| Heart_Failure | monocyte | *STXBP5* | 2.746248041 | 0.000377489 | 0.011389066 | 1 |
| Heart_Failure | monocyte | *GALNT10* | 2.792405281 | 0.000989982 | 0.015508077 | 1 |
| Heart_Failure | monocyte | *SAP30L* | 2.462273533 | 0.001583928 | 0.01864041 | 1 |
| Heart_Failure | monocyte | *SLU7* | 1.696309372 | 0.008551146 | 0.048918155 | 1 |
| Heart_Failure | monocyte | *USP49* | 2.088128006 | 0.003827462 | 0.030575929 | 1 |
| Heart_Failure | monocyte | *BRI3* | 2.872584232 | 0.000465783 | 0.011604726 | 1 |
| Heart_Failure | monocyte | *PEX2* | 2.350996175 | 0.007082543 | 0.04310547 | 1 |
| Heart_Failure | monocyte | *OXR1* | 2.535128452 | 0.001271642 | 0.016854347 | 1 |
| Heart_Failure | monocyte | *INTS1* | 4.66960014 | 0.000722341 | 0.014254661 | 1 |
| Heart_Failure | monocyte | *INTS8* | 2.286550045 | 0.003351019 | 0.028241098 | 1 |
| Heart_Failure | monocyte | *TMEM65* | 3.69579907 | 0.000178376 | 0.011389066 | 1 |
| Heart_Failure | monocyte | *TAF3* | 2.659114232 | 0.002205545 | 0.022061708 | 1 |
| Heart_Failure | monocyte | *PDZD8* | 2.431263134 | 0.001584767 | 0.01864041 | 1 |
| Heart_Failure | monocyte | *STK32C* | 3.008639552 | 0.007357675 | 0.044223425 | 1 |
| Heart_Failure | monocyte | *METTL3* | 2.281547316 | 0.006708754 | 0.041554619 | 1 |
| Heart_Failure | monocyte | *PTER* | 3.877347216 | 0.000200188 | 0.011389066 | 1 |
| Heart_Failure | monocyte | *TAF10* | 5.405680342 | 0.000117144 | 0.011389066 | 0.736482207 |
| Heart_Failure | monocyte | *RPL27A* | 4.335309254 | 3.29E-05 | 0.009395601 | 0.206703218 |
| Heart_Failure | monocyte | *TMEM41B* | 1.8698199 | 0.009009117 | 0.049751338 | 1 |
| Heart_Failure | monocyte | *ZNF143* | 3.731064858 | 0.001224629 | 0.016601601 | 1 |
| Heart_Failure | monocyte | *TMX3* | 2.137868462 | 0.005036683 | 0.03545983 | 1 |
| Heart_Failure | monocyte | *BEAN1* | 2.578642599 | 0.004551664 | 0.033430274 | 1 |
| Heart_Failure | monocyte | *AP1G1* | 2.437531622 | 0.002323921 | 0.022793274 | 1 |
| Heart_Failure | monocyte | *FAM111A* | 2.334013397 | 0.004900361 | 0.03493035 | 1 |
| Heart_Failure | monocyte | *MTMR10* | 1.940080308 | 0.002523495 | 0.023895639 | 1 |
| Heart_Failure | monocyte | *TSC22D4* | 4.156181732 | 0.000544917 | 0.012549055 | 1 |
| Heart_Failure | monocyte | *CCNDBP1* | 2.386520884 | 0.006382286 | 0.040408293 | 1 |
| Heart_Failure | monocyte | *AKTIP* | 4.990034297 | 0.000262617 | 0.011389066 | 1 |
| Heart_Failure | monocyte | *DUSP18* | 4.40750529 | 7.35E-05 | 0.011269868 | 0.462064579 |
| Heart_Failure | monocyte | *SP2* | 3.550315637 | 0.004175219 | 0.03192457 | 1 |
| Heart_Failure | monocyte | *HDHD2* | 1.327733936 | 0.001635742 | 0.018937074 | 1 |
| Heart_Failure | monocyte | *GATAD2A* | 2.305438091 | 0.005545266 | 0.037326643 | 1 |
| Heart_Failure | monocyte | *PROCA1* | 2.764501534 | 0.001962548 | 0.020744974 | 1 |
| Heart_Failure | monocyte | *RPL13* | 3.396973396 | 0.000253913 | 0.011389066 | 1 |
| Heart_Failure | monocyte | *ZNF444* | 3.590087911 | 0.003105397 | 0.027116153 | 1 |
| Heart_Failure | monocyte | *KIFC2* | 1.366608837 | 0.008795807 | 0.049330273 | 1 |
| Heart_Failure | monocyte | *MIS12* | 4.466413807 | 0.000637208 | 0.013540529 | 1 |
| Heart_Failure | monocyte | *SRP68* | 3.953794416 | 0.001062995 | 0.015840489 | 1 |
| Heart_Failure | monocyte | *TMEM68* | 2.165139575 | 0.005987638 | 0.038926702 | 1 |
| Heart_Failure | monocyte | *HRASLS5* | 0.488791241 | 0.004802264 | 0.034549195 | 1 |
| Heart_Failure | monocyte | *TRANK1* | 2.444309975 | 0.004659186 | 0.033981786 | 1 |
| Heart_Failure | monocyte | *TTC21A* | 4.831286032 | 0.001113568 | 0.016083148 | 1 |
| Heart_Failure | monocyte | *GLYCTK* | 4.0139491 | 0.000624577 | 0.013459015 | 1 |
| Heart_Failure | monocyte | *UBTD2* | 2.592436174 | 0.002133323 | 0.021667534 | 1 |
| Heart_Failure | monocyte | *RFWD3* | 1.665738982 | 0.008563507 | 0.048922662 | 1 |
| Heart_Failure | monocyte | *REEP4* | 4.265491596 | 0.005782665 | 0.037949491 | 1 |
| Heart_Failure | monocyte | *FNTA* | 4.480326919 | 7.06E-05 | 0.011101372 | 0.444054886 |
| Heart_Failure | monocyte | *TRAPPC11* | 3.58637073 | 0.000329381 | 0.011389066 | 1 |
| Heart_Failure | monocyte | *DNAJC21* | 3.774517061 | 0.000761652 | 0.014350186 | 1 |
| Heart_Failure | monocyte | *TMEM129* | 2.063328024 | 0.002431966 | 0.023462924 | 1 |

| | | | | | | |
|---|---|---|---|---|---|---|
| Heart_Failure | monocyte | *FEM1B* | 1.587372204 | 0.007514355 | 0.044996142 | 1 |
| Heart_Failure | monocyte | *XPO6* | 2.491101554 | 0.00402783 | 0.031379139 | 1 |
| Heart_Failure | monocyte | *SLC33A1* | 3.998604486 | 0.000321275 | 0.011389066 | 1 |
| Heart_Failure | monocyte | *DTWD2* | 1.87995296 | 0.005952515 | 0.038821022 | 1 |
| Heart_Failure | monocyte | *DCXR* | 3.862451802 | 0.005312206 | 0.036262586 | 1 |
| Heart_Failure | monocyte | *BRD3* | 2.817022235 | 0.004106316 | 0.031637758 | 1 |
| Heart_Failure | monocyte | *MAP3K2* | 2.909145454 | 0.000525515 | 0.012379507 | 1 |
| Heart_Failure | monocyte | *CNTROB* | 4.660240028 | 0.001324821 | 0.017175592 | 1 |
| Heart_Failure | monocyte | *CHRNB1* | 2.098169679 | 0.000513778 | 0.012314203 | 1 |
| Heart_Failure | monocyte | *PDCD6IP* | 1.671332218 | 0.006148982 | 0.039568731 | 1 |
| Heart_Failure | monocyte | *GLB1* | 2.08118808 | 0.004395219 | 0.032935327 | 1 |
| Heart_Failure | monocyte | *NFXL1* | 3.625433729 | 0.000786388 | 0.01444259 | 1 |
| Heart_Failure | monocyte | *CD14* | 2.182326451 | 0.005641885 | 0.037654428 | 1 |
| Heart_Failure | monocyte | *STAT2* | 3.129674367 | 0.000398592 | 0.011389066 | 1 |
| Heart_Failure | monocyte | *NUDCD2* | 6.014883718 | 3.03E-05 | 0.00906704 | 0.190407841 |
| Heart_Failure | monocyte | *RNF34* | 2.92535509 | 0.000756071 | 0.01431753 | 1 |
| Heart_Failure | monocyte | *AKAP13* | 2.505077454 | 0.00027641 | 0.011389066 | 1 |
| Heart_Failure | monocyte | *FOXN2* | 1.804547753 | 0.00528957 | 0.036147314 | 1 |
| Heart_Failure | monocyte | *TSEN34* | 5.952240581 | 0.001171586 | 0.016152985 | 1 |
| Heart_Failure | monocyte | *NDUFA3* | 2.934125205 | 0.007150218 | 0.043224444 | 1 |
| Heart_Failure | monocyte | *ZNF160* | 1.98557712 | 0.005488714 | 0.037144825 | 1 |
| Heart_Failure | monocyte | *C11orf24* | 3.789446376 | 0.007725855 | 0.045607934 | 1 |
| Heart_Failure | monocyte | *GRIK1* | 3.647093499 | 0.00103549 | 0.015840489 | 1 |
| Heart_Failure | monocyte | *LRRC8C* | 3.197273675 | 0.006530033 | 0.040809462 | 1 |
| Heart_Failure | monocyte | *RSL1D1* | 2.7564282 | 0.004279318 | 0.03244665 | 1 |
| Heart_Failure | monocyte | *PPID* | 3.161381947 | 0.004197912 | 0.03202946 | 1 |
| Heart_Failure | monocyte | *TBCA* | 2.380448139 | 0.000689611 | 0.01394556 | 1 |
| Heart_Failure | monocyte | *BCL2L1* | 1.948243969 | 0.003536655 | 0.029065292 | 1 |
| Heart_Failure | monocyte | *TCEA2* | 5.937115233 | 0.00044723 | 0.011600275 | 1 |
| Heart_Failure | monocyte | *TRAPPC12* | 2.650501053 | 0.003146139 | 0.027395812 | 1 |
| Heart_Failure | monocyte | *TVP23B* | 3.095331617 | 0.002192848 | 0.022058295 | 1 |
| Heart_Failure | monocyte | *SRGAP2C* | 2.797380715 | 0.004319492 | 0.0326141 | 1 |
| Heart_Failure | monocyte | *ATPAF2* | 3.194868702 | 0.002640207 | 0.02448227 | 1 |
| Heart_Failure | monocyte | *C20orf196* | 3.824061197 | 0.000569017 | 0.01286838 | 1 |
| Heart_Failure | monocyte | *MALT1* | 3.881523252 | 0.000103851 | 0.011389066 | 0.65291165 |
| Heart_Failure | monocyte | *ZNF131* | 2.772230314 | 0.001343386 | 0.01728185 | 1 |
| Heart_Failure | monocyte | *SUCLG2* | 2.591386422 | 0.001383622 | 0.017502683 | 1 |
| Heart_Failure | monocyte | *ZNF24* | 3.936319083 | 0.000348346 | 0.011389066 | 1 |
| Heart_Failure | monocyte | *MANEA* | 3.987885068 | 0.001073703 | 0.015840489 | 1 |
| Heart_Failure | monocyte | *FUT10* | 1.262622171 | 0.009013325 | 0.049751338 | 1 |
| Heart_Failure | monocyte | *CFL1* | 4.786201347 | 8.23E-05 | 0.011389066 | 0.517542126 |
| Heart_Failure | monocyte | *CBWD1* | 3.089602839 | 0.004699789 | 0.034153156 | 1 |
| Heart_Failure | monocyte | *DCP2* | 3.370607813 | 0.000606022 | 0.013396857 | 1 |
| Heart_Failure | monocyte | *NADSYN1* | 2.789891501 | 0.005141947 | 0.035529945 | 1 |
| Heart_Failure | monocyte | *HECTD4* | 1.523874049 | 0.006716805 | 0.041557112 | 1 |
| Heart_Failure | monocyte | *NOC3L* | 3.769612833 | 0.00034159 | 0.011389066 | 1 |
| Heart_Failure | monocyte | *PARP15* | 1.228237014 | 0.006144565 | 0.039568731 | 1 |
| Heart_Failure | monocyte | *THAP2* | 2.264984245 | 0.007563665 | 0.045116469 | 1 |
| Heart_Failure | monocyte | *PPP1R14B* | 3.356932191 | 0.008599723 | 0.048922662 | 1 |
| Heart_Failure | monocyte | *SMARCC1* | 1.945950121 | 0.003948801 | 0.03107148 | 1 |

| | | | | | | |
|---|---|---|---|---|---|---|
| Heart_Failure | monocyte | *AGFG1* | 1.477491952 | 0.003015075 | 0.026660729 | 1 |
| Heart_Failure | monocyte | *RNF213* | 3.122089274 | 0.000176616 | 0.011389066 | 1 |
| Heart_Failure | monocyte | *GOLIM4* | 1.592347808 | 0.002123237 | 0.021667534 | 1 |
| Heart_Failure | monocyte | *USMG5* | 2.152719246 | 0.00818005 | 0.047530474 | 1 |
| Heart_Failure | monocyte | *LEMD3* | 1.674895534 | 0.008827032 | 0.049383136 | 1 |
| Heart_Failure | monocyte | *IL20RB* | 1.494946147 | 0.006904703 | 0.042170383 | 1 |
| Heart_Failure | monocyte | *SRP72* | 1.818472487 | 0.005626816 | 0.037633819 | 1 |
| Heart_Failure | monocyte | *NDUFA11* | 3.563364567 | 0.005184684 | 0.035624159 | 1 |
| Heart_Failure | monocyte | *DHX36* | 2.016831251 | 0.005617033 | 0.037608399 | 1 |
| Heart_Failure | monocyte | *FBXW8* | 2.408257061 | 0.003005449 | 0.026660729 | 1 |
| Heart_Failure | monocyte | *ATR* | 1.616538471 | 0.004872606 | 0.03481145 | 1 |
| Heart_Failure | monocyte | *TMEM9B* | 3.0213332 | 0.003358303 | 0.028264591 | 1 |
| Heart_Failure | monocyte | *SMAD2* | 2.138950302 | 0.000689847 | 0.01394556 | 1 |
| Heart_Failure | monocyte | *CLTB* | 2.404491389 | 0.00484079 | 0.03474206 | 1 |
| Heart_Failure | monocyte | *PPP2R2D* | 2.022318209 | 0.00289991 | 0.026082599 | 1 |
| Heart_Failure | monocyte | *MCTP1* | 2.087081646 | 0.000188027 | 0.011389066 | 1 |
| Heart_Failure | monocyte | *UCP2* | 4.895164179 | 0.000102516 | 0.011389066 | 0.644519361 |
| Heart_Failure | monocyte | *RPS6KB2* | 4.84755865 | 0.000189123 | 0.011389066 | 1 |
| Heart_Failure | monocyte | *MSRA* | 2.204214728 | 0.004868058 | 0.03481145 | 1 |
| Heart_Failure | monocyte | *CTDNEP1* | 4.869528579 | 0.000557697 | 0.01265792 | 1 |
| Heart_Failure | monocyte | *IP6K1* | 3.71455199 | 9.76E-05 | 0.011389066 | 0.61341444 |
| Heart_Failure | monocyte | *TCP11L1* | 2.445100268 | 0.00084895 | 0.014523588 | 1 |
| Heart_Failure | monocyte | *COX8A* | 5.011920007 | 0.00072127 | 0.014254661 | 1 |
| Heart_Failure | monocyte | *CRLF3* | 3.505919734 | 4.53E-05 | 0.009491021 | 0.284730635 |
| Heart_Failure | monocyte | *RNPEP* | 3.936494278 | 0.000213251 | 0.011389066 | 1 |
| Heart_Failure | monocyte | *LRRC37A* | 2.108891001 | 0.006655132 | 0.041426552 | 1 |
| Heart_Failure | monocyte | *RUFY1* | 1.71366897 | 0.008033926 | 0.046898139 | 1 |
| Heart_Failure | monocyte | *LRRC37A3* | 3.338834763 | 0.002289596 | 0.022597628 | 1 |
| Heart_Failure | monocyte | *THAP4* | 2.127340108 | 0.005706987 | 0.037841897 | 1 |
| Heart_Failure | monocyte | *SEC24C* | 3.243555864 | 0.00436547 | 0.032868331 | 1 |
| Heart_Failure | monocyte | *MTHFR* | 3.510435924 | 0.00247433 | 0.02367749 | 1 |
| Heart_Failure | monocyte | *POLE* | 2.4727826 | 0.007126367 | 0.043204889 | 1 |
| Heart_Failure | monocyte | *TALDO1* | 2.121925644 | 0.003786209 | 0.030439765 | 1 |
| Heart_Failure | monocyte | *MAN1B1* | 3.3043241 | 0.002454687 | 0.023561244 | 1 |
| Heart_Failure | monocyte | *TOP3A* | 3.756976223 | 0.0002521 | 0.011389066 | 1 |
| Heart_Failure | monocyte | *NIM1K* | 3.318951437 | 0.002769352 | 0.025269836 | 1 |
| Heart_Failure | monocyte | *ATOX1* | 4.542561589 | 5.19E-05 | 0.010524778 | 0.326268103 |
| Heart_Failure | monocyte | *ZNF354C* | 2.941808926 | 0.003962085 | 0.031137035 | 1 |
| Heart_Failure | monocyte | *NDUFV2* | 7.122943814 | 4.14E-05 | 0.009491021 | 0.260523399 |
| Heart_Failure | monocyte | *LCORL* | 3.291675746 | 0.000161631 | 0.011389066 | 1 |
| Heart_Failure | monocyte | *ZC3H12D* | 6.934108596 | 0.000313909 | 0.011389066 | 1 |
| Heart_Failure | monocyte | *BEND3* | 7.42753187 | 0.002012587 | 0.021053467 | 1 |
| Heart_Failure | monocyte | *GAK* | 4.620054989 | 8.46E-06 | 0.004431396 | 0.053176749 |
| Heart_Failure | monocyte | *RMI1* | 2.861987014 | 0.001823388 | 0.020006349 | 1 |
| Heart_Failure | monocyte | *CTC1* | 2.276269134 | 0.006293959 | 0.040091306 | 1 |
| Heart_Failure | monocyte | *CALR* | 3.378532867 | 0.000381139 | 0.011389066 | 1 |
| Heart_Failure | monocyte | *HLA-DQB1* | -0.876088312 | 1.87E-07 | 0.000293539 | 0.001174158 |
| Heart_Failure | monocyte | *SERTAD2* | 2.323295826 | 0.003484939 | 0.028753037 | 1 |
| Heart_Failure | monocyte | *R3HDM2* | 2.835151636 | 0.000375373 | 0.011389066 | 1 |
| Heart_Failure | monocyte | *FGD6* | 4.127216697 | 0.000187155 | 0.011389066 | 1 |

| | | | | | | |
|---|---|---|---|---|---|---|
| Heart_Failure | monocyte | *ZNF609* | 1.895791211 | 0.002806684 | 0.025513898 | 1 |
| Heart_Failure | monocyte | *PCGF5* | 2.3396673 | 0.001966598 | 0.020744974 | 1 |
| Heart_Failure | monocyte | *ZDHHC20* | 3.662972736 | 0.00013083 | 0.011389066 | 0.822531143 |
| Heart_Failure | monocyte | *FKRP* | 1.066227881 | 0.002509567 | 0.023895639 | 1 |
| Heart_Failure | monocyte | *POLR2A* | 2.448624945 | 0.005983808 | 0.038926702 | 1 |
| Heart_Failure | monocyte | *DDX60L* | 3.479209158 | 0.000179636 | 0.011389066 | 1 |
| Heart_Failure | monocyte | *SETD2* | 1.605457504 | 0.006176577 | 0.039608989 | 1 |
| Heart_Failure | monocyte | *C3orf58* | 2.793706637 | 0.005417768 | 0.036783482 | 1 |
| Heart_Failure | monocyte | *RFX7* | 2.555994909 | 0.001230317 | 0.016601601 | 1 |
| Heart_Failure | monocyte | *ADO* | 6.922430012 | 0.000731515 | 0.014268876 | 1 |
| Heart_Failure | monocyte | *WRB* | 0.943562411 | 0.002348013 | 0.022886147 | 1 |
| Heart_Failure | monocyte | *TNRC18* | 3.187700915 | 0.000270383 | 0.011389066 | 1 |
| Heart_Failure | monocyte | *ZNF708* | 2.508676247 | 0.001517157 | 0.018237795 | 1 |
| Heart_Failure | monocyte | *IST1* | 2.078844983 | 0.003273808 | 0.028091022 | 1 |
| Heart_Failure | monocyte | *ERCC6L2* | 1.407807941 | 0.00496977 | 0.035225419 | 1 |
| Heart_Failure | monocyte | *KBTBD3* | 3.967425287 | 0.001646189 | 0.018990078 | 1 |
| Heart_Failure | monocyte | *FES* | 3.564698802 | 0.006728987 | 0.041557112 | 1 |
| Heart_Failure | monocyte | *TRAK1* | 1.690405117 | 0.008491628 | 0.048799693 | 1 |
| Heart_Failure | monocyte | *SKA2* | 3.971267806 | 0.000543164 | 0.012549055 | 1 |
| Heart_Failure | monocyte | *LENG9* | 1.789119629 | 0.008777306 | 0.049270469 | 1 |
| Heart_Failure | monocyte | *CEP63* | 1.027658908 | 0.004857779 | 0.034784574 | 1 |
| Heart_Failure | monocyte | *HMGN4* | 2.768925768 | 0.007847417 | 0.046073974 | 1 |
| Heart_Failure | monocyte | *WBP2NL* | 3.579743223 | 0.001936351 | 0.020703807 | 1 |
| Heart_Failure | monocyte | *PTTG1IP* | 2.341358884 | 0.002550484 | 0.024059456 | 1 |
| Heart_Failure | monocyte | *TANGO2* | 2.825588604 | 0.00242022 | 0.02340911 | 1 |
| Heart_Failure | monocyte | *NDUFB1* | 2.191934996 | 0.00873181 | 0.049218173 | 1 |
| Heart_Failure | monocyte | *BMP8A* | 5.307035886 | 0.000892299 | 0.014889101 | 1 |
| Heart_Failure | monocyte | *CBX6* | 3.094859434 | 0.007161883 | 0.043253369 | 1 |
| Heart_Failure | monocyte | *BTBD9* | 2.150558949 | 0.001231307 | 0.016601601 | 1 |
| Heart_Failure | monocyte | *DENND5A* | 2.593657817 | 0.002066216 | 0.021295573 | 1 |
| Heart_Failure | monocyte | *VPS33B* | 3.587677546 | 0.000640291 | 0.013540529 | 1 |
| Heart_Failure | monocyte | *TM2D3* | 3.524354806 | 0.001591172 | 0.01864041 | 1 |
| Heart_Failure | monocyte | *SS18L1* | 3.608170849 | 0.000895193 | 0.014889101 | 1 |
| Heart_Failure | monocyte | *THAP7* | 5.625759273 | 0.001108608 | 0.016083148 | 1 |
| Heart_Failure | monocyte | *TXNRD2* | 4.39473374 | 0.000738243 | 0.014268876 | 1 |
| Heart_Failure | monocyte | *SNN* | 6.095541827 | 0.001829545 | 0.0200307 | 1 |
| Heart_Failure | monocyte | *NDUFA12* | 5.051717155 | 0.000242068 | 0.011389066 | 1 |
| Heart_Failure | monocyte | *TMEM106A* | 3.897092639 | 0.001296052 | 0.017046605 | 1 |
| Heart_Failure | monocyte | *BRF1* | 2.818983091 | 0.002520922 | 0.023895639 | 1 |
| Heart_Failure | monocyte | *SP140L* | 3.084935405 | 0.001373269 | 0.017441898 | 1 |
| Heart_Failure | monocyte | *SH3BGR* | 0.497269475 | 0.003985112 | 0.031162187 | 1 |
| Heart_Failure | monocyte | *MUC1* | 2.24842498 | 0.000123184 | 0.011389066 | 0.774457225 |
| Heart_Failure | monocyte | *GSAP* | 3.418826451 | 6.75E-05 | 0.011101372 | 0.42420295 |
| Heart_Failure | monocyte | *C2orf76* | 3.569388767 | 0.000102451 | 0.011389066 | 0.644106765 |
| Heart_Failure | monocyte | *WWOX* | 1.146362261 | 0.002608058 | 0.024363837 | 1 |
| Heart_Failure | monocyte | *POLR1D* | 2.429555449 | 0.004540587 | 0.033430274 | 1 |
| Heart_Failure | monocyte | *KPNA4* | 3.545225082 | 0.000264199 | 0.011389066 | 1 |
| Heart_Failure | monocyte | *RPS23* | 1.671527639 | 0.008965592 | 0.049628243 | 1 |
| Heart_Failure | monocyte | *NF2* | 2.274481686 | 0.006322501 | 0.040153831 | 1 |
| Heart_Failure | monocyte | *RPS19BP1* | 4.013607338 | 0.001407896 | 0.017630601 | 1 |

| | | | | | | |
|---|---|---|---|---|---|---|
| Heart_Failure | monocyte | *FNBP1* | 1.491356903 | 0.00877504 | 0.049270469 | 1 |
| Heart_Failure | monocyte | *MCRS1* | 2.429357775 | 0.000452929 | 0.011604726 | 1 |
| Heart_Failure | monocyte | *ZC3H6* | 2.002977348 | 0.00770396 | 0.045562812 | 1 |
| Heart_Failure | monocyte | *LINC00265* | 4.25997396 | 0.001317311 | 0.017175592 | 1 |
| Heart_Failure | monocyte | *PLSCR1* | 2.787959727 | 0.001960604 | 0.020744974 | 1 |
| Heart_Failure | monocyte | *NBR1* | 4.090626347 | 0.000184912 | 0.011389066 | 1 |
| Heart_Failure | monocyte | *NDOR1* | 4.728957881 | 0.001713996 | 0.019518456 | 1 |
| Heart_Failure | monocyte | *SUMO2* | 3.358228562 | 0.00137001 | 0.017435739 | 1 |
| Heart_Failure | monocyte | *TMEM201* | 4.586340625 | 0.000728236 | 0.014268876 | 1 |
| Heart_Failure | monocyte | *LITAF* | 1.943218563 | 0.0041577 | 0.031877388 | 1 |
| Heart_Failure | monocyte | *SF3B3* | 4.044782097 | 0.000298123 | 0.011389066 | 1 |
| Heart_Failure | monocyte | *ZNF33A* | 2.356026242 | 0.001526952 | 0.018285615 | 1 |
| Heart_Failure | monocyte | *C15orf61* | 4.777909759 | 0.001818102 | 0.020006349 | 1 |
| Heart_Failure | monocyte | *TSPYL1* | 2.06976618 | 0.007622727 | 0.045271496 | 1 |
| Heart_Failure | monocyte | *PNRC2* | 3.889107654 | 0.001637588 | 0.018937074 | 1 |
| Heart_Failure | monocyte | *ZKSCAN3* | 2.67853339 | 0.004112036 | 0.031643051 | 1 |
| Heart_Failure | monocyte | *MPHOSPH8* | 1.832745663 | 0.003284692 | 0.028091022 | 1 |
| Heart_Failure | monocyte | *PPIA* | 2.751262268 | 0.00326226 | 0.028091022 | 1 |
| Heart_Failure | monocyte | *POM121* | 2.807312007 | 0.001730664 | 0.019604839 | 1 |
| Heart_Failure | monocyte | *LONP1* | 2.237184336 | 0.001872476 | 0.020319415 | 1 |
| Heart_Failure | monocyte | *ASB13* | 5.402804119 | 0.000441506 | 0.011517615 | 1 |
| Heart_Failure | monocyte | *EVL* | 2.108235389 | 0.002182285 | 0.02202251 | 1 |
| Heart_Failure | monocyte | *ZNF124* | 3.973799617 | 0.000420514 | 0.011479484 | 1 |
| Heart_Failure | monocyte | *PIK3R4* | 1.765546981 | 0.004049849 | 0.031448189 | 1 |
| Heart_Failure | monocyte | *ANAPC7* | 4.755424153 | 1.60E-05 | 0.006105651 | 0.100620482 |
| Heart_Failure | monocyte | *MYO18A* | 2.880452286 | 0.003288531 | 0.028091022 | 1 |
| Heart_Failure | monocyte | *HDAC2* | 4.661724989 | 4.33E-05 | 0.009491021 | 0.272197245 |
| Heart_Failure | monocyte | *TECPR2* | 1.939693489 | 0.007691815 | 0.045535255 | 1 |
| Heart_Failure | monocyte | *AMZ2* | 3.64578791 | 0.000524476 | 0.012379507 | 1 |
| Heart_Failure | monocyte | *NF1* | 1.30453543 | 0.008178638 | 0.047530474 | 1 |
| Heart_Failure | monocyte | *HLA-DQA1* | -0.605387314 | 0.001349672 | 0.01728185 | 1 |
| Heart_Failure | monocyte | *PPTC7* | 2.568787561 | 0.001143289 | 0.016135389 | 1 |
| Heart_Failure | monocyte | *NHLRC2* | 1.879183842 | 0.00348297 | 0.028753037 | 1 |
| Heart_Failure | monocyte | *ZFP28* | 4.960146707 | 0.001148466 | 0.016135389 | 1 |
| Heart_Failure | monocyte | *GMFB* | 2.488655898 | 0.008444009 | 0.048704116 | 1 |
| Heart_Failure | monocyte | *ZMYM1* | 2.642285373 | 0.002566309 | 0.024099196 | 1 |
| Heart_Failure | monocyte | *ABCB8* | 1.492561757 | 0.008284836 | 0.047975347 | 1 |
| Heart_Failure | monocyte | *SND1* | 2.00414808 | 0.001789638 | 0.019813108 | 1 |
| Heart_Failure | monocyte | *GTF2E2* | 2.02691028 | 0.005766654 | 0.037938944 | 1 |
| Heart_Failure | monocyte | *ZNF786* | 3.069380132 | 0.003812485 | 0.030495028 | 1 |
| Heart_Failure | monocyte | *SLC22A5* | 1.070218281 | 6.64E-05 | 0.011101372 | 0.417744615 |
| Heart_Failure | monocyte | *ATG7* | 2.101220951 | 0.002644133 | 0.024482565 | 1 |
| Heart_Failure | monocyte | *PIGN* | 1.572348984 | 0.002351591 | 0.022886147 | 1 |
| Heart_Failure | monocyte | *ZNF624* | 4.603545432 | 0.000733436 | 0.014268876 | 1 |
| Heart_Failure | monocyte | *PHF2* | 3.637766774 | 0.000415513 | 0.0114576 | 1 |
| Heart_Failure | monocyte | *RPS26* | 1.30908998 | 1.38E-05 | 0.00577283 | 0.086592452 |
| Heart_Failure | monocyte | *TAF13* | 2.675477609 | 0.001872125 | 0.020319415 | 1 |
| Heart_Failure | monocyte | *ZNF181* | 3.991795237 | 0.000900706 | 0.014941255 | 1 |
| Heart_Failure | monocyte | *SGTB* | 3.429583921 | 0.008962708 | 0.049628243 | 1 |
| Heart_Failure | monocyte | *NOL8* | 3.186906077 | 0.001056744 | 0.015840489 | 1 |

| | | | | | | |
|---|---|---|---|---|---|---|
| Heart_Failure | monocyte | *MRPL42* | 1.657123937 | 0.007999786 | 0.046815675 | 1 |
| Heart_Failure | monocyte | *AGAP9* | 6.246031264 | 0.000440359 | 0.011517615 | 1 |
| Heart_Failure | monocyte | *SFI1* | 3.140401556 | 0.002982607 | 0.026522845 | 1 |
| Heart_Failure | monocyte | *MIER1* | 2.91412302 | 0.000393589 | 0.011389066 | 1 |
| Heart_Failure | monocyte | *ZNF251* | 1.906062281 | 0.005217036 | 0.035768499 | 1 |
| Heart_Failure | monocyte | *HSD17B11* | 3.473581937 | 0.001291249 | 0.017046605 | 1 |
| Heart_Failure | monocyte | *QRICH1* | 1.876213598 | 0.001451801 | 0.017792349 | 1 |
| Heart_Failure | monocyte | *STYX* | 4.259440017 | 0.000463165 | 0.011604726 | 1 |
| Heart_Failure | monocyte | *ZKSCAN8* | 3.263771546 | 0.001203927 | 0.016454541 | 1 |
| Heart_Failure | monocyte | *TMA16* | 2.661147772 | 0.00327814 | 0.028091022 | 1 |
| Heart_Failure | monocyte | *TLK1* | 1.63532563 | 0.006461673 | 0.040520538 | 1 |
| Heart_Failure | monocyte | *CCDC69* | 2.083592693 | 0.00445903 | 0.03311146 | 1 |
| Heart_Failure | monocyte | *MDM4* | 2.143930042 | 0.002107927 | 0.021619147 | 1 |
| Heart_Failure | monocyte | *FAM19A2* | 3.5077689 | 0.000251903 | 0.011389066 | 1 |
| Heart_Failure | monocyte | *TUSC1* | 7.122342997 | 0.001030455 | 0.015840489 | 1 |
| Heart_Failure | monocyte | *CEP290* | 2.262999197 | 0.005733183 | 0.037861895 | 1 |
| Heart_Failure | monocyte | *ZNF652* | 3.309802291 | 0.000323547 | 0.011389066 | 1 |
| Heart_Failure | monocyte | *APCDD1L* | 3.492813932 | 0.006059794 | 0.039155114 | 1 |
| Heart_Failure | monocyte | *CAPZA2* | 3.056690409 | 0.000607301 | 0.013396857 | 1 |
| Heart_Failure | monocyte | *SMG5* | 2.627545612 | 0.005221772 | 0.035768499 | 1 |
| Heart_Failure | monocyte | *PJA2* | 1.980425098 | 0.006268274 | 0.039968644 | 1 |
| Heart_Failure | monocyte | *SYS1* | 4.177032598 | 0.006451646 | 0.040520538 | 1 |
| Heart_Failure | monocyte | *GIGYF2* | 2.767131417 | 0.000465937 | 0.011604726 | 1 |
| Heart_Failure | monocyte | *HLA-DRA* | 0.96581558 | 0.001061095 | 0.015840489 | 1 |
| Heart_Failure | monocyte | *NELFE* | 2.125950077 | 0.000167942 | 0.011389066 | 1 |
| Heart_Failure | monocyte | *BAG6* | 3.176657399 | 3.95E-07 | 0.000497085 | 0.002485426 |
| Heart_Failure | monocyte | *NFKBIL1* | 2.653603512 | 0.000847646 | 0.014523588 | 1 |
| Heart_Failure | monocyte | *MICB* | 0.8007436 | 0.000595552 | 0.013277434 | 1 |
| Heart_Failure | monocyte | *MICA* | 1.516329257 | 8.41E-07 | 0.000881049 | 0.005286296 |
| Heart_Failure | monocyte | *FAM216A* | 2.4265482 | 0.002742153 | 0.025131074 | 1 |
| Heart_Failure | monocyte | *TRIM13* | 2.947916966 | 0.000654731 | 0.013585129 | 1 |
| Heart_Failure | monocyte | *FAM205A* | -8.54838836 | 0.003621829 | 0.029323162 | 1 |
| Heart_Failure | monocyte | *NAP1L4* | 1.398940536 | 0.008287136 | 0.047975347 | 1 |
| Heart_Failure | monocyte | *DENND6B* | 6.7064956 | 0.000431641 | 0.011479484 | 1 |
| Heart_Failure | monocyte | *LIN52* | 1.552092949 | 0.003324778 | 0.028241098 | 1 |
| Heart_Failure | monocyte | *RNPS1* | 2.508102854 | 0.002747809 | 0.025146256 | 1 |
| Heart_Failure | monocyte | *HLA-A* | 1.588166576 | 0.003424826 | 0.028571324 | 1 |
| Heart_Failure | monocyte | *SACM1L* | 2.660122833 | 0.00602604 | 0.039013224 | 1 |
| Heart_Failure | monocyte | *HEXA* | 2.170392569 | 0.008660948 | 0.049055298 | 1 |
| Heart_Failure | monocyte | *PPP1CB* | 3.493833554 | 0.000128604 | 0.011389066 | 0.808535718 |
| Heart_Failure | monocyte | *ATF6B* | 3.463282107 | 2.14E-08 | 6.72E-05 | 0.000134377 |
| Heart_Failure | monocyte | *SLC35F6* | 4.684075584 | 0.001231327 | 0.016601601 | 1 |
| Heart_Failure | monocyte | *SMIM7* | 3.550389569 | 0.000403801 | 0.011389066 | 1 |
| Heart_Failure | monocyte | *PAXIP1-AS2* | 4.456729456 | 0.000352032 | 0.011389066 | 1 |
| Heart_Failure | monocyte | *LYRM4* | 1.92140802 | 0.005142877 | 0.035529945 | 1 |
| Heart_Failure | monocyte | *PHB2* | 3.417740484 | 0.001854422 | 0.020240886 | 1 |
| Heart_Failure | monocyte | *RNASEK* | 5.00293443 | 0.000327133 | 0.011389066 | 1 |
| Heart_Failure | monocyte | *UBA52* | 1.703508792 | 0.005159669 | 0.035529945 | 1 |
| Heart_Failure | monocyte | *OSER1-AS1* | 3.620638942 | 0.007799204 | 0.045868657 | 1 |
| Heart_Failure | monocyte | *LINC00863* | 2.56882669 | 0.002516962 | 0.023895639 | 1 |

| | | | | | | |
|---|---|---|---|---|---|---|
| Heart_Failure | monocyte | *PLEKHM1* | 3.379248421 | 0.001147458 | 0.016135389 | 1 |
| Heart_Failure | monocyte | *ZNF688* | 4.673807796 | 0.008000993 | 0.046815675 | 1 |
| Heart_Failure | monocyte | *RPS18* | 2.746797437 | 4.46E-05 | 0.009491021 | 0.28034679 |
| Heart_Failure | monocyte | *HLA-DQB2* | 1.48330437 | 7.03E-15 | 4.42E-11 | 4.42E-11 |
| Heart_Failure | monocyte | *TRIM26* | 2.921243829 | 0.002210733 | 0.022061708 | 1 |
| Heart_Failure | monocyte | *ZNF736* | 2.623166552 | 4.03E-05 | 0.009491021 | 0.253235491 |
| Heart_Failure | monocyte | *HLA-DQA2* | 1.025104837 | 3.31E-08 | 6.94E-05 | 0.000208275 |
| Heart_Failure | monocyte | *LRRC37A2* | 3.794598151 | 0.000251408 | 0.011389066 | 1 |
| Heart_Failure | monocyte | *PSMB9* | 2.818266601 | 1.09E-05 | 0.005272415 | 0.068541401 |
| Heart_Failure | monocyte | *NSUN6* | 1.686875839 | 0.008481709 | 0.04878729 | 1 |
| Heart_Failure | monocyte | *PI4KA* | 3.332803795 | 0.000240982 | 0.011389066 | 1 |
| Heart_Failure | monocyte | *C22orf39* | 8.19410128 | 1.65E-05 | 0.006105651 | 0.10379606 |
| Heart_Failure | monocyte | *AP5Z1* | 3.479599593 | 0.001171338 | 0.016152985 | 1 |
| Heart_Failure | monocyte | *APOBEC3C* | 6.135333347 | 0.001202817 | 0.016454541 | 1 |
| Heart_Failure | monocyte | *LINC00847* | 4.227446115 | 0.007594199 | 0.04521281 | 1 |
| Heart_Failure | monocyte | *PDCD6* | 2.225991611 | 0.008531208 | 0.048892536 | 1 |
| Heart_Failure | monocyte | *PRKDC* | 2.654253581 | 0.000979467 | 0.015508077 | 1 |
| Heart_Failure | monocyte | *FPGT* | 5.628625296 | 0.000475791 | 0.011604726 | 1 |
| Heart_Failure | monocyte | *POLR2M* | 4.571754897 | 0.001194744 | 0.016400334 | 1 |
| Heart_Failure | monocyte | *CTSO* | 2.650287702 | 0.004743825 | 0.034280954 | 1 |
| Heart_Failure | monocyte | *ZBED6* | 10.46193547 | 0.005091594 | 0.035460966 | 1 |
| Heart_Failure | monocyte | *OVCH1-AS1* | 4.041514735 | 0.000789698 | 0.01444259 | 1 |
| Heart_Failure | monocyte | *CUX1* | 1.917532086 | 0.004104921 | 0.031637758 | 1 |
| Heart_Failure | monocyte | *IQCH-AS1* | 1.743890285 | 0.003522657 | 0.028988144 | 1 |
| Heart_Failure | monocyte | *HOXB7* | 4.056374925 | 0.002812062 | 0.025513898 | 1 |
| Heart_Failure | monocyte | *AC009120.5* | 7.43560012 | 0.001150995 | 0.016135389 | 1 |
| Heart_Failure | monocyte | *GTF2H5* | 1.455629244 | 0.004998176 | 0.035307337 | 1 |
| Heart_Failure | monocyte | *POLR2J4* | 4.973031569 | 0.001137268 | 0.016135389 | 1 |
| Heart_Failure | neutrophil | *HLA-DQB1* | -5.345849814 | 5.20E-20 | 3.27E-16 | 3.27E-16 |
| Heart_Failure | nk_cell | *SEMA3F* | -0.795367277 | 1.76E-05 | 0.00921619 | 0.110594278 |
| Heart_Failure | nk_cell | *RBM6* | -0.540610826 | 4.78E-05 | 0.018767755 | 0.300284087 |
| Heart_Failure | nk_cell | *SLC17A9* | 3.480013148 | 4.70E-05 | 0.018767755 | 0.295800182 |
| Heart_Failure | nk_cell | *CHMP1A* | 1.986438083 | 3.92E-05 | 0.01759193 | 0.246287022 |
| Heart_Failure | nk_cell | *REEP2* | -0.762159889 | 0.000123341 | 0.035247419 | 0.775443208 |
| Heart_Failure | nk_cell | *NMT1* | 0.935423714 | 7.64E-05 | 0.025271246 | 0.480153681 |
| Heart_Failure | nk_cell | *RMDN3* | 2.099190795 | 7.61E-05 | 0.025271246 | 0.478470225 |
| Heart_Failure | nk_cell | *PPOX* | 1.774837697 | 0.000120157 | 0.035247419 | 0.755428515 |
| Heart_Failure | nk_cell | *EPB41* | 1.148700574 | 0.000113537 | 0.035247419 | 0.713808918 |
| Heart_Failure | nk_cell | *RNF123* | 1.165666493 | 1.39E-06 | 0.001094072 | 0.008752573 |
| Heart_Failure | nk_cell | *ORMDL3* | -0.932702062 | 7.36E-08 | 9.25E-05 | 0.000462437 |
| Heart_Failure | nk_cell | *HLA-DQB1* | -1.396057855 | 1.07E-21 | 6.70E-18 | 6.70E-18 |
| Heart_Failure | nk_cell | *BMP8A* | 3.520950246 | 0.000160477 | 0.043865942 | 1 |
| Heart_Failure | nk_cell | *HLA-DRB1* | -1.05768158 | 4.18E-06 | 0.002922715 | 0.026304436 |
| Heart_Failure | nk_cell | *HLA-DQA1* | -1.791051165 | 6.15E-16 | 1.29E-12 | 3.86E-12 |
| Heart_Failure | nk_cell | *SLC22A5* | 1.040926656 | 2.13E-07 | 0.000191065 | 0.001337455 |
| Heart_Failure | nk_cell | *RPS26* | 0.548081468 | 6.21E-05 | 0.022968082 | 0.390457395 |
| Heart_Failure | nk_cell | *HLA-DRA* | 1.34353074 | 1.29E-07 | 0.000134767 | 0.000808602 |
| Heart_Failure | nk_cell | *BAG6* | 1.477339267 | 8.51E-06 | 0.005348087 | 0.053480868 |
| Heart_Failure | nk_cell | *MICA* | 0.607625872 | 2.61E-05 | 0.012608251 | 0.163907259 |
| Heart_Failure | nk_cell | *ATF6B* | 1.29682463 | 1.18E-05 | 0.006763192 | 0.074395107 |

| | | | | | | |
|---|---|---|---|---|---|---|
| Heart_Failure | nk_cell | *HLA-DQB2* | 0.648320085 | 1.32E-13 | 2.08E-10 | 8.31E-10 |
| Heart_Failure | nk_cell | *HLA-DQA2* | 1.412974512 | 8.05E-21 | 2.53E-17 | 5.06E-17 |
| Heart_Failure | pericyte | *SCYL3* | 1.563595002 | 0.002864542 | 0.022825571 | 1 |
| Heart_Failure | pericyte | *C1orf112* | 3.272276419 | 0.001540861 | 0.015450392 | 1 |
| Heart_Failure | pericyte | *NFYA* | 2.26478563 | 0.002538419 | 0.0210264 | 1 |
| Heart_Failure | pericyte | *SEMA3F* | -0.634434157 | 0.002585061 | 0.021272619 | 1 |
| Heart_Failure | pericyte | *KRIT1* | 2.985758333 | 0.000362337 | 0.006042474 | 1 |
| Heart_Failure | pericyte | *MAD1L1* | 3.27381842 | 0.000664253 | 0.008810466 | 1 |
| Heart_Failure | pericyte | *CYP26B1* | 0.933312922 | 0.0083336 | 0.046905412 | 1 |
| Heart_Failure | pericyte | *DBNDD1* | -0.969347152 | 0.006751983 | 0.0404283 | 1 |
| Heart_Failure | pericyte | *RBM6* | -0.740442994 | 1.54E-05 | 0.001740485 | 0.097018528 |
| Heart_Failure | pericyte | *GCFC2* | 2.821420841 | 0.001137452 | 0.0123585 | 1 |
| Heart_Failure | pericyte | *SYNRG* | 3.192145734 | 0.000223288 | 0.004655112 | 1 |
| Heart_Failure | pericyte | *ARHGAP44* | 1.458982243 | 0.00198835 | 0.018169701 | 1 |
| Heart_Failure | pericyte | *TRAPPC6A* | 3.608038298 | 0.001940584 | 0.017863035 | 1 |
| Heart_Failure | pericyte | *RPUSD1* | 1.517621821 | 0.007752016 | 0.044818101 | 1 |
| Heart_Failure | pericyte | *PSMB1* | 3.546460804 | 2.90E-05 | 0.001861562 | 0.182433094 |
| Heart_Failure | pericyte | *SEC62* | 3.083790502 | 0.000516777 | 0.007443425 | 1 |
| Heart_Failure | pericyte | *TTC19* | 2.159039986 | 0.000529467 | 0.007488118 | 1 |
| Heart_Failure | pericyte | *ZBTB32* | 8.028666485 | 4.67E-05 | 0.002128526 | 0.293828618 |
| Heart_Failure | pericyte | *ERCC1* | 3.422648082 | 2.84E-05 | 0.001861562 | 0.178793479 |
| Heart_Failure | pericyte | *MDH1* | 2.531621647 | 4.76E-05 | 0.002128526 | 0.299392562 |
| Heart_Failure | pericyte | *OSBPL5* | 3.785404704 | 0.000448334 | 0.006942549 | 1 |
| Heart_Failure | pericyte | *AQR* | 2.050043262 | 0.008916747 | 0.049003134 | 1 |
| Heart_Failure | pericyte | *UBR2* | 2.183147244 | 0.003025664 | 0.023736046 | 1 |
| Heart_Failure | pericyte | *NR1H3* | 1.909877071 | 0.001247901 | 0.01311965 | 1 |
| Heart_Failure | pericyte | *VIM* | 1.812236842 | 0.004678874 | 0.031293706 | 1 |
| Heart_Failure | pericyte | *FAS* | 1.18668939 | 0.000452272 | 0.006952162 | 1 |
| Heart_Failure | pericyte | *MIPEP* | 2.680078085 | 0.006763361 | 0.040457896 | 1 |
| Heart_Failure | pericyte | *VRK2* | 2.052643613 | 0.005139678 | 0.03341588 | 1 |
| Heart_Failure | pericyte | *ARID4A* | 2.900993022 | 0.000877349 | 0.010466596 | 1 |
| Heart_Failure | pericyte | *FUT8* | 2.373818511 | 0.002495005 | 0.020886947 | 1 |
| Heart_Failure | pericyte | *ADSS* | 3.136838384 | 0.002275289 | 0.019622417 | 1 |
| Heart_Failure | pericyte | *CUL3* | 2.830585559 | 0.000382555 | 0.006247076 | 1 |
| Heart_Failure | pericyte | *MRI1* | 1.335942642 | 0.004694504 | 0.031364872 | 1 |
| Heart_Failure | pericyte | *BOD1L1* | 1.829900412 | 0.005643003 | 0.035548658 | 1 |
| Heart_Failure | pericyte | *SKIV2L2* | 2.240406011 | 0.003823523 | 0.027354305 | 1 |
| Heart_Failure | pericyte | *PHLPP2* | 1.896078635 | 0.002182937 | 0.019149613 | 1 |
| Heart_Failure | pericyte | *PSMA4* | 3.934829586 | 0.000129894 | 0.003346908 | 0.816645567 |
| Heart_Failure | pericyte | *TDP1* | 3.705286906 | 0.002752845 | 0.022183725 | 1 |
| Heart_Failure | pericyte | *CNTLN* | 2.2848734 | 0.003588406 | 0.026490062 | 1 |
| Heart_Failure | pericyte | *DSG2* | 1.013921082 | 0.005587912 | 0.035272289 | 1 |
| Heart_Failure | pericyte | *POLR2B* | 3.63123997 | 0.000114806 | 0.003204538 | 0.721784132 |
| Heart_Failure | pericyte | *CP* | 5.650112053 | 3.43E-05 | 0.001918699 | 0.215864484 |
| Heart_Failure | pericyte | *GOPC* | 3.124122802 | 0.000276292 | 0.005271845 | 1 |
| Heart_Failure | pericyte | *LMO3* | 0.668670271 | 0.003757248 | 0.027065715 | 1 |
| Heart_Failure | pericyte | *NFE2L3* | 4.072254903 | 0.002665578 | 0.02169166 | 1 |
| Heart_Failure | pericyte | *FOXN3* | 1.822891663 | 0.005971773 | 0.037172807 | 1 |
| Heart_Failure | pericyte | *AKR7A2* | 2.831414231 | 0.000482147 | 0.007183074 | 1 |
| Heart_Failure | pericyte | *MRTO4* | 2.840021372 | 0.000373293 | 0.006159824 | 1 |

| | | | | | | |
|---|---|---|---|---|---|---|
| Heart_Failure | pericyte | *ANAPC4* | 3.017214172 | 0.000747021 | 0.009449737 | 1 |
| Heart_Failure | pericyte | *SDCCAG8* | 1.681268819 | 7.77E-05 | 0.002693349 | 0.488474757 |
| Heart_Failure | pericyte | *TBC1D22A* | 1.835278111 | 0.006671874 | 0.040139781 | 1 |
| Heart_Failure | pericyte | *GINM1* | 3.61442959 | 7.65E-05 | 0.002686157 | 0.480822034 |
| Heart_Failure | pericyte | *DCBLD2* | 3.512186142 | 4.76E-05 | 0.002128526 | 0.298973026 |
| Heart_Failure | pericyte | *GDI2* | 1.938562979 | 0.006364174 | 0.038695901 | 1 |
| Heart_Failure | pericyte | *LAMC2* | -0.580224003 | 0.001007026 | 0.011387006 | 1 |
| Heart_Failure | pericyte | *CDK14* | 2.835091178 | 2.04E-05 | 0.001740485 | 0.128125419 |
| Heart_Failure | pericyte | *RIOK2* | 2.878097079 | 0.005997422 | 0.037295542 | 1 |
| Heart_Failure | pericyte | *DNAJC25* | 1.099905369 | 0.002003476 | 0.018228438 | 1 |
| Heart_Failure | pericyte | *YBX3* | 2.34303051 | 0.006908546 | 0.041052955 | 1 |
| Heart_Failure | pericyte | *QSER1* | 2.816024663 | 0.002727808 | 0.022015059 | 1 |
| Heart_Failure | pericyte | *NCKAP1* | 2.404132705 | 0.000958035 | 0.011031435 | 1 |
| Heart_Failure | pericyte | *VMP1* | 2.84673151 | 0.000300252 | 0.005471554 | 1 |
| Heart_Failure | pericyte | *ADCK1* | 3.299718052 | 0.000458297 | 0.006976539 | 1 |
| Heart_Failure | pericyte | *ATP2C2* | 2.333559722 | 0.001120356 | 0.012228605 | 1 |
| Heart_Failure | pericyte | *IPO5* | 3.389278763 | 0.001253583 | 0.013157391 | 1 |
| Heart_Failure | pericyte | *ANKRD44* | 1.753318406 | 0.006861976 | 0.040853448 | 1 |
| Heart_Failure | pericyte | *MAP2K4* | 2.29655291 | 0.005314525 | 0.034024865 | 1 |
| Heart_Failure | pericyte | *ASB1* | 3.300713886 | 0.001712756 | 0.01648789 | 1 |
| Heart_Failure | pericyte | *TBC1D1* | 2.642531167 | 5.16E-05 | 0.002162658 | 0.324398634 |
| Heart_Failure | pericyte | *GOLGA5* | 3.277642534 | 0.001946496 | 0.017871199 | 1 |
| Heart_Failure | pericyte | *SP100* | 2.915387821 | 0.000235809 | 0.004736515 | 1 |
| Heart_Failure | pericyte | *STOML1* | 2.251260404 | 0.002014958 | 0.018253657 | 1 |
| Heart_Failure | pericyte | *RASSF1* | 2.293754123 | 0.001803446 | 0.017050024 | 1 |
| Heart_Failure | pericyte | *CYFIP1* | 2.832874337 | 0.001174543 | 0.012622827 | 1 |
| Heart_Failure | pericyte | *PITX1* | 6.430113883 | 0.000872208 | 0.010425044 | 1 |
| Heart_Failure | pericyte | *DNAJA2* | 4.941746227 | 3.46E-05 | 0.001918699 | 0.217523356 |
| Heart_Failure | pericyte | *NEDD4* | 1.652652506 | 0.002588686 | 0.021274596 | 1 |
| Heart_Failure | pericyte | *TMEM260* | 2.259725417 | 0.009194667 | 0.049919579 | 1 |
| Heart_Failure | pericyte | *WIPI1* | 2.591483812 | 0.009060076 | 0.049316621 | 1 |
| Heart_Failure | pericyte | *AP3M2* | 2.770910955 | 0.008307189 | 0.046868605 | 1 |
| Heart_Failure | pericyte | *TESK2* | 2.35940352 | 0.001332532 | 0.013824469 | 1 |
| Heart_Failure | pericyte | *VASH1* | 4.212628173 | 0.000153586 | 0.00371105 | 0.965593681 |
| Heart_Failure | pericyte | *EPN2* | 2.147492117 | 0.005370624 | 0.03431414 | 1 |
| Heart_Failure | pericyte | *NFATC3* | 1.772764787 | 0.003746222 | 0.027065715 | 1 |
| Heart_Failure | pericyte | *TRNT1* | 2.552823353 | 0.007744012 | 0.044818101 | 1 |
| Heart_Failure | pericyte | *PDE8A* | 1.782019852 | 0.001172989 | 0.012622827 | 1 |
| Heart_Failure | pericyte | *PICALM* | 1.755952081 | 0.006244124 | 0.038261994 | 1 |
| Heart_Failure | pericyte | *LMAN1* | 3.386429753 | 0.000653681 | 0.008706969 | 1 |
| Heart_Failure | pericyte | *SRI* | 4.4780871 | 4.88E-05 | 0.002128526 | 0.307033037 |
| Heart_Failure | pericyte | *EIF4G3* | 2.291053151 | 0.001003975 | 0.011372954 | 1 |
| Heart_Failure | pericyte | *VPS9D1* | 4.651645717 | 0.000285955 | 0.005334716 | 1 |
| Heart_Failure | pericyte | *PLD1* | 3.649760686 | 1.38E-05 | 0.001740485 | 0.086602897 |
| Heart_Failure | pericyte | *TUBA3D* | 0.803470388 | 0.009029754 | 0.049297742 | 1 |
| Heart_Failure | pericyte | *RBMS2* | 3.556597523 | 3.66E-05 | 0.001918699 | 0.230243889 |
| Heart_Failure | pericyte | *GPATCH1* | 2.062194363 | 0.008312151 | 0.046868605 | 1 |
| Heart_Failure | pericyte | *DYNC1I2* | 2.642405814 | 0.001195274 | 0.012736757 | 1 |
| Heart_Failure | pericyte | *SIRT6* | 7.81277062 | 4.28E-05 | 0.002108725 | 0.269050012 |
| Heart_Failure | pericyte | *PIK3C3* | 2.22989706 | 0.008936157 | 0.049024099 | 1 |

| | | | | | | |
|---|---|---|---|---|---|---|
| Heart_Failure | pericyte | *SYNJ2* | 2.935722612 | 4.77E-05 | 0.002128526 | 0.299975461 |
| Heart_Failure | pericyte | *PCM1* | 2.441118111 | 0.000463478 | 0.007038378 | 1 |
| Heart_Failure | pericyte | *OSBPL6* | 3.749754002 | 0.001853838 | 0.017421646 | 1 |
| Heart_Failure | pericyte | *XRCC5* | 2.471894859 | 0.002305695 | 0.019722323 | 1 |
| Heart_Failure | pericyte | *REXO1* | 5.432277199 | 3.33E-05 | 0.001918699 | 0.209621633 |
| Heart_Failure | pericyte | *DDX1* | 3.310274816 | 0.000789594 | 0.009733684 | 1 |
| Heart_Failure | pericyte | *DNM2* | 2.182950547 | 0.009057943 | 0.049316621 | 1 |
| Heart_Failure | pericyte | *RIMS1* | 3.171827352 | 0.001083012 | 0.011945435 | 1 |
| Heart_Failure | pericyte | *PSEN1* | 1.776657463 | 0.008368585 | 0.04704352 | 1 |
| Heart_Failure | pericyte | *ZNF506* | 3.281170491 | 0.000328334 | 0.005782167 | 1 |
| Heart_Failure | pericyte | *GSK3B* | 2.133957678 | 0.004115141 | 0.028746549 | 1 |
| Heart_Failure | pericyte | *ERC1* | 2.197944258 | 0.001522175 | 0.015311861 | 1 |
| Heart_Failure | pericyte | *PLOD1* | 4.316550025 | 0.000149314 | 0.003670989 | 0.938736574 |
| Heart_Failure | pericyte | *PDS5B* | 3.245856433 | 5.67E-05 | 0.002283454 | 0.356218852 |
| Heart_Failure | pericyte | *OXCT1* | -0.995291621 | 0.00823271 | 0.046587802 | 1 |
| Heart_Failure | pericyte | *ZNF324* | 2.108476785 | 0.000893133 | 0.010614609 | 1 |
| Heart_Failure | pericyte | *GSTP1* | 4.929458397 | 9.48E-06 | 0.001740485 | 0.059569771 |
| Heart_Failure | pericyte | *WBP11* | 4.069684572 | 0.000738876 | 0.009422549 | 1 |
| Heart_Failure | pericyte | *COL16A1* | -1.19001754 | 0.001038651 | 0.011660716 | 1 |
| Heart_Failure | pericyte | *NCOA1* | 2.260400024 | 0.001202327 | 0.012790239 | 1 |
| Heart_Failure | pericyte | *CD59* | 2.15700689 | 0.006439896 | 0.039080719 | 1 |
| Heart_Failure | pericyte | *TAF9* | 6.795304746 | 4.81E-05 | 0.002128526 | 0.302390724 |
| Heart_Failure | pericyte | *OVGP1* | 1.884977289 | 0.003757262 | 0.027065715 | 1 |
| Heart_Failure | pericyte | *SLC25A24* | 2.366739297 | 0.000830337 | 0.010058438 | 1 |
| Heart_Failure | pericyte | *RRN3* | 2.617253182 | 0.001165472 | 0.012568303 | 1 |
| Heart_Failure | pericyte | *CHMP5* | 3.301428652 | 0.000621857 | 0.008336066 | 1 |
| Heart_Failure | pericyte | *PSMC5* | 2.008677319 | 8.36E-05 | 0.002749485 | 0.525458212 |
| Heart_Failure | pericyte | *KLHL42* | 2.279873828 | 0.002531804 | 0.0210264 | 1 |
| Heart_Failure | pericyte | *ERGIC2* | 3.832934867 | 6.37E-05 | 0.002397441 | 0.400372635 |
| Heart_Failure | pericyte | *ANAPC5* | 3.752384455 | 6.49E-06 | 0.001424095 | 0.040828628 |
| Heart_Failure | pericyte | *SLC23A2* | 3.705058984 | 8.72E-05 | 0.002769335 | 0.547984364 |
| Heart_Failure | pericyte | *OAS1* | 3.816064156 | 0.004880282 | 0.032151896 | 1 |
| Heart_Failure | pericyte | *KIF16B* | 2.649704489 | 0.002252892 | 0.01956344 | 1 |
| Heart_Failure | pericyte | *FUS* | 2.896011625 | 0.000350944 | 0.005952796 | 1 |
| Heart_Failure | pericyte | *FXYD5* | 5.413418256 | 4.76E-06 | 0.001108016 | 0.02991642 |
| Heart_Failure | pericyte | *KCNH4* | 1.199249787 | 0.003744812 | 0.027065715 | 1 |
| Heart_Failure | pericyte | *GPATCH2L* | 1.543733284 | 0.008008636 | 0.045565877 | 1 |
| Heart_Failure | pericyte | *PAPOLA* | 2.439783784 | 0.00320947 | 0.02478862 | 1 |
| Heart_Failure | pericyte | *MUL1* | 2.220440262 | 0.000826705 | 0.010053184 | 1 |
| Heart_Failure | pericyte | *PABPC4* | -1.239864445 | 0.006129751 | 0.037819181 | 1 |
| Heart_Failure | pericyte | *USP48* | 1.716632946 | 0.001924755 | 0.017769364 | 1 |
| Heart_Failure | pericyte | *TXNL1* | 2.727839878 | 0.003824469 | 0.027354305 | 1 |
| Heart_Failure | pericyte | *ZFHX4* | 2.346454263 | 0.003051708 | 0.023893012 | 1 |
| Heart_Failure | pericyte | *TMEM101* | 1.94206287 | 0.008636521 | 0.047797366 | 1 |
| Heart_Failure | pericyte | *PPP2R3C* | 2.927188021 | 0.00051082 | 0.007424617 | 1 |
| Heart_Failure | pericyte | *SLC22A17* | 1.639663011 | 0.001606298 | 0.015815708 | 1 |
| Heart_Failure | pericyte | *SCFD1* | 1.659165282 | 0.005253558 | 0.033841308 | 1 |
| Heart_Failure | pericyte | *DPYSL2* | 2.306018691 | 0.000125786 | 0.00330886 | 0.790817475 |
| Heart_Failure | pericyte | *MAP3K1* | 2.635393851 | 0.000616195 | 0.008295544 | 1 |
| Heart_Failure | pericyte | *PCSK5* | 2.987011139 | 3.45E-05 | 0.001918699 | 0.216653067 |

| | | | | | | |
|---|---|---|---|---|---|---|
| Heart_Failure | pericyte | *NRP1* | 2.760729037 | 4.94E-05 | 0.002128526 | 0.310764793 |
| Heart_Failure | pericyte | *PRTFDC1* | 0.878083198 | 0.006523209 | 0.039434054 | 1 |
| Heart_Failure | pericyte | *TSPAN15* | 0.890726539 | 0.002659464 | 0.02168619 | 1 |
| Heart_Failure | pericyte | *CDC34* | 4.430154807 | 0.000128486 | 0.003332666 | 0.807791333 |
| Heart_Failure | pericyte | *MTAP* | 4.996643243 | 0.00011926 | 0.003259951 | 0.749788628 |
| Heart_Failure | pericyte | *LZTR1* | 4.754793162 | 2.20E-05 | 0.001740485 | 0.138228434 |
| Heart_Failure | pericyte | *LRP5L* | 3.49831529 | 0.000529954 | 0.007488118 | 1 |
| Heart_Failure | pericyte | *SH3BP1* | 4.844553911 | 0.006947354 | 0.041205671 | 1 |
| Heart_Failure | pericyte | *POLDIP3* | 2.930573474 | 0.000115194 | 0.003204538 | 0.724225579 |
| Heart_Failure | pericyte | *TIMP3* | 2.209646633 | 0.001262807 | 0.013210092 | 1 |
| Heart_Failure | pericyte | *TTLL1* | 5.154753255 | 0.000227885 | 0.004697424 | 1 |
| Heart_Failure | pericyte | *HMGXB4* | 3.474739817 | 0.000517381 | 0.007443425 | 1 |
| Heart_Failure | pericyte | *RBFOX2* | 2.891916779 | 0.000199557 | 0.00440216 | 1 |
| Heart_Failure | pericyte | *SLC25A17* | 2.472011363 | 0.005690229 | 0.035810281 | 1 |
| Heart_Failure | pericyte | *KCTD17* | 1.934119089 | 0.005859962 | 0.036658292 | 1 |
| Heart_Failure | pericyte | *EP300* | 2.331644894 | 0.001773527 | 0.016834554 | 1 |
| Heart_Failure | pericyte | *SOS2* | 2.410330493 | 0.001867066 | 0.017493653 | 1 |
| Heart_Failure | pericyte | *CNIH1* | 2.737451036 | 0.003125226 | 0.024377538 | 1 |
| Heart_Failure | pericyte | *PCK2* | 3.183720378 | 0.000987237 | 0.011223795 | 1 |
| Heart_Failure | pericyte | *KIAA0391* | 2.595546868 | 0.002983677 | 0.023494003 | 1 |
| Heart_Failure | pericyte | *CD40* | 3.580789176 | 0.00079462 | 0.009760099 | 1 |
| Heart_Failure | pericyte | *NDRG3* | 2.868323043 | 0.005173912 | 0.033465418 | 1 |
| Heart_Failure | pericyte | *BMP7* | 3.43633051 | 0.000343866 | 0.005913159 | 1 |
| Heart_Failure | pericyte | *NELFCD* | 2.984333822 | 0.003981195 | 0.028123337 | 1 |
| Heart_Failure | pericyte | *SLC17A9* | 4.146944844 | 0.000207745 | 0.004519343 | 1 |
| Heart_Failure | pericyte | *RNF24* | 2.822964619 | 4.51E-05 | 0.002128526 | 0.283507224 |
| Heart_Failure | pericyte | *CCM2L* | 0.798797195 | 0.008552772 | 0.047507995 | 1 |
| Heart_Failure | pericyte | *MYL9* | 2.805820181 | 7.79E-06 | 0.001579196 | 0.04895508 |
| Heart_Failure | pericyte | *RBFA* | 2.760670159 | 0.000676149 | 0.008949365 | 1 |
| Heart_Failure | pericyte | *SMCHD1* | 1.986205185 | 0.004905594 | 0.032227243 | 1 |
| Heart_Failure | pericyte | *MYL12A* | 3.855405567 | 0.000228871 | 0.004702319 | 1 |
| Heart_Failure | pericyte | *STK24* | 3.277387561 | 0.000122275 | 0.00330886 | 0.768744453 |
| Heart_Failure | pericyte | *ARHGEF7* | 2.265502785 | 0.002178353 | 0.019149613 | 1 |
| Heart_Failure | pericyte | *TSC22D1* | 2.464990952 | 0.001645745 | 0.016141653 | 1 |
| Heart_Failure | pericyte | *CORO1A* | -1.744120675 | 0.005771471 | 0.036212811 | 1 |
| Heart_Failure | pericyte | *NFAT5* | 1.838171278 | 0.005078816 | 0.033209287 | 1 |
| Heart_Failure | pericyte | *LONP2* | 2.190285539 | 0.002670347 | 0.02169166 | 1 |
| Heart_Failure | pericyte | *SETD6* | 2.856419673 | 0.007107715 | 0.041841016 | 1 |
| Heart_Failure | pericyte | *NME4* | 4.478902306 | 5.53E-05 | 0.002256679 | 0.347528521 |
| Heart_Failure | pericyte | *LMF1* | 2.746712659 | 0.001797173 | 0.017016303 | 1 |
| Heart_Failure | pericyte | *SLC7A5* | 15.84498085 | 4.78E-05 | 0.002128526 | 0.300750108 |
| Heart_Failure | pericyte | *STUB1* | 2.456227575 | 0.005528027 | 0.035019283 | 1 |
| Heart_Failure | pericyte | *RRN3P2* | 2.222401724 | 0.000947127 | 0.010925847 | 1 |
| Heart_Failure | pericyte | *RBL2* | 1.169255795 | 0.007764025 | 0.044818101 | 1 |
| Heart_Failure | pericyte | *STX4* | 3.402245025 | 0.000151487 | 0.003691475 | 0.952400446 |
| Heart_Failure | pericyte | *KAT8* | 3.594399652 | 0.000208504 | 0.004520229 | 1 |
| Heart_Failure | pericyte | *RNF40* | 2.393285298 | 0.000868602 | 0.010401721 | 1 |
| Heart_Failure | pericyte | *KNOP1* | 2.212566562 | 0.000937856 | 0.010883614 | 1 |
| Heart_Failure | pericyte | *CSK* | 4.549638675 | 0.001243837 | 0.013098836 | 1 |
| Heart_Failure | pericyte | *HERC1* | 2.871516607 | 9.04E-05 | 0.002769335 | 0.568209653 |

| | | | | | | |
|---|---|---|---|---|---|---|
| Heart_Failure | pericyte | *HOMER2* | 1.999335291 | 0.004289002 | 0.029352191 | 1 |
| Heart_Failure | pericyte | *DTWD1* | 1.961948513 | 0.002841553 | 0.02269993 | 1 |
| Heart_Failure | pericyte | *DNAJC17* | 4.423588085 | 1.74E-05 | 0.001740485 | 0.109532171 |
| Heart_Failure | pericyte | *IKBKB* | 2.661028176 | 0.003286003 | 0.025132728 | 1 |
| Heart_Failure | pericyte | *GDAP1* | 2.580897079 | 0.007088894 | 0.041769334 | 1 |
| Heart_Failure | pericyte | *CHRAC1* | 1.943158188 | 0.000938272 | 0.010883614 | 1 |
| Heart_Failure | pericyte | *ERICH1* | 2.737169794 | 0.001071503 | 0.011881027 | 1 |
| Heart_Failure | pericyte | *TUSC3* | 0.707182367 | 0.004926677 | 0.032331958 | 1 |
| Heart_Failure | pericyte | *MCM4* | 1.209627107 | 0.009213221 | 0.049977153 | 1 |
| Heart_Failure | pericyte | *FCGRT* | 1.368720242 | 0.006168991 | 0.037949559 | 1 |
| Heart_Failure | pericyte | *TLE6* | 1.142418042 | 0.007791663 | 0.044818101 | 1 |
| Heart_Failure | pericyte | *AES* | 3.517925567 | 0.001054007 | 0.011770062 | 1 |
| Heart_Failure | pericyte | *C19orf53* | 1.874285098 | 0.002970659 | 0.023433542 | 1 |
| Heart_Failure | pericyte | *TIMM44* | 3.146147094 | 3.12E-05 | 0.001918699 | 0.196146258 |
| Heart_Failure | pericyte | *SYDE1* | 2.65961232 | 0.006946561 | 0.041205671 | 1 |
| Heart_Failure | pericyte | *RPS16* | 2.621646055 | 0.000596062 | 0.008101113 | 1 |
| Heart_Failure | pericyte | *PRKD2* | 1.666348159 | 0.003693882 | 0.027035435 | 1 |
| Heart_Failure | pericyte | *FZR1* | 4.76716901 | 0.001088214 | 0.011981791 | 1 |
| Heart_Failure | pericyte | *BBC3* | 2.665634802 | 0.003190234 | 0.02478862 | 1 |
| Heart_Failure | pericyte | *CYTH2* | 4.426322382 | 2.02E-05 | 0.001740485 | 0.126833236 |
| Heart_Failure | pericyte | *LIG1* | 2.655488804 | 0.000902614 | 0.010695671 | 1 |
| Heart_Failure | pericyte | *MIER2* | 3.180105753 | 0.008504916 | 0.047449452 | 1 |
| Heart_Failure | pericyte | *ZNF14* | 5.067444805 | 2.44E-05 | 0.001740485 | 0.153162723 |
| Heart_Failure | pericyte | *ZNF85* | 2.518755833 | 0.000714335 | 0.009202924 | 1 |
| Heart_Failure | pericyte | *ETHE1* | 1.736467854 | 0.008115841 | 0.046092403 | 1 |
| Heart_Failure | pericyte | *PON2* | 2.494604767 | 0.007525585 | 0.043849261 | 1 |
| Heart_Failure | pericyte | *WDR91* | 4.22070259 | 2.18E-05 | 0.001740485 | 0.137273173 |
| Heart_Failure | pericyte | *MET* | 3.370780337 | 6.17E-05 | 0.002379208 | 0.387810888 |
| Heart_Failure | pericyte | *LMBR1* | 2.560820542 | 0.000578137 | 0.007918835 | 1 |
| Heart_Failure | pericyte | *NOD1* | 2.754504478 | 0.002374687 | 0.020147987 | 1 |
| Heart_Failure | pericyte | *CHCHD2* | 1.456712532 | 0.004156345 | 0.028937919 | 1 |
| Heart_Failure | pericyte | *EIF3B* | 2.652830566 | 0.002321829 | 0.019833344 | 1 |
| Heart_Failure | pericyte | *USP42* | 3.101712902 | 0.001904887 | 0.017712797 | 1 |
| Heart_Failure | pericyte | *AP1S1* | 9.778778193 | 2.27E-05 | 0.001740485 | 0.14261621 |
| Heart_Failure | pericyte | *PLOD3* | 4.808961684 | 1.43E-05 | 0.001740485 | 0.089853563 |
| Heart_Failure | pericyte | *RPA3* | 3.258535618 | 0.000846073 | 0.010209715 | 1 |
| Heart_Failure | pericyte | *PHF14* | 2.070339472 | 0.004295215 | 0.029352191 | 1 |
| Heart_Failure | pericyte | *ZNF862* | 2.062274196 | 0.005160311 | 0.033464241 | 1 |
| Heart_Failure | pericyte | *CLIP2* | 1.189513837 | 0.001753505 | 0.016779735 | 1 |
| Heart_Failure | pericyte | *LIMK1* | 5.169328964 | 1.83E-05 | 0.001740485 | 0.114758151 |
| Heart_Failure | pericyte | *PDLIM1* | 2.095730511 | 3.18E-06 | 0.000909616 | 0.020011554 |
| Heart_Failure | pericyte | *DNMBP* | 2.692873331 | 0.003204955 | 0.02478862 | 1 |
| Heart_Failure | pericyte | *DDX50* | 3.563700979 | 0.000283587 | 0.005306292 | 1 |
| Heart_Failure | pericyte | *MAPK8* | 2.066809511 | 0.004944878 | 0.032417568 | 1 |
| Heart_Failure | pericyte | *MICU1* | 2.364677263 | 0.0015781 | 0.015614046 | 1 |
| Heart_Failure | pericyte | *CCSER2* | 2.300842723 | 0.004011253 | 0.028255829 | 1 |
| Heart_Failure | pericyte | *MLLT6* | 4.319475266 | 2.19E-05 | 0.001740485 | 0.137611381 |
| Heart_Failure | pericyte | *RPS6KB1* | 4.02666452 | 4.73E-05 | 0.002128526 | 0.297268187 |
| Heart_Failure | pericyte | *NUP88* | 2.282437598 | 0.004217721 | 0.029139352 | 1 |
| Heart_Failure | pericyte | *C1QBP* | 4.912266451 | 0.000332923 | 0.005798899 | 1 |

| | | | | | | |
|---|---|---|---|---|---|---|
| Heart_Failure | pericyte | *FTSJ3* | 0.566271345 | 0.004162928 | 0.028951692 | 1 |
| Heart_Failure | pericyte | *AKAP10* | 2.361699378 | 0.000766168 | 0.009605939 | 1 |
| Heart_Failure | pericyte | *UTP6* | 3.147467287 | 0.001366676 | 0.014085721 | 1 |
| Heart_Failure | pericyte | *PEX12* | -1.21492536 | 0.0027793 | 0.02226367 | 1 |
| Heart_Failure | pericyte | *LRRC59* | 4.30664236 | 0.007828468 | 0.044906549 | 1 |
| Heart_Failure | pericyte | *SLAIN2* | 1.828324655 | 0.00651191 | 0.039403634 | 1 |
| Heart_Failure | pericyte | *NDUFC1* | 4.654384309 | 7.85E-05 | 0.002693349 | 0.493585013 |
| Heart_Failure | pericyte | *CPE* | 2.046036277 | 4.63E-05 | 0.002128526 | 0.290795003 |
| Heart_Failure | pericyte | *WFS1* | 5.280444662 | 6.11E-05 | 0.002379208 | 0.383923736 |
| Heart_Failure | pericyte | *GRPEL1* | 2.703412355 | 0.000905423 | 0.010699987 | 1 |
| Heart_Failure | pericyte | *SLC2A9* | 3.237324427 | 0.000197745 | 0.004377544 | 1 |
| Heart_Failure | pericyte | *STIM2* | 2.801154472 | 0.000452019 | 0.006952162 | 1 |
| Heart_Failure | pericyte | *RAPGEF2* | 2.604011311 | 5.87E-05 | 0.002333071 | 0.36916593 |
| Heart_Failure | pericyte | *UFSP2* | 4.703337007 | 3.75E-06 | 0.000964138 | 0.023576088 |
| Heart_Failure | pericyte | *KLF3* | 2.584683839 | 0.003394841 | 0.025683954 | 1 |
| Heart_Failure | pericyte | *KLHL5* | 1.617963575 | 0.001030191 | 0.011607185 | 1 |
| Heart_Failure | pericyte | *DTX4* | 4.06014087 | 0.00011708 | 0.003228426 | 0.736081146 |
| Heart_Failure | pericyte | *CCDC86* | 13.19667895 | 4.99E-05 | 0.002134554 | 0.31377948 |
| Heart_Failure | pericyte | *CBL* | 2.133162106 | 0.000492133 | 0.007302207 | 1 |
| Heart_Failure | pericyte | *MADD* | 4.463150579 | 1.01E-05 | 0.001740485 | 0.063352588 |
| Heart_Failure | pericyte | *AIP* | 2.017972179 | 0.00014096 | 0.003581691 | 0.886216571 |
| Heart_Failure | pericyte | *NDUFS8* | 4.728327171 | 0.000499506 | 0.007337373 | 1 |
| Heart_Failure | pericyte | *PPFIBP1* | 1.942403862 | 0.003260846 | 0.025001141 | 1 |
| Heart_Failure | pericyte | *PRDM4* | 2.195223973 | 0.008513278 | 0.047449452 | 1 |
| Heart_Failure | pericyte | *LTA4H* | 2.881402123 | 0.007005838 | 0.041435279 | 1 |
| Heart_Failure | pericyte | *WNT5B* | 1.250066675 | 0.00357505 | 0.026490062 | 1 |
| Heart_Failure | pericyte | *VPS29* | 3.321474088 | 0.000418905 | 0.006722771 | 1 |
| Heart_Failure | pericyte | *CDK2AP1* | 0.700698913 | 0.002244654 | 0.019534361 | 1 |
| Heart_Failure | pericyte | *CNOT2* | 2.321730465 | 0.002763185 | 0.02221502 | 1 |
| Heart_Failure | pericyte | *KRR1* | 3.325266672 | 0.000221499 | 0.004655112 | 1 |
| Heart_Failure | pericyte | *GNPTAB* | 2.39757574 | 0.004275008 | 0.029352191 | 1 |
| Heart_Failure | pericyte | *PHC1* | 5.928275522 | 0.000320192 | 0.005686577 | 1 |
| Heart_Failure | pericyte | *COL12A1* | 3.131728684 | 2.70E-05 | 0.001847988 | 0.170014941 |
| Heart_Failure | pericyte | *MCM9* | 2.236078892 | 0.007778226 | 0.044818101 | 1 |
| Heart_Failure | pericyte | *MAPK14* | 2.215958813 | 0.003014314 | 0.023688737 | 1 |
| Heart_Failure | pericyte | *PHACTR1* | 2.905716456 | 0.000512037 | 0.007424617 | 1 |
| Heart_Failure | pericyte | *MDN1* | 1.974519269 | 0.002618681 | 0.021493012 | 1 |
| Heart_Failure | pericyte | *ASCC3* | 2.038384482 | 0.007015736 | 0.041454822 | 1 |
| Heart_Failure | pericyte | *PACRG* | 5.760164322 | 1.14E-05 | 0.001740485 | 0.071694591 |
| Heart_Failure | pericyte | *TFEB* | 3.427361706 | 0.002246436 | 0.019534361 | 1 |
| Heart_Failure | pericyte | *FAM120B* | 1.565906181 | 0.001910405 | 0.017714925 | 1 |
| Heart_Failure | pericyte | *GUCA1B* | 2.578500788 | 0.00381134 | 0.02732257 | 1 |
| Heart_Failure | pericyte | *EXOC2* | 2.261289143 | 0.008380586 | 0.04704352 | 1 |
| Heart_Failure | pericyte | *SENP6* | 2.955643467 | 9.25E-05 | 0.002769335 | 0.581560387 |
| Heart_Failure | pericyte | *TBX18* | 1.463138645 | 0.004730249 | 0.031536662 | 1 |
| Heart_Failure | pericyte | *DAP* | 3.461856497 | 0.000332973 | 0.005798899 | 1 |
| Heart_Failure | pericyte | *PCDHB6* | 2.183772013 | 0.003139566 | 0.024428777 | 1 |
| Heart_Failure | pericyte | *DROSHA* | 1.90849091 | 0.002537055 | 0.0210264 | 1 |
| Heart_Failure | pericyte | *SLC12A7* | 3.577135174 | 0.000173559 | 0.003996935 | 1 |
| Heart_Failure | pericyte | *IL4* | -2.22538299 | 0.003552384 | 0.02646189 | 1 |

| | | | | | | |
|---|---|---|---|---|---|---|
| Heart_Failure | pericyte | *SMAD5* | 2.263226193 | 0.0075332 | 0.043852991 | 1 |
| Heart_Failure | pericyte | *HMGXB3* | 3.595893354 | 0.000233072 | 0.004726854 | 1 |
| Heart_Failure | pericyte | *BNIP1* | 0.773198066 | 0.00181131 | 0.017098653 | 1 |
| Heart_Failure | pericyte | *PLSCR4* | 1.505184309 | 0.008694829 | 0.048077737 | 1 |
| Heart_Failure | pericyte | *HEMK1* | 2.720727257 | 2.59E-06 | 0.000909616 | 0.016284069 |
| Heart_Failure | pericyte | *ABCC5* | 2.45201799 | 0.001651157 | 0.016169511 | 1 |
| Heart_Failure | pericyte | *KLHL24* | 1.804720845 | 0.00134603 | 0.013941503 | 1 |
| Heart_Failure | pericyte | *PLCH1* | 1.865793325 | 0.001544867 | 0.015465889 | 1 |
| Heart_Failure | pericyte | *NKTR* | 2.045427527 | 0.002473913 | 0.020793437 | 1 |
| Heart_Failure | pericyte | *MOB1A* | 4.106987973 | 0.000261025 | 0.005096468 | 1 |
| Heart_Failure | pericyte | *LMAN2L* | 1.643122658 | 0.005084577 | 0.033209287 | 1 |
| Heart_Failure | pericyte | *PIKFYVE* | 2.663411749 | 0.000450734 | 0.006952162 | 1 |
| Heart_Failure | pericyte | *KCNIP3* | 3.385087183 | 0.000721795 | 0.009274224 | 1 |
| Heart_Failure | pericyte | *NCL* | 3.70123158 | 4.26E-05 | 0.002108725 | 0.268129632 |
| Heart_Failure | pericyte | *ACTR1B* | 2.739907836 | 4.14E-05 | 0.002096665 | 0.259986463 |
| Heart_Failure | pericyte | *ACTR3* | 2.053654327 | 0.009136987 | 0.049692248 | 1 |
| Heart_Failure | pericyte | *TP53I3* | 3.133285284 | 9.46E-05 | 0.002819717 | 0.594960231 |
| Heart_Failure | pericyte | *STAM2* | 3.646081848 | 9.09E-05 | 0.002769335 | 0.571218179 |
| Heart_Failure | pericyte | *TANC1* | 1.970908336 | 0.000226014 | 0.004674178 | 1 |
| Heart_Failure | pericyte | *NRBP1* | 1.891734208 | 0.00704012 | 0.041559845 | 1 |
| Heart_Failure | pericyte | *PPM1G* | 3.075363097 | 0.000509922 | 0.007424617 | 1 |
| Heart_Failure | pericyte | *GLS* | 1.79531686 | 0.008457322 | 0.047389648 | 1 |
| Heart_Failure | pericyte | *SF3B1* | 2.283172488 | 0.002098637 | 0.018715082 | 1 |
| Heart_Failure | pericyte | *TAF1B* | 3.978569514 | 3.05E-06 | 0.000909616 | 0.019144232 |
| Heart_Failure | pericyte | *STRN* | 3.00021045 | 0.000812322 | 0.00989742 | 1 |
| Heart_Failure | pericyte | *CEBPZ* | 1.871168808 | 0.002895894 | 0.023046188 | 1 |
| Heart_Failure | pericyte | *PRKD3* | 2.991033924 | 0.000160297 | 0.003831895 | 1 |
| Heart_Failure | pericyte | *RMDN2* | 3.741864576 | 0.000529302 | 0.007488118 | 1 |
| Heart_Failure | pericyte | *ORC4* | 2.208518175 | 0.002502205 | 0.020919364 | 1 |
| Heart_Failure | pericyte | *BCL9* | 2.964682967 | 0.000455832 | 0.006976539 | 1 |
| Heart_Failure | pericyte | *RALGPS2* | 3.363307457 | 3.64E-05 | 0.001918699 | 0.228653088 |
| Heart_Failure | pericyte | *FAM20B* | 3.414736998 | 0.0015793 | 0.015614046 | 1 |
| Heart_Failure | pericyte | *ICMT* | 2.132998353 | 0.003732735 | 0.027065715 | 1 |
| Heart_Failure | pericyte | *RAP1A* | 2.617963567 | 0.000761616 | 0.009576563 | 1 |
| Heart_Failure | pericyte | *DOCK7* | 3.125226181 | 0.000178839 | 0.004073766 | 1 |
| Heart_Failure | pericyte | *RGS2* | 3.720707754 | 0.000541803 | 0.00760338 | 1 |
| Heart_Failure | pericyte | *CFHR3* | -1.285155217 | 0.000212628 | 0.004578062 | 1 |
| Heart_Failure | pericyte | *TMED5* | 2.931873202 | 0.00160383 | 0.015815708 | 1 |
| Heart_Failure | pericyte | *PRRC2C* | 1.905419269 | 0.007984559 | 0.045470039 | 1 |
| Heart_Failure | pericyte | *ABCD3* | 2.44415424 | 0.00667848 | 0.040141112 | 1 |
| Heart_Failure | pericyte | *PTBP2* | 1.471749737 | 0.001568401 | 0.015577464 | 1 |
| Heart_Failure | pericyte | *DARS2* | 3.22962686 | 0.001410948 | 0.014447276 | 1 |
| Heart_Failure | pericyte | *RCOR3* | 3.25554581 | 0.000148875 | 0.003670989 | 0.935974054 |
| Heart_Failure | pericyte | *KMT2A* | 2.559265085 | 0.001716666 | 0.01648789 | 1 |
| Heart_Failure | pericyte | *FILIP1* | 1.351194842 | 0.000465451 | 0.007051295 | 1 |
| Heart_Failure | pericyte | *SGIP1* | 0.643130857 | 0.00852679 | 0.047482665 | 1 |
| Heart_Failure | pericyte | *MED28* | 2.032335098 | 0.002298258 | 0.019712344 | 1 |
| Heart_Failure | pericyte | *VAMP8* | 1.49960637 | 0.004443151 | 0.03013386 | 1 |
| Heart_Failure | pericyte | *SPP1* | 2.470751329 | 0.003200234 | 0.02478862 | 1 |
| Heart_Failure | pericyte | *UCHL3* | 1.737624297 | 0.002242633 | 0.019534361 | 1 |

| | | | | | | |
|---|---|---|---|---|---|---|
| Heart_Failure | pericyte | *SENP5* | 2.966722578 | 0.00037473 | 0.006167354 | 1 |
| Heart_Failure | pericyte | *PTBP3* | 2.627702578 | 0.002531911 | 0.0210264 | 1 |
| Heart_Failure | pericyte | *CNTRL* | 3.055634089 | 0.000534133 | 0.007529365 | 1 |
| Heart_Failure | pericyte | *TRIM32* | 4.634708924 | 1.76E-05 | 0.001740485 | 0.110705213 |
| Heart_Failure | pericyte | *INVS* | 2.123569619 | 0.001556473 | 0.015557311 | 1 |
| Heart_Failure | pericyte | *PPP4R4* | 0.443429498 | 0.007175657 | 0.042201456 | 1 |
| Heart_Failure | pericyte | *ZC2HC1C* | 1.354528492 | 0.004793236 | 0.03173585 | 1 |
| Heart_Failure | pericyte | *EIF2B2* | 1.133090853 | 0.006228325 | 0.038247407 | 1 |
| Heart_Failure | pericyte | *CNRIP1* | 2.071028576 | 0.006253682 | 0.03828325 | 1 |
| Heart_Failure | pericyte | *CUTC* | 2.252174197 | 0.003487476 | 0.026160156 | 1 |
| Heart_Failure | pericyte | *MXI1* | 2.799462898 | 0.004889567 | 0.032155553 | 1 |
| Heart_Failure | pericyte | *TCTN3* | 2.305389324 | 0.000573968 | 0.007896143 | 1 |
| Heart_Failure | pericyte | *KANSL1* | 1.540609211 | 0.006065416 | 0.037606776 | 1 |
| Heart_Failure | pericyte | *CYSTM1* | 2.760087351 | 0.00078956 | 0.009733684 | 1 |
| Heart_Failure | pericyte | *PCDHB14* | 1.641866319 | 0.003894001 | 0.027756898 | 1 |
| Heart_Failure | pericyte | *IQSEC3* | 1.253820785 | 0.006962354 | 0.041228541 | 1 |
| Heart_Failure | pericyte | *DNAJC15* | 4.025883186 | 7.14E-05 | 0.002564014 | 0.448702373 |
| Heart_Failure | pericyte | *SMAD9* | 5.133995258 | 1.98E-05 | 0.001740485 | 0.124229166 |
| Heart_Failure | pericyte | *TEX14* | 1.439564435 | 0.001538515 | 0.015450392 | 1 |
| Heart_Failure | pericyte | *LHX4* | 2.384497835 | 0.00204293 | 0.018348428 | 1 |
| Heart_Failure | pericyte | *RNF115* | 2.082556323 | 0.008179666 | 0.046360577 | 1 |
| Heart_Failure | pericyte | *LIAS* | 3.29492233 | 0.00039558 | 0.006409823 | 1 |
| Heart_Failure | pericyte | *ACVR2A* | 2.368912564 | 0.003771363 | 0.027071238 | 1 |
| Heart_Failure | pericyte | *POLK* | 2.643055731 | 0.000619614 | 0.008323748 | 1 |
| Heart_Failure | pericyte | *FLT3* | 0.790850271 | 0.006110683 | 0.037812861 | 1 |
| Heart_Failure | pericyte | *SERAC1* | 3.80849117 | 0.000499196 | 0.007337373 | 1 |
| Heart_Failure | pericyte | *LDB3* | 1.228726289 | 0.00069059 | 0.00898911 | 1 |
| Heart_Failure | pericyte | *FAM213A* | 1.428536143 | 0.000599995 | 0.008112188 | 1 |
| Heart_Failure | pericyte | *SPATA1* | 2.724503517 | 0.003350882 | 0.025473999 | 1 |
| Heart_Failure | pericyte | *CCDC18* | 2.410826197 | 0.002269161 | 0.019598651 | 1 |
| Heart_Failure | pericyte | *BBS9* | 2.101011067 | 0.002294743 | 0.019709086 | 1 |
| Heart_Failure | pericyte | *PMS2* | 3.584459345 | 0.000688467 | 0.008980062 | 1 |
| Heart_Failure | pericyte | *KLHL7* | 4.142423392 | 2.35E-05 | 0.001740485 | 0.147940989 |
| Heart_Failure | pericyte | *FAM126A* | 1.835404336 | 0.008849197 | 0.048802547 | 1 |
| Heart_Failure | pericyte | *CCZ1* | 1.862562349 | 0.001832139 | 0.017243504 | 1 |
| Heart_Failure | pericyte | *DCAF10* | 2.260851327 | 0.004251352 | 0.02930729 | 1 |
| Heart_Failure | pericyte | *PKN1* | 2.872517291 | 0.003216951 | 0.024815913 | 1 |
| Heart_Failure | pericyte | *ATF1* | 2.068058589 | 0.004814071 | 0.031792081 | 1 |
| Heart_Failure | pericyte | *SPATS2* | 3.829519066 | 9.00E-06 | 0.001740485 | 0.056613652 |
| Heart_Failure | pericyte | *PDE1B* | 0.631198091 | 0.004563561 | 0.030751456 | 1 |
| Heart_Failure | pericyte | *SARDH* | 2.099752733 | 0.007255417 | 0.042511467 | 1 |
| Heart_Failure | pericyte | *TTC21B* | 2.373365318 | 0.004787187 | 0.03173585 | 1 |
| Heart_Failure | pericyte | *PLA2G12A* | 3.404529121 | 0.001179157 | 0.012629235 | 1 |
| Heart_Failure | pericyte | *AGO2* | 3.738144487 | 0.000102599 | 0.002958887 | 0.645037408 |
| Heart_Failure | pericyte | *CDH26* | 3.889117927 | 0.003326135 | 0.025377926 | 1 |
| Heart_Failure | pericyte | *RNF114* | 3.135413622 | 8.82E-05 | 0.002769335 | 0.554293944 |
| Heart_Failure | pericyte | *SERPINB6* | 2.563357957 | 0.006360541 | 0.038695901 | 1 |
| Heart_Failure | pericyte | *GLO1* | 2.343380313 | 0.003771975 | 0.027071238 | 1 |
| Heart_Failure | pericyte | *NUP153* | 3.425313417 | 6.04E-05 | 0.002374889 | 0.379982181 |
| Heart_Failure | pericyte | *EEF1E1* | 3.346994197 | 0.003232971 | 0.024878445 | 1 |

| | | | | | | |
|---|---|---|---|---|---|---|
| Heart_Failure | pericyte | *MIF4GD* | 7.247105267 | 2.09E-05 | 0.001740485 | 0.131391095 |
| Heart_Failure | pericyte | *PPP1R12C* | 3.020538897 | 0.000319382 | 0.005686577 | 1 |
| Heart_Failure | pericyte | *SH2D3A* | 4.064242738 | 0.000560615 | 0.007797759 | 1 |
| Heart_Failure | pericyte | *SNRPD2* | 4.339482413 | 4.02E-05 | 0.002069547 | 0.252484702 |
| Heart_Failure | pericyte | *RTN2* | 1.687603317 | 0.005513971 | 0.035019283 | 1 |
| Heart_Failure | pericyte | *SYMPK* | 3.302184608 | 0.00028149 | 0.005306292 | 1 |
| Heart_Failure | pericyte | *GPCPD1* | 2.207706226 | 0.005321224 | 0.034033099 | 1 |
| Heart_Failure | pericyte | *PANK2* | 3.890683471 | 0.000106636 | 0.003039897 | 0.670420867 |
| Heart_Failure | pericyte | *PCSK2* | 3.20511542 | 0.002579779 | 0.021256974 | 1 |
| Heart_Failure | pericyte | *DSTN* | 2.63813516 | 0.002181932 | 0.019149613 | 1 |
| Heart_Failure | pericyte | *FAM110A* | 3.577365141 | 0.000475863 | 0.007140215 | 1 |
| Heart_Failure | pericyte | *UROD* | 4.177641418 | 2.84E-05 | 0.001861562 | 0.178701655 |
| Heart_Failure | pericyte | *ST3GAL3* | 2.049080591 | 0.000775743 | 0.009657612 | 1 |
| Heart_Failure | pericyte | *KLC1* | 2.940383467 | 0.000443712 | 0.006939351 | 1 |
| Heart_Failure | pericyte | *IGFLR1* | 15.60124763 | 0.000125509 | 0.00330886 | 0.789073017 |
| Heart_Failure | pericyte | *COX6B1* | 3.544437939 | 4.93E-05 | 0.002128526 | 0.310152791 |
| Heart_Failure | pericyte | *PRDX5* | 3.630578776 | 0.000684736 | 0.008968612 | 1 |
| Heart_Failure | pericyte | *BECN1* | 3.47430971 | 0.000231876 | 0.004725416 | 1 |
| Heart_Failure | pericyte | *EMG1* | 2.899466297 | 0.002514726 | 0.020968276 | 1 |
| Heart_Failure | pericyte | *RHOJ* | 0.92117054 | 0.002480997 | 0.020825135 | 1 |
| Heart_Failure | pericyte | *AIF1L* | 1.68227979 | 0.001403744 | 0.014414082 | 1 |
| Heart_Failure | pericyte | *RAP1B* | 2.440768746 | 0.000843121 | 0.010193655 | 1 |
| Heart_Failure | pericyte | *CHTF18* | 1.539455886 | 0.002081718 | 0.018595727 | 1 |
| Heart_Failure | pericyte | *HIP1* | 3.202092278 | 1.98E-05 | 0.001740485 | 0.124228155 |
| Heart_Failure | pericyte | *PTPN12* | 2.398601104 | 0.001697572 | 0.016474431 | 1 |
| Heart_Failure | pericyte | *SPECC1* | 1.569768304 | 0.000315147 | 0.005677167 | 1 |
| Heart_Failure | pericyte | *POT1* | 2.803997012 | 0.000206838 | 0.004519343 | 1 |
| Heart_Failure | pericyte | *CDHR3* | 1.595332061 | 0.000678195 | 0.008957586 | 1 |
| Heart_Failure | pericyte | *HOXD3* | 1.903390071 | 0.0009221 | 0.010795608 | 1 |
| Heart_Failure | pericyte | *GAD1* | 10.53435476 | 0.000127277 | 0.003330142 | 0.800193523 |
| Heart_Failure | pericyte | *SUMF2* | 2.369646693 | 0.007803379 | 0.044844465 | 1 |
| Heart_Failure | pericyte | *SPCS3* | 3.392501229 | 0.001047064 | 0.011713329 | 1 |
| Heart_Failure | pericyte | *SERGEF* | 3.13917564 | 0.000907557 | 0.010705083 | 1 |
| Heart_Failure | pericyte | *SLC44A2* | 2.958412071 | 0.004194362 | 0.029041799 | 1 |
| Heart_Failure | pericyte | *MTUS1* | 3.182060937 | 1.73E-05 | 0.001740485 | 0.1085696 |
| Heart_Failure | pericyte | *REEP5* | 2.188235596 | 0.008511238 | 0.047449452 | 1 |
| Heart_Failure | pericyte | *DOCK6* | 1.476583403 | 0.007856225 | 0.045024693 | 1 |
| Heart_Failure | pericyte | *RPL36* | 3.148038316 | 0.000549152 | 0.007672261 | 1 |
| Heart_Failure | pericyte | *BST2* | 4.993534879 | 6.57E-05 | 0.002428296 | 0.412810266 |
| Heart_Failure | pericyte | *ACTN4* | 3.212679649 | 0.000180961 | 0.004092442 | 1 |
| Heart_Failure | pericyte | *PAK4* | 4.584464633 | 0.000566379 | 0.007860536 | 1 |
| Heart_Failure | pericyte | *UCK1* | 1.543544052 | 0.001405413 | 0.014414082 | 1 |
| Heart_Failure | pericyte | *MAP3K10* | 2.202364698 | 0.008553004 | 0.047507995 | 1 |
| Heart_Failure | pericyte | *CHMP1A* | 1.725538747 | 0.004187565 | 0.029026708 | 1 |
| Heart_Failure | pericyte | *TBC1D5* | 2.659120892 | 7.90E-05 | 0.002693349 | 0.496498528 |
| Heart_Failure | pericyte | *RFTN1* | 2.048647383 | 0.001935111 | 0.017838769 | 1 |
| Heart_Failure | pericyte | *AOC2* | 6.963751798 | 9.54E-05 | 0.00282898 | 0.599743852 |
| Heart_Failure | pericyte | *EXOC4* | 1.886648624 | 0.000755224 | 0.009515216 | 1 |
| Heart_Failure | pericyte | *ACAP3* | 2.322171573 | 0.001206383 | 0.012804798 | 1 |
| Heart_Failure | pericyte | *NINJ1* | 4.765034976 | 0.00014243 | 0.003596208 | 0.895455863 |

| | | | | | | |
|---|---|---|---|---|---|---|
| Heart_Failure | pericyte | *NPHP4* | 5.432983993 | 3.72E-05 | 0.001934281 | 0.234047954 |
| Heart_Failure | pericyte | *MAP1B* | 0.64816872 | 0.002952626 | 0.023405834 | 1 |
| Heart_Failure | pericyte | *KHDRBS3* | 0.38048215 | 0.009148234 | 0.04971041 | 1 |
| Heart_Failure | pericyte | *LGALS3* | 2.908962459 | 0.000332465 | 0.005798899 | 1 |
| Heart_Failure | pericyte | *TRIM5* | 2.248890857 | 0.006238508 | 0.038261994 | 1 |
| Heart_Failure | pericyte | *IMMT* | 1.812766614 | 0.004143835 | 0.028882809 | 1 |
| Heart_Failure | pericyte | *XAF1* | 2.556759609 | 3.83E-06 | 0.000964138 | 0.024103459 |
| Heart_Failure | pericyte | *REEP2* | -1.082022126 | 3.41E-05 | 0.001918699 | 0.214488275 |
| Heart_Failure | pericyte | *TERF2* | 4.15609672 | 1.25E-05 | 0.001740485 | 0.078615989 |
| Heart_Failure | pericyte | *PTPRA* | 2.69196325 | 0.000575269 | 0.007896759 | 1 |
| Heart_Failure | pericyte | *MUTYH* | 4.866868305 | 1.19E-05 | 0.001740485 | 0.074703863 |
| Heart_Failure | pericyte | *CTNNBL1* | 2.996641962 | 0.001880376 | 0.01756601 | 1 |
| Heart_Failure | pericyte | *USPL1* | 2.392781212 | 0.007502174 | 0.043794028 | 1 |
| Heart_Failure | pericyte | *POMP* | 3.977836928 | 0.000141285 | 0.003581691 | 0.888259384 |
| Heart_Failure | pericyte | *ZNF414* | 3.354048842 | 0.000257773 | 0.005096295 | 1 |
| Heart_Failure | pericyte | *CNDP2* | 4.021082094 | 0.000112727 | 0.003163896 | 0.70871278 |
| Heart_Failure | pericyte | *C1QTNF6* | 4.202216767 | 0.005757766 | 0.036162912 | 1 |
| Heart_Failure | pericyte | *FAM83F* | 12.57889268 | 0.00034939 | 0.005952796 | 1 |
| Heart_Failure | pericyte | *SWAP70* | 3.549131001 | 0.000101481 | 0.002940151 | 0.638012766 |
| Heart_Failure | pericyte | *SBF2* | 1.981919208 | 0.002384889 | 0.020207273 | 1 |
| Heart_Failure | pericyte | *NARS* | 4.285038066 | 0.000106858 | 0.003039897 | 0.671817347 |
| Heart_Failure | pericyte | *FBXO18* | 3.515784107 | 0.000360186 | 0.006022578 | 1 |
| Heart_Failure | pericyte | *ZCCHC11* | 1.929089528 | 0.00629811 | 0.038480289 | 1 |
| Heart_Failure | pericyte | *TIMM10* | 1.470873353 | 0.00074267 | 0.009423995 | 1 |
| Heart_Failure | pericyte | *FADS2* | 3.500820506 | 0.000294279 | 0.005434111 | 1 |
| Heart_Failure | pericyte | *NAA35* | 2.235454655 | 0.008562197 | 0.047507995 | 1 |
| Heart_Failure | pericyte | *ADAM19* | 0.516036557 | 0.005114804 | 0.033333854 | 1 |
| Heart_Failure | pericyte | *SRPK2* | 1.836657653 | 0.00707147 | 0.041705753 | 1 |
| Heart_Failure | pericyte | *FAIM2* | 1.899179061 | 0.009059324 | 0.049316621 | 1 |
| Heart_Failure | pericyte | *PKIB* | 2.31689862 | 0.004293558 | 0.029352191 | 1 |
| Heart_Failure | pericyte | *USP15* | 2.280970812 | 0.001069017 | 0.011874397 | 1 |
| Heart_Failure | pericyte | *USP37* | 3.037939439 | 0.000193714 | 0.004349567 | 1 |
| Heart_Failure | pericyte | *GCC2* | 1.858054287 | 0.003027884 | 0.023736046 | 1 |
| Heart_Failure | pericyte | *SLC41A2* | 3.2940543 | 0.00023468 | 0.004728943 | 1 |
| Heart_Failure | pericyte | *LRCH1* | 3.200025316 | 0.000125124 | 0.00330886 | 0.786654572 |
| Heart_Failure | pericyte | *SETDB2* | 1.998903514 | 0.003240028 | 0.024902268 | 1 |
| Heart_Failure | pericyte | *ZDHHC4* | 2.383021117 | 0.000303372 | 0.005512434 | 1 |
| Heart_Failure | pericyte | *DDX56* | 3.326636516 | 0.00011184 | 0.003153093 | 0.703139801 |
| Heart_Failure | pericyte | *NMT1* | 1.331314914 | 2.20E-05 | 0.001740485 | 0.138028751 |
| Heart_Failure | pericyte | *RPS6KC1* | 2.365884856 | 0.002550308 | 0.021069368 | 1 |
| Heart_Failure | pericyte | *UGGT1* | 3.824451543 | 0.000123494 | 0.00330886 | 0.776409043 |
| Heart_Failure | pericyte | *ODF2* | 3.283722679 | 0.003994569 | 0.028186145 | 1 |
| Heart_Failure | pericyte | *ALDOB* | 1.128025824 | 0.008288047 | 0.046858772 | 1 |
| Heart_Failure | pericyte | *STX17* | 3.852154207 | 9.17E-05 | 0.002769335 | 0.576335754 |
| Heart_Failure | pericyte | *FPGS* | 2.90364823 | 0.001331233 | 0.013824469 | 1 |
| Heart_Failure | pericyte | *GARNL3* | 3.921383887 | 7.02E-05 | 0.00255121 | 0.44135936 |
| Heart_Failure | pericyte | *APTX* | 2.853249568 | 9.59E-05 | 0.002831023 | 0.60300784 |
| Heart_Failure | pericyte | *TLN1* | 3.435600174 | 3.12E-06 | 0.000909616 | 0.019628681 |
| Heart_Failure | pericyte | *TMEM8B* | 2.936517695 | 0.005517261 | 0.035019283 | 1 |
| Heart_Failure | pericyte | *KIF13A* | 2.679604077 | 0.000278392 | 0.005271845 | 1 |

| | | | | | | |
|---|---|---|---|---|---|---|
| Heart_Failure | pericyte | *TMEM63B* | 2.059039074 | 0.000370462 | 0.006145369 | 1 |
| Heart_Failure | pericyte | *FLOT1* | 2.446878146 | 0.000357781 | 0.006014363 | 1 |
| Heart_Failure | pericyte | *TPMT* | 3.113526493 | 0.001444131 | 0.014620375 | 1 |
| Heart_Failure | pericyte | *FAM8A1* | 2.773089364 | 0.000223611 | 0.004655112 | 1 |
| Heart_Failure | pericyte | *ARRB1* | 2.717034272 | 0.000974942 | 0.011144479 | 1 |
| Heart_Failure | pericyte | *CCDC90B* | 2.859687404 | 0.000567885 | 0.007864081 | 1 |
| Heart_Failure | pericyte | *RAB30* | 2.440653326 | 0.003689048 | 0.027031519 | 1 |
| Heart_Failure | pericyte | *NARS2* | 3.66730576 | 0.000167033 | 0.003947887 | 1 |
| Heart_Failure | pericyte | *SULF1* | 1.826285236 | 0.000633434 | 0.008455203 | 1 |
| Heart_Failure | pericyte | *TGS1* | 1.574270957 | 0.003335347 | 0.025417365 | 1 |
| Heart_Failure | pericyte | *NEK1* | 1.823989648 | 0.008569121 | 0.047507995 | 1 |
| Heart_Failure | pericyte | *DDX60* | 4.132005007 | 0.000296956 | 0.005443039 | 1 |
| Heart_Failure | pericyte | *RMDN3* | 3.645295904 | 6.57E-06 | 0.001424095 | 0.041298754 |
| Heart_Failure | pericyte | *UACA* | 2.199211541 | 0.002690877 | 0.021800961 | 1 |
| Heart_Failure | pericyte | *IFT172* | 3.195946088 | 0.000186513 | 0.004202893 | 1 |
| Heart_Failure | pericyte | *ADCY3* | 2.528831285 | 0.000170694 | 0.003961988 | 1 |
| Heart_Failure | pericyte | *ABCG5* | 1.877369067 | 0.000266218 | 0.005134086 | 1 |
| Heart_Failure | pericyte | *STAMBPL1* | 2.707803018 | 2.43E-05 | 0.001740485 | 0.153034564 |
| Heart_Failure | pericyte | *ATAD1* | 3.023806342 | 0.001901812 | 0.017712797 | 1 |
| Heart_Failure | pericyte | *ARL3* | 2.684788281 | 0.000530016 | 0.007488118 | 1 |
| Heart_Failure | pericyte | *DNAJC13* | 3.067633547 | 0.000597887 | 0.008101113 | 1 |
| Heart_Failure | pericyte | *RPS24* | 3.512630732 | 0.000167695 | 0.003948674 | 1 |
| Heart_Failure | pericyte | *STAT4* | 0.786430735 | 0.003381461 | 0.025660013 | 1 |
| Heart_Failure | pericyte | *CARF* | 1.487533765 | 0.001068526 | 0.011874397 | 1 |
| Heart_Failure | pericyte | *NAB1* | 4.10999083 | 5.20E-05 | 0.002166418 | 0.32712907 |
| Heart_Failure | pericyte | *SENP7* | 2.79845576 | 0.000520953 | 0.007467279 | 1 |
| Heart_Failure | pericyte | *COX17* | 4.803423147 | 1.54E-05 | 0.001740485 | 0.096847836 |
| Heart_Failure | pericyte | *USP8* | 2.120795679 | 0.002182707 | 0.019149613 | 1 |
| Heart_Failure | pericyte | *UBL7* | 3.018242744 | 0.000985001 | 0.011218665 | 1 |
| Heart_Failure | pericyte | *ARHGAP24* | 2.50621604 | 0.000150138 | 0.003672831 | 0.943917455 |
| Heart_Failure | pericyte | *HERC3* | 2.915341171 | 0.00080744 | 0.009857039 | 1 |
| Heart_Failure | pericyte | *AP1AR* | 2.443593157 | 0.008884939 | 0.048913847 | 1 |
| Heart_Failure | pericyte | *BBS7* | 4.080909532 | 0.000441129 | 0.006918249 | 1 |
| Heart_Failure | pericyte | *KIAA1109* | 2.50968943 | 0.000794842 | 0.009760099 | 1 |
| Heart_Failure | pericyte | *SCARB2* | 2.491260125 | 0.003296533 | 0.025182632 | 1 |
| Heart_Failure | pericyte | *ANXA3* | 1.539085299 | 0.001825181 | 0.017203767 | 1 |
| Heart_Failure | pericyte | *CPNE8* | 1.86864344 | 9.98E-05 | 0.002917323 | 0.627224518 |
| Heart_Failure | pericyte | *ALG10* | 2.237857158 | 0.001356534 | 0.014027184 | 1 |
| Heart_Failure | pericyte | *LGR5* | -1.666295542 | 0.002635782 | 0.021577035 | 1 |
| Heart_Failure | pericyte | *GAS2L3* | 4.819383168 | 0.000221982 | 0.004655112 | 1 |
| Heart_Failure | pericyte | *TDG* | 3.116228357 | 0.000217932 | 0.004644531 | 1 |
| Heart_Failure | pericyte | *GLTP* | 2.342082555 | 0.006964325 | 0.041228541 | 1 |
| Heart_Failure | pericyte | *WDFY2* | 1.956538445 | 0.007344426 | 0.042952935 | 1 |
| Heart_Failure | pericyte | *LPAR6* | 3.513877604 | 1.03E-05 | 0.001740485 | 0.064942381 |
| Heart_Failure | pericyte | *TMX1* | 4.483142661 | 5.12E-05 | 0.002162658 | 0.321581975 |
| Heart_Failure | pericyte | *AK7* | 1.849544945 | 0.00479546 | 0.03173585 | 1 |
| Heart_Failure | pericyte | *C14orf79* | 7.365742878 | 1.45E-06 | 0.000702609 | 0.009133923 |
| Heart_Failure | pericyte | *TCF12* | 2.363636865 | 0.000492466 | 0.007302207 | 1 |
| Heart_Failure | pericyte | *SORD* | 1.772345382 | 0.000829132 | 0.010058438 | 1 |
| Heart_Failure | pericyte | *ZSCAN29* | 4.384255396 | 3.30E-05 | 0.001918699 | 0.207446133 |

| | | | | | | |
|---|---|---|---|---|---|---|
| Heart_Failure | pericyte | *SRP14* | 2.220641 | 0.00375829 | 0.027065715 | 1 |
| Heart_Failure | pericyte | *UBE2Q2* | 3.460461463 | 0.000292661 | 0.005427606 | 1 |
| Heart_Failure | pericyte | *TPM1* | 0.663830235 | 0.003132631 | 0.02440502 | 1 |
| Heart_Failure | pericyte | *IGF1R* | 3.054021231 | 0.000134259 | 0.003445248 | 0.844085739 |
| Heart_Failure | pericyte | *SCAMP2* | 1.979345212 | 0.005167723 | 0.033464241 | 1 |
| Heart_Failure | pericyte | *MFGE8* | 1.717405431 | 0.007314343 | 0.042816827 | 1 |
| Heart_Failure | pericyte | *CRTC3* | 3.251082979 | 0.000145003 | 0.003632017 | 0.911636164 |
| Heart_Failure | pericyte | *ZFHX3* | 2.076535911 | 0.004013433 | 0.028255829 | 1 |
| Heart_Failure | pericyte | *KIFC3* | 4.391972667 | 5.29E-05 | 0.002189468 | 0.33279916 |
| Heart_Failure | pericyte | *CMTM3* | 3.603083657 | 0.004042324 | 0.02836394 | 1 |
| Heart_Failure | pericyte | *RPS2* | 2.307416643 | 0.003387595 | 0.025660013 | 1 |
| Heart_Failure | pericyte | *NDUFB10* | 2.030083749 | 0.000475164 | 0.007140215 | 1 |
| Heart_Failure | pericyte | *MYO19* | 2.449767199 | 0.00267104 | 0.02169166 | 1 |
| Heart_Failure | pericyte | *SPATA22* | 0.892965547 | 0.002961107 | 0.023405834 | 1 |
| Heart_Failure | pericyte | *ABCA8* | 0.937662641 | 0.006592576 | 0.039776894 | 1 |
| Heart_Failure | pericyte | *ASXL3* | 3.355202073 | 0.000695738 | 0.009037411 | 1 |
| Heart_Failure | pericyte | *NARF* | 2.018612514 | 0.006123734 | 0.037819181 | 1 |
| Heart_Failure | pericyte | *DPP9* | 2.601792617 | 0.005638901 | 0.035548658 | 1 |
| Heart_Failure | pericyte | *IFITM3* | 4.508109487 | 2.97E-05 | 0.001885777 | 0.186691921 |
| Heart_Failure | pericyte | *WTIP* | 2.176917921 | 0.002773091 | 0.02226367 | 1 |
| Heart_Failure | pericyte | *EVI5L* | 1.033084415 | 0.005285637 | 0.033911818 | 1 |
| Heart_Failure | pericyte | *RCN3* | 3.101735345 | 0.005872056 | 0.03669743 | 1 |
| Heart_Failure | pericyte | *RERE* | 2.126461822 | 0.000123402 | 0.00330886 | 0.775831504 |
| Heart_Failure | pericyte | *GPN2* | 4.408842972 | 0.000364544 | 0.006063203 | 1 |
| Heart_Failure | pericyte | *SERBP1* | 3.855480571 | 0.000128811 | 0.003332666 | 0.80983778 |
| Heart_Failure | pericyte | *RPS8* | 2.193161109 | 0.000445632 | 0.006942549 | 1 |
| Heart_Failure | pericyte | *MOB3C* | 3.043471721 | 0.005531119 | 0.035019283 | 1 |
| Heart_Failure | pericyte | *SYPL2* | 0.534575915 | 0.001658544 | 0.016191411 | 1 |
| Heart_Failure | pericyte | *GPR161* | 2.785944758 | 0.002183915 | 0.019149613 | 1 |
| Heart_Failure | pericyte | *TIPRL* | 3.144104131 | 0.003520352 | 0.02628557 | 1 |
| Heart_Failure | pericyte | *TMCO1* | 2.88932747 | 0.002651433 | 0.021648781 | 1 |
| Heart_Failure | pericyte | *MGST3* | 2.958444937 | 0.000434802 | 0.006903037 | 1 |
| Heart_Failure | pericyte | *RFWD2* | 2.083456709 | 0.002542673 | 0.02103393 | 1 |
| Heart_Failure | pericyte | *CERS2* | 2.738051432 | 0.002961381 | 0.023405834 | 1 |
| Heart_Failure | pericyte | *SEMA6C* | 3.435755932 | 0.000682077 | 0.008964119 | 1 |
| Heart_Failure | pericyte | *DYRK3* | 3.068766276 | 0.002293341 | 0.019709086 | 1 |
| Heart_Failure | pericyte | *TP53BP2* | 4.610594985 | 2.05E-05 | 0.001740485 | 0.12862377 |
| Heart_Failure | pericyte | *RAB13* | 4.556735826 | 0.000512531 | 0.007424617 | 1 |
| Heart_Failure | pericyte | *GATAD2B* | 2.031144312 | 0.0017753 | 0.016834554 | 1 |
| Heart_Failure | pericyte | *FLG* | -0.559135721 | 0.004748597 | 0.031625456 | 1 |
| Heart_Failure | pericyte | *CEP170* | 2.993801962 | 1.60E-05 | 0.001740485 | 0.100558396 |
| Heart_Failure | pericyte | *NVL* | 2.945081876 | 0.001689434 | 0.016441907 | 1 |
| Heart_Failure | pericyte | *GUK1* | 3.914895576 | 0.000348114 | 0.005952796 | 1 |
| Heart_Failure | pericyte | *CDC42BPA* | 1.77718898 | 0.000887847 | 0.010571774 | 1 |
| Heart_Failure | pericyte | *PYCR2* | 1.551354029 | 0.002466924 | 0.020762455 | 1 |
| Heart_Failure | pericyte | *CIAO1* | 1.938488756 | 0.007230779 | 0.04240663 | 1 |
| Heart_Failure | pericyte | *EXOC6B* | 1.687848164 | 0.007713233 | 0.044776637 | 1 |
| Heart_Failure | pericyte | *TMEM177* | 1.970763486 | 0.004668544 | 0.031257863 | 1 |
| Heart_Failure | pericyte | *METTL21A* | 3.621274142 | 0.000499039 | 0.007337373 | 1 |
| Heart_Failure | pericyte | *NBEAL1* | 1.096878998 | 0.000357366 | 0.006014363 | 1 |

| | | | | | | |
|---|---|---|---|---|---|---|
| Heart_Failure | pericyte | *FANCD2* | 5.310538074 | 0.000317256 | 0.005686577 | 1 |
| Heart_Failure | pericyte | *TAMM41* | 3.162540188 | 0.000306355 | 0.005550582 | 1 |
| Heart_Failure | pericyte | *OSBPL10* | 0.604494212 | 0.007970002 | 0.045440657 | 1 |
| Heart_Failure | pericyte | *STAC* | 0.659682274 | 0.007513276 | 0.043818153 | 1 |
| Heart_Failure | pericyte | *TMF1* | 2.492522206 | 0.005912141 | 0.036874632 | 1 |
| Heart_Failure | pericyte | *LIMD1* | 2.889291738 | 1.64E-05 | 0.001740485 | 0.103078547 |
| Heart_Failure | pericyte | *BOC* | 0.766969101 | 0.004431254 | 0.030118154 | 1 |
| Heart_Failure | pericyte | *PYURF* | 2.512429117 | 0.003615291 | 0.026646346 | 1 |
| Heart_Failure | pericyte | *CISD2* | 2.871653891 | 0.000457626 | 0.006976539 | 1 |
| Heart_Failure | pericyte | *USP53* | 1.959680024 | 0.004300112 | 0.029353752 | 1 |
| Heart_Failure | pericyte | *CBR4* | 1.965226406 | 0.008373833 | 0.04704352 | 1 |
| Heart_Failure | pericyte | *NDUFS6* | 4.083988976 | 0.000351492 | 0.005952796 | 1 |
| Heart_Failure | pericyte | *LHFPL2* | 2.967199694 | 0.002108847 | 0.018779488 | 1 |
| Heart_Failure | pericyte | *SSBP2* | 2.815254116 | 4.68E-05 | 0.002128526 | 0.294167352 |
| Heart_Failure | pericyte | *BHMT* | -0.585943671 | 0.000229853 | 0.00470712 | 1 |
| Heart_Failure | pericyte | *TSLP* | 0.822402244 | 0.004768484 | 0.031690759 | 1 |
| Heart_Failure | pericyte | *MYLK4* | 3.360486044 | 0.00513279 | 0.033405645 | 1 |
| Heart_Failure | pericyte | *FAM193B* | 2.746540876 | 0.003464873 | 0.026044671 | 1 |
| Heart_Failure | pericyte | *IRAK1BP1* | 2.689691592 | 0.0036278 | 0.026707238 | 1 |
| Heart_Failure | pericyte | *C7orf50* | 3.337024028 | 0.000372037 | 0.00615525 | 1 |
| Heart_Failure | pericyte | *TACC1* | 1.987134054 | 0.003924637 | 0.027843851 | 1 |
| Heart_Failure | pericyte | *SLC26A7* | 0.306256344 | 0.007412871 | 0.043312936 | 1 |
| Heart_Failure | pericyte | *AK3* | 2.561152694 | 0.000996576 | 0.01130952 | 1 |
| Heart_Failure | pericyte | *ZNF462* | 3.226368743 | 0.000713236 | 0.009202924 | 1 |
| Heart_Failure | pericyte | *WDR31* | 5.409231441 | 2.84E-06 | 0.000909616 | 0.017852795 |
| Heart_Failure | pericyte | *INPP5E* | 2.794422523 | 0.003070207 | 0.024007946 | 1 |
| Heart_Failure | pericyte | *DPH7* | 3.58884345 | 0.002010788 | 0.01824217 | 1 |
| Heart_Failure | pericyte | *RSU1* | 3.286608282 | 7.59E-05 | 0.002686157 | 0.477491588 |
| Heart_Failure | pericyte | *ADIRF* | 3.747516326 | 1.36E-05 | 0.001740485 | 0.085407406 |
| Heart_Failure | pericyte | *RPP30* | 3.610923506 | 0.000155281 | 0.003726156 | 0.976252777 |
| Heart_Failure | pericyte | *MTG1* | 0.88159148 | 0.006730264 | 0.040336676 | 1 |
| Heart_Failure | pericyte | *PAOX* | 3.266301678 | 0.000262971 | 0.005105204 | 1 |
| Heart_Failure | pericyte | *EIF3M* | 2.273251811 | 0.002269416 | 0.019598651 | 1 |
| Heart_Failure | pericyte | *C11orf49* | 2.221889759 | 0.003583014 | 0.026490062 | 1 |
| Heart_Failure | pericyte | *ENDOD1* | 1.393738287 | 0.001907359 | 0.017712797 | 1 |
| Heart_Failure | pericyte | *P4HA3* | 1.917342268 | 0.002148911 | 0.01905529 | 1 |
| Heart_Failure | pericyte | *ROM1* | 0.778495 | 0.002000227 | 0.018225259 | 1 |
| Heart_Failure | pericyte | *INCENP* | 1.93000538 | 0.001702139 | 0.01648789 | 1 |
| Heart_Failure | pericyte | *PLCH2* | 9.355411901 | 0.000687339 | 0.008980062 | 1 |
| Heart_Failure | pericyte | *CPSF7* | 2.446561404 | 0.008997148 | 0.049247626 | 1 |
| Heart_Failure | pericyte | *DUSP15* | -2.347033947 | 0.000169721 | 0.003961988 | 1 |
| Heart_Failure | pericyte | *COMMD7* | 4.292807842 | 0.000437289 | 0.006907621 | 1 |
| Heart_Failure | pericyte | *TBX6* | 0.784614551 | 0.006636252 | 0.039963715 | 1 |
| Heart_Failure | pericyte | *TAOK2* | 4.777583587 | 4.62E-06 | 0.001108016 | 0.029067272 |
| Heart_Failure | pericyte | *ARID5B* | 1.474003779 | 0.008907048 | 0.048992658 | 1 |
| Heart_Failure | pericyte | *SAP18* | 3.295491115 | 0.000194984 | 0.004362517 | 1 |
| Heart_Failure | pericyte | *CCDC102B* | 2.298628647 | 0.000105837 | 0.003038357 | 0.665400199 |
| Heart_Failure | pericyte | *PRSS23* | 1.248740079 | 0.006786388 | 0.040557055 | 1 |
| Heart_Failure | pericyte | *DOCK1* | 2.599564524 | 0.000175445 | 0.004025628 | 1 |
| Heart_Failure | pericyte | *DHX37* | 2.253604457 | 0.000602253 | 0.008125245 | 1 |

| | | | | | | |
|---|---|---|---|---|---|---|
| Heart_Failure | pericyte | *THRB* | 1.855297437 | 0.004867532 | 0.032111408 | 1 |
| Heart_Failure | pericyte | *BTBD11* | 5.602812582 | 1.34E-05 | 0.001740485 | 0.084438244 |
| Heart_Failure | pericyte | *FAM177A1* | 3.733980709 | 8.12E-05 | 0.002716178 | 0.510641477 |
| Heart_Failure | pericyte | *ADAMTS12* | 0.518856774 | 0.004104609 | 0.028704871 | 1 |
| Heart_Failure | pericyte | *VPS26B* | 4.387498626 | 2.67E-05 | 0.00184475 | 0.167872237 |
| Heart_Failure | pericyte | *INPP1* | 2.456881652 | 0.000496379 | 0.007337373 | 1 |
| Heart_Failure | pericyte | *ASAP2* | 3.499942227 | 1.76E-06 | 0.000789824 | 0.011057543 |
| Heart_Failure | pericyte | *BICD1* | 2.572290844 | 0.002579201 | 0.021256974 | 1 |
| Heart_Failure | pericyte | *PTPN14* | 1.696990078 | 0.003453956 | 0.02600899 | 1 |
| Heart_Failure | pericyte | *RNF219* | 3.431019051 | 0.001973157 | 0.01805712 | 1 |
| Heart_Failure | pericyte | *TGOLN2* | 2.765293637 | 0.002963424 | 0.023405834 | 1 |
| Heart_Failure | pericyte | *CWF19L2* | 2.275281019 | 0.002710379 | 0.021930697 | 1 |
| Heart_Failure | pericyte | *TMEM123* | 3.009458006 | 0.001657273 | 0.016191411 | 1 |
| Heart_Failure | pericyte | *SPARCL1* | 0.779597291 | 0.004289355 | 0.029352191 | 1 |
| Heart_Failure | pericyte | *MBNL1* | 1.974619634 | 0.00071414 | 0.009202924 | 1 |
| Heart_Failure | pericyte | *PELO* | 4.987013929 | 6.29E-05 | 0.002380436 | 0.395152323 |
| Heart_Failure | pericyte | *SRP19* | 3.745180436 | 8.38E-05 | 0.002749485 | 0.526949167 |
| Heart_Failure | pericyte | *ASAP1* | 2.208885499 | 5.85E-05 | 0.002333071 | 0.367685466 |
| Heart_Failure | pericyte | *TUBGCP5* | 3.646505931 | 2.41E-05 | 0.001740485 | 0.151679574 |
| Heart_Failure | pericyte | *CFDP1* | 2.352814214 | 0.003705518 | 0.027057845 | 1 |
| Heart_Failure | pericyte | *C7orf31* | 3.117270754 | 0.00168009 | 0.016376316 | 1 |
| Heart_Failure | pericyte | *ANKRD29* | 1.661993884 | 0.001041365 | 0.011670338 | 1 |
| Heart_Failure | pericyte | *THY1* | 1.447909796 | 0.001430365 | 0.0145711 | 1 |
| Heart_Failure | pericyte | *TBCEL* | 3.585733605 | 0.000423233 | 0.00677065 | 1 |
| Heart_Failure | pericyte | *PRKCA* | 3.054484977 | 0.000255038 | 0.005058121 | 1 |
| Heart_Failure | pericyte | *LONRF1* | 1.93593558 | 0.006899299 | 0.041036797 | 1 |
| Heart_Failure | pericyte | *ATP5G3* | 1.554147923 | 0.002256869 | 0.01957095 | 1 |
| Heart_Failure | pericyte | *CXXC1* | 3.56079131 | 0.000116063 | 0.003214493 | 0.729689954 |
| Heart_Failure | pericyte | *PPP4R1* | 2.234666938 | 0.008809212 | 0.048624685 | 1 |
| Heart_Failure | pericyte | *CDK19* | 2.830875523 | 0.000506542 | 0.007406121 | 1 |
| Heart_Failure | pericyte | *TTC39B* | 3.7153109 | 2.13E-07 | 0.000111645 | 0.001339739 |
| Heart_Failure | pericyte | *PDIA4* | 3.625858915 | 0.004765065 | 0.031690759 | 1 |
| Heart_Failure | pericyte | *DEPTOR* | 3.349638004 | 0.000984109 | 0.011218665 | 1 |
| Heart_Failure | pericyte | *TMBIM4* | 1.421398415 | 0.001234353 | 0.013020769 | 1 |
| Heart_Failure | pericyte | *DCK* | 2.669758335 | 0.001491673 | 0.015029085 | 1 |
| Heart_Failure | pericyte | *DRAM2* | 1.967903437 | 0.002860319 | 0.022820842 | 1 |
| Heart_Failure | pericyte | *CXCL13* | 6.0877631 | 0.00143186 | 0.0145711 | 1 |
| Heart_Failure | pericyte | *C14orf2* | 1.712840679 | 0.000438938 | 0.006916301 | 1 |
| Heart_Failure | pericyte | *RPUSD3* | 5.020540599 | 0.000548575 | 0.007672261 | 1 |
| Heart_Failure | pericyte | *FCHO2* | 2.50763573 | 0.001382796 | 0.014228543 | 1 |
| Heart_Failure | pericyte | *TIMP4* | 0.952634608 | 0.009000401 | 0.049247626 | 1 |
| Heart_Failure | pericyte | *IL34* | 4.476064544 | 6.77E-05 | 0.00247341 | 0.425426545 |
| Heart_Failure | pericyte | *DYRK1A* | 2.540994429 | 0.000597225 | 0.008101113 | 1 |
| Heart_Failure | pericyte | *LCA5L* | 0.647232037 | 0.004291346 | 0.029352191 | 1 |
| Heart_Failure | pericyte | *SPPL3* | 2.297659333 | 0.000447275 | 0.006942549 | 1 |
| Heart_Failure | pericyte | *C12orf43* | 1.609632237 | 0.000239628 | 0.004797912 | 1 |
| Heart_Failure | pericyte | *RADIL* | 1.920571901 | 0.001068585 | 0.011874397 | 1 |
| Heart_Failure | pericyte | *LRRC43* | 8.010764166 | 0.000387141 | 0.006290239 | 1 |
| Heart_Failure | pericyte | *AUTS2* | 2.17599975 | 0.00058821 | 0.008021857 | 1 |
| Heart_Failure | pericyte | *MITD1* | 2.217246777 | 0.001098228 | 0.012052831 | 1 |

| | | | | | | |
|---|---|---|---|---|---|---|
| Heart_Failure | pericyte | *NCF1* | 3.379704024 | 0.00072282 | 0.009274224 | 1 |
| Heart_Failure | pericyte | *DYNC1I1* | 4.325242955 | 0.000100857 | 0.002935599 | 0.634089409 |
| Heart_Failure | pericyte | *ELK4* | 4.905948148 | 2.16E-06 | 0.000849094 | 0.0135855 |
| Heart_Failure | pericyte | *RNF166* | 2.058819815 | 0.001187251 | 0.012694298 | 1 |
| Heart_Failure | pericyte | *NIT1* | 1.698030503 | 7.30E-05 | 0.002605975 | 0.458651616 |
| Heart_Failure | pericyte | *CCAR2* | 3.332100858 | 0.003938848 | 0.027855501 | 1 |
| Heart_Failure | pericyte | *ALG8* | 3.17848687 | 0.001715845 | 0.01648789 | 1 |
| Heart_Failure | pericyte | *PSMD4* | 4.656245269 | 2.21E-05 | 0.001740485 | 0.139169879 |
| Heart_Failure | pericyte | *STARD9* | 2.673666766 | 0.000118855 | 0.003259951 | 0.74724107 |
| Heart_Failure | pericyte | *RSPRY1* | 2.509965896 | 0.002927313 | 0.023237391 | 1 |
| Heart_Failure | pericyte | *SLC37A1* | 3.486917544 | 7.70E-05 | 0.002689226 | 0.484060718 |
| Heart_Failure | pericyte | *C21orf33* | -2.332724481 | 0.001740349 | 0.016679233 | 1 |
| Heart_Failure | pericyte | *FDPS* | 1.992884349 | 0.003589873 | 0.026490062 | 1 |
| Heart_Failure | pericyte | *FCRL3* | -0.881625205 | 0.000910601 | 0.010720876 | 1 |
| Heart_Failure | pericyte | *ZNF333* | 3.499764529 | 3.45E-06 | 0.000943167 | 0.021692838 |
| Heart_Failure | pericyte | *ORAI2* | 3.670339197 | 4.29E-05 | 0.002108725 | 0.269916793 |
| Heart_Failure | pericyte | *RPL8* | 2.594184977 | 0.00646418 | 0.039152503 | 1 |
| Heart_Failure | pericyte | *PGAP3* | 7.170407701 | 2.32E-05 | 0.001740485 | 0.145885202 |
| Heart_Failure | pericyte | *ALDH16A1* | 3.875634323 | 0.000754104 | 0.009515216 | 1 |
| Heart_Failure | pericyte | *JMJD8* | 1.511819243 | 0.000860798 | 0.010357221 | 1 |
| Heart_Failure | pericyte | *MEIOB* | 3.426655694 | 0.00358482 | 0.026490062 | 1 |
| Heart_Failure | pericyte | *FLYWCH2* | 1.981637396 | 0.001432311 | 0.0145711 | 1 |
| Heart_Failure | pericyte | *RPL29* | 2.593478352 | 4.59E-05 | 0.002128526 | 0.288566614 |
| Heart_Failure | pericyte | *C1orf123* | 1.628130694 | 0.002163092 | 0.019140963 | 1 |
| Heart_Failure | pericyte | *USP24* | 1.368410285 | 0.003669182 | 0.02694877 | 1 |
| Heart_Failure | pericyte | *RBP7* | 4.214368379 | 0.001451875 | 0.014675141 | 1 |
| Heart_Failure | pericyte | *CAMK2N1* | 2.099474165 | 0.002027401 | 0.018276772 | 1 |
| Heart_Failure | pericyte | *NFIA* | 1.877101753 | 0.002268078 | 0.019598651 | 1 |
| Heart_Failure | pericyte | *TM2D1* | 2.399126314 | 0.004883897 | 0.032151896 | 1 |
| Heart_Failure | pericyte | *NEXN* | 2.633600161 | 0.004626142 | 0.031060884 | 1 |
| Heart_Failure | pericyte | *B3GALT2* | 2.153773982 | 0.002028959 | 0.018276772 | 1 |
| Heart_Failure | pericyte | *ZNF496* | 4.045535048 | 0.001075264 | 0.011901729 | 1 |
| Heart_Failure | pericyte | *HAAO* | 3.239385584 | 0.000743487 | 0.009423995 | 1 |
| Heart_Failure | pericyte | *SMC6* | 3.146218437 | 7.06E-05 | 0.002552693 | 0.444168656 |
| Heart_Failure | pericyte | *CCDC74A* | 6.113114465 | 0.002164661 | 0.019140963 | 1 |
| Heart_Failure | pericyte | *NOSTRIN* | 1.087237621 | 0.000222257 | 0.004655112 | 1 |
| Heart_Failure | pericyte | *BBS5* | 1.052485434 | 0.002278718 | 0.019625072 | 1 |
| Heart_Failure | pericyte | *PDLIM5* | 2.837902038 | 0.000170781 | 0.003961988 | 1 |
| Heart_Failure | pericyte | *LYSMD1* | 1.511479785 | 0.001078333 | 0.011914723 | 1 |
| Heart_Failure | pericyte | *ERCC3* | 1.22585442 | 0.0044562 | 0.030189794 | 1 |
| Heart_Failure | pericyte | *DCAF16* | 4.406592265 | 4.92E-05 | 0.002128526 | 0.30959487 |
| Heart_Failure | pericyte | *ANTXR2* | 1.96414136 | 0.00029666 | 0.005443039 | 1 |
| Heart_Failure | pericyte | *PBXIP1* | 3.600602675 | 5.60E-05 | 0.002269684 | 0.351801021 |
| Heart_Failure | pericyte | *SLC22A15* | 1.624367935 | 0.000657617 | 0.008740887 | 1 |
| Heart_Failure | pericyte | *LRRC58* | 3.234284487 | 0.003715156 | 0.027065715 | 1 |
| Heart_Failure | pericyte | *IGFBP7* | 0.502830393 | 0.007870844 | 0.04506739 | 1 |
| Heart_Failure | pericyte | *EFHB* | 0.376111225 | 0.00058366 | 0.007977113 | 1 |
| Heart_Failure | pericyte | *GMPS* | 2.962477979 | 0.003526285 | 0.026298639 | 1 |
| Heart_Failure | pericyte | *TIPARP* | 2.358731201 | 0.003932772 | 0.027843851 | 1 |
| Heart_Failure | pericyte | *SMIM12* | 1.811877241 | 0.000425719 | 0.006793136 | 1 |

| | | | | | | |
|---|---|---|---|---|---|---|
| Heart_Failure | pericyte | *TPRA1* | 3.339293506 | 0.003510604 | 0.026243959 | 1 |
| Heart_Failure | pericyte | *MEAF6* | 3.531530139 | 0.00277986 | 0.02226367 | 1 |
| Heart_Failure | pericyte | *FAM208A* | 1.89474268 | 0.003385267 | 0.025660013 | 1 |
| Heart_Failure | pericyte | *LRPAP1* | 2.676916512 | 0.005212204 | 0.033609358 | 1 |
| Heart_Failure | pericyte | *ABLIM2* | 4.676285007 | 3.58E-05 | 0.001918699 | 0.225341526 |
| Heart_Failure | pericyte | *RNF123* | 1.712408635 | 1.22E-07 | 7.66E-05 | 0.000765709 |
| Heart_Failure | pericyte | *ANXA5* | 1.763937527 | 0.007569203 | 0.043998431 | 1 |
| Heart_Failure | pericyte | *MAP9* | 3.358686473 | 0.000219516 | 0.004655112 | 1 |
| Heart_Failure | pericyte | *CEP44* | 2.966538534 | 0.003732446 | 0.027065715 | 1 |
| Heart_Failure | pericyte | *C4orf45* | 2.873273559 | 0.000682967 | 0.008964119 | 1 |
| Heart_Failure | pericyte | *ARFIP1* | 1.812323541 | 0.008492337 | 0.047449452 | 1 |
| Heart_Failure | pericyte | *PRRC1* | 3.767621585 | 0.000234299 | 0.004728943 | 1 |
| Heart_Failure | pericyte | *NDUFS4* | 3.018387523 | 0.000680012 | 0.008962764 | 1 |
| Heart_Failure | pericyte | *TIGD6* | 3.762931742 | 0.000309321 | 0.005588228 | 1 |
| Heart_Failure | pericyte | *ERAP1* | 1.280572878 | 0.001878436 | 0.01756601 | 1 |
| Heart_Failure | pericyte | *RICTOR* | 2.117903966 | 0.005303516 | 0.033988995 | 1 |
| Heart_Failure | pericyte | *UBLCP1* | 1.888815034 | 0.000278017 | 0.005271845 | 1 |
| Heart_Failure | pericyte | *TMEM200A* | 1.199342286 | 0.001579534 | 0.015614046 | 1 |
| Heart_Failure | pericyte | *SAP30L* | 5.625223713 | 4.45E-08 | 3.21E-05 | 0.000280029 |
| Heart_Failure | pericyte | *RPS14* | 2.613029565 | 0.001457925 | 0.014712641 | 1 |
| Heart_Failure | pericyte | *KCNK5* | 1.798343866 | 0.006127666 | 0.037819181 | 1 |
| Heart_Failure | pericyte | *PEX2* | 3.005168743 | 0.003705552 | 0.027057845 | 1 |
| Heart_Failure | pericyte | *SUN1* | 1.439996752 | 0.007789149 | 0.044818101 | 1 |
| Heart_Failure | pericyte | *GPER1* | 0.662003696 | 0.003454351 | 0.02600899 | 1 |
| Heart_Failure | pericyte | *SLC4A2* | 5.346111355 | 0.000381068 | 0.006247076 | 1 |
| Heart_Failure | pericyte | *SLC25A32* | 2.85985083 | 0.002032044 | 0.018276772 | 1 |
| Heart_Failure | pericyte | *DCAF13* | 2.816667504 | 0.000149479 | 0.003670989 | 0.939773253 |
| Heart_Failure | pericyte | *FAM219A* | 4.039223107 | 6.46E-05 | 0.002414618 | 0.406273846 |
| Heart_Failure | pericyte | *CCDC171* | 2.383449506 | 0.007572181 | 0.043998431 | 1 |
| Heart_Failure | pericyte | *HNRNPK* | 3.4634709 | 0.000263096 | 0.005105204 | 1 |
| Heart_Failure | pericyte | *NOL6* | 2.617542787 | 0.002331484 | 0.019888793 | 1 |
| Heart_Failure | pericyte | *TRMT10B* | 3.36838489 | 0.000628455 | 0.00840659 | 1 |
| Heart_Failure | pericyte | *LRFN5* | 0.304996475 | 0.006165898 | 0.037949559 | 1 |
| Heart_Failure | pericyte | *CFL2* | 2.984778058 | 0.001110865 | 0.01216726 | 1 |
| Heart_Failure | pericyte | *ZCCHC24* | 4.015977067 | 2.88E-05 | 0.001861562 | 0.180784769 |
| Heart_Failure | pericyte | *PGM2L1* | 2.941858841 | 0.000331602 | 0.005798899 | 1 |
| Heart_Failure | pericyte | *ATP5C1* | 2.444254413 | 0.001138154 | 0.0123585 | 1 |
| Heart_Failure | pericyte | *TAF3* | 2.701477071 | 0.000971813 | 0.011128936 | 1 |
| Heart_Failure | pericyte | *QSOX2* | 6.665379268 | 0.000217892 | 0.004644531 | 1 |
| Heart_Failure | pericyte | *SDCCAG3* | 4.067921501 | 0.001164001 | 0.012568303 | 1 |
| Heart_Failure | pericyte | *AK8* | 2.76261875 | 0.006041249 | 0.037493913 | 1 |
| Heart_Failure | pericyte | *TSC1* | 3.42689859 | 2.80E-05 | 0.001861562 | 0.17585482 |
| Heart_Failure | pericyte | *HSPA12A* | 0.912031169 | 0.000523123 | 0.007474709 | 1 |
| Heart_Failure | pericyte | *ARHGAP42* | 0.840806363 | 0.001920225 | 0.017769364 | 1 |
| Heart_Failure | pericyte | *CLMN* | 1.197705808 | 0.001566527 | 0.015577464 | 1 |
| Heart_Failure | pericyte | *JAM3* | 1.867197118 | 0.004436811 | 0.030123361 | 1 |
| Heart_Failure | pericyte | *COX11* | 1.57437303 | 0.005719813 | 0.035960464 | 1 |
| Heart_Failure | pericyte | *ZNF202* | 2.406871109 | 0.000292409 | 0.005427606 | 1 |
| Heart_Failure | pericyte | *ILK* | 2.53876589 | 0.004502595 | 0.030465674 | 1 |
| Heart_Failure | pericyte | *TAF10* | 4.433135378 | 6.27E-05 | 0.002380436 | 0.394273956 |

| | | | | | | |
|---|---|---|---|---|---|---|
| Heart_Failure | pericyte | *WDR88* | 3.103528649 | 0.008124243 | 0.04609848 | 1 |
| Heart_Failure | pericyte | *LMO1* | 2.072146836 | 0.002519492 | 0.020980191 | 1 |
| Heart_Failure | pericyte | *RPL27A* | 3.680310792 | 7.98E-05 | 0.002697406 | 0.501717491 |
| Heart_Failure | pericyte | *PRTG* | 3.087218824 | 0.000178802 | 0.004073766 | 1 |
| Heart_Failure | pericyte | *TMEM41B* | 4.005310251 | 8.90E-05 | 0.002769335 | 0.559697156 |
| Heart_Failure | pericyte | *ZNF143* | 3.138815041 | 0.000149015 | 0.003670989 | 0.936856613 |
| Heart_Failure | pericyte | *CCDC68* | 0.899545929 | 0.004629243 | 0.031060884 | 1 |
| Heart_Failure | pericyte | *CENPV* | 2.361818825 | 0.004950177 | 0.032418503 | 1 |
| Heart_Failure | pericyte | *DUSP18* | 3.217066911 | 0.00032308 | 0.005721701 | 1 |
| Heart_Failure | pericyte | *HDHD2* | 1.097937073 | 0.000180185 | 0.004089609 | 1 |
| Heart_Failure | pericyte | *TRIM68* | 0.860578618 | 0.008861491 | 0.048827514 | 1 |
| Heart_Failure | pericyte | *ZNF646* | 1.919202835 | 8.81E-05 | 0.002769335 | 0.553994389 |
| Heart_Failure | pericyte | *GATAD2A* | 2.460353165 | 0.004283795 | 0.029352191 | 1 |
| Heart_Failure | pericyte | *RPL13* | 3.424000375 | 3.30E-05 | 0.001918699 | 0.207436757 |
| Heart_Failure | pericyte | *ZNF528* | 3.009931015 | 0.000536001 | 0.007538786 | 1 |
| Heart_Failure | pericyte | *SERTAD3* | 1.049892502 | 0.007203271 | 0.04230661 | 1 |
| Heart_Failure | pericyte | *TMC4* | 3.983223512 | 0.003437652 | 0.025976587 | 1 |
| Heart_Failure | pericyte | *TTYH1* | 4.46459162 | 0.000222792 | 0.004655112 | 1 |
| Heart_Failure | pericyte | *CDC42EP5* | 1.884525214 | 0.000470667 | 0.00709612 | 1 |
| Heart_Failure | pericyte | *PPP1R14A* | 1.967805058 | 0.002080512 | 0.018595727 | 1 |
| Heart_Failure | pericyte | *CYB5D2* | 4.291670304 | 6.49E-05 | 0.002414618 | 0.408070372 |
| Heart_Failure | pericyte | *CD320* | 5.146122255 | 0.000770064 | 0.009605939 | 1 |
| Heart_Failure | pericyte | *SPRYD3* | 6.353647401 | 4.41E-05 | 0.002128526 | 0.277174741 |
| Heart_Failure | pericyte | *NUDT8* | -0.925182728 | 0.003829411 | 0.02735853 | 1 |
| Heart_Failure | pericyte | *PRDX2* | 4.533169422 | 2.39E-05 | 0.001740485 | 0.15030114 |
| Heart_Failure | pericyte | *ATP5H* | 3.626235755 | 0.000349162 | 0.005952796 | 1 |
| Heart_Failure | pericyte | *DDB1* | 3.322669001 | 0.00156031 | 0.015570901 | 1 |
| Heart_Failure | pericyte | *KIAA1586* | 2.147989677 | 0.002009987 | 0.01824217 | 1 |
| Heart_Failure | pericyte | *PTGDR* | 1.714375811 | 5.38E-05 | 0.002212446 | 0.338504254 |
| Heart_Failure | pericyte | *GLYCTK* | 2.883909713 | 0.001178833 | 0.012629235 | 1 |
| Heart_Failure | pericyte | *DTYMK* | 2.88398541 | 0.000727547 | 0.009296927 | 1 |
| Heart_Failure | pericyte | *ZSWIM1* | 1.082661995 | 0.008465877 | 0.047395342 | 1 |
| Heart_Failure | pericyte | *NPNT* | 1.895651714 | 0.000521415 | 0.007467279 | 1 |
| Heart_Failure | pericyte | *CNNM3* | 2.025675204 | 0.006075054 | 0.037629426 | 1 |
| Heart_Failure | pericyte | *STX18* | 3.371296734 | 0.000352225 | 0.005952796 | 1 |
| Heart_Failure | pericyte | *ZNF608* | 2.759870672 | 3.27E-05 | 0.001918699 | 0.205550877 |
| Heart_Failure | pericyte | *FEM1B* | 2.101356853 | 0.003102851 | 0.024233074 | 1 |
| Heart_Failure | pericyte | *FAM110B* | 1.946319169 | 0.000210175 | 0.004540796 | 1 |
| Heart_Failure | pericyte | *PTK2* | 2.41260733 | 0.000107913 | 0.00305607 | 0.678447467 |
| Heart_Failure | pericyte | *RNASE6* | 2.210920115 | 0.002082296 | 0.018595727 | 1 |
| Heart_Failure | pericyte | *CLIC4* | 2.198183209 | 0.005492509 | 0.034986225 | 1 |
| Heart_Failure | pericyte | *SLC38A11* | 4.064281752 | 0.000943836 | 0.010907903 | 1 |
| Heart_Failure | pericyte | *HINT1* | 2.556560072 | 0.007920948 | 0.045230702 | 1 |
| Heart_Failure | pericyte | *MT1E* | 4.102271998 | 0.001207769 | 0.012804798 | 1 |
| Heart_Failure | pericyte | *CALML6* | -1.81201066 | 0.004287549 | 0.029352191 | 1 |
| Heart_Failure | pericyte | *BRD3* | 3.581926023 | 0.001923192 | 0.017769364 | 1 |
| Heart_Failure | pericyte | *MAP3K2* | 2.384437712 | 0.00240262 | 0.020302783 | 1 |
| Heart_Failure | pericyte | *CNTROB* | 5.634776539 | 6.13E-05 | 0.002379208 | 0.385586894 |
| Heart_Failure | pericyte | *ZNF212* | 2.659476277 | 0.002365794 | 0.020099657 | 1 |
| Heart_Failure | pericyte | *FAM161A* | 1.78266604 | 0.007972172 | 0.045440657 | 1 |

| | | | | | | |
|---|---|---|---|---|---|---|
| Heart_Failure | pericyte | *SEMA3E* | 2.931624821 | 0.000161057 | 0.00383548 | 1 |
| Heart_Failure | pericyte | *NFXL1* | 3.956347229 | 0.00014458 | 0.003632017 | 0.908974177 |
| Heart_Failure | pericyte | *DLGAP1* | 2.481590546 | 9.83E-05 | 0.002888384 | 0.61811407 |
| Heart_Failure | pericyte | *RNF34* | 1.99574758 | 0.006229576 | 0.038247407 | 1 |
| Heart_Failure | pericyte | *ACYP2* | 1.948751308 | 0.004030207 | 0.028310515 | 1 |
| Heart_Failure | pericyte | *TMEM133* | 4.816552615 | 0.001223973 | 0.012954746 | 1 |
| Heart_Failure | pericyte | *TTLL6* | 6.988311198 | 0.00143584 | 0.014583403 | 1 |
| Heart_Failure | pericyte | *AKAP13* | 1.892147657 | 0.00186003 | 0.017453744 | 1 |
| Heart_Failure | pericyte | *KBTBD2* | 2.388351573 | 0.004238316 | 0.029249499 | 1 |
| Heart_Failure | pericyte | *TSEN34* | 4.613304211 | 0.002627609 | 0.021538172 | 1 |
| Heart_Failure | pericyte | *SOX7* | 3.638292808 | 0.000207343 | 0.004519343 | 1 |
| Heart_Failure | pericyte | *MFN1* | 2.564201031 | 0.000317719 | 0.005686577 | 1 |
| Heart_Failure | pericyte | *ATP6V0E2* | 4.723760354 | 0.003491072 | 0.026160156 | 1 |
| Heart_Failure | pericyte | *LRRC8C* | 4.119323391 | 0.002344142 | 0.019969675 | 1 |
| Heart_Failure | pericyte | *TBCA* | 2.097825939 | 0.001991753 | 0.018174386 | 1 |
| Heart_Failure | pericyte | *BCL2L1* | 3.11198882 | 8.27E-05 | 0.002749331 | 0.519623604 |
| Heart_Failure | pericyte | *LRRC34* | 0.647996362 | 0.009032759 | 0.049297742 | 1 |
| Heart_Failure | pericyte | *ORMDL3* | -1.149786529 | 3.21E-09 | 3.37E-06 | 2.02E-05 |
| Heart_Failure | pericyte | *MTBP* | 4.131557853 | 0.00022452 | 0.004658604 | 1 |
| Heart_Failure | pericyte | *SPTLC3* | 3.670922249 | 0.000525898 | 0.007488118 | 1 |
| Heart_Failure | pericyte | *COPRS* | 4.69745377 | 2.48E-05 | 0.001753917 | 0.156098573 |
| Heart_Failure | pericyte | *SUCLG2* | 1.787873695 | 0.009033084 | 0.049297742 | 1 |
| Heart_Failure | pericyte | *RCAN2* | 2.713310253 | 0.003266199 | 0.025011683 | 1 |
| Heart_Failure | pericyte | *FIBP* | 1.497179828 | 0.007747435 | 0.044818101 | 1 |
| Heart_Failure | pericyte | *PDE3A* | 1.848345376 | 0.000123417 | 0.00330886 | 0.775920681 |
| Heart_Failure | pericyte | *ZMAT3* | 3.761854205 | 0.002454038 | 0.020681682 | 1 |
| Heart_Failure | pericyte | *CYP7B1* | 2.039501882 | 4.80E-05 | 0.002128526 | 0.302006648 |
| Heart_Failure | pericyte | *LCLAT1* | 2.556717606 | 0.002172617 | 0.019149613 | 1 |
| Heart_Failure | pericyte | *DCAKD* | -0.852945787 | 0.000387199 | 0.006290239 | 1 |
| Heart_Failure | pericyte | *ABLIM3* | 3.617768496 | 6.82E-06 | 0.001428887 | 0.042866622 |
| Heart_Failure | pericyte | *GLRX* | 3.104331482 | 0.00029474 | 0.005434111 | 1 |
| Heart_Failure | pericyte | *OLR1* | 8.143123323 | 0.004133735 | 0.028844389 | 1 |
| Heart_Failure | pericyte | *PEAK1* | 1.830393566 | 0.006332705 | 0.03857918 | 1 |
| Heart_Failure | pericyte | *CSPG4* | 0.873404505 | 0.004721768 | 0.031513538 | 1 |
| Heart_Failure | pericyte | *RNF213* | 2.445386796 | 0.001266462 | 0.013226319 | 1 |
| Heart_Failure | pericyte | *PLK3* | 4.927469233 | 0.000169844 | 0.003961988 | 1 |
| Heart_Failure | pericyte | *DPY19L1* | 3.136993436 | 0.003345282 | 0.025462212 | 1 |
| Heart_Failure | pericyte | *GOLIM4* | 3.345963401 | 1.91E-06 | 0.000799666 | 0.01199499 |
| Heart_Failure | pericyte | *USMG5* | 3.52318305 | 0.000148476 | 0.003670989 | 0.933469783 |
| Heart_Failure | pericyte | *NRROS* | -0.830488339 | 0.000555562 | 0.007744607 | 1 |
| Heart_Failure | pericyte | *BBS1* | 2.397259398 | 0.000204237 | 0.004489642 | 1 |
| Heart_Failure | pericyte | *ANKRD36C* | 2.618109787 | 0.000726853 | 0.009296927 | 1 |
| Heart_Failure | pericyte | *LARP7* | 2.772439426 | 0.003899077 | 0.027760567 | 1 |
| Heart_Failure | pericyte | *FGFBP3* | 6.958498147 | 2.01E-05 | 0.001740485 | 0.126152578 |
| Heart_Failure | pericyte | *C4orf32* | 0.639627557 | 0.006447371 | 0.039088351 | 1 |
| Heart_Failure | pericyte | *SRP72* | 1.189170474 | 0.008970033 | 0.04916704 | 1 |
| Heart_Failure | pericyte | *C4orf26* | -3.110064092 | 0.005553808 | 0.03511409 | 1 |
| Heart_Failure | pericyte | *PTDSS2* | 1.500048439 | 0.006583262 | 0.039758857 | 1 |
| Heart_Failure | pericyte | *CADM2* | 0.95519492 | 0.005154212 | 0.033464241 | 1 |
| Heart_Failure | pericyte | *ARHGAP1* | 1.796919583 | 0.007739386 | 0.044818101 | 1 |

| | | | | | | |
|---|---|---|---|---|---|---|
| Heart_Failure | pericyte | *TP53I11* | 2.732774324 | 0.004506613 | 0.030465674 | 1 |
| Heart_Failure | pericyte | *MCTP1* | 1.566233368 | 0.00034184 | 0.005904244 | 1 |
| Heart_Failure | pericyte | *UCP2* | 3.241282715 | 0.000778587 | 0.009673865 | 1 |
| Heart_Failure | pericyte | *RPS6KB2* | 1.629881097 | 0.000479246 | 0.007173851 | 1 |
| Heart_Failure | pericyte | *A2M* | 2.498258384 | 3.58E-05 | 0.001918699 | 0.225091935 |
| Heart_Failure | pericyte | *IP6K1* | 3.152734468 | 8.72E-05 | 0.002769335 | 0.547988798 |
| Heart_Failure | pericyte | *TCP11L1* | 1.680166841 | 0.004804697 | 0.031763544 | 1 |
| Heart_Failure | pericyte | *CRLF3* | 2.785718808 | 9.25E-05 | 0.002769335 | 0.581383142 |
| Heart_Failure | pericyte | *SLCO3A1* | 1.438442589 | 0.00036004 | 0.006022578 | 1 |
| Heart_Failure | pericyte | *RNF152* | 0.881418311 | 0.003683448 | 0.027021979 | 1 |
| Heart_Failure | pericyte | *BASP1* | 1.351414552 | 0.000340885 | 0.005903983 | 1 |
| Heart_Failure | pericyte | *ANKLE2* | 3.105682675 | 0.002223017 | 0.019411265 | 1 |
| Heart_Failure | pericyte | *DPP7* | 3.490384977 | 0.001299543 | 0.013526861 | 1 |
| Heart_Failure | pericyte | *SEC24C* | 3.04487698 | 0.005274437 | 0.033906324 | 1 |
| Heart_Failure | pericyte | *DSCAML1* | 3.771682701 | 4.08E-05 | 0.002084743 | 0.256423402 |
| Heart_Failure | pericyte | *FBXO39* | 0.514157327 | 0.00103457 | 0.01163567 | 1 |
| Heart_Failure | pericyte | *ZBTB38* | 3.062848956 | 3.52E-05 | 0.001918699 | 0.221142768 |
| Heart_Failure | pericyte | *TIMM22* | 4.910643901 | 0.000337756 | 0.005865949 | 1 |
| Heart_Failure | pericyte | *PAWR* | 0.677506744 | 0.004626035 | 0.031060884 | 1 |
| Heart_Failure | pericyte | *NAP1L5* | 2.054299637 | 0.001607482 | 0.015815708 | 1 |
| Heart_Failure | pericyte | *RPLP2* | 2.576132465 | 0.002492488 | 0.020886947 | 1 |
| Heart_Failure | pericyte | *ZNF354C* | 4.515614354 | 1.75E-05 | 0.001740485 | 0.110084693 |
| Heart_Failure | pericyte | *IMP3* | 4.939794387 | 8.40E-05 | 0.002749485 | 0.527901175 |
| Heart_Failure | pericyte | *MLF1* | 4.160028516 | 1.97E-05 | 0.001740485 | 0.123920854 |
| Heart_Failure | pericyte | *C2orf69* | 3.943918704 | 0.001260238 | 0.013205198 | 1 |
| Heart_Failure | pericyte | *GRAMD1C* | 1.236059688 | 0.002985797 | 0.023494003 | 1 |
| Heart_Failure | pericyte | *ZNF518B* | 2.426402047 | 0.002057232 | 0.01845052 | 1 |
| Heart_Failure | pericyte | *LCORL* | 2.225957785 | 0.004027223 | 0.028310515 | 1 |
| Heart_Failure | pericyte | *CPNE7* | -2.799905378 | 0.00074326 | 0.009423995 | 1 |
| Heart_Failure | pericyte | *TRIM73* | -2.297126982 | 0.002026338 | 0.018276772 | 1 |
| Heart_Failure | pericyte | *EFCAB13* | 1.44844664 | 0.007911423 | 0.04521738 | 1 |
| Heart_Failure | pericyte | *GAK* | 2.430131131 | 0.002132721 | 0.018938446 | 1 |
| Heart_Failure | pericyte | *KLHDC7A* | -0.582124016 | 0.003903347 | 0.027760567 | 1 |
| Heart_Failure | pericyte | *CALR* | 2.621168076 | 0.001698017 | 0.016474431 | 1 |
| Heart_Failure | pericyte | *HLA-DQB1* | -1.78626292 | 9.31E-23 | 3.45E-19 | 5.85E-19 |
| Heart_Failure | pericyte | *EGR3* | 1.170159431 | 0.005971363 | 0.037172807 | 1 |
| Heart_Failure | pericyte | *TIGD5* | 1.86448359 | 0.006629742 | 0.039962785 | 1 |
| Heart_Failure | pericyte | *PDXDC1* | 1.93878923 | 0.007825272 | 0.044906549 | 1 |
| Heart_Failure | pericyte | *SSC5D* | 0.516941869 | 0.003250688 | 0.024953696 | 1 |
| Heart_Failure | pericyte | *ZADH2* | 3.634609551 | 8.52E-05 | 0.002769335 | 0.535600226 |
| Heart_Failure | pericyte | *ZNF594* | 3.469338815 | 0.000968744 | 0.011114042 | 1 |
| Heart_Failure | pericyte | *PCGF5* | 2.2718614 | 0.005392666 | 0.03441999 | 1 |
| Heart_Failure | pericyte | *SRSF8* | 6.371266718 | 5.13E-05 | 0.002162658 | 0.322469473 |
| Heart_Failure | pericyte | *MRPL14* | 2.220483509 | 0.002920071 | 0.023209212 | 1 |
| Heart_Failure | pericyte | *ZFP82* | 3.467677012 | 0.000767506 | 0.009605939 | 1 |
| Heart_Failure | pericyte | *FKRP* | 0.920544323 | 0.0010985 | 0.012052831 | 1 |
| Heart_Failure | pericyte | *ZBTB20* | 1.737437477 | 0.002120092 | 0.018852922 | 1 |
| Heart_Failure | pericyte | *WRB* | 0.646626339 | 0.003634764 | 0.026727206 | 1 |
| Heart_Failure | pericyte | *TNRC18* | 2.963436251 | 0.000396863 | 0.006414082 | 1 |
| Heart_Failure | pericyte | *TDRKH* | -1.088767625 | 0.001717762 | 0.01648789 | 1 |

| | | | | | | |
|---|---|---|---|---|---|---|
| Heart_Failure | pericyte | *IST1* | 2.039065316 | 0.008080596 | 0.045933732 | 1 |
| Heart_Failure | pericyte | *ERCC6L2* | 1.91475431 | 0.00111774 | 0.012221271 | 1 |
| Heart_Failure | pericyte | *C8orf33* | 2.772427258 | 0.001362307 | 0.014063752 | 1 |
| Heart_Failure | pericyte | *FBXL6* | 1.637862896 | 0.007900595 | 0.045196581 | 1 |
| Heart_Failure | pericyte | *TSHZ2* | 1.528038544 | 0.001424405 | 0.014561359 | 1 |
| Heart_Failure | pericyte | *FES* | 3.925788162 | 0.000197147 | 0.004377544 | 1 |
| Heart_Failure | pericyte | *SPNS3* | 2.87379399 | 0.004176593 | 0.029014629 | 1 |
| Heart_Failure | pericyte | *TRAK1* | 1.828457396 | 0.008308454 | 0.046868605 | 1 |
| Heart_Failure | pericyte | *ACBD3* | 2.906839378 | 0.001287813 | 0.013426997 | 1 |
| Heart_Failure | pericyte | *PLCXD3* | 1.14813647 | 0.004656022 | 0.03120726 | 1 |
| Heart_Failure | pericyte | *COL18A1* | 0.407052467 | 0.003738581 | 0.027065715 | 1 |
| Heart_Failure | pericyte | *C12orf60* | 2.252726794 | 0.000481023 | 0.007183074 | 1 |
| Heart_Failure | pericyte | *WBP2NL* | 4.937409603 | 6.67E-05 | 0.002451327 | 0.419176837 |
| Heart_Failure | pericyte | *GAS6* | 2.104957799 | 0.001947156 | 0.017871199 | 1 |
| Heart_Failure | pericyte | *GPR19* | 2.137780415 | 0.005801935 | 0.036367661 | 1 |
| Heart_Failure | pericyte | *CTNNA3* | 1.537495252 | 0.000318918 | 0.005686577 | 1 |
| Heart_Failure | pericyte | *C19orf71* | 2.755498522 | 0.0035803 | 0.026490062 | 1 |
| Heart_Failure | pericyte | *GRIN2A* | 1.530975301 | 0.000937822 | 0.010883614 | 1 |
| Heart_Failure | pericyte | *PSMG1* | -0.563351151 | 0.003203858 | 0.02478862 | 1 |
| Heart_Failure | pericyte | *BMP8A* | 3.67133347 | 1.92E-05 | 0.001740485 | 0.120657404 |
| Heart_Failure | pericyte | *CBX6* | 4.900084079 | 0.000277242 | 0.005271845 | 1 |
| Heart_Failure | pericyte | *BTBD9* | 1.784204444 | 0.006150919 | 0.037912577 | 1 |
| Heart_Failure | pericyte | *DENND5A* | 3.041162659 | 6.24E-05 | 0.002380436 | 0.392121386 |
| Heart_Failure | pericyte | *TBX1* | 2.56095379 | 0.000782022 | 0.009697381 | 1 |
| Heart_Failure | pericyte | *UQCR10* | 2.630296263 | 0.000419171 | 0.006722771 | 1 |
| Heart_Failure | pericyte | *ACOT1* | -0.790801821 | 0.001770631 | 0.016834554 | 1 |
| Heart_Failure | pericyte | *CSF1* | 1.570599173 | 0.000260733 | 0.005096468 | 1 |
| Heart_Failure | pericyte | *WDR27* | 3.198823886 | 0.000124358 | 0.00330886 | 0.781841877 |
| Heart_Failure | pericyte | *SNN* | 4.399336382 | 0.000260807 | 0.005096468 | 1 |
| Heart_Failure | pericyte | *ZNF93* | 7.222156559 | 1.73E-05 | 0.001740485 | 0.108526156 |
| Heart_Failure | pericyte | *NDUFA12* | 3.956265728 | 3.57E-05 | 0.001918699 | 0.224527304 |
| Heart_Failure | pericyte | *ZFP90* | 3.714714867 | 0.000514236 | 0.007432185 | 1 |
| Heart_Failure | pericyte | *KIAA0825* | 2.95808564 | 0.000382494 | 0.006247076 | 1 |
| Heart_Failure | pericyte | *GAS2L1* | 4.252578941 | 0.000441263 | 0.006918249 | 1 |
| Heart_Failure | pericyte | *ATP6V0A2* | 2.435521716 | 0.002359152 | 0.020070349 | 1 |
| Heart_Failure | pericyte | *LSAMP* | 1.356510608 | 0.004099012 | 0.028704871 | 1 |
| Heart_Failure | pericyte | *MYBL1* | -0.523264856 | 0.003584467 | 0.026490062 | 1 |
| Heart_Failure | pericyte | *C16orf52* | 3.471228817 | 0.000153282 | 0.00371105 | 0.963682961 |
| Heart_Failure | pericyte | *POMK* | 4.974321435 | 9.03E-05 | 0.002769335 | 0.567928786 |
| Heart_Failure | pericyte | *ZNF529* | 2.06814827 | 0.002302462 | 0.0197215 | 1 |
| Heart_Failure | pericyte | *WWOX* | 1.231560169 | 0.00373071 | 0.027065715 | 1 |
| Heart_Failure | pericyte | *TOR3A* | 6.325353677 | 3.41E-05 | 0.001918699 | 0.214432265 |
| Heart_Failure | pericyte | *CYP4X1* | 1.442359732 | 0.003926987 | 0.027843851 | 1 |
| Heart_Failure | pericyte | *RPS23* | 1.85611155 | 0.003755555 | 0.027065715 | 1 |
| Heart_Failure | pericyte | *IFT140* | 2.731492917 | 0.000344237 | 0.005913159 | 1 |
| Heart_Failure | pericyte | *ISG15* | 1.491069772 | 0.005184226 | 0.033497665 | 1 |
| Heart_Failure | pericyte | *DNAH17* | 3.937057865 | 0.005504527 | 0.035019283 | 1 |
| Heart_Failure | pericyte | *TMEM220* | 3.293530983 | 0.000325391 | 0.005746433 | 1 |
| Heart_Failure | pericyte | *KLHL17* | 2.675817783 | 0.005086773 | 0.033209287 | 1 |
| Heart_Failure | pericyte | *LINC00265* | 2.572197516 | 0.000409985 | 0.006609166 | 1 |

| | | | | | | |
|---|---|---|---|---|---|---|
| Heart_Failure | pericyte | *LAMTOR4* | 1.702046268 | 0.006806678 | 0.040639681 | 1 |
| Heart_Failure | pericyte | *SRSF10* | 2.638457996 | 0.001713073 | 0.01648789 | 1 |
| Heart_Failure | pericyte | *NBR1* | 2.667816803 | 0.00552434 | 0.035019283 | 1 |
| Heart_Failure | pericyte | *NDOR1* | 4.756743821 | 3.63E-05 | 0.001918699 | 0.228047819 |
| Heart_Failure | pericyte | *DPYD* | 2.448997299 | 0.000215167 | 0.004616916 | 1 |
| Heart_Failure | pericyte | *MSL1* | -0.957543972 | 0.003766341 | 0.027071238 | 1 |
| Heart_Failure | pericyte | *FAM196A* | 2.962250026 | 0.000698004 | 0.009048145 | 1 |
| Heart_Failure | pericyte | *UTS2B* | -0.695759781 | 0.005116458 | 0.033333854 | 1 |
| Heart_Failure | pericyte | *NELFB* | 5.15913298 | 1.76E-05 | 0.001740485 | 0.110962723 |
| Heart_Failure | pericyte | *ADAT2* | 2.219590724 | 0.001191202 | 0.012714915 | 1 |
| Heart_Failure | pericyte | *S100A13* | 2.924501434 | 0.003206494 | 0.02478862 | 1 |
| Heart_Failure | pericyte | *ZKSCAN3* | 3.594640188 | 0.000139356 | 0.003561519 | 0.876133599 |
| Heart_Failure | pericyte | *HLA-DRB1* | -1.667419475 | 1.43E-07 | 8.17E-05 | 0.000898595 |
| Heart_Failure | pericyte | *MPHOSPH8* | 2.882268462 | 0.000196673 | 0.004377544 | 1 |
| Heart_Failure | pericyte | *SUPT5H* | 3.204700582 | 0.001234334 | 0.013020769 | 1 |
| Heart_Failure | pericyte | *SUPT3H* | 2.734952711 | 0.000940624 | 0.010890795 | 1 |
| Heart_Failure | pericyte | *ZKSCAN7* | 3.636633232 | 0.008185182 | 0.046360577 | 1 |
| Heart_Failure | pericyte | *WDR5* | 2.745546556 | 0.008808869 | 0.048624685 | 1 |
| Heart_Failure | pericyte | *ASB13* | 5.151678323 | 2.37E-05 | 0.001740485 | 0.149055178 |
| Heart_Failure | pericyte | *EVL* | 1.415266149 | 0.00685208 | 0.0408332 | 1 |
| Heart_Failure | pericyte | *MYL6B* | 5.488854779 | 9.29E-06 | 0.001740485 | 0.058410832 |
| Heart_Failure | pericyte | *HDAC2* | 3.83530378 | 0.00017243 | 0.003985542 | 1 |
| Heart_Failure | pericyte | *HRH1* | 2.444911657 | 0.004357282 | 0.029711746 | 1 |
| Heart_Failure | pericyte | *FAM180B* | -0.742549471 | 0.006830486 | 0.040743135 | 1 |
| Heart_Failure | pericyte | *TOMM7* | 4.563631795 | 2.39E-05 | 0.001740485 | 0.15013372 |
| Heart_Failure | pericyte | *ZNF431* | 2.140233755 | 0.006328353 | 0.03857918 | 1 |
| Heart_Failure | pericyte | *NF1* | 2.179441835 | 0.000769262 | 0.009605939 | 1 |
| Heart_Failure | pericyte | *HLA-DQA1* | -1.938244514 | 1.43E-15 | 2.99E-12 | 8.97E-12 |
| Heart_Failure | pericyte | *STRN3* | 2.259349388 | 0.002639487 | 0.021579263 | 1 |
| Heart_Failure | pericyte | *NHLRC2* | 2.703897684 | 0.000806093 | 0.009857039 | 1 |
| Heart_Failure | pericyte | *KPNA5* | 3.584530715 | 2.61E-05 | 0.001823804 | 0.164142372 |
| Heart_Failure | pericyte | *ANKRD36B* | 1.690169585 | 0.003756036 | 0.027065715 | 1 |
| Heart_Failure | pericyte | *SRGAP1* | 2.819951909 | 0.000245998 | 0.004909805 | 1 |
| Heart_Failure | pericyte | *PCDHB16* | 0.703017384 | 0.00590078 | 0.036840321 | 1 |
| Heart_Failure | pericyte | *ZMYM1* | 3.630585106 | 0.000570242 | 0.007867313 | 1 |
| Heart_Failure | pericyte | *IGF2R* | 3.113070029 | 2.07E-05 | 0.001740485 | 0.130282156 |
| Heart_Failure | pericyte | *ZNF682* | 4.800203584 | 1.11E-05 | 0.001740485 | 0.069482186 |
| Heart_Failure | pericyte | *KANK2* | 2.987036442 | 0.002392558 | 0.020244971 | 1 |
| Heart_Failure | pericyte | *GTF2E2* | 2.497478744 | 0.002673936 | 0.02169166 | 1 |
| Heart_Failure | pericyte | *ZNF517* | 6.991806762 | 0.000968208 | 0.011114042 | 1 |
| Heart_Failure | pericyte | *SLC22A5* | 1.587283629 | 1.42E-09 | 1.79E-06 | 8.94E-06 |
| Heart_Failure | pericyte | *ZNF624* | 5.421924362 | 0.000165456 | 0.003925377 | 1 |
| Heart_Failure | pericyte | *RPS26* | 0.704321533 | 7.87E-05 | 0.002693349 | 0.494787928 |
| Heart_Failure | pericyte | *PSAP* | 1.91948139 | 0.00832185 | 0.046881245 | 1 |
| Heart_Failure | pericyte | *CCDC180* | 0.959108143 | 0.008446264 | 0.047369901 | 1 |
| Heart_Failure | pericyte | *ZNF677* | 5.618183433 | 0.000124588 | 0.00330886 | 0.783285249 |
| Heart_Failure | pericyte | *NOL8* | 3.627062876 | 0.000283214 | 0.005306292 | 1 |
| Heart_Failure | pericyte | *MAK16* | 3.403253327 | 8.64E-05 | 0.002769335 | 0.543485923 |
| Heart_Failure | pericyte | *MIER1* | 2.736567899 | 0.000928927 | 0.010847621 | 1 |
| Heart_Failure | pericyte | *ZNF251* | 2.976653857 | 0.000470176 | 0.00709612 | 1 |

| | | | | | | |
|---|---|---|---|---|---|---|
| Heart_Failure | pericyte | *DDRGK1* | 4.805342386 | 0.000271079 | 0.005201888 | 1 |
| Heart_Failure | pericyte | *ZKSCAN8* | 2.499375485 | 0.002208524 | 0.019311528 | 1 |
| Heart_Failure | pericyte | *GFPT1* | 2.723822731 | 5.90E-05 | 0.002333071 | 0.370958245 |
| Heart_Failure | pericyte | *ZNF808* | 1.871265969 | 0.003929359 | 0.027843851 | 1 |
| Heart_Failure | pericyte | *ARC* | 6.094085021 | 0.00626672 | 0.038325749 | 1 |
| Heart_Failure | pericyte | *LRBA* | 2.64191833 | 0.000271388 | 0.005201888 | 1 |
| Heart_Failure | pericyte | *MDM4* | 2.704359018 | 0.000915345 | 0.010743937 | 1 |
| Heart_Failure | pericyte | *PAPSS2* | 1.704914813 | 0.007775419 | 0.044818101 | 1 |
| Heart_Failure | pericyte | *FAN1* | 4.537331473 | 4.76E-05 | 0.002128526 | 0.29924585 |
| Heart_Failure | pericyte | *ZNF652* | 2.970716365 | 0.000864068 | 0.010367169 | 1 |
| Heart_Failure | pericyte | *TMEM184B* | 2.678503759 | 0.006689087 | 0.040166464 | 1 |
| Heart_Failure | pericyte | *SFT2D1* | 3.525770173 | 0.000784846 | 0.009713245 | 1 |
| Heart_Failure | pericyte | *ITPRIPL1* | 1.162148621 | 0.004516251 | 0.030498031 | 1 |
| Heart_Failure | pericyte | *MAP3K3* | 2.858026475 | 0.001154024 | 0.012487687 | 1 |
| Heart_Failure | pericyte | *APRT* | 5.571010081 | 1.95E-05 | 0.001740485 | 0.122889481 |
| Heart_Failure | pericyte | *SMG5* | 2.629558884 | 0.002437097 | 0.02056648 | 1 |
| Heart_Failure | pericyte | *SYS1* | 4.500650443 | 8.09E-05 | 0.002716178 | 0.508465744 |
| Heart_Failure | pericyte | *NEU4* | -4.453665976 | 0.004793162 | 0.03173585 | 1 |
| Heart_Failure | pericyte | *GIGYF2* | 2.476191753 | 0.00156646 | 0.015577464 | 1 |
| Heart_Failure | pericyte | *HLA-DRA* | 1.883019467 | 4.19E-08 | 3.21E-05 | 0.000263546 |
| Heart_Failure | pericyte | *COL15A1* | 1.526446748 | 0.004104255 | 0.028704871 | 1 |
| Heart_Failure | pericyte | *PRRT1* | 1.642955416 | 1.57E-05 | 0.001740485 | 0.098421004 |
| Heart_Failure | pericyte | *DXO* | 0.872409167 | 0.002508278 | 0.020942288 | 1 |
| Heart_Failure | pericyte | *NELFE* | 1.388088345 | 0.001026106 | 0.011581915 | 1 |
| Heart_Failure | pericyte | *BAG6* | 2.68831255 | 4.59E-08 | 3.21E-05 | 0.000288817 |
| Heart_Failure | pericyte | *MICB* | 0.549046988 | 8.98E-05 | 0.002769335 | 0.564833236 |
| Heart_Failure | pericyte | *MICA* | 0.772415959 | 3.51E-05 | 0.001918699 | 0.22084828 |
| Heart_Failure | pericyte | *GABBR1* | 1.556561398 | 0.001768177 | 0.016834554 | 1 |
| Heart_Failure | pericyte | *ATXN2* | 1.671532714 | 0.008595854 | 0.047614213 | 1 |
| Heart_Failure | pericyte | *TRIM13* | 2.722239036 | 0.00042882 | 0.0068253 | 1 |
| Heart_Failure | pericyte | *FAM71F2* | 4.415382708 | 0.002755764 | 0.022183725 | 1 |
| Heart_Failure | pericyte | *FAM205A* | -5.558534439 | 0.001441672 | 0.014619024 | 1 |
| Heart_Failure | pericyte | *PRR13* | 2.209937289 | 0.004199935 | 0.029048398 | 1 |
| Heart_Failure | pericyte | *MT1A* | 3.241033613 | 0.004182548 | 0.029023932 | 1 |
| Heart_Failure | pericyte | *NAP1L4* | 1.886517566 | 0.00272268 | 0.022001912 | 1 |
| Heart_Failure | pericyte | *SERF1B* | -1.77354313 | 0.007757498 | 0.044818101 | 1 |
| Heart_Failure | pericyte | *C1QTNF9B* | 93.04988699 | 8.81E-05 | 0.002769335 | 0.554002664 |
| Heart_Failure | pericyte | *RNPS1* | 2.078332305 | 0.003497942 | 0.026180429 | 1 |
| Heart_Failure | pericyte | *NYNRIN* | 4.530402724 | 0.000861592 | 0.010357221 | 1 |
| Heart_Failure | pericyte | *SLC48A1* | 3.279678506 | 0.008482673 | 0.047447124 | 1 |
| Heart_Failure | pericyte | *RNF208* | 2.899731657 | 0.005819035 | 0.036438519 | 1 |
| Heart_Failure | pericyte | *SUPT4H1* | 3.02120235 | 0.000501508 | 0.007349601 | 1 |
| Heart_Failure | pericyte | *HEXA* | 3.678149342 | 0.000929993 | 0.010847621 | 1 |
| Heart_Failure | pericyte | *LBH* | 1.304675471 | 0.003452631 | 0.02600899 | 1 |
| Heart_Failure | pericyte | *ATF6B* | 1.308806432 | 0.000299689 | 0.005471554 | 1 |
| Heart_Failure | pericyte | *SLC35F6* | 4.335325332 | 0.002031253 | 0.018276772 | 1 |
| Heart_Failure | pericyte | *ITGA1* | 1.182603093 | 0.000570621 | 0.007867313 | 1 |
| Heart_Failure | pericyte | *ZNF726* | 4.649767879 | 0.000805808 | 0.009857039 | 1 |
| Heart_Failure | pericyte | *ZNF90* | 2.518211757 | 0.006315986 | 0.038552045 | 1 |
| Heart_Failure | pericyte | *CPNE1* | -0.647159131 | 0.002794647 | 0.022353624 | 1 |

| | | | | | | |
|---|---|---|---|---|---|---|
| Heart_Failure | pericyte | *LYRM4* | 3.816345101 | 1.73E-05 | 0.001740485 | 0.10867055 |
| Heart_Failure | pericyte | *PRCD* | 1.199788134 | 0.005195905 | 0.033538657 | 1 |
| Heart_Failure | pericyte | *COL28A1* | 3.318953774 | 0.000259378 | 0.005096468 | 1 |
| Heart_Failure | pericyte | *NPEPL1* | 4.345671149 | 7.62E-05 | 0.002686157 | 0.479296738 |
| Heart_Failure | pericyte | *CYB5RL* | 3.502963129 | 1.64E-05 | 0.001740485 | 0.103222326 |
| Heart_Failure | pericyte | *BTBD19* | 2.247529522 | 0.001131501 | 0.012328855 | 1 |
| Heart_Failure | pericyte | *VPS52* | 4.056898155 | 3.22E-05 | 0.001918699 | 0.202690315 |
| Heart_Failure | pericyte | *LINC01117* | 2.132056701 | 0.000281932 | 0.005306292 | 1 |
| Heart_Failure | pericyte | *SLC26A6* | 1.293738836 | 0.008925881 | 0.049010493 | 1 |
| Heart_Failure | pericyte | *ERCC6* | 3.750798113 | 0.00023225 | 0.004725416 | 1 |
| Heart_Failure | pericyte | *NOL7* | 3.359523022 | 0.001773607 | 0.016834554 | 1 |
| Heart_Failure | pericyte | *ELFN1* | 11.11514294 | 0.000127655 | 0.003330142 | 0.802564211 |
| Heart_Failure | pericyte | *AC064836.3* | 8.638393903 | 0.000154061 | 0.00371105 | 0.968584155 |
| Heart_Failure | pericyte | *HSBP1L1* | 1.568117424 | 0.005286079 | 0.033911818 | 1 |
| Heart_Failure | pericyte | *HCG20* | 3.040222717 | 0.005490772 | 0.034986225 | 1 |
| Heart_Failure | pericyte | *PROB1* | 1.944308287 | 0.003562246 | 0.026490062 | 1 |
| Heart_Failure | pericyte | *RPL41* | 6.711873776 | 2.11E-05 | 0.001740485 | 0.13275346 |
| Heart_Failure | pericyte | *PATL2* | 4.667221604 | 3.11E-05 | 0.001918699 | 0.195822309 |
| Heart_Failure | pericyte | *CISD3* | 3.10338255 | 0.000456556 | 0.006976539 | 1 |
| Heart_Failure | pericyte | *PPP5D1* | 0.98958512 | 0.00671028 | 0.040255279 | 1 |
| Heart_Failure | pericyte | *RPS18* | 2.279166677 | 3.18E-06 | 0.000909616 | 0.019964285 |
| Heart_Failure | pericyte | *HLA-DQB2* | 0.843734151 | 2.52E-14 | 3.96E-11 | 1.59E-10 |
| Heart_Failure | pericyte | *LINC00342* | 2.695098273 | 0.000448006 | 0.006942549 | 1 |
| Heart_Failure | pericyte | *AC123768.3* | 2.99035782 | 0.00188568 | 0.017589419 | 1 |
| Heart_Failure | pericyte | *ZNF736* | 1.145055487 | 0.005557264 | 0.03511409 | 1 |
| Heart_Failure | pericyte | *ANKRD65* | 1.621971857 | 0.00601869 | 0.037390812 | 1 |
| Heart_Failure | pericyte | *KIAA0040* | 3.562497113 | 1.60E-05 | 0.001740485 | 0.100545248 |
| Heart_Failure | pericyte | *HLA-DQA2* | 1.925835296 | 1.10E-22 | 3.45E-19 | 6.91E-19 |
| Heart_Failure | pericyte | *LRRC37A2* | 3.003841551 | 9.15E-05 | 0.002769335 | 0.574992226 |
| Heart_Failure | pericyte | *ASB14* | 1.771400399 | 0.001761001 | 0.016825856 | 1 |
| Heart_Failure | pericyte | *TNFSF12* | 1.314759093 | 7.93E-05 | 0.002693349 | 0.498269563 |
| Heart_Failure | pericyte | *PI4KA* | 2.029936546 | 0.006425529 | 0.039031206 | 1 |
| Heart_Failure | pericyte | *ARFGAP3* | 3.810232645 | 1.98E-05 | 0.001740485 | 0.124183733 |
| Heart_Failure | pericyte | *DECR2* | 2.291966767 | 0.001714315 | 0.01648789 | 1 |
| Heart_Failure | pericyte | *FGF14-IT1* | 20.08873979 | 2.84E-05 | 0.001861562 | 0.178660891 |
| Heart_Failure | pericyte | *NAT6* | 2.871420859 | 8.64E-05 | 0.002769335 | 0.543157753 |
| Heart_Failure | pericyte | *WDR92* | 1.884790386 | 0.003229105 | 0.024878445 | 1 |
| Heart_Failure | pericyte | *TTC4* | -0.528277621 | 0.000903356 | 0.010695671 | 1 |
| Heart_Failure | pericyte | *APOBEC3C* | 8.552670224 | 8.86E-05 | 0.002769335 | 0.557056046 |
| Heart_Failure | pericyte | *UBE2V1* | 6.303228568 | 2.18E-05 | 0.001740485 | 0.136972759 |
| Heart_Failure | pericyte | *N4BP2L2* | 1.803951876 | 0.003888373 | 0.027748244 | 1 |
| Heart_Failure | pericyte | *RAB30-AS1* | 1.926011706 | 0.002193636 | 0.019208062 | 1 |
| Heart_Failure | pericyte | *PGAM5* | 2.946956748 | 0.000915977 | 0.010743937 | 1 |
| Heart_Failure | pericyte | *C5orf17* | 0.961092235 | 0.003467375 | 0.026044671 | 1 |
| Heart_Failure | pericyte | *YJEFN3* | 1.32921063 | 0.004423836 | 0.03010028 | 1 |
| Heart_Failure | pericyte | *FAM218A* | 7.131284079 | 0.000264277 | 0.005112345 | 1 |
| Heart_Failure | pericyte | *TMEM200B* | 1.482516151 | 0.000437115 | 0.006907621 | 1 |
| Heart_Failure | pericyte | *ZNF260* | 2.72010999 | 0.008560689 | 0.047507995 | 1 |
| Heart_Failure | pericyte | *FPGT* | 5.485103049 | 2.25E-05 | 0.001740485 | 0.141329429 |
| Heart_Failure | pericyte | *MPV17L2* | 1.827698528 | 0.000247722 | 0.004928565 | 1 |

| | | | | | | |
|---|---|---|---|---|---|---|
| Heart_Failure | pericyte | *POLR2M* | 4.545179195 | 4.18E-05 | 0.002101368 | 0.262671018 |
| Heart_Failure | pericyte | *CTSO* | 3.796441048 | 3.26E-05 | 0.001918699 | 0.205015465 |
| Heart_Failure | pericyte | *ZNF253* | 2.85167582 | 0.004553607 | 0.030717305 | 1 |
| Heart_Failure | pericyte | *FNTB* | 2.811266683 | 0.001140314 | 0.012360614 | 1 |
| Heart_Failure | pericyte | *CEP95* | 2.51975979 | 0.001952917 | 0.017897943 | 1 |
| Heart_Failure | pericyte | *AC009120.5* | 1.693527083 | 0.004384357 | 0.029863979 | 1 |
| Heart_Failure | pericyte | *AC139100.2* | 4.223430273 | 0.005259588 | 0.033845475 | 1 |
| Heart_Failure | pericyte | *ZNF582-AS1* | 2.173312482 | 0.001642611 | 0.016136085 | 1 |
| Heart_Failure | pericyte | *YTHDF3-AS1* | 2.005976334 | 0.007206995 | 0.04230661 | 1 |
| Heart_Failure | pericyte | *CASC15* | 0.358023887 | 0.004614062 | 0.031058468 | 1 |
| Heart_Failure | pericyte | *LINC01023* | 2.24038894 | 0.005168407 | 0.033464241 | 1 |
| Heart_Failure | SMC | *SEMA3F* | -0.769649161 | 1.94E-06 | 0.001352293 | 0.01217064 |
| Heart_Failure | SMC | *RBM6* | -0.588023565 | 5.67E-07 | 0.000445764 | 0.003566109 |
| Heart_Failure | SMC | *COL16A1* | -1.31275357 | 0.000125185 | 0.041423073 | 0.787038391 |
| Heart_Failure | SMC | *PDLIM4* | 1.727947737 | 7.38E-05 | 0.029934561 | 0.464129032 |
| Heart_Failure | SMC | *REEP2* | -0.721375689 | 7.62E-05 | 0.029934561 | 0.47895298 |
| Heart_Failure | SMC | *RNF123* | 1.025275813 | 3.73E-06 | 0.002343819 | 0.023438187 |
| Heart_Failure | SMC | *ORMDL3* | -1.196635387 | 1.22E-11 | 1.54E-08 | 7.69E-08 |
| Heart_Failure | SMC | *DCAKD* | -0.656513263 | 8.38E-05 | 0.030694614 | 0.52665347 |
| Heart_Failure | SMC | *HLA-DQB1* | -1.242289119 | 6.73E-24 | 4.23E-20 | 4.23E-20 |
| Heart_Failure | SMC | *MUC1* | 1.180290802 | 2.51E-05 | 0.013133593 | 0.15760312 |
| Heart_Failure | SMC | *HLA-DRB1* | -1.353645056 | 3.57E-10 | 3.74E-07 | 2.25E-06 |
| Heart_Failure | SMC | *HLA-DQA1* | -1.477761662 | 7.31E-17 | 1.53E-13 | 4.60E-13 |
| Heart_Failure | SMC | *SLC22A5* | 0.941551662 | 1.56E-07 | 0.000140485 | 0.000983392 |
| Heart_Failure | SMC | *RPS26* | 0.465078413 | 0.000146845 | 0.046160763 | 0.923215268 |
| Heart_Failure | SMC | *HLA-DRA* | 1.070330801 | 6.77E-06 | 0.003872166 | 0.042593825 |
| Heart_Failure | SMC | *BAG6* | 1.255744617 | 8.79E-05 | 0.030694614 | 0.552503053 |
| Heart_Failure | SMC | *MICA* | 0.544822285 | 2.97E-05 | 0.014352203 | 0.186578643 |
| Heart_Failure | SMC | *ATF6B* | 1.160003096 | 6.07E-05 | 0.027281041 | 0.381934571 |
| Heart_Failure | SMC | *HLA-DQB2* | 0.517016148 | 1.37E-12 | 2.15E-09 | 8.59E-09 |
| Heart_Failure | SMC | *HLA-DQA2* | 1.330992763 | 1.13E-22 | 3.54E-19 | 7.09E-19 |
| Heart_Failure | t_cell | *RBM6* | -1.0804493 | 2.52E-06 | 0.001581608 | 0.015816083 |
| Heart_Failure | t_cell | *CDK2AP1* | 1.085242734 | 0.000173929 | 0.049457827 | 1 |
| Heart_Failure | t_cell | *PDLIM4* | 3.566491047 | 3.72E-06 | 0.001950378 | 0.023404539 |
| Heart_Failure | t_cell | *REEP2* | -1.461358087 | 5.00E-05 | 0.017477006 | 0.314586116 |
| Heart_Failure | t_cell | *NMT1* | 1.531886579 | 0.000180934 | 0.049457827 | 1 |
| Heart_Failure | t_cell | *TSLP* | 1.347170543 | 3.22E-05 | 0.012661871 | 0.202589943 |
| Heart_Failure | t_cell | *RNF123* | 1.933784599 | 7.35E-06 | 0.003555493 | 0.046221409 |
| Heart_Failure | t_cell | *C2* | 3.759818479 | 6.19E-05 | 0.020476151 | 0.389046874 |
| Heart_Failure | t_cell | *ORMDL3* | -1.464808166 | 1.73E-06 | 0.00135753 | 0.010860243 |
| Heart_Failure | t_cell | *HLA-DQB1* | -2.461290649 | 8.82E-23 | 5.54E-19 | 5.54E-19 |
| Heart_Failure | t_cell | *MUC1* | 2.430368241 | 1.48E-05 | 0.00619383 | 0.092907453 |
| Heart_Failure | t_cell | *MCRS1* | 3.478742721 | 4.46E-05 | 0.016490277 | 0.28033471 |
| Heart_Failure | t_cell | *HLA-DRB1* | -2.365097972 | 2.87E-08 | 3.60E-05 | 0.000180164 |
| Heart_Failure | t_cell | *HLA-DQA1* | -3.217217918 | 2.79E-16 | 5.84E-13 | 1.75E-12 |
| Heart_Failure | t_cell | *SLC22A5* | 1.911779805 | 4.42E-08 | 4.64E-05 | 0.000278145 |
| Heart_Failure | t_cell | *RPS26* | 0.970594754 | 6.73E-05 | 0.021162958 | 0.423259159 |
| Heart_Failure | t_cell | *HLA-DRA* | 2.113559938 | 3.84E-07 | 0.00034504 | 0.002415277 |
| Heart_Failure | t_cell | *PRRT1* | 1.830665818 | 0.000156588 | 0.046879519 | 0.984469904 |
| Heart_Failure | t_cell | *BAG6* | 2.91747236 | 2.23E-06 | 0.001555683 | 0.014001143 |

| | | | | | | |
|---|---|---|---|---|---|---|
| Heart_Failure | t_cell | *MICA* | 1.154470878 | 3.07E-06 | 0.001754649 | 0.019301135 |
| Heart_Failure | t_cell | *ATF6B* | 2.493382985 | 8.25E-06 | 0.003703732 | 0.051852248 |
| Heart_Failure | t_cell | *HLA-DQB2* | 1.078359827 | 7.91E-14 | 1.24E-10 | 4.98E-10 |
| Heart_Failure | t_cell | *HLA-DQA2* | 2.143056408 | 7.22E-20 | 2.27E-16 | 4.54E-16 |

d. Identified trait-associated genes from CTS TWAS analysis in visceral adipose tissue

| disorder | celltype | gene | Estimates | Pvalues | Pvalues_BH | Pvalue_Bonferroni |
|---|---|---|---|---|---|---|
| CAD | adipocyte | *HS3ST1* | 0.352918631 | 0.009538934 | 0.042134669 | 1 |
| CAD | adipocyte | *TMEM176A* | 1.208011339 | 0.010107857 | 0.043433303 | 1 |
| CAD | adipocyte | *ALS2* | 1.173478292 | 0.003910992 | 0.024297225 | 1 |
| CAD | adipocyte | *RBM6* | -0.19121658 | 0.009474718 | 0.042059772 | 1 |
| CAD | adipocyte | *POLR2J* | 2.095674105 | 0.000145736 | 0.009084269 | 0.707109468 |
| CAD | adipocyte | *CIAPIN1* | 0.99659324 | 0.000806741 | 0.017317396 | 1 |
| CAD | adipocyte | *GCFC2* | 1.100141694 | 0.004858673 | 0.027865581 | 1 |
| CAD | adipocyte | *SYNRG* | 1.033125546 | 0.002509407 | 0.019127952 | 1 |
| CAD | adipocyte | *ZNF263* | 1.525461246 | 0.000419506 | 0.014334098 | 1 |
| CAD | adipocyte | *BAIAP2L1* | 1.036322625 | 0.006330726 | 0.032622856 | 1 |
| CAD | adipocyte | *AGK* | 0.689822419 | 0.003716024 | 0.023439618 | 1 |
| CAD | adipocyte | *FARP2* | 1.049995077 | 0.001299888 | 0.017317396 | 1 |
| CAD | adipocyte | *VPS41* | 1.14408336 | 0.001172725 | 0.017317396 | 1 |
| CAD | adipocyte | *PAX6* | 0.237271575 | 0.008858093 | 0.040356305 | 1 |
| CAD | adipocyte | *TSR3* | 1.949809079 | 0.001641313 | 0.017317396 | 1 |
| CAD | adipocyte | *PSMB1* | 1.550810923 | 0.00119664 | 0.017317396 | 1 |
| CAD | adipocyte | *PKD1* | 1.321013349 | 0.000918484 | 0.017317396 | 1 |
| CAD | adipocyte | *MED24* | 1.351786729 | 0.00785352 | 0.037031368 | 1 |
| CAD | adipocyte | *RPS20* | 1.424750595 | 5.19E-05 | 0.006632491 | 0.252034647 |
| CAD | adipocyte | *POMT2* | 0.560515813 | 0.004769196 | 0.027515026 | 1 |
| CAD | adipocyte | *PPP5C* | 0.730196667 | 0.003657792 | 0.023229851 | 1 |
| CAD | adipocyte | *CEP68* | 0.793273113 | 0.000323021 | 0.013060813 | 1 |
| CAD | adipocyte | *MAP4K5* | 1.110543524 | 0.001537808 | 0.017317396 | 1 |
| CAD | adipocyte | *RNF14* | 0.822368468 | 0.012279603 | 0.04916762 | 1 |
| CAD | adipocyte | *ZC3H3* | 0.377095002 | 0.003530069 | 0.022806785 | 1 |
| CAD | adipocyte | *MDH1* | 0.656866186 | 0.000220226 | 0.010928681 | 1 |
| CAD | adipocyte | *NUDCD3* | 0.430717717 | 0.006364377 | 0.032746511 | 1 |
| CAD | adipocyte | *CHDH* | 1.604288387 | 0.010123535 | 0.043433303 | 1 |
| CAD | adipocyte | *RUFY3* | 0.913993802 | 0.003349872 | 0.02208367 | 1 |
| CAD | adipocyte | *WWTR1* | 1.226196161 | 0.000978723 | 0.017317396 | 1 |
| CAD | adipocyte | *TTC27* | 1.102665086 | 0.001426311 | 0.017317396 | 1 |
| CAD | adipocyte | *HGF* | 0.857816471 | 0.004341195 | 0.026165812 | 1 |
| CAD | adipocyte | *OSBPL5* | 0.853110081 | 0.011654738 | 0.047882123 | 1 |
| CAD | adipocyte | *ZDHHC6* | 1.327166728 | 0.003017779 | 0.020710417 | 1 |
| CAD | adipocyte | *SEC63* | 0.962899784 | 0.002981949 | 0.020593222 | 1 |
| CAD | adipocyte | *FAS* | 0.722856394 | 0.011341682 | 0.047034054 | 1 |
| CAD | adipocyte | *AGPAT4* | 0.763383951 | 0.009150315 | 0.040994762 | 1 |
| CAD | adipocyte | *VRK2* | 1.083737298 | 0.000694173 | 0.017317396 | 1 |
| CAD | adipocyte | *VEZT* | 1.228086037 | 0.001516023 | 0.017317396 | 1 |
| CAD | adipocyte | *BRD9* | 1.000741594 | 0.011157427 | 0.046428676 | 1 |
| CAD | adipocyte | *ANK1* | 3.029696431 | 0.001372269 | 0.017317396 | 1 |
| CAD | adipocyte | *ATP6V0A1* | 1.144031768 | 3.68E-05 | 0.00557455 | 0.178385611 |
| CAD | adipocyte | *PIAS1* | 0.866256584 | 0.001023432 | 0.017317396 | 1 |
| CAD | adipocyte | *VCL* | 1.097293988 | 0.005130096 | 0.028582558 | 1 |
| CAD | adipocyte | *ADSS* | 1.463795646 | 0.001002959 | 0.017317396 | 1 |
| CAD | adipocyte | *TBC1D23* | 1.28062039 | 0.002730083 | 0.01980024 | 1 |

| | | | | | | |
|---|---|---|---|---|---|---|
| CAD | adipocyte | *TDP1* | 1.396967235 | 0.008603145 | 0.039528846 | 1 |
| CAD | adipocyte | *AP2S1* | 1.470885588 | 0.009815076 | 0.042787737 | 1 |
| CAD | adipocyte | *CTNNA1* | 1.134198768 | 0.001248956 | 0.017317396 | 1 |
| CAD | adipocyte | *YTHDC2* | 1.26109602 | 0.001413501 | 0.017317396 | 1 |
| CAD | adipocyte | *POLR2B* | 1.172574073 | 0.002019703 | 0.018113863 | 1 |
| CAD | adipocyte | *ARAP2* | 0.80351888 | 0.000178388 | 0.010064395 | 0.865537954 |
| CAD | adipocyte | *ANO2* | 0.810548489 | 0.00647399 | 0.032926414 | 1 |
| CAD | adipocyte | *MAPK9* | 1.550053207 | 0.001878547 | 0.017489118 | 1 |
| CAD | adipocyte | *CYBA* | 1.057961288 | 0.002208423 | 0.018635253 | 1 |
| CAD | adipocyte | *THOC3* | 4.271874897 | 0.001565064 | 0.017317396 | 1 |
| CAD | adipocyte | *LY75* | 1.382315727 | 0.005282467 | 0.028895751 | 1 |
| CAD | adipocyte | *TBC1D22A* | 0.849513003 | 0.003663733 | 0.023237168 | 1 |
| CAD | adipocyte | *GINM1* | 0.730877425 | 0.008974456 | 0.040581603 | 1 |
| CAD | adipocyte | *ATP11B* | 0.900449647 | 0.000578974 | 0.016524601 | 1 |
| CAD | adipocyte | *PRDM6* | 1.078859805 | 0.009295669 | 0.041378519 | 1 |
| CAD | adipocyte | *NCKAP1* | 0.862798469 | 0.011845835 | 0.048258598 | 1 |
| CAD | adipocyte | *ADCK1* | 1.773224959 | 0.004368908 | 0.026250054 | 1 |
| CAD | adipocyte | *RNF4* | 1.555280176 | 0.001700278 | 0.017317396 | 1 |
| CAD | adipocyte | *CASP8* | 1.066731104 | 0.001644025 | 0.017317396 | 1 |
| CAD | adipocyte | *TM7SF3* | 1.087487019 | 0.002723901 | 0.019784984 | 1 |
| CAD | adipocyte | *SUGP2* | 1.168168185 | 0.002476394 | 0.019127952 | 1 |
| CAD | adipocyte | *SLC12A2* | 1.559449967 | 0.000627689 | 0.017302381 | 1 |
| CAD | adipocyte | *PMS1* | 1.055844298 | 0.004986738 | 0.028202564 | 1 |
| CAD | adipocyte | *UHRF1BP1* | 0.591760458 | 6.93E-07 | 0.001682 | 0.003364001 |
| CAD | adipocyte | *MAP2K4* | 1.036556924 | 0.004678825 | 0.027311752 | 1 |
| CAD | adipocyte | *GOLGA5* | 1.512367902 | 0.000551754 | 0.016127182 | 1 |
| CAD | adipocyte | *TRAM1* | 0.828897962 | 0.00511357 | 0.028582558 | 1 |
| CAD | adipocyte | *DHX8* | 1.719771657 | 0.001289534 | 0.017317396 | 1 |
| CAD | adipocyte | *ROGDI* | 3.571161308 | 0.001315776 | 0.017317396 | 1 |
| CAD | adipocyte | *CBFB* | 1.034572378 | 0.006756458 | 0.033829428 | 1 |
| CAD | adipocyte | *IP6K2* | 1.133672052 | 0.001253459 | 0.017317396 | 1 |
| CAD | adipocyte | *SRBD1* | 1.192244497 | 0.001256195 | 0.017317396 | 1 |
| CAD | adipocyte | *CYFIP1* | 1.001602837 | 0.004014147 | 0.024729689 | 1 |
| CAD | adipocyte | *NUCKS1* | 1.045615109 | 0.004158879 | 0.025285551 | 1 |
| CAD | adipocyte | *CLEC2D* | 0.882590512 | 0.002828241 | 0.019974713 | 1 |
| CAD | adipocyte | *NEDD4* | 0.547042503 | 0.001647441 | 0.017317396 | 1 |
| CAD | adipocyte | *GNB5* | 0.368674117 | 0.004587492 | 0.027012756 | 1 |
| CAD | adipocyte | *LRP6* | 1.205151253 | 0.002123473 | 0.018430441 | 1 |
| CAD | adipocyte | *DGCR2* | 0.903274868 | 0.003808064 | 0.023782688 | 1 |
| CAD | adipocyte | *ZXDC* | 1.483207914 | 0.000119008 | 0.008341315 | 0.577428291 |
| CAD | adipocyte | *GBA2* | 1.679080425 | 0.001200316 | 0.017317396 | 1 |
| CAD | adipocyte | *MAP4K4* | 0.890616958 | 0.004084849 | 0.025024858 | 1 |
| CAD | adipocyte | *WDR1* | 1.157153517 | 0.001545247 | 0.017317396 | 1 |
| CAD | adipocyte | *SNX13* | 1.190085147 | 0.001258854 | 0.017317396 | 1 |
| CAD | adipocyte | *RPS6KA2* | 0.683816468 | 0.003601454 | 0.023027229 | 1 |
| CAD | adipocyte | *TRIB2* | 0.345605758 | 0.012379366 | 0.049307203 | 1 |
| CAD | adipocyte | *AFF4* | 0.902801776 | 0.004016281 | 0.024729689 | 1 |
| CAD | adipocyte | *TRNT1* | 1.162443178 | 0.000886299 | 0.017317396 | 1 |

| | | | | | | |
|---|---|---|---|---|---|---|
| CAD | adipocyte | *CRMP1* | 1.362447619 | 0.00657802 | 0.03335063 | 1 |
| CAD | adipocyte | *SCARB1* | 0.856305172 | 0.000394953 | 0.013687955 | 1 |
| CAD | adipocyte | *LLGL2* | 1.65792206 | 0.00015425 | 0.009261544 | 0.748422047 |
| CAD | adipocyte | *PPP2R2C* | 1.775429995 | 0.00957241 | 0.042134669 | 1 |
| CAD | adipocyte | *ZNF532* | 1.364909201 | 0.000790402 | 0.017317396 | 1 |
| CAD | adipocyte | *ZZEF1* | 1.248378784 | 9.18E-05 | 0.007830023 | 0.44526405 |
| CAD | adipocyte | *TIPIN* | 0.313471469 | 0.000128351 | 0.008530929 | 0.622757793 |
| CAD | adipocyte | *ADD2* | 0.958292378 | 0.000600821 | 0.016753936 | 1 |
| CAD | adipocyte | *FNDC3B* | 0.999522869 | 0.000819236 | 0.017317396 | 1 |
| CAD | adipocyte | *KIFAP3* | 0.799512462 | 0.012260507 | 0.049163621 | 1 |
| CAD | adipocyte | *FMO4* | 1.634403764 | 5.45E-09 | 2.64E-05 | 2.64E-05 |
| CAD | adipocyte | *ANKRD13A* | 1.511925756 | 0.000861836 | 0.017317396 | 1 |
| CAD | adipocyte | *PAG1* | 0.77297326 | 0.007479542 | 0.035851767 | 1 |
| CAD | adipocyte | *XAB2* | 2.485992025 | 0.000155556 | 0.009261544 | 0.754757002 |
| CAD | adipocyte | *IL4R* | 0.738380863 | 0.006946737 | 0.034352818 | 1 |
| CAD | adipocyte | *DYNC1I2* | 1.227377002 | 0.001670742 | 0.017317396 | 1 |
| CAD | adipocyte | *AMPH* | 0.734143063 | 0.004471511 | 0.02662058 | 1 |
| CAD | adipocyte | *PIK3C3* | 1.266150326 | 0.000499086 | 0.01542398 | 1 |
| CAD | adipocyte | *PCM1* | 0.935597099 | 0.001227896 | 0.017317396 | 1 |
| CAD | adipocyte | *XRCC5* | 1.28827524 | 0.000132759 | 0.008657898 | 0.644146909 |
| CAD | adipocyte | *DNM2* | 1.301310968 | 0.000124425 | 0.008409031 | 0.603711165 |
| CAD | adipocyte | *STX7* | 1.032922727 | 0.00054248 | 0.016090781 | 1 |
| CAD | adipocyte | *PCDHB4* | 4.110683139 | 0.001192481 | 0.017317396 | 1 |
| CAD | adipocyte | *ME2* | 1.331301843 | 0.001699707 | 0.017317396 | 1 |
| CAD | adipocyte | *ZCCHC6* | 1.158404059 | 0.001413869 | 0.017317396 | 1 |
| CAD | adipocyte | *PIBF1* | 1.08023275 | 0.001165185 | 0.017317396 | 1 |
| CAD | adipocyte | *ZNF264* | 1.675708437 | 0.001320858 | 0.017317396 | 1 |
| CAD | adipocyte | *CHMP2B* | 1.238740403 | 0.002875698 | 0.020221576 | 1 |
| CAD | adipocyte | *WBP11* | 1.79699468 | 0.001559776 | 0.017317396 | 1 |
| CAD | adipocyte | *NCOA1* | 0.918014082 | 0.003261934 | 0.021710427 | 1 |
| CAD | adipocyte | *HACE1* | 1.386494694 | 0.001844377 | 0.017489118 | 1 |
| CAD | adipocyte | *SLC25A24* | 1.017875849 | 0.006784243 | 0.033849217 | 1 |
| CAD | adipocyte | *DDHD2* | 0.664112939 | 0.003280432 | 0.021773812 | 1 |
| CAD | adipocyte | *MAST2* | 0.672976969 | 0.007696552 | 0.036539796 | 1 |
| CAD | adipocyte | *EIF2AK1* | 1.337360794 | 0.003462815 | 0.022492071 | 1 |
| CAD | adipocyte | *TRIP6* | 1.165881988 | 0.001395579 | 0.017317396 | 1 |
| CAD | adipocyte | *SRRT* | 1.453262628 | 0.001313381 | 0.017317396 | 1 |
| CAD | adipocyte | *ADAMTS2* | 1.144413473 | 0.008152431 | 0.038070832 | 1 |
| CAD | adipocyte | *UIMC1* | 1.297757775 | 0.000693438 | 0.017317396 | 1 |
| CAD | adipocyte | *LPCAT2* | 0.739581565 | 0.011970344 | 0.048613338 | 1 |
| CAD | adipocyte | *AAMDC* | 0.382455717 | 0.004875121 | 0.027926905 | 1 |
| CAD | adipocyte | *ASAP3* | 1.743004169 | 7.09E-05 | 0.007733492 | 0.343874896 |
| CAD | adipocyte | *ANKRD10* | 0.966088141 | 0.004193647 | 0.025402713 | 1 |
| CAD | adipocyte | *DOCK3* | 1.426563787 | 0.001531886 | 0.017317396 | 1 |
| CAD | adipocyte | *ARHGAP28* | 0.897069594 | 0.001645545 | 0.017317396 | 1 |
| CAD | adipocyte | *EBF4* | 0.832506209 | 0.012006407 | 0.048613338 | 1 |
| CAD | adipocyte | *SNX5* | 1.656112183 | 0.001630442 | 0.017317396 | 1 |
| CAD | adipocyte | *RPH3A* | 1.830341434 | 0.003474468 | 0.02253759 | 1 |
| CAD | adipocyte | *KIF16B* | 1.334031672 | 0.001710725 | 0.017317396 | 1 |
| CAD | adipocyte | *ZNF302* | 0.519500223 | 0.001910394 | 0.017489118 | 1 |

| | | | | | | |
|---|---|---|---|---|---|---|
| CAD | adipocyte | *GRAMD1A* | 1.486358922 | 0.001619769 | 0.017317396 | 1 |
| CAD | adipocyte | *CDIP1* | 1.72419712 | 0.0020853 | 0.018429644 | 1 |
| CAD | adipocyte | *LYZ* | 0.535574107 | 0.004534242 | 0.026839221 | 1 |
| CAD | adipocyte | *SPG21* | 0.639070579 | 0.009858752 | 0.042862603 | 1 |
| CAD | adipocyte | *RAB11FIP3* | 0.949994515 | 0.000764144 | 0.017317396 | 1 |
| CAD | adipocyte | *CERS4* | 1.326756336 | 0.005190087 | 0.028681435 | 1 |
| CAD | adipocyte | *USP48* | 0.663754073 | 0.002196365 | 0.018565786 | 1 |
| CAD | adipocyte | *GLG1* | 1.107760539 | 0.001593401 | 0.017317396 | 1 |
| CAD | adipocyte | *DTX2* | 1.990788711 | 0.001597306 | 0.017317396 | 1 |
| CAD | adipocyte | *PPP2R3C* | 0.645720395 | 0.003241307 | 0.021655208 | 1 |
| CAD | adipocyte | *SCFD1* | 1.068563317 | 0.002983725 | 0.020593222 | 1 |
| CAD | adipocyte | *SUPT16H* | 1.54670808 | 0.001346421 | 0.017317396 | 1 |
| CAD | adipocyte | *COMT* | 1.271475829 | 0.001662494 | 0.017317396 | 1 |
| CAD | adipocyte | *LRRFIP2* | 1.071552755 | 0.001632607 | 0.017317396 | 1 |
| CAD | adipocyte | *SEC22C* | 1.540120103 | 0.002452948 | 0.019127952 | 1 |
| CAD | adipocyte | *XYLB* | 1.704406365 | 0.001630025 | 0.017317396 | 1 |
| CAD | adipocyte | *MSH2* | 1.626253583 | 0.001214021 | 0.017317396 | 1 |
| CAD | adipocyte | *PSMD5* | 1.302273301 | 0.005456278 | 0.029561594 | 1 |
| CAD | adipocyte | *WAC* | 0.960967755 | 0.002930655 | 0.020459936 | 1 |
| CAD | adipocyte | *ITPR3* | 0.609999129 | 3.49E-06 | 0.002461025 | 0.016943883 |
| CAD | adipocyte | *SCD* | -0.369181538 | 0.007783003 | 0.036806169 | 1 |
| CAD | adipocyte | *RAB18* | 1.349136886 | 0.002330675 | 0.019037772 | 1 |
| CAD | adipocyte | *MARCH2* | 0.34297207 | 0.009426717 | 0.041885012 | 1 |
| CAD | adipocyte | *MTAP* | 1.13615195 | 1.95E-05 | 0.003854781 | 0.09466903 |
| CAD | adipocyte | *MED15* | 1.055574512 | 0.005781661 | 0.030625131 | 1 |
| CAD | adipocyte | *LZTR1* | 0.812283765 | 0.00250738 | 0.019127952 | 1 |
| CAD | adipocyte | *SMARCB1* | 0.305982107 | 0.008400749 | 0.038856466 | 1 |
| CAD | adipocyte | *CABIN1* | 0.821492982 | 0.009922912 | 0.042949124 | 1 |
| CAD | adipocyte | *SUSD2* | -0.155155666 | 0.004542432 | 0.026845164 | 1 |
| CAD | adipocyte | *CCDC134* | 0.658134141 | 0.002582442 | 0.019276933 | 1 |
| CAD | adipocyte | *TTC28* | 0.915011671 | 0.001160964 | 0.017317396 | 1 |
| CAD | adipocyte | *CENPM* | -0.40464672 | 0.008579802 | 0.039491768 | 1 |
| CAD | adipocyte | *TIMP3* | 0.9191352 | 0.002947871 | 0.020491506 | 1 |
| CAD | adipocyte | *PPP6R2* | 0.903082095 | 0.011969119 | 0.048613338 | 1 |
| CAD | adipocyte | *CYB5R3* | 0.639266271 | 0.003462612 | 0.022492071 | 1 |
| CAD | adipocyte | *TSPO* | 1.52437439 | 0.002432365 | 0.019127952 | 1 |
| CAD | adipocyte | *RBM23* | 0.723172891 | 0.000350444 | 0.013592277 | 1 |
| CAD | adipocyte | *NIN* | 1.081765957 | 0.001130305 | 0.017317396 | 1 |
| CAD | adipocyte | *CNIH1* | 1.425616297 | 0.000192762 | 0.010214126 | 0.93527898 |
| CAD | adipocyte | *ATP6V1D* | 1.209867358 | 0.00515126 | 0.028582558 | 1 |
| CAD | adipocyte | *GSKIP* | 2.016293954 | 0.001863765 | 0.017489118 | 1 |
| CAD | adipocyte | *YY1* | 1.390296848 | 0.001764885 | 0.017475962 | 1 |
| CAD | adipocyte | *CCNB1IP1* | 1.313322468 | 0.01094633 | 0.045785858 | 1 |
| CAD | adipocyte | *CINP* | 1.740363135 | 0.000852969 | 0.017317396 | 1 |
| CAD | adipocyte | *TRPC4AP* | 1.029986788 | 8.22E-05 | 0.007733492 | 0.398924039 |
| CAD | adipocyte | *ABHD12* | 0.833364068 | 0.000200694 | 0.010359248 | 0.973769309 |
| CAD | adipocyte | *NDRG3* | 1.538626422 | 7.52E-05 | 0.007733492 | 0.36485552 |
| CAD | adipocyte | *CSTF1* | 1.969417797 | 0.002770008 | 0.019873367 | 1 |
| CAD | adipocyte | *CTSZ* | 1.114512577 | 0.002819514 | 0.019971215 | 1 |
| CAD | adipocyte | *DIDO1* | 1.222836582 | 0.000156264 | 0.009261544 | 0.758190863 |

| | | | | | | |
|---|---|---|---|---|---|---|
| CAD | adipocyte | *GMEB2* | 1.857038761 | 0.00227349 | 0.018849915 | 1 |
| CAD | adipocyte | *SEC23B* | 0.393975395 | 0.006932511 | 0.034323005 | 1 |
| CAD | adipocyte | *METTL4* | 1.302642232 | 0.005702962 | 0.030384947 | 1 |
| CAD | adipocyte | *LPIN2* | 1.060534273 | 0.003462524 | 0.022492071 | 1 |
| CAD | adipocyte | *MIB1* | 1.214543501 | 0.001520853 | 0.017317396 | 1 |
| CAD | adipocyte | *STK24* | 1.09456429 | 0.004700188 | 0.027311752 | 1 |
| CAD | adipocyte | *ARHGEF7* | 1.194597264 | 0.00188247 | 0.017489118 | 1 |
| CAD | adipocyte | *SUPT20H* | 1.109389864 | 0.000577949 | 0.016524601 | 1 |
| CAD | adipocyte | *MRPS31* | 1.419347289 | 0.001094344 | 0.017317396 | 1 |
| CAD | adipocyte | *KATNAL1* | 1.316486519 | 0.001570449 | 0.017317396 | 1 |
| CAD | adipocyte | *INTS6* | 1.041552399 | 0.002100257 | 0.018430441 | 1 |
| CAD | adipocyte | *CLN5* | 1.273369239 | 0.004793929 | 0.027586809 | 1 |
| CAD | adipocyte | *MGRN1* | 0.984955802 | 0.000400236 | 0.013772673 | 1 |
| CAD | adipocyte | *DHODH* | 0.609251241 | 0.002117623 | 0.018430441 | 1 |
| CAD | adipocyte | *PSMD7* | 1.738897014 | 8.64E-05 | 0.007830023 | 0.419170145 |
| CAD | adipocyte | *MON1B* | 2.027658192 | 0.002930679 | 0.020459936 | 1 |
| CAD | adipocyte | *LMF1* | 1.3606611 | 0.002742059 | 0.019834836 | 1 |
| CAD | adipocyte | *RBL2* | 0.895015957 | 0.000883386 | 0.017317396 | 1 |
| CAD | adipocyte | *KAT8* | 1.836818534 | 0.000536174 | 0.016058757 | 1 |
| CAD | adipocyte | *C16orf62* | 1.272510886 | 0.001379628 | 0.017317396 | 1 |
| CAD | adipocyte | *KNOP1* | 1.344805459 | 0.002678813 | 0.019693337 | 1 |
| CAD | adipocyte | *IQCH* | 1.496465283 | 0.002564595 | 0.019262254 | 1 |
| CAD | adipocyte | *HERC1* | 1.158023649 | 0.000383079 | 0.013616273 | 1 |
| CAD | adipocyte | *TTC23* | 1.407971722 | 0.001603387 | 0.017317396 | 1 |
| CAD | adipocyte | *SLC30A4* | 1.741227265 | 0.001256403 | 0.017317396 | 1 |
| CAD | adipocyte | *ZFAND1* | 1.304494557 | 0.002982295 | 0.020593222 | 1 |
| CAD | adipocyte | *NIPAL2* | 1.382578106 | 9.19E-05 | 0.007830023 | 0.446014983 |
| CAD | adipocyte | *RAB2A* | 0.766719565 | 0.005304022 | 0.028948387 | 1 |
| CAD | adipocyte | *CHRAC1* | 2.881949327 | 0.001482473 | 0.017317396 | 1 |
| CAD | adipocyte | *SNX16* | 1.180429661 | 0.001280075 | 0.017317396 | 1 |
| CAD | adipocyte | *GSDMD* | 1.076906932 | 0.001023183 | 0.017317396 | 1 |
| CAD | adipocyte | *ERICH1* | 0.825265741 | 0.007842728 | 0.037016454 | 1 |
| CAD | adipocyte | *TUSC3* | 0.709629797 | 0.010563657 | 0.044673236 | 1 |
| CAD | adipocyte | *SNRNP70* | 1.110843528 | 0.004672556 | 0.027311752 | 1 |
| CAD | adipocyte | *FCGRT* | 0.429556388 | 0.011122556 | 0.046363094 | 1 |
| CAD | adipocyte | *NOVA2* | 0.205958955 | 0.012356402 | 0.049280717 | 1 |
| CAD | adipocyte | *SGTA* | 1.186126836 | 0.004923454 | 0.028054408 | 1 |
| CAD | adipocyte | *MED25* | 2.000944385 | 0.000986138 | 0.017317396 | 1 |
| CAD | adipocyte | *ANKRD27* | 1.248653108 | 0.004944726 | 0.028060596 | 1 |
| CAD | adipocyte | *TGFB1* | 1.368264135 | 0.001234838 | 0.017317396 | 1 |
| CAD | adipocyte | *RABAC1* | 1.097499338 | 3.21E-05 | 0.005177026 | 0.155629163 |
| CAD | adipocyte | *GRIN2D* | 1.383996603 | 0.006978932 | 0.034352818 | 1 |
| CAD | adipocyte | *DNASE2* | 0.973635375 | 0.001006206 | 0.017317396 | 1 |
| CAD | adipocyte | *RAB3A* | 7.20135175 | 0.002427633 | 0.019127952 | 1 |
| CAD | adipocyte | *ZNF14* | 1.504388521 | 0.008185288 | 0.038114221 | 1 |
| CAD | adipocyte | *ZNF85* | 1.574644924 | 0.008266435 | 0.038418336 | 1 |
| CAD | adipocyte | *ETHE1* | 1.526657399 | 0.002350023 | 0.019062957 | 1 |
| CAD | adipocyte | *RASA4* | 15.14702786 | 0.001471167 | 0.017317396 | 1 |
| CAD | adipocyte | *DNAH11* | 1.320028595 | 0.003249988 | 0.021660636 | 1 |
| CAD | adipocyte | *STEAP1B* | 0.571115718 | 0.004899587 | 0.027991197 | 1 |

| | | | | | | |
|---|---|---|---|---|---|---|
| CAD | adipocyte | *IQCE* | 1.909357949 | 0.001272494 | 0.017317396 | 1 |
| CAD | adipocyte | *HIBADH* | 0.669310168 | 0.003395498 | 0.022293578 | 1 |
| CAD | adipocyte | *CPVL* | 0.689555362 | 0.006410321 | 0.032840444 | 1 |
| CAD | adipocyte | *CHN2* | 0.526948953 | 0.006051829 | 0.031607615 | 1 |
| CAD | adipocyte | *GRB10* | 1.343068327 | 0.000358688 | 0.013592277 | 1 |
| CAD | adipocyte | *PLEKHA8* | 1.688827222 | 0.000494862 | 0.01542398 | 1 |
| CAD | adipocyte | *CHCHD2* | 0.652220242 | 0.004903652 | 0.027991197 | 1 |
| CAD | adipocyte | *ZKSCAN1* | 1.247061129 | 0.000832179 | 0.017317396 | 1 |
| CAD | adipocyte | *EIF3B* | 1.405267677 | 0.00327316 | 0.021755303 | 1 |
| CAD | adipocyte | *C1GALT1* | 1.4375811 | 0.001142883 | 0.017317396 | 1 |
| CAD | adipocyte | *NRF1* | 0.760633647 | 0.009999206 | 0.043103145 | 1 |
| CAD | adipocyte | *TSPAN13* | 1.747080799 | 0.000195588 | 0.010214126 | 0.948990818 |
| CAD | adipocyte | *RARRES2* | 1.055990092 | 0.00103113 | 0.017317396 | 1 |
| CAD | adipocyte | *COA1* | 1.115465468 | 0.000960801 | 0.017317396 | 1 |
| CAD | adipocyte | *BLVRA* | 0.923551865 | 0.000375514 | 0.013616273 | 1 |
| CAD | adipocyte | *AEBP1* | 0.932925768 | 0.007277767 | 0.035176868 | 1 |
| CAD | adipocyte | *GCK* | 10.4011025 | 0.002357332 | 0.019062957 | 1 |
| CAD | adipocyte | *FKTN* | 1.328650858 | 0.002928558 | 0.020459936 | 1 |
| CAD | adipocyte | *MEGF9* | 1.408486603 | 0.001661593 | 0.017317396 | 1 |
| CAD | adipocyte | *CORO2A* | 0.873274679 | 0.001217849 | 0.017317396 | 1 |
| CAD | adipocyte | *ECM2* | 0.86461743 | 0.008945265 | 0.040525141 | 1 |
| CAD | adipocyte | *PTGDS* | 1.151466492 | 0.009197111 | 0.041043909 | 1 |
| CAD | adipocyte | *DNMBP* | 0.624336873 | 0.007034871 | 0.03446529 | 1 |
| CAD | adipocyte | *TRDMT1* | 1.463954077 | 0.001743913 | 0.017324379 | 1 |
| CAD | adipocyte | *ATE1* | 0.684281954 | 0.002685672 | 0.019713888 | 1 |
| CAD | adipocyte | *CDH23* | 1.126777564 | 0.00240665 | 0.019127952 | 1 |
| CAD | adipocyte | *BMPR1A* | 0.854147875 | 0.011745503 | 0.048020254 | 1 |
| CAD | adipocyte | *LIPA* | 0.510777784 | 5.10E-05 | 0.006632491 | 0.247669382 |
| CAD | adipocyte | *SFXN3* | 1.483495338 | 0.003230745 | 0.021621486 | 1 |
| CAD | adipocyte | *TNKS2* | 1.310788262 | 0.001234852 | 0.017317396 | 1 |
| CAD | adipocyte | *PITRM1* | 1.079128646 | 0.011360712 | 0.047065316 | 1 |
| CAD | adipocyte | *TFAM* | 1.896410936 | 0.001192828 | 0.017317396 | 1 |
| CAD | adipocyte | *RPL19* | 0.723692099 | 0.007977309 | 0.037469411 | 1 |
| CAD | adipocyte | *TRIM37* | 1.037277976 | 0.010160659 | 0.043512372 | 1 |
| CAD | adipocyte | *RAI1* | 1.582599099 | 0.001217921 | 0.017317396 | 1 |
| CAD | adipocyte | *GOSR1* | 1.329186272 | 0.001910366 | 0.017489118 | 1 |
| CAD | adipocyte | *ICAM2* | 1.156770055 | 0.005075472 | 0.028502532 | 1 |
| CAD | adipocyte | *RAB5C* | 0.728270587 | 0.000156522 | 0.009261544 | 0.759446631 |
| CAD | adipocyte | *NAGLU* | 1.305514555 | 0.000184847 | 0.010191792 | 0.896877702 |
| CAD | adipocyte | *CNTNAP1* | 1.136753067 | 7.92E-05 | 0.007733492 | 0.384195973 |
| CAD | adipocyte | *EFTUD2* | 1.652931971 | 0.001420576 | 0.017317396 | 1 |
| CAD | adipocyte | *FAM20A* | 0.885833325 | 0.004742653 | 0.027427118 | 1 |
| CAD | adipocyte | *DHRS7B* | 1.077369985 | 0.009875598 | 0.042866611 | 1 |
| CAD | adipocyte | *SLAIN2* | 0.94453757 | 0.012390313 | 0.049307203 | 1 |
| CAD | adipocyte | *UBE2D3* | 1.098187994 | 0.001425562 | 0.017317396 | 1 |
| CAD | adipocyte | *KLHL2* | 0.998543049 | 0.007465391 | 0.035827967 | 1 |
| CAD | adipocyte | *GAR1* | 1.747564082 | 0.004005029 | 0.024726056 | 1 |
| CAD | adipocyte | *STIM2* | 1.412976382 | 0.000120201 | 0.008341315 | 0.583216923 |
| CAD | adipocyte | *BST1* | 0.879409611 | 0.000978509 | 0.017317396 | 1 |
| CAD | adipocyte | *UFSP2* | 1.484336006 | 0.001618089 | 0.017317396 | 1 |

| | | | | | | |
|---|---|---|---|---|---|---|
| CAD | adipocyte | *KLHL5* | 1.222328604 | 0.002895074 | 0.020298987 | 1 |
| CAD | adipocyte | *ELP4* | 1.051527051 | 6.89E-05 | 0.007733492 | 0.334538349 |
| CAD | adipocyte | *DNAJC4* | 0.362101647 | 0.009784116 | 0.042729549 | 1 |
| CAD | adipocyte | *DCPS* | 1.203077037 | 0.007873304 | 0.037088614 | 1 |
| CAD | adipocyte | *CEP164* | 1.177430053 | 0.001240856 | 0.017317396 | 1 |
| CAD | adipocyte | *GALNT18* | 0.485519703 | 0.011036893 | 0.046085204 | 1 |
| CAD | adipocyte | *SLC35F2* | 1.374460207 | 0.000734172 | 0.017317396 | 1 |
| CAD | adipocyte | *TCIRG1* | 0.987134709 | 0.00247792 | 0.019127952 | 1 |
| CAD | adipocyte | *PRDM4* | 1.065611087 | 0.000333079 | 0.013356179 | 1 |
| CAD | adipocyte | *CORO1C* | 1.370510159 | 0.001461539 | 0.017317396 | 1 |
| CAD | adipocyte | *CAPRIN2* | 0.340107047 | 0.009060379 | 0.040893914 | 1 |
| CAD | adipocyte | *IL23A* | 10.10157512 | 0.004151686 | 0.025274753 | 1 |
| CAD | adipocyte | *PRR4* | 0.444382628 | 0.002452822 | 0.019127952 | 1 |
| CAD | adipocyte | *PARP11* | 1.495218506 | 0.001377637 | 0.017317396 | 1 |
| CAD | adipocyte | *GPN3* | 1.863262092 | 0.00519635 | 0.028683379 | 1 |
| CAD | adipocyte | *VPS29* | 1.101396266 | 0.006797269 | 0.033860727 | 1 |
| CAD | adipocyte | *CAND1* | 1.392460787 | 0.000804904 | 0.017317396 | 1 |
| CAD | adipocyte | *MDM1* | 1.705497818 | 0.002577624 | 0.019270618 | 1 |
| CAD | adipocyte | *GNPTAB* | 0.967665893 | 0.010121608 | 0.043433303 | 1 |
| CAD | adipocyte | *C12orf57* | 1.457768799 | 0.005489488 | 0.029561594 | 1 |
| CAD | adipocyte | *SUDS3* | 1.554065502 | 0.001666537 | 0.017317396 | 1 |
| CAD | adipocyte | *C2CD5* | 1.061330039 | 0.001582688 | 0.017317396 | 1 |
| CAD | adipocyte | *FRK* | 1.697278271 | 0.001809214 | 0.017489118 | 1 |
| CAD | adipocyte | *TMEM14C* | 1.742637602 | 0.001198627 | 0.017317396 | 1 |
| CAD | adipocyte | *SMIM8* | 0.454031322 | 0.005268632 | 0.028868018 | 1 |
| CAD | adipocyte | *RNGTT* | 1.264438352 | 0.000993948 | 0.017317396 | 1 |
| CAD | adipocyte | *CD83* | 1.31603248 | 0.012121733 | 0.048808839 | 1 |
| CAD | adipocyte | *FIG4* | 0.995120264 | 0.012062383 | 0.048650608 | 1 |
| CAD | adipocyte | *PACRG* | 1.35919235 | 0.003082169 | 0.021068977 | 1 |
| CAD | adipocyte | *QKI* | 0.903387752 | 0.003705533 | 0.023436034 | 1 |
| CAD | adipocyte | *EXOC2* | 1.308255869 | 0.000726062 | 0.017317396 | 1 |
| CAD | adipocyte | *TMEM30A* | 1.515020169 | 0.001641259 | 0.017317396 | 1 |
| CAD | adipocyte | *BTN2A1* | 1.242685084 | 8.99E-05 | 0.007830023 | 0.436137286 |
| CAD | adipocyte | *ARSB* | 1.403681294 | 0.002422714 | 0.019127952 | 1 |
| CAD | adipocyte | *SLC12A7* | 1.142240493 | 0.007527876 | 0.035950053 | 1 |
| CAD | adipocyte | *SKP1* | 1.485262569 | 0.001000081 | 0.017317396 | 1 |
| CAD | adipocyte | *PPWD1* | 1.329307733 | 0.002253289 | 0.018849915 | 1 |
| CAD | adipocyte | *TXNDC15* | 1.37304148 | 0.002310013 | 0.018932742 | 1 |
| CAD | adipocyte | *SMAD5* | 1.320947458 | 0.001360742 | 0.017317396 | 1 |
| CAD | adipocyte | *ERGIC1* | 1.293575579 | 0.001302752 | 0.017317396 | 1 |
| CAD | adipocyte | *ATP6V0E1* | 0.939014307 | 0.010815822 | 0.04551463 | 1 |
| CAD | adipocyte | *BNIP1* | 0.307867526 | 0.000496289 | 0.01542398 | 1 |
| CAD | adipocyte | *UNC5A* | 6.454702698 | 0.002793412 | 0.019873367 | 1 |
| CAD | adipocyte | *TBCCD1* | 1.994174944 | 0.001417435 | 0.017317396 | 1 |
| CAD | adipocyte | *PCCB* | 0.564765313 | 0.000259311 | 0.011672743 | 1 |
| CAD | adipocyte | *C3orf14* | 0.378649236 | 0.003622364 | 0.023095543 | 1 |
| CAD | adipocyte | *CBLB* | 0.711892455 | 0.004758648 | 0.027486858 | 1 |
| CAD | adipocyte | *PLXNA1* | 1.489268799 | 9.28E-06 | 0.002813584 | 0.045017337 |
| CAD | adipocyte | *SCAP* | 0.991089707 | 0.002779949 | 0.019873367 | 1 |
| CAD | adipocyte | *NEK11* | 0.696921058 | 0.001674952 | 0.017317396 | 1 |

| | | | | | | |
|---|---|---|---|---|---|---|
| CAD | adipocyte | *PLSCR4* | 0.78476514 | 0.007419332 | 0.0356775 | 1 |
| CAD | adipocyte | *COMMD2* | 1.464644838 | 0.000609518 | 0.016899319 | 1 |
| CAD | adipocyte | *VIPR1* | 1.13955575 | 0.00796333 | 0.037439999 | 1 |
| CAD | adipocyte | *NKTR* | 0.723112666 | 0.007665275 | 0.03645052 | 1 |
| CAD | adipocyte | *ACTR1B* | 1.282955262 | 0.008174481 | 0.038100461 | 1 |
| CAD | adipocyte | *NRBP1* | 1.164144878 | 0.011378322 | 0.047065316 | 1 |
| CAD | adipocyte | *PPM1G* | 1.041540261 | 0.006980041 | 0.034352818 | 1 |
| CAD | adipocyte | *POLE4* | 1.810727518 | 0.00022224 | 0.010928681 | 1 |
| CAD | adipocyte | *GGCX* | 1.02737708 | 5.71E-06 | 0.002461025 | 0.027726699 |
| CAD | adipocyte | *HDLBP* | 1.058398563 | 0.000388784 | 0.013669404 | 1 |
| CAD | adipocyte | *PASK* | -0.213170642 | 0.012106611 | 0.048788437 | 1 |
| CAD | adipocyte | *TPO* | 0.626153575 | 0.009756138 | 0.04264575 | 1 |
| CAD | adipocyte | *TAF1B* | 1.118544011 | 5.54E-05 | 0.006893412 | 0.268843078 |
| CAD | adipocyte | *NOL10* | 1.054317201 | 0.009877344 | 0.042866611 | 1 |
| CAD | adipocyte | *STRN* | 1.259560534 | 0.001214307 | 0.017317396 | 1 |
| CAD | adipocyte | *CEBPZ* | 1.451655848 | 0.000161602 | 0.009446908 | 0.78409334 |
| CAD | adipocyte | *PRKD3* | 1.159405076 | 0.000543876 | 0.016090781 | 1 |
| CAD | adipocyte | *SLC25A12* | 0.94647553 | 0.003106515 | 0.02116968 | 1 |
| CAD | adipocyte | *DARS* | 1.283148368 | 0.002389945 | 0.019127952 | 1 |
| CAD | adipocyte | *WIPF1* | 1.135505973 | 0.000383961 | 0.013616273 | 1 |
| CAD | adipocyte | *PNO1* | 2.20747509 | 0.004110179 | 0.025116611 | 1 |
| CAD | adipocyte | *AAK1* | 1.082069063 | 0.001937328 | 0.017702287 | 1 |
| CAD | adipocyte | *NFE2L2* | 0.787801771 | 0.001403946 | 0.017317396 | 1 |
| CAD | adipocyte | *MSH6* | 1.055649 | 0.00887534 | 0.040396951 | 1 |
| CAD | adipocyte | *ALMS1* | 1.426803424 | 0.001123575 | 0.017317396 | 1 |
| CAD | adipocyte | *DNAJC16* | 0.968774262 | 0.00166556 | 0.017317396 | 1 |
| CAD | adipocyte | *PARK7* | 0.988152503 | 0.010902524 | 0.045641973 | 1 |
| CAD | adipocyte | *EDEM3* | 1.032711115 | 0.009176354 | 0.041043909 | 1 |
| CAD | adipocyte | *CAPZA1* | 0.818766718 | 0.009591378 | 0.042134669 | 1 |
| CAD | adipocyte | *ASH1L* | 0.890406716 | 0.000167762 | 0.009671522 | 0.813980022 |
| CAD | adipocyte | *DOCK7* | 1.258713858 | 0.00085555 | 0.017317396 | 1 |
| CAD | adipocyte | *DNAJC6* | 0.512755244 | 0.009556776 | 0.042134669 | 1 |
| CAD | adipocyte | *IVNS1ABP* | 0.339197717 | 0.007802352 | 0.036861743 | 1 |
| CAD | adipocyte | *PRG4* | 1.05822798 | 0.008969305 | 0.040581603 | 1 |
| CAD | adipocyte | *CFHR3* | -0.565486452 | 0.009799441 | 0.042757993 | 1 |
| CAD | adipocyte | *TBCE* | 1.730964212 | 0.000491351 | 0.01542398 | 1 |
| CAD | adipocyte | *ZNHIT6* | 0.893898254 | 0.010246339 | 0.043803898 | 1 |
| CAD | adipocyte | *CD46* | 0.77909735 | 0.000222988 | 0.010928681 | 1 |
| CAD | adipocyte | *EBNA1BP2* | 0.885953317 | 0.010408525 | 0.04422256 | 1 |
| CAD | adipocyte | *ERI3* | 1.455769276 | 0.000288858 | 0.01225006 | 1 |
| CAD | adipocyte | *PRRC2C* | 1.352673975 | 6.09E-06 | 0.002461025 | 0.029532298 |
| CAD | adipocyte | *ABCD3* | 1.366758723 | 0.002617264 | 0.019387729 | 1 |
| CAD | adipocyte | *MTFR1L* | 1.066354057 | 0.005675337 | 0.030326801 | 1 |
| CAD | adipocyte | *RPA2* | 0.879064946 | 0.003931035 | 0.024390514 | 1 |
| CAD | adipocyte | *CD3EAP* | 1.390159963 | 4.94E-05 | 0.006632491 | 0.239728595 |
| CAD | adipocyte | *CAMSAP2* | 0.746528812 | 0.001284529 | 0.017317396 | 1 |
| CAD | adipocyte | *KLF7* | 0.706416779 | 0.003562293 | 0.022932654 | 1 |
| CAD | adipocyte | *SGIP1* | 1.017816208 | 0.007027833 | 0.03446529 | 1 |
| CAD | adipocyte | *FOXO3* | 0.903229806 | 0.004107192 | 0.025116611 | 1 |
| CAD | adipocyte | *MFSD1* | 0.90717516 | 0.01199358 | 0.048613338 | 1 |

| | | | | | | |
|---|---|---|---|---|---|---|
| CAD | adipocyte | *UBN1* | 1.22511323 | 0.002394199 | 0.019127952 | 1 |
| CAD | adipocyte | *WDR35* | 1.474466396 | 3.31E-05 | 0.005177026 | 0.160487806 |
| CAD | adipocyte | *C2orf40* | 1.404271744 | 1.33E-05 | 0.003229199 | 0.064583978 |
| CAD | adipocyte | *SENP5* | 1.118259685 | 0.005179255 | 0.028654215 | 1 |
| CAD | adipocyte | *FKBP15* | 0.446202712 | 0.001198276 | 0.017317396 | 1 |
| CAD | adipocyte | *RAB14* | 1.339150428 | 0.001590932 | 0.017317396 | 1 |
| CAD | adipocyte | *TRIM32* | 2.399766069 | 0.000144913 | 0.009084269 | 0.703116317 |
| CAD | adipocyte | *PPP6C* | 1.07833296 | 0.010838185 | 0.045524493 | 1 |
| CAD | adipocyte | *MAPKAP1* | 1.347515903 | 0.000169431 | 0.009671522 | 0.822079377 |
| CAD | adipocyte | *DENND1A* | 0.994703223 | 0.000780597 | 0.017317396 | 1 |
| CAD | adipocyte | *KDSR* | 1.427145606 | 0.001386651 | 0.017317396 | 1 |
| CAD | adipocyte | *NEK9* | 1.367265933 | 8.14E-05 | 0.007733492 | 0.39504161 |
| CAD | adipocyte | *TTLL5* | 1.30303263 | 0.000299694 | 0.012535493 | 1 |
| CAD | adipocyte | *FLVCR2* | 0.709642136 | 0.006784793 | 0.033849217 | 1 |
| CAD | adipocyte | *EIF2B2* | 0.446272185 | 6.30E-05 | 0.007638908 | 0.305556331 |
| CAD | adipocyte | *CRIPT* | 1.321919907 | 0.012319729 | 0.049197797 | 1 |
| CAD | adipocyte | *SLC17A5* | 0.881362667 | 0.001570239 | 0.017317396 | 1 |
| CAD | adipocyte | *PPP1R3C* | 1.441336411 | 0.003568262 | 0.022932654 | 1 |
| CAD | adipocyte | *MXI1* | 1.029545717 | 0.004005499 | 0.024726056 | 1 |
| CAD | adipocyte | *WDR11* | 0.940054061 | 0.003413205 | 0.022379554 | 1 |
| CAD | adipocyte | *KANSL1* | 0.903515048 | 0.00179464 | 0.017489118 | 1 |
| CAD | adipocyte | *HOXB3* | 1.402838584 | 0.001246113 | 0.017317396 | 1 |
| CAD | adipocyte | *PCDHB10* | 0.774231231 | 0.011427199 | 0.047146912 | 1 |
| CAD | adipocyte | *GORAB* | 1.46346659 | 0.001153385 | 0.017317396 | 1 |
| CAD | adipocyte | *SLC10A7* | 1.282298951 | 0.001471587 | 0.017317396 | 1 |
| CAD | adipocyte | *DNAJC15* | 1.466870194 | 0.000216765 | 0.010928681 | 1 |
| CAD | adipocyte | *SIL1* | 0.985018413 | 0.004606508 | 0.027076951 | 1 |
| CAD | adipocyte | *ZNF549* | 1.877603844 | 0.000798249 | 0.017317396 | 1 |
| CAD | adipocyte | *A1BG* | 0.877847098 | 0.003119776 | 0.021174108 | 1 |
| CAD | adipocyte | *ZSCAN18* | 0.82725806 | 0.01201681 | 0.048613338 | 1 |
| CAD | adipocyte | *LHX4* | 2.144543904 | 0.003368486 | 0.022176245 | 1 |
| CAD | adipocyte | *SEC22A* | 1.236155724 | 0.001734595 | 0.017317396 | 1 |
| CAD | adipocyte | *ZCCHC17* | 1.477938479 | 0.003120257 | 0.021174108 | 1 |
| CAD | adipocyte | *TNFSF10* | 1.209133531 | 0.002426102 | 0.019127952 | 1 |
| CAD | adipocyte | *ZNF639* | 1.601367647 | 0.004163882 | 0.025285551 | 1 |
| CAD | adipocyte | *RBBP6* | 1.099614071 | 0.003644458 | 0.023205918 | 1 |
| CAD | adipocyte | *NAA60* | 2.56663324 | 0.001525532 | 0.017317396 | 1 |
| CAD | adipocyte | *ODF2L* | 1.295887958 | 0.000748297 | 0.017317396 | 1 |
| CAD | adipocyte | *SPATA1* | 1.474108672 | 0.000266579 | 0.011758536 | 1 |
| CAD | adipocyte | *KLHL7* | 1.367357667 | 0.001781837 | 0.017489118 | 1 |
| CAD | adipocyte | *NT5C3A* | 1.027263861 | 0.004880907 | 0.027927074 | 1 |
| CAD | adipocyte | *SMU1* | 1.484563831 | 0.000384466 | 0.013616273 | 1 |
| CAD | adipocyte | *RBM19* | 1.608243591 | 0.001393167 | 0.017317396 | 1 |
| CAD | adipocyte | *DDX54* | 1.081340378 | 0.002792439 | 0.019873367 | 1 |
| CAD | adipocyte | *CCDC91* | 1.059344029 | 0.001013772 | 0.017317396 | 1 |
| CAD | adipocyte | *PFDN5* | 1.185452012 | 0.003095433 | 0.021123824 | 1 |
| CAD | adipocyte | *PLA2G12A* | 1.033605789 | 0.011836003 | 0.048258598 | 1 |
| CAD | adipocyte | *OBSL1* | 0.915968524 | 0.009590831 | 0.042134669 | 1 |
| CAD | adipocyte | *SLC12A4* | 1.506813816 | 0.003709578 | 0.023436034 | 1 |
| CAD | adipocyte | *WFDC3* | -0.121126095 | 0.001882213 | 0.017489118 | 1 |

| | | | | | | |
|---|---|---|---|---|---|---|
| CAD | adipocyte | *CSE1L* | 0.857397294 | 0.008830222 | 0.040267141 | 1 |
| CAD | adipocyte | *PTGIS* | 0.53543202 | 4.77E-06 | 0.002461025 | 0.023143532 |
| CAD | adipocyte | *RNF114* | 1.181747055 | 0.001643513 | 0.017317396 | 1 |
| CAD | adipocyte | *MTRR* | 0.785861315 | 0.008325963 | 0.038621008 | 1 |
| CAD | adipocyte | *STAMBP* | 1.429449949 | 0.002997431 | 0.020658429 | 1 |
| CAD | adipocyte | *SNRNP27* | 1.898005531 | 0.000850227 | 0.017317396 | 1 |
| CAD | adipocyte | *USP22* | 1.120974025 | 0.003727281 | 0.023454819 | 1 |
| CAD | adipocyte | *SNRPC* | -0.632559827 | 0.000316217 | 0.013060813 | 1 |
| CAD | adipocyte | *TCP11* | 0.45035157 | 0.000319774 | 0.013060813 | 1 |
| CAD | adipocyte | *GLO1* | 1.17236759 | 0.000520602 | 0.015787247 | 1 |
| CAD | adipocyte | *ATXN1* | 0.843741791 | 0.002132153 | 0.018430441 | 1 |
| CAD | adipocyte | *RUNX2* | 1.176408329 | 0.002497363 | 0.019127952 | 1 |
| CAD | adipocyte | *GOT2* | 1.31331608 | 0.000725337 | 0.017317396 | 1 |
| CAD | adipocyte | *CLYBL* | 1.339100984 | 0.002288615 | 0.018884968 | 1 |
| CAD | adipocyte | *GRK4* | 0.165246486 | 8.21E-05 | 0.007733492 | 0.398362615 |
| CAD | adipocyte | *MRPS7* | 1.481769004 | 0.005134207 | 0.028582558 | 1 |
| CAD | adipocyte | *NT5C* | 1.502790078 | 0.004535895 | 0.026839221 | 1 |
| CAD | adipocyte | *OPRL1* | 5.836928891 | 0.006235766 | 0.032255798 | 1 |
| CAD | adipocyte | *SNRPD2* | 1.943220311 | 0.001568088 | 0.017317396 | 1 |
| CAD | adipocyte | *PSMF1* | 0.588498904 | 0.005711268 | 0.030384947 | 1 |
| CAD | adipocyte | *DTD1* | 0.855396383 | 0.001725318 | 0.017317396 | 1 |
| CAD | adipocyte | *RRBP1* | 1.170309395 | 6.50E-05 | 0.007688119 | 0.315212859 |
| CAD | adipocyte | *FLRT3* | 1.589647473 | 0.001233829 | 0.017317396 | 1 |
| CAD | adipocyte | *KLC1* | 1.633297298 | 0.000146037 | 0.009084269 | 0.708572981 |
| CAD | adipocyte | *TUBGCP3* | 1.097630419 | 0.005271448 | 0.028868018 | 1 |
| CAD | adipocyte | *COX6B1* | 1.29947988 | 0.002269442 | 0.018849915 | 1 |
| CAD | adipocyte | *IRF3* | 1.395254598 | 0.002428203 | 0.019127952 | 1 |
| CAD | adipocyte | *TRAP1* | 0.221511718 | 0.00548215 | 0.029561594 | 1 |
| CAD | adipocyte | *KTN1* | 1.1200175 | 0.001214586 | 0.017317396 | 1 |
| CAD | adipocyte | *CANX* | 1.362166614 | 9.20E-05 | 0.007830023 | 0.446311296 |
| CAD | adipocyte | *ZNF484* | 1.342577823 | 0.000808471 | 0.017317396 | 1 |
| CAD | adipocyte | *HELB* | 1.019040181 | 0.011694118 | 0.047934033 | 1 |
| CAD | adipocyte | *EMC1* | 1.1203121 | 0.002782748 | 0.019873367 | 1 |
| CAD | adipocyte | *SMARCA4* | 1.340109492 | 1.17E-05 | 0.003147371 | 0.056652683 |
| CAD | adipocyte | *HIP1* | 0.965447202 | 0.001505489 | 0.017317396 | 1 |
| CAD | adipocyte | *KDR* | 0.731366574 | 0.011747742 | 0.048020254 | 1 |
| CAD | adipocyte | *SPECC1* | 1.10933295 | 0.001570059 | 0.017317396 | 1 |
| CAD | adipocyte | *MTX2* | 1.274988182 | 0.002003723 | 0.018037223 | 1 |
| CAD | adipocyte | *TWSG1* | 1.178667468 | 0.009192589 | 0.041043909 | 1 |
| CAD | adipocyte | *ARPP19* | 1.509970067 | 0.00163179 | 0.017317396 | 1 |
| CAD | adipocyte | *VPS13C* | 0.939645552 | 0.001260789 | 0.017317396 | 1 |
| CAD | adipocyte | *SERGEF* | 0.995062492 | 2.07E-05 | 0.003854781 | 0.100224294 |
| CAD | adipocyte | *PLD2* | 1.335931155 | 0.006811103 | 0.033894842 | 1 |
| CAD | adipocyte | *PHF20L1* | 0.899321858 | 0.008586936 | 0.039491768 | 1 |
| CAD | adipocyte | *MTUS1* | 0.677575434 | 0.007278973 | 0.035176868 | 1 |
| CAD | adipocyte | *MIS18BP1* | 0.615870513 | 0.006697313 | 0.033643116 | 1 |
| CAD | adipocyte | *EPB41L4A* | 1.207259051 | 0.000781539 | 0.017317396 | 1 |
| CAD | adipocyte | *REEP5* | 1.331161598 | 0.001358426 | 0.017317396 | 1 |
| CAD | adipocyte | *TMEM8A* | 1.521421098 | 0.012042802 | 0.048613338 | 1 |
| CAD | adipocyte | *PHF10* | 1.191136412 | 0.011516051 | 0.047392604 | 1 |

| | | | | | | |
|---|---|---|---|---|---|---|
| CAD | adipocyte | *XPO7* | 1.065576809 | 0.008345827 | 0.038676174 | 1 |
| CAD | adipocyte | *FAM98C* | 0.514906537 | 0.002025459 | 0.018131966 | 1 |
| CAD | adipocyte | *PLVAP* | 0.174941584 | 0.010001733 | 0.043103145 | 1 |
| CAD | adipocyte | *LSM7* | 0.480234191 | 0.00462177 | 0.027083126 | 1 |
| CAD | adipocyte | *ACTN4* | 1.384346769 | 1.75E-05 | 0.003854781 | 0.084959998 |
| CAD | adipocyte | *MAP1S* | 2.551547401 | 0.001385103 | 0.017317396 | 1 |
| CAD | adipocyte | *ATXN10* | 1.107364959 | 0.001530902 | 0.017317396 | 1 |
| CAD | adipocyte | *ZNF337* | 0.590434824 | 0.00731338 | 0.03530798 | 1 |
| CAD | adipocyte | *POMT1* | 0.848390859 | 0.006428068 | 0.032840444 | 1 |
| CAD | adipocyte | *DNMT1* | 1.369165839 | 0.002456822 | 0.019127952 | 1 |
| CAD | adipocyte | *SLC35D2* | 1.520813148 | 0.001029204 | 0.017317396 | 1 |
| CAD | adipocyte | *CHMP1A* | 1.26885503 | 0.009484285 | 0.042063757 | 1 |
| CAD | adipocyte | *KIF3A* | 1.452122254 | 0.000276936 | 0.012105358 | 1 |
| CAD | adipocyte | *EXOC4* | 0.672649557 | 0.006748094 | 0.033824125 | 1 |
| CAD | adipocyte | *C1orf159* | 1.61323524 | 0.002983412 | 0.020593222 | 1 |
| CAD | adipocyte | *MAP1B* | 0.600721283 | 0.001416014 | 0.017317396 | 1 |
| CAD | adipocyte | *KHDRBS3* | 1.715944793 | 0.001175543 | 0.017317396 | 1 |
| CAD | adipocyte | *PRKAB2* | 1.260577581 | 0.003191481 | 0.021432905 | 1 |
| CAD | adipocyte | *SPATA6* | 0.917020035 | 0.001782098 | 0.017489118 | 1 |
| CAD | adipocyte | *LRRC41* | 1.206716686 | 0.009991464 | 0.043103145 | 1 |
| CAD | adipocyte | *TRIM22* | 0.795154059 | 0.00734791 | 0.035439421 | 1 |
| CAD | adipocyte | *TIMM10B* | 0.53155071 | 0.00161995 | 0.017317396 | 1 |
| CAD | adipocyte | *SERPINF1* | 1.370058524 | 0.002707043 | 0.019782057 | 1 |
| CAD | adipocyte | *DDC* | -0.768720681 | 0.009922676 | 0.042949124 | 1 |
| CAD | adipocyte | *MUTYH* | 0.559657516 | 0.011375815 | 0.047065316 | 1 |
| CAD | adipocyte | *AP3B1* | 1.110732791 | 0.001221388 | 0.017317396 | 1 |
| CAD | adipocyte | *ALOX5AP* | 1.023336861 | 0.011069977 | 0.046183603 | 1 |
| CAD | adipocyte | *WASF3* | 1.039246999 | 0.011450048 | 0.047201046 | 1 |
| CAD | adipocyte | *MPRIP* | 1.314853311 | 0.001510984 | 0.017317396 | 1 |
| CAD | adipocyte | *DSTYK* | 1.25488487 | 0.001727359 | 0.017317396 | 1 |
| CAD | adipocyte | *EPSTI1* | 0.597446527 | 0.012291905 | 0.04916762 | 1 |
| CAD | adipocyte | *CSNK1G2* | 1.930433941 | 0.001389547 | 0.017317396 | 1 |
| CAD | adipocyte | *IPO8* | 1.429118458 | 0.00028968 | 0.01225006 | 1 |
| CAD | adipocyte | *RRAS2* | 0.76013369 | 0.001964727 | 0.017918898 | 1 |
| CAD | adipocyte | *COX16* | 4.106349136 | 0.000363521 | 0.013592277 | 1 |
| CAD | adipocyte | *ELP3* | 1.139887937 | 0.007033427 | 0.03446529 | 1 |
| CAD | adipocyte | *PEBP4* | 2.052885806 | 0.002273599 | 0.018849915 | 1 |
| CAD | adipocyte | *NAPG* | 1.078356377 | 0.005362271 | 0.029167871 | 1 |
| CAD | adipocyte | *KIDINS220* | 1.172212265 | 0.001380357 | 0.017317396 | 1 |
| CAD | adipocyte | *KIAA1468* | 1.286771329 | 0.000943945 | 0.017317396 | 1 |
| CAD | adipocyte | *IL15RA* | 0.946261673 | 0.001417219 | 0.017317396 | 1 |
| CAD | adipocyte | *KLRD1* | 1.49579423 | 0.001088376 | 0.017317396 | 1 |
| CAD | adipocyte | *ELP2* | 1.254052045 | 0.002625187 | 0.019416779 | 1 |
| CAD | adipocyte | *DHX34* | 1.665558013 | 0.001297149 | 0.017317396 | 1 |
| CAD | adipocyte | *CLOCK* | 1.22580782 | 0.001905291 | 0.017489118 | 1 |
| CAD | adipocyte | *ARGLU1* | 0.909207876 | 0.00164542 | 0.017317396 | 1 |
| CAD | adipocyte | *OSTF1* | 1.11320217 | 0.002434001 | 0.019127952 | 1 |
| CAD | adipocyte | *UBQLN1* | 1.057897689 | 0.007375891 | 0.035503794 | 1 |
| CAD | adipocyte | *DMTF1* | 1.128585474 | 0.000797702 | 0.017317396 | 1 |
| CAD | adipocyte | *SRPK2* | 0.979519857 | 0.00180559 | 0.017489118 | 1 |

| | | | | | | |
|---|---|---|---|---|---|---|
| CAD | adipocyte | *MAP3K7* | 1.292629385 | 0.001279504 | 0.017317396 | 1 |
| CAD | adipocyte | *PHF21A* | 1.065571686 | 0.001206463 | 0.017317396 | 1 |
| CAD | adipocyte | *CAPRIN1* | 0.988683543 | 0.006029018 | 0.031556416 | 1 |
| CAD | adipocyte | *ATP5G2* | 1.04655438 | 0.004551405 | 0.02686547 | 1 |
| CAD | adipocyte | *B4GALNT1* | 1.084981576 | 0.000472597 | 0.015389541 | 1 |
| CAD | adipocyte | *USP15* | 1.014915295 | 0.002314352 | 0.01893632 | 1 |
| CAD | adipocyte | *NTPCR* | 1.484640938 | 0.002353707 | 0.019062957 | 1 |
| CAD | adipocyte | *PIGC* | 1.103883281 | 0.009595797 | 0.042134669 | 1 |
| CAD | adipocyte | *SP110* | 0.748850982 | 0.007661726 | 0.03645052 | 1 |
| CAD | adipocyte | *USP37* | 0.711972557 | 0.005888944 | 0.031032029 | 1 |
| CAD | adipocyte | *GCC2* | 1.026164247 | 0.000556295 | 0.016162526 | 1 |
| CAD | adipocyte | *ANKRD36* | 1.151177364 | 0.001791734 | 0.017489118 | 1 |
| CAD | adipocyte | *EPC2* | 1.226185981 | 0.000480682 | 0.01542398 | 1 |
| CAD | adipocyte | *STAB2* | 1.895393792 | 0.001185533 | 0.017317396 | 1 |
| CAD | adipocyte | *APPL2* | 0.792217013 | 0.005488598 | 0.029561594 | 1 |
| CAD | adipocyte | *DRAM1* | 0.782790235 | 0.011483745 | 0.047299771 | 1 |
| CAD | adipocyte | *RCBTB2* | 1.461888566 | 0.001241623 | 0.017317396 | 1 |
| CAD | adipocyte | *KDELR2* | 1.586545465 | 0.001730168 | 0.017317396 | 1 |
| CAD | adipocyte | *NUPL2* | 1.470143852 | 0.001603804 | 0.017317396 | 1 |
| CAD | adipocyte | *ZNF189* | 1.785723414 | 0.00267617 | 0.019693337 | 1 |
| CAD | adipocyte | *STX17* | 1.429382091 | 0.00083121 | 0.017317396 | 1 |
| CAD | adipocyte | *POLR1E* | 0.244499632 | 0.000881146 | 0.017317396 | 1 |
| CAD | adipocyte | *DENND4C* | 0.787914827 | 0.008374576 | 0.038772369 | 1 |
| CAD | adipocyte | *TCF19* | 0.477243877 | 0.000498151 | 0.01542398 | 1 |
| CAD | adipocyte | *NRM* | 5.634638074 | 0.001514694 | 0.017317396 | 1 |
| CAD | adipocyte | *NUMA1* | 1.117903549 | 0.004062894 | 0.02492182 | 1 |
| CAD | adipocyte | *RAB30* | 1.409640316 | 0.003824251 | 0.023849957 | 1 |
| CAD | adipocyte | *NARS2* | 1.37203325 | 0.001792935 | 0.017489118 | 1 |
| CAD | adipocyte | *TGS1* | 1.608868918 | 0.000220844 | 0.010928681 | 1 |
| CAD | adipocyte | *NEK1* | 0.823956771 | 0.012202267 | 0.049012164 | 1 |
| CAD | adipocyte | *SLC37A4* | 0.629797714 | 2.99E-05 | 0.00500475 | 0.145137752 |
| CAD | adipocyte | *FDX1* | 0.726610703 | 0.003500958 | 0.022648861 | 1 |
| CAD | adipocyte | *MAP2K5* | 1.17802897 | 0.000742615 | 0.017317396 | 1 |
| CAD | adipocyte | *NUSAP1* | 0.760778104 | 0.005160412 | 0.028582558 | 1 |
| CAD | adipocyte | *PARP6* | 1.238638152 | 0.007667565 | 0.03645052 | 1 |
| CAD | adipocyte | *ADAM10* | 0.987029612 | 0.000549211 | 0.016127182 | 1 |
| CAD | adipocyte | *ZNF280D* | 1.107223405 | 0.000863149 | 0.017317396 | 1 |
| CAD | adipocyte | *RTCA* | 1.115106654 | 0.000346691 | 0.013592277 | 1 |
| CAD | adipocyte | *ACTR2* | 1.169158008 | 0.001190973 | 0.017317396 | 1 |
| CAD | adipocyte | *CENPO* | 0.814367713 | 0.002782924 | 0.019873367 | 1 |
| CAD | adipocyte | *LRPPRC* | 1.262589342 | 0.001773634 | 0.017489118 | 1 |
| CAD | adipocyte | *MYOF* | 0.272865926 | 0.003373191 | 0.022177133 | 1 |
| CAD | adipocyte | *ARL3* | 0.442355099 | 0.010551475 | 0.044673236 | 1 |
| CAD | adipocyte | *SMARCAL1* | 1.396423278 | 0.003143908 | 0.021275091 | 1 |
| CAD | adipocyte | *STAT4* | 1.311000885 | 0.002935652 | 0.020465209 | 1 |
| CAD | adipocyte | *FASTKD1* | 0.835042208 | 0.005215282 | 0.028755166 | 1 |
| CAD | adipocyte | *ITGAV* | 0.93766475 | 0.002272023 | 0.018849915 | 1 |
| CAD | adipocyte | *COX17* | 1.672595804 | 0.001893239 | 0.017489118 | 1 |
| CAD | adipocyte | *AP1AR* | 1.537345914 | 0.002153763 | 0.018430441 | 1 |
| CAD | adipocyte | *KIAA1109* | 1.051370593 | 0.001281767 | 0.017317396 | 1 |

| | | | | | | |
|---|---|---|---|---|---|---|
| CAD | adipocyte | *PRDM5* | 1.431805901 | 0.001131406 | 0.017317396 | 1 |
| CAD | adipocyte | *NUP54* | 0.644286055 | 0.000341553 | 0.013583718 | 1 |
| CAD | adipocyte | *PPA2* | 0.92829458 | 0.002809515 | 0.019938662 | 1 |
| CAD | adipocyte | *CLSTN3* | 0.903029185 | 0.002477384 | 0.019127952 | 1 |
| CAD | adipocyte | *MARCH9* | 2.314929138 | 0.001692 | 0.017317396 | 1 |
| CAD | adipocyte | *POC1B* | 1.396728104 | 0.002746715 | 0.019834836 | 1 |
| CAD | adipocyte | *RITA1* | 2.9292935 | 0.004798695 | 0.027586809 | 1 |
| CAD | adipocyte | *GLTP* | 1.705569346 | 0.002605264 | 0.019357953 | 1 |
| CAD | adipocyte | *KANSL2* | 0.592665464 | 0.008538936 | 0.039345599 | 1 |
| CAD | adipocyte | *CERS5* | 1.03919416 | 0.010767448 | 0.045350398 | 1 |
| CAD | adipocyte | *ESYT1* | 1.03234085 | 0.003135677 | 0.021249029 | 1 |
| CAD | adipocyte | *WDFY2* | 0.687006237 | 0.012360806 | 0.049280717 | 1 |
| CAD | adipocyte | *DIAPH3* | 0.246582771 | 0.004615136 | 0.027076951 | 1 |
| CAD | adipocyte | *TMX1* | 1.591758994 | 0.001225418 | 0.017317396 | 1 |
| CAD | adipocyte | *C14orf37* | 1.222302854 | 0.001497955 | 0.017317396 | 1 |
| CAD | adipocyte | *RAB15* | 2.138100981 | 0.002511136 | 0.019127952 | 1 |
| CAD | adipocyte | *WARS* | 1.055162435 | 0.000878031 | 0.017317396 | 1 |
| CAD | adipocyte | *SERF2* | 1.098308227 | 0.004478254 | 0.026628049 | 1 |
| CAD | adipocyte | *SRP14* | 0.877353745 | 0.004679251 | 0.027311752 | 1 |
| CAD | adipocyte | *UBE2Q2* | 0.871757273 | 0.006446625 | 0.032840444 | 1 |
| CAD | adipocyte | *HMG20A* | 1.423347195 | 0.001507476 | 0.017317396 | 1 |
| CAD | adipocyte | *ULK3* | 2.718658875 | 2.58E-05 | 0.00464164 | 0.125324278 |
| CAD | adipocyte | *ITGAX* | 1.264546121 | 0.008139031 | 0.038058918 | 1 |
| CAD | adipocyte | *TGFB1I1* | 1.057741348 | 0.011404254 | 0.047092292 | 1 |
| CAD | adipocyte | *DHX38* | 1.807576686 | 0.001364678 | 0.017317396 | 1 |
| CAD | adipocyte | *ZCCHC14* | 0.923684795 | 0.005981121 | 0.031339522 | 1 |
| CAD | adipocyte | *TLDC1* | 1.96857745 | 0.001153388 | 0.017317396 | 1 |
| CAD | adipocyte | *TCF25* | 1.087206915 | 0.000112918 | 0.008341315 | 0.547879255 |
| CAD | adipocyte | *NCOR1* | 1.037748142 | 0.001848971 | 0.017489118 | 1 |
| CAD | adipocyte | *KSR1* | 1.539597955 | 0.000283653 | 0.01225006 | 1 |
| CAD | adipocyte | *MYO19* | 1.399719898 | 7.35E-06 | 0.002545589 | 0.035638249 |
| CAD | adipocyte | *SSH2* | 1.020545865 | 0.001525711 | 0.017317396 | 1 |
| CAD | adipocyte | *IMPA2* | 1.381466373 | 0.000193992 | 0.010214126 | 0.941248449 |
| CAD | adipocyte | *GNAL* | 1.546062597 | 0.002480433 | 0.019127952 | 1 |
| CAD | adipocyte | *GALNT1* | 1.09124958 | 0.002112097 | 0.018430441 | 1 |
| CAD | adipocyte | *IGFBP4* | 0.581316051 | 0.002863675 | 0.020166254 | 1 |
| CAD | adipocyte | *CACNA1A* | 1.437740659 | 0.00472378 | 0.027416006 | 1 |
| CAD | adipocyte | *NFIC* | 0.881087456 | 0.003654474 | 0.023229851 | 1 |
| CAD | adipocyte | *DPP9* | 1.484224084 | 0.003584561 | 0.023005676 | 1 |
| CAD | adipocyte | *TMEM91* | 0.32011049 | 0.000119845 | 0.008341315 | 0.581488273 |
| CAD | adipocyte | *SIRT3* | 1.230232509 | 4.66E-05 | 0.006632491 | 0.226315882 |
| CAD | adipocyte | *EVI5L* | 2.129287905 | 0.002747117 | 0.019834836 | 1 |
| CAD | adipocyte | *CTU1* | 2.285793029 | 0.00362204 | 0.023095543 | 1 |
| CAD | adipocyte | *RERE* | 0.815835422 | 0.003444176 | 0.022491444 | 1 |
| CAD | adipocyte | *EVA1B* | 0.503596654 | 0.00026273 | 0.011695107 | 1 |
| CAD | adipocyte | *RPS8* | 1.047152978 | 0.00132247 | 0.017317396 | 1 |
| CAD | adipocyte | *SLC44A3* | 1.365872134 | 0.00612288 | 0.031773492 | 1 |
| CAD | adipocyte | *STRIP1* | 2.367223376 | 1.24E-05 | 0.003158083 | 0.060003573 |
| CAD | adipocyte | *TIPRL* | 0.941408702 | 0.003193002 | 0.021432905 | 1 |
| CAD | adipocyte | *MAEL* | 5.003060398 | 0.001236094 | 0.017317396 | 1 |

| | | | | | | |
|---|---|---|---|---|---|---|
| CAD | adipocyte | *RFWD2* | 1.149677712 | 0.000379129 | 0.013616273 | 1 |
| CAD | adipocyte | *SNX27* | 1.229556444 | 8.65E-06 | 0.002798143 | 0.04197214 |
| CAD | adipocyte | *SETDB1* | 1.63293843 | 9.97E-06 | 0.00284493 | 0.048363808 |
| CAD | adipocyte | *SEMA6C* | 1.241073021 | 0.001844973 | 0.017489118 | 1 |
| CAD | adipocyte | *SMYD2* | 1.356372416 | 0.002294376 | 0.018900359 | 1 |
| CAD | adipocyte | *RAB13* | 1.842715742 | 0.003244711 | 0.021655208 | 1 |
| CAD | adipocyte | *S100A8* | 1.246871909 | 0.004176482 | 0.025330363 | 1 |
| CAD | adipocyte | *C1orf43* | 1.033858182 | 0.012202534 | 0.049012164 | 1 |
| CAD | adipocyte | *CEP170* | 1.008864518 | 0.005904247 | 0.03107094 | 1 |
| CAD | adipocyte | *ACP1* | 1.321178204 | 0.007753159 | 0.036736649 | 1 |
| CAD | adipocyte | *MBOAT2* | 0.9013563 | 0.008731195 | 0.040009491 | 1 |
| CAD | adipocyte | *PLEKHA6* | 0.354652878 | 0.009532875 | 0.042134669 | 1 |
| CAD | adipocyte | *RHOB* | 1.413233143 | 0.007514297 | 0.035920558 | 1 |
| CAD | adipocyte | *GALM* | 1.529838911 | 0.00156498 | 0.017317396 | 1 |
| CAD | adipocyte | *WDPCP* | 0.625126233 | 6.63E-06 | 0.002473817 | 0.03215962 |
| CAD | adipocyte | *VPS54* | 1.125587222 | 0.002460605 | 0.019127952 | 1 |
| CAD | adipocyte | *EXOC6B* | 0.975143612 | 0.001889633 | 0.017489118 | 1 |
| CAD | adipocyte | *TEX261* | 2.567299563 | 0.001583248 | 0.017317396 | 1 |
| CAD | adipocyte | *RALB* | 1.066337186 | 0.004926289 | 0.028054408 | 1 |
| CAD | adipocyte | *METTL21A* | 1.218540197 | 0.001321702 | 0.017317396 | 1 |
| CAD | adipocyte | *SUMF1* | 0.981256691 | 0.002541593 | 0.019238394 | 1 |
| CAD | adipocyte | *ESPNL* | 16.98658149 | 0.002556977 | 0.019262254 | 1 |
| CAD | adipocyte | *TAMM41* | 1.224729443 | 0.000241108 | 0.011357815 | 1 |
| CAD | adipocyte | *EAF1* | 1.360271167 | 0.010717719 | 0.045180167 | 1 |
| CAD | adipocyte | *CNTN4* | 0.601290624 | 0.012253879 | 0.049163621 | 1 |
| CAD | adipocyte | *CTDSPL* | 0.74359576 | 0.002264594 | 0.018849915 | 1 |
| CAD | adipocyte | *ARL6IP5* | 1.360725422 | 0.001250766 | 0.017317396 | 1 |
| CAD | adipocyte | *LIMD1* | 0.676933643 | 0.002007443 | 0.018037244 | 1 |
| CAD | adipocyte | *BOC* | 0.956605306 | 0.001664486 | 0.017317396 | 1 |
| CAD | adipocyte | *AMT* | -0.175483662 | 0.001445271 | 0.017317396 | 1 |
| CAD | adipocyte | *CISD2* | 1.697173138 | 0.000840643 | 0.017317396 | 1 |
| CAD | adipocyte | *METTL14* | 1.268968228 | 0.005060064 | 0.02848194 | 1 |
| CAD | adipocyte | *CBR4* | 1.09301287 | 0.001160075 | 0.017317396 | 1 |
| CAD | adipocyte | *MARCH6* | 1.235866362 | 0.00089026 | 0.017317396 | 1 |
| CAD | adipocyte | *MYO10* | 0.737227335 | 0.007138279 | 0.034774028 | 1 |
| CAD | adipocyte | *GTF2H2* | 2.671397003 | 0.002167112 | 0.018453035 | 1 |
| CAD | adipocyte | *FEM1C* | 2.014852715 | 0.001289613 | 0.017317396 | 1 |
| CAD | adipocyte | *FBXO38* | 1.35195542 | 0.000124784 | 0.008409031 | 0.605450213 |
| CAD | adipocyte | *TBC1D7* | 0.833402704 | 0.000361966 | 0.013592277 | 1 |
| CAD | adipocyte | *IRAK1BP1* | 1.161544586 | 4.81E-06 | 0.002461025 | 0.023352535 |
| CAD | adipocyte | *PHIP* | 1.227420519 | 0.000255775 | 0.011672743 | 1 |
| CAD | adipocyte | *PNRC1* | 1.311202087 | 0.001999207 | 0.018030022 | 1 |
| CAD | adipocyte | *RNF217* | 0.990026096 | 0.003799612 | 0.023782688 | 1 |
| CAD | adipocyte | *RBAK* | 2.801319649 | 0.001358372 | 0.017317396 | 1 |
| CAD | adipocyte | *FBXO25* | 1.426341502 | 0.002113028 | 0.018430441 | 1 |
| CAD | adipocyte | *CSGALNACT1* | 0.358894065 | 0.000371499 | 0.013616273 | 1 |
| CAD | adipocyte | *BIN3* | 1.010697352 | 0.004408972 | 0.026442933 | 1 |
| CAD | adipocyte | *DOCK5* | 0.709030389 | 0.004059 | 0.02492182 | 1 |
| CAD | adipocyte | *TACC1* | 0.882704573 | 0.001722728 | 0.017317396 | 1 |
| CAD | adipocyte | *DPYS* | 0.648052574 | 0.011311143 | 0.046947531 | 1 |

| | | | | | | |
|---|---|---|---|---|---|---|
| CAD | adipocyte | *LRP12* | 1.8326334 | 0.001377006 | 0.017317396 | 1 |
| CAD | adipocyte | *SLC39A4* | 0.212940021 | 0.010430881 | 0.044278771 | 1 |
| CAD | adipocyte | *AK3* | 1.244470408 | 0.003799891 | 0.023782688 | 1 |
| CAD | adipocyte | *HAUS6* | 1.559631165 | 0.004564871 | 0.026912218 | 1 |
| CAD | adipocyte | *FBXO10* | 0.561355198 | 9.91E-05 | 0.00815332 | 0.481045899 |
| CAD | adipocyte | *REXO4* | 1.877981191 | 0.001488086 | 0.017317396 | 1 |
| CAD | adipocyte | *SH3GLB2* | 1.195899368 | 0.005810819 | 0.030746014 | 1 |
| CAD | adipocyte | *SLC39A12* | 1.242862394 | 0.010002914 | 0.043103145 | 1 |
| CAD | adipocyte | *RSU1* | 1.157935471 | 0.000955958 | 0.017317396 | 1 |
| CAD | adipocyte | *PARD3* | 0.919642588 | 0.002506213 | 0.019127952 | 1 |
| CAD | adipocyte | *ADD3* | 1.05978395 | 0.000969572 | 0.017317396 | 1 |
| CAD | adipocyte | *LRRC27* | 0.316057532 | 0.005105836 | 0.028582558 | 1 |
| CAD | adipocyte | *PAOX* | 1.997444786 | 0.008932938 | 0.040525141 | 1 |
| CAD | adipocyte | *HSD17B12* | -0.189455136 | 0.001380779 | 0.017317396 | 1 |
| CAD | adipocyte | *PTPRJ* | 0.90460343 | 0.001600629 | 0.017317396 | 1 |
| CAD | adipocyte | *TTC12* | 0.495278037 | 0.000364179 | 0.013592277 | 1 |
| CAD | adipocyte | *NPAT* | 1.215172463 | 0.009558241 | 0.042134669 | 1 |
| CAD | adipocyte | *CHEK1* | 2.672733222 | 0.00420373 | 0.025432042 | 1 |
| CAD | adipocyte | *SAP18* | 1.560255937 | 0.004450442 | 0.02662058 | 1 |
| CAD | adipocyte | *FAM124A* | 1.142960814 | 0.005159244 | 0.028582558 | 1 |
| CAD | adipocyte | *FAM173B* | 0.86805223 | 0.010249549 | 0.043803898 | 1 |
| CAD | adipocyte | *ANK3* | 0.710643636 | 0.009122777 | 0.040909163 | 1 |
| CAD | adipocyte | *TMEM18* | 2.087594646 | 0.001168325 | 0.017317396 | 1 |
| CAD | adipocyte | *NDUFC2* | 1.845954749 | 0.002947393 | 0.020491506 | 1 |
| CAD | adipocyte | *NUBPL* | 1.206819852 | 0.001835036 | 0.017489118 | 1 |
| CAD | adipocyte | *ANKRD50* | 1.238519059 | 0.00047578 | 0.015389905 | 1 |
| CAD | adipocyte | *UPF2* | 0.911004898 | 0.011789622 | 0.048110383 | 1 |
| CAD | adipocyte | *FAM160B1* | 1.152550874 | 0.001898559 | 0.017489118 | 1 |
| CAD | adipocyte | *PIGF* | 1.46872022 | 0.001903362 | 0.017489118 | 1 |
| CAD | adipocyte | *MFSD6* | 1.223153401 | 0.002587941 | 0.019288308 | 1 |
| CAD | adipocyte | *BICD1* | 0.851514153 | 0.004142741 | 0.025274753 | 1 |
| CAD | adipocyte | *TIAL1* | 1.373487397 | 0.000512464 | 0.015737195 | 1 |
| CAD | adipocyte | *PTPN14* | 1.099398172 | 0.000873262 | 0.017317396 | 1 |
| CAD | adipocyte | *EPG5* | 1.309518534 | 0.001172176 | 0.017317396 | 1 |
| CAD | adipocyte | *KCNK13* | 1.218530983 | 0.000647089 | 0.017317396 | 1 |
| CAD | adipocyte | *CWF19L2* | 0.91166524 | 0.000717494 | 0.017317396 | 1 |
| CAD | adipocyte | *SPARCL1* | 0.578996687 | 0.010124292 | 0.043433303 | 1 |
| CAD | adipocyte | *MBNL1* | 0.902423412 | 0.001303974 | 0.017317396 | 1 |
| CAD | adipocyte | *MED21* | 0.871020966 | 0.006402487 | 0.032840444 | 1 |
| CAD | adipocyte | *JAKMIP1* | 2.262783668 | 0.006280386 | 0.032452005 | 1 |
| CAD | adipocyte | *SREK1IP1* | 1.164250555 | 0.01169701 | 0.047934033 | 1 |
| CAD | adipocyte | *MR1* | 0.796747954 | 0.004467589 | 0.02662058 | 1 |
| CAD | adipocyte | *CAST* | 0.527748947 | 0.008812692 | 0.040225004 | 1 |
| CAD | adipocyte | *TMEM87B* | 1.046002734 | 0.002894391 | 0.020298987 | 1 |
| CAD | adipocyte | *PLA2R1* | 0.258421181 | 0.012140602 | 0.048844278 | 1 |
| CAD | adipocyte | *LPCAT1* | 1.031112871 | 0.010255818 | 0.043803898 | 1 |
| CAD | adipocyte | *CFDP1* | 1.208988774 | 0.001532201 | 0.017317396 | 1 |
| CAD | adipocyte | *ZDHHC7* | 1.505987988 | 0.001054356 | 0.017317396 | 1 |
| CAD | adipocyte | *FBXO36* | 1.379438208 | 0.008124018 | 0.038048008 | 1 |
| CAD | adipocyte | *SEMA3D* | 0.680547834 | 0.007256495 | 0.035138237 | 1 |

| | | | | | | |
|---|---|---|---|---|---|---|
| CAD | adipocyte | *PPP2R5E* | 1.219566991 | 0.001618175 | 0.017317396 | 1 |
| CAD | adipocyte | *ANKRD29* | 0.87956301 | 0.006695995 | 0.033643116 | 1 |
| CAD | adipocyte | *ANKH* | 0.905964215 | 0.005010542 | 0.028301688 | 1 |
| CAD | adipocyte | *ROBO4* | 0.766441959 | 0.011275307 | 0.046878999 | 1 |
| CAD | adipocyte | *ROBO3* | 0.906942245 | 5.32E-06 | 0.002461025 | 0.025800709 |
| CAD | adipocyte | *PRKCA* | 1.029723135 | 0.002305266 | 0.018925801 | 1 |
| CAD | adipocyte | *CCSAP* | 2.219935482 | 0.002035284 | 0.018152939 | 1 |
| CAD | adipocyte | *PDE1C* | 0.753504281 | 0.006090303 | 0.031773492 | 1 |
| CAD | adipocyte | *TRMT44* | 1.201530154 | 0.005121959 | 0.028582558 | 1 |
| CAD | adipocyte | *NUP205* | 1.368723362 | 0.002131068 | 0.018430441 | 1 |
| CAD | adipocyte | *PDIA4* | 1.393507556 | 0.004245528 | 0.025652926 | 1 |
| CAD | adipocyte | *C14orf2* | 0.699640494 | 0.009203581 | 0.041043909 | 1 |
| CAD | adipocyte | *RPL30* | 0.96680736 | 0.001359261 | 0.017317396 | 1 |
| CAD | adipocyte | *ZDHHC5* | 1.759959083 | 0.001166872 | 0.017317396 | 1 |
| CAD | adipocyte | *MED19* | 0.474833777 | 0.004694662 | 0.027311752 | 1 |
| CAD | adipocyte | *VPS8* | 0.883631287 | 0.000589726 | 0.016687501 | 1 |
| CAD | adipocyte | *GHRL* | 0.656348741 | 0.011146843 | 0.046424449 | 1 |
| CAD | adipocyte | *NTAN1* | 0.705947372 | 0.002510671 | 0.019127952 | 1 |
| CAD | adipocyte | *FCHO2* | 0.860834817 | 0.00011132 | 0.008341315 | 0.540123706 |
| CAD | adipocyte | *NECAP2* | 1.505039338 | 0.0015831 | 0.017317396 | 1 |
| CAD | adipocyte | *MMP14* | 1.868211993 | 0.000733424 | 0.017317396 | 1 |
| CAD | adipocyte | *DHRS4* | 2.359762735 | 0.001674524 | 0.017317396 | 1 |
| CAD | adipocyte | *AASDH* | 0.969677935 | 0.002230842 | 0.018759181 | 1 |
| CAD | adipocyte | *SLC38A10* | 1.452090682 | 0.00211316 | 0.018430441 | 1 |
| CAD | adipocyte | *EYA3* | 0.925452011 | 0.001698587 | 0.017317396 | 1 |
| CAD | adipocyte | *FANCC* | 0.875575904 | 0.010569842 | 0.044673236 | 1 |
| CAD | adipocyte | *MITD1* | 0.977180604 | 0.002423044 | 0.019127952 | 1 |
| CAD | adipocyte | *ELK4* | 1.443178489 | 0.001535524 | 0.017317396 | 1 |
| CAD | adipocyte | *ARHGAP27* | 0.935756453 | 0.009856943 | 0.042862603 | 1 |
| CAD | adipocyte | *TMEM69* | 1.534082368 | 0.007229175 | 0.035071054 | 1 |
| CAD | adipocyte | *CHCHD6* | 0.997995773 | 2.02E-05 | 0.003854781 | 0.098247197 |
| CAD | adipocyte | *SSU72* | 1.479963689 | 0.000868857 | 0.017317396 | 1 |
| CAD | adipocyte | *TAOK1* | 1.088825758 | 0.002145558 | 0.018430441 | 1 |
| CAD | adipocyte | *S100A1* | -0.801003528 | 0.0123022 | 0.049168267 | 1 |
| CAD | adipocyte | *ZBTB7B* | 1.155907259 | 0.005871979 | 0.031002003 | 1 |
| CAD | adipocyte | *IER2* | 0.915387095 | 0.005771319 | 0.030603758 | 1 |
| CAD | adipocyte | *CPSF4* | 0.971930946 | 8.29E-05 | 0.007733492 | 0.402141577 |
| CAD | adipocyte | *ZNF333* | 1.593599403 | 0.002448035 | 0.019127952 | 1 |
| CAD | adipocyte | *RPL8* | 1.084129066 | 0.001141195 | 0.017317396 | 1 |
| CAD | adipocyte | *ABCF3* | 0.340220045 | 0.009122436 | 0.040909163 | 1 |
| CAD | adipocyte | *PGAP3* | 2.469881856 | 0.001123342 | 0.017317396 | 1 |
| CAD | adipocyte | *ALDH16A1* | 0.272509998 | 0.010368012 | 0.04408904 | 1 |
| CAD | adipocyte | *ASB16* | 1.374893255 | 0.00353915 | 0.022835048 | 1 |
| CAD | adipocyte | *RPS6KA4* | 2.445803988 | 0.00137318 | 0.017317396 | 1 |
| CAD | adipocyte | *COA7* | 2.528329549 | 0.006655522 | 0.033568181 | 1 |
| CAD | adipocyte | *LAPTM5* | 0.997907456 | 0.005707487 | 0.030384947 | 1 |
| CAD | adipocyte | *TM2D1* | 1.301832921 | 0.001498284 | 0.017317396 | 1 |
| CAD | adipocyte | *NEXN* | 0.772842469 | 0.005245303 | 0.028855115 | 1 |
| CAD | adipocyte | *CNST* | 1.030120753 | 0.006929919 | 0.034323005 | 1 |
| CAD | adipocyte | *HAAO* | 1.449398773 | 0.003159273 | 0.021297167 | 1 |

| | | | | | | |
|---|---|---|---|---|---|---|
| CAD | adipocyte | *MRPL55* | 0.472478647 | 0.006303711 | 0.03253788 | 1 |
| CAD | adipocyte | *KIAA1841* | 0.997229866 | 0.010997283 | 0.045959359 | 1 |
| CAD | adipocyte | *NOSTRIN* | 0.467818636 | 0.003423886 | 0.022389079 | 1 |
| CAD | adipocyte | *PACRGL* | 0.82043817 | 0.007910747 | 0.037228852 | 1 |
| CAD | adipocyte | *RNF149* | 0.916343382 | 0.003903064 | 0.024279059 | 1 |
| CAD | adipocyte | *GPR155* | 1.390421074 | 0.002171616 | 0.018453035 | 1 |
| CAD | adipocyte | *DAPL1* | 0.893755118 | 0.001968748 | 0.017921887 | 1 |
| CAD | adipocyte | *PBXIP1* | 1.999664762 | 0.000180641 | 0.010074346 | 0.876468113 |
| CAD | adipocyte | *LRRC58* | 1.073656954 | 0.008922509 | 0.040525141 | 1 |
| CAD | adipocyte | *GTPBP8* | 1.957598434 | 0.002284509 | 0.018883196 | 1 |
| CAD | adipocyte | *WDFY3* | 0.821766874 | 0.005154114 | 0.028582558 | 1 |
| CAD | adipocyte | *C4orf36* | 1.589642834 | 0.003418548 | 0.02238434 | 1 |
| CAD | adipocyte | *TIPARP* | 1.723247651 | 4.85E-05 | 0.006632491 | 0.235312667 |
| CAD | adipocyte | *CPA3* | 1.865241498 | 0.005396567 | 0.029288753 | 1 |
| CAD | adipocyte | *SLC4A1AP* | 1.736750387 | 0.002255969 | 0.018849915 | 1 |
| CAD | adipocyte | *SNIP1* | 1.966619582 | 0.001811974 | 0.017489118 | 1 |
| CAD | adipocyte | *RPN1* | 1.415069547 | 0.001861862 | 0.017489118 | 1 |
| CAD | adipocyte | *SFMBT1* | 1.509037768 | 0.005471508 | 0.029561594 | 1 |
| CAD | adipocyte | *UVSSA* | 0.430677043 | 0.010644447 | 0.044916181 | 1 |
| CAD | adipocyte | *FAM208A* | 1.263653052 | 0.000431086 | 0.014425028 | 1 |
| CAD | adipocyte | *ABLIM2* | 0.663219574 | 0.005851274 | 0.030926342 | 1 |
| CAD | adipocyte | *RNF123* | 0.570053613 | 6.07E-06 | 0.002461025 | 0.029463648 |
| CAD | adipocyte | *HSPA4L* | 0.611687933 | 0.004730315 | 0.027421131 | 1 |
| CAD | adipocyte | *MST1R* | 0.394469022 | 0.00012034 | 0.008341315 | 0.583892078 |
| CAD | adipocyte | *ETNPPL* | 10.01394846 | 0.005137543 | 0.028582558 | 1 |
| CAD | adipocyte | *MAP9* | 1.419941514 | 0.007670235 | 0.03645052 | 1 |
| CAD | adipocyte | *ARFIP1* | 0.854177431 | 0.007100211 | 0.03465817 | 1 |
| CAD | adipocyte | *WDR41* | 0.749331322 | 0.009189395 | 0.041043909 | 1 |
| CAD | adipocyte | *NDUFS4* | 1.202022169 | 0.002220644 | 0.018705844 | 1 |
| CAD | adipocyte | *SERINC5* | 0.9352082 | 0.001743942 | 0.017324379 | 1 |
| CAD | adipocyte | *UTP15* | 1.612631148 | 0.005890457 | 0.031032029 | 1 |
| CAD | adipocyte | *PDSS2* | 1.151135728 | 0.001411855 | 0.017317396 | 1 |
| CAD | adipocyte | *GALNT10* | 0.586304632 | 0.012285821 | 0.04916762 | 1 |
| CAD | adipocyte | *SAP30L* | 1.161629126 | 0.001679008 | 0.017317396 | 1 |
| CAD | adipocyte | *RPS14* | 1.007202864 | 0.00108027 | 0.017317396 | 1 |
| CAD | adipocyte | *RP9* | 1.743412438 | 0.006953263 | 0.034352818 | 1 |
| CAD | adipocyte | *MIOS* | 1.088920602 | 0.007505675 | 0.035914728 | 1 |
| CAD | adipocyte | *BRI3* | 1.327047546 | 0.001573183 | 0.017317396 | 1 |
| CAD | adipocyte | *RAD21* | 1.198685641 | 0.00207304 | 0.018388281 | 1 |
| CAD | adipocyte | *TNFRSF11B* | 3.104923483 | 1.41E-05 | 0.003255419 | 0.068363802 |
| CAD | adipocyte | *OXR1* | 1.054146543 | 0.001328723 | 0.017317396 | 1 |
| CAD | adipocyte | *FASTK* | 1.584134406 | 0.008444536 | 0.038947614 | 1 |
| CAD | adipocyte | *INTS8* | 1.093935709 | 0.008142036 | 0.038058918 | 1 |
| CAD | adipocyte | *TMEM65* | 1.069891353 | 0.002455345 | 0.019127952 | 1 |
| CAD | adipocyte | *ZMAT4* | 2.531272218 | 0.005240341 | 0.028855115 | 1 |
| CAD | adipocyte | *AQP3* | 3.841096341 | 0.006333621 | 0.032622856 | 1 |
| CAD | adipocyte | *TRMT10B* | 1.097397823 | 0.004699954 | 0.027311752 | 1 |
| CAD | adipocyte | *PIGO* | 1.388740494 | 9.56E-05 | 0.007995986 | 0.463767204 |
| CAD | adipocyte | *WRN* | 1.276383593 | 0.000320609 | 0.013060813 | 1 |
| CAD | adipocyte | *MARCH8* | 1.392303113 | 0.001587251 | 0.017317396 | 1 |

| | | | | | | |
|---|---|---|---|---|---|---|
| CAD | adipocyte | *MICU2* | 1.03133022 | 0.004371402 | 0.026250054 | 1 |
| CAD | adipocyte | *QSOX2* | 1.17808632 | 0.00611196 | 0.031773492 | 1 |
| CAD | adipocyte | *METTL3* | 1.500954424 | 0.000689582 | 0.017317396 | 1 |
| CAD | adipocyte | *TCP11L2* | 1.437289898 | 0.001030562 | 0.017317396 | 1 |
| CAD | adipocyte | *JAM3* | 0.589960485 | 0.011715252 | 0.047968273 | 1 |
| CAD | adipocyte | *API5* | 1.53177377 | 0.001430773 | 0.017317396 | 1 |
| CAD | adipocyte | *ZNF202* | 1.648037071 | 0.000455844 | 0.014978586 | 1 |
| CAD | adipocyte | *STXBP4* | 1.079750539 | 0.001974861 | 0.017943871 | 1 |
| CAD | adipocyte | *WBP1L* | 1.005197339 | 0.001902808 | 0.017489118 | 1 |
| CAD | adipocyte | *RPL27A* | 1.173666743 | 0.001833107 | 0.017489118 | 1 |
| CAD | adipocyte | *C16orf46* | -0.84369113 | 0.003008324 | 0.020696384 | 1 |
| CAD | adipocyte | *TMEM41B* | 1.239856109 | 0.005129316 | 0.028582558 | 1 |
| CAD | adipocyte | *TMX3* | 1.130256793 | 0.000709852 | 0.017317396 | 1 |
| CAD | adipocyte | *SEC11C* | 1.394852465 | 0.004151423 | 0.025274753 | 1 |
| CAD | adipocyte | *SLFN5* | 1.130329052 | 0.002113248 | 0.018430441 | 1 |
| CAD | adipocyte | *SAAL1* | 2.285499382 | 0.001261263 | 0.017317396 | 1 |
| CAD | adipocyte | *PIP4K2C* | 1.379718202 | 0.00045689 | 0.014978586 | 1 |
| CAD | adipocyte | *MTMR10* | 0.716306788 | 0.002810809 | 0.019938662 | 1 |
| CAD | adipocyte | *MS4A14* | -0.13307101 | 0.003058989 | 0.020963582 | 1 |
| CAD | adipocyte | *EPB42* | 1.204549268 | 0.00359837 | 0.023027229 | 1 |
| CAD | adipocyte | *SMAD3* | 0.945835559 | 0.002193614 | 0.018565786 | 1 |
| CAD | adipocyte | *SP2* | 1.813942121 | 0.002572617 | 0.019265876 | 1 |
| CAD | adipocyte | *FBXO22* | 1.272294554 | 0.003731888 | 0.023454819 | 1 |
| CAD | adipocyte | *ZNF91* | 0.885824239 | 0.005481714 | 0.029561594 | 1 |
| CAD | adipocyte | *ENGASE* | 0.373801522 | 0.001826449 | 0.017489118 | 1 |
| CAD | adipocyte | *VKORC1* | 1.510748724 | 0.004469039 | 0.02662058 | 1 |
| CAD | adipocyte | *RPL13* | 0.75957282 | 0.011288371 | 0.046893129 | 1 |
| CAD | adipocyte | *ZNF641* | 1.81937604 | 4.94E-05 | 0.006632491 | 0.23958415 |
| CAD | adipocyte | *ZNF528* | 1.809306792 | 0.000650641 | 0.017317396 | 1 |
| CAD | adipocyte | *SERTAD3* | 2.631047256 | 0.002709783 | 0.019782057 | 1 |
| CAD | adipocyte | *LENG8* | 1.046874531 | 0.005497111 | 0.029569824 | 1 |
| CAD | adipocyte | *CYB5D2* | 1.279526547 | 0.002511234 | 0.019127952 | 1 |
| CAD | adipocyte | *NDUFV1* | 1.439904321 | 0.002711267 | 0.019782057 | 1 |
| CAD | adipocyte | *PRDX2* | 1.438730666 | 0.007047081 | 0.03446529 | 1 |
| CAD | adipocyte | *ATP5H* | 1.077217734 | 0.009664998 | 0.042361853 | 1 |
| CAD | adipocyte | *ATG16L2* | 1.262446961 | 0.002265558 | 0.018849915 | 1 |
| CAD | adipocyte | *FAM83B* | 3.391839483 | 0.011867434 | 0.048306033 | 1 |
| CAD | adipocyte | *COA6* | 2.579524353 | 0.001195085 | 0.017317396 | 1 |
| CAD | adipocyte | *PCMTD1* | 1.151860945 | 0.001399212 | 0.017317396 | 1 |
| CAD | adipocyte | *MLKL* | 0.319816192 | 0.001636398 | 0.017317396 | 1 |
| CAD | adipocyte | *SLC20A2* | 0.92236827 | 0.005919906 | 0.031119591 | 1 |
| CAD | adipocyte | *NBPF9* | 2.425468321 | 0.004738697 | 0.027427118 | 1 |
| CAD | adipocyte | *ADAM9* | 0.893109379 | 0.000535452 | 0.016058757 | 1 |
| CAD | adipocyte | *DNAJC21* | 1.662936646 | 0.001218023 | 0.017317396 | 1 |
| CAD | adipocyte | *SNTB2* | 1.278323452 | 0.001109927 | 0.017317396 | 1 |
| CAD | adipocyte | *STX18* | 1.389990715 | 0.001205923 | 0.017317396 | 1 |
| CAD | adipocyte | *ZBTB49* | 1.830122435 | 0.006697289 | 0.033643116 | 1 |
| CAD | adipocyte | *ATOH8* | 0.521544238 | 0.000290346 | 0.01225006 | 1 |
| CAD | adipocyte | *CEP120* | 1.255691659 | 0.003159154 | 0.021297167 | 1 |
| CAD | adipocyte | *MFF* | 1.316760898 | 0.00147541 | 0.017317396 | 1 |

| | | | | | | |
|---|---|---|---|---|---|---|
| CAD | adipocyte | *MFSD7* | 2.919022428 | 0.002380427 | 0.019127952 | 1 |
| CAD | adipocyte | *MAP2K1* | 1.384539869 | 0.001247583 | 0.017317396 | 1 |
| CAD | adipocyte | *UBE2V2* | 1.09157188 | 0.00236197 | 0.019068686 | 1 |
| CAD | adipocyte | *XPO6* | 1.071091259 | 0.003340445 | 0.022051479 | 1 |
| CAD | adipocyte | *THBS3* | 1.271238569 | 0.000428066 | 0.014423463 | 1 |
| CAD | adipocyte | *PGM2* | 0.699778069 | 0.006855553 | 0.034081088 | 1 |
| CAD | adipocyte | *HINT1* | 1.338483299 | 0.002443142 | 0.019127952 | 1 |
| CAD | adipocyte | *LUZP1* | 1.38002127 | 0.001125472 | 0.017317396 | 1 |
| CAD | adipocyte | *UGP2* | 0.546928995 | 0.010856162 | 0.045526445 | 1 |
| CAD | adipocyte | *CSGALNACT2* | 1.53668444 | 0.000105369 | 0.008341315 | 0.511251905 |
| CAD | adipocyte | *AVEN* | 1.359581904 | 0.002140298 | 0.018430441 | 1 |
| CAD | adipocyte | *TOR1AIP2* | 0.838722084 | 0.012397936 | 0.049307203 | 1 |
| CAD | adipocyte | *MAP3K2* | 0.667938818 | 0.001827291 | 0.017489118 | 1 |
| CAD | adipocyte | *MYO7B* | 0.388224573 | 0.006219089 | 0.032203865 | 1 |
| CAD | adipocyte | *HNRNPA3* | 0.940168857 | 0.000695333 | 0.017317396 | 1 |
| CAD | adipocyte | *PDCD6IP* | 1.136903467 | 0.00259522 | 0.019312893 | 1 |
| CAD | adipocyte | *CNGB3* | 0.579806138 | 0.002655051 | 0.019607776 | 1 |
| CAD | adipocyte | *STX8* | 1.147301596 | 0.000631187 | 0.017302381 | 1 |
| CAD | adipocyte | *NFRKB* | 1.521521816 | 0.00242957 | 0.019127952 | 1 |
| CAD | adipocyte | *NFXL1* | 1.059980405 | 0.001505561 | 0.017317396 | 1 |
| CAD | adipocyte | *MSANTD4* | 1.943080331 | 0.001699851 | 0.017317396 | 1 |
| CAD | adipocyte | *TSNARE1* | 0.566290197 | 0.003323317 | 0.02199827 | 1 |
| CAD | adipocyte | *FEZ2* | 0.919336836 | 0.001417532 | 0.017317396 | 1 |
| CAD | adipocyte | *MFN1* | 0.808671661 | 0.000643096 | 0.017317396 | 1 |
| CAD | adipocyte | *PDE7B* | 0.730954734 | 0.001376999 | 0.017317396 | 1 |
| CAD | adipocyte | *MCC* | 1.161384635 | 0.001337917 | 0.017317396 | 1 |
| CAD | adipocyte | *PPID* | 1.663834609 | 0.000485125 | 0.01542398 | 1 |
| CAD | adipocyte | *TBCA* | 1.175650883 | 0.002352623 | 0.019062957 | 1 |
| CAD | adipocyte | *PIK3CD* | 0.768655603 | 0.001984269 | 0.017995654 | 1 |
| CAD | adipocyte | *CAMTA1* | 0.729236999 | 0.011612992 | 0.047751049 | 1 |
| CAD | adipocyte | *NXNL1* | 0.136418901 | 0.008202238 | 0.038156528 | 1 |
| CAD | adipocyte | *TRAPPC12* | 1.347259757 | 0.000843871 | 0.017317396 | 1 |
| CAD | adipocyte | *C3AR1* | -0.23521467 | 0.007485128 | 0.035851767 | 1 |
| CAD | adipocyte | *ATPAF2* | 0.635861552 | 0.005075118 | 0.028502532 | 1 |
| CAD | adipocyte | *NME6* | 0.590146559 | 0.000856279 | 0.017317396 | 1 |
| CAD | adipocyte | *TEFM* | 1.392844542 | 2.50E-06 | 0.002461025 | 0.012117784 |
| CAD | adipocyte | *MALT1* | 1.271588095 | 0.001447286 | 0.017317396 | 1 |
| CAD | adipocyte | *MBOAT1* | 1.143717835 | 0.003340198 | 0.022051479 | 1 |
| CAD | adipocyte | *PAIP1* | 1.474977619 | 0.000670228 | 0.017317396 | 1 |
| CAD | adipocyte | *BSG* | 1.281005898 | 0.002149205 | 0.018430441 | 1 |
| CAD | adipocyte | *HINFP* | 1.03328326 | 0.010543674 | 0.044673236 | 1 |
| CAD | adipocyte | *SUCLG2* | 1.207852261 | 0.000680573 | 0.017317396 | 1 |
| CAD | adipocyte | *FGGY* | 1.152702885 | 7.33E-05 | 0.007733492 | 0.355815882 |
| CAD | adipocyte | *AFF1* | 0.99071308 | 0.00171934 | 0.017317396 | 1 |
| CAD | adipocyte | *ZNF738* | 0.268244182 | 0.004516568 | 0.026790203 | 1 |
| CAD | adipocyte | *CFL1* | 1.062310081 | 0.002573019 | 0.019265876 | 1 |
| CAD | adipocyte | *CBWD1* | 1.622273491 | 0.009529388 | 0.042134669 | 1 |
| CAD | adipocyte | *DCP2* | 1.105792745 | 0.002659583 | 0.019611396 | 1 |
| CAD | adipocyte | *ZNF621* | 1.636245954 | 0.001041836 | 0.017317396 | 1 |
| CAD | adipocyte | *GXYLT2* | 0.970446161 | 0.00644989 | 0.032840444 | 1 |

| | | | | | | |
|---|---|---|---|---|---|---|
| CAD | adipocyte | *MAP3K11* | 0.948617493 | 0.000120145 | 0.008341315 | 0.582941346 |
| CAD | adipocyte | *KCNK7* | 2.0592454 | 0.002056197 | 0.018272288 | 1 |
| CAD | adipocyte | *PPP1R14B* | 1.557077841 | 0.000253982 | 0.011672743 | 1 |
| CAD | adipocyte | *PEAK1* | 0.862073767 | 0.001233069 | 0.017317396 | 1 |
| CAD | adipocyte | *GOLIM4* | 1.224068501 | 0.00064934 | 0.017317396 | 1 |
| CAD | adipocyte | *XXYLT1* | 1.459291831 | 0.001731453 | 0.017317396 | 1 |
| CAD | adipocyte | *UBXN2A* | 0.893615307 | 0.006105646 | 0.031773492 | 1 |
| CAD | adipocyte | *SLC25A30* | 1.154962928 | 0.006698114 | 0.033643116 | 1 |
| CAD | adipocyte | *LEMD3* | 1.452165635 | 0.001228592 | 0.017317396 | 1 |
| CAD | adipocyte | *SNAPC5* | 0.760297518 | 0.005266048 | 0.028868018 | 1 |
| CAD | adipocyte | *ANKRD36C* | 0.994516429 | 0.002478271 | 0.019127952 | 1 |
| CAD | adipocyte | *MSL2* | 1.385300505 | 0.002451035 | 0.019127952 | 1 |
| CAD | adipocyte | *ANGEL2* | 1.444886132 | 0.001082324 | 0.017317396 | 1 |
| CAD | adipocyte | *SH3PXD2B* | 0.666410482 | 0.007360134 | 0.035463129 | 1 |
| CAD | adipocyte | *LARP7* | 1.329820167 | 0.00089224 | 0.017317396 | 1 |
| CAD | adipocyte | *CEP135* | 0.975628099 | 0.005385905 | 0.029263619 | 1 |
| CAD | adipocyte | *NLRP6* | -1.33603644 | 0.009121606 | 0.040909163 | 1 |
| CAD | adipocyte | *RSRC1* | 0.896562061 | 0.00376106 | 0.023607587 | 1 |
| CAD | adipocyte | *KCTD13* | 0.523166599 | 0.001702135 | 0.017317396 | 1 |
| CAD | adipocyte | *DHX36* | 1.249941193 | 0.001623149 | 0.017317396 | 1 |
| CAD | adipocyte | *TVP23C* | 3.713656408 | 0.001127494 | 0.017317396 | 1 |
| CAD | adipocyte | *ATG13* | 1.244668516 | 0.007117071 | 0.034705558 | 1 |
| CAD | adipocyte | *CCNE2* | -0.38710483 | 0.006106522 | 0.031773492 | 1 |
| CAD | adipocyte | *CLTB* | 1.720550442 | 0.001122057 | 0.017317396 | 1 |
| CAD | adipocyte | *MCTP1* | 0.733290142 | 0.004692981 | 0.027311752 | 1 |
| CAD | adipocyte | *TTLL11* | 1.267280182 | 0.001860891 | 0.017489118 | 1 |
| CAD | adipocyte | *ZDHHC21* | 0.754120266 | 0.007156014 | 0.034825455 | 1 |
| CAD | adipocyte | *A2M* | 0.854095521 | 0.002690491 | 0.019719428 | 1 |
| CAD | adipocyte | *B3GALT6* | 3.28512172 | 0.00498718 | 0.028202564 | 1 |
| CAD | adipocyte | *CRLF3* | 0.969992512 | 2.01E-06 | 0.002461025 | 0.009768363 |
| CAD | adipocyte | *WDR25* | 1.566922465 | 0.002770028 | 0.019873367 | 1 |
| CAD | adipocyte | *ANKLE2* | 1.136293061 | 0.0020958 | 0.018430441 | 1 |
| CAD | adipocyte | *DPP7* | 1.444631039 | 0.002561867 | 0.019262254 | 1 |
| CAD | adipocyte | *MAN1B1* | 1.894119852 | 0.001205096 | 0.017317396 | 1 |
| CAD | adipocyte | *SLC25A22* | -0.632593777 | 0.009098531 | 0.040909163 | 1 |
| CAD | adipocyte | *THAP5* | 1.431370513 | 0.006122007 | 0.031773492 | 1 |
| CAD | adipocyte | *NAALADL2* | 0.511259646 | 0.004975207 | 0.02820059 | 1 |
| CAD | adipocyte | *CD151* | 0.935497998 | 0.000450029 | 0.014955757 | 1 |
| CAD | adipocyte | *SLC35G5* | 12.63816543 | 0.002150057 | 0.018430441 | 1 |
| CAD | adipocyte | *PCDHB9* | 0.439164894 | 0.010645838 | 0.044916181 | 1 |
| CAD | adipocyte | *CENPBD1* | 4.593058148 | 0.00199895 | 0.018030022 | 1 |
| CAD | adipocyte | *RPS27* | 1.335871651 | 0.000595 | 0.016687501 | 1 |
| CAD | adipocyte | *ASB8* | 1.132596358 | 0.002792788 | 0.019873367 | 1 |
| CAD | adipocyte | *IMPDH2* | 0.624310675 | 0.000826427 | 0.017317396 | 1 |
| CAD | adipocyte | *NT5DC1* | 0.99546331 | 0.008312943 | 0.038597512 | 1 |
| CAD | adipocyte | *CA8* | 0.716799603 | 0.006654037 | 0.033568181 | 1 |
| CAD | adipocyte | *ERN1* | 1.123480247 | 0.002532749 | 0.019231452 | 1 |
| CAD | adipocyte | *NSUN3* | 1.008341476 | 0.009707051 | 0.042507771 | 1 |
| CAD | adipocyte | *C17orf62* | 1.82445245 | 0.001857216 | 0.017489118 | 1 |
| CAD | adipocyte | *GAK* | 1.475090918 | 0.000361561 | 0.013592277 | 1 |

| | | | | | | |
|---|---|---|---|---|---|---|
| CAD | adipocyte | *CTC1* | 0.657834606 | 0.007448789 | 0.035783687 | 1 |
| CAD | adipocyte | *SPTY2D1* | 1.669577629 | 0.001832796 | 0.017489118 | 1 |
| CAD | adipocyte | *PACS2* | 1.476012196 | 0.001284192 | 0.017317396 | 1 |
| CAD | adipocyte | *GIPC3* | 7.762446132 | 0.004614571 | 0.027076951 | 1 |
| CAD | adipocyte | *PDXDC1* | 1.352352992 | 0.000226454 | 0.010987538 | 1 |
| CAD | adipocyte | *SEPT1* | 1.779926484 | 0.007171663 | 0.034831741 | 1 |
| CAD | adipocyte | *EXOC3* | 0.976775014 | 0.009724764 | 0.042546938 | 1 |
| CAD | adipocyte | *LYNX1* | -0.278802059 | 0.006920999 | 0.034323005 | 1 |
| CAD | adipocyte | *MCFD2* | 0.798105243 | 0.005692241 | 0.030383667 | 1 |
| CAD | adipocyte | *DEFB119* | 6.232830465 | 0.000362388 | 0.013592277 | 1 |
| CAD | adipocyte | *ZDHHC20* | 1.280264238 | 0.001278824 | 0.017317396 | 1 |
| CAD | adipocyte | *CAPS2* | 1.573126538 | 1.87E-05 | 0.003854781 | 0.090697775 |
| CAD | adipocyte | *CMTR2* | 0.834919949 | 0.005350966 | 0.029139043 | 1 |
| CAD | adipocyte | *MRPL14* | 1.041521592 | 0.00013383 | 0.008657898 | 0.649342319 |
| CAD | adipocyte | *ADIPOQ* | -0.120689291 | 0.004492893 | 0.026682393 | 1 |
| CAD | adipocyte | *MUC16* | 0.797042248 | 0.00614208 | 0.031839071 | 1 |
| CAD | adipocyte | *TMEM136* | 1.180460035 | 2.93E-06 | 0.002461025 | 0.014200948 |
| CAD | adipocyte | *DDX60L* | 0.730423914 | 0.010362989 | 0.04408904 | 1 |
| CAD | adipocyte | *SGSH* | 2.672657234 | 0.001325913 | 0.017317396 | 1 |
| CAD | adipocyte | *ZBTB20* | 0.786629328 | 0.001728458 | 0.017317396 | 1 |
| CAD | adipocyte | *C3orf58* | 1.603891849 | 0.000259822 | 0.011672743 | 1 |
| CAD | adipocyte | *TMEM252* | 67.92090452 | 0.002304917 | 0.018925801 | 1 |
| CAD | adipocyte | *RELL1* | 0.714347285 | 0.003499889 | 0.022648861 | 1 |
| CAD | adipocyte | *RFX7* | 1.202907994 | 0.00119434 | 0.017317396 | 1 |
| CAD | adipocyte | *ZNF329* | 1.476947126 | 0.005350365 | 0.029139043 | 1 |
| CAD | adipocyte | *IDH2* | 1.402069489 | 0.005051486 | 0.02846668 | 1 |
| CAD | adipocyte | *ERCC6L2* | 1.318815734 | 0.000193524 | 0.010214126 | 0.938976256 |
| CAD | adipocyte | *BACE2* | 0.222231909 | 0.006734669 | 0.03379174 | 1 |
| CAD | adipocyte | *KCNA4* | 1.669756025 | 0.004781777 | 0.027554849 | 1 |
| CAD | adipocyte | *KBTBD3* | 1.634267336 | 0.00260944 | 0.019359334 | 1 |
| CAD | adipocyte | *CLN8* | 0.226175677 | 0.008943808 | 0.040525141 | 1 |
| CAD | adipocyte | *FES* | -0.580308083 | 0.002356736 | 0.019062957 | 1 |
| CAD | adipocyte | *GLRX5* | 2.037946165 | 0.002785278 | 0.019873367 | 1 |
| CAD | adipocyte | *TRAK1* | 0.613596041 | 0.002053816 | 0.018272288 | 1 |
| CAD | adipocyte | *SKA2* | 1.268895552 | 0.002506676 | 0.019127952 | 1 |
| CAD | adipocyte | *PLCXD3* | 1.26904168 | 0.001374695 | 0.017317396 | 1 |
| CAD | adipocyte | *ALG12* | 1.965770652 | 0.002978268 | 0.020593222 | 1 |
| CAD | adipocyte | *ZNF721* | 1.170621795 | 0.000232942 | 0.011080708 | 1 |
| CAD | adipocyte | *C12orf60* | 1.734864239 | 0.001746006 | 0.017324379 | 1 |
| CAD | adipocyte | *ABAT* | 1.031717155 | 0.008424352 | 0.038891491 | 1 |
| CAD | adipocyte | *ARHGEF37* | 1.806302662 | 0.003011469 | 0.020696384 | 1 |
| CAD | adipocyte | *CSMD1* | 1.947407902 | 0.001080601 | 0.017317396 | 1 |
| CAD | adipocyte | *C19orf71* | 1.092663479 | 0.006652065 | 0.033568181 | 1 |
| CAD | adipocyte | *SERHL2* | 1.74547114 | 0.006628897 | 0.033568181 | 1 |
| CAD | adipocyte | *BTBD9* | 0.861811646 | 0.005504111 | 0.029574692 | 1 |
| CAD | adipocyte | *TTC32* | 1.622748826 | 0.006425422 | 0.032840444 | 1 |
| CAD | adipocyte | *DENND5A* | 1.045260716 | 0.003719807 | 0.023439618 | 1 |
| CAD | adipocyte | *COPB2* | 1.385189166 | 0.001816568 | 0.017489118 | 1 |
| CAD | adipocyte | *TMEM255B* | 1.100237077 | 0.001192305 | 0.017317396 | 1 |
| CAD | adipocyte | *SNN* | 1.924081938 | 0.007049782 | 0.03446529 | 1 |

| | | | | | | |
|---|---|---|---|---|---|---|
| CAD | adipocyte | *ZFP90* | 1.550035001 | 0.001200619 | 0.017317396 | 1 |
| CAD | adipocyte | *TMEM106A* | 0.269497284 | 0.008937183 | 0.040525141 | 1 |
| CAD | adipocyte | *IFITM2* | 1.029737495 | 0.00644779 | 0.032840444 | 1 |
| CAD | adipocyte | *SP140L* | 0.810256893 | 0.001865462 | 0.017489118 | 1 |
| CAD | adipocyte | *AHNAK2* | 0.856496003 | 0.005030182 | 0.028379586 | 1 |
| CAD | adipocyte | *SP1* | 0.959270738 | 0.004457261 | 0.02662058 | 1 |
| CAD | adipocyte | *ZNF529* | 1.369411432 | 0.000950559 | 0.017317396 | 1 |
| CAD | adipocyte | *C15orf41* | 0.624755945 | 0.010894314 | 0.045641973 | 1 |
| CAD | adipocyte | *WWOX* | 0.894470874 | 0.001402831 | 0.017317396 | 1 |
| CAD | adipocyte | *NBPF12* | 0.19273275 | 0.011398417 | 0.047092292 | 1 |
| CAD | adipocyte | *TOR3A* | 1.204366133 | 0.003160338 | 0.021297167 | 1 |
| CAD | adipocyte | *GNG2* | 1.353285979 | 0.00114376 | 0.017317396 | 1 |
| CAD | adipocyte | *TMEM222* | 1.681656478 | 0.002031739 | 0.018152939 | 1 |
| CAD | adipocyte | *SEPT10* | 0.842758476 | 0.006575663 | 0.03335063 | 1 |
| CAD | adipocyte | *VSIG10L* | -0.536502402 | 0.009919548 | 0.042949124 | 1 |
| CAD | adipocyte | *TMPRSS6* | 9.178919235 | 0.007053593 | 0.03446529 | 1 |
| CAD | adipocyte | *GJA4* | 1.41924585 | 0.008760959 | 0.040073145 | 1 |
| CAD | adipocyte | *EXD3* | 1.571470719 | 0.001413897 | 0.017317396 | 1 |
| CAD | adipocyte | *ZFP69* | 2.41969998 | 0.002824744 | 0.019974713 | 1 |
| CAD | adipocyte | *FAM122A* | 2.800746011 | 0.00154238 | 0.017317396 | 1 |
| CAD | adipocyte | *PLSCR1* | 0.71383883 | 0.002554004 | 0.019262254 | 1 |
| CAD | adipocyte | *C22orf34* | 0.873489146 | 0.003808563 | 0.023782688 | 1 |
| CAD | adipocyte | *COL25A1* | 1.093894977 | 0.008041935 | 0.037736432 | 1 |
| CAD | adipocyte | *SRSF10* | 0.594198684 | 0.005513819 | 0.029594083 | 1 |
| CAD | adipocyte | *NBR1* | 0.939344035 | 0.004943803 | 0.028060596 | 1 |
| CAD | adipocyte | *MTF1* | 1.008485048 | 0.008732492 | 0.040009491 | 1 |
| CAD | adipocyte | *RPL14* | 1.175417529 | 0.001586183 | 0.017317396 | 1 |
| CAD | adipocyte | *FAM196A* | 1.264874244 | 0.001137561 | 0.017317396 | 1 |
| CAD | adipocyte | *NELFB* | 1.59905021 | 0.005114861 | 0.028582558 | 1 |
| CAD | adipocyte | *SF3B3* | 1.087524515 | 0.01035126 | 0.04408904 | 1 |
| CAD | adipocyte | *TSPYL1* | 1.524872291 | 0.000891846 | 0.017317396 | 1 |
| CAD | adipocyte | *PNRC2* | 1.543391471 | 0.004323635 | 0.026092382 | 1 |
| CAD | adipocyte | *FHIT* | 0.897853249 | 0.002389851 | 0.019127952 | 1 |
| CAD | adipocyte | *WDSUB1* | 0.69354847 | 0.011786171 | 0.048110383 | 1 |
| CAD | adipocyte | *SEMA4A* | 0.938471779 | 0.000975764 | 0.017317396 | 1 |
| CAD | adipocyte | *MPHOSPH8* | 0.883835744 | 0.001469034 | 0.017317396 | 1 |
| CAD | adipocyte | *PPIA* | 0.589334928 | 0.012016388 | 0.048613338 | 1 |
| CAD | adipocyte | *SUPT3H* | 0.921665461 | 0.007565744 | 0.036095369 | 1 |
| CAD | adipocyte | *CD55* | 0.887361761 | 0.003973809 | 0.024593014 | 1 |
| CAD | adipocyte | *PIK3R4* | 1.493045682 | 0.002080561 | 0.018421314 | 1 |
| CAD | adipocyte | *ZNF605* | 0.460868609 | 0.001908963 | 0.017489118 | 1 |
| CAD | adipocyte | *ZNF846* | 1.083550553 | 0.000393808 | 0.013687955 | 1 |
| CAD | adipocyte | *ZFP62* | 1.057178592 | 0.002160701 | 0.018453035 | 1 |
| CAD | adipocyte | *TOMM7* | 1.356457402 | 0.001244304 | 0.017317396 | 1 |
| CAD | adipocyte | *SLC39A10* | 1.082320771 | 0.003691959 | 0.02338562 | 1 |
| CAD | adipocyte | *WDR5B* | 0.655294532 | 0.007168244 | 0.034831741 | 1 |
| CAD | adipocyte | *ZNF138* | 0.863304383 | 0.001313923 | 0.017317396 | 1 |
| CAD | adipocyte | *ZNF470* | 1.214301101 | 0.000516817 | 0.01577103 | 1 |
| CAD | adipocyte | *GMFB* | 1.796788873 | 0.000787136 | 0.017317396 | 1 |
| CAD | adipocyte | *ZMYM1* | 1.507753664 | 0.00181146 | 0.017489118 | 1 |

| | | | | | | |
|---|---|---|---|---|---|---|
| CAD | adipocyte | *SND1* | 0.875647807 | 0.004454371 | 0.02662058 | 1 |
| CAD | adipocyte | *CCL4L2* | 1.955437271 | 0.010066467 | 0.043338507 | 1 |
| CAD | adipocyte | *ZNF720* | 1.029189008 | 0.003568457 | 0.022932654 | 1 |
| CAD | adipocyte | *MAP3K5* | 0.803070717 | 0.006961681 | 0.034352818 | 1 |
| CAD | adipocyte | *SLC2A10* | 0.755271531 | 0.003602157 | 0.023027229 | 1 |
| CAD | adipocyte | *ZNF665* | 1.533050048 | 0.005757861 | 0.030593003 | 1 |
| CAD | adipocyte | *ATG7* | 0.790840007 | 0.000994518 | 0.017317396 | 1 |
| CAD | adipocyte | *TOPORS* | 1.646238214 | 0.003193733 | 0.021432905 | 1 |
| CAD | adipocyte | *FAM114A1* | 1.406567277 | 0.000424075 | 0.014388884 | 1 |
| CAD | adipocyte | *TXNRD3* | 0.641584987 | 0.009110425 | 0.040909163 | 1 |
| CAD | adipocyte | *ADH5* | 0.922868967 | 0.006440725 | 0.032840444 | 1 |
| CAD | adipocyte | *ZNF677* | 1.897251062 | 0.000571171 | 0.016495956 | 1 |
| CAD | adipocyte | *ZNF71* | 0.517027825 | 0.001048024 | 0.017317396 | 1 |
| CAD | adipocyte | *MPZL1* | 1.014282365 | 0.002722085 | 0.019784984 | 1 |
| CAD | adipocyte | *VPS13A* | 0.848841052 | 0.006981014 | 0.034352818 | 1 |
| CAD | adipocyte | *NOL8* | 1.142455909 | 0.002515692 | 0.019131878 | 1 |
| CAD | adipocyte | *MRPL42* | 0.449766763 | 0.000594431 | 0.016687501 | 1 |
| CAD | adipocyte | *MAK16* | 1.352751722 | 0.002171508 | 0.018453035 | 1 |
| CAD | adipocyte | *HIBCH* | 0.390879873 | 0.003899233 | 0.024279059 | 1 |
| CAD | adipocyte | *ZNF544* | 2.038108153 | 0.001212024 | 0.017317396 | 1 |
| CAD | adipocyte | *SOWAHC* | 1.274669878 | 0.002350553 | 0.019062957 | 1 |
| CAD | adipocyte | *MIER1* | 1.172086365 | 0.000108555 | 0.008341315 | 0.526710031 |
| CAD | adipocyte | *MAN1A2* | 1.220466746 | 0.001464211 | 0.017317396 | 1 |
| CAD | adipocyte | *SVIP* | 1.7214861 | 0.000688408 | 0.017317396 | 1 |
| CAD | adipocyte | *DDRGK1* | 1.486743446 | 0.000368015 | 0.013616273 | 1 |
| CAD | adipocyte | *HSD17B11* | 1.049453672 | 0.004359041 | 0.02624078 | 1 |
| CAD | adipocyte | *RPS6KL1* | 0.599636634 | 0.009835144 | 0.04283673 | 1 |
| CAD | adipocyte | *STYX* | 1.286563168 | 0.008762903 | 0.040073145 | 1 |
| CAD | adipocyte | *HELZ* | 0.83689204 | 0.009571596 | 0.042134669 | 1 |
| CAD | adipocyte | *TMEM116* | -0.241351281 | 0.000195778 | 0.010214126 | 0.949913701 |
| CAD | adipocyte | *TMA16* | 1.508330952 | 0.002276597 | 0.018849915 | 1 |
| CAD | adipocyte | *HLA-DRB5* | -0.061441565 | 0.009106928 | 0.040909163 | 1 |
| CAD | adipocyte | *TTC37* | 0.753241835 | 0.009588471 | 0.042134669 | 1 |
| CAD | adipocyte | *PAPSS2* | 0.935547822 | 0.002857442 | 0.020151608 | 1 |
| CAD | adipocyte | *ANKRD13B* | 1.915831051 | 0.005483968 | 0.029561594 | 1 |
| CAD | adipocyte | *PPP1R14C* | 5.911517079 | 0.002458989 | 0.019127952 | 1 |
| CAD | adipocyte | *GPATCH3* | 1.911713122 | 0.012038026 | 0.048613338 | 1 |
| CAD | adipocyte | *RPL10A* | 1.129917824 | 0.00180689 | 0.017489118 | 1 |
| CAD | adipocyte | *RCSD1* | 0.777290498 | 0.007735616 | 0.036689356 | 1 |
| CAD | adipocyte | *HMGN2* | 0.621166726 | 0.002756297 | 0.019871547 | 1 |
| CAD | adipocyte | *NAGA* | 0.489086986 | 0.00994088 | 0.042988548 | 1 |
| CAD | adipocyte | *C6orf163* | -0.201636559 | 0.011687532 | 0.047934033 | 1 |
| CAD | adipocyte | *PCMTD2* | 1.342992518 | 0.001275962 | 0.017317396 | 1 |
| CAD | adipocyte | *RUFY2* | 1.186663045 | 0.002537308 | 0.019235969 | 1 |
| CAD | adipocyte | *AGAP6* | 0.356158993 | 0.008417922 | 0.038891491 | 1 |
| CAD | adipocyte | *LINC00619* | 5.333497806 | 0.00346043 | 0.022492071 | 1 |
| CAD | adipocyte | *BMPR2* | 1.001237297 | 0.003307018 | 0.021920292 | 1 |
| CAD | adipocyte | *TCEA3* | 0.874618135 | 0.000229011 | 0.011001618 | 1 |
| CAD | adipocyte | *BAG6* | 0.623076776 | 0.000900358 | 0.017317396 | 1 |
| CAD | adipocyte | *LILRB3* | 1.144113957 | 0.000102895 | 0.008320739 | 0.499244339 |

| | | | | | | |
|---|---|---|---|---|---|---|
| CAD | adipocyte | *DCTN1* | 1.848116619 | 0.001769558 | 0.017486546 | 1 |
| CAD | adipocyte | *FAM216A* | 1.723614877 | 0.000109311 | 0.008341315 | 0.530376714 |
| CAD | adipocyte | *SLC35B4* | 1.404301243 | 0.006450318 | 0.032840444 | 1 |
| CAD | adipocyte | *IPO7* | 1.123054406 | 0.001599185 | 0.017317396 | 1 |
| CAD | adipocyte | *LIN52* | 0.487272473 | 0.004818141 | 0.027665821 | 1 |
| CAD | adipocyte | *C1QTNF9B* | 10.89898659 | 0.006043209 | 0.031596603 | 1 |
| CAD | adipocyte | *HCG27* | 0.246155652 | 0.002562899 | 0.019262254 | 1 |
| CAD | adipocyte | *COL6A6* | 0.475688485 | 0.007235393 | 0.035071054 | 1 |
| CAD | adipocyte | *METTL6* | 0.966273425 | 0.002195932 | 0.018565786 | 1 |
| CAD | adipocyte | *SACM1L* | 1.483383798 | 0.001156268 | 0.017317396 | 1 |
| CAD | adipocyte | *TAS2R14* | 0.231138002 | 0.005608064 | 0.030033474 | 1 |
| CAD | adipocyte | *C17orf51* | 2.908947096 | 0.006763097 | 0.033829428 | 1 |
| CAD | adipocyte | *LCAT* | 1.384691723 | 0.01050375 | 0.044549122 | 1 |
| CAD | adipocyte | *PPP1CB* | 0.681213563 | 0.001106305 | 0.017317396 | 1 |
| CAD | adipocyte | *ATF6B* | 0.633029035 | 0.000923251 | 0.017317396 | 1 |
| CAD | adipocyte | *GALT* | 0.886097125 | 0.002143135 | 0.018430441 | 1 |
| CAD | adipocyte | *ZNF891* | 1.749544342 | 0.000248757 | 0.011605454 | 1 |
| CAD | adipocyte | *TSPAN4* | 0.746460823 | 0.00395923 | 0.024534076 | 1 |
| CAD | adipocyte | *PAXIP1-AS2* | 1.532044044 | 0.001096069 | 0.017317396 | 1 |
| CAD | adipocyte | *LYRM4* | 1.381003172 | 0.001706612 | 0.017317396 | 1 |
| CAD | adipocyte | *PRCD* | 1.210249942 | 0.00085568 | 0.017317396 | 1 |
| CAD | adipocyte | *SH3D21* | 0.579959061 | 0.005298229 | 0.028948387 | 1 |
| CAD | adipocyte | *ANG* | 1.755541917 | 0.009056018 | 0.040893914 | 1 |
| CAD | adipocyte | *GNAT3* | 2.317715703 | 0.001078726 | 0.017317396 | 1 |
| CAD | adipocyte | *VPS16* | 1.809630992 | 0.003083053 | 0.021068977 | 1 |
| CAD | adipocyte | *MYL5* | 0.645242612 | 0.008801099 | 0.040209919 | 1 |
| CAD | adipocyte | *PAM16* | 1.219016759 | 0.005557791 | 0.029797132 | 1 |
| CAD | adipocyte | *RNASEK* | 0.500433027 | 0.00965549 | 0.042358444 | 1 |
| CAD | adipocyte | *C5orf66* | 0.865464724 | 0.010846314 | 0.045524493 | 1 |
| CAD | adipocyte | *CYP4A22-AS1* | 2.251889545 | 0.002121924 | 0.018430441 | 1 |
| CAD | adipocyte | *ERCC6* | 1.018982202 | 0.006517497 | 0.033112978 | 1 |
| CAD | adipocyte | *ZNF717* | 0.126776831 | 0.012009258 | 0.048613338 | 1 |
| CAD | adipocyte | *SCAMP4* | 1.788793371 | 0.001215122 | 0.017317396 | 1 |
| CAD | adipocyte | *RPS18* | 1.153317863 | 8.14E-05 | 0.007733492 | 0.395094023 |
| CAD | adipocyte | *HLA-DQB2* | 0.076521322 | 0.005762985 | 0.030593003 | 1 |
| CAD | adipocyte | *GPX1* | 0.608090946 | 0.003230081 | 0.021621486 | 1 |
| CAD | adipocyte | *LINC00707* | 1.075760277 | 0.005656939 | 0.030261815 | 1 |
| CAD | adipocyte | *TXNDC5* | 5.407647764 | 0.009337847 | 0.041528171 | 1 |
| CAD | adipocyte | *LY6G5B* | -0.152230346 | 0.005928506 | 0.031131074 | 1 |
| CAD | adipocyte | *KRBOX1* | 33.98732031 | 0.003118245 | 0.021174108 | 1 |
| CAD | adipocyte | *NSUN6* | 1.300866644 | 7.71E-05 | 0.007733492 | 0.374177528 |
| CAD | adipocyte | *PI4KA* | 0.838060905 | 0.007775091 | 0.036804626 | 1 |
| CAD | adipocyte | *ARFGAP3* | 1.597889137 | 0.00132637 | 0.017317396 | 1 |
| CAD | adipocyte | *MRPL20* | 1.191034991 | 0.000939493 | 0.017317396 | 1 |
| CAD | adipocyte | *LINC00960* | -0.112908221 | 0.002720562 | 0.019784984 | 1 |
| CAD | adipocyte | *MRPL33* | 0.823530004 | 0.005267987 | 0.028868018 | 1 |
| CAD | adipocyte | *NBPF15* | 1.560287041 | 0.001990628 | 0.018019639 | 1 |
| CAD | adipocyte | *WFDC6* | -1.000792168 | 0.001282817 | 0.017317396 | 1 |
| CAD | adipocyte | *UPK3B* | 0.94994296 | 0.006787982 | 0.033849217 | 1 |
| CAD | adipocyte | *NEAT1* | 0.713697701 | 0.002114609 | 0.018430441 | 1 |

| | | | | | | |
|---|---|---|---|---|---|---|
| CAD | adipocyte | *UBAP1L* | 1.162163314 | 0.010325572 | 0.044024319 | 1 |
| CAD | adipocyte | *LINC00926* | 3.213450084 | 0.004041558 | 0.024853788 | 1 |
| CAD | adipocyte | *CDK11B* | 1.842502741 | 0.001494201 | 0.017317396 | 1 |
| CAD | adipocyte | *ABHD14A* | 1.244394722 | 0.008073838 | 0.03784953 | 1 |
| CAD | adipocyte | *C5orf17* | 1.277511766 | 0.008169703 | 0.038100461 | 1 |
| CAD | adipocyte | *TMEM150C* | 0.266823746 | 0.010318598 | 0.044024319 | 1 |
| CAD | adipocyte | *PDCD6* | 1.625533707 | 0.001520016 | 0.017317396 | 1 |
| CAD | adipocyte | *GOLGA8K* | -0.295877569 | 0.006418935 | 0.032840444 | 1 |
| CAD | adipocyte | *SMIM20* | 1.957361255 | 0.001622963 | 0.017317396 | 1 |
| CAD | adipocyte | *ZNF550* | 1.688255287 | 0.008686621 | 0.03987463 | 1 |
| CAD | adipocyte | *TAS2R20* | 0.418315061 | 0.001824634 | 0.017489118 | 1 |
| CAD | adipocyte | *HMBS* | 0.800776864 | 2.94E-05 | 0.00500475 | 0.142825614 |
| CAD | adipocyte | *CNPY2* | 1.31200188 | 0.009112806 | 0.040909163 | 1 |
| CAD | adipocyte | *HIF1A-AS2* | 1.182772097 | 0.006872018 | 0.034127976 | 1 |
| CAD | adipocyte | *MRPL46* | 0.869005268 | 0.010835068 | 0.045524493 | 1 |
| CAD | adipocyte | *IQCH-AS1* | 0.572368823 | 0.001872327 | 0.017489118 | 1 |
| CAD | adipocyte | *LINC00672* | 2.085696437 | 0.006989311 | 0.034358804 | 1 |
| CAD | adipocyte | *DYNLL2* | 2.089973273 | 0.010153034 | 0.043512372 | 1 |
| CAD | adipocyte | *MYO15B* | 0.633054718 | 0.002455244 | 0.019127952 | 1 |
| CAD | adipocyte | *ZNF582-AS1* | 3.227386281 | 0.001623815 | 0.017317396 | 1 |
| CAD | adipocyte | *ZNF224* | 1.627160877 | 0.001198282 | 0.017317396 | 1 |
| CAD | adipocyte | *TRABD2B* | 0.718098811 | 0.012043123 | 0.048613338 | 1 |
| CAD | adipocyte | *AC093157.1* | 1.146867428 | 0.006647292 | 0.033568181 | 1 |
| CAD | adipocyte | *ZNF587B* | 2.245795912 | 0.009289546 | 0.041378519 | 1 |
| CAD | adipocyte | *CASC15* | 0.848773898 | 0.00493321 | 0.028060596 | 1 |
| CAD | adipocyte | *GRIN2B* | 0.251519605 | 0.005940515 | 0.031160411 | 1 |
| CAD | ASPC | *MDH1* | 0.401797394 | 4.45E-05 | 0.022878356 | 0.215686564 |
| CAD | ASPC | *ARAP2* | 0.599785389 | 0.0001249 | 0.025876666 | 0.606016615 |
| CAD | ASPC | *UHRF1BP1* | 0.274330227 | 2.66E-06 | 0.00644652 | 0.012893041 |
| CAD | ASPC | *ZXDC* | 0.843499086 | 0.000529885 | 0.047293644 | 1 |
| CAD | ASPC | *TIPIN* | 0.14939527 | 0.000179173 | 0.026701942 | 0.869346066 |
| CAD | ASPC | *FMO4* | 0.708540516 | 5.61E-07 | 0.002721171 | 0.002721171 |
| CAD | ASPC | *RPH3A* | 1.281314835 | 0.000431345 | 0.039939376 | 1 |
| CAD | ASPC | *ITPR3* | 0.265929184 | 7.00E-05 | 0.022878356 | 0.339808812 |
| CAD | ASPC | *MTAP* | 0.508760701 | 0.000152295 | 0.025876666 | 0.738932983 |
| CAD | ASPC | *TRPC4AP* | 0.50876808 | 0.000436271 | 0.039939376 | 1 |
| CAD | ASPC | *PSMD7* | 0.914057005 | 0.000567472 | 0.049167391 | 1 |
| CAD | ASPC | *RABAC1* | 0.51724162 | 0.000379708 | 0.038382152 | 1 |
| CAD | ASPC | *LIPA* | 0.224125791 | 0.000379362 | 0.038382152 | 1 |
| CAD | ASPC | *NAGLU* | 0.704234265 | 0.000312066 | 0.036050998 | 1 |
| CAD | ASPC | *PCCB* | 0.279923424 | 0.000319885 | 0.03609497 | 1 |
| CAD | ASPC | *GGCX* | 0.463819398 | 3.92E-05 | 0.022878356 | 0.190312 |
| CAD | ASPC | *PRRC2C* | 0.681861567 | 0.000163462 | 0.025876666 | 0.793119602 |
| CAD | ASPC | *C2orf40* | 0.730664151 | 0.000112495 | 0.025876666 | 0.545824843 |
| CAD | ASPC | *NEK9* | 0.692945259 | 0.000536099 | 0.047293644 | 1 |
| CAD | ASPC | *EIF2B2* | 0.198914004 | 0.000246824 | 0.031677292 | 1 |
| CAD | ASPC | *PTGIS* | 0.243585644 | 6.79E-05 | 0.022878356 | 0.329575189 |
| CAD | ASPC | *SNRPC* | -0.346824035 | 5.51E-05 | 0.022878356 | 0.267538428 |
| CAD | ASPC | *TCP11* | 0.216519301 | 0.000329067 | 0.036287091 | 1 |
| CAD | ASPC | *GRK4* | 0.079811592 | 8.63E-05 | 0.024627088 | 0.418660503 |

| | | | | | | |
|---|---|---|---|---|---|---|
| CAD | ASPC | *SMARCA4* | 0.634302537 | 0.000204462 | 0.028344216 | 0.992047551 |
| CAD | ASPC | *SERGEF* | 0.44112421 | 0.000254638 | 0.031677292 | 1 |
| CAD | ASPC | *ACTN4* | 0.768512841 | 0.000165329 | 0.025876666 | 0.802176661 |
| CAD | ASPC | *SLC37A4* | 0.290041482 | 7.07E-05 | 0.022878356 | 0.343175346 |
| CAD | ASPC | *ULK3* | 1.453608115 | 0.000388937 | 0.038512746 | 1 |
| CAD | ASPC | *MYO19* | 0.632686066 | 0.000136649 | 0.025876666 | 0.663022652 |
| CAD | ASPC | *TMEM91* | 0.144703303 | 0.000337621 | 0.036403059 | 1 |
| CAD | ASPC | *SIRT3* | 0.619960261 | 0.000134055 | 0.025876666 | 0.650436465 |
| CAD | ASPC | *STRIP1* | 1.354906294 | 0.000141 | 0.025876666 | 0.684134018 |
| CAD | ASPC | *SNX27* | 0.550784841 | 0.000181608 | 0.026701942 | 0.881164091 |
| CAD | ASPC | *SETDB1* | 0.798849575 | 0.000165292 | 0.025876666 | 0.801995466 |
| CAD | ASPC | *ARNT* | -0.438087887 | 0.000434304 | 0.039939376 | 1 |
| CAD | ASPC | *WDPCP* | 0.316179135 | 1.12E-05 | 0.018149538 | 0.054448613 |
| CAD | ASPC | *IRAK1BP1* | 0.531079517 | 6.80E-05 | 0.022878356 | 0.329966928 |
| CAD | ASPC | *FBXO10* | 0.266060694 | 0.000132548 | 0.025876666 | 0.643125113 |
| CAD | ASPC | *ROBO3* | 0.432509009 | 7.58E-05 | 0.022973859 | 0.367581739 |
| CAD | ASPC | *CHCHD6* | 0.487956135 | 0.000151021 | 0.025876666 | 0.732754021 |
| CAD | ASPC | *ZBTB7B* | 0.939358336 | 0.00040174 | 0.038984807 | 1 |
| CAD | ASPC | *CPSF4* | 0.481902012 | 0.000137606 | 0.025876666 | 0.667662857 |
| CAD | ASPC | *RNF123* | 0.260712948 | 2.09E-05 | 0.018375272 | 0.101626327 |
| CAD | ASPC | *MST1R* | 0.18501238 | 0.000261148 | 0.031677292 | 1 |
| CAD | ASPC | *TNFRSF11B* | 1.157130878 | 0.000226266 | 0.030495641 | 1 |
| CAD | ASPC | *TEFM* | 0.640065015 | 5.56E-05 | 0.022878356 | 0.269796089 |
| CAD | ASPC | *FGGY* | 0.633164944 | 0.000127279 | 0.025876666 | 0.617556938 |
| CAD | ASPC | *CRLF3* | 0.446817802 | 2.29E-05 | 0.018375272 | 0.1109077 |
| CAD | ASPC | *CAPS2* | 0.788156084 | 0.000249068 | 0.031677292 | 1 |
| CAD | ASPC | *TMEM136* | 0.598560039 | 2.45E-05 | 0.018375272 | 0.11888687 |
| CAD | ASPC | *FES* | -0.353583062 | 0.000195529 | 0.027903069 | 0.948704348 |
| CAD | ASPC | *TMEM116* | -0.120601894 | 0.000122796 | 0.025876666 | 0.595806094 |
| CAD | ASPC | *LILRB3* | 0.583680043 | 0.000285665 | 0.033805971 | 1 |
| CAD | ASPC | *TAS2R50* | 3.387243989 | 0.00036959 | 0.038382152 | 1 |
| CAD | ASPC | *HMBS* | 0.39229664 | 2.65E-05 | 0.018375272 | 0.128626907 |
| CAD | LEC | *UHRF1BP1* | 1.74255244 | 1.06E-05 | 0.02579835 | 0.0515967 |
| CAD | LEC | *ITPR3* | 2.165682445 | 7.73E-06 | 0.02579835 | 0.037505077 |
| CAD | LEC | *RNF123* | 1.86734768 | 2.20E-05 | 0.03555378 | 0.106661341 |
| CAD | macrophage | *KRIT1* | 1.012316864 | 0.007118704 | 0.038943652 | 1 |
| CAD | macrophage | *ALS2* | 1.271933666 | 0.00386846 | 0.032549158 | 1 |
| CAD | macrophage | *POLR2J* | 2.542134151 | 0.000281005 | 0.0200505 | 1 |
| CAD | macrophage | *CIAPIN1* | 1.318658764 | 0.002956517 | 0.032549158 | 1 |
| CAD | macrophage | *GCFC2* | 1.004463363 | 0.003553067 | 0.032549158 | 1 |
| CAD | macrophage | *SYNRG* | 1.229358739 | 0.003974077 | 0.032549158 | 1 |
| CAD | macrophage | *ZNF263* | 1.843289827 | 0.000799734 | 0.028118192 | 1 |
| CAD | macrophage | *BAIAP2L1* | 1.778587365 | 0.002542087 | 0.032549158 | 1 |
| CAD | macrophage | *AGK* | 1.060630713 | 0.001302735 | 0.032549158 | 1 |
| CAD | macrophage | *FARP2* | 1.29706029 | 0.002496311 | 0.032549158 | 1 |
| CAD | macrophage | *PAX6* | 0.371908411 | 0.011301199 | 0.048268855 | 1 |
| CAD | macrophage | *PSMB1* | 1.634310737 | 0.005874314 | 0.036176476 | 1 |
| CAD | macrophage | *MGST1* | 0.330994922 | 0.002555491 | 0.032549158 | 1 |
| CAD | macrophage | *PKD1* | 1.221731722 | 0.001425266 | 0.032549158 | 1 |
| CAD | macrophage | *MED24* | 1.767502638 | 0.004425573 | 0.032821343 | 1 |

| | | | | | | |
|---|---|---|---|---|---|---|
| CAD | macrophage | *RPS20* | 1.59409122 | 0.000288052 | 0.020255464 | 1 |
| CAD | macrophage | *POMT2* | 0.933529008 | 0.00242976 | 0.032549158 | 1 |
| CAD | macrophage | *PPP5C* | 1.017365282 | 0.001439636 | 0.032549158 | 1 |
| CAD | macrophage | *CEP68* | 1.081264359 | 0.000214195 | 0.01939755 | 1 |
| CAD | macrophage | *MAP4K5* | 0.938032213 | 0.003555905 | 0.032549158 | 1 |
| CAD | macrophage | *ZC3H3* | 0.62782841 | 0.002228668 | 0.032549158 | 1 |
| CAD | macrophage | *MDH1* | 1.300379418 | 3.77E-06 | 0.002613731 | 0.01829612 |
| CAD | macrophage | *NUDCD3* | 0.713512474 | 0.002060116 | 0.032549158 | 1 |
| CAD | macrophage | *CHDH* | 1.723972974 | 0.007312407 | 0.039031065 | 1 |
| CAD | macrophage | *RUFY3* | 1.029214359 | 0.012005171 | 0.049828136 | 1 |
| CAD | macrophage | *WWTR1* | 0.613054072 | 0.007502981 | 0.039482739 | 1 |
| CAD | macrophage | *TTC27* | 1.230411007 | 0.001867245 | 0.032549158 | 1 |
| CAD | macrophage | *OSBPL5* | 0.873644591 | 0.008584834 | 0.041487662 | 1 |
| CAD | macrophage | *GCLM* | 1.639878204 | 0.003113366 | 0.032549158 | 1 |
| CAD | macrophage | *SEC63* | 1.15854896 | 0.004117813 | 0.032707875 | 1 |
| CAD | macrophage | *FAS* | 1.241145606 | 0.003614718 | 0.032549158 | 1 |
| CAD | macrophage | *VRK2* | 1.263020172 | 0.00270812 | 0.032549158 | 1 |
| CAD | macrophage | *VEZT* | 1.075623857 | 0.003045128 | 0.032549158 | 1 |
| CAD | macrophage | *BRD9* | 0.977420977 | 0.008730383 | 0.041948605 | 1 |
| CAD | macrophage | *VCL* | 1.190693205 | 0.006655761 | 0.037885063 | 1 |
| CAD | macrophage | *ADSS* | 1.700364526 | 0.003059712 | 0.032549158 | 1 |
| CAD | macrophage | *TBC1D23* | 1.262708535 | 0.002863307 | 0.032549158 | 1 |
| CAD | macrophage | *DNAH5* | 1.085175124 | 0.011924261 | 0.049567582 | 1 |
| CAD | macrophage | *CTNNA1* | 1.193628031 | 0.003951899 | 0.032549158 | 1 |
| CAD | macrophage | *POLR2B* | 1.358771372 | 0.003438906 | 0.032549158 | 1 |
| CAD | macrophage | *ANO2* | 0.834854593 | 0.0057881 | 0.036134117 | 1 |
| CAD | macrophage | *AKR7A2* | 1.313937987 | 0.007938873 | 0.040168337 | 1 |
| CAD | macrophage | *GINM1* | 1.029764494 | 0.003443761 | 0.032549158 | 1 |
| CAD | macrophage | *ATP11B* | 1.154868262 | 0.000472705 | 0.02312941 | 1 |
| CAD | macrophage | *DNAJC25* | 1.85281079 | 0.004519483 | 0.032925653 | 1 |
| CAD | macrophage | *PRDM6* | 2.026355593 | 0.003050996 | 0.032549158 | 1 |
| CAD | macrophage | *RNF4* | 1.639522268 | 0.004935246 | 0.033650257 | 1 |
| CAD | macrophage | *CASP8* | 1.390612529 | 0.002342496 | 0.032549158 | 1 |
| CAD | macrophage | *TM7SF3* | 1.264153883 | 0.007032833 | 0.038675956 | 1 |
| CAD | macrophage | *TMEM161A* | 1.324933443 | 0.005609817 | 0.035488787 | 1 |
| CAD | macrophage | *SUGP2* | 1.3664187 | 0.003572189 | 0.032549158 | 1 |
| CAD | macrophage | *SLC12A2* | 1.338138963 | 0.002100893 | 0.032549158 | 1 |
| CAD | macrophage | *PMS1* | 1.281380175 | 0.005652591 | 0.035618667 | 1 |
| CAD | macrophage | *UHRF1BP1* | 0.923683295 | 8.71E-07 | 0.002081011 | 0.004227727 |
| CAD | macrophage | *MAP2K4* | 1.143075671 | 0.006058938 | 0.036581378 | 1 |
| CAD | macrophage | *ZMYND12* | 0.23492188 | 0.007693431 | 0.039626885 | 1 |
| CAD | macrophage | *GOLGA5* | 1.710417091 | 0.000853408 | 0.028834548 | 1 |
| CAD | macrophage | *STOML1* | 3.317326104 | 0.003028586 | 0.032549158 | 1 |
| CAD | macrophage | *IP6K2* | 1.280759114 | 0.001315974 | 0.032549158 | 1 |
| CAD | macrophage | *NUCKS1* | 1.130446784 | 0.005627643 | 0.035551639 | 1 |
| CAD | macrophage | *DRD4* | 3.650600598 | 0.004481582 | 0.032923259 | 1 |
| CAD | macrophage | *PLA2G10* | 2.672218667 | 0.009370194 | 0.043423289 | 1 |
| CAD | macrophage | *NEDD4* | 0.858664694 | 0.000870669 | 0.028834548 | 1 |
| CAD | macrophage | *GNB5* | 0.602099446 | 0.001752669 | 0.032549158 | 1 |
| CAD | macrophage | *LRP6* | 0.709431991 | 0.008340019 | 0.041082002 | 1 |

| | | | | | | |
|---|---|---|---|---|---|---|
| CAD | macrophage | *CLTCL1* | 0.850462244 | 0.004338321 | 0.032821343 | 1 |
| CAD | macrophage | *DGCR2* | 1.137354413 | 0.00220846 | 0.032549158 | 1 |
| CAD | macrophage | *ZXDC* | 1.649713234 | 0.000258468 | 0.01939755 | 1 |
| CAD | macrophage | *GBA2* | 1.994251713 | 0.000643035 | 0.026896603 | 1 |
| CAD | macrophage | *EIF2B3* | 1.056569981 | 0.005521198 | 0.035310234 | 1 |
| CAD | macrophage | *MAP4K4* | 0.751498214 | 0.006827215 | 0.038393741 | 1 |
| CAD | macrophage | *TRIB2* | 0.571106256 | 0.004675816 | 0.033233594 | 1 |
| CAD | macrophage | *ALDH3A2* | 0.75027555 | 0.003072023 | 0.032549158 | 1 |
| CAD | macrophage | *AFF4* | 1.07267372 | 0.005480765 | 0.035310234 | 1 |
| CAD | macrophage | *TRNT1* | 1.444028409 | 0.001662271 | 0.032549158 | 1 |
| CAD | macrophage | *LLGL2* | 1.361286492 | 0.000736743 | 0.027706954 | 1 |
| CAD | macrophage | *ZNF532* | 1.424241131 | 0.003029822 | 0.032549158 | 1 |
| CAD | macrophage | *ZZEF1* | 1.336109821 | 0.001640784 | 0.032549158 | 1 |
| CAD | macrophage | *TIPIN* | 0.540020178 | 2.93E-05 | 0.007261836 | 0.142080652 |
| CAD | macrophage | *ADD2* | 1.332420057 | 0.000356483 | 0.021894388 | 1 |
| CAD | macrophage | *FNDC3B* | 0.999457586 | 0.001601362 | 0.032549158 | 1 |
| CAD | macrophage | *CACNG4* | 0.462167923 | 0.006719764 | 0.038044686 | 1 |
| CAD | macrophage | *KIFAP3* | 0.994250767 | 0.009663103 | 0.044002875 | 1 |
| CAD | macrophage | *FMO4* | 2.251820311 | 4.20E-08 | 0.000203798 | 0.000203798 |
| CAD | macrophage | *GPATCH1* | 1.382781992 | 0.004298649 | 0.032743483 | 1 |
| CAD | macrophage | *XAB2* | 3.273838131 | 0.00025986 | 0.01939755 | 1 |
| CAD | macrophage | *DYNC1I2* | 1.252425031 | 0.003445896 | 0.032549158 | 1 |
| CAD | macrophage | *AMPH* | 0.907337679 | 0.002739647 | 0.032549158 | 1 |
| CAD | macrophage | *PIK3C3* | 1.406771623 | 0.001908575 | 0.032549158 | 1 |
| CAD | macrophage | *PCM1* | 1.039058821 | 0.001719702 | 0.032549158 | 1 |
| CAD | macrophage | *XRCC5* | 1.391546618 | 0.001511534 | 0.032549158 | 1 |
| CAD | macrophage | *SAR1A* | 1.09601357 | 0.006244508 | 0.037040771 | 1 |
| CAD | macrophage | *FDFT1* | 0.797314162 | 0.008040266 | 0.040358159 | 1 |
| CAD | macrophage | *PGM1* | 0.933589293 | 0.004690175 | 0.03325632 | 1 |
| CAD | macrophage | *DDX1* | 0.838973475 | 0.010899597 | 0.047086962 | 1 |
| CAD | macrophage | *DNM2* | 1.177557065 | 0.007323113 | 0.039031065 | 1 |
| CAD | macrophage | *RABL2B* | 1.91023202 | 0.002737391 | 0.032549158 | 1 |
| CAD | macrophage | *PIBF1* | 1.073785686 | 0.002724673 | 0.032549158 | 1 |
| CAD | macrophage | *ZNF264* | 1.70972932 | 0.008739515 | 0.041948605 | 1 |
| CAD | macrophage | *NOA1* | 1.385375498 | 0.009084809 | 0.042588884 | 1 |
| CAD | macrophage | *NCOA1* | 0.991715359 | 0.006495818 | 0.037431956 | 1 |
| CAD | macrophage | *HACE1* | 1.216221968 | 0.002628096 | 0.032549158 | 1 |
| CAD | macrophage | *CPNE3* | 0.866438699 | 0.010851574 | 0.046968633 | 1 |
| CAD | macrophage | *RRN3* | 1.257194082 | 0.006418449 | 0.037207064 | 1 |
| CAD | macrophage | *DDHD2* | 0.911278919 | 0.000745738 | 0.027706954 | 1 |
| CAD | macrophage | *USP40* | 1.063543277 | 0.003188266 | 0.032549158 | 1 |
| CAD | macrophage | *EIF2AK1* | 1.570292628 | 0.004601509 | 0.033148387 | 1 |
| CAD | macrophage | *ALG9* | 1.74392669 | 0.004970114 | 0.033804179 | 1 |
| CAD | macrophage | *TRIP6* | 1.627293055 | 0.00101922 | 0.030318387 | 1 |
| CAD | macrophage | *SRRT* | 1.571268924 | 0.001528199 | 0.032549158 | 1 |
| CAD | macrophage | *ADAMTS2* | 1.128046622 | 0.006603269 | 0.037885063 | 1 |
| CAD | macrophage | *UIMC1* | 1.324370993 | 0.001740754 | 0.032549158 | 1 |
| CAD | macrophage | *OGFOD1* | 0.981203615 | 0.006076956 | 0.036582371 | 1 |
| CAD | macrophage | *AAMDC* | 0.625678172 | 0.001045832 | 0.030318387 | 1 |
| CAD | macrophage | *ASAP3* | 0.731623591 | 0.004086307 | 0.032707875 | 1 |

| | | | | | | |
|---|---|---|---|---|---|---|
| CAD | macrophage | *ANKRD10* | 1.046453993 | 0.004414963 | 0.032821343 | 1 |
| CAD | macrophage | *DOCK3* | 1.193526618 | 0.003991498 | 0.032549158 | 1 |
| CAD | macrophage | *ARHGAP28* | 0.585712139 | 0.005634627 | 0.035551639 | 1 |
| CAD | macrophage | *EBF4* | 1.103901699 | 0.005739747 | 0.035980948 | 1 |
| CAD | macrophage | *MAVS* | 0.841199887 | 0.004819654 | 0.033426994 | 1 |
| CAD | macrophage | *TMEM230* | 0.958236978 | 0.006881544 | 0.038407224 | 1 |
| CAD | macrophage | *RPH3A* | 3.108560756 | 0.00010645 | 0.014757048 | 0.516496696 |
| CAD | macrophage | *ZNF302* | 0.817436331 | 0.000474888 | 0.02312941 | 1 |
| CAD | macrophage | *CDIP1* | 1.96026038 | 0.002962502 | 0.032549158 | 1 |
| CAD | macrophage | *ANKRD24* | 2.023228796 | 0.004515058 | 0.032925653 | 1 |
| CAD | macrophage | *RAB11FIP3* | 1.006534459 | 0.001176334 | 0.03188587 | 1 |
| CAD | macrophage | *PABPC4* | -0.630011368 | 0.010027585 | 0.044885278 | 1 |
| CAD | macrophage | *CERS4* | 1.056871551 | 0.004756738 | 0.033426994 | 1 |
| CAD | macrophage | *USP48* | 0.958435699 | 0.001039343 | 0.030318387 | 1 |
| CAD | macrophage | *GLG1* | 1.204992886 | 0.005436357 | 0.035310234 | 1 |
| CAD | macrophage | *TF* | 1.449016695 | 0.006290115 | 0.037040771 | 1 |
| CAD | macrophage | *PPP2R3C* | 0.828246516 | 0.009684359 | 0.044002875 | 1 |
| CAD | macrophage | *SCFD1* | 1.13240034 | 0.005115028 | 0.034469609 | 1 |
| CAD | macrophage | *SUPT16H* | 1.575556682 | 0.007595477 | 0.039573543 | 1 |
| CAD | macrophage | *LRRFIP2* | 1.137696927 | 0.009589087 | 0.044002875 | 1 |
| CAD | macrophage | *SEC22C* | 1.258788967 | 0.003989959 | 0.032549158 | 1 |
| CAD | macrophage | *XYLB* | 1.792022647 | 0.003931914 | 0.032549158 | 1 |
| CAD | macrophage | *MSH2* | 1.405361633 | 0.003027005 | 0.032549158 | 1 |
| CAD | macrophage | *PSMD5* | 1.304248114 | 0.003705265 | 0.032549158 | 1 |
| CAD | macrophage | *CRAT* | 1.343171003 | 0.008238225 | 0.040820553 | 1 |
| CAD | macrophage | *WAC* | 1.074926772 | 0.006670919 | 0.037885063 | 1 |
| CAD | macrophage | *ITPR3* | 0.930074726 | 1.45E-06 | 0.002081011 | 0.007045574 |
| CAD | macrophage | *DSP* | 1.758582898 | 0.004030062 | 0.032701827 | 1 |
| CAD | macrophage | *ERMP1* | 1.081192533 | 0.002938155 | 0.032549158 | 1 |
| CAD | macrophage | *RAB18* | 1.417201296 | 0.002965356 | 0.032549158 | 1 |
| CAD | macrophage | *TSPAN15* | 1.218032468 | 0.004140652 | 0.032707875 | 1 |
| CAD | macrophage | *CIRBP* | 0.827290983 | 0.004199189 | 0.032707875 | 1 |
| CAD | macrophage | *MTAP* | 1.659636712 | 3.90E-05 | 0.008184153 | 0.189305064 |
| CAD | macrophage | *LZTR1* | 1.065047916 | 0.001279191 | 0.032549158 | 1 |
| CAD | macrophage | *SMARCB1* | 0.466839586 | 0.011443304 | 0.048576475 | 1 |
| CAD | macrophage | *BCL2L13* | 0.812863263 | 0.002747863 | 0.032549158 | 1 |
| CAD | macrophage | *CABIN1* | 1.114881492 | 0.008740734 | 0.041948605 | 1 |
| CAD | macrophage | *SUSD2* | -0.253717706 | 0.004174275 | 0.032707875 | 1 |
| CAD | macrophage | *CCDC134* | 1.017178752 | 0.003001583 | 0.032549158 | 1 |
| CAD | macrophage | *TTC28* | 0.742164818 | 0.003456059 | 0.032549158 | 1 |
| CAD | macrophage | *CENPM* | -0.72762657 | 0.002491825 | 0.032549158 | 1 |
| CAD | macrophage | *TIMP3* | 0.586665797 | 0.006001992 | 0.036447642 | 1 |
| CAD | macrophage | *PPP6R2* | 1.083014369 | 0.008940804 | 0.042322713 | 1 |
| CAD | macrophage | *CYB5R3* | 0.879767198 | 0.001382331 | 0.032549158 | 1 |
| CAD | macrophage | *CHKB* | 1.29586792 | 0.002492327 | 0.032549158 | 1 |
| CAD | macrophage | *IFT27* | 1.613490646 | 0.007018436 | 0.038675956 | 1 |
| CAD | macrophage | *POLR3H* | 1.343163259 | 0.005738054 | 0.035980948 | 1 |
| CAD | macrophage | *RBM23* | 1.110392059 | 0.000244521 | 0.01939755 | 1 |
| CAD | macrophage | *CNIH1* | 2.045349273 | 0.000218202 | 0.01939755 | 1 |
| CAD | macrophage | *ATP6V1D* | 1.488678821 | 0.003711755 | 0.032549158 | 1 |

| | | | | | | |
|---|---|---|---|---|---|---|
| CAD | macrophage | *MTHFD1* | 1.108486693 | 0.003054074 | 0.032549158 | 1 |
| CAD | macrophage | *YY1* | 1.362038434 | 0.011713158 | 0.048993314 | 1 |
| CAD | macrophage | *CCNB1IP1* | 1.406524149 | 0.003522385 | 0.032549158 | 1 |
| CAD | macrophage | *CINP* | 1.878378814 | 0.000588758 | 0.025946406 | 1 |
| CAD | macrophage | *TRPC4AP* | 1.36500238 | 7.09E-05 | 0.011104812 | 0.344249165 |
| CAD | macrophage | *NDRG3* | 2.022572899 | 0.000255413 | 0.01939755 | 1 |
| CAD | macrophage | *DOK5* | 1.190021319 | 0.009685711 | 0.044002875 | 1 |
| CAD | macrophage | *MTG2* | 1.04561351 | 0.006204268 | 0.037040771 | 1 |
| CAD | macrophage | *DIDO1* | 1.424581386 | 0.000343567 | 0.021635859 | 1 |
| CAD | macrophage | *GMEB2* | 1.916330479 | 0.003007766 | 0.032549158 | 1 |
| CAD | macrophage | *MAP1LC3A* | 0.885848826 | 0.003707176 | 0.032549158 | 1 |
| CAD | macrophage | *METTL4* | 1.45748606 | 0.006973341 | 0.038588296 | 1 |
| CAD | macrophage | *MYOM1* | 0.697346196 | 0.009348255 | 0.043423289 | 1 |
| CAD | macrophage | *MIB1* | 1.106459333 | 0.00288125 | 0.032549158 | 1 |
| CAD | macrophage | *NALCN* | 0.581716657 | 0.010046458 | 0.044885278 | 1 |
| CAD | macrophage | *STK24* | 1.254904491 | 0.002690301 | 0.032549158 | 1 |
| CAD | macrophage | *ARHGEF7* | 1.16497093 | 0.002767672 | 0.032549158 | 1 |
| CAD | macrophage | *SUPT20H* | 1.345758652 | 0.000863825 | 0.028834548 | 1 |
| CAD | macrophage | *MRPS31* | 1.329367053 | 0.002059076 | 0.032549158 | 1 |
| CAD | macrophage | *KATNAL1* | 0.742024357 | 0.006749578 | 0.03816894 | 1 |
| CAD | macrophage | *INTS6* | 1.148033013 | 0.002559842 | 0.032549158 | 1 |
| CAD | macrophage | *CLN5* | 1.421703056 | 0.00515419 | 0.034541615 | 1 |
| CAD | macrophage | *MGRN1* | 1.338528687 | 0.000342483 | 0.021635859 | 1 |
| CAD | macrophage | *DHODH* | 0.921123353 | 0.000433816 | 0.02312941 | 1 |
| CAD | macrophage | *PSMD7* | 1.987096533 | 0.000292338 | 0.020263189 | 1 |
| CAD | macrophage | *NPRL3* | 0.687788451 | 0.008276011 | 0.040820553 | 1 |
| CAD | macrophage | *LMF1* | 1.496078356 | 0.002779388 | 0.032549158 | 1 |
| CAD | macrophage | *RBL2* | 1.113216569 | 0.000633312 | 0.026720245 | 1 |
| CAD | macrophage | *KAT8* | 1.92493015 | 0.004097052 | 0.032707875 | 1 |
| CAD | macrophage | *C16orf62* | 1.308144056 | 0.006332796 | 0.037064809 | 1 |
| CAD | macrophage | *KNOP1* | 1.59501631 | 0.004203205 | 0.032707875 | 1 |
| CAD | macrophage | *IQCH* | 1.197127663 | 0.00366111 | 0.032549158 | 1 |
| CAD | macrophage | *HERC1* | 1.054131799 | 0.008522578 | 0.04146103 | 1 |
| CAD | macrophage | *TTC23* | 1.559326216 | 0.002407454 | 0.032549158 | 1 |
| CAD | macrophage | *FAH* | 0.795962851 | 0.011547796 | 0.048784846 | 1 |
| CAD | macrophage | *SLC30A4* | 1.708650945 | 0.002304108 | 0.032549158 | 1 |
| CAD | macrophage | *ZFAND1* | 1.189063484 | 0.003169955 | 0.032549158 | 1 |
| CAD | macrophage | *NIPAL2* | 1.544872468 | 0.0018904 | 0.032549158 | 1 |
| CAD | macrophage | *RAB2A* | 1.025360175 | 0.003420114 | 0.032549158 | 1 |
| CAD | macrophage | *SNX16* | 1.76287495 | 0.000347815 | 0.021635859 | 1 |
| CAD | macrophage | *GSDMD* | 1.364781376 | 0.007043595 | 0.038675956 | 1 |
| CAD | macrophage | *ERICH1* | 1.013200669 | 0.009567688 | 0.043960626 | 1 |
| CAD | macrophage | *TUSC3* | 0.763751383 | 0.007810984 | 0.039809763 | 1 |
| CAD | macrophage | *SNRNP70* | 1.112266874 | 0.004356666 | 0.032821343 | 1 |
| CAD | macrophage | *AMH* | 22.92256387 | 0.002482973 | 0.032549158 | 1 |
| CAD | macrophage | *NOVA2* | 0.355543483 | 0.006316981 | 0.037064809 | 1 |
| CAD | macrophage | *SGTA* | 1.681839611 | 0.00403044 | 0.032701827 | 1 |
| CAD | macrophage | *RASAL3* | 3.367459704 | 0.004485128 | 0.032923259 | 1 |
| CAD | macrophage | *ANKRD27* | 1.488894609 | 0.005219722 | 0.034638531 | 1 |
| CAD | macrophage | *RABAC1* | 1.714260983 | 2.67E-05 | 0.007261836 | 0.129447986 |

| | | | | | | |
|---|---|---|---|---|---|---|
| CAD | macrophage | *CAPS* | 2.207043261 | 0.003550023 | 0.032549158 | 1 |
| CAD | macrophage | *TFPT* | 2.052800308 | 0.011583736 | 0.048784846 | 1 |
| CAD | macrophage | *ZNF14* | 1.2948724 | 0.004727356 | 0.033426994 | 1 |
| CAD | macrophage | *BET1* | 1.043889355 | 0.005327533 | 0.035073527 | 1 |
| CAD | macrophage | *SP4* | 1.087174493 | 0.011420514 | 0.048527713 | 1 |
| CAD | macrophage | *DNAH11* | 1.341278232 | 0.004247814 | 0.032707875 | 1 |
| CAD | macrophage | *IQCE* | 2.009132869 | 0.00389299 | 0.032549158 | 1 |
| CAD | macrophage | *HIBADH* | 0.920208846 | 0.001260229 | 0.032549158 | 1 |
| CAD | macrophage | *GRB10* | 0.711739224 | 0.004631488 | 0.033148387 | 1 |
| CAD | macrophage | *PLEKHA8* | 1.674434138 | 0.007654221 | 0.039573543 | 1 |
| CAD | macrophage | *CHCHD2* | 1.019442536 | 0.002758722 | 0.032549158 | 1 |
| CAD | macrophage | *ZKSCAN1* | 1.337994493 | 0.00105906 | 0.030405687 | 1 |
| CAD | macrophage | *EIF3B* | 1.533451666 | 0.005923318 | 0.036271366 | 1 |
| CAD | macrophage | *NRF1* | 1.020621336 | 0.006974842 | 0.038588296 | 1 |
| CAD | macrophage | *RARRES2* | 0.908956297 | 0.001975234 | 0.032549158 | 1 |
| CAD | macrophage | *COA1* | 1.252652164 | 0.002177831 | 0.032549158 | 1 |
| CAD | macrophage | *AEBP1* | 0.84646245 | 0.008493663 | 0.04146103 | 1 |
| CAD | macrophage | *FKTN* | 1.310868249 | 0.004187163 | 0.032707875 | 1 |
| CAD | macrophage | *CORO2A* | 1.196524206 | 0.007784369 | 0.039799533 | 1 |
| CAD | macrophage | *C5* | 1.185362157 | 0.002108909 | 0.032549158 | 1 |
| CAD | macrophage | *ECM2* | 0.73305142 | 0.004280606 | 0.032707875 | 1 |
| CAD | macrophage | *PTGR1* | 0.56160894 | 0.01168608 | 0.048925163 | 1 |
| CAD | macrophage | *KANK1* | 0.480194027 | 0.00782209 | 0.039824533 | 1 |
| CAD | macrophage | *DNMBP* | 0.943051791 | 0.006642318 | 0.037885063 | 1 |
| CAD | macrophage | *TRDMT1* | 1.402917377 | 0.001279511 | 0.032549158 | 1 |
| CAD | macrophage | *ATE1* | 1.002299883 | 0.001114123 | 0.031134444 | 1 |
| CAD | macrophage | *CDH23* | 1.212551147 | 0.003766626 | 0.032549158 | 1 |
| CAD | macrophage | *BMPR1A* | 0.957875887 | 0.004386658 | 0.032821343 | 1 |
| CAD | macrophage | *TNKS2* | 1.308830654 | 0.010144305 | 0.045156119 | 1 |
| CAD | macrophage | *PITRM1* | 1.385500164 | 0.00541485 | 0.035310234 | 1 |
| CAD | macrophage | *TFAM* | 1.915384339 | 0.008361375 | 0.04110374 | 1 |
| CAD | macrophage | *RPL19* | 1.000104441 | 0.008692191 | 0.041881342 | 1 |
| CAD | macrophage | *RAPGEFL1* | 1.440482405 | 0.004439734 | 0.032821343 | 1 |
| CAD | macrophage | *WNT3* | 0.680534128 | 0.009742024 | 0.044146082 | 1 |
| CAD | macrophage | *TRIM16L* | 1.774488893 | 0.004042338 | 0.032702724 | 1 |
| CAD | macrophage | *PIGL* | 0.885812973 | 0.010319134 | 0.045508793 | 1 |
| CAD | macrophage | *RAI1* | 1.296154546 | 0.003567473 | 0.032549158 | 1 |
| CAD | macrophage | *GOSR1* | 1.406848127 | 0.003252531 | 0.032549158 | 1 |
| CAD | macrophage | *DDX5* | 0.697694095 | 0.006654068 | 0.037885063 | 1 |
| CAD | macrophage | *NAGLU* | 1.908560742 | 0.000234532 | 0.01939755 | 1 |
| CAD | macrophage | *CNTNAP1* | 1.142938088 | 0.000183439 | 0.01939755 | 0.890047789 |
| CAD | macrophage | *DHRS7B* | 1.417411607 | 0.00491227 | 0.033650257 | 1 |
| CAD | macrophage | *MYH3* | 3.248990855 | 0.00193115 | 0.032549158 | 1 |
| CAD | macrophage | *OCIAD1* | 1.045337894 | 0.010917731 | 0.047086962 | 1 |
| CAD | macrophage | *UBE2D3* | 1.15558322 | 0.0055693 | 0.035415784 | 1 |
| CAD | macrophage | *WFS1* | 1.054707463 | 0.008758501 | 0.041963024 | 1 |
| CAD | macrophage | *TRIM2* | 0.655915362 | 0.009043626 | 0.042477903 | 1 |
| CAD | macrophage | *STIM2* | 1.52059371 | 0.001380067 | 0.032549158 | 1 |
| CAD | macrophage | *BST1* | 1.503252326 | 0.000502795 | 0.023878663 | 1 |
| CAD | macrophage | *UFSP2* | 1.436291462 | 0.003369516 | 0.032549158 | 1 |

| | | | | | | |
|---|---|---|---|---|---|---|
| CAD | macrophage | *KLHL5* | 1.437442799 | 0.002992915 | 0.032549158 | 1 |
| CAD | macrophage | *ELP4* | 1.320772609 | 0.000181315 | 0.01939755 | 0.879740358 |
| CAD | macrophage | *DNAJC4* | 0.619335023 | 0.005827084 | 0.036134117 | 1 |
| CAD | macrophage | *DCPS* | 1.666902969 | 0.008250746 | 0.040820553 | 1 |
| CAD | macrophage | *CEP164* | 1.060754989 | 0.002709055 | 0.032549158 | 1 |
| CAD | macrophage | *GALNT18* | 0.682449444 | 0.00497833 | 0.033804179 | 1 |
| CAD | macrophage | *KIAA1549L* | 6.224090292 | 0.008577266 | 0.041487662 | 1 |
| CAD | macrophage | *SLC35F2* | 1.589859009 | 0.000830136 | 0.028433213 | 1 |
| CAD | macrophage | *PRDM4* | 1.293990982 | 0.003260016 | 0.032549158 | 1 |
| CAD | macrophage | *ASIC1* | 3.187947088 | 0.005523592 | 0.035310234 | 1 |
| CAD | macrophage | *PRR4* | 0.763424258 | 0.000667828 | 0.027460164 | 1 |
| CAD | macrophage | *VPS29* | 1.209382989 | 0.011581738 | 0.048784846 | 1 |
| CAD | macrophage | *CAND1* | 1.135970491 | 0.002738518 | 0.032549158 | 1 |
| CAD | macrophage | *MDM1* | 1.365407748 | 0.003673565 | 0.032549158 | 1 |
| CAD | macrophage | *C12orf57* | 1.129462363 | 0.008467306 | 0.04146103 | 1 |
| CAD | macrophage | *SUDS3* | 1.641625863 | 0.004089036 | 0.032707875 | 1 |
| CAD | macrophage | *TMEM14C* | 1.79649223 | 0.003066852 | 0.032549158 | 1 |
| CAD | macrophage | *SMIM8* | 0.809724421 | 0.000978894 | 0.029887243 | 1 |
| CAD | macrophage | *RNGTT* | 1.32838644 | 0.006918366 | 0.038407224 | 1 |
| CAD | macrophage | *ALDH5A1* | 0.494899952 | 0.009366442 | 0.043423289 | 1 |
| CAD | macrophage | *ACOT13* | 1.655972094 | 0.003467283 | 0.032549158 | 1 |
| CAD | macrophage | *HBS1L* | 0.9409252 | 0.009618503 | 0.044002875 | 1 |
| CAD | macrophage | *PACRG* | 1.420388345 | 0.003887019 | 0.032549158 | 1 |
| CAD | macrophage | *EXOC2* | 1.394026992 | 0.004227997 | 0.032707875 | 1 |
| CAD | macrophage | *NUDT12* | 1.652736574 | 0.003143411 | 0.032549158 | 1 |
| CAD | macrophage | *SPARC* | 0.667682118 | 0.0063284 | 0.037064809 | 1 |
| CAD | macrophage | *SKP1* | 1.439815134 | 0.002333579 | 0.032549158 | 1 |
| CAD | macrophage | *PPWD1* | 1.350166576 | 0.002409859 | 0.032549158 | 1 |
| CAD | macrophage | *TXNDC15* | 1.733573805 | 0.003056197 | 0.032549158 | 1 |
| CAD | macrophage | *SMAD5* | 1.289813021 | 0.002981698 | 0.032549158 | 1 |
| CAD | macrophage | *ERGIC1* | 1.258765303 | 0.002858015 | 0.032549158 | 1 |
| CAD | macrophage | *BNIP1* | 0.511915271 | 0.000321395 | 0.021635859 | 1 |
| CAD | macrophage | *CLDN16* | 0.597141563 | 0.009801794 | 0.04428284 | 1 |
| CAD | macrophage | *PCCB* | 1.00018979 | 5.70E-05 | 0.009531314 | 0.276408116 |
| CAD | macrophage | *CBLB* | 0.824117728 | 0.004822526 | 0.033426994 | 1 |
| CAD | macrophage | *IQCG* | 1.369316915 | 0.007198449 | 0.038955499 | 1 |
| CAD | macrophage | *SCAP* | 1.323871794 | 0.004545214 | 0.032925653 | 1 |
| CAD | macrophage | *NEK11* | 0.963404405 | 0.000891969 | 0.028852217 | 1 |
| CAD | macrophage | *PLSCR4* | 0.531509368 | 0.011371 | 0.048439061 | 1 |
| CAD | macrophage | *HEMK1* | 1.172533134 | 0.003609405 | 0.032549158 | 1 |
| CAD | macrophage | *COMMD2* | 1.704702253 | 0.000395082 | 0.0228207 | 1 |
| CAD | macrophage | *ARHGEF26* | 0.832287062 | 0.00809414 | 0.04048251 | 1 |
| CAD | macrophage | *NKTR* | 0.832298032 | 0.006871679 | 0.038407224 | 1 |
| CAD | macrophage | *ACTR1B* | 1.471469116 | 0.006903091 | 0.038407224 | 1 |
| CAD | macrophage | *NRBP1* | 1.420709739 | 0.005838654 | 0.036134117 | 1 |
| CAD | macrophage | *RTN4* | 0.890701259 | 0.002354354 | 0.032549158 | 1 |
| CAD | macrophage | *POLE4* | 1.671978009 | 0.000879537 | 0.028834548 | 1 |
| CAD | macrophage | *GGCX* | 1.566647984 | 3.13E-06 | 0.00252885 | 0.015173102 |
| CAD | macrophage | *HDLBP* | 1.323474372 | 0.000886058 | 0.028852217 | 1 |
| CAD | macrophage | *PASK* | -0.378892448 | 0.00469509 | 0.03325632 | 1 |

| | | | | | | |
|---|---|---|---|---|---|---|
| CAD | macrophage | *TPO* | 0.797723543 | 0.004926484 | 0.033650257 | 1 |
| CAD | macrophage | *TAF1B* | 1.491936332 | 0.00018371 | 0.01939755 | 0.89136073 |
| CAD | macrophage | *NOL10* | 1.202362116 | 0.00880954 | 0.04203875 | 1 |
| CAD | macrophage | *STRN* | 1.370337806 | 0.003343847 | 0.032549158 | 1 |
| CAD | macrophage | *CEBPZ* | 1.481169188 | 0.000446348 | 0.02312941 | 1 |
| CAD | macrophage | *PRKD3* | 1.314193762 | 0.000939324 | 0.029734791 | 1 |
| CAD | macrophage | *SLC25A12* | 0.959330928 | 0.003045118 | 0.032549158 | 1 |
| CAD | macrophage | *DARS* | 1.351892944 | 0.007771575 | 0.039776038 | 1 |
| CAD | macrophage | *PNO1* | 2.413278562 | 0.010670565 | 0.046433704 | 1 |
| CAD | macrophage | *ORC4* | 0.836161121 | 0.007740327 | 0.039741869 | 1 |
| CAD | macrophage | *NFE2L2* | 0.867842397 | 0.011159108 | 0.047915039 | 1 |
| CAD | macrophage | *MSH6* | 1.320590731 | 0.003948343 | 0.032549158 | 1 |
| CAD | macrophage | *ALMS1* | 1.440749894 | 0.002059998 | 0.032549158 | 1 |
| CAD | macrophage | *DNAJC16* | 1.294966462 | 0.002850161 | 0.032549158 | 1 |
| CAD | macrophage | *MECR* | 0.964996623 | 0.007557288 | 0.039482739 | 1 |
| CAD | macrophage | *EDEM3* | 1.227659464 | 0.009748307 | 0.044146082 | 1 |
| CAD | macrophage | *ASH1L* | 1.090458871 | 0.00020519 | 0.01939755 | 0.995579847 |
| CAD | macrophage | *DNAJC6* | 0.722438003 | 0.003765732 | 0.032549158 | 1 |
| CAD | macrophage | *LEPR* | 0.876021804 | 0.008554445 | 0.041464704 | 1 |
| CAD | macrophage | *PRG4* | 1.121569886 | 0.007653618 | 0.039573543 | 1 |
| CAD | macrophage | *C1orf109* | 1.151389894 | 0.00377833 | 0.032549158 | 1 |
| CAD | macrophage | *TBCE* | 1.747266628 | 0.003819303 | 0.032549158 | 1 |
| CAD | macrophage | *ZNHIT6* | 1.095599586 | 0.007574675 | 0.039518625 | 1 |
| CAD | macrophage | *CD46* | 0.985708574 | 0.000213619 | 0.01939755 | 1 |
| CAD | macrophage | *EBNA1BP2* | 1.046562802 | 0.006664214 | 0.037885063 | 1 |
| CAD | macrophage | *ERI3* | 1.516289973 | 0.000794001 | 0.028118192 | 1 |
| CAD | macrophage | *PRRC2C* | 1.527033953 | 9.22E-05 | 0.013557782 | 0.447406812 |
| CAD | macrophage | *ABCD3* | 1.431126905 | 0.003195123 | 0.032549158 | 1 |
| CAD | macrophage | *DARS2* | 1.162977407 | 0.005875322 | 0.036176476 | 1 |
| CAD | macrophage | *CD3EAP* | 1.966547256 | 4.15E-05 | 0.008184153 | 0.201218927 |
| CAD | macrophage | *RCN2* | 0.833025256 | 0.011756901 | 0.049101372 | 1 |
| CAD | macrophage | *CAMSAP2* | 0.972790654 | 0.000979289 | 0.029887243 | 1 |
| CAD | macrophage | *SGIP1* | 1.080647918 | 0.00615529 | 0.036870949 | 1 |
| CAD | macrophage | *TMEM5* | 1.014976571 | 0.007208625 | 0.038955499 | 1 |
| CAD | macrophage | *UBN1* | 1.467986763 | 0.002314262 | 0.032549158 | 1 |
| CAD | macrophage | *WDR35* | 1.131694369 | 0.000466295 | 0.02312941 | 1 |
| CAD | macrophage | *C2orf40* | 1.511475446 | 5.97E-05 | 0.009652182 | 0.28956545 |
| CAD | macrophage | *SENP5* | 1.243945384 | 0.007637333 | 0.039573543 | 1 |
| CAD | macrophage | *PPP6C* | 1.452258948 | 0.002916675 | 0.032549158 | 1 |
| CAD | macrophage | *MAPKAP1* | 1.09873239 | 0.011252252 | 0.048144558 | 1 |
| CAD | macrophage | *INVS* | 0.623653219 | 0.010776008 | 0.046724924 | 1 |
| CAD | macrophage | *KDSR* | 1.504578474 | 0.003012399 | 0.032549158 | 1 |
| CAD | macrophage | *NEK9* | 1.636909359 | 9.93E-05 | 0.014164811 | 0.481603561 |
| CAD | macrophage | *TTLL5* | 1.403841822 | 0.001001572 | 0.030209641 | 1 |
| CAD | macrophage | *EIF2B2* | 0.72510272 | 3.29E-05 | 0.007261836 | 0.159760394 |
| CAD | macrophage | *COQ6* | 1.582100918 | 0.007736238 | 0.039741869 | 1 |
| CAD | macrophage | *CRIPT* | 1.141682286 | 0.005425404 | 0.035310234 | 1 |
| CAD | macrophage | *PPP1R3C* | 1.38878742 | 0.00437212 | 0.032821343 | 1 |
| CAD | macrophage | *MXI1* | 1.234743416 | 0.00453647 | 0.032925653 | 1 |
| CAD | macrophage | *KANSL1* | 0.946577111 | 0.003226656 | 0.032549158 | 1 |

| | | | | | | |
|---|---|---|---|---|---|---|
| CAD | macrophage | *HOXB3* | 0.965439097 | 0.004316071 | 0.032776704 | 1 |
| CAD | macrophage | *PCDHB10* | 1.345689777 | 0.003391942 | 0.032549158 | 1 |
| CAD | macrophage | *GORAB* | 1.81540871 | 0.005476521 | 0.035310234 | 1 |
| CAD | macrophage | *SLC10A7* | 1.368210755 | 0.006455055 | 0.037296841 | 1 |
| CAD | macrophage | *IQSEC3* | 2.675057563 | 0.004627577 | 0.033148387 | 1 |
| CAD | macrophage | *DNAJC15* | 1.460359292 | 0.005394135 | 0.035275662 | 1 |
| CAD | macrophage | *SIL1* | 1.18474585 | 0.002293649 | 0.032549158 | 1 |
| CAD | macrophage | *TEX14* | 0.700244669 | 0.007534602 | 0.039482739 | 1 |
| CAD | macrophage | *ZNF549* | 1.920679978 | 0.001415228 | 0.032549158 | 1 |
| CAD | macrophage | *A1BG* | 1.616922117 | 0.002839275 | 0.032549158 | 1 |
| CAD | macrophage | *ZSCAN18* | 0.951383033 | 0.005796743 | 0.036134117 | 1 |
| CAD | macrophage | *SEC22A* | 1.246582581 | 0.003053155 | 0.032549158 | 1 |
| CAD | macrophage | *POPDC2* | -0.528126873 | 0.011604999 | 0.048784846 | 1 |
| CAD | macrophage | *ZCCHC17* | 1.670561419 | 0.003547373 | 0.032549158 | 1 |
| CAD | macrophage | *TNFSF10* | 1.02663356 | 0.003154688 | 0.032549158 | 1 |
| CAD | macrophage | *ZNF639* | 1.616485064 | 0.003022743 | 0.032549158 | 1 |
| CAD | macrophage | *RBBP6* | 1.128202984 | 0.004277829 | 0.032707875 | 1 |
| CAD | macrophage | *ODF2L* | 0.973303451 | 0.003006344 | 0.032549158 | 1 |
| CAD | macrophage | *EEPD1* | 0.581819801 | 0.007769388 | 0.039776038 | 1 |
| CAD | macrophage | *KLHL7* | 1.187482389 | 0.00328055 | 0.032549158 | 1 |
| CAD | macrophage | *NT5C3A* | 1.191079155 | 0.005468834 | 0.035310234 | 1 |
| CAD | macrophage | *SMU1* | 1.730097718 | 0.00115026 | 0.031531429 | 1 |
| CAD | macrophage | *TRIM24* | 1.143731574 | 0.007270623 | 0.038955499 | 1 |
| CAD | macrophage | *RBM19* | 1.592779788 | 0.009736597 | 0.044146082 | 1 |
| CAD | macrophage | *USP45* | 0.975714116 | 0.00387462 | 0.032549158 | 1 |
| CAD | macrophage | *PLA2G12A* | 1.228593344 | 0.006230188 | 0.037040771 | 1 |
| CAD | macrophage | *OBSL1* | 0.757283589 | 0.006988682 | 0.038620826 | 1 |
| CAD | macrophage | *SLC12A4* | 0.991754017 | 0.00926095 | 0.043219802 | 1 |
| CAD | macrophage | *WFDC3* | -0.181172229 | 0.003921024 | 0.032549158 | 1 |
| CAD | macrophage | *PTGIS* | 0.717840507 | 5.42E-06 | 0.002922343 | 0.02630109 |
| CAD | macrophage | *MTRR* | 1.149929266 | 0.008123307 | 0.040484919 | 1 |
| CAD | macrophage | *STAMBP* | 1.574495946 | 0.003208911 | 0.032549158 | 1 |
| CAD | macrophage | *SNRNP27* | 1.998895526 | 0.000730936 | 0.027706954 | 1 |
| CAD | macrophage | *ATP8A1* | 1.199446082 | 0.008545142 | 0.04146103 | 1 |
| CAD | macrophage | *USP22* | 1.263328015 | 0.004322546 | 0.032776704 | 1 |
| CAD | macrophage | *SNRPC* | -0.997707725 | 0.00018696 | 0.01939755 | 0.907128446 |
| CAD | macrophage | *NQO2* | 1.569398379 | 0.004455128 | 0.03285149 | 1 |
| CAD | macrophage | *TCP11* | 0.696144309 | 0.00056565 | 0.025412332 | 1 |
| CAD | macrophage | *GLO1* | 1.3599707 | 0.000516748 | 0.023878663 | 1 |
| CAD | macrophage | *ATXN1* | 0.905868076 | 0.011597466 | 0.048784846 | 1 |
| CAD | macrophage | *RUNX2* | 1.208081076 | 0.002259557 | 0.032549158 | 1 |
| CAD | macrophage | *TMTC4* | 1.392644124 | 0.007735064 | 0.039741869 | 1 |
| CAD | macrophage | *GRK4* | 0.283736935 | 2.79E-05 | 0.007261836 | 0.135174837 |
| CAD | macrophage | *MRPS7* | 2.11098545 | 0.001311745 | 0.032549158 | 1 |
| CAD | macrophage | *NT5C* | 1.678937083 | 0.005337612 | 0.03509227 | 1 |
| CAD | macrophage | *SNRPD2* | 2.033953768 | 0.003456522 | 0.032549158 | 1 |
| CAD | macrophage | *PSMF1* | 0.840536452 | 0.008507323 | 0.04146103 | 1 |
| CAD | macrophage | *DTD1* | 1.151775843 | 0.004406531 | 0.032821343 | 1 |
| CAD | macrophage | *RRBP1* | 1.618451151 | 0.000222388 | 0.01939755 | 1 |
| CAD | macrophage | *FLRT3* | 0.766314743 | 0.010877096 | 0.047037139 | 1 |

| | | | | | | |
|---|---|---|---|---|---|---|
| CAD | macrophage | *KLC1* | 1.671656485 | 0.000721117 | 0.027706954 | 1 |
| CAD | macrophage | *COX6B1* | 1.427805941 | 0.009973301 | 0.044767221 | 1 |
| CAD | macrophage | *IRF3* | 1.550594762 | 0.003516224 | 0.032549158 | 1 |
| CAD | macrophage | *TRAP1* | 0.372398865 | 0.002468073 | 0.032549158 | 1 |
| CAD | macrophage | *KTN1* | 1.110344557 | 0.002785278 | 0.032549158 | 1 |
| CAD | macrophage | *RHOT1* | 0.874257276 | 0.002100074 | 0.032549158 | 1 |
| CAD | macrophage | *ZNF484* | 1.829563 | 0.001049771 | 0.030318387 | 1 |
| CAD | macrophage | *ATP13A4* | 1.37170447 | 0.007029785 | 0.038675956 | 1 |
| CAD | macrophage | *EMC1* | 1.516420597 | 0.001210845 | 0.032263164 | 1 |
| CAD | macrophage | *SMARCA4* | 1.757436135 | 4.99E-05 | 0.008652153 | 0.242260288 |
| CAD | macrophage | *HIP1* | 1.162239391 | 0.002770678 | 0.032549158 | 1 |
| CAD | macrophage | *PEX1* | 1.362290385 | 0.003552483 | 0.032549158 | 1 |
| CAD | macrophage | *KDR* | 0.914056132 | 0.006353179 | 0.03707567 | 1 |
| CAD | macrophage | *ATF4* | 1.823744754 | 0.004230877 | 0.032707875 | 1 |
| CAD | macrophage | *SPECC1* | 0.883498514 | 0.004444275 | 0.032821343 | 1 |
| CAD | macrophage | *CDHR3* | 0.644483257 | 0.009658341 | 0.044002875 | 1 |
| CAD | macrophage | *MTX2* | 1.215620363 | 0.002574414 | 0.032549158 | 1 |
| CAD | macrophage | *TWSG1* | 1.334812526 | 0.006780361 | 0.038293121 | 1 |
| CAD | macrophage | *SERGEF* | 1.197934437 | 0.000481465 | 0.02312941 | 1 |
| CAD | macrophage | *PHF20L1* | 1.122634091 | 0.008127022 | 0.040484919 | 1 |
| CAD | macrophage | *MTUS1* | 0.661727156 | 0.006292444 | 0.037040771 | 1 |
| CAD | macrophage | *EPB41L4A* | 0.641542088 | 0.006049496 | 0.036581378 | 1 |
| CAD | macrophage | *PHF10* | 1.373542034 | 0.004298763 | 0.032743483 | 1 |
| CAD | macrophage | *FAM98C* | 0.854732606 | 0.001116528 | 0.031134444 | 1 |
| CAD | macrophage | *PLVAP* | 0.295576262 | 0.006298153 | 0.037040771 | 1 |
| CAD | macrophage | *LSM7* | 0.694825033 | 0.008923948 | 0.042284174 | 1 |
| CAD | macrophage | *ACTN4* | 1.330235217 | 0.000142856 | 0.017772693 | 0.693135027 |
| CAD | macrophage | *ATXN10* | 1.147984581 | 0.003327582 | 0.032549158 | 1 |
| CAD | macrophage | *PNPLA7* | 1.091245979 | 0.004393369 | 0.032821343 | 1 |
| CAD | macrophage | *ZNF337* | 0.893082912 | 0.003055332 | 0.032549158 | 1 |
| CAD | macrophage | *POMT1* | 1.263180769 | 0.002240435 | 0.032549158 | 1 |
| CAD | macrophage | *SLC35D2* | 1.5715772 | 0.006043946 | 0.036581378 | 1 |
| CAD | macrophage | *IDO1* | 3.559273037 | 0.002137194 | 0.032549158 | 1 |
| CAD | macrophage | *KIF3A* | 1.159401159 | 0.001452012 | 0.032549158 | 1 |
| CAD | macrophage | *EXOC4* | 0.82460066 | 0.008998032 | 0.042386846 | 1 |
| CAD | macrophage | *C1orf159* | 1.856036699 | 0.004203483 | 0.032707875 | 1 |
| CAD | macrophage | *MAP1B* | 0.634476164 | 0.001217121 | 0.032263164 | 1 |
| CAD | macrophage | *KHDRBS3* | 0.860652541 | 0.00793929 | 0.040168337 | 1 |
| CAD | macrophage | *SPATA6* | 0.889430721 | 0.003095401 | 0.032549158 | 1 |
| CAD | macrophage | *LRRC41* | 1.447958427 | 0.005867162 | 0.036176476 | 1 |
| CAD | macrophage | *TIMM10B* | 1.004936508 | 0.000225429 | 0.01939755 | 1 |
| CAD | macrophage | *SERPINF1* | 1.007854162 | 0.004236353 | 0.032707875 | 1 |
| CAD | macrophage | *DDC* | -1.233404257 | 0.01065654 | 0.0464143 | 1 |
| CAD | macrophage | *MUTYH* | 0.934823323 | 0.003862797 | 0.032549158 | 1 |
| CAD | macrophage | *ZBED3* | 1.363493046 | 0.002555349 | 0.032549158 | 1 |
| CAD | macrophage | *FBXO44* | 1.34569604 | 0.005945286 | 0.036330638 | 1 |
| CAD | macrophage | *WASF3* | 0.598003143 | 0.011231813 | 0.048099521 | 1 |
| CAD | macrophage | *PEMT* | 1.309560753 | 0.005714176 | 0.035960029 | 1 |
| CAD | macrophage | *SCO1* | 0.983475286 | 0.011217304 | 0.048079823 | 1 |
| CAD | macrophage | *MPRIP* | 0.981346153 | 0.003893333 | 0.032549158 | 1 |

| | | | | | | |
|---|---|---|---|---|---|---|
| CAD | macrophage | *DSTYK* | 1.556216639 | 0.004633492 | 0.033148387 | 1 |
| CAD | macrophage | *CSNK1G2* | 1.952640333 | 0.007061961 | 0.038717101 | 1 |
| CAD | macrophage | *PDZD2* | 0.527345896 | 0.008254661 | 0.040820553 | 1 |
| CAD | macrophage | *IPO8* | 1.518535974 | 0.003444198 | 0.032549158 | 1 |
| CAD | macrophage | *RRAS2* | 0.75925992 | 0.001100487 | 0.031043976 | 1 |
| CAD | macrophage | *COX16* | 2.27352453 | 0.004198252 | 0.032707875 | 1 |
| CAD | macrophage | *PEBP4* | 1.068939061 | 0.009437715 | 0.043541159 | 1 |
| CAD | macrophage | *CTIF* | 0.675170404 | 0.003717952 | 0.032549158 | 1 |
| CAD | macrophage | *NAPG* | 1.33011817 | 0.005588517 | 0.035445077 | 1 |
| CAD | macrophage | *KIDINS220* | 1.195210155 | 0.003040088 | 0.032549158 | 1 |
| CAD | macrophage | *KLRD1* | 0.779772114 | 0.007674918 | 0.039573543 | 1 |
| CAD | macrophage | *PRPF38A* | 1.420222648 | 0.011119645 | 0.047787883 | 1 |
| CAD | macrophage | *ELP2* | 1.172946325 | 0.003169617 | 0.032549158 | 1 |
| CAD | macrophage | *TPGS2* | 1.206510715 | 0.011887969 | 0.049468634 | 1 |
| CAD | macrophage | *CLOCK* | 1.293733884 | 0.005486362 | 0.035310234 | 1 |
| CAD | macrophage | *ARGLU1* | 0.854091655 | 0.003422042 | 0.032549158 | 1 |
| CAD | macrophage | *KLB* | 0.28800986 | 0.008675715 | 0.041843508 | 1 |
| CAD | macrophage | *UBQLN1* | 1.356706927 | 0.003358679 | 0.032549158 | 1 |
| CAD | macrophage | *GOLM1* | 0.416996911 | 0.008988231 | 0.042386846 | 1 |
| CAD | macrophage | *MSI1* | 0.718327251 | 0.007554003 | 0.039482739 | 1 |
| CAD | macrophage | *DMTF1* | 1.102608019 | 0.00169624 | 0.032549158 | 1 |
| CAD | macrophage | *SRPK2* | 1.03411541 | 0.008479865 | 0.04146103 | 1 |
| CAD | macrophage | *MAP3K7* | 1.426314198 | 0.005556286 | 0.035379398 | 1 |
| CAD | macrophage | *PHF21A* | 1.15048422 | 0.00361004 | 0.032549158 | 1 |
| CAD | macrophage | *CAPRIN1* | 1.237825127 | 0.007749592 | 0.039747381 | 1 |
| CAD | macrophage | *ATP5G2* | 1.293130122 | 0.006913326 | 0.038407224 | 1 |
| CAD | macrophage | *B4GALNT1* | 1.709578418 | 0.000181117 | 0.01939755 | 0.878780821 |
| CAD | macrophage | *NTPCR* | 1.674921502 | 0.003574483 | 0.032549158 | 1 |
| CAD | macrophage | *PIGC* | 1.129379271 | 0.008788249 | 0.04203875 | 1 |
| CAD | macrophage | *MRPL44* | 2.151162728 | 0.010480574 | 0.045895078 | 1 |
| CAD | macrophage | *USP37* | 1.020014777 | 0.006257546 | 0.037040771 | 1 |
| CAD | macrophage | *GCC2* | 1.242072427 | 0.001212957 | 0.032263164 | 1 |
| CAD | macrophage | *ANKRD36* | 1.14092459 | 0.003178823 | 0.032549158 | 1 |
| CAD | macrophage | *EPC2* | 1.353332502 | 0.002098252 | 0.032549158 | 1 |
| CAD | macrophage | *STAB2* | 0.891650122 | 0.008761035 | 0.041963024 | 1 |
| CAD | macrophage | *TBC1D4* | 0.898119976 | 0.0073561 | 0.039108508 | 1 |
| CAD | macrophage | *SUCLA2* | 1.336916221 | 0.002806826 | 0.032549158 | 1 |
| CAD | macrophage | *SCN2A* | 0.791946456 | 0.002442028 | 0.032549158 | 1 |
| CAD | macrophage | *STX17* | 1.396406068 | 0.00170568 | 0.032549158 | 1 |
| CAD | macrophage | *POLR1E* | 0.416901308 | 0.000427836 | 0.02312941 | 1 |
| CAD | macrophage | *APTX* | 1.12958318 | 0.008265209 | 0.040820553 | 1 |
| CAD | macrophage | *RIPK1* | 0.936229543 | 0.006380252 | 0.03707567 | 1 |
| CAD | macrophage | *TCF19* | 0.71664617 | 0.001881257 | 0.032549158 | 1 |
| CAD | macrophage | *NUMA1* | 1.31137873 | 0.002086621 | 0.032549158 | 1 |
| CAD | macrophage | *RAB30* | 0.851890274 | 0.010447264 | 0.045795852 | 1 |
| CAD | macrophage | *NARS2* | 1.440200085 | 0.003244398 | 0.032549158 | 1 |
| CAD | macrophage | *TGS1* | 1.677407523 | 0.000548709 | 0.02511638 | 1 |
| CAD | macrophage | *NEK1* | 0.94083367 | 0.007187247 | 0.038955499 | 1 |
| CAD | macrophage | *SLC37A4* | 0.722916416 | 0.00124021 | 0.032456612 | 1 |
| CAD | macrophage | *FDX1* | 1.07040827 | 0.00432339 | 0.032776704 | 1 |

| | | | | | | |
|---|---|---|---|---|---|---|
| CAD | macrophage | *MAP2K5* | 1.266236423 | 0.001779243 | 0.032549158 | 1 |
| CAD | macrophage | *NDUFAF1* | 0.881300813 | 0.004373045 | 0.032821343 | 1 |
| CAD | macrophage | *PARP6* | 1.390285445 | 0.007425809 | 0.039333376 | 1 |
| CAD | macrophage | *ADAM10* | 1.233431813 | 0.002925133 | 0.032549158 | 1 |
| CAD | macrophage | *ZNF280D* | 1.134962393 | 0.00133697 | 0.032549158 | 1 |
| CAD | macrophage | *RTCA* | 1.499648119 | 0.000721506 | 0.027706954 | 1 |
| CAD | macrophage | *DYNC2LI1* | 0.667080715 | 0.004116316 | 0.032707875 | 1 |
| CAD | macrophage | *CENPO* | 1.097974273 | 0.001473575 | 0.032549158 | 1 |
| CAD | macrophage | *LRPPRC* | 1.343492702 | 0.003229249 | 0.032549158 | 1 |
| CAD | macrophage | *BTBD16* | 0.816343403 | 0.003808132 | 0.032549158 | 1 |
| CAD | macrophage | *ARL3* | 0.694740758 | 0.010365685 | 0.045639112 | 1 |
| CAD | macrophage | *PLCE1* | 0.506823736 | 0.005983568 | 0.036447642 | 1 |
| CAD | macrophage | *RBP4* | 1.228982777 | 0.001319724 | 0.032549158 | 1 |
| CAD | macrophage | *SMARCAL1* | 1.504696913 | 0.002871035 | 0.032549158 | 1 |
| CAD | macrophage | *STAT4* | 1.346832338 | 0.003818939 | 0.032549158 | 1 |
| CAD | macrophage | *ITGAV* | 1.206105988 | 0.00504329 | 0.034080841 | 1 |
| CAD | macrophage | *AP1AR* | 1.860506915 | 0.004855332 | 0.033586837 | 1 |
| CAD | macrophage | *KIAA1109* | 1.177799254 | 0.003320118 | 0.032549158 | 1 |
| CAD | macrophage | *PRDM5* | 0.659631846 | 0.009973901 | 0.044767221 | 1 |
| CAD | macrophage | *NUP54* | 1.008276675 | 0.000126121 | 0.016538869 | 0.611938141 |
| CAD | macrophage | *PPA2* | 1.150251812 | 0.004753091 | 0.033426994 | 1 |
| CAD | macrophage | *KIF21A* | 0.595076344 | 0.009640012 | 0.044002875 | 1 |
| CAD | macrophage | *CLSTN3* | 0.861620689 | 0.003449349 | 0.032549158 | 1 |
| CAD | macrophage | *PPFIA2* | 0.763444167 | 0.009006942 | 0.042387667 | 1 |
| CAD | macrophage | *POC1B* | 1.519752876 | 0.004880198 | 0.033586837 | 1 |
| CAD | macrophage | *RITA1* | 1.712669464 | 0.008070169 | 0.040450887 | 1 |
| CAD | macrophage | *ACVRL1* | 2.15414381 | 0.006218945 | 0.037040771 | 1 |
| CAD | macrophage | *KANSL2* | 0.925027354 | 0.007286316 | 0.038955499 | 1 |
| CAD | macrophage | *CERS5* | 0.972986445 | 0.008820261 | 0.04203875 | 1 |
| CAD | macrophage | *LMBR1L* | 1.104707235 | 0.007958351 | 0.040222834 | 1 |
| CAD | macrophage | *ESYT1* | 1.592057386 | 0.000751079 | 0.027706954 | 1 |
| CAD | macrophage | *DIAPH3* | 0.406832045 | 0.003745904 | 0.032549158 | 1 |
| CAD | macrophage | *C14orf37* | 1.039501796 | 0.003356754 | 0.032549158 | 1 |
| CAD | macrophage | *RAB15* | 1.573907119 | 0.004149771 | 0.032707875 | 1 |
| CAD | macrophage | *SRP14* | 1.17628997 | 0.008036233 | 0.040358159 | 1 |
| CAD | macrophage | *UBE2Q2* | 1.172717747 | 0.008079235 | 0.040454537 | 1 |
| CAD | macrophage | *HMG20A* | 1.508942786 | 0.004808575 | 0.033426994 | 1 |
| CAD | macrophage | *BBS4* | 0.997685633 | 0.00954471 | 0.043896621 | 1 |
| CAD | macrophage | *TGFB1I1* | 1.398650304 | 0.005394588 | 0.035275662 | 1 |
| CAD | macrophage | *DHX38* | 1.832502113 | 0.002921269 | 0.032549158 | 1 |
| CAD | macrophage | *NUDT7* | 1.532525611 | 0.004159824 | 0.032707875 | 1 |
| CAD | macrophage | *ZCCHC14* | 1.179362446 | 0.005074189 | 0.034241953 | 1 |
| CAD | macrophage | *TLDC1* | 1.227345947 | 0.005432466 | 0.035310234 | 1 |
| CAD | macrophage | *TIGD7* | 1.141213777 | 0.006848566 | 0.038393741 | 1 |
| CAD | macrophage | *TCF25* | 1.28618593 | 0.000248711 | 0.01939755 | 1 |
| CAD | macrophage | *NCOR1* | 1.008129868 | 0.002694034 | 0.032549158 | 1 |
| CAD | macrophage | *KSR1* | 1.493528823 | 0.001036226 | 0.030318387 | 1 |
| CAD | macrophage | *MYO19* | 1.836523996 | 2.02E-05 | 0.006130632 | 0.098090117 |
| CAD | macrophage | *RASL10B* | 1.343526073 | 0.007120252 | 0.038943652 | 1 |
| CAD | macrophage | *IMPA2* | 1.774493406 | 0.000719588 | 0.027706954 | 1 |

| | | | | | | |
|---|---|---|---|---|---|---|
| CAD | macrophage | *GNAL* | 0.782083462 | 0.00890464 | 0.042244682 | 1 |
| CAD | macrophage | *IGFBP4* | 0.845230399 | 0.001609037 | 0.032549158 | 1 |
| CAD | macrophage | *CACNA1A* | 1.189753866 | 0.005208116 | 0.034616138 | 1 |
| CAD | macrophage | *NFIC* | 1.181553102 | 0.001295011 | 0.032549158 | 1 |
| CAD | macrophage | *TMEM91* | 0.520493669 | 0.000114202 | 0.015391918 | 0.554109047 |
| CAD | macrophage | *SIRT3* | 1.451327981 | 3.16E-05 | 0.007261836 | 0.153553053 |
| CAD | macrophage | *EVI5L* | 2.362482942 | 0.003290808 | 0.032549158 | 1 |
| CAD | macrophage | *RERE* | 0.954223108 | 0.003868745 | 0.032549158 | 1 |
| CAD | macrophage | *EVA1B* | 0.815679103 | 0.000344248 | 0.021635859 | 1 |
| CAD | macrophage | *RPS8* | 1.229446085 | 0.002301926 | 0.032549158 | 1 |
| CAD | macrophage | *CYP4B1* | 0.451347188 | 0.008359918 | 0.04110374 | 1 |
| CAD | macrophage | *SLC44A3* | 0.828360325 | 0.011851752 | 0.049402663 | 1 |
| CAD | macrophage | *STRIP1* | 2.24771213 | 0.001442012 | 0.032549158 | 1 |
| CAD | macrophage | *TIPRL* | 1.247032533 | 0.009627843 | 0.044002875 | 1 |
| CAD | macrophage | *SNX27* | 1.223819648 | 0.001124643 | 0.031181525 | 1 |
| CAD | macrophage | *SETDB1* | 1.903105698 | 8.95E-05 | 0.013557782 | 0.43429981 |
| CAD | macrophage | *SEMA6C* | 1.355323268 | 0.001663289 | 0.032549158 | 1 |
| CAD | macrophage | *RAB13* | 2.279107832 | 0.009663248 | 0.044002875 | 1 |
| CAD | macrophage | *SCCPDH* | 1.349166199 | 0.007934113 | 0.040168337 | 1 |
| CAD | macrophage | *ACP1* | 1.542553389 | 0.006256932 | 0.037040771 | 1 |
| CAD | macrophage | *MBOAT2* | 0.920600939 | 0.007659968 | 0.039573543 | 1 |
| CAD | macrophage | *PLEKHA6* | 0.534753309 | 0.003807695 | 0.032549158 | 1 |
| CAD | macrophage | *WDPCP* | 0.887636766 | 1.80E-06 | 0.002081011 | 0.008717475 |
| CAD | macrophage | *VPS54* | 1.204567704 | 0.004740782 | 0.033426994 | 1 |
| CAD | macrophage | *EXOC6B* | 1.029737317 | 0.005490078 | 0.035310234 | 1 |
| CAD | macrophage | *GALNT13* | 0.717827603 | 0.010407225 | 0.045780468 | 1 |
| CAD | macrophage | *SUMF1* | 1.066382923 | 0.006076321 | 0.036582371 | 1 |
| CAD | macrophage | *ACKR3* | 0.446913632 | 0.011636089 | 0.048797153 | 1 |
| CAD | macrophage | *TAMM41* | 1.494281319 | 0.0003659 | 0.021917844 | 1 |
| CAD | macrophage | *EAF1* | 1.758589479 | 0.003569378 | 0.032549158 | 1 |
| CAD | macrophage | *CNTN4* | 0.589536805 | 0.010526317 | 0.046012334 | 1 |
| CAD | macrophage | *CTDSPL* | 1.041774011 | 0.001796389 | 0.032549158 | 1 |
| CAD | macrophage | *ARL6IP5* | 1.328504846 | 0.010093014 | 0.045010391 | 1 |
| CAD | macrophage | *BOC* | 0.535570518 | 0.008906906 | 0.042244682 | 1 |
| CAD | macrophage | *AMT* | -0.251756559 | 0.004151087 | 0.032707875 | 1 |
| CAD | macrophage | *SCD5* | 1.365806434 | 0.004505374 | 0.032923259 | 1 |
| CAD | macrophage | *METTL14* | 1.512507198 | 0.00405077 | 0.032702724 | 1 |
| CAD | macrophage | *USP53* | 0.588096886 | 0.010277088 | 0.045435278 | 1 |
| CAD | macrophage | *CBR4* | 0.981886908 | 0.002186962 | 0.032549158 | 1 |
| CAD | macrophage | *MARCH6* | 1.238129861 | 0.001902243 | 0.032549158 | 1 |
| CAD | macrophage | *MYO10* | 0.857362098 | 0.004908287 | 0.033650257 | 1 |
| CAD | macrophage | *GTF2H2* | 3.095713762 | 0.003518188 | 0.032549158 | 1 |
| CAD | macrophage | *SPATA9* | 0.71943244 | 0.010716456 | 0.046549905 | 1 |
| CAD | macrophage | *FEM1C* | 2.121868574 | 0.004539518 | 0.032925653 | 1 |
| CAD | macrophage | *FBXO38* | 1.6094068 | 0.000665072 | 0.027460164 | 1 |
| CAD | macrophage | *TBC1D7* | 1.091649276 | 0.002184414 | 0.032549158 | 1 |
| CAD | macrophage | *FAM193B* | 1.000104445 | 0.011207706 | 0.048079823 | 1 |
| CAD | macrophage | *RPL7L1* | 1.440799098 | 0.008499486 | 0.04146103 | 1 |
| CAD | macrophage | *IRAK1BP1* | 1.248124493 | 1.74E-05 | 0.00603517 | 0.08449238 |
| CAD | macrophage | *PHIP* | 1.267100189 | 0.002122622 | 0.032549158 | 1 |

| | | | | | | |
|---|---|---|---|---|---|---|
| CAD | macrophage | *PNRC1* | 1.568778574 | 0.010799514 | 0.046785039 | 1 |
| CAD | macrophage | *RNF217* | 1.163864206 | 0.001642042 | 0.032549158 | 1 |
| CAD | macrophage | *TMEM209* | 1.751700097 | 0.003001253 | 0.032549158 | 1 |
| CAD | macrophage | *FBXO25* | 1.167608674 | 0.00380086 | 0.032549158 | 1 |
| CAD | macrophage | *CSGALNACT1* | 0.596186625 | 0.000256622 | 0.01939755 | 1 |
| CAD | macrophage | *BIN3* | 1.252726103 | 0.002712251 | 0.032549158 | 1 |
| CAD | macrophage | *TACC1* | 0.849333115 | 0.002977265 | 0.032549158 | 1 |
| CAD | macrophage | *DPYS* | 0.750913202 | 0.007279123 | 0.038955499 | 1 |
| CAD | macrophage | *LRP12* | 1.787219798 | 0.00282071 | 0.032549158 | 1 |
| CAD | macrophage | *AK3* | 1.049785363 | 0.004419671 | 0.032821343 | 1 |
| CAD | macrophage | *HAUS6* | 1.675954083 | 0.006135479 | 0.03684325 | 1 |
| CAD | macrophage | *FBXO10* | 0.760561605 | 0.000737165 | 0.027706954 | 1 |
| CAD | macrophage | *REXO4* | 1.875102427 | 0.002947323 | 0.032549158 | 1 |
| CAD | macrophage | *SH3GLB2* | 1.554471447 | 0.004256971 | 0.032707875 | 1 |
| CAD | macrophage | *INPP5E* | 1.621744171 | 0.008860597 | 0.042107363 | 1 |
| CAD | macrophage | *RSU1* | 1.322993933 | 0.004345247 | 0.032821343 | 1 |
| CAD | macrophage | *PARD3* | 0.516737565 | 0.008144606 | 0.0405309 | 1 |
| CAD | macrophage | *ADD3* | 0.877137 | 0.003059172 | 0.032549158 | 1 |
| CAD | macrophage | *GAS2* | 1.144888406 | 0.005795507 | 0.036134117 | 1 |
| CAD | macrophage | *LIN7C* | 1.768895878 | 0.006840432 | 0.038393741 | 1 |
| CAD | macrophage | *HSD17B12* | -0.296621572 | 0.001898002 | 0.032549158 | 1 |
| CAD | macrophage | *TTC12* | 0.788444513 | 0.000181978 | 0.01939755 | 0.882955023 |
| CAD | macrophage | *NUDT22* | 0.95535713 | 0.003036779 | 0.032549158 | 1 |
| CAD | macrophage | *FAM124A* | 0.856342454 | 0.00873671 | 0.041948605 | 1 |
| CAD | macrophage | *PRSS23* | 0.565944012 | 0.003634218 | 0.032549158 | 1 |
| CAD | macrophage | *FAM173B* | 1.540823618 | 0.000471033 | 0.02312941 | 1 |
| CAD | macrophage | *ANK3* | 0.898742824 | 0.003274138 | 0.032549158 | 1 |
| CAD | macrophage | *DLG5* | 0.548355662 | 0.002654047 | 0.032549158 | 1 |
| CAD | macrophage | *TMEM18* | 2.298618748 | 0.00416975 | 0.032707875 | 1 |
| CAD | macrophage | *NDUFC2* | 2.055548737 | 0.007513083 | 0.039482739 | 1 |
| CAD | macrophage | *NUBPL* | 1.282621483 | 0.00364063 | 0.032549158 | 1 |
| CAD | macrophage | *ANKRD50* | 1.580415007 | 0.002438331 | 0.032549158 | 1 |
| CAD | macrophage | *UPF2* | 1.034936815 | 0.009677132 | 0.044002875 | 1 |
| CAD | macrophage | *FAM160B1* | 1.169152559 | 0.001542646 | 0.032549158 | 1 |
| CAD | macrophage | *PIGF* | 1.514675132 | 0.007552853 | 0.039482739 | 1 |
| CAD | macrophage | *MFSD6* | 1.345079775 | 0.00345684 | 0.032549158 | 1 |
| CAD | macrophage | *RNF144A* | 0.644886348 | 0.009414688 | 0.043541159 | 1 |
| CAD | macrophage | *BICD1* | 1.385785873 | 0.004630615 | 0.033148387 | 1 |
| CAD | macrophage | *TIAL1* | 1.500141293 | 0.002048769 | 0.032549158 | 1 |
| CAD | macrophage | *PTPN14* | 0.606100965 | 0.005307195 | 0.034987108 | 1 |
| CAD | macrophage | *EPG5* | 1.458939506 | 0.003366379 | 0.032549158 | 1 |
| CAD | macrophage | *CWF19L2* | 1.132455003 | 0.000584358 | 0.025946406 | 1 |
| CAD | macrophage | *SPARCL1* | 0.628960527 | 0.007615966 | 0.039573543 | 1 |
| CAD | macrophage | *MBNL1* | 0.94482082 | 0.004014733 | 0.032683698 | 1 |
| CAD | macrophage | *GPD1L* | 0.817533209 | 0.004994832 | 0.033847661 | 1 |
| CAD | macrophage | *MED21* | 1.4757946 | 0.001002422 | 0.030209641 | 1 |
| CAD | macrophage | *JAKMIP1* | 1.676780725 | 0.008043351 | 0.040358159 | 1 |
| CAD | macrophage | *CAST* | 0.768296557 | 0.005928111 | 0.036271366 | 1 |
| CAD | macrophage | *SCOC* | 1.191321399 | 0.005136944 | 0.034519543 | 1 |
| CAD | macrophage | *TMEM87B* | 1.540429934 | 0.003168579 | 0.032549158 | 1 |

| | | | | | | |
|---|---|---|---|---|---|---|
| CAD | macrophage | *PLA2R1* | 0.412619609 | 0.002768999 | 0.032549158 | 1 |
| CAD | macrophage | *CFDP1* | 1.204512562 | 0.002876489 | 0.032549158 | 1 |
| CAD | macrophage | *FBXO36* | 1.375291617 | 0.00325612 | 0.032549158 | 1 |
| CAD | macrophage | *SEMA3D* | 0.630308863 | 0.008205295 | 0.04079108 | 1 |
| CAD | macrophage | *PPP2R5E* | 1.20852725 | 0.01023003 | 0.045329776 | 1 |
| CAD | macrophage | *ANKRD29* | 0.916059603 | 0.005969147 | 0.03643057 | 1 |
| CAD | macrophage | *ROBO4* | 0.932575413 | 0.006456996 | 0.037296841 | 1 |
| CAD | macrophage | *ROBO3* | 1.180404263 | 7.81E-06 | 0.003442744 | 0.037870189 |
| CAD | macrophage | *PRKCA* | 1.091664774 | 0.003795117 | 0.032549158 | 1 |
| CAD | macrophage | *ABCA5* | 0.6273012 | 0.001790058 | 0.032549158 | 1 |
| CAD | macrophage | *PGM5* | 0.944723843 | 0.008990354 | 0.042386846 | 1 |
| CAD | macrophage | *PDE1C* | 0.964835155 | 0.003514695 | 0.032549158 | 1 |
| CAD | macrophage | *OXNAD1* | 0.931053587 | 0.005143782 | 0.034519543 | 1 |
| CAD | macrophage | *APCDD1* | 0.533585031 | 0.000871203 | 0.028834548 | 1 |
| CAD | macrophage | *TRMT44* | 1.410672871 | 0.002190766 | 0.032549158 | 1 |
| CAD | macrophage | *ALS2CR12* | 1.091053631 | 0.003510761 | 0.032549158 | 1 |
| CAD | macrophage | *C14orf2* | 1.101858633 | 0.004586745 | 0.033148387 | 1 |
| CAD | macrophage | *RPL30* | 1.126820291 | 0.002323552 | 0.032549158 | 1 |
| CAD | macrophage | *NODAL* | 1.451279478 | 0.01178241 | 0.049155848 | 1 |
| CAD | macrophage | *ZDHHC5* | 1.820835375 | 0.005763939 | 0.036085979 | 1 |
| CAD | macrophage | *MED19* | 0.792519177 | 0.001371925 | 0.032549158 | 1 |
| CAD | macrophage | *RPUSD3* | 1.314688231 | 0.004393656 | 0.032821343 | 1 |
| CAD | macrophage | *GHRL* | 1.193782993 | 0.000942517 | 0.029734791 | 1 |
| CAD | macrophage | *CLDN12* | 1.360763096 | 0.006265109 | 0.037040771 | 1 |
| CAD | macrophage | *GATAD1* | 1.188432543 | 0.008029614 | 0.040358159 | 1 |
| CAD | macrophage | *CLEC18A* | -0.428468475 | 0.011559516 | 0.048784846 | 1 |
| CAD | macrophage | *AASDH* | 1.191586814 | 0.000959883 | 0.029887243 | 1 |
| CAD | macrophage | *AP3S2* | 1.644721814 | 0.000409431 | 0.023038679 | 1 |
| CAD | macrophage | *WDR66* | 0.482543871 | 0.009290654 | 0.043219802 | 1 |
| CAD | macrophage | *EYA3* | 1.098249697 | 0.006787322 | 0.038293121 | 1 |
| CAD | macrophage | *FANCC* | 0.832070666 | 0.00525675 | 0.034764704 | 1 |
| CAD | macrophage | *ESYT3* | 0.904921702 | 0.005474862 | 0.035310234 | 1 |
| CAD | macrophage | *MITD1* | 1.2859713 | 0.003748158 | 0.032549158 | 1 |
| CAD | macrophage | *CCAR2* | 1.29090681 | 0.007533835 | 0.039482739 | 1 |
| CAD | macrophage | *THEM4* | 0.573036821 | 0.006852634 | 0.038393741 | 1 |
| CAD | macrophage | *TMEM69* | 1.858633925 | 0.010764463 | 0.046716616 | 1 |
| CAD | macrophage | *ACE* | 1.134338639 | 0.009610624 | 0.044002875 | 1 |
| CAD | macrophage | *CHCHD6* | 1.555383914 | 1.97E-05 | 0.006130632 | 0.095365777 |
| CAD | macrophage | *SSU72* | 1.733783417 | 0.002747857 | 0.032549158 | 1 |
| CAD | macrophage | *CCDC58* | 2.157801023 | 0.003162054 | 0.032549158 | 1 |
| CAD | macrophage | *TAOK1* | 1.142671061 | 0.008481133 | 0.04146103 | 1 |
| CAD | macrophage | *IER2* | 1.277700053 | 0.007127363 | 0.038943652 | 1 |
| CAD | macrophage | *CPSF4* | 1.656968927 | 1.02E-05 | 0.004105132 | 0.049261582 |
| CAD | macrophage | *ZNF333* | 1.814826994 | 0.003678325 | 0.032549158 | 1 |
| CAD | macrophage | *RPL8* | 1.349193599 | 0.003750576 | 0.032549158 | 1 |
| CAD | macrophage | *ABCF3* | 0.535948409 | 0.010448477 | 0.045795852 | 1 |
| CAD | macrophage | *TMEM143* | 1.857069271 | 0.00273547 | 0.032549158 | 1 |
| CAD | macrophage | *ASB16* | 2.260724406 | 0.000304891 | 0.020835672 | 1 |
| CAD | macrophage | *IP6K3* | 4.21293896 | 0.01015859 | 0.045178258 | 1 |
| CAD | macrophage | *C1orf123* | 1.577168055 | 0.007478313 | 0.039482739 | 1 |

| | | | | | | |
|---|---|---|---|---|---|---|
| CAD | macrophage | *TM2D1* | 1.359621125 | 0.003423021 | 0.032549158 | 1 |
| CAD | macrophage | *NEXN* | 0.744061648 | 0.00424669 | 0.032707875 | 1 |
| CAD | macrophage | *FCRLB* | 2.403176715 | 0.007386092 | 0.039209322 | 1 |
| CAD | macrophage | *CNST* | 1.170700724 | 0.010188173 | 0.045238419 | 1 |
| CAD | macrophage | *HAAO* | 1.855383713 | 0.001552133 | 0.032549158 | 1 |
| CAD | macrophage | *MRPL55* | 0.863638237 | 0.00136872 | 0.032549158 | 1 |
| CAD | macrophage | *KIAA1841* | 1.562636152 | 0.002004923 | 0.032549158 | 1 |
| CAD | macrophage | *NOSTRIN* | 0.709999273 | 0.001244215 | 0.032456612 | 1 |
| CAD | macrophage | *INHBB* | 1.270325567 | 0.001366713 | 0.032549158 | 1 |
| CAD | macrophage | *RPRD2* | 0.882822219 | 0.006911706 | 0.038407224 | 1 |
| CAD | macrophage | *PACRGL* | 1.130211241 | 0.005732533 | 0.035980948 | 1 |
| CAD | macrophage | *C1QTNF7* | 0.675459836 | 0.003690353 | 0.032549158 | 1 |
| CAD | macrophage | *VPS72* | 1.62914524 | 0.003409625 | 0.032549158 | 1 |
| CAD | macrophage | *GNPDA2* | 0.709841497 | 0.009039925 | 0.042477903 | 1 |
| CAD | macrophage | *DAPL1* | 1.54859854 | 0.000435452 | 0.02312941 | 1 |
| CAD | macrophage | *PBXIP1* | 2.239191641 | 0.000725772 | 0.027706954 | 1 |
| CAD | macrophage | *SUCLG1* | 0.907394286 | 0.008252941 | 0.040820553 | 1 |
| CAD | macrophage | *AIM2* | 2.516463435 | 0.004546617 | 0.032925653 | 1 |
| CAD | macrophage | *GTPBP8* | 2.023711402 | 0.003878917 | 0.032549158 | 1 |
| CAD | macrophage | *C4orf36* | 1.411179125 | 0.003445039 | 0.032549158 | 1 |
| CAD | macrophage | *TIPARP* | 1.615971419 | 0.003325217 | 0.032549158 | 1 |
| CAD | macrophage | *RPP14* | 1.521371321 | 0.009285546 | 0.043219802 | 1 |
| CAD | macrophage | *CRELD1* | 1.249745901 | 0.007559659 | 0.039482739 | 1 |
| CAD | macrophage | *CPA3* | 1.58470718 | 0.00928266 | 0.043219802 | 1 |
| CAD | macrophage | *SLC4A1AP* | 1.817542672 | 0.007290106 | 0.038955499 | 1 |
| CAD | macrophage | *CLEC3B* | 2.358989274 | 0.002577042 | 0.032549158 | 1 |
| CAD | macrophage | *FYCO1* | 0.709484973 | 0.01193218 | 0.049567582 | 1 |
| CAD | macrophage | *SNIP1* | 2.035971036 | 0.003802375 | 0.032549158 | 1 |
| CAD | macrophage | *UVSSA* | 0.665604427 | 0.010909892 | 0.047086962 | 1 |
| CAD | macrophage | *FAM208A* | 1.387425115 | 0.00113596 | 0.031316348 | 1 |
| CAD | macrophage | *ABLIM2* | 0.857865749 | 0.002919188 | 0.032549158 | 1 |
| CAD | macrophage | *RNF123* | 0.922832665 | 4.49E-06 | 0.00272289 | 0.021783119 |
| CAD | macrophage | *HSPA4L* | 0.970863163 | 0.000598928 | 0.025946406 | 1 |
| CAD | macrophage | *MST1R* | 0.630576829 | 4.65E-05 | 0.008352699 | 0.225522876 |
| CAD | macrophage | *RAD54L2* | 1.007346083 | 0.00906198 | 0.042522947 | 1 |
| CAD | macrophage | *HMGB2* | 0.91404265 | 0.011421815 | 0.048527713 | 1 |
| CAD | macrophage | *MAP9* | 0.853284781 | 0.010573873 | 0.046178607 | 1 |
| CAD | macrophage | *CMBL* | 0.970242713 | 0.009426562 | 0.043541159 | 1 |
| CAD | macrophage | *C5orf63* | 0.902449282 | 0.002396182 | 0.032549158 | 1 |
| CAD | macrophage | *WDR41* | 1.032207373 | 0.005514346 | 0.035310234 | 1 |
| CAD | macrophage | *NDUFS4* | 1.255935829 | 0.002831649 | 0.032549158 | 1 |
| CAD | macrophage | *SERINC5* | 1.211067499 | 0.002265531 | 0.032549158 | 1 |
| CAD | macrophage | *UTP15* | 2.178459707 | 0.001796985 | 0.032549158 | 1 |
| CAD | macrophage | *PDSS2* | 1.190175102 | 0.006024998 | 0.036541612 | 1 |
| CAD | macrophage | *RPS14* | 1.184835034 | 0.002272924 | 0.032549158 | 1 |
| CAD | macrophage | *RP9* | 2.210280809 | 0.003172201 | 0.032549158 | 1 |
| CAD | macrophage | *USP49* | 1.959278235 | 0.000979405 | 0.029887243 | 1 |
| CAD | macrophage | *SLC13A4* | 1.080668862 | 0.004773272 | 0.033426994 | 1 |
| CAD | macrophage | *SOX17* | 0.688297538 | 0.005388625 | 0.035275662 | 1 |
| CAD | macrophage | *RAD21* | 1.309094708 | 0.002961172 | 0.032549158 | 1 |

| | | | | | | |
|---|---|---|---|---|---|---|
| CAD | macrophage | *TNFRSF11B* | 0.838724987 | 0.006377048 | 0.03707567 | 1 |
| CAD | macrophage | *OXR1* | 1.077193675 | 0.002266221 | 0.032549158 | 1 |
| CAD | macrophage | *GPER1* | 0.842619853 | 0.008539807 | 0.04146103 | 1 |
| CAD | macrophage | *FASTK* | 2.097546088 | 0.001473188 | 0.032549158 | 1 |
| CAD | macrophage | *ALDH7A1* | 0.936070008 | 0.003287829 | 0.032549158 | 1 |
| CAD | macrophage | *FZD6* | 0.532707594 | 0.003848265 | 0.032549158 | 1 |
| CAD | macrophage | *AQP7* | 0.30324296 | 0.007359041 | 0.039108508 | 1 |
| CAD | macrophage | *TRMT10B* | 1.280449231 | 0.00358468 | 0.032549158 | 1 |
| CAD | macrophage | *PIGO* | 1.997184098 | 0.000272523 | 0.019735559 | 1 |
| CAD | macrophage | *WRN* | 1.087106831 | 0.001099726 | 0.031043976 | 1 |
| CAD | macrophage | *MARCH8* | 1.481296489 | 0.007798618 | 0.039809763 | 1 |
| CAD | macrophage | *CFL2* | 1.471040169 | 0.007938413 | 0.040168337 | 1 |
| CAD | macrophage | *GJB2* | 11.54603423 | 0.002573673 | 0.032549158 | 1 |
| CAD | macrophage | *NOXRED1* | 0.855973248 | 0.007277287 | 0.038955499 | 1 |
| CAD | macrophage | *QSOX2* | 1.782151709 | 0.003701161 | 0.032549158 | 1 |
| CAD | macrophage | *METTL3* | 1.806739249 | 0.000784482 | 0.028118192 | 1 |
| CAD | macrophage | *HSPA12A* | 0.371978618 | 0.007979476 | 0.040266188 | 1 |
| CAD | macrophage | *UBTD1* | 0.992243271 | 0.005508647 | 0.035310234 | 1 |
| CAD | macrophage | *TTC7B* | 1.069296464 | 0.011318652 | 0.048300878 | 1 |
| CAD | macrophage | *SLC39A13* | 1.161557207 | 0.011584618 | 0.048784846 | 1 |
| CAD | macrophage | *IFI27L1* | 1.322708363 | 0.002231363 | 0.032549158 | 1 |
| CAD | macrophage | *TCP11L2* | 1.092347387 | 0.003627429 | 0.032549158 | 1 |
| CAD | macrophage | *JAM3* | 0.777957899 | 0.004652525 | 0.033148387 | 1 |
| CAD | macrophage | *TRMT61A* | 1.65120788 | 0.002750827 | 0.032549158 | 1 |
| CAD | macrophage | *API5* | 1.611213748 | 0.004174962 | 0.032707875 | 1 |
| CAD | macrophage | *COX11* | 1.023064651 | 0.00554165 | 0.035379061 | 1 |
| CAD | macrophage | *STXBP4* | 1.206096769 | 0.002064767 | 0.032549158 | 1 |
| CAD | macrophage | *WBP1L* | 1.200947047 | 0.005894986 | 0.036251552 | 1 |
| CAD | macrophage | *WDR88* | 1.30359964 | 0.004233576 | 0.032707875 | 1 |
| CAD | macrophage | *RPL27A* | 1.462083604 | 0.005808388 | 0.036134117 | 1 |
| CAD | macrophage | *C16orf46* | -1.364829669 | 0.002936801 | 0.032549158 | 1 |
| CAD | macrophage | *TMEM41B* | 1.552425131 | 0.002196568 | 0.032549158 | 1 |
| CAD | macrophage | *TMX3* | 1.402942375 | 0.001668661 | 0.032549158 | 1 |
| CAD | macrophage | *SLFN5* | 1.173379035 | 0.00170806 | 0.032549158 | 1 |
| CAD | macrophage | *SAAL1* | 2.301911352 | 0.007664055 | 0.039573543 | 1 |
| CAD | macrophage | *PIP4K2C* | 2.025982256 | 0.000400178 | 0.022843094 | 1 |
| CAD | macrophage | *MTMR10* | 0.966762719 | 0.00609223 | 0.036628873 | 1 |
| CAD | macrophage | *MS4A7* | -0.185128282 | 0.006484631 | 0.037411925 | 1 |
| CAD | macrophage | *MS4A14* | -0.176771032 | 0.000753775 | 0.027706954 | 1 |
| CAD | macrophage | *EPB42* | 0.995283177 | 0.003866547 | 0.032549158 | 1 |
| CAD | macrophage | *SMAD3* | 0.725753041 | 0.004678183 | 0.033233594 | 1 |
| CAD | macrophage | *CNPY4* | 0.827885336 | 0.006631371 | 0.037885063 | 1 |
| CAD | macrophage | *SP2* | 1.925378728 | 0.003006923 | 0.032549158 | 1 |
| CAD | macrophage | *FBXO22* | 1.297974616 | 0.004763621 | 0.033426994 | 1 |
| CAD | macrophage | *LOXHD1* | 0.630236619 | 0.008612161 | 0.041578314 | 1 |
| CAD | macrophage | *ZNF91* | 0.908527756 | 0.003228506 | 0.032549158 | 1 |
| CAD | macrophage | *ENGASE* | 0.546289767 | 0.003778424 | 0.032549158 | 1 |
| CAD | macrophage | *VKORC1* | 2.477039863 | 0.006509554 | 0.037466616 | 1 |
| CAD | macrophage | *SPATA33* | 1.413523221 | 0.007226853 | 0.038955499 | 1 |
| CAD | macrophage | *ZNF641* | 1.430046941 | 0.0098945 | 0.044575779 | 1 |

| | | | | | | |
|---|---|---|---|---|---|---|
| CAD | macrophage | *ZNF528* | 1.569278975 | 0.001569464 | 0.032549158 | 1 |
| CAD | macrophage | *TTYH1* | 1.287976117 | 0.01150689 | 0.048761073 | 1 |
| CAD | macrophage | *LENG8* | 1.163704117 | 0.002238957 | 0.032549158 | 1 |
| CAD | macrophage | *KIFC2* | 1.219992856 | 0.000942644 | 0.029734791 | 1 |
| CAD | macrophage | *CYB5D2* | 1.378566936 | 0.001339477 | 0.032549158 | 1 |
| CAD | macrophage | *NDUFV1* | 1.356541847 | 0.002991565 | 0.032549158 | 1 |
| CAD | macrophage | *PRDX2* | 1.499511428 | 0.003886366 | 0.032549158 | 1 |
| CAD | macrophage | *MIS12* | 1.450489357 | 0.003071524 | 0.032549158 | 1 |
| CAD | macrophage | *FAM83B* | 3.947354537 | 0.006880135 | 0.038407224 | 1 |
| CAD | macrophage | *PDHB* | 1.307458437 | 0.006253577 | 0.037040771 | 1 |
| CAD | macrophage | *PCMTD1* | 1.196763324 | 0.003304565 | 0.032549158 | 1 |
| CAD | macrophage | *SLC20A2* | 1.082263215 | 0.001948294 | 0.032549158 | 1 |
| CAD | macrophage | *SLC16A4* | 0.447361175 | 0.008828831 | 0.04203875 | 1 |
| CAD | macrophage | *DNAJC21* | 1.771041578 | 0.004264405 | 0.032707875 | 1 |
| CAD | macrophage | *SNTB2* | 1.184475007 | 0.00259825 | 0.032549158 | 1 |
| CAD | macrophage | *STX18* | 1.455545242 | 0.005180212 | 0.034616138 | 1 |
| CAD | macrophage | *ATOH8* | 0.874828933 | 0.000231478 | 0.01939755 | 1 |
| CAD | macrophage | *CEP120* | 1.431727135 | 0.008278529 | 0.040820553 | 1 |
| CAD | macrophage | *MFF* | 1.353127491 | 0.002785702 | 0.032549158 | 1 |
| CAD | macrophage | *LGALS9* | -0.25414235 | 0.004651393 | 0.033148387 | 1 |
| CAD | macrophage | *UBE2V2* | 1.481504696 | 0.001751203 | 0.032549158 | 1 |
| CAD | macrophage | *THBS3* | 1.400428964 | 0.000512166 | 0.023878663 | 1 |
| CAD | macrophage | *NPIPB3* | -0.807460011 | 0.011605145 | 0.048784846 | 1 |
| CAD | macrophage | *HINT1* | 1.532391445 | 0.005199001 | 0.034616138 | 1 |
| CAD | macrophage | *CLIC3* | 1.464268866 | 0.00896963 | 0.042381544 | 1 |
| CAD | macrophage | *DFFB* | 1.793128044 | 0.009182231 | 0.043004041 | 1 |
| CAD | macrophage | *NFU1* | 1.085915736 | 0.001463291 | 0.032549158 | 1 |
| CAD | macrophage | *UGP2* | 0.887195143 | 0.00072602 | 0.027706954 | 1 |
| CAD | macrophage | *CSGALNACT2* | 1.33015152 | 0.011634042 | 0.048797153 | 1 |
| CAD | macrophage | *AVEN* | 1.27118352 | 0.002729994 | 0.032549158 | 1 |
| CAD | macrophage | *TOR1AIP2* | 1.068984364 | 0.008970702 | 0.042381544 | 1 |
| CAD | macrophage | *MAP3K2* | 0.974104297 | 0.001606166 | 0.032549158 | 1 |
| CAD | macrophage | *MYO7B* | 0.63701218 | 0.001867618 | 0.032549158 | 1 |
| CAD | macrophage | *HNRNPA3* | 1.277398245 | 0.000413101 | 0.023038679 | 1 |
| CAD | macrophage | *CNGB3* | 0.769707829 | 0.001090391 | 0.031043976 | 1 |
| CAD | macrophage | *STX8* | 1.239139387 | 0.002714887 | 0.032549158 | 1 |
| CAD | macrophage | *NFRKB* | 1.694785699 | 0.003989078 | 0.032549158 | 1 |
| CAD | macrophage | *NFXL1* | 1.447690547 | 0.000790844 | 0.028118192 | 1 |
| CAD | macrophage | *ELOVL6* | 0.779914775 | 0.008515385 | 0.04146103 | 1 |
| CAD | macrophage | *TTLL6* | -1.164701606 | 0.010266707 | 0.045435278 | 1 |
| CAD | macrophage | *MSANTD4* | 2.203577923 | 0.004414655 | 0.032821343 | 1 |
| CAD | macrophage | *ZNF160* | 0.535575601 | 0.011331784 | 0.048314424 | 1 |
| CAD | macrophage | *ZNF415* | 0.828152752 | 0.004087545 | 0.032707875 | 1 |
| CAD | macrophage | *TSNARE1* | 0.874739018 | 0.003488409 | 0.032549158 | 1 |
| CAD | macrophage | *FEZ2* | 1.066633491 | 0.002067126 | 0.032549158 | 1 |
| CAD | macrophage | *MFN1* | 1.02409573 | 0.000447444 | 0.02312941 | 1 |
| CAD | macrophage | *PDE7B* | 1.084569877 | 0.000810137 | 0.028144167 | 1 |
| CAD | macrophage | *MCC* | 0.968293817 | 0.003532508 | 0.032549158 | 1 |
| CAD | macrophage | *PPID* | 1.801167364 | 0.004046736 | 0.032702724 | 1 |
| CAD | macrophage | *TBCA* | 1.258502623 | 0.005225764 | 0.034638531 | 1 |

| | | | | | | |
|---|---|---|---|---|---|---|
| CAD | macrophage | *RGS19* | -0.267252068 | 0.002555504 | 0.032549158 | 1 |
| CAD | macrophage | *CAMTA1* | 0.943214227 | 0.007281851 | 0.038955499 | 1 |
| CAD | macrophage | *NXNL1* | 0.23884883 | 0.004981448 | 0.033804179 | 1 |
| CAD | macrophage | *TRAPPC12* | 1.410738423 | 0.002342938 | 0.032549158 | 1 |
| CAD | macrophage | *C3AR1* | -0.224396262 | 0.004130146 | 0.032707875 | 1 |
| CAD | macrophage | *ATPAF2* | 0.931140007 | 0.007996494 | 0.040289709 | 1 |
| CAD | macrophage | *ZNF554* | 1.461228137 | 0.0016831 | 0.032549158 | 1 |
| CAD | macrophage | *ANKRD20A4* | 2.705903638 | 0.009628227 | 0.044002875 | 1 |
| CAD | macrophage | *NME6* | 0.969533828 | 0.000593609 | 0.025946406 | 1 |
| CAD | macrophage | *TEFM* | 1.720946818 | 6.78E-06 | 0.003288554 | 0.032885544 |
| CAD | macrophage | *MALT1* | 1.358711995 | 0.011759232 | 0.049101372 | 1 |
| CAD | macrophage | *PAIP1* | 1.725077687 | 0.00047044 | 0.02312941 | 1 |
| CAD | macrophage | *BSG* | 1.369566809 | 0.003330097 | 0.032549158 | 1 |
| CAD | macrophage | *HINFP* | 1.552461699 | 0.003663616 | 0.032549158 | 1 |
| CAD | macrophage | *SUCLG2* | 1.180269252 | 0.007808103 | 0.039809763 | 1 |
| CAD | macrophage | *FGGY* | 1.40147298 | 4.22E-05 | 0.008184153 | 0.204603837 |
| CAD | macrophage | *AFF1* | 1.051345018 | 0.00462423 | 0.033148387 | 1 |
| CAD | macrophage | *ZNF738* | 0.422816013 | 0.00537615 | 0.035275662 | 1 |
| CAD | macrophage | *CBWD1* | 2.245992908 | 0.002858778 | 0.032549158 | 1 |
| CAD | macrophage | *ZNF621* | 1.796077433 | 0.002570444 | 0.032549158 | 1 |
| CAD | macrophage | *GXYLT2* | 0.604407025 | 0.010039765 | 0.044885278 | 1 |
| CAD | macrophage | *MZT2A* | 0.587543452 | 0.0063805 | 0.03707567 | 1 |
| CAD | macrophage | *KCNK7* | 3.50568685 | 0.000689039 | 0.027629914 | 1 |
| CAD | macrophage | *SCAI* | 0.606467749 | 0.009237972 | 0.043219802 | 1 |
| CAD | macrophage | *UQCRH* | 1.210895488 | 0.005504751 | 0.035310234 | 1 |
| CAD | macrophage | *CD7* | 1.514944918 | 0.009275303 | 0.043219802 | 1 |
| CAD | macrophage | *XXYLT1* | 1.544590329 | 0.003600557 | 0.032549158 | 1 |
| CAD | macrophage | *UBXN2A* | 0.913619467 | 0.003583301 | 0.032549158 | 1 |
| CAD | macrophage | *LEMD3* | 1.539797798 | 0.003693175 | 0.032549158 | 1 |
| CAD | macrophage | *RALGAPA1* | 0.668333035 | 0.007670124 | 0.039573543 | 1 |
| CAD | macrophage | *SNAPC5* | 1.074441151 | 0.009930831 | 0.044697951 | 1 |
| CAD | macrophage | *ANKRD36C* | 1.268315288 | 0.002221162 | 0.032549158 | 1 |
| CAD | macrophage | *MSL2* | 1.434833416 | 0.009285248 | 0.043219802 | 1 |
| CAD | macrophage | *ANGEL2* | 1.547343023 | 0.002035927 | 0.032549158 | 1 |
| CAD | macrophage | *SH3PXD2B* | 0.673731187 | 0.007019371 | 0.038675956 | 1 |
| CAD | macrophage | *LARP7* | 1.364168561 | 0.007907596 | 0.040168337 | 1 |
| CAD | macrophage | *FGFBP3* | 2.818229255 | 0.004640159 | 0.033148387 | 1 |
| CAD | macrophage | *NLRP6* | -2.29177982 | 0.005610037 | 0.035488787 | 1 |
| CAD | macrophage | *SEZ6L2* | 0.490230833 | 0.00478083 | 0.033426994 | 1 |
| CAD | macrophage | *KCTD13* | 0.803724293 | 0.00226412 | 0.032549158 | 1 |
| CAD | macrophage | *DHX36* | 1.25129545 | 0.003328863 | 0.032549158 | 1 |
| CAD | macrophage | *TVP23C* | 1.956811573 | 0.007046485 | 0.038675956 | 1 |
| CAD | macrophage | *FAM182B* | 0.349927172 | 0.007281967 | 0.038955499 | 1 |
| CAD | macrophage | *ATG13* | 1.446765462 | 0.010592956 | 0.046220344 | 1 |
| CAD | macrophage | *CHRNA7* | 0.740735994 | 0.01019895 | 0.045238419 | 1 |
| CAD | macrophage | *CLTB* | 1.661822778 | 0.002722434 | 0.032549158 | 1 |
| CAD | macrophage | *TTLL11* | 1.294548556 | 0.00236668 | 0.032549158 | 1 |
| CAD | macrophage | *ZDHHC21* | 0.991239155 | 0.006410167 | 0.037203507 | 1 |
| CAD | macrophage | *SLC35G1* | 1.617551049 | 0.00313777 | 0.032549158 | 1 |
| CAD | macrophage | *TAC4* | 1.157796947 | 0.005998656 | 0.036447642 | 1 |

| | | | | | | |
|---|---|---|---|---|---|---|
| CAD | macrophage | *CRLF3* | 1.368007639 | 2.14E-06 | 0.002081011 | 0.010405057 |
| CAD | macrophage | *WDR25* | 1.463736463 | 0.002923439 | 0.032549158 | 1 |
| CAD | macrophage | *RNF152* | 0.575906468 | 0.006667968 | 0.037885063 | 1 |
| CAD | macrophage | *ANKLE2* | 1.419108357 | 0.004487165 | 0.032923259 | 1 |
| CAD | macrophage | *MAN1B1* | 1.954236549 | 0.008109852 | 0.04048251 | 1 |
| CAD | macrophage | *THAP5* | 1.890980457 | 0.002187355 | 0.032549158 | 1 |
| CAD | macrophage | *NAALADL2* | 0.6849304 | 0.00195098 | 0.032549158 | 1 |
| CAD | macrophage | *CD151* | 1.280898449 | 0.000389671 | 0.022779316 | 1 |
| CAD | macrophage | *PCDHB9* | 0.701950513 | 0.002226145 | 0.032549158 | 1 |
| CAD | macrophage | *RPS27* | 1.454083468 | 0.001487714 | 0.032549158 | 1 |
| CAD | macrophage | *ASB8* | 1.573779612 | 0.003203776 | 0.032549158 | 1 |
| CAD | macrophage | *IMPDH2* | 0.938878852 | 0.001527076 | 0.032549158 | 1 |
| CAD | macrophage | *SH2B1* | 1.088211369 | 0.00415363 | 0.032707875 | 1 |
| CAD | macrophage | *SH2D4B* | 3.208321751 | 0.009817558 | 0.044311435 | 1 |
| CAD | macrophage | *ERN1* | 1.497382393 | 0.002599454 | 0.032549158 | 1 |
| CAD | macrophage | *EFCAB13* | 0.59896635 | 0.009361885 | 0.043423289 | 1 |
| CAD | macrophage | *CTC1* | 0.979996297 | 0.009465133 | 0.043613318 | 1 |
| CAD | macrophage | *ZNF664* | 1.629871844 | 0.003967424 | 0.032549158 | 1 |
| CAD | macrophage | *HLA-DQB1* | -0.199180157 | 0.00493796 | 0.033650257 | 1 |
| CAD | macrophage | *PDXDC1* | 1.264895555 | 0.000679682 | 0.027629914 | 1 |
| CAD | macrophage | *SEPT1* | 1.006216684 | 0.010446016 | 0.045795852 | 1 |
| CAD | macrophage | *LYNX1* | -0.459881195 | 0.001464012 | 0.032549158 | 1 |
| CAD | macrophage | *MTURN* | 0.92297248 | 0.005195104 | 0.034616138 | 1 |
| CAD | macrophage | *CCDC66* | 0.989676099 | 0.007535284 | 0.039482739 | 1 |
| CAD | macrophage | *MCFD2* | 1.087207201 | 0.00297405 | 0.032549158 | 1 |
| CAD | macrophage | *DEFB119* | 8.930467684 | 0.000203164 | 0.01939755 | 0.985750932 |
| CAD | macrophage | *CAPS2* | 1.73239697 | 4.54E-05 | 0.008352699 | 0.220148569 |
| CAD | macrophage | *CMTR2* | 1.258862335 | 0.003982337 | 0.032549158 | 1 |
| CAD | macrophage | *MRPL14* | 1.281102199 | 0.001748165 | 0.032549158 | 1 |
| CAD | macrophage | *NQO1* | 0.601517995 | 0.009440499 | 0.043541159 | 1 |
| CAD | macrophage | *MUC16* | 0.695582544 | 0.008102231 | 0.04048251 | 1 |
| CAD | macrophage | *TMEM136* | 1.151692231 | 3.26E-05 | 0.007261836 | 0.158122459 |
| CAD | macrophage | *TMEM45A* | 0.661969339 | 0.007906311 | 0.040168337 | 1 |
| CAD | macrophage | *ZBTB20* | 0.722347239 | 0.003387534 | 0.032549158 | 1 |
| CAD | macrophage | *C3orf58* | 1.790296207 | 0.002384215 | 0.032549158 | 1 |
| CAD | macrophage | *RFX7* | 1.213776382 | 0.009753627 | 0.044146082 | 1 |
| CAD | macrophage | *ZNF329* | 1.412867463 | 0.003813312 | 0.032549158 | 1 |
| CAD | macrophage | *ERCC6L2* | 1.485759917 | 0.000832134 | 0.028433213 | 1 |
| CAD | macrophage | *MRPL41* | 2.088526459 | 0.010031963 | 0.044885278 | 1 |
| CAD | macrophage | *BACE2* | 0.37155118 | 0.003244567 | 0.032549158 | 1 |
| CAD | macrophage | *FIGN* | 0.542789275 | 0.008565075 | 0.041474793 | 1 |
| CAD | macrophage | *KBTBD3* | 1.589340883 | 0.002364296 | 0.032549158 | 1 |
| CAD | macrophage | *FES* | -0.674082664 | 0.001043886 | 0.030318387 | 1 |
| CAD | macrophage | *GLRX5* | 2.269160337 | 0.003193904 | 0.032549158 | 1 |
| CAD | macrophage | *SKA2* | 1.257012834 | 0.002660956 | 0.032549158 | 1 |
| CAD | macrophage | *TTC3* | 0.309518886 | 0.011264602 | 0.048154934 | 1 |
| CAD | macrophage | *PLCXD3* | 0.813105759 | 0.006175842 | 0.03694844 | 1 |
| CAD | macrophage | *ALG12* | 2.127633935 | 0.00350993 | 0.032549158 | 1 |
| CAD | macrophage | *ZNF721* | 1.14955135 | 0.00055757 | 0.025283455 | 1 |
| CAD | macrophage | *C12orf60* | 0.842717722 | 0.011518082 | 0.04876591 | 1 |

| | | | | | | |
|---|---|---|---|---|---|---|
| CAD | macrophage | *WBP2NL* | 1.171866841 | 0.007282332 | 0.038955499 | 1 |
| CAD | macrophage | *CSMD1* | 1.91759123 | 0.002361167 | 0.032549158 | 1 |
| CAD | macrophage | *C19orf71* | 1.718420618 | 0.001830929 | 0.032549158 | 1 |
| CAD | macrophage | *LRIT3* | 11.82024044 | 0.004436952 | 0.032821343 | 1 |
| CAD | macrophage | *SERHL2* | 1.43958925 | 0.004816862 | 0.033426994 | 1 |
| CAD | macrophage | *NDUFB1* | 1.105344708 | 0.006526139 | 0.037517564 | 1 |
| CAD | macrophage | *BTBD9* | 0.948980744 | 0.005207094 | 0.034616138 | 1 |
| CAD | macrophage | *TTC32* | 2.0123646 | 0.005553601 | 0.035379398 | 1 |
| CAD | macrophage | *PP2D1* | 2.517412011 | 0.003935723 | 0.032549158 | 1 |
| CAD | macrophage | *COPB2* | 1.432858501 | 0.003020389 | 0.032549158 | 1 |
| CAD | macrophage | *TCTE3* | 1.350371961 | 0.0039903 | 0.032549158 | 1 |
| CAD | macrophage | *UBE2G2* | 0.917630557 | 0.004799221 | 0.033426994 | 1 |
| CAD | macrophage | *BTBD6* | 0.577857545 | 0.011866878 | 0.049423254 | 1 |
| CAD | macrophage | *ZFP90* | 1.414951409 | 0.002916464 | 0.032549158 | 1 |
| CAD | macrophage | *IFITM2* | 1.1809568 | 0.005418649 | 0.035310234 | 1 |
| CAD | macrophage | *TARSL2* | 0.793078894 | 0.002792226 | 0.032549158 | 1 |
| CAD | macrophage | *AHNAK2* | 1.06577209 | 0.002770683 | 0.032549158 | 1 |
| CAD | macrophage | *SP1* | 1.776216018 | 0.000258632 | 0.01939755 | 1 |
| CAD | macrophage | *DMWD* | 0.910617452 | 0.007643189 | 0.039573543 | 1 |
| CAD | macrophage | *RTN4RL1* | 0.691499464 | 0.005820447 | 0.036134117 | 1 |
| CAD | macrophage | *RNPC3* | 0.640681758 | 0.010619457 | 0.046294343 | 1 |
| CAD | macrophage | *ZNF529* | 1.474736425 | 0.001737618 | 0.032549158 | 1 |
| CAD | macrophage | *C15orf41* | 0.924301317 | 0.006631257 | 0.037885063 | 1 |
| CAD | macrophage | *WWOX* | 0.83386936 | 0.002857897 | 0.032549158 | 1 |
| CAD | macrophage | *NBPF12* | 0.347656183 | 0.00518655 | 0.034616138 | 1 |
| CAD | macrophage | *TMEM222* | 1.918168901 | 0.006365572 | 0.03707567 | 1 |
| CAD | macrophage | *SEPT10* | 1.11829464 | 0.007260607 | 0.038955499 | 1 |
| CAD | macrophage | *MIR22HG* | 1.074420464 | 0.010505488 | 0.045962696 | 1 |
| CAD | macrophage | *VSIG10L* | -0.901789055 | 0.005991218 | 0.036447642 | 1 |
| CAD | macrophage | *WDR86* | 0.415960377 | 0.003980791 | 0.032549158 | 1 |
| CAD | macrophage | *GJA4* | 2.23305121 | 0.002966682 | 0.032549158 | 1 |
| CAD | macrophage | *SBK2* | 9.505306166 | 0.009532793 | 0.043883406 | 1 |
| CAD | macrophage | *EXD3* | 1.56585972 | 0.00278597 | 0.032549158 | 1 |
| CAD | macrophage | *C2orf88* | 0.771087338 | 0.009950328 | 0.0447442 | 1 |
| CAD | macrophage | *ZFP69* | 2.602156536 | 0.002521035 | 0.032549158 | 1 |
| CAD | macrophage | *MESP2* | 0.62325755 | 0.009802096 | 0.04428284 | 1 |
| CAD | macrophage | *AGRN* | 1.430373263 | 0.011085855 | 0.0476849 | 1 |
| CAD | macrophage | *ZNF626* | 0.960556379 | 0.010200089 | 0.045238419 | 1 |
| CAD | macrophage | *NCR3LG1* | 3.866705014 | 0.006647011 | 0.037885063 | 1 |
| CAD | macrophage | *PLSCR1* | 1.106651456 | 0.006229106 | 0.037040771 | 1 |
| CAD | macrophage | *COL25A1* | 0.752056667 | 0.008535811 | 0.04146103 | 1 |
| CAD | macrophage | *SRSF10* | 0.793512722 | 0.00301704 | 0.032549158 | 1 |
| CAD | macrophage | *RPL14* | 1.419499785 | 0.003473954 | 0.032549158 | 1 |
| CAD | macrophage | *FAM196A* | 0.975968551 | 0.003357349 | 0.032549158 | 1 |
| CAD | macrophage | *ADAT2* | 1.436740181 | 0.010433888 | 0.045795852 | 1 |
| CAD | macrophage | *SF3B3* | 1.327069322 | 0.006272135 | 0.037040771 | 1 |
| CAD | macrophage | *PLAC9* | 0.76982422 | 0.007469814 | 0.039480976 | 1 |
| CAD | macrophage | *HLA-DRB1* | -0.219785703 | 0.004500166 | 0.032923259 | 1 |
| CAD | macrophage | *AKR1C3* | 0.744971603 | 0.011613046 | 0.048784846 | 1 |
| CAD | macrophage | *WDSUB1* | 1.010552345 | 0.005259129 | 0.034764704 | 1 |

| | | | | | | |
|---|---|---|---|---|---|---|
| CAD | macrophage | *MPHOSPH8* | 0.974267319 | 0.001390948 | 0.032549158 | 1 |
| CAD | macrophage | *PPIA* | 0.921144979 | 0.010281932 | 0.045435278 | 1 |
| CAD | macrophage | *SUPT3H* | 1.030114184 | 0.007433781 | 0.039333376 | 1 |
| CAD | macrophage | *ZKSCAN7* | 0.977104703 | 0.010002618 | 0.044854623 | 1 |
| CAD | macrophage | *PIK3R4* | 1.66465135 | 0.002883875 | 0.032549158 | 1 |
| CAD | macrophage | *ZNF605* | 0.788293321 | 0.000479286 | 0.02312941 | 1 |
| CAD | macrophage | *CACNA1H* | 1.558703457 | 0.00725938 | 0.038955499 | 1 |
| CAD | macrophage | *ADH1B* | 0.513748632 | 0.006839728 | 0.038393741 | 1 |
| CAD | macrophage | *ZFP62* | 1.488640081 | 0.001041464 | 0.030318387 | 1 |
| CAD | macrophage | *TOMM7* | 1.410408596 | 0.002746077 | 0.032549158 | 1 |
| CAD | macrophage | *ANKRD36B* | 0.756023093 | 0.006438138 | 0.037276664 | 1 |
| CAD | macrophage | *PCDHB16* | 0.362957999 | 0.005453908 | 0.035310234 | 1 |
| CAD | macrophage | *WDR5B* | 1.156856629 | 0.003126564 | 0.032549158 | 1 |
| CAD | macrophage | *ZNF138* | 1.08695783 | 0.000943767 | 0.029734791 | 1 |
| CAD | macrophage | *ZNF429* | 0.95219415 | 0.01008087 | 0.044997592 | 1 |
| CAD | macrophage | *ZNF470* | 1.276884407 | 0.000440753 | 0.02312941 | 1 |
| CAD | macrophage | *GMFB* | 1.876282135 | 0.004894793 | 0.033639572 | 1 |
| CAD | macrophage | *ZMYM1* | 1.590185301 | 0.003316255 | 0.032549158 | 1 |
| CAD | macrophage | *ADAM32* | 0.989463906 | 0.011076924 | 0.0476849 | 1 |
| CAD | macrophage | *ABCB8* | 1.300663703 | 0.008242799 | 0.040820553 | 1 |
| CAD | macrophage | *SULT1A2* | 0.710834904 | 0.011485039 | 0.048711023 | 1 |
| CAD | macrophage | *RAMP2-AS1* | 0.698410567 | 0.006359139 | 0.03707567 | 1 |
| CAD | macrophage | *ZNF720* | 1.336886214 | 0.001343901 | 0.032549158 | 1 |
| CAD | macrophage | *MAP3K5* | 1.065279353 | 0.003527551 | 0.032549158 | 1 |
| CAD | macrophage | *SLC2A10* | 0.942222336 | 0.00208888 | 0.032549158 | 1 |
| CAD | macrophage | *ZNF665* | 1.165054089 | 0.004276692 | 0.032707875 | 1 |
| CAD | macrophage | *RAB40C* | 1.419198397 | 0.003621752 | 0.032549158 | 1 |
| CAD | macrophage | *TOPORS* | 1.76396211 | 0.00732838 | 0.039031065 | 1 |
| CAD | macrophage | *FAM114A1* | 1.0331258 | 0.002259166 | 0.032549158 | 1 |
| CAD | macrophage | *TXNRD3* | 0.936993171 | 0.002134049 | 0.032549158 | 1 |
| CAD | macrophage | *AC122129.1* | 2.392076593 | 0.005123373 | 0.034477958 | 1 |
| CAD | macrophage | *ADH5* | 0.96916367 | 0.002721995 | 0.032549158 | 1 |
| CAD | macrophage | *ZNF677* | 1.488325345 | 0.001648172 | 0.032549158 | 1 |
| CAD | macrophage | *ZNF71* | 0.877900311 | 0.000346478 | 0.021635859 | 1 |
| CAD | macrophage | *MPZL1* | 0.957924288 | 0.004261111 | 0.032707875 | 1 |
| CAD | macrophage | *VPS13A* | 0.880708608 | 0.004780417 | 0.033426994 | 1 |
| CAD | macrophage | *NOL8* | 1.419720809 | 0.00487178 | 0.033586837 | 1 |
| CAD | macrophage | *MRPL42* | 0.728736813 | 0.000363621 | 0.021917844 | 1 |
| CAD | macrophage | *MAK16* | 1.89015001 | 0.001664101 | 0.032549158 | 1 |
| CAD | macrophage | *SFI1* | 1.200804895 | 0.011045695 | 0.047596545 | 1 |
| CAD | macrophage | *HIBCH* | 0.622857088 | 0.000878543 | 0.028834548 | 1 |
| CAD | macrophage | *ZNF544* | 1.78539269 | 0.002183063 | 0.032549158 | 1 |
| CAD | macrophage | *MIER1* | 1.444540563 | 0.000333504 | 0.021635859 | 1 |
| CAD | macrophage | *MAN1A2* | 1.271727712 | 0.003313232 | 0.032549158 | 1 |
| CAD | macrophage | *SVIP* | 0.93765658 | 0.004817394 | 0.033426994 | 1 |
| CAD | macrophage | *DDRGK1* | 1.898247744 | 0.000619349 | 0.026360348 | 1 |
| CAD | macrophage | *RPS6KL1* | 1.382174348 | 0.000613929 | 0.026360348 | 1 |
| CAD | macrophage | *STYX* | 1.650282168 | 0.007234091 | 0.038955499 | 1 |
| CAD | macrophage | *HELZ* | 1.096721411 | 0.004793201 | 0.033426994 | 1 |
| CAD | macrophage | *TMEM116* | -0.333777015 | 0.001165837 | 0.031778892 | 1 |

| | | | | | | |
|---|---|---|---|---|---|---|
| CAD | macrophage | *GFPT1* | 0.907208843 | 0.004863969 | 0.033586837 | 1 |
| CAD | macrophage | *TMA16* | 1.36427005 | 0.003001627 | 0.032549158 | 1 |
| CAD | macrophage | *HLA-DRB5* | -0.107415384 | 0.002690643 | 0.032549158 | 1 |
| CAD | macrophage | *TTC37* | 0.935380702 | 0.007394554 | 0.039211338 | 1 |
| CAD | macrophage | *TUSC1* | 3.300523712 | 0.003401866 | 0.032549158 | 1 |
| CAD | macrophage | *RPL10A* | 1.359633999 | 0.00294019 | 0.032549158 | 1 |
| CAD | macrophage | *HMGN2* | 1.131939227 | 0.00025836 | 0.01939755 | 1 |
| CAD | macrophage | *TYW1* | 0.824628255 | 0.008825609 | 0.04203875 | 1 |
| CAD | macrophage | *ECT2L* | 1.245011443 | 0.008469074 | 0.04146103 | 1 |
| CAD | macrophage | *PCMTD2* | 1.108492851 | 0.00356394 | 0.032549158 | 1 |
| CAD | macrophage | *RUFY2* | 0.977316238 | 0.003782334 | 0.032549158 | 1 |
| CAD | macrophage | *ASAH2B* | 0.791645924 | 0.004422163 | 0.032821343 | 1 |
| CAD | macrophage | *AGAP6* | 0.582446525 | 0.007287496 | 0.038955499 | 1 |
| CAD | macrophage | *LINC00619* | 4.094366612 | 0.006888366 | 0.038407224 | 1 |
| CAD | macrophage | *BMPR2* | 0.973490773 | 0.00407127 | 0.032707875 | 1 |
| CAD | macrophage | *TCEA3* | 0.892929831 | 0.000264555 | 0.019448798 | 1 |
| CAD | macrophage | *PRRT1* | 0.338862192 | 0.003246152 | 0.032549158 | 1 |
| CAD | macrophage | *BAG6* | 0.93289324 | 0.001400662 | 0.032549158 | 1 |
| CAD | macrophage | *ZNF814* | 0.87239731 | 0.005925584 | 0.036271366 | 1 |
| CAD | macrophage | *MALRD1* | 0.817620608 | 0.007253394 | 0.038955499 | 1 |
| CAD | macrophage | *DCTN1* | 2.055762697 | 0.003391285 | 0.032549158 | 1 |
| CAD | macrophage | *FAM216A* | 1.672865987 | 0.000385351 | 0.022779316 | 1 |
| CAD | macrophage | *SCGB2B2* | 1.121823042 | 0.002633904 | 0.032549158 | 1 |
| CAD | macrophage | *IPO7* | 1.382220829 | 0.003908694 | 0.032549158 | 1 |
| CAD | macrophage | *NAP1L4* | 0.95333859 | 0.006934508 | 0.038452839 | 1 |
| CAD | macrophage | *LIN52* | 0.909506286 | 0.000514681 | 0.023878663 | 1 |
| CAD | macrophage | *HCG27* | 0.414618544 | 0.001497248 | 0.032549158 | 1 |
| CAD | macrophage | *COL6A6* | 0.648492859 | 0.002360999 | 0.032549158 | 1 |
| CAD | macrophage | *METTL6* | 1.19245498 | 0.0012235 | 0.032263164 | 1 |
| CAD | macrophage | *SLC48A1* | 1.351067955 | 0.006799335 | 0.038316343 | 1 |
| CAD | macrophage | *TAS2R50* | 9.772565249 | 0.000138617 | 0.017699256 | 0.672571725 |
| CAD | macrophage | *TAS2R14* | 0.352881336 | 0.008500556 | 0.04146103 | 1 |
| CAD | macrophage | *C17orf51* | 1.623638767 | 0.010123829 | 0.045106355 | 1 |
| CAD | macrophage | *LCAT* | 1.111684915 | 0.010714429 | 0.046549905 | 1 |
| CAD | macrophage | *SRA1* | 1.081359396 | 0.006683063 | 0.037885063 | 1 |
| CAD | macrophage | *PPP1CB* | 0.999675903 | 0.00148211 | 0.032549158 | 1 |
| CAD | macrophage | *ATF6B* | 0.978403346 | 0.000812074 | 0.028144167 | 1 |
| CAD | macrophage | *IRF9* | 1.571291203 | 0.011399565 | 0.048518147 | 1 |
| CAD | macrophage | *GALT* | 1.456130579 | 0.00047767 | 0.02312941 | 1 |
| CAD | macrophage | *REPIN1* | 2.657043744 | 0.006061712 | 0.036581378 | 1 |
| CAD | macrophage | *ZNF891* | 1.795554494 | 0.000685184 | 0.027629914 | 1 |
| CAD | macrophage | *SMIM7* | 1.327213543 | 0.005837884 | 0.036134117 | 1 |
| CAD | macrophage | *PAXIP1-AS2* | 1.823828637 | 0.002734582 | 0.032549158 | 1 |
| CAD | macrophage | *LYRM4* | 1.48790389 | 0.003636798 | 0.032549158 | 1 |
| CAD | macrophage | *PRCD* | 0.870716111 | 0.001908342 | 0.032549158 | 1 |
| CAD | macrophage | *SH3D21* | 0.901943256 | 0.001650969 | 0.032549158 | 1 |
| CAD | macrophage | *ANG* | 1.187832449 | 0.007641289 | 0.039573543 | 1 |
| CAD | macrophage | *EML6* | 1.584781486 | 0.003844263 | 0.032549158 | 1 |
| CAD | macrophage | *VPS16* | 2.055237943 | 0.005908272 | 0.036271366 | 1 |
| CAD | macrophage | *MYL5* | 1.189048047 | 0.000976323 | 0.029887243 | 1 |

| | | | | | | |
|---|---|---|---|---|---|---|
| CAD | macrophage | *NPEPL1* | 1.991315766 | 0.010292886 | 0.045442296 | 1 |
| CAD | macrophage | *PAM16* | 0.903015455 | 0.00879795 | 0.04203875 | 1 |
| CAD | macrophage | *APOL6* | 1.114879091 | 0.003442483 | 0.032549158 | 1 |
| CAD | macrophage | *AC062017.1* | 2.097448313 | 0.00316583 | 0.032549158 | 1 |
| CAD | macrophage | *LINC01117* | 0.90026125 | 0.008840372 | 0.042052437 | 1 |
| CAD | macrophage | *ERCC6* | 1.115891167 | 0.00500205 | 0.033849299 | 1 |
| CAD | macrophage | *LINC00115* | 1.492263087 | 0.005297302 | 0.034969404 | 1 |
| CAD | macrophage | *ZNF717* | 0.228220396 | 0.004505574 | 0.032923259 | 1 |
| CAD | macrophage | *AC009299.3* | 1.987801024 | 0.001371717 | 0.032549158 | 1 |
| CAD | macrophage | *SLC2A1-AS1* | 0.851487703 | 0.007233614 | 0.038955499 | 1 |
| CAD | macrophage | *RPS18* | 1.358521107 | 0.000253139 | 0.01939755 | 1 |
| CAD | macrophage | *HLA-DQB2* | 0.118354499 | 0.009433639 | 0.043541159 | 1 |
| CAD | macrophage | *LINC00959* | 1.494796755 | 0.009872554 | 0.044518245 | 1 |
| CAD | macrophage | *LINC00707* | 1.86668328 | 0.002263554 | 0.032549158 | 1 |
| CAD | macrophage | *LY6G5B* | -0.228327916 | 0.010326707 | 0.045508793 | 1 |
| CAD | macrophage | *NSUN6* | 1.344621667 | 0.000180599 | 0.01939755 | 0.876267058 |
| CAD | macrophage | *ARFGAP3* | 1.625136851 | 0.002979979 | 0.032549158 | 1 |
| CAD | macrophage | *MRPL20* | 1.615946009 | 0.000776763 | 0.028118192 | 1 |
| CAD | macrophage | *LINC00960* | -0.190208722 | 0.001779727 | 0.032549158 | 1 |
| CAD | macrophage | *DECR2* | 2.991177979 | 0.003226234 | 0.032549158 | 1 |
| CAD | macrophage | *MRPL33* | 0.963720893 | 0.003857599 | 0.032549158 | 1 |
| CAD | macrophage | *C7orf73* | 0.986559815 | 0.007194096 | 0.038955499 | 1 |
| CAD | macrophage | *NBPF15* | 1.656049875 | 0.003242968 | 0.032549158 | 1 |
| CAD | macrophage | *WFDC6* | -1.621596135 | 0.001279279 | 0.032549158 | 1 |
| CAD | macrophage | *UPK3B* | 0.99372102 | 0.0062754 | 0.037040771 | 1 |
| CAD | macrophage | *APOBEC3D* | 7.261933326 | 0.007983527 | 0.040266188 | 1 |
| CAD | macrophage | *NEAT1* | 0.803410981 | 0.005823761 | 0.036134117 | 1 |
| CAD | macrophage | *RAB30-AS1* | 0.747610739 | 0.005588074 | 0.035445077 | 1 |
| CAD | macrophage | *UBAP1L* | 1.312753109 | 0.004923045 | 0.033650257 | 1 |
| CAD | macrophage | *CDK11B* | 1.938154283 | 0.003920059 | 0.032549158 | 1 |
| CAD | macrophage | *ABHD14A* | 1.299546698 | 0.002320558 | 0.032549158 | 1 |
| CAD | macrophage | *C5orf17* | 1.686286116 | 0.003322904 | 0.032549158 | 1 |
| CAD | macrophage | *TMEM150C* | 0.465017817 | 0.003587767 | 0.032549158 | 1 |
| CAD | macrophage | *PDCD6* | 1.684271294 | 0.005862238 | 0.036176476 | 1 |
| CAD | macrophage | *PCDHA10* | 0.463254174 | 0.004609842 | 0.033148387 | 1 |
| CAD | macrophage | *ZNF550* | 1.771130549 | 0.004164218 | 0.032707875 | 1 |
| CAD | macrophage | *TMEM200B* | 0.720915242 | 0.002265698 | 0.032549158 | 1 |
| CAD | macrophage | *SNHG9* | 0.857992281 | 0.006329297 | 0.037064809 | 1 |
| CAD | macrophage | *TAS2R20* | 0.687835358 | 0.001555729 | 0.032549158 | 1 |
| CAD | macrophage | *HMBS* | 1.354206623 | 1.22E-05 | 0.004557288 | 0.05924474 |
| CAD | macrophage | *CNPY2* | 1.349762169 | 0.003774234 | 0.032549158 | 1 |
| CAD | macrophage | *SLC25A21-AS1* | 0.731426174 | 0.002680044 | 0.032549158 | 1 |
| CAD | macrophage | *MRPL46* | 1.548567744 | 0.002122143 | 0.032549158 | 1 |
| CAD | macrophage | *IQCH-AS1* | 0.95875574 | 0.000763093 | 0.027838563 | 1 |
| CAD | macrophage | *PWRN1* | 0.722765468 | 0.011686782 | 0.048925163 | 1 |
| CAD | macrophage | *MYO15B* | 0.873760644 | 0.007516665 | 0.039482739 | 1 |
| CAD | macrophage | *ILF3-AS1* | 0.78190517 | 0.007220944 | 0.038955499 | 1 |
| CAD | macrophage | *ZNF224* | 1.701861463 | 0.003324136 | 0.032549158 | 1 |
| CAD | macrophage | *TRABD2B* | 0.906543939 | 0.006148018 | 0.036870949 | 1 |
| CAD | macrophage | *AC093157.1* | 1.401461668 | 0.004879456 | 0.033586837 | 1 |

| | | | | | | |
|---|---|---|---|---|---|---|
| CAD | macrophage | *ZNF587B* | 2.804242271 | 0.002899834 | 0.032549158 | 1 |
| CAD | macrophage | *CASC15* | 0.770884049 | 0.006683762 | 0.037885063 | 1 |
| CAD | macrophage | *GRIN2B* | 0.477838156 | 0.000240368 | 0.01939755 | 1 |
| CAD | macrophage | *PAXIP1-AS1* | 1.762207469 | 0.001194253 | 0.032191757 | 1 |
| CAD | mesothelium | *NFYA* | -0.712292612 | 0.000825363 | 0.040451121 | 1 |
| CAD | mesothelium | *SEMA3F* | -0.330547109 | 0.000414274 | 0.030000834 | 1 |
| CAD | mesothelium | *RBM6* | -0.125597869 | 0.000158101 | 0.021917368 | 0.767107867 |
| CAD | mesothelium | *GGNBP2* | -0.859755825 | 0.000148572 | 0.021917368 | 0.720869515 |
| CAD | mesothelium | *PSMA4* | -1.064086775 | 0.00115266 | 0.045304084 | 1 |
| CAD | mesothelium | *DTNBP1* | -0.850609015 | 0.000677634 | 0.037362278 | 1 |
| CAD | mesothelium | *SNX29* | -0.53992012 | 0.001151557 | 0.045304084 | 1 |
| CAD | mesothelium | *UHRF1BP1* | 0.20384266 | 0.000180785 | 0.023000068 | 0.87716964 |
| CAD | mesothelium | *TIE1* | -0.310057558 | 2.97E-05 | 0.009017009 | 0.144272141 |
| CAD | mesothelium | *TRHDE* | -0.365478844 | 0.000269212 | 0.025110267 | 1 |
| CAD | mesothelium | *NDE1* | -0.650394734 | 0.000390794 | 0.029087091 | 1 |
| CAD | mesothelium | *AP1M1* | -1.033354559 | 0.000540889 | 0.03383175 | 1 |
| CAD | mesothelium | *TIPIN* | 0.11911217 | 0.001266157 | 0.046829884 | 1 |
| CAD | mesothelium | *FMO4* | 0.481537399 | 0.000446432 | 0.031854216 | 1 |
| CAD | mesothelium | *TXNDC16* | -0.355399002 | 8.58E-05 | 0.015911687 | 0.416376289 |
| CAD | mesothelium | *PABPC4* | -0.331547117 | 0.000132072 | 0.021360453 | 0.640813589 |
| CAD | mesothelium | *TOX4* | -0.738383607 | 0.000869201 | 0.041346686 | 1 |
| CAD | mesothelium | *ITPR3* | 0.219740254 | 0.000540359 | 0.03383175 | 1 |
| CAD | mesothelium | *SCD* | -0.278801815 | 3.40E-05 | 0.009693149 | 0.164783526 |
| CAD | mesothelium | *HPS4* | -0.825221016 | 0.0009008 | 0.041835582 | 1 |
| CAD | mesothelium | *PPP6R2* | -0.787009116 | 0.001253984 | 0.046802531 | 1 |
| CAD | mesothelium | *THOC5* | -0.19973046 | 0.001316668 | 0.047322032 | 1 |
| CAD | mesothelium | *ACO2* | -0.537247679 | 0.001150252 | 0.045304084 | 1 |
| CAD | mesothelium | *DNTTIP1* | -0.190841912 | 0.000495468 | 0.033783958 | 1 |
| CAD | mesothelium | *UBE2I* | -1.018166028 | 0.000557316 | 0.033984771 | 1 |
| CAD | mesothelium | *NFKBIB* | -0.324214091 | 0.00116715 | 0.045304084 | 1 |
| CAD | mesothelium | *PTN* | -0.870171125 | 0.000248774 | 0.02463368 | 1 |
| CAD | mesothelium | *OCIAD1* | -0.808711337 | 0.000982406 | 0.042942626 | 1 |
| CAD | mesothelium | *NDUFC1* | -1.092423861 | 0.00026538 | 0.025110267 | 1 |
| CAD | mesothelium | *GRPEL1* | -1.449077562 | 0.001343027 | 0.047564707 | 1 |
| CAD | mesothelium | *ST3GAL4* | -0.294076543 | 0.000560342 | 0.033984771 | 1 |
| CAD | mesothelium | *KCTD10* | -0.433951247 | 0.000150343 | 0.021917368 | 0.729463326 |
| CAD | mesothelium | *VPS29* | -0.925473016 | 0.000842092 | 0.040677866 | 1 |
| CAD | mesothelium | *ACAD10* | -0.455575618 | 0.001376187 | 0.047694723 | 1 |
| CAD | mesothelium | *NUP107* | -0.697784309 | 0.001021158 | 0.043846554 | 1 |
| CAD | mesothelium | *KRR1* | -0.704593946 | 0.000915843 | 0.041921427 | 1 |
| CAD | mesothelium | *IMPG1* | -0.760066939 | 0.000952826 | 0.04254006 | 1 |
| CAD | mesothelium | *MRPL19* | -0.801877952 | 0.001054479 | 0.044880105 | 1 |
| CAD | mesothelium | *HSDL2* | -0.16400735 | 0.001105828 | 0.045304084 | 1 |
| CAD | mesothelium | *ALG5* | -0.395756097 | 0.000997777 | 0.043225112 | 1 |
| CAD | mesothelium | *FABP3* | -1.271695895 | 8.85E-05 | 0.015911687 | 0.429615553 |
| CAD | mesothelium | *FAM35A* | -0.538994059 | 8.09E-05 | 0.015700909 | 0.392522725 |
| CAD | mesothelium | *TTC21B* | -0.6589166 | 0.001293323 | 0.046829884 | 1 |
| CAD | mesothelium | *WFDC3* | -0.055656191 | 0.001354789 | 0.047633605 | 1 |
| CAD | mesothelium | *PTGIS* | 0.191866808 | 0.000964428 | 0.04254006 | 1 |
| CAD | mesothelium | *SNRPC* | -0.40048539 | 5.51E-07 | 0.000617292 | 0.00267372 |

| | | | | | | |
|---|---|---|---|---|---|---|
| CAD | mesothelium | *TCP11* | 0.20183012 | 0.00029245 | 0.025338731 | 1 |
| CAD | mesothelium | *GRK4* | 0.070303592 | 0.000186652 | 0.023000068 | 0.905634021 |
| CAD | mesothelium | *ABHD8* | -0.264475482 | 0.000284638 | 0.025110267 | 1 |
| CAD | mesothelium | *EIF2AK4* | -0.654714224 | 3.93E-05 | 0.010395241 | 0.190899734 |
| CAD | mesothelium | *MAU2* | -0.970030785 | 1.34E-05 | 0.004987712 | 0.064840257 |
| CAD | mesothelium | *OSER1* | -0.642519825 | 7.41E-05 | 0.014983198 | 0.359596763 |
| CAD | mesothelium | *YWHAQ* | -0.822941229 | 0.00141801 | 0.048457599 | 1 |
| CAD | mesothelium | *FADS2* | -0.922711375 | 0.000543874 | 0.03383175 | 1 |
| CAD | mesothelium | *CARS2* | -0.339957529 | 0.000729925 | 0.038081682 | 1 |
| CAD | mesothelium | *WDR36* | -1.193164359 | 0.000166012 | 0.022374674 | 0.80548828 |
| CAD | mesothelium | *ODF2* | -1.021270229 | 0.001159411 | 0.045304084 | 1 |
| CAD | mesothelium | *SLC37A4* | 0.229805673 | 0.000697527 | 0.037800774 | 1 |
| CAD | mesothelium | *THUMPD2* | -0.77629821 | 0.000807016 | 0.040226772 | 1 |
| CAD | mesothelium | *ASCC1* | -0.329242452 | 5.84E-05 | 0.01348659 | 0.283218399 |
| CAD | mesothelium | *SENP7* | -0.628489609 | 0.000701169 | 0.037800774 | 1 |
| CAD | mesothelium | *MTMR6* | -0.97826887 | 0.000148417 | 0.021917368 | 0.720117946 |
| CAD | mesothelium | *SERF2* | -0.889438849 | 0.001185419 | 0.045648041 | 1 |
| CAD | mesothelium | *PCSK6* | -0.821369292 | 0.000493785 | 0.033783958 | 1 |
| CAD | mesothelium | *GFOD2* | -0.268423198 | 0.000727948 | 0.038081682 | 1 |
| CAD | mesothelium | *NPC1* | -0.72391885 | 0.000587118 | 0.034340737 | 1 |
| CAD | mesothelium | *TMEM91* | 0.12468501 | 0.000846757 | 0.040677866 | 1 |
| CAD | mesothelium | *NME7* | -0.568380537 | 6.36E-07 | 0.000617292 | 0.003086459 |
| CAD | mesothelium | *ARNT* | -0.682915057 | 8.64E-09 | 1.40E-05 | 4.19E-05 |
| CAD | mesothelium | *NBEAL1* | -0.904946702 | 1.65E-09 | 8.00E-06 | 8.00E-06 |
| CAD | mesothelium | *AMT* | -0.089795794 | 0.000263866 | 0.025110267 | 1 |
| CAD | mesothelium | *FBXO10* | 0.229685004 | 0.00035862 | 0.028064873 | 1 |
| CAD | mesothelium | *PAOX* | -1.642732766 | 0.001139448 | 0.045304084 | 1 |
| CAD | mesothelium | *HSD17B12* | -0.100329819 | 0.000154771 | 0.021917368 | 0.750948936 |
| CAD | mesothelium | *NPAT* | -0.992683048 | 0.001287493 | 0.046829884 | 1 |
| CAD | mesothelium | *CPSF7* | -0.653586995 | 0.000744458 | 0.038426703 | 1 |
| CAD | mesothelium | *GXYLT1* | -0.783834741 | 0.000176186 | 0.023000068 | 0.854853382 |
| CAD | mesothelium | *UPF2* | -0.758142866 | 0.001334035 | 0.047564707 | 1 |
| CAD | mesothelium | *ZNF827* | -0.6487742 | 7.23E-05 | 0.014983198 | 0.350713279 |
| CAD | mesothelium | *SREK1IP1* | -1.073361184 | 0.000587445 | 0.034340737 | 1 |
| CAD | mesothelium | *MIA3* | -1.154603925 | 4.87E-09 | 1.18E-05 | 2.36E-05 |
| CAD | mesothelium | *MMP16* | -0.105802127 | 0.001134585 | 0.045304084 | 1 |
| CAD | mesothelium | *NDUFAF6* | -0.538882958 | 5.54E-05 | 0.01344654 | 0.268930793 |
| CAD | mesothelium | *USP24* | -0.673841352 | 0.001083868 | 0.045304084 | 1 |
| CAD | mesothelium | *ARPC5* | -0.497999003 | 0.000714647 | 0.038081682 | 1 |
| CAD | mesothelium | *FLVCR1* | -0.716924958 | 0.000575376 | 0.034340737 | 1 |
| CAD | mesothelium | *ACP6* | -1.132958016 | 0.001253277 | 0.046802531 | 1 |
| CAD | mesothelium | *DCAF16* | -0.86854359 | 7.15E-05 | 0.014983198 | 0.346783704 |
| CAD | mesothelium | *ARPC2* | -0.710308607 | 0.000955978 | 0.04254006 | 1 |
| CAD | mesothelium | *MEAF6* | -0.843345366 | 0.000811955 | 0.040226772 | 1 |
| CAD | mesothelium | *RNF123* | 0.208299267 | 0.000239379 | 0.02463368 | 1 |
| CAD | mesothelium | *MST1R* | 0.148770845 | 0.000905345 | 0.041835582 | 1 |
| CAD | mesothelium | *INTS1* | -1.215175013 | 0.000208887 | 0.023570229 | 1 |
| CAD | mesothelium | *ATP5C1* | -0.725335922 | 0.001224134 | 0.046402337 | 1 |
| CAD | mesothelium | *VDAC2* | -1.314425808 | 1.25E-06 | 0.001009333 | 0.006055999 |
| CAD | mesothelium | *TRUB1* | -0.480463311 | 0.000661445 | 0.037317827 | 1 |

| | | | | | | |
|---|---|---|---|---|---|---|
| CAD | mesothelium | *CEP57* | -0.562760661 | 0.00024612 | 0.02463368 | 1 |
| CAD | mesothelium | *CCDC68* | -0.216010801 | 0.000936391 | 0.042461409 | 1 |
| CAD | mesothelium | *TMEM170A* | -0.520363374 | 0.000387503 | 0.029087091 | 1 |
| CAD | mesothelium | *MS4A7* | -0.142677185 | 0.000245769 | 0.02463368 | 1 |
| CAD | mesothelium | *MS4A14* | -0.068370106 | 0.000674498 | 0.037362278 | 1 |
| CAD | mesothelium | *TBC1D2B* | -0.355719616 | 0.001074263 | 0.045304084 | 1 |
| CAD | mesothelium | *ULK4* | -0.351930872 | 2.88E-05 | 0.009017009 | 0.139513006 |
| CAD | mesothelium | *PXK* | -0.615030587 | 0.000509635 | 0.03383175 | 1 |
| CAD | mesothelium | *ZEB2* | -0.406884825 | 5.22E-06 | 0.002109189 | 0.025310265 |
| CAD | mesothelium | *MGMT* | -0.68843055 | 0.000801696 | 0.040226772 | 1 |
| CAD | mesothelium | *DNAJC18* | -0.235491445 | 0.00034262 | 0.028064873 | 1 |
| CAD | mesothelium | *SCAND1* | -0.525743285 | 0.0003921 | 0.029087091 | 1 |
| CAD | mesothelium | *C3AR1* | -0.14885529 | 0.000198535 | 0.023000068 | 0.963290171 |
| CAD | mesothelium | *C11orf80* | -0.428388978 | 4.41E-06 | 0.00195644 | 0.021396401 |
| CAD | mesothelium | *CCNE2* | -0.228558839 | 0.000341384 | 0.028064873 | 1 |
| CAD | mesothelium | *RAB6A* | -0.767530394 | 0.000357557 | 0.028064873 | 1 |
| CAD | mesothelium | *MYPOP* | -0.912026695 | 3.73E-06 | 0.00195644 | 0.018121274 |
| CAD | mesothelium | *P4HTM* | -0.865274761 | 0.000620775 | 0.035851096 | 1 |
| CAD | mesothelium | *C8orf33* | -0.906348526 | 0.000279733 | 0.025110267 | 1 |
| CAD | mesothelium | *FES* | -0.421036594 | 1.68E-06 | 0.001164904 | 0.008154329 |
| CAD | mesothelium | *ADI1* | -1.013235621 | 0.000199094 | 0.023000068 | 0.966002862 |
| CAD | mesothelium | *TMED9* | -0.560805735 | 0.001288077 | 0.046829884 | 1 |
| CAD | mesothelium | *UBOX5* | -0.94893571 | 0.00089334 | 0.041835582 | 1 |
| CAD | mesothelium | *TNFAIP8L1* | -0.830479773 | 9.29E-05 | 0.016095278 | 0.450667775 |
| CAD | mesothelium | *PSMD13* | -0.395273792 | 0.000198928 | 0.023000068 | 0.965200763 |
| CAD | mesothelium | *IFT140* | -0.799747969 | 0.001165865 | 0.045304084 | 1 |
| CAD | mesothelium | *MSL1* | -0.272124302 | 0.000279276 | 0.025110267 | 1 |
| CAD | mesothelium | *HLA-DRB1* | -0.088572897 | 0.001212554 | 0.046325297 | 1 |
| CAD | mesothelium | *ZNF471* | -1.295437701 | 0.000219685 | 0.024225265 | 1 |
| CAD | mesothelium | *ZNF493* | -1.129663723 | 0.000106242 | 0.017775339 | 0.515484817 |
| CAD | mesothelium | *TPK1* | -0.57269637 | 0.001372034 | 0.047694723 | 1 |
| CAD | mesothelium | *MYO6* | -0.662727945 | 2.26E-06 | 0.001370515 | 0.010964117 |
| CAD | mesothelium | *ZNF100* | -0.245703226 | 0.000501328 | 0.033783958 | 1 |
| CAD | mesothelium | *IGF2R* | -0.86716585 | 4.44E-06 | 0.00195644 | 0.021520839 |
| CAD | mesothelium | *ENTPD6* | -0.386832551 | 0.000628059 | 0.035851096 | 1 |
| CAD | mesothelium | *ZNF334* | -0.341798671 | 0.000395661 | 0.029087091 | 1 |
| CAD | mesothelium | *TMEM116* | -0.124245344 | 1.91E-05 | 0.006631523 | 0.092841328 |
| CAD | mesothelium | *UCKL1* | -1.146870312 | 0.000517241 | 0.03383175 | 1 |
| CAD | mesothelium | *NCOA6* | -0.275259436 | 0.000812495 | 0.040226772 | 1 |
| CAD | mesothelium | *SFT2D1* | -1.063836995 | 0.000352585 | 0.028064873 | 1 |
| CAD | mesothelium | *SHISA4* | -1.031763814 | 0.000470682 | 0.033097783 | 1 |
| CAD | mesothelium | *STK19* | -0.280646721 | 0.000248514 | 0.02463368 | 1 |
| CAD | mesothelium | *TECPR1* | -1.072405841 | 4.07E-05 | 0.010395241 | 0.197509586 |
| CAD | mesothelium | *AL049840.1* | -0.457831318 | 0.00144788 | 0.049126659 | 1 |
| CAD | mesothelium | *WFDC6* | -0.440841912 | 0.001418174 | 0.048457599 | 1 |
| CAD | mesothelium | *GLI4* | -1.183736666 | 0.000541084 | 0.03383175 | 1 |
| CAD | mesothelium | *HMBS* | 0.313067461 | 0.000305348 | 0.025992059 | 1 |
| CAD | t_cell | *CEP68* | 3.793079157 | 0.000404605 | 0.041880841 | 1 |
| CAD | t_cell | *MDH1* | 4.64673073 | 4.37E-06 | 0.00530131 | 0.021205239 |
| CAD | t_cell | *UHRF1BP1* | 3.229787251 | 8.79E-07 | 0.003525752 | 0.004264727 |

| | | | | | | |
|---|---|---|---|---|---|---|
| CAD | t_cell | *ZXDC* | 4.674800253 | 0.000759904 | 0.04853336 | 1 |
| CAD | t_cell | *ZZEF1* | 4.425386779 | 0.000529688 | 0.044321117 | 1 |
| CAD | t_cell | *TIPIN* | 1.921835976 | 6.55E-05 | 0.021189717 | 0.317845762 |
| CAD | t_cell | *FMO4* | 3.576181608 | 0.000317513 | 0.037574938 | 1 |
| CAD | t_cell | *RPH3A* | 12.34612426 | 0.0004823 | 0.042562191 | 1 |
| CAD | t_cell | *ITPR3* | 3.822685651 | 1.45E-06 | 0.003525752 | 0.007051503 |
| CAD | t_cell | *MTAP* | 5.853126313 | 1.88E-05 | 0.01301134 | 0.091079381 |
| CAD | t_cell | *RBM23* | 3.780052031 | 0.000258288 | 0.033870623 | 1 |
| CAD | t_cell | *TRPC4AP* | 4.133017213 | 0.000216253 | 0.032992177 | 1 |
| CAD | t_cell | *ABHD12* | 3.732134696 | 0.000555587 | 0.044321117 | 1 |
| CAD | t_cell | *DIDO1* | 4.329001625 | 0.000801505 | 0.049857727 | 1 |
| CAD | t_cell | *PSMD7* | 5.58474629 | 0.000562989 | 0.044321117 | 1 |
| CAD | t_cell | *RABAC1* | 5.291860957 | 7.58E-05 | 0.022366212 | 0.367544151 |
| CAD | t_cell | *DNASE2* | 5.336042122 | 0.000705677 | 0.046903345 | 1 |
| CAD | t_cell | *LIPA* | 2.721748831 | 9.32E-05 | 0.022494799 | 0.452439219 |
| CAD | t_cell | *RAB5C* | 4.11890921 | 0.000174755 | 0.029238316 | 0.847911168 |
| CAD | t_cell | *ELP4* | 3.98186918 | 0.00037076 | 0.04183551 | 1 |
| CAD | t_cell | *PRDM4* | 5.130601799 | 0.000438548 | 0.042000222 | 1 |
| CAD | t_cell | *PCCB* | 3.054765384 | 0.000224265 | 0.032992177 | 1 |
| CAD | t_cell | *PLXNA1* | 5.92198711 | 0.000241006 | 0.033359546 | 1 |
| CAD | t_cell | *GGCX* | 3.296155241 | 0.000613001 | 0.045361526 | 1 |
| CAD | t_cell | *TAF1B* | 4.544892287 | 0.000282906 | 0.035196363 | 1 |
| CAD | t_cell | *ASH1L* | 3.439058339 | 0.000410587 | 0.041880841 | 1 |
| CAD | t_cell | *CD46* | 3.303233473 | 0.000482465 | 0.042562191 | 1 |
| CAD | t_cell | *PRRC2C* | 4.078093697 | 0.00023119 | 0.032992177 | 1 |
| CAD | t_cell | *C2orf40* | 5.936424136 | 0.00013056 | 0.024364555 | 0.633478417 |
| CAD | t_cell | *TRIM32* | 9.71818581 | 0.000475729 | 0.042562191 | 1 |
| CAD | t_cell | *NEK9* | 5.058250173 | 0.000294387 | 0.035709121 | 1 |
| CAD | t_cell | *EIF2B2* | 2.626693938 | 5.51E-05 | 0.020074456 | 0.267398948 |
| CAD | t_cell | *TCP11* | 2.900961646 | 0.000683105 | 0.046151458 | 1 |
| CAD | t_cell | *GRK4* | 1.026682598 | 5.04E-05 | 0.020074456 | 0.244539054 |
| CAD | t_cell | *SMARCA4* | 5.246583351 | 8.58E-05 | 0.022494799 | 0.416179396 |
| CAD | t_cell | *SERGEF* | 3.196676688 | 0.000540385 | 0.044321117 | 1 |
| CAD | t_cell | *ACTN4* | 4.455295435 | 0.000358224 | 0.041383363 | 1 |
| CAD | t_cell | *TIMM10B* | 3.588068335 | 0.000266364 | 0.034010524 | 1 |
| CAD | t_cell | *POLR1E* | 1.56225966 | 0.00041432 | 0.041880841 | 1 |
| CAD | t_cell | *SLC37A4* | 3.16141849 | 0.000128146 | 0.024364555 | 0.621762591 |
| CAD | t_cell | *RTCA* | 4.676830264 | 0.000671089 | 0.046151458 | 1 |
| CAD | t_cell | *NUP54* | 3.460827949 | 0.000247515 | 0.033359546 | 1 |
| CAD | t_cell | *TCF25* | 3.826642597 | 0.000566346 | 0.044321117 | 1 |
| CAD | t_cell | *MYO19* | 5.745615432 | 3.73E-05 | 0.020074456 | 0.181197668 |
| CAD | t_cell | *TMEM91* | 1.638853052 | 0.000406873 | 0.041880841 | 1 |
| CAD | t_cell | *SIRT3* | 5.449405001 | 9.56E-05 | 0.022494799 | 0.463994034 |
| CAD | t_cell | *EVA1B* | 3.088239003 | 0.000228313 | 0.032992177 | 1 |
| CAD | t_cell | *SNX27* | 4.846663539 | 0.000101117 | 0.022494799 | 0.490617832 |
| CAD | t_cell | *SETDB1* | 5.518721935 | 0.000156183 | 0.028066631 | 0.757799042 |
| CAD | t_cell | *WDPCP* | 3.240811028 | 3.92E-06 | 0.00530131 | 0.018999654 |
| CAD | t_cell | *TAMM41* | 4.680873814 | 0.000635649 | 0.046032355 | 1 |
| CAD | t_cell | *FBXO38* | 4.903470161 | 0.000593998 | 0.045032509 | 1 |
| CAD | t_cell | *TBC1D7* | 4.233582261 | 0.0005827 | 0.044877151 | 1 |

| | | | | | | |
|---|---|---|---|---|---|---|
| CAD | t_cell | *IRAK1BP1* | 4.469400983 | 7.84E-05 | 0.022366212 | 0.3802256 |
| CAD | t_cell | *FBXO10* | 2.630083109 | 0.00066244 | 0.046151458 | 1 |
| CAD | t_cell | *TTC12* | 2.891549402 | 0.000185607 | 0.030018794 | 0.900563808 |
| CAD | t_cell | *KCNK13* | 7.663764956 | 0.000458211 | 0.042562191 | 1 |
| CAD | t_cell | *CHCHD6* | 5.017799141 | 4.34E-05 | 0.020074456 | 0.210747854 |
| CAD | t_cell | *ZBTB7B* | 8.306060481 | 0.000164193 | 0.02845232 | 0.796664955 |
| CAD | t_cell | *CPSF4* | 5.30411172 | 4.65E-05 | 0.020074456 | 0.225772975 |
| CAD | t_cell | *TIPARP* | 5.756144916 | 0.00043428 | 0.042000222 | 1 |
| CAD | t_cell | *RNF123* | 3.129323354 | 6.15E-06 | 0.005964605 | 0.029823027 |
| CAD | t_cell | *PIGO* | 7.225940476 | 0.000129794 | 0.024364555 | 0.629762831 |
| CAD | t_cell | *ZNF202* | 7.733386426 | 0.000553095 | 0.044321117 | 1 |
| CAD | t_cell | *HNRNPA3* | 4.065603893 | 0.000771551 | 0.048617754 | 1 |
| CAD | t_cell | *MFN1* | 3.79752843 | 0.000684853 | 0.046151458 | 1 |
| CAD | t_cell | *NME6* | 3.672663034 | 0.000395396 | 0.041880841 | 1 |
| CAD | t_cell | *TEFM* | 5.048664881 | 5.79E-05 | 0.020074456 | 0.281042383 |
| CAD | t_cell | *FGGY* | 4.901721553 | 0.000101996 | 0.022494799 | 0.494885588 |
| CAD | t_cell | *CRLF3* | 4.151890598 | 9.67E-06 | 0.007816884 | 0.046901301 |
| CAD | t_cell | *MRPL14* | 5.587171262 | 0.000119911 | 0.024364555 | 0.581807932 |
| CAD | t_cell | *MRPL42* | 2.439974439 | 0.000750417 | 0.04853336 | 1 |
| CAD | t_cell | *HMGN2* | 3.676248141 | 0.000760209 | 0.04853336 | 1 |
| CAD | t_cell | *LIN52* | 3.241631781 | 0.000617036 | 0.045361526 | 1 |
| CAD | t_cell | *ATF6B* | 3.552024209 | 0.00044147 | 0.042000222 | 1 |
| CAD | t_cell | *GALT* | 4.992909307 | 0.000677949 | 0.046151458 | 1 |
| CAD | t_cell | *NSUN6* | 4.430432591 | 0.000547551 | 0.044321117 | 1 |
| CAD | t_cell | *HMBS* | 4.412186884 | 4.43E-05 | 0.020074456 | 0.214916385 |
| T2DM | adipocyte | *RBM6* | -0.285232096 | 4.30E-07 | 9.48E-05 | 0.002086538 |
| T2DM | adipocyte | *CYB561* | 0.428027847 | 0.001867431 | 0.049530135 | 1 |
| T2DM | adipocyte | *SS18L2* | 1.090621557 | 0.000252176 | 0.015488061 | 1 |
| T2DM | adipocyte | *PKD1* | 0.977320574 | 0.001360404 | 0.042949594 | 1 |
| T2DM | adipocyte | *CEP68* | 1.168143366 | 4.61E-12 | 4.51E-09 | 2.24E-08 |
| T2DM | adipocyte | *OSBPL5* | 0.882584086 | 0.000652777 | 0.028279221 | 1 |
| T2DM | adipocyte | *ATP6V0A1* | 0.759313705 | 0.000346417 | 0.01910018 | 1 |
| T2DM | adipocyte | *TUBG2* | -0.556353029 | 4.50E-05 | 0.004551982 | 0.218495119 |
| T2DM | adipocyte | *ITIH4* | 0.153364131 | 6.76E-06 | 0.001058419 | 0.032811002 |
| T2DM | adipocyte | *SLC6A16* | 1.044333533 | 0.000130778 | 0.009614129 | 0.634532532 |
| T2DM | adipocyte | *TM7SF3* | 1.142697009 | 3.89E-05 | 0.004256238 | 0.18876323 |
| T2DM | adipocyte | *CNN2* | 0.424525918 | 0.001658458 | 0.046783932 | 1 |
| T2DM | adipocyte | *BTBD1* | 1.067518378 | 0.000102931 | 0.008464731 | 0.499419137 |
| T2DM | adipocyte | *SEC61A2* | -0.591664314 | 0.001239512 | 0.041849617 | 1 |
| T2DM | adipocyte | *SDHA* | 0.562756748 | 0.000323372 | 0.018034511 | 1 |
| T2DM | adipocyte | *ZZEF1* | 1.181955198 | 1.32E-06 | 0.000255453 | 0.006386318 |
| T2DM | adipocyte | *FDFT1* | 0.609647674 | 0.000453257 | 0.021780124 | 1 |
| T2DM | adipocyte | *STK17B* | 0.613142026 | 0.000396305 | 0.020676022 | 1 |
| T2DM | adipocyte | *ZNF506* | 0.698050115 | 0.000370943 | 0.019663115 | 1 |
| T2DM | adipocyte | *CCNT2* | 0.912712388 | 4.04E-05 | 0.004256238 | 0.19578693 |
| T2DM | adipocyte | *ZNF586* | 0.75196346 | 0.000170014 | 0.01184425 | 0.824908792 |
| T2DM | adipocyte | *SLC25A24* | 0.922831068 | 0.00134564 | 0.042949594 | 1 |
| T2DM | adipocyte | *ITPKC* | 0.535947963 | 0.001055597 | 0.037659964 | 1 |
| T2DM | adipocyte | *PABPC4* | -0.838784185 | 5.54E-09 | 2.69E-06 | 2.69E-05 |
| T2DM | adipocyte | *PPP2R3C* | 0.581411405 | 0.000535426 | 0.024145282 | 1 |

| | | | | | | |
|---|---|---|---|---|---|---|
| T2DM | adipocyte | *CWF19L1* | -0.220712858 | 0.000426204 | 0.021767817 | 1 |
| T2DM | adipocyte | *SETD1A* | 0.716645496 | 0.001414715 | 0.043171054 | 1 |
| T2DM | adipocyte | *DIDO1* | 0.868706202 | 0.00045074 | 0.021780124 | 1 |
| T2DM | adipocyte | *NOMO3* | -0.293626452 | 0.001817869 | 0.049530135 | 1 |
| T2DM | adipocyte | *RBL2* | 0.932109227 | 6.08E-06 | 0.000983586 | 0.029507591 |
| T2DM | adipocyte | *CCDC9* | 1.129165115 | 0.000495888 | 0.022914764 | 1 |
| T2DM | adipocyte | *GCDH* | -0.519626762 | 0.001260891 | 0.042192029 | 1 |
| T2DM | adipocyte | *CHN2* | 1.016390276 | 4.65E-12 | 4.51E-09 | 2.25E-08 |
| T2DM | adipocyte | *CNTNAP1* | 1.234605844 | 2.15E-08 | 6.96E-06 | 0.000104433 |
| T2DM | adipocyte | *FAM149A* | 0.794440412 | 0.001735999 | 0.048408439 | 1 |
| T2DM | adipocyte | *PRR4* | 0.417582924 | 0.000200763 | 0.01298803 | 0.974102238 |
| T2DM | adipocyte | *CDK2AP1* | 0.54454137 | 3.36E-06 | 0.000626722 | 0.016294765 |
| T2DM | adipocyte | *GOLT1B* | 0.888774086 | 0.001868099 | 0.049530135 | 1 |
| T2DM | adipocyte | *BVES* | -0.170732911 | 0.000712082 | 0.029647644 | 1 |
| T2DM | adipocyte | *NPHP3* | 0.21499753 | 0.001835678 | 0.049530135 | 1 |
| T2DM | adipocyte | *PCCB* | 0.396974062 | 0.000796931 | 0.032222568 | 1 |
| T2DM | adipocyte | *NKTR* | 0.674569607 | 0.001157397 | 0.039827601 | 1 |
| T2DM | adipocyte | *POLE4* | 1.327402375 | 0.000406879 | 0.0210019 | 1 |
| T2DM | adipocyte | *GPX7* | -0.147977626 | 0.001402977 | 0.043083811 | 1 |
| T2DM | adipocyte | *ASH1L* | 0.632652905 | 0.00047836 | 0.022534 | 1 |
| T2DM | adipocyte | *ELOVL4* | 0.393497899 | 0.000898681 | 0.034883217 | 1 |
| T2DM | adipocyte | *TMEM5* | 0.87686422 | 0.00011227 | 0.008646564 | 0.544733508 |
| T2DM | adipocyte | *ADRA1A* | 0.125997578 | 2.31E-05 | 0.00287645 | 0.112181552 |
| T2DM | adipocyte | *TEX14* | 0.444102425 | 0.000681153 | 0.028738739 | 1 |
| T2DM | adipocyte | *SEC22A* | 0.9453269 | 0.001754976 | 0.048657955 | 1 |
| T2DM | adipocyte | *SSPN* | 0.398292225 | 0.000935712 | 0.035748601 | 1 |
| T2DM | adipocyte | *GLO1* | 1.062488673 | 4.00E-05 | 0.004256238 | 0.19390851 |
| T2DM | adipocyte | *KCNK17* | -0.442512109 | 0.001500782 | 0.044401179 | 1 |
| T2DM | adipocyte | *GRK4* | 0.110194472 | 0.000602824 | 0.026589997 | 1 |
| T2DM | adipocyte | *PRR12* | 0.647210505 | 0.000112052 | 0.008646564 | 0.543676569 |
| T2DM | adipocyte | *HELB* | 0.999159817 | 0.001242033 | 0.041849617 | 1 |
| T2DM | adipocyte | *SHC2* | 0.524310066 | 0.000872743 | 0.034709422 | 1 |
| T2DM | adipocyte | *STK33* | 0.38367967 | 0.000236438 | 0.014707658 | 1 |
| T2DM | adipocyte | *C12orf65* | 0.651205267 | 3.14E-05 | 0.003720115 | 0.152524724 |
| T2DM | adipocyte | *CHMP1A* | 1.198821043 | 0.00136752 | 0.042949594 | 1 |
| T2DM | adipocyte | *EXOC4* | 0.63200409 | 0.000884946 | 0.034883217 | 1 |
| T2DM | adipocyte | *OSER1* | 1.091714347 | 1.78E-05 | 0.002269359 | 0.086235625 |
| T2DM | adipocyte | *BTBD2* | 1.003362745 | 0.00113509 | 0.039621984 | 1 |
| T2DM | adipocyte | *P2RX4* | 0.384731167 | 0.000444913 | 0.021780124 | 1 |
| T2DM | adipocyte | *IRF4* | 0.136255886 | 0.000675464 | 0.028738739 | 1 |
| T2DM | adipocyte | *TCF19* | 0.622683347 | 2.96E-09 | 1.59E-06 | 1.43E-05 |
| T2DM | adipocyte | *FLOT1* | 0.496181135 | 0.000313279 | 0.017674749 | 1 |
| T2DM | adipocyte | *NUMA1* | 0.953477489 | 0.001369252 | 0.042949594 | 1 |
| T2DM | adipocyte | *SLC37A4* | 0.461634503 | 6.43E-05 | 0.006116457 | 0.311939301 |
| T2DM | adipocyte | *SPTBN5* | 0.105319466 | 5.94E-05 | 0.005759956 | 0.28799779 |
| T2DM | adipocyte | *MYOF* | 0.234161113 | 0.001016076 | 0.037067679 | 1 |
| T2DM | adipocyte | *HMG20A* | 1.131296575 | 0.000986116 | 0.037052094 | 1 |
| T2DM | adipocyte | *MAN2C1* | -0.167316629 | 4.19E-05 | 0.004326651 | 0.203352595 |
| T2DM | adipocyte | *MYO19* | 1.276347313 | 9.29E-08 | 2.37E-05 | 0.000450532 |
| T2DM | adipocyte | *C18orf8* | -0.653331535 | 0.000275196 | 0.016000108 | 1 |

| | | | | | | |
|---|---|---|---|---|---|---|
| T2DM | adipocyte | *PIP4K2B* | 0.966768284 | 0.000100917 | 0.008442249 | 0.48965043 |
| T2DM | adipocyte | *DUS3L* | 0.222503822 | 0.000537447 | 0.024145282 | 1 |
| T2DM | adipocyte | *SIRT3* | 0.879638008 | 0.000142977 | 0.010354073 | 0.693722917 |
| T2DM | adipocyte | *SNX27* | 0.693731394 | 0.001043979 | 0.037521394 | 1 |
| T2DM | adipocyte | *PLEKHA6* | 0.405795412 | 0.000106863 | 0.008575063 | 0.51850041 |
| T2DM | adipocyte | *NBEAL1* | 0.829141103 | 5.78E-05 | 0.00572649 | 0.280598011 |
| T2DM | adipocyte | *TAMM41* | 0.910637431 | 0.000362636 | 0.01955011 | 1 |
| T2DM | adipocyte | *BOC* | 0.769248755 | 0.000957248 | 0.036285691 | 1 |
| T2DM | adipocyte | *CISD2* | 1.630462742 | 2.79E-05 | 0.00338217 | 0.135286789 |
| T2DM | adipocyte | *USP53* | 0.551369447 | 0.00089574 | 0.034883217 | 1 |
| T2DM | adipocyte | *IRAK1BP1* | 0.764674674 | 8.42E-05 | 0.00742565 | 0.40841076 |
| T2DM | adipocyte | *SEC16A* | 0.889256109 | 0.000372837 | 0.019663115 | 1 |
| T2DM | adipocyte | *HSD17B12* | -0.242312232 | 9.10E-08 | 2.37E-05 | 0.000441521 |
| T2DM | adipocyte | *APIP* | -0.147207535 | 0.000467444 | 0.022235663 | 1 |
| T2DM | adipocyte | *CWF19L2* | 0.768145321 | 0.000196818 | 0.01290486 | 0.954959639 |
| T2DM | adipocyte | *PLA2R1* | 0.266921003 | 0.000714916 | 0.029647644 | 1 |
| T2DM | adipocyte | *JAZF1* | -0.830272788 | 3.31E-10 | 2.30E-07 | 1.61E-06 |
| T2DM | adipocyte | *VPS8* | 0.771603837 | 8.87E-05 | 0.007688297 | 0.430544635 |
| T2DM | adipocyte | *CLEC18A* | -0.450373809 | 1.75E-08 | 6.40E-06 | 8.51E-05 |
| T2DM | adipocyte | *AP3S2* | 0.802928059 | 0.00138243 | 0.042949594 | 1 |
| T2DM | adipocyte | *KRTCAP3* | 0.321061069 | 4.36E-06 | 0.000730232 | 0.021176735 |
| T2DM | adipocyte | *CCAR2* | 0.85259814 | 0.001472728 | 0.044239724 | 1 |
| T2DM | adipocyte | *VPS28* | 0.453442682 | 0.000547058 | 0.024351597 | 1 |
| T2DM | adipocyte | *SPTA1* | -0.233219251 | 0.000264085 | 0.015819036 | 1 |
| T2DM | adipocyte | *RFT1* | -0.459153103 | 1.51E-05 | 0.00203267 | 0.073176111 |
| T2DM | adipocyte | *RNF123* | 0.467837408 | 1.24E-06 | 0.000250138 | 0.006003314 |
| T2DM | adipocyte | *MST1R* | 0.396271552 | 4.51E-07 | 9.52E-05 | 0.002189616 |
| T2DM | adipocyte | *CAMLG* | 0.513882534 | 0.000931433 | 0.035748601 | 1 |
| T2DM | adipocyte | *GPER1* | 0.613172921 | 0.00171935 | 0.048221298 | 1 |
| T2DM | adipocyte | *FZD6* | 0.328217983 | 0.00118812 | 0.040596872 | 1 |
| T2DM | adipocyte | *NOXRED1* | 0.503517675 | 0.001839619 | 0.049530135 | 1 |
| T2DM | adipocyte | *QSOX2* | 1.070712748 | 0.001134437 | 0.039621984 | 1 |
| T2DM | adipocyte | *SLC39A13* | 0.815458331 | 0.001432227 | 0.043432282 | 1 |
| T2DM | adipocyte | *RAPSN* | -0.358016208 | 4.03E-05 | 0.004256238 | 0.195602398 |
| T2DM | adipocyte | *WBP1L* | 0.784120671 | 0.001557274 | 0.045244868 | 1 |
| T2DM | adipocyte | *ANPEP* | 0.150507879 | 0.000451128 | 0.021780124 | 1 |
| T2DM | adipocyte | *CNPY4* | 0.562217036 | 0.000732842 | 0.030133472 | 1 |
| T2DM | adipocyte | *CYB5D2* | 1.42822345 | 1.05E-05 | 0.001501798 | 0.051061128 |
| T2DM | adipocyte | *LTBP3* | 0.62135773 | 0.001843468 | 0.049530135 | 1 |
| T2DM | adipocyte | *RFWD3* | 1.067214376 | 0.000453379 | 0.021780124 | 1 |
| T2DM | adipocyte | *ZSWIM1* | 0.129910021 | 0.000432247 | 0.021780124 | 1 |
| T2DM | adipocyte | *ADAM9* | 0.671764493 | 0.0006696 | 0.028738739 | 1 |
| T2DM | adipocyte | *IRS1* | -0.175685441 | 3.51E-06 | 0.000630725 | 0.017029577 |
| T2DM | adipocyte | *INO80E* | -0.259354743 | 0.001616144 | 0.046399594 | 1 |
| T2DM | adipocyte | *SEMA3E* | 0.655086949 | 0.000107807 | 0.008575063 | 0.523078821 |
| T2DM | adipocyte | *SLC26A5* | 1.132257424 | 0.000261489 | 0.015819036 | 1 |
| T2DM | adipocyte | *TTLL6* | -1.160539547 | 2.70E-07 | 6.54E-05 | 0.001308605 |
| T2DM | adipocyte | *TRIAP1* | 1.039463466 | 0.000609304 | 0.02663371 | 1 |
| T2DM | adipocyte | *NUDT6* | 0.424022591 | 0.001486207 | 0.044239724 | 1 |
| T2DM | adipocyte | *RGS19* | -0.192512316 | 7.70E-05 | 0.007051382 | 0.373723224 |

| | | | | | | |
|---|---|---|---|---|---|---|
| T2DM | adipocyte | *C1QTNF4* | -0.169918152 | 0.000170877 | 0.01184425 | 0.829097466 |
| T2DM | adipocyte | *FGGY* | 0.703555971 | 0.00157087 | 0.045368233 | 1 |
| T2DM | adipocyte | *CTSW* | -0.056133175 | 0.000351195 | 0.019146024 | 1 |
| T2DM | adipocyte | *SNX32* | 0.163746176 | 0.001364651 | 0.042949594 | 1 |
| T2DM | adipocyte | *RNASEH2C* | -0.333698762 | 0.000824552 | 0.033063841 | 1 |
| T2DM | adipocyte | *EHBP1L1* | 0.300191796 | 0.00099274 | 0.037052094 | 1 |
| T2DM | adipocyte | *CCNE2* | -0.476750074 | 1.03E-05 | 0.001501798 | 0.049897629 |
| T2DM | adipocyte | *ADAMTSL1* | 0.534420523 | 0.000750565 | 0.030602864 | 1 |
| T2DM | adipocyte | *STX19* | 0.51733922 | 0.000116998 | 0.008869877 | 0.567672134 |
| T2DM | adipocyte | *HLA-DQB1* | -0.316128792 | 1.09E-15 | 2.65E-12 | 5.30E-12 |
| T2DM | adipocyte | *CAPS2* | 0.981059401 | 0.000489454 | 0.022834917 | 1 |
| T2DM | adipocyte | *ERCC6L2* | 0.846134437 | 0.001786039 | 0.049237839 | 1 |
| T2DM | adipocyte | *TTC3* | 0.222223188 | 0.000233643 | 0.014707658 | 1 |
| T2DM | adipocyte | *GP1BA* | 0.34037738 | 0.001002056 | 0.037067679 | 1 |
| T2DM | adipocyte | *POLR1D* | 0.914981355 | 0.000175996 | 0.012027199 | 0.853931125 |
| T2DM | adipocyte | *FFAR4* | 0.435005739 | 0.001043458 | 0.037521394 | 1 |
| T2DM | adipocyte | *NELFB* | 1.400130697 | 0.001364285 | 0.042949594 | 1 |
| T2DM | adipocyte | *HLA-DRB1* | -0.278912864 | 1.46E-09 | 8.85E-07 | 7.08E-06 |
| T2DM | adipocyte | *ZKSCAN7* | 0.767234803 | 3.40E-05 | 0.003923576 | 0.164790192 |
| T2DM | adipocyte | *ZNF34* | 1.040958107 | 0.000169523 | 0.01184425 | 0.822524684 |
| T2DM | adipocyte | *SULF2* | 0.341897948 | 0.0005274 | 0.024141011 | 1 |
| T2DM | adipocyte | *ZNF846* | 0.748217462 | 0.001389754 | 0.042949594 | 1 |
| T2DM | adipocyte | *ZKSCAN5* | 0.67624975 | 0.00155119 | 0.045244868 | 1 |
| T2DM | adipocyte | *HLA-DQA1* | -0.416884152 | 7.84E-13 | 1.27E-09 | 3.80E-09 |
| T2DM | adipocyte | *ZNF239* | 0.657187525 | 0.00154977 | 0.045244868 | 1 |
| T2DM | adipocyte | *FAM110D* | 0.45135243 | 0.00165836 | 0.046783932 | 1 |
| T2DM | adipocyte | *DDX42* | 0.849607291 | 0.00148246 | 0.044239724 | 1 |
| T2DM | adipocyte | *HLA-DRB5* | -0.124042127 | 6.08E-12 | 4.92E-09 | 2.95E-08 |
| T2DM | adipocyte | *NOTCH4* | -0.053009905 | 1.85E-08 | 6.40E-06 | 8.96E-05 |
| T2DM | adipocyte | *PRRT1* | 0.23475201 | 6.59E-05 | 0.006153343 | 0.319973813 |
| T2DM | adipocyte | *STK19* | -0.413940353 | 0.001377323 | 0.042949594 | 1 |
| T2DM | adipocyte | *GPANK1* | -0.429564129 | 0.000273119 | 0.016000108 | 1 |
| T2DM | adipocyte | *PSORS1C2* | 0.231658475 | 0.001626434 | 0.046420336 | 1 |
| T2DM | adipocyte | *DHX16* | 0.76446396 | 1.46E-05 | 0.002026004 | 0.070910125 |
| T2DM | adipocyte | *LIN52* | 0.587608546 | 8.98E-06 | 0.001361537 | 0.043569169 |
| T2DM | adipocyte | *HCG27* | 0.315935802 | 4.28E-07 | 9.48E-05 | 0.002076645 |
| T2DM | adipocyte | *TAS2R14* | 0.231081666 | 0.000297849 | 0.017001927 | 1 |
| T2DM | adipocyte | *DNLZ* | 0.13976468 | 0.000214033 | 0.013664306 | 1 |
| T2DM | adipocyte | *SUPT4H1* | 1.494946328 | 0.000180792 | 0.012183365 | 0.87720231 |
| T2DM | adipocyte | *ARHGAP19* | -0.76684195 | 0.001013176 | 0.037067679 | 1 |
| T2DM | adipocyte | *TMEM110* | 0.636051608 | 3.89E-06 | 0.000673541 | 0.018859134 |
| T2DM | adipocyte | *ATF6B* | 0.531874519 | 0.000277001 | 0.016000108 | 1 |
| T2DM | adipocyte | *STARD10* | 0.276690245 | 1.70E-08 | 6.40E-06 | 8.23E-05 |
| T2DM | adipocyte | *C4B* | 0.508911987 | 8.30E-05 | 0.00742565 | 0.402525738 |
| T2DM | adipocyte | *AC108463.2* | 0.529421551 | 0.001370345 | 0.042949594 | 1 |
| T2DM | adipocyte | *TMA7* | 0.764160958 | 9.87E-05 | 0.008401032 | 0.478858849 |
| T2DM | adipocyte | *HLA-DQB2* | 0.120629616 | 1.31E-08 | 5.76E-06 | 6.34E-05 |
| T2DM | adipocyte | *HLA-DQA2* | 0.22672142 | 1.72E-18 | 8.35E-15 | 8.35E-15 |
| T2DM | adipocyte | *PSMB9* | 0.217109468 | 1.62E-05 | 0.00211854 | 0.078385981 |
| T2DM | adipocyte | *CD302* | 0.802509169 | 0.001125746 | 0.039621984 | 1 |

| | | | | | | |
|---|---|---|---|---|---|---|
| T2DM | adipocyte | *C8orf58* | 1.168006714 | 0.001309706 | 0.042949594 | 1 |
| T2DM | adipocyte | *MRPL20* | 0.883372829 | 0.001351383 | 0.042949594 | 1 |
| T2DM | adipocyte | *IL10RB* | 0.691494199 | 0.001154926 | 0.039827601 | 1 |
| T2DM | adipocyte | *PLA2G4B* | 1.129199286 | 4.94E-08 | 1.50E-05 | 0.000239628 |
| T2DM | adipocyte | *JMJD7* | 0.341181489 | 8.98E-08 | 2.37E-05 | 0.000435555 |
| T2DM | adipocyte | *C4A* | -0.401486076 | 0.000185002 | 0.012296317 | 0.897631161 |
| T2DM | adipocyte | *TAS2R20* | 0.319831644 | 0.001849964 | 0.049530135 | 1 |
| T2DM | adipocyte | *C19orf84* | 14.8133675 | 0.000120943 | 0.009027956 | 0.586817167 |
| T2DM | ASPC | *RBM6* | -0.148531117 | 6.72E-08 | 1.81E-05 | 0.000325818 |
| T2DM | ASPC | *SS18L2* | 0.531889533 | 0.000832733 | 0.031375763 | 1 |
| T2DM | ASPC | *TRAF3IP3* | 0.157227904 | 0.001785945 | 0.049639169 | 1 |
| T2DM | ASPC | *CEP68* | 0.576281563 | 2.64E-11 | 1.60E-08 | 1.28E-07 |
| T2DM | ASPC | *ATP6V0A1* | 0.365277637 | 0.001811379 | 0.049837439 | 1 |
| T2DM | ASPC | *TUBG2* | -0.333374311 | 1.96E-06 | 0.000379931 | 0.009498285 |
| T2DM | ASPC | *ANO2* | 0.638309317 | 0.00041263 | 0.021074514 | 1 |
| T2DM | ASPC | *ITIH4* | 0.074627424 | 5.43E-06 | 0.000798267 | 0.026342817 |
| T2DM | ASPC | *TM7SF3* | 0.664565336 | 8.75E-05 | 0.007194094 | 0.424451525 |
| T2DM | ASPC | *CNN2* | 0.21313755 | 0.001387923 | 0.04455737 | 1 |
| T2DM | ASPC | *BTBD1* | 0.651876215 | 0.000135304 | 0.009946899 | 0.656495316 |
| T2DM | ASPC | *SEC61A2* | -0.363720757 | 9.02E-05 | 0.007235456 | 0.437536385 |
| T2DM | ASPC | *SDHA* | 0.245114948 | 0.001828331 | 0.049837439 | 1 |
| T2DM | ASPC | *ZZEF1* | 0.654272665 | 3.08E-06 | 0.000534162 | 0.014956524 |
| T2DM | ASPC | *MYH7B* | 0.229678117 | 0.001660057 | 0.048521675 | 1 |
| T2DM | ASPC | *FDFT1* | 0.342102777 | 0.000345127 | 0.019466808 | 1 |
| T2DM | ASPC | *STK17B* | 0.292934813 | 0.001734305 | 0.049209632 | 1 |
| T2DM | ASPC | *ZNF506* | 0.331330901 | 0.001139527 | 0.039123196 | 1 |
| T2DM | ASPC | *CCNT2* | 0.441243843 | 0.000274703 | 0.017309878 | 1 |
| T2DM | ASPC | *ZNF586* | 0.360618019 | 0.000615386 | 0.027646797 | 1 |
| T2DM | ASPC | *PABPC4* | -0.448204121 | 3.03E-10 | 1.47E-07 | 1.47E-06 |
| T2DM | ASPC | *CWF19L1* | -0.114428675 | 0.000165744 | 0.011654934 | 0.80419048 |
| T2DM | ASPC | *MTHFD1* | 0.614851363 | 0.000572082 | 0.02643562 | 1 |
| T2DM | ASPC | *RNF125* | 0.894993539 | 0.001762686 | 0.04943672 | 1 |
| T2DM | ASPC | *NOMO3* | -0.145573877 | 0.001521258 | 0.047267485 | 1 |
| T2DM | ASPC | *RBL2* | 0.460903588 | 4.07E-05 | 0.004248767 | 0.197516164 |
| T2DM | ASPC | *FCER2* | 1.757913972 | 0.001529471 | 0.047267485 | 1 |
| T2DM | ASPC | *RASAL3* | 1.171844234 | 0.000323889 | 0.01934569 | 1 |
| T2DM | ASPC | *GCDH* | -0.354294711 | 9.37E-05 | 0.007336243 | 0.45484704 |
| T2DM | ASPC | *ADAP1* | 2.459437198 | 0.001496294 | 0.046838841 | 1 |
| T2DM | ASPC | *CHN2* | 0.663583091 | 4.37E-14 | 4.24E-11 | 2.12E-10 |
| T2DM | ASPC | *CNTNAP1* | 0.489390434 | 4.67E-06 | 0.000735989 | 0.022653317 |
| T2DM | ASPC | *B3GAT1* | 2.765708782 | 0.000153924 | 0.011146845 | 0.746838622 |
| T2DM | ASPC | *IL23A* | 3.727675716 | 0.001223373 | 0.04113662 | 1 |
| T2DM | ASPC | *PRR4* | 0.192530101 | 0.000533165 | 0.025328992 | 1 |
| T2DM | ASPC | *CDK2AP1* | 0.257279877 | 5.59E-06 | 0.00079835 | 0.027143885 |
| T2DM | ASPC | *BVES* | -0.089159074 | 0.000247346 | 0.015791079 | 1 |
| T2DM | ASPC | *PCCB* | 0.187645675 | 0.001626968 | 0.048134438 | 1 |
| T2DM | ASPC | *GRB14* | 0.823538394 | 0.000537693 | 0.025328992 | 1 |
| T2DM | ASPC | *POLE4* | 0.864501978 | 0.000353067 | 0.019466808 | 1 |
| T2DM | ASPC | *GPX7* | -0.075607606 | 0.000708862 | 0.029396562 | 1 |
| T2DM | ASPC | *TMEM5* | 0.399121063 | 0.000683937 | 0.02937799 | 1 |

| | | | | | | |
|---|---|---|---|---|---|---|
| T2DM | ASPC | *MRC1* | 0.45587158 | 0.001702298 | 0.049027352 | 1 |
| T2DM | ASPC | *ADRA1A* | 0.05930567 | 4.12E-05 | 0.004248767 | 0.199692034 |
| T2DM | ASPC | *TEX14* | 0.208356909 | 0.001074692 | 0.037831583 | 1 |
| T2DM | ASPC | *PDZRN3* | -0.147364075 | 0.000376215 | 0.020282163 | 1 |
| T2DM | ASPC | *RAB38* | 0.268949517 | 0.001323788 | 0.043178403 | 1 |
| T2DM | ASPC | *GLO1* | 0.469782291 | 0.000758784 | 0.03017722 | 1 |
| T2DM | ASPC | *KCNK17* | -0.230088119 | 0.001034937 | 0.037473978 | 1 |
| T2DM | ASPC | *GRK4* | 0.050612561 | 0.00114499 | 0.039123196 | 1 |
| T2DM | ASPC | *OPRL1* | 1.902091475 | 0.001325965 | 0.043178403 | 1 |
| T2DM | ASPC | *PRR12* | 0.300287473 | 0.000347491 | 0.019466808 | 1 |
| T2DM | ASPC | *ABHD8* | -0.192803002 | 0.001372435 | 0.044393711 | 1 |
| T2DM | ASPC | *HELB* | 0.577150881 | 0.001242335 | 0.041286352 | 1 |
| T2DM | ASPC | *OPN1SW* | -0.126199088 | 0.00158122 | 0.048126408 | 1 |
| T2DM | ASPC | *STK33* | 0.220305375 | 5.63E-05 | 0.005358075 | 0.27326185 |
| T2DM | ASPC | *C12orf65* | 0.299969029 | 0.000110841 | 0.008403159 | 0.537802188 |
| T2DM | ASPC | *IDO1* | 1.565671769 | 5.86E-05 | 0.005373983 | 0.284113217 |
| T2DM | ASPC | *OSER1* | 0.542718824 | 3.42E-05 | 0.003854225 | 0.16573169 |
| T2DM | ASPC | *TTC9* | 1.07234205 | 0.001014298 | 0.03728315 | 1 |
| T2DM | ASPC | *P2RX4* | 0.177562495 | 0.001076001 | 0.037831583 | 1 |
| T2DM | ASPC | *SCN2A* | 0.340487358 | 0.000569066 | 0.02643562 | 1 |
| T2DM | ASPC | *IRF4* | 0.067134885 | 0.00051012 | 0.024570759 | 1 |
| T2DM | ASPC | *TCF19* | 0.328801094 | 1.30E-09 | 5.73E-07 | 6.31E-06 |
| T2DM | ASPC | *FLOT1* | 0.243840569 | 0.000790154 | 0.031169318 | 1 |
| T2DM | ASPC | *SLC37A4* | 0.223282761 | 6.44E-05 | 0.005684672 | 0.312656976 |
| T2DM | ASPC | *SPTBN5* | 0.051654538 | 4.44E-05 | 0.004486354 | 0.215345009 |
| T2DM | ASPC | *NEIL1* | -0.256260062 | 0.000511469 | 0.024570759 | 1 |
| T2DM | ASPC | *MAN2C1* | -0.083380336 | 2.38E-05 | 0.002890876 | 0.11563504 |
| T2DM | ASPC | *ITGAX* | 1.183001657 | 0.000287853 | 0.017905905 | 1 |
| T2DM | ASPC | *MYO19* | 0.611689229 | 1.46E-06 | 0.000294926 | 0.007078235 |
| T2DM | ASPC | *C18orf8* | -0.379383383 | 2.57E-05 | 0.003041981 | 0.124721204 |
| T2DM | ASPC | *PIP4K2B* | 0.459596361 | 0.000398356 | 0.020783047 | 1 |
| T2DM | ASPC | *DUS3L* | 0.104238278 | 0.00081044 | 0.031263342 | 1 |
| T2DM | ASPC | *SIRT3* | 0.427514815 | 0.000582051 | 0.026642541 | 1 |
| T2DM | ASPC | *SAE1* | -0.526301763 | 0.000753555 | 0.03017722 | 1 |
| T2DM | ASPC | *NBEAL1* | 0.363578615 | 0.001717776 | 0.049027352 | 1 |
| T2DM | ASPC | *CISD2* | 0.899682874 | 7.95E-05 | 0.006765417 | 0.385628772 |
| T2DM | ASPC | *IRAK1BP1* | 0.356324791 | 0.000481531 | 0.023840718 | 1 |
| T2DM | ASPC | *SEC16A* | 0.427024298 | 0.001029755 | 0.037473978 | 1 |
| T2DM | ASPC | *ST8SIA6* | 0.824859802 | 0.000109909 | 0.008403159 | 0.533280531 |
| T2DM | ASPC | *HSD17B12* | -0.11936558 | 5.18E-08 | 1.48E-05 | 0.000251217 |
| T2DM | ASPC | *APIP* | -0.072857148 | 0.000358491 | 0.019543808 | 1 |
| T2DM | ASPC | *POC5* | -0.148598158 | 0.000680543 | 0.02937799 | 1 |
| T2DM | ASPC | *CWF19L2* | 0.35708594 | 0.001616778 | 0.048126408 | 1 |
| T2DM | ASPC | *CLEC4F* | 0.318070079 | 0.001229351 | 0.04113662 | 1 |
| T2DM | ASPC | *JAZF1* | -0.5853013 | 8.48E-16 | 1.42E-12 | 4.12E-12 |
| T2DM | ASPC | *GOLGA7B* | 2.524671786 | 0.000731615 | 0.029830228 | 1 |
| T2DM | ASPC | *VPS8* | 0.387169362 | 0.000466618 | 0.023340498 | 1 |
| T2DM | ASPC | *CLEC18A* | -0.220668523 | 1.15E-08 | 4.29E-06 | 5.58E-05 |
| T2DM | ASPC | *AP3S2* | 0.500842069 | 0.001603597 | 0.048126408 | 1 |
| T2DM | ASPC | *KRTCAP3* | 0.153833168 | 5.10E-06 | 0.000773077 | 0.024738448 |

| | | | | | | |
|---|---|---|---|---|---|---|
| T2DM | ASPC | *ZBTB7B* | 0.666203631 | 0.001043929 | 0.037519588 | 1 |
| T2DM | ASPC | *ASGR2* | 2.282844454 | 0.000701622 | 0.02937799 | 1 |
| T2DM | ASPC | *ASRGL1* | 0.950888435 | 0.001182094 | 0.040108531 | 1 |
| T2DM | ASPC | *SPTA1* | -0.111651966 | 0.000296716 | 0.017995854 | 1 |
| T2DM | ASPC | *RFT1* | -0.244122857 | 2.35E-06 | 0.000439297 | 0.011421719 |
| T2DM | ASPC | *RNF123* | 0.214750237 | 4.70E-06 | 0.000735989 | 0.022815665 |
| T2DM | ASPC | *MST1R* | 0.194660256 | 5.23E-07 | 0.000120751 | 0.002535764 |
| T2DM | ASPC | *IL15* | 0.420823105 | 0.000219856 | 0.014612907 | 1 |
| T2DM | ASPC | *F2RL2* | 1.300058847 | 0.001102152 | 0.038472233 | 1 |
| T2DM | ASPC | *CHMP4C* | 1.841317705 | 0.000821011 | 0.031366504 | 1 |
| T2DM | ASPC | *FZD6* | 0.15491319 | 0.001714429 | 0.049027352 | 1 |
| T2DM | ASPC | *DCSTAMP* | 2.793795542 | 0.000173972 | 0.012058754 | 0.844112805 |
| T2DM | ASPC | *AQP3* | 2.45338439 | 0.001761224 | 0.04943672 | 1 |
| T2DM | ASPC | *GJB2* | 3.453366868 | 0.000698176 | 0.02937799 | 1 |
| T2DM | ASPC | *RAPSN* | -0.171213559 | 5.87E-05 | 0.005373983 | 0.284821084 |
| T2DM | ASPC | *CYB5D2* | 0.877462011 | 1.25E-05 | 0.001636212 | 0.060539857 |
| T2DM | ASPC | *RFWD3* | 0.526379282 | 0.001656472 | 0.048521675 | 1 |
| T2DM | ASPC | *ZSWIM1* | 0.063763478 | 0.00034519 | 0.019466808 | 1 |
| T2DM | ASPC | *ADAM9* | 0.375265102 | 0.00107223 | 0.037831583 | 1 |
| T2DM | ASPC | *IRS1* | -0.097063819 | 3.75E-07 | 9.09E-05 | 0.001817277 |
| T2DM | ASPC | *INO80E* | -0.134006885 | 0.000856873 | 0.031981128 | 1 |
| T2DM | ASPC | *SEMA3E* | 0.288434918 | 0.000688736 | 0.02937799 | 1 |
| T2DM | ASPC | *EMB* | -0.151964016 | 0.001790366 | 0.049639169 | 1 |
| T2DM | ASPC | *SLC26A5* | 0.563821942 | 0.001432491 | 0.045132766 | 1 |
| T2DM | ASPC | *TTLL6* | -0.519458742 | 1.42E-06 | 0.000294926 | 0.00686931 |
| T2DM | ASPC | *TRIAP1* | 0.506128707 | 0.000754608 | 0.03017722 | 1 |
| T2DM | ASPC | *RGS19* | -0.092754396 | 8.54E-05 | 0.007145008 | 0.414410453 |
| T2DM | ASPC | *C1QTNF4* | -0.085108524 | 9.10E-05 | 0.007235456 | 0.441362837 |
| T2DM | ASPC | *CTSW* | -0.027999195 | 0.000296357 | 0.017995854 | 1 |
| T2DM | ASPC | *RNASEH2C* | -0.18725378 | 0.00011415 | 0.008520825 | 0.553853635 |
| T2DM | ASPC | *EHBP1L1* | 0.14353221 | 0.001603142 | 0.048126408 | 1 |
| T2DM | ASPC | *SEZ6L2* | 0.145848123 | 0.001419186 | 0.04500583 | 1 |
| T2DM | ASPC | *CCNE2* | -0.246017042 | 3.00E-06 | 0.000534162 | 0.014573645 |
| T2DM | ASPC | *FAM87B* | 7.00982887 | 0.000231498 | 0.015178753 | 1 |
| T2DM | ASPC | *STX19* | 0.236746483 | 0.000350209 | 0.019466808 | 1 |
| T2DM | ASPC | *HLA-DQB1* | -0.151935135 | 8.77E-16 | 1.42E-12 | 4.26E-12 |
| T2DM | ASPC | *GIPC3* | 5.62190146 | 0.000403996 | 0.02085308 | 1 |
| T2DM | ASPC | *TMEM252* | 17.18190317 | 0.000506548 | 0.024570759 | 1 |
| T2DM | ASPC | *TTC3* | 0.100228033 | 0.000662514 | 0.02937799 | 1 |
| T2DM | ASPC | *LRIT3* | 3.394698088 | 0.001614681 | 0.048126408 | 1 |
| T2DM | ASPC | *GP1BA* | 0.173463001 | 0.00072238 | 0.029703305 | 1 |
| T2DM | ASPC | *DMWD* | -0.341122393 | 0.000796921 | 0.031182729 | 1 |
| T2DM | ASPC | *RASA3* | -0.210931253 | 0.000896144 | 0.033191534 | 1 |
| T2DM | ASPC | *POLR1D* | 0.446451938 | 0.000691515 | 0.02937799 | 1 |
| T2DM | ASPC | *FFAR4* | 1.059207395 | 1.47E-14 | 1.78E-11 | 7.11E-11 |
| T2DM | ASPC | *HLA-DRB1* | -0.144126115 | 1.99E-10 | 1.07E-07 | 9.67E-07 |
| T2DM | ASPC | *STK31* | 0.411877157 | 0.000811867 | 0.031263342 | 1 |
| T2DM | ASPC | *ZKSCAN7* | 0.35111619 | 0.000165684 | 0.011654934 | 0.803898164 |
| T2DM | ASPC | *ZNF34* | 0.560946855 | 0.000208239 | 0.014033008 | 1 |
| T2DM | ASPC | *SULF2* | 0.180349122 | 0.00045593 | 0.023043446 | 1 |

| | | | | | | |
|---|---|---|---|---|---|---|
| T2DM | ASPC | *HLA-DQA1* | -0.200951914 | 5.62E-13 | 4.54E-10 | 2.73E-09 |
| T2DM | ASPC | *FAM110D* | 0.242132435 | 0.000605601 | 0.027461463 | 1 |
| T2DM | ASPC | *HLA-DRB5* | -0.06033875 | 3.73E-12 | 2.59E-09 | 1.81E-08 |
| T2DM | ASPC | *ASAH2B* | 0.366832739 | 0.001130539 | 0.039123196 | 1 |
| T2DM | ASPC | *NOTCH4* | -0.025745472 | 1.81E-08 | 6.26E-06 | 8.77E-05 |
| T2DM | ASPC | *AGER* | 0.143450234 | 0.001818913 | 0.049837439 | 1 |
| T2DM | ASPC | *PRRT1* | 0.11835622 | 3.67E-05 | 0.003959204 | 0.178164182 |
| T2DM | ASPC | *STK19* | -0.225492597 | 0.000394011 | 0.020779821 | 1 |
| T2DM | ASPC | *GPANK1* | -0.238478553 | 5.37E-05 | 0.005210871 | 0.260543566 |
| T2DM | ASPC | *PSORS1C2* | 0.11306879 | 0.001395862 | 0.04455737 | 1 |
| T2DM | ASPC | *DHX16* | 0.358298698 | 5.23E-05 | 0.005174329 | 0.253542123 |
| T2DM | ASPC | *LIN52* | 0.305201237 | 1.04E-05 | 0.001445388 | 0.050588575 |
| T2DM | ASPC | *KLRC2* | 1.99644567 | 1.81E-05 | 0.00225692 | 0.088019865 |
| T2DM | ASPC | *HCG27* | 0.149939172 | 7.49E-07 | 0.000165138 | 0.003633044 |
| T2DM | ASPC | *TAS2R14* | 0.110742866 | 0.000326947 | 0.01934569 | 1 |
| T2DM | ASPC | *DNLZ* | 0.066833364 | 0.000237457 | 0.015361866 | 1 |
| T2DM | ASPC | *SUPT4H1* | 0.728245303 | 0.001286201 | 0.042453394 | 1 |
| T2DM | ASPC | *ARHGAP19* | -0.469275103 | 7.34E-05 | 0.006358224 | 0.356060537 |
| T2DM | ASPC | *TMEM110* | 0.304123783 | 4.68E-06 | 0.000735989 | 0.022718667 |
| T2DM | ASPC | *ATF6B* | 0.2452548 | 0.000702359 | 0.02937799 | 1 |
| T2DM | ASPC | *STARD10* | 0.132410811 | 2.16E-08 | 6.99E-06 | 0.000104891 |
| T2DM | ASPC | *EML6* | 0.523278385 | 0.000834187 | 0.031375763 | 1 |
| T2DM | ASPC | *EVPLL* | 0.301966862 | 0.001680584 | 0.048827492 | 1 |
| T2DM | ASPC | *C4B* | 0.251539036 | 6.44E-05 | 0.005684672 | 0.312462036 |
| T2DM | ASPC | *LINC00694* | 0.299241675 | 0.001562128 | 0.047971184 | 1 |
| T2DM | ASPC | *TMA7* | 0.358361615 | 0.000338834 | 0.019466808 | 1 |
| T2DM | ASPC | *HLA-DQB2* | 0.059701094 | 7.66E-09 | 3.10E-06 | 3.72E-05 |
| T2DM | ASPC | *HLA-DQA2* | 0.110263273 | 1.03E-18 | 4.99E-15 | 4.99E-15 |
| T2DM | ASPC | *LINC01143* | 3.694005182 | 1.63E-05 | 0.002084354 | 0.079205445 |
| T2DM | ASPC | *TXNDC5* | 3.249932684 | 0.00038874 | 0.02072712 | 1 |
| T2DM | ASPC | *PSMB9* | 0.111809969 | 1.21E-05 | 0.001627397 | 0.058586309 |
| T2DM | ASPC | *PLA2G4B* | 0.548248608 | 3.84E-08 | 1.16E-05 | 0.000186244 |
| T2DM | ASPC | *JMJD7* | 0.162915926 | 1.17E-07 | 2.98E-05 | 0.000565473 |
| T2DM | ASPC | *C4A* | -0.190037428 | 0.000187568 | 0.012818022 | 0.910079549 |
| T2DM | ASPC | *C19orf84* | 6.875658291 | 3.08E-05 | 0.003554771 | 0.149300398 |
| T2DM | ASPC | *NDUFA7* | 6.671741147 | 3.63E-05 | 0.003959204 | 0.176111674 |
| T2DM | b_cell | *NFYA* | -1.281542977 | 0.001248328 | 0.004980661 | 1 |
| T2DM | b_cell | *ENPP4* | -1.86117895 | 0.013216013 | 0.016556698 | 1 |
| T2DM | b_cell | *SEMA3F* | -2.197808783 | 0.00047217 | 0.004980661 | 1 |
| T2DM | b_cell | *KRIT1* | -1.028741021 | 0.002031864 | 0.004980661 | 1 |
| T2DM | b_cell | *RAD52* | -0.987774363 | 0.001747241 | 0.004980661 | 1 |
| T2DM | b_cell | *HS3ST1* | -1.775711307 | 0.00173031 | 0.004980661 | 1 |
| T2DM | b_cell | *AOC1* | 1.132909836 | 0.00263235 | 0.004980661 | 1 |
| T2DM | b_cell | *TMEM176A* | -1.628900502 | 0.001960271 | 0.004980661 | 1 |
| T2DM | b_cell | *M6PR* | -1.224265246 | 0.003862448 | 0.005966444 | 1 |
| T2DM | b_cell | *CYP26B1* | 1.460678656 | 0.003426478 | 0.005526494 | 1 |
| T2DM | b_cell | *DBNDD1* | -3.458112736 | 0.000135401 | 0.004980661 | 0.656966117 |
| T2DM | b_cell | *ALS2* | -1.918519589 | 0.020863159 | 0.025099938 | 1 |
| T2DM | b_cell | *RBM6* | -1.111595241 | 2.38E-05 | 0.003983276 | 0.115515003 |
| T2DM | b_cell | *RECQL* | -1.494683596 | 0.00188985 | 0.004980661 | 1 |

| | | | | | | |
|---|---|---|---|---|---|---|
| T2DM | b_cell | *ARHGAP33* | -2.926849863 | 0.00059917 | 0.004980661 | 1 |
| T2DM | b_cell | *CDC27* | -0.91077917 | 0.002514539 | 0.004980661 | 1 |
| T2DM | b_cell | *POLR2J* | -2.436768709 | 0.002396622 | 0.004980661 | 1 |
| T2DM | b_cell | *MEOX1* | -1.828677812 | 0.002144138 | 0.004980661 | 1 |
| T2DM | b_cell | *CIAPIN1* | -1.765867397 | 0.003795648 | 0.005891389 | 1 |
| T2DM | b_cell | *PON1* | 1.934370694 | 0.002547247 | 0.004980661 | 1 |
| T2DM | b_cell | *GCFC2* | -1.532048836 | 0.003946721 | 0.006048481 | 1 |
| T2DM | b_cell | *ZNF195* | -2.413728397 | 0.000454821 | 0.004980661 | 1 |
| T2DM | b_cell | *PDK2* | -1.295813752 | 0.000711767 | 0.004980661 | 1 |
| T2DM | b_cell | *GGNBP2* | -0.875539619 | 0.001291688 | 0.004980661 | 1 |
| T2DM | b_cell | *ITGA2B* | -1.914542297 | 0.002279641 | 0.004980661 | 1 |
| T2DM | b_cell | *TMEM98* | -1.102908596 | 0.001504083 | 0.004980661 | 1 |
| T2DM | b_cell | *ABCC8* | -2.725853425 | 0.014506427 | 0.017996723 | 1 |
| T2DM | b_cell | *SYNRG* | -1.221167762 | 0.002302277 | 0.004980661 | 1 |
| T2DM | b_cell | *ZNF263* | -1.852968266 | 0.003859128 | 0.005965584 | 1 |
| T2DM | b_cell | *SPATA20* | -1.526614517 | 0.002869927 | 0.005094011 | 1 |
| T2DM | b_cell | *MAP3K9* | -1.345090887 | 0.012710589 | 0.015964737 | 1 |
| T2DM | b_cell | *BAIAP2L1* | -2.504578751 | 0.013242886 | 0.016581802 | 1 |
| T2DM | b_cell | *AGK* | -1.00023999 | 0.002008707 | 0.004980661 | 1 |
| T2DM | b_cell | *FARP2* | -1.167802693 | 0.002037003 | 0.004980661 | 1 |
| T2DM | b_cell | *USH1C* | 2.8085276 | 0.002820886 | 0.005065458 | 1 |
| T2DM | b_cell | *IFRD1* | -1.065352362 | 0.002375979 | 0.004980661 | 1 |
| T2DM | b_cell | *VPS41* | -1.088169731 | 0.002033771 | 0.004980661 | 1 |
| T2DM | b_cell | *ELAC2* | -1.482919976 | 0.004673205 | 0.006833456 | 1 |
| T2DM | b_cell | *SCIN* | -1.468758895 | 0.001347335 | 0.004980661 | 1 |
| T2DM | b_cell | *CCDC124* | 1.713982256 | 0.00644903 | 0.008869244 | 1 |
| T2DM | b_cell | *CEACAM21* | -2.172359353 | 0.011734883 | 0.014889553 | 1 |
| T2DM | b_cell | *NOS2* | 3.26309402 | 0.006331742 | 0.008750103 | 1 |
| T2DM | b_cell | *CD79B* | 0.329149624 | 0.003254669 | 0.00536838 | 1 |
| T2DM | b_cell | *ST7L* | -1.289638267 | 0.000652435 | 0.004980661 | 1 |
| T2DM | b_cell | *PAX6* | -2.529955489 | 0.00453734 | 0.006681388 | 1 |
| T2DM | b_cell | *PSMB1* | -1.860018782 | 0.003469369 | 0.00557766 | 1 |
| T2DM | b_cell | *NADK* | -1.946779992 | 0.002924593 | 0.005134596 | 1 |
| T2DM | b_cell | *DLEC1* | -1.490192745 | 0.002091795 | 0.004980661 | 1 |
| T2DM | b_cell | *SS18L2* | -2.411645846 | 0.018121332 | 0.02205838 | 1 |
| T2DM | b_cell | *MGST1* | -0.633765229 | 0.004205884 | 0.006339414 | 1 |
| T2DM | b_cell | *MMP25* | -1.700585487 | 0.004499092 | 0.006653335 | 1 |
| T2DM | b_cell | *PKD1* | -1.089057098 | 0.002401695 | 0.004980661 | 1 |
| T2DM | b_cell | *MAPK8IP2* | -8.881414459 | 0.000913586 | 0.004980661 | 1 |
| T2DM | b_cell | *MED24* | -2.253014312 | 0.004335515 | 0.00647259 | 1 |
| T2DM | b_cell | *RPS20* | -1.571868108 | 0.0032038 | 0.005339692 | 1 |
| T2DM | b_cell | *POMT2* | -2.034889899 | 0.001467729 | 0.004980661 | 1 |
| T2DM | b_cell | *MLXIPL* | -0.828966506 | 0.001982167 | 0.004980661 | 1 |
| T2DM | b_cell | *DYRK4* | 0.745591307 | 0.001414428 | 0.004980661 | 1 |
| T2DM | b_cell | *NCAPD2* | -1.397930233 | 0.00639351 | 0.008817882 | 1 |
| T2DM | b_cell | *FUZ* | -1.958163353 | 0.00302735 | 0.005219866 | 1 |
| T2DM | b_cell | *LRRC23* | -1.221971486 | 0.003216741 | 0.005343249 | 1 |
| T2DM | b_cell | *CLCN6* | -1.574687317 | 0.001578125 | 0.004980661 | 1 |
| T2DM | b_cell | *SLC6A7* | 11.84633079 | 0.00351167 | 0.005621511 | 1 |
| T2DM | b_cell | *TSPAN9* | -0.950702858 | 0.000639951 | 0.004980661 | 1 |

| | | | | | | |
|---|---|---|---|---|---|---|
| T2DM | b_cell | *TTC19* | -0.979204773 | 0.002078384 | 0.004980661 | 1 |
| T2DM | b_cell | *LARS2* | -1.614530787 | 0.004975746 | 0.00716602 | 1 |
| T2DM | b_cell | *DCN* | -0.644599772 | 0.001650024 | 0.004980661 | 1 |
| T2DM | b_cell | *PPP5C* | -1.88018658 | 0.013491692 | 0.016854194 | 1 |
| T2DM | b_cell | *CEP68* | -1.048776237 | 0.024327293 | 0.028973006 | 1 |
| T2DM | b_cell | *ZBTB32* | -1.549476243 | 0.002519644 | 0.004980661 | 1 |
| T2DM | b_cell | *ERCC1* | -1.492168978 | 0.004310564 | 0.006457562 | 1 |
| T2DM | b_cell | *CD22* | 0.21262382 | 0.001915361 | 0.004980661 | 1 |
| T2DM | b_cell | *ALOX5* | -1.700637395 | 0.003353505 | 0.005460611 | 1 |
| T2DM | b_cell | *MAP4K5* | -1.521998197 | 0.003649788 | 0.005752881 | 1 |
| T2DM | b_cell | *EHD3* | 0.707241925 | 0.002589814 | 0.004980661 | 1 |
| T2DM | b_cell | *MAN2B2* | -2.280397777 | 0.004047368 | 0.006163369 | 1 |
| T2DM | b_cell | *SLC25A39* | -1.492541295 | 0.002835502 | 0.005067351 | 1 |
| T2DM | b_cell | *RNF14* | -1.643473209 | 0.003226457 | 0.005352058 | 1 |
| T2DM | b_cell | *DDX11* | 1.010093374 | 0.011125898 | 0.014161295 | 1 |
| T2DM | b_cell | *TACC3* | -1.60808759 | 0.001745829 | 0.004980661 | 1 |
| T2DM | b_cell | *ZC3H3* | -1.084709189 | 0.003065894 | 0.005236085 | 1 |
| T2DM | b_cell | *MDH1* | -0.99416515 | 0.001515389 | 0.004980661 | 1 |
| T2DM | b_cell | *COX15* | -1.090348314 | 0.002089176 | 0.004980661 | 1 |
| T2DM | b_cell | *CCDC88C* | -1.226051373 | 0.002298058 | 0.004980661 | 1 |
| T2DM | b_cell | *NUDCD3* | -0.87588482 | 0.00157439 | 0.004980661 | 1 |
| T2DM | b_cell | *CHDH* | 2.708226421 | 0.004051938 | 0.006164943 | 1 |
| T2DM | b_cell | *CLCA4* | -6.743894762 | 0.008357877 | 0.010983862 | 1 |
| T2DM | b_cell | *RUFY3* | -0.930059903 | 0.002282664 | 0.004980661 | 1 |
| T2DM | b_cell | *SLC11A1* | -1.427636115 | 0.001138466 | 0.004980661 | 1 |
| T2DM | b_cell | *WWTR1* | -1.045731892 | 0.002087913 | 0.004980661 | 1 |
| T2DM | b_cell | *AGPS* | -1.169178514 | 0.003241434 | 0.005362236 | 1 |
| T2DM | b_cell | *TTC27* | -0.903253482 | 0.001833954 | 0.004980661 | 1 |
| T2DM | b_cell | *MARCO* | -0.759788814 | 0.005757294 | 0.008086473 | 1 |
| T2DM | b_cell | *HGF* | -1.667036572 | 0.002432979 | 0.004980661 | 1 |
| T2DM | b_cell | *SLC7A9* | -1.892996234 | 0.012078414 | 0.015269532 | 1 |
| T2DM | b_cell | *OSBPL5* | -1.616638327 | 0.005356817 | 0.007622076 | 1 |
| T2DM | b_cell | *CPS1* | -1.473082016 | 0.00181164 | 0.004980661 | 1 |
| T2DM | b_cell | *NLRP2* | -1.26186274 | 0.038676087 | 0.044797416 | 1 |
| T2DM | b_cell | *SLC45A4* | -1.24305213 | 0.001165005 | 0.004980661 | 1 |
| T2DM | b_cell | *ZDHHC6* | -1.238524473 | 0.002180701 | 0.004980661 | 1 |
| T2DM | b_cell | *GRAMD1B* | -1.674564953 | 0.002467951 | 0.004980661 | 1 |
| T2DM | b_cell | *RNH1* | -1.171654968 | 0.003481137 | 0.005589525 | 1 |
| T2DM | b_cell | *GLRX2* | -1.635066459 | 0.001587372 | 0.004980661 | 1 |
| T2DM | b_cell | *ABCC2* | 2.436039587 | 0.016777849 | 0.020520828 | 1 |
| T2DM | b_cell | *GCLM* | -1.238938262 | 0.001811113 | 0.004980661 | 1 |
| T2DM | b_cell | *UBR2* | -1.047188916 | 0.002001899 | 0.004980661 | 1 |
| T2DM | b_cell | *RRAGD* | -1.668426589 | 0.00248077 | 0.004980661 | 1 |
| T2DM | b_cell | *HSF2* | -1.734988552 | 0.012303501 | 0.015505607 | 1 |
| T2DM | b_cell | *TYMP* | -1.346046916 | 0.003363665 | 0.005463988 | 1 |
| T2DM | b_cell | *TOMM34* | 1.384241704 | 0.006556744 | 0.008979205 | 1 |
| T2DM | b_cell | *SEC63* | -1.287488315 | 0.002484672 | 0.004980661 | 1 |
| T2DM | b_cell | *FAS* | -1.331329182 | 0.001923053 | 0.004980661 | 1 |
| T2DM | b_cell | *RNASET2* | -1.298985459 | 0.000402592 | 0.004980661 | 1 |
| T2DM | b_cell | *BTN3A1* | -1.623618806 | 0.001622323 | 0.004980661 | 1 |

| | | | | | | |
|---|---|---|---|---|---|---|
| T2DM | b_cell | *MIPEP* | -1.006845851 | 0.001499516 | 0.004980661 | 1 |
| T2DM | b_cell | *INSRR* | -2.826637742 | 0.001160721 | 0.004980661 | 1 |
| T2DM | b_cell | *B4GALT7* | -1.982578344 | 0.001800927 | 0.004980661 | 1 |
| T2DM | b_cell | *SH2D2A* | -4.488374605 | 0.005517169 | 0.007802188 | 1 |
| T2DM | b_cell | *VRK2* | -0.918541746 | 0.001875833 | 0.004980661 | 1 |
| T2DM | b_cell | *VEZT* | -1.180503135 | 0.002311129 | 0.004980661 | 1 |
| T2DM | b_cell | *BRD9* | -1.133057884 | 0.00215349 | 0.004980661 | 1 |
| T2DM | b_cell | *SNX1* | -1.190690766 | 0.002015276 | 0.004980661 | 1 |
| T2DM | b_cell | *ANK1* | -2.148226093 | 0.001893885 | 0.004980661 | 1 |
| T2DM | b_cell | *RABEP1* | -1.448062167 | 0.004189984 | 0.006324704 | 1 |
| T2DM | b_cell | *BAK1* | 3.11027328 | 0.001012065 | 0.004980661 | 1 |
| T2DM | b_cell | *IKZF2* | -0.978537231 | 0.0017872 | 0.004980661 | 1 |
| T2DM | b_cell | *GRN* | -1.420452847 | 0.005401032 | 0.007675983 | 1 |
| T2DM | b_cell | *IFT88* | -0.978168366 | 0.001418344 | 0.004980661 | 1 |
| T2DM | b_cell | *CHPF2* | -1.718300383 | 0.003208516 | 0.005339692 | 1 |
| T2DM | b_cell | *UBA6* | -1.553737852 | 0.000919358 | 0.004980661 | 1 |
| T2DM | b_cell | *ATP6V0A1* | -1.308719183 | 0.005650247 | 0.007957909 | 1 |
| T2DM | b_cell | *PIAS1* | -0.869270829 | 0.002349059 | 0.004980661 | 1 |
| T2DM | b_cell | *MYOC* | -1.428934983 | 0.003419252 | 0.005517197 | 1 |
| T2DM | b_cell | *SH3YL1* | -1.974291334 | 0.001130538 | 0.004980661 | 1 |
| T2DM | b_cell | *VCL* | -0.970159218 | 0.001787217 | 0.004980661 | 1 |
| T2DM | b_cell | *ADSS* | -1.547577778 | 0.002494982 | 0.004980661 | 1 |
| T2DM | b_cell | *TBC1D23* | -1.050634874 | 0.001905966 | 0.004980661 | 1 |
| T2DM | b_cell | *MYOM2* | -2.723731092 | 0.003511706 | 0.005621511 | 1 |
| T2DM | b_cell | *TUBG2* | -3.180773078 | 3.28E-05 | 0.004719449 | 0.158977336 |
| T2DM | b_cell | *FLT4* | -1.810730965 | 9.98E-05 | 0.004980661 | 0.484283213 |
| T2DM | b_cell | *NSUN2* | -1.480695115 | 0.001886794 | 0.004980661 | 1 |
| T2DM | b_cell | *MFAP3* | -1.104940713 | 0.002374693 | 0.004980661 | 1 |
| T2DM | b_cell | *MRI1* | -1.07967276 | 0.001545317 | 0.004980661 | 1 |
| T2DM | b_cell | *AGA* | 1.742130041 | 0.019214114 | 0.0232602 | 1 |
| T2DM | b_cell | *PI4K2B* | 0.64332819 | 0.00266014 | 0.004985914 | 1 |
| T2DM | b_cell | *BOD1L1* | -1.623522952 | 0.005513135 | 0.00780103 | 1 |
| T2DM | b_cell | *DNAH5* | -2.063820155 | 0.002209548 | 0.004980661 | 1 |
| T2DM | b_cell | *PNKP* | -1.46567917 | 0.001142147 | 0.004980661 | 1 |
| T2DM | b_cell | *PHLPP2* | -0.974567323 | 0.001136247 | 0.004980661 | 1 |
| T2DM | b_cell | *SPDL1* | -2.117207409 | 0.001911144 | 0.004980661 | 1 |
| T2DM | b_cell | *PQLC2* | -3.213323153 | 0.002141266 | 0.004980661 | 1 |
| T2DM | b_cell | *CTNS* | -1.487431897 | 0.005455723 | 0.007735583 | 1 |
| T2DM | b_cell | *INPP4A* | -1.333851711 | 0.002294645 | 0.004980661 | 1 |
| T2DM | b_cell | *PSMA4* | -1.186589939 | 0.00163509 | 0.004980661 | 1 |
| T2DM | b_cell | *LSG1* | -1.646218769 | 0.030414872 | 0.035749263 | 1 |
| T2DM | b_cell | *SPATA7* | -0.938191452 | 0.001586338 | 0.004980661 | 1 |
| T2DM | b_cell | *CAPG* | -1.725141513 | 0.000846735 | 0.004980661 | 1 |
| T2DM | b_cell | *AP2S1* | -1.903968818 | 0.002096497 | 0.004980661 | 1 |
| T2DM | b_cell | *TRIT1* | -1.488298104 | 0.001736074 | 0.004980661 | 1 |
| T2DM | b_cell | *CUL7* | -2.5955087 | 0.022080428 | 0.026459431 | 1 |
| T2DM | b_cell | *CTNNA1* | -1.163529358 | 0.002554472 | 0.004980661 | 1 |
| T2DM | b_cell | *CNTLN* | -1.051261881 | 0.001663572 | 0.004980661 | 1 |
| T2DM | b_cell | *YTHDC2* | -0.966188555 | 0.001907117 | 0.004980661 | 1 |
| T2DM | b_cell | *POLR2B* | -1.456389261 | 0.002564615 | 0.004980661 | 1 |

| | | | | | | |
|---|---|---|---|---|---|---|
| T2DM | b_cell | *ARAP2* | -0.85063408 | 0.002158997 | 0.004980661 | 1 |
| T2DM | b_cell | *DTNBP1* | -1.401923139 | 0.001844455 | 0.004980661 | 1 |
| T2DM | b_cell | *ANO2* | -0.951760862 | 0.0018782 | 0.004980661 | 1 |
| T2DM | b_cell | *RRM2B* | -1.489952799 | 0.00198611 | 0.004980661 | 1 |
| T2DM | b_cell | *SNX29* | -1.332974439 | 0.003524605 | 0.005637244 | 1 |
| T2DM | b_cell | *MRPS10* | -1.671424618 | 0.000510861 | 0.004980661 | 1 |
| T2DM | b_cell | *UTS2* | 1.27688935 | 0.004319364 | 0.006464391 | 1 |
| T2DM | b_cell | *EPN3* | -7.231618268 | 0.000809961 | 0.004980661 | 1 |
| T2DM | b_cell | *JKAMP* | -0.98869875 | 0.001601876 | 0.004980661 | 1 |
| T2DM | b_cell | *ARHGEF5* | -1.531143672 | 0.027622942 | 0.032665492 | 1 |
| T2DM | b_cell | *LAMC3* | -2.626910796 | 0.000964447 | 0.004980661 | 1 |
| T2DM | b_cell | *MAPK9* | -1.891215061 | 0.003489816 | 0.005595699 | 1 |
| T2DM | b_cell | *FAM160A2* | -1.102147398 | 0.00231445 | 0.004980661 | 1 |
| T2DM | b_cell | *THOC3* | -2.777760583 | 0.001843524 | 0.004980661 | 1 |
| T2DM | b_cell | *ALX4* | 1.528552514 | 0.00719515 | 0.009697464 | 1 |
| T2DM | b_cell | *FSTL4* | -2.327928429 | 0.002570349 | 0.004980661 | 1 |
| T2DM | b_cell | *AKR7A2* | -1.423553635 | 0.002171349 | 0.004980661 | 1 |
| T2DM | b_cell | *MRTO4* | -1.771685524 | 0.000967636 | 0.004980661 | 1 |
| T2DM | b_cell | *ANAPC4* | -0.963232179 | 0.002292587 | 0.004980661 | 1 |
| T2DM | b_cell | *LY75* | 3.348230892 | 0.008609961 | 0.011290685 | 1 |
| T2DM | b_cell | *OPN3* | 1.623444061 | 0.001509327 | 0.004980661 | 1 |
| T2DM | b_cell | *SDCCAG8* | -1.029983689 | 0.002831588 | 0.005067351 | 1 |
| T2DM | b_cell | *KIF1B* | -0.77908266 | 0.002866015 | 0.005094011 | 1 |
| T2DM | b_cell | *TBC1D22A* | -1.095444299 | 0.002900583 | 0.005113643 | 1 |
| T2DM | b_cell | *PLEKHH1* | -2.00086423 | 0.001142609 | 0.004980661 | 1 |
| T2DM | b_cell | *GALC* | -1.086218822 | 0.001516049 | 0.004980661 | 1 |
| T2DM | b_cell | *FAM114A2* | -1.240639905 | 0.002519032 | 0.004980661 | 1 |
| T2DM | b_cell | *GINM1* | -1.806399304 | 0.004622646 | 0.006788462 | 1 |
| T2DM | b_cell | *ITIH4* | 2.230367665 | 0.000946274 | 0.004980661 | 1 |
| T2DM | b_cell | *NPFFR2* | -2.567215074 | 0.001765913 | 0.004980661 | 1 |
| T2DM | b_cell | *PCGF2* | 1.99396927 | 0.010697603 | 0.01365915 | 1 |
| T2DM | b_cell | *IL17RB* | -3.419080137 | 0.001018762 | 0.004980661 | 1 |
| T2DM | b_cell | *MSH4* | -1.876602775 | 0.006137412 | 0.008532586 | 1 |
| T2DM | b_cell | *GDI2* | -1.519475611 | 0.003010277 | 0.005201518 | 1 |
| T2DM | b_cell | *PITHD1* | -1.887394178 | 0.001773578 | 0.004980661 | 1 |
| T2DM | b_cell | *ATP11B* | -1.404415427 | 0.006313702 | 0.008732634 | 1 |
| T2DM | b_cell | *LAMC2* | 1.378605759 | 0.015654895 | 0.019283461 | 1 |
| T2DM | b_cell | *RASGRF1* | -3.93662635 | 0.001216106 | 0.004980661 | 1 |
| T2DM | b_cell | *CROCC* | -1.586651611 | 0.000813685 | 0.004980661 | 1 |
| T2DM | b_cell | *FLYWCH1* | -1.059987046 | 0.001119242 | 0.004980661 | 1 |
| T2DM | b_cell | *ALDH18A1* | -1.738058124 | 0.001227498 | 0.004980661 | 1 |
| T2DM | b_cell | *DNAJC25* | -1.777399934 | 0.001822761 | 0.004980661 | 1 |
| T2DM | b_cell | *OGFR* | -1.224486391 | 0.002572242 | 0.004980661 | 1 |
| T2DM | b_cell | *GNA15* | -1.37148962 | 0.032101459 | 0.037640473 | 1 |
| T2DM | b_cell | *PIGV* | -2.096683282 | 0.000171798 | 0.004980661 | 0.833562567 |
| T2DM | b_cell | *RIMBP2* | -1.689569908 | 0.001768268 | 0.004980661 | 1 |
| T2DM | b_cell | *PRDM6* | -1.47791668 | 0.002022804 | 0.004980661 | 1 |
| T2DM | b_cell | *SPAG4* | 0.814324203 | 0.003137335 | 0.00528187 | 1 |
| T2DM | b_cell | *NCKAP1* | -0.847121985 | 0.001632431 | 0.004980661 | 1 |
| T2DM | b_cell | *LTK* | -12.79821735 | 0.001915665 | 0.004980661 | 1 |

| | | | | | | |
|---|---|---|---|---|---|---|
| T2DM | b_cell | *EIF4B* | -1.484060713 | 0.001422965 | 0.004980661 | 1 |
| T2DM | b_cell | *SLC6A16* | -1.63293473 | 0.005052335 | 0.007261234 | 1 |
| T2DM | b_cell | *AHRR* | 1.356099052 | 0.003703088 | 0.0057978 | 1 |
| T2DM | b_cell | *ADCK1* | -1.419548391 | 0.001821075 | 0.004980661 | 1 |
| T2DM | b_cell | *HAGH* | -1.013225709 | 0.0011946 | 0.004980661 | 1 |
| T2DM | b_cell | *RNF4* | -1.583937202 | 0.002532175 | 0.004980661 | 1 |
| T2DM | b_cell | *CASP8* | -1.169022533 | 0.00241356 | 0.004980661 | 1 |
| T2DM | b_cell | *TM7SF3* | -1.447705332 | 0.005716828 | 0.008037684 | 1 |
| T2DM | b_cell | *CDON* | -0.591404626 | 0.002389298 | 0.004980661 | 1 |
| T2DM | b_cell | *TMEM161A* | -1.80978505 | 0.002185669 | 0.004980661 | 1 |
| T2DM | b_cell | *LPAR2* | 2.054173003 | 0.002845392 | 0.005071948 | 1 |
| T2DM | b_cell | *SUGP2* | -1.673306741 | 0.004544414 | 0.006687745 | 1 |
| T2DM | b_cell | *SLC12A2* | -1.04578464 | 0.001902945 | 0.004980661 | 1 |
| T2DM | b_cell | *CNN2* | -1.624731007 | 0.020407801 | 0.024619257 | 1 |
| T2DM | b_cell | *ABCA7* | -1.887486915 | 0.000767662 | 0.004980661 | 1 |
| T2DM | b_cell | *BTBD1* | -1.609698316 | 0.006563868 | 0.008986425 | 1 |
| T2DM | b_cell | *FAR2* | -1.741092434 | 0.011461057 | 0.014568699 | 1 |
| T2DM | b_cell | *BCAS1* | 0.279644165 | 0.002428523 | 0.004980661 | 1 |
| T2DM | b_cell | *CHI3L2* | -4.640094 | 0.004818098 | 0.006983587 | 1 |
| T2DM | b_cell | *SBNO2* | -1.17304174 | 0.002256645 | 0.004980661 | 1 |
| T2DM | b_cell | *PMS1* | -0.895155384 | 0.002064136 | 0.004980661 | 1 |
| T2DM | b_cell | *UHRF1BP1* | -0.894644475 | 0.001784519 | 0.004980661 | 1 |
| T2DM | b_cell | *WDR18* | -2.069961615 | 0.000693831 | 0.004980661 | 1 |
| T2DM | b_cell | *ANKRD44* | -1.131069784 | 0.002999515 | 0.005195812 | 1 |
| T2DM | b_cell | *ADAT1* | -1.321525393 | 0.00469522 | 0.006851467 | 1 |
| T2DM | b_cell | *MAP2K4* | -1.315691647 | 0.002140833 | 0.004980661 | 1 |
| T2DM | b_cell | *TMEM206* | -1.186058073 | 0.002093763 | 0.004980661 | 1 |
| T2DM | b_cell | *SEC61A2* | -1.321199905 | 0.000426042 | 0.004980661 | 1 |
| T2DM | b_cell | *TIE1* | -1.19130339 | 0.003343251 | 0.005450757 | 1 |
| T2DM | b_cell | *NGEF* | -4.865738791 | 0.001921807 | 0.004980661 | 1 |
| T2DM | b_cell | *SPI1* | -1.283877463 | 0.001902738 | 0.004980661 | 1 |
| T2DM | b_cell | *ZNRD1* | -2.942931372 | 0.021012949 | 0.025261355 | 1 |
| T2DM | b_cell | *ATXN3* | -0.856194464 | 0.003324785 | 0.005435262 | 1 |
| T2DM | b_cell | *GOLGA5* | -1.068934652 | 0.001892554 | 0.004980661 | 1 |
| T2DM | b_cell | *FGFR2* | -1.633048317 | 0.000340885 | 0.004980661 | 1 |
| T2DM | b_cell | *MTFR1* | -1.500893093 | 0.001440554 | 0.004980661 | 1 |
| T2DM | b_cell | *MYO9A* | -0.931974559 | 0.002318124 | 0.004980661 | 1 |
| T2DM | b_cell | *EVI5* | -1.042354889 | 0.001705005 | 0.004980661 | 1 |
| T2DM | b_cell | *STOML1* | -2.546614576 | 0.00179049 | 0.004980661 | 1 |
| T2DM | b_cell | *METTL22* | -1.231997343 | 0.001499548 | 0.004980661 | 1 |
| T2DM | b_cell | *TP53BP1* | -1.556263771 | 0.003710289 | 0.005805329 | 1 |
| T2DM | b_cell | *RRP15* | -1.026875313 | 0.002441712 | 0.004980661 | 1 |
| T2DM | b_cell | *DHX8* | -1.120027131 | 0.001792263 | 0.004980661 | 1 |
| T2DM | b_cell | *CBFB* | -1.537831755 | 0.003959872 | 0.006063361 | 1 |
| T2DM | b_cell | *FGFR3* | -2.232442968 | 0.03582837 | 0.041698069 | 1 |
| T2DM | b_cell | *HEATR6* | -1.980877187 | 0.005075628 | 0.007285925 | 1 |
| T2DM | b_cell | *POLR1A* | -2.398922539 | 0.001057187 | 0.004980661 | 1 |
| T2DM | b_cell | *IP6K2* | -1.092502888 | 0.001421545 | 0.004980661 | 1 |
| T2DM | b_cell | *STON1-GTF2A* | -1.259955209 | 0.000625546 | 0.004980661 | 1 |
| T2DM | b_cell | *SRBD1* | -1.30719953 | 0.00260817 | 0.004980661 | 1 |

| | | | | | | |
|---|---|---|---|---|---|---|
| T2DM | b_cell | *CYFIP1* | -1.339707846 | 0.002225989 | 0.004980661 | 1 |
| T2DM | b_cell | *IFT80* | 1.416419185 | 0.016147133 | 0.019809327 | 1 |
| T2DM | b_cell | *NUCKS1* | -1.207002309 | 0.002706752 | 0.00500969 | 1 |
| T2DM | b_cell | *CLEC2D* | -1.579648471 | 0.002413858 | 0.004980661 | 1 |
| T2DM | b_cell | *DRD4* | -4.046547506 | 0.00224565 | 0.004980661 | 1 |
| T2DM | b_cell | *PLA2G10* | 0.47327157 | 0.002493444 | 0.004980661 | 1 |
| T2DM | b_cell | *NEDD4* | -0.895863714 | 0.002563695 | 0.004980661 | 1 |
| T2DM | b_cell | *PIGB* | -1.66895924 | 0.001873836 | 0.004980661 | 1 |
| T2DM | b_cell | *GNB5* | -1.169654442 | 0.00640971 | 0.008823536 | 1 |
| T2DM | b_cell | *LRP6* | -1.35732869 | 0.002341341 | 0.004980661 | 1 |
| T2DM | b_cell | *CLTCL1* | -1.463395592 | 0.001724811 | 0.004980661 | 1 |
| T2DM | b_cell | *DGCR2* | -1.570972894 | 0.002497925 | 0.004980661 | 1 |
| T2DM | b_cell | *ZXDC* | -0.857405986 | 0.001799453 | 0.004980661 | 1 |
| T2DM | b_cell | *WIPI1* | -1.372872219 | 0.000668523 | 0.004980661 | 1 |
| T2DM | b_cell | *GBA2* | -1.246615042 | 0.001662317 | 0.004980661 | 1 |
| T2DM | b_cell | *NDST1* | -1.090448124 | 0.002542397 | 0.004980661 | 1 |
| T2DM | b_cell | *ASNS* | -3.24855451 | 0.007664696 | 0.010216787 | 1 |
| T2DM | b_cell | *AP3M2* | -1.497303238 | 0.001905399 | 0.004980661 | 1 |
| T2DM | b_cell | *ST6GALNAC2* | -1.657017558 | 0.002094646 | 0.004980661 | 1 |
| T2DM | b_cell | *EIF2B3* | -1.044864823 | 0.001770653 | 0.004980661 | 1 |
| T2DM | b_cell | *MAP4K4* | -0.96520335 | 0.002007081 | 0.004980661 | 1 |
| T2DM | b_cell | *SNX13* | -1.23585204 | 0.002532261 | 0.004980661 | 1 |
| T2DM | b_cell | *RPS6KA2* | -1.415607871 | 0.003038897 | 0.005228627 | 1 |
| T2DM | b_cell | *ING3* | -2.0930002 | 0.004324286 | 0.006465773 | 1 |
| T2DM | b_cell | *VASH1* | -1.168687677 | 0.001700847 | 0.004980661 | 1 |
| T2DM | b_cell | *CYBRD1* | -1.060070535 | 0.002640503 | 0.004985914 | 1 |
| T2DM | b_cell | *RDH11* | -1.177546045 | 0.001899406 | 0.004980661 | 1 |
| T2DM | b_cell | *EPN2* | -1.116085126 | 0.00120358 | 0.004980661 | 1 |
| T2DM | b_cell | *ALDH3A2* | -1.04175672 | 0.002107652 | 0.004980661 | 1 |
| T2DM | b_cell | *AFF4* | -1.043568885 | 0.002313754 | 0.004980661 | 1 |
| T2DM | b_cell | *TRHDE* | -0.733203689 | 0.001583889 | 0.004980661 | 1 |
| T2DM | b_cell | *FCGR2B* | -1.331140717 | 0.001769265 | 0.004980661 | 1 |
| T2DM | b_cell | *NFATC3* | -1.034276352 | 0.003082151 | 0.005245002 | 1 |
| T2DM | b_cell | *TRNT1* | -1.509337036 | 0.002330029 | 0.004980661 | 1 |
| T2DM | b_cell | *CRMP1* | -1.081906392 | 0.001757185 | 0.004980661 | 1 |
| T2DM | b_cell | *NDE1* | -1.9052724 | 0.002185226 | 0.004980661 | 1 |
| T2DM | b_cell | *AP1M1* | -2.393604288 | 0.007022747 | 0.009506265 | 1 |
| T2DM | b_cell | *PVR* | -1.77253533 | 0.00073441 | 0.004980661 | 1 |
| T2DM | b_cell | *SCARB1* | -1.708058377 | 0.008896159 | 0.011606903 | 1 |
| T2DM | b_cell | *MOV10L1* | -1.856800542 | 0.002196693 | 0.004980661 | 1 |
| T2DM | b_cell | *ALPK1* | -0.912798463 | 0.002435036 | 0.004980661 | 1 |
| T2DM | b_cell | *LLGL2* | -1.352410274 | 0.001342765 | 0.004980661 | 1 |
| T2DM | b_cell | *SDHA* | -1.096306791 | 0.005416718 | 0.007693769 | 1 |
| T2DM | b_cell | *SMARCE1* | -2.04537755 | 0.034536974 | 0.040330541 | 1 |
| T2DM | b_cell | *ADAM11* | -2.446327801 | 0.009214496 | 0.0119408 | 1 |
| T2DM | b_cell | *PPP2R2C* | -2.443413864 | 0.001730592 | 0.004980661 | 1 |
| T2DM | b_cell | *ZNF532* | -1.253763667 | 0.002343503 | 0.004980661 | 1 |
| T2DM | b_cell | *ZZEF1* | -1.053057723 | 0.004079837 | 0.006201557 | 1 |
| T2DM | b_cell | *TIPIN* | -1.165834772 | 0.035009958 | 0.04083373 | 1 |
| T2DM | b_cell | *TTC38* | -1.02798462 | 0.001963582 | 0.004980661 | 1 |

| | | | | | | |
|---|---|---|---|---|---|---|
| T2DM | b_cell | *GRAMD4* | -1.132181501 | 0.00159127 | 0.004980661 | 1 |
| T2DM | b_cell | *ADD2* | -1.9185521 | 0.004942171 | 0.007126125 | 1 |
| T2DM | b_cell | *VPS9D1* | -1.204543371 | 0.002027311 | 0.004980661 | 1 |
| T2DM | b_cell | *FNDC3B* | -0.988135779 | 0.002768234 | 0.005045632 | 1 |
| T2DM | b_cell | *MOCOS* | 2.978254289 | 0.034202649 | 0.039949748 | 1 |
| T2DM | b_cell | *SEC31B* | -1.102475458 | 0.007752982 | 0.010311805 | 1 |
| T2DM | b_cell | *KIFAP3* | -1.702841869 | 0.0078842 | 0.010457665 | 1 |
| T2DM | b_cell | *MKRN2* | -2.443119863 | 0.015092189 | 0.018628161 | 1 |
| T2DM | b_cell | *MCM6* | -1.551169716 | 0.002328294 | 0.004980661 | 1 |
| T2DM | b_cell | *MLH1* | -1.478552717 | 0.001632127 | 0.004980661 | 1 |
| T2DM | b_cell | *FMO4* | -1.785241518 | 0.003901011 | 0.006006888 | 1 |
| T2DM | b_cell | *RGS11* | -1.364176089 | 0.002277484 | 0.004980661 | 1 |
| T2DM | b_cell | *SPAG5* | 2.962526856 | 0.006692468 | 0.009141851 | 1 |
| T2DM | b_cell | *ANKRD13A* | -1.50596977 | 0.002137767 | 0.004980661 | 1 |
| T2DM | b_cell | *PAG1* | -1.631175859 | 0.003486011 | 0.005593296 | 1 |
| T2DM | b_cell | *GPATCH1* | -2.055909696 | 0.002546818 | 0.004980661 | 1 |
| T2DM | b_cell | *ICAM3* | -1.962684518 | 0.002583727 | 0.004980661 | 1 |
| T2DM | b_cell | *NT5C2* | -0.994806023 | 0.005772796 | 0.008099943 | 1 |
| T2DM | b_cell | *XAB2* | -3.313603753 | 0.005888186 | 0.008240403 | 1 |
| T2DM | b_cell | *IL4R* | -1.184677591 | 0.001930839 | 0.004980661 | 1 |
| T2DM | b_cell | *DYNC1I2* | -1.16488986 | 0.002111768 | 0.004980661 | 1 |
| T2DM | b_cell | *APBB1IP* | -0.748373245 | 0.001720124 | 0.004980661 | 1 |
| T2DM | b_cell | *SIRT6* | -1.721116111 | 0.006414105 | 0.008826216 | 1 |
| T2DM | b_cell | *GPR137B* | -1.245469747 | 0.001844344 | 0.004980661 | 1 |
| T2DM | b_cell | *NAALAD2* | -1.001149146 | 0.008053446 | 0.010647226 | 1 |
| T2DM | b_cell | *AMPH* | -1.543637491 | 0.002019004 | 0.004980661 | 1 |
| T2DM | b_cell | *MCCC1* | -0.78777822 | 0.002572467 | 0.004980661 | 1 |
| T2DM | b_cell | *ACER3* | -0.790868586 | 0.000996335 | 0.004980661 | 1 |
| T2DM | b_cell | *PIK3C3* | -1.163931143 | 0.002083871 | 0.004980661 | 1 |
| T2DM | b_cell | *SYNJ2* | -1.188916025 | 0.002405655 | 0.004980661 | 1 |
| T2DM | b_cell | *RBFOX1* | -0.828311407 | 0.001316759 | 0.004980661 | 1 |
| T2DM | b_cell | *FGF20* | -13.66698208 | 0.006725516 | 0.00917666 | 1 |
| T2DM | b_cell | *PCM1* | -0.985774736 | 0.002584652 | 0.004980661 | 1 |
| T2DM | b_cell | *MYH7B* | 1.802381734 | 0.001897268 | 0.004980661 | 1 |
| T2DM | b_cell | *BPIFB2* | -17.56976566 | 0.014911228 | 0.018442334 | 1 |
| T2DM | b_cell | *CLUL1* | -3.26994214 | 0.001586614 | 0.004980661 | 1 |
| T2DM | b_cell | *XRCC5* | -1.086426179 | 0.002310124 | 0.004980661 | 1 |
| T2DM | b_cell | *SP140* | 1.363862873 | 0.027591111 | 0.032635805 | 1 |
| T2DM | b_cell | *SAR1A* | -1.033433674 | 0.001989531 | 0.004980661 | 1 |
| T2DM | b_cell | *RAPGEF3* | -1.110028016 | 0.0026764 | 0.004998428 | 1 |
| T2DM | b_cell | *FDFT1* | -1.063603038 | 0.004589628 | 0.006746099 | 1 |
| T2DM | b_cell | *SCGN* | 6.526645003 | 0.004471198 | 0.006626223 | 1 |
| T2DM | b_cell | *PGM1* | -1.024403046 | 0.001797331 | 0.004980661 | 1 |
| T2DM | b_cell | *DDX1* | -0.945584761 | 0.001937174 | 0.004980661 | 1 |
| T2DM | b_cell | *DNM2* | -1.346869338 | 0.002565682 | 0.004980661 | 1 |
| T2DM | b_cell | *STX7* | -1.156848309 | 0.001868689 | 0.004980661 | 1 |
| T2DM | b_cell | *RABL2B* | -1.243223266 | 0.00182039 | 0.004980661 | 1 |
| T2DM | b_cell | *DCT* | -2.586793534 | 9.99E-05 | 0.004980661 | 0.484901558 |
| T2DM | b_cell | *SCTR* | -1.193085249 | 0.001778523 | 0.004980661 | 1 |
| T2DM | b_cell | *RIF1* | -1.051680135 | 0.001848671 | 0.004980661 | 1 |

| | | | | | | |
|---|---|---|---|---|---|---|
| T2DM | b_cell | *KCNN2* | -3.319709481 | 0.009151109 | 0.011871973 | 1 |
| T2DM | b_cell | *PSEN1* | -1.213533256 | 0.00250275 | 0.004980661 | 1 |
| T2DM | b_cell | *MOK* | -1.61561876 | 0.002326036 | 0.004980661 | 1 |
| T2DM | b_cell | *STK17B* | -0.881623626 | 0.003548323 | 0.005652384 | 1 |
| T2DM | b_cell | *ZNF506* | -1.605993082 | 0.010314827 | 0.013208641 | 1 |
| T2DM | b_cell | *PCDHB4* | -2.69864639 | 0.001825084 | 0.004980661 | 1 |
| T2DM | b_cell | *ME2* | -0.99995774 | 0.001808485 | 0.004980661 | 1 |
| T2DM | b_cell | *CCNT2* | -1.085791912 | 0.004309703 | 0.006457562 | 1 |
| T2DM | b_cell | *SEMA5B* | -1.597695853 | 0.005664496 | 0.007971043 | 1 |
| T2DM | b_cell | *DOPEY1* | -0.849665887 | 0.003151685 | 0.00530235 | 1 |
| T2DM | b_cell | *LYRM2* | -1.991612119 | 0.019693756 | 0.023805208 | 1 |
| T2DM | b_cell | *ZCCHC6* | -1.500513898 | 0.003658948 | 0.005752881 | 1 |
| T2DM | b_cell | *ULK2* | -0.89600896 | 0.001775577 | 0.004980661 | 1 |
| T2DM | b_cell | *PLOD1* | -1.341840148 | 0.001918233 | 0.004980661 | 1 |
| T2DM | b_cell | *PIBF1* | -0.935591707 | 0.002088607 | 0.004980661 | 1 |
| T2DM | b_cell | *OXCT1* | -0.982002609 | 0.002131018 | 0.004980661 | 1 |
| T2DM | b_cell | *SLC27A5* | -1.654282987 | 0.002587873 | 0.004980661 | 1 |
| T2DM | b_cell | *ZNF586* | -0.83887535 | 0.003110791 | 0.005260913 | 1 |
| T2DM | b_cell | *RPS5* | -0.893195217 | 0.001816537 | 0.004980661 | 1 |
| T2DM | b_cell | *PPIE* | -1.246994465 | 0.00196696 | 0.004980661 | 1 |
| T2DM | b_cell | *NOA1* | -1.611336082 | 0.00175245 | 0.004980661 | 1 |
| T2DM | b_cell | *GSTP1* | -1.640168658 | 0.008649001 | 0.011329631 | 1 |
| T2DM | b_cell | *WBP11* | -1.975309379 | 0.003044528 | 0.00522879 | 1 |
| T2DM | b_cell | *NCOA1* | -0.862127645 | 0.001931341 | 0.004980661 | 1 |
| T2DM | b_cell | *CAD* | -1.387885758 | 0.001737218 | 0.004980661 | 1 |
| T2DM | b_cell | *CD82* | 1.262748421 | 0.00111696 | 0.004980661 | 1 |
| T2DM | b_cell | *HACE1* | -1.911752278 | 0.006898284 | 0.009372857 | 1 |
| T2DM | b_cell | *SEH1L* | -2.172085552 | 0.024188991 | 0.028822442 | 1 |
| T2DM | b_cell | *OVGP1* | -4.388955439 | 4.48E-05 | 0.004980661 | 0.217446438 |
| T2DM | b_cell | *SLC25A24* | -1.613042186 | 0.006020507 | 0.008403769 | 1 |
| T2DM | b_cell | *ZNF213* | -1.187013861 | 0.00339652 | 0.005502475 | 1 |
| T2DM | b_cell | *CPNE3* | -1.481215252 | 0.004638232 | 0.006799003 | 1 |
| T2DM | b_cell | *RRN3* | -1.737134976 | 0.002737206 | 0.005019995 | 1 |
| T2DM | b_cell | *MTIF2* | -1.425678923 | 0.001844907 | 0.004980661 | 1 |
| T2DM | b_cell | *DDHD2* | -0.983180372 | 0.002756308 | 0.005034338 | 1 |
| T2DM | b_cell | *EPS15* | -0.908729849 | 0.002715755 | 0.005012112 | 1 |
| T2DM | b_cell | *USP40* | -0.88273646 | 0.001835856 | 0.004980661 | 1 |
| T2DM | b_cell | *POMGNT1* | -1.280736722 | 0.002096748 | 0.004980661 | 1 |
| T2DM | b_cell | *MAST2* | -1.3967998 | 0.003057889 | 0.005233209 | 1 |
| T2DM | b_cell | *EIF2AK1* | -1.029668959 | 0.002020567 | 0.004980661 | 1 |
| T2DM | b_cell | *NME8* | 2.7485588 | 0.002319992 | 0.004980661 | 1 |
| T2DM | b_cell | *SNX10* | -1.412363455 | 0.000211604 | 0.004980661 | 1 |
| T2DM | b_cell | *ITPKC* | -1.444427232 | 0.009655866 | 0.012450242 | 1 |
| T2DM | b_cell | *TMED2* | -1.908182862 | 0.002551315 | 0.004980661 | 1 |
| T2DM | b_cell | *ALG9* | -2.095943386 | 0.001967317 | 0.004980661 | 1 |
| T2DM | b_cell | *ACOX3* | -1.152943807 | 0.002129679 | 0.004980661 | 1 |
| T2DM | b_cell | *HSD17B14* | -1.848929215 | 0.002521883 | 0.004980661 | 1 |
| T2DM | b_cell | *TRIP6* | -1.789289377 | 0.001836531 | 0.004980661 | 1 |
| T2DM | b_cell | *SRRT* | -1.17683811 | 0.001604244 | 0.004980661 | 1 |
| T2DM | b_cell | *ADAMTS2* | -0.924586416 | 0.001777544 | 0.004980661 | 1 |

| | | | | | | |
|---|---|---|---|---|---|---|
| T2DM | b_cell | *PSMC5* | -1.900854947 | 0.001083789 | 0.004980661 | 1 |
| T2DM | b_cell | *UIMC1* | -1.463841854 | 0.002748784 | 0.00503214 | 1 |
| T2DM | b_cell | *LPCAT2* | -1.066711822 | 0.001826672 | 0.004980661 | 1 |
| T2DM | b_cell | *OGFOD1* | -1.236729993 | 0.001186246 | 0.004980661 | 1 |
| T2DM | b_cell | *TXNDC16* | -0.990681701 | 0.002120501 | 0.004980661 | 1 |
| T2DM | b_cell | *SF3B2* | -1.213483626 | 0.003194113 | 0.00533738 | 1 |
| T2DM | b_cell | *PHACTR3* | 0.513800787 | 0.003874872 | 0.005974223 | 1 |
| T2DM | b_cell | *AAMDC* | -1.300604175 | 0.000913133 | 0.004980661 | 1 |
| T2DM | b_cell | *GP6* | -1.662002828 | 0.001299575 | 0.004980661 | 1 |
| T2DM | b_cell | *ASAP3* | -1.920520369 | 0.00507385 | 0.007285925 | 1 |
| T2DM | b_cell | *EDEM2* | -2.719183858 | 0.002388844 | 0.004980661 | 1 |
| T2DM | b_cell | *ANKRD10* | -1.422585515 | 0.003656535 | 0.005752881 | 1 |
| T2DM | b_cell | *DOCK3* | -1.052268606 | 0.001774073 | 0.004980661 | 1 |
| T2DM | b_cell | *ARHGAP28* | -0.893077076 | 0.001691783 | 0.004980661 | 1 |
| T2DM | b_cell | *PPP1R13B* | -1.063223411 | 0.001302315 | 0.004980661 | 1 |
| T2DM | b_cell | *ZNF343* | -2.122235163 | 0.030463404 | 0.035797636 | 1 |
| T2DM | b_cell | *EBF4* | -1.858460895 | 0.001920872 | 0.004980661 | 1 |
| T2DM | b_cell | *CPXM1* | -1.462994124 | 0.001217572 | 0.004980661 | 1 |
| T2DM | b_cell | *MAVS* | -1.180251042 | 0.001916947 | 0.004980661 | 1 |
| T2DM | b_cell | *XRN2* | -1.132767909 | 0.001875752 | 0.004980661 | 1 |
| T2DM | b_cell | *SNX5* | -1.086929715 | 0.001798527 | 0.004980661 | 1 |
| T2DM | b_cell | *ESF1* | -0.869274321 | 0.002072562 | 0.004980661 | 1 |
| T2DM | b_cell | *RBBP9* | 1.652397164 | 0.020695774 | 0.024910914 | 1 |
| T2DM | b_cell | *TMEM230* | -1.478114455 | 0.002993229 | 0.005192402 | 1 |
| T2DM | b_cell | *RPH3A* | -3.85021097 | 0.002478875 | 0.004980661 | 1 |
| T2DM | b_cell | *KIF16B* | -1.548615876 | 0.003336774 | 0.005445196 | 1 |
| T2DM | b_cell | *TBX5* | -8.916436589 | 0.001203787 | 0.004980661 | 1 |
| T2DM | b_cell | *FXYD5* | 1.464254801 | 0.010232733 | 0.013127768 | 1 |
| T2DM | b_cell | *ZNF302* | -1.229515015 | 0.001553932 | 0.004980661 | 1 |
| T2DM | b_cell | *CDIP1* | -1.519079088 | 0.00181738 | 0.004980661 | 1 |
| T2DM | b_cell | *KCNH4* | 1.088767456 | 0.000740298 | 0.004980661 | 1 |
| T2DM | b_cell | *GANAB* | -1.33222 | 0.001506293 | 0.004980661 | 1 |
| T2DM | b_cell | *MLF2* | -1.122400281 | 0.003264736 | 0.005375128 | 1 |
| T2DM | b_cell | *ANKRD24* | -2.650315428 | 0.002280045 | 0.004980661 | 1 |
| T2DM | b_cell | *DHX32* | -0.979648269 | 0.001891746 | 0.004980661 | 1 |
| T2DM | b_cell | *SPTLC1* | -1.180020815 | 0.001862391 | 0.004980661 | 1 |
| T2DM | b_cell | *YPEL3* | -1.573868224 | 0.022678246 | 0.027135598 | 1 |
| T2DM | b_cell | *MRPS33* | -1.589304258 | 0.000600584 | 0.004980661 | 1 |
| T2DM | b_cell | *STRN4* | -2.353451351 | 0.019052685 | 0.023076292 | 1 |
| T2DM | b_cell | *LYZ* | -1.621742318 | 0.022206711 | 0.026604188 | 1 |
| T2DM | b_cell | *SPG21* | -1.167247119 | 0.001317285 | 0.004980661 | 1 |
| T2DM | b_cell | *CHRD* | -2.696164912 | 0.003891683 | 0.005994428 | 1 |
| T2DM | b_cell | *RAB11FIP3* | -1.052226766 | 0.001689347 | 0.004980661 | 1 |
| T2DM | b_cell | *GNPTG* | -1.906592454 | 0.002167555 | 0.004980661 | 1 |
| T2DM | b_cell | *PABPC4* | -4.811005875 | 7.47E-06 | 0.001813013 | 0.036260263 |
| T2DM | b_cell | *CERS4* | -1.010056398 | 0.001784545 | 0.004980661 | 1 |
| T2DM | b_cell | *USP48* | -1.578439679 | 0.002596716 | 0.004980661 | 1 |
| T2DM | b_cell | *PDPR* | -1.051083646 | 0.002391492 | 0.004980661 | 1 |
| T2DM | b_cell | *GLG1* | -1.165349035 | 0.002488803 | 0.004980661 | 1 |
| T2DM | b_cell | *DTX2* | -1.454815614 | 0.001792614 | 0.004980661 | 1 |

| | | | | | | |
|---|---|---|---|---|---|---|
| T2DM | b_cell | *LAMB1* | -0.711886475 | 0.000961149 | 0.004980661 | 1 |
| T2DM | b_cell | *ABCC6* | -1.437836611 | 0.000889739 | 0.004980661 | 1 |
| T2DM | b_cell | *RAPGEF4* | -0.904031836 | 0.002145275 | 0.004980661 | 1 |
| T2DM | b_cell | *TF* | -1.585496332 | 0.001788647 | 0.004980661 | 1 |
| T2DM | b_cell | *ALKBH5* | -2.973223054 | 0.001825984 | 0.004980661 | 1 |
| T2DM | b_cell | *SPAG7* | -1.141335227 | 0.002011982 | 0.004980661 | 1 |
| T2DM | b_cell | *RGS17* | -1.285849296 | 0.000564635 | 0.004980661 | 1 |
| T2DM | b_cell | *PPP2R3C* | -1.116225659 | 0.005016745 | 0.007218638 | 1 |
| T2DM | b_cell | *HAUS4* | -1.835657138 | 0.005103489 | 0.007315078 | 1 |
| T2DM | b_cell | *OSGEP* | -1.569245951 | 0.002281439 | 0.004980661 | 1 |
| T2DM | b_cell | *SCFD1* | -1.018166712 | 0.002124022 | 0.004980661 | 1 |
| T2DM | b_cell | *SUPT16H* | -1.161920032 | 0.001881195 | 0.004980661 | 1 |
| T2DM | b_cell | *TOX4* | -1.475107923 | 0.002518863 | 0.004980661 | 1 |
| T2DM | b_cell | *CAPN3* | 1.370680765 | 0.005190812 | 0.007409773 | 1 |
| T2DM | b_cell | *TBX15* | 0.965263449 | 0.003961429 | 0.006063361 | 1 |
| T2DM | b_cell | *MFSD11* | -1.198363386 | 0.003206417 | 0.005339692 | 1 |
| T2DM | b_cell | *NUP50* | -2.30165829 | 0.005396424 | 0.007671682 | 1 |
| T2DM | b_cell | *COMT* | -1.439505126 | 0.002399578 | 0.004980661 | 1 |
| T2DM | b_cell | *LRRFIP2* | -1.04663471 | 0.002293558 | 0.004980661 | 1 |
| T2DM | b_cell | *SEC22C* | -1.063981263 | 0.001896968 | 0.004980661 | 1 |
| T2DM | b_cell | *XYLB* | -1.127424321 | 0.001772017 | 0.004980661 | 1 |
| T2DM | b_cell | *GABRP* | -3.44982555 | 0.008653345 | 0.011332262 | 1 |
| T2DM | b_cell | *CDC6* | -5.744167578 | 0.000543722 | 0.004980661 | 1 |
| T2DM | b_cell | *FMO2* | -0.679755202 | 0.004680555 | 0.006838318 | 1 |
| T2DM | b_cell | *MSH2* | -2.179766938 | 0.007860184 | 0.010434367 | 1 |
| T2DM | b_cell | *PSMD5* | -1.503951537 | 0.002308674 | 0.004980661 | 1 |
| T2DM | b_cell | *CRAT* | -1.717955248 | 0.001774397 | 0.004980661 | 1 |
| T2DM | b_cell | *PDE6C* | -1.694669087 | 0.002375347 | 0.004980661 | 1 |
| T2DM | b_cell | *CWF19L1* | -2.511091632 | 1.15E-05 | 0.00242553 | 0.055787195 |
| T2DM | b_cell | *CRTAC1* | -2.021685953 | 0.003019112 | 0.005210663 | 1 |
| T2DM | b_cell | *WAC* | -1.118726475 | 0.002036086 | 0.004980661 | 1 |
| T2DM | b_cell | *ITPR3* | -1.279483349 | 0.007498935 | 0.010034427 | 1 |
| T2DM | b_cell | *DSP* | -2.306274581 | 0.002264613 | 0.004980661 | 1 |
| T2DM | b_cell | *IFT74* | -1.201620044 | 0.002061832 | 0.004980661 | 1 |
| T2DM | b_cell | *IL12RB1* | -2.162470224 | 0.02561011 | 0.030411223 | 1 |
| T2DM | b_cell | *ACOT7* | -3.52442373 | 0.002461773 | 0.004980661 | 1 |
| T2DM | b_cell | *SCD* | -1.104048534 | 0.000450405 | 0.004980661 | 1 |
| T2DM | b_cell | *ERMP1* | -0.985521957 | 0.001777507 | 0.004980661 | 1 |
| T2DM | b_cell | *RAB18* | -1.04716662 | 0.001878749 | 0.004980661 | 1 |
| T2DM | b_cell | *PRTFDC1* | 1.314349367 | 0.026758148 | 0.031696908 | 1 |
| T2DM | b_cell | *TSPAN15* | -1.017502915 | 0.001944141 | 0.004980661 | 1 |
| T2DM | b_cell | *MAST3* | -1.385529519 | 0.001198791 | 0.004980661 | 1 |
| T2DM | b_cell | *OCEL1* | -1.529589071 | 0.00271722 | 0.005012128 | 1 |
| T2DM | b_cell | *CATSPERG* | -1.909394388 | 0.010994524 | 0.014008779 | 1 |
| T2DM | b_cell | *SETD1A* | -2.021146631 | 0.008940255 | 0.011645132 | 1 |
| T2DM | b_cell | *CIRBP* | -0.970336796 | 0.002176236 | 0.004980661 | 1 |
| T2DM | b_cell | *MTAP* | -1.282497549 | 0.001665461 | 0.004980661 | 1 |
| T2DM | b_cell | *POLR2E* | -1.222130545 | 0.001970576 | 0.004980661 | 1 |
| T2DM | b_cell | *MADCAM1* | -1.642303791 | 0.001551177 | 0.004980661 | 1 |
| T2DM | b_cell | *MED15* | -0.9587375 | 0.001678936 | 0.004980661 | 1 |

| | | | | | | |
|---|---|---|---|---|---|---|
| T2DM | b_cell | *SERPIND1* | -1.662016211 | 0.002866057 | 0.005094011 | 1 |
| T2DM | b_cell | *SNAP29* | -1.497085147 | 0.004559929 | 0.006708543 | 1 |
| T2DM | b_cell | *LZTR1* | -1.964999169 | 0.003793849 | 0.005890482 | 1 |
| T2DM | b_cell | *CECR2* | -1.27457845 | 0.00386979 | 0.005970845 | 1 |
| T2DM | b_cell | *SMARCB1* | -1.624035329 | 0.008994977 | 0.011697568 | 1 |
| T2DM | b_cell | *BCL2L13* | -0.993236352 | 0.003209308 | 0.005339692 | 1 |
| T2DM | b_cell | *DDTL* | 0.814752667 | 0.002058952 | 0.004980661 | 1 |
| T2DM | b_cell | *DDT* | -1.671655761 | 0.000937683 | 0.004980661 | 1 |
| T2DM | b_cell | *CABIN1* | -1.299139818 | 0.002240691 | 0.004980661 | 1 |
| T2DM | b_cell | *SPECC1L* | -2.865446513 | 0.015961066 | 0.019610811 | 1 |
| T2DM | b_cell | *PPIL2* | -1.548810016 | 0.004048888 | 0.006163369 | 1 |
| T2DM | b_cell | *PES1* | -1.39234902 | 0.001621059 | 0.004980661 | 1 |
| T2DM | b_cell | *SLC35E4* | -1.475460163 | 0.001120473 | 0.004980661 | 1 |
| T2DM | b_cell | *CRYBB3* | -3.735071103 | 0.030097811 | 0.035436686 | 1 |
| T2DM | b_cell | *CYTH4* | -1.261585377 | 0.001857689 | 0.004980661 | 1 |
| T2DM | b_cell | *CARD10* | -1.703136305 | 0.002204233 | 0.004980661 | 1 |
| T2DM | b_cell | *LRP5L* | -2.913187264 | 0.002718862 | 0.005012128 | 1 |
| T2DM | b_cell | *SH3BP1* | -2.226794798 | 0.001939805 | 0.004980661 | 1 |
| T2DM | b_cell | *HPS4* | -1.122398723 | 0.001993558 | 0.004980661 | 1 |
| T2DM | b_cell | *TRIOBP* | -1.564979846 | 0.004447292 | 0.006598857 | 1 |
| T2DM | b_cell | *EIF3L* | -1.323895526 | 0.000919772 | 0.004980661 | 1 |
| T2DM | b_cell | *CCDC134* | -2.036037545 | 0.003982682 | 0.006084374 | 1 |
| T2DM | b_cell | *PICK1* | -1.593918937 | 0.000600502 | 0.004980661 | 1 |
| T2DM | b_cell | *TTC28* | -0.855434186 | 0.002104284 | 0.004980661 | 1 |
| T2DM | b_cell | *CENPM* | 2.503478548 | 0.004403603 | 0.006552064 | 1 |
| T2DM | b_cell | *KDELR3* | -2.094171804 | 0.001213721 | 0.004980661 | 1 |
| T2DM | b_cell | *CYP2D6* | -3.653926468 | 0.002646211 | 0.004985914 | 1 |
| T2DM | b_cell | *CBY1* | -1.19614364 | 0.002711268 | 0.00500969 | 1 |
| T2DM | b_cell | *RTCB* | -1.854477954 | 0.004149668 | 0.006284079 | 1 |
| T2DM | b_cell | *TIMP3* | -0.750593544 | 0.001895285 | 0.004980661 | 1 |
| T2DM | b_cell | *PPP6R2* | -1.04076468 | 0.002031394 | 0.004980661 | 1 |
| T2DM | b_cell | *CYB5R3* | -1.303739741 | 0.001902586 | 0.004980661 | 1 |
| T2DM | b_cell | *RHBDD3* | -1.443667222 | 0.00105656 | 0.004980661 | 1 |
| T2DM | b_cell | *TOM1* | -1.689798215 | 0.002089253 | 0.004980661 | 1 |
| T2DM | b_cell | *CHKB* | -2.137145621 | 0.003012359 | 0.005203263 | 1 |
| T2DM | b_cell | *ARSA* | 1.782853824 | 0.012404384 | 0.01561652 | 1 |
| T2DM | b_cell | *TSPO* | -1.658641711 | 0.002613258 | 0.004980661 | 1 |
| T2DM | b_cell | *TTLL12* | -4.076139815 | 0.010247995 | 0.013143873 | 1 |
| T2DM | b_cell | *MTMR3* | -0.869444693 | 0.00149082 | 0.004980661 | 1 |
| T2DM | b_cell | *APOL4* | -1.822034458 | 0.002621231 | 0.004980661 | 1 |
| T2DM | b_cell | *SAMM50* | -2.225409263 | 0.006393404 | 0.008817882 | 1 |
| T2DM | b_cell | *FOXRED2* | -1.376361908 | 0.013440508 | 0.016803233 | 1 |
| T2DM | b_cell | *KIAA0930* | -1.812349712 | 0.003277947 | 0.005387318 | 1 |
| T2DM | b_cell | *SLC25A17* | -1.11048156 | 0.003537333 | 0.00564577 | 1 |
| T2DM | b_cell | *FAM118A* | -1.183604858 | 0.004893424 | 0.007066337 | 1 |
| T2DM | b_cell | *L3MBTL2* | -1.497454024 | 0.000959486 | 0.004980661 | 1 |
| T2DM | b_cell | *ZC3H7B* | -1.893942718 | 0.003673442 | 0.005764404 | 1 |
| T2DM | b_cell | *ACO2* | -1.275190297 | 0.00184677 | 0.004980661 | 1 |
| T2DM | b_cell | *POLR3H* | -1.331999245 | 0.002043467 | 0.004980661 | 1 |
| T2DM | b_cell | *DESI1* | -2.303124022 | 0.007960948 | 0.01054505 | 1 |

| | | | | | | |
|---|---|---|---|---|---|---|
| T2DM | b_cell | *MLC1* | 0.694855939 | 0.001882766 | 0.004980661 | 1 |
| T2DM | b_cell | *HDAC10* | -1.693653475 | 0.001920749 | 0.004980661 | 1 |
| T2DM | b_cell | *KHNYN* | -1.327028013 | 0.001555964 | 0.004980661 | 1 |
| T2DM | b_cell | *SDR39U1* | -1.295496559 | 0.000807169 | 0.004980661 | 1 |
| T2DM | b_cell | *RBM23* | -1.038496216 | 0.00333794 | 0.005445196 | 1 |
| T2DM | b_cell | *NIN* | -1.309723359 | 0.002046845 | 0.004980661 | 1 |
| T2DM | b_cell | *PYGL* | -0.710016393 | 0.003784681 | 0.005883778 | 1 |
| T2DM | b_cell | *CNIH1* | -2.451219492 | 0.011717207 | 0.014878799 | 1 |
| T2DM | b_cell | *ATP6V1D* | -1.760202258 | 0.004424663 | 0.006577349 | 1 |
| T2DM | b_cell | *PLEK2* | -6.459347222 | 0.000303183 | 0.004980661 | 1 |
| T2DM | b_cell | *GSTZ1* | -1.584063529 | 0.001773451 | 0.004980661 | 1 |
| T2DM | b_cell | *TMED8* | -1.102895609 | 0.001633277 | 0.004980661 | 1 |
| T2DM | b_cell | *LGMN* | -0.786179708 | 0.002275552 | 0.004980661 | 1 |
| T2DM | b_cell | *MTHFD1* | -0.955746889 | 0.002304066 | 0.004980661 | 1 |
| T2DM | b_cell | *GSKIP* | -1.597813234 | 0.00177179 | 0.004980661 | 1 |
| T2DM | b_cell | *PSMB5* | -1.039640462 | 0.006847095 | 0.009313738 | 1 |
| T2DM | b_cell | *CCNB1IP1* | -1.152148188 | 0.001789164 | 0.004980661 | 1 |
| T2DM | b_cell | *EFS* | -1.528915361 | 0.001778139 | 0.004980661 | 1 |
| T2DM | b_cell | *CINP* | -1.384860319 | 0.001866464 | 0.004980661 | 1 |
| T2DM | b_cell | *KIAA0391* | -3.442901525 | 0.00099733 | 0.004980661 | 1 |
| T2DM | b_cell | *EMC9* | -2.004058414 | 0.006147657 | 0.008544127 | 1 |
| T2DM | b_cell | *PLTP* | -0.769616471 | 0.016328059 | 0.019995897 | 1 |
| T2DM | b_cell | *TRPC4AP* | -1.033512113 | 0.00165591 | 0.004980661 | 1 |
| T2DM | b_cell | *PYGB* | -1.158578424 | 0.002668142 | 0.004992606 | 1 |
| T2DM | b_cell | *ABHD12* | -1.33652424 | 0.002509198 | 0.004980661 | 1 |
| T2DM | b_cell | *CD40* | -1.675861245 | 0.001359305 | 0.004980661 | 1 |
| T2DM | b_cell | *UQCC1* | -1.96226763 | 0.002863306 | 0.005094011 | 1 |
| T2DM | b_cell | *IFT52* | -1.356507238 | 0.003445694 | 0.005550633 | 1 |
| T2DM | b_cell | *NDRG3* | -2.203035999 | 0.022220246 | 0.026613832 | 1 |
| T2DM | b_cell | *PABPC1L* | 1.528641808 | 0.008933013 | 0.01164195 | 1 |
| T2DM | b_cell | *DOK5* | 1.550290731 | 0.012175638 | 0.015376418 | 1 |
| T2DM | b_cell | *CTSZ* | -1.947192797 | 0.030396952 | 0.035745519 | 1 |
| T2DM | b_cell | *MTG2* | -1.259436126 | 0.002308995 | 0.004980661 | 1 |
| T2DM | b_cell | *TCFL5* | -1.677143902 | 0.001647803 | 0.004980661 | 1 |
| T2DM | b_cell | *DIDO1* | -1.392452225 | 0.005133985 | 0.007343778 | 1 |
| T2DM | b_cell | *GID8* | -1.15181505 | 0.00225234 | 0.004980661 | 1 |
| T2DM | b_cell | *PTK6* | -4.667709682 | 0.00213824 | 0.004980661 | 1 |
| T2DM | b_cell | *GMEB2* | -1.655546171 | 0.001892826 | 0.004980661 | 1 |
| T2DM | b_cell | *C20orf27* | -1.547469481 | 0.003130561 | 0.005273382 | 1 |
| T2DM | b_cell | *RNF24* | -1.496850625 | 0.003354162 | 0.005460611 | 1 |
| T2DM | b_cell | *NDUFAF5* | -1.003148161 | 0.002802575 | 0.005065458 | 1 |
| T2DM | b_cell | *ANGPT4* | -3.03624049 | 0.002166019 | 0.004980661 | 1 |
| T2DM | b_cell | *RSPO4* | -1.931173751 | 0.007946165 | 0.010528343 | 1 |
| T2DM | b_cell | *SEC23B* | -1.408630569 | 0.003100361 | 0.005253181 | 1 |
| T2DM | b_cell | *CRNKL1* | -1.930015627 | 0.002995044 | 0.005193692 | 1 |
| T2DM | b_cell | *MROH8* | -1.394309624 | 0.000577917 | 0.004980661 | 1 |
| T2DM | b_cell | *TTI1* | -1.151281303 | 0.001631605 | 0.004980661 | 1 |
| T2DM | b_cell | *BPI* | -1.199684506 | 0.040396853 | 0.046690217 | 1 |
| T2DM | b_cell | *ACTR5* | -1.6839932 | 0.001326109 | 0.004980661 | 1 |
| T2DM | b_cell | *MAP1LC3A* | -1.376683299 | 0.002175948 | 0.004980661 | 1 |

| | | | | | | |
|---|---|---|---|---|---|---|
| T2DM | b_cell | *RBFA* | -1.422801107 | 0.002245293 | 0.004980661 | 1 |
| T2DM | b_cell | *METTL4* | -1.560191937 | 0.002023081 | 0.004980661 | 1 |
| T2DM | b_cell | *LPIN2* | -1.618659272 | 0.003984289 | 0.006084485 | 1 |
| T2DM | b_cell | *MYOM1* | -0.950636928 | 0.001789733 | 0.004980661 | 1 |
| T2DM | b_cell | *CEP192* | -1.091593342 | 0.002288646 | 0.004980661 | 1 |
| T2DM | b_cell | *LAMA1* | -2.429180166 | 0.001311913 | 0.004980661 | 1 |
| T2DM | b_cell | *RNF125* | -2.828389047 | 0.004619875 | 0.006786447 | 1 |
| T2DM | b_cell | *POLI* | -0.768694723 | 0.009301212 | 0.012041072 | 1 |
| T2DM | b_cell | *MIB1* | -1.197221766 | 0.002398368 | 0.004980661 | 1 |
| T2DM | b_cell | *NALCN* | -1.420645033 | 0.002147572 | 0.004980661 | 1 |
| T2DM | b_cell | *STK24* | -0.892106374 | 0.001839779 | 0.004980661 | 1 |
| T2DM | b_cell | *ACP5* | -1.456140257 | 0.013902126 | 0.017317873 | 1 |
| T2DM | b_cell | *DNAJC3* | -1.535764745 | 0.004193807 | 0.006325256 | 1 |
| T2DM | b_cell | *UGGT2* | -0.959991622 | 0.002323234 | 0.004980661 | 1 |
| T2DM | b_cell | *ARHGEF7* | -0.899682988 | 0.001874884 | 0.004980661 | 1 |
| T2DM | b_cell | *PARP4* | -1.072929763 | 0.002455108 | 0.004980661 | 1 |
| T2DM | b_cell | *SUPT20H* | -1.2778161 | 0.002226361 | 0.004980661 | 1 |
| T2DM | b_cell | *MRPS31* | -1.36672963 | 0.002665448 | 0.004991101 | 1 |
| T2DM | b_cell | *RGCC* | -1.03166914 | 0.003881621 | 0.005980827 | 1 |
| T2DM | b_cell | *DGKH* | -1.493701871 | 0.001107141 | 0.004980661 | 1 |
| T2DM | b_cell | *KATNAL1* | -1.14741336 | 0.002061983 | 0.004980661 | 1 |
| T2DM | b_cell | *INTS6* | -1.144881507 | 0.002153148 | 0.004980661 | 1 |
| T2DM | b_cell | *CLN5* | -1.063902265 | 0.001653251 | 0.004980661 | 1 |
| T2DM | b_cell | *MGRN1* | -1.006750799 | 0.002038373 | 0.004980661 | 1 |
| T2DM | b_cell | *ELMO3* | -2.511048763 | 0.009641643 | 0.012440254 | 1 |
| T2DM | b_cell | *LONP2* | -0.955698228 | 0.003246169 | 0.00536425 | 1 |
| T2DM | b_cell | *PLLP* | -1.235017809 | 0.001762687 | 0.004980661 | 1 |
| T2DM | b_cell | *DHODH* | -1.26476649 | 0.003797091 | 0.005891598 | 1 |
| T2DM | b_cell | *CCDC113* | 1.354346584 | 0.004788136 | 0.006951453 | 1 |
| T2DM | b_cell | *PSMD7* | -1.025321157 | 0.001954946 | 0.004980661 | 1 |
| T2DM | b_cell | *COG4* | -1.108525153 | 0.000972845 | 0.004980661 | 1 |
| T2DM | b_cell | *MON1B* | -2.527183866 | 0.003519087 | 0.005631468 | 1 |
| T2DM | b_cell | *NPRL3* | -0.948860739 | 0.001879636 | 0.004980661 | 1 |
| T2DM | b_cell | *NAGPA* | -1.575591312 | 0.001087255 | 0.004980661 | 1 |
| T2DM | b_cell | *NOMO3* | -3.177267791 | 3.31E-05 | 0.004719449 | 0.160461276 |
| T2DM | b_cell | *LMF1* | -1.837722094 | 0.005602584 | 0.007904547 | 1 |
| T2DM | b_cell | *NARFL* | -2.634064119 | 0.025653878 | 0.030455741 | 1 |
| T2DM | b_cell | *MTHFSD* | -1.234107977 | 0.001492705 | 0.004980661 | 1 |
| T2DM | b_cell | *HAGHL* | -2.162356671 | 0.00288493 | 0.005104976 | 1 |
| T2DM | b_cell | *SLC7A5* | 1.16120347 | 0.00235912 | 0.004980661 | 1 |
| T2DM | b_cell | *RHBDL1* | -6.365415976 | 0.000974829 | 0.004980661 | 1 |
| T2DM | b_cell | *NUBP1* | -1.406150243 | 0.002478793 | 0.004980661 | 1 |
| T2DM | b_cell | *UBE2I* | -1.450341717 | 0.001903437 | 0.004980661 | 1 |
| T2DM | b_cell | *CAPN15* | -1.817731566 | 0.002492388 | 0.004980661 | 1 |
| T2DM | b_cell | *EARS2* | -2.705682031 | 0.004535204 | 0.006680269 | 1 |
| T2DM | b_cell | *CPPED1* | -1.393414929 | 0.002693363 | 0.00500636 | 1 |
| T2DM | b_cell | *HMOX2* | -1.142975411 | 0.002599939 | 0.004980661 | 1 |
| T2DM | b_cell | *TOX3* | -4.899593053 | 0.004084784 | 0.006203247 | 1 |
| T2DM | b_cell | *RRN3P2* | -1.891825647 | 0.0008731 | 0.004980661 | 1 |
| T2DM | b_cell | *RBL2* | -1.033784004 | 0.004641483 | 0.006801715 | 1 |

| | | | | | | |
|---|---|---|---|---|---|---|
| T2DM | b_cell | *CDIPT* | -1.548523142 | 0.003401052 | 0.005504304 | 1 |
| T2DM | b_cell | *KAT8* | -1.619458034 | 0.001998061 | 0.004980661 | 1 |
| T2DM | b_cell | *TMC5* | -1.44409937 | 0.001639826 | 0.004980661 | 1 |
| T2DM | b_cell | *C16orf62* | -1.094130719 | 0.002043229 | 0.004980661 | 1 |
| T2DM | b_cell | *RNF40* | -1.285891046 | 0.002022274 | 0.004980661 | 1 |
| T2DM | b_cell | *IQCH* | -1.382307735 | 0.002066608 | 0.004980661 | 1 |
| T2DM | b_cell | *LACTB* | -2.511872856 | 0.009301306 | 0.012041072 | 1 |
| T2DM | b_cell | *HERC1* | -1.04878182 | 0.002647613 | 0.004985914 | 1 |
| T2DM | b_cell | *TRIP4* | -1.561959008 | 0.002720235 | 0.005012753 | 1 |
| T2DM | b_cell | *AP3B2* | -1.949078985 | 0.000405088 | 0.004980661 | 1 |
| T2DM | b_cell | *TTC23* | -0.953722406 | 0.001789499 | 0.004980661 | 1 |
| T2DM | b_cell | *FAH* | -1.182154633 | 0.001696069 | 0.004980661 | 1 |
| T2DM | b_cell | *RPAP1* | -2.310455878 | 0.000706635 | 0.004980661 | 1 |
| T2DM | b_cell | *SLC30A4* | -1.28261976 | 0.001912698 | 0.004980661 | 1 |
| T2DM | b_cell | *ZDHHC2* | -1.323979519 | 0.001393514 | 0.004980661 | 1 |
| T2DM | b_cell | *ZFAND1* | -1.422224363 | 0.001774682 | 0.004980661 | 1 |
| T2DM | b_cell | *EYA1* | -1.392636534 | 0.002171267 | 0.004980661 | 1 |
| T2DM | b_cell | *IMPAD1* | -1.112525661 | 0.001843744 | 0.004980661 | 1 |
| T2DM | b_cell | *SFRP1* | -1.236119991 | 0.001723286 | 0.004980661 | 1 |
| T2DM | b_cell | *NIPAL2* | -1.302105711 | 0.002618334 | 0.004980661 | 1 |
| T2DM | b_cell | *RAB2A* | -0.806192271 | 0.002188538 | 0.004980661 | 1 |
| T2DM | b_cell | *ARMC1* | -1.606737569 | 0.001416014 | 0.004980661 | 1 |
| T2DM | b_cell | *SPAG1* | -1.466967577 | 0.00275685 | 0.005034338 | 1 |
| T2DM | b_cell | *SNX16* | -1.238146342 | 0.001890344 | 0.004980661 | 1 |
| T2DM | b_cell | *GSDMD* | -2.394380629 | 0.006396167 | 0.008818041 | 1 |
| T2DM | b_cell | *TSTA3* | 1.510717836 | 0.001672051 | 0.004980661 | 1 |
| T2DM | b_cell | *SQLE* | 1.008289272 | 0.00608558 | 0.00847753 | 1 |
| T2DM | b_cell | *R3HCC1* | -1.578138948 | 0.004725483 | 0.006883231 | 1 |
| T2DM | b_cell | *TNFRSF10A* | -1.15525459 | 0.040465658 | 0.046758602 | 1 |
| T2DM | b_cell | *ERICH1* | -1.010496499 | 0.001999486 | 0.004980661 | 1 |
| T2DM | b_cell | *TUSC3* | -0.998744107 | 0.001817535 | 0.004980661 | 1 |
| T2DM | b_cell | *KLHDC4* | -1.589652179 | 0.002951 | 0.00515973 | 1 |
| T2DM | b_cell | *MAN2B1* | -1.694249832 | 0.003921653 | 0.006021474 | 1 |
| T2DM | b_cell | *TULP2* | 3.550043987 | 0.002825125 | 0.005065458 | 1 |
| T2DM | b_cell | *NFKBIB* | -1.831976263 | 0.002759246 | 0.005036819 | 1 |
| T2DM | b_cell | *SARS2* | 0.661017014 | 0.009485072 | 0.012252814 | 1 |
| T2DM | b_cell | *SNRNP70* | -1.021793279 | 0.001838284 | 0.004980661 | 1 |
| T2DM | b_cell | *LIN7B* | -1.682583611 | 0.001673413 | 0.004980661 | 1 |
| T2DM | b_cell | *FCGRT* | -1.55158218 | 0.001746056 | 0.004980661 | 1 |
| T2DM | b_cell | *PEX11G* | -2.017791251 | 0.00186307 | 0.004980661 | 1 |
| T2DM | b_cell | *CD37* | 1.142649139 | 0.004637143 | 0.006799003 | 1 |
| T2DM | b_cell | *AMH* | -16.11166272 | 0.001737253 | 0.004980661 | 1 |
| T2DM | b_cell | *DKKL1* | 1.734670798 | 0.002246142 | 0.004980661 | 1 |
| T2DM | b_cell | *FCER2* | 0.747736363 | 0.002559705 | 0.004980661 | 1 |
| T2DM | b_cell | *CLEC4M* | 1.237347824 | 0.002834952 | 0.005067351 | 1 |
| T2DM | b_cell | *NOVA2* | -1.316880896 | 0.008292724 | 0.010910058 | 1 |
| T2DM | b_cell | *SGTA* | -1.495447657 | 0.001643699 | 0.004980661 | 1 |
| T2DM | b_cell | *MED25* | -2.636032272 | 0.027649023 | 0.032688367 | 1 |
| T2DM | b_cell | *TIMM44* | -1.229467554 | 0.002435662 | 0.004980661 | 1 |
| T2DM | b_cell | *IL27RA* | -1.789868243 | 0.002396261 | 0.004980661 | 1 |

| | | | | | | |
|---|---|---|---|---|---|---|
| T2DM | b_cell | *FAM32A* | -1.736915608 | 0.001700297 | 0.004980661 | 1 |
| T2DM | b_cell | *C19orf44* | -1.118766436 | 0.006462779 | 0.008884636 | 1 |
| T2DM | b_cell | *OLFM2* | -1.356107751 | 0.001515889 | 0.004980661 | 1 |
| T2DM | b_cell | *RASAL3* | -2.047866228 | 0.002126039 | 0.004980661 | 1 |
| T2DM | b_cell | *PDCD5* | -1.557609602 | 0.00253998 | 0.004980661 | 1 |
| T2DM | b_cell | *ANKRD27* | -1.350198058 | 0.002185059 | 0.004980661 | 1 |
| T2DM | b_cell | *POLR2I* | -1.586218122 | 0.003460309 | 0.00556863 | 1 |
| T2DM | b_cell | *CCDC9* | -1.618536117 | 0.003541097 | 0.005648061 | 1 |
| T2DM | b_cell | *TGFB1* | -1.239100459 | 0.002110245 | 0.004980661 | 1 |
| T2DM | b_cell | *CEACAM4* | 2.724933601 | 0.001342846 | 0.004980661 | 1 |
| T2DM | b_cell | *ICAM5* | -5.563044483 | 3.49E-05 | 0.004835391 | 0.169238699 |
| T2DM | b_cell | *CD33* | -1.035485361 | 0.021867019 | 0.026229611 | 1 |
| T2DM | b_cell | *RABAC1* | -1.996397719 | 0.006996616 | 0.00947727 | 1 |
| T2DM | b_cell | *GRIN2D* | -2.394146952 | 0.00257843 | 0.004980661 | 1 |
| T2DM | b_cell | *CCDC114* | 3.02328241 | 0.002533896 | 0.004980661 | 1 |
| T2DM | b_cell | *CARD8* | -1.350028474 | 0.001884768 | 0.004980661 | 1 |
| T2DM | b_cell | *LIG1* | -2.925445749 | 0.042701736 | 0.049244982 | 1 |
| T2DM | b_cell | *CAPS* | -1.79477154 | 0.001727675 | 0.004980661 | 1 |
| T2DM | b_cell | *MIER2* | -1.873043789 | 0.004196193 | 0.006326889 | 1 |
| T2DM | b_cell | *PLEKHA4* | -1.385589426 | 0.00172631 | 0.004980661 | 1 |
| T2DM | b_cell | *TNPO2* | -1.300928438 | 0.00269118 | 0.00500636 | 1 |
| T2DM | b_cell | *GCDH* | -1.768211581 | 0.000255916 | 0.004980661 | 1 |
| T2DM | b_cell | *DNASE2* | -1.617539084 | 0.001719005 | 0.004980661 | 1 |
| T2DM | b_cell | *TFPT* | -2.240871815 | 0.002475118 | 0.004980661 | 1 |
| T2DM | b_cell | *RAB3A* | 1.612338138 | 0.002808313 | 0.005065458 | 1 |
| T2DM | b_cell | *COPE* | -2.905593939 | 0.00992543 | 0.012767281 | 1 |
| T2DM | b_cell | *ETV2* | -15.69168463 | 0.001654904 | 0.004980661 | 1 |
| T2DM | b_cell | *MAG* | 6.713367751 | 0.000971593 | 0.004980661 | 1 |
| T2DM | b_cell | *LSR* | -1.171220667 | 0.005265722 | 0.007512286 | 1 |
| T2DM | b_cell | *ZNF14* | -1.243902615 | 0.001789703 | 0.004980661 | 1 |
| T2DM | b_cell | *ZNF85* | -1.40566262 | 0.001827388 | 0.004980661 | 1 |
| T2DM | b_cell | *ETHE1* | -2.550456908 | 0.002561741 | 0.004980661 | 1 |
| T2DM | b_cell | *SMG9* | -2.445127788 | 0.003860448 | 0.005965584 | 1 |
| T2DM | b_cell | *GTPBP10* | -0.912952917 | 0.002580095 | 0.004980661 | 1 |
| T2DM | b_cell | *RASA4* | -10.05970447 | 0.001717131 | 0.004980661 | 1 |
| T2DM | b_cell | *BET1* | -1.022350723 | 0.001681281 | 0.004980661 | 1 |
| T2DM | b_cell | *SP4* | -1.095290879 | 0.002095037 | 0.004980661 | 1 |
| T2DM | b_cell | *DNAH11* | -1.231914359 | 0.001782763 | 0.004980661 | 1 |
| T2DM | b_cell | *STEAP1B* | -2.110325076 | 0.005841715 | 0.008184811 | 1 |
| T2DM | b_cell | *PTN* | -1.079149372 | 0.001782302 | 0.004980661 | 1 |
| T2DM | b_cell | *ADAP1* | 1.144895441 | 0.002749423 | 0.00503214 | 1 |
| T2DM | b_cell | *MET* | -0.887900116 | 0.001735155 | 0.004980661 | 1 |
| T2DM | b_cell | *RNF32* | -2.471811923 | 0.000559977 | 0.004980661 | 1 |
| T2DM | b_cell | *VIPR2* | -0.91782869 | 0.001801161 | 0.004980661 | 1 |
| T2DM | b_cell | *HIBADH* | -0.857097106 | 0.002406693 | 0.004980661 | 1 |
| T2DM | b_cell | *CPVL* | -1.462740872 | 0.00326961 | 0.005378257 | 1 |
| T2DM | b_cell | *CHN2* | -0.805893809 | 0.012618915 | 0.015874249 | 1 |
| T2DM | b_cell | *GRB10* | -0.836982291 | 0.001951128 | 0.004980661 | 1 |
| T2DM | b_cell | *EPHB6* | -1.316701159 | 0.000778106 | 0.004980661 | 1 |
| T2DM | b_cell | *CHCHD2* | -1.634669479 | 0.003075376 | 0.005243051 | 1 |

| | | | | | | |
|---|---|---|---|---|---|---|
| T2DM | b_cell | *ZKSCAN1* | -1.703303478 | 0.004331597 | 0.00647259 | 1 |
| T2DM | b_cell | *EIF3B* | -2.014091567 | 0.00447661 | 0.006632218 | 1 |
| T2DM | b_cell | *TAF6* | 1.070468079 | 0.002754558 | 0.005034338 | 1 |
| T2DM | b_cell | *TFR2* | -2.667082752 | 0.001459983 | 0.004980661 | 1 |
| T2DM | b_cell | *MOSPD3* | -1.943925045 | 0.000832812 | 0.004980661 | 1 |
| T2DM | b_cell | *RBM28* | -0.895622243 | 0.002558129 | 0.004980661 | 1 |
| T2DM | b_cell | *IMPDH1* | -1.601395944 | 0.001720522 | 0.004980661 | 1 |
| T2DM | b_cell | *C1GALT1* | -0.927319223 | 0.001904848 | 0.004980661 | 1 |
| T2DM | b_cell | *NRF1* | -1.095152145 | 0.002481469 | 0.004980661 | 1 |
| T2DM | b_cell | *ZNF862* | -1.146005961 | 0.001954162 | 0.004980661 | 1 |
| T2DM | b_cell | *MEST* | -0.845069911 | 0.000983512 | 0.004980661 | 1 |
| T2DM | b_cell | *ACTR3C* | -1.710354146 | 0.003007775 | 0.005199149 | 1 |
| T2DM | b_cell | *POU6F2* | -1.145786986 | 0.001697202 | 0.004980661 | 1 |
| T2DM | b_cell | *TSPAN13* | -1.666748125 | 0.001909401 | 0.004980661 | 1 |
| T2DM | b_cell | *RARRES2* | -0.987864455 | 0.002081634 | 0.004980661 | 1 |
| T2DM | b_cell | *TMEM176B* | -1.470140094 | 0.024674254 | 0.029350204 | 1 |
| T2DM | b_cell | *COA1* | -1.105331035 | 0.002418844 | 0.004980661 | 1 |
| T2DM | b_cell | *BLVRA* | -1.101477944 | 0.001355734 | 0.004980661 | 1 |
| T2DM | b_cell | *URGCP* | -1.210725841 | 0.002832893 | 0.005067351 | 1 |
| T2DM | b_cell | *AEBP1* | -1.416756171 | 0.001863181 | 0.004980661 | 1 |
| T2DM | b_cell | *GCK* | -9.177637252 | 0.001750315 | 0.004980661 | 1 |
| T2DM | b_cell | *BCL7B* | -1.650733609 | 0.003639198 | 0.005744108 | 1 |
| T2DM | b_cell | *TBL2* | -1.461210647 | 0.002516532 | 0.004980661 | 1 |
| T2DM | b_cell | *FKTN* | -1.035429294 | 0.001841999 | 0.004980661 | 1 |
| T2DM | b_cell | *NMRK1* | -1.162548492 | 0.002482309 | 0.004980661 | 1 |
| T2DM | b_cell | *MEGF9* | -0.931467585 | 0.001757707 | 0.004980661 | 1 |
| T2DM | b_cell | *TRIM14* | -1.062130855 | 0.003045542 | 0.00522879 | 1 |
| T2DM | b_cell | *C5* | -1.124028848 | 0.002169451 | 0.004980661 | 1 |
| T2DM | b_cell | *OGN* | -0.925532277 | 0.001145286 | 0.004980661 | 1 |
| T2DM | b_cell | *ECM2* | -0.774735288 | 0.001772509 | 0.004980661 | 1 |
| T2DM | b_cell | *PTGR1* | -0.878817125 | 0.001351298 | 0.004980661 | 1 |
| T2DM | b_cell | *SUSD1* | -1.09232673 | 0.005095439 | 0.007308031 | 1 |
| T2DM | b_cell | *PLGRKT* | -1.501807739 | 0.001381177 | 0.004980661 | 1 |
| T2DM | b_cell | *DOCK8* | -0.625751258 | 0.018485678 | 0.022479325 | 1 |
| T2DM | b_cell | *KANK1* | -0.863571523 | 0.001202494 | 0.004980661 | 1 |
| T2DM | b_cell | *NCS1* | -2.079458309 | 0.006337027 | 0.008754913 | 1 |
| T2DM | b_cell | *FUBP3* | -1.15550704 | 0.00177422 | 0.004980661 | 1 |
| T2DM | b_cell | *RGP1* | -2.68276473 | 0.006780915 | 0.009239258 | 1 |
| T2DM | b_cell | *PTGDS* | -1.300178534 | 0.001798952 | 0.004980661 | 1 |
| T2DM | b_cell | *ATRNL1* | -0.792556408 | 0.005315254 | 0.007574041 | 1 |
| T2DM | b_cell | *HPS1* | -1.965884325 | 0.002191482 | 0.004980661 | 1 |
| T2DM | b_cell | *PHYH* | -1.958468569 | 0.001609315 | 0.004980661 | 1 |
| T2DM | b_cell | *DNMBP* | -1.757889952 | 0.002001741 | 0.004980661 | 1 |
| T2DM | b_cell | *PKD2L1* | 1.545297262 | 0.00165628 | 0.004980661 | 1 |
| T2DM | b_cell | *CUBN* | -1.169266052 | 0.00219941 | 0.004980661 | 1 |
| T2DM | b_cell | *TRDMT1* | -1.661571861 | 0.002229672 | 0.004980661 | 1 |
| T2DM | b_cell | *ATE1* | -1.081671673 | 0.003498836 | 0.005604606 | 1 |
| T2DM | b_cell | *PLEKHA1* | -1.137938067 | 0.001783189 | 0.004980661 | 1 |
| T2DM | b_cell | *CDH23* | -0.869248683 | 0.001709487 | 0.004980661 | 1 |
| T2DM | b_cell | *SPOCK2* | -1.263171617 | 0.002490961 | 0.004980661 | 1 |

| | | | | | | |
|---|---|---|---|---|---|---|
| T2DM | b_cell | *BMPR1A* | -0.746715982 | 0.001755733 | 0.004980661 | 1 |
| T2DM | b_cell | *LIPA* | -1.139955628 | 0.004307224 | 0.006457562 | 1 |
| T2DM | b_cell | *SFXN3* | 1.40259516 | 0.010026512 | 0.012883643 | 1 |
| T2DM | b_cell | *TNKS2* | -1.428590674 | 0.002801728 | 0.005065458 | 1 |
| T2DM | b_cell | *ANKRD26* | -0.878121895 | 0.001930363 | 0.004980661 | 1 |
| T2DM | b_cell | *ACBD5* | -1.434780191 | 0.002407386 | 0.004980661 | 1 |
| T2DM | b_cell | *LHPP* | -1.282694444 | 0.000975971 | 0.004980661 | 1 |
| T2DM | b_cell | *TFAM* | -1.962990612 | 0.002587298 | 0.004980661 | 1 |
| T2DM | b_cell | *RPL28* | -0.81817647 | 0.001562483 | 0.004980661 | 1 |
| T2DM | b_cell | *PBLD* | -1.466387129 | 0.001769946 | 0.004980661 | 1 |
| T2DM | b_cell | *TSPAN14* | -1.043826486 | 0.001689872 | 0.004980661 | 1 |
| T2DM | b_cell | *KRT23* | 0.754735267 | 0.001770451 | 0.004980661 | 1 |
| T2DM | b_cell | *TADA2A* | -1.425371168 | 0.014046406 | 0.017475169 | 1 |
| T2DM | b_cell | *PSMB3* | -1.254211696 | 0.002106464 | 0.004980661 | 1 |
| T2DM | b_cell | *RPL19* | -1.321210276 | 0.004224255 | 0.006357347 | 1 |
| T2DM | b_cell | *RAPGEFL1* | 1.702927825 | 0.007767907 | 0.010328826 | 1 |
| T2DM | b_cell | *WNT3* | -2.485257961 | 0.003410963 | 0.005510766 | 1 |
| T2DM | b_cell | *ASPA* | -0.895849861 | 0.002823161 | 0.005065458 | 1 |
| T2DM | b_cell | *RAD51C* | -2.808850926 | 0.004646284 | 0.006806694 | 1 |
| T2DM | b_cell | *TRIM37* | -2.084595925 | 0.005182461 | 0.007400029 | 1 |
| T2DM | b_cell | *PNPO* | -3.025219829 | 0.000867842 | 0.004980661 | 1 |
| T2DM | b_cell | *TRIM16L* | -2.725776577 | 0.002225132 | 0.004980661 | 1 |
| T2DM | b_cell | *CDK5RAP3* | -2.966490895 | 0.004986339 | 0.007179144 | 1 |
| T2DM | b_cell | *PIGL* | -0.858827785 | 0.001714758 | 0.004980661 | 1 |
| T2DM | b_cell | *MED13* | -0.934132438 | 0.001796156 | 0.004980661 | 1 |
| T2DM | b_cell | *CHRNE* | 1.423958765 | 0.000674073 | 0.004980661 | 1 |
| T2DM | b_cell | *GOSR1* | -1.075121473 | 0.001932645 | 0.004980661 | 1 |
| T2DM | b_cell | *DRG2* | -1.446136809 | 0.001860122 | 0.004980661 | 1 |
| T2DM | b_cell | *FTSJ3* | -1.978929892 | 0.004906856 | 0.007079413 | 1 |
| T2DM | b_cell | *ICAM2* | -1.246532074 | 0.001877678 | 0.004980661 | 1 |
| T2DM | b_cell | *DDX5* | -0.909294112 | 0.003289614 | 0.005395946 | 1 |
| T2DM | b_cell | *C17orf75* | -1.597111072 | 0.003001475 | 0.005195812 | 1 |
| T2DM | b_cell | *PSMD11* | -1.860042142 | 0.00290085 | 0.005113643 | 1 |
| T2DM | b_cell | *PEX12* | 2.13053442 | 0.015551715 | 0.019161229 | 1 |
| T2DM | b_cell | *DHX58* | -2.081192972 | 0.001310305 | 0.004980661 | 1 |
| T2DM | b_cell | *RAB5C* | -1.69693418 | 0.005050341 | 0.00726052 | 1 |
| T2DM | b_cell | *NAGLU* | -2.101538287 | 0.010183777 | 0.013071875 | 1 |
| T2DM | b_cell | *HSD17B1* | 2.155039979 | 0.009006666 | 0.011703359 | 1 |
| T2DM | b_cell | *CNTNAP1* | -1.654032395 | 0.026261881 | 0.0311623 | 1 |
| T2DM | b_cell | *EZH1* | -1.680494551 | 0.005105758 | 0.007315078 | 1 |
| T2DM | b_cell | *SGCA* | 1.718252117 | 0.021518529 | 0.025830752 | 1 |
| T2DM | b_cell | *ABCC3* | -1.106987055 | 0.008134624 | 0.010731157 | 1 |
| T2DM | b_cell | *EFTUD2* | -1.829472062 | 0.00303637 | 0.005228627 | 1 |
| T2DM | b_cell | *HLF* | -1.513351501 | 0.002055749 | 0.004980661 | 1 |
| T2DM | b_cell | *EFNB3* | 0.931119548 | 0.005102403 | 0.007315078 | 1 |
| T2DM | b_cell | *FAM20A* | -1.00288864 | 0.001828842 | 0.004980661 | 1 |
| T2DM | b_cell | *DPH1* | -1.666214612 | 0.001719516 | 0.004980661 | 1 |
| T2DM | b_cell | *DHRS7B* | -1.114393058 | 0.002000227 | 0.004980661 | 1 |
| T2DM | b_cell | *MYH3* | -2.210858896 | 0.001751348 | 0.004980661 | 1 |
| T2DM | b_cell | *SLAIN2* | -1.00369991 | 0.002085371 | 0.004980661 | 1 |

| | | | | | | |
|---|---|---|---|---|---|---|
| T2DM | b_cell | *OCIAD1* | -0.820702152 | 0.001718448 | 0.004980661 | 1 |
| T2DM | b_cell | *UBE2D3* | -1.063807858 | 0.00238782 | 0.004980661 | 1 |
| T2DM | b_cell | *NDUFC1* | -1.742031256 | 0.002285462 | 0.004980661 | 1 |
| T2DM | b_cell | *KLHL2* | -1.360276119 | 0.002069086 | 0.004980661 | 1 |
| T2DM | b_cell | *CPE* | -0.736641706 | 0.003766902 | 0.005865506 | 1 |
| T2DM | b_cell | *WFS1* | -1.420357497 | 0.001796334 | 0.004980661 | 1 |
| T2DM | b_cell | *GRPEL1* | -2.594915799 | 0.002410705 | 0.004980661 | 1 |
| T2DM | b_cell | *GAR1* | -1.554950806 | 0.001909283 | 0.004980661 | 1 |
| T2DM | b_cell | *AADAT* | -2.054460601 | 0.001762235 | 0.004980661 | 1 |
| T2DM | b_cell | *TRIM2* | -0.940394748 | 0.001431984 | 0.004980661 | 1 |
| T2DM | b_cell | *STIM2* | -1.357076605 | 0.002886983 | 0.005106687 | 1 |
| T2DM | b_cell | *BST1* | -1.630286372 | 0.003392039 | 0.005497821 | 1 |
| T2DM | b_cell | *UFSP2* | -1.090040072 | 0.001818299 | 0.004980661 | 1 |
| T2DM | b_cell | *KLHL5* | -1.787309883 | 0.008188912 | 0.010791038 | 1 |
| T2DM | b_cell | *FAM149A* | -1.077650754 | 0.002671348 | 0.004994752 | 1 |
| T2DM | b_cell | *DDX25* | -1.485591797 | 0.015748975 | 0.019382121 | 1 |
| T2DM | b_cell | *HTATIP2* | -1.336849356 | 0.00504122 | 0.007249556 | 1 |
| T2DM | b_cell | *CTSC* | -0.78306765 | 0.003798211 | 0.005891598 | 1 |
| T2DM | b_cell | *ELP4* | -1.203511158 | 0.0017747 | 0.004980661 | 1 |
| T2DM | b_cell | *TECTA* | -2.599187306 | 0.001712347 | 0.004980661 | 1 |
| T2DM | b_cell | *SC5D* | -1.389061007 | 0.001386472 | 0.004980661 | 1 |
| T2DM | b_cell | *B3GAT1* | -10.91810323 | 0.003742407 | 0.005846002 | 1 |
| T2DM | b_cell | *P2RX3* | -17.03428608 | 0.001965656 | 0.004980661 | 1 |
| T2DM | b_cell | *SNX15* | -6.337286917 | 0.000666321 | 0.004980661 | 1 |
| T2DM | b_cell | *UNC93B1* | -2.841537541 | 0.026413634 | 0.031327048 | 1 |
| T2DM | b_cell | *DCPS* | -1.446539535 | 0.002000438 | 0.004980661 | 1 |
| T2DM | b_cell | *FOXRED1* | -1.30548305 | 0.004486168 | 0.006640295 | 1 |
| T2DM | b_cell | *NRXN2* | -4.340057368 | 0.001614599 | 0.004980661 | 1 |
| T2DM | b_cell | *ST3GAL4* | -1.337029671 | 0.001403863 | 0.004980661 | 1 |
| T2DM | b_cell | *CCDC86* | 0.978738538 | 0.003909567 | 0.006010413 | 1 |
| T2DM | b_cell | *PANX1* | -1.844521036 | 0.001478651 | 0.004980661 | 1 |
| T2DM | b_cell | *CEP164* | -1.06028942 | 0.001849528 | 0.004980661 | 1 |
| T2DM | b_cell | *GALNT18* | -0.871106901 | 0.001940117 | 0.004980661 | 1 |
| T2DM | b_cell | *KIAA1549L* | 0.623931424 | 0.00247353 | 0.004980661 | 1 |
| T2DM | b_cell | *ACCS* | -1.503186479 | 0.026437261 | 0.031347407 | 1 |
| T2DM | b_cell | *SLC35F2* | -1.564410097 | 0.001961095 | 0.004980661 | 1 |
| T2DM | b_cell | *C11orf21* | -9.438131714 | 1.42E-05 | 0.002875366 | 0.06900878 |
| T2DM | b_cell | *RPS13* | -1.348100918 | 0.004393271 | 0.006538697 | 1 |
| T2DM | b_cell | *NDUFS8* | -1.593558977 | 0.001563872 | 0.004980661 | 1 |
| T2DM | b_cell | *TCIRG1* | -1.439644672 | 0.001943905 | 0.004980661 | 1 |
| T2DM | b_cell | *GTF2H1* | -1.785820842 | 0.003763254 | 0.005865503 | 1 |
| T2DM | b_cell | *PRDM4* | -1.180066752 | 0.001704849 | 0.004980661 | 1 |
| T2DM | b_cell | *CORO1C* | -1.719115313 | 0.004672107 | 0.006833456 | 1 |
| T2DM | b_cell | *ASIC1* | -5.949846657 | 0.003663182 | 0.005752881 | 1 |
| T2DM | b_cell | *CAPRIN2* | -1.234985798 | 0.001986902 | 0.004980661 | 1 |
| T2DM | b_cell | *KCTD10* | -1.323826008 | 0.001499837 | 0.004980661 | 1 |
| T2DM | b_cell | *IL23A* | -9.79072408 | 0.001847939 | 0.004980661 | 1 |
| T2DM | b_cell | *ACSS3* | -0.758596227 | 0.002401906 | 0.004980661 | 1 |
| T2DM | b_cell | *LTA4H* | -1.101077296 | 0.006910236 | 0.009386468 | 1 |
| T2DM | b_cell | *PRR4* | -1.018202923 | 0.010719864 | 0.013683972 | 1 |

| | | | | | | |
|---|---|---|---|---|---|---|
| T2DM | b_cell | *PARP11* | -1.30580058 | 0.002089268 | 0.004980661 | 1 |
| T2DM | b_cell | *GPN3* | -1.677505679 | 0.001887017 | 0.004980661 | 1 |
| T2DM | b_cell | *VPS29* | -1.061373068 | 0.001835997 | 0.004980661 | 1 |
| T2DM | b_cell | *AKAP3* | -1.679146598 | 0.006515402 | 0.008932954 | 1 |
| T2DM | b_cell | *ACAD10* | -0.988033019 | 0.001373602 | 0.004980661 | 1 |
| T2DM | b_cell | *LTBR* | -1.479976621 | 0.002180119 | 0.004980661 | 1 |
| T2DM | b_cell | *DDX55* | -2.408512613 | 0.00964496 | 0.012440254 | 1 |
| T2DM | b_cell | *ENDOU* | -4.402770221 | 0.001729773 | 0.004980661 | 1 |
| T2DM | b_cell | *VDR* | -2.418865109 | 0.001742151 | 0.004980661 | 1 |
| T2DM | b_cell | *STX2* | -1.331170067 | 0.007061822 | 0.009546938 | 1 |
| T2DM | b_cell | *CAND1* | -0.908308054 | 0.001722747 | 0.004980661 | 1 |
| T2DM | b_cell | *MDM1* | -1.240496536 | 0.001715123 | 0.004980661 | 1 |
| T2DM | b_cell | *NUP107* | -1.372681878 | 0.002704467 | 0.00500969 | 1 |
| T2DM | b_cell | *KRR1* | -1.266403621 | 0.001350614 | 0.004980661 | 1 |
| T2DM | b_cell | *MRPL51* | -1.307923857 | 0.002352603 | 0.004980661 | 1 |
| T2DM | b_cell | *GNB3* | -3.843668651 | 8.07E-05 | 0.004980661 | 0.391341914 |
| T2DM | b_cell | *CDCA3* | 0.863730867 | 0.006731735 | 0.009182564 | 1 |
| T2DM | b_cell | *CHPT1* | -1.291632345 | 0.001376223 | 0.004980661 | 1 |
| T2DM | b_cell | *GNPTAB* | -0.949820421 | 0.00165597 | 0.004980661 | 1 |
| T2DM | b_cell | *SPSB2* | 1.906852465 | 0.005951493 | 0.008319402 | 1 |
| T2DM | b_cell | *ATN1* | -1.176467453 | 0.002064948 | 0.004980661 | 1 |
| T2DM | b_cell | *NT5DC3* | -1.216505832 | 0.001750408 | 0.004980661 | 1 |
| T2DM | b_cell | *SUDS3* | -1.668798316 | 0.002696532 | 0.005007108 | 1 |
| T2DM | b_cell | *GOLT1B* | -1.652211866 | 0.003706017 | 0.005800514 | 1 |
| T2DM | b_cell | *C2CD5* | -1.321800924 | 0.002721869 | 0.005013768 | 1 |
| T2DM | b_cell | *FRK* | -1.365113753 | 0.001740456 | 0.004980661 | 1 |
| T2DM | b_cell | *RWDD1* | -1.477829874 | 0.001738219 | 0.004980661 | 1 |
| T2DM | b_cell | *TMEM14C* | -1.474479501 | 0.002020192 | 0.004980661 | 1 |
| T2DM | b_cell | *SMIM8* | -0.910658616 | 0.002087659 | 0.004980661 | 1 |
| T2DM | b_cell | *FAM184A* | -1.030951069 | 0.00235143 | 0.004980661 | 1 |
| T2DM | b_cell | *RNGTT* | -1.595048804 | 0.003960951 | 0.006063361 | 1 |
| T2DM | b_cell | *GABRR2* | -2.615172039 | 0.004358806 | 0.006501361 | 1 |
| T2DM | b_cell | *HDDC2* | -1.166898609 | 0.001833091 | 0.004980661 | 1 |
| T2DM | b_cell | *UST* | -0.759618523 | 0.003414288 | 0.005511708 | 1 |
| T2DM | b_cell | *MTRF1L* | -1.14658458 | 0.006149235 | 0.008544127 | 1 |
| T2DM | b_cell | *SLC26A8* | -1.329912381 | 0.038126999 | 0.044245921 | 1 |
| T2DM | b_cell | *MRPL18* | -1.867684946 | 0.003115171 | 0.00526397 | 1 |
| T2DM | b_cell | *RNF8* | -2.203524378 | 0.039604833 | 0.045829394 | 1 |
| T2DM | b_cell | *PHACTR1* | -0.834989431 | 0.001046565 | 0.004980661 | 1 |
| T2DM | b_cell | *MDGA1* | -1.940035813 | 0.001717192 | 0.004980661 | 1 |
| T2DM | b_cell | *CD83* | -1.700793666 | 0.001142476 | 0.004980661 | 1 |
| T2DM | b_cell | *MDN1* | -0.981100577 | 0.001686283 | 0.004980661 | 1 |
| T2DM | b_cell | *GLP1R* | -11.94731702 | 0.012033681 | 0.015216945 | 1 |
| T2DM | b_cell | *BVES* | -3.159157338 | 3.00E-06 | 0.001038114 | 0.014533594 |
| T2DM | b_cell | *COL9A1* | -4.36040598 | 0.006402956 | 0.008820881 | 1 |
| T2DM | b_cell | *WASF1* | -1.052162973 | 0.001805615 | 0.004980661 | 1 |
| T2DM | b_cell | *ALDH5A1* | -1.168312172 | 0.004027704 | 0.006141553 | 1 |
| T2DM | b_cell | *VNN1* | -2.355932962 | 0.003247074 | 0.00536425 | 1 |
| T2DM | b_cell | *VNN2* | 1.280416146 | 0.001370321 | 0.004980661 | 1 |
| T2DM | b_cell | *ACOT13* | -1.223975464 | 0.001912436 | 0.004980661 | 1 |

| | | | | | | |
|---|---|---|---|---|---|---|
| T2DM | b_cell | *C6orf62* | -0.897825402 | 0.00154483 | 0.004980661 | 1 |
| T2DM | b_cell | *HBS1L* | -1.045177023 | 0.001495825 | 0.004980661 | 1 |
| T2DM | b_cell | *PEX7* | -1.608393042 | 0.005529112 | 0.007814522 | 1 |
| T2DM | b_cell | *FIG4* | -1.010155296 | 0.001962858 | 0.004980661 | 1 |
| T2DM | b_cell | *PHACTR2* | -0.868255246 | 0.001449567 | 0.004980661 | 1 |
| T2DM | b_cell | *CUTA* | -1.498099179 | 0.000611685 | 0.004980661 | 1 |
| T2DM | b_cell | *PACRG* | -1.238727806 | 0.001766116 | 0.004980661 | 1 |
| T2DM | b_cell | *QKI* | -0.712764137 | 0.001843613 | 0.004980661 | 1 |
| T2DM | b_cell | *FAM120B* | -1.120621574 | 0.003090888 | 0.005245425 | 1 |
| T2DM | b_cell | *PRPH2* | -1.76841394 | 0.000548285 | 0.004980661 | 1 |
| T2DM | b_cell | *DNPH1* | -1.250662508 | 0.008467029 | 0.011118274 | 1 |
| T2DM | b_cell | *EXOC2* | -1.042467113 | 0.002212023 | 0.004980661 | 1 |
| T2DM | b_cell | *TMEM30A* | -1.296506183 | 0.001905111 | 0.004980661 | 1 |
| T2DM | b_cell | *IMPG1* | -4.538553953 | 0.001693968 | 0.004980661 | 1 |
| T2DM | b_cell | *BTN2A1* | -1.341471101 | 0.001838311 | 0.004980661 | 1 |
| T2DM | b_cell | *LY86* | -3.333593564 | 0.023721482 | 0.02830709 | 1 |
| T2DM | b_cell | *PRSS16* | -4.720570578 | 0.038506835 | 0.044654676 | 1 |
| T2DM | b_cell | *PCDHB2* | -3.683767353 | 0.001996592 | 0.004980661 | 1 |
| T2DM | b_cell | *NUDT12* | -1.21025855 | 0.001787622 | 0.004980661 | 1 |
| T2DM | b_cell | *CEP72* | -1.834354669 | 0.001445786 | 0.004980661 | 1 |
| T2DM | b_cell | *DAP* | -2.023318715 | 0.007423372 | 0.00995528 | 1 |
| T2DM | b_cell | *NME5* | -1.21233774 | 0.008126982 | 0.010729828 | 1 |
| T2DM | b_cell | *KIF20A* | 1.218175024 | 0.003254241 | 0.00536838 | 1 |
| T2DM | b_cell | *PFDN1* | -1.12421136 | 0.002649929 | 0.004985914 | 1 |
| T2DM | b_cell | *TMCO6* | -1.674072177 | 0.001044827 | 0.004980661 | 1 |
| T2DM | b_cell | *SPARC* | -0.952547819 | 0.002236289 | 0.004980661 | 1 |
| T2DM | b_cell | *IK* | -1.470181077 | 0.00159153 | 0.004980661 | 1 |
| T2DM | b_cell | *PCDHB15* | -3.087460839 | 0.000227831 | 0.004980661 | 1 |
| T2DM | b_cell | *HAVCR1* | 0.898910679 | 0.002250221 | 0.004980661 | 1 |
| T2DM | b_cell | *RNF130* | -0.714367103 | 0.002354412 | 0.004980661 | 1 |
| T2DM | b_cell | *ARSB* | -1.772918792 | 0.002726066 | 0.005013768 | 1 |
| T2DM | b_cell | *THBS4* | -1.107762501 | 0.0028411 | 0.005070867 | 1 |
| T2DM | b_cell | *BTNL8* | -1.169136288 | 0.00463106 | 0.006792594 | 1 |
| T2DM | b_cell | *MSH3* | -0.817118617 | 0.001567131 | 0.004980661 | 1 |
| T2DM | b_cell | *RASGRF2* | -0.797198378 | 0.001160728 | 0.004980661 | 1 |
| T2DM | b_cell | *DROSHA* | -1.147691648 | 0.003171157 | 0.005315561 | 1 |
| T2DM | b_cell | *CDH6* | -1.248681381 | 0.000637516 | 0.004980661 | 1 |
| T2DM | b_cell | *LMNB1* | 0.909401644 | 0.00392669 | 0.00602349 | 1 |
| T2DM | b_cell | *SLC12A7* | -1.24462475 | 0.001778441 | 0.004980661 | 1 |
| T2DM | b_cell | *SKP1* | -1.50355535 | 0.00254732 | 0.004980661 | 1 |
| T2DM | b_cell | *PPWD1* | -1.424983277 | 0.002474306 | 0.004980661 | 1 |
| T2DM | b_cell | *TXNDC15* | -1.380778807 | 0.002018598 | 0.004980661 | 1 |
| T2DM | b_cell | *SMAD5* | -1.004162574 | 0.00188667 | 0.004980661 | 1 |
| T2DM | b_cell | *ERGIC1* | -1.726302304 | 0.004301813 | 0.006452055 | 1 |
| T2DM | b_cell | *ATP6V0E1* | -1.45595777 | 0.002625824 | 0.004980661 | 1 |
| T2DM | b_cell | *BNIP1* | -1.440856511 | 0.012359895 | 0.015568591 | 1 |
| T2DM | b_cell | *UNC5A* | -6.263283785 | 0.001761359 | 0.004980661 | 1 |
| T2DM | b_cell | *ACTR8* | 1.412105033 | 0.013361122 | 0.016712597 | 1 |
| T2DM | b_cell | *CLDN16* | -1.907045658 | 0.002056615 | 0.004980661 | 1 |
| T2DM | b_cell | *NPHP3* | -0.858240249 | 0.017287079 | 0.021106419 | 1 |

| | | | | | | |
|---|---|---|---|---|---|---|
| T2DM | b_cell | *PCCB* | -1.013603074 | 0.00735202 | 0.009875969 | 1 |
| T2DM | b_cell | *CEP70* | -1.199847509 | 0.001678747 | 0.004980661 | 1 |
| T2DM | b_cell | *HYAL1* | -1.824577288 | 0.006410339 | 0.008823536 | 1 |
| T2DM | b_cell | *CYB561D2* | -3.793808007 | 0.000891027 | 0.004980661 | 1 |
| T2DM | b_cell | *C3orf14* | -2.257798957 | 0.002401414 | 0.004980661 | 1 |
| T2DM | b_cell | *CBLB* | -0.892134409 | 0.002257949 | 0.004980661 | 1 |
| T2DM | b_cell | *IQCG* | -1.430546357 | 0.002563664 | 0.004980661 | 1 |
| T2DM | b_cell | *NCBP2* | -1.986788064 | 0.001054463 | 0.004980661 | 1 |
| T2DM | b_cell | *PLXNA1* | -1.530999105 | 0.001805978 | 0.004980661 | 1 |
| T2DM | b_cell | *SCAP* | -1.840130304 | 0.003560586 | 0.005663608 | 1 |
| T2DM | b_cell | *KIAA1257* | -3.30558343 | 0.001774689 | 0.004980661 | 1 |
| T2DM | b_cell | *NEK11* | -0.930062733 | 0.002186531 | 0.004980661 | 1 |
| T2DM | b_cell | *PLSCR4* | -0.747735822 | 0.001805372 | 0.004980661 | 1 |
| T2DM | b_cell | *HEMK1* | -1.416388078 | 0.000924269 | 0.004980661 | 1 |
| T2DM | b_cell | *COMMD2* | -1.098021307 | 0.001750727 | 0.004980661 | 1 |
| T2DM | b_cell | *AADAC* | -0.965502945 | 0.026319184 | 0.03122266 | 1 |
| T2DM | b_cell | *ABHD14B* | -1.454477191 | 0.00110439 | 0.004980661 | 1 |
| T2DM | b_cell | *ARHGEF26* | -1.049923426 | 0.001786133 | 0.004980661 | 1 |
| T2DM | b_cell | *VIPR1* | -1.919253356 | 0.002007622 | 0.004980661 | 1 |
| T2DM | b_cell | *DNAH1* | -1.159811074 | 0.004520517 | 0.006668758 | 1 |
| T2DM | b_cell | *NKTR* | -0.959274912 | 0.002987797 | 0.005187056 | 1 |
| T2DM | b_cell | *NEK4* | -2.07327444 | 0.002451506 | 0.004980661 | 1 |
| T2DM | b_cell | *ADAM23* | 0.668972398 | 0.004209495 | 0.006341033 | 1 |
| T2DM | b_cell | *LMAN2L* | -1.793484046 | 0.004168877 | 0.006301702 | 1 |
| T2DM | b_cell | *ACTR1B* | -1.420842248 | 0.002019276 | 0.004980661 | 1 |
| T2DM | b_cell | *SLC35F5* | -0.84517667 | 0.002921612 | 0.005134596 | 1 |
| T2DM | b_cell | *OTOF* | 1.108887217 | 0.002509405 | 0.004980661 | 1 |
| T2DM | b_cell | *CENPA* | 1.585899806 | 0.011388375 | 0.014483983 | 1 |
| T2DM | b_cell | *ITGB6* | -2.020944813 | 0.00050132 | 0.004980661 | 1 |
| T2DM | b_cell | *PPM1G* | -1.80457543 | 0.001767947 | 0.004980661 | 1 |
| T2DM | b_cell | *INO80B* | 1.95659894 | 0.003865852 | 0.005967901 | 1 |
| T2DM | b_cell | *MOGS* | -1.728123356 | 0.00118749 | 0.004980661 | 1 |
| T2DM | b_cell | *GRB14* | -1.391341634 | 0.001866032 | 0.004980661 | 1 |
| T2DM | b_cell | *RTN4* | -0.937388911 | 0.0019844 | 0.004980661 | 1 |
| T2DM | b_cell | *GALNT3* | 2.4022531 | 0.003923034 | 0.00602169 | 1 |
| T2DM | b_cell | *POLE4* | -1.407031773 | 0.002501435 | 0.004980661 | 1 |
| T2DM | b_cell | *ACADL* | -0.995961951 | 0.001568985 | 0.004980661 | 1 |
| T2DM | b_cell | *MRPL19* | -1.363459521 | 0.002087356 | 0.004980661 | 1 |
| T2DM | b_cell | *DNAH6* | -0.62661329 | 0.040510952 | 0.046799795 | 1 |
| T2DM | b_cell | *IGFBP2* | -1.33556666 | 0.001641275 | 0.004980661 | 1 |
| T2DM | b_cell | *GGCX* | -1.525896743 | 0.00173456 | 0.004980661 | 1 |
| T2DM | b_cell | *GNLY* | -1.053548572 | 0.002440395 | 0.004980661 | 1 |
| T2DM | b_cell | *CHMP3* | -1.371129376 | 0.000480673 | 0.004980661 | 1 |
| T2DM | b_cell | *WNT6* | -3.243102258 | 0.00030294 | 0.004980661 | 1 |
| T2DM | b_cell | *IL1RL2* | 1.028656043 | 0.002190184 | 0.004980661 | 1 |
| T2DM | b_cell | *IL18R1* | -2.616019316 | 0.006849437 | 0.009313897 | 1 |
| T2DM | b_cell | *MLPH* | -1.801756669 | 0.002565146 | 0.004980661 | 1 |
| T2DM | b_cell | *HDLBP* | -1.160988938 | 0.002256561 | 0.004980661 | 1 |
| T2DM | b_cell | *TPO* | -1.262903104 | 0.001297947 | 0.004980661 | 1 |
| T2DM | b_cell | *TAF1B* | -1.196121251 | 0.002100457 | 0.004980661 | 1 |

| | | | | | | |
|---|---|---|---|---|---|---|
| T2DM | b_cell | *NOL10* | -0.897916922 | 0.001633429 | 0.004980661 | 1 |
| T2DM | b_cell | *STRN* | -1.361244959 | 0.00266631 | 0.004991101 | 1 |
| T2DM | b_cell | *CEBPZ* | -0.944507905 | 0.001813095 | 0.004980661 | 1 |
| T2DM | b_cell | *PRKD3* | -1.470860901 | 0.003613011 | 0.005715791 | 1 |
| T2DM | b_cell | *SLC25A12* | -1.462233233 | 0.002183036 | 0.004980661 | 1 |
| T2DM | b_cell | *DARS* | -1.435705313 | 0.00281267 | 0.005065458 | 1 |
| T2DM | b_cell | *WIPF1* | -1.63938189 | 0.011333745 | 0.014422064 | 1 |
| T2DM | b_cell | *PNO1* | -1.627592846 | 0.001758648 | 0.004980661 | 1 |
| T2DM | b_cell | *ORC4* | -1.053210416 | 0.002620547 | 0.004980661 | 1 |
| T2DM | b_cell | *AAK1* | -0.844426499 | 0.001916853 | 0.004980661 | 1 |
| T2DM | b_cell | *NFE2L2* | -1.09500237 | 0.003034616 | 0.005228627 | 1 |
| T2DM | b_cell | *MSH6* | -1.77897085 | 0.002755224 | 0.005034338 | 1 |
| T2DM | b_cell | *DNAJC16* | -1.38558324 | 0.003659887 | 0.005752881 | 1 |
| T2DM | b_cell | *MARK1* | -0.924681335 | 0.001819173 | 0.004980661 | 1 |
| T2DM | b_cell | *MORN1* | -1.4335962 | 0.003155504 | 0.005303461 | 1 |
| T2DM | b_cell | *GPX7* | -2.582467697 | 4.36E-06 | 0.001410638 | 0.021159568 |
| T2DM | b_cell | *CEP104* | -1.159238476 | 0.002032353 | 0.004980661 | 1 |
| T2DM | b_cell | *FAM20B* | -1.622835797 | 0.002387149 | 0.004980661 | 1 |
| T2DM | b_cell | *WRAP73* | -2.360043532 | 0.006217041 | 0.008624423 | 1 |
| T2DM | b_cell | *PARK7* | -1.671875479 | 0.003414714 | 0.005511708 | 1 |
| T2DM | b_cell | *MECR* | -1.983314548 | 0.003398894 | 0.005502647 | 1 |
| T2DM | b_cell | *KCNC4* | -1.493595473 | 0.000837365 | 0.004980661 | 1 |
| T2DM | b_cell | *EDEM3* | -1.512679164 | 0.002251936 | 0.004980661 | 1 |
| T2DM | b_cell | *RAP1A* | -1.106088391 | 0.002290872 | 0.004980661 | 1 |
| T2DM | b_cell | *CAPZA1* | -1.33246844 | 0.002340273 | 0.004980661 | 1 |
| T2DM | b_cell | *ASH1L* | -0.831142311 | 0.003184684 | 0.005332504 | 1 |
| T2DM | b_cell | *GON4L* | -1.285355079 | 0.003167146 | 0.005314172 | 1 |
| T2DM | b_cell | *DOCK7* | -1.164440871 | 0.0023579 | 0.004980661 | 1 |
| T2DM | b_cell | *FBXO6* | 1.376050781 | 0.00402346 | 0.00613701 | 1 |
| T2DM | b_cell | *DNAJC6* | -1.461104026 | 0.001543001 | 0.004980661 | 1 |
| T2DM | b_cell | *LEPR* | -1.118116951 | 0.001910457 | 0.004980661 | 1 |
| T2DM | b_cell | *IVNS1ABP* | -1.245362891 | 0.002167466 | 0.004980661 | 1 |
| T2DM | b_cell | *MFN2* | -1.705154943 | 0.001857885 | 0.004980661 | 1 |
| T2DM | b_cell | *PRG4* | -1.198421642 | 0.00164597 | 0.004980661 | 1 |
| T2DM | b_cell | *NCF2* | -1.453098379 | 0.003855833 | 0.005965584 | 1 |
| T2DM | b_cell | *CFHR3* | -5.268351746 | 0.013341119 | 0.01669188 | 1 |
| T2DM | b_cell | *CRYZ* | -1.28697678 | 0.001076156 | 0.004980661 | 1 |
| T2DM | b_cell | *TMEM9* | -1.602795639 | 0.000817122 | 0.004980661 | 1 |
| T2DM | b_cell | *WARS2* | -1.328558665 | 0.001431934 | 0.004980661 | 1 |
| T2DM | b_cell | *OSCP1* | -0.991595831 | 0.004059009 | 0.006172326 | 1 |
| T2DM | b_cell | *MRPS15* | -1.891822896 | 0.002901455 | 0.005113643 | 1 |
| T2DM | b_cell | *EXOC8* | 1.248765469 | 0.003909824 | 0.006010413 | 1 |
| T2DM | b_cell | *GNPAT* | -1.303859271 | 0.001606607 | 0.004980661 | 1 |
| T2DM | b_cell | *C1orf109* | 1.682459136 | 0.040708282 | 0.047016563 | 1 |
| T2DM | b_cell | *TBCE* | -1.045556952 | 0.00190793 | 0.004980661 | 1 |
| T2DM | b_cell | *ACADM* | -0.834282664 | 0.002136733 | 0.004980661 | 1 |
| T2DM | b_cell | *PADI2* | -3.50520178 | 0.004373313 | 0.006512988 | 1 |
| T2DM | b_cell | *SDHB* | -1.080873069 | 0.002023003 | 0.004980661 | 1 |
| T2DM | b_cell | *CTBS* | -1.645948929 | 0.00132939 | 0.004980661 | 1 |
| T2DM | b_cell | *KLHL12* | -1.182887795 | 0.002515477 | 0.004980661 | 1 |

| | | | | | | |
|---|---|---|---|---|---|---|
| T2DM | b_cell | *ZNHIT6* | -1.180556216 | 0.001851862 | 0.004980661 | 1 |
| T2DM | b_cell | *KIF17* | -3.057750009 | 0.001827792 | 0.004980661 | 1 |
| T2DM | b_cell | *CD46* | -0.905084678 | 0.00305031 | 0.005229719 | 1 |
| T2DM | b_cell | *EBNA1BP2* | -1.33926952 | 0.001436525 | 0.004980661 | 1 |
| T2DM | b_cell | *ERI3* | -1.577563752 | 0.003355783 | 0.005460611 | 1 |
| T2DM | b_cell | *PRDX1* | -1.471670931 | 0.002741671 | 0.005023637 | 1 |
| T2DM | b_cell | *CCDC181* | 1.651479508 | 0.038154445 | 0.044267185 | 1 |
| T2DM | b_cell | *NSUN4* | -2.002179434 | 0.000940687 | 0.004980661 | 1 |
| T2DM | b_cell | *DR1* | -1.323792802 | 0.0035709 | 0.005668618 | 1 |
| T2DM | b_cell | *PRRC2C* | -1.093080362 | 0.0025402 | 0.004980661 | 1 |
| T2DM | b_cell | *ABCD3* | -1.231537531 | 0.001974948 | 0.004980661 | 1 |
| T2DM | b_cell | *VAMP4* | -1.382716573 | 0.003682208 | 0.005776293 | 1 |
| T2DM | b_cell | *PTBP2* | -0.844262918 | 0.003078886 | 0.005245002 | 1 |
| T2DM | b_cell | *DARS2* | -1.702429406 | 0.002304906 | 0.004980661 | 1 |
| T2DM | b_cell | *MTFR1L* | -2.588556248 | 0.007909619 | 0.010485648 | 1 |
| T2DM | b_cell | *RPS6KA1* | -1.814472112 | 0.001091868 | 0.004980661 | 1 |
| T2DM | b_cell | *RPA2* | -2.011757464 | 0.001957526 | 0.004980661 | 1 |
| T2DM | b_cell | *MARC2* | -0.975266605 | 0.001831032 | 0.004980661 | 1 |
| T2DM | b_cell | *SLC5A9* | -1.366093908 | 0.002286438 | 0.004980661 | 1 |
| T2DM | b_cell | *ESYT2* | -1.119208894 | 0.00238122 | 0.004980661 | 1 |
| T2DM | b_cell | *CD3EAP* | 1.271401346 | 0.009331649 | 0.012073909 | 1 |
| T2DM | b_cell | *RCN2* | -1.304122685 | 0.002364784 | 0.004980661 | 1 |
| T2DM | b_cell | *IFT46* | -2.328656743 | 0.001985818 | 0.004980661 | 1 |
| T2DM | b_cell | *APOA1* | 2.338089573 | 0.00118554 | 0.004980661 | 1 |
| T2DM | b_cell | *DDX59* | -2.006697331 | 0.001974668 | 0.004980661 | 1 |
| T2DM | b_cell | *CAMSAP2* | -0.822263147 | 0.002016557 | 0.004980661 | 1 |
| T2DM | b_cell | *KLF7* | -1.231867779 | 0.003206711 | 0.005339692 | 1 |
| T2DM | b_cell | *CASC1* | -1.662307258 | 0.00043498 | 0.004980661 | 1 |
| T2DM | b_cell | *ELOVL4* | 3.638040948 | 6.96E-05 | 0.004980661 | 0.337920835 |
| T2DM | b_cell | *CNR1* | -1.765188344 | 0.005645739 | 0.00795387 | 1 |
| T2DM | b_cell | *SGIP1* | -1.228518034 | 0.001820903 | 0.004980661 | 1 |
| T2DM | b_cell | *AKAP7* | -1.071962317 | 0.001726441 | 0.004980661 | 1 |
| T2DM | b_cell | *ALDH8A1* | -1.596695138 | 0.002000305 | 0.004980661 | 1 |
| T2DM | b_cell | *MED28* | -2.062890426 | 0.002179482 | 0.004980661 | 1 |
| T2DM | b_cell | *TMEM5* | -1.23242001 | 0.004190953 | 0.006324704 | 1 |
| T2DM | b_cell | *VAMP8* | -1.254450564 | 0.00148587 | 0.004980661 | 1 |
| T2DM | b_cell | *FOXO3* | -0.72331432 | 0.001911685 | 0.004980661 | 1 |
| T2DM | b_cell | *SPP1* | -3.520627481 | 0.010596424 | 0.013537085 | 1 |
| T2DM | b_cell | *MFSD1* | -1.234277518 | 0.002028005 | 0.004980661 | 1 |
| T2DM | b_cell | *UBN1* | -0.941999756 | 0.001877126 | 0.004980661 | 1 |
| T2DM | b_cell | *WDR35* | -1.012224157 | 0.00179855 | 0.004980661 | 1 |
| T2DM | b_cell | *DNAH7* | -1.319147351 | 0.001230195 | 0.004980661 | 1 |
| T2DM | b_cell | *C2orf40* | -1.584233219 | 0.001889082 | 0.004980661 | 1 |
| T2DM | b_cell | *CPSF3* | -1.470892514 | 0.002457301 | 0.004980661 | 1 |
| T2DM | b_cell | *PIGZ* | -1.426475234 | 0.011901505 | 0.015074955 | 1 |
| T2DM | b_cell | *SENP5* | -1.415178845 | 0.002925964 | 0.005134596 | 1 |
| T2DM | b_cell | *CCDC92* | -1.740400336 | 0.00090482 | 0.004980661 | 1 |
| T2DM | b_cell | *FKBP15* | -1.109699479 | 0.001642633 | 0.004980661 | 1 |
| T2DM | b_cell | *RAB14* | -1.592130117 | 0.002842936 | 0.005071296 | 1 |
| T2DM | b_cell | *TRIM32* | -1.946465211 | 0.001891403 | 0.004980661 | 1 |

| | | | | | | |
|---|---|---|---|---|---|---|
| T2DM | b_cell | *PPP6C* | -1.395684066 | 0.002335037 | 0.004980661 | 1 |
| T2DM | b_cell | *HDHD3* | -1.269749054 | 0.000243526 | 0.004980661 | 1 |
| T2DM | b_cell | *RBM18* | -2.706927899 | 0.006963602 | 0.009440457 | 1 |
| T2DM | b_cell | *HSDL2* | -0.734198578 | 0.002867111 | 0.005094011 | 1 |
| T2DM | b_cell | *MAPKAP1* | -1.345624318 | 0.003260304 | 0.005371476 | 1 |
| T2DM | b_cell | *INVS* | -0.89239333 | 0.003167474 | 0.005314172 | 1 |
| T2DM | b_cell | *DENND1A* | -0.800233625 | 0.002079883 | 0.004980661 | 1 |
| T2DM | b_cell | *KDSR* | -1.269389639 | 0.001798691 | 0.004980661 | 1 |
| T2DM | b_cell | *DCAF4* | -1.63273344 | 0.001414366 | 0.004980661 | 1 |
| T2DM | b_cell | *IFI27L2* | -2.382387637 | 0.003465643 | 0.005573516 | 1 |
| T2DM | b_cell | *NEK9* | -1.157869796 | 0.001832246 | 0.004980661 | 1 |
| T2DM | b_cell | *TTLL5* | -1.012716232 | 0.001716268 | 0.004980661 | 1 |
| T2DM | b_cell | *PPP4R4* | -1.688271114 | 0.004318284 | 0.006464391 | 1 |
| T2DM | b_cell | *ALDH6A1* | -1.065248095 | 0.003457268 | 0.005565581 | 1 |
| T2DM | b_cell | *ESRRB* | -2.145905969 | 0.00454256 | 0.006687045 | 1 |
| T2DM | b_cell | *EIF2B2* | -1.625378938 | 0.004135194 | 0.006267133 | 1 |
| T2DM | b_cell | *COQ6* | -2.013744526 | 0.002315997 | 0.004980661 | 1 |
| T2DM | b_cell | *DNMT3A* | -1.478928921 | 0.002827135 | 0.005065458 | 1 |
| T2DM | b_cell | *CNRIP1* | -1.057630858 | 0.00081608 | 0.004980661 | 1 |
| T2DM | b_cell | *EPCAM* | -8.794455748 | 0.001278451 | 0.004980661 | 1 |
| T2DM | b_cell | *SLC17A5* | -1.113973448 | 0.002579661 | 0.004980661 | 1 |
| T2DM | b_cell | *TECTB* | -5.721307672 | 0.003116404 | 0.00526397 | 1 |
| T2DM | b_cell | *ELOVL3* | 2.797134433 | 0.014216358 | 0.017682074 | 1 |
| T2DM | b_cell | *PPP1R3C* | -1.90094728 | 0.002070558 | 0.004980661 | 1 |
| T2DM | b_cell | *PYROXD2* | -1.121074332 | 0.002562526 | 0.004980661 | 1 |
| T2DM | b_cell | *MXI1* | -1.319981891 | 0.001694891 | 0.004980661 | 1 |
| T2DM | b_cell | *HELLS* | -2.143400406 | 0.00540832 | 0.007684091 | 1 |
| T2DM | b_cell | *TCTN3* | -1.754807238 | 0.002048799 | 0.004980661 | 1 |
| T2DM | b_cell | *WDR11* | -1.470859951 | 0.002727014 | 0.005013768 | 1 |
| T2DM | b_cell | *KANSL1* | -0.852183051 | 0.002154501 | 0.004980661 | 1 |
| T2DM | b_cell | *HOXB3* | -0.976317129 | 0.001793484 | 0.004980661 | 1 |
| T2DM | b_cell | *CAAP1* | -1.01920129 | 0.001230697 | 0.004980661 | 1 |
| T2DM | b_cell | *GORAB* | -1.326881571 | 0.002202958 | 0.004980661 | 1 |
| T2DM | b_cell | *ACAT2* | -1.550023165 | 0.00226701 | 0.004980661 | 1 |
| T2DM | b_cell | *SNX19* | -1.486395288 | 0.001772457 | 0.004980661 | 1 |
| T2DM | b_cell | *SLC10A7* | -1.317190531 | 0.002422558 | 0.004980661 | 1 |
| T2DM | b_cell | *MASTL* | -3.05105759 | 0.000452246 | 0.004980661 | 1 |
| T2DM | b_cell | *MRC1* | -0.814270789 | 0.002148157 | 0.004980661 | 1 |
| T2DM | b_cell | *IQSEC3* | 1.60294266 | 0.006126096 | 0.008522692 | 1 |
| T2DM | b_cell | *CCDC77* | -1.404307088 | 0.000893764 | 0.004980661 | 1 |
| T2DM | b_cell | *DNAJC15* | -1.628237114 | 0.002725489 | 0.005013768 | 1 |
| T2DM | b_cell | *UFM1* | -1.030649288 | 0.001740234 | 0.004980661 | 1 |
| T2DM | b_cell | *ALG5* | -1.858216909 | 0.003192283 | 0.00533738 | 1 |
| T2DM | b_cell | *SIL1* | -1.247950877 | 0.002777684 | 0.005051064 | 1 |
| T2DM | b_cell | *ZFP30* | -1.206200589 | 0.00226289 | 0.004980661 | 1 |
| T2DM | b_cell | *ARL1* | -1.305033812 | 0.001696559 | 0.004980661 | 1 |
| T2DM | b_cell | *APAF1* | -1.662588676 | 0.002194046 | 0.004980661 | 1 |
| T2DM | b_cell | *EPHX2* | -1.626457583 | 0.000831529 | 0.004980661 | 1 |
| T2DM | b_cell | *AKAP1* | -0.995547353 | 0.001636619 | 0.004980661 | 1 |
| T2DM | b_cell | *SCPEP1* | -1.075760921 | 0.00196048 | 0.004980661 | 1 |

| | | | | | | |
|---|---|---|---|---|---|---|
| T2DM | b_cell | *TBX2* | -1.532611297 | 0.01612161 | 0.019788022 | 1 |
| T2DM | b_cell | *TEX14* | -1.478759828 | 0.020664917 | 0.024879945 | 1 |
| T2DM | b_cell | *TRIM6* | -3.676507187 | 0.001459344 | 0.004980661 | 1 |
| T2DM | b_cell | *ADCY7* | -1.12293426 | 0.001354089 | 0.004980661 | 1 |
| T2DM | b_cell | *ECHDC2* | -0.758372212 | 0.000998546 | 0.004980661 | 1 |
| T2DM | b_cell | *TAS2R10* | 1.733921721 | 0.002493133 | 0.004980661 | 1 |
| T2DM | b_cell | *PYROXD1* | -1.341775312 | 0.000899969 | 0.004980661 | 1 |
| T2DM | b_cell | *ZNF549* | -1.646272251 | 0.00221571 | 0.004980661 | 1 |
| T2DM | b_cell | *A1BG* | -2.427151963 | 0.002412208 | 0.004980661 | 1 |
| T2DM | b_cell | *ZSCAN18* | -1.053473429 | 0.00182646 | 0.004980661 | 1 |
| T2DM | b_cell | *PDZRN3* | -0.765193642 | 0.000391969 | 0.004980661 | 1 |
| T2DM | b_cell | *LHX4* | -3.206532465 | 0.011793424 | 0.014952102 | 1 |
| T2DM | b_cell | *SEC22A* | -1.325999333 | 0.003109303 | 0.005260699 | 1 |
| T2DM | b_cell | *POPDC2* | 0.857644235 | 0.006746361 | 0.009199928 | 1 |
| T2DM | b_cell | *CD80* | -3.785584779 | 2.26E-05 | 0.003918133 | 0.109707712 |
| T2DM | b_cell | *MAPK8IP1* | -3.437105714 | 0.005339438 | 0.007599575 | 1 |
| T2DM | b_cell | *CAT* | -0.749042534 | 0.001609233 | 0.004980661 | 1 |
| T2DM | b_cell | *PILRB* | -0.931961959 | 0.026535496 | 0.031448516 | 1 |
| T2DM | b_cell | *ZCCHC17* | -2.096188978 | 0.034808202 | 0.040627711 | 1 |
| T2DM | b_cell | *FABP3* | -2.494059806 | 0.001465005 | 0.004980661 | 1 |
| T2DM | b_cell | *TNFSF10* | -1.012438113 | 0.002020027 | 0.004980661 | 1 |
| T2DM | b_cell | *ZNF639* | -1.827000104 | 0.001806838 | 0.004980661 | 1 |
| T2DM | b_cell | *CPXM2* | -1.041331746 | 0.001661772 | 0.004980661 | 1 |
| T2DM | b_cell | *FLT3* | 2.109682835 | 0.001942495 | 0.004980661 | 1 |
| T2DM | b_cell | *MTIF3* | -1.221485421 | 0.002823312 | 0.005065458 | 1 |
| T2DM | b_cell | *FMOD* | -2.531101628 | 0.012012303 | 0.015197835 | 1 |
| T2DM | b_cell | *CD244* | -2.950535838 | 0.003411321 | 0.005510766 | 1 |
| T2DM | b_cell | *RBBP6* | -0.931605566 | 0.001791305 | 0.004980661 | 1 |
| T2DM | b_cell | *SERAC1* | -1.447466991 | 0.001327091 | 0.004980661 | 1 |
| T2DM | b_cell | *LDB3* | -1.643744791 | 0.002626721 | 0.004980661 | 1 |
| T2DM | b_cell | *OPN4* | -9.171905722 | 0.013550836 | 0.016919366 | 1 |
| T2DM | b_cell | *FAM35A* | -1.401065989 | 0.003491528 | 0.005596596 | 1 |
| T2DM | b_cell | *NAA60* | -3.180028253 | 0.004333017 | 0.00647259 | 1 |
| T2DM | b_cell | *ODF2L* | -1.396024347 | 0.003207515 | 0.005339692 | 1 |
| T2DM | b_cell | *SPATA1* | -1.326834009 | 0.002616835 | 0.004980661 | 1 |
| T2DM | b_cell | *LRRC39* | -1.108659299 | 0.006788347 | 0.009246788 | 1 |
| T2DM | b_cell | *RWDD3* | 0.563676437 | 0.00225006 | 0.004980661 | 1 |
| T2DM | b_cell | *PMS2* | -1.435818876 | 0.004012661 | 0.006124262 | 1 |
| T2DM | b_cell | *EEPD1* | -0.73128295 | 0.001102851 | 0.004980661 | 1 |
| T2DM | b_cell | *KLHL7* | -1.043854838 | 0.001872519 | 0.004980661 | 1 |
| T2DM | b_cell | *NT5C3A* | -1.924765673 | 0.014571598 | 0.018059104 | 1 |
| T2DM | b_cell | *CCZ1* | -1.392198088 | 0.002718789 | 0.005012128 | 1 |
| T2DM | b_cell | *POLM* | -1.171791344 | 0.001315067 | 0.004980661 | 1 |
| T2DM | b_cell | *SMU1* | -1.564315261 | 0.003275828 | 0.005386079 | 1 |
| T2DM | b_cell | *GLIPR2* | -1.384954867 | 0.002414865 | 0.004980661 | 1 |
| T2DM | b_cell | *TRIM24* | -1.170911369 | 0.001581796 | 0.004980661 | 1 |
| T2DM | b_cell | *C7orf49* | -0.931895234 | 0.008232703 | 0.010836971 | 1 |
| T2DM | b_cell | *CISD1* | -2.321787216 | 0.002185463 | 0.004980661 | 1 |
| T2DM | b_cell | *RBM19* | -1.061138975 | 0.001777471 | 0.004980661 | 1 |
| T2DM | b_cell | *CCDC91* | -0.744450984 | 0.001888886 | 0.004980661 | 1 |

| | | | | | | |
|---|---|---|---|---|---|---|
| T2DM | b_cell | *EBPL* | -1.132181254 | 0.003037948 | 0.005228627 | 1 |
| T2DM | b_cell | *ATP7B* | -0.952799703 | 0.002603833 | 0.004980661 | 1 |
| T2DM | b_cell | *CENPK* | -2.288977654 | 0.001304701 | 0.004980661 | 1 |
| T2DM | b_cell | *ITIH5* | -1.171949281 | 0.001067753 | 0.004980661 | 1 |
| T2DM | b_cell | *TSFM* | -1.310656661 | 0.003543053 | 0.005649323 | 1 |
| T2DM | b_cell | *ARHGAP9* | -1.947581535 | 0.001337995 | 0.004980661 | 1 |
| T2DM | b_cell | *PFDN5* | -1.306185656 | 0.002113397 | 0.004980661 | 1 |
| T2DM | b_cell | *PDE1B* | -1.457711698 | 0.00170413 | 0.004980661 | 1 |
| T2DM | b_cell | *SMUG1* | -1.125656926 | 0.002900252 | 0.005113643 | 1 |
| T2DM | b_cell | *SARDH* | -2.524809032 | 0.006814472 | 0.009279769 | 1 |
| T2DM | b_cell | *DBH* | 3.593142717 | 0.002435644 | 0.004980661 | 1 |
| T2DM | b_cell | *NDUFAF4* | -1.750391035 | 0.004366421 | 0.006506718 | 1 |
| T2DM | b_cell | *USP45* | -0.839269591 | 0.001782546 | 0.004980661 | 1 |
| T2DM | b_cell | *TTC21B* | -1.342670104 | 0.001604525 | 0.004980661 | 1 |
| T2DM | b_cell | *ACVR1C* | -0.830986176 | 0.000670696 | 0.004980661 | 1 |
| T2DM | b_cell | *SLC36A1* | -1.232363191 | 0.006550023 | 0.008975067 | 1 |
| T2DM | b_cell | *PLA2G12A* | -1.193480246 | 0.002086209 | 0.004980661 | 1 |
| T2DM | b_cell | *C4BPA* | -2.419860202 | 0.00203176 | 0.004980661 | 1 |
| T2DM | b_cell | *C4BPB* | -2.415665832 | 0.00152513 | 0.004980661 | 1 |
| T2DM | b_cell | *RAB38* | -1.382336264 | 0.005559223 | 0.007852504 | 1 |
| T2DM | b_cell | *DAW1* | -3.163219868 | 0.004989439 | 0.007181477 | 1 |
| T2DM | b_cell | *DNPEP* | -1.420574737 | 0.002268654 | 0.004980661 | 1 |
| T2DM | b_cell | *OBSL1* | -1.210982133 | 0.00121184 | 0.004980661 | 1 |
| T2DM | b_cell | *SLC12A4* | -1.334814589 | 0.001879096 | 0.004980661 | 1 |
| T2DM | b_cell | *WFDC3* | -1.462729483 | 0.004048662 | 0.006163369 | 1 |
| T2DM | b_cell | *SLC12A5* | 1.194939544 | 0.009177418 | 0.011902922 | 1 |
| T2DM | b_cell | *MATN4* | 1.670436623 | 0.003601671 | 0.005703429 | 1 |
| T2DM | b_cell | *VAPB* | -1.291591954 | 0.000590464 | 0.004980661 | 1 |
| T2DM | b_cell | *TOX2* | -1.697716072 | 0.000945537 | 0.004980661 | 1 |
| T2DM | b_cell | *SRSF6* | -0.961496976 | 0.001796881 | 0.004980661 | 1 |
| T2DM | b_cell | *CSE1L* | -1.08643074 | 0.001547017 | 0.004980661 | 1 |
| T2DM | b_cell | *PTGIS* | -0.820163601 | 0.003287073 | 0.0053936 | 1 |
| T2DM | b_cell | *RNF114* | -1.050369917 | 0.001638376 | 0.004980661 | 1 |
| T2DM | b_cell | *NEURL2* | 3.076005274 | 0.001858982 | 0.004980661 | 1 |
| T2DM | b_cell | *MTRR* | -1.153662399 | 0.001714704 | 0.004980661 | 1 |
| T2DM | b_cell | *PEPD* | -0.98433907 | 0.001809594 | 0.004980661 | 1 |
| T2DM | b_cell | *CHST8* | -2.606926455 | 0.004049633 | 0.006163369 | 1 |
| T2DM | b_cell | *STAMBP* | -1.045715671 | 0.001800719 | 0.004980661 | 1 |
| T2DM | b_cell | *NAGK* | -1.217917839 | 0.001451706 | 0.004980661 | 1 |
| T2DM | b_cell | *ATP8A1* | -0.92032827 | 0.001842902 | 0.004980661 | 1 |
| T2DM | b_cell | *USP22* | -1.298128425 | 0.002481087 | 0.004980661 | 1 |
| T2DM | b_cell | *HIF3A* | -0.986713072 | 0.001447048 | 0.004980661 | 1 |
| T2DM | b_cell | *ZNF576* | -1.514528509 | 0.002611368 | 0.004980661 | 1 |
| T2DM | b_cell | *SNRPC* | -1.755159011 | 0.005502023 | 0.007792124 | 1 |
| T2DM | b_cell | *SERPINB6* | -1.599995274 | 0.004227801 | 0.006360711 | 1 |
| T2DM | b_cell | *PEX6* | -1.47468276 | 0.00233576 | 0.004980661 | 1 |
| T2DM | b_cell | *NQO2* | -1.128528934 | 0.00187339 | 0.004980661 | 1 |
| T2DM | b_cell | *RPS10* | 0.700723673 | 0.00427514 | 0.006423964 | 1 |
| T2DM | b_cell | *MED20* | -2.054515046 | 0.00140357 | 0.004980661 | 1 |
| T2DM | b_cell | *GNMT* | 0.97370344 | 0.001968052 | 0.004980661 | 1 |

| | | | | | | |
|---|---|---|---|---|---|---|
| T2DM | b_cell | *TREM1* | 2.589766778 | 0.004510741 | 0.006658386 | 1 |
| T2DM | b_cell | *MEA1* | -3.843009669 | 0.00073155 | 0.004980661 | 1 |
| T2DM | b_cell | *KLHL31* | -0.991071712 | 0.004954587 | 0.007137665 | 1 |
| T2DM | b_cell | *KCNK17* | -7.754866254 | 7.00E-06 | 0.00178716 | 0.033956043 |
| T2DM | b_cell | *RREB1* | -0.808877358 | 0.001623822 | 0.004980661 | 1 |
| T2DM | b_cell | *RIOK1* | -1.199802241 | 0.00438202 | 0.006523953 | 1 |
| T2DM | b_cell | *ATXN1* | -0.895688299 | 0.002102388 | 0.004980661 | 1 |
| T2DM | b_cell | *RUNX2* | -2.154872442 | 0.007543729 | 0.010080466 | 1 |
| T2DM | b_cell | *CXCL6* | -3.375448858 | 0.001400413 | 0.004980661 | 1 |
| T2DM | b_cell | *SH3TC1* | -1.386346253 | 0.000908814 | 0.004980661 | 1 |
| T2DM | b_cell | *BBS2* | -1.125845013 | 0.015790193 | 0.019425461 | 1 |
| T2DM | b_cell | *MT1G* | -5.570283836 | 0.035581354 | 0.041430461 | 1 |
| T2DM | b_cell | *GOT2* | -3.079385962 | 0.012760313 | 0.016018898 | 1 |
| T2DM | b_cell | *CLYBL* | -1.170268641 | 0.00196852 | 0.004980661 | 1 |
| T2DM | b_cell | *TMTC4* | -1.503332959 | 0.001753699 | 0.004980661 | 1 |
| T2DM | b_cell | *ABCC4* | -1.216449808 | 0.00194164 | 0.004980661 | 1 |
| T2DM | b_cell | *ATP5S* | -1.085227784 | 0.004481537 | 0.006636059 | 1 |
| T2DM | b_cell | *TEKT3* | -1.744824297 | 0.012026563 | 0.015211909 | 1 |
| T2DM | b_cell | *HS3ST3B1* | -2.651316636 | 0.00130782 | 0.004980661 | 1 |
| T2DM | b_cell | *MRPS7* | -1.259908604 | 0.001819288 | 0.004980661 | 1 |
| T2DM | b_cell | *GGA3* | -1.095286014 | 0.001841926 | 0.004980661 | 1 |
| T2DM | b_cell | *NUP85* | -1.080779871 | 0.001844999 | 0.004980661 | 1 |
| T2DM | b_cell | *NT5C* | -1.510587616 | 0.001930632 | 0.004980661 | 1 |
| T2DM | b_cell | *DDX31* | -1.156458218 | 0.00134377 | 0.004980661 | 1 |
| T2DM | b_cell | *OPRL1* | 4.672345552 | 0.007790497 | 0.010350354 | 1 |
| T2DM | b_cell | *IL1B* | 4.087512203 | 0.003109581 | 0.005260699 | 1 |
| T2DM | b_cell | *PAX8* | -1.563818399 | 0.013406177 | 0.016764632 | 1 |
| T2DM | b_cell | *POLR1B* | -1.390472845 | 0.001451962 | 0.004980661 | 1 |
| T2DM | b_cell | *CCDC93* | -1.483099285 | 0.00321313 | 0.005339692 | 1 |
| T2DM | b_cell | *PSD4* | -1.978839164 | 0.001624971 | 0.004980661 | 1 |
| T2DM | b_cell | *GTF2F1* | -1.939175898 | 0.002002446 | 0.004980661 | 1 |
| T2DM | b_cell | *TNFSF9* | -37.19915788 | 0.001094122 | 0.004980661 | 1 |
| T2DM | b_cell | *GPR108* | -1.698527051 | 0.00319358 | 0.00533738 | 1 |
| T2DM | b_cell | *SNRPD2* | -1.653888694 | 0.002062244 | 0.004980661 | 1 |
| T2DM | b_cell | *SDCBP2* | -3.571821929 | 0.003225734 | 0.005352058 | 1 |
| T2DM | b_cell | *PSMF1* | -1.625317578 | 0.001071375 | 0.004980661 | 1 |
| T2DM | b_cell | *DTD1* | -1.479336975 | 0.002526551 | 0.004980661 | 1 |
| T2DM | b_cell | *RBCK1* | -1.130742165 | 0.00148312 | 0.004980661 | 1 |
| T2DM | b_cell | *RRBP1* | -0.924465749 | 0.002149329 | 0.004980661 | 1 |
| T2DM | b_cell | *FLRT3* | -1.396467189 | 0.001700259 | 0.004980661 | 1 |
| T2DM | b_cell | *PCSK2* | -6.169594293 | 0.002552401 | 0.004980661 | 1 |
| T2DM | b_cell | *MKKS* | -1.405803957 | 0.002658382 | 0.004985914 | 1 |
| T2DM | b_cell | *LAMP5* | 3.172729044 | 0.002561446 | 0.004980661 | 1 |
| T2DM | b_cell | *MGME1* | -1.979907385 | 0.00890996 | 0.01162127 | 1 |
| T2DM | b_cell | *MCM8* | -1.608025651 | 0.006107581 | 0.008503294 | 1 |
| T2DM | b_cell | *MRPS26* | -1.363517726 | 0.002399514 | 0.004980661 | 1 |
| T2DM | b_cell | *NCLN* | 1.602950413 | 0.037312113 | 0.043362484 | 1 |
| T2DM | b_cell | *UROD* | 2.202167333 | 0.00770676 | 0.010258766 | 1 |
| T2DM | b_cell | *KLC1* | -1.416877685 | 0.002449177 | 0.004980661 | 1 |
| T2DM | b_cell | *TUBGCP3* | -1.347226387 | 0.002313759 | 0.004980661 | 1 |

| | | | | | | |
|---|---|---|---|---|---|---|
| T2DM | b_cell | *MCF2L* | -1.088564601 | 0.00568419 | 0.00799643 | 1 |
| T2DM | b_cell | *COX6B1* | -1.114684216 | 0.001742014 | 0.004980661 | 1 |
| T2DM | b_cell | *PRDX5* | -1.761277122 | 0.002291034 | 0.004980661 | 1 |
| T2DM | b_cell | *IRF3* | -1.413512967 | 0.002456147 | 0.004980661 | 1 |
| T2DM | b_cell | *PRR12* | -1.072066855 | 0.005491038 | 0.007778837 | 1 |
| T2DM | b_cell | *TRAP1* | -1.299243011 | 0.010485629 | 0.013416738 | 1 |
| T2DM | b_cell | *EMG1* | -1.257817082 | 0.001801315 | 0.004980661 | 1 |
| T2DM | b_cell | *ATG14* | -0.887274995 | 0.002125719 | 0.004980661 | 1 |
| T2DM | b_cell | *KTN1* | -1.165971512 | 0.002620431 | 0.004980661 | 1 |
| T2DM | b_cell | *ZBTB1* | -0.962624359 | 0.001834363 | 0.004980661 | 1 |
| T2DM | b_cell | *RHOT1* | -0.848962002 | 0.001233112 | 0.004980661 | 1 |
| T2DM | b_cell | *WDR60* | -0.994218079 | 0.001662821 | 0.004980661 | 1 |
| T2DM | b_cell | *AIF1L* | -1.706085007 | 0.00183686 | 0.004980661 | 1 |
| T2DM | b_cell | *CANX* | -1.636478503 | 0.004249737 | 0.00638777 | 1 |
| T2DM | b_cell | *ZNF484* | -1.326036425 | 0.001788969 | 0.004980661 | 1 |
| T2DM | b_cell | *FGD3* | -1.270423704 | 0.001843756 | 0.004980661 | 1 |
| T2DM | b_cell | *ABHD8* | -1.478126322 | 0.000294091 | 0.004980661 | 1 |
| T2DM | b_cell | *ATP13A4* | 1.891797173 | 0.003239454 | 0.005362236 | 1 |
| T2DM | b_cell | *HELB* | -2.031812779 | 0.007536878 | 0.010075596 | 1 |
| T2DM | b_cell | *RAB3IP* | -2.152999079 | 0.00175701 | 0.004980661 | 1 |
| T2DM | b_cell | *TAS2R4* | -2.186014739 | 0.000875235 | 0.004980661 | 1 |
| T2DM | b_cell | *LRRC61* | -1.408007326 | 0.004430414 | 0.006581864 | 1 |
| T2DM | b_cell | *IDUA* | -3.157402073 | 0.016776992 | 0.020520828 | 1 |
| T2DM | b_cell | *UBR4* | -1.50118895 | 0.001575247 | 0.004980661 | 1 |
| T2DM | b_cell | *GFER* | -2.738513972 | 0.002237436 | 0.004980661 | 1 |
| T2DM | b_cell | *WDR24* | 1.010482004 | 0.007157225 | 0.009657079 | 1 |
| T2DM | b_cell | *SMARCA4* | -1.139354329 | 0.001968246 | 0.004980661 | 1 |
| T2DM | b_cell | *PNKD* | -1.14609596 | 0.002440562 | 0.004980661 | 1 |
| T2DM | b_cell | *RNF6* | -1.294914552 | 0.00182993 | 0.004980661 | 1 |
| T2DM | b_cell | *HIP1* | -0.807174889 | 0.001917243 | 0.004980661 | 1 |
| T2DM | b_cell | *PTPN12* | -1.11630043 | 0.001766699 | 0.004980661 | 1 |
| T2DM | b_cell | *STYXL1* | -1.167766222 | 0.002328031 | 0.004980661 | 1 |
| T2DM | b_cell | *STEAP4* | -1.191938454 | 0.001023737 | 0.004980661 | 1 |
| T2DM | b_cell | *PEX1* | -1.844429813 | 0.0035262 | 0.005637273 | 1 |
| T2DM | b_cell | *SRD5A3* | -1.4975719 | 0.002977338 | 0.005186119 | 1 |
| T2DM | b_cell | *KDR* | -1.072089813 | 0.001830527 | 0.004980661 | 1 |
| T2DM | b_cell | *DGCR6L* | -2.103592547 | 0.004861331 | 0.007028361 | 1 |
| T2DM | b_cell | *VPREB3* | 1.489295291 | 0.002908869 | 0.00512387 | 1 |
| T2DM | b_cell | *MGAT3* | -2.04217867 | 0.003924551 | 0.006022112 | 1 |
| T2DM | b_cell | *ATF4* | -2.558045563 | 0.027484155 | 0.03251722 | 1 |
| T2DM | b_cell | *A4GALT* | -1.542683618 | 0.002221465 | 0.004980661 | 1 |
| T2DM | b_cell | *CDC42EP1* | -2.643958983 | 0.003480436 | 0.005589525 | 1 |
| T2DM | b_cell | *RIBC2* | 3.435295461 | 0.000232212 | 0.004980661 | 1 |
| T2DM | b_cell | *SPECC1* | -0.834956165 | 0.001792484 | 0.004980661 | 1 |
| T2DM | b_cell | *CDHR3* | -1.91950105 | 0.012705931 | 0.015963019 | 1 |
| T2DM | b_cell | *PRKRIP1* | -1.165244151 | 0.001776576 | 0.004980661 | 1 |
| T2DM | b_cell | *OPN1SW* | -2.810171754 | 4.30E-05 | 0.004980661 | 0.208717898 |
| T2DM | b_cell | *MTX2* | -1.426311772 | 0.002894842 | 0.005113643 | 1 |
| T2DM | b_cell | *PDE11A* | -3.296618993 | 0.001556609 | 0.004980661 | 1 |
| T2DM | b_cell | *GAD1* | 2.446998846 | 0.006607076 | 0.009040477 | 1 |

| | | | | | | |
|---|---|---|---|---|---|---|
| T2DM | b_cell | *ORMDL1* | -0.927939309 | 0.002647253 | 0.004985914 | 1 |
| T2DM | b_cell | *HOXD10* | 3.256146541 | 0.010962127 | 0.013978512 | 1 |
| T2DM | b_cell | *PSMG2* | -1.265343035 | 0.00328183 | 0.005388642 | 1 |
| T2DM | b_cell | *TWSG1* | 1.140975331 | 0.008231352 | 0.010836971 | 1 |
| T2DM | b_cell | *WDFY4* | -0.921714753 | 0.007972037 | 0.010556858 | 1 |
| T2DM | b_cell | *EIF2AK4* | -1.848589462 | 0.002563558 | 0.004980661 | 1 |
| T2DM | b_cell | *TMOD2* | -1.077126142 | 0.001801977 | 0.004980661 | 1 |
| T2DM | b_cell | *IVD* | -1.385332574 | 0.001711872 | 0.004980661 | 1 |
| T2DM | b_cell | *ARPP19* | -2.198828891 | 0.012441092 | 0.015658671 | 1 |
| T2DM | b_cell | *VPS13C* | -1.178786298 | 0.002107801 | 0.004980661 | 1 |
| T2DM | b_cell | *CALML4* | -6.023813318 | 0.001353823 | 0.004980661 | 1 |
| T2DM | b_cell | *ANAPC13* | -1.392438096 | 0.001250551 | 0.004980661 | 1 |
| T2DM | b_cell | *SUMF2* | -2.057357767 | 0.027191148 | 0.032178402 | 1 |
| T2DM | b_cell | *BBOX1* | -5.110651471 | 0.001881163 | 0.004980661 | 1 |
| T2DM | b_cell | *SERGEF* | -0.973780141 | 0.00269068 | 0.00500636 | 1 |
| T2DM | b_cell | *DCTD* | -1.348250862 | 0.000936908 | 0.004980661 | 1 |
| T2DM | b_cell | *RPAIN* | -1.185027383 | 0.000932089 | 0.004980661 | 1 |
| T2DM | b_cell | *USP6* | 1.722229491 | 0.018314962 | 0.022282898 | 1 |
| T2DM | b_cell | *PLD2* | -1.767200367 | 0.001860166 | 0.004980661 | 1 |
| T2DM | b_cell | *PHF20L1* | -1.112945568 | 0.001712498 | 0.004980661 | 1 |
| T2DM | b_cell | *PUS7L* | -1.691341967 | 0.002723779 | 0.005013768 | 1 |
| T2DM | b_cell | *KRI1* | -1.300755573 | 0.003080925 | 0.005245002 | 1 |
| T2DM | b_cell | *AP1M2* | -3.121797147 | 0.001767114 | 0.004980661 | 1 |
| T2DM | b_cell | *MTUS1* | -0.947561437 | 0.002295746 | 0.004980661 | 1 |
| T2DM | b_cell | *KLK10* | -1.482281777 | 0.004780119 | 0.006944054 | 1 |
| T2DM | b_cell | *KLK8* | -10.96865019 | 0.001905446 | 0.004980661 | 1 |
| T2DM | b_cell | *RIPK3* | 1.545709108 | 0.010523379 | 0.013457942 | 1 |
| T2DM | b_cell | *ADCY4* | -1.528454921 | 0.003364059 | 0.005463988 | 1 |
| T2DM | b_cell | *BCL2L2* | -1.200367097 | 0.001472142 | 0.004980661 | 1 |
| T2DM | b_cell | *MIS18BP1* | -1.410830056 | 0.004426762 | 0.006578454 | 1 |
| T2DM | b_cell | *EPB41L4A* | -0.922989454 | 0.001844948 | 0.004980661 | 1 |
| T2DM | b_cell | *CDO1* | -1.36660576 | 0.001229691 | 0.004980661 | 1 |
| T2DM | b_cell | *REEP5* | -1.168904694 | 0.002093575 | 0.004980661 | 1 |
| T2DM | b_cell | *RHBDF2* | 1.910757722 | 0.033316087 | 0.038961112 | 1 |
| T2DM | b_cell | *AANAT* | 3.686986651 | 0.00511251 | 0.007321693 | 1 |
| T2DM | b_cell | *TMEM8A* | -2.353179313 | 0.002462088 | 0.004980661 | 1 |
| T2DM | b_cell | *MAU2* | -1.249815798 | 0.001752892 | 0.004980661 | 1 |
| T2DM | b_cell | *SHC2* | -1.781398274 | 0.015947886 | 0.019604545 | 1 |
| T2DM | b_cell | *ERMARD* | -1.201662968 | 0.001986729 | 0.004980661 | 1 |
| T2DM | b_cell | *PHF10* | -1.508080552 | 0.001912627 | 0.004980661 | 1 |
| T2DM | b_cell | *PRKCSH* | -1.783163208 | 0.003410543 | 0.005510766 | 1 |
| T2DM | b_cell | *CDC16* | -1.234783867 | 0.002250142 | 0.004980661 | 1 |
| T2DM | b_cell | *XPO7* | -1.684853323 | 0.003196752 | 0.005339291 | 1 |
| T2DM | b_cell | *ATP8B3* | -2.979722453 | 0.003697496 | 0.00579416 | 1 |
| T2DM | b_cell | *PLVAP* | -1.412376576 | 0.002477647 | 0.004980661 | 1 |
| T2DM | b_cell | *DDA1* | -1.648582935 | 0.001536377 | 0.004980661 | 1 |
| T2DM | b_cell | *LSM7* | -1.213435737 | 0.002800919 | 0.005065458 | 1 |
| T2DM | b_cell | *TULP4* | -0.840948605 | 0.002097966 | 0.004980661 | 1 |
| T2DM | b_cell | *QRSL1* | -1.696216735 | 0.004192137 | 0.006324704 | 1 |
| T2DM | b_cell | *RSPH3* | -1.560065857 | 0.007538008 | 0.010075596 | 1 |

| | | | | | | |
|---|---|---|---|---|---|---|
| T2DM | b_cell | *ACTN4* | -1.157260923 | 0.002315373 | 0.004980661 | 1 |
| T2DM | b_cell | *STK33* | -0.884283585 | 0.009793121 | 0.01261381 | 1 |
| T2DM | b_cell | *NDUFA10* | -1.200873068 | 0.003117308 | 0.00526397 | 1 |
| T2DM | b_cell | *MAP1S* | -1.77735722 | 0.001790603 | 0.004980661 | 1 |
| T2DM | b_cell | *SSBP4* | -1.280267982 | 0.002009024 | 0.004980661 | 1 |
| T2DM | b_cell | *PGPEP1* | -1.189195248 | 0.004869119 | 0.007036292 | 1 |
| T2DM | b_cell | *KIAA1683* | -1.570274899 | 0.003293246 | 0.005398253 | 1 |
| T2DM | b_cell | *LSM4* | -1.240458916 | 0.002595779 | 0.004980661 | 1 |
| T2DM | b_cell | *TRPM4* | -1.369582349 | 0.002030714 | 0.004980661 | 1 |
| T2DM | b_cell | *UBAC1* | -1.171018972 | 0.000888256 | 0.004980661 | 1 |
| T2DM | b_cell | *ATXN10* | -0.94801612 | 0.001941413 | 0.004980661 | 1 |
| T2DM | b_cell | *CYP2E1* | 1.216629347 | 0.00298517 | 0.005187056 | 1 |
| T2DM | b_cell | *PNPLA7* | -0.982116523 | 0.00177622 | 0.004980661 | 1 |
| T2DM | b_cell | *PAK4* | -1.210030244 | 0.00152163 | 0.004980661 | 1 |
| T2DM | b_cell | *ZNF337* | -0.961165495 | 0.001853191 | 0.004980661 | 1 |
| T2DM | b_cell | *CEP85* | -1.684526148 | 0.000901662 | 0.004980661 | 1 |
| T2DM | b_cell | *POMT1* | -1.343416075 | 0.002209349 | 0.004980661 | 1 |
| T2DM | b_cell | *CHMP2A* | -2.30893027 | 0.038745127 | 0.044866672 | 1 |
| T2DM | b_cell | *YIPF2* | -1.751454061 | 0.001563911 | 0.004980661 | 1 |
| T2DM | b_cell | *NPAS1* | -2.358346142 | 0.000619432 | 0.004980661 | 1 |
| T2DM | b_cell | *MAP3K10* | -1.853309488 | 0.002156905 | 0.004980661 | 1 |
| T2DM | b_cell | *LRRC47* | -1.367067733 | 0.001503925 | 0.004980661 | 1 |
| T2DM | b_cell | *THEMIS2* | -1.291058257 | 0.010992413 | 0.014008779 | 1 |
| T2DM | b_cell | *ZNF317* | -1.412664235 | 0.002100855 | 0.004980661 | 1 |
| T2DM | b_cell | *ANGPTL6* | 0.950345552 | 0.003245164 | 0.00536425 | 1 |
| T2DM | b_cell | *DNMT1* | -2.342834681 | 0.009889658 | 0.012724641 | 1 |
| T2DM | b_cell | *C12orf65* | -1.347465371 | 0.012955781 | 0.016247467 | 1 |
| T2DM | b_cell | *NOL11* | -2.333222503 | 0.03788747 | 0.04397847 | 1 |
| T2DM | b_cell | *PKDREJ* | -6.605824009 | 0.000315069 | 0.004980661 | 1 |
| T2DM | b_cell | *HSD17B3* | -2.109304822 | 0.001705239 | 0.004980661 | 1 |
| T2DM | b_cell | *SLC35D2* | -2.068775178 | 0.007727094 | 0.010283011 | 1 |
| T2DM | b_cell | *ULBP2* | 4.898537856 | 0.002304715 | 0.004980661 | 1 |
| T2DM | b_cell | *TTLL9* | -2.626419069 | 0.00193999 | 0.004980661 | 1 |
| T2DM | b_cell | *COX4I2* | -2.586994558 | 0.001870037 | 0.004980661 | 1 |
| T2DM | b_cell | *CHMP1A* | -1.298258026 | 0.002253695 | 0.004980661 | 1 |
| T2DM | b_cell | *IDO1* | -2.684866626 | 0.001989375 | 0.004980661 | 1 |
| T2DM | b_cell | *MRPS25* | -1.687463862 | 0.001978207 | 0.004980661 | 1 |
| T2DM | b_cell | *GALNT15* | -1.09542271 | 0.001739549 | 0.004980661 | 1 |
| T2DM | b_cell | *KCNC3* | 0.727853608 | 0.002804748 | 0.005065458 | 1 |
| T2DM | b_cell | *PDLIM4* | -2.62371415 | 0.002552218 | 0.004980661 | 1 |
| T2DM | b_cell | *KIF3A* | -1.275278915 | 0.002042526 | 0.004980661 | 1 |
| T2DM | b_cell | *ACLY* | -1.410178277 | 0.001201663 | 0.004980661 | 1 |
| T2DM | b_cell | *EXOC4* | -0.881265868 | 0.003251129 | 0.00536838 | 1 |
| T2DM | b_cell | *C1orf159* | -1.185901539 | 0.001817913 | 0.004980661 | 1 |
| T2DM | b_cell | *PPFIA1* | -0.908617347 | 0.001853392 | 0.004980661 | 1 |
| T2DM | b_cell | *BARX1* | -7.285214179 | 0.000744763 | 0.004980661 | 1 |
| T2DM | b_cell | *NINJ1* | -1.530805357 | 0.003172088 | 0.005315561 | 1 |
| T2DM | b_cell | *NPHP4* | -1.220455121 | 0.00249424 | 0.004980661 | 1 |
| T2DM | b_cell | *MAP1B* | -0.834477947 | 0.002916201 | 0.005130172 | 1 |
| T2DM | b_cell | *CKMT2* | -2.204878755 | 0.013778582 | 0.017177204 | 1 |

| | | | | | | |
|---|---|---|---|---|---|---|
| T2DM | b_cell | *KHDRBS3* | -1.106162268 | 0.001787958 | 0.004980661 | 1 |
| T2DM | b_cell | *CHD1L* | -1.110106028 | 0.001416555 | 0.004980661 | 1 |
| T2DM | b_cell | *ZNF304* | -1.860987236 | 0.006851051 | 0.009313897 | 1 |
| T2DM | b_cell | *ZNF132* | 2.30360693 | 0.017012473 | 0.02079207 | 1 |
| T2DM | b_cell | *RHPN2* | -0.976405736 | 0.019302573 | 0.023361458 | 1 |
| T2DM | b_cell | *GCH1* | -1.792407726 | 0.005556628 | 0.007851124 | 1 |
| T2DM | b_cell | *MATN3* | -3.967193225 | 0.001617528 | 0.004980661 | 1 |
| T2DM | b_cell | *TRIM21* | -2.047457956 | 0.002029941 | 0.004980661 | 1 |
| T2DM | b_cell | *SPATA6* | -0.83244655 | 0.00176199 | 0.004980661 | 1 |
| T2DM | b_cell | *LRRC41* | -1.118598352 | 0.001560895 | 0.004980661 | 1 |
| T2DM | b_cell | *CCT6B* | -2.405309739 | 0.024479461 | 0.029139927 | 1 |
| T2DM | b_cell | *CNGA4* | 1.559067248 | 0.029025509 | 0.034215687 | 1 |
| T2DM | b_cell | *TRIM22* | -1.753463316 | 0.00306795 | 0.005237753 | 1 |
| T2DM | b_cell | *TIMM10B* | -1.39902856 | 0.002282138 | 0.004980661 | 1 |
| T2DM | b_cell | *IMMT* | -0.932382938 | 0.002316473 | 0.004980661 | 1 |
| T2DM | b_cell | *CARD6* | -1.348640471 | 0.002524011 | 0.004980661 | 1 |
| T2DM | b_cell | *MYBBP1A* | -1.481708116 | 0.00664844 | 0.009089385 | 1 |
| T2DM | b_cell | *RPA1* | -1.01650876 | 0.001777912 | 0.004980661 | 1 |
| T2DM | b_cell | *SERPINF1* | -0.971273572 | 0.001765514 | 0.004980661 | 1 |
| T2DM | b_cell | *TMEM128* | -1.359176992 | 0.001305356 | 0.004980661 | 1 |
| T2DM | b_cell | *SEC61G* | -2.631896923 | 0.01360045 | 0.016968214 | 1 |
| T2DM | b_cell | *FIGNL1* | -1.806968679 | 0.002450115 | 0.004980661 | 1 |
| T2DM | b_cell | *RGS22* | -1.019524722 | 0.003256213 | 0.00536838 | 1 |
| T2DM | b_cell | *PRMT7* | -1.169004974 | 0.001374449 | 0.004980661 | 1 |
| T2DM | b_cell | *IGHMBP2* | -1.64436514 | 0.000768665 | 0.004980661 | 1 |
| T2DM | b_cell | *MUTYH* | 1.969256719 | 0.008998373 | 0.011698849 | 1 |
| T2DM | b_cell | *CTNNBL1* | -1.522227417 | 0.003125021 | 0.005269748 | 1 |
| T2DM | b_cell | *OSER1* | -1.228387184 | 0.004334641 | 0.00647259 | 1 |
| T2DM | b_cell | *SERINC3* | -1.533177136 | 0.00153805 | 0.004980661 | 1 |
| T2DM | b_cell | *DMGDH* | -1.055944669 | 0.001006311 | 0.004980661 | 1 |
| T2DM | b_cell | *BHMT2* | -0.890654646 | 0.002870838 | 0.005094011 | 1 |
| T2DM | b_cell | *AP3B1* | -0.917704667 | 0.001958715 | 0.004980661 | 1 |
| T2DM | b_cell | *ZBED3* | -1.493187917 | 0.002785809 | 0.005058661 | 1 |
| T2DM | b_cell | *FBXO44* | 2.524630832 | 0.014032354 | 0.017462165 | 1 |
| T2DM | b_cell | *RSG1* | 1.307627625 | 0.002352383 | 0.004980661 | 1 |
| T2DM | b_cell | *CASP9* | -1.012477192 | 0.017969326 | 0.021895322 | 1 |
| T2DM | b_cell | *ATP8A2* | -2.187662594 | 0.002861471 | 0.005094011 | 1 |
| T2DM | b_cell | *WASF3* | -0.94255082 | 0.001783611 | 0.004980661 | 1 |
| T2DM | b_cell | *PEMT* | -1.399814861 | 0.001797554 | 0.004980661 | 1 |
| T2DM | b_cell | *SCO1* | -1.433308523 | 0.002816882 | 0.005065458 | 1 |
| T2DM | b_cell | *MPRIP* | -0.979586084 | 0.00188322 | 0.004980661 | 1 |
| T2DM | b_cell | *DSTYK* | -1.329474786 | 0.002131479 | 0.004980661 | 1 |
| T2DM | b_cell | *CCNA1* | -18.23220502 | 0.006212193 | 0.008621727 | 1 |
| T2DM | b_cell | *COG6* | -1.046823476 | 0.002126951 | 0.004980661 | 1 |
| T2DM | b_cell | *BTBD2* | -1.083891085 | 0.002352911 | 0.004980661 | 1 |
| T2DM | b_cell | *CSNK1G2* | -1.252468131 | 0.001793784 | 0.004980661 | 1 |
| T2DM | b_cell | *PDZD2* | -0.857759897 | 0.001998178 | 0.004980661 | 1 |
| T2DM | b_cell | *SLC2A11* | -1.492352217 | 0.004099624 | 0.006221975 | 1 |
| T2DM | b_cell | *FAM83F* | -8.536747522 | 0.003064126 | 0.005235984 | 1 |
| T2DM | b_cell | *GIMAP4* | -1.295019826 | 0.001329753 | 0.004980661 | 1 |

| | | | | | | |
|---|---|---|---|---|---|---|
| T2DM | b_cell | *IPO8* | -1.214117664 | 0.00174708 | 0.004980661 | 1 |
| T2DM | b_cell | *LARS* | -1.676859358 | 0.007977069 | 0.010559711 | 1 |
| T2DM | b_cell | *SPINK5* | 1.163270576 | 0.003126166 | 0.005269748 | 1 |
| T2DM | b_cell | *IMPA1* | -1.309879932 | 0.001087088 | 0.004980661 | 1 |
| T2DM | b_cell | *RRAS2* | -1.102684382 | 0.002922804 | 0.005134596 | 1 |
| T2DM | b_cell | *CATSPERB* | -1.005655454 | 0.001404859 | 0.004980661 | 1 |
| T2DM | b_cell | *COX16* | 1.587874994 | 0.003045419 | 0.00522879 | 1 |
| T2DM | b_cell | *TTC9* | 0.717207044 | 0.003119109 | 0.00526397 | 1 |
| T2DM | b_cell | *ELP3* | -1.666410268 | 0.002713322 | 0.00501115 | 1 |
| T2DM | b_cell | *PEBP4* | -2.1278022 | 0.002127685 | 0.004980661 | 1 |
| T2DM | b_cell | *CTIF* | -0.845206583 | 0.001848192 | 0.004980661 | 1 |
| T2DM | b_cell | *MRO* | -3.647103975 | 0.002914717 | 0.005129563 | 1 |
| T2DM | b_cell | *THUMPD3* | -1.668498525 | 0.001565046 | 0.004980661 | 1 |
| T2DM | b_cell | *CNTN6* | 0.616064957 | 0.00285053 | 0.005078826 | 1 |
| T2DM | b_cell | *CHL1* | -1.047203526 | 0.001744134 | 0.004980661 | 1 |
| T2DM | b_cell | *GSTM5* | -1.763878357 | 0.0041544 | 0.006288569 | 1 |
| T2DM | b_cell | *GSTM3* | -0.934857042 | 0.021345734 | 0.025644024 | 1 |
| T2DM | b_cell | *PSRC1* | -4.366218803 | 0.012200273 | 0.015403519 | 1 |
| T2DM | b_cell | *TRIM45* | -2.564531598 | 0.00186984 | 0.004980661 | 1 |
| T2DM | b_cell | *CEPT1* | -0.810440479 | 0.00178755 | 0.004980661 | 1 |
| T2DM | b_cell | *NAPG* | -1.736109436 | 0.003078925 | 0.005245002 | 1 |
| T2DM | b_cell | *PPHLN1* | -1.52089289 | 0.006354385 | 0.008773897 | 1 |
| T2DM | b_cell | *FKBP11* | 1.423534064 | 0.006942283 | 0.009419452 | 1 |
| T2DM | b_cell | *YWHAQ* | -1.565151287 | 0.00328344 | 0.005389463 | 1 |
| T2DM | b_cell | *KIDINS220* | -1.049569939 | 0.002127004 | 0.004980661 | 1 |
| T2DM | b_cell | *LPIN1* | -0.949781936 | 0.001584016 | 0.004980661 | 1 |
| T2DM | b_cell | *NARS* | -1.948440135 | 0.020563388 | 0.024788462 | 1 |
| T2DM | b_cell | *KIAA1468* | -1.037062941 | 0.002007422 | 0.004980661 | 1 |
| T2DM | b_cell | *IL15RA* | -1.214231261 | 0.001999734 | 0.004980661 | 1 |
| T2DM | b_cell | *TMEM241* | -1.204750739 | 0.002967446 | 0.005173571 | 1 |
| T2DM | b_cell | *SOX5* | -0.693061581 | 0.001977787 | 0.004980661 | 1 |
| T2DM | b_cell | *KLRD1* | -0.992103187 | 0.001887332 | 0.004980661 | 1 |
| T2DM | b_cell | *ACP2* | -1.627275198 | 0.001807715 | 0.004980661 | 1 |
| T2DM | b_cell | *HOOK1* | -1.713349471 | 0.002096151 | 0.004980661 | 1 |
| T2DM | b_cell | *ZCCHC11* | -1.005295242 | 0.002141198 | 0.004980661 | 1 |
| T2DM | b_cell | *PRPF38A* | -2.596716855 | 0.007508914 | 0.01004501 | 1 |
| T2DM | b_cell | *ELP2* | -0.927882053 | 0.001800669 | 0.004980661 | 1 |
| T2DM | b_cell | *TPGS2* | -2.159594141 | 0.007597719 | 0.010135862 | 1 |
| T2DM | b_cell | *TIMM10* | 1.017754039 | 0.00446064 | 0.006612596 | 1 |
| T2DM | b_cell | *FADS2* | -1.353448239 | 0.001844595 | 0.004980661 | 1 |
| T2DM | b_cell | *TMEM258* | -2.062924028 | 0.007601846 | 0.010138581 | 1 |
| T2DM | b_cell | *CLOCK* | -1.012798422 | 0.001974925 | 0.004980661 | 1 |
| T2DM | b_cell | *GGACT* | -3.549754922 | 0.001401677 | 0.004980661 | 1 |
| T2DM | b_cell | *CLDN10* | -2.683109336 | 0.008769362 | 0.011465627 | 1 |
| T2DM | b_cell | *ARGLU1* | -0.85180557 | 0.002077231 | 0.004980661 | 1 |
| T2DM | b_cell | *ERCC5* | -2.574659263 | 0.001362868 | 0.004980661 | 1 |
| T2DM | b_cell | *CARS2* | -1.195825157 | 0.001330015 | 0.004980661 | 1 |
| T2DM | b_cell | *KLB* | -1.051289496 | 0.000319949 | 0.004980661 | 1 |
| T2DM | b_cell | *WDR36* | -1.030648912 | 0.00178454 | 0.004980661 | 1 |
| T2DM | b_cell | *OSTF1* | -1.340899567 | 0.002167326 | 0.004980661 | 1 |

| | | | | | | |
|---|---|---|---|---|---|---|
| T2DM | b_cell | *UBQLN1* | -1.282551315 | 0.002344614 | 0.004980661 | 1 |
| T2DM | b_cell | *GOLM1* | -1.861929248 | 0.001729258 | 0.004980661 | 1 |
| T2DM | b_cell | *ADAM19* | 1.161872648 | 0.002394297 | 0.004980661 | 1 |
| T2DM | b_cell | *USP30* | -1.180721581 | 0.003306606 | 0.005412542 | 1 |
| T2DM | b_cell | *MSI1* | -3.406299725 | 0.003355721 | 0.005460611 | 1 |
| T2DM | b_cell | *P2RX4* | 2.378537887 | 0.012675683 | 0.015933268 | 1 |
| T2DM | b_cell | *DTX1* | -1.38486997 | 0.001982029 | 0.004980661 | 1 |
| T2DM | b_cell | *DMTF1* | -1.027580876 | 0.002081678 | 0.004980661 | 1 |
| T2DM | b_cell | *TMEM60* | -1.87316118 | 0.00229237 | 0.004980661 | 1 |
| T2DM | b_cell | *POM121C* | -2.161331219 | 0.001104764 | 0.004980661 | 1 |
| T2DM | b_cell | *SRPK2* | -1.022421368 | 0.002096425 | 0.004980661 | 1 |
| T2DM | b_cell | *TES* | -1.597651487 | 0.001359479 | 0.004980661 | 1 |
| T2DM | b_cell | *MTO1* | -1.217478779 | 0.001675715 | 0.004980661 | 1 |
| T2DM | b_cell | *KHDC1* | -1.108447448 | 0.002583774 | 0.004980661 | 1 |
| T2DM | b_cell | *NT5E* | -1.176738622 | 0.004406611 | 0.006554531 | 1 |
| T2DM | b_cell | *AKIRIN2* | -1.080070377 | 0.002025975 | 0.004980661 | 1 |
| T2DM | b_cell | *MAP3K7* | -1.214014912 | 0.002087305 | 0.004980661 | 1 |
| T2DM | b_cell | *PRR5L* | -1.353153357 | 0.002878354 | 0.005096998 | 1 |
| T2DM | b_cell | *PHF21A* | -0.961376851 | 0.00209188 | 0.004980661 | 1 |
| T2DM | b_cell | *CAPRIN1* | -1.349246485 | 0.002539819 | 0.004980661 | 1 |
| T2DM | b_cell | *ATP5G2* | -1.059109322 | 0.001953522 | 0.004980661 | 1 |
| T2DM | b_cell | *B4GALNT1* | -8.032638232 | 0.002503924 | 0.004980661 | 1 |
| T2DM | b_cell | *FAIM2* | -3.429481665 | 0.000644929 | 0.004980661 | 1 |
| T2DM | b_cell | *KRT7* | -2.389423213 | 0.005758195 | 0.008086473 | 1 |
| T2DM | b_cell | *SLC26A10* | -4.448768605 | 0.00132048 | 0.004980661 | 1 |
| T2DM | b_cell | *MAP7* | -1.1126566 | 0.001176759 | 0.004980661 | 1 |
| T2DM | b_cell | *NHSL1* | -1.394845557 | 0.034143238 | 0.039899565 | 1 |
| T2DM | b_cell | *AHI1* | -0.981759286 | 0.003577726 | 0.005673975 | 1 |
| T2DM | b_cell | *SMPD2* | -2.409659878 | 0.002010029 | 0.004980661 | 1 |
| T2DM | b_cell | *MICAL1* | -1.329498926 | 0.022245018 | 0.026636926 | 1 |
| T2DM | b_cell | *CCT7* | -2.414225748 | 0.007871467 | 0.010445949 | 1 |
| T2DM | b_cell | *SMYD5* | -1.972094714 | 0.001071932 | 0.004980661 | 1 |
| T2DM | b_cell | *USP15* | -1.166361389 | 0.002714201 | 0.00501115 | 1 |
| T2DM | b_cell | *MDM2* | -0.888064128 | 0.001740581 | 0.004980661 | 1 |
| T2DM | b_cell | *KLHL36* | -1.506993589 | 0.003126876 | 0.005269748 | 1 |
| T2DM | b_cell | *MPHOSPH6* | -1.184671646 | 0.002214984 | 0.004980661 | 1 |
| T2DM | b_cell | *CCDC102A* | -1.911905409 | 0.000858043 | 0.004980661 | 1 |
| T2DM | b_cell | *ZNF670* | -4.196471547 | 0.002941388 | 0.005147158 | 1 |
| T2DM | b_cell | *KCNK1* | -1.722707195 | 0.001189495 | 0.004980661 | 1 |
| T2DM | b_cell | *CAPN9* | -9.009779975 | 0.004207113 | 0.006339414 | 1 |
| T2DM | b_cell | *NTPCR* | -1.142665005 | 0.001853937 | 0.004980661 | 1 |
| T2DM | b_cell | *PIGC* | -1.051647345 | 0.001937059 | 0.004980661 | 1 |
| T2DM | b_cell | *SP110* | -2.041673318 | 0.008426293 | 0.011070775 | 1 |
| T2DM | b_cell | *MRPL44* | -2.185942211 | 0.00225869 | 0.004980661 | 1 |
| T2DM | b_cell | *USP37* | -1.139928363 | 0.002385386 | 0.004980661 | 1 |
| T2DM | b_cell | *SLC19A3* | -0.749163493 | 0.002009793 | 0.004980661 | 1 |
| T2DM | b_cell | *TMBIM1* | -1.056488775 | 0.00178774 | 0.004980661 | 1 |
| T2DM | b_cell | *TSGA10* | -0.764086263 | 0.002313255 | 0.004980661 | 1 |
| T2DM | b_cell | *GCC2* | -1.085297996 | 0.001970064 | 0.004980661 | 1 |
| T2DM | b_cell | *MRPS9* | -1.605274613 | 0.014485091 | 0.017974849 | 1 |

| | | | | | | |
|---|---|---|---|---|---|---|
| T2DM | b_cell | *ANKRD36* | -1.176409738 | 0.002105705 | 0.004980661 | 1 |
| T2DM | b_cell | *EPC2* | -1.158544811 | 0.002394852 | 0.004980661 | 1 |
| T2DM | b_cell | *STAB2* | -1.226099972 | 0.001790584 | 0.004980661 | 1 |
| T2DM | b_cell | *APPL2* | -1.064860928 | 0.001822027 | 0.004980661 | 1 |
| T2DM | b_cell | *DRAM1* | -1.327537015 | 0.002124635 | 0.004980661 | 1 |
| T2DM | b_cell | *SLC41A2* | -0.890043245 | 0.002314945 | 0.004980661 | 1 |
| T2DM | b_cell | *NEK3* | -1.16357504 | 0.002983021 | 0.005186119 | 1 |
| T2DM | b_cell | *RNASEH2B* | -1.236730733 | 0.002182723 | 0.004980661 | 1 |
| T2DM | b_cell | *CKAP2* | -1.158465945 | 0.010130393 | 0.013006792 | 1 |
| T2DM | b_cell | *TBC1D4* | -0.791707507 | 0.001502366 | 0.004980661 | 1 |
| T2DM | b_cell | *SUCLA2* | -1.357234993 | 0.002257619 | 0.004980661 | 1 |
| T2DM | b_cell | *RCBTB1* | -2.266484932 | 0.000196251 | 0.004980661 | 0.952211414 |
| T2DM | b_cell | *RCBTB2* | -1.077264887 | 0.001854203 | 0.004980661 | 1 |
| T2DM | b_cell | *SETDB2* | -1.052049407 | 0.001746122 | 0.004980661 | 1 |
| T2DM | b_cell | *SCRN1* | -1.137454844 | 0.001593063 | 0.004980661 | 1 |
| T2DM | b_cell | *C7orf25* | -10.1132865 | 0.000738474 | 0.004980661 | 1 |
| T2DM | b_cell | *CHST12* | -1.096013088 | 0.001750618 | 0.004980661 | 1 |
| T2DM | b_cell | *KDELR2* | -1.411274884 | 0.001909348 | 0.004980661 | 1 |
| T2DM | b_cell | *NUPL2* | -1.041406141 | 0.001832901 | 0.004980661 | 1 |
| T2DM | b_cell | *ZDHHC4* | 2.235831428 | 0.021655721 | 0.025989008 | 1 |
| T2DM | b_cell | *AOAH* | -0.897955984 | 0.001616111 | 0.004980661 | 1 |
| T2DM | b_cell | *TBRG4* | -1.708744914 | 0.001501019 | 0.004980661 | 1 |
| T2DM | b_cell | *NMT1* | -1.230354258 | 0.008972833 | 0.011674443 | 1 |
| T2DM | b_cell | *BRIP1* | -1.985684557 | 0.001668848 | 0.004980661 | 1 |
| T2DM | b_cell | *RTP4* | 1.666558486 | 0.019189569 | 0.023236284 | 1 |
| T2DM | b_cell | *NDUFB5* | -1.994215395 | 0.003278802 | 0.005387318 | 1 |
| T2DM | b_cell | *SCN2A* | -1.294544912 | 0.00387145 | 0.005970845 | 1 |
| T2DM | b_cell | *RPS6KC1* | -0.975086945 | 0.00248674 | 0.004980661 | 1 |
| T2DM | b_cell | *SMPD4* | -1.292652973 | 0.001467772 | 0.004980661 | 1 |
| T2DM | b_cell | *IMP4* | -1.742999537 | 0.001445164 | 0.004980661 | 1 |
| T2DM | b_cell | *NIPSNAP3A* | -1.285671766 | 0.000857135 | 0.004980661 | 1 |
| T2DM | b_cell | *ODF2* | -2.081603565 | 0.002700347 | 0.00500969 | 1 |
| T2DM | b_cell | *SLC2A8* | -3.471801734 | 0.001829954 | 0.004980661 | 1 |
| T2DM | b_cell | *ZNF189* | -2.037383904 | 0.002825814 | 0.005065458 | 1 |
| T2DM | b_cell | *STX17* | -1.532930376 | 0.002624933 | 0.004980661 | 1 |
| T2DM | b_cell | *ANP32B* | -1.565448381 | 0.001399223 | 0.004980661 | 1 |
| T2DM | b_cell | *DSCC1* | -1.726192404 | 0.001933658 | 0.004980661 | 1 |
| T2DM | b_cell | *DERL1* | -1.155318766 | 0.001579125 | 0.004980661 | 1 |
| T2DM | b_cell | *NOV* | -2.278846701 | 0.004794647 | 0.006954746 | 1 |
| T2DM | b_cell | *POLR1E* | -1.404500354 | 0.008938285 | 0.011645132 | 1 |
| T2DM | b_cell | *IL11RA* | -1.778036142 | 0.0016246 | 0.004980661 | 1 |
| T2DM | b_cell | *APTX* | -1.060998102 | 0.001376875 | 0.004980661 | 1 |
| T2DM | b_cell | *TMEM8B* | -1.288045064 | 0.002983194 | 0.005186119 | 1 |
| T2DM | b_cell | *GRHPR* | -1.244190631 | 0.001152381 | 0.004980661 | 1 |
| T2DM | b_cell | *DENND4C* | -1.110194103 | 0.002147473 | 0.004980661 | 1 |
| T2DM | b_cell | *GMPR* | -1.363271121 | 0.001302721 | 0.004980661 | 1 |
| T2DM | b_cell | *TMEM14B* | -1.217368216 | 0.001742106 | 0.004980661 | 1 |
| T2DM | b_cell | *FRS3* | -1.938004097 | 0.002142972 | 0.004980661 | 1 |
| T2DM | b_cell | *IRF4* | 2.345151487 | 2.19E-05 | 0.003918133 | 0.106121637 |
| T2DM | b_cell | *LRRC1* | -0.93698874 | 0.003004683 | 0.005197406 | 1 |

| | | | | | | |
|---|---|---|---|---|---|---|
| T2DM | b_cell | *RIPK1* | -1.68307645 | 0.004507416 | 0.006657702 | 1 |
| T2DM | b_cell | *TCF19* | 1.548316079 | 0.000136108 | 0.004980661 | 0.660395179 |
| T2DM | b_cell | *FLOT1* | -1.185954373 | 0.006012305 | 0.008397598 | 1 |
| T2DM | b_cell | *TPMT* | -1.473828528 | 0.001625033 | 0.004980661 | 1 |
| T2DM | b_cell | *NRM* | 6.391511604 | 0.043046328 | 0.049610637 | 1 |
| T2DM | b_cell | *TLR2* | -1.999384947 | 0.001673354 | 0.004980661 | 1 |
| T2DM | b_cell | *MYO7A* | -1.704090087 | 0.003777069 | 0.005875252 | 1 |
| T2DM | b_cell | *NUMA1* | -1.204090365 | 0.002726998 | 0.005013768 | 1 |
| T2DM | b_cell | *SYTL2* | -1.21627349 | 0.00048793 | 0.004980661 | 1 |
| T2DM | b_cell | *RAB30* | -1.291233292 | 0.001783834 | 0.004980661 | 1 |
| T2DM | b_cell | *PRCP* | -1.180149775 | 0.003004025 | 0.005197406 | 1 |
| T2DM | b_cell | *NARS2* | -1.654015751 | 0.003685553 | 0.005779672 | 1 |
| T2DM | b_cell | *SULF1* | -0.640370592 | 0.002433781 | 0.004980661 | 1 |
| T2DM | b_cell | *TGS1* | -0.980431312 | 0.001883819 | 0.004980661 | 1 |
| T2DM | b_cell | *NEK1* | -0.848122512 | 0.002165599 | 0.004980661 | 1 |
| T2DM | b_cell | *TRPC6* | -1.263799043 | 0.002354894 | 0.004980661 | 1 |
| T2DM | b_cell | *SLC37A4* | 3.427792646 | 0.034199161 | 0.039949748 | 1 |
| T2DM | b_cell | *PPP2R1B* | -0.740985264 | 0.003331972 | 0.005441511 | 1 |
| T2DM | b_cell | *FDX1* | -1.161972593 | 0.002269529 | 0.004980661 | 1 |
| T2DM | b_cell | *FXYD6* | -2.392543138 | 0.000937223 | 0.004980661 | 1 |
| T2DM | b_cell | *FXYD2* | -3.188846072 | 0.001717692 | 0.004980661 | 1 |
| T2DM | b_cell | *MAP2K5* | -0.918912845 | 0.002137412 | 0.004980661 | 1 |
| T2DM | b_cell | *NUSAP1* | -2.286551017 | 0.007453493 | 0.009987392 | 1 |
| T2DM | b_cell | *NDUFAF1* | -2.538009072 | 0.035341215 | 0.041190385 | 1 |
| T2DM | b_cell | *PARP6* | -1.207286446 | 0.001917289 | 0.004980661 | 1 |
| T2DM | b_cell | *ADAM10* | -1.59079921 | 0.023654304 | 0.028240817 | 1 |
| T2DM | b_cell | *ZNF280D* | -0.742435434 | 0.001862819 | 0.004980661 | 1 |
| T2DM | b_cell | *RSL24D1* | -1.715988108 | 0.00191222 | 0.004980661 | 1 |
| T2DM | b_cell | *SLC44A5* | -1.036835285 | 0.001716405 | 0.004980661 | 1 |
| T2DM | b_cell | *DBT* | -1.126967858 | 0.001791952 | 0.004980661 | 1 |
| T2DM | b_cell | *RTCA* | -2.094520715 | 0.031112386 | 0.036516037 | 1 |
| T2DM | b_cell | *KHK* | -3.147719637 | 0.000559792 | 0.004980661 | 1 |
| T2DM | b_cell | *ADCY3* | -1.168358408 | 0.002696496 | 0.005007108 | 1 |
| T2DM | b_cell | *DYNC2LI1* | -1.339168591 | 0.001462878 | 0.004980661 | 1 |
| T2DM | b_cell | *LHCGR* | -3.264906365 | 0.000565708 | 0.004980661 | 1 |
| T2DM | b_cell | *THUMPD2* | -1.091816994 | 0.001945963 | 0.004980661 | 1 |
| T2DM | b_cell | *ACTR2* | -1.251332035 | 0.002506833 | 0.004980661 | 1 |
| T2DM | b_cell | *ATRAID* | -1.977944475 | 0.023410904 | 0.027963985 | 1 |
| T2DM | b_cell | *CENPO* | -1.502580946 | 0.002981489 | 0.005186119 | 1 |
| T2DM | b_cell | *LRPPRC* | -1.016752169 | 0.001913254 | 0.004980661 | 1 |
| T2DM | b_cell | *DTNB* | -1.190602051 | 0.002232242 | 0.004980661 | 1 |
| T2DM | b_cell | *CYP2C8* | 2.258771687 | 0.003470717 | 0.00557798 | 1 |
| T2DM | b_cell | *MYOF* | -0.818861993 | 0.013655638 | 0.017032688 | 1 |
| T2DM | b_cell | *TACC2* | -0.882121736 | 0.001947431 | 0.004980661 | 1 |
| T2DM | b_cell | *CALHM2* | -2.905563267 | 0.015212173 | 0.018761938 | 1 |
| T2DM | b_cell | *ARL3* | -1.825453228 | 0.013010651 | 0.016312062 | 1 |
| T2DM | b_cell | *PLCE1* | -0.829997428 | 0.001875938 | 0.004980661 | 1 |
| T2DM | b_cell | *RBP4* | -1.249385766 | 0.002303252 | 0.004980661 | 1 |
| T2DM | b_cell | *ASCC1* | -1.23895875 | 0.001221468 | 0.004980661 | 1 |
| T2DM | b_cell | *RPS24* | -1.075137073 | 0.002098021 | 0.004980661 | 1 |

| | | | | | | |
|---|---|---|---|---|---|---|
| T2DM | b_cell | *DNA2* | -1.150930873 | 0.003305996 | 0.005412542 | 1 |
| T2DM | b_cell | *ATIC* | -3.034006142 | 0.003266819 | 0.005376733 | 1 |
| T2DM | b_cell | *SMARCAL1* | -1.227527075 | 0.001918732 | 0.004980661 | 1 |
| T2DM | b_cell | *BARD1* | -1.345828486 | 0.00155386 | 0.004980661 | 1 |
| T2DM | b_cell | *STAT4* | -1.101941626 | 0.001821993 | 0.004980661 | 1 |
| T2DM | b_cell | *SSB* | -1.340121625 | 0.002521447 | 0.004980661 | 1 |
| T2DM | b_cell | *FASTKD1* | -0.961278972 | 0.00222631 | 0.004980661 | 1 |
| T2DM | b_cell | *ITGAV* | -1.527485951 | 0.004155225 | 0.006288569 | 1 |
| T2DM | b_cell | *SLC35A5* | -1.466515355 | 0.003648586 | 0.005752881 | 1 |
| T2DM | b_cell | *SENP7* | -1.506211162 | 0.005436503 | 0.007715096 | 1 |
| T2DM | b_cell | *COX17* | -2.441351648 | 0.003916922 | 0.006016115 | 1 |
| T2DM | b_cell | *SPPL2A* | -1.056122466 | 0.00112127 | 0.004980661 | 1 |
| T2DM | b_cell | *PPCDC* | -0.985720775 | 0.001927771 | 0.004980661 | 1 |
| T2DM | b_cell | *SEMA7A* | -3.80369011 | 0.025319415 | 0.030080755 | 1 |
| T2DM | b_cell | *PCDH10* | 1.600803734 | 0.003085553 | 0.005245002 | 1 |
| T2DM | b_cell | *AP1AR* | -1.218847513 | 0.001761958 | 0.004980661 | 1 |
| T2DM | b_cell | *KIAA1109* | -0.999964732 | 0.002121082 | 0.004980661 | 1 |
| T2DM | b_cell | *BMPR1B* | -1.125372476 | 0.001777823 | 0.004980661 | 1 |
| T2DM | b_cell | *MMRN1* | -1.023719554 | 0.000678668 | 0.004980661 | 1 |
| T2DM | b_cell | *PRDM5* | -1.158046594 | 0.001935087 | 0.004980661 | 1 |
| T2DM | b_cell | *NAAA* | -1.259725446 | 0.009684114 | 0.012482617 | 1 |
| T2DM | b_cell | *NUP54* | -0.922120321 | 0.003574484 | 0.005671484 | 1 |
| T2DM | b_cell | *G3BP2* | -1.673779041 | 0.001095828 | 0.004980661 | 1 |
| T2DM | b_cell | *CNOT6L* | -1.639861191 | 0.002659266 | 0.004985914 | 1 |
| T2DM | b_cell | *ANXA3* | -0.927868478 | 0.003871162 | 0.005970845 | 1 |
| T2DM | b_cell | *PPA2* | -1.162706658 | 0.001899479 | 0.004980661 | 1 |
| T2DM | b_cell | *ENPEP* | -1.040070017 | 0.006918581 | 0.009392545 | 1 |
| T2DM | b_cell | *PAPSS1* | -1.736773559 | 0.006181079 | 0.008581 | 1 |
| T2DM | b_cell | *PARVG* | -3.025489781 | 0.002292585 | 0.004980661 | 1 |
| T2DM | b_cell | *B4GALNT3* | -2.84936752 | 0.001918298 | 0.004980661 | 1 |
| T2DM | b_cell | *KIF21A* | -0.834627888 | 0.001793241 | 0.004980661 | 1 |
| T2DM | b_cell | *ZCRB1* | -1.429342421 | 0.00267029 | 0.004994698 | 1 |
| T2DM | b_cell | *C1RL* | -0.894145826 | 0.00584458 | 0.008186461 | 1 |
| T2DM | b_cell | *CLSTN3* | -1.449650885 | 0.002538386 | 0.004980661 | 1 |
| T2DM | b_cell | *KLRG1* | -1.529235824 | 0.00140577 | 0.004980661 | 1 |
| T2DM | b_cell | *VAMP1* | 2.110184467 | 0.005039151 | 0.007248728 | 1 |
| T2DM | b_cell | *TAPBPL* | -1.295737341 | 0.001123231 | 0.004980661 | 1 |
| T2DM | b_cell | *PEX5* | -1.446960667 | 0.004360545 | 0.00650159 | 1 |
| T2DM | b_cell | *PPFIA2* | -1.027606922 | 0.001769747 | 0.004980661 | 1 |
| T2DM | b_cell | *MARCH9* | -1.7220792 | 0.001777961 | 0.004980661 | 1 |
| T2DM | b_cell | *LGR5* | -3.103738103 | 0.015982707 | 0.019632429 | 1 |
| T2DM | b_cell | *POC1B* | -1.015075475 | 0.001727211 | 0.004980661 | 1 |
| T2DM | b_cell | *AMDHD1* | -2.15287253 | 0.005067351 | 0.007280659 | 1 |
| T2DM | b_cell | *RITA1* | -2.218872979 | 0.001780491 | 0.004980661 | 1 |
| T2DM | b_cell | *SDSL* | 0.491215386 | 0.002825884 | 0.005065458 | 1 |
| T2DM | b_cell | *GLTP* | -2.70378009 | 0.004083887 | 0.006203247 | 1 |
| T2DM | b_cell | *FOXN4* | -2.270999185 | 0.003793224 | 0.005890482 | 1 |
| T2DM | b_cell | *MTMR6* | -2.066748026 | 0.003777829 | 0.005875252 | 1 |
| T2DM | b_cell | *CCDC65* | -1.043080154 | 0.002109079 | 0.004980661 | 1 |
| T2DM | b_cell | *ACVRL1* | -1.435433289 | 0.001838778 | 0.004980661 | 1 |

| | | | | | | |
|---|---|---|---|---|---|---|
| T2DM | b_cell | *BRCA2* | -3.844724549 | 0.019634359 | 0.023739325 | 1 |
| T2DM | b_cell | *KANSL2* | -1.13254967 | 0.003064756 | 0.005235984 | 1 |
| T2DM | b_cell | *CERS5* | -1.618784247 | 0.002825658 | 0.005065458 | 1 |
| T2DM | b_cell | *LMBR1L* | -1.123572625 | 0.001881773 | 0.004980661 | 1 |
| T2DM | b_cell | *ESYT1* | -1.39008883 | 0.002249194 | 0.004980661 | 1 |
| T2DM | b_cell | *ZNF740* | -1.896592741 | 0.002604218 | 0.004980661 | 1 |
| T2DM | b_cell | *WDFY2* | -1.04151526 | 0.001891325 | 0.004980661 | 1 |
| T2DM | b_cell | *HNRNPA1L2* | 0.867643808 | 0.002764297 | 0.005040349 | 1 |
| T2DM | b_cell | *RB1* | -0.796139962 | 0.001799915 | 0.004980661 | 1 |
| T2DM | b_cell | *VPS33A* | -1.58998055 | 0.004940069 | 0.007125212 | 1 |
| T2DM | b_cell | *DIAPH3* | -1.023220983 | 0.014333699 | 0.017809759 | 1 |
| T2DM | b_cell | *GRTP1* | -1.247269033 | 0.002660456 | 0.004985914 | 1 |
| T2DM | b_cell | *NOVA1* | -0.61302978 | 0.006768948 | 0.009228136 | 1 |
| T2DM | b_cell | *FITM1* | 1.361188715 | 0.001845511 | 0.004980661 | 1 |
| T2DM | b_cell | *RTN1* | -0.762710507 | 0.003210124 | 0.005339692 | 1 |
| T2DM | b_cell | *C14orf37* | -1.190646508 | 0.002283483 | 0.004980661 | 1 |
| T2DM | b_cell | *RAB15* | -1.767404383 | 0.002021689 | 0.004980661 | 1 |
| T2DM | b_cell | *EFCAB11* | 3.018871966 | 0.003968503 | 0.006071904 | 1 |
| T2DM | b_cell | *PTGR2* | -1.233380747 | 0.003529187 | 0.005639428 | 1 |
| T2DM | b_cell | *SLC24A4* | -1.831955246 | 0.001890647 | 0.004980661 | 1 |
| T2DM | b_cell | *C14orf79* | -1.541690711 | 0.002933189 | 0.005141558 | 1 |
| T2DM | b_cell | *WARS* | -1.282715338 | 0.002377603 | 0.004980661 | 1 |
| T2DM | b_cell | *DUOXA1* | -3.078044111 | 0.008710581 | 0.011391843 | 1 |
| T2DM | b_cell | *SERF2* | -1.485682293 | 0.003220791 | 0.005348144 | 1 |
| T2DM | b_cell | *ZSCAN29* | -1.331242746 | 0.024169648 | 0.028813546 | 1 |
| T2DM | b_cell | *SRP14* | -1.108651129 | 0.002044465 | 0.004980661 | 1 |
| T2DM | b_cell | *UBE2Q2* | -1.873386719 | 0.016112237 | 0.019781522 | 1 |
| T2DM | b_cell | *HMG20A* | -1.886703357 | 0.014021717 | 0.017453405 | 1 |
| T2DM | b_cell | *SCAPER* | -0.995239107 | 0.001443464 | 0.004980661 | 1 |
| T2DM | b_cell | *NEIL1* | -1.306088313 | 0.000379282 | 0.004980661 | 1 |
| T2DM | b_cell | *MAN2C1* | -1.280116229 | 1.08E-05 | 0.002384095 | 0.052450099 |
| T2DM | b_cell | *PIF1* | -2.617354077 | 0.000668246 | 0.004980661 | 1 |
| T2DM | b_cell | *CYP11A1* | -2.249218961 | 0.008943881 | 0.011646728 | 1 |
| T2DM | b_cell | *BBS4* | -0.969547674 | 0.002368242 | 0.004980661 | 1 |
| T2DM | b_cell | *ULK3* | -2.400516464 | 0.002377006 | 0.004980661 | 1 |
| T2DM | b_cell | *PCSK6* | -1.18353897 | 0.001788283 | 0.004980661 | 1 |
| T2DM | b_cell | *HAPLN3* | -2.20572035 | 0.000819874 | 0.004980661 | 1 |
| T2DM | b_cell | *POLG* | -1.87480966 | 0.002014766 | 0.004980661 | 1 |
| T2DM | b_cell | *ABHD2* | -2.036254212 | 0.018548878 | 0.022522312 | 1 |
| T2DM | b_cell | *MFGE8* | -1.096542024 | 0.002210217 | 0.004980661 | 1 |
| T2DM | b_cell | *UNC45A* | -2.32770559 | 0.007217363 | 0.009719996 | 1 |
| T2DM | b_cell | *IQGAP1* | -1.03765414 | 0.002426279 | 0.004980661 | 1 |
| T2DM | b_cell | *CRTC3* | -0.945379523 | 0.002476276 | 0.004980661 | 1 |
| T2DM | b_cell | *ITGAX* | -2.344562501 | 0.002446295 | 0.004980661 | 1 |
| T2DM | b_cell | *TGFB1I1* | -1.865164345 | 0.001893983 | 0.004980661 | 1 |
| T2DM | b_cell | *DHX38* | -2.45591367 | 0.018536437 | 0.022518475 | 1 |
| T2DM | b_cell | *NUDT7* | -1.441629579 | 0.001626398 | 0.004980661 | 1 |
| T2DM | b_cell | *GCSH* | -2.179641388 | 0.000321705 | 0.004980661 | 1 |
| T2DM | b_cell | *CMTM3* | 1.697887054 | 0.01272066 | 0.015973251 | 1 |
| T2DM | b_cell | *ZCCHC14* | -1.760953319 | 0.003397654 | 0.005502475 | 1 |

| | | | | | | |
|---|---|---|---|---|---|---|
| T2DM | b_cell | *TLDC1* | -1.286431274 | 0.001779484 | 0.004980661 | 1 |
| T2DM | b_cell | *RPL3L* | 7.211520419 | 0.000212879 | 0.004980661 | 1 |
| T2DM | b_cell | *TIGD7* | -1.674182793 | 0.004288363 | 0.006435861 | 1 |
| T2DM | b_cell | *DEF8* | -1.420749683 | 0.001648673 | 0.004980661 | 1 |
| T2DM | b_cell | *TCF25* | -1.125680618 | 0.002254662 | 0.004980661 | 1 |
| T2DM | b_cell | *GALNS* | -1.244705011 | 0.001178232 | 0.004980661 | 1 |
| T2DM | b_cell | *GAS8* | -2.00129959 | 0.003113761 | 0.00526397 | 1 |
| T2DM | b_cell | *NCOR1* | -0.946072044 | 0.002127174 | 0.004980661 | 1 |
| T2DM | b_cell | *KSR1* | -1.013070397 | 0.001763145 | 0.004980661 | 1 |
| T2DM | b_cell | *PRPSAP2* | -1.668474856 | 0.007562574 | 0.010099268 | 1 |
| T2DM | b_cell | *RASL10B* | -6.19810268 | 0.001514639 | 0.004980661 | 1 |
| T2DM | b_cell | *UNC45B* | -9.570244445 | 0.001166402 | 0.004980661 | 1 |
| T2DM | b_cell | *VPS53* | -1.090376329 | 0.000523063 | 0.004980661 | 1 |
| T2DM | b_cell | *SGSM2* | -1.215355283 | 0.001829017 | 0.004980661 | 1 |
| T2DM | b_cell | *LRRC46* | -4.888600855 | 0.000295682 | 0.004980661 | 1 |
| T2DM | b_cell | *SCRN2* | -0.967687768 | 0.001745587 | 0.004980661 | 1 |
| T2DM | b_cell | *SSH2* | -0.977560327 | 0.002309798 | 0.004980661 | 1 |
| T2DM | b_cell | *RHBDL3* | -2.425978183 | 0.001736764 | 0.004980661 | 1 |
| T2DM | b_cell | *G6PC3* | -1.915267218 | 0.001002208 | 0.004980661 | 1 |
| T2DM | b_cell | *CLTC* | -0.917319222 | 0.003211582 | 0.005339692 | 1 |
| T2DM | b_cell | *IMPA2* | -1.360641224 | 0.002632047 | 0.004980661 | 1 |
| T2DM | b_cell | *GNAL* | -1.061941917 | 0.00179587 | 0.004980661 | 1 |
| T2DM | b_cell | *RPRD1A* | -1.446242789 | 0.002137773 | 0.004980661 | 1 |
| T2DM | b_cell | *GALNT1* | -1.75460765 | 0.006152243 | 0.008545859 | 1 |
| T2DM | b_cell | *ADCYAP1* | -3.148686398 | 0.04029618 | 0.046584958 | 1 |
| T2DM | b_cell | *C18orf8* | -1.73516554 | 0.000291068 | 0.004980661 | 1 |
| T2DM | b_cell | *NPC1* | -1.342100797 | 0.001297529 | 0.004980661 | 1 |
| T2DM | b_cell | *CCDC40* | -1.124955212 | 0.003031018 | 0.005224333 | 1 |
| T2DM | b_cell | *ARHGDIA* | -1.612146731 | 0.000938914 | 0.004980661 | 1 |
| T2DM | b_cell | *TMC6* | -1.209803947 | 0.004481939 | 0.006636059 | 1 |
| T2DM | b_cell | *CARD14* | -1.499243761 | 0.001190743 | 0.004980661 | 1 |
| T2DM | b_cell | *TBCD* | -1.260138335 | 0.002264215 | 0.004980661 | 1 |
| T2DM | b_cell | *FN3KRP* | -2.141877489 | 3.30E-05 | 0.004719449 | 0.160264851 |
| T2DM | b_cell | *NARF* | -1.007390024 | 0.001809415 | 0.004980661 | 1 |
| T2DM | b_cell | *TRIM65* | -1.336655581 | 0.002625745 | 0.004980661 | 1 |
| T2DM | b_cell | *SECTM1* | -2.365820192 | 0.028288777 | 0.033395829 | 1 |
| T2DM | b_cell | *RNF157* | -1.278803805 | 0.002064159 | 0.004980661 | 1 |
| T2DM | b_cell | *ZNF750* | -4.113903332 | 0.005121615 | 0.007330406 | 1 |
| T2DM | b_cell | *WDR45B* | -1.247120261 | 0.001765755 | 0.004980661 | 1 |
| T2DM | b_cell | *RNF165* | -1.654401965 | 0.002753577 | 0.005034338 | 1 |
| T2DM | b_cell | *MAPK4* | 4.111361697 | 0.016373492 | 0.020046476 | 1 |
| T2DM | b_cell | *FBXO15* | -2.563552851 | 0.002446034 | 0.004980661 | 1 |
| T2DM | b_cell | *CBLN2* | -2.315956625 | 0.001729534 | 0.004980661 | 1 |
| T2DM | b_cell | *PIP4K2B* | -1.793660148 | 0.009706404 | 0.012505436 | 1 |
| T2DM | b_cell | *ERBB2* | -1.349966687 | 0.00176698 | 0.004980661 | 1 |
| T2DM | b_cell | *MIEN1* | 1.123707097 | 0.004449001 | 0.006599374 | 1 |
| T2DM | b_cell | *PNMT* | -5.116630988 | 0.03861748 | 0.044750898 | 1 |
| T2DM | b_cell | *IGFBP4* | -0.877304106 | 0.002113938 | 0.004980661 | 1 |
| T2DM | b_cell | *TXNL4A* | -1.131498812 | 0.001409285 | 0.004980661 | 1 |
| T2DM | b_cell | *CACNA1A* | -1.918093214 | 0.000864812 | 0.004980661 | 1 |

| | | | | | | |
|---|---|---|---|---|---|---|
| T2DM | b_cell | *NFIC* | -0.74103204 | 0.0018162 | 0.004980661 | 1 |
| T2DM | b_cell | *PRDM15* | -1.905528091 | 0.038566475 | 0.04470554 | 1 |
| T2DM | b_cell | *PFKL* | -2.47033777 | 0.003650017 | 0.005752881 | 1 |
| T2DM | b_cell | *DUS3L* | 3.564421668 | 0.000322145 | 0.004980661 | 1 |
| T2DM | b_cell | *TMEM91* | 0.836460489 | 0.022541585 | 0.02697873 | 1 |
| T2DM | b_cell | *SIRT3* | -1.329318745 | 0.00462588 | 0.006790008 | 1 |
| T2DM | b_cell | *HUNK* | -1.618212511 | 0.000253237 | 0.004980661 | 1 |
| T2DM | b_cell | *COL6A1* | -0.84195437 | 0.000483865 | 0.004980661 | 1 |
| T2DM | b_cell | *IFNAR1* | -1.769838049 | 0.00672033 | 0.009172164 | 1 |
| T2DM | b_cell | *TMEM50B* | -1.306234816 | 0.020931344 | 0.025175387 | 1 |
| T2DM | b_cell | *DOPEY2* | -1.200090339 | 0.003086779 | 0.005245002 | 1 |
| T2DM | b_cell | *URB1* | -1.070190788 | 0.003053897 | 0.00523119 | 1 |
| T2DM | b_cell | *SAE1* | -1.825743331 | 0.000722392 | 0.004980661 | 1 |
| T2DM | b_cell | *GEMIN7* | -3.349114761 | 0.000162594 | 0.004980661 | 0.78890829 |
| T2DM | b_cell | *CAPN10* | -1.480405516 | 0.002792813 | 0.005061629 | 1 |
| T2DM | b_cell | *FBN3* | -2.218731037 | 0.031668183 | 0.037159377 | 1 |
| T2DM | b_cell | *EVI5L* | -1.477685174 | 0.001832626 | 0.004980661 | 1 |
| T2DM | b_cell | *SLC47A1* | 2.334473361 | 0.00450153 | 0.006654912 | 1 |
| T2DM | b_cell | *CTU1* | -1.955720684 | 0.00192935 | 0.004980661 | 1 |
| T2DM | b_cell | *RERE* | -0.877934528 | 0.002446709 | 0.004980661 | 1 |
| T2DM | b_cell | *MMEL1* | -1.951633113 | 0.040069162 | 0.04633355 | 1 |
| T2DM | b_cell | *EPHA2* | -1.512447523 | 0.001945133 | 0.004980661 | 1 |
| T2DM | b_cell | *ARHGEF19* | -2.121200838 | 0.000222954 | 0.004980661 | 1 |
| T2DM | b_cell | *PEX14* | -0.932242511 | 0.001650738 | 0.004980661 | 1 |
| T2DM | b_cell | *ZNF593* | -6.587598404 | 0.005995549 | 0.008378572 | 1 |
| T2DM | b_cell | *C1orf216* | 2.515876167 | 0.00387759 | 0.005976514 | 1 |
| T2DM | b_cell | *PLK4* | 1.357661115 | 0.002761247 | 0.005037286 | 1 |
| T2DM | b_cell | *HSPG2* | -0.892189898 | 0.001272131 | 0.004980661 | 1 |
| T2DM | b_cell | *ITGB3BP* | -1.287748704 | 0.003663024 | 0.005752881 | 1 |
| T2DM | b_cell | *PIGK* | -1.070875003 | 0.001620398 | 0.004980661 | 1 |
| T2DM | b_cell | *RPS8* | -1.017904559 | 0.002199104 | 0.004980661 | 1 |
| T2DM | b_cell | *CYP4B1* | -1.096731313 | 0.00654847 | 0.008975067 | 1 |
| T2DM | b_cell | *SLC44A3* | -1.646800311 | 0.002343745 | 0.004980661 | 1 |
| T2DM | b_cell | *STRIP1* | -1.264484787 | 0.002100697 | 0.004980661 | 1 |
| T2DM | b_cell | *GPR161* | -1.436706725 | 0.001947398 | 0.004980661 | 1 |
| T2DM | b_cell | *ALDH9A1* | -2.190115682 | 0.004437291 | 0.00658611 | 1 |
| T2DM | b_cell | *ATP1B1* | -1.052278719 | 0.00215528 | 0.004980661 | 1 |
| T2DM | b_cell | *TIPRL* | -2.118650282 | 0.003609554 | 0.005712612 | 1 |
| T2DM | b_cell | *NME7* | -1.164244632 | 0.001115248 | 0.004980661 | 1 |
| T2DM | b_cell | *XCL2* | 0.890888708 | 0.002664269 | 0.004991101 | 1 |
| T2DM | b_cell | *MAEL* | -3.676206175 | 0.001760036 | 0.004980661 | 1 |
| T2DM | b_cell | *RFWD2* | -0.846218793 | 0.002052474 | 0.004980661 | 1 |
| T2DM | b_cell | *RGS16* | -2.634512006 | 0.000453495 | 0.004980661 | 1 |
| T2DM | b_cell | *TARS2* | -1.367283813 | 0.00358836 | 0.005684205 | 1 |
| T2DM | b_cell | *SNX27* | -1.587643368 | 0.008221502 | 0.010831042 | 1 |
| T2DM | b_cell | *SETDB1* | -1.6644886 | 0.002817094 | 0.005065458 | 1 |
| T2DM | b_cell | *SEMA6C* | 1.094006428 | 0.003657376 | 0.005752881 | 1 |
| T2DM | b_cell | *ARNT* | -1.421840307 | 0.00304847 | 0.00522879 | 1 |
| T2DM | b_cell | *POGZ* | -0.947149684 | 0.002278408 | 0.004980661 | 1 |
| T2DM | b_cell | *GABPB2* | -0.863643654 | 0.003973264 | 0.006073812 | 1 |

| | | | | | | |
|---|---|---|---|---|---|---|
| T2DM | b_cell | *DYRK3* | -1.745731537 | 0.00201576 | 0.004980661 | 1 |
| T2DM | b_cell | *EIF2D* | -1.395938586 | 0.001624253 | 0.004980661 | 1 |
| T2DM | b_cell | *TAF1A* | -1.931122934 | 0.002804579 | 0.005065458 | 1 |
| T2DM | b_cell | *ADAM15* | -1.100159994 | 0.001841633 | 0.004980661 | 1 |
| T2DM | b_cell | *S100A8* | -1.864101478 | 0.002553698 | 0.004980661 | 1 |
| T2DM | b_cell | *CREB3L4* | -2.378829957 | 0.000567922 | 0.004980661 | 1 |
| T2DM | b_cell | *KCNN3* | -1.065835342 | 0.002592002 | 0.004980661 | 1 |
| T2DM | b_cell | *C1orf43* | -1.361980902 | 0.001832331 | 0.004980661 | 1 |
| T2DM | b_cell | *RIT1* | -2.933442372 | 0.004114472 | 0.006240519 | 1 |
| T2DM | b_cell | *SCCPDH* | -1.534100832 | 0.002144168 | 0.004980661 | 1 |
| T2DM | b_cell | *CEP170* | -1.254565522 | 0.002297533 | 0.004980661 | 1 |
| T2DM | b_cell | *ACP1* | -1.098667399 | 0.001729065 | 0.004980661 | 1 |
| T2DM | b_cell | *CNIH4* | -1.932117098 | 0.003566906 | 0.005668447 | 1 |
| T2DM | b_cell | *MBOAT2* | -1.180530854 | 0.001799309 | 0.004980661 | 1 |
| T2DM | b_cell | *PYCR2* | -1.625956141 | 0.002793702 | 0.005061629 | 1 |
| T2DM | b_cell | *PLEKHA6* | -0.98029225 | 0.012227309 | 0.015425612 | 1 |
| T2DM | b_cell | *OSR1* | -1.602438612 | 0.001826425 | 0.004980661 | 1 |
| T2DM | b_cell | *RHOB* | -1.145821937 | 0.001832 | 0.004980661 | 1 |
| T2DM | b_cell | *GALM* | -1.343530644 | 0.002098601 | 0.004980661 | 1 |
| T2DM | b_cell | *WDPCP* | -0.627935012 | 0.00183118 | 0.004980661 | 1 |
| T2DM | b_cell | *VPS54* | -1.133011846 | 0.00232487 | 0.004980661 | 1 |
| T2DM | b_cell | *ABHD1* | -2.272977894 | 0.000523932 | 0.004980661 | 1 |
| T2DM | b_cell | *CIAO1* | -1.206859847 | 0.002444945 | 0.004980661 | 1 |
| T2DM | b_cell | *ZNF514* | -1.279762152 | 0.000688766 | 0.004980661 | 1 |
| T2DM | b_cell | *EXOC6B* | -0.874510215 | 0.002096428 | 0.004980661 | 1 |
| T2DM | b_cell | *TEX261* | -3.60230878 | 0.011817266 | 0.014974503 | 1 |
| T2DM | b_cell | *THNSL2* | -1.175730962 | 0.002816241 | 0.005065458 | 1 |
| T2DM | b_cell | *RALB* | -1.828436268 | 0.00278006 | 0.005051064 | 1 |
| T2DM | b_cell | *RABL2A* | -1.355231228 | 0.001250609 | 0.004980661 | 1 |
| T2DM | b_cell | *FBLN7* | -1.426540235 | 0.003764466 | 0.005865506 | 1 |
| T2DM | b_cell | *LIPT1* | -1.408404761 | 0.007368661 | 0.009895584 | 1 |
| T2DM | b_cell | *CNGA3* | -39.80259523 | 0.000162026 | 0.004980661 | 0.786148967 |
| T2DM | b_cell | *FAHD2B* | -3.207673571 | 0.010443085 | 0.013365827 | 1 |
| T2DM | b_cell | *LYG1* | -2.116994853 | 0.002066448 | 0.004980661 | 1 |
| T2DM | b_cell | *THSD7B* | -1.086019811 | 0.002002287 | 0.004980661 | 1 |
| T2DM | b_cell | *GPR17* | -0.853491901 | 0.037360437 | 0.043408247 | 1 |
| T2DM | b_cell | *POLR2D* | -1.424770101 | 0.002570958 | 0.004980661 | 1 |
| T2DM | b_cell | *GALNT13* | -1.040586553 | 0.001740975 | 0.004980661 | 1 |
| T2DM | b_cell | *PHOSPHO2* | 1.840403778 | 0.008508368 | 0.011166513 | 1 |
| T2DM | b_cell | *METTL21A* | -1.391286075 | 0.002790893 | 0.005061629 | 1 |
| T2DM | b_cell | *NBEAL1* | -0.853632652 | 0.003338718 | 0.005445196 | 1 |
| T2DM | b_cell | *SUMF1* | -0.800516461 | 0.001921893 | 0.004980661 | 1 |
| T2DM | b_cell | *ACKR3* | -1.404347296 | 0.00180326 | 0.004980661 | 1 |
| T2DM | b_cell | *ESPNL* | 2.771861823 | 0.002492438 | 0.004980661 | 1 |
| T2DM | b_cell | *ANKMY1* | -1.650625998 | 0.007043997 | 0.00952815 | 1 |
| T2DM | b_cell | *TAMM41* | -1.459606644 | 0.005302352 | 0.007557877 | 1 |
| T2DM | b_cell | *STK11IP* | -2.053844794 | 0.001500269 | 0.004980661 | 1 |
| T2DM | b_cell | *EAF1* | -1.490784411 | 0.002040221 | 0.004980661 | 1 |
| T2DM | b_cell | *CNTN4* | -0.811605584 | 0.001716202 | 0.004980661 | 1 |
| T2DM | b_cell | *SLC25A38* | 1.009941467 | 0.004701338 | 0.006854234 | 1 |

| | | | | | | |
|---|---|---|---|---|---|---|
| T2DM | b_cell | *CTDSPL* | -0.901699378 | 0.001302933 | 0.004980661 | 1 |
| T2DM | b_cell | *PTPRG* | -0.910931553 | 0.001965077 | 0.004980661 | 1 |
| T2DM | b_cell | *SLC25A26* | -0.865912859 | 0.001644083 | 0.004980661 | 1 |
| T2DM | b_cell | *ARL6IP5* | -1.398345653 | 0.00251751 | 0.004980661 | 1 |
| T2DM | b_cell | *LIMD1* | -1.790950343 | 0.02147456 | 0.025784352 | 1 |
| T2DM | b_cell | *ZNF660* | -1.249006821 | 0.020570421 | 0.02479078 | 1 |
| T2DM | b_cell | *ADPRH* | -3.857354952 | 0.003086655 | 0.005245002 | 1 |
| T2DM | b_cell | *IGSF11* | -1.230721783 | 0.008182122 | 0.010786579 | 1 |
| T2DM | b_cell | *BOC* | -0.739491034 | 0.002197498 | 0.004980661 | 1 |
| T2DM | b_cell | *MED12L* | -1.203158829 | 0.001855865 | 0.004980661 | 1 |
| T2DM | b_cell | *EIF2A* | -1.248592415 | 0.002710411 | 0.00500969 | 1 |
| T2DM | b_cell | *NCEH1* | -1.109045381 | 0.000836268 | 0.004980661 | 1 |
| T2DM | b_cell | *CCDC39* | -0.803446291 | 0.002683256 | 0.004999677 | 1 |
| T2DM | b_cell | *MUC4* | -1.95839365 | 0.001488573 | 0.004980661 | 1 |
| T2DM | b_cell | *CENPC* | -0.870354977 | 0.001317697 | 0.004980661 | 1 |
| T2DM | b_cell | *SCD5* | -1.278058883 | 0.00176859 | 0.004980661 | 1 |
| T2DM | b_cell | *TRMT10A* | -1.66200464 | 0.001609994 | 0.004980661 | 1 |
| T2DM | b_cell | *PYURF* | -2.266445876 | 0.002188302 | 0.004980661 | 1 |
| T2DM | b_cell | *CISD2* | -1.494699064 | 0.003172684 | 0.005315561 | 1 |
| T2DM | b_cell | *DDIT4L* | -4.01046599 | 0.006480572 | 0.008894974 | 1 |
| T2DM | b_cell | *METTL14* | -1.049742418 | 0.00146861 | 0.004980661 | 1 |
| T2DM | b_cell | *USP53* | -1.288375729 | 0.009835928 | 0.012662224 | 1 |
| T2DM | b_cell | *MARCH1* | -0.918203851 | 0.000916326 | 0.004980661 | 1 |
| T2DM | b_cell | *CBR4* | -0.906493825 | 0.001857448 | 0.004980661 | 1 |
| T2DM | b_cell | *CYP4V2* | -1.104491069 | 0.002193823 | 0.004980661 | 1 |
| T2DM | b_cell | *MARCH6* | -1.171078408 | 0.002505384 | 0.004980661 | 1 |
| T2DM | b_cell | *NKD2* | 1.143728993 | 0.003562517 | 0.005663608 | 1 |
| T2DM | b_cell | *MYO10* | -0.970343723 | 0.001693776 | 0.004980661 | 1 |
| T2DM | b_cell | *FAM105A* | -1.32241239 | 0.000693598 | 0.004980661 | 1 |
| T2DM | b_cell | *GTF2H2* | 0.943664622 | 0.003737278 | 0.005841905 | 1 |
| T2DM | b_cell | *SPATA9* | -1.035587366 | 0.001772357 | 0.004980661 | 1 |
| T2DM | b_cell | *FEM1C* | -1.32185406 | 0.001815814 | 0.004980661 | 1 |
| T2DM | b_cell | *COMMD10* | -0.9536636 | 0.001735196 | 0.004980661 | 1 |
| T2DM | b_cell | *YIPF5* | -2.02480889 | 0.004112801 | 0.006239935 | 1 |
| T2DM | b_cell | *CXCL14* | -1.201625413 | 0.001174003 | 0.004980661 | 1 |
| T2DM | b_cell | *FBXO38* | -1.180658422 | 0.002160088 | 0.004980661 | 1 |
| T2DM | b_cell | *PCYOX1L* | 2.018600755 | 0.008078583 | 0.010674642 | 1 |
| T2DM | b_cell | *BOD1* | -1.477382621 | 0.001971414 | 0.004980661 | 1 |
| T2DM | b_cell | *FAM50B* | -3.634744696 | 0.003849888 | 0.005958423 | 1 |
| T2DM | b_cell | *MYLK4* | 2.517864052 | 0.030205692 | 0.035546451 | 1 |
| T2DM | b_cell | *GFOD1* | -0.901888145 | 0.001706072 | 0.004980661 | 1 |
| T2DM | b_cell | *TRIM7* | 2.222502478 | 0.037805065 | 0.043893318 | 1 |
| T2DM | b_cell | *HIGD2A* | -1.398893558 | 0.001667441 | 0.004980661 | 1 |
| T2DM | b_cell | *RNF44* | 2.091971442 | 0.032906147 | 0.038514854 | 1 |
| T2DM | b_cell | *MUT* | -2.478319982 | 0.016282004 | 0.019949566 | 1 |
| T2DM | b_cell | *DOK3* | -1.285542325 | 0.013446874 | 0.016803981 | 1 |
| T2DM | b_cell | *DAAM2* | -1.161597543 | 0.002091465 | 0.004980661 | 1 |
| T2DM | b_cell | *MLIP* | -1.486119115 | 0.002377063 | 0.004980661 | 1 |
| T2DM | b_cell | *ANO7* | 0.495390316 | 0.002955986 | 0.005166587 | 1 |
| T2DM | b_cell | *TCTE1* | -5.151500072 | 0.023258276 | 0.027795358 | 1 |

| | | | | | | |
|---|---|---|---|---|---|---|
| T2DM | b_cell | *RPL7L1* | -1.357588426 | 0.001853844 | 0.004980661 | 1 |
| T2DM | b_cell | *TPBG* | -1.100601821 | 0.00160561 | 0.004980661 | 1 |
| T2DM | b_cell | *IRAK1BP1* | -0.987512912 | 0.003788389 | 0.005887656 | 1 |
| T2DM | b_cell | *PHIP* | -1.058100339 | 0.002493183 | 0.004980661 | 1 |
| T2DM | b_cell | *RNF217* | -0.79261248 | 0.001770013 | 0.004980661 | 1 |
| T2DM | b_cell | *ARHGAP18* | -0.938809507 | 0.003000778 | 0.005195812 | 1 |
| T2DM | b_cell | *ABRACL* | -2.171096186 | 0.012205968 | 0.015406701 | 1 |
| T2DM | b_cell | *SLC18B1* | -1.990422434 | 0.007978545 | 0.010559711 | 1 |
| T2DM | b_cell | *PNLDC1* | 3.405802747 | 0.002814926 | 0.005065458 | 1 |
| T2DM | b_cell | *WTAP* | -1.230512933 | 0.002268441 | 0.004980661 | 1 |
| T2DM | b_cell | *SLC22A3* | -1.066110059 | 0.001792972 | 0.004980661 | 1 |
| T2DM | b_cell | *GNA12* | -1.098104287 | 0.001148851 | 0.004980661 | 1 |
| T2DM | b_cell | *SDK1* | -0.761040653 | 0.001366109 | 0.004980661 | 1 |
| T2DM | b_cell | *CCZ1B* | -1.220728816 | 0.001187066 | 0.004980661 | 1 |
| T2DM | b_cell | *RBAK* | -3.844359629 | 0.012300973 | 0.015505607 | 1 |
| T2DM | b_cell | *POMZP3* | -1.299925457 | 0.001476622 | 0.004980661 | 1 |
| T2DM | b_cell | *PSPH* | -1.042027251 | 0.008812967 | 0.011513332 | 1 |
| T2DM | b_cell | *GIGYF1* | -2.384358067 | 0.00895598 | 0.011656227 | 1 |
| T2DM | b_cell | *TRIM4* | -1.480410536 | 0.000395681 | 0.004980661 | 1 |
| T2DM | b_cell | *TMEM209* | -1.146158387 | 0.00180145 | 0.004980661 | 1 |
| T2DM | b_cell | *EPHA1* | -4.029745311 | 0.001694048 | 0.004980661 | 1 |
| T2DM | b_cell | *NCAPG2* | 1.724476417 | 0.006715507 | 0.00916816 | 1 |
| T2DM | b_cell | *CSGALNACT1* | -0.672434093 | 0.003617203 | 0.005720557 | 1 |
| T2DM | b_cell | *BIN3* | -1.154418079 | 0.002095843 | 0.004980661 | 1 |
| T2DM | b_cell | *TACC1* | -0.849962516 | 0.001873117 | 0.004980661 | 1 |
| T2DM | b_cell | *ADHFE1* | -0.965546325 | 0.000252348 | 0.004980661 | 1 |
| T2DM | b_cell | *RPL7* | -2.398092163 | 0.027904174 | 0.032965924 | 1 |
| T2DM | b_cell | *ATP6V0D2* | -5.526917844 | 0.004041178 | 0.006158228 | 1 |
| T2DM | b_cell | *DPYS* | -0.91487388 | 0.001764608 | 0.004980661 | 1 |
| T2DM | b_cell | *LRP12* | -1.197266839 | 0.001782649 | 0.004980661 | 1 |
| T2DM | b_cell | *MAL2* | -1.057634279 | 0.004212415 | 0.006341494 | 1 |
| T2DM | b_cell | *SLC39A4* | -3.567213484 | 0.00026742 | 0.004980661 | 1 |
| T2DM | b_cell | *AK3* | -1.547739452 | 0.002945763 | 0.005152431 | 1 |
| T2DM | b_cell | *HAUS6* | -1.322072282 | 0.001897286 | 0.004980661 | 1 |
| T2DM | b_cell | *C9orf72* | -0.828957549 | 0.002273482 | 0.004980661 | 1 |
| T2DM | b_cell | *FBXO10* | -3.223061177 | 0.005630539 | 0.007937064 | 1 |
| T2DM | b_cell | *ZNF462* | -0.933391215 | 0.002506471 | 0.004980661 | 1 |
| T2DM | b_cell | *ALAD* | -1.338032615 | 0.002200278 | 0.004980661 | 1 |
| T2DM | b_cell | *WDR31* | -1.183555781 | 0.003933963 | 0.006030834 | 1 |
| T2DM | b_cell | *POLE3* | -1.588197713 | 0.006298897 | 0.008714642 | 1 |
| T2DM | b_cell | *SURF1* | -1.358197724 | 0.000830027 | 0.004980661 | 1 |
| T2DM | b_cell | *SURF6* | -1.821708732 | 0.002138303 | 0.004980661 | 1 |
| T2DM | b_cell | *MED22* | -1.915209078 | 0.000642651 | 0.004980661 | 1 |
| T2DM | b_cell | *REXO4* | -1.230059057 | 0.001814324 | 0.004980661 | 1 |
| T2DM | b_cell | *SH3GLB2* | -1.400103794 | 0.002087236 | 0.004980661 | 1 |
| T2DM | b_cell | *INPP5E* | -1.773897253 | 0.0014876 | 0.004980661 | 1 |
| T2DM | b_cell | *SEC16A* | -1.093919136 | 0.003101888 | 0.005253181 | 1 |
| T2DM | b_cell | *SLC39A12* | -3.520349991 | 0.003367837 | 0.00546696 | 1 |
| T2DM | b_cell | *TMEM236* | -1.883693786 | 0.022013882 | 0.026386204 | 1 |
| T2DM | b_cell | *RSU1* | -1.646513448 | 0.006427436 | 0.008842053 | 1 |

| | | | | | | |
|---|---|---|---|---|---|---|
| T2DM | b_cell | *ST8SIA6* | -1.418549357 | 0.002244935 | 0.004980661 | 1 |
| T2DM | b_cell | *PARD3* | -0.822427365 | 0.001983506 | 0.004980661 | 1 |
| T2DM | b_cell | *CDHR1* | -9.291207917 | 0.004123279 | 0.006251922 | 1 |
| T2DM | b_cell | *RGR* | 1.373759877 | 0.003223329 | 0.005350527 | 1 |
| T2DM | b_cell | *FRA10AC1* | -1.722468563 | 0.014277796 | 0.017744843 | 1 |
| T2DM | b_cell | *ADD3* | -1.017284029 | 0.002202951 | 0.004980661 | 1 |
| T2DM | b_cell | *MTG1* | -1.427661812 | 0.004835383 | 0.007003366 | 1 |
| T2DM | b_cell | *PAOX* | -2.450100891 | 0.002374163 | 0.004980661 | 1 |
| T2DM | b_cell | *PDCD11* | -2.305890831 | 0.006267374 | 0.008678453 | 1 |
| T2DM | b_cell | *RGS10* | -1.529091847 | 0.002128468 | 0.004980661 | 1 |
| T2DM | b_cell | *GAS2* | -1.648320446 | 0.002703993 | 0.00500969 | 1 |
| T2DM | b_cell | *LIN7C* | -1.200599404 | 0.00188247 | 0.004980661 | 1 |
| T2DM | b_cell | *ZNF215* | 1.226994805 | 0.002488784 | 0.004980661 | 1 |
| T2DM | b_cell | *HSD17B12* | -2.603409313 | 4.05E-08 | 2.81E-05 | 0.00019674 |
| T2DM | b_cell | *APIP* | -1.907635697 | 9.50E-06 | 0.00219398 | 0.046073571 |
| T2DM | b_cell | *EIF3M* | -0.99272478 | 0.001055342 | 0.004980661 | 1 |
| T2DM | b_cell | *TNKS1BP1* | -0.866666731 | 0.007075027 | 0.009562125 | 1 |
| T2DM | b_cell | *PTPRJ* | -1.362493216 | 0.003410602 | 0.005510766 | 1 |
| T2DM | b_cell | *CCDC82* | -2.139579722 | 0.006494934 | 0.008912166 | 1 |
| T2DM | b_cell | *CAPN5* | -2.452578414 | 0.001948341 | 0.004980661 | 1 |
| T2DM | b_cell | *INTS4* | -1.631950224 | 0.005359698 | 0.007623938 | 1 |
| T2DM | b_cell | *RPS3* | -1.303418789 | 0.002536471 | 0.004980661 | 1 |
| T2DM | b_cell | *TTC12* | -1.050872134 | 0.004949037 | 0.007131787 | 1 |
| T2DM | b_cell | *DRD2* | -12.53935857 | 0.001663945 | 0.004980661 | 1 |
| T2DM | b_cell | *NPAT* | -2.156186798 | 0.014380661 | 0.017858963 | 1 |
| T2DM | b_cell | *ATM* | -1.277615382 | 0.002937728 | 0.005145797 | 1 |
| T2DM | b_cell | *SLX4IP* | -1.781349684 | 0.003540583 | 0.005648061 | 1 |
| T2DM | b_cell | *FADS1* | -1.630059716 | 0.002641334 | 0.004985914 | 1 |
| T2DM | b_cell | *TMC2* | -2.81285339 | 0.001321566 | 0.004980661 | 1 |
| T2DM | b_cell | *EML3* | -1.520817882 | 0.007546059 | 0.010080804 | 1 |
| T2DM | b_cell | *INCENP* | -4.459011925 | 0.006951344 | 0.009426473 | 1 |
| T2DM | b_cell | *PLCH2* | -7.327237074 | 0.009106517 | 0.011823607 | 1 |
| T2DM | b_cell | *CPSF7* | -1.361614105 | 0.001779995 | 0.004980661 | 1 |
| T2DM | b_cell | *CCDC15* | -1.857611316 | 0.000368216 | 0.004980661 | 1 |
| T2DM | b_cell | *CHEK1* | 0.63391938 | 0.002780584 | 0.005051064 | 1 |
| T2DM | b_cell | *CABLES2* | -2.383577167 | 0.00262788 | 0.004980661 | 1 |
| T2DM | b_cell | *NUDT22* | -1.347905238 | 0.001180523 | 0.004980661 | 1 |
| T2DM | b_cell | *TBX6* | 1.977869721 | 0.000459521 | 0.004980661 | 1 |
| T2DM | b_cell | *ALDOA* | -1.616993393 | 0.002076152 | 0.004980661 | 1 |
| T2DM | b_cell | *ANXA8L1* | -0.968341287 | 0.001550119 | 0.004980661 | 1 |
| T2DM | b_cell | *CWC15* | -1.28746223 | 0.002065502 | 0.004980661 | 1 |
| T2DM | b_cell | *FCGR1A* | -3.923072165 | 0.029972319 | 0.035297498 | 1 |
| T2DM | b_cell | *TMEM218* | -2.152538666 | 0.001916246 | 0.004980661 | 1 |
| T2DM | b_cell | *FAM124A* | -1.591581685 | 0.002127434 | 0.004980661 | 1 |
| T2DM | b_cell | *LYPD6B* | 0.307004732 | 0.002361807 | 0.004980661 | 1 |
| T2DM | b_cell | *WDR17* | -1.325804251 | 0.001183468 | 0.004980661 | 1 |
| T2DM | b_cell | *CCDC102B* | -0.800263556 | 0.001608837 | 0.004980661 | 1 |
| T2DM | b_cell | *RGS18* | -1.802668302 | 0.006317388 | 0.008735243 | 1 |
| T2DM | b_cell | *PRSS23* | -0.873892346 | 0.002216075 | 0.004980661 | 1 |
| T2DM | b_cell | *PPP1R1C* | -2.36982555 | 0.001751635 | 0.004980661 | 1 |

| | | | | | | |
|---|---|---|---|---|---|---|
| T2DM | b_cell | *FAM173B* | -1.179134098 | 0.001700889 | 0.004980661 | 1 |
| T2DM | b_cell | *GPR158* | -2.209654123 | 0.002793153 | 0.005061629 | 1 |
| T2DM | b_cell | *UBE3B* | -0.98695668 | 0.001940815 | 0.004980661 | 1 |
| T2DM | b_cell | *ANK3* | -1.828528255 | 0.021920421 | 0.026287169 | 1 |
| T2DM | b_cell | *PLBD2* | -2.3908575 | 0.022726416 | 0.027186531 | 1 |
| T2DM | b_cell | *DLG5* | -1.032781173 | 0.000953831 | 0.004980661 | 1 |
| T2DM | b_cell | *GXYLT1* | -2.214958778 | 0.007395296 | 0.009920369 | 1 |
| T2DM | b_cell | *EXT2* | -0.951932878 | 0.001535526 | 0.004980661 | 1 |
| T2DM | b_cell | *TMEM18* | -2.979657353 | 0.013224133 | 0.016562595 | 1 |
| T2DM | b_cell | *NDUFC2* | -1.980869684 | 0.002317832 | 0.004980661 | 1 |
| T2DM | b_cell | *ADAMTS12* | -0.815819557 | 0.0025274 | 0.004980661 | 1 |
| T2DM | b_cell | *NUBPL* | -1.176299741 | 0.002318287 | 0.004980661 | 1 |
| T2DM | b_cell | *VIPAS39* | -1.637243501 | 0.001366586 | 0.004980661 | 1 |
| T2DM | b_cell | *ANKRD50* | -1.2788032 | 0.002340093 | 0.004980661 | 1 |
| T2DM | b_cell | *UPF2* | -1.097177896 | 0.002068443 | 0.004980661 | 1 |
| T2DM | b_cell | *C4orf33* | -1.429048917 | 0.00164384 | 0.004980661 | 1 |
| T2DM | b_cell | *PTPRO* | -1.412002861 | 0.001716748 | 0.004980661 | 1 |
| T2DM | b_cell | *EPS8* | -0.732271447 | 0.002171918 | 0.004980661 | 1 |
| T2DM | b_cell | *QDPR* | -1.111934036 | 0.001854602 | 0.004980661 | 1 |
| T2DM | b_cell | *FAM160B1* | -0.974598047 | 0.002019364 | 0.004980661 | 1 |
| T2DM | b_cell | *ZNF827* | -1.013509473 | 0.002036972 | 0.004980661 | 1 |
| T2DM | b_cell | *AKR1C2* | -1.129220299 | 0.003811663 | 0.005908686 | 1 |
| T2DM | b_cell | *PIGF* | -1.473487783 | 0.002379304 | 0.004980661 | 1 |
| T2DM | b_cell | *INPP1* | -1.1894494 | 0.002964939 | 0.005173571 | 1 |
| T2DM | b_cell | *MFSD6* | -1.431082061 | 0.002161058 | 0.004980661 | 1 |
| T2DM | b_cell | *RNF144A* | -0.87872005 | 0.001828251 | 0.004980661 | 1 |
| T2DM | b_cell | *CENPU* | -2.275718922 | 0.004724287 | 0.006883231 | 1 |
| T2DM | b_cell | *CCDC122* | -0.999866468 | 0.00159271 | 0.004980661 | 1 |
| T2DM | b_cell | *GUF1* | -1.206842707 | 0.001592761 | 0.004980661 | 1 |
| T2DM | b_cell | *GABRA2* | -4.871373858 | 0.001073078 | 0.004980661 | 1 |
| T2DM | b_cell | *PARP8* | -0.91045024 | 0.001610786 | 0.004980661 | 1 |
| T2DM | b_cell | *CACUL1* | -1.220401834 | 0.002151205 | 0.004980661 | 1 |
| T2DM | b_cell | *BEND6* | -3.832469262 | 0.003530702 | 0.005639428 | 1 |
| T2DM | b_cell | *TIAL1* | -1.603946089 | 0.003330795 | 0.005441511 | 1 |
| T2DM | b_cell | *TMEM56* | 1.032765206 | 0.003210763 | 0.005339692 | 1 |
| T2DM | b_cell | *PTPN14* | -0.912448825 | 0.002277965 | 0.004980661 | 1 |
| T2DM | b_cell | *MGAT5* | -1.413939417 | 0.002283638 | 0.004980661 | 1 |
| T2DM | b_cell | *TMEM178A* | -2.526408919 | 0.001870321 | 0.004980661 | 1 |
| T2DM | b_cell | *EPG5* | -1.479399966 | 0.002172559 | 0.004980661 | 1 |
| T2DM | b_cell | *SPC25* | -1.566660766 | 0.001333926 | 0.004980661 | 1 |
| T2DM | b_cell | *KCNK13* | -3.039261197 | 0.004514384 | 0.006661737 | 1 |
| T2DM | b_cell | *ATG10* | -0.91577655 | 0.005815212 | 0.008152387 | 1 |
| T2DM | b_cell | *POC5* | -2.783809007 | 8.18E-05 | 0.004980661 | 0.397066576 |
| T2DM | b_cell | *CWF19L2* | -1.380835012 | 0.01021547 | 0.013109088 | 1 |
| T2DM | b_cell | *ZNF547* | 1.039084999 | 0.006556521 | 0.008979205 | 1 |
| T2DM | b_cell | *NMT2* | -1.057613982 | 0.001437577 | 0.004980661 | 1 |
| T2DM | b_cell | *ZNF837* | -3.585275941 | 0.004238226 | 0.006372443 | 1 |
| T2DM | b_cell | *CCDC50* | -1.42001618 | 0.00639726 | 0.008818041 | 1 |
| T2DM | b_cell | *PLEKHH2* | -0.722385076 | 0.003305778 | 0.005412542 | 1 |
| T2DM | b_cell | *SPARCL1* | -0.706964661 | 0.001858798 | 0.004980661 | 1 |

| | | | | | | |
|---|---|---|---|---|---|---|
| T2DM | b_cell | *MBNL1* | -0.852721529 | 0.002313164 | 0.004980661 | 1 |
| T2DM | b_cell | *GPD1L* | -1.187895998 | 0.001685098 | 0.004980661 | 1 |
| T2DM | b_cell | *CCNO* | -15.38630207 | 0.001952102 | 0.004980661 | 1 |
| T2DM | b_cell | *CLEC4F* | -2.352228725 | 0.012926384 | 0.016214792 | 1 |
| T2DM | b_cell | *SLC30A6* | -0.897126833 | 0.00208901 | 0.004980661 | 1 |
| T2DM | b_cell | *GPR180* | -0.974249367 | 0.001755854 | 0.004980661 | 1 |
| T2DM | b_cell | *TCTEX1D1* | 1.276697942 | 0.002711345 | 0.00500969 | 1 |
| T2DM | b_cell | *IFIT5* | -1.296998413 | 0.026201059 | 0.031097735 | 1 |
| T2DM | b_cell | *PRDM8* | -2.664477536 | 0.001551652 | 0.004980661 | 1 |
| T2DM | b_cell | *GGPS1* | -1.361367529 | 0.00357151 | 0.005668618 | 1 |
| T2DM | b_cell | *ZNF117* | -0.504509495 | 0.018402538 | 0.022383834 | 1 |
| T2DM | b_cell | *MED21* | -1.196875954 | 0.001828924 | 0.004980661 | 1 |
| T2DM | b_cell | *JAKMIP1* | -1.889018253 | 0.002006971 | 0.004980661 | 1 |
| T2DM | b_cell | *SREK1IP1* | -1.035879686 | 0.001696696 | 0.004980661 | 1 |
| T2DM | b_cell | *MR1* | -1.07022544 | 0.002647069 | 0.004985914 | 1 |
| T2DM | b_cell | *CENPH* | -2.338872415 | 0.001466084 | 0.004980661 | 1 |
| T2DM | b_cell | *CARHSP1* | -1.312419658 | 0.001831031 | 0.004980661 | 1 |
| T2DM | b_cell | *CAST* | -1.065094527 | 0.002688569 | 0.005005732 | 1 |
| T2DM | b_cell | *SCOC* | -1.525943774 | 0.002839019 | 0.005070867 | 1 |
| T2DM | b_cell | *CLGN* | 4.512401306 | 0.014401517 | 0.017880287 | 1 |
| T2DM | b_cell | *CETN3* | -1.319929862 | 0.001072886 | 0.004980661 | 1 |
| T2DM | b_cell | *SYCP2L* | -1.967925858 | 0.000558826 | 0.004980661 | 1 |
| T2DM | b_cell | *MERTK* | -0.669447988 | 0.006369204 | 0.008791857 | 1 |
| T2DM | b_cell | *TMEM87B* | -1.576538458 | 0.002878077 | 0.005096998 | 1 |
| T2DM | b_cell | *PLA2R1* | -0.708132288 | 0.008600416 | 0.011281216 | 1 |
| T2DM | b_cell | *SLC25A27* | -1.070076122 | 0.001402067 | 0.004980661 | 1 |
| T2DM | b_cell | *FRMD1* | -2.381328619 | 0.002341069 | 0.004980661 | 1 |
| T2DM | b_cell | *INO80C* | -2.023526959 | 0.002017661 | 0.004980661 | 1 |
| T2DM | b_cell | *LPCAT1* | -1.513598127 | 0.001913969 | 0.004980661 | 1 |
| T2DM | b_cell | *NMRAL1* | -1.157872344 | 0.004325666 | 0.006465845 | 1 |
| T2DM | b_cell | *ADPRHL1* | -2.777148912 | 0.003525021 | 0.005637244 | 1 |
| T2DM | b_cell | *CFDP1* | -1.061613872 | 0.002091581 | 0.004980661 | 1 |
| T2DM | b_cell | *ZDHHC7* | -1.187574377 | 0.001868741 | 0.004980661 | 1 |
| T2DM | b_cell | *JAZF1* | -1.455994273 | 6.36E-05 | 0.004980661 | 0.308523652 |
| T2DM | b_cell | *FBXO36* | -1.232137804 | 0.001759711 | 0.004980661 | 1 |
| T2DM | b_cell | *MCOLN2* | -2.899819041 | 0.014710178 | 0.018212244 | 1 |
| T2DM | b_cell | *GDPD1* | -1.332164451 | 0.001645866 | 0.004980661 | 1 |
| T2DM | b_cell | *SEMA3D* | -0.843951811 | 0.001768488 | 0.004980661 | 1 |
| T2DM | b_cell | *PPP2R5E* | -1.222151107 | 0.002497573 | 0.004980661 | 1 |
| T2DM | b_cell | *ANKRD29* | -1.053092889 | 0.001867148 | 0.004980661 | 1 |
| T2DM | b_cell | *DNAAF1* | -3.761733654 | 0.005393068 | 0.007669158 | 1 |
| T2DM | b_cell | *ANKH* | -1.05464418 | 0.002562675 | 0.004980661 | 1 |
| T2DM | b_cell | *UBASH3B* | -1.259006177 | 0.001476616 | 0.004980661 | 1 |
| T2DM | b_cell | *ROBO4* | -1.106257981 | 0.001826256 | 0.004980661 | 1 |
| T2DM | b_cell | *ROBO3* | -1.012781243 | 0.001583528 | 0.004980661 | 1 |
| T2DM | b_cell | *CC2D1B* | -1.923529381 | 0.001745526 | 0.004980661 | 1 |
| T2DM | b_cell | *PRKCA* | -1.329868438 | 0.003022284 | 0.005212984 | 1 |
| T2DM | b_cell | *LRRK1* | -1.904589247 | 0.006344746 | 0.008763082 | 1 |
| T2DM | b_cell | *ABCA5* | -1.346506987 | 0.00556186 | 0.007853942 | 1 |
| T2DM | b_cell | *ENPP3* | -1.456360431 | 0.001516558 | 0.004980661 | 1 |

| | | | | | | |
|---|---|---|---|---|---|---|
| T2DM | b_cell | *MIA3* | -0.889549584 | 0.001959656 | 0.004980661 | 1 |
| T2DM | b_cell | *NEIL2* | 2.370585712 | 0.005155955 | 0.007369293 | 1 |
| T2DM | b_cell | *PGM5* | -0.843159878 | 0.00173706 | 0.004980661 | 1 |
| T2DM | b_cell | *CCSAP* | -1.614826822 | 0.001790294 | 0.004980661 | 1 |
| T2DM | b_cell | *GBP5* | -1.787654697 | 0.001131849 | 0.004980661 | 1 |
| T2DM | b_cell | *LY96* | -1.585390224 | 0.001439248 | 0.004980661 | 1 |
| T2DM | b_cell | *PDE1C* | -1.015341301 | 0.001776167 | 0.004980661 | 1 |
| T2DM | b_cell | *RABGEF1* | -1.466209083 | 0.004838537 | 0.007005137 | 1 |
| T2DM | b_cell | *MRPL39* | -1.03575903 | 0.006981381 | 0.009459275 | 1 |
| T2DM | b_cell | *XPC* | -0.971994885 | 0.002134808 | 0.004980661 | 1 |
| T2DM | b_cell | *FLCN* | -1.068981594 | 0.001765891 | 0.004980661 | 1 |
| T2DM | b_cell | *OXNAD1* | -0.905897587 | 0.001865525 | 0.004980661 | 1 |
| T2DM | b_cell | *APCDD1* | -0.92825453 | 0.001804642 | 0.004980661 | 1 |
| T2DM | b_cell | *MPPE1* | -1.169634972 | 0.001390969 | 0.004980661 | 1 |
| T2DM | b_cell | *RAB6B* | -2.214138044 | 0.001234972 | 0.004980661 | 1 |
| T2DM | b_cell | *ANKRD40* | -1.24632604 | 0.00126613 | 0.004980661 | 1 |
| T2DM | b_cell | *CYP2U1* | -1.199977235 | 0.003495384 | 0.005600926 | 1 |
| T2DM | b_cell | *PROM2* | -7.651587528 | 0.00142979 | 0.004980661 | 1 |
| T2DM | b_cell | *AK9* | -0.959948931 | 0.002405752 | 0.004980661 | 1 |
| T2DM | b_cell | *MARVELD1* | -1.469772784 | 0.001785429 | 0.004980661 | 1 |
| T2DM | b_cell | *GOLGA7B* | 1.911857795 | 0.002642054 | 0.004985914 | 1 |
| T2DM | b_cell | *TRMT44* | -2.192072561 | 0.004369153 | 0.006508791 | 1 |
| T2DM | b_cell | *MOV10* | -1.336393187 | 0.001228851 | 0.004980661 | 1 |
| T2DM | b_cell | *HEATR3* | -1.337207767 | 0.008894659 | 0.011606903 | 1 |
| T2DM | b_cell | *CNOT8* | -2.188858564 | 0.006946395 | 0.009422396 | 1 |
| T2DM | b_cell | *LRGUK* | -1.342693278 | 0.002811188 | 0.005065458 | 1 |
| T2DM | b_cell | *SETD9* | -2.47712601 | 0.002634043 | 0.004980661 | 1 |
| T2DM | b_cell | *NUP205* | -1.572671294 | 0.001963815 | 0.004980661 | 1 |
| T2DM | b_cell | *ZKSCAN2* | -2.170378454 | 0.006465344 | 0.008884636 | 1 |
| T2DM | b_cell | *PDIA4* | -1.620937901 | 0.002039512 | 0.004980661 | 1 |
| T2DM | b_cell | *KDM8* | -1.819385893 | 0.002118224 | 0.004980661 | 1 |
| T2DM | b_cell | *ALS2CR12* | -1.520682204 | 0.002771427 | 0.005049555 | 1 |
| T2DM | b_cell | *FZD7* | -1.378600539 | 0.002965065 | 0.005173571 | 1 |
| T2DM | b_cell | *DEPTOR* | -1.118132436 | 0.002369873 | 0.004980661 | 1 |
| T2DM | b_cell | *RRAGA* | -1.570879149 | 0.002350987 | 0.004980661 | 1 |
| T2DM | b_cell | *TMBIM4* | -0.949449466 | 0.002050939 | 0.004980661 | 1 |
| T2DM | b_cell | *PSD3* | -0.755093673 | 0.003230238 | 0.005354669 | 1 |
| T2DM | b_cell | *UGT2B4* | -1.278017431 | 0.016174483 | 0.019837865 | 1 |
| T2DM | b_cell | *MMP16* | -1.033936458 | 0.001837621 | 0.004980661 | 1 |
| T2DM | b_cell | *KCNMA1* | -0.753306689 | 0.00452762 | 0.006671124 | 1 |
| T2DM | b_cell | *NDUFAF6* | -1.2711076 | 0.000684395 | 0.004980661 | 1 |
| T2DM | b_cell | *DRAM2* | -0.861929855 | 0.002636363 | 0.004983106 | 1 |
| T2DM | b_cell | *C8orf37* | 1.639323914 | 0.03302255 | 0.038636463 | 1 |
| T2DM | b_cell | *ART3* | -3.032756164 | 0.01376096 | 0.017159645 | 1 |
| T2DM | b_cell | *WHAMM* | -1.554733036 | 0.000635343 | 0.004980661 | 1 |
| T2DM | b_cell | *N6AMT1* | -2.538534871 | 0.002706428 | 0.00500969 | 1 |
| T2DM | b_cell | *NAA11* | -3.793396532 | 0.001085228 | 0.004980661 | 1 |
| T2DM | b_cell | *SFXN2* | -1.097902201 | 0.029518178 | 0.034788001 | 1 |
| T2DM | b_cell | *C14orf2* | -2.262175208 | 0.002778952 | 0.005051064 | 1 |
| T2DM | b_cell | *TDRD9* | -4.026682137 | 0.001057534 | 0.004980661 | 1 |

| | | | | | | |
|---|---|---|---|---|---|---|
| T2DM | b_cell | *PCDH1* | -2.273320925 | 3.96E-05 | 0.004980661 | 0.192321527 |
| T2DM | b_cell | *RPL30* | -0.990444421 | 0.001897846 | 0.004980661 | 1 |
| T2DM | b_cell | *SUPV3L1* | -1.163636168 | 0.001744021 | 0.004980661 | 1 |
| T2DM | b_cell | *HKDC1* | 1.451513954 | 0.003091903 | 0.005245425 | 1 |
| T2DM | b_cell | *TYSND1* | -1.063102407 | 0.002703616 | 0.00500969 | 1 |
| T2DM | b_cell | *NODAL* | -2.288348717 | 0.001766074 | 0.004980661 | 1 |
| T2DM | b_cell | *ZDHHC5* | -1.288426384 | 0.001876913 | 0.004980661 | 1 |
| T2DM | b_cell | *MED19* | -1.368595028 | 0.002185076 | 0.004980661 | 1 |
| T2DM | b_cell | *WDYHV1* | -2.143328922 | 0.001053984 | 0.004980661 | 1 |
| T2DM | b_cell | *ATAD2* | -0.957552718 | 0.0044948 | 0.006649015 | 1 |
| T2DM | b_cell | *FBRS* | -1.212732636 | 0.00403157 | 0.006145515 | 1 |
| T2DM | b_cell | *SASS6* | -2.370481459 | 0.000662051 | 0.004980661 | 1 |
| T2DM | b_cell | *ITGAD* | -2.557077654 | 0.008089138 | 0.010685678 | 1 |
| T2DM | b_cell | *VPS8* | -1.044794049 | 0.004456546 | 0.006608546 | 1 |
| T2DM | b_cell | *MPV17L* | 2.049759733 | 0.007386518 | 0.009911334 | 1 |
| T2DM | b_cell | *GHRL* | -2.148709468 | 0.003860663 | 0.005965584 | 1 |
| T2DM | b_cell | *NTAN1* | -2.129637259 | 0.008807879 | 0.011509784 | 1 |
| T2DM | b_cell | *SLC6A1* | -4.79929557 | 0.002065008 | 0.004980661 | 1 |
| T2DM | b_cell | *FCHO2* | -1.07863023 | 0.003532197 | 0.005639428 | 1 |
| T2DM | b_cell | *TIMP4* | 2.098291992 | 0.011967409 | 0.015152889 | 1 |
| T2DM | b_cell | *NECAP2* | -1.15735187 | 0.00177838 | 0.004980661 | 1 |
| T2DM | b_cell | *STEAP2* | -0.782003318 | 0.007698034 | 0.01025559 | 1 |
| T2DM | b_cell | *CLDN12* | -2.853566164 | 0.004314117 | 0.006460523 | 1 |
| T2DM | b_cell | *MMP14* | -2.137189275 | 0.002245671 | 0.004980661 | 1 |
| T2DM | b_cell | *GATAD1* | -1.523879892 | 0.002803367 | 0.005065458 | 1 |
| T2DM | b_cell | *TMED6* | -7.472020908 | 0.028269746 | 0.033381554 | 1 |
| T2DM | b_cell | *CLEC18A* | -10.66313511 | 2.23E-09 | 2.70E-06 | 1.08E-05 |
| T2DM | b_cell | *DHRS4* | 1.90052255 | 0.018730779 | 0.022726116 | 1 |
| T2DM | b_cell | *DHRS1* | -1.372239176 | 0.005691124 | 0.008003865 | 1 |
| T2DM | b_cell | *AASDH* | -1.084364068 | 0.002701624 | 0.00500969 | 1 |
| T2DM | b_cell | *LCA5L* | -1.37495504 | 0.001142565 | 0.004980661 | 1 |
| T2DM | b_cell | *SLC38A10* | -1.713851844 | 0.003303484 | 0.005412542 | 1 |
| T2DM | b_cell | *C9orf43* | -2.447550165 | 0.008174425 | 0.010780731 | 1 |
| T2DM | b_cell | *DGKI* | -1.144249268 | 0.000847223 | 0.004980661 | 1 |
| T2DM | b_cell | *SNX22* | 0.499722046 | 0.002481895 | 0.004980661 | 1 |
| T2DM | b_cell | *SLC34A2* | -32.55651617 | 0.004747587 | 0.006902995 | 1 |
| T2DM | b_cell | *AP3S2* | -1.653885809 | 0.004703623 | 0.006855506 | 1 |
| T2DM | b_cell | *SPPL3* | -1.230149085 | 0.001898362 | 0.004980661 | 1 |
| T2DM | b_cell | *C12orf43* | -2.140689818 | 0.000555929 | 0.004980661 | 1 |
| T2DM | b_cell | *PEX10* | -1.550783495 | 0.000955268 | 0.004980661 | 1 |
| T2DM | b_cell | *LDLRAP1* | -1.601002529 | 0.000939475 | 0.004980661 | 1 |
| T2DM | b_cell | *KRTCAP3* | 4.0207391 | 5.51E-06 | 0.00157185 | 0.026721457 |
| T2DM | b_cell | *PAFAH2* | -1.556122812 | 0.001433482 | 0.004980661 | 1 |
| T2DM | b_cell | *EXTL1* | -22.91555843 | 0.001741658 | 0.004980661 | 1 |
| T2DM | b_cell | *RHPN1* | 1.986794527 | 0.015018939 | 0.018551908 | 1 |
| T2DM | b_cell | *LRRC43* | 0.546814621 | 0.002683198 | 0.004999677 | 1 |
| T2DM | b_cell | *EYA3* | -1.178317252 | 0.003043591 | 0.00522879 | 1 |
| T2DM | b_cell | *DZIP1L* | -2.180286765 | 0.008224728 | 0.01083235 | 1 |
| T2DM | b_cell | *FANCC* | -0.96501955 | 0.001834251 | 0.004980661 | 1 |
| T2DM | b_cell | *CLSTN2* | -0.903472155 | 0.001511762 | 0.004980661 | 1 |

| | | | | | | |
|---|---|---|---|---|---|---|
| T2DM | b_cell | *COLEC12* | -1.293966586 | 0.036542621 | 0.042498753 | 1 |
| T2DM | b_cell | *MITD1* | -1.690107095 | 0.002651523 | 0.004985914 | 1 |
| T2DM | b_cell | *TSPAN33* | 1.40219198 | 0.011027013 | 0.014046486 | 1 |
| T2DM | b_cell | *FAM86C1* | -3.454688533 | 0.001884339 | 0.004980661 | 1 |
| T2DM | b_cell | *PPP1R9A* | -0.809892143 | 0.001837814 | 0.004980661 | 1 |
| T2DM | b_cell | *DYNC1I1* | -0.994624837 | 0.002228354 | 0.004980661 | 1 |
| T2DM | b_cell | *COPG2* | -0.826604634 | 0.001366467 | 0.004980661 | 1 |
| T2DM | b_cell | *ELK4* | -1.122309454 | 0.001679945 | 0.004980661 | 1 |
| T2DM | b_cell | *DUSP23* | -1.734500146 | 0.001770007 | 0.004980661 | 1 |
| T2DM | b_cell | *RNF166* | -1.47801047 | 0.002932212 | 0.005141558 | 1 |
| T2DM | b_cell | *F11R* | -1.516016356 | 0.000945536 | 0.004980661 | 1 |
| T2DM | b_cell | *SPATA2L* | -2.145121396 | 0.000605895 | 0.004980661 | 1 |
| T2DM | b_cell | *CDA* | -2.187717436 | 0.004169098 | 0.006301702 | 1 |
| T2DM | b_cell | *SLC5A11* | -1.675461263 | 0.002223013 | 0.004980661 | 1 |
| T2DM | b_cell | *CCAR2* | -1.227851999 | 0.002897149 | 0.005113643 | 1 |
| T2DM | b_cell | *CDC42SE2* | -1.645085073 | 0.00592832 | 0.008291787 | 1 |
| T2DM | b_cell | *RAPGEF6* | -2.486630213 | 0.000615188 | 0.004980661 | 1 |
| T2DM | b_cell | *CLIC6* | 1.564699177 | 0.001235403 | 0.004980661 | 1 |
| T2DM | b_cell | *CBR1* | -1.220256764 | 0.007376398 | 0.009900494 | 1 |
| T2DM | b_cell | *CBR3* | -1.636565044 | 0.016216526 | 0.019874566 | 1 |
| T2DM | b_cell | *CHAF1B* | -2.437699037 | 0.000695572 | 0.004980661 | 1 |
| T2DM | b_cell | *SIM2* | 0.869936835 | 0.032647818 | 0.038253372 | 1 |
| T2DM | b_cell | *HLCS* | -0.854353111 | 0.001558529 | 0.004980661 | 1 |
| T2DM | b_cell | *ARHGAP27* | -2.700075637 | 0.008782816 | 0.011480124 | 1 |
| T2DM | b_cell | *IRX6* | -1.47507486 | 0.00065375 | 0.004980661 | 1 |
| T2DM | b_cell | *HK2* | -1.022333562 | 0.002244574 | 0.004980661 | 1 |
| T2DM | b_cell | *THEM4* | -1.73575074 | 0.031796827 | 0.03729229 | 1 |
| T2DM | b_cell | *AMFR* | -1.138849812 | 0.001726308 | 0.004980661 | 1 |
| T2DM | b_cell | *TMEM69* | -2.501395439 | 0.002658984 | 0.004985914 | 1 |
| T2DM | b_cell | *TEPP* | 3.958181407 | 0.007442194 | 0.009975007 | 1 |
| T2DM | b_cell | *CHCHD6* | -1.022995108 | 0.002235535 | 0.004980661 | 1 |
| T2DM | b_cell | *LRRC36* | -1.668952447 | 0.00112198 | 0.004980661 | 1 |
| T2DM | b_cell | *ZFYVE28* | -1.806360305 | 0.001166497 | 0.004980661 | 1 |
| T2DM | b_cell | *C16orf86* | 1.317358991 | 0.003638968 | 0.005744108 | 1 |
| T2DM | b_cell | *RGS12* | -0.990648026 | 0.001272826 | 0.004980661 | 1 |
| T2DM | b_cell | *LYPD5* | -3.550856494 | 0.000880694 | 0.004980661 | 1 |
| T2DM | b_cell | *CCDC107* | -1.256168287 | 0.001410951 | 0.004980661 | 1 |
| T2DM | b_cell | *ZNF233* | -1.714802987 | 0.002182246 | 0.004980661 | 1 |
| T2DM | b_cell | *ATAD3B* | -1.464706815 | 0.001337075 | 0.004980661 | 1 |
| T2DM | b_cell | *SSU72* | -1.385933256 | 0.00192981 | 0.004980661 | 1 |
| T2DM | b_cell | *CPAMD8* | -1.408709078 | 0.006633284 | 0.009071221 | 1 |
| T2DM | b_cell | *ANKLE1* | -9.290637337 | 0.002309959 | 0.004980661 | 1 |
| T2DM | b_cell | *CCDC58* | -2.21764858 | 0.002616183 | 0.004980661 | 1 |
| T2DM | b_cell | *CILP2* | -5.614847634 | 0.027119996 | 0.03210203 | 1 |
| T2DM | b_cell | *SLC37A1* | -2.728810544 | 0.011729555 | 0.014889553 | 1 |
| T2DM | b_cell | *PDE9A* | -1.113361394 | 0.003019865 | 0.005210663 | 1 |
| T2DM | b_cell | *NDUFV3* | -0.75151306 | 0.00516146 | 0.007372212 | 1 |
| T2DM | b_cell | *HSF2BP* | -2.856908074 | 0.000360565 | 0.004980661 | 1 |
| T2DM | b_cell | *CSTB* | -2.187049888 | 0.02709646 | 0.032081997 | 1 |
| T2DM | b_cell | *C21orf33* | 0.768461632 | 0.005458871 | 0.007737786 | 1 |

| | | | | | | |
|---|---|---|---|---|---|---|
| T2DM | b_cell | *ICOSLG* | 2.665128069 | 0.00851618 | 0.011173743 | 1 |
| T2DM | b_cell | *C21orf2* | -1.753963148 | 0.001454612 | 0.004980661 | 1 |
| T2DM | b_cell | *FTCD* | -3.435041906 | 0.001191982 | 0.004980661 | 1 |
| T2DM | b_cell | *LSS* | -1.95418912 | 0.000408451 | 0.004980661 | 1 |
| T2DM | b_cell | *VAV2* | -2.077653323 | 0.005143255 | 0.00735487 | 1 |
| T2DM | b_cell | *MCM3AP* | -1.033986109 | 0.001692786 | 0.004980661 | 1 |
| T2DM | b_cell | *DIP2A* | -2.10230187 | 0.003280268 | 0.005387901 | 1 |
| T2DM | b_cell | *S100B* | -0.916287103 | 0.003652518 | 0.005752881 | 1 |
| T2DM | b_cell | *PRMT2* | -1.154324242 | 0.001481166 | 0.004980661 | 1 |
| T2DM | b_cell | *ZNF208* | -1.614163582 | 0.001565664 | 0.004980661 | 1 |
| T2DM | b_cell | *ADAMTS13* | -1.818686461 | 0.001115684 | 0.004980661 | 1 |
| T2DM | b_cell | *CACFD1* | -3.574932743 | 0.001641304 | 0.004980661 | 1 |
| T2DM | b_cell | *GPSM1* | -1.812285164 | 0.008183306 | 0.010786579 | 1 |
| T2DM | b_cell | *TAOK1* | -1.130785252 | 0.002676861 | 0.004998428 | 1 |
| T2DM | b_cell | *TLCD1* | -6.784024991 | 0.036888145 | 0.042880039 | 1 |
| T2DM | b_cell | *S100A1* | -5.519812885 | 0.011744778 | 0.014898212 | 1 |
| T2DM | b_cell | *VPS11* | -3.114070674 | 0.007010807 | 0.009493842 | 1 |
| T2DM | b_cell | *FDPS* | -0.96933472 | 0.002680941 | 0.004999202 | 1 |
| T2DM | b_cell | *PAQR6* | -5.081094591 | 0.000378077 | 0.004980661 | 1 |
| T2DM | b_cell | *CCR5* | 5.261872145 | 0.009132514 | 0.011854189 | 1 |
| T2DM | b_cell | *NBEAL2* | -2.074187563 | 0.00260175 | 0.004980661 | 1 |
| T2DM | b_cell | *PTH1R* | -1.268883417 | 0.001092145 | 0.004980661 | 1 |
| T2DM | b_cell | *FCRL3* | 0.340577809 | 0.003061415 | 0.005233962 | 1 |
| T2DM | b_cell | *FGFR4* | -8.784561685 | 0.000597523 | 0.004980661 | 1 |
| T2DM | b_cell | *LY6K* | -2.884970062 | 0.008952573 | 0.01165492 | 1 |
| T2DM | b_cell | *IER2* | -1.661067092 | 0.001599395 | 0.004980661 | 1 |
| T2DM | b_cell | *CPSF4* | -1.385676534 | 0.001575101 | 0.004980661 | 1 |
| T2DM | b_cell | *TONSL* | 1.808466997 | 0.00177692 | 0.004980661 | 1 |
| T2DM | b_cell | *MUM1* | -1.346939094 | 0.002508663 | 0.004980661 | 1 |
| T2DM | b_cell | *ZNF333* | -1.615633561 | 0.002321451 | 0.004980661 | 1 |
| T2DM | b_cell | *SH2B2* | 1.175445364 | 0.003427292 | 0.005526494 | 1 |
| T2DM | b_cell | *RPL8* | -1.255492743 | 0.001748378 | 0.004980661 | 1 |
| T2DM | b_cell | *DVL3* | -1.299608429 | 0.000809619 | 0.004980661 | 1 |
| T2DM | b_cell | *ABCF3* | -1.632674445 | 0.007190773 | 0.009694257 | 1 |
| T2DM | b_cell | *PCYT1A* | -1.317965428 | 0.000580047 | 0.004980661 | 1 |
| T2DM | b_cell | *FBXO27* | -1.264935361 | 0.003627095 | 0.005732341 | 1 |
| T2DM | b_cell | *DMKN* | -0.653580062 | 0.025297212 | 0.030061737 | 1 |
| T2DM | b_cell | *U2AF1L4* | -1.087803923 | 0.012657722 | 0.01591894 | 1 |
| T2DM | b_cell | *BDH1* | -1.551059273 | 0.002244373 | 0.004980661 | 1 |
| T2DM | b_cell | *LRRC56* | -3.702483632 | 0.008643032 | 0.011329017 | 1 |
| T2DM | b_cell | *PGAP3* | -1.658094384 | 0.001800363 | 0.004980661 | 1 |
| T2DM | b_cell | *ZNF577* | -1.674924564 | 0.007477522 | 0.010014059 | 1 |
| T2DM | b_cell | *TMEM143* | -1.32667386 | 0.002138508 | 0.004980661 | 1 |
| T2DM | b_cell | *CCL16* | 2.786719631 | 0.036083012 | 0.041974292 | 1 |
| T2DM | b_cell | *ALDH16A1* | 1.20848891 | 0.004882951 | 0.007053313 | 1 |
| T2DM | b_cell | *DCD* | 5.192488804 | 0.007789195 | 0.010350354 | 1 |
| T2DM | b_cell | *SIGLEC11* | -1.816997875 | 0.015022851 | 0.018552017 | 1 |
| T2DM | b_cell | *ASB16* | -2.769977614 | 0.008013394 | 0.010602942 | 1 |
| T2DM | b_cell | *DBF4B* | -1.03396185 | 0.006126783 | 0.008522692 | 1 |
| T2DM | b_cell | *PLCD3* | -1.642995923 | 0.001749493 | 0.004980661 | 1 |

| | | | | | | |
|---|---|---|---|---|---|---|
| T2DM | b_cell | *IP6K3* | -24.90830324 | 0.000247462 | 0.004980661 | 1 |
| T2DM | b_cell | *LEMD2* | -2.365071368 | 0.042341235 | 0.048856046 | 1 |
| T2DM | b_cell | *CXCL16* | -1.107382839 | 0.039112403 | 0.045270367 | 1 |
| T2DM | b_cell | *ASGR2* | -10.12184381 | 0.002293715 | 0.004980661 | 1 |
| T2DM | b_cell | *POLR3K* | 2.536343802 | 0.014821048 | 0.018340149 | 1 |
| T2DM | b_cell | *WDR90* | -1.715484848 | 0.000963718 | 0.004980661 | 1 |
| T2DM | b_cell | *JMJD8* | -1.829057797 | 0.021347117 | 0.025644024 | 1 |
| T2DM | b_cell | *MEIOB* | -1.932110881 | 0.005506062 | 0.007793294 | 1 |
| T2DM | b_cell | *HS3ST6* | 1.312725873 | 0.002016114 | 0.004980661 | 1 |
| T2DM | b_cell | *SHANK2* | -1.219076481 | 0.001956017 | 0.004980661 | 1 |
| T2DM | b_cell | *ASRGL1* | -1.700274532 | 0.00186625 | 0.004980661 | 1 |
| T2DM | b_cell | *TTC9C* | -1.145382095 | 0.001417609 | 0.004980661 | 1 |
| T2DM | b_cell | *RPS6KA4* | -3.508333979 | 0.030658217 | 0.036009118 | 1 |
| T2DM | b_cell | *TPCN2* | -1.458298419 | 0.002015735 | 0.004980661 | 1 |
| T2DM | b_cell | *PDZK1IP1* | -2.49413176 | 0.001823356 | 0.004980661 | 1 |
| T2DM | b_cell | *CMPK1* | -1.302227564 | 0.001975957 | 0.004980661 | 1 |
| T2DM | b_cell | *SLC1A7* | 0.928073781 | 0.009135232 | 0.011854546 | 1 |
| T2DM | b_cell | *USP24* | -1.508862683 | 0.00356836 | 0.005668447 | 1 |
| T2DM | b_cell | *ZSWIM5* | -1.046695954 | 0.002030879 | 0.004980661 | 1 |
| T2DM | b_cell | *AK4* | -1.132999141 | 0.001731956 | 0.004980661 | 1 |
| T2DM | b_cell | *RBP7* | -1.125882484 | 0.002278998 | 0.004980661 | 1 |
| T2DM | b_cell | *SLC25A34* | -3.146715887 | 0.001367351 | 0.004980661 | 1 |
| T2DM | b_cell | *PDPN* | -1.294464341 | 0.001903987 | 0.004980661 | 1 |
| T2DM | b_cell | *LAPTM5* | -2.151516373 | 0.037789532 | 0.043885785 | 1 |
| T2DM | b_cell | *SDC3* | -2.079224127 | 0.00726284 | 0.009772407 | 1 |
| T2DM | b_cell | *SYNC* | -1.374481912 | 0.002655939 | 0.004985914 | 1 |
| T2DM | b_cell | *CAMK2N1* | 0.951668757 | 0.002244329 | 0.004980661 | 1 |
| T2DM | b_cell | *MEGF6* | -1.752669029 | 0.002076617 | 0.004980661 | 1 |
| T2DM | b_cell | *OMA1* | -1.540094567 | 0.002629549 | 0.004980661 | 1 |
| T2DM | b_cell | *TM2D1* | -1.288107723 | 0.002421526 | 0.004980661 | 1 |
| T2DM | b_cell | *NEXN* | -1.487906032 | 0.003652669 | 0.005752881 | 1 |
| T2DM | b_cell | *LRRIQ3* | -1.205561849 | 0.001768699 | 0.004980661 | 1 |
| T2DM | b_cell | *TYW3* | -1.299950165 | 0.001499617 | 0.004980661 | 1 |
| T2DM | b_cell | *WDR63* | -1.096346417 | 0.000922972 | 0.004980661 | 1 |
| T2DM | b_cell | *HFM1* | -1.1119224 | 0.001409023 | 0.004980661 | 1 |
| T2DM | b_cell | *ARPC5* | -1.487125127 | 0.003131206 | 0.005273382 | 1 |
| T2DM | b_cell | *ZNF496* | 1.484034802 | 0.020603288 | 0.024818061 | 1 |
| T2DM | b_cell | *TRIM58* | -2.229095242 | 0.03227598 | 0.037826825 | 1 |
| T2DM | b_cell | *FCRLB* | 0.429488055 | 0.0024679 | 0.004980661 | 1 |
| T2DM | b_cell | *KLHDC9* | -2.950808488 | 0.002599311 | 0.004980661 | 1 |
| T2DM | b_cell | *FLVCR1* | -1.861079497 | 0.001548878 | 0.004980661 | 1 |
| T2DM | b_cell | *TDRD5* | -3.723802318 | 0.001519207 | 0.004980661 | 1 |
| T2DM | b_cell | *BROX* | -1.181358676 | 0.002607028 | 0.004980661 | 1 |
| T2DM | b_cell | *ACP6* | -1.255830508 | 0.001694718 | 0.004980661 | 1 |
| T2DM | b_cell | *CNST* | -1.759440633 | 0.018651094 | 0.022640758 | 1 |
| T2DM | b_cell | *PM20D1* | -1.809428102 | 0.002372819 | 0.004980661 | 1 |
| T2DM | b_cell | *PKDCC* | -1.793150798 | 0.039615426 | 0.045830721 | 1 |
| T2DM | b_cell | *HAAO* | -1.285265206 | 0.001729936 | 0.004980661 | 1 |
| T2DM | b_cell | *B3GALNT2* | -1.71641527 | 0.002207384 | 0.004980661 | 1 |
| T2DM | b_cell | *KIAA1841* | -1.074995744 | 0.001683161 | 0.004980661 | 1 |

| | | | | | | |
|---|---|---|---|---|---|---|
| T2DM | b_cell | *KCNJ3* | -6.058527134 | 0.001427939 | 0.004980661 | 1 |
| T2DM | b_cell | *DUSP19* | -2.036309426 | 0.001809315 | 0.004980661 | 1 |
| T2DM | b_cell | *NUP35* | -1.523786596 | 0.001701504 | 0.004980661 | 1 |
| T2DM | b_cell | *C2orf48* | 2.286460109 | 0.00233794 | 0.004980661 | 1 |
| T2DM | b_cell | *CCDC74A* | -6.485364412 | 0.000803189 | 0.004980661 | 1 |
| T2DM | b_cell | *NOSTRIN* | -1.146943015 | 0.003202869 | 0.005339692 | 1 |
| T2DM | b_cell | *RPRD2* | -0.846694807 | 0.002120741 | 0.004980661 | 1 |
| T2DM | b_cell | *CTSS* | -0.785700551 | 0.004869717 | 0.007036292 | 1 |
| T2DM | b_cell | *PACRGL* | -1.007693139 | 0.001835822 | 0.004980661 | 1 |
| T2DM | b_cell | *C1QTNF7* | -1.425259127 | 0.00428368 | 0.006430822 | 1 |
| T2DM | b_cell | *LYSMD1* | -2.186886441 | 0.001238618 | 0.004980661 | 1 |
| T2DM | b_cell | *VPS72* | -1.333028735 | 0.00181482 | 0.004980661 | 1 |
| T2DM | b_cell | *RNF149* | -1.324311304 | 0.00297975 | 0.005186119 | 1 |
| T2DM | b_cell | *DHX57* | -0.984263119 | 0.001811513 | 0.004980661 | 1 |
| T2DM | b_cell | *S100A9* | -0.955212457 | 0.002347592 | 0.004980661 | 1 |
| T2DM | b_cell | *TDRD10* | -1.891226011 | 0.003659792 | 0.005752881 | 1 |
| T2DM | b_cell | *DCAF16* | -1.965515561 | 0.001372496 | 0.004980661 | 1 |
| T2DM | b_cell | *GNPDA2* | -1.502008832 | 0.003089671 | 0.005245425 | 1 |
| T2DM | b_cell | *NIPAL1* | -1.459373382 | 0.002546242 | 0.004980661 | 1 |
| T2DM | b_cell | *ANTXR2* | -1.208348061 | 0.002835231 | 0.005067351 | 1 |
| T2DM | b_cell | *CGGBP1* | -0.873336693 | 0.004523447 | 0.006669026 | 1 |
| T2DM | b_cell | *GPR155* | -1.075838401 | 0.001734075 | 0.004980661 | 1 |
| T2DM | b_cell | *DAPL1* | 1.163185578 | 0.001622946 | 0.004980661 | 1 |
| T2DM | b_cell | *PBXIP1* | -2.350758841 | 0.004848379 | 0.007011724 | 1 |
| T2DM | b_cell | *CLDN1* | -0.894932551 | 0.002060219 | 0.004980661 | 1 |
| T2DM | b_cell | *DCST2* | 2.454926897 | 0.001895311 | 0.004980661 | 1 |
| T2DM | b_cell | *POGLUT1* | -1.664809103 | 0.002844819 | 0.005071948 | 1 |
| T2DM | b_cell | *SLC22A15* | -1.415692165 | 0.001448714 | 0.004980661 | 1 |
| T2DM | b_cell | *LRRC58* | -1.364487391 | 0.002693385 | 0.00500636 | 1 |
| T2DM | b_cell | *LMOD1* | -1.037162689 | 0.000999385 | 0.004980661 | 1 |
| T2DM | b_cell | *TMEM169* | 1.352122266 | 0.00582055 | 0.008157512 | 1 |
| T2DM | b_cell | *ARPC2* | -1.295951729 | 0.002053498 | 0.004980661 | 1 |
| T2DM | b_cell | *CCT3* | -1.441418462 | 0.002015233 | 0.004980661 | 1 |
| T2DM | b_cell | *GLB1L* | -2.932501778 | 0.012701277 | 0.015961304 | 1 |
| T2DM | b_cell | *SERPINI1* | -1.22912937 | 0.001728462 | 0.004980661 | 1 |
| T2DM | b_cell | *SUCLG1* | -0.956450423 | 0.002481706 | 0.004980661 | 1 |
| T2DM | b_cell | *PRKCI* | -0.893755677 | 0.001530014 | 0.004980661 | 1 |
| T2DM | b_cell | *AIM2* | -2.444889443 | 0.001503005 | 0.004980661 | 1 |
| T2DM | b_cell | *EFHB* | -1.03294218 | 0.035022215 | 0.040838209 | 1 |
| T2DM | b_cell | *GTPBP8* | -1.761394406 | 0.002024733 | 0.004980661 | 1 |
| T2DM | b_cell | *WDFY3* | -0.810285182 | 0.001901484 | 0.004980661 | 1 |
| T2DM | b_cell | *PTPN13* | -0.746940046 | 0.001689411 | 0.004980661 | 1 |
| T2DM | b_cell | *C4orf36* | -2.035207532 | 0.003456484 | 0.005565581 | 1 |
| T2DM | b_cell | *PPM1K* | -1.923002387 | 0.003583695 | 0.00567867 | 1 |
| T2DM | b_cell | *TIPARP* | -1.14652162 | 0.002099559 | 0.004980661 | 1 |
| T2DM | b_cell | *RPL9* | -1.326093159 | 0.000830448 | 0.004980661 | 1 |
| T2DM | b_cell | *DNASE1L3* | -2.223922731 | 0.00919087 | 0.011917184 | 1 |
| T2DM | b_cell | *IL17RC* | -1.509934158 | 0.000865288 | 0.004980661 | 1 |
| T2DM | b_cell | *CRELD1* | -1.269337789 | 0.002037558 | 0.004980661 | 1 |
| T2DM | b_cell | *TTC14* | -0.994068488 | 0.001438463 | 0.004980661 | 1 |

| | | | | | | |
|---|---|---|---|---|---|---|
| T2DM | b_cell | *RCHY1* | -1.378585039 | 0.0013352 | 0.004980661 | 1 |
| T2DM | b_cell | *CCDC158* | -2.215205787 | 0.002728019 | 0.005013768 | 1 |
| T2DM | b_cell | *SLC4A1AP* | -1.253636981 | 0.001841921 | 0.004980661 | 1 |
| T2DM | b_cell | *CDCP1* | -1.384692844 | 0.001883486 | 0.004980661 | 1 |
| T2DM | b_cell | *CLEC3B* | -2.686503828 | 0.004233147 | 0.006366778 | 1 |
| T2DM | b_cell | *FYCO1* | -1.634091026 | 0.002629412 | 0.004980661 | 1 |
| T2DM | b_cell | *NMNAT3* | -1.199498262 | 0.003701023 | 0.005796437 | 1 |
| T2DM | b_cell | *MEAF6* | -1.423873121 | 0.002233921 | 0.004980661 | 1 |
| T2DM | b_cell | *SNIP1* | -2.294384532 | 0.003262388 | 0.005373084 | 1 |
| T2DM | b_cell | *DNALI1* | -1.908851383 | 0.018063694 | 0.021993738 | 1 |
| T2DM | b_cell | *RPN1* | 1.366159573 | 0.01887127 | 0.022873696 | 1 |
| T2DM | b_cell | *SENP2* | -1.714796744 | 0.002502338 | 0.004980661 | 1 |
| T2DM | b_cell | *IFT122* | -1.277591306 | 0.001989406 | 0.004980661 | 1 |
| T2DM | b_cell | *RFT1* | -4.457187778 | 6.54E-06 | 0.001762168 | 0.031719025 |
| T2DM | b_cell | *SFMBT1* | -2.181821451 | 0.004160356 | 0.006292409 | 1 |
| T2DM | b_cell | *UVSSA* | -1.059564641 | 0.004698027 | 0.006851467 | 1 |
| T2DM | b_cell | *FAM208A* | -1.084765045 | 0.001806354 | 0.004980661 | 1 |
| T2DM | b_cell | *PIGX* | -1.31739046 | 0.000937978 | 0.004980661 | 1 |
| T2DM | b_cell | *ABLIM2* | -1.180140545 | 0.001652936 | 0.004980661 | 1 |
| T2DM | b_cell | *ERMAP* | -1.235844333 | 0.001128656 | 0.004980661 | 1 |
| T2DM | b_cell | *BDH2* | -1.10046707 | 0.000875625 | 0.004980661 | 1 |
| T2DM | b_cell | *CDC25A* | -11.84778257 | 0.000869047 | 0.004980661 | 1 |
| T2DM | b_cell | *ZNF589* | -1.238798551 | 0.000706798 | 0.004980661 | 1 |
| T2DM | b_cell | *ATRIP* | -3.739105735 | 0.000521502 | 0.004980661 | 1 |
| T2DM | b_cell | *RNF123* | 3.369067016 | 0.001213628 | 0.004980661 | 1 |
| T2DM | b_cell | *HSPA4L* | -1.236791697 | 0.003054211 | 0.00523119 | 1 |
| T2DM | b_cell | *MFSD8* | -1.571101192 | 0.002328536 | 0.004980661 | 1 |
| T2DM | b_cell | *RAD54L2* | -0.980368742 | 0.001457202 | 0.004980661 | 1 |
| T2DM | b_cell | *TEX264* | -1.550740721 | 0.00242293 | 0.004980661 | 1 |
| T2DM | b_cell | *ETNPPL* | -11.13201037 | 0.001811452 | 0.004980661 | 1 |
| T2DM | b_cell | *PRSS12* | -1.756528801 | 0.004628184 | 0.006790429 | 1 |
| T2DM | b_cell | *HMGB2* | -1.493617436 | 0.002090002 | 0.004980661 | 1 |
| T2DM | b_cell | *ANXA5* | -1.660027511 | 0.007787868 | 0.010350354 | 1 |
| T2DM | b_cell | *MAP9* | -2.682150039 | 0.008668472 | 0.011349008 | 1 |
| T2DM | b_cell | *CEP44* | -1.517147848 | 0.001830174 | 0.004980661 | 1 |
| T2DM | b_cell | *HPGD* | 1.642501298 | 0.006386729 | 0.008813541 | 1 |
| T2DM | b_cell | *C4orf45* | -1.335252495 | 0.001652149 | 0.004980661 | 1 |
| T2DM | b_cell | *TMEM144* | -1.121877725 | 0.001591456 | 0.004980661 | 1 |
| T2DM | b_cell | *IL15* | -1.729867285 | 0.024547027 | 0.029213189 | 1 |
| T2DM | b_cell | *ARFIP1* | -1.072259353 | 0.002219048 | 0.004980661 | 1 |
| T2DM | b_cell | *LSM6* | -1.068702756 | 0.001538082 | 0.004980661 | 1 |
| T2DM | b_cell | *ITGA2* | -1.218695604 | 0.007482179 | 0.010017531 | 1 |
| T2DM | b_cell | *MOCS2* | -1.586027781 | 0.001683967 | 0.004980661 | 1 |
| T2DM | b_cell | *NDUFAF2* | -0.996361357 | 0.001478053 | 0.004980661 | 1 |
| T2DM | b_cell | *LMBRD2* | -1.997484164 | 0.003045275 | 0.00522879 | 1 |
| T2DM | b_cell | *RNF180* | -1.018801967 | 0.001493521 | 0.004980661 | 1 |
| T2DM | b_cell | *PGGT1B* | -1.430975541 | 0.002144989 | 0.004980661 | 1 |
| T2DM | b_cell | *F2RL2* | -5.083849346 | 0.002343115 | 0.004980661 | 1 |
| T2DM | b_cell | *CCDC112* | -1.473292418 | 0.001636736 | 0.004980661 | 1 |
| T2DM | b_cell | *CMBL* | -1.353291566 | 0.002053408 | 0.004980661 | 1 |

| | | | | | | |
|---|---|---|---|---|---|---|
| T2DM | b_cell | *C5orf63* | -1.329262592 | 0.00210629 | 0.004980661 | 1 |
| T2DM | b_cell | *F2RL1* | 2.389738486 | 0.007038959 | 0.00952399 | 1 |
| T2DM | b_cell | *WDR41* | -1.581431905 | 0.003859818 | 0.005965584 | 1 |
| T2DM | b_cell | *NDUFS4* | -1.122230004 | 0.001995792 | 0.004980661 | 1 |
| T2DM | b_cell | *SERINC5* | -0.798280951 | 0.001815294 | 0.004980661 | 1 |
| T2DM | b_cell | *CASP3* | -1.251910043 | 0.002230891 | 0.004980661 | 1 |
| T2DM | b_cell | *PRIMPOL* | -1.319212441 | 0.001485385 | 0.004980661 | 1 |
| T2DM | b_cell | *ERAP1* | -1.647191745 | 0.006969237 | 0.009445458 | 1 |
| T2DM | b_cell | *ERAP2* | -0.787753143 | 0.014255626 | 0.017721829 | 1 |
| T2DM | b_cell | *TLR3* | -2.17142615 | 0.001787206 | 0.004980661 | 1 |
| T2DM | b_cell | *CCDC127* | -1.798268598 | 0.002364574 | 0.004980661 | 1 |
| T2DM | b_cell | *GRIK2* | -1.045475566 | 0.00194922 | 0.004980661 | 1 |
| T2DM | b_cell | *DCBLD1* | -0.958222301 | 0.004350277 | 0.006490634 | 1 |
| T2DM | b_cell | *PDSS2* | -1.086482231 | 0.00230466 | 0.004980661 | 1 |
| T2DM | b_cell | *RAET1E* | -1.584559375 | 0.001886088 | 0.004980661 | 1 |
| T2DM | b_cell | *DAGLB* | -1.640148052 | 0.000848074 | 0.004980661 | 1 |
| T2DM | b_cell | *STK17A* | -1.017447992 | 0.001428189 | 0.004980661 | 1 |
| T2DM | b_cell | *GALNT10* | -1.700718242 | 0.002170522 | 0.004980661 | 1 |
| T2DM | b_cell | *SAP30L* | -2.014989051 | 0.003229685 | 0.005354669 | 1 |
| T2DM | b_cell | *RPS14* | -1.030782112 | 0.002176445 | 0.004980661 | 1 |
| T2DM | b_cell | *CAMLG* | -1.085348181 | 0.004774698 | 0.006938255 | 1 |
| T2DM | b_cell | *RELL2* | 0.602021409 | 0.001827739 | 0.004980661 | 1 |
| T2DM | b_cell | *MIOS* | -1.087910653 | 0.001748864 | 0.004980661 | 1 |
| T2DM | b_cell | *USP49* | -1.096170451 | 0.001736741 | 0.004980661 | 1 |
| T2DM | b_cell | *CHMP4C* | -4.612617661 | 0.001870324 | 0.004980661 | 1 |
| T2DM | b_cell | *SLC13A4* | -1.29717613 | 0.001965417 | 0.004980661 | 1 |
| T2DM | b_cell | *BRI3* | -0.940153292 | 0.001850694 | 0.004980661 | 1 |
| T2DM | b_cell | *CTSB* | -0.922701703 | 0.002737615 | 0.005019995 | 1 |
| T2DM | b_cell | *SOX17* | -1.959786111 | 0.016860765 | 0.020611849 | 1 |
| T2DM | b_cell | *RAD21* | -1.049882456 | 0.00165334 | 0.004980661 | 1 |
| T2DM | b_cell | *TNFRSF11B* | -2.366645184 | 0.002132577 | 0.004980661 | 1 |
| T2DM | b_cell | *DEFB1* | -5.894995855 | 0.035528712 | 0.04138903 | 1 |
| T2DM | b_cell | *SUN1* | -0.911264794 | 0.001904506 | 0.004980661 | 1 |
| T2DM | b_cell | *OXR1* | -0.825368025 | 0.002104777 | 0.004980661 | 1 |
| T2DM | b_cell | *INTS1* | -2.855882151 | 0.021666 | 0.025994914 | 1 |
| T2DM | b_cell | *SLC4A2* | -1.510256007 | 0.001783165 | 0.004980661 | 1 |
| T2DM | b_cell | *FASTK* | -1.46704682 | 0.002030256 | 0.004980661 | 1 |
| T2DM | b_cell | *ALDH7A1* | -1.11906374 | 0.002394307 | 0.004980661 | 1 |
| T2DM | b_cell | *YWHAZ* | -1.058229947 | 0.002045057 | 0.004980661 | 1 |
| T2DM | b_cell | *FZD6* | -1.128233301 | 0.013022225 | 0.016322355 | 1 |
| T2DM | b_cell | *SLC25A32* | -2.087663856 | 0.003766164 | 0.005865506 | 1 |
| T2DM | b_cell | *DCSTAMP* | -18.85263854 | 0.014936558 | 0.018468955 | 1 |
| T2DM | b_cell | *INTS8* | -1.132080749 | 0.00192176 | 0.004980661 | 1 |
| T2DM | b_cell | *RPP25L* | -2.240019686 | 0.000734847 | 0.004980661 | 1 |
| T2DM | b_cell | *C9orf24* | -1.477257694 | 0.002135994 | 0.004980661 | 1 |
| T2DM | b_cell | *NUDT2* | -1.193675947 | 0.005766058 | 0.00809283 | 1 |
| T2DM | b_cell | *TMEM65* | -1.443332493 | 0.002613421 | 0.004980661 | 1 |
| T2DM | b_cell | *CCDC171* | -1.02439258 | 0.001720366 | 0.004980661 | 1 |
| T2DM | b_cell | *ZMAT4* | -2.848494476 | 0.001871206 | 0.004980661 | 1 |
| T2DM | b_cell | *C8orf34* | -1.163498422 | 0.002215411 | 0.004980661 | 1 |

| | | | | | | |
|---|---|---|---|---|---|---|
| T2DM | b_cell | *RASEF* | -2.263633818 | 0.001492406 | 0.004980661 | 1 |
| T2DM | b_cell | *ANKS6* | -2.061027209 | 0.017484108 | 0.021314796 | 1 |
| T2DM | b_cell | *AQP7* | -0.725952158 | 0.002739065 | 0.005020757 | 1 |
| T2DM | b_cell | *NOL6* | -2.206109364 | 0.00061858 | 0.004980661 | 1 |
| T2DM | b_cell | *AQP3* | 4.197031014 | 0.007111415 | 0.009605954 | 1 |
| T2DM | b_cell | *TRMT10B* | -1.125809304 | 0.001790992 | 0.004980661 | 1 |
| T2DM | b_cell | *VCP* | -1.155622236 | 0.0015974 | 0.004980661 | 1 |
| T2DM | b_cell | *PIGO* | -2.087398672 | 0.00130339 | 0.004980661 | 1 |
| T2DM | b_cell | *WRN* | -0.871696067 | 0.002020374 | 0.004980661 | 1 |
| T2DM | b_cell | *MARCH8* | -1.822002576 | 0.004670193 | 0.006833456 | 1 |
| T2DM | b_cell | *TSHR* | -0.982080395 | 0.001095697 | 0.004980661 | 1 |
| T2DM | b_cell | *CFL2* | -2.34583351 | 0.003058817 | 0.005233209 | 1 |
| T2DM | b_cell | *PGM2L1* | 1.143959706 | 0.007160166 | 0.009658361 | 1 |
| T2DM | b_cell | *GJB2* | -8.734414612 | 0.001874869 | 0.004980661 | 1 |
| T2DM | b_cell | *CRYL1* | -1.420420707 | 0.002438778 | 0.004980661 | 1 |
| T2DM | b_cell | *REEP3* | -0.995027616 | 0.001278604 | 0.004980661 | 1 |
| T2DM | b_cell | *MICU2* | -0.808943284 | 0.001881504 | 0.004980661 | 1 |
| T2DM | b_cell | *LRR1* | -1.840202395 | 0.001761765 | 0.004980661 | 1 |
| T2DM | b_cell | *RPL36AL* | -1.490607623 | 0.007494997 | 0.010031925 | 1 |
| T2DM | b_cell | *AKR1E2* | -1.511808849 | 0.001747137 | 0.004980661 | 1 |
| T2DM | b_cell | *ATP5C1* | -1.753748464 | 0.003832476 | 0.005933368 | 1 |
| T2DM | b_cell | *VDAC2* | -1.270039327 | 0.001333134 | 0.004980661 | 1 |
| T2DM | b_cell | *PDZD8* | -1.480706956 | 0.009526349 | 0.01230286 | 1 |
| T2DM | b_cell | *CLEC1B* | 0.738253326 | 0.002872554 | 0.005094011 | 1 |
| T2DM | b_cell | *SNAPC4* | -1.386608075 | 0.00277406 | 0.005051064 | 1 |
| T2DM | b_cell | *STOX1* | -1.221415415 | 0.001862976 | 0.004980661 | 1 |
| T2DM | b_cell | *RET* | -2.172086679 | 0.002016375 | 0.004980661 | 1 |
| T2DM | b_cell | *CASP7* | -1.689857731 | 0.009216467 | 0.0119408 | 1 |
| T2DM | b_cell | *VWA2* | -4.104397696 | 0.002514837 | 0.004980661 | 1 |
| T2DM | b_cell | *METTL3* | -2.107499022 | 0.003194508 | 0.00533738 | 1 |
| T2DM | b_cell | *TRUB1* | 1.009919895 | 0.009573811 | 0.012360866 | 1 |
| T2DM | b_cell | *HSPA12A* | -0.868525868 | 0.002826813 | 0.005065458 | 1 |
| T2DM | b_cell | *UBTD1* | -1.03932562 | 0.001778942 | 0.004980661 | 1 |
| T2DM | b_cell | *OTOGL* | -2.941461103 | 0.0056645 | 0.007971043 | 1 |
| T2DM | b_cell | *PACSIN3* | -3.407725647 | 5.60E-05 | 0.004980661 | 0.271620383 |
| T2DM | b_cell | *TTC7B* | -0.915603929 | 0.001744599 | 0.004980661 | 1 |
| T2DM | b_cell | *SLC39A13* | 2.218544112 | 0.005281098 | 0.007532007 | 1 |
| T2DM | b_cell | *PSMC3* | -3.149209497 | 0.017477825 | 0.021312493 | 1 |
| T2DM | b_cell | *RAPSN* | -6.195212033 | 2.90E-07 | 0.000140787 | 0.001407869 |
| T2DM | b_cell | *TC2N* | -0.850323182 | 0.001795952 | 0.004980661 | 1 |
| T2DM | b_cell | *CPSF2* | -1.109249928 | 0.007292962 | 0.009807498 | 1 |
| T2DM | b_cell | *IFI27L1* | -1.279593693 | 0.001870793 | 0.004980661 | 1 |
| T2DM | b_cell | *PTER* | -1.053619674 | 0.001432895 | 0.004980661 | 1 |
| T2DM | b_cell | *C1QL3* | 2.165253554 | 0.002867502 | 0.005094011 | 1 |
| T2DM | b_cell | *ARL5B* | -1.41513018 | 0.001503387 | 0.004980661 | 1 |
| T2DM | b_cell | *LIPC* | -1.911706475 | 0.005327656 | 0.007585241 | 1 |
| T2DM | b_cell | *CEP57* | -1.817341058 | 0.001722642 | 0.004980661 | 1 |
| T2DM | b_cell | *TCP11L2* | -1.990718057 | 0.013814943 | 0.017218111 | 1 |
| T2DM | b_cell | *JAM3* | -1.162562024 | 0.002299444 | 0.004980661 | 1 |
| T2DM | b_cell | *IKBIP* | -2.012127442 | 0.004659726 | 0.006820209 | 1 |

| | | | | | | |
|---|---|---|---|---|---|---|
| T2DM | b_cell | *LARP6* | -1.260141167 | 0.002561695 | 0.004980661 | 1 |
| T2DM | b_cell | *API5* | -1.380897729 | 0.002193802 | 0.004980661 | 1 |
| T2DM | b_cell | *ASPG* | -7.387046246 | 0.01854161 | 0.022519122 | 1 |
| T2DM | b_cell | *COX11* | -1.616131675 | 0.001909794 | 0.004980661 | 1 |
| T2DM | b_cell | *STXBP4* | -1.828053636 | 0.002167105 | 0.004980661 | 1 |
| T2DM | b_cell | *WBP1L* | -0.873476587 | 0.002320728 | 0.004980661 | 1 |
| T2DM | b_cell | *APBB1* | -1.473706004 | 0.001535569 | 0.004980661 | 1 |
| T2DM | b_cell | *NUDT13* | -1.692897827 | 0.013579355 | 0.016946253 | 1 |
| T2DM | b_cell | *WDR88* | -2.822199796 | 0.012259606 | 0.015458318 | 1 |
| T2DM | b_cell | *CYB5R2* | -1.958420073 | 0.026880197 | 0.031833712 | 1 |
| T2DM | b_cell | *SERPINB8* | 0.617516694 | 0.003363152 | 0.005463988 | 1 |
| T2DM | b_cell | *PLD4* | -7.600368581 | 0.001127202 | 0.004980661 | 1 |
| T2DM | b_cell | *XRRA1* | -1.008902316 | 0.001200707 | 0.004980661 | 1 |
| T2DM | b_cell | *TRIM66* | -0.680589465 | 0.02376126 | 0.028347586 | 1 |
| T2DM | b_cell | *RPL27A* | -1.715702861 | 0.004182718 | 0.006316386 | 1 |
| T2DM | b_cell | *CDYL2* | -0.965484765 | 0.008858007 | 0.011565944 | 1 |
| T2DM | b_cell | *CENPN* | -3.113032333 | 0.002232904 | 0.004980661 | 1 |
| T2DM | b_cell | *C16orf46* | -10.89402568 | 0.000885774 | 0.004980661 | 1 |
| T2DM | b_cell | *TMEM41B* | -0.98349371 | 0.001787255 | 0.004980661 | 1 |
| T2DM | b_cell | *TMX3* | -1.026130893 | 0.001595984 | 0.004980661 | 1 |
| T2DM | b_cell | *PRKCB* | -1.731548599 | 0.010298904 | 0.013195216 | 1 |
| T2DM | b_cell | *NDST2* | -14.10126115 | 0.001615627 | 0.004980661 | 1 |
| T2DM | b_cell | *CCDC68* | -1.30109828 | 0.004738643 | 0.006892056 | 1 |
| T2DM | b_cell | *CLEC4D* | -8.927288536 | 0.002018525 | 0.004980661 | 1 |
| T2DM | b_cell | *ZSCAN21* | -1.211467526 | 0.003074218 | 0.00524292 | 1 |
| T2DM | b_cell | *RIMKLB* | -1.647926575 | 0.001400395 | 0.004980661 | 1 |
| T2DM | b_cell | *TMED3* | -1.025423342 | 0.001421614 | 0.004980661 | 1 |
| T2DM | b_cell | *SEC11C* | 1.655684615 | 0.013886649 | 0.017303036 | 1 |
| T2DM | b_cell | *GALR1* | -2.932844596 | 0.028075076 | 0.033159753 | 1 |
| T2DM | b_cell | *CENPV* | -2.114710203 | 0.002060876 | 0.004980661 | 1 |
| T2DM | b_cell | *FAM96B* | -1.221851159 | 0.0016761 | 0.004980661 | 1 |
| T2DM | b_cell | *ZNF592* | -1.226873289 | 0.00199187 | 0.004980661 | 1 |
| T2DM | b_cell | *ACSM1* | -2.085960536 | 0.002187675 | 0.004980661 | 1 |
| T2DM | b_cell | *SLFN5* | -1.709644523 | 0.006242788 | 0.008651373 | 1 |
| T2DM | b_cell | *CATSPER2* | -1.262454142 | 0.001511688 | 0.004980661 | 1 |
| T2DM | b_cell | *ZNF667-AS1* | -1.632198411 | 0.002557068 | 0.004980661 | 1 |
| T2DM | b_cell | *SAAL1* | -3.096482603 | 0.011755216 | 0.014907555 | 1 |
| T2DM | b_cell | *YPEL4* | 1.871112771 | 0.009331411 | 0.012073909 | 1 |
| T2DM | b_cell | *LDHC* | 1.878399222 | 0.004099681 | 0.006221975 | 1 |
| T2DM | b_cell | *FAM96A* | -1.185712839 | 0.001981629 | 0.004980661 | 1 |
| T2DM | b_cell | *FAM111A* | -1.361568751 | 0.003752422 | 0.005854261 | 1 |
| T2DM | b_cell | *TMEM170A* | 1.958779043 | 0.032670698 | 0.038270938 | 1 |
| T2DM | b_cell | *DCTN5* | -1.0139647 | 0.003436242 | 0.005539085 | 1 |
| T2DM | b_cell | *STAT6* | -1.312871437 | 0.002280777 | 0.004980661 | 1 |
| T2DM | b_cell | *STX3* | -1.604293905 | 0.001249682 | 0.004980661 | 1 |
| T2DM | b_cell | *PIP4K2C* | -1.57960551 | 0.001869219 | 0.004980661 | 1 |
| T2DM | b_cell | *MTMR10* | -1.078431537 | 0.002529423 | 0.004980661 | 1 |
| T2DM | b_cell | *NYAP1* | -2.74393067 | 0.00046772 | 0.004980661 | 1 |
| T2DM | b_cell | *MS4A7* | -1.013489571 | 0.001132423 | 0.004980661 | 1 |
| T2DM | b_cell | *MS4A14* | -1.475043505 | 0.001262552 | 0.004980661 | 1 |

| | | | | | | |
|---|---|---|---|---|---|---|
| T2DM | b_cell | *DIS3L* | -1.405560062 | 0.004508905 | 0.006657702 | 1 |
| T2DM | b_cell | *CCNDBP1* | -2.167997585 | 0.013448034 | 0.016803981 | 1 |
| T2DM | b_cell | *EPB42* | -1.74613944 | 0.006673063 | 0.00911791 | 1 |
| T2DM | b_cell | *SMAD3* | -1.295448405 | 0.002385354 | 0.004980661 | 1 |
| T2DM | b_cell | *MAP1A* | -3.06973309 | 0.001600356 | 0.004980661 | 1 |
| T2DM | b_cell | *RCCD1* | -3.156485873 | 0.00140947 | 0.004980661 | 1 |
| T2DM | b_cell | *AKTIP* | -2.396433759 | 0.014794504 | 0.018311973 | 1 |
| T2DM | b_cell | *CNPY4* | -1.594266074 | 0.008974788 | 0.011674443 | 1 |
| T2DM | b_cell | *MEI1* | 0.724200812 | 0.002349771 | 0.004980661 | 1 |
| T2DM | b_cell | *SAMD14* | -3.818917087 | 0.006468949 | 0.008886564 | 1 |
| T2DM | b_cell | *ACSF2* | -1.718318922 | 0.003245857 | 0.00536425 | 1 |
| T2DM | b_cell | *ISLR2* | -3.605211464 | 0.002975235 | 0.005185287 | 1 |
| T2DM | b_cell | *SP2* | -1.950546088 | 0.002517458 | 0.004980661 | 1 |
| T2DM | b_cell | *COQ7* | -2.487918631 | 0.015682936 | 0.019313098 | 1 |
| T2DM | b_cell | *GPRC5B* | -1.215701417 | 0.002479242 | 0.004980661 | 1 |
| T2DM | b_cell | *FBXO22* | -1.058407667 | 0.001589066 | 0.004980661 | 1 |
| T2DM | b_cell | *TBC1D2B* | -0.779130297 | 0.001698644 | 0.004980661 | 1 |
| T2DM | b_cell | *LOXHD1* | -1.048482363 | 0.002633413 | 0.004980661 | 1 |
| T2DM | b_cell | *KATNAL2* | -0.947200843 | 0.002010767 | 0.004980661 | 1 |
| T2DM | b_cell | *HDHD2* | 1.186061986 | 0.000807649 | 0.004980661 | 1 |
| T2DM | b_cell | *ZNF91* | -1.022189316 | 0.002630586 | 0.004980661 | 1 |
| T2DM | b_cell | *POP5* | -1.987048807 | 0.001550539 | 0.004980661 | 1 |
| T2DM | b_cell | *ENGASE* | -1.186321902 | 0.004794288 | 0.006954746 | 1 |
| T2DM | b_cell | *MYO5B* | -1.097247172 | 0.001551347 | 0.004980661 | 1 |
| T2DM | b_cell | *RRM1* | -1.317500332 | 0.001309198 | 0.004980661 | 1 |
| T2DM | b_cell | *ZNF23* | -3.649148652 | 0.003664912 | 0.005752881 | 1 |
| T2DM | b_cell | *VKORC1* | -2.48312796 | 0.002619116 | 0.004980661 | 1 |
| T2DM | b_cell | *SMG8* | -3.109629647 | 0.023166596 | 0.027692615 | 1 |
| T2DM | b_cell | *JSRP1* | 1.007101261 | 0.002522881 | 0.004980661 | 1 |
| T2DM | b_cell | *SPATA33* | -1.401857631 | 0.001805007 | 0.004980661 | 1 |
| T2DM | b_cell | *RPL13* | -1.154291525 | 0.002036027 | 0.004980661 | 1 |
| T2DM | b_cell | *ZNF641* | -1.250720958 | 0.002321848 | 0.004980661 | 1 |
| T2DM | b_cell | *TP53I13* | -1.681616433 | 0.003910732 | 0.006010413 | 1 |
| T2DM | b_cell | *TUBA1C* | -1.243957112 | 0.024844541 | 0.029538278 | 1 |
| T2DM | b_cell | *ZNF528* | -1.250665803 | 0.001794224 | 0.004980661 | 1 |
| T2DM | b_cell | *SERTAD3* | -2.820002612 | 0.00217314 | 0.004980661 | 1 |
| T2DM | b_cell | *TMC4* | -2.047797826 | 0.001404782 | 0.004980661 | 1 |
| T2DM | b_cell | *TTYH1* | -1.976128043 | 0.001770108 | 0.004980661 | 1 |
| T2DM | b_cell | *LENG8* | -1.525697602 | 0.002747381 | 0.00503214 | 1 |
| T2DM | b_cell | *CDC42EP5* | -1.769874335 | 0.001009044 | 0.004980661 | 1 |
| T2DM | b_cell | *DNAAF3* | -2.386314643 | 0.009471416 | 0.012238432 | 1 |
| T2DM | b_cell | *CHAF1A* | -1.650195304 | 0.00149537 | 0.004980661 | 1 |
| T2DM | b_cell | *FAM57A* | 1.591986962 | 0.017467137 | 0.021304814 | 1 |
| T2DM | b_cell | *KIFC2* | -2.038487551 | 0.001023535 | 0.004980661 | 1 |
| T2DM | b_cell | *SRR* | -1.145051772 | 0.014941431 | 0.018470273 | 1 |
| T2DM | b_cell | *TRPV3* | -2.08258186 | 0.0145192 | 0.018007965 | 1 |
| T2DM | b_cell | *CYB5D2* | -0.991660311 | 0.002625667 | 0.004980661 | 1 |
| T2DM | b_cell | *KLK5* | -2.57064362 | 0.001180352 | 0.004980661 | 1 |
| T2DM | b_cell | *KLK6* | -12.67576933 | 0.001771198 | 0.004980661 | 1 |
| T2DM | b_cell | *ZNF83* | -1.592878032 | 0.005081435 | 0.007290101 | 1 |

| | | | | | | |
|---|---|---|---|---|---|---|
| T2DM | b_cell | *SPRYD3* | -1.860168321 | 0.00157368 | 0.004980661 | 1 |
| T2DM | b_cell | *SOAT2* | -3.406559715 | 0.00146501 | 0.004980661 | 1 |
| T2DM | b_cell | *ZNF558* | -1.50170883 | 0.003932424 | 0.006030379 | 1 |
| T2DM | b_cell | *NDUFV1* | -1.64571213 | 0.002829334 | 0.005067351 | 1 |
| T2DM | b_cell | *ZNF232* | -1.512711516 | 0.008919195 | 0.011630189 | 1 |
| T2DM | b_cell | *MIS12* | -1.737537885 | 0.002356368 | 0.004980661 | 1 |
| T2DM | b_cell | *ATP5H* | -1.753488037 | 0.002509888 | 0.004980661 | 1 |
| T2DM | b_cell | *MGAT5B* | -4.094693111 | 0.002479145 | 0.004980661 | 1 |
| T2DM | b_cell | *GSDMA* | -2.593353216 | 0.005077022 | 0.007285925 | 1 |
| T2DM | b_cell | *TMEM99* | -1.821683932 | 0.001216536 | 0.004980661 | 1 |
| T2DM | b_cell | *ZNF598* | -3.970619666 | 0.004728468 | 0.006885513 | 1 |
| T2DM | b_cell | *MLST8* | -2.24797542 | 0.002813952 | 0.005065458 | 1 |
| T2DM | b_cell | *ECI1* | -1.16298946 | 0.00145463 | 0.004980661 | 1 |
| T2DM | b_cell | *ABCA3* | -2.470690349 | 0.001494641 | 0.004980661 | 1 |
| T2DM | b_cell | *BSCL2* | 2.879162231 | 0.008304336 | 0.010922374 | 1 |
| T2DM | b_cell | *ATG16L2* | -2.190498116 | 0.014358643 | 0.017836185 | 1 |
| T2DM | b_cell | *RPSA* | -1.481423357 | 0.008697043 | 0.011380273 | 1 |
| T2DM | b_cell | *ULK4* | -0.850671626 | 0.001336405 | 0.004980661 | 1 |
| T2DM | b_cell | *LTBP3* | -1.371714939 | 0.005442507 | 0.007721358 | 1 |
| T2DM | b_cell | *NAALADL1* | -1.894992642 | 0.002840888 | 0.005070867 | 1 |
| T2DM | b_cell | *SCARA5* | -1.121023954 | 0.002328368 | 0.004980661 | 1 |
| T2DM | b_cell | *KIAA1586* | -2.147357801 | 0.001406993 | 0.004980661 | 1 |
| T2DM | b_cell | *FAM83B* | -16.55480554 | 0.001169952 | 0.004980661 | 1 |
| T2DM | b_cell | *POLR2J3* | -1.193749893 | 0.018283084 | 0.022249692 | 1 |
| T2DM | b_cell | *NT5DC2* | 2.088583652 | 0.03863241 | 0.044757511 | 1 |
| T2DM | b_cell | *COA6* | 1.009646775 | 0.004146635 | 0.006281446 | 1 |
| T2DM | b_cell | *PDHB* | -3.083553852 | 0.001380805 | 0.004980661 | 1 |
| T2DM | b_cell | *PXK* | -1.740814061 | 0.011539153 | 0.014660375 | 1 |
| T2DM | b_cell | *PCMTD1* | -1.227911551 | 0.002775137 | 0.005051064 | 1 |
| T2DM | b_cell | *ACOX2* | -1.619068426 | 0.001560124 | 0.004980661 | 1 |
| T2DM | b_cell | *FAM107A* | -2.395636071 | 0.001326722 | 0.004980661 | 1 |
| T2DM | b_cell | *SEPT2* | -1.129906364 | 0.002359883 | 0.004980661 | 1 |
| T2DM | b_cell | *DTYMK* | -1.911161446 | 0.001678888 | 0.004980661 | 1 |
| T2DM | b_cell | *ING5* | -0.936294133 | 0.002001096 | 0.004980661 | 1 |
| T2DM | b_cell | *RFWD3* | -1.115304887 | 0.002429762 | 0.004980661 | 1 |
| T2DM | b_cell | *KLHL30* | -6.178494922 | 0.00177859 | 0.004980661 | 1 |
| T2DM | b_cell | *SERINC2* | -7.285591159 | 8.75E-05 | 0.004980661 | 0.424558198 |
| T2DM | b_cell | *SLC20A2* | -1.077620707 | 0.001697196 | 0.004980661 | 1 |
| T2DM | b_cell | *TMUB2* | -1.175839926 | 0.001746595 | 0.004980661 | 1 |
| T2DM | b_cell | *ZSWIM1* | 1.538782582 | 1.50E-05 | 0.002909763 | 0.072744083 |
| T2DM | b_cell | *NBPF9* | -1.81154772 | 0.001731416 | 0.004980661 | 1 |
| T2DM | b_cell | *ZNF30* | -1.773940978 | 0.013562035 | 0.016928993 | 1 |
| T2DM | b_cell | *SLC16A4* | -1.029914262 | 0.001934461 | 0.004980661 | 1 |
| T2DM | b_cell | *DNAJC21* | -1.507490357 | 0.00214813 | 0.004980661 | 1 |
| T2DM | b_cell | *NPNT* | -0.947175146 | 0.005106383 | 0.007315078 | 1 |
| T2DM | b_cell | *GSTM4* | -3.816794881 | 0.008469429 | 0.011118417 | 1 |
| T2DM | b_cell | *TCTN2* | -1.071575037 | 0.004361639 | 0.00650159 | 1 |
| T2DM | b_cell | *PPIP5K1* | -1.544555718 | 0.00067504 | 0.004980661 | 1 |
| T2DM | b_cell | *ADAL* | -0.910953654 | 0.008643863 | 0.011329017 | 1 |
| T2DM | b_cell | *SNTB2* | -0.997446268 | 0.001928583 | 0.004980661 | 1 |

| | | | | | | |
|---|---|---|---|---|---|---|
| T2DM | b_cell | *ZNF507* | -1.274985451 | 0.001190417 | 0.004980661 | 1 |
| T2DM | b_cell | *STX18* | -0.987732191 | 0.001828105 | 0.004980661 | 1 |
| T2DM | b_cell | *ZBTB49* | -1.448086288 | 0.00186518 | 0.004980661 | 1 |
| T2DM | b_cell | *GFM1* | -1.022132913 | 0.001825727 | 0.004980661 | 1 |
| T2DM | b_cell | *ATOH8* | -1.310662633 | 0.002925867 | 0.005134596 | 1 |
| T2DM | b_cell | *TMEM129* | -2.745529852 | 0.001530209 | 0.004980661 | 1 |
| T2DM | b_cell | *MFF* | -0.923830426 | 0.001841214 | 0.004980661 | 1 |
| T2DM | b_cell | *LGALS9* | 1.753287253 | 0.017098509 | 0.020891958 | 1 |
| T2DM | b_cell | *CPLX1* | -1.328959165 | 0.008463014 | 0.01111601 | 1 |
| T2DM | b_cell | *SIGLEC7* | 1.719220578 | 0.020817599 | 0.025051337 | 1 |
| T2DM | b_cell | *MFSD7* | 2.965699034 | 0.018700212 | 0.022694705 | 1 |
| T2DM | b_cell | *MAP2K1* | -1.71203928 | 0.004277096 | 0.006424914 | 1 |
| T2DM | b_cell | *IRS1* | -1.320801333 | 2.85E-06 | 0.001038114 | 0.013811725 |
| T2DM | b_cell | *UPF3A* | -1.234828217 | 0.001816012 | 0.004980661 | 1 |
| T2DM | b_cell | *HSPBAP1* | -1.538549478 | 0.002587344 | 0.004980661 | 1 |
| T2DM | b_cell | *AFAP1L2* | -1.073232972 | 0.019046059 | 0.023074027 | 1 |
| T2DM | b_cell | *UBE2V2* | -1.190573408 | 0.00233397 | 0.004980661 | 1 |
| T2DM | b_cell | *CPT1C* | -4.192510504 | 0.000127473 | 0.004980661 | 0.618499921 |
| T2DM | b_cell | *PCSK9* | 2.435420672 | 0.00193921 | 0.004980661 | 1 |
| T2DM | b_cell | *XPO6* | -1.363290574 | 0.001925918 | 0.004980661 | 1 |
| T2DM | b_cell | *RGS14* | -1.803870343 | 0.01286864 | 0.016150709 | 1 |
| T2DM | b_cell | *RAB24* | 1.070293461 | 0.00284165 | 0.005070867 | 1 |
| T2DM | b_cell | *THBS3* | -1.228475354 | 0.001986015 | 0.004980661 | 1 |
| T2DM | b_cell | *NPIPB3* | 0.919738956 | 0.004697288 | 0.006851467 | 1 |
| T2DM | b_cell | *SH3TC2* | -1.518621146 | 0.005327143 | 0.007585241 | 1 |
| T2DM | b_cell | *GPRIN1* | 1.225797307 | 0.001900168 | 0.004980661 | 1 |
| T2DM | b_cell | *P2RY12* | -1.592658493 | 0.001251949 | 0.004980661 | 1 |
| T2DM | b_cell | *SNUPN* | -1.3190993 | 0.000918158 | 0.004980661 | 1 |
| T2DM | b_cell | *RNASE6* | -2.149652378 | 0.001003397 | 0.004980661 | 1 |
| T2DM | b_cell | *SLC38A11* | -1.443750766 | 0.002525884 | 0.004980661 | 1 |
| T2DM | b_cell | *CCDC8* | -2.528446575 | 0.000959097 | 0.004980661 | 1 |
| T2DM | b_cell | *ZEB2* | -0.648729148 | 0.001183965 | 0.004980661 | 1 |
| T2DM | b_cell | *HINT1* | -1.301527272 | 0.001798498 | 0.004980661 | 1 |
| T2DM | b_cell | *DTWD2* | -1.096598309 | 0.001335787 | 0.004980661 | 1 |
| T2DM | b_cell | *INO80E* | -3.556172298 | 4.03E-05 | 0.004980661 | 0.195526868 |
| T2DM | b_cell | *DFFB* | -2.105716176 | 0.002830668 | 0.005067351 | 1 |
| T2DM | b_cell | *NFU1* | -1.025157522 | 0.001262724 | 0.004980661 | 1 |
| T2DM | b_cell | *C15orf40* | -1.087665576 | 0.002657266 | 0.004985914 | 1 |
| T2DM | b_cell | *PROKR1* | 2.919919634 | 0.001659262 | 0.004980661 | 1 |
| T2DM | b_cell | *LUZP1* | -0.941720058 | 0.001832924 | 0.004980661 | 1 |
| T2DM | b_cell | *LRRC45* | -2.894016244 | 0.000159776 | 0.004980661 | 0.775232539 |
| T2DM | b_cell | *CHRNA5* | -4.848402956 | 0.000986672 | 0.004980661 | 1 |
| T2DM | b_cell | *UGP2* | -0.946339931 | 0.001463766 | 0.004980661 | 1 |
| T2DM | b_cell | *HNRNPF* | -1.251304086 | 0.002388658 | 0.004980661 | 1 |
| T2DM | b_cell | *CSGALNACT2* | -1.555120814 | 0.003185004 | 0.005332504 | 1 |
| T2DM | b_cell | *AVEN* | -1.971537607 | 0.008046858 | 0.010641415 | 1 |
| T2DM | b_cell | *CTNND2* | -1.572984619 | 0.001080572 | 0.004980661 | 1 |
| T2DM | b_cell | *CALML6* | -31.18356353 | 0.001595488 | 0.004980661 | 1 |
| T2DM | b_cell | *TPST1* | -1.23386077 | 0.002025598 | 0.004980661 | 1 |
| T2DM | b_cell | *TOR1AIP2* | -0.937996454 | 0.002011872 | 0.004980661 | 1 |

| | | | | | | |
|---|---|---|---|---|---|---|
| T2DM | b_cell | *OTUD3* | -1.230481422 | 0.001669208 | 0.004980661 | 1 |
| T2DM | b_cell | *ZFPM2* | -0.577531556 | 0.001351019 | 0.004980661 | 1 |
| T2DM | b_cell | *MAP3K2* | -1.057566427 | 0.001902523 | 0.004980661 | 1 |
| T2DM | b_cell | *UBE2E3* | -1.540721501 | 0.003548467 | 0.005652384 | 1 |
| T2DM | b_cell | *NIPA1* | -1.795966597 | 0.001835255 | 0.004980661 | 1 |
| T2DM | b_cell | *HNRNPA3* | -1.170734853 | 0.002513772 | 0.004980661 | 1 |
| T2DM | b_cell | *RNF150* | -0.695917792 | 0.002698345 | 0.005008557 | 1 |
| T2DM | b_cell | *HOXD4* | 4.705606844 | 0.00060014 | 0.004980661 | 1 |
| T2DM | b_cell | *CHRNB1* | 1.290100461 | 0.001160625 | 0.004980661 | 1 |
| T2DM | b_cell | *USP38* | -2.429010693 | 0.009635663 | 0.012437413 | 1 |
| T2DM | b_cell | *ANKK1* | -6.292556016 | 0.00029624 | 0.004980661 | 1 |
| T2DM | b_cell | *PDCD6IP* | -1.473246922 | 0.004297939 | 0.006448238 | 1 |
| T2DM | b_cell | *CRTAP* | -1.051175335 | 0.002061423 | 0.004980661 | 1 |
| T2DM | b_cell | *CNGB3* | -1.084561954 | 0.002258655 | 0.004980661 | 1 |
| T2DM | b_cell | *ELP5* | -1.415562966 | 0.001535184 | 0.004980661 | 1 |
| T2DM | b_cell | *STX8* | -1.244811002 | 0.002874568 | 0.005094011 | 1 |
| T2DM | b_cell | *NFRKB* | -1.33731302 | 0.001966142 | 0.004980661 | 1 |
| T2DM | b_cell | *SEMA3E* | -1.445493216 | 0.010387773 | 0.013298542 | 1 |
| T2DM | b_cell | *ADORA2B* | 0.405741279 | 0.00565704 | 0.007965165 | 1 |
| T2DM | b_cell | *MGMT* | -1.116841547 | 0.002189833 | 0.004980661 | 1 |
| T2DM | b_cell | *METTL7B* | 0.474638974 | 0.002453068 | 0.004980661 | 1 |
| T2DM | b_cell | *NFXL1* | -1.126681679 | 0.001509544 | 0.004980661 | 1 |
| T2DM | b_cell | *DNAJC18* | -1.321979611 | 0.002501593 | 0.004980661 | 1 |
| T2DM | b_cell | *SPATA24* | 0.695508664 | 0.002286968 | 0.004980661 | 1 |
| T2DM | b_cell | *HSD17B13* | 0.640984939 | 0.002526651 | 0.004980661 | 1 |
| T2DM | b_cell | *ELOVL6* | -1.01462569 | 0.001807675 | 0.004980661 | 1 |
| T2DM | b_cell | *ARL6IP1* | -2.712233992 | 0.001191561 | 0.004980661 | 1 |
| T2DM | b_cell | *SMAGP* | -1.468820565 | 0.007140271 | 0.009639564 | 1 |
| T2DM | b_cell | *EMB* | -1.399081362 | 0.000237123 | 0.004980661 | 1 |
| T2DM | b_cell | *STAT2* | -1.603261389 | 0.004436358 | 0.00658611 | 1 |
| T2DM | b_cell | *SLC26A5* | -1.05469957 | 0.002541152 | 0.004980661 | 1 |
| T2DM | b_cell | *GTSF1* | 1.262789734 | 0.003090161 | 0.005245425 | 1 |
| T2DM | b_cell | *ARMC10* | -1.049548119 | 0.002146206 | 0.004980661 | 1 |
| T2DM | b_cell | *ACYP2* | -0.887752943 | 0.001972228 | 0.004980661 | 1 |
| T2DM | b_cell | *TRABD* | -2.208196092 | 0.010123076 | 0.013000837 | 1 |
| T2DM | b_cell | *TMEM133* | -3.463678988 | 0.02476081 | 0.029445944 | 1 |
| T2DM | b_cell | *HOXB9* | -11.08791425 | 0.001841752 | 0.004980661 | 1 |
| T2DM | b_cell | *TTLL6* | -9.054713542 | 6.85E-07 | 0.000302001 | 0.003322012 |
| T2DM | b_cell | *KCNS3* | -1.4361855 | 0.034849205 | 0.040665787 | 1 |
| T2DM | b_cell | *FOXN2* | -1.679199092 | 0.005131284 | 0.007342079 | 1 |
| T2DM | b_cell | *GPR27* | 0.704538828 | 0.00481356 | 0.006980093 | 1 |
| T2DM | b_cell | *TRIAP1* | -2.515345528 | 0.006912447 | 0.009386844 | 1 |
| T2DM | b_cell | *RPS9* | -1.27275447 | 0.004904667 | 0.007078359 | 1 |
| T2DM | b_cell | *TSEN34* | 1.934227852 | 0.02093625 | 0.025175387 | 1 |
| T2DM | b_cell | *MSANTD4* | -1.412519047 | 0.001665209 | 0.004980661 | 1 |
| T2DM | b_cell | *NDUFA3* | -1.322215823 | 0.001820713 | 0.004980661 | 1 |
| T2DM | b_cell | *PAQR8* | 1.859740666 | 0.01593562 | 0.019597852 | 1 |
| T2DM | b_cell | *NUDT6* | -1.0760159 | 0.006012625 | 0.008397598 | 1 |
| T2DM | b_cell | *ZNF160* | -1.081463006 | 0.002553747 | 0.004980661 | 1 |
| T2DM | b_cell | *S1PR1* | -1.48318908 | 0.003662673 | 0.005752881 | 1 |

| | | | | | | |
|---|---|---|---|---|---|---|
| T2DM | b_cell | *PYGO1* | -1.243705124 | 0.00174703 | 0.004980661 | 1 |
| T2DM | b_cell | *TSNARE1* | -1.52791251 | 0.010513374 | 0.013448692 | 1 |
| T2DM | b_cell | *FPR1* | -1.433483642 | 0.001952764 | 0.004980661 | 1 |
| T2DM | b_cell | *FEZ2* | -0.892752465 | 0.001778987 | 0.004980661 | 1 |
| T2DM | b_cell | *SOX7* | -2.686497145 | 0.003578393 | 0.005673975 | 1 |
| T2DM | b_cell | *TRMT61B* | -1.624518438 | 0.007696913 | 0.01025559 | 1 |
| T2DM | b_cell | *MFN1* | -0.983537103 | 0.003358368 | 0.005462555 | 1 |
| T2DM | b_cell | *GIMAP8* | -1.136832219 | 0.001372088 | 0.004980661 | 1 |
| T2DM | b_cell | *ATP6V0E2* | -2.035186548 | 0.004525103 | 0.006669442 | 1 |
| T2DM | b_cell | *SCAND1* | -2.195136086 | 0.002355478 | 0.004980661 | 1 |
| T2DM | b_cell | *SOSTDC1* | 1.283950911 | 0.001622753 | 0.004980661 | 1 |
| T2DM | b_cell | *CANT1* | -1.657109965 | 0.001474585 | 0.004980661 | 1 |
| T2DM | b_cell | *LURAP1* | -1.719618023 | 0.000306354 | 0.004980661 | 1 |
| T2DM | b_cell | *PDE7B* | -0.744825725 | 0.001875401 | 0.004980661 | 1 |
| T2DM | b_cell | *MRPL36* | 2.50682288 | 0.018495848 | 0.022486057 | 1 |
| T2DM | b_cell | *MCC* | -0.912364359 | 0.001897273 | 0.004980661 | 1 |
| T2DM | b_cell | *ZNF561* | -1.984391193 | 0.000495332 | 0.004980661 | 1 |
| T2DM | b_cell | *HOPX* | 1.661886909 | 0.006402056 | 0.008820881 | 1 |
| T2DM | b_cell | *PPID* | -1.602913003 | 0.002332643 | 0.004980661 | 1 |
| T2DM | b_cell | *ETFDH* | -0.682173949 | 0.003970765 | 0.006071904 | 1 |
| T2DM | b_cell | *TBCA* | -0.914116106 | 0.00183051 | 0.004980661 | 1 |
| T2DM | b_cell | *NMUR1* | -2.286638045 | 0.001997019 | 0.004980661 | 1 |
| T2DM | b_cell | *PIK3CD* | -1.131302599 | 0.002621864 | 0.004980661 | 1 |
| T2DM | b_cell | *RGS19* | -2.125025377 | 4.85E-06 | 0.001471621 | 0.023545932 |
| T2DM | b_cell | *ANO5* | -1.329240996 | 0.001639361 | 0.004980661 | 1 |
| T2DM | b_cell | *VAT1L* | -1.775023595 | 0.001858838 | 0.004980661 | 1 |
| T2DM | b_cell | *CAMTA1* | -0.932354972 | 0.001811859 | 0.004980661 | 1 |
| T2DM | b_cell | *LGALS4* | -3.500459501 | 0.000391212 | 0.004980661 | 1 |
| T2DM | b_cell | *LRRC34* | -1.325032828 | 0.024254388 | 0.028893271 | 1 |
| T2DM | b_cell | *PAH* | 0.773453332 | 0.002413456 | 0.004980661 | 1 |
| T2DM | b_cell | *SPATA5L1* | -1.401986231 | 0.000618975 | 0.004980661 | 1 |
| T2DM | b_cell | *METTL18* | -1.340658192 | 0.011979827 | 0.015160699 | 1 |
| T2DM | b_cell | *PWWP2B* | -2.163644732 | 0.000925342 | 0.004980661 | 1 |
| T2DM | b_cell | *EXOSC10* | -1.363872922 | 0.001127159 | 0.004980661 | 1 |
| T2DM | b_cell | *TRAPPC12* | -1.275762522 | 0.002250731 | 0.004980661 | 1 |
| T2DM | b_cell | *C3AR1* | -1.821262202 | 0.001074867 | 0.004980661 | 1 |
| T2DM | b_cell | *PRNP* | -2.096878807 | 0.006032404 | 0.008415533 | 1 |
| T2DM | b_cell | *TVP23B* | -1.828701112 | 0.002761579 | 0.005037286 | 1 |
| T2DM | b_cell | *FBXW10* | -4.019457641 | 0.001385806 | 0.004980661 | 1 |
| T2DM | b_cell | *SRGAP2C* | -1.153862756 | 0.002050152 | 0.004980661 | 1 |
| T2DM | b_cell | *ATPAF2* | -1.240779067 | 0.003830188 | 0.005931718 | 1 |
| T2DM | b_cell | *ZNF554* | -1.786406599 | 0.001212797 | 0.004980661 | 1 |
| T2DM | b_cell | *ANKRD20A4* | -4.07607961 | 0.001832587 | 0.004980661 | 1 |
| T2DM | b_cell | *SERF1A* | -12.47381189 | 0.001517062 | 0.004980661 | 1 |
| T2DM | b_cell | *EIF2AK3* | -1.506044537 | 0.001893631 | 0.004980661 | 1 |
| T2DM | b_cell | *NME6* | -2.148306604 | 0.035578219 | 0.041430461 | 1 |
| T2DM | b_cell | *MTBP* | -1.28971456 | 0.001349575 | 0.004980661 | 1 |
| T2DM | b_cell | *TEFM* | -1.061013724 | 0.002654269 | 0.004985914 | 1 |
| T2DM | b_cell | *MALT1* | -1.867242081 | 0.032910673 | 0.038514854 | 1 |
| T2DM | b_cell | *MBOAT1* | -1.107785668 | 0.001842878 | 0.004980661 | 1 |

| | | | | | | |
|---|---|---|---|---|---|---|
| T2DM | b_cell | *CXCR6* | 2.402022198 | 0.00432283 | 0.00646559 | 1 |
| T2DM | b_cell | *PAIP1* | -1.19350212 | 0.001673026 | 0.004980661 | 1 |
| T2DM | b_cell | *MACROD2* | -0.644174584 | 0.002047809 | 0.004980661 | 1 |
| T2DM | b_cell | *BSG* | -0.97254569 | 0.001738718 | 0.004980661 | 1 |
| T2DM | b_cell | *HINFP* | -1.798512916 | 0.002648063 | 0.004985914 | 1 |
| T2DM | b_cell | *TP53RK* | -4.775872037 | 0.003580976 | 0.005676215 | 1 |
| T2DM | b_cell | *SUCLG2* | -0.844291578 | 0.00186987 | 0.004980661 | 1 |
| T2DM | b_cell | *ARNT2* | -1.090174857 | 0.002060119 | 0.004980661 | 1 |
| T2DM | b_cell | *GTPBP2* | -1.771483131 | 0.004418898 | 0.006570792 | 1 |
| T2DM | b_cell | *FGGY* | -0.968736942 | 0.002680215 | 0.004999202 | 1 |
| T2DM | b_cell | *MANEA* | -2.286405157 | 0.006035979 | 0.008415681 | 1 |
| T2DM | b_cell | *AFF1* | -1.0137601 | 0.002370123 | 0.004980661 | 1 |
| T2DM | b_cell | *FIBP* | 2.697672406 | 0.001895661 | 0.004980661 | 1 |
| T2DM | b_cell | *CTSW* | -2.135017809 | 2.18E-09 | 2.70E-06 | 1.06E-05 |
| T2DM | b_cell | *SNTG2* | -0.921832182 | 0.000955183 | 0.004980661 | 1 |
| T2DM | b_cell | *SMPDL3A* | -1.448418198 | 0.000766285 | 0.004980661 | 1 |
| T2DM | b_cell | *RAD9A* | -0.874851051 | 0.002199148 | 0.004980661 | 1 |
| T2DM | b_cell | *EFEMP2* | -2.111363414 | 0.019861809 | 0.023990415 | 1 |
| T2DM | b_cell | *ZFAND4* | -2.040283846 | 0.00145338 | 0.004980661 | 1 |
| T2DM | b_cell | *ZNF738* | -1.086300087 | 0.008689008 | 0.011372826 | 1 |
| T2DM | b_cell | *SLFN11* | -1.421522403 | 0.002190222 | 0.004980661 | 1 |
| T2DM | b_cell | *FAM71D* | -1.897767039 | 0.004684275 | 0.006841693 | 1 |
| T2DM | b_cell | *FUT10* | -1.009832629 | 0.000931979 | 0.004980661 | 1 |
| T2DM | b_cell | *ZNF596* | -1.191232425 | 0.00180002 | 0.004980661 | 1 |
| T2DM | b_cell | *CFL1* | -1.434475822 | 0.001710897 | 0.004980661 | 1 |
| T2DM | b_cell | *FAM192A* | -1.11896833 | 0.002079009 | 0.004980661 | 1 |
| T2DM | b_cell | *CBWD1* | -1.403117146 | 0.001852281 | 0.004980661 | 1 |
| T2DM | b_cell | *DCP2* | -0.996367464 | 0.001606693 | 0.004980661 | 1 |
| T2DM | b_cell | *ZNF621* | -2.341707432 | 0.009937356 | 0.012779234 | 1 |
| T2DM | b_cell | *NADSYN1* | -1.121775155 | 0.001588356 | 0.004980661 | 1 |
| T2DM | b_cell | *RNASEH2C* | -1.534615009 | 0.000100705 | 0.004980661 | 0.488618999 |
| T2DM | b_cell | *MYEOV* | 1.545114028 | 0.004903936 | 0.007078359 | 1 |
| T2DM | b_cell | *ANKRD13D* | -1.164268644 | 0.001925194 | 0.004980661 | 1 |
| T2DM | b_cell | *MRGPRF* | -1.916173076 | 0.000496958 | 0.004980661 | 1 |
| T2DM | b_cell | *LCLAT1* | -1.213442612 | 0.001686434 | 0.004980661 | 1 |
| T2DM | b_cell | *SH3RF3* | -1.10285776 | 0.001481167 | 0.004980661 | 1 |
| T2DM | b_cell | *GXYLT2* | -0.850699878 | 0.001772268 | 0.004980661 | 1 |
| T2DM | b_cell | *DCAKD* | -0.99818659 | 0.001502334 | 0.004980661 | 1 |
| T2DM | b_cell | *NOC3L* | -2.087575145 | 0.002729382 | 0.005014373 | 1 |
| T2DM | b_cell | *RHOD* | -2.134906193 | 0.002600105 | 0.004980661 | 1 |
| T2DM | b_cell | *PARP14* | -1.795751091 | 0.00733042 | 0.009855139 | 1 |
| T2DM | b_cell | *PARP15* | -1.740091521 | 0.004789511 | 0.006951453 | 1 |
| T2DM | b_cell | *AHSA2* | -1.202778474 | 0.000909041 | 0.004980661 | 1 |
| T2DM | b_cell | *IQCB1* | -1.116998946 | 0.001638135 | 0.004980661 | 1 |
| T2DM | b_cell | *ZNF483* | -0.908494387 | 0.002414962 | 0.004980661 | 1 |
| T2DM | b_cell | *SLC2A14* | -2.188759959 | 0.00203403 | 0.004980661 | 1 |
| T2DM | b_cell | *MMRN2* | -1.169847854 | 0.002649144 | 0.004985914 | 1 |
| T2DM | b_cell | *MZT2A* | -1.822864972 | 0.01445527 | 0.017942433 | 1 |
| T2DM | b_cell | *MAP3K11* | -1.778480628 | 0.001113518 | 0.004980661 | 1 |
| T2DM | b_cell | *KCNK7* | -29.98559204 | 0.00020537 | 0.004980661 | 0.996453313 |

| | | | | | | |
|---|---|---|---|---|---|---|
| T2DM | b_cell | *NDNF* | -2.316985746 | 0.001827949 | 0.004980661 | 1 |
| T2DM | b_cell | *ARV1* | -1.423185425 | 0.001018248 | 0.004980661 | 1 |
| T2DM | b_cell | *CCDC36* | -1.720287916 | 0.011463993 | 0.014568699 | 1 |
| T2DM | b_cell | *PPP1R14B* | -2.030057068 | 0.001860427 | 0.004980661 | 1 |
| T2DM | b_cell | *ZNF417* | -2.353574872 | 0.003912287 | 0.006010898 | 1 |
| T2DM | b_cell | *PEAK1* | -1.085473155 | 0.002199325 | 0.004980661 | 1 |
| T2DM | b_cell | *TNFRSF10D* | -1.441472609 | 0.003569065 | 0.005668447 | 1 |
| T2DM | b_cell | *MST1* | -1.492027456 | 0.000347124 | 0.004980661 | 1 |
| T2DM | b_cell | *GMPPB* | 1.319441832 | 0.004507835 | 0.006657702 | 1 |
| T2DM | b_cell | *NUDT18* | -4.069184268 | 0.001859233 | 0.004980661 | 1 |
| T2DM | b_cell | *SULT1B1* | 1.639816516 | 0.002607376 | 0.004980661 | 1 |
| T2DM | b_cell | *NUDT4* | -1.074943671 | 0.001960884 | 0.004980661 | 1 |
| T2DM | b_cell | *SCAI* | -0.800784388 | 0.002250567 | 0.004980661 | 1 |
| T2DM | b_cell | *HSPB7* | -2.029710171 | 0.018007464 | 0.021930777 | 1 |
| T2DM | b_cell | *UQCRH* | -0.948461073 | 0.001906398 | 0.004980661 | 1 |
| T2DM | b_cell | *C11orf80* | -1.078828298 | 0.001451545 | 0.004980661 | 1 |
| T2DM | b_cell | *CD7* | -2.424094519 | 0.00308284 | 0.005245002 | 1 |
| T2DM | b_cell | *RNF213* | -1.176973679 | 0.001861193 | 0.004980661 | 1 |
| T2DM | b_cell | *TUBB8* | 1.111651492 | 0.002403797 | 0.004980661 | 1 |
| T2DM | b_cell | *GPR160* | -2.118330529 | 0.001489473 | 0.004980661 | 1 |
| T2DM | b_cell | *GOLIM4* | -1.750323952 | 0.003562068 | 0.005663608 | 1 |
| T2DM | b_cell | *HOXB2* | -1.443342344 | 0.000505189 | 0.004980661 | 1 |
| T2DM | b_cell | *XXYLT1* | -1.71751281 | 0.00282046 | 0.005065458 | 1 |
| T2DM | b_cell | *UBXN2A* | -1.448495294 | 0.003362528 | 0.005463988 | 1 |
| T2DM | b_cell | *NRROS* | -1.877626658 | 0.001074237 | 0.004980661 | 1 |
| T2DM | b_cell | *CEP19* | 0.661016567 | 0.007101997 | 0.009595904 | 1 |
| T2DM | b_cell | *SLC25A30* | -1.833299488 | 0.001943948 | 0.004980661 | 1 |
| T2DM | b_cell | *CTSF* | -1.429804398 | 0.002312482 | 0.004980661 | 1 |
| T2DM | b_cell | *LEMD3* | -0.984508954 | 0.001794495 | 0.004980661 | 1 |
| T2DM | b_cell | *CTU2* | -2.532470355 | 0.010577008 | 0.013522963 | 1 |
| T2DM | b_cell | *GLIS1* | -1.404506862 | 0.001776877 | 0.004980661 | 1 |
| T2DM | b_cell | *RALGAPA1* | -1.02919091 | 0.002068447 | 0.004980661 | 1 |
| T2DM | b_cell | *ANKRD36C* | -1.074800268 | 0.001958058 | 0.004980661 | 1 |
| T2DM | b_cell | *MSL2* | -1.432655919 | 0.002418328 | 0.004980661 | 1 |
| T2DM | b_cell | *ZNF497* | 3.589882771 | 0.002731515 | 0.005016394 | 1 |
| T2DM | b_cell | *TRAM1L1* | -4.277588208 | 0.017280277 | 0.021103424 | 1 |
| T2DM | b_cell | *ANGEL2* | -1.930789634 | 0.007700329 | 0.010255833 | 1 |
| T2DM | b_cell | *ZNF266* | -1.615508864 | 0.000234094 | 0.004980661 | 1 |
| T2DM | b_cell | *SH3PXD2B* | -0.896015334 | 0.001759644 | 0.004980661 | 1 |
| T2DM | b_cell | *KIAA1551* | -1.275774483 | 0.002063383 | 0.004980661 | 1 |
| T2DM | b_cell | *LARP7* | -1.457272409 | 0.002016813 | 0.004980661 | 1 |
| T2DM | b_cell | *FGFBP3* | 0.733380129 | 0.002818054 | 0.005065458 | 1 |
| T2DM | b_cell | *C4orf32* | -0.922967684 | 0.00180724 | 0.004980661 | 1 |
| T2DM | b_cell | *HRAS* | 1.756044197 | 0.00628242 | 0.008694324 | 1 |
| T2DM | b_cell | *SRP72* | -0.978603517 | 0.002991527 | 0.005191305 | 1 |
| T2DM | b_cell | *PCP2* | -5.771440262 | 0.00268017 | 0.004999202 | 1 |
| T2DM | b_cell | *CEP135* | -1.206233148 | 0.002159543 | 0.004980661 | 1 |
| T2DM | b_cell | *NLRP6* | -26.35939941 | 0.001667793 | 0.004980661 | 1 |
| T2DM | b_cell | *RSRC1* | -1.200321905 | 0.002423275 | 0.004980661 | 1 |
| T2DM | b_cell | *PTDSS2* | -1.305982628 | 0.001402122 | 0.004980661 | 1 |

| | | | | | | |
|---|---|---|---|---|---|---|
| T2DM | b_cell | *SEZ6L2* | -1.262304224 | 0.030702513 | 0.036052419 | 1 |
| T2DM | b_cell | *KCTD13* | -1.131340743 | 0.003638028 | 0.005744108 | 1 |
| T2DM | b_cell | *DHX36* | -1.447734571 | 0.003156948 | 0.005303461 | 1 |
| T2DM | b_cell | *FBXW8* | -1.130107143 | 0.002165464 | 0.004980661 | 1 |
| T2DM | b_cell | *SLC22A1* | -2.420260489 | 0.002494185 | 0.004980661 | 1 |
| T2DM | b_cell | *CHST2* | 0.569521878 | 0.002459795 | 0.004980661 | 1 |
| T2DM | b_cell | *TVP23C* | -2.382507295 | 0.001837959 | 0.004980661 | 1 |
| T2DM | b_cell | *PACS1* | -1.564178582 | 0.004437344 | 0.00658611 | 1 |
| T2DM | b_cell | *SH3BP5L* | -1.743927973 | 0.001779161 | 0.004980661 | 1 |
| T2DM | b_cell | *CADM2* | -0.883520811 | 0.002557349 | 0.004980661 | 1 |
| T2DM | b_cell | *PSMD2* | -1.3285152 | 0.001686294 | 0.004980661 | 1 |
| T2DM | b_cell | *FAM182B* | -1.366331587 | 0.00118992 | 0.004980661 | 1 |
| T2DM | b_cell | *ARHGAP1* | -1.670225401 | 0.001845199 | 0.004980661 | 1 |
| T2DM | b_cell | *ATG13* | -1.383214591 | 0.00179601 | 0.004980661 | 1 |
| T2DM | b_cell | *ZNF519* | -1.869508693 | 0.035462017 | 0.041321256 | 1 |
| T2DM | b_cell | *BANF1* | -2.358087238 | 0.003698375 | 0.00579416 | 1 |
| T2DM | b_cell | *CHRNA7* | -2.222286156 | 0.018820272 | 0.022823285 | 1 |
| T2DM | b_cell | *SCUBE2* | -1.504443269 | 0.001955826 | 0.004980661 | 1 |
| T2DM | b_cell | *EIF1AD* | -1.938961366 | 0.001632661 | 0.004980661 | 1 |
| T2DM | b_cell | *EIF3F* | -1.86325277 | 0.001159962 | 0.004980661 | 1 |
| T2DM | b_cell | *ZNF25* | -1.301744903 | 0.001032253 | 0.004980661 | 1 |
| T2DM | b_cell | *CLTB* | -1.584420131 | 0.002131603 | 0.004980661 | 1 |
| T2DM | b_cell | *PPP2R2D* | -1.379305347 | 0.004825166 | 0.006990655 | 1 |
| T2DM | b_cell | *MCTP1* | -0.760521493 | 0.001828266 | 0.004980661 | 1 |
| T2DM | b_cell | *UBQLNL* | -3.918057157 | 0.009467742 | 0.012236943 | 1 |
| T2DM | b_cell | *KCNE3* | -1.453798591 | 0.001722019 | 0.004980661 | 1 |
| T2DM | b_cell | *UCP2* | -1.674814276 | 0.000683475 | 0.004980661 | 1 |
| T2DM | b_cell | *C11orf68* | -2.505054292 | 0.000760738 | 0.004980661 | 1 |
| T2DM | b_cell | *PAAF1* | -0.771525229 | 0.014533897 | 0.018021587 | 1 |
| T2DM | b_cell | *MRPL48* | -1.453255681 | 0.001678168 | 0.004980661 | 1 |
| T2DM | b_cell | *RAB6A* | -1.129126774 | 0.001873289 | 0.004980661 | 1 |
| T2DM | b_cell | *TMEM70* | 2.326711898 | 0.020363282 | 0.02457166 | 1 |
| T2DM | b_cell | *RPS6KB2* | -1.64487408 | 0.00286573 | 0.005094011 | 1 |
| T2DM | b_cell | *RMI2* | -1.957703565 | 0.005805422 | 0.008141014 | 1 |
| T2DM | b_cell | *TOM1L2* | -1.100401476 | 0.001449501 | 0.004980661 | 1 |
| T2DM | b_cell | *TEX26* | -5.663541443 | 0.020153106 | 0.02433015 | 1 |
| T2DM | b_cell | *GPR156* | -2.342535951 | 0.006276774 | 0.008688989 | 1 |
| T2DM | b_cell | *TTLL11* | -1.080917573 | 0.001709654 | 0.004980661 | 1 |
| T2DM | b_cell | *PRIMA1* | -1.495278914 | 0.001673667 | 0.004980661 | 1 |
| T2DM | b_cell | *CTDNEP1* | -1.511547038 | 0.002656507 | 0.004985914 | 1 |
| T2DM | b_cell | *ZDHHC21* | -1.224857653 | 0.00184227 | 0.004980661 | 1 |
| T2DM | b_cell | *A2M* | -0.782807106 | 0.001849418 | 0.004980661 | 1 |
| T2DM | b_cell | *DOK7* | -18.86378086 | 0.003157811 | 0.005303461 | 1 |
| T2DM | b_cell | *ZNF613* | -1.705449818 | 0.004946174 | 0.007129779 | 1 |
| T2DM | b_cell | *JAKMIP2* | -1.478275899 | 0.003100445 | 0.005253181 | 1 |
| T2DM | b_cell | *TCP11L1* | -1.424805695 | 0.010581565 | 0.013525225 | 1 |
| T2DM | b_cell | *CCDC57* | -1.596700657 | 0.00507688 | 0.007285925 | 1 |
| T2DM | b_cell | *MYPOP* | -1.483688919 | 0.002818298 | 0.005065458 | 1 |
| T2DM | b_cell | *CIDEA* | -0.85747374 | 0.001348547 | 0.004980661 | 1 |
| T2DM | b_cell | *SMIM19* | -1.026561737 | 0.011974399 | 0.015157784 | 1 |

| | | | | | | |
|---|---|---|---|---|---|---|
| T2DM | b_cell | *ZNF404* | -1.348189331 | 0.00983084 | 0.012659032 | 1 |
| T2DM | b_cell | *SLC35G1* | -1.916800697 | 0.002199875 | 0.004980661 | 1 |
| T2DM | b_cell | *ZNF135* | -1.722367353 | 0.000428235 | 0.004980661 | 1 |
| T2DM | b_cell | *OR4N2* | -4.786424823 | 0.003546221 | 0.005652384 | 1 |
| T2DM | b_cell | *TAC4* | -2.144315682 | 0.003007834 | 0.005199149 | 1 |
| T2DM | b_cell | *CDC26* | -2.048061718 | 0.001586351 | 0.004980661 | 1 |
| T2DM | b_cell | *CRLF3* | -1.12576317 | 0.003903411 | 0.006008677 | 1 |
| T2DM | b_cell | *DNAJC30* | -1.830185669 | 0.001843322 | 0.004980661 | 1 |
| T2DM | b_cell | *SPRYD4* | -2.25010589 | 0.000104912 | 0.004980661 | 0.509031003 |
| T2DM | b_cell | *WDR25* | -1.639545918 | 0.002359657 | 0.004980661 | 1 |
| T2DM | b_cell | *PLA2G16* | -0.753025872 | 0.001740754 | 0.004980661 | 1 |
| T2DM | b_cell | *B3GNT5* | -1.282358291 | 0.001520921 | 0.004980661 | 1 |
| T2DM | b_cell | *RMDN1* | -0.845710848 | 0.002851344 | 0.005078826 | 1 |
| T2DM | b_cell | *RNF152* | -1.106520615 | 0.001344784 | 0.004980661 | 1 |
| T2DM | b_cell | *BDNF* | -1.995267993 | 0.001724508 | 0.004980661 | 1 |
| T2DM | b_cell | *BOK* | -1.182167203 | 0.001803504 | 0.004980661 | 1 |
| T2DM | b_cell | *PFN4* | 2.670102684 | 0.001176978 | 0.004980661 | 1 |
| T2DM | b_cell | *RUFY1* | -1.745262545 | 0.015543057 | 0.019160292 | 1 |
| T2DM | b_cell | *VSIG10* | -1.536993429 | 0.006244252 | 0.008651373 | 1 |
| T2DM | b_cell | *ANKLE2* | -1.677460901 | 0.002600214 | 0.004980661 | 1 |
| T2DM | b_cell | *C8G* | 4.37951622 | 0.00250985 | 0.004980661 | 1 |
| T2DM | b_cell | *MUC20* | 1.227941712 | 0.003235208 | 0.005361076 | 1 |
| T2DM | b_cell | *LY6H* | 2.123701022 | 0.004508364 | 0.006657702 | 1 |
| T2DM | b_cell | *SHMT1* | -0.958963046 | 0.001965622 | 0.004980661 | 1 |
| T2DM | b_cell | *DPP7* | -2.039500984 | 0.00650157 | 0.008918749 | 1 |
| T2DM | b_cell | *MTHFR* | -1.107153175 | 0.001824192 | 0.004980661 | 1 |
| T2DM | b_cell | *TMEM80* | 1.631360208 | 0.036668238 | 0.042634625 | 1 |
| T2DM | b_cell | *WDR73* | -0.926635745 | 0.001875517 | 0.004980661 | 1 |
| T2DM | b_cell | *DSCAML1* | -0.925396639 | 0.00177019 | 0.004980661 | 1 |
| T2DM | b_cell | *TALDO1* | -1.283733508 | 0.001883651 | 0.004980661 | 1 |
| T2DM | b_cell | *AP006621.1* | -1.588835806 | 0.001604326 | 0.004980661 | 1 |
| T2DM | b_cell | *MAN1B1* | -2.632653456 | 0.009430566 | 0.012192141 | 1 |
| T2DM | b_cell | *MIEF2* | -2.567014165 | 0.001983129 | 0.004980661 | 1 |
| T2DM | b_cell | *NAP1L5* | 1.901010312 | 0.007336932 | 0.0098585 | 1 |
| T2DM | b_cell | *NIM1K* | -2.714666207 | 0.007232068 | 0.009733701 | 1 |
| T2DM | b_cell | *RBM44* | 2.422067399 | 0.003461976 | 0.005569466 | 1 |
| T2DM | b_cell | *SLC25A22* | 0.823176589 | 0.006465715 | 0.008884636 | 1 |
| T2DM | b_cell | *SAMD12* | -0.685894011 | 0.016546962 | 0.02024864 | 1 |
| T2DM | b_cell | *C18orf32* | -1.464153155 | 0.003609824 | 0.005712612 | 1 |
| T2DM | b_cell | *GBA* | -2.458364672 | 0.000736016 | 0.004980661 | 1 |
| T2DM | b_cell | *ACAD9* | -0.953198353 | 0.003980987 | 0.0060837 | 1 |
| T2DM | b_cell | *IL17RA* | -0.935885536 | 0.001953368 | 0.004980661 | 1 |
| T2DM | b_cell | *MBOAT4* | -3.710851235 | 0.017190656 | 0.020999261 | 1 |
| T2DM | b_cell | *AGTRAP* | -2.371250276 | 0.008129774 | 0.01073039 | 1 |
| T2DM | b_cell | *THAP5* | -2.023459382 | 0.002340945 | 0.004980661 | 1 |
| T2DM | b_cell | *NAALADL2* | -0.74406373 | 0.002900322 | 0.005113643 | 1 |
| T2DM | b_cell | *CD151* | -1.898395948 | 0.00578532 | 0.008115169 | 1 |
| T2DM | b_cell | *SLC35G5* | -11.5650881 | 0.001874935 | 0.004980661 | 1 |
| T2DM | b_cell | *FAM87B* | 1.112946356 | 0.002447572 | 0.004980661 | 1 |
| T2DM | b_cell | *PCDHB9* | -3.275852664 | 0.004754996 | 0.006911696 | 1 |

| | | | | | | |
|---|---|---|---|---|---|---|
| T2DM | b_cell | *CENPBD1* | 2.877266744 | 0.007267332 | 0.009775741 | 1 |
| T2DM | b_cell | *RPS27* | -1.089445309 | 0.001912132 | 0.004980661 | 1 |
| T2DM | b_cell | *RIC8A* | 1.483038835 | 0.023838037 | 0.028432192 | 1 |
| T2DM | b_cell | *ASB8* | -1.198488498 | 0.002263261 | 0.004980661 | 1 |
| T2DM | b_cell | *DPY19L2* | -1.196969157 | 0.003253631 | 0.00536838 | 1 |
| T2DM | b_cell | *ADAMTSL1* | -0.781696955 | 0.003186094 | 0.005332504 | 1 |
| T2DM | b_cell | *IMPDH2* | 1.36219621 | 0.042042969 | 0.048523426 | 1 |
| T2DM | b_cell | *GRAMD1C* | -1.321716202 | 0.001172818 | 0.004980661 | 1 |
| T2DM | b_cell | *DALRD3* | 1.555203902 | 0.014222433 | 0.017685096 | 1 |
| T2DM | b_cell | *ZNF114* | -4.472235236 | 0.001496801 | 0.004980661 | 1 |
| T2DM | b_cell | *SPINK6* | 2.348850317 | 0.002988009 | 0.005187056 | 1 |
| T2DM | b_cell | *ZNF454* | -1.621718239 | 0.003308947 | 0.005413018 | 1 |
| T2DM | b_cell | *SH2B1* | -1.326771595 | 0.002120101 | 0.004980661 | 1 |
| T2DM | b_cell | *VN1R1* | -6.252235416 | 0.000336547 | 0.004980661 | 1 |
| T2DM | b_cell | *SH2D4B* | 0.957300331 | 0.002570786 | 0.004980661 | 1 |
| T2DM | b_cell | *RNF212* | -1.422639828 | 0.001232005 | 0.004980661 | 1 |
| T2DM | b_cell | *PRSS36* | -2.980260148 | 0.001165813 | 0.004980661 | 1 |
| T2DM | b_cell | *WDR6* | -1.954168862 | 0.007592805 | 0.010132092 | 1 |
| T2DM | b_cell | *GEN1* | -2.151303339 | 0.000495439 | 0.004980661 | 1 |
| T2DM | b_cell | *TMPRSS9* | -1.49860175 | 0.002156876 | 0.004980661 | 1 |
| T2DM | b_cell | *TMEM11* | -1.507026574 | 0.001146828 | 0.004980661 | 1 |
| T2DM | b_cell | *ZFAND2A* | 0.945143494 | 0.003210843 | 0.005339692 | 1 |
| T2DM | b_cell | *ZNF223* | -6.128195957 | 0.000321461 | 0.004980661 | 1 |
| T2DM | b_cell | *FAM220A* | 2.476444924 | 0.008926772 | 0.011636942 | 1 |
| T2DM | b_cell | *NT5DC1* | -1.059530968 | 0.002176737 | 0.004980661 | 1 |
| T2DM | b_cell | *GLDC* | 0.523974606 | 0.002939226 | 0.005146562 | 1 |
| T2DM | b_cell | *P4HTM* | -1.21350473 | 0.00187828 | 0.004980661 | 1 |
| T2DM | b_cell | *CA8* | -2.195370623 | 0.001242605 | 0.004980661 | 1 |
| T2DM | b_cell | *ERN1* | -1.157612421 | 0.0017909 | 0.004980661 | 1 |
| T2DM | b_cell | *ZNF713* | -1.865499612 | 0.015082927 | 0.018621466 | 1 |
| T2DM | b_cell | *PARP10* | -1.283206894 | 0.001636479 | 0.004980661 | 1 |
| T2DM | b_cell | *NSUN3* | -0.953716802 | 0.001884969 | 0.004980661 | 1 |
| T2DM | b_cell | *RPP25* | -2.866367509 | 0.00833751 | 0.010960065 | 1 |
| T2DM | b_cell | *STX19* | 1.664302885 | 0.001317824 | 0.004980661 | 1 |
| T2DM | b_cell | *MPI* | -1.604649871 | 0.001798716 | 0.004980661 | 1 |
| T2DM | b_cell | *TRIM73* | -14.68962588 | 0.000328126 | 0.004980661 | 1 |
| T2DM | b_cell | *EFCAB13* | -1.189963118 | 0.003649219 | 0.005752881 | 1 |
| T2DM | b_cell | *EXOSC4* | -8.077716979 | 0.000532594 | 0.004980661 | 1 |
| T2DM | b_cell | *HYI* | -1.41100395 | 0.001662076 | 0.004980661 | 1 |
| T2DM | b_cell | *C17orf62* | -2.224939023 | 0.002586805 | 0.004980661 | 1 |
| T2DM | b_cell | *GAK* | -2.034637406 | 0.010258517 | 0.013150416 | 1 |
| T2DM | b_cell | *TUFM* | -1.571914402 | 0.001586645 | 0.004980661 | 1 |
| T2DM | b_cell | *CTC1* | -1.246223994 | 0.003316176 | 0.005423015 | 1 |
| T2DM | b_cell | *CCDC89* | -5.121491039 | 0.002311412 | 0.004980661 | 1 |
| T2DM | b_cell | *HTR1F* | -6.121109223 | 0.000962269 | 0.004980661 | 1 |
| T2DM | b_cell | *SPTY2D1* | -1.23979053 | 0.001822592 | 0.004980661 | 1 |
| T2DM | b_cell | *ZNF664* | -1.876425007 | 0.002050071 | 0.004980661 | 1 |
| T2DM | b_cell | *CDH4* | -1.823272333 | 0.00166754 | 0.004980661 | 1 |
| T2DM | b_cell | *NSUN7* | -1.230935635 | 0.022366358 | 0.026775616 | 1 |
| T2DM | b_cell | *HLA-DQB1* | -1.033728909 | 0.028527192 | 0.033652792 | 1 |

| | | | | | | |
|---|---|---|---|---|---|---|
| T2DM | b_cell | *PACS2* | -1.060079812 | 0.001821975 | 0.004980661 | 1 |
| T2DM | b_cell | *LINC00174* | -1.297094994 | 0.000823961 | 0.004980661 | 1 |
| T2DM | b_cell | *LBX2* | -3.245055965 | 0.00483976 | 0.007005137 | 1 |
| T2DM | b_cell | *ZFPM1* | -1.616062184 | 0.000535205 | 0.004980661 | 1 |
| T2DM | b_cell | *FCER1A* | 0.761484421 | 0.003985263 | 0.006084485 | 1 |
| T2DM | b_cell | *SERTAD2* | -1.29738466 | 0.002098415 | 0.004980661 | 1 |
| T2DM | b_cell | *AKAP5* | 2.801082352 | 0.03485809 | 0.040666375 | 1 |
| T2DM | b_cell | *GIPC3* | -8.321931937 | 0.001940992 | 0.004980661 | 1 |
| T2DM | b_cell | *PDXDC1* | -1.265459406 | 0.003258265 | 0.00536994 | 1 |
| T2DM | b_cell | *C1orf194* | -4.971210325 | 0.043259353 | 0.049844308 | 1 |
| T2DM | b_cell | *B3GNT3* | 1.979754234 | 0.000640617 | 0.004980661 | 1 |
| T2DM | b_cell | *SEPHS2* | -2.458753555 | 0.01453933 | 0.018023717 | 1 |
| T2DM | b_cell | *SSC5D* | -1.217087753 | 0.00401384 | 0.006124262 | 1 |
| T2DM | b_cell | *ZADH2* | -1.358456721 | 0.001777992 | 0.004980661 | 1 |
| T2DM | b_cell | *SEPT1* | -1.437653753 | 0.001775901 | 0.004980661 | 1 |
| T2DM | b_cell | *EXOC3* | -1.136469616 | 0.001871865 | 0.004980661 | 1 |
| T2DM | b_cell | *HCLS1* | -1.286657986 | 0.000353539 | 0.004980661 | 1 |
| T2DM | b_cell | *MTURN* | -0.974362528 | 0.001792974 | 0.004980661 | 1 |
| T2DM | b_cell | *CCDC66* | -1.077559895 | 0.002150012 | 0.004980661 | 1 |
| T2DM | b_cell | *DEFB124* | -9.79658203 | 0.006605322 | 0.009040477 | 1 |
| T2DM | b_cell | *MCFD2* | -1.169631141 | 0.002316601 | 0.004980661 | 1 |
| T2DM | b_cell | *GLIPR1L2* | -1.542483662 | 0.001240732 | 0.004980661 | 1 |
| T2DM | b_cell | *DEFB119* | 23.63508103 | 0.003120916 | 0.005265189 | 1 |
| T2DM | b_cell | *MB21D2* | -1.259219633 | 0.00154635 | 0.004980661 | 1 |
| T2DM | b_cell | *SLC47A2* | -1.409673005 | 0.00245097 | 0.004980661 | 1 |
| T2DM | b_cell | *CHST13* | -14.87524078 | 0.000752454 | 0.004980661 | 1 |
| T2DM | b_cell | *SLC36A4* | -1.868348321 | 0.002216883 | 0.004980661 | 1 |
| T2DM | b_cell | *ZDHHC20* | -0.872013418 | 0.001809739 | 0.004980661 | 1 |
| T2DM | b_cell | *OR51E1* | -2.287878062 | 0.022741456 | 0.027197817 | 1 |
| T2DM | b_cell | *BHLHE22* | -2.822393826 | 0.026621544 | 0.031542792 | 1 |
| T2DM | b_cell | *ZNF443* | -5.293033132 | 0.001853103 | 0.004980661 | 1 |
| T2DM | b_cell | *CAPS2* | -1.499828539 | 0.004676823 | 0.006834924 | 1 |
| T2DM | b_cell | *KCTD2* | -1.857117925 | 0.001434484 | 0.004980661 | 1 |
| T2DM | b_cell | *D2HGDH* | -1.04549489 | 0.000810307 | 0.004980661 | 1 |
| T2DM | b_cell | *CMTR2* | -1.122984399 | 0.001904076 | 0.004980661 | 1 |
| T2DM | b_cell | *LRRC57* | -1.281120512 | 0.004733119 | 0.006886085 | 1 |
| T2DM | b_cell | *LSMEM1* | -1.416336882 | 0.012376328 | 0.015585244 | 1 |
| T2DM | b_cell | *NQO1* | -1.86318758 | 0.002967351 | 0.005173571 | 1 |
| T2DM | b_cell | *RPH3AL* | -1.032570706 | 0.00172993 | 0.004980661 | 1 |
| T2DM | b_cell | *ADIPOQ* | -0.76505071 | 0.000505503 | 0.004980661 | 1 |
| T2DM | b_cell | *F2R* | -1.787610683 | 0.013909288 | 0.017322347 | 1 |
| T2DM | b_cell | *MUC16* | -1.175120547 | 0.002138626 | 0.004980661 | 1 |
| T2DM | b_cell | *SLC25A41* | 1.67114573 | 0.00258183 | 0.004980661 | 1 |
| T2DM | b_cell | *TMEM136* | -1.418575931 | 0.002734566 | 0.005019866 | 1 |
| T2DM | b_cell | *TMEM132E* | 2.31745051 | 0.000859649 | 0.004980661 | 1 |
| T2DM | b_cell | *ZNF322* | -1.23354711 | 0.001721873 | 0.004980661 | 1 |
| T2DM | b_cell | *DDX60L* | -1.697050789 | 0.004695401 | 0.006851467 | 1 |
| T2DM | b_cell | *OGFOD3* | 1.471079536 | 0.035879117 | 0.041747117 | 1 |
| T2DM | b_cell | *TMEM45A* | -1.228932869 | 0.002153018 | 0.004980661 | 1 |
| T2DM | b_cell | *ZFP41* | -3.179591175 | 0.00609351 | 0.008486139 | 1 |

| | | | | | | |
|---|---|---|---|---|---|---|
| T2DM | b_cell | *ZBTB20* | -0.810794184 | 0.002494593 | 0.004980661 | 1 |
| T2DM | b_cell | *C3orf58* | -1.119071787 | 0.002111643 | 0.004980661 | 1 |
| T2DM | b_cell | *AMIGO1* | -3.369965087 | 0.000869542 | 0.004980661 | 1 |
| T2DM | b_cell | *TMEM252* | -62.70352215 | 0.001950047 | 0.004980661 | 1 |
| T2DM | b_cell | *ODF3L2* | 0.934330216 | 0.002406256 | 0.004980661 | 1 |
| T2DM | b_cell | *RELL1* | -1.344391792 | 0.002068979 | 0.004980661 | 1 |
| T2DM | b_cell | *RFX7* | -1.182886985 | 0.002309864 | 0.004980661 | 1 |
| T2DM | b_cell | *CLDN7* | -2.251532401 | 0.001217049 | 0.004980661 | 1 |
| T2DM | b_cell | *ZNF329* | -1.178000971 | 0.001801437 | 0.004980661 | 1 |
| T2DM | b_cell | *MGAT4C* | -1.164395399 | 0.001190296 | 0.004980661 | 1 |
| T2DM | b_cell | *IDH2* | -2.286978461 | 0.003488082 | 0.005594769 | 1 |
| T2DM | b_cell | *PTCHD3* | 1.644186204 | 0.000792688 | 0.004980661 | 1 |
| T2DM | b_cell | *WRB* | -1.573041199 | 0.000639328 | 0.004980661 | 1 |
| T2DM | b_cell | *ZNF708* | -1.340524376 | 0.001987425 | 0.004980661 | 1 |
| T2DM | b_cell | *ERCC6L2* | -1.072270129 | 0.002479992 | 0.004980661 | 1 |
| T2DM | b_cell | *MRPL41* | -2.783554743 | 0.006113722 | 0.008509403 | 1 |
| T2DM | b_cell | *HHIPL1* | -1.538541986 | 0.001689154 | 0.004980661 | 1 |
| T2DM | b_cell | *FAM153B* | -1.148104877 | 0.00219833 | 0.004980661 | 1 |
| T2DM | b_cell | *BACE2* | -0.828535532 | 0.003069646 | 0.005238805 | 1 |
| T2DM | b_cell | *SYNM* | -1.563285248 | 0.001777781 | 0.004980661 | 1 |
| T2DM | b_cell | *KCNA4* | -1.805374637 | 0.001793345 | 0.004980661 | 1 |
| T2DM | b_cell | *FIGN* | -0.707180239 | 0.001775652 | 0.004980661 | 1 |
| T2DM | b_cell | *IZUMO1* | 3.36595415 | 0.000915013 | 0.004980661 | 1 |
| T2DM | b_cell | *C8orf33* | -1.197085234 | 0.001650436 | 0.004980661 | 1 |
| T2DM | b_cell | *FBXL6* | -2.192378723 | 0.001688366 | 0.004980661 | 1 |
| T2DM | b_cell | *YBEY* | -1.043456251 | 0.011815475 | 0.014974503 | 1 |
| T2DM | b_cell | *CLN8* | -1.12046076 | 0.007750772 | 0.010311694 | 1 |
| T2DM | b_cell | *NXPH4* | -15.18091402 | 0.001864056 | 0.004980661 | 1 |
| T2DM | b_cell | *NPLOC4* | -1.692551479 | 0.001759142 | 0.004980661 | 1 |
| T2DM | b_cell | *CAPN12* | -1.607405063 | 0.005602547 | 0.007904547 | 1 |
| T2DM | b_cell | *KPNA2* | 1.717110299 | 0.013521255 | 0.016886777 | 1 |
| T2DM | b_cell | *ORAI1* | 1.200880612 | 0.00308733 | 0.005245002 | 1 |
| T2DM | b_cell | *FES* | -1.428847796 | 0.001094486 | 0.004980661 | 1 |
| T2DM | b_cell | *GLRX5* | -2.949936815 | 0.019395733 | 0.023468354 | 1 |
| T2DM | b_cell | *MXRA7* | -1.289910505 | 0.002242751 | 0.004980661 | 1 |
| T2DM | b_cell | *MFSD5* | 1.324476826 | 0.003743964 | 0.005846002 | 1 |
| T2DM | b_cell | *ADI1* | -1.1565102 | 0.001789368 | 0.004980661 | 1 |
| T2DM | b_cell | *CLEC4G* | -9.33478069 | 0.000205482 | 0.004980661 | 0.996999344 |
| T2DM | b_cell | *TRAK1* | -1.103664828 | 0.001420854 | 0.004980661 | 1 |
| T2DM | b_cell | *SKA2* | -1.10121666 | 0.001670721 | 0.004980661 | 1 |
| T2DM | b_cell | *TTC3* | -0.635122624 | 0.015959151 | 0.019610811 | 1 |
| T2DM | b_cell | *PPP1R27* | 1.15364428 | 0.003093086 | 0.005245597 | 1 |
| T2DM | b_cell | *BRICD5* | -5.78265497 | 0.003754736 | 0.005855988 | 1 |
| T2DM | b_cell | *GRID1* | -1.278196057 | 0.001044371 | 0.004980661 | 1 |
| T2DM | b_cell | *ACBD3* | -1.518580327 | 0.002314917 | 0.004980661 | 1 |
| T2DM | b_cell | *PLCXD3* | -1.0027425 | 0.001781359 | 0.004980661 | 1 |
| T2DM | b_cell | *GPIHBP1* | -1.881671465 | 0.0001132 | 0.004980661 | 0.549244949 |
| T2DM | b_cell | *ALG12* | -2.634733009 | 0.003790515 | 0.005889074 | 1 |
| T2DM | b_cell | *COL18A1* | -0.918821207 | 0.00218982 | 0.004980661 | 1 |
| T2DM | b_cell | *ZNF721* | -1.072290809 | 0.00235836 | 0.004980661 | 1 |

| | | | | | | |
|---|---|---|---|---|---|---|
| T2DM | b_cell | *C11orf54* | -1.165947581 | 0.002926043 | 0.005134596 | 1 |
| T2DM | b_cell | *CEP63* | -0.912600572 | 0.001427164 | 0.004980661 | 1 |
| T2DM | b_cell | *HMGN4* | -2.306819461 | 0.005858926 | 0.00820182 | 1 |
| T2DM | b_cell | *C12orf60* | -1.312527315 | 0.001773688 | 0.004980661 | 1 |
| T2DM | b_cell | *ABAT* | -1.138949843 | 0.00147551 | 0.004980661 | 1 |
| T2DM | b_cell | *LYSMD4* | -1.477482904 | 0.0032412 | 0.005362236 | 1 |
| T2DM | b_cell | *WBP2NL* | -1.147454906 | 0.001564948 | 0.004980661 | 1 |
| T2DM | b_cell | *ARHGEF37* | -1.691986004 | 0.00209881 | 0.004980661 | 1 |
| T2DM | b_cell | *CSMD1* | -1.314844188 | 0.001815972 | 0.004980661 | 1 |
| T2DM | b_cell | *RABIF* | 2.155147279 | 0.035700357 | 0.041559053 | 1 |
| T2DM | b_cell | *FANCF* | -2.949850444 | 0.002332148 | 0.004980661 | 1 |
| T2DM | b_cell | *SMDT1* | -1.225404599 | 0.002403546 | 0.004980661 | 1 |
| T2DM | b_cell | *RUVBL2* | -1.607033033 | 0.002316622 | 0.004980661 | 1 |
| T2DM | b_cell | *CCDC125* | -1.053628893 | 0.002152077 | 0.004980661 | 1 |
| T2DM | b_cell | *C19orf71* | 1.564074647 | 0.005159231 | 0.007371198 | 1 |
| T2DM | b_cell | *LRIT3* | -14.9314832 | 0.002239722 | 0.004980661 | 1 |
| T2DM | b_cell | *GPR132* | -2.83432675 | 0.000716944 | 0.004980661 | 1 |
| T2DM | b_cell | *PSMG1* | -2.154700177 | 0.014890391 | 0.018421259 | 1 |
| T2DM | b_cell | *SERHL2* | -1.393101084 | 0.00174283 | 0.004980661 | 1 |
| T2DM | b_cell | *PCBP3* | -1.023972872 | 0.00097844 | 0.004980661 | 1 |
| T2DM | b_cell | *SFXN4* | -1.15476642 | 0.002027239 | 0.004980661 | 1 |
| T2DM | b_cell | *MRPL54* | -1.436819467 | 0.001299386 | 0.004980661 | 1 |
| T2DM | b_cell | *HMCES* | -1.066916648 | 0.001833344 | 0.004980661 | 1 |
| T2DM | b_cell | *DGCR6* | 0.881418969 | 0.004840974 | 0.007005137 | 1 |
| T2DM | b_cell | *RP1L1* | 0.42603503 | 0.002893488 | 0.005113643 | 1 |
| T2DM | b_cell | *NDUFB1* | -1.124077093 | 0.002260876 | 0.004980661 | 1 |
| T2DM | b_cell | *FAM19A1* | -1.162585474 | 0.002296576 | 0.004980661 | 1 |
| T2DM | b_cell | *TRMT12* | -2.183253323 | 0.00320137 | 0.005339692 | 1 |
| T2DM | b_cell | *MRGPRX2* | 2.584513555 | 0.002825843 | 0.005065458 | 1 |
| T2DM | b_cell | *UPP1* | -1.622059048 | 0.001383911 | 0.004980661 | 1 |
| T2DM | b_cell | *FIGLA* | -7.739763187 | 0.002305631 | 0.004980661 | 1 |
| T2DM | b_cell | *ACSM2A* | -14.5650614 | 0.001392113 | 0.004980661 | 1 |
| T2DM | b_cell | *KREMEN1* | -1.806513837 | 0.000470775 | 0.004980661 | 1 |
| T2DM | b_cell | *ZNF703* | -2.665485372 | 0.000701378 | 0.004980661 | 1 |
| T2DM | b_cell | *NPIPA5* | -1.410244244 | 0.008823528 | 0.011524026 | 1 |
| T2DM | b_cell | *EMILIN3* | 1.781822431 | 0.000839521 | 0.004980661 | 1 |
| T2DM | b_cell | *FAM162B* | -1.399703452 | 0.000805882 | 0.004980661 | 1 |
| T2DM | b_cell | *BTBD9* | -0.759212005 | 0.001874276 | 0.004980661 | 1 |
| T2DM | b_cell | *NUDT14* | -3.300708976 | 0.003193359 | 0.00533738 | 1 |
| T2DM | b_cell | *FAM3B* | -1.325190625 | 0.001563172 | 0.004980661 | 1 |
| T2DM | b_cell | *NPIPA7* | -5.665890305 | 0.015750969 | 0.019382121 | 1 |
| T2DM | b_cell | *TTC32* | -1.413317756 | 0.001744329 | 0.004980661 | 1 |
| T2DM | b_cell | *PP2D1* | -2.521880266 | 0.002626891 | 0.004980661 | 1 |
| T2DM | b_cell | *DENND5A* | -1.366828001 | 0.002474623 | 0.004980661 | 1 |
| T2DM | b_cell | *NIPSNAP1* | -1.621641315 | 0.000761928 | 0.004980661 | 1 |
| T2DM | b_cell | *CNTN2* | -2.231093904 | 0.007704653 | 0.010258766 | 1 |
| T2DM | b_cell | *CRELD2* | -1.538629414 | 0.002579022 | 0.004980661 | 1 |
| T2DM | b_cell | *GOLGA6L4* | 2.288271768 | 0.007118343 | 0.009612636 | 1 |
| T2DM | b_cell | *PGP* | -1.20011091 | 0.001229069 | 0.004980661 | 1 |
| T2DM | b_cell | *OAF* | -1.078399039 | 0.00072663 | 0.004980661 | 1 |

| | | | | | | |
|---|---|---|---|---|---|---|
| T2DM | b_cell | *ALDH1A3* | -1.183026767 | 0.0015345 | 0.004980661 | 1 |
| T2DM | b_cell | *CLECL1* | 3.316312169 | 0.007426783 | 0.009957102 | 1 |
| T2DM | b_cell | *PKP3* | 6.072754485 | 0.001678664 | 0.004980661 | 1 |
| T2DM | b_cell | *A3GALT2* | -2.774385796 | 0.007873214 | 0.010445949 | 1 |
| T2DM | b_cell | *TOP1MT* | -1.41155224 | 0.00683075 | 0.009296718 | 1 |
| T2DM | b_cell | *COPB2* | -1.917384948 | 0.006630341 | 0.009069753 | 1 |
| T2DM | b_cell | *WDR27* | -0.810268247 | 0.002582113 | 0.004980661 | 1 |
| T2DM | b_cell | *TXNRD2* | -2.203579963 | 0.011716268 | 0.014878799 | 1 |
| T2DM | b_cell | *TMEM255B* | -1.601001037 | 0.001940959 | 0.004980661 | 1 |
| T2DM | b_cell | *HDDC3* | -1.494487588 | 0.001288916 | 0.004980661 | 1 |
| T2DM | b_cell | *DUSP8* | 1.852978084 | 0.002673322 | 0.004996518 | 1 |
| T2DM | b_cell | *FAM19A3* | 1.433333129 | 0.002615098 | 0.004980661 | 1 |
| T2DM | b_cell | *SNN* | -2.210987316 | 0.001794813 | 0.004980661 | 1 |
| T2DM | b_cell | *KRBA2* | -4.86230852 | 0.000528582 | 0.004980661 | 1 |
| T2DM | b_cell | *SEPT5* | -4.348047301 | 0.001625729 | 0.004980661 | 1 |
| T2DM | b_cell | *RNLS* | -0.826173366 | 0.001597761 | 0.004980661 | 1 |
| T2DM | b_cell | *NDUFA12* | -1.316129912 | 0.001931448 | 0.004980661 | 1 |
| T2DM | b_cell | *TCTE3* | -1.334518022 | 0.001669906 | 0.004980661 | 1 |
| T2DM | b_cell | *UBE2G2* | -1.141372462 | 0.001618869 | 0.004980661 | 1 |
| T2DM | b_cell | *TMED9* | -1.99015454 | 0.001862801 | 0.004980661 | 1 |
| T2DM | b_cell | *DRD1* | -35.5726461 | 0.001756231 | 0.004980661 | 1 |
| T2DM | b_cell | *BTBD6* | -2.099210414 | 0.000459229 | 0.004980661 | 1 |
| T2DM | b_cell | *NUTM2A* | 1.573298714 | 0.0282956 | 0.033395829 | 1 |
| T2DM | b_cell | *WT1* | -0.772739786 | 0.003531796 | 0.005639428 | 1 |
| T2DM | b_cell | *ZFP90* | -1.029883291 | 0.001796381 | 0.004980661 | 1 |
| T2DM | b_cell | *FAM227A* | -1.347472424 | 0.006044966 | 0.00842579 | 1 |
| T2DM | b_cell | *NOC4L* | -3.623655609 | 0.011451069 | 0.014559902 | 1 |
| T2DM | b_cell | *SORCS2* | -1.66944763 | 0.001205166 | 0.004980661 | 1 |
| T2DM | b_cell | *TMEM106A* | -1.138997065 | 0.0032135 | 0.005339692 | 1 |
| T2DM | b_cell | *BRI3BP* | -1.714539637 | 0.001881071 | 0.004980661 | 1 |
| T2DM | b_cell | *UBOX5* | -1.337477919 | 0.001625791 | 0.004980661 | 1 |
| T2DM | b_cell | *MAFF* | -1.736884246 | 0.004489761 | 0.006643586 | 1 |
| T2DM | b_cell | *SEMA4B* | -1.28253166 | 0.002068929 | 0.004980661 | 1 |
| T2DM | b_cell | *CIB1* | -2.326204169 | 0.005933211 | 0.008296236 | 1 |
| T2DM | b_cell | *SGCZ* | -1.097809656 | 0.001086514 | 0.004980661 | 1 |
| T2DM | b_cell | *C6orf120* | -1.58366681 | 0.002045804 | 0.004980661 | 1 |
| T2DM | b_cell | *IFITM2* | -2.211699388 | 0.016203091 | 0.01986793 | 1 |
| T2DM | b_cell | *GP1BA* | 1.096654282 | 0.000729332 | 0.004980661 | 1 |
| T2DM | b_cell | *CCDC137* | -1.180144743 | 0.005505018 | 0.007793294 | 1 |
| T2DM | b_cell | *CDK10* | -1.126024339 | 0.004787108 | 0.006951453 | 1 |
| T2DM | b_cell | *ATP6V0A2* | -1.484010072 | 0.00281386 | 0.005065458 | 1 |
| T2DM | b_cell | *C14orf80* | 1.646658895 | 0.003086231 | 0.005245002 | 1 |
| T2DM | b_cell | *TNFAIP8L1* | -2.173933727 | 0.001908502 | 0.004980661 | 1 |
| T2DM | b_cell | *SP140L* | -1.943600103 | 0.028636297 | 0.03377329 | 1 |
| T2DM | b_cell | *TARSL2* | -1.154047539 | 0.001316303 | 0.004980661 | 1 |
| T2DM | b_cell | *SH3BGR* | -1.554886796 | 0.000848523 | 0.004980661 | 1 |
| T2DM | b_cell | *FAM174B* | -0.968610126 | 0.002055411 | 0.004980661 | 1 |
| T2DM | b_cell | *SV2B* | -1.670404373 | 0.002075412 | 0.004980661 | 1 |
| T2DM | b_cell | *AHNAK2* | -1.819068716 | 0.00108321 | 0.004980661 | 1 |
| T2DM | b_cell | *SP1* | -1.607390352 | 0.003269962 | 0.005378257 | 1 |

| | | | | | | |
|---|---|---|---|---|---|---|
| T2DM | b_cell | *PCGF3* | -1.524517814 | 0.003364879 | 0.005463988 | 1 |
| T2DM | b_cell | *PMEL* | 1.347745818 | 0.01985093 | 0.023983245 | 1 |
| T2DM | b_cell | *C6orf201* | -3.415988864 | 0.001792957 | 0.004980661 | 1 |
| T2DM | b_cell | *MYBL1* | -1.776404289 | 0.001094087 | 0.004980661 | 1 |
| T2DM | b_cell | *ADARB2* | -3.225687199 | 0.002961618 | 0.005172703 | 1 |
| T2DM | b_cell | *DMWD* | -1.462538619 | 0.00049614 | 0.004980661 | 1 |
| T2DM | b_cell | *SLC52A2* | -3.151091406 | 0.008270238 | 0.010883427 | 1 |
| T2DM | b_cell | *PIGP* | -1.507835786 | 2.97E-05 | 0.004645541 | 0.14401177 |
| T2DM | b_cell | *GNB1L* | -2.040605671 | 0.007914191 | 0.010488843 | 1 |
| T2DM | b_cell | *NPIPB4* | -1.50272219 | 0.002062155 | 0.004980661 | 1 |
| T2DM | b_cell | *TRIM69* | 1.496337555 | 0.002409438 | 0.004980661 | 1 |
| T2DM | b_cell | *IFITM1* | 1.115667809 | 0.003830031 | 0.005931718 | 1 |
| T2DM | b_cell | *LINC00839* | -4.738351744 | 0.001761447 | 0.004980661 | 1 |
| T2DM | b_cell | *RTN4RL1* | -1.450508195 | 0.003762124 | 0.005865503 | 1 |
| T2DM | b_cell | *RNPC3* | -0.741404088 | 0.00257688 | 0.004980661 | 1 |
| T2DM | b_cell | *RASA3* | -1.063109856 | 0.000429903 | 0.004980661 | 1 |
| T2DM | b_cell | *ZNF529* | -1.083988938 | 0.002163421 | 0.004980661 | 1 |
| T2DM | b_cell | *C15orf41* | -1.449180531 | 0.006240607 | 0.008651373 | 1 |
| T2DM | b_cell | *WWOX* | -0.926263542 | 0.002592435 | 0.004980661 | 1 |
| T2DM | b_cell | *POLR1D* | -1.071706604 | 0.003163576 | 0.005311304 | 1 |
| T2DM | b_cell | *FFAR4* | -1.175488918 | 0.007217878 | 0.009719996 | 1 |
| T2DM | b_cell | *ZNF749* | -1.822390413 | 0.000311676 | 0.004980661 | 1 |
| T2DM | b_cell | *NBPF12* | -1.006210818 | 0.008098041 | 0.010694528 | 1 |
| T2DM | b_cell | *TOR3A* | -1.854031007 | 0.001190852 | 0.004980661 | 1 |
| T2DM | b_cell | *ZNF555* | 1.9038535 | 0.008024035 | 0.010614126 | 1 |
| T2DM | b_cell | *CCDC30* | -0.930337381 | 0.00257796 | 0.004980661 | 1 |
| T2DM | b_cell | *ZNF197* | -1.076394372 | 0.001563737 | 0.004980661 | 1 |
| T2DM | b_cell | *RPS23* | -1.123033878 | 0.001418091 | 0.004980661 | 1 |
| T2DM | b_cell | *GNG2* | -0.862772411 | 0.001895722 | 0.004980661 | 1 |
| T2DM | b_cell | *TMEM222* | -1.399671815 | 0.001612299 | 0.004980661 | 1 |
| T2DM | b_cell | *SEPT10* | -1.370763823 | 0.001539068 | 0.004980661 | 1 |
| T2DM | b_cell | *FAM86B1* | -4.97763974 | 0.003625225 | 0.005731376 | 1 |
| T2DM | b_cell | *SMYD4* | -1.080242685 | 0.002093062 | 0.004980661 | 1 |
| T2DM | b_cell | *CEACAM19* | -1.764667747 | 0.000256097 | 0.004980661 | 1 |
| T2DM | b_cell | *MIR22HG* | -1.270323681 | 0.001647711 | 0.004980661 | 1 |
| T2DM | b_cell | *CYP27C1* | -3.0085217 | 0.000137495 | 0.004980661 | 0.667126406 |
| T2DM | b_cell | *BCR* | -1.082675826 | 0.001722671 | 0.004980661 | 1 |
| T2DM | b_cell | *FOXI2* | -4.389426226 | 0.030201216 | 0.035546451 | 1 |
| T2DM | b_cell | *ZNF732* | -2.747409258 | 0.002280209 | 0.004980661 | 1 |
| T2DM | b_cell | *HYAL3* | -3.464785489 | 0.024359321 | 0.029004031 | 1 |
| T2DM | b_cell | *VSIG10L* | -4.634556681 | 0.009852662 | 0.012680402 | 1 |
| T2DM | b_cell | *QRFPR* | -6.442847759 | 0.007469482 | 0.010006054 | 1 |
| T2DM | b_cell | *MAPT* | -1.13473783 | 0.001783509 | 0.004980661 | 1 |
| T2DM | b_cell | *TNFRSF18* | 4.162348423 | 0.003392514 | 0.005497821 | 1 |
| T2DM | b_cell | *C1QL4* | -18.40639744 | 0.001718854 | 0.004980661 | 1 |
| T2DM | b_cell | *RTN4RL2* | -4.015968198 | 0.000525787 | 0.004980661 | 1 |
| T2DM | b_cell | *TMEM232* | -0.968429218 | 0.00166775 | 0.004980661 | 1 |
| T2DM | b_cell | *EFCAB6* | -0.972307943 | 0.002153963 | 0.004980661 | 1 |
| T2DM | b_cell | *RHD* | -1.290262153 | 0.006405722 | 0.008822186 | 1 |
| T2DM | b_cell | *TMPRSS6* | -10.90953804 | 0.001651952 | 0.004980661 | 1 |

| | | | | | | |
|---|---|---|---|---|---|---|
| T2DM | b_cell | *CYP4A11* | -5.046258072 | 0.01225385 | 0.015455077 | 1 |
| T2DM | b_cell | *TMEM216* | -1.617034753 | 0.028891131 | 0.034065557 | 1 |
| T2DM | b_cell | *C3orf70* | -1.557176246 | 0.013080335 | 0.016390958 | 1 |
| T2DM | b_cell | *ENTPD5* | -1.202604906 | 0.00103655 | 0.004980661 | 1 |
| T2DM | b_cell | *MRPS21* | -1.600626155 | 0.001778916 | 0.004980661 | 1 |
| T2DM | b_cell | *WDR86* | -0.866078668 | 0.008131831 | 0.01073039 | 1 |
| T2DM | b_cell | *EPOR* | -1.718154211 | 0.024962563 | 0.029671326 | 1 |
| T2DM | b_cell | *LUZP2* | -1.93056099 | 0.001781588 | 0.004980661 | 1 |
| T2DM | b_cell | *GJA4* | -2.12750782 | 0.001950455 | 0.004980661 | 1 |
| T2DM | b_cell | *IFT140* | -1.143855435 | 0.00178284 | 0.004980661 | 1 |
| T2DM | b_cell | *SBK2* | -14.05619449 | 0.001866751 | 0.004980661 | 1 |
| T2DM | b_cell | *ZNF385C* | -3.195776264 | 0.001316119 | 0.004980661 | 1 |
| T2DM | b_cell | *DHRS4L2* | -1.794283225 | 0.000500024 | 0.004980661 | 1 |
| T2DM | b_cell | *SAMD11* | -2.683883633 | 0.00458201 | 0.006736943 | 1 |
| T2DM | b_cell | *C2orf88* | -1.138029483 | 0.002395137 | 0.004980661 | 1 |
| T2DM | b_cell | *KBTBD12* | -1.180376387 | 0.005516836 | 0.007802188 | 1 |
| T2DM | b_cell | *FANCA* | -1.935280051 | 0.003334628 | 0.005444015 | 1 |
| T2DM | b_cell | *DNAH17* | -1.867106755 | 0.000639829 | 0.004980661 | 1 |
| T2DM | b_cell | *ZFP69* | 2.096759032 | 0.018906003 | 0.022910072 | 1 |
| T2DM | b_cell | *TMEM220* | -1.178137313 | 0.006860262 | 0.009323807 | 1 |
| T2DM | b_cell | *FAM122A* | -1.847440066 | 0.001767418 | 0.004980661 | 1 |
| T2DM | b_cell | *GFRAL* | 4.316756939 | 0.009967999 | 0.012811849 | 1 |
| T2DM | b_cell | *LDLRAD2* | -2.079550801 | 0.000910221 | 0.004980661 | 1 |
| T2DM | b_cell | *CPSF4L* | -7.765114352 | 0.001661049 | 0.004980661 | 1 |
| T2DM | b_cell | *KLHL17* | -2.352703083 | 0.005156379 | 0.007369293 | 1 |
| T2DM | b_cell | *OR7D2* | -9.865625149 | 0.007054794 | 0.009540095 | 1 |
| T2DM | b_cell | *S100A3* | -7.102043346 | 0.019507719 | 0.023597969 | 1 |
| T2DM | b_cell | *TMEM221* | -4.695998679 | 0.015327842 | 0.018899794 | 1 |
| T2DM | b_cell | *MESP2* | -3.984572218 | 0.001123221 | 0.004980661 | 1 |
| T2DM | b_cell | *AGRN* | -1.845885505 | 0.001752719 | 0.004980661 | 1 |
| T2DM | b_cell | *FAM166A* | -7.828909108 | 0.030735529 | 0.036082455 | 1 |
| T2DM | b_cell | *ZNF626* | -1.415440473 | 0.001805675 | 0.004980661 | 1 |
| T2DM | b_cell | *LINC00265* | -1.844251905 | 0.001725151 | 0.004980661 | 1 |
| T2DM | b_cell | *LAMTOR4* | -1.702233993 | 0.003739385 | 0.005843316 | 1 |
| T2DM | b_cell | *PRKAR1B* | -1.146072861 | 0.001575776 | 0.004980661 | 1 |
| T2DM | b_cell | *NCR3LG1* | -2.587915919 | 0.001387462 | 0.004980661 | 1 |
| T2DM | b_cell | *ZNF793* | -1.086191763 | 0.010585913 | 0.013527218 | 1 |
| T2DM | b_cell | *RUFY4* | -3.097902307 | 0.001804628 | 0.004980661 | 1 |
| T2DM | b_cell | *ZNF669* | -2.575103664 | 0.006817008 | 0.009280618 | 1 |
| T2DM | b_cell | *CENPP* | -0.758421291 | 0.001514828 | 0.004980661 | 1 |
| T2DM | b_cell | *C3orf62* | -1.128573422 | 0.003745187 | 0.005846002 | 1 |
| T2DM | b_cell | *ENO4* | -1.12984793 | 0.001644325 | 0.004980661 | 1 |
| T2DM | b_cell | *ZP3* | -1.580240885 | 0.000705519 | 0.004980661 | 1 |
| T2DM | b_cell | *H3F3C* | -50.87397439 | 0.002050118 | 0.004980661 | 1 |
| T2DM | b_cell | *PDCD1* | 2.301554938 | 0.00209456 | 0.004980661 | 1 |
| T2DM | b_cell | *SELL* | 0.387624701 | 0.00255162 | 0.004980661 | 1 |
| T2DM | b_cell | *C19orf54* | -2.189861379 | 0.002080155 | 0.004980661 | 1 |
| T2DM | b_cell | *C22orf34* | -1.076239859 | 0.001869186 | 0.004980661 | 1 |
| T2DM | b_cell | *COL25A1* | -0.890616674 | 0.001719653 | 0.004980661 | 1 |
| T2DM | b_cell | *SRSF10* | -1.044193412 | 0.002362986 | 0.004980661 | 1 |

| | | | | | | |
|---|---|---|---|---|---|---|
| T2DM | b_cell | *NBR1* | -0.938929012 | 0.001671057 | 0.004980661 | 1 |
| T2DM | b_cell | *FAM72B* | -5.388544244 | 0.004818847 | 0.006983587 | 1 |
| T2DM | b_cell | *GOLGA8M* | -1.722646766 | 0.002062324 | 0.004980661 | 1 |
| T2DM | b_cell | *RHCE* | -2.744391625 | 0.001872559 | 0.004980661 | 1 |
| T2DM | b_cell | *IDO2* | -4.138637738 | 0.004059335 | 0.006172326 | 1 |
| T2DM | b_cell | *UROS* | -1.271822627 | 0.00182433 | 0.004980661 | 1 |
| T2DM | b_cell | *ZBED6CL* | -6.989925386 | 0.000442166 | 0.004980661 | 1 |
| T2DM | b_cell | *OSTN* | -2.181902835 | 0.012916248 | 0.016206268 | 1 |
| T2DM | b_cell | *FSIP2* | -1.232303705 | 0.001607462 | 0.004980661 | 1 |
| T2DM | b_cell | *NOXA1* | -1.474038994 | 0.001834856 | 0.004980661 | 1 |
| T2DM | b_cell | *FZD9* | 2.37709817 | 0.002917178 | 0.005130172 | 1 |
| T2DM | b_cell | *SKOR1* | -6.369174012 | 0.003255272 | 0.00536838 | 1 |
| T2DM | b_cell | *MTF1* | -2.029641511 | 0.001899799 | 0.004980661 | 1 |
| T2DM | b_cell | *FAM26F* | -1.68966732 | 0.008648104 | 0.011329631 | 1 |
| T2DM | b_cell | *ENTPD8* | -204.4676792 | 0.001604787 | 0.004980661 | 1 |
| T2DM | b_cell | *RPL14* | -1.089990808 | 0.00198199 | 0.004980661 | 1 |
| T2DM | b_cell | *ASTL* | -5.1444565 | 0.001731637 | 0.004980661 | 1 |
| T2DM | b_cell | *MSL1* | -1.217394397 | 0.001490496 | 0.004980661 | 1 |
| T2DM | b_cell | *FAM196A* | -1.209868563 | 0.001914792 | 0.004980661 | 1 |
| T2DM | b_cell | *UTS2B* | -1.90689117 | 0.002271257 | 0.004980661 | 1 |
| T2DM | b_cell | *C9orf152* | -4.496576273 | 0.002312808 | 0.004980661 | 1 |
| T2DM | b_cell | *NELFB* | -2.635599403 | 0.012601958 | 0.015857028 | 1 |
| T2DM | b_cell | *SLC15A5* | -1.872909894 | 0.006515607 | 0.008932954 | 1 |
| T2DM | b_cell | *LIPI* | -1.449921435 | 0.00195929 | 0.004980661 | 1 |
| T2DM | b_cell | *ADAT2* | -2.045199042 | 0.0042829 | 0.006430822 | 1 |
| T2DM | b_cell | *LITAF* | -1.125239776 | 0.002013571 | 0.004980661 | 1 |
| T2DM | b_cell | *SF3B3* | -1.16312482 | 0.001751546 | 0.004980661 | 1 |
| T2DM | b_cell | *PLAC9* | -1.057686433 | 0.001172306 | 0.004980661 | 1 |
| T2DM | b_cell | *ZNF573* | -1.282176549 | 0.001472926 | 0.004980661 | 1 |
| T2DM | b_cell | *FAM47E* | -1.924738353 | 0.001247413 | 0.004980661 | 1 |
| T2DM | b_cell | *ZNF600* | -1.839101541 | 0.020642133 | 0.024858682 | 1 |
| T2DM | b_cell | *TSPYL1* | -1.481666853 | 0.001702523 | 0.004980661 | 1 |
| T2DM | b_cell | *PNRC2* | -1.246453115 | 0.001618064 | 0.004980661 | 1 |
| T2DM | b_cell | *FHIT* | -0.673348508 | 0.001750251 | 0.004980661 | 1 |
| T2DM | b_cell | *ZKSCAN3* | -1.636896961 | 0.000420806 | 0.004980661 | 1 |
| T2DM | b_cell | *SLC35E2B* | -1.789506868 | 0.01065157 | 0.013603953 | 1 |
| T2DM | b_cell | *KIAA0408* | -2.135214952 | 0.000990229 | 0.004980661 | 1 |
| T2DM | b_cell | *BLOC1S2* | -2.416229008 | 0.009212838 | 0.0119408 | 1 |
| T2DM | b_cell | *HLA-DRB1* | -3.067037196 | 7.15E-09 | 5.95E-06 | 3.47E-05 |
| T2DM | b_cell | *AKR1C3* | -1.395636492 | 0.001920293 | 0.004980661 | 1 |
| T2DM | b_cell | *WDSUB1* | -0.936591679 | 0.002401156 | 0.004980661 | 1 |
| T2DM | b_cell | *COLCA1* | -3.337831702 | 0.000189623 | 0.004980661 | 0.920052943 |
| T2DM | b_cell | *SEMA4A* | -2.7419136 | 0.002509272 | 0.004980661 | 1 |
| T2DM | b_cell | *HRCT1* | -2.277432306 | 0.002293916 | 0.004980661 | 1 |
| T2DM | b_cell | *MPHOSPH8* | -0.978265502 | 0.002037777 | 0.004980661 | 1 |
| T2DM | b_cell | *SIRPB2* | -1.51282554 | 0.000256541 | 0.004980661 | 1 |
| T2DM | b_cell | *SRGAP3* | -1.125383289 | 0.006241673 | 0.008651373 | 1 |
| T2DM | b_cell | *XPNPEP3* | -0.886603728 | 0.001167113 | 0.004980661 | 1 |
| T2DM | b_cell | *PPIA* | -1.826358654 | 0.001631769 | 0.004980661 | 1 |
| T2DM | b_cell | *ZNF471* | -2.353492229 | 0.029734156 | 0.035025522 | 1 |

| | | | | | | |
|---|---|---|---|---|---|---|
| T2DM | b_cell | *ZNF493* | -1.028165895 | 0.001551865 | 0.004980661 | 1 |
| T2DM | b_cell | *GTF2IRD2* | -1.401330888 | 0.0026487 | 0.004985914 | 1 |
| T2DM | b_cell | *SUPT3H* | -0.957577188 | 0.001679055 | 0.004980661 | 1 |
| T2DM | b_cell | *GIMAP5* | -2.17333805 | 0.001040434 | 0.004980661 | 1 |
| T2DM | b_cell | *STK31* | -1.983375173 | 0.00473127 | 0.006886085 | 1 |
| T2DM | b_cell | *ZKSCAN7* | -1.390294931 | 0.008896552 | 0.011606903 | 1 |
| T2DM | b_cell | *CD55* | -1.662988796 | 0.004731822 | 0.006886085 | 1 |
| T2DM | b_cell | *CPNE4* | -1.359899105 | 0.003356048 | 0.005460611 | 1 |
| T2DM | b_cell | *ZNF565* | -1.668774241 | 0.004522503 | 0.006669026 | 1 |
| T2DM | b_cell | *LONP1* | -1.569851392 | 0.008067534 | 0.010662946 | 1 |
| T2DM | b_cell | *ZNF34* | 1.562949673 | 0.002777107 | 0.005051064 | 1 |
| T2DM | b_cell | *EVL* | -0.933499839 | 0.002361702 | 0.004980661 | 1 |
| T2DM | b_cell | *ZNF124* | -1.028921285 | 0.001648136 | 0.004980661 | 1 |
| T2DM | b_cell | *PIK3R4* | -1.291126687 | 0.001948763 | 0.004980661 | 1 |
| T2DM | b_cell | *ZNF605* | -1.069966884 | 0.00562566 | 0.007932491 | 1 |
| T2DM | b_cell | *RFX8* | -1.212806087 | 0.001558943 | 0.004980661 | 1 |
| T2DM | b_cell | *C20orf96* | -1.599362641 | 0.001696016 | 0.004980661 | 1 |
| T2DM | b_cell | *SULT1A1* | -1.20546115 | 0.001071341 | 0.004980661 | 1 |
| T2DM | b_cell | *TPK1* | -1.124002786 | 0.002824668 | 0.005065458 | 1 |
| T2DM | b_cell | *MYO18A* | -1.457105207 | 0.001889474 | 0.004980661 | 1 |
| T2DM | b_cell | *SPTSSB* | -4.734521022 | 0.011734119 | 0.014889553 | 1 |
| T2DM | b_cell | *CACNA1H* | -2.674394704 | 0.002207357 | 0.004980661 | 1 |
| T2DM | b_cell | *SULF2* | -0.980185405 | 0.01209089 | 0.015281323 | 1 |
| T2DM | b_cell | *XRCC2* | -3.226208278 | 0.001740617 | 0.004980661 | 1 |
| T2DM | b_cell | *MYO6* | -1.193151644 | 0.001615851 | 0.004980661 | 1 |
| T2DM | b_cell | *ZNF846* | -1.238972479 | 0.003292425 | 0.005398253 | 1 |
| T2DM | b_cell | *ADH1B* | -0.588032929 | 0.001783094 | 0.004980661 | 1 |
| T2DM | b_cell | *ZKSCAN5* | -1.750603156 | 0.005890663 | 0.008241492 | 1 |
| T2DM | b_cell | *ZNF502* | 1.370697005 | 0.006255558 | 0.008664563 | 1 |
| T2DM | b_cell | *TRAPPC4* | -1.706271552 | 0.032792663 | 0.038395271 | 1 |
| T2DM | b_cell | *ZFP62* | -1.358652935 | 0.002035518 | 0.004980661 | 1 |
| T2DM | b_cell | *TOMM7* | -1.4241366 | 0.002452412 | 0.004980661 | 1 |
| T2DM | b_cell | *ZNF33B* | -2.193074988 | 0.010992081 | 0.014008779 | 1 |
| T2DM | b_cell | *AMZ2* | -1.742266728 | 0.001336193 | 0.004980661 | 1 |
| T2DM | b_cell | *ZNF431* | -1.163594254 | 0.001799143 | 0.004980661 | 1 |
| T2DM | b_cell | *HLA-DQA1* | -3.952548456 | 7.36E-09 | 5.95E-06 | 3.57E-05 |
| T2DM | b_cell | *COL27A1* | -1.598665639 | 0.002075044 | 0.004980661 | 1 |
| T2DM | b_cell | *GM2A* | -1.591469024 | 0.007903857 | 0.010480873 | 1 |
| T2DM | b_cell | *ZNF239* | -1.92150144 | 0.008701511 | 0.011383049 | 1 |
| T2DM | b_cell | *SPRR2B* | 2.319168269 | 0.002790307 | 0.005061629 | 1 |
| T2DM | b_cell | *ANKRD36B* | -1.383956397 | 0.001985881 | 0.004980661 | 1 |
| T2DM | b_cell | *FAM3C* | -1.245185604 | 0.001248419 | 0.004980661 | 1 |
| T2DM | b_cell | *SLC39A10* | -0.996582222 | 0.001820454 | 0.004980661 | 1 |
| T2DM | b_cell | *PCDHB16* | -1.792406095 | 0.019835917 | 0.023971076 | 1 |
| T2DM | b_cell | *FUT11* | -2.070426348 | 0.011093109 | 0.014123265 | 1 |
| T2DM | b_cell | *WDR5B* | 1.976332438 | 0.003561439 | 0.005663608 | 1 |
| T2DM | b_cell | *NPIPB9* | -6.407929188 | 0.041183777 | 0.047543109 | 1 |
| T2DM | b_cell | *ZNF138* | -1.029528604 | 0.002389113 | 0.004980661 | 1 |
| T2DM | b_cell | *ZNF429* | -1.029986994 | 0.001868663 | 0.004980661 | 1 |
| T2DM | b_cell | *ZNF470* | -2.116207852 | 0.000864155 | 0.004980661 | 1 |

| | | | | | | |
|---|---|---|---|---|---|---|
| T2DM | b_cell | *ZNF100* | -1.799912672 | 0.001373935 | 0.004980661 | 1 |
| T2DM | b_cell | *ANXA6* | -1.669133619 | 0.003555916 | 0.005660533 | 1 |
| T2DM | b_cell | *GMFB* | -1.95544709 | 0.00300162 | 0.005195812 | 1 |
| T2DM | b_cell | *ZNF420* | -1.791935404 | 0.003000754 | 0.005195812 | 1 |
| T2DM | b_cell | *ZNF763* | -2.47707825 | 0.00467441 | 0.006833456 | 1 |
| T2DM | b_cell | *ZMYM1* | -1.093377286 | 0.001815756 | 0.004980661 | 1 |
| T2DM | b_cell | *IGF2R* | -1.068600802 | 0.001654465 | 0.004980661 | 1 |
| T2DM | b_cell | *GAL3ST4* | 2.134641828 | 0.00390713 | 0.006010413 | 1 |
| T2DM | b_cell | *SLC25A29* | -0.975765977 | 0.003956056 | 0.006060873 | 1 |
| T2DM | b_cell | *ZNF772* | -2.695880752 | 0.000688962 | 0.004980661 | 1 |
| T2DM | b_cell | *ZNF257* | -1.723484965 | 0.002452445 | 0.004980661 | 1 |
| T2DM | b_cell | *ADAM32* | -1.733306286 | 0.001946382 | 0.004980661 | 1 |
| T2DM | b_cell | *LRRC8B* | -1.085025603 | 0.002342768 | 0.004980661 | 1 |
| T2DM | b_cell | *ABCB8* | -1.133781745 | 0.00132348 | 0.004980661 | 1 |
| T2DM | b_cell | *SND1* | -1.277657926 | 0.002501398 | 0.004980661 | 1 |
| T2DM | b_cell | *SULT1A2* | -1.826856848 | 0.004082302 | 0.006203247 | 1 |
| T2DM | b_cell | *NEK5* | -1.136641636 | 0.001447468 | 0.004980661 | 1 |
| T2DM | b_cell | *PIWIL2* | 1.62943866 | 0.003481357 | 0.005589525 | 1 |
| T2DM | b_cell | *ENTPD4* | -2.567457408 | 0.005849009 | 0.0081903 | 1 |
| T2DM | b_cell | *TBC1D9B* | -1.166257573 | 0.001795017 | 0.004980661 | 1 |
| T2DM | b_cell | *FAM110D* | -1.896876514 | 0.020589593 | 0.024807724 | 1 |
| T2DM | b_cell | *CCL4L2* | -3.954005635 | 0.003745923 | 0.005846002 | 1 |
| T2DM | b_cell | *ZNF165* | 0.63926516 | 0.009043945 | 0.011748654 | 1 |
| T2DM | b_cell | *RAMP2-AS1* | -1.174860495 | 0.002169932 | 0.004980661 | 1 |
| T2DM | b_cell | *ZNF720* | -1.063292103 | 0.001280271 | 0.004980661 | 1 |
| T2DM | b_cell | *ZNF98* | -1.242201833 | 0.001411345 | 0.004980661 | 1 |
| T2DM | b_cell | *ZNF786* | -1.417556341 | 0.001171749 | 0.004980661 | 1 |
| T2DM | b_cell | *ZNF517* | -2.127389755 | 0.001373656 | 0.004980661 | 1 |
| T2DM | b_cell | *ZNF675* | -2.117978858 | 0.002779304 | 0.005051064 | 1 |
| T2DM | b_cell | *SLC22A5* | -1.208743888 | 0.003072756 | 0.005242268 | 1 |
| T2DM | b_cell | *VEPH1* | -2.625317197 | 0.017447359 | 0.021286041 | 1 |
| T2DM | b_cell | *MAP3K5* | -0.843602916 | 0.001801089 | 0.004980661 | 1 |
| T2DM | b_cell | *GSTK1* | -1.088643999 | 0.001315262 | 0.004980661 | 1 |
| T2DM | b_cell | *GYPE* | 2.248577459 | 0.002357826 | 0.004980661 | 1 |
| T2DM | b_cell | *PCDHB11* | -2.576105925 | 0.000640232 | 0.004980661 | 1 |
| T2DM | b_cell | *SLC2A10* | -1.263263041 | 0.002310128 | 0.004980661 | 1 |
| T2DM | b_cell | *ZNF665* | -2.606161204 | 0.023331093 | 0.027875514 | 1 |
| T2DM | b_cell | *RPF2* | -1.442880986 | 0.002421574 | 0.004980661 | 1 |
| T2DM | b_cell | *MYO5A* | -0.747925519 | 0.001994041 | 0.004980661 | 1 |
| T2DM | b_cell | *ATG7* | -0.925439153 | 0.002633704 | 0.004980661 | 1 |
| T2DM | b_cell | *RAB40C* | -1.174251466 | 0.001807443 | 0.004980661 | 1 |
| T2DM | b_cell | *PIGN* | -1.04420632 | 0.002554143 | 0.004980661 | 1 |
| T2DM | b_cell | *HHLA3* | -2.479637581 | 0.000482705 | 0.004980661 | 1 |
| T2DM | b_cell | *TOPORS* | -1.262299348 | 0.001818544 | 0.004980661 | 1 |
| T2DM | b_cell | *BCO2* | -0.93552756 | 0.00295984 | 0.005171459 | 1 |
| T2DM | b_cell | *ENTPD6* | -2.815814003 | 0.033332109 | 0.038970456 | 1 |
| T2DM | b_cell | *ENPP1* | -1.263089297 | 0.00123438 | 0.004980661 | 1 |
| T2DM | b_cell | *MFAP5* | -0.993748549 | 0.001417474 | 0.004980661 | 1 |
| T2DM | b_cell | *PDCD1LG2* | -0.970699208 | 0.010887796 | 0.01389103 | 1 |
| T2DM | b_cell | *DNAH10* | -2.060303601 | 0.000394439 | 0.004980661 | 1 |

| | | | | | | |
|---|---|---|---|---|---|---|
| T2DM | b_cell | *NMB* | -1.505895943 | 0.006846916 | 0.009313738 | 1 |
| T2DM | b_cell | *FAM114A1* | -0.889003402 | 0.001964267 | 0.004980661 | 1 |
| T2DM | b_cell | *RPS26* | -2.212293872 | 0.00011775 | 0.004980661 | 0.571323677 |
| T2DM | b_cell | *PSAP* | -1.054056203 | 0.003411859 | 0.005510766 | 1 |
| T2DM | b_cell | *TXNRD3* | -1.196466251 | 0.00213286 | 0.004980661 | 1 |
| T2DM | b_cell | *EME2* | -2.54234253 | 0.002005561 | 0.004980661 | 1 |
| T2DM | b_cell | *ZNF780A* | -1.901908495 | 0.000440259 | 0.004980661 | 1 |
| T2DM | b_cell | *ATAD3A* | -2.805857223 | 0.009947011 | 0.012788261 | 1 |
| T2DM | b_cell | *AC122129.1* | -4.261679881 | 0.002007876 | 0.004980661 | 1 |
| T2DM | b_cell | *CCDC180* | -1.947001951 | 0.033742168 | 0.039440376 | 1 |
| T2DM | b_cell | *ZNF181* | -1.696114356 | 0.005526449 | 0.007813034 | 1 |
| T2DM | b_cell | *SGTB* | -1.947275728 | 0.000773506 | 0.004980661 | 1 |
| T2DM | b_cell | *ADH5* | -1.641580984 | 0.003664801 | 0.005752881 | 1 |
| T2DM | b_cell | *ZNF677* | -1.348122419 | 0.001853132 | 0.004980661 | 1 |
| T2DM | b_cell | *ZNF71* | -1.925903347 | 0.000781663 | 0.004980661 | 1 |
| T2DM | b_cell | *RPL12* | -1.149236908 | 0.004156636 | 0.006288742 | 1 |
| T2DM | b_cell | *MPZL1* | -0.988211934 | 0.001823626 | 0.004980661 | 1 |
| T2DM | b_cell | *VPS13A* | -1.027512881 | 0.002450647 | 0.004980661 | 1 |
| T2DM | b_cell | *NOL8* | -1.785658218 | 0.002707446 | 0.00500969 | 1 |
| T2DM | b_cell | *DLGAP2* | -0.849623426 | 0.002631824 | 0.004980661 | 1 |
| T2DM | b_cell | *MRPL42* | -1.013885519 | 0.001856412 | 0.004980661 | 1 |
| T2DM | b_cell | *ZNF560* | 2.999760084 | 0.032145699 | 0.03768324 | 1 |
| T2DM | b_cell | *AGAP9* | 1.815501687 | 0.00677265 | 0.009230589 | 1 |
| T2DM | b_cell | *ZNF273* | -1.711418032 | 0.001470012 | 0.004980661 | 1 |
| T2DM | b_cell | *MAK16* | -2.895685864 | 0.021462982 | 0.025776829 | 1 |
| T2DM | b_cell | *ZNF667* | -2.662209 | 0.007336984 | 0.0098585 | 1 |
| T2DM | b_cell | *SFI1* | -1.223098397 | 0.002114244 | 0.004980661 | 1 |
| T2DM | b_cell | *ZNF649* | -1.694922187 | 0.001657542 | 0.004980661 | 1 |
| T2DM | b_cell | *ADH4* | -1.355691282 | 0.00080903 | 0.004980661 | 1 |
| T2DM | b_cell | *HIBCH* | -1.053956906 | 0.001614082 | 0.004980661 | 1 |
| T2DM | b_cell | *ZNF544* | -1.365762334 | 0.001804544 | 0.004980661 | 1 |
| T2DM | b_cell | *SOWAHC* | 1.079322267 | 0.003058308 | 0.005233209 | 1 |
| T2DM | b_cell | *MIER1* | -1.215508961 | 0.00333173 | 0.005441511 | 1 |
| T2DM | b_cell | *MAN1A2* | -1.067766071 | 0.0020398 | 0.004980661 | 1 |
| T2DM | b_cell | *SVIP* | -1.108644203 | 0.001702424 | 0.004980661 | 1 |
| T2DM | b_cell | *DDRGK1* | -2.089508042 | 0.003550138 | 0.005653189 | 1 |
| T2DM | b_cell | *ZNF607* | -2.836840933 | 0.003156276 | 0.005303461 | 1 |
| T2DM | b_cell | *ZNF334* | -1.957658013 | 0.006479038 | 0.008894974 | 1 |
| T2DM | b_cell | *HSD17B11* | -1.8419742 | 0.005434874 | 0.007715041 | 1 |
| T2DM | b_cell | *SULT1C2* | -1.182763499 | 0.040020682 | 0.046288522 | 1 |
| T2DM | b_cell | *RPS6KL1* | -2.066852982 | 0.003240891 | 0.005362236 | 1 |
| T2DM | b_cell | *DDX42* | -1.652019126 | 0.006031104 | 0.008415533 | 1 |
| T2DM | b_cell | *HELZ* | -0.71732337 | 0.001827244 | 0.004980661 | 1 |
| T2DM | b_cell | *TMEM116* | -1.350360162 | 8.86E-05 | 0.004980661 | 0.429970024 |
| T2DM | b_cell | *UCKL1* | -1.195030831 | 0.001859809 | 0.004980661 | 1 |
| T2DM | b_cell | *ZNF442* | -2.051979966 | 0.001520754 | 0.004980661 | 1 |
| T2DM | b_cell | *GFPT1* | -0.964406394 | 0.001956905 | 0.004980661 | 1 |
| T2DM | b_cell | *ZNF568* | -1.285334493 | 0.00181291 | 0.004980661 | 1 |
| T2DM | b_cell | *ZNF587* | -2.291983599 | 0.015938401 | 0.019597852 | 1 |
| T2DM | b_cell | *SH3BGRL2* | -1.120369586 | 0.001449315 | 0.004980661 | 1 |

| | | | | | | |
|---|---|---|---|---|---|---|
| T2DM | b_cell | *TMA16* | -1.16178485 | 0.001878421 | 0.004980661 | 1 |
| T2DM | b_cell | *HLA-DRB5* | -2.266981421 | 1.12E-11 | 2.72E-08 | 5.45E-08 |
| T2DM | b_cell | *CNGA1* | -1.193254551 | 0.007566125 | 0.010099268 | 1 |
| T2DM | b_cell | *C20orf203* | -3.439938725 | 0.004345893 | 0.006486089 | 1 |
| T2DM | b_cell | *ZNF534* | -2.179739438 | 0.029534759 | 0.034799089 | 1 |
| T2DM | b_cell | *NCOA6* | -1.257448128 | 0.002181068 | 0.004980661 | 1 |
| T2DM | b_cell | *FAM19A2* | -0.883045301 | 0.001503723 | 0.004980661 | 1 |
| T2DM | b_cell | *TTC37* | -0.947258272 | 0.002627083 | 0.004980661 | 1 |
| T2DM | b_cell | *PAPSS2* | -0.910461387 | 0.00183666 | 0.004980661 | 1 |
| T2DM | b_cell | *CEP290* | -1.181091851 | 0.002161118 | 0.004980661 | 1 |
| T2DM | b_cell | *ANKRD13B* | -2.137477218 | 0.001759937 | 0.004980661 | 1 |
| T2DM | b_cell | *ECI2* | -1.824613307 | 0.002756801 | 0.005034338 | 1 |
| T2DM | b_cell | *UNC13B* | -1.020023851 | 0.002034093 | 0.004980661 | 1 |
| T2DM | b_cell | *PPP1R14C* | -5.467254585 | 0.001722177 | 0.004980661 | 1 |
| T2DM | b_cell | *GPATCH3* | 2.603953834 | 0.016216695 | 0.019874566 | 1 |
| T2DM | b_cell | *RPL10A* | -1.201572682 | 0.001965318 | 0.004980661 | 1 |
| T2DM | b_cell | *COLGALT2* | -1.37992516 | 0.00121211 | 0.004980661 | 1 |
| T2DM | b_cell | *RCSD1* | -1.134009601 | 0.001990043 | 0.004980661 | 1 |
| T2DM | b_cell | *GRIN3A* | -2.105079876 | 0.018533671 | 0.022518475 | 1 |
| T2DM | b_cell | *TMEM184B* | -1.300621894 | 0.002418885 | 0.004980661 | 1 |
| T2DM | b_cell | *SFT2D1* | -1.030815478 | 0.00176801 | 0.004980661 | 1 |
| T2DM | b_cell | *HMGN2* | -1.101004984 | 0.000901851 | 0.004980661 | 1 |
| T2DM | b_cell | *DENND4B* | -1.519792313 | 0.001580132 | 0.004980661 | 1 |
| T2DM | b_cell | *RYR3* | -1.010589211 | 0.002377821 | 0.004980661 | 1 |
| T2DM | b_cell | *TYW1* | -1.512513007 | 0.006134394 | 0.008530834 | 1 |
| T2DM | b_cell | *PRMT6* | 1.023447909 | 0.002941688 | 0.005147158 | 1 |
| T2DM | b_cell | *SHISA4* | -6.450416159 | 0.000371974 | 0.004980661 | 1 |
| T2DM | b_cell | *MAP3K3* | -1.998609227 | 0.002138002 | 0.004980661 | 1 |
| T2DM | b_cell | *C1orf174* | -1.626829184 | 0.001769605 | 0.004980661 | 1 |
| T2DM | b_cell | *DZIP3* | -0.969204635 | 0.001805968 | 0.004980661 | 1 |
| T2DM | b_cell | *KIAA0753* | -2.346895636 | 0.018506017 | 0.022492784 | 1 |
| T2DM | b_cell | *TBKBP1* | -3.066757411 | 0.000934857 | 0.004980661 | 1 |
| T2DM | b_cell | *CCDC167* | 1.127876305 | 0.003822429 | 0.005923484 | 1 |
| T2DM | b_cell | *ZFP2* | -1.319790476 | 0.001821756 | 0.004980661 | 1 |
| T2DM | b_cell | *L3MBTL3* | -2.407935761 | 0.028306816 | 0.033400941 | 1 |
| T2DM | b_cell | *MFAP3L* | -2.114358533 | 0.001763839 | 0.004980661 | 1 |
| T2DM | b_cell | *NAGA* | -2.195559588 | 0.00268475 | 0.005000541 | 1 |
| T2DM | b_cell | *SMG5* | -1.151797607 | 0.001310087 | 0.004980661 | 1 |
| T2DM | b_cell | *TGM2* | -1.182962424 | 0.00484238 | 0.007005137 | 1 |
| T2DM | b_cell | *PJA2* | -1.147516581 | 0.001938789 | 0.004980661 | 1 |
| T2DM | b_cell | *AL590822.1* | -1.854490125 | 0.001307561 | 0.004980661 | 1 |
| T2DM | b_cell | *EFCAB2* | -1.094510108 | 0.002069966 | 0.004980661 | 1 |
| T2DM | b_cell | *CHML* | 0.961143373 | 0.00315314 | 0.005302958 | 1 |
| T2DM | b_cell | *TATDN3* | -0.994757623 | 0.002889021 | 0.005108429 | 1 |
| T2DM | b_cell | *SAMD5* | -1.159575914 | 0.001933511 | 0.004980661 | 1 |
| T2DM | b_cell | *ECT2L* | -2.079766991 | 0.001899906 | 0.004980661 | 1 |
| T2DM | b_cell | *GPR52* | -38.49825725 | 0.000774538 | 0.004980661 | 1 |
| T2DM | b_cell | *FAM229B* | -1.482512834 | 0.001446579 | 0.004980661 | 1 |
| T2DM | b_cell | *DDO* | -2.54933244 | 0.010287647 | 0.013184275 | 1 |
| T2DM | b_cell | *RBM20* | 1.27940034 | 0.005607767 | 0.00790956 | 1 |

| | | | | | | |
|---|---|---|---|---|---|---|
| T2DM | b_cell | *C6orf163* | 0.808703257 | 0.003811528 | 0.005908686 | 1 |
| T2DM | b_cell | *PCMTD2* | -1.255197154 | 0.002125345 | 0.004980661 | 1 |
| T2DM | b_cell | *SAMD13* | -2.025386055 | 0.001474188 | 0.004980661 | 1 |
| T2DM | b_cell | *EFCAB7* | -1.014712884 | 0.002093929 | 0.004980661 | 1 |
| T2DM | b_cell | *IFIT1B* | -8.927438588 | 0.002022061 | 0.004980661 | 1 |
| T2DM | b_cell | *LIPN* | -4.000242144 | 0.001764874 | 0.004980661 | 1 |
| T2DM | b_cell | *INPP5B* | -0.979589977 | 0.00128964 | 0.004980661 | 1 |
| T2DM | b_cell | *NEU4* | -4.228148201 | 0.01731046 | 0.021129648 | 1 |
| T2DM | b_cell | *C2orf72* | -18.87958841 | 0.000156316 | 0.004980661 | 0.758445495 |
| T2DM | b_cell | *RUFY2* | -0.816959332 | 0.001790566 | 0.004980661 | 1 |
| T2DM | b_cell | *PHACTR4* | -1.199369261 | 0.001622927 | 0.004980661 | 1 |
| T2DM | b_cell | *ASAH2B* | -2.015592437 | 0.007150515 | 0.009650709 | 1 |
| T2DM | b_cell | *SYT15* | -3.563942645 | 0.016533423 | 0.020237177 | 1 |
| T2DM | b_cell | *LINC00619* | -16.37115612 | 0.001599646 | 0.004980661 | 1 |
| T2DM | b_cell | *DAXX* | -1.400036677 | 0.012312122 | 0.015512443 | 1 |
| T2DM | b_cell | *BMPR2* | -1.178334206 | 0.002868436 | 0.005094011 | 1 |
| T2DM | b_cell | *TCEA3* | -1.085116733 | 0.002013162 | 0.004980661 | 1 |
| T2DM | b_cell | *COL15A1* | -0.915113975 | 0.001455714 | 0.004980661 | 1 |
| T2DM | b_cell | *NOTCH4* | -0.870214254 | 9.68E-08 | 5.87E-05 | 0.000469634 |
| T2DM | b_cell | *SMIM5* | 5.27491422 | 0.030333982 | 0.035688769 | 1 |
| T2DM | b_cell | *SKIV2L* | 0.55764752 | 0.042708756 | 0.049244982 | 1 |
| T2DM | b_cell | *LAYN* | -1.821727106 | 0.001737124 | 0.004980661 | 1 |
| T2DM | b_cell | *C6orf48* | -1.828214358 | 0.001475259 | 0.004980661 | 1 |
| T2DM | b_cell | *LY6G5C* | 0.910941762 | 0.01463042 | 0.018127374 | 1 |
| T2DM | b_cell | *GPANK1* | -1.707198878 | 0.000239826 | 0.004980661 | 1 |
| T2DM | b_cell | *BAG6* | -1.683343032 | 0.007823405 | 0.010388386 | 1 |
| T2DM | b_cell | *LST1* | -1.233338858 | 0.003777985 | 0.005875252 | 1 |
| T2DM | b_cell | *ZNF814* | -1.088724846 | 0.002286571 | 0.004980661 | 1 |
| T2DM | b_cell | *MICA* | -2.421323847 | 0.021977983 | 0.026349685 | 1 |
| T2DM | b_cell | *HLA-C* | -1.761698665 | 0.000223157 | 0.004980661 | 1 |
| T2DM | b_cell | *PSORS1C2* | 3.700261753 | 0.013328458 | 0.016680341 | 1 |
| T2DM | b_cell | *DHX16* | -1.535649286 | 0.015551222 | 0.019161229 | 1 |
| T2DM | b_cell | *ABCF1* | -1.605051621 | 0.021273652 | 0.025568432 | 1 |
| T2DM | b_cell | *LILRB3* | 2.591980796 | 0.017342991 | 0.021164033 | 1 |
| T2DM | b_cell | *HLA-E* | -1.134011591 | 0.002607055 | 0.004980661 | 1 |
| T2DM | b_cell | *ZNF468* | -2.791293314 | 0.002328905 | 0.004980661 | 1 |
| T2DM | b_cell | *HLA-G* | 2.65815905 | 0.010313375 | 0.013208641 | 1 |
| T2DM | b_cell | *FAM153C* | -2.02508091 | 0.000101964 | 0.004980661 | 0.49472881 |
| T2DM | b_cell | *TRIM27* | -1.136164708 | 0.001898966 | 0.004980661 | 1 |
| T2DM | b_cell | *MALRD1* | -1.163884577 | 0.001765192 | 0.004980661 | 1 |
| T2DM | b_cell | *FAM196B* | -1.078171147 | 0.001734417 | 0.004980661 | 1 |
| T2DM | b_cell | *MROH6* | -3.584653732 | 0.011346482 | 0.014434486 | 1 |
| T2DM | b_cell | *FAM216A* | -2.30049929 | 0.038569125 | 0.04470554 | 1 |
| T2DM | b_cell | *ZBTB48* | -1.625301866 | 0.001203437 | 0.004980661 | 1 |
| T2DM | b_cell | *MZT1* | -2.387797793 | 0.00204288 | 0.004980661 | 1 |
| T2DM | b_cell | *ZNF155* | -1.482914587 | 0.001332599 | 0.004980661 | 1 |
| T2DM | b_cell | *ZNF783* | -1.178572647 | 0.001995858 | 0.004980661 | 1 |
| T2DM | b_cell | *ZNF425* | -1.774013149 | 0.002009776 | 0.004980661 | 1 |
| T2DM | b_cell | *C12orf73* | -1.938194891 | 0.001609391 | 0.004980661 | 1 |
| T2DM | b_cell | *PCDHGA1* | 1.411700557 | 0.014950322 | 0.018476557 | 1 |

| | | | | | | |
|---|---|---|---|---|---|---|
| T2DM | b_cell | *PCDHA7* | -44.01752514 | 0.004569644 | 0.006720797 | 1 |
| T2DM | b_cell | *PCDHA4* | -4.392903706 | 0.002004854 | 0.004980661 | 1 |
| T2DM | b_cell | *TRIM13* | -1.188539415 | 0.00369788 | 0.00579416 | 1 |
| T2DM | b_cell | *SPIRE2* | -1.717075333 | 0.008329199 | 0.010952107 | 1 |
| T2DM | b_cell | *PKHD1L1* | -0.751700376 | 0.000585179 | 0.004980661 | 1 |
| T2DM | b_cell | *SLFN12L* | -1.089238485 | 0.001081035 | 0.004980661 | 1 |
| T2DM | b_cell | *SLC35B4* | -2.380637428 | 0.03879633 | 0.044915245 | 1 |
| T2DM | b_cell | *SYCE1L* | 1.388397591 | 0.000302918 | 0.004980661 | 1 |
| T2DM | b_cell | *TMEM231* | -2.397869421 | 0.001728971 | 0.004980661 | 1 |
| T2DM | b_cell | *SCGB2B2* | -1.2009778 | 0.001933723 | 0.004980661 | 1 |
| T2DM | b_cell | *CCDC144NL* | -1.997163802 | 0.001815915 | 0.004980661 | 1 |
| T2DM | b_cell | *TMEM170B* | -0.977588788 | 0.00238217 | 0.004980661 | 1 |
| T2DM | b_cell | *GOLGA6L10* | -4.074287501 | 0.004171269 | 0.00630302 | 1 |
| T2DM | b_cell | *SAP25* | -11.32871063 | 0.001390493 | 0.004980661 | 1 |
| T2DM | b_cell | *IPO7* | -1.389502678 | 0.002884964 | 0.005104976 | 1 |
| T2DM | b_cell | *PRR13* | -3.950971739 | 0.006173732 | 0.008573254 | 1 |
| T2DM | b_cell | *TECPR1* | -1.157426106 | 0.001759286 | 0.004980661 | 1 |
| T2DM | b_cell | *MT1A* | -3.302712317 | 0.001593125 | 0.004980661 | 1 |
| T2DM | b_cell | *CFI* | -1.109755053 | 0.001886473 | 0.004980661 | 1 |
| T2DM | b_cell | *ATP6AP1L* | -0.881101108 | 0.006670659 | 0.009117193 | 1 |
| T2DM | b_cell | *RGL3* | -1.085880345 | 0.004135895 | 0.006267133 | 1 |
| T2DM | b_cell | *NAP1L4* | -0.933804466 | 0.002427502 | 0.004980661 | 1 |
| T2DM | b_cell | *SMN2* | 0.801115326 | 0.012105605 | 0.015295936 | 1 |
| T2DM | b_cell | *SERF1B* | -4.299844681 | 0.000379184 | 0.004980661 | 1 |
| T2DM | b_cell | *LIN52* | -1.017672694 | 0.010777429 | 0.013753836 | 1 |
| T2DM | b_cell | *C17orf107* | 1.974993659 | 0.000797887 | 0.004980661 | 1 |
| T2DM | b_cell | *KLRC2* | -2.90502175 | 0.002910155 | 0.00512387 | 1 |
| T2DM | b_cell | *RFPL3S* | 1.165810193 | 0.024037777 | 0.028663381 | 1 |
| T2DM | b_cell | *C1QTNF9B* | 1.458729436 | 0.002482196 | 0.004980661 | 1 |
| T2DM | b_cell | *HBA1* | -1.621318061 | 0.020471975 | 0.024684399 | 1 |
| T2DM | b_cell | *COL6A6* | -0.965169886 | 0.001420985 | 0.004980661 | 1 |
| T2DM | b_cell | *HLA-A* | -1.452672714 | 0.00761027 | 0.010147027 | 1 |
| T2DM | b_cell | *LNP1* | -1.81314054 | 0.005453305 | 0.007734416 | 1 |
| T2DM | b_cell | *PRSS50* | -10.32410346 | 0.000546056 | 0.004980661 | 1 |
| T2DM | b_cell | *METTL6* | -1.530190599 | 0.003101408 | 0.005253181 | 1 |
| T2DM | b_cell | *SACM1L* | -1.337836495 | 0.002155675 | 0.004980661 | 1 |
| T2DM | b_cell | *SLC48A1* | -1.200364574 | 0.001641916 | 0.004980661 | 1 |
| T2DM | b_cell | *TAS2R50* | -24.51336224 | 0.040822625 | 0.047137405 | 1 |
| T2DM | b_cell | *C17orf51* | -2.861401152 | 0.001954357 | 0.004980661 | 1 |
| T2DM | b_cell | *OR2A42* | -3.473855986 | 0.00344171 | 0.005546057 | 1 |
| T2DM | b_cell | *NUP62* | -1.798850636 | 0.001369958 | 0.004980661 | 1 |
| T2DM | b_cell | *AL021068.1* | -4.113419267 | 0.015726312 | 0.0193616 | 1 |
| T2DM | b_cell | *FAM106A* | -7.335350195 | 0.033279346 | 0.038927529 | 1 |
| T2DM | b_cell | *ZNF254* | -0.920053551 | 0.001508421 | 0.004980661 | 1 |
| T2DM | b_cell | *AC022498.1* | -8.40486876 | 0.003385327 | 0.005489842 | 1 |
| T2DM | b_cell | *ASIC3* | 5.42657037 | 0.001381375 | 0.004980661 | 1 |
| T2DM | b_cell | *GIMAP1* | -1.451691991 | 0.001271438 | 0.004980661 | 1 |
| T2DM | b_cell | *DNLZ* | 2.481622128 | 1.84E-06 | 0.000744907 | 0.008938881 |
| T2DM | b_cell | *NOTCH2NL* | 2.024262196 | 0.002212482 | 0.004980661 | 1 |
| T2DM | b_cell | *GSTM2* | -0.952243375 | 0.002910426 | 0.00512387 | 1 |

| | | | | | | |
|---|---|---|---|---|---|---|
| T2DM | b_cell | *ARHGAP19* | 1.156652796 | 0.012117121 | 0.015306501 | 1 |
| T2DM | b_cell | *LCAT* | -1.570909113 | 0.001994108 | 0.004980661 | 1 |
| T2DM | b_cell | *GPC2* | -8.658825349 | 0.007023899 | 0.009506265 | 1 |
| T2DM | b_cell | *ERV3-1* | -0.63657034 | 0.006330811 | 0.008750103 | 1 |
| T2DM | b_cell | *SRA1* | -1.289818004 | 0.001713674 | 0.004980661 | 1 |
| T2DM | b_cell | *TMEM110* | 1.222668991 | 0.000422609 | 0.004980661 | 1 |
| T2DM | b_cell | *LBH* | -1.408514093 | 0.001392416 | 0.004980661 | 1 |
| T2DM | b_cell | *PPP1CB* | -1.019288633 | 0.001654435 | 0.004980661 | 1 |
| T2DM | b_cell | *NCKIPSD* | -1.797287706 | 0.007204275 | 0.009707065 | 1 |
| T2DM | b_cell | *ATF6B* | -1.45787353 | 0.019573023 | 0.023671064 | 1 |
| T2DM | b_cell | *SLC35F6* | 1.552093927 | 0.006942103 | 0.009419452 | 1 |
| T2DM | b_cell | *EMP2* | -1.14157782 | 0.001850918 | 0.004980661 | 1 |
| T2DM | b_cell | *SLC23A3* | -1.99534031 | 0.018832511 | 0.02283242 | 1 |
| T2DM | b_cell | *LTB4R* | -3.670489118 | 0.032779181 | 0.038388748 | 1 |
| T2DM | b_cell | *IRF9* | -1.26542729 | 0.002008022 | 0.004980661 | 1 |
| T2DM | b_cell | *ZNF726* | -2.897172457 | 0.000551909 | 0.004980661 | 1 |
| T2DM | b_cell | *ZNF90* | -1.548330075 | 0.002178162 | 0.004980661 | 1 |
| T2DM | b_cell | *SMIM7* | -1.470876703 | 0.003351141 | 0.005460611 | 1 |
| T2DM | b_cell | *TSPAN4* | -1.215495781 | 0.001740151 | 0.004980661 | 1 |
| T2DM | b_cell | *CPNE1* | -1.510122545 | 0.00069292 | 0.004980661 | 1 |
| T2DM | b_cell | *ARL16* | -1.178612866 | 0.001555328 | 0.004980661 | 1 |
| T2DM | b_cell | *PAXIP1-AS2* | -1.202610286 | 0.001705479 | 0.004980661 | 1 |
| T2DM | b_cell | *LYRM4* | -0.962105645 | 0.001856573 | 0.004980661 | 1 |
| T2DM | b_cell | *PRCD* | -1.660663388 | 0.002461886 | 0.004980661 | 1 |
| T2DM | b_cell | *SH3D21* | 1.668967826 | 0.017777119 | 0.021666562 | 1 |
| T2DM | b_cell | *FIS1* | -2.022905098 | 0.015182297 | 0.018729852 | 1 |
| T2DM | b_cell | *ANG* | -1.82126433 | 0.001942249 | 0.004980661 | 1 |
| T2DM | b_cell | *COLCA2* | -2.313240559 | 0.002871988 | 0.005094011 | 1 |
| T2DM | b_cell | *MBLAC1* | 2.241048933 | 0.019963649 | 0.024107423 | 1 |
| T2DM | b_cell | *LCNL1* | -4.098435939 | 0.009372384 | 0.012120152 | 1 |
| T2DM | b_cell | *GNAT3* | -2.030954951 | 0.001936248 | 0.004980661 | 1 |
| T2DM | b_cell | *PLIN5* | -0.905856397 | 0.001176864 | 0.004980661 | 1 |
| T2DM | b_cell | *NUTM2D* | 5.563735701 | 0.001563732 | 0.004980661 | 1 |
| T2DM | b_cell | *CPEB1* | -1.378300737 | 0.000928769 | 0.004980661 | 1 |
| T2DM | b_cell | *EML6* | -1.554204412 | 0.002965991 | 0.005173571 | 1 |
| T2DM | b_cell | *ARHGEF33* | -1.768369644 | 0.007371984 | 0.009897307 | 1 |
| T2DM | b_cell | *IFRD2* | -1.69925123 | 0.000233087 | 0.004980661 | 1 |
| T2DM | b_cell | *AC005082.1* | -6.521341671 | 0.003237629 | 0.005362236 | 1 |
| T2DM | b_cell | *SMTNL1* | -1.992727541 | 0.001482891 | 0.004980661 | 1 |
| T2DM | b_cell | *UBXN2B* | -1.126965787 | 0.001993907 | 0.004980661 | 1 |
| T2DM | b_cell | *C5orf49* | 6.785125132 | 0.004599881 | 0.006759122 | 1 |
| T2DM | b_cell | *GOLGA8B* | -1.102225896 | 0.003484565 | 0.005592825 | 1 |
| T2DM | b_cell | *VPS16* | -1.378123303 | 0.001881251 | 0.004980661 | 1 |
| T2DM | b_cell | *MYL5* | -2.303979315 | 0.002986773 | 0.005187056 | 1 |
| T2DM | b_cell | *NPEPL1* | -1.476158496 | 0.001689033 | 0.004980661 | 1 |
| T2DM | b_cell | *TMEM242* | -2.114134787 | 0.027872014 | 0.032935951 | 1 |
| T2DM | b_cell | *TSTD1* | -1.397008203 | 0.001676294 | 0.004980661 | 1 |
| T2DM | b_cell | *AC009403.2* | 0.995578215 | 0.003378026 | 0.005481666 | 1 |
| T2DM | b_cell | *PAM16* | 1.340655893 | 0.005479215 | 0.007764355 | 1 |
| T2DM | b_cell | *AC016773.1* | -5.317420124 | 0.000630892 | 0.004980661 | 1 |

| | | | | | | |
|---|---|---|---|---|---|---|
| T2DM | b_cell | *AP000350.4* | -1.399585422 | 0.014660314 | 0.018159776 | 1 |
| T2DM | b_cell | *RNASEK* | -1.342584896 | 0.004211013 | 0.006341352 | 1 |
| T2DM | b_cell | *NBPF1* | -1.645691209 | 0.001384891 | 0.004980661 | 1 |
| T2DM | b_cell | *RNU6ATAC35I* | -21.64906304 | 0.00068209 | 0.004980661 | 1 |
| T2DM | b_cell | *PPP3R1* | -1.375438622 | 0.00321248 | 0.005339692 | 1 |
| T2DM | b_cell | *AP4M1* | -1.119593447 | 0.00621769 | 0.008624423 | 1 |
| T2DM | b_cell | *ZNF880* | -1.300121542 | 0.001048133 | 0.004980661 | 1 |
| T2DM | b_cell | *TIGD1* | -2.009886601 | 0.00149633 | 0.004980661 | 1 |
| T2DM | b_cell | *XKR9* | -1.281756454 | 0.001148237 | 0.004980661 | 1 |
| T2DM | b_cell | *SLC12A8* | -1.562857505 | 0.001202341 | 0.004980661 | 1 |
| T2DM | b_cell | *APOL6* | -0.868954549 | 0.00179768 | 0.004980661 | 1 |
| T2DM | b_cell | *OR2A1* | 0.579042636 | 0.002401802 | 0.004980661 | 1 |
| T2DM | b_cell | *UBA52* | -1.166142287 | 0.006034393 | 0.008415681 | 1 |
| T2DM | b_cell | *TIAF1* | 0.821365834 | 0.002244017 | 0.004980661 | 1 |
| T2DM | b_cell | *AC062017.1* | 1.943635254 | 0.022824737 | 0.027290691 | 1 |
| T2DM | b_cell | *ADRA2B* | -5.712094785 | 0.020427355 | 0.024636721 | 1 |
| T2DM | b_cell | *EXOSC6* | -1.628076165 | 0.000824416 | 0.004980661 | 1 |
| T2DM | b_cell | *ZNF844* | -1.276219716 | 0.001582979 | 0.004980661 | 1 |
| T2DM | b_cell | *LINC00571* | -1.659519435 | 0.001328222 | 0.004980661 | 1 |
| T2DM | b_cell | *C5orf66* | -1.139184132 | 0.001099435 | 0.004980661 | 1 |
| T2DM | b_cell | *LINC01133* | -1.120960074 | 0.001249747 | 0.004980661 | 1 |
| T2DM | b_cell | *C4B* | 9.38711923 | 0.001687457 | 0.004980661 | 1 |
| T2DM | b_cell | *LINC01117* | -1.54280329 | 0.001876837 | 0.004980661 | 1 |
| T2DM | b_cell | *RASAL2-AS1* | -5.220181169 | 0.002161887 | 0.004980661 | 1 |
| T2DM | b_cell | *AC069213.1* | 0.879337615 | 0.003660906 | 0.005752881 | 1 |
| T2DM | b_cell | *LINC00240* | -1.917683749 | 0.001575541 | 0.004980661 | 1 |
| T2DM | b_cell | *LINC00184* | -38.90417264 | 0.001770314 | 0.004980661 | 1 |
| T2DM | b_cell | *AL049840.1* | -1.154578989 | 0.000956801 | 0.004980661 | 1 |
| T2DM | b_cell | *PLEKHM1* | -1.493178735 | 0.0039705 | 0.006071904 | 1 |
| T2DM | b_cell | *AP000350.6* | -1.854784835 | 0.002979673 | 0.005186119 | 1 |
| T2DM | b_cell | *SFTA1P* | -4.747244545 | 0.027768424 | 0.032821533 | 1 |
| T2DM | b_cell | *CYP4A22-AS1* | -2.257288771 | 0.001923062 | 0.004980661 | 1 |
| T2DM | b_cell | *MIAT* | -2.032719498 | 0.00213073 | 0.004980661 | 1 |
| T2DM | b_cell | *ERCC6* | -1.647974024 | 0.001891854 | 0.004980661 | 1 |
| T2DM | b_cell | *LINC00115* | -2.215940425 | 0.001910828 | 0.004980661 | 1 |
| T2DM | b_cell | *HAR1A* | -21.15925357 | 0.001279302 | 0.004980661 | 1 |
| T2DM | b_cell | *LINC00937* | 1.400883701 | 0.031740378 | 0.037235086 | 1 |
| T2DM | b_cell | *FTCDNL1* | -1.175751306 | 0.012665221 | 0.015924243 | 1 |
| T2DM | b_cell | *FAM138B* | -22.43572677 | 0.001691776 | 0.004980661 | 1 |
| T2DM | b_cell | *ZNF717* | -0.89486966 | 0.002929019 | 0.005137961 | 1 |
| T2DM | b_cell | *AC017074.2* | 3.285297188 | 0.001948352 | 0.004980661 | 1 |
| T2DM | b_cell | *AC009299.3* | -1.549991766 | 0.002135187 | 0.004980661 | 1 |
| T2DM | b_cell | *SCAMP4* | -1.566798262 | 0.00207842 | 0.004980661 | 1 |
| T2DM | b_cell | *SLC2A1-AS1* | 0.78897488 | 0.00226654 | 0.004980661 | 1 |
| T2DM | b_cell | *AC005884.1* | -1.955137412 | 0.000621919 | 0.004980661 | 1 |
| T2DM | b_cell | *HCG20* | -3.467159857 | 0.024639017 | 0.029315476 | 1 |
| T2DM | b_cell | *C14orf144* | -4.035892028 | 0.007565623 | 0.010099268 | 1 |
| T2DM | b_cell | *POLR2J2* | -44.98751582 | 0.024183907 | 0.028822442 | 1 |
| T2DM | b_cell | *LINC00578* | -3.435081091 | 0.010890847 | 0.013891269 | 1 |
| T2DM | b_cell | *C19orf24* | -2.05498315 | 0.001033265 | 0.004980661 | 1 |

| | | | | | | |
|---|---|---|---|---|---|---|
| T2DM | b_cell | *ZNF883* | -1.622327156 | 0.002235184 | 0.004980661 | 1 |
| T2DM | b_cell | *ARL17B* | -1.191735283 | 0.001853043 | 0.004980661 | 1 |
| T2DM | b_cell | *DHFR* | -1.285035549 | 0.000244178 | 0.004980661 | 1 |
| T2DM | b_cell | *PATL2* | -1.189408291 | 0.002694066 | 0.00500636 | 1 |
| T2DM | b_cell | *CASC20* | -2.419568133 | 0.00146417 | 0.004980661 | 1 |
| T2DM | b_cell | *PSMG3-AS1* | -2.445428187 | 0.003109239 | 0.005260699 | 1 |
| T2DM | b_cell | *AC112229.4* | -2.554063185 | 0.004321237 | 0.006465199 | 1 |
| T2DM | b_cell | *KLHL7-AS1* | -2.943534008 | 0.00417701 | 0.006309729 | 1 |
| T2DM | b_cell | *LINC00332* | -4.061712504 | 0.000130442 | 0.004980661 | 0.632904782 |
| T2DM | b_cell | *AC092171.4* | -1.824967867 | 0.001780934 | 0.004980661 | 1 |
| T2DM | b_cell | *RPS18* | -1.22767367 | 0.002659126 | 0.004985914 | 1 |
| T2DM | b_cell | *DIRC3* | -0.691934492 | 0.001841416 | 0.004980661 | 1 |
| T2DM | b_cell | *TAPBP* | -1.453010771 | 0.009350597 | 0.0120952 | 1 |
| T2DM | b_cell | *AC107021.1* | -1.757016949 | 0.002981045 | 0.005186119 | 1 |
| T2DM | b_cell | *TMA7* | -1.754993308 | 0.03035986 | 0.035710555 | 1 |
| T2DM | b_cell | *MTRNR2L4* | 3.022878823 | 0.009288645 | 0.012031101 | 1 |
| T2DM | b_cell | *AC009495.3* | 1.27632204 | 0.002025692 | 0.004980661 | 1 |
| T2DM | b_cell | *HLA-DQB2* | 1.374830644 | 1.82E-07 | 9.81E-05 | 0.000883274 |
| T2DM | b_cell | *GOLGA8N* | -1.569828259 | 0.023705203 | 0.028294624 | 1 |
| T2DM | b_cell | *LINC00665* | -1.711554107 | 0.006018561 | 0.008403469 | 1 |
| T2DM | b_cell | *LYRM9* | 1.308395709 | 0.014671386 | 0.018168853 | 1 |
| T2DM | b_cell | *LINC01166* | 2.419071816 | 0.00968612 | 0.012482617 | 1 |
| T2DM | b_cell | *LINC00702* | -3.014690179 | 0.001643248 | 0.004980661 | 1 |
| T2DM | b_cell | *RPS28* | -1.430851319 | 0.001692125 | 0.004980661 | 1 |
| T2DM | b_cell | *TRIM26* | -1.080775705 | 0.002600465 | 0.004980661 | 1 |
| T2DM | b_cell | *ZNF736* | -0.88668231 | 0.005763192 | 0.008091148 | 1 |
| T2DM | b_cell | *PINLYP* | -3.002007174 | 0.006476673 | 0.008894655 | 1 |
| T2DM | b_cell | *ANKRD65* | -4.301741139 | 0.001787989 | 0.004980661 | 1 |
| T2DM | b_cell | *LINC00957* | 1.041473806 | 0.005327803 | 0.007585241 | 1 |
| T2DM | b_cell | *LINC00484* | -1.243051766 | 0.005728094 | 0.008051191 | 1 |
| T2DM | b_cell | *KIAA0040* | -1.504906463 | 0.00154898 | 0.004980661 | 1 |
| T2DM | b_cell | *LINC00607* | -1.161223639 | 0.002936739 | 0.005145797 | 1 |
| T2DM | b_cell | *AP001468.1* | -3.00340154 | 0.003729695 | 0.00583193 | 1 |
| T2DM | b_cell | *LINC00941* | -10.38969811 | 1.63E-05 | 0.003041201 | 0.079071228 |
| T2DM | b_cell | *AC019117.2* | -13.48309116 | 0.001071527 | 0.004980661 | 1 |
| T2DM | b_cell | *ZBTB22* | 2.927776828 | 0.026444609 | 0.031348459 | 1 |
| T2DM | b_cell | *ZBED5* | -1.141089501 | 0.001559847 | 0.004980661 | 1 |
| T2DM | b_cell | *CDKN2AIPNL* | 1.609252577 | 0.038295027 | 0.044419668 | 1 |
| T2DM | b_cell | *LINC00987* | -2.320567589 | 0.000827809 | 0.004980661 | 1 |
| T2DM | b_cell | *GS1-124K5.4* | 2.055244625 | 0.000978651 | 0.004980661 | 1 |
| T2DM | b_cell | *LINC00959* | -1.466501836 | 0.001790377 | 0.004980661 | 1 |
| T2DM | b_cell | *HLA-DQA2* | 3.471919156 | 5.50E-12 | 2.67E-08 | 2.67E-08 |
| T2DM | b_cell | *LINC01143* | -7.750671056 | 0.002058605 | 0.004980661 | 1 |
| T2DM | b_cell | *LINC00211* | -6.504950019 | 0.005745843 | 0.0080738 | 1 |
| T2DM | b_cell | *LINC00707* | 1.074307292 | 0.00205116 | 0.004980661 | 1 |
| T2DM | b_cell | *TXNDC5* | 1.091733494 | 0.002512193 | 0.004980661 | 1 |
| T2DM | b_cell | *LINC01119* | 1.007039313 | 0.002520012 | 0.004980661 | 1 |
| T2DM | b_cell | *NME1* | 0.871164235 | 0.003864898 | 0.005967901 | 1 |
| T2DM | b_cell | *TNFSF12* | -1.702612794 | 0.003169127 | 0.005315106 | 1 |
| T2DM | b_cell | *CDRT4* | -4.550194406 | 0.005594478 | 0.007897704 | 1 |

| | | | | | | |
|---|---|---|---|---|---|---|
| T2DM | b_cell | *C1orf226* | -5.217624065 | 0.011902752 | 0.015074955 | 1 |
| T2DM | b_cell | *LILRA2* | -2.659690841 | 0.006057672 | 0.008441075 | 1 |
| T2DM | b_cell | *LY6G5B* | -2.041175 | 0.013267272 | 0.01660805 | 1 |
| T2DM | b_cell | *PPIL3* | -1.043274964 | 0.003054403 | 0.00523119 | 1 |
| T2DM | b_cell | *C1QTNF9* | -6.500400955 | 0.001472441 | 0.004980661 | 1 |
| T2DM | b_cell | *PNMA2* | -2.605192962 | 0.001722153 | 0.004980661 | 1 |
| T2DM | b_cell | *KRBOX1* | -26.32102748 | 0.001681342 | 0.004980661 | 1 |
| T2DM | b_cell | *ARHGEF25* | -1.552567528 | 0.01398408 | 0.017411023 | 1 |
| T2DM | b_cell | *LINC00886* | -1.257482394 | 0.000270466 | 0.004980661 | 1 |
| T2DM | b_cell | *MIF* | 1.031341608 | 0.016293768 | 0.019958941 | 1 |
| T2DM | b_cell | *NSUN6* | -1.044621694 | 0.002094002 | 0.004980661 | 1 |
| T2DM | b_cell | *HLA-DOB* | 0.612877483 | 0.001116156 | 0.004980661 | 1 |
| T2DM | b_cell | *LINC00877* | 1.846114212 | 0.001969655 | 0.004980661 | 1 |
| T2DM | b_cell | *CD302* | -1.526388466 | 0.004310809 | 0.006457562 | 1 |
| T2DM | b_cell | *INMT* | -1.711013572 | 0.004649267 | 0.006806954 | 1 |
| T2DM | b_cell | *ATP5O* | -1.254547786 | 0.002792025 | 0.005061629 | 1 |
| T2DM | b_cell | *PLEKHO2* | -1.587428028 | 0.003060956 | 0.005233962 | 1 |
| T2DM | b_cell | *C8orf58* | -2.647717727 | 0.0052872 | 0.007538494 | 1 |
| T2DM | b_cell | *HOGA1* | -3.143070379 | 0.001706033 | 0.004980661 | 1 |
| T2DM | b_cell | *PI4KA* | -1.385943427 | 0.002548311 | 0.004980661 | 1 |
| T2DM | b_cell | *LINC00969* | -0.813747108 | 0.003383372 | 0.005488505 | 1 |
| T2DM | b_cell | *AMACR* | 1.217297717 | 0.018004631 | 0.021930777 | 1 |
| T2DM | b_cell | *ARFGAP3* | -1.290244668 | 0.001965107 | 0.004980661 | 1 |
| T2DM | b_cell | *MRPL20* | -1.082575 | 0.00256421 | 0.004980661 | 1 |
| T2DM | b_cell | *DECR2* | -2.265148845 | 0.001739714 | 0.004980661 | 1 |
| T2DM | b_cell | *ZNF709* | -12.39542712 | 0.001874412 | 0.004980661 | 1 |
| T2DM | b_cell | *ERVW-1* | -5.0920821 | 0.003307541 | 0.005412542 | 1 |
| T2DM | b_cell | *MRPL33* | -1.100240615 | 0.00203294 | 0.004980661 | 1 |
| T2DM | b_cell | *STON1* | 1.506000402 | 0.042914114 | 0.049470012 | 1 |
| T2DM | b_cell | *VSIG8* | -3.083281745 | 0.001543419 | 0.004980661 | 1 |
| T2DM | b_cell | *C7orf73* | -0.993595873 | 0.00168804 | 0.004980661 | 1 |
| T2DM | b_cell | *FGF14-IT1* | -4.865931246 | 0.001501764 | 0.004980661 | 1 |
| T2DM | b_cell | *PALM2* | -1.324479506 | 0.00304786 | 0.00522879 | 1 |
| T2DM | b_cell | *NBPF15* | -1.495417848 | 0.001938605 | 0.004980661 | 1 |
| T2DM | b_cell | *UPK3B* | -1.10235939 | 0.001688294 | 0.004980661 | 1 |
| T2DM | b_cell | *IL10RB* | 1.440303791 | 0.007800949 | 0.010361403 | 1 |
| T2DM | b_cell | *PLA2G4B* | 10.70765521 | 0.003727806 | 0.005830856 | 1 |
| T2DM | b_cell | *LEFTY1* | -27.30656088 | 0.009645556 | 0.012440254 | 1 |
| T2DM | b_cell | *TTC4* | -1.576859198 | 0.015137078 | 0.018678815 | 1 |
| T2DM | b_cell | *JMJD7* | 1.139732091 | 5.39E-05 | 0.004980661 | 0.261636779 |
| T2DM | b_cell | *APOBEC3D* | -8.812467375 | 0.00207798 | 0.004980661 | 1 |
| T2DM | b_cell | *TMEM199* | -1.878386238 | 0.004692112 | 0.006851076 | 1 |
| T2DM | b_cell | *SPRR2F* | 3.308836485 | 0.014990354 | 0.018521313 | 1 |
| T2DM | b_cell | *P2RY11* | -2.174886413 | 0.001063712 | 0.004980661 | 1 |
| T2DM | b_cell | *DBNDD2* | 2.510645538 | 0.002404733 | 0.004980661 | 1 |
| T2DM | b_cell | *SCARF2* | -1.633023016 | 0.001875534 | 0.004980661 | 1 |
| T2DM | b_cell | *APOBEC3C* | 1.182997085 | 0.003040002 | 0.005228675 | 1 |
| T2DM | b_cell | *CCDC13* | -2.309313474 | 0.00100609 | 0.004980661 | 1 |
| T2DM | b_cell | *C4A* | -9.04390116 | 6.19E-05 | 0.004980661 | 0.300377461 |
| T2DM | b_cell | *CRYBB2* | -5.86077784 | 0.00524404 | 0.007483554 | 1 |

| | | | | | | |
|---|---|---|---|---|---|---|
| T2DM | b_cell | *NEAT1* | -0.590361808 | 0.002195432 | 0.004980661 | 1 |
| T2DM | b_cell | *RAB30-AS1* | -1.048717184 | 0.001167803 | 0.004980661 | 1 |
| T2DM | b_cell | *UBAP1L* | -1.177863822 | 0.001656081 | 0.004980661 | 1 |
| T2DM | b_cell | *DNAJC3-AS1* | -1.333616831 | 0.001634349 | 0.004980661 | 1 |
| T2DM | b_cell | *LINC00926* | 0.142038098 | 0.002369006 | 0.004980661 | 1 |
| T2DM | b_cell | *DPH6-AS1* | -0.764462445 | 0.001997861 | 0.004980661 | 1 |
| T2DM | b_cell | *CDK11B* | -2.044724801 | 0.003038702 | 0.005228627 | 1 |
| T2DM | b_cell | *ABHD14A* | -1.930238696 | 0.002168192 | 0.004980661 | 1 |
| T2DM | b_cell | *SRP14-AS1* | -1.361066414 | 5.99E-05 | 0.004980661 | 0.290444174 |
| T2DM | b_cell | *GDNF-AS1* | -3.353583111 | 0.001328706 | 0.004980661 | 1 |
| T2DM | b_cell | *CCDC153* | -2.266601173 | 0.005116051 | 0.007324603 | 1 |
| T2DM | b_cell | *AL357673.1* | 2.180959168 | 0.009062373 | 0.011769442 | 1 |
| T2DM | b_cell | *C5orf17* | -1.184217219 | 0.00190663 | 0.004980661 | 1 |
| T2DM | b_cell | *TMEM150C* | -2.584800822 | 0.000301169 | 0.004980661 | 1 |
| T2DM | b_cell | *LINC00939* | -3.754245082 | 0.016787412 | 0.02052735 | 1 |
| T2DM | b_cell | *LINC01060* | -1.365675319 | 0.001954279 | 0.004980661 | 1 |
| T2DM | b_cell | *LINC00942* | 1.022082293 | 0.002629 | 0.004980661 | 1 |
| T2DM | b_cell | *PDCD6* | -1.128796226 | 0.001826922 | 0.004980661 | 1 |
| T2DM | b_cell | *TMEM158* | 3.653309058 | 0.003628199 | 0.005732341 | 1 |
| T2DM | b_cell | *DNAH10OS* | -14.6027172 | 0.00017426 | 0.004980661 | 0.845511882 |
| T2DM | b_cell | *PCDHA10* | -1.976693389 | 0.012225657 | 0.015425612 | 1 |
| T2DM | b_cell | *SMIM20* | -1.435461226 | 0.001795482 | 0.004980661 | 1 |
| T2DM | b_cell | *LINC00989* | -1.942070664 | 0.001248309 | 0.004980661 | 1 |
| T2DM | b_cell | *GLI4* | -1.981777366 | 0.001979045 | 0.004980661 | 1 |
| T2DM | b_cell | *SNHG18* | -1.730277128 | 0.00059613 | 0.004980661 | 1 |
| T2DM | b_cell | *ALG1L2* | -2.195000119 | 0.00213342 | 0.004980661 | 1 |
| T2DM | b_cell | *LINC01094* | -1.806827408 | 0.001747865 | 0.004980661 | 1 |
| T2DM | b_cell | *TMEM200B* | -2.342571387 | 0.025505596 | 0.030294529 | 1 |
| T2DM | b_cell | *PCDHGA6* | -1.254052692 | 0.030413037 | 0.035749263 | 1 |
| T2DM | b_cell | *UTP14C* | 1.990962172 | 0.01614464 | 0.019809327 | 1 |
| T2DM | b_cell | *PCDHGB2* | -8.250897349 | 0.001755208 | 0.004980661 | 1 |
| T2DM | b_cell | *LYN* | -1.344253227 | 0.005632301 | 0.007937243 | 1 |
| T2DM | b_cell | *ARMS2* | 0.631254007 | 0.00216995 | 0.004980661 | 1 |
| T2DM | b_cell | *MEX3A* | -3.050850555 | 0.01003913 | 0.012896441 | 1 |
| T2DM | b_cell | *DPP3* | -1.414921212 | 0.023535754 | 0.028106196 | 1 |
| T2DM | b_cell | *BRK1* | -1.253371417 | 0.000920873 | 0.004980661 | 1 |
| T2DM | b_cell | *SNHG9* | -1.30593999 | 0.003048691 | 0.00522879 | 1 |
| T2DM | b_cell | *TAS2R43* | 1.685290911 | 0.000886343 | 0.004980661 | 1 |
| T2DM | b_cell | *PCDHA3* | 2.646340939 | 0.037496863 | 0.043556327 | 1 |
| T2DM | b_cell | *TAS2R20* | 4.926221524 | 2.88E-05 | 0.004645541 | 0.139504023 |
| T2DM | b_cell | *ZNF432* | -1.502524335 | 0.001105772 | 0.004980661 | 1 |
| T2DM | b_cell | *HMBS* | 4.082554455 | 0.007186524 | 0.009691221 | 1 |
| T2DM | b_cell | *TAS2R31* | -3.768971955 | 0.001951902 | 0.004980661 | 1 |
| T2DM | b_cell | *AC093668.2* | -1.773658235 | 0.011087297 | 0.014119571 | 1 |
| T2DM | b_cell | *KIAA1147* | -1.248824677 | 0.002065213 | 0.004980661 | 1 |
| T2DM | b_cell | *GATC* | -2.60582331 | 0.00064352 | 0.004980661 | 1 |
| T2DM | b_cell | *ZBED6* | -3.383023067 | 0.001604555 | 0.004980661 | 1 |
| T2DM | b_cell | *LINC00592* | -8.814160706 | 0.000873795 | 0.004980661 | 1 |
| T2DM | b_cell | *CHURC1* | -1.238279956 | 0.002581229 | 0.004980661 | 1 |
| T2DM | b_cell | *RTEL1* | 2.14069924 | 0.002898553 | 0.005113643 | 1 |

| | | | | | | |
|---|---|---|---|---|---|---|
| T2DM | b_cell | *LINC00640* | -1.803525483 | 0.035284794 | 0.041134507 | 1 |
| T2DM | b_cell | *HIF1A-AS2* | -0.968941236 | 0.001717677 | 0.004980661 | 1 |
| T2DM | b_cell | *SLC25A21-AS1* | -2.145219642 | 0.003118668 | 0.00526397 | 1 |
| T2DM | b_cell | *RNASE4* | 3.21436576 | 0.003140946 | 0.005286116 | 1 |
| T2DM | b_cell | *LINC00639* | -1.21566435 | 0.001250688 | 0.004980661 | 1 |
| T2DM | b_cell | *SMIM6* | -16.78726876 | 0.009001278 | 0.011699491 | 1 |
| T2DM | b_cell | *LINC00924* | -1.105163326 | 0.018792473 | 0.022795269 | 1 |
| T2DM | b_cell | *MRPL46* | -2.904418428 | 0.007519953 | 0.010057005 | 1 |
| T2DM | b_cell | *UBE2Q2L* | 1.702743783 | 0.002253391 | 0.004980661 | 1 |
| T2DM | b_cell | *IQCH-AS1* | -1.815281284 | 0.00722798 | 0.009730898 | 1 |
| T2DM | b_cell | *AC083843.1* | 5.919634502 | 0.002677435 | 0.004998428 | 1 |
| T2DM | b_cell | *PWRN1* | -1.893380851 | 0.003015045 | 0.005206049 | 1 |
| T2DM | b_cell | *LINC01003* | 2.205299487 | 0.020362108 | 0.02457166 | 1 |
| T2DM | b_cell | *LINC00662* | -1.300089527 | 0.001930798 | 0.004980661 | 1 |
| T2DM | b_cell | *C19orf84* | -18.68442678 | 0.004626497 | 0.006790008 | 1 |
| T2DM | b_cell | *LINC00667* | -1.100168525 | 0.002348341 | 0.004980661 | 1 |
| T2DM | b_cell | *LINC00672* | -3.441744406 | 0.002064957 | 0.004980661 | 1 |
| T2DM | b_cell | *LINC00909* | -1.524543856 | 0.003906073 | 0.006010413 | 1 |
| T2DM | b_cell | *DYNLL2* | -3.984191828 | 0.01025437 | 0.013148573 | 1 |
| T2DM | b_cell | *STRADA* | -1.933066126 | 0.001495457 | 0.004980661 | 1 |
| T2DM | b_cell | *MYO15B* | -1.270015348 | 0.002873553 | 0.005094011 | 1 |
| T2DM | b_cell | *ILF3-AS1* | -0.993577649 | 0.002709869 | 0.00500969 | 1 |
| T2DM | b_cell | *ZNF582-AS1* | -2.120998712 | 0.001803346 | 0.004980661 | 1 |
| T2DM | b_cell | *S1PR2* | 0.620324221 | 0.0018588 | 0.004980661 | 1 |
| T2DM | b_cell | *ZNF224* | -1.220760855 | 0.001882219 | 0.004980661 | 1 |
| T2DM | b_cell | *LIN37* | -8.997041576 | 0.001391054 | 0.004980661 | 1 |
| T2DM | b_cell | *NDUFA7* | 0.803060812 | 0.002383085 | 0.004980661 | 1 |
| T2DM | b_cell | *SCGB1B2P* | -5.028872531 | 0.016108491 | 0.019781522 | 1 |
| T2DM | b_cell | *AC026740.1* | 1.040845279 | 0.002525388 | 0.004980661 | 1 |
| T2DM | b_cell | *ZNF728* | -1.852643862 | 0.002814612 | 0.005065458 | 1 |
| T2DM | b_cell | *TRABD2B* | -1.211543446 | 0.001954292 | 0.004980661 | 1 |
| T2DM | b_cell | *AC093157.1* | -2.180152924 | 0.005427061 | 0.007706205 | 1 |
| T2DM | b_cell | *FBXO17* | -0.938396153 | 0.003407994 | 0.005510766 | 1 |
| T2DM | b_cell | *ZNF587B* | -1.913031699 | 0.001751435 | 0.004980661 | 1 |
| T2DM | b_cell | *SNHG8* | -2.943661186 | 0.003768092 | 0.005865506 | 1 |
| T2DM | b_cell | *AP001172.2* | 1.451634106 | 0.000174056 | 0.004980661 | 0.844520598 |
| T2DM | b_cell | *HCG17* | -1.135746987 | 0.001870204 | 0.004980661 | 1 |
| T2DM | b_cell | *YTHDF3-AS1* | 0.612750446 | 0.004648317 | 0.006806954 | 1 |
| T2DM | b_cell | *MILR1* | -1.064373689 | 0.03049546 | 0.035826628 | 1 |
| T2DM | b_cell | *GTF2H5* | -1.627195003 | 0.04262129 | 0.049167498 | 1 |
| T2DM | b_cell | *FGF14-AS2* | -2.610125477 | 0.001559056 | 0.004980661 | 1 |
| T2DM | b_cell | *CASC15* | -0.91023222 | 0.001793043 | 0.004980661 | 1 |
| T2DM | b_cell | *AP000347.2* | 0.502526769 | 0.002735475 | 0.005019866 | 1 |
| T2DM | b_cell | *DOC2B* | -1.542699438 | 0.002120262 | 0.004980661 | 1 |
| T2DM | b_cell | *GRIN2B* | -0.861449378 | 0.006702821 | 0.009153416 | 1 |
| T2DM | b_cell | *AP000350.5* | -1.575914707 | 0.034683475 | 0.040491872 | 1 |
| T2DM | b_cell | *PAXIP1-AS1* | -1.264881799 | 0.001684898 | 0.004980661 | 1 |
| T2DM | dendritic_cell | *RBM6* | -2.333902841 | 2.50E-09 | 6.06E-06 | 1.21E-05 |
| T2DM | dendritic_cell | *ZNF195* | -3.00009277 | 0.000311382 | 0.025607234 | 1 |
| T2DM | dendritic_cell | *TSPAN9* | -1.536947399 | 0.000597475 | 0.037648691 | 1 |

| | | | | | | |
|---|---|---|---|---|---|---|
| T2DM | dendritic_cell | *TUBG2* | -1.727342735 | 7.23E-05 | 0.010708302 | 0.35097457 |
| T2DM | dendritic_cell | *FLT4* | -0.944248789 | 0.000999138 | 0.048967842 | 1 |
| T2DM | dendritic_cell | *ITIH4* | 0.724572129 | 1.68E-05 | 0.004080377 | 0.081607545 |
| T2DM | dendritic_cell | *PIGV* | -1.602345275 | 0.000737466 | 0.04112859 | 1 |
| T2DM | dendritic_cell | *SEC61A2* | -3.747503352 | 1.21E-05 | 0.003079247 | 0.058505686 |
| T2DM | dendritic_cell | *CD5L* | 1.848993741 | 0.000900198 | 0.045497525 | 1 |
| T2DM | dendritic_cell | *TACR2* | 10.08738194 | 0.000753033 | 0.041454274 | 1 |
| T2DM | dendritic_cell | *DCT* | -1.394932549 | 0.000707006 | 0.040357563 | 1 |
| T2DM | dendritic_cell | *PABPC4* | -5.136114056 | 2.96E-07 | 0.00019333 | 0.001433898 |
| T2DM | dendritic_cell | *ALKBH5* | -3.390002148 | 0.000216421 | 0.021001499 | 1 |
| T2DM | dendritic_cell | *CWF19L1* | -2.274275665 | 6.57E-06 | 0.002124638 | 0.031869576 |
| T2DM | dendritic_cell | *SCD* | -2.411226805 | 0.000125986 | 0.01455433 | 0.611281848 |
| T2DM | dendritic_cell | *NOMO3* | -1.974094315 | 8.37E-05 | 0.010971815 | 0.405957163 |
| T2DM | dendritic_cell | *TLE6* | 2.785261588 | 0.00035274 | 0.027363923 | 1 |
| T2DM | dendritic_cell | *GCDH* | -1.847843157 | 0.000223984 | 0.021309215 | 1 |
| T2DM | dendritic_cell | *RNF32* | -6.438750674 | 4.02E-05 | 0.006503466 | 0.195103975 |
| T2DM | dendritic_cell | *CHN2* | 1.580348783 | 0.000458312 | 0.032227978 | 1 |
| T2DM | dendritic_cell | *CNTNAP1* | 5.388307831 | 0.000266285 | 0.022884391 | 1 |
| T2DM | dendritic_cell | *CDCA3* | -3.55504156 | 0.000440192 | 0.031877803 | 1 |
| T2DM | dendritic_cell | *BVES* | -1.626434782 | 2.55E-05 | 0.00516173 | 0.123881531 |
| T2DM | dendritic_cell | *GPX7* | -1.316154738 | 3.72E-05 | 0.006453468 | 0.180697104 |
| T2DM | dendritic_cell | *MASTL* | -1.932116694 | 0.000760394 | 0.041454274 | 1 |
| T2DM | dendritic_cell | *TEX14* | 2.246756687 | 0.000898793 | 0.045497525 | 1 |
| T2DM | dendritic_cell | *PDZRN3* | -0.736118449 | 0.000831873 | 0.044354387 | 1 |
| T2DM | dendritic_cell | *RPS10* | -5.457804599 | 0.000589179 | 0.0376144 | 1 |
| T2DM | dendritic_cell | *KCNK17* | -4.099040487 | 3.62E-05 | 0.006453468 | 0.17547661 |
| T2DM | dendritic_cell | *XRCC3* | -3.221882357 | 0.000183675 | 0.019373716 | 0.891190948 |
| T2DM | dendritic_cell | *ABHD8* | -3.382616884 | 3.97E-05 | 0.006503466 | 0.192699128 |
| T2DM | dendritic_cell | *RCBTB1* | -2.014551866 | 0.00026884 | 0.022884391 | 1 |
| T2DM | dendritic_cell | *IRF4* | 0.505948484 | 0.000886931 | 0.045497525 | 1 |
| T2DM | dendritic_cell | *SYTL2* | -2.275392357 | 0.000194399 | 0.019650467 | 0.943222414 |
| T2DM | dendritic_cell | *SLC37A4* | 2.117609661 | 0.000308072 | 0.025607234 | 1 |
| T2DM | dendritic_cell | *SPTBN5* | 0.90008898 | 7.30E-05 | 0.010708302 | 0.354203106 |
| T2DM | dendritic_cell | *NEIL1* | -2.148258757 | 0.000107865 | 0.013419515 | 0.523361104 |
| T2DM | dendritic_cell | *MAN2C1* | -1.492942278 | 6.20E-07 | 0.000300773 | 0.003007728 |
| T2DM | dendritic_cell | *VPS53* | -2.742891576 | 7.50E-05 | 0.010708302 | 0.364082269 |
| T2DM | dendritic_cell | *C18orf8* | -1.941577125 | 0.000149239 | 0.01609133 | 0.724109868 |
| T2DM | dendritic_cell | *SAE1* | -2.39204503 | 0.000566862 | 0.037167783 | 1 |
| T2DM | dendritic_cell | *HSD17B12* | -1.667542539 | 7.70E-09 | 8.48E-06 | 3.74E-05 |
| T2DM | dendritic_cell | *APIP* | -1.497056113 | 1.11E-05 | 0.002993213 | 0.053877829 |
| T2DM | dendritic_cell | *POC5* | -2.495906192 | 2.01E-05 | 0.00424378 | 0.097606935 |
| T2DM | dendritic_cell | *JAZF1* | -3.769477671 | 8.01E-10 | 3.89E-06 | 3.89E-06 |
| T2DM | dendritic_cell | *NDUFAF6* | -2.142906025 | 0.000336901 | 0.026797462 | 1 |
| T2DM | dendritic_cell | *CLEC18A* | -4.27435355 | 8.74E-09 | 8.48E-06 | 4.24E-05 |
| T2DM | dendritic_cell | *SPTA1* | -2.223506927 | 4.65E-05 | 0.007278644 | 0.22563797 |
| T2DM | dendritic_cell | *RFT1* | -2.795597307 | 9.80E-07 | 0.000421727 | 0.004753819 |
| T2DM | dendritic_cell | *PACSIN3* | -1.552623613 | 0.000647065 | 0.03924449 | 1 |
| T2DM | dendritic_cell | *ANPEP* | 0.587007185 | 0.000585489 | 0.0376144 | 1 |
| T2DM | dendritic_cell | *PDHB* | -5.237319007 | 0.000619034 | 0.038019631 | 1 |
| T2DM | dendritic_cell | *SERINC2* | -4.26723478 | 0.000538239 | 0.037167783 | 1 |

| | | | | | | |
|---|---|---|---|---|---|---|
| T2DM | dendritic_cell | *IRS1* | -1.082660018 | 5.21E-07 | 0.000280925 | 0.002528326 |
| T2DM | dendritic_cell | *INO80E* | -2.046614031 | 1.89E-05 | 0.00424378 | 0.091829409 |
| T2DM | dendritic_cell | *LRRC45* | -1.570276235 | 0.000872178 | 0.045497525 | 1 |
| T2DM | dendritic_cell | *TTLL6* | -6.101647005 | 3.19E-07 | 0.00019333 | 0.001546637 |
| T2DM | dendritic_cell | *C1QTNF4* | -1.590343203 | 1.97E-05 | 0.00424378 | 0.095499309 |
| T2DM | dendritic_cell | *CTSW* | -0.546901263 | 8.10E-05 | 0.010970251 | 0.393032436 |
| T2DM | dendritic_cell | *SNX32* | 1.626996048 | 0.000117902 | 0.01395266 | 0.572059045 |
| T2DM | dendritic_cell | *RNASEH2C* | -1.185385224 | 0.000239055 | 0.021884782 | 1 |
| T2DM | dendritic_cell | *HOXB2* | -1.941651323 | 0.000552759 | 0.037167783 | 1 |
| T2DM | dendritic_cell | *CCNE2* | -4.326777239 | 1.04E-06 | 0.000421727 | 0.005060729 |
| T2DM | dendritic_cell | *C11orf68* | -5.115181225 | 0.00093089 | 0.046563694 | 1 |
| T2DM | dendritic_cell | *SPRYD4* | -1.923301275 | 0.000229606 | 0.021424048 | 1 |
| T2DM | dendritic_cell | *SLC25A22* | -2.765093612 | 0.000396329 | 0.029584436 | 1 |
| T2DM | dendritic_cell | *GEMIN4* | -4.030491195 | 0.000550262 | 0.037167783 | 1 |
| T2DM | dendritic_cell | *PIGP* | -0.999905577 | 0.000265391 | 0.022884391 | 1 |
| T2DM | dendritic_cell | *RASA3* | -1.682609949 | 0.000199277 | 0.019732507 | 0.966892844 |
| T2DM | dendritic_cell | *DNAH10* | -2.645328106 | 0.000674474 | 0.039909148 | 1 |
| T2DM | dendritic_cell | *TMEM116* | -1.186025592 | 0.000190513 | 0.019650467 | 0.924370353 |
| T2DM | dendritic_cell | *HLA-DRB5* | -0.457804293 | 0.000360942 | 0.027363923 | 1 |
| T2DM | dendritic_cell | *MAP3K3* | -2.144625719 | 0.000700484 | 0.040357563 | 1 |
| T2DM | dendritic_cell | *NOTCH4* | -0.468457229 | 5.71E-09 | 8.48E-06 | 2.77E-05 |
| T2DM | dendritic_cell | *PRRT1* | 1.87180847 | 0.000112053 | 0.013592027 | 0.54368109 |
| T2DM | dendritic_cell | *STK19* | -4.005069908 | 3.68E-05 | 0.006453468 | 0.178769836 |
| T2DM | dendritic_cell | *SKIV2L* | -1.414387422 | 0.000149213 | 0.01609133 | 0.723983159 |
| T2DM | dendritic_cell | *GPANK1* | -4.368469884 | 2.00E-06 | 0.000747573 | 0.009718449 |
| T2DM | dendritic_cell | *FAM153C* | -0.951053093 | 0.000720728 | 0.040662468 | 1 |
| T2DM | dendritic_cell | *HCG27* | 1.792131984 | 0.000561171 | 0.037167783 | 1 |
| T2DM | dendritic_cell | *DNLZ* | 1.113097301 | 0.000614102 | 0.038019631 | 1 |
| T2DM | dendritic_cell | *TMEM110* | 5.343724143 | 2.58E-06 | 0.000892583 | 0.012496168 |
| T2DM | dendritic_cell | *STARD10* | 1.652057761 | 2.76E-05 | 0.005355752 | 0.133893804 |
| T2DM | dendritic_cell | *IFRD2* | -3.211832551 | 8.14E-05 | 0.010970251 | 0.39492903 |
| T2DM | dendritic_cell | *AC016773.1* | -2.50086754 | 0.000438197 | 0.031877803 | 1 |
| T2DM | dendritic_cell | *C4B* | 4.737314335 | 7.68E-06 | 0.002328529 | 0.037256471 |
| T2DM | dendritic_cell | *AL049840.1* | -2.673442243 | 0.000953852 | 0.047225402 | 1 |
| T2DM | dendritic_cell | *AC108463.2* | 4.290993704 | 0.000252817 | 0.022716092 | 1 |
| T2DM | dendritic_cell | *HLA-DQB2* | 0.289375896 | 8.63E-05 | 0.011018612 | 0.41870725 |
| T2DM | dendritic_cell | *LINC00957* | -3.453240599 | 0.000869052 | 0.045497525 | 1 |
| T2DM | dendritic_cell | *LINC00941* | -5.250795132 | 0.000828789 | 0.044354387 | 1 |
| T2DM | dendritic_cell | *HLA-DQA2* | 0.590754172 | 2.73E-08 | 2.21E-05 | 0.000132403 |
| T2DM | dendritic_cell | *PSMB9* | 1.568851366 | 0.000451065 | 0.032184803 | 1 |
| T2DM | dendritic_cell | *PLA2G4B* | 1.485501411 | 0.000667999 | 0.039909148 | 1 |
| T2DM | dendritic_cell | *JMJD7* | 1.219538472 | 8.45E-06 | 0.002412735 | 0.041016494 |
| T2DM | dendritic_cell | *C4A* | -3.649524221 | 0.000135783 | 0.015321424 | 0.658821232 |
| T2DM | dendritic_cell | *SRP14-AS1* | -0.995714603 | 0.000360164 | 0.027363923 | 1 |
| T2DM | dendritic_cell | *DNAH10OS* | -9.47315233 | 0.000693638 | 0.040357563 | 1 |
| T2DM | dendritic_cell | *SNHG8* | -3.111850937 | 0.000317582 | 0.025681834 | 1 |
| T2DM | endometrium | *NFYA* | 2.956555709 | 3.39E-07 | 8.27E-07 | 0.001646593 |
| T2DM | endometrium | *ENPP4* | 4.634742145 | 2.47E-07 | 6.54E-07 | 0.001196355 |
| T2DM | endometrium | *SEMA3F* | 2.955486447 | 1.16E-05 | 1.85E-05 | 0.056238411 |
| T2DM | endometrium | *RAD52* | 5.390727071 | 4.13E-07 | 9.66E-07 | 0.002005469 |

| | | | | | | |
|---|---|---|---|---|---|---|
| T2DM | endometrium | *HS3ST1* | 4.124498867 | 0.000972331 | 0.001252057 | 1 |
| T2DM | endometrium | *AOC1* | 5.372419778 | 3.26E-05 | 4.89E-05 | 0.158288807 |
| T2DM | endometrium | *SNX11* | 2.53605456 | 5.11E-09 | 4.20E-07 | 2.48E-05 |
| T2DM | endometrium | *TMEM176A* | 5.623863511 | 4.74E-08 | 4.34E-07 | 0.000229995 |
| T2DM | endometrium | *M6PR* | 4.831004859 | 2.08E-08 | 4.34E-07 | 0.00010105 |
| T2DM | endometrium | *CYP26B1* | 4.077710071 | 6.01E-08 | 4.34E-07 | 0.000291543 |
| T2DM | endometrium | *ALS2* | 6.332179246 | 9.89E-06 | 1.60E-05 | 0.047998801 |
| T2DM | endometrium | *CD38* | 5.29964439 | 4.42E-08 | 4.34E-07 | 0.000214287 |
| T2DM | endometrium | *RECQL* | 6.618729159 | 1.14E-05 | 1.83E-05 | 0.055298765 |
| T2DM | endometrium | *ARHGAP33* | 2.836847943 | 2.75E-05 | 4.15E-05 | 0.133382276 |
| T2DM | endometrium | *CDC27* | 3.140809515 | 3.58E-08 | 4.34E-07 | 0.000173842 |
| T2DM | endometrium | *POLR2J* | 2.98634806 | 1.60E-07 | 4.86E-07 | 0.000774676 |
| T2DM | endometrium | *MEOX1* | -0.998723922 | 4.07E-06 | 7.13E-06 | 0.019745586 |
| T2DM | endometrium | *LIG3* | -2.128359782 | 0.000214668 | 0.000293566 | 1 |
| T2DM | endometrium | *CIAPIN1* | 2.713041832 | 6.88E-08 | 4.34E-07 | 0.000333875 |
| T2DM | endometrium | *PON1* | 10.27063736 | 4.22E-08 | 4.34E-07 | 0.000204856 |
| T2DM | endometrium | *GCFC2* | 3.778976027 | 5.29E-08 | 4.34E-07 | 0.000256764 |
| T2DM | endometrium | *ZNF195* | 3.24320788 | 0.008156121 | 0.009749569 | 1 |
| T2DM | endometrium | *PDK2* | 2.585786851 | 0.007716918 | 0.009254198 | 1 |
| T2DM | endometrium | *GGNBP2* | 4.714503857 | 0.000146586 | 0.000203268 | 0.711233217 |
| T2DM | endometrium | *ITGA2B* | 3.611236005 | 1.43E-07 | 4.78E-07 | 0.000695387 |
| T2DM | endometrium | *ASB4* | -1.68383903 | 0.001498232 | 0.001898021 | 1 |
| T2DM | endometrium | *TMEM98* | 2.5794675 | 3.78E-07 | 9.01E-07 | 0.00183352 |
| T2DM | endometrium | *ABCC8* | 4.120268032 | 8.07E-05 | 0.000114822 | 0.391773205 |
| T2DM | endometrium | *SYNRG* | 3.106535651 | 6.94E-08 | 4.34E-07 | 0.000336768 |
| T2DM | endometrium | *ZNF263* | 3.396217348 | 4.65E-08 | 4.34E-07 | 0.000225789 |
| T2DM | endometrium | *SPATA20* | 2.851107723 | 2.38E-07 | 6.36E-07 | 0.001154929 |
| T2DM | endometrium | *MAP3K9* | 1.635840597 | 0.022833755 | 0.026390991 | 1 |
| T2DM | endometrium | *BAIAP2L1* | 3.307005814 | 7.52E-08 | 4.36E-07 | 0.000364732 |
| T2DM | endometrium | *FARP2* | 2.817043676 | 6.71E-08 | 4.34E-07 | 0.000325466 |
| T2DM | endometrium | *USH1C* | -7.743826171 | 2.57E-06 | 4.69E-06 | 0.012468296 |
| T2DM | endometrium | *IFRD1* | 2.817816385 | 8.75E-07 | 1.81E-06 | 0.004243992 |
| T2DM | endometrium | *VPS41* | 4.936737756 | 1.94E-07 | 5.49E-07 | 0.000941066 |
| T2DM | endometrium | *ELAC2* | 4.874868612 | 1.58E-06 | 3.05E-06 | 0.00766815 |
| T2DM | endometrium | *CCDC124* | 3.036527683 | 3.96E-07 | 9.35E-07 | 0.001922044 |
| T2DM | endometrium | *CEACAM21* | 2.937425144 | 4.10E-05 | 6.07E-05 | 0.199152495 |
| T2DM | endometrium | *NOS2* | 7.014532243 | 0.007139395 | 0.008584968 | 1 |
| T2DM | endometrium | *CD79B* | -1.766312091 | 4.82E-07 | 1.09E-06 | 0.002339897 |
| T2DM | endometrium | *ST7L* | 2.244612483 | 9.86E-06 | 1.60E-05 | 0.047860375 |
| T2DM | endometrium | *PAX6* | 2.423876794 | 0.002874799 | 0.003558298 | 1 |
| T2DM | endometrium | *TSR3* | 3.49973926 | 1.96E-07 | 5.53E-07 | 0.00095278 |
| T2DM | endometrium | *PIGQ* | 5.742036227 | 0.000153953 | 0.000213179 | 0.746977649 |
| T2DM | endometrium | *FMO3* | 2.53123065 | 6.97E-06 | 1.16E-05 | 0.033813991 |
| T2DM | endometrium | *PSMB1* | 5.54987386 | 7.00E-08 | 4.34E-07 | 0.000339672 |
| T2DM | endometrium | *NADK* | 2.284802069 | 1.37E-07 | 4.78E-07 | 0.00066266 |
| T2DM | endometrium | *DLEC1* | -2.245781913 | 7.31E-05 | 0.000104508 | 0.354806201 |
| T2DM | endometrium | *CYB561* | 5.290930581 | 1.30E-09 | 2.63E-07 | 6.29E-06 |
| T2DM | endometrium | *SS18L2* | 4.779522281 | 9.29E-09 | 4.34E-07 | 4.51E-05 |
| T2DM | endometrium | *MGST1* | 1.964696357 | 2.46E-08 | 4.34E-07 | 0.00011956 |
| T2DM | endometrium | *MMP25* | -2.435291696 | 0.001071495 | 0.001375005 | 1 |

| | | | | | | |
|---|---|---|---|---|---|---|
| T2DM | endometrium | *IL32* | 3.002087436 | 8.58E-08 | 4.38E-07 | 0.000416208 |
| T2DM | endometrium | *PKD1* | 4.230056857 | 1.68E-08 | 4.34E-07 | 8.17E-05 |
| T2DM | endometrium | *MAPK8IP2* | 10.79504435 | 8.43E-05 | 0.000119663 | 0.409008064 |
| T2DM | endometrium | *MED24* | 2.227271772 | 1.50E-07 | 4.78E-07 | 0.000728297 |
| T2DM | endometrium | *RPS20* | 5.323472434 | 3.68E-06 | 6.49E-06 | 0.017877345 |
| T2DM | endometrium | *TRAF3IP3* | 2.948224741 | 5.34E-09 | 4.27E-07 | 2.59E-05 |
| T2DM | endometrium | *POMT2* | 2.402970368 | 8.29E-07 | 1.73E-06 | 0.004023307 |
| T2DM | endometrium | *MLXIPL* | 1.45937023 | 2.14E-06 | 3.99E-06 | 0.010364165 |
| T2DM | endometrium | *DYRK4* | 3.71035602 | 2.00E-08 | 4.34E-07 | 9.69E-05 |
| T2DM | endometrium | *FUZ* | 3.920170495 | 6.19E-08 | 4.34E-07 | 0.000300567 |
| T2DM | endometrium | *SLC6A13* | 4.326590789 | 4.53E-05 | 6.66E-05 | 0.219873446 |
| T2DM | endometrium | *ZNF200* | 3.314860229 | 5.41E-08 | 4.34E-07 | 0.000262421 |
| T2DM | endometrium | *LRRC23* | 5.23053862 | 1.03E-06 | 2.09E-06 | 0.005003532 |
| T2DM | endometrium | *CLCN6* | 2.63223449 | 3.56E-07 | 8.59E-07 | 0.001729572 |
| T2DM | endometrium | *SLC6A7* | 77.81671983 | 7.88E-06 | 1.29E-05 | 0.038233571 |
| T2DM | endometrium | *TSPAN9* | 2.611023091 | 2.31E-05 | 3.51E-05 | 0.111994568 |
| T2DM | endometrium | *UTP18* | 3.45474762 | 4.63E-08 | 4.34E-07 | 0.000224797 |
| T2DM | endometrium | *TTC19* | 4.144690312 | 1.15E-07 | 4.59E-07 | 0.00055882 |
| T2DM | endometrium | *DPF1* | 6.892779966 | 0.000231564 | 0.00031587 | 1 |
| T2DM | endometrium | *PPP5C* | 4.985537082 | 9.76E-09 | 4.34E-07 | 4.73E-05 |
| T2DM | endometrium | *CEP68* | 6.27472717 | 3.11E-06 | 5.57E-06 | 0.01510019 |
| T2DM | endometrium | *ZBTB32* | 3.022922716 | 8.08E-08 | 4.36E-07 | 0.000392058 |
| T2DM | endometrium | *ERCC1* | 3.585607751 | 4.59E-08 | 4.34E-07 | 0.000222501 |
| T2DM | endometrium | *ALOX5* | 1.977695651 | 7.73E-07 | 1.63E-06 | 0.003750641 |
| T2DM | endometrium | *MAP4K5* | 4.920439177 | 1.19E-06 | 2.36E-06 | 0.00576132 |
| T2DM | endometrium | *EHD3* | -1.841142939 | 4.90E-06 | 8.42E-06 | 0.023796446 |
| T2DM | endometrium | *MAN2B2* | 3.738384445 | 1.03E-07 | 4.47E-07 | 0.000497544 |
| T2DM | endometrium | *SLC25A39* | 2.732761695 | 3.33E-07 | 8.15E-07 | 0.001614485 |
| T2DM | endometrium | *RNF14* | 5.319943383 | 3.10E-08 | 4.34E-07 | 0.000150566 |
| T2DM | endometrium | *DDX11* | 3.086294265 | 1.38E-05 | 2.18E-05 | 0.066938771 |
| T2DM | endometrium | *TACC3* | 2.944113534 | 1.77E-07 | 5.19E-07 | 0.000859821 |
| T2DM | endometrium | *ZC3H3* | 3.843529856 | 4.97E-07 | 1.12E-06 | 0.002411807 |
| T2DM | endometrium | *MDH1* | 2.541689143 | 2.71E-07 | 6.97E-07 | 0.001314645 |
| T2DM | endometrium | *CCDC88C* | 2.094459488 | 8.13E-07 | 1.70E-06 | 0.003944967 |
| T2DM | endometrium | *NUDCD3* | 4.12830392 | 2.35E-05 | 3.58E-05 | 0.114102352 |
| T2DM | endometrium | *CHDH* | 6.357928148 | 5.80E-08 | 4.34E-07 | 0.000281524 |
| T2DM | endometrium | *CLCA4* | 9.650535352 | 7.75E-05 | 0.000110535 | 0.376260468 |
| T2DM | endometrium | *RUFY3* | 4.884912341 | 2.26E-07 | 6.09E-07 | 0.00109478 |
| T2DM | endometrium | *SLC11A1* | 1.41219727 | 0.000565068 | 0.000744019 | 1 |
| T2DM | endometrium | *WWTR1* | 4.594885699 | 1.33E-07 | 4.78E-07 | 0.000646527 |
| T2DM | endometrium | *AGPS* | 4.922815941 | 1.36E-07 | 4.78E-07 | 0.000657512 |
| T2DM | endometrium | *TTC27* | 3.180244198 | 6.28E-08 | 4.34E-07 | 0.000304515 |
| T2DM | endometrium | *MARCO* | 1.551436288 | 1.21E-07 | 4.69E-07 | 0.000588238 |
| T2DM | endometrium | *HGF* | 3.326120776 | 6.48E-08 | 4.34E-07 | 0.000314351 |
| T2DM | endometrium | *SLC7A9* | 1.655314121 | 0.011434275 | 0.013528189 | 1 |
| T2DM | endometrium | *OSBPL5* | 4.435357286 | 9.64E-09 | 4.34E-07 | 4.68E-05 |
| T2DM | endometrium | *CPS1* | 3.256233146 | 0.005501868 | 0.006665434 | 1 |
| T2DM | endometrium | *SLC45A4* | 3.040192659 | 1.75E-06 | 3.33E-06 | 0.008473472 |
| T2DM | endometrium | *ZDHHC6* | 2.477214524 | 7.52E-08 | 4.36E-07 | 0.000365058 |
| T2DM | endometrium | *GRAMD1B* | 1.893858557 | 5.47E-06 | 9.29E-06 | 0.026531867 |

| | | | | | | |
|---|---|---|---|---|---|---|
| T2DM | endometrium | *RNH1* | 5.134162056 | 1.06E-07 | 4.48E-07 | 0.000512188 |
| T2DM | endometrium | *GLRX2* | 3.113375488 | 1.92E-07 | 5.47E-07 | 0.00093339 |
| T2DM | endometrium | *ABCC2* | 3.99817264 | 2.86E-07 | 7.27E-07 | 0.001389633 |
| T2DM | endometrium | *GCLM* | 2.29457016 | 1.54E-07 | 4.79E-07 | 0.000749021 |
| T2DM | endometrium | *UBR2* | 2.302013173 | 1.04E-07 | 4.47E-07 | 0.000503328 |
| T2DM | endometrium | *RRAGD* | 3.195941489 | 1.20E-07 | 4.67E-07 | 0.000584255 |
| T2DM | endometrium | *HSF2* | 2.361342099 | 5.15E-08 | 4.34E-07 | 0.000249726 |
| T2DM | endometrium | *TYMP* | 2.032947757 | 4.28E-07 | 9.89E-07 | 0.002078519 |
| T2DM | endometrium | *TOMM34* | 3.434289486 | 2.86E-07 | 7.27E-07 | 0.001389276 |
| T2DM | endometrium | *SEC63* | 3.718412495 | 6.37E-08 | 4.34E-07 | 0.000309298 |
| T2DM | endometrium | *FAS* | 3.095348246 | 8.56E-08 | 4.38E-07 | 0.00041551 |
| T2DM | endometrium | *AGPAT4* | 2.349014885 | 9.82E-08 | 4.42E-07 | 0.000476588 |
| T2DM | endometrium | *BTN3A1* | 2.342450389 | 6.64E-05 | 9.54E-05 | 0.322164291 |
| T2DM | endometrium | *INSRR* | -4.025556334 | 1.48E-05 | 2.31E-05 | 0.071627414 |
| T2DM | endometrium | *B4GALT7* | 3.917510929 | 2.32E-07 | 6.23E-07 | 0.001125802 |
| T2DM | endometrium | *SH2D2A* | 9.603063396 | 3.90E-08 | 4.34E-07 | 0.000189158 |
| T2DM | endometrium | *VRK2* | 5.323877078 | 2.37E-06 | 4.37E-06 | 0.011486713 |
| T2DM | endometrium | *VEZT* | 4.917576852 | 2.27E-07 | 6.12E-07 | 0.001101719 |
| T2DM | endometrium | *BRD9* | 5.665667607 | 5.74E-05 | 8.32E-05 | 0.278502094 |
| T2DM | endometrium | *SNX1* | 2.135657308 | 1.13E-07 | 4.57E-07 | 0.000547815 |
| T2DM | endometrium | *ANK1* | 3.987101934 | 1.38E-07 | 4.78E-07 | 0.000671338 |
| T2DM | endometrium | *RABEP1* | 2.773120572 | 5.07E-08 | 4.34E-07 | 0.000245924 |
| T2DM | endometrium | *BAK1* | 4.672956884 | 5.20E-06 | 8.89E-06 | 0.025250188 |
| T2DM | endometrium | *IKZF2* | 5.867014277 | 3.25E-07 | 8.00E-07 | 0.001575621 |
| T2DM | endometrium | *GRN* | 1.645715762 | 1.27E-07 | 4.76E-07 | 0.000616241 |
| T2DM | endometrium | *CENPQ* | 1.238019606 | 0.00315604 | 0.003890525 | 1 |
| T2DM | endometrium | *IFT88* | 3.621335908 | 3.61E-07 | 8.69E-07 | 0.001753146 |
| T2DM | endometrium | *UBA6* | 3.591763038 | 0.000677018 | 0.000886132 | 1 |
| T2DM | endometrium | *ATP6V0A1* | 3.807369707 | 6.55E-09 | 4.34E-07 | 3.18E-05 |
| T2DM | endometrium | *PIAS1* | 3.16378488 | 2.98E-08 | 4.34E-07 | 0.000144575 |
| T2DM | endometrium | *MYOC* | 2.919456896 | 4.56E-08 | 4.34E-07 | 0.000221244 |
| T2DM | endometrium | *SH3YL1* | 2.569042017 | 9.37E-07 | 1.92E-06 | 0.004548408 |
| T2DM | endometrium | *ADSS* | 2.865659598 | 7.13E-08 | 4.34E-07 | 0.000345973 |
| T2DM | endometrium | *TBC1D23* | 5.789064365 | 1.12E-06 | 2.24E-06 | 0.005426049 |
| T2DM | endometrium | *MYOM2* | 4.817745907 | 9.08E-07 | 1.87E-06 | 0.004403803 |
| T2DM | endometrium | *TUBG2* | -4.359994146 | 7.91E-07 | 1.66E-06 | 0.00383561 |
| T2DM | endometrium | *FLT4* | 1.36964933 | 0.0028715 | 0.003555121 | 1 |
| T2DM | endometrium | *NSUN2* | 5.568646772 | 2.47E-07 | 6.55E-07 | 0.001199969 |
| T2DM | endometrium | *MFAP3* | 4.091820382 | 1.31E-06 | 2.57E-06 | 0.006359915 |
| T2DM | endometrium | *AGA* | 4.993120508 | 1.11E-07 | 4.57E-07 | 0.000538195 |
| T2DM | endometrium | *PI4K2B* | 5.923162987 | 1.39E-07 | 4.78E-07 | 0.000675802 |
| T2DM | endometrium | *BOD1L1* | 4.22515894 | 4.35E-07 | 1.00E-06 | 0.002110905 |
| T2DM | endometrium | *DNAH5* | 3.934368891 | 1.16E-07 | 4.59E-07 | 0.000560797 |
| T2DM | endometrium | *PNKP* | 2.766489571 | 1.03E-05 | 1.66E-05 | 0.050092169 |
| T2DM | endometrium | *PHLPP2* | 1.958450715 | 1.38E-06 | 2.70E-06 | 0.006703828 |
| T2DM | endometrium | *PQLC2* | -5.114167176 | 0.000657214 | 0.00086114 | 1 |
| T2DM | endometrium | *CTNS* | 3.615964727 | 5.19E-05 | 7.56E-05 | 0.251868546 |
| T2DM | endometrium | *INPP4A* | 1.959886174 | 1.71E-07 | 5.06E-07 | 0.000828094 |
| T2DM | endometrium | *PSMA4* | 3.502773858 | 1.03E-07 | 4.47E-07 | 0.000500939 |
| T2DM | endometrium | *LSG1* | 1.795984267 | 2.06E-06 | 3.87E-06 | 0.009995961 |

| | | | | | | |
|---|---|---|---|---|---|---|
| T2DM | endometrium | *TDP1* | 4.53842273 | 7.17E-08 | 4.34E-07 | 0.000347874 |
| T2DM | endometrium | *MED17* | 2.571236208 | 1.40E-07 | 4.78E-07 | 0.000679325 |
| T2DM | endometrium | *CAPG* | 2.492013913 | 6.28E-06 | 1.05E-05 | 0.030469409 |
| T2DM | endometrium | *AP2S1* | 3.766788139 | 8.90E-08 | 4.38E-07 | 0.000431762 |
| T2DM | endometrium | *TRIT1* | 2.337853707 | 8.03E-07 | 1.68E-06 | 0.003894858 |
| T2DM | endometrium | *CUL7* | 5.560532649 | 7.12E-08 | 4.34E-07 | 0.000345623 |
| T2DM | endometrium | *CTNNA1* | 3.988562566 | 7.98E-08 | 4.36E-07 | 0.000387186 |
| T2DM | endometrium | *CNTLN* | 2.902892314 | 1.51E-07 | 4.78E-07 | 0.000730515 |
| T2DM | endometrium | *YTHDC2* | 3.311711729 | 5.05E-08 | 4.34E-07 | 0.000245172 |
| T2DM | endometrium | *POLR2B* | 2.754763286 | 7.71E-08 | 4.36E-07 | 0.000373926 |
| T2DM | endometrium | *ARAP2* | 2.52939878 | 4.83E-08 | 4.34E-07 | 0.000234218 |
| T2DM | endometrium | *DTNBP1* | 4.353377463 | 0.003601021 | 0.004411046 | 1 |
| T2DM | endometrium | *ANO2* | 1.757542574 | 1.38E-07 | 4.78E-07 | 0.00066812 |
| T2DM | endometrium | *SNX29* | 1.73063712 | 1.22E-07 | 4.69E-07 | 0.000589878 |
| T2DM | endometrium | *MRPS10* | 1.539762298 | 0.000163462 | 0.000225896 | 0.793119728 |
| T2DM | endometrium | *UTS2* | 4.536590934 | 1.43E-05 | 2.25E-05 | 0.069555242 |
| T2DM | endometrium | *RCN1* | 1.977238158 | 4.57E-07 | 1.04E-06 | 0.002215277 |
| T2DM | endometrium | *JKAMP* | 4.88267968 | 0.000238958 | 0.000325589 | 1 |
| T2DM | endometrium | *ARHGEF5* | 4.288028157 | 0.000514587 | 0.000679396 | 1 |
| T2DM | endometrium | *MCUR1* | 4.036332124 | 5.78E-08 | 4.34E-07 | 0.000280664 |
| T2DM | endometrium | *LAMC3* | -0.614881084 | 1.07E-06 | 2.16E-06 | 0.005215598 |
| T2DM | endometrium | *MAPK9* | 6.570405827 | 0.000132444 | 0.000184396 | 0.642619505 |
| T2DM | endometrium | *FAM160A2* | 4.056514812 | 5.32E-08 | 4.34E-07 | 0.000258206 |
| T2DM | endometrium | *CYBA* | 2.995705452 | 5.46E-08 | 4.34E-07 | 0.000265033 |
| T2DM | endometrium | *THOC3* | 17.99322227 | 1.88E-05 | 2.91E-05 | 0.09144049 |
| T2DM | endometrium | *ALX4* | 4.224164184 | 6.83E-05 | 9.79E-05 | 0.331618737 |
| T2DM | endometrium | *FSTL4* | 3.295672192 | 2.20E-05 | 3.36E-05 | 0.106887894 |
| T2DM | endometrium | *AKR7A2* | 7.071431554 | 6.88E-08 | 4.34E-07 | 0.000333592 |
| T2DM | endometrium | *MRTO4* | 2.896125353 | 8.33E-07 | 1.74E-06 | 0.004040492 |
| T2DM | endometrium | *ANAPC4* | 3.032693619 | 3.33E-08 | 4.34E-07 | 0.000161814 |
| T2DM | endometrium | *LY75* | 5.386549263 | 8.38E-08 | 4.38E-07 | 0.000406755 |
| T2DM | endometrium | *OPN3* | 4.147681175 | 0.000414937 | 0.000552793 | 1 |
| T2DM | endometrium | *SDCCAG8* | 3.478367508 | 2.22E-07 | 6.02E-07 | 0.00107848 |
| T2DM | endometrium | *KIF1B* | 5.1210401 | 5.05E-07 | 1.13E-06 | 0.002447856 |
| T2DM | endometrium | *TBC1D22A* | 3.307376253 | 3.87E-08 | 4.34E-07 | 0.000187564 |
| T2DM | endometrium | *PLEKHH1* | -3.441858443 | 0.000836804 | 0.001085608 | 1 |
| T2DM | endometrium | *GALC* | -2.515817905 | 0.021491405 | 0.024869139 | 1 |
| T2DM | endometrium | *FAM114A2* | 4.722150142 | 4.89E-08 | 4.34E-07 | 0.000237377 |
| T2DM | endometrium | *GINM1* | 3.839143867 | 8.46E-08 | 4.38E-07 | 0.000410334 |
| T2DM | endometrium | *ITIH4* | 2.830018021 | 3.53E-08 | 4.34E-07 | 0.00017106 |
| T2DM | endometrium | *NPFFR2* | 4.737182807 | 1.60E-07 | 4.86E-07 | 0.000774862 |
| T2DM | endometrium | *PCGF2* | 2.771794528 | 2.11E-07 | 5.81E-07 | 0.001021738 |
| T2DM | endometrium | *IL17RB* | 6.058238317 | 2.22E-05 | 3.39E-05 | 0.107863951 |
| T2DM | endometrium | *MSH4* | 1.773612895 | 2.44E-05 | 3.70E-05 | 0.118274986 |
| T2DM | endometrium | *GDI2* | 3.372799729 | 5.88E-08 | 4.34E-07 | 0.000285115 |
| T2DM | endometrium | *PITHD1* | 4.402306769 | 2.96E-07 | 7.45E-07 | 0.001437234 |
| T2DM | endometrium | *ATP11B* | 3.347230175 | 2.35E-08 | 4.34E-07 | 0.000114154 |
| T2DM | endometrium | *LAMC2* | -1.922615049 | 0.01660615 | 0.019382497 | 1 |
| T2DM | endometrium | *RASGRF1* | 4.919869051 | 7.32E-05 | 0.00010452 | 0.354951409 |
| T2DM | endometrium | *CROCC* | 2.006890633 | 0.023845447 | 0.027527507 | 1 |

| | | | | | | |
|---|---|---|---|---|---|---|
| T2DM | endometrium | *ALDH18A1* | -3.148328979 | 0.000745963 | 0.000970873 | 1 |
| T2DM | endometrium | *DNAJC25* | 3.298761944 | 1.44E-07 | 4.78E-07 | 0.000698758 |
| T2DM | endometrium | *GNA15* | 2.964943027 | 1.62E-06 | 3.11E-06 | 0.007840317 |
| T2DM | endometrium | *PIGV* | -3.028521387 | 2.52E-06 | 4.61E-06 | 0.012221649 |
| T2DM | endometrium | *RIMBP2* | 3.119814638 | 1.51E-07 | 4.78E-07 | 0.000732892 |
| T2DM | endometrium | *PRDM6* | -2.27767263 | 4.91E-05 | 7.18E-05 | 0.238213151 |
| T2DM | endometrium | *SPAG4* | 5.382216617 | 0.006455051 | 0.007789084 | 1 |
| T2DM | endometrium | *NCKAP1* | 3.270525585 | 9.05E-08 | 4.38E-07 | 0.000439191 |
| T2DM | endometrium | *LTK* | 23.8335465 | 1.24E-07 | 4.71E-07 | 0.0006015 |
| T2DM | endometrium | *EIF4B* | 3.311671187 | 0.003815108 | 0.004660349 | 1 |
| T2DM | endometrium | *SLC6A16* | 2.497816574 | 3.92E-08 | 4.34E-07 | 0.000190072 |
| T2DM | endometrium | *AHRR* | 8.71403885 | 1.65E-07 | 4.97E-07 | 0.000802873 |
| T2DM | endometrium | *ADCK1* | 2.458763344 | 1.51E-07 | 4.78E-07 | 0.000733783 |
| T2DM | endometrium | *HAGH* | 3.533028615 | 5.33E-06 | 9.07E-06 | 0.025848436 |
| T2DM | endometrium | *RNF4* | 4.267818643 | 4.96E-08 | 4.34E-07 | 0.000240682 |
| T2DM | endometrium | *CASP8* | 6.164757492 | 7.62E-05 | 0.000108688 | 0.36975527 |
| T2DM | endometrium | *TM7SF3* | 2.527786295 | 2.46E-08 | 4.34E-07 | 0.000119377 |
| T2DM | endometrium | *CDON* | 1.218131143 | 1.81E-07 | 5.23E-07 | 0.00087639 |
| T2DM | endometrium | *TMEM161A* | 3.174859493 | 9.65E-08 | 4.42E-07 | 0.000467998 |
| T2DM | endometrium | *LPAR2* | 4.861253186 | 4.00E-08 | 4.34E-07 | 0.000194245 |
| T2DM | endometrium | *SUGP2* | 2.769316981 | 5.80E-08 | 4.34E-07 | 0.000281525 |
| T2DM | endometrium | *SLC12A2* | -2.458162813 | 0.00229768 | 0.002868848 | 1 |
| T2DM | endometrium | *CNN2* | 7.105201516 | 1.83E-09 | 3.07E-07 | 8.90E-06 |
| T2DM | endometrium | *ABCA7* | 2.806227376 | 3.49E-06 | 6.17E-06 | 0.016930485 |
| T2DM | endometrium | *DDX20* | 2.966234831 | 2.57E-07 | 6.72E-07 | 0.001248912 |
| T2DM | endometrium | *BTBD1* | 4.402904473 | 7.59E-09 | 4.34E-07 | 3.68E-05 |
| T2DM | endometrium | *FAR2* | 6.002754738 | 1.43E-07 | 4.78E-07 | 0.000693335 |
| T2DM | endometrium | *BCAS1* | 4.328086781 | 3.57E-07 | 8.60E-07 | 0.001731652 |
| T2DM | endometrium | *CHI3L2* | -6.37712596 | 0.000621815 | 0.000815859 | 1 |
| T2DM | endometrium | *SBNO2* | 4.153640703 | 2.72E-08 | 4.34E-07 | 0.000131977 |
| T2DM | endometrium | *PMS1* | 4.678253375 | 2.92E-08 | 4.34E-07 | 0.000141632 |
| T2DM | endometrium | *UHRF1BP1* | 2.688063562 | 9.97E-07 | 2.03E-06 | 0.00483708 |
| T2DM | endometrium | *WDR18* | 1.816985631 | 0.002104943 | 0.002636682 | 1 |
| T2DM | endometrium | *ANKRD44* | 1.606154801 | 1.05E-07 | 4.47E-07 | 0.000508534 |
| T2DM | endometrium | *MAP2K4* | 2.772239283 | 9.63E-08 | 4.42E-07 | 0.000467305 |
| T2DM | endometrium | *TMEM206* | 2.272307346 | 2.21E-07 | 6.00E-07 | 0.001069873 |
| T2DM | endometrium | *SEC61A2* | -3.483704248 | 0.019365371 | 0.0224948 | 1 |
| T2DM | endometrium | *TIE1* | 4.884511055 | 8.81E-08 | 4.38E-07 | 0.000427245 |
| T2DM | endometrium | *NGEF* | 8.044723016 | 1.53E-06 | 2.96E-06 | 0.007441356 |
| T2DM | endometrium | *SPI1* | 4.260067719 | 7.77E-06 | 1.28E-05 | 0.037717607 |
| T2DM | endometrium | *ZNRD1* | -2.107226396 | 6.93E-06 | 1.15E-05 | 0.033627109 |
| T2DM | endometrium | *ATXN3* | 3.233633711 | 2.31E-08 | 4.34E-07 | 0.000111973 |
| T2DM | endometrium | *GOLGA5* | 4.699094894 | 0.006978485 | 0.008399804 | 1 |
| T2DM | endometrium | *FGFR2* | 1.861070237 | 9.04E-05 | 0.000127685 | 0.438470356 |
| T2DM | endometrium | *MTFR1* | 4.55827556 | 4.27E-06 | 7.43E-06 | 0.020729713 |
| T2DM | endometrium | *FECH* | 1.844095168 | 0.018734028 | 0.021782292 | 1 |
| T2DM | endometrium | *MYO9A* | 3.594997487 | 7.12E-08 | 4.34E-07 | 0.000345479 |
| T2DM | endometrium | *TRAM1* | 2.909932631 | 7.24E-08 | 4.34E-07 | 0.000351288 |
| T2DM | endometrium | *EVI5* | 3.185916593 | 2.54E-07 | 6.68E-07 | 0.0012331 |
| T2DM | endometrium | *STOML1* | -6.129014607 | 0.005765509 | 0.006976123 | 1 |

| | | | | | | |
|---|---|---|---|---|---|---|
| T2DM | endometrium | *METTL22* | 6.278998035 | 3.99E-07 | 9.39E-07 | 0.00193536 |
| T2DM | endometrium | *TP53BP1* | 3.258024936 | 3.93E-08 | 4.34E-07 | 0.000190866 |
| T2DM | endometrium | *RRP15* | 3.483875737 | 1.03E-06 | 2.09E-06 | 0.005002782 |
| T2DM | endometrium | *DHX8* | 7.280329457 | 1.05E-06 | 2.12E-06 | 0.005101601 |
| T2DM | endometrium | *CBFB* | 4.727602879 | 4.88E-08 | 4.34E-07 | 0.000236922 |
| T2DM | endometrium | *FGFR3* | -1.384263456 | 1.15E-05 | 1.84E-05 | 0.055923871 |
| T2DM | endometrium | *IFI35* | 5.337229661 | 1.38E-06 | 2.70E-06 | 0.006718465 |
| T2DM | endometrium | *HEATR6* | 6.931175545 | 7.41E-07 | 1.58E-06 | 0.003596261 |
| T2DM | endometrium | *POLR1A* | 1.95339901 | 3.85E-06 | 6.75E-06 | 0.018663184 |
| T2DM | endometrium | *IP6K2* | 3.496810732 | 2.09E-07 | 5.79E-07 | 0.001016047 |
| T2DM | endometrium | *STON1-GTF2A* | -1.450193483 | 0.00025049 | 0.000340347 | 1 |
| T2DM | endometrium | *SRBD1* | 3.520729641 | 5.34E-08 | 4.34E-07 | 0.000259186 |
| T2DM | endometrium | *CYFIP1* | 3.407018837 | 9.35E-08 | 4.39E-07 | 0.00045364 |
| T2DM | endometrium | *IFT80* | 4.529742356 | 3.27E-07 | 8.03E-07 | 0.001584867 |
| T2DM | endometrium | *NUCKS1* | 3.092668613 | 3.75E-08 | 4.34E-07 | 0.000182066 |
| T2DM | endometrium | *CLEC2D* | 3.110083204 | 1.58E-07 | 4.84E-07 | 0.000764462 |
| T2DM | endometrium | *DRD4* | -2.540170781 | 7.05E-06 | 1.17E-05 | 0.034227203 |
| T2DM | endometrium | *PLA2G10* | 6.843745145 | 1.34E-07 | 4.78E-07 | 0.000648135 |
| T2DM | endometrium | *NEDD4* | 5.159414763 | 1.10E-05 | 1.76E-05 | 0.05328144 |
| T2DM | endometrium | *PIGB* | 3.419949205 | 1.63E-07 | 4.92E-07 | 0.000792484 |
| T2DM | endometrium | *GNB5* | 5.211844054 | 9.75E-06 | 1.58E-05 | 0.047300192 |
| T2DM | endometrium | *LRP6* | 3.119144176 | 6.20E-08 | 4.34E-07 | 0.000300814 |
| T2DM | endometrium | *DAPP1* | 1.357363138 | 0.005623545 | 0.006807745 | 1 |
| T2DM | endometrium | *CLTCL1* | 1.972334589 | 2.51E-07 | 6.60E-07 | 0.001216435 |
| T2DM | endometrium | *DGCR2* | 5.435545227 | 3.96E-06 | 6.94E-06 | 0.019202197 |
| T2DM | endometrium | *ZXDC* | 3.924767588 | 5.50E-08 | 4.34E-07 | 0.000266779 |
| T2DM | endometrium | *GBA2* | 6.419635972 | 0.000686797 | 0.000896996 | 1 |
| T2DM | endometrium | *NDST1* | 6.569359655 | 7.38E-06 | 1.22E-05 | 0.035804756 |
| T2DM | endometrium | *ASNS* | -2.346036498 | 1.13E-05 | 1.81E-05 | 0.054875321 |
| T2DM | endometrium | *AP3M2* | 6.537445911 | 0.000252151 | 0.000342412 | 1 |
| T2DM | endometrium | *ST6GALNAC2* | 3.027881893 | 2.46E-07 | 6.53E-07 | 0.001193267 |
| T2DM | endometrium | *EIF2B3* | 4.44470942 | 1.91E-07 | 5.45E-07 | 0.000925187 |
| T2DM | endometrium | *MAP4K4* | 4.941709948 | 1.80E-06 | 3.41E-06 | 0.008718821 |
| T2DM | endometrium | *WDR1* | 2.861274421 | 8.82E-08 | 4.38E-07 | 0.000428145 |
| T2DM | endometrium | *SNX13* | 3.285660195 | 5.08E-08 | 4.34E-07 | 0.000246248 |
| T2DM | endometrium | *RPS6KA2* | 2.706183518 | 8.48E-08 | 4.38E-07 | 0.000411451 |
| T2DM | endometrium | *ING3* | 2.725750857 | 8.99E-08 | 4.38E-07 | 0.000436222 |
| T2DM | endometrium | *VASH1* | 3.327817196 | 2.55E-07 | 6.69E-07 | 0.001238649 |
| T2DM | endometrium | *TRIB2* | 3.038529982 | 2.16E-08 | 4.34E-07 | 0.000104911 |
| T2DM | endometrium | *CPSF1* | 2.678793011 | 3.52E-07 | 8.52E-07 | 0.00171023 |
| T2DM | endometrium | *CYBRD1* | 4.458411175 | 1.11E-07 | 4.57E-07 | 0.000540066 |
| T2DM | endometrium | *RDH11* | 2.721209246 | 9.43E-08 | 4.39E-07 | 0.000457368 |
| T2DM | endometrium | *EPN2* | 2.991639655 | 9.81E-07 | 2.00E-06 | 0.004759265 |
| T2DM | endometrium | *ALDH3A2* | 3.424064796 | 1.23E-07 | 4.70E-07 | 0.000598872 |
| T2DM | endometrium | *AFF4* | 3.492107018 | 4.93E-08 | 4.34E-07 | 0.000239445 |
| T2DM | endometrium | *TRHDE* | 2.208273869 | 1.57E-07 | 4.84E-07 | 0.00076417 |
| T2DM | endometrium | *FCGR2B* | 2.455770066 | 1.55E-07 | 4.80E-07 | 0.000751579 |
| T2DM | endometrium | *NFATC3* | 2.820968668 | 2.47E-08 | 4.34E-07 | 0.000119858 |
| T2DM | endometrium | *TRNT1* | 1.789937462 | 1.78E-07 | 5.19E-07 | 0.000861582 |
| T2DM | endometrium | *NDE1* | 4.680133842 | 2.82E-07 | 7.20E-07 | 0.00136993 |

| | | | | | | |
|---|---|---|---|---|---|---|
| T2DM | endometrium | *AP1M1* | 5.991330287 | 1.12E-07 | 4.57E-07 | 0.000542479 |
| T2DM | endometrium | *PVR* | 4.22253459 | 0.010007526 | 0.011886539 | 1 |
| T2DM | endometrium | *SCARB1* | 2.528755713 | 4.81E-08 | 4.34E-07 | 0.000233302 |
| T2DM | endometrium | *MOV10L1* | -1.730133972 | 1.12E-05 | 1.79E-05 | 0.05416569 |
| T2DM | endometrium | *ALPK1* | 2.806019587 | 3.30E-08 | 4.34E-07 | 0.000159896 |
| T2DM | endometrium | *LLGL2* | 2.914303766 | 2.79E-07 | 7.14E-07 | 0.001355281 |
| T2DM | endometrium | *SDHA* | 3.546165517 | 3.12E-09 | 3.72E-07 | 1.52E-05 |
| T2DM | endometrium | *SMARCE1* | 5.091051681 | 0.002229996 | 0.002789363 | 1 |
| T2DM | endometrium | *ADAM11* | -1.695459228 | 0.040504372 | 0.045971278 | 1 |
| T2DM | endometrium | *CD5L* | 8.050187634 | 8.86E-07 | 1.83E-06 | 0.004297716 |
| T2DM | endometrium | *PPP2R2C* | -5.528839809 | 0.030307089 | 0.034738955 | 1 |
| T2DM | endometrium | *ZNF532* | 2.646850421 | 6.29E-08 | 4.34E-07 | 0.000304968 |
| T2DM | endometrium | *ZZEF1* | 5.543109686 | 2.67E-09 | 3.60E-07 | 1.29E-05 |
| T2DM | endometrium | *TACR2* | 38.93486157 | 3.44E-07 | 8.36E-07 | 0.001669657 |
| T2DM | endometrium | *TIPIN* | 3.18113549 | 0.00012063 | 0.000168625 | 0.585299075 |
| T2DM | endometrium | *TTC38* | 4.037481123 | 1.03E-07 | 4.47E-07 | 0.000498323 |
| T2DM | endometrium | *GRAMD4* | 4.464922107 | 1.64E-07 | 4.93E-07 | 0.000795826 |
| T2DM | endometrium | *ADD2* | 4.107625907 | 2.16E-08 | 4.34E-07 | 0.000104623 |
| T2DM | endometrium | *VPS9D1* | 2.272952283 | 1.18E-07 | 4.61E-07 | 0.000571425 |
| T2DM | endometrium | *FNDC3B* | 3.107610282 | 5.11E-08 | 4.34E-07 | 0.000248 |
| T2DM | endometrium | *CACNG4* | 2.373496165 | 0.016088009 | 0.018791291 | 1 |
| T2DM | endometrium | *MOCOS* | 3.659449181 | 5.06E-08 | 4.34E-07 | 0.000245402 |
| T2DM | endometrium | *SEC31B* | 4.869095395 | 4.57E-09 | 4.20E-07 | 2.22E-05 |
| T2DM | endometrium | *KIFAP3* | 3.654692882 | 4.84E-08 | 4.34E-07 | 0.000235056 |
| T2DM | endometrium | *MKRN2* | 5.6742734 | 1.16E-05 | 1.86E-05 | 0.056421046 |
| T2DM | endometrium | *FMO4* | 3.424377362 | 4.93E-08 | 4.34E-07 | 0.000239069 |
| T2DM | endometrium | *RGS11* | 2.092249151 | 8.92E-07 | 1.84E-06 | 0.004325574 |
| T2DM | endometrium | *SPAG5* | 9.522219979 | 4.77E-06 | 8.22E-06 | 0.023148309 |
| T2DM | endometrium | *PAG1* | 2.05707482 | 9.97E-08 | 4.44E-07 | 0.000483852 |
| T2DM | endometrium | *GPATCH1* | 7.074953445 | 6.31E-06 | 1.06E-05 | 0.030618427 |
| T2DM | endometrium | *ICAM3* | -1.055891096 | 4.29E-06 | 7.47E-06 | 0.020837323 |
| T2DM | endometrium | *NT5C2* | 4.445070592 | 2.97E-08 | 4.34E-07 | 0.000144231 |
| T2DM | endometrium | *XAB2* | 3.700043446 | 8.48E-08 | 4.38E-07 | 0.000411368 |
| T2DM | endometrium | *IL4R* | 5.418144266 | 7.07E-08 | 4.34E-07 | 0.000343135 |
| T2DM | endometrium | *DYNC1I2* | 4.89847422 | 1.25E-07 | 4.74E-07 | 0.00060788 |
| T2DM | endometrium | *APBB1IP* | 1.905820869 | 1.91E-07 | 5.45E-07 | 0.000924946 |
| T2DM | endometrium | *LRCH4* | 4.232074961 | 2.77E-06 | 5.02E-06 | 0.01345037 |
| T2DM | endometrium | *SIRT6* | -1.842866241 | 5.98E-05 | 8.64E-05 | 0.290067714 |
| T2DM | endometrium | *GPR137B* | 3.080066684 | 3.33E-07 | 8.15E-07 | 0.001614393 |
| T2DM | endometrium | *AMPH* | 5.104308699 | 0.024923572 | 0.028737921 | 1 |
| T2DM | endometrium | *MCCC1* | 4.157253114 | 1.92E-05 | 2.95E-05 | 0.092921355 |
| T2DM | endometrium | *ACER3* | 2.52407075 | 3.19E-06 | 5.70E-06 | 0.015478305 |
| T2DM | endometrium | *PIK3C3* | 4.080132482 | 7.86E-08 | 4.36E-07 | 0.000381573 |
| T2DM | endometrium | *SYNJ2* | 2.553444194 | 2.22E-06 | 4.12E-06 | 0.010753991 |
| T2DM | endometrium | *RBFOX1* | 1.76025517 | 1.53E-06 | 2.96E-06 | 0.007446162 |
| T2DM | endometrium | *PCM1* | 4.620176538 | 5.94E-07 | 1.31E-06 | 0.002882199 |
| T2DM | endometrium | *MYH7B* | -2.16269182 | 0.003882432 | 0.004739009 | 1 |
| T2DM | endometrium | *CLUL1* | 4.144504156 | 7.41E-05 | 0.000105848 | 0.359669987 |
| T2DM | endometrium | *FKBP7* | -1.010758555 | 5.91E-06 | 9.96E-06 | 0.028677197 |
| T2DM | endometrium | *XRCC5* | 3.848029952 | 4.78E-08 | 4.34E-07 | 0.000232138 |

| | | | | | | |
|---|---|---|---|---|---|---|
| T2DM | endometrium | *SP140* | 2.335718203 | 9.72E-07 | 1.98E-06 | 0.004716521 |
| T2DM | endometrium | *SAR1A* | 4.729944254 | 4.53E-08 | 4.34E-07 | 0.000219966 |
| T2DM | endometrium | *RAPGEF3* | 1.848037908 | 3.62E-05 | 5.40E-05 | 0.175596649 |
| T2DM | endometrium | *DUSP13* | 11.69294291 | 0.037509498 | 0.042682008 | 1 |
| T2DM | endometrium | *FDFT1* | 3.33881737 | 5.10E-09 | 4.20E-07 | 2.47E-05 |
| T2DM | endometrium | *SCGN* | 19.39227092 | 2.02E-07 | 5.64E-07 | 0.000980774 |
| T2DM | endometrium | *PGM1* | 3.71728774 | 5.51E-08 | 4.34E-07 | 0.000267557 |
| T2DM | endometrium | *DDX1* | 2.331791183 | 1.05E-07 | 4.47E-07 | 0.000507445 |
| T2DM | endometrium | *DNM2* | 2.561938821 | 6.89E-08 | 4.34E-07 | 0.000334064 |
| T2DM | endometrium | *STX7* | 3.72578669 | 1.72E-07 | 5.08E-07 | 0.000832657 |
| T2DM | endometrium | *RABL2B* | 2.764060796 | 1.05E-07 | 4.47E-07 | 0.000509186 |
| T2DM | endometrium | *DCT* | 1.768882246 | 0.006659924 | 0.008026317 | 1 |
| T2DM | endometrium | *SCTR* | 2.208189846 | 1.51E-07 | 4.78E-07 | 0.000733676 |
| T2DM | endometrium | *RIF1* | 5.190926621 | 8.03E-07 | 1.68E-06 | 0.003897255 |
| T2DM | endometrium | *KCNN2* | -4.537800164 | 0.000454725 | 0.00060381 | 1 |
| T2DM | endometrium | *PSEN1* | 2.65020463 | 4.93E-08 | 4.34E-07 | 0.000239013 |
| T2DM | endometrium | *MOK* | 4.085887751 | 6.04E-07 | 1.32E-06 | 0.002929688 |
| T2DM | endometrium | *STK17B* | 3.627259912 | 3.91E-09 | 4.12E-07 | 1.90E-05 |
| T2DM | endometrium | *ZNF506* | 5.523251354 | 2.10E-09 | 3.40E-07 | 1.02E-05 |
| T2DM | endometrium | *PCDHB4* | 4.996982025 | 1.47E-07 | 4.78E-07 | 0.000715631 |
| T2DM | endometrium | *ME2* | 2.350058738 | 9.94E-08 | 4.44E-07 | 0.000482111 |
| T2DM | endometrium | *CCNT2* | 3.402726742 | 6.23E-09 | 4.34E-07 | 3.02E-05 |
| T2DM | endometrium | *SEMA5B* | -1.655811667 | 4.22E-05 | 6.22E-05 | 0.204711266 |
| T2DM | endometrium | *DOPEY1* | 4.639588658 | 5.08E-05 | 7.42E-05 | 0.246659517 |
| T2DM | endometrium | *LYRM2* | 3.185563389 | 7.46E-08 | 4.36E-07 | 0.000361753 |
| T2DM | endometrium | *ZCCHC6* | 2.617081144 | 6.35E-08 | 4.34E-07 | 0.00030794 |
| T2DM | endometrium | *ULK2* | 5.165715175 | 1.72E-06 | 3.29E-06 | 0.008367265 |
| T2DM | endometrium | *PLOD1* | 6.919297885 | 7.69E-07 | 1.63E-06 | 0.00373123 |
| T2DM | endometrium | *PIBF1* | 2.889779777 | 4.87E-08 | 4.34E-07 | 0.000236107 |
| T2DM | endometrium | *OXCT1* | 2.894093747 | 5.13E-08 | 4.34E-07 | 0.000248912 |
| T2DM | endometrium | *SLC27A5* | 3.198769357 | 9.57E-08 | 4.42E-07 | 0.000464257 |
| T2DM | endometrium | *ZNF586* | 5.386475181 | 5.45E-09 | 4.27E-07 | 2.64E-05 |
| T2DM | endometrium | *ZNF264* | 2.54884352 | 1.03E-07 | 4.47E-07 | 0.000499562 |
| T2DM | endometrium | *RPS5* | 5.274794388 | 2.83E-05 | 4.26E-05 | 0.137147206 |
| T2DM | endometrium | *CHMP2B* | 4.06283731 | 1.11E-07 | 4.57E-07 | 0.000536239 |
| T2DM | endometrium | *PPIE* | 4.408774411 | 9.01E-06 | 1.47E-05 | 0.043732728 |
| T2DM | endometrium | *NOA1* | -1.368432115 | 6.60E-06 | 1.10E-05 | 0.032029115 |
| T2DM | endometrium | *GSTP1* | 4.802796352 | 0.001621094 | 0.002047787 | 1 |
| T2DM | endometrium | *WBP11* | 2.183674641 | 1.44E-07 | 4.78E-07 | 0.000700814 |
| T2DM | endometrium | *NCOA1* | 3.958933708 | 2.38E-07 | 6.36E-07 | 0.001156416 |
| T2DM | endometrium | *CAD* | 2.529586663 | 1.95E-07 | 5.51E-07 | 0.000947777 |
| T2DM | endometrium | *CD59* | 2.376476321 | 2.42E-08 | 4.34E-07 | 0.000117591 |
| T2DM | endometrium | *CD82* | 3.366858043 | 2.55E-08 | 4.34E-07 | 0.00012386 |
| T2DM | endometrium | *HACE1* | 4.079108507 | 4.92E-08 | 4.34E-07 | 0.000238698 |
| T2DM | endometrium | *SEH1L* | 5.261165855 | 6.17E-06 | 1.04E-05 | 0.029917484 |
| T2DM | endometrium | *SLC25A24* | 2.327506356 | 5.03E-08 | 4.34E-07 | 0.00024408 |
| T2DM | endometrium | *PILRA* | 1.452496026 | 3.12E-07 | 7.74E-07 | 0.001512169 |
| T2DM | endometrium | *CPNE3* | 4.337733987 | 1.49E-08 | 4.34E-07 | 7.22E-05 |
| T2DM | endometrium | *RRN3* | 3.521356971 | 5.72E-08 | 4.34E-07 | 0.000277369 |
| T2DM | endometrium | *MTIF2* | 7.349044485 | 1.37E-06 | 2.67E-06 | 0.006635929 |

| | | | | | | |
|---|---|---|---|---|---|---|
| T2DM | endometrium | *DDHD2* | 2.896532233 | 2.40E-08 | 4.34E-07 | 0.000116266 |
| T2DM | endometrium | *EPS15* | 3.715169492 | 1.14E-07 | 4.57E-07 | 0.000551231 |
| T2DM | endometrium | *USP40* | 3.285512217 | 5.22E-08 | 4.34E-07 | 0.000253347 |
| T2DM | endometrium | *POMGNT1* | 5.55159871 | 1.05E-07 | 4.47E-07 | 0.000508862 |
| T2DM | endometrium | *MAST2* | 4.280866635 | 6.29E-08 | 4.34E-07 | 0.000305177 |
| T2DM | endometrium | *NME8* | -4.200716426 | 1.79E-06 | 3.39E-06 | 0.008666664 |
| T2DM | endometrium | *EPDR1* | 2.90143951 | 6.81E-06 | 1.13E-05 | 0.03304967 |
| T2DM | endometrium | *SNX10* | 1.518448863 | 3.46E-05 | 5.17E-05 | 0.167775053 |
| T2DM | endometrium | *TMED2* | 5.202874748 | 0.000136924 | 0.000190433 | 0.664353782 |
| T2DM | endometrium | *ALG9* | 4.778044888 | 7.49E-08 | 4.36E-07 | 0.000363572 |
| T2DM | endometrium | *ACOX3* | 5.766846636 | 1.78E-07 | 5.20E-07 | 0.000864289 |
| T2DM | endometrium | *HSD17B14* | 3.535589547 | 1.16E-07 | 4.59E-07 | 0.000560656 |
| T2DM | endometrium | *TRIP6* | 4.369506099 | 1.45E-07 | 4.78E-07 | 0.000701475 |
| T2DM | endometrium | *ACHE* | -1.299488883 | 6.33E-07 | 1.38E-06 | 0.003069029 |
| T2DM | endometrium | *SRRT* | 2.975276759 | 1.36E-07 | 4.78E-07 | 0.000660771 |
| T2DM | endometrium | *ADAMTS2* | 3.335037687 | 0.028003247 | 0.032174226 | 1 |
| T2DM | endometrium | *PSMC5* | 2.942647918 | 6.96E-07 | 1.50E-06 | 0.003377742 |
| T2DM | endometrium | *UIMC1* | 3.214345853 | 7.80E-08 | 4.36E-07 | 0.000378379 |
| T2DM | endometrium | *LPCAT2* | 1.97357228 | 1.45E-07 | 4.78E-07 | 0.000701621 |
| T2DM | endometrium | *OGFOD1* | 3.566288657 | 6.76E-07 | 1.46E-06 | 0.003278138 |
| T2DM | endometrium | *TXNDC16* | 5.262511683 | 1.42E-05 | 2.23E-05 | 0.068977145 |
| T2DM | endometrium | *SF3B2* | 5.356090679 | 8.27E-08 | 4.37E-07 | 0.000401448 |
| T2DM | endometrium | *PHACTR3* | 3.108208082 | 0.000170317 | 0.000234966 | 0.826376748 |
| T2DM | endometrium | *AAMDC* | 2.867407488 | 3.45E-05 | 5.16E-05 | 0.167613204 |
| T2DM | endometrium | *ASAP3* | 2.204481206 | 1.10E-07 | 4.56E-07 | 0.000531933 |
| T2DM | endometrium | *EDEM2* | 3.846009901 | 5.46E-06 | 9.28E-06 | 0.026487893 |
| T2DM | endometrium | *ANKRD10* | 3.412934718 | 7.24E-08 | 4.34E-07 | 0.000351457 |
| T2DM | endometrium | *DOCK3* | 6.180733348 | 1.57E-07 | 4.84E-07 | 0.000763584 |
| T2DM | endometrium | *ARHGAP28* | 2.683121833 | 1.01E-07 | 4.47E-07 | 0.000491569 |
| T2DM | endometrium | *PPP1R13B* | 2.398529206 | 3.77E-07 | 8.99E-07 | 0.001826917 |
| T2DM | endometrium | *NSFL1C* | 2.12648907 | 7.35E-08 | 4.35E-07 | 0.000356531 |
| T2DM | endometrium | *ZNF343* | 6.8973771 | 5.30E-07 | 1.18E-06 | 0.002571373 |
| T2DM | endometrium | *EBF4* | 9.303474104 | 1.03E-06 | 2.08E-06 | 0.00497891 |
| T2DM | endometrium | *CPXM1* | -2.130926431 | 2.25E-05 | 3.42E-05 | 0.108995083 |
| T2DM | endometrium | *MAVS* | 4.277956396 | 2.23E-07 | 6.04E-07 | 0.001083374 |
| T2DM | endometrium | *XRN2* | 4.099302966 | 1.09E-07 | 4.55E-07 | 0.000529443 |
| T2DM | endometrium | *SNX5* | 6.887591512 | 4.22E-07 | 9.79E-07 | 0.002045958 |
| T2DM | endometrium | *ESF1* | 4.311130507 | 4.62E-08 | 4.34E-07 | 0.000224245 |
| T2DM | endometrium | *RBBP9* | -2.895485135 | 1.96E-05 | 3.00E-05 | 0.094878566 |
| T2DM | endometrium | *TMEM230* | 3.734247265 | 4.73E-08 | 4.34E-07 | 0.000229465 |
| T2DM | endometrium | *RPH3A* | 7.413832727 | 1.09E-07 | 4.55E-07 | 0.000529138 |
| T2DM | endometrium | *KIF16B* | 1.881135666 | 1.17E-07 | 4.61E-07 | 0.000566391 |
| T2DM | endometrium | *TBX5* | -1.217844885 | 1.28E-06 | 2.53E-06 | 0.006226055 |
| T2DM | endometrium | *FXYD5* | 2.459584927 | 1.34E-07 | 4.78E-07 | 0.000651698 |
| T2DM | endometrium | *ZNF302* | 3.677409368 | 0.000535521 | 0.000706265 | 1 |
| T2DM | endometrium | *GRAMD1A* | 4.022009347 | 5.21E-08 | 4.34E-07 | 0.000252992 |
| T2DM | endometrium | *CDIP1* | -2.498466301 | 8.54E-05 | 0.000121024 | 0.414143886 |
| T2DM | endometrium | *KCNH4* | 3.452045932 | 5.77E-05 | 8.36E-05 | 0.280113861 |
| T2DM | endometrium | *GANAB* | 5.767488549 | 2.21E-05 | 3.37E-05 | 0.10724725 |
| T2DM | endometrium | *MLF2* | 6.94366954 | 7.10E-07 | 1.52E-06 | 0.003444519 |

| | | | | | | |
|---|---|---|---|---|---|---|
| T2DM | endometrium | *ANKRD24* | -4.053523323 | 0.000377813 | 0.000505696 | 1 |
| T2DM | endometrium | *DHX32* | 4.320507075 | 1.48E-07 | 4.78E-07 | 0.000716445 |
| T2DM | endometrium | *SPTLC1* | 3.416026648 | 1.22E-07 | 4.69E-07 | 0.000594282 |
| T2DM | endometrium | *YPEL3* | 3.108077198 | 0.024438576 | 0.028198804 | 1 |
| T2DM | endometrium | *MRPS33* | 4.060300805 | 1.51E-05 | 2.36E-05 | 0.073393684 |
| T2DM | endometrium | *MAEA* | 2.455442269 | 4.16E-07 | 9.69E-07 | 0.00201631 |
| T2DM | endometrium | *STRN4* | 2.485895458 | 1.46E-07 | 4.78E-07 | 0.00071043 |
| T2DM | endometrium | *LYZ* | 1.777973222 | 6.80E-08 | 4.34E-07 | 0.000329944 |
| T2DM | endometrium | *MUL1* | 2.417574483 | 1.03E-05 | 1.66E-05 | 0.050005326 |
| T2DM | endometrium | *SPG21* | 4.38576645 | 1.16E-06 | 2.31E-06 | 0.005615404 |
| T2DM | endometrium | *CHRD* | -1.016931975 | 2.23E-06 | 4.14E-06 | 0.01080113 |
| T2DM | endometrium | *RAB11FIP3* | -2.214381047 | 0.000718161 | 0.000935691 | 1 |
| T2DM | endometrium | *GNPTG* | 3.556252304 | 1.17E-07 | 4.61E-07 | 0.000566381 |
| T2DM | endometrium | *PABPC4* | 1.810256867 | 0.000177209 | 0.000244128 | 0.859817805 |
| T2DM | endometrium | *USP48* | 3.057488298 | 3.35E-07 | 8.19E-07 | 0.001623611 |
| T2DM | endometrium | *PDPR* | -2.163047583 | 0.005985168 | 0.00723649 | 1 |
| T2DM | endometrium | *GLG1* | 3.48618886 | 4.79E-08 | 4.34E-07 | 0.000232281 |
| T2DM | endometrium | *DTX2* | 2.69468282 | 1.49E-07 | 4.78E-07 | 0.000723089 |
| T2DM | endometrium | *LAMB1* | 1.986135964 | 8.02E-06 | 1.31E-05 | 0.038901819 |
| T2DM | endometrium | *ABCC6* | 4.173145317 | 0.000125257 | 0.00017484 | 0.607745423 |
| T2DM | endometrium | *RAPGEF4* | -1.973264288 | 0.017065846 | 0.01990947 | 1 |
| T2DM | endometrium | *TF* | 4.343137184 | 7.99E-08 | 4.36E-07 | 0.000387461 |
| T2DM | endometrium | *SPAG7* | 2.001977917 | 4.87E-07 | 1.10E-06 | 0.002360809 |
| T2DM | endometrium | *RGS17* | 3.056890579 | 1.53E-05 | 2.39E-05 | 0.074402856 |
| T2DM | endometrium | *PPP2R3C* | 4.38614531 | 4.12E-09 | 4.16E-07 | 2.00E-05 |
| T2DM | endometrium | *HAUS4* | 1.286417451 | 0.02328826 | 0.026897081 | 1 |
| T2DM | endometrium | *OSGEP* | 3.064027521 | 1.19E-07 | 4.65E-07 | 0.000579432 |
| T2DM | endometrium | *SCFD1* | 4.5946044 | 1.63E-07 | 4.92E-07 | 0.000790362 |
| T2DM | endometrium | *SUPT16H* | 4.613887922 | 5.29E-08 | 4.34E-07 | 0.000256636 |
| T2DM | endometrium | *TOX4* | 2.144341655 | 1.15E-07 | 4.59E-07 | 0.000559069 |
| T2DM | endometrium | *CAPN3* | -3.098131256 | 2.43E-06 | 4.47E-06 | 0.011791348 |
| T2DM | endometrium | *UNC13D* | 2.409996969 | 1.35E-06 | 2.65E-06 | 0.00657093 |
| T2DM | endometrium | *MFSD11* | 3.98960244 | 1.62E-07 | 4.91E-07 | 0.000787558 |
| T2DM | endometrium | *NUP50* | 2.539072486 | 3.47E-07 | 8.40E-07 | 0.001681514 |
| T2DM | endometrium | *COMT* | 4.905565025 | 6.98E-08 | 4.34E-07 | 0.000338829 |
| T2DM | endometrium | *LRRFIP2* | 3.372055238 | 3.49E-08 | 4.34E-07 | 0.000169225 |
| T2DM | endometrium | *XYLB* | 5.675440648 | 6.75E-08 | 4.34E-07 | 0.000327711 |
| T2DM | endometrium | *GABRP* | 6.595827607 | 5.41E-07 | 1.20E-06 | 0.002624467 |
| T2DM | endometrium | *CDC6* | 5.334634484 | 0.001054946 | 0.001354843 | 1 |
| T2DM | endometrium | *FMO2* | 2.457736748 | 1.87E-07 | 5.38E-07 | 0.00090973 |
| T2DM | endometrium | *MSH2* | 2.39199562 | 1.07E-07 | 4.49E-07 | 0.000517501 |
| T2DM | endometrium | *CRAT* | 3.177377775 | 1.50E-07 | 4.78E-07 | 0.000730149 |
| T2DM | endometrium | *NANS* | 5.421368048 | 3.38E-08 | 4.34E-07 | 0.000164167 |
| T2DM | endometrium | *PDE6C* | -2.560934557 | 7.33E-05 | 0.0001047 | 0.355666342 |
| T2DM | endometrium | *CWF19L1* | -3.093489048 | 6.97E-05 | 9.98E-05 | 0.337964435 |
| T2DM | endometrium | *CRTAC1* | -2.235317407 | 0.000171486 | 0.000236513 | 0.832052451 |
| T2DM | endometrium | *WAC* | 3.754824961 | 1.86E-07 | 5.35E-07 | 0.000902478 |
| T2DM | endometrium | *ITPR3* | 3.130183763 | 1.61E-08 | 4.34E-07 | 7.80E-05 |
| T2DM | endometrium | *DSP* | 2.998409232 | 1.46E-07 | 4.78E-07 | 0.000708004 |
| T2DM | endometrium | *IFT74* | 3.326107192 | 9.59E-08 | 4.42E-07 | 0.000465082 |

| | | | | | | |
|---|---|---|---|---|---|---|
| T2DM | endometrium | *IL12RB1* | 2.998673446 | 8.86E-05 | 0.000125377 | 0.43004479 |
| T2DM | endometrium | *ACOT7* | 3.306953719 | 1.21E-06 | 2.39E-06 | 0.005851834 |
| T2DM | endometrium | *SCD* | 2.19299841 | 1.72E-05 | 2.66E-05 | 0.083419874 |
| T2DM | endometrium | *ERMP1* | -2.32237593 | 0.004558251 | 0.005547187 | 1 |
| T2DM | endometrium | *RAB18* | 4.329036006 | 5.57E-08 | 4.34E-07 | 0.000270148 |
| T2DM | endometrium | *PRTFDC1* | -2.305332754 | 0.03701032 | 0.042143645 | 1 |
| T2DM | endometrium | *TSPAN15* | 5.029504676 | 3.72E-08 | 4.34E-07 | 0.000180601 |
| T2DM | endometrium | *MAST3* | 1.992617181 | 2.62E-06 | 4.78E-06 | 0.012722619 |
| T2DM | endometrium | *OCEL1* | 2.641108111 | 9.25E-07 | 1.90E-06 | 0.004486511 |
| T2DM | endometrium | *CATSPERG* | 2.542635186 | 5.56E-06 | 9.42E-06 | 0.026977622 |
| T2DM | endometrium | *SETD1A* | 2.823652404 | 3.17E-08 | 4.34E-07 | 0.000153763 |
| T2DM | endometrium | *CIRBP* | 3.962567755 | 0.000908131 | 0.001173703 | 1 |
| T2DM | endometrium | *MARCH2* | 2.168792761 | 1.87E-08 | 4.34E-07 | 9.06E-05 |
| T2DM | endometrium | *MTAP* | -2.676753847 | 0.000286374 | 0.00038726 | 1 |
| T2DM | endometrium | *POLR2E* | 5.460702434 | 6.37E-08 | 4.34E-07 | 0.000309234 |
| T2DM | endometrium | *MADCAM1* | 5.693091069 | 0.003234718 | 0.003983465 | 1 |
| T2DM | endometrium | *MED15* | 4.092661921 | 0.004626245 | 0.00562429 | 1 |
| T2DM | endometrium | *SNAP29* | 3.910763387 | 6.68E-07 | 1.44E-06 | 0.003240125 |
| T2DM | endometrium | *LZTR1* | 4.013591415 | 8.93E-08 | 4.38E-07 | 0.000433069 |
| T2DM | endometrium | *MMP11* | -0.583506017 | 1.82E-06 | 3.44E-06 | 0.008811609 |
| T2DM | endometrium | *CECR2* | 5.209133802 | 9.92E-09 | 4.34E-07 | 4.81E-05 |
| T2DM | endometrium | *SMARCB1* | 2.152258414 | 2.26E-07 | 6.09E-07 | 0.001095537 |
| T2DM | endometrium | *BCL2L13* | 5.28689249 | 1.68E-07 | 5.03E-07 | 0.000816666 |
| T2DM | endometrium | *DDTL* | 3.461853302 | 0.000115103 | 0.000161085 | 0.558480008 |
| T2DM | endometrium | *DDT* | -2.917952331 | 5.97E-05 | 8.63E-05 | 0.289508902 |
| T2DM | endometrium | *OSM* | -1.930199454 | 1.19E-06 | 2.36E-06 | 0.005766521 |
| T2DM | endometrium | *CABIN1* | 4.881137249 | 9.91E-08 | 4.44E-07 | 0.000480813 |
| T2DM | endometrium | *SPECC1L* | -1.947455188 | 1.93E-05 | 2.97E-05 | 0.093638772 |
| T2DM | endometrium | *PPIL2* | 4.38706835 | 0.002527209 | 0.003142496 | 1 |
| T2DM | endometrium | *PES1* | 2.407725931 | 4.82E-07 | 1.09E-06 | 0.002340405 |
| T2DM | endometrium | *SLC35E4* | -2.345732341 | 5.58E-05 | 8.10E-05 | 0.270865875 |
| T2DM | endometrium | *CRYBB3* | -4.058444546 | 0.001607303 | 0.002031952 | 1 |
| T2DM | endometrium | *CYTH4* | 2.345506389 | 1.42E-07 | 4.78E-07 | 0.000690919 |
| T2DM | endometrium | *CARD10* | 2.834754438 | 1.52E-06 | 2.94E-06 | 0.007380499 |
| T2DM | endometrium | *LRP5L* | 2.300078532 | 2.33E-06 | 4.31E-06 | 0.011306556 |
| T2DM | endometrium | *LGALS2* | 4.082176091 | 3.37E-06 | 5.98E-06 | 0.016357635 |
| T2DM | endometrium | *SH3BP1* | 4.165169285 | 1.36E-07 | 4.78E-07 | 0.000660363 |
| T2DM | endometrium | *HPS4* | 5.050672246 | 4.77E-08 | 4.34E-07 | 0.000231548 |
| T2DM | endometrium | *TRIOBP* | 3.865942288 | 3.81E-08 | 4.34E-07 | 0.000184814 |
| T2DM | endometrium | *ANKRD54* | -2.961452501 | 0.000290137 | 0.000392129 | 1 |
| T2DM | endometrium | *EIF3L* | 2.232211135 | 7.72E-07 | 1.63E-06 | 0.003745008 |
| T2DM | endometrium | *CCDC134* | -2.799660929 | 0.015521198 | 0.018159839 | 1 |
| T2DM | endometrium | *PICK1* | 2.225526386 | 7.93E-06 | 1.30E-05 | 0.038491833 |
| T2DM | endometrium | *TTC28* | 2.538746225 | 5.50E-08 | 4.34E-07 | 0.000266826 |
| T2DM | endometrium | *CENPM* | 3.418819466 | 0.0426298 | 0.048338348 | 1 |
| T2DM | endometrium | *SLC5A4* | 0.681048758 | 0.006363793 | 0.007682787 | 1 |
| T2DM | endometrium | *KDELR3* | -2.192178005 | 8.75E-06 | 1.43E-05 | 0.042471319 |
| T2DM | endometrium | *CYP2D6* | -6.571896404 | 0.003891721 | 0.004747958 | 1 |
| T2DM | endometrium | *CBY1* | 2.677137666 | 3.69E-07 | 8.85E-07 | 0.001791338 |
| T2DM | endometrium | *RTCB* | 4.834313704 | 2.01E-07 | 5.62E-07 | 0.000975653 |

| | | | | | | |
|---|---|---|---|---|---|---|
| T2DM | endometrium | *RAB36* | 4.649514358 | 8.78E-06 | 1.43E-05 | 0.0425813 |
| T2DM | endometrium | *TIMP3* | 3.193590949 | 6.05E-08 | 4.34E-07 | 0.000293784 |
| T2DM | endometrium | *PPP6R2* | 3.352615478 | 6.21E-08 | 4.34E-07 | 0.000301335 |
| T2DM | endometrium | *RHBDD3* | 4.388942263 | 1.80E-06 | 3.42E-06 | 0.008742306 |
| T2DM | endometrium | *TOM1* | 1.846998916 | 5.27E-07 | 1.18E-06 | 0.002557173 |
| T2DM | endometrium | *CHKB* | -3.397033003 | 0.000250005 | 0.000339782 | 1 |
| T2DM | endometrium | *THOC5* | 2.754606331 | 1.43E-05 | 2.24E-05 | 0.069171654 |
| T2DM | endometrium | *ARSA* | 2.340849806 | 4.23E-07 | 9.81E-07 | 0.002053863 |
| T2DM | endometrium | *TSPO* | 4.529139194 | 5.17E-08 | 4.34E-07 | 0.000250748 |
| T2DM | endometrium | *TTLL12* | -1.954040297 | 6.65E-06 | 1.11E-05 | 0.032285272 |
| T2DM | endometrium | *MTMR3* | 4.032631675 | 1.24E-05 | 1.96E-05 | 0.059928179 |
| T2DM | endometrium | *APOL4* | 3.079886258 | 1.31E-06 | 2.57E-06 | 0.006347276 |
| T2DM | endometrium | *SAMM50* | 6.511135281 | 2.50E-08 | 4.34E-07 | 0.000121103 |
| T2DM | endometrium | *FOXRED2* | 3.189066256 | 3.87E-08 | 4.34E-07 | 0.000187656 |
| T2DM | endometrium | *IFT27* | 2.979057184 | 7.29E-08 | 4.34E-07 | 0.00035357 |
| T2DM | endometrium | *KIAA0930* | 3.611113981 | 5.24E-08 | 4.34E-07 | 0.000254057 |
| T2DM | endometrium | *SLC25A17* | 4.134872163 | 1.30E-08 | 4.34E-07 | 6.29E-05 |
| T2DM | endometrium | *FAM118A* | -1.642650529 | 0.002105216 | 0.002636682 | 1 |
| T2DM | endometrium | *ZC3H7B* | 5.847590813 | 2.19E-07 | 5.98E-07 | 0.001064303 |
| T2DM | endometrium | *ACO2* | 4.561238618 | 0.001982069 | 0.002490806 | 1 |
| T2DM | endometrium | *POLR3H* | -1.879565543 | 3.26E-05 | 4.89E-05 | 0.158317484 |
| T2DM | endometrium | *DESI1* | 3.334125829 | 3.92E-08 | 4.34E-07 | 0.000190418 |
| T2DM | endometrium | *MLC1* | 4.795272975 | 0.000511675 | 0.000675918 | 1 |
| T2DM | endometrium | *HDAC10* | 4.343078964 | 7.23E-08 | 4.34E-07 | 0.00035071 |
| T2DM | endometrium | *SDR39U1* | -1.994946003 | 0.000103348 | 0.000145177 | 0.501443081 |
| T2DM | endometrium | *RBM23* | 4.701805384 | 1.39E-08 | 4.34E-07 | 6.73E-05 |
| T2DM | endometrium | *AP4S1* | 3.772548346 | 9.55E-08 | 4.42E-07 | 0.000463241 |
| T2DM | endometrium | *NIN* | 5.555144374 | 6.59E-07 | 1.43E-06 | 0.003199741 |
| T2DM | endometrium | *PYGL* | 2.098517362 | 2.42E-06 | 4.45E-06 | 0.011738263 |
| T2DM | endometrium | *CNIH1* | 2.722016937 | 1.06E-07 | 4.48E-07 | 0.000513994 |
| T2DM | endometrium | *ATP6V1D* | 2.786416283 | 7.00E-08 | 4.34E-07 | 0.000339879 |
| T2DM | endometrium | *PLEK2* | -3.393958012 | 4.82E-07 | 1.09E-06 | 0.002340343 |
| T2DM | endometrium | *GSTZ1* | -2.064733586 | 2.94E-05 | 4.42E-05 | 0.142554377 |
| T2DM | endometrium | *TMED8* | 4.664123036 | 5.28E-07 | 1.18E-06 | 0.002563499 |
| T2DM | endometrium | *LGMN* | 1.513059022 | 8.64E-08 | 4.38E-07 | 0.00041903 |
| T2DM | endometrium | *MTHFD1* | 6.485102595 | 1.92E-05 | 2.95E-05 | 0.093108403 |
| T2DM | endometrium | *TELO2* | 8.508313007 | 2.17E-07 | 5.95E-07 | 0.001051729 |
| T2DM | endometrium | *GSKIP* | -2.147271333 | 2.08E-05 | 3.18E-05 | 0.100743476 |
| T2DM | endometrium | *PSMB5* | 6.346383005 | 1.74E-06 | 3.32E-06 | 0.008460328 |
| T2DM | endometrium | *YY1* | 2.793261432 | 6.88E-08 | 4.34E-07 | 0.000333911 |
| T2DM | endometrium | *CCNB1IP1* | 3.562711838 | 6.63E-08 | 4.34E-07 | 0.000321599 |
| T2DM | endometrium | *CINP* | 8.001489694 | 9.32E-05 | 0.000131511 | 0.452136365 |
| T2DM | endometrium | *KIAA0391* | -4.318569658 | 8.85E-06 | 1.44E-05 | 0.042939475 |
| T2DM | endometrium | *EMC9* | 3.609312961 | 1.41E-06 | 2.74E-06 | 0.006830761 |
| T2DM | endometrium | *PLTP* | 1.985619487 | 1.17E-07 | 4.61E-07 | 0.000567231 |
| T2DM | endometrium | *TRPC4AP* | 4.475595582 | 2.33E-06 | 4.31E-06 | 0.011298939 |
| T2DM | endometrium | *PYGB* | 3.940327867 | 5.21E-06 | 8.90E-06 | 0.025300007 |
| T2DM | endometrium | *ABHD12* | 2.890117094 | 1.34E-07 | 4.78E-07 | 0.000648566 |
| T2DM | endometrium | *CD40* | 2.444637912 | 2.90E-07 | 7.33E-07 | 0.001405871 |
| T2DM | endometrium | *UQCC1* | 2.56670966 | 7.73E-07 | 1.63E-06 | 0.003750104 |

| | | | | | | |
|---|---|---|---|---|---|---|
| T2DM | endometrium | *NDRG3* | 2.556744364 | 8.09E-08 | 4.36E-07 | 0.000392742 |
| T2DM | endometrium | *PABPC1L* | 6.986637319 | 5.86E-08 | 4.34E-07 | 0.000284342 |
| T2DM | endometrium | *DOK5* | 3.703842256 | 7.97E-08 | 4.36E-07 | 0.000386542 |
| T2DM | endometrium | *CSTF1* | 3.111863095 | 1.37E-07 | 4.78E-07 | 0.000664345 |
| T2DM | endometrium | *NELFCD* | 2.804193771 | 4.90E-08 | 4.34E-07 | 0.000237648 |
| T2DM | endometrium | *CTSZ* | 1.849703267 | 1.39E-07 | 4.78E-07 | 0.000672385 |
| T2DM | endometrium | *TCFL5* | -2.739186681 | 6.32E-05 | 9.11E-05 | 0.306855694 |
| T2DM | endometrium | *DIDO1* | 2.355580247 | 3.26E-08 | 4.34E-07 | 0.000158311 |
| T2DM | endometrium | *GID8* | 6.060556658 | 0.000557058 | 0.000733871 | 1 |
| T2DM | endometrium | *NKAIN4* | 2.624936254 | 0.02708973 | 0.03114677 | 1 |
| T2DM | endometrium | *PTK6* | -1.395319543 | 2.44E-06 | 4.48E-06 | 0.011818312 |
| T2DM | endometrium | *GMEB2* | 5.076732378 | 5.84E-08 | 4.34E-07 | 0.000283146 |
| T2DM | endometrium | *C20orf27* | 2.96705774 | 1.46E-07 | 4.78E-07 | 0.000707302 |
| T2DM | endometrium | *CDC25B* | 4.371937255 | 3.65E-06 | 6.44E-06 | 0.017728019 |
| T2DM | endometrium | *RNF24* | 3.008606242 | 4.72E-08 | 4.34E-07 | 0.00022904 |
| T2DM | endometrium | *NDUFAF5* | 2.569333471 | 1.40E-06 | 2.73E-06 | 0.006787955 |
| T2DM | endometrium | *TRIB3* | 4.658855649 | 1.62E-07 | 4.91E-07 | 0.000786962 |
| T2DM | endometrium | *RASSF2* | 3.05840867 | 7.32E-08 | 4.35E-07 | 0.000355229 |
| T2DM | endometrium | *ANGPT4* | 5.691631116 | 1.53E-07 | 4.78E-07 | 0.000740555 |
| T2DM | endometrium | *RSPO4* | 3.160037262 | 1.33E-05 | 2.10E-05 | 0.064300504 |
| T2DM | endometrium | *SIRPB1* | -2.062681016 | 2.20E-06 | 4.09E-06 | 0.010671174 |
| T2DM | endometrium | *CRNKL1* | 2.677817365 | 3.33E-07 | 8.15E-07 | 0.001615413 |
| T2DM | endometrium | *MANBAL* | 5.291151163 | 2.79E-08 | 4.34E-07 | 0.000135495 |
| T2DM | endometrium | *TTI1* | 2.088539953 | 2.01E-07 | 5.62E-07 | 0.000976721 |
| T2DM | endometrium | *BPI* | -0.682644972 | 6.10E-06 | 1.03E-05 | 0.029610762 |
| T2DM | endometrium | *ACTR5* | 3.653190269 | 3.83E-07 | 9.09E-07 | 0.001860139 |
| T2DM | endometrium | *DNTTIP1* | 1.929700781 | 2.38E-06 | 4.39E-06 | 0.011567613 |
| T2DM | endometrium | *MAP1LC3A* | 3.965348567 | 4.36E-08 | 4.34E-07 | 0.000211586 |
| T2DM | endometrium | *RBFA* | 2.635588317 | 1.36E-07 | 4.78E-07 | 0.000660116 |
| T2DM | endometrium | *METTL4* | 2.040012253 | 1.68E-07 | 5.03E-07 | 0.000816427 |
| T2DM | endometrium | *LPIN2* | 3.779714656 | 3.96E-08 | 4.34E-07 | 0.000192031 |
| T2DM | endometrium | *MYOM1* | 2.329171098 | 9.49E-08 | 4.41E-07 | 0.00046037 |
| T2DM | endometrium | *CEP192* | 2.56121051 | 1.92E-07 | 5.46E-07 | 0.000929173 |
| T2DM | endometrium | *LAMA1* | -1.329417199 | 2.69E-06 | 4.90E-06 | 0.013066537 |
| T2DM | endometrium | *RNF125* | 2.793799639 | 1.41E-07 | 4.78E-07 | 0.000684475 |
| T2DM | endometrium | *POLI* | 1.924508741 | 5.77E-06 | 9.76E-06 | 0.028008836 |
| T2DM | endometrium | *MIB1* | 3.299536128 | 5.09E-08 | 4.34E-07 | 0.000246925 |
| T2DM | endometrium | *NALCN* | 2.595680559 | 2.40E-07 | 6.41E-07 | 0.001166596 |
| T2DM | endometrium | *STK24* | 4.793148341 | 0.000293857 | 0.000396936 | 1 |
| T2DM | endometrium | *ACP5* | 3.425784816 | 3.52E-08 | 4.34E-07 | 0.000170994 |
| T2DM | endometrium | *DNAJC3* | 5.290846223 | 8.16E-08 | 4.36E-07 | 0.000396099 |
| T2DM | endometrium | *UGGT2* | 4.675498469 | 1.12E-07 | 4.57E-07 | 0.000541016 |
| T2DM | endometrium | *ARHGEF7* | 3.566509245 | 5.23E-08 | 4.34E-07 | 0.000253945 |
| T2DM | endometrium | *PARP4* | 4.696908281 | 6.04E-08 | 4.34E-07 | 0.000292888 |
| T2DM | endometrium | *SUPT20H* | 4.483840884 | 0.00170493 | 0.002149772 | 1 |
| T2DM | endometrium | *MRPS31* | 5.873718563 | 1.75E-07 | 5.14E-07 | 0.000848744 |
| T2DM | endometrium | *RGCC* | 3.028222136 | 3.22E-06 | 5.75E-06 | 0.015647241 |
| T2DM | endometrium | *DGKH* | 3.543289345 | 1.53E-05 | 2.39E-05 | 0.074385817 |
| T2DM | endometrium | *KATNAL1* | 4.784147377 | 9.41E-08 | 4.39E-07 | 0.00045674 |
| T2DM | endometrium | *INTS6* | 4.217642514 | 0.001427006 | 0.001813495 | 1 |

| | | | | | | |
|---|---|---|---|---|---|---|
| T2DM | endometrium | *CLN5* | 3.257402104 | 9.32E-08 | 4.39E-07 | 0.000452137 |
| T2DM | endometrium | *MGRN1* | 2.699175601 | 6.92E-08 | 4.34E-07 | 0.000335581 |
| T2DM | endometrium | *ELMO3* | -1.983506975 | 1.63E-05 | 2.52E-05 | 0.07888132 |
| T2DM | endometrium | *LONP2* | 3.594867751 | 2.46E-08 | 4.34E-07 | 0.000119242 |
| T2DM | endometrium | *PLLP* | 2.841849752 | 1.36E-06 | 2.66E-06 | 0.006578804 |
| T2DM | endometrium | *DHODH* | 3.167697037 | 3.07E-08 | 4.34E-07 | 0.00014917 |
| T2DM | endometrium | *CCDC113* | 4.10183569 | 1.28E-06 | 2.52E-06 | 0.00619998 |
| T2DM | endometrium | *PSMD7* | 2.763256827 | 6.70E-08 | 4.34E-07 | 0.000324932 |
| T2DM | endometrium | *COG4* | 4.18310511 | 7.51E-05 | 0.000107175 | 0.364286641 |
| T2DM | endometrium | *MON1B* | 3.90214087 | 7.67E-08 | 4.36E-07 | 0.000372227 |
| T2DM | endometrium | *TAF1C* | 1.778477126 | 0.000170158 | 0.000234815 | 0.825608262 |
| T2DM | endometrium | *NAGPA* | -3.192755405 | 0.001489444 | 0.001888861 | 1 |
| T2DM | endometrium | *ZNF500* | 4.383394906 | 0.003034404 | 0.003747245 | 1 |
| T2DM | endometrium | *NME4* | -3.501566961 | 0.012243497 | 0.014464438 | 1 |
| T2DM | endometrium | *NOMO3* | 2.061253037 | 0.007844576 | 0.009393357 | 1 |
| T2DM | endometrium | *LMF1* | 3.442678624 | 0.027667541 | 0.031796046 | 1 |
| T2DM | endometrium | *NARFL* | 4.623823529 | 9.78E-08 | 4.42E-07 | 0.000474732 |
| T2DM | endometrium | *MTHFSD* | 1.814075229 | 7.41E-06 | 1.22E-05 | 0.035948381 |
| T2DM | endometrium | *CLCN7* | 1.828896129 | 1.70E-05 | 2.63E-05 | 0.082345263 |
| T2DM | endometrium | *HAGHL* | 2.641438953 | 0.002087917 | 0.002618396 | 1 |
| T2DM | endometrium | *SLC7A5* | 6.099866467 | 3.39E-06 | 6.01E-06 | 0.016438816 |
| T2DM | endometrium | *RHBDL1* | -2.032611914 | 1.83E-06 | 3.47E-06 | 0.00890266 |
| T2DM | endometrium | *NUBP1* | 7.752709543 | 5.16E-07 | 1.16E-06 | 0.002504312 |
| T2DM | endometrium | *CAPN15* | -2.079656169 | 3.81E-05 | 5.66E-05 | 0.184921648 |
| T2DM | endometrium | *CLUAP1* | 4.440883956 | 1.47E-07 | 4.78E-07 | 0.000715196 |
| T2DM | endometrium | *EARS2* | -2.773125011 | 0.000788945 | 0.001024889 | 1 |
| T2DM | endometrium | *CPPED1* | 3.607236358 | 3.59E-08 | 4.34E-07 | 0.000174411 |
| T2DM | endometrium | *HMOX2* | 7.326021785 | 6.51E-08 | 4.34E-07 | 0.00031567 |
| T2DM | endometrium | *TOX3* | -4.877415531 | 0.000172014 | 0.000237105 | 0.834609728 |
| T2DM | endometrium | *RRN3P2* | 2.292062649 | 8.98E-07 | 1.85E-06 | 0.00435469 |
| T2DM | endometrium | *RBL2* | 3.892230878 | 2.37E-09 | 3.52E-07 | 1.15E-05 |
| T2DM | endometrium | *PYCARD* | 3.410140705 | 4.88E-08 | 4.34E-07 | 0.000236886 |
| T2DM | endometrium | *CDIPT* | 3.284783814 | 4.45E-08 | 4.34E-07 | 0.000215912 |
| T2DM | endometrium | *KAT8* | -2.911841445 | 0.007898429 | 0.009453176 | 1 |
| T2DM | endometrium | *TMC5* | 2.527073603 | 4.38E-07 | 1.01E-06 | 0.002124358 |
| T2DM | endometrium | *C16orf62* | 4.236983576 | 6.66E-08 | 4.34E-07 | 0.000323369 |
| T2DM | endometrium | *RNF40* | 4.497688272 | 3.76E-08 | 4.34E-07 | 0.000182542 |
| T2DM | endometrium | *KNOP1* | 1.998020578 | 2.17E-07 | 5.95E-07 | 0.001053282 |
| T2DM | endometrium | *IQCH* | 2.282929777 | 1.14E-07 | 4.57E-07 | 0.000551045 |
| T2DM | endometrium | *LACTB* | 2.784579801 | 2.50E-07 | 6.60E-07 | 0.001211922 |
| T2DM | endometrium | *HERC1* | 4.440422801 | 1.93E-07 | 5.48E-07 | 0.000937985 |
| T2DM | endometrium | *TRIP4* | 2.802925642 | 9.38E-08 | 4.39E-07 | 0.000454999 |
| T2DM | endometrium | *AP3B2* | 1.20914636 | 0.018497671 | 0.021515036 | 1 |
| T2DM | endometrium | *CTSH* | 1.888298426 | 5.46E-08 | 4.34E-07 | 0.000265139 |
| T2DM | endometrium | *TTC23* | 5.267955732 | 1.03E-07 | 4.47E-07 | 0.000498476 |
| T2DM | endometrium | *FAH* | 3.019992914 | 1.14E-07 | 4.57E-07 | 0.000552635 |
| T2DM | endometrium | *SLC30A4* | 2.399219686 | 1.29E-07 | 4.78E-07 | 0.000626579 |
| T2DM | endometrium | *ZDHHC2* | 2.342471535 | 3.98E-07 | 9.36E-07 | 0.001929903 |
| T2DM | endometrium | *ZFAND1* | -2.701390127 | 0.007262515 | 0.00872869 | 1 |
| T2DM | endometrium | *EYA1* | -1.660025594 | 2.43E-05 | 3.69E-05 | 0.118017535 |

| | | | | | | |
|---|---|---|---|---|---|---|
| T2DM | endometrium | *IMPAD1* | 6.53245905 | 1.55E-07 | 4.80E-07 | 0.000752544 |
| T2DM | endometrium | *SFRP1* | 6.035372797 | 8.72E-08 | 4.38E-07 | 0.000423286 |
| T2DM | endometrium | *NIPAL2* | 3.435927932 | 0.026253472 | 0.030199585 | 1 |
| T2DM | endometrium | *RAB2A* | 4.037281431 | 4.21E-08 | 4.34E-07 | 0.000204438 |
| T2DM | endometrium | *ARMC1* | 2.363824533 | 5.57E-07 | 1.23E-06 | 0.002700834 |
| T2DM | endometrium | *SPAG1* | 2.942290453 | 8.30E-08 | 4.37E-07 | 0.000402751 |
| T2DM | endometrium | *CHRAC1* | -1.818852884 | 8.36E-06 | 1.37E-05 | 0.040574291 |
| T2DM | endometrium | *SNX16* | 3.382495463 | 7.63E-08 | 4.36E-07 | 0.00037026 |
| T2DM | endometrium | *GSDMD* | -2.821837917 | 0.014727846 | 0.017248252 | 1 |
| T2DM | endometrium | *TSTA3* | 4.219912072 | 0.020680799 | 0.023959703 | 1 |
| T2DM | endometrium | *SQLE* | 2.474973615 | 6.02E-07 | 1.32E-06 | 0.002922597 |
| T2DM | endometrium | *R3HCC1* | 3.957811948 | 0.003147992 | 0.003882576 | 1 |
| T2DM | endometrium | *TNFRSF10A* | 3.113047681 | 4.15E-08 | 4.34E-07 | 0.000201463 |
| T2DM | endometrium | *ERICH1* | 4.709897817 | 3.88E-07 | 9.19E-07 | 0.001884574 |
| T2DM | endometrium | *TUSC3* | 1.847125466 | 1.45E-07 | 4.78E-07 | 0.000701761 |
| T2DM | endometrium | *MAN2B1* | 4.050461494 | 9.55E-07 | 1.95E-06 | 0.004632102 |
| T2DM | endometrium | *TULP2* | 15.94423602 | 1.50E-07 | 4.78E-07 | 0.000726045 |
| T2DM | endometrium | *NFKBIB* | 3.448839045 | 1.90E-07 | 5.44E-07 | 0.00092299 |
| T2DM | endometrium | *SNRNP70* | 5.203536473 | 2.67E-05 | 4.04E-05 | 0.129484732 |
| T2DM | endometrium | *LIN7B* | 9.274440514 | 8.81E-07 | 1.83E-06 | 0.004276519 |
| T2DM | endometrium | *PPP1R37* | 3.26824694 | 0.00216327 | 0.002707994 | 1 |
| T2DM | endometrium | *FCGRT* | 1.806500608 | 4.60E-07 | 1.04E-06 | 0.002229815 |
| T2DM | endometrium | *ARHGEF18* | 7.463699279 | 0.001076724 | 0.001381349 | 1 |
| T2DM | endometrium | *PEX11G* | -1.978494717 | 1.22E-05 | 1.94E-05 | 0.059247165 |
| T2DM | endometrium | *ERCC2* | 3.415553343 | 1.32E-07 | 4.78E-07 | 0.000639813 |
| T2DM | endometrium | *CD37* | 2.634177751 | 1.19E-07 | 4.65E-07 | 0.000579074 |
| T2DM | endometrium | *AMH* | 29.80444057 | 1.60E-07 | 4.87E-07 | 0.000777186 |
| T2DM | endometrium | *DKKL1* | 2.278726863 | 0.026286797 | 0.030230751 | 1 |
| T2DM | endometrium | *FCER2* | 9.213923551 | 1.47E-07 | 4.78E-07 | 0.00071411 |
| T2DM | endometrium | *CLEC4M* | 23.98071699 | 0.002675687 | 0.003316923 | 1 |
| T2DM | endometrium | *TLE6* | 7.653963002 | 3.97E-07 | 9.36E-07 | 0.001926901 |
| T2DM | endometrium | *NOVA2* | 2.479618297 | 8.93E-07 | 1.85E-06 | 0.004334587 |
| T2DM | endometrium | *SGTA* | 5.32030476 | 1.34E-07 | 4.78E-07 | 0.000651454 |
| T2DM | endometrium | *KIR3DX1* | 16.96706665 | 1.27E-06 | 2.51E-06 | 0.006164912 |
| T2DM | endometrium | *MED25* | 5.602858906 | 0.008392913 | 0.010020829 | 1 |
| T2DM | endometrium | *TIMM44* | 6.224518662 | 8.12E-08 | 4.36E-07 | 0.000394062 |
| T2DM | endometrium | *IL27RA* | 3.846659741 | 7.76E-08 | 4.36E-07 | 0.000376375 |
| T2DM | endometrium | *FAM32A* | 4.167202451 | 1.30E-07 | 4.78E-07 | 0.000630063 |
| T2DM | endometrium | *C19orf44* | -1.439984439 | 0.020158293 | 0.023365513 | 1 |
| T2DM | endometrium | *RASAL3* | 3.88220904 | 1.17E-07 | 4.61E-07 | 0.000566611 |
| T2DM | endometrium | *ZNF419* | 2.245007224 | 0.002618526 | 0.003249384 | 1 |
| T2DM | endometrium | *AURKC* | 4.288767219 | 0.000959568 | 0.001236607 | 1 |
| T2DM | endometrium | *ANKRD27* | 5.172606051 | 6.41E-08 | 4.34E-07 | 0.00031083 |
| T2DM | endometrium | *TGFB1* | 2.975054229 | 6.13E-08 | 4.34E-07 | 0.000297639 |
| T2DM | endometrium | *CEACAM4* | 5.835258542 | 0.000480662 | 0.000636336 | 1 |
| T2DM | endometrium | *MRPL4* | 4.139526872 | 3.78E-08 | 4.34E-07 | 0.000183445 |
| T2DM | endometrium | *CD33* | 1.869185877 | 1.41E-05 | 2.22E-05 | 0.068545843 |
| T2DM | endometrium | *RABAC1* | 4.163678677 | 2.97E-08 | 4.34E-07 | 0.000144162 |
| T2DM | endometrium | *GRIN2D* | -2.028958612 | 9.22E-06 | 1.50E-05 | 0.044757467 |
| T2DM | endometrium | *CCDC114* | -6.346785143 | 0.000109905 | 0.00015421 | 0.533258441 |

| | | | | | | |
|---|---|---|---|---|---|---|
| T2DM | endometrium | *CARD8* | 2.712664146 | 1.46E-07 | 4.78E-07 | 0.000709451 |
| T2DM | endometrium | *LIG1* | -3.239227053 | 0.000384127 | 0.000513723 | 1 |
| T2DM | endometrium | *ZNF175* | 2.718430376 | 2.47E-07 | 6.54E-07 | 0.001197142 |
| T2DM | endometrium | *PLA2G4C* | 2.037155179 | 0.000138597 | 0.000192685 | 0.672472383 |
| T2DM | endometrium | *CAPS* | -1.385431695 | 5.48E-06 | 9.30E-06 | 0.026594695 |
| T2DM | endometrium | *MIER2* | 2.614710115 | 8.18E-08 | 4.36E-07 | 0.000396965 |
| T2DM | endometrium | *PLEKHA4* | 4.738037653 | 6.47E-08 | 4.34E-07 | 0.000314107 |
| T2DM | endometrium | *TNPO2* | 6.914124409 | 1.06E-05 | 1.70E-05 | 0.051250178 |
| T2DM | endometrium | *GCDH* | 1.971173274 | 7.12E-06 | 1.18E-05 | 0.034566635 |
| T2DM | endometrium | *DNASE2* | -3.50026477 | 0.011681419 | 0.013813855 | 1 |
| T2DM | endometrium | *TFPT* | 3.275975495 | 1.11E-07 | 4.57E-07 | 0.000536827 |
| T2DM | endometrium | *RAB3A* | 11.2653911 | 1.34E-07 | 4.78E-07 | 0.000649211 |
| T2DM | endometrium | *UPK1A* | 8.551907207 | 4.89E-05 | 7.16E-05 | 0.23743273 |
| T2DM | endometrium | *ETV2* | 25.804213 | 1.36E-06 | 2.67E-06 | 0.00661135 |
| T2DM | endometrium | *LSR* | 3.004167387 | 1.77E-06 | 3.37E-06 | 0.008604515 |
| T2DM | endometrium | *ZNF14* | 7.301566245 | 1.46E-07 | 4.78E-07 | 0.00070807 |
| T2DM | endometrium | *ZNF85* | 6.812134661 | 6.04E-08 | 4.34E-07 | 0.000292839 |
| T2DM | endometrium | *ETHE1* | 4.051674295 | 1.30E-07 | 4.78E-07 | 0.000630104 |
| T2DM | endometrium | *SMG9* | 2.597699387 | 1.58E-07 | 4.85E-07 | 0.00076793 |
| T2DM | endometrium | *GTPBP10* | 4.832094551 | 5.99E-05 | 8.65E-05 | 0.29052553 |
| T2DM | endometrium | *RASA4* | -1.82300832 | 1.86E-06 | 3.52E-06 | 0.009042657 |
| T2DM | endometrium | *BET1* | 3.484426891 | 8.04E-08 | 4.36E-07 | 0.000390231 |
| T2DM | endometrium | *DNAH11* | 2.833763089 | 1.06E-07 | 4.48E-07 | 0.000514287 |
| T2DM | endometrium | *STEAP1B* | 5.325958296 | 5.03E-08 | 4.34E-07 | 0.000244036 |
| T2DM | endometrium | *PTN* | 2.66296246 | 9.73E-08 | 4.42E-07 | 0.000472283 |
| T2DM | endometrium | *ADAP1* | 9.116856642 | 1.46E-07 | 4.78E-07 | 0.000708328 |
| T2DM | endometrium | *MET* | 1.547627107 | 4.28E-07 | 9.89E-07 | 0.002075392 |
| T2DM | endometrium | *RNF32* | 2.683839179 | 1.38E-06 | 2.70E-06 | 0.006704953 |
| T2DM | endometrium | *IQCE* | 2.475700993 | 1.32E-07 | 4.78E-07 | 0.000640082 |
| T2DM | endometrium | *VIPR2* | 5.145094554 | 1.33E-07 | 4.78E-07 | 0.000647181 |
| T2DM | endometrium | *HIBADH* | 3.974096933 | 3.22E-08 | 4.34E-07 | 0.000156416 |
| T2DM | endometrium | *CPVL* | 1.506261467 | 1.61E-07 | 4.89E-07 | 0.000781563 |
| T2DM | endometrium | *CHN2* | 2.514027272 | 2.70E-10 | 1.09E-07 | 1.31E-06 |
| T2DM | endometrium | *GRB10* | 1.891064841 | 9.09E-08 | 4.38E-07 | 0.000441027 |
| T2DM | endometrium | *ABHD11* | 4.044192768 | 3.92E-07 | 9.27E-07 | 0.001902765 |
| T2DM | endometrium | *PLEKHA8* | 7.031926065 | 9.18E-08 | 4.38E-07 | 0.000445612 |
| T2DM | endometrium | *EPHB6* | 1.993727639 | 3.09E-05 | 4.64E-05 | 0.149719686 |
| T2DM | endometrium | *CHCHD2* | 2.757561957 | 8.97E-08 | 4.38E-07 | 0.000435384 |
| T2DM | endometrium | *CCL24* | -1.127095909 | 2.15E-06 | 4.01E-06 | 0.010431695 |
| T2DM | endometrium | *ZKSCAN1* | 2.609262214 | 9.82E-08 | 4.42E-07 | 0.000476551 |
| T2DM | endometrium | *EIF3B* | 4.225662137 | 0.013673828 | 0.016087637 | 1 |
| T2DM | endometrium | *TAF6* | 4.032203797 | 0.000110147 | 0.000154461 | 0.534434708 |
| T2DM | endometrium | *TFR2* | -4.697216637 | 5.27E-05 | 7.68E-05 | 0.255935825 |
| T2DM | endometrium | *MOSPD3* | -2.193243903 | 5.23E-06 | 8.93E-06 | 0.025385768 |
| T2DM | endometrium | *RBM28* | 4.637583293 | 0.000352062 | 0.000472925 | 1 |
| T2DM | endometrium | *IMPDH1* | -3.347223524 | 0.002290188 | 0.00286023 | 1 |
| T2DM | endometrium | *C1GALT1* | 5.158079814 | 5.64E-08 | 4.34E-07 | 0.000273519 |
| T2DM | endometrium | *NRF1* | 3.243417575 | 3.98E-08 | 4.34E-07 | 0.000193267 |
| T2DM | endometrium | *ZNF862* | 2.822605972 | 7.25E-08 | 4.34E-07 | 0.000351887 |
| T2DM | endometrium | *MEST* | 2.657450866 | 0.000414398 | 0.000552331 | 1 |

| | | | | | | |
|---|---|---|---|---|---|---|
| T2DM | endometrium | *ACTR3C* | 2.828493079 | 8.84E-08 | 4.38E-07 | 0.000428879 |
| T2DM | endometrium | *POU6F2* | -1.866071306 | 4.45E-05 | 6.55E-05 | 0.215804349 |
| T2DM | endometrium | *TSPAN13* | 5.359319478 | 6.18E-08 | 4.34E-07 | 0.000300066 |
| T2DM | endometrium | *RARRES2* | 3.854261624 | 4.96E-08 | 4.34E-07 | 0.000240521 |
| T2DM | endometrium | *TMEM176B* | 2.442015371 | 3.84E-06 | 6.74E-06 | 0.018623467 |
| T2DM | endometrium | *COA1* | 3.466520577 | 4.19E-08 | 4.34E-07 | 0.000203355 |
| T2DM | endometrium | *BLVRA* | 5.441518508 | 4.54E-06 | 7.86E-06 | 0.022032141 |
| T2DM | endometrium | *URGCP* | 5.005496797 | 1.31E-08 | 4.34E-07 | 6.34E-05 |
| T2DM | endometrium | *GCK* | 55.93392987 | 9.62E-05 | 0.000135517 | 0.466719263 |
| T2DM | endometrium | *BCL7B* | 2.91543259 | 6.93E-08 | 4.34E-07 | 0.000336402 |
| T2DM | endometrium | *TBL2* | -2.612151586 | 0.000856398 | 0.001109249 | 1 |
| T2DM | endometrium | *SPATA6L* | 3.284239595 | 5.92E-08 | 4.34E-07 | 0.000287435 |
| T2DM | endometrium | *SLC1A1* | 1.818116998 | 2.88E-08 | 4.34E-07 | 0.00013982 |
| T2DM | endometrium | *FKTN* | 4.319598699 | 5.70E-08 | 4.34E-07 | 0.000276341 |
| T2DM | endometrium | *NMRK1* | 5.552420047 | 7.12E-06 | 1.18E-05 | 0.034525837 |
| T2DM | endometrium | *MEGF9* | 3.693767576 | 5.81E-08 | 4.34E-07 | 0.00028207 |
| T2DM | endometrium | *TRIM14* | 2.071123551 | 1.03E-07 | 4.47E-07 | 0.000498808 |
| T2DM | endometrium | *CORO2A* | 3.504546521 | 3.21E-07 | 7.94E-07 | 0.001559021 |
| T2DM | endometrium | *C5* | 5.818474595 | 0.000955386 | 0.001232199 | 1 |
| T2DM | endometrium | *OGN* | 2.391062352 | 7.40E-07 | 1.58E-06 | 0.003592147 |
| T2DM | endometrium | *ECM2* | 5.059551897 | 4.56E-06 | 7.89E-06 | 0.022136472 |
| T2DM | endometrium | *PTGR1* | 4.277540374 | 2.09E-05 | 3.20E-05 | 0.101645975 |
| T2DM | endometrium | *SUSD1* | 1.806971425 | 4.17E-07 | 9.71E-07 | 0.002021407 |
| T2DM | endometrium | *PLGRKT* | -2.029006967 | 4.72E-05 | 6.93E-05 | 0.229256329 |
| T2DM | endometrium | *DOCK8* | 1.3193714 | 6.61E-08 | 4.34E-07 | 0.000320912 |
| T2DM | endometrium | *KANK1* | 3.143433336 | 1.26E-06 | 2.50E-06 | 0.006136704 |
| T2DM | endometrium | *NCS1* | 3.526272493 | 4.39E-06 | 7.63E-06 | 0.021321878 |
| T2DM | endometrium | *FUBP3* | 5.411653503 | 1.64E-06 | 3.14E-06 | 0.007934233 |
| T2DM | endometrium | *RGP1* | 2.806255905 | 1.04E-07 | 4.47E-07 | 0.000505739 |
| T2DM | endometrium | *ATRNL1* | -0.989029669 | 0.015846501 | 0.018518117 | 1 |
| T2DM | endometrium | *HPS1* | 4.424095678 | 2.45E-07 | 6.51E-07 | 0.0011881 |
| T2DM | endometrium | *PHYH* | 3.816330318 | 7.26E-05 | 0.000103824 | 0.352171178 |
| T2DM | endometrium | *DNMBP* | 2.545949398 | 3.27E-07 | 8.03E-07 | 0.001585756 |
| T2DM | endometrium | *CUBN* | 4.334034421 | 3.63E-08 | 4.34E-07 | 0.00017629 |
| T2DM | endometrium | *TRDMT1* | 7.246785695 | 1.16E-06 | 2.31E-06 | 0.00563095 |
| T2DM | endometrium | *ATE1* | 4.866975824 | 7.83E-08 | 4.36E-07 | 0.000380055 |
| T2DM | endometrium | *PLEKHA1* | 4.194491301 | 0.002526614 | 0.003142496 | 1 |
| T2DM | endometrium | *CDH23* | 2.992869107 | 7.08E-08 | 4.34E-07 | 0.000343742 |
| T2DM | endometrium | *SPOCK2* | 1.918072251 | 0.000273882 | 0.000371259 | 1 |
| T2DM | endometrium | *BMPR1A* | 4.130873232 | 0.000572897 | 0.000754122 | 1 |
| T2DM | endometrium | *LIPA* | 1.816990139 | 9.91E-08 | 4.44E-07 | 0.000481014 |
| T2DM | endometrium | *SFXN3* | 2.496666199 | 1.51E-07 | 4.78E-07 | 0.000730484 |
| T2DM | endometrium | *TNKS2* | 3.132585763 | 5.43E-08 | 4.34E-07 | 0.000263622 |
| T2DM | endometrium | *ANKRD26* | 3.938084262 | 5.65E-08 | 4.34E-07 | 0.000274072 |
| T2DM | endometrium | *ACBD5* | 4.675901138 | 0.007639672 | 0.009168362 | 1 |
| T2DM | endometrium | *PITRM1* | 6.936628725 | 4.71E-06 | 8.12E-06 | 0.022848428 |
| T2DM | endometrium | *TFAM* | 7.186495946 | 1.16E-07 | 4.60E-07 | 0.000563732 |
| T2DM | endometrium | *RPL28* | 4.349330501 | 9.02E-06 | 1.47E-05 | 0.043778507 |
| T2DM | endometrium | *PBLD* | 2.69033367 | 1.71E-07 | 5.07E-07 | 0.000829479 |
| T2DM | endometrium | *TSPAN14* | 4.247796346 | 1.09E-05 | 1.75E-05 | 0.052864176 |

| | | | | | | |
|---|---|---|---|---|---|---|
| T2DM | endometrium | *TADA2A* | 2.070240184 | 3.81E-07 | 9.06E-07 | 0.001847548 |
| T2DM | endometrium | *PSMB3* | 6.97145515 | 1.21E-07 | 4.69E-07 | 0.000588685 |
| T2DM | endometrium | *RPL19* | 5.388476574 | 7.05E-06 | 1.17E-05 | 0.034215033 |
| T2DM | endometrium | *RAPGEFL1* | 3.86843724 | 2.83E-07 | 7.22E-07 | 0.001375106 |
| T2DM | endometrium | *WNT3* | 4.387103159 | 1.10E-06 | 2.22E-06 | 0.005359018 |
| T2DM | endometrium | *RAD51C* | 2.153940804 | 2.45E-06 | 4.51E-06 | 0.011911098 |
| T2DM | endometrium | *TRIM37* | 2.716286925 | 1.15E-07 | 4.59E-07 | 0.000557837 |
| T2DM | endometrium | *PNPO* | 10.05048895 | 0.000110695 | 0.000155184 | 0.537092668 |
| T2DM | endometrium | *TRIM16L* | 3.640309316 | 1.44E-07 | 4.78E-07 | 0.000696805 |
| T2DM | endometrium | *CDK5RAP3* | -2.476160713 | 4.19E-05 | 6.19E-05 | 0.203393383 |
| T2DM | endometrium | *PIGL* | 4.520578056 | 1.25E-07 | 4.74E-07 | 0.000608114 |
| T2DM | endometrium | *MED13* | 3.48211847 | 1.11E-07 | 4.57E-07 | 0.00053685 |
| T2DM | endometrium | *ENO3* | 5.972042739 | 1.31E-07 | 4.78E-07 | 0.000633583 |
| T2DM | endometrium | *CHRNE* | 1.461874832 | 0.013168443 | 0.015515611 | 1 |
| T2DM | endometrium | *RAI1* | 2.893718353 | 8.08E-08 | 4.36E-07 | 0.000391896 |
| T2DM | endometrium | *GOSR1* | 5.709269868 | 1.73E-05 | 2.68E-05 | 0.084065182 |
| T2DM | endometrium | *DRG2* | 2.659349844 | 1.84E-07 | 5.31E-07 | 0.000894818 |
| T2DM | endometrium | *FTSJ3* | -1.528606196 | 0.007812524 | 0.009357286 | 1 |
| T2DM | endometrium | *ALDH3A1* | -6.606996032 | 0.002979236 | 0.003681929 | 1 |
| T2DM | endometrium | *ICAM2* | 6.025653167 | 4.86E-08 | 4.34E-07 | 0.000235706 |
| T2DM | endometrium | *DDX5* | 2.977946884 | 2.77E-08 | 4.34E-07 | 0.000134213 |
| T2DM | endometrium | *C17orf75* | 2.449094335 | 2.86E-07 | 7.27E-07 | 0.001389536 |
| T2DM | endometrium | *PSMD11* | 3.643695636 | 2.17E-07 | 5.95E-07 | 0.001054239 |
| T2DM | endometrium | *PEX12* | -6.16030039 | 7.93E-05 | 0.000112898 | 0.384757097 |
| T2DM | endometrium | *DHX58* | -2.606464057 | 1.34E-05 | 2.12E-05 | 0.065163266 |
| T2DM | endometrium | *RAB5C* | 2.921554732 | 6.88E-08 | 4.34E-07 | 0.00033364 |
| T2DM | endometrium | *NAGLU* | 2.911783876 | 3.54E-08 | 4.34E-07 | 0.000171916 |
| T2DM | endometrium | *HSD17B1* | 4.484430265 | 1.12E-07 | 4.57E-07 | 0.000542716 |
| T2DM | endometrium | *EZH1* | 1.94007425 | 1.80E-07 | 5.23E-07 | 0.000873565 |
| T2DM | endometrium | *ABCC3* | 3.653172936 | 1.29E-07 | 4.78E-07 | 0.000625588 |
| T2DM | endometrium | *EFTUD2* | 2.226282544 | 1.24E-07 | 4.71E-07 | 0.000601272 |
| T2DM | endometrium | *HLF* | 2.05594055 | 3.26E-07 | 8.01E-07 | 0.001579364 |
| T2DM | endometrium | *EFNB3* | -3.390578903 | 1.02E-05 | 1.64E-05 | 0.04935281 |
| T2DM | endometrium | *FAM20A* | -2.171900873 | 0.003610279 | 0.00442127 | 1 |
| T2DM | endometrium | *DPH1* | -2.027730036 | 1.50E-05 | 2.34E-05 | 0.072758783 |
| T2DM | endometrium | *DHRS7B* | 2.468006291 | 7.58E-08 | 4.36E-07 | 0.000367929 |
| T2DM | endometrium | *RCVRN* | -2.618747971 | 0.001303995 | 0.001663245 | 1 |
| T2DM | endometrium | *MYH3* | 4.069105994 | 1.67E-07 | 4.99E-07 | 0.000807937 |
| T2DM | endometrium | *SLAIN2* | 5.900354205 | 6.58E-06 | 1.10E-05 | 0.031903524 |
| T2DM | endometrium | *OCIAD1* | 5.206190833 | 1.30E-06 | 2.55E-06 | 0.006290708 |
| T2DM | endometrium | *UBE2D3* | 3.186559171 | 4.43E-08 | 4.34E-07 | 0.000214775 |
| T2DM | endometrium | *KLHL2* | 5.120968594 | 6.90E-08 | 4.34E-07 | 0.000334874 |
| T2DM | endometrium | *CPE* | 3.086661886 | 1.01E-08 | 4.34E-07 | 4.90E-05 |
| T2DM | endometrium | *WFS1* | 8.419383765 | 0.000189842 | 0.000260791 | 0.921112084 |
| T2DM | endometrium | *GAR1* | -1.917171229 | 1.74E-05 | 2.69E-05 | 0.084472555 |
| T2DM | endometrium | *AADAT* | -1.093716148 | 3.43E-06 | 6.07E-06 | 0.016626399 |
| T2DM | endometrium | *TRIM2* | 2.571839217 | 1.82E-07 | 5.25E-07 | 0.000880953 |
| T2DM | endometrium | *STIM2* | 3.165031778 | 4.91E-08 | 4.34E-07 | 0.000238002 |
| T2DM | endometrium | *BST1* | 3.328165818 | 3.51E-08 | 4.34E-07 | 0.000170396 |
| T2DM | endometrium | *UFSP2* | 3.994010035 | 5.35E-08 | 4.34E-07 | 0.000259707 |

| | | | | | | |
|---|---|---|---|---|---|---|
| T2DM | endometrium | *KLHL5* | 2.274224704 | 8.49E-08 | 4.38E-07 | 0.000412089 |
| T2DM | endometrium | *FAM149A* | -1.578498081 | 6.65E-05 | 9.54E-05 | 0.322457043 |
| T2DM | endometrium | *DDX25* | 3.500194751 | 4.36E-08 | 4.34E-07 | 0.000211524 |
| T2DM | endometrium | *CTSC* | 1.746207081 | 7.87E-08 | 4.36E-07 | 0.000381744 |
| T2DM | endometrium | *ELP4* | 4.414982304 | 4.79E-06 | 8.25E-06 | 0.02325498 |
| T2DM | endometrium | *TECTA* | 5.814331589 | 1.51E-07 | 4.78E-07 | 0.000732703 |
| T2DM | endometrium | *SC5D* | 5.158098139 | 4.20E-07 | 9.76E-07 | 0.002035847 |
| T2DM | endometrium | *B3GAT1* | -6.571021574 | 4.98E-06 | 8.53E-06 | 0.024149344 |
| T2DM | endometrium | *P2RX3* | -2.14253389 | 1.71E-06 | 3.26E-06 | 0.008289295 |
| T2DM | endometrium | *DNAJC4* | 2.983537657 | 4.29E-07 | 9.89E-07 | 0.002080385 |
| T2DM | endometrium | *UNC93B1* | 2.607256293 | 2.84E-07 | 7.23E-07 | 0.001379218 |
| T2DM | endometrium | *PUS3* | 4.421977795 | 1.13E-07 | 4.57E-07 | 0.00054951 |
| T2DM | endometrium | *DCPS* | 8.641200468 | 6.42E-05 | 9.23E-05 | 0.311420447 |
| T2DM | endometrium | *NRXN2* | -1.714283016 | 3.31E-06 | 5.88E-06 | 0.016049827 |
| T2DM | endometrium | *ST3GAL4* | 3.07677252 | 7.92E-07 | 1.66E-06 | 0.003840481 |
| T2DM | endometrium | *CCND1* | 3.544601101 | 3.91E-05 | 5.80E-05 | 0.189712445 |
| T2DM | endometrium | *CCDC86* | 5.468783128 | 9.18E-06 | 1.49E-05 | 0.044560309 |
| T2DM | endometrium | *PANX1* | -0.922743168 | 2.88E-06 | 5.21E-06 | 0.013975194 |
| T2DM | endometrium | *CEP164* | 4.630338342 | 9.42E-08 | 4.39E-07 | 0.000457091 |
| T2DM | endometrium | *GALNT18* | 1.693698857 | 1.29E-07 | 4.78E-07 | 0.000627639 |
| T2DM | endometrium | *KIAA1549L* | 9.71682356 | 1.24E-07 | 4.71E-07 | 0.000601293 |
| T2DM | endometrium | *ACCS* | 2.220436117 | 1.59E-07 | 4.86E-07 | 0.000770204 |
| T2DM | endometrium | *SLC22A18* | 1.896338896 | 0.00018725 | 0.000257449 | 0.908538647 |
| T2DM | endometrium | *SLC35F2* | -3.445698509 | 0.024595567 | 0.028373203 | 1 |
| T2DM | endometrium | *ELMOD1* | -4.181519645 | 0.010740535 | 0.012726026 | 1 |
| T2DM | endometrium | *RPS13* | 4.13157829 | 0.001574476 | 0.00199201 | 1 |
| T2DM | endometrium | *NDUFS8* | 5.568210144 | 3.75E-07 | 8.97E-07 | 0.001820035 |
| T2DM | endometrium | *TCIRG1* | -2.971152626 | 0.003816091 | 0.004660376 | 1 |
| T2DM | endometrium | *GTF2H1* | 5.422113533 | 9.83E-06 | 1.59E-05 | 0.047707007 |
| T2DM | endometrium | *PTPN5* | 2.346982922 | 0.005170251 | 0.006271515 | 1 |
| T2DM | endometrium | *PRDM4* | 5.523265608 | 0.001245893 | 0.001592485 | 1 |
| T2DM | endometrium | *COQ5* | 1.825670504 | 3.15E-08 | 4.34E-07 | 0.000152932 |
| T2DM | endometrium | *CORO1C* | 3.117352649 | 5.75E-08 | 4.34E-07 | 0.000278939 |
| T2DM | endometrium | *ASIC1* | 5.882906207 | 1.60E-07 | 4.86E-07 | 0.000774669 |
| T2DM | endometrium | *CAPRIN2* | 3.916789889 | 3.88E-05 | 5.76E-05 | 0.188187653 |
| T2DM | endometrium | *KCTD10* | 3.040958533 | 2.60E-07 | 6.77E-07 | 0.001262031 |
| T2DM | endometrium | *IL23A* | 18.14595537 | 1.43E-07 | 4.78E-07 | 0.000692811 |
| T2DM | endometrium | *LTA4H* | 1.804776234 | 3.39E-08 | 4.34E-07 | 0.000164709 |
| T2DM | endometrium | *PRR4* | 6.456027853 | 3.77E-09 | 4.07E-07 | 1.83E-05 |
| T2DM | endometrium | *PARP11* | 5.776594413 | 1.54E-07 | 4.78E-07 | 0.000746299 |
| T2DM | endometrium | *GPN3* | 8.348083939 | 1.25E-07 | 4.73E-07 | 0.000604705 |
| T2DM | endometrium | *VPS29* | 3.939468174 | 7.60E-08 | 4.36E-07 | 0.000368737 |
| T2DM | endometrium | *AKAP3* | 1.823416891 | 0.001439821 | 0.001828799 | 1 |
| T2DM | endometrium | *ACAD10* | 3.660030585 | 2.89E-07 | 7.32E-07 | 0.001401795 |
| T2DM | endometrium | *LTBR* | 3.44430361 | 2.20E-07 | 6.00E-07 | 0.001068799 |
| T2DM | endometrium | *DDX55* | 6.087746655 | 4.09E-06 | 7.15E-06 | 0.019841243 |
| T2DM | endometrium | *ENDOU* | -3.529312375 | 6.02E-06 | 1.01E-05 | 0.029193033 |
| T2DM | endometrium | *VDR* | 7.600276349 | 7.09E-08 | 4.34E-07 | 0.000344094 |
| T2DM | endometrium | *STX2* | 3.367199634 | 3.72E-08 | 4.34E-07 | 0.000180305 |
| T2DM | endometrium | *CAND1* | 3.477508744 | 6.49E-08 | 4.34E-07 | 0.000314773 |

| | | | | | | |
|---|---|---|---|---|---|---|
| T2DM | endometrium | *MDM1* | -3.012200367 | 0.007730705 | 0.009268441 | 1 |
| T2DM | endometrium | *NUP107* | 4.933311868 | 1.10E-07 | 4.56E-07 | 0.000532875 |
| T2DM | endometrium | *KRR1* | 3.567012615 | 0.032438622 | 0.037112048 | 1 |
| T2DM | endometrium | *MRPL51* | -1.83944644 | 0.000234528 | 0.000319734 | 1 |
| T2DM | endometrium | *GNB3* | -3.566867583 | 7.24E-07 | 1.55E-06 | 0.003511095 |
| T2DM | endometrium | *CDCA3* | -1.09393732 | 4.18E-07 | 9.74E-07 | 0.002029844 |
| T2DM | endometrium | *CHPT1* | 2.52173075 | 1.05E-06 | 2.12E-06 | 0.005088732 |
| T2DM | endometrium | *GNPTAB* | 4.800773152 | 3.29E-07 | 8.08E-07 | 0.001596904 |
| T2DM | endometrium | *SPSB2* | 3.559448949 | 7.95E-07 | 1.67E-06 | 0.003856904 |
| T2DM | endometrium | *C12orf57* | 8.405225737 | 6.42E-07 | 1.39E-06 | 0.003116986 |
| T2DM | endometrium | *NT5DC3* | 6.332722357 | 6.74E-05 | 9.68E-05 | 0.327223963 |
| T2DM | endometrium | *GOLT1B* | 7.552385252 | 1.97E-08 | 4.34E-07 | 9.56E-05 |
| T2DM | endometrium | *C2CD5* | 2.611525731 | 6.33E-08 | 4.34E-07 | 0.00030721 |
| T2DM | endometrium | *BTN3A3* | 3.277535389 | 4.20E-06 | 7.31E-06 | 0.020370869 |
| T2DM | endometrium | *FRK* | -3.294120402 | 0.002583897 | 0.003208053 | 1 |
| T2DM | endometrium | *RWDD1* | 4.781645552 | 8.57E-07 | 1.78E-06 | 0.004157162 |
| T2DM | endometrium | *RSPH4A* | -1.998941398 | 1.58E-05 | 2.46E-05 | 0.076853222 |
| T2DM | endometrium | *SMIM8* | 4.355283069 | 4.11E-07 | 9.62E-07 | 0.001994358 |
| T2DM | endometrium | *FAM184A* | 3.598243584 | 3.28E-08 | 4.34E-07 | 0.000159095 |
| T2DM | endometrium | *RNGTT* | 4.920662464 | 1.62E-07 | 4.91E-07 | 0.000788232 |
| T2DM | endometrium | *GABRR2* | 4.659598877 | 1.10E-06 | 2.21E-06 | 0.005335604 |
| T2DM | endometrium | *HDDC2* | 4.090543034 | 6.16E-07 | 1.35E-06 | 0.002987765 |
| T2DM | endometrium | *UST* | -1.214137187 | 0.000221488 | 0.000302466 | 1 |
| T2DM | endometrium | *SLC26A8* | 3.340641048 | 1.99E-07 | 5.58E-07 | 0.000965576 |
| T2DM | endometrium | *MRPL18* | 3.191549977 | 8.39E-08 | 4.38E-07 | 0.000406841 |
| T2DM | endometrium | *RNF8* | 2.527402446 | 1.46E-07 | 4.78E-07 | 0.00070929 |
| T2DM | endometrium | *PHACTR1* | 1.679825664 | 7.31E-07 | 1.56E-06 | 0.003548931 |
| T2DM | endometrium | *MDGA1* | 2.987663976 | 2.07E-07 | 5.76E-07 | 0.00100631 |
| T2DM | endometrium | *CD83* | 2.898254348 | 4.53E-07 | 1.03E-06 | 0.00219757 |
| T2DM | endometrium | *MDN1* | 5.017771239 | 5.87E-07 | 1.29E-06 | 0.002847783 |
| T2DM | endometrium | *GLP1R* | 10.84462328 | 0.008485403 | 0.010124974 | 1 |
| T2DM | endometrium | *COL9A1* | 9.582356432 | 2.65E-08 | 4.34E-07 | 0.000128693 |
| T2DM | endometrium | *WASF1* | 5.296711186 | 3.62E-06 | 6.38E-06 | 0.017561977 |
| T2DM | endometrium | *VNN1* | 2.261683332 | 0.002251068 | 0.00281427 | 1 |
| T2DM | endometrium | *VNN2* | 6.707144564 | 1.75E-08 | 4.34E-07 | 8.48E-05 |
| T2DM | endometrium | *ACOT13* | 3.484650925 | 5.93E-08 | 4.34E-07 | 0.000287905 |
| T2DM | endometrium | *C6orf62* | 2.677282728 | 1.32E-07 | 4.78E-07 | 0.000638118 |
| T2DM | endometrium | *HBS1L* | 5.040602829 | 5.95E-06 | 1.00E-05 | 0.028860193 |
| T2DM | endometrium | *PEX7* | 2.175042083 | 5.97E-07 | 1.31E-06 | 0.002894429 |
| T2DM | endometrium | *FIG4* | 6.341244535 | 1.52E-05 | 2.37E-05 | 0.073591056 |
| T2DM | endometrium | *PERP* | 2.864235802 | 6.25E-07 | 1.36E-06 | 0.003030653 |
| T2DM | endometrium | *PHACTR2* | 3.433409754 | 3.97E-07 | 9.36E-07 | 0.00192811 |
| T2DM | endometrium | *CUTA* | -2.649744466 | 0.039290799 | 0.044667047 | 1 |
| T2DM | endometrium | *PACRG* | 2.290729969 | 1.52E-07 | 4.78E-07 | 0.000736979 |
| T2DM | endometrium | *QKI* | 2.591839019 | 5.20E-08 | 4.34E-07 | 0.000252073 |
| T2DM | endometrium | *FAM120B* | 3.844337003 | 3.20E-08 | 4.34E-07 | 0.000155179 |
| T2DM | endometrium | *GUCA1B* | -3.249908935 | 1.49E-05 | 2.34E-05 | 0.072536657 |
| T2DM | endometrium | *PRPH2* | -1.515916934 | 0.000210593 | 0.00028848 | 1 |
| T2DM | endometrium | *DNPH1* | 2.73929341 | 5.46E-08 | 4.34E-07 | 0.000264914 |
| T2DM | endometrium | *EXOC2* | 2.798128765 | 4.48E-08 | 4.34E-07 | 0.000217445 |

| | | | | | | |
|---|---|---|---|---|---|---|
| T2DM | endometrium | *IMPG1* | -8.302489595 | 0.00028655 | 0.000387389 | 1 |
| T2DM | endometrium | *BTN2A1* | 4.793832585 | 8.94E-08 | 4.38E-07 | 0.000433958 |
| T2DM | endometrium | *LY86* | 2.792549034 | 2.63E-07 | 6.82E-07 | 0.001274648 |
| T2DM | endometrium | *PRSS16* | 5.6564012 | 0.001718123 | 0.002164719 | 1 |
| T2DM | endometrium | *PCDHB2* | 6.202312356 | 9.91E-07 | 2.01E-06 | 0.004807369 |
| T2DM | endometrium | *NUDT12* | 2.542966505 | 1.23E-07 | 4.69E-07 | 0.00059469 |
| T2DM | endometrium | *CEP72* | 6.642484946 | 5.61E-07 | 1.24E-06 | 0.002724385 |
| T2DM | endometrium | *DAP* | -2.0770857 | 0.030613158 | 0.035073209 | 1 |
| T2DM | endometrium | *NME5* | -1.505246392 | 0.000309496 | 0.000417364 | 1 |
| T2DM | endometrium | *KIF20A* | -6.127830591 | 5.21E-07 | 1.17E-06 | 0.002529215 |
| T2DM | endometrium | *PFDN1* | 3.957491678 | 3.76E-08 | 4.34E-07 | 0.00018243 |
| T2DM | endometrium | *TMCO6* | 2.630471245 | 1.79E-06 | 3.41E-06 | 0.008707404 |
| T2DM | endometrium | *SPARC* | 3.565602509 | 4.16E-08 | 4.34E-07 | 0.000201695 |
| T2DM | endometrium | *IK* | 4.859926424 | 0.000118602 | 0.000165885 | 0.575456411 |
| T2DM | endometrium | *PCDHB15* | 2.216239902 | 0.00550719 | 0.006670216 | 1 |
| T2DM | endometrium | *HAVCR1* | 5.929166807 | 5.63E-05 | 8.17E-05 | 0.273366146 |
| T2DM | endometrium | *RNF130* | 2.869931719 | 2.62E-08 | 4.34E-07 | 0.000127005 |
| T2DM | endometrium | *ARSB* | 2.129054318 | 1.43E-07 | 4.78E-07 | 0.000694337 |
| T2DM | endometrium | *THBS4* | 3.033567443 | 5.17E-08 | 4.34E-07 | 0.000251028 |
| T2DM | endometrium | *BTNL8* | 1.269220496 | 0.000916757 | 0.001183951 | 1 |
| T2DM | endometrium | *RASGRF2* | 1.309441794 | 9.32E-07 | 1.91E-06 | 0.004521958 |
| T2DM | endometrium | *CDH6* | 2.134994581 | 2.87E-06 | 5.19E-06 | 0.013904502 |
| T2DM | endometrium | *LMNB1* | 6.154982065 | 2.28E-06 | 4.24E-06 | 0.011086008 |
| T2DM | endometrium | *SLC12A7* | 2.298872856 | 1.53E-07 | 4.78E-07 | 0.000740988 |
| T2DM | endometrium | *SKP1* | 4.453117453 | 5.58E-08 | 4.34E-07 | 0.000270801 |
| T2DM | endometrium | *PPWD1* | 3.435716406 | 5.56E-08 | 4.34E-07 | 0.000269702 |
| T2DM | endometrium | *TXNDC15* | 6.822167068 | 1.23E-07 | 4.70E-07 | 0.000597763 |
| T2DM | endometrium | *SMAD5* | 3.757204128 | 5.01E-08 | 4.34E-07 | 0.000243049 |
| T2DM | endometrium | *ATP6V0E1* | 5.414376204 | 2.07E-05 | 3.17E-05 | 0.100536768 |
| T2DM | endometrium | *BNIP1* | 2.599567065 | 8.31E-07 | 1.73E-06 | 0.004031136 |
| T2DM | endometrium | *UNC5A* | 11.63608388 | 1.46E-07 | 4.78E-07 | 0.000706198 |
| T2DM | endometrium | *ACTR8* | -2.867755539 | 2.83E-05 | 4.27E-05 | 0.137366329 |
| T2DM | endometrium | *TBCCD1* | 6.373924286 | 5.83E-08 | 4.34E-07 | 0.000283047 |
| T2DM | endometrium | *CLDN16* | 3.351034794 | 4.98E-07 | 1.12E-06 | 0.002417787 |
| T2DM | endometrium | *NPHP3* | 3.728842509 | 1.72E-09 | 3.07E-07 | 8.33E-06 |
| T2DM | endometrium | *PCCB* | 2.36665118 | 8.65E-09 | 4.34E-07 | 4.20E-05 |
| T2DM | endometrium | *CEP70* | 5.786786711 | 5.49E-06 | 9.31E-06 | 0.026621793 |
| T2DM | endometrium | *CYB561D2* | 2.44555959 | 2.95E-06 | 5.33E-06 | 0.014310941 |
| T2DM | endometrium | *C3orf14* | 3.357277551 | 1.19E-05 | 1.90E-05 | 0.057913007 |
| T2DM | endometrium | *CBLB* | 3.953009047 | 0.000255254 | 0.000346528 | 1 |
| T2DM | endometrium | *IQCG* | 3.449370415 | 4.37E-08 | 4.34E-07 | 0.000212027 |
| T2DM | endometrium | *NCBP2* | 5.253419075 | 0.003172152 | 0.003909393 | 1 |
| T2DM | endometrium | *SLC41A3* | 4.737484521 | 0.004576789 | 0.00556835 | 1 |
| T2DM | endometrium | *PLXNA1* | 3.07382209 | 1.48E-07 | 4.78E-07 | 0.000717779 |
| T2DM | endometrium | *SCAP* | 6.197775907 | 5.09E-07 | 1.14E-06 | 0.002469131 |
| T2DM | endometrium | *KIAA1257* | 6.102630466 | 1.48E-07 | 4.78E-07 | 0.000718918 |
| T2DM | endometrium | *NEK11* | -1.800283765 | 0.015781476 | 0.018451017 | 1 |
| T2DM | endometrium | *PLSCR4* | 3.643816512 | 8.62E-08 | 4.38E-07 | 0.000418312 |
| T2DM | endometrium | *HEMK1* | 2.609636721 | 8.30E-07 | 1.73E-06 | 0.004024987 |
| T2DM | endometrium | *COMMD2* | 6.378534141 | 5.29E-07 | 1.18E-06 | 0.002566691 |

| | | | | | | |
|---|---|---|---|---|---|---|
| T2DM | endometrium | *AADAC* | 1.147627721 | 0.002615539 | 0.003246507 | 1 |
| T2DM | endometrium | *ABHD14B* | 4.206350046 | 7.27E-07 | 1.55E-06 | 0.003525662 |
| T2DM | endometrium | *ARHGEF26* | 3.842388566 | 5.54E-08 | 4.34E-07 | 0.000268641 |
| T2DM | endometrium | *VIPR1* | 3.949714843 | 1.12E-07 | 4.57E-07 | 0.000544033 |
| T2DM | endometrium | *DNAH1* | -1.986235192 | 0.002340679 | 0.002918781 | 1 |
| T2DM | endometrium | *NKTR* | 3.3295986 | 1.83E-08 | 4.34E-07 | 8.86E-05 |
| T2DM | endometrium | *NEK4* | 2.118284001 | 3.45E-07 | 8.37E-07 | 0.001674051 |
| T2DM | endometrium | *ADAM23* | 2.72405167 | 5.14E-07 | 1.15E-06 | 0.002495555 |
| T2DM | endometrium | *LMAN2L* | 4.152999007 | 3.55E-08 | 4.34E-07 | 0.000172166 |
| T2DM | endometrium | *ACTR1B* | 2.998049225 | 8.94E-08 | 4.38E-07 | 0.000433941 |
| T2DM | endometrium | *SLC35F5* | 4.441798029 | 8.54E-09 | 4.34E-07 | 4.14E-05 |
| T2DM | endometrium | *STEAP3* | 0.846862733 | 0.002826682 | 0.003501627 | 1 |
| T2DM | endometrium | *OTOF* | 7.588221604 | 0.002975451 | 0.003678188 | 1 |
| T2DM | endometrium | *CENPA* | 3.140213784 | 1.00E-06 | 2.03E-06 | 0.004858719 |
| T2DM | endometrium | *NRBP1* | 2.607389264 | 1.40E-07 | 4.78E-07 | 0.000677709 |
| T2DM | endometrium | *ITGB6* | -1.933152792 | 2.07E-06 | 3.89E-06 | 0.010055979 |
| T2DM | endometrium | *PPM1G* | 2.642131115 | 2.10E-07 | 5.80E-07 | 0.00101787 |
| T2DM | endometrium | *INO80B* | 5.852972949 | 5.59E-07 | 1.24E-06 | 0.002710642 |
| T2DM | endometrium | *GRB14* | 2.57382756 | 1.39E-07 | 4.78E-07 | 0.000673229 |
| T2DM | endometrium | *RTN4* | 3.199401517 | 1.48E-07 | 4.78E-07 | 0.000718679 |
| T2DM | endometrium | *GALNT3* | 3.955406664 | 2.36E-08 | 4.34E-07 | 0.00011433 |
| T2DM | endometrium | *POLE4* | 7.295273433 | 1.22E-08 | 4.34E-07 | 5.93E-05 |
| T2DM | endometrium | *ACADL* | 3.717986662 | 0.005055739 | 0.006135679 | 1 |
| T2DM | endometrium | *MRPL19* | 5.248692863 | 2.61E-07 | 6.79E-07 | 0.001268748 |
| T2DM | endometrium | *DNAH6* | 1.363452755 | 0.003394449 | 0.004170643 | 1 |
| T2DM | endometrium | *IGFBP2* | 4.308205284 | 7.69E-07 | 1.63E-06 | 0.003729538 |
| T2DM | endometrium | *GNLY* | 2.00270158 | 1.31E-07 | 4.78E-07 | 0.000634367 |
| T2DM | endometrium | *CHMP3* | 2.201520221 | 9.15E-06 | 1.49E-05 | 0.044379712 |
| T2DM | endometrium | *WNT6* | -1.583362147 | 1.16E-06 | 2.32E-06 | 0.00564133 |
| T2DM | endometrium | *IL1RL2* | -3.670746879 | 0.000149378 | 0.000206962 | 0.72478199 |
| T2DM | endometrium | *IL18R1* | 3.770525995 | 2.05E-07 | 5.71E-07 | 0.000994904 |
| T2DM | endometrium | *MLPH* | 3.338763934 | 1.98E-07 | 5.58E-07 | 0.000962983 |
| T2DM | endometrium | *HDLBP* | 4.013628017 | 6.51E-08 | 4.34E-07 | 0.000315775 |
| T2DM | endometrium | *PASK* | -2.212292965 | 1.15E-05 | 1.84E-05 | 0.055784513 |
| T2DM | endometrium | *TPO* | 2.150094814 | 4.17E-07 | 9.72E-07 | 0.002024879 |
| T2DM | endometrium | *TAF1B* | 5.777736497 | 2.05E-06 | 3.84E-06 | 0.009930619 |
| T2DM | endometrium | *NOL10* | 3.322969042 | 8.34E-08 | 4.38E-07 | 0.000404637 |
| T2DM | endometrium | *STRN* | 4.068692449 | 5.12E-08 | 4.34E-07 | 0.000248592 |
| T2DM | endometrium | *CEBPZ* | 5.136688326 | 1.01E-07 | 4.47E-07 | 0.000490031 |
| T2DM | endometrium | *PRKD3* | 2.697158917 | 5.81E-08 | 4.34E-07 | 0.000282105 |
| T2DM | endometrium | *SLC25A12* | 4.816946994 | 1.54E-06 | 2.96E-06 | 0.007448149 |
| T2DM | endometrium | *DARS* | 3.607554802 | 5.05E-08 | 4.34E-07 | 0.00024506 |
| T2DM | endometrium | *WIPF1* | 2.249354709 | 5.51E-08 | 4.34E-07 | 0.000267307 |
| T2DM | endometrium | *PNO1* | -1.184649216 | 5.07E-06 | 8.67E-06 | 0.024580711 |
| T2DM | endometrium | *ORC4* | 4.404115064 | 3.34E-08 | 4.34E-07 | 0.00016192 |
| T2DM | endometrium | *AAK1* | 3.583556725 | 5.98E-08 | 4.34E-07 | 0.000290173 |
| T2DM | endometrium | *CD207* | 7.470322207 | 0.00010746 | 0.000150823 | 0.521396346 |
| T2DM | endometrium | *GRIN3B* | 2.102969149 | 0.007063574 | 0.0084959 | 1 |
| T2DM | endometrium | *NFE2L2* | 4.598642811 | 8.92E-08 | 4.38E-07 | 0.00043295 |
| T2DM | endometrium | *MSH6* | 4.187812241 | 7.30E-08 | 4.34E-07 | 0.000354061 |

| | | | | | | |
|---|---|---|---|---|---|---|
| T2DM | endometrium | *ALMS1* | 2.497849049 | 7.64E-08 | 4.36E-07 | 0.000370726 |
| T2DM | endometrium | *DNAJC16* | 2.820166089 | 3.11E-08 | 4.34E-07 | 0.000150991 |
| T2DM | endometrium | *MARK1* | 3.58564915 | 5.38E-08 | 4.34E-07 | 0.000261153 |
| T2DM | endometrium | *MORN1* | 2.788965868 | 1.97E-07 | 5.53E-07 | 0.000953681 |
| T2DM | endometrium | *RALGPS2* | 3.525142099 | 2.66E-06 | 4.84E-06 | 0.012885305 |
| T2DM | endometrium | *CEP104* | 4.980875184 | 7.80E-08 | 4.36E-07 | 0.00037849 |
| T2DM | endometrium | *WRAP73* | 5.583587073 | 1.93E-08 | 4.34E-07 | 9.38E-05 |
| T2DM | endometrium | *ICMT* | 3.215283717 | 2.01E-07 | 5.62E-07 | 0.000975712 |
| T2DM | endometrium | *PARK7* | 5.745860533 | 4.75E-08 | 4.34E-07 | 0.000230251 |
| T2DM | endometrium | *KIAA1324* | -5.943416543 | 1.57E-05 | 2.44E-05 | 0.075985037 |
| T2DM | endometrium | *MECR* | 3.931073272 | 1.04E-07 | 4.47E-07 | 0.000505068 |
| T2DM | endometrium | *EDEM3* | 5.045869722 | 5.12E-07 | 1.15E-06 | 0.002482748 |
| T2DM | endometrium | *RAP1A* | 3.471321061 | 5.43E-08 | 4.34E-07 | 0.000263613 |
| T2DM | endometrium | *CAPZA1* | 2.750189544 | 9.21E-08 | 4.38E-07 | 0.000446628 |
| T2DM | endometrium | *ASH1L* | 4.247619716 | 2.43E-08 | 4.34E-07 | 0.000117781 |
| T2DM | endometrium | *GON4L* | 4.25567268 | 5.57E-08 | 4.34E-07 | 0.000270271 |
| T2DM | endometrium | *DOCK7* | 3.619666469 | 4.58E-08 | 4.34E-07 | 0.000222374 |
| T2DM | endometrium | *FBXO2* | 3.481532838 | 0.000184841 | 0.000254281 | 0.896849288 |
| T2DM | endometrium | *FBXO6* | 3.732353196 | 2.93E-07 | 7.39E-07 | 0.001423342 |
| T2DM | endometrium | *DNAJC6* | 2.563729263 | 3.31E-07 | 8.11E-07 | 0.001604276 |
| T2DM | endometrium | *LEPR* | 2.941736371 | 7.83E-08 | 4.36E-07 | 0.000379903 |
| T2DM | endometrium | *IVNS1ABP* | 3.057167652 | 7.23E-07 | 1.55E-06 | 0.003508392 |
| T2DM | endometrium | *MFN2* | 2.259496102 | 3.03E-07 | 7.59E-07 | 0.001472011 |
| T2DM | endometrium | *PRG4* | 2.186236111 | 1.86E-07 | 5.35E-07 | 0.000903545 |
| T2DM | endometrium | *NCF2* | 2.270208326 | 8.94E-07 | 1.85E-06 | 0.004339149 |
| T2DM | endometrium | *BCAS2* | 3.608882784 | 1.22E-08 | 4.34E-07 | 5.90E-05 |
| T2DM | endometrium | *TNNI3K* | 10.34191657 | 1.43E-08 | 4.34E-07 | 6.92E-05 |
| T2DM | endometrium | *CFHR3* | 7.152782826 | 0.000244554 | 0.000332747 | 1 |
| T2DM | endometrium | *CRYZ* | 1.723168543 | 2.31E-05 | 3.51E-05 | 0.111982681 |
| T2DM | endometrium | *TMEM9* | 2.420967424 | 2.71E-06 | 4.92E-06 | 0.013136136 |
| T2DM | endometrium | *WARS2* | 3.405895722 | 5.51E-07 | 1.22E-06 | 0.002673523 |
| T2DM | endometrium | *OSCP1* | 2.968902227 | 1.56E-08 | 4.34E-07 | 7.56E-05 |
| T2DM | endometrium | *MRPS15* | 2.383464922 | 1.15E-07 | 4.59E-07 | 0.000558046 |
| T2DM | endometrium | *EXOC8* | 4.230413091 | 1.76E-07 | 5.16E-07 | 0.0008531 |
| T2DM | endometrium | *GNPAT* | 6.060013697 | 5.65E-06 | 9.56E-06 | 0.027402953 |
| T2DM | endometrium | *C1orf109* | 4.091410549 | 4.42E-07 | 1.01E-06 | 0.002145279 |
| T2DM | endometrium | *TBCE* | 6.926171869 | 1.18E-05 | 1.88E-05 | 0.057072441 |
| T2DM | endometrium | *ACADM* | 5.227035836 | 8.91E-08 | 4.38E-07 | 0.000432426 |
| T2DM | endometrium | *SLAMF1* | 3.093522788 | 0.000144832 | 0.000201008 | 0.702722932 |
| T2DM | endometrium | *PADI2* | 3.825248636 | 0.000883376 | 0.001142361 | 1 |
| T2DM | endometrium | *SDHB* | 4.159287894 | 3.67E-08 | 4.34E-07 | 0.000178066 |
| T2DM | endometrium | *CTBS* | 2.255709464 | 3.73E-06 | 6.56E-06 | 0.018104146 |
| T2DM | endometrium | *KLHL12* | 2.313078739 | 9.10E-08 | 4.38E-07 | 0.000441411 |
| T2DM | endometrium | *ZNHIT6* | 5.015905446 | 2.63E-07 | 6.82E-07 | 0.001275709 |
| T2DM | endometrium | *GBP3* | 1.693986448 | 0.001605272 | 0.002029914 | 1 |
| T2DM | endometrium | *KIF17* | 5.310750264 | 5.00E-07 | 1.12E-06 | 0.002425302 |
| T2DM | endometrium | *CD46* | 3.517526462 | 1.44E-08 | 4.34E-07 | 6.98E-05 |
| T2DM | endometrium | *EBNA1BP2* | 3.437756451 | 2.14E-07 | 5.88E-07 | 0.001038589 |
| T2DM | endometrium | *ERI3* | 2.174863916 | 9.67E-08 | 4.42E-07 | 0.000469319 |
| T2DM | endometrium | *PRDX1* | 2.019798697 | 1.41E-07 | 4.78E-07 | 0.000684258 |

| | | | | | | |
|---|---|---|---|---|---|---|
| T2DM | endometrium | *CCDC181* | 3.246010765 | 3.25E-05 | 4.87E-05 | 0.15748958 |
| T2DM | endometrium | *NSUN4* | 4.521550157 | 1.43E-05 | 2.25E-05 | 0.069551606 |
| T2DM | endometrium | *DR1* | 6.461789665 | 9.21E-07 | 1.90E-06 | 0.004470151 |
| T2DM | endometrium | *PRRC2C* | 3.341637343 | 4.01E-08 | 4.34E-07 | 0.000194391 |
| T2DM | endometrium | *ABCD3* | 4.075108704 | 5.88E-08 | 4.34E-07 | 0.000285393 |
| T2DM | endometrium | *VAMP4* | 2.754146439 | 7.94E-08 | 4.36E-07 | 0.000385348 |
| T2DM | endometrium | *PTBP2* | 3.60304227 | 1.41E-08 | 4.34E-07 | 6.83E-05 |
| T2DM | endometrium | *DARS2* | -1.994465242 | 5.43E-05 | 7.89E-05 | 0.263324529 |
| T2DM | endometrium | *IRF6* | -1.263011345 | 0.018499738 | 0.021515036 | 1 |
| T2DM | endometrium | *SERPINC1* | -2.05082404 | 9.12E-06 | 1.48E-05 | 0.044240052 |
| T2DM | endometrium | *RPS6KA1* | 3.024444533 | 6.65E-07 | 1.44E-06 | 0.003224786 |
| T2DM | endometrium | *RPA2* | 2.112930501 | 2.92E-07 | 7.38E-07 | 0.001418438 |
| T2DM | endometrium | *SLC5A9* | 2.613134971 | 1.02E-07 | 4.47E-07 | 0.000496363 |
| T2DM | endometrium | *ESYT2* | 3.550930392 | 6.68E-08 | 4.34E-07 | 0.000324317 |
| T2DM | endometrium | *CD3EAP* | 2.738801621 | 4.20E-07 | 9.77E-07 | 0.002040153 |
| T2DM | endometrium | *RCN2* | 3.08069624 | 4.97E-08 | 4.34E-07 | 0.000241038 |
| T2DM | endometrium | *IFT46* | 3.547524832 | 1.70E-06 | 3.25E-06 | 0.008258792 |
| T2DM | endometrium | *APOA1* | 23.1632148 | 7.82E-08 | 4.36E-07 | 0.00037965 |
| T2DM | endometrium | *DDX59* | 2.395032547 | 5.56E-07 | 1.23E-06 | 0.002699822 |
| T2DM | endometrium | *CAMSAP2* | 4.422259091 | 1.27E-07 | 4.75E-07 | 0.000614139 |
| T2DM | endometrium | *KLF7* | 3.51220533 | 2.50E-08 | 4.34E-07 | 0.000121235 |
| T2DM | endometrium | *CASC1* | 1.918268193 | 9.09E-05 | 0.000128417 | 0.441113901 |
| T2DM | endometrium | *LRMP* | 2.074382767 | 4.15E-07 | 9.68E-07 | 0.002011614 |
| T2DM | endometrium | *ELOVL4* | 8.429738661 | 4.84E-09 | 4.20E-07 | 2.35E-05 |
| T2DM | endometrium | *CNR1* | 5.268727532 | 6.79E-05 | 9.73E-05 | 0.329494945 |
| T2DM | endometrium | *AKAP7* | 4.033576463 | 1.55E-07 | 4.80E-07 | 0.000751862 |
| T2DM | endometrium | *ALDH8A1* | -1.560604596 | 0.001160459 | 0.001487203 | 1 |
| T2DM | endometrium | *MED28* | 4.284103444 | 0.004826452 | 0.005861813 | 1 |
| T2DM | endometrium | *TMEM5* | 8.00363288 | 1.63E-08 | 4.34E-07 | 7.92E-05 |
| T2DM | endometrium | *VAMP8* | 6.303249478 | 3.77E-05 | 5.61E-05 | 0.182946661 |
| T2DM | endometrium | *FOXO3* | 3.841684474 | 6.79E-08 | 4.34E-07 | 0.000329368 |
| T2DM | endometrium | *SPP1* | -1.481610611 | 5.57E-06 | 9.43E-06 | 0.027006047 |
| T2DM | endometrium | *MFSD1* | 3.911753103 | 6.69E-08 | 4.34E-07 | 0.00032483 |
| T2DM | endometrium | *UBN1* | 5.736303348 | 1.69E-07 | 5.04E-07 | 0.000819402 |
| T2DM | endometrium | *WDR35* | -2.410394414 | 0.006481589 | 0.007819162 | 1 |
| T2DM | endometrium | *DNAH7* | 2.236826988 | 5.42E-07 | 1.20E-06 | 0.002629527 |
| T2DM | endometrium | *C2orf40* | 4.48077422 | 8.81E-08 | 4.38E-07 | 0.00042723 |
| T2DM | endometrium | *CPSF3* | 5.451651274 | 4.19E-06 | 7.31E-06 | 0.020348111 |
| T2DM | endometrium | *SENP5* | 3.623383578 | 5.52E-08 | 4.34E-07 | 0.000268061 |
| T2DM | endometrium | *CCDC92* | 1.656052061 | 8.71E-07 | 1.81E-06 | 0.004227329 |
| T2DM | endometrium | *FKBP15* | 3.524198241 | 3.73E-06 | 6.56E-06 | 0.018088106 |
| T2DM | endometrium | *CTNNAL1* | 2.219635831 | 1.99E-08 | 4.34E-07 | 9.65E-05 |
| T2DM | endometrium | *RAB14* | 5.192394279 | 5.85E-08 | 4.34E-07 | 0.00028366 |
| T2DM | endometrium | *TRIM32* | -2.34087296 | 1.65E-05 | 2.55E-05 | 0.079873709 |
| T2DM | endometrium | *PPP6C* | 5.151963224 | 8.50E-08 | 4.38E-07 | 0.000412479 |
| T2DM | endometrium | *HDHD3* | 1.039688892 | 0.014991674 | 0.017552993 | 1 |
| T2DM | endometrium | *RBM18* | 2.455124831 | 1.99E-07 | 5.58E-07 | 0.00096517 |
| T2DM | endometrium | *HSDL2* | 2.933874447 | 8.49E-08 | 4.38E-07 | 0.000411925 |
| T2DM | endometrium | *MAPKAP1* | 4.255456797 | 7.15E-08 | 4.34E-07 | 0.000346837 |
| T2DM | endometrium | *INVS* | 2.723657828 | 1.93E-08 | 4.34E-07 | 9.35E-05 |

| | | | | | | |
|---|---|---|---|---|---|---|
| T2DM | endometrium | *DENND1A* | 3.871682311 | 4.69E-08 | 4.34E-07 | 0.000227395 |
| T2DM | endometrium | *KDSR* | 3.039729241 | 9.50E-08 | 4.42E-07 | 0.000461139 |
| T2DM | endometrium | *DCAF4* | 6.97420356 | 3.76E-06 | 6.60E-06 | 0.018226777 |
| T2DM | endometrium | *IFI27L2* | 3.03393194 | 1.31E-07 | 4.78E-07 | 0.00063532 |
| T2DM | endometrium | *NEK9* | 3.716746426 | 0.012669437 | 0.014948175 | 1 |
| T2DM | endometrium | *ACOT2* | 1.419173936 | 0.000407962 | 0.000544548 | 1 |
| T2DM | endometrium | *TTLL5* | 5.141647168 | 1.09E-06 | 2.20E-06 | 0.005308261 |
| T2DM | endometrium | *FLVCR2* | 2.554623515 | 2.31E-07 | 6.19E-07 | 0.001118765 |
| T2DM | endometrium | *PPP4R4* | 2.971410536 | 1.31E-06 | 2.58E-06 | 0.006373055 |
| T2DM | endometrium | *ALDH6A1* | 3.621454062 | 7.41E-07 | 1.58E-06 | 0.003593557 |
| T2DM | endometrium | *ESRRB* | 3.533137572 | 4.26E-06 | 7.43E-06 | 0.02069368 |
| T2DM | endometrium | *EIF2B2* | 2.13137729 | 4.15E-07 | 9.69E-07 | 0.002015431 |
| T2DM | endometrium | *COQ6* | 3.345785238 | 1.11E-07 | 4.57E-07 | 0.000537601 |
| T2DM | endometrium | *DNMT3A* | 2.437577898 | 1.35E-07 | 4.78E-07 | 0.000656441 |
| T2DM | endometrium | *CNRIP1* | 2.117202356 | 1.95E-06 | 3.66E-06 | 0.009442424 |
| T2DM | endometrium | *CRIPT* | 5.832220148 | 5.72E-08 | 4.34E-07 | 0.0002775 |
| T2DM | endometrium | *EPCAM* | 6.866498949 | 0.000158719 | 0.000219529 | 0.770106941 |
| T2DM | endometrium | *SLC17A5* | 6.129566032 | 6.32E-08 | 4.34E-07 | 0.000306707 |
| T2DM | endometrium | *ELOVL3* | 4.824509723 | 0.003585119 | 0.004392676 | 1 |
| T2DM | endometrium | *PPP1R3C* | -4.133167228 | 0.022710469 | 0.026254752 | 1 |
| T2DM | endometrium | *MXI1* | 3.654695052 | 1.41E-07 | 4.78E-07 | 0.000682099 |
| T2DM | endometrium | *HELLS* | 6.345731514 | 5.62E-05 | 8.15E-05 | 0.272452666 |
| T2DM | endometrium | *TCTN3* | 5.473194143 | 4.08E-06 | 7.14E-06 | 0.01980265 |
| T2DM | endometrium | *WDR11* | 5.134009426 | 8.01E-07 | 1.68E-06 | 0.003885706 |
| T2DM | endometrium | *KANSL1* | 3.250559915 | 6.01E-08 | 4.34E-07 | 0.000291489 |
| T2DM | endometrium | *HOXB3* | 2.952629703 | 6.83E-08 | 4.34E-07 | 0.000331539 |
| T2DM | endometrium | *CAAP1* | 4.284100582 | 3.02E-05 | 4.54E-05 | 0.146576677 |
| T2DM | endometrium | *PCDHB10* | -1.541894522 | 2.28E-06 | 4.24E-06 | 0.011085377 |
| T2DM | endometrium | *PCDHB12* | 1.86899408 | 0.03434525 | 0.03919171 | 1 |
| T2DM | endometrium | *GORAB* | -2.322722745 | 0.000130841 | 0.000182269 | 0.634841369 |
| T2DM | endometrium | *ACAT2* | 3.615597341 | 2.41E-07 | 6.41E-07 | 0.001168614 |
| T2DM | endometrium | *SNX19* | 2.946839238 | 4.00E-07 | 9.41E-07 | 0.00194225 |
| T2DM | endometrium | *SLC10A7* | 3.408864343 | 4.87E-08 | 4.34E-07 | 0.000236365 |
| T2DM | endometrium | *MASTL* | 2.908471261 | 0.008146931 | 0.009740983 | 1 |
| T2DM | endometrium | *MRC1* | 1.554197785 | 1.00E-07 | 4.45E-07 | 0.000485529 |
| T2DM | endometrium | *IQSEC3* | 8.856541658 | 0.005580268 | 0.00675704 | 1 |
| T2DM | endometrium | *CCDC77* | -3.640331071 | 0.011183568 | 0.013234798 | 1 |
| T2DM | endometrium | *DNAJC15* | 4.449517 | 6.15E-08 | 4.34E-07 | 0.00029844 |
| T2DM | endometrium | *UFM1* | 3.025195228 | 8.74E-08 | 4.38E-07 | 0.000423947 |
| T2DM | endometrium | *ALG5* | 5.178676399 | 3.07E-06 | 5.51E-06 | 0.014913609 |
| T2DM | endometrium | *SIL1* | 5.035821235 | 7.32E-07 | 1.56E-06 | 0.003549796 |
| T2DM | endometrium | *ZFP30* | 1.959142074 | 1.35E-07 | 4.78E-07 | 0.000656289 |
| T2DM | endometrium | *ARL1* | 6.8604414 | 2.00E-06 | 3.77E-06 | 0.009722437 |
| T2DM | endometrium | *APAF1* | 6.184617145 | 1.04E-06 | 2.10E-06 | 0.00503601 |
| T2DM | endometrium | *ADRA1A* | 2.495199268 | 1.40E-10 | 6.80E-08 | 6.80E-07 |
| T2DM | endometrium | *EPHX2* | 3.998292666 | 6.75E-05 | 9.68E-05 | 0.327372546 |
| T2DM | endometrium | *AKAP1* | -1.860901656 | 8.52E-05 | 0.000120829 | 0.413357402 |
| T2DM | endometrium | *SCPEP1* | 1.86614847 | 3.91E-07 | 9.25E-07 | 0.001898762 |
| T2DM | endometrium | *TBX2* | 3.258466613 | 0.000478425 | 0.00063372 | 1 |
| T2DM | endometrium | *TEX14* | -0.937859544 | 1.61E-05 | 2.50E-05 | 0.078196648 |

| | | | | | | |
|---|---|---|---|---|---|---|
| T2DM | endometrium | *TRIM6* | 6.470194801 | 3.21E-07 | 7.93E-07 | 0.001555795 |
| T2DM | endometrium | *ADCY7* | 1.883178161 | 7.89E-07 | 1.66E-06 | 0.003827426 |
| T2DM | endometrium | *ECHDC2* | 3.234338369 | 1.49E-05 | 2.34E-05 | 0.072482722 |
| T2DM | endometrium | *PYROXD1* | 3.212894151 | 7.93E-06 | 1.30E-05 | 0.038474234 |
| T2DM | endometrium | *ZNF549* | 3.97331817 | 5.52E-08 | 4.34E-07 | 0.000267626 |
| T2DM | endometrium | *A1BG* | -2.736175367 | 2.12E-05 | 3.24E-05 | 0.102754884 |
| T2DM | endometrium | *ZSCAN18* | 3.557327786 | 6.16E-08 | 4.34E-07 | 0.000298761 |
| T2DM | endometrium | *ZNF211* | 3.503995884 | 1.09E-08 | 4.34E-07 | 5.27E-05 |
| T2DM | endometrium | *PDZRN3* | 1.736092911 | 0.000135378 | 0.000188426 | 0.656854587 |
| T2DM | endometrium | *LHX4* | 3.958843017 | 8.92E-08 | 4.38E-07 | 0.000432961 |
| T2DM | endometrium | *SEC22A* | 3.402357567 | 2.95E-08 | 4.34E-07 | 0.000143127 |
| T2DM | endometrium | *POPDC2* | 3.127868783 | 0.000358727 | 0.000481212 | 1 |
| T2DM | endometrium | *MAPK8IP1* | 7.048803272 | 9.69E-08 | 4.42E-07 | 0.000470274 |
| T2DM | endometrium | *CAT* | 2.593340307 | 3.58E-07 | 8.62E-07 | 0.001737148 |
| T2DM | endometrium | *PILRB* | 2.122865288 | 2.47E-08 | 4.34E-07 | 0.000119915 |
| T2DM | endometrium | *ZCCHC17* | 4.985755486 | 5.58E-08 | 4.34E-07 | 0.000270933 |
| T2DM | endometrium | *FABP3* | 4.415672025 | 2.93E-07 | 7.39E-07 | 0.001422811 |
| T2DM | endometrium | *TNFSF10* | 3.560642158 | 4.07E-08 | 4.34E-07 | 0.00019735 |
| T2DM | endometrium | *ZNF639* | 2.400902301 | 2.00E-07 | 5.61E-07 | 0.000971392 |
| T2DM | endometrium | *CPXM2* | 1.79672263 | 5.34E-07 | 1.19E-06 | 0.002588757 |
| T2DM | endometrium | *FLT3* | 5.076981435 | 1.40E-08 | 4.34E-07 | 6.82E-05 |
| T2DM | endometrium | *MTIF3* | 4.872705066 | 2.71E-08 | 4.34E-07 | 0.000131394 |
| T2DM | endometrium | *FMOD* | 3.316173746 | 1.04E-07 | 4.47E-07 | 0.000506898 |
| T2DM | endometrium | *CD244* | 3.710150881 | 1.57E-07 | 4.84E-07 | 0.000761406 |
| T2DM | endometrium | *SERAC1* | 2.518169185 | 3.54E-07 | 8.53E-07 | 0.001715467 |
| T2DM | endometrium | *LDB3* | 3.189751331 | 1.06E-07 | 4.48E-07 | 0.000512008 |
| T2DM | endometrium | *OPN4* | 14.90927423 | 3.09E-05 | 4.65E-05 | 0.150086431 |
| T2DM | endometrium | *FAM35A* | 4.676166636 | 7.76E-08 | 4.36E-07 | 0.000376288 |
| T2DM | endometrium | *NAA60* | -3.936771084 | 0.017479594 | 0.020382358 | 1 |
| T2DM | endometrium | *ODF2L* | 5.635112072 | 2.12E-06 | 3.96E-06 | 0.010293581 |
| T2DM | endometrium | *SPATA1* | 3.922842362 | 3.34E-08 | 4.34E-07 | 0.000162103 |
| T2DM | endometrium | *LRRC39* | -1.329758499 | 0.001675612 | 0.002113903 | 1 |
| T2DM | endometrium | *RWDD3* | 3.880058242 | 2.67E-06 | 4.86E-06 | 0.012963865 |
| T2DM | endometrium | *EEPD1* | 2.208410976 | 7.08E-07 | 1.52E-06 | 0.003433889 |
| T2DM | endometrium | *KLHL7* | 3.834004213 | 5.10E-08 | 4.34E-07 | 0.000247512 |
| T2DM | endometrium | *NT5C3A* | 3.753071504 | 9.46E-08 | 4.41E-07 | 0.000459064 |
| T2DM | endometrium | *CCZ1* | 3.605943915 | 1.02E-06 | 2.07E-06 | 0.004961546 |
| T2DM | endometrium | *POLM* | -1.473550424 | 0.037414276 | 0.042583642 | 1 |
| T2DM | endometrium | *SMU1* | 6.860566108 | 7.72E-06 | 1.27E-05 | 0.037454506 |
| T2DM | endometrium | *GLIPR2* | -2.23162315 | 9.97E-05 | 0.000140167 | 0.483577444 |
| T2DM | endometrium | *SLC25A51* | 2.600220074 | 6.76E-07 | 1.46E-06 | 0.003279601 |
| T2DM | endometrium | *TRIM24* | 5.21182488 | 8.68E-07 | 1.80E-06 | 0.004212153 |
| T2DM | endometrium | *C7orf49* | 3.638783283 | 4.92E-08 | 4.34E-07 | 0.000238594 |
| T2DM | endometrium | *RBM19* | 5.978072548 | 0.000457665 | 0.000607383 | 1 |
| T2DM | endometrium | *DDX54* | 6.694900323 | 4.99E-08 | 4.34E-07 | 0.000242243 |
| T2DM | endometrium | *CCDC91* | 4.293780546 | 0.000192321 | 0.000264121 | 0.93313914 |
| T2DM | endometrium | *ATP7B* | 0.9986486 | 0.000961208 | 0.001238391 | 1 |
| T2DM | endometrium | *CENPK* | 5.311770015 | 2.57E-06 | 4.69E-06 | 0.012451029 |
| T2DM | endometrium | *ITIH5* | 1.801406014 | 2.57E-06 | 4.69E-06 | 0.012464888 |
| T2DM | endometrium | *TSFM* | 3.537069115 | 2.67E-08 | 4.34E-07 | 0.000129365 |

| | | | | | | |
|---|---|---|---|---|---|---|
| T2DM | endometrium | *ARHGAP9* | 3.386344139 | 3.80E-07 | 9.04E-07 | 0.001842418 |
| T2DM | endometrium | *PFDN5* | 6.03076806 | 4.32E-06 | 7.51E-06 | 0.020962047 |
| T2DM | endometrium | *PDE1B* | 2.675108747 | 1.70E-07 | 5.06E-07 | 0.000826615 |
| T2DM | endometrium | *CDK2* | 3.266553132 | 2.43E-05 | 3.70E-05 | 0.118077381 |
| T2DM | endometrium | *SMUG1* | 3.5966053 | 8.54E-05 | 0.000121073 | 0.414431965 |
| T2DM | endometrium | *SARDH* | 4.187951883 | 7.13E-06 | 1.18E-05 | 0.034595305 |
| T2DM | endometrium | *DBH* | 75.52268341 | 1.38E-07 | 4.78E-07 | 0.00067031 |
| T2DM | endometrium | *NDUFAF4* | 5.372057944 | 1.82E-06 | 3.46E-06 | 0.008849148 |
| T2DM | endometrium | *USP45* | 2.918485502 | 5.89E-08 | 4.34E-07 | 0.000286017 |
| T2DM | endometrium | *TTC21B* | 4.129173532 | 3.70E-07 | 8.86E-07 | 0.001794958 |
| T2DM | endometrium | *ACVR1C* | 1.196858848 | 4.69E-06 | 8.10E-06 | 0.022766759 |
| T2DM | endometrium | *SLC36A1* | 2.470619805 | 4.43E-06 | 7.68E-06 | 0.021505882 |
| T2DM | endometrium | *PLA2G12A* | 6.709786098 | 2.96E-06 | 5.34E-06 | 0.014349958 |
| T2DM | endometrium | *C4BPA* | 3.98180333 | 1.83E-06 | 3.46E-06 | 0.008862986 |
| T2DM | endometrium | *C4BPB* | 3.157985845 | 5.10E-05 | 7.44E-05 | 0.247371048 |
| T2DM | endometrium | *RAB38* | 6.94558298 | 2.89E-09 | 3.60E-07 | 1.40E-05 |
| T2DM | endometrium | *DAW1* | 6.523859912 | 7.71E-08 | 4.36E-07 | 0.00037407 |
| T2DM | endometrium | *DNPEP* | 3.229260246 | 6.56E-08 | 4.34E-07 | 0.000318339 |
| T2DM | endometrium | *OBSL1* | 2.37510194 | 3.97E-07 | 9.36E-07 | 0.001926624 |
| T2DM | endometrium | *SLC12A4* | 4.307646324 | 5.16E-08 | 4.34E-07 | 0.000250173 |
| T2DM | endometrium | *FAM210B* | 8.451998237 | 4.41E-08 | 4.34E-07 | 0.000213751 |
| T2DM | endometrium | *SLC12A5* | -8.258190985 | 6.06E-06 | 1.02E-05 | 0.029404772 |
| T2DM | endometrium | *MATN4* | 16.96189305 | 1.15E-06 | 2.29E-06 | 0.005556765 |
| T2DM | endometrium | *VAPB* | 1.727123097 | 4.48E-06 | 7.76E-06 | 0.021749284 |
| T2DM | endometrium | *TOX2* | -1.203538447 | 6.85E-06 | 1.14E-05 | 0.033224511 |
| T2DM | endometrium | *SRSF6* | 3.456103785 | 7.06E-08 | 4.34E-07 | 0.000342688 |
| T2DM | endometrium | *CSE1L* | 3.040996964 | 1.52E-07 | 4.78E-07 | 0.000738654 |
| T2DM | endometrium | *PTGIS* | 1.835107349 | 5.41E-08 | 4.34E-07 | 0.000262561 |
| T2DM | endometrium | *RNF114* | 5.382114098 | 3.15E-07 | 7.81E-07 | 0.001527635 |
| T2DM | endometrium | *NEURL2* | 4.130251637 | 1.39E-05 | 2.19E-05 | 0.067316886 |
| T2DM | endometrium | *MTRR* | -2.605942381 | 0.016376366 | 0.019118895 | 1 |
| T2DM | endometrium | *PEPD* | 5.632807057 | 1.04E-07 | 4.47E-07 | 0.000506185 |
| T2DM | endometrium | *CHST8* | -3.807001842 | 0.001399555 | 0.001780451 | 1 |
| T2DM | endometrium | *STAMBP* | 6.549747201 | 7.88E-06 | 1.29E-05 | 0.038213922 |
| T2DM | endometrium | *NAGK* | 2.593063173 | 3.07E-07 | 7.66E-07 | 0.001491529 |
| T2DM | endometrium | *SNRNP27* | 2.714091234 | 9.58E-08 | 4.42E-07 | 0.00046486 |
| T2DM | endometrium | *ATP8A1* | 5.81914633 | 6.33E-07 | 1.38E-06 | 0.003073047 |
| T2DM | endometrium | *USP22* | 3.929616415 | 4.24E-08 | 4.34E-07 | 0.00020558 |
| T2DM | endometrium | *HIF3A* | 2.586044549 | 2.29E-07 | 6.16E-07 | 0.001110402 |
| T2DM | endometrium | *ZNF576* | -2.85058016 | 0.002482355 | 0.003090682 | 1 |
| T2DM | endometrium | *BTN2A2* | 2.886773882 | 2.21E-07 | 6.01E-07 | 0.001071923 |
| T2DM | endometrium | *MRS2* | 2.227543159 | 3.16E-08 | 4.34E-07 | 0.000153315 |
| T2DM | endometrium | *SNRPC* | 5.210141969 | 0.000677376 | 0.000886361 | 1 |
| T2DM | endometrium | *SERPINB6* | 4.031000339 | 6.23E-08 | 4.34E-07 | 0.00030241 |
| T2DM | endometrium | *PEX6* | 1.684956487 | 0.005887872 | 0.007120627 | 1 |
| T2DM | endometrium | *NQO2* | 5.231834333 | 0.001058726 | 0.001359339 | 1 |
| T2DM | endometrium | *ZNF391* | 1.579602364 | 4.13E-05 | 6.10E-05 | 0.200380499 |
| T2DM | endometrium | *RPS10* | -1.291631058 | 9.61E-07 | 1.96E-06 | 0.00466066 |
| T2DM | endometrium | *MED20* | 3.108986804 | 5.11E-06 | 8.74E-06 | 0.024811695 |
| T2DM | endometrium | *GNMT* | 4.009733452 | 0.035561244 | 0.040550683 | 1 |

| | | | | | | |
|---|---|---|---|---|---|---|
| T2DM | endometrium | *TREM1* | 5.18008241 | 6.51E-07 | 1.41E-06 | 0.003156723 |
| T2DM | endometrium | *MEA1* | 2.84014219 | 0.008066515 | 0.009649589 | 1 |
| T2DM | endometrium | *KLHL31* | 2.712849687 | 1.12E-06 | 2.25E-06 | 0.00545301 |
| T2DM | endometrium | *GLO1* | 7.581643199 | 3.20E-08 | 4.34E-07 | 0.000155179 |
| T2DM | endometrium | *KCNK17* | -2.726466667 | 8.74E-08 | 4.38E-07 | 0.000424207 |
| T2DM | endometrium | *RREB1* | 3.880734814 | 5.10E-07 | 1.14E-06 | 0.002472854 |
| T2DM | endometrium | *RIOK1* | 2.525624149 | 3.54E-08 | 4.34E-07 | 0.000171823 |
| T2DM | endometrium | *ATXN1* | 4.273344324 | 6.96E-06 | 1.16E-05 | 0.033764534 |
| T2DM | endometrium | *RUNX2* | 3.847900139 | 5.41E-08 | 4.34E-07 | 0.000262628 |
| T2DM | endometrium | *CXCL6* | 5.951757907 | 3.09E-07 | 7.70E-07 | 0.00149979 |
| T2DM | endometrium | *SH3TC1* | 1.996011744 | 2.61E-06 | 4.76E-06 | 0.012653293 |
| T2DM | endometrium | *BBS2* | 3.223036666 | 2.60E-08 | 4.34E-07 | 0.000126054 |
| T2DM | endometrium | *MT1G* | 10.05213354 | 4.84E-05 | 7.09E-05 | 0.234652457 |
| T2DM | endometrium | *GOT2* | 2.911436041 | 1.16E-07 | 4.59E-07 | 0.000560496 |
| T2DM | endometrium | *CLYBL* | 4.774728459 | 7.98E-08 | 4.36E-07 | 0.000387231 |
| T2DM | endometrium | *ABCC4* | 5.334046977 | 2.37E-07 | 6.35E-07 | 0.001151763 |
| T2DM | endometrium | *ATP5S* | 4.765139493 | 2.76E-07 | 7.06E-07 | 0.001338507 |
| T2DM | endometrium | *GRK4* | 2.063008431 | 1.03E-09 | 2.33E-07 | 5.02E-06 |
| T2DM | endometrium | *TEKT3* | 2.29815503 | 0.00035693 | 0.000478933 | 1 |
| T2DM | endometrium | *HS3ST3B1* | 4.315669645 | 1.17E-06 | 2.33E-06 | 0.005694696 |
| T2DM | endometrium | *MRPS7* | -2.946921115 | 0.014264909 | 0.016734366 | 1 |
| T2DM | endometrium | *GGA3* | 6.237198419 | 3.02E-07 | 7.58E-07 | 0.001466376 |
| T2DM | endometrium | *NUP85* | 4.009029424 | 1.03E-07 | 4.47E-07 | 0.000500173 |
| T2DM | endometrium | *NT5C* | 7.219708376 | 6.35E-07 | 1.38E-06 | 0.00308308 |
| T2DM | endometrium | *DDX31* | -2.38803485 | 0.033658405 | 0.038444111 | 1 |
| T2DM | endometrium | *OPRL1* | 9.097035743 | 1.39E-07 | 4.78E-07 | 0.000673719 |
| T2DM | endometrium | *PPDPF* | -2.296936495 | 0.003441153 | 0.004223748 | 1 |
| T2DM | endometrium | *IL1B* | 14.04130846 | 2.29E-07 | 6.16E-07 | 0.001112157 |
| T2DM | endometrium | *PAX8* | 2.707033752 | 1.81E-06 | 3.43E-06 | 0.008761817 |
| T2DM | endometrium | *POLR1B* | 2.102379166 | 4.70E-06 | 8.12E-06 | 0.022822447 |
| T2DM | endometrium | *CCDC93* | 3.735319166 | 7.62E-08 | 4.36E-07 | 0.00036975 |
| T2DM | endometrium | *PSD4* | 8.488443444 | 8.33E-07 | 1.74E-06 | 0.004040256 |
| T2DM | endometrium | *GTF2F1* | 5.080834604 | 2.60E-07 | 6.77E-07 | 0.001260337 |
| T2DM | endometrium | *TNFSF9* | 31.92267088 | 0.003047491 | 0.00376245 | 1 |
| T2DM | endometrium | *SH2D3A* | 3.589928713 | 1.38E-06 | 2.70E-06 | 0.006691835 |
| T2DM | endometrium | *GPR108* | -2.649050925 | 0.001369749 | 0.001743905 | 1 |
| T2DM | endometrium | *SNRPD2* | 4.951475886 | 5.08E-08 | 4.34E-07 | 0.000246303 |
| T2DM | endometrium | *SDCBP2* | -4.370666977 | 3.71E-05 | 5.53E-05 | 0.180229682 |
| T2DM | endometrium | *PSMF1* | 2.438449005 | 8.44E-07 | 1.76E-06 | 0.004097454 |
| T2DM | endometrium | *DTD1* | 5.041965113 | 0.000875221 | 0.001133024 | 1 |
| T2DM | endometrium | *RBCK1* | 3.390050574 | 2.22E-07 | 6.02E-07 | 0.001077875 |
| T2DM | endometrium | *RRBP1* | 3.690897898 | 2.73E-08 | 4.34E-07 | 0.000132667 |
| T2DM | endometrium | *FLRT3* | 2.768393733 | 1.49E-07 | 4.78E-07 | 0.00072113 |
| T2DM | endometrium | *PCSK2* | 11.466898 | 1.97E-07 | 5.53E-07 | 0.000954092 |
| T2DM | endometrium | *MKKS* | 6.362730884 | 1.04E-06 | 2.10E-06 | 0.005042287 |
| T2DM | endometrium | *LAMP5* | 55.29878922 | 9.70E-07 | 1.98E-06 | 0.004704609 |
| T2DM | endometrium | *MGME1* | 3.622091845 | 9.12E-08 | 4.38E-07 | 0.000442695 |
| T2DM | endometrium | *ITPA* | 3.890963286 | 1.33E-07 | 4.78E-07 | 0.000644303 |
| T2DM | endometrium | *MRPS26* | -2.185774311 | 0.012775078 | 0.015066767 | 1 |
| T2DM | endometrium | *NCLN* | 2.422653057 | 3.79E-07 | 9.03E-07 | 0.001837408 |

| | | | | | | |
|---|---|---|---|---|---|---|
| T2DM | endometrium | *UROD* | 7.591779482 | 6.47E-08 | 4.34E-07 | 0.00031392 |
| T2DM | endometrium | *KLC1* | 2.241599138 | 9.60E-08 | 4.42E-07 | 0.000465871 |
| T2DM | endometrium | *XRCC3* | 3.440763815 | 0.000630427 | 0.000826712 | 1 |
| T2DM | endometrium | *TUBGCP3* | 2.059535561 | 1.14E-07 | 4.57E-07 | 0.000552327 |
| T2DM | endometrium | *MCF2L* | 2.211039211 | 9.77E-08 | 4.42E-07 | 0.000474039 |
| T2DM | endometrium | *PROZ* | 6.027276978 | 0.009289626 | 0.011050077 | 1 |
| T2DM | endometrium | *PRDX5* | 6.343592567 | 0.000908337 | 0.001173703 | 1 |
| T2DM | endometrium | *IRF3* | 6.984036516 | 9.44E-07 | 1.93E-06 | 0.004578209 |
| T2DM | endometrium | *PRR12* | 6.006169997 | 5.75E-10 | 1.86E-07 | 2.79E-06 |
| T2DM | endometrium | *TSKS* | -1.895149421 | 7.84E-06 | 1.29E-05 | 0.038045265 |
| T2DM | endometrium | *TRAP1* | 2.477831496 | 2.44E-06 | 4.49E-06 | 0.011858126 |
| T2DM | endometrium | *EMG1* | 3.02192597 | 1.82E-07 | 5.25E-07 | 0.000881283 |
| T2DM | endometrium | *ATG14* | 3.178171921 | 5.31E-08 | 4.34E-07 | 0.000257437 |
| T2DM | endometrium | *KTN1* | 4.545331346 | 2.61E-07 | 6.78E-07 | 0.001264628 |
| T2DM | endometrium | *L3HYPDH* | 2.688594063 | 4.83E-06 | 8.32E-06 | 0.0234484 |
| T2DM | endometrium | *RHOT1* | 3.872689614 | 6.82E-06 | 1.14E-05 | 0.033081327 |
| T2DM | endometrium | *WDR60* | 4.272640817 | 3.29E-06 | 5.84E-06 | 0.015950342 |
| T2DM | endometrium | *AIF1L* | 3.142585466 | 1.41E-07 | 4.78E-07 | 0.000683373 |
| T2DM | endometrium | *CANX* | 3.69836764 | 4.66E-08 | 4.34E-07 | 0.000226027 |
| T2DM | endometrium | *FGD3* | 2.232148975 | 4.50E-07 | 1.03E-06 | 0.002184629 |
| T2DM | endometrium | *ABHD8* | 1.932818461 | 1.29E-05 | 2.05E-05 | 0.06271197 |
| T2DM | endometrium | *ATP13A4* | 5.595615995 | 8.98E-08 | 4.38E-07 | 0.000435769 |
| T2DM | endometrium | *HELB* | 2.73153801 | 4.79E-08 | 4.34E-07 | 0.000232543 |
| T2DM | endometrium | *RAB3IP* | 3.681168881 | 1.53E-07 | 4.78E-07 | 0.000743427 |
| T2DM | endometrium | *YEATS4* | -1.544904287 | 0.000407813 | 0.000544498 | 1 |
| T2DM | endometrium | *TAS2R4* | 3.180866747 | 5.88E-06 | 9.92E-06 | 0.028551082 |
| T2DM | endometrium | *LRRC61* | 2.814670871 | 1.21E-07 | 4.68E-07 | 0.00058569 |
| T2DM | endometrium | *IDUA* | 5.975284454 | 9.69E-08 | 4.42E-07 | 0.000470199 |
| T2DM | endometrium | *EMC1* | 3.520139353 | 8.05E-08 | 4.36E-07 | 0.000390623 |
| T2DM | endometrium | *PLA2G5* | 1.979459258 | 0.000106457 | 0.000149459 | 0.516529997 |
| T2DM | endometrium | *UBR4* | 3.703153222 | 4.57E-06 | 7.89E-06 | 0.022160535 |
| T2DM | endometrium | *WFIKKN1* | 4.920486444 | 0.010197296 | 0.012106014 | 1 |
| T2DM | endometrium | *WDR24* | 3.555721817 | 2.30E-05 | 3.50E-05 | 0.111432214 |
| T2DM | endometrium | *SMARCA4* | 4.871774833 | 9.10E-08 | 4.38E-07 | 0.000441461 |
| T2DM | endometrium | *PNKD* | 5.750879305 | 3.29E-08 | 4.34E-07 | 0.000159697 |
| T2DM | endometrium | *RNF6* | 7.670907602 | 7.00E-05 | 0.000100212 | 0.339519655 |
| T2DM | endometrium | *HIP1* | 4.141721024 | 8.01E-08 | 4.36E-07 | 0.000388853 |
| T2DM | endometrium | *PTPN12* | 3.808106558 | 2.98E-07 | 7.48E-07 | 0.001445706 |
| T2DM | endometrium | *STEAP4* | 2.24912707 | 1.06E-06 | 2.13E-06 | 0.005135504 |
| T2DM | endometrium | *PEX1* | 3.591604285 | 5.73E-08 | 4.34E-07 | 0.000278035 |
| T2DM | endometrium | *KDR* | 2.48729869 | 9.89E-08 | 4.44E-07 | 0.000479626 |
| T2DM | endometrium | *MGAT3* | -2.194616535 | 3.69E-05 | 5.49E-05 | 0.178805459 |
| T2DM | endometrium | *ATF4* | 3.650826591 | 7.63E-08 | 4.36E-07 | 0.000370211 |
| T2DM | endometrium | *A4GALT* | 2.883115879 | 9.43E-07 | 1.93E-06 | 0.004575262 |
| T2DM | endometrium | *CDC42EP1* | 8.831476344 | 7.54E-06 | 1.24E-05 | 0.036591802 |
| T2DM | endometrium | *SPECC1* | 4.245488261 | 6.54E-08 | 4.34E-07 | 0.000317378 |
| T2DM | endometrium | *CDHR3* | 5.294341864 | 1.26E-07 | 4.75E-07 | 0.000611936 |
| T2DM | endometrium | *PRKRIP1* | 3.5080707 | 1.09E-07 | 4.55E-07 | 0.000529156 |
| T2DM | endometrium | *IRF5* | 2.317800955 | 1.04E-05 | 1.68E-05 | 0.050560391 |
| T2DM | endometrium | *OPN1SW* | -2.051203699 | 1.43E-07 | 4.78E-07 | 0.000696045 |

| | | | | | | |
|---|---|---|---|---|---|---|
| T2DM | endometrium | *HOXD1* | -2.33346558 | 1.22E-05 | 1.93E-05 | 0.058975879 |
| T2DM | endometrium | *HOXD3* | -1.014921056 | 0.004176069 | 0.005089748 | 1 |
| T2DM | endometrium | *MTX2* | 5.651360389 | 2.39E-06 | 4.41E-06 | 0.011610885 |
| T2DM | endometrium | *GAD1* | 6.940397508 | 4.34E-06 | 7.53E-06 | 0.021038804 |
| T2DM | endometrium | *ORMDL1* | 3.692090005 | 1.74E-08 | 4.34E-07 | 8.44E-05 |
| T2DM | endometrium | *HOXD9* | 1.64168716 | 0.010175891 | 0.01208356 | 1 |
| T2DM | endometrium | *HOXD10* | -1.904569398 | 1.58E-06 | 3.05E-06 | 0.007683675 |
| T2DM | endometrium | *PSMG2* | 5.874410491 | 9.20E-08 | 4.38E-07 | 0.000446329 |
| T2DM | endometrium | *TWSG1* | 2.67595566 | 7.84E-08 | 4.36E-07 | 0.000380217 |
| T2DM | endometrium | *WDFY4* | 1.764151671 | 2.19E-06 | 4.08E-06 | 0.010637538 |
| T2DM | endometrium | *EIF2AK4* | 2.927003355 | 2.67E-07 | 6.89E-07 | 0.00129496 |
| T2DM | endometrium | *TMOD2* | 4.550787427 | 0.005218325 | 0.006328246 | 1 |
| T2DM | endometrium | *IVD* | 3.09377598 | 1.19E-06 | 2.36E-06 | 0.005759453 |
| T2DM | endometrium | *ARPP19* | 6.133014161 | 1.28E-07 | 4.78E-07 | 0.000621702 |
| T2DM | endometrium | *VPS13C* | 2.542680291 | 1.14E-07 | 4.57E-07 | 0.000551498 |
| T2DM | endometrium | *CALML4* | -2.385914048 | 2.42E-06 | 4.45E-06 | 0.011729254 |
| T2DM | endometrium | *ANAPC13* | -1.821433706 | 0.000155079 | 0.000214616 | 0.752444088 |
| T2DM | endometrium | *SUMF2* | 4.508322351 | 5.84E-08 | 4.34E-07 | 0.000283533 |
| T2DM | endometrium | *BBOX1* | -11.96002274 | 0.001503647 | 0.001904384 | 1 |
| T2DM | endometrium | *SERGEF* | 3.286867467 | 1.63E-08 | 4.34E-07 | 7.93E-05 |
| T2DM | endometrium | *DCTD* | 4.617814155 | 4.93E-05 | 7.21E-05 | 0.239111309 |
| T2DM | endometrium | *RPAIN* | 4.091431829 | 0.000362573 | 0.000486237 | 1 |
| T2DM | endometrium | *USP6* | -1.903741146 | 0.005361069 | 0.006498103 | 1 |
| T2DM | endometrium | *PLD2* | -3.856654897 | 0.00406668 | 0.004957671 | 1 |
| T2DM | endometrium | *PHF20L1* | 4.984522408 | 6.34E-05 | 9.12E-05 | 0.307510658 |
| T2DM | endometrium | *PUS7L* | 3.677528911 | 1.17E-07 | 4.61E-07 | 0.000568685 |
| T2DM | endometrium | *AP1M2* | -3.234711101 | 0.00049798 | 0.000658365 | 1 |
| T2DM | endometrium | *MTUS1* | 4.672963763 | 7.66E-08 | 4.36E-07 | 0.000371838 |
| T2DM | endometrium | *KLK8* | -17.42425365 | 4.31E-05 | 6.35E-05 | 0.209293529 |
| T2DM | endometrium | *RIPK3* | 3.001752223 | 0.000547443 | 0.0007214 | 1 |
| T2DM | endometrium | *ADCY4* | 2.950184532 | 1.35E-07 | 4.78E-07 | 0.000653226 |
| T2DM | endometrium | *BCL2L2* | 2.086317296 | 4.09E-07 | 9.58E-07 | 0.001984506 |
| T2DM | endometrium | *DTD2* | -1.983722689 | 0.00082573 | 0.001071527 | 1 |
| T2DM | endometrium | *PARP2* | 2.730907572 | 1.22E-07 | 4.69E-07 | 0.000594187 |
| T2DM | endometrium | *MIS18BP1* | 3.282354121 | 3.87E-08 | 4.34E-07 | 0.000187738 |
| T2DM | endometrium | *RNASE1* | 1.627575789 | 6.21E-08 | 4.34E-07 | 0.000301308 |
| T2DM | endometrium | *EPB41L4A* | 3.908387369 | 8.61E-08 | 4.38E-07 | 0.000417615 |
| T2DM | endometrium | *REEP5* | 5.67083729 | 1.37E-06 | 2.67E-06 | 0.006625141 |
| T2DM | endometrium | *RHBDF2* | 2.449031543 | 7.06E-08 | 4.34E-07 | 0.000342733 |
| T2DM | endometrium | *TMEM8A* | 4.437382232 | 7.21E-08 | 4.34E-07 | 0.000349895 |
| T2DM | endometrium | *MAU2* | 4.594171231 | 2.56E-07 | 6.71E-07 | 0.001244161 |
| T2DM | endometrium | *SHC2* | 9.263769459 | 1.03E-09 | 2.33E-07 | 4.99E-06 |
| T2DM | endometrium | *ERMARD* | 3.627362784 | 5.81E-08 | 4.34E-07 | 0.00028202 |
| T2DM | endometrium | *PHF10* | 2.286021575 | 1.51E-07 | 4.78E-07 | 0.000734706 |
| T2DM | endometrium | *GALNT8* | -4.294364937 | 0.000426671 | 0.000567647 | 1 |
| T2DM | endometrium | *CDC16* | 3.862558536 | 3.22E-07 | 7.94E-07 | 0.001560323 |
| T2DM | endometrium | *XPO7* | 5.257702707 | 2.70E-07 | 6.95E-07 | 0.001310575 |
| T2DM | endometrium | *FAM98C* | 3.831782172 | 0.012794478 | 0.01508598 | 1 |
| T2DM | endometrium | *PLVAP* | 1.834326901 | 0.000514426 | 0.000679368 | 1 |
| T2DM | endometrium | *DDA1* | -3.817451796 | 0.001496132 | 0.001896352 | 1 |

| | | | | | | |
|---|---|---|---|---|---|---|
| T2DM | endometrium | *MRPL34* | -1.666602837 | 0.016616322 | 0.019389705 | 1 |
| T2DM | endometrium | *TULP4* | 4.532575102 | 9.02E-08 | 4.38E-07 | 0.000437469 |
| T2DM | endometrium | *QRSL1* | 1.94323473 | 4.34E-07 | 9.98E-07 | 0.002104792 |
| T2DM | endometrium | *RSPH3* | 1.878032094 | 0.000610947 | 0.000802468 | 1 |
| T2DM | endometrium | *ACTN4* | 2.834695553 | 4.94E-08 | 4.34E-07 | 0.000239767 |
| T2DM | endometrium | *STK33* | 2.820163067 | 1.76E-09 | 3.07E-07 | 8.54E-06 |
| T2DM | endometrium | *MAP1S* | 3.290136247 | 1.49E-07 | 4.78E-07 | 0.000724286 |
| T2DM | endometrium | *PGPEP1* | 4.808249533 | 5.96E-08 | 4.34E-07 | 0.000289373 |
| T2DM | endometrium | *KIAA1683* | -1.527154943 | 1.92E-05 | 2.96E-05 | 0.093334137 |
| T2DM | endometrium | *LSM4* | -2.035743894 | 0.00217998 | 0.002728208 | 1 |
| T2DM | endometrium | *TRPM4* | 5.04942384 | 5.25E-06 | 8.96E-06 | 0.025487741 |
| T2DM | endometrium | *UBAC1* | 4.394520665 | 0.000303792 | 0.000410128 | 1 |
| T2DM | endometrium | *ATXN10* | 4.854286956 | 6.55E-07 | 1.42E-06 | 0.003179547 |
| T2DM | endometrium | *PNPLA7* | 4.151935978 | 5.34E-08 | 4.34E-07 | 0.000258934 |
| T2DM | endometrium | *PAK4* | 3.782209125 | 1.41E-07 | 4.78E-07 | 0.000684353 |
| T2DM | endometrium | *ZNF337* | 4.99205417 | 1.20E-06 | 2.39E-06 | 0.005844124 |
| T2DM | endometrium | *CEP85* | 2.370470107 | 1.05E-05 | 1.70E-05 | 0.05117181 |
| T2DM | endometrium | *POMT1* | -2.645466757 | 0.000411252 | 0.000548487 | 1 |
| T2DM | endometrium | *CHMP2A* | 5.363388806 | 4.07E-07 | 9.55E-07 | 0.001974921 |
| T2DM | endometrium | *YIPF2* | -3.255738471 | 0.001423018 | 0.001809826 | 1 |
| T2DM | endometrium | *NPAS1* | -3.960502479 | 7.97E-06 | 1.31E-05 | 0.038681999 |
| T2DM | endometrium | *MAP3K10* | -1.68167744 | 1.61E-05 | 2.50E-05 | 0.078127206 |
| T2DM | endometrium | *LRRC47* | 2.390854204 | 3.86E-07 | 9.14E-07 | 0.001872215 |
| T2DM | endometrium | *THEMIS2* | 3.011013115 | 2.56E-08 | 4.34E-07 | 0.000124094 |
| T2DM | endometrium | *ZNF317* | -2.660488531 | 0.000451074 | 0.000599291 | 1 |
| T2DM | endometrium | *DNMT1* | 4.083676967 | 4.76E-08 | 4.34E-07 | 0.000230794 |
| T2DM | endometrium | *NOL11* | -2.64322534 | 0.001446871 | 0.001836312 | 1 |
| T2DM | endometrium | *HSD17B3* | 8.680290142 | 7.47E-08 | 4.36E-07 | 0.000362443 |
| T2DM | endometrium | *SLC35D2* | 2.336673955 | 1.08E-07 | 4.54E-07 | 0.000525735 |
| T2DM | endometrium | *ULBP2* | 28.27991655 | 5.55E-06 | 9.40E-06 | 0.026914464 |
| T2DM | endometrium | *TTLL9* | 4.887608222 | 1.39E-07 | 4.78E-07 | 0.000674631 |
| T2DM | endometrium | *COX4I2* | 4.590002496 | 4.79E-07 | 1.08E-06 | 0.002323777 |
| T2DM | endometrium | *ATP6V1E1* | 3.875922116 | 1.39E-07 | 4.78E-07 | 0.000674413 |
| T2DM | endometrium | *CHMP1A* | 6.334959051 | 1.54E-08 | 4.34E-07 | 7.48E-05 |
| T2DM | endometrium | *IDO1* | 4.969897024 | 1.25E-07 | 4.73E-07 | 0.000605825 |
| T2DM | endometrium | *HAUS8* | 2.684782298 | 7.23E-06 | 1.20E-05 | 0.035098289 |
| T2DM | endometrium | *MRPS25* | 3.376285314 | 2.65E-07 | 6.86E-07 | 0.001286238 |
| T2DM | endometrium | *GALNT15* | 2.006144394 | 1.60E-07 | 4.87E-07 | 0.000778214 |
| T2DM | endometrium | *SLC6A6* | 2.230477278 | 1.56E-07 | 4.81E-07 | 0.000756406 |
| T2DM | endometrium | *KCNC3* | -3.709433174 | 1.87E-05 | 2.89E-05 | 0.090770833 |
| T2DM | endometrium | *NAPSA* | 4.855452265 | 0.002062926 | 0.002589062 | 1 |
| T2DM | endometrium | *PDLIM4* | 5.123664462 | 8.05E-08 | 4.36E-07 | 0.000390672 |
| T2DM | endometrium | *KIF3A* | 6.980135431 | 4.85E-05 | 7.11E-05 | 0.235391908 |
| T2DM | endometrium | *ACLY* | 1.992951223 | 4.29E-07 | 9.89E-07 | 0.002079771 |
| T2DM | endometrium | *NDUFA2* | -1.986154076 | 6.39E-05 | 9.19E-05 | 0.309903172 |
| T2DM | endometrium | *EXOC4* | 3.462850514 | 1.87E-08 | 4.34E-07 | 9.08E-05 |
| T2DM | endometrium | *C1orf159* | 6.320050095 | 7.88E-08 | 4.36E-07 | 0.000382472 |
| T2DM | endometrium | *PPFIA1* | 3.700619699 | 7.46E-08 | 4.36E-07 | 0.000361786 |
| T2DM | endometrium | *BARX1* | 8.999348535 | 5.96E-05 | 8.62E-05 | 0.289190431 |
| T2DM | endometrium | *NINJ1* | 2.407445949 | 4.30E-07 | 9.91E-07 | 0.002085544 |

| | | | | | | |
|---|---|---|---|---|---|---|
| T2DM | endometrium | *CKMT2* | 4.44806832 | 3.76E-07 | 8.99E-07 | 0.001826441 |
| T2DM | endometrium | *KHDRBS3* | 3.36547426 | 6.76E-08 | 4.34E-07 | 0.000328212 |
| T2DM | endometrium | *CHD1L* | 4.934999939 | 8.97E-05 | 0.000126837 | 0.435303152 |
| T2DM | endometrium | *PRKAB2* | 3.301814911 | 1.08E-07 | 4.54E-07 | 0.000525403 |
| T2DM | endometrium | *ZNF304* | 3.522178827 | 6.40E-07 | 1.39E-06 | 0.003106163 |
| T2DM | endometrium | *ZNF132* | 3.289801216 | 3.31E-06 | 5.88E-06 | 0.016074943 |
| T2DM | endometrium | *RHPN2* | 2.240675788 | 1.49E-07 | 4.78E-07 | 0.000720745 |
| T2DM | endometrium | *GCH1* | 3.002319339 | 7.23E-08 | 4.34E-07 | 0.000350925 |
| T2DM | endometrium | *MATN3* | 4.363142045 | 0.000378485 | 0.000506456 | 1 |
| T2DM | endometrium | *TRIM21* | 5.04615529 | 1.03E-07 | 4.47E-07 | 0.00049947 |
| T2DM | endometrium | *LRRC41* | -2.775927823 | 0.006344255 | 0.007661106 | 1 |
| T2DM | endometrium | *CCT6B* | 3.302608306 | 5.28E-06 | 9.01E-06 | 0.02562858 |
| T2DM | endometrium | *CNGA4* | 3.489517759 | 0.000247357 | 0.000336331 | 1 |
| T2DM | endometrium | *RRP8* | -2.738218054 | 0.001301515 | 0.001660955 | 1 |
| T2DM | endometrium | *TIMM10B* | 5.540522561 | 5.08E-06 | 8.69E-06 | 0.024665532 |
| T2DM | endometrium | *IMMT* | 4.187270945 | 2.13E-08 | 4.34E-07 | 0.000103137 |
| T2DM | endometrium | *CARD6* | 3.955685881 | 2.85E-07 | 7.25E-07 | 0.001384673 |
| T2DM | endometrium | *MYBBP1A* | 3.75964244 | 3.34E-08 | 4.34E-07 | 0.000162164 |
| T2DM | endometrium | *RPA1* | 4.345524146 | 4.46E-07 | 1.02E-06 | 0.002162628 |
| T2DM | endometrium | *TMEM128* | -2.437921622 | 0.032649204 | 0.037326564 | 1 |
| T2DM | endometrium | *SEC61G* | 5.471271887 | 8.72E-05 | 0.000123413 | 0.422936761 |
| T2DM | endometrium | *FIGNL1* | -1.730526703 | 1.89E-05 | 2.92E-05 | 0.091885243 |
| T2DM | endometrium | *DDC* | 8.778839687 | 0.004680888 | 0.005689296 | 1 |
| T2DM | endometrium | *EIF5A* | 2.964316301 | 1.51E-05 | 2.36E-05 | 0.073386394 |
| T2DM | endometrium | *RGS22* | -1.910683225 | 0.004489347 | 0.005466076 | 1 |
| T2DM | endometrium | *REEP2* | -0.801577379 | 1.62E-06 | 3.11E-06 | 0.007851455 |
| T2DM | endometrium | *PRMT7* | 2.688996822 | 2.55E-06 | 4.67E-06 | 0.012379325 |
| T2DM | endometrium | *IGHMBP2* | 2.335175107 | 0.010555461 | 0.012515908 | 1 |
| T2DM | endometrium | *MUTYH* | 2.918344204 | 5.00E-08 | 4.34E-07 | 0.000242571 |
| T2DM | endometrium | *CTNNBL1* | 4.581609934 | 3.62E-08 | 4.34E-07 | 0.000175468 |
| T2DM | endometrium | *OSER1* | 4.304052569 | 3.76E-09 | 4.07E-07 | 1.82E-05 |
| T2DM | endometrium | *SERINC3* | 2.290980819 | 4.23E-07 | 9.81E-07 | 0.002052599 |
| T2DM | endometrium | *DMGDH* | 1.719870763 | 9.53E-07 | 1.95E-06 | 0.004622603 |
| T2DM | endometrium | *AP3B1* | 4.924969066 | 4.16E-06 | 7.27E-06 | 0.020206793 |
| T2DM | endometrium | *ZBED3* | 2.806467948 | 5.68E-08 | 4.34E-07 | 0.000275402 |
| T2DM | endometrium | *FBXO44* | 3.385449748 | 4.26E-08 | 4.34E-07 | 0.000206858 |
| T2DM | endometrium | *RSG1* | 10.00028085 | 0.002255672 | 0.002819299 | 1 |
| T2DM | endometrium | *CASP9* | 2.59830347 | 3.31E-08 | 4.34E-07 | 0.000160439 |
| T2DM | endometrium | *ATP8A2* | -1.575389658 | 7.30E-06 | 1.21E-05 | 0.035415743 |
| T2DM | endometrium | *ALOX5AP* | 2.672880321 | 9.77E-08 | 4.42E-07 | 0.00047381 |
| T2DM | endometrium | *WASF3* | 3.955417544 | 0.009147601 | 0.010886475 | 1 |
| T2DM | endometrium | *GPR12* | 13.86196285 | 0.00027393 | 0.000371259 | 1 |
| T2DM | endometrium | *SCO1* | 5.344058041 | 3.93E-08 | 4.34E-07 | 0.000190871 |
| T2DM | endometrium | *MPRIP* | 4.245883623 | 5.98E-08 | 4.34E-07 | 0.00029028 |
| T2DM | endometrium | *DSTYK* | 6.015451466 | 2.84E-07 | 7.22E-07 | 0.001376299 |
| T2DM | endometrium | *COG6* | 5.428572556 | 7.00E-07 | 1.50E-06 | 0.00339655 |
| T2DM | endometrium | *EPSTI1* | 2.562875763 | 5.79E-08 | 4.34E-07 | 0.000281097 |
| T2DM | endometrium | *BTBD2* | 7.229078736 | 4.73E-08 | 4.34E-07 | 0.000229703 |
| T2DM | endometrium | *CSNK1G2* | -1.949955975 | 3.78E-05 | 5.62E-05 | 0.183590414 |
| T2DM | endometrium | *CNDP2* | 3.516174794 | 7.69E-08 | 4.36E-07 | 0.000373218 |

| | | | | | | |
|---|---|---|---|---|---|---|
| T2DM | endometrium | *PDZD2* | 4.428553603 | 6.73E-06 | 1.12E-05 | 0.032675693 |
| T2DM | endometrium | *GSTT2B* | -1.06038014 | 0.039736793 | 0.045142336 | 1 |
| T2DM | endometrium | *FAM83F* | 16.16647848 | 1.52E-07 | 4.78E-07 | 0.000736275 |
| T2DM | endometrium | *GIMAP4* | 2.176546422 | 6.45E-07 | 1.40E-06 | 0.003127608 |
| T2DM | endometrium | *ADCK2* | 2.737549962 | 1.46E-06 | 2.83E-06 | 0.00706527 |
| T2DM | endometrium | *IPO8* | 5.887129083 | 8.78E-05 | 0.000124222 | 0.425834114 |
| T2DM | endometrium | *LARS* | 3.100571302 | 0.00054714 | 0.000721196 | 1 |
| T2DM | endometrium | *SPINK5* | 5.258895329 | 1.53E-07 | 4.78E-07 | 0.000741233 |
| T2DM | endometrium | *IMPA1* | 1.978981468 | 2.76E-06 | 5.01E-06 | 0.013389178 |
| T2DM | endometrium | *LRRCC1* | 6.898265952 | 6.16E-06 | 1.04E-05 | 0.02990143 |
| T2DM | endometrium | *RRAS2* | 5.12936009 | 8.87E-08 | 4.38E-07 | 0.000430586 |
| T2DM | endometrium | *CATSPERB* | 1.821086955 | 3.11E-07 | 7.73E-07 | 0.0015094 |
| T2DM | endometrium | *TTC9* | 3.588148931 | 1.35E-07 | 4.78E-07 | 0.000657227 |
| T2DM | endometrium | *ELP3* | 4.412847061 | 1.22E-07 | 4.69E-07 | 0.000593718 |
| T2DM | endometrium | *PEBP4* | 2.750071719 | 1.59E-07 | 4.86E-07 | 0.000771385 |
| T2DM | endometrium | *CTIF* | 2.382648963 | 1.16E-07 | 4.60E-07 | 0.000562331 |
| T2DM | endometrium | *MRO* | 6.263158206 | 1.23E-06 | 2.44E-06 | 0.005976912 |
| T2DM | endometrium | *THUMPD3* | -2.096015479 | 3.56E-05 | 5.32E-05 | 0.172713623 |
| T2DM | endometrium | *CNTN6* | 3.968936424 | 1.52E-07 | 4.78E-07 | 0.000738383 |
| T2DM | endometrium | *CHL1* | 3.336586558 | 1.16E-07 | 4.60E-07 | 0.000562955 |
| T2DM | endometrium | *GSTM5* | -2.203800703 | 0.013879172 | 0.016305507 | 1 |
| T2DM | endometrium | *GSTM3* | -1.137279917 | 0.002089756 | 0.002620024 | 1 |
| T2DM | endometrium | *VAV3* | 1.552455613 | 1.15E-06 | 2.30E-06 | 0.005577232 |
| T2DM | endometrium | *PSRC1* | -3.878480823 | 6.09E-05 | 8.79E-05 | 0.295380322 |
| T2DM | endometrium | *TRIM45* | -3.849670302 | 4.83E-05 | 7.09E-05 | 0.234502169 |
| T2DM | endometrium | *CEPT1* | 4.136887597 | 7.21E-08 | 4.34E-07 | 0.00034997 |
| T2DM | endometrium | *NAPG* | 5.565131293 | 1.53E-07 | 4.78E-07 | 0.000742123 |
| T2DM | endometrium | *PPHLN1* | 3.522903339 | 3.45E-08 | 4.34E-07 | 0.000167385 |
| T2DM | endometrium | *FKBP11* | 2.787986084 | 1.15E-07 | 4.59E-07 | 0.000560004 |
| T2DM | endometrium | *YWHAQ* | 3.15153541 | 5.12E-08 | 4.34E-07 | 0.000248409 |
| T2DM | endometrium | *KIDINS220* | 4.98947119 | 1.14E-06 | 2.29E-06 | 0.00555243 |
| T2DM | endometrium | *LPIN1* | 4.024881668 | 2.72E-07 | 6.99E-07 | 0.001320594 |
| T2DM | endometrium | *NARS* | 6.192877446 | 1.38E-08 | 4.34E-07 | 6.71E-05 |
| T2DM | endometrium | *KIAA1468* | 5.083522528 | 1.83E-07 | 5.27E-07 | 0.000886558 |
| T2DM | endometrium | *IL15RA* | 4.233490404 | 6.54E-08 | 4.34E-07 | 0.000317305 |
| T2DM | endometrium | *TMEM241* | 3.546353798 | 2.68E-06 | 4.88E-06 | 0.013015732 |
| T2DM | endometrium | *SOX5* | 2.620457529 | 5.80E-08 | 4.34E-07 | 0.000281316 |
| T2DM | endometrium | *KLRD1* | 4.809332773 | 4.96E-08 | 4.34E-07 | 0.00024072 |
| T2DM | endometrium | *ACP2* | 7.866469219 | 1.93E-05 | 2.97E-05 | 0.093581628 |
| T2DM | endometrium | *HOOK1* | 1.686459305 | 0.001356801 | 0.001728328 | 1 |
| T2DM | endometrium | *ZCCHC11* | 2.838444585 | 7.12E-08 | 4.34E-07 | 0.000345327 |
| T2DM | endometrium | *PRPF38A* | 7.264560507 | 3.41E-07 | 8.31E-07 | 0.001656073 |
| T2DM | endometrium | *ELP2* | 3.967475393 | 5.87E-08 | 4.34E-07 | 0.000284834 |
| T2DM | endometrium | *TPGS2* | 5.252600845 | 6.31E-08 | 4.34E-07 | 0.000306085 |
| T2DM | endometrium | *TIMM10* | -1.884953812 | 5.84E-06 | 9.86E-06 | 0.028316126 |
| T2DM | endometrium | *DHX34* | 6.652231691 | 1.76E-07 | 5.16E-07 | 0.000853388 |
| T2DM | endometrium | *FADS2* | -2.095238413 | 3.93E-05 | 5.83E-05 | 0.190734106 |
| T2DM | endometrium | *CLOCK* | 3.897035578 | 5.53E-08 | 4.34E-07 | 0.000268525 |
| T2DM | endometrium | *GGACT* | 4.931907819 | 1.71E-05 | 2.65E-05 | 0.082962171 |
| T2DM | endometrium | *CLDN10* | 5.845779933 | 5.82E-08 | 4.34E-07 | 0.000282375 |

| | | | | | | |
|---|---|---|---|---|---|---|
| T2DM | endometrium | *ARGLU1* | 3.055917922 | 4.78E-08 | 4.34E-07 | 0.000231733 |
| T2DM | endometrium | *ERCC5* | -3.680078099 | 1.74E-05 | 2.69E-05 | 0.084532811 |
| T2DM | endometrium | *KLB* | 1.585574881 | 0.000325692 | 0.000438716 | 1 |
| T2DM | endometrium | *WDR36* | 5.796050558 | 1.25E-07 | 4.73E-07 | 0.000604695 |
| T2DM | endometrium | *OSTF1* | 3.019568146 | 6.56E-08 | 4.34E-07 | 0.000318256 |
| T2DM | endometrium | *UBQLN1* | 4.429294406 | 6.05E-08 | 4.34E-07 | 0.000293488 |
| T2DM | endometrium | *GOLM1* | 2.29688939 | 5.83E-06 | 9.86E-06 | 0.028289927 |
| T2DM | endometrium | *ADAM19* | 2.272162314 | 5.16E-08 | 4.34E-07 | 0.000250146 |
| T2DM | endometrium | *MSI1* | 5.754194029 | 1.94E-06 | 3.66E-06 | 0.009436821 |
| T2DM | endometrium | *P2RX4* | 2.478191793 | 5.42E-09 | 4.27E-07 | 2.63E-05 |
| T2DM | endometrium | *DTX1* | 2.1430018 | 1.39E-07 | 4.78E-07 | 0.000675376 |
| T2DM | endometrium | *DMTF1* | 3.75819441 | 6.52E-08 | 4.34E-07 | 0.00031643 |
| T2DM | endometrium | *TMEM60* | 3.211807824 | 1.05E-07 | 4.47E-07 | 0.000509205 |
| T2DM | endometrium | *POM121C* | 2.790168663 | 1.52E-06 | 2.93E-06 | 0.007354429 |
| T2DM | endometrium | *SRPK2* | 4.254102353 | 4.57E-07 | 1.04E-06 | 0.002215894 |
| T2DM | endometrium | *TES* | 2.598610621 | 4.29E-07 | 9.89E-07 | 0.002079144 |
| T2DM | endometrium | *MTO1* | -2.309944174 | 0.007352695 | 0.008830514 | 1 |
| T2DM | endometrium | *NT5E* | 2.430603707 | 4.66E-08 | 4.34E-07 | 0.000226027 |
| T2DM | endometrium | *AKIRIN2* | 3.692946644 | 4.01E-08 | 4.34E-07 | 0.000194696 |
| T2DM | endometrium | *MAP3K7* | 6.007869529 | 9.20E-06 | 1.49E-05 | 0.04466034 |
| T2DM | endometrium | *PRR5L* | 4.841560605 | 7.03E-05 | 0.000100685 | 0.341322171 |
| T2DM | endometrium | *PHF21A* | 4.051082755 | 1.05E-07 | 4.47E-07 | 0.000507314 |
| T2DM | endometrium | *CAPRIN1* | 5.990515749 | 5.31E-06 | 9.05E-06 | 0.025783001 |
| T2DM | endometrium | *ATP5G2* | 6.169240373 | 7.90E-07 | 1.66E-06 | 0.003833788 |
| T2DM | endometrium | *B4GALNT1* | 8.67865928 | 0.000620857 | 0.000814822 | 1 |
| T2DM | endometrium | *PAN2* | -3.014386029 | 0.000213496 | 0.000292128 | 1 |
| T2DM | endometrium | *KRT7* | 2.469894553 | 0.003464896 | 0.004249665 | 1 |
| T2DM | endometrium | *SLC26A10* | 5.792536571 | 0.008486865 | 0.010124974 | 1 |
| T2DM | endometrium | *MAP7* | 3.322816642 | 0.000304112 | 0.000410446 | 1 |
| T2DM | endometrium | *NHSL1* | 1.784510195 | 3.37E-08 | 4.34E-07 | 0.000163577 |
| T2DM | endometrium | *AHI1* | 3.100576316 | 5.11E-05 | 7.45E-05 | 0.248065903 |
| T2DM | endometrium | *SMPD2* | 2.075265819 | 0.000448529 | 0.000596236 | 1 |
| T2DM | endometrium | *MICAL1* | -1.255131363 | 0.000144455 | 0.000200543 | 0.700897641 |
| T2DM | endometrium | *CCT7* | 3.562603219 | 8.02E-08 | 4.36E-07 | 0.000389061 |
| T2DM | endometrium | *USP15* | 3.162064874 | 5.10E-08 | 4.34E-07 | 0.000247599 |
| T2DM | endometrium | *MDM2* | 5.344420223 | 1.02E-06 | 2.08E-06 | 0.004972499 |
| T2DM | endometrium | *KLHL36* | 2.087796294 | 9.73E-08 | 4.42E-07 | 0.000472327 |
| T2DM | endometrium | *MPHOSPH6* | 2.652411024 | 6.85E-08 | 4.34E-07 | 0.000332502 |
| T2DM | endometrium | *CCDC102A* | 5.393986787 | 0.000236801 | 0.000322741 | 1 |
| T2DM | endometrium | *AGT* | 3.750269822 | 5.05E-06 | 8.65E-06 | 0.024514654 |
| T2DM | endometrium | *ZNF670* | 6.041616947 | 1.90E-05 | 2.93E-05 | 0.092135096 |
| T2DM | endometrium | *KCNK1* | -2.466330623 | 0.001293002 | 0.001650959 | 1 |
| T2DM | endometrium | *CAPN9* | -1.444596162 | 1.75E-06 | 3.34E-06 | 0.008508699 |
| T2DM | endometrium | *NTPCR* | 6.975004657 | 8.48E-05 | 0.000120313 | 0.411351002 |
| T2DM | endometrium | *PIGC* | 5.76290954 | 7.29E-08 | 4.34E-07 | 0.000353638 |
| T2DM | endometrium | *SP110* | 4.302390021 | 1.19E-07 | 4.65E-07 | 0.000577888 |
| T2DM | endometrium | *MRPL44* | 3.164745368 | 1.28E-07 | 4.78E-07 | 0.000623274 |
| T2DM | endometrium | *USP37* | 4.342996853 | 5.57E-08 | 4.34E-07 | 0.000270431 |
| T2DM | endometrium | *SLC19A3* | 2.675440605 | 3.95E-08 | 4.34E-07 | 0.00019147 |
| T2DM | endometrium | *TMBIM1* | 5.924342938 | 1.04E-07 | 4.47E-07 | 0.000505164 |

| | | | | | | |
|---|---|---|---|---|---|---|
| T2DM | endometrium | *TSGA10* | 2.080391278 | 1.18E-05 | 1.89E-05 | 0.057323499 |
| T2DM | endometrium | *GCC2* | 4.07010279 | 1.17E-07 | 4.61E-07 | 0.000567682 |
| T2DM | endometrium | *MRPS9* | 4.340547813 | 4.91E-08 | 4.34E-07 | 0.000238092 |
| T2DM | endometrium | *ANKRD36* | 4.903024521 | 7.78E-08 | 4.36E-07 | 0.000377485 |
| T2DM | endometrium | *EPC2* | 4.843901415 | 1.50E-07 | 4.78E-07 | 0.000729457 |
| T2DM | endometrium | *APPL2* | 4.683704097 | 7.77E-07 | 1.64E-06 | 0.003767663 |
| T2DM | endometrium | *DRAM1* | 3.202284397 | 5.69E-08 | 4.34E-07 | 0.000276058 |
| T2DM | endometrium | *SLC41A2* | 2.435969513 | 6.21E-08 | 4.34E-07 | 0.000301129 |
| T2DM | endometrium | *NEK3* | 3.698115178 | 1.99E-05 | 3.05E-05 | 0.096320299 |
| T2DM | endometrium | *RNASEH2B* | 2.922413165 | 9.04E-08 | 4.38E-07 | 0.000438484 |
| T2DM | endometrium | *TBC1D4* | 2.967957303 | 2.36E-07 | 6.32E-07 | 0.001145169 |
| T2DM | endometrium | *SUCLA2* | 5.420865457 | 1.35E-07 | 4.78E-07 | 0.00065539 |
| T2DM | endometrium | *RCBTB1* | 1.870275062 | 1.54E-05 | 2.40E-05 | 0.074753015 |
| T2DM | endometrium | *RCBTB2* | 2.445221414 | 9.37E-08 | 4.39E-07 | 0.000454759 |
| T2DM | endometrium | *SETDB2* | 3.795450411 | 6.86E-08 | 4.34E-07 | 0.00033297 |
| T2DM | endometrium | *SCRN1* | 3.322900273 | 1.21E-07 | 4.69E-07 | 0.000587531 |
| T2DM | endometrium | *CHST12* | 3.977720303 | 8.81E-08 | 4.38E-07 | 0.000427447 |
| T2DM | endometrium | *KDELR2* | 3.496528616 | 7.16E-08 | 4.34E-07 | 0.000347245 |
| T2DM | endometrium | *NUPL2* | 4.727425754 | 5.95E-08 | 4.34E-07 | 0.000288682 |
| T2DM | endometrium | *ZDHHC4* | -2.494416879 | 8.50E-05 | 0.000120575 | 0.41236569 |
| T2DM | endometrium | *AOAH* | 1.528688507 | 6.10E-07 | 1.33E-06 | 0.002961014 |
| T2DM | endometrium | *TBRG4* | 3.038361983 | 2.74E-07 | 7.04E-07 | 0.001331156 |
| T2DM | endometrium | *MTHFS* | 3.329461656 | 7.40E-07 | 1.58E-06 | 0.003592043 |
| T2DM | endometrium | *NMT1* | 4.111663916 | 3.72E-06 | 6.55E-06 | 0.018055097 |
| T2DM | endometrium | *MYCBPAP* | 1.966460845 | 0.000871921 | 0.001129053 | 1 |
| T2DM | endometrium | *RTP4* | 4.294367588 | 0.000130279 | 0.000181589 | 0.632112927 |
| T2DM | endometrium | *NDUFB5* | 3.177932 | 9.73E-08 | 4.42E-07 | 0.000472144 |
| T2DM | endometrium | *SCN2A* | 3.982464764 | 1.02E-08 | 4.34E-07 | 4.93E-05 |
| T2DM | endometrium | *RPS6KC1* | 5.193454566 | 4.73E-07 | 1.07E-06 | 0.002295203 |
| T2DM | endometrium | *SMPD4* | 3.315382872 | 7.41E-07 | 1.58E-06 | 0.003595919 |
| T2DM | endometrium | *CCDC115* | -1.591788005 | 0.026084183 | 0.030026206 | 1 |
| T2DM | endometrium | *IMP4* | 7.442495392 | 3.91E-07 | 9.25E-07 | 0.001896775 |
| T2DM | endometrium | *NIPSNAP3A* | -3.303070919 | 0.011913903 | 0.014081914 | 1 |
| T2DM | endometrium | *ODF2* | 6.156102812 | 1.92E-07 | 5.47E-07 | 0.000933289 |
| T2DM | endometrium | *TOR1B* | 7.103376864 | 4.90E-07 | 1.11E-06 | 0.00237751 |
| T2DM | endometrium | *SLC2A8* | 3.913597056 | 5.83E-07 | 1.29E-06 | 0.002829895 |
| T2DM | endometrium | *ZNF189* | 7.986965647 | 6.05E-07 | 1.33E-06 | 0.002934241 |
| T2DM | endometrium | *STX17* | 5.783100348 | 4.30E-07 | 9.92E-07 | 0.00208842 |
| T2DM | endometrium | *PRPF4* | -1.098586553 | 0.022369998 | 0.025867309 | 1 |
| T2DM | endometrium | *ANP32B* | 5.650955667 | 0.000243541 | 0.000331461 | 1 |
| T2DM | endometrium | *DSCC1* | 1.252363872 | 0.001427025 | 0.001813495 | 1 |
| T2DM | endometrium | *DERL1* | 6.318846993 | 0.000152814 | 0.000211663 | 0.741455674 |
| T2DM | endometrium | *NOV* | 4.459897307 | 1.65E-07 | 4.96E-07 | 0.000801036 |
| T2DM | endometrium | *IL11RA* | 2.743208788 | 7.41E-07 | 1.58E-06 | 0.003596734 |
| T2DM | endometrium | *APTX* | 4.540091046 | 4.98E-07 | 1.12E-06 | 0.00241436 |
| T2DM | endometrium | *TMEM8B* | 3.730450941 | 3.10E-08 | 4.34E-07 | 0.000150358 |
| T2DM | endometrium | *GRHPR* | -1.968388419 | 2.41E-05 | 3.66E-05 | 0.116837475 |
| T2DM | endometrium | *DENND4C* | 2.478061614 | 9.29E-08 | 4.39E-07 | 0.000450932 |
| T2DM | endometrium | *GMPR* | 5.286411198 | 2.37E-06 | 4.37E-06 | 0.011483538 |
| T2DM | endometrium | *TMEM14B* | -2.727201406 | 0.002564888 | 0.003186082 | 1 |

| | | | | | | |
|---|---|---|---|---|---|---|
| T2DM | endometrium | *FRS3* | 2.952277397 | 1.27E-07 | 4.76E-07 | 0.000615956 |
| T2DM | endometrium | *CAPN11* | 2.507699828 | 0.005809671 | 0.007027804 | 1 |
| T2DM | endometrium | *IRF4* | 2.913873138 | 5.33E-08 | 4.34E-07 | 0.000258769 |
| T2DM | endometrium | *LRRC1* | 4.462294008 | 1.13E-07 | 4.57E-07 | 0.000548716 |
| T2DM | endometrium | *RIPK1* | 3.716709671 | 7.81E-08 | 4.36E-07 | 0.000379153 |
| T2DM | endometrium | *TUBB2B* | 3.414510599 | 0.000699107 | 0.000912091 | 1 |
| T2DM | endometrium | *TCF19* | 10.36645476 | 9.39E-12 | 5.69E-09 | 4.56E-08 |
| T2DM | endometrium | *FLOT1* | 3.062416211 | 1.30E-08 | 4.34E-07 | 6.31E-05 |
| T2DM | endometrium | *TPMT* | 2.166806609 | 8.66E-06 | 1.41E-05 | 0.041997475 |
| T2DM | endometrium | *NRM* | 8.696219231 | 1.16E-07 | 4.60E-07 | 0.000564806 |
| T2DM | endometrium | *VARS2* | 1.905660446 | 3.93E-05 | 5.83E-05 | 0.190743492 |
| T2DM | endometrium | *TLR2* | 3.576637689 | 2.81E-07 | 7.18E-07 | 0.001364187 |
| T2DM | endometrium | *MYO7A* | 3.094780601 | 6.78E-07 | 1.46E-06 | 0.003288105 |
| T2DM | endometrium | *NUMA1* | 3.436583925 | 2.60E-08 | 4.34E-07 | 0.000126134 |
| T2DM | endometrium | *SYTL2* | 1.723495831 | 5.87E-06 | 9.90E-06 | 0.028457603 |
| T2DM | endometrium | *RAB30* | 2.399202081 | 1.29E-07 | 4.78E-07 | 0.000628109 |
| T2DM | endometrium | *PRCP* | 5.657068368 | 1.44E-06 | 2.79E-06 | 0.006964573 |
| T2DM | endometrium | *NARS2* | 3.841959505 | 0.01043085 | 0.012377228 | 1 |
| T2DM | endometrium | *GGH* | 1.973997412 | 0.001927274 | 0.002423201 | 1 |
| T2DM | endometrium | *SULF1* | 2.34534959 | 4.79E-08 | 4.34E-07 | 0.000232491 |
| T2DM | endometrium | *NEK1* | 3.515077441 | 2.49E-08 | 4.34E-07 | 0.000120882 |
| T2DM | endometrium | *TRPC6* | 4.02213714 | 0.012605584 | 0.014877716 | 1 |
| T2DM | endometrium | *MMP27* | 8.539514127 | 0.003315396 | 0.004077643 | 1 |
| T2DM | endometrium | *SLC37A4* | -1.707231529 | 0.008925866 | 0.010630808 | 1 |
| T2DM | endometrium | *POU2F3* | -2.495906449 | 0.000454344 | 0.00060347 | 1 |
| T2DM | endometrium | *PPP2R1B* | 2.033805239 | 7.87E-08 | 4.36E-07 | 0.000381762 |
| T2DM | endometrium | *FDX1* | 4.103480768 | 4.38E-08 | 4.34E-07 | 0.000212692 |
| T2DM | endometrium | *FXYD2* | 4.885597632 | 4.36E-06 | 7.57E-06 | 0.02115674 |
| T2DM | endometrium | *MAP2K5* | 3.309462738 | 3.45E-08 | 4.34E-07 | 0.000167586 |
| T2DM | endometrium | *NUSAP1* | 11.03284825 | 1.06E-08 | 4.34E-07 | 5.15E-05 |
| T2DM | endometrium | *NDUFAF1* | -1.964699767 | 3.43E-05 | 5.12E-05 | 0.166296865 |
| T2DM | endometrium | *PARP6* | 6.243246495 | 2.83E-07 | 7.21E-07 | 0.001372343 |
| T2DM | endometrium | *ADAM10* | 2.282439101 | 5.55E-08 | 4.34E-07 | 0.00026928 |
| T2DM | endometrium | *ZNF280D* | 4.031175456 | 1.03E-07 | 4.47E-07 | 0.000499408 |
| T2DM | endometrium | *SPTBN5* | 2.087039327 | 3.77E-08 | 4.34E-07 | 0.000182889 |
| T2DM | endometrium | *SLC44A5* | 6.412773992 | 1.33E-05 | 2.10E-05 | 0.064437738 |
| T2DM | endometrium | *DBT* | 6.765868698 | 3.60E-06 | 6.35E-06 | 0.017474796 |
| T2DM | endometrium | *RTCA* | 4.257700211 | 5.01E-08 | 4.34E-07 | 0.000242867 |
| T2DM | endometrium | *KHK* | 3.525970726 | 0.000154494 | 0.000213867 | 0.749604857 |
| T2DM | endometrium | *ADCY3* | 2.491086146 | 5.08E-08 | 4.34E-07 | 0.000246309 |
| T2DM | endometrium | *DYNC2LI1* | 3.9157655 | 4.18E-06 | 7.29E-06 | 0.020278898 |
| T2DM | endometrium | *LHCGR* | 5.02997991 | 0.000699833 | 0.000912548 | 1 |
| T2DM | endometrium | *THUMPD2* | 5.45624306 | 1.92E-07 | 5.46E-07 | 0.000929797 |
| T2DM | endometrium | *ACTR2* | 3.072507558 | 5.03E-08 | 4.34E-07 | 0.000243817 |
| T2DM | endometrium | *ATRAID* | 5.331797484 | 1.11E-08 | 4.34E-07 | 5.40E-05 |
| T2DM | endometrium | *CENPO* | -1.098386979 | 8.18E-06 | 1.34E-05 | 0.039709194 |
| T2DM | endometrium | *LRPPRC* | 5.465614623 | 1.08E-05 | 1.74E-05 | 0.052581911 |
| T2DM | endometrium | *DTNB* | 5.476768934 | 2.31E-06 | 4.29E-06 | 0.011226312 |
| T2DM | endometrium | *CYP2C8* | 7.0301648 | 0.039335068 | 0.044706899 | 1 |
| T2DM | endometrium | *MYOF* | 3.890682805 | 1.30E-09 | 2.63E-07 | 6.31E-06 |

| | | | | | | |
|---|---|---|---|---|---|---|
| T2DM | endometrium | *BTBD16* | 3.831394551 | 6.75E-06 | 1.12E-05 | 0.032742306 |
| T2DM | endometrium | *TACC2* | 4.037889672 | 4.49E-08 | 4.34E-07 | 0.00021784 |
| T2DM | endometrium | *CALHM2* | 5.563484816 | 0.002345398 | 0.002923913 | 1 |
| T2DM | endometrium | *ARL3* | 2.736730272 | 4.69E-08 | 4.34E-07 | 0.000227455 |
| T2DM | endometrium | *PLCE1* | 2.092036642 | 1.79E-07 | 5.21E-07 | 0.000866722 |
| T2DM | endometrium | *RPS24* | 4.497355941 | 1.74E-07 | 5.12E-07 | 0.000841885 |
| T2DM | endometrium | *ATIC* | 2.124538058 | 8.16E-06 | 1.34E-05 | 0.039572653 |
| T2DM | endometrium | *SMARCAL1* | 2.527683621 | 9.58E-08 | 4.42E-07 | 0.000464991 |
| T2DM | endometrium | *BARD1* | 2.994938811 | 1.78E-07 | 5.20E-07 | 0.000862444 |
| T2DM | endometrium | *STAT4* | 2.035879489 | 1.60E-07 | 4.87E-07 | 0.000777968 |
| T2DM | endometrium | *SSB* | 3.535397967 | 2.14E-07 | 5.88E-07 | 0.001037939 |
| T2DM | endometrium | *FASTKD1* | 5.754416689 | 8.86E-08 | 4.38E-07 | 0.00042965 |
| T2DM | endometrium | *ITGAV* | 5.746987033 | 8.38E-06 | 1.37E-05 | 0.040674622 |
| T2DM | endometrium | *SLC35A5* | 3.304820115 | 3.60E-08 | 4.34E-07 | 0.000174508 |
| T2DM | endometrium | *SENP7* | 3.098326337 | 4.09E-08 | 4.34E-07 | 0.000198307 |
| T2DM | endometrium | *COX17* | 3.493164299 | 8.12E-08 | 4.36E-07 | 0.000393747 |
| T2DM | endometrium | *SPPL2A* | 3.956122275 | 4.56E-06 | 7.89E-06 | 0.022141424 |
| T2DM | endometrium | *PPCDC* | 4.495528944 | 9.24E-06 | 1.50E-05 | 0.044855082 |
| T2DM | endometrium | *SEMA7A* | 3.797026541 | 0.008694129 | 0.010362052 | 1 |
| T2DM | endometrium | *PCDH10* | 8.479799295 | 0.001317294 | 0.001679766 | 1 |
| T2DM | endometrium | *AP1AR* | 5.156205226 | 0.009357039 | 0.011127538 | 1 |
| T2DM | endometrium | *PRKG2* | -2.03977814 | 0.003276495 | 0.004031842 | 1 |
| T2DM | endometrium | *KIAA1109* | 4.10661579 | 9.76E-08 | 4.42E-07 | 0.000473417 |
| T2DM | endometrium | *MMRN1* | 1.472865633 | 5.40E-06 | 9.18E-06 | 0.026186062 |
| T2DM | endometrium | *PRDM5* | 6.28276656 | 1.33E-05 | 2.10E-05 | 0.06450427 |
| T2DM | endometrium | *NAAA* | 3.245535118 | 4.95E-09 | 4.20E-07 | 2.40E-05 |
| T2DM | endometrium | *NUP54* | 4.973898472 | 1.72E-08 | 4.34E-07 | 8.33E-05 |
| T2DM | endometrium | *G3BP2* | -2.383373182 | 0.034370743 | 0.039211578 | 1 |
| T2DM | endometrium | *CNOT6L* | 2.615250022 | 1.71E-07 | 5.06E-07 | 0.000827505 |
| T2DM | endometrium | *ANXA3* | 1.931697825 | 3.62E-08 | 4.34E-07 | 0.000175844 |
| T2DM | endometrium | *PPA2* | 5.04973546 | 7.15E-06 | 1.18E-05 | 0.034700229 |
| T2DM | endometrium | *ENPEP* | -1.149410457 | 9.68E-05 | 0.00013635 | 0.469725126 |
| T2DM | endometrium | *PAPSS1* | 4.794348246 | 3.34E-08 | 4.34E-07 | 0.000162192 |
| T2DM | endometrium | *FBN2* | 1.271155464 | 0.003822814 | 0.004667411 | 1 |
| T2DM | endometrium | *PARVG* | 2.742985659 | 5.30E-07 | 1.18E-06 | 0.002571636 |
| T2DM | endometrium | *B4GALNT3* | -5.614026169 | 0.000739038 | 0.000962118 | 1 |
| T2DM | endometrium | *PDE6H* | 13.02020376 | 0.000690932 | 0.000902153 | 1 |
| T2DM | endometrium | *ERP27* | 4.479672925 | 6.65E-05 | 9.55E-05 | 0.322707774 |
| T2DM | endometrium | *KIF21A* | 5.355069438 | 2.16E-05 | 3.30E-05 | 0.104772018 |
| T2DM | endometrium | *ALG10* | -3.591328973 | 0.000279145 | 0.000377799 | 1 |
| T2DM | endometrium | *C1RL* | 3.425345399 | 2.84E-06 | 5.14E-06 | 0.013772178 |
| T2DM | endometrium | *CLSTN3* | -2.523379579 | 0.000196159 | 0.000269088 | 0.951763052 |
| T2DM | endometrium | *KLRG1* | 1.470292998 | 2.53E-05 | 3.84E-05 | 0.122831995 |
| T2DM | endometrium | *VAMP1* | -2.377045404 | 0.00196571 | 0.002470887 | 1 |
| T2DM | endometrium | *PEX5* | -1.75597387 | 8.03E-05 | 0.000114217 | 0.389594449 |
| T2DM | endometrium | *LGR5* | 6.219105725 | 1.37E-06 | 2.67E-06 | 0.00662548 |
| T2DM | endometrium | *POC1B* | 6.068675007 | 3.17E-07 | 7.86E-07 | 0.00153859 |
| T2DM | endometrium | *AMDHD1* | -1.046767332 | 2.36E-06 | 4.36E-06 | 0.011464142 |
| T2DM | endometrium | *SDSL* | 4.139925352 | 5.85E-06 | 9.89E-06 | 0.028389782 |
| T2DM | endometrium | *GLTP* | 2.72889899 | 1.43E-07 | 4.78E-07 | 0.00069392 |

| | | | | | | |
|---|---|---|---|---|---|---|
| T2DM | endometrium | *FOXN4* | 4.301574599 | 2.60E-07 | 6.77E-07 | 0.001262309 |
| T2DM | endometrium | *MTMR6* | 2.375158602 | 1.29E-07 | 4.78E-07 | 0.000626328 |
| T2DM | endometrium | *CCDC65* | 6.499423863 | 2.32E-06 | 4.29E-06 | 0.011234824 |
| T2DM | endometrium | *ACVRL1* | 7.168874668 | 5.45E-08 | 4.34E-07 | 0.000264408 |
| T2DM | endometrium | *BRCA2* | 6.577047408 | 1.22E-07 | 4.69E-07 | 0.000591311 |
| T2DM | endometrium | *KANSL2* | 5.544713285 | 2.18E-08 | 4.34E-07 | 0.000105723 |
| T2DM | endometrium | *CERS5* | 3.434760281 | 8.71E-08 | 4.38E-07 | 0.000422538 |
| T2DM | endometrium | *LMBR1L* | 6.472255187 | 4.05E-05 | 6.00E-05 | 0.196643837 |
| T2DM | endometrium | *ESYT1* | 5.140287137 | 5.45E-08 | 4.34E-07 | 0.000264255 |
| T2DM | endometrium | *ZNF740* | 6.976351729 | 1.42E-06 | 2.76E-06 | 0.006869837 |
| T2DM | endometrium | *WDFY2* | 2.394589917 | 9.17E-08 | 4.38E-07 | 0.000445018 |
| T2DM | endometrium | *RB1* | 4.03560109 | 1.86E-07 | 5.35E-07 | 0.000901463 |
| T2DM | endometrium | *VPS33A* | -2.467305194 | 0.001360977 | 0.001733192 | 1 |
| T2DM | endometrium | *DIAPH3* | 3.769617502 | 2.44E-05 | 3.70E-05 | 0.118146582 |
| T2DM | endometrium | *GRTP1* | 2.394588614 | 0.003340793 | 0.004106797 | 1 |
| T2DM | endometrium | *CBLN3* | 1.180817001 | 0.000241582 | 0.000328887 | 1 |
| T2DM | endometrium | *NOVA1* | 2.45095379 | 4.42E-08 | 4.34E-07 | 0.000214466 |
| T2DM | endometrium | *FITM1* | 10.83359677 | 0.000368482 | 0.000493751 | 1 |
| T2DM | endometrium | *TMX1* | 6.027493811 | 9.14E-08 | 4.38E-07 | 0.000443575 |
| T2DM | endometrium | *RTN1* | 1.509036676 | 8.37E-08 | 4.38E-07 | 0.00040631 |
| T2DM | endometrium | *C14orf37* | 4.404794186 | 9.18E-08 | 4.38E-07 | 0.000445395 |
| T2DM | endometrium | *RAB15* | 5.663686194 | 4.29E-08 | 4.34E-07 | 0.000208081 |
| T2DM | endometrium | *EFCAB11* | 5.443437161 | 8.47E-09 | 4.34E-07 | 4.11E-05 |
| T2DM | endometrium | *PTGR2* | 1.370735431 | 0.000595622 | 0.000782974 | 1 |
| T2DM | endometrium | *SLC24A4* | 3.412277508 | 1.40E-07 | 4.78E-07 | 0.000678585 |
| T2DM | endometrium | *C14orf79* | -2.027309493 | 5.18E-05 | 7.54E-05 | 0.251281837 |
| T2DM | endometrium | *WARS* | 6.16292063 | 0.000334362 | 0.000449895 | 1 |
| T2DM | endometrium | *DUOXA1* | 2.739695658 | 0.007807244 | 0.009353271 | 1 |
| T2DM | endometrium | *SERF2* | 5.687244237 | 2.20E-06 | 4.10E-06 | 0.010690273 |
| T2DM | endometrium | *ZSCAN29* | 3.860061304 | 1.07E-08 | 4.34E-07 | 5.20E-05 |
| T2DM | endometrium | *UBE2Q2* | 3.69016437 | 2.47E-08 | 4.34E-07 | 0.000119967 |
| T2DM | endometrium | *HMG20A* | 3.277968813 | 3.33E-08 | 4.34E-07 | 0.000161591 |
| T2DM | endometrium | *SCAPER* | 2.688350948 | 0.011851127 | 0.014011128 | 1 |
| T2DM | endometrium | *MAN2C1* | -1.745662994 | 0.003029715 | 0.003742407 | 1 |
| T2DM | endometrium | *PIF1* | 4.410344716 | 0.025650983 | 0.029548568 | 1 |
| T2DM | endometrium | *CYP11A1* | 4.458185037 | 4.27E-07 | 9.88E-07 | 0.002072297 |
| T2DM | endometrium | *BBS4* | 4.360905983 | 1.84E-08 | 4.34E-07 | 8.91E-05 |
| T2DM | endometrium | *ULK3* | 4.240916642 | 9.18E-08 | 4.38E-07 | 0.000445654 |
| T2DM | endometrium | *PCSK6* | -2.805806803 | 0.006594844 | 0.007949859 | 1 |
| T2DM | endometrium | *CELF6* | 21.54805225 | 4.41E-06 | 7.65E-06 | 0.021400455 |
| T2DM | endometrium | *HAPLN3* | -1.281418362 | 1.95E-06 | 3.66E-06 | 0.009442386 |
| T2DM | endometrium | *POLG* | 5.181749633 | 3.26E-07 | 8.03E-07 | 0.001583753 |
| T2DM | endometrium | *MFGE8* | 5.353618185 | 5.17E-08 | 4.34E-07 | 0.000250811 |
| T2DM | endometrium | *UNC45A* | 2.204086184 | 1.29E-07 | 4.78E-07 | 0.000624559 |
| T2DM | endometrium | *IQGAP1* | 2.85723217 | 9.80E-08 | 4.42E-07 | 0.000475603 |
| T2DM | endometrium | *CRTC3* | 2.912639545 | 3.77E-08 | 4.34E-07 | 0.000182802 |
| T2DM | endometrium | *ITGAX* | 3.497595675 | 1.14E-07 | 4.57E-07 | 0.000551477 |
| T2DM | endometrium | *TGFB1I1* | -1.42608401 | 5.88E-06 | 9.91E-06 | 0.028509255 |
| T2DM | endometrium | *DHX38* | 2.601363704 | 1.09E-07 | 4.55E-07 | 0.000528987 |
| T2DM | endometrium | *NUDT7* | 5.559210429 | 8.45E-08 | 4.38E-07 | 0.000410039 |

| | | | | | | |
|---|---|---|---|---|---|---|
| T2DM | endometrium | *GCSH* | 2.813160195 | 1.73E-05 | 2.68E-05 | 0.083970031 |
| T2DM | endometrium | *ZCCHC14* | 3.35536206 | 8.16E-08 | 4.36E-07 | 0.000395712 |
| T2DM | endometrium | *TLDC1* | 6.374137792 | 6.29E-08 | 4.34E-07 | 0.000305363 |
| T2DM | endometrium | *RPL3L* | 12.24463314 | 1.20E-06 | 2.37E-06 | 0.005804782 |
| T2DM | endometrium | *TIGD7* | 2.779875513 | 4.57E-08 | 4.34E-07 | 0.000221746 |
| T2DM | endometrium | *DEF8* | 2.75727561 | 4.49E-07 | 1.03E-06 | 0.002180927 |
| T2DM | endometrium | *TCF25* | 2.773091134 | 7.51E-08 | 4.36E-07 | 0.000364485 |
| T2DM | endometrium | *GALNS* | 5.363557604 | 0.001376691 | 0.001752283 | 1 |
| T2DM | endometrium | *GAS8* | 5.705472512 | 3.19E-05 | 4.78E-05 | 0.154706228 |
| T2DM | endometrium | *NCOR1* | 3.906364128 | 1.04E-07 | 4.47E-07 | 0.000506387 |
| T2DM | endometrium | *KSR1* | 3.641950164 | 6.21E-08 | 4.34E-07 | 0.000301247 |
| T2DM | endometrium | *GFOD2* | 2.059918753 | 7.92E-07 | 1.67E-06 | 0.003843791 |
| T2DM | endometrium | *PRPSAP2* | 4.304779543 | 2.66E-08 | 4.34E-07 | 0.000129296 |
| T2DM | endometrium | *MYO19* | 4.334427971 | 1.06E-09 | 2.33E-07 | 5.12E-06 |
| T2DM | endometrium | *RASL10B* | 9.432702545 | 5.23E-06 | 8.93E-06 | 0.025397484 |
| T2DM | endometrium | *UNC45B* | 12.46048155 | 3.93E-05 | 5.83E-05 | 0.190864612 |
| T2DM | endometrium | *C17orf80* | 2.798066881 | 2.58E-07 | 6.72E-07 | 0.001250735 |
| T2DM | endometrium | *SGSM2* | 6.966841975 | 2.76E-06 | 5.02E-06 | 0.013411101 |
| T2DM | endometrium | *SCRN2* | 1.106271093 | 0.000269086 | 0.000364898 | 1 |
| T2DM | endometrium | *SSH2* | 3.814386744 | 1.13E-07 | 4.57E-07 | 0.000547017 |
| T2DM | endometrium | *RHBDL3* | -1.675156583 | 4.84E-06 | 8.32E-06 | 0.02348142 |
| T2DM | endometrium | *G6PC3* | -4.592198416 | 0.000276396 | 0.000374288 | 1 |
| T2DM | endometrium | *CLTC* | 4.022259256 | 2.16E-08 | 4.34E-07 | 0.000104736 |
| T2DM | endometrium | *IMPA2* | 2.254116479 | 8.74E-08 | 4.38E-07 | 0.000423938 |
| T2DM | endometrium | *GNAL* | 4.149535974 | 5.21E-08 | 4.34E-07 | 0.000253 |
| T2DM | endometrium | *RPRD1A* | 5.042101631 | 2.69E-05 | 4.07E-05 | 0.130702962 |
| T2DM | endometrium | *GALNT1* | 3.027961762 | 6.83E-08 | 4.34E-07 | 0.000331519 |
| T2DM | endometrium | *ADCYAP1* | 2.744747978 | 0.031237092 | 0.035771152 | 1 |
| T2DM | endometrium | *C18orf8* | -4.042583126 | 0.00377917 | 0.004617693 | 1 |
| T2DM | endometrium | *NPC1* | 4.522344191 | 1.50E-06 | 2.91E-06 | 0.007299288 |
| T2DM | endometrium | *SLC14A1* | 2.460344536 | 9.46E-06 | 1.53E-05 | 0.045915795 |
| T2DM | endometrium | *CCDC40* | 2.187445604 | 1.13E-07 | 4.57E-07 | 0.000549498 |
| T2DM | endometrium | *ARHGDIA* | -3.123942207 | 0.002007111 | 0.002520969 | 1 |
| T2DM | endometrium | *TMC6* | 2.549592472 | 3.80E-08 | 4.34E-07 | 0.000184171 |
| T2DM | endometrium | *CARD14* | -2.249714416 | 2.10E-05 | 3.21E-05 | 0.101901507 |
| T2DM | endometrium | *ANAPC11* | -2.986595345 | 0.033325177 | 0.038072465 | 1 |
| T2DM | endometrium | *TBCD* | 4.327491427 | 6.70E-08 | 4.34E-07 | 0.000325232 |
| T2DM | endometrium | *FN3KRP* | 1.159298412 | 0.003951114 | 0.004817995 | 1 |
| T2DM | endometrium | *SECTM1* | -1.772153612 | 5.28E-05 | 7.69E-05 | 0.25629284 |
| T2DM | endometrium | *RNF157* | 2.186527744 | 1.32E-06 | 2.58E-06 | 0.006391226 |
| T2DM | endometrium | *ZNF750* | 8.75458532 | 3.49E-08 | 4.34E-07 | 0.000169159 |
| T2DM | endometrium | *WDR45B* | 2.372828306 | 1.66E-07 | 4.97E-07 | 0.000804501 |
| T2DM | endometrium | *RNF165* | 2.85314448 | 1.29E-07 | 4.78E-07 | 0.000624448 |
| T2DM | endometrium | *MAPK4* | 6.365699845 | 1.50E-07 | 4.78E-07 | 0.000725867 |
| T2DM | endometrium | *FBXO15* | -4.49963415 | 0.000278587 | 0.00037715 | 1 |
| T2DM | endometrium | *CBLN2* | -3.995919876 | 5.98E-05 | 8.64E-05 | 0.290228917 |
| T2DM | endometrium | *NT5C3B* | -0.981223857 | 0.001587989 | 0.002008583 | 1 |
| T2DM | endometrium | *PIP4K2B* | 7.81669318 | 7.36E-09 | 4.34E-07 | 3.57E-05 |
| T2DM | endometrium | *ERBB2* | -1.477119577 | 1.20E-05 | 1.92E-05 | 0.058395019 |
| T2DM | endometrium | *MIEN1* | 5.282478948 | 2.29E-07 | 6.16E-07 | 0.001111333 |

| | | | | | | |
|---|---|---|---|---|---|---|
| T2DM | endometrium | *PNMT* | 9.324266382 | 3.69E-05 | 5.50E-05 | 0.179110658 |
| T2DM | endometrium | *IGFBP4* | 4.699342407 | 1.43E-07 | 4.78E-07 | 0.000692461 |
| T2DM | endometrium | *TXNL4A* | 4.313148755 | 0.006996991 | 0.00841999 | 1 |
| T2DM | endometrium | *CACNA1A* | 3.701452388 | 8.54E-07 | 1.78E-06 | 0.004142992 |
| T2DM | endometrium | *SAMD1* | -3.101642534 | 8.98E-05 | 0.000126942 | 0.435790404 |
| T2DM | endometrium | *NFIC* | 4.602904087 | 3.79E-07 | 9.03E-07 | 0.001838292 |
| T2DM | endometrium | *PRDM15* | 4.291511039 | 0.000145865 | 0.000202327 | 0.707738229 |
| T2DM | endometrium | *PFKL* | 2.3352438 | 2.18E-07 | 5.97E-07 | 0.001060007 |
| T2DM | endometrium | *DUS3L* | 3.363833573 | 7.15E-10 | 2.17E-07 | 3.47E-06 |
| T2DM | endometrium | *DPP9* | 3.238204266 | 7.94E-08 | 4.36E-07 | 0.000385214 |
| T2DM | endometrium | *TMEM91* | -2.691382438 | 0.001035347 | 0.00133073 | 1 |
| T2DM | endometrium | *SIRT3* | 6.371891756 | 2.85E-09 | 3.60E-07 | 1.38E-05 |
| T2DM | endometrium | *HUNK* | 1.024662666 | 0.014255908 | 0.016727852 | 1 |
| T2DM | endometrium | *IFNAR1* | 2.957105322 | 5.13E-07 | 1.15E-06 | 0.002487164 |
| T2DM | endometrium | *TRPM2* | 1.628231952 | 7.84E-07 | 1.65E-06 | 0.003804563 |
| T2DM | endometrium | *TMEM50B* | 3.582609286 | 2.64E-08 | 4.34E-07 | 0.000127895 |
| T2DM | endometrium | *DOPEY2* | 1.846761395 | 1.58E-07 | 4.84E-07 | 0.000764897 |
| T2DM | endometrium | *URB1* | 4.502260947 | 7.05E-08 | 4.34E-07 | 0.000341906 |
| T2DM | endometrium | *SAE1* | 2.977188749 | 1.29E-06 | 2.55E-06 | 0.006277188 |
| T2DM | endometrium | *GEMIN7* | -1.806884661 | 7.46E-07 | 1.59E-06 | 0.003619797 |
| T2DM | endometrium | *SLC6A3* | 4.078179747 | 6.35E-05 | 9.13E-05 | 0.307989254 |
| T2DM | endometrium | *CAPN10* | -2.64171801 | 0.002906367 | 0.003594619 | 1 |
| T2DM | endometrium | *CACNG8* | 6.21962582 | 0.000216308 | 0.000295558 | 1 |
| T2DM | endometrium | *ZNF787* | 2.831285404 | 1.10E-07 | 4.57E-07 | 0.000535846 |
| T2DM | endometrium | *FBN3* | 2.628168014 | 0.003250426 | 0.004000778 | 1 |
| T2DM | endometrium | *EVI5L* | 3.080566608 | 1.18E-07 | 4.63E-07 | 0.000574524 |
| T2DM | endometrium | *CTU1* | 3.666364527 | 1.25E-07 | 4.74E-07 | 0.000607437 |
| T2DM | endometrium | *RERE* | 3.057349987 | 3.85E-08 | 4.34E-07 | 0.000186682 |
| T2DM | endometrium | *EPHA2* | 2.763919515 | 2.20E-07 | 6.00E-07 | 0.001068708 |
| T2DM | endometrium | *ARHGEF19* | -1.623291503 | 0.000208221 | 0.000285312 | 1 |
| T2DM | endometrium | *PEX14* | 2.234878803 | 2.06E-07 | 5.72E-07 | 0.000998174 |
| T2DM | endometrium | *ZNF593* | -1.292299557 | 2.36E-06 | 4.36E-06 | 0.011450835 |
| T2DM | endometrium | *C1orf216* | 5.288411799 | 6.44E-07 | 1.40E-06 | 0.003126337 |
| T2DM | endometrium | *EVA1B* | -2.727464785 | 1.67E-06 | 3.19E-06 | 0.008099206 |
| T2DM | endometrium | *PLK4* | -1.58318232 | 8.54E-06 | 1.39E-05 | 0.041444446 |
| T2DM | endometrium | *NBPF3* | 1.211775302 | 0.006694755 | 0.008066291 | 1 |
| T2DM | endometrium | *HSPG2* | 2.186951691 | 3.60E-06 | 6.35E-06 | 0.017447224 |
| T2DM | endometrium | *ITGB3BP* | 2.994178796 | 7.69E-07 | 1.63E-06 | 0.003729629 |
| T2DM | endometrium | *PIGK* | 4.774104058 | 9.29E-07 | 1.91E-06 | 0.004506685 |
| T2DM | endometrium | *RPS8* | 4.034520225 | 4.31E-08 | 4.34E-07 | 0.000208942 |
| T2DM | endometrium | *CYP4B1* | 2.977298768 | 0.014662509 | 0.017180028 | 1 |
| T2DM | endometrium | *SLC44A3* | 2.949194153 | 9.26E-08 | 4.39E-07 | 0.000449275 |
| T2DM | endometrium | *STRIP1* | 5.787428773 | 2.36E-08 | 4.34E-07 | 0.000114741 |
| T2DM | endometrium | *GPR161* | 2.620559346 | 2.09E-07 | 5.78E-07 | 0.001012358 |
| T2DM | endometrium | *ALDH9A1* | -2.66589337 | 0.025999855 | 0.029936236 | 1 |
| T2DM | endometrium | *ATP1B1* | 2.813985153 | 1.07E-07 | 4.49E-07 | 0.000517758 |
| T2DM | endometrium | *TIPRL* | 2.668097338 | 1.70E-07 | 5.06E-07 | 0.00082464 |
| T2DM | endometrium | *NME7* | 4.095687419 | 6.03E-06 | 1.02E-05 | 0.029279178 |
| T2DM | endometrium | *XCL1* | -4.225329654 | 1.05E-05 | 1.69E-05 | 0.051027198 |
| T2DM | endometrium | *XCL2* | 4.078724115 | 0.004236455 | 0.00516205 | 1 |

| | | | | | | |
|---|---|---|---|---|---|---|
| T2DM | endometrium | *MAEL* | 6.81724584 | 1.52E-07 | 4.78E-07 | 0.000736007 |
| T2DM | endometrium | *RFWD2* | 4.192074657 | 1.08E-07 | 4.52E-07 | 0.000522805 |
| T2DM | endometrium | *RGS16* | 3.193486951 | 8.14E-06 | 1.33E-05 | 0.039476147 |
| T2DM | endometrium | *TARS2* | -2.279643231 | 0.000485188 | 0.000642153 | 1 |
| T2DM | endometrium | *SNX27* | 3.111012614 | 1.50E-08 | 4.34E-07 | 7.29E-05 |
| T2DM | endometrium | *SETDB1* | 4.412271285 | 9.81E-08 | 4.42E-07 | 0.000475819 |
| T2DM | endometrium | *SEMA6C* | 3.468152018 | 1.13E-07 | 4.57E-07 | 0.00055038 |
| T2DM | endometrium | *ARNT* | 4.133152027 | 2.24E-07 | 6.06E-07 | 0.001086583 |
| T2DM | endometrium | *POGZ* | 3.277143319 | 3.50E-08 | 4.34E-07 | 0.000170004 |
| T2DM | endometrium | *GABPB2* | 2.603568411 | 1.51E-08 | 4.34E-07 | 7.33E-05 |
| T2DM | endometrium | *DYRK3* | 3.249055726 | 1.42E-07 | 4.78E-07 | 0.000690859 |
| T2DM | endometrium | *TAF1A* | -2.418202214 | 0.000138802 | 0.000192916 | 0.673469198 |
| T2DM | endometrium | *SMYD2* | 2.838601426 | 8.95E-08 | 4.38E-07 | 0.00043432 |
| T2DM | endometrium | *ADAM15* | 3.243681877 | 0.000111242 | 0.000155906 | 0.539745562 |
| T2DM | endometrium | *RAB13* | 2.844668346 | 1.39E-07 | 4.78E-07 | 0.000673508 |
| T2DM | endometrium | *S100A8* | 2.20472607 | 1.50E-07 | 4.78E-07 | 0.00072752 |
| T2DM | endometrium | *CREB3L4* | 3.182909536 | 1.42E-05 | 2.23E-05 | 0.0687719 |
| T2DM | endometrium | *KCNN3* | 2.212858175 | 3.23E-07 | 7.96E-07 | 0.001565793 |
| T2DM | endometrium | *C1orf43* | 6.085228246 | 2.08E-06 | 3.89E-06 | 0.010077831 |
| T2DM | endometrium | *RIT1* | 2.544484434 | 2.38E-07 | 6.36E-07 | 0.001156778 |
| T2DM | endometrium | *FLG* | -1.259034116 | 2.10E-06 | 3.92E-06 | 0.01017763 |
| T2DM | endometrium | *SCCPDH* | 3.316132484 | 7.28E-08 | 4.34E-07 | 0.000353104 |
| T2DM | endometrium | *CEP170* | 2.471216675 | 7.12E-08 | 4.34E-07 | 0.000345527 |
| T2DM | endometrium | *ACP1* | 6.223739954 | 0.000330975 | 0.000445462 | 1 |
| T2DM | endometrium | *CNIH4* | 4.409043713 | 0.012484776 | 0.014738718 | 1 |
| T2DM | endometrium | *MBOAT2* | 2.214383605 | 1.44E-07 | 4.78E-07 | 0.000697151 |
| T2DM | endometrium | *PYCR2* | -3.016533687 | 0.002648817 | 0.003285291 | 1 |
| T2DM | endometrium | *PLEKHA6* | 3.991096784 | 3.04E-10 | 1.13E-07 | 1.47E-06 |
| T2DM | endometrium | *OSR1* | 3.556495609 | 1.15E-05 | 1.84E-05 | 0.055942476 |
| T2DM | endometrium | *RHOB* | 3.677001575 | 5.85E-08 | 4.34E-07 | 0.000283686 |
| T2DM | endometrium | *GALM* | 3.112923398 | 6.41E-08 | 4.34E-07 | 0.000311207 |
| T2DM | endometrium | *WDPCP* | 3.172028578 | 5.74E-07 | 1.27E-06 | 0.00278337 |
| T2DM | endometrium | *VPS54* | 3.615071971 | 5.24E-08 | 4.34E-07 | 0.000254188 |
| T2DM | endometrium | *REG3G* | 14.52695913 | 0.00371971 | 0.004551836 | 1 |
| T2DM | endometrium | *CIAO1* | 6.350306068 | 6.13E-08 | 4.34E-07 | 0.000297487 |
| T2DM | endometrium | *ZNF514* | 3.599284449 | 0.001798317 | 0.00226282 | 1 |
| T2DM | endometrium | *EXOC6B* | 4.227517193 | 1.06E-06 | 2.14E-06 | 0.00515315 |
| T2DM | endometrium | *TEX261* | 3.189377597 | 1.50E-07 | 4.78E-07 | 0.000729824 |
| T2DM | endometrium | *THNSL2* | 1.965899277 | 1.92E-06 | 3.62E-06 | 0.009320898 |
| T2DM | endometrium | *RALB* | 6.229287804 | 9.08E-08 | 4.38E-07 | 0.000440342 |
| T2DM | endometrium | *FBLN7* | 3.381641719 | 0.000307656 | 0.000415114 | 1 |
| T2DM | endometrium | *LIPT1* | -1.773707353 | 0.00078095 | 0.001015047 | 1 |
| T2DM | endometrium | *CNGA3* | 33.77417325 | 0.001463904 | 0.001857443 | 1 |
| T2DM | endometrium | *FAHD2B* | -2.123528983 | 1.07E-05 | 1.72E-05 | 0.051981095 |
| T2DM | endometrium | *LYG1* | 2.854617668 | 3.17E-06 | 5.66E-06 | 0.015356734 |
| T2DM | endometrium | *THSD7B* | -2.402961117 | 0.003741839 | 0.004576608 | 1 |
| T2DM | endometrium | *GPR17* | 1.500185281 | 5.50E-05 | 7.99E-05 | 0.266818306 |
| T2DM | endometrium | *POLR2D* | -2.757249724 | 0.000879217 | 0.00113759 | 1 |
| T2DM | endometrium | *GALNT13* | -0.862035819 | 6.17E-06 | 1.04E-05 | 0.029960113 |
| T2DM | endometrium | *PHOSPHO2* | 3.999192945 | 6.35E-07 | 1.38E-06 | 0.003081753 |

| | | | | | | |
|---|---|---|---|---|---|---|
| T2DM | endometrium | *METTL21A* | 5.307417612 | 3.94E-08 | 4.34E-07 | 0.00019105 |
| T2DM | endometrium | *NBEAL1* | 3.511407286 | 6.08E-09 | 4.34E-07 | 2.95E-05 |
| T2DM | endometrium | *SUMF1* | 2.788813832 | 5.07E-08 | 4.34E-07 | 0.000246169 |
| T2DM | endometrium | *ACKR3* | 2.439450795 | 1.12E-06 | 2.24E-06 | 0.005421181 |
| T2DM | endometrium | *ESPNL* | 37.95279307 | 1.09E-07 | 4.55E-07 | 0.000529611 |
| T2DM | endometrium | *ANKMY1* | 5.357510588 | 1.78E-07 | 5.20E-07 | 0.000863205 |
| T2DM | endometrium | *TAMM41* | 3.563732197 | 1.34E-08 | 4.34E-07 | 6.49E-05 |
| T2DM | endometrium | *STK11IP* | -1.328963871 | 3.93E-06 | 6.90E-06 | 0.019084776 |
| T2DM | endometrium | *EAF1* | 8.581548619 | 7.13E-08 | 4.34E-07 | 0.000345932 |
| T2DM | endometrium | *SLC25A38* | 3.221481308 | 3.35E-07 | 8.20E-07 | 0.001626805 |
| T2DM | endometrium | *CTDSPL* | 3.946916905 | 8.94E-07 | 1.85E-06 | 0.004336176 |
| T2DM | endometrium | *PTPRG* | 2.60623076 | 1.08E-07 | 4.52E-07 | 0.000523109 |
| T2DM | endometrium | *ARL6IP5* | 2.601536602 | 7.37E-08 | 4.35E-07 | 0.000357525 |
| T2DM | endometrium | *LIMD1* | 3.438465057 | 3.89E-08 | 4.34E-07 | 0.000188521 |
| T2DM | endometrium | *ZNF660* | -1.018221942 | 1.41E-05 | 2.22E-05 | 0.068430861 |
| T2DM | endometrium | *PLA1A* | 8.079979114 | 7.95E-06 | 1.30E-05 | 0.038566584 |
| T2DM | endometrium | *ADPRH* | 3.557003479 | 3.71E-05 | 5.53E-05 | 0.18006906 |
| T2DM | endometrium | *IGSF11* | -1.365337956 | 9.83E-05 | 0.000138341 | 0.477139591 |
| T2DM | endometrium | *BOC* | 3.988534745 | 1.63E-08 | 4.34E-07 | 7.93E-05 |
| T2DM | endometrium | *MED12L* | 2.278464292 | 1.35E-07 | 4.78E-07 | 0.000656145 |
| T2DM | endometrium | *EIF2A* | 1.960641219 | 2.42E-07 | 6.43E-07 | 0.001171875 |
| T2DM | endometrium | *NCEH1* | 1.734501489 | 1.51E-06 | 2.92E-06 | 0.007318976 |
| T2DM | endometrium | *AMT* | 1.942069614 | 1.19E-06 | 2.37E-06 | 0.005795102 |
| T2DM | endometrium | *CCDC39* | 4.278169049 | 1.05E-08 | 4.34E-07 | 5.11E-05 |
| T2DM | endometrium | *MUC4* | 3.41506747 | 3.97E-07 | 9.36E-07 | 0.001926611 |
| T2DM | endometrium | *DGKQ* | -2.578702244 | 0.000166061 | 0.000229225 | 0.805727596 |
| T2DM | endometrium | *SLC26A1* | -1.938911209 | 4.31E-05 | 6.34E-05 | 0.208949718 |
| T2DM | endometrium | *CENPC* | 3.532466219 | 5.99E-07 | 1.32E-06 | 0.002906889 |
| T2DM | endometrium | *SCD5* | 3.459878517 | 8.23E-08 | 4.37E-07 | 0.000399157 |
| T2DM | endometrium | *PYURF* | 7.316440045 | 0.000684083 | 0.000893933 | 1 |
| T2DM | endometrium | *CISD2* | 4.075396155 | 1.56E-08 | 4.34E-07 | 7.55E-05 |
| T2DM | endometrium | *DDIT4L* | -4.11252199 | 4.68E-05 | 6.88E-05 | 0.227300202 |
| T2DM | endometrium | *METTL14* | 3.402227612 | 0.043538378 | 0.04934553 | 1 |
| T2DM | endometrium | *USP53* | 4.707042307 | 1.04E-08 | 4.34E-07 | 5.06E-05 |
| T2DM | endometrium | *MARCH1* | 2.038917164 | 1.25E-06 | 2.48E-06 | 0.006085746 |
| T2DM | endometrium | *CBR4* | 4.738053023 | 1.69E-05 | 2.62E-05 | 0.081960586 |
| T2DM | endometrium | *CYP4V2* | 2.452335261 | 0.000196237 | 0.000269118 | 0.952140364 |
| T2DM | endometrium | *MARCH6* | 3.694353641 | 4.87E-08 | 4.34E-07 | 0.000236289 |
| T2DM | endometrium | *MYO10* | 3.380980225 | 7.07E-08 | 4.34E-07 | 0.000343048 |
| T2DM | endometrium | *FAM105A* | 1.736100569 | 2.05E-05 | 3.14E-05 | 0.099510142 |
| T2DM | endometrium | *GTF2H2* | 10.3043306 | 8.95E-08 | 4.38E-07 | 0.000434179 |
| T2DM | endometrium | *SPATA9* | 1.916941488 | 1.52E-07 | 4.78E-07 | 0.000737702 |
| T2DM | endometrium | *FEM1C* | 4.725254156 | 5.38E-08 | 4.34E-07 | 0.000260967 |
| T2DM | endometrium | *COMMD10* | 2.725596549 | 1.79E-07 | 5.21E-07 | 0.000869826 |
| T2DM | endometrium | *YIPF5* | 5.0753981 | 2.96E-08 | 4.34E-07 | 0.000143857 |
| T2DM | endometrium | *CXCL14* | 3.916557768 | 8.24E-06 | 1.35E-05 | 0.039969288 |
| T2DM | endometrium | *FBXO38* | 3.866349216 | 4.51E-08 | 4.34E-07 | 0.000218622 |
| T2DM | endometrium | *PCYOX1L* | -5.083474206 | 0.014700245 | 0.017220084 | 1 |
| T2DM | endometrium | *BOD1* | 2.750007644 | 1.52E-07 | 4.78E-07 | 0.000737484 |
| T2DM | endometrium | *FAM50B* | 7.440779516 | 5.17E-08 | 4.34E-07 | 0.000250789 |

| | | | | | | |
|---|---|---|---|---|---|---|
| T2DM | endometrium | *MYLK4* | -2.148914703 | 6.10E-06 | 1.03E-05 | 0.029595952 |
| T2DM | endometrium | *TBC1D7* | 2.708111037 | 1.11E-07 | 4.57E-07 | 0.000539291 |
| T2DM | endometrium | *GFOD1* | 3.550624995 | 0.014123282 | 0.016580248 | 1 |
| T2DM | endometrium | *TRIM7* | 4.269713716 | 0.000126467 | 0.000176428 | 0.613616441 |
| T2DM | endometrium | *FAM193B* | 2.372735269 | 6.16E-08 | 4.34E-07 | 0.000298838 |
| T2DM | endometrium | *MUT* | 2.081152864 | 2.75E-07 | 7.06E-07 | 0.00133614 |
| T2DM | endometrium | *DOK3* | 2.344323574 | 5.18E-06 | 8.85E-06 | 0.02511174 |
| T2DM | endometrium | *ABT1* | 4.281890943 | 4.23E-07 | 9.81E-07 | 0.002052154 |
| T2DM | endometrium | *DAAM2* | 1.882052897 | 1.22E-07 | 4.69E-07 | 0.000592972 |
| T2DM | endometrium | *MLIP* | 6.798034553 | 9.83E-05 | 0.000138282 | 0.47679482 |
| T2DM | endometrium | *ANO7* | 3.553765531 | 7.56E-05 | 0.000107853 | 0.366700065 |
| T2DM | endometrium | *TCTE1* | 5.79690184 | 0.003219105 | 0.003965245 | 1 |
| T2DM | endometrium | *RPL7L1* | 4.574312598 | 6.30E-08 | 4.34E-07 | 0.000305685 |
| T2DM | endometrium | *TPBG* | 2.050589476 | 2.01E-07 | 5.62E-07 | 0.000977316 |
| T2DM | endometrium | *IRAK1BP1* | 2.762693407 | 9.23E-09 | 4.34E-07 | 4.48E-05 |
| T2DM | endometrium | *PHIP* | 4.022284939 | 6.49E-08 | 4.34E-07 | 0.000314892 |
| T2DM | endometrium | *PNRC1* | 2.612269018 | 9.14E-08 | 4.38E-07 | 0.000443488 |
| T2DM | endometrium | *RNF217* | 3.776006553 | 7.32E-08 | 4.35E-07 | 0.000355244 |
| T2DM | endometrium | *ARHGAP18* | 2.180946894 | 3.54E-08 | 4.34E-07 | 0.000171732 |
| T2DM | endometrium | *ABRACL* | 3.601193728 | 3.21E-06 | 5.72E-06 | 0.01555214 |
| T2DM | endometrium | *SLC18B1* | 3.033707465 | 7.23E-07 | 1.55E-06 | 0.003508842 |
| T2DM | endometrium | *PNLDC1* | -5.10388233 | 0.000116041 | 0.00016235 | 0.563030508 |
| T2DM | endometrium | *WTAP* | 4.604041671 | 2.48E-06 | 4.55E-06 | 0.012029809 |
| T2DM | endometrium | *SLC22A3* | 1.975708485 | 1.50E-07 | 4.78E-07 | 0.000728957 |
| T2DM | endometrium | *SDK1* | -1.622163561 | 0.000366654 | 0.000491438 | 1 |
| T2DM | endometrium | *CCZ1B* | 3.759329844 | 0.000563471 | 0.000742118 | 1 |
| T2DM | endometrium | *RBAK* | 6.634119867 | 5.37E-08 | 4.34E-07 | 0.000260732 |
| T2DM | endometrium | *POMZP3* | 2.333280751 | 7.19E-07 | 1.54E-06 | 0.003490396 |
| T2DM | endometrium | *PSPH* | 3.619630352 | 0.000240807 | 0.000327924 | 1 |
| T2DM | endometrium | *GIGYF1* | 6.551030512 | 2.60E-06 | 4.75E-06 | 0.012629296 |
| T2DM | endometrium | *TRIM4* | 1.419736958 | 0.000129186 | 0.000180118 | 0.626812322 |
| T2DM | endometrium | *TMEM209* | 3.855454468 | 5.88E-08 | 4.34E-07 | 0.000285403 |
| T2DM | endometrium | *EPHA1* | 7.365792687 | 1.79E-07 | 5.21E-07 | 0.000870218 |
| T2DM | endometrium | *NCAPG2* | 2.715839733 | 5.06E-08 | 4.34E-07 | 0.000245635 |
| T2DM | endometrium | *FBXO25* | 6.034567232 | 1.03E-07 | 4.47E-07 | 0.000497522 |
| T2DM | endometrium | *CSGALNACT1* | 3.016096339 | 4.44E-08 | 4.34E-07 | 0.000215208 |
| T2DM | endometrium | *BIN3* | 3.408286065 | 5.87E-08 | 4.34E-07 | 0.000284703 |
| T2DM | endometrium | *CHMP7* | 4.457782843 | 5.02E-08 | 4.34E-07 | 0.000243616 |
| T2DM | endometrium | *DOCK5* | 2.545050862 | 6.90E-08 | 4.34E-07 | 0.000334557 |
| T2DM | endometrium | *TACC1* | 3.109199553 | 7.11E-08 | 4.34E-07 | 0.000344983 |
| T2DM | endometrium | *ADHFE1* | 1.193596506 | 0.002892408 | 0.003578267 | 1 |
| T2DM | endometrium | *RPL7* | -1.999433156 | 9.44E-05 | 0.000133169 | 0.458236141 |
| T2DM | endometrium | *ATP6V0D2* | 10.20592416 | 5.02E-07 | 1.13E-06 | 0.002433813 |
| T2DM | endometrium | *DPYS* | 1.69205773 | 1.52E-07 | 4.78E-07 | 0.000739641 |
| T2DM | endometrium | *LRP12* | 6.813897527 | 0.00038682 | 0.000517039 | 1 |
| T2DM | endometrium | *MAL2* | 3.345720267 | 8.16E-05 | 0.000116073 | 0.396157928 |
| T2DM | endometrium | *SLC39A4* | -1.351328968 | 3.84E-07 | 9.11E-07 | 0.001865336 |
| T2DM | endometrium | *AK3* | 2.888899329 | 6.29E-08 | 4.34E-07 | 0.000305249 |
| T2DM | endometrium | *HAUS6* | 5.245290871 | 5.08E-08 | 4.34E-07 | 0.000246486 |
| T2DM | endometrium | *C9orf72* | 1.785314339 | 7.12E-07 | 1.53E-06 | 0.003453473 |

| | | | | | | |
|---|---|---|---|---|---|---|
| T2DM | endometrium | *FBXO10* | 4.428168102 | 7.81E-05 | 0.000111312 | 0.379128283 |
| T2DM | endometrium | *ZNF462* | 2.262260925 | 3.70E-08 | 4.34E-07 | 0.000179603 |
| T2DM | endometrium | *ALAD* | 3.246891581 | 7.47E-07 | 1.59E-06 | 0.003624183 |
| T2DM | endometrium | *POLE3* | 3.192927136 | 4.59E-07 | 1.04E-06 | 0.002228894 |
| T2DM | endometrium | *SURF6* | -3.049315737 | 0.001062425 | 0.001363727 | 1 |
| T2DM | endometrium | *MED22* | 2.821898566 | 3.22E-06 | 5.74E-06 | 0.015616169 |
| T2DM | endometrium | *REXO4* | 2.859441408 | 1.01E-07 | 4.46E-07 | 0.00048851 |
| T2DM | endometrium | *SH3GLB2* | 7.734871271 | 5.40E-08 | 4.34E-07 | 0.000262192 |
| T2DM | endometrium | *INPP5E* | 9.177574118 | 0.000118853 | 0.000166188 | 0.57667366 |
| T2DM | endometrium | *SEC16A* | 7.176531592 | 3.69E-05 | 5.50E-05 | 0.179140025 |
| T2DM | endometrium | *SLC39A12* | 14.14574457 | 0.000912794 | 0.001179147 | 1 |
| T2DM | endometrium | *TMEM236* | 4.439956151 | 1.15E-07 | 4.59E-07 | 0.000557202 |
| T2DM | endometrium | *RSU1* | 2.690415529 | 5.93E-08 | 4.34E-07 | 0.000287892 |
| T2DM | endometrium | *ST8SIA6* | 8.972508462 | 7.27E-05 | 0.000103974 | 0.352782231 |
| T2DM | endometrium | *PARD3* | 3.84518417 | 2.41E-07 | 6.41E-07 | 0.001167274 |
| T2DM | endometrium | *RGR* | 12.4140064 | 0.028687797 | 0.032952934 | 1 |
| T2DM | endometrium | *FRA10AC1* | 3.847061929 | 3.98E-08 | 4.34E-07 | 0.000192934 |
| T2DM | endometrium | *ADD3* | 4.604166099 | 2.87E-07 | 7.28E-07 | 0.00139294 |
| T2DM | endometrium | *LRRC27* | 2.632651363 | 4.45E-07 | 1.02E-06 | 0.002158143 |
| T2DM | endometrium | *MTG1* | 3.508168839 | 3.78E-05 | 5.62E-05 | 0.18341943 |
| T2DM | endometrium | *PAOX* | 10.05105558 | 3.54E-07 | 8.54E-07 | 0.001716656 |
| T2DM | endometrium | *PDCD11* | 3.811648685 | 9.24E-08 | 4.38E-07 | 0.000448375 |
| T2DM | endometrium | *RGS10* | 2.454283141 | 1.31E-07 | 4.78E-07 | 0.000635458 |
| T2DM | endometrium | *LIN7C* | 2.416979115 | 1.18E-07 | 4.61E-07 | 0.000570116 |
| T2DM | endometrium | *ZNF215* | 2.329609001 | 1.32E-05 | 2.09E-05 | 0.064029591 |
| T2DM | endometrium | *APIP* | 0.841890112 | 0.026180808 | 0.03012314 | 1 |
| T2DM | endometrium | *EIF3M* | 4.357719153 | 0.001016027 | 0.001306936 | 1 |
| T2DM | endometrium | *TNKS1BP1* | 4.89703494 | 1.80E-07 | 5.23E-07 | 0.000874051 |
| T2DM | endometrium | *PTPRJ* | 1.6364753 | 1.18E-07 | 4.63E-07 | 0.000573602 |
| T2DM | endometrium | *ENDOD1* | 2.513609514 | 1.23E-07 | 4.70E-07 | 0.000597163 |
| T2DM | endometrium | *CCDC82* | 2.040462998 | 8.35E-05 | 0.000118616 | 0.405190973 |
| T2DM | endometrium | *CAPN5* | -2.092119433 | 7.36E-06 | 1.22E-05 | 0.035726376 |
| T2DM | endometrium | *RPS3* | 5.066782572 | 6.29E-05 | 9.06E-05 | 0.305123599 |
| T2DM | endometrium | *TTC12* | 5.086170573 | 5.10E-08 | 4.34E-07 | 0.000247407 |
| T2DM | endometrium | *DRD2* | 23.20297418 | 1.64E-07 | 4.93E-07 | 0.000794089 |
| T2DM | endometrium | *NPAT* | 4.347814023 | 5.42E-08 | 4.34E-07 | 0.00026308 |
| T2DM | endometrium | *ATM* | 3.220083608 | 8.08E-08 | 4.36E-07 | 0.000392283 |
| T2DM | endometrium | *SLX4IP* | 2.86918511 | 2.70E-07 | 6.96E-07 | 0.001312032 |
| T2DM | endometrium | *ST14* | 4.699816084 | 1.04E-07 | 4.47E-07 | 0.00050234 |
| T2DM | endometrium | *FADS1* | 5.063443537 | 6.37E-08 | 4.34E-07 | 0.000309219 |
| T2DM | endometrium | *TMC2* | 4.893806935 | 3.69E-07 | 8.85E-07 | 0.00179153 |
| T2DM | endometrium | *EML3* | 2.606412242 | 3.48E-08 | 4.34E-07 | 0.000168755 |
| T2DM | endometrium | *INCENP* | 3.292065144 | 5.99E-07 | 1.32E-06 | 0.002905978 |
| T2DM | endometrium | *PLCH2* | 13.02788077 | 3.90E-06 | 6.85E-06 | 0.018922887 |
| T2DM | endometrium | *CPSF7* | 4.842324105 | 0.001168993 | 0.001497348 | 1 |
| T2DM | endometrium | *CHEK1* | 4.912383342 | 1.47E-07 | 4.78E-07 | 0.000714913 |
| T2DM | endometrium | *DUSP15* | -0.972409293 | 2.96E-06 | 5.34E-06 | 0.014349934 |
| T2DM | endometrium | *SPATA25* | 6.470410247 | 5.95E-05 | 8.61E-05 | 0.28884638 |
| T2DM | endometrium | *CABLES2* | 3.622660965 | 5.15E-05 | 7.51E-05 | 0.250037964 |
| T2DM | endometrium | *TRPT1* | 2.749196888 | 2.97E-06 | 5.35E-06 | 0.014401468 |

| | | | | | | |
|---|---|---|---|---|---|---|
| T2DM | endometrium | *NUDT22* | -3.131543737 | 0.001189688 | 0.001522249 | 1 |
| T2DM | endometrium | *TBX6* | 2.06560918 | 0.000162319 | 0.00022438 | 0.787573107 |
| T2DM | endometrium | *ALDOA* | 4.662113438 | 0.002100134 | 0.002631676 | 1 |
| T2DM | endometrium | *ANXA8L1* | 1.466979365 | 4.73E-06 | 8.14E-06 | 0.022926606 |
| T2DM | endometrium | *CWC15* | 3.102181323 | 2.32E-07 | 6.23E-07 | 0.001127138 |
| T2DM | endometrium | *FCGR1A* | -2.407673703 | 2.34E-06 | 4.33E-06 | 0.011357556 |
| T2DM | endometrium | *TMEM218* | 3.293125656 | 2.61E-07 | 6.78E-07 | 0.001265826 |
| T2DM | endometrium | *SAP18* | 2.835274843 | 8.94E-08 | 4.38E-07 | 0.000433906 |
| T2DM | endometrium | *FAM124A* | 8.486817583 | 0.000576841 | 0.000759108 | 1 |
| T2DM | endometrium | *WDR17* | 2.243303601 | 5.37E-07 | 1.19E-06 | 0.002605468 |
| T2DM | endometrium | *CCDC102B* | 1.737604022 | 2.14E-07 | 5.88E-07 | 0.001037591 |
| T2DM | endometrium | *RGS18* | 2.964117354 | 1.51E-06 | 2.93E-06 | 0.007343026 |
| T2DM | endometrium | *PRSS23* | 2.579559053 | 6.44E-08 | 4.34E-07 | 0.000312472 |
| T2DM | endometrium | *PPP1R1C* | 4.372902438 | 1.54E-07 | 4.79E-07 | 0.000748972 |
| T2DM | endometrium | *FAM173B* | 2.057980075 | 1.91E-07 | 5.45E-07 | 0.000925393 |
| T2DM | endometrium | *GPR158* | 4.31440036 | 9.51E-08 | 4.42E-07 | 0.000461627 |
| T2DM | endometrium | *UBE3B* | 3.080508401 | 1.09E-06 | 2.20E-06 | 0.00531235 |
| T2DM | endometrium | *ANK3* | 2.560200029 | 3.98E-08 | 4.34E-07 | 0.000193308 |
| T2DM | endometrium | *PLBD2* | 2.634982393 | 9.81E-08 | 4.42E-07 | 0.000476201 |
| T2DM | endometrium | *DLG5* | 3.016836458 | 5.86E-06 | 9.90E-06 | 0.028444332 |
| T2DM | endometrium | *GXYLT1* | 2.649487101 | 1.43E-07 | 4.78E-07 | 0.000695456 |
| T2DM | endometrium | *EXT2* | 2.514178331 | 2.95E-07 | 7.43E-07 | 0.001432363 |
| T2DM | endometrium | *TMEM18* | 8.133050482 | 1.12E-07 | 4.57E-07 | 0.000545064 |
| T2DM | endometrium | *NDUFC2* | 3.042463119 | 1.07E-07 | 4.49E-07 | 0.000519038 |
| T2DM | endometrium | *ADAMTS12* | 2.82679542 | 0.003763487 | 0.004601925 | 1 |
| T2DM | endometrium | *NUBPL* | 3.268473121 | 5.11E-08 | 4.34E-07 | 0.000247973 |
| T2DM | endometrium | *VIPAS39* | 3.007261608 | 2.50E-07 | 6.60E-07 | 0.001213382 |
| T2DM | endometrium | *ANKRD50* | 5.905323623 | 2.89E-08 | 4.34E-07 | 0.0001404 |
| T2DM | endometrium | *UPF2* | 3.249624772 | 6.31E-08 | 4.34E-07 | 0.000306378 |
| T2DM | endometrium | *C4orf33* | 3.208502706 | 2.51E-07 | 6.61E-07 | 0.001219994 |
| T2DM | endometrium | *PTPRO* | 2.249660718 | 5.28E-07 | 1.18E-06 | 0.002562973 |
| T2DM | endometrium | *EPS8* | 3.362167844 | 4.96E-08 | 4.34E-07 | 0.000240858 |
| T2DM | endometrium | *QDPR* | 4.448260148 | 2.28E-07 | 6.15E-07 | 0.001108143 |
| T2DM | endometrium | *FAM160B1* | 3.976033286 | 0.008817419 | 0.010506413 | 1 |
| T2DM | endometrium | *ZNF827* | 2.14091692 | 9.42E-08 | 4.39E-07 | 0.000456832 |
| T2DM | endometrium | *AKR1C2* | 4.304167589 | 3.69E-08 | 4.34E-07 | 0.000178916 |
| T2DM | endometrium | *PIGF* | 6.132871775 | 3.48E-07 | 8.43E-07 | 0.001688112 |
| T2DM | endometrium | *MFSD6* | 4.05559794 | 6.11E-08 | 4.34E-07 | 0.000296528 |
| T2DM | endometrium | *CENPU* | -1.030887383 | 4.16E-06 | 7.27E-06 | 0.020207945 |
| T2DM | endometrium | *BICD1* | 2.684534461 | 6.27E-08 | 4.34E-07 | 0.000304126 |
| T2DM | endometrium | *CCDC122* | 2.286879102 | 0.003396758 | 0.004172423 | 1 |
| T2DM | endometrium | *GABRA2* | 7.399513183 | 1.68E-05 | 2.60E-05 | 0.081320246 |
| T2DM | endometrium | *PARP8* | 2.849011733 | 4.37E-06 | 7.58E-06 | 0.021199874 |
| T2DM | endometrium | *CACUL1* | 3.377231971 | 7.15E-08 | 4.34E-07 | 0.000346869 |
| T2DM | endometrium | *BEND6* | -1.078411383 | 2.12E-06 | 3.96E-06 | 0.010283322 |
| T2DM | endometrium | *TIAL1* | 3.978663557 | 7.10E-08 | 4.34E-07 | 0.000344499 |
| T2DM | endometrium | *PTPN14* | 3.413736479 | 2.56E-08 | 4.34E-07 | 0.000124052 |
| T2DM | endometrium | *MGAT5* | 2.288965051 | 1.63E-07 | 4.92E-07 | 0.000792417 |
| T2DM | endometrium | *TMEM178A* | 4.689838188 | 1.40E-07 | 4.78E-07 | 0.000681246 |
| T2DM | endometrium | *HAUS1* | 2.076362634 | 1.40E-05 | 2.20E-05 | 0.067870367 |

| | | | | | | |
|---|---|---|---|---|---|---|
| T2DM | endometrium | *SPC25* | 1.651079556 | 0.001303001 | 0.001662414 | 1 |
| T2DM | endometrium | *KCNK13* | 4.218532774 | 8.15E-08 | 4.36E-07 | 0.000395455 |
| T2DM | endometrium | *ATG10* | 2.003273271 | 0.001167787 | 0.001496198 | 1 |
| T2DM | endometrium | *POC5* | 2.033074091 | 0.004513006 | 0.005493504 | 1 |
| T2DM | endometrium | *CWF19L2* | 4.419420833 | 6.90E-09 | 4.34E-07 | 3.35E-05 |
| T2DM | endometrium | *ZNF773* | 2.099486419 | 0.000263821 | 0.000357958 | 1 |
| T2DM | endometrium | *NMT2* | 3.096305422 | 2.50E-07 | 6.60E-07 | 0.001213912 |
| T2DM | endometrium | *ZNF837* | 3.21752316 | 0.00129658 | 0.001655092 | 1 |
| T2DM | endometrium | *CCDC50* | 3.248061324 | 5.85E-05 | 8.46E-05 | 0.283745115 |
| T2DM | endometrium | *SPARCL1* | 3.003449702 | 4.76E-08 | 4.34E-07 | 0.000230963 |
| T2DM | endometrium | *MBNL1* | 2.836761143 | 5.57E-08 | 4.34E-07 | 0.000270093 |
| T2DM | endometrium | *GPD1L* | 4.782181557 | 1.21E-06 | 2.41E-06 | 0.00589493 |
| T2DM | endometrium | *CCNO* | 28.41490158 | 1.80E-07 | 5.23E-07 | 0.000874712 |
| T2DM | endometrium | *CLEC4F* | 5.717646508 | 8.76E-09 | 4.34E-07 | 4.25E-05 |
| T2DM | endometrium | *SLC30A6* | 4.966881237 | 0.000148718 | 0.000206107 | 0.721580478 |
| T2DM | endometrium | *RASGRP3* | 2.279821613 | 3.20E-07 | 7.91E-07 | 0.001551709 |
| T2DM | endometrium | *GPR180* | 2.76485521 | 2.08E-06 | 3.89E-06 | 0.010071339 |
| T2DM | endometrium | *TCTEX1D1* | 5.618387687 | 0.00058781 | 0.000773124 | 1 |
| T2DM | endometrium | *IFIT5* | 2.748853838 | 2.03E-05 | 3.11E-05 | 0.098378284 |
| T2DM | endometrium | *PRDM8* | 6.814390249 | 0.000438558 | 0.000583141 | 1 |
| T2DM | endometrium | *GGPS1* | 5.074758608 | 1.78E-08 | 4.34E-07 | 8.63E-05 |
| T2DM | endometrium | *ZNF117* | 1.298051275 | 5.53E-05 | 8.03E-05 | 0.268513642 |
| T2DM | endometrium | *MED21* | 4.41670935 | 9.04E-08 | 4.38E-07 | 0.000438454 |
| T2DM | endometrium | *JAKMIP1* | 3.540629099 | 1.39E-07 | 4.78E-07 | 0.000673291 |
| T2DM | endometrium | *SREK1IP1* | 3.962693096 | 7.60E-08 | 4.36E-07 | 0.000368614 |
| T2DM | endometrium | *MR1* | 2.559409732 | 4.41E-08 | 4.34E-07 | 0.000214047 |
| T2DM | endometrium | *CENPH* | -3.055195429 | 1.28E-05 | 2.04E-05 | 0.062320142 |
| T2DM | endometrium | *CARHSP1* | 1.615412328 | 2.58E-07 | 6.74E-07 | 0.001253698 |
| T2DM | endometrium | *CAST* | 3.487284699 | 9.97E-08 | 4.44E-07 | 0.000483932 |
| T2DM | endometrium | *SCOC* | 4.506832062 | 0.018780458 | 0.021831046 | 1 |
| T2DM | endometrium | *CLGN* | 8.523690027 | 0.000102913 | 0.000144609 | 0.499333637 |
| T2DM | endometrium | *CETN3* | 4.346964025 | 0.000322411 | 0.000434418 | 1 |
| T2DM | endometrium | *SYCP2L* | -3.531701488 | 0.000250574 | 0.000340366 | 1 |
| T2DM | endometrium | *MERTK* | 0.675471308 | 0.002022958 | 0.002540215 | 1 |
| T2DM | endometrium | *TMEM87B* | 4.400006496 | 5.68E-08 | 4.34E-07 | 0.000275548 |
| T2DM | endometrium | *SLC25A27* | 2.326626377 | 2.18E-06 | 4.06E-06 | 0.010554345 |
| T2DM | endometrium | *FRMD1* | -3.550340132 | 0.000132356 | 0.000184326 | 0.642190645 |
| T2DM | endometrium | *LPCAT1* | 2.621320619 | 1.41E-07 | 4.78E-07 | 0.000684299 |
| T2DM | endometrium | *NMRAL1* | 2.794695448 | 3.68E-08 | 4.34E-07 | 0.000178687 |
| T2DM | endometrium | *ADPRHL1* | 5.578184583 | 7.28E-08 | 4.34E-07 | 0.000353092 |
| T2DM | endometrium | *CFDP1* | 2.721380979 | 5.75E-08 | 4.34E-07 | 0.00027893 |
| T2DM | endometrium | *ZDHHC7* | 6.50543365 | 1.64E-06 | 3.14E-06 | 0.007933589 |
| T2DM | endometrium | *JAZF1* | -2.487441932 | 4.21E-07 | 9.77E-07 | 0.002040336 |
| T2DM | endometrium | *FBXO36* | 4.007115683 | 6.94E-08 | 4.34E-07 | 0.000336678 |
| T2DM | endometrium | *MCOLN2* | 4.12795446 | 4.40E-07 | 1.01E-06 | 0.002133947 |
| T2DM | endometrium | *GDPD1* | 2.420730265 | 1.93E-07 | 5.48E-07 | 0.000938305 |
| T2DM | endometrium | *SEMA3D* | 4.709654863 | 1.07E-07 | 4.49E-07 | 0.000517541 |
| T2DM | endometrium | *PPP2R5E* | 3.819590783 | 4.90E-08 | 4.34E-07 | 0.000237957 |
| T2DM | endometrium | *ANKRD29* | 1.952100701 | 1.39E-07 | 4.78E-07 | 0.000673008 |
| T2DM | endometrium | *DNAAF1* | -4.499274166 | 0.000188233 | 0.000258727 | 0.913306438 |

| | | | | | | |
|---|---|---|---|---|---|---|
| T2DM | endometrium | *ANKH* | 2.583763915 | 3.57E-08 | 4.34E-07 | 0.000173229 |
| T2DM | endometrium | *UBASH3B* | 2.560296661 | 1.69E-06 | 3.23E-06 | 0.008208633 |
| T2DM | endometrium | *ROBO4* | 3.284531962 | 6.62E-08 | 4.34E-07 | 0.000321087 |
| T2DM | endometrium | *ROBO3* | 2.12086921 | 2.91E-07 | 7.36E-07 | 0.001412572 |
| T2DM | endometrium | *CC2D1B* | 3.715081653 | 1.13E-06 | 2.26E-06 | 0.005477364 |
| T2DM | endometrium | *PRKCA* | 3.678576694 | 5.80E-08 | 4.34E-07 | 0.000281558 |
| T2DM | endometrium | *LRRK1* | 1.683273248 | 1.86E-07 | 5.35E-07 | 0.000903041 |
| T2DM | endometrium | *ABCA5* | 2.600224613 | 5.20E-08 | 4.34E-07 | 0.000252478 |
| T2DM | endometrium | *MIA3* | 4.5581895 | 5.15E-08 | 4.34E-07 | 0.000249712 |
| T2DM | endometrium | *NEIL2* | 4.350384458 | 1.23E-07 | 4.70E-07 | 0.000597194 |
| T2DM | endometrium | *PGM5* | 3.892672833 | 0.003663341 | 0.004483989 | 1 |
| T2DM | endometrium | *CCSAP* | 2.988441222 | 1.50E-07 | 4.78E-07 | 0.000729057 |
| T2DM | endometrium | *GBP5* | 2.164803657 | 9.60E-05 | 0.000135258 | 0.46569379 |
| T2DM | endometrium | *ATP5G3* | 3.822125154 | 4.58E-09 | 4.20E-07 | 2.22E-05 |
| T2DM | endometrium | *LY96* | 2.063841795 | 5.28E-07 | 1.18E-06 | 0.002561968 |
| T2DM | endometrium | *PDE1C* | 1.877639477 | 1.49E-07 | 4.78E-07 | 0.000721854 |
| T2DM | endometrium | *XPC* | 6.354155625 | 5.15E-07 | 1.15E-06 | 0.002497208 |
| T2DM | endometrium | *FLCN* | 2.303707969 | 1.80E-07 | 5.23E-07 | 0.000875634 |
| T2DM | endometrium | *OXNAD1* | 3.091648315 | 5.02E-08 | 4.34E-07 | 0.000243331 |
| T2DM | endometrium | *APCDD1* | 3.120995216 | 0.002329318 | 0.00290536 | 1 |
| T2DM | endometrium | *MPPE1* | 2.632315809 | 1.55E-05 | 2.41E-05 | 0.074988741 |
| T2DM | endometrium | *ANKRD40* | 4.500934195 | 3.40E-07 | 8.29E-07 | 0.001651211 |
| T2DM | endometrium | *CYP2U1* | 3.198939852 | 0.032572311 | 0.037247432 | 1 |
| T2DM | endometrium | *AK9* | 2.382079812 | 7.65E-08 | 4.36E-07 | 0.000371047 |
| T2DM | endometrium | *MARCKS* | 2.508653049 | 1.11E-07 | 4.57E-07 | 0.000540983 |
| T2DM | endometrium | *GOLGA7B* | 15.67281977 | 1.16E-07 | 4.59E-07 | 0.000560414 |
| T2DM | endometrium | *TRMT44* | 5.668279466 | 1.20E-07 | 4.65E-07 | 0.000581086 |
| T2DM | endometrium | *MOV10* | 6.021897144 | 0.002097224 | 0.002628709 | 1 |
| T2DM | endometrium | *HEATR3* | 1.998173708 | 2.32E-06 | 4.30E-06 | 0.011279058 |
| T2DM | endometrium | *CNOT8* | 6.662268987 | 4.09E-05 | 6.04E-05 | 0.198330255 |
| T2DM | endometrium | *LRGUK* | 2.710001473 | 0.003910182 | 0.004769282 | 1 |
| T2DM | endometrium | *SETD9* | 1.624634586 | 0.000178914 | 0.000246337 | 0.86809234 |
| T2DM | endometrium | *NUP205* | 2.581928988 | 1.36E-07 | 4.78E-07 | 0.0006617 |
| T2DM | endometrium | *ZKSCAN2* | -1.418645979 | 9.19E-06 | 1.49E-05 | 0.044588091 |
| T2DM | endometrium | *KDM8* | 3.428654437 | 1.28E-07 | 4.78E-07 | 0.000622922 |
| T2DM | endometrium | *ALS2CR12* | 5.969392229 | 1.81E-08 | 4.34E-07 | 8.78E-05 |
| T2DM | endometrium | *FZD7* | 3.761799823 | 1.30E-07 | 4.78E-07 | 0.000629178 |
| T2DM | endometrium | *DEPTOR* | 2.960595213 | 1.66E-07 | 4.97E-07 | 0.000803394 |
| T2DM | endometrium | *RRAGA* | 3.034279865 | 8.79E-08 | 4.38E-07 | 0.00042665 |
| T2DM | endometrium | *TMBIM4* | 4.967982175 | 2.93E-07 | 7.38E-07 | 0.001420757 |
| T2DM | endometrium | *PSD3* | 4.200667524 | 2.10E-06 | 3.92E-06 | 0.01016803 |
| T2DM | endometrium | *UGT2B4* | 1.134071444 | 0.012905018 | 0.015208926 | 1 |
| T2DM | endometrium | *MMP16* | 2.124874778 | 5.07E-05 | 7.40E-05 | 0.245894415 |
| T2DM | endometrium | *KCNMA1* | 1.500776371 | 1.19E-07 | 4.64E-07 | 0.00057636 |
| T2DM | endometrium | *NDUFAF6* | 3.296929874 | 8.66E-06 | 1.41E-05 | 0.041995591 |
| T2DM | endometrium | *DRAM2* | 3.026248657 | 1.04E-06 | 2.11E-06 | 0.005057389 |
| T2DM | endometrium | *C8orf37* | 2.576023727 | 2.21E-06 | 4.12E-06 | 0.010738872 |
| T2DM | endometrium | *PPEF2* | 8.841507555 | 3.25E-06 | 5.78E-06 | 0.015749979 |
| T2DM | endometrium | *ART3* | 2.417820923 | 0.020952019 | 0.024256549 | 1 |
| T2DM | endometrium | *WHAMM* | 2.084838414 | 6.25E-06 | 1.05E-05 | 0.030310397 |

| | | | | | | |
|---|---|---|---|---|---|---|
| T2DM | endometrium | *N6AMT1* | 2.829679712 | 1.26E-06 | 2.49E-06 | 0.006128646 |
| T2DM | endometrium | *RWDD2B* | 2.712018481 | 2.63E-06 | 4.79E-06 | 0.012738492 |
| T2DM | endometrium | *CCT8* | 1.878979992 | 2.56E-07 | 6.70E-07 | 0.001241886 |
| T2DM | endometrium | *NAA11* | -5.213531172 | 5.75E-05 | 8.33E-05 | 0.27919078 |
| T2DM | endometrium | *SFXN2* | 2.968109145 | 1.36E-07 | 4.78E-07 | 0.000660536 |
| T2DM | endometrium | *TDRD9* | 6.683259119 | 6.77E-07 | 1.46E-06 | 0.003284083 |
| T2DM | endometrium | *RPL30* | 3.644727033 | 1.03E-07 | 4.47E-07 | 0.000500448 |
| T2DM | endometrium | *SUPV3L1* | 7.164609553 | 1.51E-05 | 2.36E-05 | 0.073108888 |
| T2DM | endometrium | *HKDC1* | -2.970473237 | 1.61E-06 | 3.09E-06 | 0.007794771 |
| T2DM | endometrium | *NODAL* | -5.426184027 | 0.006279091 | 0.007584304 | 1 |
| T2DM | endometrium | *ZDHHC5* | 5.122523215 | 5.16E-08 | 4.34E-07 | 0.000250288 |
| T2DM | endometrium | *MED19* | -2.539860337 | 0.001233108 | 0.001576559 | 1 |
| T2DM | endometrium | *ATAD2* | 5.787509217 | 1.42E-05 | 2.23E-05 | 0.068851884 |
| T2DM | endometrium | *FBRS* | 2.556659825 | 3.01E-08 | 4.34E-07 | 0.000146074 |
| T2DM | endometrium | *SASS6* | -3.247915787 | 2.95E-05 | 4.44E-05 | 0.143005777 |
| T2DM | endometrium | *ITGAD* | 3.969154073 | 2.45E-05 | 3.71E-05 | 0.118694811 |
| T2DM | endometrium | *VPS8* | 3.507598176 | 4.82E-09 | 4.20E-07 | 2.34E-05 |
| T2DM | endometrium | *MPV17L* | 4.887462428 | 9.79E-07 | 1.99E-06 | 0.00474815 |
| T2DM | endometrium | *RPUSD3* | 3.608043995 | 1.45E-07 | 4.78E-07 | 0.00070379 |
| T2DM | endometrium | *GHRL* | 4.830737547 | 0.007876399 | 0.009429136 | 1 |
| T2DM | endometrium | *NMNAT2* | 2.51542891 | 2.64E-07 | 6.84E-07 | 0.001281511 |
| T2DM | endometrium | *SLC6A1* | 9.016263965 | 1.32E-07 | 4.78E-07 | 0.000641923 |
| T2DM | endometrium | *FCHO2* | 2.753367395 | 2.34E-08 | 4.34E-07 | 0.000113776 |
| T2DM | endometrium | *TIMP4* | 2.331634899 | 1.65E-08 | 4.34E-07 | 8.00E-05 |
| T2DM | endometrium | *NECAP2* | 2.664273649 | 1.03E-07 | 4.47E-07 | 0.000501148 |
| T2DM | endometrium | *STEAP2* | 1.771110683 | 2.10E-08 | 4.34E-07 | 0.000102125 |
| T2DM | endometrium | *CLDN12* | 3.861033528 | 8.85E-08 | 4.38E-07 | 0.000429547 |
| T2DM | endometrium | *MMP14* | 6.413486368 | 1.13E-07 | 4.57E-07 | 0.000545871 |
| T2DM | endometrium | *GATAD1* | 2.330584933 | 1.04E-07 | 4.47E-07 | 0.000505876 |
| T2DM | endometrium | *TMED6* | 16.22887431 | 9.75E-07 | 1.99E-06 | 0.004729885 |
| T2DM | endometrium | *CLEC18A* | -1.632247511 | 2.51E-09 | 3.58E-07 | 1.22E-05 |
| T2DM | endometrium | *DHRS4* | 2.931761845 | 1.44E-07 | 4.78E-07 | 0.000699074 |
| T2DM | endometrium | *AASDH* | 5.570434197 | 2.84E-08 | 4.34E-07 | 0.000137623 |
| T2DM | endometrium | *ETS2* | 2.177135355 | 7.00E-09 | 4.34E-07 | 3.40E-05 |
| T2DM | endometrium | *LCA5L* | -1.553570137 | 1.72E-05 | 2.67E-05 | 0.083579387 |
| T2DM | endometrium | *MX1* | 2.228904007 | 7.49E-06 | 1.23E-05 | 0.03635137 |
| T2DM | endometrium | *SLC38A10* | 4.364251063 | 0.00673584 | 0.008113777 | 1 |
| T2DM | endometrium | *DGKI* | 1.730578036 | 3.05E-06 | 5.48E-06 | 0.014805081 |
| T2DM | endometrium | *SNX22* | -4.095777706 | 8.39E-06 | 1.37E-05 | 0.040715417 |
| T2DM | endometrium | *SLC34A2* | -40.86609412 | 0.000486114 | 0.000643203 | 1 |
| T2DM | endometrium | *AP3S2* | 8.875922347 | 1.95E-07 | 5.51E-07 | 0.000947455 |
| T2DM | endometrium | *SPPL3* | 4.020428546 | 6.38E-07 | 1.39E-06 | 0.003094874 |
| T2DM | endometrium | *C12orf43* | -1.573861745 | 1.48E-06 | 2.86E-06 | 0.007158151 |
| T2DM | endometrium | *PEX10* | -3.750749492 | 0.001747236 | 0.002200256 | 1 |
| T2DM | endometrium | *KRTCAP3* | 4.823722459 | 3.36E-06 | 5.97E-06 | 0.016309617 |
| T2DM | endometrium | *PAFAH2* | 3.859614516 | 1.76E-05 | 2.72E-05 | 0.08550926 |
| T2DM | endometrium | *EXTL1* | 42.28593593 | 1.53E-07 | 4.78E-07 | 0.000741893 |
| T2DM | endometrium | *WDR66* | 2.776731909 | 0.014996216 | 0.017554075 | 1 |
| T2DM | endometrium | *RHPN1* | 4.357492859 | 3.57E-05 | 5.34E-05 | 0.173451792 |
| T2DM | endometrium | *LRRC43* | -6.124225242 | 0.000319554 | 0.000430807 | 1 |

| | | | | | | |
|---|---|---|---|---|---|---|
| T2DM | endometrium | *EYA3* | 3.468647647 | 2.48E-08 | 4.34E-07 | 0.000120408 |
| T2DM | endometrium | *DZIP1L* | 6.288847811 | 5.72E-06 | 9.68E-06 | 0.027751324 |
| T2DM | endometrium | *ESYT3* | -4.136267526 | 7.49E-06 | 1.23E-05 | 0.036349055 |
| T2DM | endometrium | *CLSTN2* | 1.623609463 | 2.26E-07 | 6.09E-07 | 0.00109561 |
| T2DM | endometrium | *COLEC12* | 1.337716099 | 1.41E-07 | 4.78E-07 | 0.000682832 |
| T2DM | endometrium | *MITD1* | 5.710578632 | 7.63E-07 | 1.62E-06 | 0.003703757 |
| T2DM | endometrium | *TSPAN33* | 4.002173856 | 2.50E-05 | 3.79E-05 | 0.121358973 |
| T2DM | endometrium | *FAM86C1* | -1.496508709 | 2.98E-06 | 5.37E-06 | 0.014473619 |
| T2DM | endometrium | *PPP1R9A* | 3.739099858 | 5.71E-08 | 4.34E-07 | 0.000276962 |
| T2DM | endometrium | *DYNC1I1* | 3.436379989 | 5.27E-08 | 4.34E-07 | 0.000255536 |
| T2DM | endometrium | *COPG2* | 1.598142282 | 4.54E-05 | 6.67E-05 | 0.220152635 |
| T2DM | endometrium | *ELK4* | 6.906111046 | 2.28E-05 | 3.47E-05 | 0.110425933 |
| T2DM | endometrium | *DUSP23* | 6.265092693 | 2.91E-05 | 4.38E-05 | 0.141210713 |
| T2DM | endometrium | *RNF166* | 2.480012983 | 1.80E-06 | 3.41E-06 | 0.00872456 |
| T2DM | endometrium | *F11R* | 2.190507134 | 3.00E-05 | 4.51E-05 | 0.145408695 |
| T2DM | endometrium | *SPATA2L* | -4.646559776 | 0.040011891 | 0.045444218 | 1 |
| T2DM | endometrium | *FGF17* | -1.747903992 | 2.22E-06 | 4.12E-06 | 0.010747911 |
| T2DM | endometrium | *CDA* | 1.812218427 | 0.00688707 | 0.008293886 | 1 |
| T2DM | endometrium | *CCAR2* | -2.407366143 | 0.000931132 | 0.001201556 | 1 |
| T2DM | endometrium | *CDC42SE2* | 1.648597004 | 1.67E-07 | 5.00E-07 | 0.000811087 |
| T2DM | endometrium | *RAPGEF6* | 3.36278972 | 7.98E-06 | 1.31E-05 | 0.038725379 |
| T2DM | endometrium | *IFNAR2* | 1.65128946 | 1.26E-07 | 4.75E-07 | 0.000611137 |
| T2DM | endometrium | *SV2A* | -2.417588411 | 6.32E-06 | 1.06E-05 | 0.030662911 |
| T2DM | endometrium | *CLIC6* | -2.402080291 | 2.75E-05 | 4.15E-05 | 0.133398234 |
| T2DM | endometrium | *CBR1* | -1.792758395 | 0.004315107 | 0.005256565 | 1 |
| T2DM | endometrium | *CBR3* | 3.971177236 | 5.86E-08 | 4.34E-07 | 0.000284421 |
| T2DM | endometrium | *CHAF1B* | 2.687321202 | 0.001669378 | 0.002106586 | 1 |
| T2DM | endometrium | *HLCS* | 3.006560984 | 3.50E-07 | 8.46E-07 | 0.001696362 |
| T2DM | endometrium | *ARHGAP27* | 4.568380539 | 0.025453071 | 0.029327547 | 1 |
| T2DM | endometrium | *IRX6* | 1.998015564 | 1.20E-05 | 1.91E-05 | 0.058137189 |
| T2DM | endometrium | *HK2* | 3.519163267 | 3.56E-08 | 4.34E-07 | 0.000172836 |
| T2DM | endometrium | *THEM4* | 2.167919824 | 8.28E-08 | 4.37E-07 | 0.000401742 |
| T2DM | endometrium | *AMFR* | 2.277580635 | 1.74E-07 | 5.12E-07 | 0.000843649 |
| T2DM | endometrium | *TMEM69* | 3.090905213 | 1.59E-07 | 4.86E-07 | 0.00077218 |
| T2DM | endometrium | *ACE* | 5.822496364 | 5.01E-08 | 4.34E-07 | 0.00024314 |
| T2DM | endometrium | *TEPP* | 10.17249453 | 1.66E-06 | 3.19E-06 | 0.008067972 |
| T2DM | endometrium | *CHCHD6* | 6.282425325 | 1.59E-06 | 3.06E-06 | 0.007704213 |
| T2DM | endometrium | *CTBP1* | 2.501058125 | 1.50E-07 | 4.78E-07 | 0.000729292 |
| T2DM | endometrium | *LRRC36* | 1.553285638 | 0.001468991 | 0.001863411 | 1 |
| T2DM | endometrium | *ZFYVE28* | 2.647042912 | 5.94E-07 | 1.31E-06 | 0.002884508 |
| T2DM | endometrium | *C16orf86* | 3.598573784 | 2.87E-07 | 7.28E-07 | 0.001393886 |
| T2DM | endometrium | *RGS12* | 3.42903043 | 5.70E-07 | 1.26E-06 | 0.002764518 |
| T2DM | endometrium | *LYPD5* | 4.960823039 | 1.15E-05 | 1.85E-05 | 0.056015217 |
| T2DM | endometrium | *CCDC107* | 6.122019566 | 1.66E-06 | 3.18E-06 | 0.008049689 |
| T2DM | endometrium | *ZNF233* | 5.682380904 | 1.64E-07 | 4.94E-07 | 0.000797486 |
| T2DM | endometrium | *SSU72* | 6.203611507 | 1.79E-07 | 5.21E-07 | 0.000866609 |
| T2DM | endometrium | *CPAMD8* | 5.810358868 | 1.10E-08 | 4.34E-07 | 5.35E-05 |
| T2DM | endometrium | *ANKLE1* | 7.282596215 | 0.008467303 | 0.010106606 | 1 |
| T2DM | endometrium | *CCDC58* | 7.395492016 | 0.001028677 | 0.001322857 | 1 |
| T2DM | endometrium | *SLC37A1* | 2.780691317 | 1.26E-07 | 4.75E-07 | 0.000612188 |

| | | | | | | |
|---|---|---|---|---|---|---|
| T2DM | endometrium | *PDE9A* | 2.193234399 | 7.68E-08 | 4.36E-07 | 0.00037286 |
| T2DM | endometrium | *NDUFV3* | 1.211778365 | 0.004482898 | 0.005459594 | 1 |
| T2DM | endometrium | *CSTB* | 2.142441725 | 1.68E-06 | 3.21E-06 | 0.0081295 |
| T2DM | endometrium | *C21orf2* | -1.171773845 | 2.57E-06 | 4.69E-06 | 0.01246576 |
| T2DM | endometrium | *PCNT* | 4.309771739 | 2.47E-06 | 4.53E-06 | 0.011971088 |
| T2DM | endometrium | *DIP2A* | 3.40548026 | 1.29E-05 | 2.04E-05 | 0.062561618 |
| T2DM | endometrium | *S100B* | 0.907803272 | 0.012862279 | 0.01516224 | 1 |
| T2DM | endometrium | *PRMT2* | 3.624260779 | 2.49E-06 | 4.57E-06 | 0.01209816 |
| T2DM | endometrium | *ZNF208* | 2.197635762 | 3.76E-07 | 8.98E-07 | 0.001822317 |
| T2DM | endometrium | *ADAMTS13* | -1.514221554 | 3.79E-06 | 6.67E-06 | 0.018409698 |
| T2DM | endometrium | *CACFD1* | 6.343814228 | 3.27E-07 | 8.03E-07 | 0.001586882 |
| T2DM | endometrium | *GPSM1* | 3.965738367 | 5.09E-08 | 4.34E-07 | 0.000246786 |
| T2DM | endometrium | *RDH13* | 3.825956473 | 0.00350014 | 0.004291806 | 1 |
| T2DM | endometrium | *TAOK1* | 2.567024991 | 4.32E-08 | 4.34E-07 | 0.000209584 |
| T2DM | endometrium | *TLCD1* | 6.048930278 | 0.025051296 | 0.02887147 | 1 |
| T2DM | endometrium | *S100A1* | 10.29348465 | 2.36E-06 | 4.35E-06 | 0.011431796 |
| T2DM | endometrium | *ZBTB7B* | 3.639827755 | 4.55E-08 | 4.34E-07 | 0.000220976 |
| T2DM | endometrium | *FDPS* | 3.620988561 | 1.79E-08 | 4.34E-07 | 8.67E-05 |
| T2DM | endometrium | *CCR5* | 6.059017273 | 3.20E-08 | 4.34E-07 | 0.000155408 |
| T2DM | endometrium | *NBEAL2* | 3.992177386 | 2.72E-07 | 6.99E-07 | 0.001320536 |
| T2DM | endometrium | *PTH1R* | -2.389112977 | 9.78E-05 | 0.000137647 | 0.474467787 |
| T2DM | endometrium | *FGFR4* | 8.768231667 | 0.000610619 | 0.000802254 | 1 |
| T2DM | endometrium | *LY6K* | 3.929844132 | 0.009662974 | 0.011482917 | 1 |
| T2DM | endometrium | *IER2* | 3.523689198 | 2.38E-07 | 6.36E-07 | 0.001156348 |
| T2DM | endometrium | *CPSF4* | -3.178662145 | 0.003648437 | 0.004466873 | 1 |
| T2DM | endometrium | *VPS28* | 6.585795406 | 1.12E-06 | 2.25E-06 | 0.005446053 |
| T2DM | endometrium | *TONSL* | 5.983579563 | 3.44E-08 | 4.34E-07 | 0.000167093 |
| T2DM | endometrium | *MUM1* | 3.489311829 | 5.97E-08 | 4.34E-07 | 0.000289492 |
| T2DM | endometrium | *ZNF333* | 2.388408164 | 1.13E-07 | 4.57E-07 | 0.000548482 |
| T2DM | endometrium | *SH2B2* | 4.834140619 | 2.99E-06 | 5.39E-06 | 0.014529541 |
| T2DM | endometrium | *LRWD1* | 4.364917349 | 5.29E-06 | 9.02E-06 | 0.025677666 |
| T2DM | endometrium | *DVL3* | -3.251614951 | 0.00078436 | 0.001019206 | 1 |
| T2DM | endometrium | *ABCF3* | 3.378329293 | 1.17E-07 | 4.61E-07 | 0.000569492 |
| T2DM | endometrium | *PCYT1A* | 3.009387823 | 6.43E-05 | 9.24E-05 | 0.311790972 |
| T2DM | endometrium | *FBXO27* | -1.626578313 | 0.000597885 | 0.000785736 | 1 |
| T2DM | endometrium | *DMKN* | 0.87502752 | 0.000802714 | 0.001042496 | 1 |
| T2DM | endometrium | *BDH1* | -2.956952107 | 0.028985263 | 0.033270996 | 1 |
| T2DM | endometrium | *LRRC56* | 5.25554604 | 9.36E-05 | 0.000132103 | 0.454301301 |
| T2DM | endometrium | *PGAP3* | -3.994589773 | 0.003889891 | 0.00474692 | 1 |
| T2DM | endometrium | *ZNF577* | 4.201476501 | 4.14E-08 | 4.34E-07 | 0.000200953 |
| T2DM | endometrium | *TMEM143* | 7.822808958 | 4.07E-08 | 4.34E-07 | 0.000197532 |
| T2DM | endometrium | *CCL16* | -3.23002266 | 9.35E-06 | 1.52E-05 | 0.045371912 |
| T2DM | endometrium | *ALDH16A1* | 3.216217548 | 0.002514522 | 0.003129126 | 1 |
| T2DM | endometrium | *DCD* | -10.59771317 | 2.77E-05 | 4.17E-05 | 0.134249953 |
| T2DM | endometrium | *SIGLEC11* | 1.827335658 | 0.004955255 | 0.006015236 | 1 |
| T2DM | endometrium | *ASB16* | 2.882489821 | 8.57E-08 | 4.38E-07 | 0.000416024 |
| T2DM | endometrium | *DBF4B* | 1.630500906 | 0.002883883 | 0.003568631 | 1 |
| T2DM | endometrium | *PLCD3* | 10.67949951 | 2.96E-06 | 5.34E-06 | 0.014377901 |
| T2DM | endometrium | *IP6K3* | 26.81993912 | 0.000175061 | 0.000241237 | 0.849393993 |
| T2DM | endometrium | *CXCL16* | 1.124152369 | 0.013359424 | 0.015732992 | 1 |

| | | | | | | |
|---|---|---|---|---|---|---|
| T2DM | endometrium | *ASGR2* | 19.51958623 | 9.11E-08 | 4.38E-07 | 0.000442102 |
| T2DM | endometrium | *POLR3K* | -5.339020505 | 0.003458873 | 0.00424335 | 1 |
| T2DM | endometrium | *SNRNP25* | 5.746831804 | 2.22E-08 | 4.34E-07 | 0.00010752 |
| T2DM | endometrium | *WDR90* | 2.642947594 | 2.44E-06 | 4.48E-06 | 0.011833489 |
| T2DM | endometrium | *JMJD8* | 3.075343328 | 8.61E-07 | 1.79E-06 | 0.004176301 |
| T2DM | endometrium | *MEIOB* | 3.923052329 | 1.12E-07 | 4.57E-07 | 0.000544602 |
| T2DM | endometrium | *HS3ST6* | 4.711783394 | 0.000383564 | 0.00051311 | 1 |
| T2DM | endometrium | *SHANK2* | 2.281031523 | 1.26E-07 | 4.74E-07 | 0.000610191 |
| T2DM | endometrium | *ASRGL1* | 3.148200539 | 1.41E-07 | 4.78E-07 | 0.000684639 |
| T2DM | endometrium | *TTC9C* | -2.568342983 | 0.004756271 | 0.005779471 | 1 |
| T2DM | endometrium | *RPS6KA4* | 4.591096574 | 8.19E-08 | 4.36E-07 | 0.000397417 |
| T2DM | endometrium | *TPCN2* | 4.079040693 | 1.38E-07 | 4.78E-07 | 0.000667359 |
| T2DM | endometrium | *PDZK1IP1* | 4.628051934 | 1.46E-07 | 4.78E-07 | 0.000706553 |
| T2DM | endometrium | *CMPK1* | 4.857493731 | 2.08E-07 | 5.78E-07 | 0.001011116 |
| T2DM | endometrium | *COA7* | -2.494979529 | 1.58E-05 | 2.46E-05 | 0.076639186 |
| T2DM | endometrium | *SLC1A7* | 2.262318429 | 0.000274919 | 0.000372496 | 1 |
| T2DM | endometrium | *C1orf123* | 2.543297357 | 2.76E-07 | 7.06E-07 | 0.001336777 |
| T2DM | endometrium | *USP24* | 2.702698672 | 9.05E-08 | 4.38E-07 | 0.000439178 |
| T2DM | endometrium | *ZSWIM5* | 3.464996065 | 6.63E-08 | 4.34E-07 | 0.000321858 |
| T2DM | endometrium | *AK4* | 2.694963809 | 4.26E-07 | 9.86E-07 | 0.002065777 |
| T2DM | endometrium | *RBP7* | 6.554615833 | 2.83E-08 | 4.34E-07 | 0.000137476 |
| T2DM | endometrium | *SLC25A34* | -1.112418569 | 1.83E-06 | 3.47E-06 | 0.008888257 |
| T2DM | endometrium | *PDPN* | 2.423343481 | 1.33E-07 | 4.78E-07 | 0.000647568 |
| T2DM | endometrium | *LAPTM5* | 2.007174885 | 1.46E-07 | 4.78E-07 | 0.000708821 |
| T2DM | endometrium | *SDC3* | 2.505868889 | 6.20E-08 | 4.34E-07 | 0.000300651 |
| T2DM | endometrium | *SYNC* | 2.250800685 | 1.20E-07 | 4.65E-07 | 0.000580475 |
| T2DM | endometrium | *CAMK2N1* | 4.495427903 | 5.56E-07 | 1.23E-06 | 0.002695885 |
| T2DM | endometrium | *MEGF6* | 5.508132873 | 0.000115028 | 0.000161026 | 0.558117805 |
| T2DM | endometrium | *TM2D1* | 3.376884667 | 5.13E-08 | 4.34E-07 | 0.000248816 |
| T2DM | endometrium | *NEXN* | 2.047759755 | 8.94E-08 | 4.38E-07 | 0.000433961 |
| T2DM | endometrium | *LRRIQ3* | 2.229825002 | 1.52E-07 | 4.78E-07 | 0.000737732 |
| T2DM | endometrium | *TYW3* | 3.811296693 | 1.89E-07 | 5.42E-07 | 0.000916307 |
| T2DM | endometrium | *WDR63* | 1.672887965 | 2.66E-06 | 4.84E-06 | 0.012887763 |
| T2DM | endometrium | *HFM1* | 3.14596644 | 0.043576408 | 0.049365569 | 1 |
| T2DM | endometrium | *ARPC5* | 4.473161978 | 8.26E-08 | 4.37E-07 | 0.000400632 |
| T2DM | endometrium | *ZNF496* | 5.27258867 | 0.003355215 | 0.004123481 | 1 |
| T2DM | endometrium | *TRIM58* | 2.650253652 | 0.003153012 | 0.00388778 | 1 |
| T2DM | endometrium | *FCRLB* | -2.572326684 | 2.95E-06 | 5.33E-06 | 0.014319913 |
| T2DM | endometrium | *TDRD5* | -4.93006319 | 1.87E-05 | 2.89E-05 | 0.090932718 |
| T2DM | endometrium | *BROX* | 2.386390203 | 4.43E-06 | 7.67E-06 | 0.021478648 |
| T2DM | endometrium | *CNST* | 3.439465787 | 4.98E-08 | 4.34E-07 | 0.000241786 |
| T2DM | endometrium | *PM20D1* | 2.999856007 | 1.83E-06 | 3.47E-06 | 0.008894545 |
| T2DM | endometrium | *PKDCC* | 3.151104303 | 3.45E-07 | 8.37E-07 | 0.001673726 |
| T2DM | endometrium | *B3GALNT2* | 2.997096651 | 3.98E-06 | 6.97E-06 | 0.019289247 |
| T2DM | endometrium | *MRPL55* | 5.746839648 | 1.31E-06 | 2.57E-06 | 0.006343667 |
| T2DM | endometrium | *KCNJ3* | -1.488586183 | 1.63E-06 | 3.13E-06 | 0.007912561 |
| T2DM | endometrium | *CLHC1* | 2.240928761 | 1.81E-08 | 4.34E-07 | 8.79E-05 |
| T2DM | endometrium | *DUSP19* | -3.942664728 | 0.000334955 | 0.000450569 | 1 |
| T2DM | endometrium | *NUP35* | -2.90734732 | 0.00377836 | 0.004617693 | 1 |
| T2DM | endometrium | *C2orf48* | 25.77176309 | 1.06E-07 | 4.49E-07 | 0.000516154 |

| | | | | | | |
|---|---|---|---|---|---|---|
| T2DM | endometrium | *CCDC74A* | 3.763968785 | 0.030768673 | 0.03524306 | 1 |
| T2DM | endometrium | *H3F3A* | 5.63263049 | 4.09E-06 | 7.15E-06 | 0.019829021 |
| T2DM | endometrium | *NOSTRIN* | 3.743047499 | 7.65E-08 | 4.36E-07 | 0.000371398 |
| T2DM | endometrium | *INHBB* | 4.65720665 | 2.90E-07 | 7.33E-07 | 0.00140663 |
| T2DM | endometrium | *RPRD2* | 4.205570659 | 4.65E-08 | 4.34E-07 | 0.000225459 |
| T2DM | endometrium | *CTSS* | 1.327934976 | 8.86E-07 | 1.83E-06 | 0.004296997 |
| T2DM | endometrium | *PACRGL* | 3.764121011 | 6.13E-08 | 4.34E-07 | 0.000297532 |
| T2DM | endometrium | *C1QTNF7* | 3.61423448 | 2.44E-08 | 4.34E-07 | 0.000118542 |
| T2DM | endometrium | *VPS72* | 4.918494565 | 5.27E-08 | 4.34E-07 | 0.000255924 |
| T2DM | endometrium | *ERCC3* | 2.446284633 | 4.44E-08 | 4.34E-07 | 0.000215525 |
| T2DM | endometrium | *RNF149* | 2.442758032 | 7.02E-08 | 4.34E-07 | 0.000340812 |
| T2DM | endometrium | *DHX57* | 3.49848057 | 5.96E-08 | 4.34E-07 | 0.000289286 |
| T2DM | endometrium | *S100A9* | 5.620289258 | 9.57E-07 | 1.96E-06 | 0.004645422 |
| T2DM | endometrium | *TDRD10* | 1.935687894 | 1.48E-07 | 4.78E-07 | 0.000719517 |
| T2DM | endometrium | *DCAF16* | 4.276345761 | 6.92E-07 | 1.49E-06 | 0.00335695 |
| T2DM | endometrium | *GNPDA2* | 4.757719559 | 1.73E-07 | 5.10E-07 | 0.000837157 |
| T2DM | endometrium | *NIPAL1* | 3.042903536 | 1.47E-07 | 4.78E-07 | 0.000711188 |
| T2DM | endometrium | *ANTXR2* | 4.260754016 | 5.51E-07 | 1.22E-06 | 0.002671498 |
| T2DM | endometrium | *CGGBP1* | 2.812106385 | 1.00E-06 | 2.03E-06 | 0.004857587 |
| T2DM | endometrium | *GPR155* | 2.835120327 | 9.09E-08 | 4.38E-07 | 0.000441125 |
| T2DM | endometrium | *DAPL1* | 7.382776669 | 7.63E-09 | 4.34E-07 | 3.70E-05 |
| T2DM | endometrium | *PBXIP1* | 2.424270317 | 1.22E-07 | 4.69E-07 | 0.000592288 |
| T2DM | endometrium | *CLDN1* | 1.679422187 | 1.42E-07 | 4.78E-07 | 0.000690563 |
| T2DM | endometrium | *DCST2* | 5.081754098 | 1.53E-08 | 4.34E-07 | 7.40E-05 |
| T2DM | endometrium | *LINC01116* | 10.23740619 | 1.85E-08 | 4.34E-07 | 8.98E-05 |
| T2DM | endometrium | *POGLUT1* | 2.518008332 | 2.15E-05 | 3.29E-05 | 0.104553905 |
| T2DM | endometrium | *SLC22A15* | 5.322099744 | 1.42E-05 | 2.23E-05 | 0.068733895 |
| T2DM | endometrium | *LRRC58* | 6.593342067 | 2.54E-08 | 4.34E-07 | 0.000123458 |
| T2DM | endometrium | *LMOD1* | 1.702968183 | 7.97E-07 | 1.67E-06 | 0.003865 |
| T2DM | endometrium | *TMEM169* | 3.531776421 | 1.20E-05 | 1.92E-05 | 0.058394114 |
| T2DM | endometrium | *ARPC2* | 4.726505751 | 4.78E-07 | 1.08E-06 | 0.002317693 |
| T2DM | endometrium | *GLB1L* | 2.688005876 | 1.04E-06 | 2.11E-06 | 0.005066216 |
| T2DM | endometrium | *SERPINI1* | 6.915378333 | 2.61E-07 | 6.78E-07 | 0.001266139 |
| T2DM | endometrium | *SUCLG1* | 6.191708393 | 2.22E-07 | 6.02E-07 | 0.001076056 |
| T2DM | endometrium | *SPTA1* | -2.163972715 | 4.94E-09 | 4.20E-07 | 2.39E-05 |
| T2DM | endometrium | *PRKCI* | 3.03686494 | 1.33E-06 | 2.62E-06 | 0.006474896 |
| T2DM | endometrium | *AIM2* | -5.900443169 | 0.012671444 | 0.014948175 | 1 |
| T2DM | endometrium | *EFHB* | 1.180406405 | 0.0007659 | 0.000996287 | 1 |
| T2DM | endometrium | *GTPBP8* | 4.703679002 | 6.01E-08 | 4.34E-07 | 0.000291845 |
| T2DM | endometrium | *WDFY3* | 3.149843127 | 5.43E-08 | 4.34E-07 | 0.000263368 |
| T2DM | endometrium | *PTPN13* | 2.352613856 | 1.05E-06 | 2.13E-06 | 0.005114633 |
| T2DM | endometrium | *PPM1K* | 3.706525246 | 1.38E-07 | 4.78E-07 | 0.00067104 |
| T2DM | endometrium | *TIPARP* | -2.355154141 | 0.000395425 | 0.00052825 | 1 |
| T2DM | endometrium | *DCLK3* | 7.995887233 | 0.000486272 | 0.000643236 | 1 |
| T2DM | endometrium | *RPL9* | -1.111055786 | 0.001286269 | 0.001642794 | 1 |
| T2DM | endometrium | *RPP14* | 3.516981713 | 1.45E-07 | 4.78E-07 | 0.000703026 |
| T2DM | endometrium | *DNASE1L3* | -2.70143136 | 0.000683963 | 0.000893933 | 1 |
| T2DM | endometrium | *IL17RC* | 2.493742764 | 9.75E-07 | 1.99E-06 | 0.004729017 |
| T2DM | endometrium | *CRELD1* | 2.5394789 | 9.79E-08 | 4.42E-07 | 0.000474849 |
| T2DM | endometrium | *TTC14* | 3.967350467 | 1.44E-06 | 2.80E-06 | 0.007000735 |

| | | | | | | |
|---|---|---|---|---|---|---|
| T2DM | endometrium | *RCHY1* | 2.325152278 | 8.61E-07 | 1.79E-06 | 0.004179861 |
| T2DM | endometrium | *CCDC158* | 4.855431971 | 8.16E-08 | 4.36E-07 | 0.000395745 |
| T2DM | endometrium | *CPA3* | -4.783262264 | 0.000657419 | 0.000861176 | 1 |
| T2DM | endometrium | *SLC4A1AP* | 5.06525637 | 0.00911386 | 0.010848982 | 1 |
| T2DM | endometrium | *KIAA1143* | 3.623748811 | 1.77E-06 | 3.36E-06 | 0.008571395 |
| T2DM | endometrium | *CDCP1* | 2.487660359 | 3.06E-07 | 7.63E-07 | 0.001482348 |
| T2DM | endometrium | *CLEC3B* | 5.771837409 | 1.92E-08 | 4.34E-07 | 9.33E-05 |
| T2DM | endometrium | *FYCO1* | 2.6466217 | 2.55E-07 | 6.69E-07 | 0.001239049 |
| T2DM | endometrium | *NMNAT3* | 3.570400635 | 1.63E-08 | 4.34E-07 | 7.90E-05 |
| T2DM | endometrium | *SNIP1* | 3.253431776 | 9.09E-08 | 4.38E-07 | 0.000440939 |
| T2DM | endometrium | *DNALI1* | 4.363236386 | 1.44E-07 | 4.78E-07 | 0.00069822 |
| T2DM | endometrium | *RPN1* | 2.054061637 | 1.44E-07 | 4.78E-07 | 0.000697765 |
| T2DM | endometrium | *SENP2* | 4.016444228 | 0.000355127 | 0.00047691 | 1 |
| T2DM | endometrium | *IFT122* | 3.749478016 | 1.12E-07 | 4.57E-07 | 0.000545109 |
| T2DM | endometrium | *RFT1* | 1.715009308 | 0.000232656 | 0.00031727 | 1 |
| T2DM | endometrium | *SFMBT1* | 5.406827743 | 3.35E-08 | 4.34E-07 | 0.000162322 |
| T2DM | endometrium | *UVSSA* | 4.94893768 | 1.05E-08 | 4.34E-07 | 5.07E-05 |
| T2DM | endometrium | *FAM208A* | 3.54819561 | 9.00E-08 | 4.38E-07 | 0.000436464 |
| T2DM | endometrium | *LRPAP1* | 3.808937049 | 3.43E-08 | 4.34E-07 | 0.000166234 |
| T2DM | endometrium | *PIGX* | 2.334189683 | 6.88E-06 | 1.14E-05 | 0.033372909 |
| T2DM | endometrium | *ABLIM2* | 3.03962365 | 2.17E-07 | 5.94E-07 | 0.001050507 |
| T2DM | endometrium | *ZNF691* | -2.40418466 | 0.002519223 | 0.003134172 | 1 |
| T2DM | endometrium | *METAP1* | 2.010308681 | 2.85E-07 | 7.25E-07 | 0.001383979 |
| T2DM | endometrium | *BDH2* | 3.903891569 | 0.002404372 | 0.002995895 | 1 |
| T2DM | endometrium | *CDC25A* | 8.88473248 | 0.000701739 | 0.000914542 | 1 |
| T2DM | endometrium | *ZNF589* | -2.664881247 | 0.010590212 | 0.012554046 | 1 |
| T2DM | endometrium | *ATRIP* | -1.83956946 | 9.53E-08 | 4.42E-07 | 0.000462359 |
| T2DM | endometrium | *RNF123* | 6.802212645 | 1.36E-12 | 1.65E-09 | 6.59E-09 |
| T2DM | endometrium | *HSPA4L* | -2.378007553 | 0.003191593 | 0.003932354 | 1 |
| T2DM | endometrium | *MFSD8* | 5.19132084 | 2.47E-07 | 6.54E-07 | 0.001196738 |
| T2DM | endometrium | *MST1R* | 4.755781841 | 1.72E-12 | 1.67E-09 | 8.37E-09 |
| T2DM | endometrium | *RAD54L2* | 4.942710818 | 8.47E-07 | 1.76E-06 | 0.004107908 |
| T2DM | endometrium | *TEX264* | 4.188289404 | 4.50E-07 | 1.03E-06 | 0.002183885 |
| T2DM | endometrium | *ETNPPL* | 20.77364798 | 1.48E-07 | 4.78E-07 | 0.00071942 |
| T2DM | endometrium | *HMGB2* | 6.16830116 | 4.64E-07 | 1.06E-06 | 0.002253446 |
| T2DM | endometrium | *ANXA5* | 2.565062045 | 4.80E-08 | 4.34E-07 | 0.000233082 |
| T2DM | endometrium | *MAP9* | 3.905859579 | 5.33E-08 | 4.34E-07 | 0.000258711 |
| T2DM | endometrium | *CEP44* | -2.433108099 | 0.006585324 | 0.007940355 | 1 |
| T2DM | endometrium | *HPGD* | -2.483756232 | 1.26E-05 | 1.99E-05 | 0.06092131 |
| T2DM | endometrium | *C4orf45* | 5.565606222 | 1.17E-05 | 1.86E-05 | 0.056657745 |
| T2DM | endometrium | *TMEM144* | 2.195226705 | 2.22E-07 | 6.02E-07 | 0.001077095 |
| T2DM | endometrium | *IL15* | 2.814992161 | 1.39E-08 | 4.34E-07 | 6.77E-05 |
| T2DM | endometrium | *ARFIP1* | 3.941501759 | 5.63E-08 | 4.34E-07 | 0.000272986 |
| T2DM | endometrium | *LSM6* | 3.579076419 | 3.74E-07 | 8.96E-07 | 0.001816904 |
| T2DM | endometrium | *ITGA2* | 2.582199054 | 7.36E-08 | 4.35E-07 | 0.000357168 |
| T2DM | endometrium | *MOCS2* | 3.108445027 | 1.54E-07 | 4.78E-07 | 0.000745208 |
| T2DM | endometrium | *NDUFAF2* | 3.338196529 | 3.03E-07 | 7.59E-07 | 0.001472182 |
| T2DM | endometrium | *LMBRD2* | 5.235277282 | 2.79E-07 | 7.13E-07 | 0.00135206 |
| T2DM | endometrium | *RNF180* | 2.539747063 | 1.72E-07 | 5.08E-07 | 0.000833625 |
| T2DM | endometrium | *PGGT1B* | 3.286222231 | 9.29E-08 | 4.39E-07 | 0.000450669 |

| | | | | | | |
|---|---|---|---|---|---|---|
| T2DM | endometrium | *F2RL2* | 7.063332655 | 1.34E-07 | 4.78E-07 | 0.000648062 |
| T2DM | endometrium | *CCDC112* | 2.312417882 | 2.17E-07 | 5.95E-07 | 0.001053763 |
| T2DM | endometrium | *CMBL* | -2.973995636 | 0.001169968 | 0.001498201 | 1 |
| T2DM | endometrium | *C5orf63* | 4.009049425 | 1.63E-07 | 4.92E-07 | 0.000792092 |
| T2DM | endometrium | *F2RL1* | 5.610332352 | 0.033880159 | 0.038688287 | 1 |
| T2DM | endometrium | *WDR41* | 5.35131531 | 1.43E-05 | 2.25E-05 | 0.069563639 |
| T2DM | endometrium | *SERINC5* | 2.501883981 | 7.00E-08 | 4.34E-07 | 0.000339805 |
| T2DM | endometrium | *CASP3* | 3.642984971 | 9.24E-08 | 4.38E-07 | 0.000448418 |
| T2DM | endometrium | *PRIMPOL* | 3.863894039 | 4.10E-07 | 9.60E-07 | 0.001989944 |
| T2DM | endometrium | *ERAP1* | 2.571972474 | 1.00E-07 | 4.45E-07 | 0.000486119 |
| T2DM | endometrium | *ERAP2* | 2.401313208 | 1.48E-06 | 2.87E-06 | 0.007169475 |
| T2DM | endometrium | *UTP15* | -2.174500007 | 7.93E-06 | 1.30E-05 | 0.038459606 |
| T2DM | endometrium | *TLR3* | 7.986447861 | 6.73E-07 | 1.45E-06 | 0.003265015 |
| T2DM | endometrium | *KLKB1* | 2.504281597 | 1.16E-06 | 2.31E-06 | 0.005616177 |
| T2DM | endometrium | *NSA2* | 2.304454633 | 3.19E-06 | 5.69E-06 | 0.015461279 |
| T2DM | endometrium | *CCDC127* | 5.752956546 | 5.58E-08 | 4.34E-07 | 0.000270649 |
| T2DM | endometrium | *GRIK2* | 1.555658852 | 8.99E-06 | 1.46E-05 | 0.043612098 |
| T2DM | endometrium | *PDSS2* | 2.773212069 | 5.12E-08 | 4.34E-07 | 0.000248526 |
| T2DM | endometrium | *RAET1E* | 2.961225177 | 1.33E-07 | 4.78E-07 | 0.000647608 |
| T2DM | endometrium | *DAGLB* | 1.833008141 | 0.010812636 | 0.012808327 | 1 |
| T2DM | endometrium | *STK17A* | 4.538958434 | 6.29E-07 | 1.37E-06 | 0.003053261 |
| T2DM | endometrium | *GALNT10* | 2.441433303 | 2.65E-07 | 6.85E-07 | 0.001284306 |
| T2DM | endometrium | *SAP30L* | 2.520126653 | 1.28E-07 | 4.77E-07 | 0.0006192 |
| T2DM | endometrium | *RPS14* | 4.869766988 | 6.83E-06 | 1.14E-05 | 0.033155823 |
| T2DM | endometrium | *RP9* | 3.819974215 | 7.58E-08 | 4.36E-07 | 0.000367921 |
| T2DM | endometrium | *CAMLG* | 5.978081026 | 0.000140148 | 0.000194674 | 0.679997559 |
| T2DM | endometrium | *RELL2* | 6.2503919 | 2.66E-05 | 4.02E-05 | 0.12904794 |
| T2DM | endometrium | *MIOS* | 3.18891995 | 8.21E-08 | 4.37E-07 | 0.000398175 |
| T2DM | endometrium | *CHMP4C* | 8.536861777 | 1.39E-07 | 4.78E-07 | 0.000676552 |
| T2DM | endometrium | *SLC13A4* | 2.556662183 | 9.41E-08 | 4.39E-07 | 0.000456474 |
| T2DM | endometrium | *BRI3* | 3.461600322 | 0.019508056 | 0.022655119 | 1 |
| T2DM | endometrium | *CTSB* | 1.287477971 | 7.69E-07 | 1.63E-06 | 0.003729323 |
| T2DM | endometrium | *SOX17* | 4.890637574 | 9.35E-09 | 4.34E-07 | 4.54E-05 |
| T2DM | endometrium | *TNFRSF11B* | 4.510283396 | 9.76E-08 | 4.42E-07 | 0.000473322 |
| T2DM | endometrium | *DEFB1* | 5.835808472 | 0.013332793 | 0.015705441 | 1 |
| T2DM | endometrium | *SUN1* | 4.662685493 | 1.52E-05 | 2.37E-05 | 0.073739313 |
| T2DM | endometrium | *OXR1* | 2.925612654 | 0.013484007 | 0.015872005 | 1 |
| T2DM | endometrium | *GPER1* | 7.909418309 | 1.68E-09 | 3.07E-07 | 8.13E-06 |
| T2DM | endometrium | *INTS1* | 8.502729274 | 7.09E-05 | 0.000101464 | 0.344064015 |
| T2DM | endometrium | *SLC4A2* | -3.200029834 | 0.000722594 | 0.000941216 | 1 |
| T2DM | endometrium | *ALDH7A1* | 5.19933453 | 6.95E-08 | 4.34E-07 | 0.000337123 |
| T2DM | endometrium | *YWHAZ* | 4.516348823 | 9.20E-07 | 1.89E-06 | 0.004462482 |
| T2DM | endometrium | *SLC25A32* | 4.3712582 | 2.21E-07 | 6.02E-07 | 0.001074331 |
| T2DM | endometrium | *DCSTAMP* | -4.400430055 | 3.12E-06 | 5.59E-06 | 0.015160459 |
| T2DM | endometrium | *INTS8* | -2.076836252 | 0.042811643 | 0.048533199 | 1 |
| T2DM | endometrium | *RPP25L* | -3.438740934 | 2.17E-05 | 3.31E-05 | 0.105163765 |
| T2DM | endometrium | *C9orf24* | 2.81165802 | 1.11E-07 | 4.57E-07 | 0.000536793 |
| T2DM | endometrium | *TMEM65* | 4.871261184 | 7.40E-08 | 4.36E-07 | 0.000358998 |
| T2DM | endometrium | *CCDC171* | 5.119810108 | 4.04E-05 | 5.99E-05 | 0.196164473 |
| T2DM | endometrium | *METTL2B* | 2.592816316 | 5.11E-05 | 7.45E-05 | 0.24802463 |

| | | | | | | |
|---|---|---|---|---|---|---|
| T2DM | endometrium | *ZMAT4* | 5.322249655 | 1.38E-07 | 4.78E-07 | 0.000668518 |
| T2DM | endometrium | *C8orf34* | 1.884590174 | 2.08E-07 | 5.78E-07 | 0.001011517 |
| T2DM | endometrium | *ANKS6* | 5.029416779 | 1.31E-08 | 4.34E-07 | 6.35E-05 |
| T2DM | endometrium | *AQP7* | 1.769796659 | 3.09E-07 | 7.69E-07 | 0.001498151 |
| T2DM | endometrium | *NOL6* | -3.087131593 | 5.99E-06 | 1.01E-05 | 0.029058656 |
| T2DM | endometrium | *AQP3* | 8.664350071 | 1.40E-07 | 4.78E-07 | 0.000679811 |
| T2DM | endometrium | *TRMT10B* | 3.279541103 | 7.84E-08 | 4.36E-07 | 0.000380569 |
| T2DM | endometrium | *VCP* | 3.877259579 | 3.74E-07 | 8.96E-07 | 0.001815189 |
| T2DM | endometrium | *PIGO* | 3.561541297 | 5.28E-07 | 1.18E-06 | 0.002562028 |
| T2DM | endometrium | *WRN* | 3.41472216 | 3.55E-08 | 4.34E-07 | 0.000172321 |
| T2DM | endometrium | *MARCH8* | 5.519880805 | 5.04E-08 | 4.34E-07 | 0.000244537 |
| T2DM | endometrium | *TSHR* | 3.808312345 | 3.62E-06 | 6.39E-06 | 0.017583712 |
| T2DM | endometrium | *CFL2* | 7.710337648 | 7.82E-05 | 0.000111378 | 0.379465955 |
| T2DM | endometrium | *PGM2L1* | 2.640628123 | 4.21E-07 | 9.77E-07 | 0.002041382 |
| T2DM | endometrium | *SLC16A9* | 3.719816652 | 1.51E-07 | 4.78E-07 | 0.000731837 |
| T2DM | endometrium | *GJB2* | 16.17362434 | 1.38E-07 | 4.78E-07 | 0.000669727 |
| T2DM | endometrium | *CRYL1* | 4.734597806 | 8.66E-08 | 4.38E-07 | 0.00042023 |
| T2DM | endometrium | *REEP3* | 4.658393142 | 0.000145407 | 0.000201749 | 0.705515853 |
| T2DM | endometrium | *MICU2* | 5.144016187 | 3.18E-07 | 7.88E-07 | 0.001542759 |
| T2DM | endometrium | *LRR1* | 11.17757877 | 0.000104907 | 0.000147325 | 0.509007865 |
| T2DM | endometrium | *C10orf25* | 1.37166285 | 0.020791506 | 0.024082213 | 1 |
| T2DM | endometrium | *TMEM63C* | -3.349205571 | 4.40E-07 | 1.01E-06 | 0.0021365 |
| T2DM | endometrium | *NOXRED1* | 3.720797246 | 7.36E-09 | 4.34E-07 | 3.57E-05 |
| T2DM | endometrium | *AKR1E2* | 5.271547404 | 0.008301286 | 0.009915765 | 1 |
| T2DM | endometrium | *ATP5C1* | 5.082800376 | 1.77E-07 | 5.19E-07 | 0.000859636 |
| T2DM | endometrium | *VDAC2* | 6.200554977 | 9.42E-07 | 1.93E-06 | 0.004569915 |
| T2DM | endometrium | *PDZD8* | 5.900753118 | 9.07E-08 | 4.38E-07 | 0.00044019 |
| T2DM | endometrium | *QSOX2* | 3.275170363 | 4.63E-08 | 4.34E-07 | 0.000224788 |
| T2DM | endometrium | *CLEC1B* | -2.274775649 | 1.02E-05 | 1.65E-05 | 0.049714708 |
| T2DM | endometrium | *SNAPC4* | -2.371494415 | 0.00354469 | 0.004345335 | 1 |
| T2DM | endometrium | *TSC1* | 2.60374505 | 2.65E-07 | 6.86E-07 | 0.001286298 |
| T2DM | endometrium | *STOX1* | 1.887143148 | 4.72E-06 | 8.13E-06 | 0.022886339 |
| T2DM | endometrium | *RET* | 4.106434514 | 1.13E-07 | 4.57E-07 | 0.000546489 |
| T2DM | endometrium | *CASP7* | 4.034184693 | 1.90E-07 | 5.44E-07 | 0.000922748 |
| T2DM | endometrium | *PPP1R36* | -3.866021599 | 0.001198497 | 0.001533115 | 1 |
| T2DM | endometrium | *VWA2* | 7.483945129 | 3.13E-07 | 7.76E-07 | 0.001517085 |
| T2DM | endometrium | *METTL3* | 5.896929518 | 0.001438442 | 0.001827525 | 1 |
| T2DM | endometrium | *TRUB1* | 2.18766771 | 4.48E-07 | 1.02E-06 | 0.002174424 |
| T2DM | endometrium | *HSPA12A* | 2.893817111 | 3.38E-07 | 8.25E-07 | 0.001638651 |
| T2DM | endometrium | *UBTD1* | 6.72182351 | 9.14E-07 | 1.88E-06 | 0.004432496 |
| T2DM | endometrium | *OTOGL* | 5.287876073 | 1.42E-06 | 2.76E-06 | 0.006887085 |
| T2DM | endometrium | *TTC7B* | 4.478277488 | 9.93E-08 | 4.44E-07 | 0.000481901 |
| T2DM | endometrium | *SLC39A13* | -2.96798153 | 0.003549631 | 0.004350293 | 1 |
| T2DM | endometrium | *PSMC3* | -3.492103112 | 0.003431725 | 0.004213242 | 1 |
| T2DM | endometrium | *TC2N* | 1.513668579 | 3.24E-07 | 7.98E-07 | 0.00157033 |
| T2DM | endometrium | *CPSF2* | 2.683362575 | 4.28E-05 | 6.31E-05 | 0.207801297 |
| T2DM | endometrium | *IFI27L1* | -2.637363999 | 0.000290591 | 0.000392634 | 1 |
| T2DM | endometrium | *C1QL3* | -1.905009306 | 0.00050354 | 0.000665534 | 1 |
| T2DM | endometrium | *ARL5B* | 3.01014705 | 2.66E-07 | 6.87E-07 | 0.001289555 |
| T2DM | endometrium | *LIPC* | 3.702962647 | 3.03E-07 | 7.59E-07 | 0.001472025 |

| | | | | | | |
|---|---|---|---|---|---|---|
| T2DM | endometrium | *CEP57* | 2.363093182 | 4.05E-07 | 9.49E-07 | 0.001962898 |
| T2DM | endometrium | *TCP11L2* | 4.563314222 | 4.45E-08 | 4.34E-07 | 0.000215927 |
| T2DM | endometrium | *JAM3* | 3.963188625 | 6.73E-08 | 4.34E-07 | 0.00032665 |
| T2DM | endometrium | *IKBIP* | 4.363868669 | 4.01E-08 | 4.34E-07 | 0.000194442 |
| T2DM | endometrium | *ZFYVE19* | 2.807868213 | 1.69E-06 | 3.23E-06 | 0.008204753 |
| T2DM | endometrium | *TRMT61A* | 5.039880321 | 8.69E-08 | 4.38E-07 | 0.000421562 |
| T2DM | endometrium | *LARP6* | 4.168192094 | 3.41E-08 | 4.34E-07 | 0.000165423 |
| T2DM | endometrium | *API5* | 5.267866784 | 6.79E-08 | 4.34E-07 | 0.000329625 |
| T2DM | endometrium | *ASPG* | 15.69295889 | 6.02E-07 | 1.32E-06 | 0.002919364 |
| T2DM | endometrium | *GABRB3* | 2.933037574 | 2.47E-06 | 4.53E-06 | 0.011986097 |
| T2DM | endometrium | *COX11* | 5.241092269 | 0.001054766 | 0.001354843 | 1 |
| T2DM | endometrium | *ZNF202* | -4.174462358 | 0.000216146 | 0.000295419 | 1 |
| T2DM | endometrium | *STXBP4* | 4.977052122 | 1.41E-06 | 2.75E-06 | 0.006842762 |
| T2DM | endometrium | *WBP1L* | 5.84100257 | 7.34E-08 | 4.35E-07 | 0.000355958 |
| T2DM | endometrium | *APBB1* | 6.25893512 | 7.19E-07 | 1.54E-06 | 0.003490732 |
| T2DM | endometrium | *NUDT13* | -1.717874721 | 0.024855205 | 0.028665903 | 1 |
| T2DM | endometrium | *WDR88* | -3.239934003 | 0.007407512 | 0.008894147 | 1 |
| T2DM | endometrium | *CYB5R2* | 3.936456751 | 4.88E-06 | 8.38E-06 | 0.023662199 |
| T2DM | endometrium | *SERPINB8* | 2.042664714 | 0.00144548 | 0.001835025 | 1 |
| T2DM | endometrium | *PLD4* | 4.196519665 | 0.041322769 | 0.046867245 | 1 |
| T2DM | endometrium | *XRRA1* | 2.800969348 | 0.014365025 | 0.01684774 | 1 |
| T2DM | endometrium | *RPL27A* | 4.794218437 | 0.001426605 | 0.001813495 | 1 |
| T2DM | endometrium | *CDYL2* | 3.069242599 | 1.58E-07 | 4.85E-07 | 0.000767676 |
| T2DM | endometrium | *CENPN* | 5.949581755 | 1.03E-07 | 4.47E-07 | 0.000498757 |
| T2DM | endometrium | *TMEM41B* | 3.422415412 | 5.78E-08 | 4.34E-07 | 0.000280441 |
| T2DM | endometrium | *TMX3* | 3.521979872 | 1.18E-07 | 4.61E-07 | 0.000570234 |
| T2DM | endometrium | *PRKCB* | 1.708670277 | 1.40E-07 | 4.78E-07 | 0.000680464 |
| T2DM | endometrium | *NDST2* | -3.216490755 | 1.98E-06 | 3.73E-06 | 0.009616902 |
| T2DM | endometrium | *CCDC68* | 4.396019833 | 1.12E-07 | 4.57E-07 | 0.000543493 |
| T2DM | endometrium | *CLEC4D* | 6.301181418 | 0.016185738 | 0.01890089 | 1 |
| T2DM | endometrium | *ZSCAN21* | 2.269644379 | 4.31E-07 | 9.92E-07 | 0.00208927 |
| T2DM | endometrium | *RIMKLB* | 3.442083102 | 2.02E-05 | 3.09E-05 | 0.09780285 |
| T2DM | endometrium | *TMED3* | -1.388751877 | 2.82E-05 | 4.25E-05 | 0.13678504 |
| T2DM | endometrium | *SEC11C* | 2.684085576 | 1.38E-07 | 4.78E-07 | 0.000668536 |
| T2DM | endometrium | *SPDYE6* | -1.509784664 | 0.000472409 | 0.000626436 | 1 |
| T2DM | endometrium | *ZNF592* | 3.520731312 | 2.88E-07 | 7.29E-07 | 0.001396367 |
| T2DM | endometrium | *ACSM1* | 3.673136959 | 5.41E-07 | 1.20E-06 | 0.002624957 |
| T2DM | endometrium | *SLFN5* | 4.656818387 | 3.41E-08 | 4.34E-07 | 0.000165225 |
| T2DM | endometrium | *CATSPER2* | 2.41724041 | 2.12E-07 | 5.85E-07 | 0.001029927 |
| T2DM | endometrium | *ZNF667-AS1* | 3.484917556 | 1.45E-07 | 4.78E-07 | 0.00070339 |
| T2DM | endometrium | *YPEL4* | 3.9051699 | 0.001802901 | 0.002268 | 1 |
| T2DM | endometrium | *LDHC* | 2.783329447 | 1.23E-05 | 1.95E-05 | 0.059495037 |
| T2DM | endometrium | *FAM111A* | 4.274433113 | 2.94E-07 | 7.40E-07 | 0.00142618 |
| T2DM | endometrium | *ANPEP* | 2.913392855 | 1.03E-09 | 2.33E-07 | 4.98E-06 |
| T2DM | endometrium | *DCTN5* | 2.584595859 | 2.29E-07 | 6.16E-07 | 0.001112412 |
| T2DM | endometrium | *STAT6* | 3.523773443 | 5.66E-08 | 4.34E-07 | 0.00027486 |
| T2DM | endometrium | *STX3* | 5.797235062 | 4.08E-06 | 7.14E-06 | 0.01981228 |
| T2DM | endometrium | *PIP4K2C* | 3.275623103 | 1.37E-07 | 4.78E-07 | 0.000665435 |
| T2DM | endometrium | *MS4A7* | 1.533050945 | 3.07E-06 | 5.50E-06 | 0.014877033 |
| T2DM | endometrium | *MS4A14* | 0.948930555 | 0.019839739 | 0.023018272 | 1 |

| | | | | | | |
|---|---|---|---|---|---|---|
| T2DM | endometrium | *DIS3L* | 3.543818233 | 4.42E-08 | 4.34E-07 | 0.000214689 |
| T2DM | endometrium | *CCNDBP1* | 2.388775342 | 8.92E-08 | 4.38E-07 | 0.000432899 |
| T2DM | endometrium | *EPB42* | 3.878243854 | 5.07E-08 | 4.34E-07 | 0.000246017 |
| T2DM | endometrium | *SMAD3* | 4.487162677 | 1.94E-07 | 5.50E-07 | 0.000943597 |
| T2DM | endometrium | *RCCD1* | 3.27530869 | 8.83E-07 | 1.83E-06 | 0.004284638 |
| T2DM | endometrium | *CNPY4* | 3.941892205 | 4.80E-09 | 4.20E-07 | 2.33E-05 |
| T2DM | endometrium | *MEI1* | 3.852775444 | 5.36E-08 | 4.34E-07 | 0.000259861 |
| T2DM | endometrium | *SAMD14* | 6.42140573 | 4.97E-06 | 8.52E-06 | 0.024105339 |
| T2DM | endometrium | *PIP5KL1* | 3.020697394 | 0.006504532 | 0.007844889 | 1 |
| T2DM | endometrium | *ACSF2* | 2.586082561 | 1.29E-07 | 4.78E-07 | 0.000626306 |
| T2DM | endometrium | *URM1* | 4.59889936 | 2.55E-07 | 6.69E-07 | 0.001236671 |
| T2DM | endometrium | *ENDOG* | 2.495151964 | 0.00036863 | 0.000493813 | 1 |
| T2DM | endometrium | *ISLR2* | 4.926594592 | 4.20E-05 | 6.20E-05 | 0.203894911 |
| T2DM | endometrium | *SP2* | 7.646957646 | 2.37E-07 | 6.34E-07 | 0.00114955 |
| T2DM | endometrium | *GPRC5B* | 2.308292371 | 1.36E-07 | 4.78E-07 | 0.000660128 |
| T2DM | endometrium | *FBXO22* | 1.883115614 | 1.90E-07 | 5.43E-07 | 0.000920235 |
| T2DM | endometrium | *TBC1D2B* | 3.478082243 | 0.003051364 | 0.003766273 | 1 |
| T2DM | endometrium | *LOXHD1* | 2.05124132 | 8.45E-08 | 4.38E-07 | 0.000409986 |
| T2DM | endometrium | *KATNAL2* | 2.117623793 | 2.90E-07 | 7.34E-07 | 0.001407601 |
| T2DM | endometrium | *ZNF91* | 3.552328309 | 2.13E-08 | 4.34E-07 | 0.000103246 |
| T2DM | endometrium | *POP5* | 2.220303224 | 3.53E-07 | 8.53E-07 | 0.001713413 |
| T2DM | endometrium | *MYO5B* | 1.970946213 | 2.44E-07 | 6.48E-07 | 0.00118265 |
| T2DM | endometrium | *ZNF23* | 6.802434813 | 3.49E-07 | 8.45E-07 | 0.001694397 |
| T2DM | endometrium | *ZNF229* | -1.940574839 | 0.01560794 | 0.018256925 | 1 |
| T2DM | endometrium | *SMG8* | 5.369594749 | 3.36E-05 | 5.02E-05 | 0.162837785 |
| T2DM | endometrium | *RAB8A* | 5.486815336 | 4.56E-08 | 4.34E-07 | 0.00022117 |
| T2DM | endometrium | *JSRP1* | 4.347848086 | 0.00828293 | 0.009898714 | 1 |
| T2DM | endometrium | *SPATA33* | 5.761638854 | 4.87E-08 | 4.34E-07 | 0.000236051 |
| T2DM | endometrium | *PROCA1* | -2.498505965 | 0.000462473 | 0.000613595 | 1 |
| T2DM | endometrium | *RPL13* | 3.752471826 | 1.78E-07 | 5.19E-07 | 0.000861382 |
| T2DM | endometrium | *TP53I13* | 2.379702996 | 6.70E-08 | 4.34E-07 | 0.000324883 |
| T2DM | endometrium | *TUBA1C* | 4.977192324 | 9.22E-09 | 4.34E-07 | 4.47E-05 |
| T2DM | endometrium | *ZNF528* | -2.676621968 | 0.030281368 | 0.034717674 | 1 |
| T2DM | endometrium | *TMC4* | 5.363203195 | 3.07E-07 | 7.66E-07 | 0.001490534 |
| T2DM | endometrium | *TTYH1* | 3.964114379 | 1.34E-07 | 4.78E-07 | 0.000649172 |
| T2DM | endometrium | *LENG8* | 4.6876722 | 1.22E-07 | 4.69E-07 | 0.00059421 |
| T2DM | endometrium | *CDC42EP5* | -1.928554967 | 7.24E-06 | 1.20E-05 | 0.035134792 |
| T2DM | endometrium | *C19orf33* | 4.133137654 | 4.70E-06 | 8.12E-06 | 0.022815692 |
| T2DM | endometrium | *DNAAF3* | 2.409356888 | 0.002996587 | 0.003702429 | 1 |
| T2DM | endometrium | *FAM57A* | 3.226487744 | 9.19E-06 | 1.49E-05 | 0.044601479 |
| T2DM | endometrium | *KIFC2* | -4.577110742 | 0.001717014 | 0.002163883 | 1 |
| T2DM | endometrium | *SERPINF2* | 3.769134463 | 6.29E-06 | 1.05E-05 | 0.030509271 |
| T2DM | endometrium | *SRR* | 2.310792777 | 0.000111561 | 0.000156308 | 0.541294773 |
| T2DM | endometrium | *TRPV3* | 3.3700651 | 4.20E-06 | 7.32E-06 | 0.020384251 |
| T2DM | endometrium | *CYB5D2* | 4.116038256 | 7.95E-09 | 4.34E-07 | 3.86E-05 |
| T2DM | endometrium | *C19orf48* | 2.158359228 | 1.38E-05 | 2.18E-05 | 0.067161385 |
| T2DM | endometrium | *KLK5* | -2.452252547 | 5.38E-06 | 9.15E-06 | 0.026086233 |
| T2DM | endometrium | *KLK6* | 23.41678294 | 1.57E-07 | 4.83E-07 | 0.000760279 |
| T2DM | endometrium | *ZNF83* | 3.683033268 | 7.46E-08 | 4.36E-07 | 0.000361851 |
| T2DM | endometrium | *SPRYD3* | 3.330028764 | 2.54E-07 | 6.68E-07 | 0.001234191 |

| | | | | | | |
|---|---|---|---|---|---|---|
| T2DM | endometrium | *SOAT2* | -0.534673489 | 6.10E-07 | 1.33E-06 | 0.00295791 |
| T2DM | endometrium | *ZNF558* | 5.723256254 | 3.04E-07 | 7.60E-07 | 0.001475196 |
| T2DM | endometrium | *NDUFV1* | 6.496755668 | 1.00E-06 | 2.04E-06 | 0.004870421 |
| T2DM | endometrium | *NUDT8* | -2.668742176 | 6.44E-06 | 1.08E-05 | 0.031263207 |
| T2DM | endometrium | *PRDX2* | 4.378284251 | 5.42E-08 | 4.34E-07 | 0.000263078 |
| T2DM | endometrium | *MIS12* | 3.518372436 | 6.44E-08 | 4.34E-07 | 0.000312376 |
| T2DM | endometrium | *MGAT5B* | -1.089950972 | 2.18E-06 | 4.07E-06 | 0.01059837 |
| T2DM | endometrium | *GSDMA* | 2.155815148 | 0.00856617 | 0.010214563 | 1 |
| T2DM | endometrium | *TMEM99* | 2.71673301 | 5.01E-06 | 8.59E-06 | 0.024326618 |
| T2DM | endometrium | *ZNF598* | -4.093051309 | 0.010730383 | 0.012717102 | 1 |
| T2DM | endometrium | *MLST8* | 6.536319987 | 2.80E-08 | 4.34E-07 | 0.00013592 |
| T2DM | endometrium | *ECI1* | 4.222499339 | 0.00034565 | 0.000464698 | 1 |
| T2DM | endometrium | *ATG16L2* | 2.357262625 | 1.06E-07 | 4.48E-07 | 0.000512882 |
| T2DM | endometrium | *RPSA* | 3.87711854 | 4.55E-08 | 4.34E-07 | 0.000220595 |
| T2DM | endometrium | *LTBP3* | 2.057092176 | 5.10E-08 | 4.34E-07 | 0.000247601 |
| T2DM | endometrium | *NAALADL1* | -2.769598932 | 0.000509924 | 0.000673788 | 1 |
| T2DM | endometrium | *KIAA1586* | 3.222174733 | 1.24E-06 | 2.44E-06 | 0.005993982 |
| T2DM | endometrium | *FAM83B* | -7.189064392 | 2.16E-06 | 4.03E-06 | 0.010490016 |
| T2DM | endometrium | *POLR2J3* | 2.703152595 | 1.44E-07 | 4.78E-07 | 0.00069818 |
| T2DM | endometrium | *COA6* | 3.124855162 | 1.42E-07 | 4.78E-07 | 0.000687559 |
| T2DM | endometrium | *PDHB* | 5.202295785 | 1.70E-05 | 2.63E-05 | 0.082437304 |
| T2DM | endometrium | *PXK* | 2.651852718 | 3.43E-08 | 4.34E-07 | 0.000166469 |
| T2DM | endometrium | *PCMTD1* | 2.764474685 | 5.36E-08 | 4.34E-07 | 0.000260132 |
| T2DM | endometrium | *ACOX2* | 2.366828104 | 3.41E-07 | 8.30E-07 | 0.001653577 |
| T2DM | endometrium | *SCN11A* | 2.600351039 | 1.50E-06 | 2.91E-06 | 0.007288376 |
| T2DM | endometrium | *SEPT2* | 2.345274377 | 5.06E-06 | 8.67E-06 | 0.024559484 |
| T2DM | endometrium | *DTYMK* | -3.947147891 | 0.000648081 | 0.000849632 | 1 |
| T2DM | endometrium | *ING5* | 3.664833009 | 4.55E-08 | 4.34E-07 | 0.000220726 |
| T2DM | endometrium | *MLKL* | 3.508701279 | 0.000417566 | 0.000556143 | 1 |
| T2DM | endometrium | *KLHL30* | 11.16967873 | 2.80E-07 | 7.16E-07 | 0.001359184 |
| T2DM | endometrium | *CCDC110* | 7.983357823 | 8.76E-08 | 4.38E-07 | 0.000425199 |
| T2DM | endometrium | *FNTA* | 2.905110738 | 9.66E-08 | 4.42E-07 | 0.000468889 |
| T2DM | endometrium | *SERINC2* | 6.178978089 | 0.001089428 | 0.001397278 | 1 |
| T2DM | endometrium | *SLC20A2* | 3.796437632 | 1.50E-07 | 4.78E-07 | 0.000728943 |
| T2DM | endometrium | *TMUB2* | 2.125442447 | 1.72E-07 | 5.09E-07 | 0.000834611 |
| T2DM | endometrium | *ZSWIM1* | 2.726083453 | 3.99E-07 | 9.39E-07 | 0.001937089 |
| T2DM | endometrium | *NBPF9* | 5.905869135 | 6.77E-08 | 4.34E-07 | 0.000328505 |
| T2DM | endometrium | *ADAM9* | 2.647667537 | 1.80E-08 | 4.34E-07 | 8.75E-05 |
| T2DM | endometrium | *SLC16A4* | 3.1203757 | 1.90E-06 | 3.59E-06 | 0.009232444 |
| T2DM | endometrium | *DNAJC21* | 5.029278879 | 5.28E-08 | 4.34E-07 | 0.000256326 |
| T2DM | endometrium | *NPNT* | 1.9846677 | 6.24E-08 | 4.34E-07 | 0.000302777 |
| T2DM | endometrium | *GSTM4* | 3.15037245 | 3.55E-07 | 8.56E-07 | 0.001722747 |
| T2DM | endometrium | *TCTN2* | 1.539590834 | 0.000680654 | 0.00089017 | 1 |
| T2DM | endometrium | *PPIP5K1* | 2.422388802 | 3.43E-06 | 6.07E-06 | 0.016638801 |
| T2DM | endometrium | *ADAL* | 2.475556609 | 3.65E-05 | 5.44E-05 | 0.176947125 |
| T2DM | endometrium | *SNTB2* | 4.12326679 | 6.17E-08 | 4.34E-07 | 0.000299569 |
| T2DM | endometrium | *ZNF507* | 4.070677296 | 5.70E-07 | 1.26E-06 | 0.002767889 |
| T2DM | endometrium | *ZBTB49* | 5.401577759 | 4.61E-08 | 4.34E-07 | 0.000223637 |
| T2DM | endometrium | *GFM1* | 3.945965591 | 1.91E-05 | 2.94E-05 | 0.092622988 |
| T2DM | endometrium | *ATOH8* | 2.481787334 | 1.68E-07 | 5.02E-07 | 0.000814414 |

| | | | | | | |
|---|---|---|---|---|---|---|
| T2DM | endometrium | *BTNL3* | 2.728355344 | 0.002383996 | 0.002971269 | 1 |
| T2DM | endometrium | *TMEM129* | -2.298890875 | 1.21E-05 | 1.92E-05 | 0.058606485 |
| T2DM | endometrium | *CEP120* | 3.203478074 | 1.07E-07 | 4.49E-07 | 0.00051782 |
| T2DM | endometrium | *MFF* | 2.480056654 | 7.18E-08 | 4.34E-07 | 0.000348384 |
| T2DM | endometrium | *LGALS9* | 2.667621066 | 1.41E-05 | 2.22E-05 | 0.068546472 |
| T2DM | endometrium | *CPLX1* | 2.67816375 | 2.65E-07 | 6.86E-07 | 0.001287188 |
| T2DM | endometrium | *SIGLEC7* | 2.726526243 | 0.002412408 | 0.003005136 | 1 |
| T2DM | endometrium | *MAP2K1* | 2.936978914 | 6.08E-08 | 4.34E-07 | 0.00029482 |
| T2DM | endometrium | *KLK7* | 4.486022771 | 9.42E-08 | 4.39E-07 | 0.00045688 |
| T2DM | endometrium | *IRS1* | -1.083954267 | 0.044005912 | 0.049840496 | 1 |
| T2DM | endometrium | *UPF3A* | 2.918149127 | 3.33E-07 | 8.15E-07 | 0.001613602 |
| T2DM | endometrium | *C8orf46* | 1.811730593 | 3.53E-07 | 8.52E-07 | 0.001711689 |
| T2DM | endometrium | *HSPBAP1* | 1.824096504 | 2.51E-07 | 6.60E-07 | 0.001216034 |
| T2DM | endometrium | *UBE2V2* | 5.563471784 | 3.69E-08 | 4.34E-07 | 0.000178959 |
| T2DM | endometrium | *CPT1C* | 2.862394006 | 0.000256071 | 0.000347541 | 1 |
| T2DM | endometrium | *PCSK9* | 21.62577515 | 8.94E-08 | 4.38E-07 | 0.000433695 |
| T2DM | endometrium | *XPO6* | 5.698471475 | 1.11E-06 | 2.22E-06 | 0.005372073 |
| T2DM | endometrium | *RGS14* | -1.05769086 | 7.47E-06 | 1.23E-05 | 0.036241615 |
| T2DM | endometrium | *RAB24* | 6.670578779 | 1.56E-07 | 4.81E-07 | 0.000755735 |
| T2DM | endometrium | *THBS3* | 7.565199951 | 1.90E-06 | 3.59E-06 | 0.009240649 |
| T2DM | endometrium | *NPIPB3* | 4.522420434 | 8.38E-05 | 0.000118973 | 0.406530997 |
| T2DM | endometrium | *SH3TC2* | 2.757770257 | 9.59E-07 | 1.96E-06 | 0.004652673 |
| T2DM | endometrium | *GPRIN1* | 7.803576616 | 0.009916017 | 0.011780733 | 1 |
| T2DM | endometrium | *PGM2* | 2.512678712 | 3.48E-07 | 8.43E-07 | 0.001689811 |
| T2DM | endometrium | *P2RY12* | 3.265907717 | 1.17E-06 | 2.33E-06 | 0.00568348 |
| T2DM | endometrium | *SNUPN* | 4.508290486 | 0.000147897 | 0.000205027 | 0.717595153 |
| T2DM | endometrium | *RNASE6* | -1.75835669 | 4.27E-06 | 7.43E-06 | 0.020717316 |
| T2DM | endometrium | *CD52* | 1.639859421 | 6.04E-05 | 8.72E-05 | 0.2929133 |
| T2DM | endometrium | *SLC38A11* | -1.446536765 | 1.41E-05 | 2.21E-05 | 0.068183677 |
| T2DM | endometrium | *CCDC8* | 3.631670175 | 7.32E-06 | 1.21E-05 | 0.035532536 |
| T2DM | endometrium | *ZEB2* | 2.378841888 | 7.39E-07 | 1.58E-06 | 0.003585899 |
| T2DM | endometrium | *HINT1* | 5.878684033 | 3.36E-07 | 8.20E-07 | 0.001628197 |
| T2DM | endometrium | *DTWD2* | 3.569973225 | 1.03E-06 | 2.09E-06 | 0.005002598 |
| T2DM | endometrium | *CLIC3* | -4.763489686 | 0.001613987 | 0.002039872 | 1 |
| T2DM | endometrium | *DFFB* | -2.771870856 | 0.000213398 | 0.000292076 | 1 |
| T2DM | endometrium | *NFU1* | 5.130013752 | 4.61E-05 | 6.77E-05 | 0.223743058 |
| T2DM | endometrium | *C15orf40* | 3.296460869 | 0.015066898 | 0.017632559 | 1 |
| T2DM | endometrium | *RGPD8* | 1.177464436 | 0.001202053 | 0.001537259 | 1 |
| T2DM | endometrium | *LUZP1* | 3.105523847 | 5.62E-08 | 4.34E-07 | 0.000272701 |
| T2DM | endometrium | *LRRC45* | 2.015312767 | 0.007024918 | 0.008451501 | 1 |
| T2DM | endometrium | *CHRNA5* | 7.521524442 | 2.00E-06 | 3.77E-06 | 0.009726672 |
| T2DM | endometrium | *UGP2* | 4.084934586 | 8.35E-06 | 1.36E-05 | 0.040506655 |
| T2DM | endometrium | *HNRNPF* | 5.885729605 | 8.50E-07 | 1.77E-06 | 0.00412498 |
| T2DM | endometrium | *CSGALNACT2* | 5.089236618 | 2.83E-08 | 4.34E-07 | 0.000137398 |
| T2DM | endometrium | *CTNND2* | 2.401955663 | 3.10E-06 | 5.55E-06 | 0.015038898 |
| T2DM | endometrium | *CALML6* | 54.48224637 | 4.81E-07 | 1.09E-06 | 0.002331628 |
| T2DM | endometrium | *MUC3A* | -2.140621678 | 0.000957165 | 0.001233837 | 1 |
| T2DM | endometrium | *TPST1* | 3.491306338 | 1.21E-06 | 2.40E-06 | 0.005877815 |
| T2DM | endometrium | *TOR1AIP2* | 4.014232117 | 4.38E-08 | 4.34E-07 | 0.000212583 |
| T2DM | endometrium | *ZFPM2* | 2.285736044 | 7.98E-07 | 1.67E-06 | 0.0038725 |

| | | | | | | |
|---|---|---|---|---|---|---|
| T2DM | endometrium | *MAP3K2* | 2.765934327 | 1.47E-07 | 4.78E-07 | 0.000714905 |
| T2DM | endometrium | *MYO7B* | 4.267565006 | 2.96E-07 | 7.45E-07 | 0.001438148 |
| T2DM | endometrium | *UBE2E3* | 5.052742568 | 5.36E-05 | 7.80E-05 | 0.260237469 |
| T2DM | endometrium | *NIPA1* | 2.478418095 | 1.93E-07 | 5.47E-07 | 0.00093423 |
| T2DM | endometrium | *HNRNPA3* | 4.438803647 | 5.37E-08 | 4.34E-07 | 0.000260546 |
| T2DM | endometrium | *RNF150* | 3.64334573 | 0.000898229 | 0.001161259 | 1 |
| T2DM | endometrium | *CHRNB1* | 2.051840152 | 5.46E-05 | 7.92E-05 | 0.264692451 |
| T2DM | endometrium | *USP38* | 6.590111946 | 6.71E-08 | 4.34E-07 | 0.000325773 |
| T2DM | endometrium | *ANKK1* | 3.838177714 | 0.018061417 | 0.021025431 | 1 |
| T2DM | endometrium | *PDCD6IP* | 3.325169273 | 5.02E-08 | 4.34E-07 | 0.000243792 |
| T2DM | endometrium | *CRTAP* | 5.519752958 | 8.59E-08 | 4.38E-07 | 0.000416894 |
| T2DM | endometrium | *CNGB3* | 2.040496308 | 1.48E-07 | 4.78E-07 | 0.000719331 |
| T2DM | endometrium | *STX8* | 2.38333959 | 6.35E-08 | 4.34E-07 | 0.000307889 |
| T2DM | endometrium | *NFRKB* | 4.889465316 | 4.76E-08 | 4.34E-07 | 0.000230927 |
| T2DM | endometrium | *SEMA3E* | -2.1699979 | 0.028723617 | 0.032986269 | 1 |
| T2DM | endometrium | *MGMT* | 3.74754421 | 7.64E-08 | 4.36E-07 | 0.00037072 |
| T2DM | endometrium | *METTL7B* | 5.224694193 | 0.008294498 | 0.009910098 | 1 |
| T2DM | endometrium | *NFXL1* | 6.052555671 | 5.36E-06 | 9.12E-06 | 0.026011973 |
| T2DM | endometrium | *DNAJC18* | 4.041482795 | 0.002826849 | 0.003501627 | 1 |
| T2DM | endometrium | *HSD17B13* | 6.70417963 | 1.02E-07 | 4.47E-07 | 0.00049266 |
| T2DM | endometrium | *ELOVL6* | 5.066172908 | 6.10E-08 | 4.34E-07 | 0.000295856 |
| T2DM | endometrium | *SERPINB9* | 2.01372291 | 1.98E-07 | 5.56E-07 | 0.000959748 |
| T2DM | endometrium | *SMAGP* | -1.241018933 | 0.039705218 | 0.04511703 | 1 |
| T2DM | endometrium | *CDH2* | -0.966286669 | 2.58E-06 | 4.72E-06 | 0.012534415 |
| T2DM | endometrium | *EMB* | 1.459883797 | 1.34E-05 | 2.12E-05 | 0.065218006 |
| T2DM | endometrium | *STAT2* | 5.280044678 | 3.38E-08 | 4.34E-07 | 0.000163861 |
| T2DM | endometrium | *SLC26A5* | 7.505602744 | 6.18E-06 | 1.04E-05 | 0.030006742 |
| T2DM | endometrium | *GTSF1* | 8.707259113 | 1.41E-05 | 2.21E-05 | 0.068239956 |
| T2DM | endometrium | *ARMC10* | 5.789831151 | 1.39E-06 | 2.70E-06 | 0.006723929 |
| T2DM | endometrium | *ACYP2* | 3.784029232 | 2.00E-07 | 5.61E-07 | 0.000971902 |
| T2DM | endometrium | *TRABD* | 2.65106321 | 6.60E-08 | 4.34E-07 | 0.00032 |
| T2DM | endometrium | *TMEM133* | 4.315183496 | 0.001618973 | 0.00204564 | 1 |
| T2DM | endometrium | *HOXB9* | 10.93900826 | 4.20E-05 | 6.20E-05 | 0.203829518 |
| T2DM | endometrium | *KCNS3* | -1.535349567 | 0.001690767 | 0.002132467 | 1 |
| T2DM | endometrium | *FOXN2* | 2.262816073 | 8.08E-08 | 4.36E-07 | 0.000392124 |
| T2DM | endometrium | *TRIAP1* | 5.087358366 | 1.41E-08 | 4.34E-07 | 6.83E-05 |
| T2DM | endometrium | *TSEN34* | 3.775269267 | 4.14E-07 | 9.68E-07 | 0.002010964 |
| T2DM | endometrium | *MSANTD4* | 2.577649039 | 1.82E-07 | 5.25E-07 | 0.000881926 |
| T2DM | endometrium | *NDUFA3* | 8.457258616 | 7.87E-07 | 1.66E-06 | 0.003820094 |
| T2DM | endometrium | *NUDT6* | 6.578270472 | 8.67E-09 | 4.34E-07 | 4.20E-05 |
| T2DM | endometrium | *ZNF160* | 1.971952142 | 1.36E-07 | 4.78E-07 | 0.000659581 |
| T2DM | endometrium | *ZNF415* | 7.485135793 | 1.64E-06 | 3.14E-06 | 0.007936676 |
| T2DM | endometrium | *PYGO1* | 3.006884392 | 1.28E-07 | 4.78E-07 | 0.000621469 |
| T2DM | endometrium | *TSNARE1* | 2.383380953 | 2.55E-08 | 4.34E-07 | 0.000123544 |
| T2DM | endometrium | *FPR1* | 2.668423013 | 1.46E-07 | 4.78E-07 | 0.000710391 |
| T2DM | endometrium | *FEZ2* | 3.558349826 | 9.97E-08 | 4.44E-07 | 0.000483655 |
| T2DM | endometrium | *SOX7* | -4.294460551 | 0.000419864 | 0.000558897 | 1 |
| T2DM | endometrium | *TRMT61B* | 5.497335536 | 1.38E-07 | 4.78E-07 | 0.00067161 |
| T2DM | endometrium | *MFN1* | 5.512654833 | 3.01E-05 | 4.53E-05 | 0.146185332 |
| T2DM | endometrium | *GIMAP8* | 3.278981015 | 3.85E-07 | 9.12E-07 | 0.001866771 |

| | | | | | | |
|---|---|---|---|---|---|---|
| T2DM | endometrium | *ATP6V0E2* | -2.379681637 | 5.00E-05 | 7.31E-05 | 0.242758381 |
| T2DM | endometrium | *MORN4* | 2.343300286 | 0.000163666 | 0.000226113 | 0.794108786 |
| T2DM | endometrium | *SCAND1* | 4.195114394 | 1.52E-07 | 4.78E-07 | 0.00073851 |
| T2DM | endometrium | *SOSTDC1* | 7.227306426 | 2.33E-06 | 4.32E-06 | 0.011327896 |
| T2DM | endometrium | *GAA* | 1.562997755 | 0.006259428 | 0.007562436 | 1 |
| T2DM | endometrium | *PGAM1* | 3.372845541 | 1.54E-06 | 2.97E-06 | 0.007479718 |
| T2DM | endometrium | *LURAP1* | -2.140249858 | 1.95E-05 | 2.99E-05 | 0.0944104 |
| T2DM | endometrium | *PDE7B* | 3.711734957 | 5.16E-08 | 4.34E-07 | 0.000250147 |
| T2DM | endometrium | *MCC* | 2.099247036 | 8.15E-08 | 4.36E-07 | 0.000395497 |
| T2DM | endometrium | *ZNF561* | -2.474344996 | 0.017924692 | 0.020871276 | 1 |
| T2DM | endometrium | *HOPX* | 4.204047132 | 2.19E-06 | 4.08E-06 | 0.010635905 |
| T2DM | endometrium | *PPID* | 2.572388401 | 9.71E-08 | 4.42E-07 | 0.000471333 |
| T2DM | endometrium | *ETFDH* | 2.1823645 | 0.000264833 | 0.000359231 | 1 |
| T2DM | endometrium | *TBCA* | 5.340710206 | 2.31E-06 | 4.29E-06 | 0.011230714 |
| T2DM | endometrium | *NMUR1* | 4.29874451 | 1.33E-07 | 4.78E-07 | 0.00064473 |
| T2DM | endometrium | *PIK3CD* | 3.007434637 | 3.66E-08 | 4.34E-07 | 0.000177598 |
| T2DM | endometrium | *ANO5* | 2.386259562 | 2.54E-07 | 6.68E-07 | 0.001234718 |
| T2DM | endometrium | *VAT1L* | 3.265429975 | 1.62E-07 | 4.91E-07 | 0.000786474 |
| T2DM | endometrium | *CAMTA1* | 3.795281222 | 8.65E-08 | 4.38E-07 | 0.000419581 |
| T2DM | endometrium | *LGALS4* | 2.88027049 | 0.002952468 | 0.003650707 | 1 |
| T2DM | endometrium | *LRRC34* | 2.191992983 | 1.25E-05 | 1.98E-05 | 0.060580853 |
| T2DM | endometrium | *PAH* | 14.78784046 | 1.26E-06 | 2.49E-06 | 0.006127547 |
| T2DM | endometrium | *SPATA5L1* | 2.349600182 | 0.000194711 | 0.000267328 | 0.944735628 |
| T2DM | endometrium | *NXNL1* | -1.482828279 | 0.000282802 | 0.000382536 | 1 |
| T2DM | endometrium | *RHNO1* | 5.323151473 | 4.49E-05 | 6.60E-05 | 0.217730183 |
| T2DM | endometrium | *PWWP2B* | -2.611986897 | 6.29E-06 | 1.05E-05 | 0.030512808 |
| T2DM | endometrium | *EXOSC10* | 4.935044808 | 2.37E-06 | 4.37E-06 | 0.011493838 |
| T2DM | endometrium | *TRAPPC12* | 2.844651555 | 6.55E-08 | 4.34E-07 | 0.000317853 |
| T2DM | endometrium | *C3AR1* | 1.757070291 | 8.32E-06 | 1.36E-05 | 0.040349637 |
| T2DM | endometrium | *PRNP* | 3.711895647 | 3.10E-07 | 7.72E-07 | 0.001505228 |
| T2DM | endometrium | *TVP23B* | -2.474782394 | 0.010978053 | 0.013001102 | 1 |
| T2DM | endometrium | *FBXW10* | -1.23464563 | 2.11E-06 | 3.95E-06 | 0.010238875 |
| T2DM | endometrium | *SRGAP2C* | 1.823845684 | 3.43E-07 | 8.34E-07 | 0.001665692 |
| T2DM | endometrium | *ATPAF2* | 7.505792243 | 9.20E-08 | 4.38E-07 | 0.000446291 |
| T2DM | endometrium | *ZNF554* | -2.677820042 | 1.58E-05 | 2.46E-05 | 0.076633822 |
| T2DM | endometrium | *ANKRD20A4* | 14.19204489 | 5.40E-08 | 4.34E-07 | 0.000261969 |
| T2DM | endometrium | *EIF2AK3* | 3.580342258 | 1.33E-07 | 4.78E-07 | 0.000643812 |
| T2DM | endometrium | *NME6* | 3.817884413 | 2.36E-08 | 4.34E-07 | 0.000114307 |
| T2DM | endometrium | *CD8B* | 1.826450615 | 0.032555364 | 0.037236828 | 1 |
| T2DM | endometrium | *SLFN12* | 2.191608745 | 3.82E-07 | 9.07E-07 | 0.001851409 |
| T2DM | endometrium | *MTBP* | -3.024934248 | 0.000614973 | 0.000807318 | 1 |
| T2DM | endometrium | *TEFM* | 3.702729526 | 0.01031894 | 0.012247431 | 1 |
| T2DM | endometrium | *MALT1* | 5.704411371 | 2.34E-07 | 6.28E-07 | 0.001137521 |
| T2DM | endometrium | *MBOAT1* | 6.219704671 | 3.02E-06 | 5.43E-06 | 0.014646883 |
| T2DM | endometrium | *CXCR6* | 12.69574131 | 3.15E-05 | 4.73E-05 | 0.153020877 |
| T2DM | endometrium | *PAIP1* | 4.992911981 | 1.12E-07 | 4.57E-07 | 0.000542678 |
| T2DM | endometrium | *MACROD2* | 2.41498068 | 8.16E-08 | 4.36E-07 | 0.000395733 |
| T2DM | endometrium | *TP53RK* | 9.852110802 | 0.001053423 | 0.001353604 | 1 |
| T2DM | endometrium | *CLEC12A* | 0.488546135 | 0.020007319 | 0.02319606 | 1 |
| T2DM | endometrium | *SUCLG2* | 4.509062101 | 0.000207362 | 0.000284215 | 1 |

| | | | | | | |
|---|---|---|---|---|---|---|
| T2DM | endometrium | *ARNT2* | 2.96300042 | 1.42E-05 | 2.23E-05 | 0.068979703 |
| T2DM | endometrium | *MANEA* | 2.29247606 | 1.28E-07 | 4.78E-07 | 0.000619981 |
| T2DM | endometrium | *AFF1* | 3.574820353 | 8.27E-08 | 4.37E-07 | 0.000401277 |
| T2DM | endometrium | *FIBP* | 3.941067559 | 2.72E-08 | 4.34E-07 | 0.000132042 |
| T2DM | endometrium | *SNTG2* | -2.031539635 | 0.000369891 | 0.000495365 | 1 |
| T2DM | endometrium | *SMPDL3A* | 1.925662698 | 2.00E-05 | 3.07E-05 | 0.09709754 |
| T2DM | endometrium | *RAD9A* | 3.000178727 | 0.001649259 | 0.00208228 | 1 |
| T2DM | endometrium | *EFEMP2* | 2.561305598 | 9.39E-08 | 4.39E-07 | 0.000455388 |
| T2DM | endometrium | *ZFAND4* | 4.380489115 | 0.002045288 | 0.002567591 | 1 |
| T2DM | endometrium | *SLFN11* | 2.117113433 | 1.36E-07 | 4.78E-07 | 0.000660528 |
| T2DM | endometrium | *FAM71D* | 3.781091805 | 1.32E-07 | 4.78E-07 | 0.000638401 |
| T2DM | endometrium | *FUT10* | -1.817739727 | 0.002073375 | 0.002600831 | 1 |
| T2DM | endometrium | *ZNF596* | 3.390337849 | 8.38E-08 | 4.38E-07 | 0.000406447 |
| T2DM | endometrium | *CFL1* | 6.884214009 | 2.79E-06 | 5.05E-06 | 0.013531297 |
| T2DM | endometrium | *FAM192A* | 3.53777282 | 0.014482093 | 0.016976834 | 1 |
| T2DM | endometrium | *RAB43* | -8.265698469 | 1.87E-06 | 3.53E-06 | 0.009068324 |
| T2DM | endometrium | *CBWD1* | 6.293360703 | 4.26E-08 | 4.34E-07 | 0.00020651 |
| T2DM | endometrium | *SNX32* | 2.714943251 | 5.42E-08 | 4.34E-07 | 0.000262781 |
| T2DM | endometrium | *ZNF621* | 6.692410303 | 1.70E-07 | 5.06E-07 | 0.000826022 |
| T2DM | endometrium | *NADSYN1* | -2.332053773 | 0.000239692 | 0.000326498 | 1 |
| T2DM | endometrium | *RNASEH2C* | -3.441474443 | 2.64E-06 | 4.81E-06 | 0.012820455 |
| T2DM | endometrium | *MYEOV* | -11.40591758 | 4.57E-05 | 6.71E-05 | 0.221615191 |
| T2DM | endometrium | *ANKRD13D* | 1.895842117 | 1.42E-07 | 4.78E-07 | 0.00068679 |
| T2DM | endometrium | *MRGPRF* | 2.343936291 | 5.04E-06 | 8.63E-06 | 0.024441158 |
| T2DM | endometrium | *LCLAT1* | -2.487491122 | 0.001785691 | 0.002247516 | 1 |
| T2DM | endometrium | *SH3RF3* | 2.186831076 | 2.09E-07 | 5.79E-07 | 0.001015666 |
| T2DM | endometrium | *GXYLT2* | 5.338247567 | 8.44E-07 | 1.76E-06 | 0.004097099 |
| T2DM | endometrium | *DCAKD* | 3.082566196 | 1.60E-07 | 4.87E-07 | 0.000775674 |
| T2DM | endometrium | *TRMT112* | -2.514674443 | 9.38E-05 | 0.00013237 | 0.455352683 |
| T2DM | endometrium | *NOC3L* | -2.009284531 | 4.96E-05 | 7.25E-05 | 0.240585659 |
| T2DM | endometrium | *RHOD* | -3.015939391 | 6.77E-05 | 9.71E-05 | 0.328325269 |
| T2DM | endometrium | *PARP14* | 1.798120065 | 8.98E-06 | 1.46E-05 | 0.04355711 |
| T2DM | endometrium | *PARP15* | 1.863267717 | 1.15E-05 | 1.84E-05 | 0.055668502 |
| T2DM | endometrium | *AHSA2* | 2.874216272 | 4.94E-06 | 8.47E-06 | 0.02395417 |
| T2DM | endometrium | *IQCB1* | 3.613682248 | 2.68E-07 | 6.91E-07 | 0.001300218 |
| T2DM | endometrium | *ZNF483* | 4.851697125 | 2.99E-06 | 5.39E-06 | 0.014521683 |
| T2DM | endometrium | *SLC2A14* | 2.26619099 | 0.030321144 | 0.034746856 | 1 |
| T2DM | endometrium | *MZT2A* | 3.096236448 | 5.81E-08 | 4.34E-07 | 0.000281886 |
| T2DM | endometrium | *MAP3K11* | 5.448551077 | 1.52E-06 | 2.94E-06 | 0.007389151 |
| T2DM | endometrium | *CCDC36* | 1.831959249 | 3.38E-07 | 8.25E-07 | 0.001640471 |
| T2DM | endometrium | *EHBP1L1* | 2.55006322 | 3.78E-09 | 4.07E-07 | 1.83E-05 |
| T2DM | endometrium | *PPP1R14B* | 3.737265582 | 1.76E-07 | 5.17E-07 | 0.000855527 |
| T2DM | endometrium | *ZNF417* | 4.373126751 | 4.20E-07 | 9.77E-07 | 0.002037604 |
| T2DM | endometrium | *PEAK1* | 2.657887959 | 8.80E-08 | 4.38E-07 | 0.000426803 |
| T2DM | endometrium | *TNFRSF10D* | 5.720466579 | 7.26E-07 | 1.55E-06 | 0.003523344 |
| T2DM | endometrium | *GMPPB* | 4.769371708 | 3.60E-07 | 8.66E-07 | 0.001745343 |
| T2DM | endometrium | *NUDT18* | 7.54191936 | 1.47E-07 | 4.78E-07 | 0.000711315 |
| T2DM | endometrium | *SULT1B1* | 13.91785016 | 2.05E-07 | 5.71E-07 | 0.000996683 |
| T2DM | endometrium | *NUDT4* | -2.457035678 | 0.0094067 | 0.011183854 | 1 |
| T2DM | endometrium | *SCAI* | 3.744060852 | 7.06E-08 | 4.34E-07 | 0.000342471 |

| | | | | | | |
|---|---|---|---|---|---|---|
| T2DM | endometrium | *HSPB7* | 2.849678233 | 5.08E-06 | 8.69E-06 | 0.024652166 |
| T2DM | endometrium | *UQCRH* | 6.211942559 | 4.57E-06 | 7.90E-06 | 0.022186452 |
| T2DM | endometrium | *C11orf80* | 1.758332834 | 4.40E-07 | 1.01E-06 | 0.002135917 |
| T2DM | endometrium | *CD7* | 4.812667037 | 8.03E-08 | 4.36E-07 | 0.000389663 |
| T2DM | endometrium | *RNF213* | 3.409459259 | 2.71E-07 | 6.97E-07 | 0.001316203 |
| T2DM | endometrium | *TUBB8* | 4.305175488 | 7.41E-06 | 1.22E-05 | 0.035971463 |
| T2DM | endometrium | *GPR160* | 3.441295953 | 1.21E-06 | 2.40E-06 | 0.005882436 |
| T2DM | endometrium | *GOLIM4* | 1.94741887 | 1.41E-07 | 4.78E-07 | 0.000684935 |
| T2DM | endometrium | *HOXB2* | -2.782393273 | 4.55E-06 | 7.88E-06 | 0.022096073 |
| T2DM | endometrium | *XXYLT1* | -2.373473599 | 0.021444437 | 0.024820708 | 1 |
| T2DM | endometrium | *UBXN2A* | 3.877356186 | 6.50E-08 | 4.34E-07 | 0.000315432 |
| T2DM | endometrium | *NRROS* | 2.309000454 | 7.76E-05 | 0.000110573 | 0.376500009 |
| T2DM | endometrium | *CEP19* | -2.26793178 | 0.018168665 | 0.021145206 | 1 |
| T2DM | endometrium | *SLC25A30* | 3.032627876 | 1.40E-07 | 4.78E-07 | 0.000677659 |
| T2DM | endometrium | *CTSF* | 4.550199319 | 2.85E-06 | 5.15E-06 | 0.013806353 |
| T2DM | endometrium | *LEMD3* | 4.111148994 | 5.40E-08 | 4.34E-07 | 0.000262122 |
| T2DM | endometrium | *CTU2* | 3.536608976 | 0.000157994 | 0.000218588 | 0.766587951 |
| T2DM | endometrium | *SNX31* | 1.944683591 | 0.002674302 | 0.003316053 | 1 |
| T2DM | endometrium | *GLIS1* | 2.601535896 | 1.51E-07 | 4.78E-07 | 0.000734365 |
| T2DM | endometrium | *RALGAPA1* | 2.962877035 | 1.22E-07 | 4.69E-07 | 0.000593892 |
| T2DM | endometrium | *SNAPC5* | 8.632569595 | 1.77E-06 | 3.36E-06 | 0.008572435 |
| T2DM | endometrium | *ANKRD36C* | 3.169466354 | 0.035909742 | 0.040919226 | 1 |
| T2DM | endometrium | *TMEM81* | 5.541558019 | 0.001900011 | 0.002389542 | 1 |
| T2DM | endometrium | *MSL2* | 4.791386614 | 6.34E-08 | 4.34E-07 | 0.000307585 |
| T2DM | endometrium | *ZNF497* | 20.07886124 | 2.80E-07 | 7.16E-07 | 0.001360551 |
| T2DM | endometrium | *TRAM1L1* | 7.329977849 | 0.000611632 | 0.00080315 | 1 |
| T2DM | endometrium | *ANGEL2* | 2.803188387 | 6.84E-08 | 4.34E-07 | 0.000331738 |
| T2DM | endometrium | *ZNF266* | 1.463570176 | 0.000101721 | 0.000142975 | 0.493549176 |
| T2DM | endometrium | *SH3PXD2B* | 2.63803814 | 8.33E-08 | 4.38E-07 | 0.000404349 |
| T2DM | endometrium | *KIAA1551* | 3.010804 | 5.25E-07 | 1.17E-06 | 0.002547976 |
| T2DM | endometrium | *LARP7* | 3.713733319 | 9.02E-08 | 4.38E-07 | 0.000437807 |
| T2DM | endometrium | *FGFBP3* | 5.052743052 | 1.37E-07 | 4.78E-07 | 0.000666612 |
| T2DM | endometrium | *C4orf32* | 5.310198891 | 2.96E-06 | 5.34E-06 | 0.014364247 |
| T2DM | endometrium | *HRAS* | 3.611404225 | 4.00E-07 | 9.41E-07 | 0.001943176 |
| T2DM | endometrium | *SRP72* | 4.063701305 | 0.000692868 | 0.000904438 | 1 |
| T2DM | endometrium | *NLRP6* | -10.05299536 | 2.16E-07 | 5.94E-07 | 0.001049041 |
| T2DM | endometrium | *RSRC1* | 4.38201857 | 2.82E-07 | 7.20E-07 | 0.001368843 |
| T2DM | endometrium | *PTDSS2* | 4.911037112 | 4.89E-05 | 7.16E-05 | 0.237097647 |
| T2DM | endometrium | *SEZ6L2* | 3.660918496 | 2.39E-09 | 3.52E-07 | 1.16E-05 |
| T2DM | endometrium | *KCTD13* | 2.28167701 | 7.01E-08 | 4.34E-07 | 0.0003399 |
| T2DM | endometrium | *DHX36* | 5.338065128 | 6.35E-07 | 1.38E-06 | 0.003081753 |
| T2DM | endometrium | *FBXW8* | 6.100323379 | 9.13E-06 | 1.49E-05 | 0.044322015 |
| T2DM | endometrium | *SLC22A1* | 4.723547315 | 8.30E-08 | 4.37E-07 | 0.000402846 |
| T2DM | endometrium | *CHST2* | 8.497933217 | 6.76E-06 | 1.13E-05 | 0.032778732 |
| T2DM | endometrium | *TVP23C* | 4.422153314 | 1.43E-07 | 4.78E-07 | 0.000694106 |
| T2DM | endometrium | *PACS1* | 3.488011319 | 5.44E-07 | 1.21E-06 | 0.002639058 |
| T2DM | endometrium | *SH3BP5L* | 3.222961301 | 1.54E-07 | 4.79E-07 | 0.000748422 |
| T2DM | endometrium | *CADM2* | 1.665447868 | 1.42E-07 | 4.78E-07 | 0.000691092 |
| T2DM | endometrium | *PSMD2* | 7.386227885 | 1.21E-05 | 1.92E-05 | 0.058588865 |
| T2DM | endometrium | *ARHGAP1* | 3.446486639 | 4.68E-07 | 1.06E-06 | 0.002272697 |

| | | | | | | |
|---|---|---|---|---|---|---|
| T2DM | endometrium | *ATG13* | 5.130558951 | 1.51E-07 | 4.78E-07 | 0.000733951 |
| T2DM | endometrium | *CCNE2* | -2.323042981 | 5.34E-08 | 4.34E-07 | 0.000259064 |
| T2DM | endometrium | *ZNF519* | 2.958277823 | 1.05E-07 | 4.48E-07 | 0.000511202 |
| T2DM | endometrium | *BANF1* | 4.894591931 | 1.80E-08 | 4.34E-07 | 8.74E-05 |
| T2DM | endometrium | *CHRNA7* | -1.927580591 | 5.71E-05 | 8.28E-05 | 0.277076372 |
| T2DM | endometrium | *TMEM9B* | 4.891298345 | 9.12E-08 | 4.38E-07 | 0.00044226 |
| T2DM | endometrium | *SCUBE2* | 2.796584684 | 1.44E-07 | 4.78E-07 | 0.000697589 |
| T2DM | endometrium | *EIF1AD* | 5.896879272 | 5.30E-07 | 1.18E-06 | 0.002571283 |
| T2DM | endometrium | *EIF3F* | 3.741988566 | 1.45E-06 | 2.81E-06 | 0.007028416 |
| T2DM | endometrium | *ZNF25* | 4.289216288 | 0.000881234 | 0.001139896 | 1 |
| T2DM | endometrium | *CLTB* | -2.609006902 | 0.029253939 | 0.033563517 | 1 |
| T2DM | endometrium | *PPP2R2D* | 1.849706078 | 8.58E-08 | 4.38E-07 | 0.000416403 |
| T2DM | endometrium | *MCTP1* | 1.407448901 | 1.45E-07 | 4.78E-07 | 0.000701126 |
| T2DM | endometrium | *LRRC25* | 2.934862087 | 2.02E-05 | 3.10E-05 | 0.098182267 |
| T2DM | endometrium | *UBQLNL* | -1.138085222 | 9.01E-07 | 1.86E-06 | 0.004373105 |
| T2DM | endometrium | *KCNE3* | 3.99565192 | 5.00E-07 | 1.13E-06 | 0.002427201 |
| T2DM | endometrium | *UCP2* | 1.970515755 | 3.06E-06 | 5.49E-06 | 0.014857074 |
| T2DM | endometrium | *C11orf68* | 3.967626701 | 1.00E-06 | 2.04E-06 | 0.004869058 |
| T2DM | endometrium | *PAAF1* | 2.084652995 | 0.000171608 | 0.000236613 | 0.832640184 |
| T2DM | endometrium | *MRPL48* | 4.066101603 | 1.44E-07 | 4.78E-07 | 0.000696682 |
| T2DM | endometrium | *RAB6A* | 3.594843821 | 1.13E-07 | 4.57E-07 | 0.000549436 |
| T2DM | endometrium | *TMEM70* | 3.04264915 | 5.28E-08 | 4.34E-07 | 0.000256371 |
| T2DM | endometrium | *RPS6KB2* | -1.458308099 | 1.01E-05 | 1.63E-05 | 0.048920743 |
| T2DM | endometrium | *RMI2* | -2.241787978 | 0.000136937 | 0.000190433 | 0.664419425 |
| T2DM | endometrium | *TOM1L2* | 4.357002996 | 0.000188952 | 0.000259641 | 0.916793111 |
| T2DM | endometrium | *TEX26* | 9.833384349 | 2.73E-05 | 4.12E-05 | 0.13239131 |
| T2DM | endometrium | *GPR156* | 9.626877849 | 1.45E-05 | 2.27E-05 | 0.070301742 |
| T2DM | endometrium | *TTLL11* | 5.581546903 | 0.000247396 | 0.000336331 | 1 |
| T2DM | endometrium | *PRIMA1* | -3.40777371 | 0.00058887 | 0.000774309 | 1 |
| T2DM | endometrium | *CTDNEP1* | 5.017174824 | 1.33E-07 | 4.78E-07 | 0.000643915 |
| T2DM | endometrium | *HOXD8* | 2.511173738 | 0.000432407 | 0.000575121 | 1 |
| T2DM | endometrium | *ZDHHC21* | 3.302789373 | 1.89E-07 | 5.43E-07 | 0.00091944 |
| T2DM | endometrium | *A2M* | 2.52701705 | 5.61E-08 | 4.34E-07 | 0.000272093 |
| T2DM | endometrium | *DOK7* | 19.38913783 | 8.72E-07 | 1.81E-06 | 0.004232351 |
| T2DM | endometrium | *ORAI3* | 7.269087876 | 6.56E-07 | 1.42E-06 | 0.003183637 |
| T2DM | endometrium | *B3GALT6* | 6.241268437 | 1.48E-07 | 4.78E-07 | 0.000716183 |
| T2DM | endometrium | *ZNF613* | 6.724866003 | 5.60E-06 | 9.49E-06 | 0.027190939 |
| T2DM | endometrium | *JAKMIP2* | 2.820497064 | 1.80E-07 | 5.22E-07 | 0.000871179 |
| T2DM | endometrium | *TCP11L1* | 5.622859903 | 3.54E-06 | 6.26E-06 | 0.017199691 |
| T2DM | endometrium | *CCDC57* | 2.390838652 | 1.96E-07 | 5.51E-07 | 0.000948952 |
| T2DM | endometrium | *CIDEA* | 2.172664208 | 2.51E-07 | 6.61E-07 | 0.001219109 |
| T2DM | endometrium | *SMIM19* | 3.878026787 | 3.06E-06 | 5.49E-06 | 0.014849722 |
| T2DM | endometrium | *ZNF404* | 2.114760708 | 2.25E-05 | 3.42E-05 | 0.108975267 |
| T2DM | endometrium | *SLC35G1* | -3.241144858 | 0.013668356 | 0.016085099 | 1 |
| T2DM | endometrium | *ZNF135* | 2.060938537 | 4.53E-05 | 6.66E-05 | 0.219580161 |
| T2DM | endometrium | *OR4N2* | 8.902968205 | 3.41E-07 | 8.30E-07 | 0.001655179 |
| T2DM | endometrium | *TAC4* | 6.81997084 | 7.72E-08 | 4.36E-07 | 0.00037475 |
| T2DM | endometrium | *CDC26* | -1.648384117 | 6.55E-06 | 1.09E-05 | 0.031786547 |
| T2DM | endometrium | *CRLF3* | 2.695703451 | 2.21E-08 | 4.34E-07 | 0.000107343 |
| T2DM | endometrium | *DNAJC30* | 4.089740077 | 2.02E-07 | 5.63E-07 | 0.000978242 |

| | | | | | | |
|---|---|---|---|---|---|---|
| T2DM | endometrium | *SPRYD4* | 2.034891431 | 0.000686655 | 0.000896996 | 1 |
| T2DM | endometrium | *VPS37D* | 2.440283702 | 0.009210861 | 0.010959072 | 1 |
| T2DM | endometrium | *ZNF575* | 1.488872477 | 0.003092361 | 0.003815905 | 1 |
| T2DM | endometrium | *WDR25* | -2.255813488 | 0.000228387 | 0.000311623 | 1 |
| T2DM | endometrium | *PLA2G16* | 2.748403243 | 9.60E-07 | 1.96E-06 | 0.004655509 |
| T2DM | endometrium | *B3GNT5* | -3.003364283 | 0.002544519 | 0.00316159 | 1 |
| T2DM | endometrium | *RMDN1* | 4.620680011 | 4.05E-05 | 5.99E-05 | 0.196294223 |
| T2DM | endometrium | *RNF152* | 3.320716422 | 1.98E-06 | 3.73E-06 | 0.009616358 |
| T2DM | endometrium | *LRRC37A* | -1.41852426 | 0.000817161 | 0.001060692 | 1 |
| T2DM | endometrium | *BDNF* | 3.533417742 | 3.80E-07 | 9.04E-07 | 0.001843218 |
| T2DM | endometrium | *BOK* | 2.151190125 | 1.49E-07 | 4.78E-07 | 0.000721954 |
| T2DM | endometrium | *PFN4* | 3.193347952 | 1.66E-08 | 4.34E-07 | 8.05E-05 |
| T2DM | endometrium | *RUFY1* | 3.468408969 | 1.55E-07 | 4.81E-07 | 0.000754229 |
| T2DM | endometrium | *VSIG10* | 3.276256528 | 6.46E-08 | 4.34E-07 | 0.000313429 |
| T2DM | endometrium | *C8G* | 63.42619617 | 1.32E-07 | 4.78E-07 | 0.000642066 |
| T2DM | endometrium | *MUC20* | 4.474972027 | 1.95E-07 | 5.50E-07 | 0.000944896 |
| T2DM | endometrium | *LY6H* | 5.736544158 | 3.91E-05 | 5.81E-05 | 0.189933675 |
| T2DM | endometrium | *SHMT1* | 3.655025798 | 3.93E-08 | 4.34E-07 | 0.000190737 |
| T2DM | endometrium | *DPP7* | 5.552788168 | 3.67E-08 | 4.34E-07 | 0.000178026 |
| T2DM | endometrium | *C19orf18* | 2.703922213 | 6.30E-07 | 1.37E-06 | 0.003055326 |
| T2DM | endometrium | *TMEM80* | 4.744361709 | 1.01E-07 | 4.47E-07 | 0.000491895 |
| T2DM | endometrium | *DSCAML1* | 2.455994062 | 8.37E-08 | 4.38E-07 | 0.000406173 |
| T2DM | endometrium | *TALDO1* | 2.688922683 | 9.77E-08 | 4.42E-07 | 0.000473964 |
| T2DM | endometrium | *AP006621.1* | 6.193318822 | 2.66E-05 | 4.02E-05 | 0.129056425 |
| T2DM | endometrium | *MAN1B1* | 4.235860513 | 5.92E-08 | 4.34E-07 | 0.000287356 |
| T2DM | endometrium | *FBXO39* | 2.071350187 | 0.035827056 | 0.040844191 | 1 |
| T2DM | endometrium | *SAMD9L* | 2.748123905 | 2.24E-07 | 6.07E-07 | 0.001089003 |
| T2DM | endometrium | *MIEF2* | 3.63670472 | 1.82E-07 | 5.25E-07 | 0.000881817 |
| T2DM | endometrium | *NAP1L5* | -1.534875244 | 2.07E-06 | 3.89E-06 | 0.010067746 |
| T2DM | endometrium | *NIM1K* | 5.338071969 | 3.62E-07 | 8.70E-07 | 0.001755869 |
| T2DM | endometrium | *RBM44* | 6.412041068 | 3.06E-06 | 5.49E-06 | 0.014847947 |
| T2DM | endometrium | *SLC25A22* | -1.474003337 | 6.21E-07 | 1.36E-06 | 0.003012158 |
| T2DM | endometrium | *SAMD12* | 1.166094737 | 0.003246784 | 0.003997309 | 1 |
| T2DM | endometrium | *C18orf32* | -2.333892583 | 0.005661224 | 0.006851648 | 1 |
| T2DM | endometrium | *ACAD9* | 2.45116953 | 3.93E-07 | 9.28E-07 | 0.001905705 |
| T2DM | endometrium | *IL17RA* | 2.546255037 | 9.21E-08 | 4.38E-07 | 0.000446937 |
| T2DM | endometrium | *MBOAT4* | 7.339703555 | 1.91E-06 | 3.61E-06 | 0.00927689 |
| T2DM | endometrium | *AGTRAP* | 2.69194824 | 1.33E-07 | 4.78E-07 | 0.00064619 |
| T2DM | endometrium | *THAP5* | 2.580724381 | 1.48E-07 | 4.78E-07 | 0.000718323 |
| T2DM | endometrium | *NAALADL2* | 3.885701894 | 5.06E-08 | 4.34E-07 | 0.00024538 |
| T2DM | endometrium | *CD151* | 2.755453467 | 9.89E-08 | 4.44E-07 | 0.000479726 |
| T2DM | endometrium | *SLC35G5* | 21.49474339 | 1.29E-07 | 4.78E-07 | 0.000626594 |
| T2DM | endometrium | *FAM87B* | 20.01774653 | 1.28E-07 | 4.78E-07 | 0.000620689 |
| T2DM | endometrium | *PCDHB9* | 5.081642703 | 1.20E-05 | 1.92E-05 | 0.058350187 |
| T2DM | endometrium | *CENPBD1* | 5.763731135 | 1.42E-07 | 4.78E-07 | 0.000689538 |
| T2DM | endometrium | *RPS27* | 3.073502264 | 0.038534757 | 0.043817821 | 1 |
| T2DM | endometrium | *RIC8A* | 2.928599095 | 2.43E-06 | 4.46E-06 | 0.011773029 |
| T2DM | endometrium | *ASB8* | -2.590784146 | 0.016070535 | 0.0187754 | 1 |
| T2DM | endometrium | *ODF3B* | 3.667259003 | 2.10E-07 | 5.81E-07 | 0.001021078 |
| T2DM | endometrium | *DPY19L2* | 2.769178466 | 8.18E-05 | 0.000116261 | 0.396913959 |

| | | | | | | |
|---|---|---|---|---|---|---|
| T2DM | endometrium | *ADAMTSL1* | 1.575983645 | 4.68E-08 | 4.34E-07 | 0.000227053 |
| T2DM | endometrium | *IMPDH2* | 2.672937249 | 4.33E-06 | 7.52E-06 | 0.021003465 |
| T2DM | endometrium | *GRAMD1C* | 2.448309446 | 3.57E-06 | 6.30E-06 | 0.017303667 |
| T2DM | endometrium | *DALRD3* | 3.811225446 | 6.34E-08 | 4.34E-07 | 0.00030773 |
| T2DM | endometrium | *ZNF114* | 6.674864574 | 5.48E-06 | 9.30E-06 | 0.026575627 |
| T2DM | endometrium | *SPINK6* | 10.03413911 | 3.38E-05 | 5.06E-05 | 0.164146302 |
| T2DM | endometrium | *ZNF454* | 2.114075548 | 9.11E-05 | 0.000128695 | 0.442194448 |
| T2DM | endometrium | *SH2B1* | 5.771607041 | 1.74E-07 | 5.12E-07 | 0.000842891 |
| T2DM | endometrium | *VN1R1* | 5.999744138 | 0.000586085 | 0.000771064 | 1 |
| T2DM | endometrium | *KDELC2* | 3.117286619 | 1.61E-07 | 4.89E-07 | 0.000781821 |
| T2DM | endometrium | *PLEC* | 2.723086414 | 4.28E-07 | 9.89E-07 | 0.002077601 |
| T2DM | endometrium | *SH2D4B* | 10.2079338 | 1.79E-07 | 5.21E-07 | 0.000868037 |
| T2DM | endometrium | *RNF212* | 3.83947719 | 0.020624171 | 0.023899804 | 1 |
| T2DM | endometrium | *PRSS36* | 4.840203137 | 1.19E-06 | 2.36E-06 | 0.005764264 |
| T2DM | endometrium | *TMPRSS9* | 5.333972124 | 1.50E-06 | 2.91E-06 | 0.007275808 |
| T2DM | endometrium | *TMEM11* | -3.946021857 | 0.007932808 | 0.009491981 | 1 |
| T2DM | endometrium | *ZFAND2A* | 3.796399518 | 1.51E-07 | 4.78E-07 | 0.000734531 |
| T2DM | endometrium | *FAM220A* | 4.336211291 | 1.32E-07 | 4.78E-07 | 0.000641602 |
| T2DM | endometrium | *NT5DC1* | 5.141366659 | 2.17E-07 | 5.95E-07 | 0.001055294 |
| T2DM | endometrium | *GLDC* | 4.720980764 | 7.56E-07 | 1.60E-06 | 0.003666597 |
| T2DM | endometrium | *P4HTM* | 5.275596193 | 0.007323183 | 0.008797248 | 1 |
| T2DM | endometrium | *ERN1* | 5.555615925 | 1.28E-07 | 4.78E-07 | 0.00062297 |
| T2DM | endometrium | *GPR35* | 1.748763763 | 0.014626621 | 0.017142117 | 1 |
| T2DM | endometrium | *PARP10* | 6.200026542 | 3.42E-07 | 8.32E-07 | 0.001659746 |
| T2DM | endometrium | *NSUN3* | 5.113966906 | 0.001006306 | 0.001294775 | 1 |
| T2DM | endometrium | *STX19* | 3.950896477 | 9.82E-10 | 2.33E-07 | 4.77E-06 |
| T2DM | endometrium | *MPI* | 2.732042501 | 7.75E-07 | 1.64E-06 | 0.003760353 |
| T2DM | endometrium | *TRIM73* | -6.524832795 | 0.034229909 | 0.039069282 | 1 |
| T2DM | endometrium | *EFCAB13* | 2.153611108 | 6.47E-08 | 4.34E-07 | 0.000313973 |
| T2DM | endometrium | *EXOSC4* | 6.469750006 | 0.031275204 | 0.035806345 | 1 |
| T2DM | endometrium | *ZNF852* | -4.921887983 | 0.000211138 | 0.000289146 | 1 |
| T2DM | endometrium | *HYI* | -1.788896677 | 4.06E-05 | 6.01E-05 | 0.19714296 |
| T2DM | endometrium | *C17orf62* | 2.499485396 | 1.70E-07 | 5.06E-07 | 0.000826073 |
| T2DM | endometrium | *GAK* | 2.42405976 | 9.30E-08 | 4.39E-07 | 0.000451131 |
| T2DM | endometrium | *TUFM* | 3.318528214 | 6.44E-07 | 1.40E-06 | 0.003126387 |
| T2DM | endometrium | *CTC1* | -2.372495998 | 0.005457426 | 0.006613244 | 1 |
| T2DM | endometrium | *C3orf38* | 3.862044968 | 3.59E-08 | 4.34E-07 | 0.00017433 |
| T2DM | endometrium | *CCDC89* | 9.919770006 | 8.26E-08 | 4.37E-07 | 0.000400706 |
| T2DM | endometrium | *HTR1F* | 8.844850483 | 6.65E-06 | 1.11E-05 | 0.032248372 |
| T2DM | endometrium | *SPTY2D1* | 6.956387586 | 9.88E-08 | 4.44E-07 | 0.000479482 |
| T2DM | endometrium | *CDH4* | 3.952497793 | 4.19E-06 | 7.30E-06 | 0.020323557 |
| T2DM | endometrium | *NSUN7* | 2.420636878 | 0.01378951 | 0.016211947 | 1 |
| T2DM | endometrium | *PACS2* | -2.331485513 | 0.012042188 | 0.014230076 | 1 |
| T2DM | endometrium | *LINC00174* | 2.195138075 | 0.001029813 | 0.001323968 | 1 |
| T2DM | endometrium | *GEMIN4* | 3.356727744 | 6.05E-06 | 1.02E-05 | 0.029350545 |
| T2DM | endometrium | *ZFPM1* | 2.738391013 | 2.68E-05 | 4.06E-05 | 0.130222032 |
| T2DM | endometrium | *FCER1A* | 3.329978163 | 0.000126463 | 0.000176428 | 0.61359814 |
| T2DM | endometrium | *RPRML* | 18.29179449 | 1.90E-05 | 2.93E-05 | 0.092224153 |
| T2DM | endometrium | *SERTAD2* | 2.038126167 | 1.18E-07 | 4.61E-07 | 0.000571187 |
| T2DM | endometrium | *AKAP5* | 5.781711081 | 9.62E-08 | 4.42E-07 | 0.000466567 |

| | | | | | | |
|---|---|---|---|---|---|---|
| T2DM | endometrium | *GIPC3* | 15.44911022 | 1.34E-07 | 4.78E-07 | 0.000648593 |
| T2DM | endometrium | *PDXDC1* | 3.834691067 | 3.05E-08 | 4.34E-07 | 0.000147879 |
| T2DM | endometrium | *C1orf194* | -0.988141057 | 1.60E-06 | 3.08E-06 | 0.007772194 |
| T2DM | endometrium | *B3GNT3* | 4.909961249 | 0.000418057 | 0.000556644 | 1 |
| T2DM | endometrium | *SEPT1* | 2.952470665 | 1.32E-07 | 4.78E-07 | 0.000638682 |
| T2DM | endometrium | *TDRD6* | 1.756335188 | 0.000177305 | 0.00024419 | 0.860282706 |
| T2DM | endometrium | *CSNK1A1L* | -17.82228782 | 0.000363027 | 0.000486711 | 1 |
| T2DM | endometrium | *LYNX1* | 2.807016768 | 0.000182892 | 0.000251671 | 0.887393709 |
| T2DM | endometrium | *HCLS1* | 1.667161778 | 1.69E-05 | 2.62E-05 | 0.082065347 |
| T2DM | endometrium | *MTURN* | 5.66719213 | 1.58E-05 | 2.46E-05 | 0.076641294 |
| T2DM | endometrium | *CCDC66* | 4.427745512 | 9.96E-08 | 4.44E-07 | 0.000483368 |
| T2DM | endometrium | *DEFB124* | 20.58193976 | 8.39E-08 | 4.38E-07 | 0.000406951 |
| T2DM | endometrium | *MCFD2* | -2.057022675 | 0.037369057 | 0.042542155 | 1 |
| T2DM | endometrium | *DEFB119* | 50.18963996 | 4.86E-08 | 4.34E-07 | 0.000235723 |
| T2DM | endometrium | *TSPYL5* | -1.493258043 | 2.06E-06 | 3.87E-06 | 0.010018999 |
| T2DM | endometrium | *MB21D2* | 5.772614992 | 3.28E-06 | 5.84E-06 | 0.015918265 |
| T2DM | endometrium | *SLC47A2* | 2.988839382 | 2.01E-07 | 5.62E-07 | 0.000975112 |
| T2DM | endometrium | *CHST13* | 18.69985172 | 4.91E-05 | 7.18E-05 | 0.238288939 |
| T2DM | endometrium | *SLC36A4* | 6.005238284 | 1.00E-05 | 1.62E-05 | 0.048662583 |
| T2DM | endometrium | *ZDHHC20* | 3.871031777 | 5.12E-08 | 4.34E-07 | 0.000248404 |
| T2DM | endometrium | *OR51E1* | 3.372524072 | 0.000222038 | 0.000303131 | 1 |
| T2DM | endometrium | *PSMG4* | 5.008229598 | 3.29E-06 | 5.84E-06 | 0.015945282 |
| T2DM | endometrium | *BHLHE22* | 3.75551704 | 0.00092035 | 0.001188275 | 1 |
| T2DM | endometrium | *ZNF443* | 9.364257084 | 3.80E-07 | 9.04E-07 | 0.001842902 |
| T2DM | endometrium | *CAPS2* | 2.416356429 | 3.83E-08 | 4.34E-07 | 0.000185817 |
| T2DM | endometrium | *KCTD2* | -2.531939677 | 7.97E-05 | 0.000113384 | 0.386641026 |
| T2DM | endometrium | *D2HGDH* | 3.318482039 | 0.000532771 | 0.00070302 | 1 |
| T2DM | endometrium | *ST20* | -0.997785988 | 3.96E-05 | 5.86E-05 | 0.192049229 |
| T2DM | endometrium | *LSMEM1* | 2.977116837 | 2.55E-07 | 6.69E-07 | 0.001238176 |
| T2DM | endometrium | *NQO1* | 2.280935639 | 2.64E-07 | 6.84E-07 | 0.001280168 |
| T2DM | endometrium | *FKRP* | 1.812839463 | 2.05E-05 | 3.14E-05 | 0.099482893 |
| T2DM | endometrium | *RPH3AL* | 1.866307621 | 2.27E-07 | 6.12E-07 | 0.001100306 |
| T2DM | endometrium | *ADIPOQ* | 0.99318273 | 0.009604079 | 0.011415725 | 1 |
| T2DM | endometrium | *F2R* | -1.630189994 | 6.24E-05 | 9.00E-05 | 0.302958553 |
| T2DM | endometrium | *MUC16* | 2.244186403 | 1.02E-07 | 4.47E-07 | 0.00049732 |
| T2DM | endometrium | *PENK* | 8.080080394 | 0.000878689 | 0.001137209 | 1 |
| T2DM | endometrium | *SLC25A41* | 13.31635413 | 7.78E-08 | 4.36E-07 | 0.000377516 |
| T2DM | endometrium | *TMEM136* | 3.351522512 | 5.48E-08 | 4.34E-07 | 0.00026569 |
| T2DM | endometrium | *TMEM132E* | 1.991953429 | 0.025018184 | 0.028840159 | 1 |
| T2DM | endometrium | *ZNF322* | 5.166613089 | 2.96E-05 | 4.45E-05 | 0.143402648 |
| T2DM | endometrium | *DDX60L* | 5.128428653 | 3.90E-08 | 4.34E-07 | 0.000189377 |
| T2DM | endometrium | *TMEM45A* | -1.257612601 | 1.08E-05 | 1.74E-05 | 0.05250304 |
| T2DM | endometrium | *SGSH* | 4.622801213 | 8.54E-08 | 4.38E-07 | 0.000414214 |
| T2DM | endometrium | *ZFP41* | 5.247423839 | 7.34E-06 | 1.21E-05 | 0.035602034 |
| T2DM | endometrium | *ZBTB20* | 2.738760993 | 5.61E-08 | 4.34E-07 | 0.000272267 |
| T2DM | endometrium | *C3orf58* | 4.341392257 | 2.56E-08 | 4.34E-07 | 0.000124231 |
| T2DM | endometrium | *AMIGO1* | -3.777052642 | 2.48E-05 | 3.76E-05 | 0.120435691 |
| T2DM | endometrium | *TMEM252* | -23.96449725 | 2.77E-06 | 5.02E-06 | 0.013450259 |
| T2DM | endometrium | *ODF3L2* | 9.257740553 | 1.94E-05 | 2.99E-05 | 0.094228363 |
| T2DM | endometrium | *RELL1* | 4.729407617 | 2.69E-07 | 6.94E-07 | 0.001307573 |

| | | | | | | |
|---|---|---|---|---|---|---|
| T2DM | endometrium | *RFX7* | 2.904732331 | 5.52E-08 | 4.34E-07 | 0.000267795 |
| T2DM | endometrium | *CLDN7* | -3.379482308 | 0.000308254 | 0.000415804 | 1 |
| T2DM | endometrium | *ZNF329* | -2.670065337 | 0.001495674 | 0.001896266 | 1 |
| T2DM | endometrium | *ADO* | 4.092729366 | 4.12E-08 | 4.34E-07 | 0.000199896 |
| T2DM | endometrium | *MGAT4C* | 1.670990659 | 1.14E-05 | 1.83E-05 | 0.055336667 |
| T2DM | endometrium | *IDH2* | 2.28702905 | 1.73E-07 | 5.12E-07 | 0.000841729 |
| T2DM | endometrium | *PTCHD3* | 3.365387575 | 0.006955243 | 0.008373906 | 1 |
| T2DM | endometrium | *FAM181B* | -2.107333454 | 5.49E-06 | 9.32E-06 | 0.026651974 |
| T2DM | endometrium | *TDRKH* | 3.044354578 | 1.39E-05 | 2.19E-05 | 0.067354489 |
| T2DM | endometrium | *ZNF708* | 4.02770662 | 2.54E-05 | 3.85E-05 | 0.123245624 |
| T2DM | endometrium | *ERCC6L2* | 4.207611929 | 2.44E-08 | 4.34E-07 | 0.000118502 |
| T2DM | endometrium | *MRPL41* | 4.33783436 | 6.46E-08 | 4.34E-07 | 0.00031329 |
| T2DM | endometrium | *ARL6IP4* | -1.562644758 | 0.000181297 | 0.000249547 | 0.879652737 |
| T2DM | endometrium | *HHIPL1* | 2.766607528 | 2.40E-07 | 6.40E-07 | 0.001164919 |
| T2DM | endometrium | *FAM153B* | 2.198347128 | 1.02E-07 | 4.47E-07 | 0.000492773 |
| T2DM | endometrium | *BACE2* | 2.422809641 | 0.002307896 | 0.00287938 | 1 |
| T2DM | endometrium | *SYNM* | 5.405410827 | 7.05E-08 | 4.34E-07 | 0.000342173 |
| T2DM | endometrium | *KCNA4* | 3.353053301 | 1.50E-07 | 4.78E-07 | 0.00072625 |
| T2DM | endometrium | *FIGN* | 3.106554503 | 5.50E-08 | 4.34E-07 | 0.000267055 |
| T2DM | endometrium | *C8orf33* | 5.227849154 | 1.10E-07 | 4.56E-07 | 0.00053157 |
| T2DM | endometrium | *FBXL6* | -2.75278453 | 1.49E-05 | 2.33E-05 | 0.072121931 |
| T2DM | endometrium | *KBTBD3* | 2.680379811 | 2.97E-07 | 7.47E-07 | 0.001443426 |
| T2DM | endometrium | *CLN8* | 2.289954442 | 1.93E-07 | 5.48E-07 | 0.000937294 |
| T2DM | endometrium | *NXPH4* | 28.35337586 | 1.44E-07 | 4.78E-07 | 0.000697408 |
| T2DM | endometrium | *NPLOC4* | 2.339862949 | 1.03E-06 | 2.09E-06 | 0.005002453 |
| T2DM | endometrium | *CAPN12* | 3.313845263 | 8.07E-08 | 4.36E-07 | 0.000391484 |
| T2DM | endometrium | *KPNA2* | -4.312240755 | 0.000126602 | 0.000176566 | 0.614271382 |
| T2DM | endometrium | *ORAI1* | 3.861219338 | 1.31E-07 | 4.78E-07 | 0.00063743 |
| T2DM | endometrium | *FES* | 4.836986426 | 4.75E-05 | 6.96E-05 | 0.230339372 |
| T2DM | endometrium | *GLRX5* | 7.044189065 | 5.81E-08 | 4.34E-07 | 0.000281716 |
| T2DM | endometrium | *CAV3* | 2.554351162 | 6.33E-07 | 1.38E-06 | 0.003072563 |
| T2DM | endometrium | *MXRA7* | 2.56978102 | 0.00012147 | 0.000169604 | 0.58937458 |
| T2DM | endometrium | *MFSD5* | 4.565301736 | 1.45E-05 | 2.27E-05 | 0.070441283 |
| T2DM | endometrium | *ADI1* | 3.825845159 | 6.59E-08 | 4.34E-07 | 0.000319529 |
| T2DM | endometrium | *CLEC4G* | -4.200287171 | 5.66E-07 | 1.25E-06 | 0.002743992 |
| T2DM | endometrium | *TRAK1* | 3.378543237 | 8.15E-07 | 1.71E-06 | 0.003956731 |
| T2DM | endometrium | *SKA2* | 6.246467431 | 7.66E-07 | 1.62E-06 | 0.003716097 |
| T2DM | endometrium | *TTC3* | 2.465316867 | 4.58E-10 | 1.59E-07 | 2.22E-06 |
| T2DM | endometrium | *BRICD5* | 5.873179203 | 1.49E-07 | 4.78E-07 | 0.000722039 |
| T2DM | endometrium | *GRID1* | -3.014843426 | 0.000275268 | 0.000372865 | 1 |
| T2DM | endometrium | *ACBD3* | 5.181687631 | 7.61E-05 | 0.000108573 | 0.369255222 |
| T2DM | endometrium | *PLCXD3* | 1.992575258 | 1.37E-07 | 4.78E-07 | 0.000664017 |
| T2DM | endometrium | *GPIHBP1* | 1.058909411 | 0.019921267 | 0.023101813 | 1 |
| T2DM | endometrium | *VMO1* | -2.383038523 | 0.000657674 | 0.000861278 | 1 |
| T2DM | endometrium | *ALG12* | 2.741549739 | 1.46E-07 | 4.78E-07 | 0.000708059 |
| T2DM | endometrium | *COL18A1* | 5.310709603 | 1.54E-06 | 2.97E-06 | 0.007464778 |
| T2DM | endometrium | *ZNF721* | 4.576123823 | 1.59E-07 | 4.86E-07 | 0.000773797 |
| T2DM | endometrium | *C11orf54* | 1.616524848 | 0.002442464 | 0.003041795 | 1 |
| T2DM | endometrium | *CEP63* | 2.532629695 | 0.025748206 | 0.029653524 | 1 |
| T2DM | endometrium | *ODF3L1* | 7.191962343 | 0.014435116 | 0.016925854 | 1 |

| | | | | | | |
|---|---|---|---|---|---|---|
| T2DM | endometrium | *C12orf60* | 2.573791191 | 1.37E-07 | 4.78E-07 | 0.000667127 |
| T2DM | endometrium | *ABAT* | 3.846712831 | 1.33E-07 | 4.78E-07 | 0.000647383 |
| T2DM | endometrium | *LYSMD4* | -2.410250933 | 0.008926198 | 0.010630808 | 1 |
| T2DM | endometrium | *WBP2NL* | 3.559645901 | 1.84E-07 | 5.31E-07 | 0.000894694 |
| T2DM | endometrium | *BEGAIN* | 4.323420179 | 7.32E-07 | 1.56E-06 | 0.003552046 |
| T2DM | endometrium | *ARHGEF37* | 5.731199888 | 5.29E-08 | 4.34E-07 | 0.000256788 |
| T2DM | endometrium | *CSMD1* | -2.8954274 | 0.00053418 | 0.000704687 | 1 |
| T2DM | endometrium | *RABIF* | 3.137511556 | 2.49E-07 | 6.58E-07 | 0.001207491 |
| T2DM | endometrium | *FANCF* | -2.485864165 | 1.34E-05 | 2.12E-05 | 0.064993326 |
| T2DM | endometrium | *C2CD4C* | 2.207574535 | 0.001340897 | 0.001708517 | 1 |
| T2DM | endometrium | *RUVBL2* | -2.646943644 | 0.000112696 | 0.000157852 | 0.546800542 |
| T2DM | endometrium | *PTTG1IP* | 2.238871466 | 1.92E-07 | 5.47E-07 | 0.000933559 |
| T2DM | endometrium | *PLGLB1* | 3.035178117 | 0.007753421 | 0.009293379 | 1 |
| T2DM | endometrium | *CCDC125* | 3.83753782 | 6.47E-08 | 4.34E-07 | 0.000313711 |
| T2DM | endometrium | *C10orf107* | 1.98380745 | 1.04E-05 | 1.67E-05 | 0.050278313 |
| T2DM | endometrium | *FHL3* | 3.539629393 | 1.15E-06 | 2.30E-06 | 0.005590604 |
| T2DM | endometrium | *C19orf71* | -3.215920174 | 3.30E-05 | 4.94E-05 | 0.159985391 |
| T2DM | endometrium | *LRIT3* | 20.48982317 | 1.27E-07 | 4.75E-07 | 0.00061421 |
| T2DM | endometrium | *NPIPA1* | 2.35731775 | 1.47E-06 | 2.85E-06 | 0.007120669 |
| T2DM | endometrium | *GPR132* | 2.435862858 | 2.94E-06 | 5.32E-06 | 0.014288956 |
| T2DM | endometrium | *PSMG1* | -2.227947205 | 0.041186538 | 0.046723657 | 1 |
| T2DM | endometrium | *AC131971.1* | 11.48935113 | 0.000110005 | 0.000154306 | 0.533745138 |
| T2DM | endometrium | *SERHL2* | 4.79402843 | 6.12E-08 | 4.34E-07 | 0.000296706 |
| T2DM | endometrium | *PCBP3* | 1.704347188 | 9.50E-05 | 0.000133862 | 0.46075145 |
| T2DM | endometrium | *SFXN4* | 2.733018092 | 9.57E-08 | 4.42E-07 | 0.000464547 |
| T2DM | endometrium | *MRPL54* | 2.666069635 | 0.014153805 | 0.016612061 | 1 |
| T2DM | endometrium | *RP1L1* | 4.642409093 | 0.008393374 | 0.010020829 | 1 |
| T2DM | endometrium | *NDUFB1* | 5.70733641 | 1.00E-07 | 4.45E-07 | 0.000486235 |
| T2DM | endometrium | *TRMT12* | 4.328493835 | 8.09E-08 | 4.36E-07 | 0.000392599 |
| T2DM | endometrium | *ALYREF* | -4.244634561 | 2.97E-06 | 5.35E-06 | 0.014419238 |
| T2DM | endometrium | *MRGPRX2* | 13.42722068 | 0.009019845 | 0.010739703 | 1 |
| T2DM | endometrium | *FIGLA* | 13.69586255 | 3.62E-07 | 8.71E-07 | 0.001758619 |
| T2DM | endometrium | *ACSM2A* | 25.88028943 | 3.19E-07 | 7.91E-07 | 0.001550207 |
| T2DM | endometrium | *KREMEN1* | -1.784039468 | 6.46E-06 | 1.08E-05 | 0.031335098 |
| T2DM | endometrium | *ZNF703* | 3.701042256 | 9.99E-06 | 1.62E-05 | 0.048493319 |
| T2DM | endometrium | *EMILIN3* | 5.7051008 | 0.017692902 | 0.020616225 | 1 |
| T2DM | endometrium | *FAM162B* | 3.344666647 | 2.54E-05 | 3.85E-05 | 0.123208422 |
| T2DM | endometrium | *BTBD9* | 4.313771645 | 4.23E-07 | 9.81E-07 | 0.002050591 |
| T2DM | endometrium | *NUDT14* | 7.215105657 | 2.00E-07 | 5.61E-07 | 0.000970516 |
| T2DM | endometrium | *NPIPA7* | -3.244902586 | 3.04E-07 | 7.59E-07 | 0.001473904 |
| T2DM | endometrium | *TTC32* | -1.323562746 | 7.81E-06 | 1.28E-05 | 0.037875864 |
| T2DM | endometrium | *PP2D1* | 7.662166389 | 5.93E-08 | 4.34E-07 | 0.000287673 |
| T2DM | endometrium | *DENND5A* | 1.924984284 | 1.23E-07 | 4.70E-07 | 0.000598843 |
| T2DM | endometrium | *NIPSNAP1* | -2.339236909 | 7.41E-06 | 1.22E-05 | 0.035968774 |
| T2DM | endometrium | *CNTN2* | 4.364753814 | 4.59E-07 | 1.04E-06 | 0.002227948 |
| T2DM | endometrium | *CRELD2* | -3.00064207 | 0.00070039 | 0.000913029 | 1 |
| T2DM | endometrium | *GOLGA6L4* | -1.357174363 | 5.87E-06 | 9.90E-06 | 0.028473323 |
| T2DM | endometrium | *PGP* | 3.474272518 | 4.82E-07 | 1.09E-06 | 0.002336307 |
| T2DM | endometrium | *ACOT1* | 4.221223907 | 4.00E-06 | 7.01E-06 | 0.019399317 |
| T2DM | endometrium | *OAF* | 1.573903869 | 7.02E-05 | 0.000100566 | 0.340819338 |

| | | | | | | |
|---|---|---|---|---|---|---|
| T2DM | endometrium | *ALDH1A3* | 1.786138438 | 5.32E-06 | 9.07E-06 | 0.025833648 |
| T2DM | endometrium | *CLECL1* | 4.165314674 | 7.63E-08 | 4.36E-07 | 0.000370245 |
| T2DM | endometrium | *MROH7* | 4.775284691 | 9.22E-07 | 1.90E-06 | 0.004472436 |
| T2DM | endometrium | *PKP3* | -1.366086525 | 1.70E-06 | 3.25E-06 | 0.008249597 |
| T2DM | endometrium | *SS18L1* | 4.875044087 | 4.79E-08 | 4.34E-07 | 0.000232437 |
| T2DM | endometrium | *COPB2* | 2.914970096 | 6.99E-08 | 4.34E-07 | 0.000339073 |
| T2DM | endometrium | *WDR27* | 4.955308521 | 7.00E-08 | 4.34E-07 | 0.000339787 |
| T2DM | endometrium | *TMEM255B* | 2.973762402 | 1.42E-07 | 4.78E-07 | 0.00069054 |
| T2DM | endometrium | *DUSP8* | -1.050965144 | 3.31E-06 | 5.87E-06 | 0.016038652 |
| T2DM | endometrium | *FAM19A3* | -12.39927225 | 4.98E-07 | 1.12E-06 | 0.002415935 |
| T2DM | endometrium | *SNN* | 4.093334634 | 1.49E-07 | 4.78E-07 | 0.000720608 |
| T2DM | endometrium | *KRBA2* | 4.822316358 | 0.000539305 | 0.000711062 | 1 |
| T2DM | endometrium | *SEPT5* | 5.827546857 | 3.36E-05 | 5.02E-05 | 0.162801468 |
| T2DM | endometrium | *RNLS* | 4.218332062 | 0.000843022 | 0.001093383 | 1 |
| T2DM | endometrium | *NDUFA12* | 3.424441444 | 1.20E-07 | 4.66E-07 | 0.000582101 |
| T2DM | endometrium | *TCTE3* | 6.139482613 | 8.63E-08 | 4.38E-07 | 0.000418719 |
| T2DM | endometrium | *UBE2G2* | 3.439559806 | 2.60E-07 | 6.77E-07 | 0.001259794 |
| T2DM | endometrium | *TMED9* | 5.758462666 | 8.28E-07 | 1.73E-06 | 0.004017184 |
| T2DM | endometrium | *DRD1* | 65.6950546 | 1.48E-07 | 4.78E-07 | 0.000715776 |
| T2DM | endometrium | *BTBD6* | 2.4323694 | 6.32E-06 | 1.06E-05 | 0.030664548 |
| T2DM | endometrium | *WT1* | 1.569297158 | 5.46E-08 | 4.34E-07 | 0.000264832 |
| T2DM | endometrium | *ZFP90* | 3.081039232 | 6.86E-08 | 4.34E-07 | 0.000332887 |
| T2DM | endometrium | *FAM227A* | 2.317728817 | 3.13E-06 | 5.60E-06 | 0.015175533 |
| T2DM | endometrium | *NOC4L* | 5.125143857 | 1.70E-07 | 5.06E-07 | 0.000825828 |
| T2DM | endometrium | *NDUFA6* | 2.858467635 | 4.17E-09 | 4.16E-07 | 2.02E-05 |
| T2DM | endometrium | *SORCS2* | 6.300347834 | 2.18E-06 | 4.07E-06 | 0.010591645 |
| T2DM | endometrium | *TMEM106A* | 2.180510855 | 2.92E-07 | 7.38E-07 | 0.001417908 |
| T2DM | endometrium | *UBOX5* | 2.427894905 | 2.01E-07 | 5.62E-07 | 0.00097319 |
| T2DM | endometrium | *MAFF* | -1.653471198 | 4.96E-05 | 7.25E-05 | 0.240707494 |
| T2DM | endometrium | *SEMA4B* | 5.541308856 | 3.43E-08 | 4.34E-07 | 0.0001663 |
| T2DM | endometrium | *CIB1* | -2.16386923 | 0.000186005 | 0.00025581 | 0.902497062 |
| T2DM | endometrium | *SGCZ* | 1.646392637 | 4.13E-06 | 7.21E-06 | 0.020039817 |
| T2DM | endometrium | *C6orf120* | 3.185866817 | 1.49E-07 | 4.78E-07 | 0.000724594 |
| T2DM | endometrium | *SIGIRR* | 3.703848328 | 2.80E-07 | 7.16E-07 | 0.001359163 |
| T2DM | endometrium | *IFITM2* | 3.368088214 | 7.39E-08 | 4.36E-07 | 0.000358727 |
| T2DM | endometrium | *GP1BA* | 5.214054137 | 0.000845285 | 0.001095731 | 1 |
| T2DM | endometrium | *CCDC137* | 4.911769481 | 3.52E-08 | 4.34E-07 | 0.000170841 |
| T2DM | endometrium | *CDK10* | 2.070924522 | 1.10E-06 | 2.21E-06 | 0.005353506 |
| T2DM | endometrium | *ATP6V0A2* | 6.559061366 | 5.94E-08 | 4.34E-07 | 0.000288012 |
| T2DM | endometrium | *C14orf80* | 5.150488856 | 5.74E-07 | 1.27E-06 | 0.002782776 |
| T2DM | endometrium | *TNFAIP8L1* | 8.306707071 | 0.000806818 | 0.001047546 | 1 |
| T2DM | endometrium | *SP140L* | 2.108088785 | 7.87E-08 | 4.36E-07 | 0.000382054 |
| T2DM | endometrium | *TARSL2* | 3.944479893 | 1.34E-06 | 2.63E-06 | 0.006498246 |
| T2DM | endometrium | *SH3BGR* | -1.697000017 | 0.000843367 | 0.001093537 | 1 |
| T2DM | endometrium | *FAM174B* | 4.120441686 | 3.06E-08 | 4.34E-07 | 0.000148229 |
| T2DM | endometrium | *SV2B* | 2.816389775 | 9.14E-07 | 1.88E-06 | 0.004436353 |
| T2DM | endometrium | *AHNAK2* | 3.660779153 | 5.75E-07 | 1.27E-06 | 0.002789039 |
| T2DM | endometrium | *SP1* | 2.041786188 | 1.09E-07 | 4.55E-07 | 0.000530654 |
| T2DM | endometrium | *PCGF3* | 2.323262568 | 8.06E-08 | 4.36E-07 | 0.000391192 |
| T2DM | endometrium | *C6orf201* | 2.954000365 | 5.40E-06 | 9.18E-06 | 0.026181342 |

| | | | | | | |
|---|---|---|---|---|---|---|
| T2DM | endometrium | *MYBL1* | 2.91621056 | 9.49E-07 | 1.94E-06 | 0.004605157 |
| T2DM | endometrium | *ADARB2* | 5.98186393 | 2.18E-07 | 5.97E-07 | 0.001058937 |
| T2DM | endometrium | *C11orf87* | 8.936493599 | 0.019685426 | 0.022850164 | 1 |
| T2DM | endometrium | *SLC52A2* | 3.165823628 | 1.72E-07 | 5.08E-07 | 0.000832694 |
| T2DM | endometrium | *PIGP* | -1.89240931 | 1.72E-06 | 3.28E-06 | 0.008349832 |
| T2DM | endometrium | *ARL17A* | 3.687662398 | 2.21E-08 | 4.34E-07 | 0.000107192 |
| T2DM | endometrium | *EVI2B* | 2.415229351 | 2.12E-07 | 5.83E-07 | 0.001026265 |
| T2DM | endometrium | *NPIPB4* | 2.596665517 | 6.95E-07 | 1.49E-06 | 0.003371033 |
| T2DM | endometrium | *ZNF829* | 3.119463455 | 1.59E-05 | 2.48E-05 | 0.077286883 |
| T2DM | endometrium | *TRIM69* | 2.417066224 | 0.005167783 | 0.006270089 | 1 |
| T2DM | endometrium | *IFITM1* | 5.81836025 | 3.79E-05 | 5.63E-05 | 0.183948967 |
| T2DM | endometrium | *LINC00839* | 8.433304916 | 3.24E-07 | 8.00E-07 | 0.001573417 |
| T2DM | endometrium | *RTN4RL1* | 5.285567944 | 6.13E-05 | 8.84E-05 | 0.297362921 |
| T2DM | endometrium | *RNPC3* | 3.029963053 | 1.38E-08 | 4.34E-07 | 6.67E-05 |
| T2DM | endometrium | *ZNF529* | 6.543520031 | 2.50E-05 | 3.79E-05 | 0.121297049 |
| T2DM | endometrium | *C15orf41* | 2.538010614 | 2.08E-08 | 4.34E-07 | 0.000100882 |
| T2DM | endometrium | *WWOX* | 3.680701597 | 3.82E-07 | 9.07E-07 | 0.001853335 |
| T2DM | endometrium | *CYP4Z1* | 1.775793524 | 5.33E-05 | 7.75E-05 | 0.258417511 |
| T2DM | endometrium | *FFAR4* | 2.825131814 | 4.29E-09 | 4.16E-07 | 2.08E-05 |
| T2DM | endometrium | *CYP4F12* | 2.707886925 | 2.22E-07 | 6.02E-07 | 0.001077559 |
| T2DM | endometrium | *ZNF749* | -1.362908778 | 8.23E-07 | 1.72E-06 | 0.003993788 |
| T2DM | endometrium | *NBPF12* | 2.820451416 | 0.000346546 | 0.000465773 | 1 |
| T2DM | endometrium | *GPAT2* | 4.45482998 | 0.014110431 | 0.01656917 | 1 |
| T2DM | endometrium | *TOR3A* | 2.723154057 | 4.44E-07 | 1.02E-06 | 0.002155704 |
| T2DM | endometrium | *ZNF555* | 5.55112281 | 3.78E-08 | 4.34E-07 | 0.000183412 |
| T2DM | endometrium | *GOLGA8R* | -3.885085463 | 0.000214174 | 0.000292973 | 1 |
| T2DM | endometrium | *CCDC30* | 3.835429513 | 8.21E-07 | 1.72E-06 | 0.003984395 |
| T2DM | endometrium | *GLDN* | 0.982355302 | 8.81E-05 | 0.000124662 | 0.427464653 |
| T2DM | endometrium | *ZNF197* | 4.35479181 | 3.35E-07 | 8.20E-07 | 0.001626319 |
| T2DM | endometrium | *RPS23* | 4.326189074 | 2.34E-05 | 3.57E-05 | 0.113746203 |
| T2DM | endometrium | *GNG2* | 1.883029393 | 1.04E-07 | 4.47E-07 | 0.00050423 |
| T2DM | endometrium | *BTN3A2* | 1.2921912 | 0.001170771 | 0.001498834 | 1 |
| T2DM | endometrium | *TMEM222* | 3.971366696 | 1.03E-07 | 4.47E-07 | 0.000500883 |
| T2DM | endometrium | *SEPT10* | 3.068215315 | 2.13E-07 | 5.86E-07 | 0.001032588 |
| T2DM | endometrium | *FAM86B1* | -5.590823064 | 0.000195953 | 0.000268881 | 0.950764879 |
| T2DM | endometrium | *SMYD4* | 2.955128649 | 9.36E-08 | 4.39E-07 | 0.000454141 |
| T2DM | endometrium | *CEACAM19* | -1.577344071 | 8.85E-07 | 1.83E-06 | 0.00429324 |
| T2DM | endometrium | *MIR22HG* | -2.895948322 | 0.000682026 | 0.000891724 | 1 |
| T2DM | endometrium | *BCR* | 3.898584466 | 1.47E-07 | 4.78E-07 | 0.000712295 |
| T2DM | endometrium | *FOXI2* | -4.291142333 | 0.002535469 | 0.003151959 | 1 |
| T2DM | endometrium | *ZNF732* | 5.11419816 | 1.70E-07 | 5.06E-07 | 0.0008266 |
| T2DM | endometrium | *VSIG10L* | -1.224029959 | 2.48E-06 | 4.55E-06 | 0.012050928 |
| T2DM | endometrium | *TRABD2A* | 2.506366121 | 0.000480609 | 0.000636336 | 1 |
| T2DM | endometrium | *QRFPR* | 11.94125442 | 1.29E-06 | 2.54E-06 | 0.006249081 |
| T2DM | endometrium | *MAPT* | 3.083553975 | 8.11E-08 | 4.36E-07 | 0.000393426 |
| T2DM | endometrium | *TMEM17* | -2.156097664 | 0.004894691 | 0.005943203 | 1 |
| T2DM | endometrium | *TNFRSF18* | 10.27094579 | 7.50E-07 | 1.59E-06 | 0.003639425 |
| T2DM | endometrium | *C1QL4* | 34.09904691 | 1.58E-07 | 4.84E-07 | 0.00076453 |
| T2DM | endometrium | *RTN4RL2* | 5.439031473 | 1.07E-05 | 1.73E-05 | 0.05215413 |
| T2DM | endometrium | *TMEM232* | 3.597289609 | 6.57E-06 | 1.10E-05 | 0.031886562 |

| | | | | | | |
|---|---|---|---|---|---|---|
| T2DM | endometrium | *EFCAB6* | 4.450604817 | 2.07E-08 | 4.34E-07 | 0.000100257 |
| T2DM | endometrium | *RHD* | 5.245465609 | 2.93E-07 | 7.39E-07 | 0.001424002 |
| T2DM | endometrium | *TMPRSS6* | 19.91065368 | 1.72E-07 | 5.08E-07 | 0.000832651 |
| T2DM | endometrium | *CYP4A11* | 9.749633318 | 1.63E-06 | 3.14E-06 | 0.007932826 |
| T2DM | endometrium | *TMEM216* | -0.863525012 | 1.16E-05 | 1.85E-05 | 0.056305302 |
| T2DM | endometrium | *ENTPD5* | 3.856413846 | 0.00031974 | 0.000430938 | 1 |
| T2DM | endometrium | *ANGPTL5* | 4.278645881 | 7.08E-06 | 1.17E-05 | 0.034329923 |
| T2DM | endometrium | *WDR86* | -1.065568859 | 0.000160396 | 0.000221785 | 0.778242267 |
| T2DM | endometrium | *EPOR* | -1.730860223 | 0.000280144 | 0.000379046 | 1 |
| T2DM | endometrium | *LUZP2* | 6.885231545 | 5.52E-08 | 4.34E-07 | 0.000267844 |
| T2DM | endometrium | *GJA4* | 3.959638425 | 1.33E-07 | 4.78E-07 | 0.000646416 |
| T2DM | endometrium | *IFT140* | -2.322882903 | 0.000396481 | 0.000529514 | 1 |
| T2DM | endometrium | *SBK2* | 26.33198591 | 1.42E-07 | 4.78E-07 | 0.000689183 |
| T2DM | endometrium | *ZNF385C* | 3.230370337 | 0.000767067 | 0.000997537 | 1 |
| T2DM | endometrium | *EXD3* | 2.47579843 | 9.41E-08 | 4.39E-07 | 0.00045652 |
| T2DM | endometrium | *SAMD11* | 5.441465661 | 4.74E-07 | 1.07E-06 | 0.002300991 |
| T2DM | endometrium | *TRPV2* | 2.640209107 | 3.64E-07 | 8.74E-07 | 0.001767342 |
| T2DM | endometrium | *C2orf88* | 4.043836865 | 2.51E-08 | 4.34E-07 | 0.000121823 |
| T2DM | endometrium | *FANCA* | 2.29542737 | 1.18E-05 | 1.89E-05 | 0.057436187 |
| T2DM | endometrium | *DNAH17* | 2.713128893 | 4.41E-06 | 7.66E-06 | 0.021421267 |
| T2DM | endometrium | *FANCM* | 2.830270142 | 1.87E-06 | 3.53E-06 | 0.009059318 |
| T2DM | endometrium | *ZFP69* | 3.529702598 | 1.19E-07 | 4.65E-07 | 0.000577878 |
| T2DM | endometrium | *TMEM220* | 5.476713204 | 3.74E-08 | 4.34E-07 | 0.000181627 |
| T2DM | endometrium | *P2RX2* | -3.119081019 | 4.48E-06 | 7.76E-06 | 0.021747853 |
| T2DM | endometrium | *FAM122A* | -1.958018934 | 1.03E-05 | 1.67E-05 | 0.050204869 |
| T2DM | endometrium | *GFRAL* | 6.98035375 | 1.47E-07 | 4.78E-07 | 0.00071194 |
| T2DM | endometrium | *LDLRAD2* | 3.712755793 | 0.000121184 | 0.000169329 | 0.587985859 |
| T2DM | endometrium | *CPSF4L* | 11.75395586 | 4.74E-06 | 8.17E-06 | 0.023013283 |
| T2DM | endometrium | *KLHL17* | -1.664395551 | 6.97E-06 | 1.16E-05 | 0.033808804 |
| T2DM | endometrium | *ZSCAN23* | 1.507530846 | 0.011040834 | 0.013072261 | 1 |
| T2DM | endometrium | *MAPK12* | 7.566369115 | 1.08E-05 | 1.74E-05 | 0.052487583 |
| T2DM | endometrium | *AGRN* | 6.296726 | 0.036023641 | 0.041039377 | 1 |
| T2DM | endometrium | *FAM166A* | -8.258006002 | 0.000653465 | 0.000856459 | 1 |
| T2DM | endometrium | *ZNF626* | 7.845444568 | 9.15E-08 | 4.38E-07 | 0.000443727 |
| T2DM | endometrium | *LINC00265* | 3.337122465 | 2.28E-07 | 6.14E-07 | 0.001106287 |
| T2DM | endometrium | *LAMTOR4* | 3.291924931 | 5.64E-08 | 4.34E-07 | 0.000273672 |
| T2DM | endometrium | *PRKAR1B* | 4.108403285 | 0.007212232 | 0.008670404 | 1 |
| T2DM | endometrium | *NCR3LG1* | -1.470227947 | 3.20E-06 | 5.70E-06 | 0.015504501 |
| T2DM | endometrium | *ZNF793* | 2.290021763 | 1.95E-07 | 5.51E-07 | 0.000946913 |
| T2DM | endometrium | *AGAP4* | 4.852372098 | 0.000695458 | 0.000907574 | 1 |
| T2DM | endometrium | *RUFY4* | 5.748585101 | 1.52E-07 | 4.78E-07 | 0.000736417 |
| T2DM | endometrium | *CENPP* | 2.752438365 | 0.000216614 | 0.000295892 | 1 |
| T2DM | endometrium | *PLSCR1* | 2.177639546 | 8.51E-08 | 4.38E-07 | 0.000413128 |
| T2DM | endometrium | *ENO4* | 4.916512834 | 9.36E-07 | 1.92E-06 | 0.004542419 |
| T2DM | endometrium | *ZNF559* | 2.037861176 | 3.21E-05 | 4.81E-05 | 0.155588052 |
| T2DM | endometrium | *H3F3C* | 83.01991425 | 1.96E-06 | 3.69E-06 | 0.009502778 |
| T2DM | endometrium | *JAKMIP3* | -1.355738172 | 0.000660782 | 0.000865115 | 1 |
| T2DM | endometrium | *PDCD1* | 19.18636205 | 9.35E-06 | 1.52E-05 | 0.045388839 |
| T2DM | endometrium | *SELL* | 3.556358573 | 1.92E-05 | 2.96E-05 | 0.093220573 |
| T2DM | endometrium | *C19orf54* | 4.127796441 | 1.30E-07 | 4.78E-07 | 0.000632456 |

| | | | | | | |
|---|---|---|---|---|---|---|
| T2DM | endometrium | *C22orf34* | 1.996264907 | 1.41E-07 | 4.78E-07 | 0.000683051 |
| T2DM | endometrium | *COL25A1* | -2.11272941 | 0.014072348 | 0.016528452 | 1 |
| T2DM | endometrium | *SRSF10* | 4.05575308 | 1.59E-07 | 4.86E-07 | 0.000773173 |
| T2DM | endometrium | *NBR1* | 4.346266772 | 1.13E-07 | 4.57E-07 | 0.000550183 |
| T2DM | endometrium | *NDOR1* | -2.146113034 | 2.38E-05 | 3.62E-05 | 0.115700134 |
| T2DM | endometrium | *FAM72B* | -2.352827922 | 3.52E-06 | 6.22E-06 | 0.017059602 |
| T2DM | endometrium | *GOLGA8M* | 3.232393113 | 1.33E-07 | 4.78E-07 | 0.00064761 |
| T2DM | endometrium | *RHCE* | 7.285741814 | 3.16E-07 | 7.84E-07 | 0.001533547 |
| T2DM | endometrium | *IDO2* | 8.179677931 | 1.05E-07 | 4.48E-07 | 0.000510856 |
| T2DM | endometrium | *SLC4A5* | 8.535050741 | 1.76E-07 | 5.17E-07 | 0.000855616 |
| T2DM | endometrium | *UROS* | 3.538685932 | 8.49E-08 | 4.38E-07 | 0.000412085 |
| T2DM | endometrium | *ZBED6CL* | 6.293224206 | 0.001281137 | 0.001636671 | 1 |
| T2DM | endometrium | *OSTN* | 3.150824368 | 4.65E-07 | 1.06E-06 | 0.002257011 |
| T2DM | endometrium | *FSIP2* | 2.7841232 | 1.92E-07 | 5.46E-07 | 0.000930243 |
| T2DM | endometrium | *NOXA1* | -1.626608776 | 1.40E-05 | 2.21E-05 | 0.067928411 |
| T2DM | endometrium | *FZD9* | 25.95454459 | 3.09E-06 | 5.54E-06 | 0.014997689 |
| T2DM | endometrium | *SKOR1* | 6.157592991 | 0.002261141 | 0.002825408 | 1 |
| T2DM | endometrium | *MTF1* | 2.674596211 | 2.56E-07 | 6.70E-07 | 0.001242136 |
| T2DM | endometrium | *ZDHHC11* | 3.416857481 | 0.005250272 | 0.006365397 | 1 |
| T2DM | endometrium | *FAM26F* | 2.734979515 | 1.58E-05 | 2.46E-05 | 0.076787061 |
| T2DM | endometrium | *ENTPD8* | 298.3967569 | 2.68E-07 | 6.91E-07 | 0.001298829 |
| T2DM | endometrium | *RPL14* | 4.666014412 | 0.001182012 | 0.001512825 | 1 |
| T2DM | endometrium | *ASTL* | -12.01948665 | 0.019276496 | 0.022402289 | 1 |
| T2DM | endometrium | *MSL1* | 2.112527514 | 4.38E-07 | 1.01E-06 | 0.0021256 |
| T2DM | endometrium | *FAM196A* | 2.683707338 | 9.24E-08 | 4.38E-07 | 0.00044831 |
| T2DM | endometrium | *UTS2B* | 3.381115265 | 5.36E-07 | 1.19E-06 | 0.002601365 |
| T2DM | endometrium | *C9orf152* | 7.758184995 | 7.79E-07 | 1.64E-06 | 0.003777984 |
| T2DM | endometrium | *NELFB* | 2.520596194 | 1.02E-07 | 4.47E-07 | 0.000497269 |
| T2DM | endometrium | *SLC15A5* | 3.948848967 | 7.64E-08 | 4.36E-07 | 0.000370772 |
| T2DM | endometrium | *LIPI* | -2.202136998 | 0.000355924 | 0.000477848 | 1 |
| T2DM | endometrium | *ADAT2* | 5.99086779 | 3.27E-08 | 4.34E-07 | 0.000158499 |
| T2DM | endometrium | *ALKBH2* | 5.068161669 | 2.60E-09 | 3.60E-07 | 1.26E-05 |
| T2DM | endometrium | *LITAF* | 3.727484968 | 4.51E-08 | 4.34E-07 | 0.000218725 |
| T2DM | endometrium | *VSTM1* | -0.794746646 | 3.56E-06 | 6.28E-06 | 0.017257033 |
| T2DM | endometrium | *SF3B3* | 4.894347862 | 0.00153644 | 0.001945409 | 1 |
| T2DM | endometrium | *PLAC9* | 3.628338735 | 9.15E-07 | 1.88E-06 | 0.004437335 |
| T2DM | endometrium | *ZNF573* | 3.99532408 | 5.96E-07 | 1.31E-06 | 0.002891954 |
| T2DM | endometrium | *FAM47E* | 3.234656127 | 6.38E-07 | 1.39E-06 | 0.003096569 |
| T2DM | endometrium | *ZNF600* | 1.991142242 | 5.70E-07 | 1.26E-06 | 0.00276698 |
| T2DM | endometrium | *TSPYL1* | -3.50868868 | 0.008568834 | 0.010215229 | 1 |
| T2DM | endometrium | *PNRC2* | -2.931386729 | 0.000851999 | 0.001103844 | 1 |
| T2DM | endometrium | *FHIT* | 3.560249295 | 1.32E-07 | 4.78E-07 | 0.000642737 |
| T2DM | endometrium | *ZKSCAN3* | -3.622030498 | 0.000733357 | 0.000954978 | 1 |
| T2DM | endometrium | *RRP7A* | -3.427804638 | 0.023129709 | 0.026720321 | 1 |
| T2DM | endometrium | *SLC35E2B* | 3.44202237 | 6.48E-08 | 4.34E-07 | 0.000314231 |
| T2DM | endometrium | *KIAA0408* | 1.882717778 | 0.002301891 | 0.002873366 | 1 |
| T2DM | endometrium | *HLA-DRB1* | 0.607085662 | 0.040429297 | 0.045896806 | 1 |
| T2DM | endometrium | *AKR1C3* | 3.461877919 | 6.94E-08 | 4.34E-07 | 0.000336542 |
| T2DM | endometrium | *WDSUB1* | 6.037527332 | 1.37E-05 | 2.16E-05 | 0.066289663 |
| T2DM | endometrium | *ZNF79* | -1.478235236 | 8.74E-06 | 1.43E-05 | 0.042426604 |

| | | | | | | |
|---|---|---|---|---|---|---|
| T2DM | endometrium | *COLCA1* | -2.523220457 | 5.88E-07 | 1.30E-06 | 0.002854545 |
| T2DM | endometrium | *SEMA4A* | -3.327165304 | 5.64E-05 | 8.18E-05 | 0.273667313 |
| T2DM | endometrium | *HRCT1* | 3.05679379 | 4.21E-05 | 6.21E-05 | 0.20408154 |
| T2DM | endometrium | *MPHOSPH8* | 3.301296928 | 6.87E-08 | 4.34E-07 | 0.000333201 |
| T2DM | endometrium | *SIRPB2* | 1.496178422 | 0.000413001 | 0.000550667 | 1 |
| T2DM | endometrium | *XPNPEP3* | 2.292366966 | 2.11E-05 | 3.23E-05 | 0.10241619 |
| T2DM | endometrium | *ZNF107* | 4.586042729 | 1.79E-07 | 5.21E-07 | 0.000867737 |
| T2DM | endometrium | *PPIA* | 5.347251985 | 2.93E-06 | 5.30E-06 | 0.014217032 |
| T2DM | endometrium | *ZNF471* | 2.811315154 | 1.05E-07 | 4.48E-07 | 0.000511209 |
| T2DM | endometrium | *ZNF493* | 6.265043806 | 6.94E-06 | 1.15E-05 | 0.033695911 |
| T2DM | endometrium | *GTF2IRD2* | 5.583258031 | 1.49E-08 | 4.34E-07 | 7.23E-05 |
| T2DM | endometrium | *SUPT3H* | 4.967924504 | 2.39E-06 | 4.41E-06 | 0.011606888 |
| T2DM | endometrium | *GIMAP5* | 4.462706079 | 1.53E-05 | 2.38E-05 | 0.074064313 |
| T2DM | endometrium | *STK31* | 4.272605397 | 2.54E-08 | 4.34E-07 | 0.000123334 |
| T2DM | endometrium | *ZKSCAN7* | 6.382537964 | 1.60E-10 | 7.08E-08 | 7.79E-07 |
| T2DM | endometrium | *CD55* | 2.310936567 | 8.18E-08 | 4.36E-07 | 0.00039679 |
| T2DM | endometrium | *CPNE4* | 2.603572402 | 1.76E-07 | 5.16E-07 | 0.000851936 |
| T2DM | endometrium | *ZNF565* | 2.626573934 | 2.47E-07 | 6.55E-07 | 0.001199748 |
| T2DM | endometrium | *ZNF34* | 3.470757605 | 1.90E-08 | 4.34E-07 | 9.23E-05 |
| T2DM | endometrium | *EVL* | 2.735875279 | 0.031392311 | 0.03593194 | 1 |
| T2DM | endometrium | *PIK3R4* | 5.060897991 | 0.00314415 | 0.003878824 | 1 |
| T2DM | endometrium | *ZNF605* | 5.632759371 | 2.17E-05 | 3.31E-05 | 0.105209064 |
| T2DM | endometrium | *RFX8* | 1.873828216 | 3.61E-06 | 6.36E-06 | 0.01750044 |
| T2DM | endometrium | *C20orf96* | 3.613257274 | 1.30E-07 | 4.78E-07 | 0.00062891 |
| T2DM | endometrium | *TPK1* | 4.275369748 | 1.52E-08 | 4.34E-07 | 7.35E-05 |
| T2DM | endometrium | *MYO18A* | 3.909592767 | 1.53E-07 | 4.78E-07 | 0.000742731 |
| T2DM | endometrium | *SPTSSB* | -1.574400409 | 3.46E-06 | 6.12E-06 | 0.0167742 |
| T2DM | endometrium | *CACNA1H* | 5.033687059 | 1.44E-07 | 4.78E-07 | 0.000699163 |
| T2DM | endometrium | *SULF2* | 5.733574878 | 1.48E-08 | 4.34E-07 | 7.20E-05 |
| T2DM | endometrium | *XRCC2* | -1.306675327 | 2.72E-06 | 4.93E-06 | 0.01317728 |
| T2DM | endometrium | *MYO6* | 5.090969504 | 6.77E-05 | 9.71E-05 | 0.328454232 |
| T2DM | endometrium | *ZNF846* | 5.404965155 | 1.63E-08 | 4.34E-07 | 7.92E-05 |
| T2DM | endometrium | *ADH1B* | 3.600685956 | 2.46E-07 | 6.53E-07 | 0.001193168 |
| T2DM | endometrium | *ZNF502* | 3.041766842 | 2.19E-07 | 5.97E-07 | 0.001062348 |
| T2DM | endometrium | *TRAPPC4* | 3.097040767 | 2.49E-07 | 6.59E-07 | 0.001208382 |
| T2DM | endometrium | *ZFP62* | -2.935364749 | 0.013869817 | 0.016298463 | 1 |
| T2DM | endometrium | *ZNF33B* | 2.266255635 | 2.58E-06 | 4.72E-06 | 0.012534554 |
| T2DM | endometrium | *AMZ2* | 2.890392202 | 9.63E-07 | 1.97E-06 | 0.004674255 |
| T2DM | endometrium | *ZNF431* | 5.342576936 | 1.33E-05 | 2.10E-05 | 0.064289819 |
| T2DM | endometrium | *HLA-DQA1* | 0.810471209 | 0.040153948 | 0.045594888 | 1 |
| T2DM | endometrium | *GM2A* | -1.508499304 | 0.00021503 | 0.000293977 | 1 |
| T2DM | endometrium | *ZNF239* | 4.587528782 | 7.89E-09 | 4.34E-07 | 3.83E-05 |
| T2DM | endometrium | *SPRR2B* | -6.07417544 | 1.24E-06 | 2.45E-06 | 0.006016674 |
| T2DM | endometrium | *FAM3C* | 3.860122659 | 7.99E-07 | 1.68E-06 | 0.003878074 |
| T2DM | endometrium | *SLC39A10* | 5.70969586 | 3.61E-07 | 8.69E-07 | 0.001752096 |
| T2DM | endometrium | *AP2A1* | 1.930572464 | 1.26E-07 | 4.74E-07 | 0.000609824 |
| T2DM | endometrium | *PCDHB16* | 2.583065574 | 0.001758804 | 0.002214249 | 1 |
| T2DM | endometrium | *HIST1H3E* | 2.160368827 | 0.012340849 | 0.014575901 | 1 |
| T2DM | endometrium | *FUT11* | 2.656452922 | 4.90E-08 | 4.34E-07 | 0.000237969 |
| T2DM | endometrium | *WDR5B* | 4.126976932 | 5.82E-08 | 4.34E-07 | 0.000282363 |

| | | | | | | |
|---|---|---|---|---|---|---|
| T2DM | endometrium | *ZNF138* | 5.025507824 | 2.45E-08 | 4.34E-07 | 0.000118973 |
| T2DM | endometrium | *ZNF429* | 3.570668144 | 5.31E-08 | 4.34E-07 | 0.000257733 |
| T2DM | endometrium | *ZNF100* | 2.111074503 | 8.87E-07 | 1.83E-06 | 0.004302293 |
| T2DM | endometrium | *ANXA6* | 4.517764599 | 2.52E-07 | 6.61E-07 | 0.001220337 |
| T2DM | endometrium | *GMFB* | 2.149566474 | 1.46E-07 | 4.78E-07 | 0.000708692 |
| T2DM | endometrium | *ZNF420* | 5.409835906 | 0.000971159 | 0.00125088 | 1 |
| T2DM | endometrium | *ZNF763* | -2.065322433 | 1.30E-05 | 2.05E-05 | 0.062891896 |
| T2DM | endometrium | *ZMYM1* | 6.414871234 | 1.98E-05 | 3.04E-05 | 0.096168223 |
| T2DM | endometrium | *IGF2R* | 4.254787375 | 2.21E-07 | 6.01E-07 | 0.00107298 |
| T2DM | endometrium | *SLC25A29* | 2.208868319 | 3.65E-08 | 4.34E-07 | 0.000177299 |
| T2DM | endometrium | *ZNF772* | 1.553441546 | 0.031560327 | 0.036115733 | 1 |
| T2DM | endometrium | *ZNF257* | 6.341173808 | 3.78E-05 | 5.62E-05 | 0.183200811 |
| T2DM | endometrium | *ADAM32* | -2.493590466 | 0.000101182 | 0.000142259 | 0.49093518 |
| T2DM | endometrium | *LRRC8B* | 3.66827492 | 3.89E-08 | 4.34E-07 | 0.000188517 |
| T2DM | endometrium | *SND1* | 3.353762536 | 1.00E-07 | 4.45E-07 | 0.000487234 |
| T2DM | endometrium | *NEK5* | 2.430180896 | 1.21E-05 | 1.92E-05 | 0.058484334 |
| T2DM | endometrium | *PIWIL2* | -2.657470896 | 0.008511033 | 0.010151311 | 1 |
| T2DM | endometrium | *ENTPD4* | 3.533138415 | 2.58E-07 | 6.72E-07 | 0.001250166 |
| T2DM | endometrium | *TBC1D9B* | 4.289267808 | 1.16E-06 | 2.31E-06 | 0.005632584 |
| T2DM | endometrium | *FAM110D* | 7.965873068 | 8.68E-05 | 0.000122879 | 0.420985016 |
| T2DM | endometrium | *C6orf141* | 0.92698453 | 0.019704105 | 0.022866376 | 1 |
| T2DM | endometrium | *CCL4L2* | 8.08253554 | 5.77E-08 | 4.34E-07 | 0.000280041 |
| T2DM | endometrium | *ZNF165* | -2.983527907 | 2.33E-06 | 4.31E-06 | 0.011308703 |
| T2DM | endometrium | *RAMP2-AS1* | -1.935625583 | 7.68E-05 | 0.000109439 | 0.372421676 |
| T2DM | endometrium | *ZNF720* | 2.191245197 | 2.91E-07 | 7.36E-07 | 0.001414181 |
| T2DM | endometrium | *MRPL21* | 2.281399039 | 0.0007733 | 0.001005373 | 1 |
| T2DM | endometrium | *ZNF98* | 2.051063136 | 1.08E-06 | 2.18E-06 | 0.005251546 |
| T2DM | endometrium | *ZNF517* | 3.438382767 | 3.27E-07 | 8.04E-07 | 0.001588282 |
| T2DM | endometrium | *ZNF675* | 2.990066837 | 1.53E-07 | 4.78E-07 | 0.000744678 |
| T2DM | endometrium | *SLC22A5* | -2.227879861 | 0.001114189 | 0.001428659 | 1 |
| T2DM | endometrium | *VEPH1* | 2.427046672 | 1.46E-07 | 4.78E-07 | 0.000710478 |
| T2DM | endometrium | *MAP3K5* | 4.584666656 | 6.09E-07 | 1.33E-06 | 0.002955868 |
| T2DM | endometrium | *GSTK1* | 2.73601424 | 6.52E-07 | 1.41E-06 | 0.003162787 |
| T2DM | endometrium | *GYPE* | 5.162796625 | 8.29E-08 | 4.37E-07 | 0.000402017 |
| T2DM | endometrium | *PCDHB11* | -1.074247821 | 6.61E-07 | 1.43E-06 | 0.003205861 |
| T2DM | endometrium | *SLC2A10* | 3.004283611 | 7.35E-08 | 4.35E-07 | 0.000356793 |
| T2DM | endometrium | *ZNF665* | 2.352519456 | 1.42E-07 | 4.78E-07 | 0.000687948 |
| T2DM | endometrium | *RPF2* | 5.697804906 | 4.91E-08 | 4.34E-07 | 0.000238096 |
| T2DM | endometrium | *MYO5A* | 1.748797805 | 1.56E-07 | 4.81E-07 | 0.000755235 |
| T2DM | endometrium | *ATG7* | 3.240837022 | 3.36E-08 | 4.34E-07 | 0.000163247 |
| T2DM | endometrium | *RAB40C* | 3.32494248 | 7.43E-08 | 4.36E-07 | 0.000360746 |
| T2DM | endometrium | *PIGN* | 3.906869089 | 0.000455339 | 0.000604461 | 1 |
| T2DM | endometrium | *HHLA3* | -2.903045003 | 6.53E-05 | 9.38E-05 | 0.316798233 |
| T2DM | endometrium | *TOPORS* | 5.991840181 | 0.001498091 | 0.001898021 | 1 |
| T2DM | endometrium | *BCO2* | 1.49088706 | 5.19E-06 | 8.86E-06 | 0.025164835 |
| T2DM | endometrium | *ENTPD6* | 2.254561692 | 9.89E-07 | 2.01E-06 | 0.004798072 |
| T2DM | endometrium | *ENPP1* | -0.861227523 | 2.98E-06 | 5.37E-06 | 0.014474754 |
| T2DM | endometrium | *CCDC154* | 3.084555555 | 0.001541053 | 0.001950741 | 1 |
| T2DM | endometrium | *MFAP5* | 1.725551729 | 4.49E-07 | 1.03E-06 | 0.002179813 |
| T2DM | endometrium | *PDCD1LG2* | 1.564312057 | 6.84E-07 | 1.47E-06 | 0.003318759 |

| | | | | | | |
|---|---|---|---|---|---|---|
| T2DM | endometrium | *NMB* | -2.34721559 | 0.015815983 | 0.018486906 | 1 |
| T2DM | endometrium | *FAM114A1* | 5.295429542 | 8.13E-08 | 4.36E-07 | 0.000394594 |
| T2DM | endometrium | *PSAP* | 1.622306302 | 6.00E-08 | 4.34E-07 | 0.000291114 |
| T2DM | endometrium | *TXNRD3* | 2.484424636 | 1.13E-07 | 4.57E-07 | 0.000546306 |
| T2DM | endometrium | *EME2* | -4.461516074 | 0.00170641 | 0.002151079 | 1 |
| T2DM | endometrium | *AC122129.1* | 7.972733151 | 1.47E-07 | 4.78E-07 | 0.000713954 |
| T2DM | endometrium | *CCDC180* | 4.628336865 | 0.000414476 | 0.000552331 | 1 |
| T2DM | endometrium | *OCLN* | -1.8059738 | 4.97E-05 | 7.25E-05 | 0.240935448 |
| T2DM | endometrium | *ZNF181* | 4.078724821 | 7.84E-08 | 4.36E-07 | 0.000380472 |
| T2DM | endometrium | *SGTB* | 2.564537906 | 1.22E-06 | 2.41E-06 | 0.005912707 |
| T2DM | endometrium | *ADH5* | 5.635362599 | 6.93E-07 | 1.49E-06 | 0.003360777 |
| T2DM | endometrium | *HRNR* | 0.674174577 | 0.043563117 | 0.049362038 | 1 |
| T2DM | endometrium | *ZNF677* | 2.552443749 | 1.42E-07 | 4.78E-07 | 0.000690621 |
| T2DM | endometrium | *ZNF311* | 3.096594084 | 1.67E-06 | 3.19E-06 | 0.008096647 |
| T2DM | endometrium | *ZNF71* | -3.370042803 | 5.52E-05 | 8.02E-05 | 0.268002666 |
| T2DM | endometrium | *MPZL1* | 4.850160803 | 3.65E-07 | 8.74E-07 | 0.001769004 |
| T2DM | endometrium | *VPS13A* | 2.742661939 | 3.29E-08 | 4.34E-07 | 0.00015962 |
| T2DM | endometrium | *NOL8* | 2.878838318 | 1.23E-07 | 4.69E-07 | 0.000594483 |
| T2DM | endometrium | *DLGAP2* | 4.592905205 | 1.88E-08 | 4.34E-07 | 9.14E-05 |
| T2DM | endometrium | *ZNF560* | -2.433502283 | 6.03E-06 | 1.01E-05 | 0.029261746 |
| T2DM | endometrium | *AGAP9* | 4.543019143 | 1.54E-06 | 2.97E-06 | 0.007467638 |
| T2DM | endometrium | *ZNF273* | 2.753433563 | 3.77E-07 | 9.00E-07 | 0.00182996 |
| T2DM | endometrium | *MAK16* | 2.877693177 | 8.80E-08 | 4.38E-07 | 0.000426925 |
| T2DM | endometrium | *SFI1* | 3.288402165 | 5.39E-08 | 4.34E-07 | 0.000261321 |
| T2DM | endometrium | *ZNF649* | 2.664847506 | 3.14E-06 | 5.61E-06 | 0.015214847 |
| T2DM | endometrium | *ADH4* | -2.084529124 | 0.000164226 | 0.000226821 | 0.796822185 |
| T2DM | endometrium | *HIBCH* | 2.241816863 | 5.09E-07 | 1.14E-06 | 0.002471566 |
| T2DM | endometrium | *ZNF544* | -2.979412524 | 0.000474108 | 0.000628516 | 1 |
| T2DM | endometrium | *SOWAHC* | 5.12805287 | 1.78E-07 | 5.20E-07 | 0.00086557 |
| T2DM | endometrium | *AC135178.1* | 18.96259477 | 0.013714641 | 0.016131742 | 1 |
| T2DM | endometrium | *MIER1* | 4.441597491 | 0.00092263 | 0.001190902 | 1 |
| T2DM | endometrium | *MAN1A2* | 5.234929877 | 1.62E-06 | 3.11E-06 | 0.007839466 |
| T2DM | endometrium | *SVIP* | -2.594398167 | 0.01987609 | 0.023054934 | 1 |
| T2DM | endometrium | *DDRGK1* | 7.469516697 | 2.74E-07 | 7.03E-07 | 0.001329553 |
| T2DM | endometrium | *ZNF607* | 3.601904031 | 0.000679168 | 0.000888466 | 1 |
| T2DM | endometrium | *ZNF334* | 2.226292677 | 1.03E-07 | 4.47E-07 | 0.000499013 |
| T2DM | endometrium | *HSD17B11* | 3.038618722 | 5.39E-08 | 4.34E-07 | 0.000261526 |
| T2DM | endometrium | *SULT1C2* | 2.695749116 | 1.16E-06 | 2.32E-06 | 0.005647952 |
| T2DM | endometrium | *RPS6KL1* | 9.126589291 | 1.89E-05 | 2.92E-05 | 0.091807672 |
| T2DM | endometrium | *DDX42* | 3.80330562 | 1.68E-08 | 4.34E-07 | 8.13E-05 |
| T2DM | endometrium | *STYX* | -2.529700788 | 0.029470708 | 0.033804225 | 1 |
| T2DM | endometrium | *HELZ* | 4.165105691 | 1.39E-07 | 4.78E-07 | 0.000672494 |
| T2DM | endometrium | *TMEM116* | -1.686964235 | 0.017545881 | 0.020449824 | 1 |
| T2DM | endometrium | *ZNF442* | -3.339115909 | 8.31E-05 | 0.000118019 | 0.403035259 |
| T2DM | endometrium | *GFPT1* | 5.265798957 | 3.26E-05 | 4.88E-05 | 0.15808492 |
| T2DM | endometrium | *ZNF568* | 1.967952479 | 2.07E-07 | 5.76E-07 | 0.001006626 |
| T2DM | endometrium | *ZNF587* | 5.743466966 | 1.64E-07 | 4.93E-07 | 0.000794263 |
| T2DM | endometrium | *FLVCR1-AS1* | 1.456891815 | 0.000349423 | 0.00046951 | 1 |
| T2DM | endometrium | *SH3BGRL2* | 2.459959837 | 0.007271447 | 0.008737261 | 1 |
| T2DM | endometrium | *TMA16* | 5.171487349 | 6.35E-08 | 4.34E-07 | 0.000308215 |

| | | | | | | |
|---|---|---|---|---|---|---|
| T2DM | endometrium | *HLA-DRB5* | -1.167487462 | 6.61E-05 | 9.50E-05 | 0.320815093 |
| T2DM | endometrium | *CNGA1* | 3.025060367 | 4.91E-05 | 7.18E-05 | 0.238143927 |
| T2DM | endometrium | *ZNF28* | 5.115558616 | 7.42E-08 | 4.36E-07 | 0.000360113 |
| T2DM | endometrium | *C20orf203* | 8.93771611 | 1.56E-07 | 4.82E-07 | 0.000757909 |
| T2DM | endometrium | *NCOA6* | 3.565387379 | 4.73E-07 | 1.07E-06 | 0.002297066 |
| T2DM | endometrium | *TTC37* | 4.094701834 | 1.89E-08 | 4.34E-07 | 9.17E-05 |
| T2DM | endometrium | *TUSC1* | -1.075607574 | 3.07E-06 | 5.50E-06 | 0.014876571 |
| T2DM | endometrium | *PAPSS2* | 5.312635392 | 7.96E-07 | 1.67E-06 | 0.00386441 |
| T2DM | endometrium | *CEP290* | 4.88545002 | 4.01E-07 | 9.41E-07 | 0.001944746 |
| T2DM | endometrium | *ANKRD13B* | 3.946951495 | 1.53E-07 | 4.78E-07 | 0.000742035 |
| T2DM | endometrium | *ECI2* | 2.542075679 | 1.51E-07 | 4.78E-07 | 0.000733564 |
| T2DM | endometrium | *UNC13B* | 3.293498721 | 5.74E-08 | 4.34E-07 | 0.000278401 |
| T2DM | endometrium | *PPP1R14C* | -1.373346161 | 2.09E-06 | 3.92E-06 | 0.010160974 |
| T2DM | endometrium | *GPATCH3* | 3.934510073 | 1.19E-07 | 4.65E-07 | 0.000579283 |
| T2DM | endometrium | *RPL10A* | 4.834678429 | 1.35E-07 | 4.78E-07 | 0.000657118 |
| T2DM | endometrium | *COLGALT2* | 5.723139233 | 0.000356429 | 0.000478394 | 1 |
| T2DM | endometrium | *RCSD1* | 1.743726509 | 1.44E-07 | 4.78E-07 | 0.000698996 |
| T2DM | endometrium | *GRIN3A* | -0.833870535 | 5.47E-06 | 9.28E-06 | 0.026517123 |
| T2DM | endometrium | *TMEM184B* | 3.805742657 | 8.45E-08 | 4.38E-07 | 0.000410141 |
| T2DM | endometrium | *SCAMP5* | 1.845091859 | 6.23E-05 | 8.99E-05 | 0.302498328 |
| T2DM | endometrium | *SFT2D1* | 4.780077209 | 6.61E-08 | 4.34E-07 | 0.000320584 |
| T2DM | endometrium | *HMGN2* | 3.046038565 | 2.25E-06 | 4.17E-06 | 0.01089924 |
| T2DM | endometrium | *DENND4B* | 1.995100539 | 4.12E-07 | 9.63E-07 | 0.001997545 |
| T2DM | endometrium | *RYR3* | 1.857012179 | 2.18E-07 | 5.97E-07 | 0.001060133 |
| T2DM | endometrium | *TYW1* | 3.360321261 | 3.30E-08 | 4.34E-07 | 0.000160331 |
| T2DM | endometrium | *ITPRIPL1* | 1.598414637 | 0.004596179 | 0.005590539 | 1 |
| T2DM | endometrium | *PRMT6* | 8.003338759 | 3.39E-07 | 8.26E-07 | 0.001643348 |
| T2DM | endometrium | *SHISA4* | -10.92608144 | 2.66E-06 | 4.84E-06 | 0.012910881 |
| T2DM | endometrium | *MAP3K3* | 1.560788591 | 7.19E-07 | 1.54E-06 | 0.003489333 |
| T2DM | endometrium | *C1orf174* | 2.882435945 | 3.48E-07 | 8.43E-07 | 0.001689862 |
| T2DM | endometrium | *DZIP3* | 2.83448203 | 7.79E-08 | 4.36E-07 | 0.000378047 |
| T2DM | endometrium | *KIAA0753* | 4.504198043 | 0.002503571 | 0.003116298 | 1 |
| T2DM | endometrium | *TBKBP1* | 2.371768796 | 0.000330784 | 0.000445328 | 1 |
| T2DM | endometrium | *CCDC167* | -5.547771023 | 0.023429214 | 0.027053438 | 1 |
| T2DM | endometrium | *ZFP2* | 5.670543286 | 1.11E-07 | 4.57E-07 | 0.00053648 |
| T2DM | endometrium | *L3MBTL3* | 2.477127881 | 0.036401167 | 0.041459733 | 1 |
| T2DM | endometrium | *MFAP3L* | 3.829454462 | 2.09E-07 | 5.79E-07 | 0.001015778 |
| T2DM | endometrium | *NAGA* | 3.204036518 | 3.82E-07 | 9.07E-07 | 0.001854453 |
| T2DM | endometrium | *SMG5* | 4.759578353 | 5.35E-05 | 7.78E-05 | 0.259680813 |
| T2DM | endometrium | *TGM2* | -1.393600339 | 5.41E-05 | 7.86E-05 | 0.262487124 |
| T2DM | endometrium | *PJA2* | 4.420608386 | 0.000451029 | 0.000599291 | 1 |
| T2DM | endometrium | *AL590822.1* | 3.195971783 | 4.52E-07 | 1.03E-06 | 0.002194673 |
| T2DM | endometrium | *AP001007.1* | 5.102427991 | 8.87E-05 | 0.000125501 | 0.430593911 |
| T2DM | endometrium | *EFCAB2* | 2.179906118 | 1.67E-07 | 5.01E-07 | 0.000812195 |
| T2DM | endometrium | *CHML* | 5.807616349 | 2.57E-07 | 6.71E-07 | 0.001245707 |
| T2DM | endometrium | *TATDN3* | 2.237346441 | 2.45E-07 | 6.51E-07 | 0.001188992 |
| T2DM | endometrium | *CR1* | 1.170264952 | 0.000135624 | 0.000188714 | 0.658047348 |
| T2DM | endometrium | *SAMD5* | 2.13911969 | 1.65E-07 | 4.96E-07 | 0.000800555 |
| T2DM | endometrium | *ECT2L* | 4.006587484 | 1.13E-07 | 4.57E-07 | 0.000548852 |
| T2DM | endometrium | *GPR52* | 41.91849674 | 0.017769754 | 0.020700803 | 1 |

| | | | | | | |
|---|---|---|---|---|---|---|
| T2DM | endometrium | *FAM229B* | 2.57241282 | 4.24E-07 | 9.81E-07 | 0.002055335 |
| T2DM | endometrium | *DDO* | 4.894269523 | 1.09E-06 | 2.20E-06 | 0.005297132 |
| T2DM | endometrium | *RBM20* | 3.548645517 | 4.43E-07 | 1.02E-06 | 0.002149307 |
| T2DM | endometrium | *C6orf163* | 2.43772975 | 0.000326661 | 0.000439899 | 1 |
| T2DM | endometrium | *PCMTD2* | 3.282382215 | 5.54E-08 | 4.34E-07 | 0.000268932 |
| T2DM | endometrium | *SAMD13* | -1.899058602 | 6.33E-06 | 1.06E-05 | 0.03071601 |
| T2DM | endometrium | *EFCAB7* | 6.032083322 | 1.12E-07 | 4.57E-07 | 0.000542444 |
| T2DM | endometrium | *IFIT1B* | 16.72490659 | 1.27E-07 | 4.77E-07 | 0.000617772 |
| T2DM | endometrium | *LIPN* | 7.386333959 | 1.59E-07 | 4.86E-07 | 0.000770345 |
| T2DM | endometrium | *INPP5B* | 1.945341648 | 0.000194953 | 0.000267585 | 0.945911417 |
| T2DM | endometrium | *NEU4* | -2.46045484 | 8.34E-06 | 1.36E-05 | 0.040479425 |
| T2DM | endometrium | *C2orf72* | -3.917236367 | 7.75E-07 | 1.64E-06 | 0.0037594 |
| T2DM | endometrium | *RUFY2* | 5.339043239 | 7.60E-06 | 1.25E-05 | 0.036884221 |
| T2DM | endometrium | *PHACTR4* | 3.062928148 | 4.48E-07 | 1.02E-06 | 0.00217177 |
| T2DM | endometrium | *ASAH2B* | -1.900302903 | 3.85E-05 | 5.71E-05 | 0.186618482 |
| T2DM | endometrium | *AGAP6* | 2.723801027 | 1.43E-08 | 4.34E-07 | 6.93E-05 |
| T2DM | endometrium | *SYT15* | 5.907267268 | 2.60E-05 | 3.93E-05 | 0.125957962 |
| T2DM | endometrium | *LINC00619* | 28.08454549 | 3.64E-07 | 8.73E-07 | 0.001764129 |
| T2DM | endometrium | *DAXX* | 3.161114151 | 3.32E-08 | 4.34E-07 | 0.000161132 |
| T2DM | endometrium | *BMPR2* | 3.341546449 | 4.80E-08 | 4.34E-07 | 0.000232663 |
| T2DM | endometrium | *TCEA3* | 2.33527075 | 1.37E-07 | 4.78E-07 | 0.000662988 |
| T2DM | endometrium | *HSD17B8* | 3.852670506 | 3.28E-06 | 5.84E-06 | 0.01591521 |
| T2DM | endometrium | *OXLD1* | 2.956615096 | 2.91E-08 | 4.34E-07 | 0.000141274 |
| T2DM | endometrium | *TAP2* | 1.710991945 | 0.002865437 | 0.003548519 | 1 |
| T2DM | endometrium | *COL15A1* | 3.32777896 | 5.18E-05 | 7.54E-05 | 0.251180818 |
| T2DM | endometrium | *NOTCH4* | -0.822439581 | 2.69E-07 | 6.93E-07 | 0.001303804 |
| T2DM | endometrium | *AGER* | 3.82348509 | 4.93E-08 | 4.34E-07 | 0.000239292 |
| T2DM | endometrium | *SMIM5* | 7.517426359 | 9.68E-07 | 1.98E-06 | 0.004698598 |
| T2DM | endometrium | *STK19* | 2.04328409 | 3.92E-05 | 5.81E-05 | 0.190076977 |
| T2DM | endometrium | *LAYN* | -3.261101012 | 0.000850786 | 0.001102568 | 1 |
| T2DM | endometrium | *GPANK1* | 2.183942504 | 1.42E-05 | 2.24E-05 | 0.069129058 |
| T2DM | endometrium | *BAG6* | 3.259373831 | 2.95E-08 | 4.34E-07 | 0.000143238 |
| T2DM | endometrium | *LST1* | 2.297073845 | 1.26E-07 | 4.75E-07 | 0.000613429 |
| T2DM | endometrium | *ZNF814* | 4.873596658 | 7.84E-08 | 4.36E-07 | 0.000380594 |
| T2DM | endometrium | *MICA* | 2.673387305 | 1.08E-05 | 1.73E-05 | 0.052186759 |
| T2DM | endometrium | *HLA-C* | 1.185010756 | 0.033915712 | 0.038719773 | 1 |
| T2DM | endometrium | *CCHCR1* | 1.782962094 | 1.53E-06 | 2.96E-06 | 0.007436273 |
| T2DM | endometrium | *PSORS1C2* | 4.188159574 | 0.000199735 | 0.000273838 | 0.969113845 |
| T2DM | endometrium | *DHX16* | 8.31633911 | 1.06E-09 | 2.33E-07 | 5.14E-06 |
| T2DM | endometrium | *ABCF1* | 4.546516802 | 6.45E-08 | 4.34E-07 | 0.000313054 |
| T2DM | endometrium | *LILRB3* | 3.94996565 | 2.18E-07 | 5.97E-07 | 0.001059678 |
| T2DM | endometrium | *HLA-E* | 3.700193278 | 1.07E-06 | 2.15E-06 | 0.005186027 |
| T2DM | endometrium | *ZNF468* | -3.414173466 | 2.91E-05 | 4.39E-05 | 0.141348674 |
| T2DM | endometrium | *HCG9* | 3.976107378 | 6.29E-05 | 9.07E-05 | 0.305348709 |
| T2DM | endometrium | *HLA-G* | 3.037242488 | 6.24E-07 | 1.36E-06 | 0.003025564 |
| T2DM | endometrium | *MALRD1* | 3.398651016 | 8.18E-08 | 4.36E-07 | 0.000396676 |
| T2DM | endometrium | *FAM196B* | 1.968830005 | 1.93E-07 | 5.48E-07 | 0.000936201 |
| T2DM | endometrium | *MROH6* | 2.475237332 | 0.029045685 | 0.033332465 | 1 |
| T2DM | endometrium | *DCTN1* | 8.116926593 | 0.000140722 | 0.000195416 | 0.682781855 |
| T2DM | endometrium | *ZBTB48* | 4.649174338 | 1.83E-06 | 3.47E-06 | 0.008884297 |

| | | | | | | |
|---|---|---|---|---|---|---|
| T2DM | endometrium | *MZT1* | 3.807729919 | 1.36E-07 | 4.78E-07 | 0.000661938 |
| T2DM | endometrium | *ZNF155* | 2.852197673 | 4.13E-07 | 9.65E-07 | 0.002003153 |
| T2DM | endometrium | *ZNF425* | 4.384225193 | 2.73E-06 | 4.96E-06 | 0.013255426 |
| T2DM | endometrium | *C12orf73* | 6.251054649 | 3.14E-06 | 5.61E-06 | 0.015218565 |
| T2DM | endometrium | *PCDHGA1* | -2.214772241 | 1.10E-06 | 2.21E-06 | 0.005327538 |
| T2DM | endometrium | *PCDHA4* | 6.978035361 | 2.76E-06 | 5.02E-06 | 0.013415706 |
| T2DM | endometrium | *TRIM13* | 5.008798435 | 4.90E-08 | 4.34E-07 | 0.000237755 |
| T2DM | endometrium | *SPIRE2* | -1.7806479 | 0.000341281 | 0.000458951 | 1 |
| T2DM | endometrium | *PKHD1L1* | 1.286240402 | 1.64E-05 | 2.55E-05 | 0.079713476 |
| T2DM | endometrium | *SLFN12L* | 1.759701537 | 1.14E-06 | 2.29E-06 | 0.005553593 |
| T2DM | endometrium | *SLC35B4* | 3.702660449 | 5.43E-08 | 4.34E-07 | 0.0002634 |
| T2DM | endometrium | *SYCE1L* | 2.427786668 | 2.26E-05 | 3.44E-05 | 0.109569168 |
| T2DM | endometrium | *TMEM231* | 4.021179883 | 1.01E-06 | 2.05E-06 | 0.004901001 |
| T2DM | endometrium | *LINC00654* | -2.496835354 | 0.012406498 | 0.014649873 | 1 |
| T2DM | endometrium | *SCGB2B2* | 3.397508911 | 5.96E-08 | 4.34E-07 | 0.000289322 |
| T2DM | endometrium | *CCDC144NL* | 3.710378029 | 1.46E-07 | 4.78E-07 | 0.000710386 |
| T2DM | endometrium | *TMEM170B* | 1.718798439 | 5.71E-07 | 1.26E-06 | 0.002771488 |
| T2DM | endometrium | *GOLGA6L10* | 8.207987072 | 8.42E-08 | 4.38E-07 | 0.000408704 |
| T2DM | endometrium | *NT5M* | 4.522670599 | 9.77E-08 | 4.42E-07 | 0.000474067 |
| T2DM | endometrium | *IPO7* | 5.470507389 | 0.000422915 | 0.000562805 | 1 |
| T2DM | endometrium | *PRR13* | 4.868244359 | 7.99E-06 | 1.31E-05 | 0.038745274 |
| T2DM | endometrium | *TECPR1* | 2.985098495 | 9.15E-08 | 4.38E-07 | 0.000444192 |
| T2DM | endometrium | *MT1A* | -1.404375185 | 2.49E-06 | 4.57E-06 | 0.0120883 |
| T2DM | endometrium | *C15orf59* | 3.865386152 | 0.015716519 | 0.018379501 | 1 |
| T2DM | endometrium | *MT1M* | 3.478170343 | 1.91E-07 | 5.46E-07 | 0.000928469 |
| T2DM | endometrium | *CFI* | 1.874373938 | 1.57E-07 | 4.84E-07 | 0.00076366 |
| T2DM | endometrium | *ATP6AP1L* | 3.104733276 | 4.08E-07 | 9.56E-07 | 0.001979302 |
| T2DM | endometrium | *RGL3* | 2.151452718 | 1.25E-07 | 4.73E-07 | 0.000604854 |
| T2DM | endometrium | *NAP1L4* | 3.071284318 | 2.31E-08 | 4.34E-07 | 0.000112202 |
| T2DM | endometrium | *SERF1B* | 2.614083994 | 0.017545464 | 0.020449824 | 1 |
| T2DM | endometrium | *LIN52* | 2.435974406 | 2.89E-09 | 3.60E-07 | 1.40E-05 |
| T2DM | endometrium | *C17orf107* | 3.686942976 | 0.010476664 | 0.012425513 | 1 |
| T2DM | endometrium | *KLRC2* | 5.754616885 | 8.03E-08 | 4.36E-07 | 0.000389748 |
| T2DM | endometrium | *C1QTNF9B* | -1.920075253 | 1.71E-06 | 3.27E-06 | 0.008314874 |
| T2DM | endometrium | *ZDHHC11B* | -2.275879993 | 2.07E-05 | 3.17E-05 | 0.100486227 |
| T2DM | endometrium | *HCG27* | 4.037528475 | 2.69E-13 | 4.35E-10 | 1.30E-09 |
| T2DM | endometrium | *COL6A6* | 3.895164864 | 0.001554106 | 0.001966751 | 1 |
| T2DM | endometrium | *HLA-A* | 3.605278099 | 1.86E-07 | 5.35E-07 | 0.000901276 |
| T2DM | endometrium | *LNP1* | 3.31740749 | 1.50E-07 | 4.78E-07 | 0.000725384 |
| T2DM | endometrium | *PRSS50* | 6.404733202 | 0.018426165 | 0.021439749 | 1 |
| T2DM | endometrium | *METTL6* | 5.521542546 | 7.11E-08 | 4.34E-07 | 0.000344746 |
| T2DM | endometrium | *SACM1L* | 2.835933272 | 7.27E-08 | 4.34E-07 | 0.000352921 |
| T2DM | endometrium | *SLC48A1* | 7.054444662 | 4.69E-07 | 1.06E-06 | 0.00227605 |
| T2DM | endometrium | *TAS2R50* | 70.99166783 | 7.80E-09 | 4.34E-07 | 3.79E-05 |
| T2DM | endometrium | *AC084018.1* | -2.746104463 | 9.67E-06 | 1.57E-05 | 0.046923369 |
| T2DM | endometrium | *C17orf51* | 4.270066765 | 1.50E-07 | 4.78E-07 | 0.000727708 |
| T2DM | endometrium | *OR2A42* | -1.799329381 | 1.27E-06 | 2.51E-06 | 0.006178367 |
| T2DM | endometrium | *ZNF580* | -2.506364449 | 0.002305309 | 0.002876893 | 1 |
| T2DM | endometrium | *NUP62* | 3.054235163 | 1.01E-05 | 1.63E-05 | 0.049068299 |
| T2DM | endometrium | *AL021068.1* | 4.368712878 | 0.003427902 | 0.004209612 | 1 |

| | | | | | | |
|---|---|---|---|---|---|---|
| T2DM | endometrium | *FGFR1OP* | 8.088593513 | 0.000526075 | 0.000694374 | 1 |
| T2DM | endometrium | *ZNF254* | 4.873523096 | 1.71E-06 | 3.26E-06 | 0.008294062 |
| T2DM | endometrium | *TCTEX1D2* | 7.570139689 | 2.49E-08 | 4.34E-07 | 0.000120851 |
| T2DM | endometrium | *AC022498.1* | 16.6675086 | 9.29E-08 | 4.39E-07 | 0.000450614 |
| T2DM | endometrium | *LINGO4* | 3.796810352 | 0.027394421 | 0.03148963 | 1 |
| T2DM | endometrium | *GIMAP1* | 4.966569911 | 4.97E-05 | 7.26E-05 | 0.241153195 |
| T2DM | endometrium | *ARHGEF35* | 3.585487793 | 0.023037607 | 0.026620259 | 1 |
| T2DM | endometrium | *DNLZ* | 2.476928656 | 1.90E-05 | 2.93E-05 | 0.092165169 |
| T2DM | endometrium | *SUPT4H1* | 4.291687442 | 2.53E-08 | 4.34E-07 | 0.00012263 |
| T2DM | endometrium | *MXD3* | 3.817262287 | 1.02E-07 | 4.47E-07 | 0.000493775 |
| T2DM | endometrium | *ARHGAP19* | 3.737171327 | 3.85E-05 | 5.71E-05 | 0.186600917 |
| T2DM | endometrium | *LCAT* | 2.959580091 | 1.16E-07 | 4.60E-07 | 0.000564325 |
| T2DM | endometrium | *GPC2* | -4.185083618 | 3.27E-06 | 5.82E-06 | 0.015855286 |
| T2DM | endometrium | *ERV3-1* | 1.491088567 | 3.25E-07 | 8.01E-07 | 0.001578614 |
| T2DM | endometrium | *CCL14* | 0.831190136 | 0.035053707 | 0.039981332 | 1 |
| T2DM | endometrium | *GBP7* | -0.835106318 | 2.36E-06 | 4.36E-06 | 0.011473918 |
| T2DM | endometrium | *SRA1* | 3.678660043 | 1.89E-07 | 5.43E-07 | 0.000918007 |
| T2DM | endometrium | *TMEM110* | 10.04076812 | 5.91E-12 | 4.10E-09 | 2.87E-08 |
| T2DM | endometrium | *TMX2* | 4.292059713 | 7.44E-08 | 4.36E-07 | 0.000360907 |
| T2DM | endometrium | *LBH* | 2.44187822 | 3.95E-07 | 9.31E-07 | 0.001914219 |
| T2DM | endometrium | *PPP1CB* | 3.139276473 | 1.45E-07 | 4.78E-07 | 0.000702603 |
| T2DM | endometrium | *NCKIPSD* | -2.103172242 | 0.000699823 | 0.000912548 | 1 |
| T2DM | endometrium | *ATF6B* | 7.074470417 | 4.28E-09 | 4.16E-07 | 2.07E-05 |
| T2DM | endometrium | *SLC35F6* | 2.947822362 | 9.99E-08 | 4.45E-07 | 0.000484696 |
| T2DM | endometrium | *EMP2* | 4.148225557 | 1.17E-07 | 4.61E-07 | 0.000569014 |
| T2DM | endometrium | *PPM1N* | 3.863021157 | 1.77E-08 | 4.34E-07 | 8.58E-05 |
| T2DM | endometrium | *SLC23A3* | -1.249936393 | 7.87E-06 | 1.29E-05 | 0.038166676 |
| T2DM | endometrium | *LTB4R* | 3.299364556 | 3.82E-07 | 9.07E-07 | 0.001853968 |
| T2DM | endometrium | *IRF9* | 4.681427316 | 4.02E-08 | 4.34E-07 | 0.000194825 |
| T2DM | endometrium | *GALT* | 2.509535169 | 6.82E-08 | 4.34E-07 | 0.000330901 |
| T2DM | endometrium | *ZNF90* | 2.848870597 | 2.22E-07 | 6.02E-07 | 0.001077172 |
| T2DM | endometrium | *REPIN1* | 10.6588259 | 3.59E-05 | 5.36E-05 | 0.174169173 |
| T2DM | endometrium | *ZNF891* | 4.976915176 | 0.010436914 | 0.012381396 | 1 |
| T2DM | endometrium | *SMIM7* | 4.206809559 | 1.76E-08 | 4.34E-07 | 8.55E-05 |
| T2DM | endometrium | *TSPAN4* | 3.271750941 | 1.33E-07 | 4.78E-07 | 0.000643845 |
| T2DM | endometrium | *PAXIP1-AS2* | 6.112455181 | 1.38E-07 | 4.78E-07 | 0.000671734 |
| T2DM | endometrium | *LYRM4* | 3.372180339 | 5.05E-08 | 4.34E-07 | 0.000244873 |
| T2DM | endometrium | *PRCD* | 5.330641624 | 0.028829216 | 0.033099706 | 1 |
| T2DM | endometrium | *SH3D21* | 4.062812377 | 1.27E-05 | 2.01E-05 | 0.061504681 |
| T2DM | endometrium | *FIS1* | 2.898742327 | 0.003307322 | 0.004068745 | 1 |
| T2DM | endometrium | *ANG* | -2.954875302 | 0.000165309 | 0.000228253 | 0.802080551 |
| T2DM | endometrium | *COLCA2* | 2.225511802 | 0.00223711 | 0.002797541 | 1 |
| T2DM | endometrium | *MBLAC1* | 2.275200243 | 0.002799321 | 0.003469299 | 1 |
| T2DM | endometrium | *SOGA3* | 1.608856407 | 0.008114318 | 0.00970438 | 1 |
| T2DM | endometrium | *LCNL1* | 6.319497917 | 7.32E-06 | 1.21E-05 | 0.03552853 |
| T2DM | endometrium | *GNAT3* | 3.056928967 | 1.48E-07 | 4.78E-07 | 0.000719952 |
| T2DM | endometrium | *PLIN5* | 1.855021725 | 1.08E-05 | 1.74E-05 | 0.0523816 |
| T2DM | endometrium | *SEC14L6* | 0.934087816 | 0.002640462 | 0.003275767 | 1 |
| T2DM | endometrium | *STARD10* | 4.318713538 | 3.96E-14 | 9.60E-11 | 1.92E-10 |
| T2DM | endometrium | *EML6* | 9.533926361 | 1.09E-08 | 4.34E-07 | 5.27E-05 |

| | | | | | | |
|---|---|---|---|---|---|---|
| T2DM | endometrium | *ARHGEF33* | 3.730459304 | 9.60E-08 | 4.42E-07 | 0.000465589 |
| T2DM | endometrium | *IFRD2* | 2.812660898 | 0.00256697 | 0.003187853 | 1 |
| T2DM | endometrium | *EVPLL* | 13.00753547 | 1.45E-08 | 4.34E-07 | 7.05E-05 |
| T2DM | endometrium | *AC005082.1* | 12.17242567 | 3.22E-07 | 7.96E-07 | 0.001564729 |
| T2DM | endometrium | *ACSM4* | 3.270941809 | 2.82E-05 | 4.25E-05 | 0.136651111 |
| T2DM | endometrium | *UBXN2B* | 4.140737956 | 0.003327971 | 0.004092071 | 1 |
| T2DM | endometrium | *C5orf49* | 26.82900849 | 1.37E-06 | 2.69E-06 | 0.006665735 |
| T2DM | endometrium | *GOLGA8B* | 2.069112287 | 3.08E-07 | 7.68E-07 | 0.00149617 |
| T2DM | endometrium | *VPS16* | 4.797330577 | 0.03325387 | 0.037999947 | 1 |
| T2DM | endometrium | *MYL5* | -3.883829469 | 0.000983599 | 0.001265895 | 1 |
| T2DM | endometrium | *NPEPL1* | 2.704812231 | 1.74E-07 | 5.13E-07 | 0.000845794 |
| T2DM | endometrium | *TMEM242* | 6.560703528 | 3.03E-06 | 5.44E-06 | 0.014700521 |
| T2DM | endometrium | *SLC35E2* | 2.50513003 | 6.05E-07 | 1.33E-06 | 0.002935955 |
| T2DM | endometrium | *LINC01139* | -1.04262973 | 0.000225272 | 0.00030746 | 1 |
| T2DM | endometrium | *AC009403.2* | -4.069731179 | 4.60E-05 | 6.75E-05 | 0.223012992 |
| T2DM | endometrium | *PAM16* | 10.26975932 | 1.19E-06 | 2.37E-06 | 0.005782064 |
| T2DM | endometrium | *AC016773.1* | 2.164350342 | 0.006403464 | 0.007728758 | 1 |
| T2DM | endometrium | *AP000350.4* | -1.252541078 | 3.36E-05 | 5.02E-05 | 0.162824407 |
| T2DM | endometrium | *RNASEK* | 6.518001657 | 4.89E-06 | 8.41E-06 | 0.023738151 |
| T2DM | endometrium | *FAM228B* | 2.443292547 | 3.25E-08 | 4.34E-07 | 0.000157497 |
| T2DM | endometrium | *RNU6ATAC35I* | 25.99803761 | 7.31E-05 | 0.000104508 | 0.354730501 |
| T2DM | endometrium | *PPP3R1* | -2.461305113 | 0.011120522 | 0.013163399 | 1 |
| T2DM | endometrium | *FANCG* | -2.331523198 | 2.96E-06 | 5.34E-06 | 0.014344013 |
| T2DM | endometrium | *AP4M1* | 4.612486753 | 1.01E-07 | 4.47E-07 | 0.00049196 |
| T2DM | endometrium | *ZNF880* | 1.982208296 | 4.78E-06 | 8.22E-06 | 0.023176563 |
| T2DM | endometrium | *TIGD1* | 4.065529989 | 1.38E-05 | 2.18E-05 | 0.067155966 |
| T2DM | endometrium | *FXYD7* | 3.700669961 | 6.39E-06 | 1.07E-05 | 0.030985617 |
| T2DM | endometrium | *XKR9* | -1.399865882 | 0.000392014 | 0.000523836 | 1 |
| T2DM | endometrium | *SLC12A8* | 2.375687674 | 3.52E-06 | 6.22E-06 | 0.017067461 |
| T2DM | endometrium | *APOL6* | 5.024490146 | 0.000303204 | 0.000409448 | 1 |
| T2DM | endometrium | *OR2A1* | 4.167575623 | 5.85E-05 | 8.46E-05 | 0.283822723 |
| T2DM | endometrium | *UBA52* | 5.255605639 | 1.85E-08 | 4.34E-07 | 8.98E-05 |
| T2DM | endometrium | *TIAF1* | 5.224935375 | 1.31E-06 | 2.57E-06 | 0.006335894 |
| T2DM | endometrium | *RAB6C* | 81.18185231 | 3.02E-07 | 7.58E-07 | 0.00146767 |
| T2DM | endometrium | *AC062017.1* | 2.865279178 | 1.47E-07 | 4.78E-07 | 0.000714285 |
| T2DM | endometrium | *ADRA2B* | 10.64137759 | 6.83E-06 | 1.14E-05 | 0.033130732 |
| T2DM | endometrium | *LINC00571* | -2.130044544 | 1.23E-05 | 1.95E-05 | 0.05958162 |
| T2DM | endometrium | *CERS1* | 10.65129124 | 0.024127892 | 0.027846939 | 1 |
| T2DM | endometrium | *OSER1-AS1* | 4.832824483 | 1.16E-07 | 4.60E-07 | 0.000564133 |
| T2DM | endometrium | *C5orf66* | 2.410256942 | 4.71E-07 | 1.07E-06 | 0.002284269 |
| T2DM | endometrium | *LINC01133* | 1.934439887 | 4.09E-07 | 9.59E-07 | 0.001986751 |
| T2DM | endometrium | *LINC01117* | 2.868019025 | 1.33E-07 | 4.78E-07 | 0.000644924 |
| T2DM | endometrium | *RASAL2-AS1* | -5.247680906 | 3.18E-05 | 4.77E-05 | 0.154161938 |
| T2DM | endometrium | *LINC00240* | 1.432741851 | 0.013858761 | 0.016289416 | 1 |
| T2DM | endometrium | *LINC00184* | 72.33567785 | 1.54E-07 | 4.78E-07 | 0.000745992 |
| T2DM | endometrium | *AL049840.1* | 2.413669873 | 7.19E-07 | 1.54E-06 | 0.00349101 |
| T2DM | endometrium | *PLEKHM1* | 5.19482848 | 1.29E-08 | 4.34E-07 | 6.27E-05 |
| T2DM | endometrium | *AP000350.6* | 1.928788527 | 0.001160109 | 0.001487146 | 1 |
| T2DM | endometrium | *SFTA1P* | -4.290999329 | 0.02193692 | 0.025378621 | 1 |
| T2DM | endometrium | *CYP4A22-AS1* | 3.409078945 | 1.51E-07 | 4.78E-07 | 0.000732129 |

| | | | | | | |
|---|---|---|---|---|---|---|
| T2DM | endometrium | *FAM225B* | -5.131017184 | 4.25E-05 | 6.26E-05 | 0.206057113 |
| T2DM | endometrium | *MIAT* | 3.784889321 | 1.72E-07 | 5.09E-07 | 0.000836292 |
| T2DM | endometrium | *ERCC6* | 3.541243808 | 1.39E-07 | 4.78E-07 | 0.000673683 |
| T2DM | endometrium | *LINC00694* | 10.90048887 | 3.60E-09 | 4.07E-07 | 1.75E-05 |
| T2DM | endometrium | *LINC00115* | -1.140214031 | 3.49E-06 | 6.17E-06 | 0.016923455 |
| T2DM | endometrium | *HAR1A* | -1.32481281 | 1.46E-06 | 2.83E-06 | 0.007077374 |
| T2DM | endometrium | *LINC00937* | -2.699145344 | 0.001734597 | 0.002184908 | 1 |
| T2DM | endometrium | *FTCDNL1* | 2.320852287 | 4.85E-06 | 8.33E-06 | 0.023518627 |
| T2DM | endometrium | *FAM138B* | 41.39529694 | 1.63E-07 | 4.92E-07 | 0.000790979 |
| T2DM | endometrium | *ZNF717* | 1.310332218 | 0.011622398 | 0.01374741 | 1 |
| T2DM | endometrium | *AC017074.2* | 26.63079334 | 0.001336262 | 0.001703059 | 1 |
| T2DM | endometrium | *AC009299.3* | 9.157043947 | 3.08E-08 | 4.34E-07 | 0.000149656 |
| T2DM | endometrium | *SCAMP4* | 4.447350048 | 5.30E-08 | 4.34E-07 | 0.000257289 |
| T2DM | endometrium | *SLC2A1-AS1* | -2.723917625 | 3.59E-05 | 5.36E-05 | 0.174259057 |
| T2DM | endometrium | *LINC01068* | 4.497324846 | 0.002541992 | 0.003159258 | 1 |
| T2DM | endometrium | *AC005884.1* | 4.230656249 | 0.007799723 | 0.009346568 | 1 |
| T2DM | endometrium | *AC108463.2* | 9.835795145 | 1.83E-09 | 3.07E-07 | 8.88E-06 |
| T2DM | endometrium | *C14orf144* | -2.942954508 | 1.39E-05 | 2.20E-05 | 0.067592532 |
| T2DM | endometrium | *POLR2J2* | -33.8022944 | 0.022216361 | 0.025695776 | 1 |
| T2DM | endometrium | *LINC00578* | 3.979035416 | 1.95E-07 | 5.50E-07 | 0.000944859 |
| T2DM | endometrium | *C19orf24* | -1.241165835 | 2.52E-06 | 4.62E-06 | 0.012238594 |
| T2DM | endometrium | *ZNF883* | 2.947462258 | 3.03E-07 | 7.59E-07 | 0.001471758 |
| T2DM | endometrium | *ARL17B* | 3.305726582 | 0.017453552 | 0.020356883 | 1 |
| T2DM | endometrium | *PATL2* | 2.634285876 | 7.25E-08 | 4.34E-07 | 0.000351872 |
| T2DM | endometrium | *CASC20* | 4.326219963 | 2.57E-07 | 6.71E-07 | 0.001244748 |
| T2DM | endometrium | *PSMG3-AS1* | 6.451601982 | 2.70E-07 | 6.95E-07 | 0.001309624 |
| T2DM | endometrium | *AC112229.4* | 4.560991525 | 9.46E-07 | 1.94E-06 | 0.004588291 |
| T2DM | endometrium | *KLHL7-AS1* | 5.097731931 | 1.63E-06 | 3.13E-06 | 0.007906254 |
| T2DM | endometrium | *AC092171.4* | 3.375747142 | 1.51E-07 | 4.78E-07 | 0.00073289 |
| T2DM | endometrium | *RPS18* | 4.312888933 | 2.03E-07 | 5.65E-07 | 0.000984485 |
| T2DM | endometrium | *DIRC3* | 1.601061414 | 1.01E-07 | 4.47E-07 | 0.000491521 |
| T2DM | endometrium | *TAPBP* | 3.622462619 | 1.01E-08 | 4.34E-07 | 4.91E-05 |
| T2DM | endometrium | *AC107021.1* | 4.334499583 | 9.32E-07 | 1.91E-06 | 0.004519726 |
| T2DM | endometrium | *TMA7* | 3.989661839 | 3.15E-09 | 3.72E-07 | 1.53E-05 |
| T2DM | endometrium | *MTRNR2L4* | -4.783997952 | 0.035863056 | 0.040875628 | 1 |
| T2DM | endometrium | *LINC00865* | -0.887981355 | 1.98E-06 | 3.72E-06 | 0.009603462 |
| T2DM | endometrium | *AC009495.3* | 14.37583418 | 7.96E-05 | 0.000113345 | 0.386394591 |
| T2DM | endometrium | *HLA-DQB2* | 1.836472738 | 3.91E-11 | 2.11E-08 | 1.90E-07 |
| T2DM | endometrium | *GOLGA8N* | -1.737979023 | 0.00477939 | 0.005806109 | 1 |
| T2DM | endometrium | *LYRM9* | -2.472238793 | 9.83E-06 | 1.59E-05 | 0.047710795 |
| T2DM | endometrium | *LINC01166* | 3.903837456 | 0.002269369 | 0.002834958 | 1 |
| T2DM | endometrium | *LINC00702* | 5.454486889 | 2.14E-07 | 5.88E-07 | 0.001038541 |
| T2DM | endometrium | *GPX1* | 3.022549713 | 5.81E-07 | 1.28E-06 | 0.002818877 |
| T2DM | endometrium | *RPS28* | 4.348513008 | 0.040614527 | 0.04608552 | 1 |
| T2DM | endometrium | *TRIM26* | 5.525007368 | 0.000139369 | 0.000193648 | 0.676218677 |
| T2DM | endometrium | *PINLYP* | 3.029073459 | 7.71E-07 | 1.63E-06 | 0.003739734 |
| T2DM | endometrium | *ANKRD65* | 11.27800204 | 8.81E-08 | 4.38E-07 | 0.000427358 |
| T2DM | endometrium | *ZSCAN31* | 2.254883656 | 4.95E-06 | 8.49E-06 | 0.023995647 |
| T2DM | endometrium | *AC009495.2* | 76.9045019 | 1.13E-06 | 2.26E-06 | 0.005485288 |
| T2DM | endometrium | *LINC00957* | 5.95088451 | 0.000121235 | 0.000169329 | 0.588230167 |

| | | | | | | |
|---|---|---|---|---|---|---|
| T2DM | endometrium | *LINC00484* | -1.489752735 | 0.001636735 | 0.002067007 | 1 |
| T2DM | endometrium | *KIAA0040* | 2.240927402 | 3.05E-07 | 7.62E-07 | 0.001479919 |
| T2DM | endometrium | *LINC00607* | 4.558207086 | 0.002067348 | 0.002593942 | 1 |
| T2DM | endometrium | *AC019117.2* | 20.49768885 | 3.38E-06 | 5.99E-06 | 0.01639311 |
| T2DM | endometrium | *ZBTB22* | 4.374692613 | 1.12E-06 | 2.25E-06 | 0.005456145 |
| T2DM | endometrium | *ZBED5* | 4.525187817 | 6.94E-07 | 1.49E-06 | 0.003364905 |
| T2DM | endometrium | *CDKN2AIPNL* | 3.612132685 | 0.007672813 | 0.009205858 | 1 |
| T2DM | endometrium | *LINC00987* | 3.568229484 | 1.94E-05 | 2.99E-05 | 0.094276382 |
| T2DM | endometrium | *GS1-124K5.4* | 2.191001596 | 1.02E-05 | 1.64E-05 | 0.049320561 |
| T2DM | endometrium | *LINC00959* | -1.764211191 | 1.57E-05 | 2.44E-05 | 0.075942714 |
| T2DM | endometrium | *HLA-DQA2* | 3.047285449 | 3.66E-23 | 1.78E-19 | 1.78E-19 |
| T2DM | endometrium | *IRGM* | 3.162736879 | 8.21E-07 | 1.72E-06 | 0.003981875 |
| T2DM | endometrium | *LINC01143* | -13.57371232 | 0.000113228 | 0.000158551 | 0.549380755 |
| T2DM | endometrium | *LINC00211* | -1.228300093 | 2.22E-06 | 4.12E-06 | 0.010756374 |
| T2DM | endometrium | *LRRC37A2* | 4.890136338 | 4.31E-08 | 4.34E-07 | 0.000209104 |
| T2DM | endometrium | *LINC00707* | 13.75097278 | 0.000466511 | 0.000618784 | 1 |
| T2DM | endometrium | *TXNDC5* | 14.77417791 | 1.30E-07 | 4.78E-07 | 0.000628751 |
| T2DM | endometrium | *LINC01119* | -3.460797381 | 9.69E-05 | 0.000136423 | 0.470113148 |
| T2DM | endometrium | *NME1* | 2.092346164 | 0.026091771 | 0.030027816 | 1 |
| T2DM | endometrium | *CDRT4* | 7.719875365 | 3.42E-06 | 6.07E-06 | 0.01660747 |
| T2DM | endometrium | *APOBEC3G* | 1.087825254 | 0.006160022 | 0.00744419 | 1 |
| T2DM | endometrium | *C1orf226* | 12.95122736 | 1.29E-08 | 4.34E-07 | 6.27E-05 |
| T2DM | endometrium | *PSMB9* | 3.250827649 | 1.83E-08 | 4.34E-07 | 8.86E-05 |
| T2DM | endometrium | *PPIL3* | 2.374520047 | 0.001257318 | 0.001606665 | 1 |
| T2DM | endometrium | *L1TD1* | 4.219016651 | 0.000385198 | 0.000515012 | 1 |
| T2DM | endometrium | *C1QTNF9* | 11.16127985 | 4.28E-07 | 9.89E-07 | 0.002074372 |
| T2DM | endometrium | *ISY1* | 2.864521616 | 2.02E-05 | 3.10E-05 | 0.09793751 |
| T2DM | endometrium | *PNMA2* | 4.289776074 | 1.43E-06 | 2.79E-06 | 0.006952322 |
| T2DM | endometrium | *KRBOX1* | 45.81515185 | 1.70E-07 | 5.06E-07 | 0.000824455 |
| T2DM | endometrium | *ARHGEF25* | 3.334862703 | 2.50E-07 | 6.60E-07 | 0.001213314 |
| T2DM | endometrium | *LINC00886* | 1.3907382 | 0.000121239 | 0.000169329 | 0.588250131 |
| T2DM | endometrium | *MIF* | -2.088841819 | 0.000476394 | 0.000631374 | 1 |
| T2DM | endometrium | *NSUN6* | 5.138655352 | 2.15E-07 | 5.92E-07 | 0.001045303 |
| T2DM | endometrium | *HLA-DOB* | -1.341560836 | 0.000749209 | 0.000974835 | 1 |
| T2DM | endometrium | *LINC00877* | 4.154076169 | 1.55E-07 | 4.80E-07 | 0.000750938 |
| T2DM | endometrium | *CD302* | 3.067104612 | 2.54E-08 | 4.34E-07 | 0.000123158 |
| T2DM | endometrium | *INMT* | 4.415860484 | 0.00139381 | 0.001773608 | 1 |
| T2DM | endometrium | *ATP5O* | -2.459985722 | 0.001665552 | 0.002102305 | 1 |
| T2DM | endometrium | *PLEKHO2* | 2.906781724 | 4.02E-07 | 9.44E-07 | 0.001951511 |
| T2DM | endometrium | *C8orf58* | 5.84927987 | 1.80E-08 | 4.34E-07 | 8.74E-05 |
| T2DM | endometrium | *HOGA1* | -1.800035368 | 3.68E-06 | 6.48E-06 | 0.017841378 |
| T2DM | endometrium | *PI4KA* | 2.998337738 | 7.95E-08 | 4.36E-07 | 0.000385644 |
| T2DM | endometrium | *LINC00969* | 3.148609609 | 2.03E-07 | 5.65E-07 | 0.000984396 |
| T2DM | endometrium | *AMACR* | 1.22528069 | 0.0074261 | 0.00891426 | 1 |
| T2DM | endometrium | *BGLAP* | -4.510855407 | 4.07E-06 | 7.13E-06 | 0.019752765 |
| T2DM | endometrium | *MRPL20* | 5.907999535 | 1.25E-08 | 4.34E-07 | 6.08E-05 |
| T2DM | endometrium | *DECR2* | -5.034000344 | 0.000489923 | 0.000647889 | 1 |
| T2DM | endometrium | *ZNF709* | -5.832745828 | 3.21E-06 | 5.72E-06 | 0.015560863 |
| T2DM | endometrium | *ERVW-1* | 9.866675463 | 1.23E-07 | 4.70E-07 | 0.000596794 |
| T2DM | endometrium | *MRPL33* | 3.157162969 | 6.89E-08 | 4.34E-07 | 0.000334229 |

| | | | | | | |
|---|---|---|---|---|---|---|
| T2DM | endometrium | *STON1* | 5.341575945 | 2.10E-06 | 3.92E-06 | 0.010179016 |
| T2DM | endometrium | *VSIG8* | 5.164686735 | 7.37E-07 | 1.57E-06 | 0.003577575 |
| T2DM | endometrium | *C7orf73* | 3.431924302 | 1.03E-07 | 4.47E-07 | 0.000498669 |
| T2DM | endometrium | *FGF14-IT1* | 8.765603361 | 2.07E-07 | 5.76E-07 | 0.001005552 |
| T2DM | endometrium | *PALM2* | 2.646305463 | 6.47E-08 | 4.34E-07 | 0.000314157 |
| T2DM | endometrium | *NBPF15* | 7.206810549 | 2.36E-07 | 6.32E-07 | 0.001145764 |
| T2DM | endometrium | *UPK3B* | 2.019733477 | 1.74E-07 | 5.12E-07 | 0.000841956 |
| T2DM | endometrium | *IL10RB* | 2.241623693 | 4.39E-08 | 4.34E-07 | 0.000212788 |
| T2DM | endometrium | *NME1-NME2* | 14.46778577 | 1.42E-05 | 2.24E-05 | 0.069124208 |
| T2DM | endometrium | *PLA2G4B* | 20.68874573 | 2.68E-12 | 2.16E-09 | 1.30E-08 |
| T2DM | endometrium | *LEFTY1* | -28.28506542 | 0.000213129 | 0.000291789 | 1 |
| T2DM | endometrium | *TTC4* | 3.820516987 | 2.31E-08 | 4.34E-07 | 0.000112273 |
| T2DM | endometrium | *JMJD7* | 3.006614982 | 0.000982408 | 0.001264698 | 1 |
| T2DM | endometrium | *APOBEC3D* | -13.61615496 | 4.22E-05 | 6.23E-05 | 0.204947112 |
| T2DM | endometrium | *TMEM199* | 2.681826576 | 4.66E-05 | 6.85E-05 | 0.22631433 |
| T2DM | endometrium | *SPRR2F* | 4.449901539 | 0.019363386 | 0.0224948 | 1 |
| T2DM | endometrium | *P2RY11* | 1.495030057 | 0.013718805 | 0.01613273 | 1 |
| T2DM | endometrium | *DBNDD2* | 12.83055705 | 0.000956632 | 0.001233479 | 1 |
| T2DM | endometrium | *LILRA6* | 2.519117703 | 3.34E-05 | 4.99E-05 | 0.161855218 |
| T2DM | endometrium | *SCARF2* | -0.979137526 | 4.10E-06 | 7.17E-06 | 0.019904777 |
| T2DM | endometrium | *APOBEC3C* | 4.025199058 | 6.91E-08 | 4.34E-07 | 0.000335235 |
| T2DM | endometrium | *CCDC13* | -1.909702257 | 3.21E-06 | 5.72E-06 | 0.015577074 |
| T2DM | endometrium | *C4A* | -6.511043647 | 5.87E-06 | 9.91E-06 | 0.02849172 |
| T2DM | endometrium | *NEAT1* | 2.530865697 | 3.12E-08 | 4.34E-07 | 0.000151607 |
| T2DM | endometrium | *RAB30-AS1* | 3.716120289 | 3.94E-06 | 6.91E-06 | 0.019122887 |
| T2DM | endometrium | *UBAP1L* | 3.278058033 | 1.06E-07 | 4.48E-07 | 0.000513512 |
| T2DM | endometrium | *LINC00926* | 6.648077522 | 6.30E-08 | 4.34E-07 | 0.000305616 |
| T2DM | endometrium | *DPH6-AS1* | 4.359428806 | 1.22E-07 | 4.69E-07 | 0.000589761 |
| T2DM | endometrium | *BCKDHA* | 5.072314716 | 2.63E-05 | 3.97E-05 | 0.127372944 |
| T2DM | endometrium | *CDK11B* | -2.773236438 | 0.020939836 | 0.02424823 | 1 |
| T2DM | endometrium | *ABHD14A* | 3.345824599 | 1.07E-07 | 4.49E-07 | 0.000517434 |
| T2DM | endometrium | *SRP14-AS1* | -1.891298316 | 6.31E-05 | 9.09E-05 | 0.306326344 |
| T2DM | endometrium | *GDNF-AS1* | -0.618438924 | 1.68E-06 | 3.22E-06 | 0.008163524 |
| T2DM | endometrium | *CCDC153* | -2.010565329 | 2.67E-05 | 4.04E-05 | 0.129699085 |
| T2DM | endometrium | *AL357673.1* | -2.275581363 | 0.00037515 | 0.000502271 | 1 |
| T2DM | endometrium | *C5orf17* | -2.555048487 | 0.003779237 | 0.004617693 | 1 |
| T2DM | endometrium | *TMEM150C* | 2.031376669 | 6.14E-05 | 8.86E-05 | 0.298038239 |
| T2DM | endometrium | *LINC00939* | 7.212030495 | 3.01E-06 | 5.42E-06 | 0.014611113 |
| T2DM | endometrium | *LINC01060* | 5.294949005 | 3.08E-05 | 4.63E-05 | 0.149619276 |
| T2DM | endometrium | *LINC00942* | -2.603362708 | 1.87E-06 | 3.53E-06 | 0.009074078 |
| T2DM | endometrium | *PDCD6* | 6.697868131 | 4.29E-05 | 6.33E-05 | 0.208323343 |
| T2DM | endometrium | *GOLGA8K* | 3.468002071 | 0.004615645 | 0.005612809 | 1 |
| T2DM | endometrium | *TMEM158* | 12.17233946 | 2.50E-07 | 6.60E-07 | 0.001212839 |
| T2DM | endometrium | *DNAH10OS* | 16.32345485 | 9.17E-05 | 0.000129435 | 0.44486885 |
| T2DM | endometrium | *PCDHA10* | 3.549704079 | 4.93E-06 | 8.46E-06 | 0.023910679 |
| T2DM | endometrium | *ZNF718* | 2.275022557 | 6.76E-08 | 4.34E-07 | 0.000327911 |
| T2DM | endometrium | *SMIM20* | -3.41552849 | 0.002156684 | 0.002700447 | 1 |
| T2DM | endometrium | *LINC00989* | 3.270451159 | 6.39E-07 | 1.39E-06 | 0.003098901 |
| T2DM | endometrium | *ATP6V1E2* | 2.640950545 | 2.29E-06 | 4.24E-06 | 0.011093531 |
| T2DM | endometrium | *GLI4* | -2.369029179 | 3.71E-05 | 5.53E-05 | 0.180059849 |

| | | | | | | |
|---|---|---|---|---|---|---|
| T2DM | endometrium | *SNHG18* | 2.52556881 | 4.07E-06 | 7.13E-06 | 0.019749585 |
| T2DM | endometrium | *ALG1L2* | 3.213844397 | 1.62E-07 | 4.91E-07 | 0.000786881 |
| T2DM | endometrium | *ZNF550* | 2.736619481 | 1.93E-07 | 5.48E-07 | 0.000938108 |
| T2DM | endometrium | *LINC01094* | 3.33025508 | 1.58E-07 | 4.85E-07 | 0.000767985 |
| T2DM | endometrium | *TMEM200B* | -2.59342799 | 0.000935692 | 0.001207119 | 1 |
| T2DM | endometrium | *PCDHGB6* | 5.645743874 | 8.26E-07 | 1.72E-06 | 0.004005402 |
| T2DM | endometrium | *SLC10A5* | 4.42713011 | 0.037628135 | 0.042796931 | 1 |
| T2DM | endometrium | *PCDHGA6* | -1.409388658 | 0.000622934 | 0.000817106 | 1 |
| T2DM | endometrium | *UTP14C* | 3.072978897 | 1.57E-07 | 4.84E-07 | 0.000763371 |
| T2DM | endometrium | *PCDHGB2* | 14.65889228 | 3.43E-07 | 8.34E-07 | 0.00166422 |
| T2DM | endometrium | *LYN* | 1.567563153 | 3.10E-07 | 7.72E-07 | 0.001505107 |
| T2DM | endometrium | *SIGLEC14* | 3.306303841 | 0.037622611 | 0.042796931 | 1 |
| T2DM | endometrium | *ARMS2* | 6.949290728 | 1.22E-05 | 1.93E-05 | 0.059028642 |
| T2DM | endometrium | *FPGT* | 3.023826417 | 2.23E-09 | 3.48E-07 | 1.08E-05 |
| T2DM | endometrium | *MEX3A* | -1.680566905 | 5.68E-06 | 9.61E-06 | 0.027551394 |
| T2DM | endometrium | *DPP3* | 1.999870537 | 0.001440731 | 0.001829476 | 1 |
| T2DM | endometrium | *SNHG9* | -1.863860948 | 8.55E-05 | 0.000121173 | 0.414894935 |
| T2DM | endometrium | *TAS2R43* | 2.675342248 | 0.013369524 | 0.015741065 | 1 |
| T2DM | endometrium | *TAS2R20* | 6.338251627 | 4.89E-05 | 7.16E-05 | 0.23724182 |
| T2DM | endometrium | *ZNF432* | 3.886360074 | 4.13E-06 | 7.22E-06 | 0.02006125 |
| T2DM | endometrium | *HMBS* | -1.176197338 | 1.27E-05 | 2.01E-05 | 0.061404254 |
| T2DM | endometrium | *TAS2R31* | 3.687461357 | 0.001324396 | 0.001688378 | 1 |
| T2DM | endometrium | *KIAA1147* | 3.639559384 | 8.43E-08 | 4.38E-07 | 0.000408926 |
| T2DM | endometrium | *GATC* | 2.11150041 | 0.03017135 | 0.034599714 | 1 |
| T2DM | endometrium | *ZBED6* | 6.113785483 | 2.22E-07 | 6.02E-07 | 0.001077734 |
| T2DM | endometrium | *CNPY2* | 2.326847241 | 1.15E-07 | 4.59E-07 | 0.000558831 |
| T2DM | endometrium | *CHURC1* | -1.926168105 | 0.017867188 | 0.020809312 | 1 |
| T2DM | endometrium | *PDF* | -5.433044402 | 1.31E-06 | 2.57E-06 | 0.006351352 |
| T2DM | endometrium | *LINC00640* | 2.70398432 | 0.000409721 | 0.000546595 | 1 |
| T2DM | endometrium | *HIF1A-AS2* | 4.151027907 | 6.55E-08 | 4.34E-07 | 0.000317722 |
| T2DM | endometrium | *LINC00638* | 8.194185381 | 1.82E-07 | 5.26E-07 | 0.000884551 |
| T2DM | endometrium | *SLC25A21-AS1* | 3.210459005 | 1.81E-07 | 5.25E-07 | 0.000879624 |
| T2DM | endometrium | *RNASE4* | 12.38925266 | 3.82E-07 | 9.07E-07 | 0.001854278 |
| T2DM | endometrium | *MC1R* | 8.704991669 | 9.36E-09 | 4.34E-07 | 4.54E-05 |
| T2DM | endometrium | *LINC00639* | 2.065882717 | 4.37E-07 | 1.01E-06 | 0.002122447 |
| T2DM | endometrium | *SMIM6* | -11.39294207 | 0.019535129 | 0.022681131 | 1 |
| T2DM | endometrium | *LINC00924* | 2.123624702 | 3.50E-06 | 6.19E-06 | 0.016974721 |
| T2DM | endometrium | *ITGB3* | 2.87748543 | 0.00344294 | 0.004224872 | 1 |
| T2DM | endometrium | *MRPL46* | -2.54428342 | 8.63E-05 | 0.000122314 | 0.418924226 |
| T2DM | endometrium | *IQCH-AS1* | 3.068973117 | 8.33E-08 | 4.38E-07 | 0.000404055 |
| T2DM | endometrium | *PWRN1* | 3.598049337 | 1.95E-07 | 5.51E-07 | 0.000946284 |
| T2DM | endometrium | *LINC01003* | 3.338325553 | 1.12E-07 | 4.57E-07 | 0.000542538 |
| T2DM | endometrium | *C15orf65* | 6.19920587 | 0.002003978 | 0.002517686 | 1 |
| T2DM | endometrium | *GOLGA8H* | -0.938070551 | 1.53E-05 | 2.39E-05 | 0.074297095 |
| T2DM | endometrium | *LINC00662* | 4.083447755 | 1.91E-07 | 5.46E-07 | 0.00092875 |
| T2DM | endometrium | *C19orf84* | 39.71354562 | 4.95E-08 | 4.34E-07 | 0.000240041 |
| T2DM | endometrium | *LINC00675* | -1.760632529 | 0.002184565 | 0.002733241 | 1 |
| T2DM | endometrium | *LINC00667* | 4.405875982 | 0.000130407 | 0.000181716 | 0.632736743 |
| T2DM | endometrium | *LINC00672* | 13.62038732 | 2.90E-08 | 4.34E-07 | 0.000140572 |
| T2DM | endometrium | *LINC00909* | 6.116503204 | 3.15E-07 | 7.81E-07 | 0.001528053 |

| | | | | | | |
|---|---|---|---|---|---|---|
| T2DM | endometrium | *DYNLL2* | 3.352592809 | 2.06E-07 | 5.73E-07 | 0.001000863 |
| T2DM | endometrium | *STRADA* | 3.890694783 | 1.86E-06 | 3.51E-06 | 0.009010735 |
| T2DM | endometrium | *MYO15B* | 5.163001628 | 1.31E-07 | 4.78E-07 | 0.000637107 |
| T2DM | endometrium | *LINC00663* | 2.154654221 | 1.14E-05 | 1.82E-05 | 0.055232508 |
| T2DM | endometrium | *ILF3-AS1* | 5.966265142 | 4.42E-08 | 4.34E-07 | 0.0002146 |
| T2DM | endometrium | *ZNF582-AS1* | -4.606628558 | 0.000409061 | 0.000545865 | 1 |
| T2DM | endometrium | *ZNF285* | 3.24859789 | 0.0004783 | 0.00063372 | 1 |
| T2DM | endometrium | *S1PR2* | 3.49792277 | 4.46E-05 | 6.56E-05 | 0.216236079 |
| T2DM | endometrium | *ZNF224* | 6.861430822 | 6.90E-07 | 1.49E-06 | 0.003348094 |
| T2DM | endometrium | *LIN37* | -1.432074712 | 1.41E-06 | 2.75E-06 | 0.006846213 |
| T2DM | endometrium | *NDUFA7* | 18.33499161 | 1.10E-07 | 4.56E-07 | 0.00053281 |
| T2DM | endometrium | *AC004466.1* | 4.163819949 | 0.003739439 | 0.004574826 | 1 |
| T2DM | endometrium | *SCGB1B2P* | 7.865474157 | 6.25E-05 | 9.01E-05 | 0.303230839 |
| T2DM | endometrium | *ZNF728* | 9.578716027 | 4.57E-07 | 1.04E-06 | 0.002219462 |
| T2DM | endometrium | *TRABD2B* | 2.268064816 | 1.21E-07 | 4.69E-07 | 0.000587499 |
| T2DM | endometrium | *AC093157.1* | 2.185870949 | 1.51E-07 | 4.78E-07 | 0.000734274 |
| T2DM | endometrium | *FBXO17* | 2.696575594 | 2.07E-08 | 4.34E-07 | 0.000100633 |
| T2DM | endometrium | *ZNF587B* | 3.536123572 | 1.54E-07 | 4.78E-07 | 0.000744803 |
| T2DM | endometrium | *SNHG8* | 1.872044732 | 4.66E-06 | 8.06E-06 | 0.022627573 |
| T2DM | endometrium | *AP001172.2* | 1.98840578 | 0.006058608 | 0.00732346 | 1 |
| T2DM | endometrium | *HCG17* | 2.910280715 | 1.06E-07 | 4.48E-07 | 0.00051412 |
| T2DM | endometrium | *YTHDF3-AS1* | 3.887530414 | 0.007714074 | 0.009253075 | 1 |
| T2DM | endometrium | *MILR1* | 1.86913608 | 4.13E-05 | 6.10E-05 | 0.200323734 |
| T2DM | endometrium | *GTF2H5* | 3.058130733 | 1.60E-07 | 4.87E-07 | 0.000778661 |
| T2DM | endometrium | *FGF14-AS2* | 4.631428068 | 2.97E-07 | 7.45E-07 | 0.001439567 |
| T2DM | endometrium | *CASC15* | 1.847945805 | 1.27E-07 | 4.77E-07 | 0.000618413 |
| T2DM | endometrium | *AP000347.2* | -5.348518399 | 0.000247226 | 0.000336288 | 1 |
| T2DM | endometrium | *DOC2B* | -1.150939054 | 0.032936073 | 0.03764566 | 1 |
| T2DM | endometrium | *GRIN2B* | 4.232052373 | 7.75E-07 | 1.64E-06 | 0.003760728 |
| T2DM | endometrium | *PAXIP1-AS1* | 4.013082284 | 8.84E-08 | 4.38E-07 | 0.000428774 |
| T2DM | endothelial | *NFYA* | -1.026491927 | 7.01E-06 | 5.11E-05 | 0.033992513 |
| T2DM | endothelial | *ENPP4* | 0.414777531 | 7.32E-05 | 0.000294466 | 0.355126598 |
| T2DM | endothelial | *SEMA3F* | 0.184902267 | 1.99E-05 | 0.000106436 | 0.096431114 |
| T2DM | endothelial | *KRIT1* | -0.56158477 | 0.001305034 | 0.003039858 | 1 |
| T2DM | endothelial | *RAD52* | -0.69605965 | 5.11E-05 | 0.000222374 | 0.248152028 |
| T2DM | endothelial | *HS3ST1* | -0.438010639 | 0.01785819 | 0.028483872 | 1 |
| T2DM | endothelial | *AOC1* | -0.573176666 | 0.015032502 | 0.024516874 | 1 |
| T2DM | endothelial | *TMEM176A* | -1.152642827 | 2.34E-05 | 0.000121372 | 0.113725191 |
| T2DM | endothelial | *DBNDD1* | -0.526658733 | 2.14E-06 | 2.46E-05 | 0.010366931 |
| T2DM | endothelial | *CD38* | -0.663079167 | 2.18E-06 | 2.48E-05 | 0.010565442 |
| T2DM | endothelial | *RBM6* | -0.567149434 | 1.21E-09 | 5.36E-07 | 5.89E-06 |
| T2DM | endothelial | *RECQL* | -0.5408112 | 0.021634341 | 0.033590343 | 1 |
| T2DM | endothelial | *ARHGAP33* | -0.680721976 | 0.000389398 | 0.001108776 | 1 |
| T2DM | endothelial | *POLR2J* | -0.98614231 | 0.003501665 | 0.0070265 | 1 |
| T2DM | endothelial | *MEOX1* | 0.120844343 | 2.63E-06 | 2.76E-05 | 0.012779395 |
| T2DM | endothelial | *GCFC2* | -0.586244541 | 0.000716527 | 0.001827771 | 1 |
| T2DM | endothelial | *RHBDD2* | -0.501756216 | 0.032069118 | 0.047309018 | 1 |
| T2DM | endothelial | *ZNF195* | -0.74988752 | 0.000154728 | 0.000526404 | 0.75074061 |
| T2DM | endothelial | *PDK2* | -0.609720776 | 1.39E-05 | 8.24E-05 | 0.067498204 |
| T2DM | endothelial | *GGNBP2* | -0.729749511 | 8.81E-06 | 5.97E-05 | 0.0427457 |

| | | | | | | |
|---|---|---|---|---|---|---|
| T2DM | endothelial | *ITGA2B* | -1.041871683 | 4.45E-05 | 0.000198829 | 0.215729296 |
| T2DM | endothelial | *TMEM98* | -0.607678657 | 0.000182421 | 0.000601294 | 0.885104921 |
| T2DM | endothelial | *ABCC8* | -0.459212664 | 0.003881906 | 0.0076721 | 1 |
| T2DM | endothelial | *SYNRG* | -0.558480796 | 0.000851463 | 0.002116444 | 1 |
| T2DM | endothelial | *ZNF263* | -0.645546122 | 0.005881247 | 0.010891693 | 1 |
| T2DM | endothelial | *BAIAP2L1* | 0.373857327 | 0.000127031 | 0.000451211 | 0.616354764 |
| T2DM | endothelial | *FARP2* | -0.588811877 | 0.001071911 | 0.002564553 | 1 |
| T2DM | endothelial | *IFRD1* | -0.410937298 | 0.000724558 | 0.001843502 | 1 |
| T2DM | endothelial | *SCIN* | -0.702691574 | 0.000316674 | 0.000946077 | 1 |
| T2DM | endothelial | *NOS2* | -0.978115679 | 0.017878578 | 0.028507019 | 1 |
| T2DM | endothelial | *CD79B* | 0.365452799 | 0.000794842 | 0.001992247 | 1 |
| T2DM | endothelial | *ST7L* | -0.423159003 | 0.002486847 | 0.005239332 | 1 |
| T2DM | endothelial | *BAIAP3* | 0.315150082 | 0.007800445 | 0.0138789 | 1 |
| T2DM | endothelial | *TSR3* | -1.367109619 | 0.000316854 | 0.000946077 | 1 |
| T2DM | endothelial | *PIGQ* | -0.643060858 | 5.42E-05 | 0.000231621 | 0.262889839 |
| T2DM | endothelial | *FMO3* | -0.49642889 | 6.61E-05 | 0.000270476 | 0.320514492 |
| T2DM | endothelial | *NADK* | -0.592376669 | 0.025839388 | 0.03911785 | 1 |
| T2DM | endothelial | *DLEC1* | -0.611467387 | 9.11E-06 | 6.09E-05 | 0.044222903 |
| T2DM | endothelial | *CYB561* | 0.460671677 | 3.84E-06 | 3.45E-05 | 0.018622511 |
| T2DM | endothelial | *SS18L2* | 0.55376891 | 0.019694753 | 0.030925221 | 1 |
| T2DM | endothelial | *MGST1* | -0.246968761 | 0.001020073 | 0.00245682 | 1 |
| T2DM | endothelial | *IL32* | 0.652087341 | 0.001887512 | 0.004151501 | 1 |
| T2DM | endothelial | *PKD1* | -0.728104565 | 0.000324659 | 0.000962276 | 1 |
| T2DM | endothelial | *MAPK8IP2* | -2.299792297 | 9.04E-06 | 6.06E-05 | 0.043884906 |
| T2DM | endothelial | *MED24* | -0.905706212 | 7.45E-05 | 0.000298604 | 0.36161002 |
| T2DM | endothelial | *POMT2* | -0.809568166 | 3.79E-05 | 0.000173974 | 0.183716516 |
| T2DM | endothelial | *MLXIPL* | -0.266275153 | 0.000318415 | 0.000949569 | 1 |
| T2DM | endothelial | *FUZ* | -1.075452189 | 8.36E-05 | 0.000324173 | 0.405864088 |
| T2DM | endothelial | *SLC6A13* | -0.408334089 | 0.002194582 | 0.004715728 | 1 |
| T2DM | endothelial | *LRRC23* | -0.522592422 | 0.000970366 | 0.002351894 | 1 |
| T2DM | endothelial | *CLCN6* | -0.737354487 | 0.00014767 | 0.000508874 | 0.716494295 |
| T2DM | endothelial | *SLC6A7* | 4.277470516 | 1.10E-05 | 6.92E-05 | 0.053350259 |
| T2DM | endothelial | *TSPAN9* | 0.204958934 | 0.007327179 | 0.013176973 | 1 |
| T2DM | endothelial | *UTP18* | -0.533238713 | 0.003740002 | 0.007431332 | 1 |
| T2DM | endothelial | *DCN* | -0.466561129 | 7.70E-05 | 0.00030533 | 0.373419073 |
| T2DM | endothelial | *CEP68* | 0.571873473 | 1.17E-05 | 7.28E-05 | 0.05681123 |
| T2DM | endothelial | *ZBTB32* | -0.902221629 | 0.001576344 | 0.003555752 | 1 |
| T2DM | endothelial | *ERCC1* | -0.51306457 | 0.003820559 | 0.007569356 | 1 |
| T2DM | endothelial | *ALOX5* | -0.432122031 | 0.000132585 | 0.000466501 | 0.643304632 |
| T2DM | endothelial | *MAP4K5* | -0.45520195 | 0.009350677 | 0.016242754 | 1 |
| T2DM | endothelial | *MAN2B2* | -0.872619651 | 5.82E-05 | 0.00024484 | 0.282145211 |
| T2DM | endothelial | *MDH1* | -0.64397941 | 8.40E-05 | 0.00032521 | 0.407538424 |
| T2DM | endothelial | *COX15* | -0.707295289 | 0.001026667 | 0.002469702 | 1 |
| T2DM | endothelial | *CCDC88C* | 0.283847895 | 0.000177229 | 0.000587777 | 0.859917266 |
| T2DM | endothelial | *NUDCD3* | -0.446786658 | 0.003362555 | 0.006797965 | 1 |
| T2DM | endothelial | *CHDH* | -1.915966269 | 8.93E-05 | 0.00034037 | 0.433290699 |
| T2DM | endothelial | *CLCA4* | -1.213143417 | 0.000303203 | 0.000912052 | 1 |
| T2DM | endothelial | *RUFY3* | -0.466786426 | 0.003153019 | 0.006422523 | 1 |
| T2DM | endothelial | *SLC11A1* | -0.235517887 | 0.002243338 | 0.004797126 | 1 |
| T2DM | endothelial | *AGPS* | -0.401475677 | 0.016967442 | 0.027215216 | 1 |

| | | | | | | |
|---|---|---|---|---|---|---|
| T2DM | endothelial | *TTC27* | -0.597610117 | 0.000502523 | 0.001364432 | 1 |
| T2DM | endothelial | *VSIG2* | 0.516280177 | 6.22E-06 | 4.74E-05 | 0.030187406 |
| T2DM | endothelial | *HGF* | -0.807004812 | 0.001108434 | 0.002636334 | 1 |
| T2DM | endothelial | *CPS1* | -0.525447978 | 0.000897025 | 0.002209323 | 1 |
| T2DM | endothelial | *SLC45A4* | -0.525590888 | 0.010344588 | 0.017754489 | 1 |
| T2DM | endothelial | *GRAMD1B* | -0.352026875 | 0.001844855 | 0.0040743 | 1 |
| T2DM | endothelial | *RNH1* | -0.468599617 | 0.003542216 | 0.007093204 | 1 |
| T2DM | endothelial | *ABCC2* | -1.244913496 | 1.16E-06 | 1.78E-05 | 0.005637516 |
| T2DM | endothelial | *GCLM* | -0.706020144 | 0.008171585 | 0.014422893 | 1 |
| T2DM | endothelial | *UBR2* | -0.451781675 | 0.002738006 | 0.005699187 | 1 |
| T2DM | endothelial | *RRAGD* | -0.570809064 | 1.65E-06 | 2.20E-05 | 0.007996692 |
| T2DM | endothelial | *HSF2* | -0.416679296 | 0.017733259 | 0.028337251 | 1 |
| T2DM | endothelial | *TYMP* | -0.375082956 | 0.003166411 | 0.006447094 | 1 |
| T2DM | endothelial | *TOMM34* | -0.831853544 | 0.020227196 | 0.031628216 | 1 |
| T2DM | endothelial | *SEC63* | -0.610716514 | 0.000214514 | 0.00068792 | 1 |
| T2DM | endothelial | *FAS* | -0.696953331 | 9.67E-07 | 1.61E-05 | 0.004693199 |
| T2DM | endothelial | *RNASET2* | -0.626203327 | 6.60E-07 | 1.33E-05 | 0.003200209 |
| T2DM | endothelial | *MIPEP* | -0.778373127 | 1.24E-05 | 7.55E-05 | 0.059984907 |
| T2DM | endothelial | *INSRR* | -0.896367342 | 1.40E-07 | 1.06E-05 | 0.000676973 |
| T2DM | endothelial | *SH2D2A* | -1.423327455 | 0.026733334 | 0.040320216 | 1 |
| T2DM | endothelial | *SNX1* | -0.638258733 | 7.34E-05 | 0.000294675 | 0.355967996 |
| T2DM | endothelial | *ANK1* | 0.405724117 | 1.18E-05 | 7.28E-05 | 0.057036793 |
| T2DM | endothelial | *RABEP1* | -0.514174174 | 0.002156265 | 0.004649865 | 1 |
| T2DM | endothelial | *IKZF2* | -0.738775044 | 5.16E-05 | 0.000223806 | 0.250496856 |
| T2DM | endothelial | *GRN* | -0.427854695 | 0.003131471 | 0.006392838 | 1 |
| T2DM | endothelial | *IFT88* | -0.552773092 | 0.001663507 | 0.003724658 | 1 |
| T2DM | endothelial | *CHPF2* | -0.533459482 | 0.016033338 | 0.025913977 | 1 |
| T2DM | endothelial | *UBA6* | -0.471773314 | 0.006256214 | 0.011485112 | 1 |
| T2DM | endothelial | *ATP6V0A1* | -0.467012381 | 0.005898966 | 0.010907691 | 1 |
| T2DM | endothelial | *PIAS1* | -0.388309474 | 0.009584159 | 0.016578375 | 1 |
| T2DM | endothelial | *ASTE1* | -0.817966148 | 0.000216766 | 0.000694225 | 1 |
| T2DM | endothelial | *MYOC* | -0.691285414 | 9.54E-05 | 0.000359416 | 0.462928287 |
| T2DM | endothelial | *SH3YL1* | -0.841357679 | 5.20E-06 | 4.18E-05 | 0.025230682 |
| T2DM | endothelial | *VCL* | -0.608589272 | 0.000390496 | 0.001110517 | 1 |
| T2DM | endothelial | *TBC1D23* | -0.666767852 | 0.000163876 | 0.00054988 | 0.795125955 |
| T2DM | endothelial | *MYOM2* | -0.741826331 | 5.89E-05 | 0.000247257 | 0.285829484 |
| T2DM | endothelial | *TUBG2* | -1.214299347 | 6.44E-09 | 1.49E-06 | 3.12E-05 |
| T2DM | endothelial | *NSUN2* | -0.764268579 | 0.000387206 | 0.001105131 | 1 |
| T2DM | endothelial | *MFAP3* | 0.302218443 | 0.018823284 | 0.029778472 | 1 |
| T2DM | endothelial | *MRI1* | -0.599602684 | 0.000152736 | 0.000522252 | 0.741075963 |
| T2DM | endothelial | *AGA* | -1.099313853 | 2.87E-05 | 0.000139848 | 0.139428298 |
| T2DM | endothelial | *PI4K2B* | 1.01458546 | 0.000153914 | 0.00052443 | 0.746788631 |
| T2DM | endothelial | *BOD1L1* | -0.41740639 | 0.014504373 | 0.023759357 | 1 |
| T2DM | endothelial | *DNAH5* | -0.747471607 | 8.31E-07 | 1.52E-05 | 0.004034279 |
| T2DM | endothelial | *PHLPP2* | -0.546335185 | 1.46E-05 | 8.51E-05 | 0.070640545 |
| T2DM | endothelial | *PQLC2* | -0.911863523 | 3.00E-07 | 1.24E-05 | 0.001457074 |
| T2DM | endothelial | *PSMA4* | -0.856940472 | 0.000461863 | 0.001271827 | 1 |
| T2DM | endothelial | *TDP1* | -0.959464449 | 0.000127988 | 0.000453615 | 0.620999304 |
| T2DM | endothelial | *SPATA7* | -0.676193793 | 0.000311055 | 0.000931054 | 1 |
| T2DM | endothelial | *MED17* | -0.894976702 | 0.001320535 | 0.003068599 | 1 |

| | | | | | | |
|---|---|---|---|---|---|---|
| T2DM | endothelial | *CAPG* | -0.628956832 | 1.71E-06 | 2.22E-05 | 0.008300628 |
| T2DM | endothelial | *AP2S1* | -0.964973662 | 2.25E-06 | 2.52E-05 | 0.010918617 |
| T2DM | endothelial | *TRIT1* | -0.448143231 | 0.005176239 | 0.009799107 | 1 |
| T2DM | endothelial | *CUL7* | -0.73516741 | 8.31E-05 | 0.0003225 | 0.403125024 |
| T2DM | endothelial | *YTHDC2* | -0.511655284 | 0.004643715 | 0.008930362 | 1 |
| T2DM | endothelial | *POLR2B* | -0.444982234 | 0.026902356 | 0.040537338 | 1 |
| T2DM | endothelial | *ANO2* | 0.085600235 | 3.68E-06 | 3.34E-05 | 0.017846337 |
| T2DM | endothelial | *RRM2B* | -0.871815433 | 4.91E-06 | 4.04E-05 | 0.023827782 |
| T2DM | endothelial | *SNX29* | -0.548847641 | 8.82E-05 | 0.000336933 | 0.427904985 |
| T2DM | endothelial | *MRPS10* | -0.216417046 | 0.032970561 | 0.048388736 | 1 |
| T2DM | endothelial | *UTS2* | -0.579638928 | 0.019244466 | 0.030335981 | 1 |
| T2DM | endothelial | *EPN3* | -0.995966217 | 6.41E-06 | 4.82E-05 | 0.031117416 |
| T2DM | endothelial | *JKAMP* | -0.59836265 | 0.001339013 | 0.003102621 | 1 |
| T2DM | endothelial | *ARHGEF5* | -0.301889928 | 0.019825653 | 0.031080475 | 1 |
| T2DM | endothelial | *LAMC3* | -0.630519007 | 5.59E-06 | 4.37E-05 | 0.027133268 |
| T2DM | endothelial | *MAPK9* | -0.537801029 | 0.025890515 | 0.039183025 | 1 |
| T2DM | endothelial | *CYBA* | -0.636228754 | 0.000307789 | 0.000924129 | 1 |
| T2DM | endothelial | *THOC3* | -1.349170085 | 0.029795191 | 0.044422632 | 1 |
| T2DM | endothelial | *ALX4* | -0.676532581 | 9.32E-06 | 6.19E-05 | 0.045216235 |
| T2DM | endothelial | *FSTL4* | -0.456980665 | 0.006228463 | 0.011442826 | 1 |
| T2DM | endothelial | *AKR7A2* | -0.910048508 | 0.000360279 | 0.001043628 | 1 |
| T2DM | endothelial | *MRTO4* | -1.229082679 | 2.13E-06 | 2.46E-05 | 0.01032686 |
| T2DM | endothelial | *ANAPC4* | -0.521356542 | 0.007936095 | 0.014078951 | 1 |
| T2DM | endothelial | *LY75* | 1.124659806 | 0.005402833 | 0.010148875 | 1 |
| T2DM | endothelial | *SDCCAG8* | -0.356509533 | 0.005320135 | 0.01003589 | 1 |
| T2DM | endothelial | *TBC1D22A* | -0.33079475 | 0.03382293 | 0.049475085 | 1 |
| T2DM | endothelial | *PLEKHH1* | -0.670478646 | 5.56E-06 | 4.36E-05 | 0.026963375 |
| T2DM | endothelial | *GALC* | -0.810523862 | 1.87E-05 | 0.000101569 | 0.090904436 |
| T2DM | endothelial | *ITIH4* | 0.293310229 | 3.65E-05 | 0.000168929 | 0.177037189 |
| T2DM | endothelial | *PCGF2* | 0.373991017 | 8.87E-05 | 0.000338364 | 0.430399268 |
| T2DM | endothelial | *IL17RB* | -1.231710884 | 3.19E-05 | 0.000151359 | 0.154689282 |
| T2DM | endothelial | *MSH4* | -0.302421372 | 0.00243327 | 0.005137609 | 1 |
| T2DM | endothelial | *PITHD1* | -0.767084326 | 0.001413426 | 0.003244059 | 1 |
| T2DM | endothelial | *ATP11B* | -0.431912581 | 0.00739056 | 0.01325593 | 1 |
| T2DM | endothelial | *LAMC2* | -0.339709474 | 0.006162146 | 0.011333864 | 1 |
| T2DM | endothelial | *CROCC* | -0.430225382 | 0.000683459 | 0.001753646 | 1 |
| T2DM | endothelial | *FLYWCH1* | -0.416654515 | 0.002127539 | 0.00459609 | 1 |
| T2DM | endothelial | *ALDH18A1* | -0.886372322 | 2.51E-06 | 2.67E-05 | 0.012187417 |
| T2DM | endothelial | *DNAJC25* | -0.983199421 | 0.012202654 | 0.020538637 | 1 |
| T2DM | endothelial | *OGFR* | -0.503596098 | 0.013752123 | 0.022718864 | 1 |
| T2DM | endothelial | *PIGV* | -0.585898047 | 9.78E-06 | 6.36E-05 | 0.047467327 |
| T2DM | endothelial | *RIMBP2* | 0.623654028 | 0.001729072 | 0.003850141 | 1 |
| T2DM | endothelial | *PRDM6* | -0.824932946 | 1.18E-06 | 1.79E-05 | 0.005712677 |
| T2DM | endothelial | *SPAG4* | -0.98535745 | 0.002622213 | 0.005495887 | 1 |
| T2DM | endothelial | *SLC6A16* | -0.602277886 | 0.012546699 | 0.021043964 | 1 |
| T2DM | endothelial | *AHRR* | -1.439956597 | 6.67E-07 | 1.33E-05 | 0.003234593 |
| T2DM | endothelial | *ADCK1* | -1.001860333 | 6.53E-06 | 4.87E-05 | 0.031684605 |
| T2DM | endothelial | *HAGH* | -0.519689968 | 0.002547579 | 0.005362627 | 1 |
| T2DM | endothelial | *RNF4* | -0.749448594 | 0.000322991 | 0.000958502 | 1 |
| T2DM | endothelial | *CASP8* | -0.651560942 | 0.000931951 | 0.002280296 | 1 |

| | | | | | | |
|---|---|---|---|---|---|---|
| T2DM | endothelial | *CDON* | -0.26411094 | 0.000147633 | 0.000508874 | 0.716314022 |
| T2DM | endothelial | *TMEM161A* | -0.953935332 | 2.86E-06 | 2.88E-05 | 0.013862782 |
| T2DM | endothelial | *LPAR2* | -0.776617242 | 1.81E-05 | 9.95E-05 | 0.087955815 |
| T2DM | endothelial | *SUGP2* | -0.626271833 | 0.000776524 | 0.001951163 | 1 |
| T2DM | endothelial | *CNN2* | -0.414061792 | 0.01380447 | 0.022781729 | 1 |
| T2DM | endothelial | *ABCA7* | -0.940612878 | 8.43E-06 | 5.77E-05 | 0.040884774 |
| T2DM | endothelial | *DDX20* | -0.772626681 | 0.000482552 | 0.001318324 | 1 |
| T2DM | endothelial | *FAR2* | 0.36356303 | 0.001801841 | 0.003986563 | 1 |
| T2DM | endothelial | *BCAS1* | -0.820670319 | 9.63E-06 | 6.29E-05 | 0.046711535 |
| T2DM | endothelial | *CHI3L2* | -0.92382543 | 7.40E-06 | 5.32E-05 | 0.03588228 |
| T2DM | endothelial | *SBNO2* | -0.721405371 | 4.79E-05 | 0.000210709 | 0.232201564 |
| T2DM | endothelial | *PMS1* | -0.535431808 | 0.005197323 | 0.009827519 | 1 |
| T2DM | endothelial | *UHRF1BP1* | -0.432506544 | 0.000553629 | 0.001477562 | 1 |
| T2DM | endothelial | *WDR18* | -0.390674908 | 0.000358309 | 0.001039783 | 1 |
| T2DM | endothelial | *ANKRD44* | -0.452230301 | 0.003097014 | 0.006332368 | 1 |
| T2DM | endothelial | *MAP2K4* | -0.534566183 | 0.002656806 | 0.005563583 | 1 |
| T2DM | endothelial | *SEC61A2* | -1.128002154 | 4.42E-09 | 1.19E-06 | 2.14E-05 |
| T2DM | endothelial | *TIE1* | 0.140086242 | 2.93E-06 | 2.92E-05 | 0.014211982 |
| T2DM | endothelial | *SPI1* | -0.54577071 | 1.04E-05 | 6.63E-05 | 0.050521565 |
| T2DM | endothelial | *ZNRD1* | -0.551069836 | 0.000318668 | 0.000949739 | 1 |
| T2DM | endothelial | *ATXN3* | -0.288319984 | 0.027145877 | 0.040844229 | 1 |
| T2DM | endothelial | *GOLGA5* | -0.610937538 | 0.008065469 | 0.014261536 | 1 |
| T2DM | endothelial | *FGFR2* | -0.316177696 | 0.019787351 | 0.031040487 | 1 |
| T2DM | endothelial | *MTFR1* | -0.708532296 | 7.48E-05 | 0.000299559 | 0.363064914 |
| T2DM | endothelial | *FECH* | -0.340619403 | 0.002275776 | 0.004851523 | 1 |
| T2DM | endothelial | *MYO9A* | -0.397323943 | 0.005646668 | 0.010545663 | 1 |
| T2DM | endothelial | *TRAM1* | -0.494502952 | 0.013910108 | 0.022909654 | 1 |
| T2DM | endothelial | *EVI5* | -0.543113407 | 8.09E-05 | 0.000316238 | 0.392745806 |
| T2DM | endothelial | *STOML1* | -1.84574181 | 0.0004091 | 0.001154043 | 1 |
| T2DM | endothelial | *METTL22* | -0.963082987 | 2.02E-05 | 0.000107774 | 0.097883063 |
| T2DM | endothelial | *TP53BP1* | -0.623705571 | 0.000606571 | 0.001594303 | 1 |
| T2DM | endothelial | *RRP15* | -0.350263973 | 0.01466595 | 0.023975469 | 1 |
| T2DM | endothelial | *DHX8* | -0.629127565 | 0.010892021 | 0.018549188 | 1 |
| T2DM | endothelial | *ROGDI* | -1.663583224 | 1.69E-06 | 2.21E-05 | 0.008177124 |
| T2DM | endothelial | *CBFB* | -0.510956613 | 0.007337416 | 0.013185609 | 1 |
| T2DM | endothelial | *FGFR3* | 0.493528362 | 0.001367169 | 0.003154306 | 1 |
| T2DM | endothelial | *HEATR6* | -0.667454947 | 0.001435676 | 0.003281158 | 1 |
| T2DM | endothelial | *POLR1A* | -0.671321682 | 5.16E-06 | 4.17E-05 | 0.025026861 |
| T2DM | endothelial | *STON1-GTF2A* | -0.178231892 | 0.010157382 | 0.017470264 | 1 |
| T2DM | endothelial | *SRBD1* | -0.51548482 | 0.004040741 | 0.007922118 | 1 |
| T2DM | endothelial | *CYFIP1* | -0.442593488 | 0.016448301 | 0.026478817 | 1 |
| T2DM | endothelial | *IFT80* | -1.214677332 | 8.37E-06 | 5.75E-05 | 0.040600522 |
| T2DM | endothelial | *NUCKS1* | -0.465061462 | 0.007349659 | 0.013197834 | 1 |
| T2DM | endothelial | *CLEC2D* | -0.606900727 | 0.001079 | 0.002580241 | 1 |
| T2DM | endothelial | *DRD4* | -1.422449274 | 5.26E-07 | 1.27E-05 | 0.002551605 |
| T2DM | endothelial | *PLA2G10* | -1.78417047 | 0.030574481 | 0.045477432 | 1 |
| T2DM | endothelial | *NEDD4* | 0.342168853 | 0.000716869 | 0.001827771 | 1 |
| T2DM | endothelial | *PIGB* | -0.655206997 | 0.002327718 | 0.004947037 | 1 |
| T2DM | endothelial | *DAPP1* | -0.27926249 | 0.003867642 | 0.007647026 | 1 |
| T2DM | endothelial | *CLTCL1* | -0.79832292 | 2.19E-05 | 0.000114662 | 0.10629204 |

| | | | | | | |
|---|---|---|---|---|---|---|
| T2DM | endothelial | *DGCR2* | -0.640754817 | 0.000671148 | 0.001729372 | 1 |
| T2DM | endothelial | *ZXDC* | -0.480470275 | 0.010667219 | 0.018198786 | 1 |
| T2DM | endothelial | *WIPI1* | -0.818287687 | 1.64E-06 | 2.20E-05 | 0.007971826 |
| T2DM | endothelial | *GBA2* | -0.582279015 | 0.027072672 | 0.040756006 | 1 |
| T2DM | endothelial | *NDST1* | 0.423352621 | 0.002240151 | 0.004792422 | 1 |
| T2DM | endothelial | *ASNS* | -0.684374172 | 0.003964643 | 0.007809183 | 1 |
| T2DM | endothelial | *ST6GALNAC2* | 0.528970477 | 0.005536221 | 0.010355338 | 1 |
| T2DM | endothelial | *EIF2B3* | -0.64881293 | 7.35E-06 | 5.29E-05 | 0.035685693 |
| T2DM | endothelial | *SNX13* | -0.471501977 | 0.006810562 | 0.012380984 | 1 |
| T2DM | endothelial | *RPS6KA2* | 0.29095072 | 0.013813654 | 0.022781729 | 1 |
| T2DM | endothelial | *VASH1* | 0.360234303 | 0.001068033 | 0.002557798 | 1 |
| T2DM | endothelial | *TRIB2* | 0.490988462 | 5.17E-05 | 0.000223806 | 0.251029099 |
| T2DM | endothelial | *CYBRD1* | -0.517842016 | 0.000511083 | 0.001384575 | 1 |
| T2DM | endothelial | *RDH11* | -0.773287254 | 0.00122184 | 0.002877847 | 1 |
| T2DM | endothelial | *EPN2* | -0.34276807 | 0.031152875 | 0.046153816 | 1 |
| T2DM | endothelial | *ALDH3A2* | -0.570484879 | 6.59E-05 | 0.000270407 | 0.319891744 |
| T2DM | endothelial | *AFF4* | -0.437182572 | 0.007246244 | 0.013060467 | 1 |
| T2DM | endothelial | *TRHDE* | -0.480927659 | 1.01E-06 | 1.63E-05 | 0.004878037 |
| T2DM | endothelial | *FCGR2B* | -0.920243884 | 2.49E-06 | 2.66E-05 | 0.012095148 |
| T2DM | endothelial | *NFATC3* | -0.379462485 | 0.019448472 | 0.030619292 | 1 |
| T2DM | endothelial | *CRMP1* | -0.634974463 | 5.18E-07 | 1.27E-05 | 0.002511126 |
| T2DM | endothelial | *AP1M1* | -0.720830394 | 0.00412335 | 0.008067136 | 1 |
| T2DM | endothelial | *SCARB1* | 0.363373414 | 0.000849358 | 0.002112293 | 1 |
| T2DM | endothelial | *MOV10L1* | -0.843640801 | 8.17E-06 | 5.64E-05 | 0.039618683 |
| T2DM | endothelial | *ADAM11* | -0.301795789 | 0.005439655 | 0.010206189 | 1 |
| T2DM | endothelial | *PPP2R2C* | -0.847317916 | 3.14E-07 | 1.24E-05 | 0.001522683 |
| T2DM | endothelial | *ZNF532* | -0.406171295 | 0.033234705 | 0.048746913 | 1 |
| T2DM | endothelial | *TACR2* | 4.30513469 | 8.63E-08 | 8.55E-06 | 0.000418718 |
| T2DM | endothelial | *TTC38* | -0.6434909 | 0.000217703 | 0.000696306 | 1 |
| T2DM | endothelial | *GRAMD4* | -0.8089689 | 7.84E-05 | 0.000308851 | 0.380504085 |
| T2DM | endothelial | *ADD2* | 0.236994211 | 2.13E-06 | 2.46E-05 | 0.010351542 |
| T2DM | endothelial | *VPS9D1* | -0.727005894 | 0.003767363 | 0.007476173 | 1 |
| T2DM | endothelial | *FNDC3B* | -0.428802505 | 0.000968236 | 0.002348941 | 1 |
| T2DM | endothelial | *CACNG4* | -0.537346871 | 0.000679349 | 0.001745869 | 1 |
| T2DM | endothelial | *MOCOS* | -0.649613303 | 0.004335719 | 0.008428249 | 1 |
| T2DM | endothelial | *KIFAP3* | -0.486543853 | 0.007889768 | 0.014009111 | 1 |
| T2DM | endothelial | *MKRN2* | -0.559429967 | 0.00729611 | 0.013140581 | 1 |
| T2DM | endothelial | *MCM6* | -0.766873715 | 0.000132378 | 0.000466287 | 0.642296321 |
| T2DM | endothelial | *MLH1* | -0.75169066 | 4.42E-05 | 0.000197899 | 0.214522536 |
| T2DM | endothelial | *FMO4* | -0.703416788 | 0.000232272 | 0.000735629 | 1 |
| T2DM | endothelial | *RGS11* | -0.432173938 | 0.000153653 | 0.000523911 | 0.745525111 |
| T2DM | endothelial | *SPAG5* | -2.075734308 | 5.49E-06 | 4.31E-05 | 0.026614014 |
| T2DM | endothelial | *ANKRD13A* | -0.540765055 | 0.029224417 | 0.043629807 | 1 |
| T2DM | endothelial | *PAG1* | -0.675059107 | 4.57E-06 | 3.87E-05 | 0.022165751 |
| T2DM | endothelial | *GPATCH1* | -0.832254935 | 0.000500955 | 0.001360937 | 1 |
| T2DM | endothelial | *IL4R* | -0.770678456 | 0.000116758 | 0.000421511 | 0.566511321 |
| T2DM | endothelial | *APBB1IP* | -0.454905068 | 1.41E-05 | 8.33E-05 | 0.068479386 |
| T2DM | endothelial | *LRCH4* | -0.807077717 | 8.40E-06 | 5.77E-05 | 0.04076133 |
| T2DM | endothelial | *SIRT6* | -0.661696554 | 0.002681351 | 0.005610141 | 1 |
| T2DM | endothelial | *GPR137B* | -0.473902226 | 0.005204299 | 0.009836876 | 1 |

| | | | | | | |
|---|---|---|---|---|---|---|
| T2DM | endothelial | *NAALAD2* | -0.220465339 | 0.010627288 | 0.018137039 | 1 |
| T2DM | endothelial | *AMPH* | -0.720829659 | 1.00E-06 | 1.63E-05 | 0.004854934 |
| T2DM | endothelial | *MCCC1* | -0.417576763 | 0.000194669 | 0.000634342 | 0.944535487 |
| T2DM | endothelial | *PIK3C3* | -0.439830183 | 0.018009416 | 0.028654484 | 1 |
| T2DM | endothelial | *SYNJ2* | 0.232771308 | 6.45E-05 | 0.000265927 | 0.312729816 |
| T2DM | endothelial | *RBFOX1* | -0.422507404 | 5.35E-05 | 0.000229775 | 0.259646172 |
| T2DM | endothelial | *FGF20* | -1.888382904 | 0.001221314 | 0.002877847 | 1 |
| T2DM | endothelial | *PCM1* | -0.409060408 | 0.005967932 | 0.011010041 | 1 |
| T2DM | endothelial | *BPIFB2* | -2.096754643 | 0.003844107 | 0.007609795 | 1 |
| T2DM | endothelial | *CLUL1* | -0.726411754 | 0.000619293 | 0.001621593 | 1 |
| T2DM | endothelial | *FKBP7* | -0.584439444 | 0.002213954 | 0.004751041 | 1 |
| T2DM | endothelial | *SP140* | -0.494800622 | 0.015433445 | 0.025078057 | 1 |
| T2DM | endothelial | *SAR1A* | -0.709634264 | 6.66E-05 | 0.000272277 | 0.322920286 |
| T2DM | endothelial | *RAPGEF3* | 0.157377566 | 9.67E-05 | 0.000361965 | 0.469106184 |
| T2DM | endothelial | *SCGN* | -4.60909324 | 3.18E-07 | 1.24E-05 | 0.001541753 |
| T2DM | endothelial | *PGM1* | -0.74681664 | 8.45E-06 | 5.78E-05 | 0.040979631 |
| T2DM | endothelial | *DDX1* | -0.516347777 | 0.005662031 | 0.010562159 | 1 |
| T2DM | endothelial | *DNM2* | -0.547047223 | 0.002759535 | 0.005736617 | 1 |
| T2DM | endothelial | *STX7* | -0.514308227 | 0.002070907 | 0.004495767 | 1 |
| T2DM | endothelial | *RABL2B* | -0.808257703 | 1.17E-06 | 1.79E-05 | 0.005673616 |
| T2DM | endothelial | *DCT* | -0.594622132 | 8.23E-07 | 1.51E-05 | 0.003993397 |
| T2DM | endothelial | *SCTR* | -0.647426786 | 4.26E-07 | 1.24E-05 | 0.002064829 |
| T2DM | endothelial | *RIF1* | -0.613106115 | 0.000390695 | 0.001110517 | 1 |
| T2DM | endothelial | *KCNN2* | 0.278521501 | 1.21E-06 | 1.81E-05 | 0.005880315 |
| T2DM | endothelial | *PSEN1* | -0.560153737 | 0.001399734 | 0.0032172 | 1 |
| T2DM | endothelial | *MOK* | -0.622404755 | 1.50E-05 | 8.68E-05 | 0.072684504 |
| T2DM | endothelial | *STK17B* | -0.463891924 | 0.004345268 | 0.008433297 | 1 |
| T2DM | endothelial | *PCDHB4* | 1.003444132 | 0.00146594 | 0.003337748 | 1 |
| T2DM | endothelial | *ME2* | -0.624779786 | 0.003885389 | 0.007672735 | 1 |
| T2DM | endothelial | *LYRM2* | -0.504265824 | 0.00739294 | 0.01325593 | 1 |
| T2DM | endothelial | *ZCCHC6* | -0.633447657 | 8.25E-05 | 0.000320628 | 0.400144246 |
| T2DM | endothelial | *ULK2* | -0.615362579 | 8.11E-06 | 5.62E-05 | 0.03936824 |
| T2DM | endothelial | *PLOD1* | -0.830431936 | 1.86E-05 | 0.000100843 | 0.090053153 |
| T2DM | endothelial | *OXCT1* | -0.454788888 | 0.016201679 | 0.026177337 | 1 |
| T2DM | endothelial | *ZNF264* | -0.717649512 | 0.003086481 | 0.006313493 | 1 |
| T2DM | endothelial | *CHMP2B* | -0.716292671 | 0.005672464 | 0.01056943 | 1 |
| T2DM | endothelial | *PPIE* | -0.56602421 | 0.000257044 | 0.000800499 | 1 |
| T2DM | endothelial | *NCOA1* | -0.41168762 | 0.004842178 | 0.009253346 | 1 |
| T2DM | endothelial | *CAD* | -0.927864915 | 3.16E-05 | 0.00015044 | 0.153448713 |
| T2DM | endothelial | *CD59* | 0.353771126 | 0.00035121 | 0.001021014 | 1 |
| T2DM | endothelial | *CD82* | 0.484527737 | 0.000277694 | 0.000850613 | 1 |
| T2DM | endothelial | *SEH1L* | -0.472274162 | 0.01316924 | 0.021957784 | 1 |
| T2DM | endothelial | *SLC25A24* | -0.548042715 | 0.003497769 | 0.007021587 | 1 |
| T2DM | endothelial | *ZNF213* | -0.517726222 | 0.001070445 | 0.002562309 | 1 |
| T2DM | endothelial | *CPNE3* | -0.481529931 | 0.011786466 | 0.019946959 | 1 |
| T2DM | endothelial | *RRN3* | -0.546900954 | 0.028062182 | 0.042082351 | 1 |
| T2DM | endothelial | *MTIF2* | -0.902140518 | 1.58E-05 | 8.96E-05 | 0.076425201 |
| T2DM | endothelial | *DDHD2* | -0.52619238 | 0.000756619 | 0.001912039 | 1 |
| T2DM | endothelial | *EPS15* | -0.31690717 | 0.021730793 | 0.033686201 | 1 |
| T2DM | endothelial | *USP40* | -0.621014817 | 3.62E-05 | 0.000167762 | 0.175550731 |

| | | | | | | |
|---|---|---|---|---|---|---|
| T2DM | endothelial | *EIF2AK1* | -0.696728402 | 0.001026656 | 0.002469702 | 1 |
| T2DM | endothelial | *NME8* | -6.390639898 | 1.25E-06 | 1.85E-05 | 0.006045054 |
| T2DM | endothelial | *SNX10* | -0.640339058 | 2.21E-08 | 4.12E-06 | 0.000107252 |
| T2DM | endothelial | *TMED2* | -0.509452443 | 0.013097024 | 0.021844882 | 1 |
| T2DM | endothelial | *ALG9* | -1.364369683 | 3.35E-05 | 0.000157293 | 0.162518966 |
| T2DM | endothelial | *ACOX3* | -0.596618803 | 0.00348937 | 0.007010528 | 1 |
| T2DM | endothelial | *HSD17B14* | -0.780635992 | 7.17E-06 | 5.17E-05 | 0.03477032 |
| T2DM | endothelial | *TRIP6* | -0.889463226 | 1.68E-06 | 2.21E-05 | 0.008160057 |
| T2DM | endothelial | *ACHE* | -1.003245983 | 1.20E-06 | 1.80E-05 | 0.005822713 |
| T2DM | endothelial | *SRRT* | -0.728352764 | 0.003454722 | 0.006958203 | 1 |
| T2DM | endothelial | *ADAMTS2* | -0.531469122 | 5.14E-07 | 1.27E-05 | 0.002491677 |
| T2DM | endothelial | *PSMC5* | -1.047976103 | 5.88E-05 | 0.000247188 | 0.285502591 |
| T2DM | endothelial | *LPCAT2* | 0.305803033 | 6.93E-05 | 0.000281922 | 0.336051401 |
| T2DM | endothelial | *OGFOD1* | -0.727443315 | 0.001327115 | 0.003080938 | 1 |
| T2DM | endothelial | *TXNDC16* | -0.539170601 | 0.001587404 | 0.003579036 | 1 |
| T2DM | endothelial | *SF3B2* | -0.43761321 | 0.019474688 | 0.030629233 | 1 |
| T2DM | endothelial | *AAMDC* | -0.530143653 | 0.000101572 | 0.000376492 | 0.492828483 |
| T2DM | endothelial | *GP6* | -0.379235699 | 9.12E-05 | 0.00034643 | 0.442737479 |
| T2DM | endothelial | *ASAP3* | -0.73049711 | 0.000229794 | 0.000728256 | 1 |
| T2DM | endothelial | *EDEM2* | -0.88034877 | 6.54E-06 | 4.87E-05 | 0.031727905 |
| T2DM | endothelial | *ANKRD10* | -0.430515816 | 0.011041125 | 0.018770687 | 1 |
| T2DM | endothelial | *DOCK3* | -0.6493433 | 6.75E-07 | 1.34E-05 | 0.003275023 |
| T2DM | endothelial | *PPP1R13B* | 0.261993005 | 0.00073488 | 0.001864873 | 1 |
| T2DM | endothelial | *CPXM1* | -0.727775323 | 1.81E-05 | 9.95E-05 | 0.087842909 |
| T2DM | endothelial | *MAVS* | -0.595417225 | 0.000427522 | 0.001194207 | 1 |
| T2DM | endothelial | *SNX5* | -0.822496388 | 0.000107454 | 0.00039378 | 0.521364798 |
| T2DM | endothelial | *ESF1* | -0.548969929 | 0.001611205 | 0.00362092 | 1 |
| T2DM | endothelial | *TMEM230* | -0.63818544 | 0.000373125 | 0.00107315 | 1 |
| T2DM | endothelial | *RPH3A* | -1.380508806 | 1.59E-06 | 2.16E-05 | 0.007725368 |
| T2DM | endothelial | *KIF16B* | -0.50342564 | 0.009919994 | 0.01710441 | 1 |
| T2DM | endothelial | *TBX5* | -1.481699139 | 2.18E-06 | 2.48E-05 | 0.010600501 |
| T2DM | endothelial | *FXYD5* | 0.476569995 | 0.001328673 | 0.003082701 | 1 |
| T2DM | endothelial | *ZNF302* | -0.527900837 | 0.000365565 | 0.001055386 | 1 |
| T2DM | endothelial | *CDIP1* | 0.488965024 | 0.016700039 | 0.026839546 | 1 |
| T2DM | endothelial | *KCNH4* | 0.387190183 | 1.51E-06 | 2.11E-05 | 0.007313362 |
| T2DM | endothelial | *GANAB* | -0.78363594 | 9.85E-05 | 0.000366802 | 0.477943147 |
| T2DM | endothelial | *MLF2* | -0.522275642 | 0.004470016 | 0.008640844 | 1 |
| T2DM | endothelial | *ANKRD24* | -0.732711291 | 3.81E-07 | 1.24E-05 | 0.001847343 |
| T2DM | endothelial | *DHX32* | -0.462514258 | 0.016212145 | 0.026185529 | 1 |
| T2DM | endothelial | *SPTLC1* | -0.597509342 | 0.002687353 | 0.005620274 | 1 |
| T2DM | endothelial | *MRPS33* | -1.074254196 | 1.21E-05 | 7.42E-05 | 0.058570621 |
| T2DM | endothelial | *MAEA* | -0.589329918 | 0.004906714 | 0.009365609 | 1 |
| T2DM | endothelial | *STRN4* | -0.562246975 | 0.007599739 | 0.01358156 | 1 |
| T2DM | endothelial | *LYZ* | -0.394018884 | 0.020538531 | 0.03204275 | 1 |
| T2DM | endothelial | *MUL1* | -0.463641136 | 0.003641166 | 0.007258397 | 1 |
| T2DM | endothelial | *SPG21* | -0.740751516 | 0.000170299 | 0.000567117 | 0.826289682 |
| T2DM | endothelial | *RAB11FIP3* | -0.636119259 | 0.000921754 | 0.002262191 | 1 |
| T2DM | endothelial | *GNPTG* | -0.869417226 | 0.000114383 | 0.000413859 | 0.554985301 |
| T2DM | endothelial | *PABPC4* | -1.393176209 | 7.56E-14 | 3.67E-10 | 3.67E-10 |
| T2DM | endothelial | *CERS4* | -0.64941587 | 8.58E-07 | 1.53E-05 | 0.00416161 |

| | | | | | | |
|---|---|---|---|---|---|---|
| T2DM | endothelial | *USP48* | -0.548350661 | 0.000562691 | 0.001499273 | 1 |
| T2DM | endothelial | *GLG1* | -0.421976609 | 0.012967662 | 0.021658897 | 1 |
| T2DM | endothelial | *DTX2* | -1.096558139 | 7.04E-06 | 5.11E-05 | 0.034147779 |
| T2DM | endothelial | *LAMB1* | -0.407591774 | 2.88E-05 | 0.000140037 | 0.139896528 |
| T2DM | endothelial | *ABCC6* | -0.744341603 | 6.21E-07 | 1.31E-05 | 0.003014891 |
| T2DM | endothelial | *RAPGEF4* | 0.34711955 | 0.000783844 | 0.001966501 | 1 |
| T2DM | endothelial | *TF* | -1.158203505 | 3.18E-06 | 3.05E-05 | 0.015444222 |
| T2DM | endothelial | *ALKBH5* | -1.016316719 | 4.62E-06 | 3.90E-05 | 0.022410548 |
| T2DM | endothelial | *SPAG7* | -0.393253201 | 0.00710491 | 0.012834336 | 1 |
| T2DM | endothelial | *RGS17* | -0.770427586 | 8.14E-09 | 1.79E-06 | 3.95E-05 |
| T2DM | endothelial | *SCFD1* | -0.461316917 | 0.005539763 | 0.010357968 | 1 |
| T2DM | endothelial | *SUPT16H* | -0.681634299 | 0.001722513 | 0.003841721 | 1 |
| T2DM | endothelial | *TOX4* | -0.597423217 | 0.003692986 | 0.007343594 | 1 |
| T2DM | endothelial | *CAPN3* | -1.763173235 | 4.67E-06 | 3.91E-05 | 0.022675031 |
| T2DM | endothelial | *TBX15* | -0.558312757 | 0.000211822 | 0.000680184 | 1 |
| T2DM | endothelial | *MFSD11* | -0.461169081 | 0.001551882 | 0.003507736 | 1 |
| T2DM | endothelial | *COMT* | -0.66814582 | 0.000780921 | 0.001960181 | 1 |
| T2DM | endothelial | *XYLB* | -0.85099479 | 6.50E-06 | 4.87E-05 | 0.031556063 |
| T2DM | endothelial | *GABRP* | 0.165053986 | 1.75E-06 | 2.24E-05 | 0.008505203 |
| T2DM | endothelial | *CDC6* | -1.008816014 | 6.21E-07 | 1.31E-05 | 0.003013956 |
| T2DM | endothelial | *PSMD5* | -0.719304129 | 0.000520282 | 0.001405573 | 1 |
| T2DM | endothelial | *CRAT* | -1.128730364 | 0.002163205 | 0.004660688 | 1 |
| T2DM | endothelial | *NANS* | -0.686112328 | 0.018012403 | 0.028654484 | 1 |
| T2DM | endothelial | *PDE6C* | -0.959714735 | 0.001390092 | 0.003199834 | 1 |
| T2DM | endothelial | *CWF19L1* | -0.590507937 | 1.38E-07 | 1.06E-05 | 0.000669514 |
| T2DM | endothelial | *CRTAC1* | 0.220091716 | 0.004525287 | 0.008733768 | 1 |
| T2DM | endothelial | *WAC* | -0.40438652 | 0.012258647 | 0.020623772 | 1 |
| T2DM | endothelial | *DSP* | -1.112909712 | 0.001049977 | 0.002520776 | 1 |
| T2DM | endothelial | *IFT74* | -0.632006564 | 5.06E-05 | 0.000220759 | 0.245484348 |
| T2DM | endothelial | *ACOT7* | -1.039428735 | 1.55E-05 | 8.91E-05 | 0.075241569 |
| T2DM | endothelial | *SCD* | -0.768292328 | 4.91E-07 | 1.27E-05 | 0.002383495 |
| T2DM | endothelial | *ERMP1* | -0.710760876 | 0.000100417 | 0.000372781 | 0.487224549 |
| T2DM | endothelial | *RAB18* | -0.637228073 | 0.000965601 | 0.002343719 | 1 |
| T2DM | endothelial | *PRTFDC1* | -0.540506276 | 9.03E-07 | 1.56E-05 | 0.004380947 |
| T2DM | endothelial | *MAST3* | -0.514631888 | 2.42E-05 | 0.000124381 | 0.117210127 |
| T2DM | endothelial | *OCEL1* | -0.409775972 | 0.007091029 | 0.01281403 | 1 |
| T2DM | endothelial | *SETD1A* | -0.699799095 | 0.000525868 | 0.001419085 | 1 |
| T2DM | endothelial | *CIRBP* | -0.483262977 | 0.001809659 | 0.004002035 | 1 |
| T2DM | endothelial | *MTAP* | -0.79941213 | 0.000292213 | 0.000884479 | 1 |
| T2DM | endothelial | *POLR2E* | -0.725337056 | 0.001757396 | 0.003904252 | 1 |
| T2DM | endothelial | *MADCAM1* | -0.80019958 | 8.50E-07 | 1.53E-05 | 0.004122232 |
| T2DM | endothelial | *MED15* | -0.662033978 | 0.000250155 | 0.000783571 | 1 |
| T2DM | endothelial | *SERPIND1* | -0.789739254 | 0.000370836 | 0.001067197 | 1 |
| T2DM | endothelial | *SNAP29* | -0.362439494 | 0.01739515 | 0.027872534 | 1 |
| T2DM | endothelial | *MMP11* | -0.560381864 | 0.022901491 | 0.035219661 | 1 |
| T2DM | endothelial | *BCL2L13* | -0.459510371 | 0.00179242 | 0.003969339 | 1 |
| T2DM | endothelial | *CABIN1* | -0.560416004 | 0.004779159 | 0.009158167 | 1 |
| T2DM | endothelial | *PPIL2* | -0.53636127 | 0.002892734 | 0.005985308 | 1 |
| T2DM | endothelial | *PES1* | 0.281293184 | 4.05E-05 | 0.000183952 | 0.196460918 |
| T2DM | endothelial | *TOP3B* | -0.638913276 | 0.033372916 | 0.048934841 | 1 |

| | | | | | | |
|---|---|---|---|---|---|---|
| T2DM | endothelial | *CRYBB3* | 0.433326625 | 2.94E-06 | 2.92E-05 | 0.014244036 |
| T2DM | endothelial | *CYTH4* | -0.812989752 | 9.78E-07 | 1.61E-05 | 0.004743691 |
| T2DM | endothelial | *CARD10* | 0.226940902 | 1.17E-05 | 7.27E-05 | 0.056652403 |
| T2DM | endothelial | *LRP5L* | -0.599221094 | 0.001940818 | 0.004255242 | 1 |
| T2DM | endothelial | *LGALS2* | -0.414907355 | 0.001872683 | 0.004120753 | 1 |
| T2DM | endothelial | *SH3BP1* | -0.996669175 | 4.12E-07 | 1.24E-05 | 0.001998049 |
| T2DM | endothelial | *HPS4* | -0.712596547 | 0.000374209 | 0.00107563 | 1 |
| T2DM | endothelial | *TRIOBP* | -0.446899618 | 0.019342916 | 0.030481269 | 1 |
| T2DM | endothelial | *ANKRD54* | -0.663693877 | 4.09E-05 | 0.000185218 | 0.19855403 |
| T2DM | endothelial | *EIF3L* | -1.005744722 | 6.05E-06 | 4.65E-05 | 0.029356465 |
| T2DM | endothelial | *CCDC134* | -0.694440462 | 0.001454844 | 0.003318343 | 1 |
| T2DM | endothelial | *PICK1* | -0.730339526 | 1.67E-05 | 9.35E-05 | 0.081125315 |
| T2DM | endothelial | *SLC5A4* | 0.105066336 | 0.000129746 | 0.000458841 | 0.629529944 |
| T2DM | endothelial | *KDELR3* | -1.110601914 | 5.65E-05 | 0.000240114 | 0.274104846 |
| T2DM | endothelial | *CYP2D6* | -1.358970769 | 5.70E-05 | 0.000241092 | 0.276773254 |
| T2DM | endothelial | *CBY1* | -0.352667411 | 0.014638181 | 0.023938137 | 1 |
| T2DM | endothelial | *RTCB* | -0.570731778 | 0.004591803 | 0.008847526 | 1 |
| T2DM | endothelial | *RAB36* | -0.714627887 | 8.00E-05 | 0.000314078 | 0.388106026 |
| T2DM | endothelial | *PPP6R2* | -0.533050324 | 0.00099021 | 0.002395063 | 1 |
| T2DM | endothelial | *RHBDD3* | -1.043410133 | 8.08E-06 | 5.61E-05 | 0.039200546 |
| T2DM | endothelial | *TOM1* | -0.543598007 | 1.29E-05 | 7.77E-05 | 0.062360663 |
| T2DM | endothelial | *CHKB* | -0.894112442 | 0.000111164 | 0.000404626 | 0.539366587 |
| T2DM | endothelial | *THOC5* | -0.713383167 | 9.86E-06 | 6.41E-05 | 0.047854772 |
| T2DM | endothelial | *TTLL12* | -0.93722301 | 0.004819092 | 0.009223213 | 1 |
| T2DM | endothelial | *MTMR3* | -0.525185499 | 0.000239416 | 0.000756279 | 1 |
| T2DM | endothelial | *APOL4* | -0.505050071 | 0.002732367 | 0.005689891 | 1 |
| T2DM | endothelial | *FOXRED2* | -0.334045947 | 0.002424012 | 0.00512029 | 1 |
| T2DM | endothelial | *KIAA0930* | -0.697713842 | 4.63E-06 | 3.90E-05 | 0.022477 |
| T2DM | endothelial | *L3MBTL2* | -1.059218699 | 1.03E-05 | 6.59E-05 | 0.0501195 |
| T2DM | endothelial | *ZC3H7B* | -0.724888268 | 0.000526797 | 0.001420799 | 1 |
| T2DM | endothelial | *ACO2* | -0.715737435 | 2.15E-05 | 0.000113102 | 0.104270985 |
| T2DM | endothelial | *POLR3H* | -0.918923425 | 6.82E-06 | 5.03E-05 | 0.033112293 |
| T2DM | endothelial | *DESI1* | -0.568076505 | 0.034040667 | 0.049733609 | 1 |
| T2DM | endothelial | *HDAC10* | -1.232969173 | 7.29E-05 | 0.000294079 | 0.353482753 |
| T2DM | endothelial | *SDR39U1* | -0.341574953 | 0.001891596 | 0.004156713 | 1 |
| T2DM | endothelial | *RBM23* | -0.436163287 | 0.010569091 | 0.018069496 | 1 |
| T2DM | endothelial | *AP4S1* | -0.584716188 | 0.015026852 | 0.0245159 | 1 |
| T2DM | endothelial | *PYGL* | -0.21109443 | 0.005698394 | 0.01060146 | 1 |
| T2DM | endothelial | *CNIH1* | -0.675771233 | 0.007054504 | 0.012767046 | 1 |
| T2DM | endothelial | *ATP6V1D* | -0.672767218 | 0.00039943 | 0.001132691 | 1 |
| T2DM | endothelial | *PLEK2* | -0.981115085 | 4.07E-06 | 3.57E-05 | 0.01974099 |
| T2DM | endothelial | *GSTZ1* | -0.630198141 | 0.000467411 | 0.001284309 | 1 |
| T2DM | endothelial | *TMED8* | -0.701158187 | 0.000320152 | 0.000952993 | 1 |
| T2DM | endothelial | *LGMN* | -0.486810224 | 8.57E-05 | 0.000329425 | 0.415775048 |
| T2DM | endothelial | *MTHFD1* | -0.658987827 | 3.10E-05 | 0.000148428 | 0.150505801 |
| T2DM | endothelial | *TELO2* | -0.764782338 | 0.020357758 | 0.031801623 | 1 |
| T2DM | endothelial | *YY1* | -0.571419363 | 0.004791325 | 0.009177857 | 1 |
| T2DM | endothelial | *CCNB1IP1* | -0.884411184 | 4.48E-05 | 0.000199981 | 0.2173792 |
| T2DM | endothelial | *EFS* | -0.634411972 | 3.34E-07 | 1.24E-05 | 0.001621185 |
| T2DM | endothelial | *KIAA0391* | -1.703400457 | 2.26E-06 | 2.52E-05 | 0.010955773 |

| | | | | | | |
|---|---|---|---|---|---|---|
| T2DM | endothelial | *TRPC4AP* | -0.463629832 | 0.005189493 | 0.009820367 | 1 |
| T2DM | endothelial | *PYGB* | -0.400456943 | 0.007317165 | 0.013168726 | 1 |
| T2DM | endothelial | *ABHD12* | -0.538303677 | 0.003946475 | 0.00778386 | 1 |
| T2DM | endothelial | *CD40* | 0.449633916 | 0.006316157 | 0.011577634 | 1 |
| T2DM | endothelial | *UQCC1* | -0.677208779 | 2.63E-05 | 0.000131366 | 0.127818936 |
| T2DM | endothelial | *IFT52* | -0.411681656 | 0.019468573 | 0.030629233 | 1 |
| T2DM | endothelial | *NDRG3* | -0.580578494 | 0.014274301 | 0.023429942 | 1 |
| T2DM | endothelial | *PABPC1L* | -0.880431734 | 7.95E-07 | 1.49E-05 | 0.003857884 |
| T2DM | endothelial | *DOK5* | -0.931164002 | 0.001734326 | 0.003860068 | 1 |
| T2DM | endothelial | *CSTF1* | -0.877424164 | 0.015699806 | 0.025442704 | 1 |
| T2DM | endothelial | *NELFCD* | -0.586639226 | 0.00907416 | 0.015825961 | 1 |
| T2DM | endothelial | *CTSZ* | -0.636346656 | 0.001960598 | 0.004290854 | 1 |
| T2DM | endothelial | *MTG2* | -0.667491554 | 0.000142329 | 0.000493623 | 0.690578304 |
| T2DM | endothelial | *TCFL5* | -0.902278876 | 0.000382836 | 0.001095882 | 1 |
| T2DM | endothelial | *DIDO1* | -0.440146036 | 0.014203289 | 0.023329166 | 1 |
| T2DM | endothelial | *GID8* | -0.726092239 | 0.000604174 | 0.001590588 | 1 |
| T2DM | endothelial | *NKAIN4* | -0.547275617 | 0.002991918 | 0.006138176 | 1 |
| T2DM | endothelial | *PTK6* | -2.380595565 | 1.29E-06 | 1.88E-05 | 0.006277831 |
| T2DM | endothelial | *GMEB2* | -1.02744435 | 4.20E-05 | 0.000189722 | 0.203951208 |
| T2DM | endothelial | *C20orf27* | -0.516106178 | 0.010608317 | 0.018128408 | 1 |
| T2DM | endothelial | *CDC25B* | 0.342647432 | 0.028680958 | 0.042910887 | 1 |
| T2DM | endothelial | *RNF24* | -0.63043115 | 7.78E-06 | 5.48E-05 | 0.037758595 |
| T2DM | endothelial | *NDUFAF5* | -0.333431419 | 0.002906371 | 0.006008398 | 1 |
| T2DM | endothelial | *TRIB3* | -1.532572566 | 0.004833298 | 0.009240016 | 1 |
| T2DM | endothelial | *RASSF2* | -0.463143279 | 0.00777279 | 0.013834769 | 1 |
| T2DM | endothelial | *ANGPT4* | -1.659486424 | 7.05E-06 | 5.11E-05 | 0.034191765 |
| T2DM | endothelial | *RSPO4* | -0.364252995 | 0.007996225 | 0.014159738 | 1 |
| T2DM | endothelial | *SEC23B* | -0.478181712 | 0.001942514 | 0.004257035 | 1 |
| T2DM | endothelial | *CRNKL1* | -0.431615574 | 0.031868384 | 0.047070137 | 1 |
| T2DM | endothelial | *MROH8* | -0.61655592 | 4.27E-07 | 1.24E-05 | 0.002069697 |
| T2DM | endothelial | *MANBAL* | -0.423511744 | 0.005903314 | 0.010911574 | 1 |
| T2DM | endothelial | *TTI1* | -0.808125153 | 0.000467719 | 0.001284309 | 1 |
| T2DM | endothelial | *BPI* | -0.172940137 | 0.012815618 | 0.021434463 | 1 |
| T2DM | endothelial | *DNTTIP1* | -0.459602029 | 0.003321192 | 0.006719943 | 1 |
| T2DM | endothelial | *MAP1LC3A* | -0.81812512 | 1.66E-05 | 9.29E-05 | 0.08058056 |
| T2DM | endothelial | *RBFA* | -0.671099734 | 0.005718293 | 0.01062704 | 1 |
| T2DM | endothelial | *LPIN2* | -0.678381285 | 2.65E-05 | 0.000131898 | 0.128600302 |
| T2DM | endothelial | *MYOM1* | -0.672306767 | 1.97E-06 | 2.37E-05 | 0.009568296 |
| T2DM | endothelial | *LAMA1* | -0.975198055 | 2.16E-05 | 0.000113344 | 0.104616595 |
| T2DM | endothelial | *RNF125* | 0.556205595 | 0.000985586 | 0.002386258 | 1 |
| T2DM | endothelial | *MIB1* | -0.433244872 | 0.013825747 | 0.022786184 | 1 |
| T2DM | endothelial | *STK24* | -0.577535108 | 0.000869066 | 0.00215248 | 1 |
| T2DM | endothelial | *ACP5* | -0.366333933 | 0.001285866 | 0.003000974 | 1 |
| T2DM | endothelial | *DNAJC3* | -0.478356826 | 0.012357138 | 0.020767868 | 1 |
| T2DM | endothelial | *UGGT2* | -0.381774481 | 0.019986582 | 0.031302419 | 1 |
| T2DM | endothelial | *SUPT20H* | -0.583525044 | 0.001722915 | 0.003841721 | 1 |
| T2DM | endothelial | *RGCC* | 0.27564699 | 0.002130472 | 0.004600379 | 1 |
| T2DM | endothelial | *DGKH* | 0.225591775 | 0.029827024 | 0.044447395 | 1 |
| T2DM | endothelial | *KATNAL1* | -0.605097814 | 0.000834006 | 0.00207731 | 1 |
| T2DM | endothelial | *INTS6* | -0.495316445 | 0.005016322 | 0.009529833 | 1 |

| | | | | | | |
|---|---|---|---|---|---|---|
| T2DM | endothelial | *CLN5* | -0.498924629 | 0.032797224 | 0.048236475 | 1 |
| T2DM | endothelial | *MGRN1* | -0.619100337 | 0.000287582 | 0.000874897 | 1 |
| T2DM | endothelial | *DHODH* | -0.558696784 | 0.00061023 | 0.001601318 | 1 |
| T2DM | endothelial | *PSMD7* | -0.610237863 | 0.005789187 | 0.010745652 | 1 |
| T2DM | endothelial | *COG4* | -0.577688107 | 0.00295326 | 0.00607943 | 1 |
| T2DM | endothelial | *MON1B* | -1.091735391 | 2.50E-05 | 0.000126317 | 0.121138284 |
| T2DM | endothelial | *NPRL3* | -0.571847205 | 7.60E-05 | 0.000302894 | 0.368924381 |
| T2DM | endothelial | *NAGPA* | -0.623781114 | 0.00035963 | 0.001042992 | 1 |
| T2DM | endothelial | *ZNF500* | -0.527376528 | 0.002984853 | 0.006126272 | 1 |
| T2DM | endothelial | *NME4* | -1.08962784 | 1.75E-05 | 9.69E-05 | 0.085103694 |
| T2DM | endothelial | *NOMO3* | -0.642091068 | 0.000663364 | 0.001713615 | 1 |
| T2DM | endothelial | *NARFL* | -0.640361019 | 0.005892637 | 0.010900143 | 1 |
| T2DM | endothelial | *CLCN7* | -0.518135961 | 0.000168752 | 0.000562739 | 0.818785914 |
| T2DM | endothelial | *HAGHL* | -0.362005327 | 0.00736734 | 0.013224689 | 1 |
| T2DM | endothelial | *RHBDL1* | -2.427190078 | 1.45E-07 | 1.08E-05 | 0.0007014 |
| T2DM | endothelial | *NUBP1* | -0.802149464 | 7.79E-05 | 0.000307756 | 0.377924524 |
| T2DM | endothelial | *UBE2I* | -0.732554933 | 0.0005698 | 0.001514058 | 1 |
| T2DM | endothelial | *CAPN15* | -0.774066756 | 0.005007951 | 0.009517656 | 1 |
| T2DM | endothelial | *EARS2* | -0.594966447 | 8.34E-06 | 5.74E-05 | 0.040453763 |
| T2DM | endothelial | *CPPED1* | -0.674409038 | 0.000769264 | 0.001936931 | 1 |
| T2DM | endothelial | *KAT8* | -0.859870915 | 0.000544702 | 0.001456942 | 1 |
| T2DM | endothelial | *TMC5* | -0.667364935 | 2.42E-06 | 2.63E-05 | 0.011762392 |
| T2DM | endothelial | *C16orf62* | -0.604637674 | 0.000397829 | 0.001129471 | 1 |
| T2DM | endothelial | *RNF40* | -0.928030692 | 0.000162229 | 0.000545485 | 0.787134567 |
| T2DM | endothelial | *IQCH* | -0.687092131 | 0.002227903 | 0.004776749 | 1 |
| T2DM | endothelial | *LACTB* | -0.86721214 | 2.41E-05 | 0.000124302 | 0.116968144 |
| T2DM | endothelial | *HERC1* | -0.343961742 | 0.026822788 | 0.040442564 | 1 |
| T2DM | endothelial | *TRIP4* | -0.599964525 | 0.002307174 | 0.004905525 | 1 |
| T2DM | endothelial | *AP3B2* | -0.318232899 | 0.000156177 | 0.000529539 | 0.75776977 |
| T2DM | endothelial | *CTSH* | 0.204418841 | 0.007607015 | 0.013589557 | 1 |
| T2DM | endothelial | *TTC23* | -0.563712218 | 0.006865024 | 0.012470646 | 1 |
| T2DM | endothelial | *FAH* | -0.691280451 | 5.07E-07 | 1.27E-05 | 0.002459346 |
| T2DM | endothelial | *RPAP1* | -1.091471795 | 7.76E-07 | 1.47E-05 | 0.003765669 |
| T2DM | endothelial | *ZDHHC2* | 0.318099466 | 0.000473312 | 0.001297463 | 1 |
| T2DM | endothelial | *EYA1* | -0.583035467 | 0.000458694 | 0.001264536 | 1 |
| T2DM | endothelial | *IMPAD1* | -0.767649638 | 0.00070644 | 0.001806127 | 1 |
| T2DM | endothelial | *SFRP1* | -0.879635366 | 2.04E-06 | 2.42E-05 | 0.009915019 |
| T2DM | endothelial | *RAB2A* | -0.391648953 | 0.01140415 | 0.01933366 | 1 |
| T2DM | endothelial | *ARMC1* | -0.717138733 | 4.32E-05 | 0.000194368 | 0.209723305 |
| T2DM | endothelial | *CHRAC1* | -1.355085302 | 0.000538538 | 0.001444438 | 1 |
| T2DM | endothelial | *SNX16* | -0.810428983 | 0.000338572 | 0.0009926 | 1 |
| T2DM | endothelial | *SQLE* | -0.644476487 | 0.000459777 | 0.001266801 | 1 |
| T2DM | endothelial | *TNFRSF10A* | 0.239626369 | 7.21E-07 | 1.40E-05 | 0.003498719 |
| T2DM | endothelial | *TUSC3* | 0.23894193 | 2.78E-05 | 0.000136593 | 0.134781204 |
| T2DM | endothelial | *KLHDC4* | -0.57229716 | 0.004695365 | 0.009015398 | 1 |
| T2DM | endothelial | *MAN2B1* | -0.515637059 | 9.82E-05 | 0.00036628 | 0.476530735 |
| T2DM | endothelial | *TULP2* | -2.732438195 | 5.18E-06 | 4.18E-05 | 0.025118695 |
| T2DM | endothelial | *SNRNP70* | -0.632861741 | 0.000109105 | 0.000398928 | 0.529377107 |
| T2DM | endothelial | *PPP1R37* | -0.898237347 | 7.89E-06 | 5.52E-05 | 0.038265025 |
| T2DM | endothelial | *FCGRT* | -0.441754976 | 0.004645596 | 0.00893044 | 1 |

| | | | | | | |
|---|---|---|---|---|---|---|
| T2DM | endothelial | *PEX11G* | -0.561954279 | 0.005696851 | 0.01060146 | 1 |
| T2DM | endothelial | *ERCC2* | -1.167436193 | 3.02E-06 | 2.98E-05 | 0.01464844 |
| T2DM | endothelial | *CD37* | -0.66044606 | 0.001020299 | 0.00245682 | 1 |
| T2DM | endothelial | *AMH* | -2.703008252 | 5.28E-07 | 1.27E-05 | 0.002560452 |
| T2DM | endothelial | *FCER2* | -1.239980748 | 3.78E-07 | 1.24E-05 | 0.001832463 |
| T2DM | endothelial | *CLEC4M* | -0.936267164 | 4.86E-07 | 1.27E-05 | 0.002357016 |
| T2DM | endothelial | *NOVA2* | 0.172045691 | 4.07E-06 | 3.57E-05 | 0.019766856 |
| T2DM | endothelial | *KIR3DX1* | -1.40558122 | 0.002689 | 0.005621296 | 1 |
| T2DM | endothelial | *MED25* | -0.98048122 | 1.02E-05 | 6.54E-05 | 0.049474446 |
| T2DM | endothelial | *TIMM44* | -0.686987618 | 0.000831049 | 0.002071006 | 1 |
| T2DM | endothelial | *IL27RA* | 0.444425515 | 1.66E-05 | 9.29E-05 | 0.080554095 |
| T2DM | endothelial | *C19orf44* | -0.243640417 | 0.024078567 | 0.036790847 | 1 |
| T2DM | endothelial | *OLFM2* | -0.580898525 | 1.86E-05 | 0.00010097 | 0.090267499 |
| T2DM | endothelial | *RASAL3* | -1.098245868 | 0.01641571 | 0.026435123 | 1 |
| T2DM | endothelial | *ZNF419* | 0.217480468 | 0.031753707 | 0.046943628 | 1 |
| T2DM | endothelial | *ANKRD27* | -0.633329047 | 0.002963182 | 0.006094251 | 1 |
| T2DM | endothelial | *TGFB1* | -0.473898954 | 0.022232515 | 0.034299575 | 1 |
| T2DM | endothelial | *MRPL4* | -0.609019514 | 0.007336796 | 0.013185609 | 1 |
| T2DM | endothelial | *GRIN2D* | -0.945003384 | 1.94E-06 | 2.36E-05 | 0.009424359 |
| T2DM | endothelial | *CARD8* | 0.300511444 | 0.005809719 | 0.010775983 | 1 |
| T2DM | endothelial | *LIG1* | -1.013559848 | 0.000128754 | 0.000455663 | 0.624713931 |
| T2DM | endothelial | *ZNF175* | -0.661874888 | 0.014140664 | 0.023245756 | 1 |
| T2DM | endothelial | *CAPS* | -1.264340471 | 1.55E-06 | 2.14E-05 | 0.007532775 |
| T2DM | endothelial | *MIER2* | 0.455063759 | 0.028068304 | 0.042082351 | 1 |
| T2DM | endothelial | *PLEKHA4* | -0.652474916 | 3.13E-07 | 1.24E-05 | 0.001519605 |
| T2DM | endothelial | *TNPO2* | -0.712373288 | 8.27E-05 | 0.000321365 | 0.4013845 |
| T2DM | endothelial | *GCDH* | -1.158237173 | 6.01E-10 | 3.24E-07 | 2.92E-06 |
| T2DM | endothelial | *DNASE2* | -0.982861145 | 0.000136319 | 0.000478251 | 0.661420922 |
| T2DM | endothelial | *TFPT* | -1.157955476 | 0.00105204 | 0.002523233 | 1 |
| T2DM | endothelial | *RAB3A* | 2.2818749 | 0.005842331 | 0.010831865 | 1 |
| T2DM | endothelial | *COPE* | -0.773184584 | 0.000605166 | 0.001591473 | 1 |
| T2DM | endothelial | *ETV2* | -5.854409409 | 0.000136698 | 0.000479072 | 0.663260383 |
| T2DM | endothelial | *MAG* | 1.204505061 | 7.56E-06 | 5.41E-05 | 0.036695005 |
| T2DM | endothelial | *LSR* | 0.305077571 | 0.000250595 | 0.000784444 | 1 |
| T2DM | endothelial | *ZNF14* | -0.900498922 | 0.000416619 | 0.001171196 | 1 |
| T2DM | endothelial | *ETHE1* | -0.952756161 | 0.001591549 | 0.003586715 | 1 |
| T2DM | endothelial | *GTPBP10* | -0.403247444 | 0.014094835 | 0.02318242 | 1 |
| T2DM | endothelial | *RASA4* | 1.120921838 | 4.95E-06 | 4.06E-05 | 0.024008882 |
| T2DM | endothelial | *BET1* | -0.68732007 | 0.000594951 | 0.001568859 | 1 |
| T2DM | endothelial | *DNAH11* | -0.689323877 | 4.67E-07 | 1.27E-05 | 0.002266881 |
| T2DM | endothelial | *STEAP1B* | 0.412042552 | 6.21E-06 | 4.74E-05 | 0.030119127 |
| T2DM | endothelial | *PTN* | -0.800896207 | 9.33E-06 | 6.19E-05 | 0.045276845 |
| T2DM | endothelial | *ADAP1* | -1.686304329 | 3.15E-07 | 1.24E-05 | 0.001530216 |
| T2DM | endothelial | *MET* | 0.207522638 | 8.52E-05 | 0.000328423 | 0.413484632 |
| T2DM | endothelial | *IQCE* | -0.958243239 | 1.46E-05 | 8.53E-05 | 0.070891148 |
| T2DM | endothelial | *VIPR2* | -0.572326864 | 8.69E-07 | 1.53E-05 | 0.004217263 |
| T2DM | endothelial | *HIBADH* | -0.479592464 | 0.000518951 | 0.001404322 | 1 |
| T2DM | endothelial | *CPVL* | -0.628397309 | 8.09E-06 | 5.61E-05 | 0.03923205 |
| T2DM | endothelial | *GRB10* | 0.308324163 | 0.000486469 | 0.001326783 | 1 |
| T2DM | endothelial | *PLEKHA8* | -0.655691934 | 0.007034768 | 0.012736081 | 1 |

| | | | | | | |
|---|---|---|---|---|---|---|
| T2DM | endothelial | *EPHB6* | -0.454619698 | 3.62E-05 | 0.000167762 | 0.175646419 |
| T2DM | endothelial | *CCL24* | -0.543833076 | 0.028141659 | 0.042169033 | 1 |
| T2DM | endothelial | *ZKSCAN1* | -0.619057721 | 0.000597027 | 0.001573479 | 1 |
| T2DM | endothelial | *EIF3B* | -0.763165059 | 0.000364209 | 0.001052498 | 1 |
| T2DM | endothelial | *TFR2* | -1.25988092 | 3.18E-07 | 1.24E-05 | 0.001541274 |
| T2DM | endothelial | *RBM28* | -0.484570131 | 0.002295132 | 0.004888096 | 1 |
| T2DM | endothelial | *IMPDH1* | 0.448449328 | 0.010224588 | 0.017573398 | 1 |
| T2DM | endothelial | *C1GALT1* | -0.698705544 | 3.35E-05 | 0.000157293 | 0.162452795 |
| T2DM | endothelial | *NRF1* | -0.501465825 | 0.002651863 | 0.005555629 | 1 |
| T2DM | endothelial | *ZNF862* | -0.62463731 | 0.000375398 | 0.001077132 | 1 |
| T2DM | endothelial | *MEST* | -0.434048583 | 5.79E-07 | 1.27E-05 | 0.002810238 |
| T2DM | endothelial | *ACTR3C* | -0.519008968 | 1.52E-06 | 2.12E-05 | 0.00738303 |
| T2DM | endothelial | *POU6F2* | -0.465576131 | 3.06E-07 | 1.24E-05 | 0.001483304 |
| T2DM | endothelial | *TSPAN13* | 0.204856554 | 5.28E-06 | 4.21E-05 | 0.025604909 |
| T2DM | endothelial | *RARRES2* | -0.576970604 | 0.000365644 | 0.001055386 | 1 |
| T2DM | endothelial | *COA1* | -0.430172768 | 0.010900551 | 0.0185512 | 1 |
| T2DM | endothelial | *BLVRA* | -0.80809578 | 4.80E-06 | 3.99E-05 | 0.023302022 |
| T2DM | endothelial | *URGCP* | -0.638915154 | 0.000579866 | 0.001534922 | 1 |
| T2DM | endothelial | *AEBP1* | -0.831895853 | 1.89E-06 | 2.34E-05 | 0.009180767 |
| T2DM | endothelial | *GCK* | -6.255684062 | 1.34E-06 | 1.93E-05 | 0.006516768 |
| T2DM | endothelial | *TBL2* | -0.651288505 | 0.000551087 | 0.001471586 | 1 |
| T2DM | endothelial | *SPATA6L* | 0.424544963 | 0.001619536 | 0.003637959 | 1 |
| T2DM | endothelial | *SLC1A1* | 0.251960289 | 0.00017936 | 0.000592816 | 0.870254014 |
| T2DM | endothelial | *FKTN* | -0.639981355 | 0.001304651 | 0.003039858 | 1 |
| T2DM | endothelial | *MEGF9* | -0.714057618 | 9.31E-06 | 6.19E-05 | 0.045185034 |
| T2DM | endothelial | *CORO2A* | 0.227931991 | 6.64E-06 | 4.92E-05 | 0.032211324 |
| T2DM | endothelial | *C5* | -0.688455736 | 4.53E-06 | 3.85E-05 | 0.022000632 |
| T2DM | endothelial | *OGN* | -0.624726677 | 2.48E-05 | 0.000125919 | 0.120120241 |
| T2DM | endothelial | *ECM2* | -0.549173961 | 1.98E-06 | 2.38E-05 | 0.009598373 |
| T2DM | endothelial | *PTGR1* | -0.596532029 | 3.70E-06 | 3.34E-05 | 0.017948013 |
| T2DM | endothelial | *SUSD1* | -0.245705734 | 0.01677539 | 0.026942798 | 1 |
| T2DM | endothelial | *PLGRKT* | -0.477933822 | 0.00540589 | 0.010150688 | 1 |
| T2DM | endothelial | *KANK1* | -0.569296568 | 2.15E-06 | 2.46E-05 | 0.01041552 |
| T2DM | endothelial | *NCS1* | -0.458015775 | 9.25E-05 | 0.000350222 | 0.448984789 |
| T2DM | endothelial | *FUBP3* | -0.679706461 | 0.00011312 | 0.000410214 | 0.548859632 |
| T2DM | endothelial | *RGP1* | -0.871736801 | 0.002595086 | 0.005446089 | 1 |
| T2DM | endothelial | *PTGDS* | -0.98922747 | 2.11E-05 | 0.0001119 | 0.102388193 |
| T2DM | endothelial | *ATRNL1* | -0.145092092 | 0.008936393 | 0.015608128 | 1 |
| T2DM | endothelial | *HPS1* | -0.822659819 | 3.90E-05 | 0.00017767 | 0.18903351 |
| T2DM | endothelial | *PHYH* | -0.432045948 | 0.013822474 | 0.022786184 | 1 |
| T2DM | endothelial | *DNMBP* | -0.667630839 | 0.000188748 | 0.000619205 | 0.915804668 |
| T2DM | endothelial | *ATE1* | -0.354223471 | 0.023843604 | 0.036506522 | 1 |
| T2DM | endothelial | *PLEKHA1* | -0.533186556 | 0.004500299 | 0.008695051 | 1 |
| T2DM | endothelial | *CDH23* | 0.316109064 | 0.004713134 | 0.009045937 | 1 |
| T2DM | endothelial | *BMPR1A* | -0.512332454 | 1.27E-06 | 1.86E-05 | 0.006160507 |
| T2DM | endothelial | *TNKS2* | -0.552485634 | 0.003549118 | 0.007104093 | 1 |
| T2DM | endothelial | *ANKRD26* | -0.566437097 | 0.000335603 | 0.000986278 | 1 |
| T2DM | endothelial | *ACBD5* | -0.719973804 | 0.001003268 | 0.002424231 | 1 |
| T2DM | endothelial | *LHPP* | -0.847462022 | 4.73E-07 | 1.27E-05 | 0.002296037 |
| T2DM | endothelial | *PITRM1* | -0.903959418 | 0.000120646 | 0.000432331 | 0.585376527 |

| | | | | | | |
|---|---|---|---|---|---|---|
| T2DM | endothelial | *TFAM* | -0.651437333 | 0.019053359 | 0.030103191 | 1 |
| T2DM | endothelial | *PBLD* | -0.8284123 | 1.33E-06 | 1.92E-05 | 0.006460951 |
| T2DM | endothelial | *PSMB3* | -0.674927087 | 0.00840331 | 0.014772776 | 1 |
| T2DM | endothelial | *RAPGEFL1* | -1.10186188 | 4.16E-07 | 1.24E-05 | 0.002019858 |
| T2DM | endothelial | *WNT3* | -0.614368111 | 6.53E-06 | 4.87E-05 | 0.031690247 |
| T2DM | endothelial | *ASPA* | -0.451917324 | 3.90E-05 | 0.00017767 | 0.189168919 |
| T2DM | endothelial | *RAD51C* | -0.724100312 | 5.38E-05 | 0.000230529 | 0.261188822 |
| T2DM | endothelial | *TRIM37* | -0.791814089 | 4.63E-05 | 0.000205564 | 0.224681216 |
| T2DM | endothelial | *PNPO* | 0.716066692 | 0.005401638 | 0.010148875 | 1 |
| T2DM | endothelial | *TRIM16L* | -1.400508482 | 3.68E-06 | 3.34E-05 | 0.017868769 |
| T2DM | endothelial | *CDK5RAP3* | -1.00410867 | 2.03E-05 | 0.000108012 | 0.098290838 |
| T2DM | endothelial | *PIGL* | -0.613895628 | 0.000474569 | 0.001299439 | 1 |
| T2DM | endothelial | *MED13* | -0.505092941 | 0.0013902 | 0.003199834 | 1 |
| T2DM | endothelial | *RAI1* | -0.646527923 | 0.007226758 | 0.013030185 | 1 |
| T2DM | endothelial | *GOSR1* | -0.610730243 | 0.001086397 | 0.002595876 | 1 |
| T2DM | endothelial | *DRG2* | -0.949243739 | 0.000422488 | 0.001182187 | 1 |
| T2DM | endothelial | *FTSJ3* | -0.294579922 | 9.62E-05 | 0.000361144 | 0.466959069 |
| T2DM | endothelial | *ALDH3A1* | -0.984279466 | 0.005908067 | 0.0109162 | 1 |
| T2DM | endothelial | *ICAM2* | 0.207799753 | 8.85E-06 | 5.97E-05 | 0.042936631 |
| T2DM | endothelial | *C17orf75* | -0.401080806 | 0.021535466 | 0.033458239 | 1 |
| T2DM | endothelial | *PSMD11* | -0.589023835 | 0.00287563 | 0.005952456 | 1 |
| T2DM | endothelial | *NAGLU* | -0.727967902 | 4.83E-05 | 0.000212462 | 0.234345841 |
| T2DM | endothelial | *HSD17B1* | -0.668467002 | 0.001813607 | 0.004008939 | 1 |
| T2DM | endothelial | *EZH1* | -0.456138366 | 0.006382296 | 0.011685623 | 1 |
| T2DM | endothelial | *ABCC3* | -0.273895335 | 0.02980125 | 0.044422632 | 1 |
| T2DM | endothelial | *EFTUD2* | -0.769405936 | 0.000581508 | 0.001538429 | 1 |
| T2DM | endothelial | *EFNB3* | -0.789645399 | 3.72E-05 | 0.000171791 | 0.180723936 |
| T2DM | endothelial | *FAM20A* | -0.557631119 | 0.004410038 | 0.008535104 | 1 |
| T2DM | endothelial | *DPH1* | -1.167993413 | 0.000228476 | 0.000725026 | 1 |
| T2DM | endothelial | *DHRS7B* | -0.521391187 | 0.009260099 | 0.016092408 | 1 |
| T2DM | endothelial | *MYH3* | -1.646949802 | 4.30E-06 | 3.71E-05 | 0.020849121 |
| T2DM | endothelial | *SLAIN2* | -0.476819155 | 0.014368823 | 0.023577116 | 1 |
| T2DM | endothelial | *OCIAD1* | -0.475904244 | 0.007954559 | 0.014101395 | 1 |
| T2DM | endothelial | *UBE2D3* | -0.466584685 | 0.003077002 | 0.006299416 | 1 |
| T2DM | endothelial | *NDUFC1* | -0.577062777 | 0.022886776 | 0.035215105 | 1 |
| T2DM | endothelial | *GRPEL1* | -1.175636524 | 0.000431257 | 0.001202563 | 1 |
| T2DM | endothelial | *AADAT* | -1.196183699 | 1.93E-06 | 2.35E-05 | 0.009345379 |
| T2DM | endothelial | *TRIM2* | -0.686571981 | 0.0001785 | 0.000590778 | 0.866081126 |
| T2DM | endothelial | *BST1* | -0.660871457 | 7.52E-06 | 5.39E-05 | 0.036465995 |
| T2DM | endothelial | *UFSP2* | -0.607662386 | 0.00674175 | 0.012269681 | 1 |
| T2DM | endothelial | *KLHL5* | 0.324695437 | 0.004820706 | 0.009223213 | 1 |
| T2DM | endothelial | *FAM149A* | -0.592619802 | 0.007835936 | 0.013931829 | 1 |
| T2DM | endothelial | *DDX25* | -0.357266121 | 0.002964228 | 0.006094251 | 1 |
| T2DM | endothelial | *CTSC* | -0.259972554 | 0.025421564 | 0.038545446 | 1 |
| T2DM | endothelial | *ELP4* | -0.417613168 | 0.012127492 | 0.020431456 | 1 |
| T2DM | endothelial | *TECTA* | -1.254881403 | 4.87E-07 | 1.27E-05 | 0.002363581 |
| T2DM | endothelial | *SC5D* | -0.988083221 | 1.43E-05 | 8.42E-05 | 0.069541717 |
| T2DM | endothelial | *B3GAT1* | -2.221284192 | 1.58E-06 | 2.16E-05 | 0.007673862 |
| T2DM | endothelial | *P2RX3* | 6.416584056 | 0.013300117 | 0.022107629 | 1 |
| T2DM | endothelial | *UNC93B1* | -0.845044703 | 5.95E-05 | 0.000248357 | 0.288556488 |

| | | | | | | |
|---|---|---|---|---|---|---|
| T2DM | endothelial | *DCPS* | -0.934980975 | 0.000957795 | 0.002328266 | 1 |
| T2DM | endothelial | *FOXRED1* | -0.486616681 | 0.000300534 | 0.000905147 | 1 |
| T2DM | endothelial | *NRXN2* | -0.685161108 | 2.78E-07 | 1.24E-05 | 0.001347499 |
| T2DM | endothelial | *ST3GAL4* | -0.531800067 | 0.012333185 | 0.020734793 | 1 |
| T2DM | endothelial | *CCDC86* | -1.440685125 | 2.95E-05 | 0.000142584 | 0.143089856 |
| T2DM | endothelial | *PANX1* | -1.202196483 | 3.95E-05 | 0.000179976 | 0.191853994 |
| T2DM | endothelial | *CEP164* | -0.657873938 | 0.000211175 | 0.000678558 | 1 |
| T2DM | endothelial | *GALNT18* | 0.248421989 | 6.80E-05 | 0.000277081 | 0.329726777 |
| T2DM | endothelial | *KIAA1549L* | 0.83820269 | 8.27E-06 | 5.71E-05 | 0.040141007 |
| T2DM | endothelial | *SLC35F2* | 0.580105723 | 0.008662424 | 0.015178794 | 1 |
| T2DM | endothelial | *C11orf21* | -1.438957659 | 0.000957615 | 0.002328266 | 1 |
| T2DM | endothelial | *ELMOD1* | -0.558084382 | 0.031549725 | 0.046670508 | 1 |
| T2DM | endothelial | *NDUFS8* | -0.705437401 | 0.00626322 | 0.011493624 | 1 |
| T2DM | endothelial | *GTF2H1* | -0.688095786 | 3.85E-05 | 0.000176404 | 0.186988471 |
| T2DM | endothelial | *CORO1C* | -0.668273788 | 0.000199799 | 0.000647579 | 0.969426309 |
| T2DM | endothelial | *ASIC1* | -1.037218864 | 3.04E-07 | 1.24E-05 | 0.001475177 |
| T2DM | endothelial | *IL23A* | 1.681641035 | 9.58E-06 | 6.29E-05 | 0.04646004 |
| T2DM | endothelial | *ACSS3* | -0.425225654 | 1.36E-05 | 8.10E-05 | 0.066046623 |
| T2DM | endothelial | *PARP11* | -0.565708187 | 0.008357671 | 0.014708531 | 1 |
| T2DM | endothelial | *GPN3* | -1.101411553 | 6.61E-05 | 0.000270476 | 0.320490851 |
| T2DM | endothelial | *VPS29* | -0.471112932 | 0.012547442 | 0.021043964 | 1 |
| T2DM | endothelial | *AKAP3* | -0.242428493 | 0.011068307 | 0.018810307 | 1 |
| T2DM | endothelial | *ACAD10* | -0.732724456 | 2.40E-06 | 2.61E-05 | 0.011644108 |
| T2DM | endothelial | *CDK2AP1* | 0.686948467 | 0.003484254 | 0.007006401 | 1 |
| T2DM | endothelial | *DDX55* | -0.821840931 | 3.43E-05 | 0.000160798 | 0.16658658 |
| T2DM | endothelial | *ENDOU* | -2.033575122 | 5.83E-07 | 1.27E-05 | 0.002829374 |
| T2DM | endothelial | *VDR* | -1.415056161 | 4.43E-07 | 1.24E-05 | 0.002147096 |
| T2DM | endothelial | *CAND1* | -0.661551636 | 0.000399123 | 0.001132481 | 1 |
| T2DM | endothelial | *MDM1* | -0.699111585 | 0.010164936 | 0.017477063 | 1 |
| T2DM | endothelial | *NUP107* | -0.551128937 | 0.003043965 | 0.006237043 | 1 |
| T2DM | endothelial | *KRR1* | -0.68782615 | 0.002075631 | 0.004499983 | 1 |
| T2DM | endothelial | *GNB3* | -0.253495448 | 0.033535795 | 0.049143968 | 1 |
| T2DM | endothelial | *CDCA3* | -1.15010821 | 6.59E-06 | 4.89E-05 | 0.031953251 |
| T2DM | endothelial | *CHPT1* | -0.565831401 | 4.53E-06 | 3.85E-05 | 0.021996085 |
| T2DM | endothelial | *GNPTAB* | -0.671993814 | 5.22E-05 | 0.000224985 | 0.253333067 |
| T2DM | endothelial | *NT5DC3* | -0.694167267 | 1.54E-06 | 2.13E-05 | 0.007453692 |
| T2DM | endothelial | *SUDS3* | -0.726418428 | 0.000679154 | 0.001745869 | 1 |
| T2DM | endothelial | *C2CD5* | -0.458235429 | 0.010550484 | 0.018056004 | 1 |
| T2DM | endothelial | *BTN3A3* | 0.28841462 | 0.01844237 | 0.029242607 | 1 |
| T2DM | endothelial | *FRK* | -0.729352012 | 3.79E-07 | 1.24E-05 | 0.001837853 |
| T2DM | endothelial | *RSPH4A* | -0.396087685 | 0.026720768 | 0.040313796 | 1 |
| T2DM | endothelial | *SMIM8* | -0.443521828 | 0.005136358 | 0.009731203 | 1 |
| T2DM | endothelial | *FAM184A* | 0.406415343 | 0.000883446 | 0.002182655 | 1 |
| T2DM | endothelial | *RNGTT* | -0.495245327 | 0.00823704 | 0.014522573 | 1 |
| T2DM | endothelial | *UST* | -0.346559326 | 0.001040334 | 0.002500098 | 1 |
| T2DM | endothelial | *SLC26A8* | 0.438773323 | 0.000336926 | 0.000988969 | 1 |
| T2DM | endothelial | *RNF8* | -0.736206997 | 0.001093415 | 0.002608283 | 1 |
| T2DM | endothelial | *MDGA1* | -0.696624517 | 2.94E-07 | 1.24E-05 | 0.001427648 |
| T2DM | endothelial | *CD83* | -0.853837494 | 4.79E-08 | 6.11E-06 | 0.000232209 |
| T2DM | endothelial | *MDN1* | -0.539363566 | 0.003675734 | 0.007312284 | 1 |

| | | | | | | |
|---|---|---|---|---|---|---|
| T2DM | endothelial | *GLP1R* | -1.854172304 | 0.000141672 | 0.000491698 | 0.687394306 |
| T2DM | endothelial | *BVES* | -0.370049227 | 0.00038754 | 0.001105436 | 1 |
| T2DM | endothelial | *COL9A1* | -1.445637169 | 0.004340493 | 0.008428637 | 1 |
| T2DM | endothelial | *WASF1* | -0.595706299 | 2.69E-06 | 2.76E-05 | 0.013033633 |
| T2DM | endothelial | *VNN2* | -0.492665424 | 0.030861193 | 0.0458056 | 1 |
| T2DM | endothelial | *ACOT13* | -0.783549807 | 0.000146411 | 0.00050581 | 0.710385102 |
| T2DM | endothelial | *C6orf62* | -0.628035697 | 0.000198502 | 0.000644236 | 0.963133441 |
| T2DM | endothelial | *HBS1L* | -0.553377958 | 0.003133176 | 0.006392838 | 1 |
| T2DM | endothelial | *PEX7* | -0.340182471 | 0.01162579 | 0.019688773 | 1 |
| T2DM | endothelial | *FIG4* | -0.683900404 | 0.000485737 | 0.001325532 | 1 |
| T2DM | endothelial | *PERP* | 0.357600497 | 7.66E-05 | 0.000304956 | 0.371741842 |
| T2DM | endothelial | *CUTA* | -0.567126066 | 0.004099238 | 0.008029675 | 1 |
| T2DM | endothelial | *PACRG* | -0.95302609 | 2.45E-05 | 0.000125288 | 0.118773006 |
| T2DM | endothelial | *QKI* | -0.357978218 | 0.013262017 | 0.022074547 | 1 |
| T2DM | endothelial | *GUCA1B* | -0.818021927 | 6.16E-06 | 4.71E-05 | 0.029868874 |
| T2DM | endothelial | *PRPH2* | -0.212678454 | 0.000270163 | 0.000833265 | 1 |
| T2DM | endothelial | *IMPG1* | 0.992979906 | 8.77E-05 | 0.000335639 | 0.425654746 |
| T2DM | endothelial | *LY86* | -0.857298742 | 1.37E-07 | 1.06E-05 | 0.000665285 |
| T2DM | endothelial | *PRSS16* | -0.770018927 | 0.003819816 | 0.007569356 | 1 |
| T2DM | endothelial | *NUDT12* | -0.86342084 | 2.19E-06 | 2.48E-05 | 0.010631714 |
| T2DM | endothelial | *CEP72* | -1.204499853 | 5.26E-06 | 4.21E-05 | 0.025507516 |
| T2DM | endothelial | *KIF20A* | -3.064770259 | 4.44E-08 | 5.99E-06 | 0.00021566 |
| T2DM | endothelial | *PFDN1* | -0.441831197 | 0.010487258 | 0.017973924 | 1 |
| T2DM | endothelial | *TMCO6* | -0.671653617 | 0.001643336 | 0.003686299 | 1 |
| T2DM | endothelial | *SPARC* | -0.514530676 | 0.000610054 | 0.001601318 | 1 |
| T2DM | endothelial | *IK* | -0.466798623 | 0.018977304 | 0.029992794 | 1 |
| T2DM | endothelial | *PCDHB15* | -0.565373308 | 0.000145418 | 0.000503258 | 0.705568149 |
| T2DM | endothelial | *HAVCR1* | -0.617235478 | 0.00047816 | 0.001308535 | 1 |
| T2DM | endothelial | *RNF130* | -0.421100709 | 0.001863695 | 0.004104698 | 1 |
| T2DM | endothelial | *ARSB* | -0.802453104 | 0.000170243 | 0.000567117 | 0.826019343 |
| T2DM | endothelial | *THBS4* | -0.564918938 | 0.000567025 | 0.001508335 | 1 |
| T2DM | endothelial | *BTNL8* | 0.135706621 | 0.008839119 | 0.015460492 | 1 |
| T2DM | endothelial | *MSH3* | -0.497371195 | 0.000733364 | 0.001862 | 1 |
| T2DM | endothelial | *RASGRF2* | 0.196131115 | 0.001556809 | 0.003516591 | 1 |
| T2DM | endothelial | *CDH6* | -0.759888958 | 1.52E-07 | 1.10E-05 | 0.000735106 |
| T2DM | endothelial | *LMNB1* | -0.864264626 | 1.58E-05 | 8.96E-05 | 0.076474728 |
| T2DM | endothelial | *SKP1* | -0.51339499 | 0.015955693 | 0.025805674 | 1 |
| T2DM | endothelial | *PPWD1* | -0.586422025 | 0.003212477 | 0.006535405 | 1 |
| T2DM | endothelial | *TXNDC15* | -0.548177361 | 0.031826403 | 0.04703123 | 1 |
| T2DM | endothelial | *ERGIC1* | -0.584827419 | 0.003144842 | 0.006411249 | 1 |
| T2DM | endothelial | *ATP6V0E1* | -0.475892643 | 0.012023569 | 0.020270451 | 1 |
| T2DM | endothelial | *UNC5A* | -4.002925689 | 8.84E-07 | 1.54E-05 | 0.004289927 |
| T2DM | endothelial | *ACTR8* | -0.808687903 | 0.007322019 | 0.013172575 | 1 |
| T2DM | endothelial | *TBCCD1* | -0.982547166 | 0.000160992 | 0.000542706 | 0.781132065 |
| T2DM | endothelial | *CLDN16* | -0.569319695 | 2.37E-06 | 2.60E-05 | 0.011507778 |
| T2DM | endothelial | *CEP70* | -0.63042813 | 0.001772125 | 0.003935172 | 1 |
| T2DM | endothelial | *HYAL1* | 0.361563343 | 1.65E-05 | 9.26E-05 | 0.079831211 |
| T2DM | endothelial | *CYB561D2* | -1.066800466 | 4.02E-08 | 5.74E-06 | 0.000194994 |
| T2DM | endothelial | *CBLB* | -0.415144811 | 0.004122825 | 0.008067136 | 1 |
| T2DM | endothelial | *IQCG* | -0.71389457 | 0.000210666 | 0.000677635 | 1 |

| | | | | | | |
|---|---|---|---|---|---|---|
| T2DM | endothelial | *NCBP2* | -0.855280226 | 0.000860933 | 0.002135606 | 1 |
| T2DM | endothelial | *SLC41A3* | 0.370375375 | 0.011835335 | 0.020006093 | 1 |
| T2DM | endothelial | *PLXNA1* | -0.875351894 | 1.33E-06 | 1.92E-05 | 0.006435124 |
| T2DM | endothelial | *SCAP* | -0.712455488 | 0.000521173 | 0.001407195 | 1 |
| T2DM | endothelial | *KIAA1257* | 0.885970553 | 5.03E-05 | 0.000219601 | 0.243977209 |
| T2DM | endothelial | *NEK11* | -0.543958131 | 5.99E-05 | 0.000249601 | 0.290535318 |
| T2DM | endothelial | *HEMK1* | -0.857485529 | 3.28E-08 | 5.31E-06 | 0.000159152 |
| T2DM | endothelial | *COMMD2* | -0.538778835 | 0.017702528 | 0.028300713 | 1 |
| T2DM | endothelial | *ABHD14B* | -1.04139414 | 1.97E-06 | 2.37E-05 | 0.009542161 |
| T2DM | endothelial | *ARHGEF26* | -0.805307285 | 4.73E-05 | 0.000208702 | 0.229363084 |
| T2DM | endothelial | *VIPR1* | -1.165627868 | 1.67E-06 | 2.20E-05 | 0.008094751 |
| T2DM | endothelial | *NEK4* | -0.483393568 | 0.030301633 | 0.045113078 | 1 |
| T2DM | endothelial | *ADAM23* | 0.432587516 | 0.011842607 | 0.020007078 | 1 |
| T2DM | endothelial | *LMAN2L* | -0.720566885 | 8.18E-05 | 0.000318435 | 0.396769926 |
| T2DM | endothelial | *CENPA* | -0.518268731 | 0.024678112 | 0.037570819 | 1 |
| T2DM | endothelial | *NRBP1* | -0.650582901 | 0.005530997 | 0.010349555 | 1 |
| T2DM | endothelial | *ITGB6* | -0.924245918 | 5.42E-08 | 6.74E-06 | 0.00026287 |
| T2DM | endothelial | *PPM1G* | -0.806826602 | 0.000503338 | 0.001365882 | 1 |
| T2DM | endothelial | *MOGS* | -0.554790073 | 9.60E-05 | 0.000360585 | 0.465638222 |
| T2DM | endothelial | *GRB14* | 0.164703523 | 4.99E-06 | 4.08E-05 | 0.024190579 |
| T2DM | endothelial | *RTN4* | -0.478225243 | 0.00028749 | 0.000874897 | 1 |
| T2DM | endothelial | *POLE4* | 0.545902985 | 0.002126642 | 0.00459609 | 1 |
| T2DM | endothelial | *ACADL* | -0.489555342 | 6.66E-07 | 1.33E-05 | 0.003233025 |
| T2DM | endothelial | *MRPL19* | -0.579835257 | 0.003485069 | 0.007006401 | 1 |
| T2DM | endothelial | *IGFBP2* | 0.376241815 | 0.002981974 | 0.006122953 | 1 |
| T2DM | endothelial | *GGCX* | -0.501270727 | 0.027397483 | 0.041142862 | 1 |
| T2DM | endothelial | *GNLY* | -0.581429139 | 0.000697074 | 0.001785747 | 1 |
| T2DM | endothelial | *CHMP3* | -0.501325986 | 0.010034781 | 0.017277771 | 1 |
| T2DM | endothelial | *WNT6* | -0.784986974 | 8.35E-08 | 8.44E-06 | 0.000405275 |
| T2DM | endothelial | *IL1RL2* | -0.963903302 | 0.000255324 | 0.000796675 | 1 |
| T2DM | endothelial | *MLPH* | 0.144337553 | 2.88E-06 | 2.89E-05 | 0.013989169 |
| T2DM | endothelial | *HDLBP* | -0.538308953 | 0.00161099 | 0.00362092 | 1 |
| T2DM | endothelial | *TPO* | 0.083157644 | 3.87E-06 | 3.45E-05 | 0.018758205 |
| T2DM | endothelial | *TAF1B* | -0.647478836 | 0.000275876 | 0.000846679 | 1 |
| T2DM | endothelial | *NOL10* | -0.529643207 | 0.006788691 | 0.01234585 | 1 |
| T2DM | endothelial | *CEBPZ* | -0.594598262 | 0.002952404 | 0.00607943 | 1 |
| T2DM | endothelial | *SLC25A12* | -0.555651474 | 0.002229653 | 0.004777053 | 1 |
| T2DM | endothelial | *DARS* | -0.549277545 | 0.003855219 | 0.00762868 | 1 |
| T2DM | endothelial | *WIPF1* | -0.409722437 | 0.022949348 | 0.035282078 | 1 |
| T2DM | endothelial | *ORC4* | -0.463072808 | 0.007399496 | 0.013257886 | 1 |
| T2DM | endothelial | *CD207* | 0.283313559 | 6.33E-07 | 1.32E-05 | 0.003070183 |
| T2DM | endothelial | *NFE2L2* | -0.398941903 | 0.012203811 | 0.020538637 | 1 |
| T2DM | endothelial | *MSH6* | -0.700977816 | 0.00156559 | 0.003533136 | 1 |
| T2DM | endothelial | *DNAJC16* | -0.499848287 | 0.01449817 | 0.023757217 | 1 |
| T2DM | endothelial | *MARK1* | -0.662033947 | 2.42E-05 | 0.000124381 | 0.117291605 |
| T2DM | endothelial | *MORN1* | -0.524330603 | 0.000700146 | 0.001792669 | 1 |
| T2DM | endothelial | *GPX7* | -0.367169745 | 0.000113523 | 0.000411363 | 0.550815672 |
| T2DM | endothelial | *RALGPS2* | -0.618137678 | 8.63E-07 | 1.53E-05 | 0.004188959 |
| T2DM | endothelial | *CEP104* | -0.542051758 | 0.009962141 | 0.01717098 | 1 |
| T2DM | endothelial | *FAM20B* | -0.565204851 | 0.013335465 | 0.022151207 | 1 |

| | | | | | | |
|---|---|---|---|---|---|---|
| T2DM | endothelial | *ICMT* | -1.352570762 | 8.06E-05 | 0.000315499 | 0.390903449 |
| T2DM | endothelial | *KIAA1324* | -1.71310421 | 6.17E-05 | 0.000255778 | 0.299191351 |
| T2DM | endothelial | *MECR* | -0.759812388 | 4.27E-06 | 3.70E-05 | 0.020700832 |
| T2DM | endothelial | *KCNC4* | 0.262387807 | 0.027350955 | 0.041085706 | 1 |
| T2DM | endothelial | *EDEM3* | -0.559924994 | 0.004337755 | 0.008428637 | 1 |
| T2DM | endothelial | *RAP1A* | -0.392256601 | 0.021238837 | 0.033029114 | 1 |
| T2DM | endothelial | *CAPZA1* | -0.578930434 | 0.000591452 | 0.001560482 | 1 |
| T2DM | endothelial | *ASH1L* | -0.324500351 | 0.019236772 | 0.030335981 | 1 |
| T2DM | endothelial | *DOCK7* | -0.597903785 | 7.00E-06 | 5.11E-05 | 0.033944197 |
| T2DM | endothelial | *FBXO2* | -0.469728823 | 0.000716608 | 0.001827771 | 1 |
| T2DM | endothelial | *DNAJC6* | -0.531702525 | 0.023882237 | 0.036530832 | 1 |
| T2DM | endothelial | *LEPR* | 0.306444767 | 0.008631564 | 0.015135652 | 1 |
| T2DM | endothelial | *MFN2* | -0.768460173 | 4.23E-06 | 3.67E-05 | 0.020502516 |
| T2DM | endothelial | *PRG4* | -0.795484957 | 6.45E-07 | 1.33E-05 | 0.003128759 |
| T2DM | endothelial | *NCF2* | -0.349566622 | 0.00636955 | 0.011666689 | 1 |
| T2DM | endothelial | *CFHR3* | 0.735446428 | 6.86E-05 | 0.000279403 | 0.332769077 |
| T2DM | endothelial | *CRYZ* | -0.330641429 | 0.00135299 | 0.003129031 | 1 |
| T2DM | endothelial | *TMEM9* | -0.725969657 | 9.34E-05 | 0.000353114 | 0.453240656 |
| T2DM | endothelial | *WARS2* | -0.687877455 | 1.52E-05 | 8.78E-05 | 0.073790474 |
| T2DM | endothelial | *OSCP1* | -0.407734913 | 6.22E-05 | 0.000257411 | 0.301686166 |
| T2DM | endothelial | *MRPS15* | -0.858917698 | 0.00032072 | 0.000954097 | 1 |
| T2DM | endothelial | *EXOC8* | -1.440628721 | 6.16E-05 | 0.000255778 | 0.298930566 |
| T2DM | endothelial | *C1orf109* | -1.017590071 | 6.37E-07 | 1.32E-05 | 0.003088649 |
| T2DM | endothelial | *TBCE* | -0.748394743 | 0.000428896 | 0.001196668 | 1 |
| T2DM | endothelial | *ACADM* | -0.444329044 | 0.013618625 | 0.022544376 | 1 |
| T2DM | endothelial | *SLAMF1* | -0.77061495 | 3.00E-06 | 2.97E-05 | 0.014576617 |
| T2DM | endothelial | *PADI2* | -0.588605921 | 0.00140373 | 0.003224858 | 1 |
| T2DM | endothelial | *SDHB* | -0.715194349 | 0.000276653 | 0.000847958 | 1 |
| T2DM | endothelial | *KLHL12* | -0.543210662 | 0.014796907 | 0.024181406 | 1 |
| T2DM | endothelial | *ZNHIT6* | -0.468643891 | 0.017901274 | 0.028529799 | 1 |
| T2DM | endothelial | *KIF17* | 0.409370221 | 9.90E-06 | 6.42E-05 | 0.048048572 |
| T2DM | endothelial | *EBNA1BP2* | -1.014693181 | 4.21E-06 | 3.67E-05 | 0.020444262 |
| T2DM | endothelial | *ERI3* | -0.60720338 | 0.00141648 | 0.003246463 | 1 |
| T2DM | endothelial | *CCDC181* | -0.745838834 | 0.001059018 | 0.002537459 | 1 |
| T2DM | endothelial | *NSUN4* | -0.696964917 | 0.001851381 | 0.004086853 | 1 |
| T2DM | endothelial | *PRRC2C* | -0.359848853 | 0.027340928 | 0.041083365 | 1 |
| T2DM | endothelial | *ABCD3* | -0.533489537 | 0.011287636 | 0.019156211 | 1 |
| T2DM | endothelial | *DARS2* | -0.68785566 | 0.005068776 | 0.009614426 | 1 |
| T2DM | endothelial | *IRF6* | 0.220778653 | 6.60E-07 | 1.33E-05 | 0.003203209 |
| T2DM | endothelial | *MTFR1L* | -0.870704393 | 0.000817374 | 0.002039023 | 1 |
| T2DM | endothelial | *RPS6KA1* | -1.106493054 | 1.71E-07 | 1.14E-05 | 0.000828709 |
| T2DM | endothelial | *RPA2* | -0.881000767 | 6.37E-06 | 4.81E-05 | 0.030918738 |
| T2DM | endothelial | *SLC5A9* | -0.64965111 | 0.014202683 | 0.023329166 | 1 |
| T2DM | endothelial | *CD3EAP* | -1.108614298 | 2.11E-06 | 2.46E-05 | 0.010218213 |
| T2DM | endothelial | *RCN2* | -0.540889883 | 0.012540347 | 0.021043964 | 1 |
| T2DM | endothelial | *A4GNT* | -0.428661058 | 0.000774679 | 0.001947535 | 1 |
| T2DM | endothelial | *IFT46* | -0.611430165 | 0.001725011 | 0.003842963 | 1 |
| T2DM | endothelial | *DDX59* | -0.435440664 | 0.03360264 | 0.049212197 | 1 |
| T2DM | endothelial | *CASC1* | -0.502981552 | 2.33E-05 | 0.000120727 | 0.112901303 |
| T2DM | endothelial | *LRMP* | 0.278952123 | 0.003740171 | 0.007431332 | 1 |

| | | | | | | |
|---|---|---|---|---|---|---|
| T2DM | endothelial | *CNR1* | 0.476651748 | 0.002706598 | 0.005645922 | 1 |
| T2DM | endothelial | *SGIP1* | 0.370716501 | 9.41E-05 | 0.000355219 | 0.456811048 |
| T2DM | endothelial | *AKAP7* | -0.677426267 | 2.84E-05 | 0.000138513 | 0.137958845 |
| T2DM | endothelial | *MED28* | -0.742838121 | 0.000200756 | 0.000650245 | 0.97406701 |
| T2DM | endothelial | *TMEM5* | -0.476429789 | 0.025281366 | 0.038368841 | 1 |
| T2DM | endothelial | *VAMP8* | -0.865532048 | 3.33E-06 | 3.11E-05 | 0.016133815 |
| T2DM | endothelial | *FOXO3* | -0.497145932 | 0.000415464 | 0.001169275 | 1 |
| T2DM | endothelial | *SPP1* | -0.703685017 | 0.008649662 | 0.015161907 | 1 |
| T2DM | endothelial | *MFSD1* | -0.687958029 | 6.18E-06 | 4.72E-05 | 0.029977446 |
| T2DM | endothelial | *WDR35* | -0.747760454 | 1.83E-05 | 0.000100166 | 0.088847434 |
| T2DM | endothelial | *DNAH7* | -0.723746858 | 3.22E-07 | 1.24E-05 | 0.001560909 |
| T2DM | endothelial | *CPSF3* | -0.626304752 | 0.000177633 | 0.000588713 | 0.861876441 |
| T2DM | endothelial | *SENP5* | -0.457461517 | 0.013321225 | 0.022135132 | 1 |
| T2DM | endothelial | *CCDC92* | -0.829381261 | 1.60E-06 | 2.16E-05 | 0.007782982 |
| T2DM | endothelial | *FKBP15* | -0.5269748 | 4.92E-05 | 0.000215385 | 0.238647085 |
| T2DM | endothelial | *CTNNAL1* | 0.321598414 | 0.002705317 | 0.005645677 | 1 |
| T2DM | endothelial | *PPP6C* | -0.682710973 | 0.000435434 | 0.001211424 | 1 |
| T2DM | endothelial | *HDHD3* | -0.279521899 | 0.000166135 | 0.000555157 | 0.806088265 |
| T2DM | endothelial | *HSDL2* | -0.331795549 | 0.001856392 | 0.004094187 | 1 |
| T2DM | endothelial | *MAPKAP1* | -0.420734551 | 0.015411376 | 0.02505898 | 1 |
| T2DM | endothelial | *INVS* | -0.327671255 | 0.020882869 | 0.032548565 | 1 |
| T2DM | endothelial | *DENND1A* | -0.422024333 | 0.005444259 | 0.01021088 | 1 |
| T2DM | endothelial | *KDSR* | -0.768047334 | 4.70E-05 | 0.000207601 | 0.227946171 |
| T2DM | endothelial | *NEK9* | -0.629124927 | 0.001034929 | 0.002488344 | 1 |
| T2DM | endothelial | *TTLL5* | -0.509096688 | 0.00537416 | 0.010117222 | 1 |
| T2DM | endothelial | *FLVCR2* | -0.845942658 | 4.68E-06 | 3.91E-05 | 0.022697403 |
| T2DM | endothelial | *PPP4R4* | -0.377984926 | 0.010618494 | 0.018128408 | 1 |
| T2DM | endothelial | *COQ6* | -0.905133707 | 0.000462753 | 0.00127341 | 1 |
| T2DM | endothelial | *DNMT3A* | -0.430621543 | 0.015205547 | 0.024765799 | 1 |
| T2DM | endothelial | *CRIPT* | -0.712688324 | 0.006189103 | 0.011379131 | 1 |
| T2DM | endothelial | *SLC17A5* | -0.63052161 | 0.000681578 | 0.001750671 | 1 |
| T2DM | endothelial | *TECTB* | -0.795757196 | 9.10E-07 | 1.57E-05 | 0.004416942 |
| T2DM | endothelial | *PPP1R3C* | -1.31498527 | 0.000198357 | 0.000644195 | 0.962427757 |
| T2DM | endothelial | *PYROXD2* | -0.610794694 | 0.000197008 | 0.000640671 | 0.955880698 |
| T2DM | endothelial | *MXI1* | -0.719305231 | 0.000118124 | 0.000425167 | 0.573139383 |
| T2DM | endothelial | *C10orf88* | -0.761014184 | 0.000179682 | 0.000593478 | 0.871818627 |
| T2DM | endothelial | *HELLS* | -0.539583095 | 0.015388889 | 0.025030804 | 1 |
| T2DM | endothelial | *TCTN3* | -0.6314174 | 0.002401128 | 0.005083016 | 1 |
| T2DM | endothelial | *WDR11* | -0.465943424 | 0.012301891 | 0.02068935 | 1 |
| T2DM | endothelial | *KANSL1* | -0.346600764 | 0.013472338 | 0.02233269 | 1 |
| T2DM | endothelial | *CAAP1* | -0.565108529 | 0.001100061 | 0.00262156 | 1 |
| T2DM | endothelial | *ACAT2* | -0.680832584 | 5.81E-05 | 0.00024484 | 0.281745559 |
| T2DM | endothelial | *SNX19* | -0.443169027 | 0.018760923 | 0.029689497 | 1 |
| T2DM | endothelial | *MASTL* | -0.691029383 | 0.000155569 | 0.000528216 | 0.754821356 |
| T2DM | endothelial | *MRC1* | -0.584321802 | 6.47E-05 | 0.00026669 | 0.314007107 |
| T2DM | endothelial | *CCDC77* | -0.990182333 | 3.40E-08 | 5.32E-06 | 0.000164861 |
| T2DM | endothelial | *UFM1* | -0.694661059 | 0.000457073 | 0.001260783 | 1 |
| T2DM | endothelial | *ALG5* | -0.477250212 | 0.018710336 | 0.029619103 | 1 |
| T2DM | endothelial | *SIL1* | -0.585335289 | 8.14E-05 | 0.00031767 | 0.395181056 |
| T2DM | endothelial | *ARL1* | -0.797398628 | 0.000759687 | 0.001918682 | 1 |

| | | | | | | |
|---|---|---|---|---|---|---|
| T2DM | endothelial | *APAF1* | -0.58440593 | 0.010479126 | 0.017972682 | 1 |
| T2DM | endothelial | *EPHX2* | -0.745430308 | 1.23E-05 | 7.52E-05 | 0.059660187 |
| T2DM | endothelial | *AKAP1* | -0.760407603 | 5.66E-05 | 0.000240114 | 0.274450788 |
| T2DM | endothelial | *SCPEP1* | -0.421558292 | 0.000183518 | 0.0006045 | 0.890428135 |
| T2DM | endothelial | *TRIM6* | -2.375412308 | 0.000434323 | 0.001209722 | 1 |
| T2DM | endothelial | *ADCY7* | -0.62842038 | 1.08E-05 | 6.80E-05 | 0.052167464 |
| T2DM | endothelial | *ECHDC2* | -0.569147558 | 1.69E-07 | 1.14E-05 | 0.00082174 |
| T2DM | endothelial | *PYROXD1* | -0.574965069 | 0.001366039 | 0.003153197 | 1 |
| T2DM | endothelial | *A1BG* | -0.783527913 | 2.26E-06 | 2.52E-05 | 0.010956317 |
| T2DM | endothelial | *PDZRN3* | -0.51786391 | 2.10E-09 | 6.79E-07 | 1.02E-05 |
| T2DM | endothelial | *LHX4* | -1.180471443 | 1.93E-05 | 0.000104475 | 0.093714358 |
| T2DM | endothelial | *SEC22A* | -0.493763121 | 0.007586114 | 0.013568714 | 1 |
| T2DM | endothelial | *MAPK8IP1* | -1.091237172 | 0.005718714 | 0.01062704 | 1 |
| T2DM | endothelial | *CAT* | -0.388612116 | 0.001434526 | 0.003280075 | 1 |
| T2DM | endothelial | *ZCCHC17* | -0.72867591 | 0.000893472 | 0.002201689 | 1 |
| T2DM | endothelial | *FABP3* | -1.805818388 | 4.32E-05 | 0.000194299 | 0.209453902 |
| T2DM | endothelial | *TNFSF10* | 0.327656028 | 0.001392716 | 0.003204104 | 1 |
| T2DM | endothelial | *ZNF639* | -1.04037079 | 7.88E-06 | 5.52E-05 | 0.038219052 |
| T2DM | endothelial | *CPXM2* | 0.191930538 | 3.13E-05 | 0.000149559 | 0.151951694 |
| T2DM | endothelial | *FLT3* | -0.834158212 | 0.003670455 | 0.007304778 | 1 |
| T2DM | endothelial | *FMOD* | -0.788604889 | 5.91E-05 | 0.000247945 | 0.286946956 |
| T2DM | endothelial | *CD244* | -0.98721609 | 2.24E-06 | 2.52E-05 | 0.010872219 |
| T2DM | endothelial | *RBBP6* | -0.524924597 | 0.004829463 | 0.009236325 | 1 |
| T2DM | endothelial | *SERAC1* | -0.985008319 | 0.000154647 | 0.000526404 | 0.750347792 |
| T2DM | endothelial | *LDB3* | -0.776366835 | 1.18E-05 | 7.28E-05 | 0.057029441 |
| T2DM | endothelial | *OPN4* | -1.626276184 | 0.000127944 | 0.000453615 | 0.62078562 |
| T2DM | endothelial | *FAM35A* | -0.477753254 | 0.007699153 | 0.013728157 | 1 |
| T2DM | endothelial | *NAA60* | -0.923851032 | 0.013446315 | 0.02229717 | 1 |
| T2DM | endothelial | *PMS2* | -0.540335653 | 0.028075138 | 0.042082351 | 1 |
| T2DM | endothelial | *KLHL7* | -0.678489067 | 0.00030956 | 0.000928297 | 1 |
| T2DM | endothelial | *POLM* | -0.283734106 | 0.007876333 | 0.013993397 | 1 |
| T2DM | endothelial | *SMU1* | -0.702967872 | 0.000403584 | 0.0011418 | 1 |
| T2DM | endothelial | *GLIPR2* | -0.656763853 | 0.001252654 | 0.002936172 | 1 |
| T2DM | endothelial | *CISD1* | -0.43537762 | 0.007825686 | 0.013918706 | 1 |
| T2DM | endothelial | *RBM19* | -0.802078808 | 0.000117741 | 0.000424111 | 0.571277166 |
| T2DM | endothelial | *DDX54* | -0.929210693 | 3.80E-05 | 0.000174493 | 0.184438716 |
| T2DM | endothelial | *SSPN* | -0.366786359 | 0.006155491 | 0.011325916 | 1 |
| T2DM | endothelial | *CCDC91* | -0.519686284 | 0.000369308 | 0.001064062 | 1 |
| T2DM | endothelial | *CENPK* | 0.529083776 | 0.013784626 | 0.022757062 | 1 |
| T2DM | endothelial | *ITIH5* | -0.471683514 | 2.14E-05 | 0.000112933 | 0.10367283 |
| T2DM | endothelial | *ARHGAP9* | -1.14866259 | 1.14E-07 | 9.72E-06 | 0.000552739 |
| T2DM | endothelial | *PDE1B* | -0.826514228 | 3.77E-07 | 1.24E-05 | 0.001829414 |
| T2DM | endothelial | *CDK2* | -0.759825079 | 0.000599529 | 0.001579215 | 1 |
| T2DM | endothelial | *SMUG1* | -0.379605408 | 0.002785024 | 0.005782172 | 1 |
| T2DM | endothelial | *SARDH* | -0.52237949 | 0.000183684 | 0.000604637 | 0.891235559 |
| T2DM | endothelial | *DBH* | -6.877419797 | 5.18E-07 | 1.27E-05 | 0.002515402 |
| T2DM | endothelial | *USP45* | -0.565333239 | 0.001266485 | 0.002961439 | 1 |
| T2DM | endothelial | *TTC21B* | -0.537385163 | 0.005383736 | 0.010124763 | 1 |
| T2DM | endothelial | *ACVR1C* | -0.40815234 | 5.95E-06 | 4.60E-05 | 0.028877308 |
| T2DM | endothelial | *SLC36A1* | -0.26399524 | 0.008391582 | 0.014757504 | 1 |

| | | | | | | |
|---|---|---|---|---|---|---|
| T2DM | endothelial | *PLA2G12A* | -0.632059385 | 0.003063005 | 0.006273406 | 1 |
| T2DM | endothelial | *C4BPA* | -0.80406823 | 5.07E-05 | 0.000221166 | 0.246157624 |
| T2DM | endothelial | *C4BPB* | -0.557120974 | 0.000607327 | 0.001595426 | 1 |
| T2DM | endothelial | *RAB38* | -0.554204288 | 0.000322878 | 0.000958502 | 1 |
| T2DM | endothelial | *DAW1* | -0.820328152 | 1.14E-05 | 7.11E-05 | 0.05518245 |
| T2DM | endothelial | *DNPEP* | -0.767257843 | 0.001234409 | 0.002901818 | 1 |
| T2DM | endothelial | *OBSL1* | -0.805488024 | 1.68E-07 | 1.14E-05 | 0.000814745 |
| T2DM | endothelial | *FAM210B* | -0.653263372 | 0.022095542 | 0.034120805 | 1 |
| T2DM | endothelial | *WFDC3* | -0.177035193 | 0.002260186 | 0.00482677 | 1 |
| T2DM | endothelial | *SLC12A5* | -0.973561504 | 0.000573028 | 0.001520969 | 1 |
| T2DM | endothelial | *MATN4* | -1.460765639 | 2.24E-07 | 1.24E-05 | 0.001087138 |
| T2DM | endothelial | *VAPB* | -0.509271039 | 0.000333389 | 0.000980366 | 1 |
| T2DM | endothelial | *TOX2* | 0.166057349 | 0.000683034 | 0.001753482 | 1 |
| T2DM | endothelial | *SRSF6* | -0.681885344 | 0.000266632 | 0.00082559 | 1 |
| T2DM | endothelial | *CSE1L* | -0.681111777 | 0.002033777 | 0.004425553 | 1 |
| T2DM | endothelial | *PTGIS* | 0.308858035 | 0.00075186 | 0.001901995 | 1 |
| T2DM | endothelial | *RNF114* | -0.724848061 | 0.000275886 | 0.000846679 | 1 |
| T2DM | endothelial | *PEPD* | -0.745054073 | 1.42E-05 | 8.35E-05 | 0.068778269 |
| T2DM | endothelial | *CHST8* | -0.800912386 | 0.00014594 | 0.000504703 | 0.708098547 |
| T2DM | endothelial | *STAMBP* | -0.758377688 | 0.000124254 | 0.000442645 | 0.602882332 |
| T2DM | endothelial | *NAGK* | -0.716425324 | 0.000361664 | 0.001047014 | 1 |
| T2DM | endothelial | *SNRNP27* | -0.596986377 | 0.03420675 | 0.049961213 | 1 |
| T2DM | endothelial | *ATP8A1* | 0.286804245 | 0.000117474 | 0.000423464 | 0.569982628 |
| T2DM | endothelial | *USP22* | -0.582425783 | 0.001539733 | 0.003484508 | 1 |
| T2DM | endothelial | *HIF3A* | 0.214064984 | 4.59E-05 | 0.000204368 | 0.222761651 |
| T2DM | endothelial | *ZNF576* | -0.596442736 | 0.014962798 | 0.024427825 | 1 |
| T2DM | endothelial | *BTN2A2* | 0.233404471 | 0.01774798 | 0.028345358 | 1 |
| T2DM | endothelial | *PEX6* | -0.268206955 | 0.004041064 | 0.007922118 | 1 |
| T2DM | endothelial | *NQO2* | -0.756479994 | 3.29E-05 | 0.000155306 | 0.159810383 |
| T2DM | endothelial | *MED20* | -0.726568978 | 4.66E-05 | 0.000206694 | 0.226329937 |
| T2DM | endothelial | *MEA1* | 0.337072077 | 0.006397264 | 0.011704194 | 1 |
| T2DM | endothelial | *KLHL31* | -0.29484712 | 0.000641477 | 0.001667084 | 1 |
| T2DM | endothelial | *KCNK17* | -1.328879724 | 3.29E-09 | 9.99E-07 | 1.60E-05 |
| T2DM | endothelial | *RREB1* | -0.517296723 | 0.000343566 | 0.001002394 | 1 |
| T2DM | endothelial | *ATXN1* | -0.48201189 | 0.000275546 | 0.000846679 | 1 |
| T2DM | endothelial | *RUNX2* | -0.778930175 | 0.000136804 | 0.000479072 | 0.663775073 |
| T2DM | endothelial | *CXCL6* | -0.986288557 | 1.98E-07 | 1.24E-05 | 0.000961185 |
| T2DM | endothelial | *SH3TC1* | -0.597175252 | 2.51E-05 | 0.000127016 | 0.121934928 |
| T2DM | endothelial | *MT1G* | -0.966093162 | 0.000414911 | 0.001168396 | 1 |
| T2DM | endothelial | *CLYBL* | -0.543249236 | 0.007064771 | 0.012776097 | 1 |
| T2DM | endothelial | *ABCC4* | 0.213169022 | 2.26E-05 | 0.000117423 | 0.109438671 |
| T2DM | endothelial | *GRK4* | 0.145889779 | 0.026084084 | 0.039414505 | 1 |
| T2DM | endothelial | *TEKT3* | -0.268534173 | 0.009391272 | 0.016291188 | 1 |
| T2DM | endothelial | *HS3ST3B1* | -1.210780028 | 1.79E-05 | 9.88E-05 | 0.087077745 |
| T2DM | endothelial | *MRPS7* | -0.954978633 | 1.55E-05 | 8.91E-05 | 0.07525289 |
| T2DM | endothelial | *GGA3* | -0.763598296 | 0.000186753 | 0.00061349 | 0.906124126 |
| T2DM | endothelial | *NUP85* | -0.51849821 | 0.016369408 | 0.026369312 | 1 |
| T2DM | endothelial | *NT5C* | -0.770508524 | 7.90E-07 | 1.49E-05 | 0.003834607 |
| T2DM | endothelial | *DDX31* | -0.70439591 | 0.000130192 | 0.000459747 | 0.631692008 |
| T2DM | endothelial | *OPRL1* | -2.737325017 | 4.36E-07 | 1.24E-05 | 0.002117018 |

| | | | | | | |
|---|---|---|---|---|---|---|
| T2DM | endothelial | *IL1B* | -2.511078221 | 2.80E-05 | 0.000137135 | 0.135901046 |
| T2DM | endothelial | *POLR1B* | -0.35254443 | 0.009033879 | 0.015761373 | 1 |
| T2DM | endothelial | *PSD4* | -0.912648937 | 4.05E-07 | 1.24E-05 | 0.001966769 |
| T2DM | endothelial | *GTF2F1* | -0.719167 | 0.006329973 | 0.011598576 | 1 |
| T2DM | endothelial | *TNFSF9* | -6.256055635 | 2.57E-06 | 2.72E-05 | 0.012450456 |
| T2DM | endothelial | *SH2D3A* | -0.881152935 | 2.63E-06 | 2.76E-05 | 0.012774922 |
| T2DM | endothelial | *PSMF1* | -0.451891084 | 0.015304757 | 0.024902307 | 1 |
| T2DM | endothelial | *FLRT3* | -0.69524088 | 3.14E-07 | 1.24E-05 | 0.001524714 |
| T2DM | endothelial | *MKKS* | -0.493008295 | 0.021439569 | 0.03331992 | 1 |
| T2DM | endothelial | *LAMP5* | -5.590331611 | 3.42E-07 | 1.24E-05 | 0.001657253 |
| T2DM | endothelial | *MGME1* | -0.442475566 | 0.027655436 | 0.041491705 | 1 |
| T2DM | endothelial | *ITPA* | -0.825833842 | 1.57E-05 | 8.96E-05 | 0.076367125 |
| T2DM | endothelial | *NCLN* | -0.572973945 | 0.026591949 | 0.040144411 | 1 |
| T2DM | endothelial | *TUBGCP3* | -0.452776657 | 0.025112614 | 0.038172433 | 1 |
| T2DM | endothelial | *MCF2L* | 0.147319517 | 2.61E-06 | 2.76E-05 | 0.012657723 |
| T2DM | endothelial | *COX6B1* | -0.648706979 | 0.004026685 | 0.007900314 | 1 |
| T2DM | endothelial | *IRF3* | -0.660865071 | 0.002668956 | 0.005586616 | 1 |
| T2DM | endothelial | *TSKS* | -0.779669211 | 1.03E-05 | 6.59E-05 | 0.050003929 |
| T2DM | endothelial | *TRAP1* | -0.223569268 | 0.032041359 | 0.047282443 | 1 |
| T2DM | endothelial | *ZNF384* | -0.888531119 | 0.000143728 | 0.000498122 | 0.697370341 |
| T2DM | endothelial | *EMG1* | -0.806547852 | 3.37E-05 | 0.000158186 | 0.163722155 |
| T2DM | endothelial | *ATG14* | -0.522047205 | 0.000407617 | 0.001151198 | 1 |
| T2DM | endothelial | *ZBTB1* | -0.612326549 | 0.0001344 | 0.000472544 | 0.652111118 |
| T2DM | endothelial | *RHOT1* | -0.599427197 | 6.52E-05 | 0.000268285 | 0.316576477 |
| T2DM | endothelial | *WDR60* | -0.422916174 | 0.009106176 | 0.01587039 | 1 |
| T2DM | endothelial | *AIF1L* | 0.176622953 | 4.33E-06 | 3.73E-05 | 0.020987444 |
| T2DM | endothelial | *FGD3* | -0.624366357 | 5.92E-05 | 0.000247945 | 0.287120869 |
| T2DM | endothelial | *ABHD8* | -0.939056642 | 9.68E-09 | 2.04E-06 | 4.70E-05 |
| T2DM | endothelial | *ATP13A4* | -1.009066189 | 2.17E-05 | 0.000113922 | 0.105265202 |
| T2DM | endothelial | *HELB* | -0.547123423 | 0.02130853 | 0.033126879 | 1 |
| T2DM | endothelial | *RAB3IP* | -1.220710245 | 2.24E-05 | 0.000117043 | 0.108849965 |
| T2DM | endothelial | *TAS2R4* | -0.897442929 | 1.40E-05 | 8.25E-05 | 0.067687057 |
| T2DM | endothelial | *LRRC61* | -0.454937672 | 4.41E-05 | 0.000197668 | 0.214073922 |
| T2DM | endothelial | *IDUA* | -0.897759477 | 1.17E-05 | 7.27E-05 | 0.05653637 |
| T2DM | endothelial | *EMC1* | -0.729162775 | 0.002202777 | 0.004729148 | 1 |
| T2DM | endothelial | *UBR4* | -0.415225023 | 0.013273401 | 0.022082296 | 1 |
| T2DM | endothelial | *WFIKKN1* | -0.616871669 | 0.009503079 | 0.016455724 | 1 |
| T2DM | endothelial | *WDR24* | -0.924279765 | 0.000126656 | 0.000450534 | 0.614536742 |
| T2DM | endothelial | *SMARCA4* | -0.498510551 | 0.016233043 | 0.026210558 | 1 |
| T2DM | endothelial | *PNKD* | -0.584220766 | 0.003742678 | 0.007433269 | 1 |
| T2DM | endothelial | *STYXL1* | -0.310546181 | 0.00973677 | 0.01680043 | 1 |
| T2DM | endothelial | *STEAP4* | -0.740718049 | 5.82E-05 | 0.00024484 | 0.28230063 |
| T2DM | endothelial | *PEX1* | -0.717918517 | 0.000857165 | 0.002128502 | 1 |
| T2DM | endothelial | *KDR* | 0.32108545 | 8.95E-05 | 0.000341034 | 0.434477065 |
| T2DM | endothelial | *MGAT3* | 0.626780635 | 0.006876229 | 0.012486326 | 1 |
| T2DM | endothelial | *ATF4* | -0.781626262 | 0.003865025 | 0.007644965 | 1 |
| T2DM | endothelial | *A4GALT* | 0.35579643 | 0.000499027 | 0.00135798 | 1 |
| T2DM | endothelial | *SPECC1* | -0.539530818 | 0.002697566 | 0.005634675 | 1 |
| T2DM | endothelial | *CDHR3* | -0.703531138 | 1.38E-05 | 8.20E-05 | 0.067055981 |
| T2DM | endothelial | *PRKRIP1* | -0.630044321 | 0.000268033 | 0.000828538 | 1 |

| | | | | | | |
|---|---|---|---|---|---|---|
| T2DM | endothelial | *OPN1SW* | -0.71934601 | 8.86E-06 | 5.97E-05 | 0.042972803 |
| T2DM | endothelial | *HOXD1* | 0.554595434 | 5.36E-06 | 4.25E-05 | 0.025985453 |
| T2DM | endothelial | *HOXD3* | 0.14207573 | 5.25E-07 | 1.27E-05 | 0.002548636 |
| T2DM | endothelial | *PDE11A* | -0.373234859 | 0.001982395 | 0.004332694 | 1 |
| T2DM | endothelial | *GAD1* | -0.98316258 | 5.46E-07 | 1.27E-05 | 0.002649258 |
| T2DM | endothelial | *ORMDL1* | -0.37902323 | 0.022513565 | 0.034711095 | 1 |
| T2DM | endothelial | *HOXD9* | 0.232765401 | 2.48E-05 | 0.000125956 | 0.120526475 |
| T2DM | endothelial | *HOXD10* | 0.730433889 | 0.002302466 | 0.004897661 | 1 |
| T2DM | endothelial | *TWSG1* | -0.813072819 | 2.48E-05 | 0.000125919 | 0.120165231 |
| T2DM | endothelial | *WDFY4* | -0.215069981 | 0.017767918 | 0.028367864 | 1 |
| T2DM | endothelial | *TMOD2* | -0.514833194 | 0.019175969 | 0.03027719 | 1 |
| T2DM | endothelial | *IVD* | -0.55735785 | 0.000663619 | 0.001713615 | 1 |
| T2DM | endothelial | *ARPP19* | -0.55344921 | 0.018099117 | 0.028782994 | 1 |
| T2DM | endothelial | *CALML4* | 1.057658489 | 0.00015522 | 0.0005274 | 0.7531279 |
| T2DM | endothelial | *SUMF2* | -0.634605982 | 0.003287582 | 0.006663052 | 1 |
| T2DM | endothelial | *BBOX1* | -2.251684814 | 4.07E-07 | 1.24E-05 | 0.001974843 |
| T2DM | endothelial | *DCTD* | -0.768365987 | 0.000325091 | 0.000962969 | 1 |
| T2DM | endothelial | *RPAIN* | -0.738637388 | 4.34E-05 | 0.000195046 | 0.210649725 |
| T2DM | endothelial | *PLD2* | 0.563725592 | 0.022739436 | 0.035014835 | 1 |
| T2DM | endothelial | *PHF20L1* | -0.558701424 | 0.001911747 | 0.004199094 | 1 |
| T2DM | endothelial | *PUS7L* | -0.599572594 | 0.002278379 | 0.004854938 | 1 |
| T2DM | endothelial | *KRI1* | -0.31431572 | 0.017405973 | 0.027872534 | 1 |
| T2DM | endothelial | *MTUS1* | 0.311991688 | 0.003485872 | 0.007006401 | 1 |
| T2DM | endothelial | *KLK8* | -4.930615222 | 4.50E-07 | 1.25E-05 | 0.002185255 |
| T2DM | endothelial | *RIPK3* | 0.278976785 | 0.032215769 | 0.047482051 | 1 |
| T2DM | endothelial | *ADCY4* | 0.154029599 | 2.89E-06 | 2.89E-05 | 0.014024655 |
| T2DM | endothelial | *BCL2L2* | -0.707882649 | 2.15E-05 | 0.000113102 | 0.104187812 |
| T2DM | endothelial | *PARP2* | -0.840307708 | 0.000589285 | 0.001556456 | 1 |
| T2DM | endothelial | *MIS18BP1* | -0.37454622 | 0.024429286 | 0.037252517 | 1 |
| T2DM | endothelial | *EPB41L4A* | 0.268751162 | 0.000588691 | 0.001555734 | 1 |
| T2DM | endothelial | *REEP5* | -0.54496271 | 0.003825677 | 0.007576402 | 1 |
| T2DM | endothelial | *TMEM8A* | -1.086390227 | 0.0005414 | 0.001449709 | 1 |
| T2DM | endothelial | *MAU2* | -0.650390665 | 0.00062562 | 0.00163463 | 1 |
| T2DM | endothelial | *SHC2* | 0.732823264 | 3.35E-05 | 0.000157293 | 0.162641363 |
| T2DM | endothelial | *ERMARD* | -0.756626362 | 0.000165476 | 0.000554098 | 0.802887666 |
| T2DM | endothelial | *CDC16* | -0.38200536 | 0.023203289 | 0.035627328 | 1 |
| T2DM | endothelial | *XPO7* | -0.501923864 | 0.013409145 | 0.022250743 | 1 |
| T2DM | endothelial | *FAM98C* | -0.703543285 | 0.000471382 | 0.001292904 | 1 |
| T2DM | endothelial | *ATP8B3* | -0.372774221 | 0.000809299 | 0.00202304 | 1 |
| T2DM | endothelial | *PLVAP* | 0.12283166 | 1.33E-05 | 7.97E-05 | 0.064386593 |
| T2DM | endothelial | *DDA1* | -0.896246208 | 0.004721232 | 0.009050738 | 1 |
| T2DM | endothelial | *TULP4* | -0.443112577 | 0.007951669 | 0.014101395 | 1 |
| T2DM | endothelial | *RSPH3* | -0.239384193 | 0.008539824 | 0.014991036 | 1 |
| T2DM | endothelial | *STK33* | 0.528434902 | 0.000240866 | 0.000759377 | 1 |
| T2DM | endothelial | *NDUFA10* | -0.29120673 | 0.023440128 | 0.035934124 | 1 |
| T2DM | endothelial | *MAP1S* | -1.346540213 | 7.64E-06 | 5.43E-05 | 0.037062947 |
| T2DM | endothelial | *KIAA1683* | -0.648773164 | 0.002108548 | 0.004561156 | 1 |
| T2DM | endothelial | *TRPM4* | 0.383070375 | 0.000321703 | 0.000955851 | 1 |
| T2DM | endothelial | *UBAC1* | -0.802954948 | 7.01E-05 | 0.000283999 | 0.340231241 |
| T2DM | endothelial | *ATXN10* | -0.541160281 | 0.000636352 | 0.001655538 | 1 |

| | | | | | | |
|---|---|---|---|---|---|---|
| T2DM | endothelial | *PNPLA7* | -0.721655298 | 3.52E-06 | 3.24E-05 | 0.017073908 |
| T2DM | endothelial | *PAK4* | -0.65184577 | 0.010842477 | 0.018484784 | 1 |
| T2DM | endothelial | *CEP85* | -0.603394213 | 7.02E-06 | 5.11E-05 | 0.034081241 |
| T2DM | endothelial | *YIPF2* | -0.853884594 | 0.000111172 | 0.000404626 | 0.539405029 |
| T2DM | endothelial | *NPAS1* | -1.471050472 | 7.76E-05 | 0.00030696 | 0.376640347 |
| T2DM | endothelial | *MAP3K10* | -0.575229658 | 0.021231139 | 0.033029114 | 1 |
| T2DM | endothelial | *ZNF317* | -0.647372768 | 0.008515691 | 0.014954083 | 1 |
| T2DM | endothelial | *ANGPTL6* | 0.483908939 | 0.000644297 | 0.001673518 | 1 |
| T2DM | endothelial | *PKDREJ* | -0.658998609 | 0.000389626 | 0.001108776 | 1 |
| T2DM | endothelial | *HSD17B3* | 0.757664067 | 0.001666958 | 0.003728944 | 1 |
| T2DM | endothelial | *SLC35D2* | -0.502693104 | 0.024638673 | 0.037522548 | 1 |
| T2DM | endothelial | *ULBP2* | 1.00590684 | 2.00E-06 | 2.38E-05 | 0.009691209 |
| T2DM | endothelial | *TTLL9* | 0.874510869 | 0.000243014 | 0.000763667 | 1 |
| T2DM | endothelial | *ATP6V1E1* | -0.731939351 | 0.000661103 | 0.001709846 | 1 |
| T2DM | endothelial | *CHMP1A* | -0.840412304 | 0.001263673 | 0.002956289 | 1 |
| T2DM | endothelial | *IDO1* | 0.162205585 | 2.68E-06 | 2.76E-05 | 0.013010412 |
| T2DM | endothelial | *HAUS8* | -0.832429234 | 0.000172707 | 0.000574348 | 0.83797391 |
| T2DM | endothelial | *MRPS25* | -0.751761319 | 1.71E-05 | 9.53E-05 | 0.083137543 |
| T2DM | endothelial | *GALNT15* | 0.185640674 | 1.06E-05 | 6.74E-05 | 0.051559511 |
| T2DM | endothelial | *KCNC3* | -0.863023668 | 7.93E-06 | 5.54E-05 | 0.038453193 |
| T2DM | endothelial | *PDLIM4* | -1.491056097 | 0.00223377 | 0.004780878 | 1 |
| T2DM | endothelial | *KIF3A* | -0.71794172 | 0.002003589 | 0.004371139 | 1 |
| T2DM | endothelial | *ACLY* | -0.731911486 | 0.000943871 | 0.002304811 | 1 |
| T2DM | endothelial | *C1orf159* | -0.900422106 | 1.71E-05 | 9.53E-05 | 0.08301211 |
| T2DM | endothelial | *PPFIA1* | -0.590308469 | 0.000163215 | 0.000548422 | 0.791920657 |
| T2DM | endothelial | *BARX1* | -1.506235787 | 4.79E-07 | 1.27E-05 | 0.002324431 |
| T2DM | endothelial | *NINJ1* | -0.467121049 | 0.000403106 | 0.001141351 | 1 |
| T2DM | endothelial | *NPHP4* | -0.483129595 | 0.015431344 | 0.025078057 | 1 |
| T2DM | endothelial | *MAP1B* | -0.465121631 | 0.000452573 | 0.001251929 | 1 |
| T2DM | endothelial | *CKMT2* | -0.470774256 | 0.016355646 | 0.026358428 | 1 |
| T2DM | endothelial | *KHDRBS3* | -0.764683702 | 1.53E-06 | 2.13E-05 | 0.007431224 |
| T2DM | endothelial | *CHD1L* | -0.7099488 | 3.75E-05 | 0.000172425 | 0.181736429 |
| T2DM | endothelial | *RHPN2* | -0.192384263 | 0.02289127 | 0.035215105 | 1 |
| T2DM | endothelial | *GCH1* | 0.363649387 | 3.31E-05 | 0.000155866 | 0.160688267 |
| T2DM | endothelial | *MATN3* | -0.786350947 | 1.53E-05 | 8.84E-05 | 0.074433367 |
| T2DM | endothelial | *SPATA6* | -0.545795242 | 0.000773833 | 0.001946417 | 1 |
| T2DM | endothelial | *LRRC41* | -0.83594595 | 0.000204525 | 0.00066113 | 0.992356028 |
| T2DM | endothelial | *GAS2L2* | 1.388690413 | 0.004246867 | 0.008272099 | 1 |
| T2DM | endothelial | *CCT6B* | -0.426035694 | 0.007063139 | 0.012776097 | 1 |
| T2DM | endothelial | *CNGA4* | -0.90483646 | 6.35E-08 | 7.20E-06 | 0.000307913 |
| T2DM | endothelial | *IMMT* | -0.466471661 | 0.015450291 | 0.025097025 | 1 |
| T2DM | endothelial | *RPA1* | -0.494008298 | 0.007156389 | 0.012917707 | 1 |
| T2DM | endothelial | *SERPINF1* | -0.725891781 | 6.03E-06 | 4.64E-05 | 0.029236118 |
| T2DM | endothelial | *TMEM128* | -0.674066812 | 0.000110153 | 0.000402154 | 0.534462444 |
| T2DM | endothelial | *FIGNL1* | -0.851919114 | 0.000241347 | 0.000760173 | 1 |
| T2DM | endothelial | *RGS22* | -0.429073109 | 2.46E-05 | 0.000125809 | 0.119518985 |
| T2DM | endothelial | *REEP2* | 0.427284222 | 1.58E-05 | 8.96E-05 | 0.076681936 |
| T2DM | endothelial | *PRMT7* | -0.48688893 | 0.000656601 | 0.001700923 | 1 |
| T2DM | endothelial | *IGHMBP2* | -0.421395307 | 0.002842231 | 0.005888344 | 1 |
| T2DM | endothelial | *DMGDH* | -0.683107944 | 2.57E-05 | 0.000128761 | 0.124640272 |

| | | | | | | |
|---|---|---|---|---|---|---|
| T2DM | endothelial | *BHMT2* | -0.195641706 | 0.000648945 | 0.001683787 | 1 |
| T2DM | endothelial | *AP3B1* | -0.398744213 | 0.016498672 | 0.026533496 | 1 |
| T2DM | endothelial | *ZBED3* | -0.676183537 | 0.009101174 | 0.015867372 | 1 |
| T2DM | endothelial | *FBXO44* | -0.871845603 | 3.88E-05 | 0.000177581 | 0.188413604 |
| T2DM | endothelial | *ATP8A2* | -0.979114591 | 0.003018393 | 0.006187694 | 1 |
| T2DM | endothelial | *ALOX5AP* | -0.975973864 | 0.000118209 | 0.000425167 | 0.573550346 |
| T2DM | endothelial | *WASF3* | -0.666440404 | 2.67E-06 | 2.76E-05 | 0.012965255 |
| T2DM | endothelial | *GPR12* | 1.166867291 | 8.59E-07 | 1.53E-05 | 0.004165964 |
| T2DM | endothelial | *SCO1* | -0.496000434 | 0.0245989 | 0.03747374 | 1 |
| T2DM | endothelial | *DSTYK* | -0.550292098 | 0.010022397 | 0.017264208 | 1 |
| T2DM | endothelial | *CCNA1* | -2.525195566 | 0.00019403 | 0.00063311 | 0.941434359 |
| T2DM | endothelial | *BTBD2* | -0.736521974 | 0.000139061 | 0.000484714 | 0.67472163 |
| T2DM | endothelial | *CSNK1G2* | -0.893050934 | 0.000578848 | 0.001533062 | 1 |
| T2DM | endothelial | *CNDP2* | 0.427705695 | 0.026882514 | 0.040520025 | 1 |
| T2DM | endothelial | *PDZD2* | -0.389400246 | 0.008885767 | 0.01553226 | 1 |
| T2DM | endothelial | *SLC2A11* | -0.502290193 | 0.002419452 | 0.005112884 | 1 |
| T2DM | endothelial | *FAM83F* | 0.578953564 | 2.40E-06 | 2.61E-05 | 0.011654149 |
| T2DM | endothelial | *GIMAP4* | 0.246166344 | 5.12E-05 | 0.000222374 | 0.248292237 |
| T2DM | endothelial | *LARS* | -0.314636392 | 0.017101857 | 0.02741269 | 1 |
| T2DM | endothelial | *SPINK5* | 0.390115889 | 5.83E-06 | 4.53E-05 | 0.028266948 |
| T2DM | endothelial | *RRAS2* | -0.53529019 | 2.72E-05 | 0.000134552 | 0.131995886 |
| T2DM | endothelial | *CATSPERB* | -0.647227512 | 7.72E-06 | 5.45E-05 | 0.037468701 |
| T2DM | endothelial | *COX16* | 1.502156575 | 0.000565881 | 0.001506945 | 1 |
| T2DM | endothelial | *TTC9* | -1.110583595 | 0.008934368 | 0.015608128 | 1 |
| T2DM | endothelial | *ELP3* | -0.594695313 | 0.003668674 | 0.007304229 | 1 |
| T2DM | endothelial | *PEBP4* | -1.116604082 | 5.01E-06 | 4.09E-05 | 0.02430829 |
| T2DM | endothelial | *CTIF* | -0.516668677 | 3.48E-05 | 0.000162586 | 0.169089912 |
| T2DM | endothelial | *MRO* | -0.689610552 | 2.39E-06 | 2.61E-05 | 0.011580563 |
| T2DM | endothelial | *THUMPD3* | -0.987679474 | 1.40E-05 | 8.25E-05 | 0.067717843 |
| T2DM | endothelial | *CNTN6* | -1.087536286 | 2.88E-06 | 2.89E-05 | 0.01398344 |
| T2DM | endothelial | *CHL1* | -0.600692742 | 1.80E-06 | 2.28E-05 | 0.008754213 |
| T2DM | endothelial | *GSTM5* | -0.418404713 | 0.001217926 | 0.002871418 | 1 |
| T2DM | endothelial | *PSRC1* | -0.702947876 | 0.002716948 | 0.005665076 | 1 |
| T2DM | endothelial | *TRIM45* | -1.188626125 | 3.42E-06 | 3.18E-05 | 0.016615109 |
| T2DM | endothelial | *CEPT1* | -0.61303885 | 7.95E-05 | 0.000312522 | 0.385591366 |
| T2DM | endothelial | *NAPG* | -0.618455327 | 0.003482908 | 0.007006401 | 1 |
| T2DM | endothelial | *PPHLN1* | -0.508945221 | 0.002267947 | 0.004841215 | 1 |
| T2DM | endothelial | *FKBP11* | -1.010114491 | 5.51E-06 | 4.33E-05 | 0.026746822 |
| T2DM | endothelial | *YWHAQ* | -0.456141429 | 0.024467176 | 0.037296493 | 1 |
| T2DM | endothelial | *KIDINS220* | -0.487116403 | 0.003302422 | 0.006690334 | 1 |
| T2DM | endothelial | *LPIN1* | -0.634088778 | 3.30E-06 | 3.11E-05 | 0.016032122 |
| T2DM | endothelial | *KIAA1468* | -0.428664724 | 0.020968985 | 0.032672292 | 1 |
| T2DM | endothelial | *IL15RA* | 0.442619402 | 0.001102699 | 0.002625268 | 1 |
| T2DM | endothelial | *TMEM241* | -0.430278864 | 0.000536503 | 0.001442167 | 1 |
| T2DM | endothelial | *SOX5* | -0.438299643 | 0.000695914 | 0.001783716 | 1 |
| T2DM | endothelial | *ACP2* | -0.887652082 | 5.38E-06 | 4.25E-05 | 0.026093006 |
| T2DM | endothelial | *HOOK1* | -0.288695126 | 0.00138721 | 0.003195985 | 1 |
| T2DM | endothelial | *ZCCHC11* | -0.447171925 | 0.002034003 | 0.004425553 | 1 |
| T2DM | endothelial | *PRPF38A* | -0.876197466 | 0.000557445 | 0.001486928 | 1 |
| T2DM | endothelial | *ELP2* | -0.639120739 | 0.000281723 | 0.000861322 | 1 |

| | | | | | | |
|---|---|---|---|---|---|---|
| T2DM | endothelial | *DHX34* | -0.870797378 | 7.32E-05 | 0.000294466 | 0.355026477 |
| T2DM | endothelial | *FADS2* | -0.920757947 | 3.16E-06 | 3.03E-05 | 0.015314767 |
| T2DM | endothelial | *CLOCK* | -0.390209519 | 0.032637818 | 0.048074891 | 1 |
| T2DM | endothelial | *GGACT* | -0.88765029 | 0.001132835 | 0.00268935 | 1 |
| T2DM | endothelial | *CLDN10* | 0.466925298 | 2.17E-06 | 2.47E-05 | 0.010510103 |
| T2DM | endothelial | *CARS2* | -0.668507662 | 3.18E-05 | 0.000151058 | 0.154230166 |
| T2DM | endothelial | *KLB* | -0.45639247 | 2.82E-07 | 1.24E-05 | 0.001368896 |
| T2DM | endothelial | *WDR36* | -0.77280978 | 0.000159847 | 0.000539344 | 0.775576163 |
| T2DM | endothelial | *UBQLN1* | -0.599169973 | 0.001298674 | 0.003029406 | 1 |
| T2DM | endothelial | *GOLM1* | -0.368179891 | 0.007772392 | 0.013834769 | 1 |
| T2DM | endothelial | *ADAM19* | -0.322313979 | 0.009254049 | 0.016087656 | 1 |
| T2DM | endothelial | *USP30* | -0.369313312 | 0.001522677 | 0.003454131 | 1 |
| T2DM | endothelial | *MSI1* | -0.851997532 | 0.002742949 | 0.005707028 | 1 |
| T2DM | endothelial | *DTX1* | -0.565309236 | 4.26E-07 | 1.24E-05 | 0.002068939 |
| T2DM | endothelial | *DMTF1* | -0.510391639 | 0.001472182 | 0.003348817 | 1 |
| T2DM | endothelial | *TMEM60* | -1.145269989 | 3.82E-05 | 0.000175219 | 0.185381766 |
| T2DM | endothelial | *POM121C* | -0.872742636 | 2.36E-06 | 2.60E-05 | 0.011465238 |
| T2DM | endothelial | *SRPK2* | -0.411758716 | 0.007701575 | 0.013728157 | 1 |
| T2DM | endothelial | *TES* | 0.356605447 | 0.013697228 | 0.022635882 | 1 |
| T2DM | endothelial | *MTO1* | -0.628166821 | 4.58E-05 | 0.000203999 | 0.222154882 |
| T2DM | endothelial | *KHDC1* | -0.342072543 | 0.000117039 | 0.000422211 | 0.567873777 |
| T2DM | endothelial | *AKIRIN2* | -0.748774871 | 0.000256583 | 0.000799576 | 1 |
| T2DM | endothelial | *PHF21A* | -0.418621918 | 0.00789384 | 0.014009111 | 1 |
| T2DM | endothelial | *CAPRIN1* | -0.477610867 | 0.016287975 | 0.026269439 | 1 |
| T2DM | endothelial | *ATP5G2* | -0.462922492 | 0.027202093 | 0.040912758 | 1 |
| T2DM | endothelial | *B4GALNT1* | -1.197041894 | 0.011952131 | 0.0201588 | 1 |
| T2DM | endothelial | *FAIM2* | -0.947421154 | 7.76E-07 | 1.47E-05 | 0.003767225 |
| T2DM | endothelial | *PAN2* | -0.949626793 | 1.30E-06 | 1.89E-05 | 0.006324589 |
| T2DM | endothelial | *SLC26A10* | -1.12469252 | 7.58E-05 | 0.000302527 | 0.367991486 |
| T2DM | endothelial | *MAP7* | -0.547324157 | 8.46E-06 | 5.78E-05 | 0.04105387 |
| T2DM | endothelial | *NHSL1* | -0.463380722 | 0.000712987 | 0.001821703 | 1 |
| T2DM | endothelial | *AHI1* | -0.28801524 | 0.010748617 | 0.01833121 | 1 |
| T2DM | endothelial | *SMPD2* | -0.271408416 | 0.025917955 | 0.039212322 | 1 |
| T2DM | endothelial | *MICAL1* | 0.411430599 | 0.018210163 | 0.028940619 | 1 |
| T2DM | endothelial | *SMYD5* | -0.769637936 | 0.000362661 | 0.001048648 | 1 |
| T2DM | endothelial | *USP15* | -0.357302877 | 0.027446289 | 0.041203402 | 1 |
| T2DM | endothelial | *MDM2* | -0.645579992 | 7.83E-05 | 0.000308683 | 0.379988855 |
| T2DM | endothelial | *KLHL36* | -0.694533422 | 0.000324311 | 0.000961833 | 1 |
| T2DM | endothelial | *MPHOSPH6* | -0.808659283 | 1.33E-05 | 7.98E-05 | 0.064581915 |
| T2DM | endothelial | *CCDC102A* | -0.817428717 | 0.002392272 | 0.005068691 | 1 |
| T2DM | endothelial | *AGT* | 0.312824585 | 0.021657779 | 0.033597625 | 1 |
| T2DM | endothelial | *ZNF670* | -0.912893254 | 9.11E-05 | 0.000346379 | 0.441979251 |
| T2DM | endothelial | *KCNK1* | -0.321778357 | 0.014391223 | 0.023605888 | 1 |
| T2DM | endothelial | *CAPN9* | -1.01064634 | 4.34E-07 | 1.24E-05 | 0.002105967 |
| T2DM | endothelial | *PIGC* | 0.369245375 | 0.021603394 | 0.03355303 | 1 |
| T2DM | endothelial | *MRPL44* | -1.055336033 | 1.82E-06 | 2.29E-05 | 0.008806395 |
| T2DM | endothelial | *USP37* | -0.499230216 | 0.008260889 | 0.014559329 | 1 |
| T2DM | endothelial | *SLC19A3* | -0.496332589 | 3.47E-06 | 3.20E-05 | 0.016816048 |
| T2DM | endothelial | *TMBIM1* | -0.593487939 | 0.010895545 | 0.018549188 | 1 |
| T2DM | endothelial | *TSGA10* | -0.274654773 | 0.000540709 | 0.001448659 | 1 |

| | | | | | | |
|---|---|---|---|---|---|---|
| T2DM | endothelial | *GCC2* | -0.43105067 | 0.012614562 | 0.021141918 | 1 |
| T2DM | endothelial | *EPC2* | -0.497875236 | 0.004269195 | 0.008305588 | 1 |
| T2DM | endothelial | *STAB2* | -0.749715135 | 0.00493705 | 0.009405012 | 1 |
| T2DM | endothelial | *APPL2* | -0.580423239 | 0.000385202 | 0.001101356 | 1 |
| T2DM | endothelial | *DRAM1* | -0.58613646 | 0.009744507 | 0.016807802 | 1 |
| T2DM | endothelial | *SLC41A2* | -0.554106651 | 0.000121102 | 0.000433642 | 0.587584823 |
| T2DM | endothelial | *RNASEH2B* | -0.624059905 | 0.000112668 | 0.00040918 | 0.546663879 |
| T2DM | endothelial | *TBC1D4* | -0.575806399 | 1.24E-05 | 7.55E-05 | 0.060100974 |
| T2DM | endothelial | *SUCLA2* | -0.629996814 | 0.001552166 | 0.003507736 | 1 |
| T2DM | endothelial | *RCBTB1* | -0.365432644 | 0.0314332 | 0.046512317 | 1 |
| T2DM | endothelial | *SETDB2* | -0.521999181 | 0.022632417 | 0.034861107 | 1 |
| T2DM | endothelial | *SCRN1* | -0.771198015 | 1.02E-06 | 1.64E-05 | 0.004962391 |
| T2DM | endothelial | *C7orf25* | -1.405846097 | 6.47E-05 | 0.00026669 | 0.314161053 |
| T2DM | endothelial | *NUPL2* | -0.68795312 | 0.00062258 | 0.001628442 | 1 |
| T2DM | endothelial | *ZDHHC4* | -0.945664519 | 1.02E-05 | 6.56E-05 | 0.049637016 |
| T2DM | endothelial | *AOAH* | -0.43416885 | 0.000771282 | 0.001941007 | 1 |
| T2DM | endothelial | *TBRG4* | -1.261489816 | 9.40E-06 | 6.22E-05 | 0.04562823 |
| T2DM | endothelial | *MTHFS* | -0.491808717 | 0.003369581 | 0.006809332 | 1 |
| T2DM | endothelial | *NMT1* | -0.279737811 | 0.028207555 | 0.042254726 | 1 |
| T2DM | endothelial | *MYCBPAP* | -0.252229908 | 0.020176914 | 0.031559763 | 1 |
| T2DM | endothelial | *BRIP1* | -0.675733631 | 4.73E-06 | 3.95E-05 | 0.022959999 |
| T2DM | endothelial | *RTP4* | 0.439735043 | 0.017471983 | 0.027964902 | 1 |
| T2DM | endothelial | *NDUFB5* | -0.750591171 | 0.000931248 | 0.002279724 | 1 |
| T2DM | endothelial | *SCN2A* | -0.511025813 | 0.032718758 | 0.048150262 | 1 |
| T2DM | endothelial | *RPS6KC1* | -0.437746138 | 0.007735327 | 0.013783256 | 1 |
| T2DM | endothelial | *SMPD4* | -0.60583683 | 0.002196971 | 0.004718771 | 1 |
| T2DM | endothelial | *IMP4* | -0.963382551 | 0.005344976 | 0.010067477 | 1 |
| T2DM | endothelial | *NIPSNAP3A* | -0.492324835 | 0.032949546 | 0.048372526 | 1 |
| T2DM | endothelial | *TOR1B* | -0.845827845 | 0.002178162 | 0.004684594 | 1 |
| T2DM | endothelial | *SLC2A8* | -1.33536772 | 6.15E-06 | 4.71E-05 | 0.029839717 |
| T2DM | endothelial | *ZNF189* | -0.618964441 | 0.025208807 | 0.038270692 | 1 |
| T2DM | endothelial | *DSCC1* | -0.23792249 | 0.00162934 | 0.003656595 | 1 |
| T2DM | endothelial | *DERL1* | -0.689183891 | 0.003975966 | 0.007822948 | 1 |
| T2DM | endothelial | *NOV* | 0.249658735 | 2.75E-06 | 2.79E-05 | 0.013342575 |
| T2DM | endothelial | *POLR1E* | -0.231729394 | 0.023031809 | 0.035386427 | 1 |
| T2DM | endothelial | *APTX* | -0.763569578 | 9.61E-06 | 6.29E-05 | 0.046633931 |
| T2DM | endothelial | *TMEM8B* | -0.648400481 | 0.000618509 | 0.001620412 | 1 |
| T2DM | endothelial | *GRHPR* | -0.631852969 | 0.000342334 | 0.001000002 | 1 |
| T2DM | endothelial | *DENND4C* | -0.469849293 | 0.001861088 | 0.00410268 | 1 |
| T2DM | endothelial | *GMPR* | -0.564803953 | 0.023136432 | 0.035535919 | 1 |
| T2DM | endothelial | *TMEM14B* | -0.629455282 | 0.00434113 | 0.008428637 | 1 |
| T2DM | endothelial | *CAPN11* | 0.272896925 | 1.56E-05 | 8.94E-05 | 0.075716357 |
| T2DM | endothelial | *LRRC1* | 0.312388011 | 8.59E-05 | 0.00032982 | 0.416892651 |
| T2DM | endothelial | *RIPK1* | -0.445721535 | 0.016245874 | 0.026222549 | 1 |
| T2DM | endothelial | *TCF19* | 1.110887729 | 8.91E-08 | 8.64E-06 | 0.000432208 |
| T2DM | endothelial | *FLOT1* | 0.428851153 | 0.001362932 | 0.003147524 | 1 |
| T2DM | endothelial | *TPMT* | -0.353096963 | 0.006541387 | 0.011940861 | 1 |
| T2DM | endothelial | *NRM* | 1.7430877 | 0.010023346 | 0.017264208 | 1 |
| T2DM | endothelial | *TLR2* | -1.081162494 | 1.04E-06 | 1.65E-05 | 0.005025247 |
| T2DM | endothelial | *MYO7A* | -0.494976095 | 0.003522805 | 0.007063079 | 1 |

| | | | | | | |
|---|---|---|---|---|---|---|
| T2DM | endothelial | *NUMA1* | -0.520261283 | 0.004530727 | 0.008740791 | 1 |
| T2DM | endothelial | *SYTL2* | -0.7954911 | 4.14E-08 | 5.74E-06 | 0.000201024 |
| T2DM | endothelial | *PRCP* | 0.353335453 | 0.00859342 | 0.015074214 | 1 |
| T2DM | endothelial | *NARS2* | -0.666130456 | 0.000330535 | 0.000974337 | 1 |
| T2DM | endothelial | *GGH* | -0.416950725 | 0.000731878 | 0.001859586 | 1 |
| T2DM | endothelial | *SULF1* | 0.241551771 | 0.002255132 | 0.004818097 | 1 |
| T2DM | endothelial | *TGS1* | -0.734964409 | 4.68E-05 | 0.000207014 | 0.226886866 |
| T2DM | endothelial | *NEK1* | -0.437251341 | 0.008957027 | 0.015638537 | 1 |
| T2DM | endothelial | *TRPC6* | -0.589919113 | 0.003788978 | 0.007515993 | 1 |
| T2DM | endothelial | *MMP27* | -1.070637409 | 0.002043325 | 0.004443843 | 1 |
| T2DM | endothelial | *SLC37A4* | 0.593207875 | 0.005498407 | 0.010297592 | 1 |
| T2DM | endothelial | *PPP2R1B* | -0.297104632 | 0.002929509 | 0.006048502 | 1 |
| T2DM | endothelial | *FDX1* | -0.656225041 | 1.36E-05 | 8.10E-05 | 0.066132104 |
| T2DM | endothelial | *FXYD2* | -0.912633619 | 7.70E-06 | 5.45E-05 | 0.037348462 |
| T2DM | endothelial | *MAP2K5* | -0.375303024 | 0.024848037 | 0.037805794 | 1 |
| T2DM | endothelial | *NUSAP1* | -0.67857779 | 0.000122378 | 0.00043789 | 0.59377874 |
| T2DM | endothelial | *NDUFAF1* | -0.73769545 | 0.00010263 | 0.000379253 | 0.49795919 |
| T2DM | endothelial | *PARP6* | -0.771708478 | 5.21E-05 | 0.000224667 | 0.252750534 |
| T2DM | endothelial | *ZNF280D* | -0.359255937 | 0.020839655 | 0.032502092 | 1 |
| T2DM | endothelial | *RSL24D1* | -0.530858316 | 0.033595263 | 0.049212197 | 1 |
| T2DM | endothelial | *SPTBN5* | 0.165691711 | 0.002176528 | 0.004683156 | 1 |
| T2DM | endothelial | *DBT* | -0.793772964 | 0.000111438 | 0.000405018 | 0.540699207 |
| T2DM | endothelial | *ADCY3* | -0.658742719 | 2.98E-05 | 0.000143544 | 0.144548394 |
| T2DM | endothelial | *DYNC2LI1* | -0.657834207 | 9.61E-06 | 6.29E-05 | 0.046623058 |
| T2DM | endothelial | *LHCGR* | -0.858265792 | 2.84E-07 | 1.24E-05 | 0.001378646 |
| T2DM | endothelial | *THUMPD2* | -0.657612736 | 0.00038357 | 0.001097336 | 1 |
| T2DM | endothelial | *CENPO* | -0.760021564 | 6.96E-05 | 0.00028313 | 0.337774577 |
| T2DM | endothelial | *LRPPRC* | -0.516874537 | 0.004896682 | 0.009350138 | 1 |
| T2DM | endothelial | *MYOF* | 0.339994826 | 0.00036028 | 0.001043628 | 1 |
| T2DM | endothelial | *BTBD16* | -1.119462307 | 8.26E-08 | 8.44E-06 | 0.000400888 |
| T2DM | endothelial | *TACC2* | -0.615036041 | 1.17E-05 | 7.28E-05 | 0.056919468 |
| T2DM | endothelial | *PLCE1* | -0.481940067 | 0.000138027 | 0.000482152 | 0.669708489 |
| T2DM | endothelial | *RBP4* | -0.694589258 | 0.000447532 | 0.001240107 | 1 |
| T2DM | endothelial | *ASCC1* | -0.491170166 | 0.005133548 | 0.009729677 | 1 |
| T2DM | endothelial | *ATIC* | -0.662001581 | 2.60E-07 | 1.24E-05 | 0.001259431 |
| T2DM | endothelial | *SMARCAL1* | -0.713968474 | 0.000835715 | 0.002080498 | 1 |
| T2DM | endothelial | *STAT4* | -0.834814903 | 1.23E-05 | 7.52E-05 | 0.059702991 |
| T2DM | endothelial | *SSB* | -0.494077663 | 0.002162281 | 0.004660688 | 1 |
| T2DM | endothelial | *FASTKD1* | -0.586631924 | 0.002392137 | 0.005068691 | 1 |
| T2DM | endothelial | *SLC35A5* | -0.513277256 | 0.006838 | 0.012426208 | 1 |
| T2DM | endothelial | *SPPL2A* | -0.638976673 | 0.000279839 | 0.000856642 | 1 |
| T2DM | endothelial | *SEMA7A* | 0.407461295 | 3.71E-05 | 0.00017142 | 0.180162356 |
| T2DM | endothelial | *PCDH10* | -1.251021352 | 2.49E-05 | 0.000125956 | 0.120665869 |
| T2DM | endothelial | *AP1AR* | -0.620893733 | 0.020294264 | 0.031722863 | 1 |
| T2DM | endothelial | *BMPR1B* | -0.582347079 | 2.34E-06 | 2.58E-05 | 0.011371473 |
| T2DM | endothelial | *MMRN1* | 0.174619099 | 0.030690832 | 0.045608551 | 1 |
| T2DM | endothelial | *TRPC3* | 1.143837232 | 0.000292947 | 0.000885593 | 1 |
| T2DM | endothelial | *G3BP2* | -0.441977774 | 0.021895352 | 0.033854763 | 1 |
| T2DM | endothelial | *ANXA3* | 0.287850826 | 1.00E-05 | 6.48E-05 | 0.048683512 |
| T2DM | endothelial | *ENPEP* | -0.195856608 | 0.032083707 | 0.047316154 | 1 |

| | | | | | | |
|---|---|---|---|---|---|---|
| T2DM | endothelial | *PAPSS1* | -0.532151655 | 0.006970995 | 0.012625333 | 1 |
| T2DM | endothelial | *PARVG* | -1.036305859 | 6.75E-05 | 0.000275479 | 0.327268627 |
| T2DM | endothelial | *B4GALNT3* | -0.875572151 | 7.65E-07 | 1.46E-05 | 0.003712799 |
| T2DM | endothelial | *PDE6H* | 1.286088541 | 0.015773102 | 0.025544424 | 1 |
| T2DM | endothelial | *KIF21A* | -0.575023185 | 1.54E-06 | 2.13E-05 | 0.007489593 |
| T2DM | endothelial | *ALG10* | -0.794118186 | 0.00015039 | 0.000516047 | 0.729690767 |
| T2DM | endothelial | *ZCRB1* | -0.679960665 | 0.004129522 | 0.008072699 | 1 |
| T2DM | endothelial | *PEX5* | -0.429493467 | 0.008473489 | 0.014890753 | 1 |
| T2DM | endothelial | *PPFIA2* | -0.718519273 | 1.60E-06 | 2.16E-05 | 0.007769475 |
| T2DM | endothelial | *LGR5* | -0.637817143 | 0.002079429 | 0.004504193 | 1 |
| T2DM | endothelial | *RITA1* | 0.824531324 | 0.005881335 | 0.010891693 | 1 |
| T2DM | endothelial | *SDSL* | -0.682172747 | 7.03E-06 | 5.11E-05 | 0.034103269 |
| T2DM | endothelial | *FOXN4* | -0.638183342 | 3.07E-05 | 0.000147076 | 0.148987773 |
| T2DM | endothelial | *MTMR6* | -0.547522088 | 0.03082086 | 0.045773741 | 1 |
| T2DM | endothelial | *CCDC65* | -0.696825566 | 7.35E-05 | 0.000295096 | 0.356771067 |
| T2DM | endothelial | *ACVRL1* | 0.420180365 | 7.68E-05 | 0.000305012 | 0.372420129 |
| T2DM | endothelial | *BRCA2* | -1.272628985 | 3.41E-06 | 3.18E-05 | 0.01654395 |
| T2DM | endothelial | *CERS5* | -0.611535439 | 0.001780133 | 0.003947535 | 1 |
| T2DM | endothelial | *ZNF740* | -0.71834405 | 2.56E-05 | 0.000128761 | 0.12431495 |
| T2DM | endothelial | *WDFY2* | -0.607000199 | 2.76E-05 | 0.000136232 | 0.133819944 |
| T2DM | endothelial | *GRTP1* | -0.347920157 | 0.000436245 | 0.001212822 | 1 |
| T2DM | endothelial | *NOVA1* | -0.157907974 | 0.031351756 | 0.046420116 | 1 |
| T2DM | endothelial | *FITM1* | 1.364958269 | 0.000210692 | 0.000677635 | 1 |
| T2DM | endothelial | *TMX1* | -0.552765308 | 0.021653736 | 0.033597625 | 1 |
| T2DM | endothelial | *RTN1* | -0.360415928 | 0.000339336 | 0.000993241 | 1 |
| T2DM | endothelial | *RAB15* | -1.078831746 | 4.76E-06 | 3.96E-05 | 0.023081186 |
| T2DM | endothelial | *SLC24A4* | -1.054048497 | 5.77E-07 | 1.27E-05 | 0.002798615 |
| T2DM | endothelial | *C14orf79* | -0.794318133 | 0.000304344 | 0.000914918 | 1 |
| T2DM | endothelial | *WARS* | 0.411984315 | 0.000659753 | 0.001707265 | 1 |
| T2DM | endothelial | *SRP14* | 0.359832323 | 0.003963378 | 0.007809183 | 1 |
| T2DM | endothelial | *HMG20A* | -0.646544293 | 0.00071794 | 0.00182954 | 1 |
| T2DM | endothelial | *NEIL1* | -0.573869403 | 0.004009386 | 0.007874022 | 1 |
| T2DM | endothelial | *MAN2C1* | -0.408052203 | 2.86E-07 | 1.24E-05 | 0.00138707 |
| T2DM | endothelial | *PIF1* | -0.743531636 | 9.49E-08 | 8.85E-06 | 0.000460365 |
| T2DM | endothelial | *CYP11A1* | -0.511765373 | 0.001995885 | 0.004356291 | 1 |
| T2DM | endothelial | *BBS4* | -0.617942757 | 0.000327871 | 0.000968247 | 1 |
| T2DM | endothelial | *PCSK6* | -0.797435335 | 1.69E-06 | 2.21E-05 | 0.008215912 |
| T2DM | endothelial | *HAPLN3* | 0.296388106 | 8.22E-05 | 0.000319932 | 0.398955539 |
| T2DM | endothelial | *POLG* | -0.806493221 | 0.000379013 | 0.001085579 | 1 |
| T2DM | endothelial | *ABHD2* | -0.474237312 | 0.033656981 | 0.049276907 | 1 |
| T2DM | endothelial | *MFGE8* | -0.626555542 | 0.002894707 | 0.005986836 | 1 |
| T2DM | endothelial | *UNC45A* | -0.694254361 | 0.005396915 | 0.010145614 | 1 |
| T2DM | endothelial | *IQGAP1* | -0.342939571 | 0.019239535 | 0.030335981 | 1 |
| T2DM | endothelial | *CRTC3* | -0.480760186 | 0.000755509 | 0.00191023 | 1 |
| T2DM | endothelial | *ITGAX* | -1.360048523 | 1.84E-05 | 0.00010058 | 0.0895161 |
| T2DM | endothelial | *NUDT7* | -0.978352593 | 0.001415818 | 0.003246463 | 1 |
| T2DM | endothelial | *GCSH* | -0.96377243 | 4.20E-09 | 1.19E-06 | 2.04E-05 |
| T2DM | endothelial | *CMTM3* | -0.977891007 | 3.23E-05 | 0.000152733 | 0.156704368 |
| T2DM | endothelial | *TLDC1* | -0.979557044 | 8.43E-05 | 0.000325463 | 0.409107076 |
| T2DM | endothelial | *ADAD2* | 0.287937579 | 0.029572034 | 0.04412162 | 1 |

| | | | | | | |
|---|---|---|---|---|---|---|
| T2DM | endothelial | *TIGD7* | -0.650884614 | 0.004915508 | 0.009375018 | 1 |
| T2DM | endothelial | *DEF8* | -0.730914502 | 0.000276247 | 0.000847252 | 1 |
| T2DM | endothelial | *TCF25* | -0.436898548 | 0.006616932 | 0.012068838 | 1 |
| T2DM | endothelial | *GALNS* | -0.86179798 | 2.67E-07 | 1.24E-05 | 0.001293165 |
| T2DM | endothelial | *GAS8* | -0.584133263 | 0.00502819 | 0.009544905 | 1 |
| T2DM | endothelial | *NCOR1* | -0.392951301 | 0.01033468 | 0.017743761 | 1 |
| T2DM | endothelial | *GFOD2* | 0.298360827 | 0.00349764 | 0.007021587 | 1 |
| T2DM | endothelial | *PRPSAP2* | -0.483606087 | 0.004664684 | 0.008963583 | 1 |
| T2DM | endothelial | *RASL10B* | -1.317830863 | 1.20E-06 | 1.80E-05 | 0.005830066 |
| T2DM | endothelial | *UNC45B* | -2.026813524 | 1.12E-06 | 1.73E-05 | 0.005410744 |
| T2DM | endothelial | *C17orf80* | -0.594545809 | 0.021837768 | 0.033790782 | 1 |
| T2DM | endothelial | *VPS53* | -0.696471319 | 7.03E-06 | 5.11E-05 | 0.034104243 |
| T2DM | endothelial | *SGSM2* | -0.82522296 | 9.34E-05 | 0.000353114 | 0.453398031 |
| T2DM | endothelial | *LRRC46* | -0.539826363 | 0.012990528 | 0.021682161 | 1 |
| T2DM | endothelial | *SSH2* | -0.408591142 | 0.005020062 | 0.009533205 | 1 |
| T2DM | endothelial | *RHBDL3* | 0.376430221 | 1.65E-05 | 9.27E-05 | 0.080105608 |
| T2DM | endothelial | *G6PC3* | -1.069968683 | 0.001260748 | 0.002950868 | 1 |
| T2DM | endothelial | *CLTC* | -0.35588295 | 0.017475182 | 0.027964902 | 1 |
| T2DM | endothelial | *IMPA2* | -0.709712733 | 6.28E-06 | 4.77E-05 | 0.030453559 |
| T2DM | endothelial | *RPRD1A* | -0.50295461 | 0.006426552 | 0.011753348 | 1 |
| T2DM | endothelial | *GALNT1* | -0.47062909 | 0.013294463 | 0.022105804 | 1 |
| T2DM | endothelial | *ADCYAP1* | -0.485868468 | 0.002164242 | 0.004660853 | 1 |
| T2DM | endothelial | *C18orf8* | -1.125804437 | 4.66E-09 | 1.19E-06 | 2.26E-05 |
| T2DM | endothelial | *NPC1* | -0.790505483 | 1.18E-05 | 7.32E-05 | 0.057466012 |
| T2DM | endothelial | *CCDC40* | -0.474956817 | 5.69E-05 | 0.000240518 | 0.275874334 |
| T2DM | endothelial | *ARHGDIA* | -0.506679876 | 0.01396121 | 0.022978219 | 1 |
| T2DM | endothelial | *TMC6* | 0.526899114 | 0.003973172 | 0.007820622 | 1 |
| T2DM | endothelial | *CARD14* | -0.833399144 | 7.01E-06 | 5.11E-05 | 0.034035277 |
| T2DM | endothelial | *ANAPC11* | -0.988599761 | 0.000217856 | 0.000696336 | 1 |
| T2DM | endothelial | *TBCD* | 0.34815483 | 0.000957011 | 0.002328266 | 1 |
| T2DM | endothelial | *FN3KRP* | -0.376944201 | 0.000158349 | 0.000535564 | 0.768308461 |
| T2DM | endothelial | *NARF* | -0.709906031 | 0.000298044 | 0.000898762 | 1 |
| T2DM | endothelial | *SECTM1* | -0.410480079 | 0.012770325 | 0.021373445 | 1 |
| T2DM | endothelial | *RNF157* | -0.455160832 | 3.67E-05 | 0.000169537 | 0.177844659 |
| T2DM | endothelial | *ZNF750* | 1.479067952 | 5.18E-05 | 0.000223806 | 0.251110647 |
| T2DM | endothelial | *WDR45B* | -0.611911276 | 0.005290792 | 0.0099848 | 1 |
| T2DM | endothelial | *MAPK4* | -2.441488142 | 0.000715101 | 0.001826141 | 1 |
| T2DM | endothelial | *FBXO15* | -1.018453361 | 4.40E-06 | 3.77E-05 | 0.021353141 |
| T2DM | endothelial | *CBLN2* | -0.754144206 | 3.27E-07 | 1.24E-05 | 0.001588779 |
| T2DM | endothelial | *ERBB2* | -0.806688783 | 0.001257829 | 0.002946879 | 1 |
| T2DM | endothelial | *MIEN1* | -0.957145049 | 0.001717252 | 0.003836146 | 1 |
| T2DM | endothelial | *PNMT* | 0.401789694 | 1.71E-06 | 2.22E-05 | 0.008286035 |
| T2DM | endothelial | *IGFBP4* | 0.328094034 | 0.004950295 | 0.00941915 | 1 |
| T2DM | endothelial | *CACNA1A* | -1.348592194 | 8.54E-06 | 5.82E-05 | 0.041455989 |
| T2DM | endothelial | *SAMD1* | -1.181976941 | 0.000153629 | 0.000523911 | 0.745410276 |
| T2DM | endothelial | *NFIC* | -0.556825142 | 1.60E-05 | 9.06E-05 | 0.077728344 |
| T2DM | endothelial | *PRDM15* | -0.407249372 | 0.004149313 | 0.008101597 | 1 |
| T2DM | endothelial | *PFKL* | -0.692417698 | 0.007586958 | 0.013568714 | 1 |
| T2DM | endothelial | *TMEM91* | -0.31963298 | 0.011867231 | 0.020034726 | 1 |
| T2DM | endothelial | *SIRT3* | -0.577198433 | 0.002550078 | 0.00536556 | 1 |

| | | | | | | |
|---|---|---|---|---|---|---|
| T2DM | endothelial | *HUNK* | -0.277127217 | 0.000251639 | 0.000786995 | 1 |
| T2DM | endothelial | *COL6A1* | -0.394441828 | 2.47E-06 | 2.65E-05 | 0.01197714 |
| T2DM | endothelial | *DOPEY2* | -0.412638588 | 0.00495372 | 0.009421972 | 1 |
| T2DM | endothelial | *SAE1* | -0.839597347 | 0.000233992 | 0.000739627 | 1 |
| T2DM | endothelial | *GEMIN7* | -1.17299362 | 5.75E-07 | 1.27E-05 | 0.002787874 |
| T2DM | endothelial | *CACNG8* | 0.389897473 | 1.24E-06 | 1.85E-05 | 0.006018915 |
| T2DM | endothelial | *EVI5L* | -0.966272696 | 0.002300719 | 0.004896092 | 1 |
| T2DM | endothelial | *SLC47A1* | -0.753469194 | 0.000935977 | 0.00228784 | 1 |
| T2DM | endothelial | *RERE* | -0.336668803 | 0.014467581 | 0.023715102 | 1 |
| T2DM | endothelial | *PEX14* | -0.540416572 | 0.000172262 | 0.000573263 | 0.835817285 |
| T2DM | endothelial | *CNKSR1* | -0.247811827 | 0.007424674 | 0.013298088 | 1 |
| T2DM | endothelial | *ZNF593* | -1.435361351 | 0.009353309 | 0.016242754 | 1 |
| T2DM | endothelial | *PLK4* | -0.708442941 | 0.000289602 | 0.000879319 | 1 |
| T2DM | endothelial | *NBPF3* | -0.239464675 | 0.004425037 | 0.008560717 | 1 |
| T2DM | endothelial | *ITGB3BP* | -0.281373677 | 0.031067122 | 0.046068971 | 1 |
| T2DM | endothelial | *PIGK* | -0.548666356 | 0.005554019 | 0.010376628 | 1 |
| T2DM | endothelial | *CYP4B1* | 0.403849295 | 0.004405666 | 0.008530044 | 1 |
| T2DM | endothelial | *SLC44A3* | -0.80817345 | 1.91E-06 | 2.35E-05 | 0.009277788 |
| T2DM | endothelial | *STRIP1* | -0.752076178 | 0.005889745 | 0.010900143 | 1 |
| T2DM | endothelial | *ALDH9A1* | -0.828670097 | 0.000674906 | 0.001737213 | 1 |
| T2DM | endothelial | *ATP1B1* | -0.604882852 | 1.57E-05 | 8.96E-05 | 0.076118594 |
| T2DM | endothelial | *NME7* | -0.651277803 | 0.000166076 | 0.000555157 | 0.805800712 |
| T2DM | endothelial | *XCL1* | 0.981260748 | 0.003952279 | 0.00779214 | 1 |
| T2DM | endothelial | *XCL2* | -0.560302334 | 0.001313132 | 0.003055787 | 1 |
| T2DM | endothelial | *MAEL* | -1.01236212 | 3.77E-07 | 1.24E-05 | 0.001828842 |
| T2DM | endothelial | *RGS16* | -0.70061033 | 3.88E-08 | 5.71E-06 | 0.000188496 |
| T2DM | endothelial | *TARS2* | -0.659517078 | 0.003753687 | 0.007452082 | 1 |
| T2DM | endothelial | *SNX27* | -0.5295205 | 0.003237419 | 0.006580628 | 1 |
| T2DM | endothelial | *SETDB1* | -0.661458513 | 0.000871613 | 0.002157281 | 1 |
| T2DM | endothelial | *SEMA6C* | 0.544288444 | 5.37E-05 | 0.000230342 | 0.260628902 |
| T2DM | endothelial | *ARNT* | -0.579789419 | 0.000191271 | 0.000626213 | 0.928046954 |
| T2DM | endothelial | *POGZ* | -0.412362799 | 0.008769923 | 0.015350529 | 1 |
| T2DM | endothelial | *GABPB2* | -0.379976443 | 0.000811178 | 0.002026692 | 1 |
| T2DM | endothelial | *DYRK3* | 0.343804368 | 1.21E-05 | 7.42E-05 | 0.058632634 |
| T2DM | endothelial | *EIF2D* | -0.87253468 | 7.00E-05 | 0.000283999 | 0.339509396 |
| T2DM | endothelial | *TAF1A* | -0.668553106 | 5.99E-06 | 4.62E-05 | 0.029086541 |
| T2DM | endothelial | *SMYD2* | -0.867022131 | 8.13E-05 | 0.000317546 | 0.394709603 |
| T2DM | endothelial | *ADAM15* | 0.236997707 | 0.000157202 | 0.000532644 | 0.762745724 |
| T2DM | endothelial | *RAB13* | 0.568146524 | 0.010503769 | 0.017995863 | 1 |
| T2DM | endothelial | *S100A8* | -0.908932642 | 9.35E-06 | 6.20E-05 | 0.045351727 |
| T2DM | endothelial | *KCNN3* | 0.275074761 | 9.60E-05 | 0.000360585 | 0.4658759 |
| T2DM | endothelial | *C1orf43* | -0.706281491 | 0.000644939 | 0.001674287 | 1 |
| T2DM | endothelial | *FLG* | -0.269625976 | 0.000954238 | 0.002324278 | 1 |
| T2DM | endothelial | *SCCPDH* | -0.839860544 | 1.53E-05 | 8.84E-05 | 0.07447093 |
| T2DM | endothelial | *CEP170* | -0.614499508 | 0.000368938 | 0.001063629 | 1 |
| T2DM | endothelial | *ACP1* | -0.794884829 | 0.000414421 | 0.001167695 | 1 |
| T2DM | endothelial | *CNIH4* | -0.589716685 | 0.012678443 | 0.021226986 | 1 |
| T2DM | endothelial | *MBOAT2* | 0.441667674 | 0.002787883 | 0.005785632 | 1 |
| T2DM | endothelial | *OSR1* | -0.449316269 | 0.005725762 | 0.010636064 | 1 |
| T2DM | endothelial | *GALM* | -0.773894326 | 2.52E-05 | 0.00012706 | 0.122358334 |

| | | | | | | |
|---|---|---|---|---|---|---|
| T2DM | endothelial | *WDPCP* | -0.402637223 | 0.000228274 | 0.000724859 | 1 |
| T2DM | endothelial | *VPS54* | -0.436656988 | 0.010557478 | 0.018056004 | 1 |
| T2DM | endothelial | *REG3G* | 2.240809765 | 4.47E-05 | 0.000199636 | 0.216805029 |
| T2DM | endothelial | *ABHD1* | -0.894254823 | 1.62E-08 | 3.28E-06 | 7.88E-05 |
| T2DM | endothelial | *CIAO1* | -0.560289921 | 0.010851021 | 0.018492853 | 1 |
| T2DM | endothelial | *ZNF514* | -0.802927097 | 1.45E-05 | 8.48E-05 | 0.070311883 |
| T2DM | endothelial | *EXOC6B* | -0.31329865 | 0.030710603 | 0.045623958 | 1 |
| T2DM | endothelial | *TEX261* | -1.310885974 | 9.04E-05 | 0.000344005 | 0.438606546 |
| T2DM | endothelial | *THNSL2* | -0.340674714 | 0.000491362 | 0.001339375 | 1 |
| T2DM | endothelial | *RALB* | 0.402001601 | 0.000627668 | 0.001637335 | 1 |
| T2DM | endothelial | *RABL2A* | -0.723170054 | 5.94E-05 | 0.000248341 | 0.288053082 |
| T2DM | endothelial | *FBLN7* | -0.39462993 | 0.00095195 | 0.002319871 | 1 |
| T2DM | endothelial | *CNGA3* | -5.950211858 | 5.65E-08 | 6.85E-06 | 0.000274086 |
| T2DM | endothelial | *FAHD2B* | -0.54485309 | 0.005481869 | 0.01027705 | 1 |
| T2DM | endothelial | *LYG1* | -0.570270377 | 0.000223714 | 0.000712712 | 1 |
| T2DM | endothelial | *THSD7B* | -0.572283339 | 0.00660357 | 0.012049838 | 1 |
| T2DM | endothelial | *GPR17* | 0.166256512 | 0.000610841 | 0.001602053 | 1 |
| T2DM | endothelial | *POLR2D* | -0.666306922 | 0.005196635 | 0.009827519 | 1 |
| T2DM | endothelial | *GALNT13* | -0.626891121 | 5.58E-07 | 1.27E-05 | 0.002709529 |
| T2DM | endothelial | *NBEAL1* | -0.36405343 | 0.012539589 | 0.021043964 | 1 |
| T2DM | endothelial | *SUMF1* | -0.497501834 | 0.000270488 | 0.000833274 | 1 |
| T2DM | endothelial | *ACKR3* | -0.510285914 | 0.000158395 | 0.000535564 | 0.768534672 |
| T2DM | endothelial | *ESPNL* | -8.52914933 | 5.00E-07 | 1.27E-05 | 0.002427865 |
| T2DM | endothelial | *TAMM41* | -0.469108921 | 0.015174932 | 0.024724235 | 1 |
| T2DM | endothelial | *STK11IP* | -0.993429204 | 0.000532574 | 0.001433195 | 1 |
| T2DM | endothelial | *EAF1* | -0.87939439 | 0.004921098 | 0.009381991 | 1 |
| T2DM | endothelial | *CNTN4* | -0.546932717 | 9.23E-07 | 1.58E-05 | 0.004479296 |
| T2DM | endothelial | *SLC25A38* | -1.125232998 | 1.22E-05 | 7.46E-05 | 0.058982001 |
| T2DM | endothelial | *CTDSPL* | -0.605005752 | 0.00034487 | 0.001004991 | 1 |
| T2DM | endothelial | *SLC25A26* | -0.600797019 | 0.000294373 | 0.000888798 | 1 |
| T2DM | endothelial | *ARL6IP5* | -0.483801933 | 0.015731897 | 0.025486199 | 1 |
| T2DM | endothelial | *LIMD1* | -0.589397929 | 0.000113777 | 0.000411976 | 0.552047302 |
| T2DM | endothelial | *PLA1A* | 0.261942245 | 4.39E-06 | 3.77E-05 | 0.021320604 |
| T2DM | endothelial | *ADPRH* | 0.359835482 | 0.000519948 | 0.001405452 | 1 |
| T2DM | endothelial | *IGSF11* | -0.264726256 | 0.018398537 | 0.029190378 | 1 |
| T2DM | endothelial | *BOC* | -0.519463477 | 2.13E-05 | 0.000112933 | 0.103482011 |
| T2DM | endothelial | *MED12L* | -0.689594553 | 6.90E-07 | 1.34E-05 | 0.003347625 |
| T2DM | endothelial | *EIF2A* | -0.429722613 | 0.000290717 | 0.00088105 | 1 |
| T2DM | endothelial | *NCEH1* | -0.629831472 | 0.00013108 | 0.000462547 | 0.636001754 |
| T2DM | endothelial | *CCDC39* | -0.479556758 | 3.16E-05 | 0.00015044 | 0.153413974 |
| T2DM | endothelial | *MUC4* | -0.948083047 | 5.87E-07 | 1.28E-05 | 0.002848902 |
| T2DM | endothelial | *DGKQ* | -0.42382164 | 0.022608908 | 0.034835955 | 1 |
| T2DM | endothelial | *CENPC* | -0.58537274 | 0.000196819 | 0.000640487 | 0.954966468 |
| T2DM | endothelial | *TRMT10A* | -0.97600812 | 0.000883498 | 0.002182655 | 1 |
| T2DM | endothelial | *PYURF* | -0.875396972 | 0.003083719 | 0.006310503 | 1 |
| T2DM | endothelial | *CISD2* | -0.802169772 | 0.000339405 | 0.000993241 | 1 |
| T2DM | endothelial | *DDIT4L* | 1.177119544 | 1.49E-05 | 8.66E-05 | 0.072285993 |
| T2DM | endothelial | *METTL14* | -0.811477344 | 0.00012409 | 0.000442384 | 0.602084307 |
| T2DM | endothelial | *USP53* | -0.334088477 | 0.030041172 | 0.044739032 | 1 |
| T2DM | endothelial | *MARCH1* | -0.572501323 | 2.59E-07 | 1.24E-05 | 0.001257841 |

| | | | | | | |
|---|---|---|---|---|---|---|
| T2DM | endothelial | *CBR4* | -0.460172473 | 0.004630852 | 0.00891269 | 1 |
| T2DM | endothelial | *MARCH6* | -0.481578885 | 0.004024966 | 0.007900135 | 1 |
| T2DM | endothelial | *NKD2* | -0.606311129 | 0.00972463 | 0.016785452 | 1 |
| T2DM | endothelial | *MYO10* | 0.308174956 | 0.00022566 | 0.000717968 | 1 |
| T2DM | endothelial | *FAM105A* | -0.398878342 | 0.00040683 | 0.001150313 | 1 |
| T2DM | endothelial | *GTF2H2* | -1.298009329 | 1.74E-06 | 2.24E-05 | 0.008423213 |
| T2DM | endothelial | *SPATA9* | -0.565787829 | 4.28E-07 | 1.24E-05 | 0.002077762 |
| T2DM | endothelial | *CXCL14* | -0.515293028 | 0.007396231 | 0.013256931 | 1 |
| T2DM | endothelial | *PCYOX1L* | -1.388307576 | 0.001454852 | 0.003318343 | 1 |
| T2DM | endothelial | *FAM50B* | 1.424768377 | 0.003554918 | 0.007112768 | 1 |
| T2DM | endothelial | *MYLK4* | -1.467161611 | 1.24E-05 | 7.57E-05 | 0.060311322 |
| T2DM | endothelial | *GFOD1* | 0.196488915 | 2.40E-05 | 0.000123817 | 0.116264334 |
| T2DM | endothelial | *FAM193B* | -0.603352994 | 0.000906043 | 0.002227011 | 1 |
| T2DM | endothelial | *MUT* | -0.832658542 | 5.17E-05 | 0.000223806 | 0.25108472 |
| T2DM | endothelial | *DAAM2* | -0.534444891 | 1.01E-06 | 1.63E-05 | 0.00490636 |
| T2DM | endothelial | *MLIP* | 0.462890795 | 0.021190409 | 0.03298552 | 1 |
| T2DM | endothelial | *RPL7L1* | -0.858696553 | 0.000291275 | 0.00088219 | 1 |
| T2DM | endothelial | *TPBG* | -0.528690646 | 0.019449435 | 0.030619292 | 1 |
| T2DM | endothelial | *IRAK1BP1* | -0.400168237 | 0.023320927 | 0.035785307 | 1 |
| T2DM | endothelial | *PHIP* | -0.422435591 | 0.007183575 | 0.012957139 | 1 |
| T2DM | endothelial | *RNF217* | -0.52612386 | 0.001355259 | 0.003132785 | 1 |
| T2DM | endothelial | *ARHGAP18* | -0.429002315 | 0.001053573 | 0.00252566 | 1 |
| T2DM | endothelial | *SLC18B1* | -0.495165805 | 0.000743694 | 0.001885268 | 1 |
| T2DM | endothelial | *WTAP* | -0.454645036 | 0.006888947 | 0.012504741 | 1 |
| T2DM | endothelial | *SLC22A3* | -0.580060188 | 4.40E-07 | 1.24E-05 | 0.002135864 |
| T2DM | endothelial | *GNA12* | -0.765109041 | 4.53E-06 | 3.85E-05 | 0.021999766 |
| T2DM | endothelial | *SDK1* | -0.393416466 | 4.76E-05 | 0.000210076 | 0.231083428 |
| T2DM | endothelial | *CCZ1B* | -0.521964698 | 0.002052818 | 0.004462488 | 1 |
| T2DM | endothelial | *RBAK* | -1.329258972 | 0.000474079 | 0.001298832 | 1 |
| T2DM | endothelial | *POMZP3* | -0.574990773 | 0.001526123 | 0.003459136 | 1 |
| T2DM | endothelial | *PSPH* | -0.267692683 | 0.013895792 | 0.022893848 | 1 |
| T2DM | endothelial | *TRIM4* | -0.372387272 | 9.56E-05 | 0.000359896 | 0.463906388 |
| T2DM | endothelial | *TMEM209* | -0.776989442 | 0.001393729 | 0.003204916 | 1 |
| T2DM | endothelial | *NCAPG2* | -0.70022327 | 0.000456308 | 0.001259388 | 1 |
| T2DM | endothelial | *CHMP7* | -0.79681453 | 4.10E-05 | 0.000185594 | 0.199142351 |
| T2DM | endothelial | *DOCK5* | -0.576303159 | 1.57E-05 | 8.96E-05 | 0.076326215 |
| T2DM | endothelial | *TACC1* | 0.253297735 | 0.002167905 | 0.00466667 | 1 |
| T2DM | endothelial | *ADHFE1* | -0.353763885 | 5.48E-07 | 1.27E-05 | 0.002658629 |
| T2DM | endothelial | *RPL7* | -0.54908754 | 0.001143484 | 0.002709075 | 1 |
| T2DM | endothelial | *ATP6V0D2* | -1.194958672 | 1.01E-05 | 6.48E-05 | 0.048936076 |
| T2DM | endothelial | *DPYS* | -0.580979686 | 7.63E-07 | 1.46E-05 | 0.003702439 |
| T2DM | endothelial | *MAL2* | -0.325771913 | 0.000287177 | 0.000874897 | 1 |
| T2DM | endothelial | *GSDMC* | -1.474304111 | 0.017945879 | 0.028576766 | 1 |
| T2DM | endothelial | *SLC39A4* | -0.481299622 | 0.000119123 | 0.000427503 | 0.577984523 |
| T2DM | endothelial | *HAUS6* | -0.885907765 | 5.81E-05 | 0.00024484 | 0.281971547 |
| T2DM | endothelial | *C9orf72* | -0.260254111 | 0.005809969 | 0.010775983 | 1 |
| T2DM | endothelial | *ALAD* | -0.566225826 | 0.000576873 | 0.001529503 | 1 |
| T2DM | endothelial | *POLE3* | 0.401591634 | 0.0038846 | 0.007672735 | 1 |
| T2DM | endothelial | *SURF1* | -0.473183321 | 0.003603042 | 0.007191262 | 1 |
| T2DM | endothelial | *MED22* | -0.977155236 | 1.25E-06 | 1.85E-05 | 0.006078932 |

| | | | | | | |
|---|---|---|---|---|---|---|
| T2DM | endothelial | *REXO4* | -0.788443263 | 0.002975302 | 0.006114429 | 1 |
| T2DM | endothelial | *SH3GLB2* | -0.94469906 | 0.000388542 | 0.001106991 | 1 |
| T2DM | endothelial | *INPP5E* | -0.950844911 | 0.0069304 | 0.012556498 | 1 |
| T2DM | endothelial | *SLC39A12* | -1.550776447 | 0.0001018 | 0.000377048 | 0.493933406 |
| T2DM | endothelial | *ST8SIA6* | 0.148631171 | 3.56E-06 | 3.26E-05 | 0.017253245 |
| T2DM | endothelial | *PARD3* | -0.491365374 | 2.71E-05 | 0.000134216 | 0.131263039 |
| T2DM | endothelial | *CDHR1* | -1.343463965 | 5.19E-05 | 0.000224178 | 0.251752036 |
| T2DM | endothelial | *RGR* | -3.346556981 | 1.62E-07 | 1.14E-05 | 0.000783783 |
| T2DM | endothelial | *ADD3* | -0.548956335 | 0.000161745 | 0.000544234 | 0.784784997 |
| T2DM | endothelial | *LRRC27* | -0.413735685 | 0.001230083 | 0.002894454 | 1 |
| T2DM | endothelial | *PAOX* | -1.235796065 | 9.40E-05 | 0.000355117 | 0.456324803 |
| T2DM | endothelial | *PDCD11* | -0.733334979 | 0.001161695 | 0.002748192 | 1 |
| T2DM | endothelial | *RGS10* | -0.761746821 | 1.78E-06 | 2.27E-05 | 0.008656369 |
| T2DM | endothelial | *LIN7C* | -0.85340613 | 0.000530491 | 0.001428381 | 1 |
| T2DM | endothelial | *HSD17B12* | -0.355102001 | 2.60E-05 | 0.000130064 | 0.126161715 |
| T2DM | endothelial | *APIP* | -0.422581892 | 1.40E-07 | 1.06E-05 | 0.000680006 |
| T2DM | endothelial | *EIF3M* | -0.794364914 | 2.18E-05 | 0.000114152 | 0.105704605 |
| T2DM | endothelial | *PTPRJ* | -0.519334592 | 0.002728303 | 0.005683867 | 1 |
| T2DM | endothelial | *ENDOD1* | 0.470276814 | 0.00027233 | 0.000838417 | 1 |
| T2DM | endothelial | *CCDC82* | -0.31672457 | 0.010452294 | 0.017933002 | 1 |
| T2DM | endothelial | *CAPN5* | 0.781084346 | 0.000166767 | 0.000556501 | 0.809151805 |
| T2DM | endothelial | *INTS4* | -0.589521318 | 0.001450208 | 0.003311252 | 1 |
| T2DM | endothelial | *DRD2* | -3.007008205 | 3.91E-07 | 1.24E-05 | 0.001899535 |
| T2DM | endothelial | *NPAT* | -0.702280315 | 0.001211107 | 0.002858118 | 1 |
| T2DM | endothelial | *ATM* | -0.530291653 | 0.000327751 | 0.000968247 | 1 |
| T2DM | endothelial | *SLX4IP* | -0.454216494 | 0.008276174 | 0.014580972 | 1 |
| T2DM | endothelial | *ST14* | -1.416291791 | 8.60E-05 | 0.000330006 | 0.417457416 |
| T2DM | endothelial | *FADS1* | -0.794461738 | 0.000108884 | 0.00039842 | 0.528304967 |
| T2DM | endothelial | *TMC2* | -1.419572594 | 1.23E-07 | 1.01E-05 | 0.000595138 |
| T2DM | endothelial | *INCENP* | -1.425007216 | 4.86E-05 | 0.000213398 | 0.235591862 |
| T2DM | endothelial | *PLCH2* | -1.276263407 | 1.50E-05 | 8.68E-05 | 0.072646944 |
| T2DM | endothelial | *CPSF7* | -0.733381019 | 9.21E-05 | 0.000348716 | 0.446704593 |
| T2DM | endothelial | *CCDC15* | -0.683754963 | 6.63E-08 | 7.31E-06 | 0.000321511 |
| T2DM | endothelial | *CHEK1* | -1.042795308 | 2.90E-07 | 1.24E-05 | 0.001405886 |
| T2DM | endothelial | *CABLES2* | -0.5432925 | 0.000633903 | 0.001650051 | 1 |
| T2DM | endothelial | *TRPT1* | -0.588949039 | 0.000227253 | 0.000722091 | 1 |
| T2DM | endothelial | *NUDT22* | -0.81796282 | 1.64E-06 | 2.19E-05 | 0.007933648 |
| T2DM | endothelial | *ALDOA* | -0.573629362 | 0.006029953 | 0.011111786 | 1 |
| T2DM | endothelial | *ANXA8L1* | -0.180589372 | 0.012801332 | 0.021417953 | 1 |
| T2DM | endothelial | *FCGR1A* | -0.486721045 | 0.011336212 | 0.019231924 | 1 |
| T2DM | endothelial | *TMEM218* | -0.95629218 | 3.12E-05 | 0.000148977 | 0.151212108 |
| T2DM | endothelial | *SAP18* | -0.587386181 | 0.023522547 | 0.036049084 | 1 |
| T2DM | endothelial | *FAM124A* | 0.235857692 | 5.86E-06 | 4.54E-05 | 0.028436415 |
| T2DM | endothelial | *LYPD6B* | 0.415313165 | 1.65E-05 | 9.27E-05 | 0.080014307 |
| T2DM | endothelial | *WDR17* | -0.87110958 | 3.86E-06 | 3.45E-05 | 0.018748724 |
| T2DM | endothelial | *CCDC102B* | -0.530467872 | 8.85E-06 | 5.97E-05 | 0.042949587 |
| T2DM | endothelial | *RGS18* | -0.335873055 | 0.020433694 | 0.031889444 | 1 |
| T2DM | endothelial | *PRSS23* | -0.35492143 | 0.010869376 | 0.018517631 | 1 |
| T2DM | endothelial | *PPP1R1C* | -1.789727519 | 5.21E-06 | 4.19E-05 | 0.02527976 |
| T2DM | endothelial | *FAM173B* | -0.820817334 | 0.000152583 | 0.000522097 | 0.740334208 |

| | | | | | | |
|---|---|---|---|---|---|---|
| T2DM | endothelial | *GPR158* | -0.658827814 | 2.34E-06 | 2.58E-05 | 0.01134519 |
| T2DM | endothelial | *UBE3B* | -0.349638701 | 0.019924191 | 0.031224863 | 1 |
| T2DM | endothelial | *ANK3* | -0.609020657 | 0.000126748 | 0.000450534 | 0.614979424 |
| T2DM | endothelial | *PLBD2* | -0.748622649 | 0.000293411 | 0.000886444 | 1 |
| T2DM | endothelial | *DLG5* | -0.593407538 | 1.06E-06 | 1.67E-05 | 0.00513434 |
| T2DM | endothelial | *TMEM18* | -1.046754877 | 0.000305213 | 0.000916963 | 1 |
| T2DM | endothelial | *ADAMTS12* | -0.3880033 | 1.48E-05 | 8.63E-05 | 0.072011689 |
| T2DM | endothelial | *NUBPL* | -0.597062857 | 0.000196737 | 0.000640487 | 0.954568363 |
| T2DM | endothelial | *VIPAS39* | -0.985032127 | 6.38E-05 | 0.000263579 | 0.309442175 |
| T2DM | endothelial | *ANKRD50* | 0.425351624 | 0.019357431 | 0.030494238 | 1 |
| T2DM | endothelial | *UPF2* | -0.451350058 | 0.008676793 | 0.015195281 | 1 |
| T2DM | endothelial | *PTPRO* | -0.573294097 | 3.21E-06 | 3.06E-05 | 0.015559252 |
| T2DM | endothelial | *EPS8* | -0.456495301 | 0.000419303 | 0.001175986 | 1 |
| T2DM | endothelial | *QDPR* | -0.661263736 | 0.000210017 | 0.000676628 | 1 |
| T2DM | endothelial | *FAM160B1* | -0.527838197 | 0.007961074 | 0.014107791 | 1 |
| T2DM | endothelial | *ZNF827* | -0.641387165 | 1.84E-05 | 0.00010058 | 0.089512005 |
| T2DM | endothelial | *PIGF* | -0.65436917 | 0.001751286 | 0.00389246 | 1 |
| T2DM | endothelial | *INPP1* | 0.408443883 | 4.53E-05 | 0.000202092 | 0.219876624 |
| T2DM | endothelial | *MFSD6* | 0.364200907 | 0.013224354 | 0.022026971 | 1 |
| T2DM | endothelial | *RNF144A* | 0.331928077 | 0.003891342 | 0.007681363 | 1 |
| T2DM | endothelial | *BICD1* | 0.303109055 | 0.021234178 | 0.033029114 | 1 |
| T2DM | endothelial | *CCDC122* | -0.308489185 | 0.004636905 | 0.008920802 | 1 |
| T2DM | endothelial | *GUF1* | -0.688103608 | 0.003990013 | 0.007846777 | 1 |
| T2DM | endothelial | *GABRA2* | 0.343361578 | 7.87E-06 | 5.52E-05 | 0.038197755 |
| T2DM | endothelial | *PARP8* | -0.443964544 | 3.21E-05 | 0.000152151 | 0.155802892 |
| T2DM | endothelial | *CACUL1* | -0.596845751 | 0.0003434 | 0.001002394 | 1 |
| T2DM | endothelial | *BEND6* | -0.652066608 | 1.97E-06 | 2.37E-05 | 0.009557143 |
| T2DM | endothelial | *TIAL1* | -0.544076069 | 0.00494744 | 0.009417411 | 1 |
| T2DM | endothelial | *TMEM56* | -1.238843557 | 3.55E-07 | 1.24E-05 | 0.001722827 |
| T2DM | endothelial | *MGAT5* | -0.481268508 | 0.00438276 | 0.008494848 | 1 |
| T2DM | endothelial | *TMEM178A* | 0.62592791 | 2.95E-05 | 0.000142608 | 0.143320817 |
| T2DM | endothelial | *EPG5* | -0.683117726 | 0.000317923 | 0.000948684 | 1 |
| T2DM | endothelial | *SPC25* | 0.096111828 | 8.32E-06 | 5.73E-05 | 0.040347212 |
| T2DM | endothelial | *POC5* | -0.659776699 | 1.44E-05 | 8.46E-05 | 0.07001935 |
| T2DM | endothelial | *ZNF773* | -0.490622695 | 0.000537493 | 0.001443467 | 1 |
| T2DM | endothelial | *ZNF837* | -0.426201659 | 0.01918286 | 0.030278217 | 1 |
| T2DM | endothelial | *PLEKHH2* | -0.323458217 | 0.000519469 | 0.00140494 | 1 |
| T2DM | endothelial | *SPARCL1* | 0.207883236 | 7.76E-05 | 0.00030696 | 0.376430463 |
| T2DM | endothelial | *GPD1L* | -0.420624006 | 0.033858749 | 0.049512553 | 1 |
| T2DM | endothelial | *CCNO* | 5.254561012 | 0.00589171 | 0.010900143 | 1 |
| T2DM | endothelial | *CLEC4F* | 0.157580696 | 1.06E-06 | 1.67E-05 | 0.005136591 |
| T2DM | endothelial | *SLC30A6* | -0.509042562 | 0.003465091 | 0.006976191 | 1 |
| T2DM | endothelial | *RASGRP3* | 0.30130285 | 0.000265082 | 0.000822365 | 1 |
| T2DM | endothelial | *GPR180* | -0.409419674 | 0.00019743 | 0.000641614 | 0.95792992 |
| T2DM | endothelial | *TCTEX1D1* | 0.628078197 | 9.67E-05 | 0.000361965 | 0.469064605 |
| T2DM | endothelial | *PRDM8* | -1.018517397 | 3.14E-05 | 0.000149942 | 0.152491203 |
| T2DM | endothelial | *ZNF117* | -0.087746725 | 0.027225733 | 0.040922942 | 1 |
| T2DM | endothelial | *JAKMIP1* | -1.102635442 | 9.52E-07 | 1.60E-05 | 0.004620344 |
| T2DM | endothelial | *SREK1IP1* | -0.756954823 | 7.74E-05 | 0.000306465 | 0.375419206 |
| T2DM | endothelial | *MR1* | -0.656494323 | 0.000151526 | 0.000519214 | 0.735206373 |

| | | | | | | |
|---|---|---|---|---|---|---|
| T2DM | endothelial | *CENPH* | -1.226378756 | 0.000434832 | 0.001210445 | 1 |
| T2DM | endothelial | *CLGN* | -1.267520424 | 1.57E-05 | 8.96E-05 | 0.076193305 |
| T2DM | endothelial | *CETN3* | -0.72769805 | 1.99E-06 | 2.38E-05 | 0.009661066 |
| T2DM | endothelial | *SYCP2L* | -0.745621819 | 3.03E-05 | 0.000145758 | 0.147215806 |
| T2DM | endothelial | *TMEM87B* | 0.32084709 | 0.03183235 | 0.04703123 | 1 |
| T2DM | endothelial | *SLC25A27* | -0.339346811 | 0.004909021 | 0.009366327 | 1 |
| T2DM | endothelial | *ADPRHL1* | -0.957399617 | 8.14E-06 | 5.64E-05 | 0.03951116 |
| T2DM | endothelial | *CFDP1* | -0.527562198 | 0.001833784 | 0.004051695 | 1 |
| T2DM | endothelial | *ZDHHC7* | -0.771579188 | 1.28E-05 | 7.77E-05 | 0.062305745 |
| T2DM | endothelial | *JAZF1* | -0.925680583 | 1.16E-12 | 1.40E-09 | 5.62E-09 |
| T2DM | endothelial | *FBXO36* | -0.80831821 | 8.81E-07 | 1.54E-05 | 0.004274672 |
| T2DM | endothelial | *MCOLN2* | -0.658653314 | 2.80E-05 | 0.000137135 | 0.135662696 |
| T2DM | endothelial | *GDPD1* | -0.935104895 | 0.000467461 | 0.001284309 | 1 |
| T2DM | endothelial | *SEMA3D* | -0.59415339 | 1.79E-06 | 2.27E-05 | 0.008678149 |
| T2DM | endothelial | *PPP2R5E* | -0.532216407 | 0.002104953 | 0.004555412 | 1 |
| T2DM | endothelial | *ANKRD29* | 0.180167817 | 9.42E-06 | 6.23E-05 | 0.0456942 |
| T2DM | endothelial | *DNAAF1* | -0.877172442 | 8.61E-05 | 0.000330006 | 0.417788187 |
| T2DM | endothelial | *UBASH3B* | -0.42859458 | 0.009224323 | 0.016045808 | 1 |
| T2DM | endothelial | *ROBO4* | 0.262157393 | 2.63E-05 | 0.000131293 | 0.127616441 |
| T2DM | endothelial | *ROBO3* | -0.660420849 | 9.84E-05 | 0.000366796 | 0.477568398 |
| T2DM | endothelial | *PRKCA* | -0.593122394 | 1.01E-05 | 6.48E-05 | 0.048927371 |
| T2DM | endothelial | *LRRK1* | -0.671102513 | 0.000287605 | 0.000874897 | 1 |
| T2DM | endothelial | *ABCA5* | -0.475466566 | 0.000451449 | 0.001250245 | 1 |
| T2DM | endothelial | *ENPP3* | -0.957214489 | 1.44E-06 | 2.04E-05 | 0.006984106 |
| T2DM | endothelial | *MIA3* | -0.55980497 | 0.002598474 | 0.005450841 | 1 |
| T2DM | endothelial | *CCSAP* | -1.140363104 | 1.90E-06 | 2.34E-05 | 0.009219519 |
| T2DM | endothelial | *GBP5* | -0.368649727 | 0.003440286 | 0.006932005 | 1 |
| T2DM | endothelial | *LY96* | -0.554868144 | 0.002373136 | 0.005036945 | 1 |
| T2DM | endothelial | *PDE1C* | -0.742250728 | 3.46E-06 | 3.20E-05 | 0.016803392 |
| T2DM | endothelial | *RABGEF1* | -0.195318353 | 0.025073344 | 0.038124683 | 1 |
| T2DM | endothelial | *XPC* | -0.544508946 | 0.007652236 | 0.013660282 | 1 |
| T2DM | endothelial | *FLCN* | -0.572719017 | 2.51E-06 | 2.67E-05 | 0.012162662 |
| T2DM | endothelial | *OXNAD1* | -0.594479475 | 0.000766721 | 0.001931531 | 1 |
| T2DM | endothelial | *APCDD1* | -0.455190329 | 2.57E-05 | 0.000128761 | 0.124635921 |
| T2DM | endothelial | *RAB6B* | -0.584624158 | 0.028615766 | 0.042826557 | 1 |
| T2DM | endothelial | *ANKRD40* | -0.949195896 | 0.000132429 | 0.000466287 | 0.642544099 |
| T2DM | endothelial | *CYP2U1* | -0.491415099 | 0.005690632 | 0.010595161 | 1 |
| T2DM | endothelial | *MARCKS* | -0.455955576 | 0.018918467 | 0.029919297 | 1 |
| T2DM | endothelial | *MARVELD1* | -1.005020974 | 0.001120446 | 0.002663597 | 1 |
| T2DM | endothelial | *GOLGA7B* | -4.354264984 | 6.56E-07 | 1.33E-05 | 0.003180982 |
| T2DM | endothelial | *TRMT44* | -0.78797044 | 0.000815721 | 0.002035946 | 1 |
| T2DM | endothelial | *LRGUK* | -0.36531648 | 0.001241566 | 0.002914406 | 1 |
| T2DM | endothelial | *SETD9* | -0.382032096 | 0.000337418 | 0.000989814 | 1 |
| T2DM | endothelial | *ZKSCAN2* | -0.523222912 | 0.007307645 | 0.013156473 | 1 |
| T2DM | endothelial | *PDIA4* | -0.741420534 | 0.011876069 | 0.020042674 | 1 |
| T2DM | endothelial | *KDM8* | -0.989010052 | 0.00456284 | 0.008795749 | 1 |
| T2DM | endothelial | *ALS2CR12* | -0.814977923 | 4.88E-05 | 0.000213996 | 0.236679979 |
| T2DM | endothelial | *FZD7* | -0.554815833 | 6.71E-05 | 0.000274119 | 0.325378834 |
| T2DM | endothelial | *DEPTOR* | -0.370389329 | 0.013589276 | 0.022511152 | 1 |
| T2DM | endothelial | *MMP16* | 0.165998628 | 8.41E-05 | 0.00032521 | 0.408073027 |

| | | | | | | |
|---|---|---|---|---|---|---|
| T2DM | endothelial | *KCNMA1* | -0.228810828 | 0.00944089 | 0.016353873 | 1 |
| T2DM | endothelial | *NDUFAF6* | -0.660254412 | 0.000148015 | 0.000509701 | 0.718168857 |
| T2DM | endothelial | *DRAM2* | -0.276716031 | 0.021836111 | 0.033790782 | 1 |
| T2DM | endothelial | *C8orf37* | -0.359440839 | 0.014547362 | 0.023813698 | 1 |
| T2DM | endothelial | *WHAMM* | -0.62768767 | 3.26E-05 | 0.000153848 | 0.158155308 |
| T2DM | endothelial | *NAA11* | -1.31025504 | 0.002400381 | 0.005083016 | 1 |
| T2DM | endothelial | *C14orf2* | -0.769949554 | 0.000115058 | 0.000415681 | 0.558259584 |
| T2DM | endothelial | *TDRD9* | -2.184909063 | 0.002070084 | 0.004495767 | 1 |
| T2DM | endothelial | *PCDH1* | 0.1171007 | 0.000970423 | 0.002351894 | 1 |
| T2DM | endothelial | *SUPV3L1* | -0.847562311 | 5.27E-06 | 4.21E-05 | 0.025579605 |
| T2DM | endothelial | *TYSND1* | -0.383020344 | 0.000795356 | 0.001992291 | 1 |
| T2DM | endothelial | *NODAL* | -1.681370624 | 4.81E-06 | 3.99E-05 | 0.023324468 |
| T2DM | endothelial | *ZDHHC5* | -0.672451132 | 0.00759613 | 0.013580111 | 1 |
| T2DM | endothelial | *WDYHV1* | -0.239146568 | 0.032764622 | 0.048203138 | 1 |
| T2DM | endothelial | *ATAD2* | -0.436060878 | 0.000993512 | 0.002401853 | 1 |
| T2DM | endothelial | *FBRS* | -0.501823884 | 0.019813239 | 0.031071053 | 1 |
| T2DM | endothelial | *SASS6* | -1.106355626 | 3.70E-06 | 3.34E-05 | 0.017929969 |
| T2DM | endothelial | *VPS8* | -0.366091344 | 0.021821984 | 0.033790782 | 1 |
| T2DM | endothelial | *MPV17L* | -0.7302598 | 0.018478029 | 0.029289577 | 1 |
| T2DM | endothelial | *RPUSD3* | -0.963844236 | 7.01E-05 | 0.000283999 | 0.339956196 |
| T2DM | endothelial | *GHRL* | -0.726295545 | 0.001526424 | 0.003459136 | 1 |
| T2DM | endothelial | *NMNAT2* | 0.354155651 | 0.000955242 | 0.002325558 | 1 |
| T2DM | endothelial | *SLC6A1* | -3.01012499 | 8.01E-06 | 5.58E-05 | 0.038886636 |
| T2DM | endothelial | *TIMP4* | 0.392061032 | 0.000918088 | 0.002255476 | 1 |
| T2DM | endothelial | *NECAP2* | -0.703676864 | 0.003250874 | 0.006599682 | 1 |
| T2DM | endothelial | *CLDN12* | -1.057808449 | 1.67E-06 | 2.20E-05 | 0.0080991 |
| T2DM | endothelial | *MMP14* | -0.959590457 | 1.49E-06 | 2.09E-05 | 0.007242527 |
| T2DM | endothelial | *GATAD1* | -0.591120857 | 0.00142235 | 0.003256839 | 1 |
| T2DM | endothelial | *CLEC18A* | -1.074091647 | 2.01E-09 | 6.79E-07 | 9.75E-06 |
| T2DM | endothelial | *DHRS4* | -1.202347155 | 2.91E-05 | 0.000140859 | 0.141000208 |
| T2DM | endothelial | *AASDH* | -0.610121493 | 0.000270313 | 0.000833265 | 1 |
| T2DM | endothelial | *ETS2* | 0.299243403 | 6.70E-06 | 4.95E-05 | 0.032490386 |
| T2DM | endothelial | *MX1* | 0.244687979 | 0.001270536 | 0.00296948 | 1 |
| T2DM | endothelial | *SLC38A10* | -0.739329562 | 0.000120565 | 0.000432331 | 0.584982173 |
| T2DM | endothelial | *DGKI* | -0.556840111 | 9.51E-07 | 1.60E-05 | 0.00461438 |
| T2DM | endothelial | *SNX22* | 0.952872186 | 0.002924025 | 0.006039749 | 1 |
| T2DM | endothelial | *SLC34A2* | 6.095538208 | 6.51E-05 | 0.000267724 | 0.315646547 |
| T2DM | endothelial | *AP3S2* | -0.694387221 | 0.001718998 | 0.003836513 | 1 |
| T2DM | endothelial | *SPPL3* | -0.542536179 | 0.000629202 | 0.001639574 | 1 |
| T2DM | endothelial | *C12orf43* | -0.888768731 | 0.002227735 | 0.004776749 | 1 |
| T2DM | endothelial | *PEX10* | -0.974619043 | 8.83E-05 | 0.000337096 | 0.428449222 |
| T2DM | endothelial | *KRTCAP3* | 0.4993941 | 0.000374633 | 0.001076211 | 1 |
| T2DM | endothelial | *EXTL1* | -16.0072662 | 1.59E-06 | 2.16E-05 | 0.007727316 |
| T2DM | endothelial | *WDR66* | -0.670992037 | 1.08E-06 | 1.70E-05 | 0.005238715 |
| T2DM | endothelial | *RHPN1* | 0.47157174 | 0.002939331 | 0.00605936 | 1 |
| T2DM | endothelial | *DZIP1L* | -0.606514356 | 1.92E-06 | 2.35E-05 | 0.00930031 |
| T2DM | endothelial | *ESYT3* | -1.083924616 | 1.66E-06 | 2.20E-05 | 0.008042936 |
| T2DM | endothelial | *CLSTN2* | -0.488544581 | 2.46E-07 | 1.24E-05 | 0.001192088 |
| T2DM | endothelial | *COLEC12* | -0.37793963 | 0.000203405 | 0.000657948 | 0.986921589 |
| T2DM | endothelial | *MITD1* | -0.468941902 | 0.026650761 | 0.040220682 | 1 |

| | | | | | | |
|---|---|---|---|---|---|---|
| T2DM | endothelial | *TSPAN33* | -1.236023633 | 3.31E-07 | 1.24E-05 | 0.001608014 |
| T2DM | endothelial | *FAM86C1* | -1.541337454 | 5.37E-07 | 1.27E-05 | 0.002604134 |
| T2DM | endothelial | *DYNC1I1* | -0.456974485 | 0.012945208 | 0.02162884 | 1 |
| T2DM | endothelial | *COPG2* | -0.262002634 | 0.000267786 | 0.000828538 | 1 |
| T2DM | endothelial | *DUSP23* | 0.453128711 | 0.000344337 | 0.00100404 | 1 |
| T2DM | endothelial | *SPATA2L* | -1.405406605 | 2.33E-08 | 4.12E-06 | 0.000112927 |
| T2DM | endothelial | *FGF17* | -1.829827926 | 0.000213984 | 0.000686674 | 1 |
| T2DM | endothelial | *CDA* | 0.186900679 | 0.000190448 | 0.000623939 | 0.924053305 |
| T2DM | endothelial | *SLC5A11* | -0.239057558 | 0.006007681 | 0.011079159 | 1 |
| T2DM | endothelial | *CDC42SE2* | -0.48034686 | 0.004251953 | 0.008275361 | 1 |
| T2DM | endothelial | *RAPGEF6* | -1.036030129 | 3.90E-05 | 0.00017767 | 0.189218624 |
| T2DM | endothelial | *IFNAR2* | 0.273484387 | 0.005669903 | 0.010568716 | 1 |
| T2DM | endothelial | *CHAF1B* | -0.584713118 | 7.18E-06 | 5.17E-05 | 0.034816745 |
| T2DM | endothelial | *SIM2* | -0.297903801 | 0.007623441 | 0.013613888 | 1 |
| T2DM | endothelial | *HLCS* | -0.544969425 | 4.88E-05 | 0.000214004 | 0.236902036 |
| T2DM | endothelial | *ARHGAP27* | 0.470446102 | 0.004271008 | 0.008305784 | 1 |
| T2DM | endothelial | *IRX6* | -0.486008971 | 3.95E-06 | 3.50E-05 | 0.019149689 |
| T2DM | endothelial | *HK2* | -0.540454387 | 5.59E-06 | 4.37E-05 | 0.027126257 |
| T2DM | endothelial | *THEM4* | -0.462757196 | 0.001259457 | 0.002949269 | 1 |
| T2DM | endothelial | *AMFR* | -0.692658794 | 0.001863064 | 0.004104698 | 1 |
| T2DM | endothelial | *TMEM69* | -0.931305299 | 0.00193213 | 0.004238108 | 1 |
| T2DM | endothelial | *ACE* | 0.191089202 | 4.64E-06 | 3.90E-05 | 0.02250053 |
| T2DM | endothelial | *TEPP* | -2.423060368 | 0.002874697 | 0.005952456 | 1 |
| T2DM | endothelial | *CHCHD6* | -0.538134812 | 0.006656645 | 0.012128443 | 1 |
| T2DM | endothelial | *CTBP1* | -0.777966864 | 0.000481415 | 0.0013167 | 1 |
| T2DM | endothelial | *LRRC36* | -0.232277471 | 0.012517633 | 0.021030318 | 1 |
| T2DM | endothelial | *C16orf86* | -0.569678529 | 0.007380261 | 0.013242984 | 1 |
| T2DM | endothelial | *LYPD5* | 0.582103194 | 0.001231028 | 0.002895273 | 1 |
| T2DM | endothelial | *CCDC107* | -0.596260273 | 0.019947553 | 0.03125138 | 1 |
| T2DM | endothelial | *ATAD3B* | -0.697007614 | 0.000282955 | 0.000864544 | 1 |
| T2DM | endothelial | *SSU72* | -0.681100376 | 0.004221677 | 0.008229641 | 1 |
| T2DM | endothelial | *CPAMD8* | 0.237681636 | 1.86E-06 | 2.32E-05 | 0.009018894 |
| T2DM | endothelial | *ANKLE1* | -1.461380735 | 9.81E-05 | 0.000366031 | 0.475840189 |
| T2DM | endothelial | *CCDC58* | -1.037194202 | 4.91E-06 | 4.04E-05 | 0.023839205 |
| T2DM | endothelial | *SLC37A1* | 0.471115879 | 0.001994519 | 0.004355268 | 1 |
| T2DM | endothelial | *PDE9A* | 0.253791105 | 1.01E-05 | 6.48E-05 | 0.048803826 |
| T2DM | endothelial | *HSF2BP* | -0.852093231 | 2.16E-07 | 1.24E-05 | 0.001046456 |
| T2DM | endothelial | *C21orf33* | -0.502189664 | 0.004433591 | 0.008573848 | 1 |
| T2DM | endothelial | *C21orf2* | -0.353618895 | 0.001527096 | 0.003459136 | 1 |
| T2DM | endothelial | *FTCD* | -0.365774194 | 0.001134691 | 0.002689556 | 1 |
| T2DM | endothelial | *LSS* | -0.381744486 | 1.22E-05 | 7.46E-05 | 0.059069929 |
| T2DM | endothelial | *VAV2* | -0.473803409 | 0.010061937 | 0.017318382 | 1 |
| T2DM | endothelial | *MCM3AP* | -0.387349861 | 0.012560863 | 0.021059194 | 1 |
| T2DM | endothelial | *PRMT2* | -0.392765849 | 0.01177965 | 0.01994238 | 1 |
| T2DM | endothelial | *ADAMTS13* | -0.689230694 | 1.13E-05 | 7.09E-05 | 0.054972306 |
| T2DM | endothelial | *CACFD1* | -2.118350166 | 0.000128561 | 0.000455311 | 0.623775729 |
| T2DM | endothelial | *RDH13* | -0.47850661 | 0.005546089 | 0.010365803 | 1 |
| T2DM | endothelial | *TAOK1* | -0.463838343 | 0.004534865 | 0.008745296 | 1 |
| T2DM | endothelial | *S100A1* | 1.079085577 | 1.94E-05 | 0.000104591 | 0.093922913 |
| T2DM | endothelial | *ZBTB7B* | -0.807401511 | 2.94E-06 | 2.92E-05 | 0.014260805 |

| | | | | | | |
|---|---|---|---|---|---|---|
| T2DM | endothelial | *VPS11* | -0.810230241 | 3.98E-06 | 3.51E-05 | 0.01929248 |
| T2DM | endothelial | *PAQR6* | -0.664946374 | 3.22E-05 | 0.000152523 | 0.156336313 |
| T2DM | endothelial | *PTH1R* | -0.626141712 | 2.76E-07 | 1.24E-05 | 0.001339417 |
| T2DM | endothelial | *FGFR4* | -1.466099497 | 4.24E-07 | 1.24E-05 | 0.002058225 |
| T2DM | endothelial | *IER2* | -0.776370144 | 0.000862539 | 0.002138497 | 1 |
| T2DM | endothelial | *CPSF4* | -0.694634317 | 0.004809231 | 0.00920852 | 1 |
| T2DM | endothelial | *MUM1* | -0.641320434 | 1.50E-05 | 8.71E-05 | 0.072995841 |
| T2DM | endothelial | *ZNF333* | -0.737731019 | 0.00218803 | 0.004703732 | 1 |
| T2DM | endothelial | *SH2B2* | -0.629377208 | 0.000377752 | 0.001082606 | 1 |
| T2DM | endothelial | *LRWD1* | 0.502647131 | 0.001205586 | 0.002846473 | 1 |
| T2DM | endothelial | *DVL3* | -0.825723143 | 1.03E-07 | 9.42E-06 | 0.000499237 |
| T2DM | endothelial | *PCYT1A* | -0.704103891 | 8.00E-05 | 0.000314078 | 0.388200018 |
| T2DM | endothelial | *FBXO27* | -0.314895357 | 0.00049973 | 0.00135913 | 1 |
| T2DM | endothelial | *BDH1* | -0.733482757 | 4.86E-06 | 4.01E-05 | 0.023559622 |
| T2DM | endothelial | *PGAP3* | -1.199192202 | 0.000433164 | 0.001207185 | 1 |
| T2DM | endothelial | *ZNF577* | -0.485067406 | 0.005518421 | 0.010330008 | 1 |
| T2DM | endothelial | *TMEM143* | -0.839443811 | 4.06E-06 | 3.57E-05 | 0.019716025 |
| T2DM | endothelial | *CCL16* | 0.209367834 | 5.32E-06 | 4.22E-05 | 0.025806622 |
| T2DM | endothelial | *DCD* | -1.656812315 | 0.000623103 | 0.00162893 | 1 |
| T2DM | endothelial | *ASB16* | -0.992821509 | 2.48E-05 | 0.000125919 | 0.120246927 |
| T2DM | endothelial | *AQP5* | 1.001947312 | 0.016478202 | 0.026509361 | 1 |
| T2DM | endothelial | *IP6K3* | -4.129441205 | 4.60E-08 | 6.03E-06 | 0.000223223 |
| T2DM | endothelial | *LEMD2* | -0.77405616 | 0.001522751 | 0.003454131 | 1 |
| T2DM | endothelial | *ASGR2* | -2.619132953 | 6.53E-07 | 1.33E-05 | 0.003168059 |
| T2DM | endothelial | *POLR3K* | 0.733257856 | 0.017737063 | 0.028337251 | 1 |
| T2DM | endothelial | *SNRNP25* | -0.781885712 | 0.000309046 | 0.000927329 | 1 |
| T2DM | endothelial | *WDR90* | -0.729418001 | 9.85E-07 | 1.61E-05 | 0.004778635 |
| T2DM | endothelial | *CCDC78* | -0.592086974 | 0.02595618 | 0.039246887 | 1 |
| T2DM | endothelial | *MEIOB* | -0.569757527 | 0.002149839 | 0.00463807 | 1 |
| T2DM | endothelial | *SHANK2* | 0.324889202 | 3.73E-05 | 0.000172064 | 0.181183189 |
| T2DM | endothelial | *ASRGL1* | 0.276232885 | 8.60E-06 | 5.85E-05 | 0.041715535 |
| T2DM | endothelial | *TTC9C* | -0.489685015 | 0.013694883 | 0.022635882 | 1 |
| T2DM | endothelial | *TPCN2* | -0.679565975 | 2.65E-06 | 2.76E-05 | 0.012867186 |
| T2DM | endothelial | *CYP4A22* | 1.030540101 | 0.005848863 | 0.010839833 | 1 |
| T2DM | endothelial | *COA7* | -1.449615595 | 3.09E-06 | 3.00E-05 | 0.014980703 |
| T2DM | endothelial | *SLC1A7* | -0.394620148 | 0.000349067 | 0.001016 | 1 |
| T2DM | endothelial | *USP24* | -0.459016497 | 0.008114558 | 0.014332667 | 1 |
| T2DM | endothelial | *AK4* | -0.376541979 | 0.02886552 | 0.043147105 | 1 |
| T2DM | endothelial | *RBP7* | 0.240762394 | 1.09E-05 | 6.88E-05 | 0.052922976 |
| T2DM | endothelial | *PDPN* | -0.703453605 | 6.85E-07 | 1.34E-05 | 0.003325782 |
| T2DM | endothelial | *LAPTM5* | -0.8147466 | 6.51E-06 | 4.87E-05 | 0.031576669 |
| T2DM | endothelial | *SYNC* | -0.62537018 | 2.57E-05 | 0.000128761 | 0.124475497 |
| T2DM | endothelial | *CAMK2N1* | -0.458722524 | 0.013379957 | 0.022217506 | 1 |
| T2DM | endothelial | *MEGF6* | -0.485653283 | 0.026522892 | 0.040052621 | 1 |
| T2DM | endothelial | *OMA1* | -0.623667181 | 0.014268601 | 0.023428511 | 1 |
| T2DM | endothelial | *TM2D1* | -0.543480067 | 0.003151973 | 0.006422523 | 1 |
| T2DM | endothelial | *NEXN* | -0.578070252 | 5.19E-06 | 4.18E-05 | 0.025187148 |
| T2DM | endothelial | *LRRIQ3* | -0.83050851 | 1.38E-06 | 1.96E-05 | 0.006704002 |
| T2DM | endothelial | *HFM1* | -0.763403607 | 1.27E-05 | 7.72E-05 | 0.061768528 |
| T2DM | endothelial | *ARPC5* | -0.57919993 | 0.00110492 | 0.002629266 | 1 |

| | | | | | | |
|---|---|---|---|---|---|---|
| T2DM | endothelial | *ZNF496* | -0.535847495 | 0.030556238 | 0.045464233 | 1 |
| T2DM | endothelial | *TRIM58* | 0.331640903 | 0.000658588 | 0.00170516 | 1 |
| T2DM | endothelial | *FCRLB* | -1.396539894 | 4.13E-07 | 1.24E-05 | 0.002005826 |
| T2DM | endothelial | *KLHDC9* | -0.383589634 | 0.001931583 | 0.004238108 | 1 |
| T2DM | endothelial | *FLVCR1* | -0.960047983 | 0.000310774 | 0.000930788 | 1 |
| T2DM | endothelial | *TDRD5* | -0.633415569 | 6.19E-07 | 1.31E-05 | 0.003004422 |
| T2DM | endothelial | *ACP6* | -0.973238597 | 1.47E-05 | 8.58E-05 | 0.071354154 |
| T2DM | endothelial | *PKDCC* | -0.497316214 | 4.65E-05 | 0.000206204 | 0.225587697 |
| T2DM | endothelial | *HAAO* | -0.828705565 | 9.09E-07 | 1.57E-05 | 0.004411732 |
| T2DM | endothelial | *B3GALNT2* | -0.527721377 | 9.73E-05 | 0.000363645 | 0.472011054 |
| T2DM | endothelial | *MRPL55* | -0.506984672 | 0.006896067 | 0.012508306 | 1 |
| T2DM | endothelial | *KCNJ3* | 0.901574531 | 0.018510056 | 0.029321186 | 1 |
| T2DM | endothelial | *C2orf48* | -5.526661704 | 2.09E-05 | 0.000111192 | 0.101518196 |
| T2DM | endothelial | *CCDC74A* | -0.983230104 | 2.04E-05 | 0.000108744 | 0.099065507 |
| T2DM | endothelial | *NOSTRIN* | 0.112850883 | 3.04E-06 | 2.99E-05 | 0.014766543 |
| T2DM | endothelial | *INHBB* | 0.37435087 | 1.04E-05 | 6.61E-05 | 0.050307141 |
| T2DM | endothelial | *RPRD2* | -0.47575427 | 0.002591521 | 0.005440961 | 1 |
| T2DM | endothelial | *C1QTNF7* | -0.541258984 | 4.42E-06 | 3.78E-05 | 0.021442664 |
| T2DM | endothelial | *VPS72* | -1.004465542 | 7.51E-05 | 0.000300232 | 0.364181594 |
| T2DM | endothelial | *ERCC3* | -0.623218665 | 0.001751204 | 0.00389246 | 1 |
| T2DM | endothelial | *RNF149* | -0.594218637 | 3.90E-05 | 0.00017767 | 0.189211734 |
| T2DM | endothelial | *DHX57* | -0.725679114 | 0.000151063 | 0.000517992 | 0.732959088 |
| T2DM | endothelial | *S100A9* | -0.571330096 | 0.000110316 | 0.000402445 | 0.535251478 |
| T2DM | endothelial | *DCAF16* | -0.947454238 | 3.06E-06 | 2.99E-05 | 0.014866648 |
| T2DM | endothelial | *NIPAL1* | -0.720369496 | 0.000468345 | 0.001285301 | 1 |
| T2DM | endothelial | *ANTXR2* | -0.506703816 | 1.38E-05 | 8.17E-05 | 0.066767225 |
| T2DM | endothelial | *GPR155* | -0.733996782 | 0.000240041 | 0.000757268 | 1 |
| T2DM | endothelial | *DAPL1* | -0.761983393 | 0.002945903 | 0.006069435 | 1 |
| T2DM | endothelial | *PBXIP1* | -0.935563578 | 7.31E-05 | 0.000294466 | 0.354820524 |
| T2DM | endothelial | *CLDN1* | -0.556469134 | 1.10E-05 | 6.92E-05 | 0.053300543 |
| T2DM | endothelial | *LINC01116* | 0.263916498 | 2.07E-06 | 2.44E-05 | 0.010049462 |
| T2DM | endothelial | *SLC22A15* | -0.769669315 | 1.19E-05 | 7.32E-05 | 0.057572706 |
| T2DM | endothelial | *LRRC58* | -0.685010194 | 0.00261588 | 0.005484983 | 1 |
| T2DM | endothelial | *LMOD1* | -0.706526717 | 3.83E-07 | 1.24E-05 | 0.00186034 |
| T2DM | endothelial | *TMEM169* | -0.450910034 | 0.011495924 | 0.019482439 | 1 |
| T2DM | endothelial | *ARPC2* | -0.560948369 | 0.001551138 | 0.003507736 | 1 |
| T2DM | endothelial | *GLB1L* | -0.639598234 | 8.42E-05 | 0.00032521 | 0.408302716 |
| T2DM | endothelial | *SERPINI1* | 0.409165159 | 0.024544139 | 0.037402061 | 1 |
| T2DM | endothelial | *SUCLG1* | -0.44985121 | 0.019538679 | 0.030709967 | 1 |
| T2DM | endothelial | *SPTA1* | -0.603212667 | 4.20E-06 | 3.67E-05 | 0.020396873 |
| T2DM | endothelial | *PRKCI* | -0.460394968 | 0.000386515 | 0.00110446 | 1 |
| T2DM | endothelial | *AIM2* | 0.177038239 | 3.71E-06 | 3.34E-05 | 0.01801664 |
| T2DM | endothelial | *GTPBP8* | -0.938220191 | 0.00080611 | 0.002016106 | 1 |
| T2DM | endothelial | *PTPN13* | -0.388063622 | 5.77E-05 | 0.000243702 | 0.280013252 |
| T2DM | endothelial | *PPM1K* | -0.695394524 | 0.000358036 | 0.001039611 | 1 |
| T2DM | endothelial | *TIPARP* | -0.797878189 | 0.000222979 | 0.000710836 | 1 |
| T2DM | endothelial | *RPP14* | -1.061914516 | 1.88E-06 | 2.33E-05 | 0.009137425 |
| T2DM | endothelial | *DNASE1L3* | 0.206501412 | 2.13E-06 | 2.46E-05 | 0.010336752 |
| T2DM | endothelial | *IL17RC* | -1.126849782 | 8.59E-07 | 1.53E-05 | 0.004166555 |
| T2DM | endothelial | *CRELD1* | -0.826270761 | 8.78E-05 | 0.000335639 | 0.425925896 |

| | | | | | | |
|---|---|---|---|---|---|---|
| T2DM | endothelial | *TTC14* | -0.550329558 | 0.000427305 | 0.001194207 | 1 |
| T2DM | endothelial | *CCDC158* | -0.692179507 | 2.56E-06 | 2.72E-05 | 0.012422655 |
| T2DM | endothelial | *CPA3* | -1.532739119 | 5.38E-06 | 4.25E-05 | 0.026106955 |
| T2DM | endothelial | *SLC4A1AP* | -0.884851784 | 0.000140319 | 0.0004877 | 0.680829763 |
| T2DM | endothelial | *CDCP1* | -0.625779364 | 2.23E-06 | 2.52E-05 | 0.010835176 |
| T2DM | endothelial | *CLEC3B* | 1.256054726 | 7.03E-05 | 0.00028438 | 0.340971213 |
| T2DM | endothelial | *FYCO1* | -0.511523785 | 0.006448606 | 0.011780361 | 1 |
| T2DM | endothelial | *SNIP1* | -0.720383501 | 0.013687629 | 0.022635438 | 1 |
| T2DM | endothelial | *SENP2* | -0.597558054 | 0.000141498 | 0.000491444 | 0.686547795 |
| T2DM | endothelial | *IFT122* | -0.642817586 | 0.000348131 | 0.001013885 | 1 |
| T2DM | endothelial | *RFT1* | -1.081319051 | 8.12E-10 | 3.94E-07 | 3.94E-06 |
| T2DM | endothelial | *SFMBT1* | -0.818266029 | 2.11E-06 | 2.46E-05 | 0.010239113 |
| T2DM | endothelial | *UVSSA* | -0.366116917 | 0.022557733 | 0.034768146 | 1 |
| T2DM | endothelial | *FAM208A* | -0.600135613 | 0.000743388 | 0.001885268 | 1 |
| T2DM | endothelial | *LRPAP1* | -0.532566346 | 0.014592026 | 0.023870705 | 1 |
| T2DM | endothelial | *PIGX* | -0.415794068 | 0.006490801 | 0.011852979 | 1 |
| T2DM | endothelial | *ERMAP* | -0.823631058 | 9.51E-06 | 6.26E-05 | 0.046119929 |
| T2DM | endothelial | *BDH2* | -0.821746577 | 3.90E-06 | 3.47E-05 | 0.018936476 |
| T2DM | endothelial | *CDC25A* | 0.913233826 | 0.00073203 | 0.001859586 | 1 |
| T2DM | endothelial | *ZNF589* | -0.664731962 | 5.59E-05 | 0.000238149 | 0.271251299 |
| T2DM | endothelial | *ATRIP* | -0.44526615 | 8.09E-05 | 0.000316238 | 0.392767192 |
| T2DM | endothelial | *MFSD8* | -0.578483425 | 0.005326137 | 0.01003589 | 1 |
| T2DM | endothelial | *MST1R* | 0.756570521 | 1.70E-06 | 2.22E-05 | 0.00825463 |
| T2DM | endothelial | *RAD54L2* | -0.731573488 | 8.32E-05 | 0.000322717 | 0.4037189 |
| T2DM | endothelial | *ETNPPL* | -1.86616318 | 5.83E-07 | 1.27E-05 | 0.002827641 |
| T2DM | endothelial | *ANXA5* | -0.444246035 | 0.01755912 | 0.02808996 | 1 |
| T2DM | endothelial | *MAP9* | -0.699149523 | 9.65E-07 | 1.61E-05 | 0.004681741 |
| T2DM | endothelial | *CEP44* | -0.661166702 | 0.000495099 | 0.001348804 | 1 |
| T2DM | endothelial | *HPGD* | -1.6217705 | 6.40E-05 | 0.000264105 | 0.310322919 |
| T2DM | endothelial | *TMEM144* | -0.614047433 | 0.007346318 | 0.013196719 | 1 |
| T2DM | endothelial | *IL15* | 0.406132142 | 0.000139448 | 0.000485566 | 0.676600419 |
| T2DM | endothelial | *ITGA2* | 0.318292258 | 1.34E-05 | 8.00E-05 | 0.064778046 |
| T2DM | endothelial | *NDUFAF2* | -0.593675519 | 0.000225845 | 0.000718087 | 1 |
| T2DM | endothelial | *LMBRD2* | -0.657263813 | 0.003514604 | 0.007049548 | 1 |
| T2DM | endothelial | *PGGT1B* | -0.460205714 | 0.02886401 | 0.043147105 | 1 |
| T2DM | endothelial | *F2RL2* | -1.676720442 | 4.32E-07 | 1.24E-05 | 0.002096536 |
| T2DM | endothelial | *CMBL* | -0.614732658 | 8.44E-07 | 1.53E-05 | 0.004097394 |
| T2DM | endothelial | *C5orf63* | -0.711337326 | 9.59E-06 | 6.29E-05 | 0.046544902 |
| T2DM | endothelial | *F2RL1* | -1.344163541 | 1.58E-05 | 8.96E-05 | 0.076585188 |
| T2DM | endothelial | *WDR41* | -0.592407293 | 0.000590935 | 0.001559965 | 1 |
| T2DM | endothelial | *NDUFS4* | -0.579058773 | 0.001133311 | 0.00268935 | 1 |
| T2DM | endothelial | *SERINC5* | -0.569894246 | 0.000253262 | 0.000790752 | 1 |
| T2DM | endothelial | *PRIMPOL* | -0.708908558 | 7.08E-05 | 0.00028619 | 0.343714408 |
| T2DM | endothelial | *ERAP1* | -0.504846133 | 0.001244241 | 0.002919273 | 1 |
| T2DM | endothelial | *UTP15* | -1.346938706 | 5.39E-07 | 1.27E-05 | 0.002616708 |
| T2DM | endothelial | *CCDC127* | -1.02071206 | 0.00011891 | 0.000427055 | 0.576951422 |
| T2DM | endothelial | *GRIK2* | -0.238775526 | 0.008485155 | 0.014905856 | 1 |
| T2DM | endothelial | *PDSS2* | -0.478157749 | 0.003376503 | 0.006817642 | 1 |
| T2DM | endothelial | *RAET1E* | -1.027953626 | 1.97E-06 | 2.37E-05 | 0.009570967 |
| T2DM | endothelial | *DAGLB* | -0.237494988 | 0.0339615 | 0.049647846 | 1 |

| | | | | | | |
|---|---|---|---|---|---|---|
| T2DM | endothelial | *STK17A* | -0.460817293 | 0.023882614 | 0.036530832 | 1 |
| T2DM | endothelial | *RP9* | -1.035106716 | 0.000224741 | 0.000715515 | 1 |
| T2DM | endothelial | *RELL2* | 0.627199507 | 0.006707862 | 0.012212589 | 1 |
| T2DM | endothelial | *MIOS* | -0.780046306 | 0.000292535 | 0.000884901 | 1 |
| T2DM | endothelial | *USP49* | -0.844810366 | 3.31E-05 | 0.000155866 | 0.160697706 |
| T2DM | endothelial | *CHMP4C* | 0.643402512 | 6.35E-06 | 4.81E-05 | 0.030820207 |
| T2DM | endothelial | *SLC13A4* | -0.792845008 | 0.000107613 | 0.000394068 | 0.522139678 |
| T2DM | endothelial | *CTSB* | -0.225432677 | 0.011084413 | 0.018831083 | 1 |
| T2DM | endothelial | *SOX17* | 0.134252866 | 1.05E-06 | 1.67E-05 | 0.00508646 |
| T2DM | endothelial | *RAD21* | -0.490168932 | 0.015529927 | 0.025209503 | 1 |
| T2DM | endothelial | *DEFB1* | -0.787147407 | 0.014142925 | 0.023245756 | 1 |
| T2DM | endothelial | *OXR1* | -0.502053814 | 0.000575128 | 0.001525708 | 1 |
| T2DM | endothelial | *GPER1* | 0.762536144 | 0.002936418 | 0.00605761 | 1 |
| T2DM | endothelial | *INTS1* | -1.002793967 | 0.000262103 | 0.000815207 | 1 |
| T2DM | endothelial | *SLC4A2* | -0.897931519 | 1.82E-06 | 2.29E-05 | 0.008823117 |
| T2DM | endothelial | *FASTK* | -1.024740685 | 6.55E-06 | 4.87E-05 | 0.031785552 |
| T2DM | endothelial | *ALDH7A1* | -0.496885242 | 0.008886553 | 0.01553226 | 1 |
| T2DM | endothelial | *YWHAZ* | -0.490557289 | 0.002275135 | 0.004851523 | 1 |
| T2DM | endothelial | *FZD6* | 0.351304548 | 4.62E-06 | 3.90E-05 | 0.022400154 |
| T2DM | endothelial | *SLC25A32* | -0.578769937 | 0.008387127 | 0.014755019 | 1 |
| T2DM | endothelial | *DCSTAMP* | -3.404725784 | 2.47E-05 | 0.000125919 | 0.119876194 |
| T2DM | endothelial | *INTS8* | -0.513381311 | 0.015463043 | 0.025109331 | 1 |
| T2DM | endothelial | *RPP25L* | -0.898918721 | 9.02E-06 | 6.05E-05 | 0.043752982 |
| T2DM | endothelial | *C9orf24* | -0.986188988 | 1.58E-05 | 8.96E-05 | 0.076499985 |
| T2DM | endothelial | *ZMAT4* | -2.101875518 | 0.000206127 | 0.00066542 | 1 |
| T2DM | endothelial | *C8orf34* | -0.397053144 | 2.13E-06 | 2.46E-05 | 0.01033274 |
| T2DM | endothelial | *ANKS6* | -0.536306832 | 0.009654509 | 0.016688165 | 1 |
| T2DM | endothelial | *NOL6* | -0.840150875 | 9.97E-07 | 1.63E-05 | 0.004835478 |
| T2DM | endothelial | *AQP3* | -3.54380839 | 1.18E-05 | 7.30E-05 | 0.057230354 |
| T2DM | endothelial | *TRMT10B* | -0.640728436 | 0.000533162 | 0.00143398 | 1 |
| T2DM | endothelial | *VCP* | -0.428156145 | 0.024217563 | 0.036962445 | 1 |
| T2DM | endothelial | *PIGO* | -1.329465946 | 2.02E-05 | 0.000107774 | 0.097966975 |
| T2DM | endothelial | *WRN* | -0.39664814 | 0.025121104 | 0.038173379 | 1 |
| T2DM | endothelial | *TSHR* | -0.607321209 | 1.26E-07 | 1.02E-05 | 0.000610823 |
| T2DM | endothelial | *CFL2* | -0.939024627 | 0.000421723 | 0.001181409 | 1 |
| T2DM | endothelial | *PGM2L1* | 0.378128415 | 0.001945428 | 0.004261496 | 1 |
| T2DM | endothelial | *SLC16A9* | -1.040864519 | 3.76E-07 | 1.24E-05 | 0.001822664 |
| T2DM | endothelial | *GJB2* | 1.219472271 | 6.34E-06 | 4.81E-05 | 0.030765181 |
| T2DM | endothelial | *CRYL1* | -0.4705313 | 0.021085534 | 0.032843341 | 1 |
| T2DM | endothelial | *REEP3* | -0.44312937 | 0.018330464 | 0.02910487 | 1 |
| T2DM | endothelial | *MICU2* | -0.576796409 | 0.000375227 | 0.001077132 | 1 |
| T2DM | endothelial | *LRR1* | -1.235463736 | 1.10E-06 | 1.71E-05 | 0.005330922 |
| T2DM | endothelial | *TMEM63C* | -2.545269683 | 1.17E-06 | 1.79E-05 | 0.005689826 |
| T2DM | endothelial | *AKR1E2* | 0.438947394 | 0.013657674 | 0.022593603 | 1 |
| T2DM | endothelial | *ATP5C1* | -0.49536982 | 0.014928594 | 0.024388397 | 1 |
| T2DM | endothelial | *VDAC2* | -1.034422275 | 3.31E-06 | 3.11E-05 | 0.016067126 |
| T2DM | endothelial | *QSOX2* | -0.812828838 | 0.010954851 | 0.018637075 | 1 |
| T2DM | endothelial | *TSC1* | -0.38012942 | 0.008340617 | 0.014683843 | 1 |
| T2DM | endothelial | *STOX1* | -0.372098558 | 4.05E-05 | 0.000183952 | 0.196460699 |
| T2DM | endothelial | *RET* | -0.837321757 | 5.23E-07 | 1.27E-05 | 0.002536214 |

| | | | | | | |
|---|---|---|---|---|---|---|
| T2DM | endothelial | *PPP1R36* | -0.919266504 | 0.002072 | 0.004496129 | 1 |
| T2DM | endothelial | *VWA2* | 0.545971139 | 7.07E-06 | 5.11E-05 | 0.03431566 |
| T2DM | endothelial | *METTL3* | -0.854277672 | 0.000241534 | 0.000760173 | 1 |
| T2DM | endothelial | *TRUB1* | -0.856119784 | 0.000298671 | 0.000900094 | 1 |
| T2DM | endothelial | *HSPA12A* | -0.354225185 | 0.000290181 | 0.000880105 | 1 |
| T2DM | endothelial | *UBTD1* | -0.707756183 | 1.29E-06 | 1.88E-05 | 0.006242965 |
| T2DM | endothelial | *OTOGL* | -0.691203157 | 0.000111247 | 0.000404626 | 0.539770963 |
| T2DM | endothelial | *PACSIN3* | -0.455293927 | 0.000321521 | 0.000955851 | 1 |
| T2DM | endothelial | *SLC39A13* | -0.798623296 | 0.002697722 | 0.005634675 | 1 |
| T2DM | endothelial | *RAPSN* | -0.993932491 | 5.77E-09 | 1.40E-06 | 2.80E-05 |
| T2DM | endothelial | *TC2N* | -0.392306521 | 0.000283594 | 0.000865951 | 1 |
| T2DM | endothelial | *IFI27L1* | -0.877379894 | 0.000654931 | 0.001697504 | 1 |
| T2DM | endothelial | *PTER* | -0.754344729 | 1.25E-05 | 7.61E-05 | 0.060712504 |
| T2DM | endothelial | *C1QL3* | -0.352557592 | 0.013596921 | 0.02251613 | 1 |
| T2DM | endothelial | *LIPC* | -0.521858328 | 0.003559217 | 0.007118433 | 1 |
| T2DM | endothelial | *CEP57* | -0.486597674 | 0.011013061 | 0.018729539 | 1 |
| T2DM | endothelial | *JAM3* | 0.420067611 | 0.006646577 | 0.012114647 | 1 |
| T2DM | endothelial | *TRMT61A* | -0.812928211 | 9.76E-07 | 1.61E-05 | 0.004735369 |
| T2DM | endothelial | *LARP6* | -0.725282434 | 0.000416871 | 0.001171196 | 1 |
| T2DM | endothelial | *API5* | -0.576000604 | 0.00857856 | 0.01505359 | 1 |
| T2DM | endothelial | *ASPG* | -1.428900185 | 0.000242063 | 0.000761172 | 1 |
| T2DM | endothelial | *COX11* | -0.692162046 | 0.001455366 | 0.003318343 | 1 |
| T2DM | endothelial | *STXBP4* | -0.798519005 | 5.40E-06 | 4.25E-05 | 0.026188593 |
| T2DM | endothelial | *WBP1L* | -0.487808216 | 0.00813561 | 0.014364622 | 1 |
| T2DM | endothelial | *APBB1* | -0.764568255 | 0.006232888 | 0.011446621 | 1 |
| T2DM | endothelial | *WDR88* | -0.844459597 | 1.73E-06 | 2.24E-05 | 0.008395668 |
| T2DM | endothelial | *CYB5R2* | -0.374114 | 0.000436436 | 0.001212822 | 1 |
| T2DM | endothelial | *XRRA1* | -0.51314254 | 0.000859939 | 0.002134233 | 1 |
| T2DM | endothelial | *CENPN* | -1.45657725 | 1.86E-06 | 2.32E-05 | 0.009033572 |
| T2DM | endothelial | *TMEM41B* | -0.752700446 | 4.69E-05 | 0.00020733 | 0.227440983 |
| T2DM | endothelial | *TMX3* | -0.613645219 | 0.001425252 | 0.003261844 | 1 |
| T2DM | endothelial | *PRKCB* | -0.562131863 | 1.82E-05 | 9.98E-05 | 0.088292646 |
| T2DM | endothelial | *NDST2* | -3.214563001 | 4.21E-07 | 1.24E-05 | 0.002045003 |
| T2DM | endothelial | *CCDC68* | 0.147206889 | 2.27E-06 | 2.52E-05 | 0.011033338 |
| T2DM | endothelial | *CLEC4D* | -1.215771868 | 0.005914626 | 0.01092416 | 1 |
| T2DM | endothelial | *TMED3* | -0.312205808 | 1.32E-05 | 7.94E-05 | 0.063806077 |
| T2DM | endothelial | *SEC11C* | 0.535921237 | 0.003111317 | 0.006356256 | 1 |
| T2DM | endothelial | *CENPV* | -0.857899439 | 1.45E-06 | 2.04E-05 | 0.007011213 |
| T2DM | endothelial | *ZNF592* | -0.57013913 | 0.000897817 | 0.00221015 | 1 |
| T2DM | endothelial | *ACSM1* | -0.811568966 | 5.97E-05 | 0.000249068 | 0.289666593 |
| T2DM | endothelial | *SAAL1* | -1.13206976 | 7.68E-05 | 0.000305025 | 0.372740215 |
| T2DM | endothelial | *YPEL4* | -0.666577838 | 7.01E-05 | 0.000283999 | 0.340105036 |
| T2DM | endothelial | *FAM96A* | -0.803191309 | 1.85E-05 | 0.000100816 | 0.089928138 |
| T2DM | endothelial | *STAT6* | -0.683555614 | 0.000148272 | 0.000510225 | 0.719417044 |
| T2DM | endothelial | *PIP4K2C* | -0.970526143 | 2.80E-06 | 2.83E-05 | 0.013595404 |
| T2DM | endothelial | *MTMR10* | -0.463711213 | 0.011953188 | 0.0201588 | 1 |
| T2DM | endothelial | *NYAP1* | -0.265831445 | 0.03156639 | 0.046680928 | 1 |
| T2DM | endothelial | *MS4A7* | -0.3919007 | 0.00010599 | 0.0003893 | 0.514265898 |
| T2DM | endothelial | *MS4A14* | -0.211881305 | 0.001447938 | 0.003307625 | 1 |
| T2DM | endothelial | *DIS3L* | -0.507844595 | 0.001486926 | 0.003379187 | 1 |

| | | | | | | |
|---|---|---|---|---|---|---|
| T2DM | endothelial | *CCNDBP1* | -0.642178132 | 0.002932681 | 0.006052475 | 1 |
| T2DM | endothelial | *EPB42* | -0.448427387 | 0.020333838 | 0.031774486 | 1 |
| T2DM | endothelial | *SMAD3* | -0.517527362 | 0.00276796 | 0.005751667 | 1 |
| T2DM | endothelial | *MAP1A* | -0.66677328 | 9.95E-06 | 6.44E-05 | 0.048294052 |
| T2DM | endothelial | *RCCD1* | -1.112130678 | 5.58E-07 | 1.27E-05 | 0.002709326 |
| T2DM | endothelial | *MEI1* | -0.693786902 | 0.000889386 | 0.002193849 | 1 |
| T2DM | endothelial | *SAMD14* | -0.796403558 | 0.002346059 | 0.004983835 | 1 |
| T2DM | endothelial | *ACSF2* | -0.670216051 | 0.00013685 | 0.000479072 | 0.663993875 |
| T2DM | endothelial | *ENDOG* | -0.44907086 | 0.001461851 | 0.00333 | 1 |
| T2DM | endothelial | *ISLR2* | 0.5419342 | 0.000452213 | 0.001251647 | 1 |
| T2DM | endothelial | *SP2* | -0.792837994 | 0.003379447 | 0.006820747 | 1 |
| T2DM | endothelial | *COQ7* | -0.632842364 | 0.009704911 | 0.016769312 | 1 |
| T2DM | endothelial | *GPRC5B* | 0.359950992 | 5.45E-05 | 0.000232638 | 0.264277123 |
| T2DM | endothelial | *FBXO22* | -0.737505346 | 0.000210748 | 0.000677635 | 1 |
| T2DM | endothelial | *TBC1D2B* | -0.545167739 | 8.02E-06 | 5.58E-05 | 0.038898092 |
| T2DM | endothelial | *LOXHD1* | -0.622965672 | 0.000402395 | 0.001140433 | 1 |
| T2DM | endothelial | *KATNAL2* | -0.406753785 | 0.003403431 | 0.006866298 | 1 |
| T2DM | endothelial | *ZNF91* | -0.456080926 | 0.005778048 | 0.010729081 | 1 |
| T2DM | endothelial | *POP5* | -0.979951495 | 8.01E-06 | 5.58E-05 | 0.038874891 |
| T2DM | endothelial | *ENGASE* | -0.321614004 | 0.029212229 | 0.043625035 | 1 |
| T2DM | endothelial | *MYO5B* | -0.578218923 | 4.99E-07 | 1.27E-05 | 0.002421692 |
| T2DM | endothelial | *ART5* | -0.589977522 | 0.007993875 | 0.014159738 | 1 |
| T2DM | endothelial | *ZNF23* | -0.955213887 | 0.019745147 | 0.0309843 | 1 |
| T2DM | endothelial | *ZNF229* | 0.555642449 | 3.20E-05 | 0.000151835 | 0.155327041 |
| T2DM | endothelial | *SPATA33* | -0.986122945 | 0.000706323 | 0.001806127 | 1 |
| T2DM | endothelial | *PROCA1* | -0.467181098 | 0.005052904 | 0.009588067 | 1 |
| T2DM | endothelial | *ZNF641* | -0.792858203 | 2.58E-05 | 0.000129277 | 0.125269719 |
| T2DM | endothelial | *TP53I13* | -0.725068768 | 5.01E-05 | 0.000218963 | 0.243049474 |
| T2DM | endothelial | *TMC4* | -1.115257451 | 6.29E-07 | 1.32E-05 | 0.003053974 |
| T2DM | endothelial | *TTYH1* | -0.922162252 | 3.36E-07 | 1.24E-05 | 0.001631229 |
| T2DM | endothelial | *LENG8* | -0.678309817 | 0.000137531 | 0.000480762 | 0.667298286 |
| T2DM | endothelial | *CDC42EP5* | 0.301288942 | 0.005155825 | 0.009764272 | 1 |
| T2DM | endothelial | *FAM57A* | -0.530937056 | 0.002386731 | 0.005061371 | 1 |
| T2DM | endothelial | *KIFC2* | -1.162117915 | 2.52E-05 | 0.00012706 | 0.12212151 |
| T2DM | endothelial | *SERPINF2* | -0.404871454 | 0.001380747 | 0.003184118 | 1 |
| T2DM | endothelial | *TRPV3* | -0.32461921 | 0.028972607 | 0.043293837 | 1 |
| T2DM | endothelial | *CYB5D2* | -0.573937028 | 0.006385733 | 0.011687506 | 1 |
| T2DM | endothelial | *KLK5* | -1.153156185 | 1.99E-06 | 2.38E-05 | 0.009653218 |
| T2DM | endothelial | *KLK6* | -9.872111628 | 1.95E-05 | 0.000105235 | 0.094711461 |
| T2DM | endothelial | *ZNF83* | -0.430069131 | 0.013514722 | 0.022395297 | 1 |
| T2DM | endothelial | *SPRYD3* | -1.027352441 | 0.003659799 | 0.007292544 | 1 |
| T2DM | endothelial | *SOAT2* | -0.638963251 | 5.19E-05 | 0.000224211 | 0.252012959 |
| T2DM | endothelial | *ZNF558* | -0.547399913 | 0.001285636 | 0.003000974 | 1 |
| T2DM | endothelial | *NDUFV1* | -0.533438303 | 0.015995177 | 0.025860913 | 1 |
| T2DM | endothelial | *PRDX2* | -0.742123923 | 0.003927538 | 0.00774966 | 1 |
| T2DM | endothelial | *ZNF232* | -0.188346385 | 0.032659542 | 0.048092291 | 1 |
| T2DM | endothelial | *MIS12* | -0.882660097 | 0.000251735 | 0.000786995 | 1 |
| T2DM | endothelial | *MGAT5B* | 0.563850393 | 7.68E-06 | 5.45E-05 | 0.037248864 |
| T2DM | endothelial | *GSDMA* | -0.355719659 | 0.00471728 | 0.009049408 | 1 |
| T2DM | endothelial | *MLST8* | -1.159785212 | 1.72E-05 | 9.56E-05 | 0.083442153 |

| | | | | | | |
|---|---|---|---|---|---|---|
| T2DM | endothelial | *ECI1* | -0.519933572 | 0.003356397 | 0.006788344 | 1 |
| T2DM | endothelial | *LTBP3* | -0.447236168 | 0.025409123 | 0.038538626 | 1 |
| T2DM | endothelial | *NAALADL1* | -0.414608951 | 0.023252935 | 0.035692262 | 1 |
| T2DM | endothelial | *SCARA5* | -0.492738911 | 0.000123871 | 0.000441928 | 0.601022622 |
| T2DM | endothelial | *KIAA1586* | -0.707393289 | 0.000447032 | 0.001239427 | 1 |
| T2DM | endothelial | *FAM83B* | -4.434535009 | 5.76E-07 | 1.27E-05 | 0.002796286 |
| T2DM | endothelial | *COA6* | -0.928614582 | 0.012842305 | 0.021464301 | 1 |
| T2DM | endothelial | *PDHB* | -0.824991667 | 0.001779032 | 0.003946898 | 1 |
| T2DM | endothelial | *PXK* | -0.568636487 | 0.001608122 | 0.003617343 | 1 |
| T2DM | endothelial | *ACOX2* | -0.858208593 | 5.11E-06 | 4.15E-05 | 0.024804629 |
| T2DM | endothelial | *FAM107A* | 0.211473181 | 5.85E-06 | 4.54E-05 | 0.028396522 |
| T2DM | endothelial | *SCN11A* | 0.26989836 | 1.84E-05 | 0.000100319 | 0.089082845 |
| T2DM | endothelial | *SEPT2* | -0.258483058 | 0.014448967 | 0.023692595 | 1 |
| T2DM | endothelial | *DTYMK* | -1.057816972 | 0.001713609 | 0.00382977 | 1 |
| T2DM | endothelial | *ING5* | -0.645558671 | 4.87E-05 | 0.000213809 | 0.236258493 |
| T2DM | endothelial | *MLKL* | 0.335903494 | 0.000627367 | 0.001637335 | 1 |
| T2DM | endothelial | *RFWD3* | -0.711015632 | 0.000929947 | 0.002277689 | 1 |
| T2DM | endothelial | *KLHL30* | -1.579269067 | 5.01E-07 | 1.27E-05 | 0.002432861 |
| T2DM | endothelial | *CCDC110* | -1.440033778 | 3.22E-07 | 1.24E-05 | 0.001562583 |
| T2DM | endothelial | *SLC20A2* | -0.658653686 | 2.45E-05 | 0.000125492 | 0.119091655 |
| T2DM | endothelial | *TMUB2* | -0.813498817 | 0.00017814 | 0.000589989 | 0.864334134 |
| T2DM | endothelial | *ZSWIM1* | 0.170457423 | 0.023889598 | 0.036530832 | 1 |
| T2DM | endothelial | *NBPF9* | -0.921976526 | 3.36E-07 | 1.24E-05 | 0.001631199 |
| T2DM | endothelial | *NDUFS5* | -0.651431103 | 0.002356976 | 0.005004834 | 1 |
| T2DM | endothelial | *ZNF30* | 0.370222293 | 0.005690641 | 0.010595161 | 1 |
| T2DM | endothelial | *SLC16A4* | -0.382121873 | 0.003310539 | 0.006701676 | 1 |
| T2DM | endothelial | *GSTM4* | -1.061283344 | 4.20E-07 | 1.24E-05 | 0.002038145 |
| T2DM | endothelial | *TCTN2* | -0.157531908 | 0.027556317 | 0.041355784 | 1 |
| T2DM | endothelial | *PPIP5K1* | -0.872398288 | 1.05E-05 | 6.69E-05 | 0.051014342 |
| T2DM | endothelial | *SNTB2* | -0.491463351 | 0.006894307 | 0.012508306 | 1 |
| T2DM | endothelial | *ZNF507* | -0.755133263 | 0.000410381 | 0.001156984 | 1 |
| T2DM | endothelial | *STX18* | -0.551757671 | 0.005657397 | 0.010557972 | 1 |
| T2DM | endothelial | *ZBTB49* | -1.063185748 | 0.000247084 | 0.000774952 | 1 |
| T2DM | endothelial | *ATOH8* | 0.383549662 | 0.000416666 | 0.001171196 | 1 |
| T2DM | endothelial | *BTNL3* | 0.318419959 | 1.48E-05 | 8.59E-05 | 0.071592052 |
| T2DM | endothelial | *TMEM129* | -0.950844351 | 4.03E-06 | 3.56E-05 | 0.019575115 |
| T2DM | endothelial | *CEP120* | -0.595555719 | 0.007090265 | 0.01281403 | 1 |
| T2DM | endothelial | *MFF* | -0.542034605 | 0.003562307 | 0.007121678 | 1 |
| T2DM | endothelial | *CPLX1* | 0.224292689 | 6.43E-06 | 4.83E-05 | 0.031211358 |
| T2DM | endothelial | *SIGLEC7* | -0.411122286 | 0.003133092 | 0.006392838 | 1 |
| T2DM | endothelial | *MAP2K1* | -0.525607511 | 0.009151911 | 0.015932928 | 1 |
| T2DM | endothelial | *KLK7* | -1.187553583 | 4.52E-06 | 3.85E-05 | 0.021945476 |
| T2DM | endothelial | *IRS1* | -0.467571443 | 8.90E-13 | 1.40E-09 | 4.32E-09 |
| T2DM | endothelial | *HSPBAP1* | -0.478649755 | 0.005665581 | 0.01056472 | 1 |
| T2DM | endothelial | *AFAP1L2* | 0.161430856 | 1.09E-05 | 6.86E-05 | 0.052647435 |
| T2DM | endothelial | *UBE2V2* | -0.668805378 | 0.000933404 | 0.0022827 | 1 |
| T2DM | endothelial | *PCSK9* | -1.901334674 | 9.14E-05 | 0.000346491 | 0.443273319 |
| T2DM | endothelial | *XPO6* | -0.6644625 | 0.000868649 | 0.00215248 | 1 |
| T2DM | endothelial | *RAB24* | -1.918461861 | 2.26E-05 | 0.000117761 | 0.109870586 |
| T2DM | endothelial | *THBS3* | -0.831538085 | 2.48E-05 | 0.000125956 | 0.120568406 |

| | | | | | | |
|---|---|---|---|---|---|---|
| T2DM | endothelial | *NPIPB3* | -1.051364805 | 0.000138887 | 0.000484662 | 0.673879418 |
| T2DM | endothelial | *SH3TC2* | 0.29555008 | 2.80E-05 | 0.000137195 | 0.136097755 |
| T2DM | endothelial | *P2RY12* | -0.652341308 | 4.62E-07 | 1.27E-05 | 0.002242675 |
| T2DM | endothelial | *SNUPN* | -0.858284541 | 7.65E-07 | 1.46E-05 | 0.003710292 |
| T2DM | endothelial | *RNASE6* | -0.558927621 | 0.001592673 | 0.003587581 | 1 |
| T2DM | endothelial | *CD52* | -0.33317674 | 0.000336118 | 0.000987195 | 1 |
| T2DM | endothelial | *SLC38A11* | -0.589741454 | 0.000130174 | 0.000459747 | 0.631604042 |
| T2DM | endothelial | *CCDC8* | -0.835124622 | 2.27E-06 | 2.52E-05 | 0.011001505 |
| T2DM | endothelial | *ZEB2* | -0.470117402 | 5.63E-07 | 1.27E-05 | 0.002731608 |
| T2DM | endothelial | *DTWD2* | -0.663712146 | 1.78E-06 | 2.27E-05 | 0.008637674 |
| T2DM | endothelial | *INO80E* | -0.63604347 | 2.14E-05 | 0.000113102 | 0.103974071 |
| T2DM | endothelial | *DFFB* | -0.902383451 | 0.002413007 | 0.005101486 | 1 |
| T2DM | endothelial | *NFU1* | -0.757503453 | 3.49E-05 | 0.000162863 | 0.169540204 |
| T2DM | endothelial | *C15orf40* | -0.427130285 | 0.01298541 | 0.021681077 | 1 |
| T2DM | endothelial | *PROKR1* | 0.759728766 | 9.14E-05 | 0.000346491 | 0.443508651 |
| T2DM | endothelial | *LRRC45* | -0.540296996 | 0.000453803 | 0.001253503 | 1 |
| T2DM | endothelial | *UGP2* | -0.570976362 | 1.38E-06 | 1.96E-05 | 0.006718321 |
| T2DM | endothelial | *CSGALNACT2* | -0.545837187 | 0.012526257 | 0.021037521 | 1 |
| T2DM | endothelial | *CTNND2* | -0.640597698 | 6.17E-05 | 0.000255778 | 0.299260701 |
| T2DM | endothelial | *CALML6* | -5.562012954 | 5.22E-07 | 1.27E-05 | 0.002533536 |
| T2DM | endothelial | *MUC3A* | 0.324887796 | 0.031912073 | 0.047120322 | 1 |
| T2DM | endothelial | *TOR1AIP2* | -0.615764356 | 0.000387799 | 0.001105524 | 1 |
| T2DM | endothelial | *OTUD3* | -0.511715423 | 0.004143094 | 0.00809597 | 1 |
| T2DM | endothelial | *ZFPM2* | -0.408933982 | 7.95E-05 | 0.000312522 | 0.385652677 |
| T2DM | endothelial | *MAP3K2* | -0.459800886 | 0.003589688 | 0.007170508 | 1 |
| T2DM | endothelial | *MYO7B* | 0.27510143 | 3.31E-06 | 3.11E-05 | 0.016066195 |
| T2DM | endothelial | *NIPA1* | -0.66678416 | 0.00941116 | 0.016314022 | 1 |
| T2DM | endothelial | *HNRNPA3* | -0.442466144 | 0.013940066 | 0.022951205 | 1 |
| T2DM | endothelial | *RNF150* | -0.418077689 | 4.08E-05 | 0.000184975 | 0.197967496 |
| T2DM | endothelial | *HOXD4* | 0.97748469 | 6.24E-07 | 1.31E-05 | 0.003026775 |
| T2DM | endothelial | *USP38* | -0.748052753 | 0.001787607 | 0.003962297 | 1 |
| T2DM | endothelial | *ANKK1* | -1.048525204 | 9.77E-07 | 1.61E-05 | 0.004740817 |
| T2DM | endothelial | *PDCD6IP* | -0.40100453 | 0.021764739 | 0.033728046 | 1 |
| T2DM | endothelial | *CRTAP* | -0.628738569 | 0.001350058 | 0.003123739 | 1 |
| T2DM | endothelial | *CNGB3* | -0.592117642 | 0.000325987 | 0.000965031 | 1 |
| T2DM | endothelial | *ELP5* | -0.495551928 | 0.030913764 | 0.045869598 | 1 |
| T2DM | endothelial | *STX8* | -0.399627456 | 0.015593882 | 0.025296394 | 1 |
| T2DM | endothelial | *NFRKB* | -0.859798238 | 9.00E-06 | 6.04E-05 | 0.043644195 |
| T2DM | endothelial | *SEMA3E* | 0.19752354 | 8.01E-07 | 1.49E-05 | 0.00388436 |
| T2DM | endothelial | *ADORA2B* | -0.250319017 | 0.005704015 | 0.01060785 | 1 |
| T2DM | endothelial | *MGMT* | -0.383238594 | 0.022963998 | 0.035293417 | 1 |
| T2DM | endothelial | *METTL7B* | -0.683652104 | 0.016742005 | 0.026898082 | 1 |
| T2DM | endothelial | *DNAJC18* | 0.269168318 | 2.25E-05 | 0.000117176 | 0.109090765 |
| T2DM | endothelial | *HSD17B13* | 0.558874817 | 6.37E-06 | 4.81E-05 | 0.030924715 |
| T2DM | endothelial | *ELOVL6* | -0.748462532 | 4.33E-06 | 3.73E-05 | 0.021013097 |
| T2DM | endothelial | *ARL6IP1* | 0.296000549 | 0.009437321 | 0.016353529 | 1 |
| T2DM | endothelial | *SERPINB9* | -0.744377612 | 6.57E-05 | 0.000269755 | 0.318849917 |
| T2DM | endothelial | *SMAGP* | 0.197989632 | 0.001718671 | 0.003836513 | 1 |
| T2DM | endothelial | *CDH2* | 0.681920351 | 0.002700076 | 0.005637164 | 1 |
| T2DM | endothelial | *EMB* | -0.629784911 | 1.97E-09 | 6.79E-07 | 9.56E-06 |

| | | | | | | |
|---|---|---|---|---|---|---|
| T2DM | endothelial | *STAT2* | -0.638747003 | 0.000462963 | 0.00127341 | 1 |
| T2DM | endothelial | *GTSF1* | 0.854965122 | 0.031349178 | 0.046420116 | 1 |
| T2DM | endothelial | *ARMC10* | -0.548740006 | 0.00399218 | 0.007846777 | 1 |
| T2DM | endothelial | *ACYP2* | -0.503849109 | 0.000104082 | 0.00038316 | 0.505005358 |
| T2DM | endothelial | *TRABD* | -0.557860301 | 0.019093775 | 0.030157225 | 1 |
| T2DM | endothelial | *TMEM133* | -0.539787608 | 0.004251237 | 0.008275361 | 1 |
| T2DM | endothelial | *HOXB9* | -1.190787966 | 4.47E-07 | 1.25E-05 | 0.002170288 |
| T2DM | endothelial | *TTLL6* | -2.809564902 | 2.53E-10 | 1.76E-07 | 1.23E-06 |
| T2DM | endothelial | *FOXN2* | -0.616205347 | 0.000669812 | 0.001727767 | 1 |
| T2DM | endothelial | *TRIAP1* | 1.154671148 | 0.000138946 | 0.000484662 | 0.674164569 |
| T2DM | endothelial | *TSEN34* | -1.091872091 | 0.002132359 | 0.004602404 | 1 |
| T2DM | endothelial | *ZNF160* | -0.410757402 | 0.005272662 | 0.009954457 | 1 |
| T2DM | endothelial | *ZNF415* | -0.838599515 | 0.001725057 | 0.003842963 | 1 |
| T2DM | endothelial | *S1PR1* | 0.177329134 | 2.63E-06 | 2.76E-05 | 0.01276926 |
| T2DM | endothelial | *PYGO1* | -0.622964168 | 4.77E-07 | 1.27E-05 | 0.00231551 |
| T2DM | endothelial | *FPR1* | -0.810945984 | 2.10E-06 | 2.46E-05 | 0.010198876 |
| T2DM | endothelial | *SOX7* | 0.178714709 | 2.06E-06 | 2.43E-05 | 0.009991314 |
| T2DM | endothelial | *TRMT61B* | -0.522969766 | 0.001346066 | 0.003115988 | 1 |
| T2DM | endothelial | *MFN1* | -0.373923214 | 0.033102081 | 0.048567069 | 1 |
| T2DM | endothelial | *GIMAP8* | 0.184806537 | 1.83E-05 | 0.000100166 | 0.088774927 |
| T2DM | endothelial | *ATP6V0E2* | -0.564979736 | 0.002073009 | 0.004496308 | 1 |
| T2DM | endothelial | *SOSTDC1* | -0.543927427 | 0.006078059 | 0.011191933 | 1 |
| T2DM | endothelial | *PGAM1* | -0.380664635 | 0.012820142 | 0.021434642 | 1 |
| T2DM | endothelial | *ZNF561* | -0.605743604 | 5.34E-05 | 0.000229538 | 0.259048925 |
| T2DM | endothelial | *HOPX* | -0.693959378 | 0.003612674 | 0.007207523 | 1 |
| T2DM | endothelial | *PPID* | -0.748500533 | 0.004176178 | 0.008147494 | 1 |
| T2DM | endothelial | *ETFDH* | -0.212301588 | 0.003600165 | 0.007188477 | 1 |
| T2DM | endothelial | *TBCA* | -0.521634289 | 0.00327749 | 0.006645375 | 1 |
| T2DM | endothelial | *NMUR1* | -1.11549337 | 5.37E-07 | 1.27E-05 | 0.0026035 |
| T2DM | endothelial | *PIK3CD* | -0.61281277 | 1.55E-05 | 8.91E-05 | 0.075097507 |
| T2DM | endothelial | *RGS19* | -0.464823701 | 2.48E-06 | 2.65E-05 | 0.012015216 |
| T2DM | endothelial | *ANO5* | -0.861184202 | 1.00E-05 | 6.48E-05 | 0.048729895 |
| T2DM | endothelial | *VAT1L* | 0.241117641 | 6.53E-06 | 4.87E-05 | 0.031704817 |
| T2DM | endothelial | *LGALS4* | -0.740346188 | 1.97E-05 | 0.000105937 | 0.095696352 |
| T2DM | endothelial | *LRRC34* | -0.212878027 | 0.013764995 | 0.022732388 | 1 |
| T2DM | endothelial | *SPATA5L1* | -0.508897097 | 5.87E-05 | 0.000246622 | 0.284601345 |
| T2DM | endothelial | *EXOSC10* | -0.854261982 | 0.000118373 | 0.000425443 | 0.574347577 |
| T2DM | endothelial | *TRAPPC12* | -0.637416419 | 0.000362459 | 0.001048648 | 1 |
| T2DM | endothelial | *C3AR1* | -0.448987651 | 7.05E-05 | 0.000285028 | 0.342033526 |
| T2DM | endothelial | *PRNP* | -0.609567169 | 0.000350407 | 0.00101929 | 1 |
| T2DM | endothelial | *TVP23B* | -0.446872444 | 0.020087703 | 0.031430356 | 1 |
| T2DM | endothelial | *FBXW10* | -1.323315641 | 1.85E-07 | 1.19E-05 | 0.000896005 |
| T2DM | endothelial | *SRGAP2C* | -0.470069362 | 0.000445705 | 0.001236457 | 1 |
| T2DM | endothelial | *ZNF554* | -1.394771136 | 3.07E-06 | 2.99E-05 | 0.014883878 |
| T2DM | endothelial | *ANKRD20A4* | -1.966081243 | 0.030956879 | 0.045919529 | 1 |
| T2DM | endothelial | *SERF1A* | 3.304955107 | 0.000182096 | 0.000600632 | 0.883530326 |
| T2DM | endothelial | *EIF2AK3* | -0.684623699 | 0.001315358 | 0.003059499 | 1 |
| T2DM | endothelial | *MTBP* | -0.8504783 | 0.000164354 | 0.000551102 | 0.797444829 |
| T2DM | endothelial | *TEFM* | -0.494846783 | 0.007569129 | 0.013546815 | 1 |
| T2DM | endothelial | *MBOAT1* | -0.732051498 | 9.88E-05 | 0.00036768 | 0.479454338 |

| | | | | | | |
|---|---|---|---|---|---|---|
| T2DM | endothelial | *CXCR6* | -0.958093942 | 3.32E-06 | 3.11E-05 | 0.01612054 |
| T2DM | endothelial | *PAIP1* | -0.625627872 | 0.009143992 | 0.015924856 | 1 |
| T2DM | endothelial | *C1QTNF4* | -0.464311855 | 1.47E-07 | 1.08E-05 | 0.000714429 |
| T2DM | endothelial | *MACROD2* | -0.418990433 | 4.39E-05 | 0.000196918 | 0.21286836 |
| T2DM | endothelial | *BSG* | -0.590813739 | 0.004929292 | 0.009393923 | 1 |
| T2DM | endothelial | *HINFP* | -0.649686361 | 0.013400694 | 0.022244326 | 1 |
| T2DM | endothelial | *SUCLG2* | -0.487245056 | 0.003107802 | 0.00635175 | 1 |
| T2DM | endothelial | *MANEA* | -0.787950108 | 0.000885874 | 0.00218741 | 1 |
| T2DM | endothelial | *FIBP* | 0.84974668 | 0.000499001 | 0.00135798 | 1 |
| T2DM | endothelial | *CTSW* | -0.165614677 | 4.76E-07 | 1.27E-05 | 0.002311077 |
| T2DM | endothelial | *SNTG2* | 0.187235919 | 0.000745032 | 0.001887673 | 1 |
| T2DM | endothelial | *SMPDL3A* | -0.470632381 | 0.000202261 | 0.000654684 | 0.981370981 |
| T2DM | endothelial | *RAD9A* | -0.32305244 | 0.010543895 | 0.018051863 | 1 |
| T2DM | endothelial | *EFEMP2* | -0.617102375 | 0.010483133 | 0.017973201 | 1 |
| T2DM | endothelial | *ZFAND4* | -0.721576098 | 0.000329188 | 0.000970955 | 1 |
| T2DM | endothelial | *ZNF738* | 0.319216107 | 0.007690004 | 0.013717907 | 1 |
| T2DM | endothelial | *FAM71D* | -0.571710536 | 0.006042637 | 0.011130931 | 1 |
| T2DM | endothelial | *FUT10* | -0.24693329 | 0.028400819 | 0.042517981 | 1 |
| T2DM | endothelial | *ZNF596* | -0.877701174 | 0.000104835 | 0.000385641 | 0.508659914 |
| T2DM | endothelial | *FAM192A* | -0.61932142 | 0.00032768 | 0.000968247 | 1 |
| T2DM | endothelial | *CBWD1* | -1.065334549 | 2.68E-05 | 0.000132993 | 0.129869004 |
| T2DM | endothelial | *NADSYN1* | -0.553368166 | 0.00691015 | 0.012529166 | 1 |
| T2DM | endothelial | *RNASEH2C* | -0.74843085 | 9.72E-06 | 6.33E-05 | 0.047179821 |
| T2DM | endothelial | *MYEOV* | -1.918379986 | 9.06E-06 | 6.07E-05 | 0.04397391 |
| T2DM | endothelial | *MRGPRF* | -0.72493355 | 3.85E-08 | 5.71E-06 | 0.000186898 |
| T2DM | endothelial | *GXYLT2* | -0.555499089 | 1.17E-06 | 1.79E-05 | 0.005698884 |
| T2DM | endothelial | *DCAKD* | -0.733494442 | 1.57E-06 | 2.15E-05 | 0.007615289 |
| T2DM | endothelial | *NOC3L* | -0.84985902 | 1.99E-05 | 0.000106532 | 0.096624358 |
| T2DM | endothelial | *PARP15* | -0.310342855 | 0.001182422 | 0.002794503 | 1 |
| T2DM | endothelial | *AHSA2* | -0.617058794 | 2.15E-05 | 0.000113102 | 0.104279973 |
| T2DM | endothelial | *IQCB1* | -0.54681885 | 0.001236441 | 0.002904904 | 1 |
| T2DM | endothelial | *ZNF483* | -0.502817688 | 0.000158885 | 0.000536844 | 0.770908025 |
| T2DM | endothelial | *SLC2A14* | -0.474507598 | 3.05E-05 | 0.000146198 | 0.147952872 |
| T2DM | endothelial | *MMRN2* | 0.188515314 | 5.29E-06 | 4.22E-05 | 0.025678308 |
| T2DM | endothelial | *MAP3K11* | 0.372227184 | 0.004593347 | 0.008847526 | 1 |
| T2DM | endothelial | *KCNK7* | -2.695694872 | 0.010152842 | 0.017468649 | 1 |
| T2DM | endothelial | *NDNF* | 0.216725072 | 5.13E-06 | 4.15E-05 | 0.024881206 |
| T2DM | endothelial | *ARV1* | -0.60731654 | 0.002536847 | 0.005342353 | 1 |
| T2DM | endothelial | *CCDC36* | -0.317128665 | 0.021868626 | 0.033824219 | 1 |
| T2DM | endothelial | *EHBP1L1* | 0.413652794 | 0.005657611 | 0.010557972 | 1 |
| T2DM | endothelial | *PPP1R14B* | -1.287841311 | 0.000148502 | 0.000510654 | 0.720532367 |
| T2DM | endothelial | *ZNF417* | -0.69511683 | 0.003310782 | 0.006701676 | 1 |
| T2DM | endothelial | *TNFRSF10D* | 0.335003688 | 1.97E-05 | 0.000105937 | 0.095703786 |
| T2DM | endothelial | *MST1* | -0.270613529 | 0.000988231 | 0.002391471 | 1 |
| T2DM | endothelial | *GMPPB* | -1.20429188 | 5.81E-06 | 4.53E-05 | 0.028210747 |
| T2DM | endothelial | *SULT1B1* | 0.49627453 | 3.15E-06 | 3.03E-05 | 0.015289059 |
| T2DM | endothelial | *NUDT4* | -0.745304757 | 0.00033315 | 0.000980257 | 1 |
| T2DM | endothelial | *SCAI* | -0.410099491 | 0.006282444 | 0.011520187 | 1 |
| T2DM | endothelial | *HSPB7* | -0.33486462 | 0.012096125 | 0.020385689 | 1 |
| T2DM | endothelial | *UQCRH* | -0.688447884 | 0.000206313 | 0.000665578 | 1 |

| | | | | | | |
|---|---|---|---|---|---|---|
| T2DM | endothelial | *C11orf80* | -0.421686636 | 0.024056598 | 0.036774611 | 1 |
| T2DM | endothelial | *CD7* | -1.100643198 | 1.68E-05 | 9.39E-05 | 0.081614123 |
| T2DM | endothelial | *RNF213* | -0.418202828 | 0.008015121 | 0.014188022 | 1 |
| T2DM | endothelial | *GPR160* | 0.202243506 | 7.05E-06 | 5.11E-05 | 0.034186975 |
| T2DM | endothelial | *XXYLT1* | -0.721411616 | 0.000760038 | 0.001918682 | 1 |
| T2DM | endothelial | *UBXN2A* | -0.598733596 | 0.000233904 | 0.000739627 | 1 |
| T2DM | endothelial | *CTSF* | -0.536889567 | 0.002590075 | 0.005440279 | 1 |
| T2DM | endothelial | *LEMD3* | -0.499346091 | 0.018338038 | 0.029105712 | 1 |
| T2DM | endothelial | *GLIS1* | -0.691747394 | 3.59E-07 | 1.24E-05 | 0.001741399 |
| T2DM | endothelial | *MSL2* | -0.674359124 | 0.000619812 | 0.001622075 | 1 |
| T2DM | endothelial | *ZNF497* | -1.989623061 | 1.58E-06 | 2.16E-05 | 0.007656414 |
| T2DM | endothelial | *ANGEL2* | -0.52013664 | 0.013224151 | 0.022026971 | 1 |
| T2DM | endothelial | *ZNF266* | -0.427697467 | 4.27E-05 | 0.00019267 | 0.207312503 |
| T2DM | endothelial | *LARP7* | -0.761617144 | 6.17E-05 | 0.000255819 | 0.299564239 |
| T2DM | endothelial | *C4orf32* | 0.240257897 | 5.95E-05 | 0.000248357 | 0.288591113 |
| T2DM | endothelial | *PCP2* | -0.707914002 | 0.000275258 | 0.000846357 | 1 |
| T2DM | endothelial | *NLRP6* | -3.126351629 | 0.000205912 | 0.00066517 | 0.999084673 |
| T2DM | endothelial | *RSRC1* | -0.468228922 | 0.003992929 | 0.007846777 | 1 |
| T2DM | endothelial | *PTDSS2* | -0.735237029 | 2.22E-05 | 0.000115758 | 0.107539023 |
| T2DM | endothelial | *DHX36* | -0.548001902 | 0.002245139 | 0.004798861 | 1 |
| T2DM | endothelial | *SLC22A1* | -1.191332378 | 2.65E-06 | 2.76E-05 | 0.012875386 |
| T2DM | endothelial | *CHST2* | 1.004039498 | 0.000801608 | 0.002006915 | 1 |
| T2DM | endothelial | *TVP23C* | 0.855454964 | 0.017929495 | 0.028560049 | 1 |
| T2DM | endothelial | *PACS1* | -0.489378862 | 0.001414864 | 0.003245825 | 1 |
| T2DM | endothelial | *SH3BP5L* | -1.33030257 | 1.20E-05 | 7.41E-05 | 0.058327482 |
| T2DM | endothelial | *CADM2* | 0.10392213 | 3.97E-06 | 3.51E-05 | 0.019270607 |
| T2DM | endothelial | *PSMD2* | -0.63626347 | 0.016824607 | 0.027012903 | 1 |
| T2DM | endothelial | *FAM182B* | -0.41327419 | 0.000289152 | 0.000878501 | 1 |
| T2DM | endothelial | *ARHGAP1* | -0.701483553 | 1.33E-05 | 7.97E-05 | 0.064369536 |
| T2DM | endothelial | *ATG13* | -0.814936353 | 1.66E-05 | 9.28E-05 | 0.080315048 |
| T2DM | endothelial | *CCNE2* | -1.154439127 | 5.76E-12 | 5.59E-09 | 2.79E-08 |
| T2DM | endothelial | *ZNF519* | -0.615613086 | 4.78E-05 | 0.000210709 | 0.23212576 |
| T2DM | endothelial | *BANF1* | 0.850678047 | 0.005247484 | 0.00991141 | 1 |
| T2DM | endothelial | *CHRNA7* | 0.60988616 | 1.10E-06 | 1.71E-05 | 0.005333505 |
| T2DM | endothelial | *SCUBE2* | 0.340255947 | 2.14E-05 | 0.000112933 | 0.103657663 |
| T2DM | endothelial | *EIF1AD* | -1.25849882 | 3.65E-06 | 3.33E-05 | 0.017698594 |
| T2DM | endothelial | *ZNF25* | -0.674363377 | 0.000950571 | 0.002317674 | 1 |
| T2DM | endothelial | *CLTB* | -0.88851881 | 1.71E-05 | 9.53E-05 | 0.08289523 |
| T2DM | endothelial | *PPP2R2D* | -0.400100193 | 0.022144738 | 0.034185895 | 1 |
| T2DM | endothelial | *MCTP1* | 0.198505972 | 4.08E-05 | 0.000184975 | 0.198107786 |
| T2DM | endothelial | *LRRC25* | -0.389747507 | 0.009387783 | 0.016290959 | 1 |
| T2DM | endothelial | *UCP2* | -0.67186007 | 9.69E-07 | 1.61E-05 | 0.004702823 |
| T2DM | endothelial | *MRPL48* | -0.818419037 | 0.00025767 | 0.000801933 | 1 |
| T2DM | endothelial | *TMEM70* | -0.70941308 | 0.011848877 | 0.020010703 | 1 |
| T2DM | endothelial | *TOM1L2* | -0.650509926 | 2.44E-05 | 0.000125288 | 0.118594964 |
| T2DM | endothelial | *TEX26* | -0.95608875 | 0.000144316 | 0.0004998 | 0.700219566 |
| T2DM | endothelial | *GPR156* | -0.581064787 | 1.73E-05 | 9.58E-05 | 0.08380128 |
| T2DM | endothelial | *TTLL11* | -0.583297337 | 0.006193125 | 0.011382214 | 1 |
| T2DM | endothelial | *PRIMA1* | -0.762445459 | 3.04E-07 | 1.24E-05 | 0.001474995 |
| T2DM | endothelial | *HOXD8* | 0.209747991 | 3.12E-06 | 3.02E-05 | 0.015119039 |

| | | | | | | |
|---|---|---|---|---|---|---|
| T2DM | endothelial | *ZDHHC21* | -0.362618941 | 0.033448151 | 0.049030342 | 1 |
| T2DM | endothelial | *A2M* | 0.255922105 | 0.000332579 | 0.000979171 | 1 |
| T2DM | endothelial | *DOK7* | 2.189780623 | 8.58E-05 | 0.000329425 | 0.416063303 |
| T2DM | endothelial | *ORAI3* | -0.399202337 | 0.018957963 | 0.029971989 | 1 |
| T2DM | endothelial | *B3GALT6* | 1.26197599 | 0.004625837 | 0.008906572 | 1 |
| T2DM | endothelial | *JAKMIP2* | -0.618152553 | 0.000113122 | 0.000410214 | 0.548866999 |
| T2DM | endothelial | *TCP11L1* | -0.367646053 | 0.021666729 | 0.033597625 | 1 |
| T2DM | endothelial | *CCDC57* | -0.491242327 | 0.000221927 | 0.000708415 | 1 |
| T2DM | endothelial | *MYPOP* | -0.718897692 | 0.015146834 | 0.024686745 | 1 |
| T2DM | endothelial | *CIDEA* | -0.637024703 | 5.03E-06 | 4.09E-05 | 0.024387345 |
| T2DM | endothelial | *SLC35G1* | -0.939316987 | 0.001236919 | 0.002904904 | 1 |
| T2DM | endothelial | *ZNF135* | -0.498157327 | 0.000165996 | 0.000555157 | 0.805411065 |
| T2DM | endothelial | *OR4N2* | -1.021080914 | 3.72E-06 | 3.34E-05 | 0.018028322 |
| T2DM | endothelial | *TAC4* | -0.975689441 | 3.59E-05 | 0.00016699 | 0.174337177 |
| T2DM | endothelial | *CDC26* | -0.495929009 | 0.000537582 | 0.001443467 | 1 |
| T2DM | endothelial | *CRLF3* | -0.44605044 | 0.002010789 | 0.004380938 | 1 |
| T2DM | endothelial | *DNAJC30* | -0.963509162 | 0.00028039 | 0.000857789 | 1 |
| T2DM | endothelial | *SPRYD4* | -0.664603133 | 2.64E-05 | 0.000131688 | 0.128264323 |
| T2DM | endothelial | *WDR25* | -0.72316941 | 0.002010071 | 0.004380938 | 1 |
| T2DM | endothelial | *PLA2G16* | -0.382035653 | 0.000627334 | 0.001637335 | 1 |
| T2DM | endothelial | *B3GNT5* | -0.762222657 | 0.000857191 | 0.002128502 | 1 |
| T2DM | endothelial | *RNF152* | 0.288397327 | 0.00644016 | 0.011769363 | 1 |
| T2DM | endothelial | *BDNF* | -0.870412583 | 1.38E-06 | 1.96E-05 | 0.006702549 |
| T2DM | endothelial | *BOK* | -0.88882086 | 1.56E-05 | 8.92E-05 | 0.075488475 |
| T2DM | endothelial | *RUFY1* | -0.556513464 | 0.00016629 | 0.000555293 | 0.806841337 |
| T2DM | endothelial | *ANKLE2* | -0.603257257 | 0.005247818 | 0.00991141 | 1 |
| T2DM | endothelial | *C8G* | -13.27077429 | 3.12E-07 | 1.24E-05 | 0.001515802 |
| T2DM | endothelial | *MUC20* | -0.893545722 | 1.07E-05 | 6.78E-05 | 0.051927419 |
| T2DM | endothelial | *SHMT1* | -0.697963195 | 2.82E-05 | 0.000137607 | 0.136644191 |
| T2DM | endothelial | *TMEM80* | -0.680185998 | 5.94E-05 | 0.000248341 | 0.288075635 |
| T2DM | endothelial | *WDR73* | -0.586317772 | 0.002449446 | 0.005166272 | 1 |
| T2DM | endothelial | *DSCAML1* | -0.669488553 | 2.67E-06 | 2.76E-05 | 0.012945434 |
| T2DM | endothelial | *TALDO1* | -0.830243454 | 0.000268267 | 0.000828538 | 1 |
| T2DM | endothelial | *AP006621.1* | -0.817011718 | 1.16E-05 | 7.26E-05 | 0.056404547 |
| T2DM | endothelial | *MAN1B1* | -0.978875706 | 2.76E-05 | 0.000136232 | 0.13394791 |
| T2DM | endothelial | *FBXO39* | 0.33042331 | 0.002378608 | 0.005046352 | 1 |
| T2DM | endothelial | *NAP1L5* | -2.157822178 | 1.58E-05 | 8.97E-05 | 0.076864286 |
| T2DM | endothelial | *RBM44* | 0.376538301 | 3.69E-06 | 3.34E-05 | 0.017919531 |
| T2DM | endothelial | *SLC25A22* | -0.785274567 | 0.020004732 | 0.031320735 | 1 |
| T2DM | endothelial | *SAMD12* | 0.140965317 | 0.000366113 | 0.001056112 | 1 |
| T2DM | endothelial | *C18orf32* | -0.42124105 | 0.011574709 | 0.019609108 | 1 |
| T2DM | endothelial | *GBA* | -1.169199142 | 0.000219119 | 0.00069991 | 1 |
| T2DM | endothelial | *IL17RA* | -0.609464033 | 0.00038698 | 0.001105131 | 1 |
| T2DM | endothelial | *MBOAT4* | 0.78360675 | 0.008793135 | 0.015385608 | 1 |
| T2DM | endothelial | *NAALADL2* | -0.35782724 | 0.002825591 | 0.005856372 | 1 |
| T2DM | endothelial | *SLC35G5* | 4.159500659 | 0.019532686 | 0.030709967 | 1 |
| T2DM | endothelial | *FAM87B* | -8.187298229 | 7.57E-06 | 5.41E-05 | 0.036735114 |
| T2DM | endothelial | *PCDHB9* | -0.693392934 | 0.001628306 | 0.003655966 | 1 |
| T2DM | endothelial | *ASB8* | -0.796220648 | 0.000264079 | 0.000819777 | 1 |
| T2DM | endothelial | *ODF3B* | 0.340653477 | 0.02338107 | 0.035854916 | 1 |

| | | | | | | |
|---|---|---|---|---|---|---|
| T2DM | endothelial | *ADAMTSL1* | -0.444857076 | 0.001305831 | 0.003040256 | 1 |
| T2DM | endothelial | *IMPDH2* | -0.579584277 | 0.013275775 | 0.022082296 | 1 |
| T2DM | endothelial | *GRAMD1C* | 0.264671625 | 0.001534396 | 0.00347405 | 1 |
| T2DM | endothelial | *DALRD3* | -0.741267989 | 0.000695209 | 0.001782852 | 1 |
| T2DM | endothelial | *ZNF114* | 0.456673372 | 1.51E-05 | 8.73E-05 | 0.0732064 |
| T2DM | endothelial | *SPINK6* | -1.210020621 | 6.76E-05 | 0.000275982 | 0.328142367 |
| T2DM | endothelial | *SH2B1* | -0.70761067 | 3.28E-06 | 3.10E-05 | 0.015928447 |
| T2DM | endothelial | *VN1R1* | -1.023836642 | 0.014010298 | 0.023051192 | 1 |
| T2DM | endothelial | *KDELC2* | -0.996869063 | 0.000539456 | 0.001446099 | 1 |
| T2DM | endothelial | *SH2D4B* | 0.805161077 | 7.62E-06 | 5.43E-05 | 0.036950108 |
| T2DM | endothelial | *RNF212* | -0.595667904 | 2.67E-07 | 1.24E-05 | 0.001294257 |
| T2DM | endothelial | *PRSS36* | -0.977785892 | 2.23E-07 | 1.24E-05 | 0.001080053 |
| T2DM | endothelial | *WDR6* | -0.313454271 | 0.025155894 | 0.038214276 | 1 |
| T2DM | endothelial | *GEN1* | -0.247993818 | 0.024282511 | 0.03704992 | 1 |
| T2DM | endothelial | *TMPRSS9* | -0.593193432 | 5.39E-06 | 4.25E-05 | 0.026134182 |
| T2DM | endothelial | *TMEM11* | -1.00660844 | 0.001041171 | 0.002500872 | 1 |
| T2DM | endothelial | *NT5DC1* | -0.448499376 | 0.013053095 | 0.021779097 | 1 |
| T2DM | endothelial | *GLDC* | -1.59534516 | 3.22E-06 | 3.06E-05 | 0.015629868 |
| T2DM | endothelial | *LINC00314* | 0.140406501 | 6.05E-05 | 0.000251794 | 0.293339994 |
| T2DM | endothelial | *P4HTM* | -0.880734052 | 1.72E-05 | 9.57E-05 | 0.083642764 |
| T2DM | endothelial | *CA8* | 0.157736589 | 4.85E-06 | 4.01E-05 | 0.023511999 |
| T2DM | endothelial | *ERN1* | -0.726035064 | 0.00055106 | 0.001471586 | 1 |
| T2DM | endothelial | *PARP10* | -0.920032824 | 6.98E-05 | 0.000283524 | 0.338527634 |
| T2DM | endothelial | *MPI* | -0.465977119 | 0.02535254 | 0.03846483 | 1 |
| T2DM | endothelial | *EXOSC4* | -1.329625744 | 0.00450265 | 0.008695051 | 1 |
| T2DM | endothelial | *ZNF852* | -1.133369898 | 3.73E-07 | 1.24E-05 | 0.001809842 |
| T2DM | endothelial | *GAK* | -0.473104401 | 0.03032738 | 0.045137561 | 1 |
| T2DM | endothelial | *TUFM* | -0.458044638 | 0.024430649 | 0.037252517 | 1 |
| T2DM | endothelial | *C3orf38* | -0.435694375 | 0.004061972 | 0.00795989 | 1 |
| T2DM | endothelial | *CCDC89* | -1.661244007 | 8.06E-07 | 1.49E-05 | 0.003911897 |
| T2DM | endothelial | *HTR1F* | -1.698993386 | 5.46E-07 | 1.27E-05 | 0.002650265 |
| T2DM | endothelial | *SPTY2D1* | -0.938400737 | 2.43E-05 | 0.00012514 | 0.11813244 |
| T2DM | endothelial | *ZNF664* | -1.006800415 | 2.14E-06 | 2.46E-05 | 0.010390353 |
| T2DM | endothelial | *CDH4* | -0.658331681 | 1.85E-05 | 0.000100816 | 0.089839774 |
| T2DM | endothelial | *HLA-DQB1* | -0.390800149 | 2.97E-07 | 1.24E-05 | 0.001440967 |
| T2DM | endothelial | *LINC00174* | -0.401137746 | 0.000975461 | 0.002362924 | 1 |
| T2DM | endothelial | *GEMIN4* | -1.252841948 | 6.38E-08 | 7.20E-06 | 0.000309652 |
| T2DM | endothelial | *LBX2* | -0.410424725 | 0.004228784 | 0.008240185 | 1 |
| T2DM | endothelial | *ZFPM1* | -0.68073056 | 0.000407504 | 0.001151198 | 1 |
| T2DM | endothelial | *RPRML* | 2.463485586 | 6.89E-07 | 1.34E-05 | 0.003344995 |
| T2DM | endothelial | *AKAP5* | -1.074778016 | 5.71E-07 | 1.27E-05 | 0.002769338 |
| T2DM | endothelial | *GIPC3* | 0.908435772 | 4.38E-06 | 3.76E-05 | 0.021228917 |
| T2DM | endothelial | *PDXDC1* | -0.563684067 | 0.000241588 | 0.000760173 | 1 |
| T2DM | endothelial | *B3GNT3* | 0.657958623 | 0.000186933 | 0.000613666 | 0.906998233 |
| T2DM | endothelial | *SSC5D* | -0.266965289 | 0.001457837 | 0.003322416 | 1 |
| T2DM | endothelial | *SEPT1* | -0.968581568 | 1.19E-06 | 1.80E-05 | 0.005777606 |
| T2DM | endothelial | *EXOC3* | -0.817067639 | 4.40E-05 | 0.000197134 | 0.213299278 |
| T2DM | endothelial | *CSNK1A1L* | -2.529629174 | 0.007260855 | 0.013081942 | 1 |
| T2DM | endothelial | *HCLS1* | -0.585070899 | 6.33E-05 | 0.000261838 | 0.307136495 |
| T2DM | endothelial | *MTURN* | -0.600472233 | 0.001343376 | 0.003111246 | 1 |

| | | | | | | |
|---|---|---|---|---|---|---|
| T2DM | endothelial | *CCDC66* | -0.50823135 | 0.002915229 | 0.006024144 | 1 |
| T2DM | endothelial | *DEFB124* | -1.960162219 | 1.20E-05 | 7.41E-05 | 0.058417278 |
| T2DM | endothelial | *GLIPR1L2* | -0.490582559 | 0.031413764 | 0.046497737 | 1 |
| T2DM | endothelial | *DEFB119* | -4.790525395 | 2.37E-05 | 0.000122563 | 0.114964229 |
| T2DM | endothelial | *MB21D2* | -0.750545592 | 0.000161486 | 0.000543741 | 0.78353035 |
| T2DM | endothelial | *SLC47A2* | -0.565331484 | 1.74E-05 | 9.63E-05 | 0.084432365 |
| T2DM | endothelial | *CHST13* | -2.477659633 | 2.42E-07 | 1.24E-05 | 0.001174731 |
| T2DM | endothelial | *SLC36A4* | -0.498165427 | 0.026045099 | 0.039367857 | 1 |
| T2DM | endothelial | *OR51E1* | 0.178312356 | 3.44E-06 | 3.18E-05 | 0.01667815 |
| T2DM | endothelial | *PSMG4* | -0.613202872 | 0.001279432 | 0.002988832 | 1 |
| T2DM | endothelial | *BHLHE22* | -0.435479244 | 0.005323499 | 0.01003589 | 1 |
| T2DM | endothelial | *ZNF443* | 1.063725037 | 2.97E-05 | 0.000143186 | 0.144045268 |
| T2DM | endothelial | *CAPS2* | -0.56958924 | 0.001788534 | 0.003962542 | 1 |
| T2DM | endothelial | *KCTD2* | -0.794675003 | 0.000340972 | 0.000996623 | 1 |
| T2DM | endothelial | *D2HGDH* | -0.670627459 | 1.37E-06 | 1.96E-05 | 0.006661199 |
| T2DM | endothelial | *CMTR2* | -0.719415059 | 0.000582833 | 0.001541094 | 1 |
| T2DM | endothelial | *MRPL14* | -0.898926162 | 0.000209071 | 0.000674027 | 1 |
| T2DM | endothelial | *LSMEM1* | -0.274127246 | 0.020874358 | 0.032545753 | 1 |
| T2DM | endothelial | *NQO1* | -0.570561973 | 0.002962229 | 0.006094251 | 1 |
| T2DM | endothelial | *FKRP* | -0.290830127 | 0.010280952 | 0.017657762 | 1 |
| T2DM | endothelial | *RPH3AL* | 0.322423801 | 0.013233004 | 0.022033814 | 1 |
| T2DM | endothelial | *ADIPOQ* | -0.207245852 | 0.000558231 | 0.001488207 | 1 |
| T2DM | endothelial | *F2R* | 0.254585203 | 9.70E-06 | 6.33E-05 | 0.047072188 |
| T2DM | endothelial | *MUC16* | -0.729804051 | 3.05E-06 | 2.99E-05 | 0.014776109 |
| T2DM | endothelial | *PENK* | -0.825991218 | 0.004399342 | 0.0085212 | 1 |
| T2DM | endothelial | *SLC25A41* | -1.759569236 | 8.65E-07 | 1.53E-05 | 0.004197731 |
| T2DM | endothelial | *ZNF322* | -0.582506112 | 0.005004235 | 0.009514321 | 1 |
| T2DM | endothelial | *DDX60L* | 0.366299658 | 0.002295954 | 0.004888096 | 1 |
| T2DM | endothelial | *OGFOD3* | -0.508290527 | 0.004010039 | 0.007874022 | 1 |
| T2DM | endothelial | *TMEM45A* | -0.520106159 | 1.57E-06 | 2.15E-05 | 0.007605256 |
| T2DM | endothelial | *SGSH* | -1.294146359 | 2.65E-06 | 2.76E-05 | 0.012867239 |
| T2DM | endothelial | *ZFP41* | -0.683870346 | 6.56E-05 | 0.000269504 | 0.318283755 |
| T2DM | endothelial | *TMEM252* | 2.556424699 | 2.26E-06 | 2.52E-05 | 0.01097294 |
| T2DM | endothelial | *RFX7* | -0.417817687 | 0.020030681 | 0.031351247 | 1 |
| T2DM | endothelial | *CLDN7* | -0.592899984 | 1.76E-06 | 2.25E-05 | 0.008562517 |
| T2DM | endothelial | *ADO* | 0.469055602 | 0.015845765 | 0.025644981 | 1 |
| T2DM | endothelial | *MGAT4C* | -0.354661525 | 3.55E-05 | 0.000165136 | 0.17223705 |
| T2DM | endothelial | *IDH2* | -0.957012195 | 2.88E-05 | 0.000139894 | 0.139614002 |
| T2DM | endothelial | *PTCHD3* | 0.404753228 | 0.017574008 | 0.028104512 | 1 |
| T2DM | endothelial | *FAM181B* | 1.009287732 | 0.00010271 | 0.000379262 | 0.498349837 |
| T2DM | endothelial | *ZNF708* | -0.454459991 | 0.00337257 | 0.006812536 | 1 |
| T2DM | endothelial | *ERCC6L2* | -0.395137364 | 0.032922715 | 0.048365793 | 1 |
| T2DM | endothelial | *MRPL41* | -1.058584854 | 9.81E-05 | 0.000366031 | 0.47580605 |
| T2DM | endothelial | *ARL6IP4* | 0.450994679 | 0.016521212 | 0.026560942 | 1 |
| T2DM | endothelial | *HHIPL1* | -0.922342377 | 8.02E-05 | 0.000314697 | 0.389280269 |
| T2DM | endothelial | *FAM153B* | -0.58078485 | 1.46E-06 | 2.06E-05 | 0.007107936 |
| T2DM | endothelial | *BACE2* | 0.205653647 | 0.027213175 | 0.040916742 | 1 |
| T2DM | endothelial | *SYNM* | -1.077302285 | 0.000637531 | 0.001657716 | 1 |
| T2DM | endothelial | *KCNA4* | -0.558599633 | 3.58E-07 | 1.24E-05 | 0.001738223 |
| T2DM | endothelial | *FIGN* | -0.46306039 | 9.37E-07 | 1.59E-05 | 0.004546932 |

| | | | | | | |
|---|---|---|---|---|---|---|
| T2DM | endothelial | *IZUMO1* | 0.364470197 | 8.18E-07 | 1.51E-05 | 0.003968031 |
| T2DM | endothelial | *FBXL6* | -1.116949117 | 0.003532449 | 0.007076567 | 1 |
| T2DM | endothelial | *NXPH4* | -4.122360167 | 4.13E-07 | 1.24E-05 | 0.002004346 |
| T2DM | endothelial | *NPLOC4* | -0.571803788 | 0.000146907 | 0.000506965 | 0.712792949 |
| T2DM | endothelial | *CAPN12* | 0.428231955 | 1.97E-05 | 0.000105937 | 0.095767352 |
| T2DM | endothelial | *KPNA2* | -0.772218504 | 0.019474431 | 0.030629233 | 1 |
| T2DM | endothelial | *ORAI1* | 0.3984857 | 8.91E-06 | 6.00E-05 | 0.043248655 |
| T2DM | endothelial | *CAV3* | -0.536921469 | 0.000339047 | 0.000993241 | 1 |
| T2DM | endothelial | *ADI1* | -0.868072499 | 4.29E-05 | 0.000193101 | 0.207969784 |
| T2DM | endothelial | *CLEC4G* | -1.940091565 | 2.32E-05 | 0.000120544 | 0.112588199 |
| T2DM | endothelial | *TRAK1* | -0.361903175 | 0.024089967 | 0.036790847 | 1 |
| T2DM | endothelial | *SKA2* | -0.815721206 | 2.52E-05 | 0.00012706 | 0.122319548 |
| T2DM | endothelial | *PPP1R27* | -2.703242734 | 2.37E-06 | 2.60E-05 | 0.01151471 |
| T2DM | endothelial | *GRID1* | -0.744591093 | 3.49E-07 | 1.24E-05 | 0.001694332 |
| T2DM | endothelial | *ACBD3* | -0.523501697 | 0.006746322 | 0.012273399 | 1 |
| T2DM | endothelial | *PLCXD3* | 0.231924293 | 2.60E-05 | 0.000130093 | 0.126320367 |
| T2DM | endothelial | *GPIHBP1* | 0.106413562 | 0.000426124 | 0.001191672 | 1 |
| T2DM | endothelial | *ALG12* | -1.060848925 | 3.62E-06 | 3.31E-05 | 0.017559802 |
| T2DM | endothelial | *CEP63* | -0.522025537 | 0.000194526 | 0.0006343 | 0.943838076 |
| T2DM | endothelial | *ODF3L1* | -1.29311232 | 0.000233766 | 0.000739627 | 1 |
| T2DM | endothelial | *HMGN4* | -0.605916767 | 0.000245913 | 0.00077178 | 1 |
| T2DM | endothelial | *ZNF320* | 0.172439656 | 0.030020786 | 0.044722398 | 1 |
| T2DM | endothelial | *C12orf60* | -0.87001934 | 1.00E-06 | 1.63E-05 | 0.004867248 |
| T2DM | endothelial | *ABAT* | -0.835980167 | 1.05E-06 | 1.67E-05 | 0.005113372 |
| T2DM | endothelial | *LYSMD4* | -0.550492719 | 0.007868688 | 0.013984936 | 1 |
| T2DM | endothelial | *WBP2NL* | -0.819527217 | 2.33E-05 | 0.000120727 | 0.113000931 |
| T2DM | endothelial | *BEGAIN* | -1.271436105 | 9.47E-06 | 6.25E-05 | 0.045940499 |
| T2DM | endothelial | *CSMD1* | 0.38313175 | 7.56E-05 | 0.000301973 | 0.366595046 |
| T2DM | endothelial | *C2CD4C* | 0.201962255 | 4.18E-06 | 3.66E-05 | 0.020291663 |
| T2DM | endothelial | *RUVBL2* | -1.013967997 | 0.000370592 | 0.001067129 | 1 |
| T2DM | endothelial | *ZNF623* | -0.614359877 | 0.001091572 | 0.002605592 | 1 |
| T2DM | endothelial | *CCDC125* | -0.612812171 | 0.000199232 | 0.000646172 | 0.966673088 |
| T2DM | endothelial | *LRIT3* | 4.021779647 | 0.000605053 | 0.001591473 | 1 |
| T2DM | endothelial | *GPR132* | -0.547033694 | 7.81E-08 | 8.24E-06 | 0.000379079 |
| T2DM | endothelial | *SERHL2* | -1.075120375 | 2.77E-05 | 0.000136232 | 0.134188139 |
| T2DM | endothelial | *PCBP3* | -0.324162587 | 2.78E-05 | 0.000136593 | 0.134816898 |
| T2DM | endothelial | *SFXN4* | -0.620934819 | 0.004718673 | 0.009049408 | 1 |
| T2DM | endothelial | *MRPL54* | -0.424036747 | 0.005483776 | 0.01027705 | 1 |
| T2DM | endothelial | *HMCES* | -0.698244221 | 8.54E-05 | 0.000328653 | 0.414431258 |
| T2DM | endothelial | *DGCR6* | -0.718391775 | 0.009410598 | 0.016314022 | 1 |
| T2DM | endothelial | *NDUFB1* | -0.473682837 | 0.022166581 | 0.034208732 | 1 |
| T2DM | endothelial | *FAM19A1* | -0.681353123 | 9.28E-06 | 6.19E-05 | 0.045029732 |
| T2DM | endothelial | *TRMT12* | -0.93840349 | 1.96E-05 | 0.000105777 | 0.095305385 |
| T2DM | endothelial | *ALYREF* | -0.54300693 | 0.025049948 | 0.03810105 | 1 |
| T2DM | endothelial | *MRGPRX2* | -2.110563749 | 0.000545525 | 0.00145834 | 1 |
| T2DM | endothelial | *UPP1* | 0.391549866 | 0.000326457 | 0.000965834 | 1 |
| T2DM | endothelial | *FIGLA* | 0.360018581 | 2.44E-06 | 2.63E-05 | 0.011839233 |
| T2DM | endothelial | *ACSM2A* | -2.411566637 | 4.19E-07 | 1.24E-05 | 0.002033286 |
| T2DM | endothelial | *KREMEN1* | -0.480209218 | 1.93E-05 | 0.000104268 | 0.093423812 |
| T2DM | endothelial | *ZNF703* | -1.057709257 | 2.82E-05 | 0.000137747 | 0.136921004 |

| | | | | | | |
|---|---|---|---|---|---|---|
| T2DM | endothelial | *NPIPA5* | -0.147009185 | 0.016869769 | 0.027076453 | 1 |
| T2DM | endothelial | *FAM162B* | -0.638890443 | 1.84E-06 | 2.31E-05 | 0.008939697 |
| T2DM | endothelial | *BTBD9* | -0.457629901 | 0.001515122 | 0.003440044 | 1 |
| T2DM | endothelial | *NUDT14* | 0.730956638 | 0.000571862 | 0.001518706 | 1 |
| T2DM | endothelial | *FAM3B* | -0.133228631 | 0.008299426 | 0.01461663 | 1 |
| T2DM | endothelial | *NPIPA7* | -0.712543154 | 0.002585153 | 0.005434645 | 1 |
| T2DM | endothelial | *TTC32* | -1.069813325 | 2.17E-05 | 0.000113922 | 0.105378227 |
| T2DM | endothelial | *PP2D1* | -1.096916613 | 0.000920808 | 0.002261012 | 1 |
| T2DM | endothelial | *NIPSNAP1* | -0.826230081 | 0.00027399 | 0.000842993 | 1 |
| T2DM | endothelial | *CNTN2* | -0.436609421 | 0.030830301 | 0.045773751 | 1 |
| T2DM | endothelial | *GOLGA6L4* | -0.537929582 | 0.001664569 | 0.003725318 | 1 |
| T2DM | endothelial | *OAF* | -0.345739161 | 0.000148732 | 0.000511083 | 0.72164866 |
| T2DM | endothelial | *ALDH1A3* | -0.266430864 | 0.022254632 | 0.034322783 | 1 |
| T2DM | endothelial | *SS18L1* | -0.714401205 | 0.00132183 | 0.003070138 | 1 |
| T2DM | endothelial | *COPB2* | -0.666803938 | 0.000745474 | 0.001887807 | 1 |
| T2DM | endothelial | *WDR27* | -0.381770731 | 0.017904624 | 0.028529799 | 1 |
| T2DM | endothelial | *TXNRD2* | -0.72639482 | 0.000310219 | 0.000929698 | 1 |
| T2DM | endothelial | *TMEM255B* | 0.285950248 | 1.13E-05 | 7.07E-05 | 0.054675993 |
| T2DM | endothelial | *HDDC3* | -1.019815738 | 2.48E-05 | 0.000125919 | 0.120252448 |
| T2DM | endothelial | *DUSP8* | 0.69723339 | 0.002773295 | 0.005760285 | 1 |
| T2DM | endothelial | *KRBA2* | -1.019667743 | 0.000428461 | 0.001196141 | 1 |
| T2DM | endothelial | *SEPT5* | -1.064410425 | 1.34E-05 | 8.02E-05 | 0.065046395 |
| T2DM | endothelial | *RNLS* | -0.625006846 | 7.06E-06 | 5.11E-05 | 0.034270644 |
| T2DM | endothelial | *TCTE3* | -0.979985126 | 2.21E-06 | 2.49E-05 | 0.010702192 |
| T2DM | endothelial | *UBE2G2* | -0.578495012 | 0.00072268 | 0.001839688 | 1 |
| T2DM | endothelial | *DRD1* | -5.940180475 | 5.60E-07 | 1.27E-05 | 0.002715346 |
| T2DM | endothelial | *BTBD6* | -0.697148539 | 0.001010969 | 0.002439196 | 1 |
| T2DM | endothelial | *WT1* | -0.343827445 | 0.002589727 | 0.005440279 | 1 |
| T2DM | endothelial | *ZFP90* | -0.525233737 | 0.021840043 | 0.033790782 | 1 |
| T2DM | endothelial | *FAM227A* | -0.291545486 | 0.003018605 | 0.006187694 | 1 |
| T2DM | endothelial | *SORCS2* | -0.902366083 | 2.01E-07 | 1.24E-05 | 0.000977548 |
| T2DM | endothelial | *BRI3BP* | -1.007497962 | 1.67E-06 | 2.20E-05 | 0.008100127 |
| T2DM | endothelial | *IFNE* | -1.366983866 | 0.008087551 | 0.014293516 | 1 |
| T2DM | endothelial | *UBOX5* | -1.021494994 | 9.32E-06 | 6.19E-05 | 0.045219858 |
| T2DM | endothelial | *SEMA4B* | -0.667976026 | 1.27E-06 | 1.86E-05 | 0.006172764 |
| T2DM | endothelial | *CIB1* | -0.38803109 | 0.011356851 | 0.019260203 | 1 |
| T2DM | endothelial | *SGCZ* | -0.401680694 | 3.45E-05 | 0.000161223 | 0.167187836 |
| T2DM | endothelial | *SIGIRR* | 0.477033727 | 0.001016741 | 0.002451257 | 1 |
| T2DM | endothelial | *IFITM2* | 0.375362072 | 0.012648605 | 0.021191654 | 1 |
| T2DM | endothelial | *TNFAIP8L1* | 0.567343392 | 0.000675483 | 0.001737775 | 1 |
| T2DM | endothelial | *TARSL2* | -0.634016006 | 0.000102272 | 0.00037822 | 0.496224984 |
| T2DM | endothelial | *SH3BGR* | -0.273235017 | 0.000802642 | 0.002008467 | 1 |
| T2DM | endothelial | *FAM174B* | 0.348433301 | 0.026924981 | 0.040558835 | 1 |
| T2DM | endothelial | *SV2B* | 0.171660959 | 6.30E-06 | 4.79E-05 | 0.030584181 |
| T2DM | endothelial | *AHNAK2* | 0.478289866 | 0.021097952 | 0.032852138 | 1 |
| T2DM | endothelial | *SP1* | -0.71103494 | 5.00E-05 | 0.000218639 | 0.242470111 |
| T2DM | endothelial | *PSMD13* | -0.748039921 | 0.001134568 | 0.002689556 | 1 |
| T2DM | endothelial | *C6orf201* | -0.644965931 | 0.002119117 | 0.004581977 | 1 |
| T2DM | endothelial | *MYBL1* | -1.100361737 | 2.61E-07 | 1.24E-05 | 0.001266192 |
| T2DM | endothelial | *ADARB2* | 0.267003655 | 3.17E-06 | 3.04E-05 | 0.015389775 |

| | | | | | | |
|---|---|---|---|---|---|---|
| T2DM | endothelial | *DMWD* | -0.868995174 | 0.000482111 | 0.00131786 | 1 |
| T2DM | endothelial | *SLC52A2* | -0.898142033 | 0.000511646 | 0.001385327 | 1 |
| T2DM | endothelial | *PIGP* | -0.217310167 | 0.001470284 | 0.003346069 | 1 |
| T2DM | endothelial | *ARL17A* | -0.377197817 | 0.013813304 | 0.022781729 | 1 |
| T2DM | endothelial | *GNB1L* | -0.39981797 | 0.003168999 | 0.006449657 | 1 |
| T2DM | endothelial | *ZNF829* | -0.661044773 | 0.001169484 | 0.002765272 | 1 |
| T2DM | endothelial | *IFITM1* | 0.546473084 | 0.000127756 | 0.000453455 | 0.619873251 |
| T2DM | endothelial | *LINC00839* | 1.293419473 | 0.000268242 | 0.000828538 | 1 |
| T2DM | endothelial | *RNPC3* | -0.4527803 | 0.000314345 | 0.000939744 | 1 |
| T2DM | endothelial | *RASA3* | -0.713419554 | 1.91E-09 | 6.79E-07 | 9.25E-06 |
| T2DM | endothelial | *ZNF529* | -0.529524919 | 0.015072786 | 0.024574313 | 1 |
| T2DM | endothelial | *WWOX* | -0.40084994 | 0.001183166 | 0.002794897 | 1 |
| T2DM | endothelial | *FFAR4* | -0.531607545 | 0.001927057 | 0.004230805 | 1 |
| T2DM | endothelial | *TOR3A* | -1.153403267 | 9.12E-05 | 0.000346394 | 0.44234571 |
| T2DM | endothelial | *ZNF555* | -0.626471261 | 0.018331509 | 0.02910487 | 1 |
| T2DM | endothelial | *CCDC30* | -0.395721595 | 0.003813115 | 0.007560783 | 1 |
| T2DM | endothelial | *ZNF197* | -0.670437182 | 0.000544534 | 0.001456942 | 1 |
| T2DM | endothelial | *GNG2* | -0.611050276 | 3.16E-06 | 3.03E-05 | 0.015321299 |
| T2DM | endothelial | *TMEM222* | -0.708335425 | 0.015637041 | 0.025349456 | 1 |
| T2DM | endothelial | *FAM86B1* | -0.943280326 | 9.57E-05 | 0.000359976 | 0.464369582 |
| T2DM | endothelial | *MIR22HG* | -0.728500688 | 4.04E-07 | 1.24E-05 | 0.001960458 |
| T2DM | endothelial | *CYP27C1* | -0.677664589 | 6.23E-07 | 1.31E-05 | 0.003022383 |
| T2DM | endothelial | *ZNF732* | 0.853613178 | 0.000940837 | 0.00229856 | 1 |
| T2DM | endothelial | *VSIG10L* | 0.749460261 | 0.000664437 | 0.001714814 | 1 |
| T2DM | endothelial | *TRABD2A* | 0.218163364 | 0.033808288 | 0.049468581 | 1 |
| T2DM | endothelial | *QRFPR* | -1.458633716 | 0.000153071 | 0.000523028 | 0.742700189 |
| T2DM | endothelial | *MAPT* | -0.761655745 | 1.15E-06 | 1.78E-05 | 0.005594952 |
| T2DM | endothelial | *TNFRSF18* | 1.148624965 | 5.42E-05 | 0.000231621 | 0.262881902 |
| T2DM | endothelial | *C1QL4* | -3.074266222 | 5.46E-07 | 1.27E-05 | 0.002650122 |
| T2DM | endothelial | *TMEM232* | -0.451937999 | 0.001329145 | 0.003082701 | 1 |
| T2DM | endothelial | *EFCAB6* | -0.61251671 | 3.33E-06 | 3.11E-05 | 0.01615454 |
| T2DM | endothelial | *TMPRSS6* | -5.916335641 | 2.99E-07 | 1.24E-05 | 0.001448993 |
| T2DM | endothelial | *TMEM216* | 0.457694184 | 0.032849538 | 0.048284144 | 1 |
| T2DM | endothelial | *ENTPD5* | -0.722503194 | 2.40E-05 | 0.000123897 | 0.116463103 |
| T2DM | endothelial | *MRPS21* | -0.376170066 | 0.015020647 | 0.024514019 | 1 |
| T2DM | endothelial | *ANGPTL5* | -0.449655734 | 6.08E-05 | 0.000252858 | 0.294914084 |
| T2DM | endothelial | *WDR86* | -0.297440589 | 0.001969915 | 0.004307357 | 1 |
| T2DM | endothelial | *EPOR* | -0.39685325 | 0.000483271 | 0.001319546 | 1 |
| T2DM | endothelial | *LUZP2* | 0.621752727 | 0.000184842 | 0.000607623 | 0.896852179 |
| T2DM | endothelial | *GJA4* | 0.322379709 | 6.93E-06 | 5.10E-05 | 0.033611307 |
| T2DM | endothelial | *SBK2* | -2.362844587 | 5.99E-07 | 1.30E-05 | 0.002906859 |
| T2DM | endothelial | *ZNF385C* | -0.559009498 | 0.001774527 | 0.003938704 | 1 |
| T2DM | endothelial | *DHRS4L2* | -0.357101517 | 0.007161997 | 0.012923024 | 1 |
| T2DM | endothelial | *SAMD11* | -0.760053359 | 5.55E-05 | 0.000236924 | 0.269382915 |
| T2DM | endothelial | *C2orf88* | -0.649588433 | 3.54E-05 | 0.00016476 | 0.171679605 |
| T2DM | endothelial | *KBTBD12* | -0.403518601 | 0.000353689 | 0.001027603 | 1 |
| T2DM | endothelial | *DNAH17* | -0.881279887 | 0.000175145 | 0.00058126 | 0.849802689 |
| T2DM | endothelial | *FANCM* | -0.908336592 | 2.44E-06 | 2.63E-05 | 0.011823432 |
| T2DM | endothelial | *ZFP69* | -1.223902414 | 2.95E-06 | 2.93E-05 | 0.014303821 |
| T2DM | endothelial | *TMEM220* | -0.397577984 | 0.004503434 | 0.008695051 | 1 |

| | | | | | | |
|---|---|---|---|---|---|---|
| T2DM | endothelial | *P2RX2* | -0.329566355 | 0.030630454 | 0.04553277 | 1 |
| T2DM | endothelial | *FAM122A* | -1.343997809 | 0.000375851 | 0.001077796 | 1 |
| T2DM | endothelial | *GFRAL* | -1.569642719 | 0.00105146 | 0.002523089 | 1 |
| T2DM | endothelial | *LDLRAD2* | 0.33992113 | 0.002723276 | 0.005675832 | 1 |
| T2DM | endothelial | *KLHL17* | -0.532230182 | 0.000813216 | 0.002030738 | 1 |
| T2DM | endothelial | *S100A3* | -0.686154391 | 0.026986284 | 0.040638563 | 1 |
| T2DM | endothelial | *TMEM221* | -0.692628182 | 0.000673799 | 0.001735283 | 1 |
| T2DM | endothelial | *MESP2* | -0.711841205 | 1.74E-06 | 2.24E-05 | 0.008445613 |
| T2DM | endothelial | *MAPK12* | 0.388466876 | 1.44E-05 | 8.43E-05 | 0.069712572 |
| T2DM | endothelial | *AGRN* | 0.486373332 | 5.10E-05 | 0.000222308 | 0.24765166 |
| T2DM | endothelial | *FAM166A* | -1.236345511 | 0.001155843 | 0.002735682 | 1 |
| T2DM | endothelial | *ZNF626* | -0.73971054 | 0.018653388 | 0.02953859 | 1 |
| T2DM | endothelial | *LINC00265* | -0.959944646 | 7.97E-07 | 1.49E-05 | 0.003867786 |
| T2DM | endothelial | *NCR3LG1* | -2.106265331 | 4.96E-06 | 4.07E-05 | 0.024065427 |
| T2DM | endothelial | *ZNF793* | 0.315734293 | 0.001659718 | 0.003717892 | 1 |
| T2DM | endothelial | *RUFY4* | -0.857572172 | 3.62E-07 | 1.24E-05 | 0.001757974 |
| T2DM | endothelial | *ZNF669* | 0.567482393 | 5.37E-05 | 0.000230342 | 0.260747341 |
| T2DM | endothelial | *CENPP* | -0.387628622 | 8.08E-05 | 0.000316018 | 0.391862557 |
| T2DM | endothelial | *PLSCR1* | 0.339633253 | 0.001988457 | 0.004343987 | 1 |
| T2DM | endothelial | *C3orf62* | -0.528133376 | 0.002269187 | 0.004841732 | 1 |
| T2DM | endothelial | *ENO4* | -0.560816901 | 8.58E-07 | 1.53E-05 | 0.004165038 |
| T2DM | endothelial | *ZP3* | -0.393555829 | 3.16E-05 | 0.00015044 | 0.15339675 |
| T2DM | endothelial | *PDCD1* | -1.88288664 | 0.000284474 | 0.000867547 | 1 |
| T2DM | endothelial | *C19orf54* | -1.082266417 | 1.14E-06 | 1.76E-05 | 0.005531383 |
| T2DM | endothelial | *C22orf34* | 0.268059452 | 3.04E-05 | 0.000145997 | 0.14760254 |
| T2DM | endothelial | *COL25A1* | -0.605686678 | 0.001359275 | 0.003140571 | 1 |
| T2DM | endothelial | *SRSF10* | -0.388282144 | 0.01626301 | 0.026241479 | 1 |
| T2DM | endothelial | *NBR1* | -0.635858616 | 0.000923594 | 0.002265559 | 1 |
| T2DM | endothelial | *NDOR1* | -1.161158621 | 0.000135169 | 0.000474903 | 0.655841368 |
| T2DM | endothelial | *FAM72B* | -0.88992694 | 0.012658863 | 0.02120152 | 1 |
| T2DM | endothelial | *RHCE* | -1.282943877 | 0.000670792 | 0.001729372 | 1 |
| T2DM | endothelial | *IDO2* | 1.272739213 | 0.000453114 | 0.001252712 | 1 |
| T2DM | endothelial | *SLC4A5* | -2.372663396 | 2.72E-06 | 2.78E-05 | 0.013215818 |
| T2DM | endothelial | *UROS* | -0.860632039 | 0.000173064 | 0.000575141 | 0.839705517 |
| T2DM | endothelial | *ZBED6CL* | -1.397587785 | 1.63E-06 | 2.19E-05 | 0.007920754 |
| T2DM | endothelial | *OSTN* | -0.46916648 | 0.000408556 | 0.001153177 | 1 |
| T2DM | endothelial | *FSIP2* | -0.675168201 | 0.000188964 | 0.000619495 | 0.916851946 |
| T2DM | endothelial | *NOXA1* | -0.955943568 | 1.93E-06 | 2.36E-05 | 0.009375002 |
| T2DM | endothelial | *FZD9* | -4.079785217 | 1.23E-07 | 1.01E-05 | 0.000594475 |
| T2DM | endothelial | *SKOR1* | -1.061076202 | 0.000766257 | 0.001931531 | 1 |
| T2DM | endothelial | *MTF1* | -0.850282869 | 0.000899293 | 0.00221154 | 1 |
| T2DM | endothelial | *ENTPD8* | -27.61002789 | 4.90E-07 | 1.27E-05 | 0.002375386 |
| T2DM | endothelial | *FBF1* | -0.653249633 | 1.02E-05 | 6.56E-05 | 0.049709378 |
| T2DM | endothelial | *ASTL* | -2.719993279 | 3.62E-07 | 1.24E-05 | 0.001754217 |
| T2DM | endothelial | *MSL1* | -0.537876367 | 0.001252401 | 0.002936172 | 1 |
| T2DM | endothelial | *FAM196A* | 0.347586742 | 0.00410391 | 0.008035581 | 1 |
| T2DM | endothelial | *UTS2B* | -0.656683285 | 5.31E-06 | 4.22E-05 | 0.025784734 |
| T2DM | endothelial | *C9orf152* | -1.490598978 | 0.000124403 | 0.00044285 | 0.603605178 |
| T2DM | endothelial | *NELFB* | -0.702162316 | 0.015624372 | 0.025337385 | 1 |
| T2DM | endothelial | *SLC15A5* | -0.56346423 | 0.004146696 | 0.008099746 | 1 |

| | | | | | | |
|---|---|---|---|---|---|---|
| T2DM | endothelial | *LIPI* | -0.406080329 | 0.002454614 | 0.005173669 | 1 |
| T2DM | endothelial | *ADAT2* | -0.83285303 | 6.08E-05 | 0.000252858 | 0.295084734 |
| T2DM | endothelial | *LITAF* | -0.690090068 | 0.000214674 | 0.000687977 | 1 |
| T2DM | endothelial | *SF3B3* | -0.693900032 | 0.000538085 | 0.00144402 | 1 |
| T2DM | endothelial | *PLAC9* | -0.540317793 | 0.0091632 | 0.015946861 | 1 |
| T2DM | endothelial | *ZNF573* | -0.474277045 | 0.023340734 | 0.035804375 | 1 |
| T2DM | endothelial | *TSPYL1* | -0.709389537 | 0.025182451 | 0.038242645 | 1 |
| T2DM | endothelial | *PNRC2* | -0.954434001 | 0.000110848 | 0.000404081 | 0.537832407 |
| T2DM | endothelial | *FHIT* | -0.49652999 | 2.57E-05 | 0.000128761 | 0.124489225 |
| T2DM | endothelial | *ZKSCAN3* | -0.73876197 | 5.67E-05 | 0.000240225 | 0.275220925 |
| T2DM | endothelial | *RRP7A* | -0.837253312 | 0.001407266 | 0.00323145 | 1 |
| T2DM | endothelial | *SLC35E2B* | -0.432252827 | 0.010230145 | 0.017576722 | 1 |
| T2DM | endothelial | *KIAA0408* | -0.397490317 | 1.33E-05 | 7.97E-05 | 0.06435435 |
| T2DM | endothelial | *BLOC1S2* | -0.414767906 | 0.001086608 | 0.002595876 | 1 |
| T2DM | endothelial | *HLA-DRB1* | -0.22138595 | 0.002078384 | 0.004503938 | 1 |
| T2DM | endothelial | *AKR1C3* | 0.438224527 | 0.002410138 | 0.005097641 | 1 |
| T2DM | endothelial | *COLCA1* | -0.890577707 | 1.07E-07 | 9.63E-06 | 0.000520029 |
| T2DM | endothelial | *SEMA4A* | -0.964753705 | 8.70E-06 | 5.90E-05 | 0.042207267 |
| T2DM | endothelial | *HRCT1* | 0.321093066 | 0.000527983 | 0.001423207 | 1 |
| T2DM | endothelial | *MPHOSPH8* | -0.475439034 | 0.002062987 | 0.004482584 | 1 |
| T2DM | endothelial | *SIRPB2* | -0.378394823 | 2.03E-07 | 1.24E-05 | 0.000984571 |
| T2DM | endothelial | *SRGAP3* | -0.416555854 | 0.000243383 | 0.000764334 | 1 |
| T2DM | endothelial | *XPNPEP3* | -0.357995959 | 0.000444792 | 0.001235334 | 1 |
| T2DM | endothelial | *ZNF107* | 0.32780068 | 0.023878081 | 0.036530832 | 1 |
| T2DM | endothelial | *PPIA* | -0.750030486 | 0.00040319 | 0.001141351 | 1 |
| T2DM | endothelial | *ZNF471* | -0.799454069 | 0.000945843 | 0.002308467 | 1 |
| T2DM | endothelial | *ZNF493* | -0.771831068 | 0.000229007 | 0.000726236 | 1 |
| T2DM | endothelial | *GTF2IRD2* | -0.77488244 | 2.95E-05 | 0.000142584 | 0.14315463 |
| T2DM | endothelial | *GIMAP5* | 0.208485012 | 1.34E-05 | 8.02E-05 | 0.065129773 |
| T2DM | endothelial | *STK31* | -0.871206171 | 0.002032229 | 0.004425553 | 1 |
| T2DM | endothelial | *ZKSCAN7* | -0.506561367 | 0.000222781 | 0.000710672 | 1 |
| T2DM | endothelial | *CPNE4* | 0.42763389 | 0.000779557 | 0.001957769 | 1 |
| T2DM | endothelial | *ZNF565* | -0.488747818 | 0.001872543 | 0.004120753 | 1 |
| T2DM | endothelial | *ZNF124* | -0.782722307 | 0.000103746 | 0.000382334 | 0.503373737 |
| T2DM | endothelial | *PIK3R4* | -0.723600114 | 0.001004103 | 0.002425041 | 1 |
| T2DM | endothelial | *RFX8* | -0.385521292 | 0.000417122 | 0.001171223 | 1 |
| T2DM | endothelial | *C20orf96* | -1.197672246 | 4.66E-06 | 3.91E-05 | 0.022623125 |
| T2DM | endothelial | *SULT1A1* | -0.735425053 | 7.59E-08 | 8.19E-06 | 0.000368452 |
| T2DM | endothelial | *SPTSSB* | -0.870038262 | 0.016465455 | 0.026497641 | 1 |
| T2DM | endothelial | *CACNA1H* | -1.289000429 | 3.22E-06 | 3.06E-05 | 0.015619541 |
| T2DM | endothelial | *SULF2* | 0.420407844 | 0.00028392 | 0.000866402 | 1 |
| T2DM | endothelial | *XRCC2* | -1.038413345 | 3.31E-07 | 1.24E-05 | 0.001605992 |
| T2DM | endothelial | *MYO6* | 0.292215213 | 0.022081691 | 0.034110272 | 1 |
| T2DM | endothelial | *ZNF846* | -0.598410564 | 0.000181534 | 0.000599184 | 0.880800948 |
| T2DM | endothelial | *ADH1B* | -0.436942574 | 4.68E-06 | 3.91E-05 | 0.022728706 |
| T2DM | endothelial | *AMZ2* | -0.903580335 | 5.67E-05 | 0.000240225 | 0.275298414 |
| T2DM | endothelial | *ZNF431* | -0.44098708 | 0.021780892 | 0.033742302 | 1 |
| T2DM | endothelial | *HLA-DQA1* | -0.706317875 | 1.88E-10 | 1.52E-07 | 9.11E-07 |
| T2DM | endothelial | *COL27A1* | -0.365273199 | 2.65E-06 | 2.76E-05 | 0.01284899 |
| T2DM | endothelial | *ZNF239* | -0.559424793 | 7.71E-05 | 0.0003058 | 0.37429862 |

| | | | | | | |
|---|---|---|---|---|---|---|
| T2DM | endothelial | *SPRR2B* | -3.001744999 | 0.004127717 | 0.008072423 | 1 |
| T2DM | endothelial | *FAM3C* | -0.753686706 | 8.42E-05 | 0.00032521 | 0.408463771 |
| T2DM | endothelial | *SLC39A10* | -0.438787198 | 0.0328165 | 0.048250199 | 1 |
| T2DM | endothelial | *AP2A1* | -0.464460852 | 0.015786301 | 0.025557268 | 1 |
| T2DM | endothelial | *WDR5B* | -0.60557731 | 0.032934992 | 0.048365793 | 1 |
| T2DM | endothelial | *ZNF138* | -0.521824693 | 0.004689547 | 0.009007791 | 1 |
| T2DM | endothelial | *ZNF429* | -0.705695949 | 0.000136046 | 0.000477636 | 0.660093349 |
| T2DM | endothelial | *ZNF100* | -0.525666638 | 0.001688012 | 0.003774301 | 1 |
| T2DM | endothelial | *ANXA6* | -0.635459947 | 0.000102892 | 0.000379644 | 0.499231639 |
| T2DM | endothelial | *GMFB* | -0.74596132 | 8.75E-07 | 1.53E-05 | 0.004245796 |
| T2DM | endothelial | *ZMYM1* | -0.583107803 | 0.008089447 | 0.014293516 | 1 |
| T2DM | endothelial | *IGF2R* | -0.481603162 | 0.009361897 | 0.016251851 | 1 |
| T2DM | endothelial | *GAL3ST4* | -1.01555035 | 7.56E-06 | 5.41E-05 | 0.036670431 |
| T2DM | endothelial | *ZNF257* | 0.491070801 | 0.003221218 | 0.00655044 | 1 |
| T2DM | endothelial | *ABCB8* | -0.850580257 | 4.08E-07 | 1.24E-05 | 0.001981399 |
| T2DM | endothelial | *SND1* | -0.57399202 | 0.000163568 | 0.000549226 | 0.793631091 |
| T2DM | endothelial | *SULT1A2* | -0.609435587 | 0.000256404 | 0.000799533 | 1 |
| T2DM | endothelial | *NEK5* | -0.390871351 | 0.000819039 | 0.002042126 | 1 |
| T2DM | endothelial | *TBC1D9B* | -0.47088287 | 0.009606361 | 0.016610857 | 1 |
| T2DM | endothelial | *FAM110D* | 0.156905958 | 7.89E-07 | 1.49E-05 | 0.003828141 |
| T2DM | endothelial | *C6orf141* | 0.123260392 | 1.41E-05 | 8.33E-05 | 0.068525996 |
| T2DM | endothelial | *ZNF165* | -0.457675041 | 0.000165311 | 0.000553928 | 0.802087146 |
| T2DM | endothelial | *RAMP2-AS1* | -0.716988658 | 0.004161092 | 0.008121327 | 1 |
| T2DM | endothelial | *ZNF720* | -0.815677514 | 7.42E-05 | 0.000297466 | 0.359933987 |
| T2DM | endothelial | *ZNF98* | -0.555507173 | 0.000174007 | 0.000577879 | 0.844281788 |
| T2DM | endothelial | *ZNF517* | -1.056984159 | 0.006673186 | 0.012154017 | 1 |
| T2DM | endothelial | *ZNF675* | -0.602464217 | 0.01818827 | 0.028915296 | 1 |
| T2DM | endothelial | *SLC22A5* | -0.59927861 | 8.54E-05 | 0.000328653 | 0.414420704 |
| T2DM | endothelial | *VEPH1* | -0.782339486 | 2.69E-06 | 2.76E-05 | 0.013045553 |
| T2DM | endothelial | *SHPK* | -0.349767616 | 0.010538982 | 0.01804982 | 1 |
| T2DM | endothelial | *MAP3K5* | -0.56315113 | 7.59E-05 | 0.000302527 | 0.36817537 |
| T2DM | endothelial | *GSTK1* | -0.379641368 | 0.031077317 | 0.04607001 | 1 |
| T2DM | endothelial | *SLC2A10* | -0.704265692 | 1.10E-05 | 6.94E-05 | 0.053552291 |
| T2DM | endothelial | *ZNF665* | -0.897725507 | 0.000629178 | 0.001639574 | 1 |
| T2DM | endothelial | *RPF2* | -0.73378284 | 0.000838374 | 0.002086046 | 1 |
| T2DM | endothelial | *MYO5A* | -0.420972766 | 0.000445492 | 0.001236457 | 1 |
| T2DM | endothelial | *ATG7* | -0.456493155 | 0.000762615 | 0.001924186 | 1 |
| T2DM | endothelial | *RAB40C* | -0.878413158 | 5.93E-06 | 4.59E-05 | 0.02877336 |
| T2DM | endothelial | *PIGN* | -0.375773927 | 0.012192087 | 0.020533151 | 1 |
| T2DM | endothelial | *HHLA3* | -0.455380959 | 0.00156399 | 0.003531168 | 1 |
| T2DM | endothelial | *TOPORS* | -0.795512195 | 0.001145901 | 0.002713476 | 1 |
| T2DM | endothelial | *BCO2* | -0.213461599 | 0.009723264 | 0.016785452 | 1 |
| T2DM | endothelial | *ENTPD6* | -0.576112498 | 0.016911734 | 0.027134832 | 1 |
| T2DM | endothelial | *ENPP1* | -0.597905918 | 2.57E-07 | 1.24E-05 | 0.001245058 |
| T2DM | endothelial | *MFAP5* | -0.465300661 | 0.000958982 | 0.002329985 | 1 |
| T2DM | endothelial | *PDCD1LG2* | -0.175136966 | 0.018496121 | 0.02930868 | 1 |
| T2DM | endothelial | *DNAH10* | -0.962797215 | 6.15E-08 | 7.20E-06 | 0.000298233 |
| T2DM | endothelial | *FAM114A1* | -0.63587237 | 0.00034059 | 0.000996107 | 1 |
| T2DM | endothelial | *RPS26* | -0.285392 | 0.01344341 | 0.02229717 | 1 |
| T2DM | endothelial | *PSAP* | -0.381450946 | 0.00922667 | 0.016045808 | 1 |

| | | | | | | |
|---|---|---|---|---|---|---|
| T2DM | endothelial | *TXNRD3* | -0.71879354 | 2.77E-05 | 0.000136232 | 0.134171631 |
| T2DM | endothelial | *EME2* | -1.160519067 | 6.04E-07 | 1.30E-05 | 0.002928272 |
| T2DM | endothelial | *ZNF780A* | -0.195107384 | 0.002230013 | 0.004777053 | 1 |
| T2DM | endothelial | *ATAD3A* | -0.67846244 | 1.61E-05 | 9.10E-05 | 0.07826793 |
| T2DM | endothelial | *AC122129.1* | -2.071897458 | 8.51E-07 | 1.53E-05 | 0.004129383 |
| T2DM | endothelial | *OCLN* | 0.311123324 | 2.65E-06 | 2.76E-05 | 0.012847143 |
| T2DM | endothelial | *SGTB* | -1.048238511 | 3.14E-06 | 3.03E-05 | 0.015230782 |
| T2DM | endothelial | *ADH5* | -0.677582227 | 1.60E-05 | 9.06E-05 | 0.077806055 |
| T2DM | endothelial | *ZNF311* | -0.345980944 | 0.002939764 | 0.00605936 | 1 |
| T2DM | endothelial | *FCHSD1* | -0.701976764 | 0.003437175 | 0.006928614 | 1 |
| T2DM | endothelial | *MPZL1* | -0.620752763 | 8.15E-05 | 0.000317752 | 0.395601122 |
| T2DM | endothelial | *NOL8* | -0.733626799 | 0.000421991 | 0.001181478 | 1 |
| T2DM | endothelial | *DLGAP2* | -0.456428027 | 1.43E-05 | 8.42E-05 | 0.069436145 |
| T2DM | endothelial | *MRPL42* | -0.397805452 | 0.008976951 | 0.015667686 | 1 |
| T2DM | endothelial | *AGAP9* | -1.003518736 | 2.45E-05 | 0.000125288 | 0.118654927 |
| T2DM | endothelial | *ZNF273* | -0.630863593 | 0.006132371 | 0.011287657 | 1 |
| T2DM | endothelial | *ZNF667* | -0.762759352 | 5.62E-05 | 0.000239021 | 0.272483722 |
| T2DM | endothelial | *ADH4* | -0.418870403 | 2.16E-06 | 2.47E-05 | 0.010478732 |
| T2DM | endothelial | *HIBCH* | -0.40089031 | 0.001599836 | 0.003600374 | 1 |
| T2DM | endothelial | *ZNF544* | -1.038062308 | 5.31E-05 | 0.000228724 | 0.257771778 |
| T2DM | endothelial | *SOWAHC* | -1.520554076 | 0.000421468 | 0.001181375 | 1 |
| T2DM | endothelial | *AC135178.1* | 1.886167035 | 9.63E-06 | 6.29E-05 | 0.04671069 |
| T2DM | endothelial | *MAN1A2* | -0.489451316 | 0.005324456 | 0.01003589 | 1 |
| T2DM | endothelial | *SVIP* | 0.400130821 | 0.001651824 | 0.003703629 | 1 |
| T2DM | endothelial | *DDRGK1* | -0.751845787 | 0.002980591 | 0.006122705 | 1 |
| T2DM | endothelial | *ZNF334* | -0.619885916 | 5.34E-05 | 0.000229538 | 0.259147922 |
| T2DM | endothelial | *HSD17B11* | 0.331514142 | 0.034036758 | 0.049733609 | 1 |
| T2DM | endothelial | *RPS6KL1* | -0.815392487 | 0.003138177 | 0.006400351 | 1 |
| T2DM | endothelial | *DDX42* | -0.612114047 | 0.000871894 | 0.002157281 | 1 |
| T2DM | endothelial | *STYX* | -0.657415433 | 0.009527772 | 0.016492598 | 1 |
| T2DM | endothelial | *HELZ* | -0.523518426 | 0.000191714 | 0.000627238 | 0.93019467 |
| T2DM | endothelial | *TMEM116* | -0.435058349 | 8.45E-07 | 1.53E-05 | 0.004101057 |
| T2DM | endothelial | *UCKL1* | -0.689915876 | 0.008058033 | 0.014258781 | 1 |
| T2DM | endothelial | *GFPT1* | -0.617771977 | 9.93E-06 | 6.43E-05 | 0.048166051 |
| T2DM | endothelial | *ZNF568* | -0.534697473 | 0.015247644 | 0.024826031 | 1 |
| T2DM | endothelial | *ZNF587* | -0.684178825 | 0.00021729 | 0.000695441 | 1 |
| T2DM | endothelial | *SH3BGRL2* | 0.210164012 | 0.000155703 | 0.0005283 | 0.755469086 |
| T2DM | endothelial | *TMA16* | -0.731420676 | 0.000578749 | 0.001533062 | 1 |
| T2DM | endothelial | *HLA-DRB5* | -0.172964448 | 2.71E-06 | 2.78E-05 | 0.013159842 |
| T2DM | endothelial | *CNGA1* | -0.261287238 | 0.016291167 | 0.026269439 | 1 |
| T2DM | endothelial | *ZNF28* | -0.893335697 | 0.00200975 | 0.004380938 | 1 |
| T2DM | endothelial | *C20orf203* | 0.997908586 | 0.000882247 | 0.002181785 | 1 |
| T2DM | endothelial | *NCOA6* | -0.391969655 | 0.013644181 | 0.022578979 | 1 |
| T2DM | endothelial | *FAM19A2* | -0.636102135 | 1.75E-06 | 2.24E-05 | 0.008479263 |
| T2DM | endothelial | *TUSC1* | -1.248368828 | 3.60E-07 | 1.24E-05 | 0.001744955 |
| T2DM | endothelial | *PAPSS2* | -0.500780594 | 0.005380282 | 0.010122191 | 1 |
| T2DM | endothelial | *CEP290* | -0.427497189 | 0.018221128 | 0.028948564 | 1 |
| T2DM | endothelial | *ANKRD13B* | -1.616333446 | 6.05E-06 | 4.65E-05 | 0.029339129 |
| T2DM | endothelial | *PPP1R14C* | -4.086504788 | 4.22E-06 | 3.67E-05 | 0.020478042 |
| T2DM | endothelial | *GPATCH3* | -1.074327074 | 6.86E-07 | 1.34E-05 | 0.003326548 |

| | | | | | | |
|---|---|---|---|---|---|---|
| T2DM | endothelial | *COLGALT2* | -0.68477025 | 1.14E-07 | 9.72E-06 | 0.000552504 |
| T2DM | endothelial | *RCSD1* | -0.695153796 | 0.000239793 | 0.000756979 | 1 |
| T2DM | endothelial | *GRIN3A* | -0.449836482 | 0.005956514 | 0.010993156 | 1 |
| T2DM | endothelial | *TMEM184B* | 0.331608839 | 0.026389544 | 0.039863658 | 1 |
| T2DM | endothelial | *SCAMP5* | -0.301939514 | 0.001597101 | 0.003595885 | 1 |
| T2DM | endothelial | *SFT2D1* | -0.770761015 | 0.000149472 | 0.00051326 | 0.72523704 |
| T2DM | endothelial | *HMGN2* | -0.791096888 | 1.95E-07 | 1.24E-05 | 0.000946299 |
| T2DM | endothelial | *DENND4B* | -0.664019178 | 0.000183981 | 0.000605206 | 0.892678176 |
| T2DM | endothelial | *RYR3* | 0.116184107 | 4.28E-06 | 3.70E-05 | 0.020747002 |
| T2DM | endothelial | *TYW1* | -0.534814569 | 0.000747167 | 0.001891108 | 1 |
| T2DM | endothelial | *PRMT6* | 1.09539959 | 9.64E-05 | 0.000361411 | 0.467666132 |
| T2DM | endothelial | *SHISA4* | 0.969660184 | 0.001418101 | 0.003248643 | 1 |
| T2DM | endothelial | *C1orf174* | -0.72855082 | 0.003267574 | 0.00662804 | 1 |
| T2DM | endothelial | *DZIP3* | -0.709135904 | 6.96E-06 | 5.11E-05 | 0.033791175 |
| T2DM | endothelial | *KIAA0753* | -0.688963251 | 0.001006871 | 0.002430516 | 1 |
| T2DM | endothelial | *L3MBTL3* | -0.449003559 | 0.009543498 | 0.016513928 | 1 |
| T2DM | endothelial | *NAGA* | -0.73184105 | 0.000393033 | 0.001116508 | 1 |
| T2DM | endothelial | *SMG5* | -0.679409922 | 0.000265646 | 0.000823062 | 1 |
| T2DM | endothelial | *TGM2* | 0.300124749 | 0.000123421 | 0.000441297 | 0.598840316 |
| T2DM | endothelial | *PJA2* | -0.567202399 | 0.000766363 | 0.001931531 | 1 |
| T2DM | endothelial | *AL590822.1* | -0.560713566 | 2.17E-07 | 1.24E-05 | 0.001053617 |
| T2DM | endothelial | *EFCAB2* | -0.437860952 | 0.003666067 | 0.007302034 | 1 |
| T2DM | endothelial | *TATDN3* | -0.34841287 | 0.011828541 | 0.02000421 | 1 |
| T2DM | endothelial | *SAMD5* | 0.295753069 | 4.62E-05 | 0.000205239 | 0.224120969 |
| T2DM | endothelial | *ECT2L* | -1.282611381 | 7.81E-05 | 0.000308507 | 0.379155201 |
| T2DM | endothelial | *GPR52* | -7.484049513 | 0.007765645 | 0.013832199 | 1 |
| T2DM | endothelial | *PCMTD2* | -0.61388494 | 0.001748214 | 0.003889196 | 1 |
| T2DM | endothelial | *SAMD13* | 0.5901045 | 0.032925317 | 0.048365793 | 1 |
| T2DM | endothelial | *EFCAB7* | -0.648134567 | 0.000898589 | 0.002210931 | 1 |
| T2DM | endothelial | *IFIT1B* | 2.038605278 | 1.79E-05 | 9.85E-05 | 0.086646306 |
| T2DM | endothelial | *LIPN* | -1.199910642 | 3.20E-07 | 1.24E-05 | 0.001553789 |
| T2DM | endothelial | *INPP5B* | -0.299207487 | 0.000925756 | 0.002269716 | 1 |
| T2DM | endothelial | *NEU4* | -0.799966093 | 0.000262774 | 0.000816247 | 1 |
| T2DM | endothelial | *C2orf72* | -3.681442817 | 1.71E-07 | 1.14E-05 | 0.000830737 |
| T2DM | endothelial | *RUFY2* | -0.555291818 | 0.001335841 | 0.003096751 | 1 |
| T2DM | endothelial | *ASAH2B* | 0.951721462 | 0.002408397 | 0.005096182 | 1 |
| T2DM | endothelial | *SYT15* | 0.330333947 | 2.75E-06 | 2.79E-05 | 0.013338874 |
| T2DM | endothelial | *LINC00619* | 0.494230168 | 2.26E-06 | 2.52E-05 | 0.010943502 |
| T2DM | endothelial | *TCEA3* | -0.598580989 | 3.64E-06 | 3.32E-05 | 0.017646794 |
| T2DM | endothelial | *HSD17B8* | 0.352803748 | 0.019701125 | 0.030925221 | 1 |
| T2DM | endothelial | *AGER* | 0.447658196 | 0.014943287 | 0.024404184 | 1 |
| T2DM | endothelial | *PRRT1* | 0.424949683 | 0.000312899 | 0.000935995 | 1 |
| T2DM | endothelial | *SMIM5* | -0.921252112 | 0.001482482 | 0.003370666 | 1 |
| T2DM | endothelial | *STK19* | -1.100266483 | 2.78E-07 | 1.24E-05 | 0.001350361 |
| T2DM | endothelial | *SKIV2L* | -0.378778452 | 8.55E-06 | 5.82E-05 | 0.041472754 |
| T2DM | endothelial | *GPANK1* | -0.854692051 | 9.46E-08 | 8.85E-06 | 0.000458949 |
| T2DM | endothelial | *LST1* | -0.497990626 | 0.000720416 | 0.001834885 | 1 |
| T2DM | endothelial | *ZNF814* | -0.545781169 | 0.002089614 | 0.004524233 | 1 |
| T2DM | endothelial | *MICA* | -0.425163807 | 0.0036387 | 0.007256463 | 1 |
| T2DM | endothelial | *PSORS1C2* | 0.326868459 | 0.029759638 | 0.044387877 | 1 |

| | | | | | | |
|---|---|---|---|---|---|---|
| T2DM | endothelial | *HLA-E* | 0.256453141 | 0.001133496 | 0.00268935 | 1 |
| T2DM | endothelial | *ZNF468* | -1.337746106 | 0.010556617 | 0.018056004 | 1 |
| T2DM | endothelial | *RNF39* | -0.428714743 | 0.032688592 | 0.048120463 | 1 |
| T2DM | endothelial | *HCG9* | -0.424596058 | 0.023767967 | 0.036402202 | 1 |
| T2DM | endothelial | *HLA-G* | 0.359214466 | 0.001658807 | 0.003717566 | 1 |
| T2DM | endothelial | *FAM153C* | -0.320668034 | 7.84E-06 | 5.51E-05 | 0.038023077 |
| T2DM | endothelial | *TRIM27* | -0.588688505 | 0.000965336 | 0.002343719 | 1 |
| T2DM | endothelial | *MALRD1* | -0.554919159 | 3.59E-07 | 1.24E-05 | 0.001742215 |
| T2DM | endothelial | *FAM196B* | -0.735505482 | 2.74E-06 | 2.79E-05 | 0.013312235 |
| T2DM | endothelial | *FAM216A* | -0.857311542 | 0.000102243 | 0.00037822 | 0.49608102 |
| T2DM | endothelial | *ZBTB48* | -0.863070086 | 7.68E-06 | 5.45E-05 | 0.037282916 |
| T2DM | endothelial | *ZNF155* | -0.581718587 | 0.011167169 | 0.018965034 | 1 |
| T2DM | endothelial | *ZNF783* | -0.779858872 | 0.000886702 | 0.002188342 | 1 |
| T2DM | endothelial | *ZNF425* | -0.669232971 | 0.000123759 | 0.000441852 | 0.600476434 |
| T2DM | endothelial | *C12orf73* | -0.600230753 | 0.017961372 | 0.028592053 | 1 |
| T2DM | endothelial | *PCDHA7* | -6.166063039 | 9.72E-05 | 0.000363645 | 0.471659162 |
| T2DM | endothelial | *PCDHA4* | -1.337450089 | 0.00010541 | 0.000387462 | 0.511449611 |
| T2DM | endothelial | *PKHD1L1* | -0.351933119 | 9.49E-06 | 6.26E-05 | 0.046056232 |
| T2DM | endothelial | *SLFN12L* | -0.478584484 | 0.005119911 | 0.009707623 | 1 |
| T2DM | endothelial | *SYCE1L* | 0.29676934 | 0.000161067 | 0.000542706 | 0.781497071 |
| T2DM | endothelial | *TMEM231* | 0.516681468 | 0.029539008 | 0.044085902 | 1 |
| T2DM | endothelial | *SCGB2B2* | -0.743903623 | 3.47E-05 | 0.000162331 | 0.168577941 |
| T2DM | endothelial | *CCDC144NL* | -0.899016347 | 3.50E-07 | 1.24E-05 | 0.001699892 |
| T2DM | endothelial | *TMEM170B* | -0.335877691 | 0.000794928 | 0.001992247 | 1 |
| T2DM | endothelial | *SAP25* | -1.368514188 | 0.00050083 | 0.001360937 | 1 |
| T2DM | endothelial | *NT5M* | -0.909200632 | 3.66E-07 | 1.24E-05 | 0.001776789 |
| T2DM | endothelial | *IPO7* | -0.646277156 | 0.000706519 | 0.001806127 | 1 |
| T2DM | endothelial | *PRR13* | -0.952533236 | 0.000249713 | 0.000782693 | 1 |
| T2DM | endothelial | *TECPR1* | 0.417592593 | 0.001316456 | 0.003060586 | 1 |
| T2DM | endothelial | *MT1A* | -1.442579177 | 3.90E-06 | 3.47E-05 | 0.018898945 |
| T2DM | endothelial | *MT1M* | -1.430475554 | 3.75E-05 | 0.00017263 | 0.182124228 |
| T2DM | endothelial | *CFI* | 0.331735945 | 0.003429229 | 0.006915469 | 1 |
| T2DM | endothelial | *RGL3* | -0.350490763 | 0.006433026 | 0.011760755 | 1 |
| T2DM | endothelial | *NAP1L4* | -0.438244547 | 0.009711599 | 0.016774894 | 1 |
| T2DM | endothelial | *SMN2* | -0.288786611 | 0.021964428 | 0.03395075 | 1 |
| T2DM | endothelial | *SERF1B* | -0.70651785 | 1.36E-05 | 8.09E-05 | 0.065880097 |
| T2DM | endothelial | *KLRC2* | -1.506190765 | 3.16E-06 | 3.03E-05 | 0.015315046 |
| T2DM | endothelial | *LINC00487* | 0.765380708 | 0.001961824 | 0.004291601 | 1 |
| T2DM | endothelial | *RFPL3S* | -0.293760501 | 0.021660993 | 0.033597625 | 1 |
| T2DM | endothelial | *C1QTNF9B* | 4.826617661 | 0.028763713 | 0.043021436 | 1 |
| T2DM | endothelial | *COL6A6* | -0.590450432 | 2.88E-06 | 2.89E-05 | 0.013961038 |
| T2DM | endothelial | *LNP1* | -0.521327299 | 0.000262374 | 0.000815528 | 1 |
| T2DM | endothelial | *PRSS50* | -1.538734525 | 5.38E-07 | 1.27E-05 | 0.002609352 |
| T2DM | endothelial | *METTL6* | -0.645120698 | 0.0012179 | 0.002871418 | 1 |
| T2DM | endothelial | *SACM1L* | -0.455365704 | 0.033777142 | 0.049437916 | 1 |
| T2DM | endothelial | *SLC48A1* | 0.342309398 | 0.000106997 | 0.000392402 | 0.519148116 |
| T2DM | endothelial | *TAS2R50* | -4.648433006 | 0.000193539 | 0.000631932 | 0.93905139 |
| T2DM | endothelial | *AC084018.1* | -0.306468916 | 0.008197137 | 0.014457474 | 1 |
| T2DM | endothelial | *C17orf51* | -1.257636399 | 4.40E-07 | 1.24E-05 | 0.002133718 |
| T2DM | endothelial | *OR2A42* | -0.371164733 | 0.013180073 | 0.021968297 | 1 |

| | | | | | | |
|---|---|---|---|---|---|---|
| T2DM | endothelial | *ZNF580* | -0.490290354 | 0.002812589 | 0.005834408 | 1 |
| T2DM | endothelial | *NUP62* | -0.574176571 | 0.000265356 | 0.000822688 | 1 |
| T2DM | endothelial | *AL021068.1* | -0.63503752 | 0.001224528 | 0.002882781 | 1 |
| T2DM | endothelial | *ZNF254* | -0.645434696 | 0.000159126 | 0.000537285 | 0.772079071 |
| T2DM | endothelial | *AC022498.1* | -3.373142841 | 8.05E-05 | 0.000315343 | 0.390394633 |
| T2DM | endothelial | *GIMAP1* | 0.250686795 | 4.61E-05 | 0.000205095 | 0.223758149 |
| T2DM | endothelial | *DNLZ* | 0.223419775 | 0.004179031 | 0.008149782 | 1 |
| T2DM | endothelial | *GSTM2* | -0.396536204 | 2.83E-05 | 0.000137812 | 0.137123282 |
| T2DM | endothelial | *LCAT* | -1.003953944 | 0.001800179 | 0.003984703 | 1 |
| T2DM | endothelial | *GPC2* | -1.738652861 | 5.65E-05 | 0.000240114 | 0.274289886 |
| T2DM | endothelial | *ERV3-1* | -0.155965493 | 0.00437095 | 0.008479748 | 1 |
| T2DM | endothelial | *CCL14* | 0.131626846 | 2.90E-05 | 0.000140587 | 0.140587058 |
| T2DM | endothelial | *SRA1* | -0.694808043 | 0.000569695 | 0.001514058 | 1 |
| T2DM | endothelial | *TMEM110* | 1.253072044 | 7.73E-06 | 5.45E-05 | 0.037484104 |
| T2DM | endothelial | *LBH* | -0.869845738 | 3.08E-06 | 3.00E-05 | 0.014947811 |
| T2DM | endothelial | *PPP1CB* | -0.486496266 | 0.004383856 | 0.008494848 | 1 |
| T2DM | endothelial | *NCKIPSD* | -0.465681214 | 0.002233694 | 0.004780878 | 1 |
| T2DM | endothelial | *EMP2* | 0.308418524 | 0.015870257 | 0.025676054 | 1 |
| T2DM | endothelial | *LTB4R* | -1.064282413 | 0.00031996 | 0.000952993 | 1 |
| T2DM | endothelial | *IRF9* | 0.481713088 | 0.001125676 | 0.002674721 | 1 |
| T2DM | endothelial | *GALT* | -0.632829301 | 0.006919315 | 0.012541098 | 1 |
| T2DM | endothelial | *ZNF726* | -1.574408326 | 1.01E-06 | 1.63E-05 | 0.004914176 |
| T2DM | endothelial | *ZNF90* | -0.563719839 | 0.029027644 | 0.043362725 | 1 |
| T2DM | endothelial | *REPIN1* | -1.137450354 | 0.001497079 | 0.003400669 | 1 |
| T2DM | endothelial | *SMIM7* | -0.654931639 | 0.001382781 | 0.003187293 | 1 |
| T2DM | endothelial | *TSPAN4* | -0.660024559 | 0.000157805 | 0.000534314 | 0.76567176 |
| T2DM | endothelial | *PAXIP1-AS2* | -0.689552325 | 0.007690171 | 0.013717907 | 1 |
| T2DM | endothelial | *SH3D21* | -0.892190034 | 0.00142588 | 0.003261844 | 1 |
| T2DM | endothelial | *ANG* | -0.747176705 | 0.024104907 | 0.036802079 | 1 |
| T2DM | endothelial | *COLCA2* | -0.40478023 | 0.000101455 | 0.000376347 | 0.492261952 |
| T2DM | endothelial | *SOGA3* | -0.426784667 | 1.76E-05 | 9.74E-05 | 0.08560266 |
| T2DM | endothelial | *LCNL1* | -0.757775181 | 5.14E-05 | 0.000223269 | 0.249614207 |
| T2DM | endothelial | *GNAT3* | -1.26127342 | 2.11E-05 | 0.000111803 | 0.102187776 |
| T2DM | endothelial | *PLIN5* | -0.336621302 | 2.20E-05 | 0.000115159 | 0.106867398 |
| T2DM | endothelial | *STARD10* | 0.370360503 | 0.000154818 | 0.000526404 | 0.751179018 |
| T2DM | endothelial | *NUTM2D* | -2.402277464 | 0.003965752 | 0.007809183 | 1 |
| T2DM | endothelial | *CPEB1* | -0.650150113 | 7.33E-07 | 1.42E-05 | 0.00355556 |
| T2DM | endothelial | *ARHGEF33* | -0.473269426 | 0.008061407 | 0.01425955 | 1 |
| T2DM | endothelial | *IFRD2* | -0.946240733 | 2.23E-08 | 4.12E-06 | 0.000108335 |
| T2DM | endothelial | *EVPLL* | -0.600449221 | 0.000252795 | 0.000789802 | 1 |
| T2DM | endothelial | *AC005082.1* | -2.43853052 | 0.000114501 | 0.000413978 | 0.555558139 |
| T2DM | endothelial | *ACSM4* | -0.258615807 | 0.025596071 | 0.038773692 | 1 |
| T2DM | endothelial | *UBXN2B* | -0.615858365 | 0.00010636 | 0.000390364 | 0.516061117 |
| T2DM | endothelial | *C5orf49* | -3.920145905 | 2.38E-07 | 1.24E-05 | 0.001156499 |
| T2DM | endothelial | *GOLGA8B* | -0.294108235 | 0.032451099 | 0.047814374 | 1 |
| T2DM | endothelial | *VPS16* | -1.03337361 | 3.03E-05 | 0.00014575 | 0.14691636 |
| T2DM | endothelial | *NPEPL1* | -1.135493334 | 1.13E-05 | 7.09E-05 | 0.054887526 |
| T2DM | endothelial | *TMEM242* | -0.477614822 | 0.030619578 | 0.045530552 | 1 |
| T2DM | endothelial | *SLC35E2* | -0.665907025 | 1.14E-06 | 1.76E-05 | 0.005537036 |
| T2DM | endothelial | *TSTD1* | -1.030718422 | 8.85E-06 | 5.97E-05 | 0.042950119 |

| | | | | | | |
|---|---|---|---|---|---|---|
| T2DM | endothelial | *AC009403.2* | -0.616493221 | 2.46E-06 | 2.65E-05 | 0.011938509 |
| T2DM | endothelial | *PAM16* | -1.118760769 | 0.00109175 | 0.002605592 | 1 |
| T2DM | endothelial | *AC016773.1* | -0.379389441 | 0.034232745 | 0.049984134 | 1 |
| T2DM | endothelial | *NBPF1* | -0.591746058 | 9.93E-05 | 0.000369339 | 0.481987264 |
| T2DM | endothelial | *RNU6ATAC35I* | -3.390210109 | 2.99E-07 | 1.24E-05 | 0.001450956 |
| T2DM | endothelial | *PPP3R1* | -0.541875004 | 0.006028376 | 0.011111786 | 1 |
| T2DM | endothelial | *FANCG* | -0.615264128 | 0.000139722 | 0.000485972 | 0.67793119 |
| T2DM | endothelial | *TIGD1* | -0.672729944 | 0.000153331 | 0.000523549 | 0.743963659 |
| T2DM | endothelial | *FXYD7* | -0.908533175 | 4.07E-05 | 0.000184594 | 0.197331169 |
| T2DM | endothelial | *XKR9* | -0.158837233 | 0.0220701 | 0.034103225 | 1 |
| T2DM | endothelial | *SLC12A8* | -0.55477151 | 3.69E-05 | 0.000170539 | 0.179065466 |
| T2DM | endothelial | *APOL6* | -0.665237494 | 1.62E-05 | 9.15E-05 | 0.078768496 |
| T2DM | endothelial | *OR2A1* | -0.473974228 | 0.000123654 | 0.000441804 | 0.599969725 |
| T2DM | endothelial | *TIAF1* | 0.524795957 | 0.025759579 | 0.0390092 | 1 |
| T2DM | endothelial | *RAB6C* | -7.373140426 | 1.88E-06 | 2.33E-05 | 0.009109252 |
| T2DM | endothelial | *AC062017.1* | -1.052883629 | 1.25E-06 | 1.85E-05 | 0.006078309 |
| T2DM | endothelial | *EXOSC6* | -0.606126796 | 0.019223546 | 0.030332568 | 1 |
| T2DM | endothelial | *LINC00571* | -0.733774243 | 7.72E-06 | 5.45E-05 | 0.037480112 |
| T2DM | endothelial | *OSER1-AS1* | -0.358451466 | 0.017839606 | 0.028463587 | 1 |
| T2DM | endothelial | *C5orf66* | -0.903572385 | 3.61E-05 | 0.000167672 | 0.175217334 |
| T2DM | endothelial | *LINC01133* | -0.792173311 | 2.11E-06 | 2.46E-05 | 0.010254736 |
| T2DM | endothelial | *C4B* | 0.986336117 | 0.000632092 | 0.001646222 | 1 |
| T2DM | endothelial | *LINC01117* | 0.305371197 | 1.35E-05 | 8.08E-05 | 0.065656627 |
| T2DM | endothelial | *LINC00240* | -0.325694846 | 0.000566376 | 0.001507435 | 1 |
| T2DM | endothelial | *LINC00184* | -6.637986759 | 5.46E-07 | 1.27E-05 | 0.002648968 |
| T2DM | endothelial | *AL049840.1* | -0.816047304 | 0.000110123 | 0.000402154 | 0.534317745 |
| T2DM | endothelial | *PLEKHM1* | -0.546898335 | 0.01122983 | 0.019064777 | 1 |
| T2DM | endothelial | *AP000350.6* | -0.270982262 | 0.009112368 | 0.015875479 | 1 |
| T2DM | endothelial | *SFTA1P* | -0.769596443 | 0.000508095 | 0.001377251 | 1 |
| T2DM | endothelial | *CYP4A22-AS1* | -1.208066464 | 9.34E-07 | 1.59E-05 | 0.004530993 |
| T2DM | endothelial | *MIAT* | -1.059288846 | 3.23E-06 | 3.07E-05 | 0.015689212 |
| T2DM | endothelial | *ERCC6* | -0.798496616 | 5.72E-06 | 4.46E-05 | 0.027729432 |
| T2DM | endothelial | *LINC00694* | 0.665177601 | 1.85E-07 | 1.19E-05 | 0.000896144 |
| T2DM | endothelial | *LINC00115* | -1.435846133 | 1.94E-05 | 0.000104664 | 0.094092509 |
| T2DM | endothelial | *HAR1A* | -4.248625203 | 6.17E-07 | 1.31E-05 | 0.002995036 |
| T2DM | endothelial | *LINC00937* | -0.360653929 | 0.016357219 | 0.026358428 | 1 |
| T2DM | endothelial | *FAM138B* | -8.847848039 | 2.82E-07 | 1.24E-05 | 0.001366494 |
| T2DM | endothelial | *ZNF717* | -0.170528655 | 0.017400673 | 0.027872534 | 1 |
| T2DM | endothelial | *AC009299.3* | -0.997193935 | 3.86E-06 | 3.45E-05 | 0.018715263 |
| T2DM | endothelial | *SCAMP4* | -0.907483011 | 2.06E-05 | 0.000109424 | 0.0997943 |
| T2DM | endothelial | *SLC2A1-AS1* | -0.908419807 | 0.000193052 | 0.000630767 | 0.936689054 |
| T2DM | endothelial | *AC005884.1* | -0.958538275 | 7.67E-05 | 0.000305012 | 0.372243305 |
| T2DM | endothelial | *C14orf144* | 0.610702901 | 1.26E-05 | 7.66E-05 | 0.061183996 |
| T2DM | endothelial | *POLR2J2* | 2.746640379 | 0.000662068 | 0.001711431 | 1 |
| T2DM | endothelial | *LINC00578* | -0.65110376 | 3.88E-07 | 1.24E-05 | 0.001884749 |
| T2DM | endothelial | *C19orf24* | -0.792531849 | 0.020381264 | 0.031825157 | 1 |
| T2DM | endothelial | *ZNF883* | -0.757369192 | 0.000290224 | 0.000880105 | 1 |
| T2DM | endothelial | *ARL17B* | -0.522976944 | 0.004383986 | 0.008494848 | 1 |
| T2DM | endothelial | *DHFR* | -0.20754277 | 0.001016979 | 0.002451257 | 1 |
| T2DM | endothelial | *PATL2* | -0.634301081 | 7.30E-05 | 0.000294466 | 0.3544379 |

| | | | | | | |
|---|---|---|---|---|---|---|
| T2DM | endothelial | *CASC20* | -1.001438714 | 1.68E-07 | 1.14E-05 | 0.000815058 |
| T2DM | endothelial | *PSMG3-AS1* | -0.854554802 | 8.64E-06 | 5.87E-05 | 0.041917268 |
| T2DM | endothelial | *KLHL7-AS1* | -0.627390527 | 1.29E-05 | 7.77E-05 | 0.062430388 |
| T2DM | endothelial | *LINC00332* | -0.68755074 | 9.84E-07 | 1.61E-05 | 0.004775023 |
| T2DM | endothelial | *AC092171.4* | -1.395793089 | 1.06E-05 | 6.72E-05 | 0.051307695 |
| T2DM | endothelial | *DIRC3* | -0.507725093 | 2.72E-05 | 0.000134552 | 0.131916 |
| T2DM | endothelial | *AC107021.1* | -0.65936653 | 6.65E-06 | 4.92E-05 | 0.032276947 |
| T2DM | endothelial | *MTRNR2L4* | -0.930848102 | 0.000103779 | 0.000382334 | 0.50353425 |
| T2DM | endothelial | *LINC00865* | -0.441049923 | 0.007493096 | 0.013415683 | 1 |
| T2DM | endothelial | *HLA-DQB2* | 0.237171451 | 1.14E-07 | 9.72E-06 | 0.000554264 |
| T2DM | endothelial | *LYRM9* | -0.951772832 | 3.86E-06 | 3.45E-05 | 0.018731266 |
| T2DM | endothelial | *LINC00342* | -1.022710798 | 0.005498982 | 0.010297592 | 1 |
| T2DM | endothelial | *LINC00702* | -1.391967101 | 3.32E-07 | 1.24E-05 | 0.001612439 |
| T2DM | endothelial | *GPX1* | -0.512246898 | 0.024751893 | 0.037671325 | 1 |
| T2DM | endothelial | *PINLYP* | -0.722067134 | 2.73E-06 | 2.78E-05 | 0.013232353 |
| T2DM | endothelial | *ANKRD65* | 1.554586172 | 0.011817682 | 0.019992814 | 1 |
| T2DM | endothelial | *ZSCAN31* | 0.230704651 | 0.000328707 | 0.000970124 | 1 |
| T2DM | endothelial | *AC009495.2* | -4.208822422 | 0.019582899 | 0.030759542 | 1 |
| T2DM | endothelial | *KIAA0040* | -0.814530019 | 0.000529061 | 0.00142532 | 1 |
| T2DM | endothelial | *LINC00607* | -0.553831307 | 0.000192801 | 0.000630372 | 0.935472175 |
| T2DM | endothelial | *AP001468.1* | -0.310609653 | 0.004945658 | 0.009417411 | 1 |
| T2DM | endothelial | *LINC00941* | -1.114309414 | 0.003257078 | 0.006609512 | 1 |
| T2DM | endothelial | *AC019117.2* | -3.542971292 | 4.89E-07 | 1.27E-05 | 0.002372001 |
| T2DM | endothelial | *ZBED5* | -0.688480204 | 0.000139505 | 0.000485566 | 0.676879642 |
| T2DM | endothelial | *CDKN2AIPNL* | -0.455859127 | 0.025527997 | 0.038682648 | 1 |
| T2DM | endothelial | *LINC00987* | 0.243400644 | 2.94E-05 | 0.000142487 | 0.142772125 |
| T2DM | endothelial | *GS1-124K5.4* | 0.159814083 | 0.024087206 | 0.036790847 | 1 |
| T2DM | endothelial | *LINC00959* | -1.068397864 | 3.24E-05 | 0.00015315 | 0.157285298 |
| T2DM | endothelial | *HLA-DQA2* | 0.375129222 | 5.18E-13 | 1.26E-09 | 2.51E-09 |
| T2DM | endothelial | *LINC01143* | 0.230593842 | 2.05E-06 | 2.42E-05 | 0.009929197 |
| T2DM | endothelial | *LINC00211* | 0.562239017 | 1.85E-06 | 2.32E-05 | 0.008997769 |
| T2DM | endothelial | *LRRC37A2* | -0.739183926 | 0.00089032 | 0.002195038 | 1 |
| T2DM | endothelial | *LINC00707* | -1.133266439 | 0.025464073 | 0.038597839 | 1 |
| T2DM | endothelial | *TXNDC5* | 1.833277962 | 2.44E-05 | 0.000125169 | 0.118284835 |
| T2DM | endothelial | *LINC01119* | -1.043987472 | 3.03E-05 | 0.000145758 | 0.147092644 |
| T2DM | endothelial | *TNFSF12* | 0.42857303 | 0.028248554 | 0.042303082 | 1 |
| T2DM | endothelial | *C1orf226* | 1.414673755 | 1.98E-05 | 0.000106242 | 0.096149097 |
| T2DM | endothelial | *LILRA2* | -0.277137803 | 0.007892838 | 0.014009111 | 1 |
| T2DM | endothelial | *PSMB9* | 0.363345274 | 7.59E-05 | 0.000302527 | 0.368052947 |
| T2DM | endothelial | *C1QTNF9* | 0.346111985 | 3.04E-06 | 2.99E-05 | 0.014756058 |
| T2DM | endothelial | *ISY1* | -0.525796659 | 8.49E-05 | 0.000327271 | 0.4117074 |
| T2DM | endothelial | *PNMA2* | -0.884254729 | 2.71E-06 | 2.78E-05 | 0.013148445 |
| T2DM | endothelial | *KRBOX1* | -4.163090839 | 5.10E-07 | 1.27E-05 | 0.002473775 |
| T2DM | endothelial | *LINC00886* | -0.381365747 | 0.000332022 | 0.000978125 | 1 |
| T2DM | endothelial | *MIF* | -0.284930579 | 0.005375556 | 0.010117222 | 1 |
| T2DM | endothelial | *NSUN6* | -0.521283961 | 0.003250493 | 0.006599682 | 1 |
| T2DM | endothelial | *TDGF1* | -0.524521756 | 0.00245004 | 0.005166272 | 1 |
| T2DM | endothelial | *CD302* | -0.682552928 | 0.000131524 | 0.000463775 | 0.638154518 |
| T2DM | endothelial | *PLEKHO2* | -0.422754479 | 0.01955731 | 0.030729296 | 1 |
| T2DM | endothelial | *HOGA1* | -1.021703632 | 3.13E-07 | 1.24E-05 | 0.00151629 |

| | | | | | | |
|---|---|---|---|---|---|---|
| T2DM | endothelial | *PI4KA* | -0.51833317 | 0.003531609 | 0.007076567 | 1 |
| T2DM | endothelial | *LINC00969* | -0.322757785 | 0.002749876 | 0.005718988 | 1 |
| T2DM | endothelial | *DECR2* | -1.545359012 | 1.19E-06 | 1.80E-05 | 0.005787081 |
| T2DM | endothelial | *ZNF709* | 3.805565108 | 0.000178975 | 0.000591949 | 0.868388551 |
| T2DM | endothelial | *MRPL33* | -0.455114422 | 0.023663499 | 0.036253647 | 1 |
| T2DM | endothelial | *STON1* | 0.386102101 | 0.008196253 | 0.014457474 | 1 |
| T2DM | endothelial | *VSIG8* | -1.219059363 | 1.80E-05 | 9.91E-05 | 0.087365391 |
| T2DM | endothelial | *C7orf73* | -0.539895215 | 0.006271384 | 0.011504255 | 1 |
| T2DM | endothelial | *FGF14-IT1* | -1.399286407 | 2.47E-07 | 1.24E-05 | 0.00120024 |
| T2DM | endothelial | *PALM2* | -0.534869348 | 6.41E-06 | 4.82E-05 | 0.031087456 |
| T2DM | endothelial | *UPK3B* | -0.823685188 | 3.34E-06 | 3.11E-05 | 0.016191328 |
| T2DM | endothelial | *PLA2G4B* | 2.598784777 | 3.55E-10 | 2.15E-07 | 1.72E-06 |
| T2DM | endothelial | *LEFTY1* | 3.776509217 | 0.014530075 | 0.023793427 | 1 |
| T2DM | endothelial | *JMJD7* | 0.336528209 | 0.005939894 | 0.010966653 | 1 |
| T2DM | endothelial | *APOBEC3D* | -3.873390625 | 5.75E-07 | 1.27E-05 | 0.002789154 |
| T2DM | endothelial | *TMEM199* | -0.335523698 | 0.010614996 | 0.018128408 | 1 |
| T2DM | endothelial | *SPRR2F* | -0.725978701 | 0.00484627 | 0.009257521 | 1 |
| T2DM | endothelial | *P2RY11* | -0.326444692 | 0.000929075 | 0.002276703 | 1 |
| T2DM | endothelial | *SCARF2* | -0.942402112 | 3.48E-05 | 0.000162331 | 0.168661518 |
| T2DM | endothelial | *APOBEC3C* | -0.933636133 | 3.84E-05 | 0.000176052 | 0.186438551 |
| T2DM | endothelial | *CCDC13* | -0.606186982 | 2.39E-06 | 2.61E-05 | 0.011610119 |
| T2DM | endothelial | *C4A* | -0.894821744 | 7.33E-05 | 0.000294645 | 0.355636867 |
| T2DM | endothelial | *NEAT1* | -0.321159377 | 0.002824914 | 0.005856372 | 1 |
| T2DM | endothelial | *RAB30-AS1* | -0.621788983 | 5.12E-05 | 0.000222374 | 0.248392062 |
| T2DM | endothelial | *UBAP1L* | -0.90170137 | 2.71E-05 | 0.000134479 | 0.13165449 |
| T2DM | endothelial | *DNAJC3-AS1* | -0.904084802 | 2.78E-05 | 0.00013661 | 0.134970453 |
| T2DM | endothelial | *LINC00926* | -1.566428108 | 6.42E-07 | 1.33E-05 | 0.003115835 |
| T2DM | endothelial | *DPH6-AS1* | -0.529997534 | 7.83E-05 | 0.000308683 | 0.379893492 |
| T2DM | endothelial | *BCKDHA* | 0.448254032 | 0.027148112 | 0.040844229 | 1 |
| T2DM | endothelial | *CDK11B* | -0.904072312 | 0.000146468 | 0.00050581 | 0.710663358 |
| T2DM | endothelial | *ABHD14A* | -0.983798134 | 2.68E-05 | 0.000132993 | 0.12993426 |
| T2DM | endothelial | *SRP14-AS1* | -0.322209662 | 3.52E-06 | 3.24E-05 | 0.017082712 |
| T2DM | endothelial | *GDNF-AS1* | -0.796472824 | 3.55E-07 | 1.24E-05 | 0.001722575 |
| T2DM | endothelial | *CCDC153* | 0.808578854 | 0.003567934 | 0.00712999 | 1 |
| T2DM | endothelial | *C5orf17* | -0.811130733 | 5.67E-05 | 0.000240225 | 0.275061286 |
| T2DM | endothelial | *TMEM150C* | 0.190976672 | 0.000417464 | 0.001171506 | 1 |
| T2DM | endothelial | *LINC00939* | -0.74448153 | 0.001101583 | 0.002623899 | 1 |
| T2DM | endothelial | *LINC01060* | 0.222446095 | 1.32E-05 | 7.96E-05 | 0.064082201 |
| T2DM | endothelial | *TMEM158* | -2.718586376 | 0.000453917 | 0.001253503 | 1 |
| T2DM | endothelial | *DNAH10OS* | -2.267189482 | 2.38E-08 | 4.12E-06 | 0.000115479 |
| T2DM | endothelial | *PCDHA10* | -0.406084416 | 0.000949415 | 0.002316019 | 1 |
| T2DM | endothelial | *SMIM20* | 0.518174916 | 0.014565979 | 0.023836131 | 1 |
| T2DM | endothelial | *LINC00989* | -0.788931256 | 0.017823374 | 0.028447043 | 1 |
| T2DM | endothelial | *GLI4* | -0.911949526 | 6.75E-07 | 1.34E-05 | 0.003274782 |
| T2DM | endothelial | *SNHG18* | -0.822700631 | 2.59E-08 | 4.34E-06 | 0.000125823 |
| T2DM | endothelial | *ALG1L2* | -1.062982128 | 2.98E-06 | 2.95E-05 | 0.014436306 |
| T2DM | endothelial | *LINC01094* | -1.21134576 | 9.36E-07 | 1.59E-05 | 0.00453911 |
| T2DM | endothelial | *TMEM200B* | -0.543043616 | 0.000269762 | 0.000832624 | 1 |
| T2DM | endothelial | *PCDHGB6* | 0.537310958 | 0.002442293 | 0.005154417 | 1 |
| T2DM | endothelial | *PCDHGB2* | -3.824195693 | 3.28E-06 | 3.10E-05 | 0.015918682 |

| | | | | | | |
|---|---|---|---|---|---|---|
| T2DM | endothelial | *LYN* | -0.274417299 | 0.031956674 | 0.047171823 | 1 |
| T2DM | endothelial | *ARMS2* | -0.674504617 | 0.000613771 | 0.00160887 | 1 |
| T2DM | endothelial | *MEX3A* | 0.455380134 | 0.008678097 | 0.015195281 | 1 |
| T2DM | endothelial | *BRK1* | -0.406190166 | 0.032142737 | 0.047388806 | 1 |
| T2DM | endothelial | *SNHG9* | -0.65835275 | 0.00050436 | 0.001367889 | 1 |
| T2DM | endothelial | *PCDHA3* | -1.114838982 | 3.43E-06 | 3.18E-05 | 0.016624231 |
| T2DM | endothelial | *TAS2R20* | 0.771192322 | 0.000288498 | 0.000877064 | 1 |
| T2DM | endothelial | *HMBS* | -0.485872318 | 0.018403414 | 0.029190378 | 1 |
| T2DM | endothelial | *GATC* | -0.418636214 | 0.006618957 | 0.012068838 | 1 |
| T2DM | endothelial | *ZBED6* | -1.763982651 | 3.99E-07 | 1.24E-05 | 0.001935896 |
| T2DM | endothelial | *CNPY2* | -0.813645681 | 5.03E-06 | 4.09E-05 | 0.024422268 |
| T2DM | endothelial | *LINC00592* | -1.207707216 | 7.61E-06 | 5.43E-05 | 0.036938009 |
| T2DM | endothelial | *RTEL1* | -0.438285485 | 0.010614826 | 0.018128408 | 1 |
| T2DM | endothelial | *PDF* | 1.186564817 | 8.75E-05 | 0.000335146 | 0.424630545 |
| T2DM | endothelial | *HIF1A-AS2* | -0.666215581 | 0.001094837 | 0.002610393 | 1 |
| T2DM | endothelial | *SLC25A21-AS1* | -0.64375716 | 0.020385941 | 0.031825157 | 1 |
| T2DM | endothelial | *RNASE4* | 1.466262717 | 5.59E-05 | 0.000238149 | 0.271249007 |
| T2DM | endothelial | *LINC00639* | 0.10085843 | 5.26E-06 | 4.21E-05 | 0.02550796 |
| T2DM | endothelial | *SMIM6* | -2.012087439 | 0.001888595 | 0.004152 | 1 |
| T2DM | endothelial | *ITGB3* | 0.361579481 | 0.000730022 | 0.00185643 | 1 |
| T2DM | endothelial | *MRPL46* | -0.698458744 | 0.031094789 | 0.046081831 | 1 |
| T2DM | endothelial | *UBE2Q2L* | -0.980950201 | 2.80E-05 | 0.000137135 | 0.135858955 |
| T2DM | endothelial | *AC083843.1* | -4.930719878 | 5.41E-07 | 1.27E-05 | 0.002622559 |
| T2DM | endothelial | *PWRN1* | -0.571460986 | 4.92E-06 | 4.04E-05 | 0.023865089 |
| T2DM | endothelial | *LINC00662* | -0.556786905 | 0.002560407 | 0.005384957 | 1 |
| T2DM | endothelial | *C19orf84* | -3.34448885 | 2.05E-06 | 2.42E-05 | 0.009936472 |
| T2DM | endothelial | *LINC00667* | -0.46678406 | 0.001949048 | 0.004267501 | 1 |
| T2DM | endothelial | *LINC00672* | -2.422440409 | 0.000297512 | 0.000897717 | 1 |
| T2DM | endothelial | *LINC00909* | -0.59891592 | 0.001304871 | 0.003039858 | 1 |
| T2DM | endothelial | *DYNLL2* | -1.324943858 | 0.000654375 | 0.001696969 | 1 |
| T2DM | endothelial | *STRADA* | -0.732540948 | 0.000302 | 0.000908998 | 1 |
| T2DM | endothelial | *ILF3-AS1* | -0.584998804 | 0.000137378 | 0.000480576 | 0.666558247 |
| T2DM | endothelial | *ZNF582-AS1* | 0.759736684 | 0.016976891 | 0.027221373 | 1 |
| T2DM | endothelial | *ZNF285* | 0.298514677 | 0.025956978 | 0.039246887 | 1 |
| T2DM | endothelial | *NDUFA7* | -2.945162971 | 4.77E-07 | 1.27E-05 | 0.002313268 |
| T2DM | endothelial | *AC004466.1* | 0.557561327 | 0.000152175 | 0.000521069 | 0.738355168 |
| T2DM | endothelial | *LINC00664* | 2.748703064 | 1.30E-06 | 1.88E-05 | 0.006284863 |
| T2DM | endothelial | *SCGB1B2P* | -0.774992156 | 0.015535749 | 0.02521052 | 1 |
| T2DM | endothelial | *AC026740.1* | -1.284417557 | 0.015295648 | 0.024895835 | 1 |
| T2DM | endothelial | *ZNF728* | -0.767250811 | 0.011837901 | 0.020006093 | 1 |
| T2DM | endothelial | *TRABD2B* | 0.269479416 | 1.73E-05 | 9.61E-05 | 0.084165453 |
| T2DM | endothelial | *FBXO17* | -0.426423715 | 4.11E-05 | 0.000185853 | 0.199606515 |
| T2DM | endothelial | *ZNF587B* | -0.867878316 | 3.14E-07 | 1.24E-05 | 0.001525102 |
| T2DM | endothelial | *SNHG8* | -0.81032555 | 4.29E-06 | 3.71E-05 | 0.020807458 |
| T2DM | endothelial | *HCG17* | -0.616917655 | 9.96E-05 | 0.000370189 | 0.483466595 |
| T2DM | endothelial | *GTF2H5* | -0.389762175 | 0.003239323 | 0.00658174 | 1 |
| T2DM | endothelial | *FGF14-AS2* | -1.560190347 | 5.02E-06 | 4.09E-05 | 0.024363383 |
| T2DM | endothelial | *CASC15* | -0.460880477 | 0.02276819 | 0.035047988 | 1 |
| T2DM | endothelial | *AP000347.2* | -0.764635538 | 0.001854025 | 0.004090827 | 1 |
| T2DM | endothelial | *DOC2B* | 0.117959087 | 9.48E-05 | 0.000357518 | 0.460125853 |

| | | | | | | |
|---|---|---|---|---|---|---|
| T2DM | endothelial | *GRIN2B* | -0.271499962 | 0.007148375 | 0.012908044 | 1 |
| T2DM | endothelial | *AP000350.5* | -0.215658994 | 0.005416028 | 0.01016579 | 1 |
| T2DM | endothelial | *PAXIP1-AS1* | -0.823782968 | 6.50E-07 | 1.33E-05 | 0.003154003 |
| T2DM | LEC | *RBM6* | -0.896713075 | 3.91E-06 | 0.00090412 | 0.018986526 |
| T2DM | LEC | *CEP68* | 1.586879615 | 6.68E-05 | 0.010124122 | 0.323971891 |
| T2DM | LEC | *ITIH4* | 0.559971606 | 5.10E-06 | 0.001125357 | 0.024757846 |
| T2DM | LEC | *PABPC4* | -2.167044184 | 1.88E-06 | 0.000480895 | 0.009137011 |
| T2DM | LEC | *CHN2* | 1.740018166 | 5.47E-07 | 0.000155988 | 0.002651788 |
| T2DM | LEC | *CNTNAP1* | 2.972072734 | 1.13E-05 | 0.002391194 | 0.054997469 |
| T2DM | LEC | *PRR4* | 1.344599471 | 0.000431295 | 0.041032245 | 1 |
| T2DM | LEC | *CDK2AP1* | 1.283310662 | 0.000377205 | 0.038940437 | 1 |
| T2DM | LEC | *BVES* | -0.597108323 | 0.000539368 | 0.045120963 | 1 |
| T2DM | LEC | *ADRA1A* | 0.436165504 | 1.80E-05 | 0.003365959 | 0.087514929 |
| T2DM | LEC | *GRK4* | 0.401883979 | 0.000519415 | 0.045003598 | 1 |
| T2DM | LEC | *PRR12* | 1.825019661 | 0.000347315 | 0.0366342 | 1 |
| T2DM | LEC | *C12orf65* | 1.555330465 | 0.000254879 | 0.027481591 | 1 |
| T2DM | LEC | *IRF4* | 0.469089764 | 0.000530799 | 0.045120963 | 1 |
| T2DM | LEC | *TCF19* | 2.514764431 | 4.97E-10 | 4.83E-07 | 2.41E-06 |
| T2DM | LEC | *SLC37A4* | 1.582268766 | 0.000123953 | 0.014913401 | 0.601420827 |
| T2DM | LEC | *SPTBN5* | 0.3865377 | 5.24E-05 | 0.008200233 | 0.25420721 |
| T2DM | LEC | *MAN2C1* | -0.582293786 | 7.31E-05 | 0.010749119 | 0.354720916 |
| T2DM | LEC | *MYO19* | 2.286021918 | 7.57E-05 | 0.010803347 | 0.367313808 |
| T2DM | LEC | *DUS3L* | 0.755922811 | 0.00047417 | 0.04260501 | 1 |
| T2DM | LEC | *HSD17B12* | -0.838510378 | 2.22E-07 | 8.27E-05 | 0.001075632 |
| T2DM | LEC | *JAZF1* | -1.457495667 | 3.44E-05 | 0.005752879 | 0.166833488 |
| T2DM | LEC | *CLEC18A* | -1.619167078 | 1.94E-08 | 9.39E-06 | 9.39E-05 |
| T2DM | LEC | *KRTCAP3* | 1.156423373 | 3.00E-06 | 0.000728007 | 0.014560148 |
| T2DM | LEC | *SPTA1* | -0.731842775 | 0.000408177 | 0.041032245 | 1 |
| T2DM | LEC | *RFT1* | -1.39609628 | 0.00012602 | 0.014913401 | 0.611449454 |
| T2DM | LEC | *RNF123* | 1.621360623 | 1.49E-06 | 0.000400707 | 0.007212726 |
| T2DM | LEC | *MST1R* | 1.482027608 | 3.44E-07 | 0.000119368 | 0.001671154 |
| T2DM | LEC | *RAPSN* | -1.040961739 | 0.000458297 | 0.04260501 | 1 |
| T2DM | LEC | *ANPEP* | 0.479614545 | 0.000473619 | 0.04260501 | 1 |
| T2DM | LEC | *ZSWIM1* | 0.472834758 | 0.000427694 | 0.041032245 | 1 |
| T2DM | LEC | *IRS1* | -0.561683055 | 2.63E-05 | 0.004727129 | 0.127632479 |
| T2DM | LEC | *TTLL6* | -3.793466335 | 4.62E-07 | 0.000149517 | 0.002242762 |
| T2DM | LEC | *RGS19* | -0.677789274 | 0.000125944 | 0.014913401 | 0.611081269 |
| T2DM | LEC | *C1QTNF4* | -0.624984971 | 0.000123141 | 0.014913401 | 0.597480483 |
| T2DM | LEC | *CCNE2* | -1.331001353 | 0.000203413 | 0.022952541 | 0.986959243 |
| T2DM | LEC | *HLA-DQB1* | -1.131718685 | 2.57E-15 | 6.23E-12 | 1.25E-11 |
| T2DM | LEC | *TTC3* | 0.67817715 | 0.000484374 | 0.042730587 | 1 |
| T2DM | LEC | *HLA-DRB1* | -0.920625502 | 1.43E-08 | 8.71E-06 | 6.94E-05 |
| T2DM | LEC | *ZKSCAN7* | 2.185277272 | 9.78E-05 | 0.012822348 | 0.474426878 |
| T2DM | LEC | *ZNF34* | 3.105180138 | 0.000429924 | 0.041032245 | 1 |
| T2DM | LEC | *HLA-DQA1* | -1.391918444 | 1.40E-11 | 1.70E-08 | 6.81E-08 |
| T2DM | LEC | *HLA-DRB5* | -0.456290171 | 3.98E-12 | 6.43E-09 | 1.93E-08 |
| T2DM | LEC | *NOTCH4* | -0.19611065 | 1.44E-08 | 8.71E-06 | 6.96E-05 |
| T2DM | LEC | *PRRT1* | 0.827148644 | 3.97E-05 | 0.006422297 | 0.192668903 |
| T2DM | LEC | *DHX16* | 2.234586088 | 8.07E-05 | 0.011192101 | 0.391723525 |
| T2DM | LEC | *LIN52* | 1.731591197 | 2.95E-05 | 0.005115218 | 0.143226103 |

| | | | | | | |
|---|---|---|---|---|---|---|
| T2DM | LEC | *HCG27* | 1.065022092 | 5.43E-07 | 0.000155988 | 0.002633437 |
| T2DM | LEC | *DNLZ* | 0.511821289 | 0.000193865 | 0.022396046 | 0.940633917 |
| T2DM | LEC | *TMEM110* | 1.839391337 | 1.47E-05 | 0.002862602 | 0.071565042 |
| T2DM | LEC | *STARD10* | 1.004024988 | 1.65E-08 | 8.92E-06 | 8.03E-05 |
| T2DM | LEC | *C4B* | 1.777398808 | 9.47E-05 | 0.012761911 | 0.459428795 |
| T2DM | LEC | *TMA7* | 2.012703467 | 0.000551243 | 0.045332747 | 1 |
| T2DM | LEC | *HLA-DQB2* | 0.443223177 | 7.84E-09 | 6.34E-06 | 3.80E-05 |
| T2DM | LEC | *HLA-DQA2* | 0.786183695 | 4.26E-18 | 2.07E-14 | 2.07E-14 |
| T2DM | LEC | *PSMB9* | 0.761699344 | 1.44E-05 | 0.002862602 | 0.069803563 |
| T2DM | LEC | *PLA2G4B* | 3.954704997 | 5.11E-08 | 2.25E-05 | 0.000247728 |
| T2DM | LEC | *JMJD7* | 1.211287887 | 9.31E-08 | 3.76E-05 | 0.000451716 |
| T2DM | LEC | *C4A* | -1.514035031 | 0.000239888 | 0.026453158 | 1 |
| T2DM | macrophage | *RBM6* | -0.457033998 | 2.75E-07 | 7.41E-05 | 0.001333178 |
| T2DM | macrophage | *CEP68* | 1.213728819 | 5.71E-08 | 1.75E-05 | 0.00027719 |
| T2DM | macrophage | *TUBG2* | -0.712565618 | 0.000321499 | 0.030586503 | 1 |
| T2DM | macrophage | *ITIH4* | 0.249204645 | 6.27E-06 | 0.00138385 | 0.030444699 |
| T2DM | macrophage | *PABPC4* | -1.114855098 | 2.65E-09 | 2.57E-06 | 1.29E-05 |
| T2DM | macrophage | *CWF19L1* | -0.372780375 | 0.000193244 | 0.021222239 | 0.937618493 |
| T2DM | macrophage | *NOMO3* | -0.526099165 | 0.000539887 | 0.042943138 | 1 |
| T2DM | macrophage | *CNTNAP1* | 0.955267983 | 4.43E-05 | 0.006137554 | 0.214814384 |
| T2DM | macrophage | *C11orf21* | -1.165173024 | 0.000485739 | 0.039280095 | 1 |
| T2DM | macrophage | *CDK2AP1* | 0.869551249 | 2.10E-06 | 0.000484801 | 0.010180826 |
| T2DM | macrophage | *BVES* | -0.294741786 | 0.000311485 | 0.030586503 | 1 |
| T2DM | macrophage | *ELOVL4* | 0.631094873 | 0.000475939 | 0.039280095 | 1 |
| T2DM | macrophage | *ADRA1A* | 0.197686937 | 2.93E-05 | 0.004474549 | 0.142327403 |
| T2DM | macrophage | *GRK4* | 0.179014247 | 0.000553844 | 0.043342773 | 1 |
| T2DM | macrophage | *C12orf65* | 0.797311956 | 0.000302225 | 0.030586503 | 1 |
| T2DM | macrophage | *TCF19* | 0.988102792 | 2.17E-08 | 8.76E-06 | 0.000105081 |
| T2DM | macrophage | *MAN2C1* | -0.269113498 | 3.86E-05 | 0.005503554 | 0.187120823 |
| T2DM | macrophage | *MYO19* | 1.228057784 | 0.000196826 | 0.021222239 | 0.95500074 |
| T2DM | macrophage | *C18orf8* | -1.131109336 | 3.17E-05 | 0.004658031 | 0.153715018 |
| T2DM | macrophage | *PLEKHA6* | 0.501899651 | 0.000388834 | 0.035596612 | 1 |
| T2DM | macrophage | *HSD17B12* | -0.402964561 | 3.56E-08 | 1.23E-05 | 0.000172878 |
| T2DM | macrophage | *APIP* | -0.239639177 | 0.000417184 | 0.036803247 | 1 |
| T2DM | macrophage | *POC5* | -0.500248194 | 0.000447286 | 0.03857758 | 1 |
| T2DM | macrophage | *JAZF1* | -1.354927703 | 2.36E-11 | 2.86E-08 | 1.14E-07 |
| T2DM | macrophage | *CLEC18A* | -0.75681618 | 5.66E-09 | 4.58E-06 | 2.75E-05 |
| T2DM | macrophage | *KRTCAP3* | 0.498686148 | 9.71E-06 | 0.00169841 | 0.047103295 |
| T2DM | macrophage | *SPTA1* | -0.374373794 | 0.000308916 | 0.030586503 | 1 |
| T2DM | macrophage | *RFT1* | -0.754425766 | 7.14E-06 | 0.001506922 | 0.034659195 |
| T2DM | macrophage | *RNF123* | 0.679929537 | 1.01E-05 | 0.00169841 | 0.049044448 |
| T2DM | macrophage | *MST1R* | 0.589961998 | 6.45E-07 | 0.000164784 | 0.003130887 |
| T2DM | macrophage | *RAPSN* | -0.594443892 | 8.75E-06 | 0.00169841 | 0.042465739 |
| T2DM | macrophage | *ZSWIM1* | 0.215426401 | 0.000319189 | 0.030586503 | 1 |
| T2DM | macrophage | *IRS1* | -0.269582236 | 9.55E-06 | 0.00169841 | 0.046328555 |
| T2DM | macrophage | *SEMA3E* | 0.832263441 | 0.00041053 | 0.036803247 | 1 |
| T2DM | macrophage | *EMB* | -0.464550248 | 0.000453199 | 0.03857758 | 1 |
| T2DM | macrophage | *TTLL6* | -1.884969708 | 5.76E-08 | 1.75E-05 | 0.000279473 |
| T2DM | macrophage | *RGS19* | -0.283308456 | 2.95E-05 | 0.004474549 | 0.143185567 |
| T2DM | macrophage | *C1QTNF4* | -0.274529989 | 0.000171147 | 0.019771517 | 0.830403724 |

| | | | | | | |
|---|---|---|---|---|---|---|
| T2DM | macrophage | *CTSW* | -0.087934441 | 0.00048341 | 0.039280095 | 1 |
| T2DM | macrophage | *CCNE2* | -0.753248311 | 1.02E-05 | 0.00169841 | 0.049253885 |
| T2DM | macrophage | *STX19* | 0.734624261 | 0.000374945 | 0.034985237 | 1 |
| T2DM | macrophage | *HLA-DQB1* | -0.407426502 | 5.87E-14 | 1.42E-10 | 2.85E-10 |
| T2DM | macrophage | *HLA-DRB1* | -0.342063097 | 7.69E-09 | 4.66E-06 | 3.73E-05 |
| T2DM | macrophage | *ZKSCAN7* | 1.067202832 | 0.000238109 | 0.025115324 | 1 |
| T2DM | macrophage | *HLA-DQA1* | -0.36231215 | 6.73E-09 | 4.66E-06 | 3.27E-05 |
| T2DM | macrophage | *HLA-DRB5* | -0.18978062 | 4.33E-12 | 7.00E-09 | 2.10E-08 |
| T2DM | macrophage | *NOTCH4* | -0.086521373 | 1.63E-08 | 7.91E-06 | 7.91E-05 |
| T2DM | macrophage | *PRRT1* | 0.355304539 | 5.55E-05 | 0.007475833 | 0.269130004 |
| T2DM | macrophage | *STK19* | -0.69715061 | 0.000641563 | 0.048638493 | 1 |
| T2DM | macrophage | *GPANK1* | -0.725182069 | 0.000126914 | 0.015019187 | 0.615786653 |
| T2DM | macrophage | *DHX16* | 0.929465712 | 0.000582782 | 0.044883448 | 1 |
| T2DM | macrophage | *LIN52* | 0.773182111 | 0.000115009 | 0.013950646 | 0.558025831 |
| T2DM | macrophage | *HCG27* | 0.486321749 | 1.15E-06 | 0.000277929 | 0.005558574 |
| T2DM | macrophage | *DNLZ* | 0.228764964 | 0.000183654 | 0.020722964 | 0.891087442 |
| T2DM | macrophage | *ARHGAP19* | -1.50397056 | 8.04E-05 | 0.010263389 | 0.390008779 |
| T2DM | macrophage | *TMEM110* | 0.984962917 | 9.61E-06 | 0.00169841 | 0.046642626 |
| T2DM | macrophage | *STARD10* | 0.447244745 | 1.55E-08 | 7.91E-06 | 7.54E-05 |
| T2DM | macrophage | *C4B* | 0.848780258 | 5.88E-05 | 0.00770599 | 0.285121622 |
| T2DM | macrophage | *HLA-DQB2* | 0.195495261 | 2.13E-08 | 8.76E-06 | 0.000103414 |
| T2DM | macrophage | *HLA-DQA2* | 0.347410844 | 6.47E-17 | 3.14E-13 | 3.14E-13 |
| T2DM | macrophage | *PSMB9* | 0.359503939 | 2.08E-05 | 0.003365444 | 0.100963327 |
| T2DM | macrophage | *PLA2G4B* | 1.85476628 | 2.41E-08 | 9.01E-06 | 0.000117159 |
| T2DM | macrophage | *JMJD7* | 0.557271963 | 6.66E-08 | 1.90E-05 | 0.000323196 |
| T2DM | macrophage | *C4A* | -0.656981632 | 9.52E-05 | 0.011846924 | 0.462030039 |
| T2DM | mast_cell | *NFYA* | 1.78288824 | 0.000132205 | 0.000250081 | 0.641456817 |
| T2DM | mast_cell | *ENPP4* | 2.842258999 | 0.000165378 | 0.00029952 | 0.802414698 |
| T2DM | mast_cell | *SEMA3F* | -3.624696574 | 5.12E-05 | 0.000136328 | 0.248596355 |
| T2DM | mast_cell | *KRIT1* | 2.236218177 | 3.08E-05 | 0.000134478 | 0.149512171 |
| T2DM | mast_cell | *RAD52* | 1.493352878 | 5.18E-05 | 0.000136402 | 0.251215595 |
| T2DM | mast_cell | *HS3ST1* | 2.087483933 | 0.000925936 | 0.001349521 | 1 |
| T2DM | mast_cell | *AOC1* | 2.842873859 | 0.003638053 | 0.004781104 | 1 |
| T2DM | mast_cell | *SNX11* | 1.655661317 | 1.56E-06 | 0.000102598 | 0.007592248 |
| T2DM | mast_cell | *M6PR* | 3.486919856 | 6.29E-05 | 0.000149953 | 0.304960047 |
| T2DM | mast_cell | *CYP26B1* | 4.693173591 | 1.80E-05 | 0.000134478 | 0.087332921 |
| T2DM | mast_cell | *CD38* | 1.618726768 | 3.59E-05 | 0.000134478 | 0.17419084 |
| T2DM | mast_cell | *ARHGAP33* | 1.52990737 | 0.005126105 | 0.006614856 | 1 |
| T2DM | mast_cell | *CDC27* | 2.051261169 | 1.94E-05 | 0.000134478 | 0.09404094 |
| T2DM | mast_cell | *POLR2J* | 3.391274309 | 4.25E-05 | 0.000134478 | 0.206153549 |
| T2DM | mast_cell | *MEOX1* | 2.279697409 | 3.31E-05 | 0.000134478 | 0.160407443 |
| T2DM | mast_cell | *LIG3* | 2.899530157 | 0.000299446 | 0.000490351 | 1 |
| T2DM | mast_cell | *CIAPIN1* | 5.04288106 | 1.47E-05 | 0.000134478 | 0.071083451 |
| T2DM | mast_cell | *PON1* | -1.557729192 | 0.000228779 | 0.000389896 | 1 |
| T2DM | mast_cell | *GCFC2* | 2.338873977 | 3.22E-05 | 0.000134478 | 0.156118605 |
| T2DM | mast_cell | *RHBDD2* | 1.828036786 | 4.33E-05 | 0.000134478 | 0.210330114 |
| T2DM | mast_cell | *GGNBP2* | 2.031824914 | 0.000180013 | 0.000319583 | 0.873421644 |
| T2DM | mast_cell | *ITGA2B* | -2.387055952 | 0.001869725 | 0.002577978 | 1 |
| T2DM | mast_cell | *TMEM98* | 1.227223939 | 0.000125652 | 0.000240973 | 0.60966132 |
| T2DM | mast_cell | *ABCC8* | 2.139130565 | 0.005469681 | 0.007030171 | 1 |

| | | | | | | |
|---|---|---|---|---|---|---|
| T2DM | mast_cell | *SYNRG* | 2.349038951 | 4.54E-05 | 0.000134478 | 0.220068512 |
| T2DM | mast_cell | *ZNF263* | 5.125844439 | 6.70E-05 | 0.000155401 | 0.325152276 |
| T2DM | mast_cell | *MAP3K9* | 1.141713305 | 0.002993057 | 0.003977626 | 1 |
| T2DM | mast_cell | *BAIAP2L1* | 2.41860782 | 3.34E-05 | 0.000134478 | 0.162234421 |
| T2DM | mast_cell | *AGK* | 3.218724069 | 0.000116979 | 0.000227819 | 0.567583892 |
| T2DM | mast_cell | *FARP2* | 2.012131902 | 3.75E-05 | 0.000134478 | 0.181785184 |
| T2DM | mast_cell | *USH1C* | -2.480790698 | 0.000126783 | 0.000242663 | 0.615151749 |
| T2DM | mast_cell | *IFRD1* | 1.275565323 | 0.000185167 | 0.000326977 | 0.898432379 |
| T2DM | mast_cell | *VPS41* | 3.24163669 | 6.41E-05 | 0.000151573 | 0.310875342 |
| T2DM | mast_cell | *SCIN* | -2.272266987 | 0.004312662 | 0.005623498 | 1 |
| T2DM | mast_cell | *CCDC124* | 2.378792664 | 0.010639422 | 0.013256927 | 1 |
| T2DM | mast_cell | *CEACAM21* | 1.533058516 | 0.004980757 | 0.006443884 | 1 |
| T2DM | mast_cell | *CD79B* | -3.383084642 | 1.25E-05 | 0.000134478 | 0.060564262 |
| T2DM | mast_cell | *ST7L* | 1.262033946 | 0.015308215 | 0.018794397 | 1 |
| T2DM | mast_cell | *PAX6* | -1.404552533 | 0.016432283 | 0.020118455 | 1 |
| T2DM | mast_cell | *TSR3* | 7.503790169 | 0.002649485 | 0.003551189 | 1 |
| T2DM | mast_cell | *PIGQ* | -1.832906828 | 0.01284903 | 0.01589179 | 1 |
| T2DM | mast_cell | *FMO3* | 1.414873059 | 0.001702136 | 0.002362346 | 1 |
| T2DM | mast_cell | *PSMB1* | 3.296959045 | 3.23E-05 | 0.000134478 | 0.156833097 |
| T2DM | mast_cell | *NADK* | 3.70751173 | 3.08E-05 | 0.000134478 | 0.149512176 |
| T2DM | mast_cell | *DLEC1* | 1.716502293 | 9.61E-05 | 0.000197267 | 0.466142409 |
| T2DM | mast_cell | *CYB561* | 3.592651578 | 6.52E-07 | 7.18E-05 | 0.003162095 |
| T2DM | mast_cell | *SS18L2* | 2.777920916 | 5.85E-06 | 0.000134478 | 0.028379041 |
| T2DM | mast_cell | *MGST1* | 1.241654436 | 1.60E-05 | 0.000134478 | 0.077602769 |
| T2DM | mast_cell | *MMP25* | 2.678160531 | 0.00052835 | 0.000806077 | 1 |
| T2DM | mast_cell | *IL32* | 2.420287514 | 1.93E-05 | 0.000134478 | 0.093605054 |
| T2DM | mast_cell | *PKD1* | 5.17906266 | 1.41E-05 | 0.000134478 | 0.068513621 |
| T2DM | mast_cell | *MAPK8IP2* | 5.314072835 | 0.013779496 | 0.016969064 | 1 |
| T2DM | mast_cell | *MED24* | 2.112790966 | 3.87E-05 | 0.000134478 | 0.187746677 |
| T2DM | mast_cell | *RPS20* | 1.680025576 | 5.21E-05 | 0.000136609 | 0.252863307 |
| T2DM | mast_cell | *TRAF3IP3* | 1.950681034 | 2.11E-06 | 0.000124855 | 0.010238094 |
| T2DM | mast_cell | *POMT2* | 1.499832742 | 0.000259518 | 0.000434501 | 1 |
| T2DM | mast_cell | *MLXIPL* | 0.93900589 | 0.0016136 | 0.00225106 | 1 |
| T2DM | mast_cell | *DYRK4* | 2.389392779 | 5.96E-06 | 0.000134478 | 0.028938034 |
| T2DM | mast_cell | *NCAPD2* | 1.497605554 | 0.001326114 | 0.001870982 | 1 |
| T2DM | mast_cell | *FUZ* | 3.94221977 | 1.16E-05 | 0.000134478 | 0.056431638 |
| T2DM | mast_cell | *ZNF200* | 2.067158052 | 0.008737623 | 0.010966101 | 1 |
| T2DM | mast_cell | *LRRC23* | 1.53620027 | 4.75E-05 | 0.000134478 | 0.230343194 |
| T2DM | mast_cell | *CLCN6* | 1.398700215 | 0.000128635 | 0.000245145 | 0.624138108 |
| T2DM | mast_cell | *SLC6A7* | 47.62704865 | 0.003833034 | 0.005025097 | 1 |
| T2DM | mast_cell | *TSPAN9* | 1.02928907 | 0.001162487 | 0.00166236 | 1 |
| T2DM | mast_cell | *UTP18* | 2.006672871 | 1.98E-05 | 0.000134478 | 0.096119023 |
| T2DM | mast_cell | *TTC19* | 2.343738686 | 3.97E-05 | 0.000134478 | 0.192495381 |
| T2DM | mast_cell | *DPF1* | 3.510946927 | 0.009156564 | 0.011474083 | 1 |
| T2DM | mast_cell | *LARS2* | 1.83378136 | 1.58E-05 | 0.000134478 | 0.076508923 |
| T2DM | mast_cell | *DCN* | 1.332766268 | 5.84E-05 | 0.000143793 | 0.283127563 |
| T2DM | mast_cell | *PPP5C* | 3.20975441 | 5.45E-06 | 0.000134478 | 0.026435378 |
| T2DM | mast_cell | *CEP68* | 5.947732008 | 8.07E-11 | 9.79E-08 | 3.92E-07 |
| T2DM | mast_cell | *ZBTB32* | -2.361380642 | 0.007365213 | 0.009330552 | 1 |
| T2DM | mast_cell | *ERCC1* | 1.605890894 | 2.38E-05 | 0.000134478 | 0.115367617 |

| | | | | | | |
|---|---|---|---|---|---|---|
| T2DM | mast_cell | *CD22* | -0.158554825 | 0.000198214 | 0.000345948 | 0.961736754 |
| T2DM | mast_cell | *ALOX5* | -1.092359004 | 0.001227146 | 0.001745014 | 1 |
| T2DM | mast_cell | *MAP4K5* | 2.398260287 | 5.88E-05 | 0.000144373 | 0.285173786 |
| T2DM | mast_cell | *EHD3* | 3.828181118 | 4.20E-05 | 0.000134478 | 0.203838812 |
| T2DM | mast_cell | *MAN2B2* | 3.847699212 | 0.000106122 | 0.000211747 | 0.514903132 |
| T2DM | mast_cell | *SLC25A39* | 2.51609303 | 0.000854468 | 0.001256328 | 1 |
| T2DM | mast_cell | *RNF14* | 1.817206113 | 2.44E-05 | 0.000134478 | 0.118597209 |
| T2DM | mast_cell | *DDX11* | 1.46077893 | 0.001085086 | 0.001562267 | 1 |
| T2DM | mast_cell | *TACC3* | 2.511118194 | 5.23E-05 | 0.000136633 | 0.253855662 |
| T2DM | mast_cell | *ZC3H3* | 1.686526959 | 5.53E-05 | 0.00014004 | 0.268486291 |
| T2DM | mast_cell | *MDH1* | 1.819029411 | 0.000163516 | 0.000296702 | 0.793381782 |
| T2DM | mast_cell | *MTMR11* | 1.587119832 | 0.003903636 | 0.005114891 | 1 |
| T2DM | mast_cell | *COX15* | 2.144114291 | 2.09E-05 | 0.000134478 | 0.101219242 |
| T2DM | mast_cell | *CCDC88C* | 1.343610528 | 0.000173047 | 0.000310054 | 0.839625957 |
| T2DM | mast_cell | *NUDCD3* | 1.811478177 | 0.000177094 | 0.00031544 | 0.859259647 |
| T2DM | mast_cell | *CHDH* | 4.272240363 | 1.82E-05 | 0.000134478 | 0.088107818 |
| T2DM | mast_cell | *CLCA4* | 4.980081369 | 0.006153343 | 0.007856847 | 1 |
| T2DM | mast_cell | *RUFY3* | 3.237723165 | 3.29E-05 | 0.000134478 | 0.159411398 |
| T2DM | mast_cell | *SLC11A1* | 0.67541555 | 0.031368222 | 0.037267045 | 1 |
| T2DM | mast_cell | *WWTR1* | 1.594331478 | 3.37E-05 | 0.000134478 | 0.163725159 |
| T2DM | mast_cell | *AGPS* | 3.478866255 | 0.000110095 | 0.000218082 | 0.534181454 |
| T2DM | mast_cell | *TTC27* | 1.436141145 | 4.19E-05 | 0.000134478 | 0.203405844 |
| T2DM | mast_cell | *MARCO* | 0.961734405 | 4.71E-05 | 0.000134478 | 0.22847549 |
| T2DM | mast_cell | *HGF* | 4.837886925 | 4.48E-05 | 0.000134478 | 0.217499531 |
| T2DM | mast_cell | *OSBPL5* | 2.256263225 | 8.14E-06 | 0.000134478 | 0.039490179 |
| T2DM | mast_cell | *SLC45A4* | 1.545806009 | 0.000440395 | 0.00068597 | 1 |
| T2DM | mast_cell | *ZDHHC6* | 3.708801158 | 1.89E-05 | 0.000134478 | 0.091584686 |
| T2DM | mast_cell | *GRAMD1B* | 1.105014737 | 0.001036537 | 0.001497254 | 1 |
| T2DM | mast_cell | *RNH1* | 2.166145449 | 1.76E-05 | 0.000134478 | 0.085281106 |
| T2DM | mast_cell | *ABCC2* | 5.110844802 | 0.00023828 | 0.000403678 | 1 |
| T2DM | mast_cell | *GCLM* | 3.251292798 | 3.46E-05 | 0.000134478 | 0.168042458 |
| T2DM | mast_cell | *UBR2* | 1.875451566 | 6.68E-05 | 0.000155169 | 0.32399195 |
| T2DM | mast_cell | *RRAGD* | 2.103314915 | 3.90E-05 | 0.000134478 | 0.189401498 |
| T2DM | mast_cell | *HSF2* | 1.95677754 | 1.21E-05 | 0.000134478 | 0.058467091 |
| T2DM | mast_cell | *TYMP* | 1.555615139 | 0.000368394 | 0.000587976 | 1 |
| T2DM | mast_cell | *TOMM34* | 2.214864182 | 9.57E-05 | 0.000196916 | 0.464274347 |
| T2DM | mast_cell | *SEC63* | 3.273526405 | 8.71E-05 | 0.000184883 | 0.422827949 |
| T2DM | mast_cell | *FAS* | 1.388287119 | 4.96E-05 | 0.000135163 | 0.240747397 |
| T2DM | mast_cell | *RNASET2* | 0.935205817 | 0.002600185 | 0.003490896 | 1 |
| T2DM | mast_cell | *AGPAT4* | 4.630789284 | 0.000188605 | 0.000331639 | 0.915113586 |
| T2DM | mast_cell | *BTN3A1* | 1.150973971 | 0.012859075 | 0.01589611 | 1 |
| T2DM | mast_cell | *MIPEP* | 1.832182602 | 8.82E-05 | 0.000186205 | 0.42789893 |
| T2DM | mast_cell | *INSRR* | 3.014889178 | 0.000189415 | 0.000332745 | 0.91904199 |
| T2DM | mast_cell | *VRK2* | 1.904352577 | 3.47E-05 | 0.000134478 | 0.168420132 |
| T2DM | mast_cell | *VEZT* | 2.666896786 | 3.58E-05 | 0.000134478 | 0.173887277 |
| T2DM | mast_cell | *BRD9* | 2.784826818 | 2.33E-05 | 0.000134478 | 0.112924679 |
| T2DM | mast_cell | *SNX1* | 1.941781956 | 4.99E-05 | 0.000135379 | 0.242051895 |
| T2DM | mast_cell | *ANK1* | 2.648720043 | 4.18E-05 | 0.000134478 | 0.202944878 |
| T2DM | mast_cell | *RABEP1* | 2.334600795 | 2.71E-05 | 0.000134478 | 0.131720855 |
| T2DM | mast_cell | *BAK1* | 2.653765357 | 0.000331163 | 0.000535245 | 1 |

| | | | | | | |
|---|---|---|---|---|---|---|
| T2DM | mast_cell | *IKZF2* | 2.132085994 | 3.74E-05 | 0.000134478 | 0.181421779 |
| T2DM | mast_cell | *GRN* | 1.425253132 | 3.83E-05 | 0.000134478 | 0.186027441 |
| T2DM | mast_cell | *IFT88* | 1.962448247 | 0.00015027 | 0.000276179 | 0.729111419 |
| T2DM | mast_cell | *CHPF2* | 2.073912246 | 6.99E-05 | 0.000159617 | 0.339026602 |
| T2DM | mast_cell | *UBA6* | 1.619428708 | 0.001441953 | 0.002022652 | 1 |
| T2DM | mast_cell | *ATP6V0A1* | 2.027909285 | 6.57E-06 | 0.000134478 | 0.031862176 |
| T2DM | mast_cell | *PIAS1* | 2.214137545 | 1.87E-05 | 0.000134478 | 0.090927454 |
| T2DM | mast_cell | *ASTE1* | 1.863100377 | 4.12E-05 | 0.000134478 | 0.200128454 |
| T2DM | mast_cell | *MYOC* | 1.955487878 | 1.45E-05 | 0.000134478 | 0.070435297 |
| T2DM | mast_cell | *SH3YL1* | 2.197604183 | 0.001891242 | 0.002604279 | 1 |
| T2DM | mast_cell | *VCL* | 2.203238757 | 4.21E-05 | 0.000134478 | 0.204414973 |
| T2DM | mast_cell | *ADSS* | 4.03361064 | 4.24E-05 | 0.000134478 | 0.205482397 |
| T2DM | mast_cell | *TBC1D23* | 1.683907653 | 3.59E-05 | 0.000134478 | 0.174064837 |
| T2DM | mast_cell | *MYOM2* | -1.062553623 | 0.000348658 | 0.000560347 | 1 |
| T2DM | mast_cell | *TUBG2* | 1.011981532 | 0.00803203 | 0.010119815 | 1 |
| T2DM | mast_cell | *NSUN2* | 3.587611138 | 0.000136049 | 0.000255757 | 0.660108768 |
| T2DM | mast_cell | *MFAP3* | 1.926070394 | 0.000133932 | 0.000252736 | 0.649837958 |
| T2DM | mast_cell | *MRI1* | 1.90100475 | 0.000523935 | 0.000800595 | 1 |
| T2DM | mast_cell | *AGA* | 1.880997973 | 5.98E-05 | 0.000145429 | 0.290131138 |
| T2DM | mast_cell | *BOD1L1* | 2.909761704 | 0.001570728 | 0.002195038 | 1 |
| T2DM | mast_cell | *DNAH5* | 2.605926092 | 3.70E-05 | 0.000134478 | 0.179478921 |
| T2DM | mast_cell | *PNKP* | 1.498168565 | 0.005425829 | 0.006975655 | 1 |
| T2DM | mast_cell | *PHLPP2* | 1.349268762 | 0.000935045 | 0.001361188 | 1 |
| T2DM | mast_cell | *SPDL1* | -1.776025813 | 0.001301255 | 0.001840726 | 1 |
| T2DM | mast_cell | *PQLC2* | -4.90692096 | 0.032010292 | 0.037992647 | 1 |
| T2DM | mast_cell | *CTNS* | 1.512282665 | 0.000138967 | 0.000260134 | 0.674268221 |
| T2DM | mast_cell | *INPP4A* | 1.662455735 | 7.80E-05 | 0.000171028 | 0.378313716 |
| T2DM | mast_cell | *PSMA4* | 2.309177672 | 5.24E-05 | 0.000136633 | 0.254321225 |
| T2DM | mast_cell | *LSG1* | 1.164459793 | 0.018684306 | 0.022726561 | 1 |
| T2DM | mast_cell | *TDP1* | 1.537711105 | 5.30E-05 | 0.000137292 | 0.256981532 |
| T2DM | mast_cell | *SPATA7* | 1.217356624 | 6.30E-05 | 0.000150165 | 0.305736275 |
| T2DM | mast_cell | *MED17* | 2.574717208 | 3.33E-05 | 0.000134478 | 0.161733029 |
| T2DM | mast_cell | *CAPG* | -2.084550724 | 0.000500424 | 0.000769346 | 1 |
| T2DM | mast_cell | *AP2S1* | 1.993945941 | 4.74E-05 | 0.000134478 | 0.230088386 |
| T2DM | mast_cell | *CUL7* | 1.843624786 | 3.48E-05 | 0.000134478 | 0.168782012 |
| T2DM | mast_cell | *CTNNA1* | 1.868089028 | 3.20E-05 | 0.000134478 | 0.15536703 |
| T2DM | mast_cell | *CNTLN* | 2.433825069 | 0.000179919 | 0.000319534 | 0.872967949 |
| T2DM | mast_cell | *YTHDC2* | 2.479050489 | 3.21E-05 | 0.000134478 | 0.155701459 |
| T2DM | mast_cell | *POLR2B* | 3.652419123 | 0.000101361 | 0.000205174 | 0.491801225 |
| T2DM | mast_cell | *ARAP2* | 1.080332117 | 3.26E-05 | 0.000134478 | 0.15830531 |
| T2DM | mast_cell | *DTNBP1* | 3.54514659 | 0.000293585 | 0.000482872 | 1 |
| T2DM | mast_cell | *ANO2* | 1.164717473 | 4.21E-05 | 0.000134478 | 0.204374386 |
| T2DM | mast_cell | *RRM2B* | 3.444264796 | 4.32E-05 | 0.000134478 | 0.209600427 |
| T2DM | mast_cell | *SNX29* | 2.538246333 | 8.04E-05 | 0.000174893 | 0.390012166 |
| T2DM | mast_cell | *EPN3* | -2.296343187 | 3.66E-05 | 0.000134478 | 0.177378317 |
| T2DM | mast_cell | *RCN1* | 2.299530469 | 0.000343761 | 0.00055321 | 1 |
| T2DM | mast_cell | *JKAMP* | 1.66250887 | 0.000101569 | 0.000205424 | 0.492812091 |
| T2DM | mast_cell | *MCUR1* | 2.371177056 | 3.15E-05 | 0.000134478 | 0.152964473 |
| T2DM | mast_cell | *LAMC3* | -1.722718404 | 0.000131789 | 0.000249488 | 0.639437996 |
| T2DM | mast_cell | *MAPK9* | 4.963406218 | 8.80E-05 | 0.000186041 | 0.427185375 |

| | | | | | | |
|---|---|---|---|---|---|---|
| T2DM | mast_cell | *FAM160A2* | 1.605306547 | 3.30E-05 | 0.000134478 | 0.159994682 |
| T2DM | mast_cell | *CYBA* | 1.957958539 | 3.05E-05 | 0.000134478 | 0.147993946 |
| T2DM | mast_cell | *THOC3* | -3.836800075 | 0.000900153 | 0.001313942 | 1 |
| T2DM | mast_cell | *ALX4* | 2.198560555 | 0.0074631 | 0.009449625 | 1 |
| T2DM | mast_cell | *FSTL4* | 1.77577739 | 0.003254225 | 0.004303489 | 1 |
| T2DM | mast_cell | *AKR7A2* | 1.782167326 | 4.03E-05 | 0.000134478 | 0.195532274 |
| T2DM | mast_cell | *MRTO4* | -3.505341259 | 0.035818442 | 0.042264368 | 1 |
| T2DM | mast_cell | *ANAPC4* | 1.815999643 | 1.91E-05 | 0.000134478 | 0.092441863 |
| T2DM | mast_cell | *LY75* | 8.473639996 | 1.30E-05 | 0.000134478 | 0.063101935 |
| T2DM | mast_cell | *OPN3* | 2.01269877 | 3.79E-06 | 0.000134478 | 0.018382602 |
| T2DM | mast_cell | *SDCCAG8* | 1.753977655 | 4.07E-05 | 0.000134478 | 0.197696521 |
| T2DM | mast_cell | *KIF1B* | 2.532863114 | 7.19E-06 | 0.000134478 | 0.034894676 |
| T2DM | mast_cell | *TBC1D22A* | 1.669478488 | 2.50E-05 | 0.000134478 | 0.121193673 |
| T2DM | mast_cell | *PLEKHH1* | 1.656269701 | 0.001822084 | 0.002515866 | 1 |
| T2DM | mast_cell | *GALC* | 3.421675398 | 0.016666522 | 0.020379527 | 1 |
| T2DM | mast_cell | *FAM114A2* | 1.584028339 | 3.43E-05 | 0.000134478 | 0.166663245 |
| T2DM | mast_cell | *GINM1* | 2.005696137 | 4.54E-05 | 0.000134478 | 0.220275917 |
| T2DM | mast_cell | *ITIH4* | 1.741814131 | 3.31E-07 | 5.73E-05 | 0.001603691 |
| T2DM | mast_cell | *NPFFR2* | -5.235283159 | 0.005885821 | 0.007531119 | 1 |
| T2DM | mast_cell | *PCGF2* | 1.740014282 | 6.50E-05 | 0.000152782 | 0.315437182 |
| T2DM | mast_cell | *IL17RB* | 2.721464426 | 0.001599993 | 0.002234005 | 1 |
| T2DM | mast_cell | *GDI2* | 3.416238112 | 6.92E-05 | 0.000158597 | 0.335750015 |
| T2DM | mast_cell | *PITHD1* | 3.705213111 | 0.003531312 | 0.004648379 | 1 |
| T2DM | mast_cell | *ATP11B* | 2.364192974 | 1.44E-05 | 0.000134478 | 0.069662663 |
| T2DM | mast_cell | *LAMC2* | 1.423567069 | 0.001847198 | 0.002547642 | 1 |
| T2DM | mast_cell | *RASGRF1* | 2.571803138 | 0.008538973 | 0.010727887 | 1 |
| T2DM | mast_cell | *FLYWCH1* | 1.276049165 | 0.001713798 | 0.002377172 | 1 |
| T2DM | mast_cell | *ALDH18A1* | 1.971284662 | 0.000273146 | 0.000453871 | 1 |
| T2DM | mast_cell | *DNAJC25* | 2.184019415 | 4.53E-05 | 0.000134478 | 0.219888548 |
| T2DM | mast_cell | *OGFR* | 2.377150192 | 1.96E-05 | 0.000134478 | 0.094975675 |
| T2DM | mast_cell | *GNA15* | 1.272191302 | 0.036798163 | 0.043367668 | 1 |
| T2DM | mast_cell | *RIMBP2* | -2.804482734 | 0.001456163 | 0.002040815 | 1 |
| T2DM | mast_cell | *PRDM6* | 6.576795379 | 3.46E-05 | 0.000134478 | 0.167800566 |
| T2DM | mast_cell | *NCKAP1* | 1.737676736 | 5.23E-05 | 0.000136633 | 0.253884909 |
| T2DM | mast_cell | *LTK* | 15.94508313 | 3.60E-05 | 0.000134478 | 0.17450252 |
| T2DM | mast_cell | *EIF4B* | 1.950344616 | 0.002204818 | 0.003003307 | 1 |
| T2DM | mast_cell | *SLC6A16* | 1.698399163 | 1.17E-05 | 0.000134478 | 0.0566607 |
| T2DM | mast_cell | *AHRR* | 3.470386495 | 7.29E-05 | 0.000163513 | 0.353515858 |
| T2DM | mast_cell | *ADCK1* | 1.62608753 | 4.79E-05 | 0.000134478 | 0.232620373 |
| T2DM | mast_cell | *HAGH* | 1.934305874 | 0.021119428 | 0.025515803 | 1 |
| T2DM | mast_cell | *RNF4* | 3.950813889 | 4.56E-05 | 0.000134478 | 0.221395878 |
| T2DM | mast_cell | *CASP8* | 1.274921915 | 3.03E-05 | 0.000134478 | 0.146994775 |
| T2DM | mast_cell | *TM7SF3* | 3.546973751 | 3.69E-06 | 0.000134478 | 0.017904387 |
| T2DM | mast_cell | *CDON* | 0.890755006 | 8.01E-05 | 0.000174583 | 0.388825689 |
| T2DM | mast_cell | *TMEM161A* | 1.740066908 | 4.13E-05 | 0.000134478 | 0.200257025 |
| T2DM | mast_cell | *LPAR2* | 3.246740193 | 1.29E-05 | 0.000134478 | 0.062752173 |
| T2DM | mast_cell | *SUGP2* | 1.858619471 | 2.84E-05 | 0.000134478 | 0.13802823 |
| T2DM | mast_cell | *SLC12A2* | 2.00768879 | 2.94E-05 | 0.000134478 | 0.142494085 |
| T2DM | mast_cell | *CNN2* | 3.14141219 | 9.16E-07 | 8.78E-05 | 0.004442113 |
| T2DM | mast_cell | *ABCA7* | 1.882591478 | 0.002386392 | 0.003227983 | 1 |

| | | | | | | |
|---|---|---|---|---|---|---|
| T2DM | mast_cell | *DDX20* | 2.682738467 | 0.00013009 | 0.000247141 | 0.631198386 |
| T2DM | mast_cell | *BTBD1* | 3.991033231 | 3.27E-06 | 0.000134478 | 0.015848314 |
| T2DM | mast_cell | *FAR2* | 3.292875062 | 1.11E-05 | 0.000134478 | 0.053965581 |
| T2DM | mast_cell | *BCAS1* | 2.712227636 | 0.000119673 | 0.000231553 | 0.580654111 |
| T2DM | mast_cell | *CHI3L2* | -2.044373741 | 0.000379111 | 0.000602123 | 1 |
| T2DM | mast_cell | *SBNO2* | 1.727073546 | 2.47E-05 | 0.000134478 | 0.119732058 |
| T2DM | mast_cell | *PMS1* | 4.595110189 | 0.000230077 | 0.000391834 | 1 |
| T2DM | mast_cell | *UHRF1BP1* | 1.645400675 | 0.00052405 | 0.000800595 | 1 |
| T2DM | mast_cell | *ANKRD44* | 1.41185279 | 2.76E-05 | 0.000134478 | 0.13395636 |
| T2DM | mast_cell | *ADAT1* | -0.949889869 | 0.003202562 | 0.004242104 | 1 |
| T2DM | mast_cell | *MAP2K4* | 1.690528605 | 5.55E-05 | 0.00014008 | 0.269093424 |
| T2DM | mast_cell | *TMEM206* | 1.577378109 | 7.99E-05 | 0.000174442 | 0.38778512 |
| T2DM | mast_cell | *SEC61A2* | 1.518933913 | 0.006395169 | 0.008148467 | 1 |
| T2DM | mast_cell | *TIE1* | 1.543183789 | 2.81E-05 | 0.000134478 | 0.13623218 |
| T2DM | mast_cell | *ZMYND12* | 1.240835888 | 0.000782833 | 0.001158373 | 1 |
| T2DM | mast_cell | *NGEF* | -5.579151665 | 0.011671563 | 0.014490896 | 1 |
| T2DM | mast_cell | *SPI1* | 1.420921473 | 0.000144487 | 0.000267241 | 0.701049369 |
| T2DM | mast_cell | *ZNRD1* | 1.30351881 | 0.007540589 | 0.009537784 | 1 |
| T2DM | mast_cell | *ATXN3* | 2.053862495 | 1.57E-05 | 0.000134478 | 0.076394808 |
| T2DM | mast_cell | *GOLGA5* | 5.019116222 | 0.000594084 | 0.000898814 | 1 |
| T2DM | mast_cell | *FGFR2* | 0.923870494 | 0.015424203 | 0.018927221 | 1 |
| T2DM | mast_cell | *MTFR1* | 2.555004045 | 0.000705917 | 0.001052908 | 1 |
| T2DM | mast_cell | *FECH* | 0.980065784 | 0.003222284 | 0.004263573 | 1 |
| T2DM | mast_cell | *MYO9A* | 1.995325899 | 3.43E-05 | 0.000134478 | 0.166376505 |
| T2DM | mast_cell | *TRAM1* | 3.30964075 | 6.88E-05 | 0.000158046 | 0.333951151 |
| T2DM | mast_cell | *EVI5* | 1.376144694 | 9.11E-05 | 0.00019059 | 0.441788 |
| T2DM | mast_cell | *STOML1* | 3.123536804 | 4.66E-05 | 0.000134478 | 0.226071306 |
| T2DM | mast_cell | *METTL22* | 1.974900468 | 7.32E-05 | 0.000164124 | 0.354999412 |
| T2DM | mast_cell | *TP53BP1* | 3.122833751 | 2.24E-05 | 0.000134478 | 0.108825424 |
| T2DM | mast_cell | *RRP15* | 1.810120932 | 0.000176517 | 0.000314644 | 0.85646012 |
| T2DM | mast_cell | *DHX8* | 2.513158396 | 3.30E-05 | 0.000134478 | 0.160201763 |
| T2DM | mast_cell | *ROGDI* | -4.439542048 | 0.002713986 | 0.003634629 | 1 |
| T2DM | mast_cell | *CBFB* | 3.265503865 | 2.45E-05 | 0.000134478 | 0.118736345 |
| T2DM | mast_cell | *IFI35* | 2.406159489 | 0.000264763 | 0.000441697 | 1 |
| T2DM | mast_cell | *HEATR6* | 2.311050216 | 2.44E-05 | 0.000134478 | 0.118358091 |
| T2DM | mast_cell | *POLR1A* | 1.281185115 | 0.002545517 | 0.003422236 | 1 |
| T2DM | mast_cell | *IP6K2* | 2.922943642 | 0.00028713 | 0.000473863 | 1 |
| T2DM | mast_cell | *SRBD1* | 3.505231504 | 7.94E-05 | 0.00017358 | 0.385346613 |
| T2DM | mast_cell | *CYFIP1* | 1.919445211 | 5.46E-05 | 0.000139259 | 0.265009517 |
| T2DM | mast_cell | *NUCKS1* | 3.3828657 | 3.78E-05 | 0.000134478 | 0.183356461 |
| T2DM | mast_cell | *CLEC2D* | 1.09559597 | 7.39E-05 | 0.000165254 | 0.358766758 |
| T2DM | mast_cell | *DRD4* | 7.899138385 | 5.96E-05 | 0.000145187 | 0.289109144 |
| T2DM | mast_cell | *PLA2G10* | 13.64900712 | 2.94E-05 | 0.000134478 | 0.142554708 |
| T2DM | mast_cell | *NEDD4* | 3.262654203 | 0.000499084 | 0.000767773 | 1 |
| T2DM | mast_cell | *PIGB* | 1.482624311 | 7.91E-05 | 0.000173038 | 0.383626062 |
| T2DM | mast_cell | *GNB5* | 3.121392931 | 2.83E-05 | 0.000134478 | 0.137508456 |
| T2DM | mast_cell | *LRP6* | 1.749630589 | 3.80E-05 | 0.000134478 | 0.184427251 |
| T2DM | mast_cell | *CLTCL1* | 1.421812764 | 8.02E-05 | 0.000174583 | 0.388970929 |
| T2DM | mast_cell | *DGCR2* | 2.712309075 | 0.000112774 | 0.000222703 | 0.54718187 |
| T2DM | mast_cell | *ZXDC* | 2.880460466 | 3.82E-05 | 0.000134478 | 0.185464024 |

| | | | | | | |
|---|---|---|---|---|---|---|
| T2DM | mast_cell | *GBA2* | 2.692413129 | 4.90E-05 | 0.000135021 | 0.237747216 |
| T2DM | mast_cell | *NDST1* | 3.150647466 | 2.10E-05 | 0.000134478 | 0.102064462 |
| T2DM | mast_cell | *ASNS* | 3.284678897 | 0.00084302 | 0.001240623 | 1 |
| T2DM | mast_cell | *AP3M2* | 2.552118784 | 4.95E-05 | 0.000135163 | 0.240133199 |
| T2DM | mast_cell | *ST6GALNAC2* | 3.406400009 | 0.000207 | 0.00035883 | 1 |
| T2DM | mast_cell | *EIF2B3* | 2.024326124 | 5.30E-05 | 0.000137292 | 0.256938208 |
| T2DM | mast_cell | *MAP4K4* | 2.766378015 | 4.32E-05 | 0.000134478 | 0.209712936 |
| T2DM | mast_cell | *WDR1* | 2.786773497 | 3.06E-05 | 0.000134478 | 0.148449005 |
| T2DM | mast_cell | *SNX13* | 1.587021179 | 3.43E-05 | 0.000134478 | 0.166536896 |
| T2DM | mast_cell | *RPS6KA2* | 1.954724416 | 4.45E-05 | 0.000134478 | 0.215705737 |
| T2DM | mast_cell | *ING3* | 5.230639996 | 0.002158637 | 0.002944533 | 1 |
| T2DM | mast_cell | *VASH1* | 2.871203231 | 0.016613799 | 0.02032018 | 1 |
| T2DM | mast_cell | *TRIB2* | 2.701371867 | 6.11E-06 | 0.000134478 | 0.029642028 |
| T2DM | mast_cell | *CPSF1* | 1.953809048 | 0.000181099 | 0.000321277 | 0.878691591 |
| T2DM | mast_cell | *CYBRD1* | 1.920217981 | 2.62E-05 | 0.000134478 | 0.126948345 |
| T2DM | mast_cell | *RDH11* | 3.282643582 | 2.60E-05 | 0.000134478 | 0.12637746 |
| T2DM | mast_cell | *EPN2* | 1.785902218 | 0.000563439 | 0.000855384 | 1 |
| T2DM | mast_cell | *ALDH3A2* | 1.489942393 | 5.47E-05 | 0.000139371 | 0.265500935 |
| T2DM | mast_cell | *AFF4* | 2.204651572 | 3.02E-05 | 0.000134478 | 0.146672934 |
| T2DM | mast_cell | *TRHDE* | 1.616706288 | 9.25E-05 | 0.000192537 | 0.448610902 |
| T2DM | mast_cell | *FCGR2B* | 1.61829758 | 5.07E-05 | 0.000136098 | 0.245990999 |
| T2DM | mast_cell | *NFATC3* | 2.082249941 | 1.28E-05 | 0.000134478 | 0.061934205 |
| T2DM | mast_cell | *TRNT1* | 1.845790101 | 4.79E-05 | 0.000134478 | 0.232188994 |
| T2DM | mast_cell | *CRMP1* | 1.749253361 | 4.17E-05 | 0.000134478 | 0.20233773 |
| T2DM | mast_cell | *NDE1* | 1.335296095 | 8.58E-05 | 0.000183071 | 0.416120173 |
| T2DM | mast_cell | *AP1M1* | 1.759821687 | 4.83E-05 | 0.000134478 | 0.234529521 |
| T2DM | mast_cell | *PVR* | 2.133752221 | 0.004460802 | 0.005807302 | 1 |
| T2DM | mast_cell | *SCARB1* | 1.302328962 | 2.72E-05 | 0.000134478 | 0.131866481 |
| T2DM | mast_cell | *MOV10L1* | 3.526262375 | 6.60E-05 | 0.000154113 | 0.320228973 |
| T2DM | mast_cell | *ALPK1* | 2.424880028 | 1.96E-05 | 0.000134478 | 0.095235123 |
| T2DM | mast_cell | *LLGL2* | 2.075563584 | 0.000114355 | 0.000224693 | 0.554848958 |
| T2DM | mast_cell | *SDHA* | 2.701548675 | 1.26E-06 | 9.43E-05 | 0.006129515 |
| T2DM | mast_cell | *SMARCE1* | 2.132108516 | 1.86E-05 | 0.000134478 | 0.090377641 |
| T2DM | mast_cell | *ADAM11* | -1.100863529 | 0.000213432 | 0.000368006 | 1 |
| T2DM | mast_cell | *CD5L* | 4.616976398 | 3.82E-05 | 0.000134478 | 0.185520183 |
| T2DM | mast_cell | *PPP2R2C* | 2.974731578 | 5.20E-05 | 0.000136581 | 0.252538611 |
| T2DM | mast_cell | *ZNF532* | 1.819442284 | 3.22E-05 | 0.000134478 | 0.156465642 |
| T2DM | mast_cell | *ZZEF1* | 2.297123599 | 2.49E-06 | 0.000128766 | 0.012104011 |
| T2DM | mast_cell | *TACR2* | 22.95265755 | 1.78E-05 | 0.000134478 | 0.086451356 |
| T2DM | mast_cell | *TIPIN* | 1.736729294 | 4.93E-05 | 0.000135163 | 0.239172603 |
| T2DM | mast_cell | *TTC38* | 1.774544185 | 4.85E-05 | 0.000134706 | 0.235465816 |
| T2DM | mast_cell | *GRAMD4* | 3.546807366 | 0.000288346 | 0.000475223 | 1 |
| T2DM | mast_cell | *ADD2* | 2.551123501 | 8.16E-06 | 0.000134478 | 0.039608757 |
| T2DM | mast_cell | *VPS9D1* | 1.872914482 | 2.95E-05 | 0.000134478 | 0.142932755 |
| T2DM | mast_cell | *FNDC3B* | 2.115929742 | 3.04E-05 | 0.000134478 | 0.147418721 |
| T2DM | mast_cell | *MOCOS* | 2.447382905 | 1.33E-05 | 0.000134478 | 0.064643315 |
| T2DM | mast_cell | *SEC31B* | 3.226510367 | 3.11E-06 | 0.000134478 | 0.015098821 |
| T2DM | mast_cell | *KIFAP3* | 2.28428816 | 2.95E-05 | 0.000134478 | 0.143158356 |
| T2DM | mast_cell | *MKRN2* | 1.478025441 | 8.21E-05 | 0.000177654 | 0.398484629 |
| T2DM | mast_cell | *MLH1* | 2.779546853 | 0.031092346 | 0.036957389 | 1 |

| | | | | | | |
|---|---|---|---|---|---|---|
| T2DM | mast_cell | *FMO4* | 3.866819815 | 6.43E-05 | 0.000151762 | 0.312021815 |
| T2DM | mast_cell | *RGS11* | 1.26864178 | 0.000287027 | 0.000473853 | 1 |
| T2DM | mast_cell | *SPAG5* | 5.214223464 | 0.001082874 | 0.001560007 | 1 |
| T2DM | mast_cell | *ANKRD13A* | 2.355687365 | 3.09E-05 | 0.000134478 | 0.14985377 |
| T2DM | mast_cell | *GPATCH1* | 1.539371188 | 6.46E-05 | 0.000152326 | 0.313487506 |
| T2DM | mast_cell | *ICAM3* | 3.859076805 | 8.00E-05 | 0.000174521 | 0.388308707 |
| T2DM | mast_cell | *NT5C2* | 2.45328898 | 4.67E-06 | 0.000134478 | 0.02263933 |
| T2DM | mast_cell | *XAB2* | 2.475781638 | 2.64E-05 | 0.000134478 | 0.127975214 |
| T2DM | mast_cell | *IL4R* | 2.429629203 | 3.21E-05 | 0.000134478 | 0.155888462 |
| T2DM | mast_cell | *DYNC1I2* | 2.576672458 | 3.42E-05 | 0.000134478 | 0.166008405 |
| T2DM | mast_cell | *APBB1IP* | 1.364748137 | 0.000103667 | 0.000208193 | 0.502994104 |
| T2DM | mast_cell | *LRCH4* | 2.937340562 | 0.011094198 | 0.013812946 | 1 |
| T2DM | mast_cell | *GPR137B* | 1.555401121 | 0.000120147 | 0.000232252 | 0.582951817 |
| T2DM | mast_cell | *NAALAD2* | 1.434131585 | 0.001102657 | 0.001583805 | 1 |
| T2DM | mast_cell | *AMPH* | 1.897619272 | 4.74E-05 | 0.000134478 | 0.229923659 |
| T2DM | mast_cell | *MCCC1* | 1.606789697 | 2.54E-05 | 0.000134478 | 0.123359917 |
| T2DM | mast_cell | *PIK3C3* | 1.91767852 | 4.00E-05 | 0.000134478 | 0.193876864 |
| T2DM | mast_cell | *RBFOX1* | 1.025247023 | 0.000418649 | 0.000653566 | 1 |
| T2DM | mast_cell | *PCM1* | 1.995408935 | 2.05E-05 | 0.000134478 | 0.099255029 |
| T2DM | mast_cell | *MYH7B* | 2.787141395 | 1.55E-06 | 0.000102598 | 0.007497038 |
| T2DM | mast_cell | *FKBP7* | 1.78424747 | 8.26E-05 | 0.000178055 | 0.4006248 |
| T2DM | mast_cell | *XRCC5* | 2.32978925 | 2.83E-05 | 0.000134478 | 0.137273883 |
| T2DM | mast_cell | *SP140* | 1.533737271 | 0.000321459 | 0.000521456 | 1 |
| T2DM | mast_cell | *SAR1A* | 1.514977417 | 3.38E-05 | 0.000134478 | 0.16404159 |
| T2DM | mast_cell | *FDFT1* | 2.077220242 | 3.16E-06 | 0.000134478 | 0.015355975 |
| T2DM | mast_cell | *SCGN* | 12.79915583 | 5.91E-05 | 0.000144683 | 0.286617187 |
| T2DM | mast_cell | *PGM1* | 2.424563513 | 3.38E-05 | 0.000134478 | 0.164040792 |
| T2DM | mast_cell | *DDX1* | 2.446912708 | 5.61E-05 | 0.000140825 | 0.271967471 |
| T2DM | mast_cell | *DNM2* | 2.140919697 | 3.26E-05 | 0.000134478 | 0.158332706 |
| T2DM | mast_cell | *STX7* | 1.958383144 | 6.85E-05 | 0.000157661 | 0.332256595 |
| T2DM | mast_cell | *RABL2B* | 2.771065218 | 3.14E-05 | 0.000134478 | 0.152513401 |
| T2DM | mast_cell | *SCTR* | 1.459035003 | 4.83E-05 | 0.000134478 | 0.234512455 |
| T2DM | mast_cell | *RIF1* | 3.200311778 | 0.000104461 | 0.000209439 | 0.506842698 |
| T2DM | mast_cell | *KCNN2* | 5.064709998 | 6.07E-06 | 0.000134478 | 0.029466903 |
| T2DM | mast_cell | *PSEN1* | 2.06820162 | 2.24E-05 | 0.000134478 | 0.108527791 |
| T2DM | mast_cell | *MOK* | 1.801239947 | 0.000123841 | 0.000237782 | 0.600874706 |
| T2DM | mast_cell | *STK17B* | 2.695334714 | 1.79E-06 | 0.000112139 | 0.008670667 |
| T2DM | mast_cell | *ZNF506* | 4.641349761 | 1.23E-06 | 9.40E-05 | 0.005948967 |
| T2DM | mast_cell | *PCDHB4* | 4.649644841 | 3.65E-05 | 0.000134478 | 0.177009596 |
| T2DM | mast_cell | *ME2* | 2.226981903 | 3.25E-05 | 0.000134478 | 0.157523039 |
| T2DM | mast_cell | *CCNT2* | 3.499166829 | 1.80E-06 | 0.000112139 | 0.008746833 |
| T2DM | mast_cell | *SEMA5B* | -1.21444508 | 0.017746295 | 0.021623561 | 1 |
| T2DM | mast_cell | *DOPEY1* | 1.981251864 | 1.03E-05 | 0.000134478 | 0.050183268 |
| T2DM | mast_cell | *LYRM2* | 2.996280302 | 7.72E-05 | 0.000169753 | 0.374813645 |
| T2DM | mast_cell | *ZCCHC6* | 1.923079653 | 3.21E-05 | 0.000134478 | 0.155880667 |
| T2DM | mast_cell | *ULK2* | 1.740064548 | 4.27E-05 | 0.000134478 | 0.206962121 |
| T2DM | mast_cell | *PLOD1* | 2.685252935 | 5.69E-05 | 0.000141911 | 0.276016269 |
| T2DM | mast_cell | *PIBF1* | 1.859464897 | 3.03E-05 | 0.000134478 | 0.147006211 |
| T2DM | mast_cell | *OXCT1* | 1.190474708 | 3.63E-05 | 0.000134478 | 0.175979088 |
| T2DM | mast_cell | *SLC27A5* | 2.123707157 | 2.88E-05 | 0.000134478 | 0.139661707 |

| | | | | | | |
|---|---|---|---|---|---|---|
| T2DM | mast_cell | *ZNF586* | 3.134194362 | 2.14E-06 | 0.000125054 | 0.010379444 |
| T2DM | mast_cell | *ZNF264* | 2.315226951 | 3.27E-05 | 0.000134478 | 0.15855469 |
| T2DM | mast_cell | *RPS5* | 2.234749835 | 4.25E-05 | 0.000134478 | 0.206001914 |
| T2DM | mast_cell | *CHMP2B* | 2.666962424 | 6.54E-05 | 0.000153213 | 0.317309691 |
| T2DM | mast_cell | *PPIE* | 1.72316134 | 0.000201575 | 0.000350804 | 0.978041822 |
| T2DM | mast_cell | *NOA1* | 3.296295719 | 7.37E-05 | 0.00016487 | 0.357439069 |
| T2DM | mast_cell | *GSTP1* | 3.335199104 | 8.10E-06 | 0.000134478 | 0.039293677 |
| T2DM | mast_cell | *WBP11* | 1.447393712 | 4.50E-05 | 0.000134478 | 0.218123348 |
| T2DM | mast_cell | *NCOA1* | 2.392288504 | 5.76E-05 | 0.000142772 | 0.279261468 |
| T2DM | mast_cell | *CAD* | 4.467321819 | 0.000500186 | 0.000769225 | 1 |
| T2DM | mast_cell | *CD59* | 1.978605499 | 8.83E-06 | 0.000134478 | 0.042849827 |
| T2DM | mast_cell | *CD82* | -0.725664578 | 0.003536109 | 0.004653432 | 1 |
| T2DM | mast_cell | *HACE1* | 2.167730584 | 3.17E-05 | 0.000134478 | 0.154001542 |
| T2DM | mast_cell | *SEH1L* | 2.909299696 | 0.000746743 | 0.001108012 | 1 |
| T2DM | mast_cell | *SLC25A24* | 2.705673445 | 1.25E-05 | 0.000134478 | 0.060742805 |
| T2DM | mast_cell | *PILRA* | 0.810856078 | 0.00013955 | 0.000260924 | 0.677098863 |
| T2DM | mast_cell | *ZNF213* | 2.222690115 | 1.73E-05 | 0.000134478 | 0.083824545 |
| T2DM | mast_cell | *CPNE3* | 2.13703512 | 1.20E-05 | 0.000134478 | 0.057987136 |
| T2DM | mast_cell | *RRN3* | 1.53905163 | 3.63E-05 | 0.000134478 | 0.176351577 |
| T2DM | mast_cell | *MTIF2* | 4.600766452 | 9.47E-05 | 0.000195667 | 0.459426152 |
| T2DM | mast_cell | *DDHD2* | 3.783862047 | 2.96E-05 | 0.000134478 | 0.143476938 |
| T2DM | mast_cell | *EPS15* | 2.148113126 | 3.74E-05 | 0.000134478 | 0.181257019 |
| T2DM | mast_cell | *USP40* | 1.741040925 | 3.41E-05 | 0.000134478 | 0.165242333 |
| T2DM | mast_cell | *POMGNT1* | 2.171580158 | 4.07E-05 | 0.000134478 | 0.197655456 |
| T2DM | mast_cell | *MAST2* | 2.402849733 | 3.18E-05 | 0.000134478 | 0.154401876 |
| T2DM | mast_cell | *EIF2AK1* | 4.980596299 | 5.94E-05 | 0.000144951 | 0.288275566 |
| T2DM | mast_cell | *NME8* | 49.57741527 | 1.67E-05 | 0.000134478 | 0.081187775 |
| T2DM | mast_cell | *EPDR1* | 1.519198334 | 3.43E-05 | 0.000134478 | 0.166311701 |
| T2DM | mast_cell | *SNX10* | 0.81204352 | 0.007581673 | 0.00957976 | 1 |
| T2DM | mast_cell | *ITPKC* | 1.606403269 | 6.61E-06 | 0.000134478 | 0.032069633 |
| T2DM | mast_cell | *TMED2* | 2.400818672 | 0.000227786 | 0.000388476 | 1 |
| T2DM | mast_cell | *ALG9* | 2.214765118 | 4.32E-05 | 0.000134478 | 0.209715447 |
| T2DM | mast_cell | *ACOX3* | 3.117278692 | 4.20E-05 | 0.000134478 | 0.203875727 |
| T2DM | mast_cell | *HSD17B14* | 2.323246412 | 3.65E-05 | 0.000134478 | 0.177177608 |
| T2DM | mast_cell | *TRIP6* | 1.789786085 | 7.01E-05 | 0.000159886 | 0.339917699 |
| T2DM | mast_cell | *ACHE* | -2.521764908 | 2.25E-05 | 0.000134478 | 0.109149614 |
| T2DM | mast_cell | *SRRT* | 2.060379131 | 5.99E-05 | 0.000145495 | 0.290407393 |
| T2DM | mast_cell | *ADAMTS2* | 1.533111783 | 3.99E-05 | 0.000134478 | 0.193762944 |
| T2DM | mast_cell | *PSMC5* | 1.564484503 | 0.000209511 | 0.000362406 | 1 |
| T2DM | mast_cell | *UIMC1* | 2.342859695 | 5.16E-05 | 0.000136402 | 0.250504513 |
| T2DM | mast_cell | *LPCAT2* | 1.553865294 | 4.01E-05 | 0.000134478 | 0.19437176 |
| T2DM | mast_cell | *OGFOD1* | 1.836210988 | 0.000248546 | 0.000419028 | 1 |
| T2DM | mast_cell | *TXNDC16* | 1.922468729 | 4.51E-05 | 0.000134478 | 0.218656797 |
| T2DM | mast_cell | *SF3B2* | 1.732521278 | 1.96E-05 | 0.000134478 | 0.095088644 |
| T2DM | mast_cell | *PHACTR3* | 1.489211133 | 0.019482182 | 0.023649624 | 1 |
| T2DM | mast_cell | *AAMDC* | 1.266704397 | 0.001604192 | 0.002239223 | 1 |
| T2DM | mast_cell | *GP6* | -1.243141098 | 0.02106988 | 0.025468624 | 1 |
| T2DM | mast_cell | *ASAP3* | 1.220249015 | 4.66E-05 | 0.000134478 | 0.226089677 |
| T2DM | mast_cell | *ANKRD10* | 2.404666039 | 4.99E-05 | 0.000135379 | 0.242166049 |
| T2DM | mast_cell | *DOCK3* | 2.169490321 | 3.64E-05 | 0.000134478 | 0.176816081 |

| | | | | | | |
|---|---|---|---|---|---|---|
| T2DM | mast_cell | *ARHGAP28* | 1.366863997 | 5.76E-05 | 0.000142813 | 0.27948581 |
| T2DM | mast_cell | *PPP1R13B* | 1.443090442 | 0.000170433 | 0.000306609 | 0.826940012 |
| T2DM | mast_cell | *NSFL1C* | 2.36642481 | 1.56E-05 | 0.000134478 | 0.075800353 |
| T2DM | mast_cell | *ZNF343* | 2.250237381 | 1.60E-05 | 0.000134478 | 0.077871181 |
| T2DM | mast_cell | *EBF4* | 5.628899657 | 0.000112138 | 0.000221627 | 0.54409333 |
| T2DM | mast_cell | *MAVS* | 2.030236485 | 6.84E-05 | 0.000157548 | 0.331717156 |
| T2DM | mast_cell | *XRN2* | 2.623454301 | 6.28E-05 | 0.000149953 | 0.304851389 |
| T2DM | mast_cell | *SNX5* | 5.15396542 | 0.001070214 | 0.001542685 | 1 |
| T2DM | mast_cell | *ESF1* | 3.47396504 | 5.62E-05 | 0.000140825 | 0.272642964 |
| T2DM | mast_cell | *TMEM230* | 2.503572612 | 2.97E-05 | 0.000134478 | 0.144181772 |
| T2DM | mast_cell | *RPH3A* | 4.917936399 | 3.32E-05 | 0.000134478 | 0.161044889 |
| T2DM | mast_cell | *KIF16B* | 1.99355079 | 3.18E-05 | 0.000134478 | 0.154128252 |
| T2DM | mast_cell | *FXYD5* | -2.281044713 | 0.02433376 | 0.029289854 | 1 |
| T2DM | mast_cell | *ZNF302* | 2.168104593 | 0.0047641 | 0.006182245 | 1 |
| T2DM | mast_cell | *GRAMD1A* | 2.697857036 | 3.21E-05 | 0.000134478 | 0.1559769 |
| T2DM | mast_cell | *CDIP1* | -2.953293989 | 0.010593783 | 0.013203452 | 1 |
| T2DM | mast_cell | *KCNH4* | 1.888981635 | 0.001297645 | 0.001836691 | 1 |
| T2DM | mast_cell | *GANAB* | 3.088293898 | 0.000328798 | 0.00053213 | 1 |
| T2DM | mast_cell | *MLF2* | 4.303515268 | 4.54E-05 | 0.000134478 | 0.220460698 |
| T2DM | mast_cell | *ANKRD24* | 2.345370018 | 4.40E-05 | 0.000134478 | 0.213392366 |
| T2DM | mast_cell | *DHX32* | 1.966248875 | 4.90E-05 | 0.000135021 | 0.237663928 |
| T2DM | mast_cell | *SPTLC1* | 2.784290904 | 0.000109677 | 0.000217471 | 0.532152746 |
| T2DM | mast_cell | *YPEL3* | 2.18021273 | 4.13E-06 | 0.000134478 | 0.020030771 |
| T2DM | mast_cell | *STRN4* | 2.601376091 | 5.81E-05 | 0.000143474 | 0.282069285 |
| T2DM | mast_cell | *LYZ* | 1.436055983 | 1.54E-05 | 0.000134478 | 0.074848105 |
| T2DM | mast_cell | *MUL1* | 1.436042975 | 0.004893682 | 0.006343613 | 1 |
| T2DM | mast_cell | *SPG21* | 1.985751808 | 0.000207204 | 0.000359054 | 1 |
| T2DM | mast_cell | *CHRD* | -1.667354392 | 0.001632948 | 0.002274781 | 1 |
| T2DM | mast_cell | *RAB11FIP3* | 2.970179558 | 0.000117753 | 0.000228639 | 0.571335919 |
| T2DM | mast_cell | *GNPTG* | 2.842117022 | 7.26E-05 | 0.000163044 | 0.352098202 |
| T2DM | mast_cell | *CERS4* | 2.688890295 | 3.31E-05 | 0.000134478 | 0.160697451 |
| T2DM | mast_cell | *USP48* | 2.478687082 | 0.001684698 | 0.002341494 | 1 |
| T2DM | mast_cell | *PDPR* | 2.316800092 | 1.26E-05 | 0.000134478 | 0.061088078 |
| T2DM | mast_cell | *GLG1* | 2.248973476 | 2.96E-05 | 0.000134478 | 0.143674237 |
| T2DM | mast_cell | *DTX2* | 4.224610832 | 3.38E-05 | 0.000134478 | 0.163821419 |
| T2DM | mast_cell | *LAMB1* | 0.923167321 | 0.00078548 | 0.001161936 | 1 |
| T2DM | mast_cell | *ABCC6* | 1.362730095 | 0.000581667 | 0.000881127 | 1 |
| T2DM | mast_cell | *RAPGEF4* | 1.146565578 | 3.26E-05 | 0.000134478 | 0.158416333 |
| T2DM | mast_cell | *TF* | 6.954346536 | 0.000103786 | 0.000208304 | 0.503567376 |
| T2DM | mast_cell | *ALKBH5* | 2.340357814 | 0.025137978 | 0.030161627 | 1 |
| T2DM | mast_cell | *SPAG7* | 1.707911723 | 0.000501054 | 0.000770071 | 1 |
| T2DM | mast_cell | *RGS17* | -3.412843719 | 0.000965176 | 0.001402106 | 1 |
| T2DM | mast_cell | *PPP2R3C* | 2.688902936 | 2.29E-06 | 0.000126109 | 0.011097561 |
| T2DM | mast_cell | *SCFD1* | 2.140266807 | 3.00E-05 | 0.000134478 | 0.145341043 |
| T2DM | mast_cell | *SUPT16H* | 2.023407569 | 3.46E-05 | 0.000134478 | 0.168008981 |
| T2DM | mast_cell | *TOX4* | 1.920191145 | 3.62E-05 | 0.000134478 | 0.175593877 |
| T2DM | mast_cell | *CAPN3* | -4.519444547 | 0.000145091 | 0.000267878 | 0.703982815 |
| T2DM | mast_cell | *TBX15* | 1.895398437 | 0.001454921 | 0.002039665 | 1 |
| T2DM | mast_cell | *UNC13D* | -0.908915318 | 0.001889416 | 0.002602909 | 1 |
| T2DM | mast_cell | *MFSD11* | 2.700973145 | 0.000188066 | 0.000330854 | 0.912496508 |

| | | | | | | |
|---|---|---|---|---|---|---|
| T2DM | mast_cell | *NUP50* | 2.606622893 | 0.001250646 | 0.00177379 | 1 |
| T2DM | mast_cell | *COMT* | 3.628595635 | 4.97E-05 | 0.000135163 | 0.240944674 |
| T2DM | mast_cell | *LRRFIP2* | 2.061550224 | 2.27E-05 | 0.000134478 | 0.109986316 |
| T2DM | mast_cell | *SEC22C* | 3.309450995 | 2.33E-05 | 0.000134478 | 0.113149873 |
| T2DM | mast_cell | *XYLB* | 2.18708083 | 3.68E-05 | 0.000134478 | 0.178657393 |
| T2DM | mast_cell | *GABRP* | 3.911753064 | 0.000160087 | 0.00029157 | 0.776742893 |
| T2DM | mast_cell | *FMO2* | 1.312017873 | 9.29E-05 | 0.000193112 | 0.45072914 |
| T2DM | mast_cell | *MSH2* | 3.119354261 | 3.14E-05 | 0.000134478 | 0.152457135 |
| T2DM | mast_cell | *PSMD5* | 2.6463345 | 3.51E-05 | 0.000134478 | 0.170363683 |
| T2DM | mast_cell | *CRAT* | -3.692202415 | 0.033381849 | 0.039516647 | 1 |
| T2DM | mast_cell | *NANS* | 1.677991503 | 3.70E-05 | 0.000134478 | 0.179338979 |
| T2DM | mast_cell | *PDE6C* | 6.019063833 | 0.004045591 | 0.005289466 | 1 |
| T2DM | mast_cell | *WAC* | 2.847147393 | 0.000216178 | 0.000371554 | 1 |
| T2DM | mast_cell | *ITPR3* | 3.212204701 | 0.000303405 | 0.00049583 | 1 |
| T2DM | mast_cell | *DSP* | 2.560596758 | 3.86E-05 | 0.000134478 | 0.187333487 |
| T2DM | mast_cell | *IFT74* | 2.727894992 | 0.000101137 | 0.000204978 | 0.490717984 |
| T2DM | mast_cell | *IL12RB1* | 1.528912783 | 0.0074959 | 0.009488679 | 1 |
| T2DM | mast_cell | *ACOT7* | -2.638441663 | 0.000562947 | 0.000855035 | 1 |
| T2DM | mast_cell | *CDC7* | 1.035431724 | 0.028394754 | 0.033875423 | 1 |
| T2DM | mast_cell | *ERMP1* | 2.413427231 | 4.86E-05 | 0.000134775 | 0.23585299 |
| T2DM | mast_cell | *RAB18* | 2.834224121 | 3.37E-05 | 0.000134478 | 0.163324946 |
| T2DM | mast_cell | *TSPAN15* | 1.532716838 | 3.30E-05 | 0.000134478 | 0.160139824 |
| T2DM | mast_cell | *MAST3* | 1.303012328 | 0.000922934 | 0.00134598 | 1 |
| T2DM | mast_cell | *OCEL1* | 1.989777318 | 0.002434645 | 0.00328501 | 1 |
| T2DM | mast_cell | *CATSPERG* | 1.440380798 | 0.014719991 | 0.018104282 | 1 |
| T2DM | mast_cell | *SETD1A* | 3.361744924 | 2.88E-06 | 0.000134478 | 0.013978035 |
| T2DM | mast_cell | *CIRBP* | 1.524618826 | 3.52E-05 | 0.000134478 | 0.170796392 |
| T2DM | mast_cell | *MARCH2* | 3.139508869 | 7.12E-06 | 0.000134478 | 0.034544347 |
| T2DM | mast_cell | *MTAP* | 1.849909873 | 9.26E-05 | 0.000192728 | 0.449440836 |
| T2DM | mast_cell | *POLR2E* | 3.742617715 | 4.66E-05 | 0.000134478 | 0.226335739 |
| T2DM | mast_cell | *MADCAM1* | 1.645927477 | 8.77E-05 | 0.000185825 | 0.425538936 |
| T2DM | mast_cell | *MED15* | 2.742734244 | 6.23E-05 | 0.000149337 | 0.302501435 |
| T2DM | mast_cell | *SERPIND1* | 2.136540395 | 2.99E-05 | 0.000134478 | 0.144876121 |
| T2DM | mast_cell | *SNAP29* | 1.825859959 | 7.78E-05 | 0.000170845 | 0.377568124 |
| T2DM | mast_cell | *LZTR1* | 4.052367009 | 0.003737619 | 0.004909292 | 1 |
| T2DM | mast_cell | *MMP11* | 6.133472222 | 1.61E-05 | 0.000134478 | 0.078333109 |
| T2DM | mast_cell | *CECR2* | 1.418360241 | 1.73E-05 | 0.000134478 | 0.083736254 |
| T2DM | mast_cell | *SMARCB1* | 1.902725137 | 0.018665452 | 0.022709322 | 1 |
| T2DM | mast_cell | *BCL2L13* | 2.473337266 | 8.05E-06 | 0.000134478 | 0.039035825 |
| T2DM | mast_cell | *DDT* | 1.905254629 | 0.000373047 | 0.000593839 | 1 |
| T2DM | mast_cell | *OSM* | -2.335101513 | 0.000363481 | 0.000581473 | 1 |
| T2DM | mast_cell | *CABIN1* | 2.845684573 | 3.98E-05 | 0.000134478 | 0.192981853 |
| T2DM | mast_cell | *PPIL2* | 1.603976911 | 3.20E-05 | 0.000134478 | 0.155432495 |
| T2DM | mast_cell | *PES1* | 1.53547735 | 0.0001383 | 0.000259138 | 0.671031098 |
| T2DM | mast_cell | *SLC35E4* | 1.733111424 | 0.008081551 | 0.010176924 | 1 |
| T2DM | mast_cell | *TOP3B* | 1.860691781 | 8.28E-05 | 0.000178199 | 0.401611101 |
| T2DM | mast_cell | *CRYBB3* | 3.606240109 | 0.002256373 | 0.003068246 | 1 |
| T2DM | mast_cell | *CYTH4* | 1.55800666 | 4.35E-05 | 0.000134478 | 0.211146784 |
| T2DM | mast_cell | *CARD10* | 1.716846878 | 0.000388493 | 0.000614396 | 1 |
| T2DM | mast_cell | *LRP5L* | 1.466266271 | 0.000815344 | 0.001202081 | 1 |

| | | | | | | |
|---|---|---|---|---|---|---|
| T2DM | mast_cell | *LGALS2* | 1.672978622 | 0.000101187 | 0.000204994 | 0.490960834 |
| T2DM | mast_cell | *SH3BP1* | 2.779757752 | 4.00E-05 | 0.000134478 | 0.194218007 |
| T2DM | mast_cell | *HPS4* | 1.669730203 | 3.30E-05 | 0.000134478 | 0.160310079 |
| T2DM | mast_cell | *TRIOBP* | 1.67514166 | 2.60E-05 | 0.000134478 | 0.126175226 |
| T2DM | mast_cell | *ANKRD54* | 1.368221764 | 0.001091184 | 0.001569649 | 1 |
| T2DM | mast_cell | *EIF3L* | 1.784143978 | 0.00039023 | 0.000616541 | 1 |
| T2DM | mast_cell | *CCDC134* | 1.906404918 | 3.87E-05 | 0.000134478 | 0.187874624 |
| T2DM | mast_cell | *PICK1* | 1.262715102 | 0.002246423 | 0.003056547 | 1 |
| T2DM | mast_cell | *TTC28* | 1.947307548 | 3.76E-05 | 0.000134478 | 0.182469551 |
| T2DM | mast_cell | *SLC5A4* | 0.362850074 | 0.03076949 | 0.036609506 | 1 |
| T2DM | mast_cell | *KDELR3* | -3.424975258 | 0.015184049 | 0.018646674 | 1 |
| T2DM | mast_cell | *CYP2D6* | 4.109382027 | 0.000152741 | 0.000280294 | 0.741097564 |
| T2DM | mast_cell | *CBY1* | 2.041066985 | 0.002826337 | 0.003769486 | 1 |
| T2DM | mast_cell | *RTCB* | 2.608948302 | 6.95E-05 | 0.000159011 | 0.336997843 |
| T2DM | mast_cell | *RAB36* | 2.526117706 | 0.001985794 | 0.002725502 | 1 |
| T2DM | mast_cell | *TIMP3* | 1.934801795 | 3.27E-05 | 0.000134478 | 0.158869221 |
| T2DM | mast_cell | *PPP6R2* | 2.601298925 | 4.80E-05 | 0.000134478 | 0.232793742 |
| T2DM | mast_cell | *CYB5R3* | 1.631606563 | 6.25E-05 | 0.00014957 | 0.303476603 |
| T2DM | mast_cell | *RHBDD3* | 1.515108729 | 0.000221844 | 0.000379678 | 1 |
| T2DM | mast_cell | *TOM1* | 1.391340876 | 0.000255655 | 0.00042892 | 1 |
| T2DM | mast_cell | *CHKB* | 3.340677861 | 3.51E-05 | 0.000134478 | 0.17035098 |
| T2DM | mast_cell | *ARSA* | 1.422840783 | 0.00028345 | 0.000468905 | 1 |
| T2DM | mast_cell | *TSPO* | 2.410208933 | 3.23E-05 | 0.000134478 | 0.156725292 |
| T2DM | mast_cell | *TTLL12* | 2.322581445 | 0.000167257 | 0.000302358 | 0.811529697 |
| T2DM | mast_cell | *MTMR3* | 2.099721094 | 0.000352964 | 0.000566518 | 1 |
| T2DM | mast_cell | *APOL4* | 2.115918157 | 0.001201485 | 0.001711067 | 1 |
| T2DM | mast_cell | *FOXRED2* | 2.040469963 | 1.20E-05 | 0.000134478 | 0.058083521 |
| T2DM | mast_cell | *IFT27* | 1.721492497 | 2.99E-05 | 0.000134478 | 0.145317076 |
| T2DM | mast_cell | *SLC25A17* | 2.154424195 | 8.49E-06 | 0.000134478 | 0.041202952 |
| T2DM | mast_cell | *FAM118A* | 1.530072249 | 0.000759516 | 0.001124899 | 1 |
| T2DM | mast_cell | *ZC3H7B* | 3.321683092 | 5.70E-05 | 0.000142008 | 0.276507517 |
| T2DM | mast_cell | *ACO2* | 2.596756419 | 7.67E-05 | 0.000168814 | 0.372066845 |
| T2DM | mast_cell | *POLR3H* | 2.475409972 | 2.38E-05 | 0.000134478 | 0.11529333 |
| T2DM | mast_cell | *DESI1* | 6.006046646 | 3.15E-05 | 0.000134478 | 0.152816594 |
| T2DM | mast_cell | *MLC1* | 2.312871765 | 0.016069386 | 0.019684085 | 1 |
| T2DM | mast_cell | *HDAC10* | 4.811334126 | 2.63E-05 | 0.000134478 | 0.127772736 |
| T2DM | mast_cell | *KHNYN* | 1.960488349 | 0.000546542 | 0.000831814 | 1 |
| T2DM | mast_cell | *SDR39U1* | 0.930806224 | 0.011649714 | 0.014471175 | 1 |
| T2DM | mast_cell | *RBM23* | 3.136967181 | 1.13E-05 | 0.000134478 | 0.054712831 |
| T2DM | mast_cell | *AP4S1* | 4.095372303 | 0.000252856 | 0.000425254 | 1 |
| T2DM | mast_cell | *NIN* | 2.714324946 | 5.16E-05 | 0.000136402 | 0.250484247 |
| T2DM | mast_cell | *PYGL* | 0.987716541 | 0.000175027 | 0.000312908 | 0.849232381 |
| T2DM | mast_cell | *CNIH1* | 3.32523176 | 4.09E-05 | 0.000134478 | 0.198372072 |
| T2DM | mast_cell | *ATP6V1D* | 4.000412141 | 9.62E-05 | 0.000197405 | 0.466664766 |
| T2DM | mast_cell | *GSTZ1* | 2.381803808 | 0.011523203 | 0.014321063 | 1 |
| T2DM | mast_cell | *TMED8* | 2.868077112 | 0.000298534 | 0.000489024 | 1 |
| T2DM | mast_cell | *LGMN* | 1.005479706 | 2.83E-05 | 0.000134478 | 0.137213763 |
| T2DM | mast_cell | *MTHFD1* | 2.275278794 | 1.20E-05 | 0.000134478 | 0.05815105 |
| T2DM | mast_cell | *TELO2* | 2.03433758 | 4.78E-05 | 0.000134478 | 0.231782415 |
| T2DM | mast_cell | *PSMB5* | 3.19030301 | 9.75E-07 | 9.10E-05 | 0.004730354 |

| | | | | | | |
|---|---|---|---|---|---|---|
| T2DM | mast_cell | *YY1* | 4.061718132 | 9.37E-05 | 0.000194208 | 0.454447771 |
| T2DM | mast_cell | *CCNB1IP1* | 2.688129634 | 3.30E-05 | 0.000134478 | 0.15991098 |
| T2DM | mast_cell | *EFS* | 2.148752572 | 4.20E-05 | 0.000134478 | 0.203978548 |
| T2DM | mast_cell | *CINP* | 1.632071789 | 4.37E-05 | 0.000134478 | 0.211800021 |
| T2DM | mast_cell | *KIAA0391* | 3.276818992 | 0.000548058 | 0.000833701 | 1 |
| T2DM | mast_cell | *EMC9* | -1.490014729 | 0.011426366 | 0.014208285 | 1 |
| T2DM | mast_cell | *PLTP* | 1.243294022 | 3.84E-05 | 0.000134478 | 0.186097871 |
| T2DM | mast_cell | *TRPC4AP* | 2.550805053 | 0.000215837 | 0.000371144 | 1 |
| T2DM | mast_cell | *PYGB* | 1.795588483 | 0.00021276 | 0.000367109 | 1 |
| T2DM | mast_cell | *ABHD12* | 2.654560341 | 0.000121132 | 0.000233505 | 0.587731663 |
| T2DM | mast_cell | *CD40* | 2.409293948 | 0.000107264 | 0.000213472 | 0.520445575 |
| T2DM | mast_cell | *UQCC1* | 1.865236147 | 0.000743102 | 0.001102946 | 1 |
| T2DM | mast_cell | *IFT52* | 1.303239271 | 3.82E-05 | 0.000134478 | 0.185226535 |
| T2DM | mast_cell | *NDRG3* | 1.649034287 | 3.26E-05 | 0.000134478 | 0.158143079 |
| T2DM | mast_cell | *PABPC1L* | 1.850519369 | 4.60E-05 | 0.000134478 | 0.222976454 |
| T2DM | mast_cell | *DOK5* | 6.029968925 | 0.00558634 | 0.007166822 | 1 |
| T2DM | mast_cell | *CSTF1* | 2.062401434 | 4.29E-05 | 0.000134478 | 0.207932937 |
| T2DM | mast_cell | *NELFCD* | 4.330473153 | 1.66E-05 | 0.000134478 | 0.080378873 |
| T2DM | mast_cell | *CTSZ* | 1.48160561 | 4.01E-05 | 0.000134478 | 0.194636935 |
| T2DM | mast_cell | *MTG2* | 1.58361166 | 3.47E-05 | 0.000134478 | 0.168545792 |
| T2DM | mast_cell | *TCFL5* | -2.500750005 | 0.002262272 | 0.003073801 | 1 |
| T2DM | mast_cell | *DIDO1* | 3.938075566 | 8.75E-06 | 0.000134478 | 0.04246061 |
| T2DM | mast_cell | *GID8* | 3.816040523 | 2.58E-05 | 0.000134478 | 0.125214851 |
| T2DM | mast_cell | *BIRC7* | -0.844597324 | 6.07E-05 | 0.00014641 | 0.294723591 |
| T2DM | mast_cell | *NKAIN4* | -2.317447839 | 0.00467192 | 0.00606587 | 1 |
| T2DM | mast_cell | *PTK6* | 6.194454093 | 2.98E-05 | 0.000134478 | 0.144695428 |
| T2DM | mast_cell | *GMEB2* | 4.049989343 | 3.88E-05 | 0.000134478 | 0.188115723 |
| T2DM | mast_cell | *CDC25B* | 2.619661403 | 0.002829995 | 0.003773327 | 1 |
| T2DM | mast_cell | *RNF24* | 1.50289923 | 3.35E-05 | 0.000134478 | 0.162321397 |
| T2DM | mast_cell | *NDUFAF5* | 1.464506004 | 0.000444548 | 0.000690665 | 1 |
| T2DM | mast_cell | *TRIB3* | 3.080504542 | 5.05E-05 | 0.000135795 | 0.244974032 |
| T2DM | mast_cell | *RASSF2* | 2.286510059 | 2.38E-05 | 0.000134478 | 0.115451328 |
| T2DM | mast_cell | *ANGPT4* | 3.785255659 | 4.18E-05 | 0.000134478 | 0.202899946 |
| T2DM | mast_cell | *RSPO4* | -1.262714156 | 0.035297276 | 0.041679821 | 1 |
| T2DM | mast_cell | *SIRPB1* | -2.449052084 | 0.024566205 | 0.029540329 | 1 |
| T2DM | mast_cell | *SEC23B* | 2.407701607 | 0.000263756 | 0.000440531 | 1 |
| T2DM | mast_cell | *CRNKL1* | 2.723325053 | 0.006746192 | 0.008575459 | 1 |
| T2DM | mast_cell | *MROH8* | 1.159407427 | 0.002895887 | 0.003853769 | 1 |
| T2DM | mast_cell | *MANBAL* | 2.37469765 | 0.028986814 | 0.034556271 | 1 |
| T2DM | mast_cell | *TTI1* | 1.84084106 | 5.88E-05 | 0.000144373 | 0.285457876 |
| T2DM | mast_cell | *BPI* | 0.735227423 | 0.03772261 | 0.044392458 | 1 |
| T2DM | mast_cell | *ACTR5* | 1.871187396 | 0.000121366 | 0.000233862 | 0.588865613 |
| T2DM | mast_cell | *MAP1LC3A* | 3.91632751 | 2.53E-05 | 0.000134478 | 0.122755538 |
| T2DM | mast_cell | *RBFA* | 4.477270803 | 0.000371139 | 0.000591385 | 1 |
| T2DM | mast_cell | *METTL4* | 1.789143524 | 4.57E-05 | 0.000134478 | 0.221735841 |
| T2DM | mast_cell | *LPIN2* | 2.854718911 | 2.36E-05 | 0.000134478 | 0.114707031 |
| T2DM | mast_cell | *MYOM1* | 1.20933345 | 4.56E-05 | 0.000134478 | 0.221283212 |
| T2DM | mast_cell | *CEP192* | 1.618924315 | 9.51E-05 | 0.000196364 | 0.461651377 |
| T2DM | mast_cell | *LAMA1* | 2.299947365 | 0.000603895 | 0.000913087 | 1 |
| T2DM | mast_cell | *RNF125* | 1.854649756 | 4.33E-05 | 0.000134478 | 0.209903182 |

| | | | | | | |
|---|---|---|---|---|---|---|
| T2DM | mast_cell | *POLI* | 1.026568256 | 0.00057353 | 0.000869616 | 1 |
| T2DM | mast_cell | *MIB1* | 3.123754384 | 5.22E-05 | 0.000136624 | 0.253116213 |
| T2DM | mast_cell | *NALCN* | 1.668842527 | 7.62E-05 | 0.000168292 | 0.36994011 |
| T2DM | mast_cell | *STK24* | 2.175456485 | 3.21E-05 | 0.000134478 | 0.155593516 |
| T2DM | mast_cell | *ACP5* | 2.189624448 | 1.29E-05 | 0.000134478 | 0.062553843 |
| T2DM | mast_cell | *DNAJC3* | 2.258261284 | 2.00E-05 | 0.000134478 | 0.096966772 |
| T2DM | mast_cell | *UGGT2* | 2.084091512 | 1.98E-05 | 0.000134478 | 0.096149577 |
| T2DM | mast_cell | *ARHGEF7* | 1.730853194 | 3.28E-05 | 0.000134478 | 0.159136056 |
| T2DM | mast_cell | *PARP4* | 2.881581153 | 2.28E-05 | 0.000134478 | 0.110435402 |
| T2DM | mast_cell | *SUPT20H* | 3.709536581 | 0.000179758 | 0.000319366 | 0.87218788 |
| T2DM | mast_cell | *MRPS31* | 2.823968358 | 2.19E-05 | 0.000134478 | 0.106433512 |
| T2DM | mast_cell | *RGCC* | 1.219868975 | 8.27E-05 | 0.000178199 | 0.401297459 |
| T2DM | mast_cell | *DGKH* | 1.524984685 | 0.000855849 | 0.001257975 | 1 |
| T2DM | mast_cell | *KATNAL1* | 2.148411624 | 3.26E-05 | 0.000134478 | 0.158135022 |
| T2DM | mast_cell | *INTS6* | 2.191908336 | 3.60E-05 | 0.000134478 | 0.174507106 |
| T2DM | mast_cell | *CLN5* | 2.546054511 | 5.10E-05 | 0.000136328 | 0.247392262 |
| T2DM | mast_cell | *MGRN1* | 2.922463542 | 5.14E-05 | 0.000136328 | 0.249504376 |
| T2DM | mast_cell | *LONP2* | 2.681355602 | 1.66E-05 | 0.000134478 | 0.080585714 |
| T2DM | mast_cell | *PLLP* | 1.70155515 | 0.000642018 | 0.000965017 | 1 |
| T2DM | mast_cell | *DHODH* | 3.409508394 | 1.22E-05 | 0.000134478 | 0.059104056 |
| T2DM | mast_cell | *PSMD7* | 2.341144315 | 2.47E-05 | 0.000134478 | 0.120028142 |
| T2DM | mast_cell | *COG4* | 2.081436504 | 0.005942077 | 0.00759709 | 1 |
| T2DM | mast_cell | *MON1B* | 1.772334033 | 4.53E-05 | 0.000134478 | 0.219942032 |
| T2DM | mast_cell | *NPRL3* | 1.6946012 | 6.49E-05 | 0.000152782 | 0.315061015 |
| T2DM | mast_cell | *NME4* | 2.589711166 | 3.59E-05 | 0.000134478 | 0.17400699 |
| T2DM | mast_cell | *LMF1* | 2.088400152 | 2.82E-05 | 0.000134478 | 0.136768965 |
| T2DM | mast_cell | *NARFL* | 1.777420216 | 4.28E-05 | 0.000134478 | 0.207676637 |
| T2DM | mast_cell | *MTHFSD* | 1.100342169 | 0.006384501 | 0.00813701 | 1 |
| T2DM | mast_cell | *SLC7A5* | 3.54336332 | 0.004103282 | 0.005360562 | 1 |
| T2DM | mast_cell | *RHBDL1* | 6.5005937 | 0.000443166 | 0.000688739 | 1 |
| T2DM | mast_cell | *NUBP1* | 3.30369485 | 1.90E-05 | 0.000134478 | 0.092354082 |
| T2DM | mast_cell | *UBE2I* | 1.81857593 | 5.54E-05 | 0.00014004 | 0.268608925 |
| T2DM | mast_cell | *CAPN15* | 5.123604881 | 0.000126039 | 0.000241621 | 0.6115435 |
| T2DM | mast_cell | *CLUAP1* | 2.254631439 | 4.67E-06 | 0.000134478 | 0.022665844 |
| T2DM | mast_cell | *EARS2* | 1.46635094 | 0.001174538 | 0.001676628 | 1 |
| T2DM | mast_cell | *CPPED1* | 3.78462077 | 2.30E-05 | 0.000134478 | 0.111511922 |
| T2DM | mast_cell | *HMOX2* | 1.750127321 | 1.78E-05 | 0.000134478 | 0.086373958 |
| T2DM | mast_cell | *TOX3* | -1.600969553 | 0.000243979 | 0.000412325 | 1 |
| T2DM | mast_cell | *RBL2* | 2.524590707 | 1.49E-06 | 0.000100592 | 0.007242606 |
| T2DM | mast_cell | *PYCARD* | 2.862346714 | 9.39E-06 | 0.000134478 | 0.045547658 |
| T2DM | mast_cell | *CDIPT* | 3.885522682 | 1.33E-05 | 0.000134478 | 0.064400735 |
| T2DM | mast_cell | *KAT8* | 2.3012604 | 4.09E-05 | 0.000134478 | 0.198665806 |
| T2DM | mast_cell | *TMC5* | 1.602797771 | 0.000141893 | 0.000264035 | 0.688464824 |
| T2DM | mast_cell | *C16orf62* | 2.101783425 | 3.27E-05 | 0.000134478 | 0.158538813 |
| T2DM | mast_cell | *RNF40* | 1.617378145 | 3.58E-05 | 0.000134478 | 0.173925874 |
| T2DM | mast_cell | *KNOP1* | 4.223539633 | 0.007175091 | 0.009094447 | 1 |
| T2DM | mast_cell | *LACTB* | 3.669082822 | 0.002788376 | 0.00371988 | 1 |
| T2DM | mast_cell | *HERC1* | 2.443107975 | 2.55E-05 | 0.000134478 | 0.123770157 |
| T2DM | mast_cell | *TRIP4* | 2.088554826 | 4.72E-05 | 0.000134478 | 0.228920531 |
| T2DM | mast_cell | *AP3B2* | -1.282502119 | 0.000656117 | 0.000984074 | 1 |

| | | | | | | |
|---|---|---|---|---|---|---|
| T2DM | mast_cell | *TTC23* | 1.636115484 | 3.95E-05 | 0.000134478 | 0.191467019 |
| T2DM | mast_cell | *FAH* | 1.548327113 | 5.84E-05 | 0.000143838 | 0.283360222 |
| T2DM | mast_cell | *RPAP1* | 1.426182765 | 0.000478726 | 0.000739974 | 1 |
| T2DM | mast_cell | *SLC30A4* | 3.231005875 | 2.53E-05 | 0.000134478 | 0.122570188 |
| T2DM | mast_cell | *ZDHHC2* | 1.872364675 | 0.000208734 | 0.000361319 | 1 |
| T2DM | mast_cell | *ZFAND1* | 3.452787436 | 8.89E-05 | 0.000187353 | 0.431392499 |
| T2DM | mast_cell | *EYA1* | 1.622273007 | 8.59E-05 | 0.00018324 | 0.41687088 |
| T2DM | mast_cell | *IMPAD1* | 3.422796517 | 3.37E-05 | 0.000134478 | 0.163654289 |
| T2DM | mast_cell | *SFRP1* | 1.498429621 | 5.24E-05 | 0.000136633 | 0.254064195 |
| T2DM | mast_cell | *NIPAL2* | 1.478239803 | 2.81E-05 | 0.000134478 | 0.136389745 |
| T2DM | mast_cell | *RAB2A* | 2.1060281 | 1.69E-05 | 0.000134478 | 0.081781686 |
| T2DM | mast_cell | *ARMC1* | 1.8081399 | 0.000398161 | 0.000626216 | 1 |
| T2DM | mast_cell | *SPAG1* | 2.231226195 | 4.48E-05 | 0.000134478 | 0.217409315 |
| T2DM | mast_cell | *CHRAC1* | 2.314672735 | 4.74E-05 | 0.000134478 | 0.229890619 |
| T2DM | mast_cell | *SNX16* | 2.839227157 | 3.92E-05 | 0.000134478 | 0.1899568 |
| T2DM | mast_cell | *GSDMD* | 2.531752663 | 2.56E-05 | 0.000134478 | 0.124102544 |
| T2DM | mast_cell | *TSTA3* | 2.834513142 | 6.60E-06 | 0.000134478 | 0.032000925 |
| T2DM | mast_cell | *SQLE* | 1.848711367 | 0.000298255 | 0.000488731 | 1 |
| T2DM | mast_cell | *TNFRSF10A* | 1.940953234 | 1.24E-05 | 0.000134478 | 0.059930639 |
| T2DM | mast_cell | *ERICH1* | 1.921496737 | 3.69E-05 | 0.000134478 | 0.178931874 |
| T2DM | mast_cell | *TUSC3* | 2.137154997 | 3.18E-05 | 0.000134478 | 0.154526125 |
| T2DM | mast_cell | *MAN2B1* | 2.319617735 | 0.000558744 | 0.000849319 | 1 |
| T2DM | mast_cell | *TULP2* | 10.48530906 | 3.91E-05 | 0.000134478 | 0.189881251 |
| T2DM | mast_cell | *NFKBIB* | 3.889578485 | 0.000297811 | 0.000488283 | 1 |
| T2DM | mast_cell | *SARS2* | -1.476882648 | 0.000414424 | 0.000647388 | 1 |
| T2DM | mast_cell | *SNRNP70* | 2.637953011 | 4.92E-05 | 0.000135162 | 0.238826602 |
| T2DM | mast_cell | *LIN7B* | 2.465089249 | 5.90E-05 | 0.000144683 | 0.28649872 |
| T2DM | mast_cell | *PPP1R37* | -3.154360051 | 6.70E-05 | 0.000155401 | 0.325197789 |
| T2DM | mast_cell | *FCGRT* | 1.501415355 | 0.000328787 | 0.00053213 | 1 |
| T2DM | mast_cell | *ARHGEF18* | 3.647752087 | 0.023304462 | 0.028099714 | 1 |
| T2DM | mast_cell | *PEX11G* | 1.935569294 | 0.000524762 | 0.000801178 | 1 |
| T2DM | mast_cell | *ERCC2* | 2.258070732 | 4.26E-05 | 0.000134478 | 0.206609729 |
| T2DM | mast_cell | *CD37* | -1.873452481 | 0.028641843 | 0.034153409 | 1 |
| T2DM | mast_cell | *AMH* | 19.66614745 | 5.19E-05 | 0.000136402 | 0.2518763 |
| T2DM | mast_cell | *FCER2* | 6.07530683 | 4.76E-05 | 0.000134478 | 0.230948017 |
| T2DM | mast_cell | *CLEC4M* | 18.24399614 | 5.14E-05 | 0.000136328 | 0.249161693 |
| T2DM | mast_cell | *NOVA2* | 1.491069265 | 0.000176222 | 0.000314348 | 0.855027206 |
| T2DM | mast_cell | *SGTA* | 4.728026193 | 0.000222615 | 0.000380863 | 1 |
| T2DM | mast_cell | *KIR3DX1* | 10.11467039 | 0.000158436 | 0.000289432 | 0.768731499 |
| T2DM | mast_cell | *MED25* | 2.938462351 | 3.29E-05 | 0.000134478 | 0.159525033 |
| T2DM | mast_cell | *TIMM44* | 2.25690618 | 2.36E-05 | 0.000134478 | 0.114637759 |
| T2DM | mast_cell | *IL27RA* | 2.305948697 | 2.80E-05 | 0.000134478 | 0.135674845 |
| T2DM | mast_cell | *FAM32A* | 6.965266662 | 0.000606392 | 0.000916292 | 1 |
| T2DM | mast_cell | *C19orf44* | 1.33753411 | 0.000103306 | 0.00020762 | 0.501238306 |
| T2DM | mast_cell | *OLFM2* | 1.373344298 | 0.000347355 | 0.000558611 | 1 |
| T2DM | mast_cell | *PDCD5* | 1.720316634 | 3.67E-05 | 0.000134478 | 0.178277798 |
| T2DM | mast_cell | *ANKRD27* | 4.510702934 | 0.000113901 | 0.000224288 | 0.552646091 |
| T2DM | mast_cell | *POLR2I* | 2.130381589 | 1.88E-05 | 0.000134478 | 0.091348669 |
| T2DM | mast_cell | *CCDC9* | 7.982787701 | 0.000387187 | 0.00061273 | 1 |
| T2DM | mast_cell | *TGFB1* | 3.169793591 | 3.47E-05 | 0.000134478 | 0.16855963 |

| | | | | | | |
|---|---|---|---|---|---|---|
| T2DM | mast_cell | *CEACAM4* | 3.011004183 | 0.008732379 | 0.010962355 | 1 |
| T2DM | mast_cell | *MRPL4* | 2.471763833 | 2.02E-05 | 0.000134478 | 0.097997945 |
| T2DM | mast_cell | *ICAM5* | -2.84706468 | 4.15E-06 | 0.000134478 | 0.020131118 |
| T2DM | mast_cell | *CD33* | 0.97613923 | 0.002346062 | 0.003179634 | 1 |
| T2DM | mast_cell | *RABAC1* | 1.750242755 | 2.52E-05 | 0.000134478 | 0.122086188 |
| T2DM | mast_cell | *GRIN2D* | 3.10369856 | 2.82E-05 | 0.000134478 | 0.136711088 |
| T2DM | mast_cell | *CCDC114* | 4.599036964 | 0.025839155 | 0.030955946 | 1 |
| T2DM | mast_cell | *CARD8* | 3.162645167 | 0.000488111 | 0.00075328 | 1 |
| T2DM | mast_cell | *ZNF175* | 1.888986492 | 8.45E-05 | 0.000181073 | 0.410129797 |
| T2DM | mast_cell | *PLA2G4C* | 1.09699393 | 0.001652103 | 0.002298826 | 1 |
| T2DM | mast_cell | *CAPS* | 3.044077467 | 4.19E-05 | 0.000134478 | 0.203353237 |
| T2DM | mast_cell | *MIER2* | 3.431996524 | 1.78E-05 | 0.000134478 | 0.086240187 |
| T2DM | mast_cell | *PLEKHA4* | 1.68409943 | 5.15E-05 | 0.000136328 | 0.249704437 |
| T2DM | mast_cell | *TNPO2* | 1.679979547 | 2.21E-05 | 0.000134478 | 0.107299771 |
| T2DM | mast_cell | *GCDH* | 1.426604311 | 0.007567692 | 0.009568627 | 1 |
| T2DM | mast_cell | *TFPT* | 3.880665445 | 2.06E-05 | 0.000134478 | 0.099788945 |
| T2DM | mast_cell | *RAB3A* | 7.449388256 | 4.28E-05 | 0.000134478 | 0.20768228 |
| T2DM | mast_cell | *UPK1A* | -1.073255698 | 0.000493497 | 0.000760625 | 1 |
| T2DM | mast_cell | *ETV2* | 15.64820114 | 0.000372143 | 0.000592595 | 1 |
| T2DM | mast_cell | *MAG* | -1.981606766 | 0.001267083 | 0.001795527 | 1 |
| T2DM | mast_cell | *LSR* | -0.967117639 | 0.005003437 | 0.006466882 | 1 |
| T2DM | mast_cell | *ZNF14* | 5.078418809 | 6.33E-05 | 0.000150641 | 0.307005856 |
| T2DM | mast_cell | *ZNF85* | 1.726067402 | 4.55E-05 | 0.000134478 | 0.220780358 |
| T2DM | mast_cell | *ETHE1* | 2.630845028 | 6.41E-05 | 0.000151578 | 0.311037705 |
| T2DM | mast_cell | *GTPBP10* | 2.594605084 | 3.47E-05 | 0.000134478 | 0.168423649 |
| T2DM | mast_cell | *RASA4* | 12.15575289 | 5.05E-05 | 0.000135777 | 0.244806214 |
| T2DM | mast_cell | *BET1* | 3.848375487 | 0.000173302 | 0.000310281 | 0.84086223 |
| T2DM | mast_cell | *SP4* | 1.67384189 | 2.82E-05 | 0.000134478 | 0.136984201 |
| T2DM | mast_cell | *DNAH11* | 3.146377877 | 3.37E-05 | 0.000134478 | 0.16366392 |
| T2DM | mast_cell | *STEAP1B* | 3.119165713 | 2.31E-05 | 0.000134478 | 0.111866335 |
| T2DM | mast_cell | *PTN* | 1.707736396 | 4.22E-05 | 0.000134478 | 0.204553691 |
| T2DM | mast_cell | *ADAP1* | 6.025228981 | 4.63E-05 | 0.000134478 | 0.224872331 |
| T2DM | mast_cell | *MET* | 1.919345109 | 0.003344176 | 0.004416424 | 1 |
| T2DM | mast_cell | *RNF32* | 2.420509641 | 0.001225857 | 0.001743729 | 1 |
| T2DM | mast_cell | *IQCE* | 2.405838229 | 3.50E-05 | 0.000134478 | 0.170030104 |
| T2DM | mast_cell | *VIPR2* | 1.758822434 | 4.14E-05 | 0.000134478 | 0.200891456 |
| T2DM | mast_cell | *HIBADH* | 1.850640761 | 1.52E-05 | 0.000134478 | 0.073565034 |
| T2DM | mast_cell | *CPVL* | 1.335733633 | 4.33E-05 | 0.000134478 | 0.210329959 |
| T2DM | mast_cell | *CHN2* | 2.390249684 | 1.56E-08 | 5.04E-06 | 7.55E-05 |
| T2DM | mast_cell | *GRB10* | 1.340051392 | 3.20E-05 | 0.000134478 | 0.155043039 |
| T2DM | mast_cell | *ABHD11* | 2.383619278 | 6.93E-05 | 0.000158841 | 0.336426253 |
| T2DM | mast_cell | *PLEKHA8* | 1.405041676 | 3.61E-05 | 0.000134478 | 0.175300368 |
| T2DM | mast_cell | *CRHR2* | 2.613768501 | 0.005094959 | 0.006578165 | 1 |
| T2DM | mast_cell | *EPHB6* | 1.044888672 | 0.025339113 | 0.030386895 | 1 |
| T2DM | mast_cell | *CHCHD2* | 2.776466981 | 4.96E-05 | 0.000135163 | 0.240898732 |
| T2DM | mast_cell | *ZKSCAN1* | 1.903333296 | 5.14E-05 | 0.000136328 | 0.249188013 |
| T2DM | mast_cell | *EIF3B* | 1.818405802 | 4.08E-05 | 0.000134478 | 0.198082917 |
| T2DM | mast_cell | *TAF6* | 1.8869586 | 0.000156554 | 0.000286426 | 0.75960182 |
| T2DM | mast_cell | *TFR2* | 2.991764597 | 0.000123977 | 0.000237949 | 0.601534551 |
| T2DM | mast_cell | *MOSPD3* | 1.751552255 | 0.000786688 | 0.001163014 | 1 |

| | | | | | | |
|---|---|---|---|---|---|---|
| T2DM | mast_cell | *RBM28* | 2.41187716 | 2.11E-05 | 0.000134478 | 0.102325362 |
| T2DM | mast_cell | *IMPDH1* | 2.359516493 | 0.000131482 | 0.000249102 | 0.637949118 |
| T2DM | mast_cell | *C1GALT1* | 2.068103631 | 2.54E-05 | 0.000134478 | 0.123383976 |
| T2DM | mast_cell | *NRF1* | 2.378327291 | 2.64E-05 | 0.000134478 | 0.128316777 |
| T2DM | mast_cell | *ZNF862* | 2.710230248 | 5.07E-05 | 0.000136098 | 0.246202022 |
| T2DM | mast_cell | *MEST* | 0.918956906 | 0.00051072 | 0.000783438 | 1 |
| T2DM | mast_cell | *ACTR3C* | 3.892411378 | 3.16E-05 | 0.000134478 | 0.153426905 |
| T2DM | mast_cell | *POU6F2* | 1.81535531 | 5.02E-05 | 0.0001354 | 0.243584738 |
| T2DM | mast_cell | *TSPAN13* | 2.03662789 | 4.35E-05 | 0.000134478 | 0.211131675 |
| T2DM | mast_cell | *RARRES2* | 2.79348093 | 3.81E-05 | 0.000134478 | 0.184680494 |
| T2DM | mast_cell | *TMEM176B* | 1.366308661 | 0.00692253 | 0.008792701 | 1 |
| T2DM | mast_cell | *COA1* | 2.08400957 | 2.59E-05 | 0.000134478 | 0.125728515 |
| T2DM | mast_cell | *BLVRA* | 3.179495833 | 0.005901638 | 0.007549366 | 1 |
| T2DM | mast_cell | *URGCP* | 2.263549054 | 1.06E-05 | 0.000134478 | 0.051431349 |
| T2DM | mast_cell | *AEBP1* | 2.68411058 | 3.95E-05 | 0.000134478 | 0.191790785 |
| T2DM | mast_cell | *GCK* | 11.21321804 | 4.77E-05 | 0.000134478 | 0.231610721 |
| T2DM | mast_cell | *SPATA6L* | 1.469281906 | 3.53E-05 | 0.000134478 | 0.171197917 |
| T2DM | mast_cell | *SLC1A1* | 1.141627562 | 7.94E-06 | 0.000134478 | 0.038521083 |
| T2DM | mast_cell | *FKTN* | 2.167855523 | 3.42E-05 | 0.000134478 | 0.165955193 |
| T2DM | mast_cell | *NMRK1* | 1.454973518 | 3.57E-05 | 0.000134478 | 0.173401732 |
| T2DM | mast_cell | *MEGF9* | 2.44764858 | 3.57E-05 | 0.000134478 | 0.173170176 |
| T2DM | mast_cell | *TRIM14* | 1.362970904 | 3.20E-05 | 0.000134478 | 0.1553655 |
| T2DM | mast_cell | *CORO2A* | 2.259175414 | 9.60E-05 | 0.000197267 | 0.465627891 |
| T2DM | mast_cell | *C5* | 2.122108355 | 2.64E-05 | 0.000134478 | 0.128236191 |
| T2DM | mast_cell | *OGN* | 1.457079622 | 0.000371468 | 0.000591715 | 1 |
| T2DM | mast_cell | *ECM2* | 2.143123071 | 3.51E-05 | 0.000134478 | 0.170290419 |
| T2DM | mast_cell | *PTGR1* | 1.254846955 | 0.00011816 | 0.00022905 | 0.57331339 |
| T2DM | mast_cell | *SUSD1* | 1.463605624 | 0.000708546 | 0.001056179 | 1 |
| T2DM | mast_cell | *PLGRKT* | 1.865019448 | 0.004484369 | 0.00583485 | 1 |
| T2DM | mast_cell | *DOCK8* | 1.220553378 | 8.28E-05 | 0.000178199 | 0.401661047 |
| T2DM | mast_cell | *KANK1* | 1.231768214 | 0.000180313 | 0.00032 | 0.874879147 |
| T2DM | mast_cell | *NCS1* | 1.976424696 | 0.001010107 | 0.001462124 | 1 |
| T2DM | mast_cell | *FUBP3* | 1.912512515 | 5.61E-05 | 0.000140825 | 0.272000996 |
| T2DM | mast_cell | *PTGDS* | 2.978942768 | 3.32E-05 | 0.000134478 | 0.161278267 |
| T2DM | mast_cell | *HPS1* | 1.977585904 | 8.22E-05 | 0.000177654 | 0.398654824 |
| T2DM | mast_cell | *PHYH* | 1.852109886 | 0.007690559 | 0.009709756 | 1 |
| T2DM | mast_cell | *DNMBP* | 1.162392743 | 0.000117844 | 0.000228639 | 0.571780657 |
| T2DM | mast_cell | *PKD2L1* | -1.538146555 | 0.002949093 | 0.003920273 | 1 |
| T2DM | mast_cell | *CUBN* | 4.474841004 | 7.62E-05 | 0.000168279 | 0.369616812 |
| T2DM | mast_cell | *TRDMT1* | 4.126943872 | 5.94E-05 | 0.000144951 | 0.28830788 |
| T2DM | mast_cell | *ATE1* | 2.92813858 | 9.20E-06 | 0.000134478 | 0.04466196 |
| T2DM | mast_cell | *PLEKHA1* | 1.736469034 | 6.77E-05 | 0.000156404 | 0.328316103 |
| T2DM | mast_cell | *CDH23* | 3.070387053 | 6.35E-05 | 0.000150963 | 0.308192388 |
| T2DM | mast_cell | *SPOCK2* | 0.919686639 | 0.00716253 | 0.009083271 | 1 |
| T2DM | mast_cell | *BMPR1A* | 1.715357868 | 3.66E-05 | 0.000134478 | 0.177795368 |
| T2DM | mast_cell | *LIPA* | 1.407423133 | 3.32E-05 | 0.000134478 | 0.161074351 |
| T2DM | mast_cell | *SFXN3* | 2.673991089 | 3.54E-05 | 0.000134478 | 0.171910308 |
| T2DM | mast_cell | *TNKS2* | 1.494501313 | 3.71E-05 | 0.000134478 | 0.179777368 |
| T2DM | mast_cell | *ANKRD26* | 3.356102259 | 0.000116733 | 0.000227587 | 0.566388801 |
| T2DM | mast_cell | *ACBD5* | 2.831785748 | 2.36E-05 | 0.000134478 | 0.114471929 |

| | | | | | | |
|---|---|---|---|---|---|---|
| T2DM | mast_cell | *LHPP* | 2.392792823 | 0.011732977 | 0.014559694 | 1 |
| T2DM | mast_cell | *PITRM1* | 4.176684057 | 0.011229029 | 0.013973647 | 1 |
| T2DM | mast_cell | *TFAM* | 2.876627055 | 3.20E-05 | 0.000134478 | 0.155186918 |
| T2DM | mast_cell | *RPL28* | 1.622264364 | 0.000100785 | 0.000204434 | 0.48900652 |
| T2DM | mast_cell | *DNAJC12* | 4.629387392 | 4.11E-05 | 0.000134478 | 0.199651037 |
| T2DM | mast_cell | *PBLD* | 1.757494455 | 5.94E-05 | 0.000144951 | 0.288188151 |
| T2DM | mast_cell | *TSPAN14* | 2.325754858 | 0.000559178 | 0.000849713 | 1 |
| T2DM | mast_cell | *TADA2A* | 1.526612709 | 0.000757539 | 0.001122657 | 1 |
| T2DM | mast_cell | *RPL19* | 1.981407806 | 1.50E-05 | 0.000134478 | 0.072703181 |
| T2DM | mast_cell | *RAPGEFL1* | 2.418219036 | 9.14E-05 | 0.000191197 | 0.443576636 |
| T2DM | mast_cell | *WNT3* | 2.66629601 | 0.000276865 | 0.000459422 | 1 |
| T2DM | mast_cell | *ASPA* | 1.621852719 | 2.34E-05 | 0.000134478 | 0.113771456 |
| T2DM | mast_cell | *RAD51C* | 1.579014634 | 0.002379707 | 0.003220737 | 1 |
| T2DM | mast_cell | *TRIM37* | 2.831761939 | 5.74E-05 | 0.000142483 | 0.278479258 |
| T2DM | mast_cell | *PNPO* | 2.978831415 | 0.000370169 | 0.00059042 | 1 |
| T2DM | mast_cell | *TRIM16L* | 2.428148977 | 4.58E-05 | 0.000134478 | 0.222390573 |
| T2DM | mast_cell | *CDK5RAP3* | 2.267499548 | 3.75E-05 | 0.000134478 | 0.181798208 |
| T2DM | mast_cell | *PIGL* | 1.746576318 | 4.31E-05 | 0.000134478 | 0.20911142 |
| T2DM | mast_cell | *MED13* | 1.453449422 | 4.92E-05 | 0.000135162 | 0.238787335 |
| T2DM | mast_cell | *ENO3* | 3.633130987 | 2.52E-05 | 0.000134478 | 0.122203479 |
| T2DM | mast_cell | *RAI1* | 3.855141075 | 3.60E-05 | 0.000134478 | 0.174883141 |
| T2DM | mast_cell | *GOSR1* | 2.669345091 | 3.37E-05 | 0.000134478 | 0.163710281 |
| T2DM | mast_cell | *DRG2* | 1.747744439 | 5.70E-05 | 0.000142008 | 0.276578974 |
| T2DM | mast_cell | *ALDH3A1* | 5.253395292 | 0.011525917 | 0.014321063 | 1 |
| T2DM | mast_cell | *ICAM2* | 2.203540797 | 3.22E-05 | 0.000134478 | 0.156246942 |
| T2DM | mast_cell | *DDX5* | 2.19360301 | 1.88E-05 | 0.000134478 | 0.091148351 |
| T2DM | mast_cell | *C17orf75* | 1.578534083 | 9.68E-05 | 0.000198277 | 0.469497271 |
| T2DM | mast_cell | *PSMD11* | 2.707600172 | 0.000230875 | 0.000392808 | 1 |
| T2DM | mast_cell | *PEX12* | -4.201315861 | 0.013706923 | 0.016888265 | 1 |
| T2DM | mast_cell | *DHX58* | 2.090950972 | 0.000339545 | 0.00054697 | 1 |
| T2DM | mast_cell | *RAB5C* | 1.891024289 | 2.23E-05 | 0.000134478 | 0.108137969 |
| T2DM | mast_cell | *NAGLU* | 2.67830276 | 8.27E-06 | 0.000134478 | 0.04013961 |
| T2DM | mast_cell | *HSD17B1* | 2.540674295 | 3.48E-05 | 0.000134478 | 0.168651965 |
| T2DM | mast_cell | *CNTNAP1* | 3.948758292 | 5.05E-08 | 1.36E-05 | 0.000245145 |
| T2DM | mast_cell | *EZH1* | 2.238403136 | 0.000163393 | 0.00029659 | 0.792784251 |
| T2DM | mast_cell | *ABCC3* | 1.543076136 | 2.26E-05 | 0.000134478 | 0.109638796 |
| T2DM | mast_cell | *EFTUD2* | 3.553914747 | 3.58E-05 | 0.000134478 | 0.173561446 |
| T2DM | mast_cell | *HLF* | 2.626902171 | 0.002086817 | 0.002853787 | 1 |
| T2DM | mast_cell | *EFNB3* | 1.966555562 | 0.006576347 | 0.008368328 | 1 |
| T2DM | mast_cell | *FAM20A* | 1.545161051 | 3.96E-05 | 0.000134478 | 0.192184506 |
| T2DM | mast_cell | *DPH1* | 6.22969052 | 0.000331789 | 0.000535898 | 1 |
| T2DM | mast_cell | *DHRS7B* | 2.25083989 | 2.95E-05 | 0.000134478 | 0.143234001 |
| T2DM | mast_cell | *MYH3* | 2.689651152 | 5.17E-05 | 0.000136402 | 0.250759909 |
| T2DM | mast_cell | *TMEM97* | -1.019483817 | 0.001071431 | 0.001543981 | 1 |
| T2DM | mast_cell | *GNRHR* | 2.462470964 | 3.27E-06 | 0.000134478 | 0.015888809 |
| T2DM | mast_cell | *SLAIN2* | 3.09220973 | 2.48E-05 | 0.000134478 | 0.120486137 |
| T2DM | mast_cell | *OCIAD1* | 3.006706925 | 6.38E-05 | 0.000151327 | 0.309766785 |
| T2DM | mast_cell | *UBE2D3* | 2.058496468 | 2.78E-05 | 0.000134478 | 0.134837948 |
| T2DM | mast_cell | *NDUFC1* | 1.892199083 | 4.49E-05 | 0.000134478 | 0.218084251 |
| T2DM | mast_cell | *KLHL2* | 3.258450487 | 3.87E-05 | 0.000134478 | 0.187947442 |

| | | | | | | |
|---|---|---|---|---|---|---|
| T2DM | mast_cell | *CPE* | 1.688428472 | 6.18E-06 | 0.000134478 | 0.029989358 |
| T2DM | mast_cell | *WFS1* | 2.011215782 | 4.22E-05 | 0.000134478 | 0.204591704 |
| T2DM | mast_cell | *GRPEL1* | 6.542689768 | 0.008846521 | 0.011097032 | 1 |
| T2DM | mast_cell | *GAR1* | 2.709212768 | 3.18E-05 | 0.000134478 | 0.154367308 |
| T2DM | mast_cell | *AADAT* | 2.449826998 | 6.54E-05 | 0.000153213 | 0.317456818 |
| T2DM | mast_cell | *TRIM2* | 2.295057855 | 0.000141897 | 0.000264035 | 0.688483955 |
| T2DM | mast_cell | *STIM2* | 1.980872188 | 3.01E-05 | 0.000134478 | 0.146097523 |
| T2DM | mast_cell | *BST1* | 4.134133519 | 1.32E-05 | 0.000134478 | 0.064165647 |
| T2DM | mast_cell | *UFSP2* | 4.039406034 | 5.67E-05 | 0.000141687 | 0.275298699 |
| T2DM | mast_cell | *KLHL5* | 3.505444644 | 5.81E-05 | 0.000143427 | 0.281687099 |
| T2DM | mast_cell | *FAM149A* | 2.111706701 | 1.03E-05 | 0.000134478 | 0.050111226 |
| T2DM | mast_cell | *DDX25* | 2.203867909 | 1.58E-05 | 0.000134478 | 0.07650648 |
| T2DM | mast_cell | *HTATIP2* | 1.614027768 | 7.56E-05 | 0.000167152 | 0.366730921 |
| T2DM | mast_cell | *CTSC* | 1.356596853 | 3.00E-05 | 0.000134478 | 0.145671953 |
| T2DM | mast_cell | *ELP4* | 1.668733523 | 9.68E-05 | 0.000198277 | 0.469519642 |
| T2DM | mast_cell | *SC5D* | 3.218443537 | 0.000272966 | 0.000453727 | 1 |
| T2DM | mast_cell | *B3GAT1* | 16.05838707 | 9.78E-06 | 0.000134478 | 0.047428903 |
| T2DM | mast_cell | *P2RX3* | 21.49287119 | 3.88E-05 | 0.000134478 | 0.188427269 |
| T2DM | mast_cell | *DNAJC4* | 1.825148735 | 0.000154581 | 0.000283244 | 0.750029126 |
| T2DM | mast_cell | *UNC93B1* | 2.478882048 | 0.000139313 | 0.000260682 | 0.675947133 |
| T2DM | mast_cell | *FOXRED1* | 1.681255269 | 2.36E-05 | 0.000134478 | 0.114683301 |
| T2DM | mast_cell | *NRXN2* | -5.110522411 | 0.000696236 | 0.001038787 | 1 |
| T2DM | mast_cell | *CCND1* | 1.886332129 | 0.026301765 | 0.031479073 | 1 |
| T2DM | mast_cell | *CCDC86* | 3.063254659 | 0.002370971 | 0.003210704 | 1 |
| T2DM | mast_cell | *PANX1* | 3.321567481 | 0.000185463 | 0.000327224 | 0.899866272 |
| T2DM | mast_cell | *CEP164* | 2.067624782 | 3.78E-05 | 0.000134478 | 0.183383486 |
| T2DM | mast_cell | *GALNT18* | 1.071381832 | 4.37E-05 | 0.000134478 | 0.212147841 |
| T2DM | mast_cell | *KIAA1549L* | -3.321815678 | 0.000275991 | 0.000458128 | 1 |
| T2DM | mast_cell | *ACCS* | 1.63779305 | 9.41E-05 | 0.00019472 | 0.456619139 |
| T2DM | mast_cell | *C11orf21* | -3.739785771 | 2.92E-06 | 0.000134478 | 0.014167112 |
| T2DM | mast_cell | *ELMOD1* | -2.073742271 | 0.001793552 | 0.002479729 | 1 |
| T2DM | mast_cell | *RPS13* | 2.686696371 | 1.43E-05 | 0.000134478 | 0.069278471 |
| T2DM | mast_cell | *NDUFS8* | 1.451298706 | 8.31E-05 | 0.000178688 | 0.403298479 |
| T2DM | mast_cell | *TCIRG1* | 1.995051064 | 4.70E-05 | 0.000134478 | 0.228097524 |
| T2DM | mast_cell | *GTF2H1* | 1.886171311 | 6.88E-05 | 0.000158046 | 0.333797893 |
| T2DM | mast_cell | *PRDM4* | 3.674112672 | 0.000256026 | 0.000429246 | 1 |
| T2DM | mast_cell | *COQ5* | 1.120379398 | 8.18E-06 | 0.000134478 | 0.039689298 |
| T2DM | mast_cell | *CORO1C* | 2.873512072 | 3.58E-05 | 0.000134478 | 0.173476545 |
| T2DM | mast_cell | *ASIC1* | 3.87200417 | 5.24E-05 | 0.000136633 | 0.254385366 |
| T2DM | mast_cell | *CAPRIN2* | 1.923708775 | 0.000617471 | 0.000930714 | 1 |
| T2DM | mast_cell | *KCTD10* | 1.503473356 | 9.88E-05 | 0.000201328 | 0.479199878 |
| T2DM | mast_cell | *IL23A* | -3.377238871 | 0.00017253 | 0.000309356 | 0.837116172 |
| T2DM | mast_cell | *ACSS3* | 1.054993854 | 3.00E-05 | 0.000134478 | 0.145331987 |
| T2DM | mast_cell | *LTA4H* | 1.322623573 | 9.19E-06 | 0.000134478 | 0.044576957 |
| T2DM | mast_cell | *PRR4* | 4.300226713 | 1.33E-06 | 9.63E-05 | 0.006468573 |
| T2DM | mast_cell | *PARP11* | 2.38634185 | 3.20E-05 | 0.000134478 | 0.155452415 |
| T2DM | mast_cell | *GPN3* | 1.768675115 | 4.66E-05 | 0.000134478 | 0.225970062 |
| T2DM | mast_cell | *VPS29* | 2.46804866 | 4.45E-05 | 0.000134478 | 0.216143984 |
| T2DM | mast_cell | *ACAD10* | 1.282879468 | 9.57E-05 | 0.000196916 | 0.464112163 |
| T2DM | mast_cell | *LTBR* | 1.945184631 | 9.60E-05 | 0.000197267 | 0.465805224 |

| | | | | | | |
|---|---|---|---|---|---|---|
| T2DM | mast_cell | *DDX55* | 2.142522248 | 9.04E-05 | 0.000189794 | 0.438615048 |
| T2DM | mast_cell | *ENDOU* | 5.234963027 | 6.53E-05 | 0.000153213 | 0.317023282 |
| T2DM | mast_cell | *VDR* | 4.273044612 | 4.16E-05 | 0.000134478 | 0.202024347 |
| T2DM | mast_cell | *STX2* | 1.848158966 | 1.81E-05 | 0.000134478 | 0.087669901 |
| T2DM | mast_cell | *CAND1* | 1.992360412 | 3.99E-05 | 0.000134478 | 0.193646193 |
| T2DM | mast_cell | *MDM1* | 5.348368789 | 0.000213757 | 0.000368324 | 1 |
| T2DM | mast_cell | *NUP107* | 1.862578344 | 3.19E-05 | 0.000134478 | 0.154676693 |
| T2DM | mast_cell | *KRR1* | 2.729585018 | 0.000254729 | 0.000427885 | 1 |
| T2DM | mast_cell | *MRPL51* | 1.238128649 | 0.000648486 | 0.000973832 | 1 |
| T2DM | mast_cell | *GNB3* | -1.927761555 | 0.000827365 | 0.001218692 | 1 |
| T2DM | mast_cell | *CHPT1* | 1.746265534 | 0.001381113 | 0.00194462 | 1 |
| T2DM | mast_cell | *SPSB2* | 2.12899561 | 0.000239359 | 0.000405365 | 1 |
| T2DM | mast_cell | *ATN1* | 2.900270617 | 0.000625284 | 0.000941614 | 1 |
| T2DM | mast_cell | *NT5DC3* | 2.030951285 | 7.14E-05 | 0.000161624 | 0.346199321 |
| T2DM | mast_cell | *SUDS3* | 3.430101603 | 3.87E-05 | 0.000134478 | 0.187781076 |
| T2DM | mast_cell | *GOLT1B* | 5.863955382 | 1.79E-05 | 0.000134478 | 0.086808606 |
| T2DM | mast_cell | *C2CD5* | 1.438280605 | 3.76E-05 | 0.000134478 | 0.182244395 |
| T2DM | mast_cell | *BTN3A3* | 1.355800032 | 0.000259101 | 0.000434102 | 1 |
| T2DM | mast_cell | *FRK* | 1.665841835 | 5.00E-05 | 0.000135379 | 0.242640992 |
| T2DM | mast_cell | *RWDD1* | 1.91213654 | 0.000115025 | 0.000225313 | 0.558100494 |
| T2DM | mast_cell | *RSPH4A* | 1.985087119 | 0.033936621 | 0.040141512 | 1 |
| T2DM | mast_cell | *TMEM14C* | 5.365791623 | 0.000295375 | 0.000485171 | 1 |
| T2DM | mast_cell | *SMIM8* | 2.163600541 | 6.57E-05 | 0.000153604 | 0.318727985 |
| T2DM | mast_cell | *FAM184A* | 1.325782609 | 2.75E-05 | 0.000134478 | 0.133288156 |
| T2DM | mast_cell | *RNGTT* | 2.44102939 | 3.78E-05 | 0.000134478 | 0.18324275 |
| T2DM | mast_cell | *GABRR2* | 2.916463714 | 0.000329269 | 0.000532715 | 1 |
| T2DM | mast_cell | *HDDC2* | 1.83397004 | 0.000135932 | 0.000255715 | 0.659543822 |
| T2DM | mast_cell | *UST* | 1.011560793 | 2.49E-05 | 0.000134478 | 0.120705909 |
| T2DM | mast_cell | *MTRF1L* | 1.781927285 | 7.50E-06 | 0.000134478 | 0.036374934 |
| T2DM | mast_cell | *SLC26A8* | 2.023277312 | 4.61E-05 | 0.000134478 | 0.223894849 |
| T2DM | mast_cell | *MRPL18* | 3.732242767 | 6.07E-05 | 0.00014641 | 0.294444823 |
| T2DM | mast_cell | *RNF8* | 2.68883618 | 6.42E-05 | 0.000151618 | 0.311429158 |
| T2DM | mast_cell | *PHACTR1* | 1.515372943 | 0.000724981 | 0.001079352 | 1 |
| T2DM | mast_cell | *MDGA1* | 1.950590379 | 6.95E-05 | 0.000159011 | 0.337102876 |
| T2DM | mast_cell | *CD83* | 1.866679691 | 0.000140547 | 0.000262484 | 0.681932225 |
| T2DM | mast_cell | *MDN1* | 2.016262403 | 5.79E-05 | 0.000143246 | 0.28104832 |
| T2DM | mast_cell | *BVES* | -1.388327914 | 0.006073569 | 0.007761116 | 1 |
| T2DM | mast_cell | *COL9A1* | 6.394834691 | 8.52E-06 | 0.000134478 | 0.041345267 |
| T2DM | mast_cell | *WASF1* | 1.24192532 | 7.65E-05 | 0.000168736 | 0.371417643 |
| T2DM | mast_cell | *VNN2* | 4.259401339 | 5.74E-06 | 0.000134478 | 0.027859025 |
| T2DM | mast_cell | *ACOT13* | 4.576832619 | 9.29E-05 | 0.000193112 | 0.450915776 |
| T2DM | mast_cell | *C6orf62* | 1.836845025 | 7.55E-05 | 0.000167051 | 0.366120961 |
| T2DM | mast_cell | *HBS1L* | 2.654602597 | 0.000239527 | 0.000405492 | 1 |
| T2DM | mast_cell | *PEX7* | 1.744985291 | 0.001050203 | 0.001515641 | 1 |
| T2DM | mast_cell | *FIG4* | 2.254720564 | 2.54E-05 | 0.000134478 | 0.123157378 |
| T2DM | mast_cell | *PERP* | 2.124523769 | 0.000404026 | 0.000633387 | 1 |
| T2DM | mast_cell | *PHACTR2* | 2.480439095 | 0.000515765 | 0.00078943 | 1 |
| T2DM | mast_cell | *CUTA* | 1.28746376 | 0.001254583 | 0.001778853 | 1 |
| T2DM | mast_cell | *PACRG* | 5.592247401 | 0.000184435 | 0.00032624 | 0.894876932 |
| T2DM | mast_cell | *QKI* | 1.746217066 | 3.21E-05 | 0.000134478 | 0.155604257 |

| | | | | | | |
|---|---|---|---|---|---|---|
| T2DM | mast_cell | *FAM120B* | 2.145430666 | 1.87E-05 | 0.000134478 | 0.090878522 |
| T2DM | mast_cell | *GUCA1B* | -2.748970798 | 0.000613347 | 0.000925937 | 1 |
| T2DM | mast_cell | *DNPH1* | 1.766832684 | 1.85E-05 | 0.000134478 | 0.089833951 |
| T2DM | mast_cell | *EXOC2* | 3.259696134 | 2.27E-05 | 0.000134478 | 0.110109954 |
| T2DM | mast_cell | *TMEM30A* | 3.164618168 | 3.75E-05 | 0.000134478 | 0.181783343 |
| T2DM | mast_cell | *IMPG1* | 4.898178461 | 0.000176295 | 0.000314364 | 0.855383451 |
| T2DM | mast_cell | *BTN2A1* | 3.233507795 | 7.26E-05 | 0.000163044 | 0.352175175 |
| T2DM | mast_cell | *LY86* | 1.823633652 | 8.26E-05 | 0.000178055 | 0.400619548 |
| T2DM | mast_cell | *PCDHB2* | 3.694901205 | 0.00038523 | 0.00061043 | 1 |
| T2DM | mast_cell | *NUDT12* | 3.282503178 | 3.35E-05 | 0.000134478 | 0.162425094 |
| T2DM | mast_cell | *CEP72* | 2.080475471 | 0.000104716 | 0.000209703 | 0.508083097 |
| T2DM | mast_cell | *DAP* | 2.675747449 | 0.00016736 | 0.000302432 | 0.812030482 |
| T2DM | mast_cell | *NME5* | 1.489512025 | 9.53E-05 | 0.000196519 | 0.462224182 |
| T2DM | mast_cell | *PFDN1* | 2.196459928 | 2.06E-05 | 0.000134478 | 0.099850363 |
| T2DM | mast_cell | *TMCO6* | 2.148147049 | 0.012333809 | 0.01527013 | 1 |
| T2DM | mast_cell | *SPARC* | 1.488782961 | 2.83E-05 | 0.000134478 | 0.137268517 |
| T2DM | mast_cell | *IK* | 1.358408202 | 0.000127493 | 0.000243708 | 0.618598441 |
| T2DM | mast_cell | *HAVCR1* | 2.92706782 | 0.006455076 | 0.00822048 | 1 |
| T2DM | mast_cell | *RNF130* | 2.061234189 | 1.61E-05 | 0.000134478 | 0.078240825 |
| T2DM | mast_cell | *ARSB* | 1.550590929 | 4.61E-05 | 0.000134478 | 0.223838585 |
| T2DM | mast_cell | *THBS4* | 1.442763904 | 2.60E-05 | 0.000134478 | 0.126290876 |
| T2DM | mast_cell | *BTNL8* | 0.582424883 | 0.036995792 | 0.043589992 | 1 |
| T2DM | mast_cell | *MSH3* | 1.379061116 | 0.00012267 | 0.000235815 | 0.595196344 |
| T2DM | mast_cell | *RASGRF2* | 1.206669011 | 0.001169665 | 0.001671329 | 1 |
| T2DM | mast_cell | *DROSHA* | 2.145039973 | 1.35E-05 | 0.000134478 | 0.065639172 |
| T2DM | mast_cell | *CDH6* | 1.129194578 | 0.000793506 | 0.001172655 | 1 |
| T2DM | mast_cell | *LMNB1* | 3.018348661 | 0.039350817 | 0.046230064 | 1 |
| T2DM | mast_cell | *SLC12A7* | 2.683224375 | 3.53E-05 | 0.000134478 | 0.171219194 |
| T2DM | mast_cell | *SKP1* | 2.174728779 | 3.35E-05 | 0.000134478 | 0.162328229 |
| T2DM | mast_cell | *PPWD1* | 1.747439501 | 3.67E-05 | 0.000134478 | 0.177909335 |
| T2DM | mast_cell | *TXNDC15* | 1.736891764 | 3.71E-05 | 0.000134478 | 0.180082948 |
| T2DM | mast_cell | *SMAD5* | 2.976906015 | 3.55E-05 | 0.000134478 | 0.172207721 |
| T2DM | mast_cell | *ERGIC1* | 3.553247251 | 5.12E-05 | 0.000136328 | 0.248358116 |
| T2DM | mast_cell | *ATP6V0E1* | 1.671682966 | 4.37E-05 | 0.000134478 | 0.211845644 |
| T2DM | mast_cell | *UNC5A* | 7.744721008 | 4.48E-05 | 0.000134478 | 0.21759153 |
| T2DM | mast_cell | *ACTR8* | 3.382100273 | 0.001170137 | 0.001671329 | 1 |
| T2DM | mast_cell | *TBCCD1* | 6.0972777 | 0.000200382 | 0.000349479 | 0.972251907 |
| T2DM | mast_cell | *CLDN16* | 2.089345762 | 0.000158612 | 0.000289535 | 0.769583503 |
| T2DM | mast_cell | *NPHP3* | 2.241389331 | 8.11E-07 | 8.37E-05 | 0.003935631 |
| T2DM | mast_cell | *PCCB* | 3.576338227 | 4.03E-07 | 6.51E-05 | 0.001953464 |
| T2DM | mast_cell | *CEP70* | 1.653807461 | 6.13E-05 | 0.0001477 | 0.297616434 |
| T2DM | mast_cell | *C3orf14* | 1.870153349 | 0.002123358 | 0.002901305 | 1 |
| T2DM | mast_cell | *CBLB* | 1.600180658 | 2.52E-05 | 0.000134478 | 0.122127924 |
| T2DM | mast_cell | *IQCG* | 2.572167147 | 2.34E-05 | 0.000134478 | 0.113708617 |
| T2DM | mast_cell | *NCBP2* | 2.649803217 | 0.000741675 | 0.001101165 | 1 |
| T2DM | mast_cell | *PLXNA1* | 4.711596961 | 0.00012758 | 0.000243708 | 0.619018171 |
| T2DM | mast_cell | *SCAP* | 2.578922095 | 3.95E-05 | 0.000134478 | 0.191868186 |
| T2DM | mast_cell | *KIAA1257* | 5.293670926 | 3.91E-05 | 0.000134478 | 0.189837221 |
| T2DM | mast_cell | *NEK11* | 1.755695276 | 2.09E-05 | 0.000134478 | 0.101526745 |
| T2DM | mast_cell | *PLSCR4* | 1.309664219 | 3.83E-05 | 0.000134478 | 0.185692218 |

| | | | | | | |
|---|---|---|---|---|---|---|
| T2DM | mast_cell | *HEMK1* | 2.418135268 | 0.000967237 | 0.00140468 | 1 |
| T2DM | mast_cell | *COMMD2* | 4.545863321 | 0.002463331 | 0.003320945 | 1 |
| T2DM | mast_cell | *ABHD14B* | 1.552675122 | 0.000183183 | 0.000324617 | 0.888802671 |
| T2DM | mast_cell | *ARHGEF26* | 1.790551335 | 3.86E-05 | 0.000134478 | 0.187347237 |
| T2DM | mast_cell | *VIPR1* | 2.389963278 | 4.08E-05 | 0.000134478 | 0.197726944 |
| T2DM | mast_cell | *DNAH1* | 3.973928217 | 8.86E-06 | 0.000134478 | 0.042978764 |
| T2DM | mast_cell | *NKTR* | 2.133619977 | 1.02E-05 | 0.000134478 | 0.049333155 |
| T2DM | mast_cell | *NEK4* | 3.042751947 | 0.001106258 | 0.001588036 | 1 |
| T2DM | mast_cell | *ADAM23* | 1.732440751 | 0.000165786 | 0.000300119 | 0.804392091 |
| T2DM | mast_cell | *ACTR1B* | 1.726276127 | 3.99E-05 | 0.000134478 | 0.193550195 |
| T2DM | mast_cell | *SLC35F5* | 2.762199024 | 5.13E-06 | 0.000134478 | 0.024885056 |
| T2DM | mast_cell | *CENPA* | -1.578341344 | 0.000464518 | 0.000719618 | 1 |
| T2DM | mast_cell | *NRBP1* | 2.405226273 | 5.97E-05 | 0.000145324 | 0.289777023 |
| T2DM | mast_cell | *PPM1G* | 2.292541385 | 0.000102143 | 0.000206134 | 0.495597157 |
| T2DM | mast_cell | *INO80B* | 3.177555467 | 0.000103317 | 0.00020762 | 0.501295193 |
| T2DM | mast_cell | *MOGS* | 1.537993087 | 0.004968559 | 0.006430367 | 1 |
| T2DM | mast_cell | *RTN4* | 1.941858239 | 8.29E-05 | 0.00017822 | 0.402065103 |
| T2DM | mast_cell | *GALNT3* | -0.85705433 | 0.000528469 | 0.000806077 | 1 |
| T2DM | mast_cell | *ACADL* | 1.251047539 | 9.44E-05 | 0.000195212 | 0.457967011 |
| T2DM | mast_cell | *MRPL19* | 1.397823204 | 5.01E-05 | 0.000135379 | 0.24289907 |
| T2DM | mast_cell | *DNAH6* | 0.896155372 | 9.22E-05 | 0.000192375 | 0.447463832 |
| T2DM | mast_cell | *IGFBP2* | 2.11856783 | 0.000212208 | 0.000366548 | 1 |
| T2DM | mast_cell | *GGCX* | 1.725401499 | 9.96E-05 | 0.000202664 | 0.483354134 |
| T2DM | mast_cell | *GNLY* | -1.572804884 | 0.032427692 | 0.038441037 | 1 |
| T2DM | mast_cell | *CHMP3* | 1.440376202 | 0.012521767 | 0.015494928 | 1 |
| T2DM | mast_cell | *WNT6* | 1.910168917 | 0.035864935 | 0.042308938 | 1 |
| T2DM | mast_cell | *IL1RL2* | 4.348176408 | 2.18E-05 | 0.000134478 | 0.105722165 |
| T2DM | mast_cell | *IL18R1* | -0.187901341 | 0.000135519 | 0.000255156 | 0.657537096 |
| T2DM | mast_cell | *MLPH* | -0.320523179 | 0.000142985 | 0.000265505 | 0.693764429 |
| T2DM | mast_cell | *HDLBP* | 2.609436965 | 4.05E-05 | 0.000134478 | 0.196386962 |
| T2DM | mast_cell | *TPO* | 1.379352745 | 0.000128589 | 0.000245145 | 0.623914104 |
| T2DM | mast_cell | *TAF1B* | 2.643115054 | 5.12E-05 | 0.000136328 | 0.248502382 |
| T2DM | mast_cell | *NOL10* | 1.902563543 | 4.91E-05 | 0.000135162 | 0.238428324 |
| T2DM | mast_cell | *STRN* | 3.688635535 | 6.91E-05 | 0.000158505 | 0.335237048 |
| T2DM | mast_cell | *CEBPZ* | 3.504613745 | 5.48E-05 | 0.000139536 | 0.265955362 |
| T2DM | mast_cell | *PRKD3* | 3.429350978 | 5.66E-05 | 0.00014153 | 0.274709565 |
| T2DM | mast_cell | *SLC25A12* | 1.478462227 | 7.41E-05 | 0.000165367 | 0.35967388 |
| T2DM | mast_cell | *DARS* | 1.737814003 | 3.42E-05 | 0.000134478 | 0.165945538 |
| T2DM | mast_cell | *WIPF1* | 2.525131151 | 1.98E-05 | 0.000134478 | 0.095832646 |
| T2DM | mast_cell | *PNO1* | -3.1662525 | 0.003403497 | 0.004491098 | 1 |
| T2DM | mast_cell | *ORC4* | 1.936479862 | 1.91E-05 | 0.000134478 | 0.092605325 |
| T2DM | mast_cell | *AAK1* | 1.880759738 | 3.10E-05 | 0.000134478 | 0.150324524 |
| T2DM | mast_cell | *CD207* | 4.186319156 | 0.001028073 | 0.001486356 | 1 |
| T2DM | mast_cell | *GRIN3B* | 1.0957966 | 0.036674527 | 0.04323246 | 1 |
| T2DM | mast_cell | *NFE2L2* | 1.829576754 | 1.81E-05 | 0.000134478 | 0.087609791 |
| T2DM | mast_cell | *MSH6* | 2.715388926 | 4.59E-05 | 0.000134478 | 0.222683764 |
| T2DM | mast_cell | *ALMS1* | 4.311280966 | 0.000236949 | 0.000402267 | 1 |
| T2DM | mast_cell | *DNAJC16* | 2.265879072 | 1.16E-05 | 0.000134478 | 0.056268232 |
| T2DM | mast_cell | *MARK1* | 1.18819887 | 4.29E-05 | 0.000134478 | 0.20804629 |
| T2DM | mast_cell | *MORN1* | 1.701233492 | 6.60E-05 | 0.000154113 | 0.32039993 |

| | | | | | | |
|---|---|---|---|---|---|---|
| T2DM | mast_cell | *GPX7* | -1.675220093 | 0.000274614 | 0.000456154 | 1 |
| T2DM | mast_cell | *RALGPS2* | 1.958163627 | 0.001061569 | 0.001530678 | 1 |
| T2DM | mast_cell | *CEP104* | 1.983771418 | 3.56E-05 | 0.000134478 | 0.172925328 |
| T2DM | mast_cell | *FAM20B* | 1.985733799 | 4.15E-05 | 0.000134478 | 0.201129067 |
| T2DM | mast_cell | *WRAP73* | 1.999388411 | 2.09E-05 | 0.000134478 | 0.101202963 |
| T2DM | mast_cell | *ICMT* | 3.482897019 | 6.07E-05 | 0.00014641 | 0.294510124 |
| T2DM | mast_cell | *PARK7* | 3.407637867 | 2.00E-05 | 0.000134478 | 0.097048064 |
| T2DM | mast_cell | *KIAA1324* | 3.394750182 | 0.000733694 | 0.001091654 | 1 |
| T2DM | mast_cell | *MECR* | 3.379488514 | 0.000114384 | 0.000224693 | 0.554991673 |
| T2DM | mast_cell | *KCNC4* | 1.286181032 | 0.001129674 | 0.001618314 | 1 |
| T2DM | mast_cell | *EDEM3* | 2.731458168 | 9.33E-05 | 0.000193584 | 0.452600118 |
| T2DM | mast_cell | *RAP1A* | 2.450708588 | 3.61E-05 | 0.000134478 | 0.175282913 |
| T2DM | mast_cell | *CAPZA1* | 1.754370183 | 5.54E-05 | 0.00014004 | 0.26864064 |
| T2DM | mast_cell | *ASH1L* | 2.432588675 | 5.50E-06 | 0.000134478 | 0.026692143 |
| T2DM | mast_cell | *GON4L* | 2.320782513 | 2.25E-05 | 0.000134478 | 0.109081923 |
| T2DM | mast_cell | *DOCK7* | 2.08782325 | 2.83E-05 | 0.000134478 | 0.137287981 |
| T2DM | mast_cell | *FBXO2* | 1.706625411 | 0.013417968 | 0.016557472 | 1 |
| T2DM | mast_cell | *FBXO6* | 2.682896595 | 0.000151467 | 0.000278167 | 0.734918056 |
| T2DM | mast_cell | *DNAJC6* | 1.747865368 | 0.000112845 | 0.000222752 | 0.547525589 |
| T2DM | mast_cell | *LEPR* | 1.76820663 | 4.84E-05 | 0.000134478 | 0.234749367 |
| T2DM | mast_cell | *IVNS1ABP* | 1.083611376 | 0.000102302 | 0.000206134 | 0.496371479 |
| T2DM | mast_cell | *MFN2* | 1.837784144 | 9.88E-05 | 0.000201328 | 0.479361479 |
| T2DM | mast_cell | *PRG4* | 2.024044834 | 5.31E-05 | 0.000137311 | 0.257870376 |
| T2DM | mast_cell | *NCF2* | 1.441139734 | 0.00016161 | 0.000293792 | 0.78413048 |
| T2DM | mast_cell | *BCAS2* | 3.180871724 | 4.93E-06 | 0.000134478 | 0.023917126 |
| T2DM | mast_cell | *TNNI3K* | 6.229940466 | 2.10E-06 | 0.000124855 | 0.010209431 |
| T2DM | mast_cell | *CFHR3* | 3.495112781 | 0.013693965 | 0.016876586 | 1 |
| T2DM | mast_cell | *TMEM9* | 1.87228184 | 0.005158025 | 0.006648974 | 1 |
| T2DM | mast_cell | *WARS2* | 1.8529594 | 0.000226406 | 0.00038694 | 1 |
| T2DM | mast_cell | *OSCP1* | 1.43468549 | 1.11E-05 | 0.000134478 | 0.053782305 |
| T2DM | mast_cell | *MRPS15* | 5.593761081 | 0.001690163 | 0.002347744 | 1 |
| T2DM | mast_cell | *EXOC8* | 2.76267016 | 5.81E-05 | 0.000143427 | 0.281793133 |
| T2DM | mast_cell | *GNPAT* | 1.870317041 | 6.92E-05 | 0.000158597 | 0.335671625 |
| T2DM | mast_cell | *C1orf109* | 3.194807867 | 0.000489591 | 0.000755084 | 1 |
| T2DM | mast_cell | *TBCE* | 1.538201751 | 3.50E-05 | 0.000134478 | 0.16984256 |
| T2DM | mast_cell | *ACADM* | 2.184703484 | 1.72E-05 | 0.000134478 | 0.083614878 |
| T2DM | mast_cell | *SLAMF1* | 1.328771474 | 0.030593682 | 0.036409258 | 1 |
| T2DM | mast_cell | *PADI2* | -0.57870965 | 0.000117507 | 0.000228423 | 0.570144266 |
| T2DM | mast_cell | *SDHB* | 3.660422651 | 2.57E-05 | 0.000134478 | 0.124908713 |
| T2DM | mast_cell | *CTBS* | 1.131590087 | 0.000741273 | 0.001100905 | 1 |
| T2DM | mast_cell | *KLHL12* | 2.367484864 | 2.17E-05 | 0.000134478 | 0.105051999 |
| T2DM | mast_cell | *ZNHIT6* | 2.249333833 | 5.19E-05 | 0.000136402 | 0.251724089 |
| T2DM | mast_cell | *KIF17* | 3.326988553 | 0.000171229 | 0.000307739 | 0.830803549 |
| T2DM | mast_cell | *CD46* | 2.281659413 | 7.72E-06 | 0.000134478 | 0.037441689 |
| T2DM | mast_cell | *EBNA1BP2* | 3.1650716 | 0.000153235 | 0.000280882 | 0.743495346 |
| T2DM | mast_cell | *ERI3* | 2.071870471 | 3.35E-05 | 0.000134478 | 0.162330254 |
| T2DM | mast_cell | *PRDX1* | 2.038118807 | 4.30E-05 | 0.000134478 | 0.208777927 |
| T2DM | mast_cell | *CCDC181* | -2.910820254 | 0.006093443 | 0.007784461 | 1 |
| T2DM | mast_cell | *NSUN4* | 2.40229684 | 0.014770946 | 0.018162349 | 1 |
| T2DM | mast_cell | *DR1* | 2.212275335 | 1.46E-05 | 0.000134478 | 0.070701261 |

| | | | | | | |
|---|---|---|---|---|---|---|
| T2DM | mast_cell | *PRRC2C* | 2.678353374 | 3.10E-05 | 0.000134478 | 0.15043734 |
| T2DM | mast_cell | *ABCD3* | 1.949347603 | 3.77E-05 | 0.000134478 | 0.182874803 |
| T2DM | mast_cell | *VAMP4* | 2.123081022 | 3.78E-05 | 0.000134478 | 0.183593046 |
| T2DM | mast_cell | *PTBP2* | 2.513079716 | 8.52E-06 | 0.000134478 | 0.041330845 |
| T2DM | mast_cell | *DARS2* | 3.387018173 | 5.50E-05 | 0.000139632 | 0.266836387 |
| T2DM | mast_cell | *MTFR1L* | 4.834899904 | 1.94E-05 | 0.000134478 | 0.094138535 |
| T2DM | mast_cell | *RPS6KA1* | 1.917506076 | 0.000209924 | 0.000362991 | 1 |
| T2DM | mast_cell | *RPA2* | 2.668561271 | 0.000179294 | 0.000318658 | 0.869935013 |
| T2DM | mast_cell | *MARC2* | 2.295135314 | 2.87E-05 | 0.000134478 | 0.139175073 |
| T2DM | mast_cell | *SLC5A9* | 2.810687619 | 2.52E-05 | 0.000134478 | 0.12211711 |
| T2DM | mast_cell | *ESYT2* | 1.812434685 | 3.77E-05 | 0.000134478 | 0.18306026 |
| T2DM | mast_cell | *CD3EAP* | 1.759754128 | 0.000130512 | 0.000247846 | 0.633245882 |
| T2DM | mast_cell | *RCN2* | 2.740141919 | 2.25E-05 | 0.000134478 | 0.109181891 |
| T2DM | mast_cell | *APOA1* | 13.37225904 | 1.75E-05 | 0.000134478 | 0.084918459 |
| T2DM | mast_cell | *DDX59* | 2.174879365 | 0.000563033 | 0.000855035 | 1 |
| T2DM | mast_cell | *CAMSAP2* | 1.655785857 | 2.80E-05 | 0.000134478 | 0.135861336 |
| T2DM | mast_cell | *KLF7* | 1.74487362 | 1.83E-05 | 0.000134478 | 0.08893559 |
| T2DM | mast_cell | *LRMP* | 1.584854904 | 0.00117341 | 0.001675511 | 1 |
| T2DM | mast_cell | *ELOVL4* | 5.026999659 | 1.10E-06 | 9.21E-05 | 0.00533429 |
| T2DM | mast_cell | *CNR1* | 2.799130126 | 0.000264909 | 0.000441697 | 1 |
| T2DM | mast_cell | *SGIP1* | 3.008035872 | 2.94E-05 | 0.000134478 | 0.142646484 |
| T2DM | mast_cell | *AKAP7* | 2.028496026 | 5.62E-05 | 0.000140825 | 0.272501993 |
| T2DM | mast_cell | *MED28* | 1.632941828 | 0.00015543 | 0.000284691 | 0.75414629 |
| T2DM | mast_cell | *TMEM5* | 4.252042465 | 8.88E-07 | 8.78E-05 | 0.004309511 |
| T2DM | mast_cell | *VAMP8* | 2.021073635 | 0.000101325 | 0.000205174 | 0.491626917 |
| T2DM | mast_cell | *FOXO3* | 1.557580332 | 2.77E-05 | 0.000134478 | 0.134437808 |
| T2DM | mast_cell | *SPP1* | 2.503343579 | 0.00011969 | 0.000231553 | 0.580735891 |
| T2DM | mast_cell | *MFSD1* | 1.356479289 | 4.68E-05 | 0.000134478 | 0.227180314 |
| T2DM | mast_cell | *UBN1* | 1.469336654 | 3.60E-05 | 0.000134478 | 0.174551803 |
| T2DM | mast_cell | *WDR35* | 1.232670024 | 5.17E-05 | 0.000136402 | 0.251053772 |
| T2DM | mast_cell | *DNAH7* | 1.418305543 | 0.00016995 | 0.00030586 | 0.824598636 |
| T2DM | mast_cell | *C2orf40* | 2.054016369 | 5.50E-05 | 0.000139632 | 0.266805434 |
| T2DM | mast_cell | *CPSF3* | 2.432626765 | 0.000107815 | 0.000214393 | 0.523119243 |
| T2DM | mast_cell | *PIGZ* | 0.98828913 | 0.023521534 | 0.028333287 | 1 |
| T2DM | mast_cell | *SENP5* | 2.098419168 | 3.40E-05 | 0.000134478 | 0.164862519 |
| T2DM | mast_cell | *CCDC92* | 1.726838052 | 0.001016648 | 0.001470276 | 1 |
| T2DM | mast_cell | *FKBP15* | 1.35466922 | 0.000183475 | 0.000324898 | 0.890219299 |
| T2DM | mast_cell | *CTNNAL1* | 1.656170192 | 6.62E-06 | 0.000134478 | 0.032116418 |
| T2DM | mast_cell | *RAB14* | 3.426499564 | 3.05E-05 | 0.000134478 | 0.14794325 |
| T2DM | mast_cell | *TRIM32* | 2.403800895 | 4.31E-05 | 0.000134478 | 0.209220057 |
| T2DM | mast_cell | *PPP6C* | 1.753938681 | 3.57E-05 | 0.000134478 | 0.173420881 |
| T2DM | mast_cell | *RBM18* | 2.375133787 | 5.96E-05 | 0.000145187 | 0.289356987 |
| T2DM | mast_cell | *HSDL2* | 1.613370473 | 3.72E-05 | 0.000134478 | 0.180402616 |
| T2DM | mast_cell | *MAPKAP1* | 1.887734667 | 2.92E-05 | 0.000134478 | 0.141542585 |
| T2DM | mast_cell | *INVS* | 2.381604858 | 1.12E-05 | 0.000134478 | 0.054112424 |
| T2DM | mast_cell | *DENND1A* | 1.546032323 | 2.32E-05 | 0.000134478 | 0.112587159 |
| T2DM | mast_cell | *KDSR* | 2.125371637 | 5.13E-05 | 0.000136328 | 0.248998741 |
| T2DM | mast_cell | *DCAF4* | 2.227962189 | 0.000132344 | 0.000250246 | 0.64213085 |
| T2DM | mast_cell | *IFI27L2* | 1.981014225 | 4.74E-05 | 0.000134478 | 0.229793699 |
| T2DM | mast_cell | *NEK9* | 2.702983724 | 5.36E-05 | 0.000138012 | 0.259920638 |

| | | | | | | |
|---|---|---|---|---|---|---|
| T2DM | mast_cell | *ACOT2* | 0.688329252 | 0.011665397 | 0.014486948 | 1 |
| T2DM | mast_cell | *TTLL5* | 1.586404033 | 5.00E-05 | 0.000135379 | 0.242753592 |
| T2DM | mast_cell | *FLVCR2* | 1.667739278 | 6.86E-05 | 0.000157889 | 0.332987683 |
| T2DM | mast_cell | *PPP4R4* | 1.725541799 | 0.000426728 | 0.000665751 | 1 |
| T2DM | mast_cell | *ALDH6A1* | 1.317356591 | 5.72E-05 | 0.000142446 | 0.277769058 |
| T2DM | mast_cell | *ESRRB* | 2.004169228 | 0.000878916 | 0.001287785 | 1 |
| T2DM | mast_cell | *EIF2B2* | 1.409698452 | 0.000133957 | 0.000252736 | 0.649958033 |
| T2DM | mast_cell | *COQ6* | 2.246570774 | 5.12E-05 | 0.000136328 | 0.248254399 |
| T2DM | mast_cell | *DNMT3A* | 2.835606795 | 0.000227588 | 0.000388412 | 1 |
| T2DM | mast_cell | *CNRIP1* | 1.545571934 | 0.002471355 | 0.003329913 | 1 |
| T2DM | mast_cell | *CRIPT* | 2.1656636 | 3.55E-05 | 0.000134478 | 0.17240777 |
| T2DM | mast_cell | *EPCAM* | -1.53322438 | 6.29E-05 | 0.000149953 | 0.304982722 |
| T2DM | mast_cell | *SLC17A5* | 1.623591389 | 2.06E-05 | 0.000134478 | 0.099961721 |
| T2DM | mast_cell | *PPP1R3C* | 2.401522157 | 3.33E-05 | 0.000134478 | 0.161541843 |
| T2DM | mast_cell | *PYROXD2* | 1.474537987 | 3.28E-05 | 0.000134478 | 0.159215913 |
| T2DM | mast_cell | *MXI1* | 2.337570953 | 8.89E-05 | 0.000187353 | 0.431568801 |
| T2DM | mast_cell | *C10orf88* | 1.889383903 | 0.002532869 | 0.003406176 | 1 |
| T2DM | mast_cell | *HELLS* | 1.694234855 | 4.43E-05 | 0.000134478 | 0.214975936 |
| T2DM | mast_cell | *TCTN3* | 2.437499649 | 0.000171879 | 0.000308529 | 0.833955222 |
| T2DM | mast_cell | *WDR11* | 2.026424702 | 5.12E-05 | 0.000136328 | 0.248328604 |
| T2DM | mast_cell | *KANSL1* | 1.923724702 | 3.08E-05 | 0.000134478 | 0.149220638 |
| T2DM | mast_cell | *HOXB3* | 2.544607175 | 3.63E-05 | 0.000134478 | 0.175947363 |
| T2DM | mast_cell | *CAAP1* | 2.061831757 | 0.000731063 | 0.001088072 | 1 |
| T2DM | mast_cell | *PCDHB10* | 4.783196396 | 0.001790154 | 0.002476711 | 1 |
| T2DM | mast_cell | *GORAB* | 3.104257816 | 1.54E-05 | 0.000134478 | 0.074732673 |
| T2DM | mast_cell | *ACAT2* | 1.738027662 | 7.21E-05 | 0.000162581 | 0.350037642 |
| T2DM | mast_cell | *SNX19* | 2.128198306 | 0.000337084 | 0.000543727 | 1 |
| T2DM | mast_cell | *SLC10A7* | 3.466975568 | 4.76E-05 | 0.000134478 | 0.23091409 |
| T2DM | mast_cell | *MASTL* | 1.158287541 | 0.002324028 | 0.003151353 | 1 |
| T2DM | mast_cell | *MRC1* | 1.039149836 | 3.04E-05 | 0.000134478 | 0.147285603 |
| T2DM | mast_cell | *IQSEC3* | 2.4348051 | 5.62E-05 | 0.000140825 | 0.272918708 |
| T2DM | mast_cell | *CCDC77* | 2.529358638 | 0.000869242 | 0.001275344 | 1 |
| T2DM | mast_cell | *DNAJC15* | 1.969781352 | 3.75E-05 | 0.000134478 | 0.181773402 |
| T2DM | mast_cell | *UFM1* | 3.419716202 | 8.05E-05 | 0.000175007 | 0.390579581 |
| T2DM | mast_cell | *ALG5* | 1.443657108 | 9.21E-05 | 0.000192375 | 0.4471079 |
| T2DM | mast_cell | *SIL1* | 2.081908431 | 3.06E-05 | 0.000134478 | 0.14859809 |
| T2DM | mast_cell | *ZFP30* | 1.409415933 | 4.36E-05 | 0.000134478 | 0.211424554 |
| T2DM | mast_cell | *ARL1* | 2.327545498 | 6.44E-05 | 0.000151874 | 0.312404219 |
| T2DM | mast_cell | *APAF1* | 3.645552709 | 0.00014804 | 0.000272644 | 0.718292034 |
| T2DM | mast_cell | *ADRA1A* | 1.408236983 | 2.10E-06 | 0.000124855 | 0.010168832 |
| T2DM | mast_cell | *EPHX2* | 1.899452178 | 0.003204321 | 0.004243276 | 1 |
| T2DM | mast_cell | *AKAP1* | 1.757597428 | 5.29E-05 | 0.000137292 | 0.256474468 |
| T2DM | mast_cell | *SCPEP1* | 1.627610199 | 0.00052973 | 0.000807747 | 1 |
| T2DM | mast_cell | *TBX2* | 1.767126938 | 0.000277906 | 0.000460519 | 1 |
| T2DM | mast_cell | *TEX14* | 3.090273419 | 0.011910522 | 0.014768682 | 1 |
| T2DM | mast_cell | *TRIM6* | 4.177796099 | 0.000104991 | 0.000210155 | 0.509415056 |
| T2DM | mast_cell | *ADCY7* | 1.959014166 | 0.001986268 | 0.002725502 | 1 |
| T2DM | mast_cell | *ECHDC2* | 1.065261349 | 0.000320007 | 0.000519462 | 1 |
| T2DM | mast_cell | *PYROXD1* | 1.529745896 | 0.000940015 | 0.001367603 | 1 |
| T2DM | mast_cell | *ZNF549* | 4.001397484 | 2.96E-05 | 0.000134478 | 0.143467433 |

| | | | | | | |
|---|---|---|---|---|---|---|
| T2DM | mast_cell | *A1BG* | -1.832467094 | 0.000508464 | 0.000780223 | 1 |
| T2DM | mast_cell | *ZSCAN18* | 2.669905597 | 3.71E-05 | 0.000134478 | 0.179806493 |
| T2DM | mast_cell | *ZNF211* | 2.335860357 | 0.030844618 | 0.036680903 | 1 |
| T2DM | mast_cell | *PDZRN3* | 0.830200547 | 0.017898784 | 0.021803892 | 1 |
| T2DM | mast_cell | *LHX4* | 1.970629196 | 4.68E-05 | 0.000134478 | 0.227047614 |
| T2DM | mast_cell | *SEC22A* | 2.384183901 | 1.73E-05 | 0.000134478 | 0.083996616 |
| T2DM | mast_cell | *POPDC2* | 1.455154477 | 0.031461184 | 0.037359193 | 1 |
| T2DM | mast_cell | *MAPK8IP1* | 4.602366873 | 2.61E-05 | 0.000134478 | 0.126630568 |
| T2DM | mast_cell | *CAT* | 1.472028224 | 0.000199669 | 0.000348362 | 0.968795425 |
| T2DM | mast_cell | *PILRB* | 1.545630033 | 0.000184605 | 0.000326422 | 0.895702347 |
| T2DM | mast_cell | *ZCCHC17* | 2.856543218 | 3.16E-05 | 0.000134478 | 0.15353056 |
| T2DM | mast_cell | *FABP3* | -3.287660135 | 0.000433188 | 0.000675178 | 1 |
| T2DM | mast_cell | *TNFSF10* | 1.811428011 | 2.78E-05 | 0.000134478 | 0.135047154 |
| T2DM | mast_cell | *ZNF639* | 5.439170918 | 0.002612704 | 0.003505763 | 1 |
| T2DM | mast_cell | *CPXM2* | 1.264122215 | 0.000230549 | 0.0003925 | 1 |
| T2DM | mast_cell | *FLT3* | 3.514362402 | 3.33E-06 | 0.000134478 | 0.016155517 |
| T2DM | mast_cell | *MTIF3* | 3.244446222 | 1.45E-05 | 0.000134478 | 0.070452218 |
| T2DM | mast_cell | *FMOD* | 1.619898478 | 4.60E-05 | 0.000134478 | 0.223280434 |
| T2DM | mast_cell | *CD244* | 2.474100156 | 4.23E-05 | 0.000134478 | 0.205121997 |
| T2DM | mast_cell | *RBBP6* | 2.12566702 | 3.78E-05 | 0.000134478 | 0.18352387 |
| T2DM | mast_cell | *SERAC1* | 1.623517408 | 0.000114067 | 0.000224319 | 0.55345498 |
| T2DM | mast_cell | *LDB3* | 2.08514948 | 3.73E-05 | 0.000134478 | 0.180740035 |
| T2DM | mast_cell | *OPN4* | 8.135529516 | 0.002433614 | 0.003284533 | 1 |
| T2DM | mast_cell | *FAM35A* | 2.589479709 | 3.00E-05 | 0.000134478 | 0.145777677 |
| T2DM | mast_cell | *NAA60* | 2.070850387 | 4.66E-05 | 0.000134478 | 0.226337032 |
| T2DM | mast_cell | *ODF2L* | 2.883442893 | 2.58E-05 | 0.000134478 | 0.125033792 |
| T2DM | mast_cell | *SPATA1* | 2.164633385 | 2.22E-05 | 0.000134478 | 0.107770229 |
| T2DM | mast_cell | *LRRC39* | 1.052058887 | 0.000946283 | 0.001376309 | 1 |
| T2DM | mast_cell | *PMS2* | 1.879088464 | 1.03E-05 | 0.000134478 | 0.050022509 |
| T2DM | mast_cell | *EEPD1* | 1.508282704 | 0.000749807 | 0.001112217 | 1 |
| T2DM | mast_cell | *KLHL7* | 1.917646504 | 3.43E-05 | 0.000134478 | 0.166329693 |
| T2DM | mast_cell | *NT5C3A* | 2.760543705 | 6.78E-05 | 0.000156533 | 0.328876783 |
| T2DM | mast_cell | *CCZ1* | 2.25275036 | 0.002463089 | 0.003320945 | 1 |
| T2DM | mast_cell | *POLM* | 0.96660806 | 0.026672948 | 0.03190758 | 1 |
| T2DM | mast_cell | *SMU1* | 2.665881847 | 1.52E-05 | 0.000134478 | 0.073793327 |
| T2DM | mast_cell | *SLC25A51* | 1.499051069 | 8.64E-05 | 0.000183795 | 0.419301965 |
| T2DM | mast_cell | *TRIM24* | 3.474463323 | 0.000563995 | 0.000855961 | 1 |
| T2DM | mast_cell | *C7orf49* | 2.132426569 | 2.72E-05 | 0.000134478 | 0.131916076 |
| T2DM | mast_cell | *CISD1* | 1.249138471 | 0.000400507 | 0.000628815 | 1 |
| T2DM | mast_cell | *RBM19* | 2.401847741 | 3.42E-05 | 0.000134478 | 0.166011823 |
| T2DM | mast_cell | *DDX54* | 1.86809007 | 3.50E-05 | 0.000134478 | 0.17004467 |
| T2DM | mast_cell | *SSPN* | 3.122658477 | 9.23E-07 | 8.78E-05 | 0.004477767 |
| T2DM | mast_cell | *CCDC91* | 1.575425813 | 2.93E-05 | 0.000134478 | 0.142299295 |
| T2DM | mast_cell | *EBPL* | 1.951960852 | 2.13E-05 | 0.000134478 | 0.103331797 |
| T2DM | mast_cell | *CENPK* | 2.329757031 | 0.000312647 | 0.000508707 | 1 |
| T2DM | mast_cell | *ITIH5* | 1.193938882 | 0.00206493 | 0.002827043 | 1 |
| T2DM | mast_cell | *TSFM* | 1.930000403 | 1.54E-05 | 0.000134478 | 0.074810989 |
| T2DM | mast_cell | *ARHGAP9* | 2.196539706 | 0.000115411 | 0.000225797 | 0.55997628 |
| T2DM | mast_cell | *PFDN5* | 2.438356042 | 3.59E-05 | 0.000134478 | 0.174362308 |
| T2DM | mast_cell | *PDE1B* | 1.768539418 | 5.25E-05 | 0.000136633 | 0.254814828 |

| | | | | | | |
|---|---|---|---|---|---|---|
| T2DM | mast_cell | *CDK2* | -1.881790498 | 0.000108308 | 0.000215284 | 0.525508459 |
| T2DM | mast_cell | *SMUG1* | 1.319753153 | 0.00010512 | 0.000210327 | 0.51004283 |
| T2DM | mast_cell | *SARDH* | 2.345022964 | 0.00111175 | 0.001594976 | 1 |
| T2DM | mast_cell | *DBH* | 49.87021663 | 4.47E-05 | 0.000134478 | 0.217014854 |
| T2DM | mast_cell | *NDUFAF4* | 2.44648517 | 9.07E-05 | 0.000190296 | 0.439964448 |
| T2DM | mast_cell | *USP45* | 2.504856896 | 3.53E-05 | 0.000134478 | 0.171499567 |
| T2DM | mast_cell | *TTC21B* | 1.635145703 | 9.53E-05 | 0.000196519 | 0.462410299 |
| T2DM | mast_cell | *ACVR1C* | 0.842898566 | 0.007262652 | 0.009203026 | 1 |
| T2DM | mast_cell | *PLA2G12A* | 1.741463011 | 3.06E-05 | 0.000134478 | 0.148581387 |
| T2DM | mast_cell | *C4BPA* | 2.3714897 | 0.000514908 | 0.000788615 | 1 |
| T2DM | mast_cell | *C4BPB* | 1.659492735 | 0.006744471 | 0.008575459 | 1 |
| T2DM | mast_cell | *RAB38* | -1.372170972 | 0.009835671 | 0.012286992 | 1 |
| T2DM | mast_cell | *DAW1* | 4.238962597 | 2.56E-05 | 0.000134478 | 0.124124156 |
| T2DM | mast_cell | *DNPEP* | 2.875369042 | 2.43E-05 | 0.000134478 | 0.117894377 |
| T2DM | mast_cell | *OBSL1* | 3.545824015 | 0.00444505 | 0.005789902 | 1 |
| T2DM | mast_cell | *SLC12A4* | 2.393499327 | 3.14E-05 | 0.000134478 | 0.152294853 |
| T2DM | mast_cell | *FAM210B* | 5.57210902 | 0.005855777 | 0.00749663 | 1 |
| T2DM | mast_cell | *SLC12A5* | -2.751131866 | 6.38E-06 | 0.000134478 | 0.030945005 |
| T2DM | mast_cell | *MATN4* | 5.169739703 | 0.000135325 | 0.000255088 | 0.656596454 |
| T2DM | mast_cell | *VAPB* | 1.082010313 | 0.001948772 | 0.002678595 | 1 |
| T2DM | mast_cell | *SRSF6* | 1.562646411 | 4.34E-05 | 0.000134478 | 0.21059243 |
| T2DM | mast_cell | *CSE1L* | 1.874675359 | 7.06E-05 | 0.000160722 | 0.342498576 |
| T2DM | mast_cell | *PTGIS* | 2.354811618 | 3.92E-05 | 0.000134478 | 0.190176103 |
| T2DM | mast_cell | *RNF114* | 1.90503456 | 5.72E-05 | 0.000142294 | 0.277331645 |
| T2DM | mast_cell | *NEURL2* | 2.105278964 | 2.44E-06 | 0.000128679 | 0.011838433 |
| T2DM | mast_cell | *MTRR* | 1.750438187 | 6.49E-05 | 0.000152689 | 0.314692769 |
| T2DM | mast_cell | *PEPD* | 3.594671753 | 4.09E-05 | 0.000134478 | 0.1984608 |
| T2DM | mast_cell | *CHST8* | 3.157333515 | 6.97E-05 | 0.000159406 | 0.338419806 |
| T2DM | mast_cell | *STAMBP* | 2.902537028 | 3.38E-05 | 0.000134478 | 0.164068449 |
| T2DM | mast_cell | *NAGK* | 1.948031025 | 0.000145283 | 0.000268129 | 0.704911878 |
| T2DM | mast_cell | *SNRNP27* | 2.372852444 | 2.70E-05 | 0.000134478 | 0.130900528 |
| T2DM | mast_cell | *ATP8A1* | 1.575968085 | 3.71E-05 | 0.000134478 | 0.17981187 |
| T2DM | mast_cell | *USP22* | 2.115352887 | 2.62E-05 | 0.000134478 | 0.127223566 |
| T2DM | mast_cell | *HIF3A* | 1.127722507 | 8.62E-05 | 0.000183584 | 0.418204972 |
| T2DM | mast_cell | *ZNF576* | 1.995363965 | 3.79E-05 | 0.000134478 | 0.183973568 |
| T2DM | mast_cell | *MRS2* | 1.80419779 | 1.02E-05 | 0.000134478 | 0.049311282 |
| T2DM | mast_cell | *SNRPC* | 2.352142472 | 2.83E-05 | 0.000134478 | 0.137335182 |
| T2DM | mast_cell | *SERPINB6* | 1.722593682 | 3.68E-05 | 0.000134478 | 0.178320896 |
| T2DM | mast_cell | *PEX6* | 0.813274483 | 0.003054595 | 0.004054964 | 1 |
| T2DM | mast_cell | *NQO2* | 1.602612191 | 3.94E-05 | 0.000134478 | 0.191293007 |
| T2DM | mast_cell | *ZNF391* | 0.886924544 | 0.00199246 | 0.002733224 | 1 |
| T2DM | mast_cell | *RPS10* | 2.444780747 | 0.013286954 | 0.016404148 | 1 |
| T2DM | mast_cell | *MED20* | 1.796567416 | 0.001272608 | 0.001802305 | 1 |
| T2DM | mast_cell | *TCP11* | -2.0855247 | 0.001185802 | 0.00169022 | 1 |
| T2DM | mast_cell | *TREM1* | 3.053248016 | 0.016054167 | 0.019670409 | 1 |
| T2DM | mast_cell | *KLHL31* | 1.314090216 | 0.000106614 | 0.000212439 | 0.517293053 |
| T2DM | mast_cell | *GLO1* | 2.578911528 | 3.64E-06 | 0.000134478 | 0.017674585 |
| T2DM | mast_cell | *RREB1* | 1.60095319 | 7.12E-05 | 0.000161512 | 0.34544594 |
| T2DM | mast_cell | *RIOK1* | 1.995653714 | 8.63E-06 | 0.000134478 | 0.041880873 |
| T2DM | mast_cell | *ATXN1* | 2.418753018 | 5.34E-05 | 0.000137618 | 0.25898432 |

| | | | | | | |
|---|---|---|---|---|---|---|
| T2DM | mast_cell | *RUNX2* | 1.344882796 | 4.26E-05 | 0.000134478 | 0.206553124 |
| T2DM | mast_cell | *CXCL6* | 3.85700609 | 9.78E-05 | 0.000199936 | 0.474754553 |
| T2DM | mast_cell | *SH3TC1* | 1.182808923 | 0.000804615 | 0.001187707 | 1 |
| T2DM | mast_cell | *BBS2* | 2.039816331 | 1.40E-05 | 0.000134478 | 0.068121949 |
| T2DM | mast_cell | *MT1G* | -3.007902248 | 0.002198098 | 0.002994994 | 1 |
| T2DM | mast_cell | *GOT2* | 3.663227435 | 2.18E-05 | 0.000134478 | 0.105536158 |
| T2DM | mast_cell | *CLYBL* | 2.554052444 | 3.50E-05 | 0.000134478 | 0.169817704 |
| T2DM | mast_cell | *TMTC4* | 1.509512959 | 6.03E-05 | 0.000146092 | 0.292475693 |
| T2DM | mast_cell | *ABCC4* | 3.975300983 | 0.001308375 | 0.001850258 | 1 |
| T2DM | mast_cell | *ATP5S* | 2.492354246 | 4.33E-05 | 0.000134478 | 0.210168233 |
| T2DM | mast_cell | *GRK4* | 1.377799623 | 6.37E-07 | 7.18E-05 | 0.003090853 |
| T2DM | mast_cell | *HS3ST3B1* | 2.670390029 | 0.000324415 | 0.000525564 | 1 |
| T2DM | mast_cell | *MRPS7* | 1.696473209 | 4.27E-05 | 0.000134478 | 0.2072341 |
| T2DM | mast_cell | *GGA3* | 1.987657949 | 4.03E-05 | 0.000134478 | 0.195642853 |
| T2DM | mast_cell | *NUP85* | 1.442898515 | 5.55E-05 | 0.000140094 | 0.269261572 |
| T2DM | mast_cell | *NT5C* | 4.90063717 | 0.000143853 | 0.000266607 | 0.697976624 |
| T2DM | mast_cell | *DDX31* | 2.092885452 | 0.000569328 | 0.000863784 | 1 |
| T2DM | mast_cell | *OPRL1* | 6.045844126 | 4.25E-05 | 0.000134478 | 0.206365095 |
| T2DM | mast_cell | *PPDPF* | 2.121086002 | 1.43E-05 | 0.000134478 | 0.06955126 |
| T2DM | mast_cell | *IL1B* | 8.654835812 | 8.61E-05 | 0.000183456 | 0.417729869 |
| T2DM | mast_cell | *PAX8* | 1.472814795 | 0.000331501 | 0.000535612 | 1 |
| T2DM | mast_cell | *POLR1B* | 1.308004512 | 0.004570616 | 0.005940699 | 1 |
| T2DM | mast_cell | *CCDC93* | 1.764726312 | 3.95E-05 | 0.000134478 | 0.191684467 |
| T2DM | mast_cell | *PSD4* | 2.279853196 | 9.98E-05 | 0.000203034 | 0.484438733 |
| T2DM | mast_cell | *SH2D3A* | 3.00745526 | 0.012298799 | 0.015230672 | 1 |
| T2DM | mast_cell | *GPR108* | 2.46347899 | 0.000100324 | 0.000203585 | 0.486770709 |
| T2DM | mast_cell | *SNRPD2* | 4.766323296 | 4.20E-05 | 0.000134478 | 0.204018175 |
| T2DM | mast_cell | *SDCBP2* | -2.096695564 | 0.000406834 | 0.000637172 | 1 |
| T2DM | mast_cell | *DTD1* | 1.918578831 | 3.72E-05 | 0.000134478 | 0.180686099 |
| T2DM | mast_cell | *RBCK1* | 3.10263613 | 0.000313967 | 0.000510342 | 1 |
| T2DM | mast_cell | *RRBP1* | 2.250169706 | 1.83E-05 | 0.000134478 | 0.088657504 |
| T2DM | mast_cell | *FLRT3* | -2.925130742 | 0.004698136 | 0.006098276 | 1 |
| T2DM | mast_cell | *PCSK2* | -1.96042857 | 0.000214399 | 0.000369019 | 1 |
| T2DM | mast_cell | *MKKS* | 2.719794347 | 2.46E-05 | 0.000134478 | 0.119572585 |
| T2DM | mast_cell | *LAMP5* | 33.91437896 | 0.000317925 | 0.000516255 | 1 |
| T2DM | mast_cell | *MGME1* | 2.388052937 | 4.87E-05 | 0.000134902 | 0.236415002 |
| T2DM | mast_cell | *ITPA* | 2.342797354 | 8.00E-05 | 0.000174493 | 0.388071886 |
| T2DM | mast_cell | *NCLN* | 3.418515144 | 0.001786115 | 0.002473239 | 1 |
| T2DM | mast_cell | *UROD* | 4.406006816 | 1.29E-05 | 0.000134478 | 0.062718773 |
| T2DM | mast_cell | *KLC1* | 3.065809955 | 3.37E-05 | 0.000134478 | 0.163593957 |
| T2DM | mast_cell | *TUBGCP3* | 2.531126556 | 3.74E-05 | 0.000134478 | 0.181286274 |
| T2DM | mast_cell | *MCF2L* | 1.443749942 | 2.14E-05 | 0.000134478 | 0.103966579 |
| T2DM | mast_cell | *PROZ* | -1.827893836 | 0.000827679 | 0.001218785 | 1 |
| T2DM | mast_cell | *IGFLR1* | -1.602625877 | 0.002175356 | 0.002966506 | 1 |
| T2DM | mast_cell | *COX6B1* | 2.682023108 | 4.63E-05 | 0.000134478 | 0.224431929 |
| T2DM | mast_cell | *PRDX5* | 3.466820351 | 5.22E-05 | 0.000136624 | 0.253309217 |
| T2DM | mast_cell | *IRF3* | 4.135591012 | 2.81E-05 | 0.000134478 | 0.136229288 |
| T2DM | mast_cell | *PRR12* | 5.542670032 | 5.02E-07 | 6.81E-05 | 0.002434082 |
| T2DM | mast_cell | *TRAP1* | 1.358022174 | 0.000765709 | 0.00113338 | 1 |
| T2DM | mast_cell | *ZNF384* | 1.684358837 | 0.000147049 | 0.000270977 | 0.713483509 |

| | | | | | | |
|---|---|---|---|---|---|---|
| T2DM | mast_cell | *EMG1* | 1.997187566 | 7.66E-05 | 0.000168736 | 0.371557468 |
| T2DM | mast_cell | *ATG14* | 2.135515277 | 3.17E-05 | 0.000134478 | 0.153998542 |
| T2DM | mast_cell | *KTN1* | 2.153243613 | 3.21E-05 | 0.000134478 | 0.155816616 |
| T2DM | mast_cell | *L3HYPDH* | 1.492300313 | 0.000888568 | 0.001299377 | 1 |
| T2DM | mast_cell | *ZBTB1* | 2.128582652 | 4.88E-05 | 0.000134902 | 0.236843095 |
| T2DM | mast_cell | *RHOT1* | 1.369323751 | 0.000185392 | 0.000327219 | 0.899524002 |
| T2DM | mast_cell | *WDR60* | 1.996600443 | 0.00017707 | 0.00031544 | 0.859145747 |
| T2DM | mast_cell | *AIF1L* | 3.608570662 | 3.04E-05 | 0.000134478 | 0.147446458 |
| T2DM | mast_cell | *CANX* | 2.063548197 | 2.93E-05 | 0.000134478 | 0.142194284 |
| T2DM | mast_cell | *ZNF484* | 3.98134156 | 7.24E-05 | 0.000162881 | 0.35133503 |
| T2DM | mast_cell | *FGD3* | 1.414084385 | 0.000142793 | 0.000265249 | 0.692830155 |
| T2DM | mast_cell | *HELB* | 1.936338995 | 1.73E-05 | 0.000134478 | 0.083785059 |
| T2DM | mast_cell | *RAB3IP* | 1.845485102 | 6.75E-05 | 0.000156274 | 0.327549898 |
| T2DM | mast_cell | *YEATS4* | 1.857184083 | 3.11E-05 | 0.000134478 | 0.15104612 |
| T2DM | mast_cell | *TAS2R4* | 1.820126034 | 0.001651524 | 0.002298679 | 1 |
| T2DM | mast_cell | *LRRC61* | 1.79144737 | 4.76E-05 | 0.000134478 | 0.230769874 |
| T2DM | mast_cell | *IDUA* | 2.801270576 | 4.35E-05 | 0.000134478 | 0.210879374 |
| T2DM | mast_cell | *EMC1* | 1.895552974 | 4.47E-05 | 0.000134478 | 0.216880827 |
| T2DM | mast_cell | *UBR4* | 1.808773863 | 0.000571606 | 0.000866968 | 1 |
| T2DM | mast_cell | *GFER* | 2.734568907 | 0.00038609 | 0.000611593 | 1 |
| T2DM | mast_cell | *WDR24* | 1.899140151 | 0.005263883 | 0.006780026 | 1 |
| T2DM | mast_cell | *SMARCA4* | 3.191062906 | 5.05E-05 | 0.000135809 | 0.245135121 |
| T2DM | mast_cell | *PNKD* | 1.493560045 | 2.27E-05 | 0.000134478 | 0.110170127 |
| T2DM | mast_cell | *RNF6* | 1.541366597 | 4.53E-05 | 0.000134478 | 0.219749338 |
| T2DM | mast_cell | *HIP1* | 1.822515541 | 2.75E-05 | 0.000134478 | 0.133222078 |
| T2DM | mast_cell | *PTPN12* | 1.899734749 | 9.40E-05 | 0.000194647 | 0.456058242 |
| T2DM | mast_cell | *STEAP4* | 1.222243415 | 0.000280513 | 0.000464522 | 1 |
| T2DM | mast_cell | *PEX1* | 2.830263305 | 3.24E-05 | 0.000134478 | 0.157358568 |
| T2DM | mast_cell | *SRD5A3* | 2.593267231 | 0.013791147 | 0.016979103 | 1 |
| T2DM | mast_cell | *KDR* | 1.316993724 | 4.49E-05 | 0.000134478 | 0.217688927 |
| T2DM | mast_cell | *DGCR6L* | -1.256773783 | 0.000810696 | 0.001195955 | 1 |
| T2DM | mast_cell | *MGAT3* | 2.604357722 | 4.03E-05 | 0.000134478 | 0.195738964 |
| T2DM | mast_cell | *ATF4* | 4.321182019 | 4.61E-05 | 0.000134478 | 0.223662358 |
| T2DM | mast_cell | *A4GALT* | 1.579195143 | 0.000264827 | 0.000441697 | 1 |
| T2DM | mast_cell | *CDC42EP1* | 3.079888278 | 5.23E-05 | 0.000136624 | 0.25357415 |
| T2DM | mast_cell | *RIBC2* | 2.540694856 | 0.002887102 | 0.003843133 | 1 |
| T2DM | mast_cell | *SPECC1* | 3.180996062 | 4.74E-05 | 0.000134478 | 0.229904252 |
| T2DM | mast_cell | *CDHR3* | 1.124406205 | 4.69E-05 | 0.000134478 | 0.227767117 |
| T2DM | mast_cell | *PRKRIP1* | 2.205674727 | 6.89E-05 | 0.000158111 | 0.334247044 |
| T2DM | mast_cell | *IRF5* | 1.02572702 | 0.028433468 | 0.033913272 | 1 |
| T2DM | mast_cell | *OPN1SW* | -2.965817225 | 0.000117204 | 0.000228108 | 0.568674081 |
| T2DM | mast_cell | *HOXD1* | 2.552952677 | 0.042270901 | 0.049540679 | 1 |
| T2DM | mast_cell | *HOXD3* | 1.396258587 | 3.37E-05 | 0.000134478 | 0.163736474 |
| T2DM | mast_cell | *MTX2* | 1.946166018 | 3.17E-05 | 0.000134478 | 0.153749982 |
| T2DM | mast_cell | *GAD1* | 4.055353241 | 0.000995626 | 0.001443315 | 1 |
| T2DM | mast_cell | *ORMDL1* | 2.691123652 | 1.06E-05 | 0.000134478 | 0.051555242 |
| T2DM | mast_cell | *HOXD10* | 4.522066917 | 0.000382412 | 0.000607154 | 1 |
| T2DM | mast_cell | *PSMG2* | 2.619898389 | 1.32E-05 | 0.000134478 | 0.063929358 |
| T2DM | mast_cell | *WDFY4* | 1.119664846 | 0.0002875 | 0.000474311 | 1 |
| T2DM | mast_cell | *EIF2AK4* | 1.48964406 | 0.000123772 | 0.000237744 | 0.600541199 |

| | | | | | | |
|---|---|---|---|---|---|---|
| T2DM | mast_cell | *TMOD2* | 1.725878123 | 3.98E-05 | 0.000134478 | 0.19294389 |
| T2DM | mast_cell | *IVD* | 1.692729535 | 0.000383501 | 0.000608484 | 1 |
| T2DM | mast_cell | *KNSTRN* | 0.744059908 | 0.020756681 | 0.025127599 | 1 |
| T2DM | mast_cell | *ARPP19* | 3.136737816 | 3.20E-05 | 0.000134478 | 0.155482267 |
| T2DM | mast_cell | *VPS13C* | 2.809161952 | 0.000309199 | 0.000503942 | 1 |
| T2DM | mast_cell | *CALML4* | -3.887838105 | 0.000246751 | 0.000416574 | 1 |
| T2DM | mast_cell | *SUMF2* | 2.037751674 | 3.63E-05 | 0.000134478 | 0.176087626 |
| T2DM | mast_cell | *BBOX1* | 6.347941009 | 4.24E-05 | 0.000134478 | 0.205958586 |
| T2DM | mast_cell | *SERGEF* | 1.564565867 | 1.31E-05 | 0.000134478 | 0.063454091 |
| T2DM | mast_cell | *DCTD* | 2.37598153 | 0.006413566 | 0.008169762 | 1 |
| T2DM | mast_cell | *RPAIN* | 1.623872511 | 0.000926467 | 0.001349521 | 1 |
| T2DM | mast_cell | *PLD2* | 4.371747738 | 3.17E-05 | 0.000134478 | 0.15361258 |
| T2DM | mast_cell | *PHF20L1* | 2.042367515 | 7.00E-05 | 0.000159858 | 0.339697919 |
| T2DM | mast_cell | *PUS7L* | 2.7081007 | 0.000101855 | 0.000205891 | 0.494198359 |
| T2DM | mast_cell | *KRI1* | 1.354408278 | 0.000306907 | 0.000500878 | 1 |
| T2DM | mast_cell | *MTUS1* | 1.317538654 | 2.77E-05 | 0.000134478 | 0.134265252 |
| T2DM | mast_cell | *KLK10* | 1.726898487 | 0.005046606 | 0.006520941 | 1 |
| T2DM | mast_cell | *KLK8* | 13.64581777 | 4.33E-05 | 0.000134478 | 0.210321988 |
| T2DM | mast_cell | *ADCY4* | 1.925157948 | 3.99E-05 | 0.000134478 | 0.19339775 |
| T2DM | mast_cell | *BCL2L2* | 2.110286298 | 0.000289144 | 0.000476214 | 1 |
| T2DM | mast_cell | *MIS18BP1* | 2.0719698 | 2.23E-05 | 0.000134478 | 0.108041349 |
| T2DM | mast_cell | *RNASE1* | 1.519680023 | 1.41E-05 | 0.000134478 | 0.068328116 |
| T2DM | mast_cell | *EPB41L4A* | 1.969263278 | 3.84E-05 | 0.000134478 | 0.186208411 |
| T2DM | mast_cell | *CDO1* | 1.161701444 | 0.001321305 | 0.001865824 | 1 |
| T2DM | mast_cell | *REEP5* | 3.606766956 | 6.77E-05 | 0.000156404 | 0.328448175 |
| T2DM | mast_cell | *RHBDF2* | 1.557363916 | 2.31E-05 | 0.000134478 | 0.112098296 |
| T2DM | mast_cell | *AANAT* | -1.41920067 | 0.001882848 | 0.002594598 | 1 |
| T2DM | mast_cell | *TMEM8A* | 2.564651535 | 3.68E-05 | 0.000134478 | 0.178367555 |
| T2DM | mast_cell | *MAU2* | 1.563781919 | 6.87E-05 | 0.000157925 | 0.333369678 |
| T2DM | mast_cell | *SHC2* | 2.260246934 | 2.33E-06 | 0.000126966 | 0.011299977 |
| T2DM | mast_cell | *ERMARD* | 2.103253319 | 3.56E-05 | 0.000134478 | 0.172540734 |
| T2DM | mast_cell | *PHF10* | 3.295357075 | 5.21E-05 | 0.000136609 | 0.252830939 |
| T2DM | mast_cell | *GALNT8* | 2.88269187 | 0.000398379 | 0.000626356 | 1 |
| T2DM | mast_cell | *PRKCSH* | 4.605754892 | 2.52E-05 | 0.000134478 | 0.122349619 |
| T2DM | mast_cell | *CDC16* | 2.360027868 | 0.000172071 | 0.00030876 | 0.834886083 |
| T2DM | mast_cell | *XPO7* | 2.132714814 | 4.69E-05 | 0.000134478 | 0.227473882 |
| T2DM | mast_cell | *FAM98C* | 2.255073858 | 0.00223748 | 0.003045232 | 1 |
| T2DM | mast_cell | *ATP8B3* | -1.561804387 | 0.000183594 | 0.00032499 | 0.890799796 |
| T2DM | mast_cell | *PLVAP* | 0.883470412 | 0.013041583 | 0.016113512 | 1 |
| T2DM | mast_cell | *DDA1* | 3.427373252 | 0.000118555 | 0.000229632 | 0.5752271 |
| T2DM | mast_cell | *MRPL34* | 0.929482935 | 0.007007505 | 0.008898054 | 1 |
| T2DM | mast_cell | *LSM7* | 1.750776087 | 3.76E-05 | 0.000134478 | 0.182664037 |
| T2DM | mast_cell | *TULP4* | 2.004452916 | 2.48E-05 | 0.000134478 | 0.120516675 |
| T2DM | mast_cell | *QRSL1* | 1.674770496 | 0.000992013 | 0.001438508 | 1 |
| T2DM | mast_cell | *RSPH3* | 0.918637321 | 0.007944841 | 0.010015165 | 1 |
| T2DM | mast_cell | *ACTN4* | 2.727970951 | 2.57E-05 | 0.000134478 | 0.124804872 |
| T2DM | mast_cell | *STK33* | 1.963020874 | 6.81E-07 | 7.18E-05 | 0.003304389 |
| T2DM | mast_cell | *NDUFA10* | 1.737506757 | 0.000441115 | 0.00068665 | 1 |
| T2DM | mast_cell | *MAP1S* | 5.750432192 | 3.60E-05 | 0.000134478 | 0.174722533 |
| T2DM | mast_cell | *SSBP4* | 3.201028033 | 7.13E-05 | 0.000161597 | 0.345979963 |

| | | | | | | |
|---|---|---|---|---|---|---|
| T2DM | mast_cell | *KIAA1683* | 4.117790863 | 0.000250518 | 0.000421908 | 1 |
| T2DM | mast_cell | *LSM4* | 1.672025123 | 0.003039906 | 0.00403878 | 1 |
| T2DM | mast_cell | *TRPM4* | 1.561365675 | 0.000112387 | 0.000222028 | 0.54529981 |
| T2DM | mast_cell | *UBAC1* | 1.537794598 | 0.000616597 | 0.000929975 | 1 |
| T2DM | mast_cell | *ATXN10* | 2.094195696 | 3.48E-05 | 0.000134478 | 0.168923731 |
| T2DM | mast_cell | *PNPLA7* | 2.227360896 | 3.44E-05 | 0.000134478 | 0.167100781 |
| T2DM | mast_cell | *PAK4* | 4.435364504 | 0.004898228 | 0.006347811 | 1 |
| T2DM | mast_cell | *ZNF337* | 2.213055603 | 8.24E-05 | 0.000178055 | 0.399993751 |
| T2DM | mast_cell | *POMT1* | 2.342929914 | 3.51E-05 | 0.000134478 | 0.17039613 |
| T2DM | mast_cell | *CHMP2A* | 2.012989542 | 6.50E-05 | 0.000152782 | 0.315495455 |
| T2DM | mast_cell | *YIPF2* | 3.053374399 | 0.001140617 | 0.001633011 | 1 |
| T2DM | mast_cell | *NPAS1* | 2.166694522 | 0.000675347 | 0.001010417 | 1 |
| T2DM | mast_cell | *MAP3K10* | 1.730547261 | 8.91E-05 | 0.000187451 | 0.432125763 |
| T2DM | mast_cell | *LRRC47* | 3.122287591 | 0.000873506 | 0.001280825 | 1 |
| T2DM | mast_cell | *THEMIS2* | 2.003508056 | 6.67E-06 | 0.000134478 | 0.032375044 |
| T2DM | mast_cell | *ZNF317* | 1.738612315 | 4.82E-05 | 0.000134478 | 0.234027574 |
| T2DM | mast_cell | *DNMT1* | 3.015381238 | 2.63E-05 | 0.000134478 | 0.127606138 |
| T2DM | mast_cell | *C12orf65* | 2.141674113 | 1.07E-06 | 9.21E-05 | 0.005187747 |
| T2DM | mast_cell | *NOL11* | 3.609196285 | 2.55E-05 | 0.000134478 | 0.123488039 |
| T2DM | mast_cell | *PKDREJ* | -2.542672188 | 0.005407637 | 0.006954109 | 1 |
| T2DM | mast_cell | *HSD17B3* | 2.546269672 | 5.39E-05 | 0.000138255 | 0.261747391 |
| T2DM | mast_cell | *SLC35D2* | 2.659937651 | 3.56E-05 | 0.000134478 | 0.172682079 |
| T2DM | mast_cell | *ULBP2* | 16.20913885 | 0.000659476 | 0.0009885 | 1 |
| T2DM | mast_cell | *TTLL9* | 3.235656197 | 4.22E-05 | 0.000134478 | 0.204970163 |
| T2DM | mast_cell | *COX4I2* | 2.863491462 | 0.000169535 | 0.000305226 | 0.822584594 |
| T2DM | mast_cell | *ATP6V1E1* | 3.025027447 | 0.000116484 | 0.000227345 | 0.565179698 |
| T2DM | mast_cell | *CHMP1A* | 6.129245207 | 0.002780687 | 0.003712684 | 1 |
| T2DM | mast_cell | *IDO1* | 3.305440116 | 3.68E-05 | 0.000134478 | 0.178553798 |
| T2DM | mast_cell | *HAUS8* | -2.567172617 | 0.030250257 | 0.036018221 | 1 |
| T2DM | mast_cell | *MRPS25* | 1.9852499 | 0.000135476 | 0.000255156 | 0.65732737 |
| T2DM | mast_cell | *GALNT15* | 1.324268489 | 5.01E-05 | 0.000135379 | 0.243248821 |
| T2DM | mast_cell | *SLC6A6* | 1.473552629 | 4.90E-05 | 0.000135021 | 0.237921721 |
| T2DM | mast_cell | *KCNC3* | -3.122132713 | 0.018081918 | 0.022010403 | 1 |
| T2DM | mast_cell | *NAPSA* | 2.325419965 | 0.033802907 | 0.039993101 | 1 |
| T2DM | mast_cell | *PDLIM4* | 3.425472846 | 2.47E-05 | 0.000134478 | 0.12008593 |
| T2DM | mast_cell | *ACLY* | 3.06529864 | 0.0027166 | 0.003637125 | 1 |
| T2DM | mast_cell | *NDUFA2* | 2.631708757 | 0.000181259 | 0.000321444 | 0.879470302 |
| T2DM | mast_cell | *EXOC4* | 2.031831797 | 9.79E-06 | 0.000134478 | 0.047523928 |
| T2DM | mast_cell | *C1orf159* | -2.276527242 | 0.003771119 | 0.004949275 | 1 |
| T2DM | mast_cell | *PPFIA1* | 2.548242602 | 4.79E-05 | 0.000134478 | 0.232466931 |
| T2DM | mast_cell | *BARX1* | -3.68495136 | 7.55E-05 | 0.000167051 | 0.36617609 |
| T2DM | mast_cell | *NINJ1* | 1.463475672 | 0.000157665 | 0.000288132 | 0.764989741 |
| T2DM | mast_cell | *NPHP4* | 1.566684952 | 3.76E-05 | 0.000134478 | 0.182565841 |
| T2DM | mast_cell | *MAP1B* | 1.729046943 | 9.88E-06 | 0.000134478 | 0.047924431 |
| T2DM | mast_cell | *CKMT2* | 1.791042987 | 7.62E-05 | 0.000168279 | 0.369708523 |
| T2DM | mast_cell | *KHDRBS3* | 2.088325242 | 3.59E-05 | 0.000134478 | 0.17433482 |
| T2DM | mast_cell | *CHD1L* | 1.705364907 | 0.000158759 | 0.000289695 | 0.770300255 |
| T2DM | mast_cell | *PRKAB2* | 2.557141558 | 4.74E-05 | 0.000134478 | 0.230224649 |
| T2DM | mast_cell | *ZNF304* | 2.108104831 | 0.000172653 | 0.000309461 | 0.83771143 |
| T2DM | mast_cell | *ZNF132* | 1.82713556 | 0.000796819 | 0.001176915 | 1 |

| | | | | | | |
|---|---|---|---|---|---|---|
| T2DM | mast_cell | *RHPN2* | 1.379272595 | 4.02E-05 | 0.000134478 | 0.195233578 |
| T2DM | mast_cell | *GCH1* | 1.732926143 | 2.84E-05 | 0.000134478 | 0.13792037 |
| T2DM | mast_cell | *TRIM21* | 2.490231439 | 4.89E-05 | 0.000134955 | 0.237251611 |
| T2DM | mast_cell | *SPATA6* | 1.850539208 | 3.87E-05 | 0.000134478 | 0.187972683 |
| T2DM | mast_cell | *LRRC41* | 2.178702487 | 6.17E-05 | 0.000148384 | 0.299587692 |
| T2DM | mast_cell | *CCT6B* | 1.716723751 | 0.022784035 | 0.02747903 | 1 |
| T2DM | mast_cell | *CNGA4* | 1.674778375 | 0.02810388 | 0.033553155 | 1 |
| T2DM | mast_cell | *TRIM22* | 1.598705179 | 4.39E-05 | 0.000134478 | 0.213177734 |
| T2DM | mast_cell | *RRP8* | 4.089360182 | 3.98E-05 | 0.000134478 | 0.19298418 |
| T2DM | mast_cell | *TIMM10B* | 2.498511175 | 0.000104722 | 0.000209703 | 0.508109193 |
| T2DM | mast_cell | *IMMT* | 2.490634652 | 1.24E-05 | 0.000134478 | 0.060060795 |
| T2DM | mast_cell | *CARD6* | 2.584363413 | 0.015539322 | 0.019058845 | 1 |
| T2DM | mast_cell | *MYBBP1A* | 2.351829301 | 1.18E-05 | 0.000134478 | 0.057239174 |
| T2DM | mast_cell | *RPA1* | 1.431643899 | 6.96E-05 | 0.000159066 | 0.337466317 |
| T2DM | mast_cell | *SERPINF1* | 3.560096133 | 5.27E-05 | 0.000136843 | 0.255484977 |
| T2DM | mast_cell | *TMEM128* | 1.732033098 | 0.000506965 | 0.000778264 | 1 |
| T2DM | mast_cell | *SEC61G* | 2.222379185 | 0.000192289 | 0.000337062 | 0.932986338 |
| T2DM | mast_cell | *FIGNL1* | 2.538978687 | 5.01E-05 | 0.000135379 | 0.243276237 |
| T2DM | mast_cell | *EIF5A* | 1.567153113 | 0.039718703 | 0.046628393 | 1 |
| T2DM | mast_cell | *RGS22* | 1.773940327 | 3.00E-05 | 0.000134478 | 0.145736849 |
| T2DM | mast_cell | *REEP2* | -3.063393072 | 0.009760564 | 0.012196306 | 1 |
| T2DM | mast_cell | *PRMT7* | 1.663528302 | 0.002617244 | 0.003510885 | 1 |
| T2DM | mast_cell | *CTNNBL1* | 3.281694211 | 0.033663687 | 0.039838099 | 1 |
| T2DM | mast_cell | *OSER1* | 6.272605391 | 4.21E-07 | 6.59E-05 | 0.002042706 |
| T2DM | mast_cell | *SERINC3* | 1.834428643 | 0.000376614 | 0.000599125 | 1 |
| T2DM | mast_cell | *DMGDH* | 1.92575122 | 0.0041002 | 0.005357977 | 1 |
| T2DM | mast_cell | *AP3B1* | 1.848828987 | 3.17E-05 | 0.000134478 | 0.1537525 |
| T2DM | mast_cell | *ZBED3* | 5.708112393 | 1.39E-05 | 0.000134478 | 0.067283675 |
| T2DM | mast_cell | *FBXO44* | 2.286379599 | 1.34E-05 | 0.000134478 | 0.064810868 |
| T2DM | mast_cell | *CASP9* | 1.56028497 | 1.19E-05 | 0.000134478 | 0.05789363 |
| T2DM | mast_cell | *ATP8A2* | 4.695277571 | 3.74E-05 | 0.000134478 | 0.181423407 |
| T2DM | mast_cell | *ALOX5AP* | 3.633558731 | 1.38E-05 | 0.000134478 | 0.066865137 |
| T2DM | mast_cell | *WASF3* | 2.469064234 | 3.42E-05 | 0.000134478 | 0.165841044 |
| T2DM | mast_cell | *GPR12* | 7.453229925 | 0.004245179 | 0.005541461 | 1 |
| T2DM | mast_cell | *PEMT* | 1.71708056 | 4.65E-05 | 0.000134478 | 0.225468256 |
| T2DM | mast_cell | *SCO1* | 3.412762465 | 2.61E-05 | 0.000134478 | 0.126854262 |
| T2DM | mast_cell | *MPRIP* | 1.994402765 | 3.20E-05 | 0.000134478 | 0.155101725 |
| T2DM | mast_cell | *COG6* | 2.965278183 | 4.88E-05 | 0.000134902 | 0.236687136 |
| T2DM | mast_cell | *EPSTI1* | 2.295782797 | 2.40E-05 | 0.000134478 | 0.11644819 |
| T2DM | mast_cell | *BTBD2* | 1.412111997 | 2.44E-05 | 0.000134478 | 0.118356138 |
| T2DM | mast_cell | *CSNK1G2* | 2.806314338 | 3.29E-05 | 0.000134478 | 0.159564631 |
| T2DM | mast_cell | *CNDP2* | 2.232414604 | 3.68E-05 | 0.000134478 | 0.17839757 |
| T2DM | mast_cell | *PDZD2* | 0.951994582 | 4.27E-05 | 0.000134478 | 0.207218797 |
| T2DM | mast_cell | *FAM83F* | -4.380579079 | 0.000373887 | 0.000594982 | 1 |
| T2DM | mast_cell | *GIMAP4* | 1.590516037 | 0.000226201 | 0.000386726 | 1 |
| T2DM | mast_cell | *ADCK2* | 1.639735743 | 0.000407582 | 0.000637932 | 1 |
| T2DM | mast_cell | *IPO8* | 2.600402345 | 5.74E-05 | 0.000142483 | 0.278555162 |
| T2DM | mast_cell | *LARS* | 1.467562723 | 0.000399894 | 0.000628127 | 1 |
| T2DM | mast_cell | *SPINK5* | 3.470665645 | 4.33E-05 | 0.000134478 | 0.210007987 |
| T2DM | mast_cell | *IMPA1* | 1.432720186 | 0.003944602 | 0.005164385 | 1 |

| | | | | | | |
|---|---|---|---|---|---|---|
| T2DM | mast_cell | *LRRCC1* | 1.858789231 | 4.97E-05 | 0.000135163 | 0.240995286 |
| T2DM | mast_cell | *RRAS2* | 1.483208112 | 2.17E-05 | 0.000134478 | 0.105135286 |
| T2DM | mast_cell | *CATSPERB* | 2.50056784 | 0.001086347 | 0.001563618 | 1 |
| T2DM | mast_cell | *COX16* | -1.806151913 | 0.000208398 | 0.000360866 | 1 |
| T2DM | mast_cell | *TTC9* | 2.381503914 | 4.20E-05 | 0.000134478 | 0.203962068 |
| T2DM | mast_cell | *ELP3* | 1.796240758 | 5.37E-05 | 0.000138064 | 0.260647107 |
| T2DM | mast_cell | *PEBP4* | 3.161482286 | 4.03E-05 | 0.000134478 | 0.195537772 |
| T2DM | mast_cell | *CTIF* | 1.621550311 | 6.64E-05 | 0.000154584 | 0.322153132 |
| T2DM | mast_cell | *MRO* | 3.756249699 | 0.000321557 | 0.000521456 | 1 |
| T2DM | mast_cell | *THUMPD3* | 3.921303716 | 0.00020236 | 0.000352044 | 0.981851942 |
| T2DM | mast_cell | *CNTN6* | 2.588035953 | 5.30E-05 | 0.000137292 | 0.256986429 |
| T2DM | mast_cell | *CHL1* | 1.246068616 | 6.41E-05 | 0.000151607 | 0.311249395 |
| T2DM | mast_cell | *GSTM5* | 1.594853947 | 0.001320736 | 0.001865564 | 1 |
| T2DM | mast_cell | *GSTM3* | 1.286645637 | 5.88E-05 | 0.000144373 | 0.285363552 |
| T2DM | mast_cell | *VAV3* | 1.492129009 | 0.003970525 | 0.005196921 | 1 |
| T2DM | mast_cell | *PSRC1* | -0.837319871 | 0.000185138 | 0.000326977 | 0.898289411 |
| T2DM | mast_cell | *TRIM45* | 5.020601953 | 0.03304593 | 0.039135673 | 1 |
| T2DM | mast_cell | *CEPT1* | 1.493668886 | 3.70E-05 | 0.000134478 | 0.179579774 |
| T2DM | mast_cell | *NAPG* | 1.826834635 | 3.93E-05 | 0.000134478 | 0.190514814 |
| T2DM | mast_cell | *PPHLN1* | 2.255411079 | 2.18E-05 | 0.000134478 | 0.105833491 |
| T2DM | mast_cell | *FKBP11* | 7.14499303 | 0.000879049 | 0.001287785 | 1 |
| T2DM | mast_cell | *YWHAQ* | 2.483502223 | 2.79E-05 | 0.000134478 | 0.135502705 |
| T2DM | mast_cell | *KIDINS220* | 2.316998653 | 3.37E-05 | 0.000134478 | 0.163432698 |
| T2DM | mast_cell | *LPIN1* | 2.284847334 | 0.00011531 | 0.00022578 | 0.559482562 |
| T2DM | mast_cell | *NARS* | 2.572204878 | 7.53E-06 | 0.000134478 | 0.036542558 |
| T2DM | mast_cell | *KIAA1468* | 1.764382586 | 3.17E-05 | 0.000134478 | 0.153631782 |
| T2DM | mast_cell | *IL15RA* | 2.070725425 | 3.51E-05 | 0.000134478 | 0.170337542 |
| T2DM | mast_cell | *TMEM241* | 1.599556349 | 0.000131633 | 0.000249292 | 0.638684962 |
| T2DM | mast_cell | *SOX5* | 1.878523724 | 3.58E-05 | 0.000134478 | 0.173839661 |
| T2DM | mast_cell | *KLRD1* | 1.980056839 | 2.98E-05 | 0.000134478 | 0.144537845 |
| T2DM | mast_cell | *ACP2* | 3.203148336 | 0.000106146 | 0.000211747 | 0.515020151 |
| T2DM | mast_cell | *ZCCHC11* | 2.006025495 | 5.31E-05 | 0.000137292 | 0.257587683 |
| T2DM | mast_cell | *PRPF38A* | 4.57489043 | 9.58E-05 | 0.000197031 | 0.464796524 |
| T2DM | mast_cell | *ELP2* | 1.864736646 | 3.55E-05 | 0.000134478 | 0.172041563 |
| T2DM | mast_cell | *TPGS2* | 4.502754649 | 8.01E-05 | 0.000174583 | 0.388885761 |
| T2DM | mast_cell | *TIMM10* | 2.600223005 | 0.00017352 | 0.000310443 | 0.841921177 |
| T2DM | mast_cell | *DHX34* | 2.452080959 | 3.65E-05 | 0.000134478 | 0.176925956 |
| T2DM | mast_cell | *FADS2* | 5.987651327 | 0.000477917 | 0.000738958 | 1 |
| T2DM | mast_cell | *TMEM258* | 2.434551962 | 5.61E-05 | 0.000140825 | 0.272059171 |
| T2DM | mast_cell | *CLOCK* | 2.040489484 | 3.15E-05 | 0.000134478 | 0.153037155 |
| T2DM | mast_cell | *GGACT* | 2.686931413 | 0.003156076 | 0.004182814 | 1 |
| T2DM | mast_cell | *ARGLU1* | 2.18911728 | 3.13E-05 | 0.000134478 | 0.151749798 |
| T2DM | mast_cell | *ERCC5* | -3.687397673 | 0.039562396 | 0.046456134 | 1 |
| T2DM | mast_cell | *CARS2* | 2.149300967 | 0.000400624 | 0.000628815 | 1 |
| T2DM | mast_cell | *KLB* | 0.7221294 | 0.016586602 | 0.020297149 | 1 |
| T2DM | mast_cell | *WDR36* | 1.760751312 | 4.01E-05 | 0.000134478 | 0.194678369 |
| T2DM | mast_cell | *OSTF1* | 1.763367616 | 3.69E-05 | 0.000134478 | 0.179083936 |
| T2DM | mast_cell | *UBQLN1* | 1.790243271 | 3.33E-05 | 0.000134478 | 0.161489334 |
| T2DM | mast_cell | *GOLM1* | 1.241251457 | 0.001423572 | 0.001999181 | 1 |
| T2DM | mast_cell | *ADAM19* | 1.75238778 | 1.36E-05 | 0.000134478 | 0.066125238 |

| | | | | | | |
|---|---|---|---|---|---|---|
| T2DM | mast_cell | *USP30* | 1.540003866 | 0.000160325 | 0.000291894 | 0.777896438 |
| T2DM | mast_cell | *MSI1* | 3.452063876 | 0.000370943 | 0.000591267 | 1 |
| T2DM | mast_cell | *P2RX4* | 2.429745234 | 5.33E-07 | 6.99E-05 | 0.002584767 |
| T2DM | mast_cell | *DTX1* | 5.468616843 | 0.000195448 | 0.000341488 | 0.948312348 |
| T2DM | mast_cell | *DMTF1* | 2.857357069 | 5.24E-05 | 0.000136633 | 0.254472982 |
| T2DM | mast_cell | *POM121C* | 2.106943843 | 0.00219019 | 0.002985057 | 1 |
| T2DM | mast_cell | *SRPK2* | 1.977901754 | 4.76E-05 | 0.000134478 | 0.230867435 |
| T2DM | mast_cell | *TES* | 1.89518519 | 0.000191115 | 0.000335246 | 0.927289789 |
| T2DM | mast_cell | *MTO1* | 1.841511188 | 0.000175298 | 0.000313276 | 0.850543614 |
| T2DM | mast_cell | *KHDC1* | 1.21801327 | 0.00063505 | 0.000955431 | 1 |
| T2DM | mast_cell | *NT5E* | 2.516287099 | 2.54E-05 | 0.000134478 | 0.123289554 |
| T2DM | mast_cell | *AKIRIN2* | 3.409196935 | 2.07E-05 | 0.000134478 | 0.100354168 |
| T2DM | mast_cell | *MAP3K7* | 2.339037952 | 3.18E-05 | 0.000134478 | 0.154331981 |
| T2DM | mast_cell | *PRR5L* | 2.217611051 | 0.000100962 | 0.000204708 | 0.489865669 |
| T2DM | mast_cell | *PHF21A* | 2.00873339 | 3.21E-05 | 0.000134478 | 0.155964231 |
| T2DM | mast_cell | *CAPRIN1* | 2.276680375 | 2.55E-05 | 0.000134478 | 0.123772196 |
| T2DM | mast_cell | *ATP5G2* | 2.296714401 | 3.08E-05 | 0.000134478 | 0.149313672 |
| T2DM | mast_cell | *B4GALNT1* | -4.703243802 | 0.028094105 | 0.033549741 | 1 |
| T2DM | mast_cell | *FAIM2* | -1.915355687 | 7.10E-05 | 0.000161307 | 0.344324727 |
| T2DM | mast_cell | *PAN2* | 2.700267087 | 0.002880432 | 0.003835307 | 1 |
| T2DM | mast_cell | *MAP7* | 1.103759619 | 0.000395425 | 0.000623064 | 1 |
| T2DM | mast_cell | *NHSL1* | 2.39600151 | 3.83E-06 | 0.000134478 | 0.0185817 |
| T2DM | mast_cell | *AHI1* | 1.5903914 | 0.000239726 | 0.000405562 | 1 |
| T2DM | mast_cell | *SMPD2* | 0.970102888 | 0.030505385 | 0.036313083 | 1 |
| T2DM | mast_cell | *MICAL1* | 2.368207161 | 1.05E-06 | 9.21E-05 | 0.005116224 |
| T2DM | mast_cell | *CCT7* | 2.568258421 | 4.12E-05 | 0.000134478 | 0.200133198 |
| T2DM | mast_cell | *SMYD5* | 1.772458012 | 0.000884534 | 0.001294443 | 1 |
| T2DM | mast_cell | *USP15* | 1.750981079 | 3.15E-05 | 0.000134478 | 0.152808636 |
| T2DM | mast_cell | *MDM2* | 1.878646964 | 4.25E-05 | 0.000134478 | 0.206070377 |
| T2DM | mast_cell | *KLHL36* | 2.812677909 | 1.63E-05 | 0.000134478 | 0.07915948 |
| T2DM | mast_cell | *CCDC102A* | 1.787698352 | 0.000621147 | 0.000935674 | 1 |
| T2DM | mast_cell | *AGT* | 2.011088538 | 0.000662204 | 0.000992284 | 1 |
| T2DM | mast_cell | *KCNK1* | -1.429894927 | 0.000428209 | 0.000667847 | 1 |
| T2DM | mast_cell | *CAPN9* | 4.646188765 | 0.001128542 | 0.001617633 | 1 |
| T2DM | mast_cell | *NTPCR* | 2.062428538 | 3.49E-05 | 0.000134478 | 0.1693767 |
| T2DM | mast_cell | *PIGC* | 1.7828581 | 3.22E-05 | 0.000134478 | 0.156109483 |
| T2DM | mast_cell | *SP110* | 2.409460246 | 6.02E-05 | 0.000145946 | 0.292038526 |
| T2DM | mast_cell | *MRPL44* | 2.102932475 | 3.97E-05 | 0.000134478 | 0.19285215 |
| T2DM | mast_cell | *USP37* | 3.562158078 | 8.90E-05 | 0.000187353 | 0.431662267 |
| T2DM | mast_cell | *SLC19A3* | 1.279905171 | 2.97E-05 | 0.000134478 | 0.144233887 |
| T2DM | mast_cell | *TMBIM1* | 2.36705203 | 3.36E-05 | 0.000134478 | 0.162837188 |
| T2DM | mast_cell | *TSGA10* | 0.890202061 | 0.000336728 | 0.000543333 | 1 |
| T2DM | mast_cell | *GCC2* | 2.360303868 | 5.34E-05 | 0.000137618 | 0.258921851 |
| T2DM | mast_cell | *MRPS9* | 2.570911059 | 2.40E-05 | 0.000134478 | 0.116455109 |
| T2DM | mast_cell | *ANKRD36* | 2.297579851 | 2.97E-05 | 0.000134478 | 0.143948766 |
| T2DM | mast_cell | *EPC2* | 2.734712848 | 3.38E-05 | 0.000134478 | 0.164078015 |
| T2DM | mast_cell | *STAB2* | 1.510961929 | 4.57E-05 | 0.000134478 | 0.221581487 |
| T2DM | mast_cell | *APPL2* | 2.17098711 | 7.16E-05 | 0.000161829 | 0.347480307 |
| T2DM | mast_cell | *DRAM1* | 1.789282586 | 3.33E-05 | 0.000134478 | 0.161647474 |
| T2DM | mast_cell | *SLC41A2* | 2.099405574 | 2.27E-05 | 0.000134478 | 0.110128462 |

| | | | | | | |
|---|---|---|---|---|---|---|
| T2DM | mast_cell | *NEK3* | 1.766782328 | 0.000283668 | 0.000468946 | 1 |
| T2DM | mast_cell | *RNASEH2B* | 1.730105783 | 4.84E-05 | 0.000134478 | 0.234725291 |
| T2DM | mast_cell | *CKAP2* | 0.770587334 | 0.019895944 | 0.024127748 | 1 |
| T2DM | mast_cell | *TBC1D4* | 1.916007719 | 0.000158565 | 0.000289535 | 0.769357345 |
| T2DM | mast_cell | *SUCLA2* | 1.96043651 | 3.46E-05 | 0.000134478 | 0.167689837 |
| T2DM | mast_cell | *RCBTB1* | 1.071061555 | 0.004397935 | 0.00573007 | 1 |
| T2DM | mast_cell | *RCBTB2* | 1.88191951 | 3.47E-05 | 0.000134478 | 0.168293749 |
| T2DM | mast_cell | *SETDB2* | 2.385344701 | 4.04E-05 | 0.000134478 | 0.196059412 |
| T2DM | mast_cell | *SCRN1* | 2.024097438 | 6.22E-05 | 0.000149115 | 0.301808371 |
| T2DM | mast_cell | *CHST12* | 2.40156628 | 4.98E-05 | 0.000135363 | 0.241488434 |
| T2DM | mast_cell | *KDELR2* | 3.822463448 | 3.29E-05 | 0.000134478 | 0.159774437 |
| T2DM | mast_cell | *NUPL2* | 1.664731627 | 3.78E-05 | 0.000134478 | 0.183242929 |
| T2DM | mast_cell | *ZDHHC4* | -2.551111298 | 0.0090526 | 0.011346737 | 1 |
| T2DM | mast_cell | *AOAH* | 0.998414789 | 0.000133908 | 0.000252736 | 0.649720782 |
| T2DM | mast_cell | *TBRG4* | 1.977145957 | 8.68E-05 | 0.000184458 | 0.421302029 |
| T2DM | mast_cell | *MTHFS* | 2.009168029 | 0.000217021 | 0.000372607 | 1 |
| T2DM | mast_cell | *NMT1* | 2.229804129 | 5.47E-05 | 0.00013935 | 0.265322281 |
| T2DM | mast_cell | *BRIP1* | 1.681648595 | 0.000302511 | 0.000494686 | 1 |
| T2DM | mast_cell | *RTP4* | 2.116301038 | 0.019383255 | 0.023535424 | 1 |
| T2DM | mast_cell | *NDUFB5* | 4.426130641 | 0.000289395 | 0.000476466 | 1 |
| T2DM | mast_cell | *SCN2A* | 6.153319199 | 1.40E-05 | 0.000134478 | 0.067788788 |
| T2DM | mast_cell | *RPS6KC1* | 1.962297117 | 1.68E-05 | 0.000134478 | 0.081271364 |
| T2DM | mast_cell | *SMPD4* | 1.675047088 | 0.000226842 | 0.000387412 | 1 |
| T2DM | mast_cell | *CCDC115* | 1.681252747 | 0.000242078 | 0.000409397 | 1 |
| T2DM | mast_cell | *IMP4* | 2.83470044 | 9.57E-05 | 0.000196916 | 0.464102032 |
| T2DM | mast_cell | *NIPSNAP3A* | 2.285117964 | 0.004854138 | 0.006294035 | 1 |
| T2DM | mast_cell | *ODF2* | 3.30998484 | 6.40E-05 | 0.000151548 | 0.310672988 |
| T2DM | mast_cell | *TOR1B* | 1.960501443 | 6.65E-05 | 0.000154763 | 0.322836238 |
| T2DM | mast_cell | *SLC2A8* | 2.246319086 | 0.000169207 | 0.000304748 | 0.820990746 |
| T2DM | mast_cell | *ZNF189* | 1.780069559 | 4.14E-05 | 0.000134478 | 0.20110657 |
| T2DM | mast_cell | *STX17* | 2.020268335 | 3.80E-05 | 0.000134478 | 0.184158469 |
| T2DM | mast_cell | *ANP32B* | 2.803504618 | 0.000242507 | 0.00040998 | 1 |
| T2DM | mast_cell | *DERL1* | 2.231857208 | 7.05E-05 | 0.000160662 | 0.342048449 |
| T2DM | mast_cell | *NOV* | 2.831240753 | 5.38E-05 | 0.000138108 | 0.261162083 |
| T2DM | mast_cell | *POLR1E* | 1.306671821 | 0.001109316 | 0.001591955 | 1 |
| T2DM | mast_cell | *IL11RA* | 2.336672467 | 0.000737917 | 0.001096928 | 1 |
| T2DM | mast_cell | *APTX* | 2.741740148 | 0.000213919 | 0.000368324 | 1 |
| T2DM | mast_cell | *TMEM8B* | 1.761009749 | 1.86E-05 | 0.000134478 | 0.090077714 |
| T2DM | mast_cell | *DENND4C* | 2.259351942 | 7.84E-05 | 0.00017179 | 0.380514659 |
| T2DM | mast_cell | *GMPR* | -2.518208959 | 0.001196121 | 0.001703928 | 1 |
| T2DM | mast_cell | *FRS3* | 2.149485202 | 3.94E-05 | 0.000134478 | 0.190937128 |
| T2DM | mast_cell | *IRF4* | 1.756864399 | 2.41E-06 | 0.000128421 | 0.011686323 |
| T2DM | mast_cell | *LRRC1* | 1.414984536 | 2.54E-05 | 0.000134478 | 0.12312426 |
| T2DM | mast_cell | *RIPK1* | 2.354561566 | 5.06E-05 | 0.000136053 | 0.245711145 |
| T2DM | mast_cell | *TUBB2B* | 1.692912282 | 0.015990615 | 0.01959749 | 1 |
| T2DM | mast_cell | *TCF19* | 3.461090585 | 1.25E-09 | 6.08E-07 | 6.08E-06 |
| T2DM | mast_cell | *FLOT1* | 2.142215608 | 3.76E-06 | 0.000134478 | 0.018253925 |
| T2DM | mast_cell | *NRM* | 5.791651822 | 3.60E-05 | 0.000134478 | 0.174462468 |
| T2DM | mast_cell | *VARS2* | 0.973951983 | 0.002735958 | 0.003661023 | 1 |
| T2DM | mast_cell | *TLR2* | 2.318170881 | 8.83E-05 | 0.000186449 | 0.428646972 |

| | | | | | | |
|---|---|---|---|---|---|---|
| T2DM | mast_cell | *MYO7A* | 1.90328636 | 0.000179157 | 0.00031853 | 0.869269667 |
| T2DM | mast_cell | *NUMA1* | 2.889948132 | 1.41E-05 | 0.000134478 | 0.068489676 |
| T2DM | mast_cell | *RAB30* | 1.425858679 | 5.18E-05 | 0.000136402 | 0.251371719 |
| T2DM | mast_cell | *PRCP* | 2.240190515 | 1.90E-05 | 0.000134478 | 0.092022081 |
| T2DM | mast_cell | *NARS2* | 2.177645035 | 3.28E-05 | 0.000134478 | 0.159261811 |
| T2DM | mast_cell | *SULF1* | 1.97812021 | 4.77E-05 | 0.000134478 | 0.231679629 |
| T2DM | mast_cell | *TGS1* | 3.776316033 | 3.54E-05 | 0.000134478 | 0.171723984 |
| T2DM | mast_cell | *NEK1* | 2.251589396 | 1.63E-05 | 0.000134478 | 0.079055861 |
| T2DM | mast_cell | *TRPC6* | 1.566051213 | 4.71E-05 | 0.000134478 | 0.228302611 |
| T2DM | mast_cell | *SLC37A4* | 5.244100942 | 1.55E-07 | 3.14E-05 | 0.000753663 |
| T2DM | mast_cell | *POU2F3* | 2.432347338 | 0.003101304 | 0.004113594 | 1 |
| T2DM | mast_cell | *PPP2R1B* | 1.359437953 | 4.41E-05 | 0.000134478 | 0.214073785 |
| T2DM | mast_cell | *FDX1* | 1.656352484 | 3.26E-05 | 0.000134478 | 0.158018261 |
| T2DM | mast_cell | *FXYD6* | 1.20290253 | 0.002780213 | 0.003712684 | 1 |
| T2DM | mast_cell | *FXYD2* | 2.783529789 | 0.001183985 | 0.001688127 | 1 |
| T2DM | mast_cell | *MAP2K5* | 1.981073406 | 2.24E-05 | 0.000134478 | 0.108618756 |
| T2DM | mast_cell | *NUSAP1* | 6.648903284 | 3.60E-06 | 0.000134478 | 0.017447658 |
| T2DM | mast_cell | *NDUFAF1* | 1.519756538 | 0.000116443 | 0.000227345 | 0.564983305 |
| T2DM | mast_cell | *PARP6* | 1.90900223 | 3.62E-05 | 0.000134478 | 0.175819503 |
| T2DM | mast_cell | *ADAM10* | 3.117706455 | 2.98E-05 | 0.000134478 | 0.144532376 |
| T2DM | mast_cell | *ZNF280D* | 2.277811138 | 3.16E-05 | 0.000134478 | 0.153280182 |
| T2DM | mast_cell | *RSL24D1* | 1.744599802 | 8.05E-05 | 0.000175007 | 0.390617077 |
| T2DM | mast_cell | *SPTBN5* | 1.260687109 | 8.64E-07 | 8.73E-05 | 0.004192383 |
| T2DM | mast_cell | *SLC44A5* | 1.987978963 | 4.43E-05 | 0.000134478 | 0.215041091 |
| T2DM | mast_cell | *DBT* | 1.503969276 | 4.68E-05 | 0.000134478 | 0.227277661 |
| T2DM | mast_cell | *RTCA* | 4.393640781 | 0.000271922 | 0.000452151 | 1 |
| T2DM | mast_cell | *KHK* | -2.13303937 | 0.000105513 | 0.000211026 | 0.511948955 |
| T2DM | mast_cell | *ADCY3* | 2.435607289 | 1.26E-05 | 0.000134478 | 0.061232874 |
| T2DM | mast_cell | *DYNC2LI1* | 1.828102752 | 0.000536808 | 0.000817512 | 1 |
| T2DM | mast_cell | *LHCGR* | 2.288462202 | 0.019582106 | 0.023759034 | 1 |
| T2DM | mast_cell | *THUMPD2* | 1.572410265 | 3.75E-05 | 0.000134478 | 0.181916478 |
| T2DM | mast_cell | *ACTR2* | 2.58968447 | 3.11E-05 | 0.000134478 | 0.150772945 |
| T2DM | mast_cell | *ATRAID* | 4.241442095 | 8.74E-06 | 0.000134478 | 0.042414883 |
| T2DM | mast_cell | *CENPO* | 2.409079674 | 1.60E-05 | 0.000134478 | 0.077777002 |
| T2DM | mast_cell | *LRPPRC* | 2.574158792 | 3.55E-05 | 0.000134478 | 0.172076582 |
| T2DM | mast_cell | *DTNB* | 1.820370287 | 3.35E-05 | 0.000134478 | 0.162784 |
| T2DM | mast_cell | *MYOF* | 1.709432489 | 5.06E-07 | 6.81E-05 | 0.002453216 |
| T2DM | mast_cell | *BTBD16* | 2.240388181 | 0.001546197 | 0.002163874 | 1 |
| T2DM | mast_cell | *TACC2* | 2.211943307 | 2.63E-05 | 0.000134478 | 0.127570229 |
| T2DM | mast_cell | *CALHM2* | 1.858904532 | 7.39E-05 | 0.000165254 | 0.358750005 |
| T2DM | mast_cell | *ARL3* | 2.327947454 | 1.88E-05 | 0.000134478 | 0.091230134 |
| T2DM | mast_cell | *PLCE1* | 1.267315868 | 7.36E-05 | 0.000164696 | 0.356895434 |
| T2DM | mast_cell | *RBP4* | 2.426655431 | 2.14E-05 | 0.000134478 | 0.104026582 |
| T2DM | mast_cell | *ASCC1* | 2.219110239 | 0.005247043 | 0.006760131 | 1 |
| T2DM | mast_cell | *RPS24* | 2.950171148 | 7.40E-05 | 0.000165359 | 0.359202727 |
| T2DM | mast_cell | *SMARCAL1* | 2.647052878 | 3.48E-05 | 0.000134478 | 0.168956504 |
| T2DM | mast_cell | *BARD1* | 2.031269781 | 7.45E-05 | 0.000165912 | 0.361688895 |
| T2DM | mast_cell | *STAT4* | 1.630004397 | 4.21E-05 | 0.000134478 | 0.204172882 |
| T2DM | mast_cell | *SSB* | 2.71164075 | 0.001596421 | 0.002229659 | 1 |
| T2DM | mast_cell | *FASTKD1* | 1.938163721 | 2.60E-05 | 0.000134478 | 0.126087281 |

| | | | | | | |
|---|---|---|---|---|---|---|
| T2DM | mast_cell | *ITGAV* | 4.078544856 | 0.000178817 | 0.000318043 | 0.867621519 |
| T2DM | mast_cell | *SLC35A5* | 4.14172641 | 0.000206163 | 0.000357634 | 1 |
| T2DM | mast_cell | *SENP7* | 2.585756485 | 2.76E-05 | 0.000134478 | 0.133804611 |
| T2DM | mast_cell | *COX17* | 1.857929612 | 4.10E-05 | 0.000134478 | 0.198732505 |
| T2DM | mast_cell | *SPPL2A* | 2.301607649 | 0.0016853 | 0.00234166 | 1 |
| T2DM | mast_cell | *PPCDC* | 1.670740564 | 7.97E-05 | 0.000174007 | 0.386643449 |
| T2DM | mast_cell | *SEMA7A* | -0.55227341 | 0.000131473 | 0.000249102 | 0.637906129 |
| T2DM | mast_cell | *AP1AR* | 5.47188291 | 0.000378896 | 0.000602123 | 1 |
| T2DM | mast_cell | *PRKG2* | -0.29095341 | 0.000215863 | 0.000371144 | 1 |
| T2DM | mast_cell | *KIAA1109* | 2.155914363 | 3.25E-05 | 0.000134478 | 0.157891212 |
| T2DM | mast_cell | *BMPR1B* | 1.311694596 | 8.49E-05 | 0.000181573 | 0.411808097 |
| T2DM | mast_cell | *MMRN1* | 0.858924783 | 0.001569686 | 0.002194213 | 1 |
| T2DM | mast_cell | *PRDM5* | 1.762825285 | 3.56E-05 | 0.000134478 | 0.172830617 |
| T2DM | mast_cell | *TRPC3* | 4.84024783 | 4.61E-07 | 6.78E-05 | 0.002236397 |
| T2DM | mast_cell | *NAAA* | 3.306719867 | 1.21E-06 | 9.40E-05 | 0.005886841 |
| T2DM | mast_cell | *NUP54* | 2.119947767 | 6.16E-06 | 0.000134478 | 0.029885218 |
| T2DM | mast_cell | *CNOT6L* | 2.317858833 | 0.000143045 | 0.000265514 | 0.694052393 |
| T2DM | mast_cell | *ANXA3* | 2.957329235 | 3.66E-06 | 0.000134478 | 0.017745046 |
| T2DM | mast_cell | *PPA2* | 3.307953479 | 0.000359384 | 0.000575679 | 1 |
| T2DM | mast_cell | *ENPEP* | 1.106671716 | 0.00039074 | 0.000616744 | 1 |
| T2DM | mast_cell | *PAPSS1* | 1.458436936 | 2.70E-05 | 0.000134478 | 0.131219066 |
| T2DM | mast_cell | *PARVG* | 1.755015416 | 0.000156972 | 0.000287081 | 0.761626884 |
| T2DM | mast_cell | *B4GALNT3* | 3.356612834 | 7.72E-05 | 0.000169753 | 0.374649487 |
| T2DM | mast_cell | *PDE6H* | 6.876017723 | 0.00757089 | 0.009568627 | 1 |
| T2DM | mast_cell | *ERP27* | -2.49465298 | 0.022306649 | 0.026916653 | 1 |
| T2DM | mast_cell | *KIF21A* | 1.385704551 | 3.88E-05 | 0.000134478 | 0.188484927 |
| T2DM | mast_cell | *ALG10* | 2.093582478 | 0.00046736 | 0.000723558 | 1 |
| T2DM | mast_cell | *ZCRB1* | 2.650308265 | 1.52E-05 | 0.000134478 | 0.073560916 |
| T2DM | mast_cell | *C1RL* | 1.285300604 | 2.11E-05 | 0.000134478 | 0.102205381 |
| T2DM | mast_cell | *CLSTN3* | 2.988527263 | 1.80E-05 | 0.000134478 | 0.087406667 |
| T2DM | mast_cell | *VAMP1* | 3.634464135 | 0.001011577 | 0.001463815 | 1 |
| T2DM | mast_cell | *TAPBPL* | 0.942438334 | 0.009375251 | 0.011739024 | 1 |
| T2DM | mast_cell | *PEX5* | -1.346775975 | 0.005867671 | 0.007509876 | 1 |
| T2DM | mast_cell | *PPFIA2* | 1.442718196 | 4.38E-05 | 0.000134478 | 0.212311956 |
| T2DM | mast_cell | *MARCH9* | 8.168358272 | 0.000671473 | 0.001005552 | 1 |
| T2DM | mast_cell | *LGR5* | -1.15823609 | 0.000517565 | 0.000791935 | 1 |
| T2DM | mast_cell | *POC1B* | 2.185106257 | 3.95E-05 | 0.000134478 | 0.19176956 |
| T2DM | mast_cell | *AMDHD1* | -1.166346444 | 0.003981145 | 0.005209416 | 1 |
| T2DM | mast_cell | *RITA1* | 2.71071434 | 4.73E-05 | 0.000134478 | 0.22934002 |
| T2DM | mast_cell | *FOXN4* | -1.613347209 | 0.001433454 | 0.002011894 | 1 |
| T2DM | mast_cell | *MTMR6* | 3.764125315 | 4.18E-05 | 0.000134478 | 0.202942792 |
| T2DM | mast_cell | *CCDC65* | 2.216652957 | 2.63E-05 | 0.000134478 | 0.127649559 |
| T2DM | mast_cell | *ACVRL1* | 6.594507618 | 0.000118216 | 0.000229066 | 0.573582406 |
| T2DM | mast_cell | *BRCA2* | 2.214028862 | 6.51E-05 | 0.000152815 | 0.315868103 |
| T2DM | mast_cell | *KANSL2* | 3.450348239 | 1.35E-05 | 0.000134478 | 0.065596667 |
| T2DM | mast_cell | *CERS5* | 3.812484137 | 0.00029418 | 0.000483524 | 1 |
| T2DM | mast_cell | *LMBR1L* | 3.244648642 | 5.93E-05 | 0.000144951 | 0.287916722 |
| T2DM | mast_cell | *ESYT1* | 3.72823911 | 3.56E-05 | 0.000134478 | 0.172757224 |
| T2DM | mast_cell | *ZNF740* | 3.472253296 | 0.001247397 | 0.001769699 | 1 |
| T2DM | mast_cell | *WDFY2* | 2.22742036 | 5.60E-05 | 0.000140825 | 0.271714621 |

| | | | | | | |
|---|---|---|---|---|---|---|
| T2DM | mast_cell | *HNRNPA1L2* | -4.161276099 | 0.000402566 | 0.000631507 | 1 |
| T2DM | mast_cell | *RB1* | 2.081626223 | 4.58E-05 | 0.000134478 | 0.222251339 |
| T2DM | mast_cell | *VPS33A* | 3.218519649 | 1.54E-05 | 0.000134478 | 0.07448531 |
| T2DM | mast_cell | *DIAPH3* | 1.730836241 | 8.69E-06 | 0.000134478 | 0.04217308 |
| T2DM | mast_cell | *GRTP1* | 1.342198431 | 0.002517921 | 0.003387012 | 1 |
| T2DM | mast_cell | *CBLN3* | 0.612893912 | 0.006070448 | 0.007759171 | 1 |
| T2DM | mast_cell | *NOVA1* | 1.18205095 | 6.92E-06 | 0.000134478 | 0.033588214 |
| T2DM | mast_cell | *FITM1* | 5.557135094 | 0.008805783 | 0.011048786 | 1 |
| T2DM | mast_cell | *TMX1* | 3.397697249 | 3.29E-05 | 0.000134478 | 0.159618817 |
| T2DM | mast_cell | *RTN1* | 1.705172508 | 1.66E-05 | 0.000134478 | 0.080470656 |
| T2DM | mast_cell | *C14orf37* | 2.677666294 | 3.86E-05 | 0.000134478 | 0.187359647 |
| T2DM | mast_cell | *RAB15* | 1.858127691 | 4.09E-05 | 0.000134478 | 0.198588942 |
| T2DM | mast_cell | *EFCAB11* | 2.53287574 | 5.43E-06 | 0.000134478 | 0.026362773 |
| T2DM | mast_cell | *SLC24A4* | 2.268734142 | 4.24E-05 | 0.000134478 | 0.205515971 |
| T2DM | mast_cell | *C14orf79* | 3.583002935 | 1.48E-05 | 0.000134478 | 0.071626847 |
| T2DM | mast_cell | *WARS* | 4.985671883 | 0.002626412 | 0.003521235 | 1 |
| T2DM | mast_cell | *SERF2* | 3.380157142 | 4.67E-05 | 0.000134478 | 0.2267621 |
| T2DM | mast_cell | *ZSCAN29* | 2.147446772 | 5.00E-06 | 0.000134478 | 0.024278634 |
| T2DM | mast_cell | *SRP14* | 2.999347969 | 3.20E-05 | 0.000134478 | 0.155344825 |
| T2DM | mast_cell | *UBE2Q2* | 2.77459048 | 1.37E-05 | 0.000134478 | 0.066352098 |
| T2DM | mast_cell | *HMG20A* | 2.920272585 | 1.53E-05 | 0.000134478 | 0.074313463 |
| T2DM | mast_cell | *SCAPER* | 1.627450216 | 0.000203343 | 0.000353374 | 0.986620496 |
| T2DM | mast_cell | *PIF1* | 2.137121887 | 0.001793868 | 0.002479729 | 1 |
| T2DM | mast_cell | *BBS4* | 2.211429622 | 1.42E-05 | 0.000134478 | 0.068924446 |
| T2DM | mast_cell | *ULK3* | 1.907920635 | 5.22E-05 | 0.000136624 | 0.253495234 |
| T2DM | mast_cell | *PCSK6* | 1.771745441 | 4.22E-05 | 0.000134478 | 0.204910622 |
| T2DM | mast_cell | *CELF6* | 12.11685141 | 0.000637833 | 0.00095932 | 1 |
| T2DM | mast_cell | *HAPLN3* | 1.751904593 | 0.002147842 | 0.002933107 | 1 |
| T2DM | mast_cell | *POLG* | 2.680368988 | 0.000117775 | 0.000228639 | 0.571444119 |
| T2DM | mast_cell | *ABHD2* | 4.305052913 | 2.33E-05 | 0.000134478 | 0.112833221 |
| T2DM | mast_cell | *MFGE8* | 4.463714905 | 0.000157367 | 0.000287696 | 0.763546487 |
| T2DM | mast_cell | *IQGAP1* | 1.486657902 | 4.81E-05 | 0.000134478 | 0.233413833 |
| T2DM | mast_cell | *CRTC3* | 2.609397395 | 2.74E-05 | 0.000134478 | 0.133125429 |
| T2DM | mast_cell | *ITGAX* | -1.289073997 | 0.000334613 | 0.0005401 | 1 |
| T2DM | mast_cell | *TGFB1I1* | 2.294874919 | 4.60E-05 | 0.000134478 | 0.223175535 |
| T2DM | mast_cell | *DHX38* | 2.182578297 | 3.46E-05 | 0.000134478 | 0.167719543 |
| T2DM | mast_cell | *NUDT7* | 1.727785121 | 5.88E-05 | 0.000144373 | 0.285161207 |
| T2DM | mast_cell | *GCSH* | 1.548385432 | 0.004561315 | 0.005930199 | 1 |
| T2DM | mast_cell | *CMTM3* | 5.517525858 | 0.000514163 | 0.000787722 | 1 |
| T2DM | mast_cell | *ZCCHC14* | 3.892977082 | 0.000279847 | 0.000463577 | 1 |
| T2DM | mast_cell | *TLDC1* | 2.646233241 | 3.50E-05 | 0.000134478 | 0.169654743 |
| T2DM | mast_cell | *RPL3L* | 7.051613141 | 6.20E-05 | 0.000148752 | 0.300628484 |
| T2DM | mast_cell | *TIGD7* | 1.650025983 | 1.84E-05 | 0.000134478 | 0.08929978 |
| T2DM | mast_cell | *DEF8* | 2.148916296 | 0.000309316 | 0.000503963 | 1 |
| T2DM | mast_cell | *TCF25* | 2.302457452 | 5.67E-05 | 0.000141687 | 0.275230508 |
| T2DM | mast_cell | *GALNS* | 1.450087901 | 0.000148937 | 0.000273936 | 0.722642151 |
| T2DM | mast_cell | *GAS8* | 2.781807196 | 0.000337779 | 0.000544667 | 1 |
| T2DM | mast_cell | *NCOR1* | 1.648341415 | 3.28E-05 | 0.000134478 | 0.15904141 |
| T2DM | mast_cell | *KSR1* | 3.893282629 | 8.32E-05 | 0.0001787 | 0.403504305 |
| T2DM | mast_cell | *GFOD2* | 2.172436427 | 0.008083838 | 0.010177162 | 1 |

| | | | | | | |
|---|---|---|---|---|---|---|
| T2DM | mast_cell | *PRPSAP2* | 2.903450285 | 0.013557844 | 0.016721571 | 1 |
| T2DM | mast_cell | *MYO19* | 6.281367839 | 1.03E-07 | 2.38E-05 | 0.000500691 |
| T2DM | mast_cell | *UNC45B* | -8.157346367 | 0.020766392 | 0.025133084 | 1 |
| T2DM | mast_cell | *C17orf80* | 3.370670972 | 0.0003585 | 0.000574452 | 1 |
| T2DM | mast_cell | *VPS53* | 1.369401972 | 0.03875254 | 0.045560292 | 1 |
| T2DM | mast_cell | *SGSM2* | 2.032631301 | 4.62E-05 | 0.000134478 | 0.224404531 |
| T2DM | mast_cell | *SSH2* | 1.946000305 | 3.23E-05 | 0.000134478 | 0.156547998 |
| T2DM | mast_cell | *RHBDL3* | -1.50051642 | 0.000284364 | 0.000469936 | 1 |
| T2DM | mast_cell | *G6PC3* | 2.197511158 | 0.000317126 | 0.00051513 | 1 |
| T2DM | mast_cell | *CLTC* | 1.938282853 | 1.16E-05 | 0.000134478 | 0.056399155 |
| T2DM | mast_cell | *IMPA2* | 3.278049614 | 1.15E-05 | 0.000134478 | 0.055871155 |
| T2DM | mast_cell | *GNAL* | 4.04818041 | 4.80E-05 | 0.000134478 | 0.232967132 |
| T2DM | mast_cell | *RPRD1A* | 2.08726021 | 8.46E-05 | 0.000181193 | 0.410583603 |
| T2DM | mast_cell | *GALNT1* | 2.776309841 | 4.73E-05 | 0.000134478 | 0.229403149 |
| T2DM | mast_cell | *ADCYAP1* | -1.847933206 | 0.003808542 | 0.004994337 | 1 |
| T2DM | mast_cell | *C18orf8* | 1.079230719 | 0.001733437 | 0.002403039 | 1 |
| T2DM | mast_cell | *NPC1* | 2.169944337 | 0.000255282 | 0.000428533 | 1 |
| T2DM | mast_cell | *SLC14A1* | 1.363464558 | 0.002407893 | 0.003254344 | 1 |
| T2DM | mast_cell | *CCDC40* | 1.4234679 | 3.88E-05 | 0.000134478 | 0.188421256 |
| T2DM | mast_cell | *ARHGDIA* | 2.055297345 | 0.005143638 | 0.006635717 | 1 |
| T2DM | mast_cell | *CARD14* | -2.598316044 | 0.015012505 | 0.018450019 | 1 |
| T2DM | mast_cell | *ANAPC11* | 3.287876688 | 3.66E-05 | 0.000134478 | 0.177384825 |
| T2DM | mast_cell | *TBCD* | 2.908123111 | 4.17E-05 | 0.000134478 | 0.202194694 |
| T2DM | mast_cell | *NARF* | 3.844869089 | 9.09E-05 | 0.000190496 | 0.441245483 |
| T2DM | mast_cell | *TRIM65* | 2.332876736 | 1.91E-05 | 0.000134478 | 0.092818722 |
| T2DM | mast_cell | *SECTM1* | 1.995690765 | 0.000128115 | 0.000244538 | 0.621616257 |
| T2DM | mast_cell | *RNF157* | 1.318877432 | 0.000421973 | 0.000658544 | 1 |
| T2DM | mast_cell | *ZNF750* | 9.223683328 | 7.66E-06 | 0.000134478 | 0.037182546 |
| T2DM | mast_cell | *WDR45B* | 3.142078569 | 0.000106179 | 0.000211747 | 0.515181436 |
| T2DM | mast_cell | *RNF165* | 2.463379134 | 7.50E-05 | 0.000166518 | 0.363676376 |
| T2DM | mast_cell | *MAPK4* | 4.226127266 | 4.55E-05 | 0.000134478 | 0.220766838 |
| T2DM | mast_cell | *FBXO15* | -3.371232914 | 0.004346974 | 0.005666717 | 1 |
| T2DM | mast_cell | *CBLN2* | 2.822913461 | 5.17E-05 | 0.000136402 | 0.250951268 |
| T2DM | mast_cell | *PIP4K2B* | 3.764080491 | 1.79E-06 | 0.000112139 | 0.008708693 |
| T2DM | mast_cell | *ERBB2* | 2.161913047 | 5.49E-05 | 0.000139632 | 0.266423004 |
| T2DM | mast_cell | *MIEN1* | 2.203866466 | 8.10E-05 | 0.000175806 | 0.392927131 |
| T2DM | mast_cell | *PNMT* | 4.99456687 | 0.00246032 | 0.00331873 | 1 |
| T2DM | mast_cell | *IGFBP4* | 1.929438193 | 3.17E-05 | 0.000134478 | 0.153630768 |
| T2DM | mast_cell | *TXNL4A* | 2.60254608 | 0.000338024 | 0.0005447 | 1 |
| T2DM | mast_cell | *CACNA1A* | 1.663867666 | 0.000237908 | 0.00040333 | 1 |
| T2DM | mast_cell | *SAMD1* | 7.517421194 | 9.10E-05 | 0.000190496 | 0.441378167 |
| T2DM | mast_cell | *NFIC* | 1.627451879 | 3.46E-05 | 0.000134478 | 0.167803866 |
| T2DM | mast_cell | *PFKL* | -2.741746655 | 0.019186998 | 0.023308792 | 1 |
| T2DM | mast_cell | *MVB12A* | 2.662665864 | 0.026093095 | 0.031252456 | 1 |
| T2DM | mast_cell | *DUS3L* | 2.217989106 | 1.79E-07 | 3.34E-05 | 0.000867946 |
| T2DM | mast_cell | *DPP9* | 1.637962603 | 4.45E-05 | 0.000134478 | 0.215815513 |
| T2DM | mast_cell | *SIRT3* | 2.463791686 | 3.61E-06 | 0.000134478 | 0.017501861 |
| T2DM | mast_cell | *IFNAR1* | 1.632478856 | 0.000244986 | 0.000413883 | 1 |
| T2DM | mast_cell | *TMEM50B* | 1.729530376 | 1.07E-05 | 0.000134478 | 0.051882028 |
| T2DM | mast_cell | *DOPEY2* | 1.960227094 | 9.11E-05 | 0.000190661 | 0.442142933 |

| | | | | | | |
|---|---|---|---|---|---|---|
| T2DM | mast_cell | *URB1* | 2.496040364 | 2.85E-05 | 0.000134478 | 0.138437514 |
| T2DM | mast_cell | *SAE1* | 1.647317556 | 0.000507027 | 0.000778264 | 1 |
| T2DM | mast_cell | *GEMIN7* | 1.655971029 | 0.027028352 | 0.032316798 | 1 |
| T2DM | mast_cell | *SLC6A3* | 2.11850171 | 0.00344191 | 0.004538083 | 1 |
| T2DM | mast_cell | *CAPN10* | 2.000725863 | 8.33E-05 | 0.00017893 | 0.404201811 |
| T2DM | mast_cell | *CACNG8* | 3.305535601 | 0.004956099 | 0.006415954 | 1 |
| T2DM | mast_cell | *ZNF787* | 4.662933222 | 0.000142196 | 0.000264342 | 0.689933725 |
| T2DM | mast_cell | *SLC47A1* | 2.76170961 | 5.60E-05 | 0.000140825 | 0.271925029 |
| T2DM | mast_cell | *CTU1* | -3.848214248 | 0.01058974 | 0.013201803 | 1 |
| T2DM | mast_cell | *RERE* | 1.919207999 | 2.26E-05 | 0.000134478 | 0.109712046 |
| T2DM | mast_cell | *EPHA2* | 2.143215809 | 7.16E-05 | 0.000161829 | 0.347250822 |
| T2DM | mast_cell | *ARHGEF19* | -1.007467643 | 9.52E-06 | 0.000134478 | 0.046194851 |
| T2DM | mast_cell | *PEX14* | 2.062094725 | 0.000250408 | 0.000421869 | 1 |
| T2DM | mast_cell | *ZNF593* | 3.243565629 | 0.000159053 | 0.000290121 | 0.771723106 |
| T2DM | mast_cell | *C1orf216* | 2.216560557 | 1.16E-05 | 0.000134478 | 0.056463219 |
| T2DM | mast_cell | *PLK4* | 3.086534062 | 6.04E-05 | 0.000146177 | 0.292939696 |
| T2DM | mast_cell | *HSPG2* | 1.242247806 | 0.002151086 | 0.002935059 | 1 |
| T2DM | mast_cell | *ITGB3BP* | 1.695489427 | 0.000254198 | 0.000427215 | 1 |
| T2DM | mast_cell | *PIGK* | 2.352091601 | 0.000150711 | 0.000276885 | 0.731252071 |
| T2DM | mast_cell | *RPS8* | 3.232674524 | 3.35E-05 | 0.000134478 | 0.162442607 |
| T2DM | mast_cell | *CYP4B1* | 1.72541626 | 9.98E-06 | 0.000134478 | 0.048405824 |
| T2DM | mast_cell | *SLC44A3* | 1.968954161 | 2.94E-05 | 0.000134478 | 0.142597394 |
| T2DM | mast_cell | *STRIP1* | 6.635992887 | 0.000456137 | 0.000707764 | 1 |
| T2DM | mast_cell | *GPR161* | 1.698306659 | 6.61E-05 | 0.000154234 | 0.320805838 |
| T2DM | mast_cell | *ALDH9A1* | 2.875227172 | 2.89E-05 | 0.000134478 | 0.140227436 |
| T2DM | mast_cell | *ATP1B1* | 1.292004727 | 4.91E-05 | 0.000135021 | 0.238041887 |
| T2DM | mast_cell | *NME7* | 2.169475006 | 0.00100205 | 0.001451761 | 1 |
| T2DM | mast_cell | *XCL1* | 4.760157289 | 0.002293835 | 0.003114071 | 1 |
| T2DM | mast_cell | *MAEL* | 4.499297924 | 4.96E-05 | 0.000135163 | 0.240541427 |
| T2DM | mast_cell | *RFWD2* | 2.800309945 | 3.71E-05 | 0.000134478 | 0.180011054 |
| T2DM | mast_cell | *RGS16* | -3.119454468 | 0.026281263 | 0.031462296 | 1 |
| T2DM | mast_cell | *TARS2* | 3.587884401 | 3.98E-06 | 0.000134478 | 0.019321938 |
| T2DM | mast_cell | *SNX27* | 2.373023314 | 5.98E-06 | 0.000134478 | 0.028996317 |
| T2DM | mast_cell | *SETDB1* | 2.279078892 | 4.96E-05 | 0.000135163 | 0.240775991 |
| T2DM | mast_cell | *SEMA6C* | 6.980168322 | 0.002745403 | 0.003670624 | 1 |
| T2DM | mast_cell | *ARNT* | 2.015732269 | 6.42E-05 | 0.000151618 | 0.311575256 |
| T2DM | mast_cell | *POGZ* | 1.967846215 | 2.26E-05 | 0.000134478 | 0.109414233 |
| T2DM | mast_cell | *GABPB2* | 1.762794286 | 7.58E-06 | 0.000134478 | 0.036761619 |
| T2DM | mast_cell | *DYRK3* | 2.265523051 | 4.14E-05 | 0.000134478 | 0.200787158 |
| T2DM | mast_cell | *EIF2D* | 1.950379365 | 6.37E-05 | 0.000151168 | 0.309290046 |
| T2DM | mast_cell | *TAF1A* | 3.12624812 | 0.000127562 | 0.000243708 | 0.618929939 |
| T2DM | mast_cell | *SMYD2* | 4.035491983 | 3.68E-05 | 0.000134478 | 0.178519953 |
| T2DM | mast_cell | *ADAM15* | 1.233673029 | 0.000214621 | 0.00036927 | 1 |
| T2DM | mast_cell | *RAB13* | 1.885874721 | 4.33E-05 | 0.000134478 | 0.210292976 |
| T2DM | mast_cell | *S100A8* | 1.617903032 | 4.43E-05 | 0.000134478 | 0.215013534 |
| T2DM | mast_cell | *CREB3L4* | 1.785282487 | 0.003071129 | 0.004075798 | 1 |
| T2DM | mast_cell | *KCNN3* | 1.242209549 | 9.31E-05 | 0.000193411 | 0.451950637 |
| T2DM | mast_cell | *C1orf43* | 2.471002077 | 5.62E-05 | 0.000140825 | 0.272841594 |
| T2DM | mast_cell | *RIT1* | 3.427568564 | 9.79E-05 | 0.000199936 | 0.47496633 |
| T2DM | mast_cell | *SCCPDH* | 2.582809145 | 3.60E-05 | 0.000134478 | 0.174461262 |

| | | | | | | |
|---|---|---|---|---|---|---|
| T2DM | mast_cell | *CEP170* | 2.587959463 | 3.26E-05 | 0.000134478 | 0.158148941 |
| T2DM | mast_cell | *ACP1* | 2.472725291 | 4.13E-05 | 0.000134478 | 0.200368728 |
| T2DM | mast_cell | *CNIH4* | 1.456471832 | 4.57E-05 | 0.000134478 | 0.221923724 |
| T2DM | mast_cell | *MBOAT2* | 2.518019187 | 3.30E-05 | 0.000134478 | 0.160331394 |
| T2DM | mast_cell | *PYCR2* | 2.124124128 | 4.80E-05 | 0.000134478 | 0.233110043 |
| T2DM | mast_cell | *PLEKHA6* | 1.759396641 | 6.72E-07 | 7.18E-05 | 0.003260914 |
| T2DM | mast_cell | *RHOB* | 1.609534012 | 4.09E-05 | 0.000134478 | 0.198252709 |
| T2DM | mast_cell | *GALM* | 1.237720497 | 4.59E-05 | 0.000134478 | 0.222718167 |
| T2DM | mast_cell | *WDPCP* | 1.242040029 | 6.47E-05 | 0.000152379 | 0.313747936 |
| T2DM | mast_cell | *VPS54* | 2.630852414 | 3.54E-05 | 0.000134478 | 0.171734214 |
| T2DM | mast_cell | *REG3G* | 8.070840507 | 0.015335095 | 0.018822636 | 1 |
| T2DM | mast_cell | *ABHD1* | 1.920194117 | 0.001356568 | 0.00191228 | 1 |
| T2DM | mast_cell | *CIAO1* | 4.430287194 | 0.003925414 | 0.00514065 | 1 |
| T2DM | mast_cell | *ZNF514* | 1.589407761 | 0.004807861 | 0.006237363 | 1 |
| T2DM | mast_cell | *EXOC6B* | 2.01694897 | 3.41E-05 | 0.000134478 | 0.165251838 |
| T2DM | mast_cell | *TEX261* | -1.945093682 | 0.000503696 | 0.000773886 | 1 |
| T2DM | mast_cell | *THNSL2* | 1.16379858 | 0.000476986 | 0.000737755 | 1 |
| T2DM | mast_cell | *RALB* | 4.715835036 | 6.68E-05 | 0.000155169 | 0.323982783 |
| T2DM | mast_cell | *FBLN7* | 1.494142062 | 0.000239601 | 0.000405492 | 1 |
| T2DM | mast_cell | *LYG1* | 1.681584094 | 0.001005922 | 0.001456501 | 1 |
| T2DM | mast_cell | *THSD7B* | 1.347986412 | 4.07E-05 | 0.000134478 | 0.197304411 |
| T2DM | mast_cell | *GPR17* | 0.732378496 | 0.009339582 | 0.011697381 | 1 |
| T2DM | mast_cell | *POLR2D* | 4.258950904 | 0.000227718 | 0.000388476 | 1 |
| T2DM | mast_cell | *GALNT13* | 1.26847218 | 5.03E-05 | 0.00013555 | 0.243989144 |
| T2DM | mast_cell | *PHOSPHO2* | 2.537160801 | 0.000185188 | 0.000326977 | 0.898533787 |
| T2DM | mast_cell | *METTL21A* | 2.051567584 | 2.12E-05 | 0.000134478 | 0.102799937 |
| T2DM | mast_cell | *NBEAL1* | 2.270145207 | 3.90E-06 | 0.000134478 | 0.018925007 |
| T2DM | mast_cell | *SUMF1* | 1.583136693 | 3.21E-05 | 0.000134478 | 0.155948527 |
| T2DM | mast_cell | *ESPNL* | 25.20859587 | 3.45E-05 | 0.000134478 | 0.167622702 |
| T2DM | mast_cell | *TAMM41* | 3.27960286 | 5.65E-06 | 0.000134478 | 0.02741226 |
| T2DM | mast_cell | *STK11IP* | 3.947739153 | 0.00532753 | 0.006856545 | 1 |
| T2DM | mast_cell | *EAF1* | 2.307445004 | 3.15E-05 | 0.000134478 | 0.152821888 |
| T2DM | mast_cell | *CNTN4* | 2.520046534 | 4.57E-05 | 0.000134478 | 0.221859873 |
| T2DM | mast_cell | *SLC25A38* | 2.716976831 | 0.000131321 | 0.000248992 | 0.63716982 |
| T2DM | mast_cell | *CTDSPL* | 1.352727956 | 0.000124317 | 0.000238507 | 0.603184958 |
| T2DM | mast_cell | *PTPRG* | 1.942646315 | 8.93E-05 | 0.000187874 | 0.433425993 |
| T2DM | mast_cell | *SLC25A26* | 1.403512363 | 6.55E-05 | 0.000153219 | 0.317622934 |
| T2DM | mast_cell | *ARL6IP5* | 2.019170719 | 3.31E-05 | 0.000134478 | 0.160806569 |
| T2DM | mast_cell | *LIMD1* | 2.137771199 | 2.32E-05 | 0.000134478 | 0.112330352 |
| T2DM | mast_cell | *PLA1A* | 4.582530045 | 0.001821298 | 0.002515496 | 1 |
| T2DM | mast_cell | *ADPRH* | 1.821761635 | 0.007042741 | 0.008938367 | 1 |
| T2DM | mast_cell | *IGSF11* | 1.490458479 | 0.000113644 | 0.000224055 | 0.55139974 |
| T2DM | mast_cell | *BOC* | 1.270279004 | 1.99E-05 | 0.000134478 | 0.096394095 |
| T2DM | mast_cell | *EIF2A* | 1.887850488 | 0.006893026 | 0.008757519 | 1 |
| T2DM | mast_cell | *NCEH1* | 1.076636015 | 0.000518896 | 0.000793721 | 1 |
| T2DM | mast_cell | *CCDC39* | 2.174871726 | 7.04E-06 | 0.000134478 | 0.034146467 |
| T2DM | mast_cell | *MUC4* | 2.621235249 | 0.000151736 | 0.000278556 | 0.736223894 |
| T2DM | mast_cell | *CENPC* | 1.594270053 | 0.00014478 | 0.000267405 | 0.702473428 |
| T2DM | mast_cell | *SCD5* | -2.289828686 | 0.00209418 | 0.00286305 | 1 |
| T2DM | mast_cell | *TRMT10A* | 2.532003167 | 0.000100282 | 0.000203585 | 0.486569565 |

| | | | | | | |
|---|---|---|---|---|---|---|
| T2DM | mast_cell | *PYURF* | 4.582898727 | 0.024465123 | 0.029433369 | 1 |
| T2DM | mast_cell | *CISD2* | 3.30085273 | 5.85E-06 | 0.000134478 | 0.028405997 |
| T2DM | mast_cell | *DDIT4L* | 5.549123423 | 1.81E-05 | 0.000134478 | 0.087626459 |
| T2DM | mast_cell | *METTL14* | 2.775490354 | 0.000109999 | 0.000218021 | 0.533714781 |
| T2DM | mast_cell | *USP53* | 1.875481356 | 5.81E-06 | 0.000134478 | 0.028166575 |
| T2DM | mast_cell | *MARCH1* | 0.849212538 | 0.000256432 | 0.000429778 | 1 |
| T2DM | mast_cell | *CBR4* | 1.84180965 | 4.05E-05 | 0.000134478 | 0.196649052 |
| T2DM | mast_cell | *CYP4V2* | 1.188748199 | 0.005804478 | 0.007434881 | 1 |
| T2DM | mast_cell | *MARCH6* | 2.805106731 | 3.46E-05 | 0.000134478 | 0.168010558 |
| T2DM | mast_cell | *MYO10* | 3.3137197 | 6.29E-05 | 0.000149953 | 0.305004279 |
| T2DM | mast_cell | *FAM105A* | 0.942588018 | 0.004188888 | 0.005469452 | 1 |
| T2DM | mast_cell | *BHMT* | -1.602973881 | 1.16E-05 | 0.000134478 | 0.056325836 |
| T2DM | mast_cell | *GTF2H2* | 2.269455322 | 4.61E-05 | 0.000134478 | 0.22362542 |
| T2DM | mast_cell | *SPATA9* | 3.177915113 | 3.70E-05 | 0.000134478 | 0.17961841 |
| T2DM | mast_cell | *FEM1C* | 2.476502655 | 3.46E-05 | 0.000134478 | 0.167692962 |
| T2DM | mast_cell | *COMMD10* | 1.740836907 | 0.000123306 | 0.000236942 | 0.598278638 |
| T2DM | mast_cell | *YIPF5* | 2.922550097 | 1.85E-05 | 0.000134478 | 0.089667115 |
| T2DM | mast_cell | *CXCL14* | 1.580292248 | 0.000404843 | 0.000634463 | 1 |
| T2DM | mast_cell | *FBXO38* | 2.901203269 | 3.00E-05 | 0.000134478 | 0.145675939 |
| T2DM | mast_cell | *PCYOX1L* | 7.770521288 | 0.000628174 | 0.000945673 | 1 |
| T2DM | mast_cell | *BOD1* | 2.806283862 | 4.13E-05 | 0.000134478 | 0.200155145 |
| T2DM | mast_cell | *FAM50B* | 4.973235659 | 1.57E-05 | 0.000134478 | 0.076347162 |
| T2DM | mast_cell | *MYLK4* | -4.169597847 | 0.002611059 | 0.003504525 | 1 |
| T2DM | mast_cell | *GFOD1* | 1.503449633 | 4.99E-05 | 0.000135379 | 0.242163318 |
| T2DM | mast_cell | *HIGD2A* | 1.642380263 | 7.48E-05 | 0.000166327 | 0.362925979 |
| T2DM | mast_cell | *FAM193B* | 4.296507849 | 0.001245104 | 0.001767997 | 1 |
| T2DM | mast_cell | *RNF44* | 2.466727974 | 6.15E-05 | 0.000147853 | 0.29822026 |
| T2DM | mast_cell | *MUT* | 3.524015427 | 0.000882449 | 0.001292376 | 1 |
| T2DM | mast_cell | *DAAM2* | 1.574998768 | 3.79E-05 | 0.000134478 | 0.183784084 |
| T2DM | mast_cell | *MLIP* | 3.996987808 | 0.002870207 | 0.003822741 | 1 |
| T2DM | mast_cell | *ANO7* | 1.651629825 | 0.029133035 | 0.034722055 | 1 |
| T2DM | mast_cell | *TCTE1* | -2.661115586 | 0.002098938 | 0.002868746 | 1 |
| T2DM | mast_cell | *RPL7L1* | 4.457518031 | 8.18E-05 | 0.000177121 | 0.397104705 |
| T2DM | mast_cell | *TPBG* | 1.302529366 | 6.55E-05 | 0.000153235 | 0.317808779 |
| T2DM | mast_cell | *IRAK1BP1* | 3.030743872 | 1.43E-06 | 9.89E-05 | 0.006924569 |
| T2DM | mast_cell | *PHIP* | 2.432450303 | 2.60E-05 | 0.000134478 | 0.126155697 |
| T2DM | mast_cell | *PNRC1* | 4.306060291 | 5.89E-05 | 0.000144373 | 0.285579037 |
| T2DM | mast_cell | *RNF217* | 2.038504663 | 3.79E-05 | 0.000134478 | 0.184000319 |
| T2DM | mast_cell | *ARHGAP18* | 1.981709556 | 1.58E-05 | 0.000134478 | 0.076524541 |
| T2DM | mast_cell | *ABRACL* | -1.410569813 | 0.012146335 | 0.015049545 | 1 |
| T2DM | mast_cell | *SLC18B1* | 1.200554079 | 0.000120718 | 0.000233077 | 0.585723273 |
| T2DM | mast_cell | *WTAP* | 2.789999501 | 0.000386238 | 0.000611628 | 1 |
| T2DM | mast_cell | *SLC22A3* | 1.309860783 | 4.67E-05 | 0.000134478 | 0.226451527 |
| T2DM | mast_cell | *GNA12* | 1.892662121 | 0.000378725 | 0.000602089 | 1 |
| T2DM | mast_cell | *SDK1* | 1.067914709 | 0.000396212 | 0.000623757 | 1 |
| T2DM | mast_cell | *CCZ1B* | 1.806155775 | 0.00200702 | 0.002751643 | 1 |
| T2DM | mast_cell | *RBAK* | -3.928130169 | 0.034418537 | 0.040681788 | 1 |
| T2DM | mast_cell | *PSPH* | 2.166060641 | 0.000191902 | 0.000336504 | 0.931107516 |
| T2DM | mast_cell | *GIGYF1* | 3.220291002 | 0.000109663 | 0.000217471 | 0.532084224 |
| T2DM | mast_cell | *TMEM209* | 2.224745673 | 3.49E-05 | 0.000134478 | 0.16918571 |

| | | | | | | |
|---|---|---|---|---|---|---|
| T2DM | mast_cell | *EPHA1* | 4.865503649 | 5.46E-05 | 0.000139259 | 0.264812494 |
| T2DM | mast_cell | *NCAPG2* | 5.212503391 | 2.96E-05 | 0.000134478 | 0.143553768 |
| T2DM | mast_cell | *FBXO25* | 1.594561063 | 3.52E-05 | 0.000134478 | 0.170984017 |
| T2DM | mast_cell | *CSGALNACT1* | 1.377025964 | 1.63E-05 | 0.000134478 | 0.07905839 |
| T2DM | mast_cell | *BIN3* | 2.780339988 | 2.75E-05 | 0.000134478 | 0.133400013 |
| T2DM | mast_cell | *CHMP7* | 1.869438166 | 3.08E-05 | 0.000134478 | 0.149410159 |
| T2DM | mast_cell | *DOCK5* | 1.786664721 | 3.78E-05 | 0.000134478 | 0.183507846 |
| T2DM | mast_cell | *TACC1* | 1.540627411 | 3.96E-05 | 0.000134478 | 0.192361827 |
| T2DM | mast_cell | *RPL7* | 2.767595828 | 2.92E-05 | 0.000134478 | 0.14172057 |
| T2DM | mast_cell | *ATP6V0D2* | -1.188666769 | 0.000193014 | 0.000337845 | 0.936506015 |
| T2DM | mast_cell | *DPYS* | 1.115872064 | 4.96E-05 | 0.000135163 | 0.240658832 |
| T2DM | mast_cell | *LRP12* | 3.405226967 | 3.37E-05 | 0.000134478 | 0.163513466 |
| T2DM | mast_cell | *MAL2* | 1.278796104 | 7.83E-05 | 0.000171612 | 0.379920721 |
| T2DM | mast_cell | *GSDMC* | -7.557259618 | 0.003051545 | 0.004052025 | 1 |
| T2DM | mast_cell | *AK3* | 1.784031146 | 3.32E-05 | 0.000134478 | 0.161046827 |
| T2DM | mast_cell | *HAUS6* | 1.42795828 | 4.58E-05 | 0.000134478 | 0.222104037 |
| T2DM | mast_cell | *C9orf72* | 1.247312659 | 0.001102413 | 0.001583805 | 1 |
| T2DM | mast_cell | *FBXO10* | -1.702592601 | 0.000864232 | 0.001269144 | 1 |
| T2DM | mast_cell | *ZNF462* | 1.740906676 | 1.34E-05 | 0.000134478 | 0.065085773 |
| T2DM | mast_cell | *ALAD* | 2.37197306 | 0.002307451 | 0.003130804 | 1 |
| T2DM | mast_cell | *WDR31* | 1.511531688 | 3.61E-05 | 0.000134478 | 0.175396862 |
| T2DM | mast_cell | *POLE3* | 2.244279377 | 0.002831229 | 0.003773934 | 1 |
| T2DM | mast_cell | *SURF1* | 1.259277455 | 0.002572484 | 0.003456574 | 1 |
| T2DM | mast_cell | *SURF6* | 2.325732455 | 4.35E-05 | 0.000134478 | 0.211242674 |
| T2DM | mast_cell | *MED22* | -3.046892883 | 0.001979492 | 0.002717741 | 1 |
| T2DM | mast_cell | *REXO4* | 2.975812996 | 3.10E-05 | 0.000134478 | 0.150195771 |
| T2DM | mast_cell | *SH3GLB2* | 1.97294503 | 3.18E-05 | 0.000134478 | 0.154324798 |
| T2DM | mast_cell | *INPP5E* | 2.058115478 | 8.03E-05 | 0.000174788 | 0.38960258 |
| T2DM | mast_cell | *SEC16A* | 1.860257496 | 9.98E-06 | 0.000134478 | 0.048440182 |
| T2DM | mast_cell | *SLC39A12* | 4.569543297 | 3.13E-05 | 0.000134478 | 0.151979909 |
| T2DM | mast_cell | *RSU1* | 3.056597114 | 4.16E-05 | 0.000134478 | 0.202014272 |
| T2DM | mast_cell | *PARD3* | 1.370537771 | 3.61E-05 | 0.000134478 | 0.175029662 |
| T2DM | mast_cell | *FRA10AC1* | 3.601100455 | 0.000174831 | 0.000312672 | 0.848279235 |
| T2DM | mast_cell | *ADD3* | 2.230321017 | 3.08E-05 | 0.000134478 | 0.14944494 |
| T2DM | mast_cell | *LRRC27* | 1.401065855 | 0.000114871 | 0.000225103 | 0.557355563 |
| T2DM | mast_cell | *PAOX* | 4.819398447 | 3.61E-05 | 0.000134478 | 0.175326418 |
| T2DM | mast_cell | *PDCD11* | 4.092691875 | 0.006138851 | 0.007840406 | 1 |
| T2DM | mast_cell | *RGS10* | 3.497167702 | 3.35E-05 | 0.000134478 | 0.162776614 |
| T2DM | mast_cell | *GAS2* | 6.541081966 | 0.000161378 | 0.00029348 | 0.783005061 |
| T2DM | mast_cell | *LIN7C* | 3.564143259 | 2.56E-05 | 0.000134478 | 0.124118088 |
| T2DM | mast_cell | *ZNF215* | -0.652987326 | 0.032172446 | 0.038166432 | 1 |
| T2DM | mast_cell | *EIF3M* | 1.86928899 | 0.000481269 | 0.000743432 | 1 |
| T2DM | mast_cell | *TNKS1BP1* | 2.102540476 | 2.48E-06 | 0.000128766 | 0.012027665 |
| T2DM | mast_cell | *PTPRJ* | 1.659468988 | 2.87E-05 | 0.000134478 | 0.13903404 |
| T2DM | mast_cell | *ENDOD1* | 2.260528189 | 2.92E-05 | 0.000134478 | 0.141546439 |
| T2DM | mast_cell | *CCDC82* | 0.984951642 | 0.020545889 | 0.024884836 | 1 |
| T2DM | mast_cell | *CAPN5* | 3.007210236 | 4.55E-05 | 0.000134478 | 0.220983149 |
| T2DM | mast_cell | *INTS4* | 2.693077329 | 1.72E-05 | 0.000134478 | 0.083264754 |
| T2DM | mast_cell | *RPS3* | 1.829662048 | 3.67E-05 | 0.000134478 | 0.178141159 |
| T2DM | mast_cell | *TTC12* | 1.945498895 | 7.97E-06 | 0.000134478 | 0.038694364 |

| | | | | | | |
|---|---|---|---|---|---|---|
| T2DM | mast_cell | *DRD2* | -1.38633078 | 0.000129385 | 0.000246187 | 0.627777003 |
| T2DM | mast_cell | *NPAT* | 3.878024587 | 5.14E-05 | 0.000136328 | 0.24957587 |
| T2DM | mast_cell | *ATM* | 1.559542509 | 4.32E-05 | 0.000134478 | 0.209823063 |
| T2DM | mast_cell | *SLX4IP* | 2.029058725 | 0.000220099 | 0.000376957 | 1 |
| T2DM | mast_cell | *ST14* | -3.075625911 | 0.002079428 | 0.002844484 | 1 |
| T2DM | mast_cell | *FADS1* | 4.666615351 | 0.000386499 | 0.000611841 | 1 |
| T2DM | mast_cell | *TMC2* | 3.158269199 | 0.000117854 | 0.000228639 | 0.571826925 |
| T2DM | mast_cell | *EML3* | 1.956882948 | 9.45E-06 | 0.000134478 | 0.045850563 |
| T2DM | mast_cell | *INCENP* | -2.324767932 | 0.000173181 | 0.000310178 | 0.840272412 |
| T2DM | mast_cell | *PLCH2* | 7.591965625 | 0.000619842 | 0.000933999 | 1 |
| T2DM | mast_cell | *CPSF7* | 1.656459167 | 6.83E-05 | 0.000157548 | 0.331415032 |
| T2DM | mast_cell | *CCDC15* | 1.200735079 | 0.008489206 | 0.010668124 | 1 |
| T2DM | mast_cell | *CHEK1* | 3.24251191 | 4.75E-05 | 0.000134478 | 0.230405918 |
| T2DM | mast_cell | *DUSP15* | 5.998221782 | 0.002126724 | 0.002905085 | 1 |
| T2DM | mast_cell | *CABLES2* | -0.976439241 | 0.00021387 | 0.000368324 | 1 |
| T2DM | mast_cell | *TRPT1* | 1.621432147 | 0.000734438 | 0.001092426 | 1 |
| T2DM | mast_cell | *NUDT22* | 1.482643142 | 0.000187849 | 0.000330592 | 0.911441598 |
| T2DM | mast_cell | *TBX6* | 1.166348935 | 0.001633514 | 0.002274917 | 1 |
| T2DM | mast_cell | *ALDOA* | 2.499689559 | 0.000148796 | 0.000273781 | 0.721959841 |
| T2DM | mast_cell | *ANXA8L1* | 0.858755613 | 0.00438149 | 0.005710178 | 1 |
| T2DM | mast_cell | *CWC15* | 1.431751335 | 8.79E-05 | 0.000186018 | 0.426442329 |
| T2DM | mast_cell | *TMEM218* | 1.549957311 | 9.60E-05 | 0.000197267 | 0.46594366 |
| T2DM | mast_cell | *SAP18* | 3.858336861 | 3.14E-05 | 0.000134478 | 0.152363638 |
| T2DM | mast_cell | *FAM124A* | 3.763072024 | 1.91E-05 | 0.000134478 | 0.092673467 |
| T2DM | mast_cell | *LYPD6B* | 2.615104109 | 0.000797604 | 0.001177717 | 1 |
| T2DM | mast_cell | *WDR17* | 2.786712195 | 0.001630962 | 0.002272667 | 1 |
| T2DM | mast_cell | *CCDC102B* | 1.238915561 | 0.000102156 | 0.000206134 | 0.495661352 |
| T2DM | mast_cell | *RGS18* | 1.743320771 | 0.000364238 | 0.000582492 | 1 |
| T2DM | mast_cell | *PRSS23* | 1.928040745 | 4.48E-05 | 0.000134478 | 0.217148042 |
| T2DM | mast_cell | *PPP1R1C* | 7.473653531 | 4.16E-05 | 0.000134478 | 0.201971727 |
| T2DM | mast_cell | *FAM173B* | 2.560645912 | 5.61E-05 | 0.000140825 | 0.27240543 |
| T2DM | mast_cell | *GPR158* | 5.548793393 | 3.36E-05 | 0.000134478 | 0.16299055 |
| T2DM | mast_cell | *UBE3B* | 1.687525527 | 0.000299928 | 0.00049081 | 1 |
| T2DM | mast_cell | *ANK3* | 1.445665773 | 1.95E-05 | 0.000134478 | 0.094785453 |
| T2DM | mast_cell | *PLBD2* | 2.212317827 | 2.75E-05 | 0.000134478 | 0.133469215 |
| T2DM | mast_cell | *DLG5* | 1.476688678 | 0.001269243 | 0.001798063 | 1 |
| T2DM | mast_cell | *GXYLT1* | 2.475098397 | 7.46E-05 | 0.000166011 | 0.362071031 |
| T2DM | mast_cell | *EXT2* | 2.297997014 | 0.000498626 | 0.000767518 | 1 |
| T2DM | mast_cell | *TMEM18* | 5.12938564 | 4.12E-05 | 0.000134478 | 0.199748713 |
| T2DM | mast_cell | *NDUFC2* | 2.757650387 | 3.20E-05 | 0.000134478 | 0.155177856 |
| T2DM | mast_cell | *ADAMTS12* | 1.303066527 | 4.39E-05 | 0.000134478 | 0.213044478 |
| T2DM | mast_cell | *NUBPL* | 1.558380381 | 3.49E-05 | 0.000134478 | 0.169548668 |
| T2DM | mast_cell | *VIPAS39* | 3.088541092 | 0.000190864 | 0.000334926 | 0.926071402 |
| T2DM | mast_cell | *ANKRD50* | 2.426608855 | 1.87E-05 | 0.000134478 | 0.090589729 |
| T2DM | mast_cell | *UPF2* | 3.591680215 | 0.000649135 | 0.000974506 | 1 |
| T2DM | mast_cell | *C4orf33* | 2.228607817 | 0.000113527 | 0.000223916 | 0.55083422 |
| T2DM | mast_cell | *PTPRO* | 1.411605172 | 0.000178234 | 0.000317238 | 0.864790871 |
| T2DM | mast_cell | *EPS8* | 1.501891384 | 2.97E-05 | 0.000134478 | 0.144225076 |
| T2DM | mast_cell | *QDPR* | 2.122804164 | 6.85E-05 | 0.000157661 | 0.332349686 |
| T2DM | mast_cell | *FAM160B1* | 1.793898833 | 2.61E-05 | 0.000134478 | 0.126450556 |

| | | | | | | |
|---|---|---|---|---|---|---|
| T2DM | mast_cell | *ZNF827* | 1.858402586 | 3.10E-05 | 0.000134478 | 0.150463929 |
| T2DM | mast_cell | *AKR1C2* | 1.802824263 | 1.77E-05 | 0.000134478 | 0.085960125 |
| T2DM | mast_cell | *PIGF* | 2.456840089 | 3.26E-05 | 0.000134478 | 0.15815772 |
| T2DM | mast_cell | *INPP1* | 2.250045501 | 1.21E-05 | 0.000134478 | 0.058489277 |
| T2DM | mast_cell | *MFSD6* | 2.677108005 | 3.61E-05 | 0.000134478 | 0.175211671 |
| T2DM | mast_cell | *RNF144A* | 1.755598497 | 3.22E-05 | 0.000134478 | 0.156410996 |
| T2DM | mast_cell | *CENPU* | 3.024149909 | 0.002516071 | 0.003385462 | 1 |
| T2DM | mast_cell | *BICD1* | 2.311614376 | 3.04E-05 | 0.000134478 | 0.147640711 |
| T2DM | mast_cell | *CCDC122* | 1.201821985 | 0.020822489 | 0.025188411 | 1 |
| T2DM | mast_cell | *GUF1* | 1.568464418 | 7.09E-05 | 0.000161268 | 0.343967853 |
| T2DM | mast_cell | *GABRA2* | -3.666600107 | 0.000218195 | 0.000374225 | 1 |
| T2DM | mast_cell | *PARP8* | 0.887564906 | 0.000146816 | 0.000270649 | 0.712348921 |
| T2DM | mast_cell | *CACUL1* | 1.961505754 | 4.44E-05 | 0.000134478 | 0.215366942 |
| T2DM | mast_cell | *BEND6* | 1.721490421 | 0.003459773 | 0.004560396 | 1 |
| T2DM | mast_cell | *TIAL1* | 2.081411016 | 4.05E-05 | 0.000134478 | 0.196345836 |
| T2DM | mast_cell | *TMEM56* | 3.291335032 | 4.59E-05 | 0.000134478 | 0.222928916 |
| T2DM | mast_cell | *PTPN14* | 1.065174433 | 2.95E-05 | 0.000134478 | 0.143306984 |
| T2DM | mast_cell | *MGAT5* | 2.066278698 | 0.000117472 | 0.000228423 | 0.569975694 |
| T2DM | mast_cell | *EPG5* | 2.553138517 | 6.34E-05 | 0.000150843 | 0.307711088 |
| T2DM | mast_cell | *HAUS1* | 1.176370701 | 0.033383911 | 0.039516647 | 1 |
| T2DM | mast_cell | *SPC25* | 0.846821171 | 0.017201649 | 0.021002115 | 1 |
| T2DM | mast_cell | *KCNK13* | 2.809225349 | 2.57E-05 | 0.000134478 | 0.12491648 |
| T2DM | mast_cell | *ATG10* | 1.115723775 | 0.001404511 | 0.001975271 | 1 |
| T2DM | mast_cell | *CWF19L2* | 2.433438676 | 4.23E-06 | 0.000134478 | 0.020517903 |
| T2DM | mast_cell | *NMT2* | 2.283063331 | 0.000247346 | 0.000417434 | 1 |
| T2DM | mast_cell | *ZNF837* | -1.839343189 | 0.022284887 | 0.026897082 | 1 |
| T2DM | mast_cell | *CCDC50* | 1.57959219 | 0.000443166 | 0.000688739 | 1 |
| T2DM | mast_cell | *PLEKHH2* | 1.644094879 | 8.43E-06 | 0.000134478 | 0.040913575 |
| T2DM | mast_cell | *SPARCL1* | 1.348732106 | 3.32E-05 | 0.000134478 | 0.161157653 |
| T2DM | mast_cell | *MBNL1* | 1.760638076 | 3.13E-05 | 0.000134478 | 0.151908226 |
| T2DM | mast_cell | *GPD1L* | 1.405491831 | 8.59E-05 | 0.00018324 | 0.416707441 |
| T2DM | mast_cell | *CCNO* | -3.712440606 | 0.000155892 | 0.000285322 | 0.756388136 |
| T2DM | mast_cell | *CLEC4F* | 3.87859143 | 2.28E-06 | 0.000126109 | 0.011042782 |
| T2DM | mast_cell | *SLC30A6* | 1.482177084 | 3.61E-05 | 0.000134478 | 0.175102469 |
| T2DM | mast_cell | *RASGRP3* | 1.256681086 | 0.000102223 | 0.000206134 | 0.495985889 |
| T2DM | mast_cell | *GPR180* | 1.620416482 | 0.001566642 | 0.00219059 | 1 |
| T2DM | mast_cell | *TCTEX1D1* | -0.538574036 | 0.000141135 | 0.00026338 | 0.684788522 |
| T2DM | mast_cell | *IFIT5* | 1.486235949 | 0.000530088 | 0.000808039 | 1 |
| T2DM | mast_cell | *PRDM8* | 2.621312696 | 0.000429637 | 0.000669858 | 1 |
| T2DM | mast_cell | *GGPS1* | 1.955443783 | 1.47E-05 | 0.000134478 | 0.071409346 |
| T2DM | mast_cell | *ZNF117* | 0.76039014 | 4.71E-05 | 0.000134478 | 0.228305282 |
| T2DM | mast_cell | *MED21* | 3.303327849 | 7.52E-05 | 0.000166869 | 0.365109709 |
| T2DM | mast_cell | *JAKMIP1* | 2.376843828 | 3.82E-05 | 0.000134478 | 0.185162847 |
| T2DM | mast_cell | *SREK1IP1* | 1.485467611 | 5.01E-05 | 0.000135379 | 0.243034577 |
| T2DM | mast_cell | *MR1* | 2.203283424 | 1.20E-05 | 0.000134478 | 0.058354867 |
| T2DM | mast_cell | *CENPH* | 3.737654481 | 0.000521049 | 0.000796512 | 1 |
| T2DM | mast_cell | *CARHSP1* | 1.544754814 | 9.08E-05 | 0.00019042 | 0.440635137 |
| T2DM | mast_cell | *CAST* | 2.302552651 | 6.76E-05 | 0.000156404 | 0.328020503 |
| T2DM | mast_cell | *SCOC* | 2.892108754 | 1.76E-05 | 0.000134478 | 0.085409583 |
| T2DM | mast_cell | *CLGN* | 4.264249295 | 0.010765486 | 0.013410561 | 1 |

| | | | | | | |
|---|---|---|---|---|---|---|
| T2DM | mast_cell | *CETN3* | 2.036801481 | 0.002268639 | 0.003081589 | 1 |
| T2DM | mast_cell | *SYCP2L* | -0.699581942 | 7.09E-05 | 0.000161268 | 0.343985698 |
| T2DM | mast_cell | *TMEM87B* | 2.059450064 | 3.41E-05 | 0.000134478 | 0.165564313 |
| T2DM | mast_cell | *PLA2R1* | 2.00641155 | 5.69E-07 | 7.18E-05 | 0.002761828 |
| T2DM | mast_cell | *SLC25A27* | 1.191084002 | 0.000550411 | 0.000836915 | 1 |
| T2DM | mast_cell | *FRMD1* | 2.57978135 | 0.000194775 | 0.000340557 | 0.945046865 |
| T2DM | mast_cell | *INO80C* | 3.01532043 | 6.29E-05 | 0.000150057 | 0.305366435 |
| T2DM | mast_cell | *LPCAT1* | 6.856302317 | 0.000898182 | 0.001311854 | 1 |
| T2DM | mast_cell | *NMRAL1* | 3.38753772 | 0.000263079 | 0.000439553 | 1 |
| T2DM | mast_cell | *ADPRHL1* | -3.159797507 | 0.016494443 | 0.020189464 | 1 |
| T2DM | mast_cell | *CFDP1* | 1.878678491 | 3.23E-05 | 0.000134478 | 0.156585321 |
| T2DM | mast_cell | *ZDHHC7* | 4.446599776 | 0.000144526 | 0.000267241 | 0.701241444 |
| T2DM | mast_cell | *FBXO36* | 3.436710298 | 3.91E-05 | 0.000134478 | 0.189896469 |
| T2DM | mast_cell | *MCOLN2* | 1.967660513 | 0.000116169 | 0.000227096 | 0.563651201 |
| T2DM | mast_cell | *GDPD1* | 1.594058208 | 5.89E-05 | 0.000144373 | 0.285713561 |
| T2DM | mast_cell | *SEMA3D* | 1.272107718 | 4.28E-05 | 0.000134478 | 0.207510355 |
| T2DM | mast_cell | *PPP2R5E* | 2.716580902 | 3.14E-05 | 0.000134478 | 0.152166378 |
| T2DM | mast_cell | *ANKRD29* | 1.294467166 | 4.28E-05 | 0.000134478 | 0.207782442 |
| T2DM | mast_cell | *DNAAF1* | 3.896584762 | 0.000409562 | 0.000640824 | 1 |
| T2DM | mast_cell | *ANKH* | 1.914371877 | 1.71E-05 | 0.000134478 | 0.082729406 |
| T2DM | mast_cell | *UBASH3B* | 1.284877773 | 0.000297016 | 0.000487359 | 1 |
| T2DM | mast_cell | *ROBO4* | 1.423033399 | 4.33E-05 | 0.000134478 | 0.210330336 |
| T2DM | mast_cell | *ROBO3* | 1.287120515 | 0.000110574 | 0.000218891 | 0.536502901 |
| T2DM | mast_cell | *CC2D1B* | 2.388274417 | 0.001373692 | 0.001935294 | 1 |
| T2DM | mast_cell | *LRRK1* | 1.538160723 | 5.33E-05 | 0.000137613 | 0.258575277 |
| T2DM | mast_cell | *ABCA5* | 2.304512423 | 3.61E-05 | 0.000134478 | 0.175101655 |
| T2DM | mast_cell | *ENPP3* | -0.639043578 | 0.000164261 | 0.00029783 | 0.796993799 |
| T2DM | mast_cell | *MIA3* | 2.297835457 | 2.67E-05 | 0.000134478 | 0.129329502 |
| T2DM | mast_cell | *PGM5* | 1.022880472 | 5.18E-05 | 0.000136402 | 0.251530768 |
| T2DM | mast_cell | *CCSAP* | 1.976595015 | 4.75E-05 | 0.000134478 | 0.23025784 |
| T2DM | mast_cell | *GBP5* | 1.125467796 | 0.0096708 | 0.012090369 | 1 |
| T2DM | mast_cell | *ATP5G3* | 2.282140474 | 1.87E-05 | 0.000134478 | 0.09049429 |
| T2DM | mast_cell | *LY96* | 1.298621881 | 0.000175454 | 0.000313439 | 0.851301164 |
| T2DM | mast_cell | *PDE1C* | 1.24142912 | 4.75E-05 | 0.000134478 | 0.23054683 |
| T2DM | mast_cell | *MRPL39* | 1.579117581 | 0.030808172 | 0.036646543 | 1 |
| T2DM | mast_cell | *XPC* | 2.422221587 | 2.07E-05 | 0.000134478 | 0.100484374 |
| T2DM | mast_cell | *FLCN* | 3.107856138 | 0.001189012 | 0.001694298 | 1 |
| T2DM | mast_cell | *OXNAD1* | 1.613018241 | 3.30E-05 | 0.000134478 | 0.15996799 |
| T2DM | mast_cell | *APCDD1* | 1.606084013 | 0.000293843 | 0.000483133 | 1 |
| T2DM | mast_cell | *MPPE1* | 1.363880061 | 0.005934957 | 0.007589987 | 1 |
| T2DM | mast_cell | *RAB6B* | 2.135614959 | 0.000464465 | 0.000719618 | 1 |
| T2DM | mast_cell | *ANKRD40* | 1.689265263 | 0.000117849 | 0.000228639 | 0.571802945 |
| T2DM | mast_cell | *CYP2U1* | 2.32787779 | 2.23E-05 | 0.000134478 | 0.108421615 |
| T2DM | mast_cell | *PROM2* | -3.56774208 | 0.000102425 | 0.000206295 | 0.496964056 |
| T2DM | mast_cell | *AK9* | 2.289923207 | 2.96E-05 | 0.000134478 | 0.143779269 |
| T2DM | mast_cell | *MARCKS* | 2.634907108 | 7.51E-05 | 0.000166733 | 0.364478495 |
| T2DM | mast_cell | *MARVELD1* | 2.196867267 | 4.30E-05 | 0.000134478 | 0.20878057 |
| T2DM | mast_cell | *GOLGA7B* | 10.59887454 | 3.09E-05 | 0.000134478 | 0.149899432 |
| T2DM | mast_cell | *TRMT44* | 2.009723105 | 4.79E-05 | 0.000134478 | 0.232604004 |
| T2DM | mast_cell | *MOV10* | 2.054287888 | 0.000146617 | 0.000270385 | 0.71138357 |

| | | | | | | |
|---|---|---|---|---|---|---|
| T2DM | mast_cell | *HEATR3* | 1.19740598 | 0.001327098 | 0.001871825 | 1 |
| T2DM | mast_cell | *CNOT8* | 2.286392075 | 4.05E-05 | 0.000134478 | 0.19670566 |
| T2DM | mast_cell | *LRGUK* | 1.331626287 | 0.00043826 | 0.000682864 | 1 |
| T2DM | mast_cell | *NUP205* | 4.153574889 | 0.000176164 | 0.000314348 | 0.8547489 |
| T2DM | mast_cell | *PDIA4* | 2.511990031 | 3.22E-05 | 0.000134478 | 0.156447321 |
| T2DM | mast_cell | *KDM8* | 2.268708323 | 4.01E-05 | 0.000134478 | 0.194595451 |
| T2DM | mast_cell | *ALS2CR12* | 2.258517438 | 1.77E-05 | 0.000134478 | 0.086059223 |
| T2DM | mast_cell | *FZD7* | 2.992875142 | 0.000205439 | 0.000356633 | 0.99678921 |
| T2DM | mast_cell | *DEPTOR* | 1.988332536 | 0.000133973 | 0.000252736 | 0.650035984 |
| T2DM | mast_cell | *RRAGA* | 2.127246892 | 2.55E-05 | 0.000134478 | 0.123652663 |
| T2DM | mast_cell | *TMBIM4* | 2.797686021 | 8.13E-05 | 0.000176186 | 0.394259997 |
| T2DM | mast_cell | *PSD3* | 2.181142697 | 2.02E-05 | 0.000134478 | 0.097912573 |
| T2DM | mast_cell | *MMP16* | 0.933842131 | 0.000927658 | 0.001350839 | 1 |
| T2DM | mast_cell | *KCNMA1* | 0.929374859 | 5.70E-05 | 0.000142008 | 0.276632497 |
| T2DM | mast_cell | *NDUFAF6* | 1.779602377 | 0.002787666 | 0.00371988 | 1 |
| T2DM | mast_cell | *DRAM2* | 1.258601865 | 8.54E-05 | 0.000182432 | 0.414302911 |
| T2DM | mast_cell | *PPEF2* | 4.885612573 | 0.000393859 | 0.000621262 | 1 |
| T2DM | mast_cell | *WHAMM* | 1.272829698 | 0.003211558 | 0.00425054 | 1 |
| T2DM | mast_cell | *N6AMT1* | 2.217318693 | 0.004450828 | 0.005795872 | 1 |
| T2DM | mast_cell | *RWDD2B* | 1.930874841 | 0.013777235 | 0.016969064 | 1 |
| T2DM | mast_cell | *CCT8* | 1.282968088 | 0.025162183 | 0.030182178 | 1 |
| T2DM | mast_cell | *NAA11* | 2.764686183 | 0.000287759 | 0.000474578 | 1 |
| T2DM | mast_cell | *SFXN2* | 1.51526716 | 1.22E-05 | 0.000134478 | 0.059026244 |
| T2DM | mast_cell | *C14orf2* | 2.048320813 | 0.000277451 | 0.00046008 | 1 |
| T2DM | mast_cell | *TDRD9* | 4.236409472 | 0.000206786 | 0.000358586 | 1 |
| T2DM | mast_cell | *RPL30* | 2.402220832 | 6.05E-05 | 0.000146324 | 0.293611357 |
| T2DM | mast_cell | *SUPV3L1* | 1.853367251 | 4.77E-05 | 0.000134478 | 0.2312583 |
| T2DM | mast_cell | *HKDC1* | 7.959503753 | 0.001695791 | 0.002354888 | 1 |
| T2DM | mast_cell | *TYSND1* | 1.365398564 | 0.000187724 | 0.000330492 | 0.910834683 |
| T2DM | mast_cell | *NODAL* | 2.74087918 | 4.93E-05 | 0.000135163 | 0.23907215 |
| T2DM | mast_cell | *ZDHHC5* | 5.174719849 | 0.000114729 | 0.000225007 | 0.556667411 |
| T2DM | mast_cell | *MED19* | 2.026072864 | 9.67E-05 | 0.000198277 | 0.469345136 |
| T2DM | mast_cell | *ATAD2* | 1.534378377 | 5.93E-06 | 0.000134478 | 0.028772712 |
| T2DM | mast_cell | *FBRS* | 1.732424957 | 9.44E-06 | 0.000134478 | 0.045809133 |
| T2DM | mast_cell | *SASS6* | 2.39857974 | 0.003845281 | 0.00503979 | 1 |
| T2DM | mast_cell | *ITGAD* | 2.086932298 | 0.003546069 | 0.004665273 | 1 |
| T2DM | mast_cell | *VPS8* | 2.577479553 | 2.62E-06 | 0.000133575 | 0.0126896 |
| T2DM | mast_cell | *MPV17L* | -2.345415299 | 0.001004539 | 0.001454932 | 1 |
| T2DM | mast_cell | *RPUSD3* | 4.454832942 | 0.000302603 | 0.000494686 | 1 |
| T2DM | mast_cell | *NTAN1* | 4.125110425 | 0.000130617 | 0.000247947 | 0.633752125 |
| T2DM | mast_cell | *NMNAT2* | 1.373602491 | 1.00E-04 | 0.000203167 | 0.485163198 |
| T2DM | mast_cell | *SLC6A1* | 6.015906991 | 3.79E-05 | 0.000134478 | 0.183811943 |
| T2DM | mast_cell | *FCHO2* | 1.853699897 | 1.37E-05 | 0.000134478 | 0.066293571 |
| T2DM | mast_cell | *TIMP4* | 2.239868393 | 6.10E-06 | 0.000134478 | 0.029575213 |
| T2DM | mast_cell | *NECAP2* | 2.111455249 | 3.88E-05 | 0.000134478 | 0.188177781 |
| T2DM | mast_cell | *STEAP2* | 1.19230322 | 6.03E-06 | 0.000134478 | 0.029254751 |
| T2DM | mast_cell | *CLDN12* | 1.857319054 | 4.70E-05 | 0.000134478 | 0.228108563 |
| T2DM | mast_cell | *MMP14* | 3.081572826 | 5.31E-05 | 0.000137292 | 0.257696657 |
| T2DM | mast_cell | *GATAD1* | 3.044382673 | 7.66E-05 | 0.000168736 | 0.37148682 |
| T2DM | mast_cell | *CLEC18A* | -4.683666118 | 7.68E-08 | 1.96E-05 | 0.000372717 |

| | | | | | | |
|---|---|---|---|---|---|---|
| T2DM | mast_cell | *DHRS4* | -2.624468091 | 0.001659336 | 0.002307566 | 1 |
| T2DM | mast_cell | *AASDH* | 2.841626831 | 9.88E-06 | 0.000134478 | 0.047924214 |
| T2DM | mast_cell | *ETS2* | 1.752481344 | 2.87E-06 | 0.000134478 | 0.013911781 |
| T2DM | mast_cell | *LCA5L* | 1.072355076 | 0.002379478 | 0.003220737 | 1 |
| T2DM | mast_cell | *MX1* | 1.15369158 | 0.000851247 | 0.001252351 | 1 |
| T2DM | mast_cell | *SLC38A10* | 2.129160915 | 3.34E-05 | 0.000134478 | 0.162276106 |
| T2DM | mast_cell | *C9orf43* | -1.369848506 | 0.000288453 | 0.000475238 | 1 |
| T2DM | mast_cell | *DGKI* | 1.234715537 | 0.002754254 | 0.003681444 | 1 |
| T2DM | mast_cell | *SLC34A2* | 34.19869868 | 0.000297351 | 0.000487745 | 1 |
| T2DM | mast_cell | *AP3S2* | 3.899112666 | 2.70E-06 | 0.000134478 | 0.013100924 |
| T2DM | mast_cell | *SPPL3* | 2.166905219 | 0.000134651 | 0.000253916 | 0.653326184 |
| T2DM | mast_cell | *C12orf43* | 1.740679706 | 0.002008236 | 0.002752531 | 1 |
| T2DM | mast_cell | *PEX10* | 2.060501578 | 0.000462927 | 0.00071761 | 1 |
| T2DM | mast_cell | *LDLRAP1* | -2.230080603 | 0.000970122 | 0.001408027 | 1 |
| T2DM | mast_cell | *KRTCAP3* | 2.91142003 | 2.36E-09 | 8.82E-07 | 1.15E-05 |
| T2DM | mast_cell | *EXTL1* | 27.80336333 | 5.10E-05 | 0.000136328 | 0.247627216 |
| T2DM | mast_cell | *WDR66* | 1.320181899 | 0.003531027 | 0.004648379 | 1 |
| T2DM | mast_cell | *LRRC43* | 3.559606527 | 0.000917345 | 0.001338231 | 1 |
| T2DM | mast_cell | *EYA3* | 1.790637081 | 1.73E-05 | 0.000134478 | 0.083733947 |
| T2DM | mast_cell | *DZIP1L* | 1.631153734 | 4.56E-05 | 0.000134478 | 0.221252968 |
| T2DM | mast_cell | *FANCC* | 1.758971791 | 3.83E-05 | 0.000134478 | 0.185835789 |
| T2DM | mast_cell | *ESYT3* | 3.062472347 | 0.001495781 | 0.00209513 | 1 |
| T2DM | mast_cell | *CLSTN2* | 3.155549768 | 0.006708086 | 0.008531489 | 1 |
| T2DM | mast_cell | *COLEC12* | 1.362742125 | 5.37E-05 | 0.000138027 | 0.260318975 |
| T2DM | mast_cell | *MITD1* | 2.013686134 | 5.31E-05 | 0.000137292 | 0.257672409 |
| T2DM | mast_cell | *TSPAN33* | 2.168994049 | 0.005283134 | 0.006801212 | 1 |
| T2DM | mast_cell | *FAM86C1* | -4.440183772 | 0.000821221 | 0.001210378 | 1 |
| T2DM | mast_cell | *PPP1R9A* | 1.336833618 | 3.86E-05 | 0.000134478 | 0.187456434 |
| T2DM | mast_cell | *DYNC1I1* | 1.249864635 | 3.59E-05 | 0.000134478 | 0.17423561 |
| T2DM | mast_cell | *COPG2* | 0.758205907 | 0.023402969 | 0.028204472 | 1 |
| T2DM | mast_cell | *DUSP23* | 1.909782538 | 0.000144526 | 0.000267241 | 0.701239686 |
| T2DM | mast_cell | *RNF166* | 1.458867462 | 0.000512501 | 0.000785672 | 1 |
| T2DM | mast_cell | *SPATA2L* | 2.617722057 | 0.00811187 | 0.010207156 | 1 |
| T2DM | mast_cell | *FGF17* | 8.842514735 | 0.000857835 | 0.001260514 | 1 |
| T2DM | mast_cell | *CCAR2* | 2.463575143 | 9.12E-06 | 0.000134478 | 0.044239261 |
| T2DM | mast_cell | *CDC42SE2* | 1.91994623 | 6.36E-05 | 0.000150963 | 0.308407698 |
| T2DM | mast_cell | *RAPGEF6* | 1.923858969 | 0.00201129 | 0.002755939 | 1 |
| T2DM | mast_cell | *IFNAR2* | 1.979780176 | 0.000173393 | 0.000310329 | 0.841303161 |
| T2DM | mast_cell | *SV2A* | -2.86375497 | 0.03495489 | 0.041285572 | 1 |
| T2DM | mast_cell | *CLIC6* | -0.321917878 | 0.000176067 | 0.000314303 | 0.854276063 |
| T2DM | mast_cell | *CBR1* | 2.34459841 | 3.16E-05 | 0.000134478 | 0.153117119 |
| T2DM | mast_cell | *CBR3* | 1.659476797 | 1.96E-05 | 0.000134478 | 0.095219149 |
| T2DM | mast_cell | *SIM2* | -1.433840288 | 3.27E-05 | 0.000134478 | 0.158662609 |
| T2DM | mast_cell | *HLCS* | 1.92092999 | 0.000229402 | 0.000390821 | 1 |
| T2DM | mast_cell | *ARHGAP27* | 3.681027867 | 7.45E-05 | 0.00016578 | 0.361235701 |
| T2DM | mast_cell | *IRX6* | 1.079589707 | 0.003521227 | 0.004637621 | 1 |
| T2DM | mast_cell | *HK2* | 1.293963401 | 3.35E-05 | 0.000134478 | 0.162735651 |
| T2DM | mast_cell | *THEM4* | 1.569876912 | 2.80E-05 | 0.000134478 | 0.135763005 |
| T2DM | mast_cell | *AMFR* | 2.397262633 | 5.87E-05 | 0.000144373 | 0.285039191 |
| T2DM | mast_cell | *TMEM69* | 2.380999644 | 5.94E-05 | 0.000144951 | 0.287967139 |

| | | | | | | |
|---|---|---|---|---|---|---|
| T2DM | mast_cell | *ACE* | 2.100467586 | 4.25E-05 | 0.000134478 | 0.206190602 |
| T2DM | mast_cell | *TEPP* | -1.749264114 | 0.000120577 | 0.000232897 | 0.585037935 |
| T2DM | mast_cell | *CHCHD6* | 2.133043136 | 2.39E-05 | 0.000134478 | 0.115859042 |
| T2DM | mast_cell | *CTBP1* | 3.483410588 | 0.000117008 | 0.000227819 | 0.567724494 |
| T2DM | mast_cell | *ZFYVE28* | 1.756063764 | 0.000281919 | 0.000466532 | 1 |
| T2DM | mast_cell | *C16orf86* | 3.016863992 | 0.012823291 | 0.015864 | 1 |
| T2DM | mast_cell | *RGS12* | 1.811683551 | 0.000204023 | 0.000354429 | 0.989920864 |
| T2DM | mast_cell | *LYPD5* | -1.194744773 | 8.36E-05 | 0.000179257 | 0.405479914 |
| T2DM | mast_cell | *CCDC107* | 1.4346871 | 9.78E-05 | 0.000199906 | 0.474377737 |
| T2DM | mast_cell | *ATAD3B* | 2.434756499 | 0.003775058 | 0.004952641 | 1 |
| T2DM | mast_cell | *SSU72* | 4.784635605 | 0.000235142 | 0.000399478 | 1 |
| T2DM | mast_cell | *CPAMD8* | -1.374064146 | 0.016858513 | 0.020603905 | 1 |
| T2DM | mast_cell | *CCDC58* | 2.690816729 | 3.64E-05 | 0.000134478 | 0.176486132 |
| T2DM | mast_cell | *SLC37A1* | 1.891434338 | 4.47E-05 | 0.000134478 | 0.216858109 |
| T2DM | mast_cell | *PDE9A* | 1.455618052 | 2.45E-05 | 0.000134478 | 0.119008677 |
| T2DM | mast_cell | *NDUFV3* | 0.593952441 | 0.040231159 | 0.047218574 | 1 |
| T2DM | mast_cell | *HSF2BP* | -3.400261596 | 0.001141817 | 0.001634246 | 1 |
| T2DM | mast_cell | *CSTB* | 1.627358246 | 0.010567246 | 0.013177147 | 1 |
| T2DM | mast_cell | *FTCD* | -1.673714631 | 1.21E-05 | 0.000134478 | 0.058859723 |
| T2DM | mast_cell | *LSS* | 0.62194108 | 0.025820061 | 0.03094071 | 1 |
| T2DM | mast_cell | *VAV2* | 2.297595244 | 0.02122587 | 0.025638018 | 1 |
| T2DM | mast_cell | *MCM3AP* | 1.615146661 | 0.000230892 | 0.000392808 | 1 |
| T2DM | mast_cell | *PCNT* | 1.785173543 | 4.40E-05 | 0.000134478 | 0.213649729 |
| T2DM | mast_cell | *DIP2A* | 1.862332978 | 0.008094366 | 0.010187773 | 1 |
| T2DM | mast_cell | *PRMT2* | 1.923165489 | 0.000397773 | 0.000625809 | 1 |
| T2DM | mast_cell | *ZNF208* | 1.797763014 | 0.000248549 | 0.000419028 | 1 |
| T2DM | mast_cell | *ADAMTS13* | 1.621225935 | 0.000949495 | 0.001380566 | 1 |
| T2DM | mast_cell | *CACFD1* | 4.056960233 | 0.000114871 | 0.000225103 | 0.557352099 |
| T2DM | mast_cell | *GPSM1* | 3.910626511 | 0.000261301 | 0.000437184 | 1 |
| T2DM | mast_cell | *TAOK1* | 2.337581927 | 2.13E-05 | 0.000134478 | 0.103157061 |
| T2DM | mast_cell | *S100A1* | 6.00394641 | 0.000367964 | 0.000587718 | 1 |
| T2DM | mast_cell | *ZBTB7B* | 2.342104718 | 1.93E-05 | 0.000134478 | 0.093757754 |
| T2DM | mast_cell | *VPS11* | 2.376441254 | 0.002623116 | 0.003517788 | 1 |
| T2DM | mast_cell | *FDPS* | 1.95245068 | 1.34E-05 | 0.000134478 | 0.06484569 |
| T2DM | mast_cell | *CCR5* | 3.83875972 | 6.80E-06 | 0.000134478 | 0.03299655 |
| T2DM | mast_cell | *NBEAL2* | 2.865497799 | 0.000160818 | 0.000292572 | 0.780289843 |
| T2DM | mast_cell | *PTH1R* | 1.29723446 | 0.000255335 | 0.000428533 | 1 |
| T2DM | mast_cell | *LY6K* | 2.312498266 | 0.004613832 | 0.005993659 | 1 |
| T2DM | mast_cell | *IER2* | 2.075309673 | 0.000116073 | 0.000226999 | 0.563184175 |
| T2DM | mast_cell | *CPSF4* | 1.922156325 | 0.000120479 | 0.0002328 | 0.584562054 |
| T2DM | mast_cell | *VPS28* | 3.956445229 | 6.45E-07 | 7.18E-05 | 0.003131252 |
| T2DM | mast_cell | *TONSL* | 3.943793927 | 9.75E-06 | 0.000134478 | 0.047293938 |
| T2DM | mast_cell | *MUM1* | 2.596578022 | 4.07E-05 | 0.000134478 | 0.197623778 |
| T2DM | mast_cell | *ZNF333* | -2.357478996 | 0.036032099 | 0.042495806 | 1 |
| T2DM | mast_cell | *SH2B2* | 2.461942766 | 0.000297881 | 0.000488283 | 1 |
| T2DM | mast_cell | *RPL8* | 1.903495893 | 7.55E-05 | 0.000167138 | 0.366533581 |
| T2DM | mast_cell | *LRWD1* | 2.457862356 | 0.000468331 | 0.000724831 | 1 |
| T2DM | mast_cell | *DVL3* | 2.146800037 | 0.025138833 | 0.030161627 | 1 |
| T2DM | mast_cell | *ABCF3* | 2.269979345 | 4.40E-05 | 0.000134478 | 0.213374883 |
| T2DM | mast_cell | *PCYT1A* | 1.153800828 | 0.001380996 | 0.00194462 | 1 |

| | | | | | | |
|---|---|---|---|---|---|---|
| T2DM | mast_cell | *FBXO27* | 1.209203953 | 0.000672798 | 0.001007225 | 1 |
| T2DM | mast_cell | *BDH1* | 2.201609881 | 5.15E-05 | 0.000136328 | 0.250025913 |
| T2DM | mast_cell | *LRRC56* | -1.423255473 | 0.000337934 | 0.0005447 | 1 |
| T2DM | mast_cell | *PGAP3* | 4.471506215 | 3.17E-05 | 0.000134478 | 0.153666884 |
| T2DM | mast_cell | *ZNF577* | 3.436536339 | 7.53E-05 | 0.000166984 | 0.365528164 |
| T2DM | mast_cell | *TMEM143* | 2.402595904 | 2.19E-05 | 0.000134478 | 0.10627611 |
| T2DM | mast_cell | *ASB16* | 2.586078056 | 1.96E-05 | 0.000134478 | 0.094906192 |
| T2DM | mast_cell | *DBF4B* | 0.901203168 | 0.005543308 | 0.007115378 | 1 |
| T2DM | mast_cell | *IP6K3* | 12.93744481 | 0.025070548 | 0.030094582 | 1 |
| T2DM | mast_cell | *LEMD2* | 3.26086147 | 5.42E-05 | 0.000138726 | 0.263047875 |
| T2DM | mast_cell | *ASGR2* | 13.14995216 | 2.65E-05 | 0.000134478 | 0.128578911 |
| T2DM | mast_cell | *POLR3K* | -2.853163163 | 0.000684465 | 0.001023113 | 1 |
| T2DM | mast_cell | *SNRNP25* | 3.478142665 | 1.25E-05 | 0.000134478 | 0.060697823 |
| T2DM | mast_cell | *WDR90* | 1.573863971 | 0.000750693 | 0.001113191 | 1 |
| T2DM | mast_cell | *JMJD8* | 2.018231997 | 0.001177486 | 0.001679848 | 1 |
| T2DM | mast_cell | *MEIOB* | 2.532241654 | 2.75E-05 | 0.000134478 | 0.133207927 |
| T2DM | mast_cell | *SHANK2* | 4.267514392 | 2.98E-05 | 0.000134478 | 0.144474535 |
| T2DM | mast_cell | *ASRGL1* | 2.087119506 | 4.34E-05 | 0.000134478 | 0.210688828 |
| T2DM | mast_cell | *TTC9C* | 2.043786588 | 0.000251778 | 0.000423735 | 1 |
| T2DM | mast_cell | *RPS6KA4* | 6.452957222 | 4.26E-05 | 0.000134478 | 0.206879206 |
| T2DM | mast_cell | *TPCN2* | 2.182169752 | 6.36E-05 | 0.000150963 | 0.308416628 |
| T2DM | mast_cell | *PDZK1IP1* | -2.393342305 | 0.000396136 | 0.000623757 | 1 |
| T2DM | mast_cell | *CMPK1* | 3.286803841 | 0.000143473 | 0.000266004 | 0.696132014 |
| T2DM | mast_cell | *COA7* | -3.972731997 | 0.00492334 | 0.006376948 | 1 |
| T2DM | mast_cell | *C1orf123* | 2.189502321 | 9.57E-05 | 0.000196916 | 0.46432897 |
| T2DM | mast_cell | *USP24* | 2.913993734 | 0.000121087 | 0.000233505 | 0.587512703 |
| T2DM | mast_cell | *ZSWIM5* | 2.720802222 | 4.72E-05 | 0.000134478 | 0.229240645 |
| T2DM | mast_cell | *AK4* | 1.274482739 | 0.000113794 | 0.00022426 | 0.552127096 |
| T2DM | mast_cell | *RBP7* | 1.447080691 | 2.63E-05 | 0.000134478 | 0.12746316 |
| T2DM | mast_cell | *SLC25A34* | 2.804822581 | 0.001049022 | 0.001514387 | 1 |
| T2DM | mast_cell | *PDPN* | 1.607619455 | 4.17E-05 | 0.000134478 | 0.202511074 |
| T2DM | mast_cell | *LAPTM5* | 2.33118816 | 3.21E-05 | 0.000134478 | 0.155724018 |
| T2DM | mast_cell | *SDC3* | 1.681072958 | 1.91E-05 | 0.000134478 | 0.092658237 |
| T2DM | mast_cell | *SYNC* | 2.994030436 | 0.000116749 | 0.000227587 | 0.566465021 |
| T2DM | mast_cell | *CAMK2N1* | 2.710504817 | 0.000121643 | 0.000234211 | 0.590211802 |
| T2DM | mast_cell | *MEGF6* | 2.455890913 | 0.000201493 | 0.000350788 | 0.977645884 |
| T2DM | mast_cell | *OMA1* | 1.975597369 | 2.83E-05 | 0.000134478 | 0.137372393 |
| T2DM | mast_cell | *TM2D1* | 1.996562162 | 3.22E-05 | 0.000134478 | 0.156021117 |
| T2DM | mast_cell | *NEXN* | 1.930141137 | 2.67E-05 | 0.000134478 | 0.129404862 |
| T2DM | mast_cell | *LRRIQ3* | 1.857081931 | 4.23E-05 | 0.000134478 | 0.205060609 |
| T2DM | mast_cell | *TYW3* | 1.672924466 | 8.45E-05 | 0.000181073 | 0.410047103 |
| T2DM | mast_cell | *HFM1* | 1.440376773 | 0.000135883 | 0.000255715 | 0.659302471 |
| T2DM | mast_cell | *ARPC5* | 2.23830675 | 3.52E-05 | 0.000134478 | 0.170872371 |
| T2DM | mast_cell | *ZNF496* | 3.127578794 | 0.000148463 | 0.000273272 | 0.720344248 |
| T2DM | mast_cell | *FCRLB* | -8.127305308 | 0.008245004 | 0.0103693 | 1 |
| T2DM | mast_cell | *KLHDC9* | -1.61394427 | 0.000785796 | 0.001162049 | 1 |
| T2DM | mast_cell | *TDRD5* | -3.364437339 | 0.000384587 | 0.000609691 | 1 |
| T2DM | mast_cell | *BROX* | 1.357658795 | 0.017536844 | 0.021389836 | 1 |
| T2DM | mast_cell | *ACP6* | 2.415058714 | 4.52E-05 | 0.000134478 | 0.219540991 |
| T2DM | mast_cell | *CNST* | 2.561378529 | 3.24E-05 | 0.000134478 | 0.157234815 |

| | | | | | | |
|---|---|---|---|---|---|---|
| T2DM | mast_cell | *PM20D1* | 2.165529084 | 0.007409295 | 0.009383947 | 1 |
| T2DM | mast_cell | *PKDCC* | 1.18907431 | 8.77E-05 | 0.000185825 | 0.425531884 |
| T2DM | mast_cell | *B3GALNT2* | 1.423226282 | 0.000530716 | 0.000808742 | 1 |
| T2DM | mast_cell | *MRPL55* | 1.746097122 | 3.81E-05 | 0.000134478 | 0.184657446 |
| T2DM | mast_cell | *KIAA1841* | 3.619642532 | 7.96E-05 | 0.000173977 | 0.386402595 |
| T2DM | mast_cell | *KCNJ3* | 4.593187964 | 0.003366831 | 0.004445133 | 1 |
| T2DM | mast_cell | *CLHC1* | 1.31694877 | 4.58E-06 | 0.000134478 | 0.022210495 |
| T2DM | mast_cell | *DUSP19* | -3.162922672 | 0.010240979 | 0.012776866 | 1 |
| T2DM | mast_cell | *NUP35* | 2.199968614 | 6.22E-05 | 0.000149115 | 0.301727364 |
| T2DM | mast_cell | *C2orf48* | 17.48993581 | 2.20E-05 | 0.000134478 | 0.106541253 |
| T2DM | mast_cell | *H3F3A* | 1.478800862 | 2.18E-05 | 0.000134478 | 0.1059776 |
| T2DM | mast_cell | *NOSTRIN* | 1.450960442 | 3.78E-05 | 0.000134478 | 0.183429854 |
| T2DM | mast_cell | *INHBB* | 2.990182426 | 9.24E-05 | 0.000192537 | 0.448452498 |
| T2DM | mast_cell | *RPRD2* | 2.409593238 | 1.99E-05 | 0.000134478 | 0.096407355 |
| T2DM | mast_cell | *CTSS* | 0.814644406 | 0.000307819 | 0.000502029 | 1 |
| T2DM | mast_cell | *PACRGL* | 1.514742633 | 4.10E-05 | 0.000134478 | 0.198940413 |
| T2DM | mast_cell | *C1QTNF7* | 2.1904501 | 1.60E-05 | 0.000134478 | 0.077685149 |
| T2DM | mast_cell | *LYSMD1* | -1.622753981 | 0.004298577 | 0.005606639 | 1 |
| T2DM | mast_cell | *VPS72* | 2.058104279 | 3.92E-05 | 0.000134478 | 0.190234561 |
| T2DM | mast_cell | *ERCC3* | 4.168869297 | 7.97E-06 | 0.000134478 | 0.03865501 |
| T2DM | mast_cell | *RNF149* | 1.666520856 | 3.60E-05 | 0.000134478 | 0.174648746 |
| T2DM | mast_cell | *DHX57* | 2.10882088 | 3.77E-05 | 0.000134478 | 0.182947963 |
| T2DM | mast_cell | *S100A9* | 1.556591185 | 2.69E-05 | 0.000134478 | 0.130427664 |
| T2DM | mast_cell | *TDRD10* | 1.295097756 | 4.65E-05 | 0.000134478 | 0.225852973 |
| T2DM | mast_cell | *DCAF16* | 2.415147245 | 0.000329669 | 0.000533185 | 1 |
| T2DM | mast_cell | *GNPDA2* | 2.040240462 | 4.74E-05 | 0.000134478 | 0.230024002 |
| T2DM | mast_cell | *NIPAL1* | 1.816433183 | 4.63E-05 | 0.000134478 | 0.224427169 |
| T2DM | mast_cell | *ANTXR2* | 2.618146093 | 0.000119826 | 0.000231723 | 0.581393524 |
| T2DM | mast_cell | *CGGBP1* | 1.584279186 | 0.000200565 | 0.000349548 | 0.973140615 |
| T2DM | mast_cell | *GPR155* | 1.187465549 | 5.51E-05 | 0.000139817 | 0.267550193 |
| T2DM | mast_cell | *DAPL1* | -1.346319543 | 0.001175045 | 0.001676858 | 1 |
| T2DM | mast_cell | *PBXIP1* | 4.658025114 | 2.32E-05 | 0.000134478 | 0.112610881 |
| T2DM | mast_cell | *CLDN1* | 1.270762878 | 4.40E-05 | 0.000134478 | 0.213459226 |
| T2DM | mast_cell | *DCST2* | 3.349931725 | 5.35E-06 | 0.000134478 | 0.025946941 |
| T2DM | mast_cell | *LINC01116* | 3.663643918 | 1.11E-05 | 0.000134478 | 0.053950522 |
| T2DM | mast_cell | *POGLUT1* | 1.283013086 | 0.01508549 | 0.018530329 | 1 |
| T2DM | mast_cell | *SLC22A15* | -2.106878394 | 0.001519243 | 0.002127378 | 1 |
| T2DM | mast_cell | *LRRC58* | 3.776614553 | 0.032593173 | 0.038627766 | 1 |
| T2DM | mast_cell | *LMOD1* | 1.139219096 | 0.000292074 | 0.000480714 | 1 |
| T2DM | mast_cell | *TMEM169* | 1.990792288 | 0.001610381 | 0.002247215 | 1 |
| T2DM | mast_cell | *ARPC2* | 2.44004713 | 7.49E-05 | 0.000166394 | 0.363238708 |
| T2DM | mast_cell | *CCT3* | 1.497915611 | 6.37E-05 | 0.00015104 | 0.308877691 |
| T2DM | mast_cell | *GLB1L* | 2.022928074 | 0.03604482 | 0.042500478 | 1 |
| T2DM | mast_cell | *SERPINI1* | 2.023885881 | 4.68E-05 | 0.000134478 | 0.22700145 |
| T2DM | mast_cell | *SUCLG1* | 1.959847586 | 1.58E-05 | 0.000134478 | 0.076456769 |
| T2DM | mast_cell | *SPTA1* | -2.408599676 | 0.000209448 | 0.000362406 | 1 |
| T2DM | mast_cell | *PRKCI* | 1.297140598 | 0.000186052 | 0.000328024 | 0.902722561 |
| T2DM | mast_cell | *AIM2* | 2.825483334 | 7.25E-05 | 0.000162972 | 0.351692755 |
| T2DM | mast_cell | *GTPBP8* | 2.431773711 | 3.81E-05 | 0.000134478 | 0.18488801 |
| T2DM | mast_cell | *WDFY3* | 1.905310556 | 3.30E-05 | 0.000134478 | 0.160052579 |

| | | | | | | |
|---|---|---|---|---|---|---|
| T2DM | mast_cell | *PTPN13* | 1.143613437 | 0.000186888 | 0.00032926 | 0.906782671 |
| T2DM | mast_cell | *C4orf36* | 4.862275126 | 8.61E-05 | 0.000183456 | 0.417606843 |
| T2DM | mast_cell | *PPM1K* | 2.614121318 | 0.000103015 | 0.000207225 | 0.499827739 |
| T2DM | mast_cell | *TIPARP* | 1.450667526 | 3.38E-05 | 0.000134478 | 0.164110424 |
| T2DM | mast_cell | *DCLK3* | 4.198742026 | 0.007686055 | 0.009706595 | 1 |
| T2DM | mast_cell | *RPP14* | 5.477875026 | 0.009972916 | 0.01245203 | 1 |
| T2DM | mast_cell | *DNASE1L3* | 2.524819364 | 0.000197596 | 0.000345118 | 0.958736687 |
| T2DM | mast_cell | *IL17RC* | 1.600276045 | 0.000361056 | 0.000578166 | 1 |
| T2DM | mast_cell | *CRELD1* | 1.573753917 | 3.80E-05 | 0.000134478 | 0.184287559 |
| T2DM | mast_cell | *TTC14* | 1.727519872 | 0.00015309 | 0.000280723 | 0.742792697 |
| T2DM | mast_cell | *RCHY1* | 1.750160387 | 0.000759294 | 0.001124899 | 1 |
| T2DM | mast_cell | *CCDC158* | 5.610638386 | 6.36E-05 | 0.000151009 | 0.308661931 |
| T2DM | mast_cell | *CPA3* | -0.072804023 | 0.000120823 | 0.000233188 | 0.586234952 |
| T2DM | mast_cell | *SLC4A1AP* | 1.800698782 | 3.98E-05 | 0.000134478 | 0.192929784 |
| T2DM | mast_cell | *KIAA1143* | 1.530405161 | 0.000168006 | 0.000302922 | 0.815162863 |
| T2DM | mast_cell | *CDCP1* | 1.607314607 | 8.86E-05 | 0.000186835 | 0.42972117 |
| T2DM | mast_cell | *CLEC3B* | 3.951661446 | 5.94E-06 | 0.000134478 | 0.028839221 |
| T2DM | mast_cell | *FYCO1* | 2.433432652 | 0.029840802 | 0.035548139 | 1 |
| T2DM | mast_cell | *NMNAT3* | 1.870913311 | 1.31E-05 | 0.000134478 | 0.063468333 |
| T2DM | mast_cell | *TPRA1* | 4.787909063 | 0.000265341 | 0.000442266 | 1 |
| T2DM | mast_cell | *MEAF6* | 1.599404974 | 3.61E-05 | 0.000134478 | 0.175125302 |
| T2DM | mast_cell | *SNIP1* | 4.726738539 | 3.86E-05 | 0.000134478 | 0.187489663 |
| T2DM | mast_cell | *DNALI1* | 2.730887778 | 3.21E-05 | 0.000134478 | 0.155740281 |
| T2DM | mast_cell | *RPN1* | 2.44930782 | 3.10E-05 | 0.000134478 | 0.150553671 |
| T2DM | mast_cell | *SENP2* | 2.21509261 | 0.002078163 | 0.002843555 | 1 |
| T2DM | mast_cell | *IFT122* | 2.425832707 | 8.25E-05 | 0.000178055 | 0.40053259 |
| T2DM | mast_cell | *RFT1* | 0.806608799 | 0.037416732 | 0.04406456 | 1 |
| T2DM | mast_cell | *SFMBT1* | 2.371679635 | 2.64E-05 | 0.000134478 | 0.128212785 |
| T2DM | mast_cell | *UVSSA* | 1.955224093 | 6.88E-06 | 0.000134478 | 0.033382486 |
| T2DM | mast_cell | *FAM208A* | 2.834804099 | 8.71E-05 | 0.000184846 | 0.422558661 |
| T2DM | mast_cell | *LRPAP1* | 4.314464338 | 4.31E-05 | 0.000134478 | 0.209131998 |
| T2DM | mast_cell | *PIGX* | 1.349688185 | 0.002299663 | 0.00312111 | 1 |
| T2DM | mast_cell | *ABLIM2* | 1.80221759 | 8.94E-05 | 0.000188007 | 0.4339206 |
| T2DM | mast_cell | *ERMAP* | 1.450786187 | 0.000216745 | 0.000372264 | 1 |
| T2DM | mast_cell | *ZNF691* | -1.379163891 | 0.02098857 | 0.025376661 | 1 |
| T2DM | mast_cell | *METAP1* | 3.335876368 | 0.000875321 | 0.001283099 | 1 |
| T2DM | mast_cell | *BDH2* | 1.317987106 | 0.000460672 | 0.000714344 | 1 |
| T2DM | mast_cell | *ZNF589* | 1.505822624 | 0.032281247 | 0.038276786 | 1 |
| T2DM | mast_cell | *RNF123* | 4.235797429 | 7.31E-10 | 4.43E-07 | 3.55E-06 |
| T2DM | mast_cell | *MFSD8* | 3.761657935 | 0.000306103 | 0.000499734 | 1 |
| T2DM | mast_cell | *MST1R* | 3.354461248 | 2.33E-10 | 1.63E-07 | 1.13E-06 |
| T2DM | mast_cell | *RAD54L2* | 1.954784444 | 9.79E-05 | 0.000199936 | 0.475110597 |
| T2DM | mast_cell | *TEX264* | 2.227761212 | 0.000144062 | 0.000266822 | 0.698987727 |
| T2DM | mast_cell | *ETNPPL* | 13.7322445 | 4.82E-05 | 0.000134478 | 0.233762468 |
| T2DM | mast_cell | *PRSS12* | -0.666117791 | 0.000245162 | 0.000414037 | 1 |
| T2DM | mast_cell | *HMGB2* | 3.259629949 | 9.71E-05 | 0.000198686 | 0.470940695 |
| T2DM | mast_cell | *ANXA5* | 1.8385376 | 2.19E-05 | 0.000134478 | 0.106462372 |
| T2DM | mast_cell | *MAP9* | 2.295287292 | 2.72E-05 | 0.000134478 | 0.131829269 |
| T2DM | mast_cell | *CEP44* | 1.836658221 | 0.000102546 | 0.000206453 | 0.497551097 |
| T2DM | mast_cell | *HPGD* | -0.102147338 | 0.000122159 | 0.000235018 | 0.59271567 |

| | | | | | | |
|---|---|---|---|---|---|---|
| T2DM | mast_cell | *TMEM144* | 2.442147605 | 9.86E-05 | 0.000201139 | 0.478510601 |
| T2DM | mast_cell | *IL15* | 2.06952059 | 4.91E-06 | 0.000134478 | 0.023838614 |
| T2DM | mast_cell | *ARFIP1* | 2.290585477 | 2.96E-05 | 0.000134478 | 0.143595791 |
| T2DM | mast_cell | *LSM6* | 1.861241757 | 0.000129903 | 0.00024692 | 0.630290176 |
| T2DM | mast_cell | *ITGA2* | 1.680138833 | 2.67E-05 | 0.000134478 | 0.12955801 |
| T2DM | mast_cell | *MOCS2* | 5.094076791 | 0.038104189 | 0.044819763 | 1 |
| T2DM | mast_cell | *NDUFAF2* | 1.682094712 | 0.000129923 | 0.00024692 | 0.630385912 |
| T2DM | mast_cell | *LMBRD2* | 3.805020447 | 0.006164469 | 0.007868982 | 1 |
| T2DM | mast_cell | *RNF180* | 1.194057487 | 7.23E-05 | 0.000162735 | 0.350693492 |
| T2DM | mast_cell | *PGGT1B* | 3.421173726 | 0.00011911 | 0.000230614 | 0.577919696 |
| T2DM | mast_cell | *F2RL2* | 4.709931679 | 3.96E-05 | 0.000134478 | 0.192236762 |
| T2DM | mast_cell | *CCDC112* | 2.480011205 | 6.60E-05 | 0.000154113 | 0.32034318 |
| T2DM | mast_cell | *CMBL* | 1.681361623 | 4.43E-05 | 0.000134478 | 0.215159665 |
| T2DM | mast_cell | *C5orf63* | 1.606441581 | 6.04E-05 | 0.000146277 | 0.293285849 |
| T2DM | mast_cell | *WDR41* | 1.422664783 | 4.44E-05 | 0.000134478 | 0.215352092 |
| T2DM | mast_cell | *NDUFS4* | 1.578562591 | 4.04E-05 | 0.000134478 | 0.196077801 |
| T2DM | mast_cell | *SERINC5* | 1.733265768 | 3.45E-05 | 0.000134478 | 0.167265461 |
| T2DM | mast_cell | *CASP3* | 3.515699372 | 0.017609755 | 0.021473368 | 1 |
| T2DM | mast_cell | *PRIMPOL* | 2.962626473 | 0.000973754 | 0.001412853 | 1 |
| T2DM | mast_cell | *ERAP1* | 2.648746969 | 0.000155811 | 0.000285281 | 0.755995399 |
| T2DM | mast_cell | *ERAP2* | 1.254931198 | 5.85E-05 | 0.000144108 | 0.284036259 |
| T2DM | mast_cell | *UTP15* | 2.408578571 | 8.50E-05 | 0.000181679 | 0.412353105 |
| T2DM | mast_cell | *TLR3* | 5.00702733 | 0.000314257 | 0.000510641 | 1 |
| T2DM | mast_cell | *NSA2* | 1.092672417 | 0.00019259 | 0.000337224 | 0.934448144 |
| T2DM | mast_cell | *CCDC127* | 2.265264656 | 3.77E-05 | 0.000134478 | 0.182993406 |
| T2DM | mast_cell | *GRIK2* | 0.925643591 | 0.009872788 | 0.012330185 | 1 |
| T2DM | mast_cell | *DCBLD1* | 1.59432844 | 9.48E-06 | 0.000134478 | 0.046003138 |
| T2DM | mast_cell | *PDSS2* | 2.202376419 | 3.04E-05 | 0.000134478 | 0.147283017 |
| T2DM | mast_cell | *RAET1E* | 1.957584371 | 4.35E-05 | 0.000134478 | 0.210903002 |
| T2DM | mast_cell | *STK17A* | 2.707894718 | 0.000271924 | 0.000452151 | 1 |
| T2DM | mast_cell | *GALNT10* | 2.283063292 | 0.000269101 | 0.000447916 | 1 |
| T2DM | mast_cell | *SAP30L* | 2.160898597 | 3.21E-05 | 0.000134478 | 0.155670649 |
| T2DM | mast_cell | *RPS14* | 2.378732691 | 3.58E-05 | 0.000134478 | 0.173555868 |
| T2DM | mast_cell | *RP9* | 1.718467932 | 4.49E-05 | 0.000134478 | 0.217634838 |
| T2DM | mast_cell | *CAMLG* | 2.154434993 | 2.95E-06 | 0.000134478 | 0.014297377 |
| T2DM | mast_cell | *MIOS* | 1.963937194 | 4.31E-05 | 0.000134478 | 0.209005052 |
| T2DM | mast_cell | *USP49* | 2.747947076 | 3.77E-05 | 0.000134478 | 0.182991258 |
| T2DM | mast_cell | *CHMP4C* | 5.686710992 | 4.10E-05 | 0.000134478 | 0.199101119 |
| T2DM | mast_cell | *SLC13A4* | 4.43231325 | 7.42E-05 | 0.000165438 | 0.360104705 |
| T2DM | mast_cell | *BRI3* | 1.958642306 | 3.12E-05 | 0.000134478 | 0.151343504 |
| T2DM | mast_cell | *CTSB* | 1.008071005 | 0.000680177 | 0.001017329 | 1 |
| T2DM | mast_cell | *SOX17* | -1.406330487 | 0.013478151 | 0.016627508 | 1 |
| T2DM | mast_cell | *RAD21* | 2.449950406 | 6.24E-05 | 0.000149337 | 0.302707761 |
| T2DM | mast_cell | *TNFRSF11B* | 4.517308146 | 2.36E-05 | 0.000134478 | 0.11431553 |
| T2DM | mast_cell | *SUN1* | 2.372682308 | 7.63E-05 | 0.000168292 | 0.370074185 |
| T2DM | mast_cell | *OXR1* | 1.95236306 | 2.10E-05 | 0.000134478 | 0.101929252 |
| T2DM | mast_cell | *GPER1* | 2.596280759 | 3.55E-06 | 0.000134478 | 0.017207571 |
| T2DM | mast_cell | *INTS1* | 1.71199832 | 4.44E-05 | 0.000134478 | 0.215450542 |
| T2DM | mast_cell | *SLC4A2* | 3.344508579 | 6.72E-05 | 0.00015582 | 0.326286406 |
| T2DM | mast_cell | *FASTK* | 1.850958015 | 3.50E-05 | 0.000134478 | 0.169996164 |

| | | | | | | |
|---|---|---|---|---|---|---|
| T2DM | mast_cell | *ALDH7A1* | 2.09359678 | 2.18E-05 | 0.000134478 | 0.105707419 |
| T2DM | mast_cell | *YWHAZ* | 2.003373507 | 6.16E-05 | 0.000148224 | 0.299116365 |
| T2DM | mast_cell | *FZD6* | 1.774377212 | 1.63E-06 | 0.000105732 | 0.007929937 |
| T2DM | mast_cell | *SLC25A32* | 1.921345793 | 8.50E-05 | 0.000181679 | 0.412411634 |
| T2DM | mast_cell | *DCSTAMP* | 33.39908574 | 1.35E-06 | 9.63E-05 | 0.006567709 |
| T2DM | mast_cell | *INTS8* | 3.304625697 | 3.85E-05 | 0.000134478 | 0.186644827 |
| T2DM | mast_cell | *C9orf24* | 3.380883445 | 2.29E-05 | 0.000134478 | 0.111083579 |
| T2DM | mast_cell | *NUDT2* | 1.101629045 | 0.000615281 | 0.000928278 | 1 |
| T2DM | mast_cell | *TMEM65* | 2.956332917 | 3.36E-05 | 0.000134478 | 0.162824269 |
| T2DM | mast_cell | *CCDC171* | 1.582546547 | 6.62E-05 | 0.000154268 | 0.321032033 |
| T2DM | mast_cell | *ZMAT4* | 3.551149498 | 4.13E-05 | 0.000134478 | 0.200223618 |
| T2DM | mast_cell | *C8orf34* | 1.113716983 | 6.63E-05 | 0.000154419 | 0.321499353 |
| T2DM | mast_cell | *ANKS6* | 1.610364348 | 1.63E-05 | 0.000134478 | 0.079226958 |
| T2DM | mast_cell | *AQP7* | 1.086498183 | 0.000197791 | 0.000345334 | 0.95968182 |
| T2DM | mast_cell | *NOL6* | 1.635904214 | 0.003620509 | 0.004759337 | 1 |
| T2DM | mast_cell | *AQP3* | -8.123858211 | 0.002149465 | 0.002934498 | 1 |
| T2DM | mast_cell | *TRMT10B* | 2.058402017 | 4.80E-05 | 0.000134478 | 0.2328403 |
| T2DM | mast_cell | *VCP* | 2.082080307 | 0.00014142 | 0.000263507 | 0.686171572 |
| T2DM | mast_cell | *WRN* | 1.804358755 | 2.43E-05 | 0.000134478 | 0.117899802 |
| T2DM | mast_cell | *MARCH8* | 4.737423235 | 0.001820004 | 0.002514425 | 1 |
| T2DM | mast_cell | *TSHR* | 1.519257601 | 0.000311067 | 0.000506475 | 1 |
| T2DM | mast_cell | *CFL2* | 2.940118439 | 5.00E-05 | 0.000135379 | 0.242593736 |
| T2DM | mast_cell | *PGM2L1* | -1.594242487 | 0.001659896 | 0.002307683 | 1 |
| T2DM | mast_cell | *SLC16A9* | 2.460265053 | 4.77E-05 | 0.000134478 | 0.231651289 |
| T2DM | mast_cell | *GJB2* | 10.72684514 | 4.23E-05 | 0.000134478 | 0.205136491 |
| T2DM | mast_cell | *CRYL1* | 1.714348742 | 3.91E-05 | 0.000134478 | 0.189549034 |
| T2DM | mast_cell | *REEP3* | 2.681641139 | 0.000692088 | 0.001034188 | 1 |
| T2DM | mast_cell | *MICU2* | 1.864846261 | 2.72E-05 | 0.000134478 | 0.131768696 |
| T2DM | mast_cell | *LRR1* | -2.839570548 | 0.001098173 | 0.001578299 | 1 |
| T2DM | mast_cell | *RPL36AL* | 1.739660284 | 2.74E-05 | 0.000134478 | 0.133025055 |
| T2DM | mast_cell | *C10orf25* | 0.867371865 | 0.025069405 | 0.030094582 | 1 |
| T2DM | mast_cell | *NOXRED1* | 2.675511275 | 2.19E-06 | 0.000126109 | 0.010648791 |
| T2DM | mast_cell | *AKR1E2* | 2.615641584 | 7.34E-05 | 0.000164447 | 0.356094717 |
| T2DM | mast_cell | *ATP5C1* | 3.149563441 | 6.60E-05 | 0.000154113 | 0.320016783 |
| T2DM | mast_cell | *VDAC2* | 1.545576513 | 0.000101885 | 0.000205891 | 0.494345303 |
| T2DM | mast_cell | *PDZD8* | 2.345595233 | 6.20E-06 | 0.000134478 | 0.030096943 |
| T2DM | mast_cell | *QSOX2* | 2.211519491 | 1.47E-05 | 0.000134478 | 0.071479606 |
| T2DM | mast_cell | *SNAPC4* | 2.540927779 | 0.001427282 | 0.002003811 | 1 |
| T2DM | mast_cell | *TSC1* | 1.86720302 | 0.000213223 | 0.000367777 | 1 |
| T2DM | mast_cell | *STOX1* | 1.069416235 | 0.001348917 | 0.001902047 | 1 |
| T2DM | mast_cell | *RET* | 3.07075581 | 3.31E-05 | 0.000134478 | 0.160536388 |
| T2DM | mast_cell | *CASP7* | 1.83575122 | 0.037650941 | 0.044318866 | 1 |
| T2DM | mast_cell | *PPP1R36* | 5.282838561 | 0.002407203 | 0.003254318 | 1 |
| T2DM | mast_cell | *VWA2* | 6.703099914 | 0.003270344 | 0.004322448 | 1 |
| T2DM | mast_cell | *METTL3* | 2.922585916 | 4.68E-05 | 0.000134478 | 0.226974626 |
| T2DM | mast_cell | *TRUB1* | 2.865142672 | 0.000763189 | 0.001129994 | 1 |
| T2DM | mast_cell | *HSPA12A* | 1.530181441 | 8.79E-05 | 0.000186018 | 0.426643075 |
| T2DM | mast_cell | *UBTD1* | 1.492874316 | 4.31E-05 | 0.000134478 | 0.20892464 |
| T2DM | mast_cell | *OTOGL* | 3.13907454 | 0.000320819 | 0.000520606 | 1 |
| T2DM | mast_cell | *PACSIN3* | -2.174806768 | 0.000159949 | 0.000291428 | 0.776074003 |

| | | | | | | |
|---|---|---|---|---|---|---|
| T2DM | mast_cell | *TTC7B* | 3.797079788 | 0.002581711 | 0.003468013 | 1 |
| T2DM | mast_cell | *SLC39A13* | -2.023126748 | 0.023501224 | 0.028315853 | 1 |
| T2DM | mast_cell | *PSMC3* | 1.936897236 | 4.27E-05 | 0.000134478 | 0.207348592 |
| T2DM | mast_cell | *RAPSN* | -3.592933678 | 5.94E-05 | 0.000144951 | 0.288065322 |
| T2DM | mast_cell | *TC2N* | 1.361318663 | 0.000281223 | 0.000465539 | 1 |
| T2DM | mast_cell | *CPSF2* | 1.353932941 | 0.005201545 | 0.006703292 | 1 |
| T2DM | mast_cell | *IFI27L1* | 1.578258669 | 4.43E-05 | 0.000134478 | 0.214729667 |
| T2DM | mast_cell | *PTER* | 3.03182362 | 0.001029166 | 0.001487493 | 1 |
| T2DM | mast_cell | *C1QL3* | 2.767371011 | 9.67E-05 | 0.000198277 | 0.469012671 |
| T2DM | mast_cell | *ARL5B* | 2.223428567 | 0.000109275 | 0.000216852 | 0.530203134 |
| T2DM | mast_cell | *LIPC* | -1.561039819 | 0.005847271 | 0.007487717 | 1 |
| T2DM | mast_cell | *CEP57* | 2.649885019 | 0.012003529 | 0.014880205 | 1 |
| T2DM | mast_cell | *TCP11L2* | 2.363094005 | 2.72E-05 | 0.000134478 | 0.132182367 |
| T2DM | mast_cell | *JAM3* | 2.274642102 | 3.68E-05 | 0.000134478 | 0.178346479 |
| T2DM | mast_cell | *IKBIP* | 1.771421153 | 3.12E-05 | 0.000134478 | 0.151194859 |
| T2DM | mast_cell | *TRMT61A* | 2.306334567 | 4.49E-05 | 0.000134478 | 0.217765755 |
| T2DM | mast_cell | *LARP6* | 1.635840224 | 2.64E-05 | 0.000134478 | 0.128010482 |
| T2DM | mast_cell | *API5* | 3.663974274 | 3.88E-05 | 0.000134478 | 0.188174011 |
| T2DM | mast_cell | *ASPG* | 9.280207803 | 0.000132156 | 0.000250081 | 0.641219558 |
| T2DM | mast_cell | *GABRB3* | 1.53313883 | 0.029929888 | 0.035645512 | 1 |
| T2DM | mast_cell | *COX11* | 1.543177822 | 7.14E-05 | 0.000161643 | 0.346400613 |
| T2DM | mast_cell | *ZNF202* | 2.101550412 | 0.000113855 | 0.000224288 | 0.552425305 |
| T2DM | mast_cell | *STXBP4* | 1.556820726 | 0.000111703 | 0.000220857 | 0.541984129 |
| T2DM | mast_cell | *WBP1L* | 2.670199127 | 9.51E-06 | 0.000134478 | 0.046118757 |
| T2DM | mast_cell | *NUDT13* | 1.425338281 | 5.15E-05 | 0.000136328 | 0.249753086 |
| T2DM | mast_cell | *WDR88* | 1.792605142 | 3.29E-05 | 0.000134478 | 0.159727202 |
| T2DM | mast_cell | *SERPINB8* | 0.946093612 | 0.039164446 | 0.046022256 | 1 |
| T2DM | mast_cell | *XRRA1* | 1.326261912 | 0.000212416 | 0.000366776 | 1 |
| T2DM | mast_cell | *TRIM66* | 1.057131806 | 3.11E-05 | 0.000134478 | 0.150801003 |
| T2DM | mast_cell | *RPL27A* | 2.103345848 | 3.16E-05 | 0.000134478 | 0.153185528 |
| T2DM | mast_cell | *CDYL2* | 1.43698903 | 2.44E-05 | 0.000134478 | 0.118315516 |
| T2DM | mast_cell | *CENPN* | -2.843939184 | 0.00064503 | 0.000969243 | 1 |
| T2DM | mast_cell | *C16orf46* | -2.294005323 | 3.88E-05 | 0.000134478 | 0.188026197 |
| T2DM | mast_cell | *TMEM41B* | 2.929463514 | 3.41E-05 | 0.000134478 | 0.165279551 |
| T2DM | mast_cell | *TMX3* | 1.652218908 | 6.54E-05 | 0.000153213 | 0.317234072 |
| T2DM | mast_cell | *PRKCB* | 2.474470337 | 4.41E-05 | 0.000134478 | 0.214024209 |
| T2DM | mast_cell | *NDST2* | -0.669391938 | 0.000122066 | 0.000234932 | 0.592263603 |
| T2DM | mast_cell | *CCDC68* | 1.719600851 | 2.54E-05 | 0.000134478 | 0.123108237 |
| T2DM | mast_cell | *ZSCAN21* | 1.878641441 | 0.001718369 | 0.002382832 | 1 |
| T2DM | mast_cell | *RIMKLB* | 1.456456517 | 0.001044748 | 0.001508665 | 1 |
| T2DM | mast_cell | *TMED3* | 0.886004222 | 0.001317367 | 0.001861889 | 1 |
| T2DM | mast_cell | *SEC11C* | 4.8887679 | 3.19E-05 | 0.000134478 | 0.154921001 |
| T2DM | mast_cell | *CENPV* | 1.654538038 | 6.70E-05 | 0.000155401 | 0.32525433 |
| T2DM | mast_cell | *FAM96B* | 2.164581395 | 0.00012731 | 0.00024348 | 0.617707586 |
| T2DM | mast_cell | *ZNF592* | 2.022493084 | 0.000141565 | 0.000263675 | 0.686872135 |
| T2DM | mast_cell | *ACSM1* | 2.277634237 | 0.000177811 | 0.000316602 | 0.862741121 |
| T2DM | mast_cell | *SLFN5* | 2.849230133 | 1.79E-05 | 0.000134478 | 0.087077079 |
| T2DM | mast_cell | *CATSPER2* | 1.82371747 | 7.12E-05 | 0.000161512 | 0.345474329 |
| T2DM | mast_cell | *ZNF667-AS1* | 3.723225549 | 0.002786857 | 0.00371988 | 1 |
| T2DM | mast_cell | *SAAL1* | 1.949330237 | 4.36E-05 | 0.000134478 | 0.211510025 |

| | | | | | | |
|---|---|---|---|---|---|---|
| T2DM | mast_cell | *LDHC* | 1.534680867 | 0.001059509 | 0.001528163 | 1 |
| T2DM | mast_cell | *FAM96A* | 2.713317699 | 3.29E-05 | 0.000134478 | 0.159802188 |
| T2DM | mast_cell | *FAM111A* | 2.649419745 | 0.00399549 | 0.00522576 | 1 |
| T2DM | mast_cell | *TMEM170A* | 1.797043474 | 4.88E-05 | 0.000134902 | 0.236695758 |
| T2DM | mast_cell | *ANPEP* | 1.444791365 | 0.000882871 | 0.001292604 | 1 |
| T2DM | mast_cell | *DCTN5* | 1.879894745 | 0.000403972 | 0.000633387 | 1 |
| T2DM | mast_cell | *STAT6* | 2.375658292 | 3.29E-05 | 0.000134478 | 0.159657875 |
| T2DM | mast_cell | *STX3* | -0.460974666 | 0.000133162 | 0.000251598 | 0.646103991 |
| T2DM | mast_cell | *PIP4K2C* | 2.453627886 | 4.82E-05 | 0.000134478 | 0.2340376 |
| T2DM | mast_cell | *MTMR10* | 2.149646301 | 1.76E-05 | 0.000134478 | 0.08528476 |
| T2DM | mast_cell | *MS4A7* | 0.924594114 | 0.000709193 | 0.00105682 | 1 |
| T2DM | mast_cell | *DIS3L* | 1.56973015 | 2.44E-05 | 0.000134478 | 0.118300252 |
| T2DM | mast_cell | *CCNDBP1* | 2.415381209 | 2.46E-05 | 0.000134478 | 0.119383997 |
| T2DM | mast_cell | *EPB42* | 3.617990561 | 6.24E-05 | 0.000149337 | 0.302847453 |
| T2DM | mast_cell | *SMAD3* | 1.964117664 | 4.73E-05 | 0.000134478 | 0.229321372 |
| T2DM | mast_cell | *AKTIP* | 1.950249105 | 1.43E-05 | 0.000134478 | 0.069540999 |
| T2DM | mast_cell | *CNPY4* | 2.319213983 | 2.27E-06 | 0.000126109 | 0.01099773 |
| T2DM | mast_cell | *MEI1* | 4.064609144 | 1.89E-05 | 0.000134478 | 0.091481844 |
| T2DM | mast_cell | *PIP5KL1* | -1.116579817 | 0.007952642 | 0.010022395 | 1 |
| T2DM | mast_cell | *ACSF2* | 1.402140383 | 4.78E-05 | 0.000134478 | 0.231770583 |
| T2DM | mast_cell | *URM1* | 1.806971639 | 7.41E-05 | 0.000165359 | 0.359491356 |
| T2DM | mast_cell | *ISLR2* | 2.637205462 | 0.004289193 | 0.005597409 | 1 |
| T2DM | mast_cell | *SP2* | 5.892900667 | 0.000309992 | 0.000504894 | 1 |
| T2DM | mast_cell | *COQ7* | 2.986135748 | 8.12E-05 | 0.000176186 | 0.394136629 |
| T2DM | mast_cell | *GPRC5B* | 1.999687699 | 3.66E-05 | 0.000134478 | 0.17780497 |
| T2DM | mast_cell | *FBXO22* | 2.369152436 | 6.28E-05 | 0.000149953 | 0.304915313 |
| T2DM | mast_cell | *TBC1D2B* | 1.429364004 | 5.69E-05 | 0.000141911 | 0.275941366 |
| T2DM | mast_cell | *LOXHD1* | 1.365921605 | 2.73E-05 | 0.000134478 | 0.132456828 |
| T2DM | mast_cell | *KATNAL2* | 1.797175878 | 0.000296845 | 0.000487325 | 1 |
| T2DM | mast_cell | *ZNF91* | 2.265684319 | 1.29E-05 | 0.000134478 | 0.062776988 |
| T2DM | mast_cell | *CCL23* | -0.660425619 | 0.001217542 | 0.001732409 | 1 |
| T2DM | mast_cell | *POP5* | 3.808804154 | 0.003775725 | 0.004952641 | 1 |
| T2DM | mast_cell | *ENGASE* | 1.629839461 | 3.70E-05 | 0.000134478 | 0.179565438 |
| T2DM | mast_cell | *MYO5B* | 2.336716781 | 0.000138288 | 0.000259138 | 0.670974707 |
| T2DM | mast_cell | *RRM1* | 0.952236011 | 0.026405608 | 0.031595563 | 1 |
| T2DM | mast_cell | *ZNF23* | 4.242497632 | 0.000102203 | 0.000206134 | 0.495888144 |
| T2DM | mast_cell | *VKORC1* | 1.963315533 | 4.53E-05 | 0.000134478 | 0.219983068 |
| T2DM | mast_cell | *SMG8* | -1.959895457 | 0.013582593 | 0.016747838 | 1 |
| T2DM | mast_cell | *RAB8A* | 3.482882749 | 3.32E-05 | 0.000134478 | 0.16117676 |
| T2DM | mast_cell | *RPL13* | 2.175488442 | 7.51E-05 | 0.000166733 | 0.364475951 |
| T2DM | mast_cell | *ZNF641* | 2.690379202 | 1.51E-05 | 0.000134478 | 0.073357194 |
| T2DM | mast_cell | *TP53I13* | 2.327800527 | 1.49E-05 | 0.000134478 | 0.072195408 |
| T2DM | mast_cell | *TUBA1C* | 1.719513602 | 2.86E-06 | 0.000134478 | 0.01386135 |
| T2DM | mast_cell | *ZNF528* | 4.159954057 | 4.66E-05 | 0.000134478 | 0.225867247 |
| T2DM | mast_cell | *SERTAD3* | -4.502278511 | 0.020337227 | 0.024644412 | 1 |
| T2DM | mast_cell | *TMC4* | 2.061118112 | 0.000107731 | 0.000214313 | 0.522708413 |
| T2DM | mast_cell | *TTYH1* | 2.419113142 | 4.84E-05 | 0.000134478 | 0.23479864 |
| T2DM | mast_cell | *LENG8* | 3.640942828 | 0.000126864 | 0.000242722 | 0.61554325 |
| T2DM | mast_cell | *CDC42EP5* | 1.439417484 | 0.001788322 | 0.002474883 | 1 |
| T2DM | mast_cell | *FAM57A* | 1.767099228 | 0.003325797 | 0.004393348 | 1 |

| | | | | | | |
|---|---|---|---|---|---|---|
| T2DM | mast_cell | *KIFC2* | 2.010758261 | 0.00039949 | 0.0006279 | 1 |
| T2DM | mast_cell | *SERPINF2* | 2.102952522 | 0.001119317 | 0.001605358 | 1 |
| T2DM | mast_cell | *TRPV3* | 1.876056038 | 0.000907051 | 0.001323613 | 1 |
| T2DM | mast_cell | *CYB5D2* | 1.928637963 | 9.40E-06 | 0.000134478 | 0.045598074 |
| T2DM | mast_cell | *KLK5* | 2.820210234 | 0.000575067 | 0.000871673 | 1 |
| T2DM | mast_cell | *KLK6* | 58.96494297 | 0.00122748 | 0.001745014 | 1 |
| T2DM | mast_cell | *ZNF83* | 3.065413963 | 0.000120948 | 0.000233337 | 0.586842058 |
| T2DM | mast_cell | *SOAT2* | -2.09990763 | 0.002431335 | 0.003283284 | 1 |
| T2DM | mast_cell | *ZNF558* | 2.723905403 | 4.15E-05 | 0.000134478 | 0.201434376 |
| T2DM | mast_cell | *NDUFV1* | 2.243987928 | 3.26E-05 | 0.000134478 | 0.15831187 |
| T2DM | mast_cell | *NUDT8* | -1.831925657 | 0.001629078 | 0.002270694 | 1 |
| T2DM | mast_cell | *PRDX2* | 1.841494785 | 4.03E-05 | 0.000134478 | 0.195293907 |
| T2DM | mast_cell | *ZNF232* | -0.896089507 | 0.009475381 | 0.011858279 | 1 |
| T2DM | mast_cell | *MIS12* | 2.668262338 | 2.93E-05 | 0.000134478 | 0.14218677 |
| T2DM | mast_cell | *ATP5H* | 3.153811567 | 3.75E-05 | 0.000134478 | 0.182146606 |
| T2DM | mast_cell | *MGAT5B* | 4.500655967 | 0.000167809 | 0.000302856 | 0.814208961 |
| T2DM | mast_cell | *TMEM99* | 1.571547162 | 0.001614603 | 0.002251812 | 1 |
| T2DM | mast_cell | *ZNF598* | 3.205890086 | 0.002858756 | 0.003808534 | 1 |
| T2DM | mast_cell | *ECI1* | 1.681089194 | 0.000253931 | 0.000426914 | 1 |
| T2DM | mast_cell | *ABCA3* | 1.522675276 | 0.002280123 | 0.003096321 | 1 |
| T2DM | mast_cell | *ATG16L2* | 1.743575977 | 3.70E-05 | 0.000134478 | 0.179328243 |
| T2DM | mast_cell | *RPSA* | 1.992994965 | 2.05E-05 | 0.000134478 | 0.099586646 |
| T2DM | mast_cell | *ULK4* | 1.556243292 | 0.000261191 | 0.000437151 | 1 |
| T2DM | mast_cell | *LTBP3* | 2.64788688 | 5.30E-06 | 0.000134478 | 0.025714948 |
| T2DM | mast_cell | *NAALADL1* | -1.64369743 | 0.032220102 | 0.038213624 | 1 |
| T2DM | mast_cell | *SCARA5* | 1.307495198 | 8.35E-05 | 0.000179166 | 0.404916289 |
| T2DM | mast_cell | *KIAA1586* | 2.266704333 | 0.022461188 | 0.027096391 | 1 |
| T2DM | mast_cell | *FAM83B* | 15.53487188 | 0.000658435 | 0.000987245 | 1 |
| T2DM | mast_cell | *POLR2J3* | 1.398371965 | 0.025047381 | 0.030081656 | 1 |
| T2DM | mast_cell | *COA6* | 5.950996972 | 3.32E-05 | 0.000134478 | 0.161199614 |
| T2DM | mast_cell | *PDHB* | 1.736038782 | 0.000451071 | 0.000700351 | 1 |
| T2DM | mast_cell | *PXK* | 4.18478746 | 1.82E-05 | 0.000134478 | 0.0881162 |
| T2DM | mast_cell | *PCMTD1* | 3.270357353 | 8.69E-05 | 0.000184595 | 0.421799831 |
| T2DM | mast_cell | *ACOX2* | 2.030113001 | 0.000140948 | 0.000263132 | 0.683879868 |
| T2DM | mast_cell | *FAM107A* | -0.739440843 | 0.000136519 | 0.000256443 | 0.662392534 |
| T2DM | mast_cell | *SCN11A* | 1.520509447 | 0.000262855 | 0.000439329 | 1 |
| T2DM | mast_cell | *SEPT2* | 1.276374767 | 0.006777633 | 0.008613168 | 1 |
| T2DM | mast_cell | *DTYMK* | 3.59261315 | 8.77E-05 | 0.00018584 | 0.425758362 |
| T2DM | mast_cell | *ING5* | 2.621521878 | 2.70E-05 | 0.000134478 | 0.130780251 |
| T2DM | mast_cell | *MLKL* | 1.36947488 | 5.25E-05 | 0.000136633 | 0.254548194 |
| T2DM | mast_cell | *RFWD3* | 1.755871663 | 1.57E-05 | 0.000134478 | 0.076276943 |
| T2DM | mast_cell | *KLHL30* | -2.098625303 | 0.000163808 | 0.00029712 | 0.794795724 |
| T2DM | mast_cell | *CCDC110* | -5.558681637 | 0.002150862 | 0.002935059 | 1 |
| T2DM | mast_cell | *FNTA* | 2.587913588 | 4.53E-05 | 0.000134478 | 0.219916617 |
| T2DM | mast_cell | *SLC20A2* | 1.825539134 | 6.69E-05 | 0.000155293 | 0.32456178 |
| T2DM | mast_cell | *TMUB2* | 1.951353729 | 4.88E-05 | 0.000134902 | 0.236768789 |
| T2DM | mast_cell | *ZSWIM1* | 1.619116844 | 7.80E-06 | 0.000134478 | 0.037831725 |
| T2DM | mast_cell | *NBPF9* | 2.605120518 | 4.65E-05 | 0.000134478 | 0.225826639 |
| T2DM | mast_cell | *ADAM9* | 2.028750824 | 5.92E-06 | 0.000134478 | 0.028723681 |
| T2DM | mast_cell | *NDUFS5* | 2.594836796 | 3.53E-05 | 0.000134478 | 0.171064701 |

| | | | | | | |
|---|---|---|---|---|---|---|
| T2DM | mast_cell | *ZNF30* | 2.209349819 | 0.009554995 | 0.011954831 | 1 |
| T2DM | mast_cell | *SLC16A4* | 1.627869788 | 0.000390612 | 0.000616744 | 1 |
| T2DM | mast_cell | *DNAJC21* | 3.096930169 | 3.17E-05 | 0.000134478 | 0.153598149 |
| T2DM | mast_cell | *NPNT* | 2.154538126 | 0.000465493 | 0.000720898 | 1 |
| T2DM | mast_cell | *GSTM4* | 4.729954285 | 0.000858385 | 0.00126094 | 1 |
| T2DM | mast_cell | *PPIP5K1* | 1.372040254 | 0.000924608 | 0.001348016 | 1 |
| T2DM | mast_cell | *ADAL* | 1.272166917 | 0.000167826 | 0.000302856 | 0.814291322 |
| T2DM | mast_cell | *SNTB2* | 2.290929849 | 3.18E-05 | 0.000134478 | 0.154116236 |
| T2DM | mast_cell | *ZNF507* | 2.239871189 | 0.000261402 | 0.000437201 | 1 |
| T2DM | mast_cell | *STX18* | 3.394520222 | 4.61E-05 | 0.000134478 | 0.223584339 |
| T2DM | mast_cell | *ZBTB49* | 3.538940499 | 2.76E-05 | 0.000134478 | 0.133867401 |
| T2DM | mast_cell | *GFM1* | 2.124536795 | 0.001827724 | 0.002522935 | 1 |
| T2DM | mast_cell | *ATOH8* | 1.606428342 | 4.94E-05 | 0.000135163 | 0.239852073 |
| T2DM | mast_cell | *BTNL3* | 1.425196173 | 0.017635569 | 0.021499443 | 1 |
| T2DM | mast_cell | *TMEM129* | 1.719789134 | 0.000262774 | 0.000439329 | 1 |
| T2DM | mast_cell | *CEP120* | 2.783805111 | 7.17E-05 | 0.000161829 | 0.34769157 |
| T2DM | mast_cell | *MFF* | 2.006396869 | 3.22E-05 | 0.000134478 | 0.156120424 |
| T2DM | mast_cell | *LGALS9* | 1.477787405 | 0.001872907 | 0.002581632 | 1 |
| T2DM | mast_cell | *CPLX1* | 1.653186007 | 6.80E-05 | 0.000156958 | 0.329926133 |
| T2DM | mast_cell | *MFSD7* | -4.658155684 | 0.003042756 | 0.00404146 | 1 |
| T2DM | mast_cell | *MAP2K1* | 2.929081028 | 3.27E-05 | 0.000134478 | 0.158553106 |
| T2DM | mast_cell | *KLK7* | 2.982549718 | 2.97E-05 | 0.000134478 | 0.144205484 |
| T2DM | mast_cell | *UPF3A* | 1.894557387 | 0.000220795 | 0.000378017 | 1 |
| T2DM | mast_cell | *C8orf46* | 1.052776722 | 2.61E-05 | 0.000134478 | 0.126706282 |
| T2DM | mast_cell | *HSPBAP1* | 1.661411872 | 9.39E-05 | 0.000194614 | 0.455785939 |
| T2DM | mast_cell | *UBE2V2* | 2.681492984 | 1.67E-05 | 0.000134478 | 0.080826311 |
| T2DM | mast_cell | *CPT1C* | 1.383238083 | 0.028301331 | 0.03378058 | 1 |
| T2DM | mast_cell | *PCSK9* | 13.71102472 | 2.00E-05 | 0.000134478 | 0.097123769 |
| T2DM | mast_cell | *XPO6* | 3.512495001 | 0.000141335 | 0.000263507 | 0.685755663 |
| T2DM | mast_cell | *RGS14* | 1.41153935 | 0.005541715 | 0.007115216 | 1 |
| T2DM | mast_cell | *RAB24* | -1.456880424 | 0.000217604 | 0.000373476 | 1 |
| T2DM | mast_cell | *THBS3* | 2.356306428 | 3.60E-05 | 0.000134478 | 0.174807966 |
| T2DM | mast_cell | *NPIPB3* | -1.518183611 | 7.10E-05 | 0.000161307 | 0.344551268 |
| T2DM | mast_cell | *SH3TC2* | 1.858676733 | 0.000649453 | 0.000974682 | 1 |
| T2DM | mast_cell | *GPRIN1* | -2.208306764 | 0.000939681 | 0.001367527 | 1 |
| T2DM | mast_cell | *P2RY12* | 2.561119986 | 0.004132729 | 0.005397578 | 1 |
| T2DM | mast_cell | *SNUPN* | 1.575482414 | 0.000582041 | 0.000881182 | 1 |
| T2DM | mast_cell | *CD52* | 0.86804431 | 0.007071325 | 0.008972298 | 1 |
| T2DM | mast_cell | *SLC38A11* | 1.595466187 | 0.000187714 | 0.000330492 | 0.910789326 |
| T2DM | mast_cell | *CCDC8* | 2.04597992 | 0.001925141 | 0.002649117 | 1 |
| T2DM | mast_cell | *ZEB2* | 1.324340645 | 0.000288325 | 0.000475223 | 1 |
| T2DM | mast_cell | *HINT1* | 2.165138271 | 4.94E-05 | 0.000135163 | 0.239681329 |
| T2DM | mast_cell | *DTWD2* | 1.50115813 | 0.000218596 | 0.000374649 | 1 |
| T2DM | mast_cell | *CLIC3* | 2.504009126 | 4.39E-05 | 0.000134478 | 0.213191106 |
| T2DM | mast_cell | *DFFB* | 3.564818964 | 2.17E-05 | 0.000134478 | 0.105238559 |
| T2DM | mast_cell | *NFU1* | 2.142540885 | 0.000219968 | 0.000376866 | 1 |
| T2DM | mast_cell | *C15orf40* | 2.843754674 | 0.000695717 | 0.001038333 | 1 |
| T2DM | mast_cell | *LUZP1* | 3.170338505 | 3.78E-05 | 0.000134478 | 0.183193064 |
| T2DM | mast_cell | *CHRNA5* | -3.128776725 | 0.000140115 | 0.00026188 | 0.679839838 |
| T2DM | mast_cell | *UGP2* | 1.395317831 | 0.000108571 | 0.000215719 | 0.526785511 |

| | | | | | | |
|---|---|---|---|---|---|---|
| T2DM | mast_cell | *HNRNPF* | 3.617058525 | 7.92E-05 | 0.000173118 | 0.384149056 |
| T2DM | mast_cell | *CSGALNACT2* | 2.140006413 | 1.99E-05 | 0.000134478 | 0.096772695 |
| T2DM | mast_cell | *AVEN* | 2.29109189 | 2.68E-05 | 0.000134478 | 0.130137706 |
| T2DM | mast_cell | *CTNND2* | 1.593428327 | 0.002039669 | 0.002793248 | 1 |
| T2DM | mast_cell | *CALML6* | 34.48935899 | 0.000160451 | 0.000292014 | 0.778510023 |
| T2DM | mast_cell | *MUC3A* | 1.592104651 | 0.02914313 | 0.034725557 | 1 |
| T2DM | mast_cell | *TPST1* | 1.223148976 | 0.000129009 | 0.000245664 | 0.625953123 |
| T2DM | mast_cell | *TOR1AIP2* | 1.837500909 | 2.78E-05 | 0.000134478 | 0.134705992 |
| T2DM | mast_cell | *OTUD3* | 1.443108591 | 0.000161788 | 0.000294006 | 0.784996812 |
| T2DM | mast_cell | *ZFPM2* | 0.972544252 | 0.000167509 | 0.000302588 | 0.812751842 |
| T2DM | mast_cell | *MAP3K2* | 1.591099223 | 7.87E-05 | 0.000172297 | 0.381810349 |
| T2DM | mast_cell | *MYO7B* | 2.79168841 | 0.004644204 | 0.006031498 | 1 |
| T2DM | mast_cell | *UBE2E3* | 1.584176166 | 4.58E-05 | 0.000134478 | 0.222296491 |
| T2DM | mast_cell | *NIPA1* | 2.200072147 | 6.84E-05 | 0.000157548 | 0.331758997 |
| T2DM | mast_cell | *HNRNPA3* | 2.800144879 | 2.56E-05 | 0.000134478 | 0.124276227 |
| T2DM | mast_cell | *RNF150* | 1.586979937 | 9.32E-06 | 0.000134478 | 0.045216708 |
| T2DM | mast_cell | *CHRNB1* | 1.088200048 | 0.002432821 | 0.003284376 | 1 |
| T2DM | mast_cell | *USP38* | 3.999548235 | 0.020073199 | 0.024330543 | 1 |
| T2DM | mast_cell | *PDCD6IP* | 2.486828168 | 3.34E-05 | 0.000134478 | 0.162191601 |
| T2DM | mast_cell | *CRTAP* | 1.956640473 | 2.70E-05 | 0.000134478 | 0.131077815 |
| T2DM | mast_cell | *CNGB3* | 2.680588544 | 8.13E-05 | 0.000176186 | 0.394481309 |
| T2DM | mast_cell | *STX8* | 2.517842343 | 3.88E-05 | 0.000134478 | 0.188344525 |
| T2DM | mast_cell | *NFRKB* | 1.943165476 | 3.58E-05 | 0.000134478 | 0.173831366 |
| T2DM | mast_cell | *ADORA2B* | -1.111389862 | 0.007830264 | 0.009875863 | 1 |
| T2DM | mast_cell | *MGMT* | 2.473604562 | 4.61E-05 | 0.000134478 | 0.223897739 |
| T2DM | mast_cell | *NFXL1* | 3.75129827 | 0.000899726 | 0.001313714 | 1 |
| T2DM | mast_cell | *DNAJC18* | 1.571985602 | 6.06E-05 | 0.000146324 | 0.293964194 |
| T2DM | mast_cell | *SPATA24* | 1.39268595 | 0.03411292 | 0.040330382 | 1 |
| T2DM | mast_cell | *HSD17B13* | -5.950570452 | 0.016605746 | 0.020315451 | 1 |
| T2DM | mast_cell | *ELOVL6* | 1.385114895 | 4.31E-05 | 0.000134478 | 0.20916548 |
| T2DM | mast_cell | *ARL6IP1* | 2.013336441 | 0.009031343 | 0.011323017 | 1 |
| T2DM | mast_cell | *SERPINB9* | 3.118037929 | 0.000126415 | 0.000242055 | 0.613367231 |
| T2DM | mast_cell | *SMAGP* | 0.749132789 | 0.041859365 | 0.049082078 | 1 |
| T2DM | mast_cell | *CDH2* | -2.533633647 | 0.000811114 | 0.001196209 | 1 |
| T2DM | mast_cell | *EMB* | 0.775634179 | 0.002482538 | 0.003342195 | 1 |
| T2DM | mast_cell | *STAT2* | 2.714801851 | 1.39E-05 | 0.000134478 | 0.067534488 |
| T2DM | mast_cell | *SLC26A5* | 4.073630407 | 4.82E-06 | 0.000134478 | 0.02340427 |
| T2DM | mast_cell | *GTSF1* | 4.673179807 | 0.002842313 | 0.003787669 | 1 |
| T2DM | mast_cell | *ARMC10* | 1.44798673 | 4.53E-05 | 0.000134478 | 0.219653406 |
| T2DM | mast_cell | *ACYP2* | 1.753913826 | 4.60E-05 | 0.000134478 | 0.223281064 |
| T2DM | mast_cell | *TRABD* | 4.69140677 | 3.14E-05 | 0.000134478 | 0.152518611 |
| T2DM | mast_cell | *HOXB9* | -2.549589116 | 9.23E-05 | 0.000192477 | 0.44797894 |
| T2DM | mast_cell | *KCNS3* | 1.550076798 | 5.56E-05 | 0.000140218 | 0.269779172 |
| T2DM | mast_cell | *FOXN2* | 2.435095311 | 2.88E-05 | 0.000134478 | 0.139514564 |
| T2DM | mast_cell | *TRIAP1* | 3.491931327 | 4.09E-06 | 0.000134478 | 0.019842981 |
| T2DM | mast_cell | *RPS9* | 1.587619513 | 5.43E-05 | 0.000138727 | 0.263303611 |
| T2DM | mast_cell | *TSEN34* | -3.908097985 | 0.002068537 | 0.002831183 | 1 |
| T2DM | mast_cell | *MSANTD4* | 5.214846185 | 9.99E-05 | 0.000203073 | 0.484734821 |
| T2DM | mast_cell | *NDUFA3* | 2.298406087 | 3.57E-05 | 0.000134478 | 0.173437519 |
| T2DM | mast_cell | *NUDT6* | 1.728835901 | 3.78E-06 | 0.000134478 | 0.018363796 |

| | | | | | | |
|---|---|---|---|---|---|---|
| T2DM | mast_cell | *ZNF160* | 1.501612896 | 5.44E-05 | 0.000139088 | 0.264127186 |
| T2DM | mast_cell | *ZNF415* | 2.0557656 | 7.42E-05 | 0.000165438 | 0.360159377 |
| T2DM | mast_cell | *S1PR1* | 1.994201514 | 1.89E-05 | 0.000134478 | 0.091822117 |
| T2DM | mast_cell | *PYGO1* | 1.541567422 | 5.59E-05 | 0.000140825 | 0.271449797 |
| T2DM | mast_cell | *TSNARE1* | 2.609678805 | 2.84E-06 | 0.000134478 | 0.013800716 |
| T2DM | mast_cell | *FPR1* | 1.75182156 | 4.91E-05 | 0.000135021 | 0.238036212 |
| T2DM | mast_cell | *FEZ2* | 2.00369103 | 4.83E-05 | 0.000134478 | 0.23436094 |
| T2DM | mast_cell | *SOX7* | 3.571892403 | 1.86E-05 | 0.000134478 | 0.090097554 |
| T2DM | mast_cell | *TRMT61B* | 2.106291863 | 1.78E-05 | 0.000134478 | 0.086502101 |
| T2DM | mast_cell | *MFN1* | 3.183110789 | 4.68E-06 | 0.000134478 | 0.022723762 |
| T2DM | mast_cell | *GIMAP8* | 1.271011425 | 0.000115402 | 0.000225797 | 0.559930222 |
| T2DM | mast_cell | *ATP6V0E2* | 2.261367086 | 0.000226518 | 0.000386995 | 1 |
| T2DM | mast_cell | *SCAND1* | 2.586052969 | 7.17E-05 | 0.000161829 | 0.347771454 |
| T2DM | mast_cell | *SOSTDC1* | 4.079957756 | 0.000286995 | 0.000473853 | 1 |
| T2DM | mast_cell | *GAA* | 0.844963602 | 0.007826259 | 0.009873378 | 1 |
| T2DM | mast_cell | *CANT1* | -1.412502674 | 0.007079767 | 0.008980661 | 1 |
| T2DM | mast_cell | *PGAM1* | 1.99986333 | 0.007633551 | 0.009642798 | 1 |
| T2DM | mast_cell | *LURAP1* | -1.174418642 | 0.017297454 | 0.021113772 | 1 |
| T2DM | mast_cell | *PDE7B* | 3.148903375 | 0.006180391 | 0.007887232 | 1 |
| T2DM | mast_cell | *MRPL36* | 1.982232893 | 9.40E-06 | 0.000134478 | 0.045625129 |
| T2DM | mast_cell | *MCC* | 1.838624971 | 3.20E-05 | 0.000134478 | 0.155495712 |
| T2DM | mast_cell | *ZNF561* | 1.173606719 | 0.025038009 | 0.030077846 | 1 |
| T2DM | mast_cell | *HOPX* | 2.511260087 | 0.000481822 | 0.000744049 | 1 |
| T2DM | mast_cell | *PPID* | 3.813356292 | 1.99E-05 | 0.000134478 | 0.096659797 |
| T2DM | mast_cell | *ETFDH* | 1.233210019 | 0.000548126 | 0.000833701 | 1 |
| T2DM | mast_cell | *TBCA* | 1.982764574 | 3.46E-05 | 0.000134478 | 0.167840673 |
| T2DM | mast_cell | *ZNF584* | 1.610439863 | 8.79E-06 | 0.000134478 | 0.042634065 |
| T2DM | mast_cell | *NMUR1* | 2.878114751 | 3.80E-05 | 0.000134478 | 0.184616907 |
| T2DM | mast_cell | *PIK3CD* | 1.494572214 | 2.34E-05 | 0.000134478 | 0.113509806 |
| T2DM | mast_cell | *RGS19* | -2.211277689 | 4.37E-06 | 0.000134478 | 0.021221451 |
| T2DM | mast_cell | *ANO5* | 3.657891162 | 0.000714363 | 0.00106387 | 1 |
| T2DM | mast_cell | *VAT1L* | -1.025015393 | 0.00023729 | 0.000402663 | 1 |
| T2DM | mast_cell | *CAMTA1* | 2.474790699 | 5.11E-05 | 0.000136328 | 0.248152657 |
| T2DM | mast_cell | *LRRC34* | 1.142556339 | 0.003084998 | 0.004093084 | 1 |
| T2DM | mast_cell | *PAH* | 8.823629834 | 0.000388925 | 0.000614879 | 1 |
| T2DM | mast_cell | *RHNO1* | 2.811810255 | 0.005337208 | 0.006867179 | 1 |
| T2DM | mast_cell | *METTL18* | -0.699764253 | 0.016692687 | 0.020406379 | 1 |
| T2DM | mast_cell | *EXOSC10* | 2.767349529 | 0.00088466 | 0.001294443 | 1 |
| T2DM | mast_cell | *TRAPPC12* | 2.079050667 | 3.39E-05 | 0.000134478 | 0.164400025 |
| T2DM | mast_cell | *PRNP* | 1.475982297 | 8.23E-05 | 0.000177899 | 0.399382595 |
| T2DM | mast_cell | *TVP23B* | 2.191540727 | 0.00061736 | 0.000930714 | 1 |
| T2DM | mast_cell | *FBXW10* | 4.592907939 | 0.00010334 | 0.00020762 | 0.501403312 |
| T2DM | mast_cell | *SRGAP2C* | 1.161320005 | 0.000108799 | 0.000216048 | 0.527894909 |
| T2DM | mast_cell | *ATPAF2* | 1.732667845 | 1.28E-05 | 0.000134478 | 0.062177917 |
| T2DM | mast_cell | *ZNF554* | 1.979885144 | 0.000130717 | 0.000248041 | 0.634241061 |
| T2DM | mast_cell | *ANKRD20A4* | 12.37436137 | 2.96E-05 | 0.000134478 | 0.143757895 |
| T2DM | mast_cell | *SERF1A* | 14.36618749 | 8.86E-05 | 0.000186903 | 0.430064136 |
| T2DM | mast_cell | *EIF2AK3* | 2.381479273 | 8.28E-05 | 0.00017822 | 0.401926801 |
| T2DM | mast_cell | *NME6* | 2.549303641 | 6.82E-06 | 0.000134478 | 0.033096796 |
| T2DM | mast_cell | *CD8B* | -1.141923231 | 0.007503822 | 0.00949623 | 1 |

| | | | | | | |
|---|---|---|---|---|---|---|
| T2DM | mast_cell | *MTBP* | -1.695114319 | 0.00036576 | 0.000584733 | 1 |
| T2DM | mast_cell | *TEFM* | 1.984440336 | 1.23E-05 | 0.000134478 | 0.059815458 |
| T2DM | mast_cell | *MALT1* | 4.106160712 | 9.54E-05 | 0.000196671 | 0.46296351 |
| T2DM | mast_cell | *MBOAT1* | 1.886672232 | 3.94E-05 | 0.000134478 | 0.191261859 |
| T2DM | mast_cell | *CXCR6* | 3.904135762 | 6.96E-05 | 0.000159066 | 0.337538485 |
| T2DM | mast_cell | *PAIP1* | 3.035921209 | 6.65E-05 | 0.000154713 | 0.322575785 |
| T2DM | mast_cell | *C1QTNF4* | -1.076693081 | 0.014162774 | 0.017432212 | 1 |
| T2DM | mast_cell | *MACROD2* | 1.15726015 | 4.60E-05 | 0.000134478 | 0.223066323 |
| T2DM | mast_cell | *BSG* | 1.542832066 | 4.47E-05 | 0.000134478 | 0.21678405 |
| T2DM | mast_cell | *HINFP* | 4.600570797 | 4.81E-05 | 0.000134478 | 0.233531607 |
| T2DM | mast_cell | *TP53RK* | 2.582037565 | 0.000146136 | 0.000269602 | 0.709052318 |
| T2DM | mast_cell | *SUCLG2* | 1.862613524 | 3.10E-05 | 0.000134478 | 0.150465396 |
| T2DM | mast_cell | *ARNT2* | 1.558927519 | 0.006354347 | 0.008102836 | 1 |
| T2DM | mast_cell | *GTPBP2* | 2.451031714 | 0.000113397 | 0.000223751 | 0.550204041 |
| T2DM | mast_cell | *FGGY* | 2.311999428 | 8.05E-06 | 0.000134478 | 0.039037531 |
| T2DM | mast_cell | *MANEA* | 2.346849124 | 3.03E-05 | 0.000134478 | 0.147138843 |
| T2DM | mast_cell | *AFF1* | 2.060283104 | 3.59E-05 | 0.000134478 | 0.174006574 |
| T2DM | mast_cell | *FIBP* | 2.771568302 | 4.63E-06 | 0.000134478 | 0.022487166 |
| T2DM | mast_cell | *CTSW* | -0.394157542 | 0.012085199 | 0.014977621 | 1 |
| T2DM | mast_cell | *SNTG2* | 0.833664882 | 0.00088789 | 0.001298777 | 1 |
| T2DM | mast_cell | *SMPDL3A* | 1.044782088 | 0.004292421 | 0.005600114 | 1 |
| T2DM | mast_cell | *RAD9A* | 1.885303148 | 0.001126989 | 0.001615883 | 1 |
| T2DM | mast_cell | *EFEMP2* | 1.763972151 | 1.94E-05 | 0.000134478 | 0.093955253 |
| T2DM | mast_cell | *ZFAND4* | 2.375319848 | 0.001936584 | 0.002663352 | 1 |
| T2DM | mast_cell | *ZNF738* | 1.907040798 | 3.37E-05 | 0.000134478 | 0.163625206 |
| T2DM | mast_cell | *SLFN11* | 1.402970821 | 4.26E-05 | 0.000134478 | 0.206756512 |
| T2DM | mast_cell | *FAM71D* | -1.213529169 | 0.001058336 | 0.001526925 | 1 |
| T2DM | mast_cell | *FUT10* | 0.972841948 | 0.033964932 | 0.040165208 | 1 |
| T2DM | mast_cell | *ZNF596* | 2.101828356 | 4.17E-05 | 0.000134478 | 0.202107301 |
| T2DM | mast_cell | *CFL1* | 2.666597253 | 4.54E-05 | 0.000134478 | 0.220048167 |
| T2DM | mast_cell | *FAM192A* | 1.523006498 | 4.75E-05 | 0.000134478 | 0.230435318 |
| T2DM | mast_cell | *RAB43* | -2.916218079 | 0.000139477 | 0.000260888 | 0.676743236 |
| T2DM | mast_cell | *CBWD1* | 2.533527079 | 3.26E-05 | 0.000134478 | 0.158055049 |
| T2DM | mast_cell | *DCP2* | 1.574910549 | 6.00E-05 | 0.000145631 | 0.291107081 |
| T2DM | mast_cell | *SNX32* | 1.881757054 | 1.32E-06 | 9.63E-05 | 0.006410549 |
| T2DM | mast_cell | *ZNF621* | 1.516574159 | 4.40E-05 | 0.000134478 | 0.213695622 |
| T2DM | mast_cell | *NADSYN1* | 2.346507767 | 0.000188985 | 0.000332111 | 0.916957424 |
| T2DM | mast_cell | *ANKRD13D* | 3.798614321 | 8.64E-05 | 0.000183795 | 0.419420232 |
| T2DM | mast_cell | *MRGPRF* | 1.562663097 | 0.003679098 | 0.004833735 | 1 |
| T2DM | mast_cell | *LCLAT1* | 1.41495281 | 9.28E-05 | 0.00019295 | 0.450151968 |
| T2DM | mast_cell | *SH3RF3* | 1.556958385 | 0.000104702 | 0.000209703 | 0.508016168 |
| T2DM | mast_cell | *GXYLT2* | 1.037748186 | 4.96E-05 | 0.000135163 | 0.240766557 |
| T2DM | mast_cell | *DCAKD* | 3.061154514 | 0.000185683 | 0.000327494 | 0.900935615 |
| T2DM | mast_cell | *TRMT112* | 1.581228544 | 0.000395366 | 0.000623064 | 1 |
| T2DM | mast_cell | *NOC3L* | 2.237362601 | 0.000114047 | 0.000224319 | 0.553354874 |
| T2DM | mast_cell | *RHOD* | 2.681982679 | 3.84E-05 | 0.000134478 | 0.18617198 |
| T2DM | mast_cell | *PARP14* | 1.030797689 | 0.014637833 | 0.018007801 | 1 |
| T2DM | mast_cell | *PARP15* | 1.014054659 | 0.002551177 | 0.003428895 | 1 |
| T2DM | mast_cell | *AHSA2* | 1.510252138 | 0.001388853 | 0.001954384 | 1 |
| T2DM | mast_cell | *VANGL1* | 2.519246829 | 2.42E-05 | 0.000134478 | 0.117656739 |

| | | | | | | |
|---|---|---|---|---|---|---|
| T2DM | mast_cell | *IQCB1* | 2.069974123 | 0.000142532 | 0.000264866 | 0.69156425 |
| T2DM | mast_cell | *ZNF483* | 1.780762696 | 3.57E-05 | 0.000134478 | 0.173200188 |
| T2DM | mast_cell | *MMRN2* | 2.183211564 | 2.98E-05 | 0.000134478 | 0.144568461 |
| T2DM | mast_cell | *MAP3K11* | 3.237539419 | 0.002701554 | 0.003618979 | 1 |
| T2DM | mast_cell | *NDNF* | 3.456126324 | 0.000269031 | 0.000447916 | 1 |
| T2DM | mast_cell | *ARV1* | 1.540509798 | 0.00069542 | 0.001038208 | 1 |
| T2DM | mast_cell | *CCDC36* | 1.179754234 | 9.07E-05 | 0.000190321 | 0.440212979 |
| T2DM | mast_cell | *EHBP1L1* | 1.719750715 | 1.20E-06 | 9.40E-05 | 0.005822115 |
| T2DM | mast_cell | *PPP1R14B* | -2.049603961 | 0.000531618 | 0.000809862 | 1 |
| T2DM | mast_cell | *PEAK1* | 1.757899229 | 5.30E-05 | 0.000137292 | 0.257296016 |
| T2DM | mast_cell | *TNFRSF10D* | 1.850597206 | 2.91E-05 | 0.000134478 | 0.141179508 |
| T2DM | mast_cell | *GMPPB* | 2.89771423 | 0.000162813 | 0.000295647 | 0.789969696 |
| T2DM | mast_cell | *NUDT18* | 5.103756523 | 4.78E-05 | 0.000134478 | 0.232131736 |
| T2DM | mast_cell | *SULT1B1* | 4.829676947 | 2.82E-05 | 0.000134478 | 0.136819745 |
| T2DM | mast_cell | *NUDT4* | 2.201989695 | 2.65E-05 | 0.000134478 | 0.12868721 |
| T2DM | mast_cell | *SCAI* | 1.865031601 | 2.57E-05 | 0.000134478 | 0.12450304 |
| T2DM | mast_cell | *UQCRH* | 1.779764622 | 3.26E-05 | 0.000134478 | 0.158164525 |
| T2DM | mast_cell | *C11orf80* | 2.493767853 | 0.001891483 | 0.002604279 | 1 |
| T2DM | mast_cell | *CD7* | 8.294906569 | 0.001103268 | 0.001584213 | 1 |
| T2DM | mast_cell | *RNF213* | 1.803933272 | 0.000103809 | 0.000208304 | 0.503679987 |
| T2DM | mast_cell | *TUBB8* | 2.321291659 | 0.001246517 | 0.001768968 | 1 |
| T2DM | mast_cell | *GPR160* | 2.113809035 | 0.000313113 | 0.000509295 | 1 |
| T2DM | mast_cell | *GOLIM4* | 2.01433984 | 4.09E-05 | 0.000134478 | 0.198559892 |
| T2DM | mast_cell | *HOXB2* | 1.608561961 | 0.005801472 | 0.007432992 | 1 |
| T2DM | mast_cell | *XXYLT1* | 3.519593 | 4.53E-05 | 0.000134478 | 0.21991977 |
| T2DM | mast_cell | *UBXN2A* | 2.625887764 | 4.12E-05 | 0.000134478 | 0.200120102 |
| T2DM | mast_cell | *NRROS* | 1.14607505 | 0.012853035 | 0.015892692 | 1 |
| T2DM | mast_cell | *SLC25A30* | 2.956552289 | 4.10E-05 | 0.000134478 | 0.198773854 |
| T2DM | mast_cell | *CTSF* | 2.500434965 | 0.010831666 | 0.013489539 | 1 |
| T2DM | mast_cell | *LEMD3* | 3.347871334 | 4.28E-05 | 0.000134478 | 0.207522808 |
| T2DM | mast_cell | *CTU2* | -1.487865681 | 0.032713585 | 0.038761005 | 1 |
| T2DM | mast_cell | *SNX31* | 0.94790539 | 0.034452397 | 0.040711892 | 1 |
| T2DM | mast_cell | *GLIS1* | 1.721625885 | 4.79E-05 | 0.000134478 | 0.232279607 |
| T2DM | mast_cell | *RALGAPA1* | 2.387479835 | 0.000183288 | 0.000324685 | 0.889311849 |
| T2DM | mast_cell | *ANKRD36C* | 2.839079621 | 2.93E-05 | 0.000134478 | 0.142107104 |
| T2DM | mast_cell | *MSL2* | 3.150246593 | 3.56E-05 | 0.000134478 | 0.172592746 |
| T2DM | mast_cell | *ZNF497* | -6.818753604 | 0.000490867 | 0.000756812 | 1 |
| T2DM | mast_cell | *TRAM1L1* | -1.357368113 | 0.000377778 | 0.00060078 | 1 |
| T2DM | mast_cell | *ANGEL2* | 2.515607819 | 3.16E-05 | 0.000134478 | 0.153362328 |
| T2DM | mast_cell | *SH3PXD2B* | 1.725384677 | 4.17E-05 | 0.000134478 | 0.202312643 |
| T2DM | mast_cell | *KIAA1551* | 1.432234118 | 0.000159809 | 0.000291282 | 0.775393344 |
| T2DM | mast_cell | *LARP7* | 2.45469622 | 5.52E-05 | 0.000139846 | 0.267804744 |
| T2DM | mast_cell | *FGFBP3* | 8.321313878 | 2.59E-05 | 0.000134478 | 0.125865599 |
| T2DM | mast_cell | *C4orf32* | 1.586307369 | 4.32E-05 | 0.000134478 | 0.209401942 |
| T2DM | mast_cell | *HRAS* | 2.483309308 | 0.000213839 | 0.000368324 | 1 |
| T2DM | mast_cell | *SRP72* | 1.933543459 | 4.80E-05 | 0.000134478 | 0.233081056 |
| T2DM | mast_cell | *CEP135* | 4.295494648 | 3.15E-05 | 0.000134478 | 0.152759889 |
| T2DM | mast_cell | *NLRP6* | -12.41873765 | 0.00013625 | 0.000256036 | 0.661084148 |
| T2DM | mast_cell | *RSRC1* | 2.16954316 | 4.74E-05 | 0.000134478 | 0.229801793 |
| T2DM | mast_cell | *PTDSS2* | 2.310737581 | 0.027154448 | 0.032459567 | 1 |

| | | | | | | |
|---|---|---|---|---|---|---|
| T2DM | mast_cell | *SEZ6L2* | 3.426198288 | 0.000194119 | 0.000339533 | 0.941865813 |
| T2DM | mast_cell | *KCTD13* | 1.512073177 | 2.22E-05 | 0.000134478 | 0.107778515 |
| T2DM | mast_cell | *DHX36* | 2.217386169 | 3.32E-05 | 0.000134478 | 0.161067447 |
| T2DM | mast_cell | *FBXW8* | 2.051668587 | 2.86E-05 | 0.000134478 | 0.138927065 |
| T2DM | mast_cell | *SLC22A1* | -1.179115991 | 0.000270856 | 0.000450684 | 1 |
| T2DM | mast_cell | *CHST2* | -1.558004184 | 0.000200464 | 0.000349497 | 0.972650737 |
| T2DM | mast_cell | *TVP23C* | 6.728887231 | 2.91E-05 | 0.000134478 | 0.141300493 |
| T2DM | mast_cell | *PACS1* | 2.201202707 | 0.000311902 | 0.000507665 | 1 |
| T2DM | mast_cell | *SH3BP5L* | 6.31277118 | 4.75E-05 | 0.000134478 | 0.230296575 |
| T2DM | mast_cell | *CADM2* | 1.095410592 | 3.98E-05 | 0.000134478 | 0.193008256 |
| T2DM | mast_cell | *PSMD2* | 2.037133889 | 6.34E-05 | 0.000150843 | 0.307720235 |
| T2DM | mast_cell | *FAM182B* | 1.163515141 | 0.005154208 | 0.006645818 | 1 |
| T2DM | mast_cell | *ARHGAP1* | 2.690234608 | 0.002355391 | 0.003191387 | 1 |
| T2DM | mast_cell | *ATG13* | 2.068484445 | 5.52E-05 | 0.000139817 | 0.267609689 |
| T2DM | mast_cell | *ZNF519* | 3.12134805 | 0.000482444 | 0.000744772 | 1 |
| T2DM | mast_cell | *BANF1* | 3.190175894 | 9.01E-06 | 0.000134478 | 0.043720152 |
| T2DM | mast_cell | *CHRNA7* | 3.738795074 | 3.04E-06 | 0.000134478 | 0.014726141 |
| T2DM | mast_cell | *TMEM9B* | 1.602823107 | 5.51E-05 | 0.000139817 | 0.267562294 |
| T2DM | mast_cell | *SCUBE2* | 4.060272774 | 4.12E-05 | 0.000134478 | 0.199839772 |
| T2DM | mast_cell | *EIF1AD* | 5.102396176 | 0.003241688 | 0.004288078 | 1 |
| T2DM | mast_cell | *EIF3F* | 2.629466789 | 0.006473872 | 0.008242253 | 1 |
| T2DM | mast_cell | *ZNF25* | 2.190619624 | 0.003995789 | 0.00522576 | 1 |
| T2DM | mast_cell | *CLTB* | 1.544094726 | 4.44E-05 | 0.000134478 | 0.215391268 |
| T2DM | mast_cell | *PPP2R2D* | 2.621348482 | 1.97E-05 | 0.000134478 | 0.09551178 |
| T2DM | mast_cell | *MCTP1* | 1.350001346 | 3.48E-05 | 0.000134478 | 0.16907226 |
| T2DM | mast_cell | *KCNE3* | 2.965607912 | 0.000739467 | 0.00109856 | 1 |
| T2DM | mast_cell | *UCP2* | 1.550912833 | 0.004473641 | 0.005822454 | 1 |
| T2DM | mast_cell | *C11orf68* | 2.451847262 | 0.000351904 | 0.000565189 | 1 |
| T2DM | mast_cell | *PAAF1* | 1.15982046 | 0.000459259 | 0.000712379 | 1 |
| T2DM | mast_cell | *MRPL48* | 2.183785505 | 7.22E-05 | 0.000162717 | 0.350491951 |
| T2DM | mast_cell | *RAB6A* | 2.519050274 | 8.27E-05 | 0.000178199 | 0.401480547 |
| T2DM | mast_cell | *TMEM70* | 2.051192662 | 1.56E-05 | 0.000134478 | 0.07558614 |
| T2DM | mast_cell | *RPS6KB2* | 3.298877761 | 2.94E-05 | 0.000134478 | 0.142496271 |
| T2DM | mast_cell | *TOM1L2* | 1.906411569 | 0.00015914 | 0.000290171 | 0.77214544 |
| T2DM | mast_cell | *TEX26* | -1.362410242 | 0.00023408 | 0.000397813 | 1 |
| T2DM | mast_cell | *GPR156* | 3.896931286 | 2.05E-05 | 0.000134478 | 0.099299923 |
| T2DM | mast_cell | *TTLL11* | 3.251656114 | 7.57E-05 | 0.000167424 | 0.36749677 |
| T2DM | mast_cell | *HOXD8* | 1.452408493 | 0.006647668 | 0.008456865 | 1 |
| T2DM | mast_cell | *ZDHHC21* | 1.824798805 | 9.09E-05 | 0.00019042 | 0.440822769 |
| T2DM | mast_cell | *A2M* | 1.968622537 | 2.96E-05 | 0.000134478 | 0.143763374 |
| T2DM | mast_cell | *DOK7* | 11.96164746 | 0.000264034 | 0.000440845 | 1 |
| T2DM | mast_cell | *ORAI3* | 3.939912663 | 0.000344036 | 0.000553468 | 1 |
| T2DM | mast_cell | *B3GALT6* | -4.886621287 | 0.000969442 | 0.00140746 | 1 |
| T2DM | mast_cell | *ZNF613* | 2.259479333 | 2.66E-05 | 0.000134478 | 0.128998078 |
| T2DM | mast_cell | *JAKMIP2* | 2.7318423 | 0.024221087 | 0.029161467 | 1 |
| T2DM | mast_cell | *TCP11L1* | 1.967249796 | 7.28E-06 | 0.000134478 | 0.035336955 |
| T2DM | mast_cell | *CCDC57* | 1.178295693 | 7.15E-05 | 0.000161829 | 0.347132617 |
| T2DM | mast_cell | *MYPOP* | 3.276511261 | 1.08E-05 | 0.000134478 | 0.052414359 |
| T2DM | mast_cell | *CIDEA* | 1.477854224 | 0.000144603 | 0.000267244 | 0.701611903 |
| T2DM | mast_cell | *SMIM19* | 1.498613872 | 1.09E-05 | 0.000134478 | 0.053126351 |

| | | | | | | |
|---|---|---|---|---|---|---|
| T2DM | mast_cell | *ZNF404* | 1.115741509 | 0.0027421 | 0.00366814 | 1 |
| T2DM | mast_cell | *SLC35G1* | 3.786270089 | 3.13E-05 | 0.000134478 | 0.151681137 |
| T2DM | mast_cell | *OR4N2* | 5.499609852 | 0.000116626 | 0.000227531 | 0.565868862 |
| T2DM | mast_cell | *TAC4* | 2.036949255 | 3.86E-05 | 0.000134478 | 0.187272871 |
| T2DM | mast_cell | *CDC26* | 1.437980291 | 0.004580637 | 0.00595213 | 1 |
| T2DM | mast_cell | *CRLF3* | 2.451241422 | 9.84E-06 | 0.000134478 | 0.047721947 |
| T2DM | mast_cell | *WDR25* | 2.679812797 | 3.30E-05 | 0.000134478 | 0.160114231 |
| T2DM | mast_cell | *PLA2G16* | 1.654214314 | 0.001136275 | 0.001627275 | 1 |
| T2DM | mast_cell | *RMDN1* | 1.84163556 | 1.50E-05 | 0.000134478 | 0.072765057 |
| T2DM | mast_cell | *RNF152* | 1.188122584 | 0.000167844 | 0.000302856 | 0.814381114 |
| T2DM | mast_cell | *LRRC37A* | 2.04733021 | 2.17E-05 | 0.000134478 | 0.10541231 |
| T2DM | mast_cell | *BDNF* | 4.474656532 | 0.001291311 | 0.001828258 | 1 |
| T2DM | mast_cell | *BOK* | 1.909828343 | 3.88E-05 | 0.000134478 | 0.188343003 |
| T2DM | mast_cell | *PFN4* | 2.026829685 | 4.41E-06 | 0.000134478 | 0.021378985 |
| T2DM | mast_cell | *RUFY1* | 1.937788586 | 9.24E-05 | 0.000192477 | 0.448087616 |
| T2DM | mast_cell | *VSIG10* | 2.852584352 | 0.000141922 | 0.000264035 | 0.688604473 |
| T2DM | mast_cell | *ANKLE2* | 2.532609251 | 4.68E-05 | 0.000134478 | 0.227010063 |
| T2DM | mast_cell | *C8G* | 42.09303173 | 4.13E-05 | 0.000134478 | 0.200460741 |
| T2DM | mast_cell | *MUC20* | -2.247041041 | 0.001167669 | 0.001669279 | 1 |
| T2DM | mast_cell | *LY6H* | 2.957285269 | 0.013344155 | 0.016470578 | 1 |
| T2DM | mast_cell | *SHMT1* | 1.378580935 | 3.44E-05 | 0.000134478 | 0.166749854 |
| T2DM | mast_cell | *DPP7* | 1.806078748 | 2.80E-05 | 0.000134478 | 0.135625255 |
| T2DM | mast_cell | *MTHFR* | 2.004428164 | 4.40E-05 | 0.000134478 | 0.213665225 |
| T2DM | mast_cell | *C19orf18* | -2.036390848 | 0.018329218 | 0.022305835 | 1 |
| T2DM | mast_cell | *TMEM80* | 1.737446348 | 3.92E-05 | 0.000134478 | 0.190032194 |
| T2DM | mast_cell | *WDR73* | 2.514377983 | 2.86E-05 | 0.000134478 | 0.138741451 |
| T2DM | mast_cell | *DSCAML1* | 2.222339589 | 3.45E-05 | 0.000134478 | 0.167491559 |
| T2DM | mast_cell | *TALDO1* | 4.884877462 | 8.62E-05 | 0.000183587 | 0.4184795 |
| T2DM | mast_cell | *AP006621.1* | 2.57587823 | 0.000299782 | 0.000490736 | 1 |
| T2DM | mast_cell | *MAN1B1* | 3.396688627 | 3.18E-05 | 0.000134478 | 0.154182516 |
| T2DM | mast_cell | *SAMD9L* | 1.743424297 | 0.00011643 | 0.000227345 | 0.564917235 |
| T2DM | mast_cell | *MIEF2* | 3.055705319 | 5.38E-05 | 0.000138064 | 0.260940309 |
| T2DM | mast_cell | *NAP1L5* | 3.333861063 | 0.000379119 | 0.000602123 | 1 |
| T2DM | mast_cell | *RBM44* | 4.017968602 | 0.00241783 | 0.003265955 | 1 |
| T2DM | mast_cell | *SLC25A22* | -6.109297763 | 2.19E-05 | 0.000134478 | 0.106066284 |
| T2DM | mast_cell | *SAMD12* | 0.569076831 | 0.034495977 | 0.040753465 | 1 |
| T2DM | mast_cell | *GBA* | 2.314910005 | 0.000515427 | 0.000789161 | 1 |
| T2DM | mast_cell | *ACAD9* | 1.132077467 | 7.34E-05 | 0.000164447 | 0.356193034 |
| T2DM | mast_cell | *IL17RA* | 1.879352148 | 4.43E-05 | 0.000134478 | 0.21511348 |
| T2DM | mast_cell | *MBOAT4* | 4.198336249 | 0.00038452 | 0.000609691 | 1 |
| T2DM | mast_cell | *AGTRAP* | 2.313543155 | 4.41E-05 | 0.000134478 | 0.213920727 |
| T2DM | mast_cell | *NAALADL2* | 1.90694895 | 9.75E-06 | 0.000134478 | 0.047288355 |
| T2DM | mast_cell | *CD151* | 1.661264208 | 4.69E-05 | 0.000134478 | 0.227563365 |
| T2DM | mast_cell | *SLC35G5* | 14.30084795 | 3.95E-05 | 0.000134478 | 0.191442125 |
| T2DM | mast_cell | *FAM87B* | 13.28572972 | 3.96E-05 | 0.000134478 | 0.191996132 |
| T2DM | mast_cell | *PCDHB9* | -2.15135329 | 0.002189487 | 0.002984937 | 1 |
| T2DM | mast_cell | *CENPBD1* | -3.480964084 | 0.000541267 | 0.000824043 | 1 |
| T2DM | mast_cell | *RPS27* | 1.949176348 | 3.48E-05 | 0.000134478 | 0.168921731 |
| T2DM | mast_cell | *RIC8A* | 1.774765802 | 0.000736853 | 0.001095683 | 1 |
| T2DM | mast_cell | *ASB8* | 5.11965062 | 3.05E-05 | 0.000134478 | 0.147810199 |

| | | | | | | |
|---|---|---|---|---|---|---|
| T2DM | mast_cell | *ODF3B* | 1.755168879 | 3.93E-05 | 0.000134478 | 0.190911548 |
| T2DM | mast_cell | *DPY19L2* | 1.441172378 | 0.003398197 | 0.004485325 | 1 |
| T2DM | mast_cell | *ADAMTSL1* | 1.29887238 | 9.28E-06 | 0.000134478 | 0.045042385 |
| T2DM | mast_cell | *IMPDH2* | 1.491389318 | 0.001091556 | 0.001569719 | 1 |
| T2DM | mast_cell | *GRAMD1C* | 1.38853545 | 0.000807484 | 0.001191579 | 1 |
| T2DM | mast_cell | *DALRD3* | 1.570917698 | 3.71E-05 | 0.000134478 | 0.180184678 |
| T2DM | mast_cell | *ZNF114* | 4.328360861 | 0.010047653 | 0.012542118 | 1 |
| T2DM | mast_cell | *SPINK6* | 5.401985355 | 0.003115157 | 0.00413084 | 1 |
| T2DM | mast_cell | *ZNF454* | 1.067308984 | 0.009659632 | 0.012079519 | 1 |
| T2DM | mast_cell | *SH2B1* | 3.474869015 | 4.28E-05 | 0.000134478 | 0.207601636 |
| T2DM | mast_cell | *KDELC2* | 2.591745993 | 6.02E-05 | 0.000145946 | 0.291949369 |
| T2DM | mast_cell | *PLEC* | 2.145226116 | 0.001540826 | 0.002156979 | 1 |
| T2DM | mast_cell | *SH2D4B* | 13.35075992 | 0.020431537 | 0.024752514 | 1 |
| T2DM | mast_cell | *RNF212* | 1.524189968 | 0.000168377 | 0.000303478 | 0.816963181 |
| T2DM | mast_cell | *PRSS36* | 2.955428486 | 0.000397742 | 0.000625809 | 1 |
| T2DM | mast_cell | *WDR6* | 1.312948297 | 0.000497399 | 0.000766153 | 1 |
| T2DM | mast_cell | *GEN1* | 0.794882078 | 0.019265611 | 0.023398434 | 1 |
| T2DM | mast_cell | *TMPRSS9* | 2.97566355 | 0.000711166 | 0.001059434 | 1 |
| T2DM | mast_cell | *TMEM11* | 4.326474991 | 0.011762023 | 0.014592006 | 1 |
| T2DM | mast_cell | *ZFAND2A* | 3.294485025 | 4.71E-05 | 0.000134478 | 0.228416184 |
| T2DM | mast_cell | *FAM220A* | 4.818505525 | 4.98E-05 | 0.000135379 | 0.241848026 |
| T2DM | mast_cell | *NT5DC1* | 1.713876372 | 2.90E-05 | 0.000134478 | 0.140868511 |
| T2DM | mast_cell | *GLDC* | 2.948578958 | 0.000238217 | 0.000403678 | 1 |
| T2DM | mast_cell | *P4HTM* | 2.276891787 | 3.31E-05 | 0.000134478 | 0.160731322 |
| T2DM | mast_cell | *ERN1* | 2.612147144 | 4.00E-05 | 0.000134478 | 0.193856539 |
| T2DM | mast_cell | *PARP10* | 2.265057794 | 7.04E-05 | 0.000160469 | 0.341477399 |
| T2DM | mast_cell | *NSUN3* | 1.996247315 | 2.82E-05 | 0.000134478 | 0.136736008 |
| T2DM | mast_cell | *STX19* | 4.154549953 | 9.53E-08 | 2.31E-05 | 0.000462308 |
| T2DM | mast_cell | *MPI* | 1.720372021 | 0.000183661 | 0.00032499 | 0.891122287 |
| T2DM | mast_cell | *TRIM73* | -8.019403491 | 2.24E-05 | 0.000134478 | 0.108449217 |
| T2DM | mast_cell | *EFCAB13* | 2.842689636 | 9.90E-05 | 0.000201516 | 0.480166837 |
| T2DM | mast_cell | *EXOSC4* | -3.324335402 | 6.03E-05 | 0.000146151 | 0.292739813 |
| T2DM | mast_cell | *ZNF852* | 1.601403574 | 0.024513648 | 0.029484438 | 1 |
| T2DM | mast_cell | *HYI* | 1.17177216 | 0.001801222 | 0.002489185 | 1 |
| T2DM | mast_cell | *C17orf62* | 1.651752359 | 5.22E-05 | 0.000136624 | 0.253465768 |
| T2DM | mast_cell | *GAK* | 2.837045184 | 3.35E-05 | 0.000134478 | 0.16238079 |
| T2DM | mast_cell | *TUFM* | 1.860715144 | 0.000190302 | 0.000334183 | 0.923346258 |
| T2DM | mast_cell | *CTC1* | 2.809956451 | 6.42E-06 | 0.000134478 | 0.03113902 |
| T2DM | mast_cell | *C3orf38* | 2.679696546 | 2.37E-05 | 0.000134478 | 0.114794628 |
| T2DM | mast_cell | *CCDC89* | 6.606021739 | 2.67E-05 | 0.000134478 | 0.129329137 |
| T2DM | mast_cell | *HTR1F* | 5.071321015 | 0.001553472 | 0.002173428 | 1 |
| T2DM | mast_cell | *SPTY2D1* | 1.653640816 | 4.31E-05 | 0.000134478 | 0.209035211 |
| T2DM | mast_cell | *ZNF664* | 2.847408183 | 3.52E-05 | 0.000134478 | 0.170925648 |
| T2DM | mast_cell | *CDH4* | 1.803931024 | 0.000390647 | 0.000616744 | 1 |
| T2DM | mast_cell | *NSUN7* | 1.898011407 | 3.25E-05 | 0.000134478 | 0.157453163 |
| T2DM | mast_cell | *HLA-DQB1* | -2.588529704 | 2.36E-10 | 1.63E-07 | 1.14E-06 |
| T2DM | mast_cell | *PACS2* | 1.977177759 | 3.54E-05 | 0.000134478 | 0.171657243 |
| T2DM | mast_cell | *LINC00174* | 0.94694485 | 0.01804039 | 0.021965363 | 1 |
| T2DM | mast_cell | *ZFPM1* | 1.279462124 | 0.002236005 | 0.003044079 | 1 |
| T2DM | mast_cell | *FCER1A* | -0.940660165 | 0.000203183 | 0.000353223 | 0.985845284 |

| | | | | | | |
|---|---|---|---|---|---|---|
| T2DM | mast_cell | *RPRML* | 11.11130662 | 0.000106059 | 0.000211747 | 0.514596028 |
| T2DM | mast_cell | *SERTAD2* | 4.144884104 | 0.00050579 | 0.000776857 | 1 |
| T2DM | mast_cell | *AKAP5* | -4.295810023 | 0.019834504 | 0.024059253 | 1 |
| T2DM | mast_cell | *GIPC3* | 10.31038263 | 3.86E-05 | 0.000134478 | 0.187134885 |
| T2DM | mast_cell | *PDXDC1* | 3.649389068 | 4.42E-05 | 0.000134478 | 0.214234059 |
| T2DM | mast_cell | *SEPHS2* | 2.402810432 | 7.33E-06 | 0.000134478 | 0.035577747 |
| T2DM | mast_cell | *SSC5D* | 1.083958666 | 0.001922598 | 0.002646367 | 1 |
| T2DM | mast_cell | *ZADH2* | 2.448214353 | 5.82E-05 | 0.000143571 | 0.282404334 |
| T2DM | mast_cell | *SEPT1* | 3.131046235 | 3.59E-05 | 0.000134478 | 0.174332578 |
| T2DM | mast_cell | *EXOC3* | 1.899945216 | 3.71E-05 | 0.000134478 | 0.180246488 |
| T2DM | mast_cell | *TDRD6* | 0.899722289 | 0.00716933 | 0.009089518 | 1 |
| T2DM | mast_cell | *LYNX1* | 1.356415123 | 0.017390766 | 0.021216997 | 1 |
| T2DM | mast_cell | *HCLS1* | 0.896146285 | 0.004918652 | 0.006372576 | 1 |
| T2DM | mast_cell | *MTURN* | 1.477455429 | 4.02E-05 | 0.000134478 | 0.194964345 |
| T2DM | mast_cell | *CCDC66* | 1.675435407 | 3.44E-05 | 0.000134478 | 0.16696494 |
| T2DM | mast_cell | *MCFD2* | 1.930624338 | 2.39E-05 | 0.000134478 | 0.115834532 |
| T2DM | mast_cell | *GLIPR1L2* | 2.226084698 | 0.00175557 | 0.002433027 | 1 |
| T2DM | mast_cell | *DEFB119* | -0.33974853 | 0.000122393 | 0.000235375 | 0.593851367 |
| T2DM | mast_cell | *TSPYL5* | 1.715077207 | 0.007782136 | 0.009820266 | 1 |
| T2DM | mast_cell | *MB21D2* | 1.585250768 | 7.54E-05 | 0.000167051 | 0.366073627 |
| T2DM | mast_cell | *SLC47A2* | 2.21984382 | 9.17E-05 | 0.00019169 | 0.444912709 |
| T2DM | mast_cell | *CHST13* | 9.783209134 | 0.008152647 | 0.010255806 | 1 |
| T2DM | mast_cell | *SLC36A4* | 2.302534303 | 9.21E-05 | 0.000192339 | 0.446668683 |
| T2DM | mast_cell | *ZDHHC20* | 1.867854471 | 3.26E-05 | 0.000134478 | 0.158228886 |
| T2DM | mast_cell | *OR51E1* | 1.679523765 | 0.011397174 | 0.014175618 | 1 |
| T2DM | mast_cell | *PSMG4* | 1.977229938 | 0.000141321 | 0.000263507 | 0.685689442 |
| T2DM | mast_cell | *BHLHE22* | 1.726810763 | 0.031905877 | 0.037877982 | 1 |
| T2DM | mast_cell | *CAPS2* | 4.430053722 | 1.55E-05 | 0.000134478 | 0.07515489 |
| T2DM | mast_cell | *KCTD2* | 1.412192234 | 0.000142046 | 0.000264165 | 0.68920706 |
| T2DM | mast_cell | *D2HGDH* | 1.116845015 | 0.0006308 | 0.00094933 | 1 |
| T2DM | mast_cell | *CMTR2* | 3.199765212 | 5.79E-05 | 0.0001432 | 0.280815548 |
| T2DM | mast_cell | *LRRC57* | 2.089237424 | 1.14E-05 | 0.000134478 | 0.055238069 |
| T2DM | mast_cell | *MRPL14* | 1.738559009 | 8.80E-05 | 0.000186018 | 0.426789987 |
| T2DM | mast_cell | *LSMEM1* | 1.822855589 | 6.21E-05 | 0.000149016 | 0.301310048 |
| T2DM | mast_cell | *NQO1* | 1.46684913 | 8.63E-05 | 0.000183587 | 0.418578888 |
| T2DM | mast_cell | *FKRP* | 0.954481613 | 0.042182889 | 0.049449475 | 1 |
| T2DM | mast_cell | *RPH3AL* | 1.668476121 | 8.14E-05 | 0.000176401 | 0.395138643 |
| T2DM | mast_cell | *F2R* | 1.746678294 | 0.001151629 | 0.001647805 | 1 |
| T2DM | mast_cell | *PENK* | -2.896530321 | 0.007726725 | 0.00975288 | 1 |
| T2DM | mast_cell | *SLC25A41* | 8.918899244 | 2.37E-05 | 0.000134478 | 0.114833727 |
| T2DM | mast_cell | *TMEM136* | 4.690308601 | 0.005367892 | 0.006904828 | 1 |
| T2DM | mast_cell | *ZNF322* | 1.846258091 | 0.000135974 | 0.000255715 | 0.659745703 |
| T2DM | mast_cell | *DDX60L* | 2.577929891 | 1.85E-05 | 0.000134478 | 0.089871818 |
| T2DM | mast_cell | *OGFOD3* | 1.961176006 | 0.003268824 | 0.004321617 | 1 |
| T2DM | mast_cell | *TMEM45A* | 2.560627683 | 7.66E-05 | 0.000168797 | 0.371860487 |
| T2DM | mast_cell | *SGSH* | 3.382701697 | 3.56E-05 | 0.000134478 | 0.172803117 |
| T2DM | mast_cell | *ZFP41* | 2.946743469 | 0.00124344 | 0.00176615 | 1 |
| T2DM | mast_cell | *ZBTB20* | 1.898303737 | 3.32E-05 | 0.000134478 | 0.161172966 |
| T2DM | mast_cell | *TMEM252* | 77.18450289 | 3.64E-05 | 0.000134478 | 0.176505864 |
| T2DM | mast_cell | *RELL1* | 1.792507552 | 5.40E-05 | 0.000138255 | 0.261992957 |

| | | | | | | |
|---|---|---|---|---|---|---|
| T2DM | mast_cell | *RFX7* | 2.247425782 | 3.21E-05 | 0.000134478 | 0.15587283 |
| T2DM | mast_cell | *CLDN7* | -1.33665667 | 0.000171856 | 0.000308529 | 0.833844756 |
| T2DM | mast_cell | *ZNF329* | 4.012548766 | 4.45E-05 | 0.000134478 | 0.216020452 |
| T2DM | mast_cell | *MGAT4C* | 0.934953276 | 0.002320548 | 0.003147693 | 1 |
| T2DM | mast_cell | *IDH2* | 1.743489443 | 5.07E-05 | 0.000136098 | 0.246105867 |
| T2DM | mast_cell | *PTCHD3* | 1.883799861 | 0.02337825 | 0.028181682 | 1 |
| T2DM | mast_cell | *WRB* | 1.01082521 | 0.016984229 | 0.020752325 | 1 |
| T2DM | mast_cell | *FAM181B* | -2.527693551 | 0.003281966 | 0.004336628 | 1 |
| T2DM | mast_cell | *TDRKH* | 1.705197134 | 0.002482232 | 0.003342195 | 1 |
| T2DM | mast_cell | *ZNF708* | 2.115101034 | 0.001439441 | 0.002019713 | 1 |
| T2DM | mast_cell | *ERCC6L2* | 1.646443212 | 2.03E-05 | 0.000134478 | 0.098365944 |
| T2DM | mast_cell | *MRPL41* | 4.294544845 | 3.34E-05 | 0.000134478 | 0.162131916 |
| T2DM | mast_cell | *HHIPL1* | 2.169165959 | 9.69E-05 | 0.000198366 | 0.469928395 |
| T2DM | mast_cell | *FAM153B* | 1.984931398 | 2.50E-05 | 0.000134478 | 0.121188364 |
| T2DM | mast_cell | *SYNM* | 5.214302959 | 5.13E-05 | 0.000136328 | 0.249110461 |
| T2DM | mast_cell | *KCNA4* | 2.216013537 | 4.82E-05 | 0.000134478 | 0.23381404 |
| T2DM | mast_cell | *FIGN* | 1.243214054 | 3.87E-05 | 0.000134478 | 0.187711036 |
| T2DM | mast_cell | *IZUMO1* | -0.981311018 | 0.003411631 | 0.004499384 | 1 |
| T2DM | mast_cell | *C8orf33* | 1.431717295 | 6.00E-05 | 0.000145631 | 0.29111558 |
| T2DM | mast_cell | *FBXL6* | -3.930567732 | 0.02088077 | 0.025252617 | 1 |
| T2DM | mast_cell | *KBTBD3* | 1.742407513 | 9.39E-05 | 0.000194614 | 0.455656784 |
| T2DM | mast_cell | *CLN8* | 1.416176152 | 6.40E-05 | 0.000151548 | 0.310644198 |
| T2DM | mast_cell | *NXPH4* | 18.87737106 | 4.35E-05 | 0.000134478 | 0.211255944 |
| T2DM | mast_cell | *NPLOC4* | 1.760978694 | 0.025724074 | 0.030840921 | 1 |
| T2DM | mast_cell | *CAPN12* | 2.166952838 | 2.09E-05 | 0.000134478 | 0.10137363 |
| T2DM | mast_cell | *KPNA2* | 2.206599867 | 0.000324077 | 0.000525191 | 1 |
| T2DM | mast_cell | *ORAI1* | 3.129872033 | 5.92E-05 | 0.000144951 | 0.287339195 |
| T2DM | mast_cell | *FES* | 1.827606641 | 0.000497725 | 0.000766411 | 1 |
| T2DM | mast_cell | *GLRX5* | 1.743856685 | 4.56E-05 | 0.000134478 | 0.221136348 |
| T2DM | mast_cell | *CAV3* | 1.838721227 | 0.00040072 | 0.000628815 | 1 |
| T2DM | mast_cell | *MXRA7* | 1.238827707 | 0.005624171 | 0.007211542 | 1 |
| T2DM | mast_cell | *MFSD5* | -1.593582511 | 0.002665524 | 0.003571699 | 1 |
| T2DM | mast_cell | *ADI1* | 1.414164855 | 4.80E-05 | 0.000134478 | 0.233037682 |
| T2DM | mast_cell | *TRAK1* | 1.892504085 | 0.000362919 | 0.000580767 | 1 |
| T2DM | mast_cell | *SKA2* | 3.943838857 | 0.000171824 | 0.000308529 | 0.833692135 |
| T2DM | mast_cell | *TTC3* | 1.858131401 | 1.73E-07 | 3.34E-05 | 0.000838841 |
| T2DM | mast_cell | *PPP1R27* | -3.877002776 | 4.03E-05 | 0.000134478 | 0.195705864 |
| T2DM | mast_cell | *BRICD5* | 3.889055575 | 4.65E-05 | 0.000134478 | 0.225752575 |
| T2DM | mast_cell | *GRID1* | 1.274246629 | 0.000394135 | 0.000621496 | 1 |
| T2DM | mast_cell | *ACBD3* | 3.135566739 | 0.000295382 | 0.000485171 | 1 |
| T2DM | mast_cell | *PLCXD3* | 2.755651735 | 3.51E-05 | 0.000134478 | 0.170080877 |
| T2DM | mast_cell | *VMO1* | -0.857302283 | 0.000753396 | 0.001116858 | 1 |
| T2DM | mast_cell | *ALG12* | 3.337927054 | 3.29E-05 | 0.000134478 | 0.159805022 |
| T2DM | mast_cell | *COL18A1* | 2.578216783 | 3.62E-05 | 0.000134478 | 0.175459134 |
| T2DM | mast_cell | *ZNF721* | 1.708536549 | 2.87E-05 | 0.000134478 | 0.139359752 |
| T2DM | mast_cell | *C11orf54* | 0.837767052 | 0.005488515 | 0.007050642 | 1 |
| T2DM | mast_cell | *CEP63* | 1.441909238 | 0.000204188 | 0.000354588 | 0.990719562 |
| T2DM | mast_cell | *HMGN4* | 2.485358317 | 0.001084782 | 0.001562267 | 1 |
| T2DM | mast_cell | *ZNF320* | 1.56192952 | 2.17E-05 | 0.000134478 | 0.105227958 |
| T2DM | mast_cell | *C12orf60* | 2.220779566 | 3.94E-05 | 0.000134478 | 0.191115104 |

| | | | | | | |
|---|---|---|---|---|---|---|
| T2DM | mast_cell | *ABAT* | 2.471060445 | 8.25E-05 | 0.000178055 | 0.400250625 |
| T2DM | mast_cell | *LYSMD4* | 1.570242893 | 3.47E-05 | 0.000134478 | 0.168409564 |
| T2DM | mast_cell | *WBP2NL* | 1.432964295 | 8.18E-05 | 0.000177103 | 0.396887857 |
| T2DM | mast_cell | *ARHGEF37* | 2.006568707 | 3.93E-05 | 0.000134478 | 0.190635241 |
| T2DM | mast_cell | *CSMD1* | 1.764590609 | 4.20E-05 | 0.000134478 | 0.203779264 |
| T2DM | mast_cell | *FANCF* | -3.124499044 | 0.001202734 | 0.001712344 | 1 |
| T2DM | mast_cell | *SMDT1* | 1.350509039 | 0.001318087 | 0.001862364 | 1 |
| T2DM | mast_cell | *C2CD4C* | 1.092685382 | 0.022097622 | 0.026677696 | 1 |
| T2DM | mast_cell | *RUVBL2* | 2.074564089 | 2.66E-05 | 0.000134478 | 0.128842861 |
| T2DM | mast_cell | *GDPGP1* | 2.482906848 | 0.001635689 | 0.002277292 | 1 |
| T2DM | mast_cell | *PTTG1IP* | 2.063022654 | 0.000172148 | 0.000308785 | 0.83526447 |
| T2DM | mast_cell | *ZNF623* | 2.807975667 | 0.008671362 | 0.010888574 | 1 |
| T2DM | mast_cell | *CCDC125* | 1.343879967 | 3.88E-05 | 0.000134478 | 0.188444988 |
| T2DM | mast_cell | *C10orf107* | 1.075572776 | 0.001397375 | 0.001965806 | 1 |
| T2DM | mast_cell | *FHL3* | -1.048738099 | 0.009747197 | 0.012182741 | 1 |
| T2DM | mast_cell | *C19orf71* | 2.694334754 | 6.54E-05 | 0.000153213 | 0.317294801 |
| T2DM | mast_cell | *LRIT3* | -1.391847959 | 0.000135507 | 0.000255156 | 0.657480601 |
| T2DM | mast_cell | *NPIPA1* | 1.314466706 | 0.000283559 | 0.000468925 | 1 |
| T2DM | mast_cell | *SF3A3* | 2.749207367 | 7.23E-05 | 0.000162818 | 0.351034916 |
| T2DM | mast_cell | *GPR132* | 1.450121287 | 0.000870116 | 0.001276241 | 1 |
| T2DM | mast_cell | *PSMG1* | 3.461379752 | 4.44E-05 | 0.000134478 | 0.215433685 |
| T2DM | mast_cell | *AC131971.1* | 5.763429514 | 0.009415972 | 0.01178697 | 1 |
| T2DM | mast_cell | *PCBP3* | 0.801690502 | 0.002258456 | 0.003069476 | 1 |
| T2DM | mast_cell | *SFXN4* | 3.892668768 | 0.000117402 | 0.000228402 | 0.56963492 |
| T2DM | mast_cell | *MRPL54* | 1.493238925 | 0.001791996 | 0.002478553 | 1 |
| T2DM | mast_cell | *HMCES* | 1.356207171 | 5.73E-05 | 0.000142472 | 0.278161858 |
| T2DM | mast_cell | *DGCR6* | -2.755603345 | 0.027555636 | 0.032922912 | 1 |
| T2DM | mast_cell | *NDUFB1* | 2.125078046 | 2.41E-05 | 0.000134478 | 0.116789072 |
| T2DM | mast_cell | *FAM19A1* | 1.487788343 | 3.04E-05 | 0.000134478 | 0.147367518 |
| T2DM | mast_cell | *TRMT12* | 2.862766285 | 2.59E-05 | 0.000134478 | 0.125582023 |
| T2DM | mast_cell | *ALYREF* | 1.588613131 | 0.0412125 | 0.048346966 | 1 |
| T2DM | mast_cell | *MRGPRX2* | -0.39459259 | 0.000117952 | 0.000228738 | 0.572303226 |
| T2DM | mast_cell | *UPP1* | 1.764245382 | 0.000144198 | 0.000266939 | 0.699647854 |
| T2DM | mast_cell | *FIGLA* | -3.740958939 | 0.00025594 | 0.000429246 | 1 |
| T2DM | mast_cell | *ACSM2A* | -4.145526523 | 0.00013704 | 0.000257222 | 0.66491855 |
| T2DM | mast_cell | *KREMEN1* | -0.758319758 | 6.24E-05 | 0.000149337 | 0.302856396 |
| T2DM | mast_cell | *ZNF703* | 2.218941836 | 0.005149321 | 0.006641283 | 1 |
| T2DM | mast_cell | *NPIPA5* | -0.793746823 | 0.000511552 | 0.000784466 | 1 |
| T2DM | mast_cell | *EMILIN3* | 5.211495812 | 0.00034034 | 0.000548068 | 1 |
| T2DM | mast_cell | *FAM162B* | 1.274238631 | 0.000708446 | 0.001056179 | 1 |
| T2DM | mast_cell | *BTBD9* | 2.029640264 | 3.36E-05 | 0.000134478 | 0.163072926 |
| T2DM | mast_cell | *NUDT14* | 2.837419187 | 6.14E-05 | 0.000147753 | 0.297870215 |
| T2DM | mast_cell | *TTC32* | 3.443873466 | 3.83E-05 | 0.000134478 | 0.185830371 |
| T2DM | mast_cell | *DENND5A* | 3.032543536 | 5.46E-05 | 0.000139259 | 0.264890737 |
| T2DM | mast_cell | *NIPSNAP1* | 2.014814073 | 0.003745166 | 0.004916544 | 1 |
| T2DM | mast_cell | *CNTN2* | -1.650850915 | 0.017332859 | 0.021151668 | 1 |
| T2DM | mast_cell | *CRELD2* | 1.960570071 | 3.52E-05 | 0.000134478 | 0.170746934 |
| T2DM | mast_cell | *PGP* | 2.151045946 | 0.000252054 | 0.000424052 | 1 |
| T2DM | mast_cell | *ACOT1* | 1.607705749 | 0.017131461 | 0.020921684 | 1 |
| T2DM | mast_cell | *OAF* | 0.778889603 | 0.017081605 | 0.020866049 | 1 |

| | | | | | | |
|---|---|---|---|---|---|---|
| T2DM | mast_cell | *ALDH1A3* | 1.03021525 | 0.001182731 | 0.001686834 | 1 |
| T2DM | mast_cell | *CLECL1* | 2.645062448 | 1.84E-05 | 0.000134478 | 0.089133249 |
| T2DM | mast_cell | *MROH7* | -0.922485108 | 0.014327781 | 0.017630838 | 1 |
| T2DM | mast_cell | *PKP3* | 16.793052 | 0.01312219 | 0.016204852 | 1 |
| T2DM | mast_cell | *SS18L1* | 3.966488413 | 3.99E-05 | 0.000134478 | 0.193769635 |
| T2DM | mast_cell | *TOP1MT* | 1.494158815 | 0.002406193 | 0.00325386 | 1 |
| T2DM | mast_cell | *COPB2* | 4.071498989 | 0.000105964 | 0.000211747 | 0.514135041 |
| T2DM | mast_cell | *WDR27* | 2.561604542 | 1.11E-05 | 0.000134478 | 0.053619664 |
| T2DM | mast_cell | *TXNRD2* | 3.451515838 | 1.15E-05 | 0.000134478 | 0.055995884 |
| T2DM | mast_cell | *TMEM255B* | 2.060433403 | 4.46E-05 | 0.000134478 | 0.2161748 |
| T2DM | mast_cell | *ZFP1* | 2.789340757 | 0.021678018 | 0.026177636 | 1 |
| T2DM | mast_cell | *DUSP8* | 3.389083313 | 0.000393359 | 0.000620676 | 1 |
| T2DM | mast_cell | *SNN* | 5.460828963 | 3.23E-05 | 0.000134478 | 0.156513933 |
| T2DM | mast_cell | *RNLS* | 1.310311057 | 6.05E-05 | 0.000146324 | 0.293712406 |
| T2DM | mast_cell | *NDUFA12* | 4.021480041 | 0.000448455 | 0.000696512 | 1 |
| T2DM | mast_cell | *TCTE3* | 1.754015148 | 5.38E-05 | 0.000138064 | 0.26080996 |
| T2DM | mast_cell | *UBE2G2* | 1.659471307 | 9.71E-05 | 0.000198686 | 0.471083491 |
| T2DM | mast_cell | *TMED9* | 2.145488011 | 0.000111055 | 0.000219664 | 0.53883667 |
| T2DM | mast_cell | *DRD1* | 44.72532191 | 3.65E-05 | 0.000134478 | 0.176955079 |
| T2DM | mast_cell | *BTBD6* | 1.404059897 | 0.001763974 | 0.002443975 | 1 |
| T2DM | mast_cell | *WT1* | 1.317817972 | 1.38E-05 | 0.000134478 | 0.066912252 |
| T2DM | mast_cell | *ZFP90* | 3.84313317 | 4.45E-05 | 0.000134478 | 0.215863253 |
| T2DM | mast_cell | *FAM227A* | 1.15329062 | 0.040749259 | 0.047815091 | 1 |
| T2DM | mast_cell | *NOC4L* | 3.2503743 | 5.65E-05 | 0.000141307 | 0.2741355 |
| T2DM | mast_cell | *NDUFA6* | 1.756575265 | 1.49E-06 | 0.000100592 | 0.007231837 |
| T2DM | mast_cell | *TMEM106A* | 1.887760298 | 0.000897347 | 0.00131103 | 1 |
| T2DM | mast_cell | *BRI3BP* | 1.852585365 | 5.09E-05 | 0.000136231 | 0.246987571 |
| T2DM | mast_cell | *MAFF* | 1.637482825 | 0.002020829 | 0.002768227 | 1 |
| T2DM | mast_cell | *SEMA4B* | 1.611736436 | 3.69E-05 | 0.000134478 | 0.179023138 |
| T2DM | mast_cell | *CIB1* | 1.726079826 | 0.004981659 | 0.006443884 | 1 |
| T2DM | mast_cell | *SGCZ* | 0.95232004 | 0.001156775 | 0.001654679 | 1 |
| T2DM | mast_cell | *C6orf120* | 3.233584765 | 7.20E-05 | 0.000162496 | 0.349547363 |
| T2DM | mast_cell | *SIGIRR* | 3.538818964 | 0.000331933 | 0.000535954 | 1 |
| T2DM | mast_cell | *IFITM2* | 3.119109885 | 4.18E-05 | 0.000134478 | 0.202649923 |
| T2DM | mast_cell | *GP1BA* | 3.673140573 | 2.61E-07 | 4.69E-05 | 0.001265871 |
| T2DM | mast_cell | *CCDC137* | 1.609478345 | 1.05E-05 | 0.000134478 | 0.051163712 |
| T2DM | mast_cell | *CDK10* | 1.212918799 | 0.023760693 | 0.028614267 | 1 |
| T2DM | mast_cell | *ATP6V0A2* | -0.999911543 | 0.000582067 | 0.000881182 | 1 |
| T2DM | mast_cell | *C14orf80* | 3.601559565 | 0.000369276 | 0.000589191 | 1 |
| T2DM | mast_cell | *TNFAIP8L1* | 5.890531099 | 0.001973473 | 0.002710244 | 1 |
| T2DM | mast_cell | *SP140L* | 2.770301022 | 1.54E-05 | 0.000134478 | 0.074798516 |
| T2DM | mast_cell | *TARSL2* | 1.958625162 | 0.000277091 | 0.00045964 | 1 |
| T2DM | mast_cell | *FAM174B* | 1.598904454 | 2.79E-05 | 0.000134478 | 0.135355528 |
| T2DM | mast_cell | *SV2B* | 1.726037576 | 0.000254774 | 0.000427885 | 1 |
| T2DM | mast_cell | *AHNAK2* | -1.717137284 | 0.000162772 | 0.000295647 | 0.78977174 |
| T2DM | mast_cell | *SP1* | 2.089738387 | 2.94E-05 | 0.000134478 | 0.142427818 |
| T2DM | mast_cell | *PCGF3* | 2.078158816 | 3.28E-05 | 0.000134478 | 0.159213022 |
| T2DM | mast_cell | *PMEL* | -1.308987914 | 0.001364683 | 0.001923161 | 1 |
| T2DM | mast_cell | *MYBL1* | 1.853625628 | 0.000259486 | 0.000434501 | 1 |
| T2DM | mast_cell | *ADARB2* | 5.36192634 | 0.014879741 | 0.018291488 | 1 |

| | | | | | | |
|---|---|---|---|---|---|---|
| T2DM | mast_cell | *DMWD* | 2.025622179 | 0.024979595 | 0.030015106 | 1 |
| T2DM | mast_cell | *SLC52A2* | 4.487631722 | 0.003145671 | 0.004170163 | 1 |
| T2DM | mast_cell | *ARL17A* | 2.854420913 | 1.24E-05 | 0.000134478 | 0.060098144 |
| T2DM | mast_cell | *GNB1L* | -1.352537129 | 0.015529816 | 0.019052002 | 1 |
| T2DM | mast_cell | *EVI2B* | 1.433953317 | 2.41E-05 | 0.000134478 | 0.116794851 |
| T2DM | mast_cell | *TRIM69* | 1.524875386 | 7.74E-05 | 0.000169981 | 0.375486943 |
| T2DM | mast_cell | *IFITM1* | -1.256043852 | 0.000110639 | 0.000218931 | 0.536820027 |
| T2DM | mast_cell | *LINC00839* | 5.402862491 | 0.00010602 | 0.000211747 | 0.514406724 |
| T2DM | mast_cell | *RTN4RL1* | 2.460727533 | 6.07E-05 | 0.00014641 | 0.294605948 |
| T2DM | mast_cell | *RNPC3* | 2.048815948 | 9.09E-06 | 0.000134478 | 0.044084531 |
| T2DM | mast_cell | *RASA3* | 1.108203701 | 0.003466975 | 0.004567408 | 1 |
| T2DM | mast_cell | *ZNF529* | 3.544973814 | 1.54E-05 | 0.000134478 | 0.074495889 |
| T2DM | mast_cell | *C15orf41* | 2.567993062 | 4.85E-06 | 0.000134478 | 0.023546031 |
| T2DM | mast_cell | *WWOX* | 1.998399422 | 3.83E-05 | 0.000134478 | 0.18566365 |
| T2DM | mast_cell | *POLR1D* | 2.637324213 | 4.42E-06 | 0.000134478 | 0.021459979 |
| T2DM | mast_cell | *FFAR4* | 1.993677213 | 1.14E-06 | 9.23E-05 | 0.005540905 |
| T2DM | mast_cell | *NBPF12* | 1.554085307 | 0.000168756 | 0.00030405 | 0.818805803 |
| T2DM | mast_cell | *TOR3A* | 2.349506091 | 0.000168002 | 0.000302922 | 0.81514419 |
| T2DM | mast_cell | *ZNF555* | 2.022767493 | 2.56E-05 | 0.000134478 | 0.124159938 |
| T2DM | mast_cell | *GOLGA8R* | 4.021014032 | 0.011521963 | 0.014321063 | 1 |
| T2DM | mast_cell | *CCDC30* | 1.483679539 | 5.73E-05 | 0.000142472 | 0.278152997 |
| T2DM | mast_cell | *GLDN* | 0.367015701 | 0.024598433 | 0.029571754 | 1 |
| T2DM | mast_cell | *ZNF197* | 1.770441406 | 9.46E-05 | 0.000195596 | 0.459064825 |
| T2DM | mast_cell | *RPS23* | 1.846126165 | 0.000164545 | 0.000298235 | 0.798374477 |
| T2DM | mast_cell | *GNG2* | 2.426242673 | 2.50E-05 | 0.000134478 | 0.121285578 |
| T2DM | mast_cell | *TMEM222* | 5.159096167 | 0.000188649 | 0.000331639 | 0.915323499 |
| T2DM | mast_cell | *SEPT10* | 1.2089989 | 9.50E-05 | 0.000196088 | 0.460770153 |
| T2DM | mast_cell | *FAM86B1* | 2.994651482 | 0.016180659 | 0.019815386 | 1 |
| T2DM | mast_cell | *SMYD4* | 2.506578738 | 7.39E-05 | 0.000165254 | 0.358668451 |
| T2DM | mast_cell | *MIR22HG* | 1.960441732 | 5.86E-05 | 0.000144186 | 0.284335454 |
| T2DM | mast_cell | *BCR* | 1.731090789 | 6.47E-05 | 0.00015243 | 0.314005652 |
| T2DM | mast_cell | *ZNF732* | 3.337840082 | 5.18E-05 | 0.000136402 | 0.25109809 |
| T2DM | mast_cell | *VSIG10L* | 3.543187445 | 0.005081706 | 0.006563115 | 1 |
| T2DM | mast_cell | *TRABD2A* | 1.327658445 | 0.001409151 | 0.001980076 | 1 |
| T2DM | mast_cell | *QRFPR* | 6.912939922 | 0.013610407 | 0.016777869 | 1 |
| T2DM | mast_cell | *MAPT* | 1.971956237 | 3.83E-05 | 0.000134478 | 0.185717933 |
| T2DM | mast_cell | *TMEM17* | 1.741170144 | 0.03283654 | 0.038897191 | 1 |
| T2DM | mast_cell | *TNFRSF18* | 6.15762184 | 0.000170493 | 0.000306609 | 0.827231608 |
| T2DM | mast_cell | *C1QL4* | 22.40032776 | 5.37E-05 | 0.000138064 | 0.260775488 |
| T2DM | mast_cell | *RTN4RL2* | -2.714437557 | 7.11E-05 | 0.000161326 | 0.344753459 |
| T2DM | mast_cell | *TMEM232* | 1.351864362 | 0.000186128 | 0.000328039 | 0.903091104 |
| T2DM | mast_cell | *EFCAB6* | 1.24159736 | 3.09E-05 | 0.000134478 | 0.149749912 |
| T2DM | mast_cell | *RHD* | 1.581649099 | 2.13E-05 | 0.000134478 | 0.103274844 |
| T2DM | mast_cell | *TMPRSS6* | 13.11599833 | 5.50E-05 | 0.000139632 | 0.26662063 |
| T2DM | mast_cell | *TMEM216* | 3.851405964 | 3.06E-06 | 0.000134478 | 0.014847265 |
| T2DM | mast_cell | *C3orf70* | 1.763328329 | 1.94E-05 | 0.000134478 | 0.094075555 |
| T2DM | mast_cell | *ENTPD5* | 1.572221294 | 0.001324284 | 0.001869487 | 1 |
| T2DM | mast_cell | *MRPS21* | 1.259519666 | 0.000442764 | 0.000688555 | 1 |
| T2DM | mast_cell | *ANGPTL5* | -1.381241935 | 0.002414962 | 0.00326299 | 1 |
| T2DM | mast_cell | *WDR86* | 1.783279715 | 3.67E-06 | 0.000134478 | 0.017785779 |

| | | | | | | |
|---|---|---|---|---|---|---|
| T2DM | mast_cell | *EPOR* | 3.600447046 | 2.73E-05 | 0.000134478 | 0.132304445 |
| T2DM | mast_cell | *LUZP2* | 3.383632506 | 3.82E-05 | 0.000134478 | 0.185174166 |
| T2DM | mast_cell | *GJA4* | 3.237327596 | 3.40E-05 | 0.000134478 | 0.16481802 |
| T2DM | mast_cell | *IFT140* | 2.211334825 | 4.27E-05 | 0.000134478 | 0.20711706 |
| T2DM | mast_cell | *SBK2* | 17.50040016 | 4.42E-05 | 0.000134478 | 0.214510715 |
| T2DM | mast_cell | *EXD3* | 4.041263542 | 4.93E-05 | 0.000135163 | 0.239382263 |
| T2DM | mast_cell | *C17orf97* | 0.415490289 | 0.03904321 | 0.045890904 | 1 |
| T2DM | mast_cell | *C2orf88* | 2.195307373 | 1.81E-05 | 0.000134478 | 0.087739639 |
| T2DM | mast_cell | *KBTBD12* | 1.65578403 | 1.35E-05 | 0.000134478 | 0.065475724 |
| T2DM | mast_cell | *FANCA* | 1.298247353 | 0.008895994 | 0.011156207 | 1 |
| T2DM | mast_cell | *FANCM* | 2.144739673 | 0.003464856 | 0.004565856 | 1 |
| T2DM | mast_cell | *ZFP69* | 2.608940572 | 3.83E-05 | 0.000134478 | 0.185837726 |
| T2DM | mast_cell | *TMEM220* | 3.057865039 | 3.24E-06 | 0.000134478 | 0.015698963 |
| T2DM | mast_cell | *FAM122A* | 2.259700022 | 4.82E-05 | 0.000134478 | 0.233956438 |
| T2DM | mast_cell | *GFRAL* | 4.510648388 | 4.94E-05 | 0.000135163 | 0.239599192 |
| T2DM | mast_cell | *LDLRAD2* | 1.616742974 | 0.002596045 | 0.003486303 | 1 |
| T2DM | mast_cell | *CPSF4L* | 6.628391518 | 0.001309507 | 0.001851319 | 1 |
| T2DM | mast_cell | *ZSCAN23* | 0.883632344 | 0.024804387 | 0.029811961 | 1 |
| T2DM | mast_cell | *MESP2* | 1.992448412 | 0.031116139 | 0.036976611 | 1 |
| T2DM | mast_cell | *MAPK12* | 3.185219688 | 0.000384638 | 0.000609691 | 1 |
| T2DM | mast_cell | *AGRN* | 2.243075992 | 4.93E-05 | 0.000135163 | 0.239389323 |
| T2DM | mast_cell | *FAM166A* | -4.315475797 | 0.00246912 | 0.003327825 | 1 |
| T2DM | mast_cell | *ZNF626* | 3.356870887 | 3.18E-05 | 0.000134478 | 0.154398622 |
| T2DM | mast_cell | *LINC00265* | 2.244634276 | 7.21E-05 | 0.000162496 | 0.349691612 |
| T2DM | mast_cell | *LAMTOR4* | 2.383649384 | 2.92E-05 | 0.000134478 | 0.141561828 |
| T2DM | mast_cell | *PRKAR1B* | 2.850207416 | 0.000526089 | 0.000802951 | 1 |
| T2DM | mast_cell | *NCR3LG1* | -2.60064521 | 0.000247621 | 0.000417752 | 1 |
| T2DM | mast_cell | *ZNF793* | 1.442540522 | 4.32E-05 | 0.000134478 | 0.209736619 |
| T2DM | mast_cell | *AGAP4* | 2.06756084 | 7.01E-05 | 0.000159988 | 0.34029477 |
| T2DM | mast_cell | *RUFY4* | 3.814299032 | 4.68E-05 | 0.000134478 | 0.227109647 |
| T2DM | mast_cell | *ZNF669* | 2.92590013 | 0.000192491 | 0.000337172 | 0.933966598 |
| T2DM | mast_cell | *CENPP* | 1.134967791 | 0.00031323 | 0.000509314 | 1 |
| T2DM | mast_cell | *PLSCR1* | 1.935419458 | 2.74E-05 | 0.000134478 | 0.133076292 |
| T2DM | mast_cell | *C3orf62* | 1.903766847 | 9.20E-06 | 0.000134478 | 0.044649855 |
| T2DM | mast_cell | *ENO4* | 1.489930064 | 8.36E-05 | 0.000179257 | 0.405413323 |
| T2DM | mast_cell | *ZP3* | 0.849512519 | 0.024389043 | 0.029349116 | 1 |
| T2DM | mast_cell | *H3F3C* | -7.572513516 | 0.000128417 | 0.000245018 | 0.623080064 |
| T2DM | mast_cell | *JAKMIP3* | 1.341147316 | 0.001417157 | 0.001990748 | 1 |
| T2DM | mast_cell | *PDCD1* | 10.57716384 | 0.001089912 | 0.001568283 | 1 |
| T2DM | mast_cell | *SELL* | 1.923324706 | 0.002385238 | 0.003227321 | 1 |
| T2DM | mast_cell | *C19orf54* | 2.735272943 | 4.03E-05 | 0.000134478 | 0.19533736 |
| T2DM | mast_cell | *C22orf34* | 1.32449074 | 4.30E-05 | 0.000134478 | 0.208693827 |
| T2DM | mast_cell | *COL25A1* | 1.082709854 | 5.08E-05 | 0.000136174 | 0.246474529 |
| T2DM | mast_cell | *SRSF10* | 1.948113337 | 4.00E-05 | 0.000134478 | 0.193863403 |
| T2DM | mast_cell | *NBR1* | 1.883203695 | 5.02E-05 | 0.0001354 | 0.243500909 |
| T2DM | mast_cell | *NDOR1* | 3.085839614 | 3.65E-05 | 0.000134478 | 0.177070152 |
| T2DM | mast_cell | *FAM72B* | 4.097241096 | 0.004093488 | 0.005350648 | 1 |
| T2DM | mast_cell | *GOLGA8M* | 2.137021794 | 4.15E-05 | 0.000134478 | 0.201572568 |
| T2DM | mast_cell | *RHCE* | 6.014732257 | 0.001767041 | 0.002447526 | 1 |
| T2DM | mast_cell | *IDO2* | 5.276967179 | 3.38E-05 | 0.000134478 | 0.164138485 |

| | | | | | | |
|---|---|---|---|---|---|---|
| T2DM | mast_cell | *SLC4A5* | 5.559341071 | 5.99E-05 | 0.000145532 | 0.290626478 |
| T2DM | mast_cell | *UROS* | 2.402746153 | 3.97E-05 | 0.000134478 | 0.192656407 |
| T2DM | mast_cell | *ZBED6CL* | -4.019243728 | 9.26E-05 | 0.000192728 | 0.449398529 |
| T2DM | mast_cell | *OSTN* | 2.158725118 | 0.000178493 | 0.000317582 | 0.866046181 |
| T2DM | mast_cell | *FSIP2* | 1.61353187 | 9.35E-05 | 0.00019387 | 0.453461465 |
| T2DM | mast_cell | *NOXA1* | 1.688855688 | 4.74E-05 | 0.000134478 | 0.230055512 |
| T2DM | mast_cell | *FZD9* | -2.478545907 | 0.000102669 | 0.000206615 | 0.498149482 |
| T2DM | mast_cell | *SKOR1* | -2.138923358 | 0.000126189 | 0.000241813 | 0.612269689 |
| T2DM | mast_cell | *FAM26F* | 1.500041832 | 0.001998119 | 0.002740213 | 1 |
| T2DM | mast_cell | *ENTPD8* | 193.3625817 | 8.68E-05 | 0.000184458 | 0.421119113 |
| T2DM | mast_cell | *RPL14* | 2.550228991 | 3.12E-05 | 0.000134478 | 0.151199339 |
| T2DM | mast_cell | *FBF1* | 2.033278781 | 0.00021254 | 0.00036686 | 1 |
| T2DM | mast_cell | *ASTL* | -5.286094629 | 0.000410611 | 0.000642051 | 1 |
| T2DM | mast_cell | *MSL1* | 2.20309243 | 0.005108396 | 0.006593758 | 1 |
| T2DM | mast_cell | *LRRK2* | 1.847650995 | 9.52E-06 | 0.000134478 | 0.046178797 |
| T2DM | mast_cell | *FAM196A* | 2.135550711 | 4.40E-05 | 0.000134478 | 0.213516343 |
| T2DM | mast_cell | *UTS2B* | 2.091625304 | 0.000193735 | 0.000338983 | 0.93999987 |
| T2DM | mast_cell | *C9orf152* | -2.547586369 | 0.00036799 | 0.000587718 | 1 |
| T2DM | mast_cell | *SLC15A5* | 2.538571371 | 2.47E-05 | 0.000134478 | 0.119643544 |
| T2DM | mast_cell | *ALKBH2* | 3.697917712 | 1.37E-06 | 9.63E-05 | 0.006647176 |
| T2DM | mast_cell | *LITAF* | 2.749630589 | 2.74E-05 | 0.000134478 | 0.132964218 |
| T2DM | mast_cell | *VSTM1* | -1.234668111 | 0.026170336 | 0.031337234 | 1 |
| T2DM | mast_cell | *SF3B3* | 1.988994027 | 5.20E-05 | 0.00013644 | 0.252140739 |
| T2DM | mast_cell | *PLAC9* | 2.574143119 | 0.001595467 | 0.002228968 | 1 |
| T2DM | mast_cell | *ZNF573* | 2.139011219 | 0.000227333 | 0.000388114 | 1 |
| T2DM | mast_cell | *FAM47E* | 3.299613499 | 0.035416384 | 0.04180012 | 1 |
| T2DM | mast_cell | *ZNF600* | 1.63930295 | 0.012384389 | 0.015328841 | 1 |
| T2DM | mast_cell | *TSPYL1* | 6.08047455 | 0.001787633 | 0.002474635 | 1 |
| T2DM | mast_cell | *PNRC2* | 1.492602231 | 5.93E-05 | 0.000144951 | 0.287879074 |
| T2DM | mast_cell | *FHIT* | 1.479310709 | 3.98E-05 | 0.000134478 | 0.192930717 |
| T2DM | mast_cell | *ZKSCAN3* | 1.474930909 | 0.031394751 | 0.037289433 | 1 |
| T2DM | mast_cell | *SLC35E2B* | 1.763103959 | 3.05E-05 | 0.000134478 | 0.147992481 |
| T2DM | mast_cell | *BLOC1S2* | 1.104344238 | 0.00803994 | 0.010127152 | 1 |
| T2DM | mast_cell | *AKR1C3* | 4.6445828 | 5.34E-05 | 0.000137618 | 0.258996842 |
| T2DM | mast_cell | *WDSUB1* | 2.630684695 | 9.34E-06 | 0.000134478 | 0.045302634 |
| T2DM | mast_cell | *COLCA1* | -3.589900641 | 0.000667499 | 0.000999908 | 1 |
| T2DM | mast_cell | *SEMA4A* | 3.517567704 | 8.80E-05 | 0.000186018 | 0.426911586 |
| T2DM | mast_cell | *MPHOSPH8* | 1.514554989 | 3.98E-05 | 0.000134478 | 0.193141495 |
| T2DM | mast_cell | *SRGAP3* | 3.141259838 | 3.59E-06 | 0.000134478 | 0.017441621 |
| T2DM | mast_cell | *XPNPEP3* | 1.118400662 | 0.020718142 | 0.025087203 | 1 |
| T2DM | mast_cell | *PPIA* | 2.316927179 | 0.000201338 | 0.000350644 | 0.976894348 |
| T2DM | mast_cell | *ZNF471* | 2.56383818 | 3.96E-05 | 0.000134478 | 0.192163613 |
| T2DM | mast_cell | *ZNF493* | 2.256303404 | 6.51E-05 | 0.0001528 | 0.315685032 |
| T2DM | mast_cell | *GTF2IRD2* | 2.469206176 | 1.34E-05 | 0.000134478 | 0.065125324 |
| T2DM | mast_cell | *SUPT3H* | 2.563729025 | 7.68E-05 | 0.000168895 | 0.37241354 |
| T2DM | mast_cell | *GIMAP5* | 1.930506213 | 0.000853098 | 0.001254692 | 1 |
| T2DM | mast_cell | *STK31* | 2.891758051 | 8.08E-06 | 0.000134478 | 0.039221112 |
| T2DM | mast_cell | *ZKSCAN7* | 2.305691127 | 1.24E-06 | 9.40E-05 | 0.006017205 |
| T2DM | mast_cell | *CD55* | 1.463553529 | 3.81E-05 | 0.000134478 | 0.184760216 |
| T2DM | mast_cell | *ZNF565* | 2.429892262 | 0.001169807 | 0.001671329 | 1 |

| | | | | | | |
|---|---|---|---|---|---|---|
| T2DM | mast_cell | *LONP1* | 2.235604164 | 8.91E-05 | 0.000187451 | 0.432263051 |
| T2DM | mast_cell | *ZNF34* | 3.355270463 | 2.40E-06 | 0.000128421 | 0.011668653 |
| T2DM | mast_cell | *ZNF781* | 0.912409566 | 0.019521653 | 0.023691611 | 1 |
| T2DM | mast_cell | *EVL* | 2.284776254 | 3.55E-05 | 0.000134478 | 0.17243505 |
| T2DM | mast_cell | *ZNF124* | 2.1311461 | 5.39E-05 | 0.000138255 | 0.261580656 |
| T2DM | mast_cell | *PIK3R4* | 2.811641707 | 3.52E-05 | 0.000134478 | 0.170935928 |
| T2DM | mast_cell | *ZNF605* | 1.596136123 | 5.74E-06 | 0.000134478 | 0.027834732 |
| T2DM | mast_cell | *RFX8* | 1.156287694 | 0.0014063 | 0.001977215 | 1 |
| T2DM | mast_cell | *C20orf96* | 1.927006779 | 5.64E-05 | 0.000141146 | 0.273682529 |
| T2DM | mast_cell | *SULT1A1* | 2.271099408 | 0.000612316 | 0.000924955 | 1 |
| T2DM | mast_cell | *TPK1* | 1.716835757 | 1.51E-05 | 0.000134478 | 0.073344068 |
| T2DM | mast_cell | *MYO18A* | 3.714013887 | 0.000694835 | 0.001037654 | 1 |
| T2DM | mast_cell | *SPTSSB* | 4.947675175 | 0.000347463 | 0.000558611 | 1 |
| T2DM | mast_cell | *CACNA1H* | 5.30667021 | 4.59E-05 | 0.000134478 | 0.222477451 |
| T2DM | mast_cell | *SULF2* | 1.721798623 | 1.04E-06 | 9.21E-05 | 0.005057671 |
| T2DM | mast_cell | *XRCC2* | 3.946424637 | 5.10E-05 | 0.000136328 | 0.247473524 |
| T2DM | mast_cell | *MYO6* | 2.42738841 | 0.000138893 | 0.000260096 | 0.673910023 |
| T2DM | mast_cell | *ZNF846* | 1.733893076 | 1.32E-05 | 0.000134478 | 0.064100981 |
| T2DM | mast_cell | *ADH1B* | 1.317268924 | 3.41E-05 | 0.000134478 | 0.165258634 |
| T2DM | mast_cell | *ZKSCAN5* | 3.696216252 | 1.89E-06 | 0.000115875 | 0.009154156 |
| T2DM | mast_cell | *ZNF502* | 3.427318063 | 0.000166343 | 0.000300931 | 0.807097808 |
| T2DM | mast_cell | *TRAPPC4* | 2.182406403 | 0.001029593 | 0.001487667 | 1 |
| T2DM | mast_cell | *ZFP62* | 3.4330363 | 3.32E-05 | 0.000134478 | 0.161005209 |
| T2DM | mast_cell | *TOMM7* | 2.289196437 | 3.15E-05 | 0.000134478 | 0.152599359 |
| T2DM | mast_cell | *ZNF33B* | 1.59896872 | 0.015983408 | 0.019593606 | 1 |
| T2DM | mast_cell | *AMZ2* | 2.66500442 | 0.035414485 | 0.04180012 | 1 |
| T2DM | mast_cell | *ZNF431* | 2.256457878 | 6.84E-05 | 0.000157548 | 0.331795281 |
| T2DM | mast_cell | *HLA-DQA1* | -3.851803881 | 4.26E-11 | 6.89E-08 | 2.07E-07 |
| T2DM | mast_cell | *COL27A1* | 1.294858683 | 0.003410768 | 0.004499384 | 1 |
| T2DM | mast_cell | *ZNF239* | 4.17682249 | 1.14E-06 | 9.23E-05 | 0.005517883 |
| T2DM | mast_cell | *ANKRD36B* | 2.533966839 | 0.000113989 | 0.000224319 | 0.553074562 |
| T2DM | mast_cell | *FAM3C* | 2.749520072 | 0.003570819 | 0.004696561 | 1 |
| T2DM | mast_cell | *SLC39A10* | 2.653572596 | 3.51E-05 | 0.000134478 | 0.170283838 |
| T2DM | mast_cell | *AP2A1* | 1.824808765 | 3.09E-05 | 0.000134478 | 0.15002207 |
| T2DM | mast_cell | *PCDHB16* | 1.377011985 | 0.008247328 | 0.010369535 | 1 |
| T2DM | mast_cell | *FUT11* | 4.79272561 | 2.03E-05 | 0.000134478 | 0.098306288 |
| T2DM | mast_cell | *WDR5B* | -1.803563225 | 0.003205747 | 0.004244007 | 1 |
| T2DM | mast_cell | *ZNF138* | 4.473570808 | 0.000413003 | 0.000645375 | 1 |
| T2DM | mast_cell | *ZNF429* | 2.144998357 | 3.29E-05 | 0.000134478 | 0.159499127 |
| T2DM | mast_cell | *ZNF470* | -2.897247462 | 0.00012627 | 0.000241872 | 0.612661096 |
| T2DM | mast_cell | *ANXA6* | 2.471420826 | 8.05E-05 | 0.000175007 | 0.390791378 |
| T2DM | mast_cell | *GMFB* | 3.705227213 | 3.37E-05 | 0.000134478 | 0.163496993 |
| T2DM | mast_cell | *ZNF420* | 2.274003609 | 4.10E-05 | 0.000134478 | 0.199033901 |
| T2DM | mast_cell | *ZMYM1* | 2.803477788 | 3.49E-05 | 0.000134478 | 0.169557295 |
| T2DM | mast_cell | *IGF2R* | 1.653399386 | 6.39E-05 | 0.000151412 | 0.310091675 |
| T2DM | mast_cell | *GAL3ST4* | 3.587779095 | 9.21E-05 | 0.000192339 | 0.446802504 |
| T2DM | mast_cell | *SLC25A29* | 2.999720502 | 2.00E-05 | 0.000134478 | 0.097170746 |
| T2DM | mast_cell | *ZNF257* | 3.227082489 | 0.000615073 | 0.000928254 | 1 |
| T2DM | mast_cell | *ADAM32* | 2.727218093 | 3.83E-05 | 0.000134478 | 0.185691318 |
| T2DM | mast_cell | *LRRC8B* | 2.59286217 | 2.00E-05 | 0.000134478 | 0.097097943 |

| | | | | | | |
|---|---|---|---|---|---|---|
| T2DM | mast_cell | *ABCB8* | 1.602882427 | 0.000101498 | 0.000205367 | 0.492469732 |
| T2DM | mast_cell | *SND1* | 2.060509663 | 5.77E-05 | 0.000142926 | 0.279925019 |
| T2DM | mast_cell | *SULT1A2* | 3.676864943 | 0.000308118 | 0.000502349 | 1 |
| T2DM | mast_cell | *NEK5* | 1.16163358 | 0.000960348 | 0.00139551 | 1 |
| T2DM | mast_cell | *ENTPD4* | 1.619749256 | 0.000106853 | 0.000212742 | 0.518452263 |
| T2DM | mast_cell | *TBC1D9B* | 2.235935097 | 0.000293436 | 0.000482791 | 1 |
| T2DM | mast_cell | *FAM110D* | 5.101283949 | 5.90E-07 | 7.18E-05 | 0.002860708 |
| T2DM | mast_cell | *CCL4L2* | 5.508152967 | 1.54E-05 | 0.000134478 | 0.074600868 |
| T2DM | mast_cell | *ZNF165* | -1.917116892 | 0.001958364 | 0.002691017 | 1 |
| T2DM | mast_cell | *RAMP2-AS1* | 1.500480979 | 3.09E-05 | 0.000134478 | 0.150065181 |
| T2DM | mast_cell | *ZNF720* | 1.92851361 | 0.000136631 | 0.000256554 | 0.662936017 |
| T2DM | mast_cell | *MRPL21* | 1.339895561 | 9.90E-05 | 0.000201516 | 0.480213583 |
| T2DM | mast_cell | *ZNF98* | 1.794720309 | 0.001959884 | 0.002692343 | 1 |
| T2DM | mast_cell | *ZNF517* | 5.549535291 | 0.000675217 | 0.001010417 | 1 |
| T2DM | mast_cell | *ZNF675* | 3.933171181 | 0.000205734 | 0.000357018 | 0.998222268 |
| T2DM | mast_cell | *SLC22A5* | 2.628434471 | 2.10E-05 | 0.000134478 | 0.101652709 |
| T2DM | mast_cell | *VEPH1* | 1.67651321 | 4.51E-05 | 0.000134478 | 0.218899386 |
| T2DM | mast_cell | *SHPK* | 2.021851921 | 0.000866543 | 0.001271768 | 1 |
| T2DM | mast_cell | *MAP3K5* | 1.440882315 | 4.06E-05 | 0.000134478 | 0.197174624 |
| T2DM | mast_cell | *GSTK1* | 1.264105529 | 0.000192485 | 0.000337172 | 0.933937595 |
| T2DM | mast_cell | *GYPE* | 3.254510435 | 3.33E-05 | 0.000134478 | 0.161779198 |
| T2DM | mast_cell | *PCDHB11* | -1.04674129 | 0.03204949 | 0.038029866 | 1 |
| T2DM | mast_cell | *ZNF665* | 4.91063068 | 4.25E-05 | 0.000134478 | 0.206424971 |
| T2DM | mast_cell | *RPF2* | 5.041830764 | 0.00089227 | 0.001304397 | 1 |
| T2DM | mast_cell | *MYO5A* | 1.127119367 | 6.68E-05 | 0.00015526 | 0.324338661 |
| T2DM | mast_cell | *ATG7* | 1.383424958 | 2.41E-05 | 0.000134478 | 0.116976905 |
| T2DM | mast_cell | *RAB40C* | 3.046092522 | 3.14E-05 | 0.000134478 | 0.152407367 |
| T2DM | mast_cell | *PIGN* | 1.96455413 | 5.09E-05 | 0.000136231 | 0.246945417 |
| T2DM | mast_cell | *HHLA3* | 1.085412304 | 0.015065182 | 0.01851007 | 1 |
| T2DM | mast_cell | *TOPORS* | 5.439024768 | 0.00035847 | 0.000574452 | 1 |
| T2DM | mast_cell | *BCO2* | 0.912832474 | 0.003197971 | 0.00423718 | 1 |
| T2DM | mast_cell | *ENPP1* | 1.346284446 | 0.000195309 | 0.000341369 | 0.947640788 |
| T2DM | mast_cell | *CCDC154* | 2.207955075 | 1.59E-05 | 0.000134478 | 0.077115474 |
| T2DM | mast_cell | *MFAP5* | 1.050713155 | 0.000211461 | 0.000365387 | 1 |
| T2DM | mast_cell | *PDCD1LG2* | 1.072801901 | 0.001246386 | 0.001768968 | 1 |
| T2DM | mast_cell | *DNAH10* | -3.542703677 | 0.000138183 | 0.000259138 | 0.67046576 |
| T2DM | mast_cell | *NMB* | 4.418072339 | 1.93E-05 | 0.000134478 | 0.093416116 |
| T2DM | mast_cell | *FAM114A1* | 2.180356473 | 2.36E-05 | 0.000134478 | 0.114368696 |
| T2DM | mast_cell | *PSAP* | 1.341181363 | 1.79E-05 | 0.000134478 | 0.086622609 |
| T2DM | mast_cell | *TXNRD3* | 1.482466217 | 4.26E-05 | 0.000134478 | 0.206681021 |
| T2DM | mast_cell | *EME2* | -3.041367208 | 0.001265379 | 0.001793637 | 1 |
| T2DM | mast_cell | *AC122129.1* | 14.22056378 | 5.78E-05 | 0.000143116 | 0.280507275 |
| T2DM | mast_cell | *OCLN* | -1.099600675 | 0.001696354 | 0.002354996 | 1 |
| T2DM | mast_cell | *ZNF181* | 1.870115169 | 2.78E-05 | 0.000134478 | 0.134821712 |
| T2DM | mast_cell | *SGTB* | 1.400577376 | 0.000368343 | 0.000587976 | 1 |
| T2DM | mast_cell | *ADH5* | 2.218891998 | 3.73E-05 | 0.000134478 | 0.180946081 |
| T2DM | mast_cell | *ZNF677* | 1.656136941 | 4.54E-05 | 0.000134478 | 0.220085541 |
| T2DM | mast_cell | *ZNF71* | 1.522571016 | 0.001388047 | 0.001953816 | 1 |
| T2DM | mast_cell | *RPL12* | 2.072270467 | 1.93E-05 | 0.000134478 | 0.093428799 |
| T2DM | mast_cell | *MPZL1* | 1.269235065 | 4.66E-05 | 0.000134478 | 0.225938275 |

| | | | | | | |
|---|---|---|---|---|---|---|
| T2DM | mast_cell | *VPS13A* | 1.840238186 | 1.78E-05 | 0.000134478 | 0.086156545 |
| T2DM | mast_cell | *NOL8* | 3.368715921 | 0.00011012 | 0.000218082 | 0.534300985 |
| T2DM | mast_cell | *DLGAP2* | 1.24465658 | 1.76E-05 | 0.000134478 | 0.085535825 |
| T2DM | mast_cell | *AGAP9* | 2.715634944 | 0.000475782 | 0.000736127 | 1 |
| T2DM | mast_cell | *MAK16* | 5.966549206 | 0.005473356 | 0.007033031 | 1 |
| T2DM | mast_cell | *ZNF667* | 2.110626799 | 0.000106657 | 0.000212439 | 0.517501416 |
| T2DM | mast_cell | *SFI1* | 2.881468817 | 2.22E-05 | 0.000134478 | 0.107593651 |
| T2DM | mast_cell | *ADH4* | 1.076259896 | 0.001927717 | 0.002651908 | 1 |
| T2DM | mast_cell | *HIBCH* | 1.411975261 | 0.000343606 | 0.000553144 | 1 |
| T2DM | mast_cell | *ZNF544* | 2.278167181 | 3.71E-05 | 0.000134478 | 0.179863203 |
| T2DM | mast_cell | *SOWAHC* | 3.332544391 | 5.96E-05 | 0.000145187 | 0.289069258 |
| T2DM | mast_cell | *AC135178.1* | -5.89480768 | 0.00028601 | 0.000472496 | 1 |
| T2DM | mast_cell | *MIER1* | 2.4999344 | 1.11E-05 | 0.000134478 | 0.053925985 |
| T2DM | mast_cell | *MAN1A2* | 2.162574505 | 3.37E-05 | 0.000134478 | 0.163731494 |
| T2DM | mast_cell | *SVIP* | 2.249405438 | 4.27E-05 | 0.000134478 | 0.206942701 |
| T2DM | mast_cell | *DDRGK1* | 1.885129143 | 3.67E-05 | 0.000134478 | 0.178136511 |
| T2DM | mast_cell | *ZNF607* | 1.82031702 | 0.010120266 | 0.012629509 | 1 |
| T2DM | mast_cell | *ZNF334* | 2.57746318 | 2.57E-05 | 0.000134478 | 0.124476186 |
| T2DM | mast_cell | *HSD17B11* | 3.005065715 | 2.48E-05 | 0.000134478 | 0.120191827 |
| T2DM | mast_cell | *SULT1C2* | 1.543657435 | 0.000210455 | 0.000363779 | 1 |
| T2DM | mast_cell | *RPS6KL1* | 4.228039133 | 4.32E-05 | 0.000134478 | 0.209385603 |
| T2DM | mast_cell | *DDX42* | 1.863390334 | 1.48E-05 | 0.000134478 | 0.071841368 |
| T2DM | mast_cell | *STYX* | 2.933493041 | 1.79E-05 | 0.000134478 | 0.087085498 |
| T2DM | mast_cell | *HELZ* | 1.707059885 | 3.09E-05 | 0.000134478 | 0.149921475 |
| T2DM | mast_cell | *UCKL1* | 3.317766508 | 2.70E-05 | 0.000134478 | 0.130994258 |
| T2DM | mast_cell | *ZNF442* | -1.698551247 | 0.000304458 | 0.000497384 | 1 |
| T2DM | mast_cell | *GFPT1* | 2.381913021 | 2.88E-05 | 0.000134478 | 0.139548401 |
| T2DM | mast_cell | *ZNF568* | 1.79615073 | 6.32E-05 | 0.000150474 | 0.306514595 |
| T2DM | mast_cell | *ZNF587* | 2.787436841 | 4.21E-05 | 0.000134478 | 0.204105787 |
| T2DM | mast_cell | *FLVCR1-AS1* | 0.702072658 | 0.015815555 | 0.01939274 | 1 |
| T2DM | mast_cell | *SH3BGRL2* | 1.064026387 | 0.000613345 | 0.000925937 | 1 |
| T2DM | mast_cell | *TMA16* | 2.177875824 | 3.35E-05 | 0.000134478 | 0.162518695 |
| T2DM | mast_cell | *HLA-DRB5* | -1.283215334 | 6.08E-12 | 1.48E-08 | 2.95E-08 |
| T2DM | mast_cell | *CNGA1* | 1.364846816 | 0.000217809 | 0.000373695 | 1 |
| T2DM | mast_cell | *ZNF28* | 4.517742021 | 6.73E-05 | 0.000155911 | 0.326633741 |
| T2DM | mast_cell | *C20orf203* | -1.280336744 | 0.000352535 | 0.000566016 | 1 |
| T2DM | mast_cell | *NCOA6* | 1.923073077 | 0.000149738 | 0.000275304 | 0.72652691 |
| T2DM | mast_cell | *FAM19A2* | 2.228559922 | 0.000108825 | 0.000216048 | 0.528021292 |
| T2DM | mast_cell | *TTC37* | 2.848707901 | 1.12E-05 | 0.000134478 | 0.054487249 |
| T2DM | mast_cell | *TUSC1* | 2.773909907 | 5.19E-05 | 0.000136402 | 0.25193396 |
| T2DM | mast_cell | *PAPSS2* | 1.390775809 | 3.93E-05 | 0.000134478 | 0.190587422 |
| T2DM | mast_cell | *CEP290* | 2.77663201 | 7.69E-05 | 0.000169168 | 0.373183631 |
| T2DM | mast_cell | *ANKRD13B* | 3.236758483 | 4.36E-05 | 0.000134478 | 0.211362217 |
| T2DM | mast_cell | *ECI2* | 3.669343616 | 0.000514105 | 0.000787722 | 1 |
| T2DM | mast_cell | *UNC13B* | 1.611365881 | 3.84E-05 | 0.000134478 | 0.186334053 |
| T2DM | mast_cell | *PPP1R14C* | 6.691831992 | 4.89E-05 | 0.000134955 | 0.237174821 |
| T2DM | mast_cell | *GPATCH3* | -3.934374912 | 0.008595271 | 0.01079582 | 1 |
| T2DM | mast_cell | *RPL10A* | 2.075054746 | 4.19E-05 | 0.000134478 | 0.203479467 |
| T2DM | mast_cell | *COLGALT2* | 2.704546419 | 0.00068289 | 0.001021073 | 1 |
| T2DM | mast_cell | *RCSD1* | 1.20420513 | 4.31E-05 | 0.000134478 | 0.208933061 |

| | | | | | | |
|---|---|---|---|---|---|---|
| T2DM | mast_cell | *TMEM184B* | 1.606928057 | 4.22E-05 | 0.000134478 | 0.204530516 |
| T2DM | mast_cell | *SFT2D1* | 2.725216547 | 3.85E-05 | 0.000134478 | 0.186574746 |
| T2DM | mast_cell | *HMGN2* | 1.404698346 | 0.000479686 | 0.000741222 | 1 |
| T2DM | mast_cell | *RYR3* | -1.210179649 | 0.0027558 | 0.003681481 | 1 |
| T2DM | mast_cell | *TYW1* | 2.013170791 | 2.07E-05 | 0.000134478 | 0.100330271 |
| T2DM | mast_cell | *PRMT6* | -2.162448804 | 0.000148066 | 0.000272644 | 0.718417629 |
| T2DM | mast_cell | *SHISA4* | -1.833385967 | 5.83E-05 | 0.000143677 | 0.282755811 |
| T2DM | mast_cell | *MAP3K3* | 2.170882839 | 0.005501641 | 0.007065633 | 1 |
| T2DM | mast_cell | *C1orf174* | 2.416682 | 0.000132602 | 0.000250638 | 0.643387007 |
| T2DM | mast_cell | *DZIP3* | 1.432027987 | 4.40E-05 | 0.000134478 | 0.213547543 |
| T2DM | mast_cell | *KIAA0753* | 1.257936699 | 0.000153016 | 0.000280694 | 0.742434932 |
| T2DM | mast_cell | *CCDC167* | 6.29713987 | 0.000116868 | 0.000227729 | 0.567044686 |
| T2DM | mast_cell | *ZFP2* | 2.63105199 | 5.25E-05 | 0.000136633 | 0.254820752 |
| T2DM | mast_cell | *MFAP3L* | 2.473988323 | 7.27E-05 | 0.000163182 | 0.3526357 |
| T2DM | mast_cell | *NAGA* | 1.778183802 | 0.000114472 | 0.000224769 | 0.555420328 |
| T2DM | mast_cell | *TGM2* | 1.499925019 | 0.000128084 | 0.000244538 | 0.621464735 |
| T2DM | mast_cell | *PJA2* | 1.805134079 | 5.96E-05 | 0.000145187 | 0.289257212 |
| T2DM | mast_cell | *AL590822.1* | -1.669315481 | 0.000218323 | 0.000374312 | 1 |
| T2DM | mast_cell | *AP001007.1* | 2.625225981 | 0.005569691 | 0.007147353 | 1 |
| T2DM | mast_cell | *EFCAB2* | 1.669681737 | 0.000106447 | 0.000212194 | 0.516481218 |
| T2DM | mast_cell | *TATDN3* | 1.801030024 | 0.000175545 | 0.000313488 | 0.851746341 |
| T2DM | mast_cell | *CR1* | 0.604493542 | 0.006502782 | 0.008276888 | 1 |
| T2DM | mast_cell | *SAMD5* | 2.38835529 | 5.60E-05 | 0.000140825 | 0.271723425 |
| T2DM | mast_cell | *ECT2L* | 2.779009297 | 4.29E-05 | 0.000134478 | 0.208041382 |
| T2DM | mast_cell | *GPR52* | -32.93287512 | 0.012959895 | 0.016016661 | 1 |
| T2DM | mast_cell | *DDO* | 2.819902674 | 0.000326743 | 0.000529158 | 1 |
| T2DM | mast_cell | *RBM20* | 3.086624308 | 0.000207319 | 0.000359126 | 1 |
| T2DM | mast_cell | *C6orf163* | 1.170203035 | 0.018915268 | 0.022995961 | 1 |
| T2DM | mast_cell | *PCMTD2* | 3.14449429 | 3.82E-05 | 0.000134478 | 0.185211104 |
| T2DM | mast_cell | *SAMD13* | 2.322099163 | 9.50E-05 | 0.000196088 | 0.460807593 |
| T2DM | mast_cell | *EFCAB7* | 3.43535408 | 2.43E-05 | 0.000134478 | 0.117874464 |
| T2DM | mast_cell | *IFIT1B* | 11.12710705 | 3.80E-05 | 0.000134478 | 0.18420224 |
| T2DM | mast_cell | *LIPN* | 4.877052279 | 5.04E-05 | 0.000135719 | 0.244566118 |
| T2DM | mast_cell | *INPP5B* | 0.900794522 | 0.01887479 | 0.022952501 | 1 |
| T2DM | mast_cell | *NEU4* | 3.87553265 | 0.002499427 | 0.003364 | 1 |
| T2DM | mast_cell | *RUFY2* | 2.004663926 | 3.24E-05 | 0.000134478 | 0.15705215 |
| T2DM | mast_cell | *PHACTR4* | 2.083945135 | 0.000399644 | 0.000627938 | 1 |
| T2DM | mast_cell | *ASAH2B* | 3.196192938 | 3.01E-06 | 0.000134478 | 0.014587237 |
| T2DM | mast_cell | *AGAP6* | 2.518659581 | 2.91E-06 | 0.000134478 | 0.014118335 |
| T2DM | mast_cell | *LINC00619* | -2.026872767 | 0.000128721 | 0.000245211 | 0.624552394 |
| T2DM | mast_cell | *DAXX* | 2.197224651 | 0.020033018 | 0.024287907 | 1 |
| T2DM | mast_cell | *BMPR2* | 2.754308514 | 4.00E-05 | 0.000134478 | 0.194064367 |
| T2DM | mast_cell | *TCEA3* | 2.024648246 | 4.82E-05 | 0.000134478 | 0.233784007 |
| T2DM | mast_cell | *HSD17B8* | -0.977159881 | 0.002731924 | 0.003656633 | 1 |
| T2DM | mast_cell | *OXLD1* | 2.030516989 | 1.04E-05 | 0.000134478 | 0.050389574 |
| T2DM | mast_cell | *TAP2* | 0.881014774 | 0.010406552 | 0.0129801 | 1 |
| T2DM | mast_cell | *COL15A1* | 0.984022807 | 0.000171248 | 0.000307739 | 0.830895303 |
| T2DM | mast_cell | *NOTCH4* | -0.560001018 | 4.67E-08 | 1.33E-05 | 0.000226664 |
| T2DM | mast_cell | *PRRT1* | 2.29236123 | 3.43E-08 | 1.04E-05 | 0.000166231 |
| T2DM | mast_cell | *STK19* | -4.443474598 | 1.57E-05 | 0.000134478 | 0.076127061 |

| | | | | | | |
|---|---|---|---|---|---|---|
| T2DM | mast_cell | *LAYN* | 1.919342093 | 0.000228087 | 0.000388854 | 1 |
| T2DM | mast_cell | *C6orf48* | 1.764948738 | 0.000488775 | 0.000754065 | 1 |
| T2DM | mast_cell | *GPANK1* | 1.175160074 | 0.006323482 | 0.008065598 | 1 |
| T2DM | mast_cell | *LST1* | 1.511704279 | 2.93E-05 | 0.000134478 | 0.142351398 |
| T2DM | mast_cell | *ZNF814* | 3.092481944 | 3.62E-05 | 0.000134478 | 0.175701919 |
| T2DM | mast_cell | *CCHCR1* | 1.047932635 | 5.73E-05 | 0.000142472 | 0.278248193 |
| T2DM | mast_cell | *PSORS1C2* | 2.589770183 | 0.000495966 | 0.000764188 | 1 |
| T2DM | mast_cell | *PSORS1C1* | 0.449327435 | 0.002742789 | 0.00366814 | 1 |
| T2DM | mast_cell | *DHX16* | 2.765038522 | 6.54E-07 | 7.18E-05 | 0.00317522 |
| T2DM | mast_cell | *ABCF1* | 1.509844015 | 1.80E-05 | 0.000134478 | 0.087334327 |
| T2DM | mast_cell | *LILRB3* | 2.551257893 | 7.32E-05 | 0.00016414 | 0.355198694 |
| T2DM | mast_cell | *HLA-E* | 1.494649223 | 8.81E-05 | 0.000186041 | 0.427335213 |
| T2DM | mast_cell | *ZNF468* | -3.935074193 | 0.004992233 | 0.00645584 | 1 |
| T2DM | mast_cell | *HCG9* | -0.961601282 | 0.000203006 | 0.000353041 | 0.984984883 |
| T2DM | mast_cell | *HLA-G* | 1.815603881 | 8.57E-05 | 0.000183057 | 0.415904583 |
| T2DM | mast_cell | *TRIM27* | 2.17018693 | 9.41E-05 | 0.00019472 | 0.456432963 |
| T2DM | mast_cell | *MALRD1* | 3.56058006 | 8.40E-05 | 0.000180125 | 0.407622805 |
| T2DM | mast_cell | *FAM196B* | 2.942123963 | 7.83E-05 | 0.000171612 | 0.37994973 |
| T2DM | mast_cell | *FAM216A* | 2.484071719 | 3.53E-05 | 0.000134478 | 0.171368991 |
| T2DM | mast_cell | *ZBTB48* | 2.352804357 | 0.000440781 | 0.000686351 | 1 |
| T2DM | mast_cell | *MZT1* | 2.406836941 | 5.62E-05 | 0.000140825 | 0.272612893 |
| T2DM | mast_cell | *ZNF155* | 1.490719269 | 0.000138328 | 0.000259138 | 0.671167878 |
| T2DM | mast_cell | *ZNF783* | 2.47356776 | 2.87E-05 | 0.000134478 | 0.139436109 |
| T2DM | mast_cell | *C12orf73* | 3.707037491 | 0.00959313 | 0.01199945 | 1 |
| T2DM | mast_cell | *PCDHGA1* | -2.473914105 | 0.002926074 | 0.00389074 | 1 |
| T2DM | mast_cell | *PCDHA4* | 3.988783927 | 0.000896379 | 0.001310009 | 1 |
| T2DM | mast_cell | *TRIM13* | 2.419356263 | 9.36E-06 | 0.000134478 | 0.045431473 |
| T2DM | mast_cell | *SPIRE2* | 1.377789909 | 0.013117052 | 0.016202632 | 1 |
| T2DM | mast_cell | *PKHD1L1* | 0.630593055 | 0.001704345 | 0.002364737 | 1 |
| T2DM | mast_cell | *SLFN12L* | 1.078291008 | 0.00038291 | 0.000607746 | 1 |
| T2DM | mast_cell | *SLC35B4* | 3.227527058 | 2.36E-05 | 0.000134478 | 0.114414665 |
| T2DM | mast_cell | *SYCE1L* | 1.391837209 | 0.00040735 | 0.000637774 | 1 |
| T2DM | mast_cell | *TMEM231* | 2.467240898 | 0.000307749 | 0.000502029 | 1 |
| T2DM | mast_cell | *LINC00654* | 2.991993608 | 0.000302313 | 0.000494546 | 1 |
| T2DM | mast_cell | *SCGB2B2* | 2.819055945 | 3.62E-05 | 0.000134478 | 0.175634525 |
| T2DM | mast_cell | *CCDC144NL* | 4.13888437 | 3.28E-05 | 0.000134478 | 0.159151482 |
| T2DM | mast_cell | *TMEM170B* | 1.348297806 | 0.00039022 | 0.000616541 | 1 |
| T2DM | mast_cell | *GOLGA6L10* | -3.806214045 | 0.004997765 | 0.006461273 | 1 |
| T2DM | mast_cell | *SAP25* | -5.818870309 | 3.83E-05 | 0.000134478 | 0.185720329 |
| T2DM | mast_cell | *IPO7* | 3.875865681 | 2.12E-05 | 0.000134478 | 0.103008065 |
| T2DM | mast_cell | *PRR13* | 1.918863394 | 0.000412478 | 0.000644763 | 1 |
| T2DM | mast_cell | *TECPR1* | 1.740244733 | 4.26E-05 | 0.000134478 | 0.20691724 |
| T2DM | mast_cell | *MT1A* | -3.108824398 | 0.000358164 | 0.000574405 | 1 |
| T2DM | mast_cell | *C15orf59* | -1.46104692 | 0.006231545 | 0.007950422 | 1 |
| T2DM | mast_cell | *CFI* | 1.4075958 | 5.30E-05 | 0.000137292 | 0.257362813 |
| T2DM | mast_cell | *ATP6AP1L* | 1.191360018 | 3.49E-05 | 0.000134478 | 0.169457414 |
| T2DM | mast_cell | *RGL3* | 1.411321332 | 2.93E-05 | 0.000134478 | 0.142180337 |
| T2DM | mast_cell | *NAP1L4* | 2.755977595 | 1.19E-05 | 0.000134478 | 0.057734802 |
| T2DM | mast_cell | *SMN2* | -1.35181517 | 0.000144638 | 0.000267244 | 0.701781905 |
| T2DM | mast_cell | *LIN52* | 2.809067195 | 1.25E-07 | 2.75E-05 | 0.000605168 |

| | | | | | | |
|---|---|---|---|---|---|---|
| T2DM | mast_cell | *C17orf107* | 2.092994418 | 0.028391712 | 0.033875423 | 1 |
| T2DM | mast_cell | *KLRC2* | 3.993734878 | 1.70E-05 | 0.000134478 | 0.082310947 |
| T2DM | mast_cell | *C1QTNF9B* | -1.550338006 | 0.000129543 | 0.000246391 | 0.628543032 |
| T2DM | mast_cell | *GOLGA8O* | -2.783840504 | 0.004533862 | 0.005896087 | 1 |
| T2DM | mast_cell | *HBA1* | 1.699789477 | 0.000926475 | 0.001349521 | 1 |
| T2DM | mast_cell | *HCG27* | 1.944322964 | 2.24E-09 | 8.82E-07 | 1.09E-05 |
| T2DM | mast_cell | *COL6A6* | 1.425815527 | 0.00013791 | 0.000258755 | 0.669141235 |
| T2DM | mast_cell | *HLA-A* | 2.010467894 | 6.76E-05 | 0.000156404 | 0.328232623 |
| T2DM | mast_cell | *METTL6* | 2.080457694 | 2.53E-05 | 0.000134478 | 0.122737827 |
| T2DM | mast_cell | *SACM1L* | 2.904850825 | 3.43E-05 | 0.000134478 | 0.166636271 |
| T2DM | mast_cell | *SLC48A1* | 1.663203883 | 5.46E-05 | 0.000139259 | 0.264792977 |
| T2DM | mast_cell | *TAS2R50* | 45.32319937 | 2.89E-06 | 0.000134478 | 0.014042794 |
| T2DM | mast_cell | *TAS2R14* | 2.733657392 | 1.47E-07 | 3.11E-05 | 0.000714671 |
| T2DM | mast_cell | *C17orf51* | 7.145106236 | 3.60E-05 | 0.000134478 | 0.174633403 |
| T2DM | mast_cell | *ZNF580* | 1.910103297 | 0.002360916 | 0.003197979 | 1 |
| T2DM | mast_cell | *NUP62* | 1.68759875 | 0.00358398 | 0.004712594 | 1 |
| T2DM | mast_cell | *FGFR1OP* | 5.716300346 | 0.000114626 | 0.000224895 | 0.55616652 |
| T2DM | mast_cell | *ZNF254* | 1.674724737 | 7.20E-05 | 0.000162466 | 0.349302885 |
| T2DM | mast_cell | *TCTEX1D2* | 4.640316258 | 9.03E-06 | 0.000134478 | 0.043806763 |
| T2DM | mast_cell | *ASIC3* | 3.518944736 | 0.000165832 | 0.000300119 | 0.804618377 |
| T2DM | mast_cell | *GIMAP1* | 1.508821652 | 0.00023235 | 0.000395012 | 1 |
| T2DM | mast_cell | *DNLZ* | 1.530817988 | 6.06E-05 | 0.000146324 | 0.293904416 |
| T2DM | mast_cell | *SUPT4H1* | 4.599466764 | 3.96E-06 | 0.000134478 | 0.019189988 |
| T2DM | mast_cell | *MXD3* | 2.729434061 | 5.50E-05 | 0.000139632 | 0.26666 |
| T2DM | mast_cell | *GSTM2* | 1.297168222 | 5.13E-05 | 0.000136328 | 0.249038387 |
| T2DM | mast_cell | *ARHGAP19* | -4.458925392 | 0.038196696 | 0.044917685 | 1 |
| T2DM | mast_cell | *LCAT* | 1.965831874 | 3.70E-05 | 0.000134478 | 0.179750561 |
| T2DM | mast_cell | *GPC2* | 6.833651374 | 0.005274074 | 0.006791351 | 1 |
| T2DM | mast_cell | *ERV3-1* | 1.052890214 | 0.000116319 | 0.000227297 | 0.564378381 |
| T2DM | mast_cell | *SRA1* | 2.474608918 | 0.000106022 | 0.000211747 | 0.514417918 |
| T2DM | mast_cell | *TMEM110* | 6.562794446 | 1.51E-09 | 6.68E-07 | 7.34E-06 |
| T2DM | mast_cell | *TMX2* | 3.197010122 | 3.02E-05 | 0.000134478 | 0.14642224 |
| T2DM | mast_cell | *LBH* | 1.587252857 | 0.000104257 | 0.000209118 | 0.505857367 |
| T2DM | mast_cell | *PPP1CB* | 2.151754798 | 0.000100151 | 0.000203404 | 0.485933099 |
| T2DM | mast_cell | *NCKIPSD* | 2.208725594 | 5.56E-05 | 0.000140218 | 0.269729318 |
| T2DM | mast_cell | *ATF6B* | 3.09098769 | 5.05E-07 | 6.81E-05 | 0.002448725 |
| T2DM | mast_cell | *SLC35F6* | 3.068089155 | 2.12E-05 | 0.000134478 | 0.102993356 |
| T2DM | mast_cell | *EMP2* | 1.994028946 | 5.54E-05 | 0.000140046 | 0.268888975 |
| T2DM | mast_cell | *PPM1N* | 2.038218972 | 0.007510447 | 0.009502136 | 1 |
| T2DM | mast_cell | *SLC23A3* | 1.496742537 | 0.007009143 | 0.008898054 | 1 |
| T2DM | mast_cell | *LTB4R* | 2.103361818 | 0.000129252 | 0.00024603 | 0.627130838 |
| T2DM | mast_cell | *IRF9* | 1.583151872 | 3.71E-05 | 0.000134478 | 0.180121828 |
| T2DM | mast_cell | *GALT* | 3.666230762 | 0.028038763 | 0.033491896 | 1 |
| T2DM | mast_cell | *ZNF726* | -4.721287085 | 0.000235931 | 0.000400678 | 1 |
| T2DM | mast_cell | *ZNF90* | 1.954077658 | 7.79E-05 | 0.000170933 | 0.37793392 |
| T2DM | mast_cell | *REPIN1* | 2.708334828 | 3.89E-05 | 0.000134478 | 0.188810415 |
| T2DM | mast_cell | *SMIM7* | 3.018307039 | 9.87E-06 | 0.000134478 | 0.047906366 |
| T2DM | mast_cell | *TSPAN4* | 3.026542243 | 0.000277831 | 0.000460519 | 1 |
| T2DM | mast_cell | *CPNE1* | 0.678740922 | 0.038082108 | 0.044804652 | 1 |
| T2DM | mast_cell | *PAXIP1-AS2* | 2.497992537 | 5.00E-05 | 0.000135379 | 0.242505095 |

| | | | | | | |
|---|---|---|---|---|---|---|
| T2DM | mast_cell | *LYRM4* | 2.198622374 | 2.96E-05 | 0.000134478 | 0.14381674 |
| T2DM | mast_cell | *PRCD* | 2.250510953 | 2.93E-05 | 0.000134478 | 0.141940738 |
| T2DM | mast_cell | *SH3D21* | 2.071712842 | 0.001838339 | 0.002536865 | 1 |
| T2DM | mast_cell | *ANG* | 1.823248449 | 4.70E-05 | 0.000134478 | 0.227840497 |
| T2DM | mast_cell | *MBLAC1* | 1.508868929 | 0.001238928 | 0.001760257 | 1 |
| T2DM | mast_cell | *LCNL1* | 3.452824775 | 0.001465746 | 0.002053653 | 1 |
| T2DM | mast_cell | *GNAT3* | 2.03104325 | 4.60E-05 | 0.000134478 | 0.223106163 |
| T2DM | mast_cell | *PLIN5* | 1.015603435 | 0.002324545 | 0.003151353 | 1 |
| T2DM | mast_cell | *STARD10* | 1.98115305 | 2.70E-05 | 0.000134478 | 0.131131105 |
| T2DM | mast_cell | *NUTM2D* | 16.3869477 | 0.001558882 | 0.002180367 | 1 |
| T2DM | mast_cell | *CPEB1* | 1.286471061 | 0.00063858 | 0.000960146 | 1 |
| T2DM | mast_cell | *EML6* | 5.72815785 | 4.37E-06 | 0.000134478 | 0.021195839 |
| T2DM | mast_cell | *ARHGEF33* | 2.387265149 | 2.89E-05 | 0.000134478 | 0.140284143 |
| T2DM | mast_cell | *IFRD2* | 1.19205473 | 0.03753516 | 0.044193302 | 1 |
| T2DM | mast_cell | *EVPLL* | 6.547199616 | 1.10E-06 | 9.21E-05 | 0.005339982 |
| T2DM | mast_cell | *AC005082.1* | 7.704554473 | 9.77E-05 | 0.000199875 | 0.474104655 |
| T2DM | mast_cell | *ACSM4* | 1.739717581 | 0.002256448 | 0.003068246 | 1 |
| T2DM | mast_cell | *UBXN2B* | 1.79306104 | 8.13E-05 | 0.000176186 | 0.394452188 |
| T2DM | mast_cell | *C5orf49* | 16.35914541 | 0.000414923 | 0.000647958 | 1 |
| T2DM | mast_cell | *GOLGA8B* | 1.568768235 | 0.000143426 | 0.000266004 | 0.695902407 |
| T2DM | mast_cell | *VPS16* | 2.191273695 | 3.44E-05 | 0.000134478 | 0.166735606 |
| T2DM | mast_cell | *MYL5* | 2.763737477 | 7.10E-05 | 0.000161307 | 0.344538452 |
| T2DM | mast_cell | *NPEPL1* | -2.885324749 | 0.002755257 | 0.003681481 | 1 |
| T2DM | mast_cell | *TMEM242* | 1.620054278 | 3.35E-05 | 0.000134478 | 0.162593499 |
| T2DM | mast_cell | *SLC35E2* | 2.573401832 | 0.001001975 | 0.001451761 | 1 |
| T2DM | mast_cell | *TSTD1* | 2.799367265 | 5.40E-05 | 0.000138255 | 0.261988776 |
| T2DM | mast_cell | *AC009403.2* | -1.905171405 | 0.000305415 | 0.000498779 | 1 |
| T2DM | mast_cell | *PAM16* | -2.073981302 | 0.000652512 | 0.00097897 | 1 |
| T2DM | mast_cell | *RNASEK* | 1.551739815 | 1.64E-05 | 0.000134478 | 0.079663662 |
| T2DM | mast_cell | *FAM228B* | 2.081871308 | 8.06E-05 | 0.000175067 | 0.3910989 |
| T2DM | mast_cell | *RNU6ATAC35I* | -4.438583252 | 8.99E-05 | 0.000188906 | 0.436366785 |
| T2DM | mast_cell | *PPP3R1* | 2.008486051 | 5.09E-05 | 0.000136231 | 0.246825627 |
| T2DM | mast_cell | *FANCG* | -2.187167283 | 0.000453104 | 0.000703283 | 1 |
| T2DM | mast_cell | *AP4M1* | 2.161266069 | 0.011248495 | 0.013994281 | 1 |
| T2DM | mast_cell | *ZNF880* | 1.308395816 | 0.017680893 | 0.021549282 | 1 |
| T2DM | mast_cell | *FXYD7* | 2.145820801 | 0.001325461 | 0.001870603 | 1 |
| T2DM | mast_cell | *SLC12A8* | 1.407722598 | 0.00097403 | 0.001412853 | 1 |
| T2DM | mast_cell | *APOL6* | 1.521385263 | 3.72E-05 | 0.000134478 | 0.180691508 |
| T2DM | mast_cell | *OR2A1* | 2.158653687 | 0.005081951 | 0.006563115 | 1 |
| T2DM | mast_cell | *UBA52* | 3.130341899 | 3.70E-06 | 0.000134478 | 0.017964264 |
| T2DM | mast_cell | *TIAF1* | 3.057748917 | 0.000296894 | 0.000487325 | 1 |
| T2DM | mast_cell | *RAB6C* | 50.13834613 | 0.000114101 | 0.000224319 | 0.553620236 |
| T2DM | mast_cell | *AC062017.1* | 1.894827838 | 4.69E-05 | 0.000134478 | 0.227340506 |
| T2DM | mast_cell | *ADRA2B* | 5.749399424 | 0.001234526 | 0.001754517 | 1 |
| T2DM | mast_cell | *EXOSC6* | 1.571561338 | 0.000395514 | 0.000623064 | 1 |
| T2DM | mast_cell | *ZNF844* | 1.464970836 | 9.22E-05 | 0.000192375 | 0.447440659 |
| T2DM | mast_cell | *LINC00571* | 1.649896332 | 0.000405765 | 0.000635703 | 1 |
| T2DM | mast_cell | *OSER1-AS1* | 1.760874449 | 1.36E-05 | 0.000134478 | 0.06579356 |
| T2DM | mast_cell | *C5orf66* | 2.479038522 | 0.00049876 | 0.000767518 | 1 |
| T2DM | mast_cell | *LINC01133* | 1.254219754 | 0.000140228 | 0.00026199 | 0.680387967 |

| | | | | | | |
|---|---|---|---|---|---|---|
| T2DM | mast_cell | *C4B* | 5.266524907 | 1.56E-05 | 0.000134478 | 0.075735568 |
| T2DM | mast_cell | *LINC01117* | 1.900854399 | 4.22E-05 | 0.000134478 | 0.204751341 |
| T2DM | mast_cell | *LINC00240* | 0.73195985 | 0.006371422 | 0.008122475 | 1 |
| T2DM | mast_cell | *LINC00184* | 47.91148393 | 4.88E-05 | 0.000134902 | 0.236887419 |
| T2DM | mast_cell | *AL049840.1* | 2.347902998 | 0.001445926 | 0.00202764 | 1 |
| T2DM | mast_cell | *PLEKHM1* | 1.4794381 | 1.97E-05 | 0.000134478 | 0.095737668 |
| T2DM | mast_cell | *SFTA1P* | 2.723831267 | 0.041280357 | 0.048414864 | 1 |
| T2DM | mast_cell | *CYP4A22-AS1* | 5.040615198 | 3.48E-05 | 0.000134478 | 0.168900069 |
| T2DM | mast_cell | *MIAT* | 2.486598638 | 5.31E-05 | 0.000137292 | 0.257624051 |
| T2DM | mast_cell | *ERCC6* | 2.652998555 | 0.000114516 | 0.000224769 | 0.555630189 |
| T2DM | mast_cell | *LINC00694* | 6.872460607 | 1.07E-06 | 9.21E-05 | 0.005186524 |
| T2DM | mast_cell | *LINC00115* | 7.629628557 | 0.000232042 | 0.000394625 | 1 |
| T2DM | mast_cell | *LINC00937* | -1.447150437 | 0.00037055 | 0.000590835 | 1 |
| T2DM | mast_cell | *FTCDNL1* | 1.294231428 | 0.0029071 | 0.003867631 | 1 |
| T2DM | mast_cell | *FAM138B* | -4.444002038 | 0.000141386 | 0.000263507 | 0.686006684 |
| T2DM | mast_cell | *ZNF717* | 0.71524341 | 0.025779575 | 0.030899827 | 1 |
| T2DM | mast_cell | *AC017074.2* | 13.32901621 | 0.019044324 | 0.023147059 | 1 |
| T2DM | mast_cell | *AC009299.3* | 1.979960029 | 3.02E-05 | 0.000134478 | 0.146324201 |
| T2DM | mast_cell | *SCAMP4* | 1.774177092 | 4.08E-05 | 0.000134478 | 0.198097738 |
| T2DM | mast_cell | *SLC2A1-AS1* | 3.782206862 | 1.68E-05 | 0.000134478 | 0.081598708 |
| T2DM | mast_cell | *AC005884.1* | 2.168651258 | 0.01169651 | 0.014518155 | 1 |
| T2DM | mast_cell | *AC108463.2* | 6.338134215 | 3.89E-07 | 6.50E-05 | 0.001885767 |
| T2DM | mast_cell | *C14orf144* | 4.274400478 | 0.001653382 | 0.002299946 | 1 |
| T2DM | mast_cell | *POLR2J2* | -9.953814688 | 1.92E-05 | 0.000134478 | 0.093059858 |
| T2DM | mast_cell | *LINC00578* | 2.88203922 | 9.79E-05 | 0.000199936 | 0.475247392 |
| T2DM | mast_cell | *C19orf24* | 3.013977822 | 0.000840164 | 0.001236795 | 1 |
| T2DM | mast_cell | *ZNF883* | 1.879583865 | 9.83E-05 | 0.000200595 | 0.477015679 |
| T2DM | mast_cell | *ARL17B* | 2.332092129 | 7.92E-05 | 0.000173118 | 0.38413027 |
| T2DM | mast_cell | *PATL2* | 2.116258105 | 2.49E-05 | 0.000134478 | 0.12078386 |
| T2DM | mast_cell | *CASC20* | -1.191160451 | 0.00016472 | 0.00029844 | 0.799221793 |
| T2DM | mast_cell | *PSMG3-AS1* | 1.863642215 | 5.36E-05 | 0.000138027 | 0.26021881 |
| T2DM | mast_cell | *AC112229.4* | 2.74531422 | 0.000237349 | 0.000402663 | 1 |
| T2DM | mast_cell | *KLHL7-AS1* | -2.167165086 | 0.004513472 | 0.005871144 | 1 |
| T2DM | mast_cell | *AC092171.4* | 2.576884389 | 4.36E-05 | 0.000134478 | 0.211374924 |
| T2DM | mast_cell | *RPS18* | 2.508537937 | 5.15E-05 | 0.000136328 | 0.249981668 |
| T2DM | mast_cell | *DIRC3* | 1.44910315 | 3.29E-05 | 0.000134478 | 0.159420712 |
| T2DM | mast_cell | *TAPBP* | 2.62414699 | 4.90E-06 | 0.000134478 | 0.023756109 |
| T2DM | mast_cell | *AC107021.1* | 1.84336232 | 0.000128527 | 0.00024513 | 0.623611916 |
| T2DM | mast_cell | *TMA7* | 3.878623522 | 6.48E-07 | 7.18E-05 | 0.003145515 |
| T2DM | mast_cell | *LINC00865* | 1.493863074 | 0.020801431 | 0.025169213 | 1 |
| T2DM | mast_cell | *AC009495.3* | 6.01316939 | 5.42E-05 | 0.000138726 | 0.263162516 |
| T2DM | mast_cell | *HLA-DQB2* | 1.158885342 | 1.17E-09 | 6.08E-07 | 5.66E-06 |
| T2DM | mast_cell | *GOLGA8N* | 1.727495819 | 0.000592831 | 0.000897198 | 1 |
| T2DM | mast_cell | *LINC00665* | 1.795497448 | 3.50E-05 | 0.000134478 | 0.169651906 |
| T2DM | mast_cell | *LYRM9* | 1.680059813 | 0.000520002 | 0.000795162 | 1 |
| T2DM | mast_cell | *LINC00342* | 2.177243793 | 8.47E-05 | 0.000181363 | 0.411150541 |
| T2DM | mast_cell | *LINC00702* | 3.583352968 | 6.63E-05 | 0.000154467 | 0.32175428 |
| T2DM | mast_cell | *RPS28* | 2.659428693 | 6.19E-05 | 0.000148752 | 0.300534655 |
| T2DM | mast_cell | *TRIM26* | 2.56044472 | 2.87E-05 | 0.000134478 | 0.139020858 |
| T2DM | mast_cell | *ZNF736* | 1.461119096 | 1.24E-05 | 0.000134478 | 0.060243149 |

| | | | | | | |
|---|---|---|---|---|---|---|
| T2DM | mast_cell | *PINLYP* | 1.632787009 | 0.000190489 | 0.00033439 | 0.924253653 |
| T2DM | mast_cell | *ANKRD65* | -4.479068642 | 0.000442377 | 0.000688395 | 1 |
| T2DM | mast_cell | *ZSCAN31* | 1.203823875 | 2.14E-05 | 0.000134478 | 0.103763636 |
| T2DM | mast_cell | *AC009495.2* | 44.19474344 | 8.99E-05 | 0.000188906 | 0.436372782 |
| T2DM | mast_cell | *LINC00957* | 2.90346562 | 0.021102466 | 0.02550166 | 1 |
| T2DM | mast_cell | *LINC00484* | 0.879581804 | 0.008481182 | 0.010660802 | 1 |
| T2DM | mast_cell | *KIAA0040* | 3.49378668 | 0.000738876 | 0.001098018 | 1 |
| T2DM | mast_cell | *LINC00607* | 1.654397887 | 2.82E-05 | 0.000134478 | 0.136994876 |
| T2DM | mast_cell | *AP001468.1* | -1.203015191 | 0.027235569 | 0.032548517 | 1 |
| T2DM | mast_cell | *LINC00941* | -6.669416097 | 5.31E-06 | 0.000134478 | 0.025752263 |
| T2DM | mast_cell | *AC019117.2* | -4.398732187 | 0.000121443 | 0.000233919 | 0.589241757 |
| T2DM | mast_cell | *ZBTB22* | -1.553674405 | 0.001014796 | 0.001468035 | 1 |
| T2DM | mast_cell | *ZBED5* | 2.589439426 | 0.000250691 | 0.000422052 | 1 |
| T2DM | mast_cell | *CDKN2AIPNL* | 1.649140579 | 0.000694654 | 0.001037654 | 1 |
| T2DM | mast_cell | *LINC00959* | -2.153458421 | 0.001207614 | 0.001718786 | 1 |
| T2DM | mast_cell | *HLA-DQA2* | 2.349034414 | 3.03E-20 | 1.47E-16 | 1.47E-16 |
| T2DM | mast_cell | *LINC01143* | -7.654053074 | 0.000598081 | 0.000904578 | 1 |
| T2DM | mast_cell | *LINC00211* | 10.83113205 | 3.26E-05 | 0.000134478 | 0.157985668 |
| T2DM | mast_cell | *LRRC37A2* | 3.053172807 | 2.69E-05 | 0.000134478 | 0.130480897 |
| T2DM | mast_cell | *LINC00707* | 6.894655877 | 0.012189414 | 0.015099065 | 1 |
| T2DM | mast_cell | *TXNDC5* | 9.8078397 | 3.97E-05 | 0.000134478 | 0.192859311 |
| T2DM | mast_cell | *LINC01119* | 3.330238216 | 1.67E-05 | 0.000134478 | 0.081269758 |
| T2DM | mast_cell | *CDRT4* | 4.346520055 | 0.000793693 | 0.001172655 | 1 |
| T2DM | mast_cell | *C1orf226* | -2.794481154 | 0.00184529 | 0.002545734 | 1 |
| T2DM | mast_cell | *LILRA2* | -1.486599936 | 0.000357921 | 0.000574283 | 1 |
| T2DM | mast_cell | *PSMB9* | 1.84563917 | 4.38E-07 | 6.64E-05 | 0.002124326 |
| T2DM | mast_cell | *L1TD1* | 2.140569425 | 0.009257666 | 0.011597778 | 1 |
| T2DM | mast_cell | *C1QTNF9* | -4.734495 | 0.000200854 | 0.000349926 | 0.974543985 |
| T2DM | mast_cell | *ISY1* | 1.356772252 | 0.001622356 | 0.002261974 | 1 |
| T2DM | mast_cell | *PNMA2* | 2.599256337 | 0.000410014 | 0.000641325 | 1 |
| T2DM | mast_cell | *KRBOX1* | 30.03017059 | 5.77E-05 | 0.000142926 | 0.279992253 |
| T2DM | mast_cell | *ARHGEF25* | 2.287835149 | 0.000237469 | 0.000402727 | 1 |
| T2DM | mast_cell | *NSUN6* | 3.871520293 | 0.000248986 | 0.000419617 | 1 |
| T2DM | mast_cell | *LINC00877* | 2.719331251 | 1.88E-05 | 0.000134478 | 0.091199534 |
| T2DM | mast_cell | *CD302* | 2.082756648 | 1.00E-05 | 0.000134478 | 0.048554473 |
| T2DM | mast_cell | *INMT* | 2.83224228 | 0.000866471 | 0.001271768 | 1 |
| T2DM | mast_cell | *ATP5O* | 2.068385432 | 1.53E-05 | 0.000134478 | 0.0742268 |
| T2DM | mast_cell | *PLEKHO2* | -1.412053002 | 0.002914096 | 0.003875876 | 1 |
| T2DM | mast_cell | *C8orf58* | -1.148052075 | 0.000442633 | 0.000688555 | 1 |
| T2DM | mast_cell | *HOGA1* | 3.826700512 | 5.26E-05 | 0.000136775 | 0.255221986 |
| T2DM | mast_cell | *PI4KA* | 1.72060161 | 4.71E-05 | 0.000134478 | 0.22828988 |
| T2DM | mast_cell | *LINC00969* | 1.664513247 | 4.82E-05 | 0.000134478 | 0.233646325 |
| T2DM | mast_cell | *ARFGAP3* | 4.923553921 | 0.00033037 | 0.00053414 | 1 |
| T2DM | mast_cell | *MRPL20* | 4.759997578 | 9.18E-06 | 0.000134478 | 0.044543185 |
| T2DM | mast_cell | *DECR2* | 2.759779656 | 5.01E-05 | 0.000135379 | 0.243272246 |
| T2DM | mast_cell | *ERVW-1* | -2.37483422 | 0.000387878 | 0.000613624 | 1 |
| T2DM | mast_cell | *MRPL33* | 2.341849569 | 4.16E-05 | 0.000134478 | 0.201866412 |
| T2DM | mast_cell | *STON1* | 2.283053035 | 0.000266124 | 0.000443419 | 1 |
| T2DM | mast_cell | *VSIG8* | 3.212605956 | 0.000223191 | 0.000381715 | 1 |
| T2DM | mast_cell | *C7orf73* | 2.701314543 | 8.74E-05 | 0.000185249 | 0.423849686 |

| | | | | | | |
|---|---|---|---|---|---|---|
| T2DM | mast_cell | *FGF14-IT1* | 5.704503913 | 7.16E-05 | 0.000161829 | 0.34742617 |
| T2DM | mast_cell | *PALM2* | 2.214272652 | 1.64E-05 | 0.000134478 | 0.079578977 |
| T2DM | mast_cell | *NBPF15* | 2.051216623 | 3.96E-05 | 0.000134478 | 0.192192125 |
| T2DM | mast_cell | *UPK3B* | 3.030572903 | 5.03E-05 | 0.000135576 | 0.244172732 |
| T2DM | mast_cell | *IL10RB* | 5.092871863 | 5.56E-06 | 0.000134478 | 0.026957302 |
| T2DM | mast_cell | *NME1-NME2* | 8.531054271 | 0.000144079 | 0.000266822 | 0.699072944 |
| T2DM | mast_cell | *PLA2G4B* | 12.79464995 | 1.82E-10 | 1.63E-07 | 8.85E-07 |
| T2DM | mast_cell | *LEFTY1* | 18.60361144 | 0.011131917 | 0.013856352 | 1 |
| T2DM | mast_cell | *TTC4* | 2.526686053 | 5.37E-06 | 0.000134478 | 0.026060555 |
| T2DM | mast_cell | *JMJD7* | 3.746340528 | 4.71E-09 | 1.63E-06 | 2.28E-05 |
| T2DM | mast_cell | *APOBEC3D* | -12.61131131 | 0.001408305 | 0.001979459 | 1 |
| T2DM | mast_cell | *TMEM199* | 1.405697049 | 0.00563672 | 0.007225724 | 1 |
| T2DM | mast_cell | *DBNDD2* | 6.46452256 | 0.019127014 | 0.023241741 | 1 |
| T2DM | mast_cell | *LILRA6* | 1.337411254 | 0.002256919 | 0.003068246 | 1 |
| T2DM | mast_cell | *SCARF2* | 2.210402317 | 7.06E-05 | 0.000160722 | 0.342413557 |
| T2DM | mast_cell | *APOBEC3C* | -1.553565573 | 0.000821915 | 0.001211034 | 1 |
| T2DM | mast_cell | *CCDC13* | 1.531414423 | 0.00788513 | 0.009942477 | 1 |
| T2DM | mast_cell | *C4A* | -4.132884465 | 0.000366556 | 0.000585814 | 1 |
| T2DM | mast_cell | *CRYBB2* | -2.39928761 | 2.99E-05 | 0.000134478 | 0.144962795 |
| T2DM | mast_cell | *NEAT1* | 1.660375768 | 1.77E-05 | 0.000134478 | 0.085977244 |
| T2DM | mast_cell | *RAB30-AS1* | 1.501563451 | 0.000268496 | 0.000447217 | 1 |
| T2DM | mast_cell | *UBAP1L* | 1.778419555 | 5.36E-05 | 0.000138012 | 0.260014605 |
| T2DM | mast_cell | *DNAJC3-AS1* | 1.680480476 | 7.43E-05 | 0.000165609 | 0.360697332 |
| T2DM | mast_cell | *LINC00926* | 3.013956677 | 3.99E-05 | 0.000134478 | 0.193757092 |
| T2DM | mast_cell | *DPH6-AS1* | 1.483657073 | 2.97E-05 | 0.000134478 | 0.14421012 |
| T2DM | mast_cell | *BCKDHA* | 2.711042696 | 0.001946874 | 0.002676745 | 1 |
| T2DM | mast_cell | *CDK11B* | 1.611302021 | 4.40E-05 | 0.000134478 | 0.213407778 |
| T2DM | mast_cell | *ABHD14A* | 1.548216532 | 5.61E-05 | 0.000140825 | 0.272092716 |
| T2DM | mast_cell | *GDNF-AS1* | 2.928050248 | 0.000361765 | 0.00057911 | 1 |
| T2DM | mast_cell | *CCDC153* | 3.168469892 | 1.24E-05 | 0.000134478 | 0.06015519 |
| T2DM | mast_cell | *AL357673.1* | 1.856977659 | 0.003744022 | 0.004916372 | 1 |
| T2DM | mast_cell | *C5orf17* | 1.338036921 | 4.34E-05 | 0.000134478 | 0.210634293 |
| T2DM | mast_cell | *TMEM150C* | 1.041723858 | 0.011838485 | 0.01468311 | 1 |
| T2DM | mast_cell | *LINC00939* | 4.013934225 | 0.000647863 | 0.000973199 | 1 |
| T2DM | mast_cell | *LINC01060* | 1.55443045 | 0.000102271 | 0.000206134 | 0.496219252 |
| T2DM | mast_cell | *LINC00942* | -5.329204044 | 0.000143265 | 0.000265821 | 0.695121687 |
| T2DM | mast_cell | *PDCD6* | 2.18450715 | 3.40E-05 | 0.000134478 | 0.164794718 |
| T2DM | mast_cell | *GOLGA8K* | 1.830919779 | 0.026926766 | 0.032203271 | 1 |
| T2DM | mast_cell | *TMEM158* | -5.868303802 | 0.001094094 | 0.001572902 | 1 |
| T2DM | mast_cell | *DNAH10OS* | 8.124437294 | 0.017953469 | 0.021865018 | 1 |
| T2DM | mast_cell | *PCDHA10* | 1.959034831 | 0.000953323 | 0.001385716 | 1 |
| T2DM | mast_cell | *ZNF718* | 2.602688475 | 1.66E-05 | 0.000134478 | 0.080559114 |
| T2DM | mast_cell | *SMIM20* | 2.70609006 | 3.57E-05 | 0.000134478 | 0.173276309 |
| T2DM | mast_cell | *LINC00989* | 2.082532716 | 0.000184841 | 0.000326721 | 0.896847863 |
| T2DM | mast_cell | *ATP6V1E2* | 1.341582161 | 9.32E-05 | 0.000193411 | 0.452000778 |
| T2DM | mast_cell | *GLI4* | 1.889034318 | 4.79E-05 | 0.000134478 | 0.232182729 |
| T2DM | mast_cell | *SNHG18* | 1.498295833 | 0.001129684 | 0.001618314 | 1 |
| T2DM | mast_cell | *ALG1L2* | 2.386889608 | 4.86E-05 | 0.000134775 | 0.235856667 |
| T2DM | mast_cell | *ZNF550* | -3.250113916 | 0.004947321 | 0.006406299 | 1 |
| T2DM | mast_cell | *LINC01094* | 2.192661288 | 5.15E-05 | 0.000136328 | 0.249772536 |

| | | | | | | |
|---|---|---|---|---|---|---|
| T2DM | mast_cell | *TMEM200B* | 2.732163531 | 7.36E-06 | 0.000134478 | 0.035694503 |
| T2DM | mast_cell | *PCDHGB6* | 2.916316192 | 1.11E-05 | 0.000134478 | 0.053665264 |
| T2DM | mast_cell | *SLC10A5* | -2.178730043 | 0.000524664 | 0.000801178 | 1 |
| T2DM | mast_cell | *PCDHGA6* | 1.857233807 | 0.000102094 | 0.000206134 | 0.495361769 |
| T2DM | mast_cell | *UTP14C* | 3.761632465 | 5.54E-05 | 0.00014004 | 0.268735985 |
| T2DM | mast_cell | *PCDHGB2* | -6.904369853 | 0.000350425 | 0.000563001 | 1 |
| T2DM | mast_cell | *LYN* | 1.488821156 | 0.000167211 | 0.000302358 | 0.811306012 |
| T2DM | mast_cell | *FPGT* | 2.82433983 | 2.44E-05 | 0.000134478 | 0.118441785 |
| T2DM | mast_cell | *MEX3A* | 2.219641846 | 0.007569148 | 0.009568627 | 1 |
| T2DM | mast_cell | *DPP3* | 0.928711649 | 0.037370897 | 0.044021266 | 1 |
| T2DM | mast_cell | *BRK1* | 1.818301775 | 0.005733102 | 0.007347335 | 1 |
| T2DM | mast_cell | *SNHG9* | 2.496669816 | 1.66E-05 | 0.000134478 | 0.080684234 |
| T2DM | mast_cell | *TAS2R20* | 3.757495725 | 0.000275801 | 0.00045797 | 1 |
| T2DM | mast_cell | *ZNF432* | 1.834993428 | 0.000604367 | 0.000913516 | 1 |
| T2DM | mast_cell | *HMBS* | 3.119536408 | 3.12E-06 | 0.000134478 | 0.01514999 |
| T2DM | mast_cell | *AC093668.2* | 2.947135724 | 2.23E-06 | 0.000126109 | 0.010827962 |
| T2DM | mast_cell | *KIAA1147* | 2.296062861 | 4.74E-05 | 0.000134478 | 0.229942964 |
| T2DM | mast_cell | *ZBED6* | -3.201320646 | 0.000323013 | 0.000523642 | 1 |
| T2DM | mast_cell | *CNPY2* | 3.917051506 | 3.62E-05 | 0.000134478 | 0.17583848 |
| T2DM | mast_cell | *LINC00592* | -2.812196346 | 2.48E-05 | 0.000134478 | 0.120244429 |
| T2DM | mast_cell | *CHURC1* | 1.844886314 | 7.41E-05 | 0.000165359 | 0.359452327 |
| T2DM | mast_cell | *RTEL1* | 2.38478303 | 0.005599 | 0.007181166 | 1 |
| T2DM | mast_cell | *HIF1A-AS2* | 3.129556619 | 4.83E-05 | 0.000134478 | 0.234432755 |
| T2DM | mast_cell | *LINC00638* | 4.879760159 | 4.12E-05 | 0.000134478 | 0.199729071 |
| T2DM | mast_cell | *RNASE4* | -5.80083046 | 0.00222379 | 0.003028299 | 1 |
| T2DM | mast_cell | *MC1R* | -0.770403503 | 0.000577544 | 0.000875154 | 1 |
| T2DM | mast_cell | *LINC00639* | 1.329979862 | 0.000130947 | 0.00024838 | 0.635355414 |
| T2DM | mast_cell | *LINC00924* | 1.336541127 | 0.000264776 | 0.000441697 | 1 |
| T2DM | mast_cell | *MRPL46* | 2.367186408 | 2.13E-05 | 0.000134478 | 0.103218416 |
| T2DM | mast_cell | *UBE2Q2L* | -3.024152065 | 0.001300745 | 0.001840541 | 1 |
| T2DM | mast_cell | *IQCH-AS1* | 2.375979437 | 4.23E-05 | 0.000134478 | 0.205223724 |
| T2DM | mast_cell | *AC083843.1* | 36.12459729 | 4.57E-05 | 0.000134478 | 0.221714856 |
| T2DM | mast_cell | *PWRN1* | 2.317144722 | 6.09E-05 | 0.000146709 | 0.29547139 |
| T2DM | mast_cell | *LINC01003* | 2.02983843 | 3.93E-05 | 0.000134478 | 0.190629036 |
| T2DM | mast_cell | *C15orf65* | 2.259153589 | 0.000181431 | 0.000321631 | 0.880304692 |
| T2DM | mast_cell | *LINC00662* | 1.662044475 | 7.13E-05 | 0.000161567 | 0.345752954 |
| T2DM | mast_cell | *C19orf84* | 28.73237412 | 6.81E-06 | 0.000134478 | 0.033034017 |
| T2DM | mast_cell | *LINC00675* | 3.377095831 | 2.53E-05 | 0.000134478 | 0.122823599 |
| T2DM | mast_cell | *LINC00667* | 1.617251781 | 6.87E-05 | 0.000157925 | 0.33337976 |
| T2DM | mast_cell | *LINC00672* | 4.35525424 | 3.31E-05 | 0.000134478 | 0.160706855 |
| T2DM | mast_cell | *LINC00909* | 3.457249175 | 2.37E-05 | 0.000134478 | 0.115214094 |
| T2DM | mast_cell | *DYNLL2* | 2.20263009 | 6.50E-05 | 0.000152782 | 0.31549135 |
| T2DM | mast_cell | *STRADA* | 1.844060366 | 0.000358234 | 0.000574405 | 1 |
| T2DM | mast_cell | *MYO15B* | 2.79324086 | 3.47E-05 | 0.000134478 | 0.168545263 |
| T2DM | mast_cell | *LINC00663* | 1.026509547 | 0.030995874 | 0.036851747 | 1 |
| T2DM | mast_cell | *ILF3-AS1* | 1.718295655 | 1.48E-05 | 0.000134478 | 0.071601131 |
| T2DM | mast_cell | *ZNF582-AS1* | 4.352851887 | 3.35E-05 | 0.000134478 | 0.162720718 |
| T2DM | mast_cell | *ZNF285* | 1.817392315 | 0.003917402 | 0.005131543 | 1 |
| T2DM | mast_cell | *S1PR2* | 1.868927995 | 0.002480743 | 0.003341634 | 1 |
| T2DM | mast_cell | *ZNF224* | 2.457356828 | 3.26E-05 | 0.000134478 | 0.1580775 |

| | | | | | | |
|---|---|---|---|---|---|---|
| T2DM | mast_cell | *LIN37* | -5.114523886 | 0.000216404 | 0.00037181 | 1 |
| T2DM | mast_cell | *NDUFA7* | 12.29747178 | 3.13E-05 | 0.000134478 | 0.152091084 |
| T2DM | mast_cell | *AC004466.1* | 2.013501871 | 0.039448752 | 0.046333901 | 1 |
| T2DM | mast_cell | *ZNF728* | 4.290758532 | 2.31E-05 | 0.000134478 | 0.1123214 |
| T2DM | mast_cell | *TRABD2B* | 1.871491595 | 3.10E-05 | 0.000134478 | 0.150616755 |
| T2DM | mast_cell | *AC093157.1* | 3.858447347 | 7.52E-05 | 0.000166834 | 0.364866123 |
| T2DM | mast_cell | *FBXO17* | 1.651251185 | 1.06E-05 | 0.000134478 | 0.051276763 |
| T2DM | mast_cell | *ZNF587B* | 2.336199317 | 4.92E-05 | 0.000135162 | 0.238831585 |
| T2DM | mast_cell | *AP001172.2* | 1.824878184 | 4.30E-05 | 0.000134478 | 0.208582581 |
| T2DM | mast_cell | *HCG17* | 1.9084317 | 5.81E-05 | 0.000143427 | 0.281834232 |
| T2DM | mast_cell | *MILR1* | 0.939484864 | 0.004847814 | 0.006287515 | 1 |
| T2DM | mast_cell | *GTF2H5* | 1.541650768 | 4.85E-05 | 0.000134672 | 0.23527263 |
| T2DM | mast_cell | *FGF14-AS2* | 2.980834123 | 9.92E-05 | 0.00020188 | 0.48128149 |
| T2DM | mast_cell | *CASC15* | 1.850329048 | 3.44E-05 | 0.000134478 | 0.167061721 |
| T2DM | mast_cell | *GRIN2B* | 1.715727253 | 6.03E-06 | 0.000134478 | 0.029255309 |
| T2DM | mast_cell | *PAXIP1-AS1* | 1.898568159 | 5.18E-05 | 0.000136402 | 0.25151437 |
| T2DM | mesothelium | *RBM6* | -0.149124974 | 4.65E-09 | 2.05E-06 | 2.25E-05 |
| T2DM | mesothelium | *GGNBP2* | -0.716292474 | 3.66E-05 | 0.003346524 | 0.177365756 |
| T2DM | mesothelium | *TSPAN9* | -0.189815267 | 0.00084676 | 0.03830539 | 1 |
| T2DM | mesothelium | *CEP68* | 0.467284753 | 8.08E-09 | 3.02E-06 | 3.92E-05 |
| T2DM | mesothelium | *SH3YL1* | -0.351521596 | 0.000498403 | 0.026247212 | 1 |
| T2DM | mesothelium | *TUBG2* | -0.350484234 | 7.34E-08 | 1.78E-05 | 0.000356356 |
| T2DM | mesothelium | *ITIH4* | 0.068135327 | 7.23E-06 | 0.000938756 | 0.035072724 |
| T2DM | mesothelium | *SEC61A2* | -0.444317523 | 3.40E-07 | 6.34E-05 | 0.001651751 |
| T2DM | mesothelium | *IFT80* | -1.037648124 | 1.00E-05 | 0.001183871 | 0.04853872 |
| T2DM | mesothelium | *PABPC4* | -0.484703008 | 2.88E-13 | 2.80E-10 | 1.40E-09 |
| T2DM | mesothelium | *ALKBH5* | -0.46605312 | 2.74E-05 | 0.002713294 | 0.132951403 |
| T2DM | mesothelium | *RGS17* | -0.226739657 | 0.001056286 | 0.045759822 | 1 |
| T2DM | mesothelium | *CWF19L1* | -0.118648089 | 2.52E-05 | 0.002552219 | 0.122506505 |
| T2DM | mesothelium | *SCD* | -0.167508942 | 0.001141624 | 0.048539725 | 1 |
| T2DM | mesothelium | *NOMO3* | -0.148424496 | 0.000481246 | 0.025659399 | 1 |
| T2DM | mesothelium | *GCDH* | -0.425445345 | 5.81E-07 | 9.72E-05 | 0.002818201 |
| T2DM | mesothelium | *RNF32* | -0.837772739 | 1.55E-06 | 0.000250486 | 0.007514572 |
| T2DM | mesothelium | *CHN2* | 0.523695896 | 3.89E-10 | 2.09E-07 | 1.89E-06 |
| T2DM | mesothelium | *CNTNAP1* | 0.480979039 | 2.67E-06 | 0.000404781 | 0.012953005 |
| T2DM | mesothelium | *C11orf21* | -0.336299552 | 0.000444664 | 0.024517149 | 1 |
| T2DM | mesothelium | *CDK2AP1* | 0.239756674 | 4.76E-06 | 0.000660151 | 0.0231053 |
| T2DM | mesothelium | *BVES* | -0.07894209 | 0.000458875 | 0.025016427 | 1 |
| T2DM | mesothelium | *GPX7* | -0.070948571 | 0.000597272 | 0.030504865 | 1 |
| T2DM | mesothelium | *CCDC92* | -0.37067017 | 0.000142663 | 0.009354049 | 0.692199609 |
| T2DM | mesothelium | *ADRA1A* | 0.052901529 | 7.72E-05 | 0.00604113 | 0.37455008 |
| T2DM | mesothelium | *ECHDC2* | -0.309193902 | 0.000131958 | 0.009074366 | 0.640260153 |
| T2DM | mesothelium | *PDZRN3* | -0.151962487 | 8.50E-05 | 0.006543739 | 0.412255536 |
| T2DM | mesothelium | *OBSL1* | -0.42063618 | 0.001033583 | 0.045179698 | 1 |
| T2DM | mesothelium | *KCNK17* | -0.219246074 | 0.000710077 | 0.034452945 | 1 |
| T2DM | mesothelium | *ABHD8* | -0.215693288 | 0.000110719 | 0.00801802 | 0.537207355 |
| T2DM | mesothelium | *NPAS1* | -0.386611786 | 0.000355498 | 0.020514651 | 1 |
| T2DM | mesothelium | *APTX* | -0.575632672 | 0.000768626 | 0.035957762 | 1 |
| T2DM | mesothelium | *IRF4* | 0.060005792 | 0.000784781 | 0.036264371 | 1 |
| T2DM | mesothelium | *TCF19* | 0.295385828 | 3.33E-09 | 1.62E-06 | 1.62E-05 |

| | | | | | | |
|---|---|---|---|---|---|---|
| T2DM | mesothelium | *SLC37A4* | 0.184732536 | 0.000370644 | 0.020911239 | 1 |
| T2DM | mesothelium | *SPTBN5* | 0.046687453 | 6.63E-05 | 0.005456217 | 0.321916823 |
| T2DM | mesothelium | *NEIL1* | -0.315925763 | 4.38E-06 | 0.000625333 | 0.021261321 |
| T2DM | mesothelium | *MAN2C1* | -0.081899746 | 7.35E-06 | 0.000938756 | 0.035672714 |
| T2DM | mesothelium | *MYO19* | 0.434304673 | 0.000359387 | 0.020514651 | 1 |
| T2DM | mesothelium | *VPS53* | -0.256272173 | 6.80E-05 | 0.005496136 | 0.329768165 |
| T2DM | mesothelium | *C18orf8* | -0.449337402 | 1.06E-07 | 2.33E-05 | 0.000513334 |
| T2DM | mesothelium | *NPC1* | -0.629447577 | 9.46E-05 | 0.007168222 | 0.458766199 |
| T2DM | mesothelium | *CACNA1A* | -0.897916122 | 9.44E-06 | 0.001144531 | 0.045781234 |
| T2DM | mesothelium | *SAE1* | -0.832715952 | 4.22E-08 | 1.14E-05 | 0.000204911 |
| T2DM | mesothelium | *HSD17B12* | -0.117232105 | 7.72E-09 | 3.02E-06 | 3.75E-05 |
| T2DM | mesothelium | *APIP* | -0.072513799 | 0.000123176 | 0.008733954 | 0.597650251 |
| T2DM | mesothelium | *EIF3M* | -0.455210731 | 5.21E-05 | 0.004436711 | 0.252892505 |
| T2DM | mesothelium | *POC5* | -0.154456754 | 0.000139872 | 0.009354049 | 0.678656606 |
| T2DM | mesothelium | *JAZF1* | -0.649408482 | 3.78E-21 | 1.84E-17 | 1.84E-17 |
| T2DM | mesothelium | *MOV10* | -0.715401017 | 0.000166489 | 0.010770713 | 0.807803489 |
| T2DM | mesothelium | *NDUFAF6* | -0.514030087 | 5.08E-07 | 8.81E-05 | 0.002466366 |
| T2DM | mesothelium | *CLEC18A* | -0.204036468 | 1.17E-08 | 4.04E-06 | 5.66E-05 |
| T2DM | mesothelium | *KRTCAP3* | 0.126991714 | 4.33E-05 | 0.003822041 | 0.210212232 |
| T2DM | mesothelium | *SPATA2L* | -0.40131862 | 0.000232607 | 0.014286189 | 1 |
| T2DM | mesothelium | *DCAF16* | -0.662914143 | 7.52E-05 | 0.005979422 | 0.364744729 |
| T2DM | mesothelium | *SPTA1* | -0.104288419 | 0.000259294 | 0.015726191 | 1 |
| T2DM | mesothelium | *RFT1* | -0.250969158 | 1.67E-07 | 3.38E-05 | 0.000810354 |
| T2DM | mesothelium | *RNF123* | 0.180059423 | 3.36E-05 | 0.003194922 | 0.16294104 |
| T2DM | mesothelium | *MST1R* | 0.142161009 | 3.44E-05 | 0.003212765 | 0.167063798 |
| T2DM | mesothelium | *RAPSN* | -0.168994413 | 1.88E-05 | 0.002029055 | 0.091307475 |
| T2DM | mesothelium | *CEP57* | -0.397115245 | 0.000726612 | 0.034906158 | 1 |
| T2DM | mesothelium | *STXBP4* | -0.514434138 | 0.000770735 | 0.035957762 | 1 |
| T2DM | mesothelium | *ANPEP* | 0.065859663 | 0.000626836 | 0.031354727 | 1 |
| T2DM | mesothelium | *PDHB* | -0.542271243 | 4.05E-05 | 0.003637231 | 0.196410497 |
| T2DM | mesothelium | *ZSWIM1* | 0.056391212 | 0.000623691 | 0.031354727 | 1 |
| T2DM | mesothelium | *ZNF507* | -0.455315063 | 0.000852634 | 0.03830539 | 1 |
| T2DM | mesothelium | *IRS1* | -0.090825856 | 2.77E-07 | 5.38E-05 | 0.001345244 |
| T2DM | mesothelium | *INO80E* | -0.143069914 | 0.000124205 | 0.008733954 | 0.602642829 |
| T2DM | mesothelium | *SEMA3E* | 0.26528613 | 0.000641336 | 0.031476242 | 1 |
| T2DM | mesothelium | *EMB* | -0.193709206 | 1.71E-05 | 0.001889742 | 0.083148642 |
| T2DM | mesothelium | *TTLL6* | -0.569556616 | 1.38E-08 | 4.12E-06 | 6.69E-05 |
| T2DM | mesothelium | *RGS19* | -0.090785776 | 3.26E-05 | 0.003165278 | 0.158263907 |
| T2DM | mesothelium | *EXOSC10* | -0.493229041 | 0.000474894 | 0.02560208 | 1 |
| T2DM | mesothelium | *C1QTNF4* | -0.078212344 | 0.000103509 | 0.007609485 | 0.502225992 |
| T2DM | mesothelium | *CTSW* | -0.026128578 | 0.000222441 | 0.014201097 | 1 |
| T2DM | mesothelium | *RNASEH2C* | -0.192697715 | 2.13E-05 | 0.002248719 | 0.103441091 |
| T2DM | mesothelium | *HOXB2* | -0.273948011 | 0.000269742 | 0.016157851 | 1 |
| T2DM | mesothelium | *CCNE2* | -0.248804147 | 3.53E-07 | 6.34E-05 | 0.001710943 |
| T2DM | mesothelium | *SLC25A22* | -0.290416162 | 0.000431978 | 0.02409148 | 1 |
| T2DM | mesothelium | *ZNF852* | -0.229602531 | 0.000762694 | 0.035957762 | 1 |
| T2DM | mesothelium | *HLA-DQB1* | -0.145764116 | 1.13E-16 | 1.83E-13 | 5.50E-13 |
| T2DM | mesothelium | *PSMD13* | -0.26827017 | 0.000973108 | 0.042922926 | 1 |
| T2DM | mesothelium | *DMWD* | -0.402856074 | 2.41E-05 | 0.002490289 | 0.117043592 |
| T2DM | mesothelium | *RASA3* | -0.240005688 | 4.98E-05 | 0.004315176 | 0.241649858 |

| | | | | | | |
|---|---|---|---|---|---|---|
| T2DM | mesothelium | *FFAR4* | 0.818204518 | 3.54E-10 | 2.09E-07 | 1.72E-06 |
| T2DM | mesothelium | *HLA-DRB1* | -0.139564231 | 2.73E-11 | 1.89E-08 | 1.33E-07 |
| T2DM | mesothelium | *HLA-DQA1* | -0.199091678 | 2.32E-14 | 2.81E-11 | 1.12E-10 |
| T2DM | mesothelium | *ABCB8* | -0.657148761 | 0.000326887 | 0.019342118 | 1 |
| T2DM | mesothelium | *HLA-DRB5* | -0.056331712 | 2.39E-12 | 1.93E-09 | 1.16E-08 |
| T2DM | mesothelium | *SHISA4* | -0.785865776 | 0.00050309 | 0.026247212 | 1 |
| T2DM | mesothelium | *MAP3K3* | -0.50597346 | 8.06E-06 | 0.001002903 | 0.039113218 |
| T2DM | mesothelium | *NOTCH4* | -0.023947409 | 1.44E-08 | 4.12E-06 | 7.00E-05 |
| T2DM | mesothelium | *PRRT1* | 0.10177651 | 0.000132786 | 0.009074366 | 0.644279989 |
| T2DM | mesothelium | *STK19* | -0.256092046 | 1.26E-05 | 0.00145522 | 0.061119234 |
| T2DM | mesothelium | *GPANK1* | -0.255029071 | 3.49E-06 | 0.000513804 | 0.01695552 |
| T2DM | mesothelium | *LIN52* | 0.231417901 | 0.000349038 | 0.020404006 | 1 |
| T2DM | mesothelium | *HCG27* | 0.13206281 | 2.59E-06 | 0.000404781 | 0.012576036 |
| T2DM | mesothelium | *TAS2R14* | 0.097309607 | 0.00064224 | 0.031476242 | 1 |
| T2DM | mesothelium | *DNLZ* | 0.061938682 | 0.000231052 | 0.014286189 | 1 |
| T2DM | mesothelium | *ARHGAP19* | -0.478161515 | 1.39E-05 | 0.001570585 | 0.067535174 |
| T2DM | mesothelium | *TMEM110* | 0.280088871 | 5.06E-06 | 0.000682179 | 0.024558434 |
| T2DM | mesothelium | *STARD10* | 0.118676049 | 5.74E-08 | 1.47E-05 | 0.0002785 |
| T2DM | mesothelium | *IFRD2* | -0.168162254 | 0.001150468 | 0.048539725 | 1 |
| T2DM | mesothelium | *C5orf66* | -0.46078582 | 0.000840144 | 0.03830539 | 1 |
| T2DM | mesothelium | *C4B* | 0.229292871 | 9.73E-05 | 0.007263852 | 0.472150369 |
| T2DM | mesothelium | *AL049840.1* | -0.358464117 | 0.001124836 | 0.048298284 | 1 |
| T2DM | mesothelium | *HLA-DQB2* | 0.053886957 | 1.36E-08 | 4.12E-06 | 6.58E-05 |
| T2DM | mesothelium | *HLA-DQA2* | 0.100336541 | 3.36E-18 | 8.16E-15 | 1.63E-14 |
| T2DM | mesothelium | *PSMB9* | 0.094552195 | 6.07E-05 | 0.005080735 | 0.294682604 |
| T2DM | mesothelium | *PLA2G4B* | 0.493888875 | 8.78E-08 | 2.03E-05 | 0.000425884 |
| T2DM | mesothelium | *JMJD7* | 0.149458254 | 1.48E-07 | 3.13E-05 | 0.000719145 |
| T2DM | mesothelium | *C4A* | -0.175214146 | 0.000141929 | 0.009354049 | 0.688641581 |
| T2DM | mesothelium | *DNAH10OS* | -1.261581355 | 0.000232596 | 0.014286189 | 1 |
| T2DM | mesothelium | *C19orf84* | 5.090707917 | 0.000904243 | 0.040251269 | 1 |
| T2DM | mesothelium | *SNHG8* | -0.195813661 | 0.000543448 | 0.028051163 | 1 |
| T2DM | monocyte | *RBM6* | -1.216133805 | 8.91E-05 | 0.02400781 | 0.432140574 |
| T2DM | monocyte | *ITIH4* | 1.228085966 | 4.23E-05 | 0.014352358 | 0.205244346 |
| T2DM | monocyte | *PABPC4* | -2.834386071 | 6.08E-05 | 0.017355402 | 0.295041829 |
| T2DM | monocyte | *HSD17B12* | -1.296905904 | 8.00E-06 | 0.003526773 | 0.038794506 |
| T2DM | monocyte | *JAZF1* | -2.397561667 | 5.77E-05 | 0.017355402 | 0.280135853 |
| T2DM | monocyte | *CLEC18A* | -4.224204087 | 5.66E-08 | 5.49E-05 | 0.000274427 |
| T2DM | monocyte | *KRTCAP3* | 2.360447354 | 1.16E-05 | 0.004324221 | 0.056214872 |
| T2DM | monocyte | *TTLL6* | -7.126216839 | 3.63E-06 | 0.001954811 | 0.0175933 |
| T2DM | monocyte | *HLA-DQB1* | -2.585010535 | 9.15E-15 | 2.22E-11 | 4.44E-11 |
| T2DM | monocyte | *HLA-DRB1* | -1.951700246 | 1.28E-07 | 0.000103126 | 0.000618753 |
| T2DM | monocyte | *HLA-DRB5* | -0.952478389 | 6.53E-10 | 1.06E-06 | 3.17E-06 |
| T2DM | monocyte | *NOTCH4* | -0.480100391 | 2.57E-08 | 3.12E-05 | 0.00012487 |
| T2DM | monocyte | *STARD10* | 2.215604157 | 3.22E-07 | 0.000223107 | 0.001561748 |
| T2DM | monocyte | *C4B* | 4.570605305 | 0.000158222 | 0.040404852 | 0.76769218 |
| T2DM | monocyte | *HLA-DQB2* | 0.928926284 | 5.07E-07 | 0.000307596 | 0.002460769 |
| T2DM | monocyte | *HLA-DQA2* | 1.774624866 | 7.07E-17 | 3.43E-13 | 3.43E-13 |
| T2DM | monocyte | *PSMB9* | 1.699046612 | 4.44E-05 | 0.014352358 | 0.215285376 |
| T2DM | monocyte | *PLA2G4B* | 7.52781166 | 9.03E-06 | 0.003649297 | 0.043791565 |
| T2DM | monocyte | *JMJD7* | 2.475949481 | 4.74E-06 | 0.00229772 | 0.022977205 |

| | | | | | | |
|---|---|---|---|---|---|---|
| T2DM | nk_cell | *NFYA* | -1.732423089 | 3.54E-06 | 0.000122264 | 0.017165033 |
| T2DM | nk_cell | *ENPP4* | -0.877801459 | 0.001901485 | 0.002879395 | 1 |
| T2DM | nk_cell | *SEMA3F* | -2.041189596 | 6.24E-07 | 5.68E-05 | 0.003025865 |
| T2DM | nk_cell | *KRIT1* | -1.164782939 | 3.28E-05 | 0.000127909 | 0.159167696 |
| T2DM | nk_cell | *RAD52* | -1.301481716 | 4.24E-05 | 0.000141957 | 0.205527035 |
| T2DM | nk_cell | *HS3ST1* | -1.248938902 | 0.001868232 | 0.002830937 | 1 |
| T2DM | nk_cell | *AOC1* | -1.167082239 | 0.000236236 | 0.000460144 | 1 |
| T2DM | nk_cell | *SNX11* | -0.354334511 | 0.038378642 | 0.047178406 | 1 |
| T2DM | nk_cell | *TMEM176A* | -1.762543666 | 3.34E-05 | 0.000128061 | 0.16212478 |
| T2DM | nk_cell | *CYP26B1* | -0.628646042 | 6.99E-05 | 0.00018653 | 0.339111558 |
| T2DM | nk_cell | *DBNDD1* | -1.047102632 | 1.34E-06 | 9.04E-05 | 0.006492738 |
| T2DM | nk_cell | *ALS2* | -1.19217771 | 4.51E-05 | 0.000147008 | 0.218894557 |
| T2DM | nk_cell | *CD38* | 0.379693031 | 9.84E-05 | 0.000233629 | 0.477537733 |
| T2DM | nk_cell | *RBM6* | -1.337109523 | 5.65E-12 | 5.48E-09 | 2.74E-08 |
| T2DM | nk_cell | *RECQL* | -1.924071542 | 0.000395997 | 0.000714533 | 1 |
| T2DM | nk_cell | *ARHGAP33* | -1.428422907 | 0.0020265 | 0.003048861 | 1 |
| T2DM | nk_cell | *SLC22A16* | 0.611622205 | 0.00074705 | 0.001227842 | 1 |
| T2DM | nk_cell | *CDC27* | -0.939002309 | 0.000373436 | 0.000678619 | 1 |
| T2DM | nk_cell | *POLR2J* | 1.172782178 | 0.001299655 | 0.002024374 | 1 |
| T2DM | nk_cell | *MEOX1* | -0.847214235 | 3.57E-05 | 0.000131704 | 0.173318895 |
| T2DM | nk_cell | *LIG3* | -0.974929899 | 0.000306611 | 0.000573287 | 1 |
| T2DM | nk_cell | *PON1* | -2.871231554 | 8.30E-05 | 0.000208464 | 0.402805581 |
| T2DM | nk_cell | *GCFC2* | -1.282480631 | 4.55E-05 | 0.000147697 | 0.220966138 |
| T2DM | nk_cell | *RHBDD2* | -0.919864427 | 7.40E-05 | 0.000194104 | 0.359128336 |
| T2DM | nk_cell | *ZNF195* | -2.092289338 | 5.20E-06 | 0.000122264 | 0.025241286 |
| T2DM | nk_cell | *PDK2* | -1.147061511 | 6.31E-06 | 0.000122264 | 0.03062518 |
| T2DM | nk_cell | *GGNBP2* | -1.385025539 | 4.66E-06 | 0.000122264 | 0.022594989 |
| T2DM | nk_cell | *ITGA2B* | -0.683517425 | 3.41E-05 | 0.000129429 | 0.165410726 |
| T2DM | nk_cell | *TMEM98* | -0.905445643 | 2.48E-05 | 0.000122264 | 0.120131075 |
| T2DM | nk_cell | *ABCC8* | -0.867083152 | 0.001132199 | 0.001792894 | 1 |
| T2DM | nk_cell | *SYNRG* | -1.066891251 | 0.025247587 | 0.031804856 | 1 |
| T2DM | nk_cell | *ZNF263* | -1.703524437 | 0.003820938 | 0.005486591 | 1 |
| T2DM | nk_cell | *SPATA20* | -1.174554588 | 0.008945618 | 0.012073473 | 1 |
| T2DM | nk_cell | *MAP3K9* | -0.477201892 | 0.019583426 | 0.025123951 | 1 |
| T2DM | nk_cell | *BAIAP2L1* | -0.950254944 | 1.84E-05 | 0.000122264 | 0.089466415 |
| T2DM | nk_cell | *FARP2* | -0.902736195 | 2.08E-05 | 0.000122264 | 0.100884196 |
| T2DM | nk_cell | *USH1C* | -6.245311958 | 5.28E-05 | 0.00015824 | 0.25606517 |
| T2DM | nk_cell | *IFRD1* | -0.814362744 | 6.12E-05 | 0.00017138 | 0.297173026 |
| T2DM | nk_cell | *VPS41* | -1.152726137 | 2.64E-05 | 0.000122391 | 0.128166955 |
| T2DM | nk_cell | *ELAC2* | -0.979704686 | 0.000549608 | 0.000941298 | 1 |
| T2DM | nk_cell | *SCIN* | -0.889702928 | 1.27E-05 | 0.000122264 | 0.061763627 |
| T2DM | nk_cell | *CCDC124* | -0.986671819 | 0.006299258 | 0.008722602 | 1 |
| T2DM | nk_cell | *CEACAM21* | 0.16192811 | 6.31E-05 | 0.000173832 | 0.306079482 |
| T2DM | nk_cell | *CD79B* | 0.74035433 | 0.010819864 | 0.014418561 | 1 |
| T2DM | nk_cell | *ST7L* | -1.245046544 | 7.59E-06 | 0.000122264 | 0.03682242 |
| T2DM | nk_cell | *PIGQ* | -1.289225788 | 0.000738229 | 0.001217344 | 1 |
| T2DM | nk_cell | *FMO3* | -0.647587509 | 3.57E-05 | 0.000131704 | 0.173402597 |
| T2DM | nk_cell | *NADK* | -1.054337536 | 2.19E-05 | 0.000122264 | 0.106267553 |
| T2DM | nk_cell | *DLEC1* | -0.887018025 | 5.85E-05 | 0.000167232 | 0.283693384 |
| T2DM | nk_cell | *CYB561* | -0.727363473 | 0.001077174 | 0.001713027 | 1 |

| | | | | | | |
|---|---|---|---|---|---|---|
| T2DM | nk_cell | *SS18L2* | -1.314059796 | 0.000272717 | 0.000518912 | 1 |
| T2DM | nk_cell | *MGST1* | -0.462839778 | 0.001160118 | 0.001831563 | 1 |
| T2DM | nk_cell | *MMP25* | 0.827340666 | 0.000161256 | 0.000337612 | 0.782412049 |
| T2DM | nk_cell | *IL32* | 0.114486454 | 5.46E-05 | 0.000160881 | 0.265017649 |
| T2DM | nk_cell | *PKD1* | -0.82801261 | 3.95E-05 | 0.000137131 | 0.191708557 |
| T2DM | nk_cell | *MAPK8IP2* | 2.230420262 | 0.015137812 | 0.019744264 | 1 |
| T2DM | nk_cell | *MED24* | -1.838761492 | 5.33E-05 | 0.000159278 | 0.258668249 |
| T2DM | nk_cell | *TRAF3IP3* | 0.131820829 | 3.04E-05 | 0.000125051 | 0.147669603 |
| T2DM | nk_cell | *POMT2* | -0.593267382 | 1.31E-05 | 0.000122264 | 0.063409263 |
| T2DM | nk_cell | *MLXIPL* | -0.506535067 | 8.58E-05 | 0.000212706 | 0.416266169 |
| T2DM | nk_cell | *DYRK4* | -0.625449266 | 0.00679871 | 0.009350153 | 1 |
| T2DM | nk_cell | *NCAPD2* | 0.587337413 | 0.015041134 | 0.019634002 | 1 |
| T2DM | nk_cell | *FUZ* | 1.985119618 | 0.008116532 | 0.010997323 | 1 |
| T2DM | nk_cell | *SLC6A13* | 0.886944963 | 0.029383666 | 0.036725798 | 1 |
| T2DM | nk_cell | *LRRC23* | -1.083207813 | 0.000832958 | 0.001355757 | 1 |
| T2DM | nk_cell | *CLCN6* | -1.768980485 | 0.000163449 | 0.000340659 | 0.793053792 |
| T2DM | nk_cell | *SLC6A7* | -16.89161954 | 1.81E-05 | 0.000122264 | 0.087636711 |
| T2DM | nk_cell | *TSPAN9* | -1.268936173 | 3.09E-07 | 3.95E-05 | 0.001499403 |
| T2DM | nk_cell | *UTP18* | -1.13227732 | 0.001037982 | 0.001657218 | 1 |
| T2DM | nk_cell | *TTC19* | -1.252552899 | 0.000161296 | 0.000337612 | 0.782607292 |
| T2DM | nk_cell | *LARS2* | -1.416013919 | 0.000986174 | 0.001585984 | 1 |
| T2DM | nk_cell | *DCN* | -0.770768907 | 1.83E-05 | 0.000122264 | 0.088553877 |
| T2DM | nk_cell | *CEP68* | -0.600996888 | 0.010189364 | 0.013638288 | 1 |
| T2DM | nk_cell | *ZBTB32* | -1.832240551 | 0.018767808 | 0.024160628 | 1 |
| T2DM | nk_cell | *ERCC1* | -1.114590573 | 0.002953685 | 0.004323161 | 1 |
| T2DM | nk_cell | *CD22* | 0.401733118 | 0.000404541 | 0.000726437 | 1 |
| T2DM | nk_cell | *MAP4K5* | -1.356512423 | 5.36E-05 | 0.000159514 | 0.260189858 |
| T2DM | nk_cell | *EHD3* | -1.846705765 | 7.07E-05 | 0.000188051 | 0.342817245 |
| T2DM | nk_cell | *MAN2B2* | -1.747730699 | 4.63E-05 | 0.000148275 | 0.224635999 |
| T2DM | nk_cell | *SLC25A39* | 0.835513612 | 0.025809141 | 0.03247561 | 1 |
| T2DM | nk_cell | *RNF14* | -1.392411171 | 0.000117076 | 0.000263449 | 0.56805468 |
| T2DM | nk_cell | *DDX11* | -0.71282961 | 1.41E-05 | 0.000122264 | 0.068472645 |
| T2DM | nk_cell | *TACC3* | -1.948685006 | 2.76E-05 | 0.000122472 | 0.13386135 |
| T2DM | nk_cell | *ZC3H3* | -0.88223974 | 0.006773213 | 0.009320372 | 1 |
| T2DM | nk_cell | *MDH1* | -1.398796487 | 4.85E-05 | 0.000151023 | 0.235188838 |
| T2DM | nk_cell | *MTMR11* | -0.514743025 | 0.003286386 | 0.004782706 | 1 |
| T2DM | nk_cell | *COX15* | -1.156725774 | 3.61E-05 | 0.000132186 | 0.175284065 |
| T2DM | nk_cell | *CCDC88C* | 0.178201303 | 7.41E-05 | 0.000194164 | 0.359592017 |
| T2DM | nk_cell | *NUDCD3* | -1.132397949 | 0.000426733 | 0.00075815 | 1 |
| T2DM | nk_cell | *CHDH* | -2.070484675 | 7.37E-05 | 0.000193525 | 0.357391532 |
| T2DM | nk_cell | *CLCA4* | -2.139951819 | 0.00047624 | 0.000832091 | 1 |
| T2DM | nk_cell | *RUFY3* | -1.104778625 | 8.23E-05 | 0.000207015 | 0.399538102 |
| T2DM | nk_cell | *SLC11A1* | -0.47356333 | 3.39E-05 | 0.000128918 | 0.164381934 |
| T2DM | nk_cell | *WWTR1* | -0.911955491 | 1.99E-05 | 0.000122264 | 0.09634586 |
| T2DM | nk_cell | *AGPS* | -1.129221679 | 0.000977836 | 0.001574141 | 1 |
| T2DM | nk_cell | *TTC27* | -1.156509065 | 2.70E-05 | 0.000122391 | 0.13096863 |
| T2DM | nk_cell | *VSIG2* | -0.722512278 | 0.000118039 | 0.000264906 | 0.572727225 |
| T2DM | nk_cell | *HGF* | -1.91399274 | 0.000317792 | 0.00059123 | 1 |
| T2DM | nk_cell | *SLC7A9* | -0.578875633 | 0.001349806 | 0.002098449 | 1 |
| T2DM | nk_cell | *OSBPL5* | 0.763099631 | 0.000350932 | 0.000642537 | 1 |

| | | | | | | |
|---|---|---|---|---|---|---|
| T2DM | nk_cell | *CPS1* | -1.135403892 | 7.52E-05 | 0.000196007 | 0.364811705 |
| T2DM | nk_cell | *NLRP2* | 0.398441014 | 0.001956194 | 0.002954079 | 1 |
| T2DM | nk_cell | *SLC45A4* | -1.771060913 | 0.000161761 | 0.000338158 | 0.78486518 |
| T2DM | nk_cell | *RNH1* | -1.096031561 | 0.001286294 | 0.00200743 | 1 |
| T2DM | nk_cell | *GLRX2* | -1.726797075 | 1.00E-05 | 0.000122264 | 0.048595523 |
| T2DM | nk_cell | *ABCC2* | -1.584029085 | 1.65E-05 | 0.000122264 | 0.079856486 |
| T2DM | nk_cell | *GCLM* | -1.856234308 | 0.008106595 | 0.010989998 | 1 |
| T2DM | nk_cell | *UBR2* | -1.137092684 | 6.21E-05 | 0.00017315 | 0.30133629 |
| T2DM | nk_cell | *RRAGD* | -0.797880618 | 3.96E-05 | 0.000137211 | 0.19223262 |
| T2DM | nk_cell | *HSF2* | -0.953013836 | 0.009007579 | 0.012143588 | 1 |
| T2DM | nk_cell | *TYMP* | -0.837454913 | 0.004436331 | 0.006299408 | 1 |
| T2DM | nk_cell | *TOMM34* | 0.714443741 | 0.00021062 | 0.00041831 | 1 |
| T2DM | nk_cell | *SEC63* | -1.350344863 | 6.96E-05 | 0.00018614 | 0.337844096 |
| T2DM | nk_cell | *FAS* | -1.841525614 | 0.000176565 | 0.000362851 | 0.856691163 |
| T2DM | nk_cell | *RNASET2* | -1.354068522 | 1.92E-05 | 0.000122264 | 0.093235131 |
| T2DM | nk_cell | *AGPAT4* | -1.447638598 | 3.65E-05 | 0.000132621 | 0.177181792 |
| T2DM | nk_cell | *BTN3A1* | 0.442568279 | 0.03330178 | 0.041282636 | 1 |
| T2DM | nk_cell | *MIPEP* | -1.296193922 | 1.05E-05 | 0.000122264 | 0.050866326 |
| T2DM | nk_cell | *INSRR* | 1.659893862 | 0.020484538 | 0.026189981 | 1 |
| T2DM | nk_cell | *B4GALT7* | -0.866020874 | 2.35E-05 | 0.000122264 | 0.114123968 |
| T2DM | nk_cell | *SH2D2A* | 0.148159477 | 4.70E-05 | 0.000148778 | 0.227927421 |
| T2DM | nk_cell | *VRK2* | -0.995821041 | 2.37E-05 | 0.000122264 | 0.114992641 |
| T2DM | nk_cell | *VEZT* | -1.23094152 | 4.28E-05 | 0.000142857 | 0.207719417 |
| T2DM | nk_cell | *BRD9* | -1.435988713 | 0.000104103 | 0.000242723 | 0.50510677 |
| T2DM | nk_cell | *SNX1* | -1.353997165 | 0.004013866 | 0.005743225 | 1 |
| T2DM | nk_cell | *ANK1* | 0.719686267 | 0.000123652 | 0.00027383 | 0.599961853 |
| T2DM | nk_cell | *RABEP1* | -1.301865726 | 0.000352298 | 0.000644794 | 1 |
| T2DM | nk_cell | *BAK1* | 0.930409698 | 2.50E-05 | 0.000122264 | 0.121339035 |
| T2DM | nk_cell | *IKZF2* | 0.769639437 | 0.002427829 | 0.003606806 | 1 |
| T2DM | nk_cell | *GRN* | -1.00815114 | 0.00222189 | 0.003319154 | 1 |
| T2DM | nk_cell | *IFT88* | -1.298032962 | 9.00E-06 | 0.000122264 | 0.043690911 |
| T2DM | nk_cell | *CHPF2* | 0.749526702 | 0.000240864 | 0.000467468 | 1 |
| T2DM | nk_cell | *UBA6* | -1.52662542 | 0.000452657 | 0.000796623 | 1 |
| T2DM | nk_cell | *ATP6V0A1* | -0.959198342 | 0.000183836 | 0.000375251 | 0.891972387 |
| T2DM | nk_cell | *PIAS1* | -1.055492528 | 0.000144949 | 0.000310916 | 0.703292907 |
| T2DM | nk_cell | *ASTE1* | -1.475620959 | 0.000107151 | 0.000247923 | 0.519894938 |
| T2DM | nk_cell | *MYOC* | -0.550330867 | 7.53E-05 | 0.000196013 | 0.3651714 |
| T2DM | nk_cell | *SH3YL1* | -2.018591775 | 1.09E-05 | 0.000122264 | 0.052659096 |
| T2DM | nk_cell | *VCL* | -1.37570923 | 4.70E-05 | 0.000148778 | 0.227856416 |
| T2DM | nk_cell | *TBC1D23* | -1.347935109 | 3.93E-05 | 0.000136619 | 0.190536735 |
| T2DM | nk_cell | *MYOM2* | 1.237211508 | 0.000333227 | 0.000616165 | 1 |
| T2DM | nk_cell | *TUBG2* | -1.045136225 | 4.86E-08 | 1.04E-05 | 0.00023584 |
| T2DM | nk_cell | *NSUN2* | -1.712222365 | 7.79E-05 | 0.000200111 | 0.378210596 |
| T2DM | nk_cell | *MFAP3* | -1.006905461 | 0.009755009 | 0.013089409 | 1 |
| T2DM | nk_cell | *MRI1* | -1.270829192 | 4.72E-05 | 0.000149114 | 0.229034219 |
| T2DM | nk_cell | *AGA* | -2.118851401 | 3.31E-05 | 0.000127948 | 0.160473483 |
| T2DM | nk_cell | *PI4K2B* | -4.867273711 | 8.34E-05 | 0.000208918 | 0.404465726 |
| T2DM | nk_cell | *BOD1L1* | -1.010357402 | 3.66E-05 | 0.000132841 | 0.177609074 |
| T2DM | nk_cell | *DNAH5* | -2.357582104 | 0.030959527 | 0.03856627 | 1 |
| T2DM | nk_cell | *PNKP* | -0.941918285 | 0.010638085 | 0.014199722 | 1 |

| | | | | | | |
|---|---|---|---|---|---|---|
| T2DM | nk_cell | *PHLPP2* | -1.127857655 | 4.98E-05 | 0.00015287 | 0.241697975 |
| T2DM | nk_cell | *SPDL1* | 0.621578969 | 0.00050101 | 0.000869109 | 1 |
| T2DM | nk_cell | *PQLC2* | -1.618672386 | 1.53E-05 | 0.000122264 | 0.074177239 |
| T2DM | nk_cell | *INPP4A* | 0.610231541 | 0.003335509 | 0.004849833 | 1 |
| T2DM | nk_cell | *PSMA4* | 0.97441732 | 0.009725772 | 0.013061015 | 1 |
| T2DM | nk_cell | *TDP1* | -2.193355107 | 0.000152188 | 0.000323491 | 0.738413804 |
| T2DM | nk_cell | *SPATA7* | -1.046818135 | 1.31E-05 | 0.000122264 | 0.063634614 |
| T2DM | nk_cell | *CAPG* | -0.630714007 | 1.06E-05 | 0.000122264 | 0.051451933 |
| T2DM | nk_cell | *AP2S1* | -2.326704732 | 0.000306076 | 0.000572506 | 1 |
| T2DM | nk_cell | *TRIT1* | -1.124105636 | 0.00010263 | 0.000240564 | 0.49796172 |
| T2DM | nk_cell | *CTNNA1* | -0.92620972 | 2.15E-05 | 0.000122264 | 0.104367661 |
| T2DM | nk_cell | *CNTLN* | -0.963992588 | 1.19E-05 | 0.000122264 | 0.057689665 |
| T2DM | nk_cell | *YTHDC2* | -1.415452166 | 0.000128984 | 0.000283694 | 0.625828555 |
| T2DM | nk_cell | *POLR2B* | -1.495567529 | 0.003399057 | 0.00493927 | 1 |
| T2DM | nk_cell | *ARAP2* | 0.535175432 | 0.000333417 | 0.000616281 | 1 |
| T2DM | nk_cell | *DTNBP1* | -1.734768632 | 0.000425698 | 0.000756866 | 1 |
| T2DM | nk_cell | *ANO2* | -0.653371587 | 2.28E-05 | 0.000122264 | 0.110829017 |
| T2DM | nk_cell | *SNX29* | -1.15138529 | 4.25E-05 | 0.000142336 | 0.206448414 |
| T2DM | nk_cell | *EPN3* | -2.098149346 | 5.08E-05 | 0.000154353 | 0.246655689 |
| T2DM | nk_cell | *RCN1* | -1.922496376 | 1.10E-05 | 0.000122264 | 0.05327413 |
| T2DM | nk_cell | *JKAMP* | -1.453907895 | 4.72E-05 | 0.000149114 | 0.229039825 |
| T2DM | nk_cell | *ARHGEF5* | -0.558107455 | 0.000741362 | 0.001221588 | 1 |
| T2DM | nk_cell | *MCUR1* | -0.934575438 | 3.27E-05 | 0.000127909 | 0.158560544 |
| T2DM | nk_cell | *MAPK9* | -1.858214585 | 0.001084331 | 0.00172328 | 1 |
| T2DM | nk_cell | *FAM160A2* | -1.187816422 | 0.00018757 | 0.000381181 | 0.91008995 |
| T2DM | nk_cell | *CYBA* | 0.536301712 | 0.000492195 | 0.000855347 | 1 |
| T2DM | nk_cell | *THOC3* | 1.381914419 | 0.000265603 | 0.000507565 | 1 |
| T2DM | nk_cell | *FSTL4* | -0.72859462 | 7.37E-05 | 0.000193525 | 0.357439987 |
| T2DM | nk_cell | *LY75* | 0.582557018 | 6.85E-05 | 0.00018443 | 0.332526017 |
| T2DM | nk_cell | *SDCCAG8* | -0.712053382 | 9.55E-05 | 0.000228386 | 0.463394451 |
| T2DM | nk_cell | *KIF1B* | -0.830147932 | 0.000197349 | 0.000396653 | 0.957538143 |
| T2DM | nk_cell | *TBC1D22A* | -1.181730104 | 0.00084654 | 0.001376017 | 1 |
| T2DM | nk_cell | *GALC* | -1.470341538 | 1.77E-05 | 0.000122264 | 0.085916132 |
| T2DM | nk_cell | *FAM114A2* | -1.533697281 | 0.002568641 | 0.003793926 | 1 |
| T2DM | nk_cell | *GINM1* | -1.343423867 | 0.00966976 | 0.012992987 | 1 |
| T2DM | nk_cell | *ITIH4* | 0.457039174 | 0.00491926 | 0.006934412 | 1 |
| T2DM | nk_cell | *NPFFR2* | -0.821899999 | 2.57E-05 | 0.000122264 | 0.124568781 |
| T2DM | nk_cell | *PCGF2* | -0.841520846 | 0.000879851 | 0.001426817 | 1 |
| T2DM | nk_cell | *MSH4* | -0.700753256 | 0.002009792 | 0.003025601 | 1 |
| T2DM | nk_cell | *GDI2* | -1.408885403 | 0.000182855 | 0.000373721 | 0.887214827 |
| T2DM | nk_cell | *PITHD1* | -0.728060751 | 1.13E-05 | 0.000122264 | 0.05492354 |
| T2DM | nk_cell | *ATP11B* | -1.091204829 | 0.006757452 | 0.009306518 | 1 |
| T2DM | nk_cell | *LAMC2* | -0.716626959 | 0.002049489 | 0.003078675 | 1 |
| T2DM | nk_cell | *RASGRF1* | -1.935938712 | 5.39E-05 | 0.00015958 | 0.261342372 |
| T2DM | nk_cell | *CROCC* | -1.061538868 | 8.00E-05 | 0.000203408 | 0.38806238 |
| T2DM | nk_cell | *FLYWCH1* | -1.02003543 | 0.00196509 | 0.002965667 | 1 |
| T2DM | nk_cell | *ALDH18A1* | -1.398263154 | 8.02E-06 | 0.000122264 | 0.038927454 |
| T2DM | nk_cell | *DNAJC25* | -0.642198644 | 2.47E-05 | 0.000122264 | 0.119915821 |
| T2DM | nk_cell | *OGFR* | -1.397982999 | 0.000746456 | 0.001227314 | 1 |
| T2DM | nk_cell | *GNA15* | -0.472619906 | 0.004462028 | 0.00633219 | 1 |

| | | | | | | |
|---|---|---|---|---|---|---|
| T2DM | nk_cell | *PIGV* | -1.255917328 | 8.81E-05 | 0.000216672 | 0.427493602 |
| T2DM | nk_cell | *RIMBP2* | -0.733246726 | 2.21E-05 | 0.000122264 | 0.107013365 |
| T2DM | nk_cell | *PRDM6* | -1.069238554 | 2.52E-05 | 0.000122264 | 0.122190382 |
| T2DM | nk_cell | *SPAG4* | -2.056940283 | 5.39E-05 | 0.00015958 | 0.261551595 |
| T2DM | nk_cell | *NCKAP1* | -0.931369601 | 1.35E-05 | 0.000122264 | 0.065362458 |
| T2DM | nk_cell | *LTK* | -4.09342352 | 2.88E-05 | 0.000123207 | 0.139976221 |
| T2DM | nk_cell | *EIF4B* | -1.315925527 | 0.000366774 | 0.000668514 | 1 |
| T2DM | nk_cell | *AHRR* | -0.941051853 | 2.55E-05 | 0.000122264 | 0.12374737 |
| T2DM | nk_cell | *ADCK1* | -1.072785322 | 1.61E-05 | 0.000122264 | 0.078274141 |
| T2DM | nk_cell | *HAGH* | -1.452463547 | 0.000664415 | 0.001110103 | 1 |
| T2DM | nk_cell | *CASP8* | 0.675644421 | 0.000423125 | 0.000753671 | 1 |
| T2DM | nk_cell | *TM7SF3* | -1.260052242 | 0.003529841 | 0.005101814 | 1 |
| T2DM | nk_cell | *CDON* | -0.349423096 | 6.56E-05 | 0.000178701 | 0.3184459 |
| T2DM | nk_cell | *TMEM161A* | -2.288207217 | 0.000226347 | 0.000444269 | 1 |
| T2DM | nk_cell | *LPAR2* | 1.02549673 | 6.63E-05 | 0.000179849 | 0.321570693 |
| T2DM | nk_cell | *SUGP2* | -1.367861869 | 0.007558158 | 0.010289614 | 1 |
| T2DM | nk_cell | *CNN2* | 0.579502961 | 1.82E-05 | 0.000122264 | 0.088495558 |
| T2DM | nk_cell | *ABCA7* | -2.015876886 | 0.000241087 | 0.000467714 | 1 |
| T2DM | nk_cell | *DDX20* | -1.656127729 | 0.002471837 | 0.003668814 | 1 |
| T2DM | nk_cell | *FAR2* | -1.25923136 | 0.000541706 | 0.000930064 | 1 |
| T2DM | nk_cell | *BCAS1* | -1.264068252 | 8.31E-05 | 0.000208464 | 0.402960671 |
| T2DM | nk_cell | *CHI3L2* | -1.837118282 | 0.000158789 | 0.000333525 | 0.7704428 |
| T2DM | nk_cell | *SBNO2* | -1.326655508 | 0.02144354 | 0.027358416 | 1 |
| T2DM | nk_cell | *PMS1* | -1.434427368 | 0.000508168 | 0.000879954 | 1 |
| T2DM | nk_cell | *UHRF1BP1* | -0.953308425 | 0.000350257 | 0.000641786 | 1 |
| T2DM | nk_cell | *ANKRD44* | 0.429363008 | 0.00029436 | 0.000554018 | 1 |
| T2DM | nk_cell | *ADAT1* | -0.622450877 | 0.017368711 | 0.022460817 | 1 |
| T2DM | nk_cell | *MAP2K4* | -1.069292569 | 1.28E-05 | 0.000122264 | 0.062011617 |
| T2DM | nk_cell | *TMEM206* | -1.024341817 | 0.000113128 | 0.000257941 | 0.548897826 |
| T2DM | nk_cell | *SEC61A2* | -1.347879686 | 8.96E-08 | 1.74E-05 | 0.000434622 |
| T2DM | nk_cell | *TIE1* | -1.016088199 | 0.02163492 | 0.027580828 | 1 |
| T2DM | nk_cell | *NGEF* | -1.569592173 | 2.62E-05 | 0.000122295 | 0.127354911 |
| T2DM | nk_cell | *SPI1* | -0.971108595 | 5.65E-05 | 0.00016385 | 0.274121487 |
| T2DM | nk_cell | *ZNRD1* | -1.484163694 | 4.03E-05 | 0.000138652 | 0.195361017 |
| T2DM | nk_cell | *ATXN3* | -0.762230039 | 0.001432321 | 0.002214666 | 1 |
| T2DM | nk_cell | *GOLGA5* | -1.417366206 | 3.58E-05 | 0.000131743 | 0.173753946 |
| T2DM | nk_cell | *FGFR2* | -0.896167502 | 2.70E-06 | 0.000122264 | 0.013105324 |
| T2DM | nk_cell | *MTFR1* | -1.379883234 | 0.007223605 | 0.009878504 | 1 |
| T2DM | nk_cell | *FECH* | -0.609322145 | 2.64E-05 | 0.000122391 | 0.12819019 |
| T2DM | nk_cell | *MYO9A* | -0.838897453 | 3.04E-05 | 0.000125051 | 0.147535564 |
| T2DM | nk_cell | *TRAM1* | -1.410529332 | 0.005858391 | 0.00815636 | 1 |
| T2DM | nk_cell | *EVI5* | -0.971852604 | 1.30E-05 | 0.000122264 | 0.063215076 |
| T2DM | nk_cell | *STOML1* | -2.94417375 | 1.86E-05 | 0.000122264 | 0.09029957 |
| T2DM | nk_cell | *METTL22* | -0.773635002 | 1.17E-05 | 0.000122264 | 0.056617848 |
| T2DM | nk_cell | *TP53BP1* | -1.180255954 | 4.47E-05 | 0.000146229 | 0.21671095 |
| T2DM | nk_cell | *RRP15* | -0.865848382 | 0.005639477 | 0.007876438 | 1 |
| T2DM | nk_cell | *DHX8* | -1.503186974 | 2.46E-05 | 0.000122264 | 0.119268114 |
| T2DM | nk_cell | *CBFB* | -1.448317245 | 0.000485836 | 0.000847028 | 1 |
| T2DM | nk_cell | *IFI35* | -1.831156232 | 2.06E-05 | 0.000122264 | 0.099922907 |
| T2DM | nk_cell | *HEATR6* | -0.969944941 | 8.15E-05 | 0.000205631 | 0.395428555 |

| | | | | | | |
|---|---|---|---|---|---|---|
| T2DM | nk_cell | *POLR1A* | -1.333818905 | 3.02E-06 | 0.000122264 | 0.014663272 |
| T2DM | nk_cell | *IP6K2* | -1.520862777 | 1.35E-05 | 0.000122264 | 0.065723685 |
| T2DM | nk_cell | *STON1-GTF2A* | -0.470485208 | 1.88E-05 | 0.000122264 | 0.091073466 |
| T2DM | nk_cell | *SRBD1* | -1.3435217 | 0.004043535 | 0.005777158 | 1 |
| T2DM | nk_cell | *CYFIP1* | -1.164811893 | 1.51E-05 | 0.000122264 | 0.07314107 |
| T2DM | nk_cell | *IFT80* | -1.902751708 | 3.47E-06 | 0.000122264 | 0.016845169 |
| T2DM | nk_cell | *NUCKS1* | -1.305250202 | 0.002399164 | 0.003566405 | 1 |
| T2DM | nk_cell | *CLEC2D* | 0.544629594 | 0.0009397 | 0.001516774 | 1 |
| T2DM | nk_cell | *DRD4* | -2.645783172 | 1.80E-05 | 0.000122264 | 0.087439897 |
| T2DM | nk_cell | *PLA2G10* | 1.400427394 | 0.000136895 | 0.000296944 | 0.664212394 |
| T2DM | nk_cell | *NEDD4* | -1.050513995 | 0.000569827 | 0.000968752 | 1 |
| T2DM | nk_cell | *GNB5* | -0.757111707 | 0.009804004 | 0.013147879 | 1 |
| T2DM | nk_cell | *LRP6* | -1.037889531 | 1.84E-05 | 0.000122264 | 0.089406657 |
| T2DM | nk_cell | *DAPP1* | -0.501476351 | 0.03226513 | 0.040048711 | 1 |
| T2DM | nk_cell | *CLTCL1* | -1.441312207 | 1.55E-05 | 0.000122264 | 0.075192395 |
| T2DM | nk_cell | *ZXDC* | -1.430880737 | 9.25E-05 | 0.000223822 | 0.448697554 |
| T2DM | nk_cell | *GBA2* | -1.539736629 | 1.43E-05 | 0.000122264 | 0.069361655 |
| T2DM | nk_cell | *NDST1* | -0.691404846 | 4.40E-05 | 0.000144733 | 0.213481145 |
| T2DM | nk_cell | *ASNS* | -1.08409519 | 0.000196382 | 0.000395208 | 0.952846678 |
| T2DM | nk_cell | *AP3M2* | -1.9916563 | 0.000127665 | 0.000281176 | 0.619430751 |
| T2DM | nk_cell | *ST6GALNAC2* | -0.791656073 | 2.54E-05 | 0.000122264 | 0.123100729 |
| T2DM | nk_cell | *EIF2B3* | -1.321266242 | 2.56E-05 | 0.000122264 | 0.124243199 |
| T2DM | nk_cell | *MAP4K4* | -1.279815668 | 0.000139942 | 0.000301777 | 0.678998844 |
| T2DM | nk_cell | *SNX13* | -1.286906917 | 6.65E-05 | 0.000180243 | 0.322454491 |
| T2DM | nk_cell | *RPS6KA2* | -1.177342353 | 4.79E-05 | 0.000149989 | 0.232483124 |
| T2DM | nk_cell | *ING3* | -1.189793561 | 2.21E-05 | 0.000122264 | 0.107329634 |
| T2DM | nk_cell | *VASH1* | -1.616403505 | 0.000163571 | 0.000340768 | 0.793648781 |
| T2DM | nk_cell | *TRIB2* | 1.207333691 | 0.000672682 | 0.001121985 | 1 |
| T2DM | nk_cell | *CPSF1* | -0.873698848 | 0.005804772 | 0.008088672 | 1 |
| T2DM | nk_cell | *CYBRD1* | -0.880791722 | 3.55E-05 | 0.000131522 | 0.17216201 |
| T2DM | nk_cell | *RDH11* | -1.836737117 | 0.00014443 | 0.000310078 | 0.700776541 |
| T2DM | nk_cell | *EPN2* | -1.079259874 | 4.87E-06 | 0.000122264 | 0.023630514 |
| T2DM | nk_cell | *ALDH3A2* | -0.891178578 | 4.65E-05 | 0.000148578 | 0.225385496 |
| T2DM | nk_cell | *AFF4* | -1.074333584 | 4.48E-05 | 0.000146556 | 0.217342702 |
| T2DM | nk_cell | *TRHDE* | -0.710014115 | 1.12E-05 | 0.000122264 | 0.054296827 |
| T2DM | nk_cell | *FCGR2B* | -1.404021612 | 1.88E-05 | 0.000122264 | 0.091210983 |
| T2DM | nk_cell | *TRNT1* | -1.299900332 | 0.020360909 | 0.026059385 | 1 |
| T2DM | nk_cell | *CRMP1* | -0.877881631 | 1.55E-05 | 0.000122264 | 0.075105809 |
| T2DM | nk_cell | *NDE1* | -1.540079457 | 1.07E-05 | 0.000122264 | 0.052020457 |
| T2DM | nk_cell | *PVR* | -1.974884904 | 3.61E-06 | 0.000122264 | 0.017514919 |
| T2DM | nk_cell | *SCARB1* | -0.992901632 | 8.10E-05 | 0.00020495 | 0.393196784 |
| T2DM | nk_cell | *MOV10L1* | -0.594176339 | 3.82E-05 | 0.000135392 | 0.185216635 |
| T2DM | nk_cell | *ALPK1* | -1.174626967 | 0.000433233 | 0.000766611 | 1 |
| T2DM | nk_cell | *LLGL2* | 0.307050724 | 0.000132706 | 0.000289649 | 0.64389083 |
| T2DM | nk_cell | *ADAM11* | -0.74667307 | 0.000922053 | 0.001489778 | 1 |
| T2DM | nk_cell | *PPP2R2C* | -0.860162532 | 2.32E-05 | 0.000122264 | 0.112479613 |
| T2DM | nk_cell | *ZNF532* | -1.140977822 | 2.67E-05 | 0.000122391 | 0.129450766 |
| T2DM | nk_cell | *ZZEF1* | -1.062346802 | 0.006532011 | 0.009021724 | 1 |
| T2DM | nk_cell | *TTC38* | 0.598916559 | 0.000461634 | 0.000809325 | 1 |
| T2DM | nk_cell | *GRAMD4* | -1.819860104 | 0.000100251 | 0.0002367 | 0.486417642 |

| | | | | | | |
|---|---|---|---|---|---|---|
| T2DM | nk_cell | *ADD2* | -1.537248504 | 0.023687788 | 0.030000822 | 1 |
| T2DM | nk_cell | *FNDC3B* | -0.931455772 | 4.81E-05 | 0.000150261 | 0.233356068 |
| T2DM | nk_cell | *CACNG4* | -1.21303374 | 0.001227054 | 0.001923019 | 1 |
| T2DM | nk_cell | *MOCOS* | -0.631429039 | 0.00010275 | 0.00024061 | 0.498543436 |
| T2DM | nk_cell | *SEC31B* | -0.769125053 | 0.014899107 | 0.019459076 | 1 |
| T2DM | nk_cell | *KIFAP3* | -1.251486043 | 5.16E-05 | 0.000155956 | 0.250465002 |
| T2DM | nk_cell | *MKRN2* | -1.327892216 | 9.01E-05 | 0.000219625 | 0.43727434 |
| T2DM | nk_cell | *MCM6* | -1.489953773 | 0.000114518 | 0.000260132 | 0.555641852 |
| T2DM | nk_cell | *MLH1* | -1.703816195 | 0.000414638 | 0.000740457 | 1 |
| T2DM | nk_cell | *FMO4* | -1.187974598 | 6.31E-05 | 0.000173832 | 0.30611809 |
| T2DM | nk_cell | *RGS11* | -0.88645839 | 0.00070092 | 0.001164281 | 1 |
| T2DM | nk_cell | *SPAG5* | 1.324849601 | 0.000247829 | 0.000478879 | 1 |
| T2DM | nk_cell | *ANKRD13A* | -0.954093317 | 1.98E-05 | 0.000122264 | 0.096220921 |
| T2DM | nk_cell | *GPATCH1* | -1.772695637 | 3.21E-05 | 0.000127628 | 0.155892389 |
| T2DM | nk_cell | *ICAM3* | 0.235077987 | 6.05E-05 | 0.000169923 | 0.293627163 |
| T2DM | nk_cell | *NT5C2* | -0.808497712 | 0.002478163 | 0.003675955 | 1 |
| T2DM | nk_cell | *XAB2* | -1.942860217 | 6.18E-05 | 0.000172731 | 0.30003441 |
| T2DM | nk_cell | *IL4R* | -1.28036332 | 2.27E-05 | 0.000122264 | 0.110353534 |
| T2DM | nk_cell | *DYNC1I2* | -1.149333496 | 2.52E-05 | 0.000122264 | 0.122363441 |
| T2DM | nk_cell | *LRCH4* | -1.561932554 | 5.02E-05 | 0.000153221 | 0.24363246 |
| T2DM | nk_cell | *SIRT6* | -1.186092238 | 0.038540831 | 0.047353789 | 1 |
| T2DM | nk_cell | *GPR137B* | -1.168165861 | 0.000123458 | 0.000273525 | 0.599019062 |
| T2DM | nk_cell | *NAALAD2* | -0.455233011 | 0.013808091 | 0.018107259 | 1 |
| T2DM | nk_cell | *AMPH* | 1.085643375 | 0.000935346 | 0.00151025 | 1 |
| T2DM | nk_cell | *MCCC1* | -0.740295274 | 0.000190742 | 0.000385617 | 0.925480673 |
| T2DM | nk_cell | *ACER3* | -0.882327643 | 3.08E-06 | 0.000122264 | 0.014960724 |
| T2DM | nk_cell | *PIK3C3* | -1.44393269 | 8.70E-05 | 0.000214575 | 0.422284013 |
| T2DM | nk_cell | *RBFOX1* | -0.884800153 | 5.15E-05 | 0.000155956 | 0.249878886 |
| T2DM | nk_cell | *PCM1* | -1.017884756 | 8.10E-05 | 0.00020495 | 0.393056296 |
| T2DM | nk_cell | *MYH7B* | -0.635645888 | 0.000455083 | 0.000800313 | 1 |
| T2DM | nk_cell | *BPIFB2* | -4.956872638 | 0.004372399 | 0.006215904 | 1 |
| T2DM | nk_cell | *CLUL1* | -1.025148158 | 3.30E-05 | 0.000127909 | 0.159939093 |
| T2DM | nk_cell | *FKBP7* | -1.020662273 | 0.000162334 | 0.000339209 | 0.787642396 |
| T2DM | nk_cell | *XRCC5* | -1.251469152 | 0.004560505 | 0.006464379 | 1 |
| T2DM | nk_cell | *SP140* | 0.325861065 | 0.0001852 | 0.000377399 | 0.898588045 |
| T2DM | nk_cell | *SAR1A* | -1.214924923 | 3.33E-05 | 0.000128061 | 0.161762791 |
| T2DM | nk_cell | *RAPGEF3* | -0.496425645 | 0.004708698 | 0.006656936 | 1 |
| T2DM | nk_cell | *SCGN* | 1.31194339 | 7.14E-05 | 0.000189116 | 0.346358713 |
| T2DM | nk_cell | *PGM1* | -1.017448658 | 1.95E-05 | 0.000122264 | 0.094678246 |
| T2DM | nk_cell | *DDX1* | -1.448887608 | 0.0004391 | 0.000776144 | 1 |
| T2DM | nk_cell | *STX7* | -0.990095116 | 1.29E-05 | 0.000122264 | 0.062589877 |
| T2DM | nk_cell | *RABL2B* | -1.935071106 | 5.96E-05 | 0.000168718 | 0.289324227 |
| T2DM | nk_cell | *DCT* | -0.796765292 | 1.20E-06 | 8.57E-05 | 0.005824823 |
| T2DM | nk_cell | *SCTR* | -0.805388569 | 1.69E-05 | 0.000122264 | 0.082028769 |
| T2DM | nk_cell | *RIF1* | -1.061228971 | 1.83E-05 | 0.000122264 | 0.088679467 |
| T2DM | nk_cell | *KCNN2* | 1.032108399 | 2.89E-05 | 0.000123207 | 0.140332804 |
| T2DM | nk_cell | *PSEN1* | -1.189421825 | 7.29E-05 | 0.000192063 | 0.353637416 |
| T2DM | nk_cell | *MOK* | -0.853618168 | 3.32E-05 | 0.000127948 | 0.161026187 |
| T2DM | nk_cell | *STK17B* | 0.636235898 | 0.000101529 | 0.000238787 | 0.492618027 |
| T2DM | nk_cell | *ZNF506* | -1.015202589 | 0.035978378 | 0.044407808 | 1 |

| | | | | | | |
|---|---|---|---|---|---|---|
| T2DM | nk_cell | *PCDHB4* | 0.906817079 | 0.000133106 | 0.000290392 | 0.645831495 |
| T2DM | nk_cell | *CCNT2* | -1.070497164 | 0.007192408 | 0.009841388 | 1 |
| T2DM | nk_cell | *SEMA5B* | -0.695409794 | 0.000508884 | 0.000880565 | 1 |
| T2DM | nk_cell | *DOPEY1* | -0.888793169 | 0.001368841 | 0.002124638 | 1 |
| T2DM | nk_cell | *LYRM2* | -1.14168284 | 0.000131982 | 0.000288458 | 0.640376074 |
| T2DM | nk_cell | *ZCCHC6* | -1.456025576 | 0.000743868 | 0.001224102 | 1 |
| T2DM | nk_cell | *ULK2* | -0.80206707 | 1.60E-05 | 0.000122264 | 0.077837436 |
| T2DM | nk_cell | *PIBF1* | -1.234274406 | 0.000224964 | 0.000442272 | 1 |
| T2DM | nk_cell | *OXCT1* | -1.261556286 | 0.000121969 | 0.000270936 | 0.591794061 |
| T2DM | nk_cell | *SLC27A5* | 1.278989792 | 0.001056859 | 0.001684585 | 1 |
| T2DM | nk_cell | *ZNF586* | -0.994461726 | 0.015615658 | 0.020334722 | 1 |
| T2DM | nk_cell | *ZNF264* | -1.288074101 | 2.04E-05 | 0.000122264 | 0.099013281 |
| T2DM | nk_cell | *CHMP2B* | -2.099995815 | 0.000321099 | 0.000596696 | 1 |
| T2DM | nk_cell | *PPIE* | -1.138146668 | 0.00635715 | 0.008792728 | 1 |
| T2DM | nk_cell | *NOA1* | -1.808311077 | 2.69E-05 | 0.000122391 | 0.130700179 |
| T2DM | nk_cell | *WBP11* | 1.029827654 | 0.026745553 | 0.033584219 | 1 |
| T2DM | nk_cell | *NCOA1* | -1.190775139 | 0.000772746 | 0.001265395 | 1 |
| T2DM | nk_cell | *CAD* | -1.899996628 | 0.004864011 | 0.006860517 | 1 |
| T2DM | nk_cell | *CD59* | -0.904408966 | 0.003403685 | 0.004941556 | 1 |
| T2DM | nk_cell | *HACE1* | -0.948013714 | 1.81E-05 | 0.000122264 | 0.087779181 |
| T2DM | nk_cell | *SEH1L* | -1.021103162 | 0.000123404 | 0.000273525 | 0.59875786 |
| T2DM | nk_cell | *OVGP1* | -1.373633661 | 1.49E-06 | 9.27E-05 | 0.007228061 |
| T2DM | nk_cell | *SLC25A24* | -1.177921694 | 7.57E-05 | 0.000196566 | 0.367382057 |
| T2DM | nk_cell | *ZNF213* | -1.101675164 | 0.000340696 | 0.000627109 | 1 |
| T2DM | nk_cell | *CPNE3* | -1.203009963 | 0.000270941 | 0.000516818 | 1 |
| T2DM | nk_cell | *RRN3* | -1.020964407 | 2.92E-05 | 0.000123467 | 0.141733843 |
| T2DM | nk_cell | *MTIF2* | -1.719066892 | 0.012124108 | 0.016046419 | 1 |
| T2DM | nk_cell | *DDHD2* | -1.017615565 | 0.000209278 | 0.000415983 | 1 |
| T2DM | nk_cell | *EPS15* | -0.958289803 | 0.000514045 | 0.000888152 | 1 |
| T2DM | nk_cell | *USP40* | -0.914380501 | 1.79E-05 | 0.000122264 | 0.086689247 |
| T2DM | nk_cell | *POMGNT1* | -0.905352005 | 3.44E-05 | 0.00012999 | 0.167036655 |
| T2DM | nk_cell | *MAST2* | -1.341890355 | 0.000195831 | 0.000394425 | 0.950170817 |
| T2DM | nk_cell | *EIF2AK1* | -1.230861314 | 3.56E-05 | 0.000131704 | 0.172724294 |
| T2DM | nk_cell | *NME8* | 2.029364316 | 4.94E-05 | 0.00015216 | 0.239605535 |
| T2DM | nk_cell | *EPDR1* | 0.642642968 | 0.002632812 | 0.003883979 | 1 |
| T2DM | nk_cell | *ITPKC* | -0.633623034 | 0.000200364 | 0.000401389 | 0.972165321 |
| T2DM | nk_cell | *TMED2* | -1.538753422 | 0.000137295 | 0.000297524 | 0.66615573 |
| T2DM | nk_cell | *ALG9* | -1.554842749 | 1.83E-05 | 0.000122264 | 0.088948982 |
| T2DM | nk_cell | *ACOX3* | -1.453936018 | 0.00017451 | 0.000359543 | 0.846724433 |
| T2DM | nk_cell | *HSD17B14* | -0.705669889 | 4.83E-05 | 0.000150747 | 0.234562021 |
| T2DM | nk_cell | *TRIP6* | -1.692366082 | 2.19E-05 | 0.000122264 | 0.106338396 |
| T2DM | nk_cell | *ACHE* | -2.103100316 | 1.31E-05 | 0.000122264 | 0.063702831 |
| T2DM | nk_cell | *SRRT* | -1.057369261 | 1.15E-05 | 0.000122264 | 0.05565148 |
| T2DM | nk_cell | *ADAMTS2* | -0.899368433 | 1.60E-05 | 0.000122264 | 0.077623782 |
| T2DM | nk_cell | *PSMC5* | 0.877370691 | 0.025838237 | 0.032503792 | 1 |
| T2DM | nk_cell | *UIMC1* | -1.536568284 | 0.000249463 | 0.000481079 | 1 |
| T2DM | nk_cell | *LPCAT2* | -1.755247848 | 0.00011129 | 0.000255069 | 0.539980639 |
| T2DM | nk_cell | *OGFOD1* | -2.028228712 | 3.29E-05 | 0.000127909 | 0.159759066 |
| T2DM | nk_cell | *TXNDC16* | -1.241853616 | 0.002797168 | 0.004103979 | 1 |
| T2DM | nk_cell | *AAMDC* | -1.099764422 | 4.91E-06 | 0.000122264 | 0.023804197 |

| | | | | | | |
|---|---|---|---|---|---|---|
| T2DM | nk_cell | *ASAP3* | -0.614273342 | 1.93E-05 | 0.000122264 | 0.093518503 |
| T2DM | nk_cell | *EDEM2* | -1.233882637 | 2.05E-06 | 0.000112031 | 0.009970765 |
| T2DM | nk_cell | *ANKRD10* | -1.013744857 | 2.08E-05 | 0.000122264 | 0.100966061 |
| T2DM | nk_cell | *DOCK3* | -0.99269603 | 1.55E-05 | 0.000122264 | 0.075105261 |
| T2DM | nk_cell | *ARHGAP28* | -0.812124224 | 1.38E-05 | 0.000122264 | 0.067040712 |
| T2DM | nk_cell | *PPP1R13B* | -1.481738354 | 0.000246114 | 0.000476135 | 1 |
| T2DM | nk_cell | *NSFL1C* | -1.037810639 | 0.000156912 | 0.000330728 | 0.761335695 |
| T2DM | nk_cell | *ZNF343* | -0.959955725 | 0.000887972 | 0.001439024 | 1 |
| T2DM | nk_cell | *EBF4* | -1.491754443 | 1.52E-05 | 0.000122264 | 0.073687341 |
| T2DM | nk_cell | *CPXM1* | -0.923104741 | 1.14E-05 | 0.000122264 | 0.055422694 |
| T2DM | nk_cell | *MAVS* | -1.265631395 | 0.000160573 | 0.00033662 | 0.779099243 |
| T2DM | nk_cell | *XRN2* | -1.551372864 | 0.000235425 | 0.000458932 | 1 |
| T2DM | nk_cell | *ESF1* | -1.004829325 | 4.15E-05 | 0.000140757 | 0.201282478 |
| T2DM | nk_cell | *RBBP9* | -1.503284431 | 0.000134741 | 0.000293037 | 0.653764979 |
| T2DM | nk_cell | *TMEM230* | -1.283367941 | 6.40E-05 | 0.000175533 | 0.31034176 |
| T2DM | nk_cell | *KIF16B* | -1.44461355 | 7.54E-05 | 0.000196203 | 0.36607249 |
| T2DM | nk_cell | *TBX5* | -2.708404319 | 3.70E-05 | 0.000133757 | 0.179501397 |
| T2DM | nk_cell | *FXYD5* | 0.832669947 | 0.000488182 | 0.000850506 | 1 |
| T2DM | nk_cell | *ZNF302* | -1.23695583 | 0.000567729 | 0.000966214 | 1 |
| T2DM | nk_cell | *GRAMD1A* | -1.312206505 | 2.26E-05 | 0.000122264 | 0.109689374 |
| T2DM | nk_cell | *CDIP1* | -1.56358456 | 1.77E-05 | 0.000122264 | 0.085961576 |
| T2DM | nk_cell | *KCNH4* | 0.943672184 | 0.000832423 | 0.00135534 | 1 |
| T2DM | nk_cell | *GANAB* | -1.320894717 | 8.43E-06 | 0.000122264 | 0.040888963 |
| T2DM | nk_cell | *MLF2* | -0.725720941 | 0.000125909 | 0.000278193 | 0.610911415 |
| T2DM | nk_cell | *ANKRD24* | -0.971267817 | 2.05E-05 | 0.000122264 | 0.099503332 |
| T2DM | nk_cell | *DHX32* | -0.851006512 | 2.88E-05 | 0.000123207 | 0.139632792 |
| T2DM | nk_cell | *SPTLC1* | -1.462045721 | 0.005379166 | 0.007538912 | 1 |
| T2DM | nk_cell | *YPEL3* | -0.842231079 | 0.035750942 | 0.044160787 | 1 |
| T2DM | nk_cell | *MRPS33* | -0.955133415 | 2.15E-06 | 0.000114775 | 0.010444538 |
| T2DM | nk_cell | *MAEA* | -1.441152833 | 0.007781753 | 0.010576209 | 1 |
| T2DM | nk_cell | *MUL1* | -1.197711347 | 0.000299569 | 0.000562938 | 1 |
| T2DM | nk_cell | *SPG21* | -1.447280801 | 1.13E-05 | 0.000122264 | 0.055043898 |
| T2DM | nk_cell | *CHRD* | -0.834208935 | 0.000178014 | 0.000365057 | 0.863725301 |
| T2DM | nk_cell | *RAB11FIP3* | -1.525519733 | 0.000106731 | 0.000247425 | 0.517860038 |
| T2DM | nk_cell | *GNPTG* | -1.214051784 | 1.10E-05 | 0.000122264 | 0.053246211 |
| T2DM | nk_cell | *PABPC4* | -2.949261577 | 1.17E-12 | 1.88E-09 | 5.65E-09 |
| T2DM | nk_cell | *CERS4* | -1.261218748 | 0.039431302 | 0.048386615 | 1 |
| T2DM | nk_cell | *USP48* | -1.283715193 | 1.71E-05 | 0.000122264 | 0.083211321 |
| T2DM | nk_cell | *PDPR* | -1.03679525 | 0.000106344 | 0.000246645 | 0.51598124 |
| T2DM | nk_cell | *GLG1* | -1.328626177 | 0.000235777 | 0.000459434 | 1 |
| T2DM | nk_cell | *DTX2* | -2.346418472 | 5.88E-05 | 0.000167252 | 0.285354452 |
| T2DM | nk_cell | *LAMB1* | -0.728243636 | 8.72E-06 | 0.000122264 | 0.042325979 |
| T2DM | nk_cell | *ABCC6* | -0.517303462 | 1.05E-05 | 0.000122264 | 0.050761589 |
| T2DM | nk_cell | *RAPGEF4* | -0.529758798 | 2.91E-05 | 0.000123467 | 0.141248639 |
| T2DM | nk_cell | *TF* | -1.586426303 | 1.60E-05 | 0.000122264 | 0.077393179 |
| T2DM | nk_cell | *ALKBH5* | -1.569917923 | 0.01117836 | 0.014871785 | 1 |
| T2DM | nk_cell | *SPAG7* | -0.84647073 | 0.020167381 | 0.025838957 | 1 |
| T2DM | nk_cell | *RGS17* | -1.350276836 | 3.54E-07 | 4.05E-05 | 0.001715671 |
| T2DM | nk_cell | *PPP2R3C* | -0.888747422 | 0.024299732 | 0.030719723 | 1 |
| T2DM | nk_cell | *OSGEP* | -1.661174154 | 0.000741462 | 0.001221588 | 1 |

| | | | | | | |
|---|---|---|---|---|---|---|
| T2DM | nk_cell | *SCFD1* | -1.26911228 | 0.000106944 | 0.000247756 | 0.518890497 |
| T2DM | nk_cell | *SUPT16H* | -1.755132459 | 0.000147765 | 0.000315701 | 0.716956644 |
| T2DM | nk_cell | *CAPN3* | 1.260807438 | 0.00250543 | 0.003706203 | 1 |
| T2DM | nk_cell | *UNC13D* | 0.624030012 | 0.001470461 | 0.002265696 | 1 |
| T2DM | nk_cell | *MFSD11* | -1.063694092 | 0.001054133 | 0.001680793 | 1 |
| T2DM | nk_cell | *NUP50* | -1.502797019 | 0.001099662 | 0.00174593 | 1 |
| T2DM | nk_cell | *COMT* | -1.396055921 | 3.35E-05 | 0.000128164 | 0.162701413 |
| T2DM | nk_cell | *LRRFIP2* | -1.149913994 | 6.61E-05 | 0.000179706 | 0.320774325 |
| T2DM | nk_cell | *SEC22C* | -1.472703745 | 4.07E-05 | 0.000139476 | 0.197521656 |
| T2DM | nk_cell | *XYLB* | -0.999632991 | 1.58E-05 | 0.000122264 | 0.076847037 |
| T2DM | nk_cell | *GABRP* | 1.797566201 | 0.003329815 | 0.004843005 | 1 |
| T2DM | nk_cell | *CDC6* | -1.773345889 | 7.79E-06 | 0.000122264 | 0.037817503 |
| T2DM | nk_cell | *FMO2* | -0.460638935 | 0.003841484 | 0.005511914 | 1 |
| T2DM | nk_cell | *MSH2* | -1.31364955 | 2.21E-05 | 0.000122264 | 0.107337792 |
| T2DM | nk_cell | *PSMD5* | -1.488612208 | 3.16E-05 | 0.000127109 | 0.153475165 |
| T2DM | nk_cell | *CRAT* | -1.155592121 | 1.72E-05 | 0.000122264 | 0.083326656 |
| T2DM | nk_cell | *NANS* | -1.911241395 | 0.010420635 | 0.013936307 | 1 |
| T2DM | nk_cell | *PDE6C* | -2.198374548 | 0.002323905 | 0.003460893 | 1 |
| T2DM | nk_cell | *CWF19L1* | -1.382221868 | 1.95E-08 | 5.90E-06 | 9.44E-05 |
| T2DM | nk_cell | *CRTAC1* | -0.737209219 | 0.001020649 | 0.001632231 | 1 |
| T2DM | nk_cell | *WAC* | -1.162986795 | 2.16E-05 | 0.000122264 | 0.104749193 |
| T2DM | nk_cell | *IFT74* | -0.814677037 | 2.30E-05 | 0.000122264 | 0.111768186 |
| T2DM | nk_cell | *IL12RB1* | 0.223597549 | 6.99E-05 | 0.00018653 | 0.338926397 |
| T2DM | nk_cell | *SCD* | -1.658537278 | 2.32E-07 | 3.38E-05 | 0.001123692 |
| T2DM | nk_cell | *ERMP1* | -1.583825413 | 0.000118758 | 0.000266027 | 0.576213607 |
| T2DM | nk_cell | *RAB18* | -1.589974599 | 0.000161361 | 0.000337612 | 0.782923358 |
| T2DM | nk_cell | *PRTFDC1* | -1.429251283 | 0.000475928 | 0.000831846 | 1 |
| T2DM | nk_cell | *TSPAN15* | -1.06032238 | 2.44E-05 | 0.000122264 | 0.118201645 |
| T2DM | nk_cell | *CATSPERG* | -0.736389354 | 0.015281736 | 0.019915924 | 1 |
| T2DM | nk_cell | *CIRBP* | -0.962211639 | 4.14E-05 | 0.000140573 | 0.200738739 |
| T2DM | nk_cell | *MTAP* | -1.500778854 | 2.58E-05 | 0.000122264 | 0.125140615 |
| T2DM | nk_cell | *POLR2E* | -0.959512179 | 3.28E-05 | 0.000127909 | 0.159176984 |
| T2DM | nk_cell | *MADCAM1* | -0.553723987 | 1.95E-05 | 0.000122264 | 0.09454988 |
| T2DM | nk_cell | *MED15* | -1.34628498 | 0.009835342 | 0.013184717 | 1 |
| T2DM | nk_cell | *SERPIND1* | -1.338483688 | 0.00023059 | 0.000450774 | 1 |
| T2DM | nk_cell | *SNAP29* | -0.93053978 | 0.003448734 | 0.004995749 | 1 |
| T2DM | nk_cell | *LZTR1* | -1.550241149 | 0.004784919 | 0.006756819 | 1 |
| T2DM | nk_cell | *MMP11* | 1.457865529 | 0.032046646 | 0.039823125 | 1 |
| T2DM | nk_cell | *CECR2* | -0.49065857 | 6.00E-05 | 0.000169532 | 0.291317532 |
| T2DM | nk_cell | *BCL2L13* | -0.810678266 | 0.000114799 | 0.000260568 | 0.557005263 |
| T2DM | nk_cell | *DDTL* | 0.718550281 | 0.000202782 | 0.00040523 | 0.983898663 |
| T2DM | nk_cell | *DDT* | -1.789411942 | 0.00712735 | 0.009763382 | 1 |
| T2DM | nk_cell | *SUSD2* | -0.432902393 | 0.0253344 | 0.031902201 | 1 |
| T2DM | nk_cell | *SPECC1L* | -1.161082106 | 0.000137398 | 0.000297615 | 0.666656815 |
| T2DM | nk_cell | *PPIL2* | -1.184677571 | 0.010392584 | 0.013902623 | 1 |
| T2DM | nk_cell | *SLC35E4* | -0.814879161 | 1.19E-05 | 0.000122264 | 0.057828968 |
| T2DM | nk_cell | *TOP3B* | -2.009673264 | 0.006340064 | 0.008771596 | 1 |
| T2DM | nk_cell | *CRYBB3* | 1.542083921 | 0.01361156 | 0.017859191 | 1 |
| T2DM | nk_cell | *CYTH4* | 0.497235228 | 0.000166898 | 0.000346359 | 0.80978811 |
| T2DM | nk_cell | *CARD10* | -0.948113082 | 7.88E-05 | 0.000201551 | 0.382159342 |

| | | | | | | |
|---|---|---|---|---|---|---|
| T2DM | nk_cell | *LRP5L* | -1.458912765 | 9.81E-06 | 0.000122264 | 0.047620569 |
| T2DM | nk_cell | *LGALS2* | 0.853217042 | 0.012692596 | 0.01672128 | 1 |
| T2DM | nk_cell | *SH3BP1* | 0.30871155 | 6.94E-05 | 0.000185742 | 0.336936346 |
| T2DM | nk_cell | *HPS4* | -0.838820243 | 2.02E-05 | 0.000122264 | 0.098143801 |
| T2DM | nk_cell | *TRIOBP* | -1.028536908 | 6.29E-05 | 0.000173739 | 0.305379588 |
| T2DM | nk_cell | *ANKRD54* | -0.702869574 | 8.45E-06 | 0.000122264 | 0.04101875 |
| T2DM | nk_cell | *EIF3L* | -1.640716376 | 1.41E-06 | 9.04E-05 | 0.006860854 |
| T2DM | nk_cell | *CCDC134* | -1.505997781 | 0.000981448 | 0.001579432 | 1 |
| T2DM | nk_cell | *PICK1* | -1.08823875 | 0.019013591 | 0.024438131 | 1 |
| T2DM | nk_cell | *TTC28* | -0.834658784 | 2.38E-05 | 0.000122264 | 0.115627019 |
| T2DM | nk_cell | *CENPM* | 0.817577003 | 0.000217263 | 0.000429743 | 1 |
| T2DM | nk_cell | *CYP2D6* | -1.175934137 | 4.78E-05 | 0.000149597 | 0.231725333 |
| T2DM | nk_cell | *CBY1* | -0.854562321 | 0.006988138 | 0.009586244 | 1 |
| T2DM | nk_cell | *RTCB* | -1.360309281 | 5.85E-05 | 0.000167232 | 0.283878018 |
| T2DM | nk_cell | *RAB36* | -0.979471176 | 5.88E-05 | 0.000167252 | 0.285421855 |
| T2DM | nk_cell | *TIMP3* | -0.849503094 | 2.39E-05 | 0.000122264 | 0.116048573 |
| T2DM | nk_cell | *PPP6R2* | -1.327098973 | 0.000113124 | 0.000257941 | 0.548877505 |
| T2DM | nk_cell | *CYB5R3* | -1.183232146 | 3.85E-05 | 0.000135801 | 0.186725958 |
| T2DM | nk_cell | *RHBDD3* | -1.94068475 | 4.03E-06 | 0.000122264 | 0.019537684 |
| T2DM | nk_cell | *TOM1* | -1.147349478 | 0.000325032 | 0.00060262 | 1 |
| T2DM | nk_cell | *CHKB* | -1.074717659 | 7.56E-05 | 0.000196393 | 0.366861565 |
| T2DM | nk_cell | *THOC5* | -1.518122407 | 5.58E-05 | 0.000163082 | 0.270715698 |
| T2DM | nk_cell | *TTLL12* | -1.829062046 | 1.48E-05 | 0.000122264 | 0.071796887 |
| T2DM | nk_cell | *MTMR3* | -1.146061855 | 2.77E-05 | 0.000122493 | 0.134505419 |
| T2DM | nk_cell | *APOL4* | -0.733292366 | 0.000105566 | 0.000244957 | 0.512204448 |
| T2DM | nk_cell | *SAMM50* | -1.399020486 | 6.56E-05 | 0.000178701 | 0.318379841 |
| T2DM | nk_cell | *FOXRED2* | -0.517292684 | 0.001085781 | 0.00172502 | 1 |
| T2DM | nk_cell | *IFT27* | -2.086170034 | 0.001287664 | 0.002008519 | 1 |
| T2DM | nk_cell | *KIAA0930* | -1.654456289 | 0.001498407 | 0.002305827 | 1 |
| T2DM | nk_cell | *SLC25A17* | -1.091380201 | 0.001031258 | 0.001647567 | 1 |
| T2DM | nk_cell | *FAM118A* | -0.567359845 | 0.026670209 | 0.033498279 | 1 |
| T2DM | nk_cell | *L3MBTL2* | 0.815391109 | 0.038893213 | 0.047750473 | 1 |
| T2DM | nk_cell | *ZC3H7B* | -1.592781787 | 4.90E-05 | 0.000151719 | 0.237743194 |
| T2DM | nk_cell | *ACO2* | -0.911740827 | 1.38E-05 | 0.000122264 | 0.06680665 |
| T2DM | nk_cell | *DESI1* | -1.479948489 | 0.000158262 | 0.000332708 | 0.767889128 |
| T2DM | nk_cell | *MLC1* | 0.231020481 | 4.36E-05 | 0.000144302 | 0.211422712 |
| T2DM | nk_cell | *HDAC10* | -1.197455304 | 2.27E-05 | 0.000122264 | 0.110215888 |
| T2DM | nk_cell | *KHNYN* | -1.421336425 | 6.00E-05 | 0.000169532 | 0.291224985 |
| T2DM | nk_cell | *SDR39U1* | -0.786453 | 2.49E-05 | 0.000122264 | 0.120787456 |
| T2DM | nk_cell | *AP4S1* | -1.048747544 | 2.76E-05 | 0.000122472 | 0.133723102 |
| T2DM | nk_cell | *NIN* | 0.77616885 | 0.018089042 | 0.023323951 | 1 |
| T2DM | nk_cell | *PYGL* | -0.447077313 | 0.001952296 | 0.002949109 | 1 |
| T2DM | nk_cell | *CNIH1* | -1.117619432 | 1.68E-05 | 0.000122264 | 0.081369803 |
| T2DM | nk_cell | *ATP6V1D* | -0.99927699 | 1.71E-05 | 0.000122264 | 0.082974761 |
| T2DM | nk_cell | *PLEK2* | -1.793259956 | 6.95E-06 | 0.000122264 | 0.033721139 |
| T2DM | nk_cell | *GSTZ1* | -0.556797999 | 3.55E-05 | 0.0001316 | 0.172396172 |
| T2DM | nk_cell | *TMED8* | -0.901162774 | 1.25E-05 | 0.000122264 | 0.060824432 |
| T2DM | nk_cell | *LGMN* | -0.947416333 | 0.000248001 | 0.000479021 | 1 |
| T2DM | nk_cell | *MTHFD1* | -0.728403586 | 3.63E-05 | 0.000132391 | 0.176208957 |
| T2DM | nk_cell | *TELO2* | -0.853451438 | 2.71E-05 | 0.000122391 | 0.131325313 |

| | | | | | | |
|---|---|---|---|---|---|---|
| T2DM | nk_cell | *GSKIP* | -1.600520059 | 1.61E-05 | 0.000122264 | 0.078080312 |
| T2DM | nk_cell | *PSMB5* | -0.76688521 | 0.024195275 | 0.030595641 | 1 |
| T2DM | nk_cell | *YY1* | -1.595910805 | 0.000149415 | 0.000318928 | 0.724960306 |
| T2DM | nk_cell | *CCNB1IP1* | -1.311811291 | 1.80E-05 | 0.000122264 | 0.087432346 |
| T2DM | nk_cell | *EFS* | -1.499190552 | 1.77E-05 | 0.000122264 | 0.086042388 |
| T2DM | nk_cell | *CINP* | -0.810574026 | 2.12E-05 | 0.000122264 | 0.102898892 |
| T2DM | nk_cell | *KIAA0391* | 1.525519282 | 0.011438139 | 0.015189436 | 1 |
| T2DM | nk_cell | *EMC9* | -0.826423583 | 0.000308412 | 0.000576209 | 1 |
| T2DM | nk_cell | *TRPC4AP* | -1.11259721 | 1.46E-05 | 0.000122264 | 0.070729745 |
| T2DM | nk_cell | *PYGB* | -0.957932932 | 0.00340585 | 0.004943219 | 1 |
| T2DM | nk_cell | *ABHD12* | -1.267787337 | 0.008917977 | 0.012039517 | 1 |
| T2DM | nk_cell | *CD40* | -0.866692151 | 8.89E-06 | 0.000122264 | 0.04311416 |
| T2DM | nk_cell | *UQCC1* | -1.255960236 | 3.59E-06 | 0.000122264 | 0.017433092 |
| T2DM | nk_cell | *IFT52* | -0.99538562 | 0.018722772 | 0.024109047 | 1 |
| T2DM | nk_cell | *NDRG3* | -1.74274006 | 0.000355271 | 0.000649746 | 1 |
| T2DM | nk_cell | *PABPC1L* | -1.179038266 | 2.01E-05 | 0.000122264 | 0.09736084 |
| T2DM | nk_cell | *DOK5* | -0.821251252 | 2.08E-05 | 0.000122264 | 0.101049273 |
| T2DM | nk_cell | *NELFCD* | -1.624038697 | 0.001247462 | 0.00195185 | 1 |
| T2DM | nk_cell | *CTSZ* | -1.256826386 | 2.43E-05 | 0.000122264 | 0.117834418 |
| T2DM | nk_cell | *MTG2* | -1.168606032 | 4.83E-05 | 0.000150747 | 0.234478919 |
| T2DM | nk_cell | *TCFL5* | -1.537754888 | 3.84E-05 | 0.000135693 | 0.186391097 |
| T2DM | nk_cell | *DIDO1* | -1.229727776 | 0.000985238 | 0.001585005 | 1 |
| T2DM | nk_cell | *NKAIN4* | -1.200304995 | 0.004358908 | 0.006200357 | 1 |
| T2DM | nk_cell | *PTK6* | -1.501471455 | 3.30E-05 | 0.000127909 | 0.160121703 |
| T2DM | nk_cell | *GMEB2* | -1.095210526 | 1.59E-05 | 0.000122264 | 0.076908034 |
| T2DM | nk_cell | *CDC25B* | 0.790174657 | 0.014848322 | 0.01939797 | 1 |
| T2DM | nk_cell | *RNF24* | -1.314063392 | 8.89E-05 | 0.000217502 | 0.431306415 |
| T2DM | nk_cell | *NDUFAF5* | -0.715392595 | 0.004730339 | 0.006683636 | 1 |
| T2DM | nk_cell | *TRIB3* | 2.15780415 | 0.009238306 | 0.012437364 | 1 |
| T2DM | nk_cell | *RASSF2* | -0.998984168 | 0.011309108 | 0.015037487 | 1 |
| T2DM | nk_cell | *ANGPT4* | -0.980433746 | 3.17E-05 | 0.000127109 | 0.153683454 |
| T2DM | nk_cell | *RSPO4* | -0.635098234 | 0.000254791 | 0.00048999 | 1 |
| T2DM | nk_cell | *CRNKL1* | -1.373265708 | 0.000670235 | 0.001119058 | 1 |
| T2DM | nk_cell | *MROH8* | -1.377394446 | 5.60E-05 | 0.000163342 | 0.271474026 |
| T2DM | nk_cell | *MANBAL* | -0.771484034 | 0.000800712 | 0.001308099 | 1 |
| T2DM | nk_cell | *DNTTIP1* | -0.890515626 | 0.02399145 | 0.030361636 | 1 |
| T2DM | nk_cell | *MAP1LC3A* | -1.191090243 | 2.67E-05 | 0.000122391 | 0.129698462 |
| T2DM | nk_cell | *METTL4* | -1.552424934 | 2.25E-05 | 0.000122264 | 0.109364017 |
| T2DM | nk_cell | *MYOM1* | -0.846633761 | 1.59E-05 | 0.000122264 | 0.076991723 |
| T2DM | nk_cell | *LAMA1* | -0.773328765 | 1.89E-05 | 0.000122264 | 0.091569315 |
| T2DM | nk_cell | *MIB1* | -1.208997339 | 4.16E-05 | 0.000140997 | 0.201766037 |
| T2DM | nk_cell | *NALCN* | -0.964083 | 8.17E-05 | 0.000205997 | 0.396544175 |
| T2DM | nk_cell | *STK24* | -1.288896908 | 4.73E-05 | 0.000149118 | 0.229641167 |
| T2DM | nk_cell | *DNAJC3* | -1.372373427 | 0.000272602 | 0.000518895 | 1 |
| T2DM | nk_cell | *UGGT2* | -0.920516085 | 4.57E-05 | 0.000147697 | 0.221881296 |
| T2DM | nk_cell | *ARHGEF7* | -1.086522051 | 2.65E-05 | 0.000122391 | 0.128677506 |
| T2DM | nk_cell | *PARP4* | -1.265533384 | 0.000275249 | 0.000522908 | 1 |
| T2DM | nk_cell | *SUPT20H* | -1.267126253 | 3.72E-05 | 0.000134067 | 0.18058838 |
| T2DM | nk_cell | *MRPS31* | -1.171142243 | 3.34E-05 | 0.000128061 | 0.161926701 |
| T2DM | nk_cell | *DGKH* | -1.4557702 | 1.08E-05 | 0.000122264 | 0.052587904 |

| | | | | | | |
|---|---|---|---|---|---|---|
| T2DM | nk_cell | *KATNAL1* | -1.18677263 | 2.62E-05 | 0.000122295 | 0.127006105 |
| T2DM | nk_cell | *INTS6* | -1.323361474 | 0.000102662 | 0.000240564 | 0.49811767 |
| T2DM | nk_cell | *CLN5* | -1.86200947 | 8.48E-05 | 0.00021115 | 0.411593925 |
| T2DM | nk_cell | *MGRN1* | -1.356746651 | 0.000103926 | 0.000242428 | 0.50425059 |
| T2DM | nk_cell | *LONP2* | -0.918383665 | 0.000337313 | 0.000621588 | 1 |
| T2DM | nk_cell | *PLLP* | -0.696820791 | 3.22E-05 | 0.000127628 | 0.156000226 |
| T2DM | nk_cell | *DHODH* | -0.875852915 | 0.000220309 | 0.000434881 | 1 |
| T2DM | nk_cell | *CCDC113* | 1.012579019 | 0.002456237 | 0.003647891 | 1 |
| T2DM | nk_cell | *COG4* | -1.795807495 | 1.41E-05 | 0.000122264 | 0.068344325 |
| T2DM | nk_cell | *MON1B* | -2.347614626 | 8.50E-05 | 0.000211192 | 0.412247474 |
| T2DM | nk_cell | *NPRL3* | -1.28081505 | 0.000816101 | 0.001330552 | 1 |
| T2DM | nk_cell | *NAGPA* | -1.174817293 | 1.28E-05 | 0.000122264 | 0.061979025 |
| T2DM | nk_cell | *ZNF500* | -0.906871421 | 0.000189333 | 0.000383567 | 0.918642463 |
| T2DM | nk_cell | *NME4* | -1.287309166 | 2.22E-05 | 0.000122264 | 0.107864908 |
| T2DM | nk_cell | *LMF1* | -1.244645612 | 3.02E-05 | 0.000124985 | 0.146740007 |
| T2DM | nk_cell | *NARFL* | -1.203244692 | 0.000141665 | 0.000305086 | 0.687359711 |
| T2DM | nk_cell | *MTHFSD* | -0.594733141 | 0.027169173 | 0.034080876 | 1 |
| T2DM | nk_cell | *HAGHL* | -0.758859161 | 0.014257527 | 0.01865629 | 1 |
| T2DM | nk_cell | *SLC7A5* | 1.316858455 | 0.00038134 | 0.000690913 | 1 |
| T2DM | nk_cell | *RHBDL1* | -2.00130643 | 1.32E-05 | 0.000122264 | 0.064136523 |
| T2DM | nk_cell | *NUBP1* | -1.434085105 | 7.75E-05 | 0.00019931 | 0.376098411 |
| T2DM | nk_cell | *UBE2I* | -1.432172182 | 0.018177462 | 0.023425511 | 1 |
| T2DM | nk_cell | *EARS2* | -1.118753368 | 0.006251511 | 0.008661431 | 1 |
| T2DM | nk_cell | *CPPED1* | -0.974478417 | 3.53E-05 | 0.000131127 | 0.17112034 |
| T2DM | nk_cell | *HMOX2* | 1.000778745 | 0.00064256 | 0.001076555 | 1 |
| T2DM | nk_cell | *TOX3* | -1.646870652 | 0.000291213 | 0.000549364 | 1 |
| T2DM | nk_cell | *PYCARD* | 0.664459874 | 6.35E-05 | 0.00017461 | 0.308186521 |
| T2DM | nk_cell | *CDIPT* | 1.141487009 | 0.001014279 | 0.001624186 | 1 |
| T2DM | nk_cell | *KAT8* | -1.062049628 | 1.49E-05 | 0.000122264 | 0.072442526 |
| T2DM | nk_cell | *TMC5* | -0.727786849 | 2.28E-05 | 0.000122264 | 0.110496857 |
| T2DM | nk_cell | *C16orf62* | -1.002290495 | 1.96E-05 | 0.000122264 | 0.095105144 |
| T2DM | nk_cell | *RNF40* | -1.69451836 | 5.47E-05 | 0.000160927 | 0.265367969 |
| T2DM | nk_cell | *KNOP1* | -1.602989574 | 1.48E-05 | 0.000122264 | 0.071806874 |
| T2DM | nk_cell | *IQCH* | -1.273988999 | 2.05E-05 | 0.000122264 | 0.099433187 |
| T2DM | nk_cell | *LACTB* | -1.246613784 | 8.36E-06 | 0.000122264 | 0.040547841 |
| T2DM | nk_cell | *HERC1* | -1.195943139 | 0.000457653 | 0.000803376 | 1 |
| T2DM | nk_cell | *TRIP4* | -1.418398265 | 4.71E-05 | 0.000149114 | 0.228614071 |
| T2DM | nk_cell | *TTC23* | -0.994269584 | 1.66E-05 | 0.000122264 | 0.080385083 |
| T2DM | nk_cell | *FAH* | -0.879145977 | 1.30E-05 | 0.000122264 | 0.06303486 |
| T2DM | nk_cell | *SLC30A4* | -1.938436748 | 7.68E-05 | 0.000198198 | 0.372612354 |
| T2DM | nk_cell | *ZDHHC2* | -1.419922697 | 0.010644685 | 0.014200718 | 1 |
| T2DM | nk_cell | *ZFAND1* | -1.484451529 | 1.72E-05 | 0.000122264 | 0.083630252 |
| T2DM | nk_cell | *EYA1* | -0.505810598 | 4.33E-05 | 0.000143917 | 0.209974682 |
| T2DM | nk_cell | *IMPAD1* | -0.834036949 | 1.89E-05 | 0.000122264 | 0.091854467 |
| T2DM | nk_cell | *SFRP1* | -1.924811558 | 4.77E-05 | 0.000149597 | 0.231674919 |
| T2DM | nk_cell | *NIPAL2* | -1.486828515 | 0.002488461 | 0.003688974 | 1 |
| T2DM | nk_cell | *RAB2A* | -1.051522853 | 0.000155072 | 0.000327561 | 0.75240845 |
| T2DM | nk_cell | *ARMC1* | -1.352250695 | 6.64E-06 | 0.000122264 | 0.032239984 |
| T2DM | nk_cell | *SPAG1* | -1.36011406 | 0.000751074 | 0.001233236 | 1 |
| T2DM | nk_cell | *CHRAC1* | -3.318977184 | 0.000267076 | 0.000510179 | 1 |

| | | | | | | |
|---|---|---|---|---|---|---|
| T2DM | nk_cell | *SNX16* | -1.820373417 | 0.000121518 | 0.000270222 | 0.589605536 |
| T2DM | nk_cell | *GSDMD* | -1.445167877 | 0.034316005 | 0.042474811 | 1 |
| T2DM | nk_cell | *TSTA3* | -0.610442907 | 0.000399507 | 0.000719528 | 1 |
| T2DM | nk_cell | *SQLE* | -1.176119288 | 0.020401408 | 0.026097451 | 1 |
| T2DM | nk_cell | *ERICH1* | -1.002633857 | 2.39E-05 | 0.000122264 | 0.116124116 |
| T2DM | nk_cell | *TUSC3* | -0.760759232 | 1.87E-05 | 0.000122264 | 0.090663848 |
| T2DM | nk_cell | *KLHDC4* | 0.843376905 | 0.006686546 | 0.009224658 | 1 |
| T2DM | nk_cell | *MAN2B1* | -0.989878318 | 0.012242058 | 0.016176052 | 1 |
| T2DM | nk_cell | *TULP2* | 3.536760395 | 0.000238045 | 0.000463029 | 1 |
| T2DM | nk_cell | *NFKBIB* | 0.673445305 | 0.000135275 | 0.000294065 | 0.656353658 |
| T2DM | nk_cell | *SARS2* | -0.770975871 | 3.79E-05 | 0.000134881 | 0.183957781 |
| T2DM | nk_cell | *SNRNP70* | -1.009093426 | 1.56E-05 | 0.000122264 | 0.075518394 |
| T2DM | nk_cell | *LIN7B* | -0.880868794 | 1.80E-05 | 0.000122264 | 0.087223805 |
| T2DM | nk_cell | *PPP1R37* | -1.934601639 | 0.000196667 | 0.000395618 | 0.954230205 |
| T2DM | nk_cell | *FCGRT* | -1.117781336 | 1.16E-05 | 0.000122264 | 0.056261991 |
| T2DM | nk_cell | *ARHGEF18* | -1.617061039 | 0.010159272 | 0.013601762 | 1 |
| T2DM | nk_cell | *PEX11G* | -1.014317313 | 6.90E-05 | 0.000185105 | 0.334748846 |
| T2DM | nk_cell | *ERCC2* | -2.272022698 | 4.65E-05 | 0.000148578 | 0.225690162 |
| T2DM | nk_cell | *CD37* | 0.516447755 | 0.000125284 | 0.000277064 | 0.607878019 |
| T2DM | nk_cell | *AMH* | -12.9949642 | 1.38E-05 | 0.000122264 | 0.06713838 |
| T2DM | nk_cell | *FCER2* | -4.997726891 | 1.95E-05 | 0.000122264 | 0.094677394 |
| T2DM | nk_cell | *CLEC4M* | -1.555796877 | 2.52E-05 | 0.000122264 | 0.122444252 |
| T2DM | nk_cell | *NOVA2* | -0.530703449 | 0.00172273 | 0.002629344 | 1 |
| T2DM | nk_cell | *SGTA* | -2.460021077 | 0.000628295 | 0.001055205 | 1 |
| T2DM | nk_cell | *KIR3DX1* | 0.108649592 | 4.65E-05 | 0.000148578 | 0.225655242 |
| T2DM | nk_cell | *MED25* | -0.975425957 | 1.71E-05 | 0.000122264 | 0.082975552 |
| T2DM | nk_cell | *TIMM44* | -1.616055589 | 0.001219345 | 0.001912172 | 1 |
| T2DM | nk_cell | *IL27RA* | 0.274336031 | 6.43E-05 | 0.000176247 | 0.311956814 |
| T2DM | nk_cell | *FAM32A* | 1.391966837 | 0.022608592 | 0.02871785 | 1 |
| T2DM | nk_cell | *C19orf44* | -0.566159968 | 0.001995752 | 0.003008198 | 1 |
| T2DM | nk_cell | *OLFM2* | -0.993989325 | 3.75E-05 | 0.00013436 | 0.182072196 |
| T2DM | nk_cell | *RASAL3* | 0.138359541 | 5.64E-05 | 0.00016385 | 0.273741416 |
| T2DM | nk_cell | *EPHX3* | -1.311002275 | 0.025249857 | 0.031804856 | 1 |
| T2DM | nk_cell | *PDCD5* | -0.66574822 | 4.01E-05 | 0.000138284 | 0.194566042 |
| T2DM | nk_cell | *ANKRD27* | -1.709069805 | 0.000703907 | 0.001168042 | 1 |
| T2DM | nk_cell | *POLR2I* | -1.359549368 | 0.021734387 | 0.02769308 | 1 |
| T2DM | nk_cell | *CCDC9* | -1.612763535 | 0.000512969 | 0.000886685 | 1 |
| T2DM | nk_cell | *ICAM5* | -1.596662678 | 6.48E-07 | 5.68E-05 | 0.003142958 |
| T2DM | nk_cell | *RABAC1* | 1.088864111 | 0.008009959 | 0.010868099 | 1 |
| T2DM | nk_cell | *GRIN2D* | -2.333813679 | 0.000107876 | 0.000248889 | 0.523413969 |
| T2DM | nk_cell | *CCDC114* | -2.057211662 | 0.012180215 | 0.016103107 | 1 |
| T2DM | nk_cell | *LIG1* | 1.01976318 | 0.002109613 | 0.003165072 | 1 |
| T2DM | nk_cell | *ZNF175* | -1.080182723 | 1.22E-05 | 0.000122264 | 0.059054603 |
| T2DM | nk_cell | *CAPS* | -0.805524575 | 2.02E-05 | 0.000122264 | 0.097907623 |
| T2DM | nk_cell | *MIER2* | -1.265229625 | 0.000187807 | 0.000381431 | 0.911237657 |
| T2DM | nk_cell | *PLEKHA4* | -0.567072212 | 2.15E-05 | 0.000122264 | 0.104498534 |
| T2DM | nk_cell | *TNPO2* | -1.166224681 | 5.26E-05 | 0.000158074 | 0.255317248 |
| T2DM | nk_cell | *GCDH* | -2.546190367 | 7.11E-09 | 2.46E-06 | 3.45E-05 |
| T2DM | nk_cell | *DNASE2* | -1.985066092 | 0.005307057 | 0.007442746 | 1 |
| T2DM | nk_cell | *TFPT* | -2.671197137 | 0.000149125 | 0.000318466 | 0.723554045 |

| | | | | | | |
|---|---|---|---|---|---|---|
| T2DM | nk_cell | *RAB3A* | -4.451490376 | 2.22E-05 | 0.000122264 | 0.10752054 |
| T2DM | nk_cell | *ETV2* | -9.801874971 | 0.0251521 | 0.031698179 | 1 |
| T2DM | nk_cell | *LSR* | -0.508141674 | 0.000211739 | 0.000420359 | 1 |
| T2DM | nk_cell | *ZNF14* | -1.968127419 | 4.96E-05 | 0.000152576 | 0.240765528 |
| T2DM | nk_cell | *ETHE1* | -1.948424811 | 0.02005889 | 0.025713537 | 1 |
| T2DM | nk_cell | *SMG9* | -2.214126645 | 0.00030959 | 0.000578187 | 1 |
| T2DM | nk_cell | *GTPBP10* | -0.897067902 | 0.029028664 | 0.036310151 | 1 |
| T2DM | nk_cell | *RASA4* | -3.196362191 | 2.62E-05 | 0.000122295 | 0.127235079 |
| T2DM | nk_cell | *BET1* | -1.009141133 | 1.23E-05 | 0.000122264 | 0.0596843 |
| T2DM | nk_cell | *SP4* | 0.944618167 | 0.002378443 | 0.00353777 | 1 |
| T2DM | nk_cell | *DNAH11* | -1.949610188 | 4.52E-05 | 0.000147139 | 0.219236822 |
| T2DM | nk_cell | *STEAP1B* | -1.30625634 | 0.001135521 | 0.001797569 | 1 |
| T2DM | nk_cell | *PTN* | -1.253023755 | 2.15E-05 | 0.000122264 | 0.104422818 |
| T2DM | nk_cell | *ADAP1* | -5.987620137 | 2.30E-05 | 0.000122264 | 0.111782765 |
| T2DM | nk_cell | *MET* | -0.646809653 | 2.65E-05 | 0.000122391 | 0.12876135 |
| T2DM | nk_cell | *RNF32* | -1.747201949 | 4.31E-07 | 4.54E-05 | 0.00208881 |
| T2DM | nk_cell | *IQCE* | -1.176524187 | 1.71E-05 | 0.000122264 | 0.08277289 |
| T2DM | nk_cell | *VIPR2* | -0.691167008 | 1.60E-05 | 0.000122264 | 0.077396432 |
| T2DM | nk_cell | *HIBADH* | -0.881057062 | 9.72E-05 | 0.000231443 | 0.471593316 |
| T2DM | nk_cell | *CPVL* | -1.275162283 | 4.09E-05 | 0.00013951 | 0.198232374 |
| T2DM | nk_cell | *GRB10* | -1.124236052 | 4.36E-05 | 0.000144302 | 0.21141651 |
| T2DM | nk_cell | *ABHD11* | -0.567571749 | 0.010471069 | 0.013999897 | 1 |
| T2DM | nk_cell | *PLEKHA8* | -0.748408952 | 2.40E-05 | 0.000122264 | 0.116288747 |
| T2DM | nk_cell | *CRHR2* | 0.958433834 | 0.000104975 | 0.000244287 | 0.509339106 |
| T2DM | nk_cell | *EPHB6* | -0.860337648 | 0.004363896 | 0.006205634 | 1 |
| T2DM | nk_cell | *CCL24* | 1.420442566 | 0.002635672 | 0.003887016 | 1 |
| T2DM | nk_cell | *ZKSCAN1* | -1.352323634 | 2.56E-05 | 0.000122264 | 0.124030103 |
| T2DM | nk_cell | *EIF3B* | -1.670141088 | 4.73E-05 | 0.000149118 | 0.229287604 |
| T2DM | nk_cell | *TAF6* | -0.71248159 | 0.01263475 | 0.016654118 | 1 |
| T2DM | nk_cell | *TFR2* | 0.380898211 | 8.77E-05 | 0.000215957 | 0.425650264 |
| T2DM | nk_cell | *MOSPD3* | -0.725634497 | 9.36E-06 | 0.000122264 | 0.045409635 |
| T2DM | nk_cell | *RBM28* | -1.051784471 | 0.000543848 | 0.00093275 | 1 |
| T2DM | nk_cell | *IMPDH1* | -1.320392128 | 3.34E-05 | 0.000128061 | 0.162111534 |
| T2DM | nk_cell | *C1GALT1* | -1.473926507 | 9.59E-05 | 0.000229078 | 0.465257536 |
| T2DM | nk_cell | *NRF1* | -1.287066344 | 0.000977218 | 0.001573668 | 1 |
| T2DM | nk_cell | *ZNF862* | -1.399082485 | 0.000103844 | 0.000242428 | 0.503851617 |
| T2DM | nk_cell | *MEST* | -0.66403665 | 6.92E-06 | 0.000122264 | 0.033588731 |
| T2DM | nk_cell | *POU6F2* | -0.587498107 | 1.76E-05 | 0.000122264 | 0.085386577 |
| T2DM | nk_cell | *TSPAN13* | -0.703385891 | 2.70E-05 | 0.000122391 | 0.130909452 |
| T2DM | nk_cell | *RARRES2* | -1.371934376 | 0.000444768 | 0.00078486 | 1 |
| T2DM | nk_cell | *COA1* | -1.297675867 | 0.000316402 | 0.000589549 | 1 |
| T2DM | nk_cell | *BLVRA* | -1.596139379 | 2.17E-05 | 0.000122264 | 0.105494471 |
| T2DM | nk_cell | *URGCP* | -1.350635434 | 0.000333189 | 0.000616165 | 1 |
| T2DM | nk_cell | *AEBP1* | -1.757322389 | 3.14E-05 | 0.000127109 | 0.152398764 |
| T2DM | nk_cell | *GCK* | -4.216447483 | 2.13E-05 | 0.000122264 | 0.103320667 |
| T2DM | nk_cell | *TBL2* | -1.265000182 | 0.012483647 | 0.016472846 | 1 |
| T2DM | nk_cell | *SPATA6L* | -0.863524998 | 3.37E-05 | 0.00012843 | 0.163620321 |
| T2DM | nk_cell | *FKTN* | -1.38634815 | 3.26E-05 | 0.000127909 | 0.158173765 |
| T2DM | nk_cell | *NMRK1* | -1.095612234 | 0.000101223 | 0.000238183 | 0.491132375 |
| T2DM | nk_cell | *MEGF9* | -1.627732791 | 0.000136575 | 0.000296493 | 0.66266092 |

| | | | | | | |
|---|---|---|---|---|---|---|
| T2DM | nk_cell | *TRIM14* | 0.711131239 | 0.000435238 | 0.000769878 | 1 |
| T2DM | nk_cell | *CORO2A* | 0.411176464 | 9.67E-05 | 0.00023082 | 0.469025678 |
| T2DM | nk_cell | *C5* | -0.75526901 | 2.78E-05 | 0.000122493 | 0.134892744 |
| T2DM | nk_cell | *OGN* | -1.03643087 | 6.76E-06 | 0.000122264 | 0.032785059 |
| T2DM | nk_cell | *ECM2* | -1.206940162 | 4.13E-05 | 0.000140573 | 0.200549669 |
| T2DM | nk_cell | *PTGR1* | -1.067919626 | 1.79E-05 | 0.000122264 | 0.086814563 |
| T2DM | nk_cell | *SUSD1* | 0.519246359 | 0.001110411 | 0.001760691 | 1 |
| T2DM | nk_cell | *PLGRKT* | -1.284923073 | 0.002161459 | 0.003233857 | 1 |
| T2DM | nk_cell | *KANK1* | -1.049563773 | 7.03E-06 | 0.000122264 | 0.034102763 |
| T2DM | nk_cell | *NCS1* | -0.884312162 | 0.022766877 | 0.028887261 | 1 |
| T2DM | nk_cell | *FUBP3* | -1.125570463 | 1.66E-05 | 0.000122264 | 0.08034968 |
| T2DM | nk_cell | *RGP1* | -2.139244672 | 0.002213831 | 0.003308134 | 1 |
| T2DM | nk_cell | *ATRNL1* | -0.32019212 | 0.014038603 | 0.018394626 | 1 |
| T2DM | nk_cell | *HPS1* | -1.677186259 | 4.75E-05 | 0.000149363 | 0.230587307 |
| T2DM | nk_cell | *PHYH* | -1.077863502 | 3.02E-05 | 0.000124985 | 0.146488204 |
| T2DM | nk_cell | *DNMBP* | -0.932054061 | 7.98E-06 | 0.000122264 | 0.038698861 |
| T2DM | nk_cell | *PKD2L1* | 1.127977587 | 4.36E-05 | 0.000144302 | 0.211478882 |
| T2DM | nk_cell | *CUBN* | -1.611473879 | 0.000444068 | 0.000784068 | 1 |
| T2DM | nk_cell | *TRDMT1* | -1.217663404 | 1.72E-05 | 0.000122264 | 0.083472088 |
| T2DM | nk_cell | *ATE1* | -0.955501501 | 0.000335979 | 0.000619837 | 1 |
| T2DM | nk_cell | *PLEKHA1* | -1.529746056 | 0.00042732 | 0.000758685 | 1 |
| T2DM | nk_cell | *CDH23* | -0.823539191 | 1.46E-05 | 0.000122264 | 0.070613174 |
| T2DM | nk_cell | *SPOCK2* | 0.392306049 | 0.003096531 | 0.00451589 | 1 |
| T2DM | nk_cell | *BMPR1A* | -0.851556771 | 1.61E-05 | 0.000122264 | 0.078297664 |
| T2DM | nk_cell | *LIPA* | -0.835459856 | 0.00418878 | 0.005967105 | 1 |
| T2DM | nk_cell | *SFXN3* | -1.301955396 | 1.75E-05 | 0.000122264 | 0.084950889 |
| T2DM | nk_cell | *TNKS2* | -1.131904415 | 0.031828656 | 0.039577816 | 1 |
| T2DM | nk_cell | *ANKRD26* | -1.047588426 | 3.47E-05 | 0.000130294 | 0.168359256 |
| T2DM | nk_cell | *ACBD5* | -1.352336443 | 0.030594866 | 0.038146328 | 1 |
| T2DM | nk_cell | *LHPP* | -1.949690516 | 1.59E-05 | 0.000122264 | 0.076947485 |
| T2DM | nk_cell | *PITRM1* | -1.130562103 | 9.28E-06 | 0.000122264 | 0.045046016 |
| T2DM | nk_cell | *TFAM* | -1.682115151 | 0.024904864 | 0.031435588 | 1 |
| T2DM | nk_cell | *RPL28* | 0.593986124 | 0.028689492 | 0.035905422 | 1 |
| T2DM | nk_cell | *DNAJC12* | -1.17545991 | 0.000168393 | 0.000348567 | 0.817040865 |
| T2DM | nk_cell | *PBLD* | -1.57800607 | 3.28E-05 | 0.000127909 | 0.159316881 |
| T2DM | nk_cell | *TSPAN14* | -1.235685613 | 0.000380997 | 0.000690549 | 1 |
| T2DM | nk_cell | *RPL19* | 0.749552035 | 0.028655484 | 0.035880363 | 1 |
| T2DM | nk_cell | *RAPGEFL1* | -0.944753182 | 1.81E-05 | 0.000122264 | 0.088025076 |
| T2DM | nk_cell | *WNT3* | -0.813553208 | 5.65E-05 | 0.00016385 | 0.273940808 |
| T2DM | nk_cell | *ASPA* | -0.587459495 | 7.73E-05 | 0.000199059 | 0.375000047 |
| T2DM | nk_cell | *RAD51C* | -1.674091791 | 8.36E-05 | 0.000209406 | 0.405831524 |
| T2DM | nk_cell | *TRIM37* | -1.774066942 | 0.000115926 | 0.000262225 | 0.562472461 |
| T2DM | nk_cell | *TRIM16L* | -0.629596103 | 2.60E-05 | 0.000122264 | 0.126073084 |
| T2DM | nk_cell | *CDK5RAP3* | -1.975491438 | 0.000182182 | 0.000372502 | 0.883948262 |
| T2DM | nk_cell | *PIGL* | -1.425537218 | 7.11E-05 | 0.000189011 | 0.345134225 |
| T2DM | nk_cell | *MED13* | -1.193068045 | 4.06E-05 | 0.000139271 | 0.197068238 |
| T2DM | nk_cell | *CHRNE* | 0.608961494 | 8.74E-05 | 0.000215323 | 0.424185659 |
| T2DM | nk_cell | *GOSR1* | -1.18497703 | 2.39E-05 | 0.000122264 | 0.116171747 |
| T2DM | nk_cell | *DRG2* | -1.084649811 | 1.95E-05 | 0.000122264 | 0.094800695 |
| T2DM | nk_cell | *FTSJ3* | -0.591631624 | 0.001942514 | 0.002937076 | 1 |

| | | | | | | |
|---|---|---|---|---|---|---|
| T2DM | nk_cell | *ICAM2* | 0.783842271 | 0.000450738 | 0.000793534 | 1 |
| T2DM | nk_cell | *DDX5* | -0.936798634 | 0.00079926 | 0.001306607 | 1 |
| T2DM | nk_cell | *C17orf75* | -0.975019438 | 0.000542807 | 0.0009315 | 1 |
| T2DM | nk_cell | *PSMD11* | -1.486694766 | 4.86E-05 | 0.000151135 | 0.235713729 |
| T2DM | nk_cell | *PEX12* | 0.736713479 | 0.000408973 | 0.000732227 | 1 |
| T2DM | nk_cell | *DHX58* | -0.912243088 | 1.64E-05 | 0.000122264 | 0.079536862 |
| T2DM | nk_cell | *RAB5C* | 1.205184685 | 0.001668141 | 0.002555674 | 1 |
| T2DM | nk_cell | *NAGLU* | -1.54558439 | 0.000801477 | 0.001308908 | 1 |
| T2DM | nk_cell | *CNTNAP1* | -0.63921388 | 0.000567741 | 0.000966214 | 1 |
| T2DM | nk_cell | *EZH1* | -0.990820671 | 1.57E-05 | 0.000122264 | 0.076323348 |
| T2DM | nk_cell | *EFTUD2* | -1.377223696 | 2.23E-05 | 0.000122264 | 0.108190093 |
| T2DM | nk_cell | *HLF* | -1.02072194 | 4.88E-05 | 0.000151402 | 0.236697594 |
| T2DM | nk_cell | *EFNB3* | -1.235048154 | 2.79E-05 | 0.000122493 | 0.135409391 |
| T2DM | nk_cell | *FAM20A* | -0.999415156 | 1.75E-05 | 0.000122264 | 0.08491175 |
| T2DM | nk_cell | *DPH1* | -0.718544291 | 1.99E-05 | 0.000122264 | 0.096694708 |
| T2DM | nk_cell | *DHRS7B* | -1.574642891 | 0.000224552 | 0.00044164 | 1 |
| T2DM | nk_cell | *MYH3* | 1.880367503 | 0.018154737 | 0.02340244 | 1 |
| T2DM | nk_cell | *GNRHR* | 0.867140661 | 0.021348896 | 0.02724483 | 1 |
| T2DM | nk_cell | *SLAIN2* | -1.107296439 | 0.036141143 | 0.044574689 | 1 |
| T2DM | nk_cell | *OCIAD1* | -1.380633923 | 7.76E-05 | 0.000199359 | 0.3763889 |
| T2DM | nk_cell | *UBE2D3* | -1.114676302 | 5.15E-05 | 0.000155956 | 0.250048412 |
| T2DM | nk_cell | *KLHL2* | -1.692052576 | 9.16E-05 | 0.000222312 | 0.444401401 |
| T2DM | nk_cell | *CPE* | -0.668098776 | 0.001385207 | 0.002146606 | 1 |
| T2DM | nk_cell | *WFS1* | -2.276346726 | 5.46E-05 | 0.000160881 | 0.26480981 |
| T2DM | nk_cell | *AADAT* | -0.754333887 | 2.61E-05 | 0.000122264 | 0.126543056 |
| T2DM | nk_cell | *TRIM2* | -1.24843291 | 1.04E-05 | 0.000122264 | 0.050291211 |
| T2DM | nk_cell | *BST1* | -1.083247334 | 7.45E-05 | 0.000194744 | 0.361640158 |
| T2DM | nk_cell | *UFSP2* | -1.298892957 | 0.035834967 | 0.044253312 | 1 |
| T2DM | nk_cell | *KLHL5* | -1.074455152 | 2.14E-05 | 0.000122264 | 0.103787349 |
| T2DM | nk_cell | *FAM149A* | -0.968652277 | 7.43E-05 | 0.000194408 | 0.360432169 |
| T2DM | nk_cell | *DDX25* | -0.656160326 | 0.003489552 | 0.005046589 | 1 |
| T2DM | nk_cell | *ELP4* | -1.301995408 | 2.76E-05 | 0.000122472 | 0.133791971 |
| T2DM | nk_cell | *TECTA* | -0.833649479 | 2.38E-05 | 0.000122264 | 0.115592455 |
| T2DM | nk_cell | *B3GAT1* | 0.143482075 | 4.94E-05 | 0.00015216 | 0.239656097 |
| T2DM | nk_cell | *P2RX3* | 0.925282167 | 5.57E-05 | 0.00016298 | 0.270384473 |
| T2DM | nk_cell | *DNAJC4* | 0.920177352 | 0.006772906 | 0.009320372 | 1 |
| T2DM | nk_cell | *SNX15* | -1.499920776 | 0.000174187 | 0.00035903 | 0.84515611 |
| T2DM | nk_cell | *UNC93B1* | -1.471806053 | 5.01E-05 | 0.000153185 | 0.242884081 |
| T2DM | nk_cell | *PUS3* | -0.66003296 | 0.014112397 | 0.018476349 | 1 |
| T2DM | nk_cell | *FOXRED1* | -0.605680213 | 0.000111525 | 0.000255366 | 0.541119831 |
| T2DM | nk_cell | *NRXN2* | -3.700694072 | 7.55E-06 | 0.000122264 | 0.036651517 |
| T2DM | nk_cell | *CCND1* | -2.05052788 | 5.42E-07 | 5.26E-05 | 0.002631725 |
| T2DM | nk_cell | *CCDC86* | -2.906724797 | 4.58E-05 | 0.000147697 | 0.222232654 |
| T2DM | nk_cell | *PANX1* | -1.791687364 | 1.33E-05 | 0.000122264 | 0.064595688 |
| T2DM | nk_cell | *CEP164* | -1.175238569 | 2.30E-05 | 0.000122264 | 0.111821886 |
| T2DM | nk_cell | *GALNT18* | -1.141234852 | 0.011871978 | 0.01572559 | 1 |
| T2DM | nk_cell | *KIAA1549L* | -1.655751214 | 3.23E-05 | 0.000127896 | 0.156928849 |
| T2DM | nk_cell | *ACCS* | -0.715174467 | 0.001299072 | 0.002024116 | 1 |
| T2DM | nk_cell | *SLC35F2* | -1.577957327 | 2.63E-05 | 0.000122391 | 0.127844314 |
| T2DM | nk_cell | *C11orf21* | 0.421489917 | 0.000262614 | 0.000502842 | 1 |

| | | | | | | |
|---|---|---|---|---|---|---|
| T2DM | nk_cell | *ELMOD1* | 0.86717527 | 9.32E-05 | 0.000224526 | 0.452418898 |
| T2DM | nk_cell | *NDUFS8* | -2.109088749 | 0.000718282 | 0.001189455 | 1 |
| T2DM | nk_cell | *GTF2H1* | -1.266631906 | 2.61E-05 | 0.000122295 | 0.126823889 |
| T2DM | nk_cell | *PRDM4* | -1.445351658 | 2.16E-05 | 0.000122264 | 0.104749597 |
| T2DM | nk_cell | *CORO1C* | -0.823403264 | 1.64E-05 | 0.000122264 | 0.079561467 |
| T2DM | nk_cell | *ASIC1* | -1.021487438 | 2.48E-05 | 0.000122264 | 0.120173705 |
| T2DM | nk_cell | *CAPRIN2* | -0.880469417 | 0.019666252 | 0.025216874 | 1 |
| T2DM | nk_cell | *KCTD10* | -0.968661357 | 1.47E-05 | 0.000122264 | 0.071381397 |
| T2DM | nk_cell | *IL23A* | 6.528095613 | 0.000617524 | 0.001039634 | 1 |
| T2DM | nk_cell | *ACSS3* | -0.801567329 | 0.000109763 | 0.000252403 | 0.532571221 |
| T2DM | nk_cell | *LTA4H* | -0.731754875 | 0.008094652 | 0.010976872 | 1 |
| T2DM | nk_cell | *PARP11* | -1.423996483 | 3.35E-05 | 0.000128164 | 0.162768229 |
| T2DM | nk_cell | *GPN3* | -1.152769983 | 1.66E-05 | 0.000122264 | 0.080662068 |
| T2DM | nk_cell | *VPS29* | -1.417810932 | 5.16E-05 | 0.000155956 | 0.250380747 |
| T2DM | nk_cell | *AKAP3* | -0.54919792 | 0.000202518 | 0.00040487 | 0.982618317 |
| T2DM | nk_cell | *ACAD10* | -1.642723438 | 4.61E-05 | 0.000147867 | 0.223647967 |
| T2DM | nk_cell | *LTBR* | -1.122851001 | 2.52E-05 | 0.000122264 | 0.122199787 |
| T2DM | nk_cell | *CDK2AP1* | 1.951385057 | 1.48E-08 | 4.80E-06 | 7.20E-05 |
| T2DM | nk_cell | *ENDOU* | -1.431695008 | 2.61E-05 | 0.000122264 | 0.126431754 |
| T2DM | nk_cell | *VDR* | -0.88325868 | 2.30E-05 | 0.000122264 | 0.111381506 |
| T2DM | nk_cell | *STX2* | -0.858261204 | 0.001189617 | 0.001871479 | 1 |
| T2DM | nk_cell | *CAND1* | -1.58214146 | 0.000102681 | 0.000240564 | 0.498207879 |
| T2DM | nk_cell | *MDM1* | -1.765698035 | 2.22E-05 | 0.000122264 | 0.107565084 |
| T2DM | nk_cell | *NUP107* | -1.312591638 | 6.05E-05 | 0.000169923 | 0.293574435 |
| T2DM | nk_cell | *KRR1* | -1.704883543 | 7.26E-06 | 0.000122264 | 0.035229657 |
| T2DM | nk_cell | *GNB3* | -1.149564641 | 3.32E-06 | 0.000122264 | 0.016126271 |
| T2DM | nk_cell | *CDCA3* | -2.32993395 | 4.84E-07 | 4.79E-05 | 0.002347319 |
| T2DM | nk_cell | *CHPT1* | -1.236717522 | 2.50E-05 | 0.000122264 | 0.121521514 |
| T2DM | nk_cell | *SPSB2* | -0.816749712 | 0.000258139 | 0.000495644 | 1 |
| T2DM | nk_cell | *ATN1* | -0.988684351 | 9.96E-05 | 0.000235947 | 0.483454912 |
| T2DM | nk_cell | *C12orf57* | 1.163346678 | 0.00934477 | 0.012570231 | 1 |
| T2DM | nk_cell | *NT5DC3* | -1.770965552 | 0.000186075 | 0.000378866 | 0.902837052 |
| T2DM | nk_cell | *SUDS3* | -1.059587886 | 1.67E-05 | 0.000122264 | 0.080945731 |
| T2DM | nk_cell | *GOLT1B* | 1.166374965 | 0.028872263 | 0.036123831 | 1 |
| T2DM | nk_cell | *C2CD5* | -1.29564302 | 5.64E-05 | 0.00016385 | 0.273797404 |
| T2DM | nk_cell | *BTN3A3* | 0.720920446 | 0.026347154 | 0.033118236 | 1 |
| T2DM | nk_cell | *FRK* | -0.967408209 | 1.57E-05 | 0.000122264 | 0.076068295 |
| T2DM | nk_cell | *RSPH4A* | -0.871243738 | 0.024656921 | 0.031146935 | 1 |
| T2DM | nk_cell | *TMEM14C* | -1.59181259 | 2.79E-05 | 0.000122493 | 0.135150344 |
| T2DM | nk_cell | *SMIM8* | -0.875049389 | 8.11E-05 | 0.00020495 | 0.39364325 |
| T2DM | nk_cell | *FAM184A* | -0.822025948 | 5.39E-05 | 0.00015958 | 0.261494819 |
| T2DM | nk_cell | *RNGTT* | -1.319171798 | 3.79E-05 | 0.000134881 | 0.183816299 |
| T2DM | nk_cell | *UST* | -0.545776153 | 0.000143484 | 0.000308592 | 0.69618395 |
| T2DM | nk_cell | *TULP1* | -0.965636543 | 0.02985042 | 0.037270777 | 1 |
| T2DM | nk_cell | *RNF8* | -1.658712475 | 3.50E-05 | 0.000130831 | 0.170036884 |
| T2DM | nk_cell | *PHACTR1* | -1.430928786 | 2.56E-05 | 0.000122264 | 0.124171047 |
| T2DM | nk_cell | *MDGA1* | -1.307794302 | 1.22E-05 | 0.000122264 | 0.059092727 |
| T2DM | nk_cell | *CD83* | -1.40347494 | 4.18E-06 | 0.000122264 | 0.020294586 |
| T2DM | nk_cell | *MDN1* | -0.987920357 | 1.54E-05 | 0.000122264 | 0.074747348 |
| T2DM | nk_cell | *GLP1R* | -3.689541771 | 0.001218634 | 0.001911675 | 1 |

| | | | | | | |
|---|---|---|---|---|---|---|
| T2DM | nk_cell | *BVES* | -0.871164538 | 6.89E-08 | 1.39E-05 | 0.000334226 |
| T2DM | nk_cell | *COL9A1* | -1.440502567 | 0.000155415 | 0.000328143 | 0.754073003 |
| T2DM | nk_cell | *WASF1* | -0.84742557 | 1.75E-05 | 0.000122264 | 0.084773284 |
| T2DM | nk_cell | *VNN1* | -0.777404453 | 0.004010332 | 0.005739861 | 1 |
| T2DM | nk_cell | *VNN2* | 1.034612667 | 1.29E-05 | 0.000122264 | 0.062805005 |
| T2DM | nk_cell | *ACOT13* | -1.326883294 | 0.03542707 | 0.043771872 | 1 |
| T2DM | nk_cell | *C6orf62* | -1.473906898 | 6.20E-05 | 0.000172988 | 0.300825814 |
| T2DM | nk_cell | *HBS1L* | -1.240877279 | 1.51E-05 | 0.000122264 | 0.073393823 |
| T2DM | nk_cell | *PEX7* | -0.784968701 | 0.014599995 | 0.019094117 | 1 |
| T2DM | nk_cell | *FIG4* | -1.573418512 | 0.00018013 | 0.000368772 | 0.873989839 |
| T2DM | nk_cell | *PERP* | -0.899782261 | 0.00463241 | 0.006556725 | 1 |
| T2DM | nk_cell | *PHACTR2* | -1.407057354 | 5.27E-05 | 0.0001582 | 0.255808684 |
| T2DM | nk_cell | *CUTA* | 0.543668572 | 0.005992179 | 0.008333062 | 1 |
| T2DM | nk_cell | *PACRG* | -0.61564354 | 1.99E-05 | 0.000122264 | 0.096418754 |
| T2DM | nk_cell | *QKI* | -0.957024013 | 3.34E-05 | 0.000128061 | 0.161878142 |
| T2DM | nk_cell | *FAM120B* | -1.114683412 | 0.00041532 | 0.000741401 | 1 |
| T2DM | nk_cell | *GUCA1B* | -0.861909339 | 5.97E-06 | 0.000122264 | 0.028946322 |
| T2DM | nk_cell | *TMEM30A* | -1.230556017 | 1.74E-05 | 0.000122264 | 0.084242947 |
| T2DM | nk_cell | *IMPG1* | -1.436526711 | 2.89E-05 | 0.000123207 | 0.140161383 |
| T2DM | nk_cell | *LY86* | -2.990477455 | 8.79E-05 | 0.000216172 | 0.426291141 |
| T2DM | nk_cell | *PRSS16* | -1.440593749 | 0.015768688 | 0.020522981 | 1 |
| T2DM | nk_cell | *PCDHB2* | -1.184921886 | 2.92E-05 | 0.000123467 | 0.141879586 |
| T2DM | nk_cell | *NUDT12* | -1.669192958 | 2.48E-05 | 0.000122264 | 0.12048512 |
| T2DM | nk_cell | *NME5* | -0.531866919 | 0.001243572 | 0.001946391 | 1 |
| T2DM | nk_cell | *KIF20A* | 1.800931005 | 0.000804503 | 0.001313409 | 1 |
| T2DM | nk_cell | *PFDN1* | -1.029500073 | 7.94E-05 | 0.000202355 | 0.385081891 |
| T2DM | nk_cell | *TMCO6* | -1.723795536 | 0.000149927 | 0.000319616 | 0.727445367 |
| T2DM | nk_cell | *SPARC* | -1.138238463 | 7.88E-05 | 0.000201551 | 0.382467915 |
| T2DM | nk_cell | *HAVCR1* | 1.300331366 | 0.021931219 | 0.027921878 | 1 |
| T2DM | nk_cell | *RNF130* | -0.998329733 | 0.000528423 | 0.00091048 | 1 |
| T2DM | nk_cell | *THBS4* | -0.630681793 | 6.08E-05 | 0.00017062 | 0.295171811 |
| T2DM | nk_cell | *BTNL8* | -0.40263398 | 0.00303343 | 0.004429191 | 1 |
| T2DM | nk_cell | *MSH3* | -1.145068156 | 3.62E-05 | 0.000132186 | 0.175637629 |
| T2DM | nk_cell | *RASGRF2* | -0.588900705 | 9.70E-06 | 0.000122264 | 0.047041745 |
| T2DM | nk_cell | *DROSHA* | -1.037275625 | 0.000395318 | 0.000713573 | 1 |
| T2DM | nk_cell | *CDH6* | -0.483485286 | 5.74E-06 | 0.000122264 | 0.027841311 |
| T2DM | nk_cell | *LMNB1* | -1.628843212 | 0.012912843 | 0.016992979 | 1 |
| T2DM | nk_cell | *SLC12A7* | -2.136605873 | 0.00146008 | 0.002253601 | 1 |
| T2DM | nk_cell | *SKP1* | -1.273442736 | 2.35E-05 | 0.000122264 | 0.114240256 |
| T2DM | nk_cell | *PPWD1* | -1.584619966 | 0.000130692 | 0.000286802 | 0.634118462 |
| T2DM | nk_cell | *TXNDC15* | -1.263963592 | 2.14E-05 | 0.000122264 | 0.103619231 |
| T2DM | nk_cell | *SMAD5* | -1.5550763 | 0.000550179 | 0.000941944 | 1 |
| T2DM | nk_cell | *ERGIC1* | -1.587048349 | 0.000176641 | 0.000362853 | 0.857059871 |
| T2DM | nk_cell | *ATP6V0E1* | -1.4556629 | 7.45E-05 | 0.000194744 | 0.361373256 |
| T2DM | nk_cell | *BNIP1* | 0.704909418 | 0.000855378 | 0.001389452 | 1 |
| T2DM | nk_cell | *UNC5A* | -2.012320891 | 2.46E-05 | 0.000122264 | 0.119539351 |
| T2DM | nk_cell | *ACTR8* | -1.862480403 | 3.80E-06 | 0.000122264 | 0.018450962 |
| T2DM | nk_cell | *TBCCD1* | -2.113855796 | 6.54E-05 | 0.000178359 | 0.317478941 |
| T2DM | nk_cell | *CLDN16* | -0.609865949 | 3.46E-05 | 0.000130101 | 0.167792682 |
| T2DM | nk_cell | *PCCB* | -0.694878576 | 0.022378682 | 0.028454237 | 1 |

| | | | | | | |
|---|---|---|---|---|---|---|
| T2DM | nk_cell | *CEP70* | -1.642501854 | 0.000184777 | 0.000376854 | 0.896536684 |
| T2DM | nk_cell | *CYB561D2* | -2.073269266 | 0.001414859 | 0.002188364 | 1 |
| T2DM | nk_cell | *C3orf14* | -1.211929995 | 0.000857653 | 0.001392682 | 1 |
| T2DM | nk_cell | *IQCG* | -1.336883226 | 4.02E-05 | 0.000138637 | 0.195201583 |
| T2DM | nk_cell | *NCBP2* | -2.329835969 | 0.000284299 | 0.000537786 | 1 |
| T2DM | nk_cell | *SLC41A3* | -0.857422951 | 1.13E-05 | 0.000122264 | 0.054993894 |
| T2DM | nk_cell | *PLXNA1* | -1.304648882 | 1.95E-05 | 0.000122264 | 0.094582346 |
| T2DM | nk_cell | *SCAP* | -1.518929483 | 9.14E-05 | 0.000221911 | 0.443378424 |
| T2DM | nk_cell | *KIAA1257* | -1.421275365 | 2.25E-05 | 0.000122264 | 0.109202251 |
| T2DM | nk_cell | *NEK11* | -0.746919912 | 5.24E-05 | 0.000157634 | 0.254420814 |
| T2DM | nk_cell | *PLSCR4* | -0.808080194 | 1.82E-05 | 0.000122264 | 0.088164913 |
| T2DM | nk_cell | *HEMK1* | -2.020400381 | 1.10E-06 | 7.99E-05 | 0.005353501 |
| T2DM | nk_cell | *COMMD2* | -1.662979933 | 0.004396503 | 0.00624834 | 1 |
| T2DM | nk_cell | *AADAC* | -0.330552921 | 0.003024883 | 0.004419371 | 1 |
| T2DM | nk_cell | *ABHD14B* | -2.248729451 | 1.18E-05 | 0.000122264 | 0.057234644 |
| T2DM | nk_cell | *ARHGEF26* | -1.118695069 | 1.70E-05 | 0.000122264 | 0.082631586 |
| T2DM | nk_cell | *VIPR1* | -0.900253177 | 2.50E-05 | 0.000122264 | 0.121481471 |
| T2DM | nk_cell | *DNAH1* | -0.846066102 | 0.000574722 | 0.000976042 | 1 |
| T2DM | nk_cell | *NKTR* | -1.021534705 | 0.000407396 | 0.000729943 | 1 |
| T2DM | nk_cell | *NEK4* | -1.065221171 | 1.71E-05 | 0.000122264 | 0.083187828 |
| T2DM | nk_cell | *ADAM23* | -1.763316392 | 8.62E-06 | 0.000122264 | 0.041816056 |
| T2DM | nk_cell | *LMAN2L* | -0.916364047 | 4.37E-05 | 0.000144321 | 0.21206162 |
| T2DM | nk_cell | *ACTR1B* | -2.101652996 | 0.000743998 | 0.001224102 | 1 |
| T2DM | nk_cell | *SLC35F5* | -0.965372828 | 0.000324596 | 0.000602041 | 1 |
| T2DM | nk_cell | *OTOF* | 1.855033148 | 0.009906824 | 0.013274761 | 1 |
| T2DM | nk_cell | *CENPA* | -0.794270978 | 3.02E-05 | 0.000124985 | 0.146712599 |
| T2DM | nk_cell | *NRBP1* | -1.887259185 | 6.36E-05 | 0.000174859 | 0.308800172 |
| T2DM | nk_cell | *ITGB6* | -0.764312254 | 4.52E-06 | 0.000122264 | 0.021923653 |
| T2DM | nk_cell | *PPM1G* | 0.841866857 | 0.010698429 | 0.014268492 | 1 |
| T2DM | nk_cell | *MOGS* | -1.122105255 | 2.26E-05 | 0.000122264 | 0.109423265 |
| T2DM | nk_cell | *GRB14* | -1.309251104 | 2.22E-05 | 0.000122264 | 0.107510833 |
| T2DM | nk_cell | *RTN4* | -0.946757694 | 2.13E-05 | 0.000122264 | 0.103424372 |
| T2DM | nk_cell | *GALNT3* | 0.327575845 | 3.74E-05 | 0.000134293 | 0.181295682 |
| T2DM | nk_cell | *ACADL* | -1.275215453 | 6.04E-05 | 0.000169923 | 0.293076519 |
| T2DM | nk_cell | *MRPL19* | -1.101832336 | 1.42E-05 | 0.000122264 | 0.068830772 |
| T2DM | nk_cell | *IGFBP2* | -1.514525788 | 0.000163895 | 0.000341144 | 0.795218018 |
| T2DM | nk_cell | *GNLY* | 0.045710277 | 5.23E-05 | 0.000157457 | 0.253821325 |
| T2DM | nk_cell | *CHMP3* | -2.028736229 | 3.72E-06 | 0.000122264 | 0.018048863 |
| T2DM | nk_cell | *WNT6* | -0.963555609 | 6.37E-06 | 0.000122264 | 0.030885417 |
| T2DM | nk_cell | *IL1RL2* | -1.108282815 | 0.000172154 | 0.000355359 | 0.835290697 |
| T2DM | nk_cell | *IL18R1* | 0.582055525 | 0.000230547 | 0.000450774 | 1 |
| T2DM | nk_cell | *MLPH* | -1.001347251 | 7.25E-05 | 0.000191287 | 0.351585178 |
| T2DM | nk_cell | *HDLBP* | -1.300686246 | 0.000119358 | 0.000267001 | 0.579124697 |
| T2DM | nk_cell | *PASK* | -0.859779769 | 0.000397152 | 0.00071635 | 1 |
| T2DM | nk_cell | *TPO* | -1.433370568 | 9.58E-06 | 0.000122264 | 0.046472445 |
| T2DM | nk_cell | *TAF1B* | -1.250379793 | 3.91E-05 | 0.000136598 | 0.189734987 |
| T2DM | nk_cell | *NOL10* | -1.404622965 | 2.59E-05 | 0.000122264 | 0.125653099 |
| T2DM | nk_cell | *STRN* | -1.186580737 | 3.05E-05 | 0.000125102 | 0.147902143 |
| T2DM | nk_cell | *CEBPZ* | -1.583219871 | 0.000163965 | 0.000341144 | 0.795556361 |
| T2DM | nk_cell | *PRKD3* | -1.378876225 | 0.000151994 | 0.000323312 | 0.737474322 |

| | | | | | | |
|---|---|---|---|---|---|---|
| T2DM | nk_cell | *SLC25A12* | -1.499553974 | 3.57E-05 | 0.000131704 | 0.173454592 |
| T2DM | nk_cell | *DARS* | -1.538539178 | 0.000480372 | 0.000838405 | 1 |
| T2DM | nk_cell | *WIPF1* | 0.711196026 | 0.002499074 | 0.003699057 | 1 |
| T2DM | nk_cell | *PNO1* | -2.074399663 | 1.94E-05 | 0.000122264 | 0.094239919 |
| T2DM | nk_cell | *ORC4* | -1.102303088 | 9.98E-05 | 0.000236122 | 0.48428582 |
| T2DM | nk_cell | *GRIN3B* | 0.479044451 | 5.38E-05 | 0.000159514 | 0.260881915 |
| T2DM | nk_cell | *NFE2L2* | -0.953055243 | 9.51E-05 | 0.000227998 | 0.461608522 |
| T2DM | nk_cell | *MSH6* | -1.267139114 | 1.61E-05 | 0.000122264 | 0.07805015 |
| T2DM | nk_cell | *ALMS1* | -1.221505188 | 2.77E-05 | 0.000122493 | 0.134215186 |
| T2DM | nk_cell | *DNAJC16* | -0.882301138 | 6.73E-05 | 0.000181892 | 0.326379376 |
| T2DM | nk_cell | *MARK1* | -1.030983654 | 3.12E-05 | 0.000126743 | 0.151273252 |
| T2DM | nk_cell | *MORN1* | -0.774664593 | 0.000112103 | 0.000256447 | 0.54392427 |
| T2DM | nk_cell | *GPX7* | 0.211086087 | 0.015689102 | 0.020424879 | 1 |
| T2DM | nk_cell | *RALGPS2* | -0.549786783 | 6.66E-06 | 0.000122264 | 0.032329197 |
| T2DM | nk_cell | *CEP104* | -1.474646647 | 0.000152688 | 0.000323794 | 0.740840958 |
| T2DM | nk_cell | *FAM20B* | -1.668045137 | 7.25E-05 | 0.000191375 | 0.351938087 |
| T2DM | nk_cell | *WRAP73* | 1.302154371 | 0.000697116 | 0.001158756 | 1 |
| T2DM | nk_cell | *ICMT* | -2.230546394 | 1.44E-05 | 0.000122264 | 0.070028156 |
| T2DM | nk_cell | *PARK7* | -1.595770357 | 0.00024518 | 0.000474516 | 1 |
| T2DM | nk_cell | *KIAA1324* | 0.507999766 | 0.000125455 | 0.000277315 | 0.608705331 |
| T2DM | nk_cell | *MECR* | -0.979072458 | 1.49E-05 | 0.000122264 | 0.072487388 |
| T2DM | nk_cell | *KCNC4* | -1.119125152 | 2.94E-05 | 0.00012351 | 0.142530656 |
| T2DM | nk_cell | *EDEM3* | -1.670959595 | 0.000109033 | 0.000250962 | 0.529028158 |
| T2DM | nk_cell | *RAP1A* | -1.073605332 | 0.017596676 | 0.022737436 | 1 |
| T2DM | nk_cell | *CAPZA1* | -1.392945726 | 5.56E-05 | 0.000162627 | 0.269636166 |
| T2DM | nk_cell | *ASH1L* | -0.887690209 | 0.000443191 | 0.000782804 | 1 |
| T2DM | nk_cell | *GON4L* | -1.028412692 | 9.29E-05 | 0.000223847 | 0.450780598 |
| T2DM | nk_cell | *DOCK7* | -1.029289712 | 2.57E-05 | 0.000122264 | 0.124458545 |
| T2DM | nk_cell | *FBXO2* | 0.582109684 | 0.000275895 | 0.00052393 | 1 |
| T2DM | nk_cell | *LEPR* | -1.142053219 | 2.58E-05 | 0.000122264 | 0.125014922 |
| T2DM | nk_cell | *MFN2* | -1.681780146 | 0.002828539 | 0.004147498 | 1 |
| T2DM | nk_cell | *PRG4* | -0.895466779 | 1.29E-05 | 0.000122264 | 0.062666302 |
| T2DM | nk_cell | *BCAS2* | -0.871252633 | 0.001183132 | 0.001863817 | 1 |
| T2DM | nk_cell | *TNNI3K* | 1.56789911 | 0.03657454 | 0.0450634 | 1 |
| T2DM | nk_cell | *CFHR3* | -1.67230524 | 0.000976402 | 0.001572875 | 1 |
| T2DM | nk_cell | *CRYZ* | -0.722732984 | 0.000535854 | 0.000921319 | 1 |
| T2DM | nk_cell | *TMEM9* | -1.234297621 | 1.23E-05 | 0.000122264 | 0.05967159 |
| T2DM | nk_cell | *WARS2* | -1.118769852 | 1.25E-05 | 0.000122264 | 0.060792488 |
| T2DM | nk_cell | *OSCP1* | -0.736108314 | 0.000456102 | 0.000801814 | 1 |
| T2DM | nk_cell | *MRPS15* | -1.478247919 | 4.73E-05 | 0.000149118 | 0.229453346 |
| T2DM | nk_cell | *GNPAT* | -1.836791436 | 0.000370668 | 0.000675106 | 1 |
| T2DM | nk_cell | *C1orf109* | -0.862401395 | 9.54E-06 | 0.000122264 | 0.046304054 |
| T2DM | nk_cell | *TBCE* | -1.43313525 | 3.84E-05 | 0.000135693 | 0.186334885 |
| T2DM | nk_cell | *ACADM* | -1.114574551 | 7.11E-05 | 0.00018898 | 0.344888967 |
| T2DM | nk_cell | *SLAMF1* | -1.07432105 | 0.036805501 | 0.045336454 | 1 |
| T2DM | nk_cell | *PADI2* | 0.85757636 | 0.002755814 | 0.004051029 | 1 |
| T2DM | nk_cell | *SDHB* | -1.555584494 | 0.00010549 | 0.000244931 | 0.511838745 |
| T2DM | nk_cell | *CTBS* | -0.849674032 | 0.016213314 | 0.021039583 | 1 |
| T2DM | nk_cell | *KLHL12* | -1.448496531 | 0.001462134 | 0.002254718 | 1 |
| T2DM | nk_cell | *ZNHIT6* | -1.518982502 | 8.00E-05 | 0.000203408 | 0.38805702 |

| | | | | | | |
|---|---|---|---|---|---|---|
| T2DM | nk_cell | *KIF17* | -2.579551397 | 7.75E-05 | 0.00019931 | 0.375908561 |
| T2DM | nk_cell | *CD46* | -0.970368325 | 0.002496321 | 0.003696109 | 1 |
| T2DM | nk_cell | *EBNA1BP2* | -0.825180131 | 1.22E-05 | 0.000122264 | 0.059324243 |
| T2DM | nk_cell | *ERI3* | -1.293535717 | 3.25E-05 | 0.000127909 | 0.157740365 |
| T2DM | nk_cell | *NSUN4* | -1.856422635 | 0.001050642 | 0.001675777 | 1 |
| T2DM | nk_cell | *PRRC2C* | -1.221044344 | 0.001389824 | 0.002153074 | 1 |
| T2DM | nk_cell | *ABCD3* | -1.341581772 | 2.53E-05 | 0.000122264 | 0.122758955 |
| T2DM | nk_cell | *PTBP2* | -0.90560189 | 0.000615136 | 0.001036693 | 1 |
| T2DM | nk_cell | *DARS2* | -1.485900037 | 0.021991164 | 0.027990852 | 1 |
| T2DM | nk_cell | *SERPINC1* | -1.350329147 | 0.029187986 | 0.036500028 | 1 |
| T2DM | nk_cell | *MTFR1L* | -0.980995536 | 4.36E-05 | 0.000144302 | 0.211668331 |
| T2DM | nk_cell | *RPS6KA1* | 0.381722417 | 0.00015703 | 0.000330833 | 0.761907384 |
| T2DM | nk_cell | *RPA2* | -1.33030774 | 0.037288505 | 0.045919753 | 1 |
| T2DM | nk_cell | *MARC2* | -0.84816857 | 2.02E-05 | 0.000122264 | 0.097930062 |
| T2DM | nk_cell | *SLC5A9* | -0.617256914 | 3.16E-05 | 0.000127109 | 0.153119433 |
| T2DM | nk_cell | *ESYT2* | -1.285008934 | 0.00024025 | 0.000466464 | 1 |
| T2DM | nk_cell | *CD3EAP* | -1.039869799 | 9.07E-06 | 0.000122264 | 0.043987903 |
| T2DM | nk_cell | *RCN2* | -1.366726592 | 5.77E-05 | 0.000165907 | 0.279719573 |
| T2DM | nk_cell | *A4GNT* | -0.948663442 | 0.001767218 | 0.002690474 | 1 |
| T2DM | nk_cell | *IFT46* | -0.920795882 | 1.75E-05 | 0.000122264 | 0.084827042 |
| T2DM | nk_cell | *DDX59* | -1.477258916 | 3.57E-05 | 0.000131704 | 0.173417215 |
| T2DM | nk_cell | *CAMSAP2* | -0.868680617 | 4.08E-05 | 0.000139476 | 0.197916886 |
| T2DM | nk_cell | *KLF7* | -1.090522468 | 0.00010802 | 0.000249102 | 0.524111329 |
| T2DM | nk_cell | *CASC1* | -1.0182767 | 2.29E-05 | 0.000122264 | 0.110880089 |
| T2DM | nk_cell | *LRMP* | 0.555829759 | 0.029678771 | 0.037085088 | 1 |
| T2DM | nk_cell | *CNR1* | -0.565907826 | 0.000189298 | 0.000383567 | 0.918475756 |
| T2DM | nk_cell | *SGIP1* | -2.037598058 | 0.00289063 | 0.004234704 | 1 |
| T2DM | nk_cell | *AKAP7* | -1.383589203 | 4.42E-05 | 0.000145325 | 0.21464446 |
| T2DM | nk_cell | *ALDH8A1* | -0.470754964 | 0.006729898 | 0.009278988 | 1 |
| T2DM | nk_cell | *MED28* | -1.119776828 | 6.86E-06 | 0.000122264 | 0.033305178 |
| T2DM | nk_cell | *FOXO3* | -1.062049867 | 6.93E-05 | 0.000185455 | 0.336230322 |
| T2DM | nk_cell | *SPP1* | 0.867633456 | 0.000207833 | 0.00041362 | 1 |
| T2DM | nk_cell | *MFSD1* | -1.214771977 | 2.02E-05 | 0.000122264 | 0.097960265 |
| T2DM | nk_cell | *UBN1* | -1.518595785 | 0.000120225 | 0.000268413 | 0.58333368 |
| T2DM | nk_cell | *WDR35* | -1.320205415 | 2.31E-05 | 0.000122264 | 0.112306228 |
| T2DM | nk_cell | *DNAH7* | -1.399645774 | 1.20E-05 | 0.000122264 | 0.058461593 |
| T2DM | nk_cell | *C2orf40* | -1.812002772 | 6.25E-05 | 0.000173501 | 0.303453702 |
| T2DM | nk_cell | *CPSF3* | -1.344696937 | 0.000524788 | 0.000904858 | 1 |
| T2DM | nk_cell | *PIGZ* | -0.557863552 | 0.00224487 | 0.003351419 | 1 |
| T2DM | nk_cell | *SENP5* | -1.007421675 | 1.92E-05 | 0.000122264 | 0.093234581 |
| T2DM | nk_cell | *CCDC92* | -1.540833812 | 6.55E-07 | 5.68E-05 | 0.003179123 |
| T2DM | nk_cell | *FKBP15* | -1.069901584 | 9.38E-05 | 0.000225551 | 0.455162836 |
| T2DM | nk_cell | *RAB14* | -1.703506555 | 0.0006038 | 0.001021848 | 1 |
| T2DM | nk_cell | *TRIM32* | 1.670103488 | 0.020598157 | 0.026328308 | 1 |
| T2DM | nk_cell | *PPP6C* | -1.177311651 | 2.12E-05 | 0.000122264 | 0.102637321 |
| T2DM | nk_cell | *HDHD3* | -0.612503006 | 0.000564797 | 0.000963232 | 1 |
| T2DM | nk_cell | *RBM18* | -1.962056846 | 0.004006216 | 0.005735662 | 1 |
| T2DM | nk_cell | *HSDL2* | -0.601390719 | 0.000307669 | 0.000575042 | 1 |
| T2DM | nk_cell | *MAPKAP1* | -1.298310161 | 0.000119829 | 0.000267809 | 0.581412581 |
| T2DM | nk_cell | *INVS* | -0.885116242 | 0.002377721 | 0.00353777 | 1 |

| | | | | | | |
|---|---|---|---|---|---|---|
| T2DM | nk_cell | *DENND1A* | -1.048192077 | 7.61E-05 | 0.000197207 | 0.369369024 |
| T2DM | nk_cell | *KDSR* | -1.125331639 | 1.18E-05 | 0.000122264 | 0.057438704 |
| T2DM | nk_cell | *DCAF4* | -1.204127975 | 1.60E-05 | 0.000122264 | 0.077559273 |
| T2DM | nk_cell | *IFI27L2* | 1.279836982 | 0.014995726 | 0.019579995 | 1 |
| T2DM | nk_cell | *NEK9* | -1.170300932 | 2.01E-05 | 0.000122264 | 0.097613331 |
| T2DM | nk_cell | *TTLL5* | -1.427263147 | 4.61E-05 | 0.000147867 | 0.223739729 |
| T2DM | nk_cell | *FLVCR2* | -1.402232331 | 3.54E-05 | 0.000131318 | 0.171632287 |
| T2DM | nk_cell | *PPP4R4* | -0.70188314 | 0.000315423 | 0.000588377 | 1 |
| T2DM | nk_cell | *ALDH6A1* | -0.807078565 | 0.000600935 | 0.001017355 | 1 |
| T2DM | nk_cell | *COQ6* | -2.258926476 | 0.000410126 | 0.000734021 | 1 |
| T2DM | nk_cell | *DNMT3A* | -1.243774077 | 4.99E-05 | 0.00015287 | 0.241965402 |
| T2DM | nk_cell | *CNRIP1* | -1.373128263 | 0.000800343 | 0.001307936 | 1 |
| T2DM | nk_cell | *CRIPT* | -2.016463357 | 0.001906748 | 0.002885696 | 1 |
| T2DM | nk_cell | *EPCAM* | -1.493660142 | 6.27E-06 | 0.000122264 | 0.030416079 |
| T2DM | nk_cell | *TECTB* | -1.69808096 | 7.31E-06 | 0.000122264 | 0.035492217 |
| T2DM | nk_cell | *ELOVL3* | -1.586697212 | 0.000332276 | 0.000615109 | 1 |
| T2DM | nk_cell | *PPP1R3C* | -0.871972172 | 2.93E-05 | 0.000123467 | 0.142268021 |
| T2DM | nk_cell | *PYROXD2* | -0.772859951 | 6.60E-05 | 0.000179489 | 0.320207975 |
| T2DM | nk_cell | *MXI1* | -1.052856707 | 9.04E-06 | 0.000122264 | 0.043853507 |
| T2DM | nk_cell | *C10orf88* | -1.881579866 | 0.000202189 | 0.000404379 | 0.981023054 |
| T2DM | nk_cell | *HELLS* | 1.078151051 | 0.003459404 | 0.005008962 | 1 |
| T2DM | nk_cell | *TCTN3* | -1.05423288 | 1.42E-05 | 0.000122264 | 0.068957226 |
| T2DM | nk_cell | *WDR11* | -1.213424492 | 2.30E-05 | 0.000122264 | 0.111382686 |
| T2DM | nk_cell | *KANSL1* | -1.101767439 | 0.000491039 | 0.000854256 | 1 |
| T2DM | nk_cell | *HOXB3* | -1.10834613 | 1.83E-05 | 0.000122264 | 0.088811389 |
| T2DM | nk_cell | *CAAP1* | -1.129180376 | 0.014052052 | 0.018407277 | 1 |
| T2DM | nk_cell | *PCDHB10* | -1.868864435 | 9.08E-05 | 0.000220956 | 0.440691067 |
| T2DM | nk_cell | *GORAB* | -1.497073186 | 5.88E-05 | 0.000167252 | 0.285146771 |
| T2DM | nk_cell | *SNX19* | -1.149747903 | 2.40E-05 | 0.000122264 | 0.116672376 |
| T2DM | nk_cell | *SLC10A7* | -1.311590535 | 4.19E-05 | 0.000141779 | 0.203429969 |
| T2DM | nk_cell | *MASTL* | -1.60952005 | 1.60E-06 | 9.70E-05 | 0.007785543 |
| T2DM | nk_cell | *MRC1* | -1.236170124 | 0.000212837 | 0.000422021 | 1 |
| T2DM | nk_cell | *IQSEC3* | -0.648192509 | 2.42E-05 | 0.000122264 | 0.117554682 |
| T2DM | nk_cell | *CCDC77* | -2.531408437 | 3.88E-06 | 0.000122264 | 0.018829704 |
| T2DM | nk_cell | *DNAJC15* | -1.71224165 | 0.000632854 | 0.001062126 | 1 |
| T2DM | nk_cell | *UFM1* | -1.43033611 | 3.59E-05 | 0.000131838 | 0.174290192 |
| T2DM | nk_cell | *ALG5* | -1.494293304 | 3.75E-05 | 0.00013436 | 0.181799238 |
| T2DM | nk_cell | *SIL1* | -0.881677217 | 2.07E-05 | 0.000122264 | 0.100558823 |
| T2DM | nk_cell | *ZFP30* | -1.064449142 | 0.01520879 | 0.019826182 | 1 |
| T2DM | nk_cell | *ARL1* | -1.775427577 | 8.33E-05 | 0.000208835 | 0.404095754 |
| T2DM | nk_cell | *APAF1* | -1.80471394 | 0.000111271 | 0.000255069 | 0.539887177 |
| T2DM | nk_cell | *ADRA1A* | 0.295821822 | 0.032835193 | 0.040714632 | 1 |
| T2DM | nk_cell | *EPHX2* | -1.231260388 | 4.72E-06 | 0.000122264 | 0.02288576 |
| T2DM | nk_cell | *AKAP1* | -0.771290103 | 1.29E-05 | 0.000122264 | 0.062780775 |
| T2DM | nk_cell | *SCPEP1* | -0.866578788 | 0.001706796 | 0.002609128 | 1 |
| T2DM | nk_cell | *TBX2* | -0.505624997 | 0.000805999 | 0.001315407 | 1 |
| T2DM | nk_cell | *TEX14* | -0.694306964 | 0.002848577 | 0.004175619 | 1 |
| T2DM | nk_cell | *ADCY7* | 0.456118543 | 0.000808137 | 0.00131801 | 1 |
| T2DM | nk_cell | *ECHDC2* | -1.069676502 | 1.41E-06 | 9.04E-05 | 0.00683505 |
| T2DM | nk_cell | *ZNF549* | -2.09754647 | 0.000346517 | 0.000636132 | 1 |

| | | | | | | |
|---|---|---|---|---|---|---|
| T2DM | nk_cell | *A1BG* | 1.153576652 | 0.000234785 | 0.000457868 | 1 |
| T2DM | nk_cell | *ZSCAN18* | -1.319496508 | 4.89E-05 | 0.000151564 | 0.237256868 |
| T2DM | nk_cell | *PDZRN3* | -1.081642161 | 4.50E-08 | 1.04E-05 | 0.000218228 |
| T2DM | nk_cell | *LHX4* | -1.172243085 | 1.64E-05 | 0.000122264 | 0.079544194 |
| T2DM | nk_cell | *SEC22A* | -1.214018369 | 7.03E-05 | 0.000187406 | 0.341079527 |
| T2DM | nk_cell | *POPDC2* | -0.91647117 | 1.15E-05 | 0.000122264 | 0.056020205 |
| T2DM | nk_cell | *CD80* | -1.233017389 | 7.45E-07 | 6.24E-05 | 0.003616395 |
| T2DM | nk_cell | *MAPK8IP1* | 0.824432119 | 5.32E-05 | 0.000159179 | 0.258267003 |
| T2DM | nk_cell | *CAT* | -0.717727785 | 3.20E-05 | 0.000127628 | 0.155373513 |
| T2DM | nk_cell | *PILRB* | 0.639010814 | 0.000324479 | 0.000602041 | 1 |
| T2DM | nk_cell | *ZCCHC17* | -1.419129992 | 2.66E-05 | 0.000122391 | 0.128981937 |
| T2DM | nk_cell | *FABP3* | -1.170475638 | 1.47E-05 | 0.000122264 | 0.071424872 |
| T2DM | nk_cell | *TNFSF10* | -1.248788704 | 5.19E-05 | 0.000156496 | 0.251718754 |
| T2DM | nk_cell | *ZNF639* | -1.787552238 | 1.44E-05 | 0.000122264 | 0.070111006 |
| T2DM | nk_cell | *CPXM2* | -0.539067289 | 1.65E-05 | 0.000122264 | 0.080092354 |
| T2DM | nk_cell | *FLT3* | 1.088814983 | 2.74E-05 | 0.000122472 | 0.132821651 |
| T2DM | nk_cell | *MTIF3* | -1.192692711 | 0.000138139 | 0.000298685 | 0.670248719 |
| T2DM | nk_cell | *FMOD* | -0.955211098 | 2.91E-05 | 0.000123467 | 0.141103415 |
| T2DM | nk_cell | *CD244* | 0.116067872 | 5.50E-05 | 0.000161243 | 0.266857739 |
| T2DM | nk_cell | *RBBP6* | -1.305462226 | 3.76E-05 | 0.00013436 | 0.182332277 |
| T2DM | nk_cell | *SERAC1* | 0.955542494 | 0.030819524 | 0.038401728 | 1 |
| T2DM | nk_cell | *LDB3* | -0.533733308 | 4.23E-05 | 0.000141957 | 0.205358828 |
| T2DM | nk_cell | *OPN4* | -3.039436076 | 0.000644073 | 0.001078345 | 1 |
| T2DM | nk_cell | *FAM35A* | -1.265836062 | 0.000142456 | 0.000306518 | 0.691198602 |
| T2DM | nk_cell | *NAA60* | -2.634747956 | 0.006988159 | 0.009586244 | 1 |
| T2DM | nk_cell | *ODF2L* | -1.194845212 | 4.67E-05 | 0.000148657 | 0.226824387 |
| T2DM | nk_cell | *SPATA1* | -1.193136451 | 4.57E-05 | 0.000147697 | 0.221962544 |
| T2DM | nk_cell | *LRRC39* | -0.465494513 | 0.011083767 | 0.014758078 | 1 |
| T2DM | nk_cell | *PMS2* | -1.321568077 | 0.001861948 | 0.002822638 | 1 |
| T2DM | nk_cell | *EEPD1* | -0.825998503 | 3.36E-06 | 0.000122264 | 0.016322116 |
| T2DM | nk_cell | *KLHL7* | -1.177345001 | 2.16E-05 | 0.000122264 | 0.1048238 |
| T2DM | nk_cell | *NT5C3A* | -1.600673168 | 0.000907433 | 0.001468111 | 1 |
| T2DM | nk_cell | *CCZ1* | -0.958729708 | 0.000440323 | 0.000778021 | 1 |
| T2DM | nk_cell | *POLM* | -0.722406146 | 0.000605985 | 0.001023761 | 1 |
| T2DM | nk_cell | *SMU1* | -1.045326794 | 3.76E-05 | 0.00013436 | 0.182282146 |
| T2DM | nk_cell | *GLIPR2* | 0.964674444 | 0.00181552 | 0.002758817 | 1 |
| T2DM | nk_cell | *SLC25A51* | 0.624497671 | 2.78E-05 | 0.000122493 | 0.135035727 |
| T2DM | nk_cell | *TRIM24* | -1.573286444 | 2.36E-05 | 0.000122264 | 0.114564564 |
| T2DM | nk_cell | *CISD1* | -1.072995715 | 3.25E-05 | 0.000127909 | 0.157808083 |
| T2DM | nk_cell | *RBM19* | -1.046099112 | 1.62E-05 | 0.000122264 | 0.078552884 |
| T2DM | nk_cell | *SSPN* | -0.600844195 | 0.000717136 | 0.001187962 | 1 |
| T2DM | nk_cell | *CCDC91* | -1.18961643 | 0.000114448 | 0.000260095 | 0.555302431 |
| T2DM | nk_cell | *EBPL* | -0.966298294 | 0.001225393 | 0.001921036 | 1 |
| T2DM | nk_cell | *ATP7B* | -0.358324865 | 0.007535544 | 0.010261706 | 1 |
| T2DM | nk_cell | *CENPK* | 0.282731733 | 9.27E-05 | 0.000223847 | 0.449872197 |
| T2DM | nk_cell | *ITIH5* | -0.986173273 | 2.33E-05 | 0.000122264 | 0.113051657 |
| T2DM | nk_cell | *TSFM* | -1.044046518 | 0.000567095 | 0.000966021 | 1 |
| T2DM | nk_cell | *ARHGAP9* | 0.304653628 | 0.000101212 | 0.000238183 | 0.491079742 |
| T2DM | nk_cell | *PDE1B* | -1.862760193 | 1.69E-05 | 0.000122264 | 0.082159492 |
| T2DM | nk_cell | *CDK2* | -1.064873291 | 8.03E-06 | 0.000122264 | 0.038950593 |

| | | | | | | |
|---|---|---|---|---|---|---|
| T2DM | nk_cell | *SMUG1* | -0.699924709 | 0.029886201 | 0.037305853 | 1 |
| T2DM | nk_cell | *SARDH* | -0.887311047 | 0.000404077 | 0.000725873 | 1 |
| T2DM | nk_cell | *DBH* | -12.903354 | 2.93E-05 | 0.000123467 | 0.141991218 |
| T2DM | nk_cell | *NDUFAF4* | 0.913786227 | 0.011421757 | 0.015174798 | 1 |
| T2DM | nk_cell | *USP45* | -1.109931503 | 2.44E-05 | 0.000122264 | 0.118367092 |
| T2DM | nk_cell | *TTC21B* | -1.358961866 | 7.77E-06 | 0.000122264 | 0.037676384 |
| T2DM | nk_cell | *ACVR1C* | -0.606917985 | 2.44E-06 | 0.000122264 | 0.011819803 |
| T2DM | nk_cell | *PLA2G12A* | -1.563654108 | 0.00011953 | 0.000267263 | 0.579961151 |
| T2DM | nk_cell | *C4BPA* | -0.78005726 | 3.04E-05 | 0.000125051 | 0.147385129 |
| T2DM | nk_cell | *C4BPB* | -0.765116256 | 3.02E-05 | 0.000124985 | 0.146605274 |
| T2DM | nk_cell | *RAB38* | -0.920140845 | 0.000783177 | 0.001281179 | 1 |
| T2DM | nk_cell | *DAW1* | -1.045616409 | 9.95E-05 | 0.000235619 | 0.482548717 |
| T2DM | nk_cell | *OBSL1* | -1.720159482 | 5.41E-06 | 0.000122264 | 0.026239915 |
| T2DM | nk_cell | *SLC12A4* | -1.868277078 | 4.65E-05 | 0.000148578 | 0.22550424 |
| T2DM | nk_cell | *FAM210B* | -1.508167174 | 0.000107651 | 0.000248764 | 0.522323718 |
| T2DM | nk_cell | *WFDC3* | -0.40474194 | 0.000915187 | 0.001479669 | 1 |
| T2DM | nk_cell | *MATN4* | 1.4728705 | 0.000220163 | 0.00043477 | 1 |
| T2DM | nk_cell | *VAPB* | -1.298051188 | 0.000147307 | 0.000315137 | 0.714731337 |
| T2DM | nk_cell | *SRSF6* | -1.267546365 | 2.99E-05 | 0.000124686 | 0.145258928 |
| T2DM | nk_cell | *CSE1L* | -1.775381389 | 4.67E-05 | 0.000148657 | 0.226613187 |
| T2DM | nk_cell | *PTGIS* | -0.635297964 | 0.000300792 | 0.000564292 | 1 |
| T2DM | nk_cell | *RNF114* | -1.208578964 | 1.24E-05 | 0.000122264 | 0.060180873 |
| T2DM | nk_cell | *NEURL2* | -0.435580049 | 0.012523255 | 0.016516128 | 1 |
| T2DM | nk_cell | *MTRR* | -1.219009753 | 1.92E-05 | 0.000122264 | 0.093064197 |
| T2DM | nk_cell | *PEPD* | -1.138894233 | 1.94E-05 | 0.000122264 | 0.094210071 |
| T2DM | nk_cell | *CHST8* | -0.876711029 | 8.72E-05 | 0.000214956 | 0.423247799 |
| T2DM | nk_cell | *STAMBP* | -1.103267269 | 1.72E-05 | 0.000122264 | 0.083612735 |
| T2DM | nk_cell | *SNRNP27* | -2.115888873 | 0.000467626 | 0.000818514 | 1 |
| T2DM | nk_cell | *ATP8A1* | 0.459838753 | 0.000264288 | 0.000505849 | 1 |
| T2DM | nk_cell | *USP22* | -1.471920417 | 0.000358235 | 0.000654919 | 1 |
| T2DM | nk_cell | *HIF3A* | -0.69213472 | 1.69E-05 | 0.000122264 | 0.081987911 |
| T2DM | nk_cell | *ZNF576* | -0.70300413 | 5.01E-05 | 0.000153185 | 0.242992538 |
| T2DM | nk_cell | *SNRPC* | 1.042713734 | 0.007901865 | 0.010730436 | 1 |
| T2DM | nk_cell | *SERPINB6* | -0.99871255 | 2.11E-05 | 0.000122264 | 0.102451949 |
| T2DM | nk_cell | *PEX6* | -0.691761538 | 0.000293732 | 0.000553901 | 1 |
| T2DM | nk_cell | *NQO2* | -1.165545103 | 1.88E-05 | 0.000122264 | 0.091286057 |
| T2DM | nk_cell | *RPS10* | -1.271055343 | 2.84E-06 | 0.000122264 | 0.013788382 |
| T2DM | nk_cell | *MED20* | -1.397863235 | 0.006855112 | 0.009419711 | 1 |
| T2DM | nk_cell | *GNMT* | 0.802246936 | 3.93E-05 | 0.000136619 | 0.190719459 |
| T2DM | nk_cell | *TREM1* | -0.911071966 | 0.000168119 | 0.000348298 | 0.815714362 |
| T2DM | nk_cell | *MEA1* | 0.592498386 | 0.003762689 | 0.005410956 | 1 |
| T2DM | nk_cell | *KLHL31* | -0.588847042 | 0.008256448 | 0.011180655 | 1 |
| T2DM | nk_cell | *GLO1* | -1.272963106 | 0.006122769 | 0.008497619 | 1 |
| T2DM | nk_cell | *KCNK17* | -2.776557019 | 4.93E-08 | 1.04E-05 | 0.000239242 |
| T2DM | nk_cell | *RREB1* | -0.890572793 | 1.53E-05 | 0.000122264 | 0.074372809 |
| T2DM | nk_cell | *ATXN1* | -1.136792451 | 0.000199554 | 0.000400594 | 0.968235386 |
| T2DM | nk_cell | *CXCL6* | -1.48029725 | 1.42E-05 | 0.000122264 | 0.068669626 |
| T2DM | nk_cell | *SH3TC1* | -1.051140085 | 1.10E-05 | 0.000122264 | 0.053415284 |
| T2DM | nk_cell | *BBS2* | -0.525772186 | 0.003796939 | 0.005455359 | 1 |
| T2DM | nk_cell | *CLYBL* | -1.633187508 | 0.000429625 | 0.000761583 | 1 |

| | | | | | | |
|---|---|---|---|---|---|---|
| T2DM | nk_cell | *TMTC4* | -1.420308955 | 1.40E-05 | 0.000122264 | 0.067822842 |
| T2DM | nk_cell | *ABCC4* | -1.359824479 | 0.011145321 | 0.014835966 | 1 |
| T2DM | nk_cell | *TEKT3* | -0.578171406 | 0.000500369 | 0.000868309 | 1 |
| T2DM | nk_cell | *HS3ST3B1* | 0.202196294 | 6.91E-05 | 0.000185105 | 0.335097268 |
| T2DM | nk_cell | *GGA3* | -1.721680073 | 0.000159678 | 0.000335249 | 0.774759464 |
| T2DM | nk_cell | *NUP85* | -0.916881823 | 2.34E-05 | 0.000122264 | 0.113582673 |
| T2DM | nk_cell | *DDX31* | -1.318670644 | 1.43E-05 | 0.000122264 | 0.069287516 |
| T2DM | nk_cell | *OPRL1* | -1.834389094 | 2.52E-05 | 0.000122264 | 0.122102232 |
| T2DM | nk_cell | *IL1B* | -2.3947965 | 7.58E-05 | 0.000196701 | 0.367958601 |
| T2DM | nk_cell | *PAX8* | -0.617875033 | 0.00051454 | 0.000888451 | 1 |
| T2DM | nk_cell | *CCDC93* | -1.250297676 | 6.62E-05 | 0.00017972 | 0.320979509 |
| T2DM | nk_cell | *PSD4* | 0.326451754 | 9.29E-05 | 0.000223847 | 0.45082717 |
| T2DM | nk_cell | *GTF2F1* | -1.874359126 | 8.19E-05 | 0.000206226 | 0.397398388 |
| T2DM | nk_cell | *TNFSF9* | -11.42733938 | 2.70E-05 | 0.000122391 | 0.13122016 |
| T2DM | nk_cell | *SH2D3A* | -1.770517026 | 3.51E-05 | 0.000130901 | 0.170359223 |
| T2DM | nk_cell | *GPR108* | -1.388155974 | 0.000375403 | 0.000681427 | 1 |
| T2DM | nk_cell | *SNRPD2* | 1.097932982 | 0.031477188 | 0.039180943 | 1 |
| T2DM | nk_cell | *SDCBP2* | 1.058253841 | 0.000141478 | 0.000304955 | 0.686453528 |
| T2DM | nk_cell | *PSMF1* | -1.566353146 | 5.53E-06 | 0.000122264 | 0.026817165 |
| T2DM | nk_cell | *RRBP1* | -1.082424807 | 6.01E-05 | 0.000169557 | 0.291637208 |
| T2DM | nk_cell | *FLRT3* | -1.447996339 | 1.36E-05 | 0.000122264 | 0.065902005 |
| T2DM | nk_cell | *PCSK2* | -4.638400607 | 0.006725864 | 0.009276263 | 1 |
| T2DM | nk_cell | *MKKS* | -1.501724982 | 0.001160393 | 0.001831563 | 1 |
| T2DM | nk_cell | *LAMP5* | 7.508050577 | 0.000177787 | 0.000364744 | 0.862620379 |
| T2DM | nk_cell | *MGME1* | -1.083662038 | 0.000134147 | 0.000292268 | 0.650880357 |
| T2DM | nk_cell | *ITPA* | -1.507953108 | 2.69E-05 | 0.000122391 | 0.130694094 |
| T2DM | nk_cell | *MCM8* | -0.623498197 | 0.022758903 | 0.028884697 | 1 |
| T2DM | nk_cell | *MRPS26* | -1.018135863 | 0.007969954 | 0.010817238 | 1 |
| T2DM | nk_cell | *NCLN* | -1.867599136 | 1.72E-05 | 0.000122264 | 0.083626261 |
| T2DM | nk_cell | *KLC1* | -1.636358414 | 0.000300942 | 0.000564292 | 1 |
| T2DM | nk_cell | *XRCC3* | -1.13059375 | 6.21E-07 | 5.68E-05 | 0.003011679 |
| T2DM | nk_cell | *TUBGCP3* | -1.576584301 | 0.001186639 | 0.001867523 | 1 |
| T2DM | nk_cell | *MCF2L* | -0.593328526 | 0.011475869 | 0.01523001 | 1 |
| T2DM | nk_cell | *TSKS* | -0.636010968 | 2.32E-05 | 0.000122264 | 0.112671901 |
| T2DM | nk_cell | *TRAP1* | -0.509348179 | 0.040184658 | 0.04924879 | 1 |
| T2DM | nk_cell | *ZNF384* | -1.947170392 | 3.95E-05 | 0.000137145 | 0.191866129 |
| T2DM | nk_cell | *EMG1* | -1.717838139 | 0.003401884 | 0.00494042 | 1 |
| T2DM | nk_cell | *ATG14* | -1.018319471 | 0.000206777 | 0.000411856 | 1 |
| T2DM | nk_cell | *KTN1* | -1.221132383 | 8.86E-05 | 0.00021713 | 0.42970087 |
| T2DM | nk_cell | *L3HYPDH* | -0.502573076 | 0.000390474 | 0.000705616 | 1 |
| T2DM | nk_cell | *ZBTB1* | -1.252252783 | 3.54E-05 | 0.000131465 | 0.171956499 |
| T2DM | nk_cell | *RHOT1* | -1.065500097 | 7.53E-06 | 0.000122264 | 0.036528833 |
| T2DM | nk_cell | *WDR60* | -1.140970908 | 2.20E-05 | 0.000122264 | 0.106539909 |
| T2DM | nk_cell | *AIF1L* | -0.72046435 | 2.58E-05 | 0.000122264 | 0.125210776 |
| T2DM | nk_cell | *CANX* | -1.293560524 | 0.011149387 | 0.014837308 | 1 |
| T2DM | nk_cell | *ZNF484* | -2.125288513 | 0.001277557 | 0.001995077 | 1 |
| T2DM | nk_cell | *FGD3* | 0.297121115 | 0.000131419 | 0.000287875 | 0.637644111 |
| T2DM | nk_cell | *ABHD8* | -0.935389602 | 3.59E-07 | 4.05E-05 | 0.001741657 |
| T2DM | nk_cell | *ATP13A4* | 1.543577042 | 0.000177736 | 0.000364744 | 0.86237693 |
| T2DM | nk_cell | *HELB* | 1.046237231 | 0.035200898 | 0.043514588 | 1 |

| | | | | | | |
|---|---|---|---|---|---|---|
| T2DM | nk_cell | *RAB3IP* | 1.124482846 | 0.002471068 | 0.003668794 | 1 |
| T2DM | nk_cell | *YEATS4* | -0.566609958 | 0.00459289 | 0.006502685 | 1 |
| T2DM | nk_cell | *TAS2R4* | -0.692275424 | 9.48E-06 | 0.000122264 | 0.046012224 |
| T2DM | nk_cell | *LRRC61* | -0.9766848 | 0.001254498 | 0.001961594 | 1 |
| T2DM | nk_cell | *IDUA* | -1.289632418 | 4.78E-05 | 0.000149597 | 0.231703169 |
| T2DM | nk_cell | *EMC1* | -1.894448693 | 0.000193541 | 0.0003903 | 0.939062743 |
| T2DM | nk_cell | *PLA2G5* | -0.353625157 | 0.010234788 | 0.013695309 | 1 |
| T2DM | nk_cell | *UBR4* | -1.299835761 | 1.52E-05 | 0.000122264 | 0.073727788 |
| T2DM | nk_cell | *WFIKKN1* | -1.183586987 | 0.035072336 | 0.043377765 | 1 |
| T2DM | nk_cell | *WDR24* | -0.868686543 | 9.57E-06 | 0.000122264 | 0.046415067 |
| T2DM | nk_cell | *SMARCA4* | -1.44327245 | 0.00600445 | 0.008345343 | 1 |
| T2DM | nk_cell | *PNKD* | -1.103763603 | 5.79E-05 | 0.000166463 | 0.281008456 |
| T2DM | nk_cell | *RNF6* | -0.816769808 | 1.94E-05 | 0.000122264 | 0.093999374 |
| T2DM | nk_cell | *HIP1* | -1.267337161 | 0.00038682 | 0.000699599 | 1 |
| T2DM | nk_cell | *PTPN12* | -1.264633467 | 0.004552076 | 0.006454317 | 1 |
| T2DM | nk_cell | *STYXL1* | -0.755984903 | 0.000615356 | 0.001036704 | 1 |
| T2DM | nk_cell | *STEAP4* | -0.832999344 | 3.83E-06 | 0.000122264 | 0.018602308 |
| T2DM | nk_cell | *PEX1* | -1.572164192 | 4.38E-05 | 0.000144434 | 0.21260704 |
| T2DM | nk_cell | *SRD5A3* | -1.149273495 | 0.001716427 | 0.002620548 | 1 |
| T2DM | nk_cell | *KDR* | -1.839600597 | 0.001208731 | 0.001897982 | 1 |
| T2DM | nk_cell | *DGCR6L* | -0.737904329 | 0.000223506 | 0.000440118 | 1 |
| T2DM | nk_cell | *MGAT3* | -1.11028934 | 0.000210553 | 0.00041831 | 1 |
| T2DM | nk_cell | *ATF4* | -1.777199899 | 0.016505266 | 0.02140127 | 1 |
| T2DM | nk_cell | *A4GALT* | -0.861491963 | 7.14E-05 | 0.000189116 | 0.346461297 |
| T2DM | nk_cell | *CDC42EP1* | -2.062322422 | 2.88E-05 | 0.000123207 | 0.139957149 |
| T2DM | nk_cell | *RIBC2* | 0.990853578 | 0.021118852 | 0.026972537 | 1 |
| T2DM | nk_cell | *SPECC1* | -1.469112593 | 0.000497731 | 0.00086404 | 1 |
| T2DM | nk_cell | *CDHR3* | -1.031067743 | 1.82E-05 | 0.000122264 | 0.088257473 |
| T2DM | nk_cell | *PRKRIP1* | -1.461391281 | 4.55E-05 | 0.000147697 | 0.220922731 |
| T2DM | nk_cell | *IRF5* | -0.40176899 | 0.005762553 | 0.008036766 | 1 |
| T2DM | nk_cell | *OPN1SW* | -0.826370584 | 4.57E-07 | 4.72E-05 | 0.002216618 |
| T2DM | nk_cell | *MTX2* | -1.008803876 | 2.01E-05 | 0.000122264 | 0.097431355 |
| T2DM | nk_cell | *GAD1* | -1.451813395 | 1.99E-05 | 0.000122264 | 0.096471541 |
| T2DM | nk_cell | *ORMDL1* | -1.078833489 | 0.000584662 | 0.00099188 | 1 |
| T2DM | nk_cell | *HOXD9* | 0.492041062 | 0.007134599 | 0.009770555 | 1 |
| T2DM | nk_cell | *HOXD10* | -1.429567683 | 4.87E-05 | 0.000151351 | 0.236258213 |
| T2DM | nk_cell | *PSMG2* | -1.130349474 | 0.000296952 | 0.000558671 | 1 |
| T2DM | nk_cell | *TWSG1* | -1.694381903 | 0.000106976 | 0.000247756 | 0.51904782 |
| T2DM | nk_cell | *EIF2AK4* | -1.490214976 | 7.29E-06 | 0.000122264 | 0.035387578 |
| T2DM | nk_cell | *TMOD2* | -1.257457599 | 3.12E-05 | 0.000126743 | 0.15133076 |
| T2DM | nk_cell | *IVD* | -1.291192592 | 0.000107852 | 0.000248889 | 0.523300229 |
| T2DM | nk_cell | *VPS13C* | -1.06730458 | 0.012520108 | 0.016516128 | 1 |
| T2DM | nk_cell | *CALML4* | -1.876811269 | 2.63E-05 | 0.000122295 | 0.12743173 |
| T2DM | nk_cell | *ANAPC13* | -0.711416486 | 0.000229221 | 0.000448641 | 1 |
| T2DM | nk_cell | *BBOX1* | 1.070448123 | 8.63E-05 | 0.000213928 | 0.418870274 |
| T2DM | nk_cell | *DCTD* | -1.79494298 | 0.000649207 | 0.00108619 | 1 |
| T2DM | nk_cell | *RPAIN* | -1.150709517 | 1.85E-06 | 0.000107137 | 0.008999541 |
| T2DM | nk_cell | *USP6* | -0.634019088 | 0.002723942 | 0.004006235 | 1 |
| T2DM | nk_cell | *PLD2* | -2.140214428 | 2.75E-05 | 0.000122472 | 0.133190493 |
| T2DM | nk_cell | *PHF20L1* | -1.530868753 | 0.000572573 | 0.000972732 | 1 |

| | | | | | | |
|---|---|---|---|---|---|---|
| T2DM | nk_cell | *PUS7L* | -1.550795642 | 0.000693537 | 0.001153992 | 1 |
| T2DM | nk_cell | *AP1M2* | -1.052158815 | 0.001707456 | 0.002609316 | 1 |
| T2DM | nk_cell | *MTUS1* | -0.957380938 | 4.87E-05 | 0.000151402 | 0.236512578 |
| T2DM | nk_cell | *KLK10* | -0.665771821 | 0.013182494 | 0.017324339 | 1 |
| T2DM | nk_cell | *KLK8* | 1.432730792 | 6.85E-05 | 0.00018443 | 0.332193017 |
| T2DM | nk_cell | *RIPK3* | 0.651345802 | 0.030598827 | 0.038146328 | 1 |
| T2DM | nk_cell | *ADCY4* | -1.222424572 | 0.000514183 | 0.000888152 | 1 |
| T2DM | nk_cell | *BCL2L2* | -1.469069702 | 8.10E-05 | 0.00020495 | 0.393125263 |
| T2DM | nk_cell | *DTD2* | -0.593687514 | 0.00040567 | 0.000727905 | 1 |
| T2DM | nk_cell | *MIS18BP1* | -0.975778477 | 7.64E-05 | 0.000197624 | 0.37074228 |
| T2DM | nk_cell | *RNASE1* | -1.063371175 | 0.000563272 | 0.000960969 | 1 |
| T2DM | nk_cell | *EPB41L4A* | -0.907935901 | 1.71E-05 | 0.000122264 | 0.08312797 |
| T2DM | nk_cell | *CDO1* | -0.853023252 | 6.47E-05 | 0.000176937 | 0.313886171 |
| T2DM | nk_cell | *REEP5* | -1.318860847 | 4.00E-05 | 0.000138284 | 0.193994152 |
| T2DM | nk_cell | *RHBDF2* | 0.800018228 | 0.000610852 | 0.001030547 | 1 |
| T2DM | nk_cell | *TMEM8A* | -2.332405315 | 0.007144697 | 0.009781622 | 1 |
| T2DM | nk_cell | *MAU2* | -1.417290688 | 1.49E-05 | 0.000122264 | 0.072155636 |
| T2DM | nk_cell | *SHC2* | -0.845914606 | 0.00131033 | 0.002039692 | 1 |
| T2DM | nk_cell | *ERMARD* | -1.272662301 | 3.21E-05 | 0.000127628 | 0.155923011 |
| T2DM | nk_cell | *PHF10* | -1.228444703 | 1.50E-05 | 0.000122264 | 0.072913135 |
| T2DM | nk_cell | *GALNT8* | -0.847981482 | 1.75E-05 | 0.000122264 | 0.084733732 |
| T2DM | nk_cell | *PRKCSH* | 1.18632796 | 0.007529619 | 0.010259396 | 1 |
| T2DM | nk_cell | *CDC16* | -1.163517585 | 0.002695783 | 0.003968429 | 1 |
| T2DM | nk_cell | *XPO7* | -1.614329739 | 9.28E-05 | 0.000223847 | 0.450487488 |
| T2DM | nk_cell | *ATP8B3* | -0.804374492 | 2.23E-05 | 0.000122264 | 0.108128457 |
| T2DM | nk_cell | *PLVAP* | -0.566090387 | 0.017216522 | 0.022269945 | 1 |
| T2DM | nk_cell | *DDA1* | -2.145757175 | 0.009750913 | 0.013087533 | 1 |
| T2DM | nk_cell | *MRPL34* | -0.698741192 | 0.000203933 | 0.000407195 | 0.98948364 |
| T2DM | nk_cell | *TULP4* | -0.96961674 | 8.49E-05 | 0.00021115 | 0.411864979 |
| T2DM | nk_cell | *QRSL1* | -0.791218282 | 0.000257902 | 0.000495384 | 1 |
| T2DM | nk_cell | *RSPH3* | -0.605178407 | 0.001185249 | 0.001865941 | 1 |
| T2DM | nk_cell | *STK33* | -0.566016812 | 0.001869107 | 0.002831379 | 1 |
| T2DM | nk_cell | *NDUFA10* | -0.805186587 | 0.001362582 | 0.002116955 | 1 |
| T2DM | nk_cell | *MAP1S* | -1.900180893 | 1.71E-05 | 0.000122264 | 0.082844859 |
| T2DM | nk_cell | *SSBP4* | -1.613919392 | 0.001683871 | 0.002578145 | 1 |
| T2DM | nk_cell | *PGPEP1* | 1.02505868 | 0.006142883 | 0.008520659 | 1 |
| T2DM | nk_cell | *KIAA1683* | -1.194664804 | 0.000556821 | 0.000951636 | 1 |
| T2DM | nk_cell | *TRPM4* | -0.879747166 | 4.21E-05 | 0.000141813 | 0.204298975 |
| T2DM | nk_cell | *ATXN10* | -1.130795179 | 3.28E-05 | 0.000127909 | 0.158954529 |
| T2DM | nk_cell | *CYP2E1* | -0.553986079 | 0.021606508 | 0.027551847 | 1 |
| T2DM | nk_cell | *PNPLA7* | -0.920432587 | 1.59E-05 | 0.000122264 | 0.077187842 |
| T2DM | nk_cell | *PAK4* | -1.945874763 | 0.003009512 | 0.004399564 | 1 |
| T2DM | nk_cell | *ZNF337* | -1.081563174 | 3.19E-05 | 0.000127545 | 0.154712226 |
| T2DM | nk_cell | *CEP85* | -0.964982551 | 1.43E-05 | 0.000122264 | 0.06937938 |
| T2DM | nk_cell | *POMT1* | -1.363377022 | 6.29E-05 | 0.000173739 | 0.305362631 |
| T2DM | nk_cell | *CHMP2A* | -1.380533851 | 0.010902902 | 0.014525228 | 1 |
| T2DM | nk_cell | *MAP3K10* | -1.115837086 | 1.68E-05 | 0.000122264 | 0.081562308 |
| T2DM | nk_cell | *LRRC47* | -0.953173431 | 1.44E-05 | 0.000122264 | 0.069941213 |
| T2DM | nk_cell | *THEMIS2* | 0.675210125 | 3.59E-05 | 0.000131838 | 0.174191225 |
| T2DM | nk_cell | *ZNF317* | -1.691587783 | 0.000429034 | 0.000760845 | 1 |

| | | | | | | |
|---|---|---|---|---|---|---|
| T2DM | nk_cell | *ANGPTL6* | 0.771493473 | 0.000555705 | 0.000950399 | 1 |
| T2DM | nk_cell | *DNMT1* | 0.932430786 | 0.001986514 | 0.002996135 | 1 |
| T2DM | nk_cell | *NOL11* | -1.677127702 | 0.000238167 | 0.000463029 | 1 |
| T2DM | nk_cell | *PKDREJ* | -1.745296317 | 7.88E-05 | 0.000201551 | 0.382543347 |
| T2DM | nk_cell | *HSD17B3* | -3.556370717 | 8.44E-05 | 0.000210701 | 0.409743586 |
| T2DM | nk_cell | *SLC35D2* | -1.570241855 | 4.44E-05 | 0.000145721 | 0.21552139 |
| T2DM | nk_cell | *ULBP2* | -5.095279923 | 0.000774469 | 0.001267787 | 1 |
| T2DM | nk_cell | *TTLL9* | -1.25979129 | 2.71E-05 | 0.000122391 | 0.131315607 |
| T2DM | nk_cell | *ATP6V1E1* | -1.826042078 | 0.000199949 | 0.000400889 | 0.970151034 |
| T2DM | nk_cell | *IDO1* | 2.254404808 | 0.001076761 | 0.001712932 | 1 |
| T2DM | nk_cell | *HAUS8* | -1.501857596 | 0.0086723 | 0.011720892 | 1 |
| T2DM | nk_cell | *MRPS25* | -1.019150238 | 6.97E-06 | 0.000122264 | 0.033821202 |
| T2DM | nk_cell | *GALNT15* | -0.617868736 | 1.97E-05 | 0.000122264 | 0.095351833 |
| T2DM | nk_cell | *KCNC3* | -1.241717623 | 4.93E-05 | 0.00015216 | 0.239000771 |
| T2DM | nk_cell | *KIF3A* | -1.737522366 | 0.000130867 | 0.000287056 | 0.634968842 |
| T2DM | nk_cell | *NDUFA2* | -1.258975887 | 0.000199831 | 0.000400818 | 0.969579351 |
| T2DM | nk_cell | *EXOC4* | -0.919523268 | 0.000372382 | 0.000676956 | 1 |
| T2DM | nk_cell | *PPFIA1* | -1.025555397 | 2.21E-05 | 0.000122264 | 0.107224156 |
| T2DM | nk_cell | *BARX1* | -2.264039579 | 1.12E-05 | 0.000122264 | 0.054230122 |
| T2DM | nk_cell | *NINJ1* | -0.625378787 | 6.78E-05 | 0.000183006 | 0.328861938 |
| T2DM | nk_cell | *NPHP4* | -0.627363045 | 5.89E-05 | 0.000167252 | 0.285558735 |
| T2DM | nk_cell | *MAP1B* | -0.802271065 | 0.000344552 | 0.000633004 | 1 |
| T2DM | nk_cell | *CKMT2* | -1.260418658 | 0.005547046 | 0.007754038 | 1 |
| T2DM | nk_cell | *KHDRBS3* | -1.234351242 | 1.73E-05 | 0.000122264 | 0.083951032 |
| T2DM | nk_cell | *CHD1L* | -1.539627875 | 0.000635985 | 0.001066274 | 1 |
| T2DM | nk_cell | *ZNF304* | -0.868270767 | 0.002483028 | 0.003682045 | 1 |
| T2DM | nk_cell | *ZNF132* | -0.689168373 | 0.020779358 | 0.026552922 | 1 |
| T2DM | nk_cell | *MATN3* | -1.233340562 | 4.04E-05 | 0.000138868 | 0.195807318 |
| T2DM | nk_cell | *TRIM21* | 1.117731193 | 0.000341885 | 0.000628934 | 1 |
| T2DM | nk_cell | *SPATA6* | -0.71255412 | 1.50E-05 | 0.000122264 | 0.072733377 |
| T2DM | nk_cell | *LRRC41* | -1.478822274 | 1.29E-05 | 0.000122264 | 0.062447326 |
| T2DM | nk_cell | *GAS2L2* | 2.388772446 | 2.58E-05 | 0.000122264 | 0.125402949 |
| T2DM | nk_cell | *CCT6B* | -1.061361436 | 0.002115019 | 0.003171222 | 1 |
| T2DM | nk_cell | *TRIM22* | -1.25928564 | 0.035407522 | 0.043758863 | 1 |
| T2DM | nk_cell | *IMMT* | -1.058595517 | 0.035846782 | 0.044256638 | 1 |
| T2DM | nk_cell | *CARD6* | -1.195222973 | 0.006196279 | 0.008589813 | 1 |
| T2DM | nk_cell | *MYBBP1A* | -0.83414245 | 0.006042533 | 0.008393464 | 1 |
| T2DM | nk_cell | *RPA1* | -1.282439066 | 0.003178767 | 0.004633037 | 1 |
| T2DM | nk_cell | *SERPINF1* | -1.631082767 | 8.67E-05 | 0.000213966 | 0.420871673 |
| T2DM | nk_cell | *TMEM128* | -1.337995106 | 4.00E-05 | 0.000138284 | 0.194131404 |
| T2DM | nk_cell | *FIGNL1* | -1.294668853 | 4.04E-05 | 0.000138934 | 0.19617436 |
| T2DM | nk_cell | *DDC* | -2.069892722 | 0.020174331 | 0.025841038 | 1 |
| T2DM | nk_cell | *EIF5A* | 0.623057556 | 0.003215931 | 0.004685794 | 1 |
| T2DM | nk_cell | *RGS22* | -0.858533551 | 0.000232137 | 0.00045325 | 1 |
| T2DM | nk_cell | *REEP2* | -1.576913636 | 0.000120266 | 0.000268413 | 0.583530008 |
| T2DM | nk_cell | *PRMT7* | -1.122582215 | 0.000268334 | 0.00051238 | 1 |
| T2DM | nk_cell | *IGHMBP2* | -1.078790128 | 2.72E-05 | 0.000122437 | 0.132211006 |
| T2DM | nk_cell | *MUTYH* | 1.27195517 | 0.005307482 | 0.007442746 | 1 |
| T2DM | nk_cell | *OSER1* | -1.208268424 | 0.031525057 | 0.039230463 | 1 |
| T2DM | nk_cell | *DMGDH* | -1.543228082 | 0.000162635 | 0.000339693 | 0.789106864 |

| | | | | | | |
|---|---|---|---|---|---|---|
| T2DM | nk_cell | *BHMT2* | -0.414098712 | 0.001171723 | 0.001848845 | 1 |
| T2DM | nk_cell | *AP3B1* | -0.94199952 | 0.037816923 | 0.046546856 | 1 |
| T2DM | nk_cell | *ZBED3* | -1.005301345 | 5.49E-05 | 0.000161068 | 0.266219619 |
| T2DM | nk_cell | *FBXO44* | -0.901276311 | 8.17E-05 | 0.000205997 | 0.396509484 |
| T2DM | nk_cell | *RSG1* | -2.581947055 | 0.004024901 | 0.00575562 | 1 |
| T2DM | nk_cell | *ATP8A2* | -2.103361436 | 0.007407604 | 0.010110181 | 1 |
| T2DM | nk_cell | *ALOX5AP* | 0.471104537 | 0.000105097 | 0.000244455 | 0.50993283 |
| T2DM | nk_cell | *WASF3* | -1.041082234 | 1.71E-05 | 0.000122264 | 0.083008012 |
| T2DM | nk_cell | *PEMT* | -1.938688503 | 2.72E-05 | 0.000122437 | 0.132062255 |
| T2DM | nk_cell | *MPRIP* | -1.512328057 | 0.000505918 | 0.00087637 | 1 |
| T2DM | nk_cell | *DSTYK* | -1.624436012 | 9.27E-05 | 0.000223847 | 0.44980739 |
| T2DM | nk_cell | *CCNA1* | -5.492621172 | 0.000610345 | 0.00103005 | 1 |
| T2DM | nk_cell | *COG6* | -1.360058549 | 0.000190585 | 0.00038546 | 0.924718938 |
| T2DM | nk_cell | *EPSTI1* | -1.394468447 | 0.000131587 | 0.000288019 | 0.63845788 |
| T2DM | nk_cell | *BTBD2* | -1.346796988 | 0.0001214 | 0.000270198 | 0.589030679 |
| T2DM | nk_cell | *CNDP2* | -1.660091759 | 4.28E-05 | 0.000142857 | 0.207856473 |
| T2DM | nk_cell | *PDZD2* | -0.877962566 | 2.32E-05 | 0.000122264 | 0.112374543 |
| T2DM | nk_cell | *GSTT2B* | -0.334319553 | 0.033823148 | 0.041886145 | 1 |
| T2DM | nk_cell | *SLC2A11* | 1.078893128 | 0.000898283 | 0.001454762 | 1 |
| T2DM | nk_cell | *GIMAP4* | 0.416132685 | 0.000258859 | 0.000496632 | 1 |
| T2DM | nk_cell | *ADCK2* | -1.088626438 | 0.000322082 | 0.000598064 | 1 |
| T2DM | nk_cell | *SFTPD* | -0.277208501 | 0.011835987 | 0.015682198 | 1 |
| T2DM | nk_cell | *IPO8* | -1.710205029 | 6.38E-05 | 0.000175261 | 0.309685802 |
| T2DM | nk_cell | *LARS* | -0.841370163 | 0.000152478 | 0.000323491 | 0.739824363 |
| T2DM | nk_cell | *SPINK5* | -0.905477988 | 3.42E-05 | 0.000129717 | 0.166145301 |
| T2DM | nk_cell | *IMPA1* | -0.979076308 | 0.0072593 | 0.009918931 | 1 |
| T2DM | nk_cell | *CATSPERB* | -1.43998346 | 0.000254741 | 0.00048999 | 1 |
| T2DM | nk_cell | *COX16* | 0.496102423 | 6.29E-05 | 0.000173739 | 0.305315801 |
| T2DM | nk_cell | *TTC9* | 1.755685071 | 0.009408009 | 0.01265179 | 1 |
| T2DM | nk_cell | *ELP3* | -1.708954842 | 0.000122092 | 0.000270992 | 0.592388337 |
| T2DM | nk_cell | *PEBP4* | -0.876626404 | 1.80E-05 | 0.000122264 | 0.087183301 |
| T2DM | nk_cell | *CTIF* | -0.72167844 | 1.71E-05 | 0.000122264 | 0.083152545 |
| T2DM | nk_cell | *MRO* | -2.047063864 | 0.010047861 | 0.013456314 | 1 |
| T2DM | nk_cell | *THUMPD3* | -2.14475441 | 2.89E-05 | 0.000123207 | 0.140246211 |
| T2DM | nk_cell | *CNTN6* | -1.142269033 | 3.21E-05 | 0.000127628 | 0.155564776 |
| T2DM | nk_cell | *CHL1* | -0.719269118 | 2.20E-05 | 0.000122264 | 0.106716545 |
| T2DM | nk_cell | *GSTM5* | -0.705615268 | 0.000167356 | 0.000347161 | 0.812010223 |
| T2DM | nk_cell | *GSTM3* | -0.391005443 | 0.013123273 | 0.017251184 | 1 |
| T2DM | nk_cell | *VAV3* | 0.407671796 | 0.001540758 | 0.002368927 | 1 |
| T2DM | nk_cell | *PSRC1* | -1.372262993 | 0.000992603 | 0.001594738 | 1 |
| T2DM | nk_cell | *TRIM45* | -2.568340381 | 7.89E-05 | 0.000201568 | 0.382978365 |
| T2DM | nk_cell | *CEPT1* | -1.000427344 | 2.13E-05 | 0.000122264 | 0.10344212 |
| T2DM | nk_cell | *NAPG* | -1.267419762 | 2.73E-05 | 0.000122437 | 0.132353888 |
| T2DM | nk_cell | *PPHLN1* | -1.04232759 | 4.14E-05 | 0.000140573 | 0.200659395 |
| T2DM | nk_cell | *FKBP11* | 1.373371845 | 0.009731458 | 0.013065035 | 1 |
| T2DM | nk_cell | *YWHAQ* | -1.200819633 | 3.10E-05 | 0.000126367 | 0.150442342 |
| T2DM | nk_cell | *KIDINS220* | -1.213168434 | 5.28E-05 | 0.00015824 | 0.256189938 |
| T2DM | nk_cell | *LPIN1* | -1.457057333 | 0.000504421 | 0.000874403 | 1 |
| T2DM | nk_cell | *KIAA1468* | -1.451215063 | 0.000838367 | 0.001364009 | 1 |
| T2DM | nk_cell | *IL15RA* | -1.511899462 | 8.18E-05 | 0.000206154 | 0.397052171 |

| | | | | | | |
|---|---|---|---|---|---|---|
| T2DM | nk_cell | *TMEM241* | -0.672608328 | 3.72E-05 | 0.000134067 | 0.180549033 |
| T2DM | nk_cell | *SOX5* | -0.702907106 | 4.39E-05 | 0.000144631 | 0.21318609 |
| T2DM | nk_cell | *KLRD1* | 0.190490638 | 8.14E-05 | 0.000205476 | 0.394924851 |
| T2DM | nk_cell | *HOOK1* | -0.654409742 | 0.000201877 | 0.00040392 | 0.979506183 |
| T2DM | nk_cell | *ZCCHC11* | -1.061395402 | 9.86E-05 | 0.00023397 | 0.478475067 |
| T2DM | nk_cell | *ELP2* | -1.395743802 | 5.34E-05 | 0.000159458 | 0.259119827 |
| T2DM | nk_cell | *TIMM10* | -2.759459934 | 0.000956116 | 0.001541221 | 1 |
| T2DM | nk_cell | *DHX34* | -1.830949246 | 5.64E-05 | 0.00016385 | 0.273861709 |
| T2DM | nk_cell | *FADS2* | -0.85248207 | 1.92E-05 | 0.000122264 | 0.092983047 |
| T2DM | nk_cell | *CLOCK* | -1.336519238 | 7.22E-05 | 0.000190817 | 0.350340593 |
| T2DM | nk_cell | *GGACT* | -1.111385377 | 2.75E-05 | 0.000122472 | 0.133269935 |
| T2DM | nk_cell | *CLDN10* | -1.064445525 | 0.000475665 | 0.000831685 | 1 |
| T2DM | nk_cell | *ARGLU1* | -1.122420472 | 0.00015717 | 0.000330841 | 0.762588456 |
| T2DM | nk_cell | *ERCC5* | -1.985090487 | 1.48E-05 | 0.000122264 | 0.071637301 |
| T2DM | nk_cell | *CARS2* | -1.44668678 | 3.25E-05 | 0.000127909 | 0.15789496 |
| T2DM | nk_cell | *KLB* | -0.562237495 | 1.27E-06 | 8.96E-05 | 0.006179382 |
| T2DM | nk_cell | *WDR36* | -1.464035342 | 2.66E-05 | 0.000122391 | 0.129172508 |
| T2DM | nk_cell | *OSTF1* | -1.337604222 | 0.018891766 | 0.024294421 | 1 |
| T2DM | nk_cell | *UBQLN1* | -1.439138243 | 8.64E-05 | 0.000213956 | 0.419434943 |
| T2DM | nk_cell | *GOLM1* | -0.987678697 | 0.001586461 | 0.00243438 | 1 |
| T2DM | nk_cell | *USP30* | -0.695003009 | 0.000491974 | 0.00085527 | 1 |
| T2DM | nk_cell | *MSI1* | 1.505009616 | 0.031568656 | 0.039274646 | 1 |
| T2DM | nk_cell | *DTX1* | -0.645805868 | 1.95E-05 | 0.000122264 | 0.094715306 |
| T2DM | nk_cell | *DMTF1* | -1.152886694 | 4.21E-05 | 0.000141813 | 0.204417177 |
| T2DM | nk_cell | *TMEM60* | 1.511721415 | 0.025003686 | 0.031535712 | 1 |
| T2DM | nk_cell | *SRPK2* | -0.999313814 | 0.017027423 | 0.022042971 | 1 |
| T2DM | nk_cell | *TES* | 0.697255058 | 0.001901991 | 0.002879395 | 1 |
| T2DM | nk_cell | *MTO1* | -1.153046587 | 4.97E-05 | 0.000152658 | 0.24104693 |
| T2DM | nk_cell | *NT5E* | -0.780960271 | 0.001546383 | 0.002376639 | 1 |
| T2DM | nk_cell | *AKIRIN2* | -1.458894431 | 8.83E-05 | 0.0002169 | 0.428619637 |
| T2DM | nk_cell | *MAP3K7* | -1.051050576 | 2.02E-05 | 0.000122264 | 0.09791005 |
| T2DM | nk_cell | *PRR5L* | 0.661420813 | 0.000743146 | 0.001223945 | 1 |
| T2DM | nk_cell | *PHF21A* | -1.219413325 | 0.000118824 | 0.000266052 | 0.576534378 |
| T2DM | nk_cell | *CAPRIN1* | -1.460358337 | 0.001693367 | 0.002591049 | 1 |
| T2DM | nk_cell | *B4GALNT1* | -2.526735416 | 7.04E-05 | 0.000187526 | 0.341485732 |
| T2DM | nk_cell | *FAIM2* | -1.068934049 | 8.09E-06 | 0.000122264 | 0.039247928 |
| T2DM | nk_cell | *PAN2* | -1.361187576 | 0.029277269 | 0.036602244 | 1 |
| T2DM | nk_cell | *KRT7* | -0.740501244 | 0.000326585 | 0.000605036 | 1 |
| T2DM | nk_cell | *SLC26A10* | -1.398479877 | 2.21E-05 | 0.000122264 | 0.107127425 |
| T2DM | nk_cell | *MAP7* | -1.154932706 | 0.001127506 | 0.001786046 | 1 |
| T2DM | nk_cell | *NHSL1* | -0.775357534 | 0.000151138 | 0.000321773 | 0.733320521 |
| T2DM | nk_cell | *AHI1* | -0.745006861 | 0.000361527 | 0.000660067 | 1 |
| T2DM | nk_cell | *SMPD2* | -0.683586635 | 9.50E-05 | 0.000227802 | 0.460843247 |
| T2DM | nk_cell | *SMYD5* | -1.359071408 | 1.47E-05 | 0.000122264 | 0.071480781 |
| T2DM | nk_cell | *USP15* | -1.228557642 | 0.002054902 | 0.003085851 | 1 |
| T2DM | nk_cell | *MDM2* | -1.372714248 | 0.00010008 | 0.000236412 | 0.485589956 |
| T2DM | nk_cell | *KLHL36* | -1.405224994 | 0.008827186 | 0.011923582 | 1 |
| T2DM | nk_cell | *MPHOSPH6* | 1.159582569 | 0.006106994 | 0.00847815 | 1 |
| T2DM | nk_cell | *KCNK1* | -0.571081845 | 1.53E-05 | 0.000122264 | 0.074309874 |
| T2DM | nk_cell | *CAPN9* | 1.837469558 | 0.004229654 | 0.006021797 | 1 |

| | | | | | | |
|---|---|---|---|---|---|---|
| T2DM | nk_cell | *NTPCR* | -1.648261119 | 4.72E-05 | 0.000149114 | 0.228875603 |
| T2DM | nk_cell | *PIGC* | -1.539519551 | 6.29E-05 | 0.000173739 | 0.305433099 |
| T2DM | nk_cell | *MRPL44* | 1.092805697 | 0.000489002 | 0.000851018 | 1 |
| T2DM | nk_cell | *USP37* | -1.244576131 | 0.000103886 | 0.000242428 | 0.504054022 |
| T2DM | nk_cell | *SLC19A3* | -0.486009358 | 2.31E-05 | 0.000122264 | 0.112312443 |
| T2DM | nk_cell | *TMBIM1* | -1.090943506 | 1.67E-05 | 0.000122264 | 0.081234148 |
| T2DM | nk_cell | *TSGA10* | -0.535053346 | 0.000299911 | 0.000563145 | 1 |
| T2DM | nk_cell | *GCC2* | -1.258253743 | 4.58E-05 | 0.000147697 | 0.222139317 |
| T2DM | nk_cell | *ANKRD36* | -1.445106739 | 7.59E-05 | 0.000196701 | 0.368028212 |
| T2DM | nk_cell | *EPC2* | -1.211447627 | 4.68E-05 | 0.000148778 | 0.227186761 |
| T2DM | nk_cell | *STAB2* | -1.417070826 | 1.75E-05 | 0.000122264 | 0.084955188 |
| T2DM | nk_cell | *APPL2* | -1.239957056 | 3.31E-05 | 0.000127948 | 0.160576804 |
| T2DM | nk_cell | *DRAM1* | -1.223783653 | 2.38E-05 | 0.000122264 | 0.115635585 |
| T2DM | nk_cell | *SLC41A2* | -0.793669868 | 4.34E-05 | 0.000144292 | 0.210665859 |
| T2DM | nk_cell | *NEK3* | -0.877764017 | 0.000335829 | 0.000619796 | 1 |
| T2DM | nk_cell | *RNASEH2B* | -1.028352341 | 0.038155819 | 0.046928272 | 1 |
| T2DM | nk_cell | *TBC1D4* | -0.887171937 | 7.93E-06 | 0.000122264 | 0.038485071 |
| T2DM | nk_cell | *SUCLA2* | -0.967765544 | 1.68E-05 | 0.000122264 | 0.081704012 |
| T2DM | nk_cell | *RCBTB1* | -1.369753418 | 0.001801291 | 0.002738053 | 1 |
| T2DM | nk_cell | *SETDB2* | -1.699777671 | 0.003448064 | 0.004995749 | 1 |
| T2DM | nk_cell | *SCRN1* | -1.797675526 | 5.06E-05 | 0.000153902 | 0.24562794 |
| T2DM | nk_cell | *C7orf25* | -2.979903473 | 3.30E-05 | 0.000127909 | 0.160073083 |
| T2DM | nk_cell | *CHST12* | 0.722346003 | 0.000998078 | 0.001602474 | 1 |
| T2DM | nk_cell | *KDELR2* | -1.804885858 | 0.01136859 | 0.015108299 | 1 |
| T2DM | nk_cell | *NUPL2* | -1.504891535 | 4.61E-05 | 0.000147867 | 0.223460791 |
| T2DM | nk_cell | *AOAH* | 0.134615127 | 8.58E-05 | 0.000212706 | 0.416204598 |
| T2DM | nk_cell | *TBRG4* | -2.882003213 | 0.000559341 | 0.000955606 | 1 |
| T2DM | nk_cell | *MTHFS* | -1.005230938 | 0.014384195 | 0.018816962 | 1 |
| T2DM | nk_cell | *NMT1* | -0.6835801 | 0.01761976 | 0.022761202 | 1 |
| T2DM | nk_cell | *BRIP1* | 0.552977701 | 0.000415472 | 0.000741401 | 1 |
| T2DM | nk_cell | *RTP4* | -1.636195912 | 1.64E-06 | 9.70E-05 | 0.007937039 |
| T2DM | nk_cell | *NDUFB5* | -1.753176652 | 4.69E-05 | 0.000148778 | 0.227476948 |
| T2DM | nk_cell | *SCN2A* | -1.032126518 | 0.000341947 | 0.000628934 | 1 |
| T2DM | nk_cell | *RPS6KC1* | -1.202062458 | 0.000437847 | 0.000774211 | 1 |
| T2DM | nk_cell | *SMPD4* | -1.423840761 | 0.004034029 | 0.005766973 | 1 |
| T2DM | nk_cell | *CCDC115* | 0.754204121 | 0.000968693 | 0.001560976 | 1 |
| T2DM | nk_cell | *IMP4* | -0.794691906 | 1.63E-05 | 0.000122264 | 0.079075496 |
| T2DM | nk_cell | *NIPSNAP3A* | -1.022311429 | 1.94E-06 | 0.00010951 | 0.009417848 |
| T2DM | nk_cell | *ODF2* | -1.876977279 | 2.64E-05 | 0.000122391 | 0.128270239 |
| T2DM | nk_cell | *TOR1B* | -1.801363863 | 1.73E-05 | 0.000122264 | 0.084180102 |
| T2DM | nk_cell | *SLC2A8* | -0.862923723 | 1.29E-05 | 0.000122264 | 0.062648798 |
| T2DM | nk_cell | *ZNF189* | -2.167780743 | 0.000685453 | 0.001141323 | 1 |
| T2DM | nk_cell | *STX17* | -1.154309848 | 1.92E-05 | 0.000122264 | 0.093204448 |
| T2DM | nk_cell | *ANP32B* | -1.629202847 | 6.48E-06 | 0.000122264 | 0.031442611 |
| T2DM | nk_cell | *DERL1* | -1.768573259 | 0.005191198 | 0.00729655 | 1 |
| T2DM | nk_cell | *NOV* | -1.062690924 | 0.00021235 | 0.000421399 | 1 |
| T2DM | nk_cell | *IL11RA* | -1.423331827 | 0.000584242 | 0.000991516 | 1 |
| T2DM | nk_cell | *APTX* | -1.582877948 | 1.23E-05 | 0.000122264 | 0.059879509 |
| T2DM | nk_cell | *TMEM8B* | -1.027439724 | 0.000248296 | 0.000479401 | 1 |
| T2DM | nk_cell | *GRHPR* | -1.154927656 | 1.97E-05 | 0.000122264 | 0.095498749 |

| | | | | | | |
|---|---|---|---|---|---|---|
| T2DM | nk_cell | *DENND4C* | -1.090765688 | 2.31E-05 | 0.000122264 | 0.111903977 |
| T2DM | nk_cell | *GMPR* | -0.764972624 | 9.08E-06 | 0.000122264 | 0.044034514 |
| T2DM | nk_cell | *TMEM14B* | -0.839357203 | 2.06E-05 | 0.000122264 | 0.099872162 |
| T2DM | nk_cell | *FRS3* | -1.642618066 | 2.47E-05 | 0.000122264 | 0.120073065 |
| T2DM | nk_cell | *CAPN11* | -0.588101512 | 0.03948845 | 0.04844449 | 1 |
| T2DM | nk_cell | *IRF4* | 0.722812563 | 0.000271805 | 0.000517791 | 1 |
| T2DM | nk_cell | *LRRC1* | -0.762035221 | 0.000143864 | 0.000309273 | 0.698028906 |
| T2DM | nk_cell | *RIPK1* | -1.241123834 | 0.000113673 | 0.000258939 | 0.551540505 |
| T2DM | nk_cell | *TCF19* | 2.207307641 | 2.24E-10 | 1.55E-07 | 1.09E-06 |
| T2DM | nk_cell | *NRM* | 4.051884303 | 0.002621612 | 0.003868631 | 1 |
| T2DM | nk_cell | *TLR2* | 1.380413049 | 0.015839872 | 0.020610099 | 1 |
| T2DM | nk_cell | *NUMA1* | -1.092313054 | 5.16E-05 | 0.000155956 | 0.250339165 |
| T2DM | nk_cell | *RAB30* | -0.966072675 | 1.41E-05 | 0.000122264 | 0.068217976 |
| T2DM | nk_cell | *PRCP* | -0.972383883 | 0.000145697 | 0.000311968 | 0.706919801 |
| T2DM | nk_cell | *NARS2* | -1.086718974 | 1.96E-05 | 0.000122264 | 0.095044683 |
| T2DM | nk_cell | *GGH* | -0.812221553 | 0.006577956 | 0.009082596 | 1 |
| T2DM | nk_cell | *SULF1* | -0.64169186 | 0.000117995 | 0.000264906 | 0.5725108 |
| T2DM | nk_cell | *TGS1* | -1.36589993 | 0.017626903 | 0.022764368 | 1 |
| T2DM | nk_cell | *NEK1* | -1.188489738 | 0.000198541 | 0.000398725 | 0.963318598 |
| T2DM | nk_cell | *TRPC6* | -0.703089971 | 3.93E-05 | 0.000136619 | 0.190584577 |
| T2DM | nk_cell | *MMP27* | -2.198608831 | 0.006731664 | 0.009278988 | 1 |
| T2DM | nk_cell | *POU2F3* | -0.820760772 | 0.006882457 | 0.00945461 | 1 |
| T2DM | nk_cell | *PPP2R1B* | -0.484952064 | 0.000134338 | 0.000292552 | 0.651806302 |
| T2DM | nk_cell | *FXYD6* | -1.064178231 | 2.51E-05 | 0.000122264 | 0.121916832 |
| T2DM | nk_cell | *MAP2K5* | -0.998697438 | 3.85E-05 | 0.000135882 | 0.186973444 |
| T2DM | nk_cell | *NUSAP1* | 1.390507046 | 2.68E-05 | 0.000122391 | 0.12979475 |
| T2DM | nk_cell | *NDUFAF1* | -1.144819593 | 3.37E-05 | 0.00012843 | 0.163561222 |
| T2DM | nk_cell | *PARP6* | -1.554772404 | 4.69E-05 | 0.000148778 | 0.227539618 |
| T2DM | nk_cell | *ZNF280D* | -1.025813139 | 3.82E-05 | 0.000135483 | 0.185476636 |
| T2DM | nk_cell | *RSL24D1* | -1.654801404 | 0.010612337 | 0.014177054 | 1 |
| T2DM | nk_cell | *SPTBN5* | 0.620468183 | 2.04E-07 | 3.10E-05 | 0.00099102 |
| T2DM | nk_cell | *SLC44A5* | -0.732592358 | 1.55E-05 | 0.000122264 | 0.075249302 |
| T2DM | nk_cell | *DBT* | -1.561031379 | 3.32E-05 | 0.000127948 | 0.161086981 |
| T2DM | nk_cell | *RTCA* | -1.590254227 | 0.001297368 | 0.002022162 | 1 |
| T2DM | nk_cell | *KHK* | -0.963610419 | 9.73E-06 | 0.000122264 | 0.047198186 |
| T2DM | nk_cell | *DYNC2LI1* | -1.286084153 | 8.04E-06 | 0.000122264 | 0.039011862 |
| T2DM | nk_cell | *LHCGR* | -0.993413285 | 1.10E-05 | 0.000122264 | 0.053304922 |
| T2DM | nk_cell | *THUMPD2* | -1.143566038 | 2.40E-05 | 0.000122264 | 0.11624166 |
| T2DM | nk_cell | *ATRAID* | 1.079359427 | 0.01203201 | 0.015933219 | 1 |
| T2DM | nk_cell | *CENPO* | -1.061711969 | 0.000114818 | 0.000260568 | 0.557094765 |
| T2DM | nk_cell | *LRPPRC* | -1.288490891 | 3.88E-05 | 0.000136418 | 0.188467232 |
| T2DM | nk_cell | *DTNB* | -1.384584828 | 0.000551405 | 0.00094371 | 1 |
| T2DM | nk_cell | *CYP2C8* | -3.124006409 | 0.001233352 | 0.001932264 | 1 |
| T2DM | nk_cell | *MYOF* | -0.432517253 | 0.022595586 | 0.028714978 | 1 |
| T2DM | nk_cell | *BTBD16* | -1.234963352 | 4.76E-06 | 0.000122264 | 0.023099802 |
| T2DM | nk_cell | *TACC2* | -0.962304338 | 3.35E-05 | 0.000128164 | 0.162650184 |
| T2DM | nk_cell | *CALHM2* | -1.572574441 | 0.00013187 | 0.000288342 | 0.639831679 |
| T2DM | nk_cell | *PLCE1* | -0.971276803 | 0.000130971 | 0.000287153 | 0.635469206 |
| T2DM | nk_cell | *RBP4* | -1.254141768 | 4.76E-05 | 0.000149363 | 0.230915307 |
| T2DM | nk_cell | *ASCC1* | -0.861379261 | 9.09E-06 | 0.000122264 | 0.0440871 |

| | | | | | | |
|---|---|---|---|---|---|---|
| T2DM | nk_cell | *DNA2* | -0.411524855 | 0.004931286 | 0.006949346 | 1 |
| T2DM | nk_cell | *ATIC* | -1.040529844 | 2.00E-06 | 0.000111758 | 0.009722952 |
| T2DM | nk_cell | *SMARCAL1* | -1.791906717 | 0.000542927 | 0.0009315 | 1 |
| T2DM | nk_cell | *BARD1* | 0.910981496 | 0.003165994 | 0.004615807 | 1 |
| T2DM | nk_cell | *STAT4* | 0.169265615 | 7.55E-05 | 0.000196203 | 0.366311126 |
| T2DM | nk_cell | *SSB* | -1.025678245 | 6.67E-05 | 0.000180685 | 0.323425361 |
| T2DM | nk_cell | *FASTKD1* | -1.237514833 | 9.82E-05 | 0.000233238 | 0.476271297 |
| T2DM | nk_cell | *ITGAV* | -1.103383082 | 4.31E-05 | 0.000143655 | 0.209210468 |
| T2DM | nk_cell | *SLC35A5* | -1.107363116 | 0.000214308 | 0.000424417 | 1 |
| T2DM | nk_cell | *SENP7* | -1.048278657 | 4.26E-05 | 0.000142336 | 0.206529937 |
| T2DM | nk_cell | *COX17* | 1.199025657 | 0.016143393 | 0.020954453 | 1 |
| T2DM | nk_cell | *SPPL2A* | -1.692852128 | 0.000129395 | 0.000284212 | 0.627823624 |
| T2DM | nk_cell | *PPCDC* | -1.119672598 | 0.00016839 | 0.000348567 | 0.817025871 |
| T2DM | nk_cell | *SEMA7A* | -1.058152184 | 0.032050557 | 0.039823125 | 1 |
| T2DM | nk_cell | *PCDH10* | 1.043875699 | 0.000163323 | 0.000340544 | 0.792445608 |
| T2DM | nk_cell | *AP1AR* | -1.852381133 | 4.08E-05 | 0.000139476 | 0.197826473 |
| T2DM | nk_cell | *PRKG2* | 0.789061188 | 0.000149473 | 0.000318928 | 0.725242888 |
| T2DM | nk_cell | *KIAA1109* | -1.281388107 | 0.000221206 | 0.000436298 | 1 |
| T2DM | nk_cell | *BMPR1B* | -0.596533371 | 2.12E-05 | 0.000122264 | 0.102826145 |
| T2DM | nk_cell | *PRDM5* | -0.825920409 | 1.72E-05 | 0.000122264 | 0.083353934 |
| T2DM | nk_cell | *TRPC3* | 0.187872989 | 3.05E-05 | 0.000125102 | 0.148121277 |
| T2DM | nk_cell | *NUP54* | -0.90684371 | 0.012420709 | 0.016400822 | 1 |
| T2DM | nk_cell | *G3BP2* | -1.696476897 | 0.000280399 | 0.000530822 | 1 |
| T2DM | nk_cell | *ANXA3* | -0.8397427 | 0.000703556 | 0.001167952 | 1 |
| T2DM | nk_cell | *PPA2* | -1.250950031 | 2.93E-05 | 0.000123467 | 0.142343167 |
| T2DM | nk_cell | *ENPEP* | -0.475556699 | 0.015135394 | 0.019744264 | 1 |
| T2DM | nk_cell | *PAPSS1* | -0.896844272 | 4.07E-05 | 0.000139476 | 0.197699918 |
| T2DM | nk_cell | *FBN2* | -0.314705437 | 0.010739535 | 0.01431938 | 1 |
| T2DM | nk_cell | *PARVG* | 0.499725414 | 0.000243274 | 0.000471581 | 1 |
| T2DM | nk_cell | *B4GALNT3* | 0.727340089 | 0.000112208 | 0.000256566 | 0.544432939 |
| T2DM | nk_cell | *ERP27* | 0.246129271 | 7.72E-05 | 0.000199059 | 0.374686237 |
| T2DM | nk_cell | *KIF21A* | -1.473723651 | 0.000643796 | 0.001078253 | 1 |
| T2DM | nk_cell | *ALG10* | -0.975479985 | 3.74E-05 | 0.00013433 | 0.181614609 |
| T2DM | nk_cell | *ZCRB1* | -1.717395206 | 0.000903032 | 0.001461966 | 1 |
| T2DM | nk_cell | *CLSTN3* | 0.981226789 | 0.000337871 | 0.000622381 | 1 |
| T2DM | nk_cell | *KLRG1* | 0.275820732 | 0.001216707 | 0.00190927 | 1 |
| T2DM | nk_cell | *VAMP1* | 1.054381509 | 0.000183245 | 0.00037436 | 0.889104913 |
| T2DM | nk_cell | *PEX5* | -0.771702947 | 0.000250549 | 0.000482789 | 1 |
| T2DM | nk_cell | *PPFIA2* | -0.750918302 | 1.61E-05 | 0.000122264 | 0.078162789 |
| T2DM | nk_cell | *MARCH9* | -1.845679642 | 1.69E-05 | 0.000122264 | 0.082061248 |
| T2DM | nk_cell | *POC1B* | -1.520730639 | 0.010575924 | 0.014132301 | 1 |
| T2DM | nk_cell | *AMDHD1* | -0.595789655 | 0.009095697 | 0.012248771 | 1 |
| T2DM | nk_cell | *RITA1* | -0.939696846 | 2.25E-05 | 0.000122264 | 0.109270023 |
| T2DM | nk_cell | *SDSL* | 1.00078725 | 0.018578995 | 0.023930258 | 1 |
| T2DM | nk_cell | *GLTP* | 1.0080136 | 0.000737866 | 0.001217344 | 1 |
| T2DM | nk_cell | *FOXN4* | -0.728832537 | 0.000101828 | 0.000239374 | 0.494068511 |
| T2DM | nk_cell | *MTMR6* | 0.997001408 | 0.022635289 | 0.028742848 | 1 |
| T2DM | nk_cell | *CCDC65* | -1.362357172 | 8.66E-05 | 0.000213966 | 0.419991608 |
| T2DM | nk_cell | *BRCA2* | -2.962123536 | 0.000244486 | 0.000473717 | 1 |
| T2DM | nk_cell | *KANSL2* | -1.039588628 | 0.000278074 | 0.000527073 | 1 |

| | | | | | | |
|---|---|---|---|---|---|---|
| T2DM | nk_cell | *CERS5* | -1.515810121 | 3.62E-05 | 0.000132186 | 0.175661884 |
| T2DM | nk_cell | *LMBR1L* | -1.72185272 | 0.000430372 | 0.000762105 | 1 |
| T2DM | nk_cell | *ESYT1* | -1.275613454 | 0.039950143 | 0.048973747 | 1 |
| T2DM | nk_cell | *ZNF740* | 0.945151509 | 0.031114658 | 0.038739625 | 1 |
| T2DM | nk_cell | *WDFY2* | -1.144417132 | 2.59E-05 | 0.000122264 | 0.125828291 |
| T2DM | nk_cell | *HNRNPA1L2* | -1.674683953 | 3.68E-05 | 0.000133329 | 0.178394676 |
| T2DM | nk_cell | *RB1* | -1.192509349 | 0.000102197 | 0.000239777 | 0.495859588 |
| T2DM | nk_cell | *VPS33A* | -1.055762158 | 0.000944426 | 0.00152339 | 1 |
| T2DM | nk_cell | *NOVA1* | -0.36268295 | 0.005102993 | 0.007176731 | 1 |
| T2DM | nk_cell | *FITM1* | 0.483560375 | 3.52E-05 | 0.000130929 | 0.170731858 |
| T2DM | nk_cell | *TMX1* | -1.692564415 | 0.006609112 | 0.009123019 | 1 |
| T2DM | nk_cell | *RTN1* | -0.651292201 | 0.000730673 | 0.001207091 | 1 |
| T2DM | nk_cell | *C14orf37* | -1.301664018 | 5.69E-05 | 0.000164236 | 0.275947237 |
| T2DM | nk_cell | *RAB15* | -2.566275609 | 0.000531664 | 0.000914881 | 1 |
| T2DM | nk_cell | *EFCAB11* | -0.797543875 | 0.000488664 | 0.000850734 | 1 |
| T2DM | nk_cell | *SLC24A4* | 0.96731654 | 0.000289868 | 0.000547465 | 1 |
| T2DM | nk_cell | *C14orf79* | -1.529113096 | 0.000807742 | 0.001317809 | 1 |
| T2DM | nk_cell | *WARS* | -1.34615306 | 7.14E-05 | 0.000189116 | 0.346307227 |
| T2DM | nk_cell | *DUOXA1* | -0.968080088 | 0.000661342 | 0.00110535 | 1 |
| T2DM | nk_cell | *DISP2* | -0.294518434 | 0.033413679 | 0.041403317 | 1 |
| T2DM | nk_cell | *UBE2Q2* | -1.322683856 | 0.008677846 | 0.011725121 | 1 |
| T2DM | nk_cell | *HMG20A* | -1.531498643 | 0.000397813 | 0.000717275 | 1 |
| T2DM | nk_cell | *SCAPER* | -1.116788894 | 1.46E-05 | 0.000122264 | 0.070809653 |
| T2DM | nk_cell | *NEIL1* | -1.604883907 | 3.98E-08 | 1.02E-05 | 0.000193104 |
| T2DM | nk_cell | *MAN2C1* | -0.853680949 | 1.14E-09 | 6.28E-07 | 5.52E-06 |
| T2DM | nk_cell | *PIF1* | 0.669521706 | 0.000737937 | 0.001217344 | 1 |
| T2DM | nk_cell | *CYP11A1* | -0.729415156 | 0.000365815 | 0.000667017 | 1 |
| T2DM | nk_cell | *BBS4* | -1.307345953 | 0.000728861 | 0.001204919 | 1 |
| T2DM | nk_cell | *PCSK6* | -1.035764221 | 1.80E-05 | 0.000122264 | 0.087124555 |
| T2DM | nk_cell | *CELF6* | 3.473216661 | 7.89E-05 | 0.000201568 | 0.38290277 |
| T2DM | nk_cell | *HAPLN3* | -0.702285555 | 1.54E-05 | 0.000122264 | 0.074804409 |
| T2DM | nk_cell | *MFGE8* | -1.51819287 | 0.000729512 | 0.001205584 | 1 |
| T2DM | nk_cell | *UNC45A* | -1.051896861 | 2.43E-05 | 0.000122264 | 0.117960395 |
| T2DM | nk_cell | *CRTC3* | -0.958573602 | 0.000118684 | 0.000265983 | 0.575852756 |
| T2DM | nk_cell | *ITGAX* | 0.144435907 | 5.75E-05 | 0.000165634 | 0.278762222 |
| T2DM | nk_cell | *TGFB1I1* | -2.888189633 | 0.000152804 | 0.000323899 | 0.741404109 |
| T2DM | nk_cell | *NUDT7* | -1.299275795 | 1.18E-05 | 0.000122264 | 0.05732331 |
| T2DM | nk_cell | *GCSH* | -0.862514364 | 1.42E-06 | 9.04E-05 | 0.006869666 |
| T2DM | nk_cell | *CMTM3* | -1.226438951 | 3.14E-05 | 0.000127109 | 0.152480661 |
| T2DM | nk_cell | *ZCCHC14* | -1.158017076 | 2.20E-05 | 0.000122264 | 0.10698265 |
| T2DM | nk_cell | *TLDC1* | -1.073463636 | 1.61E-05 | 0.000122264 | 0.078337803 |
| T2DM | nk_cell | *RPL3L* | 3.293334817 | 0.00017226 | 0.000355359 | 0.835803783 |
| T2DM | nk_cell | *TIGD7* | -1.035714501 | 9.52E-05 | 0.000227998 | 0.461695124 |
| T2DM | nk_cell | *DEF8* | -1.525081888 | 5.36E-05 | 0.000159514 | 0.260017509 |
| T2DM | nk_cell | *TCF25* | -1.295119575 | 0.000414475 | 0.000740439 | 1 |
| T2DM | nk_cell | *GALNS* | -1.593655247 | 4.86E-06 | 0.000122264 | 0.023602758 |
| T2DM | nk_cell | *GAS8* | -0.980036829 | 3.64E-05 | 0.000132553 | 0.176826291 |
| T2DM | nk_cell | *NCOR1* | -1.188184516 | 0.000123028 | 0.000272821 | 0.596933322 |
| T2DM | nk_cell | *KSR1* | -1.432169498 | 3.11E-05 | 0.000126604 | 0.15091188 |
| T2DM | nk_cell | *GFOD2* | -1.678739718 | 0.000126391 | 0.000278749 | 0.613247523 |

| | | | | | | |
|---|---|---|---|---|---|---|
| T2DM | nk_cell | *PRPSAP2* | -1.067816129 | 0.0074261 | 0.010129726 | 1 |
| T2DM | nk_cell | *RASL10B* | -1.965297964 | 2.57E-05 | 0.000122264 | 0.124572572 |
| T2DM | nk_cell | *UNC45B* | -3.023101514 | 1.74E-05 | 0.000122264 | 0.084459543 |
| T2DM | nk_cell | *C17orf80* | -0.915297718 | 1.42E-05 | 0.000122264 | 0.068901914 |
| T2DM | nk_cell | *VPS53* | -1.408031958 | 2.37E-07 | 3.38E-05 | 0.001150626 |
| T2DM | nk_cell | *SGSM2* | -1.336394099 | 2.33E-05 | 0.000122264 | 0.11300871 |
| T2DM | nk_cell | *LRRC46* | -1.457932425 | 6.46E-06 | 0.000122264 | 0.031364976 |
| T2DM | nk_cell | *SCRN2* | -0.38386714 | 0.008660449 | 0.011708135 | 1 |
| T2DM | nk_cell | *SSH2* | -1.1077657 | 8.35E-05 | 0.000209146 | 0.405115746 |
| T2DM | nk_cell | *RHBDL3* | 1.223116351 | 0.001011557 | 0.001621304 | 1 |
| T2DM | nk_cell | *CLTC* | -0.943959394 | 0.001734125 | 0.002644241 | 1 |
| T2DM | nk_cell | *IMPA2* | -1.455807214 | 0.000103296 | 0.000241421 | 0.501190945 |
| T2DM | nk_cell | *GNAL* | -1.743853596 | 7.39E-05 | 0.000193958 | 0.358627681 |
| T2DM | nk_cell | *RPRD1A* | -1.34692209 | 3.42E-05 | 0.000129717 | 0.16617102 |
| T2DM | nk_cell | *GALNT1* | -1.275818501 | 3.03E-05 | 0.000124985 | 0.147000602 |
| T2DM | nk_cell | *C18orf8* | -2.36895928 | 3.63E-05 | 0.000132391 | 0.176212104 |
| T2DM | nk_cell | *SLC14A1* | -0.977809202 | 3.62E-05 | 0.000132186 | 0.175675055 |
| T2DM | nk_cell | *CCDC40* | -0.676632906 | 0.000104885 | 0.000244195 | 0.508901783 |
| T2DM | nk_cell | *TMC6* | 0.518705533 | 6.27E-05 | 0.000173708 | 0.30433661 |
| T2DM | nk_cell | *CARD14* | -0.900628407 | 1.49E-05 | 0.000122264 | 0.072169423 |
| T2DM | nk_cell | *TBCD* | -1.507330979 | 0.001981261 | 0.002989142 | 1 |
| T2DM | nk_cell | *FN3KRP* | -0.895806517 | 0.000221912 | 0.000437512 | 1 |
| T2DM | nk_cell | *NARF* | -1.678084337 | 0.000279799 | 0.000530099 | 1 |
| T2DM | nk_cell | *CBX8* | -1.04735851 | 0.007021342 | 0.009626321 | 1 |
| T2DM | nk_cell | *SECTM1* | -0.555527761 | 0.000134556 | 0.000292764 | 0.652863548 |
| T2DM | nk_cell | *ZNF750* | 0.627922924 | 4.32E-05 | 0.000143865 | 0.209754901 |
| T2DM | nk_cell | *WDR45B* | -1.677030569 | 6.86E-05 | 0.00018443 | 0.332711122 |
| T2DM | nk_cell | *RNF165* | 0.954576225 | 0.00849873 | 0.011499118 | 1 |
| T2DM | nk_cell | *MAPK4* | -2.967090925 | 1.68E-05 | 0.000122264 | 0.081707024 |
| T2DM | nk_cell | *FBXO15* | -1.561959182 | 4.95E-05 | 0.000152245 | 0.240089965 |
| T2DM | nk_cell | *CBLN2* | -0.798440303 | 2.34E-05 | 0.000122264 | 0.113377615 |
| T2DM | nk_cell | *PIP4K2B* | -1.289552482 | 0.001828567 | 0.002776035 | 1 |
| T2DM | nk_cell | *ERBB2* | -0.546585377 | 2.46E-05 | 0.000122264 | 0.119435319 |
| T2DM | nk_cell | *PNMT* | -1.669158333 | 0.004472132 | 0.006342819 | 1 |
| T2DM | nk_cell | *IGFBP4* | -0.833157506 | 4.42E-05 | 0.000145325 | 0.214557408 |
| T2DM | nk_cell | *TXNL4A* | -0.833394065 | 1.02E-05 | 0.000122264 | 0.049392831 |
| T2DM | nk_cell | *CACNA1A* | -0.790195845 | 5.57E-06 | 0.000122264 | 0.027040211 |
| T2DM | nk_cell | *SAMD1* | -2.285693294 | 0.000445014 | 0.00078488 | 1 |
| T2DM | nk_cell | *NFIC* | -0.874948712 | 2.01E-05 | 0.000122264 | 0.097534683 |
| T2DM | nk_cell | *PRDM15* | -0.881576689 | 0.00108849 | 0.001728757 | 1 |
| T2DM | nk_cell | *PFKL* | -1.995995936 | 0.002659859 | 0.003919112 | 1 |
| T2DM | nk_cell | *MVB12A* | -1.255405548 | 0.001724976 | 0.002631117 | 1 |
| T2DM | nk_cell | *DUS3L* | 1.180935096 | 9.74E-06 | 0.000122264 | 0.04724106 |
| T2DM | nk_cell | *TMEM91* | -0.62593531 | 5.02E-06 | 0.000122264 | 0.024375578 |
| T2DM | nk_cell | *SIRT3* | -1.233341892 | 0.007102528 | 0.009734877 | 1 |
| T2DM | nk_cell | *HUNK* | -0.648367879 | 1.01E-05 | 0.000122264 | 0.049197783 |
| T2DM | nk_cell | *COL6A1* | -0.741725826 | 2.41E-06 | 0.000122264 | 0.011707937 |
| T2DM | nk_cell | *IFNAR1* | -1.007214309 | 2.86E-05 | 0.000123207 | 0.138884651 |
| T2DM | nk_cell | *DOPEY2* | -0.924933315 | 0.005305396 | 0.007442746 | 1 |
| T2DM | nk_cell | *URB1* | -0.9563578 | 0.001321932 | 0.002057093 | 1 |

| | | | | | | |
|---|---|---|---|---|---|---|
| T2DM | nk_cell | *SAE1* | -2.131760172 | 0.000560841 | 0.000957441 | 1 |
| T2DM | nk_cell | *GEMIN7* | -1.871730159 | 9.24E-07 | 7.35E-05 | 0.004483955 |
| T2DM | nk_cell | *CACNG8* | 1.275522315 | 6.05E-05 | 0.000169923 | 0.293554248 |
| T2DM | nk_cell | *ZNF787* | -1.700497752 | 0.000606726 | 0.001024655 | 1 |
| T2DM | nk_cell | *FBN3* | -0.714793784 | 0.005093247 | 0.007165102 | 1 |
| T2DM | nk_cell | *EVI5L* | 1.307054863 | 0.017769109 | 0.022935812 | 1 |
| T2DM | nk_cell | *SLC47A1* | 1.48941863 | 0.00229344 | 0.00341657 | 1 |
| T2DM | nk_cell | *CTU1* | -0.627398967 | 2.88E-05 | 0.000123207 | 0.139953664 |
| T2DM | nk_cell | *RERE* | -0.82079633 | 3.75E-05 | 0.00013436 | 0.181989816 |
| T2DM | nk_cell | *MMEL1* | -0.582900162 | 0.010641985 | 0.014200718 | 1 |
| T2DM | nk_cell | *EPHA2* | -1.015337359 | 2.92E-05 | 0.000123467 | 0.141480906 |
| T2DM | nk_cell | *ARHGEF19* | -0.357211865 | 0.015855674 | 0.020619601 | 1 |
| T2DM | nk_cell | *PEX14* | -1.021184232 | 3.15E-05 | 0.000127109 | 0.152796788 |
| T2DM | nk_cell | *CNKSR1* | -0.569095141 | 0.009602226 | 0.012905818 | 1 |
| T2DM | nk_cell | *ZNF593* | -1.449713487 | 1.02E-05 | 0.000122264 | 0.049395263 |
| T2DM | nk_cell | *EVA1B* | -1.667208095 | 0.000671203 | 0.001119902 | 1 |
| T2DM | nk_cell | *NBPF3* | -0.62737387 | 0.001302927 | 0.002028819 | 1 |
| T2DM | nk_cell | *HSPG2* | -0.816712731 | 2.31E-05 | 0.000122264 | 0.111969126 |
| T2DM | nk_cell | *ITGB3BP* | -0.750540101 | 0.00018643 | 0.000379429 | 0.904558662 |
| T2DM | nk_cell | *PIGK* | -1.375770002 | 4.94E-05 | 0.00015216 | 0.239738816 |
| T2DM | nk_cell | *CYP4B1* | -0.522086997 | 0.000423854 | 0.000754694 | 1 |
| T2DM | nk_cell | *SLC44A3* | -1.447259864 | 4.05E-05 | 0.000139129 | 0.196728218 |
| T2DM | nk_cell | *STRIP1* | -1.49941457 | 4.61E-05 | 0.000147867 | 0.223870204 |
| T2DM | nk_cell | *GPR161* | -1.288560841 | 8.32E-05 | 0.000208812 | 0.403842572 |
| T2DM | nk_cell | *ALDH9A1* | -1.53016315 | 2.81E-05 | 0.000122662 | 0.136154731 |
| T2DM | nk_cell | *ATP1B1* | -1.332724175 | 0.000767231 | 0.001257213 | 1 |
| T2DM | nk_cell | *NME7* | -0.77134767 | 3.84E-06 | 0.000122264 | 0.018642031 |
| T2DM | nk_cell | *XCL1* | 0.761694911 | 7.35E-05 | 0.000193315 | 0.356666824 |
| T2DM | nk_cell | *XCL2* | 0.231033524 | 5.94E-05 | 0.000168356 | 0.28822515 |
| T2DM | nk_cell | *MAEL* | -1.432416631 | 2.22E-05 | 0.000122264 | 0.10750492 |
| T2DM | nk_cell | *RFWD2* | -1.205479269 | 0.00057003 | 0.000968752 | 1 |
| T2DM | nk_cell | *TARS2* | -1.281539914 | 0.000680778 | 0.001133929 | 1 |
| T2DM | nk_cell | *SNX27* | -1.053760948 | 0.00025716 | 0.000494155 | 1 |
| T2DM | nk_cell | *SETDB1* | -1.381155735 | 2.43E-05 | 0.000122264 | 0.117908294 |
| T2DM | nk_cell | *SEMA6C* | -0.586805847 | 3.53E-05 | 0.000131165 | 0.171301879 |
| T2DM | nk_cell | *ARNT* | -1.010830532 | 2.84E-05 | 0.00012318 | 0.137857525 |
| T2DM | nk_cell | *POGZ* | -1.013310274 | 6.92E-05 | 0.000185314 | 0.335788949 |
| T2DM | nk_cell | *GABPB2* | -0.716085253 | 0.023897664 | 0.030258733 | 1 |
| T2DM | nk_cell | *DYRK3* | -0.836773517 | 3.20E-05 | 0.000127628 | 0.155029537 |
| T2DM | nk_cell | *EIF2D* | -1.508800368 | 1.48E-05 | 0.000122264 | 0.072039643 |
| T2DM | nk_cell | *TAF1A* | -1.208057796 | 8.09E-05 | 0.00020495 | 0.392574364 |
| T2DM | nk_cell | *RAB13* | 1.356248641 | 0.028065764 | 0.035157205 | 1 |
| T2DM | nk_cell | *S100A8* | -1.9279128 | 6.19E-05 | 0.000172781 | 0.300292787 |
| T2DM | nk_cell | *CREB3L4* | -1.933860253 | 1.94E-05 | 0.000122264 | 0.094085042 |
| T2DM | nk_cell | *KCNN3* | -0.758171754 | 0.00014166 | 0.000305086 | 0.68733308 |
| T2DM | nk_cell | *C1orf43* | -1.439173063 | 1.91E-05 | 0.000122264 | 0.092830206 |
| T2DM | nk_cell | *RIT1* | -1.474692362 | 8.10E-06 | 0.000122264 | 0.03930496 |
| T2DM | nk_cell | *FLG* | -0.578423245 | 0.000713158 | 0.001182181 | 1 |
| T2DM | nk_cell | *SCCPDH* | -1.193672736 | 1.57E-05 | 0.000122264 | 0.076354145 |
| T2DM | nk_cell | *CEP170* | -1.163563713 | 3.42E-05 | 0.000129717 | 0.165984629 |

| | | | | | | |
|---|---|---|---|---|---|---|
| T2DM | nk_cell | *ACP1* | -1.504286809 | 2.03E-05 | 0.000122264 | 0.098590111 |
| T2DM | nk_cell | *CNIH4* | -1.152781583 | 1.89E-05 | 0.000122264 | 0.091704657 |
| T2DM | nk_cell | *MBOAT2* | -0.968322095 | 1.75E-05 | 0.000122264 | 0.084862229 |
| T2DM | nk_cell | *PYCR2* | 1.11326513 | 0.001001964 | 0.001607649 | 1 |
| T2DM | nk_cell | *PLEKHA6* | -0.329527905 | 0.00039355 | 0.00071091 | 1 |
| T2DM | nk_cell | *OSR1* | -0.565791547 | 4.19E-05 | 0.000141779 | 0.203453082 |
| T2DM | nk_cell | *RHOB* | -1.246485596 | 2.01E-05 | 0.000122264 | 0.097293473 |
| T2DM | nk_cell | *GALM* | -1.300971775 | 2.35E-05 | 0.000122264 | 0.113893333 |
| T2DM | nk_cell | *WDPCP* | -0.813657863 | 5.26E-05 | 0.000158074 | 0.255448263 |
| T2DM | nk_cell | *VPS54* | -1.041237903 | 2.66E-05 | 0.000122391 | 0.128932711 |
| T2DM | nk_cell | *REG3G* | 4.682351188 | 0.002340147 | 0.003484012 | 1 |
| T2DM | nk_cell | *ABHD1* | -2.62700806 | 1.06E-06 | 7.89E-05 | 0.005133854 |
| T2DM | nk_cell | *CIAO1* | -1.382352106 | 0.000115091 | 0.000260823 | 0.558422871 |
| T2DM | nk_cell | *ZNF514* | -1.877693432 | 2.15E-06 | 0.000114775 | 0.010429976 |
| T2DM | nk_cell | *EXOC6B* | -0.941876028 | 3.23E-05 | 0.000127896 | 0.156773543 |
| T2DM | nk_cell | *TEX261* | -2.994920712 | 0.000163158 | 0.000340409 | 0.791642109 |
| T2DM | nk_cell | *RALB* | -1.891200433 | 0.000125988 | 0.000278239 | 0.611291528 |
| T2DM | nk_cell | *RABL2A* | -1.303696347 | 2.68E-05 | 0.000122391 | 0.130041031 |
| T2DM | nk_cell | *FBLN7* | -0.660586328 | 0.000406544 | 0.000728954 | 1 |
| T2DM | nk_cell | *CNGA3* | -11.7094068 | 3.22E-06 | 0.000122264 | 0.015625271 |
| T2DM | nk_cell | *FAHD2B* | -0.934565292 | 6.01E-05 | 0.000169532 | 0.291425747 |
| T2DM | nk_cell | *THSD7B* | -0.517113596 | 2.76E-05 | 0.000122493 | 0.134157792 |
| T2DM | nk_cell | *GPR17* | -0.280312009 | 0.004121955 | 0.005880543 | 1 |
| T2DM | nk_cell | *POLR2D* | -1.579286379 | 0.001565808 | 0.00240497 | 1 |
| T2DM | nk_cell | *GALNT13* | -1.122773118 | 1.57E-05 | 0.000122264 | 0.076046785 |
| T2DM | nk_cell | *PHOSPHO2* | -2.893710509 | 0.001800688 | 0.002737994 | 1 |
| T2DM | nk_cell | *METTL21A* | -1.503538594 | 0.000317772 | 0.00059123 | 1 |
| T2DM | nk_cell | *NBEAL1* | -0.941095785 | 0.000482557 | 0.000841613 | 1 |
| T2DM | nk_cell | *SUMF1* | -1.166389464 | 0.000160609 | 0.00033662 | 0.779275525 |
| T2DM | nk_cell | *ACKR3* | -1.013388731 | 5.48E-05 | 0.000161002 | 0.265653084 |
| T2DM | nk_cell | *ESPNL* | -6.396280519 | 3.33E-05 | 0.000128061 | 0.161673425 |
| T2DM | nk_cell | *ANKMY1* | -1.146927315 | 0.008599313 | 0.011628726 | 1 |
| T2DM | nk_cell | *TAMM41* | -1.283934153 | 0.000747283 | 0.001227842 | 1 |
| T2DM | nk_cell | *STK11IP* | -1.707052435 | 1.82E-05 | 0.000122264 | 0.088074625 |
| T2DM | nk_cell | *EAF1* | 0.669006373 | 0.00017558 | 0.000361286 | 0.851913346 |
| T2DM | nk_cell | *CNTN4* | -0.654870145 | 1.44E-05 | 0.000122264 | 0.069948731 |
| T2DM | nk_cell | *SLC25A38* | 0.653271301 | 0.000272381 | 0.000518678 | 1 |
| T2DM | nk_cell | *CTDSPL* | -1.082802052 | 5.26E-06 | 0.000122264 | 0.025536424 |
| T2DM | nk_cell | *PTPRG* | -0.882287908 | 1.95E-05 | 0.000122264 | 0.094617816 |
| T2DM | nk_cell | *SLC25A26* | -1.44333424 | 0.000280297 | 0.000530822 | 1 |
| T2DM | nk_cell | *LIMD1* | -1.133357747 | 0.000144298 | 0.00030993 | 0.700131899 |
| T2DM | nk_cell | *PLA1A* | -1.750508165 | 2.11E-05 | 0.000122264 | 0.102412746 |
| T2DM | nk_cell | *ADPRH* | 0.78442204 | 0.027827873 | 0.034889105 | 1 |
| T2DM | nk_cell | *IGSF11* | -0.430301017 | 0.000200698 | 0.000401892 | 0.973785403 |
| T2DM | nk_cell | *BOC* | -0.540595918 | 3.26E-05 | 0.000127909 | 0.15807143 |
| T2DM | nk_cell | *MED12L* | -1.288853532 | 2.10E-05 | 0.000122264 | 0.101711708 |
| T2DM | nk_cell | *EIF2A* | -0.910608002 | 0.000342607 | 0.000629908 | 1 |
| T2DM | nk_cell | *NCEH1* | -1.096777185 | 2.98E-06 | 0.000122264 | 0.014442426 |
| T2DM | nk_cell | *CCDC39* | -0.925526271 | 0.000204201 | 0.000407394 | 0.990781408 |
| T2DM | nk_cell | *MUC4* | -1.154408963 | 1.58E-05 | 0.000122264 | 0.076833909 |

| | | | | | | |
|---|---|---|---|---|---|---|
| T2DM | nk_cell | *DGKQ* | 0.655537008 | 0.000733396 | 0.001211177 | 1 |
| T2DM | nk_cell | *SLC26A1* | -0.569988806 | 0.012963825 | 0.017050821 | 1 |
| T2DM | nk_cell | *CENPC* | -1.201807078 | 7.45E-06 | 0.000122264 | 0.036165794 |
| T2DM | nk_cell | *SCD5* | -1.267583575 | 1.64E-05 | 0.000122264 | 0.07974347 |
| T2DM | nk_cell | *TRMT10A* | -0.673358676 | 1.85E-05 | 0.000122264 | 0.089595472 |
| T2DM | nk_cell | *CISD2* | -1.673565106 | 0.009038519 | 0.012178532 | 1 |
| T2DM | nk_cell | *DDIT4L* | -1.326666101 | 0.000149723 | 0.000319322 | 0.726456462 |
| T2DM | nk_cell | *METTL14* | -1.9792448 | 6.31E-05 | 0.000173881 | 0.306378983 |
| T2DM | nk_cell | *USP53* | -0.831981757 | 0.000290012 | 0.000547525 | 1 |
| T2DM | nk_cell | *MARCH1* | -1.269801656 | 6.47E-06 | 0.000122264 | 0.031385836 |
| T2DM | nk_cell | *CBR4* | -1.248478769 | 6.88E-05 | 0.000184893 | 0.333916093 |
| T2DM | nk_cell | *MARCH6* | -1.185736049 | 5.37E-05 | 0.000159514 | 0.260399852 |
| T2DM | nk_cell | *NKD2* | 1.12506167 | 0.00675931 | 0.009306518 | 1 |
| T2DM | nk_cell | *MYO10* | -0.516266394 | 1.90E-05 | 0.000122264 | 0.092408117 |
| T2DM | nk_cell | *BHMT* | -1.027291143 | 5.74E-06 | 0.000122264 | 0.027840218 |
| T2DM | nk_cell | *GTF2H2* | -1.889251925 | 1.89E-05 | 0.000122264 | 0.091934587 |
| T2DM | nk_cell | *SPATA9* | -0.571950019 | 1.84E-05 | 0.000122264 | 0.08917967 |
| T2DM | nk_cell | *TSLP* | -0.419654094 | 0.024837201 | 0.03135834 | 1 |
| T2DM | nk_cell | *COMMD10* | -1.203662118 | 0.000100791 | 0.000237513 | 0.489039801 |
| T2DM | nk_cell | *YIPF5* | -1.856934529 | 0.00069831 | 0.001160342 | 1 |
| T2DM | nk_cell | *CXCL14* | -0.792565998 | 1.23E-05 | 0.000122264 | 0.059820527 |
| T2DM | nk_cell | *PCYOX1L* | -1.13375012 | 1.84E-05 | 0.000122264 | 0.089188805 |
| T2DM | nk_cell | *BOD1* | 1.253255427 | 0.013590295 | 0.017836113 | 1 |
| T2DM | nk_cell | *MYLK4* | -2.369491527 | 0.028690297 | 0.035905422 | 1 |
| T2DM | nk_cell | *TBC1D7* | 1.117243869 | 0.020827203 | 0.026607054 | 1 |
| T2DM | nk_cell | *GFOD1* | -1.411688569 | 0.005038265 | 0.007091866 | 1 |
| T2DM | nk_cell | *TRIM7* | -1.134360816 | 1.77E-05 | 0.000122264 | 0.085731465 |
| T2DM | nk_cell | *FAM193B* | -1.281744896 | 9.24E-05 | 0.000223814 | 0.448298966 |
| T2DM | nk_cell | *RNF44* | 1.156931806 | 0.002072427 | 0.003111205 | 1 |
| T2DM | nk_cell | *MUT* | -1.475861053 | 1.87E-05 | 0.000122264 | 0.090749024 |
| T2DM | nk_cell | *DOK3* | -0.436150493 | 0.00045003 | 0.000792576 | 1 |
| T2DM | nk_cell | *ABT1* | 0.779980268 | 0.000111216 | 0.000255069 | 0.539618044 |
| T2DM | nk_cell | *DAAM2* | -1.173192545 | 3.73E-05 | 0.000134293 | 0.181139897 |
| T2DM | nk_cell | *MLIP* | -0.618199753 | 4.45E-05 | 0.000145873 | 0.215891671 |
| T2DM | nk_cell | *ANO7* | -0.821018077 | 0.00020034 | 0.000401389 | 0.972048886 |
| T2DM | nk_cell | *TCTE1* | 1.047251579 | 0.000206666 | 0.000411804 | 1 |
| T2DM | nk_cell | *RPL7L1* | -1.714703248 | 3.64E-05 | 0.000132426 | 0.176504327 |
| T2DM | nk_cell | *TPBG* | -0.899464676 | 1.67E-05 | 0.000122264 | 0.080989066 |
| T2DM | nk_cell | *IRAK1BP1* | -0.923364843 | 0.000529497 | 0.000911681 | 1 |
| T2DM | nk_cell | *PHIP* | -1.211656771 | 0.00020768 | 0.000413485 | 1 |
| T2DM | nk_cell | *PNRC1* | -1.166728161 | 1.89E-05 | 0.000122264 | 0.091881819 |
| T2DM | nk_cell | *RNF217* | -0.683481967 | 1.51E-05 | 0.000122264 | 0.073260301 |
| T2DM | nk_cell | *ARHGAP18* | -0.936286018 | 0.001008569 | 0.001617176 | 1 |
| T2DM | nk_cell | *ABRACL* | 0.462043105 | 0.000110625 | 0.000254143 | 0.536750669 |
| T2DM | nk_cell | *SLC18B1* | -0.969195362 | 0.000118927 | 0.00026616 | 0.577035014 |
| T2DM | nk_cell | *PNLDC1* | -1.573439774 | 0.027538385 | 0.034535085 | 1 |
| T2DM | nk_cell | *WTAP* | -1.218418562 | 7.49E-05 | 0.000195374 | 0.3633957 |
| T2DM | nk_cell | *SLC22A3* | -1.117411584 | 1.60E-05 | 0.000122264 | 0.07766556 |
| T2DM | nk_cell | *GNA12* | -1.655473935 | 4.73E-06 | 0.000122264 | 0.022928713 |
| T2DM | nk_cell | *SDK1* | -0.812074058 | 5.66E-05 | 0.000163891 | 0.274844908 |

| | | | | | | |
|---|---|---|---|---|---|---|
| T2DM | nk_cell | *RBAK* | 1.369344074 | 0.001182948 | 0.001863817 | 1 |
| T2DM | nk_cell | *POMZP3* | -1.406016021 | 0.000317519 | 0.000591175 | 1 |
| T2DM | nk_cell | *PSPH* | -0.537208455 | 0.005227397 | 0.007345303 | 1 |
| T2DM | nk_cell | *GIGYF1* | -1.242246065 | 1.98E-05 | 0.000122264 | 0.09594052 |
| T2DM | nk_cell | *TRIM4* | -0.783897067 | 0.001074991 | 0.001710698 | 1 |
| T2DM | nk_cell | *TMEM209* | -2.001275419 | 0.000208027 | 0.000413837 | 1 |
| T2DM | nk_cell | *EPHA1* | 1.877900459 | 0.000270978 | 0.000516818 | 1 |
| T2DM | nk_cell | *NCAPG2* | 1.226477157 | 0.026595611 | 0.033413233 | 1 |
| T2DM | nk_cell | *FBXO25* | -1.403205092 | 3.89E-05 | 0.000136418 | 0.1888026 |
| T2DM | nk_cell | *CSGALNACT1* | -0.644768011 | 0.000655635 | 0.001096189 | 1 |
| T2DM | nk_cell | *CHMP7* | 1.227116547 | 0.016098662 | 0.020907577 | 1 |
| T2DM | nk_cell | *DOCK5* | -1.27364106 | 8.65E-05 | 0.000213956 | 0.419568138 |
| T2DM | nk_cell | *TACC1* | -0.945466022 | 2.45E-05 | 0.000122264 | 0.118998549 |
| T2DM | nk_cell | *ADHFE1* | -0.724765562 | 5.40E-06 | 0.000122264 | 0.026222002 |
| T2DM | nk_cell | *RPL7* | -0.977106891 | 0.00135057 | 0.002098964 | 1 |
| T2DM | nk_cell | *ATP6V0D2* | -1.791380621 | 9.53E-05 | 0.000228207 | 0.462347625 |
| T2DM | nk_cell | *DPYS* | -0.471781844 | 1.94E-05 | 0.000122264 | 0.094344425 |
| T2DM | nk_cell | *LRP12* | -1.245235512 | 1.69E-05 | 0.000122264 | 0.081986623 |
| T2DM | nk_cell | *GSDMC* | -3.366780305 | 0.023463139 | 0.029755135 | 1 |
| T2DM | nk_cell | *SLC39A4* | -1.032037321 | 1.11E-05 | 0.000122264 | 0.053965649 |
| T2DM | nk_cell | *AK3* | -1.414135179 | 4.98E-05 | 0.00015287 | 0.241567516 |
| T2DM | nk_cell | *HAUS6* | -1.923056878 | 0.003024271 | 0.004419371 | 1 |
| T2DM | nk_cell | *C9orf72* | -0.567132026 | 0.000605603 | 0.001023761 | 1 |
| T2DM | nk_cell | *FBXO10* | -1.018792199 | 0.000248503 | 0.00047961 | 1 |
| T2DM | nk_cell | *ZNF462* | -0.645906539 | 4.27E-05 | 0.000142711 | 0.207358628 |
| T2DM | nk_cell | *WDR31* | -0.737795868 | 0.001122313 | 0.0017784 | 1 |
| T2DM | nk_cell | *POLE3* | 0.557143298 | 0.000145762 | 0.000311971 | 0.70723799 |
| T2DM | nk_cell | *SURF6* | -0.799530914 | 2.55E-05 | 0.000122264 | 0.123525797 |
| T2DM | nk_cell | *MED22* | 0.713194702 | 0.00488032 | 0.006881521 | 1 |
| T2DM | nk_cell | *REXO4* | 1.082477979 | 0.01889102 | 0.024294421 | 1 |
| T2DM | nk_cell | *SH3GLB2* | -2.11588483 | 0.000380955 | 0.000690549 | 1 |
| T2DM | nk_cell | *INPP5E* | -1.219865174 | 1.46E-05 | 0.000122264 | 0.070929239 |
| T2DM | nk_cell | *SLC39A12* | -1.227036179 | 5.37E-05 | 0.000159514 | 0.260571659 |
| T2DM | nk_cell | *ST8SIA6* | 0.196883396 | 6.30E-05 | 0.000173782 | 0.305682779 |
| T2DM | nk_cell | *PARD3* | -0.726399575 | 1.71E-05 | 0.000122264 | 0.083195964 |
| T2DM | nk_cell | *CDHR1* | 0.340136562 | 8.49E-05 | 0.00021115 | 0.411759005 |
| T2DM | nk_cell | *RGR* | -6.642557126 | 5.91E-06 | 0.000122264 | 0.02867354 |
| T2DM | nk_cell | *FRA10AC1* | -1.156334053 | 0.000117119 | 0.000263449 | 0.568259151 |
| T2DM | nk_cell | *ADD3* | -1.244482396 | 0.000131274 | 0.000287687 | 0.636940092 |
| T2DM | nk_cell | *LRRC27* | -0.617643248 | 0.000136133 | 0.000295666 | 0.660517712 |
| T2DM | nk_cell | *GAS2* | -1.559340361 | 8.11E-05 | 0.00020495 | 0.393434542 |
| T2DM | nk_cell | *LIN7C* | -2.047754896 | 0.001140654 | 0.001803928 | 1 |
| T2DM | nk_cell | *ZNF215* | -0.38553818 | 0.005457285 | 0.007639569 | 1 |
| T2DM | nk_cell | *HSD17B12* | -1.238959895 | 1.61E-12 | 1.96E-09 | 7.83E-09 |
| T2DM | nk_cell | *APIP* | -1.007994821 | 1.03E-07 | 1.86E-05 | 0.000500925 |
| T2DM | nk_cell | *EIF3M* | -2.002812624 | 3.02E-05 | 0.000124985 | 0.146564147 |
| T2DM | nk_cell | *TNKS1BP1* | -0.592226719 | 0.014157592 | 0.018530519 | 1 |
| T2DM | nk_cell | *PTPRJ* | 0.708969424 | 0.004102186 | 0.005856363 | 1 |
| T2DM | nk_cell | *CCDC82* | -1.024668975 | 0.000305445 | 0.000571546 | 1 |
| T2DM | nk_cell | *CAPN5* | -0.785772813 | 2.88E-05 | 0.000123207 | 0.13981144 |

| | | | | | | |
|---|---|---|---|---|---|---|
| T2DM | nk_cell | *INTS4* | -1.338099991 | 0.000155815 | 0.000328702 | 0.75601463 |
| T2DM | nk_cell | *RPS3* | 0.716749735 | 0.006434103 | 0.008896628 | 1 |
| T2DM | nk_cell | *DRD2* | -4.039674334 | 2.21E-05 | 0.000122264 | 0.107118764 |
| T2DM | nk_cell | *SLX4IP* | -1.144023824 | 2.64E-05 | 0.000122391 | 0.128292399 |
| T2DM | nk_cell | *ST14* | -1.481539587 | 4.23E-05 | 0.000141926 | 0.205083133 |
| T2DM | nk_cell | *FADS1* | 1.048308137 | 0.0018237 | 0.002769513 | 1 |
| T2DM | nk_cell | *TMC2* | -1.900392504 | 7.62E-06 | 0.000122264 | 0.036995845 |
| T2DM | nk_cell | *ROM1* | -0.698674799 | 0.000410906 | 0.000734874 | 1 |
| T2DM | nk_cell | *EML3* | -0.903309528 | 0.001635602 | 0.002506615 | 1 |
| T2DM | nk_cell | *PLCH2* | 0.138676949 | 4.86E-05 | 0.000151135 | 0.235771033 |
| T2DM | nk_cell | *CPSF7* | -1.696825123 | 6.67E-05 | 0.000180692 | 0.323620197 |
| T2DM | nk_cell | *CCDC15* | -0.716871437 | 2.49E-06 | 0.000122264 | 0.012075315 |
| T2DM | nk_cell | *CHEK1* | -0.847984644 | 2.66E-05 | 0.000122391 | 0.129283489 |
| T2DM | nk_cell | *SCN2B* | -0.690922358 | 0.021770078 | 0.027731273 | 1 |
| T2DM | nk_cell | *CABLES2* | -1.010638054 | 0.016847561 | 0.021821774 | 1 |
| T2DM | nk_cell | *TRPT1* | -1.175082794 | 0.000104172 | 0.000242767 | 0.5054405 |
| T2DM | nk_cell | *NUDT22* | -1.81882974 | 6.62E-05 | 0.000179817 | 0.321332226 |
| T2DM | nk_cell | *ANXA8L1* | -0.419010921 | 0.012206714 | 0.016133744 | 1 |
| T2DM | nk_cell | *FCGR1A* | -1.077177402 | 0.0180848 | 0.023323951 | 1 |
| T2DM | nk_cell | *TMEM218* | -1.768710652 | 1.68E-05 | 0.000122264 | 0.081453969 |
| T2DM | nk_cell | *SAP18* | -1.565184422 | 0.024632016 | 0.031123578 | 1 |
| T2DM | nk_cell | *FAM124A* | -1.160521288 | 3.84E-05 | 0.000135693 | 0.186096651 |
| T2DM | nk_cell | *LYPD6B* | -0.837102217 | 0.000353758 | 0.000647222 | 1 |
| T2DM | nk_cell | *WDR17* | -1.613614746 | 1.79E-05 | 0.000122264 | 0.087038345 |
| T2DM | nk_cell | *CCDC102B* | -0.822961198 | 1.92E-05 | 0.000122264 | 0.093202767 |
| T2DM | nk_cell | *PRSS23* | -0.810250235 | 2.24E-05 | 0.000122264 | 0.108461506 |
| T2DM | nk_cell | *PPP1R1C* | -1.055212379 | 2.12E-05 | 0.000122264 | 0.102749691 |
| T2DM | nk_cell | *FAM173B* | -1.951546299 | 0.000274049 | 0.000521037 | 1 |
| T2DM | nk_cell | *GPR158* | -1.19874622 | 7.05E-05 | 0.000187698 | 0.341986305 |
| T2DM | nk_cell | *UBE3B* | -0.951198889 | 0.006813143 | 0.009364694 | 1 |
| T2DM | nk_cell | *ANK3* | 0.652971174 | 0.00030033 | 0.000563713 | 1 |
| T2DM | nk_cell | *PLBD2* | -1.48266441 | 0.000294365 | 0.000554018 | 1 |
| T2DM | nk_cell | *DLG5* | -1.109143135 | 6.97E-06 | 0.000122264 | 0.033834346 |
| T2DM | nk_cell | *GXYLT1* | -1.799231452 | 7.52E-05 | 0.000196007 | 0.364965267 |
| T2DM | nk_cell | *EXT2* | -1.365487433 | 8.43E-05 | 0.000210467 | 0.408936599 |
| T2DM | nk_cell | *TMEM18* | -2.308488694 | 6.54E-05 | 0.000178359 | 0.317359448 |
| T2DM | nk_cell | *NDUFC2* | -2.295751579 | 0.002190142 | 0.003273743 | 1 |
| T2DM | nk_cell | *ADAMTS12* | -0.763328591 | 9.23E-05 | 0.000223745 | 0.447713617 |
| T2DM | nk_cell | *NUBPL* | -1.305899556 | 6.85E-05 | 0.00018443 | 0.332163401 |
| T2DM | nk_cell | *VIPAS39* | -1.021365393 | 6.68E-06 | 0.000122264 | 0.032402615 |
| T2DM | nk_cell | *ANKRD50* | -1.31901181 | 6.98E-05 | 0.000186464 | 0.338618065 |
| T2DM | nk_cell | *UPF2* | -1.360591693 | 0.00013243 | 0.000289306 | 0.642548478 |
| T2DM | nk_cell | *PTPRO* | -1.043009829 | 3.48E-05 | 0.000130559 | 0.169074515 |
| T2DM | nk_cell | *EPS8* | -0.97931908 | 9.18E-05 | 0.000222758 | 0.445515129 |
| T2DM | nk_cell | *QDPR* | -0.75893602 | 2.54E-05 | 0.000122264 | 0.123438661 |
| T2DM | nk_cell | *FAM160B1* | -1.103310727 | 3.87E-05 | 0.00013613 | 0.187589248 |
| T2DM | nk_cell | *ZNF827* | -1.374727623 | 0.004669638 | 0.006603638 | 1 |
| T2DM | nk_cell | *AKR1C2* | -0.619474914 | 0.000112944 | 0.000257763 | 0.548004236 |
| T2DM | nk_cell | *DPYSL4* | 0.46037023 | 0.006987631 | 0.009586244 | 1 |
| T2DM | nk_cell | *PIGF* | -1.316525778 | 2.47E-05 | 0.000122264 | 0.120057684 |

| | | | | | | |
|---|---|---|---|---|---|---|
| T2DM | nk_cell | *INPP1* | -1.192870906 | 0.000294146 | 0.000554018 | 1 |
| T2DM | nk_cell | *RNF144A* | -0.789364086 | 1.79E-05 | 0.000122264 | 0.086672765 |
| T2DM | nk_cell | *CENPU* | -1.068585819 | 0.038045812 | 0.046804838 | 1 |
| T2DM | nk_cell | *BICD1* | -1.272760933 | 4.12E-05 | 0.000140276 | 0.199753237 |
| T2DM | nk_cell | *CCDC122* | -0.695985057 | 0.003613667 | 0.005218307 | 1 |
| T2DM | nk_cell | *GUF1* | -1.133647945 | 1.57E-05 | 0.000122264 | 0.076119474 |
| T2DM | nk_cell | *GABRA2* | 1.106437772 | 0.000339396 | 0.000624952 | 1 |
| T2DM | nk_cell | *PARP8* | 0.424590616 | 0.003419911 | 0.004960661 | 1 |
| T2DM | nk_cell | *CACUL1* | -1.42946126 | 0.000544213 | 0.000933047 | 1 |
| T2DM | nk_cell | *BEND6* | -0.714902686 | 1.98E-05 | 0.000122264 | 0.096053507 |
| T2DM | nk_cell | *TIAL1* | -1.550010067 | 0.000117087 | 0.000263449 | 0.568105678 |
| T2DM | nk_cell | *TMEM56* | -2.094867247 | 1.88E-05 | 0.000122264 | 0.091285491 |
| T2DM | nk_cell | *PTPN14* | -0.920098285 | 4.67E-05 | 0.000148657 | 0.226355676 |
| T2DM | nk_cell | *MGAT5* | -1.369345877 | 3.87E-05 | 0.00013613 | 0.187858715 |
| T2DM | nk_cell | *TMEM178A* | 0.977636607 | 0.00015347 | 0.000325168 | 0.744634529 |
| T2DM | nk_cell | *EPG5* | -1.654466481 | 0.000111409 | 0.000255221 | 0.540557518 |
| T2DM | nk_cell | *KCNK13* | -2.273876012 | 0.011459805 | 0.015212852 | 1 |
| T2DM | nk_cell | *ATG10* | -0.486048321 | 0.003663785 | 0.00528596 | 1 |
| T2DM | nk_cell | *POC5* | -1.781982638 | 8.08E-07 | 6.65E-05 | 0.003921055 |
| T2DM | nk_cell | *CWF19L2* | -1.01184423 | 0.000755052 | 0.001239125 | 1 |
| T2DM | nk_cell | *ZNF773* | -1.063211823 | 0.002113774 | 0.003170334 | 1 |
| T2DM | nk_cell | *NMT2* | -1.455787909 | 5.04E-05 | 0.000153489 | 0.244721709 |
| T2DM | nk_cell | *ZNF837* | -0.983964597 | 0.000105504 | 0.000244931 | 0.511904994 |
| T2DM | nk_cell | *CCDC50* | -0.722391014 | 8.40E-05 | 0.000209939 | 0.407491989 |
| T2DM | nk_cell | *PLEKHH2* | -0.522623803 | 0.000566282 | 0.000965386 | 1 |
| T2DM | nk_cell | *SPARCL1* | -0.924900756 | 3.49E-05 | 0.000130739 | 0.1695689 |
| T2DM | nk_cell | *GPD1L* | -1.325608355 | 0.007970245 | 0.010817238 | 1 |
| T2DM | nk_cell | *CCNO* | -4.934045087 | 3.00E-05 | 0.000124725 | 0.145554364 |
| T2DM | nk_cell | *CLEC4F* | -1.043841093 | 0.00127523 | 0.001992085 | 1 |
| T2DM | nk_cell | *SLC30A6* | -1.129466558 | 8.82E-05 | 0.000216718 | 0.427801442 |
| T2DM | nk_cell | *RASGRP3* | -1.858671022 | 0.000116744 | 0.000263375 | 0.566442888 |
| T2DM | nk_cell | *GPR180* | -0.747629478 | 0.000278093 | 0.000527073 | 1 |
| T2DM | nk_cell | *TCTEX1D1* | -1.404269832 | 0.001075004 | 0.001710698 | 1 |
| T2DM | nk_cell | *IFIT5* | -0.479396145 | 0.030638105 | 0.038185483 | 1 |
| T2DM | nk_cell | *PRDM8* | 0.810882167 | 0.000373886 | 0.000679182 | 1 |
| T2DM | nk_cell | *ZNF117* | -0.199122935 | 0.004142249 | 0.005906021 | 1 |
| T2DM | nk_cell | *MED21* | -1.720961514 | 0.000152335 | 0.000323491 | 0.739129267 |
| T2DM | nk_cell | *JAKMIP1* | 0.115226569 | 5.66E-05 | 0.000163891 | 0.274386777 |
| T2DM | nk_cell | *SREK1IP1* | -1.538202449 | 2.89E-05 | 0.000123207 | 0.140041742 |
| T2DM | nk_cell | *CENPH* | -1.485280805 | 1.22E-05 | 0.000122264 | 0.059345417 |
| T2DM | nk_cell | *CARHSP1* | -1.374622807 | 9.10E-05 | 0.00022117 | 0.441455656 |
| T2DM | nk_cell | *CAST* | -1.023837252 | 0.00016284 | 0.000339975 | 0.790102051 |
| T2DM | nk_cell | *SCOC* | -1.586126745 | 0.000608054 | 0.00102654 | 1 |
| T2DM | nk_cell | *CLGN* | 0.600453253 | 8.98E-05 | 0.000219081 | 0.435532602 |
| T2DM | nk_cell | *CETN3* | -1.192409642 | 0.019278745 | 0.024765811 | 1 |
| T2DM | nk_cell | *SYCP2L* | -0.613606483 | 6.36E-06 | 0.000122264 | 0.030872368 |
| T2DM | nk_cell | *MERTK* | -0.200004219 | 0.023659065 | 0.029987926 | 1 |
| T2DM | nk_cell | *TMEM87B* | -1.483225118 | 5.12E-05 | 0.000155307 | 0.248646301 |
| T2DM | nk_cell | *PLA2R1* | -0.474640839 | 0.003945879 | 0.005650946 | 1 |
| T2DM | nk_cell | *SLC25A27* | -0.811175205 | 9.28E-05 | 0.000223847 | 0.450108976 |

| | | | | | | |
|---|---|---|---|---|---|---|
| T2DM | nk_cell | *FRMD1* | -0.768056704 | 3.76E-05 | 0.00013436 | 0.1824613 |
| T2DM | nk_cell | *INO80C* | -1.816443846 | 6.23E-05 | 0.000173265 | 0.30241557 |
| T2DM | nk_cell | *LPCAT1* | 0.399431428 | 0.000109517 | 0.000251956 | 0.531374735 |
| T2DM | nk_cell | *NMRAL1* | 1.035892341 | 0.004039506 | 0.005773102 | 1 |
| T2DM | nk_cell | *ADPRHL1* | -1.146749732 | 8.37E-05 | 0.000209406 | 0.406037408 |
| T2DM | nk_cell | *CFDP1* | -1.217419992 | 4.38E-05 | 0.000144323 | 0.212298462 |
| T2DM | nk_cell | *ZDHHC7* | -0.952191695 | 1.54E-05 | 0.000122264 | 0.074542074 |
| T2DM | nk_cell | *JAZF1* | -1.692079535 | 7.44E-06 | 0.000122264 | 0.036118475 |
| T2DM | nk_cell | *FBXO36* | -1.237347289 | 1.44E-05 | 0.000122264 | 0.069652228 |
| T2DM | nk_cell | *MCOLN2* | 0.200260241 | 6.87E-05 | 0.000184665 | 0.333319874 |
| T2DM | nk_cell | *GDPD1* | 1.049191434 | 0.03656488 | 0.04506294 | 1 |
| T2DM | nk_cell | *SEMA3D* | -0.681245063 | 1.58E-05 | 0.000122264 | 0.076715682 |
| T2DM | nk_cell | *PPP2R5E* | -1.335474864 | 0.003049388 | 0.004451152 | 1 |
| T2DM | nk_cell | *ANKRD29* | -1.156671339 | 2.41E-05 | 0.000122264 | 0.116874448 |
| T2DM | nk_cell | *DNAAF1* | -1.21615352 | 0.000166691 | 0.000346226 | 0.80878417 |
| T2DM | nk_cell | *ANKH* | -1.187964553 | 0.000129242 | 0.000284134 | 0.62708299 |
| T2DM | nk_cell | *UBASH3B* | 0.508673401 | 0.000675776 | 0.001125985 | 1 |
| T2DM | nk_cell | *ROBO4* | -1.333360172 | 2.39E-05 | 0.000122264 | 0.116067466 |
| T2DM | nk_cell | *ROBO3* | -0.521960819 | 1.60E-05 | 0.000122264 | 0.077796716 |
| T2DM | nk_cell | *CC2D1B* | -1.542880526 | 8.41E-05 | 0.000210014 | 0.407846862 |
| T2DM | nk_cell | *PRKCA* | -1.209590768 | 4.58E-05 | 0.000147697 | 0.222197192 |
| T2DM | nk_cell | *LRRK1* | -1.057477864 | 1.55E-05 | 0.000122264 | 0.07502997 |
| T2DM | nk_cell | *ABCA5* | -0.835948253 | 0.038011185 | 0.046774098 | 1 |
| T2DM | nk_cell | *ENPP3* | -0.697506645 | 1.43E-05 | 0.000122264 | 0.069426044 |
| T2DM | nk_cell | *MIA3* | -1.317568951 | 7.81E-05 | 0.000200157 | 0.378797994 |
| T2DM | nk_cell | *NEIL2* | -0.752796403 | 0.001793142 | 0.002727375 | 1 |
| T2DM | nk_cell | *PGM5* | -1.218738494 | 3.03E-05 | 0.000124985 | 0.146845627 |
| T2DM | nk_cell | *CCSAP* | -2.302782338 | 2.86E-05 | 0.000123207 | 0.138812609 |
| T2DM | nk_cell | *GBP5* | 0.159190794 | 0.000105288 | 0.000244782 | 0.510859738 |
| T2DM | nk_cell | *ATP5G3* | 0.852938047 | 0.005298926 | 0.007437196 | 1 |
| T2DM | nk_cell | *LY96* | -1.07727874 | 2.50E-05 | 0.000122264 | 0.121142683 |
| T2DM | nk_cell | *PDE1C* | -0.466982681 | 2.11E-05 | 0.000122264 | 0.102340948 |
| T2DM | nk_cell | *RABGEF1* | -0.54873651 | 0.000337074 | 0.000621384 | 1 |
| T2DM | nk_cell | *MRPL39* | -0.606277636 | 0.019385652 | 0.024896554 | 1 |
| T2DM | nk_cell | *XPC* | -1.440317581 | 0.000224026 | 0.000440785 | 1 |
| T2DM | nk_cell | *FLCN* | -1.198020031 | 5.86E-05 | 0.000167232 | 0.284167153 |
| T2DM | nk_cell | *OXNAD1* | -1.342281207 | 6.26E-05 | 0.000173589 | 0.303954098 |
| T2DM | nk_cell | *APCDD1* | -0.435195131 | 3.03E-05 | 0.000124985 | 0.146963707 |
| T2DM | nk_cell | *ANKRD40* | -0.922461378 | 6.55E-06 | 0.000122264 | 0.03177973 |
| T2DM | nk_cell | *CYP2U1* | -0.69256875 | 0.000111069 | 0.000254922 | 0.538905492 |
| T2DM | nk_cell | *AK9* | -0.991035805 | 0.031054291 | 0.038674389 | 1 |
| T2DM | nk_cell | *MARVELD1* | -0.677226011 | 2.12E-05 | 0.000122264 | 0.102937432 |
| T2DM | nk_cell | *GOLGA7B* | 0.83353211 | 6.07E-05 | 0.000170412 | 0.294641795 |
| T2DM | nk_cell | *TRMT44* | -2.002714036 | 0.000362282 | 0.000661072 | 1 |
| T2DM | nk_cell | *MOV10* | -1.708501707 | 3.68E-06 | 0.000122264 | 0.017846108 |
| T2DM | nk_cell | *CNOT8* | -1.794489624 | 9.70E-05 | 0.000231207 | 0.470505302 |
| T2DM | nk_cell | *LRGUK* | -0.43031124 | 5.73E-05 | 0.000165252 | 0.277954509 |
| T2DM | nk_cell | *SETD9* | -0.473164783 | 4.54E-06 | 0.000122264 | 0.022048142 |
| T2DM | nk_cell | *PDIA4* | 1.080322815 | 0.001856802 | 0.002816255 | 1 |
| T2DM | nk_cell | *KDM8* | 1.490434309 | 0.036044243 | 0.044477789 | 1 |

| | | | | | | |
|---|---|---|---|---|---|---|
| T2DM | nk_cell | *ALS2CR12* | -1.715659866 | 0.000605904 | 0.001023761 | 1 |
| T2DM | nk_cell | *FZD7* | -0.44425578 | 5.79E-05 | 0.000166463 | 0.281156605 |
| T2DM | nk_cell | *DEPTOR* | -0.655377501 | 3.30E-05 | 0.000127909 | 0.16006504 |
| T2DM | nk_cell | *RRAGA* | -2.262586043 | 0.003794754 | 0.005453835 | 1 |
| T2DM | nk_cell | *TMBIM4* | -1.171012049 | 7.29E-05 | 0.000192063 | 0.35378047 |
| T2DM | nk_cell | *PSD3* | -0.728343692 | 0.000627354 | 0.001053989 | 1 |
| T2DM | nk_cell | *UGT2B4* | -0.405095709 | 0.001486646 | 0.002288454 | 1 |
| T2DM | nk_cell | *MMP16* | -0.639921069 | 0.000137624 | 0.000297848 | 0.667753623 |
| T2DM | nk_cell | *KCNMA1* | -0.508085977 | 0.001572234 | 0.002414076 | 1 |
| T2DM | nk_cell | *NDUFAF6* | -1.473814612 | 3.97E-07 | 4.28E-05 | 0.0019263 |
| T2DM | nk_cell | *DRAM2* | -0.719677806 | 0.004718963 | 0.006669505 | 1 |
| T2DM | nk_cell | *C8orf37* | -0.673934423 | 0.000126775 | 0.000279342 | 0.615112025 |
| T2DM | nk_cell | *PPEF2* | -1.241638816 | 0.011593764 | 0.015382265 | 1 |
| T2DM | nk_cell | *ART3* | -0.905536746 | 0.002131014 | 0.003192244 | 1 |
| T2DM | nk_cell | *WHAMM* | -0.991019659 | 0.015419345 | 0.020089866 | 1 |
| T2DM | nk_cell | *N6AMT1* | -1.601913849 | 0.000278067 | 0.000527073 | 1 |
| T2DM | nk_cell | *RWDD2B* | -1.226344394 | 0.000348499 | 0.000639047 | 1 |
| T2DM | nk_cell | *NAA11* | -0.849456553 | 1.56E-05 | 0.000122264 | 0.075778656 |
| T2DM | nk_cell | *C14orf2* | -1.601958566 | 9.42E-06 | 0.000122264 | 0.045683462 |
| T2DM | nk_cell | *TDRD9* | 2.201600051 | 0.006127557 | 0.008501832 | 1 |
| T2DM | nk_cell | *PCDH1* | -0.681048611 | 0.002002939 | 0.003017157 | 1 |
| T2DM | nk_cell | *SUPV3L1* | -1.755235834 | 0.007500151 | 0.010222116 | 1 |
| T2DM | nk_cell | *HKDC1* | -3.036821703 | 9.50E-06 | 0.000122264 | 0.046101609 |
| T2DM | nk_cell | *TYSND1* | -0.702433318 | 0.000133688 | 0.00029153 | 0.648653813 |
| T2DM | nk_cell | *NODAL* | -0.980179373 | 2.12E-05 | 0.000122264 | 0.102654642 |
| T2DM | nk_cell | *MED19* | -0.920499362 | 0.000151616 | 0.00032265 | 0.735642796 |
| T2DM | nk_cell | *WDYHV1* | -0.7960078 | 0.002276471 | 0.003393375 | 1 |
| T2DM | nk_cell | *FBRS* | -1.197064101 | 0.006811072 | 0.0093645 | 1 |
| T2DM | nk_cell | *SASS6* | -0.664787447 | 5.68E-06 | 0.000122264 | 0.027554473 |
| T2DM | nk_cell | *ITGAD* | 0.144018546 | 5.28E-05 | 0.000158242 | 0.256352495 |
| T2DM | nk_cell | *VPS8* | -0.910152807 | 0.023672856 | 0.02999757 | 1 |
| T2DM | nk_cell | *MPV17L* | -0.924363885 | 4.37E-05 | 0.000144302 | 0.211795025 |
| T2DM | nk_cell | *RPUSD3* | -1.425939536 | 1.61E-05 | 0.000122264 | 0.078063456 |
| T2DM | nk_cell | *GHRL* | -1.633946236 | 0.000105495 | 0.000244931 | 0.511860703 |
| T2DM | nk_cell | *NTAN1* | -1.406276786 | 0.005826291 | 0.008116326 | 1 |
| T2DM | nk_cell | *NMNAT2* | -2.125312942 | 0.000103131 | 0.000241152 | 0.500390251 |
| T2DM | nk_cell | *SLC6A1* | -1.543394545 | 3.17E-05 | 0.000127109 | 0.153663682 |
| T2DM | nk_cell | *FCHO2* | -0.973335935 | 0.000265322 | 0.000507471 | 1 |
| T2DM | nk_cell | *TIMP4* | 0.676472877 | 0.039871555 | 0.048889761 | 1 |
| T2DM | nk_cell | *NECAP2* | -1.658078417 | 3.20E-05 | 0.000127628 | 0.155321058 |
| T2DM | nk_cell | *STEAP2* | -0.491754526 | 0.004829466 | 0.006815756 | 1 |
| T2DM | nk_cell | *CLDN12* | -2.116969598 | 2.75E-05 | 0.000122472 | 0.133392901 |
| T2DM | nk_cell | *MMP14* | -0.708414793 | 1.70E-05 | 0.000122264 | 0.0824894 |
| T2DM | nk_cell | *GATAD1* | -1.250762384 | 1.97E-05 | 0.000122264 | 0.095629039 |
| T2DM | nk_cell | *TMED6* | -2.454810657 | 0.002561102 | 0.003785096 | 1 |
| T2DM | nk_cell | *CLEC18A* | -2.556546835 | 2.35E-09 | 9.85E-07 | 1.14E-05 |
| T2DM | nk_cell | *DHRS4* | -1.267381969 | 1.69E-05 | 0.000122264 | 0.082160264 |
| T2DM | nk_cell | *DHRS1* | -0.611179005 | 0.000845645 | 0.001375023 | 1 |
| T2DM | nk_cell | *AASDH* | -1.136004136 | 0.000131839 | 0.000288342 | 0.639683227 |
| T2DM | nk_cell | *ETS2* | 0.54611159 | 0.02998397 | 0.037408645 | 1 |

| | | | | | | |
|---|---|---|---|---|---|---|
| T2DM | nk_cell | *LCA5L* | -0.777769048 | 0.000138305 | 0.000298911 | 0.671055691 |
| T2DM | nk_cell | *MX1* | -0.768185284 | 7.79E-05 | 0.000199993 | 0.377786298 |
| T2DM | nk_cell | *SLC38A10* | -1.641722247 | 9.48E-05 | 0.000227532 | 0.460069205 |
| T2DM | nk_cell | *C9orf43* | -0.820296489 | 0.000647962 | 0.001084481 | 1 |
| T2DM | nk_cell | *DGKI* | -0.541219727 | 5.60E-06 | 0.000122264 | 0.027166148 |
| T2DM | nk_cell | *SNX22* | 1.451909266 | 0.000192463 | 0.000388772 | 0.933829425 |
| T2DM | nk_cell | *SLC34A2* | 4.098010374 | 5.89E-05 | 0.000167252 | 0.285666267 |
| T2DM | nk_cell | *SPPL3* | -1.338731568 | 2.78E-05 | 0.000122493 | 0.135092614 |
| T2DM | nk_cell | *C12orf43* | -0.894235622 | 2.73E-06 | 0.000122264 | 0.013236197 |
| T2DM | nk_cell | *PEX10* | -2.290366872 | 0.000121572 | 0.000270222 | 0.589867862 |
| T2DM | nk_cell | *PAFAH2* | 0.698498805 | 0.026955147 | 0.033838648 | 1 |
| T2DM | nk_cell | *EXTL1* | -7.319436383 | 2.56E-05 | 0.000122264 | 0.124118341 |
| T2DM | nk_cell | *WDR66* | -0.768671361 | 7.55E-06 | 0.000122264 | 0.036634352 |
| T2DM | nk_cell | *RHPN1* | -1.011561394 | 4.17E-05 | 0.000141263 | 0.202429563 |
| T2DM | nk_cell | *DZIP1L* | -0.950631816 | 2.86E-05 | 0.000123207 | 0.139005323 |
| T2DM | nk_cell | *FANCC* | -0.845748732 | 1.62E-05 | 0.000122264 | 0.078773856 |
| T2DM | nk_cell | *ESYT3* | -1.136032993 | 1.79E-05 | 0.000122264 | 0.086794739 |
| T2DM | nk_cell | *CLSTN2* | -1.049459039 | 1.05E-05 | 0.000122264 | 0.051026479 |
| T2DM | nk_cell | *COLEC12* | -0.580666774 | 8.92E-05 | 0.000217825 | 0.432601346 |
| T2DM | nk_cell | *MITD1* | -1.399619414 | 0.011255004 | 0.014969649 | 1 |
| T2DM | nk_cell | *TSPAN33* | -1.818574491 | 2.66E-06 | 0.000122264 | 0.012894995 |
| T2DM | nk_cell | *FAM86C1* | -5.677442992 | 0.000412099 | 0.000736738 | 1 |
| T2DM | nk_cell | *PPP1R9A* | -1.205833251 | 5.76E-05 | 0.000165907 | 0.279564316 |
| T2DM | nk_cell | *DYNC1I1* | -0.916818813 | 9.25E-05 | 0.000223822 | 0.448762644 |
| T2DM | nk_cell | *COPG2* | -0.517043648 | 0.000186711 | 0.000379683 | 0.905922708 |
| T2DM | nk_cell | *DUSP23* | -0.710400545 | 2.85E-05 | 0.00012318 | 0.138331119 |
| T2DM | nk_cell | *RNF166* | 0.424414355 | 0.000225834 | 0.000443621 | 1 |
| T2DM | nk_cell | *SPATA2L* | 0.875998667 | 0.014084988 | 0.018445442 | 1 |
| T2DM | nk_cell | *FGF17* | -2.962524511 | 0.000298409 | 0.000560976 | 1 |
| T2DM | nk_cell | *CDA* | -0.661067357 | 0.000287492 | 0.000543614 | 1 |
| T2DM | nk_cell | *SLC5A11* | -0.521803177 | 0.000108438 | 0.000249949 | 0.526141786 |
| T2DM | nk_cell | *CCAR2* | -1.565820794 | 0.002393109 | 0.003558494 | 1 |
| T2DM | nk_cell | *CDC42SE2* | 0.63712069 | 0.006217495 | 0.008616762 | 1 |
| T2DM | nk_cell | *IFNAR2* | 0.644796937 | 0.012842591 | 0.016909554 | 1 |
| T2DM | nk_cell | *SV2A* | -1.576536497 | 0.032223475 | 0.040017482 | 1 |
| T2DM | nk_cell | *CLIC6* | -1.138076959 | 0.005593611 | 0.007816879 | 1 |
| T2DM | nk_cell | *CBR3* | -0.763008492 | 0.000394697 | 0.000712716 | 1 |
| T2DM | nk_cell | *CHAF1B* | -1.300583332 | 0.000569948 | 0.000968752 | 1 |
| T2DM | nk_cell | *SIM2* | 0.328227493 | 0.002247682 | 0.003353552 | 1 |
| T2DM | nk_cell | *HLCS* | -0.985950558 | 0.013362803 | 0.017551792 | 1 |
| T2DM | nk_cell | *ARHGAP27* | -2.012709992 | 5.78E-05 | 0.000166383 | 0.280688151 |
| T2DM | nk_cell | *IRX6* | -0.456095932 | 1.00E-05 | 0.000122264 | 0.048506977 |
| T2DM | nk_cell | *HK2* | -0.87324101 | 6.51E-05 | 0.000177893 | 0.315937403 |
| T2DM | nk_cell | *TMEM69* | -1.880426382 | 1.43E-05 | 0.000122264 | 0.069287863 |
| T2DM | nk_cell | *ACE* | -0.868082342 | 2.52E-05 | 0.000122264 | 0.122184887 |
| T2DM | nk_cell | *TEPP* | 0.69994303 | 8.92E-05 | 0.0002179 | 0.432967818 |
| T2DM | nk_cell | *CHCHD6* | -1.191424219 | 9.47E-05 | 0.000227317 | 0.459407763 |
| T2DM | nk_cell | *CTBP1* | -1.858746616 | 0.000372126 | 0.000676744 | 1 |
| T2DM | nk_cell | *LRRC36* | -0.56124195 | 3.78E-05 | 0.000134804 | 0.183333058 |
| T2DM | nk_cell | *ZFYVE28* | 0.548414339 | 0.001158084 | 0.001829109 | 1 |

| | | | | | | |
|---|---|---|---|---|---|---|
| T2DM | nk_cell | *C16orf86* | -0.891826532 | 0.000165872 | 0.000344672 | 0.804808534 |
| T2DM | nk_cell | *RGS12* | -1.118290377 | 9.18E-06 | 0.000122264 | 0.044526849 |
| T2DM | nk_cell | *LYPD5* | -1.126812228 | 8.98E-06 | 0.000122264 | 0.043560194 |
| T2DM | nk_cell | *CCDC107* | -2.062586402 | 0.001259429 | 0.001968669 | 1 |
| T2DM | nk_cell | *ZNF233* | -1.730262631 | 0.000385382 | 0.00069746 | 1 |
| T2DM | nk_cell | *CPAMD8* | -0.542014622 | 0.000151135 | 0.000321773 | 0.733306817 |
| T2DM | nk_cell | *ANKLE1* | -3.055981433 | 0.001460289 | 0.002253601 | 1 |
| T2DM | nk_cell | *CCDC58* | -1.229072144 | 1.58E-05 | 0.000122264 | 0.076858411 |
| T2DM | nk_cell | *CILP2* | 1.276197574 | 0.009568217 | 0.012863671 | 1 |
| T2DM | nk_cell | *PDE9A* | -1.077480999 | 0.000432165 | 0.000765 | 1 |
| T2DM | nk_cell | *HSF2BP* | -1.208131058 | 3.22E-06 | 0.000122264 | 0.015626502 |
| T2DM | nk_cell | *CSTB* | -1.214444652 | 0.00011778 | 0.000264569 | 0.571468573 |
| T2DM | nk_cell | *C21orf2* | -0.554815121 | 2.34E-05 | 0.000122264 | 0.113698174 |
| T2DM | nk_cell | *FTCD* | -0.952241925 | 0.000188333 | 0.00038234 | 0.913792459 |
| T2DM | nk_cell | *SPATC1L* | -0.319694981 | 0.002148269 | 0.003216107 | 1 |
| T2DM | nk_cell | *LSS* | -0.818203577 | 3.40E-06 | 0.000122264 | 0.016497272 |
| T2DM | nk_cell | *VAV2* | -0.755067673 | 2.07E-05 | 0.000122264 | 0.100284796 |
| T2DM | nk_cell | *MCM3AP* | -0.981585203 | 4.91E-05 | 0.000151955 | 0.23841694 |
| T2DM | nk_cell | *S100B* | -0.309766815 | 0.012572893 | 0.016577086 | 1 |
| T2DM | nk_cell | *PRMT2* | -1.310449431 | 0.000156773 | 0.000330578 | 0.760660359 |
| T2DM | nk_cell | *ZNF208* | -0.965292892 | 1.13E-05 | 0.000122264 | 0.054810877 |
| T2DM | nk_cell | *ADAMTS13* | -0.674188893 | 1.52E-05 | 0.000122264 | 0.073795664 |
| T2DM | nk_cell | *CACFD1* | -3.434306864 | 2.60E-05 | 0.000122264 | 0.126335382 |
| T2DM | nk_cell | *RDH13* | 0.727953496 | 0.02982922 | 0.037253894 | 1 |
| T2DM | nk_cell | *TAOK1* | -1.133897354 | 7.40E-05 | 0.000194104 | 0.359286342 |
| T2DM | nk_cell | *TLCD1* | -2.261489253 | 0.003697766 | 0.005328191 | 1 |
| T2DM | nk_cell | *S100A1* | -1.788494463 | 0.000596327 | 0.001010259 | 1 |
| T2DM | nk_cell | *ZBTB7B* | -0.978232508 | 3.93E-05 | 0.000136619 | 0.190714497 |
| T2DM | nk_cell | *FDPS* | -1.232170761 | 0.01081568 | 0.014416944 | 1 |
| T2DM | nk_cell | *CCR5* | 0.298334106 | 2.40E-05 | 0.000122264 | 0.116270401 |
| T2DM | nk_cell | *NBEAL2* | 0.76895814 | 0.000406977 | 0.000729461 | 1 |
| T2DM | nk_cell | *PTH1R* | -1.441607301 | 3.35E-05 | 0.000128164 | 0.162590599 |
| T2DM | nk_cell | *FCRL3* | 0.115766934 | 8.00E-05 | 0.000203408 | 0.388103142 |
| T2DM | nk_cell | *FGFR4* | 1.716208205 | 0.003006481 | 0.004396458 | 1 |
| T2DM | nk_cell | *LY6K* | -1.238124334 | 0.000429764 | 0.000761583 | 1 |
| T2DM | nk_cell | *IER2* | -1.889128406 | 0.0022561 | 0.003365077 | 1 |
| T2DM | nk_cell | *CPSF4* | 0.862748242 | 0.002968454 | 0.004342158 | 1 |
| T2DM | nk_cell | *TONSL* | -1.523924996 | 0.016034221 | 0.020829462 | 1 |
| T2DM | nk_cell | *MUM1* | -0.924804662 | 2.39E-05 | 0.000122264 | 0.115846297 |
| T2DM | nk_cell | *SH2B2* | -1.242347931 | 0.001205111 | 0.001894137 | 1 |
| T2DM | nk_cell | *LRWD1* | 0.806261971 | 4.15E-05 | 0.000140757 | 0.201148045 |
| T2DM | nk_cell | *DVL3* | -1.418273457 | 2.51E-06 | 0.000122264 | 0.01218973 |
| T2DM | nk_cell | *FBXO27* | -0.451657184 | 0.000116489 | 0.000263067 | 0.565206892 |
| T2DM | nk_cell | *U2AF1L4* | -0.276696669 | 0.022567857 | 0.028687253 | 1 |
| T2DM | nk_cell | *BDH1* | -1.793487226 | 0.000368978 | 0.00067228 | 1 |
| T2DM | nk_cell | *LRRC56* | -1.211621324 | 0.000320488 | 0.000595789 | 1 |
| T2DM | nk_cell | *ZNF577* | -0.836977433 | 8.64E-05 | 0.000213956 | 0.419264329 |
| T2DM | nk_cell | *TMEM143* | -1.211321322 | 3.52E-05 | 0.000130901 | 0.170563706 |
| T2DM | nk_cell | *CCL16* | -1.310864888 | 0.000508699 | 0.00088056 | 1 |
| T2DM | nk_cell | *DCD* | -3.398416023 | 0.002766095 | 0.004062075 | 1 |

| | | | | | | |
|---|---|---|---|---|---|---|
| T2DM | nk_cell | *SIGLEC11* | -0.567092192 | 0.001540877 | 0.002368927 | 1 |
| T2DM | nk_cell | *ASB16* | -0.945816618 | 3.62E-05 | 0.000132186 | 0.175487643 |
| T2DM | nk_cell | *PLCD3* | -0.801858932 | 1.98E-05 | 0.000122264 | 0.095985137 |
| T2DM | nk_cell | *IP6K3* | -7.815908099 | 1.64E-06 | 9.70E-05 | 0.007951894 |
| T2DM | nk_cell | *LEMD2* | -1.410592296 | 2.97E-05 | 0.000124182 | 0.144092808 |
| T2DM | nk_cell | *ASGR2* | 3.60634525 | 0.000107671 | 0.000248764 | 0.522421816 |
| T2DM | nk_cell | *SNRNP25* | -1.291793552 | 0.000456329 | 0.000801923 | 1 |
| T2DM | nk_cell | *WDR90* | -0.968822638 | 1.01E-05 | 0.000122264 | 0.048968461 |
| T2DM | nk_cell | *JMJD8* | 0.787056029 | 0.005626116 | 0.007860039 | 1 |
| T2DM | nk_cell | *MEIOB* | -0.625217551 | 0.000163189 | 0.000340409 | 0.791792029 |
| T2DM | nk_cell | *HS3ST6* | -0.887623416 | 0.018806915 | 0.02420455 | 1 |
| T2DM | nk_cell | *SHANK2* | -0.576380578 | 2.72E-05 | 0.000122437 | 0.131758166 |
| T2DM | nk_cell | *ASRGL1* | -2.368321643 | 0.018182372 | 0.023425616 | 1 |
| T2DM | nk_cell | *TTC9C* | -1.500013251 | 0.00087974 | 0.001426817 | 1 |
| T2DM | nk_cell | *TPCN2* | -0.816439379 | 1.97E-05 | 0.000122264 | 0.095692839 |
| T2DM | nk_cell | *PDZK1IP1* | -1.572957431 | 1.85E-05 | 0.000122264 | 0.089589607 |
| T2DM | nk_cell | *CMPK1* | -1.073565152 | 1.71E-05 | 0.000122264 | 0.083176937 |
| T2DM | nk_cell | *COA7* | -3.103229591 | 5.86E-05 | 0.000167232 | 0.284359199 |
| T2DM | nk_cell | *SLC1A7* | -0.671683883 | 4.58E-05 | 0.000147711 | 0.222452456 |
| T2DM | nk_cell | *C1orf123* | -2.468019715 | 1.59E-05 | 0.000122264 | 0.076912702 |
| T2DM | nk_cell | *USP24* | -1.400599364 | 0.00117983 | 0.001859824 | 1 |
| T2DM | nk_cell | *ZSWIM5* | -1.1257292 | 4.94E-05 | 0.00015216 | 0.239804737 |
| T2DM | nk_cell | *AK4* | -0.967386 | 7.61E-05 | 0.000197146 | 0.369056593 |
| T2DM | nk_cell | *RBP7* | -1.465812248 | 0.00020483 | 0.000408481 | 0.993834623 |
| T2DM | nk_cell | *SLC25A34* | -1.011268068 | 1.54E-05 | 0.000122264 | 0.074784521 |
| T2DM | nk_cell | *PDPN* | -1.092525118 | 1.98E-05 | 0.000122264 | 0.095832261 |
| T2DM | nk_cell | *LAPTM5* | 0.91874075 | 0.039525316 | 0.048477461 | 1 |
| T2DM | nk_cell | *SDC3* | -1.203112196 | 0.0002125 | 0.000421526 | 1 |
| T2DM | nk_cell | *SYNC* | -0.982630408 | 3.17E-05 | 0.000127109 | 0.15396989 |
| T2DM | nk_cell | *CAMK2N1* | -0.732451035 | 0.000559666 | 0.000955826 | 1 |
| T2DM | nk_cell | *MEGF6* | -0.634176471 | 2.52E-05 | 0.000122264 | 0.12226509 |
| T2DM | nk_cell | *OMA1* | -0.93186882 | 0.000114443 | 0.000260095 | 0.555279824 |
| T2DM | nk_cell | *TM2D1* | -1.371140874 | 6.28E-05 | 0.000173739 | 0.304804964 |
| T2DM | nk_cell | *NEXN* | -1.323637024 | 0.000255707 | 0.000491558 | 1 |
| T2DM | nk_cell | *LRRIQ3* | -0.386876645 | 2.51E-05 | 0.000122264 | 0.121801968 |
| T2DM | nk_cell | *TYW3* | -1.171799869 | 8.82E-06 | 0.000122264 | 0.042784694 |
| T2DM | nk_cell | *WDR63* | -0.860228047 | 1.43E-05 | 0.000122264 | 0.069524855 |
| T2DM | nk_cell | *HFM1* | -1.563359822 | 2.47E-05 | 0.000122264 | 0.119641405 |
| T2DM | nk_cell | *ARPC5* | -1.376669194 | 0.002126289 | 0.00318615 | 1 |
| T2DM | nk_cell | *ZNF496* | -1.669240589 | 4.59E-05 | 0.000147852 | 0.22291683 |
| T2DM | nk_cell | *TRIM58* | -0.728484686 | 0.00340012 | 0.004939336 | 1 |
| T2DM | nk_cell | *FCRLB* | -1.648575381 | 2.82E-05 | 0.000122868 | 0.13688968 |
| T2DM | nk_cell | *KLHDC9* | -0.686058334 | 0.027136483 | 0.034057479 | 1 |
| T2DM | nk_cell | *FLVCR1* | -1.466867883 | 8.85E-06 | 0.000122264 | 0.042922735 |
| T2DM | nk_cell | *TDRD5* | -1.183438438 | 2.43E-05 | 0.000122264 | 0.117893752 |
| T2DM | nk_cell | *BROX* | -0.800385632 | 0.004102592 | 0.005856363 | 1 |
| T2DM | nk_cell | *ACP6* | -1.223711825 | 1.29E-05 | 0.000122264 | 0.062592542 |
| T2DM | nk_cell | *CNST* | -1.155521043 | 2.75E-05 | 0.000122472 | 0.133276853 |
| T2DM | nk_cell | *PKDCC* | -0.637884463 | 4.66E-05 | 0.000148595 | 0.225864213 |
| T2DM | nk_cell | *HAAO* | -1.992469098 | 4.73E-05 | 0.000149118 | 0.229541659 |

| | | | | | | |
|---|---|---|---|---|---|---|
| T2DM | nk_cell | *B3GALNT2* | -1.061120524 | 4.21E-05 | 0.000141813 | 0.20420224 |
| T2DM | nk_cell | *KIAA1841* | -1.623688472 | 3.43E-05 | 0.000129854 | 0.166665223 |
| T2DM | nk_cell | *KCNJ3* | -1.900905733 | 2.69E-05 | 0.000122391 | 0.130376734 |
| T2DM | nk_cell | *DUSP19* | -1.116994863 | 2.92E-05 | 0.000123467 | 0.141554511 |
| T2DM | nk_cell | *NUP35* | -0.909398836 | 1.19E-05 | 0.000122264 | 0.057839016 |
| T2DM | nk_cell | *C2orf48* | 0.372196466 | 5.01E-05 | 0.000153185 | 0.243257704 |
| T2DM | nk_cell | *CCDC74A* | -1.884746819 | 4.49E-05 | 0.000146788 | 0.217832835 |
| T2DM | nk_cell | *NOSTRIN* | -0.656534966 | 0.000111957 | 0.000256233 | 0.543213126 |
| T2DM | nk_cell | *INHBB* | -0.832090159 | 2.85E-05 | 0.00012318 | 0.138190432 |
| T2DM | nk_cell | *RPRD2* | -1.180950565 | 0.000155572 | 0.000328333 | 0.754837541 |
| T2DM | nk_cell | *PACRGL* | -1.537955038 | 0.000228003 | 0.000446798 | 1 |
| T2DM | nk_cell | *C1QTNF7* | -0.870035515 | 3.79E-05 | 0.000134881 | 0.183978333 |
| T2DM | nk_cell | *LYSMD1* | -0.932952741 | 0.002166082 | 0.003239775 | 1 |
| T2DM | nk_cell | *ERCC3* | -1.349058639 | 0.000461898 | 0.000809364 | 1 |
| T2DM | nk_cell | *RNF149* | -1.31661687 | 0.000117703 | 0.000264518 | 0.571095089 |
| T2DM | nk_cell | *DHX57* | -1.470460628 | 3.91E-05 | 0.0001366 | 0.189874256 |
| T2DM | nk_cell | *S100A9* | -0.95970746 | 0.000100667 | 0.000237337 | 0.48843852 |
| T2DM | nk_cell | *TDRD10* | -1.043982832 | 1.78E-05 | 0.000122264 | 0.086215534 |
| T2DM | nk_cell | *DCAF16* | -1.516583935 | 0.01747402 | 0.022590979 | 1 |
| T2DM | nk_cell | *GNPDA2* | -1.247115447 | 0.000154697 | 0.000327302 | 0.750589818 |
| T2DM | nk_cell | *NIPAL1* | -0.970578044 | 5.35E-05 | 0.000159514 | 0.259666535 |
| T2DM | nk_cell | *ANTXR2* | -0.892270888 | 4.11E-05 | 0.000140092 | 0.199351081 |
| T2DM | nk_cell | *CGGBP1* | -0.550750464 | 0.023680669 | 0.029999635 | 1 |
| T2DM | nk_cell | *GPR155* | -1.713496534 | 7.82E-05 | 0.000200251 | 0.379274502 |
| T2DM | nk_cell | *DAPL1* | -0.978019902 | 0.000630328 | 0.001058253 | 1 |
| T2DM | nk_cell | *PBXIP1* | -2.082059055 | 0.000343347 | 0.00063103 | 1 |
| T2DM | nk_cell | *CLDN1* | -0.986133724 | 3.96E-05 | 0.000137211 | 0.192126196 |
| T2DM | nk_cell | *DCST2* | -0.724584916 | 0.000315979 | 0.000588986 | 1 |
| T2DM | nk_cell | *LINC01116* | 2.417125487 | 0.007369829 | 0.010067024 | 1 |
| T2DM | nk_cell | *POGLUT1* | -0.847762884 | 0.000618136 | 0.001040304 | 1 |
| T2DM | nk_cell | *SLC22A15* | -1.160449754 | 1.77E-05 | 0.000122264 | 0.08596949 |
| T2DM | nk_cell | *LRRC58* | -1.119322268 | 4.94E-05 | 0.00015216 | 0.239580396 |
| T2DM | nk_cell | *LMOD1* | -0.528083423 | 6.56E-06 | 0.000122264 | 0.03181777 |
| T2DM | nk_cell | *TMEM169* | 0.6723628 | 0.00034876 | 0.000639284 | 1 |
| T2DM | nk_cell | *ARPC2* | 0.667665957 | 0.00765614 | 0.01041716 | 1 |
| T2DM | nk_cell | *CCT3* | -1.378845456 | 0.007742888 | 0.010532238 | 1 |
| T2DM | nk_cell | *SERPINI1* | -1.082965383 | 1.53E-05 | 0.000122264 | 0.074194262 |
| T2DM | nk_cell | *SUCLG1* | -1.071400305 | 0.000194035 | 0.000391133 | 0.941456824 |
| T2DM | nk_cell | *SPTA1* | -1.419507067 | 3.22E-06 | 0.000122264 | 0.015618055 |
| T2DM | nk_cell | *PRKCI* | -0.784314912 | 3.50E-05 | 0.000130812 | 0.169794102 |
| T2DM | nk_cell | *AIM2* | -1.101185006 | 1.79E-05 | 0.000122264 | 0.086641616 |
| T2DM | nk_cell | *EFHB* | -0.359729674 | 0.001715987 | 0.002620548 | 1 |
| T2DM | nk_cell | *GTPBP8* | -2.068310681 | 4.01E-05 | 0.000138284 | 0.194471486 |
| T2DM | nk_cell | *WDFY3* | -1.008053327 | 3.51E-05 | 0.000130831 | 0.170080034 |
| T2DM | nk_cell | *PTPN13* | -0.731821134 | 6.47E-05 | 0.000176937 | 0.313722891 |
| T2DM | nk_cell | *C4orf36* | -0.886557475 | 1.69E-05 | 0.000122264 | 0.081818163 |
| T2DM | nk_cell | *TIPARP* | -1.5324293 | 7.73E-05 | 0.000199059 | 0.375026309 |
| T2DM | nk_cell | *RPL9* | -0.282730387 | 0.016827074 | 0.021801058 | 1 |
| T2DM | nk_cell | *DNASE1L3* | -0.838458429 | 0.000544475 | 0.000933166 | 1 |
| T2DM | nk_cell | *IL17RC* | -1.695029788 | 2.05E-06 | 0.000112031 | 0.009944625 |

| | | | | | | |
|---|---|---|---|---|---|---|
| T2DM | nk_cell | *CRELD1* | -1.383607898 | 2.85E-05 | 0.00012318 | 0.13808724 |
| T2DM | nk_cell | *TTC14* | -1.358783166 | 2.03E-05 | 0.000122264 | 0.098665406 |
| T2DM | nk_cell | *CCDC158* | -1.869859623 | 0.000171261 | 0.00035378 | 0.830957686 |
| T2DM | nk_cell | *CPA3* | -2.105759795 | 9.02E-06 | 0.000122264 | 0.043747704 |
| T2DM | nk_cell | *SLC4A1AP* | -1.521440324 | 2.32E-05 | 0.000122264 | 0.112397552 |
| T2DM | nk_cell | *KIAA1143* | -0.760997832 | 9.87E-05 | 0.00023397 | 0.478702618 |
| T2DM | nk_cell | *CLEC3B* | -1.342618705 | 0.000115874 | 0.000262225 | 0.562222903 |
| T2DM | nk_cell | *NMNAT3* | -0.787762706 | 0.000122809 | 0.00027246 | 0.595870732 |
| T2DM | nk_cell | *TPRA1* | -1.46988944 | 0.000112309 | 0.000256676 | 0.544922108 |
| T2DM | nk_cell | *MEAF6* | -1.71270032 | 9.27E-05 | 0.000223847 | 0.449646079 |
| T2DM | nk_cell | *SNIP1* | -1.687842832 | 2.34E-05 | 0.000122264 | 0.113334967 |
| T2DM | nk_cell | *DNALI1* | -0.665799255 | 0.000540206 | 0.000928144 | 1 |
| T2DM | nk_cell | *SENP2* | -1.110364232 | 1.03E-05 | 0.000122264 | 0.049785503 |
| T2DM | nk_cell | *IFT122* | -1.293139162 | 4.01E-05 | 0.000138284 | 0.194333946 |
| T2DM | nk_cell | *RFT1* | -2.469596566 | 1.16E-09 | 6.28E-07 | 5.65E-06 |
| T2DM | nk_cell | *SFMBT1* | -1.146649149 | 2.19E-05 | 0.000122264 | 0.106442423 |
| T2DM | nk_cell | *UVSSA* | -0.80776975 | 0.032502296 | 0.04032246 | 1 |
| T2DM | nk_cell | *FAM208A* | -1.474390681 | 0.001849981 | 0.002806788 | 1 |
| T2DM | nk_cell | *LRPAP1* | -1.411159413 | 0.000116991 | 0.000263449 | 0.56764028 |
| T2DM | nk_cell | *ABLIM2* | -1.72228998 | 0.000152428 | 0.000323491 | 0.739580797 |
| T2DM | nk_cell | *ERMAP* | -0.676818576 | 9.30E-06 | 0.000122264 | 0.045115935 |
| T2DM | nk_cell | *BDH2* | -0.924723768 | 1.88E-06 | 0.000107428 | 0.009131369 |
| T2DM | nk_cell | *CDC25A* | -2.750113146 | 1.46E-05 | 0.000122264 | 0.070764945 |
| T2DM | nk_cell | *ZNF589* | -1.419661487 | 9.50E-06 | 0.000122264 | 0.04610935 |
| T2DM | nk_cell | *ATRIP* | -0.751953781 | 0.007125457 | 0.009763382 | 1 |
| T2DM | nk_cell | *RNF123* | -0.427515245 | 0.036082082 | 0.044513161 | 1 |
| T2DM | nk_cell | *HSPA4L* | -1.363090744 | 0.003087937 | 0.004504712 | 1 |
| T2DM | nk_cell | *MFSD8* | -1.31479312 | 2.44E-05 | 0.000122264 | 0.118277867 |
| T2DM | nk_cell | *RAD54L2* | -1.60154188 | 2.81E-05 | 0.000122832 | 0.136567273 |
| T2DM | nk_cell | *TEX264* | -1.325162401 | 0.001402813 | 0.002170424 | 1 |
| T2DM | nk_cell | *ETNPPL* | 4.475927572 | 0.000189136 | 0.000383488 | 0.917686799 |
| T2DM | nk_cell | *PRSS12* | -0.598969449 | 0.000154941 | 0.000327428 | 0.751775481 |
| T2DM | nk_cell | *HMGB2* | -1.320802721 | 3.84E-05 | 0.000135693 | 0.186442082 |
| T2DM | nk_cell | *ANXA5* | -1.314824697 | 0.000501753 | 0.000870087 | 1 |
| T2DM | nk_cell | *MAP9* | 1.147220688 | 0.0013822 | 0.00214263 | 1 |
| T2DM | nk_cell | *CEP44* | -1.190644081 | 1.92E-05 | 0.000122264 | 0.093011543 |
| T2DM | nk_cell | *HPGD* | -1.393696565 | 1.22E-05 | 0.000122264 | 0.058987143 |
| T2DM | nk_cell | *C4orf45* | -0.78554497 | 1.84E-05 | 0.000122264 | 0.089307536 |
| T2DM | nk_cell | *TMEM144* | -1.544028853 | 1.66E-05 | 0.000122264 | 0.08072263 |
| T2DM | nk_cell | *ARFIP1* | -1.20813431 | 6.81E-05 | 0.000183747 | 0.330377454 |
| T2DM | nk_cell | *LSM6* | -1.104144966 | 2.58E-05 | 0.000122264 | 0.125301363 |
| T2DM | nk_cell | *ITGA2* | -0.631550602 | 0.005653767 | 0.007894123 | 1 |
| T2DM | nk_cell | *MOCS2* | -2.112011836 | 2.14E-05 | 0.000122264 | 0.103794126 |
| T2DM | nk_cell | *NDUFAF2* | -1.407399036 | 0.000625141 | 0.001051 | 1 |
| T2DM | nk_cell | *LMBRD2* | -1.52936253 | 4.57E-05 | 0.000147697 | 0.221859346 |
| T2DM | nk_cell | *RNF180* | -1.286495848 | 1.09E-05 | 0.000122264 | 0.053058158 |
| T2DM | nk_cell | *PGGT1B* | -1.251407172 | 0.025340289 | 0.031902201 | 1 |
| T2DM | nk_cell | *F2RL2* | 0.513787781 | 6.85E-05 | 0.00018443 | 0.332577328 |
| T2DM | nk_cell | *CCDC112* | -2.01879424 | 0.003867508 | 0.005546896 | 1 |
| T2DM | nk_cell | *CMBL* | -1.090803362 | 2.38E-05 | 0.000122264 | 0.115597133 |

| | | | | | | |
|---|---|---|---|---|---|---|
| T2DM | nk_cell | *C5orf63* | -1.672876389 | 0.000524692 | 0.000904858 | 1 |
| T2DM | nk_cell | *WDR41* | -1.084647992 | 3.01E-05 | 0.000124875 | 0.145854366 |
| T2DM | nk_cell | *NDUFS4* | -1.024116302 | 1.74E-05 | 0.000122264 | 0.084332697 |
| T2DM | nk_cell | *SERINC5* | -1.097844134 | 3.72E-05 | 0.000134067 | 0.18048813 |
| T2DM | nk_cell | *PRIMPOL* | -1.507732636 | 9.67E-06 | 0.000122264 | 0.046906113 |
| T2DM | nk_cell | *ERAP1* | -1.185080332 | 0.002924057 | 0.004281088 | 1 |
| T2DM | nk_cell | *ERAP2* | 0.526347973 | 0.005683039 | 0.007932712 | 1 |
| T2DM | nk_cell | *UTP15* | -1.317412083 | 1.26E-05 | 0.000122264 | 0.061066384 |
| T2DM | nk_cell | *TLR3* | -1.466224196 | 1.25E-05 | 0.000122264 | 0.060637618 |
| T2DM | nk_cell | *KLKB1* | 0.526629594 | 0.010622558 | 0.014186802 | 1 |
| T2DM | nk_cell | *GRIK2* | -0.497997073 | 9.70E-05 | 0.000231235 | 0.470793944 |
| T2DM | nk_cell | *DCBLD1* | -0.827522603 | 0.001208604 | 0.001897982 | 1 |
| T2DM | nk_cell | *PDSS2* | -1.313903304 | 0.000265345 | 0.000507471 | 1 |
| T2DM | nk_cell | *RAET1E* | -0.788748888 | 2.32E-05 | 0.000122264 | 0.112648578 |
| T2DM | nk_cell | *DAGLB* | -0.741445542 | 7.43E-05 | 0.000194408 | 0.360371707 |
| T2DM | nk_cell | *STK17A* | 0.296884819 | 0.000160395 | 0.000336461 | 0.778234599 |
| T2DM | nk_cell | *RP9* | 1.140493454 | 0.002349903 | 0.003497463 | 1 |
| T2DM | nk_cell | *CAMLG* | -0.901936174 | 0.002521736 | 0.00372805 | 1 |
| T2DM | nk_cell | *RELL2* | 0.422165113 | 2.80E-05 | 0.000122557 | 0.135876209 |
| T2DM | nk_cell | *MIOS* | -1.234417525 | 1.94E-05 | 0.000122264 | 0.094254551 |
| T2DM | nk_cell | *USP49* | -1.223250971 | 1.62E-05 | 0.000122264 | 0.078402224 |
| T2DM | nk_cell | *CHMP4C* | -7.661333638 | 0.001912463 | 0.002893443 | 1 |
| T2DM | nk_cell | *SLC13A4* | -1.3119035 | 2.11E-05 | 0.000122264 | 0.10216734 |
| T2DM | nk_cell | *BRI3* | -1.454492706 | 7.34E-05 | 0.000193189 | 0.356241187 |
| T2DM | nk_cell | *CTSB* | -0.551938011 | 0.003460837 | 0.005009541 | 1 |
| T2DM | nk_cell | *SOX17* | -0.942283999 | 0.004469708 | 0.006341235 | 1 |
| T2DM | nk_cell | *RAD21* | -1.597239456 | 5.66E-05 | 0.000163891 | 0.274787772 |
| T2DM | nk_cell | *TNFRSF11B* | -0.758710899 | 3.45E-05 | 0.000130101 | 0.167576604 |
| T2DM | nk_cell | *DEFB1* | -1.82990862 | 0.006335052 | 0.008767162 | 1 |
| T2DM | nk_cell | *SUN1* | -0.930686279 | 4.60E-05 | 0.000147852 | 0.223164907 |
| T2DM | nk_cell | *OXR1* | -0.891673734 | 3.47E-05 | 0.000130294 | 0.168600103 |
| T2DM | nk_cell | *INTS1* | -2.095035732 | 0.007385394 | 0.010082705 | 1 |
| T2DM | nk_cell | *FASTK* | -1.568440671 | 2.83E-05 | 0.00012294 | 0.137201412 |
| T2DM | nk_cell | *ALDH7A1* | -1.180573441 | 6.83E-05 | 0.00018416 | 0.331304223 |
| T2DM | nk_cell | *YWHAZ* | -1.247263526 | 9.09E-05 | 0.000220956 | 0.440807257 |
| T2DM | nk_cell | *FZD6* | -0.605885883 | 0.008248358 | 0.011172817 | 1 |
| T2DM | nk_cell | *SLC25A32* | -1.851002427 | 0.000399277 | 0.00071938 | 1 |
| T2DM | nk_cell | *DCSTAMP* | -6.571221611 | 0.00037955 | 0.000688698 | 1 |
| T2DM | nk_cell | *INTS8* | -1.465582537 | 5.10E-05 | 0.000154585 | 0.247335448 |
| T2DM | nk_cell | *RPP25L* | -1.354424452 | 8.16E-06 | 0.000122264 | 0.039616288 |
| T2DM | nk_cell | *C9orf24* | 1.346420488 | 0.020421754 | 0.026116592 | 1 |
| T2DM | nk_cell | *TMEM65* | -1.252682676 | 0.021922021 | 0.027917492 | 1 |
| T2DM | nk_cell | *CCDC171* | -1.419629619 | 6.12E-05 | 0.000171318 | 0.296722274 |
| T2DM | nk_cell | *METTL2B* | -0.427857877 | 0.040737446 | 0.049875874 | 1 |
| T2DM | nk_cell | *ZMAT4* | 0.740627882 | 0.000102868 | 0.000240654 | 0.499116672 |
| T2DM | nk_cell | *C8orf34* | -0.709928866 | 6.05E-05 | 0.000169923 | 0.293491426 |
| T2DM | nk_cell | *RASEF* | -0.63805513 | 0.005259355 | 0.007388069 | 1 |
| T2DM | nk_cell | *ANKS6* | -0.789256291 | 0.000124135 | 0.000274774 | 0.602304046 |
| T2DM | nk_cell | *AQP7* | -0.483456706 | 0.000122011 | 0.000270936 | 0.591995691 |
| T2DM | nk_cell | *NOL6* | -0.931788631 | 5.35E-06 | 0.000122264 | 0.025965817 |

| | | | | | | |
|---|---|---|---|---|---|---|
| T2DM | nk_cell | *AQP3* | -1.491713906 | 2.69E-05 | 0.000122391 | 0.130562625 |
| T2DM | nk_cell | *TRMT10B* | -1.496956501 | 0.002805958 | 0.004115632 | 1 |
| T2DM | nk_cell | *PIGO* | 0.80267594 | 0.000454334 | 0.000799286 | 1 |
| T2DM | nk_cell | *WRN* | -1.17476693 | 7.00E-05 | 0.000186641 | 0.339499421 |
| T2DM | nk_cell | *TSHR* | -1.168953835 | 2.65E-06 | 0.000122264 | 0.012868116 |
| T2DM | nk_cell | *CFL2* | -1.706179652 | 1.50E-05 | 0.000122264 | 0.073011619 |
| T2DM | nk_cell | *PGM2L1* | -1.282845812 | 0.004127877 | 0.00588726 | 1 |
| T2DM | nk_cell | *SLC16A9* | -2.136009843 | 1.73E-05 | 0.000122264 | 0.083780255 |
| T2DM | nk_cell | *GJB2* | -8.534921555 | 2.26E-05 | 0.000122264 | 0.109473887 |
| T2DM | nk_cell | *CRYL1* | -1.31502042 | 7.75E-05 | 0.00019931 | 0.37591466 |
| T2DM | nk_cell | *REEP3* | -1.252968164 | 4.21E-06 | 0.000122264 | 0.020441242 |
| T2DM | nk_cell | *LRR1* | -1.290754286 | 1.61E-05 | 0.000122264 | 0.07830952 |
| T2DM | nk_cell | *NOXRED1* | -0.990904932 | 0.005777559 | 0.008055378 | 1 |
| T2DM | nk_cell | *AKR1E2* | -0.781840222 | 1.51E-05 | 0.000122264 | 0.073215147 |
| T2DM | nk_cell | *OXGR1* | 1.150673922 | 7.63E-05 | 0.000197316 | 0.369966803 |
| T2DM | nk_cell | *ATP5C1* | -1.56148415 | 6.91E-05 | 0.000185105 | 0.335225722 |
| T2DM | nk_cell | *VDAC2* | -2.182337301 | 0.002282708 | 0.003401628 | 1 |
| T2DM | nk_cell | *CLEC1B* | -0.860025162 | 0.03494017 | 0.04322532 | 1 |
| T2DM | nk_cell | *SNAPC4* | -0.661553957 | 8.49E-05 | 0.00021115 | 0.411842727 |
| T2DM | nk_cell | *TSC1* | -0.935428186 | 0.000749782 | 0.001231531 | 1 |
| T2DM | nk_cell | *STOX1* | -0.716888409 | 0.000100505 | 0.000237068 | 0.487649897 |
| T2DM | nk_cell | *RET* | -1.03496922 | 2.58E-05 | 0.000122264 | 0.125399733 |
| T2DM | nk_cell | *CASP7* | -0.831013156 | 0.012161926 | 0.016087695 | 1 |
| T2DM | nk_cell | *PPP1R36* | -0.806174378 | 4.53E-05 | 0.00014727 | 0.219726243 |
| T2DM | nk_cell | *VWA2* | -1.293666792 | 5.84E-05 | 0.000167232 | 0.283483232 |
| T2DM | nk_cell | *METTL3* | -1.506122483 | 2.31E-05 | 0.000122264 | 0.111913799 |
| T2DM | nk_cell | *TRUB1* | -1.948446998 | 1.23E-05 | 0.000122264 | 0.059882687 |
| T2DM | nk_cell | *HSPA12A* | -0.659480524 | 0.000611954 | 0.001032047 | 1 |
| T2DM | nk_cell | *UBTD1* | -0.816797732 | 1.59E-05 | 0.000122264 | 0.077202782 |
| T2DM | nk_cell | *PACSIN3* | -0.68300043 | 0.026167182 | 0.032900535 | 1 |
| T2DM | nk_cell | *TTC7B* | -1.346210996 | 3.47E-05 | 0.000130294 | 0.168478054 |
| T2DM | nk_cell | *PSMC3* | -1.514634151 | 4.00E-05 | 0.000138284 | 0.194192532 |
| T2DM | nk_cell | *RAPSN* | -1.677921724 | 5.78E-09 | 2.16E-06 | 2.80E-05 |
| T2DM | nk_cell | *TC2N* | 0.508278971 | 0.025077007 | 0.031611753 | 1 |
| T2DM | nk_cell | *CPSF2* | -0.523957655 | 0.003479835 | 0.005034037 | 1 |
| T2DM | nk_cell | *IFI27L1* | -1.027380795 | 2.02E-05 | 0.000122264 | 0.098076001 |
| T2DM | nk_cell | *PTER* | -1.660500988 | 0.001074284 | 0.001710675 | 1 |
| T2DM | nk_cell | *C1QL3* | -0.701481568 | 0.025849075 | 0.032508998 | 1 |
| T2DM | nk_cell | *LIPC* | -1.019610504 | 0.001140527 | 0.001803928 | 1 |
| T2DM | nk_cell | *CEP57* | -1.805139005 | 2.29E-05 | 0.000122264 | 0.111029881 |
| T2DM | nk_cell | *TCP11L2* | -0.825908198 | 1.91E-05 | 0.000122264 | 0.092704418 |
| T2DM | nk_cell | *JAM3* | -1.046055421 | 7.12E-05 | 0.000189077 | 0.345443317 |
| T2DM | nk_cell | *ZFYVE19* | -1.240209512 | 0.000169057 | 0.000349793 | 0.820264637 |
| T2DM | nk_cell | *TRMT61A* | -0.730647139 | 2.82E-05 | 0.000122868 | 0.136997623 |
| T2DM | nk_cell | *LARP6* | -1.03239808 | 8.91E-05 | 0.000217825 | 0.432261506 |
| T2DM | nk_cell | *API5* | -1.617621162 | 7.47E-05 | 0.000195056 | 0.362453886 |
| T2DM | nk_cell | *ASPG* | 3.962625245 | 0.000226802 | 0.000444983 | 1 |
| T2DM | nk_cell | *COX11* | -1.637968336 | 1.78E-05 | 0.000122264 | 0.08643355 |
| T2DM | nk_cell | *ZNF202* | -1.347511015 | 8.95E-06 | 0.000122264 | 0.043439122 |
| T2DM | nk_cell | *STXBP4* | -1.517597318 | 7.14E-06 | 0.000122264 | 0.03462974 |

| | | | | | | |
|---|---|---|---|---|---|---|
| T2DM | nk_cell | *WBP1L* | -1.221969302 | 0.000302067 | 0.000566098 | 1 |
| T2DM | nk_cell | *APBB1* | -1.700065779 | 1.59E-05 | 0.000122264 | 0.076998606 |
| T2DM | nk_cell | *NUDT13* | -0.722837451 | 0.021709437 | 0.027668555 | 1 |
| T2DM | nk_cell | *WDR88* | -1.235385379 | 2.95E-05 | 0.000123723 | 0.143183202 |
| T2DM | nk_cell | *CYB5R2* | -0.706564983 | 0.000849383 | 0.001380176 | 1 |
| T2DM | nk_cell | *SERPINB8* | -0.529985775 | 0.017896862 | 0.023088427 | 1 |
| T2DM | nk_cell | *XRRA1* | -1.067795641 | 9.54E-06 | 0.000122264 | 0.046298566 |
| T2DM | nk_cell | *CENPN* | -1.725400767 | 3.81E-05 | 0.000135266 | 0.18478958 |
| T2DM | nk_cell | *C16orf46* | -3.151599551 | 5.67E-05 | 0.00016393 | 0.275074534 |
| T2DM | nk_cell | *TMEM41B* | -1.486882114 | 3.86E-05 | 0.000136122 | 0.187440118 |
| T2DM | nk_cell | *TMX3* | -1.590652337 | 4.75E-05 | 0.000149363 | 0.230672633 |
| T2DM | nk_cell | *PRKCB* | 0.251117607 | 0.000100324 | 0.000236756 | 0.486771322 |
| T2DM | nk_cell | *NDST2* | 1.944100401 | 7.98E-05 | 0.000203233 | 0.386954708 |
| T2DM | nk_cell | *CCDC68* | -0.822248671 | 0.034540481 | 0.042741753 | 1 |
| T2DM | nk_cell | *CLEC4D* | -2.899361056 | 2.99E-05 | 0.000124686 | 0.145102138 |
| T2DM | nk_cell | *RIMKLB* | -0.980535718 | 5.10E-06 | 0.000122264 | 0.024723652 |
| T2DM | nk_cell | *TMED3* | -0.661121103 | 0.002594349 | 0.003829565 | 1 |
| T2DM | nk_cell | *SEC11C* | 1.237424316 | 0.033416198 | 0.041403317 | 1 |
| T2DM | nk_cell | *GALR1* | -0.82041075 | 0.010481844 | 0.014010443 | 1 |
| T2DM | nk_cell | *CENPV* | -1.524788543 | 2.73E-05 | 0.000122437 | 0.132348671 |
| T2DM | nk_cell | *FAM96B* | -1.523115254 | 0.003705767 | 0.005335425 | 1 |
| T2DM | nk_cell | *ZNF592* | -1.089084926 | 2.61E-05 | 0.000122264 | 0.126521805 |
| T2DM | nk_cell | *ACSM1* | -0.985772869 | 4.43E-05 | 0.000145494 | 0.215040696 |
| T2DM | nk_cell | *YPEL4* | -1.271651123 | 0.00858811 | 0.011616814 | 1 |
| T2DM | nk_cell | *LDHC* | 0.716722324 | 0.00098705 | 0.001586867 | 1 |
| T2DM | nk_cell | *FAM96A* | -0.85310811 | 2.45E-05 | 0.000122264 | 0.118675885 |
| T2DM | nk_cell | *FAM111A* | -1.009813814 | 0.005026723 | 0.007077672 | 1 |
| T2DM | nk_cell | *TMEM170A* | -1.275825265 | 3.25E-05 | 0.000127909 | 0.157715495 |
| T2DM | nk_cell | *STAT6* | -1.565284525 | 0.00082806 | 0.001349143 | 1 |
| T2DM | nk_cell | *PIP4K2C* | 1.242051394 | 0.007870206 | 0.010690436 | 1 |
| T2DM | nk_cell | *MTMR10* | -1.100278833 | 0.000143982 | 0.00030939 | 0.698602629 |
| T2DM | nk_cell | *NYAP1* | -0.861287751 | 6.90E-06 | 0.000122264 | 0.033472002 |
| T2DM | nk_cell | *MS4A7* | -0.636624855 | 0.027966045 | 0.035053281 | 1 |
| T2DM | nk_cell | *MS4A14* | -0.500868299 | 0.000153643 | 0.000325392 | 0.745473476 |
| T2DM | nk_cell | *SMAD3* | -1.425967022 | 0.000265481 | 0.000507531 | 1 |
| T2DM | nk_cell | *MAP1A* | -0.975951447 | 3.37E-05 | 0.00012843 | 0.163484471 |
| T2DM | nk_cell | *RCCD1* | -1.177464634 | 4.95E-06 | 0.000122264 | 0.0240093 |
| T2DM | nk_cell | *AKTIP* | 0.920894817 | 0.000136905 | 0.000296944 | 0.664263969 |
| T2DM | nk_cell | *MEI1* | 0.578809405 | 5.40E-05 | 0.000159625 | 0.261785699 |
| T2DM | nk_cell | *SAMD14* | 0.924184897 | 9.82E-05 | 0.000233264 | 0.476558633 |
| T2DM | nk_cell | *ACSF2* | -1.324519514 | 0.007750091 | 0.010539081 | 1 |
| T2DM | nk_cell | *URM1* | -1.585596147 | 0.003746855 | 0.005391299 | 1 |
| T2DM | nk_cell | *ENDOG* | -1.028945784 | 0.000828819 | 0.001349926 | 1 |
| T2DM | nk_cell | *ISLR2* | -1.0714636 | 0.00021557 | 0.000426743 | 1 |
| T2DM | nk_cell | *SP2* | -1.607091385 | 2.21E-05 | 0.000122264 | 0.10737089 |
| T2DM | nk_cell | *COQ7* | -1.328267707 | 1.82E-05 | 0.000122264 | 0.088538714 |
| T2DM | nk_cell | *GPRC5B* | -0.933051378 | 8.23E-05 | 0.000207015 | 0.399446823 |
| T2DM | nk_cell | *FBXO22* | -1.498796026 | 0.008989782 | 0.012129705 | 1 |
| T2DM | nk_cell | *TBC1D2B* | -0.69569639 | 1.87E-05 | 0.000122264 | 0.090902599 |
| T2DM | nk_cell | *LOXHD1* | -1.24996751 | 0.000246959 | 0.000477579 | 1 |

| | | | | | | |
|---|---|---|---|---|---|---|
| T2DM | nk_cell | *KATNAL2* | -0.913256966 | 0.000300986 | 0.000564292 | 1 |
| T2DM | nk_cell | *HDHD2* | 0.739608089 | 0.004073387 | 0.005818097 | 1 |
| T2DM | nk_cell | *ZNF91* | -1.044449487 | 9.67E-05 | 0.000230845 | 0.469307861 |
| T2DM | nk_cell | *ENGASE* | -0.702236195 | 0.000405307 | 0.000727543 | 1 |
| T2DM | nk_cell | *MYO5B* | -0.902988089 | 1.25E-05 | 0.000122264 | 0.060590701 |
| T2DM | nk_cell | *RRM1* | -0.589255007 | 0.019485251 | 0.025011227 | 1 |
| T2DM | nk_cell | *ZNF23* | -1.185392782 | 7.28E-05 | 0.000192012 | 0.353301177 |
| T2DM | nk_cell | *SMG8* | -1.043771877 | 0.001377379 | 0.002135839 | 1 |
| T2DM | nk_cell | *RAB8A* | 0.871955925 | 0.00011676 | 0.000263375 | 0.566519127 |
| T2DM | nk_cell | *JSRP1* | -1.231404585 | 0.008993768 | 0.01213171 | 1 |
| T2DM | nk_cell | *ZNF641* | -1.4351056 | 0.029961319 | 0.03739 | 1 |
| T2DM | nk_cell | *TP53I13* | -1.448172338 | 0.000282469 | 0.000534532 | 1 |
| T2DM | nk_cell | *TUBA1C* | -0.573120509 | 0.037349028 | 0.045982614 | 1 |
| T2DM | nk_cell | *ZNF528* | -0.943124429 | 1.74E-05 | 0.000122264 | 0.084522722 |
| T2DM | nk_cell | *SERTAD3* | -2.347493477 | 2.07E-05 | 0.000122264 | 0.100217032 |
| T2DM | nk_cell | *TMC4* | -1.313895449 | 1.15E-05 | 0.000122264 | 0.055738971 |
| T2DM | nk_cell | *TTYH1* | -1.337346213 | 1.65E-05 | 0.000122264 | 0.080107908 |
| T2DM | nk_cell | *LENG8* | -0.950987847 | 1.49E-05 | 0.000122264 | 0.072436173 |
| T2DM | nk_cell | *CDC42EP5* | -1.089199801 | 3.16E-05 | 0.000127109 | 0.153160827 |
| T2DM | nk_cell | *KIR3DL1* | 1.316978347 | 7.55E-05 | 0.000196203 | 0.366228576 |
| T2DM | nk_cell | *C19orf33* | -0.689492237 | 0.002513141 | 0.003716477 | 1 |
| T2DM | nk_cell | *DNAAF3* | -0.753636817 | 0.000668621 | 0.001116746 | 1 |
| T2DM | nk_cell | *FAM57A* | -1.149008517 | 0.000237283 | 0.000461813 | 1 |
| T2DM | nk_cell | *KIFC2* | -0.716451069 | 1.06E-05 | 0.000122264 | 0.051424478 |
| T2DM | nk_cell | *TRPV3* | -0.78886874 | 0.008428129 | 0.011409956 | 1 |
| T2DM | nk_cell | *CYB5D2* | -1.316249667 | 0.00033505 | 0.000618593 | 1 |
| T2DM | nk_cell | *KLK5* | -0.81229672 | 1.67E-05 | 0.000122264 | 0.08091571 |
| T2DM | nk_cell | *KLK6* | -4.078077781 | 2.47E-05 | 0.000122264 | 0.119728418 |
| T2DM | nk_cell | *ZNF83* | -1.218223872 | 4.67E-05 | 0.000148657 | 0.226581931 |
| T2DM | nk_cell | *SPRYD3* | -2.313187712 | 2.53E-05 | 0.000122264 | 0.122573199 |
| T2DM | nk_cell | *SOAT2* | -1.336682213 | 4.35E-05 | 0.000144302 | 0.211160355 |
| T2DM | nk_cell | *NDUFV1* | -1.724200173 | 0.00017076 | 0.000353016 | 0.828527754 |
| T2DM | nk_cell | *PRDX2* | -1.98399474 | 0.002246775 | 0.00335323 | 1 |
| T2DM | nk_cell | *MIS12* | -2.036850062 | 0.000171275 | 0.00035378 | 0.831028537 |
| T2DM | nk_cell | *MGAT5B* | -1.306376426 | 4.70E-05 | 0.000148778 | 0.227921218 |
| T2DM | nk_cell | *GSDMA* | -0.799967722 | 0.000305122 | 0.000571162 | 1 |
| T2DM | nk_cell | *TMEM99* | -1.247053703 | 4.74E-05 | 0.000149166 | 0.229865304 |
| T2DM | nk_cell | *ZNF598* | -1.139430796 | 1.32E-05 | 0.000122264 | 0.063857853 |
| T2DM | nk_cell | *MLST8* | -0.726015624 | 5.01E-05 | 0.000153185 | 0.243049273 |
| T2DM | nk_cell | *ECI1* | -1.30604895 | 0.00083859 | 0.001364009 | 1 |
| T2DM | nk_cell | *ABCA3* | -1.535847299 | 1.70E-05 | 0.000122264 | 0.082692918 |
| T2DM | nk_cell | *ATG16L2* | -1.833360762 | 0.000457502 | 0.000803376 | 1 |
| T2DM | nk_cell | *RPSA* | 0.827280899 | 0.019394771 | 0.024901674 | 1 |
| T2DM | nk_cell | *ULK4* | -1.076561911 | 1.34E-05 | 0.000122264 | 0.064791714 |
| T2DM | nk_cell | *NAALADL1* | 0.744483172 | 0.00072304 | 0.001196518 | 1 |
| T2DM | nk_cell | *SCARA5* | -0.425794649 | 4.81E-05 | 0.000150261 | 0.233278449 |
| T2DM | nk_cell | *KIAA1586* | -1.263485278 | 8.74E-06 | 0.000122264 | 0.042422988 |
| T2DM | nk_cell | *FAM83B* | -5.171656707 | 2.11E-05 | 0.000122264 | 0.102437928 |
| T2DM | nk_cell | *COA6* | 1.157372088 | 0.000703612 | 0.001167952 | 1 |
| T2DM | nk_cell | *PDHB* | -2.78641343 | 6.53E-06 | 0.000122264 | 0.031683112 |

| | | | | | | |
|---|---|---|---|---|---|---|
| T2DM | nk_cell | *PXK* | -1.044784918 | 8.06E-05 | 0.000204734 | 0.391042627 |
| T2DM | nk_cell | *PCMTD1* | -1.279662909 | 0.000126058 | 0.00027826 | 0.611635269 |
| T2DM | nk_cell | *ACOX2* | -0.744832328 | 1.37E-05 | 0.000122264 | 0.066388408 |
| T2DM | nk_cell | *SCN11A* | -0.51230608 | 0.002016587 | 0.003034888 | 1 |
| T2DM | nk_cell | *SEPT2* | -0.716935271 | 0.001610494 | 0.002468916 | 1 |
| T2DM | nk_cell | *DTYMK* | -2.632226204 | 9.34E-05 | 0.000224675 | 0.452945583 |
| T2DM | nk_cell | *ING5* | -1.01131492 | 3.91E-05 | 0.000136598 | 0.189719215 |
| T2DM | nk_cell | *RFWD3* | -1.519879236 | 0.000634053 | 0.001063402 | 1 |
| T2DM | nk_cell | *KLHL30* | -4.276557027 | 1.80E-05 | 0.000122264 | 0.087284388 |
| T2DM | nk_cell | *CCDC110* | -1.016502163 | 2.42E-05 | 0.000122264 | 0.117491184 |
| T2DM | nk_cell | *FNTA* | -0.930032174 | 0.000101074 | 0.000238064 | 0.490411097 |
| T2DM | nk_cell | *SERINC2* | -3.142606175 | 3.19E-07 | 3.97E-05 | 0.001549221 |
| T2DM | nk_cell | *SLC20A2* | -1.522131936 | 7.30E-05 | 0.000192269 | 0.354352 |
| T2DM | nk_cell | *TMUB2* | -1.066950081 | 1.87E-05 | 0.000122264 | 0.090956945 |
| T2DM | nk_cell | *ZSWIM1* | 0.744359105 | 8.75E-07 | 7.07E-05 | 0.004243373 |
| T2DM | nk_cell | *NBPF9* | -0.67814652 | 2.27E-05 | 0.000122264 | 0.110175177 |
| T2DM | nk_cell | *ADAM9* | -0.975410554 | 0.000271808 | 0.000517791 | 1 |
| T2DM | nk_cell | *ZNF30* | -0.723541217 | 0.000360236 | 0.00065825 | 1 |
| T2DM | nk_cell | *SLC16A4* | -0.890178863 | 0.001524851 | 0.002345776 | 1 |
| T2DM | nk_cell | *NPNT* | -0.625251622 | 0.004584145 | 0.006492198 | 1 |
| T2DM | nk_cell | *TCTN2* | -0.377059057 | 0.000446432 | 0.00078681 | 1 |
| T2DM | nk_cell | *PPIP5K1* | -1.952468109 | 4.68E-05 | 0.000148657 | 0.226850834 |
| T2DM | nk_cell | *ADAL* | -0.384766939 | 0.040554558 | 0.04967703 | 1 |
| T2DM | nk_cell | *SNTB2* | -1.227622704 | 3.60E-05 | 0.000132133 | 0.174913345 |
| T2DM | nk_cell | *ZNF507* | -2.004678186 | 2.32E-05 | 0.000122264 | 0.11244124 |
| T2DM | nk_cell | *STX18* | -1.382009358 | 3.81E-05 | 0.000135266 | 0.184893064 |
| T2DM | nk_cell | *ZBTB49* | -1.703671399 | 2.19E-05 | 0.000122264 | 0.106136003 |
| T2DM | nk_cell | *GFM1* | -0.951262182 | 5.96E-05 | 0.000168718 | 0.289351959 |
| T2DM | nk_cell | *TMEM129* | -2.124978475 | 7.39E-05 | 0.000193958 | 0.358452562 |
| T2DM | nk_cell | *CEP120* | -1.540969471 | 1.73E-05 | 0.000122264 | 0.083976679 |
| T2DM | nk_cell | *MFF* | -1.371751432 | 5.84E-05 | 0.000167232 | 0.283415432 |
| T2DM | nk_cell | *LGALS9* | -0.690730314 | 0.01668453 | 0.021622152 | 1 |
| T2DM | nk_cell | *CPLX1* | -0.594557087 | 0.001014894 | 0.001624633 | 1 |
| T2DM | nk_cell | *SIGLEC7* | -0.824624909 | 0.015176519 | 0.019789431 | 1 |
| T2DM | nk_cell | *MFSD7* | -2.055098339 | 1.63E-05 | 0.000122264 | 0.078903874 |
| T2DM | nk_cell | *MAP2K1* | -1.247889911 | 0.022737961 | 0.028865669 | 1 |
| T2DM | nk_cell | *KLK7* | 1.002066379 | 0.000120943 | 0.000269553 | 0.586816195 |
| T2DM | nk_cell | *IRS1* | -1.080823487 | 2.40E-11 | 1.94E-08 | 1.16E-07 |
| T2DM | nk_cell | *UPF3A* | -1.138475069 | 6.72E-05 | 0.000181721 | 0.325825298 |
| T2DM | nk_cell | *HSPBAP1* | -1.273271856 | 0.00053875 | 0.000925971 | 1 |
| T2DM | nk_cell | *UBE2V2* | -1.347866565 | 8.03E-05 | 0.000204174 | 0.389768775 |
| T2DM | nk_cell | *PCSK9* | 4.402882147 | 2.60E-05 | 0.000122264 | 0.126336083 |
| T2DM | nk_cell | *XPO6* | -1.517479519 | 2.87E-05 | 0.000123207 | 0.139209814 |
| T2DM | nk_cell | *RGS14* | 0.337738973 | 9.13E-05 | 0.000221806 | 0.442947355 |
| T2DM | nk_cell | *RAB24* | -1.149325604 | 3.33E-05 | 0.000128061 | 0.161776413 |
| T2DM | nk_cell | *THBS3* | -1.007232628 | 2.52E-05 | 0.000122264 | 0.122240081 |
| T2DM | nk_cell | *PGM2* | -2.086409492 | 0.001068517 | 0.001702051 | 1 |
| T2DM | nk_cell | *P2RY12* | -1.688526046 | 0.001951431 | 0.002948721 | 1 |
| T2DM | nk_cell | *RNASE6* | -1.336606168 | 0.000385385 | 0.00069746 | 1 |
| T2DM | nk_cell | *CD52* | 0.154767195 | 0.000158639 | 0.000333354 | 0.769714542 |

| | | | | | | |
|---|---|---|---|---|---|---|
| T2DM | nk_cell | *SLC38A11* | -0.568230073 | 4.39E-05 | 0.000144447 | 0.212770144 |
| T2DM | nk_cell | *CCDC8* | -0.845844717 | 1.38E-05 | 0.000122264 | 0.067152319 |
| T2DM | nk_cell | *ZEB2* | -1.091099813 | 2.30E-05 | 0.000122264 | 0.11180444 |
| T2DM | nk_cell | *DTWD2* | -1.370343443 | 1.85E-05 | 0.000122264 | 0.089994715 |
| T2DM | nk_cell | *CLIC3* | 0.637766163 | 0.000120418 | 0.000268629 | 0.584267493 |
| T2DM | nk_cell | *INO80E* | -1.561058291 | 2.42E-05 | 0.000122264 | 0.117506424 |
| T2DM | nk_cell | *NFU1* | -1.520256031 | 1.20E-05 | 0.000122264 | 0.058398968 |
| T2DM | nk_cell | *C15orf40* | -1.028524619 | 0.000361595 | 0.000660067 | 1 |
| T2DM | nk_cell | *LUZP1* | -1.469578753 | 0.003591662 | 0.00518962 | 1 |
| T2DM | nk_cell | *LRRC45* | 0.407624772 | 0.031706706 | 0.039436282 | 1 |
| T2DM | nk_cell | *UGP2* | -1.012427113 | 6.78E-06 | 0.000122264 | 0.032913207 |
| T2DM | nk_cell | *CSGALNACT2* | -1.460086602 | 0.009836908 | 0.013184717 | 1 |
| T2DM | nk_cell | *AVEN* | -1.085552887 | 2.41E-05 | 0.000122264 | 0.117083308 |
| T2DM | nk_cell | *CTNND2* | -0.789353458 | 1.34E-05 | 0.000122264 | 0.065186447 |
| T2DM | nk_cell | *CALML6* | -10.01388005 | 2.13E-05 | 0.000122264 | 0.103393357 |
| T2DM | nk_cell | *TPST1* | -0.633490358 | 1.96E-05 | 0.000122264 | 0.095098816 |
| T2DM | nk_cell | *TOR1AIP2* | -1.168863536 | 4.75E-05 | 0.000149363 | 0.230541817 |
| T2DM | nk_cell | *OTUD3* | -1.262567615 | 0.000173114 | 0.000356969 | 0.83994871 |
| T2DM | nk_cell | *ZFPM2* | -0.630008549 | 5.75E-06 | 0.000122264 | 0.027875923 |
| T2DM | nk_cell | *MAP3K2* | -1.182111772 | 5.64E-05 | 0.00016385 | 0.273611106 |
| T2DM | nk_cell | *MYO7B* | -0.696937502 | 0.000336511 | 0.000620581 | 1 |
| T2DM | nk_cell | *UBE2E3* | -1.226188719 | 3.43E-05 | 0.000129717 | 0.166297216 |
| T2DM | nk_cell | *HNRNPA3* | -1.264857407 | 0.000386856 | 0.000699599 | 1 |
| T2DM | nk_cell | *RNF150* | -0.768920992 | 0.000175305 | 0.000360875 | 0.850582207 |
| T2DM | nk_cell | *HOXD4* | 2.501933371 | 1.85E-05 | 0.000122264 | 0.089754671 |
| T2DM | nk_cell | *ANKK1* | -2.036903138 | 2.35E-05 | 0.000122264 | 0.114199462 |
| T2DM | nk_cell | *PDCD6IP* | -1.100814134 | 2.95E-05 | 0.000123723 | 0.143027559 |
| T2DM | nk_cell | *CRTAP* | -1.395627507 | 0.000185444 | 0.000377739 | 0.899773457 |
| T2DM | nk_cell | *CNGB3* | -1.191103383 | 0.00022372 | 0.000440361 | 1 |
| T2DM | nk_cell | *ELP5* | -1.525439025 | 5.32E-05 | 0.000159179 | 0.258346722 |
| T2DM | nk_cell | *STX8* | -1.201238657 | 0.004567388 | 0.006472245 | 1 |
| T2DM | nk_cell | *NFRKB* | -1.889219228 | 0.000107719 | 0.000248764 | 0.522652962 |
| T2DM | nk_cell | *SEMA3E* | -0.534755571 | 0.000331473 | 0.000613858 | 1 |
| T2DM | nk_cell | *ADORA2B* | -0.637257874 | 0.001109159 | 0.00175928 | 1 |
| T2DM | nk_cell | *MGMT* | -1.319282714 | 0.000126657 | 0.00027921 | 0.614541899 |
| T2DM | nk_cell | *NFXL1* | -1.402160394 | 1.25E-05 | 0.000122264 | 0.060730242 |
| T2DM | nk_cell | *DNAJC18* | -0.957307328 | 0.000427503 | 0.000758685 | 1 |
| T2DM | nk_cell | *SPATA24* | -0.659138815 | 0.013638823 | 0.017890124 | 1 |
| T2DM | nk_cell | *HSD17B13* | -4.107948916 | 0.000193322 | 0.000390031 | 0.937996123 |
| T2DM | nk_cell | *ELOVL6* | 0.892057009 | 0.017043987 | 0.022058528 | 1 |
| T2DM | nk_cell | *ARL6IP1* | -1.444623481 | 0.006275723 | 0.008692494 | 1 |
| T2DM | nk_cell | *CDH2* | -0.812512043 | 3.18E-05 | 0.000127109 | 0.154056326 |
| T2DM | nk_cell | *STAT2* | -1.409397289 | 0.000242742 | 0.000470737 | 1 |
| T2DM | nk_cell | *SLC26A5* | -1.527209245 | 0.001102501 | 0.001749864 | 1 |
| T2DM | nk_cell | *GTSF1* | -1.890180841 | 5.59E-05 | 0.000163255 | 0.271167311 |
| T2DM | nk_cell | *ARMC10* | -1.383041949 | 0.000534423 | 0.000919185 | 1 |
| T2DM | nk_cell | *ACYP2* | -1.008287497 | 4.58E-05 | 0.000147697 | 0.222284226 |
| T2DM | nk_cell | *TRABD* | 1.054870685 | 0.001742626 | 0.002655535 | 1 |
| T2DM | nk_cell | *TMEM133* | -1.224865454 | 0.001691983 | 0.002589749 | 1 |
| T2DM | nk_cell | *HOXB9* | -2.378024676 | 1.10E-05 | 0.000122264 | 0.053238101 |

| | | | | | | |
|---|---|---|---|---|---|---|
| T2DM | nk_cell | *TTLL6* | -2.592738799 | 2.44E-09 | 9.85E-07 | 1.18E-05 |
| T2DM | nk_cell | *KCNS3* | -0.407607825 | 0.00034849 | 0.000639047 | 1 |
| T2DM | nk_cell | *FOXN2* | -1.519814649 | 0.000402956 | 0.000724642 | 1 |
| T2DM | nk_cell | *GPR27* | -0.971521026 | 0.001779716 | 0.002708652 | 1 |
| T2DM | nk_cell | *TRIAP1* | -1.091131772 | 0.000236776 | 0.00046101 | 1 |
| T2DM | nk_cell | *TSEN34* | -2.576466046 | 1.11E-05 | 0.000122264 | 0.053943119 |
| T2DM | nk_cell | *MSANTD4* | 1.148242289 | 0.031916339 | 0.039676679 | 1 |
| T2DM | nk_cell | *NDUFA3* | 0.91918541 | 0.000891484 | 0.001444234 | 1 |
| T2DM | nk_cell | *PAQR8* | -1.269690358 | 0.002665052 | 0.003925572 | 1 |
| T2DM | nk_cell | *NUDT6* | -0.780755992 | 0.001752352 | 0.002669517 | 1 |
| T2DM | nk_cell | *ZNF160* | -0.880024223 | 0.000112509 | 0.000256916 | 0.545892435 |
| T2DM | nk_cell | *ZNF415* | -1.761881006 | 7.98E-05 | 0.000203354 | 0.387390287 |
| T2DM | nk_cell | *S1PR1* | -1.232172349 | 0.000597991 | 0.001012724 | 1 |
| T2DM | nk_cell | *PYGO1* | -1.442564337 | 2.36E-05 | 0.000122264 | 0.114453196 |
| T2DM | nk_cell | *FPR1* | -1.436860458 | 2.90E-05 | 0.000123467 | 0.140757365 |
| T2DM | nk_cell | *FEZ2* | -1.243786282 | 7.80E-05 | 0.000200157 | 0.37859654 |
| T2DM | nk_cell | *SOX7* | -2.377873488 | 0.006734742 | 0.009280593 | 1 |
| T2DM | nk_cell | *TRMT61B* | -1.119603974 | 0.002179957 | 0.003259523 | 1 |
| T2DM | nk_cell | *MFN1* | -0.879705468 | 0.000196355 | 0.000395208 | 0.952713036 |
| T2DM | nk_cell | *GIMAP8* | -1.634485202 | 0.004203877 | 0.005986854 | 1 |
| T2DM | nk_cell | *ATP6V0E2* | 0.966284015 | 0.00060412 | 0.001022033 | 1 |
| T2DM | nk_cell | *MORN4* | -0.518793817 | 0.003729893 | 0.005368568 | 1 |
| T2DM | nk_cell | *SCAND1* | 1.128209798 | 0.007446708 | 0.010154983 | 1 |
| T2DM | nk_cell | *SOSTDC1* | -1.019606743 | 0.008998324 | 0.012134482 | 1 |
| T2DM | nk_cell | *GAA* | -0.488488473 | 0.008460817 | 0.011451014 | 1 |
| T2DM | nk_cell | *CANT1* | -0.856170589 | 0.00344925 | 0.004995749 | 1 |
| T2DM | nk_cell | *PGAM1* | -0.839643684 | 0.000402745 | 0.000724553 | 1 |
| T2DM | nk_cell | *LURAP1* | -0.785360538 | 5.49E-05 | 0.000161068 | 0.266406836 |
| T2DM | nk_cell | *PDE7B* | -0.898896396 | 2.63E-05 | 0.000122295 | 0.127427178 |
| T2DM | nk_cell | *MRPL36* | 0.917507888 | 0.000405809 | 0.000727905 | 1 |
| T2DM | nk_cell | *MCC* | -1.141730484 | 3.46E-05 | 0.000130101 | 0.167773455 |
| T2DM | nk_cell | *ZNF561* | -1.458923265 | 9.08E-06 | 0.000122264 | 0.044034569 |
| T2DM | nk_cell | *HOPX* | 0.157900929 | 5.84E-05 | 0.000167232 | 0.283298682 |
| T2DM | nk_cell | *PPID* | -1.926469948 | 0.000216976 | 0.00042935 | 1 |
| T2DM | nk_cell | *ETFDH* | -0.41033389 | 0.024114128 | 0.030500977 | 1 |
| T2DM | nk_cell | *TBCA* | -1.338251414 | 6.26E-05 | 0.000173589 | 0.303889634 |
| T2DM | nk_cell | *ZNF584* | -0.360651025 | 0.006623088 | 0.009139711 | 1 |
| T2DM | nk_cell | *NMUR1* | 0.201083097 | 6.04E-05 | 0.000169923 | 0.293027394 |
| T2DM | nk_cell | *PIK3CD* | 0.514434158 | 0.000176097 | 0.000362197 | 0.854423206 |
| T2DM | nk_cell | *ANO5* | -0.712780056 | 1.70E-05 | 0.000122264 | 0.082467062 |
| T2DM | nk_cell | *VAT1L* | -1.631594425 | 3.08E-05 | 0.00012591 | 0.149455011 |
| T2DM | nk_cell | *CAMTA1* | -1.398752621 | 0.000350832 | 0.000642537 | 1 |
| T2DM | nk_cell | *LGALS4* | -1.426338811 | 2.79E-05 | 0.000122493 | 0.135380397 |
| T2DM | nk_cell | *PAH* | -2.810235267 | 4.27E-05 | 0.000142711 | 0.207344608 |
| T2DM | nk_cell | *RHNO1* | 0.404960358 | 8.67E-05 | 0.000213966 | 0.420695811 |
| T2DM | nk_cell | *PWWP2B* | -0.88017898 | 1.03E-05 | 0.000122264 | 0.049760741 |
| T2DM | nk_cell | *EXOSC10* | -1.645972561 | 2.84E-06 | 0.000122264 | 0.013793063 |
| T2DM | nk_cell | *TRAPPC12* | -1.169067435 | 0.025722006 | 0.032374364 | 1 |
| T2DM | nk_cell | *C3AR1* | -0.813822742 | 0.007590398 | 0.010330606 | 1 |
| T2DM | nk_cell | *PRNP* | -1.302835465 | 0.00020915 | 0.0004159 | 1 |

| | | | | | | |
|---|---|---|---|---|---|---|
| T2DM | nk_cell | *TVP23B* | -1.411851812 | 0.000716047 | 0.001186564 | 1 |
| T2DM | nk_cell | *FBXW10* | -1.287414847 | 1.72E-05 | 0.000122264 | 0.083525635 |
| T2DM | nk_cell | *SRGAP2C* | -0.638984508 | 4.50E-05 | 0.00014683 | 0.21812351 |
| T2DM | nk_cell | *ATPAF2* | -1.132515116 | 0.001023616 | 0.001636436 | 1 |
| T2DM | nk_cell | *ZNF554* | 1.151828656 | 0.008109544 | 0.010990924 | 1 |
| T2DM | nk_cell | *ANKRD20A4* | -5.749646936 | 0.017833075 | 0.023012255 | 1 |
| T2DM | nk_cell | *SERF1A* | -3.985622042 | 2.10E-05 | 0.000122264 | 0.101749924 |
| T2DM | nk_cell | *CD8B* | 0.246897327 | 7.17E-05 | 0.000189735 | 0.347974079 |
| T2DM | nk_cell | *SLFN12* | -0.967220285 | 0.036257472 | 0.044706799 | 1 |
| T2DM | nk_cell | *MTBP* | -1.357416933 | 6.46E-06 | 0.000122264 | 0.031335691 |
| T2DM | nk_cell | *TEFM* | -0.982994209 | 9.36E-05 | 0.000225227 | 0.454282728 |
| T2DM | nk_cell | *MALT1* | -1.618050545 | 0.000410571 | 0.000734547 | 1 |
| T2DM | nk_cell | *MBOAT1* | -1.161035922 | 1.81E-05 | 0.000122264 | 0.087626318 |
| T2DM | nk_cell | *CXCR6* | 0.625377663 | 7.73E-05 | 0.000199059 | 0.374904589 |
| T2DM | nk_cell | *PAIP1* | -1.760194144 | 3.10E-05 | 0.000126367 | 0.150503461 |
| T2DM | nk_cell | *C1QTNF4* | -1.01787201 | 1.56E-07 | 2.52E-05 | 0.0007552 |
| T2DM | nk_cell | *MACROD2* | -0.754169055 | 4.54E-05 | 0.000147579 | 0.220334826 |
| T2DM | nk_cell | *BSG* | -1.553061924 | 4.99E-05 | 0.00015287 | 0.24199381 |
| T2DM | nk_cell | *HINFP* | -0.846127228 | 2.41E-05 | 0.000122264 | 0.116908182 |
| T2DM | nk_cell | *TP53RK* | 0.577379601 | 0.000172223 | 0.000355359 | 0.835623742 |
| T2DM | nk_cell | *SUCLG2* | -1.327266649 | 0.000934031 | 0.001508628 | 1 |
| T2DM | nk_cell | *ARNT2* | -0.673907806 | 4.24E-05 | 0.000141957 | 0.205553744 |
| T2DM | nk_cell | *GTPBP2* | -1.32296466 | 0.000694856 | 0.00115579 | 1 |
| T2DM | nk_cell | *FGGY* | -1.199741151 | 0.000821038 | 0.001338151 | 1 |
| T2DM | nk_cell | *MANEA* | -1.544884923 | 6.22E-05 | 0.00017316 | 0.301778339 |
| T2DM | nk_cell | *AFF1* | -0.971361223 | 3.10E-05 | 0.000126367 | 0.150366935 |
| T2DM | nk_cell | *FIBP* | 0.721348357 | 0.000110179 | 0.000253239 | 0.534586476 |
| T2DM | nk_cell | *CTSW* | 0.0743474 | 6.43E-05 | 0.000176235 | 0.311759398 |
| T2DM | nk_cell | *SNTG2* | -0.817246952 | 7.43E-06 | 0.000122264 | 0.036053039 |
| T2DM | nk_cell | *SMPDL3A* | -1.018892296 | 0.001141766 | 0.0018051 | 1 |
| T2DM | nk_cell | *RAD9A* | -0.753559325 | 0.000743591 | 0.001224102 | 1 |
| T2DM | nk_cell | *EFEMP2* | -0.650028856 | 1.77E-05 | 0.000122264 | 0.085805454 |
| T2DM | nk_cell | *ZFAND4* | -1.460166265 | 6.38E-06 | 0.000122264 | 0.030931622 |
| T2DM | nk_cell | *SLFN11* | -1.77135219 | 0.000374817 | 0.000680618 | 1 |
| T2DM | nk_cell | *FUT10* | -0.804420973 | 0.000186608 | 0.000379633 | 0.905423915 |
| T2DM | nk_cell | *ZNF596* | -1.855716844 | 0.005013568 | 0.007061199 | 1 |
| T2DM | nk_cell | *CFL1* | 0.703379431 | 0.000418908 | 0.000746709 | 1 |
| T2DM | nk_cell | *FAM192A* | -1.397024246 | 0.000398851 | 0.000718879 | 1 |
| T2DM | nk_cell | *CBWD1* | -1.12304455 | 1.85E-05 | 0.000122264 | 0.089886503 |
| T2DM | nk_cell | *DCP2* | -1.4641479 | 2.50E-05 | 0.000122264 | 0.121244739 |
| T2DM | nk_cell | *ZNF621* | -1.81379105 | 0.004358694 | 0.006200357 | 1 |
| T2DM | nk_cell | *NADSYN1* | -1.5772323 | 0.001016426 | 0.00162655 | 1 |
| T2DM | nk_cell | *RNASEH2C* | -1.67724733 | 1.96E-07 | 3.06E-05 | 0.000949301 |
| T2DM | nk_cell | *MYEOV* | -2.799613562 | 7.64E-06 | 0.000122264 | 0.037045042 |
| T2DM | nk_cell | *MRGPRF* | -0.620492296 | 4.68E-06 | 0.000122264 | 0.022719796 |
| T2DM | nk_cell | *LCLAT1* | -1.62532162 | 0.000126112 | 0.00027826 | 0.611894425 |
| T2DM | nk_cell | *SH3RF3* | -1.197846072 | 1.03E-05 | 0.000122264 | 0.049959987 |
| T2DM | nk_cell | *GXYLT2* | -0.830190554 | 1.81E-05 | 0.000122264 | 0.087946835 |
| T2DM | nk_cell | *DCAKD* | -1.753774696 | 6.45E-05 | 0.000176728 | 0.313162327 |
| T2DM | nk_cell | *TRMT112* | -1.245537071 | 0.004300427 | 0.00612076 | 1 |

| | | | | | | |
|---|---|---|---|---|---|---|
| T2DM | nk_cell | *NOC3L* | -1.429158439 | 9.35E-06 | 0.000122264 | 0.045389291 |
| T2DM | nk_cell | *RHOD* | -0.722030579 | 5.04E-05 | 0.000153489 | 0.244546135 |
| T2DM | nk_cell | *PARP14* | -0.6231977 | 0.024995023 | 0.03153328 | 1 |
| T2DM | nk_cell | *PARP15* | 0.165355057 | 0.000113579 | 0.000258846 | 0.551082919 |
| T2DM | nk_cell | *AHSA2* | -1.093816904 | 3.54E-06 | 0.000122264 | 0.017155577 |
| T2DM | nk_cell | *VANGL1* | -0.998972065 | 7.24E-05 | 0.00019113 | 0.351106398 |
| T2DM | nk_cell | *IQCB1* | -1.074358947 | 1.49E-05 | 0.000122264 | 0.072365638 |
| T2DM | nk_cell | *ZNF483* | -0.861307909 | 0.000104526 | 0.000243476 | 0.507161307 |
| T2DM | nk_cell | *SLC2A14* | -0.701973327 | 3.44E-05 | 0.000129854 | 0.166732968 |
| T2DM | nk_cell | *MMRN2* | -1.241894341 | 0.001063445 | 0.001694527 | 1 |
| T2DM | nk_cell | *MAP3K11* | -1.531880032 | 3.65E-05 | 0.000132621 | 0.177119043 |
| T2DM | nk_cell | *KCNK7* | 1.596079107 | 0.000103519 | 0.000241827 | 0.502275455 |
| T2DM | nk_cell | *NDNF* | -0.927078808 | 2.82E-05 | 0.000122832 | 0.136589516 |
| T2DM | nk_cell | *ARV1* | -1.018327143 | 8.62E-06 | 0.000122264 | 0.041809698 |
| T2DM | nk_cell | *CCDC36* | -0.692969791 | 0.000227821 | 0.000446621 | 1 |
| T2DM | nk_cell | *ZNF417* | -1.394232172 | 0.019048398 | 0.024476384 | 1 |
| T2DM | nk_cell | *PEAK1* | -1.008457859 | 2.06E-05 | 0.000122264 | 0.099713233 |
| T2DM | nk_cell | *MST1* | -0.654412996 | 2.69E-05 | 0.000122391 | 0.130733209 |
| T2DM | nk_cell | *GMPPB* | -1.733100386 | 3.04E-05 | 0.000125051 | 0.147685584 |
| T2DM | nk_cell | *NUDT18* | -4.077946599 | 3.29E-05 | 0.000127909 | 0.159681525 |
| T2DM | nk_cell | *SULT1B1* | 3.019961671 | 0.004659023 | 0.006590548 | 1 |
| T2DM | nk_cell | *NUDT4* | -0.862039782 | 2.58E-05 | 0.000122264 | 0.125135726 |
| T2DM | nk_cell | *SCAI* | -1.017964177 | 0.000418905 | 0.000746709 | 1 |
| T2DM | nk_cell | *HSPB7* | 0.71529759 | 0.022249957 | 0.028305399 | 1 |
| T2DM | nk_cell | *UQCRH* | -1.499384995 | 9.24E-05 | 0.000223814 | 0.448274243 |
| T2DM | nk_cell | *C11orf80* | -1.607832458 | 0.000371657 | 0.000676146 | 1 |
| T2DM | nk_cell | *CD7* | 0.090873014 | 5.02E-05 | 0.000153221 | 0.243775401 |
| T2DM | nk_cell | *GOLIM4* | -1.146503066 | 1.72E-05 | 0.000122264 | 0.08345941 |
| T2DM | nk_cell | *HOXB2* | -1.547404874 | 2.44E-07 | 3.38E-05 | 0.001181984 |
| T2DM | nk_cell | *XXYLT1* | -0.995789216 | 1.60E-05 | 0.000122264 | 0.077720829 |
| T2DM | nk_cell | *UBXN2A* | -1.281601458 | 7.65E-05 | 0.000197625 | 0.370942651 |
| T2DM | nk_cell | *NRROS* | 0.497680849 | 0.010971317 | 0.01461236 | 1 |
| T2DM | nk_cell | *CEP19* | -0.966493454 | 0.001460958 | 0.002253917 | 1 |
| T2DM | nk_cell | *SLC25A30* | -2.359420548 | 0.000139298 | 0.000300656 | 0.675875401 |
| T2DM | nk_cell | *CTSF* | -1.292642869 | 0.000734407 | 0.001212434 | 1 |
| T2DM | nk_cell | *LEMD3* | -1.510137585 | 0.005994612 | 0.008334057 | 1 |
| T2DM | nk_cell | *CTU2* | 0.730836255 | 0.000697076 | 0.001158756 | 1 |
| T2DM | nk_cell | *GLIS1* | -0.673715119 | 1.99E-05 | 0.000122264 | 0.096523165 |
| T2DM | nk_cell | *RALGAPA1* | -1.094222602 | 8.67E-05 | 0.000213966 | 0.420574715 |
| T2DM | nk_cell | *ANKRD36C* | -1.406799946 | 5.54E-05 | 0.000162218 | 0.268794627 |
| T2DM | nk_cell | *TMEM81* | 1.617688793 | 0.001697432 | 0.002596451 | 1 |
| T2DM | nk_cell | *MSL2* | -1.666082255 | 0.000650967 | 0.00108876 | 1 |
| T2DM | nk_cell | *ZNF497* | 3.297944621 | 0.000128751 | 0.00028331 | 0.624698868 |
| T2DM | nk_cell | *TRAM1L1* | -1.420684067 | 0.000905254 | 0.001465074 | 1 |
| T2DM | nk_cell | *ANGEL2* | -1.595957607 | 0.000116972 | 0.000263449 | 0.567549393 |
| T2DM | nk_cell | *ZNF266* | -0.840166748 | 0.00171464 | 0.00261947 | 1 |
| T2DM | nk_cell | *SH3PXD2B* | -0.633213247 | 1.79E-05 | 0.000122264 | 0.086899161 |
| T2DM | nk_cell | *LARP7* | -1.740501885 | 9.58E-05 | 0.000228964 | 0.464797696 |
| T2DM | nk_cell | *FGFBP3* | 1.227048065 | 0.000189805 | 0.000384203 | 0.920935032 |
| T2DM | nk_cell | *C4orf32* | -0.840264512 | 2.66E-05 | 0.000122391 | 0.129129316 |

| | | | | | | |
|---|---|---|---|---|---|---|
| T2DM | nk_cell | *HRAS* | 1.080697752 | 0.003595624 | 0.005193798 | 1 |
| T2DM | nk_cell | *CEP135* | -1.655601732 | 0.001466405 | 0.002260164 | 1 |
| T2DM | nk_cell | *RSRC1* | -1.103683891 | 3.00E-05 | 0.000124725 | 0.145484676 |
| T2DM | nk_cell | *PTDSS2* | -1.106750013 | 1.40E-05 | 0.000122264 | 0.06781836 |
| T2DM | nk_cell | *SEZ6L2* | 1.435809511 | 0.000424641 | 0.00075535 | 1 |
| T2DM | nk_cell | *DHX36* | -1.471874127 | 0.000310796 | 0.000579993 | 1 |
| T2DM | nk_cell | *FBXW8* | -1.440975963 | 0.004402505 | 0.006255038 | 1 |
| T2DM | nk_cell | *SLC22A1* | -1.700314983 | 4.20E-05 | 0.000141813 | 0.203748998 |
| T2DM | nk_cell | *CHST2* | 0.251428188 | 5.36E-05 | 0.000159514 | 0.260132009 |
| T2DM | nk_cell | *TVP23C* | 1.896526137 | 0.001787335 | 0.002719395 | 1 |
| T2DM | nk_cell | *PACS1* | -1.110654895 | 0.007534167 | 0.010261706 | 1 |
| T2DM | nk_cell | *CADM2* | -0.801019612 | 0.000613199 | 0.001033788 | 1 |
| T2DM | nk_cell | *PSMD2* | -1.022158643 | 1.53E-05 | 0.000122264 | 0.074041794 |
| T2DM | nk_cell | *ARHGAP1* | -1.325893382 | 0.006983952 | 0.009586244 | 1 |
| T2DM | nk_cell | *ATG13* | -1.806338056 | 5.45E-05 | 0.000160881 | 0.264240053 |
| T2DM | nk_cell | *CCNE2* | -0.935047495 | 3.14E-08 | 8.47E-06 | 0.000152387 |
| T2DM | nk_cell | *ZNF519* | -0.886721915 | 3.17E-05 | 0.000127109 | 0.153735131 |
| T2DM | nk_cell | *BANF1* | 2.855909871 | 0.00158809 | 0.002436108 | 1 |
| T2DM | nk_cell | *CHRNA7* | -0.955284071 | 0.002473255 | 0.003669795 | 1 |
| T2DM | nk_cell | *TMEM9B* | -1.346525646 | 2.65E-05 | 0.000122391 | 0.128448314 |
| T2DM | nk_cell | *SCUBE2* | -0.947642677 | 2.58E-05 | 0.000122264 | 0.125417135 |
| T2DM | nk_cell | *EIF1AD* | -1.148427943 | 6.00E-06 | 0.000122264 | 0.029105412 |
| T2DM | nk_cell | *ZNF25* | -1.803231644 | 1.05E-05 | 0.000122264 | 0.051153816 |
| T2DM | nk_cell | *CLTB* | -2.030224195 | 0.000250468 | 0.000482789 | 1 |
| T2DM | nk_cell | *PPP2R2D* | -0.953459319 | 0.03862303 | 0.047442771 | 1 |
| T2DM | nk_cell | *MCTP1* | -0.995944529 | 2.80E-05 | 0.000122557 | 0.135835629 |
| T2DM | nk_cell | *LRRC25* | -0.977665137 | 0.001034918 | 0.001652871 | 1 |
| T2DM | nk_cell | *UBQLNL* | -1.202931728 | 0.000921323 | 0.001489094 | 1 |
| T2DM | nk_cell | *KCNE3* | -1.660578744 | 0.001724586 | 0.002631117 | 1 |
| T2DM | nk_cell | *C11orf68* | 1.327026965 | 0.007770724 | 0.010564178 | 1 |
| T2DM | nk_cell | *MRPL48* | -1.776174749 | 2.68E-05 | 0.000122391 | 0.129994352 |
| T2DM | nk_cell | *RAB6A* | -1.385312574 | 5.38E-05 | 0.000159514 | 0.260887255 |
| T2DM | nk_cell | *TMEM70* | -1.221156563 | 0.000161058 | 0.000337416 | 0.781454546 |
| T2DM | nk_cell | *RPS6KB2* | 0.818650027 | 0.000213198 | 0.000422563 | 1 |
| T2DM | nk_cell | *TOM1L2* | -1.045870167 | 6.11E-06 | 0.000122264 | 0.029622793 |
| T2DM | nk_cell | *TEX26* | -1.887661904 | 0.001199342 | 0.00188568 | 1 |
| T2DM | nk_cell | *GPR156* | -1.495583057 | 0.007147472 | 0.009782661 | 1 |
| T2DM | nk_cell | *TTLL11* | -1.554309783 | 3.45E-05 | 0.000130101 | 0.167437278 |
| T2DM | nk_cell | *PRIMA1* | -0.574170991 | 2.14E-05 | 0.000122264 | 0.10400691 |
| T2DM | nk_cell | *CTDNEP1* | 1.158729079 | 0.004422823 | 0.006282066 | 1 |
| T2DM | nk_cell | *ZDHHC21* | -1.386387805 | 5.63E-05 | 0.00016385 | 0.273101421 |
| T2DM | nk_cell | *A2M* | -1.162532704 | 0.006526758 | 0.009017036 | 1 |
| T2DM | nk_cell | *DOK7* | 2.221432241 | 0.000124272 | 0.000274951 | 0.602967752 |
| T2DM | nk_cell | *ORAI3* | 0.793641087 | 3.58E-05 | 0.000131758 | 0.173920709 |
| T2DM | nk_cell | *ZNF613* | -0.863288283 | 0.000424323 | 0.000755251 | 1 |
| T2DM | nk_cell | *JAKMIP2* | 0.114518193 | 5.46E-05 | 0.000160881 | 0.264799445 |
| T2DM | nk_cell | *TCP11L1* | -0.910699417 | 0.003681775 | 0.005310336 | 1 |
| T2DM | nk_cell | *CCDC57* | -0.958707938 | 0.000121277 | 0.000270105 | 0.588434165 |
| T2DM | nk_cell | *MYPOP* | -0.9052435 | 5.38E-05 | 0.000159514 | 0.260964249 |
| T2DM | nk_cell | *CIDEA* | -0.574417828 | 9.84E-06 | 0.000122264 | 0.047753456 |

| | | | | | | |
|---|---|---|---|---|---|---|
| T2DM | nk_cell | *SLC35G1* | -1.386991588 | 1.74E-05 | 0.000122264 | 0.084642834 |
| T2DM | nk_cell | *ZNF135* | -1.174158224 | 0.000161445 | 0.000337644 | 0.783333306 |
| T2DM | nk_cell | *OR4N2* | -1.542334015 | 8.11E-05 | 0.00020495 | 0.393291856 |
| T2DM | nk_cell | *TAC4* | -1.827904226 | 3.20E-05 | 0.000127628 | 0.155159105 |
| T2DM | nk_cell | *CDC26* | -0.878174004 | 3.08E-05 | 0.00012591 | 0.149372484 |
| T2DM | nk_cell | *CRLF3* | -0.973338677 | 0.007371833 | 0.010067024 | 1 |
| T2DM | nk_cell | *DNAJC30* | -2.005505263 | 0.004115458 | 0.005873 | 1 |
| T2DM | nk_cell | *SPRYD4* | -1.266368165 | 3.46E-07 | 4.05E-05 | 0.001680704 |
| T2DM | nk_cell | *VPS37D* | -0.613073359 | 0.015934525 | 0.020711041 | 1 |
| T2DM | nk_cell | *WDR25* | -1.488728577 | 2.50E-05 | 0.000122264 | 0.121140435 |
| T2DM | nk_cell | *PLA2G16* | -0.66276139 | 5.60E-05 | 0.000163351 | 0.271653231 |
| T2DM | nk_cell | *B3GNT5* | -1.011448672 | 1.11E-05 | 0.000122264 | 0.054089835 |
| T2DM | nk_cell | *RMDN1* | -0.924635412 | 0.000594988 | 0.001008342 | 1 |
| T2DM | nk_cell | *RNF152* | -0.903257645 | 2.20E-05 | 0.000122264 | 0.106698609 |
| T2DM | nk_cell | *BDNF* | -1.160826775 | 2.20E-05 | 0.000122264 | 0.106686519 |
| T2DM | nk_cell | *BOK* | -0.881714491 | 1.64E-05 | 0.000122264 | 0.079762078 |
| T2DM | nk_cell | *RUFY1* | -1.046337451 | 2.95E-05 | 0.000123723 | 0.143250845 |
| T2DM | nk_cell | *VSIG10* | -0.791588072 | 0.000101978 | 0.000239427 | 0.494795474 |
| T2DM | nk_cell | *ANKLE2* | -1.408794681 | 2.59E-05 | 0.000122264 | 0.125782419 |
| T2DM | nk_cell | *C8G* | -10.83852528 | 2.93E-05 | 0.000123467 | 0.142124969 |
| T2DM | nk_cell | *MUC20* | 1.418425805 | 0.000370914 | 0.0006753 | 1 |
| T2DM | nk_cell | *LY6H* | -1.270559239 | 0.000188473 | 0.000382463 | 0.914468646 |
| T2DM | nk_cell | *SHMT1* | -1.553894099 | 0.000427501 | 0.000758685 | 1 |
| T2DM | nk_cell | *DPP7* | -1.756518423 | 0.001038659 | 0.001657755 | 1 |
| T2DM | nk_cell | *MTHFR* | -1.383255463 | 3.46E-05 | 0.000130101 | 0.167960621 |
| T2DM | nk_cell | *C19orf18* | -1.074794902 | 5.03E-05 | 0.000153297 | 0.244048746 |
| T2DM | nk_cell | *TMEM80* | -1.271844701 | 0.00014503 | 0.000310953 | 0.703687514 |
| T2DM | nk_cell | *WDR73* | -1.193262724 | 3.73E-05 | 0.000134293 | 0.181179686 |
| T2DM | nk_cell | *DSCAML1* | -0.567218616 | 1.78E-05 | 0.000122264 | 0.086582439 |
| T2DM | nk_cell | *TALDO1* | -1.880275772 | 8.53E-05 | 0.000211954 | 0.413946453 |
| T2DM | nk_cell | *AP006621.1* | -0.652302435 | 2.12E-05 | 0.000122264 | 0.102958628 |
| T2DM | nk_cell | *MAN1B1* | -1.906199356 | 3.89E-05 | 0.000136418 | 0.188770508 |
| T2DM | nk_cell | *FBXO39* | -0.616727037 | 0.038450793 | 0.047255128 | 1 |
| T2DM | nk_cell | *MIEF2* | -1.254591174 | 1.82E-05 | 0.000122264 | 0.088264418 |
| T2DM | nk_cell | *NAP1L5* | -1.066221832 | 1.02E-05 | 0.000122264 | 0.049562163 |
| T2DM | nk_cell | *SLC25A22* | 0.741131987 | 0.004148377 | 0.005913021 | 1 |
| T2DM | nk_cell | *GBA* | -0.927904629 | 5.20E-06 | 0.000122264 | 0.025246155 |
| T2DM | nk_cell | *ACAD9* | -0.548284597 | 0.007194559 | 0.009841556 | 1 |
| T2DM | nk_cell | *IL17RA* | -1.322376672 | 9.68E-05 | 0.000230933 | 0.469718221 |
| T2DM | nk_cell | *MBOAT4* | -1.298971108 | 0.000672978 | 0.001122093 | 1 |
| T2DM | nk_cell | *AGTRAP* | 0.817160812 | 0.000290592 | 0.000548407 | 1 |
| T2DM | nk_cell | *THAP5* | -2.298900294 | 0.001208444 | 0.001897982 | 1 |
| T2DM | nk_cell | *NAALADL2* | -0.750085523 | 0.000187605 | 0.000381181 | 0.91026074 |
| T2DM | nk_cell | *CD151* | -1.422342405 | 0.000131603 | 0.000288019 | 0.638537104 |
| T2DM | nk_cell | *FAM87B* | -4.483747611 | 2.78E-05 | 0.000122493 | 0.134774795 |
| T2DM | nk_cell | *CENPBD1* | 2.019582631 | 0.000567229 | 0.000966021 | 1 |
| T2DM | nk_cell | *RPS27* | 0.640514766 | 0.000995971 | 0.00159962 | 1 |
| T2DM | nk_cell | *RIC8A* | 0.712735926 | 0.002134698 | 0.003196776 | 1 |
| T2DM | nk_cell | *ODF3B* | -0.518649022 | 0.000231515 | 0.000452218 | 1 |
| T2DM | nk_cell | *DPY19L2* | -0.477605159 | 6.53E-05 | 0.00017822 | 0.316697459 |

| | | | | | | |
|---|---|---|---|---|---|---|
| T2DM | nk_cell | *ADAMTSL1* | -0.562579349 | 9.94E-05 | 0.000235619 | 0.482442626 |
| T2DM | nk_cell | *GRAMD1C* | -0.954456498 | 3.29E-05 | 0.000127909 | 0.159393156 |
| T2DM | nk_cell | *DALRD3* | -1.214728208 | 8.55E-05 | 0.000212227 | 0.414691099 |
| T2DM | nk_cell | *ZNF114* | -2.92222868 | 0.000380263 | 0.000689734 | 1 |
| T2DM | nk_cell | *SPINK6* | -2.143051976 | 0.000323264 | 0.000600029 | 1 |
| T2DM | nk_cell | *SH2B1* | -1.312335833 | 2.41E-05 | 0.000122264 | 0.116902611 |
| T2DM | nk_cell | *VN1R1* | -3.12323771 | 5.74E-06 | 0.000122264 | 0.027869053 |
| T2DM | nk_cell | *KDELC2* | -2.441303411 | 0.000309849 | 0.000578449 | 1 |
| T2DM | nk_cell | *PLEC* | -1.006119771 | 0.000707398 | 0.001173435 | 1 |
| T2DM | nk_cell | *SH2D4B* | -1.773394761 | 3.31E-05 | 0.000127948 | 0.160766314 |
| T2DM | nk_cell | *RNF212* | -1.700849823 | 0.001297401 | 0.002022162 | 1 |
| T2DM | nk_cell | *PRSS36* | -3.364195985 | 2.30E-05 | 0.000122264 | 0.111714479 |
| T2DM | nk_cell | *WDR6* | -0.954685736 | 0.000197021 | 0.000396165 | 0.955946111 |
| T2DM | nk_cell | *TMPRSS9* | -0.61382139 | 4.21E-05 | 0.000141813 | 0.204026744 |
| T2DM | nk_cell | *TMEM11* | -0.793357477 | 7.85E-06 | 0.000122264 | 0.03807723 |
| T2DM | nk_cell | *ZFAND2A* | -0.940767053 | 4.50E-05 | 0.00014683 | 0.218482859 |
| T2DM | nk_cell | *ZNF223* | -1.738102623 | 2.07E-05 | 0.000122264 | 0.100403018 |
| T2DM | nk_cell | *FAM220A* | -1.558489355 | 5.19E-05 | 0.000156496 | 0.251746831 |
| T2DM | nk_cell | *NT5DC1* | -1.392548865 | 0.000239241 | 0.00046469 | 1 |
| T2DM | nk_cell | *GLDC* | -0.892670413 | 1.86E-05 | 0.000122264 | 0.090265738 |
| T2DM | nk_cell | *P4HTM* | -1.084128283 | 2.19E-05 | 0.000122264 | 0.106224717 |
| T2DM | nk_cell | *CA8* | -1.496775481 | 1.63E-05 | 0.000122264 | 0.079278161 |
| T2DM | nk_cell | *ERN1* | -1.661455224 | 0.003702554 | 0.005332381 | 1 |
| T2DM | nk_cell | *ZNF713* | -0.597170293 | 0.001154858 | 0.001824607 | 1 |
| T2DM | nk_cell | *PARP10* | -1.072100297 | 1.38E-05 | 0.000122264 | 0.066721593 |
| T2DM | nk_cell | *NSUN3* | -1.310512458 | 4.05E-05 | 0.000139095 | 0.196540542 |
| T2DM | nk_cell | *RPP25* | 0.499167571 | 0.006302801 | 0.008725018 | 1 |
| T2DM | nk_cell | *STX19* | -0.669681498 | 0.00258396 | 0.00381539 | 1 |
| T2DM | nk_cell | *MPI* | -1.318159241 | 0.003652607 | 0.005271401 | 1 |
| T2DM | nk_cell | *TRIM73* | -2.190869496 | 0.028070695 | 0.035157205 | 1 |
| T2DM | nk_cell | *EFCAB13* | -0.880436946 | 0.000203286 | 0.000406071 | 0.986345503 |
| T2DM | nk_cell | *EXOSC4* | -3.763186731 | 1.10E-05 | 0.000122264 | 0.053328253 |
| T2DM | nk_cell | *ZNF852* | -2.546005354 | 9.77E-06 | 0.000122264 | 0.047408753 |
| T2DM | nk_cell | *HYI* | -0.822007856 | 0.000177093 | 0.00036363 | 0.859257639 |
| T2DM | nk_cell | *GAK* | -1.691146926 | 0.000299789 | 0.000563134 | 1 |
| T2DM | nk_cell | *CTC1* | 1.271410904 | 0.012845964 | 0.016909554 | 1 |
| T2DM | nk_cell | *C3orf38* | -0.893439703 | 0.001237983 | 0.001938268 | 1 |
| T2DM | nk_cell | *CCDC89* | -1.64825974 | 3.71E-05 | 0.000134067 | 0.180056033 |
| T2DM | nk_cell | *HTR1F* | -1.92044966 | 1.40E-05 | 0.000122264 | 0.067940387 |
| T2DM | nk_cell | *ZNF664* | -1.500921169 | 1.69E-05 | 0.000122264 | 0.081770792 |
| T2DM | nk_cell | *CDH4* | -1.035208722 | 3.29E-05 | 0.000127909 | 0.159590789 |
| T2DM | nk_cell | *NSUN7* | -0.504485572 | 0.007818277 | 0.010622873 | 1 |
| T2DM | nk_cell | *HLA-DQB1* | -1.456818771 | 1.19E-14 | 2.88E-11 | 5.77E-11 |
| T2DM | nk_cell | *PACS2* | -1.201611461 | 1.99E-05 | 0.000122264 | 0.096611472 |
| T2DM | nk_cell | *LINC00174* | -1.039779317 | 0.000164033 | 0.000341144 | 0.795888165 |
| T2DM | nk_cell | *GEMIN4* | -2.412185114 | 6.73E-07 | 5.73E-05 | 0.003267298 |
| T2DM | nk_cell | *LBX2* | 0.55638252 | 0.002679508 | 0.003945666 | 1 |
| T2DM | nk_cell | *ZFPM1* | 0.485083352 | 0.002489297 | 0.003689086 | 1 |
| T2DM | nk_cell | *FCER1A* | -0.810455956 | 0.000147427 | 0.000315256 | 0.71531599 |
| T2DM | nk_cell | *RPRML* | 0.772190518 | 1.09E-05 | 0.000122264 | 0.05291131 |

| | | | | | | |
|---|---|---|---|---|---|---|
| T2DM | nk_cell | *SERTAD2* | -1.580865511 | 0.000413352 | 0.000738706 | 1 |
| T2DM | nk_cell | *AKAP5* | 0.43714729 | 8.91E-05 | 0.000217825 | 0.432511124 |
| T2DM | nk_cell | *GIPC3* | 2.758494294 | 0.000116515 | 0.000263067 | 0.565331096 |
| T2DM | nk_cell | *PDXDC1* | -0.948099221 | 3.57E-05 | 0.000131704 | 0.17321094 |
| T2DM | nk_cell | *SEPHS2* | -1.039431825 | 0.000294324 | 0.000554018 | 1 |
| T2DM | nk_cell | *ZADH2* | -1.493819758 | 2.23E-05 | 0.000122264 | 0.108319874 |
| T2DM | nk_cell | *SEPT1* | 1.189454993 | 0.003792204 | 0.005451785 | 1 |
| T2DM | nk_cell | *CSNK1A1L* | 3.706531954 | 0.00015228 | 0.000323491 | 0.738864381 |
| T2DM | nk_cell | *LYNX1* | -0.738717314 | 4.76E-05 | 0.000149363 | 0.230899054 |
| T2DM | nk_cell | *FAHD1* | -0.45748532 | 0.001862173 | 0.002822638 | 1 |
| T2DM | nk_cell | *HCLS1* | 0.373954355 | 0.009998479 | 0.013393876 | 1 |
| T2DM | nk_cell | *MTURN* | -0.874795458 | 1.71E-05 | 0.000122264 | 0.082734018 |
| T2DM | nk_cell | *CCDC66* | -1.072182544 | 2.97E-05 | 0.000124182 | 0.144079301 |
| T2DM | nk_cell | *DEFB124* | -4.867275618 | 0.00091054 | 0.001472647 | 1 |
| T2DM | nk_cell | *GLIPR1L2* | -0.686447473 | 1.29E-05 | 0.000122264 | 0.062760942 |
| T2DM | nk_cell | *DEFB119* | -7.617877205 | 0.000137901 | 0.000298303 | 0.669093519 |
| T2DM | nk_cell | *TSPYL5* | -1.111139453 | 3.29E-05 | 0.000127909 | 0.159683307 |
| T2DM | nk_cell | *MB21D2* | -0.992227708 | 9.84E-06 | 0.000122264 | 0.047725091 |
| T2DM | nk_cell | *SLC47A2* | -1.216109451 | 0.000360328 | 0.00065825 | 1 |
| T2DM | nk_cell | *CHST13* | -4.737447205 | 7.18E-06 | 0.000122264 | 0.034852093 |
| T2DM | nk_cell | *SLC36A4* | -1.304905828 | 1.13E-05 | 0.000122264 | 0.054964742 |
| T2DM | nk_cell | *OR51E1* | -0.743006247 | 0.0018346 | 0.002784322 | 1 |
| T2DM | nk_cell | *PSMG4* | -1.173787014 | 8.92E-06 | 0.000122264 | 0.043272133 |
| T2DM | nk_cell | *ZNF443* | -1.69501483 | 2.72E-05 | 0.000122437 | 0.131868458 |
| T2DM | nk_cell | *CAPS2* | -1.207642217 | 0.000158023 | 0.000332349 | 0.766728752 |
| T2DM | nk_cell | *KCTD2* | -0.814833864 | 1.17E-05 | 0.000122264 | 0.056744574 |
| T2DM | nk_cell | *D2HGDH* | -1.283565919 | 2.65E-06 | 0.000122264 | 0.012856827 |
| T2DM | nk_cell | *CMTR2* | -1.662154797 | 0.000332734 | 0.000615722 | 1 |
| T2DM | nk_cell | *ST20* | 0.313554703 | 0.01383015 | 0.018131286 | 1 |
| T2DM | nk_cell | *MRPL14* | -1.976464963 | 0.000244669 | 0.000473717 | 1 |
| T2DM | nk_cell | *NQO1* | -1.369272222 | 0.000988023 | 0.001587905 | 1 |
| T2DM | nk_cell | *FKRP* | -0.803312474 | 0.00024732 | 0.000478086 | 1 |
| T2DM | nk_cell | *RPH3AL* | -1.376750724 | 0.000231077 | 0.000451545 | 1 |
| T2DM | nk_cell | *ADIPOQ* | -0.440243097 | 5.09E-05 | 0.000154523 | 0.247082424 |
| T2DM | nk_cell | *F2R* | 0.253547602 | 4.94E-05 | 0.00015216 | 0.239631889 |
| T2DM | nk_cell | *MUC16* | -1.554043916 | 8.25E-05 | 0.000207398 | 0.400485367 |
| T2DM | nk_cell | *PENK* | -1.6331194 | 0.015944505 | 0.020718462 | 1 |
| T2DM | nk_cell | *SLC25A41* | 6.172072699 | 0.001248762 | 0.001953253 | 1 |
| T2DM | nk_cell | *TMEM136* | -1.05947227 | 8.85E-05 | 0.00021713 | 0.429519524 |
| T2DM | nk_cell | *DDX60L* | -1.498682476 | 0.000217862 | 0.000430753 | 1 |
| T2DM | nk_cell | *OGFOD3* | -1.327661732 | 0.000457961 | 0.000803626 | 1 |
| T2DM | nk_cell | *TMEM45A* | -1.060917722 | 3.21E-05 | 0.000127628 | 0.155905233 |
| T2DM | nk_cell | *SGSH* | -2.067533197 | 1.93E-05 | 0.000122264 | 0.093422838 |
| T2DM | nk_cell | *ZFP41* | 0.93134188 | 0.000383746 | 0.000695012 | 1 |
| T2DM | nk_cell | *ZBTB20* | -0.862582581 | 7.87E-05 | 0.000201405 | 0.381662892 |
| T2DM | nk_cell | *C3orf58* | -1.543156809 | 0.014615825 | 0.019109669 | 1 |
| T2DM | nk_cell | *AMIGO1* | -1.185818382 | 2.16E-05 | 0.000122264 | 0.104671694 |
| T2DM | nk_cell | *TMEM252* | -19.85055371 | 3.26E-05 | 0.000127909 | 0.157940033 |
| T2DM | nk_cell | *ODF3L2* | 1.437727901 | 0.000129336 | 0.000284211 | 0.627538376 |
| T2DM | nk_cell | *RELL1* | -1.239084036 | 6.45E-05 | 0.000176728 | 0.313148791 |

| | | | | | | |
|---|---|---|---|---|---|---|
| T2DM | nk_cell | *RFX7* | -1.417697272 | 0.000298315 | 0.000560976 | 1 |
| T2DM | nk_cell | *CLDN7* | -0.721695755 | 1.36E-05 | 0.000122264 | 0.065875695 |
| T2DM | nk_cell | *ZNF329* | -0.886826292 | 1.66E-05 | 0.000122264 | 0.080472326 |
| T2DM | nk_cell | *ADO* | 1.223129057 | 0.006949485 | 0.009543986 | 1 |
| T2DM | nk_cell | *MGAT4C* | -0.653928608 | 0.009328169 | 0.012551379 | 1 |
| T2DM | nk_cell | *IDH2* | -2.201399294 | 0.000390074 | 0.000705157 | 1 |
| T2DM | nk_cell | *WRB* | -0.916265143 | 4.60E-05 | 0.000147852 | 0.223127717 |
| T2DM | nk_cell | *FAM181B* | -1.628554114 | 0.003698547 | 0.005328191 | 1 |
| T2DM | nk_cell | *TDRKH* | -1.113575092 | 3.69E-05 | 0.00013348 | 0.178863263 |
| T2DM | nk_cell | *ZNF708* | -1.140734692 | 0.000107333 | 0.000248226 | 0.520777886 |
| T2DM | nk_cell | *ERCC6L2* | -1.225051172 | 0.000142026 | 0.000305727 | 0.689109032 |
| T2DM | nk_cell | *MRPL41* | -1.579247477 | 1.88E-05 | 0.000122264 | 0.090995985 |
| T2DM | nk_cell | *HHIPL1* | -1.422489498 | 2.92E-05 | 0.000123467 | 0.141483042 |
| T2DM | nk_cell | *FAM153B* | -1.083871401 | 3.84E-05 | 0.000135693 | 0.18634482 |
| T2DM | nk_cell | *BACE2* | -0.648007143 | 0.00079347 | 0.001297578 | 1 |
| T2DM | nk_cell | *SYNM* | -2.656651935 | 0.00087223 | 0.001415405 | 1 |
| T2DM | nk_cell | *KCNA4* | -3.083475698 | 0.002098015 | 0.003148644 | 1 |
| T2DM | nk_cell | *FIGN* | -1.12952296 | 4.88E-05 | 0.000151402 | 0.236792687 |
| T2DM | nk_cell | *IZUMO1* | 1.197916049 | 7.17E-05 | 0.000189735 | 0.347822842 |
| T2DM | nk_cell | *CLN8* | -0.573208004 | 0.020345392 | 0.026046396 | 1 |
| T2DM | nk_cell | *NXPH4* | 6.796719841 | 0.000222892 | 0.000439088 | 1 |
| T2DM | nk_cell | *NPLOC4* | -1.314270689 | 3.74E-05 | 0.0001343 | 0.18143911 |
| T2DM | nk_cell | *CAPN12* | 0.449994043 | 5.91E-05 | 0.000167611 | 0.286782637 |
| T2DM | nk_cell | *KPNA2* | -1.651428762 | 1.22E-05 | 0.000122264 | 0.059367193 |
| T2DM | nk_cell | *ORAI1* | 0.889347829 | 0.000115028 | 0.000260823 | 0.55811345 |
| T2DM | nk_cell | *FES* | -1.395260922 | 8.37E-06 | 0.000122264 | 0.040632009 |
| T2DM | nk_cell | *GLRX5* | -2.474661125 | 0.000745226 | 0.001225707 | 1 |
| T2DM | nk_cell | *CAV3* | -0.678850076 | 5.41E-05 | 0.000159936 | 0.262454571 |
| T2DM | nk_cell | *MFSD5* | 0.766813242 | 0.000469295 | 0.00082114 | 1 |
| T2DM | nk_cell | *ADI1* | -1.974577967 | 0.000273942 | 0.000521037 | 1 |
| T2DM | nk_cell | *CLEC4G* | -2.774890558 | 3.82E-06 | 0.000122264 | 0.018535796 |
| T2DM | nk_cell | *TRAK1* | -1.383212471 | 7.62E-05 | 0.000197291 | 0.369723702 |
| T2DM | nk_cell | *SKA2* | -1.835279256 | 0.001282633 | 0.00200236 | 1 |
| T2DM | nk_cell | *PPP1R27* | 1.180131263 | 0.000228519 | 0.000447628 | 1 |
| T2DM | nk_cell | *BRICD5* | -1.182935584 | 2.39E-05 | 0.000122264 | 0.115951795 |
| T2DM | nk_cell | *GRID1* | -0.668269204 | 5.97E-06 | 0.000122264 | 0.028958485 |
| T2DM | nk_cell | *ACBD3* | -1.583146491 | 0.000118394 | 0.000265456 | 0.57444686 |
| T2DM | nk_cell | *PLCXD3* | -1.596590624 | 6.23E-05 | 0.000173265 | 0.302520994 |
| T2DM | nk_cell | *GPIHBP1* | -0.74032968 | 7.09E-05 | 0.000188482 | 0.343790733 |
| T2DM | nk_cell | *VMO1* | 1.086027945 | 0.005532061 | 0.007735321 | 1 |
| T2DM | nk_cell | *ALG12* | 1.05068901 | 0.001029811 | 0.001645799 | 1 |
| T2DM | nk_cell | *COL18A1* | -1.037632815 | 0.000101986 | 0.000239427 | 0.494837792 |
| T2DM | nk_cell | *ZNF721* | -1.285331537 | 0.000479767 | 0.000837922 | 1 |
| T2DM | nk_cell | *CEP63* | -0.786409655 | 1.80E-05 | 0.000122264 | 0.087442106 |
| T2DM | nk_cell | *HMGN4* | -1.181874191 | 3.37E-05 | 0.00012843 | 0.163535215 |
| T2DM | nk_cell | *C12orf60* | -1.504629192 | 1.79E-05 | 0.000122264 | 0.08675912 |
| T2DM | nk_cell | *ABAT* | -2.0159288 | 0.000145347 | 0.000311422 | 0.705223462 |
| T2DM | nk_cell | *LYSMD4* | -0.890081722 | 7.81E-05 | 0.000200157 | 0.378897989 |
| T2DM | nk_cell | *WBP2NL* | -1.283893552 | 1.54E-05 | 0.000122264 | 0.07474482 |
| T2DM | nk_cell | *BEGAIN* | -1.077099785 | 1.66E-05 | 0.000122264 | 0.080544491 |

| | | | | | | |
|---|---|---|---|---|---|---|
| T2DM | nk_cell | *ARHGEF37* | -2.020835517 | 5.87E-05 | 0.000167252 | 0.284773891 |
| T2DM | nk_cell | *CSMD1* | -1.025048127 | 1.85E-05 | 0.000122264 | 0.089996028 |
| T2DM | nk_cell | *RABIF* | 1.237346679 | 0.004165793 | 0.005936102 | 1 |
| T2DM | nk_cell | *FANCF* | 1.70505068 | 0.001048406 | 0.001672761 | 1 |
| T2DM | nk_cell | *RUVBL2* | 1.266094792 | 0.000633774 | 0.001063303 | 1 |
| T2DM | nk_cell | *PTTG1IP* | -1.164827969 | 0.000719611 | 0.00119125 | 1 |
| T2DM | nk_cell | *ZNF623* | -0.776157055 | 3.09E-05 | 0.000126354 | 0.150108875 |
| T2DM | nk_cell | *CCDC125* | -0.848797597 | 3.61E-05 | 0.000132133 | 0.174943709 |
| T2DM | nk_cell | *C10orf107* | -0.481998557 | 0.001014132 | 0.001624186 | 1 |
| T2DM | nk_cell | *C19orf71* | -0.72993804 | 3.03E-05 | 0.000124985 | 0.147107129 |
| T2DM | nk_cell | *LRIT3* | -3.507480454 | 2.73E-05 | 0.000122437 | 0.132301127 |
| T2DM | nk_cell | *NPIPA1* | -0.340720715 | 0.006158375 | 0.008539707 | 1 |
| T2DM | nk_cell | *SF3A3* | -0.960917687 | 0.012341454 | 0.016302949 | 1 |
| T2DM | nk_cell | *PSMG1* | -1.133355859 | 0.006459695 | 0.00892947 | 1 |
| T2DM | nk_cell | *AC131971.1* | -2.607684908 | 0.000445179 | 0.000784887 | 1 |
| T2DM | nk_cell | *SERHL2* | -0.445928278 | 2.51E-05 | 0.000122264 | 0.121751786 |
| T2DM | nk_cell | *PCBP3* | -0.60466421 | 5.90E-05 | 0.000167488 | 0.286236856 |
| T2DM | nk_cell | *HMCES* | -1.546108006 | 0.001818564 | 0.002762578 | 1 |
| T2DM | nk_cell | *DGCR6* | -1.58219284 | 0.016551059 | 0.021454913 | 1 |
| T2DM | nk_cell | *RP1L1* | -1.740221232 | 0.023918933 | 0.030277763 | 1 |
| T2DM | nk_cell | *NDUFB1* | -1.249899048 | 0.00010712 | 0.000247923 | 0.51974729 |
| T2DM | nk_cell | *FAM19A1* | 0.437269444 | 0.000118118 | 0.000264959 | 0.573107225 |
| T2DM | nk_cell | *MRGPRX2* | 1.421784952 | 9.55E-05 | 0.000228386 | 0.463279073 |
| T2DM | nk_cell | *UPP1* | 0.599543996 | 0.000316826 | 0.000590112 | 1 |
| T2DM | nk_cell | *FIGLA* | -2.443928815 | 4.66E-05 | 0.000148627 | 0.22606238 |
| T2DM | nk_cell | *ACSM2A* | -4.689183206 | 1.71E-05 | 0.000122264 | 0.082998938 |
| T2DM | nk_cell | *KREMEN1* | -0.781157264 | 7.11E-06 | 0.000122264 | 0.034488549 |
| T2DM | nk_cell | *FAM162B* | -1.341822155 | 3.26E-05 | 0.000127909 | 0.158386037 |
| T2DM | nk_cell | *BTBD9* | -1.125512845 | 8.11E-05 | 0.00020495 | 0.393708743 |
| T2DM | nk_cell | *NUDT14* | 1.711435283 | 0.007489499 | 0.010210466 | 1 |
| T2DM | nk_cell | *FAM3B* | -0.386901766 | 0.000448879 | 0.000790836 | 1 |
| T2DM | nk_cell | *NPIPA7* | -1.747592411 | 0.001928687 | 0.002917078 | 1 |
| T2DM | nk_cell | *TTC32* | -1.723421476 | 0.04060433 | 0.049725445 | 1 |
| T2DM | nk_cell | *PP2D1* | -1.529503698 | 1.64E-05 | 0.000122264 | 0.079490187 |
| T2DM | nk_cell | *DENND5A* | -1.196829446 | 2.29E-05 | 0.000122264 | 0.111324833 |
| T2DM | nk_cell | *CNTN2* | -0.726758988 | 0.000277968 | 0.000527073 | 1 |
| T2DM | nk_cell | *CRELD2* | 1.219010227 | 0.002041511 | 0.003069542 | 1 |
| T2DM | nk_cell | *GOLGA6L4* | -0.857592778 | 0.00018895 | 0.000383272 | 0.916786309 |
| T2DM | nk_cell | *PGP* | -2.009538913 | 0.000114415 | 0.000260095 | 0.555142677 |
| T2DM | nk_cell | *OAF* | -0.670416062 | 2.10E-05 | 0.000122264 | 0.102113156 |
| T2DM | nk_cell | *ALDH1A3* | -0.444057892 | 4.04E-05 | 0.000138868 | 0.195942854 |
| T2DM | nk_cell | *CLECL1* | -0.613657872 | 0.000590928 | 0.001002162 | 1 |
| T2DM | nk_cell | *PKP3* | 5.888742464 | 4.50E-05 | 0.00014683 | 0.218384045 |
| T2DM | nk_cell | *A3GALT2* | -0.872110592 | 0.000479923 | 0.000837922 | 1 |
| T2DM | nk_cell | *SS18L1* | -0.880491394 | 3.17E-05 | 0.000127109 | 0.154040931 |
| T2DM | nk_cell | *TOP1MT* | -0.528009738 | 0.001005648 | 0.001613026 | 1 |
| T2DM | nk_cell | *COPB2* | -1.545505705 | 0.003841981 | 0.005511914 | 1 |
| T2DM | nk_cell | *WDR27* | -0.993822956 | 0.00042469 | 0.00075535 | 1 |
| T2DM | nk_cell | *TXNRD2* | -0.943545813 | 6.23E-05 | 0.000173265 | 0.302401609 |
| T2DM | nk_cell | *HDDC3* | -1.545808936 | 6.99E-06 | 0.000122264 | 0.03391781 |

| | | | | | | |
|---|---|---|---|---|---|---|
| T2DM | nk_cell | *ZFP1* | 0.893262401 | 0.009072959 | 0.012221542 | 1 |
| T2DM | nk_cell | *DUSP8* | -1.038635578 | 0.000334057 | 0.000617229 | 1 |
| T2DM | nk_cell | *SNN* | 1.776327058 | 0.002404571 | 0.003573347 | 1 |
| T2DM | nk_cell | *KRBA2* | -1.477560622 | 1.03E-05 | 0.000122264 | 0.049923403 |
| T2DM | nk_cell | *SEPT5* | -2.350752244 | 0.000428122 | 0.000759506 | 1 |
| T2DM | nk_cell | *RNLS* | -0.8954263 | 1.22E-05 | 0.000122264 | 0.059173303 |
| T2DM | nk_cell | *TCTE3* | -1.238225357 | 1.32E-05 | 0.000122264 | 0.064038319 |
| T2DM | nk_cell | *UBE2G2* | -1.460335375 | 0.000138853 | 0.000299829 | 0.673716247 |
| T2DM | nk_cell | *DRD1* | -11.39210589 | 2.48E-05 | 0.000122264 | 0.120118364 |
| T2DM | nk_cell | *BTBD6* | -1.94803645 | 1.65E-05 | 0.000122264 | 0.080297978 |
| T2DM | nk_cell | *WT1* | -0.490692663 | 0.000157612 | 0.000331628 | 0.764733706 |
| T2DM | nk_cell | *ZFP90* | -1.727799026 | 0.00250173 | 0.003701859 | 1 |
| T2DM | nk_cell | *FAM227A* | -0.558282292 | 0.022609684 | 0.02871785 | 1 |
| T2DM | nk_cell | *NOC4L* | 1.486672082 | 0.000465424 | 0.000814954 | 1 |
| T2DM | nk_cell | *NDUFA6* | 0.521779826 | 0.021289128 | 0.027175703 | 1 |
| T2DM | nk_cell | *SORCS2* | -0.835387087 | 1.14E-05 | 0.000122264 | 0.055506931 |
| T2DM | nk_cell | *TMEM106A* | -0.784068233 | 0.005485401 | 0.007672287 | 1 |
| T2DM | nk_cell | *BRI3BP* | -2.352636787 | 0.003747909 | 0.005391299 | 1 |
| T2DM | nk_cell | *UBOX5* | -2.019718849 | 0.008061408 | 0.010934848 | 1 |
| T2DM | nk_cell | *MAFF* | -0.58045344 | 8.67E-05 | 0.000213966 | 0.420848641 |
| T2DM | nk_cell | *SEMA4B* | -1.75780915 | 9.76E-05 | 0.000232202 | 0.47369157 |
| T2DM | nk_cell | *SGCZ* | -0.668723825 | 3.72E-05 | 0.000134067 | 0.180556422 |
| T2DM | nk_cell | *C6orf120* | -1.696143556 | 0.012701994 | 0.016729119 | 1 |
| T2DM | nk_cell | *SIGIRR* | 0.499248956 | 0.000154023 | 0.000326056 | 0.747320499 |
| T2DM | nk_cell | *IFITM2* | 0.752113771 | 0.001176132 | 0.001854596 | 1 |
| T2DM | nk_cell | *GP1BA* | -0.541008714 | 0.013317832 | 0.01749746 | 1 |
| T2DM | nk_cell | *ATP6V0A2* | 1.088313665 | 0.007234211 | 0.00989022 | 1 |
| T2DM | nk_cell | *C14orf80* | 1.161168467 | 0.000189558 | 0.000383863 | 0.919735487 |
| T2DM | nk_cell | *TNFAIP8L1* | -2.582897119 | 9.28E-05 | 0.000223847 | 0.450247464 |
| T2DM | nk_cell | *TARSL2* | -1.270706196 | 1.45E-05 | 0.000122264 | 0.070509783 |
| T2DM | nk_cell | *SH3BGR* | -0.483908833 | 0.01710028 | 0.022125482 | 1 |
| T2DM | nk_cell | *FAM174B* | -1.026927341 | 3.51E-05 | 0.000130901 | 0.1705207 |
| T2DM | nk_cell | *SV2B* | -0.750553078 | 6.90E-05 | 0.000185105 | 0.334692078 |
| T2DM | nk_cell | *AHNAK2* | -2.867864334 | 1.49E-05 | 0.000122264 | 0.072341875 |
| T2DM | nk_cell | *SP1* | -1.114144868 | 2.53E-05 | 0.000122264 | 0.122986764 |
| T2DM | nk_cell | *PCGF3* | -1.304396811 | 0.000185117 | 0.000377389 | 0.898186551 |
| T2DM | nk_cell | *PSMD13* | -2.145073443 | 6.15E-07 | 5.68E-05 | 0.002982051 |
| T2DM | nk_cell | *PMEL* | -0.643262077 | 6.12E-05 | 0.00017138 | 0.297025226 |
| T2DM | nk_cell | *C6orf201* | -1.40011843 | 8.87E-05 | 0.000217186 | 0.430188717 |
| T2DM | nk_cell | *MYBL1* | 0.071523707 | 6.04E-05 | 0.000169923 | 0.292978883 |
| T2DM | nk_cell | *ADARB2* | 1.087317077 | 0.000108893 | 0.000250759 | 0.528348749 |
| T2DM | nk_cell | *DMWD* | -1.193266192 | 4.83E-07 | 4.79E-05 | 0.00234113 |
| T2DM | nk_cell | *SLC52A2* | 1.063442098 | 0.000886614 | 0.001437305 | 1 |
| T2DM | nk_cell | *PIGP* | -0.687662343 | 5.40E-06 | 0.000122264 | 0.026224971 |
| T2DM | nk_cell | *ARL17A* | 0.914403952 | 0.033897448 | 0.041967445 | 1 |
| T2DM | nk_cell | *EVI2B* | 0.650737233 | 2.46E-05 | 0.000122264 | 0.1192217 |
| T2DM | nk_cell | *ZNF829* | -1.556719567 | 1.47E-05 | 0.000122264 | 0.071199868 |
| T2DM | nk_cell | *IFITM1* | 0.38685286 | 9.73E-05 | 0.000231443 | 0.471912508 |
| T2DM | nk_cell | *LINC00839* | -1.517135839 | 2.51E-05 | 0.000122264 | 0.121948346 |
| T2DM | nk_cell | *RTN4RL1* | -0.625793934 | 7.54E-05 | 0.000196203 | 0.365772669 |

| | | | | | | |
|---|---|---|---|---|---|---|
| T2DM | nk_cell | *RNPC3* | -0.667699267 | 5.91E-05 | 0.000167611 | 0.286689491 |
| T2DM | nk_cell | *ZNF529* | -1.20446351 | 4.85E-05 | 0.000151023 | 0.235293763 |
| T2DM | nk_cell | *WWOX* | -0.842118969 | 3.13E-05 | 0.000127033 | 0.151804647 |
| T2DM | nk_cell | *CYP4Z1* | -0.336437311 | 0.003274384 | 0.004766669 | 1 |
| T2DM | nk_cell | *POLR1D* | -1.144651284 | 0.000403267 | 0.000724685 | 1 |
| T2DM | nk_cell | *FFAR4* | -0.501140905 | 0.000254644 | 0.00048999 | 1 |
| T2DM | nk_cell | *CYP4F12* | -1.108232896 | 2.35E-05 | 0.000122264 | 0.113960633 |
| T2DM | nk_cell | *ZNF749* | -0.575343342 | 0.000639601 | 0.001071966 | 1 |
| T2DM | nk_cell | *GPAT2* | -1.246316575 | 0.018873333 | 0.024283588 | 1 |
| T2DM | nk_cell | *TOR3A* | -1.874073719 | 4.67E-06 | 0.000122264 | 0.022657552 |
| T2DM | nk_cell | *ZNF555* | -0.975540713 | 0.000108577 | 0.00025015 | 0.526815164 |
| T2DM | nk_cell | *GOLGA8R* | 2.273506844 | 0.000121577 | 0.000270222 | 0.589893959 |
| T2DM | nk_cell | *CCDC30* | -0.782754751 | 0.000190234 | 0.000384911 | 0.923015643 |
| T2DM | nk_cell | *ZNF197* | -1.290911604 | 1.38E-05 | 0.000122264 | 0.066937074 |
| T2DM | nk_cell | *FAM228A* | 0.34461122 | 0.024084131 | 0.030470978 | 1 |
| T2DM | nk_cell | *GNG2* | 0.368220127 | 0.000183975 | 0.000375377 | 0.892646305 |
| T2DM | nk_cell | *TMEM222* | -2.406974567 | 0.00014772 | 0.000315701 | 0.716737043 |
| T2DM | nk_cell | *SEPT10* | -1.199497888 | 6.31E-06 | 0.000122264 | 0.03062135 |
| T2DM | nk_cell | *FAM86B1* | -1.900524194 | 0.000540726 | 0.000928709 | 1 |
| T2DM | nk_cell | *SMYD4* | -0.972895008 | 8.49E-05 | 0.00021115 | 0.411954581 |
| T2DM | nk_cell | *CEACAM19* | -0.810021383 | 3.46E-06 | 0.000122264 | 0.016772561 |
| T2DM | nk_cell | *MIR22HG* | -1.020922797 | 1.29E-05 | 0.000122264 | 0.062820407 |
| T2DM | nk_cell | *CYP27C1* | -0.896384596 | 1.81E-06 | 0.000106072 | 0.008803937 |
| T2DM | nk_cell | *BCR* | -1.579673521 | 0.000133821 | 0.000291689 | 0.649299839 |
| T2DM | nk_cell | *FOXI2* | -1.198051336 | 0.014028595 | 0.018386479 | 1 |
| T2DM | nk_cell | *ZNF732* | -0.883025458 | 3.64E-05 | 0.000132426 | 0.176523843 |
| T2DM | nk_cell | *HYAL3* | -0.956927409 | 0.00976763 | 0.013102721 | 1 |
| T2DM | nk_cell | *VSIG10L* | -1.448056933 | 0.000755172 | 0.001239125 | 1 |
| T2DM | nk_cell | *TRABD2A* | -0.492166005 | 0.001763957 | 0.002686353 | 1 |
| T2DM | nk_cell | *QRFPR* | -2.175300678 | 0.000152405 | 0.000323491 | 0.739471028 |
| T2DM | nk_cell | *MAPT* | -0.815688003 | 1.64E-05 | 0.000122264 | 0.079585226 |
| T2DM | nk_cell | *TMEM17* | -0.777198425 | 0.003239186 | 0.004718261 | 1 |
| T2DM | nk_cell | *TNFRSF18* | 0.310357982 | 4.58E-05 | 0.000147697 | 0.222138118 |
| T2DM | nk_cell | *C1QL4* | -5.911929785 | 2.39E-05 | 0.000122264 | 0.116183196 |
| T2DM | nk_cell | *RTN4RL2* | -1.229748976 | 8.14E-06 | 0.000122264 | 0.039517369 |
| T2DM | nk_cell | *TMEM232* | -0.759435974 | 4.22E-05 | 0.000141822 | 0.204648889 |
| T2DM | nk_cell | *EFCAB6* | -1.320625242 | 8.47E-05 | 0.00021115 | 0.410997795 |
| T2DM | nk_cell | *RHD* | -0.891411295 | 0.012109254 | 0.016031132 | 1 |
| T2DM | nk_cell | *TMPRSS6* | -5.24276855 | 1.88E-05 | 0.000122264 | 0.091449288 |
| T2DM | nk_cell | *CYP4A11* | 1.17550232 | 6.17E-05 | 0.000172546 | 0.299539354 |
| T2DM | nk_cell | *TMEM216* | 1.156372983 | 1.09E-05 | 0.000122264 | 0.053095512 |
| T2DM | nk_cell | *C3orf70* | -0.513521355 | 0.000200835 | 0.000402001 | 0.974450424 |
| T2DM | nk_cell | *ENTPD5* | -1.476756499 | 7.24E-06 | 0.000122264 | 0.035123574 |
| T2DM | nk_cell | *MRPS21* | -1.142952015 | 0.002049361 | 0.003078675 | 1 |
| T2DM | nk_cell | *ANGPTL5* | 0.913040747 | 0.029727941 | 0.037136964 | 1 |
| T2DM | nk_cell | *WDR86* | -0.524063884 | 0.00100103 | 0.001606682 | 1 |
| T2DM | nk_cell | *EPOR* | -0.512191376 | 0.00032516 | 0.000602626 | 1 |
| T2DM | nk_cell | *LUZP2* | -0.616148825 | 2.65E-05 | 0.000122391 | 0.128792556 |
| T2DM | nk_cell | *GJA4* | -2.629342523 | 5.05E-05 | 0.000153489 | 0.244815693 |
| T2DM | nk_cell | *IFT140* | -0.890919329 | 1.51E-05 | 0.000122264 | 0.073071511 |

| | | | | | | |
|---|---|---|---|---|---|---|
| T2DM | nk_cell | *SBK2* | 2.68308634 | 8.45E-05 | 0.000210701 | 0.409813902 |
| T2DM | nk_cell | *ZNF385C* | 0.581225011 | 0.001183693 | 0.001864095 | 1 |
| T2DM | nk_cell | *EXD3* | -1.546502133 | 4.35E-05 | 0.000144302 | 0.211229736 |
| T2DM | nk_cell | *DHRS4L2* | -0.740482189 | 0.023641383 | 0.029973345 | 1 |
| T2DM | nk_cell | *SAMD11* | -0.76166061 | 6.25E-05 | 0.000173467 | 0.303047188 |
| T2DM | nk_cell | *TRPV2* | 0.564216105 | 0.000193328 | 0.000390031 | 0.938025122 |
| T2DM | nk_cell | *C2orf88* | -0.788436732 | 2.62E-05 | 0.000122295 | 0.127192214 |
| T2DM | nk_cell | *KBTBD12* | -0.743388781 | 0.00362746 | 0.005236667 | 1 |
| T2DM | nk_cell | *FANCA* | 0.411382248 | 0.000495456 | 0.000860569 | 1 |
| T2DM | nk_cell | *DNAH17* | -0.63401427 | 7.43E-06 | 0.000122264 | 0.036068962 |
| T2DM | nk_cell | *FANCM* | -2.007920338 | 2.58E-06 | 0.000122264 | 0.012511277 |
| T2DM | nk_cell | *ZFP69* | 0.855325323 | 0.000270326 | 0.00051598 | 1 |
| T2DM | nk_cell | *TMEM220* | -0.739810773 | 0.000486493 | 0.000847867 | 1 |
| T2DM | nk_cell | *FAM122A* | -1.837338523 | 1.59E-05 | 0.000122264 | 0.077095609 |
| T2DM | nk_cell | *GFRAL* | -1.17903792 | 5.22E-05 | 0.000157285 | 0.25338611 |
| T2DM | nk_cell | *LDLRAD2* | -0.730472404 | 9.91E-06 | 0.000122264 | 0.048096181 |
| T2DM | nk_cell | *CPSF4L* | -2.441928884 | 3.17E-05 | 0.000127109 | 0.15396139 |
| T2DM | nk_cell | *KLHL17* | -1.048576381 | 0.000482451 | 0.000841613 | 1 |
| T2DM | nk_cell | *OR7D2* | 2.233141083 | 0.010629729 | 0.014192472 | 1 |
| T2DM | nk_cell | *S100A3* | -1.807726486 | 0.013399838 | 0.017595674 | 1 |
| T2DM | nk_cell | *TMEM221* | -1.424886959 | 0.002152739 | 0.003221804 | 1 |
| T2DM | nk_cell | *MESP2* | 0.845609267 | 0.003468438 | 0.005019046 | 1 |
| T2DM | nk_cell | *MAPK12* | -2.566031561 | 0.001400281 | 0.002167805 | 1 |
| T2DM | nk_cell | *FAM166A* | -2.422891493 | 0.005268174 | 0.007397386 | 1 |
| T2DM | nk_cell | *LINC00265* | 1.280007072 | 0.005269037 | 0.007397386 | 1 |
| T2DM | nk_cell | *LAMTOR4* | -1.132493939 | 0.040201283 | 0.049256723 | 1 |
| T2DM | nk_cell | *PRKAR1B* | -1.280168571 | 2.50E-05 | 0.000122264 | 0.12150954 |
| T2DM | nk_cell | *NCR3LG1* | -1.338245552 | 1.21E-05 | 0.000122264 | 0.058740154 |
| T2DM | nk_cell | *AGAP4* | 1.026499505 | 0.013580666 | 0.017828299 | 1 |
| T2DM | nk_cell | *RUFY4* | -2.298043433 | 1.54E-05 | 0.000122264 | 0.074803468 |
| T2DM | nk_cell | *CENPP* | -0.738554084 | 7.93E-05 | 0.000202333 | 0.384635548 |
| T2DM | nk_cell | *PLSCR1* | -1.295420931 | 0.005982149 | 0.008321499 | 1 |
| T2DM | nk_cell | *C3orf62* | -1.055429816 | 0.001738876 | 0.002650653 | 1 |
| T2DM | nk_cell | *ENO4* | -0.807405642 | 2.30E-05 | 0.000122264 | 0.111609665 |
| T2DM | nk_cell | *ZP3* | -0.845337775 | 0.000194127 | 0.000391156 | 0.941904798 |
| T2DM | nk_cell | *H3F3C* | -16.05733036 | 4.22E-05 | 0.000141838 | 0.204814691 |
| T2DM | nk_cell | *PDCD1* | 0.517472021 | 4.31E-05 | 0.000143655 | 0.209305048 |
| T2DM | nk_cell | *SELL* | 0.279034009 | 5.68E-05 | 0.000164073 | 0.275478148 |
| T2DM | nk_cell | *C19orf54* | -1.550122662 | 2.82E-05 | 0.000122868 | 0.136965599 |
| T2DM | nk_cell | *C22orf34* | 0.815275225 | 0.001209468 | 0.001898525 | 1 |
| T2DM | nk_cell | *COL25A1* | -0.526618123 | 1.71E-05 | 0.000122264 | 0.083033597 |
| T2DM | nk_cell | *SRSF10* | -1.210391937 | 0.000288058 | 0.000544449 | 1 |
| T2DM | nk_cell | *NBR1* | -1.636582673 | 0.000510782 | 0.000883535 | 1 |
| T2DM | nk_cell | *NDOR1* | -0.683812392 | 2.12E-05 | 0.000122264 | 0.102669897 |
| T2DM | nk_cell | *FAM72B* | -1.679247424 | 0.000238194 | 0.000463029 | 1 |
| T2DM | nk_cell | *GOLGA8M* | -1.498694606 | 3.23E-05 | 0.000127896 | 0.156891315 |
| T2DM | nk_cell | *RHCE* | -0.86601603 | 3.69E-05 | 0.000133524 | 0.17905608 |
| T2DM | nk_cell | *IDO2* | -1.337505826 | 8.87E-05 | 0.000217186 | 0.430246308 |
| T2DM | nk_cell | *SLC4A5* | -1.486747173 | 2.87E-05 | 0.000123207 | 0.139350699 |
| T2DM | nk_cell | *UROS* | 0.950041512 | 0.002717496 | 0.003997965 | 1 |

| | | | | | | |
|---|---|---|---|---|---|---|
| T2DM | nk_cell | *ZBED6CL* | -2.084170978 | 1.12E-05 | 0.000122264 | 0.054527182 |
| T2DM | nk_cell | *OSTN* | -0.547623503 | 0.000114201 | 0.000259899 | 0.55410406 |
| T2DM | nk_cell | *FSIP2* | -0.847318785 | 1.51E-05 | 0.000122264 | 0.073444888 |
| T2DM | nk_cell | *NOXA1* | -2.400621753 | 0.000860357 | 0.001396604 | 1 |
| T2DM | nk_cell | *FZD9* | -18.50119415 | 0.000321484 | 0.000597182 | 1 |
| T2DM | nk_cell | *MTF1* | 0.905073593 | 0.003429831 | 0.004972076 | 1 |
| T2DM | nk_cell | *ZDHHC11* | -2.018208051 | 1.28E-05 | 0.000122264 | 0.062321522 |
| T2DM | nk_cell | *ENTPD8* | -53.2833552 | 2.06E-05 | 0.000122264 | 0.100189169 |
| T2DM | nk_cell | *FBF1* | -1.165438982 | 0.016995501 | 0.022007519 | 1 |
| T2DM | nk_cell | *ASTL* | -1.652769896 | 2.41E-05 | 0.000122264 | 0.116750122 |
| T2DM | nk_cell | *MSL1* | -1.372581768 | 0.000137215 | 0.000297482 | 0.665765507 |
| T2DM | nk_cell | *LRRK2* | -0.822122229 | 0.001234432 | 0.001933332 | 1 |
| T2DM | nk_cell | *FAM196A* | -1.263350072 | 1.93E-05 | 0.000122264 | 0.093427503 |
| T2DM | nk_cell | *UTS2B* | -0.826729578 | 3.07E-05 | 0.000125754 | 0.149018735 |
| T2DM | nk_cell | *C9orf152* | -2.135765237 | 6.00E-05 | 0.000169532 | 0.291043171 |
| T2DM | nk_cell | *NELFB* | 1.289180034 | 0.00891073 | 0.012033082 | 1 |
| T2DM | nk_cell | *SLC15A5* | -0.627371835 | 0.000128244 | 0.000282323 | 0.622239545 |
| T2DM | nk_cell | *LIPI* | -0.717146033 | 6.73E-05 | 0.000181892 | 0.326496983 |
| T2DM | nk_cell | *ADAT2* | -1.431424778 | 5.63E-05 | 0.00016385 | 0.273359311 |
| T2DM | nk_cell | *ALKBH2* | 1.432694162 | 0.001366795 | 0.00212214 | 1 |
| T2DM | nk_cell | *SF3B3* | -1.732583532 | 9.41E-05 | 0.000225942 | 0.456403557 |
| T2DM | nk_cell | *PLAC9* | -1.225565627 | 3.44E-06 | 0.000122264 | 0.0166848 |
| T2DM | nk_cell | *ZNF573* | -1.466928775 | 4.21E-05 | 0.000141813 | 0.204494252 |
| T2DM | nk_cell | *FAM47E* | -0.612874854 | 1.57E-05 | 0.000122264 | 0.075971027 |
| T2DM | nk_cell | *ZNF600* | 0.612855315 | 0.005787107 | 0.008066373 | 1 |
| T2DM | nk_cell | *TSPYL1* | -1.91842585 | 2.60E-05 | 0.000122264 | 0.126144279 |
| T2DM | nk_cell | *PNRC2* | -1.401547463 | 1.15E-05 | 0.000122264 | 0.055859471 |
| T2DM | nk_cell | *FHIT* | -0.812172243 | 1.79E-05 | 0.000122264 | 0.087033355 |
| T2DM | nk_cell | *ZKSCAN3* | -1.281685794 | 3.00E-07 | 3.93E-05 | 0.001454706 |
| T2DM | nk_cell | *KIAA0408* | -0.740991308 | 2.16E-05 | 0.000122264 | 0.104644378 |
| T2DM | nk_cell | *HLA-DRB1* | -1.297552118 | 1.47E-09 | 7.15E-07 | 7.15E-06 |
| T2DM | nk_cell | *AKR1C3* | -0.722948076 | 1.87E-05 | 0.000122264 | 0.090623323 |
| T2DM | nk_cell | *WDSUB1* | -1.212620679 | 0.00070944 | 0.00117642 | 1 |
| T2DM | nk_cell | *COLCA1* | -1.957173104 | 0.000271451 | 0.000517518 | 1 |
| T2DM | nk_cell | *HRCT1* | -1.03876969 | 0.00145635 | 0.002250386 | 1 |
| T2DM | nk_cell | *MPHOSPH8* | -1.115558391 | 5.46E-05 | 0.000160881 | 0.26474723 |
| T2DM | nk_cell | *SIRPB2* | -0.900133986 | 1.20E-05 | 0.000122264 | 0.058464145 |
| T2DM | nk_cell | *SRGAP3* | -0.716217339 | 0.000584149 | 0.000991516 | 1 |
| T2DM | nk_cell | *XPNPEP3* | -0.628153133 | 5.37E-05 | 0.000159514 | 0.260533384 |
| T2DM | nk_cell | *PPIA* | -1.716102125 | 5.87E-05 | 0.000167252 | 0.284834933 |
| T2DM | nk_cell | *ZNF471* | -0.975304681 | 1.53E-05 | 0.000122264 | 0.074300834 |
| T2DM | nk_cell | *ZNF493* | -1.328720989 | 9.93E-06 | 0.000122264 | 0.048202046 |
| T2DM | nk_cell | *GTF2IRD2* | -0.866171017 | 3.93E-05 | 0.000136619 | 0.190693768 |
| T2DM | nk_cell | *SUPT3H* | -1.334021461 | 3.30E-05 | 0.000127932 | 0.160299328 |
| T2DM | nk_cell | *GIMAP5* | 0.581252578 | 0.000561007 | 0.000957441 | 1 |
| T2DM | nk_cell | *STK31* | -1.021168618 | 0.000167493 | 0.000347297 | 0.81267542 |
| T2DM | nk_cell | *ZKSCAN7* | 1.907569593 | 8.99E-05 | 0.000219093 | 0.435994613 |
| T2DM | nk_cell | *CD55* | -1.133619593 | 2.41E-05 | 0.000122264 | 0.117158484 |
| T2DM | nk_cell | *CPNE4* | -0.466716216 | 6.10E-05 | 0.000170908 | 0.295841876 |
| T2DM | nk_cell | *ZNF565* | -1.255060359 | 0.000252173 | 0.000485702 | 1 |

| | | | | | | |
|---|---|---|---|---|---|---|
| T2DM | nk_cell | *ZNF34* | -1.524503018 | 0.001573998 | 0.00241602 | 1 |
| T2DM | nk_cell | *ZNF781* | -0.531663019 | 0.000154747 | 0.000327302 | 0.750830773 |
| T2DM | nk_cell | *EVL* | 0.709583973 | 0.001189924 | 0.001871479 | 1 |
| T2DM | nk_cell | *ZNF124* | -1.83311308 | 0.000115083 | 0.000260823 | 0.5583812 |
| T2DM | nk_cell | *PIK3R4* | -1.390652877 | 2.19E-05 | 0.000122264 | 0.106353435 |
| T2DM | nk_cell | *ZNF605* | -0.730760559 | 0.002493653 | 0.003693286 | 1 |
| T2DM | nk_cell | *RFX8* | -0.722949107 | 0.000120865 | 0.000269503 | 0.586438533 |
| T2DM | nk_cell | *C20orf96* | -1.847403132 | 1.45E-05 | 0.000122264 | 0.07052834 |
| T2DM | nk_cell | *SULT1A1* | -0.886079689 | 4.49E-06 | 0.000122264 | 0.021778396 |
| T2DM | nk_cell | *TPK1* | -1.249178177 | 0.006011551 | 0.008352819 | 1 |
| T2DM | nk_cell | *MYO18A* | -1.241080466 | 0.036378724 | 0.04484491 | 1 |
| T2DM | nk_cell | *SPTSSB* | -1.510407969 | 0.000319671 | 0.000594498 | 1 |
| T2DM | nk_cell | *CACNA1H* | -0.862432504 | 3.40E-05 | 0.000129287 | 0.165099348 |
| T2DM | nk_cell | *XRCC2* | 1.749200469 | 0.000400034 | 0.000719942 | 1 |
| T2DM | nk_cell | *MYO6* | -1.178726358 | 1.24E-05 | 0.000122264 | 0.060192757 |
| T2DM | nk_cell | *ZNF846* | -1.15257652 | 0.00022888 | 0.000448154 | 1 |
| T2DM | nk_cell | *ADH1B* | -0.747807486 | 2.08E-05 | 0.000122264 | 0.100698105 |
| T2DM | nk_cell | *ZKSCAN5* | -0.89399791 | 0.000227632 | 0.000446431 | 1 |
| T2DM | nk_cell | *ZNF502* | -1.508170232 | 4.50E-05 | 0.00014683 | 0.218452941 |
| T2DM | nk_cell | *TRAPPC4* | -0.684624481 | 0.032683445 | 0.040536829 | 1 |
| T2DM | nk_cell | *ZFP62* | -1.69098079 | 0.000132642 | 0.000289639 | 0.643576844 |
| T2DM | nk_cell | *ZNF33B* | -1.269234166 | 5.77E-06 | 0.000122264 | 0.027973717 |
| T2DM | nk_cell | *AMZ2* | 0.659272709 | 0.000622149 | 0.001046332 | 1 |
| T2DM | nk_cell | *ZNF431* | -1.552608187 | 0.000334594 | 0.000617986 | 1 |
| T2DM | nk_cell | *HLA-DQA1* | -2.081600612 | 2.21E-16 | 1.07E-12 | 1.07E-12 |
| T2DM | nk_cell | *COL27A1* | -0.740796804 | 0.011637111 | 0.015435556 | 1 |
| T2DM | nk_cell | *GM2A* | -0.590262279 | 0.0021171 | 0.003173361 | 1 |
| T2DM | nk_cell | *SPRR2B* | -6.452916841 | 0.009010895 | 0.012144684 | 1 |
| T2DM | nk_cell | *ANKRD36B* | -1.441073072 | 0.000512009 | 0.000885341 | 1 |
| T2DM | nk_cell | *AP2A1* | -1.229081999 | 0.005381986 | 0.007540686 | 1 |
| T2DM | nk_cell | *HIST1H3E* | 0.668897731 | 0.003941271 | 0.005646013 | 1 |
| T2DM | nk_cell | *FUT11* | 1.001680547 | 0.000197427 | 0.000396653 | 0.957916863 |
| T2DM | nk_cell | *WDR5B* | -1.136258084 | 0.000303358 | 0.000568299 | 1 |
| T2DM | nk_cell | *NPIPB9* | -2.012287761 | 0.007413947 | 0.010115993 | 1 |
| T2DM | nk_cell | *ZNF138* | -1.208595033 | 0.000178196 | 0.000365276 | 0.864608152 |
| T2DM | nk_cell | *ZNF429* | -1.563168959 | 0.003026475 | 0.004420366 | 1 |
| T2DM | nk_cell | *ZNF100* | -1.223192195 | 1.66E-05 | 0.000122264 | 0.080664523 |
| T2DM | nk_cell | *ANXA6* | 0.763683904 | 0.005841815 | 0.008135616 | 1 |
| T2DM | nk_cell | *ZNF420* | -1.419190315 | 3.90E-05 | 0.000136502 | 0.189191353 |
| T2DM | nk_cell | *ZNF763* | -0.788033356 | 0.00016679 | 0.000346284 | 0.809265345 |
| T2DM | nk_cell | *ZMYM1* | -1.616197854 | 5.46E-05 | 0.000160881 | 0.264846273 |
| T2DM | nk_cell | *GAL3ST4* | -1.876062065 | 0.000137629 | 0.000297848 | 0.667774877 |
| T2DM | nk_cell | *SLC25A29* | -0.814482607 | 0.000764078 | 0.001252469 | 1 |
| T2DM | nk_cell | *ZNF772* | -0.953638995 | 5.82E-05 | 0.000166869 | 0.282343187 |
| T2DM | nk_cell | *ZNF257* | -1.542471725 | 0.000778874 | 0.001274568 | 1 |
| T2DM | nk_cell | *ADAM32* | -0.796538188 | 1.75E-05 | 0.000122264 | 0.085112458 |
| T2DM | nk_cell | *LRRC8B* | -1.024639555 | 5.80E-05 | 0.000166593 | 0.281541572 |
| T2DM | nk_cell | *ABCB8* | -1.500445305 | 4.82E-06 | 0.000122264 | 0.023363179 |
| T2DM | nk_cell | *SND1* | -1.255086725 | 4.09E-05 | 0.000139516 | 0.198391614 |
| T2DM | nk_cell | *NEK5* | -0.862042213 | 0.000949098 | 0.001530417 | 1 |

| | | | | | | |
|---|---|---|---|---|---|---|
| T2DM | nk_cell | *PIWIL2* | -1.056574776 | 0.00380792 | 0.005469517 | 1 |
| T2DM | nk_cell | *ENTPD4* | -1.683345769 | 0.009279664 | 0.012489579 | 1 |
| T2DM | nk_cell | *TBC1D9B* | -1.252097227 | 8.56E-05 | 0.000212483 | 0.415403997 |
| T2DM | nk_cell | *FAM110D* | 1.427982438 | 5.20E-06 | 0.000122264 | 0.025220826 |
| T2DM | nk_cell | *CCL4L2* | 0.158581711 | 5.01E-05 | 0.000153185 | 0.243241617 |
| T2DM | nk_cell | *RAMP2-AS1* | -0.699088276 | 2.84E-05 | 0.00012318 | 0.137595329 |
| T2DM | nk_cell | *ZNF720* | -1.666293255 | 0.004739556 | 0.00669471 | 1 |
| T2DM | nk_cell | *ZNF786* | 0.682449392 | 0.013047279 | 0.017155934 | 1 |
| T2DM | nk_cell | *ZNF517* | -0.830816416 | 1.59E-05 | 0.000122264 | 0.077290096 |
| T2DM | nk_cell | *ZNF675* | -2.019980293 | 0.000408549 | 0.000731738 | 1 |
| T2DM | nk_cell | *SLC22A5* | -1.081946059 | 0.000258761 | 0.000496632 | 1 |
| T2DM | nk_cell | *VEPH1* | -0.564521972 | 2.59E-05 | 0.000122264 | 0.125650139 |
| T2DM | nk_cell | *SHPK* | -0.677674289 | 0.000527287 | 0.000908844 | 1 |
| T2DM | nk_cell | *MAP3K5* | -1.270475108 | 0.001018208 | 0.001628865 | 1 |
| T2DM | nk_cell | *GSTK1* | -1.424965288 | 0.000604494 | 0.001022309 | 1 |
| T2DM | nk_cell | *GYPE* | -1.260620764 | 0.001589526 | 0.002437541 | 1 |
| T2DM | nk_cell | *PCDHB11* | -0.70155583 | 0.001462407 | 0.002254718 | 1 |
| T2DM | nk_cell | *SLC2A10* | -0.656369739 | 4.12E-05 | 0.000140378 | 0.200039265 |
| T2DM | nk_cell | *ZNF665* | -2.175227795 | 0.000218429 | 0.000431698 | 1 |
| T2DM | nk_cell | *RPF2* | -1.014354445 | 2.75E-05 | 0.000122472 | 0.133603818 |
| T2DM | nk_cell | *ATG7* | -0.970157897 | 8.98E-05 | 0.000219084 | 0.43575787 |
| T2DM | nk_cell | *PIGN* | -0.831711152 | 0.000130618 | 0.000286768 | 0.633758116 |
| T2DM | nk_cell | *HHLA3* | -0.768468445 | 1.14E-05 | 0.000122264 | 0.05534168 |
| T2DM | nk_cell | *TOPORS* | -2.04854562 | 0.000293929 | 0.000554018 | 1 |
| T2DM | nk_cell | *BCO2* | -0.510974513 | 0.005471721 | 0.007657569 | 1 |
| T2DM | nk_cell | *ENTPD6* | -1.12662302 | 2.42E-06 | 0.000122264 | 0.011748186 |
| T2DM | nk_cell | *ENPP1* | -1.01257796 | 8.77E-06 | 0.000122264 | 0.042550871 |
| T2DM | nk_cell | *MFAP5* | -0.712090717 | 1.81E-05 | 0.000122264 | 0.088044172 |
| T2DM | nk_cell | *DNAH10* | -2.203426359 | 1.36E-06 | 9.04E-05 | 0.006614356 |
| T2DM | nk_cell | *NMB* | 1.196074816 | 0.00012012 | 0.000268335 | 0.58282422 |
| T2DM | nk_cell | *FAM114A1* | -0.972487416 | 2.60E-05 | 0.000122264 | 0.125968962 |
| T2DM | nk_cell | *PSAP* | -1.00634046 | 0.000261025 | 0.000500195 | 1 |
| T2DM | nk_cell | *TXNRD3* | -0.723776798 | 3.15E-05 | 0.000127109 | 0.152681923 |
| T2DM | nk_cell | *EME2* | -0.60717246 | 2.45E-05 | 0.000122264 | 0.119085706 |
| T2DM | nk_cell | *ZNF780A* | -0.511948201 | 0.000463252 | 0.000811443 | 1 |
| T2DM | nk_cell | *AC122129.1* | -1.905450476 | 2.59E-05 | 0.000122264 | 0.125456381 |
| T2DM | nk_cell | *CCDC180* | -0.686491017 | 0.001699031 | 0.002598077 | 1 |
| T2DM | nk_cell | *OCLN* | -0.64877825 | 0.000371125 | 0.00067543 | 1 |
| T2DM | nk_cell | *ADH5* | -1.440234083 | 8.10E-05 | 0.00020495 | 0.39319478 |
| T2DM | nk_cell | *ZNF677* | -1.228952753 | 1.96E-05 | 0.000122264 | 0.095078168 |
| T2DM | nk_cell | *ZNF311* | -0.692406108 | 0.002702208 | 0.003976679 | 1 |
| T2DM | nk_cell | *ZNF71* | -1.733211029 | 0.000160189 | 0.000336176 | 0.777238117 |
| T2DM | nk_cell | *MPZL1* | -0.86258826 | 1.54E-05 | 0.000122264 | 0.074516572 |
| T2DM | nk_cell | *VPS13A* | -1.24538413 | 0.000495555 | 0.000860569 | 1 |
| T2DM | nk_cell | *NOL8* | -1.573915194 | 3.29E-05 | 0.000127909 | 0.15939662 |
| T2DM | nk_cell | *DLGAP2* | -0.450517405 | 5.46E-05 | 0.000160881 | 0.265131831 |
| T2DM | nk_cell | *MRPL42* | -0.901804017 | 9.80E-05 | 0.000233035 | 0.475623941 |
| T2DM | nk_cell | *ZNF560* | -1.005490941 | 0.018982576 | 0.024404732 | 1 |
| T2DM | nk_cell | *AGAP9* | -0.883921258 | 2.85E-05 | 0.00012318 | 0.138282731 |
| T2DM | nk_cell | *ZNF273* | -1.695491593 | 1.32E-05 | 0.000122264 | 0.064286488 |

| | | | | | | |
|---|---|---|---|---|---|---|
| T2DM | nk_cell | *MAK16* | -1.347443493 | 5.86E-05 | 0.000167232 | 0.284423988 |
| T2DM | nk_cell | *ZNF667* | -0.607571077 | 2.68E-05 | 0.000122391 | 0.130003327 |
| T2DM | nk_cell | *SFI1* | 0.703039089 | 0.00044484 | 0.00078486 | 1 |
| T2DM | nk_cell | *ADH4* | -0.549663361 | 2.19E-05 | 0.000122264 | 0.106146301 |
| T2DM | nk_cell | *HIBCH* | -0.862032107 | 2.25E-05 | 0.000122264 | 0.109012997 |
| T2DM | nk_cell | *ZNF544* | -1.112189418 | 1.72E-05 | 0.000122264 | 0.083525343 |
| T2DM | nk_cell | *SOWAHC* | -2.6869753 | 9.03E-05 | 0.000219847 | 0.437935048 |
| T2DM | nk_cell | *AC135178.1* | 4.673709712 | 0.000403093 | 0.000724642 | 1 |
| T2DM | nk_cell | *MIER1* | -1.212465878 | 0.000399682 | 0.000719577 | 1 |
| T2DM | nk_cell | *MAN1A2* | -1.423711772 | 0.000488582 | 0.000850734 | 1 |
| T2DM | nk_cell | *SVIP* | -1.915058244 | 9.54E-05 | 0.000228386 | 0.463105989 |
| T2DM | nk_cell | *DDRGK1* | -1.378229138 | 2.95E-05 | 0.000123723 | 0.143271518 |
| T2DM | nk_cell | *ZNF607* | -1.001227725 | 0.01213688 | 0.016058942 | 1 |
| T2DM | nk_cell | *ZNF334* | -0.802128037 | 7.66E-05 | 0.000197717 | 0.371509534 |
| T2DM | nk_cell | *HSD17B11* | -1.416611246 | 6.54E-05 | 0.000178359 | 0.317463392 |
| T2DM | nk_cell | *SULT1C2* | 0.673926042 | 0.000420005 | 0.000748389 | 1 |
| T2DM | nk_cell | *RPS6KL1* | -1.703989654 | 0.003071997 | 0.004482807 | 1 |
| T2DM | nk_cell | *DDX42* | -1.025796053 | 6.48E-05 | 0.000177129 | 0.31440381 |
| T2DM | nk_cell | *HELZ* | -1.217304523 | 0.000178471 | 0.000365685 | 0.865943025 |
| T2DM | nk_cell | *TMEM116* | -0.943835989 | 1.61E-06 | 9.70E-05 | 0.007804344 |
| T2DM | nk_cell | *UCKL1* | -1.135538623 | 2.02E-05 | 0.000122264 | 0.098119752 |
| T2DM | nk_cell | *ZNF442* | -1.311812936 | 0.032253897 | 0.040045012 | 1 |
| T2DM | nk_cell | *GFPT1* | -1.212448786 | 4.37E-05 | 0.000144321 | 0.212151705 |
| T2DM | nk_cell | *ZNF568* | -1.677999027 | 0.000101998 | 0.000239427 | 0.494896601 |
| T2DM | nk_cell | *ZNF587* | -1.446032582 | 0.000415913 | 0.000741915 | 1 |
| T2DM | nk_cell | *SH3BGRL2* | -0.719669153 | 3.39E-05 | 0.000128918 | 0.164499317 |
| T2DM | nk_cell | *TMA16* | -1.522659646 | 0.011653693 | 0.015453326 | 1 |
| T2DM | nk_cell | *HLA-DRB5* | -0.482810243 | 2.87E-08 | 8.20E-06 | 0.000139459 |
| T2DM | nk_cell | *CNGA1* | -0.623181953 | 0.005943124 | 0.008271955 | 1 |
| T2DM | nk_cell | *ZNF28* | 1.143565729 | 0.004850962 | 0.006844102 | 1 |
| T2DM | nk_cell | *C20orf203* | -1.104415308 | 0.000121302 | 0.000270105 | 0.588558367 |
| T2DM | nk_cell | *ZNF534* | -0.698663027 | 0.003687053 | 0.00531637 | 1 |
| T2DM | nk_cell | *NCOA6* | -1.102183423 | 0.005073191 | 0.007138957 | 1 |
| T2DM | nk_cell | *FAM19A2* | -0.690492372 | 9.80E-06 | 0.000122264 | 0.047539929 |
| T2DM | nk_cell | *TTC37* | -1.172803323 | 0.002901728 | 0.004249678 | 1 |
| T2DM | nk_cell | *TUSC1* | -0.923438045 | 2.13E-05 | 0.000122264 | 0.103232416 |
| T2DM | nk_cell | *PAPSS2* | -0.838916088 | 1.68E-05 | 0.000122264 | 0.08129931 |
| T2DM | nk_cell | *CEP290* | -1.25790889 | 6.75E-05 | 0.00018242 | 0.327625499 |
| T2DM | nk_cell | *ANKRD13B* | -1.084687893 | 1.99E-05 | 0.000122264 | 0.09643316 |
| T2DM | nk_cell | *ECI2* | -1.569380253 | 0.000225455 | 0.000443057 | 1 |
| T2DM | nk_cell | *UNC13B* | -1.042674179 | 2.45E-05 | 0.000122264 | 0.118983569 |
| T2DM | nk_cell | *PPP1R14C* | -5.078296152 | 1.40E-05 | 0.000122264 | 0.067791098 |
| T2DM | nk_cell | *GPATCH3* | -3.050353721 | 8.84E-05 | 0.0002169 | 0.428811396 |
| T2DM | nk_cell | *COLGALT2* | 0.646698949 | 0.001325753 | 0.002062377 | 1 |
| T2DM | nk_cell | *RCSD1* | 0.615949622 | 0.000625396 | 0.001051064 | 1 |
| T2DM | nk_cell | *GRIN3A* | -0.745681188 | 0.000547651 | 0.000938278 | 1 |
| T2DM | nk_cell | *TMEM184B* | -1.342234921 | 0.000491821 | 0.00085527 | 1 |
| T2DM | nk_cell | *SCAMP5* | -0.570789538 | 0.00011252 | 0.000256916 | 0.545946425 |
| T2DM | nk_cell | *PNP* | 0.307758782 | 0.005425765 | 0.00759983 | 1 |
| T2DM | nk_cell | *SFT2D1* | -1.575479143 | 3.94E-05 | 0.000136823 | 0.19114225 |

| | | | | | | |
|---|---|---|---|---|---|---|
| T2DM | nk_cell | *HMGN2* | -1.153669452 | 1.44E-06 | 9.05E-05 | 0.006971249 |
| T2DM | nk_cell | *DENND4B* | -1.04179685 | 0.039255206 | 0.048182712 | 1 |
| T2DM | nk_cell | *RYR3* | -0.869029026 | 0.000531732 | 0.000914881 | 1 |
| T2DM | nk_cell | *TYW1* | -1.091298683 | 9.72E-05 | 0.000231443 | 0.471836145 |
| T2DM | nk_cell | *PRMT6* | 0.730282819 | 9.05E-05 | 0.000220274 | 0.43900706 |
| T2DM | nk_cell | *SHISA4* | -2.186526817 | 2.61E-06 | 0.000122264 | 0.012649377 |
| T2DM | nk_cell | *MAP3K3* | -1.862438937 | 3.23E-05 | 0.000127896 | 0.156752457 |
| T2DM | nk_cell | *C1orf174* | -1.135951741 | 3.69E-05 | 0.00013348 | 0.178814024 |
| T2DM | nk_cell | *DZIP3* | -1.473561741 | 5.66E-05 | 0.000163891 | 0.274764847 |
| T2DM | nk_cell | *KIAA0753* | -1.285507375 | 4.12E-06 | 0.000122264 | 0.019999764 |
| T2DM | nk_cell | *TBKBP1* | -0.967650235 | 0.006798395 | 0.009350153 | 1 |
| T2DM | nk_cell | *ZFP2* | -1.708545445 | 2.20E-05 | 0.000122264 | 0.106557842 |
| T2DM | nk_cell | *MFAP3L* | -2.252160752 | 0.000116247 | 0.000262828 | 0.564028117 |
| T2DM | nk_cell | *NAGA* | -1.435830488 | 0.000347223 | 0.000637188 | 1 |
| T2DM | nk_cell | *SMG5* | -1.476372605 | 2.68E-05 | 0.000122391 | 0.130138216 |
| T2DM | nk_cell | *PJA2* | -1.316816714 | 2.93E-05 | 0.000123467 | 0.142357884 |
| T2DM | nk_cell | *AL590822.1* | -0.839243003 | 1.34E-05 | 0.000122264 | 0.064884016 |
| T2DM | nk_cell | *AP001007.1* | -1.075148393 | 0.001458217 | 0.002251983 | 1 |
| T2DM | nk_cell | *EFCAB2* | -0.886332535 | 2.30E-05 | 0.000122264 | 0.111388077 |
| T2DM | nk_cell | *CHML* | -1.102114521 | 0.000566458 | 0.000965386 | 1 |
| T2DM | nk_cell | *TATDN3* | -0.887050091 | 0.001400673 | 0.002167805 | 1 |
| T2DM | nk_cell | *SAMD5* | -1.432371842 | 0.000113851 | 0.000259222 | 0.552403045 |
| T2DM | nk_cell | *ECT2L* | -0.862425016 | 2.10E-05 | 0.000122264 | 0.101885564 |
| T2DM | nk_cell | *GPR52* | 5.378324264 | 0.000461709 | 0.000809325 | 1 |
| T2DM | nk_cell | *FAM229B* | -1.323641597 | 1.19E-05 | 0.000122264 | 0.057560503 |
| T2DM | nk_cell | *DDO* | -1.03371169 | 0.001172696 | 0.001849779 | 1 |
| T2DM | nk_cell | *RBM20* | -0.65538292 | 1.81E-05 | 0.000122264 | 0.087944484 |
| T2DM | nk_cell | *PCMTD2* | -1.521460586 | 8.10E-05 | 0.00020495 | 0.392838279 |
| T2DM | nk_cell | *SAMD13* | -0.998993601 | 1.35E-05 | 0.000122264 | 0.065456861 |
| T2DM | nk_cell | *EFCAB7* | -1.001051316 | 4.17E-05 | 0.000141162 | 0.202144585 |
| T2DM | nk_cell | *IFIT1B* | 1.518199162 | 7.13E-05 | 0.000189116 | 0.346163711 |
| T2DM | nk_cell | *LIPN* | -1.881644286 | 1.94E-05 | 0.000122264 | 0.094039254 |
| T2DM | nk_cell | *INPP5B* | -0.651874476 | 0.003895695 | 0.005584021 | 1 |
| T2DM | nk_cell | *NEU4* | -1.466845824 | 0.000555372 | 0.000950164 | 1 |
| T2DM | nk_cell | *C2orf72* | -5.636817643 | 2.55E-06 | 0.000122264 | 0.012356232 |
| T2DM | nk_cell | *RUFY2* | -1.034931629 | 2.15E-05 | 0.000122264 | 0.104293188 |
| T2DM | nk_cell | *PHACTR4* | -1.132965292 | 1.36E-05 | 0.000122264 | 0.066103216 |
| T2DM | nk_cell | *ASAH2B* | -1.187768038 | 0.001079764 | 0.001716585 | 1 |
| T2DM | nk_cell | *AGAP6* | -0.680683374 | 0.004021489 | 0.005752437 | 1 |
| T2DM | nk_cell | *SYT15* | -1.119651616 | 0.001447545 | 0.002237492 | 1 |
| T2DM | nk_cell | *LINC00619* | -5.118176643 | 2.74E-05 | 0.000122472 | 0.132702333 |
| T2DM | nk_cell | *BMPR2* | -0.945257515 | 2.79E-05 | 0.000122493 | 0.135476791 |
| T2DM | nk_cell | *TCEA3* | -1.236520757 | 6.34E-05 | 0.000174311 | 0.307484329 |
| T2DM | nk_cell | *HSD17B8* | -0.623887903 | 0.005479475 | 0.007666209 | 1 |
| T2DM | nk_cell | *TAP2* | 0.444298272 | 0.011346863 | 0.015083556 | 1 |
| T2DM | nk_cell | *COL15A1* | -0.666960157 | 2.59E-05 | 0.000122264 | 0.125700596 |
| T2DM | nk_cell | *NOTCH4* | -0.259294787 | 4.23E-08 | 1.03E-05 | 0.000205111 |
| T2DM | nk_cell | *AGER* | 1.155937629 | 0.011439035 | 0.015189436 | 1 |
| T2DM | nk_cell | *PRRT1* | 0.646834468 | 0.022265253 | 0.028317433 | 1 |
| T2DM | nk_cell | *SMIM5* | -1.313636282 | 0.00024466 | 0.000473717 | 1 |

| | | | | | | |
|---|---|---|---|---|---|---|
| T2DM | nk_cell | *STK19* | -0.659210711 | 9.53E-07 | 7.46E-05 | 0.004624593 |
| T2DM | nk_cell | *SKIV2L* | -0.638539444 | 2.58E-07 | 3.47E-05 | 0.001250271 |
| T2DM | nk_cell | *LAYN* | -1.601344968 | 0.001370921 | 0.002126977 | 1 |
| T2DM | nk_cell | *GPANK1* | -2.4732422 | 3.37E-07 | 4.05E-05 | 0.001632866 |
| T2DM | nk_cell | *LST1* | 0.974208593 | 0.005351634 | 0.007502493 | 1 |
| T2DM | nk_cell | *ZNF814* | -1.303850229 | 0.00015489 | 0.000327428 | 0.751524143 |
| T2DM | nk_cell | *MICA* | -1.028682573 | 0.00061991 | 0.001042927 | 1 |
| T2DM | nk_cell | *CCHCR1* | 0.413088636 | 0.022117964 | 0.028144863 | 1 |
| T2DM | nk_cell | *PSORS1C2* | 0.736796969 | 0.040784251 | 0.049920581 | 1 |
| T2DM | nk_cell | *DHX16* | -0.906240237 | 0.024449021 | 0.030900404 | 1 |
| T2DM | nk_cell | *ZNF468* | -0.940017686 | 3.90E-05 | 0.000136502 | 0.189173839 |
| T2DM | nk_cell | *HLA-G* | 0.84172167 | 0.000111019 | 0.000254922 | 0.53866533 |
| T2DM | nk_cell | *FAM153C* | -0.602378041 | 1.34E-06 | 9.04E-05 | 0.006513452 |
| T2DM | nk_cell | *TRIM27* | -1.414951062 | 0.001475543 | 0.002272804 | 1 |
| T2DM | nk_cell | *MALRD1* | -1.165236058 | 1.37E-05 | 0.000122264 | 0.066303889 |
| T2DM | nk_cell | *FAM196B* | 0.749288242 | 0.001485926 | 0.002288072 | 1 |
| T2DM | nk_cell | *MROH6* | -1.014980122 | 8.82E-05 | 0.000216729 | 0.428039413 |
| T2DM | nk_cell | *DCTN1* | -2.340395087 | 0.000182177 | 0.000372502 | 0.883924931 |
| T2DM | nk_cell | *FAM216A* | -1.877229675 | 9.98E-05 | 0.000236122 | 0.484135978 |
| T2DM | nk_cell | *ZBTB48* | -1.900767938 | 6.25E-05 | 0.000173492 | 0.303264482 |
| T2DM | nk_cell | *MZT1* | 1.337234883 | 0.012171339 | 0.016095759 | 1 |
| T2DM | nk_cell | *ZNF155* | -1.13215692 | 1.12E-05 | 0.000122264 | 0.054448413 |
| T2DM | nk_cell | *ZNF783* | -1.783036269 | 9.40E-05 | 0.000225942 | 0.456239159 |
| T2DM | nk_cell | *ZNF425* | -1.116011737 | 6.22E-05 | 0.00017316 | 0.301818349 |
| T2DM | nk_cell | *C12orf73* | -1.115913195 | 1.05E-05 | 0.000122264 | 0.050732198 |
| T2DM | nk_cell | *PCDHGA1* | -1.186385126 | 3.25E-05 | 0.000127909 | 0.157819338 |
| T2DM | nk_cell | *PCDHA7* | -12.65072975 | 0.000675572 | 0.001125985 | 1 |
| T2DM | nk_cell | *PCDHA4* | -1.409509938 | 3.14E-05 | 0.000127109 | 0.152554142 |
| T2DM | nk_cell | *TRIM13* | -1.087987881 | 0.000474813 | 0.000830495 | 1 |
| T2DM | nk_cell | *SPIRE2* | -0.700507471 | 0.005428251 | 0.007601118 | 1 |
| T2DM | nk_cell | *PKHD1L1* | -0.740176397 | 1.10E-05 | 0.000122264 | 0.053493725 |
| T2DM | nk_cell | *SLFN12L* | 0.160492992 | 0.00011638 | 0.000263006 | 0.564674347 |
| T2DM | nk_cell | *SLC35B4* | -1.69171094 | 0.016503514 | 0.02140127 | 1 |
| T2DM | nk_cell | *SYCE1L* | 0.578528766 | 9.46E-06 | 0.000122264 | 0.045909279 |
| T2DM | nk_cell | *TMEM231* | -0.817191904 | 2.09E-05 | 0.000122264 | 0.101579198 |
| T2DM | nk_cell | *SCGB2B2* | -0.844458162 | 1.57E-05 | 0.000122264 | 0.076285649 |
| T2DM | nk_cell | *TMEM170B* | -0.561723893 | 0.000179804 | 0.000368261 | 0.872410208 |
| T2DM | nk_cell | *GOLGA6L10* | -1.474928593 | 0.000100041 | 0.000236412 | 0.485400957 |
| T2DM | nk_cell | *SAP25* | -2.066498088 | 0.038327347 | 0.04712729 | 1 |
| T2DM | nk_cell | *NT5M* | -0.689456101 | 2.20E-05 | 0.000122264 | 0.106617357 |
| T2DM | nk_cell | *IPO7* | -1.133862742 | 5.69E-05 | 0.000164236 | 0.276080558 |
| T2DM | nk_cell | *TECPR1* | -1.771778881 | 5.20E-05 | 0.00015684 | 0.252512206 |
| T2DM | nk_cell | *MT1M* | 1.277350744 | 0.001011811 | 0.001621304 | 1 |
| T2DM | nk_cell | *CFI* | -0.792223587 | 2.00E-05 | 0.000122264 | 0.096830242 |
| T2DM | nk_cell | *ATP6AP1L* | -0.547769854 | 0.001961136 | 0.002960619 | 1 |
| T2DM | nk_cell | *RGL3* | -0.546564751 | 0.000232957 | 0.000454669 | 1 |
| T2DM | nk_cell | *SERF1B* | -1.191856405 | 3.57E-05 | 0.000131704 | 0.173335917 |
| T2DM | nk_cell | *KLRC2* | 0.0601709 | 5.02E-05 | 0.000153221 | 0.243468019 |
| T2DM | nk_cell | *LINC00487* | 1.498048929 | 0.012932297 | 0.017013965 | 1 |
| T2DM | nk_cell | *RFPL3S* | 0.443567579 | 0.024995259 | 0.03153328 | 1 |

| | | | | | | |
|---|---|---|---|---|---|---|
| T2DM | nk_cell | *C1QTNF9B* | -4.641303967 | 2.52E-05 | 0.000122264 | 0.122282014 |
| T2DM | nk_cell | *ZDHHC11B* | -1.036749388 | 1.32E-05 | 0.000122264 | 0.064024013 |
| T2DM | nk_cell | *HBA1* | 0.404679283 | 5.76E-05 | 0.000165883 | 0.279347238 |
| T2DM | nk_cell | *COL6A6* | -0.98211603 | 2.34E-05 | 0.000122264 | 0.11375497 |
| T2DM | nk_cell | *HLA-A* | 0.561288906 | 0.003494433 | 0.005052143 | 1 |
| T2DM | nk_cell | *LNP1* | -0.956175917 | 0.038725342 | 0.047556405 | 1 |
| T2DM | nk_cell | *PRSS50* | -3.133588229 | 1.19E-05 | 0.000122264 | 0.057952758 |
| T2DM | nk_cell | *METTL6* | -1.532302223 | 0.001366225 | 0.002121934 | 1 |
| T2DM | nk_cell | *SLC48A1* | -1.048336635 | 1.28E-05 | 0.000122264 | 0.062187122 |
| T2DM | nk_cell | *TAS2R50* | -8.830284009 | 0.002044884 | 0.00307366 | 1 |
| T2DM | nk_cell | *TAS2R14* | 1.202378527 | 1.07E-06 | 7.89E-05 | 0.005206015 |
| T2DM | nk_cell | *AC084018.1* | -0.82076117 | 7.87E-06 | 0.000122264 | 0.038201988 |
| T2DM | nk_cell | *OR2A42* | -0.990228644 | 0.000504884 | 0.000874892 | 1 |
| T2DM | nk_cell | *ZNF580* | -0.896446903 | 4.89E-05 | 0.000151564 | 0.237349409 |
| T2DM | nk_cell | *AL021068.1* | -1.329031235 | 0.001103825 | 0.001751392 | 1 |
| T2DM | nk_cell | *FGFR1OP* | 1.37963744 | 0.027158693 | 0.03407654 | 1 |
| T2DM | nk_cell | *FAM106A* | -2.278971204 | 0.005959897 | 0.008292922 | 1 |
| T2DM | nk_cell | *ZNF254* | -1.366893513 | 2.22E-05 | 0.000122264 | 0.107559506 |
| T2DM | nk_cell | *TCTEX1D2* | -0.943086174 | 0.003692284 | 0.00532233 | 1 |
| T2DM | nk_cell | *AC022498.1* | 1.83428356 | 6.17E-05 | 0.000172519 | 0.299320921 |
| T2DM | nk_cell | *GIMAP1* | 0.777079248 | 0.006466354 | 0.00893613 | 1 |
| T2DM | nk_cell | *DNLZ* | 0.587584935 | 9.84E-08 | 1.84E-05 | 0.000477577 |
| T2DM | nk_cell | *NOTCH2NL* | 0.607471944 | 5.96E-05 | 0.000168718 | 0.289040349 |
| T2DM | nk_cell | *SUPT4H1* | 0.916458011 | 0.000116896 | 0.000263449 | 0.567177358 |
| T2DM | nk_cell | *MXD3* | 1.089430586 | 0.000167713 | 0.000347604 | 0.813741206 |
| T2DM | nk_cell | *GSTM2* | -0.776755372 | 0.000204147 | 0.000407394 | 0.990520555 |
| T2DM | nk_cell | *ARHGAP19* | -1.274442214 | 6.43E-07 | 5.68E-05 | 0.003120313 |
| T2DM | nk_cell | *LCAT* | -2.434066246 | 0.001996682 | 0.003008665 | 1 |
| T2DM | nk_cell | *GPC2* | -2.959147448 | 0.000181151 | 0.000370706 | 0.878943455 |
| T2DM | nk_cell | *ERV3-1* | -0.316673759 | 0.001137398 | 0.001799953 | 1 |
| T2DM | nk_cell | *GBP7* | -0.563108855 | 0.021477549 | 0.027394603 | 1 |
| T2DM | nk_cell | *SRA1* | -1.629241294 | 9.26E-05 | 0.000223847 | 0.449059373 |
| T2DM | nk_cell | *LBH* | 0.479544158 | 0.000259796 | 0.000498037 | 1 |
| T2DM | nk_cell | *PPP1CB* | -1.33188119 | 0.000176368 | 0.0003626 | 0.855735186 |
| T2DM | nk_cell | *NCKIPSD* | -0.917541111 | 0.005739883 | 0.00800745 | 1 |
| T2DM | nk_cell | *ATF6B* | -0.859543385 | 0.003415706 | 0.004956043 | 1 |
| T2DM | nk_cell | *SLC35F6* | -1.825646614 | 0.011816582 | 0.015665042 | 1 |
| T2DM | nk_cell | *EMP2* | -1.251928601 | 2.31E-05 | 0.000122264 | 0.112129382 |
| T2DM | nk_cell | *PPM1N* | 1.064373629 | 2.80E-05 | 0.000122557 | 0.135916204 |
| T2DM | nk_cell | *SLC23A3* | -0.649319975 | 0.000616249 | 0.001037848 | 1 |
| T2DM | nk_cell | *LTB4R* | -1.963499953 | 0.014804491 | 0.019350344 | 1 |
| T2DM | nk_cell | *ZNF726* | -3.155569069 | 3.65E-06 | 0.000122264 | 0.017732149 |
| T2DM | nk_cell | *ZNF90* | -0.945421347 | 2.98E-05 | 0.000124249 | 0.144377257 |
| T2DM | nk_cell | *REPIN1* | -1.260236533 | 1.76E-05 | 0.000122264 | 0.085411254 |
| T2DM | nk_cell | *ZNF891* | -1.34181233 | 1.58E-05 | 0.000122264 | 0.076772298 |
| T2DM | nk_cell | *SMIM7* | -1.389835691 | 0.012422305 | 0.016400822 | 1 |
| T2DM | nk_cell | *TSPAN4* | -1.337826513 | 2.22E-05 | 0.000122264 | 0.10777204 |
| T2DM | nk_cell | *CPNE1* | -0.667604712 | 0.002863454 | 0.004196158 | 1 |
| T2DM | nk_cell | *PAXIP1-AS2* | -0.976789143 | 1.29E-05 | 0.000122264 | 0.062395484 |
| T2DM | nk_cell | *LYRM4* | -1.390688586 | 3.89E-05 | 0.000136418 | 0.188672809 |

| | | | | | | |
|---|---|---|---|---|---|---|
| T2DM | nk_cell | *PRCD* | -1.347699037 | 5.49E-05 | 0.000161068 | 0.26636153 |
| T2DM | nk_cell | *SH3D21* | -0.79043712 | 1.06E-05 | 0.000122264 | 0.051395248 |
| T2DM | nk_cell | *FIS1* | -0.859123591 | 6.59E-05 | 0.000179342 | 0.319766756 |
| T2DM | nk_cell | *ANG* | -1.909345657 | 2.22E-05 | 0.000122264 | 0.107503812 |
| T2DM | nk_cell | *COLCA2* | -0.728945481 | 8.22E-05 | 0.000206771 | 0.398655371 |
| T2DM | nk_cell | *SOGA3* | -0.652754077 | 0.020222347 | 0.025895705 | 1 |
| T2DM | nk_cell | *LCNL1* | -1.285194379 | 0.00023916 | 0.00046469 | 1 |
| T2DM | nk_cell | *GNAT3* | -1.15974851 | 1.58E-05 | 0.000122264 | 0.076449065 |
| T2DM | nk_cell | *PLIN5* | -0.361756025 | 2.78E-05 | 0.000122493 | 0.134850168 |
| T2DM | nk_cell | *NUTM2D* | -4.598787155 | 0.017668894 | 0.022812526 | 1 |
| T2DM | nk_cell | *CPEB1* | -0.432300635 | 1.27E-05 | 0.000122264 | 0.061530141 |
| T2DM | nk_cell | *EML6* | -1.779326022 | 0.000523064 | 0.000902526 | 1 |
| T2DM | nk_cell | *ARHGEF33* | -0.954519823 | 0.025037662 | 0.031570357 | 1 |
| T2DM | nk_cell | *IFRD2* | -1.441243445 | 1.47E-07 | 2.52E-05 | 0.000714901 |
| T2DM | nk_cell | *EVPLL* | -1.023835015 | 0.001949752 | 0.002947102 | 1 |
| T2DM | nk_cell | *AC005082.1* | -2.14684642 | 4.60E-05 | 0.000147852 | 0.223255801 |
| T2DM | nk_cell | *SMTNL1* | -0.586672009 | 0.000220679 | 0.000435435 | 1 |
| T2DM | nk_cell | *UBXN2B* | -1.090791648 | 0.02310472 | 0.029308262 | 1 |
| T2DM | nk_cell | *C5orf49* | -5.275201032 | 1.57E-05 | 0.000122264 | 0.076296045 |
| T2DM | nk_cell | *VPS16* | -1.439161953 | 2.32E-05 | 0.000122264 | 0.112377925 |
| T2DM | nk_cell | *NPEPL1* | -1.630702383 | 1.35E-05 | 0.000122264 | 0.065488853 |
| T2DM | nk_cell | *TMEM242* | -1.267732593 | 0.026508328 | 0.033312201 | 1 |
| T2DM | nk_cell | *SLC35E2* | -1.464181338 | 3.46E-05 | 0.000130101 | 0.167850826 |
| T2DM | nk_cell | *LINC01139* | -0.292596618 | 0.021251004 | 0.027134177 | 1 |
| T2DM | nk_cell | *AC009403.2* | -1.18550027 | 5.24E-05 | 0.00015751 | 0.254063033 |
| T2DM | nk_cell | *AP000350.4* | -0.427029173 | 0.001675631 | 0.002566339 | 1 |
| T2DM | nk_cell | *RNASEK* | -1.074420281 | 0.00128782 | 0.002008519 | 1 |
| T2DM | nk_cell | *RNU6ATAC35I* | -6.625715415 | 1.36E-05 | 0.000122264 | 0.065924985 |
| T2DM | nk_cell | *PPP3R1* | -1.02109767 | 0.000144892 | 0.000310916 | 0.703017999 |
| T2DM | nk_cell | *FANCG* | -1.295047844 | 6.68E-05 | 0.000180893 | 0.324159451 |
| T2DM | nk_cell | *AP4M1* | -0.665996386 | 0.003264538 | 0.004753762 | 1 |
| T2DM | nk_cell | *ZNF880* | -1.029249436 | 7.18E-05 | 0.000189848 | 0.348371828 |
| T2DM | nk_cell | *FXYD7* | -0.850271242 | 1.80E-05 | 0.000122264 | 0.087256532 |
| T2DM | nk_cell | *XKR9* | -0.452711793 | 0.00226609 | 0.003378939 | 1 |
| T2DM | nk_cell | *SLC12A8* | -0.849303674 | 3.15E-05 | 0.000127109 | 0.152797175 |
| T2DM | nk_cell | *APOL6* | -1.530550706 | 0.000259048 | 0.0004968 | 1 |
| T2DM | nk_cell | *OR2A1* | -0.954345782 | 0.00198987 | 0.003000264 | 1 |
| T2DM | nk_cell | *RAB6C* | -14.17573725 | 6.02E-05 | 0.000169714 | 0.292076939 |
| T2DM | nk_cell | *AC062017.1* | -0.639114968 | 2.28E-05 | 0.000122264 | 0.110734355 |
| T2DM | nk_cell | *ADRA2B* | -2.153401795 | 0.002782514 | 0.004083713 | 1 |
| T2DM | nk_cell | *EXOSC6* | -1.780971773 | 3.07E-06 | 0.000122264 | 0.014894042 |
| T2DM | nk_cell | *ZNF844* | -1.107857454 | 2.56E-05 | 0.000122264 | 0.123971217 |
| T2DM | nk_cell | *LINC00571* | -1.485790428 | 7.66E-05 | 0.000197717 | 0.371435547 |
| T2DM | nk_cell | *OSER1-AS1* | -0.762810142 | 0.015103967 | 0.019710717 | 1 |
| T2DM | nk_cell | *C5orf66* | -1.85316088 | 2.93E-06 | 0.000122264 | 0.014219598 |
| T2DM | nk_cell | *LINC01133* | -0.492765073 | 1.18E-05 | 0.000122264 | 0.0573832 |
| T2DM | nk_cell | *C4B* | 1.651192858 | 0.009710735 | 0.013044431 | 1 |
| T2DM | nk_cell | *LINC01117* | -2.137626108 | 6.28E-05 | 0.000173739 | 0.304739216 |
| T2DM | nk_cell | *RASAL2-AS1* | -1.456173604 | 0.003353999 | 0.004875256 | 1 |
| T2DM | nk_cell | *LINC00240* | -0.56038026 | 4.09E-05 | 0.00013951 | 0.198243266 |

| | | | | | | |
|---|---|---|---|---|---|---|
| T2DM | nk_cell | *LINC00184* | -12.57522948 | 2.36E-05 | 0.000122264 | 0.114618464 |
| T2DM | nk_cell | *AL049840.1* | -2.174784478 | 6.94E-06 | 0.000122264 | 0.033687628 |
| T2DM | nk_cell | *PLEKHM1* | -1.243591813 | 0.000345186 | 0.000633929 | 1 |
| T2DM | nk_cell | *LINC00092* | 0.501870265 | 0.005702361 | 0.007957392 | 1 |
| T2DM | nk_cell | *AP000350.6* | -0.589669694 | 0.000135751 | 0.000294968 | 0.658664021 |
| T2DM | nk_cell | *SFTA1P* | 0.898911904 | 0.000134456 | 0.000292679 | 0.652381612 |
| T2DM | nk_cell | *CYP4A22-AS1* | -1.51543844 | 1.60E-05 | 0.000122264 | 0.077756324 |
| T2DM | nk_cell | *MIAT* | 0.40857847 | 8.67E-05 | 0.000213966 | 0.420468887 |
| T2DM | nk_cell | *ERCC6* | -1.818966879 | 0.000234275 | 0.000457058 | 1 |
| T2DM | nk_cell | *LINC00694* | -1.121799874 | 0.012643442 | 0.016661048 | 1 |
| T2DM | nk_cell | *LINC00115* | 1.25721284 | 0.000457585 | 0.000803376 | 1 |
| T2DM | nk_cell | *HAR1A* | 7.019038519 | 0.002493365 | 0.003693286 | 1 |
| T2DM | nk_cell | *LINC00937* | -0.642602094 | 2.50E-05 | 0.000122264 | 0.121230427 |
| T2DM | nk_cell | *FAM138B* | -7.208225033 | 2.32E-05 | 0.000122264 | 0.112620567 |
| T2DM | nk_cell | *ZNF717* | -0.464207264 | 0.002241192 | 0.003346957 | 1 |
| T2DM | nk_cell | *AC009299.3* | -1.224458262 | 3.16E-05 | 0.000127109 | 0.153462895 |
| T2DM | nk_cell | *SCAMP4* | -2.070545423 | 0.000769846 | 0.001261071 | 1 |
| T2DM | nk_cell | *SLC2A1-AS1* | -0.8909433 | 0.000139871 | 0.000301758 | 0.678652793 |
| T2DM | nk_cell | *LINC01068* | -0.909519348 | 0.035095888 | 0.043395833 | 1 |
| T2DM | nk_cell | *AC005884.1* | -2.513211085 | 5.05E-06 | 0.000122264 | 0.024523622 |
| T2DM | nk_cell | *AC108463.2* | 3.069619132 | 0.000150619 | 0.00032095 | 0.730802014 |
| T2DM | nk_cell | *C14orf144* | -1.305620979 | 0.000277087 | 0.000525988 | 1 |
| T2DM | nk_cell | *POLR2J2* | 1.270268545 | 0.000102809 | 0.000240631 | 0.498828411 |
| T2DM | nk_cell | *LINC00578* | -0.548076276 | 2.27E-05 | 0.000122264 | 0.110152382 |
| T2DM | nk_cell | *C19orf24* | -1.335064298 | 5.07E-06 | 0.000122264 | 0.024616618 |
| T2DM | nk_cell | *ARL17B* | -1.105812242 | 2.47E-05 | 0.000122264 | 0.11987996 |
| T2DM | nk_cell | *DHFR* | -0.500493921 | 5.30E-05 | 0.000158648 | 0.257167689 |
| T2DM | nk_cell | *PATL2* | 0.609567512 | 0.000163646 | 0.000340778 | 0.794012378 |
| T2DM | nk_cell | *CASC20* | -1.659346257 | 1.04E-05 | 0.000122264 | 0.050332394 |
| T2DM | nk_cell | *PSMG3-AS1* | -1.072549428 | 3.30E-05 | 0.000127909 | 0.160142396 |
| T2DM | nk_cell | *AC112229.4* | -1.27272064 | 0.015478435 | 0.02016144 | 1 |
| T2DM | nk_cell | *KLHL7-AS1* | -0.944116957 | 0.000115152 | 0.00026084 | 0.558718979 |
| T2DM | nk_cell | *LINC00332* | -1.47004247 | 2.74E-06 | 0.000122264 | 0.013288687 |
| T2DM | nk_cell | *AC092171.4* | -1.191420471 | 1.73E-05 | 0.000122264 | 0.084045138 |
| T2DM | nk_cell | *DIRC3* | -0.606530276 | 1.87E-05 | 0.000122264 | 0.090490151 |
| T2DM | nk_cell | *TAPBP* | 0.931670177 | 0.001559103 | 0.002395431 | 1 |
| T2DM | nk_cell | *AC107021.1* | -0.922656214 | 1.12E-05 | 0.000122264 | 0.054258318 |
| T2DM | nk_cell | *TMA7* | 1.087530954 | 0.000165232 | 0.00034349 | 0.801704716 |
| T2DM | nk_cell | *MTRNR2L4* | -1.834942638 | 0.000691719 | 0.001151362 | 1 |
| T2DM | nk_cell | *LINC00865* | -0.839906865 | 6.21E-05 | 0.00017315 | 0.301453834 |
| T2DM | nk_cell | *AC009495.3* | 0.252389774 | 4.53E-05 | 0.00014727 | 0.219600915 |
| T2DM | nk_cell | *HLA-DQB2* | 0.250304032 | 0.007020598 | 0.009626321 | 1 |
| T2DM | nk_cell | *GOLGA8N* | -0.540074205 | 0.024826494 | 0.03135298 | 1 |
| T2DM | nk_cell | *LINC00665* | -1.090388618 | 0.002756069 | 0.004051029 | 1 |
| T2DM | nk_cell | *LYRM9* | -1.040994429 | 8.09E-06 | 0.000122264 | 0.039233558 |
| T2DM | nk_cell | *LINC00702* | -0.96850519 | 2.18E-05 | 0.000122264 | 0.105612607 |
| T2DM | nk_cell | *GPX1* | -1.672500526 | 7.14E-05 | 0.000189116 | 0.346320945 |
| T2DM | nk_cell | *RPS28* | -2.277855921 | 9.99E-05 | 0.000236294 | 0.484875265 |
| T2DM | nk_cell | *TRIM26* | -1.105316819 | 0.000261779 | 0.000501442 | 1 |
| T2DM | nk_cell | *ZNF736* | -0.579456594 | 0.012466601 | 0.016454828 | 1 |

| | | | | | | |
|---|---|---|---|---|---|---|
| T2DM | nk_cell | *PINLYP* | -0.98576975 | 2.18E-05 | 0.000122264 | 0.105675933 |
| T2DM | nk_cell | *ANKRD65* | -4.676293976 | 1.72E-05 | 0.000122264 | 0.083497725 |
| T2DM | nk_cell | *LINC00957* | -2.635961035 | 9.76E-07 | 7.51E-05 | 0.004733149 |
| T2DM | nk_cell | *LINC00484* | 0.37073435 | 0.003428557 | 0.004971715 | 1 |
| T2DM | nk_cell | *KIAA0040* | 0.794400727 | 0.002762473 | 0.004057983 | 1 |
| T2DM | nk_cell | *LINC00607* | -0.63349142 | 8.88E-05 | 0.000217461 | 0.431008022 |
| T2DM | nk_cell | *AP001468.1* | -0.859644428 | 0.000556641 | 0.000951636 | 1 |
| T2DM | nk_cell | *LINC00941* | 0.609839955 | 0.000943642 | 0.001522631 | 1 |
| T2DM | nk_cell | *AC019117.2* | -4.247488211 | 1.55E-05 | 0.000122264 | 0.074988887 |
| T2DM | nk_cell | *ZBTB22* | -1.124320137 | 0.032112015 | 0.039889272 | 1 |
| T2DM | nk_cell | *ZBED5* | -1.166664406 | 1.18E-05 | 0.000122264 | 0.057188039 |
| T2DM | nk_cell | *LINC00987* | 0.548601347 | 0.000528755 | 0.000910727 | 1 |
| T2DM | nk_cell | *GS1-124K5.4* | 0.611654742 | 2.72E-05 | 0.000122437 | 0.132126254 |
| T2DM | nk_cell | *LINC00959* | -1.748256468 | 2.87E-05 | 0.000123207 | 0.139077115 |
| T2DM | nk_cell | *HLA-DQA2* | 0.414878354 | 4.57E-05 | 0.000147697 | 0.221607268 |
| T2DM | nk_cell | *IRGM* | 0.759843951 | 0.000138834 | 0.000299829 | 0.673622075 |
| T2DM | nk_cell | *LINC01143* | -3.124981947 | 3.81E-05 | 0.000135266 | 0.184909075 |
| T2DM | nk_cell | *LINC00211* | 1.760312012 | 4.81E-05 | 0.000150335 | 0.233621303 |
| T2DM | nk_cell | *LRRC37A2* | -1.1713452 | 3.87E-05 | 0.00013613 | 0.187749205 |
| T2DM | nk_cell | *TXNDC5* | -2.529959894 | 3.05E-05 | 0.000125102 | 0.148047757 |
| T2DM | nk_cell | *LINC01119* | -1.034318603 | 8.07E-05 | 0.000204944 | 0.391647689 |
| T2DM | nk_cell | *NME1* | 0.599729046 | 0.002965048 | 0.004338484 | 1 |
| T2DM | nk_cell | *CDRT4* | -1.484134124 | 0.000288159 | 0.000544449 | 1 |
| T2DM | nk_cell | *APOBEC3G* | 0.141056569 | 0.000183536 | 0.000374797 | 0.89051814 |
| T2DM | nk_cell | *C1orf226* | 2.002310093 | 3.58E-05 | 0.000131743 | 0.173769664 |
| T2DM | nk_cell | *LILRA2* | -0.791192933 | 0.000430111 | 0.000761921 | 1 |
| T2DM | nk_cell | *LY6G5B* | -0.554883394 | 0.005177157 | 0.007278924 | 1 |
| T2DM | nk_cell | *PSMB9* | 0.831725415 | 1.55E-07 | 2.52E-05 | 0.000752857 |
| T2DM | nk_cell | *C1QTNF9* | 0.465555046 | 6.34E-05 | 0.000174311 | 0.307400319 |
| T2DM | nk_cell | *ISY1* | -0.518535734 | 1.65E-05 | 0.000122264 | 0.080228511 |
| T2DM | nk_cell | *PNMA2* | 1.230640258 | 0.015855091 | 0.020619601 | 1 |
| T2DM | nk_cell | *KRBOX1* | -7.984219525 | 2.33E-05 | 0.000122264 | 0.113226187 |
| T2DM | nk_cell | *ARHGEF25* | -0.517584242 | 0.000568782 | 0.000967647 | 1 |
| T2DM | nk_cell | *LINC00886* | -0.939354199 | 0.000175102 | 0.00036061 | 0.849596129 |
| T2DM | nk_cell | *NSUN6* | -1.37708638 | 0.000213616 | 0.000423219 | 1 |
| T2DM | nk_cell | *LINC00877* | -0.638420233 | 0.000724763 | 0.001198552 | 1 |
| T2DM | nk_cell | *TDGF1* | -1.533956936 | 2.76E-05 | 0.000122472 | 0.133689356 |
| T2DM | nk_cell | *CD302* | -0.799815345 | 7.45E-05 | 0.000194744 | 0.3614505 |
| T2DM | nk_cell | *INMT* | -1.155310907 | 0.001113295 | 0.001764687 | 1 |
| T2DM | nk_cell | *ATP5O* | -1.398062544 | 0.001458312 | 0.002251983 | 1 |
| T2DM | nk_cell | *C8orf58* | -1.590485662 | 0.000226341 | 0.000444269 | 1 |
| T2DM | nk_cell | *HOGA1* | -1.008863514 | 2.38E-05 | 0.000122264 | 0.115341347 |
| T2DM | nk_cell | *PI4KA* | -1.337032548 | 0.003931472 | 0.005633639 | 1 |
| T2DM | nk_cell | *LINC00969* | -0.607335232 | 0.000193216 | 0.000390031 | 0.937483613 |
| T2DM | nk_cell | *ARFGAP3* | -1.738755938 | 0.002646932 | 0.003902435 | 1 |
| T2DM | nk_cell | *MRPL20* | -1.35780141 | 0.000205343 | 0.000409336 | 0.996324061 |
| T2DM | nk_cell | *DECR2* | -0.72562248 | 2.49E-05 | 0.000122264 | 0.120725563 |
| T2DM | nk_cell | *ZNF709* | 5.79051365 | 0.000274623 | 0.000521923 | 1 |
| T2DM | nk_cell | *ERVW-1* | 2.259283122 | 0.000157148 | 0.000330841 | 0.762480401 |
| T2DM | nk_cell | *MRPL33* | -1.354060311 | 5.65E-05 | 0.00016385 | 0.273998395 |

| | | | | | | |
|---|---|---|---|---|---|---|
| T2DM | nk_cell | *STON1* | -2.107651379 | 5.07E-05 | 0.000154085 | 0.246073112 |
| T2DM | nk_cell | *VSIG8* | -1.467629514 | 3.93E-05 | 0.000136619 | 0.190512886 |
| T2DM | nk_cell | *C7orf73* | -1.381332776 | 0.007256988 | 0.009918565 | 1 |
| T2DM | nk_cell | *FGF14-IT1* | -1.543287994 | 2.22E-05 | 0.000122264 | 0.10764857 |
| T2DM | nk_cell | *PALM2* | -0.447187094 | 5.35E-05 | 0.000159514 | 0.259639607 |
| T2DM | nk_cell | *NBPF15* | -1.224851574 | 1.63E-05 | 0.000122264 | 0.079238558 |
| T2DM | nk_cell | *WFDC6* | -2.711958145 | 0.020397587 | 0.026097451 | 1 |
| T2DM | nk_cell | *UPK3B* | -1.532482835 | 2.01E-05 | 0.000122264 | 0.097708491 |
| T2DM | nk_cell | *PLA2G4B* | 2.132522517 | 0.015875733 | 0.020640155 | 1 |
| T2DM | nk_cell | *LEFTY1* | 3.824315076 | 8.38E-05 | 0.000209686 | 0.406791091 |
| T2DM | nk_cell | *TTC4* | 1.413420443 | 0.002005829 | 0.003020571 | 1 |
| T2DM | nk_cell | *JMJD7* | 0.843256316 | 0.003300791 | 0.004802231 | 1 |
| T2DM | nk_cell | *APOBEC3D* | 0.484701892 | 5.51E-05 | 0.000161492 | 0.267431259 |
| T2DM | nk_cell | *TMEM199* | 0.526722441 | 0.000593494 | 0.001006161 | 1 |
| T2DM | nk_cell | *SPRR2F* | -1.620131002 | 0.002761189 | 0.004057326 | 1 |
| T2DM | nk_cell | *P2RY11* | 0.322897043 | 0.001396452 | 0.00216265 | 1 |
| T2DM | nk_cell | *DBNDD2* | -2.983763518 | 0.004438988 | 0.006301337 | 1 |
| T2DM | nk_cell | *LILRA6* | -0.406738916 | 0.016478303 | 0.021377734 | 1 |
| T2DM | nk_cell | *SCARF2* | -1.259500334 | 2.97E-05 | 0.000124182 | 0.144175757 |
| T2DM | nk_cell | *APOBEC3C* | 0.396383158 | 5.86E-05 | 0.000167232 | 0.284462038 |
| T2DM | nk_cell | *C4A* | -2.291720591 | 3.91E-05 | 0.000136598 | 0.189677656 |
| T2DM | nk_cell | *CRYBB2* | -1.521747893 | 0.002562409 | 0.003785874 | 1 |
| T2DM | nk_cell | *NEAT1* | -0.65772407 | 4.21E-05 | 0.000141813 | 0.204280618 |
| T2DM | nk_cell | *RAB30-AS1* | -1.163749779 | 4.84E-06 | 0.000122264 | 0.023501892 |
| T2DM | nk_cell | *DNAJC3-AS1* | -1.469062175 | 2.05E-05 | 0.000122264 | 0.099533512 |
| T2DM | nk_cell | *LINC00926* | -1.19917641 | 2.99E-05 | 0.000124686 | 0.145224565 |
| T2DM | nk_cell | *DPH6-AS1* | -0.834026848 | 2.92E-05 | 0.000123467 | 0.141443305 |
| T2DM | nk_cell | *BCKDHA* | -0.894556438 | 0.027982658 | 0.035065045 | 1 |
| T2DM | nk_cell | *CDK11B* | -1.800388789 | 3.77E-05 | 0.000134709 | 0.183069894 |
| T2DM | nk_cell | *ABHD14A* | -2.244171324 | 0.000199648 | 0.000400618 | 0.968694024 |
| T2DM | nk_cell | *SRP14-AS1* | -0.666174602 | 0.000145377 | 0.000311422 | 0.705370366 |
| T2DM | nk_cell | *GDNF-AS1* | -0.921281455 | 1.72E-05 | 0.000122264 | 0.083652115 |
| T2DM | nk_cell | *CCDC153* | -0.744642507 | 0.000115305 | 0.000261064 | 0.559459117 |
| T2DM | nk_cell | *C5orf17* | -1.402947195 | 2.78E-05 | 0.000122493 | 0.134714417 |
| T2DM | nk_cell | *TMEM150C* | -1.217703835 | 2.58E-06 | 0.000122264 | 0.012519969 |
| T2DM | nk_cell | *LINC00939* | -1.264300363 | 0.000723852 | 0.001197453 | 1 |
| T2DM | nk_cell | *LINC01060* | -1.005035514 | 6.90E-05 | 0.000185105 | 0.334893246 |
| T2DM | nk_cell | *LINC00942* | -3.031099737 | 1.85E-05 | 0.000122264 | 0.09000206 |
| T2DM | nk_cell | *PDCD6* | -1.85654271 | 0.000219843 | 0.000434316 | 1 |
| T2DM | nk_cell | *TMEM158* | -2.128432787 | 4.24E-05 | 0.000141986 | 0.20573765 |
| T2DM | nk_cell | *DNAH10OS* | -4.487806085 | 1.37E-06 | 9.04E-05 | 0.00666223 |
| T2DM | nk_cell | *PCDHA10* | -0.66307037 | 0.000426594 | 0.00075815 | 1 |
| T2DM | nk_cell | *SMIM20* | -2.492251234 | 0.000230374 | 0.000450715 | 1 |
| T2DM | nk_cell | *LINC00989* | -0.613704565 | 1.79E-05 | 0.000122264 | 0.086901261 |
| T2DM | nk_cell | *GLI4* | -0.71028552 | 2.22E-05 | 0.000122264 | 0.107798648 |
| T2DM | nk_cell | *SNHG18* | -1.529605434 | 1.03E-06 | 7.81E-05 | 0.004996453 |
| T2DM | nk_cell | *ALG1L2* | -1.805607974 | 2.75E-05 | 0.000122472 | 0.133216408 |
| T2DM | nk_cell | *LINC01094* | -0.824936763 | 2.03E-05 | 0.000122264 | 0.098634881 |
| T2DM | nk_cell | *TMEM200B* | 1.610324216 | 0.00609432 | 0.008462977 | 1 |
| T2DM | nk_cell | *SLC10A5* | -1.537002391 | 0.014807879 | 0.019350344 | 1 |

| | | | | | | |
|---|---|---|---|---|---|---|
| T2DM | nk_cell | *PCDHGA6* | -0.450033152 | 0.001144995 | 0.001809614 | 1 |
| T2DM | nk_cell | *UTP14C* | 1.312276603 | 0.016136391 | 0.020950969 | 1 |
| T2DM | nk_cell | *PCDHGB2* | -2.648250193 | 2.41E-05 | 0.000122264 | 0.116899096 |
| T2DM | nk_cell | *LYN* | 0.548157615 | 0.00465421 | 0.006585659 | 1 |
| T2DM | nk_cell | *SIGLEC14* | -1.003730699 | 0.033800607 | 0.041868916 | 1 |
| T2DM | nk_cell | *MEX3A* | -1.079986185 | 0.001335024 | 0.002076134 | 1 |
| T2DM | nk_cell | *SNHG9* | -1.045552013 | 0.000170156 | 0.000351917 | 0.825596891 |
| T2DM | nk_cell | *PCDHA3* | -2.288775389 | 4.56E-05 | 0.000147697 | 0.221231102 |
| T2DM | nk_cell | *ZNF432* | -1.04243892 | 6.87E-06 | 0.000122264 | 0.033348836 |
| T2DM | nk_cell | *HMBS* | -0.639116283 | 0.000459292 | 0.000805671 | 1 |
| T2DM | nk_cell | *TAS2R31* | 0.681294501 | 0.00076011 | 0.001246385 | 1 |
| T2DM | nk_cell | *AC093668.2* | -0.72878853 | 0.000517759 | 0.000893691 | 1 |
| T2DM | nk_cell | *KIAA1147* | -1.117559175 | 2.86E-05 | 0.000123207 | 0.138974396 |
| T2DM | nk_cell | *ZBED6* | -1.079689508 | 2.31E-05 | 0.000122264 | 0.111860162 |
| T2DM | nk_cell | *CNPY2* | -1.785855182 | 7.93E-05 | 0.000202355 | 0.384942974 |
| T2DM | nk_cell | *LINC00592* | -2.584755875 | 4.80E-05 | 0.000150037 | 0.232706821 |
| T2DM | nk_cell | *CHURC1* | -0.957906105 | 0.017489027 | 0.022604358 | 1 |
| T2DM | nk_cell | *RTEL1* | 1.084043599 | 0.004799595 | 0.006775571 | 1 |
| T2DM | nk_cell | *LINC00640* | -0.591352391 | 0.006752832 | 0.009302879 | 1 |
| T2DM | nk_cell | *HIF1A-AS2* | -1.723432779 | 0.000252261 | 0.000485702 | 1 |
| T2DM | nk_cell | *LINC00638* | -1.107452262 | 0.003868817 | 0.005547134 | 1 |
| T2DM | nk_cell | *RNASE4* | -2.167582986 | 7.47E-05 | 0.000195056 | 0.362608618 |
| T2DM | nk_cell | *MC1R* | 2.809378818 | 0.000192211 | 0.000388425 | 0.932608514 |
| T2DM | nk_cell | *LINC00639* | 0.729212248 | 0.002776405 | 0.00407598 | 1 |
| T2DM | nk_cell | *LINC00924* | -0.41412637 | 0.032478402 | 0.040303122 | 1 |
| T2DM | nk_cell | *ITGB3* | -0.658532212 | 0.020084651 | 0.025739759 | 1 |
| T2DM | nk_cell | *MRPL46* | 1.47640975 | 0.000670975 | 0.001119902 | 1 |
| T2DM | nk_cell | *UBE2Q2L* | -1.82180709 | 0.000364655 | 0.000665152 | 1 |
| T2DM | nk_cell | *IQCH-AS1* | -1.067794255 | 0.019600162 | 0.025138775 | 1 |
| T2DM | nk_cell | *AC083843.1* | -9.41017671 | 2.56E-05 | 0.000122264 | 0.124091893 |
| T2DM | nk_cell | *PWRN1* | -0.615240977 | 5.19E-05 | 0.000156496 | 0.251802186 |
| T2DM | nk_cell | *LINC01003* | 1.283529617 | 0.002653057 | 0.003910277 | 1 |
| T2DM | nk_cell | *C15orf65* | -1.706697084 | 0.00024864 | 0.000479682 | 1 |
| T2DM | nk_cell | *GOLGA8H* | -0.395184695 | 0.004580904 | 0.006489503 | 1 |
| T2DM | nk_cell | *LINC00662* | -1.216544631 | 2.84E-05 | 0.00012318 | 0.137958751 |
| T2DM | nk_cell | *C19orf84* | 0.186247214 | 4.90E-05 | 0.000151762 | 0.237962582 |
| T2DM | nk_cell | *LINC00667* | -0.936075108 | 0.000315531 | 0.000588377 | 1 |
| T2DM | nk_cell | *LINC00672* | -5.229348126 | 0.004969423 | 0.007001058 | 1 |
| T2DM | nk_cell | *LINC00909* | -1.412780057 | 0.000757922 | 0.001243218 | 1 |
| T2DM | nk_cell | *DYNLL2* | 1.495463711 | 0.011835859 | 0.015682198 | 1 |
| T2DM | nk_cell | *STRADA* | -1.750008146 | 3.22E-05 | 0.000127628 | 0.1560892 |
| T2DM | nk_cell | *MYO15B* | -1.048376152 | 0.000117202 | 0.000263515 | 0.56866546 |
| T2DM | nk_cell | *ILF3-AS1* | -1.214262983 | 0.003822602 | 0.005487356 | 1 |
| T2DM | nk_cell | *ZNF285* | 0.741330545 | 0.019508673 | 0.025034668 | 1 |
| T2DM | nk_cell | *ZNF224* | -1.635506627 | 4.37E-05 | 0.000144302 | 0.211835131 |
| T2DM | nk_cell | *LIN37* | -2.773195128 | 3.49E-05 | 0.000130739 | 0.169503601 |
| T2DM | nk_cell | *NDUFA7* | 0.642692409 | 5.48E-05 | 0.000161068 | 0.265922618 |
| T2DM | nk_cell | *LINC00664* | 1.241200153 | 4.45E-05 | 0.00014589 | 0.216062983 |
| T2DM | nk_cell | *SCGB1B2P* | -1.610827133 | 0.001262281 | 0.001972492 | 1 |
| T2DM | nk_cell | *AC026740.1* | -2.701977344 | 0.030387348 | 0.037902163 | 1 |

| | | | | | | |
|---|---|---|---|---|---|---|
| T2DM | nk_cell | *ZNF728* | -0.75572044 | 3.78E-05 | 0.000134863 | 0.183548015 |
| T2DM | nk_cell | *TRABD2B* | -1.628041414 | 5.61E-05 | 0.000163673 | 0.272351904 |
| T2DM | nk_cell | *AC093157.1* | -0.885309738 | 1.58E-05 | 0.000122264 | 0.07641908 |
| T2DM | nk_cell | *FBXO17* | -0.745017982 | 0.000222594 | 0.000438678 | 1 |
| T2DM | nk_cell | *ZNF587B* | -0.646079732 | 2.42E-05 | 0.000122264 | 0.117336623 |
| T2DM | nk_cell | *SNHG8* | -1.714276794 | 3.88E-07 | 4.27E-05 | 0.001880599 |
| T2DM | nk_cell | *HCG17* | -1.1682376 | 5.81E-05 | 0.000166801 | 0.282060694 |
| T2DM | nk_cell | *GTF2H5* | -0.776439428 | 0.000738385 | 0.001217344 | 1 |
| T2DM | nk_cell | *FGF14-AS2* | -0.831444512 | 2.29E-05 | 0.000122264 | 0.110928356 |
| T2DM | nk_cell | *CASC15* | -0.808345615 | 1.66E-05 | 0.000122264 | 0.080325977 |
| T2DM | nk_cell | *AP000347.2* | -1.59600674 | 0.000304107 | 0.000569482 | 1 |
| T2DM | nk_cell | *DOC2B* | -0.495437077 | 0.002036455 | 0.003062889 | 1 |
| T2DM | nk_cell | *GRIN2B* | -0.481160302 | 0.001371225 | 0.002126977 | 1 |
| T2DM | nk_cell | *PAXIP1-AS1* | -2.019738123 | 3.32E-05 | 0.000127948 | 0.160862146 |
| T2DM | pericyte | *RBM6* | -2.444738722 | 1.04E-07 | 3.59E-05 | 0.000502256 |
| T2DM | pericyte | *CEP68* | 5.493048985 | 3.13E-06 | 0.000689544 | 0.015169977 |
| T2DM | pericyte | *TUBG2* | -4.263678903 | 6.35E-05 | 0.009942694 | 0.308223513 |
| T2DM | pericyte | *ITIH4* | 1.250195898 | 1.31E-05 | 0.002435774 | 0.063330122 |
| T2DM | pericyte | *SEC61A2* | -4.819529818 | 0.000218026 | 0.024042323 | 1 |
| T2DM | pericyte | *PABPC4* | -6.612322948 | 1.38E-09 | 9.56E-07 | 6.69E-06 |
| T2DM | pericyte | *CWF19L1* | -1.961443241 | 9.85E-05 | 0.012921129 | 0.478081757 |
| T2DM | pericyte | *GCDH* | -5.165926571 | 7.63E-05 | 0.010583692 | 0.370429226 |
| T2DM | pericyte | *CHN2* | 5.379407731 | 1.01E-06 | 0.000270988 | 0.004877783 |
| T2DM | pericyte | *CNTNAP1* | 7.005637043 | 9.32E-06 | 0.001884046 | 0.045217114 |
| T2DM | pericyte | *CDK2AP1* | 3.240351099 | 0.000261465 | 0.026992051 | 1 |
| T2DM | pericyte | *ADRA1A* | 0.858656888 | 0.000338998 | 0.032896366 | 1 |
| T2DM | pericyte | *TCF19* | 4.567511579 | 1.72E-08 | 9.98E-06 | 8.37E-05 |
| T2DM | pericyte | *SPTBN5* | 0.881086895 | 7.36E-05 | 0.010506325 | 0.357215036 |
| T2DM | pericyte | *NEIL1* | -3.798746238 | 0.000394526 | 0.037534144 | 1 |
| T2DM | pericyte | *MAN2C1* | -1.489151789 | 1.20E-05 | 0.002323236 | 0.058080906 |
| T2DM | pericyte | *C18orf8* | -4.987319195 | 7.17E-05 | 0.010506325 | 0.348104546 |
| T2DM | pericyte | *HSD17B12* | -2.098594496 | 3.30E-08 | 1.36E-05 | 0.000159937 |
| T2DM | pericyte | *APIP* | -1.304732556 | 0.000228112 | 0.024595548 | 1 |
| T2DM | pericyte | *JAZF1* | -6.093665909 | 1.41E-10 | 1.37E-07 | 6.84E-07 |
| T2DM | pericyte | *CLEC18A* | -3.728336752 | 3.64E-08 | 1.36E-05 | 0.000176773 |
| T2DM | pericyte | *KRTCAP3* | 2.336234643 | 4.77E-05 | 0.007975626 | 0.231293141 |
| T2DM | pericyte | *SPTA1* | -2.00936989 | 0.000195526 | 0.023304888 | 0.948690433 |
| T2DM | pericyte | *RFT1* | -3.988945442 | 3.78E-06 | 0.000796616 | 0.018322168 |
| T2DM | pericyte | *RNF123* | 3.03043741 | 8.02E-05 | 0.010807645 | 0.389075223 |
| T2DM | pericyte | *MST1R* | 2.089802158 | 0.000243673 | 0.025702244 | 1 |
| T2DM | pericyte | *RAPSN* | -2.971092141 | 2.06E-05 | 0.003696482 | 0.099805011 |
| T2DM | pericyte | *ANPEP* | 1.153432645 | 0.00055351 | 0.048829662 | 1 |
| T2DM | pericyte | *IRS1* | -1.482247428 | 8.34E-07 | 0.000238114 | 0.004047944 |
| T2DM | pericyte | *INO80E* | -2.140739171 | 0.000512859 | 0.046950793 | 1 |
| T2DM | pericyte | *TTLL6* | -6.985638153 | 2.38E-06 | 0.000570836 | 0.011540772 |
| T2DM | pericyte | *RGS19* | -1.687751881 | 2.55E-05 | 0.00441325 | 0.123571 |
| T2DM | pericyte | *CTSW* | -0.493460328 | 0.000212423 | 0.023969226 | 1 |
| T2DM | pericyte | *RNASEH2C* | -2.784456409 | 0.000538665 | 0.048400031 | 1 |
| T2DM | pericyte | *CCNE2* | -4.066366468 | 2.47E-06 | 0.000570836 | 0.01198755 |
| T2DM | pericyte | *HLA-DQB1* | -2.679401203 | 4.24E-16 | 1.03E-12 | 2.06E-12 |

| | | | | | | |
|---|---|---|---|---|---|---|
| T2DM | pericyte | *HLA-DRB1* | -2.453555341 | 2.15E-10 | 1.73E-07 | 1.04E-06 |
| T2DM | pericyte | *HLA-DQA1* | -3.504156581 | 2.95E-13 | 4.77E-10 | 1.43E-09 |
| T2DM | pericyte | *HLA-DRB5* | -1.056244357 | 4.27E-12 | 5.18E-09 | 2.07E-08 |
| T2DM | pericyte | *NOTCH4* | -0.445298547 | 1.85E-08 | 9.98E-06 | 8.98E-05 |
| T2DM | pericyte | *PRRT1* | 1.81581039 | 0.000279368 | 0.028239441 | 1 |
| T2DM | pericyte | *GPANK1* | -3.519992591 | 0.000205511 | 0.023741451 | 0.997140956 |
| T2DM | pericyte | *LIN52* | 3.642373238 | 0.000449807 | 0.041970494 | 1 |
| T2DM | pericyte | *HCG27* | 2.38813192 | 2.17E-06 | 0.000553319 | 0.010513064 |
| T2DM | pericyte | *DNLZ* | 1.15665006 | 0.000291823 | 0.02889647 | 1 |
| T2DM | pericyte | *ARHGAP19* | -7.501512782 | 0.000122204 | 0.015203407 | 0.592932884 |
| T2DM | pericyte | *TMEM110* | 4.476383048 | 5.56E-05 | 0.008990251 | 0.269707534 |
| T2DM | pericyte | *STARD10* | 2.279645804 | 3.62E-08 | 1.36E-05 | 0.000175435 |
| T2DM | pericyte | *C4B* | 4.238934177 | 0.000118161 | 0.015087337 | 0.573318815 |
| T2DM | pericyte | *HLA-DQB2* | 0.993032067 | 3.07E-08 | 1.36E-05 | 0.000148945 |
| T2DM | pericyte | *HLA-DQA2* | 1.874630953 | 9.30E-18 | 4.51E-14 | 4.51E-14 |
| T2DM | pericyte | *PSMB9* | 1.72566841 | 6.62E-05 | 0.010039226 | 0.321255217 |
| T2DM | pericyte | *PLA2G4B* | 8.75246279 | 3.47E-07 | 0.000105237 | 0.001683789 |
| T2DM | pericyte | *JMJD7* | 2.833108296 | 1.58E-07 | 5.10E-05 | 0.000764851 |
| T2DM | pericyte | *C4A* | -3.540130707 | 0.000196929 | 0.023304888 | 0.955500415 |
| T2DM | SMC | *RBM6* | -2.405745399 | 3.88E-09 | 2.09E-06 | 1.88E-05 |
| T2DM | SMC | *TUBG2* | -3.094995055 | 0.00015553 | 0.017150734 | 0.754632308 |
| T2DM | SMC | *ITIH4* | 0.996544294 | 7.92E-05 | 0.010107425 | 0.38408216 |
| T2DM | SMC | *SEC61A2* | -5.727321364 | 9.11E-07 | 0.000232565 | 0.004418743 |
| T2DM | SMC | *PABPC4* | -6.53300056 | 2.32E-11 | 2.25E-08 | 1.13E-07 |
| T2DM | SMC | *ALKBH5* | -4.88198208 | 5.26E-05 | 0.007292382 | 0.255233357 |
| T2DM | SMC | *CWF19L1* | -1.700715504 | 0.00012802 | 0.015528853 | 0.62115411 |
| T2DM | SMC | *RNF32* | -6.988029862 | 0.000153454 | 0.017150734 | 0.744560562 |
| T2DM | SMC | *CDK2AP1* | 3.055909672 | 3.35E-05 | 0.005246143 | 0.162630442 |
| T2DM | SMC | *BVES* | -1.276948951 | 0.000167342 | 0.018043167 | 0.811942497 |
| T2DM | SMC | *GPX7* | -1.296870852 | 0.000224722 | 0.021280021 | 1 |
| T2DM | SMC | *CCDC92* | -3.903836452 | 0.000460949 | 0.035641497 | 1 |
| T2DM | SMC | *ADRA1A* | 0.770035724 | 0.000612473 | 0.045026026 | 1 |
| T2DM | SMC | *PDZRN3* | -2.037642459 | 0.000181326 | 0.018719022 | 0.879794012 |
| T2DM | SMC | *ABHD8* | -2.896406162 | 0.000105162 | 0.013083171 | 0.510243673 |
| T2DM | SMC | *RCBTB1* | -2.179515638 | 0.000639915 | 0.046341324 | 1 |
| T2DM | SMC | *TCF19* | 4.318349502 | 4.70E-08 | 1.75E-05 | 0.000228038 |
| T2DM | SMC | *SPTBN5* | 0.797472054 | 4.69E-05 | 0.007117317 | 0.22775414 |
| T2DM | SMC | *MAN2C1* | -1.434263935 | 3.64E-06 | 0.000801848 | 0.017640655 |
| T2DM | SMC | *VPS53* | -3.14769516 | 0.000188612 | 0.019065515 | 0.915144728 |
| T2DM | SMC | *C18orf8* | -5.355245657 | 3.29E-07 | 8.87E-05 | 0.001597477 |
| T2DM | SMC | *SAE1* | -6.057814808 | 1.62E-05 | 0.002904215 | 0.078413816 |
| T2DM | SMC | *HSD17B12* | -2.077661599 | 1.64E-09 | 9.96E-07 | 7.97E-06 |
| T2DM | SMC | *APIP* | -1.194775114 | 0.000144685 | 0.016714523 | 0.702009977 |
| T2DM | SMC | *POC5* | -2.228849283 | 0.000382091 | 0.030391895 | 1 |
| T2DM | SMC | *JAZF1* | -5.676509545 | 1.85E-12 | 2.25E-09 | 8.99E-09 |
| T2DM | SMC | *CLEC18A* | -2.108517092 | 1.61E-06 | 0.000371639 | 0.007804428 |
| T2DM | SMC | *KRTCAP3* | 1.79809792 | 6.48E-05 | 0.008731754 | 0.314343135 |
| T2DM | SMC | *SPTA1* | -1.800429328 | 0.000265896 | 0.023891264 | 1 |
| T2DM | SMC | *RFT1* | -4.110732972 | 1.15E-07 | 3.29E-05 | 0.000559572 |
| T2DM | SMC | *RNF123* | 2.550289232 | 0.000199618 | 0.019766301 | 0.968548727 |

| | | | | | | |
|---|---|---|---|---|---|---|
| T2DM | SMC | *MST1R* | 2.256093395 | 1.30E-05 | 0.00249898 | 0.062914948 |
| T2DM | SMC | *RAPSN* | -2.701299409 | 5.00E-05 | 0.00713686 | 0.242653237 |
| T2DM | SMC | *PDHB* | -5.466145369 | 0.000311447 | 0.026054165 | 1 |
| T2DM | SMC | *ZSWIM1* | 0.993504733 | 0.000327673 | 0.026946914 | 1 |
| T2DM | SMC | *IRS1* | -1.278641587 | 4.38E-06 | 0.000923766 | 0.021246609 |
| T2DM | SMC | *INO80E* | -2.363547598 | 3.10E-05 | 0.005129967 | 0.150592292 |
| T2DM | SMC | *TTLL6* | -9.79198055 | 7.05E-09 | 3.42E-06 | 3.42E-05 |
| T2DM | SMC | *RGS19* | -1.243673152 | 0.000278114 | 0.024096589 | 1 |
| T2DM | SMC | *C1QTNF4* | -1.329429576 | 0.00013548 | 0.016032853 | 0.657346987 |
| T2DM | SMC | *RNASEH2C* | -3.21006986 | 1.05E-05 | 0.002123629 | 0.050967085 |
| T2DM | SMC | *HOXB2* | -4.024607701 | 7.26E-05 | 0.009519352 | 0.352216027 |
| T2DM | SMC | *CCNE2* | -2.85964097 | 4.98E-05 | 0.00713686 | 0.24174506 |
| T2DM | SMC | *ZNF852* | -3.726702093 | 0.000462781 | 0.035641497 | 1 |
| T2DM | SMC | *HLA-DQB1* | -2.380725619 | 8.18E-16 | 1.98E-12 | 3.97E-12 |
| T2DM | SMC | *PSMD13* | -3.99682891 | 0.000548561 | 0.041587749 | 1 |
| T2DM | SMC | *DMWD* | -4.698103906 | 3.17E-05 | 0.005129967 | 0.153899004 |
| T2DM | SMC | *HLA-DRB1* | -2.19558688 | 1.65E-10 | 1.14E-07 | 7.99E-07 |
| T2DM | SMC | *HLA-DQA1* | -2.595524109 | 6.39E-11 | 5.17E-08 | 3.10E-07 |
| T2DM | SMC | *HLA-DRB5* | -0.987504041 | 9.78E-13 | 1.58E-09 | 4.74E-09 |
| T2DM | SMC | *NOTCH4* | -0.407500542 | 1.58E-08 | 6.98E-06 | 7.67E-05 |
| T2DM | SMC | *PRRT1* | 1.487127222 | 0.000293654 | 0.024996615 | 1 |
| T2DM | SMC | *SKIV2L* | -1.180964487 | 0.000588591 | 0.043936038 | 1 |
| T2DM | SMC | *GPANK1* | -4.188115753 | 1.22E-06 | 0.000296927 | 0.005938531 |
| T2DM | SMC | *HCG27* | 1.920610033 | 3.01E-05 | 0.005129967 | 0.145889494 |
| T2DM | SMC | *TAS2R14* | 1.621428754 | 0.00069006 | 0.049237821 | 1 |
| T2DM | SMC | *DNLZ* | 1.068901198 | 0.000238557 | 0.02183918 | 1 |
| T2DM | SMC | *TMEM110* | 4.385285765 | 1.34E-05 | 0.00249898 | 0.06497348 |
| T2DM | SMC | *STARD10* | 1.92329792 | 9.56E-08 | 2.90E-05 | 0.000464084 |
| T2DM | SMC | *IFRD2* | -3.051051901 | 0.000272251 | 0.024017489 | 1 |
| T2DM | SMC | *C4B* | 3.659378215 | 0.000228063 | 0.021280021 | 1 |
| T2DM | SMC | *AL049840.1* | -4.382583104 | 0.000356729 | 0.028847523 | 1 |
| T2DM | SMC | *HLA-DQB2* | 0.911040124 | 3.52E-08 | 1.42E-05 | 0.000170893 |
| T2DM | SMC | *HLA-DQA2* | 1.614480796 | 5.46E-17 | 2.65E-13 | 2.65E-13 |
| T2DM | SMC | *PSMB9* | 1.415024348 | 0.000175358 | 0.018496421 | 0.850835365 |
| T2DM | SMC | *PLA2G4B* | 8.330776204 | 5.93E-08 | 2.05E-05 | 0.000287515 |
| T2DM | SMC | *JMJD7* | 2.587641933 | 7.02E-08 | 2.27E-05 | 0.000340744 |
| T2DM | SMC | *C4A* | -3.207716347 | 0.000220817 | 0.021280021 | 1 |
| T2DM | t_cell | *SEMA3F* | -2.248023969 | 1.49E-05 | 0.001201364 | 0.072081852 |
| T2DM | t_cell | *SNX11* | 0.987306137 | 0.005353696 | 0.024858025 | 1 |
| T2DM | t_cell | *M6PR* | 2.233822281 | 0.001558995 | 0.010166995 | 1 |
| T2DM | t_cell | *CYP26B1* | -1.583700849 | 0.004872109 | 0.02303124 | 1 |
| T2DM | t_cell | *DBNDD1* | -1.264904884 | 0.000356902 | 0.003764538 | 1 |
| T2DM | t_cell | *RBM6* | -1.58265328 | 1.11E-06 | 0.000213354 | 0.005406834 |
| T2DM | t_cell | *MEOX1* | -1.47791273 | 0.000456619 | 0.004344147 | 1 |
| T2DM | t_cell | *PON1* | -3.759637575 | 0.000973903 | 0.007337545 | 1 |
| T2DM | t_cell | *ITGA2B* | -2.005906164 | 0.002244755 | 0.013282382 | 1 |
| T2DM | t_cell | *SYNRG* | 2.09663507 | 0.000237975 | 0.003062745 | 1 |
| T2DM | t_cell | *USH1C* | -9.534619374 | 0.002219933 | 0.013222396 | 1 |
| T2DM | t_cell | *CEACAM21* | 0.550846851 | 0.00015047 | 0.002594722 | 0.730080103 |
| T2DM | t_cell | *ST7L* | -1.262993718 | 0.008972871 | 0.036159777 | 1 |

| | | | | | | |
|---|---|---|---|---|---|---|
| T2DM | t_cell | *TSR3* | 4.519501363 | 0.011810645 | 0.044285355 | 1 |
| T2DM | t_cell | *FMO3* | -0.972780542 | 0.000551428 | 0.004927308 | 1 |
| T2DM | t_cell | *CYB561* | 3.176852509 | 1.90E-06 | 0.000317201 | 0.009198827 |
| T2DM | t_cell | *SS18L2* | 4.509572644 | 5.03E-06 | 0.000596779 | 0.024413585 |
| T2DM | t_cell | *MMP25* | 1.109942293 | 6.46E-05 | 0.001782154 | 0.313659124 |
| T2DM | t_cell | *IL32* | 0.712605283 | 4.00E-05 | 0.001670757 | 0.1940199 |
| T2DM | t_cell | *MAPK8IP2* | -4.965815724 | 0.00019872 | 0.002828058 | 0.96418806 |
| T2DM | t_cell | *RPS20* | 2.291225321 | 0.010310863 | 0.04021568 | 1 |
| T2DM | t_cell | *TRAF3IP3* | 0.454623806 | 8.69E-06 | 0.000878186 | 0.042152906 |
| T2DM | t_cell | *SLC6A7* | -34.1402836 | 0.000148478 | 0.002594722 | 0.72041545 |
| T2DM | t_cell | *TSPAN9* | -2.062146989 | 0.000113517 | 0.002285408 | 0.550783332 |
| T2DM | t_cell | *PPP5C* | 3.17286151 | 0.000227572 | 0.003033461 | 1 |
| T2DM | t_cell | *CEP68* | 3.881220471 | 2.27E-06 | 0.000367525 | 0.011025758 |
| T2DM | t_cell | *TACC3* | 2.67092245 | 0.000126491 | 0.002417859 | 0.61373593 |
| T2DM | t_cell | *CCDC88C* | 0.658481834 | 8.19E-05 | 0.002006241 | 0.397235679 |
| T2DM | t_cell | *CHDH* | -3.634557706 | 0.006941428 | 0.030206108 | 1 |
| T2DM | t_cell | *CPS1* | -1.209291342 | 0.004938868 | 0.023265424 | 1 |
| T2DM | t_cell | *ABCC2* | -2.571387668 | 0.00044581 | 0.004281236 | 1 |
| T2DM | t_cell | *GCLM* | 3.560653868 | 0.000475511 | 0.004445429 | 1 |
| T2DM | t_cell | *INSRR* | -2.717381417 | 9.25E-05 | 0.002106122 | 0.448604008 |
| T2DM | t_cell | *SH2D2A* | 0.885294951 | 4.16E-05 | 0.001670757 | 0.201725802 |
| T2DM | t_cell | *ANK1* | -3.189476335 | 0.001188433 | 0.00847982 | 1 |
| T2DM | t_cell | *IKZF2* | 2.632038522 | 0.000295578 | 0.003358649 | 1 |
| T2DM | t_cell | *CENPQ* | -0.728903609 | 0.00377866 | 0.019044812 | 1 |
| T2DM | t_cell | *SH3YL1* | -2.357979626 | 0.007410624 | 0.03145787 | 1 |
| T2DM | t_cell | *TUBG2* | -3.753745311 | 3.77E-08 | 1.31E-05 | 0.000182704 |
| T2DM | t_cell | *FLT4* | -0.98660594 | 0.000157361 | 0.002614785 | 0.763517341 |
| T2DM | t_cell | *PI4K2B* | -3.534941418 | 0.000398029 | 0.004048711 | 1 |
| T2DM | t_cell | *DNAH5* | -2.595279017 | 0.002167345 | 0.013047097 | 1 |
| T2DM | t_cell | *PQLC2* | -4.130819165 | 0.001960811 | 0.012150514 | 1 |
| T2DM | t_cell | *INPP4A* | 1.529702397 | 0.000328434 | 0.003565777 | 1 |
| T2DM | t_cell | *TDP1* | 3.17411199 | 0.000216206 | 0.002946715 | 1 |
| T2DM | t_cell | *ARAP2* | 1.212681924 | 2.57E-05 | 0.00151791 | 0.124468603 |
| T2DM | t_cell | *ANO2* | -0.909320642 | 0.000340483 | 0.003638727 | 1 |
| T2DM | t_cell | *RCN1* | -2.301652247 | 0.012811265 | 0.046942469 | 1 |
| T2DM | t_cell | *LAMC3* | -1.533468898 | 0.000581701 | 0.005113072 | 1 |
| T2DM | t_cell | *CYBA* | 2.758532138 | 0.000422364 | 0.004123191 | 1 |
| T2DM | t_cell | *THOC3* | 1.685751757 | 5.13E-05 | 0.00172419 | 0.248830942 |
| T2DM | t_cell | *ALX4* | -1.620509042 | 0.001849052 | 0.011591213 | 1 |
| T2DM | t_cell | *AKR7A2* | 3.3156657 | 0.00799417 | 0.033351428 | 1 |
| T2DM | t_cell | *MRTO4* | -2.428130928 | 0.000111602 | 0.002275183 | 0.541493612 |
| T2DM | t_cell | *LY75* | 3.591468895 | 1.85E-05 | 0.001374162 | 0.089862708 |
| T2DM | t_cell | *PLEKHH1* | -1.526695639 | 0.004802673 | 0.022823282 | 1 |
| T2DM | t_cell | *ITIH4* | 0.821482384 | 6.32E-05 | 0.001756372 | 0.306644198 |
| T2DM | t_cell | *NPFFR2* | -1.834787162 | 0.000169351 | 0.002681745 | 0.821690009 |
| T2DM | t_cell | *ATP11B* | 2.378363798 | 0.002396051 | 0.013883495 | 1 |
| T2DM | t_cell | *LAMC2* | -0.784756005 | 0.005425642 | 0.025143472 | 1 |
| T2DM | t_cell | *DNAJC25* | 5.741830777 | 0.001937705 | 0.012053517 | 1 |
| T2DM | t_cell | *RIMBP2* | -1.860771296 | 0.000322476 | 0.00354797 | 1 |
| T2DM | t_cell | *PRDM6* | -2.276646777 | 0.003065876 | 0.016510133 | 1 |

| | | | | | | |
|---|---|---|---|---|---|---|
| T2DM | t_cell | *SPAG4* | -2.596472634 | 0.003255278 | 0.017129689 | 1 |
| T2DM | t_cell | *LTK* | 0.797574141 | 7.15E-05 | 0.001903045 | 0.346816888 |
| T2DM | t_cell | *AHRR* | -2.815746972 | 0.000307266 | 0.003445826 | 1 |
| T2DM | t_cell | *CASP8* | 1.535786577 | 2.33E-05 | 0.001454217 | 0.113127891 |
| T2DM | t_cell | *LPAR2* | 2.172923487 | 1.27E-05 | 0.00105876 | 0.061408078 |
| T2DM | t_cell | *SLC12A2* | 3.389871877 | 0.002722201 | 0.015234278 | 1 |
| T2DM | t_cell | *CNN2* | 2.108496223 | 9.36E-07 | 0.000197442 | 0.004541165 |
| T2DM | t_cell | *ABCA7* | 1.188904842 | 0.00381559 | 0.019184707 | 1 |
| T2DM | t_cell | *BTBD1* | 3.695352643 | 0.00068238 | 0.005708462 | 1 |
| T2DM | t_cell | *FAR2* | 2.730711342 | 5.78E-05 | 0.00172419 | 0.280314523 |
| T2DM | t_cell | *BCAS1* | -1.665635418 | 0.001000848 | 0.007470988 | 1 |
| T2DM | t_cell | *CHI3L2* | 3.350547182 | 0.000294487 | 0.003354647 | 1 |
| T2DM | t_cell | *ANKRD44* | 0.941128021 | 3.57E-05 | 0.001630473 | 0.173299759 |
| T2DM | t_cell | *SEC61A2* | -3.440682291 | 0.000233361 | 0.003043725 | 1 |
| T2DM | t_cell | *NGEF* | -3.73708334 | 0.0053538 | 0.024858025 | 1 |
| T2DM | t_cell | *FGFR2* | -1.06823959 | 0.00880868 | 0.035825413 | 1 |
| T2DM | t_cell | *MTFR1* | 1.594611755 | 0.001782549 | 0.011217807 | 1 |
| T2DM | t_cell | *STOML1* | -4.219722459 | 0.001344104 | 0.0092768 | 1 |
| T2DM | t_cell | *ROGDI* | -3.370633972 | 0.000594721 | 0.0051899 | 1 |
| T2DM | t_cell | *HEATR6* | 2.4393214 | 0.007102445 | 0.030632058 | 1 |
| T2DM | t_cell | *POLR1A* | -1.542341641 | 0.003779916 | 0.019044812 | 1 |
| T2DM | t_cell | *STON1-GTF2A* | -0.533481785 | 0.005868671 | 0.02663685 | 1 |
| T2DM | t_cell | *IFT80* | -3.674272256 | 0.002782181 | 0.015480668 | 1 |
| T2DM | t_cell | *CLEC2D* | 1.10297409 | 0.000106037 | 0.002217626 | 0.514489142 |
| T2DM | t_cell | *DRD4* | -3.991711799 | 0.000747982 | 0.006089276 | 1 |
| T2DM | t_cell | *PLA2G10* | 6.457073597 | 3.60E-05 | 0.001630473 | 0.174460645 |
| T2DM | t_cell | *GNB5* | 1.897990233 | 8.87E-05 | 0.002066904 | 0.43042106 |
| T2DM | t_cell | *CLTCL1* | -2.012246091 | 0.011348601 | 0.043051925 | 1 |
| T2DM | t_cell | *VASH1* | -1.654265704 | 0.003087911 | 0.016573613 | 1 |
| T2DM | t_cell | *TRIB2* | 1.834888804 | 0.004052 | 0.020123136 | 1 |
| T2DM | t_cell | *NFATC3* | 2.436013289 | 0.00016201 | 0.002646713 | 0.786073821 |
| T2DM | t_cell | *CRMP1* | -1.927546899 | 0.001840417 | 0.011552008 | 1 |
| T2DM | t_cell | *PVR* | -1.991792745 | 5.86E-05 | 0.00172419 | 0.284223948 |
| T2DM | t_cell | *MOV10L1* | -1.637506808 | 0.001521244 | 0.01004228 | 1 |
| T2DM | t_cell | *LLGL2* | 1.559279445 | 0.000201748 | 0.00285945 | 0.978879666 |
| T2DM | t_cell | *SDHA* | 3.024859114 | 9.97E-05 | 0.002149929 | 0.483734007 |
| T2DM | t_cell | *PPP2R2C* | -1.757157406 | 0.000154479 | 0.00260406 | 0.749532591 |
| T2DM | t_cell | *ZZEF1* | 3.08650389 | 0.001594268 | 0.010354591 | 1 |
| T2DM | t_cell | *VPS9D1* | 3.654847163 | 0.002220268 | 0.013222396 | 1 |
| T2DM | t_cell | *MCM6* | 2.23208782 | 0.002943917 | 0.016103593 | 1 |
| T2DM | t_cell | *SPAG5* | -3.862048223 | 0.000270953 | 0.003258353 | 1 |
| T2DM | t_cell | *PAG1* | 1.318403633 | 3.36E-05 | 0.001630473 | 0.163259508 |
| T2DM | t_cell | *ICAM3* | 1.382071981 | 5.50E-05 | 0.00172419 | 0.26665496 |
| T2DM | t_cell | *XAB2* | 6.403163528 | 8.14E-05 | 0.002006241 | 0.394962739 |
| T2DM | t_cell | *APBB1IP* | 1.220894353 | 0.004214926 | 0.020846911 | 1 |
| T2DM | t_cell | *CLUL1* | -1.689417744 | 0.004094146 | 0.020311654 | 1 |
| T2DM | t_cell | *SP140* | 0.666583287 | 0.000198096 | 0.002828058 | 0.961160555 |
| T2DM | t_cell | *FDFT1* | 2.490745437 | 0.006197544 | 0.027612931 | 1 |
| T2DM | t_cell | *SCGN* | -7.6650394 | 0.000155968 | 0.002609502 | 0.756755509 |
| T2DM | t_cell | *DNM2* | 2.385727858 | 0.009925596 | 0.039125083 | 1 |

| | | | | | | |
|---|---|---|---|---|---|---|
| T2DM | t_cell | *DCT* | -1.275679149 | 0.001374717 | 0.009445267 | 1 |
| T2DM | t_cell | *KCNN2* | -1.861885956 | 0.008644118 | 0.035363626 | 1 |
| T2DM | t_cell | *STK17B* | 1.407899193 | 3.51E-06 | 0.000470882 | 0.017030558 |
| T2DM | t_cell | *ZNF506* | 2.957853242 | 0.002866312 | 0.015789646 | 1 |
| T2DM | t_cell | *PCDHB4* | -2.64695188 | 0.000276077 | 0.003278465 | 1 |
| T2DM | t_cell | *SEMA5B* | -0.775875545 | 0.013642554 | 0.049398264 | 1 |
| T2DM | t_cell | *OXCT1* | 2.363015187 | 0.006952968 | 0.030229214 | 1 |
| T2DM | t_cell | *RPS5* | 2.068583008 | 0.010853334 | 0.041661691 | 1 |
| T2DM | t_cell | *CD82* | 1.505018626 | 1.59E-05 | 0.00124438 | 0.077197255 |
| T2DM | t_cell | *OVGP1* | -1.686346725 | 3.51E-05 | 0.001630473 | 0.170324851 |
| T2DM | t_cell | *NME8* | -24.90323847 | 0.000671652 | 0.005667572 | 1 |
| T2DM | t_cell | *SNX10* | -1.148854341 | 0.012202847 | 0.04526622 | 1 |
| T2DM | t_cell | *HSD17B14* | -1.661961233 | 0.002582682 | 0.014537323 | 1 |
| T2DM | t_cell | *ACHE* | -2.626852263 | 0.000544236 | 0.004890063 | 1 |
| T2DM | t_cell | *ADAMTS2* | -1.80314117 | 0.010849257 | 0.041661691 | 1 |
| T2DM | t_cell | *LPCAT2* | -1.918977129 | 0.003856237 | 0.019328988 | 1 |
| T2DM | t_cell | *ASAP3* | -1.794816801 | 0.003761334 | 0.018990628 | 1 |
| T2DM | t_cell | *DOCK3* | -2.082187954 | 0.013299272 | 0.048371864 | 1 |
| T2DM | t_cell | *EBF4* | -2.252155184 | 0.000825284 | 0.006521623 | 1 |
| T2DM | t_cell | *TBX5* | -3.921537928 | 0.001330146 | 0.009219809 | 1 |
| T2DM | t_cell | *FXYD5* | 2.46817794 | 8.62E-05 | 0.002066904 | 0.418411566 |
| T2DM | t_cell | *GRAMD1A* | 3.057081184 | 0.009386868 | 0.037424062 | 1 |
| T2DM | t_cell | *YPEL3* | 2.051876337 | 0.006780533 | 0.029585565 | 1 |
| T2DM | t_cell | *LYZ* | 2.211366368 | 0.002987055 | 0.016193512 | 1 |
| T2DM | t_cell | *PABPC4* | -4.703725009 | 2.40E-09 | 1.45E-06 | 1.16E-05 |
| T2DM | t_cell | *CERS4* | 3.168973437 | 0.002595352 | 0.014574822 | 1 |
| T2DM | t_cell | *GLG1* | 2.32277869 | 0.00797557 | 0.033351428 | 1 |
| T2DM | t_cell | *ABCC6* | -1.913126559 | 0.002553549 | 0.014442943 | 1 |
| T2DM | t_cell | *TF* | -2.946039358 | 0.002937362 | 0.016085869 | 1 |
| T2DM | t_cell | *ALKBH5* | -3.019950792 | 0.003864799 | 0.019331962 | 1 |
| T2DM | t_cell | *RGS17* | -2.554400131 | 2.71E-05 | 0.001541761 | 0.131258749 |
| T2DM | t_cell | *PPP2R3C* | 2.652002455 | 0.000844675 | 0.006642401 | 1 |
| T2DM | t_cell | *TBX15* | -1.20696613 | 0.00299725 | 0.01619911 | 1 |
| T2DM | t_cell | *UNC13D* | 0.926993745 | 0.002862116 | 0.015789646 | 1 |
| T2DM | t_cell | *CRAT* | -3.331391647 | 0.006210407 | 0.027644856 | 1 |
| T2DM | t_cell | *NANS* | 4.329667997 | 0.000563686 | 0.005000006 | 1 |
| T2DM | t_cell | *CWF19L1* | -1.385481424 | 0.000171098 | 0.002686624 | 0.830166966 |
| T2DM | t_cell | *IL12RB1* | 0.700114127 | 0.000263105 | 0.003231864 | 1 |
| T2DM | t_cell | *SETD1A* | 3.744356258 | 0.001072043 | 0.00784548 | 1 |
| T2DM | t_cell | *SERPIND1* | 2.817230307 | 0.000383665 | 0.003952322 | 1 |
| T2DM | t_cell | *CYTH4* | 1.424936917 | 4.20E-05 | 0.001670757 | 0.203589935 |
| T2DM | t_cell | *SH3BP1* | 1.310033621 | 4.89E-05 | 0.00172419 | 0.237343689 |
| T2DM | t_cell | *ANKRD54* | -1.430765045 | 0.007038187 | 0.030463231 | 1 |
| T2DM | t_cell | *PICK1* | -1.61569366 | 0.004473616 | 0.021597996 | 1 |
| T2DM | t_cell | *KDELR3* | -2.581460802 | 0.009239659 | 0.037050268 | 1 |
| T2DM | t_cell | *RAB36* | -1.535536311 | 0.00247598 | 0.014150126 | 1 |
| T2DM | t_cell | *RHBDD3* | -2.7254497 | 0.001541281 | 0.010119481 | 1 |
| T2DM | t_cell | *L3MBTL2* | -2.652620787 | 0.01011697 | 0.039721865 | 1 |
| T2DM | t_cell | *NIN* | 2.327856493 | 0.000270383 | 0.003258353 | 1 |
| T2DM | t_cell | *PLEK2* | -2.390654409 | 0.000524168 | 0.004807683 | 1 |

| | | | | | | |
|---|---|---|---|---|---|---|
| T2DM | t_cell | *GSKIP* | 4.863105593 | 0.000414961 | 0.004112304 | 1 |
| T2DM | t_cell | *PSMB5* | 2.158828424 | 0.001327672 | 0.009215829 | 1 |
| T2DM | t_cell | *EFS* | -2.820733956 | 0.004618389 | 0.022099036 | 1 |
| T2DM | t_cell | *CD40* | -3.162573586 | 0.001071887 | 0.00784548 | 1 |
| T2DM | t_cell | *DOK5* | -2.077405984 | 0.000931486 | 0.007125997 | 1 |
| T2DM | t_cell | *NKAIN4* | -1.399530162 | 0.006131136 | 0.027417614 | 1 |
| T2DM | t_cell | *PTK6* | 4.06969692 | 3.11E-05 | 0.001619427 | 0.151036992 |
| T2DM | t_cell | *GMEB2* | 3.931992761 | 0.00640054 | 0.028283624 | 1 |
| T2DM | t_cell | *TRIB3* | 7.316767963 | 0.000869911 | 0.006764112 | 1 |
| T2DM | t_cell | *RASSF2* | 1.323669654 | 3.05E-05 | 0.001619427 | 0.148048835 |
| T2DM | t_cell | *ANGPT4* | -3.698543722 | 0.003128648 | 0.016681537 | 1 |
| T2DM | t_cell | *MROH8* | -1.319910828 | 0.007561478 | 0.032042175 | 1 |
| T2DM | t_cell | *MAP1LC3A* | -1.960457272 | 0.013231824 | 0.048162648 | 1 |
| T2DM | t_cell | *LPIN2* | 2.911795945 | 0.000337151 | 0.003627254 | 1 |
| T2DM | t_cell | *LAMA1* | -1.869401336 | 0.000395287 | 0.004029273 | 1 |
| T2DM | t_cell | *RNF125* | 3.123173541 | 4.61E-05 | 0.001701031 | 0.223454906 |
| T2DM | t_cell | *PARP4* | 2.582915898 | 0.001599585 | 0.010354591 | 1 |
| T2DM | t_cell | *NOMO3* | -2.055411962 | 0.000287785 | 0.003332537 | 1 |
| T2DM | t_cell | *SLC7A5* | 1.993484997 | 0.010358071 | 0.04033496 | 1 |
| T2DM | t_cell | *RHBDL1* | -4.925705554 | 7.11E-05 | 0.001903045 | 0.344895611 |
| T2DM | t_cell | *HMOX2* | 2.854195581 | 1.63E-05 | 0.00124438 | 0.079023175 |
| T2DM | t_cell | *RBL2* | 2.462289779 | 1.14E-06 | 0.000213354 | 0.005547191 |
| T2DM | t_cell | *PYCARD* | 3.936624541 | 9.29E-06 | 0.000919876 | 0.045073918 |
| T2DM | t_cell | *CDIPT* | 2.789171355 | 0.011705454 | 0.044000262 | 1 |
| T2DM | t_cell | *TMC5* | -1.678231339 | 0.005331847 | 0.024858025 | 1 |
| T2DM | t_cell | *KNOP1* | -2.33567769 | 0.012509998 | 0.046041178 | 1 |
| T2DM | t_cell | *AP3B2* | -0.751441761 | 0.003155444 | 0.016732472 | 1 |
| T2DM | t_cell | *CHRAC1* | 5.477044555 | 0.011505913 | 0.043464472 | 1 |
| T2DM | t_cell | *TNFRSF10A* | 1.090870648 | 9.88E-06 | 0.000931216 | 0.047918448 |
| T2DM | t_cell | *TUSC3* | -1.713217112 | 0.002359727 | 0.013744774 | 1 |
| T2DM | t_cell | *TULP2* | -5.595699885 | 0.000590066 | 0.005167872 | 1 |
| T2DM | t_cell | *NFKBIB* | 2.261280231 | 0.008030765 | 0.03347532 | 1 |
| T2DM | t_cell | *PPP1R37* | -1.921017078 | 0.004638551 | 0.022173644 | 1 |
| T2DM | t_cell | *ARHGEF18* | 2.359632927 | 0.010794148 | 0.041533073 | 1 |
| T2DM | t_cell | *CD37* | 1.488738129 | 6.14E-05 | 0.001745457 | 0.298065063 |
| T2DM | t_cell | *AMH* | -11.63357102 | 0.000150382 | 0.002594722 | 0.729655457 |
| T2DM | t_cell | *FCER2* | -3.765361162 | 0.000182086 | 0.002738946 | 0.883482305 |
| T2DM | t_cell | *CLEC4M* | -4.53363814 | 0.00021254 | 0.002921367 | 1 |
| T2DM | t_cell | *TLE6* | 1.932541357 | 0.003472852 | 0.017868212 | 1 |
| T2DM | t_cell | *IL27RA* | 1.10787975 | 3.50E-05 | 0.001630473 | 0.170036392 |
| T2DM | t_cell | *RASAL3* | 0.52589696 | 5.83E-05 | 0.00172419 | 0.282706865 |
| T2DM | t_cell | *ZNF419* | 0.748745289 | 0.011511098 | 0.043464472 | 1 |
| T2DM | t_cell | *AURKC* | 1.353035939 | 0.010511713 | 0.040802263 | 1 |
| T2DM | t_cell | *POLR2I* | 3.066466683 | 0.000653765 | 0.005590288 | 1 |
| T2DM | t_cell | *TGFB1* | 2.922291365 | 0.000407173 | 0.004078878 | 1 |
| T2DM | t_cell | *ICAM5* | -2.238085414 | 2.20E-05 | 0.001405861 | 0.106611788 |
| T2DM | t_cell | *RABAC1* | 2.626710177 | 0.010291953 | 0.040206569 | 1 |
| T2DM | t_cell | *CARD8* | 2.05953568 | 0.005929785 | 0.026813904 | 1 |
| T2DM | t_cell | *LIG1* | 2.878555448 | 0.000731893 | 0.00599753 | 1 |
| T2DM | t_cell | *PLEKHA4* | -1.859045321 | 0.000461616 | 0.004358847 | 1 |

| | | | | | | |
|---|---|---|---|---|---|---|
| T2DM | t_cell | *GCDH* | -3.719819946 | 9.50E-05 | 0.00214103 | 0.460863477 |
| T2DM | t_cell | *ETV2* | -11.09936111 | 0.000988166 | 0.00741048 | 1 |
| T2DM | t_cell | *ZNF85* | -2.621419258 | 0.012286001 | 0.045505098 | 1 |
| T2DM | t_cell | *RASA4* | -12.1829896 | 0.000399782 | 0.004056357 | 1 |
| T2DM | t_cell | *SP4* | 2.348436705 | 4.15E-05 | 0.001670757 | 0.201286348 |
| T2DM | t_cell | *DNAH11* | -2.416447477 | 0.007244901 | 0.031027367 | 1 |
| T2DM | t_cell | *PTN* | -1.993771361 | 0.006680888 | 0.029229639 | 1 |
| T2DM | t_cell | *ADAP1* | 7.118158472 | 4.48E-05 | 0.001701031 | 0.2175812 |
| T2DM | t_cell | *MET* | -1.035604277 | 0.004248162 | 0.02092597 | 1 |
| T2DM | t_cell | *VIPR2* | -1.573793955 | 0.001888395 | 0.011807333 | 1 |
| T2DM | t_cell | *CHN2* | 4.731044796 | 1.38E-10 | 1.67E-07 | 6.67E-07 |
| T2DM | t_cell | *ABHD11* | 1.274472859 | 0.005916175 | 0.026777317 | 1 |
| T2DM | t_cell | *TFR2* | 2.131060501 | 0.000172789 | 0.002692473 | 0.8383706 |
| T2DM | t_cell | *NRF1* | 2.266915495 | 0.007987776 | 0.033351428 | 1 |
| T2DM | t_cell | *ACTR3C* | 2.654229737 | 0.008883297 | 0.035948087 | 1 |
| T2DM | t_cell | *POU6F2* | -2.041091084 | 0.002031455 | 0.012457418 | 1 |
| T2DM | t_cell | *TSPAN13* | -2.131768841 | 0.001232012 | 0.008701194 | 1 |
| T2DM | t_cell | *COA1* | 2.396231446 | 0.004435481 | 0.021478 | 1 |
| T2DM | t_cell | *AEBP1* | -2.146669362 | 0.002237015 | 0.013252742 | 1 |
| T2DM | t_cell | *GCK* | -6.554413121 | 0.000168493 | 0.002681745 | 0.817529319 |
| T2DM | t_cell | *TRIM14* | 1.697763019 | 0.000308978 | 0.003445826 | 1 |
| T2DM | t_cell | *CORO2A* | 1.193556655 | 9.12E-05 | 0.002099002 | 0.442647556 |
| T2DM | t_cell | *DOCK8* | 0.708681786 | 0.001021528 | 0.007578675 | 1 |
| T2DM | t_cell | *NCS1* | -1.003276866 | 0.007245295 | 0.031027367 | 1 |
| T2DM | t_cell | *CUBN* | 2.879873049 | 0.000270435 | 0.003258353 | 1 |
| T2DM | t_cell | *SPOCK2* | 0.669309203 | 9.59E-05 | 0.00214103 | 0.465356265 |
| T2DM | t_cell | *SFXN3* | 4.300760274 | 0.002223914 | 0.013222396 | 1 |
| T2DM | t_cell | *ACBD5* | 3.073011173 | 0.006842662 | 0.029829828 | 1 |
| T2DM | t_cell | *LHPP* | -2.116355165 | 0.009777325 | 0.038702965 | 1 |
| T2DM | t_cell | *TFAM* | 3.625311489 | 0.012435339 | 0.045848227 | 1 |
| T2DM | t_cell | *DNAJC12* | -1.919596247 | 0.006461112 | 0.028499379 | 1 |
| T2DM | t_cell | *RPL19* | 2.650576926 | 0.000132789 | 0.002459123 | 0.644290272 |
| T2DM | t_cell | *WNT3* | -1.597860129 | 0.003405968 | 0.017636878 | 1 |
| T2DM | t_cell | *PNPO* | -4.096961798 | 8.84E-05 | 0.002066904 | 0.428906607 |
| T2DM | t_cell | *TRIM16L* | -2.655522106 | 0.000689655 | 0.00573963 | 1 |
| T2DM | t_cell | *DRG2* | 3.340324423 | 0.001662725 | 0.010671353 | 1 |
| T2DM | t_cell | *FTSJ3* | -0.685525831 | 0.003241981 | 0.017079363 | 1 |
| T2DM | t_cell | *ICAM2* | 4.083069245 | 0.003605186 | 0.018374332 | 1 |
| T2DM | t_cell | *EFNB3* | -1.772195893 | 0.000663163 | 0.005615478 | 1 |
| T2DM | t_cell | *GAR1* | 4.875228532 | 0.004504233 | 0.021650583 | 1 |
| T2DM | t_cell | *AADAT* | -1.990715916 | 0.000354676 | 0.003757396 | 1 |
| T2DM | t_cell | *TRIM2* | -1.926193001 | 0.005406943 | 0.02508077 | 1 |
| T2DM | t_cell | *STIM2* | 2.677933612 | 0.000670553 | 0.005667572 | 1 |
| T2DM | t_cell | *TECTA* | -2.93199137 | 0.000401421 | 0.004057698 | 1 |
| T2DM | t_cell | *B3GAT1* | 1.696118144 | 5.71E-05 | 0.00172419 | 0.27724203 |
| T2DM | t_cell | *P2RX3* | -12.41200791 | 0.000176454 | 0.002709345 | 0.856153148 |
| T2DM | t_cell | *DCPS* | 3.882705907 | 0.000211302 | 0.002912608 | 1 |
| T2DM | t_cell | *NRXN2* | -2.91607096 | 0.000119906 | 0.002365307 | 0.58178515 |
| T2DM | t_cell | *CCND1* | -2.961278125 | 0.000432703 | 0.004190569 | 1 |
| T2DM | t_cell | *CCDC86* | -2.557085262 | 9.61E-05 | 0.00214103 | 0.46642482 |

| | | | | | | |
|---|---|---|---|---|---|---|
| T2DM | t_cell | *GALNT18* | -1.307056958 | 0.005831727 | 0.026527417 | 1 |
| T2DM | t_cell | *KIAA1549L* | -4.143000413 | 0.000266912 | 0.003258353 | 1 |
| T2DM | t_cell | *SLC35F2* | 4.581599026 | 0.000570248 | 0.005039783 | 1 |
| T2DM | t_cell | *C11orf21* | 1.15735979 | 0.006435505 | 0.028412258 | 1 |
| T2DM | t_cell | *RPS13* | 2.452917273 | 0.001103736 | 0.008028975 | 1 |
| T2DM | t_cell | *KCTD10* | -1.855013807 | 0.012400182 | 0.045784395 | 1 |
| T2DM | t_cell | *IL23A* | -17.87208908 | 0.004337184 | 0.021226528 | 1 |
| T2DM | t_cell | *PRR4* | 2.295401854 | 0.000225089 | 0.003025299 | 1 |
| T2DM | t_cell | *PARP11* | 3.125152232 | 0.001046317 | 0.007715393 | 1 |
| T2DM | t_cell | *VDR* | -3.15320837 | 0.000431368 | 0.004190569 | 1 |
| T2DM | t_cell | *KRR1* | 2.2032756 | 0.012265473 | 0.045463771 | 1 |
| T2DM | t_cell | *GNB3* | -1.109857226 | 0.001406698 | 0.009586091 | 1 |
| T2DM | t_cell | *GOLT1B* | 3.533496249 | 0.004292125 | 0.02107833 | 1 |
| T2DM | t_cell | *FRK* | -2.394637795 | 0.001482304 | 0.009879312 | 1 |
| T2DM | t_cell | *RNGTT* | 2.514336783 | 0.000360228 | 0.003777851 | 1 |
| T2DM | t_cell | *PHACTR1* | -1.690239008 | 0.000859964 | 0.006709329 | 1 |
| T2DM | t_cell | *MDGA1* | -2.980028457 | 0.003685754 | 0.018686813 | 1 |
| T2DM | t_cell | *CD83* | 2.721066261 | 0.001943516 | 0.012069293 | 1 |
| T2DM | t_cell | *BVES* | -1.306778185 | 5.84E-06 | 0.00067502 | 0.028350843 |
| T2DM | t_cell | *COL9A1* | 7.321110991 | 0.000386743 | 0.003967181 | 1 |
| T2DM | t_cell | *WASF1* | -1.530729293 | 0.005276951 | 0.024761864 | 1 |
| T2DM | t_cell | *VNN2* | 2.736475295 | 6.78E-06 | 0.000758992 | 0.032894328 |
| T2DM | t_cell | *PHACTR2* | 1.691656246 | 0.003526556 | 0.01808758 | 1 |
| T2DM | t_cell | *PACRG* | -2.225002038 | 0.002175163 | 0.013077933 | 1 |
| T2DM | t_cell | *EXOC2* | 2.99695788 | 0.000696601 | 0.005777617 | 1 |
| T2DM | t_cell | *IMPG1* | -3.979162462 | 0.000943453 | 0.007152552 | 1 |
| T2DM | t_cell | *BTN2A1* | 2.743969494 | 0.002956442 | 0.016135621 | 1 |
| T2DM | t_cell | *PCDHB2* | -2.646456528 | 0.001326465 | 0.009215829 | 1 |
| T2DM | t_cell | *CEP72* | 2.867022267 | 0.003122652 | 0.016667887 | 1 |
| T2DM | t_cell | *KIF20A* | -9.114074037 | 0.000711418 | 0.005870409 | 1 |
| T2DM | t_cell | *PCDHB15* | -1.237883827 | 0.005051123 | 0.023732447 | 1 |
| T2DM | t_cell | *CDH6* | -1.954820559 | 0.00053327 | 0.004827283 | 1 |
| T2DM | t_cell | *LMNB1* | 1.525541274 | 0.000214162 | 0.002935354 | 1 |
| T2DM | t_cell | *SKP1* | 2.761202488 | 0.012516141 | 0.046041178 | 1 |
| T2DM | t_cell | *TXNDC15* | 3.370865752 | 0.011073988 | 0.042341207 | 1 |
| T2DM | t_cell | *UNC5A* | -11.02501085 | 0.001724543 | 0.0109522 | 1 |
| T2DM | t_cell | *ACTR8* | -2.837115836 | 0.01379423 | 0.049687903 | 1 |
| T2DM | t_cell | *CLDN16* | -1.281211113 | 0.000530022 | 0.004823326 | 1 |
| T2DM | t_cell | *CYB561D2* | -2.964944135 | 0.00011749 | 0.002345932 | 0.57006147 |
| T2DM | t_cell | *CBLB* | 1.989792594 | 0.000154569 | 0.00260406 | 0.749969201 |
| T2DM | t_cell | *KIAA1257* | -5.759651082 | 0.00224835 | 0.013283765 | 1 |
| T2DM | t_cell | *ABHD14B* | -2.642760482 | 0.001462813 | 0.009774098 | 1 |
| T2DM | t_cell | *VIPR1* | 5.583779846 | 0.001485454 | 0.009886725 | 1 |
| T2DM | t_cell | *NKTR* | 2.089762503 | 0.009693713 | 0.038457804 | 1 |
| T2DM | t_cell | *ADAM23* | 1.174522381 | 0.000224417 | 0.003025299 | 1 |
| T2DM | t_cell | *ACTR1B* | 3.812804684 | 0.001001223 | 0.007470988 | 1 |
| T2DM | t_cell | *ITGB6* | -2.375667944 | 0.003365079 | 0.017481118 | 1 |
| T2DM | t_cell | *GRB14* | -1.680543272 | 0.0005509 | 0.004927308 | 1 |
| T2DM | t_cell | *GALNT3* | 1.33884798 | 9.93E-06 | 0.000931216 | 0.048202431 |
| T2DM | t_cell | *IGFBP2* | -1.581724914 | 0.00250181 | 0.014233653 | 1 |

| | | | | | | |
|---|---|---|---|---|---|---|
| T2DM | t_cell | *GNLY* | 1.48541183 | 9.66E-05 | 0.00214103 | 0.468679477 |
| T2DM | t_cell | *WNT6* | -2.417529887 | 0.001454154 | 0.009745242 | 1 |
| T2DM | t_cell | *IL1RL2* | -1.771940082 | 0.005769761 | 0.026331008 | 1 |
| T2DM | t_cell | *IL18R1* | 1.451186982 | 0.000122974 | 0.002401423 | 0.596671566 |
| T2DM | t_cell | *MLPH* | -1.418828797 | 0.001304378 | 0.009158518 | 1 |
| T2DM | t_cell | *TPO* | -1.018524824 | 0.000108989 | 0.002260662 | 0.528815448 |
| T2DM | t_cell | *WIPF1* | 1.903151791 | 3.40E-05 | 0.001630473 | 0.165165881 |
| T2DM | t_cell | *AAK1* | 2.182895241 | 0.000264733 | 0.003243646 | 1 |
| T2DM | t_cell | *RALGPS2* | -1.726217775 | 0.004857508 | 0.022993784 | 1 |
| T2DM | t_cell | *KIAA1324* | 1.85043087 | 0.000497122 | 0.004594355 | 1 |
| T2DM | t_cell | *MECR* | -1.902938481 | 0.006982716 | 0.03030424 | 1 |
| T2DM | t_cell | *RAP1A* | 2.356564629 | 0.000644851 | 0.005527942 | 1 |
| T2DM | t_cell | *CAPZA1* | 2.068152431 | 0.010170322 | 0.039827606 | 1 |
| T2DM | t_cell | *PRG4* | -2.33079329 | 0.002200362 | 0.013164187 | 1 |
| T2DM | t_cell | *EXOC8* | 4.232161609 | 0.006393969 | 0.028280343 | 1 |
| T2DM | t_cell | *SLAMF1* | 0.263739819 | 5.01E-05 | 0.00172419 | 0.242888277 |
| T2DM | t_cell | *KIF17* | -2.261665231 | 0.000341974 | 0.003638727 | 1 |
| T2DM | t_cell | *EBNA1BP2* | -2.589580951 | 0.002062376 | 0.012578458 | 1 |
| T2DM | t_cell | *NSUN4* | -2.17431866 | 0.008944619 | 0.036075887 | 1 |
| T2DM | t_cell | *DR1* | 2.273841167 | 0.001716396 | 0.010919944 | 1 |
| T2DM | t_cell | *RPS6KA1* | 1.402273871 | 0.000337158 | 0.003627254 | 1 |
| T2DM | t_cell | *SLC5A9* | -1.749273873 | 0.007531361 | 0.031942451 | 1 |
| T2DM | t_cell | *CD3EAP* | -2.605894447 | 0.000733004 | 0.00599753 | 1 |
| T2DM | t_cell | *APOA1* | 6.464243373 | 2.78E-06 | 0.000412234 | 0.013500281 |
| T2DM | t_cell | *CASC1* | -1.158380684 | 0.001553458 | 0.010149299 | 1 |
| T2DM | t_cell | *LRMP* | 0.868787071 | 0.009284318 | 0.037106681 | 1 |
| T2DM | t_cell | *ELOVL4* | 2.818473668 | 3.86E-07 | 9.72E-05 | 0.001870571 |
| T2DM | t_cell | *CNRIP1* | -1.551842561 | 0.00549037 | 0.025394922 | 1 |
| T2DM | t_cell | *EPCAM* | -3.447575497 | 0.007720549 | 0.032574004 | 1 |
| T2DM | t_cell | *TECTB* | -1.66407481 | 0.004594226 | 0.022005117 | 1 |
| T2DM | t_cell | *PPP1R3C* | -2.583919536 | 0.002116213 | 0.012818809 | 1 |
| T2DM | t_cell | *C10orf88* | -1.852905946 | 0.011647952 | 0.043903902 | 1 |
| T2DM | t_cell | *MASTL* | -1.616375868 | 0.009820151 | 0.038832414 | 1 |
| T2DM | t_cell | *MRC1* | 2.681134215 | 0.003684897 | 0.018686813 | 1 |
| T2DM | t_cell | *ADRA1A* | 0.554454705 | 0.00155419 | 0.010149299 | 1 |
| T2DM | t_cell | *TEX14* | 2.036500682 | 0.007894122 | 0.033104823 | 1 |
| T2DM | t_cell | *ECHDC2* | -1.742820705 | 0.006748095 | 0.029478532 | 1 |
| T2DM | t_cell | *ZNF549* | 3.937613274 | 0.000531181 | 0.004823326 | 1 |
| T2DM | t_cell | *PDZRN3* | -1.689164808 | 3.65E-05 | 0.001641967 | 0.177332388 |
| T2DM | t_cell | *LHX4* | 4.736287516 | 0.00055267 | 0.00492933 | 1 |
| T2DM | t_cell | *CD80* | -1.666692306 | 8.90E-05 | 0.002066904 | 0.431982861 |
| T2DM | t_cell | *PILRB* | 0.787492877 | 0.003654287 | 0.018566074 | 1 |
| T2DM | t_cell | *FABP3* | -2.727437741 | 0.000182837 | 0.002738946 | 0.887126742 |
| T2DM | t_cell | *CPXM2* | -1.137444564 | 0.000933629 | 0.007125997 | 1 |
| T2DM | t_cell | *CD244* | 1.146255237 | 4.91E-05 | 0.00172419 | 0.238013149 |
| T2DM | t_cell | *OPN4* | 4.54648682 | 0.000660742 | 0.005604753 | 1 |
| T2DM | t_cell | *PMS2* | 3.588874245 | 0.001100865 | 0.008028975 | 1 |
| T2DM | t_cell | *GLIPR2* | 1.853905312 | 0.000192652 | 0.002815504 | 0.934747338 |
| T2DM | t_cell | *CCDC91* | 2.327173526 | 0.001425801 | 0.009661989 | 1 |
| T2DM | t_cell | *CENPK* | 0.887923923 | 0.000151467 | 0.002594722 | 0.7349163 |

| | | | | | | |
|---|---|---|---|---|---|---|
| T2DM | t_cell | *ARHGAP9* | 1.516527748 | 0.000146749 | 0.002594722 | 0.712026925 |
| T2DM | t_cell | *CDK2* | -1.750133646 | 0.008301954 | 0.034310972 | 1 |
| T2DM | t_cell | *DBH* | -29.10902088 | 0.00018219 | 0.002738946 | 0.883983875 |
| T2DM | t_cell | *C4BPA* | -1.733782993 | 0.006085571 | 0.027264257 | 1 |
| T2DM | t_cell | *DAW1* | -2.082010795 | 0.007360317 | 0.031354047 | 1 |
| T2DM | t_cell | *OBSL1* | -2.704595846 | 0.002461858 | 0.014104187 | 1 |
| T2DM | t_cell | *SLC12A4* | -2.308123916 | 0.00447339 | 0.021597996 | 1 |
| T2DM | t_cell | *SLC12A5* | -2.382572742 | 0.004330814 | 0.021225365 | 1 |
| T2DM | t_cell | *MATN4* | -3.336578348 | 9.96E-05 | 0.002149929 | 0.483177843 |
| T2DM | t_cell | *ATP8A1* | 1.956177447 | 5.62E-05 | 0.00172419 | 0.272745117 |
| T2DM | t_cell | *HIF3A* | -1.43925818 | 0.002216166 | 0.013222396 | 1 |
| T2DM | t_cell | *RIOK1* | 2.219506692 | 0.004574546 | 0.021932506 | 1 |
| T2DM | t_cell | *ATXN1* | 1.982525704 | 0.000930497 | 0.007125997 | 1 |
| T2DM | t_cell | *RUNX2* | 1.378436698 | 3.97E-05 | 0.001670757 | 0.192783415 |
| T2DM | t_cell | *SH3TC1* | -1.340331321 | 0.000857652 | 0.006709329 | 1 |
| T2DM | t_cell | *GOT2* | 4.537920703 | 0.000159464 | 0.002631693 | 0.77371787 |
| T2DM | t_cell | *CLYBL* | 2.862369579 | 0.000768032 | 0.006179917 | 1 |
| T2DM | t_cell | *GRK4* | 0.567635 | 0.003416383 | 0.017666275 | 1 |
| T2DM | t_cell | *HS3ST3B1* | 0.502632345 | 9.67E-05 | 0.00214103 | 0.469063476 |
| T2DM | t_cell | *MRPS7* | -2.400803137 | 0.013885416 | 0.04979456 | 1 |
| T2DM | t_cell | *OPRL1* | -3.502478039 | 0.000185943 | 0.002767476 | 0.902197313 |
| T2DM | t_cell | *IL1B* | 7.444351427 | 0.000224934 | 0.003025299 | 1 |
| T2DM | t_cell | *PSD4* | 1.029389432 | 8.73E-05 | 0.002066904 | 0.423348329 |
| T2DM | t_cell | *TNFSF9* | -15.62683864 | 0.001324137 | 0.009215829 | 1 |
| T2DM | t_cell | *SH2D3A* | 0.902793252 | 0.000182017 | 0.002738946 | 0.883148593 |
| T2DM | t_cell | *SDCBP2* | -2.298565689 | 0.00196356 | 0.012152034 | 1 |
| T2DM | t_cell | *DTD1* | 2.443580652 | 0.000744593 | 0.006082095 | 1 |
| T2DM | t_cell | *FLRT3* | -2.476155627 | 0.001316801 | 0.009206221 | 1 |
| T2DM | t_cell | *XRCC3* | -2.2822593 | 0.003224166 | 0.017033989 | 1 |
| T2DM | t_cell | *PRR12* | 2.242394784 | 0.011113589 | 0.042459162 | 1 |
| T2DM | t_cell | *TSKS* | -1.686106636 | 0.002440661 | 0.014065311 | 1 |
| T2DM | t_cell | *AIF1L* | -1.595919423 | 0.000313491 | 0.003472735 | 1 |
| T2DM | t_cell | *FGD3* | 0.805005372 | 0.000126121 | 0.002417859 | 0.611939747 |
| T2DM | t_cell | *ABHD8* | -2.575182444 | 6.74E-05 | 0.001838153 | 0.327191309 |
| T2DM | t_cell | *ATP13A4* | 1.588773161 | 3.48E-05 | 0.001630473 | 0.169059387 |
| T2DM | t_cell | *HELB* | 1.103571804 | 2.20E-05 | 0.001405861 | 0.106538079 |
| T2DM | t_cell | *RAB3IP* | 2.09028267 | 9.75E-05 | 0.00214103 | 0.473056337 |
| T2DM | t_cell | *YEATS4* | 1.161022974 | 0.008882963 | 0.035948087 | 1 |
| T2DM | t_cell | *KDR* | -2.041392462 | 0.013466809 | 0.048871322 | 1 |
| T2DM | t_cell | *CDC42EP1* | -2.388002442 | 0.001230265 | 0.008701194 | 1 |
| T2DM | t_cell | *OPN1SW* | -1.616960536 | 0.001109769 | 0.008036716 | 1 |
| T2DM | t_cell | *GAD1* | -2.801719928 | 0.000269777 | 0.003258353 | 1 |
| T2DM | t_cell | *CALML4* | -4.627223871 | 0.001405756 | 0.009586091 | 1 |
| T2DM | t_cell | *BBOX1* | 16.75134651 | 0.000813576 | 0.006460669 | 1 |
| T2DM | t_cell | *SERGEF* | 2.778137537 | 8.58E-05 | 0.002066904 | 0.41618522 |
| T2DM | t_cell | *PLD2* | -2.625157818 | 0.001423455 | 0.009659582 | 1 |
| T2DM | t_cell | *KLK8* | -7.923581581 | 0.000177708 | 0.002719991 | 0.862237056 |
| T2DM | t_cell | *ADCY4* | -1.264681524 | 0.013387473 | 0.048619775 | 1 |
| T2DM | t_cell | *SYT5* | 4.160217807 | 0.002440409 | 0.014065311 | 1 |
| T2DM | t_cell | *PLVAP* | -0.662089627 | 0.011911758 | 0.044561178 | 1 |

| | | | | | | |
|---|---|---|---|---|---|---|
| T2DM | t_cell | *ACTN4* | 2.398307403 | 0.01208777 | 0.044942423 | 1 |
| T2DM | t_cell | *SSBP4* | 2.155273034 | 0.000204111 | 0.002870576 | 0.990348798 |
| T2DM | t_cell | *TRPM4* | -1.405972506 | 0.010207535 | 0.039908914 | 1 |
| T2DM | t_cell | *PAK4* | -2.414337159 | 0.005550946 | 0.025626252 | 1 |
| T2DM | t_cell | *NPAS1* | -4.499265147 | 0.000207504 | 0.00289313 | 1 |
| T2DM | t_cell | *THEMIS2* | 1.710885917 | 0.003842176 | 0.019297555 | 1 |
| T2DM | t_cell | *DNMT1* | 3.457309031 | 0.000151163 | 0.002594722 | 0.733441515 |
| T2DM | t_cell | *C12orf65* | 3.49642266 | 2.22E-07 | 5.98E-05 | 0.001077202 |
| T2DM | t_cell | *PKDREJ* | -1.71099771 | 0.001689887 | 0.010804673 | 1 |
| T2DM | t_cell | *HSD17B3* | -3.759153929 | 0.001690179 | 0.010804673 | 1 |
| T2DM | t_cell | *ULBP2* | 9.34768671 | 0.01011876 | 0.039721865 | 1 |
| T2DM | t_cell | *TTLL9* | -3.784974062 | 0.004501472 | 0.021650583 | 1 |
| T2DM | t_cell | *IDO1* | -1.884757152 | 0.000253668 | 0.003155887 | 1 |
| T2DM | t_cell | *HAUS8* | -2.063508705 | 0.00347642 | 0.017868212 | 1 |
| T2DM | t_cell | *GALNT15* | -2.062513431 | 0.005661693 | 0.025940069 | 1 |
| T2DM | t_cell | *SLC6A6* | 2.899421121 | 0.000105602 | 0.002217626 | 0.512383197 |
| T2DM | t_cell | *PDLIM4* | -2.287517598 | 0.000720665 | 0.005916522 | 1 |
| T2DM | t_cell | *BARX1* | -4.260175789 | 0.000123147 | 0.002401423 | 0.597511006 |
| T2DM | t_cell | *PRKAB2* | 3.400092907 | 0.004277331 | 0.021026958 | 1 |
| T2DM | t_cell | *GCH1* | 2.015737656 | 0.00017369 | 0.002692473 | 0.842743958 |
| T2DM | t_cell | *MATN3* | -1.815707997 | 0.001446435 | 0.00970692 | 1 |
| T2DM | t_cell | *TRIM21* | 2.630760637 | 5.18E-05 | 0.00172419 | 0.251302339 |
| T2DM | t_cell | *TRIM22* | 2.656531599 | 0.00019521 | 0.002825716 | 0.947159429 |
| T2DM | t_cell | *REEP2* | -1.935977973 | 0.009577767 | 0.038060055 | 1 |
| T2DM | t_cell | *IGHMBP2* | -1.060696823 | 0.013659215 | 0.049411483 | 1 |
| T2DM | t_cell | *DMGDH* | -1.663893008 | 0.005633326 | 0.025834498 | 1 |
| T2DM | t_cell | *ALOX5AP* | 1.629427762 | 2.47E-05 | 0.001498556 | 0.119884519 |
| T2DM | t_cell | *WASF3* | -1.772588819 | 0.004657832 | 0.022222029 | 1 |
| T2DM | t_cell | *CSNK1G2* | 3.754136281 | 0.000285902 | 0.003332537 | 1 |
| T2DM | t_cell | *FAM83F* | -5.29362669 | 0.001203383 | 0.008561311 | 1 |
| T2DM | t_cell | *GIMAP4* | 1.246848902 | 0.000704471 | 0.005832921 | 1 |
| T2DM | t_cell | *SPINK5* | -3.334955733 | 0.003155334 | 0.016732472 | 1 |
| T2DM | t_cell | *CATSPERB* | 1.722719423 | 0.001544416 | 0.010126363 | 1 |
| T2DM | t_cell | *COX16* | -2.971291264 | 0.000235351 | 0.003053273 | 1 |
| T2DM | t_cell | *TTC9* | 1.227315673 | 4.74E-05 | 0.001717271 | 0.23011435 |
| T2DM | t_cell | *PEBP4* | -2.731403397 | 0.002458408 | 0.014104187 | 1 |
| T2DM | t_cell | *MRO* | -2.161243525 | 0.00431952 | 0.021191415 | 1 |
| T2DM | t_cell | *CNTN6* | -1.806323719 | 0.000459063 | 0.004354585 | 1 |
| T2DM | t_cell | *CHL1* | -1.479426026 | 0.002557709 | 0.014447035 | 1 |
| T2DM | t_cell | *TRIM45* | -2.911483739 | 0.012385139 | 0.045784395 | 1 |
| T2DM | t_cell | *FKBP11* | 4.725577019 | 0.000245353 | 0.003100135 | 1 |
| T2DM | t_cell | *KLRD1* | 2.366520981 | 6.01E-05 | 0.001745457 | 0.2914525 |
| T2DM | t_cell | *TIMM10* | 2.743656484 | 0.005585309 | 0.025703381 | 1 |
| T2DM | t_cell | *FADS2* | -2.286466354 | 0.003103753 | 0.016634892 | 1 |
| T2DM | t_cell | *KLB* | -0.996781481 | 0.002011573 | 0.012354622 | 1 |
| T2DM | t_cell | *OSTF1* | 2.821857141 | 0.00024992 | 0.003134276 | 1 |
| T2DM | t_cell | *ADAM19* | 0.844393008 | 9.98E-06 | 0.000931216 | 0.048423252 |
| T2DM | t_cell | *MSI1* | -1.775938852 | 0.003273821 | 0.017191102 | 1 |
| T2DM | t_cell | *P2RX4* | 2.072915598 | 0.001119785 | 0.008061126 | 1 |
| T2DM | t_cell | *DTX1* | -1.916288381 | 0.001600392 | 0.010354591 | 1 |

| | | | | | | |
|---|---|---|---|---|---|---|
| T2DM | t_cell | *TMEM60* | 4.35807354 | 6.96E-05 | 0.001874811 | 0.337465909 |
| T2DM | t_cell | *POM121C* | -2.250807479 | 0.006614094 | 0.028989687 | 1 |
| T2DM | t_cell | *B4GALNT1* | -3.252931431 | 0.007059971 | 0.030503097 | 1 |
| T2DM | t_cell | *FAIM2* | -2.307386572 | 0.000226543 | 0.003033461 | 1 |
| T2DM | t_cell | *PAN2* | -2.465122102 | 0.007206993 | 0.030950274 | 1 |
| T2DM | t_cell | *MICAL1* | 1.971893861 | 3.17E-06 | 0.000452339 | 0.015379512 |
| T2DM | t_cell | *SMYD5* | -1.790095811 | 0.002983163 | 0.016190502 | 1 |
| T2DM | t_cell | *USP15* | 2.277933897 | 0.003587841 | 0.018305159 | 1 |
| T2DM | t_cell | *CAPN9* | -3.291770329 | 0.000208913 | 0.002904432 | 1 |
| T2DM | t_cell | *SP110* | 2.386475733 | 0.000717545 | 0.005910914 | 1 |
| T2DM | t_cell | *MRPL44* | 4.560128812 | 0.000113012 | 0.002285106 | 0.548334725 |
| T2DM | t_cell | *ANKRD36* | 2.681558216 | 0.008944459 | 0.036075887 | 1 |
| T2DM | t_cell | *STAB2* | -2.394045791 | 0.007399553 | 0.031438382 | 1 |
| T2DM | t_cell | *RNASEH2B* | 2.041483498 | 0.009146481 | 0.036706968 | 1 |
| T2DM | t_cell | *SETDB2* | 3.026921195 | 0.001071366 | 0.00784548 | 1 |
| T2DM | t_cell | *SCRN1* | -2.006722001 | 0.002195509 | 0.013164187 | 1 |
| T2DM | t_cell | *CHST12* | 2.697961885 | 0.000623773 | 0.005394914 | 1 |
| T2DM | t_cell | *AOAH* | 0.726740687 | 0.000236086 | 0.003054636 | 1 |
| T2DM | t_cell | *TBRG4* | -3.020080556 | 0.001445774 | 0.00970692 | 1 |
| T2DM | t_cell | *BRIP1* | 1.409621766 | 0.010192201 | 0.039881096 | 1 |
| T2DM | t_cell | *RTP4* | -3.329151108 | 0.000441888 | 0.004262504 | 1 |
| T2DM | t_cell | *IMP4* | -3.073464664 | 0.012806486 | 0.046942469 | 1 |
| T2DM | t_cell | *GMPR* | -2.237900793 | 0.002763047 | 0.015409544 | 1 |
| T2DM | t_cell | *FRS3* | -2.028771774 | 0.000860099 | 0.006709329 | 1 |
| T2DM | t_cell | *IRF4* | 1.022678683 | 1.05E-05 | 0.000939573 | 0.050736928 |
| T2DM | t_cell | *TCF19* | 2.442292973 | 7.51E-05 | 0.00194078 | 0.36418045 |
| T2DM | t_cell | *NRM* | -4.154007238 | 0.000330358 | 0.003569928 | 1 |
| T2DM | t_cell | *SYTL2* | 0.767372519 | 0.013833486 | 0.049784376 | 1 |
| T2DM | t_cell | *RAB30* | -2.198912912 | 0.007208122 | 0.030950274 | 1 |
| T2DM | t_cell | *GGH* | -0.938481106 | 0.007928772 | 0.03322142 | 1 |
| T2DM | t_cell | *FXYD6* | -1.154496435 | 0.00930129 | 0.037113371 | 1 |
| T2DM | t_cell | *NUSAP1* | 3.123982967 | 3.54E-06 | 0.000470882 | 0.017161783 |
| T2DM | t_cell | *SPTBN5* | 0.556902667 | 0.000434804 | 0.004202526 | 1 |
| T2DM | t_cell | *KHK* | -1.899349196 | 0.000247704 | 0.00312172 | 1 |
| T2DM | t_cell | *DYNC2LI1* | -1.615034116 | 0.010308095 | 0.04021568 | 1 |
| T2DM | t_cell | *LHCGR* | -2.403747003 | 0.00028936 | 0.003334858 | 1 |
| T2DM | t_cell | *ACTR2* | 2.649665273 | 0.002149695 | 0.012983945 | 1 |
| T2DM | t_cell | *BTBD16* | -3.179611431 | 0.000419432 | 0.004119601 | 1 |
| T2DM | t_cell | *STAT4* | 0.554454107 | 5.69E-05 | 0.00172419 | 0.276203888 |
| T2DM | t_cell | *SENP7* | 2.203354794 | 0.012987868 | 0.047488421 | 1 |
| T2DM | t_cell | *PRKG2* | 1.242841451 | 0.000567891 | 0.005028115 | 1 |
| T2DM | t_cell | *BMPR1B* | -1.469079479 | 0.010943368 | 0.041874779 | 1 |
| T2DM | t_cell | *MMRN1* | -1.206348722 | 0.000987076 | 0.00741048 | 1 |
| T2DM | t_cell | *PRDM5* | -1.867320301 | 0.013104606 | 0.047735398 | 1 |
| T2DM | t_cell | *TRPC3* | 2.318964255 | 1.11E-06 | 0.000213354 | 0.005361633 |
| T2DM | t_cell | *NAAA* | 2.330720908 | 0.000309338 | 0.003445826 | 1 |
| T2DM | t_cell | *CNOT6L* | 1.941699407 | 0.00450682 | 0.021650583 | 1 |
| T2DM | t_cell | *PARVG* | 1.294122422 | 0.000223543 | 0.003025299 | 1 |
| T2DM | t_cell | *B4GALNT3* | 1.089987015 | 6.85E-05 | 0.001857146 | 0.332429119 |
| T2DM | t_cell | *PDE6H* | 2.967915313 | 1.93E-05 | 0.001374162 | 0.093443034 |

| | | | | | | |
|---|---|---|---|---|---|---|
| T2DM | t_cell | *ERP27* | 0.998951609 | 0.000423196 | 0.004123191 | 1 |
| T2DM | t_cell | *KIF21A* | 2.537992364 | 0.006017045 | 0.027132621 | 1 |
| T2DM | t_cell | *ALG10* | -1.711919929 | 0.003899362 | 0.019484763 | 1 |
| T2DM | t_cell | *CLSTN3* | 3.725135172 | 0.000658306 | 0.005604753 | 1 |
| T2DM | t_cell | *KLRG1* | 0.521908832 | 0.003154881 | 0.016732472 | 1 |
| T2DM | t_cell | *VAMP1* | 2.211923842 | 0.000315576 | 0.003479946 | 1 |
| T2DM | t_cell | *PPFIA2* | -2.035726574 | 0.011721262 | 0.044018239 | 1 |
| T2DM | t_cell | *MARCH9* | 4.585195459 | 0.000128275 | 0.002427073 | 0.622390034 |
| T2DM | t_cell | *POC1B* | 3.067809243 | 0.001627086 | 0.010470317 | 1 |
| T2DM | t_cell | *RITA1* | 6.7891216 | 0.00602275 | 0.027133133 | 1 |
| T2DM | t_cell | *GLTP* | 4.545029888 | 0.006312216 | 0.02801018 | 1 |
| T2DM | t_cell | *FOXN4* | -1.365306836 | 0.006610368 | 0.028989687 | 1 |
| T2DM | t_cell | *MTMR6* | 3.106846246 | 0.012402642 | 0.045784395 | 1 |
| T2DM | t_cell | *KANSL2* | 2.278115588 | 0.010712141 | 0.041381613 | 1 |
| T2DM | t_cell | *HNRNPA1L2* | -3.241087059 | 0.000454895 | 0.004336245 | 1 |
| T2DM | t_cell | *RAB15* | 4.651790324 | 0.006904892 | 0.03007409 | 1 |
| T2DM | t_cell | *NEIL1* | -3.05240789 | 3.59E-06 | 0.000470882 | 0.017422626 |
| T2DM | t_cell | *MAN2C1* | -1.117917425 | 3.69E-06 | 0.000471074 | 0.017900794 |
| T2DM | t_cell | *PIF1* | -2.586190523 | 0.000341816 | 0.003638727 | 1 |
| T2DM | t_cell | *ULK3* | 4.654911168 | 0.001982089 | 0.012235491 | 1 |
| T2DM | t_cell | *PCSK6* | -2.017816767 | 0.002387057 | 0.013854068 | 1 |
| T2DM | t_cell | *POLG* | 2.339953163 | 0.008892716 | 0.035956215 | 1 |
| T2DM | t_cell | *ITGAX* | 3.198186237 | 2.50E-05 | 0.001500229 | 0.121518518 |
| T2DM | t_cell | *TGFB1I1* | -3.140290268 | 0.011270149 | 0.042922106 | 1 |
| T2DM | t_cell | *GCSH* | -2.904494498 | 6.88E-06 | 0.000758992 | 0.033395638 |
| T2DM | t_cell | *CMTM3* | 3.34996796 | 0.000593974 | 0.0051899 | 1 |
| T2DM | t_cell | *TLDC1* | 2.98537805 | 7.87E-05 | 0.00197835 | 0.381821539 |
| T2DM | t_cell | *MYO19* | 3.868218747 | 0.00028777 | 0.003332537 | 1 |
| T2DM | t_cell | *RASL10B* | -3.970237174 | 0.000485495 | 0.004530042 | 1 |
| T2DM | t_cell | *UNC45B* | 2.449775535 | 0.000326095 | 0.003565777 | 1 |
| T2DM | t_cell | *LRRC46* | -1.985054133 | 0.000276575 | 0.003278465 | 1 |
| T2DM | t_cell | *SSH2* | 2.081388065 | 0.004266771 | 0.020996323 | 1 |
| T2DM | t_cell | *RHBDL3* | -1.832513838 | 0.000792 | 0.006330781 | 1 |
| T2DM | t_cell | *G6PC3* | -3.387390737 | 0.001150573 | 0.008233896 | 1 |
| T2DM | t_cell | *GNAL* | -1.984843094 | 0.003845988 | 0.019297555 | 1 |
| T2DM | t_cell | *SLC14A1* | 0.862553892 | 0.009458554 | 0.037617135 | 1 |
| T2DM | t_cell | *TMC6* | 1.200580006 | 8.68E-06 | 0.000878186 | 0.042115999 |
| T2DM | t_cell | *TBCD* | 2.571694587 | 0.004355243 | 0.021254145 | 1 |
| T2DM | t_cell | *FN3KRP* | -0.965636049 | 0.001464503 | 0.009774098 | 1 |
| T2DM | t_cell | *RNF157* | 0.965164164 | 0.002443746 | 0.014065311 | 1 |
| T2DM | t_cell | *ZNF750* | -2.621265469 | 0.003518288 | 0.018064268 | 1 |
| T2DM | t_cell | *MAPK4* | -2.494859961 | 0.000173259 | 0.002692473 | 0.840651382 |
| T2DM | t_cell | *FBXO15* | -1.749170927 | 0.000452393 | 0.00432089 | 1 |
| T2DM | t_cell | *ERBB2* | -2.016671884 | 0.012554435 | 0.046127496 | 1 |
| T2DM | t_cell | *CACNA1A* | -4.653989835 | 0.00117174 | 0.008373023 | 1 |
| T2DM | t_cell | *TMEM91* | -1.058525246 | 0.002855427 | 0.015786085 | 1 |
| T2DM | t_cell | *HUNK* | -0.718979585 | 0.000819016 | 0.006493248 | 1 |
| T2DM | t_cell | *COL6A1* | -0.898171933 | 0.002104247 | 0.012762975 | 1 |
| T2DM | t_cell | *SLC47A1* | -1.4555163 | 0.003563254 | 0.018237244 | 1 |
| T2DM | t_cell | *ARHGEF19* | -0.456773174 | 0.011258539 | 0.042911571 | 1 |

| | | | | | | |
|---|---|---|---|---|---|---|
| T2DM | t_cell | *ZNF593* | -4.500237708 | 0.000271306 | 0.003258353 | 1 |
| T2DM | t_cell | *EVA1B* | -1.614222592 | 0.011860909 | 0.044439482 | 1 |
| T2DM | t_cell | *PLK4* | 2.138243688 | 0.012130577 | 0.045067045 | 1 |
| T2DM | t_cell | *RPS8* | 2.618153856 | 0.002372878 | 0.01378827 | 1 |
| T2DM | t_cell | *XCL1* | 0.704316859 | 7.64E-05 | 0.00194078 | 0.370688984 |
| T2DM | t_cell | *XCL2* | 1.197020484 | 0.000696256 | 0.005777617 | 1 |
| T2DM | t_cell | *MAEL* | -4.702371724 | 0.000406925 | 0.004078878 | 1 |
| T2DM | t_cell | *SEMA6C* | -2.473869362 | 0.013878546 | 0.04979456 | 1 |
| T2DM | t_cell | *DYRK3* | -2.354419476 | 0.004229965 | 0.020866389 | 1 |
| T2DM | t_cell | *SMYD2* | 3.418286253 | 0.000258476 | 0.003195379 | 1 |
| T2DM | t_cell | *CREB3L4* | -2.009283726 | 0.000106022 | 0.002217626 | 0.514416634 |
| T2DM | t_cell | *PYCR2* | 2.279307979 | 0.00373174 | 0.018860836 | 1 |
| T2DM | t_cell | *OSR1* | -1.100156039 | 0.008270918 | 0.034211843 | 1 |
| T2DM | t_cell | *GALM* | 3.029791554 | 0.000232633 | 0.003042414 | 1 |
| T2DM | t_cell | *ABHD1* | -3.277629091 | 4.20E-05 | 0.001670757 | 0.203832305 |
| T2DM | t_cell | *ZNF514* | -2.036532128 | 0.003418927 | 0.017666275 | 1 |
| T2DM | t_cell | *TEX261* | -2.946308905 | 0.001500921 | 0.009948729 | 1 |
| T2DM | t_cell | *CNGA3* | -21.14318185 | 3.18E-05 | 0.001619427 | 0.15444769 |
| T2DM | t_cell | *GALNT13* | -2.05617394 | 0.0058893 | 0.0267055 | 1 |
| T2DM | t_cell | *PHOSPHO2* | -2.464771438 | 0.000258339 | 0.003195379 | 1 |
| T2DM | t_cell | *METTL21A* | 3.226499981 | 0.001419877 | 0.009648798 | 1 |
| T2DM | t_cell | *ESPNL* | 22.49952695 | 2.80E-05 | 0.001541761 | 0.135674993 |
| T2DM | t_cell | *ANKMY1* | 2.271117056 | 0.002340221 | 0.013698845 | 1 |
| T2DM | t_cell | *TAMM41* | 2.634686118 | 0.012023585 | 0.044807806 | 1 |
| T2DM | t_cell | *STK11IP* | -2.388407209 | 0.012392659 | 0.045784395 | 1 |
| T2DM | t_cell | *EAF1* | 3.94166024 | 8.23E-05 | 0.002006827 | 0.399358563 |
| T2DM | t_cell | *CNTN4* | -1.641349314 | 0.011683649 | 0.043979103 | 1 |
| T2DM | t_cell | *CTDSPL* | -1.871217047 | 0.005344742 | 0.024858025 | 1 |
| T2DM | t_cell | *ADPRH* | -1.492413659 | 0.001325539 | 0.009215829 | 1 |
| T2DM | t_cell | *MED12L* | 3.470502751 | 0.010084456 | 0.03965136 | 1 |
| T2DM | t_cell | *MUC4* | -2.253256121 | 0.000528219 | 0.004823326 | 1 |
| T2DM | t_cell | *CISD2* | 4.339896774 | 0.002791925 | 0.01551709 | 1 |
| T2DM | t_cell | *DDIT4L* | -2.265342347 | 0.00533775 | 0.024858025 | 1 |
| T2DM | t_cell | *MYO10* | -1.7612939 | 0.002441191 | 0.014065311 | 1 |
| T2DM | t_cell | *BHMT* | -1.16895696 | 7.58E-05 | 0.00194078 | 0.367877156 |
| T2DM | t_cell | *SPATA9* | -2.028596554 | 0.005588843 | 0.025703381 | 1 |
| T2DM | t_cell | *GFOD1* | -1.733965819 | 0.005336451 | 0.024858025 | 1 |
| T2DM | t_cell | *TRIM7* | -1.784474598 | 0.010734582 | 0.041435314 | 1 |
| T2DM | t_cell | *RNF44* | 2.593208178 | 0.000130939 | 0.00244943 | 0.635316603 |
| T2DM | t_cell | *MLIP* | -1.434060405 | 0.003310816 | 0.017347816 | 1 |
| T2DM | t_cell | *TPBG* | -1.673259504 | 0.001135852 | 0.008140555 | 1 |
| T2DM | t_cell | *ABRACL* | 1.225619759 | 0.003932426 | 0.01960959 | 1 |
| T2DM | t_cell | *EPHA1* | 8.557491067 | 0.000113031 | 0.002285106 | 0.548425551 |
| T2DM | t_cell | *ADHFE1* | -0.735431878 | 0.003360541 | 0.017476255 | 1 |
| T2DM | t_cell | *DPYS* | -1.535536512 | 0.001332069 | 0.009219968 | 1 |
| T2DM | t_cell | *SEC16A* | 3.664204575 | 0.00106807 | 0.00784548 | 1 |
| T2DM | t_cell | *SLC39A12* | -2.653576835 | 0.001969453 | 0.012172976 | 1 |
| T2DM | t_cell | *ST8SIA6* | 4.906078374 | 5.46E-05 | 0.00172419 | 0.264931235 |
| T2DM | t_cell | *CDHR1* | 1.080081971 | 0.000381408 | 0.003943314 | 1 |
| T2DM | t_cell | *RGR* | -9.292880847 | 0.000231344 | 0.003040666 | 1 |

| | | | | | | |
|---|---|---|---|---|---|---|
| T2DM | t_cell | *PAOX* | 5.030537614 | 0.00075638 | 0.006116593 | 1 |
| T2DM | t_cell | *RGS10* | 3.276179643 | 0.000323571 | 0.00355196 | 1 |
| T2DM | t_cell | *HSD17B12* | -1.628073353 | 9.48E-10 | 7.67E-07 | 4.60E-06 |
| T2DM | t_cell | *APIP* | -0.905060158 | 0.000352386 | 0.003741311 | 1 |
| T2DM | t_cell | *PTPRJ* | 2.373459351 | 0.000124245 | 0.002401749 | 0.602838987 |
| T2DM | t_cell | *RPS3* | 2.245422555 | 0.006044608 | 0.02718053 | 1 |
| T2DM | t_cell | *TTC12* | 1.59777538 | 0.006978635 | 0.03030424 | 1 |
| T2DM | t_cell | *DRD2* | -18.01122306 | 0.000339631 | 0.003638727 | 1 |
| T2DM | t_cell | *NPAT* | 3.066023248 | 0.006265069 | 0.027837101 | 1 |
| T2DM | t_cell | *ATM* | 2.035080145 | 0.001190712 | 0.008483604 | 1 |
| T2DM | t_cell | *TMC2* | -5.05599655 | 0.00058796 | 0.005158734 | 1 |
| T2DM | t_cell | *INCENP* | -3.875899337 | 0.008231031 | 0.034105004 | 1 |
| T2DM | t_cell | *PLCH2* | 0.904764295 | 6.33E-05 | 0.001756372 | 0.306892457 |
| T2DM | t_cell | *CCDC15* | -1.929111748 | 0.000296418 | 0.003360332 | 1 |
| T2DM | t_cell | *CHEK1* | 3.843677313 | 4.60E-05 | 0.001701031 | 0.22324274 |
| T2DM | t_cell | *FAM124A* | -2.057115809 | 0.002816173 | 0.015633949 | 1 |
| T2DM | t_cell | *WDR17* | -1.96497896 | 0.000611783 | 0.005329212 | 1 |
| T2DM | t_cell | *PPP1R1C* | -2.483762094 | 0.000277034 | 0.003278465 | 1 |
| T2DM | t_cell | *GPR158* | -1.476746791 | 0.000613371 | 0.005333471 | 1 |
| T2DM | t_cell | *ANK3* | 0.596721701 | 2.84E-05 | 0.001547767 | 0.137751271 |
| T2DM | t_cell | *MFSD6* | 2.769504215 | 0.00020733 | 0.00289313 | 1 |
| T2DM | t_cell | *PARP8* | 0.830607955 | 0.000559283 | 0.004970041 | 1 |
| T2DM | t_cell | *BEND6* | -1.505987434 | 0.002009165 | 0.012354622 | 1 |
| T2DM | t_cell | *TMEM56* | -4.78553497 | 0.003226347 | 0.017033989 | 1 |
| T2DM | t_cell | *MGAT5* | 1.868140629 | 0.00572802 | 0.026169824 | 1 |
| T2DM | t_cell | *POC5* | -1.53636462 | 0.003109924 | 0.016634892 | 1 |
| T2DM | t_cell | *CWF19L2* | 2.2191612 | 0.010398131 | 0.040458485 | 1 |
| T2DM | t_cell | *MBNL1* | 1.797149339 | 0.000287593 | 0.003332537 | 1 |
| T2DM | t_cell | *CCNO* | 7.072879361 | 6.07E-05 | 0.001745457 | 0.294441059 |
| T2DM | t_cell | *PRDM8* | 1.265502548 | 0.00024308 | 0.003087496 | 1 |
| T2DM | t_cell | *GGPS1* | 3.151962966 | 0.000686544 | 0.005725595 | 1 |
| T2DM | t_cell | *JAKMIP1* | 0.49651703 | 6.13E-05 | 0.001745457 | 0.297594024 |
| T2DM | t_cell | *MR1* | 3.187426392 | 0.000418209 | 0.004115925 | 1 |
| T2DM | t_cell | *SYCP2L* | -1.790693698 | 0.000539249 | 0.004872321 | 1 |
| T2DM | t_cell | *TMEM87B* | 2.777631704 | 0.010537834 | 0.04087096 | 1 |
| T2DM | t_cell | *LPCAT1* | 3.372999248 | 6.15E-05 | 0.001745457 | 0.298473189 |
| T2DM | t_cell | *NMRAL1* | 1.952385076 | 0.001005473 | 0.007470988 | 1 |
| T2DM | t_cell | *JAZF1* | -2.85421505 | 1.18E-05 | 0.001006325 | 0.057360506 |
| T2DM | t_cell | *MCOLN2* | 0.836311157 | 8.27E-05 | 0.002006828 | 0.401365683 |
| T2DM | t_cell | *SEMA3D* | -1.553571037 | 0.002688831 | 0.015064908 | 1 |
| T2DM | t_cell | *ANKRD29* | -1.852088349 | 0.004231766 | 0.020866389 | 1 |
| T2DM | t_cell | *ANKH* | 2.857715619 | 0.001258497 | 0.008860305 | 1 |
| T2DM | t_cell | *UBASH3B* | 1.012466088 | 0.001371585 | 0.009439616 | 1 |
| T2DM | t_cell | *ROBO4* | -2.003316435 | 0.00419368 | 0.020762994 | 1 |
| T2DM | t_cell | *PRKCA* | 2.366637971 | 0.005503094 | 0.025429535 | 1 |
| T2DM | t_cell | *PGM5* | -1.578533185 | 0.01240859 | 0.045784395 | 1 |
| T2DM | t_cell | *CCSAP* | 2.947745818 | 5.28E-05 | 0.00172419 | 0.256122596 |
| T2DM | t_cell | *GBP5* | 0.432200856 | 0.000362222 | 0.003779571 | 1 |
| T2DM | t_cell | *PDE1C* | -1.910381618 | 0.004874914 | 0.02303124 | 1 |
| T2DM | t_cell | *OXNAD1* | 1.851975069 | 5.72E-05 | 0.00172419 | 0.277292207 |

| | | | | | | |
|---|---|---|---|---|---|---|
| T2DM | t_cell | *PROM2* | -2.532296795 | 0.004035691 | 0.020062676 | 1 |
| T2DM | t_cell | *MARVELD1* | -2.635984537 | 0.002866999 | 0.015789646 | 1 |
| T2DM | t_cell | *GOLGA7B* | 2.605181069 | 5.58E-05 | 0.00172419 | 0.270843206 |
| T2DM | t_cell | *MOV10* | -3.228461231 | 0.00439734 | 0.021400096 | 1 |
| T2DM | t_cell | *SETD9* | -0.895598802 | 0.006335533 | 0.028073066 | 1 |
| T2DM | t_cell | *KDM8* | 3.995829461 | 0.007864665 | 0.033038402 | 1 |
| T2DM | t_cell | *RRAGA* | 5.129494871 | 0.000827158 | 0.006525809 | 1 |
| T2DM | t_cell | *NAA11* | -2.852376035 | 0.000143143 | 0.002562837 | 0.694528761 |
| T2DM | t_cell | *TDRD9* | -6.627095989 | 0.000413718 | 0.004112304 | 1 |
| T2DM | t_cell | *PCDH1* | -0.823482154 | 0.002066165 | 0.012578458 | 1 |
| T2DM | t_cell | *RPL30* | 2.10038144 | 0.007025213 | 0.030434225 | 1 |
| T2DM | t_cell | *HKDC1* | 1.26101727 | 0.000123239 | 0.002401423 | 0.597954302 |
| T2DM | t_cell | *NODAL* | -2.205569711 | 0.000239623 | 0.003068607 | 1 |
| T2DM | t_cell | *ATAD2* | 2.576234845 | 4.61E-05 | 0.001701031 | 0.223655223 |
| T2DM | t_cell | *ITGAD* | 0.75069287 | 0.000128557 | 0.002427073 | 0.623757735 |
| T2DM | t_cell | *VPS8* | 2.652127121 | 0.001735936 | 0.010981438 | 1 |
| T2DM | t_cell | *MPV17L* | -1.936211701 | 0.013789674 | 0.049687903 | 1 |
| T2DM | t_cell | *NMNAT2* | -2.502255014 | 0.00759108 | 0.032136075 | 1 |
| T2DM | t_cell | *SLC6A1* | -4.592960211 | 0.000521127 | 0.004797926 | 1 |
| T2DM | t_cell | *MMP14* | -2.729725142 | 0.008574808 | 0.035198788 | 1 |
| T2DM | t_cell | *CLEC18A* | -2.287027772 | 1.71E-06 | 0.000296183 | 0.008293137 |
| T2DM | t_cell | *SLC34A2* | 7.963442585 | 6.25E-05 | 0.001756372 | 0.303090924 |
| T2DM | t_cell | *C12orf43* | -3.019780171 | 0.001697316 | 0.010836023 | 1 |
| T2DM | t_cell | *PEX10* | -2.541412496 | 0.002962811 | 0.016135621 | 1 |
| T2DM | t_cell | *KRTCAP3* | 1.814897239 | 1.64E-05 | 0.00124438 | 0.079640316 |
| T2DM | t_cell | *EXTL1* | -16.36037207 | 0.000167042 | 0.002679155 | 0.810485538 |
| T2DM | t_cell | *WDR66* | -1.425821896 | 0.00761755 | 0.032195429 | 1 |
| T2DM | t_cell | *RHPN1* | 1.397740844 | 0.011178637 | 0.042674074 | 1 |
| T2DM | t_cell | *LRRC43* | -2.747729452 | 0.002345859 | 0.013698845 | 1 |
| T2DM | t_cell | *CLSTN2* | -1.697868617 | 0.002058259 | 0.012577675 | 1 |
| T2DM | t_cell | *RNF166* | 0.971695025 | 0.001460667 | 0.009774098 | 1 |
| T2DM | t_cell | *CDC42SE2* | 1.107978351 | 8.15E-05 | 0.002006241 | 0.395360418 |
| T2DM | t_cell | *IFNAR2* | 1.086331099 | 0.013722081 | 0.049538347 | 1 |
| T2DM | t_cell | *ACE* | -2.31461261 | 0.00096896 | 0.007311654 | 1 |
| T2DM | t_cell | *TEPP* | 4.925469885 | 0.006749928 | 0.029478532 | 1 |
| T2DM | t_cell | *ZFYVE28* | 1.157652106 | 0.000464338 | 0.004358847 | 1 |
| T2DM | t_cell | *C16orf86* | -1.266317834 | 0.011345572 | 0.043051925 | 1 |
| T2DM | t_cell | *LYPD5* | -2.294522672 | 7.50E-05 | 0.00194078 | 0.363983057 |
| T2DM | t_cell | *CCDC107* | 2.406942439 | 0.002726597 | 0.015241301 | 1 |
| T2DM | t_cell | *SLC37A1* | 3.525240412 | 0.000942942 | 0.007152552 | 1 |
| T2DM | t_cell | *HSF2BP* | -2.025352915 | 0.004924626 | 0.023220882 | 1 |
| T2DM | t_cell | *CACFD1* | -3.568806045 | 0.000328504 | 0.003565777 | 1 |
| T2DM | t_cell | *PAQR6* | -1.24686951 | 0.013868057 | 0.04979456 | 1 |
| T2DM | t_cell | *CCR5* | 1.062695023 | 8.61E-06 | 0.000878186 | 0.041764353 |
| T2DM | t_cell | *NBEAL2* | 1.86370488 | 0.000355774 | 0.003760818 | 1 |
| T2DM | t_cell | *FCRL3* | 0.640745292 | 0.003811044 | 0.019181728 | 1 |
| T2DM | t_cell | *FGFR4* | -4.59524125 | 0.000163808 | 0.00266157 | 0.794798342 |
| T2DM | t_cell | *VPS28* | 2.405955826 | 0.000937152 | 0.007138241 | 1 |
| T2DM | t_cell | *DVL3* | -2.174531049 | 0.005014763 | 0.02360003 | 1 |
| T2DM | t_cell | *BDH1* | 2.613121677 | 0.000875361 | 0.006795599 | 1 |

| | | | | | | |
|---|---|---|---|---|---|---|
| T2DM | t_cell | *PLCD3* | -2.595543777 | 0.000887884 | 0.006881812 | 1 |
| T2DM | t_cell | *IP6K3* | -15.60001011 | 1.03E-05 | 0.000939573 | 0.050184101 |
| T2DM | t_cell | *ASGR2* | -7.192557599 | 0.000288907 | 0.003334858 | 1 |
| T2DM | t_cell | *POLR3K* | 4.116453911 | 0.000540552 | 0.004875015 | 1 |
| T2DM | t_cell | *WDR90* | -1.734666009 | 0.005694721 | 0.026066779 | 1 |
| T2DM | t_cell | *SHANK2* | -1.794122623 | 0.002471837 | 0.014143107 | 1 |
| T2DM | t_cell | *TPCN2* | -1.66484769 | 0.008768997 | 0.035731935 | 1 |
| T2DM | t_cell | *PDZK1IP1* | 6.951169742 | 0.004341764 | 0.021226528 | 1 |
| T2DM | t_cell | *SLC1A7* | -0.843505643 | 0.011483011 | 0.043426007 | 1 |
| T2DM | t_cell | *C1orf123* | -3.152653924 | 0.005598593 | 0.025720366 | 1 |
| T2DM | t_cell | *ZSWIM5* | 2.388496938 | 0.002349019 | 0.013698845 | 1 |
| T2DM | t_cell | *SLC25A34* | -2.102379929 | 0.000273312 | 0.003274345 | 1 |
| T2DM | t_cell | *PDPN* | -1.684880897 | 0.000674515 | 0.005681849 | 1 |
| T2DM | t_cell | *LAPTM5* | 3.388053947 | 0.000375084 | 0.003888694 | 1 |
| T2DM | t_cell | *OMA1* | 2.60198119 | 0.009282699 | 0.037106681 | 1 |
| T2DM | t_cell | *HFM1* | -1.666481571 | 0.000686788 | 0.005725595 | 1 |
| T2DM | t_cell | *FCRLB* | 4.823407719 | 0.000413857 | 0.004112304 | 1 |
| T2DM | t_cell | *KLHDC9* | -0.900532979 | 0.010147254 | 0.039769368 | 1 |
| T2DM | t_cell | *TDRD5* | -2.748385665 | 0.000299515 | 0.003387523 | 1 |
| T2DM | t_cell | *HAAO* | -2.401751354 | 0.006171406 | 0.027521749 | 1 |
| T2DM | t_cell | *KCNJ3* | -3.499777412 | 0.009672079 | 0.038403377 | 1 |
| T2DM | t_cell | *DUSP19* | -1.964269463 | 0.000720584 | 0.005916522 | 1 |
| T2DM | t_cell | *C2orf48* | -8.875446404 | 0.000785168 | 0.00628653 | 1 |
| T2DM | t_cell | *INHBB* | -2.78119655 | 0.001442985 | 0.00970692 | 1 |
| T2DM | t_cell | *LYSMD1* | -0.95281753 | 0.013879973 | 0.04979456 | 1 |
| T2DM | t_cell | *RNF149* | 2.232340488 | 0.007764361 | 0.032701978 | 1 |
| T2DM | t_cell | *TDRD10* | -1.87955544 | 0.002856592 | 0.015786085 | 1 |
| T2DM | t_cell | *NIPAL1* | -1.446113126 | 0.003643155 | 0.018548364 | 1 |
| T2DM | t_cell | *PBXIP1* | 2.019776183 | 3.13E-05 | 0.001619427 | 0.15176192 |
| T2DM | t_cell | *CLDN1* | -1.285182133 | 0.013594851 | 0.049262297 | 1 |
| T2DM | t_cell | *DCST2* | 3.174387682 | 0.000765621 | 0.006171919 | 1 |
| T2DM | t_cell | *LINC01116* | -1.854802169 | 0.007596883 | 0.032136075 | 1 |
| T2DM | t_cell | *SLC22A15* | -1.725886275 | 0.011290822 | 0.042967114 | 1 |
| T2DM | t_cell | *LRRC58* | 3.815645602 | 0.000941008 | 0.007152552 | 1 |
| T2DM | t_cell | *LMOD1* | -1.75067311 | 0.000202142 | 0.00285945 | 0.980791251 |
| T2DM | t_cell | *ARPC2* | 2.231913199 | 0.003231902 | 0.017044771 | 1 |
| T2DM | t_cell | *SPTA1* | -1.614219586 | 2.63E-05 | 0.001535034 | 0.127407792 |
| T2DM | t_cell | *GTPBP8* | 4.188637241 | 0.000868859 | 0.006764112 | 1 |
| T2DM | t_cell | *PPM1K* | 2.116581949 | 0.012053748 | 0.044884717 | 1 |
| T2DM | t_cell | *TIPARP* | 3.295197476 | 0.008516617 | 0.035019175 | 1 |
| T2DM | t_cell | *DCLK3* | 2.41781852 | 0.010785059 | 0.041531037 | 1 |
| T2DM | t_cell | *IL17RC* | -2.623083021 | 0.000103232 | 0.002196851 | 0.500882015 |
| T2DM | t_cell | *CPA3* | -2.815669418 | 0.000210063 | 0.002912079 | 1 |
| T2DM | t_cell | *CDCP1* | -1.56329721 | 0.005812634 | 0.026481595 | 1 |
| T2DM | t_cell | *CLEC3B* | -2.202746339 | 0.00443386 | 0.021478 | 1 |
| T2DM | t_cell | *FYCO1* | 1.737845763 | 0.002007852 | 0.012354622 | 1 |
| T2DM | t_cell | *RFT1* | -2.342965421 | 0.000135995 | 0.002475971 | 0.659849065 |
| T2DM | t_cell | *SFMBT1* | 3.129133088 | 4.66E-05 | 0.001701031 | 0.226237147 |
| T2DM | t_cell | *UVSSA* | 1.761583451 | 0.011971575 | 0.04475045 | 1 |
| T2DM | t_cell | *BDH2* | -2.277069298 | 0.000381978 | 0.003943314 | 1 |

| | | | | | | |
|---|---|---|---|---|---|---|
| T2DM | t_cell | *CDC25A* | -4.564764574 | 0.000367861 | 0.003821977 | 1 |
| T2DM | t_cell | *ATRIP* | -1.136140073 | 0.000531838 | 0.004823326 | 1 |
| T2DM | t_cell | *RNF123* | 2.952013935 | 2.53E-08 | 9.44E-06 | 0.000122763 |
| T2DM | t_cell | *ETNPPL* | -11.46460221 | 0.000250704 | 0.003135094 | 1 |
| T2DM | t_cell | *IL15* | 2.932556807 | 0.003447093 | 0.017766793 | 1 |
| T2DM | t_cell | *MOCS2* | 4.233704238 | 0.000644266 | 0.005527942 | 1 |
| T2DM | t_cell | *RNF180* | -2.122210418 | 0.006055679 | 0.02718053 | 1 |
| T2DM | t_cell | *F2RL2* | 1.423809381 | 5.52E-05 | 0.00172419 | 0.267794972 |
| T2DM | t_cell | *CMBL* | -1.995668202 | 0.004344176 | 0.021226528 | 1 |
| T2DM | t_cell | *SERINC5* | 2.271932565 | 0.003586096 | 0.018305159 | 1 |
| T2DM | t_cell | *CASP3* | 2.553039416 | 0.003079288 | 0.016545634 | 1 |
| T2DM | t_cell | *ERAP2* | 0.722541812 | 0.005348436 | 0.024858025 | 1 |
| T2DM | t_cell | *TLR3* | -2.03979224 | 0.000360781 | 0.003777851 | 1 |
| T2DM | t_cell | *RAET1E* | -2.411752859 | 0.001600565 | 0.010354591 | 1 |
| T2DM | t_cell | *STK17A* | 0.981991885 | 0.000119923 | 0.002365307 | 0.581865506 |
| T2DM | t_cell | *SAP30L* | 3.185135425 | 0.010141239 | 0.039769368 | 1 |
| T2DM | t_cell | *RPS14* | 2.312316209 | 0.003355263 | 0.01746753 | 1 |
| T2DM | t_cell | *CAMLG* | 2.738568408 | 0.001003605 | 0.007470988 | 1 |
| T2DM | t_cell | *CHMP4C* | -4.963906251 | 0.000402721 | 0.004062371 | 1 |
| T2DM | t_cell | *TNFRSF11B* | -2.965903985 | 0.001901221 | 0.011856972 | 1 |
| T2DM | t_cell | *RPP25L* | -1.814319456 | 0.012009013 | 0.044807806 | 1 |
| T2DM | t_cell | *TMEM65* | 2.937558438 | 0.000850225 | 0.006675232 | 1 |
| T2DM | t_cell | *ZMAT4* | -4.087654607 | 0.000751631 | 0.006108734 | 1 |
| T2DM | t_cell | *RASEF* | -0.708369919 | 0.011204141 | 0.042737807 | 1 |
| T2DM | t_cell | *AQP3* | 4.081593346 | 4.50E-05 | 0.001701031 | 0.218522321 |
| T2DM | t_cell | *GJB2* | 7.080785578 | 4.05E-05 | 0.001670757 | 0.196358792 |
| T2DM | t_cell | *MICU2* | 2.419202121 | 0.008246201 | 0.03413871 | 1 |
| T2DM | t_cell | *TMEM63C* | -6.185051389 | 0.000245274 | 0.003100135 | 1 |
| T2DM | t_cell | *AKR1E2* | -2.230028269 | 0.005899985 | 0.026728972 | 1 |
| T2DM | t_cell | *PDZD8* | 2.623557696 | 0.000203597 | 0.002870576 | 0.98785287 |
| T2DM | t_cell | *RET* | -1.751101838 | 0.000294534 | 0.003354647 | 1 |
| T2DM | t_cell | *PPP1R36* | -1.976894441 | 0.011412667 | 0.043227371 | 1 |
| T2DM | t_cell | *VWA2* | -2.867557622 | 0.00250233 | 0.014233653 | 1 |
| T2DM | t_cell | *UBTD1* | -2.051863003 | 0.010352614 | 0.04033496 | 1 |
| T2DM | t_cell | *OTOGL* | -1.451614313 | 0.008050583 | 0.033529123 | 1 |
| T2DM | t_cell | *PACSIN3* | -1.312428032 | 0.000237727 | 0.003062745 | 1 |
| T2DM | t_cell | *RAPSN* | -2.040528459 | 0.000135349 | 0.002475971 | 0.656711405 |
| T2DM | t_cell | *TC2N* | 0.848537393 | 8.71E-05 | 0.002066904 | 0.422667822 |
| T2DM | t_cell | *IFI27L1* | -2.252065923 | 0.007388795 | 0.031420188 | 1 |
| T2DM | t_cell | *ARL5B* | -2.360085856 | 0.008080722 | 0.033625784 | 1 |
| T2DM | t_cell | *TRMT61A* | -2.331665915 | 0.002250663 | 0.013283765 | 1 |
| T2DM | t_cell | *RPL27A* | 2.83639469 | 0.006008467 | 0.027119145 | 1 |
| T2DM | t_cell | *CENPN* | -3.856886049 | 0.006044622 | 0.02718053 | 1 |
| T2DM | t_cell | *C16orf46* | -3.633113249 | 0.002447816 | 0.014072039 | 1 |
| T2DM | t_cell | *PRKCB* | 0.915308281 | 4.19E-05 | 0.001670757 | 0.203318769 |
| T2DM | t_cell | *NDST2* | 24.69952246 | 0.000215117 | 0.002940128 | 1 |
| T2DM | t_cell | *CLEC4D* | -3.49099026 | 0.002562867 | 0.014459335 | 1 |
| T2DM | t_cell | *RIMKLB* | -1.350373897 | 0.01371751 | 0.049538347 | 1 |
| T2DM | t_cell | *CENPV* | -2.135189399 | 0.004842246 | 0.022943924 | 1 |
| T2DM | t_cell | *SLFN5* | 2.345751331 | 4.08E-05 | 0.001670757 | 0.19818495 |

| | | | | | | |
|---|---|---|---|---|---|---|
| T2DM | t_cell | *YPEL4* | -1.461367408 | 0.00883603 | 0.035846504 | 1 |
| T2DM | t_cell | *TMEM170A* | 2.942964857 | 0.002971945 | 0.01616578 | 1 |
| T2DM | t_cell | *ANPEP* | 0.756273984 | 0.003435545 | 0.017733259 | 1 |
| T2DM | t_cell | *STX3* | 1.953934117 | 0.002751297 | 0.01536167 | 1 |
| T2DM | t_cell | *PIP4K2C* | 3.502509801 | 0.000571865 | 0.005044439 | 1 |
| T2DM | t_cell | *NYAP1* | -1.034240657 | 0.002346684 | 0.013698845 | 1 |
| T2DM | t_cell | *SMAD3* | 2.241526226 | 0.001338983 | 0.009254624 | 1 |
| T2DM | t_cell | *AKTIP* | 1.904518543 | 1.10E-05 | 0.000971545 | 0.053434996 |
| T2DM | t_cell | *CNPY4* | 3.762329445 | 2.04E-05 | 0.001401991 | 0.099022973 |
| T2DM | t_cell | *MEI1* | 1.279491105 | 2.06E-05 | 0.001401991 | 0.099987422 |
| T2DM | t_cell | *LOXHD1* | -1.289739773 | 0.009253246 | 0.03707411 | 1 |
| T2DM | t_cell | *MYO5B* | -1.664793398 | 0.009458307 | 0.037617135 | 1 |
| T2DM | t_cell | *RAB8A* | 2.286485235 | 0.000709895 | 0.005867817 | 1 |
| T2DM | t_cell | *RPL13* | 1.946554047 | 0.010835407 | 0.041658791 | 1 |
| T2DM | t_cell | *TP53I13* | 3.472390504 | 0.00075394 | 0.006113817 | 1 |
| T2DM | t_cell | *TTYH1* | -3.912771943 | 0.008500733 | 0.034983509 | 1 |
| T2DM | t_cell | *CDC42EP5* | -1.118885491 | 0.000239695 | 0.003068607 | 1 |
| T2DM | t_cell | *KIFC2* | -2.638096863 | 0.005565593 | 0.02564507 | 1 |
| T2DM | t_cell | *KLK5* | -2.700009358 | 0.000838618 | 0.006605479 | 1 |
| T2DM | t_cell | *KLK6* | -9.126007522 | 0.000161455 | 0.002646545 | 0.78337736 |
| T2DM | t_cell | *NUDT8* | -1.150835563 | 0.000286042 | 0.003332537 | 1 |
| T2DM | t_cell | *ZNF598* | 1.882766744 | 0.005774154 | 0.026331008 | 1 |
| T2DM | t_cell | *ATG16L2* | 3.40185319 | 0.001945216 | 0.012069293 | 1 |
| T2DM | t_cell | *RPSA* | 1.813684434 | 0.001117626 | 0.008057533 | 1 |
| T2DM | t_cell | *NAALADL1* | -1.113851244 | 0.0121866 | 0.045240537 | 1 |
| T2DM | t_cell | *SCARA5* | -0.988572627 | 0.008995251 | 0.036189851 | 1 |
| T2DM | t_cell | *FAM83B* | -10.76948354 | 0.0001603 | 0.002636521 | 0.777773584 |
| T2DM | t_cell | *ACOX2* | -2.07014635 | 0.005994179 | 0.027079846 | 1 |
| T2DM | t_cell | *FAM107A* | -2.461217989 | 0.000303469 | 0.003424258 | 1 |
| T2DM | t_cell | *DTYMK* | 3.536083661 | 0.003572183 | 0.018263678 | 1 |
| T2DM | t_cell | *CCDC110* | -2.167711445 | 0.000156942 | 0.002614785 | 0.761483897 |
| T2DM | t_cell | *SERINC2* | -5.007772534 | 1.33E-05 | 0.001091132 | 0.064376765 |
| T2DM | t_cell | *ZSWIM1* | 0.931048074 | 2.66E-05 | 0.001535307 | 0.128965757 |
| T2DM | t_cell | *NBPF9* | -3.581990018 | 0.006315568 | 0.02801018 | 1 |
| T2DM | t_cell | *MFSD7* | -3.157856049 | 0.000451618 | 0.00432089 | 1 |
| T2DM | t_cell | *KLK7* | -2.585881336 | 0.00314065 | 0.016727152 | 1 |
| T2DM | t_cell | *IRS1* | -1.316789818 | 5.23E-09 | 2.54E-06 | 2.54E-05 |
| T2DM | t_cell | *CPT1C* | -2.424666405 | 0.000134694 | 0.002475971 | 0.653533375 |
| T2DM | t_cell | *XPO6* | 2.512984308 | 0.00079742 | 0.006353174 | 1 |
| T2DM | t_cell | *RGS14* | 0.830971353 | 0.001412345 | 0.009611076 | 1 |
| T2DM | t_cell | *RAB24* | 3.490621629 | 7.61E-05 | 0.00194078 | 0.369350304 |
| T2DM | t_cell | *GPRIN1* | 3.14290281 | 0.002095285 | 0.012739751 | 1 |
| T2DM | t_cell | *CD52* | 0.504762844 | 0.012011566 | 0.044807806 | 1 |
| T2DM | t_cell | *CCDC8* | -1.800439404 | 0.000231184 | 0.003040666 | 1 |
| T2DM | t_cell | *HINT1* | 2.725577682 | 0.013783965 | 0.049687903 | 1 |
| T2DM | t_cell | *CLIC3* | 6.45680322 | 0.000309641 | 0.003445826 | 1 |
| T2DM | t_cell | *INO80E* | -1.691924946 | 0.000490986 | 0.004563727 | 1 |
| T2DM | t_cell | *LUZP1* | 2.872114529 | 0.002198339 | 0.013164187 | 1 |
| T2DM | t_cell | *LRRC45* | -1.556877838 | 0.000197338 | 0.002828058 | 0.957483001 |
| T2DM | t_cell | *CSGALNACT2* | 3.058966854 | 5.71E-05 | 0.00172419 | 0.277057914 |

| | | | | | | |
|---|---|---|---|---|---|---|
| T2DM | t_cell | *CTNND2* | -1.393675324 | 0.004382493 | 0.021349254 | 1 |
| T2DM | t_cell | *CALML6* | -21.75698399 | 0.000151363 | 0.002594722 | 0.734412955 |
| T2DM | t_cell | *USP38* | 2.944903003 | 0.00816736 | 0.033928109 | 1 |
| T2DM | t_cell | *HSD17B13* | -2.448644319 | 0.000341161 | 0.003638727 | 1 |
| T2DM | t_cell | *ELOVL6* | 2.903516297 | 0.000173321 | 0.002692473 | 0.840951079 |
| T2DM | t_cell | *ARL6IP1* | -1.855686014 | 0.008614588 | 0.035294969 | 1 |
| T2DM | t_cell | *SERPINB9* | 2.320217123 | 0.002151503 | 0.012983945 | 1 |
| T2DM | t_cell | *CDH2* | -2.165344975 | 0.001500017 | 0.009948729 | 1 |
| T2DM | t_cell | *GTSF1* | 1.17343498 | 0.000127 | 0.002417859 | 0.616205811 |
| T2DM | t_cell | *TRABD* | 3.205592124 | 0.000258818 | 0.003195379 | 1 |
| T2DM | t_cell | *HOXB9* | -4.567554988 | 0.000142732 | 0.002562837 | 0.692536023 |
| T2DM | t_cell | *TTLL6* | -7.608327257 | 2.49E-10 | 2.42E-07 | 1.21E-06 |
| T2DM | t_cell | *NDUFA3* | 4.013254942 | 0.002554008 | 0.014442943 | 1 |
| T2DM | t_cell | *PAQR8* | 1.182626502 | 0.001729513 | 0.010955087 | 1 |
| T2DM | t_cell | *PYGO1* | -2.041399758 | 0.007729894 | 0.032585095 | 1 |
| T2DM | t_cell | *TSNARE1* | 2.055544996 | 0.010885882 | 0.041753596 | 1 |
| T2DM | t_cell | *FPR1* | -2.106669757 | 0.011707407 | 0.044000262 | 1 |
| T2DM | t_cell | *SOX7* | -1.828237509 | 0.001619408 | 0.010448629 | 1 |
| T2DM | t_cell | *LURAP1* | -0.824527805 | 0.004229487 | 0.020866389 | 1 |
| T2DM | t_cell | *ZNF561* | -1.291151225 | 0.013318361 | 0.04840501 | 1 |
| T2DM | t_cell | *HOPX* | 1.179261797 | 0.000231727 | 0.003040666 | 1 |
| T2DM | t_cell | *PPID* | 3.549578882 | 0.009281567 | 0.037106681 | 1 |
| T2DM | t_cell | *NMUR1* | 6.029620053 | 6.54E-05 | 0.001792929 | 0.31734838 |
| T2DM | t_cell | *PIK3CD* | 1.534374742 | 1.92E-05 | 0.001374162 | 0.093341745 |
| T2DM | t_cell | *RGS19* | -0.6873245 | 0.013666573 | 0.049411483 | 1 |
| T2DM | t_cell | *ANO5* | -1.841767502 | 0.00132108 | 0.009215829 | 1 |
| T2DM | t_cell | *VAT1L* | -1.556916887 | 0.000312043 | 0.003464608 | 1 |
| T2DM | t_cell | *LGALS4* | -1.675843639 | 0.002848374 | 0.015786085 | 1 |
| T2DM | t_cell | *PWWP2B* | -2.422812066 | 0.002963075 | 0.016135621 | 1 |
| T2DM | t_cell | *ANKRD20A4* | -3.194241452 | 0.000194494 | 0.002825402 | 0.943684144 |
| T2DM | t_cell | *SERF1A* | -8.943075306 | 0.000123736 | 0.002401474 | 0.600368613 |
| T2DM | t_cell | *CD8B* | 0.55742555 | 0.000294353 | 0.003354647 | 1 |
| T2DM | t_cell | *MALT1* | 2.792014792 | 0.008229116 | 0.034105004 | 1 |
| T2DM | t_cell | *MBOAT1* | 2.771176754 | 0.007989832 | 0.033351428 | 1 |
| T2DM | t_cell | *CXCR6* | 0.551742677 | 6.33E-05 | 0.001756372 | 0.307365135 |
| T2DM | t_cell | *C1QTNF4* | -1.134402562 | 3.21E-05 | 0.001619427 | 0.155624964 |
| T2DM | t_cell | *TP53RK* | 3.39740547 | 0.002462128 | 0.014104187 | 1 |
| T2DM | t_cell | *FIBP* | 3.540608448 | 0.000169809 | 0.002681745 | 0.823912907 |
| T2DM | t_cell | *SMPDL3A* | -1.005907254 | 0.011318671 | 0.043012736 | 1 |
| T2DM | t_cell | *ZNF596* | -2.208623004 | 0.010748357 | 0.041453911 | 1 |
| T2DM | t_cell | *CFL1* | 3.210440485 | 0.00239785 | 0.013883495 | 1 |
| T2DM | t_cell | *SNX32* | 0.83938092 | 0.007790875 | 0.03276853 | 1 |
| T2DM | t_cell | *RNASEH2C* | -2.29441542 | 2.74E-05 | 0.001541761 | 0.13275123 |
| T2DM | t_cell | *MYEOV* | -4.38039804 | 0.000193737 | 0.002822856 | 0.940011104 |
| T2DM | t_cell | *ANKRD13D* | 2.646698919 | 0.003720852 | 0.018838985 | 1 |
| T2DM | t_cell | *MRGPRF* | -2.113254752 | 9.80E-05 | 0.00214103 | 0.47530868 |
| T2DM | t_cell | *LCLAT1* | 1.84791509 | 0.013071551 | 0.047735398 | 1 |
| T2DM | t_cell | *PARP15* | 0.550581991 | 0.001208834 | 0.008574949 | 1 |
| T2DM | t_cell | *MZT2A* | 1.66765823 | 0.007183606 | 0.030899697 | 1 |
| T2DM | t_cell | *NDNF* | -2.071362101 | 0.001520774 | 0.01004228 | 1 |

| | | | | | | |
|---|---|---|---|---|---|---|
| T2DM | t_cell | *EHBP1L1* | 2.052365004 | 8.16E-05 | 0.002006241 | 0.395785384 |
| T2DM | t_cell | *ZNF417* | -1.486300518 | 0.008589608 | 0.035229736 | 1 |
| T2DM | t_cell | *NUDT18* | -3.228696236 | 0.000249993 | 0.003134276 | 1 |
| T2DM | t_cell | *SULT1B1* | -2.41893477 | 0.002065703 | 0.012578458 | 1 |
| T2DM | t_cell | *CD7* | 1.072444648 | 3.98E-05 | 0.001670757 | 0.193244119 |
| T2DM | t_cell | *RNF213* | 1.670771207 | 0.008143016 | 0.033855966 | 1 |
| T2DM | t_cell | *GPR160* | -1.702855186 | 0.003144903 | 0.016731437 | 1 |
| T2DM | t_cell | *HOXB2* | -3.022015368 | 9.94E-05 | 0.002149929 | 0.482281342 |
| T2DM | t_cell | *SLC25A30* | -2.172623679 | 0.001116138 | 0.008057533 | 1 |
| T2DM | t_cell | *GLIS1* | -2.56067268 | 0.002199073 | 0.013164187 | 1 |
| T2DM | t_cell | *SNAPC5* | 3.390746934 | 0.000746068 | 0.006083905 | 1 |
| T2DM | t_cell | *ANKRD36C* | 2.70594197 | 0.01069493 | 0.041348048 | 1 |
| T2DM | t_cell | *ZNF497* | -6.777445264 | 0.000896812 | 0.006939921 | 1 |
| T2DM | t_cell | *SH3PXD2B* | -1.63181538 | 0.005841613 | 0.026538863 | 1 |
| T2DM | t_cell | *KIAA1551* | 1.232793455 | 0.001622728 | 0.010456142 | 1 |
| T2DM | t_cell | *FGFBP3* | -5.081869938 | 0.009422329 | 0.0375346 | 1 |
| T2DM | t_cell | *PCP2* | -1.537174321 | 0.010500797 | 0.040792527 | 1 |
| T2DM | t_cell | *NLRP6* | 2.31062471 | 0.001774188 | 0.011179691 | 1 |
| T2DM | t_cell | *SEZ6L2* | 1.658563498 | 0.003348792 | 0.017452565 | 1 |
| T2DM | t_cell | *KCTD13* | 2.046494541 | 0.001386236 | 0.009500026 | 1 |
| T2DM | t_cell | *SLC22A1* | -2.854912615 | 0.002225776 | 0.013222396 | 1 |
| T2DM | t_cell | *TVP23C* | -4.355742857 | 0.003990871 | 0.019860213 | 1 |
| T2DM | t_cell | *PACS1* | 1.557833394 | 0.002253203 | 0.013283765 | 1 |
| T2DM | t_cell | *FAM182B* | -0.890490611 | 0.007793669 | 0.03276853 | 1 |
| T2DM | t_cell | *CCNE2* | -3.299153205 | 5.28E-08 | 1.60E-05 | 0.000256021 |
| T2DM | t_cell | *BANF1* | 3.980203523 | 0.002937142 | 0.016085869 | 1 |
| T2DM | t_cell | *SCUBE2* | -2.333950063 | 0.006487373 | 0.02856328 | 1 |
| T2DM | t_cell | *ZNF25* | -2.04125655 | 0.00339646 | 0.017625268 | 1 |
| T2DM | t_cell | *C11orf68* | -4.45217537 | 0.000151513 | 0.002594722 | 0.735138965 |
| T2DM | t_cell | *GPR156* | 2.596463858 | 0.012923443 | 0.047288497 | 1 |
| T2DM | t_cell | *PRIMA1* | -1.757730492 | 0.000285079 | 0.003332537 | 1 |
| T2DM | t_cell | *A2M* | 1.966619365 | 8.83E-05 | 0.002066904 | 0.428411236 |
| T2DM | t_cell | *DOK7* | -7.531556322 | 0.000269008 | 0.003258353 | 1 |
| T2DM | t_cell | *B3GALT6* | -5.126847969 | 0.000934075 | 0.007125997 | 1 |
| T2DM | t_cell | *JAKMIP2* | 0.460169542 | 5.21E-05 | 0.00172419 | 0.252799499 |
| T2DM | t_cell | *ZNF135* | -1.239925181 | 0.002123895 | 0.012849299 | 1 |
| T2DM | t_cell | *OR4N2* | -2.710187126 | 0.002368951 | 0.013781953 | 1 |
| T2DM | t_cell | *CRLF3* | 1.990082835 | 0.005603138 | 0.025720366 | 1 |
| T2DM | t_cell | *SPRYD4* | -1.469882652 | 0.002918236 | 0.016035424 | 1 |
| T2DM | t_cell | *WDR25* | -2.147159953 | 0.010597612 | 0.041053568 | 1 |
| T2DM | t_cell | *B3GNT5* | -2.01046523 | 0.006372611 | 0.028211595 | 1 |
| T2DM | t_cell | *BDNF* | -2.37650371 | 0.00449828 | 0.021650583 | 1 |
| T2DM | t_cell | *C8G* | -24.33193816 | 0.000191928 | 0.002813391 | 0.9312325 |
| T2DM | t_cell | *MUC20* | -2.05483786 | 0.005309872 | 0.024844262 | 1 |
| T2DM | t_cell | *C19orf18* | -1.452232972 | 0.007384631 | 0.031420188 | 1 |
| T2DM | t_cell | *DSCAML1* | -1.813701568 | 0.010908227 | 0.041773259 | 1 |
| T2DM | t_cell | *MIEF2* | -2.773719538 | 0.000898977 | 0.006945597 | 1 |
| T2DM | t_cell | *NIM1K* | 2.172463195 | 0.007323566 | 0.031252369 | 1 |
| T2DM | t_cell | *RBM44* | -1.858923712 | 0.011320541 | 0.043012736 | 1 |
| T2DM | t_cell | *SLC25A22* | -3.418765994 | 1.35E-06 | 0.000243413 | 0.006572153 |

| | | | | | | |
|---|---|---|---|---|---|---|
| T2DM | t_cell | *GBA* | -3.636173038 | 0.001539109 | 0.01011891 | 1 |
| T2DM | t_cell | *IL17RA* | 2.11042244 | 0.006136754 | 0.027417614 | 1 |
| T2DM | t_cell | *MBOAT4* | 1.614283647 | 5.47E-05 | 0.00172419 | 0.265279677 |
| T2DM | t_cell | *SLC35G5* | 14.73553116 | 3.24E-05 | 0.001619427 | 0.157084434 |
| T2DM | t_cell | *FAM87B* | 5.022046074 | 5.04E-05 | 0.00172419 | 0.24464208 |
| T2DM | t_cell | *CENPBD1* | -2.22938395 | 0.000179591 | 0.002737442 | 0.871376233 |
| T2DM | t_cell | *RPS27* | 2.822768991 | 0.003113042 | 0.016634892 | 1 |
| T2DM | t_cell | *IMPDH2* | -2.287900468 | 0.001038204 | 0.007678914 | 1 |
| T2DM | t_cell | *ZNF114* | -2.644100455 | 0.000392081 | 0.00401345 | 1 |
| T2DM | t_cell | *VN1R1* | -3.524656131 | 7.94E-05 | 0.001985749 | 0.385235239 |
| T2DM | t_cell | *SH2D4B* | -3.807881878 | 0.000304391 | 0.003426692 | 1 |
| T2DM | t_cell | *RNF212* | -1.84967549 | 0.004687956 | 0.022321848 | 1 |
| T2DM | t_cell | *ZNF223* | -1.89337648 | 0.000908976 | 0.00699124 | 1 |
| T2DM | t_cell | *FAM220A* | 3.994800579 | 0.002575519 | 0.014513842 | 1 |
| T2DM | t_cell | *GLDC* | -3.613235623 | 0.003107378 | 0.016634892 | 1 |
| T2DM | t_cell | *CA8* | -2.402992173 | 0.000771533 | 0.006197812 | 1 |
| T2DM | t_cell | *ERN1* | 1.830845564 | 7.26E-05 | 0.001910617 | 0.352030349 |
| T2DM | t_cell | *NSUN3* | 2.760529845 | 0.013511738 | 0.048997723 | 1 |
| T2DM | t_cell | *TRIM73* | -4.212001916 | 0.000572854 | 0.005044439 | 1 |
| T2DM | t_cell | *EXOSC4* | -4.250392197 | 0.000105184 | 0.002217626 | 0.510352086 |
| T2DM | t_cell | *ZNF852* | -3.049664162 | 4.92E-05 | 0.00172419 | 0.238939437 |
| T2DM | t_cell | *C17orf62* | 3.927983192 | 0.000464453 | 0.004358847 | 1 |
| T2DM | t_cell | *GAK* | 2.757287907 | 0.01255862 | 0.046127496 | 1 |
| T2DM | t_cell | *CTC1* | 2.966324366 | 2.05E-05 | 0.001401991 | 0.099311752 |
| T2DM | t_cell | *CCDC89* | -4.680871839 | 0.000676561 | 0.005685979 | 1 |
| T2DM | t_cell | *HTR1F* | -4.671688756 | 0.002500238 | 0.014233653 | 1 |
| T2DM | t_cell | *CDH4* | -1.42790302 | 0.003024819 | 0.016307138 | 1 |
| T2DM | t_cell | *HLA-DQB1* | -1.813933773 | 1.35E-14 | 6.54E-11 | 6.54E-11 |
| T2DM | t_cell | *GEMIN4* | -2.776006938 | 2.09E-05 | 0.001401991 | 0.101643087 |
| T2DM | t_cell | *GIPC3* | -10.06781528 | 0.000630343 | 0.005442035 | 1 |
| T2DM | t_cell | *B3GNT3* | 1.423567041 | 0.013098813 | 0.047735398 | 1 |
| T2DM | t_cell | *SEPT1* | 1.953151761 | 5.07E-05 | 0.00172419 | 0.24580115 |
| T2DM | t_cell | *GLIPR1L2* | -1.803465769 | 0.005559514 | 0.025641407 | 1 |
| T2DM | t_cell | *DEFB119* | -13.33268502 | 0.003969962 | 0.019776445 | 1 |
| T2DM | t_cell | *CHST13* | -8.974085047 | 9.24E-05 | 0.002106122 | 0.44822673 |
| T2DM | t_cell | *ZDHHC20* | 2.815974355 | 0.001129969 | 0.00811037 | 1 |
| T2DM | t_cell | *ZNF443* | -3.604017477 | 0.000241022 | 0.003077472 | 1 |
| T2DM | t_cell | *KCTD2* | -2.014859704 | 0.012023859 | 0.044807806 | 1 |
| T2DM | t_cell | *F2R* | 0.894050832 | 0.000176204 | 0.002709345 | 0.854941967 |
| T2DM | t_cell | *MUC16* | -1.843787749 | 0.007085551 | 0.030586384 | 1 |
| T2DM | t_cell | *SLC25A41* | -5.021031816 | 0.000432659 | 0.004190569 | 1 |
| T2DM | t_cell | *TMEM45A* | -1.585482462 | 0.008982422 | 0.036168226 | 1 |
| T2DM | t_cell | *ZFP41* | -1.53334963 | 0.006568389 | 0.028841468 | 1 |
| T2DM | t_cell | *TMEM252* | -43.8882186 | 0.000251479 | 0.003136697 | 1 |
| T2DM | t_cell | *MGAT4C* | -0.730836737 | 0.011654622 | 0.043903902 | 1 |
| T2DM | t_cell | *IDH2* | 2.855445934 | 0.000158257 | 0.002620688 | 0.767861537 |
| T2DM | t_cell | *ARL6IP4* | 1.432587267 | 0.00908997 | 0.036510375 | 1 |
| T2DM | t_cell | *HHIPL1* | -1.869668258 | 0.001205757 | 0.008565642 | 1 |
| T2DM | t_cell | *KCNA4* | -2.344809764 | 0.000446477 | 0.004281236 | 1 |
| T2DM | t_cell | *NXPH4* | -11.01954018 | 0.000164969 | 0.00266157 | 0.800429593 |

| | | | | | | |
|---|---|---|---|---|---|---|
| T2DM | t_cell | *CAPN12* | 1.561811225 | 5.26E-05 | 0.00172419 | 0.255408123 |
| T2DM | t_cell | *ORAI1* | 2.115996466 | 0.000111208 | 0.002275183 | 0.53958308 |
| T2DM | t_cell | *FES* | -1.863246486 | 0.006078516 | 0.027257817 | 1 |
| T2DM | t_cell | *TTC3* | 1.059851285 | 0.00227693 | 0.013391111 | 1 |
| T2DM | t_cell | *PPP1R27* | -6.911905012 | 0.001102295 | 0.008028975 | 1 |
| T2DM | t_cell | *BRICD5* | 7.42885782 | 7.28E-05 | 0.001910617 | 0.35346416 |
| T2DM | t_cell | *GRID1* | -1.971951226 | 0.004841025 | 0.022943924 | 1 |
| T2DM | t_cell | *PLCXD3* | -1.806467279 | 0.002960868 | 0.016135621 | 1 |
| T2DM | t_cell | *GPIHBP1* | -0.812249428 | 0.000549208 | 0.004925614 | 1 |
| T2DM | t_cell | *ZNF320* | 0.618914455 | 0.00833724 | 0.034427481 | 1 |
| T2DM | t_cell | *BEGAIN* | -1.975443206 | 0.000182897 | 0.002738946 | 0.887418345 |
| T2DM | t_cell | *ARHGEF37* | -2.457053119 | 0.009840831 | 0.038882502 | 1 |
| T2DM | t_cell | *RABIF* | 2.714270813 | 0.000995057 | 0.007450642 | 1 |
| T2DM | t_cell | *FANCF* | 6.259603568 | 0.000100567 | 0.002159079 | 0.48795185 |
| T2DM | t_cell | *C19orf71* | -2.831111284 | 0.009934481 | 0.039125083 | 1 |
| T2DM | t_cell | *LRIT3* | -7.835560954 | 0.000184801 | 0.002758937 | 0.896654684 |
| T2DM | t_cell | *GPR132* | 1.073193563 | 0.007218289 | 0.030966525 | 1 |
| T2DM | t_cell | *RP1L1* | -2.594936776 | 0.001063619 | 0.007831078 | 1 |
| T2DM | t_cell | *FAM19A1* | 1.398602734 | 3.11E-05 | 0.001619427 | 0.150879214 |
| T2DM | t_cell | *FIGLA* | -4.887688534 | 0.000643174 | 0.005527942 | 1 |
| T2DM | t_cell | *ACSM2A* | -25.8031434 | 0.000555219 | 0.004942973 | 1 |
| T2DM | t_cell | *ZNF703* | -2.283322135 | 0.008537758 | 0.035076378 | 1 |
| T2DM | t_cell | *FAM162B* | -1.510453491 | 0.001211781 | 0.008583302 | 1 |
| T2DM | t_cell | *FAM3B* | -0.387913575 | 0.007120377 | 0.030663274 | 1 |
| T2DM | t_cell | *ACOT1* | 1.047007196 | 0.009724628 | 0.038548933 | 1 |
| T2DM | t_cell | *CLECL1* | 1.617726559 | 2.11E-05 | 0.001401991 | 0.10234535 |
| T2DM | t_cell | *PKP3* | 11.12099233 | 0.002256207 | 0.013285338 | 1 |
| T2DM | t_cell | *ZFP1* | -2.723710845 | 0.001376299 | 0.009445267 | 1 |
| T2DM | t_cell | *SNN* | -4.083942164 | 0.003341446 | 0.017433008 | 1 |
| T2DM | t_cell | *TCTE3* | 3.848745452 | 0.00172886 | 0.010955087 | 1 |
| T2DM | t_cell | *DRD1* | -25.41207052 | 0.000169139 | 0.002681745 | 0.820664123 |
| T2DM | t_cell | *NOC4L* | 2.780643085 | 0.00040772 | 0.004078878 | 1 |
| T2DM | t_cell | *NDUFA6* | 0.876567229 | 0.00187389 | 0.01173176 | 1 |
| T2DM | t_cell | *SORCS2* | -2.306596554 | 0.000279153 | 0.003295505 | 1 |
| T2DM | t_cell | *SGCZ* | -0.849472404 | 0.009294376 | 0.037113371 | 1 |
| T2DM | t_cell | *SIGIRR* | 1.637053194 | 0.000153915 | 0.00260406 | 0.746797262 |
| T2DM | t_cell | *GP1BA* | 2.654961987 | 1.13E-05 | 0.000977311 | 0.054729439 |
| T2DM | t_cell | *SP140L* | 2.897316344 | 0.000286203 | 0.003332537 | 1 |
| T2DM | t_cell | *SV2B* | -1.161085547 | 0.002485304 | 0.014186699 | 1 |
| T2DM | t_cell | *AHNAK2* | -2.498400688 | 0.000109958 | 0.002260662 | 0.533516225 |
| T2DM | t_cell | *MYBL1* | 0.401405828 | 0.000109383 | 0.002260662 | 0.53072753 |
| T2DM | t_cell | *ADARB2* | 3.006133152 | 0.000151876 | 0.002594722 | 0.73690091 |
| T2DM | t_cell | *C11orf87* | 4.212012418 | 0.001596759 | 0.010354591 | 1 |
| T2DM | t_cell | *DMWD* | -3.374431243 | 0.000958504 | 0.007244021 | 1 |
| T2DM | t_cell | *PIGP* | -0.550617864 | 0.006501546 | 0.02859973 | 1 |
| T2DM | t_cell | *EVI2B* | 0.870159774 | 7.59E-05 | 0.00194078 | 0.368307254 |
| T2DM | t_cell | *ZNF829* | -1.838626478 | 0.001551391 | 0.010149299 | 1 |
| T2DM | t_cell | *IFITM1* | -2.370476385 | 0.000618482 | 0.005358705 | 1 |
| T2DM | t_cell | *FFAR4* | 3.5806661 | 0.002983065 | 0.016190502 | 1 |
| T2DM | t_cell | *CYP4F12* | -2.136046661 | 0.004647431 | 0.022194227 | 1 |

| | | | | | | |
|---|---|---|---|---|---|---|
| T2DM | t_cell | *ZNF749* | -0.563034546 | 0.008857822 | 0.03590489 | 1 |
| T2DM | t_cell | *TOR3A* | -3.241154289 | 0.004125686 | 0.020447221 | 1 |
| T2DM | t_cell | *GOLGA8R* | 2.715285221 | 0.002288387 | 0.013442194 | 1 |
| T2DM | t_cell | *GNG2* | 1.813652127 | 4.54E-05 | 0.001701031 | 0.220424771 |
| T2DM | t_cell | *SEPT10* | -2.047949571 | 0.010897781 | 0.041766221 | 1 |
| T2DM | t_cell | *CEACAM19* | -0.911621548 | 0.002851197 | 0.015786085 | 1 |
| T2DM | t_cell | *MIR22HG* | -2.198126544 | 0.001260019 | 0.008860305 | 1 |
| T2DM | t_cell | *CYP27C1* | -1.584057568 | 3.35E-05 | 0.001630473 | 0.162655962 |
| T2DM | t_cell | *TRABD2A* | 0.784602348 | 0.000824984 | 0.006521623 | 1 |
| T2DM | t_cell | *TNFRSF18* | 0.923454538 | 5.13E-05 | 0.00172419 | 0.24882954 |
| T2DM | t_cell | *C1QL4* | -37.44269581 | 0.010461529 | 0.040672547 | 1 |
| T2DM | t_cell | *RTN4RL2* | -3.776944922 | 0.000217951 | 0.002962178 | 1 |
| T2DM | t_cell | *TMPRSS6* | -7.796383368 | 0.00015075 | 0.002594722 | 0.731436924 |
| T2DM | t_cell | *LUZP2* | -2.657671669 | 0.000659757 | 0.005604753 | 1 |
| T2DM | t_cell | *GJA4* | -3.301470682 | 0.003861673 | 0.019331962 | 1 |
| T2DM | t_cell | *SBK2* | -10.17464245 | 0.000167309 | 0.002679155 | 0.811784053 |
| T2DM | t_cell | *C17orf97* | 0.293885041 | 0.009930381 | 0.039125083 | 1 |
| T2DM | t_cell | *SAMD11* | -1.413421774 | 0.001366444 | 0.009417597 | 1 |
| T2DM | t_cell | *TRPV2* | 1.317232785 | 0.000415299 | 0.004112304 | 1 |
| T2DM | t_cell | *DNAH17* | -2.053750654 | 0.008749961 | 0.035706317 | 1 |
| T2DM | t_cell | *FAM122A* | -2.979016883 | 0.001044261 | 0.007711953 | 1 |
| T2DM | t_cell | *GFRAL* | -2.453361154 | 0.000983811 | 0.007400701 | 1 |
| T2DM | t_cell | *LDLRAD2* | -1.632964357 | 0.001432227 | 0.009678502 | 1 |
| T2DM | t_cell | *CPSF4L* | -4.362545395 | 0.000677761 | 0.005685979 | 1 |
| T2DM | t_cell | *MAPK12* | -2.059629312 | 0.000282418 | 0.003325951 | 1 |
| T2DM | t_cell | *AGRN* | -3.565230308 | 0.008359774 | 0.034491176 | 1 |
| T2DM | t_cell | *ZNF626* | -2.438560056 | 0.001762585 | 0.011121018 | 1 |
| T2DM | t_cell | *NCR3LG1* | -3.907964865 | 0.000187183 | 0.002770691 | 0.90821154 |
| T2DM | t_cell | *RUFY4* | -5.416056004 | 0.001500137 | 0.009948729 | 1 |
| T2DM | t_cell | *ZNF669* | 1.991136905 | 0.001111633 | 0.008038215 | 1 |
| T2DM | t_cell | *H3F3C* | -34.33647141 | 0.002926207 | 0.016061037 | 1 |
| T2DM | t_cell | *PDCD1* | 0.853882694 | 5.86E-05 | 0.00172419 | 0.284489604 |
| T2DM | t_cell | *SELL* | 0.852030346 | 0.000198757 | 0.002828058 | 0.964367862 |
| T2DM | t_cell | *C19orf54* | -2.769286537 | 0.001436343 | 0.009692819 | 1 |
| T2DM | t_cell | *C22orf34* | 2.34773182 | 4.96E-05 | 0.00172419 | 0.240656516 |
| T2DM | t_cell | *NDOR1* | 4.550015187 | 0.007960681 | 0.033326335 | 1 |
| T2DM | t_cell | *GOLGA8M* | -2.058004251 | 0.001398663 | 0.009571672 | 1 |
| T2DM | t_cell | *RHCE* | -2.051191877 | 0.000529301 | 0.004823326 | 1 |
| T2DM | t_cell | *FSIP2* | -1.622071566 | 0.008667247 | 0.035428376 | 1 |
| T2DM | t_cell | *FZD9* | -11.99749559 | 4.20E-05 | 0.001670757 | 0.203595447 |
| T2DM | t_cell | *SKOR1* | -2.493396629 | 0.006468719 | 0.028507016 | 1 |
| T2DM | t_cell | *MTF1* | -2.496345243 | 0.003019723 | 0.016297771 | 1 |
| T2DM | t_cell | *ZDHHC11* | -2.06722442 | 3.35E-05 | 0.001630473 | 0.162399837 |
| T2DM | t_cell | *ENTPD8* | -118.646616 | 0.000129833 | 0.002441662 | 0.62994888 |
| T2DM | t_cell | *RPL14* | 2.572064186 | 0.012086205 | 0.044942423 | 1 |
| T2DM | t_cell | *FAM196A* | -1.861913127 | 0.006556451 | 0.028815127 | 1 |
| T2DM | t_cell | *UTS2B* | -1.627661424 | 0.002874743 | 0.015814346 | 1 |
| T2DM | t_cell | *C9orf152* | -2.695268626 | 0.000811975 | 0.006458526 | 1 |
| T2DM | t_cell | *SLC15A5* | -1.099852697 | 0.008770968 | 0.035731935 | 1 |
| T2DM | t_cell | *LIPI* | -0.943193873 | 0.007312007 | 0.03123051 | 1 |

| | | | | | | |
|---|---|---|---|---|---|---|
| T2DM | t_cell | *LITAF* | 3.051399936 | 0.002009357 | 0.012354622 | 1 |
| T2DM | t_cell | *PLAC9* | -2.083562219 | 0.012819203 | 0.046942469 | 1 |
| T2DM | t_cell | *FAM47E* | -2.486556047 | 0.000654428 | 0.005590288 | 1 |
| T2DM | t_cell | *ZKSCAN3* | -1.607144615 | 0.011873543 | 0.044452493 | 1 |
| T2DM | t_cell | *HLA-DRB1* | -1.479349207 | 5.83E-08 | 1.67E-05 | 0.000283065 |
| T2DM | t_cell | *COLCA1* | -2.13682275 | 2.79E-05 | 0.001541761 | 0.135228509 |
| T2DM | t_cell | *HRCT1* | -1.121789272 | 0.003337341 | 0.01743033 | 1 |
| T2DM | t_cell | *SIRPB2* | -1.012929803 | 0.000543094 | 0.00488885 | 1 |
| T2DM | t_cell | *ZNF107* | 1.779747317 | 0.000492371 | 0.004567849 | 1 |
| T2DM | t_cell | *ZNF471* | -2.092289127 | 0.002104365 | 0.012762975 | 1 |
| T2DM | t_cell | *SUPT3H* | 2.150484722 | 0.008829721 | 0.035846504 | 1 |
| T2DM | t_cell | *GIMAP5* | 1.345186309 | 0.006251381 | 0.027801744 | 1 |
| T2DM | t_cell | *STK31* | -1.521858968 | 0.005721965 | 0.026166798 | 1 |
| T2DM | t_cell | *EVL* | 1.467885815 | 5.23E-05 | 0.00172419 | 0.253845948 |
| T2DM | t_cell | *C20orf96* | -3.096363041 | 0.005833626 | 0.026527417 | 1 |
| T2DM | t_cell | *SULT1A1* | -2.580478524 | 0.001107534 | 0.008036716 | 1 |
| T2DM | t_cell | *TPK1* | 3.140544359 | 0.000361278 | 0.003777851 | 1 |
| T2DM | t_cell | *CACNA1H* | -3.142493658 | 0.004500293 | 0.021650583 | 1 |
| T2DM | t_cell | *XRCC2* | -3.805574435 | 0.000291304 | 0.003341384 | 1 |
| T2DM | t_cell | *TOMM7* | 2.954555938 | 0.003449365 | 0.017766793 | 1 |
| T2DM | t_cell | *HLA-DQA1* | -2.374735177 | 4.15E-12 | 6.71E-09 | 2.01E-08 |
| T2DM | t_cell | *ZNF239* | 3.187239539 | 0.007294326 | 0.031182439 | 1 |
| T2DM | t_cell | *SLC39A10* | 2.731224474 | 0.000328102 | 0.003565777 | 1 |
| T2DM | t_cell | *ZNF470* | -3.839200611 | 0.000473146 | 0.00443186 | 1 |
| T2DM | t_cell | *ANXA6* | 2.031719823 | 0.002593716 | 0.014574822 | 1 |
| T2DM | t_cell | *GAL3ST4* | 2.737604701 | 0.000101412 | 0.002167631 | 0.492052292 |
| T2DM | t_cell | *ZNF772* | -0.942726243 | 0.007671639 | 0.032395816 | 1 |
| T2DM | t_cell | *ZNF257* | 1.906700193 | 0.008924495 | 0.036054661 | 1 |
| T2DM | t_cell | *ADAM32* | -2.412478641 | 0.002058014 | 0.012577675 | 1 |
| T2DM | t_cell | *LRRC8B* | 2.460874593 | 0.013092112 | 0.047735398 | 1 |
| T2DM | t_cell | *CCL4L2* | 4.078137276 | 1.59E-05 | 0.00124438 | 0.077378503 |
| T2DM | t_cell | *ZNF165* | -1.222355102 | 0.001586251 | 0.01033086 | 1 |
| T2DM | t_cell | *RAMP2-AS1* | -1.880502693 | 0.007872732 | 0.03304368 | 1 |
| T2DM | t_cell | *ZNF98* | -1.193861701 | 0.007251656 | 0.031027367 | 1 |
| T2DM | t_cell | *MAP3K5* | 2.132385409 | 0.003334479 | 0.01743033 | 1 |
| T2DM | t_cell | *PCDHB11* | -0.746636891 | 0.008620062 | 0.035294969 | 1 |
| T2DM | t_cell | *HHLA3* | -1.140847674 | 0.006135158 | 0.027417614 | 1 |
| T2DM | t_cell | *ENPP1* | -1.806976201 | 0.001914793 | 0.011926285 | 1 |
| T2DM | t_cell | *CCDC154* | 1.03758586 | 0.008180351 | 0.033953005 | 1 |
| T2DM | t_cell | *DNAH10* | -2.309660228 | 0.003214713 | 0.017009585 | 1 |
| T2DM | t_cell | *EME2* | 5.760526913 | 0.002412027 | 0.013948932 | 1 |
| T2DM | t_cell | *ZNF677* | -2.124347116 | 0.001717213 | 0.010919944 | 1 |
| T2DM | t_cell | *ZNF71* | -1.796835728 | 0.002779429 | 0.015480668 | 1 |
| T2DM | t_cell | *VPS13A* | 2.624959945 | 0.002234429 | 0.013252742 | 1 |
| T2DM | t_cell | *ZNF273* | -2.047709414 | 0.007337648 | 0.031284946 | 1 |
| T2DM | t_cell | *ZNF667* | -1.499167054 | 0.002544677 | 0.014423802 | 1 |
| T2DM | t_cell | *SFI1* | 1.81094861 | 3.59E-05 | 0.001630473 | 0.174213739 |
| T2DM | t_cell | *SOWAHC* | -3.228952614 | 0.010601839 | 0.041053568 | 1 |
| T2DM | t_cell | *MIER1* | 2.839401263 | 0.001004868 | 0.007470988 | 1 |
| T2DM | t_cell | *MAN1A2* | 2.5022079 | 0.00332587 | 0.017407899 | 1 |

| | | | | | | |
|---|---|---|---|---|---|---|
| T2DM | t_cell | *STYX* | 3.815663521 | 0.000119392 | 0.002365307 | 0.579291377 |
| T2DM | t_cell | *ZNF442* | -1.764320929 | 0.001900184 | 0.011856972 | 1 |
| T2DM | t_cell | *SH3BGRL2* | -0.937742447 | 0.011538269 | 0.043533188 | 1 |
| T2DM | t_cell | *HLA-DRB5* | -0.652732628 | 1.38E-09 | 9.53E-07 | 6.67E-06 |
| T2DM | t_cell | *C20orf203* | -2.034363747 | 0.006142794 | 0.027419353 | 1 |
| T2DM | t_cell | *TUSC1* | -4.539028546 | 0.005052683 | 0.023732447 | 1 |
| T2DM | t_cell | *ANKRD13B* | -2.623111927 | 0.000400452 | 0.004056357 | 1 |
| T2DM | t_cell | *PPP1R14C* | -3.92962449 | 0.000154483 | 0.00260406 | 0.749552912 |
| T2DM | t_cell | *RPL10A* | 2.648758323 | 0.003297412 | 0.017296263 | 1 |
| T2DM | t_cell | *COLGALT2* | 1.530187118 | 0.000660395 | 0.005604753 | 1 |
| T2DM | t_cell | *RCSD1* | 1.158909306 | 4.25E-05 | 0.001675348 | 0.20606785 |
| T2DM | t_cell | *PRMT6* | -3.481869438 | 8.83E-05 | 0.002066904 | 0.428552312 |
| T2DM | t_cell | *SHISA4* | -4.719890039 | 5.04E-06 | 0.000596779 | 0.024467935 |
| T2DM | t_cell | *KIAA0753* | -2.140696546 | 0.008824023 | 0.035846504 | 1 |
| T2DM | t_cell | *CCDC167* | 2.918812157 | 0.000116429 | 0.002334345 | 0.564911599 |
| T2DM | t_cell | *MFAP3L* | -2.437084322 | 0.004829005 | 0.022925962 | 1 |
| T2DM | t_cell | *AL590822.1* | -2.430679615 | 0.000421287 | 0.004123035 | 1 |
| T2DM | t_cell | *SAMD5* | -1.539785728 | 0.003558062 | 0.018229899 | 1 |
| T2DM | t_cell | *GPR52* | 6.744667242 | 0.000308194 | 0.003445826 | 1 |
| T2DM | t_cell | *FAM229B* | -2.074232569 | 0.004440456 | 0.021480653 | 1 |
| T2DM | t_cell | *RBM20* | -2.838962205 | 0.000417366 | 0.004115925 | 1 |
| T2DM | t_cell | *SAMD13* | -3.714939817 | 0.007122323 | 0.030663274 | 1 |
| T2DM | t_cell | *LIPN* | 12.37211242 | 0.001097863 | 0.00802234 | 1 |
| T2DM | t_cell | *C2orf72* | -9.485154965 | 3.85E-05 | 0.001670757 | 0.186912687 |
| T2DM | t_cell | *ASAH2B* | 5.033670934 | 2.47E-05 | 0.001498556 | 0.119765765 |
| T2DM | t_cell | *LINC00619* | -13.96736968 | 0.000395102 | 0.004029273 | 1 |
| T2DM | t_cell | *DAXX* | 1.889467187 | 0.000326469 | 0.003565777 | 1 |
| T2DM | t_cell | *COL15A1* | -1.012492051 | 0.008701585 | 0.035538798 | 1 |
| T2DM | t_cell | *NOTCH4* | -0.332327108 | 3.53E-09 | 1.90E-06 | 1.71E-05 |
| T2DM | t_cell | *AGER* | 2.142741314 | 7.62E-06 | 0.000821846 | 0.036983062 |
| T2DM | t_cell | *STK19* | -2.967586313 | 5.85E-07 | 0.000129024 | 0.00283853 |
| T2DM | t_cell | *SKIV2L* | -0.931814234 | 0.000754777 | 0.006113817 | 1 |
| T2DM | t_cell | *GPANK1* | -2.764864683 | 4.64E-05 | 0.001701031 | 0.22533092 |
| T2DM | t_cell | *LST1* | 1.520692607 | 0.009027966 | 0.036291374 | 1 |
| T2DM | t_cell | *PSORS1C2* | 1.167641152 | 0.008390941 | 0.034590355 | 1 |
| T2DM | t_cell | *DHX16* | 4.080160261 | 2.46E-06 | 0.000384687 | 0.011925308 |
| T2DM | t_cell | *LILRB3* | -1.974979565 | 0.001036164 | 0.007675524 | 1 |
| T2DM | t_cell | *HLA-G* | 1.042988613 | 0.006667458 | 0.029197207 | 1 |
| T2DM | t_cell | *FAM153C* | -0.804256776 | 0.000231873 | 0.003040666 | 1 |
| T2DM | t_cell | *MALRD1* | -2.039299589 | 0.007044814 | 0.030464739 | 1 |
| T2DM | t_cell | *FAM196B* | 1.589433765 | 9.40E-05 | 0.002130478 | 0.455922338 |
| T2DM | t_cell | *MROH6* | -1.251460915 | 0.004716123 | 0.02243395 | 1 |
| T2DM | t_cell | *C12orf73* | -2.326063901 | 0.013103334 | 0.047735398 | 1 |
| T2DM | t_cell | *PCDHGA1* | -1.469754578 | 0.003649432 | 0.018560843 | 1 |
| T2DM | t_cell | *PCDHA4* | -2.542359011 | 0.000678521 | 0.005685979 | 1 |
| T2DM | t_cell | *PKHD1L1* | -0.685075076 | 0.013841534 | 0.049784376 | 1 |
| T2DM | t_cell | *SLFN12L* | 0.492128056 | 0.000150285 | 0.002594722 | 0.729183601 |
| T2DM | t_cell | *TMEM231* | -2.201006149 | 0.002518518 | 0.014308955 | 1 |
| T2DM | t_cell | *GOLGA6L10* | -2.452921024 | 0.002165133 | 0.013047097 | 1 |
| T2DM | t_cell | *SAP25* | -3.443739276 | 0.002998104 | 0.01619911 | 1 |

| | | | | | | |
|---|---|---|---|---|---|---|
| T2DM | t_cell | *MT1A* | -2.243347932 | 0.000179976 | 0.002737442 | 0.873243905 |
| T2DM | t_cell | *MT1M* | -3.170565791 | 0.001306202 | 0.009158518 | 1 |
| T2DM | t_cell | *CFI* | -1.511569609 | 0.011429379 | 0.043256902 | 1 |
| T2DM | t_cell | *NAP1L4* | 2.757884862 | 0.000421481 | 0.004123035 | 1 |
| T2DM | t_cell | *SMN2* | -0.905537154 | 0.010653567 | 0.041220978 | 1 |
| T2DM | t_cell | *SERF1B* | -1.602919786 | 0.000957632 | 0.007244021 | 1 |
| T2DM | t_cell | *LIN52* | 2.741279373 | 0.000155468 | 0.002609502 | 0.754332256 |
| T2DM | t_cell | *KLRC2* | 0.602360191 | 5.51E-05 | 0.00172419 | 0.267472822 |
| T2DM | t_cell | *C1QTNF9B* | 41.30439007 | 0.001243226 | 0.008767636 | 1 |
| T2DM | t_cell | *ZDHHC11B* | -2.017547019 | 0.000314915 | 0.003479946 | 1 |
| T2DM | t_cell | *HCG27* | 1.391448335 | 0.000109617 | 0.002260662 | 0.531860272 |
| T2DM | t_cell | *HLA-A* | 1.497519185 | 0.00492051 | 0.023220882 | 1 |
| T2DM | t_cell | *SACM1L* | 2.982720822 | 0.000509808 | 0.00470264 | 1 |
| T2DM | t_cell | *SLC48A1* | -2.261509351 | 0.001815448 | 0.011410045 | 1 |
| T2DM | t_cell | *TAS2R14* | 1.444737503 | 0.000175251 | 0.002708013 | 0.850315964 |
| T2DM | t_cell | *AC084018.1* | -1.040929354 | 0.000463941 | 0.004358847 | 1 |
| T2DM | t_cell | *C17orf51* | -4.090418436 | 0.002033445 | 0.012457418 | 1 |
| T2DM | t_cell | *FGFR1OP* | 3.500726008 | 0.003403842 | 0.017636878 | 1 |
| T2DM | t_cell | *AC022498.1* | -6.010248373 | 0.001615275 | 0.010435835 | 1 |
| T2DM | t_cell | *DNLZ* | 0.809903814 | 0.000417736 | 0.004115925 | 1 |
| T2DM | t_cell | *SUPT4H1* | 6.368290585 | 1.91E-05 | 0.001374162 | 0.092478048 |
| T2DM | t_cell | *ARHGAP19* | -5.746692696 | 5.15E-07 | 0.000119004 | 0.002499077 |
| T2DM | t_cell | *LCAT* | -2.135793641 | 0.001528444 | 0.010076104 | 1 |
| T2DM | t_cell | *GPC2* | -3.64534117 | 0.012476097 | 0.045963571 | 1 |
| T2DM | t_cell | *TMEM110* | 4.427282138 | 2.50E-08 | 9.44E-06 | 0.000121279 |
| T2DM | t_cell | *LBH* | 1.042843605 | 0.000150334 | 0.002594722 | 0.729422729 |
| T2DM | t_cell | *ATF6B* | 1.949023127 | 0.011783547 | 0.04421792 | 1 |
| T2DM | t_cell | *PPM1N* | 1.473810975 | 0.000773131 | 0.006200382 | 1 |
| T2DM | t_cell | *LTB4R* | 2.186457068 | 0.000794712 | 0.006342014 | 1 |
| T2DM | t_cell | *PRCD* | -1.958295471 | 0.005472056 | 0.025334365 | 1 |
| T2DM | t_cell | *SH3D21* | -2.260940519 | 0.00013625 | 0.002475971 | 0.661084208 |
| T2DM | t_cell | *ANG* | 4.458744909 | 0.000274676 | 0.003278465 | 1 |
| T2DM | t_cell | *COLCA2* | -0.912182068 | 0.005309137 | 0.024844262 | 1 |
| T2DM | t_cell | *LCNL1* | -1.709205619 | 0.010756487 | 0.041453911 | 1 |
| T2DM | t_cell | *GNAT3* | -1.578746184 | 0.000228424 | 0.003036473 | 1 |
| T2DM | t_cell | *STARD10* | 1.624904267 | 4.38E-08 | 1.42E-05 | 0.000212505 |
| T2DM | t_cell | *CPEB1* | -1.571911857 | 0.000488874 | 0.004552813 | 1 |
| T2DM | t_cell | *ARHGEF33* | -0.961216871 | 0.006999223 | 0.030348732 | 1 |
| T2DM | t_cell | *IFRD2* | -2.166164167 | 0.000922687 | 0.007083669 | 1 |
| T2DM | t_cell | *AC005082.1* | -4.138157787 | 0.000853403 | 0.006689353 | 1 |
| T2DM | t_cell | *C5orf49* | -17.07797876 | 0.000909207 | 0.00699124 | 1 |
| T2DM | t_cell | *GOLGA8B* | 1.093869221 | 0.008468257 | 0.034879444 | 1 |
| T2DM | t_cell | *TSTD1* | 2.630708879 | 0.000165114 | 0.00266157 | 0.80113245 |
| T2DM | t_cell | *PAM16* | -1.852365908 | 0.000384738 | 0.003954974 | 1 |
| T2DM | t_cell | *RNU6ATAC35I* | -11.99949932 | 0.000170234 | 0.002681745 | 0.825977496 |
| T2DM | t_cell | *FANCG* | -1.476747983 | 0.001654091 | 0.01063 | 1 |
| T2DM | t_cell | *FXYD7* | -1.929952613 | 0.002314649 | 0.013580022 | 1 |
| T2DM | t_cell | *UBA52* | 2.356320648 | 0.000187301 | 0.002770691 | 0.908786566 |
| T2DM | t_cell | *RAB6C* | -28.64516053 | 0.012715955 | 0.046670054 | 1 |
| T2DM | t_cell | *AC062017.1* | -2.836829968 | 0.003073502 | 0.016532853 | 1 |

| | | | | | | |
|---|---|---|---|---|---|---|
| T2DM | t_cell | *LINC00571* | -1.676505458 | 0.00166612 | 0.010679017 | 1 |
| T2DM | t_cell | *LINC01133* | -1.818586936 | 0.00046421 | 0.004358847 | 1 |
| T2DM | t_cell | *C4B* | 3.123571681 | 7.87E-05 | 0.00197835 | 0.381730812 |
| T2DM | t_cell | *LINC01117* | -2.281420864 | 0.001740263 | 0.010994473 | 1 |
| T2DM | t_cell | *RASAL2-AS1* | -1.67072887 | 0.005307864 | 0.024844262 | 1 |
| T2DM | t_cell | *LINC00184* | -37.87820761 | 0.000206687 | 0.00289313 | 1 |
| T2DM | t_cell | *CYP4A22-AS1* | -3.41213895 | 0.002992855 | 0.01619911 | 1 |
| T2DM | t_cell | *MIAT* | 0.853591792 | 5.86E-05 | 0.00172419 | 0.284491309 |
| T2DM | t_cell | *LINC00115* | 4.732959473 | 0.004409095 | 0.021435801 | 1 |
| T2DM | t_cell | *HAR1A* | 5.5514746 | 0.000190094 | 0.002803451 | 0.922335398 |
| T2DM | t_cell | *FAM138B* | -16.24868693 | 0.000139938 | 0.002533496 | 0.678976888 |
| T2DM | t_cell | *SLC2A1-AS1* | 4.06821627 | 0.000443807 | 0.004272522 | 1 |
| T2DM | t_cell | *LINC00578* | -1.93311993 | 0.000527144 | 0.004823326 | 1 |
| T2DM | t_cell | *PATL2* | 1.783604263 | 4.50E-05 | 0.001701031 | 0.218515115 |
| T2DM | t_cell | *CASC20* | -2.604051355 | 0.000146475 | 0.002594722 | 0.710695779 |
| T2DM | t_cell | *AC112229.4* | -1.346253766 | 0.003922351 | 0.019579471 | 1 |
| T2DM | t_cell | *KLHL7-AS1* | -1.497623644 | 0.004424606 | 0.021478 | 1 |
| T2DM | t_cell | *LINC00332* | -1.435687551 | 0.004432781 | 0.021478 | 1 |
| T2DM | t_cell | *RPS18* | 2.203367293 | 0.002226442 | 0.013222396 | 1 |
| T2DM | t_cell | *TAPBP* | 2.590076253 | 0.000211064 | 0.002912608 | 1 |
| T2DM | t_cell | *TMA7* | 3.708831367 | 0.00013176 | 0.00244943 | 0.63930117 |
| T2DM | t_cell | *AC009495.3* | 1.203527003 | 3.80E-05 | 0.001670757 | 0.184489047 |
| T2DM | t_cell | *HLA-DQB2* | 0.651857004 | 4.01E-07 | 9.72E-05 | 0.001943908 |
| T2DM | t_cell | *LYRM9* | -2.203911595 | 0.00110854 | 0.008036716 | 1 |
| T2DM | t_cell | *LINC00342* | 3.569187617 | 0.003258595 | 0.017129689 | 1 |
| T2DM | t_cell | *LINC00702* | -3.058311631 | 0.000260248 | 0.003204879 | 1 |
| T2DM | t_cell | *PINLYP* | -1.552854549 | 0.000765766 | 0.006171919 | 1 |
| T2DM | t_cell | *ANKRD65* | -3.501112935 | 0.000191734 | 0.002813391 | 0.930294521 |
| T2DM | t_cell | *LINC00957* | -3.626764379 | 4.38E-06 | 0.000545353 | 0.021268777 |
| T2DM | t_cell | *KIAA0040* | 2.056102212 | 0.001123092 | 0.008072949 | 1 |
| T2DM | t_cell | *AP001468.1* | -0.828213294 | 0.00988846 | 0.039038898 | 1 |
| T2DM | t_cell | *LINC00941* | -4.331093869 | 2.34E-05 | 0.001454217 | 0.113428947 |
| T2DM | t_cell | *AC019117.2* | -8.508276045 | 0.000164062 | 0.00266157 | 0.796027306 |
| T2DM | t_cell | *LINC00987* | 1.078964518 | 0.001534058 | 0.010099388 | 1 |
| T2DM | t_cell | *GS1-124K5.4* | 0.576904109 | 0.005327794 | 0.024858025 | 1 |
| T2DM | t_cell | *HLA-DQA2* | 1.151665163 | 2.90E-14 | 7.04E-11 | 1.41E-10 |
| T2DM | t_cell | *IRGM* | 1.072157249 | 0.000413135 | 0.004112304 | 1 |
| T2DM | t_cell | *LINC01143* | -5.322596538 | 0.000329867 | 0.003569928 | 1 |
| T2DM | t_cell | *LINC00211* | -3.9635924 | 0.011374702 | 0.04311723 | 1 |
| T2DM | t_cell | *TXNDC5* | -8.680184345 | 0.000459511 | 0.004354585 | 1 |
| T2DM | t_cell | *C1orf226* | -2.366757744 | 0.004564007 | 0.02190362 | 1 |
| T2DM | t_cell | *LILRA2* | -0.806704139 | 0.006051601 | 0.02718053 | 1 |
| T2DM | t_cell | *PSMB9* | 1.579448198 | 2.09E-08 | 9.23E-06 | 0.000101528 |
| T2DM | t_cell | *C1QTNF9* | -5.012377814 | 0.000135431 | 0.002475971 | 0.657109744 |
| T2DM | t_cell | *PNMA2* | -1.758992841 | 0.000360469 | 0.003777851 | 1 |
| T2DM | t_cell | *KRBOX1* | -17.94270461 | 0.000142096 | 0.002562837 | 0.689447939 |
| T2DM | t_cell | *TDGF1* | -1.695207578 | 0.000523528 | 0.004807683 | 1 |
| T2DM | t_cell | *HOGA1* | -3.616071643 | 0.000271003 | 0.003258353 | 1 |
| T2DM | t_cell | *MRPL20* | 3.68877165 | 0.000407623 | 0.004078878 | 1 |
| T2DM | t_cell | *DECR2* | -3.944206935 | 0.001513731 | 0.010019949 | 1 |

| | | | | | | |
|---|---|---|---|---|---|---|
| T2DM | t_cell | *ERVW-1* | -3.148648911 | 0.00131119 | 0.009180224 | 1 |
| T2DM | t_cell | *FGF14-IT1* | -3.760779377 | 0.000131301 | 0.00244943 | 0.637073514 |
| T2DM | t_cell | *IL10RB* | 3.65983174 | 7.18E-05 | 0.001903045 | 0.348257246 |
| T2DM | t_cell | *NME1-NME2* | 5.217292883 | 5.58E-05 | 0.00172419 | 0.270940801 |
| T2DM | t_cell | *PLA2G4B* | 4.92099633 | 4.47E-05 | 0.001701031 | 0.217072572 |
| T2DM | t_cell | *JMJD7* | 1.601017762 | 2.20E-05 | 0.001405861 | 0.106845398 |
| T2DM | t_cell | *APOBEC3D* | 2.138715965 | 6.03E-05 | 0.001745457 | 0.292690046 |
| T2DM | t_cell | *APOBEC3C* | 3.283181896 | 3.76E-05 | 0.001670757 | 0.182635974 |
| T2DM | t_cell | *CCDC13* | -1.38226229 | 0.000363849 | 0.0037884 | 1 |
| T2DM | t_cell | *C4A* | -2.665662315 | 9.13E-05 | 0.002099002 | 0.442889479 |
| T2DM | t_cell | *LINC00926* | -3.272228691 | 0.000908921 | 0.00699124 | 1 |
| T2DM | t_cell | *CDK11B* | 3.54105219 | 0.010052264 | 0.03955684 | 1 |
| T2DM | t_cell | *SRP14-AS1* | -0.697750819 | 0.001440855 | 0.00970692 | 1 |
| T2DM | t_cell | *GDNF-AS1* | -2.664816995 | 0.000290826 | 0.003341384 | 1 |
| T2DM | t_cell | *TMEM150C* | -1.47358978 | 0.00022928 | 0.003039531 | 1 |
| T2DM | t_cell | *LINC00942* | -7.290861942 | 9.78E-05 | 0.00214103 | 0.474693212 |
| T2DM | t_cell | *ZNF718* | 3.149226643 | 0.002529295 | 0.014353381 | 1 |
| T2DM | t_cell | *LINC00989* | -2.07586934 | 0.000227237 | 0.003033461 | 1 |
| T2DM | t_cell | *GLI4* | -2.791568995 | 0.004673124 | 0.022273083 | 1 |
| T2DM | t_cell | *SNHG18* | -2.563794878 | 0.000496133 | 0.004593965 | 1 |
| T2DM | t_cell | *LINC01094* | -3.091758052 | 0.001432042 | 0.009678502 | 1 |
| T2DM | t_cell | *PCDHGB2* | -9.097687259 | 0.00319452 | 0.016921193 | 1 |
| T2DM | t_cell | *SNHG9* | 2.575419378 | 0.002616166 | 0.014674723 | 1 |
| T2DM | t_cell | *PCDHA3* | -2.601907013 | 0.002344196 | 0.013698845 | 1 |
| T2DM | t_cell | *ZBED6* | -3.608204065 | 0.000241778 | 0.003079026 | 1 |
| T2DM | t_cell | *LINC00639* | -1.351471635 | 0.000127072 | 0.002417859 | 0.616554138 |
| T2DM | t_cell | *AC083843.1* | -25.67073781 | 0.000195817 | 0.002825716 | 0.950104282 |
| T2DM | t_cell | *PWRN1* | -1.545684963 | 0.009779438 | 0.038702965 | 1 |
| T2DM | t_cell | *C15orf65* | -1.577766075 | 0.000616824 | 0.0053539 | 1 |
| T2DM | t_cell | *C19orf84* | -14.17588815 | 0.000637248 | 0.005491879 | 1 |
| T2DM | t_cell | *LINC00672* | 3.23270036 | 3.15E-05 | 0.001619427 | 0.152643362 |
| T2DM | t_cell | *ZNF582-AS1* | -2.121794126 | 0.000275357 | 0.003278465 | 1 |
| T2DM | t_cell | *LIN37* | -4.446034421 | 0.004358589 | 0.021254145 | 1 |
| T2DM | t_cell | *NDUFA7* | -6.982095069 | 0.000234117 | 0.003045405 | 1 |
| T2DM | t_cell | *LINC00664* | 22.21154058 | 0.00140266 | 0.009585502 | 1 |
| T2DM | t_cell | *ZNF728* | -1.841461097 | 0.003723534 | 0.018838985 | 1 |
| T2DM | t_cell | *TRABD2B* | -1.752063593 | 0.001709242 | 0.010897822 | 1 |
| T2DM | t_cell | *YTHDF3-AS1* | -3.671933883 | 2.80E-06 | 0.000412234 | 0.013603709 |
| T2DM | t_cell | *FGF14-AS2* | -2.176397884 | 0.000196263 | 0.002825716 | 0.95226642 |
| T2DM | t_cell | *CASC15* | -1.762847752 | 0.008789569 | 0.035777675 | 1 |
| HTN | adipocyte | *RBM6* | -0.430279684 | 9.04E-06 | 0.00073078 | 0.043846773 |
| HTN | adipocyte | *CD79B* | -0.831666809 | 0.000722816 | 0.023073042 | 1 |
| HTN | adipocyte | *CEP68* | 1.576602307 | 5.50E-08 | 1.11E-05 | 0.000266763 |
| HTN | adipocyte | *RNASET2* | -0.435547882 | 0.001982085 | 0.04668484 | 1 |
| HTN | adipocyte | *ANAPC4* | 1.509840654 | 5.53E-05 | 0.003049418 | 0.268131936 |
| HTN | adipocyte | *SDCCAG8* | 1.164583607 | 7.27E-09 | 2.07E-06 | 3.53E-05 |
| HTN | adipocyte | *KIF1B* | 0.847188619 | 0.002093412 | 0.048367783 | 1 |
| HTN | adipocyte | *ITIH4* | 0.20234114 | 0.000546661 | 0.018548238 | 1 |
| HTN | adipocyte | *PIGV* | -0.544378601 | 1.21E-05 | 0.000921884 | 0.058709215 |
| HTN | adipocyte | *UHRF1BP1* | 0.790042572 | 4.70E-07 | 6.19E-05 | 0.002278719 |

| | | | | | | |
|---|---|---|---|---|---|---|
| HTN | adipocyte | *TIE1* | -0.755068594 | 0.000316117 | 0.012572135 | 1 |
| HTN | adipocyte | *POLR1A* | -0.611336198 | 0.000514537 | 0.017589608 | 1 |
| HTN | adipocyte | *FDFT1* | 1.451305976 | 1.18E-06 | 0.000135772 | 0.005702426 |
| HTN | adipocyte | *PLOD1* | 1.157098957 | 0.001692542 | 0.041686364 | 1 |
| HTN | adipocyte | *CAD* | 1.491179354 | 0.000427049 | 0.01557927 | 1 |
| HTN | adipocyte | *DDHD2* | 0.993610212 | 0.000823598 | 0.025612514 | 1 |
| HTN | adipocyte | *SNX10* | -0.624345376 | 3.47E-06 | 0.000348754 | 0.016831774 |
| HTN | adipocyte | *AAMDC* | -0.673727584 | 0.000162767 | 0.007312438 | 0.789743305 |
| HTN | adipocyte | *YPEL3* | 0.962708878 | 6.94E-05 | 0.003620308 | 0.336688616 |
| HTN | adipocyte | *PABPC4* | -1.085488095 | 1.12E-05 | 0.000894219 | 0.054547381 |
| HTN | adipocyte | *RGS17* | 0.632529179 | 0.002194041 | 0.049949907 | 1 |
| HTN | adipocyte | *SCD* | -0.758398107 | 3.22E-05 | 0.00220388 | 0.156475484 |
| HTN | adipocyte | *SETD1A* | 1.682839411 | 1.28E-05 | 0.000958991 | 0.062334387 |
| HTN | adipocyte | *MARCH2* | 0.829091272 | 1.82E-06 | 0.000201166 | 0.008851301 |
| HTN | adipocyte | *BIRC7* | -0.676001742 | 0.000939647 | 0.027750807 | 1 |
| HTN | adipocyte | *MANBAL* | 0.689782649 | 0.000825584 | 0.025612514 | 1 |
| HTN | adipocyte | *DNTTIP1* | -0.512030709 | 0.001271203 | 0.034459843 | 1 |
| HTN | adipocyte | *POLI* | -0.245597206 | 0.000194816 | 0.008439726 | 0.945249315 |
| HTN | adipocyte | *MGRN1* | 1.441021264 | 8.22E-05 | 0.004198022 | 0.398812097 |
| HTN | adipocyte | *MTHFSD* | 0.384253763 | 0.000173432 | 0.007649914 | 0.841490528 |
| HTN | adipocyte | *PYCARD* | 1.261887039 | 0.001222944 | 0.03378731 | 1 |
| HTN | adipocyte | *SNX16* | 2.190373018 | 5.53E-06 | 0.000504521 | 0.026821588 |
| HTN | adipocyte | *LIN7B* | -2.279949537 | 0.000683844 | 0.022268522 | 1 |
| HTN | adipocyte | *CD37* | -0.9715911 | 0.000301041 | 0.012071488 | 1 |
| HTN | adipocyte | *RABAC1* | 1.108594375 | 0.001404772 | 0.035919249 | 1 |
| HTN | adipocyte | *TAF6* | -0.466911588 | 0.000861848 | 0.026466361 | 1 |
| HTN | adipocyte | *NAGLU* | 1.900115873 | 3.51E-05 | 0.002269704 | 0.170227829 |
| HTN | adipocyte | *CCND1* | -1.13718233 | 8.92E-06 | 0.00073078 | 0.043256696 |
| HTN | adipocyte | *RNF130* | 0.846794456 | 0.000828764 | 0.025612514 | 1 |
| HTN | adipocyte | *PCCB* | 1.101938039 | 5.98E-08 | 1.12E-05 | 0.000290265 |
| HTN | adipocyte | *ACOT2* | 0.181205958 | 0.001361692 | 0.035713127 | 1 |
| HTN | adipocyte | *ZFP30* | 0.968657453 | 5.33E-05 | 0.003015043 | 0.258401264 |
| HTN | adipocyte | *TBX2* | -0.653842983 | 5.31E-08 | 1.11E-05 | 0.000257517 |
| HTN | adipocyte | *TEX14* | 1.216862152 | 6.02E-08 | 1.12E-05 | 0.000291871 |
| HTN | adipocyte | *MAPK8IP1* | -1.554117963 | 0.000347068 | 0.013259627 | 1 |
| HTN | adipocyte | *WFDC3* | -0.183517704 | 0.000342667 | 0.013195383 | 1 |
| HTN | adipocyte | *BTN2A2* | 0.416710464 | 5.07E-06 | 0.000473105 | 0.024601467 |
| HTN | adipocyte | *SNRPC* | -0.899528665 | 9.85E-05 | 0.004925908 | 0.477813051 |
| HTN | adipocyte | *ZNF391* | -0.247819853 | 6.75E-05 | 0.003582481 | 0.327330856 |
| HTN | adipocyte | *GRK4* | 0.176527636 | 0.001379858 | 0.035786011 | 1 |
| HTN | adipocyte | *GPR108* | 0.805241211 | 0.002035837 | 0.047489825 | 1 |
| HTN | adipocyte | *TSKS* | -0.739472621 | 0.001225591 | 0.03378731 | 1 |
| HTN | adipocyte | *TRAP1* | 0.527337813 | 4.98E-07 | 6.19E-05 | 0.002414631 |
| HTN | adipocyte | *ABHD8* | -0.862226394 | 3.72E-05 | 0.002374182 | 0.180437844 |
| HTN | adipocyte | *ADCY4* | 1.074697576 | 7.86E-06 | 0.000668961 | 0.038130758 |
| HTN | adipocyte | *CDC16* | 0.878175333 | 0.000236038 | 0.009872883 | 1 |
| HTN | adipocyte | *ATP8B3* | -0.334957931 | 0.000628711 | 0.021037986 | 1 |
| HTN | adipocyte | *PGPEP1* | 0.673201205 | 0.000356285 | 0.013505434 | 1 |
| HTN | adipocyte | *TRPM4* | -0.741459283 | 0.001012557 | 0.029243608 | 1 |
| HTN | adipocyte | *HAUS8* | -1.212183343 | 6.49E-08 | 1.17E-05 | 0.000314905 |

| | | | | | | |
|---|---|---|---|---|---|---|
| HTN | adipocyte | *PDLIM4* | 2.539277634 | 0.001824148 | 0.044220058 | 1 |
| HTN | adipocyte | *EXOC4* | 1.014834234 | 0.0018856 | 0.045054802 | 1 |
| HTN | adipocyte | *IMPA1* | 0.513260523 | 0.000650935 | 0.021632453 | 1 |
| HTN | adipocyte | *ACP2* | 2.623355891 | 5.06E-12 | 4.91E-09 | 2.46E-08 |
| HTN | adipocyte | *AGT* | 0.414811244 | 0.000374854 | 0.013990713 | 1 |
| HTN | adipocyte | *NMT1* | 0.795287488 | 9.46E-08 | 1.53E-05 | 0.00045918 |
| HTN | adipocyte | *SLC37A4* | 0.821443691 | 3.48E-05 | 0.002269704 | 0.168801493 |
| HTN | adipocyte | *NUSAP1* | 1.482637713 | 3.41E-05 | 0.002269645 | 0.165290913 |
| HTN | adipocyte | *DYNC2LI1* | 0.849005652 | 0.001058922 | 0.029871444 | 1 |
| HTN | adipocyte | *ARL3* | 0.708749563 | 0.001837304 | 0.044220058 | 1 |
| HTN | adipocyte | *G3BP2* | 0.850215834 | 0.000820896 | 0.025612514 | 1 |
| HTN | adipocyte | *FBN2* | 0.231588643 | 5.10E-05 | 0.002978618 | 0.247225296 |
| HTN | adipocyte | *FITM1* | 1.208798412 | 0.002203067 | 0.049949907 | 1 |
| HTN | adipocyte | *RTN1* | 0.564999181 | 0.000327578 | 0.012817823 | 1 |
| HTN | adipocyte | *ULK3* | 3.443343736 | 5.08E-05 | 0.002978618 | 0.246497453 |
| HTN | adipocyte | *DEF8* | 1.1844541 | 2.61E-06 | 0.000269163 | 0.01265068 |
| HTN | adipocyte | *MYO19* | 1.391029188 | 0.000704493 | 0.022637096 | 1 |
| HTN | adipocyte | *SCRN2* | -0.178710241 | 0.000987177 | 0.028681324 | 1 |
| HTN | adipocyte | *C18orf8* | -1.337190418 | 1.47E-05 | 0.001060958 | 0.071084211 |
| HTN | adipocyte | *SNX27* | 1.346445401 | 0.000212295 | 0.009115545 | 1 |
| HTN | adipocyte | *CREB3L4* | -1.110870035 | 1.89E-06 | 0.000203891 | 0.009175091 |
| HTN | adipocyte | *CEP170* | 1.579308292 | 0.001049457 | 0.029871444 | 1 |
| HTN | adipocyte | *PLEKHA6* | 0.662530593 | 0.000231204 | 0.009754805 | 1 |
| HTN | adipocyte | *CIAO1* | 1.419469395 | 0.001237533 | 0.033923788 | 1 |
| HTN | adipocyte | *CISD2* | 2.825313418 | 2.37E-05 | 0.001645115 | 0.11515803 |
| HTN | adipocyte | *ABT1* | 0.414664187 | 0.001647402 | 0.041202043 | 1 |
| HTN | adipocyte | *IRAK1BP1* | 1.754836589 | 1.50E-07 | 2.20E-05 | 0.000726449 |
| HTN | adipocyte | *SLC18B1* | -0.536829346 | 0.001924821 | 0.045557232 | 1 |
| HTN | adipocyte | *ADHFE1* | -0.233588651 | 0.001386597 | 0.035786011 | 1 |
| HTN | adipocyte | *HSD17B12* | -0.466781263 | 2.07E-09 | 8.36E-07 | 1.00E-05 |
| HTN | adipocyte | *TNKS1BP1* | 0.973723888 | 9.07E-11 | 6.29E-08 | 4.40E-07 |
| HTN | adipocyte | *TTC12* | 0.603971891 | 0.000948636 | 0.027750807 | 1 |
| HTN | adipocyte | *TBX6* | 0.212969619 | 0.001106046 | 0.030842167 | 1 |
| HTN | adipocyte | *CWF19L2* | 1.450035773 | 4.29E-05 | 0.002571941 | 0.208327252 |
| HTN | adipocyte | *MED21* | 1.53997767 | 0.00024715 | 0.010162461 | 1 |
| HTN | adipocyte | *ABCA5* | 0.78064338 | 0.000949353 | 0.027750807 | 1 |
| HTN | adipocyte | *NEIL2* | 1.003498379 | 4.50E-09 | 1.47E-06 | 2.18E-05 |
| HTN | adipocyte | *NDUFAF6* | -1.339298061 | 7.71E-05 | 0.00398031 | 0.374149131 |
| HTN | adipocyte | *SFXN2* | 0.553513456 | 1.73E-08 | 4.20E-06 | 8.41E-05 |
| HTN | adipocyte | *MED19* | 0.698672991 | 0.00156083 | 0.03944348 | 1 |
| HTN | adipocyte | *CLEC18A* | -0.636825747 | 3.52E-06 | 0.000348754 | 0.017088944 |
| HTN | adipocyte | *SLC5A11* | -0.384712331 | 8.22E-07 | 9.97E-05 | 0.003987232 |
| HTN | adipocyte | *S100B* | -0.194498639 | 5.23E-05 | 0.003015043 | 0.253679344 |
| HTN | adipocyte | *ZNF208* | -0.830853748 | 0.001379017 | 0.035786011 | 1 |
| HTN | adipocyte | *ABCF3* | 0.943629355 | 3.82E-08 | 8.43E-06 | 0.00018542 |
| HTN | adipocyte | *MRPL55* | 1.578506543 | 3.95E-12 | 4.79E-09 | 1.92E-08 |
| HTN | adipocyte | *LYSMD1* | -0.475378661 | 5.34E-05 | 0.003015043 | 0.259293741 |
| HTN | adipocyte | *DCAF16* | -1.990601174 | 0.000142052 | 0.00656416 | 0.689236795 |
| HTN | adipocyte | *RFT1* | -0.652829828 | 0.000341476 | 0.013195383 | 1 |
| HTN | adipocyte | *CDC25A* | -2.855707835 | 4.54E-09 | 1.47E-06 | 2.20E-05 |

| | | | | | | |
|---|---|---|---|---|---|---|
| HTN | adipocyte | *ZNF589* | -1.107254303 | 5.23E-09 | 1.59E-06 | 2.54E-05 |
| HTN | adipocyte | *RNF123* | 1.076308141 | 8.33E-11 | 6.29E-08 | 4.04E-07 |
| HTN | adipocyte | *MST1R* | 0.555525994 | 3.83E-05 | 0.002411424 | 0.185679658 |
| HTN | adipocyte | *ERAP1* | -1.311628437 | 3.71E-06 | 0.00035663 | 0.01801479 |
| HTN | adipocyte | *ERAP2* | 0.250188121 | 0.001271293 | 0.034459843 | 1 |
| HTN | adipocyte | *RPP25L* | -0.74395845 | 0.000931595 | 0.027750807 | 1 |
| HTN | adipocyte | *ARL5B* | -1.703189701 | 5.89E-05 | 0.003210764 | 0.285758023 |
| HTN | adipocyte | *CYB5R2* | 0.45797957 | 0.00201894 | 0.047323165 | 1 |
| HTN | adipocyte | *TRIM66* | -0.275700474 | 4.00E-05 | 0.002487836 | 0.194051205 |
| HTN | adipocyte | *RIMKLB* | -1.00134121 | 2.21E-07 | 3.15E-05 | 0.001071967 |
| HTN | adipocyte | *CNPY4* | 0.892862259 | 0.001797223 | 0.043819732 | 1 |
| HTN | adipocyte | *ZNF641* | 2.082929431 | 0.000410934 | 0.015104922 | 1 |
| HTN | adipocyte | *SRR* | 0.323973939 | 0.000116029 | 0.005574004 | 0.562974418 |
| HTN | adipocyte | *ZNF598* | -0.970835363 | 0.001786875 | 0.043787462 | 1 |
| HTN | adipocyte | *PDHB* | 1.427553753 | 0.001840983 | 0.044220058 | 1 |
| HTN | adipocyte | *UPF3A* | 0.747194491 | 0.001283215 | 0.034589772 | 1 |
| HTN | adipocyte | *C8orf46* | 0.156337437 | 0.000868226 | 0.026494544 | 1 |
| HTN | adipocyte | *CPT1C* | 1.059882832 | 1.48E-07 | 2.20E-05 | 0.000720208 |
| HTN | adipocyte | *RAB24* | -2.418312678 | 0.000676041 | 0.022163168 | 1 |
| HTN | adipocyte | *INO80E* | -0.834617022 | 3.50E-09 | 1.31E-06 | 1.70E-05 |
| HTN | adipocyte | *RGPD8* | -0.150827538 | 0.00094943 | 0.027750807 | 1 |
| HTN | adipocyte | *MYO7B* | 0.773312933 | 3.41E-05 | 0.002269645 | 0.165684074 |
| HTN | adipocyte | *DNAJC18* | -0.666259968 | 0.000326504 | 0.012817823 | 1 |
| HTN | adipocyte | *TTLL6* | -2.396293933 | 6.32E-10 | 3.41E-07 | 3.07E-06 |
| HTN | adipocyte | *PAQR8* | -0.576803692 | 0.001406566 | 0.035919249 | 1 |
| HTN | adipocyte | *MRPL36* | 0.35647405 | 0.001338708 | 0.035494047 | 1 |
| HTN | adipocyte | *RGS19* | -0.470189217 | 1.90E-08 | 4.39E-06 | 9.22E-05 |
| HTN | adipocyte | *NME6* | 1.481679584 | 1.96E-10 | 1.19E-07 | 9.50E-07 |
| HTN | adipocyte | *CTSW* | -0.110577264 | 4.16E-05 | 0.002549595 | 0.20166394 |
| HTN | adipocyte | *ZNF738* | 0.392015151 | 0.001602891 | 0.040296516 | 1 |
| HTN | adipocyte | *NOC3L* | -1.281272929 | 0.001661419 | 0.04133952 | 1 |
| HTN | adipocyte | *PPP1R14B* | 2.071574865 | 0.000215063 | 0.009153398 | 1 |
| HTN | adipocyte | *MST1* | -0.339958433 | 2.49E-06 | 0.000262775 | 0.012087663 |
| HTN | adipocyte | *C11orf80* | -1.424613893 | 1.05E-07 | 1.64E-05 | 0.000508799 |
| HTN | adipocyte | *SNX31* | -0.37419367 | 5.62E-06 | 0.000504521 | 0.027244116 |
| HTN | adipocyte | *SEZ6L2* | 0.615615017 | 0.00010563 | 0.005229766 | 0.512517094 |
| HTN | adipocyte | *CCNE2* | -1.0542256 | 1.36E-08 | 3.48E-06 | 6.61E-05 |
| HTN | adipocyte | *TMEM9B* | 1.702470357 | 0.000154716 | 0.007019612 | 0.750680794 |
| HTN | adipocyte | *CTDNEP1* | 1.127862246 | 4.20E-05 | 0.002549595 | 0.203967632 |
| HTN | adipocyte | *ORAI3* | 1.068315812 | 2.43E-07 | 3.37E-05 | 0.0011806 |
| HTN | adipocyte | *CRLF3* | 0.888804824 | 0.000930202 | 0.027750807 | 1 |
| HTN | adipocyte | *MTHFR* | -1.552887026 | 5.53E-05 | 0.003049418 | 0.268348764 |
| HTN | adipocyte | *ASB8* | 1.731365919 | 0.000510859 | 0.017589608 | 1 |
| HTN | adipocyte | *ODF3B* | 0.485015005 | 0.001686612 | 0.041686364 | 1 |
| HTN | adipocyte | *GEN1* | -0.29842535 | 0.002121232 | 0.04854819 | 1 |
| HTN | adipocyte | *RPP25* | 0.550108526 | 1.37E-06 | 0.000155141 | 0.006671055 |
| HTN | adipocyte | *MPI* | -1.647379936 | 6.22E-15 | 1.51E-11 | 3.02E-11 |
| HTN | adipocyte | *TRIM73* | -1.178057423 | 0.000692319 | 0.0223942 | 1 |
| HTN | adipocyte | *HYI* | -0.56710084 | 1.12E-06 | 0.000131973 | 0.005410899 |
| HTN | adipocyte | *CTC1* | 1.242156177 | 0.000121946 | 0.005744498 | 0.591683302 |

| | | | | | | |
|---|---|---|---|---|---|---|
| HTN | adipocyte | *CLDN7* | -0.822231658 | 3.75E-06 | 0.00035663 | 0.018188145 |
| HTN | adipocyte | *ADO* | 1.092904485 | 8.66E-10 | 4.20E-07 | 4.20E-06 |
| HTN | adipocyte | *CLN8* | 0.499226603 | 1.15E-05 | 0.000897643 | 0.055653897 |
| HTN | adipocyte | *FES* | -2.07816945 | 1.22E-16 | 5.92E-13 | 5.92E-13 |
| HTN | adipocyte | *ZNF320* | 0.256059614 | 0.000125086 | 0.005835752 | 0.606918232 |
| HTN | adipocyte | *A3GALT2* | -0.409385193 | 6.79E-05 | 0.003582481 | 0.329583984 |
| HTN | adipocyte | *NUTM2A* | -0.580215932 | 0.000118993 | 0.005660332 | 0.577353819 |
| HTN | adipocyte | *CDK10* | -0.383487511 | 0.000170898 | 0.007607324 | 0.829198315 |
| HTN | adipocyte | *CCDC30* | 0.552237263 | 0.001300774 | 0.034678554 | 1 |
| HTN | adipocyte | *KLHL17* | 0.568259711 | 0.001357594 | 0.035713127 | 1 |
| HTN | adipocyte | *FBF1* | -1.029350136 | 5.83E-06 | 0.00051424 | 0.028283212 |
| HTN | adipocyte | *HLA-DRB1* | -0.276310413 | 0.0004864 | 0.017101559 | 1 |
| HTN | adipocyte | *COLCA1* | -0.70260194 | 0.000461879 | 0.016724154 | 1 |
| HTN | adipocyte | *HRCT1* | 0.505996562 | 0.0005813 | 0.01958659 | 1 |
| HTN | adipocyte | *HIST1H3E* | 0.546709484 | 7.70E-08 | 1.33E-05 | 0.000373573 |
| HTN | adipocyte | *WDR5B* | 1.236442285 | 0.000114434 | 0.00555233 | 0.55523299 |
| HTN | adipocyte | *ZNF100* | -0.789895905 | 9.53E-05 | 0.004815131 | 0.462252567 |
| HTN | adipocyte | *SULT1A2* | 1.01763742 | 1.22E-05 | 0.000921884 | 0.059000556 |
| HTN | adipocyte | *MFAP5* | -0.536742548 | 0.000655531 | 0.02163699 | 1 |
| HTN | adipocyte | *RPS26* | -0.42546912 | 1.32E-05 | 0.000970569 | 0.064057538 |
| HTN | adipocyte | *TMEM116* | -0.620032132 | 3.46E-13 | 5.59E-10 | 1.68E-09 |
| HTN | adipocyte | *HLA-DRB5* | -0.131666004 | 2.14E-05 | 0.001506392 | 0.103941033 |
| HTN | adipocyte | *DENND4B* | -0.830989161 | 0.000500668 | 0.017476555 | 1 |
| HTN | adipocyte | *SHISA4* | -4.793374347 | 9.18E-09 | 2.47E-06 | 4.45E-05 |
| HTN | adipocyte | *HSD17B8* | -0.536581614 | 0.000154802 | 0.007019612 | 0.751098483 |
| HTN | adipocyte | *PRRT1* | 0.508377095 | 4.89E-07 | 6.19E-05 | 0.002374831 |
| HTN | adipocyte | *HLA-C* | -0.459831828 | 4.86E-07 | 6.19E-05 | 0.002360018 |
| HTN | adipocyte | *POU5F1* | -0.117407272 | 0.001097675 | 0.030785668 | 1 |
| HTN | adipocyte | *SUPT4H1* | 3.065505908 | 7.84E-06 | 0.000668961 | 0.038026153 |
| HTN | adipocyte | *MXD3* | -0.495829536 | 0.002070223 | 0.04806086 | 1 |
| HTN | adipocyte | *TMEM110* | 0.856843303 | 0.00029373 | 0.011876467 | 1 |
| HTN | adipocyte | *ATF6B* | 1.520691736 | 1.44E-09 | 6.34E-07 | 6.97E-06 |
| HTN | adipocyte | *PPM1N* | 0.402728425 | 0.001894307 | 0.045054802 | 1 |
| HTN | adipocyte | *LTB4R* | -1.307426926 | 0.000514782 | 0.017589608 | 1 |
| HTN | adipocyte | *GALT* | 1.328188372 | 0.000467855 | 0.016815072 | 1 |
| HTN | adipocyte | *UBXN2B* | 0.744734191 | 0.00147336 | 0.037427979 | 1 |
| HTN | adipocyte | *AC016773.1* | 0.56894646 | 0.000262099 | 0.010686596 | 1 |
| HTN | adipocyte | *AP4M1* | -0.513210756 | 0.000948453 | 0.027750807 | 1 |
| HTN | adipocyte | *SLC12A8* | 0.540227445 | 0.000481697 | 0.017099817 | 1 |
| HTN | adipocyte | *TIAF1* | 0.864671797 | 6.79E-05 | 0.003582481 | 0.329588237 |
| HTN | adipocyte | *EXOSC6* | -1.203174582 | 0.00010934 | 0.005358779 | 0.530519142 |
| HTN | adipocyte | *OSER1-AS1* | 0.525963838 | 0.002116718 | 0.04854819 | 1 |
| HTN | adipocyte | *AL049840.1* | -1.337937994 | 0.000482827 | 0.017099817 | 1 |
| HTN | adipocyte | *TMA7* | 1.235952685 | 0.000246395 | 0.010162461 | 1 |
| HTN | adipocyte | *HLA-DQA2* | 0.189411625 | 1.98E-05 | 0.001414286 | 0.096171454 |
| HTN | adipocyte | *TNFSF12* | 1.188759883 | 9.20E-08 | 1.53E-05 | 0.000446342 |
| HTN | adipocyte | *NSUN6* | 1.417745479 | 0.001053329 | 0.029871444 | 1 |
| HTN | adipocyte | *ZNF709* | -15.21049849 | 8.37E-06 | 0.000699916 | 0.040595103 |
| HTN | adipocyte | *WFDC6* | -1.448323828 | 0.000395649 | 0.014654102 | 1 |
| HTN | adipocyte | *PLA2G4B* | 1.332727853 | 0.000179204 | 0.007833326 | 0.869499177 |

| | | | | | | |
|---|---|---|---|---|---|---|
| HTN | adipocyte | *JMJD7* | 0.35937901 | 0.001045187 | 0.029871444 | 1 |
| HTN | adipocyte | *HMBS* | 0.810662086 | 0.001300803 | 0.034678554 | 1 |
| HTN | adipocyte | *LINC00638* | 0.929051445 | 0.000810884 | 0.025612514 | 1 |
| HTN | adipocyte | *C19orf84* | 23.61844076 | 0.000359939 | 0.013538156 | 1 |
| HTN | ASPC | *DBNDD1* | 0.15485111 | 0.002614344 | 0.017095412 | 1 |
| HTN | ASPC | *RBM6* | -0.202626691 | 1.81E-05 | 0.00035999 | 0.087837636 |
| HTN | ASPC | *ARHGAP33* | -0.242494637 | 0.005259544 | 0.028673381 | 1 |
| HTN | ASPC | *POLR2J* | 1.481864621 | 0.000231261 | 0.002784319 | 1 |
| HTN | ASPC | *CIAPIN1* | 0.742984678 | 0.000238314 | 0.002817365 | 1 |
| HTN | ASPC | *RHBDD2* | 0.383994094 | 0.004090455 | 0.023825795 | 1 |
| HTN | ASPC | *BAIAP2L1* | 1.834505551 | 0.000107195 | 0.001552564 | 0.520108847 |
| HTN | ASPC | *AGK* | 0.448674277 | 0.005118127 | 0.028283771 | 1 |
| HTN | ASPC | *SCIN* | -0.317638164 | 0.003048928 | 0.019162436 | 1 |
| HTN | ASPC | *CD79B* | -0.313330654 | 0.008948675 | 0.041077551 | 1 |
| HTN | ASPC | *TSR3* | 1.354016642 | 0.00383179 | 0.022756238 | 1 |
| HTN | ASPC | *PIGQ* | 0.30742891 | 0.007121722 | 0.035170054 | 1 |
| HTN | ASPC | *PSMB1* | 1.084952319 | 0.006821098 | 0.034014353 | 1 |
| HTN | ASPC | *IL32* | 0.962300034 | 0.009250615 | 0.041947648 | 1 |
| HTN | ASPC | *MED24* | 1.209018592 | 0.002982603 | 0.01891711 | 1 |
| HTN | ASPC | *TRAF3IP3* | 0.236512484 | 0.006233616 | 0.031971992 | 1 |
| HTN | ASPC | *DCN* | -0.323297545 | 0.003859141 | 0.022779261 | 1 |
| HTN | ASPC | *CEP68* | 0.924815839 | 4.78E-10 | 1.22E-07 | 2.32E-06 |
| HTN | ASPC | *TACC3* | 1.474103836 | 1.00E-06 | 3.53E-05 | 0.004865491 |
| HTN | ASPC | *NUDCD3* | 0.325903163 | 0.001636425 | 0.012103558 | 1 |
| HTN | ASPC | *CHDH* | 3.512627172 | 3.61E-09 | 3.89E-07 | 1.75E-05 |
| HTN | ASPC | *RNH1* | 0.422388653 | 0.001313874 | 0.010088942 | 1 |
| HTN | ASPC | *GCLM* | 1.379910563 | 0.000156604 | 0.002087477 | 0.759841488 |
| HTN | ASPC | *RNASET2* | -0.190947621 | 0.006283852 | 0.032195616 | 1 |
| HTN | ASPC | *INSRR* | -0.859074987 | 7.88E-08 | 3.98E-06 | 0.000382121 |
| HTN | ASPC | *VEZT* | 0.903127054 | 0.006621999 | 0.033399104 | 1 |
| HTN | ASPC | *SNX1* | 1.090826184 | 1.19E-05 | 0.000263104 | 0.057619803 |
| HTN | ASPC | *PIAS1* | 0.517588116 | 0.011103464 | 0.047973293 | 1 |
| HTN | ASPC | *ADSS* | 1.31504125 | 0.001723755 | 0.012576931 | 1 |
| HTN | ASPC | *AGA* | 1.274522359 | 4.88E-05 | 0.000828806 | 0.236982018 |
| HTN | ASPC | *AP2S1* | 1.211115257 | 0.010500593 | 0.046149344 | 1 |
| HTN | ASPC | *ARAP2* | 0.980824366 | 1.84E-06 | 5.83E-05 | 0.008934547 |
| HTN | ASPC | *ANO2* | 1.627417153 | 1.59E-07 | 7.02E-06 | 0.000771703 |
| HTN | ASPC | *RRM2B* | 0.865869194 | 0.005183542 | 0.028380408 | 1 |
| HTN | ASPC | *MAPK9* | 1.282515578 | 0.003179839 | 0.019882191 | 1 |
| HTN | ASPC | *CYBA* | 1.153247796 | 0.001216559 | 0.009520557 | 1 |
| HTN | ASPC | *THOC3* | 4.856899754 | 2.00E-07 | 8.51E-06 | 0.000970101 |
| HTN | ASPC | *ANAPC4* | 1.077509606 | 1.36E-07 | 6.21E-06 | 0.000658644 |
| HTN | ASPC | *LY75* | 1.42682871 | 0.003485562 | 0.021246167 | 1 |
| HTN | ASPC | *SDCCAG8* | 0.655711775 | 1.25E-10 | 4.05E-08 | 6.07E-07 |
| HTN | ASPC | *KIF1B* | 0.537850353 | 0.000172212 | 0.00226442 | 0.835570929 |
| HTN | ASPC | *TBC1D22A* | 0.828778843 | 0.001126482 | 0.009079223 | 1 |
| HTN | ASPC | *PLEKHH1* | 0.288245517 | 0.001769858 | 0.012816943 | 1 |
| HTN | ASPC | *ITIH4* | 0.101291567 | 0.000327017 | 0.003622576 | 1 |
| HTN | ASPC | *NPFFR2* | 2.262624735 | 1.86E-06 | 5.83E-05 | 0.009015786 |
| HTN | ASPC | *DNAJC25* | 2.798231253 | 7.57E-06 | 0.000177149 | 0.036734339 |

| | | | | | | |
|---|---|---|---|---|---|---|
| HTN | ASPC | *PIGV* | -0.254563119 | 2.42E-05 | 0.000461691 | 0.117269494 |
| HTN | ASPC | *EIF4B* | 0.286500529 | 0.003005649 | 0.019008969 | 1 |
| HTN | ASPC | *ADCK1* | 2.109254656 | 2.26E-05 | 0.000444214 | 0.109720962 |
| HTN | ASPC | *HAGH* | 0.412376744 | 0.000190691 | 0.00242059 | 0.925231188 |
| HTN | ASPC | *RNF4* | 1.149564885 | 0.006738463 | 0.033796734 | 1 |
| HTN | ASPC | *CASP8* | 0.745043745 | 0.002544098 | 0.016794507 | 1 |
| HTN | ASPC | *TMEM161A* | 1.20153638 | 0.007682161 | 0.037014742 | 1 |
| HTN | ASPC | *SLC12A2* | 1.667901905 | 1.66E-05 | 0.000336732 | 0.080437815 |
| HTN | ASPC | *DDX20* | 0.453447554 | 0.001391985 | 0.010602683 | 1 |
| HTN | ASPC | *BTBD1* | 0.884735448 | 0.002567572 | 0.016926441 | 1 |
| HTN | ASPC | *PMS1* | 0.756232714 | 0.009532523 | 0.042865433 | 1 |
| HTN | ASPC | *UHRF1BP1* | 0.437236698 | 1.27E-08 | 9.39E-07 | 6.14E-05 |
| HTN | ASPC | *ANKRD44* | 0.43691746 | 0.002390663 | 0.016134206 | 1 |
| HTN | ASPC | *TMEM206* | 0.272133012 | 0.010822324 | 0.046980056 | 1 |
| HTN | ASPC | *TIE1* | -0.29031554 | 0.006462242 | 0.032832248 | 1 |
| HTN | ASPC | *STOML1* | 3.510109276 | 0.000669134 | 0.006137314 | 1 |
| HTN | ASPC | *RRP15* | 0.212770389 | 0.006723725 | 0.033771753 | 1 |
| HTN | ASPC | *DHX8* | 1.296808945 | 0.005053931 | 0.02810033 | 1 |
| HTN | ASPC | *ROGDI* | 3.861004462 | 2.42E-05 | 0.000461691 | 0.117250042 |
| HTN | ASPC | *HEATR6* | 0.451561503 | 0.004581333 | 0.026099689 | 1 |
| HTN | ASPC | *POLR1A* | -0.270490549 | 0.001694572 | 0.012438828 | 1 |
| HTN | ASPC | *SRBD1* | 1.008632203 | 0.000839409 | 0.007338399 | 1 |
| HTN | ASPC | *IFT80* | -1.20803153 | 0.004328959 | 0.024915909 | 1 |
| HTN | ASPC | *DRD4* | 2.09803725 | 0.001987417 | 0.014077297 | 1 |
| HTN | ASPC | *PLA2G10* | 5.110448036 | 0.000267446 | 0.003082301 | 1 |
| HTN | ASPC | *ZXDC* | 1.05909339 | 0.000938986 | 0.007937214 | 1 |
| HTN | ASPC | *GBA2* | 1.463686314 | 0.001000445 | 0.008354838 | 1 |
| HTN | ASPC | *WDR1* | 1.101438774 | 0.001363539 | 0.010418723 | 1 |
| HTN | ASPC | *SNX13* | 0.891015951 | 0.006775867 | 0.033928285 | 1 |
| HTN | ASPC | *RPS6KA2* | 0.813251211 | 3.10E-05 | 0.000568036 | 0.150529661 |
| HTN | ASPC | *ING3* | 1.000158198 | 0.008211758 | 0.038720552 | 1 |
| HTN | ASPC | *FCGR2B* | 0.826247699 | 0.010906499 | 0.047248514 | 1 |
| HTN | ASPC | *TRNT1* | 0.690909035 | 0.007891708 | 0.037724698 | 1 |
| HTN | ASPC | *MOV10L1* | 0.454989826 | 0.007645969 | 0.036876982 | 1 |
| HTN | ASPC | *LLGL2* | 1.205820856 | 1.46E-05 | 0.000313582 | 0.071049547 |
| HTN | ASPC | *SMARCE1* | 0.528455621 | 0.00372017 | 0.022299659 | 1 |
| HTN | ASPC | *PPP2R2C* | 3.562630992 | 5.86E-08 | 3.23E-06 | 0.000284305 |
| HTN | ASPC | *ZZEF1* | 0.850705706 | 0.000414037 | 0.004338894 | 1 |
| HTN | ASPC | *TIPIN* | 0.138720433 | 0.008162835 | 0.038592365 | 1 |
| HTN | ASPC | *GRAMD4* | 0.585956378 | 0.008250219 | 0.03875553 | 1 |
| HTN | ASPC | *ADD2* | 0.794862987 | 0.000125954 | 0.001757101 | 0.611128904 |
| HTN | ASPC | *VPS9D1* | 1.181391845 | 0.000148036 | 0.002006345 | 0.718271356 |
| HTN | ASPC | *ANKRD13A* | 1.66200391 | 1.16E-05 | 0.000258862 | 0.056431898 |
| HTN | ASPC | *XAB2* | 1.363478107 | 0.003717186 | 0.022299659 | 1 |
| HTN | ASPC | *AMPH* | -0.398113522 | 0.003907581 | 0.022953489 | 1 |
| HTN | ASPC | *FDFT1* | 1.059824688 | 1.09E-10 | 3.87E-08 | 5.29E-07 |
| HTN | ASPC | *SCGN* | 5.88796919 | 0.00259156 | 0.01699223 | 1 |
| HTN | ASPC | *DNM2* | 1.020767387 | 0.000133024 | 0.001833617 | 0.645433177 |
| HTN | ASPC | *SCTR* | 1.978998866 | 5.53E-06 | 0.000141321 | 0.026850921 |
| HTN | ASPC | *PSEN1* | 0.650865561 | 0.009449828 | 0.04265169 | 1 |

| | | | | | | |
|---|---|---|---|---|---|---|
| HTN | ASPC | *ZNF506* | 0.572833487 | 0.001058855 | 0.008707735 | 1 |
| HTN | ASPC | *PCDHB4* | -1.996499811 | 0.001488606 | 0.011128992 | 1 |
| HTN | ASPC | *ME2* | 1.455965061 | 0.000293179 | 0.003300477 | 1 |
| HTN | ASPC | *ZCCHC6* | 0.962346316 | 0.005140368 | 0.028374362 | 1 |
| HTN | ASPC | *PLOD1* | 0.707246383 | 0.000176643 | 0.002297783 | 0.857073029 |
| HTN | ASPC | *WBP11* | 1.418005532 | 0.003550169 | 0.021504895 | 1 |
| HTN | ASPC | *CAD* | 0.905443097 | 3.29E-05 | 0.000600086 | 0.159622959 |
| HTN | ASPC | *MTIF2* | 1.35508074 | 0.003227116 | 0.020054426 | 1 |
| HTN | ASPC | *DDHD2* | 0.563349097 | 0.000305303 | 0.0034132 | 1 |
| HTN | ASPC | *EIF2AK1* | 1.04792942 | 0.00443276 | 0.025452961 | 1 |
| HTN | ASPC | *NME8* | -7.864996389 | 0.010575024 | 0.046191181 | 1 |
| HTN | ASPC | *SNX10* | -0.253806578 | 9.63E-05 | 0.00141995 | 0.467163437 |
| HTN | ASPC | *ALG9* | 2.521840743 | 0.000351859 | 0.003797577 | 1 |
| HTN | ASPC | *TRIP6* | 0.695783584 | 0.005881027 | 0.030748227 | 1 |
| HTN | ASPC | *LPCAT2* | 1.151410901 | 0.009233331 | 0.041908441 | 1 |
| HTN | ASPC | *AAMDC* | -0.326838237 | 0.000222115 | 0.002714617 | 1 |
| HTN | ASPC | *DOCK3* | 1.751740041 | 1.55E-06 | 5.00E-05 | 0.007496625 |
| HTN | ASPC | *NSFL1C* | 0.41676051 | 0.005313335 | 0.028872849 | 1 |
| HTN | ASPC | *SNX5* | 1.225839139 | 0.006328118 | 0.032388218 | 1 |
| HTN | ASPC | *RPH3A* | 1.849106617 | 0.00011218 | 0.001615123 | 0.544296385 |
| HTN | ASPC | *KIF16B* | 0.93145145 | 0.011191878 | 0.048312269 | 1 |
| HTN | ASPC | *TBX5* | -0.586115487 | 0.006617722 | 0.033399104 | 1 |
| HTN | ASPC | *GRAMD1A* | 1.502293899 | 0.000221316 | 0.002711682 | 1 |
| HTN | ASPC | *MLF2* | 0.435683039 | 0.001249764 | 0.009717714 | 1 |
| HTN | ASPC | *ANKRD24* | 2.637027529 | 4.48E-07 | 1.71E-05 | 0.00217241 |
| HTN | ASPC | *YPEL3* | 0.548089702 | 5.18E-06 | 0.000136649 | 0.025143413 |
| HTN | ASPC | *LYZ* | 0.379052996 | 0.003798875 | 0.022616123 | 1 |
| HTN | ASPC | *RAB11FIP3* | 0.534296057 | 0.011507312 | 0.049248514 | 1 |
| HTN | ASPC | *GNPTG* | 0.831169667 | 0.009946231 | 0.044306862 | 1 |
| HTN | ASPC | *PABPC4* | -0.483667788 | 7.63E-05 | 0.001175435 | 0.370262063 |
| HTN | ASPC | *DTX2* | 1.592503628 | 0.008065666 | 0.038292181 | 1 |
| HTN | ASPC | *TF* | 2.479527676 | 5.07E-05 | 0.000847588 | 0.246047162 |
| HTN | ASPC | *RGS17* | 0.564886319 | 9.40E-06 | 0.000213052 | 0.045593222 |
| HTN | ASPC | *PPP2R3C* | 0.391477775 | 0.008168664 | 0.038592365 | 1 |
| HTN | ASPC | *SUPT16H* | 1.099802603 | 0.009193283 | 0.041771198 | 1 |
| HTN | ASPC | *LRRFIP2* | 0.92880204 | 0.001235626 | 0.009654197 | 1 |
| HTN | ASPC | *GABRP* | 0.356729709 | 0.010768309 | 0.046848446 | 1 |
| HTN | ASPC | *MSH2* | 1.118886502 | 0.011510267 | 0.049248514 | 1 |
| HTN | ASPC | *NANS* | 1.485753584 | 0.000172799 | 0.002265797 | 0.838419122 |
| HTN | ASPC | *WAC* | 0.753426508 | 0.00580509 | 0.030504952 | 1 |
| HTN | ASPC | *DSP* | 2.712935716 | 6.95E-08 | 3.71E-06 | 0.000337242 |
| HTN | ASPC | *SCD* | -0.379822827 | 6.93E-05 | 0.00108825 | 0.336269357 |
| HTN | ASPC | *TSPAN15* | 1.568937729 | 2.66E-05 | 0.000504351 | 0.129113967 |
| HTN | ASPC | *SETD1A* | 1.002150741 | 2.72E-07 | 1.12E-05 | 0.001321184 |
| HTN | ASPC | *MARCH2* | 0.465455569 | 3.90E-08 | 2.31E-06 | 0.000189045 |
| HTN | ASPC | *MTAP* | -0.48201126 | 0.006366633 | 0.032543904 | 1 |
| HTN | ASPC | *CYTH4* | 2.237609236 | 3.55E-08 | 2.19E-06 | 0.000172279 |
| HTN | ASPC | *SH3BP1* | 4.129911058 | 1.75E-07 | 7.64E-06 | 0.000847984 |
| HTN | ASPC | *TRIOBP* | -0.48844284 | 0.007125343 | 0.035170054 | 1 |
| HTN | ASPC | *CHKB* | 0.483564411 | 0.007342539 | 0.035769075 | 1 |

| | | | | | | |
|---|---|---|---|---|---|---|
| HTN | ASPC | *THOC5* | -0.235577933 | 0.007531046 | 0.036540636 | 1 |
| HTN | ASPC | *TSPO* | 1.31930293 | 0.001764716 | 0.012798811 | 1 |
| HTN | ASPC | *POLR3H* | 1.533831711 | 0.000431613 | 0.004465218 | 1 |
| HTN | ASPC | *DESI1* | 0.665110555 | 0.00552864 | 0.029739426 | 1 |
| HTN | ASPC | *CNIH1* | 0.898885598 | 0.008332269 | 0.039019047 | 1 |
| HTN | ASPC | *ATP6V1D* | 1.209465013 | 0.001337087 | 0.010232722 | 1 |
| HTN | ASPC | *LGMN* | 0.463300169 | 0.000160319 | 0.002131146 | 0.777868366 |
| HTN | ASPC | *MTHFD1* | 1.480547874 | 1.38E-06 | 4.55E-05 | 0.006684949 |
| HTN | ASPC | *GSKIP* | 2.380147655 | 0.000169376 | 0.002233181 | 0.821810466 |
| HTN | ASPC | *YY1* | 1.018852445 | 0.007795385 | 0.037374713 | 1 |
| HTN | ASPC | *TRPC4AP* | 0.512029298 | 0.007115494 | 0.035170054 | 1 |
| HTN | ASPC | *NDRG3* | 0.973161232 | 0.002625358 | 0.017121288 | 1 |
| HTN | ASPC | *PABPC1L* | 1.83657797 | 1.77E-05 | 0.000354704 | 0.085838428 |
| HTN | ASPC | *DOK5* | -1.010412085 | 0.001801784 | 0.012989982 | 1 |
| HTN | ASPC | *CTSZ* | 1.426722145 | 0.000216493 | 0.00267283 | 1 |
| HTN | ASPC | *DIDO1* | 0.666291716 | 0.006140484 | 0.031679437 | 1 |
| HTN | ASPC | *BIRC7* | -0.32103787 | 0.001112035 | 0.008986213 | 1 |
| HTN | ASPC | *TRIB3* | 2.76277882 | 0.000472467 | 0.004795834 | 1 |
| HTN | ASPC | *SEC23B* | 0.240205372 | 0.010459877 | 0.046053831 | 1 |
| HTN | ASPC | *MANBAL* | 0.370978876 | 0.000238085 | 0.002817365 | 1 |
| HTN | ASPC | *DNTTIP1* | -0.222273499 | 0.004195941 | 0.024236556 | 1 |
| HTN | ASPC | *MYOM1* | 1.397126644 | 0.000224955 | 0.002735549 | 1 |
| HTN | ASPC | *RNF125* | 2.601232455 | 1.21E-07 | 5.77E-06 | 0.000589 |
| HTN | ASPC | *POLI* | -0.118021101 | 0.000238355 | 0.002817365 | 1 |
| HTN | ASPC | *ARHGEF7* | 0.895020127 | 0.006535909 | 0.033102538 | 1 |
| HTN | ASPC | *MGRN1* | 0.999875079 | 2.92E-07 | 1.18E-05 | 0.001417114 |
| HTN | ASPC | *PSMD7* | 1.278805079 | 0.00024589 | 0.002888763 | 1 |
| HTN | ASPC | *MON1B* | 2.159861635 | 0.000132596 | 0.001832926 | 0.643357168 |
| HTN | ASPC | *NARFL* | 0.441271396 | 0.002390869 | 0.016134206 | 1 |
| HTN | ASPC | *MTHFSD* | 0.193127369 | 9.89E-05 | 0.001450121 | 0.47999016 |
| HTN | ASPC | *HAGHL* | 0.168852659 | 0.004264543 | 0.024574303 | 1 |
| HTN | ASPC | *PYCARD* | 1.078027824 | 3.05E-06 | 8.72E-05 | 0.014815463 |
| HTN | ASPC | *KAT8* | 1.369879271 | 0.001918261 | 0.013667255 | 1 |
| HTN | ASPC | *IQCH* | 1.195754969 | 0.004748217 | 0.026913962 | 1 |
| HTN | ASPC | *TRIP4* | 0.956694561 | 9.21E-05 | 0.00136261 | 0.446936075 |
| HTN | ASPC | *FAH* | 0.831397149 | 0.00362251 | 0.021806972 | 1 |
| HTN | ASPC | *ZFAND1* | 1.665759488 | 7.33E-07 | 2.67E-05 | 0.003557186 |
| HTN | ASPC | *SFRP1* | -0.731351915 | 0.00585724 | 0.030690418 | 1 |
| HTN | ASPC | *NIPAL2* | 0.786250458 | 0.00752801 | 0.036540636 | 1 |
| HTN | ASPC | *SNX16* | 1.649506895 | 7.62E-10 | 1.71E-07 | 3.70E-06 |
| HTN | ASPC | *TSTA3* | 0.30404066 | 0.005739566 | 0.030408456 | 1 |
| HTN | ASPC | *SQLE* | 0.291684543 | 0.005705586 | 0.03032147 | 1 |
| HTN | ASPC | *TUSC3* | 1.073628366 | 0.009328442 | 0.042261064 | 1 |
| HTN | ASPC | *TULP2* | 1.766787763 | 0.0091341 | 0.04161376 | 1 |
| HTN | ASPC | *PEX11G* | 0.246405246 | 0.010040828 | 0.044532084 | 1 |
| HTN | ASPC | *CD37* | -0.409649717 | 0.002309078 | 0.015713386 | 1 |
| HTN | ASPC | *FCER2* | 5.690082319 | 2.36E-09 | 3.27E-07 | 1.14E-05 |
| HTN | ASPC | *CLEC4M* | 4.506501006 | 5.25E-09 | 5.10E-07 | 2.55E-05 |
| HTN | ASPC | *MED25* | 1.756979896 | 0.000746722 | 0.00670578 | 1 |
| HTN | ASPC | *IL27RA* | 1.412472766 | 4.96E-05 | 0.00083632 | 0.240860089 |

| | | | | | | |
|---|---|---|---|---|---|---|
| HTN | ASPC | *RASAL3* | 3.419165945 | 1.02E-09 | 1.83E-07 | 4.94E-06 |
| HTN | ASPC | *TGFB1* | 1.249770305 | 0.001037901 | 0.008579036 | 1 |
| HTN | ASPC | *ICAM5* | -0.252537913 | 0.002490278 | 0.016551823 | 1 |
| HTN | ASPC | *RABAC1* | 0.777623069 | 4.86E-05 | 0.000828806 | 0.235631649 |
| HTN | ASPC | *GRIN2D* | -0.848498775 | 0.010617827 | 0.046328865 | 1 |
| HTN | ASPC | *PLA2G4C* | -0.093949598 | 0.005059286 | 0.02810033 | 1 |
| HTN | ASPC | *PLEKHA4* | -1.075311447 | 0.005222409 | 0.028535056 | 1 |
| HTN | ASPC | *DNASE2* | 0.588134962 | 0.002451505 | 0.016348144 | 1 |
| HTN | ASPC | *TFPT* | 1.901485805 | 0.000149378 | 0.002018886 | 0.724780153 |
| HTN | ASPC | *RAB3A* | 8.406430292 | 0.000397144 | 0.004198131 | 1 |
| HTN | ASPC | *ZNF14* | 1.332493215 | 0.009616833 | 0.04317785 | 1 |
| HTN | ASPC | *ETHE1* | 1.461450246 | 0.000270251 | 0.003107248 | 1 |
| HTN | ASPC | *RASA4* | 12.02828602 | 1.28E-08 | 9.39E-07 | 6.20E-05 |
| HTN | ASPC | *SP4* | 0.837301406 | 0.000547722 | 0.005368781 | 1 |
| HTN | ASPC | *PTN* | -0.742687287 | 0.00017325 | 0.002265797 | 0.840610652 |
| HTN | ASPC | *ADAP1* | 7.591322757 | 1.16E-08 | 8.80E-07 | 5.63E-05 |
| HTN | ASPC | *IQCE* | 1.705455675 | 0.00078348 | 0.006936939 | 1 |
| HTN | ASPC | *HIBADH* | 0.426346003 | 0.007918619 | 0.037746111 | 1 |
| HTN | ASPC | *CHN2* | 0.519977199 | 0.000574316 | 0.005539918 | 1 |
| HTN | ASPC | *ZKSCAN1* | 1.379484387 | 3.39E-06 | 9.56E-05 | 0.016448309 |
| HTN | ASPC | *TAF6* | -0.227148034 | 0.000793446 | 0.007012382 | 1 |
| HTN | ASPC | *RBM28* | 0.363413385 | 0.005539389 | 0.029764245 | 1 |
| HTN | ASPC | *C1GALT1* | 1.570181444 | 1.53E-05 | 0.000321542 | 0.074445294 |
| HTN | ASPC | *MEST* | -0.204956179 | 0.003545337 | 0.021502472 | 1 |
| HTN | ASPC | *ACTR3C* | 0.657760524 | 0.000180408 | 0.002334213 | 0.875340277 |
| HTN | ASPC | *TSPAN13* | 1.510540704 | 7.09E-06 | 0.000169378 | 0.034383702 |
| HTN | ASPC | *GCK* | -6.943380031 | 0.002185998 | 0.015066001 | 1 |
| HTN | ASPC | *SPATA6L* | 0.49423385 | 0.009396421 | 0.042489687 | 1 |
| HTN | ASPC | *SLC1A1* | 0.094333009 | 0.003727334 | 0.022299659 | 1 |
| HTN | ASPC | *MEGF9* | 1.418357606 | 0.000105321 | 0.001534583 | 0.511016158 |
| HTN | ASPC | *CORO2A* | 0.730681175 | 6.66E-05 | 0.001059323 | 0.3230936 |
| HTN | ASPC | *C5* | 0.718972573 | 0.00229841 | 0.015684791 | 1 |
| HTN | ASPC | *RGP1* | 0.824661275 | 0.007405874 | 0.036005311 | 1 |
| HTN | ASPC | *PTGDS* | -0.963269864 | 0.000304279 | 0.003409614 | 1 |
| HTN | ASPC | *TRDMT1* | 1.293024481 | 0.000709852 | 0.006461917 | 1 |
| HTN | ASPC | *ATE1* | 0.551113843 | 0.000604735 | 0.005753285 | 1 |
| HTN | ASPC | *TRIM16L* | 1.987115301 | 0.000143607 | 0.001957257 | 0.696783475 |
| HTN | ASPC | *PIGL* | 0.783504826 | 0.007553173 | 0.036611382 | 1 |
| HTN | ASPC | *GOSR1* | 0.957062331 | 0.00907596 | 0.041543922 | 1 |
| HTN | ASPC | *ICAM2* | 2.182745382 | 1.86E-06 | 5.83E-05 | 0.00903206 |
| HTN | ASPC | *DDX5* | 0.468364411 | 0.000826211 | 0.007262272 | 1 |
| HTN | ASPC | *NAGLU* | 1.325687408 | 2.47E-07 | 1.02E-05 | 0.001197586 |
| HTN | ASPC | *SGCA* | 0.153828662 | 0.00509329 | 0.028210779 | 1 |
| HTN | ASPC | *EFTUD2* | 1.22128231 | 0.006155616 | 0.031679437 | 1 |
| HTN | ASPC | *MYH3* | 2.360477015 | 0.003393271 | 0.020893591 | 1 |
| HTN | ASPC | *WFS1* | 2.265369127 | 3.80E-05 | 0.000680915 | 0.184527868 |
| HTN | ASPC | *GAR1* | 1.823030487 | 0.001082328 | 0.008833468 | 1 |
| HTN | ASPC | *AADAT* | -0.775943866 | 0.003239676 | 0.020075233 | 1 |
| HTN | ASPC | *STIM2* | 1.09110775 | 0.000571831 | 0.00552694 | 1 |
| HTN | ASPC | *KLHL5* | 1.266263939 | 0.000254285 | 0.002965838 | 1 |

| | | | | | | |
|---|---|---|---|---|---|---|
| HTN | ASPC | *B3GAT1* | 8.097621928 | 1.12E-10 | 3.87E-08 | 5.41E-07 |
| HTN | ASPC | *P2RX3* | 25.35338477 | 2.98E-09 | 3.53E-07 | 1.45E-05 |
| HTN | ASPC | *NRXN2* | -2.147907549 | 2.61E-06 | 7.67E-05 | 0.012659589 |
| HTN | ASPC | *CCND1* | -0.582395996 | 1.49E-05 | 0.000314869 | 0.07210494 |
| HTN | ASPC | *KIAA1549L* | 4.613865218 | 0.001008537 | 0.008393522 | 1 |
| HTN | ASPC | *PRDM4* | 0.792602541 | 0.000146986 | 0.001997693 | 0.713176352 |
| HTN | ASPC | *CORO1C* | 1.00096824 | 0.006929722 | 0.034449807 | 1 |
| HTN | ASPC | *ASIC1* | -1.976306153 | 9.73E-06 | 0.000218579 | 0.047209754 |
| HTN | ASPC | *IL23A* | 11.79656716 | 2.58E-09 | 3.38E-07 | 1.25E-05 |
| HTN | ASPC | *PRR4* | 0.274627221 | 0.004029698 | 0.023528395 | 1 |
| HTN | ASPC | *PARP11* | 1.115571181 | 0.005688761 | 0.030298426 | 1 |
| HTN | ASPC | *GPN3* | 2.016617552 | 0.000385013 | 0.004089703 | 1 |
| HTN | ASPC | *VDR* | 4.066745005 | 5.46E-06 | 0.000140237 | 0.026504881 |
| HTN | ASPC | *NT5DC3* | 0.469093973 | 0.009890542 | 0.044148029 | 1 |
| HTN | ASPC | *FRK* | 2.326131752 | 5.22E-06 | 0.000136909 | 0.025328148 |
| HTN | ASPC | *HDDC2* | 0.296058956 | 0.009194485 | 0.041771198 | 1 |
| HTN | ASPC | *UST* | -0.201540225 | 0.004885275 | 0.027402723 | 1 |
| HTN | ASPC | *MDGA1* | -0.771775772 | 0.009517292 | 0.042865433 | 1 |
| HTN | ASPC | *CD83* | 1.265647682 | 0.000467353 | 0.004753867 | 1 |
| HTN | ASPC | *ACOT13* | 1.134918072 | 0.008604082 | 0.039818416 | 1 |
| HTN | ASPC | *FIG4* | 0.935921131 | 0.004824691 | 0.027188618 | 1 |
| HTN | ASPC | *PACRG* | 1.820607189 | 0.000182185 | 0.002336468 | 0.883960007 |
| HTN | ASPC | *QKI* | 0.73502005 | 0.006556656 | 0.033172986 | 1 |
| HTN | ASPC | *EXOC2* | 1.03613606 | 0.000860323 | 0.007467418 | 1 |
| HTN | ASPC | *BTN2A1* | 0.755339365 | 0.00117525 | 0.009268434 | 1 |
| HTN | ASPC | *LY86* | 1.553028139 | 8.50E-05 | 0.001273348 | 0.412564677 |
| HTN | ASPC | *NME5* | 0.196728904 | 0.000514295 | 0.005092568 | 1 |
| HTN | ASPC | *IK* | 0.352289934 | 0.002039431 | 0.014302228 | 1 |
| HTN | ASPC | *RNF130* | 0.595341934 | 5.35E-06 | 0.000138861 | 0.025967062 |
| HTN | ASPC | *ARSB* | 1.711543026 | 1.46E-05 | 0.000313582 | 0.070817335 |
| HTN | ASPC | *SKP1* | 0.977441372 | 0.011463439 | 0.049221774 | 1 |
| HTN | ASPC | *BNIP1* | 0.149910566 | 0.007600925 | 0.036784522 | 1 |
| HTN | ASPC | *UNC5A* | 8.835537714 | 5.65E-09 | 5.38E-07 | 2.74E-05 |
| HTN | ASPC | *CLDN16* | 0.381059243 | 0.001815296 | 0.013048614 | 1 |
| HTN | ASPC | *NPHP3* | 0.166609393 | 0.003744671 | 0.022348267 | 1 |
| HTN | ASPC | *PCCB* | 0.607559602 | 2.87E-09 | 3.48E-07 | 1.39E-05 |
| HTN | ASPC | *IQCG* | 1.031229048 | 0.006530224 | 0.033102538 | 1 |
| HTN | ASPC | *SCAP* | 0.805083344 | 0.00060911 | 0.005768672 | 1 |
| HTN | ASPC | *KIAA1257* | -2.425728862 | 0.000747781 | 0.00670578 | 1 |
| HTN | ASPC | *VIPR1* | 1.771196731 | 4.63E-05 | 0.000798601 | 0.224626418 |
| HTN | ASPC | *DNAH1* | 0.33129096 | 0.005183555 | 0.028380408 | 1 |
| HTN | ASPC | *ACTR1B* | 1.360147791 | 0.000445358 | 0.004578131 | 1 |
| HTN | ASPC | *NRBP1* | 0.91898376 | 0.011698658 | 0.049875767 | 1 |
| HTN | ASPC | *PPM1G* | 1.311867222 | 5.77E-06 | 0.0001436 | 0.028001981 |
| HTN | ASPC | *GRB14* | 2.248269891 | 3.79E-08 | 2.30E-06 | 0.000183756 |
| HTN | ASPC | *GALNT3* | 0.520790487 | 0.002501689 | 0.016604919 | 1 |
| HTN | ASPC | *POLE4* | 1.446739195 | 0.000500429 | 0.004985793 | 1 |
| HTN | ASPC | *MLPH* | 0.523006339 | 0.000618168 | 0.005779093 | 1 |
| HTN | ASPC | *STRN* | 1.087082814 | 0.001693876 | 0.012438828 | 1 |
| HTN | ASPC | *CEBPZ* | 1.062101792 | 0.000339422 | 0.003709185 | 1 |

| | | | | | | |
|---|---|---|---|---|---|---|
| HTN | ASPC | *DARS* | 0.97264263 | 0.007329518 | 0.035769075 | 1 |
| HTN | ASPC | *WIPF1* | 0.982003852 | 0.001190584 | 0.009347432 | 1 |
| HTN | ASPC | *PNO1* | 2.58346891 | 2.88E-05 | 0.000535474 | 0.139758708 |
| HTN | ASPC | *ORC4* | 0.525990903 | 0.002841561 | 0.018309763 | 1 |
| HTN | ASPC | *AAK1* | 0.952381044 | 0.001263686 | 0.00979458 | 1 |
| HTN | ASPC | *ALMS1* | 1.004626982 | 0.007301775 | 0.035713923 | 1 |
| HTN | ASPC | *MORN1* | -0.264074798 | 0.006156989 | 0.031679437 | 1 |
| HTN | ASPC | *KIAA1324* | 0.827533491 | 0.000431165 | 0.004465218 | 1 |
| HTN | ASPC | *PRG4* | 1.236633125 | 0.00409995 | 0.023852469 | 1 |
| HTN | ASPC | *MRPS15* | 0.90836159 | 0.00244228 | 0.01634475 | 1 |
| HTN | ASPC | *TBCE* | 1.184127692 | 0.00483996 | 0.027243026 | 1 |
| HTN | ASPC | *ACADM* | 0.643287187 | 0.006796287 | 0.033960439 | 1 |
| HTN | ASPC | *SDHB* | 0.99234527 | 0.005895843 | 0.030759817 | 1 |
| HTN | ASPC | *CD46* | 0.494042764 | 0.000694901 | 0.006337709 | 1 |
| HTN | ASPC | *PRRC2C* | 0.648483281 | 0.00641125 | 0.032675827 | 1 |
| HTN | ASPC | *IRF6* | -0.116222521 | 0.006079421 | 0.031447071 | 1 |
| HTN | ASPC | *RPA2* | 0.753305821 | 0.000287249 | 0.003241239 | 1 |
| HTN | ASPC | *RCN2* | 0.801364579 | 0.006389697 | 0.032600223 | 1 |
| HTN | ASPC | *CAMSAP2* | 0.42383262 | 0.009747702 | 0.043691931 | 1 |
| HTN | ASPC | *KLF7* | 0.732617502 | 0.002306606 | 0.015713386 | 1 |
| HTN | ASPC | *SGIP1* | 1.906879422 | 4.91E-05 | 0.00083073 | 0.238419508 |
| HTN | ASPC | *AKAP7* | 0.638167434 | 0.005159368 | 0.028380408 | 1 |
| HTN | ASPC | *FOXO3* | 0.831545655 | 0.001475595 | 0.011081811 | 1 |
| HTN | ASPC | *UBN1* | 1.279815605 | 6.40E-05 | 0.001027989 | 0.310452735 |
| HTN | ASPC | *FKBP15* | 0.309105989 | 0.00067203 | 0.006152242 | 1 |
| HTN | ASPC | *RAB14* | 0.962586301 | 0.009089236 | 0.041554897 | 1 |
| HTN | ASPC | *PPP6C* | 1.307624175 | 0.000632832 | 0.005882183 | 1 |
| HTN | ASPC | *ACOT2* | 0.090513582 | 0.00110975 | 0.008986213 | 1 |
| HTN | ASPC | *COQ6* | 1.237738414 | 0.004124938 | 0.023935357 | 1 |
| HTN | ASPC | *GORAB* | 0.911740551 | 0.009102286 | 0.041554897 | 1 |
| HTN | ASPC | *MRC1* | 0.87695432 | 0.000441979 | 0.004553038 | 1 |
| HTN | ASPC | *IQSEC3* | 1.748705511 | 0.000614586 | 0.005779014 | 1 |
| HTN | ASPC | *ZFP30* | 0.532209956 | 7.59E-06 | 0.000177149 | 0.036846978 |
| HTN | ASPC | *ADRA1A* | 0.063538164 | 0.010542752 | 0.046191181 | 1 |
| HTN | ASPC | *AKAP1* | 0.737389028 | 0.010986348 | 0.047509589 | 1 |
| HTN | ASPC | *TBX2* | -0.329767478 | 1.12E-08 | 8.76E-07 | 5.44E-05 |
| HTN | ASPC | *TEX14* | 0.631896757 | 7.78E-09 | 6.62E-07 | 3.77E-05 |
| HTN | ASPC | *ZNF549* | 1.467907207 | 0.001257067 | 0.009758863 | 1 |
| HTN | ASPC | *LHX4* | 1.58706177 | 0.011708264 | 0.049875767 | 1 |
| HTN | ASPC | *MAPK8IP1* | -0.790005585 | 0.000448449 | 0.004600162 | 1 |
| HTN | ASPC | *TNFSF10* | 1.069646005 | 0.001007861 | 0.008393522 | 1 |
| HTN | ASPC | *FLT3* | 0.596792848 | 0.003021463 | 0.019051685 | 1 |
| HTN | ASPC | *CD244* | 0.971340236 | 0.000150909 | 0.002022682 | 0.732210871 |
| HTN | ASPC | *LDB3* | 0.504152905 | 0.005377215 | 0.029086115 | 1 |
| HTN | ASPC | *NAA60* | 2.138633739 | 0.002043431 | 0.014306966 | 1 |
| HTN | ASPC | *SPATA1* | 1.130486292 | 0.00056096 | 0.005476414 | 1 |
| HTN | ASPC | *SMU1* | 0.82928699 | 0.010150859 | 0.044896963 | 1 |
| HTN | ASPC | *DDX54* | 0.764443212 | 0.001961052 | 0.013931223 | 1 |
| HTN | ASPC | *CCDC91* | 0.820149758 | 0.001403808 | 0.010642618 | 1 |
| HTN | ASPC | *PDE1B* | 1.606324148 | 0.003726629 | 0.022299659 | 1 |

| | | | | | | |
|---|---|---|---|---|---|---|
| HTN | ASPC | *CDK2* | -0.279999472 | 0.005566418 | 0.029825258 | 1 |
| HTN | ASPC | *DBH* | -18.51191333 | 1.02E-06 | 3.55E-05 | 0.004931197 |
| HTN | ASPC | *NDUFAF4* | 0.336012109 | 0.001721665 | 0.012576931 | 1 |
| HTN | ASPC | *WFDC3* | -0.081974822 | 0.000893675 | 0.007674535 | 1 |
| HTN | ASPC | *MATN4* | -1.231080861 | 0.005466833 | 0.029439596 | 1 |
| HTN | ASPC | *PEPD* | 1.296122009 | 0.000229267 | 0.002774071 | 1 |
| HTN | ASPC | *BTN2A2* | 0.217086255 | 9.42E-07 | 3.35E-05 | 0.004572021 |
| HTN | ASPC | *SNRPC* | -0.404943601 | 0.000343441 | 0.003736268 | 1 |
| HTN | ASPC | *ZNF391* | -0.115431704 | 0.000115367 | 0.001651216 | 0.559762094 |
| HTN | ASPC | *TCP11* | 0.216310259 | 0.006371952 | 0.032543904 | 1 |
| HTN | ASPC | *ATXN1* | 0.657515207 | 0.003308467 | 0.020434449 | 1 |
| HTN | ASPC | *GOT2* | 1.500677597 | 7.33E-05 | 0.001138447 | 0.355523998 |
| HTN | ASPC | *CLYBL* | 1.322017372 | 0.000334669 | 0.003673788 | 1 |
| HTN | ASPC | *GRK4* | 0.081516614 | 0.002295277 | 0.015684791 | 1 |
| HTN | ASPC | *MRPS7* | 1.595458543 | 0.000721647 | 0.006556988 | 1 |
| HTN | ASPC | *OPRL1* | 6.131732165 | 1.70E-09 | 2.84E-07 | 8.24E-06 |
| HTN | ASPC | *IL1B* | 1.749723687 | 1.71E-05 | 0.00034526 | 0.082862478 |
| HTN | ASPC | *PSD4* | 0.949334502 | 0.000220668 | 0.002711682 | 1 |
| HTN | ASPC | *TNFSF9* | -1.965813053 | 0.008979729 | 0.041181137 | 1 |
| HTN | ASPC | *GPR108* | 0.438881555 | 0.000666016 | 0.006127655 | 1 |
| HTN | ASPC | *SNRPD2* | 1.568617544 | 0.00280547 | 0.018125354 | 1 |
| HTN | ASPC | *MKKS* | 0.536410325 | 0.007289035 | 0.035687588 | 1 |
| HTN | ASPC | *KLC1* | 0.982853553 | 0.008470614 | 0.039420451 | 1 |
| HTN | ASPC | *COX6B1* | 0.911913756 | 0.005569185 | 0.029825258 | 1 |
| HTN | ASPC | *PRDX5* | 0.806460561 | 0.001062706 | 0.008709881 | 1 |
| HTN | ASPC | *TSKS* | -0.478573666 | 7.21E-06 | 0.000171489 | 0.034983842 |
| HTN | ASPC | *TRAP1* | 0.275184183 | 6.41E-08 | 3.49E-06 | 0.000310896 |
| HTN | ASPC | *KTN1* | 0.811053485 | 0.007731535 | 0.037179914 | 1 |
| HTN | ASPC | *RHOT1* | 0.361665084 | 0.005882391 | 0.030748227 | 1 |
| HTN | ASPC | *ZNF484* | 0.750123963 | 0.011488476 | 0.049248514 | 1 |
| HTN | ASPC | *ABHD8* | -0.433236753 | 2.84E-05 | 0.000529425 | 0.137650614 |
| HTN | ASPC | *RAB3IP* | 1.705600785 | 4.22E-05 | 0.000743968 | 0.204591291 |
| HTN | ASPC | *GFER* | -0.347118592 | 0.008233153 | 0.038746127 | 1 |
| HTN | ASPC | *SMARCA4* | 0.594713587 | 0.008113762 | 0.038482868 | 1 |
| HTN | ASPC | *RNF6* | 1.327297975 | 0.001212981 | 0.009507889 | 1 |
| HTN | ASPC | *HIP1* | 1.043336981 | 0.000450628 | 0.004612759 | 1 |
| HTN | ASPC | *PEX1* | 1.177508801 | 0.004786126 | 0.027065598 | 1 |
| HTN | ASPC | *KDR* | 1.856826752 | 4.91E-07 | 1.86E-05 | 0.002380798 |
| HTN | ASPC | *A4GALT* | -0.236694642 | 0.006651184 | 0.033476705 | 1 |
| HTN | ASPC | *GAD1* | -0.620767368 | 0.003232185 | 0.020054426 | 1 |
| HTN | ASPC | *ORMDL1* | 0.41536049 | 0.008166346 | 0.038592365 | 1 |
| HTN | ASPC | *TWSG1* | 0.907604956 | 0.010106546 | 0.044782613 | 1 |
| HTN | ASPC | *ARPP19* | 1.27569125 | 0.002751466 | 0.017823913 | 1 |
| HTN | ASPC | *BBOX1* | 8.235629434 | 2.38E-05 | 0.000458071 | 0.115433971 |
| HTN | ASPC | *SERGEF* | 0.609597749 | 0.000121455 | 0.001718072 | 0.589298666 |
| HTN | ASPC | *PLD2* | 1.909521033 | 0.001748449 | 0.012718854 | 1 |
| HTN | ASPC | *PHF20L1* | 0.758139832 | 0.00397217 | 0.02327653 | 1 |
| HTN | ASPC | *KLK8* | 12.28848556 | 3.32E-09 | 3.75E-07 | 1.61E-05 |
| HTN | ASPC | *RIPK3* | 0.188675618 | 0.002958559 | 0.018835499 | 1 |
| HTN | ASPC | *ADCY4* | 0.718919187 | 3.97E-09 | 4.19E-07 | 1.93E-05 |

| | | | | | | |
|---|---|---|---|---|---|---|
| HTN | ASPC | *RNASE1* | 0.54303956 | 0.00570121 | 0.03032147 | 1 |
| HTN | ASPC | *EPB41L4A* | 1.073149043 | 0.000422348 | 0.00438952 | 1 |
| HTN | ASPC | *RHBDF2* | 0.166658951 | 0.004817289 | 0.027178471 | 1 |
| HTN | ASPC | *TMEM8A* | 1.67915401 | 0.000131732 | 0.001826183 | 0.639164216 |
| HTN | ASPC | *CDC16* | 0.502193368 | 2.95E-05 | 0.000546693 | 0.143233476 |
| HTN | ASPC | *ATP8B3* | -0.154498947 | 0.001104106 | 0.008986213 | 1 |
| HTN | ASPC | *DDA1* | 0.716573075 | 0.005794433 | 0.030493044 | 1 |
| HTN | ASPC | *LSM7* | 0.307453204 | 0.005405005 | 0.029171394 | 1 |
| HTN | ASPC | *ACTN4* | 0.808150161 | 0.002596609 | 0.017002359 | 1 |
| HTN | ASPC | *MAP1S* | 2.709082608 | 0.000126025 | 0.001757101 | 0.611471316 |
| HTN | ASPC | *PGPEP1* | 0.334851093 | 0.000279559 | 0.003176624 | 1 |
| HTN | ASPC | *TRPM4* | -0.32444882 | 0.005072439 | 0.028127399 | 1 |
| HTN | ASPC | *THEMIS2* | 0.228079978 | 0.007196224 | 0.035461838 | 1 |
| HTN | ASPC | *HSD17B3* | 2.777480272 | 3.48E-05 | 0.000629421 | 0.16868493 |
| HTN | ASPC | *TTLL9* | 2.08130134 | 3.05E-06 | 8.72E-05 | 0.014819048 |
| HTN | ASPC | *IDO1* | 3.932280856 | 4.22E-09 | 4.27E-07 | 2.05E-05 |
| HTN | ASPC | *HAUS8* | -0.572341657 | 1.96E-07 | 8.43E-06 | 0.000949992 |
| HTN | ASPC | *GALNT15* | 1.080918916 | 0.006786395 | 0.033945968 | 1 |
| HTN | ASPC | *SLC6A6* | 1.488270152 | 6.57E-05 | 0.001052585 | 0.318933325 |
| HTN | ASPC | *PDLIM4* | 1.372636447 | 0.002115188 | 0.014640362 | 1 |
| HTN | ASPC | *EXOC4* | 0.883797862 | 3.76E-06 | 0.000101806 | 0.018223279 |
| HTN | ASPC | *C1orf159* | 1.302292071 | 0.003432399 | 0.02102778 | 1 |
| HTN | ASPC | *PPFIA1* | 0.84531185 | 0.000341022 | 0.003718286 | 1 |
| HTN | ASPC | *KHDRBS3* | 1.253678032 | 0.006009604 | 0.031185667 | 1 |
| HTN | ASPC | *GCH1* | 0.456316757 | 5.29E-05 | 0.000876007 | 0.256670036 |
| HTN | ASPC | *TIMM10B* | 0.289277274 | 0.009752342 | 0.043691931 | 1 |
| HTN | ASPC | *MUTYH* | 0.47585988 | 0.000867432 | 0.007502451 | 1 |
| HTN | ASPC | *ZBED3* | 0.750337545 | 0.004667818 | 0.026520201 | 1 |
| HTN | ASPC | *ALOX5AP* | 1.061816008 | 0.005147112 | 0.028379304 | 1 |
| HTN | ASPC | *PEMT* | 1.956813867 | 0.000539072 | 0.005294694 | 1 |
| HTN | ASPC | *DSTYK* | 1.140088134 | 0.004348461 | 0.024998498 | 1 |
| HTN | ASPC | *EPSTI1* | 0.724642025 | 0.010495174 | 0.046149344 | 1 |
| HTN | ASPC | *CSNK1G2* | 1.824669639 | 0.000276152 | 0.003160118 | 1 |
| HTN | ASPC | *CNDP2* | 0.827551213 | 0.003519941 | 0.021401948 | 1 |
| HTN | ASPC | *PDZD2* | 0.826018484 | 0.002945463 | 0.018804458 | 1 |
| HTN | ASPC | *SLC2A11* | 0.321352719 | 0.001176701 | 0.009268434 | 1 |
| HTN | ASPC | *FAM83F* | 1.708343616 | 0.002852701 | 0.018357171 | 1 |
| HTN | ASPC | *IMPA1* | 0.266459994 | 0.00028434 | 0.003223401 | 1 |
| HTN | ASPC | *COX16* | 2.592871963 | 0.003040522 | 0.019134388 | 1 |
| HTN | ASPC | *TTC9* | 3.112351666 | 2.81E-08 | 1.80E-06 | 0.000136535 |
| HTN | ASPC | *CTIF* | 0.349106361 | 0.008456427 | 0.039414585 | 1 |
| HTN | ASPC | *CNTN6* | 0.682593171 | 0.008297739 | 0.038936779 | 1 |
| HTN | ASPC | *GSTM5* | -0.183484162 | 0.00517978 | 0.028380408 | 1 |
| HTN | ASPC | *CEPT1* | 1.075675647 | 0.000169239 | 0.002233181 | 0.821146411 |
| HTN | ASPC | *KIAA1468* | 0.930246492 | 0.007033743 | 0.034824207 | 1 |
| HTN | ASPC | *KLRD1* | 1.28325611 | 0.001397051 | 0.010624591 | 1 |
| HTN | ASPC | *ACP2* | 1.514585631 | 7.08E-16 | 1.72E-12 | 3.44E-12 |
| HTN | ASPC | *DHX34* | 1.218158874 | 0.00389243 | 0.022905499 | 1 |
| HTN | ASPC | *CLOCK* | 0.913341664 | 0.006904144 | 0.034393128 | 1 |
| HTN | ASPC | *CLDN10* | 0.497094135 | 0.001083247 | 0.008833468 | 1 |

| | | | | | | |
|---|---|---|---|---|---|---|
| HTN | ASPC | *WDR36* | 1.094894752 | 0.007279885 | 0.035678788 | 1 |
| HTN | ASPC | *OSTF1* | 0.960253818 | 0.008473935 | 0.039420451 | 1 |
| HTN | ASPC | *UBQLN1* | 1.091559374 | 0.00114673 | 0.009196584 | 1 |
| HTN | ASPC | *MSI1* | 0.417201213 | 0.003413221 | 0.020963227 | 1 |
| HTN | ASPC | *TMEM60* | 1.542635516 | 4.38E-05 | 0.000769276 | 0.212526481 |
| HTN | ASPC | *AKIRIN2* | 0.711465176 | 0.00995352 | 0.044306862 | 1 |
| HTN | ASPC | *MAP3K7* | 0.980936113 | 0.00415215 | 0.024012195 | 1 |
| HTN | ASPC | *CAPRIN1* | 0.739379819 | 0.006570493 | 0.033208365 | 1 |
| HTN | ASPC | *FAIM2* | -0.434187804 | 0.000212907 | 0.002648781 | 1 |
| HTN | ASPC | *USP15* | 0.864523896 | 0.002760913 | 0.017861265 | 1 |
| HTN | ASPC | *MDM2* | 0.589996721 | 0.005610329 | 0.029980198 | 1 |
| HTN | ASPC | *AGT* | 0.20728487 | 0.000258274 | 0.002997951 | 1 |
| HTN | ASPC | *MRPL44* | 1.442433057 | 0.00205862 | 0.014371831 | 1 |
| HTN | ASPC | *USP37* | 0.548229037 | 0.002050032 | 0.014332503 | 1 |
| HTN | ASPC | *TMBIM1* | 1.372934753 | 0.001488384 | 0.011128992 | 1 |
| HTN | ASPC | *GCC2* | 0.835325552 | 0.000226232 | 0.00274419 | 1 |
| HTN | ASPC | *MRPS9* | 0.261919807 | 0.009124486 | 0.041609027 | 1 |
| HTN | ASPC | *STAB2* | 2.069572364 | 1.27E-06 | 4.26E-05 | 0.00614074 |
| HTN | ASPC | *SUCLA2* | 1.160783392 | 0.002521966 | 0.016716639 | 1 |
| HTN | ASPC | *RCBTB2* | 1.369649703 | 0.000888625 | 0.0076447 | 1 |
| HTN | ASPC | *TBRG4* | 0.917096284 | 0.002961972 | 0.018835499 | 1 |
| HTN | ASPC | *NMT1* | 0.410938558 | 1.99E-08 | 1.33E-06 | 9.68E-05 |
| HTN | ASPC | *SCN2A* | 0.757273034 | 8.15E-06 | 0.000189105 | 0.039523021 |
| HTN | ASPC | *SLC2A8* | 0.630432951 | 0.005923956 | 0.030840166 | 1 |
| HTN | ASPC | *ZNF189* | 1.339600685 | 0.008591429 | 0.039818416 | 1 |
| HTN | ASPC | *STX17* | 0.904229059 | 0.010560591 | 0.046191181 | 1 |
| HTN | ASPC | *DERL1* | 0.772488738 | 0.003607487 | 0.021787647 | 1 |
| HTN | ASPC | *IL11RA* | -0.298873935 | 0.004110428 | 0.023884788 | 1 |
| HTN | ASPC | *TMEM8B* | 0.464518389 | 0.001614785 | 0.01199078 | 1 |
| HTN | ASPC | *FRS3* | 1.174841224 | 0.006189661 | 0.031780142 | 1 |
| HTN | ASPC | *TCF19* | 0.238104264 | 0.010539666 | 0.046191181 | 1 |
| HTN | ASPC | *NRM* | 5.176594415 | 5.26E-05 | 0.000873675 | 0.255113201 |
| HTN | ASPC | *SLC37A4* | 0.44707912 | 3.20E-06 | 9.08E-05 | 0.015531694 |
| HTN | ASPC | *PPP2R1B* | -0.19241199 | 0.001700817 | 0.012465807 | 1 |
| HTN | ASPC | *FDX1* | 0.431275663 | 0.01014606 | 0.044896963 | 1 |
| HTN | ASPC | *MAP2K5* | 1.029971248 | 0.000376171 | 0.004020225 | 1 |
| HTN | ASPC | *NUSAP1* | 0.748295489 | 1.76E-05 | 0.000354259 | 0.085376345 |
| HTN | ASPC | *ADAM10* | 0.862262505 | 0.000493731 | 0.004939344 | 1 |
| HTN | ASPC | *SLC44A5* | 1.010118358 | 0.000258037 | 0.002997951 | 1 |
| HTN | ASPC | *RTCA* | 0.5512083 | 0.010764085 | 0.046848446 | 1 |
| HTN | ASPC | *DYNC2LI1* | 0.432422514 | 0.000907977 | 0.007728957 | 1 |
| HTN | ASPC | *ACTR2* | 1.083415778 | 0.001314141 | 0.010088942 | 1 |
| HTN | ASPC | *DTNB* | 0.724992015 | 0.000230581 | 0.002783032 | 1 |
| HTN | ASPC | *ARL3* | 0.366606459 | 0.00115553 | 0.009236628 | 1 |
| HTN | ASPC | *RBP4* | 0.768643194 | 0.008035683 | 0.038242827 | 1 |
| HTN | ASPC | *STAT4* | 1.267697177 | 0.000674867 | 0.006166585 | 1 |
| HTN | ASPC | *AP1AR* | 1.255954424 | 0.001788759 | 0.012934512 | 1 |
| HTN | ASPC | *G3BP2* | 0.514106165 | 6.15E-05 | 0.000991813 | 0.298354109 |
| HTN | ASPC | *CNOT6L* | 0.622244006 | 0.002104729 | 0.014588776 | 1 |
| HTN | ASPC | *PPA2* | 0.675026706 | 0.003480697 | 0.021246167 | 1 |

| | | | | | | |
|---|---|---|---|---|---|---|
| HTN | ASPC | *ENPEP* | -0.144600422 | 0.000436832 | 0.004509591 | 1 |
| HTN | ASPC | *FBN2* | 0.117701157 | 1.93E-05 | 0.000381395 | 0.093441717 |
| HTN | ASPC | *KIF21A* | 1.16371768 | 0.000525261 | 0.005180009 | 1 |
| HTN | ASPC | *PPFIA2* | 1.115756151 | 0.008616928 | 0.039818416 | 1 |
| HTN | ASPC | *POC1B* | 1.45338095 | 9.80E-05 | 0.001440208 | 0.475268615 |
| HTN | ASPC | *RITA1* | 3.327335282 | 0.000138117 | 0.001893061 | 0.670143718 |
| HTN | ASPC | *ESYT1* | 0.94815567 | 0.000190621 | 0.00242059 | 0.924893116 |
| HTN | ASPC | *RB1* | 0.642959613 | 0.00298129 | 0.01891711 | 1 |
| HTN | ASPC | *FITM1* | 0.647256535 | 0.000666818 | 0.006127655 | 1 |
| HTN | ASPC | *RTN1* | 0.365343265 | 2.86E-06 | 8.31E-05 | 0.01387068 |
| HTN | ASPC | *SLC24A4* | 2.985806079 | 7.71E-08 | 3.94E-06 | 0.000374093 |
| HTN | ASPC | *WARS* | 1.661273349 | 9.12E-08 | 4.47E-06 | 0.000442427 |
| HTN | ASPC | *UBE2Q2* | 0.762827803 | 0.002027984 | 0.014260545 | 1 |
| HTN | ASPC | *HMG20A* | 1.141300478 | 0.002685342 | 0.017481082 | 1 |
| HTN | ASPC | *ULK3* | 3.532275384 | 5.58E-11 | 3.01E-08 | 2.71E-07 |
| HTN | ASPC | *ITGAX* | 3.311236773 | 3.46E-09 | 3.82E-07 | 1.68E-05 |
| HTN | ASPC | *DHX38* | 1.321223803 | 0.006962878 | 0.034543847 | 1 |
| HTN | ASPC | *NUDT7* | 2.126305141 | 4.72E-05 | 0.000809829 | 0.229181475 |
| HTN | ASPC | *RPL3L* | 0.42513201 | 0.007633916 | 0.036855481 | 1 |
| HTN | ASPC | *DEF8* | 0.665070248 | 1.01E-07 | 4.88E-06 | 0.000487666 |
| HTN | ASPC | *TCF25* | 0.840070039 | 5.84E-05 | 0.000954738 | 0.283557234 |
| HTN | ASPC | *NCOR1* | 0.743718527 | 0.008902223 | 0.04094179 | 1 |
| HTN | ASPC | *KSR1* | 0.985573887 | 0.007860254 | 0.03764852 | 1 |
| HTN | ASPC | *MYO19* | 0.894342685 | 4.14E-05 | 0.000733176 | 0.200890292 |
| HTN | ASPC | *SCRN2* | -0.086142063 | 0.001266255 | 0.009798838 | 1 |
| HTN | ASPC | *RPRD1A* | 0.672220424 | 0.00110904 | 0.008986213 | 1 |
| HTN | ASPC | *GALNT1* | 1.028053111 | 0.000803951 | 0.007092309 | 1 |
| HTN | ASPC | *C18orf8* | -0.592338883 | 0.000130816 | 0.001818685 | 0.634721215 |
| HTN | ASPC | *NARF* | 0.742697749 | 0.004852533 | 0.027282144 | 1 |
| HTN | ASPC | *MAPK4* | 5.807916641 | 6.23E-06 | 0.00015271 | 0.030236563 |
| HTN | ASPC | *CBLN2* | 2.932797662 | 1.45E-08 | 1.03E-06 | 7.03E-05 |
| HTN | ASPC | *SIRT3* | 0.614859665 | 0.003979113 | 0.023289092 | 1 |
| HTN | ASPC | *EVI5L* | 2.053486297 | 0.000617478 | 0.005779093 | 1 |
| HTN | ASPC | *RERE* | 0.837117455 | 0.000198467 | 0.002494726 | 0.962964104 |
| HTN | ASPC | *SLC44A3* | 1.181978836 | 0.001106313 | 0.008986213 | 1 |
| HTN | ASPC | *STRIP1* | 1.41095434 | 0.002578798 | 0.016950191 | 1 |
| HTN | ASPC | *ALDH9A1* | 1.219832922 | 0.007214555 | 0.03550205 | 1 |
| HTN | ASPC | *MAEL* | 3.957952689 | 6.35E-09 | 5.92E-07 | 3.08E-05 |
| HTN | ASPC | *SNX27* | 0.836062089 | 1.56E-05 | 0.000323165 | 0.075545328 |
| HTN | ASPC | *ARNT* | -0.417574525 | 0.010775529 | 0.046848446 | 1 |
| HTN | ASPC | *SMYD2* | 1.679032011 | 0.000247947 | 0.002905892 | 1 |
| HTN | ASPC | *S100A8* | 1.978356694 | 1.50E-05 | 0.000315726 | 0.072617031 |
| HTN | ASPC | *CREB3L4* | -0.459676241 | 4.41E-05 | 0.000770178 | 0.21410951 |
| HTN | ASPC | *CEP170* | 1.52413751 | 4.05E-07 | 1.57E-05 | 0.001962849 |
| HTN | ASPC | *RHOB* | 1.620787515 | 0.000271398 | 0.003113056 | 1 |
| HTN | ASPC | *GALM* | 1.263959625 | 0.002348187 | 0.015934829 | 1 |
| HTN | ASPC | *CIAO1* | 1.015748564 | 1.59E-05 | 0.000327514 | 0.076965741 |
| HTN | ASPC | *RALB* | 1.342565756 | 0.00385288 | 0.022771806 | 1 |
| HTN | ASPC | *RABL2A* | -0.304975779 | 0.005809269 | 0.030504952 | 1 |
| HTN | ASPC | *METTL21A* | 1.437979744 | 6.91E-07 | 2.54E-05 | 0.00335465 |

| | | | | | | |
|---|---|---|---|---|---|---|
| HTN | ASPC | *EAF1* | 1.548470118 | 4.39E-05 | 0.000769276 | 0.213089413 |
| HTN | ASPC | *MED12L* | 1.374225843 | 0.001045456 | 0.008612143 | 1 |
| HTN | ASPC | *AMT* | -0.089153337 | 0.010859198 | 0.047085638 | 1 |
| HTN | ASPC | *MUC4* | -0.521848174 | 0.007731767 | 0.037179914 | 1 |
| HTN | ASPC | *CISD2* | 2.534252848 | 9.78E-11 | 3.87E-08 | 4.75E-07 |
| HTN | ASPC | *MYO10* | -0.65066423 | 0.008875953 | 0.040898502 | 1 |
| HTN | ASPC | *SPATA9* | 1.495482481 | 0.000260606 | 0.003017806 | 1 |
| HTN | ASPC | *FEM1C* | 1.577912012 | 0.003848017 | 0.022771806 | 1 |
| HTN | ASPC | *BOD1* | 0.758673368 | 0.007199075 | 0.035461838 | 1 |
| HTN | ASPC | *TBC1D7* | 0.417412895 | 0.005922283 | 0.030840166 | 1 |
| HTN | ASPC | *GFOD1* | 1.283615654 | 9.47E-07 | 3.35E-05 | 0.004594576 |
| HTN | ASPC | *ABT1* | 0.207573664 | 0.001168561 | 0.009268434 | 1 |
| HTN | ASPC | *IRAK1BP1* | 1.119996772 | 1.69E-10 | 4.82E-08 | 8.20E-07 |
| HTN | ASPC | *PHIP* | 0.88703279 | 0.002452895 | 0.016348144 | 1 |
| HTN | ASPC | *PNRC1* | 1.161159044 | 0.008039506 | 0.038242827 | 1 |
| HTN | ASPC | *SLC18B1* | -0.244628167 | 0.00511354 | 0.028283771 | 1 |
| HTN | ASPC | *SLC22A3* | 1.765641107 | 5.31E-06 | 0.000138571 | 0.025774156 |
| HTN | ASPC | *TMEM209* | 1.7855125 | 6.92E-05 | 0.00108825 | 0.33596742 |
| HTN | ASPC | *EPHA1* | 5.10901195 | 0.000234106 | 0.002804648 | 1 |
| HTN | ASPC | *NCAPG2* | 0.457131718 | 0.008220518 | 0.038724225 | 1 |
| HTN | ASPC | *FBXO25* | 1.229394824 | 0.001292359 | 0.009969037 | 1 |
| HTN | ASPC | *CSGALNACT1* | 0.229932746 | 0.001020365 | 0.008463575 | 1 |
| HTN | ASPC | *DOCK5* | 0.762404941 | 0.002432203 | 0.016299793 | 1 |
| HTN | ASPC | *ADHFE1* | -0.108229365 | 0.002876837 | 0.018487964 | 1 |
| HTN | ASPC | *DPYS* | 1.124587302 | 0.002383797 | 0.016131358 | 1 |
| HTN | ASPC | *HAUS6* | 1.235185436 | 0.007267887 | 0.035656004 | 1 |
| HTN | ASPC | *ZNF462* | 0.602577866 | 0.004974259 | 0.02780542 | 1 |
| HTN | ASPC | *SH3GLB2* | 0.840429197 | 0.009347527 | 0.042308025 | 1 |
| HTN | ASPC | *SLC39A12* | 1.135563835 | 0.000199935 | 0.002506673 | 0.970082563 |
| HTN | ASPC | *ST8SIA6* | 1.877401581 | 2.99E-07 | 1.20E-05 | 0.001450055 |
| HTN | ASPC | *RGR* | -1.260237857 | 0.001475737 | 0.011081811 | 1 |
| HTN | ASPC | *PAOX* | 1.838770721 | 0.003818586 | 0.022705614 | 1 |
| HTN | ASPC | *RGS10* | 0.665893883 | 0.009104051 | 0.041554897 | 1 |
| HTN | ASPC | *LIN7C* | 1.207784178 | 0.011545902 | 0.049357458 | 1 |
| HTN | ASPC | *ZNF215* | 0.1025095 | 0.007868986 | 0.037653176 | 1 |
| HTN | ASPC | *HSD17B12* | -0.217263346 | 7.96E-09 | 6.66E-07 | 3.86E-05 |
| HTN | ASPC | *TNKS1BP1* | 0.494987621 | 1.73E-11 | 1.20E-08 | 8.41E-08 |
| HTN | ASPC | *TTC12* | 0.31120841 | 0.000513776 | 0.005092568 | 1 |
| HTN | ASPC | *DRD2* | 15.92178523 | 3.83E-08 | 2.30E-06 | 0.000185996 |
| HTN | ASPC | *NPAT* | 1.035757978 | 0.008139193 | 0.038565786 | 1 |
| HTN | ASPC | *ST14* | 0.7894653 | 0.010513413 | 0.046163874 | 1 |
| HTN | ASPC | *EML3* | 0.282533657 | 0.007390286 | 0.035965567 | 1 |
| HTN | ASPC | *DUSP15* | 0.463154775 | 0.005780891 | 0.030493044 | 1 |
| HTN | ASPC | *TRPT1* | -0.233601414 | 0.005313985 | 0.028872849 | 1 |
| HTN | ASPC | *TBX6* | 0.109220491 | 0.000521301 | 0.005151435 | 1 |
| HTN | ASPC | *SAP18* | 1.37767045 | 0.003218339 | 0.020045416 | 1 |
| HTN | ASPC | *FAM124A* | 1.182362024 | 0.000249786 | 0.002920395 | 1 |
| HTN | ASPC | *PPP1R1C* | -1.790199994 | 0.000612297 | 0.005768672 | 1 |
| HTN | ASPC | *FAM173B* | 0.889000796 | 0.001899509 | 0.013553555 | 1 |
| HTN | ASPC | *GXYLT1* | 0.719767325 | 0.009619797 | 0.04317785 | 1 |

| | | | | | | |
|---|---|---|---|---|---|---|
| HTN | ASPC | *TMEM18* | 1.649487147 | 0.005061766 | 0.02810033 | 1 |
| HTN | ASPC | *NUBPL* | 0.988014205 | 0.002716218 | 0.017642689 | 1 |
| HTN | ASPC | *ANKRD50* | 0.843188436 | 0.001421163 | 0.010723923 | 1 |
| HTN | ASPC | *MFSD6* | 1.02241007 | 0.004575397 | 0.026099689 | 1 |
| HTN | ASPC | *RNF144A* | 1.025122187 | 0.003412287 | 0.020963227 | 1 |
| HTN | ASPC | *BICD1* | 1.111152726 | 0.001624245 | 0.012031812 | 1 |
| HTN | ASPC | *PTPN14* | 0.955638026 | 0.000224659 | 0.002735549 | 1 |
| HTN | ASPC | *KCNK13* | 0.924780545 | 0.000328486 | 0.003626698 | 1 |
| HTN | ASPC | *CWF19L2* | 1.033766152 | 1.08E-07 | 5.17E-06 | 0.000522498 |
| HTN | ASPC | *PLEKHH2* | -0.205340619 | 0.003572856 | 0.021615333 | 1 |
| HTN | ASPC | *CCNO* | 6.037177235 | 6.71E-05 | 0.001063291 | 0.325366978 |
| HTN | ASPC | *CLEC4F* | 0.440686556 | 0.009144266 | 0.041620995 | 1 |
| HTN | ASPC | *RASGRP3* | 0.493939372 | 0.007331339 | 0.035769075 | 1 |
| HTN | ASPC | *PRDM8* | -0.637771954 | 6.42E-06 | 0.000156497 | 0.031142931 |
| HTN | ASPC | *MED21* | 1.245226237 | 1.44E-07 | 6.39E-06 | 0.000696343 |
| HTN | ASPC | *JAKMIP1* | 2.31931494 | 9.73E-06 | 0.000218579 | 0.0472131 |
| HTN | ASPC | *TMEM87B* | 1.329907627 | 0.000463285 | 0.00472861 | 1 |
| HTN | ASPC | *FBXO36* | 1.23475897 | 0.005786918 | 0.030493044 | 1 |
| HTN | ASPC | *GDPD1* | 0.80911368 | 0.01030863 | 0.045511804 | 1 |
| HTN | ASPC | *ANKRD29* | 1.596130191 | 7.74E-05 | 0.001184233 | 0.375560961 |
| HTN | ASPC | *ROBO4* | 1.648150116 | 0.000196234 | 0.00247306 | 0.952128215 |
| HTN | ASPC | *ABCA5* | 0.556327154 | 2.10E-05 | 0.00041439 | 0.101939899 |
| HTN | ASPC | *NEIL2* | 0.538891258 | 1.45E-10 | 4.40E-08 | 7.04E-07 |
| HTN | ASPC | *CCSAP* | 2.220226832 | 0.000734981 | 0.006653223 | 1 |
| HTN | ASPC | *OXNAD1* | 0.999706877 | 0.003617347 | 0.021802941 | 1 |
| HTN | ASPC | *ANKRD40* | 1.1195209 | 0.000487011 | 0.004901747 | 1 |
| HTN | ASPC | *PROM2* | -0.341841698 | 0.009791213 | 0.043744905 | 1 |
| HTN | ASPC | *AK9* | 0.377166432 | 0.003778366 | 0.022521661 | 1 |
| HTN | ASPC | *GOLGA7B* | 7.893798739 | 7.89E-10 | 1.71E-07 | 3.83E-06 |
| HTN | ASPC | *MOV10* | 1.57835329 | 3.69E-06 | 0.00010079 | 0.017926901 |
| HTN | ASPC | *PDIA4* | 1.586235964 | 0.002098958 | 0.014569588 | 1 |
| HTN | ASPC | *MMP16* | -0.138582569 | 0.002581655 | 0.016950191 | 1 |
| HTN | ASPC | *NDUFAF6* | -0.591375239 | 0.001160387 | 0.009260194 | 1 |
| HTN | ASPC | *ART3* | -0.205490949 | 0.001715177 | 0.012552092 | 1 |
| HTN | ASPC | *NAA11* | -0.881518009 | 0.000190793 | 0.00242059 | 0.925727629 |
| HTN | ASPC | *SFXN2* | 0.285216068 | 1.82E-09 | 2.88E-07 | 8.85E-06 |
| HTN | ASPC | *HKDC1* | 1.51833076 | 0.001994105 | 0.01410408 | 1 |
| HTN | ASPC | *NODAL* | 3.385650333 | 9.12E-05 | 0.001353423 | 0.442569454 |
| HTN | ASPC | *ZDHHC5* | 1.520394367 | 0.001187884 | 0.009341353 | 1 |
| HTN | ASPC | *MED19* | 0.386126026 | 0.000347701 | 0.003765723 | 1 |
| HTN | ASPC | *VPS8* | 0.795741387 | 2.83E-05 | 0.000529425 | 0.137486261 |
| HTN | ASPC | *NTAN1* | 0.513541965 | 0.001756568 | 0.012758782 | 1 |
| HTN | ASPC | *FCHO2* | 0.449930117 | 0.005740755 | 0.030408456 | 1 |
| HTN | ASPC | *GATAD1* | 0.909836466 | 0.005444876 | 0.02935393 | 1 |
| HTN | ASPC | *CLEC18A* | -0.310794648 | 2.88E-06 | 8.33E-05 | 0.013994352 |
| HTN | ASPC | *ETS2* | 0.117515792 | 0.007773797 | 0.037308075 | 1 |
| HTN | ASPC | *SLC38A10* | 1.073139407 | 0.008047617 | 0.038243917 | 1 |
| HTN | ASPC | *EXTL1* | 27.58597836 | 0.002812705 | 0.018147931 | 1 |
| HTN | ASPC | *RHPN1* | 0.282724501 | 0.00894633 | 0.041077551 | 1 |
| HTN | ASPC | *FANCC* | 0.861075679 | 0.001171731 | 0.009268434 | 1 |

| | | | | | | |
|---|---|---|---|---|---|---|
| HTN | ASPC | *ELK4* | 1.601059243 | 1.81E-05 | 0.00035999 | 0.087657799 |
| HTN | ASPC | *SPATA2L* | 0.69224367 | 0.000743669 | 0.006694398 | 1 |
| HTN | ASPC | *SLC5A11* | -0.177712566 | 2.31E-06 | 6.97E-05 | 0.01122952 |
| HTN | ASPC | *ARHGAP27* | 0.89509123 | 0.000567767 | 0.005498614 | 1 |
| HTN | ASPC | *AMFR* | 0.69961332 | 0.002229757 | 0.015324049 | 1 |
| HTN | ASPC | *TMEM69* | 1.274694598 | 0.003481671 | 0.021246167 | 1 |
| HTN | ASPC | *ACE* | -1.208495531 | 0.000502041 | 0.004991607 | 1 |
| HTN | ASPC | *CHCHD6* | 0.487225609 | 0.004015302 | 0.023472585 | 1 |
| HTN | ASPC | *ATAD3B* | 0.359218786 | 0.00111309 | 0.008986213 | 1 |
| HTN | ASPC | *CCDC58* | 1.864955957 | 0.000912799 | 0.007742831 | 1 |
| HTN | ASPC | *SLC37A1* | 0.873008046 | 0.005632058 | 0.030029391 | 1 |
| HTN | ASPC | *S100B* | -0.092055764 | 8.17E-05 | 0.00123217 | 0.396596628 |
| HTN | ASPC | *ZNF208* | -0.412353876 | 0.002459544 | 0.016369968 | 1 |
| HTN | ASPC | *TAOK1* | 0.884939135 | 0.002929145 | 0.018774387 | 1 |
| HTN | ASPC | *ZBTB7B* | 1.2840054 | 0.000234962 | 0.002807973 | 1 |
| HTN | ASPC | *FDPS* | 0.811503051 | 0.00016901 | 0.002233181 | 0.820038471 |
| HTN | ASPC | *CCR5* | 0.340737063 | 0.000979664 | 0.00820955 | 1 |
| HTN | ASPC | *PTH1R* | -0.267292381 | 0.010964387 | 0.04745692 | 1 |
| HTN | ASPC | *LY6K* | 0.260079912 | 0.004797993 | 0.027101122 | 1 |
| HTN | ASPC | *IER2* | 0.608686159 | 0.010039782 | 0.044532084 | 1 |
| HTN | ASPC | *CPSF4* | 0.590066187 | 0.000386044 | 0.004089703 | 1 |
| HTN | ASPC | *ABCF3* | 0.474589285 | 1.14E-08 | 8.76E-07 | 5.52E-05 |
| HTN | ASPC | *PGAP3* | 2.659782209 | 3.58E-05 | 0.000646506 | 0.17391012 |
| HTN | ASPC | *ASGR2* | 7.108814864 | 8.09E-10 | 1.71E-07 | 3.92E-06 |
| HTN | ASPC | *CCDC78* | 0.33740132 | 0.002882488 | 0.018499776 | 1 |
| HTN | ASPC | *SHANK2* | 0.861282209 | 0.003469933 | 0.021230913 | 1 |
| HTN | ASPC | *ASRGL1* | 2.573638338 | 3.23E-07 | 1.27E-05 | 0.00156592 |
| HTN | ASPC | *RPS6KA4* | 2.794665563 | 3.66E-05 | 0.000657602 | 0.177552487 |
| HTN | ASPC | *CMPK1* | 0.615950898 | 0.003008839 | 0.019008969 | 1 |
| HTN | ASPC | *COA7* | 2.400079761 | 0.001406411 | 0.010645719 | 1 |
| HTN | ASPC | *LAPTM5* | 1.85548856 | 5.90E-06 | 0.000146124 | 0.028640277 |
| HTN | ASPC | *LRRIQ3* | 1.921983373 | 2.33E-07 | 9.80E-06 | 0.00113062 |
| HTN | ASPC | *ACP6* | 1.389376419 | 0.000723582 | 0.006562276 | 1 |
| HTN | ASPC | *CNST* | 0.971856839 | 0.001811111 | 0.01303785 | 1 |
| HTN | ASPC | *MRPL55* | 0.848691324 | 3.28E-14 | 5.30E-11 | 1.59E-10 |
| HTN | ASPC | *C2orf48* | 4.217486525 | 0.000776763 | 0.006890048 | 1 |
| HTN | ASPC | *INHBB* | 0.787565454 | 0.001428087 | 0.010759441 | 1 |
| HTN | ASPC | *LYSMD1* | -0.237838909 | 3.44E-05 | 0.000625021 | 0.166880531 |
| HTN | ASPC | *VPS72* | 1.858884019 | 0.000487952 | 0.004901747 | 1 |
| HTN | ASPC | *ERCC3* | 0.573151157 | 0.010825165 | 0.046980056 | 1 |
| HTN | ASPC | *DCAF16* | -1.235155526 | 2.30E-05 | 0.00044751 | 0.111374567 |
| HTN | ASPC | *GPR155* | 1.267404723 | 0.000554961 | 0.005428772 | 1 |
| HTN | ASPC | *DAPL1* | 0.485748615 | 0.00844218 | 0.039414585 | 1 |
| HTN | ASPC | *PBXIP1* | 1.797242269 | 8.93E-06 | 0.000204286 | 0.043308595 |
| HTN | ASPC | *LMOD1* | -0.450224245 | 0.000232544 | 0.002792834 | 1 |
| HTN | ASPC | *CRELD1* | 1.023003473 | 0.00331028 | 0.020434449 | 1 |
| HTN | ASPC | *CCDC158* | 0.792546515 | 0.000122397 | 0.001726369 | 0.59387093 |
| HTN | ASPC | *RPN1* | 1.376724266 | 0.002579963 | 0.016950191 | 1 |
| HTN | ASPC | *RFT1* | -0.313633011 | 0.000415367 | 0.00434345 | 1 |
| HTN | ASPC | *SFMBT1* | 1.973898917 | 7.15E-05 | 0.001116249 | 0.347153552 |

| | | | | | | |
|---|---|---|---|---|---|---|
| HTN | ASPC | *FAM208A* | 1.32886825 | 3.69E-06 | 0.00010079 | 0.017888073 |
| HTN | ASPC | *ABLIM2* | 0.499106034 | 0.001977909 | 0.014030427 | 1 |
| HTN | ASPC | *BDH2* | -0.432759404 | 0.001516628 | 0.011303653 | 1 |
| HTN | ASPC | *CDC25A* | -1.324403331 | 2.07E-08 | 1.36E-06 | 0.000100515 |
| HTN | ASPC | *ZNF589* | -0.55269629 | 2.30E-09 | 3.27E-07 | 1.12E-05 |
| HTN | ASPC | *RNF123* | 0.537253014 | 2.62E-11 | 1.59E-08 | 1.27E-07 |
| HTN | ASPC | *MST1R* | 0.289614811 | 1.39E-05 | 0.00030074 | 0.067365763 |
| HTN | ASPC | *ETNPPL* | 12.87978242 | 7.34E-09 | 6.60E-07 | 3.56E-05 |
| HTN | ASPC | *HMGB2* | 0.486226058 | 0.006441671 | 0.032796422 | 1 |
| HTN | ASPC | *HPGD* | 3.081659911 | 9.35E-06 | 0.000213052 | 0.045389965 |
| HTN | ASPC | *IL15* | 0.511862629 | 0.008891209 | 0.040929928 | 1 |
| HTN | ASPC | *RNF180* | 0.845706335 | 0.001834181 | 0.013164863 | 1 |
| HTN | ASPC | *F2RL2* | 4.247448655 | 5.44E-10 | 1.32E-07 | 2.64E-06 |
| HTN | ASPC | *CCDC112* | 1.050702702 | 0.000749181 | 0.00670578 | 1 |
| HTN | ASPC | *ERAP1* | -0.705526339 | 1.90E-06 | 5.88E-05 | 0.00922547 |
| HTN | ASPC | *ERAP2* | 0.124063258 | 0.000962976 | 0.008097679 | 1 |
| HTN | ASPC | *RAET1E* | 1.478033055 | 0.000991033 | 0.008290502 | 1 |
| HTN | ASPC | *SAP30L* | 0.715571893 | 0.004963904 | 0.027779544 | 1 |
| HTN | ASPC | *RP9* | 1.761163409 | 0.002194249 | 0.015101416 | 1 |
| HTN | ASPC | *USP49* | 1.167910488 | 0.006152179 | 0.031679437 | 1 |
| HTN | ASPC | *CHMP4C* | 5.658725728 | 2.16E-09 | 3.23E-07 | 1.05E-05 |
| HTN | ASPC | *BRI3* | 0.963251862 | 0.008211392 | 0.038720552 | 1 |
| HTN | ASPC | *SOX17* | 0.394510413 | 0.001170566 | 0.009268434 | 1 |
| HTN | ASPC | *RAD21* | 0.945900885 | 0.002451957 | 0.016348144 | 1 |
| HTN | ASPC | *TNFRSF11B* | 1.943880384 | 2.47E-06 | 7.40E-05 | 0.011991375 |
| HTN | ASPC | *FASTK* | 1.733855069 | 0.001305553 | 0.010054827 | 1 |
| HTN | ASPC | *ALDH7A1* | 0.776145941 | 0.000626714 | 0.005847723 | 1 |
| HTN | ASPC | *DCSTAMP* | 6.844040685 | 8.63E-08 | 4.32E-06 | 0.000418873 |
| HTN | ASPC | *RPP25L* | -0.369990179 | 0.000904657 | 0.007727813 | 1 |
| HTN | ASPC | *C9orf24* | 1.060281378 | 0.001639072 | 0.01210468 | 1 |
| HTN | ASPC | *TMEM65* | 1.026375132 | 0.0067121 | 0.0337483 | 1 |
| HTN | ASPC | *ZMAT4* | 4.838755572 | 1.74E-08 | 1.19E-06 | 8.43E-05 |
| HTN | ASPC | *AQP7* | 0.181135894 | 0.000346677 | 0.003763039 | 1 |
| HTN | ASPC | *AQP3* | 7.280520528 | 6.55E-08 | 3.53E-06 | 0.000317843 |
| HTN | ASPC | *WRN* | 1.097658825 | 0.000123329 | 0.001734474 | 0.598393619 |
| HTN | ASPC | *SLC16A9* | -1.508940384 | 0.002408566 | 0.016186096 | 1 |
| HTN | ASPC | *GJB2* | 10.43475024 | 2.47E-09 | 3.33E-07 | 1.20E-05 |
| HTN | ASPC | *MICU2* | 0.921803633 | 0.002537692 | 0.016775042 | 1 |
| HTN | ASPC | *LRR1* | 2.792784098 | 0.000104616 | 0.00152891 | 0.507598002 |
| HTN | ASPC | *ATP5C1* | 1.050751791 | 0.000328884 | 0.003626698 | 1 |
| HTN | ASPC | *STOX1* | 0.161091133 | 0.002532852 | 0.01676589 | 1 |
| HTN | ASPC | *RET* | -0.833160294 | 1.36E-06 | 4.52E-05 | 0.006602633 |
| HTN | ASPC | *UBTD1* | 1.597729164 | 8.18E-05 | 0.00123217 | 0.396758731 |
| HTN | ASPC | *IFI27L1* | 1.468535819 | 4.55E-05 | 0.000791532 | 0.220837539 |
| HTN | ASPC | *ARL5B* | -0.719086091 | 0.000848592 | 0.007405337 | 1 |
| HTN | ASPC | *API5* | 1.08465572 | 0.008452607 | 0.039414585 | 1 |
| HTN | ASPC | *WBP1L* | 1.005103136 | 2.38E-05 | 0.000458071 | 0.115264522 |
| HTN | ASPC | *CYB5R2* | 0.241449836 | 0.000740136 | 0.006674984 | 1 |
| HTN | ASPC | *TRIM66* | -0.125076549 | 0.000116069 | 0.001656378 | 0.563168646 |
| HTN | ASPC | *PRKCB* | 0.377001592 | 0.008392083 | 0.039227731 | 1 |

| | | | | | | |
|---|---|---|---|---|---|---|
| HTN | ASPC | *NDST2* | 18.25667389 | 1.96E-07 | 8.43E-06 | 0.00095219 |
| HTN | ASPC | *RIMKLB* | -0.447419341 | 2.28E-06 | 6.90E-05 | 0.01104749 |
| HTN | ASPC | *SEC11C* | 2.471271474 | 1.31E-06 | 4.37E-05 | 0.006340274 |
| HTN | ASPC | *CENPV* | 0.863917169 | 5.80E-05 | 0.000953804 | 0.281372275 |
| HTN | ASPC | *CATSPER2* | 0.87355917 | 0.005823014 | 0.030544067 | 1 |
| HTN | ASPC | *SAAL1* | 1.576816634 | 0.0105581 | 0.046191181 | 1 |
| HTN | ASPC | *NYAP1* | 0.153155342 | 0.00202599 | 0.014260545 | 1 |
| HTN | ASPC | *CNPY4* | 0.392502146 | 0.004866507 | 0.027329039 | 1 |
| HTN | ASPC | *FBXO22* | 1.076656646 | 0.002957041 | 0.018835499 | 1 |
| HTN | ASPC | *LOXHD1* | 0.462751239 | 0.00505361 | 0.02810033 | 1 |
| HTN | ASPC | *ZNF23* | -0.514535117 | 0.010576752 | 0.046191181 | 1 |
| HTN | ASPC | *ZNF641* | 1.958324525 | 7.62E-09 | 6.62E-07 | 3.70E-05 |
| HTN | ASPC | *LENG8* | 0.924077771 | 0.003218088 | 0.020045416 | 1 |
| HTN | ASPC | *SRR* | 0.159333881 | 8.66E-05 | 0.001288609 | 0.420086459 |
| HTN | ASPC | *CYB5D2* | 1.363377854 | 7.76E-05 | 0.001184233 | 0.376586202 |
| HTN | ASPC | *KLK6* | 10.66160015 | 2.19E-09 | 3.23E-07 | 1.07E-05 |
| HTN | ASPC | *PRDX2* | 1.439250717 | 0.001169102 | 0.009268434 | 1 |
| HTN | ASPC | *MIS12* | 1.526009235 | 0.001248909 | 0.009717714 | 1 |
| HTN | ASPC | *ATP5H* | 0.95146811 | 0.002136584 | 0.01474638 | 1 |
| HTN | ASPC | *ZNF598* | -0.428207294 | 0.004925686 | 0.027597492 | 1 |
| HTN | ASPC | *ATG16L2* | 1.445935331 | 0.000750461 | 0.00670578 | 1 |
| HTN | ASPC | *SCARA5* | -0.262665922 | 0.000591605 | 0.005675775 | 1 |
| HTN | ASPC | *NT5DC2* | -0.216383774 | 0.001799943 | 0.012989982 | 1 |
| HTN | ASPC | *PDHB* | 1.001073128 | 2.74E-05 | 0.000514978 | 0.132864323 |
| HTN | ASPC | *RFWD3* | 0.789337834 | 0.006033571 | 0.031276587 | 1 |
| HTN | ASPC | *KLHL30* | 1.739505656 | 0.000631433 | 0.00588045 | 1 |
| HTN | ASPC | *CCDC110* | 4.741681428 | 1.56E-05 | 0.000323165 | 0.075620596 |
| HTN | ASPC | *NBPF9* | -1.330166598 | 0.006634305 | 0.033426427 | 1 |
| HTN | ASPC | *ADAM9* | 0.558823157 | 0.004578325 | 0.026099689 | 1 |
| HTN | ASPC | *ZBTB49* | 2.258173164 | 7.09E-05 | 0.001109507 | 0.343947222 |
| HTN | ASPC | *TMEM129* | 0.480035497 | 0.001892141 | 0.013520864 | 1 |
| HTN | ASPC | *MAP2K1* | 1.028442816 | 0.00674263 | 0.033796734 | 1 |
| HTN | ASPC | *UPF3A* | 0.441325778 | 0.000180927 | 0.002334213 | 0.877858117 |
| HTN | ASPC | *C8orf46* | 0.079960823 | 0.000485702 | 0.004899435 | 1 |
| HTN | ASPC | *CPT1C* | 0.520313338 | 1.34E-07 | 6.21E-06 | 0.000652458 |
| HTN | ASPC | *RAB24* | -1.151413666 | 0.00208652 | 0.014545683 | 1 |
| HTN | ASPC | *PGM2* | 0.500866291 | 0.008544325 | 0.039671833 | 1 |
| HTN | ASPC | *CLIC3* | 3.399756244 | 7.26E-06 | 0.000171738 | 0.035206351 |
| HTN | ASPC | *INO80E* | -0.391652859 | 1.41E-08 | 1.02E-06 | 6.86E-05 |
| HTN | ASPC | *NFU1* | 0.66435264 | 0.000182506 | 0.002336468 | 0.885521223 |
| HTN | ASPC | *RGPD8* | -0.073523739 | 0.000872633 | 0.007533837 | 1 |
| HTN | ASPC | *LUZP1* | 1.108712107 | 0.00232825 | 0.015821664 | 1 |
| HTN | ASPC | *CALML6* | 11.42157711 | 2.34E-05 | 0.000454661 | 0.113665304 |
| HTN | ASPC | *MYO7B* | 0.406242914 | 6.53E-06 | 0.000158462 | 0.031692439 |
| HTN | ASPC | *HNRNPA3* | 0.569607322 | 0.003929617 | 0.023055018 | 1 |
| HTN | ASPC | *CNGB3* | 0.349994884 | 0.005763747 | 0.030463727 | 1 |
| HTN | ASPC | *STX8* | 0.719505452 | 0.009766983 | 0.043717162 | 1 |
| HTN | ASPC | *NFRKB* | 1.074385257 | 0.00833431 | 0.039019047 | 1 |
| HTN | ASPC | *DNAJC18* | -0.328610787 | 0.0004202 | 0.004384535 | 1 |
| HTN | ASPC | *TTLL6* | -0.973693877 | 1.42E-07 | 6.39E-06 | 0.000690494 |

| | | | | | | |
|---|---|---|---|---|---|---|
| HTN | ASPC | *FOXN2* | 1.181817643 | 0.000858604 | 0.007465852 | 1 |
| HTN | ASPC | *MSANTD4* | 1.540010108 | 0.0019448 | 0.013836028 | 1 |
| HTN | ASPC | *PAQR8* | -0.259288864 | 0.003698768 | 0.022238443 | 1 |
| HTN | ASPC | *TSNARE1* | 0.338047461 | 0.007018512 | 0.03478429 | 1 |
| HTN | ASPC | *FEZ2* | 0.912237142 | 8.10E-05 | 0.001231847 | 0.392959293 |
| HTN | ASPC | *SOX7* | 0.886401636 | 0.000772015 | 0.006860473 | 1 |
| HTN | ASPC | *GIMAP8* | 0.875150998 | 4.42E-07 | 1.70E-05 | 0.002145647 |
| HTN | ASPC | *MRPL36* | 0.177266747 | 0.000957248 | 0.008063489 | 1 |
| HTN | ASPC | *PPID* | 1.186141181 | 0.000919754 | 0.007788212 | 1 |
| HTN | ASPC | *NMUR1* | 3.971723006 | 3.70E-06 | 0.00010079 | 0.017940703 |
| HTN | ASPC | *PIK3CD* | 0.636440317 | 0.003509911 | 0.021367741 | 1 |
| HTN | ASPC | *RGS19* | -0.213514108 | 1.41E-07 | 6.38E-06 | 0.000682644 |
| HTN | ASPC | *ANO5* | 0.511932274 | 0.00239778 | 0.016135962 | 1 |
| HTN | ASPC | *VAT1L* | 0.999175813 | 0.000595297 | 0.005697002 | 1 |
| HTN | ASPC | *CAMTA1* | 0.783706266 | 0.000279043 | 0.003176624 | 1 |
| HTN | ASPC | *FBXW10* | 2.260932579 | 0.000195084 | 0.002464962 | 0.946545417 |
| HTN | ASPC | *ATPAF2* | 0.412013728 | 0.005368813 | 0.02907308 | 1 |
| HTN | ASPC | *ZNF554* | 0.945816237 | 0.003853185 | 0.022771806 | 1 |
| HTN | ASPC | *ANKRD20A4* | 6.976994076 | 5.33E-07 | 2.01E-05 | 0.002587838 |
| HTN | ASPC | *SERF1A* | 5.336078306 | 0.00186261 | 0.013329474 | 1 |
| HTN | ASPC | *NME6* | 0.735865941 | 7.50E-11 | 3.64E-08 | 3.64E-07 |
| HTN | ASPC | *TEFM* | 0.568234802 | 0.006511794 | 0.033049396 | 1 |
| HTN | ASPC | *MALT1* | 1.158295839 | 0.002380583 | 0.016131358 | 1 |
| HTN | ASPC | *MACROD2* | 0.305780598 | 0.004583045 | 0.026099689 | 1 |
| HTN | ASPC | *TP53RK* | 1.09570135 | 0.002005731 | 0.014165656 | 1 |
| HTN | ASPC | *FGGY* | 1.017552805 | 2.84E-06 | 8.30E-05 | 0.013773438 |
| HTN | ASPC | *AFF1* | 0.73462028 | 0.007604055 | 0.036784522 | 1 |
| HTN | ASPC | *FIBP* | 0.400845828 | 0.011249786 | 0.048475986 | 1 |
| HTN | ASPC | *CTSW* | -0.053412907 | 5.87E-05 | 0.000955008 | 0.284592486 |
| HTN | ASPC | *ZNF738* | 0.199789295 | 0.000907511 | 0.007728957 | 1 |
| HTN | ASPC | *RAB43* | -4.191043985 | 0.000298681 | 0.003354631 | 1 |
| HTN | ASPC | *CBWD1* | 1.800118867 | 0.001411713 | 0.010669212 | 1 |
| HTN | ASPC | *NOC3L* | -0.621222759 | 0.005887284 | 0.030748227 | 1 |
| HTN | ASPC | *RHOD* | 0.597723113 | 0.004052415 | 0.023632592 | 1 |
| HTN | ASPC | *AHSA2* | 0.280579066 | 0.005326106 | 0.028906339 | 1 |
| HTN | ASPC | *MAP3K11* | 0.690776651 | 4.63E-05 | 0.000798601 | 0.224636978 |
| HTN | ASPC | *PPP1R14B* | 1.331607586 | 5.46E-06 | 0.000140237 | 0.026492017 |
| HTN | ASPC | *MST1* | -0.157380733 | 6.75E-06 | 0.000162184 | 0.032761106 |
| HTN | ASPC | *NUDT18* | 2.112200082 | 0.002098504 | 0.014569588 | 1 |
| HTN | ASPC | *UQCRH* | 0.791552815 | 0.01001742 | 0.044509634 | 1 |
| HTN | ASPC | *C11orf80* | -0.635769688 | 2.50E-06 | 7.44E-05 | 0.01212115 |
| HTN | ASPC | *GPR160* | 0.455267416 | 0.000244754 | 0.00288239 | 1 |
| HTN | ASPC | *CTU2* | -0.213896333 | 0.002991133 | 0.018946445 | 1 |
| HTN | ASPC | *SNX31* | -0.172025191 | 1.48E-05 | 0.000314446 | 0.071693726 |
| HTN | ASPC | *GLIS1* | 2.316193451 | 1.63E-06 | 5.24E-05 | 0.007911857 |
| HTN | ASPC | *SNAPC5* | 0.510141248 | 0.004145389 | 0.024001701 | 1 |
| HTN | ASPC | *MSL2* | 1.130965741 | 0.003851455 | 0.022771806 | 1 |
| HTN | ASPC | *CEP135* | 1.21264393 | 0.00075648 | 0.006747131 | 1 |
| HTN | ASPC | *SEZ6L2* | 0.363812007 | 3.60E-06 | 0.000100465 | 0.017480833 |
| HTN | ASPC | *KCTD13* | 0.342311368 | 0.001545504 | 0.011501203 | 1 |

| | | | | | | |
|---|---|---|---|---|---|---|
| HTN | ASPC | *TVP23C* | 2.52024825 | 0.010694551 | 0.046621709 | 1 |
| HTN | ASPC | *SH3BP5L* | 2.162590262 | 0.001378559 | 0.010516932 | 1 |
| HTN | ASPC | *ARHGAP1* | 0.382738231 | 0.005169763 | 0.028380408 | 1 |
| HTN | ASPC | *CCNE2* | -0.494684165 | 4.58E-08 | 2.58E-06 | 0.000222288 |
| HTN | ASPC | *TMEM9B* | 1.110548733 | 2.05E-06 | 6.29E-05 | 0.009940406 |
| HTN | ASPC | *CLTB* | 1.394971361 | 0.003356783 | 0.020695184 | 1 |
| HTN | ASPC | *MCTP1* | 1.318302648 | 2.57E-06 | 7.61E-05 | 0.012482592 |
| HTN | ASPC | *PRIMA1* | -1.072311017 | 0.000238651 | 0.002817365 | 1 |
| HTN | ASPC | *CTDNEP1* | 0.648408229 | 1.52E-06 | 4.96E-05 | 0.007396772 |
| HTN | ASPC | *A2M* | 1.081476285 | 0.000326035 | 0.003619962 | 1 |
| HTN | ASPC | *ORAI3* | 0.53597536 | 1.27E-07 | 5.97E-06 | 0.000614533 |
| HTN | ASPC | *B3GALT6* | 4.485956346 | 0.001172905 | 0.009268434 | 1 |
| HTN | ASPC | *SLC35G1* | 2.05475338 | 0.00013405 | 0.001842526 | 0.650411757 |
| HTN | ASPC | *TAC4* | 1.68172614 | 0.001172903 | 0.009268434 | 1 |
| HTN | ASPC | *CRLF3* | 0.550162647 | 7.34E-05 | 0.001138447 | 0.356333769 |
| HTN | ASPC | *WDR25* | 1.46000702 | 0.001020443 | 0.008463575 | 1 |
| HTN | ASPC | *BOK* | 1.947377317 | 1.16E-06 | 3.97E-05 | 0.005642937 |
| HTN | ASPC | *C8G* | -26.01004951 | 0.001145696 | 0.009196584 | 1 |
| HTN | ASPC | *LY6H* | 0.403302622 | 0.000855998 | 0.007456559 | 1 |
| HTN | ASPC | *SHMT1* | 1.169620555 | 2.44E-05 | 0.000464068 | 0.118337259 |
| HTN | ASPC | *DPP7* | 0.990157021 | 0.011310672 | 0.048695102 | 1 |
| HTN | ASPC | *MTHFR* | -0.860481922 | 8.65E-05 | 0.001288609 | 0.41988469 |
| HTN | ASPC | *TALDO1* | 1.685810553 | 0.00017379 | 0.002266746 | 0.843229656 |
| HTN | ASPC | *CD151* | 0.660052144 | 0.000479398 | 0.004845919 | 1 |
| HTN | ASPC | *FAM87B* | 16.73652565 | 3.10E-07 | 1.23E-05 | 0.001504403 |
| HTN | ASPC | *CENPBD1* | 4.346908551 | 0.000563392 | 0.005489113 | 1 |
| HTN | ASPC | *RPS27* | 0.983315649 | 0.007629894 | 0.036855481 | 1 |
| HTN | ASPC | *ASB8* | 1.317382115 | 8.28E-07 | 3.00E-05 | 0.004018862 |
| HTN | ASPC | *ODF3B* | 0.265033224 | 0.000413141 | 0.004338873 | 1 |
| HTN | ASPC | *ADAMTSL1* | -0.34900531 | 0.00293585 | 0.018792537 | 1 |
| HTN | ASPC | *SH2B1* | 1.102349531 | 0.005762251 | 0.030463727 | 1 |
| HTN | ASPC | *SH2D4B* | 1.603175091 | 0.008339397 | 0.039019047 | 1 |
| HTN | ASPC | *WDR6* | 0.183248807 | 0.006186417 | 0.031780142 | 1 |
| HTN | ASPC | *GEN1* | -0.125975916 | 0.007238545 | 0.03558401 | 1 |
| HTN | ASPC | *NT5DC1* | 1.097909053 | 0.000279431 | 0.003176624 | 1 |
| HTN | ASPC | *ERN1* | 1.105453834 | 0.001041931 | 0.008597701 | 1 |
| HTN | ASPC | *RPP25* | 0.252972686 | 4.09E-06 | 0.000109626 | 0.019842243 |
| HTN | ASPC | *MPI* | -0.764043526 | 1.22E-13 | 1.48E-10 | 5.91E-10 |
| HTN | ASPC | *TRIM73* | -0.56187184 | 0.000828692 | 0.007270914 | 1 |
| HTN | ASPC | *HYI* | -0.278012548 | 1.17E-06 | 3.97E-05 | 0.005678193 |
| HTN | ASPC | *C17orf62* | 2.243931732 | 5.96E-06 | 0.000146842 | 0.028927847 |
| HTN | ASPC | *GAK* | 1.179869318 | 0.000493589 | 0.004939344 | 1 |
| HTN | ASPC | *CTC1* | 0.733125363 | 5.59E-06 | 0.000141351 | 0.027139346 |
| HTN | ASPC | *CCDC89* | -2.505853969 | 0.000463895 | 0.00472861 | 1 |
| HTN | ASPC | *ZNF664* | 1.715928137 | 0.003137523 | 0.019642916 | 1 |
| HTN | ASPC | *HLA-DQB1* | -0.089885995 | 0.005610474 | 0.029980198 | 1 |
| HTN | ASPC | *GIPC3* | 14.68925042 | 7.43E-08 | 3.88E-06 | 0.00036064 |
| HTN | ASPC | *SEPHS2* | 0.796897146 | 6.15E-05 | 0.000991813 | 0.298535765 |
| HTN | ASPC | *ZDHHC20* | 1.197038547 | 0.000760578 | 0.006771236 | 1 |
| HTN | ASPC | *MUC16* | 1.15625637 | 0.000181368 | 0.002334213 | 0.879998432 |

| | | | | | | |
|---|---|---|---|---|---|---|
| HTN | ASPC | *SLC25A41* | 4.116060664 | 0.000867452 | 0.007502451 | 1 |
| HTN | ASPC | *SGSH* | 2.688872729 | 0.000150775 | 0.002022682 | 0.731562185 |
| HTN | ASPC | *ZFP41* | 0.392136625 | 0.000652793 | 0.006033053 | 1 |
| HTN | ASPC | *TMEM252* | 51.9687004 | 9.22E-10 | 1.83E-07 | 4.47E-06 |
| HTN | ASPC | *CLDN7* | -0.346952228 | 5.84E-05 | 0.000954738 | 0.283546811 |
| HTN | ASPC | *ADO* | 0.541471 | 3.28E-10 | 8.84E-08 | 1.59E-06 |
| HTN | ASPC | *ERCC6L2* | 0.982353473 | 0.000478835 | 0.004845919 | 1 |
| HTN | ASPC | *MRPL41* | 1.490131541 | 0.009983916 | 0.044401432 | 1 |
| HTN | ASPC | *FAM153B* | 1.048658255 | 4.64E-05 | 0.000798601 | 0.225205571 |
| HTN | ASPC | *SYNM* | 1.158181465 | 0.004253088 | 0.024537435 | 1 |
| HTN | ASPC | *KCNA4* | 2.647008397 | 1.91E-08 | 1.29E-06 | 9.27E-05 |
| HTN | ASPC | *KBTBD3* | 1.253220721 | 0.001616235 | 0.01199078 | 1 |
| HTN | ASPC | *CLN8* | 0.271122476 | 1.08E-06 | 3.76E-05 | 0.005264025 |
| HTN | ASPC | *NXPH4* | 15.85400013 | 4.98E-09 | 4.93E-07 | 2.42E-05 |
| HTN | ASPC | *CAPN12* | 0.266169216 | 0.005973851 | 0.031033323 | 1 |
| HTN | ASPC | *FES* | -1.022065947 | 2.68E-16 | 1.30E-12 | 1.30E-12 |
| HTN | ASPC | *GLRX5* | 1.832568112 | 0.001665121 | 0.012278367 | 1 |
| HTN | ASPC | *BRICD5* | 5.266999845 | 4.90E-08 | 2.73E-06 | 0.000237644 |
| HTN | ASPC | *GRID1* | -0.439898352 | 0.000105836 | 0.001537478 | 0.513517738 |
| HTN | ASPC | *PLCXD3* | -0.692201673 | 0.008518945 | 0.039591877 | 1 |
| HTN | ASPC | *COL18A1* | 0.402534724 | 0.004128997 | 0.023935357 | 1 |
| HTN | ASPC | *ZNF721* | 0.888181939 | 0.000325899 | 0.003619962 | 1 |
| HTN | ASPC | *ZNF320* | 0.122171541 | 0.000149794 | 0.00201889 | 0.72680039 |
| HTN | ASPC | *C12orf60* | -0.987246752 | 0.001514966 | 0.011303653 | 1 |
| HTN | ASPC | *ARHGEF37* | 2.147535924 | 3.65E-06 | 0.00010079 | 0.017710759 |
| HTN | ASPC | *CSMD1* | 1.964813386 | 0.000109297 | 0.001578295 | 0.530307092 |
| HTN | ASPC | *FANCF* | 2.33347959 | 0.000402122 | 0.004241518 | 1 |
| HTN | ASPC | *LRIT3* | 10.60982428 | 9.65E-09 | 7.78E-07 | 4.68E-05 |
| HTN | ASPC | *ACSM2A* | 11.7344692 | 4.38E-08 | 2.50E-06 | 0.000212459 |
| HTN | ASPC | *KREMEN1* | -0.165617394 | 0.00294417 | 0.018804458 | 1 |
| HTN | ASPC | *TTC32* | 1.246865812 | 0.007915884 | 0.037746111 | 1 |
| HTN | ASPC | *PP2D1* | 2.174025045 | 0.000657169 | 0.006061945 | 1 |
| HTN | ASPC | *ACOT1* | 0.152393311 | 0.005791619 | 0.030493044 | 1 |
| HTN | ASPC | *ALDH1A3* | -0.17831351 | 0.000591909 | 0.005675775 | 1 |
| HTN | ASPC | *A3GALT2* | -0.197642698 | 6.65E-05 | 0.001059323 | 0.322711395 |
| HTN | ASPC | *TMEM255B* | 1.257154673 | 4.01E-06 | 0.000108177 | 0.019471943 |
| HTN | ASPC | *SNN* | 3.821332499 | 6.13E-07 | 2.29E-05 | 0.002974034 |
| HTN | ASPC | *DRD1* | 23.99294559 | 2.83E-09 | 3.48E-07 | 1.38E-05 |
| HTN | ASPC | *NUTM2A* | -0.28015061 | 0.000119534 | 0.001700823 | 0.579980489 |
| HTN | ASPC | *SORCS2* | -0.510386726 | 0.003136681 | 0.019642916 | 1 |
| HTN | ASPC | *TMEM106A* | 0.180836254 | 0.0069117 | 0.034395453 | 1 |
| HTN | ASPC | *UBOX5* | 1.074767224 | 0.004551095 | 0.026070733 | 1 |
| HTN | ASPC | *SEMA4B* | 0.823486545 | 0.001324432 | 0.010151886 | 1 |
| HTN | ASPC | *IFITM2* | 1.094368777 | 0.00901386 | 0.04129863 | 1 |
| HTN | ASPC | *CDK10* | -0.179186333 | 0.000285573 | 0.003229842 | 1 |
| HTN | ASPC | *SP1* | 1.284834904 | 4.00E-05 | 0.00071286 | 0.193897819 |
| HTN | ASPC | *SLC52A2* | 0.518195239 | 0.002617906 | 0.017095663 | 1 |
| HTN | ASPC | *ZNF829* | 0.295422484 | 0.002687734 | 0.017481082 | 1 |
| HTN | ASPC | *RNPC3* | 0.399678642 | 0.007340743 | 0.035769075 | 1 |
| HTN | ASPC | *ZNF529* | 1.108081002 | 0.000832445 | 0.007290653 | 1 |

| | | | | | | |
|---|---|---|---|---|---|---|
| HTN | ASPC | *C15orf41* | 0.475035784 | 0.009526209 | 0.042865433 | 1 |
| HTN | ASPC | *POLR1D* | 0.640638888 | 0.004595685 | 0.026140991 | 1 |
| HTN | ASPC | *FFAR4* | 1.445257027 | 1.01E-09 | 1.83E-07 | 4.89E-06 |
| HTN | ASPC | *CCDC30* | 0.313522537 | 0.000215034 | 0.002661588 | 1 |
| HTN | ASPC | *GNG2* | 0.911301162 | 0.010346419 | 0.045595662 | 1 |
| HTN | ASPC | *CYP27C1* | -0.205312255 | 0.001477727 | 0.011081811 | 1 |
| HTN | ASPC | *FOXI2* | -0.215968798 | 0.011214691 | 0.048367718 | 1 |
| HTN | ASPC | *QRFPR* | 0.86786017 | 0.000647932 | 0.005999551 | 1 |
| HTN | ASPC | *MAPT* | 1.851151011 | 1.08E-05 | 0.000241005 | 0.052298011 |
| HTN | ASPC | *C1QL4* | 26.99202971 | 2.34E-07 | 9.80E-06 | 0.001136402 |
| HTN | ASPC | *EFCAB6* | 0.965919608 | 0.002395145 | 0.016135962 | 1 |
| HTN | ASPC | *TMPRSS6* | -7.373466875 | 0.000124929 | 0.001751896 | 0.606156063 |
| HTN | ASPC | *LUZP2* | -1.446768888 | 0.001150935 | 0.009215074 | 1 |
| HTN | ASPC | *GJA4* | 1.351116341 | 0.010343837 | 0.045595662 | 1 |
| HTN | ASPC | *SBK2* | 12.181605 | 1.84E-09 | 2.88E-07 | 8.93E-06 |
| HTN | ASPC | *EXD3* | 1.351918874 | 0.00127706 | 0.009866709 | 1 |
| HTN | ASPC | *DNAH17* | 0.373391118 | 0.00212152 | 0.014663267 | 1 |
| HTN | ASPC | *FAM122A* | 2.56765465 | 0.000566593 | 0.005498216 | 1 |
| HTN | ASPC | *GFRAL* | -0.772697455 | 8.28E-05 | 0.001244132 | 0.401854542 |
| HTN | ASPC | *KLHL17* | 0.322193879 | 0.000180885 | 0.002334213 | 0.877652581 |
| HTN | ASPC | *AGRN* | 2.611999288 | 3.28E-08 | 2.07E-06 | 0.000159048 |
| HTN | ASPC | *NCR3LG1* | 2.975084604 | 1.61E-05 | 0.000329906 | 0.078187818 |
| HTN | ASPC | *RUFY4* | 4.827335405 | 3.57E-08 | 2.19E-06 | 0.000173101 |
| HTN | ASPC | *ZNF559* | -0.112291365 | 0.003118014 | 0.019571285 | 1 |
| HTN | ASPC | *H3F3C* | -7.983229369 | 0.000364628 | 0.003914104 | 1 |
| HTN | ASPC | *C19orf54* | 1.370530282 | 0.00061222 | 0.005768672 | 1 |
| HTN | ASPC | *C22orf34* | 1.878025125 | 3.25E-09 | 3.75E-07 | 1.58E-05 |
| HTN | ASPC | *COL25A1* | 1.449834244 | 1.54E-05 | 0.000321542 | 0.074597676 |
| HTN | ASPC | *SRSF10* | 0.416699742 | 0.005056957 | 0.02810033 | 1 |
| HTN | ASPC | *NDOR1* | 2.444346355 | 3.02E-05 | 0.000557606 | 0.146650505 |
| HTN | ASPC | *GOLGA8M* | 0.683468697 | 0.007267096 | 0.035656004 | 1 |
| HTN | ASPC | *NOXA1* | 1.98466161 | 5.57E-06 | 0.000141351 | 0.027008054 |
| HTN | ASPC | *FZD9* | 4.728326111 | 5.71E-06 | 0.000143454 | 0.02768657 |
| HTN | ASPC | *FBF1* | -0.482492346 | 1.61E-05 | 0.000329906 | 0.077973907 |
| HTN | ASPC | *ASTL* | 8.309041998 | 2.91E-07 | 1.18E-05 | 0.001410929 |
| HTN | ASPC | *MSL1* | -0.266828996 | 0.011490234 | 0.049248514 | 1 |
| HTN | ASPC | *LITAF* | 1.315764118 | 0.000374471 | 0.004010889 | 1 |
| HTN | ASPC | *SF3B3* | 1.242352543 | 0.000317968 | 0.00354662 | 1 |
| HTN | ASPC | *FAM47E* | 0.86166723 | 1.50E-06 | 4.92E-05 | 0.007287936 |
| HTN | ASPC | *TSPYL1* | 1.274997483 | 0.000588226 | 0.005662843 | 1 |
| HTN | ASPC | *FHIT* | 0.863193984 | 0.000385357 | 0.004089703 | 1 |
| HTN | ASPC | *HLA-DRB1* | -0.129230184 | 0.000898726 | 0.007693964 | 1 |
| HTN | ASPC | *AKR1C3* | 1.483763793 | 0.002039807 | 0.014302228 | 1 |
| HTN | ASPC | *COLCA1* | -0.457758598 | 1.09E-06 | 3.76E-05 | 0.005297076 |
| HTN | ASPC | *HRCT1* | 0.250790733 | 0.000422487 | 0.00438952 | 1 |
| HTN | ASPC | *SRGAP3* | 0.283660683 | 0.010237993 | 0.045241113 | 1 |
| HTN | ASPC | *STK31* | 0.732236907 | 0.000529276 | 0.005209021 | 1 |
| HTN | ASPC | *ZKSCAN7* | 0.454314288 | 0.004557557 | 0.026076967 | 1 |
| HTN | ASPC | *ZNF34* | 0.792922795 | 0.00227539 | 0.01559349 | 1 |
| HTN | ASPC | *ZNF124* | 1.170297991 | 0.000377393 | 0.004024418 | 1 |

| | | | | | | |
|---|---|---|---|---|---|---|
| HTN | ASPC | *PIK3R4* | 1.0898717 | 0.006944283 | 0.034486857 | 1 |
| HTN | ASPC | *ZNF605* | 0.310699981 | 0.00124452 | 0.009708056 | 1 |
| HTN | ASPC | *CACNA1H* | 0.825342387 | 0.00860881 | 0.039818416 | 1 |
| HTN | ASPC | *SULF2* | 0.289350993 | 0.001061202 | 0.008709881 | 1 |
| HTN | ASPC | *XRCC2* | 5.016667078 | 4.02E-05 | 0.000713965 | 0.19491242 |
| HTN | ASPC | *ZFP62* | 0.733457008 | 0.001671079 | 0.012303605 | 1 |
| HTN | ASPC | *HIST1H3E* | 0.272091732 | 2.79E-08 | 1.80E-06 | 0.000135575 |
| HTN | ASPC | *WDR5B* | 0.720886334 | 5.03E-06 | 0.000133234 | 0.024381902 |
| HTN | ASPC | *ZNF138* | 0.495844209 | 0.007919526 | 0.037746111 | 1 |
| HTN | ASPC | *ZNF100* | -0.36812882 | 0.000235805 | 0.002811121 | 1 |
| HTN | ASPC | *LRRC8B* | 0.469177811 | 0.01132855 | 0.048728835 | 1 |
| HTN | ASPC | *SND1* | 0.993125999 | 5.32E-05 | 0.000878108 | 0.258163667 |
| HTN | ASPC | *SULT1A2* | 0.486702672 | 1.66E-05 | 0.000336732 | 0.080478882 |
| HTN | ASPC | *C6orf141* | -0.06048166 | 0.008731975 | 0.040311649 | 1 |
| HTN | ASPC | *CCL4L2* | 2.442797268 | 1.57E-08 | 1.09E-06 | 7.61E-05 |
| HTN | ASPC | *PCDHB11* | -0.119632545 | 0.003194386 | 0.019947442 | 1 |
| HTN | ASPC | *ATG7* | 1.011368761 | 3.65E-07 | 1.43E-05 | 0.001768726 |
| HTN | ASPC | *RAB40C* | 1.673898824 | 0.00040317 | 0.004243345 | 1 |
| HTN | ASPC | *MFAP5* | -0.428180543 | 4.17E-08 | 2.44E-06 | 0.000202149 |
| HTN | ASPC | *RPS26* | -0.204414439 | 1.47E-05 | 0.000313582 | 0.071183174 |
| HTN | ASPC | *PSAP* | 0.627666315 | 0.00185739 | 0.013311753 | 1 |
| HTN | ASPC | *ATAD3A* | 0.423148333 | 0.004715676 | 0.02676077 | 1 |
| HTN | ASPC | *ZNF311* | 0.209335558 | 0.000973347 | 0.008170729 | 1 |
| HTN | ASPC | *VPS13A* | 0.594004139 | 0.01137128 | 0.04886931 | 1 |
| HTN | ASPC | *NOL8* | 1.110121703 | 6.92E-05 | 0.00108825 | 0.335567323 |
| HTN | ASPC | *AGAP9* | -0.424207305 | 0.004782318 | 0.027065598 | 1 |
| HTN | ASPC | *MAK16* | 0.844533696 | 0.005725033 | 0.03039153 | 1 |
| HTN | ASPC | *MAN1A2* | 0.899555972 | 0.006127949 | 0.031664333 | 1 |
| HTN | ASPC | *SVIP* | 1.554914513 | 1.23E-05 | 0.000270977 | 0.059885984 |
| HTN | ASPC | *HSD17B11* | 1.313501481 | 7.61E-05 | 0.001175229 | 0.369022036 |
| HTN | ASPC | *RPS6KL1* | 0.747520593 | 1.89E-06 | 5.87E-05 | 0.009156358 |
| HTN | ASPC | *HELZ* | 0.878184999 | 0.002269767 | 0.015576961 | 1 |
| HTN | ASPC | *TMEM116* | -0.295294199 | 8.67E-13 | 8.42E-10 | 4.21E-09 |
| HTN | ASPC | *ZNF442* | 0.34873571 | 0.003230861 | 0.020054426 | 1 |
| HTN | ASPC | *HLA-DRB5* | -0.06524901 | 1.23E-05 | 0.000270399 | 0.059487839 |
| HTN | ASPC | *PAPSS2* | 0.927021479 | 0.006450674 | 0.032807829 | 1 |
| HTN | ASPC | *ANKRD13B* | -1.62319358 | 0.000610354 | 0.005768672 | 1 |
| HTN | ASPC | *PPP1R14C* | 9.14174567 | 2.23E-06 | 6.81E-05 | 0.010820788 |
| HTN | ASPC | *RCSD1* | 1.646329229 | 1.84E-06 | 5.83E-05 | 0.008913014 |
| HTN | ASPC | *PNP* | -0.069882927 | 0.005616846 | 0.029981226 | 1 |
| HTN | ASPC | *DENND4B* | -0.347137161 | 0.00389469 | 0.022905499 | 1 |
| HTN | ASPC | *SHISA4* | -2.213267194 | 1.28E-07 | 5.98E-06 | 0.000622226 |
| HTN | ASPC | *MFAP3L* | 0.909665812 | 1.37E-05 | 0.000298684 | 0.066606522 |
| HTN | ASPC | *TGM2* | 0.215969349 | 0.000362399 | 0.0038988 | 1 |
| HTN | ASPC | *AL590822.1* | -0.598194631 | 0.005402314 | 0.029171394 | 1 |
| HTN | ASPC | *SAMD5* | 0.498068371 | 0.002735146 | 0.017741885 | 1 |
| HTN | ASPC | *IFIT1B* | -4.774635835 | 0.000909741 | 0.007730406 | 1 |
| HTN | ASPC | *LIPN* | 3.902855876 | 1.30E-09 | 2.25E-07 | 6.30E-06 |
| HTN | ASPC | *ASAH2B* | 0.54351178 | 0.00498413 | 0.027828536 | 1 |
| HTN | ASPC | *LINC00619* | 4.217436324 | 0.00173662 | 0.012651773 | 1 |

| | | | | | | |
|---|---|---|---|---|---|---|
| HTN | ASPC | *HSD17B8* | -0.237281068 | 0.000499838 | 0.004985793 | 1 |
| HTN | ASPC | *PRRT1* | 0.269907272 | 4.28E-08 | 2.47E-06 | 0.000207793 |
| HTN | ASPC | *C6orf48* | 0.282936956 | 0.006059474 | 0.031377342 | 1 |
| HTN | ASPC | *BAG6* | 0.322037887 | 0.007754868 | 0.037254077 | 1 |
| HTN | ASPC | *HLA-C* | -0.227094246 | 6.71E-07 | 2.49E-05 | 0.003255587 |
| HTN | ASPC | *POU5F1* | -0.056899452 | 0.001028402 | 0.008515029 | 1 |
| HTN | ASPC | *PSORS1C2* | 0.187719138 | 0.002013665 | 0.014201022 | 1 |
| HTN | ASPC | *ZNF468* | 1.239928108 | 0.002295594 | 0.015684791 | 1 |
| HTN | ASPC | *FAM196B* | 0.714091835 | 0.000736605 | 0.006655506 | 1 |
| HTN | ASPC | *MZT1* | 1.135009478 | 0.011692205 | 0.049875767 | 1 |
| HTN | ASPC | *CCDC144NL* | 3.13632254 | 5.08E-05 | 0.000847588 | 0.246648247 |
| HTN | ASPC | *TMEM170B* | 0.19715568 | 0.00033021 | 0.003633058 | 1 |
| HTN | ASPC | *TECPR1* | 0.908734339 | 0.008251126 | 0.03875553 | 1 |
| HTN | ASPC | *KLRC2* | 4.88860717 | 9.96E-10 | 1.83E-07 | 4.83E-06 |
| HTN | ASPC | *SACM1L* | 1.122036121 | 0.003889174 | 0.022905499 | 1 |
| HTN | ASPC | *SLC48A1* | 1.599891812 | 0.000352207 | 0.003797577 | 1 |
| HTN | ASPC | *TAS2R50* | 3.794022968 | 0.0024238 | 0.016265943 | 1 |
| HTN | ASPC | *TAS2R14* | 0.133667845 | 0.011588626 | 0.049496491 | 1 |
| HTN | ASPC | *SUPT4H1* | 2.197492008 | 1.56E-08 | 1.09E-06 | 7.59E-05 |
| HTN | ASPC | *MXD3* | -0.229299444 | 0.003540231 | 0.021498376 | 1 |
| HTN | ASPC | *TMEM110* | 0.42569118 | 0.000191073 | 0.00242059 | 0.927085892 |
| HTN | ASPC | *ATF6B* | 0.840832909 | 1.35E-11 | 1.09E-08 | 6.57E-08 |
| HTN | ASPC | *PPM1N* | 0.230060024 | 0.000263834 | 0.003047912 | 1 |
| HTN | ASPC | *IRF9* | 1.311631323 | 0.000336548 | 0.003686075 | 1 |
| HTN | ASPC | *GALT* | 0.86371545 | 8.56E-06 | 0.000197684 | 0.041513678 |
| HTN | ASPC | *REPIN1* | 2.699833188 | 2.69E-05 | 0.0005085 | 0.13068462 |
| HTN | ASPC | *GNAT3* | 2.765906194 | 8.90E-08 | 4.41E-06 | 0.000431904 |
| HTN | ASPC | *EML6* | 0.859396941 | 0.001402117 | 0.010642618 | 1 |
| HTN | ASPC | *EVPLL* | 0.48057244 | 0.003610319 | 0.021787647 | 1 |
| HTN | ASPC | *UBXN2B* | 0.478604669 | 3.10E-05 | 0.000568036 | 0.150347308 |
| HTN | ASPC | *GOLGA8B* | 0.202849475 | 0.003023454 | 0.019051685 | 1 |
| HTN | ASPC | *VPS16* | 1.364684205 | 0.005559654 | 0.029825258 | 1 |
| HTN | ASPC | *AC016773.1* | 0.287309242 | 0.000153832 | 0.002056178 | 0.746392659 |
| HTN | ASPC | *AP4M1* | -0.245210949 | 0.001134681 | 0.00913014 | 1 |
| HTN | ASPC | *SLC12A8* | 0.301246449 | 5.99E-05 | 0.000971967 | 0.290618066 |
| HTN | ASPC | *APOL6* | 1.365347087 | 5.02E-05 | 0.000842531 | 0.243491498 |
| HTN | ASPC | *TIAF1* | 0.50760249 | 3.50E-06 | 9.82E-05 | 0.016985085 |
| HTN | ASPC | *RAB6C* | 6.581768888 | 0.004500701 | 0.025812532 | 1 |
| HTN | ASPC | *AC062017.1* | 2.513862808 | 1.35E-05 | 0.000294662 | 0.065414921 |
| HTN | ASPC | *EXOSC6* | -0.593052223 | 0.00011501 | 0.001650969 | 0.558027512 |
| HTN | ASPC | *OSER1-AS1* | 0.256562085 | 0.002093743 | 0.014569588 | 1 |
| HTN | ASPC | *LINC01117* | 1.5585836 | 7.68E-05 | 0.001178807 | 0.372502978 |
| HTN | ASPC | *LINC00184* | 26.80362307 | 7.73E-09 | 6.62E-07 | 3.75E-05 |
| HTN | ASPC | *AL049840.1* | -0.68711508 | 0.000610229 | 0.005768672 | 1 |
| HTN | ASPC | *HAR1A* | -2.889793434 | 0.000617815 | 0.005779093 | 1 |
| HTN | ASPC | *ZNF717* | 0.089748634 | 0.005173657 | 0.028380408 | 1 |
| HTN | ASPC | *SCAMP4* | 1.361180393 | 0.005297486 | 0.028847812 | 1 |
| HTN | ASPC | *AC108463.2* | -0.515663235 | 0.000220994 | 0.002711682 | 1 |
| HTN | ASPC | *LINC00578* | -0.753484295 | 0.000883614 | 0.007615093 | 1 |
| HTN | ASPC | *CASC20* | 1.857966083 | 2.30E-05 | 0.00044751 | 0.111430055 |

| | | | | | | |
|---|---|---|---|---|---|---|
| HTN | ASPC | *DIRC3* | 0.461807985 | 0.00876758 | 0.040437548 | 1 |
| HTN | ASPC | *TMA7* | 0.774183609 | 6.59E-06 | 0.000159005 | 0.031960054 |
| HTN | ASPC | *AC009495.3* | 0.764196621 | 0.000822821 | 0.007245607 | 1 |
| HTN | ASPC | *LINC00702* | 1.613136332 | 0.000565247 | 0.005496148 | 1 |
| HTN | ASPC | *GPX1* | 0.444388701 | 0.000956758 | 0.008063489 | 1 |
| HTN | ASPC | *RPS28* | 1.213272069 | 0.000206046 | 0.002576641 | 0.99973668 |
| HTN | ASPC | *ANKRD65* | 5.912607918 | 0.000141782 | 0.001937821 | 0.687926312 |
| HTN | ASPC | *HLA-DQA2* | 0.097280506 | 5.74E-06 | 0.000143588 | 0.027856132 |
| HTN | ASPC | *LINC01143* | 8.659217111 | 4.07E-09 | 4.21E-07 | 1.98E-05 |
| HTN | ASPC | *TXNDC5* | 9.353236199 | 2.79E-09 | 3.48E-07 | 1.36E-05 |
| HTN | ASPC | *TNFSF12* | 0.592154963 | 7.52E-08 | 3.88E-06 | 0.000364728 |
| HTN | ASPC | *C1QTNF9* | 1.937672682 | 0.005332735 | 0.028909979 | 1 |
| HTN | ASPC | *PNMA2* | -0.433199533 | 0.003735417 | 0.022320498 | 1 |
| HTN | ASPC | *KRBOX1* | -14.18308038 | 4.53E-06 | 0.000120668 | 0.021961572 |
| HTN | ASPC | *NSUN6* | 1.102748559 | 8.62E-06 | 0.000198251 | 0.041830949 |
| HTN | ASPC | *HOGA1* | 4.484743392 | 7.45E-08 | 3.88E-06 | 0.000361244 |
| HTN | ASPC | *MRPL20* | 0.883459049 | 0.00063787 | 0.005917682 | 1 |
| HTN | ASPC | *LINC00960* | -0.062681352 | 0.008616799 | 0.039818416 | 1 |
| HTN | ASPC | *DECR2* | 3.665753639 | 7.40E-06 | 0.000174394 | 0.0359252 |
| HTN | ASPC | *FGF14-IT1* | -1.51311161 | 8.71E-07 | 3.13E-05 | 0.004226258 |
| HTN | ASPC | *WFDC6* | -0.675851391 | 0.000598097 | 0.005709161 | 1 |
| HTN | ASPC | *PLA2G4B* | 0.674906667 | 8.15E-05 | 0.00123217 | 0.39560531 |
| HTN | ASPC | *JMJD7* | 0.175374638 | 0.000899109 | 0.007693964 | 1 |
| HTN | ASPC | *APOBEC3D* | -5.617421713 | 0.000207512 | 0.002588304 | 1 |
| HTN | ASPC | *NEAT1* | 0.810527961 | 4.89E-05 | 0.000828806 | 0.237038406 |
| HTN | ASPC | *CDK11B* | 1.454221526 | 0.003302777 | 0.020434449 | 1 |
| HTN | ASPC | *C5orf17* | 1.195061478 | 0.005257005 | 0.028673381 | 1 |
| HTN | ASPC | *ZNF718* | 0.773120264 | 0.00941256 | 0.042523035 | 1 |
| HTN | ASPC | *SMIM20* | 1.536156679 | 0.009787442 | 0.043744905 | 1 |
| HTN | ASPC | *GLI4* | 1.270351539 | 0.005935774 | 0.03086857 | 1 |
| HTN | ASPC | *LINC01094* | 2.682141172 | 7.22E-09 | 6.60E-07 | 3.51E-05 |
| HTN | ASPC | *PCDHGB2* | -1.906075051 | 0.006819286 | 0.034014353 | 1 |
| HTN | ASPC | *HMBS* | 0.47196366 | 0.000121327 | 0.001718072 | 0.58867664 |
| HTN | ASPC | *LINC00638* | 0.460061107 | 0.000598921 | 0.005709161 | 1 |
| HTN | ASPC | *AC083843.1* | 22.92194117 | 8.15E-09 | 6.70E-07 | 3.95E-05 |
| HTN | ASPC | *C19orf84* | 18.37746334 | 8.91E-11 | 3.87E-08 | 4.32E-07 |
| HTN | ASPC | *LINC00672* | 2.809184776 | 0.009459383 | 0.042655137 | 1 |
| HTN | ASPC | *DYNLL2* | 2.16708666 | 0.00107824 | 0.008822295 | 1 |
| HTN | ASPC | *NDUFA7* | 15.91469167 | 9.78E-09 | 7.78E-07 | 4.74E-05 |
| HTN | ASPC | *AC026740.1* | 0.592518443 | 0.009931774 | 0.044291329 | 1 |
| HTN | ASPC | *ZNF587B* | 2.193255103 | 0.005188257 | 0.028380408 | 1 |
| HTN | ASPC | *CASC15* | 1.341822761 | 0.000214294 | 0.002659213 | 1 |
| HTN | ASPC | *GRIN2B* | 0.186805452 | 0.00342528 | 0.021010695 | 1 |
| HTN | b_cell | *ITIH4* | 4.393141307 | 0.000150284 | 0.040509768 | 0.729175817 |
| HTN | b_cell | *BTN2A2* | 4.428594011 | 3.44E-05 | 0.013917413 | 0.167008957 |
| HTN | b_cell | *TRAP1* | 3.322349021 | 0.000139117 | 0.039705681 | 0.674996583 |
| HTN | b_cell | *FBN2* | 3.761470913 | 2.11E-05 | 0.011348787 | 0.102139081 |
| HTN | b_cell | *SFXN2* | 3.470146589 | 6.22E-05 | 0.02010477 | 0.301571546 |
| HTN | b_cell | *CLEC18A* | -12.90927133 | 2.51E-05 | 0.012157087 | 0.121570866 |
| HTN | b_cell | *MRPL55* | 11.80603331 | 8.97E-07 | 0.00126676 | 0.004354512 |

| | | | | | | |
|---|---|---|---|---|---|---|
| HTN | b_cell | *CPT1C* | 7.497292779 | 6.66E-05 | 0.020189176 | 0.323026811 |
| HTN | b_cell | *MYO7B* | 13.09189894 | 1.46E-05 | 0.01011915 | 0.070834052 |
| HTN | b_cell | *NME6* | 7.106081427 | 5.19E-05 | 0.019378899 | 0.251925681 |
| HTN | b_cell | *SNX31* | -6.66759604 | 1.81E-05 | 0.0109777 | 0.087821598 |
| HTN | b_cell | *RPP25* | 9.03206063 | 1.31E-06 | 0.00126676 | 0.006333802 |
| HTN | b_cell | *ADO* | 15.23793544 | 7.13E-08 | 0.000172894 | 0.000345789 |
| HTN | b_cell | *HIST1H3E* | 10.49937593 | 1.79E-08 | 8.71E-05 | 8.71E-05 |
| HTN | b_cell | *PRRT1* | 6.152694183 | 2.93E-06 | 0.0023672 | 0.014203201 |
| HTN | b_cell | *AC016773.1* | 11.18520503 | 2.85E-05 | 0.01256826 | 0.138250858 |
| HTN | b_cell | *HLA-DQA2* | 4.187646013 | 1.31E-06 | 0.00126676 | 0.00633298 |
| HTN | b_cell | *PLA2G4B* | 25.41856229 | 6.13E-05 | 0.02010477 | 0.297365991 |
| HTN | dendritic_cell | *RBM6* | -2.641781817 | 8.57E-05 | 0.013407341 | 0.415627573 |
| HTN | dendritic_cell | *CD79B* | -7.186291605 | 0.000593771 | 0.049671999 | 1 |
| HTN | dendritic_cell | *PIGV* | -2.842391173 | 0.000491642 | 0.045717685 | 1 |
| HTN | dendritic_cell | *PABPC4* | -6.335237499 | 0.000232605 | 0.025650017 | 1 |
| HTN | dendritic_cell | *MARCH2* | 5.387572659 | 5.30E-05 | 0.009522104 | 0.257096812 |
| HTN | dendritic_cell | *POLI* | -1.964707013 | 0.000228386 | 0.025650017 | 1 |
| HTN | dendritic_cell | *MTHFSD* | 2.895955867 | 0.000507147 | 0.045717685 | 1 |
| HTN | dendritic_cell | *TBX2* | -4.0874811 | 1.66E-06 | 0.000892574 | 0.008033169 |
| HTN | dendritic_cell | *TEX14* | 3.994125909 | 0.000590226 | 0.049671999 | 1 |
| HTN | dendritic_cell | *ZNF391* | -1.829437892 | 0.000141118 | 0.020138358 | 0.684704156 |
| HTN | dendritic_cell | *ABHD8* | -5.096790229 | 0.000313261 | 0.033776483 | 1 |
| HTN | dendritic_cell | *HAUS8* | -9.712885669 | 1.07E-07 | 0.000103822 | 0.000519112 |
| HTN | dendritic_cell | *ACP2* | 7.405445509 | 0.00017915 | 0.023492918 | 0.869237961 |
| HTN | dendritic_cell | *NMT1* | 3.857354979 | 0.000133526 | 0.019632433 | 0.647870304 |
| HTN | dendritic_cell | *FBN2* | 1.552331579 | 0.000470202 | 0.045717685 | 1 |
| HTN | dendritic_cell | *HSD17B12* | -2.350030974 | 2.16E-06 | 0.001050161 | 0.010501614 |
| HTN | dendritic_cell | *SFXN2* | 1.998066069 | 0.000531164 | 0.046021576 | 1 |
| HTN | dendritic_cell | *CLEC18A* | -5.802330588 | 5.46E-06 | 0.002085489 | 0.026474879 |
| HTN | dendritic_cell | *SLC5A11* | -2.250991371 | 2.69E-05 | 0.005924676 | 0.130342879 |
| HTN | dendritic_cell | *S100B* | -1.650138064 | 4.94E-05 | 0.00921891 | 0.239691659 |
| HTN | dendritic_cell | *ABCF3* | 4.663106487 | 7.83E-05 | 0.012667982 | 0.380039455 |
| HTN | dendritic_cell | *MRPL55* | 4.291587874 | 0.000225817 | 0.025650017 | 1 |
| HTN | dendritic_cell | *RNF123* | 3.996039241 | 4.64E-05 | 0.009009368 | 0.225234194 |
| HTN | dendritic_cell | *TRIM66* | -1.759413319 | 0.000221341 | 0.025650017 | 1 |
| HTN | dendritic_cell | *SRR* | 2.452116448 | 0.000426681 | 0.044047974 | 1 |
| HTN | dendritic_cell | *CPT1C* | 6.893341436 | 4.69E-06 | 0.002067241 | 0.022739655 |
| HTN | dendritic_cell | *INO80E* | -3.584790331 | 1.29E-05 | 0.00392567 | 0.062810721 |
| HTN | dendritic_cell | *TTLL6* | -8.355530093 | 4.60E-05 | 0.009009368 | 0.223354233 |
| HTN | dendritic_cell | *RGS19* | -3.140192723 | 1.25E-06 | 0.000761148 | 0.006089182 |
| HTN | dendritic_cell | *NME6* | 10.87431052 | 6.26E-09 | 1.65E-05 | 3.04E-05 |
| HTN | dendritic_cell | *CTSW* | -1.007511878 | 2.37E-05 | 0.005480702 | 0.115094738 |
| HTN | dendritic_cell | *MST1* | -1.708107895 | 0.000175254 | 0.023492918 | 0.850334219 |
| HTN | dendritic_cell | *SNX31* | -2.84632188 | 1.21E-05 | 0.003906434 | 0.058596505 |
| HTN | dendritic_cell | *CCNE2* | -8.371876605 | 3.79E-08 | 4.60E-05 | 0.000184065 |
| HTN | dendritic_cell | *ORAI3* | 5.434476807 | 7.30E-05 | 0.012212214 | 0.354154192 |
| HTN | dendritic_cell | *RPP25* | 3.917002438 | 1.57E-05 | 0.004494129 | 0.076400193 |
| HTN | dendritic_cell | *MPI* | -5.67962535 | 2.88E-07 | 0.000232916 | 0.001397499 |
| HTN | dendritic_cell | *TRIM73* | -10.38718305 | 0.000508812 | 0.045717685 | 1 |
| HTN | dendritic_cell | *HYI* | -2.616136398 | 0.000189529 | 0.024199808 | 0.919592722 |

| | | | | | | |
|---|---|---|---|---|---|---|
| HTN | dendritic_cell | *CLDN7* | -6.812003847 | 5.59E-06 | 0.002085489 | 0.027111361 |
| HTN | dendritic_cell | *ADO* | 8.542398154 | 9.95E-09 | 1.65E-05 | 4.83E-05 |
| HTN | dendritic_cell | *CLN8* | 3.172298943 | 0.000228274 | 0.025650017 | 1 |
| HTN | dendritic_cell | *FES* | -4.524889323 | 4.00E-05 | 0.008445221 | 0.194240074 |
| HTN | dendritic_cell | *ZNF320* | 2.127090852 | 0.000148497 | 0.020585866 | 0.720505299 |
| HTN | dendritic_cell | *A3GALT2* | -3.211684721 | 0.00012925 | 0.019597506 | 0.627120184 |
| HTN | dendritic_cell | *NUTM2A* | -4.434206581 | 0.000372996 | 0.039342935 | 1 |
| HTN | dendritic_cell | *CDK10* | -2.968796864 | 0.000222562 | 0.025650017 | 1 |
| HTN | dendritic_cell | *HIST1H3E* | 2.747328891 | 2.34E-05 | 0.005480702 | 0.113425673 |
| HTN | dendritic_cell | *ZNF100* | -4.764248417 | 0.000519227 | 0.0458053 | 1 |
| HTN | dendritic_cell | *SULT1A2* | 6.110709619 | 0.000493165 | 0.045717685 | 1 |
| HTN | dendritic_cell | *RPS26* | -3.555597609 | 1.95E-05 | 0.004991489 | 0.094838285 |
| HTN | dendritic_cell | *TMEM116* | -3.127264941 | 1.02E-08 | 1.65E-05 | 4.96E-05 |
| HTN | dendritic_cell | *PRRT1* | 4.06035075 | 1.07E-06 | 0.000743822 | 0.005206756 |
| HTN | dendritic_cell | *HLA-C* | -2.82542543 | 1.07E-05 | 0.003707602 | 0.051906428 |
| HTN | dendritic_cell | *TMEM110* | 6.791361989 | 0.0005041 | 0.045717685 | 1 |
| HTN | dendritic_cell | *ATF6B* | 6.548311049 | 6.21E-05 | 0.01075541 | 0.301151468 |
| HTN | dendritic_cell | *TNFSF12* | 7.130668126 | 1.78E-05 | 0.004804892 | 0.086488058 |
| HTN | dendritic_cell | *WFDC6* | -12.3200221 | 0.000456 | 0.045717685 | 1 |
| HTN | endometrium | *NFYA* | 3.188707991 | 0.001365144 | 0.00325328 | 1 |
| HTN | endometrium | *ENPP4* | 5.120135508 | 0.000905368 | 0.003087532 | 1 |
| HTN | endometrium | *SEMA3F* | 2.649502106 | 0.022118566 | 0.030307619 | 1 |
| HTN | endometrium | *RAD52* | 4.896498805 | 0.007432455 | 0.011735091 | 1 |
| HTN | endometrium | *HS3ST1* | 4.959954431 | 0.020947165 | 0.028873763 | 1 |
| HTN | endometrium | *AOC1* | 5.184478767 | 0.019614083 | 0.027260822 | 1 |
| HTN | endometrium | *SNX11* | 2.213665719 | 0.002987075 | 0.005593704 | 1 |
| HTN | endometrium | *TMEM176A* | 6.156606748 | 0.000501707 | 0.003073153 | 1 |
| HTN | endometrium | *CYP26B1* | 4.785040295 | 0.000214531 | 0.003073153 | 1 |
| HTN | endometrium | *DBNDD1* | 5.420518748 | 6.61E-05 | 0.002410623 | 0.320612885 |
| HTN | endometrium | *ALS2* | 6.95781338 | 0.004701818 | 0.008029997 | 1 |
| HTN | endometrium | *CD38* | 5.642498494 | 0.000693484 | 0.003073153 | 1 |
| HTN | endometrium | *RECQL* | 7.246247014 | 0.005158765 | 0.008652032 | 1 |
| HTN | endometrium | *CDC27* | 3.551682092 | 0.000286592 | 0.003073153 | 1 |
| HTN | endometrium | *POLR2J* | 3.318048842 | 0.000699736 | 0.003073153 | 1 |
| HTN | endometrium | *MEOX1* | -1.051944495 | 0.004726062 | 0.008060003 | 1 |
| HTN | endometrium | *LIG3* | -2.294478368 | 0.020202148 | 0.027965998 | 1 |
| HTN | endometrium | *CIAPIN1* | 3.094008681 | 0.000342397 | 0.003073153 | 1 |
| HTN | endometrium | *PON1* | 10.57108246 | 0.001023454 | 0.003095606 | 1 |
| HTN | endometrium | *GCFC2* | 4.105039035 | 0.000580388 | 0.003073153 | 1 |
| HTN | endometrium | *ZNF195* | 6.498845677 | 0.002029583 | 0.00418332 | 1 |
| HTN | endometrium | *PDK2* | 4.095372747 | 0.014039981 | 0.02034707 | 1 |
| HTN | endometrium | *GGNBP2* | 5.411151869 | 0.011190238 | 0.016685629 | 1 |
| HTN | endometrium | *ITGA2B* | 3.759036001 | 0.001433937 | 0.003335313 | 1 |
| HTN | endometrium | *ASB4* | -1.950692654 | 0.032266419 | 0.042751683 | 1 |
| HTN | endometrium | *TMEM98* | 2.955338209 | 0.000704519 | 0.003073153 | 1 |
| HTN | endometrium | *ABCC8* | 4.302971054 | 0.016552193 | 0.023539873 | 1 |
| HTN | endometrium | *SYNRG* | 3.323539437 | 0.000784047 | 0.003073153 | 1 |
| HTN | endometrium | *ZNF263* | 3.514944095 | 0.000995413 | 0.003095606 | 1 |
| HTN | endometrium | *SPATA20* | 3.013873524 | 0.001476079 | 0.003392182 | 1 |
| HTN | endometrium | *MAP3K9* | 3.083383323 | 0.012508436 | 0.018374487 | 1 |

| | | | | | | |
|---|---|---|---|---|---|---|
| HTN | endometrium | *BAIAP2L1* | 3.523264226 | 0.000851557 | 0.0030808 | 1 |
| HTN | endometrium | *FARP2* | 3.103935312 | 0.000534765 | 0.003073153 | 1 |
| HTN | endometrium | *USH1C* | -8.191178376 | 0.003785858 | 0.006738439 | 1 |
| HTN | endometrium | *IFRD1* | 3.223015442 | 0.001058926 | 0.003095606 | 1 |
| HTN | endometrium | *VPS41* | 5.482057941 | 0.000766714 | 0.003073153 | 1 |
| HTN | endometrium | *ELAC2* | 3.826130345 | 0.02828251 | 0.037949873 | 1 |
| HTN | endometrium | *CCDC124* | 3.359195455 | 0.001093188 | 0.003095606 | 1 |
| HTN | endometrium | *CEACAM21* | 3.90910767 | 0.001486648 | 0.003397652 | 1 |
| HTN | endometrium | *CD79B* | -2.021248657 | 0.000800189 | 0.003073153 | 1 |
| HTN | endometrium | *ST7L* | 1.907818204 | 0.028749754 | 0.0385342 | 1 |
| HTN | endometrium | *PAX6* | 4.203413033 | 0.002620714 | 0.005074104 | 1 |
| HTN | endometrium | *TSR3* | 3.853843483 | 0.000853162 | 0.0030808 | 1 |
| HTN | endometrium | *PIGQ* | 10.23882149 | 8.55E-05 | 0.002714492 | 0.41505303 |
| HTN | endometrium | *FMO3* | 2.493455208 | 0.009957996 | 0.015127175 | 1 |
| HTN | endometrium | *PSMB1* | 6.196862046 | 0.000458267 | 0.003073153 | 1 |
| HTN | endometrium | *NADK* | 2.277167591 | 0.002233423 | 0.004507724 | 1 |
| HTN | endometrium | *DLEC1* | -2.640247507 | 0.006646813 | 0.010661268 | 1 |
| HTN | endometrium | *CYB561* | 4.395350019 | 0.003343636 | 0.00611048 | 1 |
| HTN | endometrium | *SS18L2* | 4.79430765 | 0.000797946 | 0.003073153 | 1 |
| HTN | endometrium | *MGST1* | 2.281088877 | 0.000164333 | 0.003073153 | 0.797345552 |
| HTN | endometrium | *MMP25* | -3.706544393 | 0.003754879 | 0.006689764 | 1 |
| HTN | endometrium | *IL32* | 3.11521805 | 0.001219305 | 0.003095606 | 1 |
| HTN | endometrium | *PKD1* | 4.404997017 | 0.000626662 | 0.003073153 | 1 |
| HTN | endometrium | *MAPK8IP2* | 10.75974376 | 0.022533619 | 0.03082411 | 1 |
| HTN | endometrium | *MED24* | 2.383990797 | 0.001065723 | 0.003095606 | 1 |
| HTN | endometrium | *RPS20* | 6.221675424 | 0.001639437 | 0.003610779 | 1 |
| HTN | endometrium | *TRAF3IP3* | 3.244910092 | 0.000184661 | 0.003073153 | 0.895975146 |
| HTN | endometrium | *POMT2* | 2.963048409 | 0.000403988 | 0.003073153 | 1 |
| HTN | endometrium | *MLXIPL* | 1.177684038 | 0.025971682 | 0.035062493 | 1 |
| HTN | endometrium | *DYRK4* | 3.432288792 | 0.002510701 | 0.00490613 | 1 |
| HTN | endometrium | *FUZ* | 4.041635826 | 0.001159793 | 0.003095606 | 1 |
| HTN | endometrium | *SLC6A13* | 4.227863461 | 0.020342978 | 0.028128849 | 1 |
| HTN | endometrium | *ZNF200* | 2.301855022 | 0.027961115 | 0.037570571 | 1 |
| HTN | endometrium | *LRRC23* | 4.236285268 | 0.021261346 | 0.029252874 | 1 |
| HTN | endometrium | *CLCN6* | 2.418230345 | 0.006479897 | 0.010448807 | 1 |
| HTN | endometrium | *SLC6A7* | 91.85142378 | 0.002139183 | 0.004340995 | 1 |
| HTN | endometrium | *TSPAN9* | 3.366017607 | 0.001491308 | 0.0034037 | 1 |
| HTN | endometrium | *UTP18* | 3.618345486 | 0.000863098 | 0.003084413 | 1 |
| HTN | endometrium | *TTC19* | 4.20174472 | 0.001758894 | 0.003774504 | 1 |
| HTN | endometrium | *PPP5C* | 5.152294268 | 0.000560689 | 0.003073153 | 1 |
| HTN | endometrium | *CEP68* | 9.103608305 | 8.21E-05 | 0.002714492 | 0.398142825 |
| HTN | endometrium | *ZBTB32* | 3.158473182 | 0.001101617 | 0.003095606 | 1 |
| HTN | endometrium | *ERCC1* | 3.631791994 | 0.001267974 | 0.003128219 | 1 |
| HTN | endometrium | *ALOX5* | 1.7707469 | 0.010002464 | 0.01518997 | 1 |
| HTN | endometrium | *MAP4K5* | 5.631887423 | 0.001209857 | 0.003095606 | 1 |
| HTN | endometrium | *EHD3* | -1.926922808 | 0.0053787 | 0.00895896 | 1 |
| HTN | endometrium | *MAN2B2* | 4.24194087 | 0.000439206 | 0.003073153 | 1 |
| HTN | endometrium | *SLC25A39* | 3.294035978 | 0.000342045 | 0.003073153 | 1 |
| HTN | endometrium | *RNF14* | 5.634444354 | 0.000643204 | 0.003073153 | 1 |
| HTN | endometrium | *DDX11* | 3.06409682 | 0.01199665 | 0.017703085 | 1 |

| | | | | | | |
|---|---|---|---|---|---|---|
| HTN | endometrium | *TACC3* | 3.346792341 | 0.000549595 | 0.003073153 | 1 |
| HTN | endometrium | *ZC3H3* | 3.876773189 | 0.003159427 | 0.005835379 | 1 |
| HTN | endometrium | *MDH1* | 2.720677695 | 0.001353812 | 0.003239618 | 1 |
| HTN | endometrium | *CCDC88C* | 1.90368086 | 0.00906687 | 0.013913124 | 1 |
| HTN | endometrium | *NUDCD3* | 6.821637093 | 4.75E-05 | 0.002211831 | 0.230700675 |
| HTN | endometrium | *CHDH* | 7.313849734 | 0.000280636 | 0.003073153 | 1 |
| HTN | endometrium | *RUFY3* | 5.502701874 | 0.000687362 | 0.003073153 | 1 |
| HTN | endometrium | *WWTR1* | 5.09017971 | 0.000671088 | 0.003073153 | 1 |
| HTN | endometrium | *AGPS* | 5.76069464 | 0.000329873 | 0.003073153 | 1 |
| HTN | endometrium | *TTC27* | 3.406158275 | 0.000742635 | 0.003073153 | 1 |
| HTN | endometrium | *MARCO* | 1.433574479 | 0.004423819 | 0.007637895 | 1 |
| HTN | endometrium | *HGF* | 3.558233254 | 0.000763094 | 0.003073153 | 1 |
| HTN | endometrium | *OSBPL5* | 4.710959263 | 0.00038964 | 0.003073153 | 1 |
| HTN | endometrium | *SLC45A4* | 3.807929213 | 0.00049095 | 0.003073153 | 1 |
| HTN | endometrium | *ZDHHC6* | 2.578382842 | 0.00112004 | 0.003095606 | 1 |
| HTN | endometrium | *GRAMD1B* | 2.28742523 | 0.001392812 | 0.003285331 | 1 |
| HTN | endometrium | *RNH1* | 6.850011914 | 3.64E-05 | 0.00200615 | 0.176541207 |
| HTN | endometrium | *GLRX2* | 3.328563858 | 0.001194384 | 0.003095606 | 1 |
| HTN | endometrium | *ABCC2* | 4.387529356 | 0.00104407 | 0.003095606 | 1 |
| HTN | endometrium | *GCLM* | 2.469232531 | 0.001012833 | 0.003095606 | 1 |
| HTN | endometrium | *UBR2* | 2.597867959 | 0.000474404 | 0.003073153 | 1 |
| HTN | endometrium | *RRAGD* | 3.460280501 | 0.000850519 | 0.0030808 | 1 |
| HTN | endometrium | *HSF2* | 2.253808168 | 0.002479463 | 0.00486075 | 1 |
| HTN | endometrium | *TYMP* | 2.624400294 | 0.00014512 | 0.003073153 | 0.704123303 |
| HTN | endometrium | *TOMM34* | 3.811511024 | 0.000914409 | 0.003087532 | 1 |
| HTN | endometrium | *SEC63* | 4.186227353 | 0.000394071 | 0.003073153 | 1 |
| HTN | endometrium | *FAS* | 3.305205762 | 0.000873541 | 0.003084413 | 1 |
| HTN | endometrium | *AGPAT4* | 2.502719704 | 0.000947755 | 0.003095606 | 1 |
| HTN | endometrium | *BTN3A1* | 2.840191724 | 0.004873317 | 0.008247413 | 1 |
| HTN | endometrium | *INSRR* | -4.402988213 | 0.005811641 | 0.009532821 | 1 |
| HTN | endometrium | *B4GALT7* | 3.943926804 | 0.00243971 | 0.004802221 | 1 |
| HTN | endometrium | *SH2D2A* | 9.235317224 | 0.002095447 | 0.004271894 | 1 |
| HTN | endometrium | *VRK2* | 6.166361025 | 0.00146371 | 0.003373718 | 1 |
| HTN | endometrium | *VEZT* | 5.462743591 | 0.00081758 | 0.0030808 | 1 |
| HTN | endometrium | *BRD9* | 6.225597331 | 0.010070556 | 0.01526948 | 1 |
| HTN | endometrium | *SNX1* | 2.452772575 | 0.000390645 | 0.003073153 | 1 |
| HTN | endometrium | *ANK1* | 4.219737679 | 0.001174256 | 0.003095606 | 1 |
| HTN | endometrium | *RABEP1* | 2.930813111 | 0.000801834 | 0.003073153 | 1 |
| HTN | endometrium | *BAK1* | 3.66205006 | 0.037662877 | 0.049216342 | 1 |
| HTN | endometrium | *IKZF2* | 6.469260272 | 0.001042117 | 0.003095606 | 1 |
| HTN | endometrium | *GRN* | 1.568819815 | 0.003372165 | 0.006153346 | 1 |
| HTN | endometrium | *IFT88* | 4.058527002 | 0.000901881 | 0.003087532 | 1 |
| HTN | endometrium | *UBA6* | 4.041026363 | 0.026022975 | 0.03511618 | 1 |
| HTN | endometrium | *ATP6V0A1* | 4.069732002 | 0.000306155 | 0.003073153 | 1 |
| HTN | endometrium | *PIAS1* | 3.453821232 | 0.000428087 | 0.003073153 | 1 |
| HTN | endometrium | *MYOC* | 3.319788377 | 0.000295554 | 0.003073153 | 1 |
| HTN | endometrium | *SH3YL1* | 2.636091588 | 0.003396975 | 0.006186984 | 1 |
| HTN | endometrium | *ADSS* | 3.067666682 | 0.000786933 | 0.003073153 | 1 |
| HTN | endometrium | *TBC1D23* | 6.509490055 | 0.001435845 | 0.003336552 | 1 |
| HTN | endometrium | *MYOM2* | 5.665954699 | 0.000774419 | 0.003073153 | 1 |

| | | | | | | |
|---|---|---|---|---|---|---|
| HTN | endometrium | *FLT4* | 2.369027819 | 0.002685883 | 0.005169046 | 1 |
| HTN | endometrium | *NSUN2* | 6.538752169 | 0.000420754 | 0.003073153 | 1 |
| HTN | endometrium | *MFAP3* | 4.936776958 | 0.000679224 | 0.003073153 | 1 |
| HTN | endometrium | *AGA* | 5.836115189 | 0.000304612 | 0.003073153 | 1 |
| HTN | endometrium | *PI4K2B* | 6.238210036 | 0.001244419 | 0.003115541 | 1 |
| HTN | endometrium | *BOD1L1* | 4.773089172 | 0.000891315 | 0.003084413 | 1 |
| HTN | endometrium | *DNAH5* | 4.183172654 | 0.001036429 | 0.003095606 | 1 |
| HTN | endometrium | *PNKP* | 4.227430014 | 8.75E-05 | 0.002714492 | 0.424399038 |
| HTN | endometrium | *PHLPP2* | 1.93801857 | 0.005421687 | 0.009018179 | 1 |
| HTN | endometrium | *INPP4A* | 1.939575366 | 0.002594424 | 0.00503929 | 1 |
| HTN | endometrium | *PSMA4* | 3.810684168 | 0.000752838 | 0.003073153 | 1 |
| HTN | endometrium | *LSG1* | 1.655312872 | 0.010864399 | 0.016275533 | 1 |
| HTN | endometrium | *TDP1* | 4.857723168 | 0.000789912 | 0.003073153 | 1 |
| HTN | endometrium | *MED17* | 2.657163048 | 0.00153852 | 0.003470431 | 1 |
| HTN | endometrium | *CAPG* | 3.380462334 | 0.000361682 | 0.003073153 | 1 |
| HTN | endometrium | *AP2S1* | 4.022755301 | 0.000885626 | 0.003084413 | 1 |
| HTN | endometrium | *TRIT1* | 2.629714241 | 0.001233142 | 0.003103164 | 1 |
| HTN | endometrium | *CUL7* | 5.274094405 | 0.002931731 | 0.005509202 | 1 |
| HTN | endometrium | *CTNNA1* | 4.376082151 | 0.000607963 | 0.003073153 | 1 |
| HTN | endometrium | *CNTLN* | 3.289026907 | 0.000532665 | 0.003073153 | 1 |
| HTN | endometrium | *YTHDC2* | 3.585959384 | 0.000592985 | 0.003073153 | 1 |
| HTN | endometrium | *POLR2B* | 3.005191042 | 0.000643707 | 0.003073153 | 1 |
| HTN | endometrium | *ARAP2* | 2.834648705 | 0.000370529 | 0.003073153 | 1 |
| HTN | endometrium | *ANO2* | 1.852075195 | 0.001230911 | 0.003103164 | 1 |
| HTN | endometrium | *SNX29* | 1.85557552 | 0.000958932 | 0.003095606 | 1 |
| HTN | endometrium | *UTS2* | 6.10553042 | 0.000676362 | 0.003073153 | 1 |
| HTN | endometrium | *RCN1* | 2.046626091 | 0.00237378 | 0.004702313 | 1 |
| HTN | endometrium | *JKAMP* | 4.938133536 | 0.03055471 | 0.040683714 | 1 |
| HTN | endometrium | *MCUR1* | 4.384159004 | 0.000603061 | 0.003073153 | 1 |
| HTN | endometrium | *LAMC3* | -0.640049611 | 0.003123142 | 0.00578819 | 1 |
| HTN | endometrium | *MAPK9* | 7.377568791 | 0.012488798 | 0.018351195 | 1 |
| HTN | endometrium | *FAM160A2* | 4.180534979 | 0.001100204 | 0.003095606 | 1 |
| HTN | endometrium | *CYBA* | 3.233107353 | 0.000638328 | 0.003073153 | 1 |
| HTN | endometrium | *THOC3* | 21.05100557 | 0.003573752 | 0.006419531 | 1 |
| HTN | endometrium | *ALX4* | 4.203465941 | 0.021079659 | 0.029039894 | 1 |
| HTN | endometrium | *FSTL4* | 4.452816547 | 0.00084608 | 0.0030808 | 1 |
| HTN | endometrium | *AKR7A2* | 7.493026012 | 0.000876956 | 0.003084413 | 1 |
| HTN | endometrium | *MRTO4* | 3.388250456 | 0.000791575 | 0.003073153 | 1 |
| HTN | endometrium | *ANAPC4* | 3.698264541 | 8.84E-05 | 0.002714492 | 0.428889814 |
| HTN | endometrium | *LY75* | 5.515542224 | 0.001403238 | 0.003300297 | 1 |
| HTN | endometrium | *OPN3* | 5.29953454 | 0.008645396 | 0.013346313 | 1 |
| HTN | endometrium | *SDCCAG8* | 6.010748849 | 1.89E-07 | 5.08E-05 | 0.000915111 |
| HTN | endometrium | *KIF1B* | 6.913015495 | 7.87E-05 | 0.002708518 | 0.381901054 |
| HTN | endometrium | *TBC1D22A* | 3.643126563 | 0.000424193 | 0.003073153 | 1 |
| HTN | endometrium | *FAM114A2* | 5.270063127 | 0.000394239 | 0.003073153 | 1 |
| HTN | endometrium | *GINM1* | 4.235404693 | 0.000581216 | 0.003073153 | 1 |
| HTN | endometrium | *ITIH4* | 3.519388279 | 6.58E-05 | 0.002410623 | 0.319350875 |
| HTN | endometrium | *NPFFR2* | 4.964876566 | 0.001386581 | 0.003278602 | 1 |
| HTN | endometrium | *PCGF2* | 3.142094399 | 0.000615837 | 0.003073153 | 1 |
| HTN | endometrium | *IL17RB* | 5.505578861 | 0.024857939 | 0.033682579 | 1 |

| | | | | | | |
|---|---|---|---|---|---|---|
| HTN | endometrium | *MSH4* | 1.910354128 | 0.008140903 | 0.012664206 | 1 |
| HTN | endometrium | *GDI2* | 3.705291894 | 0.000525094 | 0.003073153 | 1 |
| HTN | endometrium | *PITHD1* | 5.240816257 | 0.000382206 | 0.003073153 | 1 |
| HTN | endometrium | *ATP11B* | 3.593412104 | 0.000484098 | 0.003073153 | 1 |
| HTN | endometrium | *RASGRF1* | 5.442201775 | 0.010662188 | 0.016026312 | 1 |
| HTN | endometrium | *CROCC* | 3.24407403 | 0.033487474 | 0.044176515 | 1 |
| HTN | endometrium | *DNAJC25* | 3.485021922 | 0.001217443 | 0.003095606 | 1 |
| HTN | endometrium | *GNA15* | 2.461230537 | 0.020469563 | 0.028287759 | 1 |
| HTN | endometrium | *PIGV* | -5.055889262 | 4.79E-06 | 0.000553371 | 0.023241568 |
| HTN | endometrium | *RIMBP2* | 3.302383131 | 0.001213672 | 0.003095606 | 1 |
| HTN | endometrium | *PRDM6* | -2.308681643 | 0.016600222 | 0.023599261 | 1 |
| HTN | endometrium | *NCKAP1* | 3.418174416 | 0.001147319 | 0.003095606 | 1 |
| HTN | endometrium | *LTK* | 25.15737287 | 0.001158768 | 0.003095606 | 1 |
| HTN | endometrium | *EIF4B* | 7.738374545 | 8.31E-05 | 0.002714492 | 0.40339553 |
| HTN | endometrium | *SLC6A16* | 2.49171074 | 0.001420038 | 0.003317298 | 1 |
| HTN | endometrium | *AHRR* | 9.063945753 | 0.001527929 | 0.003456182 | 1 |
| HTN | endometrium | *ADCK1* | 2.613022594 | 0.00116053 | 0.003095606 | 1 |
| HTN | endometrium | *HAGH* | 5.633579909 | 2.39E-05 | 0.001488735 | 0.116121298 |
| HTN | endometrium | *RNF4* | 4.629861216 | 0.000574695 | 0.003073153 | 1 |
| HTN | endometrium | *CASP8* | 7.379402594 | 0.005841668 | 0.009569134 | 1 |
| HTN | endometrium | *TM7SF3* | 2.608395278 | 0.000810091 | 0.003073153 | 1 |
| HTN | endometrium | *CDON* | 1.416423202 | 0.000412415 | 0.003073153 | 1 |
| HTN | endometrium | *TMEM161A* | 3.384233835 | 0.000935507 | 0.003095606 | 1 |
| HTN | endometrium | *LPAR2* | 5.204143535 | 0.000622591 | 0.003073153 | 1 |
| HTN | endometrium | *SUGP2* | 2.964257745 | 0.000724712 | 0.003073153 | 1 |
| HTN | endometrium | *CNN2* | 5.353712779 | 0.008369172 | 0.01296527 | 1 |
| HTN | endometrium | *ABCA7* | 2.759884192 | 0.00790351 | 0.012358309 | 1 |
| HTN | endometrium | *DDX20* | 3.732696894 | 0.000160597 | 0.003073153 | 0.779214872 |
| HTN | endometrium | *BTBD1* | 4.654984903 | 0.000377168 | 0.003073153 | 1 |
| HTN | endometrium | *FAR2* | 6.614499079 | 0.000738599 | 0.003073153 | 1 |
| HTN | endometrium | *BCAS1* | 4.347188672 | 0.002917217 | 0.005498965 | 1 |
| HTN | endometrium | *SBNO2* | 4.417765897 | 0.00057874 | 0.003073153 | 1 |
| HTN | endometrium | *PMS1* | 5.233537599 | 0.000304246 | 0.003073153 | 1 |
| HTN | endometrium | *UHRF1BP1* | 4.736578414 | 5.22E-07 | 0.000105527 | 0.00253265 |
| HTN | endometrium | *MCM10* | 2.112758713 | 0.036746552 | 0.048148601 | 1 |
| HTN | endometrium | *ANKRD44* | 1.762902662 | 0.00067922 | 0.003073153 | 1 |
| HTN | endometrium | *MAP2K4* | 2.992238041 | 0.000805092 | 0.003073153 | 1 |
| HTN | endometrium | *TMEM206* | 2.624795291 | 0.000494333 | 0.003073153 | 1 |
| HTN | endometrium | *NGEF* | 7.231383404 | 0.011898162 | 0.017568437 | 1 |
| HTN | endometrium | *SPI1* | 4.093327015 | 0.012388141 | 0.018219842 | 1 |
| HTN | endometrium | *ZNRD1* | -2.263337241 | 0.004940374 | 0.008340535 | 1 |
| HTN | endometrium | *ATXN3* | 2.470020889 | 0.012981685 | 0.019003116 | 1 |
| HTN | endometrium | *FGFR2* | 1.70168813 | 0.037177296 | 0.048634198 | 1 |
| HTN | endometrium | *MTFR1* | 4.678587984 | 0.00601525 | 0.009813717 | 1 |
| HTN | endometrium | *MYO9A* | 3.769574729 | 0.001006425 | 0.003095606 | 1 |
| HTN | endometrium | *TRAM1* | 3.04098335 | 0.001053471 | 0.003095606 | 1 |
| HTN | endometrium | *EVI5* | 3.589201743 | 0.000724001 | 0.003073153 | 1 |
| HTN | endometrium | *METTL22* | 7.022231147 | 0.000965971 | 0.003095606 | 1 |
| HTN | endometrium | *TP53BP1* | 3.352294583 | 0.001000205 | 0.003095606 | 1 |
| HTN | endometrium | *RRP15* | 5.031183895 | 4.01E-05 | 0.002058697 | 0.194468598 |

| | | | | | | |
|---|---|---|---|---|---|---|
| HTN | endometrium | *DHX8* | 8.203279876 | 0.001365123 | 0.00325328 | 1 |
| HTN | endometrium | *CBFB* | 4.944264176 | 0.000896334 | 0.003087532 | 1 |
| HTN | endometrium | *FGFR3* | -1.327844097 | 0.014315676 | 0.020678672 | 1 |
| HTN | endometrium | *IFI35* | 6.724127242 | 0.000399992 | 0.003073153 | 1 |
| HTN | endometrium | *HEATR6* | 9.536225991 | 7.35E-05 | 0.002585212 | 0.356759207 |
| HTN | endometrium | *POLR1A* | 1.57926718 | 0.029705607 | 0.039651061 | 1 |
| HTN | endometrium | *IP6K2* | 3.64595659 | 0.001630286 | 0.003600432 | 1 |
| HTN | endometrium | *SRBD1* | 3.955826282 | 0.000374173 | 0.003073153 | 1 |
| HTN | endometrium | *CYFIP1* | 3.673862117 | 0.00080462 | 0.003073153 | 1 |
| HTN | endometrium | *IFT80* | 4.4413472 | 0.00355695 | 0.00640383 | 1 |
| HTN | endometrium | *NUCKS1* | 3.249198319 | 0.000765747 | 0.003073153 | 1 |
| HTN | endometrium | *CLEC2D* | 3.48927658 | 0.000616186 | 0.003073153 | 1 |
| HTN | endometrium | *DRD4* | -2.567008607 | 0.008230813 | 0.012799969 | 1 |
| HTN | endometrium | *PLA2G10* | 7.160452681 | 0.001318223 | 0.003202813 | 1 |
| HTN | endometrium | *NEDD4* | 7.401762699 | 0.000240837 | 0.003073153 | 1 |
| HTN | endometrium | *PIGB* | 3.689309391 | 0.001007326 | 0.003095606 | 1 |
| HTN | endometrium | *LRP6* | 3.32876974 | 0.000772037 | 0.003073153 | 1 |
| HTN | endometrium | *CLTCL1* | 2.086891062 | 0.001491399 | 0.0034037 | 1 |
| HTN | endometrium | *DGCR2* | 5.76127102 | 0.004420064 | 0.007637517 | 1 |
| HTN | endometrium | *ZXDC* | 4.401253244 | 0.000389041 | 0.003073153 | 1 |
| HTN | endometrium | *GBA2* | 8.070256056 | 0.012983374 | 0.019003116 | 1 |
| HTN | endometrium | *NDST1* | 8.792289734 | 0.000479142 | 0.003073153 | 1 |
| HTN | endometrium | *ASNS* | -2.037675935 | 0.026433288 | 0.035626199 | 1 |
| HTN | endometrium | *AP3M2* | 6.451707907 | 0.035501486 | 0.046643166 | 1 |
| HTN | endometrium | *ST6GALNAC2* | 3.263342835 | 0.001204981 | 0.003095606 | 1 |
| HTN | endometrium | *EIF2B3* | 4.992646006 | 0.000660578 | 0.003073153 | 1 |
| HTN | endometrium | *MAP4K4* | 5.289203172 | 0.002929949 | 0.005509202 | 1 |
| HTN | endometrium | *WDR1* | 3.07144526 | 0.000829918 | 0.0030808 | 1 |
| HTN | endometrium | *SNX13* | 3.559919867 | 0.000589486 | 0.003073153 | 1 |
| HTN | endometrium | *RPS6KA2* | 3.109030019 | 0.000340654 | 0.003073153 | 1 |
| HTN | endometrium | *ING3* | 2.885061062 | 0.000988418 | 0.003095606 | 1 |
| HTN | endometrium | *VASH1* | 3.947930454 | 0.000372195 | 0.003073153 | 1 |
| HTN | endometrium | *TRIB2* | 3.027474429 | 0.001166199 | 0.003095606 | 1 |
| HTN | endometrium | *CPSF1* | 2.15114743 | 0.017280985 | 0.024438163 | 1 |
| HTN | endometrium | *CYBRD1* | 4.837132198 | 0.000802995 | 0.003073153 | 1 |
| HTN | endometrium | *RDH11* | 3.003875716 | 0.00060433 | 0.003073153 | 1 |
| HTN | endometrium | *EPN2* | 2.834712749 | 0.006930699 | 0.011054488 | 1 |
| HTN | endometrium | *ALDH3A2* | 4.075624082 | 0.000248186 | 0.003073153 | 1 |
| HTN | endometrium | *AFF4* | 3.779692821 | 0.000590378 | 0.003073153 | 1 |
| HTN | endometrium | *TRHDE* | 2.133586683 | 0.003187529 | 0.005883014 | 1 |
| HTN | endometrium | *FCGR2B* | 2.578399884 | 0.001343258 | 0.003234314 | 1 |
| HTN | endometrium | *NFATC3* | 2.842227641 | 0.001075985 | 0.003095606 | 1 |
| HTN | endometrium | *TRNT1* | 1.943545623 | 0.000966114 | 0.003095606 | 1 |
| HTN | endometrium | *NDE1* | 5.506205465 | 0.000437128 | 0.003073153 | 1 |
| HTN | endometrium | *AP1M1* | 6.154507006 | 0.001508792 | 0.00342567 | 1 |
| HTN | endometrium | *SCARB1* | 2.549289152 | 0.00135982 | 0.003248571 | 1 |
| HTN | endometrium | *MOV10L1* | -1.722286601 | 0.010904173 | 0.016319261 | 1 |
| HTN | endometrium | *ALPK1* | 2.885462234 | 0.000942564 | 0.003095606 | 1 |
| HTN | endometrium | *LLGL2* | 3.269404892 | 0.000794988 | 0.003073153 | 1 |
| HTN | endometrium | *SDHA* | 3.803065009 | 0.000216683 | 0.003073153 | 1 |

| | | | | | | |
|---|---|---|---|---|---|---|
| HTN | endometrium | *SMARCE1* | 8.374079421 | 0.003414241 | 0.006206781 | 1 |
| HTN | endometrium | *ADAM11* | -3.559397088 | 0.012298395 | 0.018104312 | 1 |
| HTN | endometrium | *ZNF532* | 2.838250459 | 0.000732312 | 0.003073153 | 1 |
| HTN | endometrium | *ZZEF1* | 5.941075842 | 0.000205122 | 0.003073153 | 0.995249876 |
| HTN | endometrium | *TACR2* | 34.48764695 | 0.008580227 | 0.013266813 | 1 |
| HTN | endometrium | *TIPIN* | 5.24726626 | 0.000222729 | 0.003073153 | 1 |
| HTN | endometrium | *TTC38* | 4.370637486 | 0.000798006 | 0.003073153 | 1 |
| HTN | endometrium | *GRAMD4* | 5.163763747 | 0.000423508 | 0.003073153 | 1 |
| HTN | endometrium | *ADD2* | 4.642111545 | 0.000230285 | 0.003073153 | 1 |
| HTN | endometrium | *VPS9D1* | 2.470085878 | 0.000805907 | 0.003073153 | 1 |
| HTN | endometrium | *FNDC3B* | 3.393134222 | 0.00053564 | 0.003073153 | 1 |
| HTN | endometrium | *CACNG4* | 4.896949496 | 0.003844131 | 0.006817151 | 1 |
| HTN | endometrium | *MOCOS* | 3.57879374 | 0.001921511 | 0.00403077 | 1 |
| HTN | endometrium | *SEC31B* | 3.955241446 | 0.005574586 | 0.009218777 | 1 |
| HTN | endometrium | *KIFAP3* | 3.964865171 | 0.000568821 | 0.003073153 | 1 |
| HTN | endometrium | *MKRN2* | 6.701155905 | 0.002577706 | 0.00501083 | 1 |
| HTN | endometrium | *FMO4* | 3.893486598 | 0.000306627 | 0.003073153 | 1 |
| HTN | endometrium | *RGS11* | 2.032207008 | 0.005461703 | 0.009075405 | 1 |
| HTN | endometrium | *SPAG5* | 8.210846398 | 0.02166658 | 0.029747098 | 1 |
| HTN | endometrium | *PAG1* | 2.190149701 | 0.000961395 | 0.003095606 | 1 |
| HTN | endometrium | *GPATCH1* | 9.050820599 | 0.000771802 | 0.003073153 | 1 |
| HTN | endometrium | *ICAM3* | -1.083115205 | 0.006057201 | 0.009870223 | 1 |
| HTN | endometrium | *NT5C2* | 5.018621617 | 0.000269545 | 0.003073153 | 1 |
| HTN | endometrium | *XAB2* | 3.934060008 | 0.000915411 | 0.003087532 | 1 |
| HTN | endometrium | *IL4R* | 6.139279699 | 0.00037845 | 0.003073153 | 1 |
| HTN | endometrium | *DYNC1I2* | 5.441697551 | 0.000630926 | 0.003073153 | 1 |
| HTN | endometrium | *APBB1IP* | 2.136887848 | 0.000675588 | 0.003073153 | 1 |
| HTN | endometrium | *LRCH4* | 5.532172158 | 0.000361823 | 0.003073153 | 1 |
| HTN | endometrium | *SIRT6* | -1.873039136 | 0.017567219 | 0.024761449 | 1 |
| HTN | endometrium | *GPR137B* | 3.776163668 | 0.000270075 | 0.003073153 | 1 |
| HTN | endometrium | *AMPH* | 9.441871793 | 0.015746095 | 0.022536889 | 1 |
| HTN | endometrium | *MCCC1* | 3.531671282 | 0.034532263 | 0.04546826 | 1 |
| HTN | endometrium | *ACER3* | 2.932172703 | 0.001632982 | 0.003601469 | 1 |
| HTN | endometrium | *PIK3C3* | 4.456323698 | 0.000639641 | 0.003073153 | 1 |
| HTN | endometrium | *SYNJ2* | 2.189330438 | 0.018175455 | 0.025524547 | 1 |
| HTN | endometrium | *RBFOX1* | 2.230313533 | 0.000392401 | 0.003073153 | 1 |
| HTN | endometrium | *PCM1* | 4.820660933 | 0.002424256 | 0.004777617 | 1 |
| HTN | endometrium | *MYH7B* | -3.393235613 | 0.008358086 | 0.012952231 | 1 |
| HTN | endometrium | *CLUL1* | 4.186708919 | 0.019798445 | 0.027477705 | 1 |
| HTN | endometrium | *FKBP7* | -1.09416874 | 0.004313803 | 0.007488576 | 1 |
| HTN | endometrium | *XRCC5* | 4.235267945 | 0.000469544 | 0.003073153 | 1 |
| HTN | endometrium | *SP140* | 2.364928057 | 0.003897787 | 0.006894663 | 1 |
| HTN | endometrium | *SAR1A* | 4.999452862 | 0.000766236 | 0.003073153 | 1 |
| HTN | endometrium | *RAPGEF3* | 2.245230635 | 0.003487184 | 0.006294576 | 1 |
| HTN | endometrium | *DUSP13* | 27.84651465 | 0.003930396 | 0.006937171 | 1 |
| HTN | endometrium | *FDFT1* | 4.175647366 | 2.10E-05 | 0.001374309 | 0.101698843 |
| HTN | endometrium | *SCGN* | 21.20465262 | 0.000939194 | 0.003095606 | 1 |
| HTN | endometrium | *PGM1* | 3.869516181 | 0.000992699 | 0.003095606 | 1 |
| HTN | endometrium | *DDX1* | 2.385887381 | 0.001536162 | 0.003470431 | 1 |
| HTN | endometrium | *DNM2* | 2.860060647 | 0.00045607 | 0.003073153 | 1 |

| | | | | | | |
|---|---|---|---|---|---|---|
| HTN | endometrium | *STX7* | 3.990667661 | 0.001116475 | 0.003095606 | 1 |
| HTN | endometrium | *RABL2B* | 2.910275557 | 0.001116759 | 0.003095606 | 1 |
| HTN | endometrium | *DCT* | 3.252798567 | 0.003678371 | 0.006574791 | 1 |
| HTN | endometrium | *SCTR* | 2.342405842 | 0.001185108 | 0.003095606 | 1 |
| HTN | endometrium | *RIF1* | 6.075161462 | 0.000774578 | 0.003073153 | 1 |
| HTN | endometrium | *KCNN2* | -5.90835297 | 0.007876199 | 0.012323546 | 1 |
| HTN | endometrium | *PSEN1* | 2.880813185 | 0.000558512 | 0.003073153 | 1 |
| HTN | endometrium | *MOK* | 5.077237393 | 0.000306857 | 0.003073153 | 1 |
| HTN | endometrium | *STK17B* | 3.897183856 | 0.000231038 | 0.003073153 | 1 |
| HTN | endometrium | *ZNF506* | 6.326205091 | 6.51E-05 | 0.002410623 | 0.315960202 |
| HTN | endometrium | *PCDHB4* | 5.293587482 | 0.00119147 | 0.003095606 | 1 |
| HTN | endometrium | *ME2* | 2.511055776 | 0.000920278 | 0.003087532 | 1 |
| HTN | endometrium | *CCNT2* | 3.151917962 | 0.00173088 | 0.003742526 | 1 |
| HTN | endometrium | *SEMA5B* | -1.884582601 | 0.00666485 | 0.010686667 | 1 |
| HTN | endometrium | *DOPEY1* | 4.507714112 | 0.021935012 | 0.030081594 | 1 |
| HTN | endometrium | *LYRM2* | 2.781017481 | 0.006258918 | 0.010139657 | 1 |
| HTN | endometrium | *ZCCHC6* | 2.812841354 | 0.000714541 | 0.003073153 | 1 |
| HTN | endometrium | *ULK2* | 5.249158836 | 0.00466451 | 0.007971893 | 1 |
| HTN | endometrium | *PLOD1* | 10.31490052 | 1.79E-05 | 0.001270304 | 0.086912668 |
| HTN | endometrium | *PIBF1* | 3.114595598 | 0.000619119 | 0.003073153 | 1 |
| HTN | endometrium | *OXCT1* | 2.816480417 | 0.002031986 | 0.00418415 | 1 |
| HTN | endometrium | *SLC27A5* | 3.499068139 | 0.000681589 | 0.003073153 | 1 |
| HTN | endometrium | *ZNF586* | 5.485029008 | 0.000545424 | 0.003073153 | 1 |
| HTN | endometrium | *ZNF264* | 2.709982085 | 0.000989729 | 0.003095606 | 1 |
| HTN | endometrium | *RPS5* | 5.773758494 | 0.007634639 | 0.012007543 | 1 |
| HTN | endometrium | *CHMP2B* | 4.196217782 | 0.001415193 | 0.003314023 | 1 |
| HTN | endometrium | *PPIE* | 5.389665649 | 0.001581699 | 0.003533335 | 1 |
| HTN | endometrium | *NOA1* | -1.442813461 | 0.005681529 | 0.009354184 | 1 |
| HTN | endometrium | *WBP11* | 2.317466315 | 0.001157537 | 0.003095606 | 1 |
| HTN | endometrium | *NCOA1* | 4.412889488 | 0.000801245 | 0.003073153 | 1 |
| HTN | endometrium | *CAD* | 3.024392844 | 0.000292781 | 0.003073153 | 1 |
| HTN | endometrium | *CD59* | 2.534385674 | 0.000533948 | 0.003073153 | 1 |
| HTN | endometrium | *CD82* | 3.456605343 | 0.000873147 | 0.003084413 | 1 |
| HTN | endometrium | *HACE1* | 4.400627547 | 0.0006145 | 0.003073153 | 1 |
| HTN | endometrium | *SEH1L* | 4.949196799 | 0.013308013 | 0.019454799 | 1 |
| HTN | endometrium | *SLC25A24* | 2.451032381 | 0.000834914 | 0.0030808 | 1 |
| HTN | endometrium | *PILRA* | 1.323140072 | 0.006668875 | 0.010689588 | 1 |
| HTN | endometrium | *CPNE3* | 4.163399873 | 0.001556862 | 0.003498794 | 1 |
| HTN | endometrium | *RRN3* | 3.586299556 | 0.001292769 | 0.003159958 | 1 |
| HTN | endometrium | *MTIF2* | 8.355460294 | 0.001391772 | 0.003285301 | 1 |
| HTN | endometrium | *DDHD2* | 3.485531592 | 9.27E-05 | 0.00279224 | 0.449550625 |
| HTN | endometrium | *EPS15* | 4.000651068 | 0.000886131 | 0.003084413 | 1 |
| HTN | endometrium | *USP40* | 3.556775484 | 0.000602828 | 0.003073153 | 1 |
| HTN | endometrium | *POMGNT1* | 6.04878335 | 0.000743854 | 0.003073153 | 1 |
| HTN | endometrium | *MAST2* | 5.015100997 | 0.000224659 | 0.003073153 | 1 |
| HTN | endometrium | *NME8* | -4.357839649 | 0.003923666 | 0.006927811 | 1 |
| HTN | endometrium | *TMED2* | 7.222443472 | 0.002058574 | 0.004223341 | 1 |
| HTN | endometrium | *ALG9* | 5.097176379 | 0.000837183 | 0.0030808 | 1 |
| HTN | endometrium | *ACOX3* | 5.989826953 | 0.001596484 | 0.003554047 | 1 |
| HTN | endometrium | *HSD17B14* | 3.956754586 | 0.000554602 | 0.003073153 | 1 |

| | | | | | | |
|---|---|---|---|---|---|---|
| HTN | endometrium | *TRIP6* | 5.164774816 | 0.000296126 | 0.003073153 | 1 |
| HTN | endometrium | *ACHE* | -1.218820964 | 0.00653888 | 0.010529919 | 1 |
| HTN | endometrium | *SRRT* | 3.273446628 | 0.000737343 | 0.003073153 | 1 |
| HTN | endometrium | *PSMC5* | 3.269022538 | 0.001331875 | 0.003214407 | 1 |
| HTN | endometrium | *UIMC1* | 3.534576123 | 0.000585156 | 0.003073153 | 1 |
| HTN | endometrium | *LPCAT2* | 2.092813891 | 0.001168707 | 0.003095606 | 1 |
| HTN | endometrium | *OGFOD1* | 3.616086124 | 0.003363551 | 0.006142246 | 1 |
| HTN | endometrium | *TXNDC16* | 4.602767277 | 0.027115293 | 0.036494702 | 1 |
| HTN | endometrium | *SF3B2* | 4.700652908 | 0.006166771 | 0.010017132 | 1 |
| HTN | endometrium | *PHACTR3* | 3.070613655 | 0.030626565 | 0.04076502 | 1 |
| HTN | endometrium | *ASAP3* | 2.329964378 | 0.00108737 | 0.003095606 | 1 |
| HTN | endometrium | *EDEM2* | 4.560708541 | 0.001699866 | 0.003705189 | 1 |
| HTN | endometrium | *ANKRD10* | 3.764244827 | 0.000545648 | 0.003073153 | 1 |
| HTN | endometrium | *DOCK3* | 6.853225032 | 0.000713131 | 0.003073153 | 1 |
| HTN | endometrium | *ARHGAP28* | 2.890825913 | 0.000840114 | 0.0030808 | 1 |
| HTN | endometrium | *PPP1R13B* | 2.292249341 | 0.00470902 | 0.008039467 | 1 |
| HTN | endometrium | *NSFL1C* | 2.3739876 | 0.000469146 | 0.003073153 | 1 |
| HTN | endometrium | *ZNF343* | 7.114620762 | 0.002601652 | 0.005047268 | 1 |
| HTN | endometrium | *EBF4* | 11.13003028 | 0.000666678 | 0.003073153 | 1 |
| HTN | endometrium | *CPXM1* | -1.968303505 | 0.022658051 | 0.030959409 | 1 |
| HTN | endometrium | *MAVS* | 3.6341104 | 0.010440556 | 0.015746838 | 1 |
| HTN | endometrium | *XRN2* | 4.526420256 | 0.000641224 | 0.003073153 | 1 |
| HTN | endometrium | *SNX5* | 7.707412598 | 0.000983141 | 0.003095606 | 1 |
| HTN | endometrium | *ESF1* | 4.607780121 | 0.000673246 | 0.003073153 | 1 |
| HTN | endometrium | *RBBP9* | -3.521186626 | 0.002505786 | 0.004898499 | 1 |
| HTN | endometrium | *TMEM230* | 3.908359646 | 0.000877284 | 0.003084413 | 1 |
| HTN | endometrium | *RPH3A* | 8.875105406 | 0.000214881 | 0.003073153 | 1 |
| HTN | endometrium | *KIF16B* | 2.001499718 | 0.001032102 | 0.003095606 | 1 |
| HTN | endometrium | *TBX5* | -1.306564309 | 0.002493956 | 0.004881273 | 1 |
| HTN | endometrium | *FXYD5* | 2.557795658 | 0.001413459 | 0.003313092 | 1 |
| HTN | endometrium | *ZNF302* | 4.33302837 | 0.017560795 | 0.024761449 | 1 |
| HTN | endometrium | *GRAMD1A* | 4.33272799 | 0.000640936 | 0.003073153 | 1 |
| HTN | endometrium | *CDIP1* | -2.5157553 | 0.021289293 | 0.029278812 | 1 |
| HTN | endometrium | *GANAB* | 6.117058945 | 0.008812252 | 0.013577976 | 1 |
| HTN | endometrium | *MLF2* | 9.985021424 | 3.31E-05 | 0.00189203 | 0.16082255 |
| HTN | endometrium | *ANKRD24* | -4.117515791 | 0.035543391 | 0.04668558 | 1 |
| HTN | endometrium | *DHX32* | 4.539176827 | 0.001308848 | 0.003186417 | 1 |
| HTN | endometrium | *SPTLC1* | 3.384829145 | 0.002280484 | 0.004570187 | 1 |
| HTN | endometrium | *YPEL3* | 7.483357391 | 0.001612108 | 0.003579584 | 1 |
| HTN | endometrium | *MRPS33* | 5.408821142 | 0.000793153 | 0.003073153 | 1 |
| HTN | endometrium | *MAEA* | 2.661912218 | 0.001402358 | 0.003299826 | 1 |
| HTN | endometrium | *STRN4* | 2.702372382 | 0.000879173 | 0.003084413 | 1 |
| HTN | endometrium | *LYZ* | 1.949898641 | 0.000571106 | 0.003073153 | 1 |
| HTN | endometrium | *SPG21* | 4.858880421 | 0.001712693 | 0.003713132 | 1 |
| HTN | endometrium | *CHRD* | -1.118814182 | 0.002443156 | 0.004805104 | 1 |
| HTN | endometrium | *GNPTG* | 3.97184069 | 0.000571658 | 0.003073153 | 1 |
| HTN | endometrium | *PABPC4* | 1.808985996 | 0.02918021 | 0.039001558 | 1 |
| HTN | endometrium | *USP48* | 3.33158326 | 0.001209463 | 0.003095606 | 1 |
| HTN | endometrium | *GLG1* | 3.796438817 | 0.000539173 | 0.003073153 | 1 |
| HTN | endometrium | *DTX2* | 2.861549967 | 0.001163867 | 0.003095606 | 1 |

| | | | | | | |
|---|---|---|---|---|---|---|
| HTN | endometrium | *LAMB1* | 1.947552777 | 0.010821002 | 0.016224814 | 1 |
| HTN | endometrium | *TF* | 4.646179958 | 0.000830573 | 0.0030808 | 1 |
| HTN | endometrium | *SPAG7* | 2.058379421 | 0.002600683 | 0.005047268 | 1 |
| HTN | endometrium | *RGS17* | 5.251655593 | 1.53E-05 | 0.001180045 | 0.074342842 |
| HTN | endometrium | *PPP2R3C* | 4.896471256 | 0.000133197 | 0.003073153 | 0.646272518 |
| HTN | endometrium | *HAUS4* | 3.112014425 | 0.001399619 | 0.003296578 | 1 |
| HTN | endometrium | *OSGEP* | 3.364489042 | 0.000714197 | 0.003073153 | 1 |
| HTN | endometrium | *SCFD1* | 4.997678653 | 0.00091277 | 0.003087532 | 1 |
| HTN | endometrium | *SUPT16H* | 5.019847654 | 0.000568808 | 0.003073153 | 1 |
| HTN | endometrium | *TOX4* | 2.223988115 | 0.001372983 | 0.003265547 | 1 |
| HTN | endometrium | *CAPN3* | -3.015478987 | 0.007567956 | 0.011925859 | 1 |
| HTN | endometrium | *TBX15* | 7.163235928 | 0.00360557 | 0.006467366 | 1 |
| HTN | endometrium | *UNC13D* | 2.947359145 | 0.00058257 | 0.003073153 | 1 |
| HTN | endometrium | *MFSD11* | 3.620031216 | 0.005664434 | 0.009329204 | 1 |
| HTN | endometrium | *NUP50* | 2.872015688 | 0.000792257 | 0.003073153 | 1 |
| HTN | endometrium | *COMT* | 5.376295947 | 0.000582547 | 0.003073153 | 1 |
| HTN | endometrium | *LRRFIP2* | 3.753696074 | 0.000352086 | 0.003073153 | 1 |
| HTN | endometrium | *XYLB* | 6.203365494 | 0.000594308 | 0.003073153 | 1 |
| HTN | endometrium | *GABRP* | 7.996551403 | 0.000405459 | 0.003073153 | 1 |
| HTN | endometrium | *FMO2* | 3.480182255 | 1.74E-05 | 0.001262767 | 0.08456769 |
| HTN | endometrium | *MSH2* | 2.54444099 | 0.000998183 | 0.003095606 | 1 |
| HTN | endometrium | *CRAT* | 3.369048102 | 0.001188652 | 0.003095606 | 1 |
| HTN | endometrium | *NANS* | 5.910563786 | 0.000459451 | 0.003073153 | 1 |
| HTN | endometrium | *PDE6C* | -2.801190181 | 0.011577782 | 0.017200061 | 1 |
| HTN | endometrium | *WAC* | 4.389163999 | 0.000389631 | 0.003073153 | 1 |
| HTN | endometrium | *ITPR3* | 3.987739415 | 2.79E-05 | 0.001652545 | 0.135508698 |
| HTN | endometrium | *DSP* | 3.194429859 | 0.001111972 | 0.003095606 | 1 |
| HTN | endometrium | *IFT74* | 3.81480491 | 0.000368882 | 0.003073153 | 1 |
| HTN | endometrium | *ACOT7* | 3.602160746 | 0.002082878 | 0.004257002 | 1 |
| HTN | endometrium | *RAB18* | 4.723255594 | 0.000560572 | 0.003073153 | 1 |
| HTN | endometrium | *TSPAN15* | 5.417755371 | 0.000558686 | 0.003073153 | 1 |
| HTN | endometrium | *MAST3* | 1.867733578 | 0.01036133 | 0.015646802 | 1 |
| HTN | endometrium | *OCEL1* | 2.560942767 | 0.005611092 | 0.009272992 | 1 |
| HTN | endometrium | *CATSPERG* | 2.562999361 | 0.007690822 | 0.012084154 | 1 |
| HTN | endometrium | *SETD1A* | 3.341565211 | 0.00013857 | 0.003073153 | 0.67234172 |
| HTN | endometrium | *CIRBP* | 4.381559973 | 0.032732596 | 0.043343859 | 1 |
| HTN | endometrium | *MARCH2* | 2.826758786 | 1.98E-05 | 0.001324109 | 0.09617546 |
| HTN | endometrium | *MTAP* | -3.458525077 | 0.006368969 | 0.010296196 | 1 |
| HTN | endometrium | *POLR2E* | 5.707235037 | 0.001001462 | 0.003095606 | 1 |
| HTN | endometrium | *SNAP29* | 4.647040012 | 0.00058589 | 0.003073153 | 1 |
| HTN | endometrium | *LZTR1* | 4.594744162 | 0.000366089 | 0.003073153 | 1 |
| HTN | endometrium | *MMP11* | -0.613745702 | 0.00347476 | 0.006279157 | 1 |
| HTN | endometrium | *CECR2* | 4.744080593 | 0.002375787 | 0.004703108 | 1 |
| HTN | endometrium | *SMARCB1* | 2.361409651 | 0.00094564 | 0.003095606 | 1 |
| HTN | endometrium | *BCL2L13* | 5.88850361 | 0.000694698 | 0.003073153 | 1 |
| HTN | endometrium | *DDT* | -2.799387007 | 0.024981876 | 0.033829769 | 1 |
| HTN | endometrium | *OSM* | -1.994384603 | 0.003482291 | 0.006290424 | 1 |
| HTN | endometrium | *CABIN1* | 5.396053739 | 0.000606157 | 0.003073153 | 1 |
| HTN | endometrium | *SPECC1L* | -2.299134094 | 0.003320555 | 0.006077455 | 1 |
| HTN | endometrium | *PES1* | 2.345367537 | 0.004317986 | 0.007493157 | 1 |

| | | | | | | |
|---|---|---|---|---|---|---|
| HTN | endometrium | *SLC35E4* | -2.822296229 | 0.004767711 | 0.008108532 | 1 |
| HTN | endometrium | *CRYBB3* | -4.613748712 | 0.036838874 | 0.048243513 | 1 |
| HTN | endometrium | *CYTH4* | 2.504281394 | 0.001073822 | 0.003095606 | 1 |
| HTN | endometrium | *CARD10* | 3.072033636 | 0.002419073 | 0.004769338 | 1 |
| HTN | endometrium | *LRP5L* | 2.610610376 | 0.001808059 | 0.003856134 | 1 |
| HTN | endometrium | *SH3BP1* | 4.464860834 | 0.001006643 | 0.003095606 | 1 |
| HTN | endometrium | *HPS4* | 5.552467141 | 0.000476243 | 0.003073153 | 1 |
| HTN | endometrium | *TRIOBP* | 3.502219066 | 0.003730943 | 0.006657792 | 1 |
| HTN | endometrium | *EIF3L* | 2.342336407 | 0.002537103 | 0.004949747 | 1 |
| HTN | endometrium | *PICK1* | 2.994161242 | 0.000468247 | 0.003073153 | 1 |
| HTN | endometrium | *TTC28* | 2.744008912 | 0.000628278 | 0.003073153 | 1 |
| HTN | endometrium | *SLC5A4* | 0.938479197 | 0.028628601 | 0.038382419 | 1 |
| HTN | endometrium | *KDELR3* | -2.212127836 | 0.009014511 | 0.013841268 | 1 |
| HTN | endometrium | *CYP2D6* | -9.07669305 | 0.020292465 | 0.028067002 | 1 |
| HTN | endometrium | *CBY1* | 2.787309777 | 0.002060974 | 0.004226477 | 1 |
| HTN | endometrium | *RTCB* | 5.267041255 | 0.000977894 | 0.003095606 | 1 |
| HTN | endometrium | *RAB36* | 3.92290859 | 0.029017945 | 0.038818601 | 1 |
| HTN | endometrium | *TIMP3* | 3.486867765 | 0.000575478 | 0.003073153 | 1 |
| HTN | endometrium | *PPP6R2* | 3.607726767 | 0.000697809 | 0.003073153 | 1 |
| HTN | endometrium | *RHBDD3* | 5.243523059 | 0.000898956 | 0.003087532 | 1 |
| HTN | endometrium | *TOM1* | 1.847423179 | 0.003493556 | 0.006303254 | 1 |
| HTN | endometrium | *ARSA* | 2.70422608 | 0.00067035 | 0.003073153 | 1 |
| HTN | endometrium | *TSPO* | 4.907859143 | 0.000592905 | 0.003073153 | 1 |
| HTN | endometrium | *TTLL12* | -2.07995403 | 0.005254295 | 0.00879098 | 1 |
| HTN | endometrium | *MTMR3* | 5.390458966 | 0.000670621 | 0.003073153 | 1 |
| HTN | endometrium | *APOL4* | 2.751548557 | 0.011861519 | 0.017519662 | 1 |
| HTN | endometrium | *SAMM50* | 6.669185647 | 0.000890182 | 0.003084413 | 1 |
| HTN | endometrium | *FOXRED2* | 3.161769614 | 0.001512412 | 0.003430353 | 1 |
| HTN | endometrium | *IFT27* | 3.084679183 | 0.001174061 | 0.003095606 | 1 |
| HTN | endometrium | *KIAA0930* | 3.949388864 | 0.00052977 | 0.003073153 | 1 |
| HTN | endometrium | *SLC25A17* | 4.200686948 | 0.000771555 | 0.003073153 | 1 |
| HTN | endometrium | *ZC3H7B* | 6.976321105 | 0.000319695 | 0.003073153 | 1 |
| HTN | endometrium | *POLR3H* | -1.906796922 | 0.014156874 | 0.020479772 | 1 |
| HTN | endometrium | *DESI1* | 3.645694027 | 0.00047044 | 0.003073153 | 1 |
| HTN | endometrium | *MLC1* | 5.831173363 | 0.013913752 | 0.020176188 | 1 |
| HTN | endometrium | *HDAC10* | 4.672120355 | 0.000745304 | 0.003073153 | 1 |
| HTN | endometrium | *SDR39U1* | -2.457763148 | 0.005362768 | 0.008942212 | 1 |
| HTN | endometrium | *RBM23* | 4.168709728 | 0.003401659 | 0.006190699 | 1 |
| HTN | endometrium | *AP4S1* | 3.651952832 | 0.002644448 | 0.005111898 | 1 |
| HTN | endometrium | *NIN* | 6.214489439 | 0.001202313 | 0.003095606 | 1 |
| HTN | endometrium | *CNIH1* | 2.992549715 | 0.000668592 | 0.003073153 | 1 |
| HTN | endometrium | *ATP6V1D* | 2.987605446 | 0.000766239 | 0.003073153 | 1 |
| HTN | endometrium | *PLEK2* | -2.628742919 | 0.023252814 | 0.031727406 | 1 |
| HTN | endometrium | *GSTZ1* | -2.039102532 | 0.016307734 | 0.0232311 | 1 |
| HTN | endometrium | *TMED8* | 5.621276132 | 0.000433467 | 0.003073153 | 1 |
| HTN | endometrium | *LGMN* | 1.703828573 | 0.000449706 | 0.003073153 | 1 |
| HTN | endometrium | *MTHFD1* | 8.111761348 | 0.001858035 | 0.003928693 | 1 |
| HTN | endometrium | *TELO2* | 8.025041283 | 0.004421654 | 0.007637545 | 1 |
| HTN | endometrium | *GSKIP* | -2.183852218 | 0.011737529 | 0.017405407 | 1 |
| HTN | endometrium | *PSMB5* | 7.69680992 | 0.000737439 | 0.003073153 | 1 |

| | | | | | | |
|---|---|---|---|---|---|---|
| HTN | endometrium | *YY1* | 2.9865786 | 0.000787099 | 0.003073153 | 1 |
| HTN | endometrium | *CCNB1IP1* | 3.825409506 | 0.000736226 | 0.003073153 | 1 |
| HTN | endometrium | *CINP* | 8.833353726 | 0.01203367 | 0.01774692 | 1 |
| HTN | endometrium | *KIAA0391* | -3.896393053 | 0.019610483 | 0.027260822 | 1 |
| HTN | endometrium | *EMC9* | 3.20884616 | 0.012544954 | 0.01841698 | 1 |
| HTN | endometrium | *PLTP* | 1.418975985 | 0.02752518 | 0.037005313 | 1 |
| HTN | endometrium | *TRPC4AP* | 5.720816768 | 0.000441627 | 0.003073153 | 1 |
| HTN | endometrium | *PYGB* | 3.36290088 | 0.023611156 | 0.032180148 | 1 |
| HTN | endometrium | *ABHD12* | 3.096512879 | 0.001005165 | 0.003095606 | 1 |
| HTN | endometrium | *CD40* | 2.669978833 | 0.001108514 | 0.003095606 | 1 |
| HTN | endometrium | *UQCC1* | 2.900392925 | 0.001150843 | 0.003095606 | 1 |
| HTN | endometrium | *NDRG3* | 2.826852975 | 0.00055459 | 0.003073153 | 1 |
| HTN | endometrium | *PABPC1L* | 7.761591728 | 0.000453394 | 0.003073153 | 1 |
| HTN | endometrium | *DOK5* | 3.930063526 | 0.000914828 | 0.003087532 | 1 |
| HTN | endometrium | *CSTF1* | 3.286626678 | 0.001197236 | 0.003095606 | 1 |
| HTN | endometrium | *NELFCD* | 2.820962547 | 0.001401575 | 0.003299583 | 1 |
| HTN | endometrium | *CTSZ* | 1.965635981 | 0.00112138 | 0.003095606 | 1 |
| HTN | endometrium | *TCFL5* | -2.786955333 | 0.017824985 | 0.025090463 | 1 |
| HTN | endometrium | *DIDO1* | 2.527750282 | 0.000555956 | 0.003073153 | 1 |
| HTN | endometrium | *GID8* | 6.512392477 | 0.030846736 | 0.041027511 | 1 |
| HTN | endometrium | *BIRC7* | -9.707200403 | 0.002009612 | 0.004154512 | 1 |
| HTN | endometrium | *PTK6* | -1.447046779 | 0.004435439 | 0.007647743 | 1 |
| HTN | endometrium | *GMEB2* | 5.319302789 | 0.000939566 | 0.003095606 | 1 |
| HTN | endometrium | *C20orf27* | 2.757624765 | 0.004447581 | 0.007660512 | 1 |
| HTN | endometrium | *CDC25B* | 5.265808598 | 0.001169217 | 0.003095606 | 1 |
| HTN | endometrium | *RNF24* | 3.217564892 | 0.00067396 | 0.003073153 | 1 |
| HTN | endometrium | *NDUFAF5* | 2.059842755 | 0.024337607 | 0.03304031 | 1 |
| HTN | endometrium | *TRIB3* | 4.987747516 | 0.001097031 | 0.003095606 | 1 |
| HTN | endometrium | *RASSF2* | 3.419066454 | 0.000460017 | 0.003073153 | 1 |
| HTN | endometrium | *ANGPT4* | 5.6704388 | 0.002332428 | 0.004639337 | 1 |
| HTN | endometrium | *RSPO4* | 4.254174831 | 0.00064133 | 0.003073153 | 1 |
| HTN | endometrium | *SIRPB1* | -2.510950064 | 0.00079458 | 0.003073153 | 1 |
| HTN | endometrium | *CRNKL1* | 2.570150654 | 0.004351922 | 0.007543322 | 1 |
| HTN | endometrium | *MANBAL* | 7.43900964 | 5.48E-06 | 0.000597955 | 0.026609918 |
| HTN | endometrium | *TTI1* | 2.215661003 | 0.001327333 | 0.003212251 | 1 |
| HTN | endometrium | *BPI* | -0.801800778 | 0.001986306 | 0.004120376 | 1 |
| HTN | endometrium | *ACTR5* | 4.368971167 | 0.000408706 | 0.003073153 | 1 |
| HTN | endometrium | *DNTTIP1* | 1.498823374 | 0.032922027 | 0.043548985 | 1 |
| HTN | endometrium | *MAP1LC3A* | 4.253111588 | 0.000629356 | 0.003073153 | 1 |
| HTN | endometrium | *RBFA* | 2.859357418 | 0.000873338 | 0.003084413 | 1 |
| HTN | endometrium | *METTL4* | 2.16005901 | 0.001262487 | 0.003126895 | 1 |
| HTN | endometrium | *LPIN2* | 4.086735014 | 0.000546168 | 0.003073153 | 1 |
| HTN | endometrium | *MYOM1* | 2.486680186 | 0.000912192 | 0.003087532 | 1 |
| HTN | endometrium | *CEP192* | 2.537917517 | 0.00266753 | 0.005146264 | 1 |
| HTN | endometrium | *LAMA1* | -1.226909888 | 0.011697963 | 0.017357344 | 1 |
| HTN | endometrium | *RNF125* | 2.963215434 | 0.001154484 | 0.003095606 | 1 |
| HTN | endometrium | *MIB1* | 3.57357682 | 0.000593047 | 0.003073153 | 1 |
| HTN | endometrium | *NALCN* | 2.976067084 | 0.000566534 | 0.003073153 | 1 |
| HTN | endometrium | *STK24* | 5.72296531 | 0.01185452 | 0.017514657 | 1 |
| HTN | endometrium | *ACP5* | 3.358614538 | 0.00165356 | 0.003628707 | 1 |

| | | | | | | |
|---|---|---|---|---|---|---|
| HTN | endometrium | *DNAJC3* | 5.791417617 | 0.000632079 | 0.003073153 | 1 |
| HTN | endometrium | *UGGT2* | 5.090850302 | 0.000769395 | 0.003073153 | 1 |
| HTN | endometrium | *ARHGEF7* | 3.88060388 | 0.00056577 | 0.003073153 | 1 |
| HTN | endometrium | *PARP4* | 4.35369968 | 0.003464451 | 0.00626753 | 1 |
| HTN | endometrium | *SUPT20H* | 5.813758432 | 0.017888191 | 0.025157537 | 1 |
| HTN | endometrium | *MRPS31* | 6.379484153 | 0.000957082 | 0.003095606 | 1 |
| HTN | endometrium | *RGCC* | 2.455332877 | 0.027984819 | 0.037592011 | 1 |
| HTN | endometrium | *DGKH* | 4.603928574 | 0.001074117 | 0.003095606 | 1 |
| HTN | endometrium | *KATNAL1* | 5.25675624 | 0.000640138 | 0.003073153 | 1 |
| HTN | endometrium | *INTS6* | 5.377258258 | 0.017941941 | 0.025225818 | 1 |
| HTN | endometrium | *CLN5* | 3.560864238 | 0.000679546 | 0.003073153 | 1 |
| HTN | endometrium | *MGRN1* | 3.323073251 | 0.00011103 | 0.002992864 | 0.538715529 |
| HTN | endometrium | *ELMO3* | -1.924313759 | 0.01491208 | 0.021457121 | 1 |
| HTN | endometrium | *LONP2* | 3.730088074 | 0.000757083 | 0.003073153 | 1 |
| HTN | endometrium | *PLLP* | 3.007197347 | 0.002920516 | 0.005499273 | 1 |
| HTN | endometrium | *DHODH* | 3.4475097 | 0.000451685 | 0.003073153 | 1 |
| HTN | endometrium | *CCDC113* | 3.448465296 | 0.017781053 | 0.025035888 | 1 |
| HTN | endometrium | *PSMD7* | 3.004300877 | 0.000633193 | 0.003073153 | 1 |
| HTN | endometrium | *COG4* | 4.477440637 | 0.013622632 | 0.019831087 | 1 |
| HTN | endometrium | *MON1B* | 4.177892731 | 0.000809011 | 0.003073153 | 1 |
| HTN | endometrium | *TAF1C* | 2.757904027 | 0.000689462 | 0.003073153 | 1 |
| HTN | endometrium | *NOMO3* | 3.025986129 | 0.023087886 | 0.031511231 | 1 |
| HTN | endometrium | *NARFL* | 5.967356206 | 6.21E-05 | 0.002410623 | 0.301378045 |
| HTN | endometrium | *MTHFSD* | 3.182945817 | 4.71E-06 | 0.000553371 | 0.022855777 |
| HTN | endometrium | *CLCN7* | 2.098060737 | 0.004074159 | 0.007151888 | 1 |
| HTN | endometrium | *HAGHL* | 6.103251674 | 3.49E-05 | 0.0019512 | 0.169128502 |
| HTN | endometrium | *RHBDL1* | -2.031811063 | 0.005503231 | 0.009125658 | 1 |
| HTN | endometrium | *NUBP1* | 7.066978503 | 0.007726582 | 0.012120716 | 1 |
| HTN | endometrium | *CAPN15* | -2.206841367 | 0.010956374 | 0.01639233 | 1 |
| HTN | endometrium | *CLUAP1* | 4.761334336 | 0.001037516 | 0.003095606 | 1 |
| HTN | endometrium | *CPPED1* | 3.807964975 | 0.00071008 | 0.003073153 | 1 |
| HTN | endometrium | *HMOX2* | 7.956118704 | 0.000634515 | 0.003073153 | 1 |
| HTN | endometrium | *RRN3P2* | 2.458213119 | 0.002161512 | 0.004382639 | 1 |
| HTN | endometrium | *RBL2* | 3.9548016 | 0.000413975 | 0.003073153 | 1 |
| HTN | endometrium | *PYCARD* | 4.017725832 | 0.000182934 | 0.003073153 | 0.88759639 |
| HTN | endometrium | *CDIPT* | 3.508166081 | 0.000669287 | 0.003073153 | 1 |
| HTN | endometrium | *TMC5* | 2.600009342 | 0.002482357 | 0.004864456 | 1 |
| HTN | endometrium | *C16orf62* | 4.635851569 | 0.000583342 | 0.003073153 | 1 |
| HTN | endometrium | *RNF40* | 4.857107979 | 0.000543317 | 0.003073153 | 1 |
| HTN | endometrium | *KNOP1* | 2.102249358 | 0.001497982 | 0.003412305 | 1 |
| HTN | endometrium | *IQCH* | 2.452978364 | 0.000909393 | 0.003087532 | 1 |
| HTN | endometrium | *LACTB* | 2.75307476 | 0.0029933 | 0.005601888 | 1 |
| HTN | endometrium | *HERC1* | 4.957631508 | 0.000716562 | 0.003073153 | 1 |
| HTN | endometrium | *TRIP4* | 3.297577006 | 0.000256302 | 0.003073153 | 1 |
| HTN | endometrium | *CTSH* | 2.069580544 | 0.000524829 | 0.003073153 | 1 |
| HTN | endometrium | *TTC23* | 5.765741386 | 0.000697514 | 0.003073153 | 1 |
| HTN | endometrium | *FAH* | 3.240775494 | 0.000924443 | 0.003087532 | 1 |
| HTN | endometrium | *SLC30A4* | 2.596526972 | 0.000880109 | 0.003084413 | 1 |
| HTN | endometrium | *ZDHHC2* | 2.467006778 | 0.001882512 | 0.003967831 | 1 |
| HTN | endometrium | *EYA1* | -1.561252615 | 0.020841314 | 0.028760539 | 1 |

| | | | | | | |
|---|---|---|---|---|---|---|
| HTN | endometrium | *IMPAD1* | 6.68844089 | 0.001766894 | 0.003786648 | 1 |
| HTN | endometrium | *SFRP1* | 6.698197664 | 0.000545807 | 0.003073153 | 1 |
| HTN | endometrium | *RAB2A* | 4.38318284 | 0.000531695 | 0.003073153 | 1 |
| HTN | endometrium | *ARMC1* | 2.453991736 | 0.002486687 | 0.004870975 | 1 |
| HTN | endometrium | *SPAG1* | 3.057478387 | 0.001184923 | 0.003095606 | 1 |
| HTN | endometrium | *CHRAC1* | -1.866744423 | 0.007768796 | 0.012177817 | 1 |
| HTN | endometrium | *SNX16* | 4.254180362 | 8.30E-05 | 0.002714492 | 0.402715957 |
| HTN | endometrium | *TSTA3* | 7.28910722 | 0.019995306 | 0.027711289 | 1 |
| HTN | endometrium | *SQLE* | 3.154428866 | 0.000213522 | 0.003073153 | 1 |
| HTN | endometrium | *TNFRSF10A* | 3.043862377 | 0.001799741 | 0.00384346 | 1 |
| HTN | endometrium | *ERICH1* | 5.281699996 | 0.000924605 | 0.003087532 | 1 |
| HTN | endometrium | *TUSC3* | 1.956158693 | 0.001186401 | 0.003095606 | 1 |
| HTN | endometrium | *MAN2B1* | 3.059154208 | 0.031196794 | 0.041436311 | 1 |
| HTN | endometrium | *TULP2* | 17.88731628 | 0.000602649 | 0.003073153 | 1 |
| HTN | endometrium | *NFKBIB* | 3.736826948 | 0.001019358 | 0.003095606 | 1 |
| HTN | endometrium | *SNRNP70* | 5.954757228 | 0.005145045 | 0.008637978 | 1 |
| HTN | endometrium | *PPP1R37* | 4.2166153 | 0.021264403 | 0.029252874 | 1 |
| HTN | endometrium | *FCGRT* | 2.068679243 | 0.000775998 | 0.003073153 | 1 |
| HTN | endometrium | *ARHGEF18* | 9.248032837 | 0.018358658 | 0.025759459 | 1 |
| HTN | endometrium | *ERCC2* | 3.615487727 | 0.001149679 | 0.003095606 | 1 |
| HTN | endometrium | *CD37* | 2.060239801 | 0.015933951 | 0.022778883 | 1 |
| HTN | endometrium | *AMH* | 31.83397617 | 0.001121237 | 0.003095606 | 1 |
| HTN | endometrium | *FCER2* | 9.76067235 | 0.001190685 | 0.003095606 | 1 |
| HTN | endometrium | *CLEC4M* | 29.22028888 | 0.033187931 | 0.043840959 | 1 |
| HTN | endometrium | *TLE6* | 5.593701461 | 0.031007414 | 0.041212839 | 1 |
| HTN | endometrium | *NOVA2* | 2.463990308 | 0.004479109 | 0.007706609 | 1 |
| HTN | endometrium | *SGTA* | 5.916141851 | 0.000642233 | 0.003073153 | 1 |
| HTN | endometrium | *KIR3DX1* | 15.49645425 | 0.010010768 | 0.01519307 | 1 |
| HTN | endometrium | *TIMM44* | 7.091192473 | 0.00037454 | 0.003073153 | 1 |
| HTN | endometrium | *IL27RA* | 4.110412819 | 0.000832128 | 0.0030808 | 1 |
| HTN | endometrium | *FAM32A* | 4.330666571 | 0.001405719 | 0.00330293 | 1 |
| HTN | endometrium | *RASAL3* | 4.173256346 | 0.000914916 | 0.003087532 | 1 |
| HTN | endometrium | *ANKRD27* | 5.458811989 | 0.000894998 | 0.003087532 | 1 |
| HTN | endometrium | *TGFB1* | 3.203144451 | 0.000689722 | 0.003073153 | 1 |
| HTN | endometrium | *CEACAM4* | 8.288557139 | 0.003892559 | 0.006890441 | 1 |
| HTN | endometrium | *MRPL4* | 4.673011036 | 0.000300724 | 0.003073153 | 1 |
| HTN | endometrium | *RABAC1* | 5.17104597 | 6.14E-05 | 0.002410623 | 0.29796723 |
| HTN | endometrium | *GRIN2D* | -2.182674566 | 0.005487097 | 0.009105129 | 1 |
| HTN | endometrium | *CCDC114* | -6.144242269 | 0.029294477 | 0.039134582 | 1 |
| HTN | endometrium | *CARD8* | 2.950572333 | 0.000872696 | 0.003084413 | 1 |
| HTN | endometrium | *LIG1* | -3.345606643 | 0.032778649 | 0.04338298 | 1 |
| HTN | endometrium | *ZNF175* | 2.968816179 | 0.001036603 | 0.003095606 | 1 |
| HTN | endometrium | *CAPS* | -1.425829545 | 0.006467661 | 0.010432544 | 1 |
| HTN | endometrium | *MIER2* | 2.529497142 | 0.002526869 | 0.004931765 | 1 |
| HTN | endometrium | *PLEKHA4* | 5.048456827 | 0.000800846 | 0.003073153 | 1 |
| HTN | endometrium | *GCDH* | 2.237429948 | 0.003010754 | 0.005627011 | 1 |
| HTN | endometrium | *TFPT* | 3.462586314 | 0.001090827 | 0.003095606 | 1 |
| HTN | endometrium | *RAB3A* | 11.83708687 | 0.00125743 | 0.003122339 | 1 |
| HTN | endometrium | *UPK1A* | 12.24255227 | 0.000714882 | 0.003073153 | 1 |
| HTN | endometrium | *ETV2* | 27.28594157 | 0.002946767 | 0.005526754 | 1 |

| | | | | | | |
|---|---|---|---|---|---|---|
| HTN | endometrium | *LSR* | 3.770394423 | 0.00048226 | 0.003073153 | 1 |
| HTN | endometrium | *ZNF14* | 8.083215874 | 0.000703809 | 0.003073153 | 1 |
| HTN | endometrium | *ZNF85* | 7.235025818 | 0.000809729 | 0.003073153 | 1 |
| HTN | endometrium | *ETHE1* | 4.496566613 | 0.000648793 | 0.003073153 | 1 |
| HTN | endometrium | *SMG9* | 2.854917337 | 0.000796704 | 0.003073153 | 1 |
| HTN | endometrium | *RASA4* | -1.894634882 | 0.003922778 | 0.006927811 | 1 |
| HTN | endometrium | *BET1* | 3.79318034 | 0.000672794 | 0.003073153 | 1 |
| HTN | endometrium | *DNAH11* | 3.027422036 | 0.000946375 | 0.003095606 | 1 |
| HTN | endometrium | *STEAP1B* | 6.042063208 | 0.000319232 | 0.003073153 | 1 |
| HTN | endometrium | *PTN* | 2.788157378 | 0.001156299 | 0.003095606 | 1 |
| HTN | endometrium | *ADAP1* | 9.650588279 | 0.001197024 | 0.003095606 | 1 |
| HTN | endometrium | *MET* | 1.600730258 | 0.002333055 | 0.004639337 | 1 |
| HTN | endometrium | *RNF32* | 2.942386557 | 0.002063995 | 0.004229098 | 1 |
| HTN | endometrium | *IQCE* | 2.628259976 | 0.001112121 | 0.003095606 | 1 |
| HTN | endometrium | *VIPR2* | 5.852312861 | 0.000479286 | 0.003073153 | 1 |
| HTN | endometrium | *HIBADH* | 4.762870643 | 0.000114737 | 0.003017974 | 0.556705581 |
| HTN | endometrium | *CPVL* | 1.608313409 | 0.001127939 | 0.003095606 | 1 |
| HTN | endometrium | *CHN2* | 2.522826622 | 0.000225282 | 0.003073153 | 1 |
| HTN | endometrium | *GRB10* | 1.987146299 | 0.001079973 | 0.003095606 | 1 |
| HTN | endometrium | *ABHD11* | 4.364567684 | 0.001438786 | 0.003339619 | 1 |
| HTN | endometrium | *PLEKHA8* | 7.573126423 | 0.000811014 | 0.003074195 | 1 |
| HTN | endometrium | *EPHB6* | 2.577683748 | 0.001712025 | 0.003713132 | 1 |
| HTN | endometrium | *CHCHD2* | 2.962712008 | 0.000826829 | 0.0030808 | 1 |
| HTN | endometrium | *CCL24* | -1.143330519 | 0.00513904 | 0.008630884 | 1 |
| HTN | endometrium | *ZKSCAN1* | 3.018662837 | 0.000331444 | 0.003073153 | 1 |
| HTN | endometrium | *TFR2* | -4.441417266 | 0.026058985 | 0.035141243 | 1 |
| HTN | endometrium | *MOSPD3* | -2.248448438 | 0.006560519 | 0.010554257 | 1 |
| HTN | endometrium | *RBM28* | 6.779882794 | 0.002356581 | 0.004672714 | 1 |
| HTN | endometrium | *IMPDH1* | -4.45964103 | 0.018016441 | 0.02531589 | 1 |
| HTN | endometrium | *C1GALT1* | 5.754797414 | 0.000421207 | 0.003073153 | 1 |
| HTN | endometrium | *NRF1* | 3.523334771 | 0.000515564 | 0.003073153 | 1 |
| HTN | endometrium | *ZNF862* | 3.079426097 | 0.000626921 | 0.003073153 | 1 |
| HTN | endometrium | *ACTR3C* | 3.228936249 | 0.000378644 | 0.003073153 | 1 |
| HTN | endometrium | *POU6F2* | -2.06181748 | 0.008640215 | 0.01334256 | 1 |
| HTN | endometrium | *TSPAN13* | 6.055204266 | 0.000370438 | 0.003073153 | 1 |
| HTN | endometrium | *RARRES2* | 4.014932246 | 0.000944986 | 0.003095606 | 1 |
| HTN | endometrium | *TMEM176B* | 2.177740568 | 0.016473937 | 0.023440335 | 1 |
| HTN | endometrium | *COA1* | 3.766398405 | 0.00052532 | 0.003073153 | 1 |
| HTN | endometrium | *BLVRA* | 6.386273281 | 0.001734504 | 0.003747095 | 1 |
| HTN | endometrium | *URGCP* | 5.011230233 | 0.000922419 | 0.003087532 | 1 |
| HTN | endometrium | *GCK* | 63.94187563 | 0.009452949 | 0.014441344 | 1 |
| HTN | endometrium | *BCL7B* | 3.080137156 | 0.00091137 | 0.003087532 | 1 |
| HTN | endometrium | *SPATA6L* | 3.519011154 | 0.000721737 | 0.003073153 | 1 |
| HTN | endometrium | *SLC1A1* | 2.263373097 | 5.80E-05 | 0.002410623 | 0.28148779 |
| HTN | endometrium | *FKTN* | 4.69043064 | 0.000601452 | 0.003073153 | 1 |
| HTN | endometrium | *NMRK1* | 6.616841637 | 0.001840577 | 0.003903182 | 1 |
| HTN | endometrium | *MEGF9* | 4.034967512 | 0.000562166 | 0.003073153 | 1 |
| HTN | endometrium | *TRIM14* | 2.212768443 | 0.00093424 | 0.003095606 | 1 |
| HTN | endometrium | *CORO2A* | 4.189879492 | 0.00037573 | 0.003073153 | 1 |
| HTN | endometrium | *C5* | 7.467134406 | 0.013596931 | 0.019805557 | 1 |

| | | | | | | |
|---|---|---|---|---|---|---|
| HTN | endometrium | *OGN* | 2.213066695 | 0.007649714 | 0.012023457 | 1 |
| HTN | endometrium | *ECM2* | 5.932069539 | 0.001757069 | 0.003774504 | 1 |
| HTN | endometrium | *PTGR1* | 4.950411939 | 0.004155082 | 0.007270048 | 1 |
| HTN | endometrium | *SUSD1* | 1.498096417 | 0.01458478 | 0.02104233 | 1 |
| HTN | endometrium | *PLGRKT* | -2.489253074 | 0.003664014 | 0.006557653 | 1 |
| HTN | endometrium | *DOCK8* | 1.368207105 | 0.001112145 | 0.003095606 | 1 |
| HTN | endometrium | *KANK1* | 3.748834047 | 0.00076935 | 0.003073153 | 1 |
| HTN | endometrium | *NCS1* | 4.303743282 | 0.001105458 | 0.003095606 | 1 |
| HTN | endometrium | *FUBP3* | 5.717622666 | 0.003194838 | 0.005891811 | 1 |
| HTN | endometrium | *RGP1* | 2.94862363 | 0.00114041 | 0.003095606 | 1 |
| HTN | endometrium | *HPS1* | 4.709637199 | 0.001381533 | 0.00327409 | 1 |
| HTN | endometrium | *PHYH* | 4.600487256 | 0.005366915 | 0.008942823 | 1 |
| HTN | endometrium | *DNMBP* | 2.905890312 | 0.000690651 | 0.003073153 | 1 |
| HTN | endometrium | *CUBN* | 4.655515505 | 0.000573372 | 0.003073153 | 1 |
| HTN | endometrium | *TRDMT1* | 8.848663065 | 0.000548329 | 0.003073153 | 1 |
| HTN | endometrium | *ATE1* | 6.042549585 | 0.000103777 | 0.002934745 | 0.503525521 |
| HTN | endometrium | *PLEKHA1* | 5.00463774 | 0.03594151 | 0.047170194 | 1 |
| HTN | endometrium | *CDH23* | 3.248560163 | 0.000661773 | 0.003073153 | 1 |
| HTN | endometrium | *BMPR1A* | 4.802887434 | 0.019753896 | 0.02742372 | 1 |
| HTN | endometrium | *LIPA* | 1.689748449 | 0.003921637 | 0.006927811 | 1 |
| HTN | endometrium | *SFXN3* | 2.647268703 | 0.001188894 | 0.003095606 | 1 |
| HTN | endometrium | *TNKS2* | 3.377437909 | 0.000645002 | 0.003073153 | 1 |
| HTN | endometrium | *ANKRD26* | 4.173552982 | 0.000809468 | 0.003073153 | 1 |
| HTN | endometrium | *PITRM1* | 7.369839781 | 0.004641945 | 0.00793892 | 1 |
| HTN | endometrium | *TFAM* | 7.937726698 | 0.000656223 | 0.003073153 | 1 |
| HTN | endometrium | *RPL28* | 3.608645026 | 0.032026847 | 0.042462597 | 1 |
| HTN | endometrium | *PBLD* | 2.908721291 | 0.00100001 | 0.003095606 | 1 |
| HTN | endometrium | *TSPAN14* | 3.769640022 | 0.023073773 | 0.031500829 | 1 |
| HTN | endometrium | *TADA2A* | 1.918359787 | 0.006158491 | 0.010007033 | 1 |
| HTN | endometrium | *PSMB3* | 7.432645914 | 0.00102367 | 0.003095606 | 1 |
| HTN | endometrium | *RPL19* | 4.734941933 | 0.021577371 | 0.029633004 | 1 |
| HTN | endometrium | *RAPGEFL1* | 4.124477656 | 0.001440544 | 0.003339619 | 1 |
| HTN | endometrium | *WNT3* | 4.307212155 | 0.005363116 | 0.008942212 | 1 |
| HTN | endometrium | *RAD51C* | 1.969630495 | 0.012139264 | 0.017891771 | 1 |
| HTN | endometrium | *TRIM37* | 2.888767241 | 0.001031296 | 0.003095606 | 1 |
| HTN | endometrium | *TRIM16L* | 3.804879585 | 0.001371368 | 0.003263303 | 1 |
| HTN | endometrium | *CDK5RAP3* | -2.648310191 | 0.010758403 | 0.016145924 | 1 |
| HTN | endometrium | *PIGL* | 5.205248602 | 0.000396043 | 0.003073153 | 1 |
| HTN | endometrium | *MED13* | 3.710720933 | 0.000991856 | 0.003095606 | 1 |
| HTN | endometrium | *ENO3* | 6.364054025 | 0.00106032 | 0.003095606 | 1 |
| HTN | endometrium | *RAI1* | 3.092214944 | 0.000845548 | 0.0030808 | 1 |
| HTN | endometrium | *GOSR1* | 6.567308963 | 0.004013548 | 0.007063887 | 1 |
| HTN | endometrium | *DRG2* | 2.894113491 | 0.000955709 | 0.003095606 | 1 |
| HTN | endometrium | *ALDH3A1* | -11.57908237 | 0.002447411 | 0.004811522 | 1 |
| HTN | endometrium | *ICAM2* | 6.550468748 | 0.000554747 | 0.003073153 | 1 |
| HTN | endometrium | *DDX5* | 3.460792514 | 0.000171082 | 0.003073153 | 0.830090115 |
| HTN | endometrium | *C17orf75* | 2.378473389 | 0.003716121 | 0.006633782 | 1 |
| HTN | endometrium | *PSMD11* | 4.009083177 | 0.000899669 | 0.003087532 | 1 |
| HTN | endometrium | *PEX12* | -7.274141449 | 0.006674885 | 0.010695688 | 1 |
| HTN | endometrium | *DHX58* | -2.402324932 | 0.019521546 | 0.027155544 | 1 |

| | | | | | | |
|---|---|---|---|---|---|---|
| HTN | endometrium | *RAB5C* | 2.869964232 | 0.002038178 | 0.004193909 | 1 |
| HTN | endometrium | *NAGLU* | 3.399059112 | 0.000179943 | 0.003073153 | 0.873084053 |
| HTN | endometrium | *HSD17B1* | 4.231177272 | 0.003564019 | 0.006414176 | 1 |
| HTN | endometrium | *EZH1* | 2.080196775 | 0.001124759 | 0.003095606 | 1 |
| HTN | endometrium | *SGCA* | 6.056123092 | 0.000460651 | 0.003073153 | 1 |
| HTN | endometrium | *ABCC3* | 3.365719598 | 0.004628178 | 0.007923754 | 1 |
| HTN | endometrium | *EFTUD2* | 2.368724569 | 0.001057174 | 0.003095606 | 1 |
| HTN | endometrium | *HLF* | 2.318479747 | 0.000799135 | 0.003073153 | 1 |
| HTN | endometrium | *EFNB3* | -3.778437994 | 0.00419664 | 0.007319229 | 1 |
| HTN | endometrium | *DPH1* | -2.149114599 | 0.007571262 | 0.011927196 | 1 |
| HTN | endometrium | *DHRS7B* | 2.621164512 | 0.000887199 | 0.003084413 | 1 |
| HTN | endometrium | *RCVRN* | -3.037342546 | 0.029955783 | 0.039963008 | 1 |
| HTN | endometrium | *MYH3* | 4.309773994 | 0.001253252 | 0.003122339 | 1 |
| HTN | endometrium | *GNRHR* | -3.24791329 | 0.030071435 | 0.040106268 | 1 |
| HTN | endometrium | *SLAIN2* | 7.008792985 | 0.001830834 | 0.003891344 | 1 |
| HTN | endometrium | *OCIAD1* | 5.603325216 | 0.002425686 | 0.004778493 | 1 |
| HTN | endometrium | *UBE2D3* | 3.408145923 | 0.000656029 | 0.003073153 | 1 |
| HTN | endometrium | *KLHL2* | 5.488148551 | 0.000766 | 0.003073153 | 1 |
| HTN | endometrium | *CPE* | 2.882781104 | 0.00184168 | 0.003903814 | 1 |
| HTN | endometrium | *WFS1* | 9.955357142 | 0.010203936 | 0.015457227 | 1 |
| HTN | endometrium | *GAR1* | -1.955102482 | 0.010771024 | 0.016159867 | 1 |
| HTN | endometrium | *AADAT* | -1.142016531 | 0.004767857 | 0.008108532 | 1 |
| HTN | endometrium | *TRIM2* | 2.725325574 | 0.00128985 | 0.003157593 | 1 |
| HTN | endometrium | *STIM2* | 3.400818392 | 0.000645521 | 0.003073153 | 1 |
| HTN | endometrium | *BST1* | 3.532103247 | 0.000658629 | 0.003073153 | 1 |
| HTN | endometrium | *UFSP2* | 4.315873135 | 0.000623235 | 0.003073153 | 1 |
| HTN | endometrium | *KLHL5* | 2.428079269 | 0.000872059 | 0.003084413 | 1 |
| HTN | endometrium | *FAM149A* | -1.6548562 | 0.01492891 | 0.021474969 | 1 |
| HTN | endometrium | *DDX25* | 3.276547344 | 0.002847846 | 0.005397557 | 1 |
| HTN | endometrium | *CTSC* | 1.594761366 | 0.004305824 | 0.007477401 | 1 |
| HTN | endometrium | *ELP4* | 5.312647367 | 0.001356987 | 0.003244998 | 1 |
| HTN | endometrium | *TECTA* | 6.283697427 | 0.00095466 | 0.003095606 | 1 |
| HTN | endometrium | *SC5D* | 5.527869204 | 0.001596831 | 0.003554047 | 1 |
| HTN | endometrium | *B3GAT1* | -6.637718414 | 0.00725943 | 0.011514467 | 1 |
| HTN | endometrium | *P2RX3* | -2.223520763 | 0.003843352 | 0.006817151 | 1 |
| HTN | endometrium | *DNAJC4* | 3.424158685 | 0.000731305 | 0.003073153 | 1 |
| HTN | endometrium | *UNC93B1* | 2.935240442 | 0.000767701 | 0.003073153 | 1 |
| HTN | endometrium | *PUS3* | 4.215019149 | 0.003250595 | 0.005971938 | 1 |
| HTN | endometrium | *DCPS* | 8.700634244 | 0.019150304 | 0.026688032 | 1 |
| HTN | endometrium | *NRXN2* | -1.776770587 | 0.005018156 | 0.008445401 | 1 |
| HTN | endometrium | *ST3GAL4* | 3.8313947 | 0.0003449 | 0.003073153 | 1 |
| HTN | endometrium | *CCDC86* | 5.062444169 | 0.016844035 | 0.023903848 | 1 |
| HTN | endometrium | *PANX1* | -0.938300984 | 0.005611192 | 0.009272992 | 1 |
| HTN | endometrium | *CEP164* | 5.051561383 | 0.000699851 | 0.003073153 | 1 |
| HTN | endometrium | *GALNT18* | 1.751563386 | 0.001481185 | 0.003394073 | 1 |
| HTN | endometrium | *KIAA1549L* | 10.02662449 | 0.001492959 | 0.003405658 | 1 |
| HTN | endometrium | *ACCS* | 2.445331745 | 0.000777999 | 0.003073153 | 1 |
| HTN | endometrium | *SLC22A18* | 3.002380829 | 0.000575687 | 0.003073153 | 1 |
| HTN | endometrium | *RPS13* | 4.908936369 | 0.028819151 | 0.038595231 | 1 |
| HTN | endometrium | *NDUFS8* | 6.082277102 | 0.001234358 | 0.003103164 | 1 |

| | | | | | | |
|---|---|---|---|---|---|---|
| HTN | endometrium | *GTF2H1* | 5.683502212 | 0.006987771 | 0.011134536 | 1 |
| HTN | endometrium | *PRDM4* | 10.46473939 | 0.000370397 | 0.003073153 | 1 |
| HTN | endometrium | *COQ5* | 1.327921844 | 0.019152286 | 0.026688032 | 1 |
| HTN | endometrium | *CORO1C* | 3.358828903 | 0.000665501 | 0.003073153 | 1 |
| HTN | endometrium | *ASIC1* | 6.216811329 | 0.001264223 | 0.003127478 | 1 |
| HTN | endometrium | *CAPRIN2* | 4.358272741 | 0.007696922 | 0.012089824 | 1 |
| HTN | endometrium | *KCTD10* | 2.794339431 | 0.005872087 | 0.009612471 | 1 |
| HTN | endometrium | *IL23A* | 19.26265544 | 0.001148883 | 0.003095606 | 1 |
| HTN | endometrium | *LTA4H* | 1.905230995 | 0.000694125 | 0.003073153 | 1 |
| HTN | endometrium | *PRR4* | 7.660045335 | 4.68E-05 | 0.002206299 | 0.227248768 |
| HTN | endometrium | *PARP11* | 6.395219109 | 0.000719674 | 0.003073153 | 1 |
| HTN | endometrium | *GPN3* | 9.379409916 | 0.000545235 | 0.003073153 | 1 |
| HTN | endometrium | *VPS29* | 4.205824161 | 0.000834198 | 0.0030808 | 1 |
| HTN | endometrium | *ACAD10* | 3.368990653 | 0.005977641 | 0.009762206 | 1 |
| HTN | endometrium | *LTBR* | 3.702865143 | 0.001184931 | 0.003095606 | 1 |
| HTN | endometrium | *DDX55* | 7.897547661 | 0.0005036 | 0.003073153 | 1 |
| HTN | endometrium | *ENDOU* | -3.648415251 | 0.006461763 | 0.010426496 | 1 |
| HTN | endometrium | *VDR* | 8.225188823 | 0.000686898 | 0.003073153 | 1 |
| HTN | endometrium | *STX2* | 3.609663999 | 0.000593692 | 0.003073153 | 1 |
| HTN | endometrium | *CAND1* | 3.75786347 | 0.000674392 | 0.003073153 | 1 |
| HTN | endometrium | *NUP107* | 5.255036235 | 0.000993644 | 0.003095606 | 1 |
| HTN | endometrium | *GNB3* | -3.621942005 | 0.003402843 | 0.006190699 | 1 |
| HTN | endometrium | *CDCA3* | -1.134697306 | 0.002247433 | 0.004523136 | 1 |
| HTN | endometrium | *CHPT1* | 2.767438233 | 0.001815422 | 0.003865041 | 1 |
| HTN | endometrium | *GNPTAB* | 5.470492346 | 0.000707105 | 0.003073153 | 1 |
| HTN | endometrium | *SPSB2* | 3.427525 | 0.005655753 | 0.009327688 | 1 |
| HTN | endometrium | *C12orf57* | 9.728755027 | 0.000796543 | 0.003073153 | 1 |
| HTN | endometrium | *NT5DC3* | 8.960059569 | 0.00103003 | 0.003095606 | 1 |
| HTN | endometrium | *GOLT1B* | 7.892645905 | 0.000636546 | 0.003073153 | 1 |
| HTN | endometrium | *C2CD5* | 2.806221715 | 0.000715885 | 0.003073153 | 1 |
| HTN | endometrium | *RWDD1* | 5.073709866 | 0.002365154 | 0.004687797 | 1 |
| HTN | endometrium | *RSPH4A* | -1.718727285 | 0.03073371 | 0.040888391 | 1 |
| HTN | endometrium | *SMIM8* | 5.33169587 | 0.000308019 | 0.003073153 | 1 |
| HTN | endometrium | *FAM184A* | 3.967187888 | 0.00039052 | 0.003073153 | 1 |
| HTN | endometrium | *RNGTT* | 5.481903943 | 0.000681999 | 0.003073153 | 1 |
| HTN | endometrium | *GABRR2* | 5.492481373 | 0.000827156 | 0.0030808 | 1 |
| HTN | endometrium | *HDDC2* | 5.483037823 | 0.000100023 | 0.002934745 | 0.485311809 |
| HTN | endometrium | *UST* | -1.479082355 | 0.008820864 | 0.013586931 | 1 |
| HTN | endometrium | *SLC26A8* | 3.685211396 | 0.000839886 | 0.0030808 | 1 |
| HTN | endometrium | *MRPL18* | 3.393840888 | 0.00090998 | 0.003087532 | 1 |
| HTN | endometrium | *RNF8* | 2.585410646 | 0.001745013 | 0.003751898 | 1 |
| HTN | endometrium | *PHACTR1* | 1.820193896 | 0.001784381 | 0.003815698 | 1 |
| HTN | endometrium | *MDGA1* | 3.130017321 | 0.001542221 | 0.003473936 | 1 |
| HTN | endometrium | *CD83* | 3.218193261 | 0.001110023 | 0.003095606 | 1 |
| HTN | endometrium | *MDN1* | 5.626600535 | 0.001111406 | 0.003095606 | 1 |
| HTN | endometrium | *COL9A1* | 8.669840089 | 0.003391561 | 0.006181764 | 1 |
| HTN | endometrium | *WASF1* | 5.059397314 | 0.010008109 | 0.01519307 | 1 |
| HTN | endometrium | *VNN1* | 3.251993444 | 0.010559622 | 0.015901703 | 1 |
| HTN | endometrium | *VNN2* | 6.810098327 | 0.000866531 | 0.003084413 | 1 |
| HTN | endometrium | *ACOT13* | 3.752814406 | 0.000678139 | 0.003073153 | 1 |

| | | | | | | |
|---|---|---|---|---|---|---|
| HTN | endometrium | *C6orf62* | 2.655865452 | 0.002311333 | 0.004616956 | 1 |
| HTN | endometrium | *HBS1L* | 5.48903718 | 0.004099688 | 0.007188901 | 1 |
| HTN | endometrium | *PEX7* | 2.202113747 | 0.003258799 | 0.005980217 | 1 |
| HTN | endometrium | *FIG4* | 7.585547408 | 0.00259129 | 0.005035218 | 1 |
| HTN | endometrium | *PERP* | 2.923225558 | 0.003070304 | 0.005707362 | 1 |
| HTN | endometrium | *PHACTR2* | 3.818911624 | 0.001027998 | 0.003095606 | 1 |
| HTN | endometrium | *PACRG* | 2.429600607 | 0.001189249 | 0.003095606 | 1 |
| HTN | endometrium | *QKI* | 2.806627485 | 0.000599416 | 0.003073153 | 1 |
| HTN | endometrium | *FAM120B* | 4.191176091 | 0.000448823 | 0.003073153 | 1 |
| HTN | endometrium | *GUCA1B* | -3.348809907 | 0.009408035 | 0.014386318 | 1 |
| HTN | endometrium | *DNPH1* | 2.30695074 | 0.007705848 | 0.012099927 | 1 |
| HTN | endometrium | *EXOC2* | 3.011405167 | 0.000609738 | 0.003073153 | 1 |
| HTN | endometrium | *IMPG1* | -11.30609679 | 0.004037368 | 0.007092437 | 1 |
| HTN | endometrium | *BTN2A1* | 5.993326678 | 9.97E-05 | 0.002934745 | 0.483846316 |
| HTN | endometrium | *LY86* | 2.999263387 | 0.00128759 | 0.003155246 | 1 |
| HTN | endometrium | *PCDHB2* | 5.936394725 | 0.006403769 | 0.010339796 | 1 |
| HTN | endometrium | *NUDT12* | 2.706633606 | 0.001048179 | 0.003095606 | 1 |
| HTN | endometrium | *CEP72* | 8.144713298 | 0.000354814 | 0.003073153 | 1 |
| HTN | endometrium | *KIF20A* | -6.195812719 | 0.003141224 | 0.005812822 | 1 |
| HTN | endometrium | *PFDN1* | 4.225362092 | 0.0006277 | 0.003073153 | 1 |
| HTN | endometrium | *TMCO6* | 3.28033895 | 0.000527413 | 0.003073153 | 1 |
| HTN | endometrium | *SPARC* | 3.730663244 | 0.000837657 | 0.0030808 | 1 |
| HTN | endometrium | *IK* | 8.950917522 | 3.68E-05 | 0.002007522 | 0.178669457 |
| HTN | endometrium | *HAVCR1* | 6.117006048 | 0.015573108 | 0.022302455 | 1 |
| HTN | endometrium | *RNF130* | 3.567783841 | 5.64E-05 | 0.00240165 | 0.273788145 |
| HTN | endometrium | *ARSB* | 2.269632042 | 0.001095523 | 0.003095606 | 1 |
| HTN | endometrium | *THBS4* | 3.240769308 | 0.000708759 | 0.003073153 | 1 |
| HTN | endometrium | *RASGRF2* | 1.436066451 | 0.001738473 | 0.003747697 | 1 |
| HTN | endometrium | *CDH6* | 2.451198938 | 0.001760874 | 0.003777083 | 1 |
| HTN | endometrium | *LMNB1* | 6.319984796 | 0.004727693 | 0.008060003 | 1 |
| HTN | endometrium | *SLC12A7* | 2.432105484 | 0.001226352 | 0.003095606 | 1 |
| HTN | endometrium | *SKP1* | 4.89240692 | 0.000513313 | 0.003073153 | 1 |
| HTN | endometrium | *PPWD1* | 3.730428141 | 0.000595885 | 0.003073153 | 1 |
| HTN | endometrium | *TXNDC15* | 7.605853713 | 0.00059873 | 0.003073153 | 1 |
| HTN | endometrium | *SMAD5* | 4.051032546 | 0.00062355 | 0.003073153 | 1 |
| HTN | endometrium | *ATP6V0E1* | 5.964471594 | 0.006338064 | 0.010250762 | 1 |
| HTN | endometrium | *BNIP1* | 3.656407027 | 5.46E-05 | 0.00235392 | 0.265149545 |
| HTN | endometrium | *UNC5A* | 12.48255893 | 0.001025631 | 0.003095606 | 1 |
| HTN | endometrium | *ACTR8* | -3.065989567 | 0.00917365 | 0.014058923 | 1 |
| HTN | endometrium | *TBCCD1* | 6.947400026 | 0.000578988 | 0.003073153 | 1 |
| HTN | endometrium | *CLDN16* | 4.406774767 | 0.000119035 | 0.003039787 | 0.577559561 |
| HTN | endometrium | *NPHP3* | 4.74567933 | 8.13E-06 | 0.00073048 | 0.039445896 |
| HTN | endometrium | *PCCB* | 3.083709461 | 1.27E-05 | 0.001044798 | 0.061643102 |
| HTN | endometrium | *CEP70* | 7.433983391 | 0.000676499 | 0.003073153 | 1 |
| HTN | endometrium | *HYAL1* | -6.189791278 | 0.003204307 | 0.005904785 | 1 |
| HTN | endometrium | *CYB561D2* | 2.983508437 | 0.000901971 | 0.003087532 | 1 |
| HTN | endometrium | *C3orf14* | 3.62206518 | 0.005961146 | 0.009741825 | 1 |
| HTN | endometrium | *CBLB* | 4.823151285 | 0.009396363 | 0.014377532 | 1 |
| HTN | endometrium | *IQCG* | 3.69536857 | 0.000639191 | 0.003073153 | 1 |
| HTN | endometrium | *NCBP2* | 8.828485574 | 0.003897495 | 0.006894663 | 1 |

| | | | | | | |
|---|---|---|---|---|---|---|
| HTN | endometrium | *PLXNA1* | 3.55328344 | 0.000405997 | 0.003073153 | 1 |
| HTN | endometrium | *SCAP* | 7.874892214 | 0.000203162 | 0.003073153 | 0.985741279 |
| HTN | endometrium | *KIAA1257* | 6.472165217 | 0.001178414 | 0.003095606 | 1 |
| HTN | endometrium | *PLSCR4* | 4.084704404 | 0.0004769 | 0.003073153 | 1 |
| HTN | endometrium | *HEMK1* | 2.969829431 | 0.001095733 | 0.003095606 | 1 |
| HTN | endometrium | *COMMD2* | 7.005411494 | 0.001343854 | 0.003234314 | 1 |
| HTN | endometrium | *ABHD14B* | 4.558049471 | 0.001778616 | 0.003808404 | 1 |
| HTN | endometrium | *ARHGEF26* | 4.135077474 | 0.000665169 | 0.003073153 | 1 |
| HTN | endometrium | *VIPR1* | 4.265924353 | 0.000850467 | 0.0030808 | 1 |
| HTN | endometrium | *NKTR* | 3.485302481 | 0.000605685 | 0.003073153 | 1 |
| HTN | endometrium | *NEK4* | 2.005503492 | 0.004969343 | 0.008377781 | 1 |
| HTN | endometrium | *ADAM23* | 2.801415442 | 0.002650012 | 0.005120612 | 1 |
| HTN | endometrium | *LMAN2L* | 4.578046002 | 0.000404852 | 0.003073153 | 1 |
| HTN | endometrium | *ACTR1B* | 3.310738513 | 0.000587978 | 0.003073153 | 1 |
| HTN | endometrium | *SLC35F5* | 3.715905475 | 0.0050521 | 0.008496633 | 1 |
| HTN | endometrium | *CENPA* | 3.499715006 | 0.00150751 | 0.003424364 | 1 |
| HTN | endometrium | *NRBP1* | 2.835077949 | 0.000859728 | 0.0030808 | 1 |
| HTN | endometrium | *ITGB6* | -1.833707133 | 0.008776141 | 0.013535231 | 1 |
| HTN | endometrium | *PPM1G* | 3.096772876 | 0.000398212 | 0.003073153 | 1 |
| HTN | endometrium | *INO80B* | 6.684743435 | 0.000876499 | 0.003084413 | 1 |
| HTN | endometrium | *GRB14* | 2.714077663 | 0.00122532 | 0.003095606 | 1 |
| HTN | endometrium | *RTN4* | 3.454405118 | 0.000957689 | 0.003095606 | 1 |
| HTN | endometrium | *GALNT3* | 4.238633878 | 0.000495972 | 0.003073153 | 1 |
| HTN | endometrium | *POLE4* | 7.916330133 | 0.000320311 | 0.003073153 | 1 |
| HTN | endometrium | *ACADL* | 6.408558917 | 0.004910258 | 0.008301244 | 1 |
| HTN | endometrium | *MRPL19* | 5.590043771 | 0.001411279 | 0.003309766 | 1 |
| HTN | endometrium | *IGFBP2* | 4.710953857 | 0.001652874 | 0.003628707 | 1 |
| HTN | endometrium | *GNLY* | 2.09504653 | 0.001309598 | 0.003186644 | 1 |
| HTN | endometrium | *CHMP3* | 2.31680487 | 0.006574137 | 0.010569157 | 1 |
| HTN | endometrium | *WNT6* | -1.701728645 | 0.002352602 | 0.004672054 | 1 |
| HTN | endometrium | *IL1RL2* | -4.134559442 | 0.012907724 | 0.018903796 | 1 |
| HTN | endometrium | *IL18R1* | 3.488490004 | 0.005147598 | 0.008639275 | 1 |
| HTN | endometrium | *MLPH* | 3.876550805 | 0.000439757 | 0.003073153 | 1 |
| HTN | endometrium | *HDLBP* | 4.444496497 | 0.000494866 | 0.003073153 | 1 |
| HTN | endometrium | *PASK* | -2.439036637 | 0.004872311 | 0.008247413 | 1 |
| HTN | endometrium | *TPO* | 2.273568147 | 0.001838724 | 0.003900956 | 1 |
| HTN | endometrium | *TAF1B* | 6.930503239 | 0.000913872 | 0.003087532 | 1 |
| HTN | endometrium | *NOL10* | 3.644533655 | 0.000622207 | 0.003073153 | 1 |
| HTN | endometrium | *STRN* | 4.501008226 | 0.000452202 | 0.003073153 | 1 |
| HTN | endometrium | *CEBPZ* | 6.301229567 | 0.000143321 | 0.003073153 | 0.695393426 |
| HTN | endometrium | *PRKD3* | 3.004771533 | 0.000435339 | 0.003073153 | 1 |
| HTN | endometrium | *SLC25A12* | 5.342621721 | 0.001913908 | 0.004021775 | 1 |
| HTN | endometrium | *DARS* | 3.906901963 | 0.000591719 | 0.003073153 | 1 |
| HTN | endometrium | *WIPF1* | 2.469706497 | 0.000514628 | 0.003073153 | 1 |
| HTN | endometrium | *PNO1* | -1.219880129 | 0.006246755 | 0.010123332 | 1 |
| HTN | endometrium | *ORC4* | 5.192665104 | 0.000150402 | 0.003073153 | 0.729748683 |
| HTN | endometrium | *AAK1* | 3.921633674 | 0.000555964 | 0.003073153 | 1 |
| HTN | endometrium | *CD207* | 8.823646808 | 0.007745307 | 0.012146163 | 1 |
| HTN | endometrium | *NFE2L2* | 5.070252231 | 0.000599282 | 0.003073153 | 1 |
| HTN | endometrium | *MSH6* | 4.580781609 | 0.000608228 | 0.003073153 | 1 |

| | | | | | | |
|---|---|---|---|---|---|---|
| HTN | endometrium | *ALMS1* | 2.69478092 | 0.000736219 | 0.003073153 | 1 |
| HTN | endometrium | *DNAJC16* | 2.75291276 | 0.001660325 | 0.003638616 | 1 |
| HTN | endometrium | *MARK1* | 3.867680791 | 0.000638847 | 0.003073153 | 1 |
| HTN | endometrium | *MORN1* | 2.106040527 | 0.022204513 | 0.030408213 | 1 |
| HTN | endometrium | *RALGPS2* | 3.970521696 | 0.002078477 | 0.004249797 | 1 |
| HTN | endometrium | *CEP104* | 5.488412425 | 0.000570061 | 0.003073153 | 1 |
| HTN | endometrium | *WRAP73* | 4.899957302 | 0.004107787 | 0.007200499 | 1 |
| HTN | endometrium | *ICMT* | 3.449070902 | 0.00117053 | 0.003095606 | 1 |
| HTN | endometrium | *PARK7* | 6.083877173 | 0.000763773 | 0.003073153 | 1 |
| HTN | endometrium | *MECR* | 3.778117398 | 0.00292078 | 0.005499273 | 1 |
| HTN | endometrium | *EDEM3* | 5.507337257 | 0.001419874 | 0.003317298 | 1 |
| HTN | endometrium | *RAP1A* | 3.886247343 | 0.000395807 | 0.003073153 | 1 |
| HTN | endometrium | *CAPZA1* | 2.644000179 | 0.002795506 | 0.005326775 | 1 |
| HTN | endometrium | *ASH1L* | 4.503224564 | 0.000576186 | 0.003073153 | 1 |
| HTN | endometrium | *GON4L* | 4.615099877 | 0.000605658 | 0.003073153 | 1 |
| HTN | endometrium | *DOCK7* | 3.885232723 | 0.00063624 | 0.003073153 | 1 |
| HTN | endometrium | *FBXO2* | 5.410236585 | 0.0007193 | 0.003073153 | 1 |
| HTN | endometrium | *FBXO6* | 3.746891881 | 0.00273149 | 0.00523221 | 1 |
| HTN | endometrium | *DNAJC6* | 2.574688224 | 0.002841586 | 0.005389904 | 1 |
| HTN | endometrium | *LEPR* | 3.126250747 | 0.000891943 | 0.003084413 | 1 |
| HTN | endometrium | *IVNS1ABP* | 3.372293241 | 0.001464355 | 0.003373718 | 1 |
| HTN | endometrium | *MFN2* | 2.431131663 | 0.001338553 | 0.003227963 | 1 |
| HTN | endometrium | *PRG4* | 2.323508586 | 0.001260742 | 0.003126646 | 1 |
| HTN | endometrium | *NCF2* | 2.680517589 | 0.000732433 | 0.003073153 | 1 |
| HTN | endometrium | *BCAS2* | 4.053396762 | 0.000195337 | 0.003073153 | 0.947776646 |
| HTN | endometrium | *TNNI3K* | 11.38733824 | 0.000278691 | 0.003073153 | 1 |
| HTN | endometrium | *CFHR3* | 8.107723582 | 0.015514456 | 0.022238151 | 1 |
| HTN | endometrium | *CRYZ* | 2.451490759 | 0.000456015 | 0.003073153 | 1 |
| HTN | endometrium | *TMEM9* | 2.274248082 | 0.010299264 | 0.01557751 | 1 |
| HTN | endometrium | *WARS2* | 3.614986257 | 0.001975431 | 0.004106134 | 1 |
| HTN | endometrium | *OSCP1* | 2.912714571 | 0.00124022 | 0.003110503 | 1 |
| HTN | endometrium | *MRPS15* | 2.551535757 | 0.000955004 | 0.003095606 | 1 |
| HTN | endometrium | *EXOC8* | 4.633999223 | 0.000866873 | 0.003084413 | 1 |
| HTN | endometrium | *GNPAT* | 6.379622561 | 0.005409032 | 0.009000214 | 1 |
| HTN | endometrium | *C1orf109* | 4.545421472 | 0.00109188 | 0.003095606 | 1 |
| HTN | endometrium | *TBCE* | 7.978998819 | 0.003298396 | 0.006043738 | 1 |
| HTN | endometrium | *ACADM* | 5.942047757 | 0.00040217 | 0.003073153 | 1 |
| HTN | endometrium | *SLAMF1* | 4.141016476 | 0.003068281 | 0.005707362 | 1 |
| HTN | endometrium | *SDHB* | 4.453027379 | 0.000600444 | 0.003073153 | 1 |
| HTN | endometrium | *CTBS* | 2.633670624 | 0.001667255 | 0.003648858 | 1 |
| HTN | endometrium | *KLHL12* | 2.343121604 | 0.001626699 | 0.003599062 | 1 |
| HTN | endometrium | *ZNHIT6* | 5.561252453 | 0.000891751 | 0.003084413 | 1 |
| HTN | endometrium | *KIF17* | 5.35283918 | 0.003187643 | 0.005883014 | 1 |
| HTN | endometrium | *CD46* | 4.118517472 | 0.000111771 | 0.002996205 | 0.542313118 |
| HTN | endometrium | *EBNA1BP2* | 3.852739835 | 0.000715325 | 0.003073153 | 1 |
| HTN | endometrium | *ERI3* | 2.336875789 | 0.000851627 | 0.0030808 | 1 |
| HTN | endometrium | *PRDX1* | 2.070831434 | 0.001680626 | 0.003668195 | 1 |
| HTN | endometrium | *CCDC181* | 3.16131288 | 0.018482711 | 0.025911041 | 1 |
| HTN | endometrium | *NSUN4* | 6.127467058 | 0.000620356 | 0.003073153 | 1 |
| HTN | endometrium | *DR1* | 5.950172405 | 0.0085232 | 0.013199671 | 1 |

| | | | | | | |
|---|---|---|---|---|---|---|
| HTN | endometrium | *PRRC2C* | 3.776841816 | 0.000303486 | 0.003073153 | 1 |
| HTN | endometrium | *ABCD3* | 4.457907754 | 0.000554705 | 0.003073153 | 1 |
| HTN | endometrium | *VAMP4* | 2.805150459 | 0.001458389 | 0.003363763 | 1 |
| HTN | endometrium | *PTBP2* | 3.582858798 | 0.001025256 | 0.003095606 | 1 |
| HTN | endometrium | *DARS2* | -2.015624756 | 0.01757083 | 0.024761449 | 1 |
| HTN | endometrium | *IRF6* | -3.744404739 | 4.80E-05 | 0.002211831 | 0.233101424 |
| HTN | endometrium | *SERPINC1* | -2.081189834 | 0.008765031 | 0.013522394 | 1 |
| HTN | endometrium | *RPS6KA1* | 3.209048894 | 0.002136981 | 0.00433834 | 1 |
| HTN | endometrium | *RPA2* | 2.443434009 | 0.000556153 | 0.003073153 | 1 |
| HTN | endometrium | *SLC5A9* | 2.812177166 | 0.00085522 | 0.0030808 | 1 |
| HTN | endometrium | *ESYT2* | 4.016380575 | 0.000377964 | 0.003073153 | 1 |
| HTN | endometrium | *CD3EAP* | 3.246471496 | 0.000481178 | 0.003073153 | 1 |
| HTN | endometrium | *RCN2* | 3.396498294 | 0.000466603 | 0.003073153 | 1 |
| HTN | endometrium | *IFT46* | 3.33236874 | 0.008872768 | 0.013653875 | 1 |
| HTN | endometrium | *APOA1* | 21.76840728 | 0.003300598 | 0.006045489 | 1 |
| HTN | endometrium | *DDX59* | 2.214109276 | 0.007065642 | 0.011243849 | 1 |
| HTN | endometrium | *CAMSAP2* | 5.394706855 | 0.000175426 | 0.003073153 | 0.851168089 |
| HTN | endometrium | *KLF7* | 4.034527821 | 0.00019398 | 0.003073153 | 0.941190316 |
| HTN | endometrium | *CASC1* | 1.856538635 | 0.02746602 | 0.036936012 | 1 |
| HTN | endometrium | *LRMP* | 2.366151204 | 0.000776242 | 0.003073153 | 1 |
| HTN | endometrium | *ELOVL4* | 9.372118979 | 0.0001521 | 0.003073153 | 0.737991151 |
| HTN | endometrium | *CNR1* | 5.429218225 | 0.01687434 | 0.023939854 | 1 |
| HTN | endometrium | *AKAP7* | 4.623672892 | 0.000463823 | 0.003073153 | 1 |
| HTN | endometrium | *ALDH8A1* | -2.137847844 | 0.009588017 | 0.014616747 | 1 |
| HTN | endometrium | *MED28* | 5.456401143 | 0.036659957 | 0.048061094 | 1 |
| HTN | endometrium | *TMEM5* | 7.415606962 | 0.002321712 | 0.004630766 | 1 |
| HTN | endometrium | *VAMP8* | 7.332124922 | 0.005263 | 0.00880143 | 1 |
| HTN | endometrium | *FOXO3* | 4.272837362 | 0.000475888 | 0.003073153 | 1 |
| HTN | endometrium | *SPP1* | -1.515300747 | 0.006847601 | 0.010943531 | 1 |
| HTN | endometrium | *MFSD1* | 4.231215813 | 0.000675018 | 0.003073153 | 1 |
| HTN | endometrium | *UBN1* | 7.002918713 | 0.000201522 | 0.003073153 | 0.977782909 |
| HTN | endometrium | *DNAH7* | 2.305844366 | 0.002640074 | 0.005105475 | 1 |
| HTN | endometrium | *C2orf40* | 4.988749985 | 0.000525988 | 0.003073153 | 1 |
| HTN | endometrium | *CPSF3* | 4.616178324 | 0.023325149 | 0.031817155 | 1 |
| HTN | endometrium | *SENP5* | 3.916876269 | 0.000628396 | 0.003073153 | 1 |
| HTN | endometrium | *CCDC92* | 1.796414264 | 0.001897341 | 0.003995442 | 1 |
| HTN | endometrium | *FKBP15* | 5.582787341 | 1.99E-05 | 0.001324109 | 0.096659975 |
| HTN | endometrium | *CTNNAL1* | 1.92916075 | 0.004515638 | 0.007758455 | 1 |
| HTN | endometrium | *RAB14* | 5.724851109 | 0.000500067 | 0.003073153 | 1 |
| HTN | endometrium | *TRIM32* | -2.462890635 | 0.008327266 | 0.012916847 | 1 |
| HTN | endometrium | *PPP6C* | 5.660564601 | 0.000613423 | 0.003073153 | 1 |
| HTN | endometrium | *RBM18* | 2.654630655 | 0.001063718 | 0.003095606 | 1 |
| HTN | endometrium | *HSDL2* | 3.51034575 | 0.00019102 | 0.003073153 | 0.926829065 |
| HTN | endometrium | *MAPKAP1* | 4.572270926 | 0.000753104 | 0.003073153 | 1 |
| HTN | endometrium | *INVS* | 2.922934868 | 0.000448564 | 0.003073153 | 1 |
| HTN | endometrium | *DENND1A* | 4.288830116 | 0.000427475 | 0.003073153 | 1 |
| HTN | endometrium | *KDSR* | 3.306190997 | 0.000729252 | 0.003073153 | 1 |
| HTN | endometrium | *DCAF4* | 5.60892752 | 0.030394547 | 0.040503802 | 1 |
| HTN | endometrium | *IFI27L2* | 3.049316439 | 0.002017633 | 0.004162227 | 1 |
| HTN | endometrium | *NEK9* | 5.474831589 | 0.032552273 | 0.043118654 | 1 |

| | | | | | | |
|---|---|---|---|---|---|---|
| HTN | endometrium | *ACOT2* | 3.106009101 | 6.69E-06 | 0.000662056 | 0.032440727 |
| HTN | endometrium | *TTLL5* | 5.873059704 | 0.001192348 | 0.003095606 | 1 |
| HTN | endometrium | *FLVCR2* | 2.673400456 | 0.00162654 | 0.003599062 | 1 |
| HTN | endometrium | *PPP4R4* | 2.889065771 | 0.006181597 | 0.010034497 | 1 |
| HTN | endometrium | *ALDH6A1* | 4.383388142 | 0.000486809 | 0.003073153 | 1 |
| HTN | endometrium | *ESRRB* | 4.076599294 | 0.002013579 | 0.004159168 | 1 |
| HTN | endometrium | *EIF2B2* | 2.622146432 | 0.000289057 | 0.003073153 | 1 |
| HTN | endometrium | *COQ6* | 3.656258299 | 0.000734031 | 0.003073153 | 1 |
| HTN | endometrium | *DNMT3A* | 2.547734294 | 0.00134021 | 0.003228749 | 1 |
| HTN | endometrium | *CNRIP1* | 2.272743587 | 0.002942331 | 0.00552193 | 1 |
| HTN | endometrium | *CRIPT* | 6.345761937 | 0.000587221 | 0.003073153 | 1 |
| HTN | endometrium | *EPCAM* | 10.19962099 | 0.001091308 | 0.003095606 | 1 |
| HTN | endometrium | *SLC17A5* | 6.339765202 | 0.001126631 | 0.003095606 | 1 |
| HTN | endometrium | *MXI1* | 4.06005695 | 0.000663202 | 0.003073153 | 1 |
| HTN | endometrium | *TCTN3* | 5.937615288 | 0.003620983 | 0.006492612 | 1 |
| HTN | endometrium | *WDR11* | 6.075530228 | 0.000674851 | 0.003073153 | 1 |
| HTN | endometrium | *KANSL1* | 3.407180431 | 0.000945954 | 0.003095606 | 1 |
| HTN | endometrium | *HOXB3* | 3.142394295 | 0.000830042 | 0.0030808 | 1 |
| HTN | endometrium | *CAAP1* | 5.380251297 | 0.002289768 | 0.004581424 | 1 |
| HTN | endometrium | *PCDHB10* | -1.505094621 | 0.007239736 | 0.011498265 | 1 |
| HTN | endometrium | *GORAB* | -2.207614253 | 0.034334 | 0.045231759 | 1 |
| HTN | endometrium | *ACAT2* | 4.063673123 | 0.000727665 | 0.003073153 | 1 |
| HTN | endometrium | *SNX19* | 2.61645175 | 0.008799385 | 0.013562457 | 1 |
| HTN | endometrium | *SLC10A7* | 3.631994748 | 0.000714948 | 0.003073153 | 1 |
| HTN | endometrium | *MRC1* | 1.632540783 | 0.001126331 | 0.003095606 | 1 |
| HTN | endometrium | *IQSEC3* | 12.27974342 | 0.025296007 | 0.034236046 | 1 |
| HTN | endometrium | *DNAJC15* | 4.843359492 | 0.000602101 | 0.003073153 | 1 |
| HTN | endometrium | *UFM1* | 3.075695721 | 0.00154059 | 0.003471872 | 1 |
| HTN | endometrium | *ALG5* | 4.940611242 | 0.009566134 | 0.014591287 | 1 |
| HTN | endometrium | *SIL1* | 5.607851103 | 0.001325019 | 0.003211626 | 1 |
| HTN | endometrium | *ZFP30* | 2.461276266 | 0.000115693 | 0.003017974 | 0.561343088 |
| HTN | endometrium | *ARL1* | 6.330316342 | 0.010681388 | 0.016048483 | 1 |
| HTN | endometrium | *APAF1* | 6.743288847 | 0.001935268 | 0.004050808 | 1 |
| HTN | endometrium | *ADRA1A* | 2.684781397 | 5.86E-05 | 0.002410623 | 0.284366907 |
| HTN | endometrium | *EPHX2* | 3.730039259 | 0.030459697 | 0.040568337 | 1 |
| HTN | endometrium | *AKAP1* | -1.920931804 | 0.018223821 | 0.025585063 | 1 |
| HTN | endometrium | *SCPEP1* | 2.088561134 | 0.000949679 | 0.003095606 | 1 |
| HTN | endometrium | *TBX2* | -4.857070032 | 0.002442011 | 0.0048048 | 1 |
| HTN | endometrium | *TEX14* | -0.831507101 | 0.026025924 | 0.03511618 | 1 |
| HTN | endometrium | *TRIM6* | 6.821057677 | 0.001710481 | 0.003713132 | 1 |
| HTN | endometrium | *ADCY7* | 2.023038512 | 0.002015355 | 0.004161064 | 1 |
| HTN | endometrium | *ECHDC2* | 3.930585937 | 0.002192796 | 0.004436801 | 1 |
| HTN | endometrium | *PYROXD1* | 3.89390359 | 0.001624576 | 0.003598884 | 1 |
| HTN | endometrium | *ZNF549* | 4.283097351 | 0.000650497 | 0.003073153 | 1 |
| HTN | endometrium | *A1BG* | -2.784121326 | 0.011784751 | 0.017459424 | 1 |
| HTN | endometrium | *ZSCAN18* | 3.676237049 | 0.001126237 | 0.003095606 | 1 |
| HTN | endometrium | *ZNF211* | 3.299933031 | 0.001725438 | 0.003735754 | 1 |
| HTN | endometrium | *PDZRN3* | 2.891652774 | 0.000215354 | 0.003073153 | 1 |
| HTN | endometrium | *LHX4* | 4.222288363 | 0.00090065 | 0.003087532 | 1 |
| HTN | endometrium | *SEC22A* | 3.623528048 | 0.0005878 | 0.003073153 | 1 |

| | | | | | | |
|---|---|---|---|---|---|---|
| HTN | endometrium | *CAT* | 2.634767881 | 0.002610478 | 0.005058322 | 1 |
| HTN | endometrium | *PILRB* | 2.068972597 | 0.001561686 | 0.003499546 | 1 |
| HTN | endometrium | *ZCCHC17* | 5.427369551 | 0.000577839 | 0.003073153 | 1 |
| HTN | endometrium | *FABP3* | 4.382798956 | 0.00305017 | 0.005683343 | 1 |
| HTN | endometrium | *TNFSF10* | 3.828140652 | 0.000593653 | 0.003073153 | 1 |
| HTN | endometrium | *ZNF639* | 2.570786302 | 0.001192907 | 0.003095606 | 1 |
| HTN | endometrium | *CPXM2* | 1.839691902 | 0.002804821 | 0.005334767 | 1 |
| HTN | endometrium | *FLT3* | 5.451000944 | 0.000392033 | 0.003073153 | 1 |
| HTN | endometrium | *MTIF3* | 4.654142311 | 0.001995308 | 0.004130219 | 1 |
| HTN | endometrium | *FMOD* | 3.558668169 | 0.000892294 | 0.003084413 | 1 |
| HTN | endometrium | *CD244* | 4.069895498 | 0.000812006 | 0.003074195 | 1 |
| HTN | endometrium | *SERAC1* | 2.6091953 | 0.002130288 | 0.004328375 | 1 |
| HTN | endometrium | *LDB3* | 3.496297028 | 0.000692748 | 0.003073153 | 1 |
| HTN | endometrium | *OPN4* | 16.55157939 | 0.007092306 | 0.011282579 | 1 |
| HTN | endometrium | *FAM35A* | 4.481795106 | 0.002722348 | 0.005216758 | 1 |
| HTN | endometrium | *ODF2L* | 6.047658226 | 0.003055597 | 0.005691269 | 1 |
| HTN | endometrium | *SPATA1* | 4.352780151 | 0.000361083 | 0.003073153 | 1 |
| HTN | endometrium | *RWDD3* | 3.343942632 | 0.018516742 | 0.025951251 | 1 |
| HTN | endometrium | *EEPD1* | 2.306780942 | 0.00256676 | 0.00499756 | 1 |
| HTN | endometrium | *KLHL7* | 4.180814028 | 0.00054424 | 0.003073153 | 1 |
| HTN | endometrium | *NT5C3A* | 4.135722736 | 0.000618721 | 0.003073153 | 1 |
| HTN | endometrium | *CCZ1* | 4.440730071 | 0.00045924 | 0.003073153 | 1 |
| HTN | endometrium | *SMU1* | 7.97788824 | 0.002464459 | 0.004837198 | 1 |
| HTN | endometrium | *GLIPR2* | -2.141316519 | 0.029740466 | 0.039686672 | 1 |
| HTN | endometrium | *TRIM24* | 5.612090894 | 0.002045774 | 0.004202411 | 1 |
| HTN | endometrium | *RBM19* | 7.063423236 | 0.015940694 | 0.022781811 | 1 |
| HTN | endometrium | *DDX54* | 8.100400169 | 0.000123251 | 0.003066741 | 0.598014458 |
| HTN | endometrium | *CCDC91* | 5.137159162 | 0.009405596 | 0.014386318 | 1 |
| HTN | endometrium | *ATP7B* | 1.318613409 | 0.011159294 | 0.016644603 | 1 |
| HTN | endometrium | *CENPK* | 6.716063342 | 0.000537698 | 0.003073153 | 1 |
| HTN | endometrium | *ITIH5* | 2.296682046 | 0.000483038 | 0.003073153 | 1 |
| HTN | endometrium | *TSFM* | 3.522610896 | 0.001262234 | 0.003126895 | 1 |
| HTN | endometrium | *ARHGAP9* | 3.569564947 | 0.001830985 | 0.003891344 | 1 |
| HTN | endometrium | *PFDN5* | 6.509797952 | 0.003883994 | 0.006882812 | 1 |
| HTN | endometrium | *PDE1B* | 2.860530999 | 0.001134631 | 0.003095606 | 1 |
| HTN | endometrium | *SMUG1* | 3.669035569 | 0.019650974 | 0.027304274 | 1 |
| HTN | endometrium | *SARDH* | 5.195843938 | 0.001184934 | 0.003095606 | 1 |
| HTN | endometrium | *DBH* | 78.41321684 | 0.001454147 | 0.003358174 | 1 |
| HTN | endometrium | *NDUFAF4* | 8.367477512 | 1.51E-05 | 0.001180045 | 0.073437969 |
| HTN | endometrium | *USP45* | 3.120586969 | 0.000739012 | 0.003073153 | 1 |
| HTN | endometrium | *TTC21B* | 4.287912089 | 0.002118823 | 0.004308689 | 1 |
| HTN | endometrium | *ACVR1C* | 1.413132821 | 0.00165252 | 0.003628707 | 1 |
| HTN | endometrium | *SLC36A1* | 3.183977314 | 0.000574632 | 0.003073153 | 1 |
| HTN | endometrium | *PLA2G12A* | 7.299709584 | 0.0030785 | 0.005718561 | 1 |
| HTN | endometrium | *C4BPA* | 5.192592743 | 0.000292055 | 0.003073153 | 1 |
| HTN | endometrium | *C4BPB* | 3.809095355 | 0.004450363 | 0.007662584 | 1 |
| HTN | endometrium | *RAB38* | 6.676091661 | 0.000892522 | 0.003084413 | 1 |
| HTN | endometrium | *DAW1* | 6.076548171 | 0.00357283 | 0.006419531 | 1 |
| HTN | endometrium | *DNPEP* | 3.558848924 | 0.000528354 | 0.003073153 | 1 |
| HTN | endometrium | *OBSL1* | 2.461583124 | 0.002221353 | 0.004488965 | 1 |

| | | | | | | |
|---|---|---|---|---|---|---|
| HTN | endometrium | *SLC12A4* | 4.448306577 | 0.001062111 | 0.003095606 | 1 |
| HTN | endometrium | *FAM210B* | 8.922425577 | 0.000769249 | 0.003073153 | 1 |
| HTN | endometrium | *MATN4* | 19.28518538 | 0.001283084 | 0.003148975 | 1 |
| HTN | endometrium | *VAPB* | 1.860735709 | 0.004013837 | 0.007063887 | 1 |
| HTN | endometrium | *TOX2* | -1.562438508 | 0.000675423 | 0.003073153 | 1 |
| HTN | endometrium | *SRSF6* | 3.850596042 | 0.000473411 | 0.003073153 | 1 |
| HTN | endometrium | *CSE1L* | 3.429130333 | 0.000569013 | 0.003073153 | 1 |
| HTN | endometrium | *PTGIS* | 2.091387434 | 0.00030973 | 0.003073153 | 1 |
| HTN | endometrium | *RNF114* | 5.659682682 | 0.00174455 | 0.003751898 | 1 |
| HTN | endometrium | *PEPD* | 6.265659138 | 0.000573031 | 0.003073153 | 1 |
| HTN | endometrium | *STAMBP* | 7.478627256 | 0.002977694 | 0.005578289 | 1 |
| HTN | endometrium | *NAGK* | 2.921167234 | 0.000788757 | 0.003073153 | 1 |
| HTN | endometrium | *SNRNP27* | 2.879161471 | 0.000987476 | 0.003095606 | 1 |
| HTN | endometrium | *ATP8A1* | 6.070898544 | 0.002488519 | 0.004872597 | 1 |
| HTN | endometrium | *USP22* | 4.328721771 | 0.000440882 | 0.003073153 | 1 |
| HTN | endometrium | *HIF3A* | 2.756523934 | 0.00132516 | 0.003211626 | 1 |
| HTN | endometrium | *BTN2A2* | 5.342974016 | 2.38E-08 | 8.25E-06 | 0.00011553 |
| HTN | endometrium | *MRS2* | 2.146168891 | 0.001917462 | 0.004025868 | 1 |
| HTN | endometrium | *SERPINB6* | 4.455577253 | 0.000497425 | 0.003073153 | 1 |
| HTN | endometrium | *NQO2* | 6.237860556 | 0.023051672 | 0.031479514 | 1 |
| HTN | endometrium | *RPS10* | -1.290035581 | 0.004392481 | 0.007597974 | 1 |
| HTN | endometrium | *MED20* | 3.327407749 | 0.00449853 | 0.007734538 | 1 |
| HTN | endometrium | *TCP11* | 8.139826225 | 0.004278652 | 0.007440867 | 1 |
| HTN | endometrium | *TREM1* | 5.084157781 | 0.004474156 | 0.007700818 | 1 |
| HTN | endometrium | *MEA1* | 6.113764894 | 0.000901169 | 0.003087532 | 1 |
| HTN | endometrium | *KLHL31* | 2.743388691 | 0.004157952 | 0.007270048 | 1 |
| HTN | endometrium | *GLO1* | 8.3076935 | 0.00041963 | 0.003073153 | 1 |
| HTN | endometrium | *KCNK17* | -2.605796474 | 0.002910821 | 0.00549105 | 1 |
| HTN | endometrium | *RREB1* | 4.308980797 | 0.001169083 | 0.003095606 | 1 |
| HTN | endometrium | *RIOK1* | 2.522670583 | 0.001350782 | 0.00323814 | 1 |
| HTN | endometrium | *ATXN1* | 5.154187492 | 0.001600339 | 0.00355859 | 1 |
| HTN | endometrium | *RUNX2* | 4.079346281 | 0.000792628 | 0.003073153 | 1 |
| HTN | endometrium | *CXCL6* | 6.294325539 | 0.001629656 | 0.003600432 | 1 |
| HTN | endometrium | *SH3TC1* | 2.383256776 | 0.001089531 | 0.003095606 | 1 |
| HTN | endometrium | *BBS2* | 3.679046759 | 0.000216659 | 0.003073153 | 1 |
| HTN | endometrium | *GOT2* | 3.215850317 | 0.000654419 | 0.003073153 | 1 |
| HTN | endometrium | *CLYBL* | 5.332902036 | 0.000483539 | 0.003073153 | 1 |
| HTN | endometrium | *ABCC4* | 5.955446394 | 0.000784064 | 0.003073153 | 1 |
| HTN | endometrium | *GRK4* | 2.621816717 | 6.32E-06 | 0.00065196 | 0.030642114 |
| HTN | endometrium | *HS3ST3B1* | 4.602321654 | 0.002553442 | 0.004975623 | 1 |
| HTN | endometrium | *GGA3* | 6.674401766 | 0.001420039 | 0.003317298 | 1 |
| HTN | endometrium | *NUP85* | 3.986264975 | 0.002070844 | 0.004237763 | 1 |
| HTN | endometrium | *NT5C* | 8.164700555 | 0.001044043 | 0.003095606 | 1 |
| HTN | endometrium | *OPRL1* | 9.705376491 | 0.001072449 | 0.003095606 | 1 |
| HTN | endometrium | *IL1B* | 17.60672248 | 0.000159042 | 0.003073153 | 0.771670547 |
| HTN | endometrium | *PAX8* | 2.284363933 | 0.019024055 | 0.026547229 | 1 |
| HTN | endometrium | *POLR1B* | 2.50533958 | 0.001496574 | 0.003410699 | 1 |
| HTN | endometrium | *CCDC93* | 3.88622339 | 0.00113103 | 0.003095606 | 1 |
| HTN | endometrium | *PSD4* | 11.26799198 | 0.000140337 | 0.003073153 | 0.680915689 |
| HTN | endometrium | *GTF2F1* | 5.537452766 | 0.001084759 | 0.003095606 | 1 |

| | | | | | | |
|---|---|---|---|---|---|---|
| HTN | endometrium | *SH2D3A* | 3.687952695 | 0.003887934 | 0.006887278 | 1 |
| HTN | endometrium | *SNRPD2* | 5.356998337 | 0.000600488 | 0.003073153 | 1 |
| HTN | endometrium | *SDCBP2* | -4.878755636 | 0.007361829 | 0.011650227 | 1 |
| HTN | endometrium | *PSMF1* | 2.652048884 | 0.001821795 | 0.003876908 | 1 |
| HTN | endometrium | *RBCK1* | 3.551324108 | 0.001585353 | 0.003539868 | 1 |
| HTN | endometrium | *RRBP1* | 4.080795995 | 0.000347749 | 0.003073153 | 1 |
| HTN | endometrium | *FLRT3* | 2.944379638 | 0.001142215 | 0.003095606 | 1 |
| HTN | endometrium | *PCSK2* | 12.75713261 | 0.000754093 | 0.003073153 | 1 |
| HTN | endometrium | *MKKS* | 8.02198681 | 0.00033783 | 0.003073153 | 1 |
| HTN | endometrium | *LAMP5* | 61.98765867 | 0.001394397 | 0.003287471 | 1 |
| HTN | endometrium | *MGME1* | 3.765650161 | 0.001222211 | 0.003095606 | 1 |
| HTN | endometrium | *ITPA* | 4.36820895 | 0.000566492 | 0.003073153 | 1 |
| HTN | endometrium | *NCLN* | 2.563320949 | 0.001757729 | 0.003774504 | 1 |
| HTN | endometrium | *KLC1* | 2.420515226 | 0.00079995 | 0.003073153 | 1 |
| HTN | endometrium | *XRCC3* | 3.653959292 | 0.034599327 | 0.045531851 | 1 |
| HTN | endometrium | *TUBGCP3* | 2.206120377 | 0.000944149 | 0.003095606 | 1 |
| HTN | endometrium | *MCF2L* | 2.122502625 | 0.002892997 | 0.005468182 | 1 |
| HTN | endometrium | *PRDX5* | 10.30563693 | 0.001705411 | 0.003709537 | 1 |
| HTN | endometrium | *IRF3* | 7.586234408 | 0.001934124 | 0.004050808 | 1 |
| HTN | endometrium | *PRR12* | 4.115811718 | 0.01343606 | 0.019606546 | 1 |
| HTN | endometrium | *TSKS* | -2.393646075 | 0.001016074 | 0.003095606 | 1 |
| HTN | endometrium | *TRAP1* | 5.028964027 | 2.58E-08 | 8.34E-06 | 0.000125161 |
| HTN | endometrium | *EMG1* | 3.144120893 | 0.001579079 | 0.00353065 | 1 |
| HTN | endometrium | *ATG14* | 3.512217215 | 0.00046537 | 0.003073153 | 1 |
| HTN | endometrium | *KTN1* | 5.051743966 | 0.000863033 | 0.003084413 | 1 |
| HTN | endometrium | *L3HYPDH* | 3.538531007 | 0.000460663 | 0.003073153 | 1 |
| HTN | endometrium | *RHOT1* | 5.596267988 | 0.000153902 | 0.003073153 | 0.746732319 |
| HTN | endometrium | *WDR60* | 5.243623667 | 0.000889492 | 0.003084413 | 1 |
| HTN | endometrium | *AIF1L* | 3.302654614 | 0.001280393 | 0.00314396 | 1 |
| HTN | endometrium | *CANX* | 4.000280903 | 0.000581365 | 0.003073153 | 1 |
| HTN | endometrium | *FGD3* | 2.624987981 | 0.000551353 | 0.003073153 | 1 |
| HTN | endometrium | *ABHD8* | 1.650073512 | 0.030206432 | 0.040275243 | 1 |
| HTN | endometrium | *ATP13A4* | 6.492042286 | 0.000305503 | 0.003073153 | 1 |
| HTN | endometrium | *HELB* | 2.808999394 | 0.001084271 | 0.003095606 | 1 |
| HTN | endometrium | *RAB3IP* | 4.006302971 | 0.000883904 | 0.003084413 | 1 |
| HTN | endometrium | *TAS2R4* | 4.410268281 | 0.000255681 | 0.003073153 | 1 |
| HTN | endometrium | *LRRC61* | 2.401849668 | 0.00856693 | 0.013254701 | 1 |
| HTN | endometrium | *IDUA* | 5.983705358 | 0.001881168 | 0.003966721 | 1 |
| HTN | endometrium | *EMC1* | 3.666982827 | 0.001138565 | 0.003095606 | 1 |
| HTN | endometrium | *UBR4* | 4.326644099 | 0.00182427 | 0.003880473 | 1 |
| HTN | endometrium | *WFIKKN1* | 10.45437181 | 0.001486183 | 0.003397652 | 1 |
| HTN | endometrium | *WDR24* | 5.596033716 | 0.000104982 | 0.002934745 | 0.50937186 |
| HTN | endometrium | *SMARCA4* | 5.779373077 | 0.000224098 | 0.003073153 | 1 |
| HTN | endometrium | *PNKD* | 5.919368505 | 0.000931507 | 0.003093547 | 1 |
| HTN | endometrium | *RNF6* | 9.385195329 | 0.004626737 | 0.007923754 | 1 |
| HTN | endometrium | *HIP1* | 4.631729484 | 0.000476512 | 0.003073153 | 1 |
| HTN | endometrium | *PTPN12* | 4.406745851 | 0.000556323 | 0.003073153 | 1 |
| HTN | endometrium | *STEAP4* | 2.605876959 | 0.000996419 | 0.003095606 | 1 |
| HTN | endometrium | *PEX1* | 3.869961628 | 0.000664306 | 0.003073153 | 1 |
| HTN | endometrium | *KDR* | 2.65218573 | 0.000941197 | 0.003095606 | 1 |

| | | | | | | |
|---|---|---|---|---|---|---|
| HTN | endometrium | *MGAT3* | -2.429783173 | 0.007828896 | 0.012257439 | 1 |
| HTN | endometrium | *ATF4* | 3.906305321 | 0.000813534 | 0.003074195 | 1 |
| HTN | endometrium | *A4GALT* | 2.332651409 | 0.020932016 | 0.028861081 | 1 |
| HTN | endometrium | *CDC42EP1* | 10.94680205 | 0.001234185 | 0.003103164 | 1 |
| HTN | endometrium | *SPECC1* | 4.648441841 | 0.000573572 | 0.003073153 | 1 |
| HTN | endometrium | *CDHR3* | 5.800587593 | 0.000751189 | 0.003073153 | 1 |
| HTN | endometrium | *PRKRIP1* | 3.878417261 | 0.00063113 | 0.003073153 | 1 |
| HTN | endometrium | *OPN1SW* | -2.03349677 | 0.002398535 | 0.004738474 | 1 |
| HTN | endometrium | *HOXD1* | -2.869275573 | 0.001740223 | 0.003747697 | 1 |
| HTN | endometrium | *HOXD3* | -1.440348421 | 0.017960979 | 0.02524527 | 1 |
| HTN | endometrium | *MTX2* | 6.377244034 | 0.001945181 | 0.004061109 | 1 |
| HTN | endometrium | *ORMDL1* | 4.204758159 | 0.00018675 | 0.003073153 | 0.906109786 |
| HTN | endometrium | *HOXD10* | -1.798412879 | 0.008325952 | 0.012916847 | 1 |
| HTN | endometrium | *PSMG2* | 6.516098096 | 0.000562304 | 0.003073153 | 1 |
| HTN | endometrium | *TWSG1* | 2.894675923 | 0.000719831 | 0.003073153 | 1 |
| HTN | endometrium | *WDFY4* | 1.741526134 | 0.006512427 | 0.010490802 | 1 |
| HTN | endometrium | *EIF2AK4* | 2.745751265 | 0.004958962 | 0.008363185 | 1 |
| HTN | endometrium | *TMOD2* | 6.143176883 | 0.028170567 | 0.037810122 | 1 |
| HTN | endometrium | *IVD* | 4.049723109 | 0.000214351 | 0.003073153 | 1 |
| HTN | endometrium | *KNSTRN* | -2.221796639 | 0.0017397 | 0.003747697 | 1 |
| HTN | endometrium | *ARPP19* | 6.894168368 | 0.000548431 | 0.003073153 | 1 |
| HTN | endometrium | *VPS13C* | 2.876218736 | 0.000479296 | 0.003073153 | 1 |
| HTN | endometrium | *CALML4* | -2.525710923 | 0.003667781 | 0.00656014 | 1 |
| HTN | endometrium | *ANAPC13* | -2.417773447 | 0.003467807 | 0.006271264 | 1 |
| HTN | endometrium | *SUMF2* | 4.736687286 | 0.00091007 | 0.003087532 | 1 |
| HTN | endometrium | *SERGEF* | 3.858031798 | 0.000114194 | 0.003017974 | 0.554067335 |
| HTN | endometrium | *DCTD* | 5.308616267 | 0.006603243 | 0.010608919 | 1 |
| HTN | endometrium | *RPAIN* | 4.5448782 | 0.021122535 | 0.029090701 | 1 |
| HTN | endometrium | *PHF20L1* | 6.750899017 | 0.001613616 | 0.003579911 | 1 |
| HTN | endometrium | *PUS7L* | 3.814383795 | 0.001380915 | 0.00327409 | 1 |
| HTN | endometrium | *AP1M2* | -3.866696551 | 0.015402343 | 0.022097033 | 1 |
| HTN | endometrium | *MTUS1* | 4.773879543 | 0.001392123 | 0.003285301 | 1 |
| HTN | endometrium | *KLK8* | -17.50543633 | 0.016761623 | 0.023800818 | 1 |
| HTN | endometrium | *RIPK3* | 6.282604109 | 2.54E-05 | 0.001541561 | 0.123324872 |
| HTN | endometrium | *ADCY4* | 4.073619053 | 2.26E-05 | 0.001451296 | 0.109515812 |
| HTN | endometrium | *BCL2L2* | 2.114754854 | 0.002804807 | 0.005334767 | 1 |
| HTN | endometrium | *DTD2* | -2.665654924 | 0.008903719 | 0.013697161 | 1 |
| HTN | endometrium | *PARP2* | 2.910563426 | 0.00103165 | 0.003095606 | 1 |
| HTN | endometrium | *MIS18BP1* | 3.29658519 | 0.001311246 | 0.003189055 | 1 |
| HTN | endometrium | *RNASE1* | 1.730841072 | 0.000806353 | 0.003073153 | 1 |
| HTN | endometrium | *EPB41L4A* | 4.313011379 | 0.000583523 | 0.003073153 | 1 |
| HTN | endometrium | *REEP5* | 6.396865408 | 0.001516558 | 0.003433662 | 1 |
| HTN | endometrium | *RHBDF2* | 3.027409454 | 0.000105244 | 0.002934745 | 0.510645571 |
| HTN | endometrium | *AANAT* | 6.346601396 | 0.017072617 | 0.024184416 | 1 |
| HTN | endometrium | *TMEM8A* | 4.857958401 | 0.000598633 | 0.003073153 | 1 |
| HTN | endometrium | *MAU2* | 4.751983776 | 0.001918493 | 0.004026181 | 1 |
| HTN | endometrium | *SHC2* | 7.543582129 | 0.003807082 | 0.006763808 | 1 |
| HTN | endometrium | *ERMARD* | 3.821554418 | 0.000879033 | 0.003084413 | 1 |
| HTN | endometrium | *PHF10* | 2.435902839 | 0.001126358 | 0.003095606 | 1 |
| HTN | endometrium | *GALNT8* | -5.324706842 | 0.010997011 | 0.016442989 | 1 |

| | | | | | | |
|---|---|---|---|---|---|---|
| HTN | endometrium | *CDC16* | 5.818234523 | 7.43E-06 | 0.000685047 | 0.036063212 |
| HTN | endometrium | *XPO7* | 5.789530671 | 0.000978315 | 0.003095606 | 1 |
| HTN | endometrium | *FAM98C* | 8.212368574 | 0.001897744 | 0.003995442 | 1 |
| HTN | endometrium | *PLVAP* | 2.755705093 | 0.002387619 | 0.004724603 | 1 |
| HTN | endometrium | *TULP4* | 5.110610828 | 0.000450731 | 0.003073153 | 1 |
| HTN | endometrium | *QRSL1* | 2.274561199 | 0.000574867 | 0.003073153 | 1 |
| HTN | endometrium | *RSPH3* | 2.823317437 | 0.002711243 | 0.00520269 | 1 |
| HTN | endometrium | *ACTN4* | 3.079885085 | 0.000562672 | 0.003073153 | 1 |
| HTN | endometrium | *STK33* | 2.836043527 | 0.000426492 | 0.003073153 | 1 |
| HTN | endometrium | *MAP1S* | 3.498196245 | 0.001148313 | 0.003095606 | 1 |
| HTN | endometrium | *PGPEP1* | 7.266961483 | 1.86E-06 | 0.000300614 | 0.009018414 |
| HTN | endometrium | *KIAA1683* | -1.732148847 | 0.004778727 | 0.008124171 | 1 |
| HTN | endometrium | *LSM4* | -2.651483756 | 0.020155049 | 0.02790876 | 1 |
| HTN | endometrium | *UBAC1* | 7.627752849 | 0.000262696 | 0.003073153 | 1 |
| HTN | endometrium | *ATXN10* | 5.463335521 | 0.001119053 | 0.003095606 | 1 |
| HTN | endometrium | *CYP2E1* | -3.947652734 | 0.010604034 | 0.015950715 | 1 |
| HTN | endometrium | *PNPLA7* | 4.491630327 | 0.000613617 | 0.003073153 | 1 |
| HTN | endometrium | *PAK4* | 4.078449731 | 0.000952812 | 0.003095606 | 1 |
| HTN | endometrium | *ZNF337* | 6.244728437 | 0.000407472 | 0.003073153 | 1 |
| HTN | endometrium | *CEP85* | 3.173986999 | 0.000595082 | 0.003073153 | 1 |
| HTN | endometrium | *CHMP2A* | 6.694202756 | 0.000232887 | 0.003073153 | 1 |
| HTN | endometrium | *NPAS1* | -3.396369875 | 0.025792 | 0.034839305 | 1 |
| HTN | endometrium | *MAP3K10* | -1.782578745 | 0.007784631 | 0.012196006 | 1 |
| HTN | endometrium | *LRRC47* | 2.595386894 | 0.001339452 | 0.003228526 | 1 |
| HTN | endometrium | *THEMIS2* | 3.42120545 | 0.000230019 | 0.003073153 | 1 |
| HTN | endometrium | *DNMT1* | 4.412934001 | 0.000593362 | 0.003073153 | 1 |
| HTN | endometrium | *HSD17B3* | 9.460079965 | 0.000643213 | 0.003073153 | 1 |
| HTN | endometrium | *SLC35D2* | 2.504358683 | 0.000919417 | 0.003087532 | 1 |
| HTN | endometrium | *ULBP2* | 35.7907922 | 0.000817471 | 0.0030808 | 1 |
| HTN | endometrium | *TTLL9* | 5.340454661 | 0.000809066 | 0.003073153 | 1 |
| HTN | endometrium | *COX4I2* | 4.377124455 | 0.005194742 | 0.00870034 | 1 |
| HTN | endometrium | *ATP6V1E1* | 4.180841504 | 0.000943521 | 0.003095606 | 1 |
| HTN | endometrium | *CHMP1A* | 6.456072702 | 0.000791332 | 0.003073153 | 1 |
| HTN | endometrium | *IDO1* | 5.221275727 | 0.001226015 | 0.003095606 | 1 |
| HTN | endometrium | *MRPS25* | 3.356954139 | 0.002894299 | 0.005468511 | 1 |
| HTN | endometrium | *GALNT15* | 2.107868256 | 0.001350626 | 0.00323814 | 1 |
| HTN | endometrium | *SLC6A6* | 2.374758537 | 0.00115043 | 0.003095606 | 1 |
| HTN | endometrium | *KCNC3* | -4.757721109 | 0.001396826 | 0.003291598 | 1 |
| HTN | endometrium | *NAPSA* | 9.120188778 | 0.000755059 | 0.003073153 | 1 |
| HTN | endometrium | *PDLIM4* | 5.913942947 | 0.000311784 | 0.003073153 | 1 |
| HTN | endometrium | *KIF3A* | 8.842565216 | 0.002736448 | 0.005238428 | 1 |
| HTN | endometrium | *ACLY* | 2.2142118 | 0.001076596 | 0.003095606 | 1 |
| HTN | endometrium | *NDUFA2* | -2.438854644 | 0.004267021 | 0.0074233 | 1 |
| HTN | endometrium | *EXOC4* | 4.161447148 | 8.36E-05 | 0.002714492 | 0.405577136 |
| HTN | endometrium | *C1orf159* | 7.027944764 | 0.000509206 | 0.003073153 | 1 |
| HTN | endometrium | *PPFIA1* | 4.400296129 | 0.000196338 | 0.003073153 | 0.952633437 |
| HTN | endometrium | *BARX1* | 11.99878181 | 0.00183558 | 0.003897694 | 1 |
| HTN | endometrium | *NINJ1* | 2.449486845 | 0.00275302 | 0.005260989 | 1 |
| HTN | endometrium | *CKMT2* | 4.310602262 | 0.004157384 | 0.007270048 | 1 |
| HTN | endometrium | *KHDRBS3* | 3.618765099 | 0.000729531 | 0.003073153 | 1 |

| | | | | | | |
|---|---|---|---|---|---|---|
| HTN | endometrium | *CHD1L* | 5.573160573 | 0.010028668 | 0.015215477 | 1 |
| HTN | endometrium | *PRKAB2* | 3.568641867 | 0.000831327 | 0.0030808 | 1 |
| HTN | endometrium | *ZNF304* | 3.575915015 | 0.003257749 | 0.005980217 | 1 |
| HTN | endometrium | *ZNF132* | 3.526998544 | 0.003705804 | 0.0066178 | 1 |
| HTN | endometrium | *RHPN2* | 2.03485595 | 0.005476063 | 0.009096151 | 1 |
| HTN | endometrium | *GCH1* | 3.756154862 | 8.79E-05 | 0.002714492 | 0.42629548 |
| HTN | endometrium | *TRIM21* | 5.133700389 | 0.001625135 | 0.003598884 | 1 |
| HTN | endometrium | *CCT6B* | 2.597167679 | 0.037131525 | 0.048587422 | 1 |
| HTN | endometrium | *CNGA4* | 4.619082278 | 0.004743063 | 0.008077266 | 1 |
| HTN | endometrium | *TIMM10B* | 8.303450731 | 6.91E-05 | 0.002466751 | 0.335478175 |
| HTN | endometrium | *IMMT* | 3.854069514 | 0.002689751 | 0.005170632 | 1 |
| HTN | endometrium | *CARD6* | 4.478616514 | 0.000717451 | 0.003073153 | 1 |
| HTN | endometrium | *MYBBP1A* | 4.048272836 | 0.000537309 | 0.003073153 | 1 |
| HTN | endometrium | *RPA1* | 4.15491104 | 0.004958387 | 0.008363185 | 1 |
| HTN | endometrium | *SEC61G* | 5.539940418 | 0.020738465 | 0.028626752 | 1 |
| HTN | endometrium | *FIGNL1* | -1.696176621 | 0.014676467 | 0.021162026 | 1 |
| HTN | endometrium | *EIF5A* | 2.646980295 | 0.024509837 | 0.03324622 | 1 |
| HTN | endometrium | *REEP2* | -0.859170628 | 0.002768115 | 0.005283594 | 1 |
| HTN | endometrium | *PRMT7* | 2.469567778 | 0.01191182 | 0.017583253 | 1 |
| HTN | endometrium | *MUTYH* | 3.450135433 | 0.000175808 | 0.003073153 | 0.853020889 |
| HTN | endometrium | *CTNNBL1* | 4.825966543 | 0.000731239 | 0.003073153 | 1 |
| HTN | endometrium | *OSER1* | 4.047037161 | 0.001254074 | 0.003122339 | 1 |
| HTN | endometrium | *SERINC3* | 2.429533179 | 0.001793208 | 0.003832883 | 1 |
| HTN | endometrium | *DMGDH* | 1.873936593 | 0.001880147 | 0.003966721 | 1 |
| HTN | endometrium | *AP3B1* | 5.607221264 | 0.002283949 | 0.004572854 | 1 |
| HTN | endometrium | *ZBED3* | 2.94395778 | 0.000916867 | 0.003087532 | 1 |
| HTN | endometrium | *FBXO44* | 3.446696235 | 0.001164789 | 0.003095606 | 1 |
| HTN | endometrium | *CASP9* | 1.702636716 | 0.035098142 | 0.046137506 | 1 |
| HTN | endometrium | *ATP8A2* | -1.678057921 | 0.005423764 | 0.009018541 | 1 |
| HTN | endometrium | *ALOX5AP* | 2.788488094 | 0.00120617 | 0.003095606 | 1 |
| HTN | endometrium | *SCO1* | 5.943902593 | 0.000375269 | 0.003073153 | 1 |
| HTN | endometrium | *MPRIP* | 4.642842438 | 0.000561593 | 0.003073153 | 1 |
| HTN | endometrium | *DSTYK* | 6.703220509 | 0.000867872 | 0.003084413 | 1 |
| HTN | endometrium | *CCNA1* | 26.57959652 | 0.004823531 | 0.008188864 | 1 |
| HTN | endometrium | *COG6* | 5.38254301 | 0.004189639 | 0.007314412 | 1 |
| HTN | endometrium | *EPSTI1* | 2.781995549 | 0.000608215 | 0.003073153 | 1 |
| HTN | endometrium | *BTBD2* | 7.903337153 | 0.000510188 | 0.003073153 | 1 |
| HTN | endometrium | *CSNK1G2* | -1.971092408 | 0.015332362 | 0.022003142 | 1 |
| HTN | endometrium | *CNDP2* | 3.755237629 | 0.000834715 | 0.0030808 | 1 |
| HTN | endometrium | *PDZD2* | 5.025183435 | 0.002943057 | 0.00552193 | 1 |
| HTN | endometrium | *GSTT2B* | -1.936338502 | 0.028817618 | 0.038595231 | 1 |
| HTN | endometrium | *FAM83F* | 16.95131109 | 0.001352282 | 0.003239618 | 1 |
| HTN | endometrium | *GIMAP4* | 2.316963434 | 0.002040752 | 0.004197427 | 1 |
| HTN | endometrium | *ADCK2* | 3.59027328 | 0.000235506 | 0.003073153 | 1 |
| HTN | endometrium | *IPO8* | 5.928974962 | 0.021486953 | 0.029517184 | 1 |
| HTN | endometrium | *SPINK5* | 5.47051256 | 0.001480344 | 0.003394073 | 1 |
| HTN | endometrium | *IMPA1* | 3.022882378 | 3.07E-05 | 0.00179363 | 0.148871287 |
| HTN | endometrium | *LRRCC1* | 6.487016705 | 0.013337081 | 0.019485552 | 1 |
| HTN | endometrium | *RRAS2* | 4.927322748 | 0.00278373 | 0.005309221 | 1 |
| HTN | endometrium | *CATSPERB* | 1.978873505 | 0.001210322 | 0.003095606 | 1 |

| | | | | | | |
|---|---|---|---|---|---|---|
| HTN | endometrium | *TTC9* | 3.799505262 | 0.001157415 | 0.003095606 | 1 |
| HTN | endometrium | *ELP3* | 4.731954223 | 0.000960191 | 0.003095606 | 1 |
| HTN | endometrium | *PEBP4* | 2.954450015 | 0.001045307 | 0.003095606 | 1 |
| HTN | endometrium | *CTIF* | 2.74264034 | 0.000383533 | 0.003073153 | 1 |
| HTN | endometrium | *MRO* | 5.888357103 | 0.007942962 | 0.012415997 | 1 |
| HTN | endometrium | *THUMPD3* | -2.138258243 | 0.01408397 | 0.020398634 | 1 |
| HTN | endometrium | *CNTN6* | 4.254711606 | 0.001052859 | 0.003095606 | 1 |
| HTN | endometrium | *CHL1* | 4.006372581 | 0.000212059 | 0.003073153 | 1 |
| HTN | endometrium | *GSTM5* | -4.031306873 | 0.008796812 | 0.013562457 | 1 |
| HTN | endometrium | *GSTM3* | -1.424373799 | 0.024870359 | 0.033688157 | 1 |
| HTN | endometrium | *VAV3* | 1.506867984 | 0.005990843 | 0.009777185 | 1 |
| HTN | endometrium | *PSRC1* | -4.019972653 | 0.015562719 | 0.022294158 | 1 |
| HTN | endometrium | *TRIM45* | -4.115499771 | 0.011447236 | 0.017021756 | 1 |
| HTN | endometrium | *CEPT1* | 4.650343425 | 0.000424795 | 0.003073153 | 1 |
| HTN | endometrium | *NAPG* | 6.123748294 | 0.00077345 | 0.003073153 | 1 |
| HTN | endometrium | *PPHLN1* | 3.775604061 | 0.000577949 | 0.003073153 | 1 |
| HTN | endometrium | *FKBP11* | 2.978229885 | 0.000980618 | 0.003095606 | 1 |
| HTN | endometrium | *YWHAQ* | 3.34978953 | 0.000751082 | 0.003073153 | 1 |
| HTN | endometrium | *KIDINS220* | 5.609535025 | 0.001453144 | 0.003357454 | 1 |
| HTN | endometrium | *LPIN1* | 4.150283582 | 0.002026828 | 0.004179419 | 1 |
| HTN | endometrium | *NARS* | 6.195485483 | 0.000949702 | 0.003095606 | 1 |
| HTN | endometrium | *KIAA1468* | 5.651673092 | 0.000736424 | 0.003073153 | 1 |
| HTN | endometrium | *IL15RA* | 4.319135633 | 0.00133227 | 0.003214407 | 1 |
| HTN | endometrium | *SOX5* | 2.93742167 | 0.000400408 | 0.003073153 | 1 |
| HTN | endometrium | *KLRD1* | 5.284161828 | 0.000487767 | 0.003073153 | 1 |
| HTN | endometrium | *ACP2* | 20.07237111 | 2.20E-10 | 2.13E-07 | 1.07E-06 |
| HTN | endometrium | *ZCCHC11* | 3.240401149 | 0.000342773 | 0.003073153 | 1 |
| HTN | endometrium | *PRPF38A* | 8.691225358 | 0.00038364 | 0.003073153 | 1 |
| HTN | endometrium | *ELP2* | 4.337279064 | 0.00055898 | 0.003073153 | 1 |
| HTN | endometrium | *TPGS2* | 5.996898823 | 0.000324031 | 0.003073153 | 1 |
| HTN | endometrium | *TIMM10* | -1.885766814 | 0.008306173 | 0.012900625 | 1 |
| HTN | endometrium | *DHX34* | 7.386013367 | 0.000736048 | 0.003073153 | 1 |
| HTN | endometrium | *FADS2* | -2.156797787 | 0.013753919 | 0.019980244 | 1 |
| HTN | endometrium | *CLOCK* | 4.243553445 | 0.000573381 | 0.003073153 | 1 |
| HTN | endometrium | *GGACT* | 6.618557362 | 0.00078264 | 0.003073153 | 1 |
| HTN | endometrium | *CLDN10* | 7.313013527 | 7.82E-05 | 0.002708518 | 0.379259268 |
| HTN | endometrium | *ARGLU1* | 3.342989461 | 0.000508037 | 0.003073153 | 1 |
| HTN | endometrium | *ERCC5* | -3.811536093 | 0.009603275 | 0.014629542 | 1 |
| HTN | endometrium | *KLB* | 1.9506503 | 0.01006038 | 0.01525882 | 1 |
| HTN | endometrium | *WDR36* | 6.469394483 | 0.000592951 | 0.003073153 | 1 |
| HTN | endometrium | *OSTF1* | 3.255339125 | 0.000697501 | 0.003073153 | 1 |
| HTN | endometrium | *UBQLN1* | 4.895941352 | 0.000490904 | 0.003073153 | 1 |
| HTN | endometrium | *GOLM1* | 2.345482438 | 0.007056689 | 0.011233286 | 1 |
| HTN | endometrium | *ADAM19* | 2.499348625 | 0.000488475 | 0.003073153 | 1 |
| HTN | endometrium | *MSI1* | 7.453270363 | 0.0003329 | 0.003073153 | 1 |
| HTN | endometrium | *P2RX4* | 2.368744804 | 0.001171014 | 0.003095606 | 1 |
| HTN | endometrium | *DTX1* | 2.273289903 | 0.001146394 | 0.003095606 | 1 |
| HTN | endometrium | *DMTF1* | 4.006298548 | 0.000798664 | 0.003073153 | 1 |
| HTN | endometrium | *TMEM60* | 3.5453499 | 0.000633043 | 0.003073153 | 1 |
| HTN | endometrium | *POM121C* | 2.682707333 | 0.007111226 | 0.011305265 | 1 |

| | | | | | | |
|---|---|---|---|---|---|---|
| HTN | endometrium | *SRPK2* | 4.777122102 | 0.000977657 | 0.003095606 | 1 |
| HTN | endometrium | *TES* | 2.608701988 | 0.003132459 | 0.005798815 | 1 |
| HTN | endometrium | *NT5E* | 2.66664011 | 0.000484134 | 0.003073153 | 1 |
| HTN | endometrium | *AKIRIN2* | 3.957033914 | 0.000616067 | 0.003073153 | 1 |
| HTN | endometrium | *MAP3K7* | 7.104191756 | 0.002267442 | 0.004549888 | 1 |
| HTN | endometrium | *PRR5L* | 6.597163695 | 0.001616126 | 0.003583842 | 1 |
| HTN | endometrium | *PHF21A* | 4.465523337 | 0.00064323 | 0.003073153 | 1 |
| HTN | endometrium | *CAPRIN1* | 6.932531244 | 0.00216654 | 0.004390999 | 1 |
| HTN | endometrium | *ATP5G2* | 6.945552025 | 0.001212261 | 0.003095606 | 1 |
| HTN | endometrium | *B4GALNT1* | 15.501899 | 0.0003731 | 0.003073153 | 1 |
| HTN | endometrium | *KRT7* | 4.615694147 | 0.00147323 | 0.003389336 | 1 |
| HTN | endometrium | *SLC26A10* | 10.81419765 | 0.004231378 | 0.007371866 | 1 |
| HTN | endometrium | *MAP7* | 4.218468107 | 0.007604915 | 0.011968552 | 1 |
| HTN | endometrium | *NHSL1* | 1.724281366 | 0.001900961 | 0.003999768 | 1 |
| HTN | endometrium | *AHI1* | 3.646136696 | 0.005560761 | 0.009202188 | 1 |
| HTN | endometrium | *MICAL1* | -1.208157149 | 0.033218321 | 0.043869159 | 1 |
| HTN | endometrium | *CCT7* | 3.432913355 | 0.00261044 | 0.005058322 | 1 |
| HTN | endometrium | *USP15* | 3.467471089 | 0.000506349 | 0.003073153 | 1 |
| HTN | endometrium | *MDM2* | 6.618542237 | 0.000427038 | 0.003073153 | 1 |
| HTN | endometrium | *KLHL36* | 2.221169595 | 0.0009607 | 0.003095606 | 1 |
| HTN | endometrium | *MPHOSPH6* | 2.796402401 | 0.000929949 | 0.003093357 | 1 |
| HTN | endometrium | *CCDC102A* | 8.239012718 | 0.001080954 | 0.003095606 | 1 |
| HTN | endometrium | *AGT* | 6.771321163 | 1.62E-06 | 0.000271627 | 0.007877188 |
| HTN | endometrium | *ZNF670* | 7.508736808 | 0.001975861 | 0.004106134 | 1 |
| HTN | endometrium | *CAPN9* | -1.556872659 | 0.00271179 | 0.00520269 | 1 |
| HTN | endometrium | *NTPCR* | 8.388216354 | 0.005932307 | 0.009697963 | 1 |
| HTN | endometrium | *PIGC* | 6.138414185 | 0.000843292 | 0.0030808 | 1 |
| HTN | endometrium | *SP110* | 4.965937045 | 0.000374405 | 0.003073153 | 1 |
| HTN | endometrium | *MRPL44* | 3.379942918 | 0.001026636 | 0.003095606 | 1 |
| HTN | endometrium | *USP37* | 5.38590259 | 8.81E-05 | 0.002714492 | 0.427416265 |
| HTN | endometrium | *SLC19A3* | 2.708328175 | 0.001208275 | 0.003095606 | 1 |
| HTN | endometrium | *TMBIM1* | 6.532755835 | 0.000639196 | 0.003073153 | 1 |
| HTN | endometrium | *TSGA10* | 1.816792531 | 0.025943776 | 0.035034568 | 1 |
| HTN | endometrium | *GCC2* | 5.04918801 | 0.000130281 | 0.003073153 | 0.632123107 |
| HTN | endometrium | *MRPS9* | 5.46668219 | 6.36E-05 | 0.002410623 | 0.308725863 |
| HTN | endometrium | *ANKRD36* | 5.340813351 | 0.000658367 | 0.003073153 | 1 |
| HTN | endometrium | *EPC2* | 5.457307661 | 0.000572262 | 0.003073153 | 1 |
| HTN | endometrium | *APPL2* | 5.129607315 | 0.001632179 | 0.003601334 | 1 |
| HTN | endometrium | *DRAM1* | 3.410135802 | 0.000765706 | 0.003073153 | 1 |
| HTN | endometrium | *SLC41A2* | 2.388122852 | 0.002009166 | 0.004154512 | 1 |
| HTN | endometrium | *NEK3* | 4.123154952 | 0.005622583 | 0.009285491 | 1 |
| HTN | endometrium | *RNASEH2B* | 2.856647225 | 0.00235515 | 0.004672314 | 1 |
| HTN | endometrium | *TBC1D4* | 3.046127088 | 0.002016403 | 0.004161458 | 1 |
| HTN | endometrium | *SUCLA2* | 5.993541384 | 0.000691521 | 0.003073153 | 1 |
| HTN | endometrium | *RCBTB1* | 2.031424113 | 0.006273763 | 0.010160313 | 1 |
| HTN | endometrium | *RCBTB2* | 2.614331889 | 0.000892311 | 0.003084413 | 1 |
| HTN | endometrium | *SETDB2* | 4.033669572 | 0.000845749 | 0.0030808 | 1 |
| HTN | endometrium | *SCRN1* | 3.637156451 | 0.000747048 | 0.003073153 | 1 |
| HTN | endometrium | *CHST12* | 3.890288791 | 0.002322064 | 0.004630766 | 1 |
| HTN | endometrium | *KDELR2* | 3.745057806 | 0.000782878 | 0.003073153 | 1 |

| | | | | | | |
|---|---|---|---|---|---|---|
| HTN | endometrium | *NUPL2* | 5.163685411 | 0.000568264 | 0.003073153 | 1 |
| HTN | endometrium | *ZDHHC4* | -2.593592578 | 0.017381164 | 0.02455836 | 1 |
| HTN | endometrium | *AOAH* | 1.687091627 | 0.001353707 | 0.003239618 | 1 |
| HTN | endometrium | *TBRG4* | 3.375214311 | 0.000887742 | 0.003084413 | 1 |
| HTN | endometrium | *MTHFS* | 3.528200065 | 0.00226049 | 0.004541573 | 1 |
| HTN | endometrium | *NMT1* | 8.94794877 | 4.61E-09 | 2.03E-06 | 2.24E-05 |
| HTN | endometrium | *MYCBPAP* | 3.016563479 | 0.002953453 | 0.005537153 | 1 |
| HTN | endometrium | *RTP4* | 4.749898119 | 0.013765485 | 0.019989471 | 1 |
| HTN | endometrium | *NDUFB5* | 3.222196528 | 0.001650694 | 0.00362734 | 1 |
| HTN | endometrium | *SCN2A* | 4.413644635 | 0.000219662 | 0.003073153 | 1 |
| HTN | endometrium | *RPS6KC1* | 4.955170409 | 0.005150365 | 0.008640931 | 1 |
| HTN | endometrium | *SMPD4* | 4.261895444 | 0.000212042 | 0.003073153 | 1 |
| HTN | endometrium | *IMP4* | 7.179185631 | 0.004389818 | 0.007596076 | 1 |
| HTN | endometrium | *ODF2* | 6.69835073 | 0.000974764 | 0.003095606 | 1 |
| HTN | endometrium | *TOR1B* | 8.726753141 | 0.0003216 | 0.003073153 | 1 |
| HTN | endometrium | *SLC2A8* | 4.642255187 | 0.00056035 | 0.003073153 | 1 |
| HTN | endometrium | *ZNF189* | 9.062675041 | 0.000981787 | 0.003095606 | 1 |
| HTN | endometrium | *STX17* | 6.725719487 | 0.000621329 | 0.003073153 | 1 |
| HTN | endometrium | *ANP32B* | 7.255485883 | 0.006104272 | 0.009940781 | 1 |
| HTN | endometrium | *DERL1* | 9.081402097 | 0.00153635 | 0.003470431 | 1 |
| HTN | endometrium | *NOV* | 4.681583386 | 0.001380535 | 0.00327409 | 1 |
| HTN | endometrium | *IL11RA* | 2.222628539 | 0.019559409 | 0.027200416 | 1 |
| HTN | endometrium | *APTX* | 5.19184232 | 0.00081853 | 0.0030808 | 1 |
| HTN | endometrium | *TMEM8B* | 4.384462421 | 0.000152292 | 0.003073153 | 0.738922274 |
| HTN | endometrium | *GRHPR* | -1.845368994 | 0.021182277 | 0.029156428 | 1 |
| HTN | endometrium | *DENND4C* | 2.575894232 | 0.001232963 | 0.003103164 | 1 |
| HTN | endometrium | *GMPR* | 6.632468608 | 0.00056771 | 0.003073153 | 1 |
| HTN | endometrium | *FRS3* | 3.135652422 | 0.001089362 | 0.003095606 | 1 |
| HTN | endometrium | *CAPN11* | 4.461026226 | 0.004285745 | 0.007447863 | 1 |
| HTN | endometrium | *LRRC1* | 3.68081862 | 0.010868245 | 0.016275533 | 1 |
| HTN | endometrium | *RIPK1* | 4.235594892 | 0.000366384 | 0.003073153 | 1 |
| HTN | endometrium | *TCF19* | 7.962436369 | 0.002309304 | 0.004614804 | 1 |
| HTN | endometrium | *FLOT1* | 2.773540903 | 0.002720773 | 0.0052158 | 1 |
| HTN | endometrium | *TPMT* | 2.844595432 | 0.000675517 | 0.003073153 | 1 |
| HTN | endometrium | *NRM* | 9.193223747 | 0.001110724 | 0.003095606 | 1 |
| HTN | endometrium | *VARS2* | 3.171270436 | 6.81E-05 | 0.00244793 | 0.330470529 |
| HTN | endometrium | *TLR2* | 3.600999336 | 0.00261395 | 0.005063028 | 1 |
| HTN | endometrium | *MYO7A* | 3.146052733 | 0.003286569 | 0.006027506 | 1 |
| HTN | endometrium | *NUMA1* | 3.553421292 | 0.00080696 | 0.003073153 | 1 |
| HTN | endometrium | *SYTL2* | 1.801776499 | 0.005826054 | 0.009550005 | 1 |
| HTN | endometrium | *RAB30* | 2.543350072 | 0.001122348 | 0.003095606 | 1 |
| HTN | endometrium | *PRCP* | 4.960610674 | 0.013881514 | 0.02014148 | 1 |
| HTN | endometrium | *SULF1* | 2.22583476 | 0.002564036 | 0.00499426 | 1 |
| HTN | endometrium | *NEK1* | 3.691032863 | 0.000656905 | 0.003073153 | 1 |
| HTN | endometrium | *TRPC6* | 6.893421676 | 0.012815097 | 0.018790828 | 1 |
| HTN | endometrium | *MMP27* | 10.90116913 | 0.029083199 | 0.038895171 | 1 |
| HTN | endometrium | *POU2F3* | -3.485183194 | 0.004371477 | 0.007572441 | 1 |
| HTN | endometrium | *FDX1* | 4.664563256 | 0.000291724 | 0.003073153 | 1 |
| HTN | endometrium | *FXYD2* | 5.777609606 | 0.001566748 | 0.003508011 | 1 |
| HTN | endometrium | *MAP2K5* | 3.689484494 | 0.000343559 | 0.003073153 | 1 |

| | | | | | | |
|---|---|---|---|---|---|---|
| HTN | endometrium | *NUSAP1* | 17.20964848 | 2.05E-07 | 5.25E-05 | 0.000996688 |
| HTN | endometrium | *NDUFAF1* | -1.943312108 | 0.017059852 | 0.02418183 | 1 |
| HTN | endometrium | *PARP6* | 6.521135733 | 0.001797195 | 0.003839714 | 1 |
| HTN | endometrium | *ADAM10* | 2.526320235 | 0.00046474 | 0.003073153 | 1 |
| HTN | endometrium | *ZNF280D* | 4.461635641 | 0.000607134 | 0.003073153 | 1 |
| HTN | endometrium | *SPTBN5* | 2.041127261 | 0.001736948 | 0.003747526 | 1 |
| HTN | endometrium | *SLC44A5* | 7.594263241 | 0.002678514 | 0.005161298 | 1 |
| HTN | endometrium | *DBT* | 7.882669577 | 0.001677227 | 0.003664172 | 1 |
| HTN | endometrium | *RTCA* | 4.844239263 | 0.000305937 | 0.003073153 | 1 |
| HTN | endometrium | *ADCY3* | 2.828985055 | 0.000315961 | 0.003073153 | 1 |
| HTN | endometrium | *DYNC2LI1* | 6.281764756 | 1.73E-05 | 0.001262767 | 0.083735165 |
| HTN | endometrium | *THUMPD2* | 5.781307595 | 0.001319321 | 0.003203877 | 1 |
| HTN | endometrium | *ACTR2* | 3.325510006 | 0.000594873 | 0.003073153 | 1 |
| HTN | endometrium | *ATRAID* | 4.742776938 | 0.003097303 | 0.005744693 | 1 |
| HTN | endometrium | *CENPO* | -1.153827112 | 0.006383512 | 0.010313952 | 1 |
| HTN | endometrium | *LRPPRC* | 6.126966435 | 0.004091657 | 0.007180007 | 1 |
| HTN | endometrium | *DTNB* | 7.340746897 | 0.000228111 | 0.003073153 | 1 |
| HTN | endometrium | *CYP2C8* | 12.24247807 | 0.036718891 | 0.048125354 | 1 |
| HTN | endometrium | *MYOF* | 2.486039596 | 0.024023776 | 0.032650801 | 1 |
| HTN | endometrium | *BTBD16* | 4.364304405 | 0.002837817 | 0.00538486 | 1 |
| HTN | endometrium | *TACC2* | 3.972850153 | 0.001730077 | 0.003742526 | 1 |
| HTN | endometrium | *ARL3* | 3.375269014 | 8.79E-05 | 0.002714492 | 0.426418489 |
| HTN | endometrium | *PLCE1* | 2.147791038 | 0.001810215 | 0.003859035 | 1 |
| HTN | endometrium | *RPS24* | 4.8984349 | 0.000922516 | 0.003087532 | 1 |
| HTN | endometrium | *ATIC* | 2.787137494 | 0.000657597 | 0.003073153 | 1 |
| HTN | endometrium | *SMARCAL1* | 2.721461229 | 0.000828082 | 0.0030808 | 1 |
| HTN | endometrium | *BARD1* | 3.103734239 | 0.001634509 | 0.003601562 | 1 |
| HTN | endometrium | *STAT4* | 2.195836491 | 0.001001773 | 0.003095606 | 1 |
| HTN | endometrium | *SSB* | 3.549970288 | 0.002431886 | 0.004788762 | 1 |
| HTN | endometrium | *FASTKD1* | 5.708126424 | 0.002011873 | 0.004157414 | 1 |
| HTN | endometrium | *ITGAV* | 6.101296399 | 0.005900869 | 0.009656329 | 1 |
| HTN | endometrium | *SLC35A5* | 3.378390972 | 0.001043502 | 0.003095606 | 1 |
| HTN | endometrium | *SENP7* | 3.168104763 | 0.001091016 | 0.003095606 | 1 |
| HTN | endometrium | *COX17* | 3.734876198 | 0.000841466 | 0.0030808 | 1 |
| HTN | endometrium | *SPPL2A* | 3.597666921 | 0.01524378 | 0.021882492 | 1 |
| HTN | endometrium | *PPCDC* | 6.341199068 | 0.000271775 | 0.003073153 | 1 |
| HTN | endometrium | *SEMA7A* | 6.878539514 | 0.005659173 | 0.009329204 | 1 |
| HTN | endometrium | *KIAA1109* | 4.5228842 | 0.000631173 | 0.003073153 | 1 |
| HTN | endometrium | *MMRN1* | 1.561671489 | 0.004993169 | 0.008412103 | 1 |
| HTN | endometrium | *PRDM5* | 7.181888074 | 0.003755754 | 0.006689764 | 1 |
| HTN | endometrium | *NAAA* | 3.023204161 | 0.00151748 | 0.003434147 | 1 |
| HTN | endometrium | *NUP54* | 5.188629317 | 0.000617448 | 0.003073153 | 1 |
| HTN | endometrium | *CNOT6L* | 3.004273584 | 0.000471423 | 0.003073153 | 1 |
| HTN | endometrium | *ANXA3* | 2.012359266 | 0.000837067 | 0.0030808 | 1 |
| HTN | endometrium | *PPA2* | 6.495228588 | 0.000776211 | 0.003073153 | 1 |
| HTN | endometrium | *ENPEP* | -1.716611819 | 0.000701121 | 0.003073153 | 1 |
| HTN | endometrium | *PAPSS1* | 5.342591798 | 0.00034068 | 0.003073153 | 1 |
| HTN | endometrium | *FBN2* | 3.838445644 | 3.69E-07 | 7.82E-05 | 0.001791088 |
| HTN | endometrium | *PARVG* | 2.715538506 | 0.003851332 | 0.006827425 | 1 |
| HTN | endometrium | *KIF21A* | 6.163467232 | 0.00442881 | 0.007639028 | 1 |

| | | | | | | |
|---|---|---|---|---|---|---|
| HTN | endometrium | *ALG10* | -4.625515683 | 0.006440621 | 0.010395839 | 1 |
| HTN | endometrium | *C1RL* | 3.31640311 | 0.008319646 | 0.012913282 | 1 |
| HTN | endometrium | *CLSTN3* | -2.752752452 | 0.018042645 | 0.025345372 | 1 |
| HTN | endometrium | *KLRG1* | 1.698008493 | 0.00463268 | 0.007925868 | 1 |
| HTN | endometrium | *PEX5* | -1.961925603 | 0.010324564 | 0.015599253 | 1 |
| HTN | endometrium | *LGR5* | 4.662451475 | 0.035048708 | 0.046085727 | 1 |
| HTN | endometrium | *POC1B* | 6.941970201 | 0.000662691 | 0.003073153 | 1 |
| HTN | endometrium | *AMDHD1* | -1.255485599 | 0.000983874 | 0.003095606 | 1 |
| HTN | endometrium | *SDSL* | 4.964716274 | 0.001559069 | 0.003499546 | 1 |
| HTN | endometrium | *GLTP* | 2.896126573 | 0.001152837 | 0.003095606 | 1 |
| HTN | endometrium | *FOXN4* | 3.580201636 | 0.012588906 | 0.018470326 | 1 |
| HTN | endometrium | *MTMR6* | 2.540329261 | 0.001011501 | 0.003095606 | 1 |
| HTN | endometrium | *CCDC65* | 8.052942594 | 0.000656954 | 0.003073153 | 1 |
| HTN | endometrium | *ACVRL1* | 7.844617163 | 0.000535073 | 0.003073153 | 1 |
| HTN | endometrium | *BRCA2* | 6.618268805 | 0.001941755 | 0.004057448 | 1 |
| HTN | endometrium | *KANSL2* | 5.544383194 | 0.001122376 | 0.003095606 | 1 |
| HTN | endometrium | *CERS5* | 3.828688674 | 0.0005155 | 0.003073153 | 1 |
| HTN | endometrium | *LMBR1L* | 5.863427752 | 0.030429141 | 0.040538768 | 1 |
| HTN | endometrium | *ESYT1* | 5.994276842 | 0.000224198 | 0.003073153 | 1 |
| HTN | endometrium | *ZNF740* | 6.316267261 | 0.011031303 | 0.016484369 | 1 |
| HTN | endometrium | *WDFY2* | 2.567712083 | 0.000854072 | 0.0030808 | 1 |
| HTN | endometrium | *RB1* | 4.748978065 | 0.000355736 | 0.003073153 | 1 |
| HTN | endometrium | *VPS33A* | -3.33858837 | 0.011644791 | 0.017283734 | 1 |
| HTN | endometrium | *DIAPH3* | 4.120563038 | 0.007239194 | 0.011498265 | 1 |
| HTN | endometrium | *GRTP1* | 4.106821637 | 0.003393758 | 0.006183445 | 1 |
| HTN | endometrium | *NOVA1* | 3.256718926 | 2.30E-05 | 0.001451296 | 0.111710359 |
| HTN | endometrium | *FITM1* | 21.70967433 | 3.26E-05 | 0.001881463 | 0.158042887 |
| HTN | endometrium | *TMX1* | 6.617688193 | 0.000638525 | 0.003073153 | 1 |
| HTN | endometrium | *RTN1* | 1.852515232 | 0.00012844 | 0.003073153 | 0.623191701 |
| HTN | endometrium | *C14orf37* | 4.834689523 | 0.000641978 | 0.003073153 | 1 |
| HTN | endometrium | *RAB15* | 6.051677141 | 0.000655557 | 0.003073153 | 1 |
| HTN | endometrium | *EFCAB11* | 4.337109106 | 0.007564938 | 0.011924977 | 1 |
| HTN | endometrium | *PTGR2* | 2.361030415 | 0.000575937 | 0.003073153 | 1 |
| HTN | endometrium | *SLC24A4* | 3.645678561 | 0.001057894 | 0.003095606 | 1 |
| HTN | endometrium | *C14orf79* | -2.168492173 | 0.011732996 | 0.017404004 | 1 |
| HTN | endometrium | *WARS* | 8.486486293 | 0.004036139 | 0.007092437 | 1 |
| HTN | endometrium | *SERF2* | 6.463048806 | 0.00173941 | 0.003747697 | 1 |
| HTN | endometrium | *ZSCAN29* | 3.423406474 | 0.003153332 | 0.005828559 | 1 |
| HTN | endometrium | *UBE2Q2* | 4.189269444 | 0.000229368 | 0.003073153 | 1 |
| HTN | endometrium | *HMG20A* | 3.503227951 | 0.00059051 | 0.003073153 | 1 |
| HTN | endometrium | *PIF1* | 9.952217036 | 0.003377112 | 0.006160056 | 1 |
| HTN | endometrium | *CYP11A1* | 3.65459832 | 0.015829472 | 0.022642864 | 1 |
| HTN | endometrium | *BBS4* | 4.388302518 | 0.000981047 | 0.003095606 | 1 |
| HTN | endometrium | *ULK3* | 5.036889314 | 0.000221344 | 0.003073153 | 1 |
| HTN | endometrium | *CELF6* | 26.91333595 | 0.000844244 | 0.0030808 | 1 |
| HTN | endometrium | *HAPLN3* | -1.463133499 | 0.0015614 | 0.003499546 | 1 |
| HTN | endometrium | *POLG* | 5.101334351 | 0.003423049 | 0.006218133 | 1 |
| HTN | endometrium | *MFGE8* | 5.999780383 | 0.00038182 | 0.003073153 | 1 |
| HTN | endometrium | *UNC45A* | 2.240405484 | 0.001780948 | 0.003810036 | 1 |
| HTN | endometrium | *IQGAP1* | 2.952314259 | 0.001345699 | 0.00323715 | 1 |

| | | | | | | |
|---|---|---|---|---|---|---|
| HTN | endometrium | *CRTC3* | 3.004589955 | 0.00095497 | 0.003095606 | 1 |
| HTN | endometrium | *ITGAX* | 3.760060846 | 0.000904125 | 0.003087532 | 1 |
| HTN | endometrium | *TGFB1I1* | -1.504810916 | 0.00538607 | 0.008968157 | 1 |
| HTN | endometrium | *DHX38* | 2.768118524 | 0.001002905 | 0.003095606 | 1 |
| HTN | endometrium | *NUDT7* | 6.174693064 | 0.000532804 | 0.003073153 | 1 |
| HTN | endometrium | *GCSH* | 2.924799639 | 0.009306014 | 0.014248274 | 1 |
| HTN | endometrium | *ZCCHC14* | 3.735861763 | 0.000508498 | 0.003073153 | 1 |
| HTN | endometrium | *TLDC1* | 6.956974149 | 0.0005876 | 0.003073153 | 1 |
| HTN | endometrium | *RPL3L* | 16.45443857 | 0.000145448 | 0.003073153 | 0.705714066 |
| HTN | endometrium | *TIGD7* | 2.659340651 | 0.002330669 | 0.004638394 | 1 |
| HTN | endometrium | *DEF8* | 4.022264568 | 1.82E-05 | 0.001270304 | 0.08847776 |
| HTN | endometrium | *TCF25* | 3.32372679 | 0.000175094 | 0.003073153 | 0.849557212 |
| HTN | endometrium | *GALNS* | 7.299525442 | 0.011254914 | 0.016766608 | 1 |
| HTN | endometrium | *GAS8* | 6.514398075 | 0.005699492 | 0.009377393 | 1 |
| HTN | endometrium | *NCOR1* | 4.366512757 | 0.000538422 | 0.003073153 | 1 |
| HTN | endometrium | *KSR1* | 3.964452833 | 0.00060419 | 0.003073153 | 1 |
| HTN | endometrium | *GFOD2* | 2.17449287 | 0.002414574 | 0.004764341 | 1 |
| HTN | endometrium | *PRPSAP2* | 4.435288701 | 0.000850301 | 0.0030808 | 1 |
| HTN | endometrium | *MYO19* | 4.807595226 | 8.18E-05 | 0.002714492 | 0.397045309 |
| HTN | endometrium | *RASL10B* | 8.447584049 | 0.017565834 | 0.024761449 | 1 |
| HTN | endometrium | *UNC45B* | 15.95635567 | 0.002179196 | 0.004414806 | 1 |
| HTN | endometrium | *C17orf80* | 2.904820111 | 0.001849608 | 0.003913779 | 1 |
| HTN | endometrium | *SGSM2* | 7.861505991 | 0.002076421 | 0.004247384 | 1 |
| HTN | endometrium | *SSH2* | 4.212624826 | 0.000648732 | 0.003073153 | 1 |
| HTN | endometrium | *RHBDL3* | -1.823863931 | 0.003762854 | 0.006699951 | 1 |
| HTN | endometrium | *CLTC* | 3.917725492 | 0.001501742 | 0.003419265 | 1 |
| HTN | endometrium | *IMPA2* | 2.407393771 | 0.00087867 | 0.003084413 | 1 |
| HTN | endometrium | *GNAL* | 4.484106353 | 0.000616249 | 0.003073153 | 1 |
| HTN | endometrium | *RPRD1A* | 7.198612921 | 0.00048526 | 0.003073153 | 1 |
| HTN | endometrium | *GALNT1* | 3.338302422 | 0.000534923 | 0.003073153 | 1 |
| HTN | endometrium | *C18orf8* | -5.765985045 | 0.016196156 | 0.023106072 | 1 |
| HTN | endometrium | *NPC1* | 4.642896332 | 0.004041149 | 0.00709651 | 1 |
| HTN | endometrium | *SLC14A1* | 2.637828636 | 0.005706891 | 0.009386385 | 1 |
| HTN | endometrium | *CCDC40* | 2.01192803 | 0.004514799 | 0.007758455 | 1 |
| HTN | endometrium | *TMC6* | 2.593880953 | 0.001125097 | 0.003095606 | 1 |
| HTN | endometrium | *CARD14* | -2.125488481 | 0.019309353 | 0.026883496 | 1 |
| HTN | endometrium | *TBCD* | 4.842877539 | 0.000436423 | 0.003073153 | 1 |
| HTN | endometrium | *FN3KRP* | 1.86001547 | 0.007109449 | 0.011305265 | 1 |
| HTN | endometrium | *CBX8* | 4.811311575 | 0.033364216 | 0.04403786 | 1 |
| HTN | endometrium | *SECTM1* | -1.923114743 | 0.010656172 | 0.016022233 | 1 |
| HTN | endometrium | *RNF157* | 1.954427151 | 0.011842256 | 0.017514657 | 1 |
| HTN | endometrium | *ZNF750* | 9.143318356 | 0.000799783 | 0.003073153 | 1 |
| HTN | endometrium | *WDR45B* | 2.495495881 | 0.00135407 | 0.003239618 | 1 |
| HTN | endometrium | *RNF165* | 2.640660353 | 0.004441672 | 0.007653051 | 1 |
| HTN | endometrium | *MAPK4* | 6.772827292 | 0.001140461 | 0.003095606 | 1 |
| HTN | endometrium | *CBLN2* | -4.019413538 | 0.018772269 | 0.026263854 | 1 |
| HTN | endometrium | *NT5C3B* | -1.361765382 | 0.010732221 | 0.016111614 | 1 |
| HTN | endometrium | *PIP4K2B* | 7.706864081 | 0.000904292 | 0.003087532 | 1 |
| HTN | endometrium | *ERBB2* | -1.475954668 | 0.010902176 | 0.016319261 | 1 |
| HTN | endometrium | *MIEN1* | 4.499862777 | 0.010299393 | 0.01557751 | 1 |

| | | | | | | |
|---|---|---|---|---|---|---|
| HTN | endometrium | *IGFBP4* | 5.865419243 | 0.000131914 | 0.003073153 | 0.640048581 |
| HTN | endometrium | *CACNA1A* | 4.140275499 | 0.001349761 | 0.00323814 | 1 |
| HTN | endometrium | *SAMD1* | -3.486116176 | 0.010393567 | 0.015685718 | 1 |
| HTN | endometrium | *NFIC* | 5.370037944 | 0.000561797 | 0.003073153 | 1 |
| HTN | endometrium | *PFKL* | 2.456590015 | 0.001504911 | 0.00342122 | 1 |
| HTN | endometrium | *DUS3L* | 3.533920935 | 0.000164129 | 0.003073153 | 0.796352865 |
| HTN | endometrium | *DPP9* | 3.480646708 | 0.000782327 | 0.003073153 | 1 |
| HTN | endometrium | *SIRT3* | 7.079716168 | 0.000122144 | 0.003057499 | 0.59264496 |
| HTN | endometrium | *IFNAR1* | 3.190474357 | 0.001612044 | 0.003579584 | 1 |
| HTN | endometrium | *TRPM2* | 1.692869012 | 0.002796228 | 0.005326775 | 1 |
| HTN | endometrium | *TMEM50B* | 3.536969566 | 0.001385932 | 0.003278602 | 1 |
| HTN | endometrium | *DOPEY2* | 1.999068902 | 0.000953479 | 0.003095606 | 1 |
| HTN | endometrium | *URB1* | 4.574404185 | 0.001433706 | 0.003335313 | 1 |
| HTN | endometrium | *SAE1* | 3.213649637 | 0.00235336 | 0.004672054 | 1 |
| HTN | endometrium | *GEMIN7* | -1.925880435 | 0.002136831 | 0.00433834 | 1 |
| HTN | endometrium | *SLC6A3* | 4.537344792 | 0.00959083 | 0.014616747 | 1 |
| HTN | endometrium | *ZNF787* | 2.909635673 | 0.001494854 | 0.003408379 | 1 |
| HTN | endometrium | *EVI5L* | 3.2792177 | 0.001032359 | 0.003095606 | 1 |
| HTN | endometrium | *CTU1* | 3.854768692 | 0.00121667 | 0.003095606 | 1 |
| HTN | endometrium | *RERE* | 3.462523534 | 0.000289717 | 0.003073153 | 1 |
| HTN | endometrium | *EPHA2* | 2.774656948 | 0.00246347 | 0.004837198 | 1 |
| HTN | endometrium | *PEX14* | 2.378987216 | 0.001288901 | 0.003156863 | 1 |
| HTN | endometrium | *ZNF593* | -1.336375914 | 0.004495197 | 0.007731547 | 1 |
| HTN | endometrium | *PLK4* | -1.705017727 | 0.005264169 | 0.00880143 | 1 |
| HTN | endometrium | *NBPF3* | 2.0603885 | 0.007277269 | 0.011535221 | 1 |
| HTN | endometrium | *HSPG2* | 2.817437323 | 0.000511372 | 0.003073153 | 1 |
| HTN | endometrium | *ITGB3BP* | 3.126883635 | 0.002656296 | 0.005130083 | 1 |
| HTN | endometrium | *PIGK* | 4.54029915 | 0.006607447 | 0.010612159 | 1 |
| HTN | endometrium | *RPS8* | 4.289462532 | 0.00069862 | 0.003073153 | 1 |
| HTN | endometrium | *CYP4B1* | 5.251368247 | 0.012216771 | 0.018000539 | 1 |
| HTN | endometrium | *SLC44A3* | 3.204304309 | 0.000730865 | 0.003073153 | 1 |
| HTN | endometrium | *STRIP1* | 6.241949653 | 0.000456509 | 0.003073153 | 1 |
| HTN | endometrium | *GPR161* | 2.738446422 | 0.001589124 | 0.003546655 | 1 |
| HTN | endometrium | *ATP1B1* | 2.931858418 | 0.001266771 | 0.003128219 | 1 |
| HTN | endometrium | *TIPRL* | 2.989110704 | 0.000648786 | 0.003073153 | 1 |
| HTN | endometrium | *NME7* | 3.651121868 | 0.018864841 | 0.026378158 | 1 |
| HTN | endometrium | *XCL1* | -4.473968602 | 0.00661106 | 0.010614449 | 1 |
| HTN | endometrium | *MAEL* | 7.245434706 | 0.00116105 | 0.003095606 | 1 |
| HTN | endometrium | *RFWD2* | 4.5539808 | 0.000780032 | 0.003073153 | 1 |
| HTN | endometrium | *RGS16* | 3.937350325 | 0.0013646 | 0.00325328 | 1 |
| HTN | endometrium | *SNX27* | 3.674212666 | 9.94E-05 | 0.002934745 | 0.482368797 |
| HTN | endometrium | *SETDB1* | 4.885759764 | 0.000590995 | 0.003073153 | 1 |
| HTN | endometrium | *SEMA6C* | 3.911545306 | 0.000497619 | 0.003073153 | 1 |
| HTN | endometrium | *ARNT* | 3.45268155 | 0.011802021 | 0.017479672 | 1 |
| HTN | endometrium | *POGZ* | 3.163143904 | 0.001947322 | 0.004063832 | 1 |
| HTN | endometrium | *GABPB2* | 2.536423626 | 0.001327831 | 0.003212251 | 1 |
| HTN | endometrium | *DYRK3* | 3.226438485 | 0.002351923 | 0.004672054 | 1 |
| HTN | endometrium | *TAF1A* | -2.372721472 | 0.029535433 | 0.039434761 | 1 |
| HTN | endometrium | *SMYD2* | 3.039863672 | 0.000858973 | 0.0030808 | 1 |
| HTN | endometrium | *ADAM15* | 3.378200004 | 0.019133238 | 0.026684239 | 1 |

| | | | | | | |
|---|---|---|---|---|---|---|
| HTN | endometrium | *RAB13* | 2.99997754 | 0.001224131 | 0.003095606 | 1 |
| HTN | endometrium | *S100A8* | 2.344707263 | 0.001147228 | 0.003095606 | 1 |
| HTN | endometrium | *KCNN3* | 2.11222705 | 0.004524353 | 0.007767927 | 1 |
| HTN | endometrium | *C1orf43* | 5.832607244 | 0.00810082 | 0.012613985 | 1 |
| HTN | endometrium | *RIT1* | 2.665223023 | 0.001631716 | 0.003601334 | 1 |
| HTN | endometrium | *FLG* | -1.425003275 | 0.001775789 | 0.00380403 | 1 |
| HTN | endometrium | *SCCPDH* | 3.544948903 | 0.000806978 | 0.003073153 | 1 |
| HTN | endometrium | *CEP170* | 2.856162776 | 0.000288996 | 0.003073153 | 1 |
| HTN | endometrium | *ACP1* | 7.171164719 | 0.016052882 | 0.022935389 | 1 |
| HTN | endometrium | *MBOAT2* | 2.348272743 | 0.001165178 | 0.003095606 | 1 |
| HTN | endometrium | *PLEKHA6* | 4.946674651 | 5.55E-06 | 0.000597955 | 0.026907958 |
| HTN | endometrium | *OSR1* | 5.872303104 | 2.49E-05 | 0.001528309 | 0.1207364 |
| HTN | endometrium | *RHOB* | 3.945054979 | 0.000706309 | 0.003073153 | 1 |
| HTN | endometrium | *GALM* | 3.345703161 | 0.00071781 | 0.003073153 | 1 |
| HTN | endometrium | *WDPCP* | 3.297833032 | 0.002479242 | 0.00486075 | 1 |
| HTN | endometrium | *VPS54* | 3.923732511 | 0.000584093 | 0.003073153 | 1 |
| HTN | endometrium | *CIAO1* | 8.261916034 | 4.12E-05 | 0.002058697 | 0.199693584 |
| HTN | endometrium | *EXOC6B* | 4.74641096 | 0.001426797 | 0.003328251 | 1 |
| HTN | endometrium | *TEX261* | 3.384935982 | 0.001175856 | 0.003095606 | 1 |
| HTN | endometrium | *THNSL2* | 1.484369299 | 0.036367233 | 0.047703112 | 1 |
| HTN | endometrium | *RALB* | 6.846617477 | 0.000627987 | 0.003073153 | 1 |
| HTN | endometrium | *FBLN7* | 4.263636361 | 0.008083298 | 0.012598831 | 1 |
| HTN | endometrium | *LIPT1* | -2.304886135 | 0.011047539 | 0.016503281 | 1 |
| HTN | endometrium | *CNGA3* | 65.39979646 | 0.000335269 | 0.003073153 | 1 |
| HTN | endometrium | *FAHD2B* | -2.409102067 | 0.00364711 | 0.006532217 | 1 |
| HTN | endometrium | *LYG1* | 2.932130192 | 0.005334101 | 0.00891221 | 1 |
| HTN | endometrium | *GPR17* | 2.406091472 | 0.00016616 | 0.003073153 | 0.806210369 |
| HTN | endometrium | *GALNT13* | -0.887731522 | 0.006734113 | 0.010783471 | 1 |
| HTN | endometrium | *PHOSPHO2* | 3.746193861 | 0.006615899 | 0.010618704 | 1 |
| HTN | endometrium | *METTL21A* | 6.360026452 | 0.000127071 | 0.003073153 | 0.616548447 |
| HTN | endometrium | *NBEAL1* | 3.442493988 | 0.000906146 | 0.003087532 | 1 |
| HTN | endometrium | *SUMF1* | 3.025669086 | 0.000579421 | 0.003073153 | 1 |
| HTN | endometrium | *ACKR3* | 2.69891204 | 0.00171155 | 0.003713132 | 1 |
| HTN | endometrium | *ESPNL* | 39.54031879 | 0.001278695 | 0.003143367 | 1 |
| HTN | endometrium | *ANKMY1* | 5.731528265 | 0.001148592 | 0.003095606 | 1 |
| HTN | endometrium | *TAMM41* | 3.857022856 | 0.000344719 | 0.003073153 | 1 |
| HTN | endometrium | *STK11IP* | -1.337047754 | 0.00687866 | 0.010982316 | 1 |
| HTN | endometrium | *EAF1* | 9.989920427 | 0.000261223 | 0.003073153 | 1 |
| HTN | endometrium | *SLC25A38* | 3.219574671 | 0.002993754 | 0.005601888 | 1 |
| HTN | endometrium | *CTDSPL* | 3.893246117 | 0.004781777 | 0.008126508 | 1 |
| HTN | endometrium | *PTPRG* | 2.756212015 | 0.00107283 | 0.003095606 | 1 |
| HTN | endometrium | *ARL6IP5* | 2.775294037 | 0.000831556 | 0.0030808 | 1 |
| HTN | endometrium | *LIMD1* | 3.869406244 | 0.000318013 | 0.003073153 | 1 |
| HTN | endometrium | *ZNF660* | -1.018963022 | 0.011423653 | 0.016991896 | 1 |
| HTN | endometrium | *PLA1A* | 8.532286984 | 0.00604117 | 0.00984938 | 1 |
| HTN | endometrium | *ADPRH* | 4.406277122 | 0.002936511 | 0.005516049 | 1 |
| HTN | endometrium | *IGSF11* | -1.361284563 | 0.023799117 | 0.032381749 | 1 |
| HTN | endometrium | *BOC* | 4.232592518 | 0.000486402 | 0.003073153 | 1 |
| HTN | endometrium | *MED12L* | 2.446655121 | 0.000983787 | 0.003095606 | 1 |
| HTN | endometrium | *EIF2A* | 1.742085675 | 0.007563375 | 0.011924977 | 1 |

| | | | | | | |
|---|---|---|---|---|---|---|
| HTN | endometrium | *NCEH1* | 1.730816937 | 0.005205685 | 0.008712654 | 1 |
| HTN | endometrium | *CCDC39* | 3.762001184 | 0.003401454 | 0.006190699 | 1 |
| HTN | endometrium | *MUC4* | 3.472844929 | 0.002687514 | 0.005169046 | 1 |
| HTN | endometrium | *CENPC* | 3.874703096 | 0.001437278 | 0.003338282 | 1 |
| HTN | endometrium | *SCD5* | 3.694452915 | 0.000859498 | 0.0030808 | 1 |
| HTN | endometrium | *PYURF* | 8.547067029 | 0.020930057 | 0.028861081 | 1 |
| HTN | endometrium | *CISD2* | 4.701769393 | 0.000145896 | 0.003073153 | 0.707889719 |
| HTN | endometrium | *DDIT4L* | -4.159102038 | 0.016553596 | 0.023539873 | 1 |
| HTN | endometrium | *USP53* | 4.970702189 | 0.000434289 | 0.003073153 | 1 |
| HTN | endometrium | *MARCH1* | 2.338071182 | 0.00121505 | 0.003095606 | 1 |
| HTN | endometrium | *CBR4* | 5.520935837 | 0.003519623 | 0.006346047 | 1 |
| HTN | endometrium | *MARCH6* | 4.033101637 | 0.000526014 | 0.003073153 | 1 |
| HTN | endometrium | *MYO10* | 3.60414938 | 0.00082511 | 0.0030808 | 1 |
| HTN | endometrium | *FAM105A* | 2.468176261 | 0.000423767 | 0.003073153 | 1 |
| HTN | endometrium | *BHMT* | 2.700527475 | 0.02013341 | 0.027886755 | 1 |
| HTN | endometrium | *GTF2H2* | 11.32357821 | 0.000625563 | 0.003073153 | 1 |
| HTN | endometrium | *SPATA9* | 2.031480318 | 0.001200886 | 0.003095606 | 1 |
| HTN | endometrium | *FEM1C* | 5.093550641 | 0.000643977 | 0.003073153 | 1 |
| HTN | endometrium | *COMMD10* | 3.082309671 | 0.000590368 | 0.003073153 | 1 |
| HTN | endometrium | *YIPF5* | 5.388508025 | 0.000612681 | 0.003073153 | 1 |
| HTN | endometrium | *CXCL14* | 4.080097699 | 0.006852709 | 0.010944484 | 1 |
| HTN | endometrium | *FBXO38* | 4.214350597 | 0.000519408 | 0.003073153 | 1 |
| HTN | endometrium | *BOD1* | 3.081253198 | 0.000616576 | 0.003073153 | 1 |
| HTN | endometrium | *FAM50B* | 7.215849524 | 0.002112477 | 0.004302581 | 1 |
| HTN | endometrium | *MYLK4* | -2.103834176 | 0.009950199 | 0.015125567 | 1 |
| HTN | endometrium | *TBC1D7* | 3.086207473 | 0.000430193 | 0.003073153 | 1 |
| HTN | endometrium | *GFOD1* | 5.740460678 | 0.020910016 | 0.028847142 | 1 |
| HTN | endometrium | *FAM193B* | 2.384765474 | 0.001537367 | 0.003470431 | 1 |
| HTN | endometrium | *MUT* | 2.218113729 | 0.001429497 | 0.003329773 | 1 |
| HTN | endometrium | *ABT1* | 7.001975943 | 1.47E-06 | 0.000255124 | 0.007143472 |
| HTN | endometrium | *DAAM2* | 1.991933327 | 0.001117324 | 0.003095606 | 1 |
| HTN | endometrium | *MLIP* | 8.120883405 | 0.006761436 | 0.010823652 | 1 |
| HTN | endometrium | *RPL7L1* | 4.69314478 | 0.001233549 | 0.003103164 | 1 |
| HTN | endometrium | *TPBG* | 2.172281077 | 0.001349075 | 0.00323814 | 1 |
| HTN | endometrium | *IRAK1BP1* | 3.375118938 | 4.41E-05 | 0.002160348 | 0.213874438 |
| HTN | endometrium | *PHIP* | 4.533033794 | 0.000391724 | 0.003073153 | 1 |
| HTN | endometrium | *PNRC1* | 2.785761808 | 0.000910736 | 0.003087532 | 1 |
| HTN | endometrium | *RNF217* | 4.141198619 | 0.000589167 | 0.003073153 | 1 |
| HTN | endometrium | *ARHGAP18* | 2.272066531 | 0.000829266 | 0.0030808 | 1 |
| HTN | endometrium | *ABRACL* | 4.337938254 | 0.001092372 | 0.003095606 | 1 |
| HTN | endometrium | *PNLDC1* | -7.284178186 | 0.001364441 | 0.00325328 | 1 |
| HTN | endometrium | *WTAP* | 4.81736117 | 0.004122693 | 0.007221409 | 1 |
| HTN | endometrium | *SLC22A3* | 2.097856292 | 0.001168887 | 0.003095606 | 1 |
| HTN | endometrium | *RBAK* | 7.147971726 | 0.000647384 | 0.003073153 | 1 |
| HTN | endometrium | *POMZP3* | 2.721605339 | 0.000765261 | 0.003073153 | 1 |
| HTN | endometrium | *PSPH* | 4.395549425 | 0.009443342 | 0.014432916 | 1 |
| HTN | endometrium | *GIGYF1* | 9.049669899 | 0.00015727 | 0.003073153 | 0.763072336 |
| HTN | endometrium | *TRIM4* | 1.938555525 | 0.002345534 | 0.00466224 | 1 |
| HTN | endometrium | *TMEM209* | 4.150588346 | 0.000678891 | 0.003073153 | 1 |
| HTN | endometrium | *EPHA1* | 7.862866021 | 0.001181363 | 0.003095606 | 1 |

| | | | | | | |
|---|---|---|---|---|---|---|
| HTN | endometrium | *NCAPG2* | 2.988144176 | 0.000483103 | 0.003073153 | 1 |
| HTN | endometrium | *FBXO25* | 6.863251426 | 0.000425942 | 0.003073153 | 1 |
| HTN | endometrium | *CSGALNACT1* | 3.827566958 | 5.28E-05 | 0.002317582 | 0.256414026 |
| HTN | endometrium | *BIN3* | 3.565464431 | 0.000959387 | 0.003095606 | 1 |
| HTN | endometrium | *CHMP7* | 5.158807747 | 0.000240919 | 0.003073153 | 1 |
| HTN | endometrium | *DOCK5* | 2.768212755 | 0.000637614 | 0.003073153 | 1 |
| HTN | endometrium | *TACC1* | 3.514256931 | 0.000392326 | 0.003073153 | 1 |
| HTN | endometrium | *RPL7* | -2.143828316 | 0.014812874 | 0.021333353 | 1 |
| HTN | endometrium | *ATP6V0D2* | 11.62108777 | 0.000865003 | 0.003084413 | 1 |
| HTN | endometrium | *DPYS* | 1.793841798 | 0.001196961 | 0.003095606 | 1 |
| HTN | endometrium | *LRP12* | 8.025073544 | 0.014972655 | 0.021531513 | 1 |
| HTN | endometrium | *MAL2* | 4.402734501 | 0.002547064 | 0.004965189 | 1 |
| HTN | endometrium | *SLC39A4* | -1.278369906 | 0.005182152 | 0.008685252 | 1 |
| HTN | endometrium | *AK3* | 3.081530651 | 0.000781275 | 0.003073153 | 1 |
| HTN | endometrium | *HAUS6* | 5.644457786 | 0.000642713 | 0.003073153 | 1 |
| HTN | endometrium | *C9orf72* | 2.194390337 | 0.000388987 | 0.003073153 | 1 |
| HTN | endometrium | *FBXO10* | 6.348074958 | 0.000979858 | 0.003095606 | 1 |
| HTN | endometrium | *ZNF462* | 2.546376274 | 0.00031024 | 0.003073153 | 1 |
| HTN | endometrium | *ALAD* | 3.795577646 | 0.000758618 | 0.003073153 | 1 |
| HTN | endometrium | *POLE3* | 4.14455588 | 0.000138902 | 0.003073153 | 0.673954692 |
| HTN | endometrium | *MED22* | 3.086315334 | 0.003032252 | 0.005656473 | 1 |
| HTN | endometrium | *REXO4* | 3.050396934 | 0.000943059 | 0.003095606 | 1 |
| HTN | endometrium | *SH3GLB2* | 9.086921478 | 0.000200464 | 0.003073153 | 0.972650553 |
| HTN | endometrium | *INPP5E* | 11.31001312 | 0.005767179 | 0.00946629 | 1 |
| HTN | endometrium | *SEC16A* | 7.47997759 | 0.012304959 | 0.018108481 | 1 |
| HTN | endometrium | *SLC39A12* | 23.02295027 | 0.001679665 | 0.003667747 | 1 |
| HTN | endometrium | *TMEM236* | 4.22921074 | 0.003287053 | 0.006027506 | 1 |
| HTN | endometrium | *RSU1* | 2.888329801 | 0.000705155 | 0.003073153 | 1 |
| HTN | endometrium | *ST8SIA6* | 9.918736358 | 0.010683553 | 0.016048483 | 1 |
| HTN | endometrium | *PARD3* | 4.217332388 | 0.000975622 | 0.003095606 | 1 |
| HTN | endometrium | *FRA10AC1* | 4.155093436 | 0.000554798 | 0.003073153 | 1 |
| HTN | endometrium | *ADD3* | 5.130156342 | 0.000873311 | 0.003084413 | 1 |
| HTN | endometrium | *LRRC27* | 2.62157952 | 0.00342802 | 0.006222504 | 1 |
| HTN | endometrium | *MTG1* | 5.226002458 | 0.000352825 | 0.003073153 | 1 |
| HTN | endometrium | *PAOX* | 11.28440431 | 0.000874883 | 0.003084413 | 1 |
| HTN | endometrium | *PDCD11* | 4.153024418 | 0.000705395 | 0.003073153 | 1 |
| HTN | endometrium | *RGS10* | 2.619270088 | 0.001043457 | 0.003095606 | 1 |
| HTN | endometrium | *LIN7C* | 2.555306752 | 0.001113883 | 0.003095606 | 1 |
| HTN | endometrium | *ZNF215* | 3.726719235 | 4.97E-05 | 0.002232309 | 0.241089372 |
| HTN | endometrium | *APIP* | 1.686304391 | 0.009527356 | 0.014541506 | 1 |
| HTN | endometrium | *TNKS1BP1* | 10.98166703 | 9.59E-12 | 2.77E-08 | 4.65E-08 |
| HTN | endometrium | *PTPRJ* | 1.700313033 | 0.001359112 | 0.003248477 | 1 |
| HTN | endometrium | *ENDOD1* | 2.689741093 | 0.000986406 | 0.003095606 | 1 |
| HTN | endometrium | *CCDC82* | 2.11596154 | 0.0175594 | 0.024761449 | 1 |
| HTN | endometrium | *CAPN5* | -2.076644793 | 0.009591864 | 0.014616747 | 1 |
| HTN | endometrium | *RPS3* | 5.884304472 | 0.006825205 | 0.010914929 | 1 |
| HTN | endometrium | *TTC12* | 7.491437929 | 3.00E-06 | 0.000419407 | 0.01455317 |
| HTN | endometrium | *DRD2* | 24.78347088 | 0.001130778 | 0.003095606 | 1 |
| HTN | endometrium | *NPAT* | 4.742964707 | 0.000555535 | 0.003073153 | 1 |
| HTN | endometrium | *ATM* | 3.440934885 | 0.000845982 | 0.0030808 | 1 |

| | | | | | | |
|---|---|---|---|---|---|---|
| HTN | endometrium | *SLX4IP* | 2.839773753 | 0.003046149 | 0.005678031 | 1 |
| HTN | endometrium | *ST14* | 4.841997767 | 0.001419268 | 0.003317298 | 1 |
| HTN | endometrium | *FADS1* | 5.222818991 | 0.001165939 | 0.003095606 | 1 |
| HTN | endometrium | *TMC2* | 5.148526506 | 0.001848586 | 0.003913325 | 1 |
| HTN | endometrium | *EML3* | 2.951368622 | 0.000277458 | 0.003073153 | 1 |
| HTN | endometrium | *INCENP* | 3.339779873 | 0.003201428 | 0.00590172 | 1 |
| HTN | endometrium | *PLCH2* | 14.37396778 | 0.003026628 | 0.005648153 | 1 |
| HTN | endometrium | *CPSF7* | 6.377721232 | 0.01281416 | 0.018790828 | 1 |
| HTN | endometrium | *CHEK1* | 5.178324435 | 0.001259473 | 0.003125811 | 1 |
| HTN | endometrium | *DUSP15* | -0.985326376 | 0.005838212 | 0.009566701 | 1 |
| HTN | endometrium | *TBX6* | 3.952679213 | 2.66E-05 | 0.001591925 | 0.12894593 |
| HTN | endometrium | *ANXA8L1* | 1.508375059 | 0.006158152 | 0.010007033 | 1 |
| HTN | endometrium | *CWC15* | 3.340844533 | 0.001187406 | 0.003095606 | 1 |
| HTN | endometrium | *FCGR1A* | -2.527669564 | 0.003909365 | 0.006912624 | 1 |
| HTN | endometrium | *TMEM218* | 3.606030743 | 0.001028984 | 0.003095606 | 1 |
| HTN | endometrium | *SAP18* | 3.026910316 | 0.00089101 | 0.003084413 | 1 |
| HTN | endometrium | *FAM124A* | 10.56875138 | 0.012584648 | 0.018469665 | 1 |
| HTN | endometrium | *WDR17* | 2.46463828 | 0.001347215 | 0.003237588 | 1 |
| HTN | endometrium | *CCDC102B* | 1.818366169 | 0.00157977 | 0.00353065 | 1 |
| HTN | endometrium | *RGS18* | 2.950407476 | 0.005325764 | 0.008901346 | 1 |
| HTN | endometrium | *PRSS23* | 2.86295794 | 0.000478271 | 0.003073153 | 1 |
| HTN | endometrium | *PPP1R1C* | 4.634408341 | 0.001207507 | 0.003095606 | 1 |
| HTN | endometrium | *FAM173B* | 2.252736933 | 0.000904785 | 0.003087532 | 1 |
| HTN | endometrium | *GPR158* | 4.479338546 | 0.001261098 | 0.003126646 | 1 |
| HTN | endometrium | *UBE3B* | 3.375218272 | 0.001880756 | 0.003966721 | 1 |
| HTN | endometrium | *ANK3* | 2.686539674 | 0.000795436 | 0.003073153 | 1 |
| HTN | endometrium | *PLBD2* | 2.489980527 | 0.003368691 | 0.006149319 | 1 |
| HTN | endometrium | *DLG5* | 3.725376876 | 0.001125307 | 0.003095606 | 1 |
| HTN | endometrium | *GXYLT1* | 2.891318079 | 0.000832061 | 0.0030808 | 1 |
| HTN | endometrium | *EXT2* | 2.68044275 | 0.001465307 | 0.003374309 | 1 |
| HTN | endometrium | *TMEM18* | 8.978091995 | 0.000651428 | 0.003073153 | 1 |
| HTN | endometrium | *NDUFC2* | 3.207067487 | 0.001110531 | 0.003095606 | 1 |
| HTN | endometrium | *NUBPL* | 3.540467436 | 0.000592954 | 0.003073153 | 1 |
| HTN | endometrium | *VIPAS39* | 3.318486164 | 0.000923094 | 0.003087532 | 1 |
| HTN | endometrium | *ANKRD50* | 6.797720073 | 0.000201317 | 0.003073153 | 0.976788701 |
| HTN | endometrium | *UPF2* | 3.496072848 | 0.000704617 | 0.003073153 | 1 |
| HTN | endometrium | *C4orf33* | 3.681604927 | 0.000572581 | 0.003073153 | 1 |
| HTN | endometrium | *PTPRO* | 2.353128724 | 0.002258398 | 0.004540399 | 1 |
| HTN | endometrium | *EPS8* | 3.459128933 | 0.001092771 | 0.003095606 | 1 |
| HTN | endometrium | *QDPR* | 5.196282879 | 0.000433717 | 0.003073153 | 1 |
| HTN | endometrium | *ZNF827* | 2.158008625 | 0.001737054 | 0.003747526 | 1 |
| HTN | endometrium | *AKR1C2* | 4.375105196 | 0.001124421 | 0.003095606 | 1 |
| HTN | endometrium | *PIGF* | 6.827973898 | 0.000959167 | 0.003095606 | 1 |
| HTN | endometrium | *MFSD6* | 4.346659464 | 0.000728544 | 0.003073153 | 1 |
| HTN | endometrium | *CENPU* | -1.002670364 | 0.009159039 | 0.014045404 | 1 |
| HTN | endometrium | *BICD1* | 2.878099587 | 0.000733239 | 0.003073153 | 1 |
| HTN | endometrium | *CCDC122* | 3.919730718 | 0.003470031 | 0.006272947 | 1 |
| HTN | endometrium | *GABRA2* | 6.667927935 | 0.023960805 | 0.032574342 | 1 |
| HTN | endometrium | *PARP8* | 2.525720035 | 0.017778471 | 0.025035888 | 1 |
| HTN | endometrium | *CACUL1* | 3.615763464 | 0.000786535 | 0.003073153 | 1 |

| | | | | | | |
|---|---|---|---|---|---|---|
| HTN | endometrium | *BEND6* | -1.145128252 | 0.003379778 | 0.006162602 | 1 |
| HTN | endometrium | *TIAL1* | 4.394847495 | 0.00053057 | 0.003073153 | 1 |
| HTN | endometrium | *PTPN14* | 3.720223508 | 0.000410894 | 0.003073153 | 1 |
| HTN | endometrium | *MGAT5* | 2.44858485 | 0.00111034 | 0.003095606 | 1 |
| HTN | endometrium | *TMEM178A* | 4.956682119 | 0.001199652 | 0.003095606 | 1 |
| HTN | endometrium | *HAUS1* | 2.338126837 | 0.004406879 | 0.007620163 | 1 |
| HTN | endometrium | *SPC25* | 2.254287714 | 0.010605319 | 0.015950715 | 1 |
| HTN | endometrium | *KCNK13* | 4.60688383 | 0.000650393 | 0.003073153 | 1 |
| HTN | endometrium | *POC5* | 4.446068775 | 0.000300176 | 0.003073153 | 1 |
| HTN | endometrium | *CWF19L2* | 5.547765742 | 2.30E-05 | 0.001451296 | 0.111749773 |
| HTN | endometrium | *ZNF773* | 2.853760998 | 0.003892029 | 0.006890441 | 1 |
| HTN | endometrium | *NMT2* | 3.206096391 | 0.001879304 | 0.003966721 | 1 |
| HTN | endometrium | *CCDC50* | 3.035201738 | 0.028810002 | 0.038595231 | 1 |
| HTN | endometrium | *SPARCL1* | 3.282883065 | 0.000512777 | 0.003073153 | 1 |
| HTN | endometrium | *MBNL1* | 3.096939811 | 0.000556253 | 0.003073153 | 1 |
| HTN | endometrium | *GPD1L* | 5.436487563 | 0.001320108 | 0.003204185 | 1 |
| HTN | endometrium | *CCNO* | 29.72880088 | 0.001481854 | 0.003394073 | 1 |
| HTN | endometrium | *CLEC4F* | 5.953125103 | 0.000488702 | 0.003073153 | 1 |
| HTN | endometrium | *RASGRP3* | 2.527700594 | 0.00097085 | 0.003095606 | 1 |
| HTN | endometrium | *GPR180* | 2.470355452 | 0.013574251 | 0.019778459 | 1 |
| HTN | endometrium | *IFIT5* | 2.718443578 | 0.014152306 | 0.020479772 | 1 |
| HTN | endometrium | *GGPS1* | 5.533841354 | 0.000350087 | 0.003073153 | 1 |
| HTN | endometrium | *ZNF117* | 1.505207908 | 0.006498685 | 0.010472141 | 1 |
| HTN | endometrium | *MED21* | 5.788678374 | 4.54E-05 | 0.002205049 | 0.220504874 |
| HTN | endometrium | *JAKMIP1* | 3.748934375 | 0.001169479 | 0.003095606 | 1 |
| HTN | endometrium | *SREK1IP1* | 4.361852664 | 0.000571325 | 0.003073153 | 1 |
| HTN | endometrium | *MR1* | 2.629927299 | 0.001060644 | 0.003095606 | 1 |
| HTN | endometrium | *CENPH* | -3.323730607 | 0.005731242 | 0.009413672 | 1 |
| HTN | endometrium | *CARHSP1* | 1.694594501 | 0.001656979 | 0.003632924 | 1 |
| HTN | endometrium | *CAST* | 2.926084036 | 0.009270167 | 0.01419787 | 1 |
| HTN | endometrium | *CLGN* | 9.182373731 | 0.014876911 | 0.021412866 | 1 |
| HTN | endometrium | *CETN3* | 4.425926532 | 0.03303832 | 0.04366772 | 1 |
| HTN | endometrium | *TMEM87B* | 4.808447832 | 0.000553699 | 0.003073153 | 1 |
| HTN | endometrium | *SLC25A27* | 2.927327118 | 0.000522558 | 0.003073153 | 1 |
| HTN | endometrium | *FRMD1* | -3.706030921 | 0.020212563 | 0.027972434 | 1 |
| HTN | endometrium | *LPCAT1* | 2.79174056 | 0.001101143 | 0.003095606 | 1 |
| HTN | endometrium | *NMRAL1* | 2.927127937 | 0.000789163 | 0.003073153 | 1 |
| HTN | endometrium | *ADPRHL1* | 5.736500396 | 0.00126818 | 0.003128219 | 1 |
| HTN | endometrium | *CFDP1* | 2.929225463 | 0.000673907 | 0.003073153 | 1 |
| HTN | endometrium | *ZDHHC7* | 7.373316809 | 0.001562246 | 0.003499546 | 1 |
| HTN | endometrium | *JAZF1* | -1.98084074 | 0.019012675 | 0.026546618 | 1 |
| HTN | endometrium | *FBXO36* | 4.439294397 | 0.000505773 | 0.003073153 | 1 |
| HTN | endometrium | *MCOLN2* | 4.526162988 | 0.001265301 | 0.003127478 | 1 |
| HTN | endometrium | *GDPD1* | 2.530593192 | 0.001536529 | 0.003470431 | 1 |
| HTN | endometrium | *SEMA3D* | 5.213535975 | 0.000615109 | 0.003073153 | 1 |
| HTN | endometrium | *PPP2R5E* | 4.154511722 | 0.000553106 | 0.003073153 | 1 |
| HTN | endometrium | *ANKRD29* | 2.054891267 | 0.001249503 | 0.003121827 | 1 |
| HTN | endometrium | *DNAAF1* | -6.484732588 | 0.001730302 | 0.003742526 | 1 |
| HTN | endometrium | *ANKH* | 2.669806593 | 0.000917374 | 0.003087532 | 1 |
| HTN | endometrium | *UBASH3B* | 2.798573166 | 0.002319593 | 0.004629644 | 1 |

| | | | | | | |
|---|---|---|---|---|---|---|
| HTN | endometrium | *ROBO4* | 3.513907609 | 0.000769227 | 0.003073153 | 1 |
| HTN | endometrium | *ROBO3* | 2.412461649 | 0.000683429 | 0.003073153 | 1 |
| HTN | endometrium | *CC2D1B* | 4.594219426 | 0.000458263 | 0.003073153 | 1 |
| HTN | endometrium | *PRKCA* | 4.003020463 | 0.000589789 | 0.003073153 | 1 |
| HTN | endometrium | *LRRK1* | 1.741763436 | 0.001690945 | 0.0036874 | 1 |
| HTN | endometrium | *ABCA5* | 3.269853376 | 6.74E-05 | 0.002441268 | 0.327129933 |
| HTN | endometrium | *MIA3* | 4.825145025 | 0.000791124 | 0.003073153 | 1 |
| HTN | endometrium | *NEIL2* | 8.305610008 | 4.16E-09 | 2.02E-06 | 2.02E-05 |
| HTN | endometrium | *CCSAP* | 3.17586318 | 0.001157817 | 0.003095606 | 1 |
| HTN | endometrium | *GBP5* | 2.736777636 | 0.00409928 | 0.007188901 | 1 |
| HTN | endometrium | *ATP5G3* | 4.214226204 | 0.000168455 | 0.003073153 | 0.817345232 |
| HTN | endometrium | *LY96* | 2.21407543 | 0.001734663 | 0.003747095 | 1 |
| HTN | endometrium | *PDE1C* | 2.004693253 | 0.001093302 | 0.003095606 | 1 |
| HTN | endometrium | *XPC* | 5.765027225 | 0.008010792 | 0.012501886 | 1 |
| HTN | endometrium | *FLCN* | 2.337989015 | 0.002050374 | 0.004209317 | 1 |
| HTN | endometrium | *OXNAD1* | 3.342160015 | 0.000603601 | 0.003073153 | 1 |
| HTN | endometrium | *APCDD1* | 4.341452379 | 0.013686835 | 0.019894704 | 1 |
| HTN | endometrium | *MPPE1* | 2.36673787 | 0.02369331 | 0.032273986 | 1 |
| HTN | endometrium | *ANKRD40* | 5.440097185 | 0.000333341 | 0.003073153 | 1 |
| HTN | endometrium | *AK9* | 2.708046375 | 0.00037496 | 0.003073153 | 1 |
| HTN | endometrium | *MARCKS* | 2.7503022 | 0.000707328 | 0.003073153 | 1 |
| HTN | endometrium | *GOLGA7B* | 16.82317938 | 0.000926742 | 0.003091039 | 1 |
| HTN | endometrium | *TRMT44* | 5.933258182 | 0.001257044 | 0.003122339 | 1 |
| HTN | endometrium | *MOV10* | 11.98529426 | 0.000365459 | 0.003073153 | 1 |
| HTN | endometrium | *HEATR3* | 1.793388788 | 0.013607279 | 0.01981468 | 1 |
| HTN | endometrium | *CNOT8* | 6.943178552 | 0.012819379 | 0.018791428 | 1 |
| HTN | endometrium | *SETD9* | 2.76373634 | 0.000206841 | 0.003073153 | 1 |
| HTN | endometrium | *NUP205* | 2.790322394 | 0.000915426 | 0.003087532 | 1 |
| HTN | endometrium | *ZKSCAN2* | -1.599925802 | 0.003593736 | 0.006448524 | 1 |
| HTN | endometrium | *KDM8* | 3.570938407 | 0.001366085 | 0.003253925 | 1 |
| HTN | endometrium | *ALS2CR12* | 6.223721183 | 0.000634537 | 0.003073153 | 1 |
| HTN | endometrium | *FZD7* | 4.264164237 | 0.000494943 | 0.003073153 | 1 |
| HTN | endometrium | *DEPTOR* | 3.528270237 | 0.000282175 | 0.003073153 | 1 |
| HTN | endometrium | *RRAGA* | 3.134805729 | 0.001293422 | 0.003159962 | 1 |
| HTN | endometrium | *TMBIM4* | 5.269567529 | 0.001544311 | 0.003477029 | 1 |
| HTN | endometrium | *PSD3* | 3.439144236 | 0.023768397 | 0.032358098 | 1 |
| HTN | endometrium | *KCNMA1* | 1.257210848 | 0.009818323 | 0.014943069 | 1 |
| HTN | endometrium | *NDUFAF6* | 2.912244908 | 0.022182229 | 0.030386273 | 1 |
| HTN | endometrium | *DRAM2* | 3.505399494 | 0.000992083 | 0.003095606 | 1 |
| HTN | endometrium | *C8orf37* | 2.145675776 | 0.021746127 | 0.029839426 | 1 |
| HTN | endometrium | *WHAMM* | 2.531445474 | 0.001407005 | 0.003304351 | 1 |
| HTN | endometrium | *N6AMT1* | 2.855233428 | 0.004426898 | 0.007638445 | 1 |
| HTN | endometrium | *RWDD2B* | 2.94277937 | 0.003002397 | 0.005614518 | 1 |
| HTN | endometrium | *CCT8* | 1.576327538 | 0.011850913 | 0.017514657 | 1 |
| HTN | endometrium | *NAA11* | -6.126296043 | 0.005931061 | 0.009697963 | 1 |
| HTN | endometrium | *SFXN2* | 5.948400273 | 7.82E-10 | 5.57E-07 | 3.80E-06 |
| HTN | endometrium | *TDRD9* | 6.646888906 | 0.004026557 | 0.007083703 | 1 |
| HTN | endometrium | *RPL30* | 4.172937861 | 0.000390672 | 0.003073153 | 1 |
| HTN | endometrium | *SUPV3L1* | 8.77084936 | 0.00204282 | 0.0041999 | 1 |
| HTN | endometrium | *HKDC1* | -2.876828661 | 0.006838775 | 0.010933026 | 1 |

| | | | | | | |
|---|---|---|---|---|---|---|
| HTN | endometrium | *ZDHHC5* | 5.584058937 | 0.000549091 | 0.003073153 | 1 |
| HTN | endometrium | *WDYHV1* | 3.251364944 | 0.011520408 | 0.017125312 | 1 |
| HTN | endometrium | *FBRS* | 2.549467293 | 0.001298854 | 0.003166855 | 1 |
| HTN | endometrium | *SASS6* | -2.958440534 | 0.026760367 | 0.036026998 | 1 |
| HTN | endometrium | *ITGAD* | 5.375207002 | 0.000879657 | 0.003084413 | 1 |
| HTN | endometrium | *VPS8* | 4.020271238 | 9.42E-05 | 0.002820722 | 0.456956928 |
| HTN | endometrium | *MPV17L* | 6.599901792 | 0.000118869 | 0.003039787 | 0.576751333 |
| HTN | endometrium | *RPUSD3* | 3.897227392 | 0.00094484 | 0.003095606 | 1 |
| HTN | endometrium | *GHRL* | 7.539908047 | 0.015763807 | 0.022555586 | 1 |
| HTN | endometrium | *NMNAT2* | 2.632290308 | 0.001715583 | 0.003716761 | 1 |
| HTN | endometrium | *SLC6A1* | 9.291868078 | 0.001551817 | 0.003489071 | 1 |
| HTN | endometrium | *FCHO2* | 3.047615898 | 0.000320574 | 0.003073153 | 1 |
| HTN | endometrium | *TIMP4* | 2.433709454 | 0.000603489 | 0.003073153 | 1 |
| HTN | endometrium | *NECAP2* | 2.849497747 | 0.000924364 | 0.003087532 | 1 |
| HTN | endometrium | *STEAP2* | 1.503648962 | 0.005622103 | 0.009285491 | 1 |
| HTN | endometrium | *CLDN12* | 4.121734013 | 0.000888071 | 0.003084413 | 1 |
| HTN | endometrium | *MMP14* | 7.439207846 | 0.000340836 | 0.003073153 | 1 |
| HTN | endometrium | *GATAD1* | 2.535262726 | 0.000756629 | 0.003073153 | 1 |
| HTN | endometrium | *CLEC18A* | -1.828667787 | 0.000101323 | 0.002934745 | 0.491618935 |
| HTN | endometrium | *DHRS4* | 3.108481161 | 0.001168793 | 0.003095606 | 1 |
| HTN | endometrium | *AASDH* | 5.874515462 | 0.000654954 | 0.003073153 | 1 |
| HTN | endometrium | *ETS2* | 2.528678425 | 9.03E-05 | 0.002739749 | 0.438359869 |
| HTN | endometrium | *LCA5L* | -1.592538463 | 0.010326628 | 0.015599253 | 1 |
| HTN | endometrium | *MX1* | 2.901652759 | 0.00068795 | 0.003073153 | 1 |
| HTN | endometrium | *DGKI* | 1.882360101 | 0.003125208 | 0.005789808 | 1 |
| HTN | endometrium | *SNX22* | -4.046085318 | 0.010414126 | 0.015711859 | 1 |
| HTN | endometrium | *SLC34A2* | -46.2767953 | 0.021482639 | 0.029517184 | 1 |
| HTN | endometrium | *AP3S2* | 10.03999629 | 0.00061165 | 0.003073153 | 1 |
| HTN | endometrium | *SPPL3* | 4.757902452 | 0.000603165 | 0.003073153 | 1 |
| HTN | endometrium | *C12orf43* | -1.608795226 | 0.004173246 | 0.007294161 | 1 |
| HTN | endometrium | *PAFAH2* | 3.331135709 | 0.031020109 | 0.041212915 | 1 |
| HTN | endometrium | *EXTL1* | 44.75801762 | 0.001220331 | 0.003095606 | 1 |
| HTN | endometrium | *WDR66* | 5.436820553 | 0.005563954 | 0.009204332 | 1 |
| HTN | endometrium | *RHPN1* | 6.436483934 | 0.000379428 | 0.003073153 | 1 |
| HTN | endometrium | *LRRC43* | -9.919148256 | 0.000691089 | 0.003073153 | 1 |
| HTN | endometrium | *EYA3* | 3.456900519 | 0.001220886 | 0.003095606 | 1 |
| HTN | endometrium | *DZIP1L* | 7.209377059 | 0.002467999 | 0.004842189 | 1 |
| HTN | endometrium | *ESYT3* | -4.338213311 | 0.006244444 | 0.010123332 | 1 |
| HTN | endometrium | *CLSTN2* | 1.711963321 | 0.001486225 | 0.003397652 | 1 |
| HTN | endometrium | *COLEC12* | 1.372573125 | 0.001665468 | 0.003646592 | 1 |
| HTN | endometrium | *MITD1* | 6.935042062 | 0.000473432 | 0.003073153 | 1 |
| HTN | endometrium | *TSPAN33* | 3.526548603 | 0.030632577 | 0.04076502 | 1 |
| HTN | endometrium | *FAM86C1* | -1.538839314 | 0.005166035 | 0.008661231 | 1 |
| HTN | endometrium | *PPP1R9A* | 4.074139118 | 0.000576212 | 0.003073153 | 1 |
| HTN | endometrium | *DYNC1I1* | 3.644117229 | 0.000781322 | 0.003073153 | 1 |
| HTN | endometrium | *COPG2* | 1.664334749 | 0.013425024 | 0.019596335 | 1 |
| HTN | endometrium | *ELK4* | 9.005785132 | 0.001302509 | 0.003172577 | 1 |
| HTN | endometrium | *DUSP23* | 6.206238786 | 0.015928873 | 0.022778336 | 1 |
| HTN | endometrium | *FGF17* | -1.868010705 | 0.003237546 | 0.005954727 | 1 |
| HTN | endometrium | *SLC5A11* | -4.160305257 | 0.000257172 | 0.003073153 | 1 |

| | | | | | | |
|---|---|---|---|---|---|---|
| HTN | endometrium | *CDC42SE2* | 1.631585654 | 0.002574448 | 0.005006502 | 1 |
| HTN | endometrium | *RAPGEF6* | 3.980132037 | 0.002093067 | 0.004270631 | 1 |
| HTN | endometrium | *IFNAR2* | 1.553976201 | 0.003792814 | 0.006743399 | 1 |
| HTN | endometrium | *SV2A* | -2.327741407 | 0.011382261 | 0.016940715 | 1 |
| HTN | endometrium | *CLIC6* | -2.675462594 | 0.006553345 | 0.010547607 | 1 |
| HTN | endometrium | *CBR3* | 5.112051838 | 4.83E-05 | 0.002211831 | 0.234454091 |
| HTN | endometrium | *HLCS* | 3.541160201 | 0.00047793 | 0.003073153 | 1 |
| HTN | endometrium | *IRX6* | 2.016785406 | 0.010101653 | 0.015311847 | 1 |
| HTN | endometrium | *HK2* | 3.557754652 | 0.001181546 | 0.003095606 | 1 |
| HTN | endometrium | *THEM4* | 2.398120521 | 0.000556544 | 0.003073153 | 1 |
| HTN | endometrium | *AMFR* | 2.538886513 | 0.000697361 | 0.003073153 | 1 |
| HTN | endometrium | *TMEM69* | 3.457561967 | 0.000642259 | 0.003073153 | 1 |
| HTN | endometrium | *ACE* | 6.157757786 | 0.000791709 | 0.003073153 | 1 |
| HTN | endometrium | *TEPP* | 9.187964729 | 0.011778244 | 0.017459424 | 1 |
| HTN | endometrium | *CHCHD6* | 7.829520729 | 0.000497596 | 0.003073153 | 1 |
| HTN | endometrium | *CTBP1* | 2.367865277 | 0.003797895 | 0.006749959 | 1 |
| HTN | endometrium | *ZFYVE28* | 2.937909025 | 0.00125623 | 0.003122339 | 1 |
| HTN | endometrium | *C16orf86* | 4.236683733 | 0.000436787 | 0.003073153 | 1 |
| HTN | endometrium | *RGS12* | 4.088378919 | 0.000518565 | 0.003073153 | 1 |
| HTN | endometrium | *LYPD5* | 4.17301315 | 0.031741331 | 0.042124983 | 1 |
| HTN | endometrium | *CCDC107* | 7.800826842 | 0.000379902 | 0.003073153 | 1 |
| HTN | endometrium | *ZNF233* | 5.621561031 | 0.002569127 | 0.005000162 | 1 |
| HTN | endometrium | *SSU72* | 6.983760544 | 0.000624142 | 0.003073153 | 1 |
| HTN | endometrium | *CPAMD8* | 6.160260298 | 0.000420966 | 0.003073153 | 1 |
| HTN | endometrium | *CCDC58* | 8.821287277 | 0.022656447 | 0.030959409 | 1 |
| HTN | endometrium | *SLC37A1* | 3.01327732 | 0.000858251 | 0.0030808 | 1 |
| HTN | endometrium | *PDE9A* | 2.268223695 | 0.001214767 | 0.003095606 | 1 |
| HTN | endometrium | *NDUFV3* | 2.286158056 | 0.001801589 | 0.003845716 | 1 |
| HTN | endometrium | *HSF2BP* | -6.55572658 | 0.031254523 | 0.041490272 | 1 |
| HTN | endometrium | *CSTB* | 2.170353137 | 0.004742186 | 0.008077266 | 1 |
| HTN | endometrium | *C21orf2* | -1.358525552 | 0.001505421 | 0.00342122 | 1 |
| HTN | endometrium | *SPATC1L* | 1.23838885 | 0.032086425 | 0.042524811 | 1 |
| HTN | endometrium | *LSS* | 3.345878831 | 0.001057746 | 0.003095606 | 1 |
| HTN | endometrium | *DIP2A* | 4.882525393 | 0.00027228 | 0.003073153 | 1 |
| HTN | endometrium | *PRMT2* | 4.922336248 | 0.000197162 | 0.003073153 | 0.956628927 |
| HTN | endometrium | *ZNF208* | 1.941785601 | 0.008967685 | 0.013786822 | 1 |
| HTN | endometrium | *ADAMTS13* | -1.39154457 | 0.013410344 | 0.019580798 | 1 |
| HTN | endometrium | *CACFD1* | 6.610570534 | 0.001949093 | 0.004065779 | 1 |
| HTN | endometrium | *GPSM1* | 4.388955574 | 0.000448296 | 0.003073153 | 1 |
| HTN | endometrium | *RDH13* | 5.284878329 | 0.01887921 | 0.026390644 | 1 |
| HTN | endometrium | *TAOK1* | 2.782828157 | 0.000547549 | 0.003073153 | 1 |
| HTN | endometrium | *S100A1* | 10.61222532 | 0.004615438 | 0.007910317 | 1 |
| HTN | endometrium | *ZBTB7B* | 3.820974414 | 0.000833994 | 0.0030808 | 1 |
| HTN | endometrium | *FDPS* | 4.115484842 | 0.000194701 | 0.003073153 | 0.944690198 |
| HTN | endometrium | *CCR5* | 7.528491876 | 6.34E-05 | 0.002410623 | 0.307740051 |
| HTN | endometrium | *NBEAL2* | 4.699643321 | 0.000425994 | 0.003073153 | 1 |
| HTN | endometrium | *PTH1R* | -2.770404276 | 0.008542885 | 0.013221716 | 1 |
| HTN | endometrium | *FCRL3* | 4.308432404 | 0.007368749 | 0.011657375 | 1 |
| HTN | endometrium | *FGFR4* | 13.96969637 | 0.001482284 | 0.003394073 | 1 |
| HTN | endometrium | *LY6K* | 10.13586198 | 0.00010231 | 0.002934745 | 0.496407963 |

| | | | | | | |
|---|---|---|---|---|---|---|
| HTN | endometrium | *IER2* | 4.019988932 | 0.000601116 | 0.003073153 | 1 |
| HTN | endometrium | *VPS28* | 6.132571327 | 0.008312098 | 0.012905695 | 1 |
| HTN | endometrium | *TONSL* | 6.22871172 | 0.000828335 | 0.0030808 | 1 |
| HTN | endometrium | *MUM1* | 3.705878928 | 0.000806145 | 0.003073153 | 1 |
| HTN | endometrium | *ZNF333* | 2.533547966 | 0.001055395 | 0.003095606 | 1 |
| HTN | endometrium | *SH2B2* | 3.717323166 | 0.036535616 | 0.04791103 | 1 |
| HTN | endometrium | *ABCF3* | 5.729449694 | 1.70E-07 | 4.84E-05 | 0.00082327 |
| HTN | endometrium | *PCYT1A* | 4.236427211 | 0.0010566 | 0.003095606 | 1 |
| HTN | endometrium | *FBXO27* | -2.073805478 | 0.010849991 | 0.016263255 | 1 |
| HTN | endometrium | *DMKN* | 1.068620762 | 0.017181495 | 0.024318732 | 1 |
| HTN | endometrium | *U2AF1L4* | -1.900602647 | 0.004251323 | 0.007401299 | 1 |
| HTN | endometrium | *LRRC56* | 7.838461928 | 0.000694858 | 0.003073153 | 1 |
| HTN | endometrium | *ZNF577* | 4.119660657 | 0.001745267 | 0.003751898 | 1 |
| HTN | endometrium | *TMEM143* | 8.716219483 | 0.000371939 | 0.003073153 | 1 |
| HTN | endometrium | *CCL16* | -3.785269856 | 0.002501761 | 0.004892602 | 1 |
| HTN | endometrium | *ALDH16A1* | 6.014258807 | 0.001005169 | 0.003095606 | 1 |
| HTN | endometrium | *ASB16* | 3.009900252 | 0.00113517 | 0.003095606 | 1 |
| HTN | endometrium | *PLCD3* | 11.99869099 | 0.002240142 | 0.004517375 | 1 |
| HTN | endometrium | *IP6K3* | 36.86482302 | 0.002677985 | 0.005161298 | 1 |
| HTN | endometrium | *ASGR2* | 21.08204861 | 0.000780851 | 0.003073153 | 1 |
| HTN | endometrium | *SNRNP25* | 6.078313558 | 0.000572685 | 0.003073153 | 1 |
| HTN | endometrium | *WDR90* | 3.370620026 | 0.000467148 | 0.003073153 | 1 |
| HTN | endometrium | *JMJD8* | 3.809035607 | 0.000388216 | 0.003073153 | 1 |
| HTN | endometrium | *CCDC78* | 12.79943897 | 0.00021767 | 0.003073153 | 1 |
| HTN | endometrium | *MEIOB* | 4.253166725 | 0.000812994 | 0.003074195 | 1 |
| HTN | endometrium | *HS3ST6* | 6.932325732 | 0.002355416 | 0.004672314 | 1 |
| HTN | endometrium | *SHANK2* | 2.386992576 | 0.001285231 | 0.003152649 | 1 |
| HTN | endometrium | *ASRGL1* | 3.316880964 | 0.001245573 | 0.003116824 | 1 |
| HTN | endometrium | *TTC9C* | -3.335932908 | 0.032804229 | 0.043404996 | 1 |
| HTN | endometrium | *RPS6KA4* | 4.92207143 | 0.00081757 | 0.0030808 | 1 |
| HTN | endometrium | *TPCN2* | 4.62735007 | 0.000503215 | 0.003073153 | 1 |
| HTN | endometrium | *PDZK1IP1* | 4.853198577 | 0.001329261 | 0.003213539 | 1 |
| HTN | endometrium | *CMPK1* | 6.021369565 | 0.000179536 | 0.003073153 | 0.871107211 |
| HTN | endometrium | *COA7* | -2.559540049 | 0.00993356 | 0.015108976 | 1 |
| HTN | endometrium | *C1orf123* | 2.720055811 | 0.001376133 | 0.003271434 | 1 |
| HTN | endometrium | *USP24* | 2.818598645 | 0.001176042 | 0.003095606 | 1 |
| HTN | endometrium | *ZSWIM5* | 3.7630878 | 0.000639505 | 0.003073153 | 1 |
| HTN | endometrium | *AK4* | 2.983495141 | 0.001118507 | 0.003095606 | 1 |
| HTN | endometrium | *RBP7* | 6.448668591 | 0.001476564 | 0.003392182 | 1 |
| HTN | endometrium | *SLC25A34* | -1.171362144 | 0.003450719 | 0.006252012 | 1 |
| HTN | endometrium | *PDPN* | 2.553457688 | 0.001217088 | 0.003095606 | 1 |
| HTN | endometrium | *LAPTM5* | 2.136464707 | 0.001124081 | 0.003095606 | 1 |
| HTN | endometrium | *SDC3* | 2.528711379 | 0.001474534 | 0.003390728 | 1 |
| HTN | endometrium | *SYNC* | 2.187685067 | 0.002742196 | 0.005244437 | 1 |
| HTN | endometrium | *CAMK2N1* | 5.477080132 | 0.000384724 | 0.003073153 | 1 |
| HTN | endometrium | *MEGF6* | 6.331629416 | 0.00986595 | 0.015010846 | 1 |
| HTN | endometrium | *TM2D1* | 3.652458606 | 0.00060508 | 0.003073153 | 1 |
| HTN | endometrium | *NEXN* | 2.263459242 | 0.000581022 | 0.003073153 | 1 |
| HTN | endometrium | *LRRIQ3* | 2.359353904 | 0.001222461 | 0.003095606 | 1 |
| HTN | endometrium | *TYW3* | 4.263298551 | 0.000692816 | 0.003073153 | 1 |

| | | | | | | |
|---|---|---|---|---|---|---|
| HTN | endometrium | *WDR63* | 1.945583233 | 0.001477947 | 0.003392273 | 1 |
| HTN | endometrium | *ARPC5* | 4.487639827 | 0.001742232 | 0.00375036 | 1 |
| HTN | endometrium | *FCRLB* | -2.671132701 | 0.004722832 | 0.008060003 | 1 |
| HTN | endometrium | *TDRD5* | -5.017192251 | 0.011242814 | 0.016753726 | 1 |
| HTN | endometrium | *BROX* | 3.056704598 | 0.000620371 | 0.003073153 | 1 |
| HTN | endometrium | *CNST* | 3.721126368 | 0.000595974 | 0.003073153 | 1 |
| HTN | endometrium | *PM20D1* | 3.108776014 | 0.004001581 | 0.007049989 | 1 |
| HTN | endometrium | *PKDCC* | 3.671038192 | 0.000547105 | 0.003073153 | 1 |
| HTN | endometrium | *B3GALNT2* | 2.723522745 | 0.014689596 | 0.021174664 | 1 |
| HTN | endometrium | *MRPL55* | 13.66451262 | 2.12E-11 | 3.44E-08 | 1.03E-07 |
| HTN | endometrium | *KCNJ3* | -1.508045719 | 0.004693236 | 0.008018162 | 1 |
| HTN | endometrium | *CLHC1* | 2.013149829 | 0.003244115 | 0.005964549 | 1 |
| HTN | endometrium | *DUSP19* | -3.988068732 | 0.034700053 | 0.045639647 | 1 |
| HTN | endometrium | *C2orf48* | 29.59116473 | 0.000381305 | 0.003073153 | 1 |
| HTN | endometrium | *CCDC74A* | 7.047927323 | 0.018552362 | 0.025993664 | 1 |
| HTN | endometrium | *NOSTRIN* | 2.935991156 | 0.014119518 | 0.020444017 | 1 |
| HTN | endometrium | *INHBB* | 5.456772797 | 0.000467386 | 0.003073153 | 1 |
| HTN | endometrium | *RPRD2* | 4.439246455 | 0.00078646 | 0.003073153 | 1 |
| HTN | endometrium | *CTSS* | 1.168233296 | 0.011829305 | 0.017504053 | 1 |
| HTN | endometrium | *PACRGL* | 4.086340393 | 0.000621995 | 0.003073153 | 1 |
| HTN | endometrium | *C1QTNF7* | 3.906671333 | 0.000449431 | 0.003073153 | 1 |
| HTN | endometrium | *LYSMD1* | -6.967968265 | 6.44E-05 | 0.002410623 | 0.312561879 |
| HTN | endometrium | *VPS72* | 5.319301052 | 0.000612988 | 0.003073153 | 1 |
| HTN | endometrium | *ERCC3* | 2.632479152 | 0.0006083 | 0.003073153 | 1 |
| HTN | endometrium | *RNF149* | 2.652551192 | 0.000656017 | 0.003073153 | 1 |
| HTN | endometrium | *DHX57* | 3.739895347 | 0.000744485 | 0.003073153 | 1 |
| HTN | endometrium | *S100A9* | 4.977094625 | 0.011537962 | 0.017146154 | 1 |
| HTN | endometrium | *TDRD10* | 2.050822872 | 0.001192464 | 0.003095606 | 1 |
| HTN | endometrium | *DCAF16* | 3.873296845 | 0.008870415 | 0.013653875 | 1 |
| HTN | endometrium | *GNPDA2* | 4.845382245 | 0.001944797 | 0.004061109 | 1 |
| HTN | endometrium | *NIPAL1* | 3.37245734 | 0.000695377 | 0.003073153 | 1 |
| HTN | endometrium | *ANTXR2* | 5.018400328 | 0.000596084 | 0.003073153 | 1 |
| HTN | endometrium | *CGGBP1* | 3.09838779 | 0.00170593 | 0.003709537 | 1 |
| HTN | endometrium | *GPR155* | 3.022412354 | 0.000912361 | 0.003087532 | 1 |
| HTN | endometrium | *DAPL1* | 8.294325705 | 0.000158291 | 0.003073153 | 0.768027492 |
| HTN | endometrium | *PBXIP1* | 2.617022958 | 0.000886168 | 0.003084413 | 1 |
| HTN | endometrium | *CLDN1* | 1.77273909 | 0.001223507 | 0.003095606 | 1 |
| HTN | endometrium | *DCST2* | 5.547248946 | 0.000323632 | 0.003073153 | 1 |
| HTN | endometrium | *SLC22A15* | 6.615226119 | 0.001683298 | 0.003672375 | 1 |
| HTN | endometrium | *LRRC58* | 6.683445513 | 0.001013618 | 0.003095606 | 1 |
| HTN | endometrium | *LMOD1* | 1.607400358 | 0.006685509 | 0.010709174 | 1 |
| HTN | endometrium | *TMEM169* | 4.48308068 | 0.001220227 | 0.003095606 | 1 |
| HTN | endometrium | *ARPC2* | 5.264995535 | 0.001095272 | 0.003095606 | 1 |
| HTN | endometrium | *GLB1L* | 2.735206902 | 0.003823273 | 0.006787603 | 1 |
| HTN | endometrium | *SERPINI1* | 6.81171426 | 0.003150239 | 0.00582506 | 1 |
| HTN | endometrium | *SUCLG1* | 6.026620104 | 0.003336188 | 0.006099166 | 1 |
| HTN | endometrium | *SPTA1* | -1.693299513 | 0.007714895 | 0.012110214 | 1 |
| HTN | endometrium | *PRKCI* | 3.375766982 | 0.001758506 | 0.003774504 | 1 |
| HTN | endometrium | *EFHB* | 1.48577143 | 0.013686708 | 0.019894704 | 1 |
| HTN | endometrium | *GTPBP8* | 5.064674038 | 0.000683564 | 0.003073153 | 1 |

| | | | | | | |
|---|---|---|---|---|---|---|
| HTN | endometrium | *WDFY3* | 3.477267255 | 0.00047644 | 0.003073153 | 1 |
| HTN | endometrium | *PTPN13* | 3.312279094 | 6.32E-05 | 0.002410623 | 0.306769387 |
| HTN | endometrium | *PPM1K* | 3.934528198 | 0.001134638 | 0.003095606 | 1 |
| HTN | endometrium | *TIPARP* | -2.381593427 | 0.037013738 | 0.048459432 | 1 |
| HTN | endometrium | *RPP14* | 3.62121741 | 0.001621827 | 0.00359484 | 1 |
| HTN | endometrium | *IL17RC* | 2.551790954 | 0.00353739 | 0.006373345 | 1 |
| HTN | endometrium | *CRELD1* | 2.692480416 | 0.001002168 | 0.003095606 | 1 |
| HTN | endometrium | *TTC14* | 4.191662838 | 0.003039239 | 0.005667328 | 1 |
| HTN | endometrium | *RCHY1* | 2.39725126 | 0.003144157 | 0.005816031 | 1 |
| HTN | endometrium | *CCDC158* | 5.710838635 | 0.000240447 | 0.003073153 | 1 |
| HTN | endometrium | *KIAA1143* | 3.895846519 | 0.002786161 | 0.005311769 | 1 |
| HTN | endometrium | *CDCP1* | 2.733070611 | 0.001058943 | 0.003095606 | 1 |
| HTN | endometrium | *CLEC3B* | 6.163673465 | 0.000478059 | 0.003073153 | 1 |
| HTN | endometrium | *FYCO1* | 2.678649207 | 0.002395131 | 0.004734383 | 1 |
| HTN | endometrium | *NMNAT3* | 3.848989698 | 0.000394223 | 0.003073153 | 1 |
| HTN | endometrium | *SNIP1* | 3.46701678 | 0.000916401 | 0.003087532 | 1 |
| HTN | endometrium | *RPN1* | 2.177511649 | 0.001170147 | 0.003095606 | 1 |
| HTN | endometrium | *SENP2* | 6.257945723 | 0.001199048 | 0.003095606 | 1 |
| HTN | endometrium | *IFT122* | 4.194460154 | 0.000550557 | 0.003073153 | 1 |
| HTN | endometrium | *SFMBT1* | 5.950988135 | 0.00040307 | 0.003073153 | 1 |
| HTN | endometrium | *UVSSA* | 4.690180716 | 0.001592573 | 0.003551086 | 1 |
| HTN | endometrium | *FAM208A* | 4.338717196 | 0.000141701 | 0.003073153 | 0.687534326 |
| HTN | endometrium | *LRPAP1* | 3.867389176 | 0.001108643 | 0.003095606 | 1 |
| HTN | endometrium | *PIGX* | 2.464349682 | 0.00571079 | 0.009389615 | 1 |
| HTN | endometrium | *ABLIM2* | 3.452612799 | 0.000608073 | 0.003073153 | 1 |
| HTN | endometrium | *METAP1* | 2.014027892 | 0.002758724 | 0.005269814 | 1 |
| HTN | endometrium | *ZNF589* | -7.917754806 | 9.84E-06 | 0.000852897 | 0.047762218 |
| HTN | endometrium | *ATRIP* | -1.981914007 | 0.00081955 | 0.0030808 | 1 |
| HTN | endometrium | *RNF123* | 11.18994335 | 1.14E-11 | 2.77E-08 | 5.53E-08 |
| HTN | endometrium | *MFSD8* | 5.638458817 | 0.001104373 | 0.003095606 | 1 |
| HTN | endometrium | *MST1R* | 5.794815429 | 5.61E-07 | 0.000108973 | 0.002724315 |
| HTN | endometrium | *RAD54L2* | 5.631006538 | 0.001092308 | 0.003095606 | 1 |
| HTN | endometrium | *TEX264* | 4.19801932 | 0.003236205 | 0.005954519 | 1 |
| HTN | endometrium | *ETNPPL* | 22.14786606 | 0.00110981 | 0.003095606 | 1 |
| HTN | endometrium | *HMGB2* | 7.784421874 | 0.000213241 | 0.003073153 | 1 |
| HTN | endometrium | *ANXA5* | 2.704299949 | 0.000808639 | 0.003073153 | 1 |
| HTN | endometrium | *MAP9* | 4.186398904 | 0.000688838 | 0.003073153 | 1 |
| HTN | endometrium | *HPGD* | -2.509943637 | 0.010191779 | 0.015443633 | 1 |
| HTN | endometrium | *C4orf45* | 7.820988631 | 0.000336183 | 0.003073153 | 1 |
| HTN | endometrium | *TMEM144* | 2.222869875 | 0.002264178 | 0.004545218 | 1 |
| HTN | endometrium | *IL15* | 2.916477756 | 0.000621777 | 0.003073153 | 1 |
| HTN | endometrium | *ARFIP1* | 4.44870579 | 0.000360078 | 0.003073153 | 1 |
| HTN | endometrium | *LSM6* | 4.211982963 | 0.000499515 | 0.003073153 | 1 |
| HTN | endometrium | *ITGA2* | 2.336387102 | 0.00458702 | 0.00786995 | 1 |
| HTN | endometrium | *MOCS2* | 3.337403262 | 0.001038063 | 0.003095606 | 1 |
| HTN | endometrium | *NDUFAF2* | 3.858849412 | 0.00056878 | 0.003073153 | 1 |
| HTN | endometrium | *LMBRD2* | 5.119249506 | 0.003453797 | 0.006255253 | 1 |
| HTN | endometrium | *RNF180* | 2.803441007 | 0.000782611 | 0.003073153 | 1 |
| HTN | endometrium | *PGGT1B* | 3.582094932 | 0.000703126 | 0.003073153 | 1 |
| HTN | endometrium | *F2RL2* | 7.571220394 | 0.000999484 | 0.003095606 | 1 |

| | | | | | | |
|---|---|---|---|---|---|---|
| HTN | endometrium | *CCDC112* | 2.545362294 | 0.000895062 | 0.003087532 | 1 |
| HTN | endometrium | *C5orf63* | 4.138428577 | 0.001650634 | 0.00362734 | 1 |
| HTN | endometrium | *F2RL1* | 10.53060458 | 0.02045076 | 0.028269825 | 1 |
| HTN | endometrium | *WDR41* | 6.267969768 | 0.00309502 | 0.005742653 | 1 |
| HTN | endometrium | *SERINC5* | 2.602502135 | 0.001097134 | 0.003095606 | 1 |
| HTN | endometrium | *CASP3* | 3.716563113 | 0.001514939 | 0.003433202 | 1 |
| HTN | endometrium | *PRIMPOL* | 4.338273053 | 0.000933661 | 0.003095606 | 1 |
| HTN | endometrium | *ERAP1* | 1.984594277 | 0.016738526 | 0.023774979 | 1 |
| HTN | endometrium | *ERAP2* | 4.000849041 | 3.03E-06 | 0.000419407 | 0.014679237 |
| HTN | endometrium | *UTP15* | -2.214123578 | 0.008103607 | 0.012614276 | 1 |
| HTN | endometrium | *TLR3* | 8.812179715 | 0.001415222 | 0.003314023 | 1 |
| HTN | endometrium | *NSA2* | 2.160013276 | 0.011031481 | 0.016484369 | 1 |
| HTN | endometrium | *CCDC127* | 4.62201856 | 0.011076177 | 0.01653588 | 1 |
| HTN | endometrium | *PDSS2* | 2.999281253 | 0.000605405 | 0.003073153 | 1 |
| HTN | endometrium | *RAET1E* | 3.118953436 | 0.001222724 | 0.003095606 | 1 |
| HTN | endometrium | *STK17A* | 5.22350022 | 0.000845661 | 0.0030808 | 1 |
| HTN | endometrium | *GALNT10* | 2.78423428 | 0.000633744 | 0.003073153 | 1 |
| HTN | endometrium | *SAP30L* | 2.819251059 | 0.000582249 | 0.003073153 | 1 |
| HTN | endometrium | *RPS14* | 5.032467897 | 0.006804603 | 0.01088916 | 1 |
| HTN | endometrium | *RP9* | 4.08652796 | 0.000813322 | 0.003074195 | 1 |
| HTN | endometrium | *MIOS* | 3.368626338 | 0.000975715 | 0.003095606 | 1 |
| HTN | endometrium | *CHMP4C* | 9.065247721 | 0.001133685 | 0.003095606 | 1 |
| HTN | endometrium | *SLC13A4* | 2.727600031 | 0.000916832 | 0.003087532 | 1 |
| HTN | endometrium | *CTSB* | 1.145099026 | 0.010490929 | 0.015817896 | 1 |
| HTN | endometrium | *SOX17* | 5.627435644 | 0.000120053 | 0.00304009 | 0.582494905 |
| HTN | endometrium | *TNFRSF11B* | 4.816467844 | 0.000919789 | 0.003087532 | 1 |
| HTN | endometrium | *SUN1* | 4.975738961 | 0.00720709 | 0.011453915 | 1 |
| HTN | endometrium | *GPER1* | 8.74663916 | 0.000104757 | 0.002934745 | 0.508282461 |
| HTN | endometrium | *INTS1* | 9.057917803 | 0.013746198 | 0.019975008 | 1 |
| HTN | endometrium | *ALDH7A1* | 6.382020195 | 0.000116677 | 0.003027371 | 0.566118292 |
| HTN | endometrium | *YWHAZ* | 4.476168708 | 0.004631886 | 0.007925868 | 1 |
| HTN | endometrium | *SLC25A32* | 4.510940571 | 0.001859085 | 0.003928693 | 1 |
| HTN | endometrium | *DCSTAMP* | -4.486483624 | 0.005655824 | 0.009327688 | 1 |
| HTN | endometrium | *RPP25L* | -5.013845806 | 0.000312759 | 0.003073153 | 1 |
| HTN | endometrium | *C9orf24* | 3.035934367 | 0.000848533 | 0.0030808 | 1 |
| HTN | endometrium | *TMEM65* | 5.352607597 | 0.000577596 | 0.003073153 | 1 |
| HTN | endometrium | *CCDC171* | 6.343470629 | 0.003069904 | 0.005707362 | 1 |
| HTN | endometrium | *METTL2B* | 3.716836868 | 0.000725077 | 0.003073153 | 1 |
| HTN | endometrium | *ZMAT4* | 5.701804893 | 0.001018724 | 0.003095606 | 1 |
| HTN | endometrium | *C8orf34* | 2.015868639 | 0.001226886 | 0.003095606 | 1 |
| HTN | endometrium | *ANKS6* | 5.23022687 | 0.000578995 | 0.003073153 | 1 |
| HTN | endometrium | *AQP7* | 2.419951795 | 4.63E-05 | 0.002206299 | 0.224555948 |
| HTN | endometrium | *NOL6* | -2.957242449 | 0.011597053 | 0.017218146 | 1 |
| HTN | endometrium | *AQP3* | 9.149262215 | 0.001210711 | 0.003095606 | 1 |
| HTN | endometrium | *TRMT10B* | 3.60779599 | 0.000583447 | 0.003073153 | 1 |
| HTN | endometrium | *VCP* | 4.293863515 | 0.00105331 | 0.003095606 | 1 |
| HTN | endometrium | *PIGO* | 4.184304611 | 0.000603002 | 0.003073153 | 1 |
| HTN | endometrium | *WRN* | 3.924169259 | 0.000226826 | 0.003073153 | 1 |
| HTN | endometrium | *MARCH8* | 5.820849047 | 0.000821772 | 0.0030808 | 1 |
| HTN | endometrium | *TSHR* | 4.467663974 | 0.001561413 | 0.003499546 | 1 |

| | | | | | | |
|---|---|---|---|---|---|---|
| HTN | endometrium | *CFL2* | 8.934161862 | 0.007718412 | 0.012111816 | 1 |
| HTN | endometrium | *PGM2L1* | 2.551970987 | 0.004425005 | 0.007637895 | 1 |
| HTN | endometrium | *SLC16A9* | 3.940284848 | 0.001203277 | 0.003095606 | 1 |
| HTN | endometrium | *GJB2* | 17.1081325 | 0.001180324 | 0.003095606 | 1 |
| HTN | endometrium | *CRYL1* | 5.2049374 | 0.000613915 | 0.003073153 | 1 |
| HTN | endometrium | *MICU2* | 6.049159997 | 0.0004659 | 0.003073153 | 1 |
| HTN | endometrium | *LRR1* | 13.06961737 | 0.008286811 | 0.012874673 | 1 |
| HTN | endometrium | *TMEM63C* | -3.05053826 | 0.0074107 | 0.011708472 | 1 |
| HTN | endometrium | *NOXRED1* | 3.449839859 | 0.001802959 | 0.003846947 | 1 |
| HTN | endometrium | *ATP5C1* | 6.045740336 | 0.000299783 | 0.003073153 | 1 |
| HTN | endometrium | *VDAC2* | 6.464573657 | 0.002922117 | 0.005499655 | 1 |
| HTN | endometrium | *PDZD8* | 5.045314811 | 0.007814937 | 0.012239533 | 1 |
| HTN | endometrium | *QSOX2* | 3.49657843 | 0.000683439 | 0.003073153 | 1 |
| HTN | endometrium | *CLEC1B* | -2.491147047 | 0.004916402 | 0.008308736 | 1 |
| HTN | endometrium | *TSC1* | 2.939292602 | 0.000719644 | 0.003073153 | 1 |
| HTN | endometrium | *STOX1* | 2.738213149 | 0.000110709 | 0.002992864 | 0.537157767 |
| HTN | endometrium | *RET* | 4.268636577 | 0.001327088 | 0.003212251 | 1 |
| HTN | endometrium | *VWA2* | 7.735041356 | 0.002085855 | 0.004259498 | 1 |
| HTN | endometrium | *METTL3* | 6.779224426 | 0.032956323 | 0.043582469 | 1 |
| HTN | endometrium | *TRUB1* | 2.208034797 | 0.003023146 | 0.005643826 | 1 |
| HTN | endometrium | *HSPA12A* | 3.838578872 | 8.19E-05 | 0.002714492 | 0.397383822 |
| HTN | endometrium | *UBTD1* | 7.577604719 | 0.001274757 | 0.003141249 | 1 |
| HTN | endometrium | *OTOGL* | 4.0389067 | 0.03203073 | 0.042462597 | 1 |
| HTN | endometrium | *TTC7B* | 4.908604624 | 0.000675265 | 0.003073153 | 1 |
| HTN | endometrium | *RAPSN* | 4.319837945 | 0.019866432 | 0.027556297 | 1 |
| HTN | endometrium | *TC2N* | 1.596585291 | 0.001707213 | 0.003709537 | 1 |
| HTN | endometrium | *CPSF2* | 2.38075823 | 0.034585894 | 0.045526521 | 1 |
| HTN | endometrium | *C1QL3* | -2.148790075 | 0.022359635 | 0.030612006 | 1 |
| HTN | endometrium | *ARL5B* | 2.61543674 | 0.009246301 | 0.014165789 | 1 |
| HTN | endometrium | *LIPC* | 3.867577632 | 0.001847855 | 0.003913325 | 1 |
| HTN | endometrium | *CEP57* | 2.392599845 | 0.002824547 | 0.005365967 | 1 |
| HTN | endometrium | *TCP11L2* | 4.957380602 | 0.000539568 | 0.003073153 | 1 |
| HTN | endometrium | *JAM3* | 3.760971733 | 0.002864262 | 0.005424434 | 1 |
| HTN | endometrium | *IKBIP* | 4.85827506 | 0.000373505 | 0.003073153 | 1 |
| HTN | endometrium | *ZFYVE19* | 2.560290763 | 0.01105546 | 0.016510031 | 1 |
| HTN | endometrium | *TRMT61A* | 5.134422957 | 0.00150371 | 0.00342122 | 1 |
| HTN | endometrium | *LARP6* | 4.416017089 | 0.000665001 | 0.003073153 | 1 |
| HTN | endometrium | *API5* | 5.754349823 | 0.00060041 | 0.003073153 | 1 |
| HTN | endometrium | *ASPG* | 12.6750941 | 0.018955501 | 0.026482031 | 1 |
| HTN | endometrium | *GABRB3* | 3.747822307 | 0.000458751 | 0.003073153 | 1 |
| HTN | endometrium | *COX11* | 6.116147176 | 0.026080057 | 0.035159888 | 1 |
| HTN | endometrium | *STXBP4* | 5.921188052 | 0.000836503 | 0.0030808 | 1 |
| HTN | endometrium | *WBP1L* | 7.107415775 | 0.000137492 | 0.003073153 | 0.667109503 |
| HTN | endometrium | *APBB1* | 7.277447358 | 0.000795435 | 0.003073153 | 1 |
| HTN | endometrium | *NUDT13* | -3.553102081 | 0.006906941 | 0.011023841 | 1 |
| HTN | endometrium | *CYB5R2* | 6.633779048 | 7.36E-06 | 0.000685047 | 0.035700518 |
| HTN | endometrium | *TRIM66* | -3.84597167 | 3.88E-05 | 0.002046851 | 0.188310315 |
| HTN | endometrium | *RPL27A* | 5.727115844 | 0.026575541 | 0.035798036 | 1 |
| HTN | endometrium | *CDYL2* | 2.630293304 | 0.008913996 | 0.013708624 | 1 |
| HTN | endometrium | *CENPN* | 6.299593755 | 0.001038032 | 0.003095606 | 1 |

| | | | | | | |
|---|---|---|---|---|---|---|
| HTN | endometrium | *C16orf46* | 13.50329697 | 0.005355892 | 0.008939384 | 1 |
| HTN | endometrium | *TMEM41B* | 3.668850604 | 0.000710282 | 0.003073153 | 1 |
| HTN | endometrium | *TMX3* | 3.967384217 | 0.000513674 | 0.003073153 | 1 |
| HTN | endometrium | *PRKCB* | 1.833318786 | 0.001007984 | 0.003095606 | 1 |
| HTN | endometrium | *NDST2* | -3.327471365 | 0.004189353 | 0.007314412 | 1 |
| HTN | endometrium | *CCDC68* | 4.214976251 | 0.003061859 | 0.005698628 | 1 |
| HTN | endometrium | *CLEC4D* | 11.05484938 | 0.014057054 | 0.020365729 | 1 |
| HTN | endometrium | *ZSCAN21* | 1.910502124 | 0.013254674 | 0.019382664 | 1 |
| HTN | endometrium | *TMED3* | -1.667651704 | 0.003420775 | 0.00621633 | 1 |
| HTN | endometrium | *SEC11C* | 2.848683631 | 0.001135274 | 0.003095606 | 1 |
| HTN | endometrium | *SPDYE6* | -1.581338103 | 0.033053155 | 0.043674811 | 1 |
| HTN | endometrium | *ZNF592* | 4.011758429 | 0.000665297 | 0.003073153 | 1 |
| HTN | endometrium | *ACSM1* | 4.239272902 | 0.000761006 | 0.003073153 | 1 |
| HTN | endometrium | *SLFN5* | 4.953753824 | 0.000631794 | 0.003073153 | 1 |
| HTN | endometrium | *CATSPER2* | 2.601975771 | 0.001150422 | 0.003095606 | 1 |
| HTN | endometrium | *ZNF667-AS1* | 3.526028271 | 0.001953051 | 0.004068787 | 1 |
| HTN | endometrium | *YPEL4* | 6.457782205 | 0.002662928 | 0.00513943 | 1 |
| HTN | endometrium | *LDHC* | 3.089231398 | 0.004724826 | 0.008060003 | 1 |
| HTN | endometrium | *FAM111A* | 3.505589475 | 0.014374981 | 0.020758157 | 1 |
| HTN | endometrium | *ANPEP* | 2.773121004 | 0.000717587 | 0.003073153 | 1 |
| HTN | endometrium | *DCTN5* | 2.522150177 | 0.003293201 | 0.006036498 | 1 |
| HTN | endometrium | *STAT6* | 3.689904384 | 0.000935454 | 0.003095606 | 1 |
| HTN | endometrium | *STX3* | 6.700461652 | 0.001937619 | 0.004052297 | 1 |
| HTN | endometrium | *PIP4K2C* | 3.510099491 | 0.001013855 | 0.003095606 | 1 |
| HTN | endometrium | *NYAP1* | 5.263542165 | 1.86E-05 | 0.001272617 | 0.090355827 |
| HTN | endometrium | *MS4A7* | 1.338977198 | 0.017672236 | 0.024897122 | 1 |
| HTN | endometrium | *DIS3L* | 3.44296075 | 0.001967063 | 0.004094461 | 1 |
| HTN | endometrium | *CCNDBP1* | 2.552625896 | 0.000880271 | 0.003084413 | 1 |
| HTN | endometrium | *EPB42* | 4.210979165 | 0.000573427 | 0.003073153 | 1 |
| HTN | endometrium | *SMAD3* | 4.809211193 | 0.001166281 | 0.003095606 | 1 |
| HTN | endometrium | *RCCD1* | 3.496338164 | 0.002252468 | 0.004531083 | 1 |
| HTN | endometrium | *CNPY4* | 4.559379678 | 8.10E-05 | 0.002714492 | 0.393051786 |
| HTN | endometrium | *MEI1* | 4.024462667 | 0.000942432 | 0.003095606 | 1 |
| HTN | endometrium | *SAMD14* | 7.124253185 | 0.003189337 | 0.005883902 | 1 |
| HTN | endometrium | *ACSF2* | 2.747889837 | 0.001091239 | 0.003095606 | 1 |
| HTN | endometrium | *URM1* | 4.564915915 | 0.002901168 | 0.005479356 | 1 |
| HTN | endometrium | *ENDOG* | 2.719399562 | 0.023850295 | 0.032442285 | 1 |
| HTN | endometrium | *SP2* | 8.480332204 | 0.000849977 | 0.0030808 | 1 |
| HTN | endometrium | *GPRC5B* | 2.27506637 | 0.00249625 | 0.004883793 | 1 |
| HTN | endometrium | *FBXO22* | 2.031461461 | 0.00107112 | 0.003095606 | 1 |
| HTN | endometrium | *LOXHD1* | 2.153276028 | 0.001061961 | 0.003095606 | 1 |
| HTN | endometrium | *KATNAL2* | 2.411063351 | 0.000674314 | 0.003073153 | 1 |
| HTN | endometrium | *ZNF91* | 3.533868151 | 0.001180506 | 0.003095606 | 1 |
| HTN | endometrium | *POP5* | 2.386093317 | 0.00144367 | 0.003341453 | 1 |
| HTN | endometrium | *MYO5B* | 2.057460284 | 0.001707055 | 0.003709537 | 1 |
| HTN | endometrium | *ZNF229* | -3.234311363 | 0.018983433 | 0.026513419 | 1 |
| HTN | endometrium | *RAB8A* | 6.336342644 | 0.00023733 | 0.003073153 | 1 |
| HTN | endometrium | *JSRP1* | 6.896993258 | 0.014766807 | 0.021273322 | 1 |
| HTN | endometrium | *SPATA33* | 6.04639133 | 0.000859138 | 0.0030808 | 1 |
| HTN | endometrium | *PROCA1* | -2.554922466 | 0.03713138 | 0.048587422 | 1 |

| | | | | | | |
|---|---|---|---|---|---|---|
| HTN | endometrium | *RPL13* | 4.141651969 | 0.000794706 | 0.003073153 | 1 |
| HTN | endometrium | *TP53I13* | 2.493190112 | 0.000992222 | 0.003095606 | 1 |
| HTN | endometrium | *TUBA1C* | 4.869497586 | 0.001069689 | 0.003095606 | 1 |
| HTN | endometrium | *TMC4* | 5.84905314 | 0.001155294 | 0.003095606 | 1 |
| HTN | endometrium | *TTYH1* | 4.21340343 | 0.001103206 | 0.003095606 | 1 |
| HTN | endometrium | *LENG8* | 5.401688234 | 0.00038788 | 0.003073153 | 1 |
| HTN | endometrium | *CDC42EP5* | -2.022048233 | 0.006177913 | 0.010031872 | 1 |
| HTN | endometrium | *C19orf33* | 5.435557056 | 0.000457741 | 0.003073153 | 1 |
| HTN | endometrium | *CHAF1A* | -3.320784865 | 0.006126792 | 0.009968878 | 1 |
| HTN | endometrium | *FAM57A* | 3.144972411 | 0.011849555 | 0.017514657 | 1 |
| HTN | endometrium | *SERPINF2* | 4.809469565 | 0.000794506 | 0.003073153 | 1 |
| HTN | endometrium | *SRR* | 5.222556026 | 3.71E-07 | 7.82E-05 | 0.001798268 |
| HTN | endometrium | *TRPV3* | 4.842904159 | 0.000118651 | 0.003039787 | 0.575696963 |
| HTN | endometrium | *CYB5D2* | 4.440101429 | 0.000291358 | 0.003073153 | 1 |
| HTN | endometrium | *KLK5* | -2.607258287 | 0.004867262 | 0.008242917 | 1 |
| HTN | endometrium | *KLK6* | 25.13681342 | 0.001048892 | 0.003095606 | 1 |
| HTN | endometrium | *ZNF83* | 4.140496019 | 0.000430673 | 0.003073153 | 1 |
| HTN | endometrium | *SPRYD3* | 3.362385909 | 0.002449072 | 0.004812837 | 1 |
| HTN | endometrium | *SOAT2* | -0.502830744 | 0.006320522 | 0.010225799 | 1 |
| HTN | endometrium | *ZNF558* | 6.259073374 | 0.001114246 | 0.003095606 | 1 |
| HTN | endometrium | *NDUFV1* | 7.285259613 | 0.001410319 | 0.003309766 | 1 |
| HTN | endometrium | *NUDT8* | -2.797211674 | 0.005915234 | 0.009676573 | 1 |
| HTN | endometrium | *PRDX2* | 4.796048894 | 0.000526717 | 0.003073153 | 1 |
| HTN | endometrium | *MIS12* | 3.735718988 | 0.000833746 | 0.0030808 | 1 |
| HTN | endometrium | *MGAT5B* | -1.143536284 | 0.003826684 | 0.006791174 | 1 |
| HTN | endometrium | *TMEM99* | 3.461216076 | 0.000712018 | 0.003073153 | 1 |
| HTN | endometrium | *ZNF598* | -8.290423563 | 0.002628215 | 0.005086597 | 1 |
| HTN | endometrium | *MLST8* | 6.750200909 | 0.000842559 | 0.0030808 | 1 |
| HTN | endometrium | *ATG16L2* | 2.511801231 | 0.00097479 | 0.003095606 | 1 |
| HTN | endometrium | *RPSA* | 4.627194973 | 0.000145377 | 0.003073153 | 0.705368623 |
| HTN | endometrium | *LTBP3* | 2.066435956 | 0.001445061 | 0.003341962 | 1 |
| HTN | endometrium | *KIAA1586* | 3.281951668 | 0.004033981 | 0.007091621 | 1 |
| HTN | endometrium | *FAM83B* | -7.402060147 | 0.004518374 | 0.007760407 | 1 |
| HTN | endometrium | *POLR2J3* | 2.38711024 | 0.00685169 | 0.010944484 | 1 |
| HTN | endometrium | *NT5DC2* | -5.719748283 | 0.008861369 | 0.013644989 | 1 |
| HTN | endometrium | *COA6* | 3.310552912 | 0.00117177 | 0.003095606 | 1 |
| HTN | endometrium | *PDHB* | 8.497244432 | 4.32E-05 | 0.002140353 | 0.209754582 |
| HTN | endometrium | *PXK* | 2.641045283 | 0.001379676 | 0.00327409 | 1 |
| HTN | endometrium | *PCMTD1* | 2.980862181 | 0.000640653 | 0.003073153 | 1 |
| HTN | endometrium | *ACOX2* | 2.442631987 | 0.002188314 | 0.004429579 | 1 |
| HTN | endometrium | *SCN11A* | 2.72330987 | 0.003358336 | 0.006135033 | 1 |
| HTN | endometrium | *SEPT2* | 2.582720378 | 0.003449618 | 0.006252012 | 1 |
| HTN | endometrium | *ING5* | 3.976104146 | 0.000553736 | 0.003073153 | 1 |
| HTN | endometrium | *MLKL* | 5.586844364 | 0.001072859 | 0.003095606 | 1 |
| HTN | endometrium | *KLHL30* | 11.48248066 | 0.002114933 | 0.004302581 | 1 |
| HTN | endometrium | *CCDC110* | 8.521936124 | 0.00088501 | 0.003084413 | 1 |
| HTN | endometrium | *FNTA* | 3.037262658 | 0.00117239 | 0.003095606 | 1 |
| HTN | endometrium | *SLC20A2* | 4.031868374 | 0.00116651 | 0.003095606 | 1 |
| HTN | endometrium | *TMUB2* | 2.176047632 | 0.001837881 | 0.003900873 | 1 |
| HTN | endometrium | *NBPF9* | 6.330307817 | 0.000758615 | 0.003073153 | 1 |

| | | | | | | |
|---|---|---|---|---|---|---|
| HTN | endometrium | *ADAM9* | 2.839555579 | 0.000440393 | 0.003073153 | 1 |
| HTN | endometrium | *SLC16A4* | 4.111359259 | 0.000258746 | 0.003073153 | 1 |
| HTN | endometrium | *DNAJC21* | 5.476568955 | 0.000562217 | 0.003073153 | 1 |
| HTN | endometrium | *NPNT* | 2.137149284 | 0.000693278 | 0.003073153 | 1 |
| HTN | endometrium | *GSTM4* | 3.172063589 | 0.002842962 | 0.005390407 | 1 |
| HTN | endometrium | *PPIP5K1* | 2.761311534 | 0.002062906 | 0.004228653 | 1 |
| HTN | endometrium | *SNTB2* | 4.605820716 | 0.000431369 | 0.003073153 | 1 |
| HTN | endometrium | *ZNF507* | 4.634253934 | 0.000919409 | 0.003087532 | 1 |
| HTN | endometrium | *ZBTB49* | 5.854410716 | 0.000564029 | 0.003073153 | 1 |
| HTN | endometrium | *GFM1* | 5.538968343 | 0.000477025 | 0.003073153 | 1 |
| HTN | endometrium | *ATOH8* | 2.87686931 | 0.000415155 | 0.003073153 | 1 |
| HTN | endometrium | *BTNL3* | 3.94815797 | 0.010503143 | 0.015826475 | 1 |
| HTN | endometrium | *CEP120* | 3.365946416 | 0.001151205 | 0.003095606 | 1 |
| HTN | endometrium | *MFF* | 2.661239322 | 0.000766518 | 0.003073153 | 1 |
| HTN | endometrium | *LGALS9* | 3.9623173 | 0.000173986 | 0.003073153 | 0.844178393 |
| HTN | endometrium | *CPLX1* | 2.927646177 | 0.001057009 | 0.003095606 | 1 |
| HTN | endometrium | *SIGLEC7* | 4.290902118 | 0.005443646 | 0.009048499 | 1 |
| HTN | endometrium | *MAP2K1* | 3.157594133 | 0.000699414 | 0.003073153 | 1 |
| HTN | endometrium | *KLK7* | 4.58007835 | 0.001512975 | 0.003430353 | 1 |
| HTN | endometrium | *UPF3A* | 4.045886929 | 3.80E-05 | 0.002046851 | 0.184447179 |
| HTN | endometrium | *C8orf46* | 2.788074393 | 5.06E-06 | 0.00057133 | 0.024567202 |
| HTN | endometrium | *HSPBAP1* | 1.909222218 | 0.001677272 | 0.003664172 | 1 |
| HTN | endometrium | *UBE2V2* | 5.973380339 | 0.000580301 | 0.003073153 | 1 |
| HTN | endometrium | *CPT1C* | 6.961200781 | 2.27E-07 | 5.51E-05 | 0.001101021 |
| HTN | endometrium | *PCSK9* | 20.7044363 | 0.002883179 | 0.005453873 | 1 |
| HTN | endometrium | *XPO6* | 5.982715917 | 0.002909036 | 0.005489943 | 1 |
| HTN | endometrium | *RGS14* | -1.304753093 | 0.001295997 | 0.003163068 | 1 |
| HTN | endometrium | *RAB24* | 5.367388274 | 0.014012933 | 0.020313938 | 1 |
| HTN | endometrium | *THBS3* | 9.212664288 | 0.000734035 | 0.003073153 | 1 |
| HTN | endometrium | *NPIPB3* | 6.316317479 | 0.001384819 | 0.003277629 | 1 |
| HTN | endometrium | *SH3TC2* | 2.321139115 | 0.016366537 | 0.023308024 | 1 |
| HTN | endometrium | *PGM2* | 2.861025647 | 0.000732438 | 0.003073153 | 1 |
| HTN | endometrium | *P2RY12* | 4.47126189 | 0.000107299 | 0.002958035 | 0.520614117 |
| HTN | endometrium | *SNUPN* | 4.25118569 | 0.037263772 | 0.048734183 | 1 |
| HTN | endometrium | *RNASE6* | -1.611798062 | 0.014155048 | 0.020479772 | 1 |
| HTN | endometrium | *CD52* | 1.599663016 | 0.022739375 | 0.031061781 | 1 |
| HTN | endometrium | *SLC38A11* | -1.531898367 | 0.007422387 | 0.011723119 | 1 |
| HTN | endometrium | *CCDC8* | 4.466000533 | 0.001328066 | 0.003212251 | 1 |
| HTN | endometrium | *ZEB2* | 2.349818892 | 0.004417391 | 0.007635619 | 1 |
| HTN | endometrium | *HINT1* | 6.507063081 | 0.001010752 | 0.003095606 | 1 |
| HTN | endometrium | *DTWD2* | 3.882072685 | 0.001984448 | 0.004118281 | 1 |
| HTN | endometrium | *INO80E* | -7.393510705 | 8.61E-05 | 0.002714492 | 0.417985376 |
| HTN | endometrium | *DFFB* | -3.093004967 | 0.016172591 | 0.023079238 | 1 |
| HTN | endometrium | *NFU1* | 8.667792487 | 6.14E-05 | 0.002410623 | 0.297834969 |
| HTN | endometrium | *PROKR1* | 8.041237769 | 0.021181379 | 0.029156428 | 1 |
| HTN | endometrium | *LUZP1* | 3.326705057 | 0.000708631 | 0.003073153 | 1 |
| HTN | endometrium | *CHRNA5* | 6.599650254 | 0.015199279 | 0.021825067 | 1 |
| HTN | endometrium | *UGP2* | 4.515588784 | 0.004139479 | 0.007248197 | 1 |
| HTN | endometrium | *HNRNPF* | 6.320991799 | 0.002084582 | 0.004258692 | 1 |
| HTN | endometrium | *CSGALNACT2* | 5.433847411 | 0.000559831 | 0.003073153 | 1 |

| | | | | | | |
|---|---|---|---|---|---|---|
| HTN | endometrium | *CTNND2* | 2.308648212 | 0.009067044 | 0.013913124 | 1 |
| HTN | endometrium | *CALML6* | 66.08607161 | 0.000378917 | 0.003073153 | 1 |
| HTN | endometrium | *MUC3A* | -4.25408321 | 0.000133022 | 0.003073153 | 0.645421494 |
| HTN | endometrium | *TPST1* | 4.364220443 | 0.000412741 | 0.003073153 | 1 |
| HTN | endometrium | *TOR1AIP2* | 4.290836171 | 0.000658156 | 0.003073153 | 1 |
| HTN | endometrium | *ZFPM2* | 2.413819235 | 0.002412169 | 0.004761531 | 1 |
| HTN | endometrium | *MAP3K2* | 3.057106931 | 0.000720907 | 0.003073153 | 1 |
| HTN | endometrium | *MYO7B* | 6.930039147 | 1.26E-06 | 0.000227302 | 0.00613715 |
| HTN | endometrium | *UBE2E3* | 5.007198541 | 0.019804569 | 0.027478344 | 1 |
| HTN | endometrium | *NIPA1* | 2.587246021 | 0.00155767 | 0.003498988 | 1 |
| HTN | endometrium | *HNRNPA3* | 5.199990853 | 0.000208322 | 0.003073153 | 1 |
| HTN | endometrium | *CHRNB1* | 1.855599267 | 0.033649168 | 0.044377756 | 1 |
| HTN | endometrium | *USP38* | 6.821882102 | 0.001141622 | 0.003095606 | 1 |
| HTN | endometrium | *PDCD6IP* | 3.591832045 | 0.000609985 | 0.003073153 | 1 |
| HTN | endometrium | *CRTAP* | 6.372196928 | 0.000320639 | 0.003073153 | 1 |
| HTN | endometrium | *CNGB3* | 2.356087634 | 0.000412655 | 0.003073153 | 1 |
| HTN | endometrium | *STX8* | 2.594367294 | 0.000609789 | 0.003073153 | 1 |
| HTN | endometrium | *NFRKB* | 5.332443575 | 0.000527757 | 0.003073153 | 1 |
| HTN | endometrium | *ADORA2B* | 3.166305244 | 0.00681483 | 0.010901931 | 1 |
| HTN | endometrium | *MGMT* | 4.085941542 | 0.000645882 | 0.003073153 | 1 |
| HTN | endometrium | *NFXL1* | 7.772358094 | 0.000670794 | 0.003073153 | 1 |
| HTN | endometrium | *HSD17B13* | 6.851996772 | 0.001538084 | 0.003470431 | 1 |
| HTN | endometrium | *ELOVL6* | 5.532670736 | 0.000575514 | 0.003073153 | 1 |
| HTN | endometrium | *SERPINB9* | 2.183619851 | 0.001026435 | 0.003095606 | 1 |
| HTN | endometrium | *SMAGP* | -3.004819873 | 0.003744594 | 0.00667724 | 1 |
| HTN | endometrium | *CDH2* | -1.009610804 | 0.004244232 | 0.007391605 | 1 |
| HTN | endometrium | *EMB* | 1.543389883 | 0.007390845 | 0.011681323 | 1 |
| HTN | endometrium | *STAT2* | 5.37373735 | 0.001073198 | 0.003095606 | 1 |
| HTN | endometrium | *SLC26A5* | 7.593444847 | 0.007770512 | 0.012177817 | 1 |
| HTN | endometrium | *GTSF1* | 7.294709199 | 0.034185718 | 0.045060882 | 1 |
| HTN | endometrium | *ARMC10* | 5.467250347 | 0.007971609 | 0.012452751 | 1 |
| HTN | endometrium | *ACYP2* | 4.117152638 | 0.000991976 | 0.003095606 | 1 |
| HTN | endometrium | *TRABD* | 2.835245604 | 0.000771331 | 0.003073153 | 1 |
| HTN | endometrium | *HOXB9* | 12.59176068 | 0.006058022 | 0.009870223 | 1 |
| HTN | endometrium | *FOXN2* | 2.415314764 | 0.000857246 | 0.0030808 | 1 |
| HTN | endometrium | *TRIAP1* | 4.868856996 | 0.00157728 | 0.003528506 | 1 |
| HTN | endometrium | *TSEN34* | 4.1815381 | 0.001099244 | 0.003095606 | 1 |
| HTN | endometrium | *MSANTD4* | 2.770887471 | 0.001096464 | 0.003095606 | 1 |
| HTN | endometrium | *NDUFA3* | 9.305111881 | 0.001562119 | 0.003499546 | 1 |
| HTN | endometrium | *NUDT6* | 6.66933785 | 0.000682954 | 0.003073153 | 1 |
| HTN | endometrium | *ZNF160* | 1.922991535 | 0.002772746 | 0.005290352 | 1 |
| HTN | endometrium | *ZNF415* | 8.513080748 | 0.001504781 | 0.00342122 | 1 |
| HTN | endometrium | *PYGO1* | 3.319438865 | 0.000688864 | 0.003073153 | 1 |
| HTN | endometrium | *TSNARE1* | 2.640810116 | 0.000327461 | 0.003073153 | 1 |
| HTN | endometrium | *FPR1* | 2.740273937 | 0.001675083 | 0.003662686 | 1 |
| HTN | endometrium | *FEZ2* | 4.155470779 | 0.000293007 | 0.003073153 | 1 |
| HTN | endometrium | *TRMT61B* | 5.062208459 | 0.004751401 | 0.008086214 | 1 |
| HTN | endometrium | *MFN1* | 6.589216296 | 0.003694429 | 0.006599915 | 1 |
| HTN | endometrium | *GIMAP8* | 4.230032323 | 0.000137832 | 0.003073153 | 0.668762524 |
| HTN | endometrium | *ATP6V0E2* | -2.706361758 | 0.007257834 | 0.011514467 | 1 |

| | | | | | | |
|---|---|---|---|---|---|---|
| HTN | endometrium | *SCAND1* | 3.926664645 | 0.004231165 | 0.007371866 | 1 |
| HTN | endometrium | *SOSTDC1* | 6.407088244 | 0.014819328 | 0.021336314 | 1 |
| HTN | endometrium | *PGAM1* | 3.4002567 | 0.004799719 | 0.008154145 | 1 |
| HTN | endometrium | *PDE7B* | 3.954357683 | 0.000732149 | 0.003073153 | 1 |
| HTN | endometrium | *MCC* | 2.239541209 | 0.000865658 | 0.003084413 | 1 |
| HTN | endometrium | *HOPX* | 3.640102586 | 0.017012269 | 0.024121429 | 1 |
| HTN | endometrium | *PPID* | 2.79548189 | 0.000743731 | 0.003073153 | 1 |
| HTN | endometrium | *ETFDH* | 2.423777594 | 0.018370062 | 0.025768009 | 1 |
| HTN | endometrium | *TBCA* | 6.20031661 | 0.001411357 | 0.003309766 | 1 |
| HTN | endometrium | *NMUR1* | 4.606979878 | 0.000999584 | 0.003095606 | 1 |
| HTN | endometrium | *PIK3CD* | 3.235446199 | 0.000564023 | 0.003073153 | 1 |
| HTN | endometrium | *ANO5* | 2.607781318 | 0.001042141 | 0.003095606 | 1 |
| HTN | endometrium | *VAT1L* | 3.415190003 | 0.001427471 | 0.003328251 | 1 |
| HTN | endometrium | *CAMTA1* | 4.51756933 | 0.000208045 | 0.003073153 | 1 |
| HTN | endometrium | *LGALS4* | 4.691176108 | 0.004827336 | 0.008192457 | 1 |
| HTN | endometrium | *LRRC34* | 2.889652529 | 0.000800575 | 0.003073153 | 1 |
| HTN | endometrium | *PAH* | 14.9729254 | 0.004291841 | 0.007455786 | 1 |
| HTN | endometrium | *SPATA5L1* | 3.018155471 | 0.005337962 | 0.008912523 | 1 |
| HTN | endometrium | *RHNO1* | 7.081339497 | 0.001577356 | 0.003528506 | 1 |
| HTN | endometrium | *PWWP2B* | -2.406446211 | 0.015429231 | 0.022129066 | 1 |
| HTN | endometrium | *EXOSC10* | 6.110429469 | 0.000670774 | 0.003073153 | 1 |
| HTN | endometrium | *TRAPPC12* | 3.016131727 | 0.000853854 | 0.0030808 | 1 |
| HTN | endometrium | *C3AR1* | 1.71387152 | 0.011387677 | 0.016943578 | 1 |
| HTN | endometrium | *PRNP* | 3.970751408 | 0.00144053 | 0.003339619 | 1 |
| HTN | endometrium | *FBXW10* | -1.275216698 | 0.004353113 | 0.007543322 | 1 |
| HTN | endometrium | *SRGAP2C* | 1.828638637 | 0.002926712 | 0.005506168 | 1 |
| HTN | endometrium | *ATPAF2* | 9.282333988 | 0.0001203 | 0.00304009 | 0.583697213 |
| HTN | endometrium | *ZNF554* | -2.524037505 | 0.017846477 | 0.025106148 | 1 |
| HTN | endometrium | *ANKRD20A4* | 15.35883304 | 0.000613722 | 0.003073153 | 1 |
| HTN | endometrium | *EIF2AK3* | 3.869285188 | 0.000905218 | 0.003087532 | 1 |
| HTN | endometrium | *NME6* | 5.992388051 | 3.37E-07 | 7.79E-05 | 0.001635587 |
| HTN | endometrium | *SLFN12* | 2.446527782 | 0.000967954 | 0.003095606 | 1 |
| HTN | endometrium | *TEFM* | 5.501651361 | 0.026523394 | 0.035737713 | 1 |
| HTN | endometrium | *MALT1* | 6.320616076 | 0.00085493 | 0.0030808 | 1 |
| HTN | endometrium | *MBOAT1* | 6.832828425 | 0.002825648 | 0.005365967 | 1 |
| HTN | endometrium | *CXCR6* | 14.76530771 | 0.004837054 | 0.008206079 | 1 |
| HTN | endometrium | *PAIP1* | 5.459958192 | 0.000730794 | 0.003073153 | 1 |
| HTN | endometrium | *MACROD2* | 2.830482048 | 0.000252851 | 0.003073153 | 1 |
| HTN | endometrium | *TP53RK* | 16.69136905 | 0.001235123 | 0.003103477 | 1 |
| HTN | endometrium | *SUCLG2* | 5.351717355 | 0.010374128 | 0.015661254 | 1 |
| HTN | endometrium | *ARNT2* | 4.086820269 | 0.000492292 | 0.003073153 | 1 |
| HTN | endometrium | *MANEA* | 2.378996563 | 0.00141931 | 0.003317298 | 1 |
| HTN | endometrium | *AFF1* | 3.926133139 | 0.000609391 | 0.003073153 | 1 |
| HTN | endometrium | *FIBP* | 4.443160355 | 0.000264491 | 0.003073153 | 1 |
| HTN | endometrium | *CTSW* | -1.187463416 | 0.004388888 | 0.007596076 | 1 |
| HTN | endometrium | *SMPDL3A* | 2.297160965 | 0.003061897 | 0.005698628 | 1 |
| HTN | endometrium | *RAD9A* | 5.220933576 | 0.001433036 | 0.003335313 | 1 |
| HTN | endometrium | *EFEMP2* | 2.836721092 | 0.000578448 | 0.003073153 | 1 |
| HTN | endometrium | *SLFN11* | 2.161456842 | 0.001735309 | 0.003747095 | 1 |
| HTN | endometrium | *FAM71D* | 3.403782905 | 0.005691847 | 0.009367993 | 1 |

| | | | | | | |
|---|---|---|---|---|---|---|
| HTN | endometrium | *ZNF596* | 3.586597375 | 0.000967054 | 0.003095606 | 1 |
| HTN | endometrium | *CFL1* | 6.964339537 | 0.005791644 | 0.009503232 | 1 |
| HTN | endometrium | *RAB43* | -8.773848652 | 0.003224049 | 0.005938908 | 1 |
| HTN | endometrium | *CBWD1* | 6.779349352 | 0.000590016 | 0.003073153 | 1 |
| HTN | endometrium | *SNX32* | 2.892431942 | 0.000746698 | 0.003073153 | 1 |
| HTN | endometrium | *ZNF621* | 7.613696095 | 0.000534284 | 0.003073153 | 1 |
| HTN | endometrium | *RNASEH2C* | -3.077249715 | 0.01449831 | 0.020923795 | 1 |
| HTN | endometrium | *MYEOV* | -18.2056597 | 0.00015198 | 0.003073153 | 0.737408233 |
| HTN | endometrium | *ANKRD13D* | 1.99641769 | 0.001254202 | 0.003122339 | 1 |
| HTN | endometrium | *MRGPRF* | 2.447026992 | 0.005552716 | 0.009192008 | 1 |
| HTN | endometrium | *SH3RF3* | 2.301867966 | 0.001469968 | 0.003383438 | 1 |
| HTN | endometrium | *GXYLT2* | 6.290844784 | 0.000729347 | 0.003073153 | 1 |
| HTN | endometrium | *DCAKD* | 2.935746992 | 0.003668108 | 0.00656014 | 1 |
| HTN | endometrium | *TRMT112* | -2.472695386 | 0.025370287 | 0.034317434 | 1 |
| HTN | endometrium | *NOC3L* | -2.386263254 | 0.005030625 | 0.008463452 | 1 |
| HTN | endometrium | *RHOD* | -2.792494476 | 0.031756678 | 0.042133826 | 1 |
| HTN | endometrium | *PARP14* | 1.943794011 | 0.005202849 | 0.008710912 | 1 |
| HTN | endometrium | *PARP15* | 2.641410224 | 0.000294074 | 0.003073153 | 1 |
| HTN | endometrium | *AHSA2* | 4.333632072 | 6.10E-05 | 0.002410623 | 0.296197042 |
| HTN | endometrium | *IQCB1* | 4.188851934 | 0.000517796 | 0.003073153 | 1 |
| HTN | endometrium | *ZNF483* | 4.733300606 | 0.007982381 | 0.012461555 | 1 |
| HTN | endometrium | *MZT2A* | 3.364603513 | 0.000600637 | 0.003073153 | 1 |
| HTN | endometrium | *MAP3K11* | 8.370789032 | 1.71E-05 | 0.001262767 | 0.082869496 |
| HTN | endometrium | *CCDC36* | 1.521271559 | 0.013676371 | 0.019894704 | 1 |
| HTN | endometrium | *EHBP1L1* | 2.182721895 | 0.003319548 | 0.006077455 | 1 |
| HTN | endometrium | *PPP1R14B* | 4.566127341 | 0.000203688 | 0.003073153 | 0.988296037 |
| HTN | endometrium | *ZNF417* | 4.94271981 | 0.00087202 | 0.003084413 | 1 |
| HTN | endometrium | *PEAK1* | 2.814138293 | 0.000976192 | 0.003095606 | 1 |
| HTN | endometrium | *TNFRSF10D* | 6.398688501 | 0.001256314 | 0.003122339 | 1 |
| HTN | endometrium | *MST1* | -3.041251686 | 0.00323583 | 0.005954519 | 1 |
| HTN | endometrium | *GMPPB* | 4.220041799 | 0.008762709 | 0.013522394 | 1 |
| HTN | endometrium | *NUDT18* | 7.909987718 | 0.001330559 | 0.003214407 | 1 |
| HTN | endometrium | *SULT1B1* | 10.70360763 | 0.02005351 | 0.027784019 | 1 |
| HTN | endometrium | *SCAI* | 3.861813989 | 0.001212166 | 0.003095606 | 1 |
| HTN | endometrium | *HSPB7* | 4.0340889 | 0.000170726 | 0.003073153 | 0.828361633 |
| HTN | endometrium | *UQCRH* | 6.958647563 | 0.002800986 | 0.005331654 | 1 |
| HTN | endometrium | *C11orf80* | 1.436808193 | 0.016287801 | 0.023209518 | 1 |
| HTN | endometrium | *CD7* | 5.056416349 | 0.001030694 | 0.003095606 | 1 |
| HTN | endometrium | *RNF213* | 3.859989986 | 0.000701639 | 0.003073153 | 1 |
| HTN | endometrium | *TUBB8* | 4.953530872 | 0.002686327 | 0.005169046 | 1 |
| HTN | endometrium | *GPR160* | 4.809740677 | 7.85E-05 | 0.002708518 | 0.381114802 |
| HTN | endometrium | *GOLIM4* | 2.07183862 | 0.001115011 | 0.003095606 | 1 |
| HTN | endometrium | *HOXB2* | -2.289623209 | 0.028095985 | 0.037720453 | 1 |
| HTN | endometrium | *UBXN2A* | 4.358154735 | 0.000406193 | 0.003073153 | 1 |
| HTN | endometrium | *NRROS* | 2.386987364 | 0.017407368 | 0.024588224 | 1 |
| HTN | endometrium | *SLC25A30* | 3.203628602 | 0.001203742 | 0.003095606 | 1 |
| HTN | endometrium | *CTSF* | 5.199247025 | 0.001846105 | 0.003911485 | 1 |
| HTN | endometrium | *LEMD3* | 4.442592069 | 0.000625368 | 0.003073153 | 1 |
| HTN | endometrium | *SNX31* | -2.725883893 | 0.01427837 | 0.020630926 | 1 |
| HTN | endometrium | *GLIS1* | 2.75772067 | 0.001195066 | 0.003095606 | 1 |

| | | | | | | |
|---|---|---|---|---|---|---|
| HTN | endometrium | *RALGAPA1* | 2.884644585 | 0.002718219 | 0.005212964 | 1 |
| HTN | endometrium | *SNAPC5* | 11.30546286 | 0.000269718 | 0.003073153 | 1 |
| HTN | endometrium | *TMEM81* | 7.517416615 | 0.014197763 | 0.020520568 | 1 |
| HTN | endometrium | *MSL2* | 5.247096098 | 0.000564728 | 0.003073153 | 1 |
| HTN | endometrium | *ZNF497* | 23.69127913 | 0.000419256 | 0.003073153 | 1 |
| HTN | endometrium | *ANGEL2* | 3.031729329 | 0.000681995 | 0.003073153 | 1 |
| HTN | endometrium | *ZNF266* | 1.947811587 | 0.00260532 | 0.005052363 | 1 |
| HTN | endometrium | *SH3PXD2B* | 2.862591211 | 0.000710532 | 0.003073153 | 1 |
| HTN | endometrium | *KIAA1551* | 3.097947516 | 0.002657028 | 0.005130083 | 1 |
| HTN | endometrium | *LARP7* | 4.02653976 | 0.000741854 | 0.003073153 | 1 |
| HTN | endometrium | *FGFBP3* | 5.328484752 | 0.001219483 | 0.003095606 | 1 |
| HTN | endometrium | *C4orf32* | 5.61838926 | 0.003997431 | 0.007047797 | 1 |
| HTN | endometrium | *HRAS* | 3.643697979 | 0.002911886 | 0.00549105 | 1 |
| HTN | endometrium | *SRP72* | 4.64782905 | 0.023910103 | 0.032514524 | 1 |
| HTN | endometrium | *NLRP6* | -10.00114329 | 0.002677506 | 0.005161298 | 1 |
| HTN | endometrium | *RSRC1* | 3.870889533 | 0.008279948 | 0.012868132 | 1 |
| HTN | endometrium | *PTDSS2* | 6.99780306 | 0.000756129 | 0.003073153 | 1 |
| HTN | endometrium | *SEZ6L2* | 4.851461906 | 4.14E-06 | 0.00050228 | 0.020091217 |
| HTN | endometrium | *KCTD13* | 2.697039532 | 0.000207754 | 0.003073153 | 1 |
| HTN | endometrium | *DHX36* | 5.994337939 | 0.001132355 | 0.003095606 | 1 |
| HTN | endometrium | *FBXW8* | 6.883600134 | 0.003565811 | 0.006415022 | 1 |
| HTN | endometrium | *SLC22A1* | 4.876608302 | 0.001275608 | 0.003141752 | 1 |
| HTN | endometrium | *TVP23C* | 4.692355264 | 0.001155178 | 0.003095606 | 1 |
| HTN | endometrium | *PACS1* | 3.993152942 | 0.000841511 | 0.0030808 | 1 |
| HTN | endometrium | *SH3BP5L* | 3.422522182 | 0.001180082 | 0.003095606 | 1 |
| HTN | endometrium | *CADM2* | 1.460082856 | 0.007245387 | 0.011503145 | 1 |
| HTN | endometrium | *PSMD2* | 6.817983433 | 0.018701575 | 0.026180047 | 1 |
| HTN | endometrium | *ARHGAP1* | 4.565273698 | 0.000102226 | 0.002934745 | 0.496002022 |
| HTN | endometrium | *ATG13* | 5.624335858 | 0.000805813 | 0.003073153 | 1 |
| HTN | endometrium | *CCNE2* | -2.929903717 | 6.51E-05 | 0.002410623 | 0.315770038 |
| HTN | endometrium | *ZNF519* | 3.093133922 | 0.001211091 | 0.003095606 | 1 |
| HTN | endometrium | *BANF1* | 4.211608739 | 0.004803635 | 0.008157941 | 1 |
| HTN | endometrium | *CHRNA7* | -2.13565772 | 0.009444458 | 0.014432916 | 1 |
| HTN | endometrium | *TMEM9B* | 6.452410239 | 4.07E-05 | 0.002058697 | 0.197683334 |
| HTN | endometrium | *SCUBE2* | 3.001749311 | 0.001013698 | 0.003095606 | 1 |
| HTN | endometrium | *EIF1AD* | 6.505017423 | 0.001280189 | 0.00314396 | 1 |
| HTN | endometrium | *EIF3F* | 4.406668127 | 0.000956997 | 0.003095606 | 1 |
| HTN | endometrium | *ZNF25* | 5.305101062 | 0.016639397 | 0.023648024 | 1 |
| HTN | endometrium | *PPP2R2D* | 1.68852609 | 0.004438044 | 0.007649517 | 1 |
| HTN | endometrium | *MCTP1* | 1.493620844 | 0.001158284 | 0.003095606 | 1 |
| HTN | endometrium | *LRRC25* | 3.639211638 | 0.002094265 | 0.00427128 | 1 |
| HTN | endometrium | *UBQLNL* | -1.119198066 | 0.004925694 | 0.008318644 | 1 |
| HTN | endometrium | *KCNE3* | 4.496740778 | 0.000992365 | 0.003095606 | 1 |
| HTN | endometrium | *UCP2* | 2.316818751 | 0.001403949 | 0.003300368 | 1 |
| HTN | endometrium | *C11orf68* | 4.500097997 | 0.001241765 | 0.003110503 | 1 |
| HTN | endometrium | *MRPL48* | 4.363978452 | 0.001014306 | 0.003095606 | 1 |
| HTN | endometrium | *RAB6A* | 3.947708367 | 0.000697461 | 0.003073153 | 1 |
| HTN | endometrium | *TMEM70* | 3.112903844 | 0.001192688 | 0.003095606 | 1 |
| HTN | endometrium | *RPS6KB2* | -1.521719203 | 0.007319054 | 0.011597665 | 1 |
| HTN | endometrium | *RMI2* | -2.429735154 | 0.016126413 | 0.023020111 | 1 |

| | | | | | | |
|---|---|---|---|---|---|---|
| HTN | endometrium | *TOM1L2* | 5.117033101 | 0.010701985 | 0.016071196 | 1 |
| HTN | endometrium | *TEX26* | 9.323724412 | 0.020593322 | 0.02843449 | 1 |
| HTN | endometrium | *TTLL11* | 6.58624029 | 0.011822905 | 0.017501232 | 1 |
| HTN | endometrium | *CTDNEP1* | 7.592786324 | 3.37E-06 | 0.000441729 | 0.016343956 |
| HTN | endometrium | *ZDHHC21* | 3.709956932 | 0.00065874 | 0.003073153 | 1 |
| HTN | endometrium | *A2M* | 2.718560919 | 0.00067156 | 0.003073153 | 1 |
| HTN | endometrium | *DOK7* | 22.8084046 | 0.000757329 | 0.003073153 | 1 |
| HTN | endometrium | *ORAI3* | 15.12382222 | 1.70E-09 | 1.03E-06 | 8.26E-06 |
| HTN | endometrium | *B3GALT6* | 6.623900236 | 0.001167137 | 0.003095606 | 1 |
| HTN | endometrium | *ZNF613* | 6.068326055 | 0.017072075 | 0.024184416 | 1 |
| HTN | endometrium | *JAKMIP2* | 2.540856151 | 0.006201022 | 0.010059726 | 1 |
| HTN | endometrium | *TCP11L1* | 6.410387255 | 0.002091868 | 0.00426998 | 1 |
| HTN | endometrium | *CCDC57* | 2.486162908 | 0.001634276 | 0.003601562 | 1 |
| HTN | endometrium | *CIDEA* | 2.370833269 | 0.001054589 | 0.003095606 | 1 |
| HTN | endometrium | *SMIM19* | 4.547965698 | 0.001443614 | 0.003341453 | 1 |
| HTN | endometrium | *ZNF404* | 2.120725678 | 0.013344451 | 0.01949045 | 1 |
| HTN | endometrium | *ZNF135* | 2.701136135 | 0.001859083 | 0.003928693 | 1 |
| HTN | endometrium | *OR4N2* | 10.44384371 | 0.000497754 | 0.003073153 | 1 |
| HTN | endometrium | *TAC4* | 7.477348894 | 0.000604927 | 0.003073153 | 1 |
| HTN | endometrium | *CDC26* | -1.684153873 | 0.007342016 | 0.011626456 | 1 |
| HTN | endometrium | *CRLF3* | 3.249180682 | 8.67E-05 | 0.002714492 | 0.420452723 |
| HTN | endometrium | *DNAJC30* | 4.53389202 | 0.000795781 | 0.003073153 | 1 |
| HTN | endometrium | *SPRYD4* | 2.756269751 | 0.007434804 | 0.011735091 | 1 |
| HTN | endometrium | *WDR25* | -2.241469271 | 0.033038788 | 0.04366772 | 1 |
| HTN | endometrium | *PLA2G16* | 3.486088015 | 0.000297234 | 0.003073153 | 1 |
| HTN | endometrium | *RNF152* | 2.857725237 | 0.017214099 | 0.024357775 | 1 |
| HTN | endometrium | *LRRC37A* | -1.648057299 | 0.023604897 | 0.032180148 | 1 |
| HTN | endometrium | *BDNF* | 3.471382809 | 0.00367901 | 0.006574791 | 1 |
| HTN | endometrium | *BOK* | 2.281138354 | 0.001182245 | 0.003095606 | 1 |
| HTN | endometrium | *PFN4* | 2.191695344 | 0.024136142 | 0.032794333 | 1 |
| HTN | endometrium | *RUFY1* | 3.709409434 | 0.001091265 | 0.003095606 | 1 |
| HTN | endometrium | *VSIG10* | 3.349791067 | 0.001294429 | 0.00316083 | 1 |
| HTN | endometrium | *C8G* | 66.32773051 | 0.001320876 | 0.003204446 | 1 |
| HTN | endometrium | *MUC20* | 5.092193913 | 0.000566305 | 0.003073153 | 1 |
| HTN | endometrium | *LY6H* | 10.42508143 | 1.36E-05 | 0.001098678 | 0.065920695 |
| HTN | endometrium | *SHMT1* | 4.046650109 | 0.000399117 | 0.003073153 | 1 |
| HTN | endometrium | *DPP7* | 5.962703874 | 0.000578088 | 0.003073153 | 1 |
| HTN | endometrium | *C19orf18* | 2.970993847 | 0.001440607 | 0.003339619 | 1 |
| HTN | endometrium | *TMEM80* | 4.264855835 | 0.005336346 | 0.008912523 | 1 |
| HTN | endometrium | *DSCAML1* | 2.622538594 | 0.000865435 | 0.003084413 | 1 |
| HTN | endometrium | *TALDO1* | 2.871072605 | 0.000921774 | 0.003087532 | 1 |
| HTN | endometrium | *AP006621.1* | 7.380359693 | 0.003568688 | 0.006417818 | 1 |
| HTN | endometrium | *MAN1B1* | 4.553355708 | 0.000693424 | 0.003073153 | 1 |
| HTN | endometrium | *SAMD9L* | 2.978827193 | 0.0010867 | 0.003095606 | 1 |
| HTN | endometrium | *MIEF2* | 3.829986716 | 0.001382503 | 0.00327409 | 1 |
| HTN | endometrium | *NAP1L5* | -1.542552134 | 0.005495473 | 0.009115909 | 1 |
| HTN | endometrium | *NIM1K* | 6.690616806 | 0.000205499 | 0.003073153 | 0.99708304 |
| HTN | endometrium | *RBM44* | 7.944645125 | 0.000763163 | 0.003073153 | 1 |
| HTN | endometrium | *SLC25A22* | -1.546736823 | 0.002328888 | 0.004636752 | 1 |
| HTN | endometrium | *ACAD9* | 2.019300879 | 0.014997765 | 0.021561231 | 1 |

| | | | | | | |
|---|---|---|---|---|---|---|
| HTN | endometrium | *IL17RA* | 2.698856375 | 0.000981724 | 0.003095606 | 1 |
| HTN | endometrium | *MBOAT4* | 7.326280748 | 0.005654138 | 0.009327688 | 1 |
| HTN | endometrium | *AGTRAP* | 2.902980681 | 0.000929877 | 0.003093357 | 1 |
| HTN | endometrium | *THAP5* | 2.690619505 | 0.001426781 | 0.003328251 | 1 |
| HTN | endometrium | *NAALADL2* | 4.081946367 | 0.000861538 | 0.003084413 | 1 |
| HTN | endometrium | *CD151* | 3.238375847 | 0.000266842 | 0.003073153 | 1 |
| HTN | endometrium | *SLC35G5* | 22.72778221 | 0.001154719 | 0.003095606 | 1 |
| HTN | endometrium | *FAM87B* | 20.89307873 | 0.001332088 | 0.003214407 | 1 |
| HTN | endometrium | *PCDHB9* | 7.545700963 | 0.000154691 | 0.003073153 | 0.750562355 |
| HTN | endometrium | *CENPBD1* | 6.123468168 | 0.001135957 | 0.003095606 | 1 |
| HTN | endometrium | *RIC8A* | 3.166906101 | 0.003002818 | 0.005614518 | 1 |
| HTN | endometrium | *ODF3B* | 5.437655005 | 7.48E-06 | 0.000685047 | 0.036307503 |
| HTN | endometrium | *DPY19L2* | 2.821727826 | 0.019468169 | 0.027096832 | 1 |
| HTN | endometrium | *ADAMTSL1* | 1.593621061 | 0.001301531 | 0.003171787 | 1 |
| HTN | endometrium | *IMPDH2* | 3.402305373 | 0.000662662 | 0.003073153 | 1 |
| HTN | endometrium | *GRAMD1C* | 2.827458114 | 0.001833321 | 0.003894604 | 1 |
| HTN | endometrium | *DALRD3* | 3.509693134 | 0.003738788 | 0.006669337 | 1 |
| HTN | endometrium | *ZNF114* | 8.45997681 | 0.000797487 | 0.003073153 | 1 |
| HTN | endometrium | *SPINK6* | 11.78239478 | 0.004598404 | 0.007883907 | 1 |
| HTN | endometrium | *ZNF454* | 2.997014504 | 0.001241707 | 0.003110503 | 1 |
| HTN | endometrium | *SH2B1* | 6.393435621 | 0.000753157 | 0.003073153 | 1 |
| HTN | endometrium | *VN1R1* | 6.726236706 | 0.024859298 | 0.033682579 | 1 |
| HTN | endometrium | *KDELC2* | 3.226096487 | 0.001597913 | 0.003554824 | 1 |
| HTN | endometrium | *PLEC* | 2.291892772 | 0.013247268 | 0.019377674 | 1 |
| HTN | endometrium | *SH2D4B* | 11.23301167 | 0.000826662 | 0.0030808 | 1 |
| HTN | endometrium | *RNF212* | 6.41506688 | 0.024363372 | 0.033066037 | 1 |
| HTN | endometrium | *PRSS36* | 5.389588442 | 0.001640667 | 0.003611849 | 1 |
| HTN | endometrium | *WDR6* | 4.6300602 | 0.009337349 | 0.014291741 | 1 |
| HTN | endometrium | *TMPRSS9* | 4.886472259 | 0.010297008 | 0.01557751 | 1 |
| HTN | endometrium | *ZFAND2A* | 3.371812735 | 0.006633896 | 0.010644068 | 1 |
| HTN | endometrium | *FAM220A* | 4.677953625 | 0.000922916 | 0.003087532 | 1 |
| HTN | endometrium | *NT5DC1* | 5.605908495 | 0.001001564 | 0.003095606 | 1 |
| HTN | endometrium | *GLDC* | 5.219633909 | 0.001455162 | 0.003358918 | 1 |
| HTN | endometrium | *ERN1* | 6.058744546 | 0.000800724 | 0.003073153 | 1 |
| HTN | endometrium | *PARP10* | 6.382760302 | 0.002247585 | 0.004523136 | 1 |
| HTN | endometrium | *NSUN3* | 5.920903594 | 0.026659221 | 0.035900788 | 1 |
| HTN | endometrium | *RPP25* | 8.96618947 | 8.00E-09 | 3.24E-06 | 3.88E-05 |
| HTN | endometrium | *STX19* | 3.549301686 | 0.001392045 | 0.003285301 | 1 |
| HTN | endometrium | *TRIM73* | -15.12138617 | 0.004284857 | 0.007447863 | 1 |
| HTN | endometrium | *EFCAB13* | 2.122166609 | 0.001932167 | 0.004049621 | 1 |
| HTN | endometrium | *EXOSC4* | 13.91282122 | 0.007022267 | 0.011185831 | 1 |
| HTN | endometrium | *ZNF852* | -5.078737136 | 0.026040926 | 0.035126653 | 1 |
| HTN | endometrium | *HYI* | -3.282657076 | 1.17E-05 | 0.000976361 | 0.056628926 |
| HTN | endometrium | *C17orf62* | 2.698354567 | 0.001015851 | 0.003095606 | 1 |
| HTN | endometrium | *GAK* | 2.643923476 | 0.000698134 | 0.003073153 | 1 |
| HTN | endometrium | *TUFM* | 4.23335482 | 0.000218941 | 0.003073153 | 1 |
| HTN | endometrium | *C3orf38* | 3.250389457 | 0.006948932 | 0.011079928 | 1 |
| HTN | endometrium | *CCDC89* | 10.2792515 | 0.001220872 | 0.003095606 | 1 |
| HTN | endometrium | *SPTY2D1* | 7.602245983 | 0.000699086 | 0.003073153 | 1 |
| HTN | endometrium | *CDH4* | 3.150623929 | 0.032740157 | 0.043343859 | 1 |

| | | | | | | |
|---|---|---|---|---|---|---|
| HTN | endometrium | *GEMIN4* | 3.526102204 | 0.005662498 | 0.009329204 | 1 |
| HTN | endometrium | *ZFPM1* | 3.156699223 | 0.004840541 | 0.008206257 | 1 |
| HTN | endometrium | *RPRML* | 17.02574733 | 0.020508158 | 0.028333025 | 1 |
| HTN | endometrium | *SERTAD2* | 2.205996906 | 0.000845822 | 0.0030808 | 1 |
| HTN | endometrium | *AKAP5* | 5.761197316 | 0.001976056 | 0.004106134 | 1 |
| HTN | endometrium | *GIPC3* | 16.38152359 | 0.001133452 | 0.003095606 | 1 |
| HTN | endometrium | *PDXDC1* | 4.198541706 | 0.000415607 | 0.003073153 | 1 |
| HTN | endometrium | *C1orf194* | -1.067858442 | 0.002542018 | 0.004957343 | 1 |
| HTN | endometrium | *SEPHS2* | 8.311701407 | 0.031829554 | 0.042218971 | 1 |
| HTN | endometrium | *SEPT1* | 3.119458826 | 0.001173663 | 0.003095606 | 1 |
| HTN | endometrium | *CSNK1A1L* | -18.76003621 | 0.028906933 | 0.038702107 | 1 |
| HTN | endometrium | *LYNX1* | 3.650295756 | 0.004621921 | 0.00791863 | 1 |
| HTN | endometrium | *HCLS1* | 1.839617565 | 0.005721128 | 0.009400242 | 1 |
| HTN | endometrium | *MTURN* | 6.708476267 | 0.002931238 | 0.005509202 | 1 |
| HTN | endometrium | *CCDC66* | 4.570625837 | 0.001368778 | 0.003258739 | 1 |
| HTN | endometrium | *DEFB124* | 20.46366753 | 0.001926665 | 0.004039836 | 1 |
| HTN | endometrium | *DEFB119* | 52.97906122 | 0.000800257 | 0.003073153 | 1 |
| HTN | endometrium | *TSPYL5* | -1.652103104 | 0.002234518 | 0.00450806 | 1 |
| HTN | endometrium | *MB21D2* | 7.059295997 | 0.000926929 | 0.003091039 | 1 |
| HTN | endometrium | *SLC47A2* | 2.95032855 | 0.002817116 | 0.005356053 | 1 |
| HTN | endometrium | *CHST13* | 22.56794947 | 0.004343578 | 0.00753218 | 1 |
| HTN | endometrium | *SLC36A4* | 6.929432089 | 0.00301182 | 0.005627011 | 1 |
| HTN | endometrium | *ZDHHC20* | 4.21505117 | 0.000555324 | 0.003073153 | 1 |
| HTN | endometrium | *PSMG4* | 5.703213962 | 0.002044067 | 0.004200683 | 1 |
| HTN | endometrium | *ZNF443* | 9.266728771 | 0.003438142 | 0.006236709 | 1 |
| HTN | endometrium | *CAPS2* | 2.594912723 | 0.000587744 | 0.003073153 | 1 |
| HTN | endometrium | *KCTD2* | -2.595349443 | 0.018567271 | 0.026007044 | 1 |
| HTN | endometrium | *ST20* | -0.97075081 | 0.019935285 | 0.027636001 | 1 |
| HTN | endometrium | *NQO1* | 2.322432968 | 0.002281326 | 0.004570187 | 1 |
| HTN | endometrium | *FKRP* | 2.139894687 | 0.003426076 | 0.006221303 | 1 |
| HTN | endometrium | *RPH3AL* | 2.036690967 | 0.001009106 | 0.003095606 | 1 |
| HTN | endometrium | *MUC16* | 2.368790306 | 0.001075135 | 0.003095606 | 1 |
| HTN | endometrium | *PENK* | 13.95885646 | 0.000821496 | 0.0030808 | 1 |
| HTN | endometrium | *SLC25A41* | 14.01688558 | 0.000996078 | 0.003095606 | 1 |
| HTN | endometrium | *TMEM136* | 3.832500231 | 0.000297092 | 0.003073153 | 1 |
| HTN | endometrium | *ZNF322* | 5.965544535 | 0.004995347 | 0.008412126 | 1 |
| HTN | endometrium | *DDX60L* | 5.718438629 | 0.000361434 | 0.003073153 | 1 |
| HTN | endometrium | *TMEM45A* | -1.26888571 | 0.009758976 | 0.014857405 | 1 |
| HTN | endometrium | *SGSH* | 4.939598833 | 0.000865323 | 0.003084413 | 1 |
| HTN | endometrium | *ZFP41* | 8.613880053 | 1.83E-05 | 0.001270304 | 0.088921286 |
| HTN | endometrium | *ZBTB20* | 2.923094985 | 0.000740805 | 0.003073153 | 1 |
| HTN | endometrium | *C3orf58* | 4.48714932 | 0.000806323 | 0.003073153 | 1 |
| HTN | endometrium | *TMEM252* | -24.8103999 | 0.004733603 | 0.008067244 | 1 |
| HTN | endometrium | *ODF3L2* | 9.456546892 | 0.011091145 | 0.016553134 | 1 |
| HTN | endometrium | *RELL1* | 5.546170003 | 0.000446112 | 0.003073153 | 1 |
| HTN | endometrium | *RFX7* | 3.127624139 | 0.000660014 | 0.003073153 | 1 |
| HTN | endometrium | *CLDN7* | -7.291571697 | 5.85E-06 | 0.000616821 | 0.028373744 |
| HTN | endometrium | *ADO* | 7.253356955 | 1.52E-08 | 5.68E-06 | 7.38E-05 |
| HTN | endometrium | *MGAT4C* | 1.50605204 | 0.021307872 | 0.029296059 | 1 |
| HTN | endometrium | *IDH2* | 2.37101284 | 0.001612731 | 0.003579584 | 1 |

| | | | | | | |
|---|---|---|---|---|---|---|
| HTN | endometrium | *FAM181B* | -2.081029863 | 0.008984623 | 0.013804113 | 1 |
| HTN | endometrium | *TDRKH* | 3.833853711 | 0.001444152 | 0.003341453 | 1 |
| HTN | endometrium | *ERCC6L2* | 4.7466952 | 0.000249747 | 0.003073153 | 1 |
| HTN | endometrium | *MRPL41* | 4.648679836 | 0.000745917 | 0.003073153 | 1 |
| HTN | endometrium | *ARL6IP4* | -1.724063317 | 0.016200927 | 0.023106084 | 1 |
| HTN | endometrium | *HHIPL1* | 2.963497061 | 0.001278856 | 0.003143367 | 1 |
| HTN | endometrium | *FAM153B* | 2.369450537 | 0.000837014 | 0.0030808 | 1 |
| HTN | endometrium | *SYNM* | 5.743397786 | 0.000857304 | 0.0030808 | 1 |
| HTN | endometrium | *KCNA4* | 3.568690467 | 0.001136362 | 0.003095606 | 1 |
| HTN | endometrium | *FIGN* | 3.387149187 | 0.000562732 | 0.003073153 | 1 |
| HTN | endometrium | *C8orf33* | 5.886490759 | 0.000500576 | 0.003073153 | 1 |
| HTN | endometrium | *FBXL6* | -2.677819861 | 0.014196588 | 0.020520568 | 1 |
| HTN | endometrium | *KBTBD3* | 2.967047223 | 0.000958796 | 0.003095606 | 1 |
| HTN | endometrium | *CLN8* | 3.592634291 | 1.99E-06 | 0.00030216 | 0.00966913 |
| HTN | endometrium | *NXPH4* | 30.18011749 | 0.00111661 | 0.003095606 | 1 |
| HTN | endometrium | *NPLOC4* | 2.597877797 | 0.001591141 | 0.003549524 | 1 |
| HTN | endometrium | *CAPN12* | 3.844853257 | 0.000290317 | 0.003073153 | 1 |
| HTN | endometrium | *ORAI1* | 4.576200625 | 0.000272125 | 0.003073153 | 1 |
| HTN | endometrium | *GLRX5* | 7.682149936 | 0.000573856 | 0.003073153 | 1 |
| HTN | endometrium | *CAV3* | 2.063471834 | 0.019173405 | 0.026709549 | 1 |
| HTN | endometrium | *ADI1* | 4.106696053 | 0.000737037 | 0.003073153 | 1 |
| HTN | endometrium | *CLEC4G* | -4.43469006 | 0.002108252 | 0.004296194 | 1 |
| HTN | endometrium | *TRAK1* | 3.529502561 | 0.002708938 | 0.005201332 | 1 |
| HTN | endometrium | *SKA2* | 6.711143206 | 0.001988734 | 0.004123411 | 1 |
| HTN | endometrium | *TTC3* | 2.383021568 | 0.000453168 | 0.003073153 | 1 |
| HTN | endometrium | *PPP1R27* | 20.90193429 | 0.013029591 | 0.019065011 | 1 |
| HTN | endometrium | *BRICD5* | 6.247354212 | 0.001141078 | 0.003095606 | 1 |
| HTN | endometrium | *GRID1* | -3.609706666 | 0.011238096 | 0.01675184 | 1 |
| HTN | endometrium | *ACBD3* | 5.964873229 | 0.008023275 | 0.012513317 | 1 |
| HTN | endometrium | *PLCXD3* | 2.112486197 | 0.001146285 | 0.003095606 | 1 |
| HTN | endometrium | *VMO1* | -2.620903255 | 0.029186862 | 0.039001558 | 1 |
| HTN | endometrium | *ALG12* | 2.897765132 | 0.001217072 | 0.003095606 | 1 |
| HTN | endometrium | *COL18A1* | 7.593690807 | 6.32E-05 | 0.002410623 | 0.30686689 |
| HTN | endometrium | *ZNF721* | 5.554546487 | 0.000212838 | 0.003073153 | 1 |
| HTN | endometrium | *C12orf60* | 2.732044659 | 0.001132326 | 0.003095606 | 1 |
| HTN | endometrium | *ABAT* | 4.094523104 | 0.001083804 | 0.003095606 | 1 |
| HTN | endometrium | *LYSMD4* | -3.477512709 | 0.028082912 | 0.037713338 | 1 |
| HTN | endometrium | *WBP2NL* | 3.450657928 | 0.003257039 | 0.005980217 | 1 |
| HTN | endometrium | *BEGAIN* | 4.763922441 | 0.001489709 | 0.003403045 | 1 |
| HTN | endometrium | *ARHGEF37* | 6.236319629 | 0.000567958 | 0.003073153 | 1 |
| HTN | endometrium | *RABIF* | 3.535550467 | 0.00071504 | 0.003073153 | 1 |
| HTN | endometrium | *FANCF* | -2.506918661 | 0.010598323 | 0.015950082 | 1 |
| HTN | endometrium | *RUVBL2* | -2.751101514 | 0.019478955 | 0.02710407 | 1 |
| HTN | endometrium | *PTTG1IP* | 2.384363264 | 0.001248025 | 0.003119971 | 1 |
| HTN | endometrium | *CCDC125* | 3.722046825 | 0.002274747 | 0.004562659 | 1 |
| HTN | endometrium | *C10orf107* | 2.395508889 | 0.001938833 | 0.004053087 | 1 |
| HTN | endometrium | *FHL3* | 3.195795838 | 0.010584165 | 0.015933716 | 1 |
| HTN | endometrium | *C19orf71* | -3.309107397 | 0.012887633 | 0.018880072 | 1 |
| HTN | endometrium | *LRIT3* | 21.65346391 | 0.001152745 | 0.003095606 | 1 |
| HTN | endometrium | *NPIPA1* | 3.37626734 | 5.95E-05 | 0.002410623 | 0.288616324 |

| | | | | | | |
|---|---|---|---|---|---|---|
| HTN | endometrium | *GPR132* | 2.780171912 | 0.001898082 | 0.003995442 | 1 |
| HTN | endometrium | *SERHL2* | 5.146874289 | 0.000713553 | 0.003073153 | 1 |
| HTN | endometrium | *PCBP3* | 2.32348017 | 0.001952468 | 0.004068787 | 1 |
| HTN | endometrium | *SFXN4* | 2.860352522 | 0.001154204 | 0.003095606 | 1 |
| HTN | endometrium | *MRPL54* | 5.341211872 | 0.004222236 | 0.007361226 | 1 |
| HTN | endometrium | *NDUFB1* | 6.052114853 | 0.001009884 | 0.003095606 | 1 |
| HTN | endometrium | *TRMT12* | 4.572595299 | 0.00096985 | 0.003095606 | 1 |
| HTN | endometrium | *MRGPRX2* | 21.22746105 | 0.016257155 | 0.023179464 | 1 |
| HTN | endometrium | *FIGLA* | 11.87020329 | 0.010265132 | 0.015545075 | 1 |
| HTN | endometrium | *ACSM2A* | 30.14252389 | 0.000529294 | 0.003073153 | 1 |
| HTN | endometrium | *KREMEN1* | -2.191718796 | 0.001256329 | 0.003122339 | 1 |
| HTN | endometrium | *ZNF703* | 3.384721547 | 0.018695584 | 0.026179213 | 1 |
| HTN | endometrium | *FAM162B* | 4.490637464 | 0.00099722 | 0.003095606 | 1 |
| HTN | endometrium | *BTBD9* | 4.747987203 | 0.001191395 | 0.003095606 | 1 |
| HTN | endometrium | *NUDT14* | 5.559205395 | 0.01970511 | 0.027371655 | 1 |
| HTN | endometrium | *NPIPA7* | -3.247014871 | 0.00285505 | 0.005409098 | 1 |
| HTN | endometrium | *TTC32* | -1.336215266 | 0.008611107 | 0.013306081 | 1 |
| HTN | endometrium | *PP2D1* | 8.360368032 | 0.00057516 | 0.003073153 | 1 |
| HTN | endometrium | *DENND5A* | 2.018345138 | 0.001248117 | 0.003119971 | 1 |
| HTN | endometrium | *NIPSNAP1* | -2.058392579 | 0.021705973 | 0.029792752 | 1 |
| HTN | endometrium | *CNTN2* | 4.64614778 | 0.00178074 | 0.003810036 | 1 |
| HTN | endometrium | *GOLGA6L4* | -1.623037559 | 0.001609214 | 0.003576687 | 1 |
| HTN | endometrium | *PGP* | 4.133921702 | 0.000489672 | 0.003073153 | 1 |
| HTN | endometrium | *ACOT1* | 6.161055477 | 8.93E-05 | 0.002725423 | 0.433342308 |
| HTN | endometrium | *OAF* | 2.121603709 | 0.00181208 | 0.003859619 | 1 |
| HTN | endometrium | *CLECL1* | 3.070594023 | 0.021068416 | 0.029032648 | 1 |
| HTN | endometrium | *MROH7* | 4.250637458 | 0.010994381 | 0.016442989 | 1 |
| HTN | endometrium | *PKP3* | -1.432392006 | 0.003486441 | 0.006294576 | 1 |
| HTN | endometrium | *A3GALT2* | -3.398302199 | 0.019084875 | 0.026624443 | 1 |
| HTN | endometrium | *SS18L1* | 4.843380881 | 0.001594058 | 0.003552764 | 1 |
| HTN | endometrium | *COPB2* | 3.154333886 | 0.000683573 | 0.003073153 | 1 |
| HTN | endometrium | *WDR27* | 5.214083482 | 0.000959862 | 0.003095606 | 1 |
| HTN | endometrium | *TMEM255B* | 3.271698351 | 0.000751585 | 0.003073153 | 1 |
| HTN | endometrium | *DUSP8* | -1.072201113 | 0.005745327 | 0.009433614 | 1 |
| HTN | endometrium | *FAM19A3* | -12.93778194 | 0.002262836 | 0.004544403 | 1 |
| HTN | endometrium | *SNN* | 4.367121836 | 0.001101786 | 0.003095606 | 1 |
| HTN | endometrium | *KRBA2* | 6.546615754 | 0.006245796 | 0.010123332 | 1 |
| HTN | endometrium | *SEPT5* | 6.03405879 | 0.012417028 | 0.018251263 | 1 |
| HTN | endometrium | *RNLS* | 5.177467038 | 0.01707663 | 0.024184416 | 1 |
| HTN | endometrium | *NDUFA12* | 3.865013782 | 0.000505534 | 0.003073153 | 1 |
| HTN | endometrium | *TCTE3* | 6.710636881 | 0.000658883 | 0.003073153 | 1 |
| HTN | endometrium | *UBE2G2* | 4.011524158 | 0.000470981 | 0.003073153 | 1 |
| HTN | endometrium | *TMED9* | 7.006871181 | 0.000481279 | 0.003073153 | 1 |
| HTN | endometrium | *DRD1* | 71.28562562 | 0.000901322 | 0.003087532 | 1 |
| HTN | endometrium | *BTBD6* | 3.365230529 | 0.000276394 | 0.003073153 | 1 |
| HTN | endometrium | *NUTM2A* | -7.609891486 | 0.001106229 | 0.003095606 | 1 |
| HTN | endometrium | *WT1* | 1.515815303 | 0.002240998 | 0.004517375 | 1 |
| HTN | endometrium | *ZFP90* | 3.302894341 | 0.0007615 | 0.003073153 | 1 |
| HTN | endometrium | *FAM227A* | 3.077763158 | 0.000313527 | 0.003073153 | 1 |
| HTN | endometrium | *NOC4L* | 5.479367231 | 0.001136615 | 0.003095606 | 1 |

| | | | | | | |
|---|---|---|---|---|---|---|
| HTN | endometrium | *NDUFA6* | 2.4029029 | 0.004028166 | 0.007083965 | 1 |
| HTN | endometrium | *SORCS2* | 6.971937408 | 0.002284542 | 0.004572854 | 1 |
| HTN | endometrium | *TMEM106A* | 2.805996598 | 0.00012225 | 0.003057499 | 0.593154728 |
| HTN | endometrium | *UBOX5* | 2.629277288 | 0.001048284 | 0.003095606 | 1 |
| HTN | endometrium | *MAFF* | -2.071409613 | 0.003087884 | 0.005731605 | 1 |
| HTN | endometrium | *SEMA4B* | 5.938463389 | 0.000576832 | 0.003073153 | 1 |
| HTN | endometrium | *CIB1* | -2.638727271 | 0.00798023 | 0.012461555 | 1 |
| HTN | endometrium | *SGCZ* | 1.835817092 | 0.002800683 | 0.005331654 | 1 |
| HTN | endometrium | *C6orf120* | 3.234212252 | 0.001906703 | 0.004008371 | 1 |
| HTN | endometrium | *SIGIRR* | 4.222700347 | 0.000652709 | 0.003073153 | 1 |
| HTN | endometrium | *IFITM2* | 3.635564448 | 0.000721587 | 0.003073153 | 1 |
| HTN | endometrium | *CCDC137* | 5.917497008 | 0.000110432 | 0.002992864 | 0.535814163 |
| HTN | endometrium | *ATP6V0A2* | 6.755593336 | 0.00115427 | 0.003095606 | 1 |
| HTN | endometrium | *C14orf80* | 4.157191336 | 0.018815963 | 0.026317397 | 1 |
| HTN | endometrium | *SP140L* | 2.222444506 | 0.00098269 | 0.003095606 | 1 |
| HTN | endometrium | *TARSL2* | 4.177203858 | 0.002884449 | 0.00545415 | 1 |
| HTN | endometrium | *SH3BGR* | -2.398486518 | 0.006025456 | 0.009827063 | 1 |
| HTN | endometrium | *FAM174B* | 4.290218939 | 0.000788934 | 0.003073153 | 1 |
| HTN | endometrium | *SV2B* | 2.733619644 | 0.005542568 | 0.009178341 | 1 |
| HTN | endometrium | *AHNAK2* | 4.283619757 | 0.000660796 | 0.003073153 | 1 |
| HTN | endometrium | *SP1* | 2.239973357 | 0.000695744 | 0.003073153 | 1 |
| HTN | endometrium | *PCGF3* | 2.512630687 | 0.000730218 | 0.003073153 | 1 |
| HTN | endometrium | *C6orf201* | 2.793133536 | 0.012291955 | 0.018100323 | 1 |
| HTN | endometrium | *MYBL1* | 2.809899102 | 0.005970849 | 0.009754398 | 1 |
| HTN | endometrium | *ADARB2* | 6.13250725 | 0.001981086 | 0.004113063 | 1 |
| HTN | endometrium | *SLC52A2* | 3.679375248 | 0.00040517 | 0.003073153 | 1 |
| HTN | endometrium | *PIGP* | -1.630812678 | 0.016409891 | 0.023362908 | 1 |
| HTN | endometrium | *ARL17A* | 2.935593228 | 0.009527504 | 0.014541506 | 1 |
| HTN | endometrium | *EVI2B* | 2.746208027 | 0.000593875 | 0.003073153 | 1 |
| HTN | endometrium | *NPIPB4* | 2.593286353 | 0.003912897 | 0.006916349 | 1 |
| HTN | endometrium | *ZNF829* | 5.009526173 | 5.48E-05 | 0.00235392 | 0.26599293 |
| HTN | endometrium | *TRIM69* | 3.446844463 | 0.020270721 | 0.028044921 | 1 |
| HTN | endometrium | *LINC00839* | 8.454612478 | 0.002870062 | 0.005431178 | 1 |
| HTN | endometrium | *RTN4RL1* | 6.65192725 | 0.003324321 | 0.006082054 | 1 |
| HTN | endometrium | *RNPC3* | 3.288029057 | 0.000336205 | 0.003073153 | 1 |
| HTN | endometrium | *ZNF529* | 8.039788333 | 0.002574327 | 0.005006502 | 1 |
| HTN | endometrium | *C15orf41* | 2.747430017 | 0.000412338 | 0.003073153 | 1 |
| HTN | endometrium | *WWOX* | 4.101342256 | 0.000989338 | 0.003095606 | 1 |
| HTN | endometrium | *CYP4Z1* | 2.531284202 | 0.000799697 | 0.003073153 | 1 |
| HTN | endometrium | *FFAR4* | 2.921297361 | 0.000408008 | 0.003073153 | 1 |
| HTN | endometrium | *CYP4F12* | 2.947303498 | 0.001031582 | 0.003095606 | 1 |
| HTN | endometrium | *ZNF749* | -1.319139591 | 0.005478167 | 0.009096532 | 1 |
| HTN | endometrium | *TOR3A* | 3.082943073 | 0.000876944 | 0.003084413 | 1 |
| HTN | endometrium | *ZNF555* | 5.490444783 | 0.001540062 | 0.003471872 | 1 |
| HTN | endometrium | *GOLGA8R* | -4.780173512 | 0.008020658 | 0.012513259 | 1 |
| HTN | endometrium | *CCDC30* | 6.001049634 | 7.11E-06 | 0.000685047 | 0.034494402 |
| HTN | endometrium | *GLDN* | 1.140462714 | 0.008050948 | 0.012552441 | 1 |
| HTN | endometrium | *ZNF197* | 4.849988233 | 0.000939917 | 0.003095606 | 1 |
| HTN | endometrium | *RPS23* | 4.710236856 | 0.007352868 | 0.011639842 | 1 |
| HTN | endometrium | *GNG2* | 2.002953631 | 0.00098833 | 0.003095606 | 1 |

| | | | | | | |
|---|---|---|---|---|---|---|
| HTN | endometrium | *BTN3A2* | 2.110146895 | 0.002032212 | 0.00418415 | 1 |
| HTN | endometrium | *TMEM222* | 4.16007291 | 0.001176597 | 0.003095606 | 1 |
| HTN | endometrium | *SEPT10* | 3.275899485 | 0.001263183 | 0.003127023 | 1 |
| HTN | endometrium | *SMYD4* | 2.516356484 | 0.008138996 | 0.012664206 | 1 |
| HTN | endometrium | *CEACAM19* | -1.470543446 | 0.007638082 | 0.012009065 | 1 |
| HTN | endometrium | *BCR* | 4.104984463 | 0.001273806 | 0.003140501 | 1 |
| HTN | endometrium | *FOXI2* | -8.241294041 | 0.00073764 | 0.003073153 | 1 |
| HTN | endometrium | *ZNF732* | 5.060489254 | 0.002596388 | 0.005041086 | 1 |
| HTN | endometrium | *VSIG10L* | -1.321469393 | 0.003080253 | 0.005719628 | 1 |
| HTN | endometrium | *TRABD2A* | 4.386664817 | 0.00037512 | 0.003073153 | 1 |
| HTN | endometrium | *QRFPR* | 15.89706415 | 0.000175472 | 0.003073153 | 0.851388212 |
| HTN | endometrium | *MAPT* | 3.296089347 | 0.000843747 | 0.0030808 | 1 |
| HTN | endometrium | *TNFRSF18* | 9.994238154 | 0.005069343 | 0.008522679 | 1 |
| HTN | endometrium | *C1QL4* | 36.21697662 | 0.001187722 | 0.003095606 | 1 |
| HTN | endometrium | *RTN4RL2* | 4.806801727 | 0.023554793 | 0.032121376 | 1 |
| HTN | endometrium | *TMEM232* | 4.074182417 | 0.002964421 | 0.005555569 | 1 |
| HTN | endometrium | *EFCAB6* | 4.858771258 | 0.000366989 | 0.003073153 | 1 |
| HTN | endometrium | *RHD* | 7.232801185 | 3.86E-05 | 0.002046851 | 0.187360959 |
| HTN | endometrium | *TMPRSS6* | 21.19551435 | 0.001197681 | 0.003095606 | 1 |
| HTN | endometrium | *CYP4A11* | 10.22937669 | 0.003412588 | 0.0062061 | 1 |
| HTN | endometrium | *TMEM216* | -0.951369695 | 0.004921808 | 0.008314977 | 1 |
| HTN | endometrium | *ENTPD5* | 6.441386295 | 0.000466924 | 0.003073153 | 1 |
| HTN | endometrium | *ANGPTL5* | 4.118677031 | 0.011845807 | 0.017514657 | 1 |
| HTN | endometrium | *WDR86* | -1.17748459 | 0.015188876 | 0.021816585 | 1 |
| HTN | endometrium | *EPOR* | -1.831198315 | 0.025261917 | 0.034199448 | 1 |
| HTN | endometrium | *LUZP2* | 7.341309333 | 0.000745001 | 0.003073153 | 1 |
| HTN | endometrium | *GJA4* | 4.05923501 | 0.00164716 | 0.0036245 | 1 |
| HTN | endometrium | *SBK2* | 28.28389151 | 0.001000341 | 0.003095606 | 1 |
| HTN | endometrium | *ZNF385C* | 6.072974432 | 0.000231591 | 0.003073153 | 1 |
| HTN | endometrium | *EXD3* | 2.637966067 | 0.000930812 | 0.003093357 | 1 |
| HTN | endometrium | *SAMD11* | 7.515226765 | 5.14E-05 | 0.002289397 | 0.249544223 |
| HTN | endometrium | *TRPV2* | 2.952026049 | 0.000930612 | 0.003093357 | 1 |
| HTN | endometrium | *C2orf88* | 4.13465461 | 0.000911165 | 0.003087532 | 1 |
| HTN | endometrium | *KBTBD12* | -3.997478885 | 0.016272005 | 0.023193822 | 1 |
| HTN | endometrium | *FANCA* | 3.344549223 | 0.000202895 | 0.003073153 | 0.984448358 |
| HTN | endometrium | *DNAH17* | 3.815413836 | 0.000171245 | 0.003073153 | 0.830882132 |
| HTN | endometrium | *FANCM* | 3.01942198 | 0.003071295 | 0.005707362 | 1 |
| HTN | endometrium | *ZFP69* | 3.75504557 | 0.001042072 | 0.003095606 | 1 |
| HTN | endometrium | *TMEM220* | 5.84673874 | 0.000627498 | 0.003073153 | 1 |
| HTN | endometrium | *P2RX2* | -3.600610275 | 0.002050871 | 0.004209317 | 1 |
| HTN | endometrium | *FAM122A* | -2.003744123 | 0.00861695 | 0.01331087 | 1 |
| HTN | endometrium | *GFRAL* | 7.066708665 | 0.001950098 | 0.004066125 | 1 |
| HTN | endometrium | *LDLRAD2* | 5.274346567 | 0.001480315 | 0.003394073 | 1 |
| HTN | endometrium | *CPSF4L* | 11.70952411 | 0.007965357 | 0.012446992 | 1 |
| HTN | endometrium | *MAPK12* | 6.65417178 | 0.024288308 | 0.032982611 | 1 |
| HTN | endometrium | *ZNF626* | 8.649098065 | 0.000606451 | 0.003073153 | 1 |
| HTN | endometrium | *LINC00265* | 3.625176668 | 0.001066602 | 0.003095606 | 1 |
| HTN | endometrium | *LAMTOR4* | 3.309774561 | 0.001483734 | 0.003395791 | 1 |
| HTN | endometrium | *PRKAR1B* | 5.860486483 | 0.025677478 | 0.034713604 | 1 |
| HTN | endometrium | *NCR3LG1* | -1.518127296 | 0.005115546 | 0.008597378 | 1 |

| | | | | | | |
|---|---|---|---|---|---|---|
| HTN | endometrium | *RUFY4* | 6.136453657 | 0.001104475 | 0.003095606 | 1 |
| HTN | endometrium | *PLSCR1* | 2.364772696 | 0.00071032 | 0.003073153 | 1 |
| HTN | endometrium | *ENO4* | 5.641793703 | 0.00105238 | 0.003095606 | 1 |
| HTN | endometrium | *H3F3C* | 68.62413665 | 0.022067225 | 0.030245812 | 1 |
| HTN | endometrium | *JAKMIP3* | -1.811196534 | 0.008091901 | 0.012608189 | 1 |
| HTN | endometrium | *PDCD1* | 16.04246596 | 0.031011557 | 0.041212839 | 1 |
| HTN | endometrium | *SELL* | 3.852837667 | 0.007034337 | 0.01120138 | 1 |
| HTN | endometrium | *C19orf54* | 4.406318518 | 0.001038627 | 0.003095606 | 1 |
| HTN | endometrium | *C22orf34* | 2.161813127 | 0.000905248 | 0.003087532 | 1 |
| HTN | endometrium | *SRSF10* | 5.147129887 | 0.000107925 | 0.002958475 | 0.52365009 |
| HTN | endometrium | *NBR1* | 4.531249751 | 0.001287434 | 0.003155246 | 1 |
| HTN | endometrium | *NDOR1* | -2.181891767 | 0.012396541 | 0.018226671 | 1 |
| HTN | endometrium | *FAM72B* | -2.275979148 | 0.009010169 | 0.013838981 | 1 |
| HTN | endometrium | *GOLGA8M* | 3.396841141 | 0.001254503 | 0.003122339 | 1 |
| HTN | endometrium | *RHCE* | 7.231635214 | 0.003131505 | 0.005798815 | 1 |
| HTN | endometrium | *IDO2* | 8.979063871 | 0.000679488 | 0.003073153 | 1 |
| HTN | endometrium | *SLC4A5* | 9.156019656 | 0.001108971 | 0.003095606 | 1 |
| HTN | endometrium | *UROS* | 3.775804019 | 0.000878364 | 0.003084413 | 1 |
| HTN | endometrium | *ZBED6CL* | 8.287159825 | 0.013572583 | 0.019778459 | 1 |
| HTN | endometrium | *OSTN* | 3.175346203 | 0.003109453 | 0.005765023 | 1 |
| HTN | endometrium | *FSIP2* | 2.913853232 | 0.00151214 | 0.003430353 | 1 |
| HTN | endometrium | *NOXA1* | -1.667144457 | 0.009553612 | 0.014576769 | 1 |
| HTN | endometrium | *FZD9* | 37.16239467 | 0.000101127 | 0.002934745 | 0.490670079 |
| HTN | endometrium | *SKOR1* | 9.136964203 | 0.008350247 | 0.012944217 | 1 |
| HTN | endometrium | *MTF1* | 3.001359032 | 0.000762125 | 0.003073153 | 1 |
| HTN | endometrium | *ZDHHC11* | 5.043636715 | 0.016467563 | 0.023438139 | 1 |
| HTN | endometrium | *ENTPD8* | 313.9093855 | 0.001629752 | 0.003600432 | 1 |
| HTN | endometrium | *RPL14* | 5.592317994 | 0.0236571 | 0.032233712 | 1 |
| HTN | endometrium | *FBF1* | -6.80389963 | 0.001649698 | 0.00362734 | 1 |
| HTN | endometrium | *MSL1* | 1.872073469 | 0.009164851 | 0.014049876 | 1 |
| HTN | endometrium | *FAM196A* | 2.952192135 | 0.000626033 | 0.003073153 | 1 |
| HTN | endometrium | *UTS2B* | 3.207319356 | 0.005639784 | 0.009310728 | 1 |
| HTN | endometrium | *C9orf152* | 8.217019172 | 0.002318749 | 0.004629644 | 1 |
| HTN | endometrium | *NELFB* | 2.616980376 | 0.00129741 | 0.003164924 | 1 |
| HTN | endometrium | *SLC15A5* | 4.075964278 | 0.001239034 | 0.003110085 | 1 |
| HTN | endometrium | *ADAT2* | 6.317507135 | 0.000692891 | 0.003073153 | 1 |
| HTN | endometrium | *ALKBH2* | 4.820188914 | 0.000978105 | 0.003095606 | 1 |
| HTN | endometrium | *LITAF* | 4.047354174 | 0.000545949 | 0.003073153 | 1 |
| HTN | endometrium | *VSTM1* | -0.819975417 | 0.005367168 | 0.008942823 | 1 |
| HTN | endometrium | *SF3B3* | 7.086640941 | 0.00758779 | 0.011949353 | 1 |
| HTN | endometrium | *PLAC9* | 4.158240754 | 0.001056653 | 0.003095606 | 1 |
| HTN | endometrium | *ZNF573* | 4.5378713 | 0.000964168 | 0.003095606 | 1 |
| HTN | endometrium | *FAM47E* | 4.302863505 | 0.000115431 | 0.003017974 | 0.560072403 |
| HTN | endometrium | *ZNF600* | 2.477562127 | 0.000292039 | 0.003073153 | 1 |
| HTN | endometrium | *FHIT* | 4.104991067 | 0.000399116 | 0.003073153 | 1 |
| HTN | endometrium | *SLC35E2B* | 3.692494103 | 0.000737414 | 0.003073153 | 1 |
| HTN | endometrium | *KIAA0408* | 2.228542053 | 0.035701054 | 0.046879977 | 1 |
| HTN | endometrium | *AKR1C3* | 3.698212718 | 0.000798074 | 0.003073153 | 1 |
| HTN | endometrium | *WDSUB1* | 5.874313517 | 0.01376851 | 0.019989471 | 1 |
| HTN | endometrium | *COLCA1* | -3.142276987 | 0.000293503 | 0.003073153 | 1 |

| | | | | | | |
|---|---|---|---|---|---|---|
| HTN | endometrium | *SEMA4A* | -3.732727372 | 0.008535797 | 0.01321496 | 1 |
| HTN | endometrium | *HRCT1* | 5.906576245 | 4.09E-06 | 0.00050228 | 0.01982286 |
| HTN | endometrium | *MPHOSPH8* | 3.811291447 | 0.000288612 | 0.003073153 | 1 |
| HTN | endometrium | *SIRPB2* | 1.875912137 | 0.009950703 | 0.015125567 | 1 |
| HTN | endometrium | *XPNPEP3* | 2.242782088 | 0.015433878 | 0.022129189 | 1 |
| HTN | endometrium | *ZNF107* | 4.776951189 | 0.001550276 | 0.003487223 | 1 |
| HTN | endometrium | *PPIA* | 6.412143898 | 0.001099557 | 0.003095606 | 1 |
| HTN | endometrium | *ZNF471* | 3.000200075 | 0.000956175 | 0.003095606 | 1 |
| HTN | endometrium | *ZNF493* | 6.999579115 | 0.003460486 | 0.006262693 | 1 |
| HTN | endometrium | *GTF2IRD2* | 5.836486156 | 0.00056953 | 0.003073153 | 1 |
| HTN | endometrium | *SUPT3H* | 5.849034018 | 0.001225667 | 0.003095606 | 1 |
| HTN | endometrium | *GIMAP5* | 4.962277982 | 0.005121161 | 0.008603835 | 1 |
| HTN | endometrium | *STK31* | 4.438509787 | 0.000756009 | 0.003073153 | 1 |
| HTN | endometrium | *ZKSCAN7* | 7.095361952 | 3.50E-05 | 0.0019512 | 0.169754429 |
| HTN | endometrium | *CD55* | 2.471712595 | 0.00084053 | 0.0030808 | 1 |
| HTN | endometrium | *CPNE4* | 2.465576661 | 0.003983933 | 0.007026552 | 1 |
| HTN | endometrium | *ZNF565* | 2.655472157 | 0.002393096 | 0.004733512 | 1 |
| HTN | endometrium | *ZNF34* | 3.727671918 | 0.000441425 | 0.003073153 | 1 |
| HTN | endometrium | *EVL* | 4.977685363 | 0.022653448 | 0.030959409 | 1 |
| HTN | endometrium | *PIK3R4* | 6.262724985 | 0.033390091 | 0.044060028 | 1 |
| HTN | endometrium | *ZNF605* | 8.427837992 | 0.00021664 | 0.003073153 | 1 |
| HTN | endometrium | *RFX8* | 1.909962653 | 0.005981665 | 0.009765491 | 1 |
| HTN | endometrium | *C20orf96* | 3.827990138 | 0.001130878 | 0.003095606 | 1 |
| HTN | endometrium | *TPK1* | 4.290992153 | 0.000944263 | 0.003095606 | 1 |
| HTN | endometrium | *MYO18A* | 4.07420845 | 0.001452626 | 0.003357454 | 1 |
| HTN | endometrium | *SPTSSB* | -1.693495754 | 0.003657694 | 0.006548757 | 1 |
| HTN | endometrium | *CACNA1H* | 5.298844491 | 0.001268109 | 0.003128219 | 1 |
| HTN | endometrium | *SULF2* | 7.029712429 | 5.30E-05 | 0.002317582 | 0.257251651 |
| HTN | endometrium | *XRCC2* | -1.351236611 | 0.004744478 | 0.008077266 | 1 |
| HTN | endometrium | *MYO6* | 4.963350549 | 0.023753065 | 0.032346301 | 1 |
| HTN | endometrium | *ZNF846* | 5.407213563 | 0.001007324 | 0.003095606 | 1 |
| HTN | endometrium | *ADH1B* | 3.987882448 | 0.00087742 | 0.003084413 | 1 |
| HTN | endometrium | *ZNF502* | 3.321793745 | 0.000986152 | 0.003095606 | 1 |
| HTN | endometrium | *TRAPPC4* | 2.81378277 | 0.0063704 | 0.010296196 | 1 |
| HTN | endometrium | *ZNF33B* | 2.660959892 | 0.001311934 | 0.00318913 | 1 |
| HTN | endometrium | *AMZ2* | 2.896544854 | 0.004266326 | 0.0074233 | 1 |
| HTN | endometrium | *ZNF431* | 6.611424401 | 0.001702226 | 0.003705679 | 1 |
| HTN | endometrium | *HLA-DQA1* | 1.592157155 | 0.018939354 | 0.026467093 | 1 |
| HTN | endometrium | *GM2A* | -1.478924119 | 0.034691944 | 0.045639647 | 1 |
| HTN | endometrium | *ZNF239* | 4.096379647 | 0.00270428 | 0.005194444 | 1 |
| HTN | endometrium | *SPRR2B* | -6.169945002 | 0.00414139 | 0.007248927 | 1 |
| HTN | endometrium | *FAM3C* | 4.032593345 | 0.002687861 | 0.005169046 | 1 |
| HTN | endometrium | *SLC39A10* | 6.306352275 | 0.001069865 | 0.003095606 | 1 |
| HTN | endometrium | *AP2A1* | 1.797629505 | 0.004175835 | 0.007296057 | 1 |
| HTN | endometrium | *PCDHB16* | 5.326198571 | 0.000173527 | 0.003073153 | 0.841953469 |
| HTN | endometrium | *HIST1H3E* | 9.112870268 | 8.04E-10 | 5.57E-07 | 3.90E-06 |
| HTN | endometrium | *FUT11* | 2.571476395 | 0.002114734 | 0.004302581 | 1 |
| HTN | endometrium | *WDR5B* | 5.374158712 | 3.93E-05 | 0.002049405 | 0.190594701 |
| HTN | endometrium | *ZNF138* | 5.650759036 | 0.000262203 | 0.003073153 | 1 |
| HTN | endometrium | *ZNF429* | 3.801677496 | 0.000746712 | 0.003073153 | 1 |

| | | | | | | |
|---|---|---|---|---|---|---|
| HTN | endometrium | *ZNF470* | 9.48877168 | 0.035107104 | 0.046137506 | 1 |
| HTN | endometrium | *ZNF100* | 1.660859472 | 0.024386697 | 0.033088437 | 1 |
| HTN | endometrium | *ANXA6* | 5.401307526 | 0.000332355 | 0.003073153 | 1 |
| HTN | endometrium | *GMFB* | 2.290674157 | 0.001109171 | 0.003095606 | 1 |
| HTN | endometrium | *ZNF420* | 6.735414563 | 0.016814998 | 0.023869623 | 1 |
| HTN | endometrium | *ZNF763* | -2.516862853 | 0.001978977 | 0.004110444 | 1 |
| HTN | endometrium | *ZMYM1* | 7.432443764 | 0.004004976 | 0.00705341 | 1 |
| HTN | endometrium | *IGF2R* | 4.385689932 | 0.001880889 | 0.003966721 | 1 |
| HTN | endometrium | *SLC25A29* | 2.311715687 | 0.000794344 | 0.003073153 | 1 |
| HTN | endometrium | *ADAM32* | -2.527878117 | 0.021780153 | 0.029877665 | 1 |
| HTN | endometrium | *LRRC8B* | 3.91533644 | 0.000638566 | 0.003073153 | 1 |
| HTN | endometrium | *SND1* | 3.914250781 | 0.000296467 | 0.003073153 | 1 |
| HTN | endometrium | *NEK5* | 3.130258041 | 0.001032368 | 0.003095606 | 1 |
| HTN | endometrium | *PIWIL2* | -3.737838518 | 0.031223137 | 0.04145995 | 1 |
| HTN | endometrium | *ENTPD4* | 3.97930386 | 0.000730881 | 0.003073153 | 1 |
| HTN | endometrium | *TBC1D9B* | 4.614895272 | 0.002325578 | 0.004635869 | 1 |
| HTN | endometrium | *CCL4L2* | 9.321124423 | 0.0002702 | 0.003073153 | 1 |
| HTN | endometrium | *RAMP2-AS1* | -2.00238975 | 0.017256022 | 0.024409976 | 1 |
| HTN | endometrium | *ZNF720* | 2.399731429 | 0.001077053 | 0.003095606 | 1 |
| HTN | endometrium | *MRPL21* | 2.620847067 | 0.024554715 | 0.033297786 | 1 |
| HTN | endometrium | *ZNF98* | 2.450233453 | 0.000697036 | 0.003073153 | 1 |
| HTN | endometrium | *ZNF517* | 3.414185162 | 0.003158729 | 0.005835379 | 1 |
| HTN | endometrium | *ZNF675* | 3.137601497 | 0.001346882 | 0.003237588 | 1 |
| HTN | endometrium | *VEPH1* | 2.564976099 | 0.001220686 | 0.003095606 | 1 |
| HTN | endometrium | *MAP3K5* | 5.196985183 | 0.000996581 | 0.003095606 | 1 |
| HTN | endometrium | *GSTK1* | 2.423252114 | 0.010317647 | 0.015595396 | 1 |
| HTN | endometrium | *GYPE* | 5.453725682 | 0.000979451 | 0.003095606 | 1 |
| HTN | endometrium | *PCDHB11* | -1.312464257 | 0.000405609 | 0.003073153 | 1 |
| HTN | endometrium | *SLC2A10* | 3.195858357 | 0.000859678 | 0.0030808 | 1 |
| HTN | endometrium | *ZNF665* | 2.496409141 | 0.001150303 | 0.003095606 | 1 |
| HTN | endometrium | *RPF2* | 5.984345361 | 0.000853539 | 0.0030808 | 1 |
| HTN | endometrium | *MYO5A* | 1.657076818 | 0.003811055 | 0.006768388 | 1 |
| HTN | endometrium | *ATG7* | 3.960618203 | 8.57E-05 | 0.002714492 | 0.415977876 |
| HTN | endometrium | *RAB40C* | 3.567445273 | 0.000778545 | 0.003073153 | 1 |
| HTN | endometrium | *HHLA3* | -2.922725558 | 0.019284499 | 0.026856598 | 1 |
| HTN | endometrium | *TOPORS* | 7.159734579 | 0.027212748 | 0.036605559 | 1 |
| HTN | endometrium | *BCO2* | 1.224837882 | 0.029308156 | 0.039142079 | 1 |
| HTN | endometrium | *ENTPD6* | 2.356490584 | 0.002906128 | 0.005486588 | 1 |
| HTN | endometrium | *ENPP1* | -0.827513375 | 0.008972621 | 0.013790041 | 1 |
| HTN | endometrium | *MFAP5* | 1.386221966 | 0.018273233 | 0.025647013 | 1 |
| HTN | endometrium | *PDCD1LG2* | 1.927259058 | 0.000369013 | 0.003073153 | 1 |
| HTN | endometrium | *NMB* | -3.676471256 | 0.027793115 | 0.037355179 | 1 |
| HTN | endometrium | *FAM114A1* | 5.928960551 | 0.000472208 | 0.003073153 | 1 |
| HTN | endometrium | *PSAP* | 1.740529186 | 0.000713992 | 0.003073153 | 1 |
| HTN | endometrium | *TXNRD3* | 2.630398947 | 0.00107715 | 0.003095606 | 1 |
| HTN | endometrium | *AC122129.1* | 7.873488191 | 0.002515374 | 0.004913282 | 1 |
| HTN | endometrium | *OCLN* | -2.227710919 | 0.003576909 | 0.006420704 | 1 |
| HTN | endometrium | *ZNF181* | 3.557254931 | 0.006399081 | 0.010335666 | 1 |
| HTN | endometrium | *SGTB* | 2.672126932 | 0.003248404 | 0.005970172 | 1 |
| HTN | endometrium | *ADH5* | 6.335641017 | 0.001161237 | 0.003095606 | 1 |

| | | | | | | |
|---|---|---|---|---|---|---|
| HTN | endometrium | *ZNF677* | 2.665035417 | 0.001382649 | 0.00327409 | 1 |
| HTN | endometrium | *ZNF311* | 4.770485761 | 1.74E-05 | 0.001262767 | 0.084605412 |
| HTN | endometrium | *MPZL1* | 5.591699648 | 0.000641131 | 0.003073153 | 1 |
| HTN | endometrium | *VPS13A* | 2.908291446 | 0.000648467 | 0.003073153 | 1 |
| HTN | endometrium | *NOL8* | 3.346904477 | 0.000343904 | 0.003073153 | 1 |
| HTN | endometrium | *DLGAP2* | 5.312774483 | 0.000153295 | 0.003073153 | 0.743789663 |
| HTN | endometrium | *ZNF560* | -2.253583238 | 0.014710549 | 0.02119857 | 1 |
| HTN | endometrium | *ZNF273* | 2.888972628 | 0.001917515 | 0.004025868 | 1 |
| HTN | endometrium | *MAK16* | 3.099897159 | 0.00079463 | 0.003073153 | 1 |
| HTN | endometrium | *SFI1* | 3.220220224 | 0.001936072 | 0.004050808 | 1 |
| HTN | endometrium | *ZNF649* | 3.09987022 | 0.001596336 | 0.003554047 | 1 |
| HTN | endometrium | *ADH4* | -2.919152623 | 0.002126415 | 0.004322316 | 1 |
| HTN | endometrium | *HIBCH* | 2.781637606 | 0.000285877 | 0.003073153 | 1 |
| HTN | endometrium | *SOWAHC* | 5.445646795 | 0.001250569 | 0.003122339 | 1 |
| HTN | endometrium | *AC135178.1* | 27.70706724 | 0.036057253 | 0.0473093 | 1 |
| HTN | endometrium | *MAN1A2* | 5.997716347 | 0.001380053 | 0.00327409 | 1 |
| HTN | endometrium | *DDRGK1* | 8.482053202 | 0.000679039 | 0.003073153 | 1 |
| HTN | endometrium | *ZNF607* | 7.39585392 | 4.88E-05 | 0.002213026 | 0.236793828 |
| HTN | endometrium | *ZNF334* | 2.254190304 | 0.001709591 | 0.003713043 | 1 |
| HTN | endometrium | *HSD17B11* | 3.423667473 | 0.000361722 | 0.003073153 | 1 |
| HTN | endometrium | *SULT1C2* | 2.181516249 | 0.022013482 | 0.030180677 | 1 |
| HTN | endometrium | *RPS6KL1* | 16.12052039 | 1.09E-05 | 0.000930785 | 0.053054728 |
| HTN | endometrium | *DDX42* | 4.03519429 | 0.000492534 | 0.003073153 | 1 |
| HTN | endometrium | *HELZ* | 4.639183368 | 0.000637936 | 0.003073153 | 1 |
| HTN | endometrium | *TMEM116* | -8.051386889 | 4.15E-11 | 5.04E-08 | 2.01E-07 |
| HTN | endometrium | *GFPT1* | 6.041658828 | 0.005524433 | 0.009154559 | 1 |
| HTN | endometrium | *ZNF568* | 1.944091575 | 0.002827288 | 0.005366979 | 1 |
| HTN | endometrium | *ZNF587* | 5.782330573 | 0.002149491 | 0.004360089 | 1 |
| HTN | endometrium | *SH3BGRL2* | 3.930935632 | 0.012533535 | 0.018405784 | 1 |
| HTN | endometrium | *TMA16* | 5.663466988 | 0.000565065 | 0.003073153 | 1 |
| HTN | endometrium | *HLA-DRB5* | -1.22742412 | 0.014617374 | 0.021083086 | 1 |
| HTN | endometrium | *ZNF28* | 5.758296926 | 0.000422523 | 0.003073153 | 1 |
| HTN | endometrium | *C20orf203* | 9.15509502 | 0.001763449 | 0.003780934 | 1 |
| HTN | endometrium | *NCOA6* | 2.880790066 | 0.017838885 | 0.025102746 | 1 |
| HTN | endometrium | *TTC37* | 4.122882896 | 0.000984311 | 0.003095606 | 1 |
| HTN | endometrium | *TUSC1* | -1.114979422 | 0.004866936 | 0.008242917 | 1 |
| HTN | endometrium | *PAPSS2* | 5.955451607 | 0.001277175 | 0.003143251 | 1 |
| HTN | endometrium | *CEP290* | 5.049650112 | 0.00229161 | 0.004583221 | 1 |
| HTN | endometrium | *ANKRD13B* | 4.177962163 | 0.001219539 | 0.003095606 | 1 |
| HTN | endometrium | *ECI2* | 2.581848428 | 0.001906056 | 0.004008371 | 1 |
| HTN | endometrium | *UNC13B* | 3.438623547 | 0.00097333 | 0.003095606 | 1 |
| HTN | endometrium | *PPP1R14C* | -1.423957866 | 0.004192189 | 0.007314412 | 1 |
| HTN | endometrium | *GPATCH3* | 4.083671854 | 0.001380376 | 0.00327409 | 1 |
| HTN | endometrium | *RPL10A* | 5.357587872 | 0.000673056 | 0.003073153 | 1 |
| HTN | endometrium | *COLGALT2* | 9.056258631 | 0.001006272 | 0.003095606 | 1 |
| HTN | endometrium | *RCSD1* | 1.856809278 | 0.001112532 | 0.003095606 | 1 |
| HTN | endometrium | *GRIN3A* | -0.844081553 | 0.007391109 | 0.011681323 | 1 |
| HTN | endometrium | *TMEM184B* | 4.301290555 | 0.000424334 | 0.003073153 | 1 |
| HTN | endometrium | *SCAMP5* | 2.660963565 | 0.000776166 | 0.003073153 | 1 |
| HTN | endometrium | *PNP* | -2.640094514 | 0.000704859 | 0.003073153 | 1 |

| | | | | | | |
|---|---|---|---|---|---|---|
| HTN | endometrium | *SFT2D1* | 5.258466083 | 0.000542368 | 0.003073153 | 1 |
| HTN | endometrium | *HMGN2* | 3.485675382 | 0.001629003 | 0.003600432 | 1 |
| HTN | endometrium | *DENND4B* | 1.703980961 | 0.011823792 | 0.017501232 | 1 |
| HTN | endometrium | *RYR3* | 2.069186253 | 0.000774762 | 0.003073153 | 1 |
| HTN | endometrium | *TYW1* | 3.632953172 | 0.000507604 | 0.003073153 | 1 |
| HTN | endometrium | *PRMT6* | 8.232100528 | 0.00225897 | 0.004540399 | 1 |
| HTN | endometrium | *SHISA4* | -16.28138429 | 4.64E-05 | 0.002206299 | 0.225326275 |
| HTN | endometrium | *MAP3K3* | 1.666669637 | 0.002065348 | 0.004230085 | 1 |
| HTN | endometrium | *C1orf174* | 3.303629249 | 0.000675471 | 0.003073153 | 1 |
| HTN | endometrium | *DZIP3* | 3.113087866 | 0.000594012 | 0.003073153 | 1 |
| HTN | endometrium | *ZFP2* | 5.920394929 | 0.001254488 | 0.003122339 | 1 |
| HTN | endometrium | *L3MBTL3* | 6.218194402 | 0.002231894 | 0.004506513 | 1 |
| HTN | endometrium | *MFAP3L* | 4.601289462 | 0.000282763 | 0.003073153 | 1 |
| HTN | endometrium | *NAGA* | 3.619331504 | 0.000841447 | 0.0030808 | 1 |
| HTN | endometrium | *PJA2* | 5.089199676 | 0.018720865 | 0.026199491 | 1 |
| HTN | endometrium | *AL590822.1* | 2.917715711 | 0.007334108 | 0.011617726 | 1 |
| HTN | endometrium | *EFCAB2* | 2.181314218 | 0.00230599 | 0.004610081 | 1 |
| HTN | endometrium | *CHML* | 5.91680862 | 0.002244685 | 0.004521051 | 1 |
| HTN | endometrium | *TATDN3* | 2.334878425 | 0.001715899 | 0.003716761 | 1 |
| HTN | endometrium | *CR1* | 1.175039245 | 0.025722621 | 0.034764946 | 1 |
| HTN | endometrium | *SAMD5* | 2.32919388 | 0.000906451 | 0.003087532 | 1 |
| HTN | endometrium | *ECT2L* | 4.397336201 | 0.000702125 | 0.003073153 | 1 |
| HTN | endometrium | *FAM229B* | 2.621486975 | 0.002695808 | 0.005180221 | 1 |
| HTN | endometrium | *DDO* | 3.590653206 | 0.03737854 | 0.048857941 | 1 |
| HTN | endometrium | *RBM20* | 3.944454407 | 0.001086314 | 0.003095606 | 1 |
| HTN | endometrium | *PCMTD2* | 3.535407759 | 0.00065847 | 0.003073153 | 1 |
| HTN | endometrium | *SAMD13* | -1.928800916 | 0.007599355 | 0.011963683 | 1 |
| HTN | endometrium | *EFCAB7* | 6.454370698 | 0.000949388 | 0.003095606 | 1 |
| HTN | endometrium | *IFIT1B* | 17.7572595 | 0.001095245 | 0.003095606 | 1 |
| HTN | endometrium | *LIPN* | 7.932879906 | 0.001048387 | 0.003095606 | 1 |
| HTN | endometrium | *NEU4* | -2.502492867 | 0.008332908 | 0.012921466 | 1 |
| HTN | endometrium | *C2orf72* | -3.833074478 | 0.004882143 | 0.008259469 | 1 |
| HTN | endometrium | *RUFY2* | 6.077422966 | 0.003018093 | 0.005636562 | 1 |
| HTN | endometrium | *PHACTR4* | 3.800319311 | 0.000266853 | 0.003073153 | 1 |
| HTN | endometrium | *ASAH2B* | -1.940390531 | 0.014413517 | 0.020807613 | 1 |
| HTN | endometrium | *AGAP6* | 2.973045926 | 0.000315092 | 0.003073153 | 1 |
| HTN | endometrium | *LINC00619* | 29.32197002 | 0.001990319 | 0.004123411 | 1 |
| HTN | endometrium | *DAXX* | 2.505974524 | 0.010805725 | 0.016206918 | 1 |
| HTN | endometrium | *BMPR2* | 3.638577835 | 0.000540204 | 0.003073153 | 1 |
| HTN | endometrium | *TCEA3* | 2.437961618 | 0.001362943 | 0.00325328 | 1 |
| HTN | endometrium | *OXLD1* | 3.124648118 | 0.000644386 | 0.003073153 | 1 |
| HTN | endometrium | *TAP2* | 2.177723237 | 0.027160296 | 0.036545135 | 1 |
| HTN | endometrium | *COL15A1* | 5.28430046 | 0.000183084 | 0.003073153 | 0.888323658 |
| HTN | endometrium | *AGER* | 3.546281722 | 0.003235165 | 0.005954519 | 1 |
| HTN | endometrium | *PRRT1* | 3.539468408 | 0.004944744 | 0.008345008 | 1 |
| HTN | endometrium | *SMIM5* | 6.379404649 | 0.015542392 | 0.022271615 | 1 |
| HTN | endometrium | *STK19* | 2.753868228 | 0.001254223 | 0.003122339 | 1 |
| HTN | endometrium | *LAYN* | -5.356057745 | 0.001427026 | 0.003328251 | 1 |
| HTN | endometrium | *GPANK1* | 2.801958339 | 0.001190115 | 0.003095606 | 1 |
| HTN | endometrium | *BAG6* | 3.830850495 | 0.000148739 | 0.003073153 | 0.721679658 |

| | | | | | | |
|---|---|---|---|---|---|---|
| HTN | endometrium | *LST1* | 2.181111063 | 0.003494591 | 0.006303254 | 1 |
| HTN | endometrium | *ZNF814* | 5.4444598 | 0.000478582 | 0.003073153 | 1 |
| HTN | endometrium | *MICA* | 2.20608378 | 0.034497115 | 0.045434311 | 1 |
| HTN | endometrium | *POU5F1* | -1.513908843 | 0.005720494 | 0.009400242 | 1 |
| HTN | endometrium | *PSORS1C2* | 6.984268218 | 0.000305618 | 0.003073153 | 1 |
| HTN | endometrium | *DHX16* | 7.251563085 | 0.001959381 | 0.004080221 | 1 |
| HTN | endometrium | *ABCF1* | 4.224782751 | 0.003455279 | 0.006255602 | 1 |
| HTN | endometrium | *LILRB3* | 4.353519983 | 0.000883437 | 0.003084413 | 1 |
| HTN | endometrium | *HLA-E* | 3.298411883 | 0.011359777 | 0.016912439 | 1 |
| HTN | endometrium | *ZNF468* | -3.162565541 | 0.024199308 | 0.032870953 | 1 |
| HTN | endometrium | *HCG9* | 5.187380583 | 0.002374416 | 0.004702313 | 1 |
| HTN | endometrium | *HLA-G* | 2.947981799 | 0.004865662 | 0.008242917 | 1 |
| HTN | endometrium | *ZFP57* | -2.649479467 | 0.004990901 | 0.008411203 | 1 |
| HTN | endometrium | *FAM153C* | 1.914114393 | 0.017466039 | 0.024663917 | 1 |
| HTN | endometrium | *MALRD1* | 3.56989748 | 0.001040915 | 0.003095606 | 1 |
| HTN | endometrium | *FAM196B* | 2.121894735 | 0.001090733 | 0.003095606 | 1 |
| HTN | endometrium | *DCTN1* | 9.625520891 | 0.008592192 | 0.013281081 | 1 |
| HTN | endometrium | *ZBTB48* | 6.006069501 | 0.000333313 | 0.003073153 | 1 |
| HTN | endometrium | *MZT1* | 4.104706066 | 0.000943268 | 0.003095606 | 1 |
| HTN | endometrium | *ZNF155* | 2.885455647 | 0.002868394 | 0.005430139 | 1 |
| HTN | endometrium | *ZNF425* | 3.813502944 | 0.017564807 | 0.024761449 | 1 |
| HTN | endometrium | *C12orf73* | 6.762581977 | 0.003328859 | 0.00608806 | 1 |
| HTN | endometrium | *PCDHGA1* | -1.868044764 | 0.01672934 | 0.02376889 | 1 |
| HTN | endometrium | *PCDHA4* | 7.121387556 | 0.005358036 | 0.008939886 | 1 |
| HTN | endometrium | *TRIM13* | 5.284874581 | 0.00080721 | 0.003073153 | 1 |
| HTN | endometrium | *PKHD1L1* | 1.689390313 | 0.000987181 | 0.003095606 | 1 |
| HTN | endometrium | *SLFN12L* | 1.697440953 | 0.006300494 | 0.010196797 | 1 |
| HTN | endometrium | *SLC35B4* | 3.933743509 | 0.00077327 | 0.003073153 | 1 |
| HTN | endometrium | *TMEM231* | 4.669800425 | 0.000948457 | 0.003095606 | 1 |
| HTN | endometrium | *LINC00654* | -4.45035954 | 0.009479979 | 0.014478079 | 1 |
| HTN | endometrium | *SCGB2B2* | 3.668649124 | 0.000657511 | 0.003073153 | 1 |
| HTN | endometrium | *CCDC144NL* | 3.947553169 | 0.001131056 | 0.003095606 | 1 |
| HTN | endometrium | *TMEM170B* | 2.367441288 | 6.09E-05 | 0.002410623 | 0.295289864 |
| HTN | endometrium | *GOLGA6L10* | 7.064618525 | 0.007267444 | 0.011523411 | 1 |
| HTN | endometrium | *NT5M* | 4.719598785 | 0.001202619 | 0.003095606 | 1 |
| HTN | endometrium | *IPO7* | 6.076610617 | 0.022638297 | 0.030958573 | 1 |
| HTN | endometrium | *TECPR1* | 3.117163775 | 0.00116381 | 0.003095606 | 1 |
| HTN | endometrium | *MT1A* | -1.442067099 | 0.004884077 | 0.008259861 | 1 |
| HTN | endometrium | *MT1M* | 3.716455511 | 0.001199565 | 0.003095606 | 1 |
| HTN | endometrium | *CFI* | 1.957398539 | 0.001434791 | 0.003335699 | 1 |
| HTN | endometrium | *ATP6AP1L* | 3.611715584 | 0.000603729 | 0.003073153 | 1 |
| HTN | endometrium | *RGL3* | 2.193739414 | 0.001702382 | 0.003705679 | 1 |
| HTN | endometrium | *NAP1L4* | 3.30516538 | 0.000465707 | 0.003073153 | 1 |
| HTN | endometrium | *SERF1B* | 5.251139393 | 0.00548231 | 0.009100297 | 1 |
| HTN | endometrium | *LIN52* | 2.45950635 | 0.000483709 | 0.003073153 | 1 |
| HTN | endometrium | *KLRC2* | 6.277714308 | 0.000655056 | 0.003073153 | 1 |
| HTN | endometrium | *C1QTNF9B* | -1.995998341 | 0.003788942 | 0.006741455 | 1 |
| HTN | endometrium | *ZDHHC11B* | -2.356014033 | 0.010308243 | 0.015586038 | 1 |
| HTN | endometrium | *HCG27* | 3.89467968 | 4.06E-05 | 0.002058697 | 0.196919137 |
| HTN | endometrium | *COL6A6* | 5.41122038 | 0.010499283 | 0.015825574 | 1 |

| | | | | | | |
|---|---|---|---|---|---|---|
| HTN | endometrium | *HLA-A* | 3.905065562 | 0.001013146 | 0.003095606 | 1 |
| HTN | endometrium | *LNP1* | 3.503228189 | 0.001241566 | 0.003110503 | 1 |
| HTN | endometrium | *PRSS50* | 17.38689661 | 0.000195668 | 0.003073153 | 0.949382589 |
| HTN | endometrium | *METTL6* | 6.112978785 | 0.000515405 | 0.003073153 | 1 |
| HTN | endometrium | *SACM1L* | 3.035296745 | 0.000795037 | 0.003073153 | 1 |
| HTN | endometrium | *SLC48A1* | 7.996678421 | 0.0008852 | 0.003084413 | 1 |
| HTN | endometrium | *TAS2R50* | 77.36537499 | 0.00025031 | 0.003073153 | 1 |
| HTN | endometrium | *AC084018.1* | -2.879994941 | 0.006912674 | 0.011029363 | 1 |
| HTN | endometrium | *C17orf51* | 4.524975605 | 0.001195072 | 0.003095606 | 1 |
| HTN | endometrium | *OR2A42* | -1.82721274 | 0.00419237 | 0.007314412 | 1 |
| HTN | endometrium | *NUP62* | 2.51745838 | 0.034159588 | 0.045038674 | 1 |
| HTN | endometrium | *AL021068.1* | 7.115123003 | 0.005529174 | 0.009159287 | 1 |
| HTN | endometrium | *FGFR1OP* | 10.65319828 | 0.007857624 | 0.012298448 | 1 |
| HTN | endometrium | *ZNF254* | 5.50468421 | 0.001654537 | 0.003629211 | 1 |
| HTN | endometrium | *TCTEX1D2* | 6.013371667 | 0.009956562 | 0.015127175 | 1 |
| HTN | endometrium | *AC022498.1* | 17.6348958 | 0.001005737 | 0.003095606 | 1 |
| HTN | endometrium | *GIMAP1* | 6.381142519 | 0.002411451 | 0.004761531 | 1 |
| HTN | endometrium | *SUPT4H1* | 5.022262171 | 0.000147542 | 0.003073153 | 0.715873931 |
| HTN | endometrium | *ARHGAP19* | 4.474478511 | 0.004117557 | 0.007215018 | 1 |
| HTN | endometrium | *LCAT* | 3.037486749 | 0.001546219 | 0.003479709 | 1 |
| HTN | endometrium | *GPC2* | -4.080877525 | 0.008261185 | 0.012843085 | 1 |
| HTN | endometrium | *ERV3-1* | 1.743788454 | 0.000506354 | 0.003073153 | 1 |
| HTN | endometrium | *GBP7* | -0.830487834 | 0.006295015 | 0.010191329 | 1 |
| HTN | endometrium | *SRA1* | 3.918298282 | 0.001237539 | 0.003107939 | 1 |
| HTN | endometrium | *TMEM110* | 11.92732386 | 1.95E-06 | 0.00030216 | 0.009464657 |
| HTN | endometrium | *TMX2* | 4.75918642 | 0.000515255 | 0.003073153 | 1 |
| HTN | endometrium | *LBH* | 2.527672625 | 0.002243912 | 0.004521051 | 1 |
| HTN | endometrium | *PPP1CB* | 3.454075637 | 0.000756415 | 0.003073153 | 1 |
| HTN | endometrium | *NCKIPSD* | -2.801613942 | 0.008579189 | 0.013266813 | 1 |
| HTN | endometrium | *ATF6B* | 12.31558696 | 2.65E-09 | 1.43E-06 | 1.29E-05 |
| HTN | endometrium | *SLC35F6* | 2.922181313 | 0.002113353 | 0.004302581 | 1 |
| HTN | endometrium | *EMP2* | 4.370441877 | 0.001158182 | 0.003095606 | 1 |
| HTN | endometrium | *PPM1N* | 5.217806577 | 9.46E-06 | 0.000834404 | 0.045892235 |
| HTN | endometrium | *SLC23A3* | -1.074401 | 0.025353375 | 0.03430412 | 1 |
| HTN | endometrium | *LTB4R* | 2.762740029 | 0.013324551 | 0.019473109 | 1 |
| HTN | endometrium | *IRF9* | 5.114722477 | 0.000480093 | 0.003073153 | 1 |
| HTN | endometrium | *GALT* | 2.921039987 | 0.000256214 | 0.003073153 | 1 |
| HTN | endometrium | *ZNF90* | 3.079793143 | 0.001115584 | 0.003095606 | 1 |
| HTN | endometrium | *REPIN1* | 12.15019531 | 0.006105426 | 0.009940781 | 1 |
| HTN | endometrium | *ZNF891* | 7.231808663 | 0.030295879 | 0.040383407 | 1 |
| HTN | endometrium | *SMIM7* | 4.53038316 | 0.000412966 | 0.003073153 | 1 |
| HTN | endometrium | *TSPAN4* | 3.648367589 | 0.000616856 | 0.003073153 | 1 |
| HTN | endometrium | *ARL16* | 2.901153223 | 0.002451627 | 0.004815909 | 1 |
| HTN | endometrium | *PAXIP1-AS2* | 7.008946859 | 0.000438443 | 0.003073153 | 1 |
| HTN | endometrium | *LYRM4* | 3.609373079 | 0.000685356 | 0.003073153 | 1 |
| HTN | endometrium | *PRCD* | 10.57696201 | 0.011582245 | 0.017201424 | 1 |
| HTN | endometrium | *SH3D21* | 5.659718616 | 0.000399991 | 0.003073153 | 1 |
| HTN | endometrium | *ANG* | -2.954387613 | 0.028354934 | 0.038036533 | 1 |
| HTN | endometrium | *LCNL1* | 5.961024746 | 0.013811364 | 0.020045688 | 1 |
| HTN | endometrium | *GNAT3* | 3.274815139 | 0.001050321 | 0.003095606 | 1 |

| | | | | | | |
|---|---|---|---|---|---|---|
| HTN | endometrium | *PLIN5* | 2.110086689 | 0.003570493 | 0.006418686 | 1 |
| HTN | endometrium | *STARD10* | 3.529392512 | 0.000321315 | 0.003073153 | 1 |
| HTN | endometrium | *EML6* | 9.78036289 | 0.000640704 | 0.003073153 | 1 |
| HTN | endometrium | *ARHGEF33* | 3.263860806 | 0.006594249 | 0.010597978 | 1 |
| HTN | endometrium | *IFRD2* | 5.726921359 | 0.000351713 | 0.003073153 | 1 |
| HTN | endometrium | *EVPLL* | 14.41445768 | 0.000256562 | 0.003073153 | 1 |
| HTN | endometrium | *AC005082.1* | 9.77350381 | 0.016930172 | 0.024012042 | 1 |
| HTN | endometrium | *ACSM4* | 3.49978167 | 0.009099572 | 0.013958622 | 1 |
| HTN | endometrium | *UBXN2B* | 9.526741288 | 8.43E-05 | 0.002714492 | 0.409240246 |
| HTN | endometrium | *C5orf49* | 29.53503786 | 0.001972533 | 0.004104087 | 1 |
| HTN | endometrium | *GOLGA8B* | 2.590347437 | 0.000191567 | 0.003073153 | 0.92948547 |
| HTN | endometrium | *NPEPL1* | 2.901286955 | 0.00110514 | 0.003095606 | 1 |
| HTN | endometrium | *TMEM242* | 7.436464525 | 0.002066631 | 0.004230925 | 1 |
| HTN | endometrium | *SLC35E2* | 2.581297495 | 0.002764795 | 0.005279334 | 1 |
| HTN | endometrium | *LINC01139* | -1.197267732 | 0.013672086 | 0.019894704 | 1 |
| HTN | endometrium | *AC009403.2* | -3.849165422 | 0.024854951 | 0.033682579 | 1 |
| HTN | endometrium | *PAM16* | 9.732326025 | 0.007377308 | 0.011667112 | 1 |
| HTN | endometrium | *AC016773.1* | 6.388770892 | 2.80E-06 | 0.000412274 | 0.013605027 |
| HTN | endometrium | *RNASEK* | 6.612369373 | 0.006968796 | 0.01110795 | 1 |
| HTN | endometrium | *FAM228B* | 1.616181965 | 0.033305749 | 0.043972652 | 1 |
| HTN | endometrium | *RNU6ATAC35I* | 31.86311855 | 0.004663702 | 0.007971893 | 1 |
| HTN | endometrium | *FANCG* | -2.329340347 | 0.006565899 | 0.01055941 | 1 |
| HTN | endometrium | *ZNF880* | 1.864760558 | 0.012246143 | 0.018038338 | 1 |
| HTN | endometrium | *TIGD1* | 4.685824857 | 0.003544948 | 0.006384591 | 1 |
| HTN | endometrium | *FXYD7* | 3.859705309 | 0.006143336 | 0.009989097 | 1 |
| HTN | endometrium | *SLC12A8* | 3.814991237 | 1.45E-05 | 0.00115683 | 0.070566636 |
| HTN | endometrium | *APOL6* | 5.904199306 | 0.013471487 | 0.019646424 | 1 |
| HTN | endometrium | *OR2A1* | 5.32100238 | 0.002819153 | 0.005357826 | 1 |
| HTN | endometrium | *UBA52* | 5.963720307 | 0.000202567 | 0.003073153 | 0.982856825 |
| HTN | endometrium | *TIAF1* | 9.087128681 | 9.64E-07 | 0.00017993 | 0.004678171 |
| HTN | endometrium | *RAB6C* | 91.61751566 | 0.000766233 | 0.003073153 | 1 |
| HTN | endometrium | *AC062017.1* | 3.029809443 | 0.00121554 | 0.003095606 | 1 |
| HTN | endometrium | *EXOSC6* | -7.015424728 | 0.023797069 | 0.032381749 | 1 |
| HTN | endometrium | *LINC00571* | -2.339307537 | 0.005184642 | 0.008686423 | 1 |
| HTN | endometrium | *OSER1-AS1* | 7.060225409 | 6.59E-06 | 0.000662056 | 0.031987105 |
| HTN | endometrium | *C5orf66* | 2.612799993 | 0.001478002 | 0.003392273 | 1 |
| HTN | endometrium | *LINC01133* | 2.062548842 | 0.001670633 | 0.003654604 | 1 |
| HTN | endometrium | *C4B* | -7.590892376 | 0.0043844 | 0.007592115 | 1 |
| HTN | endometrium | *LINC01117* | 3.04274621 | 0.001123956 | 0.003095606 | 1 |
| HTN | endometrium | *LINC00184* | 76.99590268 | 0.001147257 | 0.003095606 | 1 |
| HTN | endometrium | *AL049840.1* | 2.293794316 | 0.006113636 | 0.009950808 | 1 |
| HTN | endometrium | *PLEKHM1* | 3.874894768 | 0.01353932 | 0.019739417 | 1 |
| HTN | endometrium | *AP000350.6* | 3.147131524 | 0.002032572 | 0.00418415 | 1 |
| HTN | endometrium | *CYP4A22-AS1* | 3.614751058 | 0.001189843 | 0.003095606 | 1 |
| HTN | endometrium | *FAM225B* | -6.865966253 | 0.001429416 | 0.003329773 | 1 |
| HTN | endometrium | *MIAT* | 3.72658853 | 0.002737975 | 0.005238428 | 1 |
| HTN | endometrium | *ERCC6* | 3.970283476 | 0.000587709 | 0.003073153 | 1 |
| HTN | endometrium | *LINC00694* | 10.65855828 | 0.000782315 | 0.003073153 | 1 |
| HTN | endometrium | *LINC00115* | -1.196514081 | 0.004594421 | 0.007879862 | 1 |
| HTN | endometrium | *HAR1A* | -1.382306408 | 0.003438416 | 0.006236709 | 1 |

| | | | | | | |
|---|---|---|---|---|---|---|
| HTN | endometrium | *LINC00937* | -3.405446114 | 0.021424302 | 0.029447794 | 1 |
| HTN | endometrium | *FTCDNL1* | 2.196636803 | 0.011784367 | 0.017459424 | 1 |
| HTN | endometrium | *FAM138B* | 44.15316786 | 0.001147224 | 0.003095606 | 1 |
| HTN | endometrium | *ZNF717* | 3.455950946 | 0.000106996 | 0.002958035 | 0.519144649 |
| HTN | endometrium | *AC009299.3* | 9.851043702 | 0.000526118 | 0.003073153 | 1 |
| HTN | endometrium | *SCAMP4* | 4.810427317 | 0.000613267 | 0.003073153 | 1 |
| HTN | endometrium | *SLC2A1-AS1* | -2.998531229 | 0.008098718 | 0.012613985 | 1 |
| HTN | endometrium | *LINC01068* | 5.59117571 | 0.028940503 | 0.038725681 | 1 |
| HTN | endometrium | *AC005884.1* | 6.399010622 | 0.019152458 | 0.026688032 | 1 |
| HTN | endometrium | *HCG20* | 6.169029596 | 0.015005125 | 0.021565422 | 1 |
| HTN | endometrium | *C14orf144* | -2.9457214 | 0.011355028 | 0.016910557 | 1 |
| HTN | endometrium | *LINC00578* | 3.727844794 | 0.004537238 | 0.007787293 | 1 |
| HTN | endometrium | *C19orf24* | -1.216565944 | 0.007247551 | 0.011503145 | 1 |
| HTN | endometrium | *ZNF883* | 3.448253382 | 0.000487253 | 0.003073153 | 1 |
| HTN | endometrium | *PATL2* | 2.849998615 | 0.000695945 | 0.003073153 | 1 |
| HTN | endometrium | *CASC20* | 4.641053681 | 0.001291938 | 0.003159517 | 1 |
| HTN | endometrium | *PSMG3-AS1* | 6.28591081 | 0.003533706 | 0.006369072 | 1 |
| HTN | endometrium | *AC112229.4* | 4.940061266 | 0.001994791 | 0.004130219 | 1 |
| HTN | endometrium | *KLHL7-AS1* | 6.695760586 | 0.000246472 | 0.003073153 | 1 |
| HTN | endometrium | *AC092171.4* | 3.583672687 | 0.001174312 | 0.003095606 | 1 |
| HTN | endometrium | *RPS18* | 4.791992389 | 0.000776575 | 0.003073153 | 1 |
| HTN | endometrium | *DIRC3* | 1.6837352 | 0.001116657 | 0.003095606 | 1 |
| HTN | endometrium | *TAPBP* | 4.046446335 | 0.000195323 | 0.003073153 | 0.94770665 |
| HTN | endometrium | *AC107021.1* | 5.206870171 | 0.000603006 | 0.003073153 | 1 |
| HTN | endometrium | *TMA7* | 4.654007441 | 5.76E-05 | 0.002410623 | 0.279531675 |
| HTN | endometrium | *LINC00865* | -0.872098844 | 0.006554212 | 0.010547607 | 1 |
| HTN | endometrium | *AC009495.3* | 21.83798011 | 0.000485116 | 0.003073153 | 1 |
| HTN | endometrium | *HLA-DQB2* | 1.133862248 | 0.017565003 | 0.024761449 | 1 |
| HTN | endometrium | *LYRM9* | -2.252912199 | 0.019024017 | 0.026547229 | 1 |
| HTN | endometrium | *LINC01166* | 4.815731033 | 0.028384089 | 0.038065119 | 1 |
| HTN | endometrium | *LINC00702* | 5.923736552 | 0.001042031 | 0.003095606 | 1 |
| HTN | endometrium | *GPX1* | 3.978620156 | 0.000128572 | 0.003073153 | 0.623830141 |
| HTN | endometrium | *RPS28* | 8.784105495 | 0.016061101 | 0.022940377 | 1 |
| HTN | endometrium | *TRIM26* | 5.802405423 | 0.019873637 | 0.027558412 | 1 |
| HTN | endometrium | *PINLYP* | 3.26379039 | 0.001934881 | 0.004050808 | 1 |
| HTN | endometrium | *ANKRD65* | 12.31835567 | 0.00067054 | 0.003073153 | 1 |
| HTN | endometrium | *AC009495.2* | 59.29021492 | 0.028933912 | 0.038725681 | 1 |
| HTN | endometrium | *LINC00957* | 8.656282478 | 0.001135735 | 0.003095606 | 1 |
| HTN | endometrium | *KIAA0040* | 2.384583766 | 0.001515952 | 0.003433662 | 1 |
| HTN | endometrium | *LINC00607* | 6.503536632 | 0.010516819 | 0.015842162 | 1 |
| HTN | endometrium | *AC019117.2* | 17.67137459 | 0.019715794 | 0.027378659 | 1 |
| HTN | endometrium | *ZBTB22* | 5.991115022 | 0.000103987 | 0.002934745 | 0.504543038 |
| HTN | endometrium | *ZBED5* | 5.700452755 | 0.000273262 | 0.003073153 | 1 |
| HTN | endometrium | *LINC00987* | 4.645078667 | 0.001209156 | 0.003095606 | 1 |
| HTN | endometrium | *LINC00959* | -1.800661827 | 0.010281214 | 0.015564571 | 1 |
| HTN | endometrium | *HLA-DQA2* | 2.790440637 | 1.27E-07 | 3.84E-05 | 0.000614149 |
| HTN | endometrium | *IRGM* | 2.76711628 | 0.012039719 | 0.017750445 | 1 |
| HTN | endometrium | *LINC01143* | -14.23844349 | 0.018415251 | 0.02582393 | 1 |
| HTN | endometrium | *LINC00211* | -1.326962096 | 0.002919232 | 0.005499273 | 1 |
| HTN | endometrium | *LRRC37A2* | 4.633289324 | 0.002518813 | 0.00491802 | 1 |

| | | | | | | |
|---|---|---|---|---|---|---|
| HTN | endometrium | *TXNDC5* | 15.56792795 | 0.001202596 | 0.003095606 | 1 |
| HTN | endometrium | *LINC01119* | -3.707064626 | 0.015071618 | 0.021654572 | 1 |
| HTN | endometrium | *NME1* | 3.525500525 | 0.029097157 | 0.038903116 | 1 |
| HTN | endometrium | *TNFSF12* | 9.046193381 | 0.000514408 | 0.003073153 | 1 |
| HTN | endometrium | *CDRT4* | 10.57330358 | 0.000213729 | 0.003073153 | 1 |
| HTN | endometrium | *C1orf226* | 10.8237076 | 0.005663173 | 0.009329204 | 1 |
| HTN | endometrium | *LY6G5B* | 2.293769774 | 0.030970818 | 0.041181257 | 1 |
| HTN | endometrium | *PSMB9* | 2.441347992 | 0.013886279 | 0.020142369 | 1 |
| HTN | endometrium | *PPIL3* | 4.460753846 | 0.000419879 | 0.003073153 | 1 |
| HTN | endometrium | *C1QTNF9* | 11.61832725 | 0.002185572 | 0.004425874 | 1 |
| HTN | endometrium | *ISY1* | 3.088334707 | 0.007466605 | 0.011777623 | 1 |
| HTN | endometrium | *PNMA2* | 3.416409749 | 0.025438911 | 0.034400668 | 1 |
| HTN | endometrium | *KRBOX1* | 48.52040813 | 0.001264705 | 0.003127478 | 1 |
| HTN | endometrium | *ARHGEF25* | 3.389298654 | 0.002278701 | 0.004568702 | 1 |
| HTN | endometrium | *LINC00886* | 1.901238066 | 0.00222333 | 0.004491089 | 1 |
| HTN | endometrium | *NSUN6* | 6.771220478 | 6.97E-05 | 0.0024668 | 0.337951603 |
| HTN | endometrium | *HLA-DOB* | -1.76845208 | 0.009692631 | 0.014761032 | 1 |
| HTN | endometrium | *LINC00877* | 3.628834606 | 0.00763072 | 0.012005271 | 1 |
| HTN | endometrium | *CD302* | 3.162832484 | 0.000826205 | 0.0030808 | 1 |
| HTN | endometrium | *ATP5O* | -3.296016547 | 0.014196695 | 0.020520568 | 1 |
| HTN | endometrium | *PLEKHO2* | 2.429386034 | 0.013681101 | 0.019894704 | 1 |
| HTN | endometrium | *C8orf58* | 5.79487559 | 0.001167115 | 0.003095606 | 1 |
| HTN | endometrium | *HOGA1* | -1.859006647 | 0.005390375 | 0.008972247 | 1 |
| HTN | endometrium | *PI4KA* | 3.21914634 | 0.000793602 | 0.003073153 | 1 |
| HTN | endometrium | *LINC00969* | 2.833487499 | 0.006484252 | 0.010452355 | 1 |
| HTN | endometrium | *BGLAP* | -5.195848755 | 0.0020052 | 0.004148925 | 1 |
| HTN | endometrium | *MRPL20* | 6.765934661 | 0.000147821 | 0.003073153 | 0.717226003 |
| HTN | endometrium | *ZNF709* | -6.469662272 | 0.002639091 | 0.005105475 | 1 |
| HTN | endometrium | *ERVW-1* | 9.596111779 | 0.002750849 | 0.005258913 | 1 |
| HTN | endometrium | *MRPL33* | 3.474978003 | 0.000548345 | 0.003073153 | 1 |
| HTN | endometrium | *STON1* | 5.919987444 | 0.002210715 | 0.004469328 | 1 |
| HTN | endometrium | *VSIG8* | 4.48164429 | 0.012385726 | 0.018219842 | 1 |
| HTN | endometrium | *C7orf73* | 3.873337046 | 0.000471886 | 0.003073153 | 1 |
| HTN | endometrium | *FGF14-IT1* | 9.396015924 | 0.001194997 | 0.003095606 | 1 |
| HTN | endometrium | *PALM2* | 2.679825894 | 0.00144155 | 0.003340211 | 1 |
| HTN | endometrium | *NBPF15* | 7.989116312 | 0.000852721 | 0.0030808 | 1 |
| HTN | endometrium | *WFDC6* | -19.49473326 | 0.001811074 | 0.00385917 | 1 |
| HTN | endometrium | *UPK3B* | 2.113304985 | 0.001458648 | 0.003363763 | 1 |
| HTN | endometrium | *IL10RB* | 2.212023821 | 0.001662756 | 0.003642298 | 1 |
| HTN | endometrium | *PLA2G4B* | 23.66760219 | 3.21E-06 | 0.00043202 | 0.015552726 |
| HTN | endometrium | *LEFTY1* | -44.69478608 | 0.000659205 | 0.003073153 | 1 |
| HTN | endometrium | *TTC4* | 3.526875364 | 0.002681451 | 0.005164908 | 1 |
| HTN | endometrium | *JMJD7* | 3.262673437 | 0.037369334 | 0.048857941 | 1 |
| HTN | endometrium | *APOBEC3D* | -13.62033935 | 0.017108533 | 0.024222528 | 1 |
| HTN | endometrium | *P2RY11* | 2.654219071 | 0.010866085 | 0.016275533 | 1 |
| HTN | endometrium | *LILRA6* | 3.175699587 | 0.002328338 | 0.004636752 | 1 |
| HTN | endometrium | *SCARF2* | -0.999482635 | 0.006201286 | 0.010059726 | 1 |
| HTN | endometrium | *APOBEC3C* | 4.245229343 | 0.000929023 | 0.003093357 | 1 |
| HTN | endometrium | *CCDC13* | -1.885150252 | 0.007452429 | 0.011759084 | 1 |
| HTN | endometrium | *NEAT1* | 2.855342628 | 0.000278473 | 0.003073153 | 1 |

| | | | | | | |
|---|---|---|---|---|---|---|
| HTN | endometrium | *RAB30-AS1* | 3.790896441 | 0.00614148 | 0.009989097 | 1 |
| HTN | endometrium | *UBAP1L* | 3.519974281 | 0.000890339 | 0.003084413 | 1 |
| HTN | endometrium | *LINC00926* | 6.858258956 | 0.001158992 | 0.003095606 | 1 |
| HTN | endometrium | *DPH6-AS1* | 4.582318046 | 0.001206024 | 0.003095606 | 1 |
| HTN | endometrium | *BCKDHA* | 7.078307115 | 0.00063836 | 0.003073153 | 1 |
| HTN | endometrium | *ABHD14A* | 3.56539337 | 0.000977547 | 0.003095606 | 1 |
| HTN | endometrium | *SRP14-AS1* | -1.932148725 | 0.017351947 | 0.02452422 | 1 |
| HTN | endometrium | *GDNF-AS1* | -0.638596131 | 0.004000974 | 0.007049989 | 1 |
| HTN | endometrium | *CCDC153* | -2.268096315 | 0.005819807 | 0.009542989 | 1 |
| HTN | endometrium | *AL357673.1* | -2.450177545 | 0.025788054 | 0.034839305 | 1 |
| HTN | endometrium | *TMEM150C* | 2.813559166 | 0.001234318 | 0.003103164 | 1 |
| HTN | endometrium | *LINC00939* | 7.474794554 | 0.00484019 | 0.008206257 | 1 |
| HTN | endometrium | *LINC01060* | 7.150486634 | 0.001053613 | 0.003095606 | 1 |
| HTN | endometrium | *LINC00942* | -2.73336126 | 0.003574933 | 0.006419531 | 1 |
| HTN | endometrium | *PDCD6* | 7.749940173 | 0.005860299 | 0.009596413 | 1 |
| HTN | endometrium | *TMEM158* | 12.41153969 | 0.002203694 | 0.004456993 | 1 |
| HTN | endometrium | *DNAH10OS* | 20.97070227 | 0.003442388 | 0.006241579 | 1 |
| HTN | endometrium | *PCDHA10* | 5.33592584 | 6.42E-05 | 0.002410623 | 0.311332077 |
| HTN | endometrium | *ZNF718* | 2.441859553 | 0.000745483 | 0.003073153 | 1 |
| HTN | endometrium | *LINC00989* | 3.616798727 | 0.001349206 | 0.00323814 | 1 |
| HTN | endometrium | *GLI4* | -2.37419846 | 0.016116084 | 0.023012136 | 1 |
| HTN | endometrium | *SNHG18* | 2.85895317 | 0.002395488 | 0.004734383 | 1 |
| HTN | endometrium | *ALG1L2* | 3.394971218 | 0.001277513 | 0.003143251 | 1 |
| HTN | endometrium | *ZNF550* | 2.886622151 | 0.001392071 | 0.003285301 | 1 |
| HTN | endometrium | *LINC01094* | 3.546256198 | 0.001155317 | 0.003095606 | 1 |
| HTN | endometrium | *PCDHGB6* | 4.166668864 | 0.034208685 | 0.045078907 | 1 |
| HTN | endometrium | *PCDHGA6* | -1.477269894 | 0.036838717 | 0.048243513 | 1 |
| HTN | endometrium | *UTP14C* | 3.379592152 | 0.000788049 | 0.003073153 | 1 |
| HTN | endometrium | *PCDHGB2* | 14.31772376 | 0.003748775 | 0.00668224 | 1 |
| HTN | endometrium | *LYN* | 1.765846595 | 0.000792301 | 0.003073153 | 1 |
| HTN | endometrium | *FPGT* | 3.167281121 | 0.000265997 | 0.003073153 | 1 |
| HTN | endometrium | *MEX3A* | -1.690294022 | 0.007886767 | 0.012336104 | 1 |
| HTN | endometrium | *SNHG9* | -1.860259531 | 0.022470666 | 0.030746664 | 1 |
| HTN | endometrium | *TAS2R20* | 6.734145793 | 0.012022835 | 0.017736332 | 1 |
| HTN | endometrium | *ZNF432* | 4.179596193 | 0.003945095 | 0.006960582 | 1 |
| HTN | endometrium | *HMBS* | -1.118330231 | 0.015674522 | 0.022441069 | 1 |
| HTN | endometrium | *KIAA1147* | 4.092194796 | 0.000453885 | 0.003073153 | 1 |
| HTN | endometrium | *ZBED6* | 6.269131386 | 0.00198963 | 0.004123411 | 1 |
| HTN | endometrium | *CNPY2* | 2.472757607 | 0.001041003 | 0.003095606 | 1 |
| HTN | endometrium | *PDF* | -5.502626122 | 0.004337754 | 0.00752477 | 1 |
| HTN | endometrium | *LINC00640* | 3.270459233 | 0.01284862 | 0.018828602 | 1 |
| HTN | endometrium | *HIF1A-AS2* | 4.482936746 | 0.000682156 | 0.003073153 | 1 |
| HTN | endometrium | *LINC00638* | 12.48436666 | 3.72E-06 | 0.000475096 | 0.018053655 |
| HTN | endometrium | *SLC25A21-AS1* | 3.427831511 | 0.001183681 | 0.003095606 | 1 |
| HTN | endometrium | *RNASE4* | 12.19465251 | 0.003622801 | 0.006493473 | 1 |
| HTN | endometrium | *MC1R* | 9.180208919 | 0.000423786 | 0.003073153 | 1 |
| HTN | endometrium | *LINC00639* | 2.089255379 | 0.002939619 | 0.00551975 | 1 |
| HTN | endometrium | *LINC00924* | 1.650299752 | 0.035855905 | 0.047070577 | 1 |
| HTN | endometrium | *MRPL46* | -2.649501896 | 0.01731764 | 0.024482864 | 1 |
| HTN | endometrium | *IQCH-AS1* | 2.946822107 | 0.002737232 | 0.005238428 | 1 |

| | | | | | | |
|---|---|---|---|---|---|---|
| HTN | endometrium | *PWRN1* | 2.935976329 | 0.01344189 | 0.019609155 | 1 |
| HTN | endometrium | *LINC01003* | 3.290875052 | 0.002327242 | 0.004636752 | 1 |
| HTN | endometrium | *C15orf65* | 8.753943202 | 0.011095825 | 0.016555025 | 1 |
| HTN | endometrium | *LINC00662* | 4.680446302 | 0.000511894 | 0.003073153 | 1 |
| HTN | endometrium | *C19orf84* | 44.86166742 | 0.000336052 | 0.003073153 | 1 |
| HTN | endometrium | *LINC00667* | 5.727409232 | 0.003792628 | 0.006743399 | 1 |
| HTN | endometrium | *LINC00672* | 14.68042527 | 0.000500401 | 0.003073153 | 1 |
| HTN | endometrium | *LINC00909* | 5.976241793 | 0.003628691 | 0.006501627 | 1 |
| HTN | endometrium | *DYNLL2* | 3.585844897 | 0.00122273 | 0.003095606 | 1 |
| HTN | endometrium | *STRADA* | 4.882336262 | 0.000495573 | 0.003073153 | 1 |
| HTN | endometrium | *MYO15B* | 6.128534758 | 0.000266145 | 0.003073153 | 1 |
| HTN | endometrium | *LINC00663* | 1.953717143 | 0.020524267 | 0.028347208 | 1 |
| HTN | endometrium | *ILF3-AS1* | 7.23131082 | 0.000112742 | 0.003005626 | 0.547023941 |
| HTN | endometrium | *ZNF224* | 7.694263437 | 0.001193729 | 0.003095606 | 1 |
| HTN | endometrium | *LIN37* | -1.415404307 | 0.005519402 | 0.009149347 | 1 |
| HTN | endometrium | *NDUFA7* | 19.29718868 | 0.001142057 | 0.003095606 | 1 |
| HTN | endometrium | *SCGB1B2P* | 10.86122134 | 0.001291766 | 0.003159517 | 1 |
| HTN | endometrium | *AC026740.1* | 22.77774414 | 0.001117762 | 0.003095606 | 1 |
| HTN | endometrium | *ZNF728* | 10.23796601 | 0.001702253 | 0.003705679 | 1 |
| HTN | endometrium | *TRABD2B* | 2.384486481 | 0.001201575 | 0.003095606 | 1 |
| HTN | endometrium | *AC093157.1* | 2.335570723 | 0.001091182 | 0.003095606 | 1 |
| HTN | endometrium | *FBXO17* | 2.846300044 | 0.000572629 | 0.003073153 | 1 |
| HTN | endometrium | *ZNF587B* | 3.755715238 | 0.001175512 | 0.003095606 | 1 |
| HTN | endometrium | *SNHG8* | 2.272224033 | 0.001214317 | 0.003095606 | 1 |
| HTN | endometrium | *HCG17* | 3.045162124 | 0.001203801 | 0.003095606 | 1 |
| HTN | endometrium | *YTHDF3-AS1* | 5.724589774 | 0.022382544 | 0.030634725 | 1 |
| HTN | endometrium | *MILR1* | 2.375506315 | 0.002418932 | 0.004769338 | 1 |
| HTN | endometrium | *GTF2H5* | 2.814489684 | 0.00499665 | 0.008412126 | 1 |
| HTN | endometrium | *FGF14-AS2* | 5.082561558 | 0.001059624 | 0.003095606 | 1 |
| HTN | endometrium | *CASC15* | 1.965389606 | 0.001074153 | 0.003095606 | 1 |
| HTN | endometrium | *GRIN2B* | 5.484738511 | 0.000193069 | 0.003073153 | 0.936771863 |
| HTN | endometrium | *AP000350.5* | 2.131794522 | 0.038156952 | 0.049848554 | 1 |
| HTN | endometrium | *PAXIP1-AS1* | 4.3889685 | 0.000660702 | 0.003073153 | 1 |
| HTN | endothelial | *ITIH4* | 0.419100773 | 0.000594726 | 0.044394041 | 1 |
| HTN | endothelial | *PIGV* | -0.898793567 | 7.86E-05 | 0.008474028 | 0.381331243 |
| HTN | endothelial | *SNX10* | -0.721339921 | 0.000243871 | 0.021513885 | 1 |
| HTN | endothelial | *MARCH2* | 1.266660514 | 6.94E-06 | 0.001402179 | 0.033652301 |
| HTN | endothelial | *POLI* | -0.450407471 | 0.00046604 | 0.039670649 | 1 |
| HTN | endothelial | *MTHFSD* | 0.684676565 | 0.000513107 | 0.041700827 | 1 |
| HTN | endothelial | *PCCB* | 0.971053616 | 0.000495656 | 0.041464211 | 1 |
| HTN | endothelial | *TBX2* | -1.232902789 | 1.53E-07 | 6.73E-05 | 0.000740538 |
| HTN | endothelial | *TEX14* | 1.502590382 | 3.34E-05 | 0.004503468 | 0.162124848 |
| HTN | endothelial | *WFDC3* | -0.362491877 | 0.000272907 | 0.023645431 | 1 |
| HTN | endothelial | *BTN2A2* | 0.807719434 | 1.79E-06 | 0.000510541 | 0.008679195 |
| HTN | endothelial | *ZNF391* | -0.480034149 | 0.000148873 | 0.014741441 | 0.7223306 |
| HTN | endothelial | *TRAP1* | 0.748990046 | 2.90E-05 | 0.004038708 | 0.140870012 |
| HTN | endothelial | *ATP8B3* | -0.660573283 | 0.000549952 | 0.042355018 | 1 |
| HTN | endothelial | *HAUS8* | -1.602072134 | 2.58E-05 | 0.003791757 | 0.125127985 |
| HTN | endothelial | *AGT* | 0.867049558 | 0.000211462 | 0.019706819 | 1 |
| HTN | endothelial | *NMT1* | 0.841582552 | 0.000121573 | 0.012550516 | 0.589874253 |

| | | | | | | |
|---|---|---|---|---|---|---|
| HTN | endothelial | *SLC37A4* | 1.551883688 | 2.36E-05 | 0.003577294 | 0.114473417 |
| HTN | endothelial | *FBN2* | 0.472481703 | 5.91E-05 | 0.006995222 | 0.286804099 |
| HTN | endothelial | *HSD17B12* | -0.67187522 | 3.62E-06 | 0.00079745 | 0.017543908 |
| HTN | endothelial | *TNKS1BP1* | 1.192717022 | 5.22E-07 | 0.000194681 | 0.002530854 |
| HTN | endothelial | *NEIL2* | 1.720397322 | 5.96E-08 | 3.61E-05 | 0.000289114 |
| HTN | endothelial | *SFXN2* | 0.841106116 | 2.58E-06 | 0.00062561 | 0.012512209 |
| HTN | endothelial | *CLEC18A* | -1.341676725 | 1.30E-05 | 0.002170332 | 0.062939619 |
| HTN | endothelial | *SLC5A11* | -0.715192268 | 1.71E-06 | 0.000510541 | 0.008314893 |
| HTN | endothelial | *S100B* | -0.365376481 | 0.000215264 | 0.019706819 | 1 |
| HTN | endothelial | *ABCF3* | 1.400775924 | 6.32E-06 | 0.001332852 | 0.030655585 |
| HTN | endothelial | *MRPL55* | 1.494583589 | 3.54E-06 | 0.00079745 | 0.017196533 |
| HTN | endothelial | *LYSMD1* | -0.81473663 | 0.000568837 | 0.043124943 | 1 |
| HTN | endothelial | *ZNF589* | -1.159038415 | 4.32E-05 | 0.005237556 | 0.209502226 |
| HTN | endothelial | *RNF123* | 1.59595151 | 3.41E-08 | 2.37E-05 | 0.000165555 |
| HTN | endothelial | *MST1R* | 1.135183121 | 2.91E-05 | 0.004038708 | 0.141354793 |
| HTN | endothelial | *ERAP2* | 0.520452647 | 0.000532863 | 0.041700827 | 1 |
| HTN | endothelial | *TRIM66* | -0.46887534 | 0.0001924 | 0.018304427 | 0.933525793 |
| HTN | endothelial | *SRR* | 0.675302339 | 7.04E-05 | 0.00813547 | 0.341689741 |
| HTN | endothelial | *CPT1C* | 1.665739712 | 4.64E-07 | 0.000187472 | 0.002249668 |
| HTN | endothelial | *INO80E* | -1.219802923 | 2.10E-06 | 0.000549305 | 0.010170143 |
| HTN | endothelial | *TTLL6* | -4.404823509 | 7.82E-09 | 9.49E-06 | 3.80E-05 |
| HTN | endothelial | *RGS19* | -0.893730297 | 1.35E-07 | 6.56E-05 | 0.000655935 |
| HTN | endothelial | *NME6* | 2.347731208 | 1.38E-08 | 1.34E-05 | 6.70E-05 |
| HTN | endothelial | *CTSW* | -0.242678831 | 1.75E-05 | 0.002731491 | 0.084676212 |
| HTN | endothelial | *MST1* | -0.627757071 | 8.67E-06 | 0.001558636 | 0.042083185 |
| HTN | endothelial | *SNX31* | -0.757328264 | 8.33E-06 | 0.001554028 | 0.040404739 |
| HTN | endothelial | *CCNE2* | -1.283845722 | 8.30E-06 | 0.001554028 | 0.040260806 |
| HTN | endothelial | *CTDNEP1* | 1.453239181 | 0.000530877 | 0.041700827 | 1 |
| HTN | endothelial | *RPP25* | 1.147876845 | 1.31E-06 | 0.000423188 | 0.006347827 |
| HTN | endothelial | *MPI* | -2.136833492 | 2.39E-09 | 3.87E-06 | 1.16E-05 |
| HTN | endothelial | *HYI* | -0.981908195 | 1.11E-05 | 0.001929032 | 0.054012906 |
| HTN | endothelial | *ADO* | 2.047010038 | 8.86E-10 | 2.15E-06 | 4.30E-06 |
| HTN | endothelial | *CLN8* | 1.035527931 | 2.15E-06 | 0.000549305 | 0.010436795 |
| HTN | endothelial | *FES* | -1.034693151 | 0.000528115 | 0.041700827 | 1 |
| HTN | endothelial | *ZNF320* | 0.502851377 | 0.000230297 | 0.020692581 | 1 |
| HTN | endothelial | *NUTM2A* | -1.244808429 | 7.85E-05 | 0.008474028 | 0.381040436 |
| HTN | endothelial | *HIST1H3E* | 1.156184509 | 3.35E-08 | 2.37E-05 | 0.000162442 |
| HTN | endothelial | *RPS26* | -0.817664699 | 3.75E-05 | 0.004686318 | 0.181916867 |
| HTN | endothelial | *TMEM116* | -0.968018371 | 1.79E-10 | 8.68E-07 | 8.68E-07 |
| HTN | endothelial | *HLA-DRB5* | -0.238441731 | 0.000166718 | 0.016178283 | 0.808914169 |
| HTN | endothelial | *PRRT1* | 1.068123873 | 1.34E-07 | 6.56E-05 | 0.000648537 |
| HTN | endothelial | *TMEM110* | 1.83030883 | 0.000143017 | 0.01445667 | 0.693920164 |
| HTN | endothelial | *ATF6B* | 1.343156045 | 3.77E-05 | 0.004686318 | 0.182766406 |
| HTN | endothelial | *AC016773.1* | 1.218159915 | 7.58E-05 | 0.008474028 | 0.3676427 |
| HTN | endothelial | *TIAF1* | 1.744053667 | 1.61E-05 | 0.002597331 | 0.077919933 |
| HTN | endothelial | *HLA-DQA2* | 0.368723048 | 3.61E-05 | 0.004686318 | 0.17510842 |
| HTN | endothelial | *TNFSF12* | 1.641854386 | 9.96E-07 | 0.00034535 | 0.004834901 |
| HTN | endothelial | *PLA2G4B* | 2.778323634 | 9.48E-05 | 0.010001577 | 0.460072562 |
| HTN | LEC | *SEMA3F* | -3.214500942 | 0.000534917 | 0.014105517 | 1 |
| HTN | LEC | *RAD52* | -2.529899451 | 0.005343839 | 0.039635608 | 1 |

| | | | | | | |
|---|---|---|---|---|---|---|
| HTN | LEC | *HS3ST1* | 0.351432153 | 0.004388322 | 0.037953901 | 1 |
| HTN | LEC | *M6PR* | -2.419206193 | 0.002615522 | 0.034391635 | 1 |
| HTN | LEC | *RBM6* | -1.644804345 | 8.28E-07 | 0.000121773 | 0.004018508 |
| HTN | LEC | *ARHGAP33* | -1.805284016 | 0.005018697 | 0.038529615 | 1 |
| HTN | LEC | *PON1* | -3.009964213 | 0.006457393 | 0.042255533 | 1 |
| HTN | LEC | *RHBDD2* | 1.886132531 | 0.001008304 | 0.020877179 | 1 |
| HTN | LEC | *SCIN* | -1.922301971 | 0.001021008 | 0.020986119 | 1 |
| HTN | LEC | *CD79B* | -3.440150247 | 0.000118576 | 0.005218385 | 0.575330871 |
| HTN | LEC | *ST7L* | -1.658315183 | 0.001338106 | 0.024600852 | 1 |
| HTN | LEC | *TSR3* | 3.788262056 | 0.003977057 | 0.03770947 | 1 |
| HTN | LEC | *NADK* | -3.625605557 | 0.003302991 | 0.037708496 | 1 |
| HTN | LEC | *DLEC1* | -1.463197683 | 0.001976297 | 0.030076364 | 1 |
| HTN | LEC | *SS18L2* | 1.234490793 | 0.005074659 | 0.038653449 | 1 |
| HTN | LEC | *MAPK8IP2* | -3.851252237 | 0.005726102 | 0.041022318 | 1 |
| HTN | LEC | *MLXIPL* | -0.795695323 | 0.00099704 | 0.020762403 | 1 |
| HTN | LEC | *FUZ* | -1.808322493 | 0.009257284 | 0.048822112 | 1 |
| HTN | LEC | *CLCN6* | -2.962460734 | 0.001336257 | 0.024600852 | 1 |
| HTN | LEC | *CEP68* | 3.506047645 | 2.92E-07 | 4.89E-05 | 0.001418057 |
| HTN | LEC | *EHD3* | 1.100852585 | 0.004710785 | 0.038101652 | 1 |
| HTN | LEC | *SLC11A1* | -0.639445388 | 0.004348651 | 0.037880891 | 1 |
| HTN | LEC | *WWTR1* | 1.159374149 | 0.006555865 | 0.042352283 | 1 |
| HTN | LEC | *OSBPL5* | 1.824550316 | 0.005313425 | 0.039535978 | 1 |
| HTN | LEC | *RNASET2* | -1.880330592 | 6.31E-05 | 0.003645071 | 0.306185976 |
| HTN | LEC | *INSRR* | -2.429196181 | 0.000965241 | 0.020451311 | 1 |
| HTN | LEC | *ANK1* | -2.092571634 | 0.005994387 | 0.041329827 | 1 |
| HTN | LEC | *CENPQ* | -0.81700248 | 0.001467769 | 0.025617315 | 1 |
| HTN | LEC | *SH3YL1* | -2.604606454 | 0.001611635 | 0.027151576 | 1 |
| HTN | LEC | *FLT4* | 0.208151331 | 0.001714437 | 0.02782089 | 1 |
| HTN | LEC | *DNAH5* | -1.420909371 | 0.006133915 | 0.041637045 | 1 |
| HTN | LEC | *SPATA7* | -2.09707233 | 0.005158706 | 0.039042822 | 1 |
| HTN | LEC | *ANO2* | 1.677373159 | 0.008468343 | 0.047375361 | 1 |
| HTN | LEC | *MRPS10* | -0.826911008 | 0.008217506 | 0.046470093 | 1 |
| HTN | LEC | *ANAPC4* | 2.880338254 | 0.007368607 | 0.043868076 | 1 |
| HTN | LEC | *LY75* | -2.751795519 | 0.005900819 | 0.041317869 | 1 |
| HTN | LEC | *SDCCAG8* | 2.02208524 | 0.000578636 | 0.014933739 | 1 |
| HTN | LEC | *ITIH4* | 0.617577752 | 0.003413858 | 0.03770947 | 1 |
| HTN | LEC | *NPFFR2* | -1.873350924 | 0.003515876 | 0.03770947 | 1 |
| HTN | LEC | *IL17RB* | -2.778476544 | 0.007069629 | 0.043229876 | 1 |
| HTN | LEC | *PIGV* | -2.16549512 | 2.25E-07 | 3.89E-05 | 0.001089745 |
| HTN | LEC | *RIMBP2* | -1.459719805 | 0.004658227 | 0.038101652 | 1 |
| HTN | LEC | *PRDM6* | -1.575281816 | 0.008129675 | 0.046277049 | 1 |
| HTN | LEC | *LTK* | -9.711926219 | 0.005059877 | 0.038653449 | 1 |
| HTN | LEC | *AHRR* | -1.87054593 | 0.003319846 | 0.03770947 | 1 |
| HTN | LEC | *ABCA7* | -1.647817274 | 0.000930203 | 0.019970552 | 1 |
| HTN | LEC | *FAR2* | 1.568696961 | 0.0069616 | 0.043126036 | 1 |
| HTN | LEC | *BCAS1* | -1.287665286 | 0.005527222 | 0.04048889 | 1 |
| HTN | LEC | *UHRF1BP1* | 1.942159608 | 0.000189923 | 0.007313544 | 0.921506545 |
| HTN | LEC | *SLC9A3* | -0.757529586 | 0.003958002 | 0.03770947 | 1 |
| HTN | LEC | *NGEF* | -4.63282238 | 0.002128978 | 0.031234415 | 1 |
| HTN | LEC | *FGFR2* | -0.986260177 | 0.000719919 | 0.017122786 | 1 |

| | | | | | | |
|---|---|---|---|---|---|---|
| HTN | LEC | *ROGDI* | 3.391455864 | 0.006535922 | 0.04230219 | 1 |
| HTN | LEC | *POLR1A* | -2.328338863 | 8.90E-06 | 0.000744373 | 0.04317361 |
| HTN | LEC | *IFT80* | -4.008788744 | 0.00022221 | 0.007962365 | 1 |
| HTN | LEC | *DRD4* | -2.52413454 | 0.00847523 | 0.047375361 | 1 |
| HTN | LEC | *PLA2G10* | -2.779886872 | 0.004195156 | 0.03770947 | 1 |
| HTN | LEC | *NEDD4* | 1.893396428 | 0.0027023 | 0.034856249 | 1 |
| HTN | LEC | *RPS6KA2* | 1.105229372 | 0.001984274 | 0.030086557 | 1 |
| HTN | LEC | *TRHDE* | -1.262525712 | 0.000656022 | 0.015995081 | 1 |
| HTN | LEC | *SCARB1* | -1.616188249 | 0.007396414 | 0.04397966 | 1 |
| HTN | LEC | *MOV10L1* | 0.548601964 | 0.00265999 | 0.034728081 | 1 |
| HTN | LEC | *ADAM11* | -0.915780982 | 0.006738517 | 0.042882211 | 1 |
| HTN | LEC | *PPP2R2C* | -1.714152809 | 0.004139929 | 0.03770947 | 1 |
| HTN | LEC | *TACR2* | 7.051964038 | 0.003948682 | 0.03770947 | 1 |
| HTN | LEC | *SPAG5* | -5.157921398 | 0.000991105 | 0.020727767 | 1 |
| HTN | LEC | *ANKRD13A* | 3.467469147 | 0.006936448 | 0.043126036 | 1 |
| HTN | LEC | *XAB2* | 3.227832562 | 0.004067318 | 0.03770947 | 1 |
| HTN | LEC | *SIRT6* | 2.730387811 | 0.006762924 | 0.042893733 | 1 |
| HTN | LEC | *SCGN* | 4.716733301 | 0.003020419 | 0.036053579 | 1 |
| HTN | LEC | *SCTR* | -0.991431757 | 0.004659987 | 0.038101652 | 1 |
| HTN | LEC | *OXCT1* | -2.262706608 | 0.006348734 | 0.041983118 | 1 |
| HTN | LEC | *NOA1* | -2.898294416 | 0.009047368 | 0.048505888 | 1 |
| HTN | LEC | *CD59* | 1.368180492 | 0.007905637 | 0.045448045 | 1 |
| HTN | LEC | *SNX10* | -2.384616771 | 8.99E-08 | 1.98E-05 | 0.000436343 |
| HTN | LEC | *ITPKC* | -1.953983024 | 0.002929677 | 0.035805527 | 1 |
| HTN | LEC | *ADAMTS2* | -1.668965868 | 0.008135681 | 0.046277049 | 1 |
| HTN | LEC | *AAMDC* | -2.461837081 | 4.67E-06 | 0.000487091 | 0.022682321 |
| HTN | LEC | *ASAP3* | -1.571698917 | 0.008830641 | 0.048396858 | 1 |
| HTN | LEC | *DOCK3* | -1.587989462 | 0.00895192 | 0.048458848 | 1 |
| HTN | LEC | *ARHGAP28* | -1.192818136 | 0.00865865 | 0.048033815 | 1 |
| HTN | LEC | *EBF4* | 2.092567861 | 0.004293327 | 0.037874946 | 1 |
| HTN | LEC | *MAVS* | -2.149123045 | 0.008255719 | 0.046543498 | 1 |
| HTN | LEC | *RPH3A* | 1.821489614 | 0.000849684 | 0.019175186 | 1 |
| HTN | LEC | *TBX5* | -4.7327036 | 0.000506832 | 0.013586467 | 1 |
| HTN | LEC | *ANKRD24* | -1.370468119 | 0.003819176 | 0.03770947 | 1 |
| HTN | LEC | *MUL1* | -1.94428736 | 8.16E-05 | 0.004397219 | 0.395749703 |
| HTN | LEC | *RAB11FIP3* | 1.537358396 | 0.004221345 | 0.03770947 | 1 |
| HTN | LEC | *PABPC4* | -3.869936587 | 7.27E-07 | 0.000117603 | 0.003528083 |
| HTN | LEC | *ABCC6* | -1.965302316 | 0.001277967 | 0.024235407 | 1 |
| HTN | LEC | *RAPGEF4* | 0.736882871 | 0.008318169 | 0.046821064 | 1 |
| HTN | LEC | *TF* | -2.123336738 | 0.007083213 | 0.043229876 | 1 |
| HTN | LEC | *RGS17* | 3.146253686 | 0.000119382 | 0.005218385 | 0.57924073 |
| HTN | LEC | *CRTAC1* | 0.310078637 | 0.004089895 | 0.03770947 | 1 |
| HTN | LEC | *ITPR3* | 1.932717351 | 0.002406313 | 0.033395663 | 1 |
| HTN | LEC | *DSP* | 0.454639151 | 0.003599866 | 0.03770947 | 1 |
| HTN | LEC | *SCD* | -2.944620047 | 2.22E-07 | 3.89E-05 | 0.001076158 |
| HTN | LEC | *MAST3* | -1.229891756 | 0.009207603 | 0.048822112 | 1 |
| HTN | LEC | *MARCH2* | 2.838343849 | 2.02E-06 | 0.000244795 | 0.009791811 |
| HTN | LEC | *MTAP* | -3.531478857 | 0.000415121 | 0.011918143 | 1 |
| HTN | LEC | *MADCAM1* | -2.239693749 | 0.002455339 | 0.033395663 | 1 |
| HTN | LEC | *CECR2* | -1.366382386 | 0.0040825 | 0.03770947 | 1 |

| | | | | | | |
|---|---|---|---|---|---|---|
| HTN | LEC | *CRYBB3* | 0.758290637 | 0.005013246 | 0.038529615 | 1 |
| HTN | LEC | *CYTH4* | -1.42889974 | 0.007768761 | 0.045059762 | 1 |
| HTN | LEC | *CARD10* | 0.36166719 | 0.004135427 | 0.03770947 | 1 |
| HTN | LEC | *LGALS2* | -1.403259074 | 0.002463873 | 0.033395663 | 1 |
| HTN | LEC | *SH3BP1* | -1.612130557 | 0.005834942 | 0.041169714 | 1 |
| HTN | LEC | *SLC5A4* | 0.22752443 | 0.004014273 | 0.03770947 | 1 |
| HTN | LEC | *THOC5* | -2.015157346 | 0.001370516 | 0.02473249 | 1 |
| HTN | LEC | *TTLL12* | -2.691728047 | 0.002538556 | 0.033816145 | 1 |
| HTN | LEC | *APOL4* | -1.302830418 | 0.007415653 | 0.043986247 | 1 |
| HTN | LEC | *ACO2* | -2.514663603 | 0.006978949 | 0.043126036 | 1 |
| HTN | LEC | *PYGL* | -0.748931986 | 0.002966096 | 0.035978744 | 1 |
| HTN | LEC | *EFS* | -2.304555892 | 0.0065143 | 0.04230219 | 1 |
| HTN | LEC | *NDRG3* | 2.572866227 | 0.002403861 | 0.033395663 | 1 |
| HTN | LEC | *DOK5* | -1.807404289 | 0.006427973 | 0.042146654 | 1 |
| HTN | LEC | *BIRC7* | -2.552579722 | 0.000603606 | 0.015096369 | 1 |
| HTN | LEC | *PTK6* | -4.662523714 | 0.008201297 | 0.046470093 | 1 |
| HTN | LEC | *CDC25B* | 1.105821411 | 0.007323867 | 0.043816771 | 1 |
| HTN | LEC | *TRIB3* | 3.682345328 | 0.005521041 | 0.04048889 | 1 |
| HTN | LEC | *ANGPT4* | -2.920730086 | 0.002029913 | 0.030587379 | 1 |
| HTN | LEC | *SIRPB1* | -2.155515273 | 0.006865478 | 0.043025132 | 1 |
| HTN | LEC | *CRNKL1* | -1.740776886 | 0.008179417 | 0.046470093 | 1 |
| HTN | LEC | *DNTTIP1* | -2.103593455 | 4.02E-05 | 0.002501052 | 0.195082052 |
| HTN | LEC | *POLI* | -1.017019546 | 1.15E-05 | 0.000914545 | 0.05578727 |
| HTN | LEC | *NALCN* | 0.196439081 | 0.00268368 | 0.034851979 | 1 |
| HTN | LEC | *DHODH* | 1.450025376 | 0.002152078 | 0.031357001 | 1 |
| HTN | LEC | *CCDC113* | -2.51879263 | 0.002219813 | 0.031775401 | 1 |
| HTN | LEC | *NME4* | -2.217515144 | 0.000954717 | 0.020317047 | 1 |
| HTN | LEC | *SLC7A5* | 0.368385475 | 0.008067175 | 0.046157941 | 1 |
| HTN | LEC | *NUBP1* | -1.85874647 | 0.002843655 | 0.035259216 | 1 |
| HTN | LEC | *TMC5* | -1.171969755 | 0.00379488 | 0.03770947 | 1 |
| HTN | LEC | *RPAP1* | -2.81865483 | 0.000899786 | 0.019754583 | 1 |
| HTN | LEC | *EYA1* | 0.222982106 | 0.002789492 | 0.035003553 | 1 |
| HTN | LEC | *NIPAL2* | 1.656874737 | 0.002939272 | 0.035822654 | 1 |
| HTN | LEC | *SNX16* | 3.397219154 | 2.67E-05 | 0.001797128 | 0.129393249 |
| HTN | LEC | *TSTA3* | 1.308155088 | 0.000500473 | 0.013519085 | 1 |
| HTN | LEC | *R3HCC1* | -1.512938647 | 0.009591254 | 0.04965591 | 1 |
| HTN | LEC | *MAN2B1* | -1.494132449 | 0.003097691 | 0.036398387 | 1 |
| HTN | LEC | *TULP2* | 1.705949827 | 0.002536175 | 0.033816145 | 1 |
| HTN | LEC | *LIN7B* | -3.963646346 | 6.62E-05 | 0.003779341 | 0.321243989 |
| HTN | LEC | *CD37* | -3.598354825 | 5.74E-06 | 0.000552257 | 0.027857918 |
| HTN | LEC | *AMH* | -9.26579205 | 0.004182332 | 0.03770947 | 1 |
| HTN | LEC | *FCER2* | -7.297001491 | 0.009661001 | 0.04965591 | 1 |
| HTN | LEC | *CLEC4M* | 1.216089645 | 0.003750762 | 0.03770947 | 1 |
| HTN | LEC | *NOVA2* | 0.480458466 | 0.007069661 | 0.043229876 | 1 |
| HTN | LEC | *RASAL3* | 1.675083488 | 0.002709958 | 0.034856249 | 1 |
| HTN | LEC | *CEACAM4* | 1.896593481 | 0.005689458 | 0.041016712 | 1 |
| HTN | LEC | *GRIN2D* | -2.243082154 | 0.002159065 | 0.031364627 | 1 |
| HTN | LEC | *TNPO2* | -2.836989179 | 0.004491003 | 0.038101652 | 1 |
| HTN | LEC | *GCDH* | -2.330561414 | 0.006890051 | 0.043025132 | 1 |
| HTN | LEC | *ETV2* | 4.480317233 | 0.007068058 | 0.043229876 | 1 |

| | | | | | | |
|---|---|---|---|---|---|---|
| HTN | LEC | *RASA4* | -5.776206705 | 0.003848189 | 0.03770947 | 1 |
| HTN | LEC | *ADAP1* | -3.780105791 | 0.00470745 | 0.038101652 | 1 |
| HTN | LEC | *MET* | -0.901359453 | 0.007063376 | 0.043229876 | 1 |
| HTN | LEC | *TAF6* | -1.878781269 | 0.000166284 | 0.006895827 | 0.80681174 |
| HTN | LEC | *TFR2* | -1.635181134 | 0.007442802 | 0.04399196 | 1 |
| HTN | LEC | *MEST* | -1.56073321 | 8.53E-06 | 0.000726275 | 0.041397649 |
| HTN | LEC | *ACTR3C* | 2.976448308 | 0.006050506 | 0.041463538 | 1 |
| HTN | LEC | *POU6F2* | -2.132257251 | 0.00067064 | 0.016269728 | 1 |
| HTN | LEC | *GCK* | 10.66634587 | 0.006195356 | 0.041807885 | 1 |
| HTN | LEC | *SPATA6L* | 0.797371358 | 0.003614791 | 0.03770947 | 1 |
| HTN | LEC | *SLC1A1* | 0.747615237 | 0.000395011 | 0.011545758 | 1 |
| HTN | LEC | *OGN* | -1.889614119 | 0.000222318 | 0.007962365 | 1 |
| HTN | LEC | *PLGRKT* | -1.831098261 | 0.00065102 | 0.015953274 | 1 |
| HTN | LEC | *DNMBP* | 1.255503611 | 0.005831151 | 0.041169714 | 1 |
| HTN | LEC | *LHPP* | -1.845278204 | 0.008129615 | 0.046277049 | 1 |
| HTN | LEC | *RPL28* | -2.065714412 | 0.004956937 | 0.038529615 | 1 |
| HTN | LEC | *RAD51C* | -2.228500038 | 0.000104219 | 0.004957552 | 0.505670332 |
| HTN | LEC | *PNPO* | -5.800398214 | 3.19E-05 | 0.002073289 | 0.154695908 |
| HTN | LEC | *TRIM16L* | -1.852692479 | 0.003835563 | 0.03770947 | 1 |
| HTN | LEC | *PEX12* | -2.136320429 | 0.001025085 | 0.020986119 | 1 |
| HTN | LEC | *EFNB3* | -1.407639573 | 0.003276326 | 0.037580928 | 1 |
| HTN | LEC | *FAM20A* | -1.364609575 | 0.009029289 | 0.048462513 | 1 |
| HTN | LEC | *DPH1* | -3.266890345 | 0.003395024 | 0.03770947 | 1 |
| HTN | LEC | *MYH3* | -3.310082585 | 0.007135367 | 0.04338446 | 1 |
| HTN | LEC | *GNRHR* | -0.943851534 | 0.009161787 | 0.048802678 | 1 |
| HTN | LEC | *WFS1* | 0.6373079 | 0.003734957 | 0.03770947 | 1 |
| HTN | LEC | *AADAT* | -3.040340671 | 0.009273382 | 0.048853907 | 1 |
| HTN | LEC | *FAM149A* | -1.115182294 | 0.00759532 | 0.044594598 | 1 |
| HTN | LEC | *TECTA* | -1.443013561 | 0.005065335 | 0.038653449 | 1 |
| HTN | LEC | *SC5D* | -1.735614634 | 0.004462401 | 0.038101652 | 1 |
| HTN | LEC | *P2RX3* | -9.722256969 | 0.005224579 | 0.039240957 | 1 |
| HTN | LEC | *SNX15* | -1.997636963 | 0.001644582 | 0.027492515 | 1 |
| HTN | LEC | *NRXN2* | -1.751480683 | 0.002768509 | 0.03496719 | 1 |
| HTN | LEC | *CCDC86* | -3.072076104 | 0.001425853 | 0.025249047 | 1 |
| HTN | LEC | *GALNT18* | 0.619835724 | 0.006040857 | 0.041459167 | 1 |
| HTN | LEC | *C11orf21* | -4.16064441 | 0.000925073 | 0.019970552 | 1 |
| HTN | LEC | *ASIC1* | -1.975844783 | 0.003845269 | 0.03770947 | 1 |
| HTN | LEC | *IL23A* | -5.581975064 | 0.004485221 | 0.038101652 | 1 |
| HTN | LEC | *ACAD10* | -3.020140249 | 0.000635592 | 0.01573415 | 1 |
| HTN | LEC | *CDK2AP1* | 1.66845361 | 0.007127211 | 0.04338446 | 1 |
| HTN | LEC | *BTN3A3* | -1.603057224 | 0.004280574 | 0.037831232 | 1 |
| HTN | LEC | *RSPH4A* | 1.668608684 | 0.003497269 | 0.03770947 | 1 |
| HTN | LEC | *UST* | -1.102433413 | 0.004168498 | 0.03770947 | 1 |
| HTN | LEC | *TULP1* | -2.030165313 | 0.001354014 | 0.024698021 | 1 |
| HTN | LEC | *PHACTR1* | -1.276468103 | 0.009240549 | 0.048822112 | 1 |
| HTN | LEC | *CD83* | -1.877513244 | 0.009609227 | 0.04965591 | 1 |
| HTN | LEC | *C6orf62* | -2.3490961 | 0.00213079 | 0.031234415 | 1 |
| HTN | LEC | *PACRG* | -1.79908373 | 0.007966722 | 0.04574501 | 1 |
| HTN | LEC | *LY86* | -2.23768148 | 0.00699673 | 0.043136128 | 1 |
| HTN | LEC | *PCDHB2* | -2.120023467 | 0.002428026 | 0.033395663 | 1 |

| | | | | | | |
|---|---|---|---|---|---|---|
| HTN | LEC | *CDH6* | -0.896537881 | 0.008121298 | 0.046277049 | 1 |
| HTN | LEC | *UNC5A* | 0.463089391 | 0.003624767 | 0.03770947 | 1 |
| HTN | LEC | *NPHP3* | 1.140880942 | 0.006609028 | 0.042480568 | 1 |
| HTN | LEC | *PCCB* | 1.8017998 | 0.003525571 | 0.03770947 | 1 |
| HTN | LEC | *KIAA1257* | 0.616442077 | 0.003654914 | 0.03770947 | 1 |
| HTN | LEC | *PLSCR4* | 1.274509834 | 0.008080444 | 0.046179402 | 1 |
| HTN | LEC | *ABHD14B* | -1.951910139 | 0.009369613 | 0.049233755 | 1 |
| HTN | LEC | *ARHGEF26* | -1.463326028 | 0.007443817 | 0.04399196 | 1 |
| HTN | LEC | *VIPR1* | -1.076333775 | 0.005901336 | 0.041317869 | 1 |
| HTN | LEC | *ADAM23* | 0.511715147 | 0.006827409 | 0.043025132 | 1 |
| HTN | LEC | *GRB14* | -1.469821321 | 0.00598081 | 0.041329827 | 1 |
| HTN | LEC | *WNT6* | -2.291179086 | 0.00137418 | 0.02473249 | 1 |
| HTN | LEC | *IL18R1* | -1.999327683 | 0.001778078 | 0.028199053 | 1 |
| HTN | LEC | *TPO* | -1.582242681 | 0.007408187 | 0.043986247 | 1 |
| HTN | LEC | *PNO1* | -2.256790727 | 0.007294851 | 0.043739036 | 1 |
| HTN | LEC | *MORN1* | -2.336315322 | 0.000307755 | 0.010021653 | 1 |
| HTN | LEC | *WRAP73* | -3.093407498 | 0.009502264 | 0.049415847 | 1 |
| HTN | LEC | *MECR* | -2.806499564 | 0.001977403 | 0.030076364 | 1 |
| HTN | LEC | *DNAJC6* | -1.741526431 | 0.004912103 | 0.038441165 | 1 |
| HTN | LEC | *TMEM9* | -1.822434186 | 0.005538366 | 0.04048889 | 1 |
| HTN | LEC | *GBP3* | -0.854856359 | 0.002715801 | 0.034856249 | 1 |
| HTN | LEC | *KIF17* | 0.878941173 | 0.00690195 | 0.043044039 | 1 |
| HTN | LEC | *IRF6* | -0.839137162 | 0.00870764 | 0.048138645 | 1 |
| HTN | LEC | *RPS6KA1* | -2.781325328 | 0.006331607 | 0.041983118 | 1 |
| HTN | LEC | *CASC1* | -1.041728062 | 0.00585472 | 0.041169714 | 1 |
| HTN | LEC | *SGIP1* | 0.260897615 | 0.003623992 | 0.03770947 | 1 |
| HTN | LEC | *DNAH7* | -1.695657362 | 0.00499934 | 0.038529615 | 1 |
| HTN | LEC | *DCAF4* | -2.223983151 | 0.000846532 | 0.019175186 | 1 |
| HTN | LEC | *HELLS* | -2.799313773 | 0.00158979 | 0.026970839 | 1 |
| HTN | LEC | *PCDHB10* | 3.775638911 | 0.006312545 | 0.041983118 | 1 |
| HTN | LEC | *SNX19* | -2.394271138 | 0.001264733 | 0.024235407 | 1 |
| HTN | LEC | *MRC1* | 1.436219207 | 0.009608411 | 0.04965591 | 1 |
| HTN | LEC | *CCDC77* | -2.866255763 | 0.003885721 | 0.03770947 | 1 |
| HTN | LEC | *UFM1* | -2.343736535 | 0.006731225 | 0.042882211 | 1 |
| HTN | LEC | *ZFP30* | 2.008425402 | 0.008474152 | 0.047375361 | 1 |
| HTN | LEC | *EPHX2* | -1.738781729 | 0.00125665 | 0.024195498 | 1 |
| HTN | LEC | *AKAP1* | -2.116603347 | 0.008259265 | 0.046543498 | 1 |
| HTN | LEC | *TBX2* | -2.02307245 | 1.23E-09 | 6.66E-07 | 5.99E-06 |
| HTN | LEC | *TEX14* | 2.043971399 | 0.003010524 | 0.036053579 | 1 |
| HTN | LEC | *TRIM6* | -2.850289693 | 0.002981109 | 0.035986396 | 1 |
| HTN | LEC | *ECHDC2* | -1.266440771 | 0.009612402 | 0.04965591 | 1 |
| HTN | LEC | *POPDC2* | -1.597974767 | 0.003296551 | 0.037708496 | 1 |
| HTN | LEC | *MAPK8IP1* | -5.934251426 | 1.36E-06 | 0.000183261 | 0.00659741 |
| HTN | LEC | *FABP3* | -2.79922558 | 0.000774267 | 0.018004378 | 1 |
| HTN | LEC | *CPXM2* | -1.064758004 | 0.006610228 | 0.042480568 | 1 |
| HTN | LEC | *SERAC1* | -2.56393341 | 0.00771936 | 0.045044321 | 1 |
| HTN | LEC | *EEPD1* | -1.364033503 | 0.003166606 | 0.037022583 | 1 |
| HTN | LEC | *C7orf49* | -1.198584897 | 0.004715784 | 0.038101652 | 1 |
| HTN | LEC | *CENPK* | 1.959982697 | 0.004767415 | 0.038101652 | 1 |
| HTN | LEC | *ARHGAP9* | -2.989551192 | 0.003181629 | 0.037108807 | 1 |

| | | | | | | |
|---|---|---|---|---|---|---|
| HTN | LEC | *PDE1B* | -1.901406324 | 0.008411645 | 0.047237619 | 1 |
| HTN | LEC | *CDK2* | -2.529486812 | 0.000488964 | 0.01339246 | 1 |
| HTN | LEC | *DBH* | -23.82424765 | 0.003345675 | 0.03770947 | 1 |
| HTN | LEC | *C4BPA* | 2.07691854 | 0.00636842 | 0.041983118 | 1 |
| HTN | LEC | *DAW1* | -2.479198813 | 0.005802426 | 0.041169714 | 1 |
| HTN | LEC | *OBSL1* | -2.860359118 | 0.004887024 | 0.038306686 | 1 |
| HTN | LEC | *WFDC3* | -0.686791773 | 0.000233251 | 0.008260845 | 1 |
| HTN | LEC | *SLC12A5* | 1.912368902 | 0.002686447 | 0.034851979 | 1 |
| HTN | LEC | *CHST8* | 0.328468737 | 0.003650653 | 0.03770947 | 1 |
| HTN | LEC | *HIF3A* | 0.767053062 | 0.007518421 | 0.044302485 | 1 |
| HTN | LEC | *BTN2A2* | 1.076451784 | 0.000599673 | 0.015075726 | 1 |
| HTN | LEC | *SNRPC* | -3.065268275 | 1.29E-06 | 0.000178219 | 0.006237657 |
| HTN | LEC | *ZNF391* | -0.953752919 | 2.24E-05 | 0.001575029 | 0.108676967 |
| HTN | LEC | *TCP11* | 2.003651623 | 0.000681733 | 0.016393911 | 1 |
| HTN | LEC | *MEA1* | 1.423886388 | 0.007849375 | 0.045235126 | 1 |
| HTN | LEC | *CXCL6* | -2.113368315 | 0.003105716 | 0.036398387 | 1 |
| HTN | LEC | *OPRL1* | -4.267739473 | 0.005672729 | 0.04099621 | 1 |
| HTN | LEC | *FLRT3* | -1.775273248 | 0.005995307 | 0.041329827 | 1 |
| HTN | LEC | *UROD* | -2.851386844 | 9.44E-05 | 0.004722281 | 0.458061297 |
| HTN | LEC | *PRR12* | -2.302167615 | 0.008621661 | 0.047917866 | 1 |
| HTN | LEC | *TSKS* | -3.023497477 | 1.81E-05 | 0.001327752 | 0.087631648 |
| HTN | LEC | *TRAP1* | 1.279403774 | 0.00030449 | 0.009982347 | 1 |
| HTN | LEC | *AIF1L* | -1.908985259 | 0.005970785 | 0.041329827 | 1 |
| HTN | LEC | *ABHD8* | -3.232189153 | 5.80E-06 | 0.000552257 | 0.0281651 |
| HTN | LEC | *LRRC61* | -1.518131297 | 0.001744231 | 0.028192068 | 1 |
| HTN | LEC | *IDUA* | -2.058316446 | 0.008002608 | 0.045842566 | 1 |
| HTN | LEC | *GFER* | -3.21552129 | 0.000188757 | 0.007313544 | 0.915848042 |
| HTN | LEC | *STYXL1* | -1.264447362 | 0.001889854 | 0.02924751 | 1 |
| HTN | LEC | *KDR* | 0.637204434 | 0.003731329 | 0.03770947 | 1 |
| HTN | LEC | *CDHR3* | -1.47685952 | 0.00965887 | 0.04965591 | 1 |
| HTN | LEC | *GAD1* | -3.302222039 | 0.000110717 | 0.005116202 | 0.537201255 |
| HTN | LEC | *HOXD10* | 0.281891193 | 0.002581446 | 0.03412855 | 1 |
| HTN | LEC | *EIF2AK4* | -3.341472987 | 0.000413864 | 0.011918143 | 1 |
| HTN | LEC | *KNSTRN* | -0.579470981 | 0.004865669 | 0.038211862 | 1 |
| HTN | LEC | *BBOX1* | -2.94375861 | 0.004345923 | 0.037880891 | 1 |
| HTN | LEC | *KRI1* | -1.139732005 | 0.002826675 | 0.03516674 | 1 |
| HTN | LEC | *KLK8* | -6.652134447 | 0.004663237 | 0.038101652 | 1 |
| HTN | LEC | *ADCY4* | 0.758912595 | 0.000132985 | 0.005710101 | 0.645241388 |
| HTN | LEC | *RHBDF2* | 1.161875896 | 0.007240234 | 0.043639272 | 1 |
| HTN | LEC | *SHC2* | 0.824500625 | 0.004541149 | 0.038101652 | 1 |
| HTN | LEC | *GALNT8* | -2.700878656 | 0.001229827 | 0.023964337 | 1 |
| HTN | LEC | *ATP8B3* | -1.401502531 | 9.15E-06 | 0.000752599 | 0.044403363 |
| HTN | LEC | *STK33* | 1.270740434 | 0.003874039 | 0.03770947 | 1 |
| HTN | LEC | *MAP1S* | -2.575979524 | 0.009173133 | 0.048802678 | 1 |
| HTN | LEC | *PGPEP1* | 1.684142663 | 0.009148726 | 0.048802678 | 1 |
| HTN | LEC | *KIAA1683* | -1.700317141 | 0.003227661 | 0.037376156 | 1 |
| HTN | LEC | *ULBP2* | 9.36019618 | 0.008206381 | 0.046470093 | 1 |
| HTN | LEC | *TTLL9* | 5.51210229 | 0.009008632 | 0.048458848 | 1 |
| HTN | LEC | *IDO1* | -1.514494074 | 0.004801827 | 0.038101652 | 1 |
| HTN | LEC | *HAUS8* | -4.092613268 | 8.38E-10 | 5.08E-07 | 4.07E-06 |

| | | | | | | |
|---|---|---|---|---|---|---|
| HTN | LEC | *MRPS25* | -2.524914789 | 0.001770354 | 0.028199053 | 1 |
| HTN | LEC | *PDLIM4* | 2.778591547 | 0.000425671 | 0.012078114 | 1 |
| HTN | LEC | *NDUFA2* | -2.388270144 | 0.00176417 | 0.028199053 | 1 |
| HTN | LEC | *MATN3* | -1.792112581 | 0.002484714 | 0.033395663 | 1 |
| HTN | LEC | *TRIM21* | -2.08347431 | 0.00298156 | 0.035986396 | 1 |
| HTN | LEC | *CCT6B* | -1.409689562 | 0.00599653 | 0.041329827 | 1 |
| HTN | LEC | *CARD6* | 1.199410103 | 0.00876403 | 0.048214523 | 1 |
| HTN | LEC | *REEP2* | -2.25708346 | 0.003595164 | 0.03770947 | 1 |
| HTN | LEC | *ATP8A2* | -2.099952553 | 0.006863798 | 0.043025132 | 1 |
| HTN | LEC | *SPINK5* | -1.575195551 | 0.005297272 | 0.039535978 | 1 |
| HTN | LEC | *IMPA1* | 1.43640887 | 0.006352679 | 0.041983118 | 1 |
| HTN | LEC | *RRAS2* | -1.488835007 | 0.007904077 | 0.045448045 | 1 |
| HTN | LEC | *CATSPERB* | 0.873153223 | 0.006932787 | 0.043126036 | 1 |
| HTN | LEC | *TTC9* | 2.157859693 | 0.004612651 | 0.038101652 | 1 |
| HTN | LEC | *MRO* | -1.919624096 | 0.00478712 | 0.038101652 | 1 |
| HTN | LEC | *TMEM241* | -1.638633661 | 0.004957542 | 0.038529615 | 1 |
| HTN | LEC | *HOOK1* | -0.746786275 | 0.008510953 | 0.04746568 | 1 |
| HTN | LEC | *FADS2* | -2.014585281 | 0.006363205 | 0.041983118 | 1 |
| HTN | LEC | *CLDN10* | 2.764827975 | 0.000101825 | 0.004891615 | 0.494053111 |
| HTN | LEC | *ADAM19* | 0.518293388 | 0.001776543 | 0.028199053 | 1 |
| HTN | LEC | *USP30* | -1.490789715 | 0.000596296 | 0.015068898 | 1 |
| HTN | LEC | *DTX1* | 0.809461247 | 0.004096554 | 0.03770947 | 1 |
| HTN | LEC | *POM121C* | -2.864314597 | 0.001409545 | 0.025143795 | 1 |
| HTN | LEC | *NT5E* | 1.223548719 | 0.00246984 | 0.033395663 | 1 |
| HTN | LEC | *PRR5L* | 0.553553394 | 0.002428532 | 0.033395663 | 1 |
| HTN | LEC | *FAIM2* | -2.676902142 | 0.001243057 | 0.02412525 | 1 |
| HTN | LEC | *SMPD2* | -0.932496921 | 0.004709024 | 0.038101652 | 1 |
| HTN | LEC | *AGT* | 1.529275865 | 0.000356765 | 0.010619775 | 1 |
| HTN | LEC | *SLC19A3* | -1.1128932 | 0.00305528 | 0.036245037 | 1 |
| HTN | LEC | *STAB2* | 0.182059453 | 0.003615538 | 0.03770947 | 1 |
| HTN | LEC | *SLC41A2* | -1.504909451 | 0.004973885 | 0.038529615 | 1 |
| HTN | LEC | *RNASEH2B* | -2.066679188 | 0.005311554 | 0.039535978 | 1 |
| HTN | LEC | *NMT1* | 1.647735546 | 0.000547731 | 0.014288133 | 1 |
| HTN | LEC | *NDUFB5* | -2.65805661 | 0.005754293 | 0.041022318 | 1 |
| HTN | LEC | *SCN2A* | 2.447390178 | 0.000889512 | 0.019617773 | 1 |
| HTN | LEC | *IMP4* | -3.876980489 | 0.002460746 | 0.033395663 | 1 |
| HTN | LEC | *IL11RA* | -2.284768352 | 0.000214483 | 0.007824586 | 1 |
| HTN | LEC | *TCF19* | 1.822349086 | 0.008698392 | 0.048138645 | 1 |
| HTN | LEC | *TLR2* | -2.615447353 | 0.002117598 | 0.031234415 | 1 |
| HTN | LEC | *SYTL2* | -1.576679235 | 0.00077925 | 0.018004378 | 1 |
| HTN | LEC | *SULF1* | -0.885623298 | 0.00365649 | 0.03770947 | 1 |
| HTN | LEC | *TRPC6* | 1.05707174 | 0.001294937 | 0.024359331 | 1 |
| HTN | LEC | *SLC37A4* | 2.762586365 | 9.58E-05 | 0.004745381 | 0.465047375 |
| HTN | LEC | *PPP2R1B* | -1.680363843 | 6.76E-06 | 0.000618856 | 0.032799394 |
| HTN | LEC | *FXYD2* | 0.820010444 | 0.003086909 | 0.036398387 | 1 |
| HTN | LEC | *NUSAP1* | 3.386121593 | 0.004943404 | 0.038529615 | 1 |
| HTN | LEC | *ADAM10* | 1.878271369 | 0.004231709 | 0.03770947 | 1 |
| HTN | LEC | *KHK* | -1.949067349 | 0.000941978 | 0.020134253 | 1 |
| HTN | LEC | *LHCGR* | -2.553270936 | 0.001829083 | 0.028628097 | 1 |
| HTN | LEC | *TACC2* | -1.7021185 | 0.003376673 | 0.03770947 | 1 |

| | | | | | | |
|---|---|---|---|---|---|---|
| HTN | LEC | *SPPL2A* | -2.780596335 | 0.001420936 | 0.025249047 | 1 |
| HTN | LEC | *MMRN1* | 0.140264295 | 0.004999622 | 0.038529615 | 1 |
| HTN | LEC | *ENPEP* | -0.994561169 | 0.000246505 | 0.008604636 | 1 |
| HTN | LEC | *FBN2* | 0.716971127 | 0.000478165 | 0.013274713 | 1 |
| HTN | LEC | *PARVG* | -2.888862972 | 0.001154915 | 0.022965772 | 1 |
| HTN | LEC | *ERP27* | -2.09800831 | 0.00549977 | 0.040431645 | 1 |
| HTN | LEC | *CLSTN3* | -1.579411519 | 0.005270141 | 0.039460994 | 1 |
| HTN | LEC | *MARCH9* | 2.213177802 | 0.005821562 | 0.041169714 | 1 |
| HTN | LEC | *AMDHD1* | -0.908742391 | 0.002242683 | 0.032004407 | 1 |
| HTN | LEC | *RITA1* | -3.061365783 | 0.007273524 | 0.043731277 | 1 |
| HTN | LEC | *FOXN4* | -1.509039358 | 0.001453137 | 0.025453509 | 1 |
| HTN | LEC | *ACVRL1* | 2.642935108 | 0.009656767 | 0.04965591 | 1 |
| HTN | LEC | *SLC24A4* | -1.360989333 | 0.005444522 | 0.040110336 | 1 |
| HTN | LEC | *WARS* | 1.54209679 | 0.001380283 | 0.02473249 | 1 |
| HTN | LEC | *PIF1* | 1.799714226 | 0.004456753 | 0.038101652 | 1 |
| HTN | LEC | *CYP11A1* | -1.228791523 | 0.006404724 | 0.042146654 | 1 |
| HTN | LEC | *PCSK6* | 0.480569614 | 0.003592074 | 0.03770947 | 1 |
| HTN | LEC | *UNC45A* | -2.943945134 | 0.009123961 | 0.048802678 | 1 |
| HTN | LEC | *ITGAX* | -1.69232439 | 0.008371378 | 0.047065963 | 1 |
| HTN | LEC | *GCSH* | -2.070930071 | 0.000809207 | 0.018516165 | 1 |
| HTN | LEC | *TIGD7* | -2.682660063 | 0.00718451 | 0.043522435 | 1 |
| HTN | LEC | *DEF8* | 3.060704125 | 0.000315079 | 0.010191745 | 1 |
| HTN | LEC | *VPS53* | -2.353664695 | 0.000350023 | 0.010542716 | 1 |
| HTN | LEC | *LRRC46* | -2.241002527 | 7.40E-05 | 0.004080647 | 0.359096908 |
| HTN | LEC | *SCRN2* | -0.781534943 | 6.99E-05 | 0.003941031 | 0.338928663 |
| HTN | LEC | *RHBDL3* | -1.937719707 | 0.00156513 | 0.026713076 | 1 |
| HTN | LEC | *C18orf8* | -3.697434136 | 8.16E-07 | 0.000121773 | 0.003959736 |
| HTN | LEC | *NPC1* | -2.830747487 | 0.005557626 | 0.04048889 | 1 |
| HTN | LEC | *TBCD* | 1.594585338 | 0.006743431 | 0.042882211 | 1 |
| HTN | LEC | *NARF* | 1.577620725 | 0.006292712 | 0.041983118 | 1 |
| HTN | LEC | *RNF157* | -1.234278678 | 0.006517515 | 0.04230219 | 1 |
| HTN | LEC | *MAPK4* | -2.287828664 | 0.004603741 | 0.038101652 | 1 |
| HTN | LEC | *MIEN1* | -3.527697326 | 0.005677958 | 0.04099621 | 1 |
| HTN | LEC | *TXNL4A* | -2.472895226 | 0.008875234 | 0.04843941 | 1 |
| HTN | LEC | *GEMIN7* | -2.352229103 | 0.00131646 | 0.024600852 | 1 |
| HTN | LEC | *CTU1* | -2.58801296 | 0.007301859 | 0.043739036 | 1 |
| HTN | LEC | *ZNF593* | -3.586445277 | 0.00595995 | 0.041329827 | 1 |
| HTN | LEC | *SLC44A3* | 1.161333228 | 0.002098129 | 0.031227369 | 1 |
| HTN | LEC | *NME7* | -2.48865492 | 0.000321752 | 0.010213202 | 1 |
| HTN | LEC | *MAEL* | -2.595356293 | 0.004272754 | 0.037831024 | 1 |
| HTN | LEC | *SEMA6C* | 4.640405923 | 0.000973973 | 0.020546599 | 1 |
| HTN | LEC | *ARNT* | -2.520266755 | 0.000474946 | 0.013274713 | 1 |
| HTN | LEC | *DYRK3* | -2.509030961 | 0.002003938 | 0.030290057 | 1 |
| HTN | LEC | *SMYD2* | 0.379834089 | 0.003636882 | 0.03770947 | 1 |
| HTN | LEC | *CREB3L4* | -3.087738861 | 0.000112273 | 0.005120678 | 0.544748213 |
| HTN | LEC | *C1orf43* | -2.887275322 | 0.007077939 | 0.043229876 | 1 |
| HTN | LEC | *ABHD1* | -2.752399652 | 0.00046743 | 0.013185873 | 1 |
| HTN | LEC | *ZNF514* | -2.470951642 | 0.000369804 | 0.010940791 | 1 |
| HTN | LEC | *THNSL2* | -1.062523901 | 0.001295282 | 0.024359331 | 1 |
| HTN | LEC | *RABL2A* | -2.535857862 | 0.000209575 | 0.007716819 | 1 |

| | | | | | | |
|---|---|---|---|---|---|---|
| HTN | LEC | *FAHD2B* | -1.405497837 | 0.002820315 | 0.03516674 | 1 |
| HTN | LEC | *GALNT13* | -1.447120742 | 0.006479414 | 0.042255533 | 1 |
| HTN | LEC | *ESPNL* | -11.66971489 | 0.003975669 | 0.03770947 | 1 |
| HTN | LEC | *SLC25A38* | -2.778111572 | 0.005549432 | 0.04048889 | 1 |
| HTN | LEC | *EIF2A* | -1.356661371 | 0.002730435 | 0.034856249 | 1 |
| HTN | LEC | *NCEH1* | -1.735795428 | 0.002147504 | 0.031357001 | 1 |
| HTN | LEC | *AMT* | -0.834879399 | 0.001037225 | 0.021137669 | 1 |
| HTN | LEC | *MUC4* | -1.85978603 | 0.002319614 | 0.032717343 | 1 |
| HTN | LEC | *SPATA9* | -1.077034058 | 0.005058577 | 0.038653449 | 1 |
| HTN | LEC | *TSLP* | -0.763590775 | 0.00300922 | 0.036053579 | 1 |
| HTN | LEC | *CXCL14* | -1.339408785 | 0.004229173 | 0.03770947 | 1 |
| HTN | LEC | *DOK3* | -0.990368466 | 0.001566031 | 0.026713076 | 1 |
| HTN | LEC | *ABT1* | 1.696427688 | 0.00027407 | 0.00943112 | 1 |
| HTN | LEC | *IRAK1BP1* | 2.55745227 | 0.005243626 | 0.03932314 | 1 |
| HTN | LEC | *SLC18B1* | -1.926467324 | 0.001708874 | 0.02782089 | 1 |
| HTN | LEC | *SLC22A3* | -1.368067289 | 0.006753142 | 0.042887753 | 1 |
| HTN | LEC | *RBAK* | -1.348736509 | 0.004597632 | 0.038101652 | 1 |
| HTN | LEC | *CSGALNACT1* | 1.136623655 | 0.000491314 | 0.01339246 | 1 |
| HTN | LEC | *DOCK5* | 0.604525508 | 0.003973932 | 0.03770947 | 1 |
| HTN | LEC | *ADHFE1* | -1.05145001 | 1.54E-05 | 0.001167203 | 0.074700979 |
| HTN | LEC | *ATP6V0D2* | 2.835725365 | 0.004011107 | 0.03770947 | 1 |
| HTN | LEC | *DPYS* | -1.133000002 | 0.006177041 | 0.041742342 | 1 |
| HTN | LEC | *SLC39A12* | 5.665314405 | 0.004552486 | 0.038101652 | 1 |
| HTN | LEC | *RGR* | -8.185298418 | 0.00359089 | 0.03770947 | 1 |
| HTN | LEC | *HSD17B12* | -1.744702992 | 3.51E-10 | 2.84E-07 | 1.70E-06 |
| HTN | LEC | *TNKS1BP1* | 1.673092738 | 0.000223183 | 0.007962365 | 1 |
| HTN | LEC | *ENDOD1* | 1.984356882 | 0.003890442 | 0.03770947 | 1 |
| HTN | LEC | *DRD2* | -7.530707359 | 0.003743349 | 0.03770947 | 1 |
| HTN | LEC | *DUSP15* | 3.059546171 | 0.000290293 | 0.009647269 | 1 |
| HTN | LEC | *SPATA25* | -2.106255366 | 0.002473816 | 0.033395663 | 1 |
| HTN | LEC | *TRPT1* | -2.215085829 | 0.000189261 | 0.007313544 | 0.918292601 |
| HTN | LEC | *TBX6* | 0.723978463 | 0.002326601 | 0.032720781 | 1 |
| HTN | LEC | *PPP1R1C* | -2.165959921 | 0.004728014 | 0.038101652 | 1 |
| HTN | LEC | *PLBD2* | -2.59365176 | 0.001278702 | 0.024235407 | 1 |
| HTN | LEC | *GABRA2* | -3.440369791 | 0.003631001 | 0.03770947 | 1 |
| HTN | LEC | *PARP8* | -1.181950779 | 0.008503405 | 0.04746568 | 1 |
| HTN | LEC | *BEND6* | -1.18432707 | 0.004082416 | 0.03770947 | 1 |
| HTN | LEC | *TMEM56* | -1.904745749 | 0.004646561 | 0.038101652 | 1 |
| HTN | LEC | *PTPN14* | 1.26293555 | 0.0047228 | 0.038101652 | 1 |
| HTN | LEC | *TMEM178A* | -1.447570433 | 0.004302632 | 0.037880891 | 1 |
| HTN | LEC | *PLEKHH2* | -1.47512025 | 0.000323201 | 0.010213202 | 1 |
| HTN | LEC | *CCNO* | -8.733960727 | 0.00443854 | 0.038049106 | 1 |
| HTN | LEC | *RASGRP3* | 0.250237314 | 0.003469934 | 0.03770947 | 1 |
| HTN | LEC | *JAKMIP1* | -1.505071316 | 0.004983917 | 0.038529615 | 1 |
| HTN | LEC | *CAST* | -2.146582477 | 0.002105417 | 0.031234415 | 1 |
| HTN | LEC | *FRMD1* | -1.330035275 | 0.007757189 | 0.045059762 | 1 |
| HTN | LEC | *INO80C* | 1.003468688 | 0.005845052 | 0.041169714 | 1 |
| HTN | LEC | *JAZF1* | -1.799503602 | 0.002910351 | 0.035749424 | 1 |
| HTN | LEC | *GDPD1* | -2.524237893 | 0.007849954 | 0.045235126 | 1 |
| HTN | LEC | *SEMA3D* | 0.752183324 | 0.004419027 | 0.038016166 | 1 |

| | | | | | | |
|---|---|---|---|---|---|---|
| HTN | LEC | *ROBO4* | 1.021011457 | 0.00423114 | 0.03770947 | 1 |
| HTN | LEC | *LRRK1* | -1.99457812 | 0.004867051 | 0.038211862 | 1 |
| HTN | LEC | *ENPP3* | 0.567100126 | 0.004982178 | 0.038529615 | 1 |
| HTN | LEC | *NEIL2* | 2.844467234 | 2.23E-06 | 0.000263653 | 0.010809792 |
| HTN | LEC | *PGM5* | 0.37977726 | 0.003852296 | 0.03770947 | 1 |
| HTN | LEC | *PDE1C* | -1.30703902 | 0.008928889 | 0.048458848 | 1 |
| HTN | LEC | *RAB6B* | 0.497203577 | 0.009143087 | 0.048802678 | 1 |
| HTN | LEC | *PROM2* | -3.068377695 | 0.002202225 | 0.031775401 | 1 |
| HTN | LEC | *MARVELD1* | -2.093176092 | 0.006676959 | 0.042739585 | 1 |
| HTN | LEC | *GOLGA7B* | 1.27032261 | 0.003398073 | 0.03770947 | 1 |
| HTN | LEC | *ALS2CR12* | -1.326280895 | 0.00916923 | 0.048802678 | 1 |
| HTN | LEC | *KCNMA1* | -0.791279775 | 0.009237081 | 0.048822112 | 1 |
| HTN | LEC | *NDUFAF6* | -3.510676444 | 1.63E-05 | 0.001216026 | 0.079041713 |
| HTN | LEC | *C8orf37* | -1.203234182 | 0.00593367 | 0.041329827 | 1 |
| HTN | LEC | *NAA11* | -4.259125706 | 0.000478787 | 0.013274713 | 1 |
| HTN | LEC | *SFXN2* | 1.945908978 | 3.83E-08 | 9.79E-06 | 0.000185961 |
| HTN | LEC | *TDRD9* | -7.597834204 | 0.000748965 | 0.017726723 | 1 |
| HTN | LEC | *NODAL* | -1.392018012 | 0.003985422 | 0.03770947 | 1 |
| HTN | LEC | *NTAN1* | 1.80860415 | 0.000195533 | 0.007415698 | 0.948725411 |
| HTN | LEC | *SLC6A1* | -2.847415215 | 0.003376073 | 0.03770947 | 1 |
| HTN | LEC | *STEAP2* | -0.898643727 | 0.002604629 | 0.034341469 | 1 |
| HTN | LEC | *CLEC18A* | -2.007019502 | 5.05E-05 | 0.003024153 | 0.244956413 |
| HTN | LEC | *C12orf43* | -1.853970412 | 0.001880706 | 0.02924739 | 1 |
| HTN | LEC | *EXTL1* | -13.15739075 | 0.003999236 | 0.03770947 | 1 |
| HTN | LEC | *RHPN1* | 1.05601699 | 0.001190604 | 0.023387907 | 1 |
| HTN | LEC | *LRRC43* | -2.990142255 | 0.000108086 | 0.00507612 | 0.524432322 |
| HTN | LEC | *CLSTN2* | -1.686387364 | 0.008784224 | 0.048231719 | 1 |
| HTN | LEC | *TSPAN33* | -2.472908901 | 0.000197209 | 0.00741752 | 0.956860058 |
| HTN | LEC | *DYNC1I1* | 1.055351972 | 0.006464222 | 0.042255533 | 1 |
| HTN | LEC | *ELK4* | 2.333660071 | 0.004833894 | 0.038136673 | 1 |
| HTN | LEC | *DUSP23* | -1.560981902 | 0.007600934 | 0.044594598 | 1 |
| HTN | LEC | *FGF17* | -4.869963277 | 0.004181257 | 0.03770947 | 1 |
| HTN | LEC | *SLC5A11* | -1.51230531 | 2.78E-08 | 8.09E-06 | 0.000134854 |
| HTN | LEC | *ARHGAP27* | 0.803128298 | 0.002735019 | 0.034856249 | 1 |
| HTN | LEC | *HK2* | -1.09853982 | 0.006174872 | 0.041742342 | 1 |
| HTN | LEC | *ACE* | -1.12895883 | 0.004538631 | 0.038101652 | 1 |
| HTN | LEC | *TEPP* | -5.493499534 | 0.000326267 | 0.010213202 | 1 |
| HTN | LEC | *CTBP1* | -3.098702266 | 0.000175996 | 0.007116091 | 0.853930923 |
| HTN | LEC | *LRRC36* | -0.892540138 | 0.000117859 | 0.005218385 | 0.57185389 |
| HTN | LEC | *ZFYVE28* | -1.343356877 | 0.007751595 | 0.045059762 | 1 |
| HTN | LEC | *C16orf86* | 1.91568899 | 0.006832705 | 0.043025132 | 1 |
| HTN | LEC | *LYPD5* | -3.519490108 | 0.002482943 | 0.033395663 | 1 |
| HTN | LEC | *HSF2BP* | -1.768282507 | 9.77E-05 | 0.004788567 | 0.474068176 |
| HTN | LEC | *PCNT* | -1.313491529 | 0.009636264 | 0.04965591 | 1 |
| HTN | LEC | *S100B* | -0.789646895 | 4.72E-06 | 0.000487091 | 0.022893298 |
| HTN | LEC | *GPSM1* | 1.160321996 | 0.001256296 | 0.024195498 | 1 |
| HTN | LEC | *ZBTB7B* | -1.456389729 | 0.007819911 | 0.045169296 | 1 |
| HTN | LEC | *FDPS* | 2.620131427 | 0.003712071 | 0.03770947 | 1 |
| HTN | LEC | *PAQR6* | -1.387227355 | 0.005320897 | 0.039535978 | 1 |
| HTN | LEC | *NBEAL2* | 1.966360558 | 0.007981134 | 0.045773594 | 1 |

| | | | | | | |
|---|---|---|---|---|---|---|
| HTN | LEC | *PTH1R* | -1.747843356 | 0.001968744 | 0.030076364 | 1 |
| HTN | LEC | *SH2B2* | -1.730421487 | 0.005072214 | 0.038653449 | 1 |
| HTN | LEC | *ABCF3* | 2.411794156 | 3.56E-05 | 0.002244143 | 0.172799011 |
| HTN | LEC | *ASGR2* | 12.96399741 | 0.00221438 | 0.031775401 | 1 |
| HTN | LEC | *TTC9C* | -1.992673655 | 0.003009593 | 0.036053579 | 1 |
| HTN | LEC | *PDZK1IP1* | -3.639663426 | 0.006972979 | 0.043126036 | 1 |
| HTN | LEC | *COA7* | -2.067470561 | 0.005170532 | 0.039042822 | 1 |
| HTN | LEC | *SLC25A34* | -1.934661787 | 0.002762927 | 0.03496719 | 1 |
| HTN | LEC | *PDPN* | 0.813249691 | 0.00424692 | 0.03770947 | 1 |
| HTN | LEC | *SYNC* | -2.202776284 | 0.006031392 | 0.041459167 | 1 |
| HTN | LEC | *MEGF6* | 1.02737891 | 0.008904947 | 0.048458848 | 1 |
| HTN | LEC | *LRRIQ3* | -0.915088666 | 0.004223521 | 0.03770947 | 1 |
| HTN | LEC | *MRPL55* | 1.830585666 | 0.002355079 | 0.033025553 | 1 |
| HTN | LEC | *NOSTRIN* | -1.620855443 | 0.001189825 | 0.023387907 | 1 |
| HTN | LEC | *LYSMD1* | -1.795035202 | 3.20E-05 | 0.002073289 | 0.1554967 |
| HTN | LEC | *S100A9* | -1.476559277 | 0.005943074 | 0.041329827 | 1 |
| HTN | LEC | *DCAF16* | -4.763227264 | 0.000139571 | 0.00594034 | 0.677198751 |
| HTN | LEC | *NIPAL1* | 1.746276727 | 0.003815242 | 0.03770947 | 1 |
| HTN | LEC | *CLDN1* | -0.976327592 | 0.008989854 | 0.048458848 | 1 |
| HTN | LEC | *LINC01116* | 0.424790361 | 0.008230578 | 0.046489831 | 1 |
| HTN | LEC | *LMOD1* | -1.700350938 | 7.77E-05 | 0.004234271 | 0.376850092 |
| HTN | LEC | *IL17RC* | -2.649549387 | 0.000868729 | 0.019335188 | 1 |
| HTN | LEC | *CPA3* | -1.840458878 | 0.004324889 | 0.037880891 | 1 |
| HTN | LEC | *RFT1* | -2.561764847 | 4.22E-05 | 0.002589016 | 0.204532297 |
| HTN | LEC | *ABLIM2* | 1.462392623 | 0.003959909 | 0.03770947 | 1 |
| HTN | LEC | *BDH2* | -2.29532831 | 0.000812849 | 0.018516165 | 1 |
| HTN | LEC | *CDC25A* | -9.307387194 | 9.14E-11 | 1.11E-07 | 4.44E-07 |
| HTN | LEC | *ZNF589* | -3.147426735 | 4.36E-10 | 3.02E-07 | 2.11E-06 |
| HTN | LEC | *RNF123* | 3.284506564 | 1.38E-08 | 5.71E-06 | 6.70E-05 |
| HTN | LEC | *MST1R* | 2.165863536 | 1.45E-05 | 0.00111594 | 0.070304203 |
| HTN | LEC | *ETNPPL* | -6.424244222 | 0.004224828 | 0.03770947 | 1 |
| HTN | LEC | *CEP44* | -2.461083799 | 0.002074648 | 0.030972892 | 1 |
| HTN | LEC | *TMEM144* | -2.632860617 | 0.005647793 | 0.040990555 | 1 |
| HTN | LEC | *LMBRD2* | -2.019503659 | 0.003568732 | 0.03770947 | 1 |
| HTN | LEC | *F2RL2* | -3.599687118 | 0.007769686 | 0.045059762 | 1 |
| HTN | LEC | *CMBL* | 2.013903331 | 0.001706067 | 0.02782089 | 1 |
| HTN | LEC | *ERAP1* | -3.742839792 | 1.77E-06 | 0.000225694 | 0.008576364 |
| HTN | LEC | *CCDC127* | -4.024480508 | 8.97E-05 | 0.004532562 | 0.435125976 |
| HTN | LEC | *GRIK2* | -0.769969503 | 0.000646641 | 0.015926408 | 1 |
| HTN | LEC | *RAET1E* | -1.489135639 | 0.005207733 | 0.039175073 | 1 |
| HTN | LEC | *DAGLB* | -0.933673906 | 0.007480803 | 0.044156762 | 1 |
| HTN | LEC | *TNFRSF11B* | -1.52033726 | 0.00716308 | 0.043498451 | 1 |
| HTN | LEC | *DEFB1* | -3.095856164 | 0.002415577 | 0.033395663 | 1 |
| HTN | LEC | *GPER1* | 2.954983036 | 0.003203173 | 0.037181326 | 1 |
| HTN | LEC | *INTS1* | -2.467070271 | 0.008890888 | 0.048458848 | 1 |
| HTN | LEC | *FZD6* | 1.073814574 | 0.001698035 | 0.02782089 | 1 |
| HTN | LEC | *RPP25L* | -2.92036429 | 7.22E-06 | 0.00062871 | 0.035009376 |
| HTN | LEC | *NUDT2* | -0.933484358 | 0.003564824 | 0.03770947 | 1 |
| HTN | LEC | *TMEM65* | 1.582568758 | 0.009406367 | 0.049233755 | 1 |
| HTN | LEC | *ZMAT4* | 0.353548842 | 0.003555374 | 0.03770947 | 1 |

| | | | | | | |
|---|---|---|---|---|---|---|
| HTN | LEC | *AQP3* | -2.692278613 | 0.004000341 | 0.03770947 | 1 |
| HTN | LEC | *SLC16A9* | -2.58953068 | 0.006474198 | 0.042255533 | 1 |
| HTN | LEC | *GJB2* | -5.682041237 | 0.004767591 | 0.038101652 | 1 |
| HTN | LEC | *TMEM63C* | 1.257853867 | 0.001790047 | 0.028199053 | 1 |
| HTN | LEC | *RET* | 1.88309955 | 0.005725509 | 0.041022318 | 1 |
| HTN | LEC | *CASP7* | -1.645908586 | 0.004143422 | 0.03770947 | 1 |
| HTN | LEC | *PPP1R36* | 1.489838495 | 0.006303012 | 0.041983118 | 1 |
| HTN | LEC | *UBTD1* | 1.706483979 | 0.009084889 | 0.048653293 | 1 |
| HTN | LEC | *OTOGL* | -1.756321996 | 0.001904825 | 0.02924751 | 1 |
| HTN | LEC | *PACSIN3* | -1.0493564 | 0.00558583 | 0.040633356 | 1 |
| HTN | LEC | *ARL5B* | -5.23736489 | 7.11E-06 | 0.00062871 | 0.034477617 |
| HTN | LEC | *LIPC* | 0.744035864 | 0.004569252 | 0.038101652 | 1 |
| HTN | LEC | *JAM3* | -2.04324156 | 0.00902746 | 0.048462513 | 1 |
| HTN | LEC | *TRMT61A* | -2.330445335 | 0.006335683 | 0.041983118 | 1 |
| HTN | LEC | *WDR88* | -2.220485499 | 0.008983326 | 0.048458848 | 1 |
| HTN | LEC | *XRRA1* | -2.160738543 | 0.001121359 | 0.022390266 | 1 |
| HTN | LEC | *TRIM66* | -1.116997689 | 1.47E-06 | 0.000192941 | 0.007138828 |
| HTN | LEC | *NDST2* | -8.268341779 | 0.003809465 | 0.03770947 | 1 |
| HTN | LEC | *RIMKLB* | -3.442935962 | 5.72E-08 | 1.32E-05 | 0.000277712 |
| HTN | LEC | *NYAP1* | 0.795153274 | 8.53E-05 | 0.004451906 | 0.414027217 |
| HTN | LEC | *MS4A7* | -1.122927528 | 0.00133778 | 0.024600852 | 1 |
| HTN | LEC | *MS4A14* | -0.622558412 | 0.001478384 | 0.025710096 | 1 |
| HTN | LEC | *RCCD1* | -2.414758434 | 0.006135715 | 0.041637045 | 1 |
| HTN | LEC | *ISLR2* | 0.328321952 | 0.008607537 | 0.047894234 | 1 |
| HTN | LEC | *TBC1D2B* | -1.594430151 | 0.008662322 | 0.048033815 | 1 |
| HTN | LEC | *LOXHD1* | 1.500658302 | 0.008840814 | 0.048396858 | 1 |
| HTN | LEC | *MYO5B* | -1.035941043 | 0.003933488 | 0.03770947 | 1 |
| HTN | LEC | *ZNF23* | -3.451291495 | 0.000108804 | 0.00507612 | 0.527916469 |
| HTN | LEC | *SMG8* | -1.757416686 | 0.007193939 | 0.043522435 | 1 |
| HTN | LEC | *CHAF1A* | -0.832033166 | 0.007014865 | 0.043193049 | 1 |
| HTN | LEC | *SRR* | 1.058522803 | 0.000501532 | 0.013519085 | 1 |
| HTN | LEC | *KLK5* | -1.776060483 | 0.001648871 | 0.027492515 | 1 |
| HTN | LEC | *KLK6* | -12.921935 | 0.006855097 | 0.043025132 | 1 |
| HTN | LEC | *SPRYD3* | -2.836616115 | 0.001701336 | 0.02782089 | 1 |
| HTN | LEC | *ZNF598* | -3.617461658 | 2.08E-05 | 0.00148624 | 0.10106429 |
| HTN | LEC | *ECI1* | -1.940711343 | 0.001691308 | 0.02782089 | 1 |
| HTN | LEC | *SCARA5* | -0.767916751 | 0.004235625 | 0.03770947 | 1 |
| HTN | LEC | *FAM83B* | -8.218922355 | 0.006058872 | 0.041463538 | 1 |
| HTN | LEC | *NT5DC2* | -1.751977092 | 0.000682517 | 0.016393911 | 1 |
| HTN | LEC | *COA6* | -1.966263675 | 0.00659817 | 0.042480568 | 1 |
| HTN | LEC | *MLKL* | 1.268118044 | 0.004211383 | 0.03770947 | 1 |
| HTN | LEC | *TMUB2* | -2.486807169 | 0.009254278 | 0.048822112 | 1 |
| HTN | LEC | *NBPF9* | -2.293437784 | 0.005894305 | 0.041317869 | 1 |
| HTN | LEC | *ADAM9* | 2.063581884 | 0.003193935 | 0.037163001 | 1 |
| HTN | LEC | *GSTM4* | -3.790240601 | 0.002737063 | 0.034856249 | 1 |
| HTN | LEC | *SNTB2* | 1.314133438 | 0.005316224 | 0.039535978 | 1 |
| HTN | LEC | *LGALS9* | 1.142482797 | 0.007435077 | 0.04399196 | 1 |
| HTN | LEC | *KLK7* | -1.973440967 | 0.004700278 | 0.038101652 | 1 |
| HTN | LEC | *IRS1* | -0.630273982 | 0.006041144 | 0.041459167 | 1 |
| HTN | LEC | *UPF3A* | 1.982531122 | 0.005659564 | 0.040990555 | 1 |

| | | | | | | |
|---|---|---|---|---|---|---|
| HTN | LEC | *C8orf46* | 0.535874119 | 0.001897596 | 0.02924751 | 1 |
| HTN | LEC | *CPT1C* | 4.036925298 | 2.18E-08 | 7.53E-06 | 0.000105598 |
| HTN | LEC | *RAB24* | -6.581304155 | 9.58E-06 | 0.000774918 | 0.046495091 |
| HTN | LEC | *ZEB2* | -1.473898309 | 0.00086717 | 0.019335188 | 1 |
| HTN | LEC | *CLIC3* | -2.491344339 | 0.007234271 | 0.043639272 | 1 |
| HTN | LEC | *INO80E* | -2.808243237 | 2.33E-08 | 7.53E-06 | 0.000112955 |
| HTN | LEC | *DFFB* | -3.257894444 | 0.002127312 | 0.031234415 | 1 |
| HTN | LEC | *RGPD8* | -0.602697025 | 0.000296568 | 0.009788752 | 1 |
| HTN | LEC | *LRRC45* | -1.08289478 | 0.006636682 | 0.042537887 | 1 |
| HTN | LEC | *MYO7B* | 2.488651404 | 0.000209938 | 0.007716819 | 1 |
| HTN | LEC | *RNF150* | -1.311435521 | 0.007622898 | 0.044629148 | 1 |
| HTN | LEC | *ANKK1* | -2.007847015 | 0.007625219 | 0.044629148 | 1 |
| HTN | LEC | *ELP5* | -2.23655443 | 0.004365041 | 0.03788762 | 1 |
| HTN | LEC | *EMB* | -1.55146195 | 0.000589689 | 0.014979953 | 1 |
| HTN | LEC | *SLC26A5* | 1.07639071 | 0.004653935 | 0.038101652 | 1 |
| HTN | LEC | *TTLL6* | -5.957278925 | 4.06E-06 | 0.000447239 | 0.019678518 |
| HTN | LEC | *KCNS3* | 1.238704534 | 0.001205543 | 0.02358587 | 1 |
| HTN | LEC | *S1PR1* | 1.27317016 | 0.005712504 | 0.041022318 | 1 |
| HTN | LEC | *FPR1* | -1.371127637 | 0.003098634 | 0.036398387 | 1 |
| HTN | LEC | *SOX7* | 1.681289417 | 0.000981072 | 0.020606749 | 1 |
| HTN | LEC | *GIMAP8* | 0.89989781 | 0.000796318 | 0.018311538 | 1 |
| HTN | LEC | *ATP6V0E2* | -1.193346113 | 0.00656408 | 0.042352283 | 1 |
| HTN | LEC | *PDE7B* | 0.900012802 | 0.007807055 | 0.045169296 | 1 |
| HTN | LEC | *MRPL36* | 1.231806938 | 0.002220087 | 0.031775401 | 1 |
| HTN | LEC | *HOPX* | -1.866027053 | 0.00609153 | 0.041544972 | 1 |
| HTN | LEC | *NMUR1* | -2.100898203 | 0.007188288 | 0.043522435 | 1 |
| HTN | LEC | *RGS19* | -1.780812051 | 4.51E-09 | 2.19E-06 | 2.19E-05 |
| HTN | LEC | *VAT1L* | -1.102644218 | 0.003870328 | 0.03770947 | 1 |
| HTN | LEC | *PAH* | -4.342825629 | 0.007724006 | 0.045044321 | 1 |
| HTN | LEC | *FBXW10* | -3.608609974 | 0.005164367 | 0.039042822 | 1 |
| HTN | LEC | *ANKRD20A4* | -4.005360832 | 0.005756931 | 0.041022318 | 1 |
| HTN | LEC | *NME6* | 4.492654328 | 3.39E-08 | 9.15E-06 | 0.00016471 |
| HTN | LEC | *TP53RK* | 4.342749209 | 0.004335541 | 0.037880891 | 1 |
| HTN | LEC | *FIBP* | 2.866720863 | 0.000347521 | 0.010538571 | 1 |
| HTN | LEC | *CTSW* | -0.449279526 | 3.24E-06 | 0.000365675 | 0.015724037 |
| HTN | LEC | *SNTG2* | 0.2530431 | 0.004621343 | 0.038101652 | 1 |
| HTN | LEC | *ZFAND4* | -2.411912658 | 0.000881644 | 0.019533042 | 1 |
| HTN | LEC | *ZNF738* | 1.604856247 | 0.000184634 | 0.007313544 | 0.895844484 |
| HTN | LEC | *SLFN11* | -2.51141269 | 0.006355298 | 0.041983118 | 1 |
| HTN | LEC | *FAM71D* | -1.444020503 | 0.00591581 | 0.041329827 | 1 |
| HTN | LEC | *CFL1* | -2.499656104 | 0.006959764 | 0.043126036 | 1 |
| HTN | LEC | *RAB43* | 5.459411686 | 0.005757667 | 0.041022318 | 1 |
| HTN | LEC | *MYEOV* | -5.365380971 | 0.00011398 | 0.005120678 | 0.553033172 |
| HTN | LEC | *MRGPRF* | -1.805953213 | 0.00725807 | 0.043692502 | 1 |
| HTN | LEC | *DCAKD* | -3.427305223 | 0.000323779 | 0.010213202 | 1 |
| HTN | LEC | *NOC3L* | -3.680992972 | 6.07E-05 | 0.003549508 | 0.294609129 |
| HTN | LEC | *RHOD* | 1.421067344 | 0.001367169 | 0.02473249 | 1 |
| HTN | LEC | *MAP3K11* | 2.860217536 | 0.000172561 | 0.007095466 | 0.837264994 |
| HTN | LEC | *NDNF* | 0.456441858 | 0.003494089 | 0.03770947 | 1 |
| HTN | LEC | *CCDC36* | -1.226938494 | 0.000329144 | 0.01023721 | 1 |

| | | | | | | |
|---|---|---|---|---|---|---|
| HTN | LEC | *MST1* | -1.373438339 | 2.24E-08 | 7.53E-06 | 0.000108738 |
| HTN | LEC | *GMPPB* | -2.89378434 | 0.000288268 | 0.009646036 | 1 |
| HTN | LEC | *C11orf80* | -4.076871949 | 1.41E-08 | 5.71E-06 | 6.85E-05 |
| HTN | LEC | *CTU2* | -1.664736254 | 0.002168993 | 0.031414785 | 1 |
| HTN | LEC | *SNX31* | -1.503167052 | 1.64E-07 | 3.06E-05 | 0.00079597 |
| HTN | LEC | *RSRC1* | -2.65402339 | 0.002067896 | 0.030967386 | 1 |
| HTN | LEC | *SLC22A1* | -1.432388405 | 0.004108931 | 0.03770947 | 1 |
| HTN | LEC | *CHST2* | -3.244221821 | 0.000775432 | 0.018004378 | 1 |
| HTN | LEC | *CADM2* | -1.478176375 | 0.003941836 | 0.03770947 | 1 |
| HTN | LEC | *PSMD2* | -2.514652023 | 0.004544779 | 0.038101652 | 1 |
| HTN | LEC | *CCNE2* | -3.89567697 | 2.47E-10 | 2.39E-07 | 1.20E-06 |
| HTN | LEC | *PPP2R2D* | -2.870107432 | 0.000284413 | 0.009646036 | 1 |
| HTN | LEC | *C11orf68* | -1.731204752 | 0.00631219 | 0.041983118 | 1 |
| HTN | LEC | *PRIMA1* | -1.657704504 | 0.00518892 | 0.03909416 | 1 |
| HTN | LEC | *DOK7* | 3.06291725 | 0.002469034 | 0.033395663 | 1 |
| HTN | LEC | *ORAI3* | 3.601513201 | 1.16E-06 | 0.000164831 | 0.005604266 |
| HTN | LEC | *B3GALT6* | -4.02611104 | 0.006883639 | 0.043025132 | 1 |
| HTN | LEC | *JAKMIP2* | -1.515406207 | 0.001270052 | 0.024235407 | 1 |
| HTN | LEC | *MYPOP* | -2.960729746 | 0.002791916 | 0.035003553 | 1 |
| HTN | LEC | *B3GNT5* | 0.894306368 | 0.008864353 | 0.048434505 | 1 |
| HTN | LEC | *BDNF* | -2.80777524 | 0.003089616 | 0.036398387 | 1 |
| HTN | LEC | *C8G* | 23.36874703 | 0.004750149 | 0.038101652 | 1 |
| HTN | LEC | *MTHFR* | -4.262759977 | 0.000326013 | 0.010213202 | 1 |
| HTN | LEC | *NAP1L5* | -1.636540219 | 0.008458332 | 0.047375361 | 1 |
| HTN | LEC | *SLC25A22* | -3.197795157 | 0.000487191 | 0.01339246 | 1 |
| HTN | LEC | *SLC35G5* | -6.625073431 | 0.004336639 | 0.037880891 | 1 |
| HTN | LEC | *FAM87B* | -6.214673411 | 0.003896852 | 0.03770947 | 1 |
| HTN | LEC | *CENPBD1* | -1.777892951 | 0.004405346 | 0.038016166 | 1 |
| HTN | LEC | *DALRD3* | -2.773757284 | 0.005854315 | 0.041169714 | 1 |
| HTN | LEC | *VN1R1* | -2.501612793 | 0.005985548 | 0.041329827 | 1 |
| HTN | LEC | *TMPRSS9* | -1.46325351 | 0.006724976 | 0.042882211 | 1 |
| HTN | LEC | *ZFAND2A* | -3.710031323 | 0.000776516 | 0.018004378 | 1 |
| HTN | LEC | *P4HTM* | -1.78480319 | 0.001087234 | 0.021798602 | 1 |
| HTN | LEC | *RPP25* | 2.135773265 | 1.26E-07 | 2.56E-05 | 0.000613372 |
| HTN | LEC | *MPI* | -4.958152136 | 1.34E-15 | 3.26E-12 | 6.52E-12 |
| HTN | LEC | *TRIM73* | -4.735463228 | 0.000175368 | 0.007116091 | 0.850884701 |
| HTN | LEC | *ZNF852* | -2.283984544 | 0.0010412 | 0.021137669 | 1 |
| HTN | LEC | *HYI* | -2.188774772 | 2.83E-08 | 8.09E-06 | 0.000137483 |
| HTN | LEC | *CTC1* | 3.04557871 | 0.003515808 | 0.03770947 | 1 |
| HTN | LEC | *CCDC89* | -2.966384897 | 0.004565041 | 0.038101652 | 1 |
| HTN | LEC | *HTR1F* | -6.859070014 | 0.000286845 | 0.009646036 | 1 |
| HTN | LEC | *ZNF664* | -2.230575152 | 0.009585672 | 0.04965591 | 1 |
| HTN | LEC | *HLA-DQB1* | -0.788156491 | 0.001342645 | 0.024600852 | 1 |
| HTN | LEC | *LINC00174* | -1.043086298 | 0.009449955 | 0.049355417 | 1 |
| HTN | LEC | *GEMIN4* | -2.220269533 | 0.002471715 | 0.033395663 | 1 |
| HTN | LEC | *SERTAD2* | 1.772806715 | 0.007523753 | 0.044302485 | 1 |
| HTN | LEC | *AKAP5* | -4.188027654 | 0.00190259 | 0.02924751 | 1 |
| HTN | LEC | *SEPT1* | -2.19034257 | 0.006780713 | 0.042950414 | 1 |
| HTN | LEC | *D2HGDH* | -1.882202518 | 0.001011158 | 0.020877179 | 1 |
| HTN | LEC | *LSMEM1* | -1.048284968 | 0.00458262 | 0.038101652 | 1 |

| | | | | | | |
|---|---|---|---|---|---|---|
| HTN | LEC | *MUC16* | -1.088008258 | 0.006245973 | 0.041983118 | 1 |
| HTN | LEC | *SLC25A41* | -3.88248127 | 0.005697705 | 0.041016712 | 1 |
| HTN | LEC | *TMEM252* | 6.62360824 | 0.003472853 | 0.03770947 | 1 |
| HTN | LEC | *CLDN7* | -3.011350636 | 4.18E-08 | 1.01E-05 | 0.00020276 |
| HTN | LEC | *ADO* | 2.217309873 | 8.80E-05 | 0.004498209 | 0.42696869 |
| HTN | LEC | *SYNM* | 0.540691837 | 0.004103237 | 0.03770947 | 1 |
| HTN | LEC | *KCNA4* | -1.367581563 | 0.00463022 | 0.038101652 | 1 |
| HTN | LEC | *CLN8* | 1.906085124 | 3.16E-06 | 0.000365182 | 0.015337637 |
| HTN | LEC | *NXPH4* | -8.784643842 | 0.004251253 | 0.03770947 | 1 |
| HTN | LEC | *CAPN12* | 1.105173455 | 0.000904547 | 0.01976964 | 1 |
| HTN | LEC | *FES* | -6.562300951 | 2.35E-17 | 1.14E-13 | 1.14E-13 |
| HTN | LEC | *BRICD5* | 5.082609995 | 0.007209294 | 0.043561014 | 1 |
| HTN | LEC | *GRID1* | -1.785264416 | 0.000374349 | 0.011008123 | 1 |
| HTN | LEC | *PLCXD3* | -1.47268859 | 0.008919169 | 0.048458848 | 1 |
| HTN | LEC | *CEP63* | -1.438656173 | 0.009489629 | 0.04940309 | 1 |
| HTN | LEC | *ODF3L1* | -4.790075992 | 0.000181324 | 0.007270927 | 0.879782214 |
| HTN | LEC | *ZNF320* | 0.808433571 | 0.000766293 | 0.018004378 | 1 |
| HTN | LEC | *LYSMD4* | -2.070239475 | 0.004782033 | 0.038101652 | 1 |
| HTN | LEC | *WBP2NL* | -2.165859595 | 0.000927592 | 0.019970552 | 1 |
| HTN | LEC | *BEGAIN* | -1.562061298 | 0.00900077 | 0.048458848 | 1 |
| HTN | LEC | *CSMD1* | 1.640759042 | 0.004807367 | 0.038101652 | 1 |
| HTN | LEC | *LRIT3* | -6.352602976 | 0.00398784 | 0.03770947 | 1 |
| HTN | LEC | *NPIPA1* | 0.773277061 | 0.006221994 | 0.041929328 | 1 |
| HTN | LEC | *AC131971.1* | -4.910374432 | 0.00051868 | 0.013827677 | 1 |
| HTN | LEC | *FAM19A1* | -1.30639846 | 0.006961228 | 0.043126036 | 1 |
| HTN | LEC | *ACSM2A* | 1.09814068 | 0.003247455 | 0.037426728 | 1 |
| HTN | LEC | *KREMEN1* | -1.162293572 | 0.004651226 | 0.038101652 | 1 |
| HTN | LEC | *ZNF703* | -2.207061288 | 0.00054702 | 0.014288133 | 1 |
| HTN | LEC | *EMILIN3* | -2.79798315 | 0.002626725 | 0.034445588 | 1 |
| HTN | LEC | *ALDH1A3* | -1.31730523 | 3.37E-05 | 0.002154464 | 0.163739261 |
| HTN | LEC | *CLECL1* | -1.30984818 | 0.003440442 | 0.03770947 | 1 |
| HTN | LEC | *A3GALT2* | -1.644659443 | 6.66E-06 | 0.000618856 | 0.032298927 |
| HTN | LEC | *TMEM255B* | 0.389416426 | 0.002816196 | 0.03516674 | 1 |
| HTN | LEC | *HDDC3* | -2.290734017 | 0.004194608 | 0.03770947 | 1 |
| HTN | LEC | *SNN* | 3.405833674 | 0.005786737 | 0.041168987 | 1 |
| HTN | LEC | *DRD1* | -20.16786044 | 0.005018004 | 0.038529615 | 1 |
| HTN | LEC | *NUTM2A* | -2.164443243 | 8.52E-05 | 0.004451906 | 0.413417311 |
| HTN | LEC | *WT1* | -0.930928694 | 0.009203272 | 0.048822112 | 1 |
| HTN | LEC | *SORCS2* | -1.764309895 | 0.004815596 | 0.038101652 | 1 |
| HTN | LEC | *BRI3BP* | -2.698065417 | 0.004128867 | 0.03770947 | 1 |
| HTN | LEC | *CDK10* | -1.475112121 | 5.16E-05 | 0.003055555 | 0.250555504 |
| HTN | LEC | *C14orf80* | -2.10968948 | 0.004657611 | 0.038101652 | 1 |
| HTN | LEC | *TNFAIP8L1* | 0.63485966 | 0.006357301 | 0.041983118 | 1 |
| HTN | LEC | *SV2B* | 0.375506156 | 0.00730116 | 0.043739036 | 1 |
| HTN | LEC | *PSMD13* | -2.429700782 | 0.003369507 | 0.03770947 | 1 |
| HTN | LEC | *C6orf201* | -1.72988942 | 0.00362192 | 0.03770947 | 1 |
| HTN | LEC | *MYBL1* | -2.002404977 | 0.000253835 | 0.008797192 | 1 |
| HTN | LEC | *C11orf87* | 2.051674955 | 0.001076825 | 0.021719723 | 1 |
| HTN | LEC | *IFITM1* | -2.909415532 | 0.001530603 | 0.026428776 | 1 |
| HTN | LEC | *LINC00839* | 1.07705393 | 0.005735908 | 0.041022318 | 1 |

| | | | | | | |
|---|---|---|---|---|---|---|
| HTN | LEC | *RASA3* | -1.508943767 | 0.008977939 | 0.048458848 | 1 |
| HTN | LEC | *SMYD4* | -2.255705474 | 0.002774602 | 0.03496719 | 1 |
| HTN | LEC | *MIR22HG* | -1.602712434 | 0.003273298 | 0.037580928 | 1 |
| HTN | LEC | *CYP27C1* | -1.097067074 | 0.004373636 | 0.037894435 | 1 |
| HTN | LEC | *FOXI2* | -1.831548306 | 0.004013555 | 0.03770947 | 1 |
| HTN | LEC | *MAPT* | -1.633368838 | 0.007819253 | 0.045169296 | 1 |
| HTN | LEC | *C1QL4* | -10.64156217 | 0.003878987 | 0.03770947 | 1 |
| HTN | LEC | *RTN4RL2* | -3.343523835 | 0.000241899 | 0.008505017 | 1 |
| HTN | LEC | *RHD* | 1.873694228 | 0.001875488 | 0.02924739 | 1 |
| HTN | LEC | *TMPRSS6* | -6.221191964 | 0.004059049 | 0.03770947 | 1 |
| HTN | LEC | *LUZP2* | -1.128416655 | 0.004013288 | 0.03770947 | 1 |
| HTN | LEC | *GJA4* | 1.48536779 | 0.006728034 | 0.042882211 | 1 |
| HTN | LEC | *SBK2* | -8.341384585 | 0.004813046 | 0.038101652 | 1 |
| HTN | LEC | *ZNF385C* | 1.590311617 | 0.005099131 | 0.038778971 | 1 |
| HTN | LEC | *KBTBD12* | -1.415346811 | 0.000925205 | 0.019970552 | 1 |
| HTN | LEC | *ZFP69* | 2.938316233 | 0.008960393 | 0.048458848 | 1 |
| HTN | LEC | *ZSCAN23* | -0.613899999 | 0.005361585 | 0.039656112 | 1 |
| HTN | LEC | *PRKAR1B* | 0.613325258 | 0.003580991 | 0.03770947 | 1 |
| HTN | LEC | *NCR3LG1* | -1.91989104 | 0.003026899 | 0.036053579 | 1 |
| HTN | LEC | *RUFY4* | -1.780926267 | 0.004344283 | 0.037880891 | 1 |
| HTN | LEC | *CENPP* | -1.29352718 | 0.000339182 | 0.010350392 | 1 |
| HTN | LEC | *C3orf62* | -1.738199313 | 0.002736119 | 0.034856249 | 1 |
| HTN | LEC | *ZNF559* | -0.953295496 | 0.000715341 | 0.017097713 | 1 |
| HTN | LEC | *H3F3C* | -45.63095126 | 0.001711232 | 0.02782089 | 1 |
| HTN | LEC | *C19orf54* | -1.701816622 | 0.007367457 | 0.043868076 | 1 |
| HTN | LEC | *C22orf34* | 2.025809441 | 0.002945845 | 0.035822654 | 1 |
| HTN | LEC | *COL25A1* | -1.079415692 | 0.008587211 | 0.047835991 | 1 |
| HTN | LEC | *GOLGA8M* | -1.382643195 | 0.006418403 | 0.042146654 | 1 |
| HTN | LEC | *SLC4A5* | -2.484509612 | 0.00687159 | 0.043025132 | 1 |
| HTN | LEC | *FZD9* | 4.919380389 | 0.000861091 | 0.019335188 | 1 |
| HTN | LEC | *FAM26F* | -1.269988977 | 0.002745138 | 0.03486757 | 1 |
| HTN | LEC | *ENTPD8* | -96.19786763 | 0.003244481 | 0.037426728 | 1 |
| HTN | LEC | *FBF1* | -3.509763819 | 1.62E-07 | 3.06E-05 | 0.000787395 |
| HTN | LEC | *ASTL* | 7.985016162 | 0.008847488 | 0.048396858 | 1 |
| HTN | LEC | *MSL1* | -1.83151831 | 0.009473045 | 0.049369727 | 1 |
| HTN | LEC | *UTS2B* | -1.92055667 | 0.008942876 | 0.048458848 | 1 |
| HTN | LEC | *HLA-DRB1* | -1.105744711 | 7.37E-05 | 0.004080647 | 0.357768319 |
| HTN | LEC | *COLCA1* | -2.231640113 | 5.58E-06 | 0.000552257 | 0.027081066 |
| HTN | LEC | *HRCT1* | 1.909526476 | 0.000335249 | 0.010350392 | 1 |
| HTN | LEC | *SRGAP3* | 1.032144859 | 0.001343616 | 0.024600852 | 1 |
| HTN | LEC | *GIMAP5* | 0.851779044 | 0.009652727 | 0.04965591 | 1 |
| HTN | LEC | *CPNE4* | -1.146647431 | 0.004821177 | 0.038101652 | 1 |
| HTN | LEC | *C20orf96* | -2.302126171 | 0.006525284 | 0.04230219 | 1 |
| HTN | LEC | *SULT1A1* | -2.080665127 | 0.000616828 | 0.015347934 | 1 |
| HTN | LEC | *CACNA1H* | -1.521432654 | 0.004752719 | 0.038101652 | 1 |
| HTN | LEC | *SULF2* | 1.319180776 | 0.000286001 | 0.009646036 | 1 |
| HTN | LEC | *HLA-DQA1* | -1.079869911 | 0.00227538 | 0.032281125 | 1 |
| HTN | LEC | *AP2A1* | -2.449336231 | 0.001441647 | 0.025435888 | 1 |
| HTN | LEC | *PCDHB16* | 1.026141916 | 0.008971015 | 0.048458848 | 1 |
| HTN | LEC | *HIST1H3E* | 1.615633726 | 7.26E-06 | 0.00062871 | 0.035207754 |

| | | | | | | |
|---|---|---|---|---|---|---|
| HTN | LEC | *ZNF100* | -2.658257347 | 8.81E-05 | 0.004498209 | 0.427329819 |
| HTN | LEC | *ZNF420* | 1.503178495 | 0.006105022 | 0.041544972 | 1 |
| HTN | LEC | *PIWIL2* | -1.169879625 | 0.007347206 | 0.043848273 | 1 |
| HTN | LEC | *ZNF517* | -4.283665986 | 0.000588985 | 0.014979953 | 1 |
| HTN | LEC | *GSTK1* | -2.305847405 | 0.001078824 | 0.021719723 | 1 |
| HTN | LEC | *MFAP5* | -2.0687862 | 4.37E-06 | 0.000471516 | 0.021218221 |
| HTN | LEC | *RPS26* | -1.554787949 | 1.17E-05 | 0.000919414 | 0.05700364 |
| HTN | LEC | *AC122129.1* | -3.228457818 | 0.001449018 | 0.025453509 | 1 |
| HTN | LEC | *SGTB* | -2.704528244 | 0.003031711 | 0.036053579 | 1 |
| HTN | LEC | *DLGAP2* | 1.268392715 | 0.001787425 | 0.028199053 | 1 |
| HTN | LEC | *AGAP9* | -3.964828227 | 0.000159389 | 0.006666868 | 0.773356711 |
| HTN | LEC | *ADH4* | -1.169054326 | 0.000407793 | 0.011847983 | 1 |
| HTN | LEC | *AC135178.1* | 4.966882469 | 0.004146315 | 0.03770947 | 1 |
| HTN | LEC | *SVIP* | 1.454605193 | 0.007080367 | 0.043229876 | 1 |
| HTN | LEC | *RPS6KL1* | 2.203749903 | 9.94E-05 | 0.004821914 | 0.482191395 |
| HTN | LEC | *TMEM116* | -2.310538951 | 3.43E-15 | 5.54E-12 | 1.66E-11 |
| HTN | LEC | *SH3BGRL2* | 0.671306336 | 0.006364455 | 0.041983118 | 1 |
| HTN | LEC | *HLA-DRB5* | -0.474096426 | 2.71E-05 | 0.001804175 | 0.131704756 |
| HTN | LEC | *TUSC1* | -4.160403528 | 0.008216399 | 0.046470093 | 1 |
| HTN | LEC | *ANKRD13B* | -2.034584807 | 0.00477421 | 0.038101652 | 1 |
| HTN | LEC | *PPP1R14C* | -3.154830875 | 0.004001568 | 0.03770947 | 1 |
| HTN | LEC | *GPATCH3* | -2.065500116 | 0.003862232 | 0.03770947 | 1 |
| HTN | LEC | *PNP* | -0.596149031 | 0.001748931 | 0.028192068 | 1 |
| HTN | LEC | *HMGN2* | -1.36727252 | 0.007720328 | 0.045044321 | 1 |
| HTN | LEC | *DENND4B* | -3.197816935 | 1.88E-05 | 0.001359283 | 0.091071974 |
| HTN | LEC | *ITPRIPL1* | -0.795732436 | 0.003997032 | 0.03770947 | 1 |
| HTN | LEC | *PRMT6* | -3.557547267 | 0.001786584 | 0.028199053 | 1 |
| HTN | LEC | *SHISA4* | -5.174762717 | 1.82E-06 | 0.000226628 | 0.008838487 |
| HTN | LEC | *MFAP3L* | 4.094468632 | 0.004009912 | 0.03770947 | 1 |
| HTN | LEC | *SMG5* | -2.272462127 | 0.004071762 | 0.03770947 | 1 |
| HTN | LEC | *TGM2* | 1.203136414 | 0.00681959 | 0.043025132 | 1 |
| HTN | LEC | *AL590822.1* | -2.125598271 | 0.000113445 | 0.005120678 | 0.550436327 |
| HTN | LEC | *SAMD5* | -0.738557233 | 0.009693637 | 0.049770928 | 1 |
| HTN | LEC | *GPR52* | -15.36006495 | 0.00156909 | 0.026713076 | 1 |
| HTN | LEC | *RBM20* | 0.928063101 | 0.003428726 | 0.03770947 | 1 |
| HTN | LEC | *IFIT1B* | -8.684964451 | 0.007773087 | 0.045059762 | 1 |
| HTN | LEC | *LIPN* | -2.27097582 | 0.004774847 | 0.038101652 | 1 |
| HTN | LEC | *INPP5B* | -0.842870937 | 0.006538879 | 0.04230219 | 1 |
| HTN | LEC | *C2orf72* | 0.947126824 | 0.003402119 | 0.03770947 | 1 |
| HTN | LEC | *LINC00619* | -8.791933372 | 0.004709481 | 0.038101652 | 1 |
| HTN | LEC | *TCEA3* | -1.290712992 | 0.009392746 | 0.049233755 | 1 |
| HTN | LEC | *PRRT1* | 1.193424166 | 0.000558179 | 0.01448281 | 1 |
| HTN | LEC | *SMIM5* | -2.087349104 | 0.00754972 | 0.044401503 | 1 |
| HTN | LEC | *SKIV2L* | -0.771554615 | 0.006297116 | 0.041983118 | 1 |
| HTN | LEC | *MICB* | -0.363306461 | 0.006875522 | 0.043025132 | 1 |
| HTN | LEC | *HLA-C* | -1.715812032 | 1.25E-07 | 2.56E-05 | 0.000606224 |
| HTN | LEC | *POU5F1* | -0.395893232 | 0.002662582 | 0.034728081 | 1 |
| HTN | LEC | *PSORS1C2* | 1.2586058 | 0.00617332 | 0.041742342 | 1 |
| HTN | LEC | *MALRD1* | -1.844014057 | 0.005350644 | 0.039635608 | 1 |
| HTN | LEC | *PCDHA4* | -2.143512386 | 0.00923407 | 0.048822112 | 1 |

| | | | | | | |
|---|---|---|---|---|---|---|
| HTN | LEC | *PKHD1L1* | 0.454250929 | 0.004094517 | 0.03770947 | 1 |
| HTN | LEC | *SLFN12L* | -1.570948664 | 0.000419531 | 0.011973896 | 1 |
| HTN | LEC | *GOLGA6L10* | -3.813638309 | 0.001323192 | 0.024600852 | 1 |
| HTN | LEC | *MT1A* | -1.70817214 | 0.006633859 | 0.042537887 | 1 |
| HTN | LEC | *MT1M* | -2.073417392 | 0.006986204 | 0.043126036 | 1 |
| HTN | LEC | *KLRC2* | -1.614486363 | 0.009235084 | 0.048822112 | 1 |
| HTN | LEC | *C1QTNF9B* | 7.688859231 | 0.004743879 | 0.038101652 | 1 |
| HTN | LEC | *TAS2R14* | 1.216753682 | 0.000582657 | 0.014957952 | 1 |
| HTN | LEC | *C17orf51* | -1.98359514 | 0.004651959 | 0.038101652 | 1 |
| HTN | LEC | *AC022498.1* | -4.5565718 | 0.008758636 | 0.048214523 | 1 |
| HTN | LEC | *GIMAP1* | 1.081855266 | 0.006802373 | 0.043025132 | 1 |
| HTN | LEC | *SUPT4H1* | 5.885773292 | 0.002383651 | 0.033329898 | 1 |
| HTN | LEC | *ARHGAP19* | -2.546441804 | 0.004438129 | 0.038049106 | 1 |
| HTN | LEC | *LBH* | -1.426978115 | 0.004106744 | 0.03770947 | 1 |
| HTN | LEC | *NCKIPSD* | -1.324022924 | 0.005174059 | 0.039042822 | 1 |
| HTN | LEC | *ATF6B* | 3.381739805 | 2.29E-05 | 0.001587564 | 0.111129499 |
| HTN | LEC | *SLC23A3* | 1.105019312 | 0.007135195 | 0.04338446 | 1 |
| HTN | LEC | *LTB4R* | -4.731555091 | 5.29E-06 | 0.000534371 | 0.025649815 |
| HTN | LEC | *REPIN1* | -2.13125693 | 0.006502158 | 0.04230219 | 1 |
| HTN | LEC | *FIS1* | -1.450878653 | 0.002892211 | 0.035707402 | 1 |
| HTN | LEC | *GNAT3* | 0.142187409 | 0.003618338 | 0.03770947 | 1 |
| HTN | LEC | *CPEB1* | -1.007147381 | 0.009443787 | 0.049355417 | 1 |
| HTN | LEC | *EML6* | -1.323644898 | 0.00876447 | 0.048214523 | 1 |
| HTN | LEC | *AC005082.1* | -8.044241365 | 0.000195633 | 0.007415698 | 0.949209379 |
| HTN | LEC | *C5orf49* | -8.393112521 | 0.006103428 | 0.041544972 | 1 |
| HTN | LEC | *TSTD1* | -2.185732209 | 0.002578649 | 0.03412855 | 1 |
| HTN | LEC | *PAM16* | -1.920322656 | 0.002252722 | 0.032053396 | 1 |
| HTN | LEC | *FAM228B* | -1.23278713 | 0.002049082 | 0.030780632 | 1 |
| HTN | LEC | *AP4M1* | -2.084431569 | 8.38E-05 | 0.004451906 | 0.406443049 |
| HTN | LEC | *APOL6* | -1.32968132 | 0.009390872 | 0.049233755 | 1 |
| HTN | LEC | *TIAF1* | 3.452583829 | 2.45E-05 | 0.001675863 | 0.118986241 |
| HTN | LEC | *ADRA2B* | -3.4597159 | 0.002306918 | 0.032633141 | 1 |
| HTN | LEC | *EXOSC6* | -4.202817861 | 4.46E-05 | 0.002707102 | 0.216568176 |
| HTN | LEC | *LINC01133* | -1.308145708 | 0.004229385 | 0.03770947 | 1 |
| HTN | LEC | *LINC01117* | 0.444725508 | 0.003501839 | 0.03770947 | 1 |
| HTN | LEC | *LINC00184* | -22.58622806 | 0.003966236 | 0.03770947 | 1 |
| HTN | LEC | *HAR1A* | -11.58624822 | 0.001797867 | 0.028230591 | 1 |
| HTN | LEC | *LINC00937* | -1.041167961 | 0.005996743 | 0.041329827 | 1 |
| HTN | LEC | *FAM138B* | -12.97648029 | 0.003912544 | 0.03770947 | 1 |
| HTN | LEC | *LINC00578* | -3.530949318 | 0.000337127 | 0.010350392 | 1 |
| HTN | LEC | *CASC20* | -1.382768269 | 0.003687934 | 0.03770947 | 1 |
| HTN | LEC | *LINC00342* | 3.412787298 | 0.003899577 | 0.03770947 | 1 |
| HTN | LEC | *LINC00702* | -1.660623793 | 0.00541019 | 0.039954704 | 1 |
| HTN | LEC | *AC019117.2* | -8.473175726 | 0.000352003 | 0.010542716 | 1 |
| HTN | LEC | *LINC00987* | 1.377567872 | 0.008744221 | 0.048214523 | 1 |
| HTN | LEC | *HLA-DQA2* | 0.505065583 | 0.001183945 | 0.023387907 | 1 |
| HTN | LEC | *LINC01143* | -4.328693967 | 0.004821602 | 0.038101652 | 1 |
| HTN | LEC | *LINC00211* | -5.083470746 | 0.000186845 | 0.007313544 | 0.906573868 |
| HTN | LEC | *TXNDC5* | 3.294325859 | 0.003620264 | 0.03770947 | 1 |
| HTN | LEC | *TNFSF12* | 3.937874469 | 7.94E-07 | 0.000121773 | 0.003852307 |

| | | | | | | |
|---|---|---|---|---|---|---|
| HTN | LEC | *CDRT4* | 3.150742795 | 0.002543877 | 0.033816145 | 1 |
| HTN | LEC | *C1orf226* | -2.951934079 | 0.002908253 | 0.035749424 | 1 |
| HTN | LEC | *C1QTNF9* | 2.760958362 | 0.006317936 | 0.041983118 | 1 |
| HTN | LEC | *PNMA2* | -1.799720092 | 0.000201351 | 0.007515032 | 0.976954107 |
| HTN | LEC | *KRBOX1* | -14.40377481 | 0.003696437 | 0.03770947 | 1 |
| HTN | LEC | *TDGF1* | -1.507052179 | 0.002848642 | 0.035259216 | 1 |
| HTN | LEC | *ATP5O* | -2.56344684 | 0.00138139 | 0.02473249 | 1 |
| HTN | LEC | *C8orf58* | 1.385272101 | 0.003881626 | 0.03770947 | 1 |
| HTN | LEC | *HOGA1* | -1.853037248 | 0.003814211 | 0.03770947 | 1 |
| HTN | LEC | *LINC00960* | -0.536685507 | 0.002928494 | 0.035805527 | 1 |
| HTN | LEC | *DECR2* | -3.769774964 | 0.009719483 | 0.049850881 | 1 |
| HTN | LEC | *ERVW-1* | -2.918589066 | 0.004413073 | 0.038016166 | 1 |
| HTN | LEC | *STON1* | -1.616558287 | 0.009471648 | 0.049369727 | 1 |
| HTN | LEC | *VSIG8* | -1.996172998 | 0.000528055 | 0.014000666 | 1 |
| HTN | LEC | *FGF14-IT1* | -2.736065371 | 0.004152299 | 0.03770947 | 1 |
| HTN | LEC | *WFDC6* | -5.679667168 | 0.000129631 | 0.005615814 | 0.628971154 |
| HTN | LEC | *UPK3B* | -1.607935631 | 0.003948464 | 0.03770947 | 1 |
| HTN | LEC | *PLA2G4B* | 3.46614548 | 0.005447797 | 0.040110336 | 1 |
| HTN | LEC | *LEFTY1* | -16.2085186 | 0.001689178 | 0.02782089 | 1 |
| HTN | LEC | *APOBEC3D* | -5.662777404 | 0.005660279 | 0.040990555 | 1 |
| HTN | LEC | *RAB30-AS1* | -1.836739512 | 0.005542927 | 0.04048889 | 1 |
| HTN | LEC | *LINC00926* | -3.087432351 | 0.004997595 | 0.038529615 | 1 |
| HTN | LEC | *CCDC153* | -1.363650897 | 0.004362469 | 0.03788762 | 1 |
| HTN | LEC | *AL357673.1* | 0.258072784 | 0.003844185 | 0.03770947 | 1 |
| HTN | LEC | *LINC01060* | 0.402280238 | 0.002497888 | 0.033479985 | 1 |
| HTN | LEC | *TMEM158* | -3.556120764 | 0.006104884 | 0.041544972 | 1 |
| HTN | LEC | *SMIM20* | -2.111322957 | 0.008787477 | 0.048231719 | 1 |
| HTN | LEC | *LINC00989* | -1.033374053 | 0.005113237 | 0.038825391 | 1 |
| HTN | LEC | *LINC01094* | -1.378990075 | 0.004857155 | 0.038211862 | 1 |
| HTN | LEC | *PCDHGB2* | -7.182719817 | 0.001595998 | 0.026981824 | 1 |
| HTN | LEC | *LYN* | 0.877425828 | 0.005853919 | 0.041169714 | 1 |
| HTN | LEC | *TAS2R31* | -1.734607698 | 0.001536061 | 0.026428964 | 1 |
| HTN | LEC | *LINC00639* | -2.432789068 | 0.001625651 | 0.027292929 | 1 |
| HTN | LEC | *ITGB3* | 0.736812089 | 0.009397567 | 0.049233755 | 1 |
| HTN | LEC | *AC083843.1* | -22.3343898 | 0.004088154 | 0.03770947 | 1 |
| HTN | LEC | *PWRN1* | -2.875973276 | 0.000154702 | 0.006527091 | 0.750615452 |
| HTN | LEC | *LINC00672* | 6.677074143 | 0.00955274 | 0.049625152 | 1 |
| HTN | LEC | *DYNLL2* | -2.870382017 | 0.009287498 | 0.048875208 | 1 |
| HTN | LEC | *NDUFA7* | -5.54967064 | 0.004805837 | 0.038101652 | 1 |
| HTN | LEC | *SCGB1B2P* | 2.490213225 | 0.007346637 | 0.043848273 | 1 |
| HTN | LEC | *TRABD2B* | 1.700312477 | 0.006424906 | 0.042146654 | 1 |
| HTN | LEC | *AP000347.2* | 1.804376209 | 0.008710991 | 0.048138645 | 1 |
| HTN | LEC | *DOC2B* | 0.476198627 | 0.004237664 | 0.03770947 | 1 |
| HTN | LEC | *GRIN2B* | 1.278425302 | 0.001526545 | 0.026428776 | 1 |
| HTN | macrophage | *SEMA3F* | -1.172269385 | 0.000589144 | 0.021988661 | 1 |
| HTN | macrophage | *RBM6* | -0.715922249 | 2.77E-06 | 0.000345199 | 0.013462774 |
| HTN | macrophage | *CD79B* | -1.456328008 | 0.000146035 | 0.007961359 | 0.70856093 |
| HTN | macrophage | *CEP68* | 1.333576651 | 0.000517843 | 0.02059488 | 1 |
| HTN | macrophage | *SDCCAG8* | 1.480679742 | 7.23E-07 | 0.000116902 | 0.003507071 |
| HTN | macrophage | *ITIH4* | 0.325386495 | 0.000596747 | 0.0221024 | 1 |

| | | | | | | |
|---|---|---|---|---|---|---|
| HTN | macrophage | *PIGV* | -0.890709122 | 9.62E-06 | 0.000952174 | 0.046656544 |
| HTN | macrophage | *UHRF1BP1* | 1.208001097 | 1.00E-06 | 0.000152022 | 0.004864691 |
| HTN | macrophage | *TIE1* | -1.221854969 | 3.68E-05 | 0.002697588 | 0.178536954 |
| HTN | macrophage | *POLR1A* | -1.017031724 | 0.000220667 | 0.010626898 | 1 |
| HTN | macrophage | *PLOD1* | 1.811180617 | 0.000726106 | 0.025164774 | 1 |
| HTN | macrophage | *AAMDC* | -1.054352879 | 2.67E-05 | 0.002039261 | 0.129510548 |
| HTN | macrophage | *YPEL3* | 1.150764506 | 0.001510931 | 0.043871959 | 1 |
| HTN | macrophage | *MUL1* | -0.759922279 | 0.001555398 | 0.044392897 | 1 |
| HTN | macrophage | *SCD* | -1.313752359 | 1.42E-05 | 0.001353025 | 0.069004273 |
| HTN | macrophage | *SETD1A* | 2.344877845 | 6.90E-05 | 0.004236389 | 0.334674763 |
| HTN | macrophage | *MARCH2* | 1.329720589 | 7.66E-07 | 0.00011989 | 0.003716586 |
| HTN | macrophage | *BIRC7* | -1.119610842 | 0.000714498 | 0.02494062 | 1 |
| HTN | macrophage | *MANBAL* | 1.06061134 | 0.001146051 | 0.035418081 | 1 |
| HTN | macrophage | *DNTTIP1* | -0.832925594 | 0.001040494 | 0.033139232 | 1 |
| HTN | macrophage | *POLI* | -0.413417334 | 0.000112333 | 0.006488553 | 0.545038487 |
| HTN | macrophage | *MTHFSD* | 0.575265678 | 0.000466288 | 0.01885358 | 1 |
| HTN | macrophage | *PYCARD* | 0.790680763 | 0.00190561 | 0.049709794 | 1 |
| HTN | macrophage | *ICAM5* | -0.894508602 | 0.001234066 | 0.037190614 | 1 |
| HTN | macrophage | *TAF6* | -0.754408313 | 0.000834822 | 0.028128875 | 1 |
| HTN | macrophage | *MEST* | -0.720862512 | 0.000103102 | 0.00602714 | 0.500252629 |
| HTN | macrophage | *RAD51C* | -1.01136361 | 0.001547194 | 0.044392897 | 1 |
| HTN | macrophage | *NAGLU* | 2.578465294 | 0.000157591 | 0.008243172 | 0.764632114 |
| HTN | macrophage | *RNF130* | 1.05278333 | 0.000421224 | 0.017468185 | 1 |
| HTN | macrophage | *PCCB* | 1.669484348 | 3.25E-07 | 6.30E-05 | 0.001576094 |
| HTN | macrophage | *MORN1* | -0.940665938 | 0.00162711 | 0.045899644 | 1 |
| HTN | macrophage | *FKBP15* | 0.741331222 | 0.001034516 | 0.033139232 | 1 |
| HTN | macrophage | *TBX2* | -1.083773822 | 1.63E-08 | 5.26E-06 | 7.89E-05 |
| HTN | macrophage | *TEX14* | 1.551111134 | 6.77E-06 | 0.000687149 | 0.032848708 |
| HTN | macrophage | *MAPK8IP1* | -2.607828124 | 1.90E-05 | 0.001558519 | 0.091952637 |
| HTN | macrophage | *WFDC3* | -0.310182127 | 0.000173367 | 0.008671929 | 0.841177091 |
| HTN | macrophage | *BTN2A2* | 0.699180108 | 1.19E-06 | 0.000175111 | 0.005778671 |
| HTN | macrophage | *SNRPC* | -1.114938108 | 0.001501079 | 0.043871959 | 1 |
| HTN | macrophage | *ZNF391* | -0.418879368 | 1.71E-05 | 0.001502302 | 0.083110603 |
| HTN | macrophage | *TSKS* | -1.254640261 | 0.000566262 | 0.021298466 | 1 |
| HTN | macrophage | *TRAP1* | 0.682794256 | 2.44E-05 | 0.001940448 | 0.118367326 |
| HTN | macrophage | *ABHD8* | -1.425378762 | 1.65E-05 | 0.001480593 | 0.079951999 |
| HTN | macrophage | *RIPK3* | 0.640189158 | 0.000506223 | 0.02029911 | 1 |
| HTN | macrophage | *ADCY4* | 1.314604056 | 0.000404323 | 0.016911848 | 1 |
| HTN | macrophage | *ATP8B3* | -0.513722783 | 0.001207212 | 0.036608694 | 1 |
| HTN | macrophage | *PGPEP1* | 1.079420581 | 0.000304893 | 0.013327409 | 1 |
| HTN | macrophage | *HAUS8* | -1.785260868 | 5.19E-07 | 8.95E-05 | 0.002518676 |
| HTN | macrophage | *GCH1* | 0.776538143 | 0.00075874 | 0.026109251 | 1 |
| HTN | macrophage | *IMPA1* | 0.743458256 | 0.001879522 | 0.04929427 | 1 |
| HTN | macrophage | *ACP2* | 0.906357539 | 2.56E-05 | 0.002000644 | 0.124039956 |
| HTN | macrophage | *NMT1* | 1.069569694 | 3.56E-06 | 0.00042111 | 0.017265525 |
| HTN | macrophage | *IL11RA* | -0.956319904 | 0.001818332 | 0.049246394 | 1 |
| HTN | macrophage | *SLC37A4* | 1.338765769 | 5.43E-06 | 0.000598823 | 0.02634821 |
| HTN | macrophage | *PPP2R1B* | -0.705730653 | 6.07E-05 | 0.003929231 | 0.294692332 |
| HTN | macrophage | *NUSAP1* | 2.271154386 | 6.80E-06 | 0.000687149 | 0.032983129 |
| HTN | macrophage | *G3BP2* | 1.297553206 | 0.000924144 | 0.030296953 | 1 |

| | | | | | | |
|---|---|---|---|---|---|---|
| HTN | macrophage | *FBN2* | 0.391602527 | 6.04E-06 | 0.000636739 | 0.029289986 |
| HTN | macrophage | *FITM1* | 2.146662732 | 0.000436337 | 0.017941587 | 1 |
| HTN | macrophage | *RTN1* | 0.866959632 | 6.52E-05 | 0.004054021 | 0.316213615 |
| HTN | macrophage | *ULK3* | 1.880878672 | 0.000880186 | 0.029251127 | 1 |
| HTN | macrophage | *HAPLN3* | -1.048584224 | 0.001519062 | 0.043871959 | 1 |
| HTN | macrophage | *DEF8* | 1.511600737 | 1.49E-06 | 0.000206343 | 0.007222017 |
| HTN | macrophage | *LRRC46* | -0.900311345 | 0.001835647 | 0.049246394 | 1 |
| HTN | macrophage | *SCRN2* | -0.322749199 | 0.000250801 | 0.011589391 | 1 |
| HTN | macrophage | *C18orf8* | -1.908956888 | 4.35E-05 | 0.003061681 | 0.211255988 |
| HTN | macrophage | *SNX27* | 1.818125805 | 0.000233713 | 0.011009454 | 1 |
| HTN | macrophage | *CREB3L4* | -1.833356344 | 3.85E-07 | 7.18E-05 | 0.001867761 |
| HTN | macrophage | *CEP170* | 1.473127328 | 0.001596422 | 0.045297315 | 1 |
| HTN | macrophage | *CISD2* | 1.422225115 | 0.000904271 | 0.029847105 | 1 |
| HTN | macrophage | *ADHFE1* | -0.433823553 | 0.000172112 | 0.008671929 | 0.835086474 |
| HTN | macrophage | *HSD17B12* | -0.758295128 | 1.57E-09 | 8.38E-07 | 7.62E-06 |
| HTN | macrophage | *TNKS1BP1* | 0.865072481 | 1.88E-05 | 0.001558519 | 0.091164092 |
| HTN | macrophage | *TRPT1* | -0.914242389 | 0.000523114 | 0.020635347 | 1 |
| HTN | macrophage | *TBX6* | 0.330701491 | 0.001747481 | 0.048356958 | 1 |
| HTN | macrophage | *PRDM8* | -1.546534506 | 0.000151661 | 0.008086367 | 0.735859431 |
| HTN | macrophage | *NEIL2* | 1.694284919 | 9.92E-10 | 7.37E-07 | 4.81E-06 |
| HTN | macrophage | *PROM2* | -1.476014927 | 0.000562082 | 0.021298466 | 1 |
| HTN | macrophage | *NDUFAF6* | -1.634278839 | 5.85E-05 | 0.003892979 | 0.283627645 |
| HTN | macrophage | *SFXN2* | 0.868400432 | 2.69E-09 | 1.19E-06 | 1.31E-05 |
| HTN | macrophage | *VPS8* | 1.268166327 | 0.001199413 | 0.036600961 | 1 |
| HTN | macrophage | *NTAN1* | 1.431442872 | 0.000562227 | 0.021298466 | 1 |
| HTN | macrophage | *CLEC18A* | -1.026717072 | 4.20E-06 | 0.00048537 | 0.020385523 |
| HTN | macrophage | *SPATA2L* | 1.735496248 | 0.001754086 | 0.048356958 | 1 |
| HTN | macrophage | *SLC5A11* | -0.63459618 | 4.81E-07 | 8.65E-05 | 0.002335151 |
| HTN | macrophage | *S100B* | -0.336821806 | 1.53E-05 | 0.001430971 | 0.074410479 |
| HTN | macrophage | *ZNF208* | -1.006964196 | 0.000338767 | 0.014596875 | 1 |
| HTN | macrophage | *ABCF3* | 1.541523062 | 2.14E-08 | 6.50E-06 | 0.00010395 |
| HTN | macrophage | *MRPL55* | 2.197818503 | 5.85E-10 | 7.10E-07 | 2.84E-06 |
| HTN | macrophage | *LYSMD1* | -0.693142997 | 0.000258323 | 0.011605403 | 1 |
| HTN | macrophage | *DCAF16* | -2.38225682 | 0.000171352 | 0.008671929 | 0.831397735 |
| HTN | macrophage | *LINC01116* | -1.645720428 | 0.001783881 | 0.048625798 | 1 |
| HTN | macrophage | *LMOD1* | -1.009315344 | 0.00062445 | 0.022610697 | 1 |
| HTN | macrophage | *RFT1* | -1.111976183 | 0.000117247 | 0.006692732 | 0.568882243 |
| HTN | macrophage | *CDC25A* | -2.754867256 | 4.94E-05 | 0.00342117 | 0.239481912 |
| HTN | macrophage | *ZNF589* | -1.771729458 | 1.73E-09 | 8.38E-07 | 8.38E-06 |
| HTN | macrophage | *RNF123* | 1.681427562 | 2.08E-10 | 3.36E-07 | 1.01E-06 |
| HTN | macrophage | *MST1R* | 0.633061611 | 0.001878127 | 0.04929427 | 1 |
| HTN | macrophage | *ERAP1* | -1.752669361 | 2.48E-06 | 0.000325076 | 0.012027808 |
| HTN | macrophage | *ERAP2* | 0.410487879 | 0.001044992 | 0.033139232 | 1 |
| HTN | macrophage | *RPP25L* | -1.344097621 | 0.000221211 | 0.010626898 | 1 |
| HTN | macrophage | *ARL5B* | -2.459534117 | 9.64E-05 | 0.00570572 | 0.46786906 |
| HTN | macrophage | *TRIM66* | -0.465158763 | 1.57E-05 | 0.001440335 | 0.076337762 |
| HTN | macrophage | *RIMKLB* | -1.558820505 | 1.43E-07 | 3.15E-05 | 0.00069368 |
| HTN | macrophage | *ZNF641* | 2.428805459 | 0.000864865 | 0.028940186 | 1 |
| HTN | macrophage | *SRR* | 0.544513362 | 5.86E-05 | 0.003892979 | 0.284187471 |
| HTN | macrophage | *C8orf46* | 0.27570765 | 0.000289888 | 0.01290401 | 1 |

| | | | | | | |
|---|---|---|---|---|---|---|
| HTN | macrophage | *CPT1C* | 0.957784317 | 0.000613895 | 0.0225653 | 1 |
| HTN | macrophage | *RAB24* | -4.101919284 | 3.73E-05 | 0.002697588 | 0.180738385 |
| HTN | macrophage | *INO80E* | -1.388007287 | 8.19E-10 | 7.37E-07 | 3.97E-06 |
| HTN | macrophage | *RGPD8* | -0.245052088 | 0.000978475 | 0.031862829 | 1 |
| HTN | macrophage | *DNAJC18* | -1.143670414 | 2.69E-05 | 0.002039261 | 0.130512729 |
| HTN | macrophage | *TTLL6* | -3.196764312 | 8.48E-08 | 2.06E-05 | 0.000411294 |
| HTN | macrophage | *MRPL36* | 0.559128781 | 0.001841913 | 0.049246394 | 1 |
| HTN | macrophage | *RGS19* | -0.442988584 | 0.000143492 | 0.007911615 | 0.696222085 |
| HTN | macrophage | *NME6* | 2.264488498 | 1.06E-09 | 7.37E-07 | 5.16E-06 |
| HTN | macrophage | *CTSW* | -0.185989548 | 1.73E-05 | 0.001502302 | 0.084128916 |
| HTN | macrophage | *MYEOV* | -2.19000899 | 0.001121228 | 0.03509804 | 1 |
| HTN | macrophage | *DCAKD* | -1.625988342 | 0.001854941 | 0.049246394 | 1 |
| HTN | macrophage | *NOC3L* | -1.959087605 | 0.000663407 | 0.023709307 | 1 |
| HTN | macrophage | *MAP3K11* | 1.289933039 | 0.000664564 | 0.023709307 | 1 |
| HTN | macrophage | *PPP1R14B* | 2.626241168 | 7.31E-05 | 0.004434087 | 0.354726985 |
| HTN | macrophage | *MST1* | -0.582305571 | 5.35E-07 | 8.95E-05 | 0.002595511 |
| HTN | macrophage | *C11orf80* | -1.796776883 | 9.48E-08 | 2.19E-05 | 0.000460148 |
| HTN | macrophage | *GPR160* | 1.186689819 | 0.00081587 | 0.027682531 | 1 |
| HTN | macrophage | *SNX31* | -0.605182498 | 5.80E-06 | 0.000625468 | 0.028146038 |
| HTN | macrophage | *CCNE2* | -1.724986441 | 4.00E-09 | 1.62E-06 | 1.94E-05 |
| HTN | macrophage | *TMEM9B* | 2.221265313 | 9.58E-05 | 0.00570572 | 0.464882237 |
| HTN | macrophage | *CTDNEP1* | 1.782341599 | 1.99E-05 | 0.001608292 | 0.096497546 |
| HTN | macrophage | *ORAI3* | 1.754057127 | 4.33E-08 | 1.12E-05 | 0.000209971 |
| HTN | macrophage | *MTHFR* | -1.892295673 | 0.001095464 | 0.034514234 | 1 |
| HTN | macrophage | *RPP25* | 0.907961683 | 1.80E-07 | 3.64E-05 | 0.000874141 |
| HTN | macrophage | *MPI* | -2.643024769 | 8.83E-16 | 4.28E-12 | 4.28E-12 |
| HTN | macrophage | *TRIM73* | -2.033798176 | 0.000303541 | 0.013327409 | 1 |
| HTN | macrophage | *HYI* | -0.985696434 | 1.55E-07 | 3.27E-05 | 0.000751136 |
| HTN | macrophage | *CTC1* | 1.743322837 | 0.000448585 | 0.01829021 | 1 |
| HTN | macrophage | *SEPHS2* | 1.545517412 | 0.000681381 | 0.023956964 | 1 |
| HTN | macrophage | *ADO* | 1.737711339 | 1.39E-09 | 8.38E-07 | 6.74E-06 |
| HTN | macrophage | *CLN8* | 0.722197819 | 2.12E-06 | 0.000285457 | 0.010276445 |
| HTN | macrophage | *FES* | -1.112729651 | 3.87E-05 | 0.00276379 | 0.187937708 |
| HTN | macrophage | *ODF3L1* | -1.921894814 | 0.001253271 | 0.037536243 | 1 |
| HTN | macrophage | *ZNF320* | 0.406491946 | 0.000168456 | 0.008671929 | 0.817349418 |
| HTN | macrophage | *ALDH1A3* | -0.57070471 | 0.000555228 | 0.021298466 | 1 |
| HTN | macrophage | *A3GALT2* | -0.684481774 | 3.52E-05 | 0.00262821 | 0.170833669 |
| HTN | macrophage | *NUTM2A* | -0.986614682 | 6.31E-05 | 0.004029095 | 0.306211233 |
| HTN | macrophage | *CDK10* | -0.62068007 | 0.000158 | 0.008243172 | 0.766615035 |
| HTN | macrophage | *KLHL17* | 0.979108699 | 0.000619769 | 0.022609907 | 1 |
| HTN | macrophage | *FBF1* | -1.506232392 | 1.47E-06 | 0.000206343 | 0.007144237 |
| HTN | macrophage | *COLCA1* | -1.199416383 | 5.51E-05 | 0.003767845 | 0.267517024 |
| HTN | macrophage | *HIST1H3E* | 0.899848525 | 4.32E-08 | 1.12E-05 | 0.000209637 |
| HTN | macrophage | *WDR5B* | 1.888358467 | 0.000244837 | 0.011422607 | 1 |
| HTN | macrophage | *ZNF100* | -1.035376766 | 0.00080277 | 0.027429868 | 1 |
| HTN | macrophage | *SULT1A2* | 1.484689339 | 5.97E-05 | 0.003912135 | 0.289498004 |
| HTN | macrophage | *PCDHB11* | -0.426602785 | 0.001285719 | 0.03827183 | 1 |
| HTN | macrophage | *MFAP5* | -0.909815937 | 6.46E-05 | 0.004054021 | 0.313198514 |
| HTN | macrophage | *RPS26* | -0.675740048 | 1.89E-05 | 0.001558519 | 0.091598938 |
| HTN | macrophage | *ZNF311* | 0.681511198 | 0.001022245 | 0.033066228 | 1 |

| | | | | | | |
|---|---|---|---|---|---|---|
| HTN | macrophage | *TMEM116* | -1.017168823 | 5.30E-14 | 1.29E-10 | 2.57E-10 |
| HTN | macrophage | *HLA-DRB5* | -0.162733335 | 0.000546241 | 0.021298466 | 1 |
| HTN | macrophage | *SHISA4* | -7.738089045 | 1.37E-08 | 5.12E-06 | 6.66E-05 |
| HTN | macrophage | *TGM2* | 0.676700812 | 0.000255789 | 0.011598944 | 1 |
| HTN | macrophage | *AL590822.1* | -1.14832173 | 0.00114312 | 0.035418081 | 1 |
| HTN | macrophage | *HSD17B8* | -0.86716624 | 0.000150358 | 0.008086367 | 0.729538827 |
| HTN | macrophage | *PRRT1* | 0.579239185 | 0.000130433 | 0.007358843 | 0.632860456 |
| HTN | macrophage | *LAYN* | -1.118214861 | 0.000670596 | 0.02374987 | 1 |
| HTN | macrophage | *HLA-C* | -0.690081021 | 2.98E-06 | 0.000361769 | 0.014470758 |
| HTN | macrophage | *POU5F1* | -0.189409175 | 0.001158551 | 0.035577775 | 1 |
| HTN | macrophage | *SUPT4H1* | 4.214537789 | 1.29E-05 | 0.001252143 | 0.062607164 |
| HTN | macrophage | *TMEM110* | 1.424242839 | 0.000195183 | 0.009565923 | 0.947026356 |
| HTN | macrophage | *ATF6B* | 2.10353685 | 4.39E-08 | 1.12E-05 | 0.000213216 |
| HTN | macrophage | *PPM1N* | 0.530284126 | 0.000339952 | 0.014596875 | 1 |
| HTN | macrophage | *UBXN2B* | 1.160476309 | 0.001326225 | 0.039236843 | 1 |
| HTN | macrophage | *AC016773.1* | 0.934883289 | 0.000232258 | 0.011009454 | 1 |
| HTN | macrophage | *AP4M1* | -0.872534977 | 0.000254931 | 0.011598944 | 1 |
| HTN | macrophage | *TIAF1* | 1.631107264 | 4.62E-06 | 0.000521466 | 0.022423059 |
| HTN | macrophage | *EXOSC6* | -1.728272966 | 0.00035041 | 0.014913946 | 1 |
| HTN | macrophage | *AL049840.1* | -1.681215004 | 0.000194305 | 0.009565923 | 0.942766613 |
| HTN | macrophage | *GPX1* | 0.767275656 | 0.001921719 | 0.049861931 | 1 |
| HTN | macrophage | *HLA-DQA2* | 0.335257084 | 2.68E-06 | 0.000342126 | 0.013000803 |
| HTN | macrophage | *TNFSF12* | 1.952731919 | 1.48E-08 | 5.14E-06 | 7.19E-05 |
| HTN | macrophage | *PNMA2* | -1.151763229 | 0.001734138 | 0.04835655 | 1 |
| HTN | macrophage | *ZNF709* | -5.135026043 | 0.001359928 | 0.039990117 | 1 |
| HTN | macrophage | *WFDC6* | -2.348300811 | 0.000390827 | 0.016489503 | 1 |
| HTN | macrophage | *PLA2G4B* | 1.969116796 | 0.000564865 | 0.021298466 | 1 |
| HTN | macrophage | *JMJD7* | 0.554375979 | 0.001765705 | 0.048402261 | 1 |
| HTN | macrophage | *HMBS* | 1.279304136 | 0.001679805 | 0.047112206 | 1 |
| HTN | macrophage | *LINC00638* | 1.626893678 | 0.000135797 | 0.00757342 | 0.658887499 |
| HTN | macrophage | *PWRN1* | -1.173236664 | 0.001857397 | 0.049246394 | 1 |
| HTN | mast_cell | *SCD* | -5.386805604 | 0.000324049 | 0.047644997 | 1 |
| HTN | mast_cell | *TBX2* | -3.00868535 | 0.000314978 | 0.047644997 | 1 |
| HTN | mast_cell | *TEX14* | 9.839984213 | 3.14E-06 | 0.001906117 | 0.01524894 |
| HTN | mast_cell | *BTN2A2* | 4.066035049 | 1.05E-05 | 0.003655256 | 0.051173579 |
| HTN | mast_cell | *HAUS8* | -10.31075865 | 4.06E-07 | 0.000492947 | 0.001971787 |
| HTN | mast_cell | *FBN2* | 2.240781106 | 9.48E-05 | 0.020909906 | 0.460017942 |
| HTN | mast_cell | *HSD17B12* | -2.714062093 | 5.72E-06 | 0.002523803 | 0.027761835 |
| HTN | mast_cell | *NEIL2* | 7.237114764 | 1.51E-06 | 0.00104914 | 0.00734398 |
| HTN | mast_cell | *CLEC18A* | -6.858180523 | 4.63E-06 | 0.002495232 | 0.022457085 |
| HTN | mast_cell | *SLC5A11* | -3.921966542 | 6.13E-07 | 0.000594655 | 0.002973274 |
| HTN | mast_cell | *S100B* | -1.905852904 | 6.39E-05 | 0.01550102 | 0.310020392 |
| HTN | mast_cell | *CDC25A* | -13.87455268 | 5.17E-05 | 0.013200561 | 0.250810659 |
| HTN | mast_cell | *ZNF589* | -4.527441109 | 0.000178978 | 0.033399985 | 0.8683996 |
| HTN | mast_cell | *RNF123* | 5.367628032 | 5.54E-06 | 0.002523803 | 0.026875148 |
| HTN | mast_cell | *SRR* | 3.028142225 | 0.00027133 | 0.042467516 | 1 |
| HTN | mast_cell | *MYO7B* | 6.253883138 | 0.000223077 | 0.037323057 | 1 |
| HTN | mast_cell | *TTLL6* | -9.514603631 | 0.000127121 | 0.025699612 | 0.616790694 |
| HTN | mast_cell | *RGS19* | -4.597277509 | 2.73E-08 | 9.40E-05 | 0.000132224 |
| HTN | mast_cell | *CTSW* | -1.105605182 | 4.17E-05 | 0.011244905 | 0.202408296 |

| | | | | | | |
|---|---|---|---|---|---|---|
| HTN | mast_cell | *SNX31* | -3.230568847 | 2.72E-05 | 0.008810403 | 0.13215605 |
| HTN | mast_cell | *SEZ6L2* | 5.869606761 | 0.000202202 | 0.035038664 | 0.98108258 |
| HTN | mast_cell | *ORAI3* | 8.502656649 | 6.90E-06 | 0.002787891 | 0.03345469 |
| HTN | mast_cell | *RPP25* | 4.772005383 | 8.50E-06 | 0.003172473 | 0.041242144 |
| HTN | mast_cell | *ADO* | 4.82965967 | 7.79E-05 | 0.017992775 | 0.377848275 |
| HTN | mast_cell | *A3GALT2* | -3.681935373 | 0.000193303 | 0.034737242 | 0.937905537 |
| HTN | mast_cell | *NUTM2A* | -5.840751399 | 0.000147657 | 0.028657236 | 0.716430889 |
| HTN | mast_cell | *HIST1H3E* | 5.149952618 | 2.37E-07 | 0.000383936 | 0.001151807 |
| HTN | mast_cell | *RPS26* | -2.930222395 | 0.000347747 | 0.049625479 | 1 |
| HTN | mast_cell | *TMEM116* | -3.563303477 | 3.87E-08 | 9.40E-05 | 0.000187949 |
| HTN | mast_cell | *HLA-DRB5* | -1.339401244 | 2.93E-05 | 0.008886585 | 0.142185362 |
| HTN | mast_cell | *HLA-DQA2* | 1.803615469 | 3.79E-05 | 0.010823559 | 0.184000506 |
| HTN | mast_cell | *TNFSF12* | 10.47868331 | 9.39E-07 | 0.000759379 | 0.004556272 |
| HTN | mast_cell | *ZNF709* | -103.8528429 | 0.000106799 | 0.022530049 | 0.518191136 |
| HTN | mast_cell | *PLA2G4B* | 12.59770248 | 0.000258124 | 0.041747334 | 1 |
| HTN | mesothelium | *SEMA3F* | -0.393133594 | 0.001408041 | 0.027999241 | 1 |
| HTN | mesothelium | *RAD52* | -0.562382099 | 0.002324899 | 0.038951294 | 1 |
| HTN | mesothelium | *M6PR* | -0.416819313 | 0.00018651 | 0.006198269 | 0.904947219 |
| HTN | mesothelium | *RBM6* | -0.217529127 | 6.52E-07 | 5.65E-05 | 0.003161783 |
| HTN | mesothelium | *GGNBP2* | -1.028233652 | 0.000561452 | 0.014646055 | 1 |
| HTN | mesothelium | *CD79B* | -0.399518908 | 0.000323138 | 0.009444976 | 1 |
| HTN | mesothelium | *ST7L* | -0.230831024 | 0.001274402 | 0.026312336 | 1 |
| HTN | mesothelium | *NADK* | -1.316162953 | 5.10E-05 | 0.002115051 | 0.247460997 |
| HTN | mesothelium | *CLCN6* | -0.551365573 | 0.000167697 | 0.005770665 | 0.813663769 |
| HTN | mesothelium | *CEP68* | 0.654331854 | 2.59E-06 | 0.000187869 | 0.012587207 |
| HTN | mesothelium | *RNASET2* | -0.235426272 | 0.000283989 | 0.008443706 | 1 |
| HTN | mesothelium | *SH3YL1* | -0.585339282 | 0.000738912 | 0.01774853 | 1 |
| HTN | mesothelium | *SPATA7* | -0.780561573 | 0.00248906 | 0.040256402 | 1 |
| HTN | mesothelium | *ANAPC4* | 0.653732004 | 0.000697475 | 0.016920744 | 1 |
| HTN | mesothelium | *SDCCAG8* | 0.493110274 | 2.70E-07 | 2.79E-05 | 0.001311268 |
| HTN | mesothelium | *ITIH4* | 0.089701758 | 0.000584938 | 0.014808844 | 1 |
| HTN | mesothelium | *PIGV* | -0.258339509 | 3.71E-06 | 0.000257334 | 0.018013389 |
| HTN | mesothelium | *NCKAP1* | -0.893710536 | 0.001297977 | 0.026685528 | 1 |
| HTN | mesothelium | *UHRF1BP1* | 0.328795488 | 4.38E-06 | 0.00028733 | 0.021262426 |
| HTN | mesothelium | *TIE1* | -0.426646192 | 1.25E-05 | 0.000713533 | 0.060650322 |
| HTN | mesothelium | *ATXN3* | -0.25231651 | 0.003652479 | 0.049227298 | 1 |
| HTN | mesothelium | *MTFR1* | -0.639177243 | 0.001262636 | 0.026312336 | 1 |
| HTN | mesothelium | *POLR1A* | -0.326784916 | 4.47E-05 | 0.00195604 | 0.217120471 |
| HTN | mesothelium | *IP6K2* | -0.81902682 | 0.001536966 | 0.029411751 | 1 |
| HTN | mesothelium | *IFT80* | -2.242933746 | 2.73E-08 | 4.73E-06 | 0.000132518 |
| HTN | mesothelium | *EPN2* | -0.401145312 | 0.001018164 | 0.02235354 | 1 |
| HTN | mesothelium | *TRHDE* | -0.452825196 | 0.000598676 | 0.014973072 | 1 |
| HTN | mesothelium | *AP1M1* | -1.266491294 | 0.001263948 | 0.026312336 | 1 |
| HTN | mesothelium | *FDFT1* | 0.685090569 | 9.73E-06 | 0.000589967 | 0.047197361 |
| HTN | mesothelium | *CCNT2* | -0.584730406 | 0.003573491 | 0.048431777 | 1 |
| HTN | mesothelium | *LYRM2* | -0.399143223 | 0.002361699 | 0.039109087 | 1 |
| HTN | mesothelium | *OXCT1* | -0.555268899 | 0.003103287 | 0.044285724 | 1 |
| HTN | mesothelium | *SNX10* | -0.31457652 | 1.97E-07 | 2.08E-05 | 0.000955691 |
| HTN | mesothelium | *AAMDC* | -0.383544437 | 3.35E-06 | 0.00023534 | 0.016238431 |
| HTN | mesothelium | *PPP1R13B* | -0.359761988 | 0.002965357 | 0.043468014 | 1 |

| | | | | | | |
|---|---|---|---|---|---|---|
| HTN | mesothelium | *MAVS* | -0.39854189 | 0.002566033 | 0.041090409 | 1 |
| HTN | mesothelium | *YPEL3* | 0.382322441 | 0.000660704 | 0.016109219 | 1 |
| HTN | mesothelium | *MUL1* | -0.22321249 | 0.000906913 | 0.020756341 | 1 |
| HTN | mesothelium | *PABPC4* | -0.599187757 | 1.50E-07 | 1.77E-05 | 0.000726801 |
| HTN | mesothelium | *RGS17* | 0.365119914 | 0.00214089 | 0.036966544 | 1 |
| HTN | mesothelium | *SCD* | -0.464293889 | 1.53E-07 | 1.77E-05 | 0.000743069 |
| HTN | mesothelium | *SETD1A* | 0.690806912 | 0.000155354 | 0.005422856 | 0.753776933 |
| HTN | mesothelium | *MARCH2* | 0.336457507 | 1.85E-05 | 0.001006728 | 0.089598753 |
| HTN | mesothelium | *MTAP* | -0.615276631 | 0.000212356 | 0.006818931 | 1 |
| HTN | mesothelium | *TRIOBP* | -0.594767486 | 0.000955252 | 0.021358906 | 1 |
| HTN | mesothelium | *THOC5* | -0.262607053 | 0.001320782 | 0.027039801 | 1 |
| HTN | mesothelium | *ACO2* | -0.713439036 | 0.001028828 | 0.022385084 | 1 |
| HTN | mesothelium | *BIRC7* | -0.305450297 | 0.000797928 | 0.018793911 | 1 |
| HTN | mesothelium | *GMEB2* | -1.540968299 | 0.002882893 | 0.04271014 | 1 |
| HTN | mesothelium | *DNTTIP1* | -0.26548947 | 0.00022923 | 0.00717563 | 1 |
| HTN | mesothelium | *POLI* | -0.118734354 | 6.50E-05 | 0.00254341 | 0.315382812 |
| HTN | mesothelium | *PARP4* | -0.696346453 | 0.000978042 | 0.021768168 | 1 |
| HTN | mesothelium | *MGRN1* | 0.558507001 | 0.002789797 | 0.042432893 | 1 |
| HTN | mesothelium | *NME4* | -1.142043836 | 0.000425216 | 0.01192572 | 1 |
| HTN | mesothelium | *MTHFSD* | 0.162003324 | 0.000419898 | 0.011845028 | 1 |
| HTN | mesothelium | *PYCARD* | 0.683633993 | 0.001509362 | 0.029289748 | 1 |
| HTN | mesothelium | *LACTB* | -0.705795452 | 0.000778759 | 0.018431888 | 1 |
| HTN | mesothelium | *SNX16* | 0.951321721 | 0.000191893 | 0.006311896 | 0.931066653 |
| HTN | mesothelium | *LIN7B* | -1.945474075 | 4.10E-08 | 6.63E-06 | 0.000198919 |
| HTN | mesothelium | *CD37* | -0.548433779 | 1.13E-05 | 0.000661425 | 0.054898292 |
| HTN | mesothelium | *ICAM5* | -0.240758478 | 0.001817244 | 0.033316233 | 1 |
| HTN | mesothelium | *TNPO2* | -0.906658994 | 0.000481559 | 0.013053218 | 1 |
| HTN | mesothelium | *TAF6* | -0.222019232 | 0.000395775 | 0.011229829 | 1 |
| HTN | mesothelium | *MEST* | -0.269863001 | 3.78E-05 | 0.001696489 | 0.183220777 |
| HTN | mesothelium | *POU6F2* | -1.127469849 | 6.25E-05 | 0.002484054 | 0.303054559 |
| HTN | mesothelium | *OGN* | -0.295109765 | 0.002810132 | 0.042434092 | 1 |
| HTN | mesothelium | *RPL28* | -0.431046887 | 0.001382059 | 0.027824692 | 1 |
| HTN | mesothelium | *RAD51C* | -0.335456281 | 0.000726569 | 0.017538876 | 1 |
| HTN | mesothelium | *PNPO* | -1.065059548 | 0.000122463 | 0.004401412 | 0.594190617 |
| HTN | mesothelium | *NAGLU* | 0.755437601 | 0.001430522 | 0.028215017 | 1 |
| HTN | mesothelium | *NRXN2* | -3.196280918 | 0.000647469 | 0.01586625 | 1 |
| HTN | mesothelium | *CCND1* | -0.659834404 | 9.31E-08 | 1.22E-05 | 0.000451624 |
| HTN | mesothelium | *KCTD10* | -0.462636762 | 0.002121745 | 0.036898593 | 1 |
| HTN | mesothelium | *VPS29* | -1.079123937 | 0.003076849 | 0.044285724 | 1 |
| HTN | mesothelium | *ACAD10* | -0.788737636 | 2.53E-05 | 0.001240229 | 0.122825493 |
| HTN | mesothelium | *C6orf62* | -0.762887699 | 0.00018559 | 0.006198269 | 0.900483396 |
| HTN | mesothelium | *CUTA* | -0.290915023 | 0.002995921 | 0.043783761 | 1 |
| HTN | mesothelium | *PCCB* | 0.479879105 | 4.34E-07 | 3.83E-05 | 0.002107553 |
| HTN | mesothelium | *NEK4* | -0.472757403 | 0.001039246 | 0.022410765 | 1 |
| HTN | mesothelium | *MORN1* | -0.272510065 | 0.00207162 | 0.03655091 | 1 |
| HTN | mesothelium | *MECR* | -0.656872416 | 0.00157502 | 0.029968614 | 1 |
| HTN | mesothelium | *CAPZA1* | -1.200407483 | 3.85E-07 | 3.73E-05 | 0.001867344 |
| HTN | mesothelium | *DDX59* | -0.351631655 | 0.002706007 | 0.041709335 | 1 |
| HTN | mesothelium | *ACOT2* | 0.08029761 | 0.001771135 | 0.032799801 | 1 |
| HTN | mesothelium | *HELLS* | -0.377879966 | 0.002872948 | 0.04271014 | 1 |

| | | | | | | |
|---|---|---|---|---|---|---|
| HTN | mesothelium | *KANSL1* | -0.735073625 | 0.003457825 | 0.04753618 | 1 |
| HTN | mesothelium | *SNX19* | -0.361969845 | 0.001752269 | 0.032700029 | 1 |
| HTN | mesothelium | *UFM1* | -0.820516902 | 0.002130967 | 0.03692662 | 1 |
| HTN | mesothelium | *ZFP30* | 0.386618736 | 0.000492954 | 0.013214444 | 1 |
| HTN | mesothelium | *TBX2* | -0.302445884 | 1.58E-08 | 2.94E-06 | 7.64E-05 |
| HTN | mesothelium | *TEX14* | 0.517585738 | 3.36E-07 | 3.32E-05 | 0.001629131 |
| HTN | mesothelium | *MAPK8IP1* | -0.87644397 | 2.56E-05 | 0.001240229 | 0.124022919 |
| HTN | mesothelium | *FABP3* | -1.47905577 | 0.000527068 | 0.013898563 | 1 |
| HTN | mesothelium | *CDK2* | -0.27918163 | 0.002608657 | 0.041408821 | 1 |
| HTN | mesothelium | *TTC21B* | -1.019034948 | 0.000154664 | 0.005422856 | 0.750428815 |
| HTN | mesothelium | *WFDC3* | -0.082885713 | 0.000285401 | 0.008443706 | 1 |
| HTN | mesothelium | *BTN2A2* | 0.181674577 | 9.88E-06 | 0.000591948 | 0.047947771 |
| HTN | mesothelium | *SNRPC* | -0.479608182 | 5.12E-06 | 0.000331216 | 0.024841186 |
| HTN | mesothelium | *ZNF391* | -0.112871983 | 4.57E-05 | 0.001970945 | 0.221838302 |
| HTN | mesothelium | *GRK4* | 0.075627078 | 0.002242114 | 0.038037545 | 1 |
| HTN | mesothelium | *UROD* | -0.404197157 | 0.001017854 | 0.02235354 | 1 |
| HTN | mesothelium | *TSKS* | -0.378147318 | 0.000213619 | 0.006818931 | 1 |
| HTN | mesothelium | *TRAP1* | 0.222187614 | 2.51E-06 | 0.000187537 | 0.012189894 |
| HTN | mesothelium | *ABHD8* | -0.451549198 | 2.47E-06 | 0.000186885 | 0.011960657 |
| HTN | mesothelium | *GFER* | -0.382363889 | 0.001669654 | 0.031522026 | 1 |
| HTN | mesothelium | *GAD1* | -0.573158938 | 0.003463656 | 0.04753618 | 1 |
| HTN | mesothelium | *EIF2AK4* | -1.102756066 | 1.40E-07 | 1.77E-05 | 0.000678982 |
| HTN | mesothelium | *ADCY4* | 0.46874554 | 3.13E-05 | 0.001445757 | 0.151804517 |
| HTN | mesothelium | *MAU2* | -1.025142273 | 0.000467503 | 0.012743402 | 1 |
| HTN | mesothelium | *PRKCSH* | -0.626516691 | 0.001360711 | 0.027587831 | 1 |
| HTN | mesothelium | *CDC16* | 0.339335848 | 0.002637124 | 0.041408821 | 1 |
| HTN | mesothelium | *ATP8B3* | -0.153326204 | 0.000461327 | 0.012717957 | 1 |
| HTN | mesothelium | *PGPEP1* | 0.268309553 | 0.001712955 | 0.032089791 | 1 |
| HTN | mesothelium | *TRPM4* | -0.435504836 | 4.84E-05 | 0.002060466 | 0.234893154 |
| HTN | mesothelium | *HAUS8* | -0.606614313 | 2.63E-09 | 5.70E-07 | 1.28E-05 |
| HTN | mesothelium | *MRPS25* | -0.76292415 | 0.0003271 | 0.009503538 | 1 |
| HTN | mesothelium | *IPO8* | -0.961289077 | 0.001539692 | 0.029411751 | 1 |
| HTN | mesothelium | *IMPA1* | 0.209059281 | 0.002115132 | 0.036898593 | 1 |
| HTN | mesothelium | *TMEM241* | -0.280887873 | 0.0008241 | 0.019102676 | 1 |
| HTN | mesothelium | *ACP2* | 1.106888575 | 2.82E-10 | 1.24E-07 | 1.37E-06 |
| HTN | mesothelium | *POM121C* | -0.505758405 | 0.000459116 | 0.012717957 | 1 |
| HTN | mesothelium | *TES* | -0.543955549 | 0.002410127 | 0.039606735 | 1 |
| HTN | mesothelium | *CCT7* | -0.66847513 | 0.00258809 | 0.041307273 | 1 |
| HTN | mesothelium | *AGT* | 0.180394205 | 0.000576959 | 0.014779764 | 1 |
| HTN | mesothelium | *TBC1D4* | -0.470082942 | 0.003507011 | 0.047797801 | 1 |
| HTN | mesothelium | *CHST12* | -0.814143348 | 0.001025511 | 0.022385084 | 1 |
| HTN | mesothelium | *NMT1* | 0.332079897 | 1.06E-06 | 8.68E-05 | 0.005119111 |
| HTN | mesothelium | *NDUFB5* | -0.909917741 | 0.001400269 | 0.02797436 | 1 |
| HTN | mesothelium | *RPS6KC1* | -0.75950861 | 0.002086337 | 0.036677203 | 1 |
| HTN | mesothelium | *IL11RA* | -0.287730596 | 0.003071287 | 0.044285724 | 1 |
| HTN | mesothelium | *DENND4C* | -0.857202552 | 0.000645072 | 0.01586625 | 1 |
| HTN | mesothelium | *SLC37A4* | 0.341208813 | 0.000129304 | 0.004579443 | 0.627383683 |
| HTN | mesothelium | *PPP2R1B* | -0.235786529 | 3.41E-05 | 0.00156211 | 0.165583688 |
| HTN | mesothelium | *NUSAP1* | 0.58360966 | 0.000258848 | 0.007898938 | 1 |
| HTN | mesothelium | *PARP6* | -1.18684096 | 0.002720865 | 0.041709335 | 1 |

| | | | | | | |
|---|---|---|---|---|---|---|
| HTN | mesothelium | *SPPL2A* | -0.726396402 | 1.17E-05 | 0.000677619 | 0.056920028 |
| HTN | mesothelium | *FBN2* | 0.10191655 | 6.35E-05 | 0.002504975 | 0.308111955 |
| HTN | mesothelium | *FITM1* | 0.528396002 | 0.002722971 | 0.041709335 | 1 |
| HTN | mesothelium | *RTN1* | 0.22173917 | 0.002406624 | 0.039606735 | 1 |
| HTN | mesothelium | *SCAPER* | -0.38168743 | 0.003091598 | 0.044285724 | 1 |
| HTN | mesothelium | *ULK3* | 1.584044923 | 0.002268373 | 0.038215784 | 1 |
| HTN | mesothelium | *HAPLN3* | -0.295479042 | 0.002054123 | 0.036374467 | 1 |
| HTN | mesothelium | *UNC45A* | -1.010076828 | 0.001784218 | 0.032916453 | 1 |
| HTN | mesothelium | *DEF8* | 0.493061846 | 2.19E-05 | 0.001128177 | 0.106048676 |
| HTN | mesothelium | *VPS53* | -0.42799346 | 0.00010777 | 0.003991619 | 0.52290208 |
| HTN | mesothelium | *LRRC46* | -0.236930686 | 0.003519673 | 0.047835998 | 1 |
| HTN | mesothelium | *SCRN2* | -0.087815288 | 0.000382552 | 0.010918498 | 1 |
| HTN | mesothelium | *C18orf8* | -0.799723519 | 3.63E-08 | 6.07E-06 | 0.000176109 |
| HTN | mesothelium | *NPC1* | -1.250653305 | 6.32E-06 | 0.000390262 | 0.030645652 |
| HTN | mesothelium | *SAE1* | -1.024861931 | 8.61E-05 | 0.003313919 | 0.417553797 |
| HTN | mesothelium | *NME7* | -0.583853844 | 0.000100422 | 0.003777127 | 0.487249378 |
| HTN | mesothelium | *ARNT* | -0.636009429 | 4.59E-05 | 0.001970945 | 0.222716773 |
| HTN | mesothelium | *POGZ* | -0.674657213 | 0.000805091 | 0.018871024 | 1 |
| HTN | mesothelium | *CREB3L4* | -0.53364844 | 3.31E-07 | 3.32E-05 | 0.001604932 |
| HTN | mesothelium | *C1orf43* | -1.37416324 | 1.98E-05 | 0.001069097 | 0.096218717 |
| HTN | mesothelium | *PLEKHA6* | 0.29767681 | 0.000466657 | 0.012743402 | 1 |
| HTN | mesothelium | *ZNF514* | -0.337822774 | 0.001345779 | 0.027435789 | 1 |
| HTN | mesothelium | *RABL2A* | -0.325694197 | 0.001454543 | 0.028572639 | 1 |
| HTN | mesothelium | *PHOSPHO2* | -0.655574937 | 0.002728745 | 0.041709335 | 1 |
| HTN | mesothelium | *CTDSPL* | -0.617885834 | 0.000593678 | 0.014925011 | 1 |
| HTN | mesothelium | *PTPRG* | -0.595228061 | 0.003249816 | 0.045572567 | 1 |
| HTN | mesothelium | *CISD2* | 1.143670897 | 0.002455738 | 0.039850297 | 1 |
| HTN | mesothelium | *FAM193B* | -0.842790061 | 0.001590452 | 0.030144032 | 1 |
| HTN | mesothelium | *ABT1* | 0.174426389 | 0.003223908 | 0.045383606 | 1 |
| HTN | mesothelium | *IRAK1BP1* | 0.755653592 | 4.19E-06 | 0.000282542 | 0.020343001 |
| HTN | mesothelium | *SLC18B1* | -0.300947595 | 0.000202488 | 0.006566261 | 0.982472573 |
| HTN | mesothelium | *GNA12* | -0.601368376 | 0.000756261 | 0.018075758 | 1 |
| HTN | mesothelium | *ADHFE1* | -0.123251339 | 0.000248654 | 0.007635887 | 1 |
| HTN | mesothelium | *HSD17B12* | -0.218391255 | 3.81E-10 | 1.54E-07 | 1.85E-06 |
| HTN | mesothelium | *EIF3M* | -0.662541287 | 0.000608472 | 0.015140041 | 1 |
| HTN | mesothelium | *TNKS1BP1* | 0.416796288 | 1.06E-09 | 2.70E-07 | 5.12E-06 |
| HTN | mesothelium | *INCENP* | -0.8489251 | 0.002328317 | 0.038951294 | 1 |
| HTN | mesothelium | *TRPT1* | -0.258403878 | 0.000866485 | 0.019925039 | 1 |
| HTN | mesothelium | *TBX6* | 0.093948566 | 0.001257685 | 0.026312336 | 1 |
| HTN | mesothelium | *ADAMTS12* | -0.24594614 | 0.002816112 | 0.042434092 | 1 |
| HTN | mesothelium | *CWF19L2* | 0.585973546 | 0.001826488 | 0.033316233 | 1 |
| HTN | mesothelium | *PLEKHH2* | -0.200359194 | 0.002416239 | 0.039606735 | 1 |
| HTN | mesothelium | *PRDM8* | -0.488553129 | 0.000278468 | 0.008340279 | 1 |
| HTN | mesothelium | *CAST* | -0.514360305 | 0.000103089 | 0.003847585 | 0.500186106 |
| HTN | mesothelium | *JAZF1* | -0.36823157 | 0.001838122 | 0.033402883 | 1 |
| HTN | mesothelium | *LRRK1* | -0.838355181 | 0.002172274 | 0.037243372 | 1 |
| HTN | mesothelium | *NEIL2* | 0.442643178 | 1.40E-08 | 2.72E-06 | 6.79E-05 |
| HTN | mesothelium | *PROM2* | -0.376228981 | 0.002161869 | 0.037196421 | 1 |
| HTN | mesothelium | *MMP16* | -0.126176006 | 0.003163403 | 0.04487963 | 1 |
| HTN | mesothelium | *NDUFAF6* | -1.081016475 | 7.78E-10 | 2.52E-07 | 3.78E-06 |

| | | | | | | |
|---|---|---|---|---|---|---|
| HTN | mesothelium | *ART3* | -0.185535207 | 0.002215286 | 0.037847077 | 1 |
| HTN | mesothelium | *SFXN2* | 0.238393639 | 5.87E-08 | 8.38E-06 | 0.000284808 |
| HTN | mesothelium | *CLEC18A* | -0.286374169 | 3.15E-06 | 0.000224804 | 0.015286648 |
| HTN | mesothelium | *SLC5A11* | -0.176284584 | 4.06E-07 | 3.78E-05 | 0.001968259 |
| HTN | mesothelium | *CDC42SE2* | -0.814937807 | 2.36E-05 | 0.001178293 | 0.114379645 |
| HTN | mesothelium | *CTBP1* | -0.92885209 | 1.67E-05 | 0.000921957 | 0.081132226 |
| HTN | mesothelium | *S100B* | -0.091647842 | 2.25E-05 | 0.001147753 | 0.10903649 |
| HTN | mesothelium | *ZNF208* | -0.533961021 | 2.02E-05 | 0.001078332 | 0.098128214 |
| HTN | mesothelium | *ABCF3* | 0.404193897 | 1.50E-07 | 1.77E-05 | 0.000727069 |
| HTN | mesothelium | *USP24* | -0.878637467 | 0.001195946 | 0.025562688 | 1 |
| HTN | mesothelium | *HFM1* | -0.441773194 | 0.00280999 | 0.042434092 | 1 |
| HTN | mesothelium | *MRPL55* | 0.687432306 | 3.92E-11 | 3.81E-08 | 1.90E-07 |
| HTN | mesothelium | *LYSMD1* | -0.226149646 | 2.08E-05 | 0.001087753 | 0.101161055 |
| HTN | mesothelium | *DCAF16* | -1.907657488 | 3.30E-11 | 3.81E-08 | 1.60E-07 |
| HTN | mesothelium | *LINC01116* | -0.652975772 | 0.001885741 | 0.034104515 | 1 |
| HTN | mesothelium | *LMOD1* | -0.427448765 | 0.000183992 | 0.006198269 | 0.892730607 |
| HTN | mesothelium | *SERPINI1* | -0.929421291 | 0.002673012 | 0.041568762 | 1 |
| HTN | mesothelium | *TPRA1* | -0.543033877 | 0.002614804 | 0.041408821 | 1 |
| HTN | mesothelium | *RFT1* | -0.33325325 | 5.24E-05 | 0.002154551 | 0.254236975 |
| HTN | mesothelium | *METAP1* | -0.510917048 | 0.003589283 | 0.048510315 | 1 |
| HTN | mesothelium | *BDH2* | -0.403470693 | 0.0025623 | 0.041090409 | 1 |
| HTN | mesothelium | *CDC25A* | -1.295212891 | 1.65E-09 | 3.82E-07 | 8.03E-06 |
| HTN | mesothelium | *ZNF589* | -0.557068382 | 8.03E-11 | 6.50E-08 | 3.90E-07 |
| HTN | mesothelium | *RNF123* | 0.462112111 | 5.77E-10 | 2.00E-07 | 2.80E-06 |
| HTN | mesothelium | *MST1R* | 0.234495695 | 6.98E-05 | 0.002708133 | 0.338516608 |
| HTN | mesothelium | *CEP44* | -0.463051029 | 0.003094474 | 0.044285724 | 1 |
| HTN | mesothelium | *ERAP1* | -0.908578123 | 1.10E-10 | 7.65E-08 | 5.36E-07 |
| HTN | mesothelium | *ERAP2* | 0.103799193 | 0.002851654 | 0.042573003 | 1 |
| HTN | mesothelium | *CCDC127* | -0.799477164 | 0.000264001 | 0.008005818 | 1 |
| HTN | mesothelium | *RPS14* | -0.886066255 | 0.001897819 | 0.034104515 | 1 |
| HTN | mesothelium | *SLC25A32* | -0.741285828 | 0.003320117 | 0.046158188 | 1 |
| HTN | mesothelium | *RPP25L* | -0.397562775 | 0.000119036 | 0.00437547 | 0.577562049 |
| HTN | mesothelium | *REEP3* | -1.04301497 | 4.35E-06 | 0.00028733 | 0.021087959 |
| HTN | mesothelium | *VDAC2* | -1.324836858 | 0.000202997 | 0.006566261 | 0.984939099 |
| HTN | mesothelium | *CASP7* | -0.224020273 | 0.002887248 | 0.04271014 | 1 |
| HTN | mesothelium | *TRUB1* | -0.55826877 | 0.002626806 | 0.041408821 | 1 |
| HTN | mesothelium | *ARL5B* | -1.052310786 | 1.65E-07 | 1.86E-05 | 0.000800427 |
| HTN | mesothelium | *CEP57* | -0.81491542 | 5.42E-05 | 0.002191523 | 0.26298274 |
| HTN | mesothelium | *CYB5R2* | 0.19578226 | 0.003094607 | 0.044285724 | 1 |
| HTN | mesothelium | *XRRA1* | -0.418835985 | 0.000368209 | 0.01062985 | 1 |
| HTN | mesothelium | *TRIM66* | -0.131136365 | 1.29E-05 | 0.000729257 | 0.062716118 |
| HTN | mesothelium | *RIMKLB* | -0.547920216 | 1.01E-09 | 2.70E-07 | 4.90E-06 |
| HTN | mesothelium | *ZNF91* | -0.743889651 | 0.002909523 | 0.042778811 | 1 |
| HTN | mesothelium | *ZNF23* | -0.555887868 | 0.002828328 | 0.042486221 | 1 |
| HTN | mesothelium | *SRR* | 0.138527004 | 0.000226069 | 0.007122645 | 1 |
| HTN | mesothelium | *SOAT2* | 0.206998547 | 0.003412094 | 0.047301376 | 1 |
| HTN | mesothelium | *ZNF598* | -0.510178949 | 0.000298255 | 0.008770503 | 1 |
| HTN | mesothelium | *NT5DC2* | -0.198537261 | 0.001969471 | 0.035261532 | 1 |
| HTN | mesothelium | *SERINC2* | -0.601955748 | 0.001249278 | 0.026312336 | 1 |
| HTN | mesothelium | *SLC20A2* | -0.812262551 | 0.003226988 | 0.045383606 | 1 |

| | | | | | | |
|---|---|---|---|---|---|---|
| HTN | mesothelium | *CEP120* | -1.130224369 | 0.001507844 | 0.029289748 | 1 |
| HTN | mesothelium | *C8orf46* | 0.069765874 | 0.001002366 | 0.022207676 | 1 |
| HTN | mesothelium | *CPT1C* | 0.461934717 | 4.27E-07 | 3.83E-05 | 0.002073118 |
| HTN | mesothelium | *XPO6* | -0.976096376 | 0.002101966 | 0.036818555 | 1 |
| HTN | mesothelium | *RAB24* | -1.571871095 | 6.32E-06 | 0.000390262 | 0.030680819 |
| HTN | mesothelium | *ZEB2* | -0.427296896 | 0.000274909 | 0.008284837 | 1 |
| HTN | mesothelium | *INO80E* | -0.407943233 | 1.89E-10 | 1.02E-07 | 9.18E-07 |
| HTN | mesothelium | *DFFB* | -1.595493284 | 0.000158917 | 0.005507593 | 0.771062957 |
| HTN | mesothelium | *RGPD8* | -0.070422437 | 0.000568133 | 0.014707701 | 1 |
| HTN | mesothelium | *MYO7B* | 0.318723748 | 0.000124001 | 0.004423906 | 0.601651271 |
| HTN | mesothelium | *DNAJC18* | -0.361889195 | 2.86E-05 | 0.001359688 | 0.138975423 |
| HTN | mesothelium | *TTLL6* | -1.094265569 | 2.18E-10 | 1.06E-07 | 1.06E-06 |
| HTN | mesothelium | *PAQR8* | -0.281677585 | 0.000615464 | 0.015235872 | 1 |
| HTN | mesothelium | *MRPL36* | 0.150653097 | 0.002424449 | 0.039607495 | 1 |
| HTN | mesothelium | *RGS19* | -0.218948858 | 5.47E-09 | 1.11E-06 | 2.65E-05 |
| HTN | mesothelium | *NME6* | 0.636142579 | 1.28E-09 | 3.12E-07 | 6.23E-06 |
| HTN | mesothelium | *CTSW* | -0.051447079 | 2.32E-05 | 0.001172127 | 0.11252422 |
| HTN | mesothelium | *ZFAND4* | -0.406470233 | 0.003468221 | 0.04753618 | 1 |
| HTN | mesothelium | *ZNF738* | 0.162081273 | 0.003678887 | 0.049445878 | 1 |
| HTN | mesothelium | *CFL1* | -1.140415471 | 0.000927049 | 0.02101889 | 1 |
| HTN | mesothelium | *DCP2* | -1.306195882 | 3.56E-05 | 0.001613962 | 0.17269397 |
| HTN | mesothelium | *MYEOV* | -0.569621718 | 0.003184409 | 0.045045929 | 1 |
| HTN | mesothelium | *DCAKD* | -1.416002144 | 6.11E-08 | 8.47E-06 | 0.000296604 |
| HTN | mesothelium | *NOC3L* | -0.984912848 | 1.98E-06 | 0.000155003 | 0.009610176 |
| HTN | mesothelium | *PPP1R14B* | 0.871798067 | 0.001538286 | 0.029411751 | 1 |
| HTN | mesothelium | *MST1* | -0.172356618 | 4.13E-07 | 3.78E-05 | 0.002005857 |
| HTN | mesothelium | *C11orf80* | -0.76306584 | 5.03E-10 | 1.88E-07 | 2.44E-06 |
| HTN | mesothelium | *CTU2* | -0.198828253 | 0.002733629 | 0.041709335 | 1 |
| HTN | mesothelium | *SNX31* | -0.169456453 | 3.97E-06 | 0.000271609 | 0.019284244 |
| HTN | mesothelium | *RSRC1* | -1.327978682 | 4.58E-08 | 6.94E-06 | 0.000222175 |
| HTN | mesothelium | *SEZ6L2* | 0.260877903 | 0.000184391 | 0.006198269 | 0.894663862 |
| HTN | mesothelium | *CCNE2* | -0.514757015 | 8.59E-10 | 2.61E-07 | 4.17E-06 |
| HTN | mesothelium | *TMEM9B* | 0.663786332 | 0.002655508 | 0.041429336 | 1 |
| HTN | mesothelium | *PPP2R2D* | -0.63148777 | 2.08E-05 | 0.001087753 | 0.100964526 |
| HTN | mesothelium | *CTDNEP1* | 0.446852649 | 0.000370248 | 0.01062985 | 1 |
| HTN | mesothelium | *ORAI3* | 0.458954074 | 1.05E-06 | 8.68E-05 | 0.005079721 |
| HTN | mesothelium | *MTHFR* | -1.270474033 | 9.24E-10 | 2.64E-07 | 4.48E-06 |
| HTN | mesothelium | *GEN1* | -0.143880043 | 0.000932564 | 0.021045588 | 1 |
| HTN | mesothelium | *P4HTM* | -1.295147804 | 9.82E-05 | 0.003722007 | 0.476416839 |
| HTN | mesothelium | *RPP25* | 0.242148303 | 1.92E-06 | 0.000152399 | 0.00929636 |
| HTN | mesothelium | *MPI* | -0.799578961 | 6.55E-17 | 1.59E-13 | 3.18E-13 |
| HTN | mesothelium | *TRIM73* | -0.535705989 | 0.000569878 | 0.014707701 | 1 |
| HTN | mesothelium | *HYI* | -0.278221214 | 1.47E-07 | 1.77E-05 | 0.00071306 |
| HTN | mesothelium | *CTC1* | 0.487615651 | 0.001237462 | 0.026222367 | 1 |
| HTN | mesothelium | *HLA-DQB1* | -0.094853489 | 0.00168896 | 0.03176293 | 1 |
| HTN | mesothelium | *AKAP5* | -1.410976555 | 0.002518866 | 0.040603115 | 1 |
| HTN | mesothelium | *CLDN7* | -0.394275229 | 8.97E-07 | 7.64E-05 | 0.004352481 |
| HTN | mesothelium | *ADO* | 0.474489361 | 2.70E-09 | 5.70E-07 | 1.31E-05 |
| HTN | mesothelium | *IDH2* | -1.165663133 | 0.002026409 | 0.036063454 | 1 |
| HTN | mesothelium | *CLN8* | 0.216492375 | 2.59E-05 | 0.001245276 | 0.12577286 |

| | | | | | | |
|---|---|---|---|---|---|---|
| HTN | mesothelium | *FES* | -1.067698597 | 2.62E-20 | 1.27E-16 | 1.27E-16 |
| HTN | mesothelium | *ZNF320* | 0.110070657 | 0.000222216 | 0.007047007 | 1 |
| HTN | mesothelium | *WBP2NL* | -0.606427806 | 0.001825803 | 0.033316233 | 1 |
| HTN | mesothelium | *ALDH1A3* | -0.147952898 | 0.001761308 | 0.032742779 | 1 |
| HTN | mesothelium | *A3GALT2* | -0.186050978 | 4.95E-05 | 0.002068815 | 0.239982534 |
| HTN | mesothelium | *NUTM2A* | -0.258770375 | 0.00012178 | 0.004401412 | 0.590874915 |
| HTN | mesothelium | *CDK10* | -0.184717082 | 5.38E-05 | 0.002191523 | 0.26090975 |
| HTN | mesothelium | *MYBL1* | -0.603135401 | 0.000923007 | 0.02101889 | 1 |
| HTN | mesothelium | *SMYD4* | -0.333248534 | 0.002908389 | 0.042778811 | 1 |
| HTN | mesothelium | *KLHL17* | 0.243169779 | 0.002336114 | 0.038951294 | 1 |
| HTN | mesothelium | *CENPP* | -0.169094647 | 0.003308393 | 0.046127358 | 1 |
| HTN | mesothelium | *ZNF559* | -0.10922367 | 0.001897427 | 0.034104515 | 1 |
| HTN | mesothelium | *FBF1* | -0.539707099 | 1.85E-07 | 2.00E-05 | 0.000899645 |
| HTN | mesothelium | *MSL1* | -0.329316146 | 0.000826785 | 0.019102676 | 1 |
| HTN | mesothelium | *HLA-DRB1* | -0.138419212 | 0.000120494 | 0.004395774 | 0.584637989 |
| HTN | mesothelium | *COLCA1* | -0.340597542 | 0.00018249 | 0.006198269 | 0.885439793 |
| HTN | mesothelium | *HRCT1* | 0.215025551 | 0.001086018 | 0.023315745 | 1 |
| HTN | mesothelium | *SULT1A1* | -0.548009623 | 0.002434797 | 0.039643076 | 1 |
| HTN | mesothelium | *AP2A1* | -0.403851275 | 0.002228824 | 0.037944756 | 1 |
| HTN | mesothelium | *HIST1H3E* | 0.242471085 | 8.87E-08 | 1.19E-05 | 0.000430156 |
| HTN | mesothelium | *WDR5B* | 0.512444625 | 0.000498726 | 0.013267119 | 1 |
| HTN | mesothelium | *ZNF100* | -0.43793852 | 2.40E-06 | 0.000184923 | 0.011650166 |
| HTN | mesothelium | *IGF2R* | -0.795636053 | 0.001364608 | 0.027587831 | 1 |
| HTN | mesothelium | *SULT1A2* | 0.424820589 | 4.23E-05 | 0.001865838 | 0.205242176 |
| HTN | mesothelium | *ZNF517* | -1.145192169 | 0.000500388 | 0.013267119 | 1 |
| HTN | mesothelium | *GSTK1* | -0.368622812 | 0.000586005 | 0.014808844 | 1 |
| HTN | mesothelium | *PCDHB11* | -0.109621837 | 0.003501215 | 0.047797801 | 1 |
| HTN | mesothelium | *MFAP5* | -0.310905896 | 2.89E-05 | 0.001359688 | 0.140047883 |
| HTN | mesothelium | *RPS26* | -0.197605121 | 5.97E-06 | 0.000381088 | 0.028962682 |
| HTN | mesothelium | *AC122129.1* | -2.333758742 | 0.003007997 | 0.043828232 | 1 |
| HTN | mesothelium | *AGAP9* | -0.458164456 | 0.000949864 | 0.021336749 | 1 |
| HTN | mesothelium | *SFI1* | -0.96198007 | 0.000578762 | 0.014779764 | 1 |
| HTN | mesothelium | *MIER1* | -0.700856339 | 0.002029127 | 0.036063454 | 1 |
| HTN | mesothelium | *TMEM116* | -0.289771965 | 3.44E-14 | 5.56E-11 | 1.67E-10 |
| HTN | mesothelium | *UCKL1* | -1.785684523 | 3.96E-05 | 0.00176092 | 0.191940247 |
| HTN | mesothelium | *ZNF568* | -0.595710654 | 0.001506796 | 0.029289748 | 1 |
| HTN | mesothelium | *HLA-DRB5* | -0.05973263 | 1.51E-05 | 0.000844885 | 0.073505037 |
| HTN | mesothelium | *NCOA6* | -0.361893271 | 0.000815777 | 0.019029578 | 1 |
| HTN | mesothelium | *PNP* | -0.07103612 | 0.002350086 | 0.039050052 | 1 |
| HTN | mesothelium | *DENND4B* | -0.490442436 | 1.09E-05 | 0.000644544 | 0.052852574 |
| HTN | mesothelium | *PRMT6* | -3.132480954 | 0.001401024 | 0.02797436 | 1 |
| HTN | mesothelium | *SHISA4* | -2.482940587 | 1.57E-10 | 9.50E-08 | 7.60E-07 |
| HTN | mesothelium | *MAP3K3* | -0.627448353 | 0.001272841 | 0.026312336 | 1 |
| HTN | mesothelium | *AL590822.1* | -0.809358902 | 5.60E-05 | 0.002243634 | 0.271479664 |
| HTN | mesothelium | *HSD17B8* | -0.248132021 | 8.83E-05 | 0.003371933 | 0.428235523 |
| HTN | mesothelium | *PRRT1* | 0.223095116 | 1.08E-06 | 8.76E-05 | 0.005256944 |
| HTN | mesothelium | *LAYN* | -0.317817875 | 0.002264214 | 0.038215784 | 1 |
| HTN | mesothelium | *HLA-C* | -0.236261629 | 2.20E-08 | 3.96E-06 | 0.000106986 |
| HTN | mesothelium | *POU5F1* | -0.052520166 | 0.001033582 | 0.022388117 | 1 |
| HTN | mesothelium | *PSORS1C2* | 0.167816974 | 0.002845152 | 0.042573003 | 1 |

| | | | | | | |
|---|---|---|---|---|---|---|
| HTN | mesothelium | *PRR13* | -0.405039239 | 0.002717627 | 0.041709335 | 1 |
| HTN | mesothelium | *SUPT4H1* | 1.364926687 | 0.000192531 | 0.006311896 | 0.934160555 |
| HTN | mesothelium | *MXD3* | -0.244943413 | 0.00076379 | 0.018166218 | 1 |
| HTN | mesothelium | *TMEM110* | 0.364783307 | 0.000542754 | 0.014234826 | 1 |
| HTN | mesothelium | *ATF6B* | 0.633957536 | 5.21E-08 | 7.66E-06 | 0.000252878 |
| HTN | mesothelium | *PPM1N* | 0.175374178 | 0.002635805 | 0.041408821 | 1 |
| HTN | mesothelium | *LTB4R* | -0.805960754 | 6.35E-06 | 0.000390262 | 0.030830706 |
| HTN | mesothelium | *AC016773.1* | 0.244744288 | 0.000486622 | 0.01311716 | 1 |
| HTN | mesothelium | *AP4M1* | -0.25640009 | 0.00023508 | 0.007311588 | 1 |
| HTN | mesothelium | *SLC12A8* | 0.218367995 | 0.001414611 | 0.028015076 | 1 |
| HTN | mesothelium | *TIAF1* | 0.411185972 | 4.90E-05 | 0.002066702 | 0.237670728 |
| HTN | mesothelium | *EXOSC6* | -0.672735198 | 2.56E-06 | 0.000187869 | 0.01243248 |
| HTN | mesothelium | *AL049840.1* | -0.987919465 | 1.74E-07 | 1.91E-05 | 0.00084223 |
| HTN | mesothelium | *PLEKHM1* | -0.746294346 | 2.38E-05 | 0.001178293 | 0.11547274 |
| HTN | mesothelium | *TMA7* | 0.473171044 | 0.003302305 | 0.046127358 | 1 |
| HTN | mesothelium | *HLA-DQA2* | 0.08251939 | 3.12E-05 | 0.001445757 | 0.151594839 |
| HTN | mesothelium | *LINC00211* | -1.129384007 | 0.003143183 | 0.044723527 | 1 |
| HTN | mesothelium | *LRRC37A2* | -0.70217192 | 0.003454625 | 0.04753618 | 1 |
| HTN | mesothelium | *TNFSF12* | 0.517061483 | 4.02E-07 | 3.78E-05 | 0.001951215 |
| HTN | mesothelium | *ZNF709* | -8.669500468 | 4.47E-08 | 6.94E-06 | 0.000217069 |
| HTN | mesothelium | *WFDC6* | -0.639824877 | 0.000429315 | 0.011971474 | 1 |
| HTN | mesothelium | *PLA2G4B* | 0.582775182 | 0.000237878 | 0.007351487 | 1 |
| HTN | mesothelium | *JMJD7* | 0.157826719 | 0.001237618 | 0.026222367 | 1 |
| HTN | mesothelium | *HMBS* | 0.33765213 | 0.003066229 | 0.044285724 | 1 |
| HTN | mesothelium | *LINC00638* | 0.393014697 | 0.001515195 | 0.029289748 | 1 |
| HTN | mesothelium | *PWRN1* | -0.392878544 | 0.002655285 | 0.041429336 | 1 |
| HTN | monocyte | *NFYA* | -2.038418615 | 0.006007389 | 0.023595115 | 1 |
| HTN | monocyte | *SEMA3F* | -1.998200577 | 0.000632445 | 0.018265624 | 1 |
| HTN | monocyte | *KRIT1* | -1.84623365 | 0.022213638 | 0.041647859 | 1 |
| HTN | monocyte | *RAD52* | -2.391783622 | 0.004548004 | 0.023595115 | 1 |
| HTN | monocyte | *HS3ST1* | -1.072419663 | 0.005407746 | 0.023595115 | 1 |
| HTN | monocyte | *AOC1* | -2.752103127 | 0.01906426 | 0.037504578 | 1 |
| HTN | monocyte | *TMEM176A* | 3.502476016 | 0.002786969 | 0.023595115 | 1 |
| HTN | monocyte | *ALS2* | -1.447400198 | 0.009065596 | 0.026214745 | 1 |
| HTN | monocyte | *RBM6* | -2.483584769 | 3.19E-06 | 0.00055237 | 0.01546635 |
| HTN | monocyte | *ARHGAP33* | -2.09801056 | 0.000402462 | 0.01501035 | 1 |
| HTN | monocyte | *POLR2J* | 4.983469587 | 0.001696851 | 0.023595115 | 1 |
| HTN | monocyte | *MEOX1* | -3.420779955 | 0.007126938 | 0.024238452 | 1 |
| HTN | monocyte | *PON1* | -4.378758807 | 0.006325724 | 0.02374425 | 1 |
| HTN | monocyte | *GCFC2* | -1.484100401 | 0.011002245 | 0.027963799 | 1 |
| HTN | monocyte | *RHBDD2* | 2.797869052 | 0.000753731 | 0.020093968 | 1 |
| HTN | monocyte | *ZNF195* | -1.652378632 | 0.0289237 | 0.049698576 | 1 |
| HTN | monocyte | *GGNBP2* | -2.160791091 | 0.01341679 | 0.030706728 | 1 |
| HTN | monocyte | *ITGA2B* | -1.647235147 | 0.00513434 | 0.023595115 | 1 |
| HTN | monocyte | *TMEM98* | -1.456851954 | 0.023026419 | 0.042480679 | 1 |
| HTN | monocyte | *ZNF263* | -2.181198564 | 0.010336112 | 0.027285536 | 1 |
| HTN | monocyte | *SPATA20* | -1.655412416 | 0.012163232 | 0.029071921 | 1 |
| HTN | monocyte | *BAIAP2L1* | -1.881745279 | 0.008688712 | 0.025839644 | 1 |
| HTN | monocyte | *USH1C* | 2.237635187 | 0.003981384 | 0.023595115 | 1 |
| HTN | monocyte | *ELAC2* | -2.061865935 | 0.004158472 | 0.023595115 | 1 |

| | | | | | | |
|---|---|---|---|---|---|---|
| HTN | monocyte | *SCIN* | -1.341858891 | 0.001703491 | 0.023595115 | 1 |
| HTN | monocyte | *CCDC124* | -1.26842524 | 0.015080868 | 0.032773807 | 1 |
| HTN | monocyte | *NOS2* | -5.613048941 | 0.02246005 | 0.041849524 | 1 |
| HTN | monocyte | *CD79B* | 0.510767188 | 0.009038837 | 0.026214249 | 1 |
| HTN | monocyte | *ST7L* | -2.982469403 | 0.00164006 | 0.023595115 | 1 |
| HTN | monocyte | *BAIAP3* | -2.936903878 | 0.01624413 | 0.034075452 | 1 |
| HTN | monocyte | *TSR3* | -3.318843542 | 0.027275621 | 0.047742177 | 1 |
| HTN | monocyte | *FMO3* | -1.444780402 | 0.003342447 | 0.023595115 | 1 |
| HTN | monocyte | *NADK* | 1.929602434 | 0.009830246 | 0.026779763 | 1 |
| HTN | monocyte | *DLEC1* | -1.370295666 | 0.002525291 | 0.023595115 | 1 |
| HTN | monocyte | *CYB561* | -2.327286847 | 0.008723631 | 0.025839644 | 1 |
| HTN | monocyte | *MGST1* | -0.556731984 | 0.019727435 | 0.038431229 | 1 |
| HTN | monocyte | *PKD1* | -2.358722259 | 0.021859951 | 0.041172245 | 1 |
| HTN | monocyte | *MAPK8IP2* | -7.114248148 | 0.0034241 | 0.023595115 | 1 |
| HTN | monocyte | *RPS20* | -1.92498869 | 0.028147855 | 0.048791054 | 1 |
| HTN | monocyte | *TRAF3IP3* | 0.397269496 | 0.002026792 | 0.023595115 | 1 |
| HTN | monocyte | *MLXIPL* | -0.82184231 | 0.000881322 | 0.020511604 | 1 |
| HTN | monocyte | *FUZ* | -1.886861713 | 0.005914502 | 0.023595115 | 1 |
| HTN | monocyte | *ZNF200* | -2.447610956 | 0.008423208 | 0.025487024 | 1 |
| HTN | monocyte | *SLC6A7* | 9.867206271 | 0.003448504 | 0.023595115 | 1 |
| HTN | monocyte | *TSPAN9* | -1.721640411 | 0.023009313 | 0.042480467 | 1 |
| HTN | monocyte | *UTP18* | -1.891553245 | 0.027292217 | 0.047753998 | 1 |
| HTN | monocyte | *TTC19* | -1.586578765 | 0.008000042 | 0.025091616 | 1 |
| HTN | monocyte | *DCN* | -0.999343457 | 0.005959986 | 0.023595115 | 1 |
| HTN | monocyte | *ZBTB32* | -2.62763176 | 0.013459465 | 0.030775364 | 1 |
| HTN | monocyte | *ALOX5* | 0.45183654 | 0.006855552 | 0.024118623 | 1 |
| HTN | monocyte | *EHD3* | 4.022482604 | 0.005062648 | 0.023595115 | 1 |
| HTN | monocyte | *MAN2B2* | 4.452576658 | 0.009568099 | 0.026650066 | 1 |
| HTN | monocyte | *RNF14* | -2.21756943 | 0.020277089 | 0.03899502 | 1 |
| HTN | monocyte | *DDX11* | -1.52438925 | 0.002547157 | 0.023595115 | 1 |
| HTN | monocyte | *ZC3H3* | -1.568433917 | 0.016747753 | 0.034534678 | 1 |
| HTN | monocyte | *MDH1* | -1.611197093 | 0.00899027 | 0.026120233 | 1 |
| HTN | monocyte | *MTMR11* | 1.009103931 | 0.000858179 | 0.020511604 | 1 |
| HTN | monocyte | *CCDC88C* | 0.832623965 | 0.011087444 | 0.028056466 | 1 |
| HTN | monocyte | *CHDH* | -2.455247425 | 0.014320286 | 0.031973856 | 1 |
| HTN | monocyte | *CLCA4* | -6.659210809 | 0.002900262 | 0.023595115 | 1 |
| HTN | monocyte | *RUFY3* | -1.29993968 | 0.014073913 | 0.031684476 | 1 |
| HTN | monocyte | *SLC11A1* | 0.280273836 | 0.005432651 | 0.023595115 | 1 |
| HTN | monocyte | *WWTR1* | -1.688749366 | 0.019818873 | 0.038510682 | 1 |
| HTN | monocyte | *VSIG2* | -1.839302453 | 0.012352693 | 0.029302872 | 1 |
| HTN | monocyte | *MARCO* | -0.935459459 | 0.004452373 | 0.023595115 | 1 |
| HTN | monocyte | *HGF* | -2.428250379 | 0.020478236 | 0.039272886 | 1 |
| HTN | monocyte | *OSBPL5* | -1.746295184 | 0.017075365 | 0.034942924 | 1 |
| HTN | monocyte | *CPS1* | -1.206769995 | 0.005863792 | 0.023595115 | 1 |
| HTN | monocyte | *NLRP2* | 1.089106234 | 0.023404465 | 0.042977291 | 1 |
| HTN | monocyte | *ZDHHC6* | -2.128735828 | 0.011445371 | 0.028463835 | 1 |
| HTN | monocyte | *GRAMD1B* | 1.481735529 | 0.00370914 | 0.023595115 | 1 |
| HTN | monocyte | *ABCC2* | 3.861324069 | 0.002951682 | 0.023595115 | 1 |
| HTN | monocyte | *TYMP* | 2.193176155 | 0.000369416 | 0.01501035 | 1 |
| HTN | monocyte | *TOMM34* | -2.181128339 | 0.011049066 | 0.028009441 | 1 |

| | | | | | | |
|---|---|---|---|---|---|---|
| HTN | monocyte | *FAS* | -2.401455633 | 0.01977379 | 0.038484729 | 1 |
| HTN | monocyte | *AGPAT4* | -1.968326095 | 0.014502318 | 0.03215962 | 1 |
| HTN | monocyte | *INSRR* | -2.477959564 | 0.002387031 | 0.023595115 | 1 |
| HTN | monocyte | *B4GALT7* | 2.45524597 | 0.007948221 | 0.025074621 | 1 |
| HTN | monocyte | *SH2D2A* | 4.133456817 | 0.004595534 | 0.023595115 | 1 |
| HTN | monocyte | *VEZT* | -1.72210026 | 0.018763642 | 0.037068889 | 1 |
| HTN | monocyte | *BRD9* | -2.049884808 | 0.020290481 | 0.039005315 | 1 |
| HTN | monocyte | *BAK1* | 1.568758666 | 0.008430889 | 0.025487024 | 1 |
| HTN | monocyte | *IKZF2* | -1.957320823 | 0.01420771 | 0.031826319 | 1 |
| HTN | monocyte | *GRN* | -1.921120838 | 0.004342877 | 0.023595115 | 1 |
| HTN | monocyte | *CENPQ* | -1.502352072 | 0.000333753 | 0.014588925 | 1 |
| HTN | monocyte | *IFT88* | -1.597901316 | 0.011113468 | 0.028084661 | 1 |
| HTN | monocyte | *CHPF2* | -2.022296041 | 0.012905362 | 0.030176778 | 1 |
| HTN | monocyte | *UBA6* | -2.569941694 | 0.013715028 | 0.031168765 | 1 |
| HTN | monocyte | *ATP6V0A1* | -1.339029133 | 0.015362683 | 0.033143504 | 1 |
| HTN | monocyte | *SH3YL1* | -1.59395289 | 0.002462294 | 0.023595115 | 1 |
| HTN | monocyte | *VCL* | -2.049700754 | 0.023985046 | 0.043824751 | 1 |
| HTN | monocyte | *NSUN2* | -1.762566002 | 0.015323829 | 0.033089106 | 1 |
| HTN | monocyte | *AGA* | 4.907579376 | 0.002508393 | 0.023595115 | 1 |
| HTN | monocyte | *PI4K2B* | 10.7702916 | 0.026838747 | 0.047215953 | 1 |
| HTN | monocyte | *BOD1L1* | -1.762243457 | 0.028969153 | 0.049738022 | 1 |
| HTN | monocyte | *PHLPP2* | -1.930129539 | 0.007012918 | 0.024238452 | 1 |
| HTN | monocyte | *SPDL1* | -2.99141113 | 0.011138614 | 0.028107124 | 1 |
| HTN | monocyte | *PQLC2* | -4.797941469 | 0.018013627 | 0.035970978 | 1 |
| HTN | monocyte | *PSMA4* | 3.92203926 | 0.013402137 | 0.030691157 | 1 |
| HTN | monocyte | *LSG1* | -1.402454791 | 0.004486357 | 0.023595115 | 1 |
| HTN | monocyte | *TDP1* | 3.928703044 | 0.016047415 | 0.033794297 | 1 |
| HTN | monocyte | *SPATA7* | -1.718483829 | 0.005792775 | 0.023595115 | 1 |
| HTN | monocyte | *AP2S1* | 3.796671068 | 0.009068389 | 0.026214745 | 1 |
| HTN | monocyte | *TRIT1* | 2.246751395 | 0.022757466 | 0.042176939 | 1 |
| HTN | monocyte | *CUL7* | -2.198895701 | 0.006473641 | 0.023864166 | 1 |
| HTN | monocyte | *CTNNA1* | -1.716295617 | 0.028120598 | 0.048791054 | 1 |
| HTN | monocyte | *YTHDC2* | -1.89676631 | 0.026981828 | 0.047401239 | 1 |
| HTN | monocyte | *ARAP2* | 1.424668682 | 0.001728668 | 0.023595115 | 1 |
| HTN | monocyte | *DTNBP1* | -2.104142718 | 0.015534109 | 0.033364983 | 1 |
| HTN | monocyte | *ANO2* | -0.924451548 | 0.005534064 | 0.023595115 | 1 |
| HTN | monocyte | *MRPS10* | -2.076434384 | 0.002544832 | 0.023595115 | 1 |
| HTN | monocyte | *UTS2* | 4.253637516 | 0.006062173 | 0.023595115 | 1 |
| HTN | monocyte | *EPN3* | -4.294897356 | 0.015650833 | 0.033438063 | 1 |
| HTN | monocyte | *RCN1* | -2.547998594 | 0.004828789 | 0.023595115 | 1 |
| HTN | monocyte | *JKAMP* | -1.558176209 | 0.005934783 | 0.023595115 | 1 |
| HTN | monocyte | *MCUR1* | 3.016454666 | 0.003118149 | 0.023595115 | 1 |
| HTN | monocyte | *CYBA* | 2.972218056 | 0.014090582 | 0.031686282 | 1 |
| HTN | monocyte | *THOC3* | 3.391411825 | 0.00304404 | 0.023595115 | 1 |
| HTN | monocyte | *ALX4* | 2.014298198 | 0.022789371 | 0.042203053 | 1 |
| HTN | monocyte | *AKR7A2* | -1.458844162 | 0.005939947 | 0.023595115 | 1 |
| HTN | monocyte | *MRTO4* | -1.419307821 | 0.008292467 | 0.025373581 | 1 |
| HTN | monocyte | *LY75* | -2.853663215 | 0.005470149 | 0.023595115 | 1 |
| HTN | monocyte | *GALC* | -2.15863544 | 0.011724149 | 0.028672163 | 1 |
| HTN | monocyte | *GINM1* | -1.664031591 | 0.014054327 | 0.03166001 | 1 |

| | | | | | | |
|---|---|---|---|---|---|---|
| HTN | monocyte | *ITIH4* | 1.323098923 | 0.010233468 | 0.027124512 | 1 |
| HTN | monocyte | *NPFFR2* | -2.187589726 | 0.004610413 | 0.023595115 | 1 |
| HTN | monocyte | *IL17RB* | -2.892490207 | 0.001590941 | 0.023595115 | 1 |
| HTN | monocyte | *RASGRF1* | -2.973981583 | 0.006949638 | 0.024187912 | 1 |
| HTN | monocyte | *ALDH18A1* | -2.470902999 | 0.020164641 | 0.038917597 | 1 |
| HTN | monocyte | *DNAJC25* | 3.298347141 | 0.003888258 | 0.023595115 | 1 |
| HTN | monocyte | *PIGV* | -5.01393789 | 3.58E-06 | 0.000579456 | 0.017383691 |
| HTN | monocyte | *RIMBP2* | -2.213646032 | 0.006811675 | 0.02405092 | 1 |
| HTN | monocyte | *PRDM6* | -1.242373477 | 0.005808191 | 0.023595115 | 1 |
| HTN | monocyte | *SPAG4* | -5.832830407 | 0.006611234 | 0.02395053 | 1 |
| HTN | monocyte | *NCKAP1* | -1.41776753 | 0.006416873 | 0.023819881 | 1 |
| HTN | monocyte | *LTK* | 25.39136601 | 0.003549886 | 0.023595115 | 1 |
| HTN | monocyte | *AHRR* | -2.50761674 | 0.004500572 | 0.023595115 | 1 |
| HTN | monocyte | *RNF4* | -2.32612811 | 0.026243206 | 0.046482276 | 1 |
| HTN | monocyte | *CDON* | -0.597520435 | 0.028768202 | 0.049515188 | 1 |
| HTN | monocyte | *TMEM161A* | -2.13074819 | 0.009421366 | 0.026481384 | 1 |
| HTN | monocyte | *LPAR2* | -1.893145484 | 0.013083312 | 0.030344279 | 1 |
| HTN | monocyte | *SUGP2* | -1.987907877 | 0.024957245 | 0.044932302 | 1 |
| HTN | monocyte | *SLC12A2* | -1.542321202 | 0.010962823 | 0.02789645 | 1 |
| HTN | monocyte | *CNN2* | -2.033067034 | 0.008072488 | 0.025188238 | 1 |
| HTN | monocyte | *ABCA7* | 1.230870819 | 0.013309446 | 0.030634456 | 1 |
| HTN | monocyte | *FAR2* | -1.786914156 | 0.02702473 | 0.047439938 | 1 |
| HTN | monocyte | *BCAS1* | -2.060704602 | 0.005050882 | 0.023595115 | 1 |
| HTN | monocyte | *CHI3L2* | 2.158228542 | 0.001666579 | 0.023595115 | 1 |
| HTN | monocyte | *SBNO2* | -2.133789395 | 0.022564829 | 0.041986971 | 1 |
| HTN | monocyte | *ANKRD44* | 1.802690968 | 0.001862879 | 0.023595115 | 1 |
| HTN | monocyte | *TIE1* | -3.250633377 | 6.94E-05 | 0.005633811 | 0.33660065 |
| HTN | monocyte | *NGEF* | 3.158399924 | 0.016471243 | 0.034355995 | 1 |
| HTN | monocyte | *SPI1* | 0.585932513 | 0.004438155 | 0.023595115 | 1 |
| HTN | monocyte | *ZNRD1* | -2.740233959 | 0.00467078 | 0.023595115 | 1 |
| HTN | monocyte | *ATXN3* | -1.829294191 | 0.002090762 | 0.023595115 | 1 |
| HTN | monocyte | *FGFR2* | -1.392325713 | 0.000857658 | 0.020511604 | 1 |
| HTN | monocyte | *MTFR1* | -2.001949207 | 0.005007343 | 0.023595115 | 1 |
| HTN | monocyte | *FECH* | -1.07609826 | 0.0048186 | 0.023595115 | 1 |
| HTN | monocyte | *MYO9A* | -1.428112366 | 0.011921086 | 0.028877239 | 1 |
| HTN | monocyte | *STOML1* | 6.842978198 | 0.005478862 | 0.023595115 | 1 |
| HTN | monocyte | *METTL22* | -2.685779953 | 0.018465589 | 0.03659928 | 1 |
| HTN | monocyte | *TP53BP1* | -1.6090574 | 0.007467416 | 0.024580667 | 1 |
| HTN | monocyte | *ROGDI* | -2.013487104 | 0.0052174 | 0.023595115 | 1 |
| HTN | monocyte | *CBFB* | -1.947299233 | 0.014777172 | 0.03242824 | 1 |
| HTN | monocyte | *IFI35* | -1.498806786 | 0.010724216 | 0.027636231 | 1 |
| HTN | monocyte | *HEATR6* | 5.084714131 | 0.002223711 | 0.023595115 | 1 |
| HTN | monocyte | *POLR1A* | -2.582728672 | 9.28E-05 | 0.006642646 | 0.450454113 |
| HTN | monocyte | *CYFIP1* | -1.622095041 | 0.010725273 | 0.027636231 | 1 |
| HTN | monocyte | *IFT80* | -1.394417535 | 0.002481487 | 0.023595115 | 1 |
| HTN | monocyte | *NUCKS1* | -1.778939511 | 0.016543672 | 0.034411074 | 1 |
| HTN | monocyte | *DRD4* | -2.424241386 | 0.006284908 | 0.023704653 | 1 |
| HTN | monocyte | *PLA2G10* | -3.595549243 | 0.00518714 | 0.023595115 | 1 |
| HTN | monocyte | *PIGB* | -1.263356872 | 0.005596156 | 0.023595115 | 1 |
| HTN | monocyte | *GNB5* | -1.744644106 | 0.001590071 | 0.023595115 | 1 |

| | | | | | | |
|---|---|---|---|---|---|---|
| HTN | monocyte | *LRP6* | -1.699944181 | 0.01086586 | 0.027878663 | 1 |
| HTN | monocyte | *DAPP1* | 0.337437072 | 0.007637117 | 0.024886026 | 1 |
| HTN | monocyte | *CLTCL1* | -2.180213211 | 0.013216457 | 0.030491729 | 1 |
| HTN | monocyte | *DGCR2* | -1.566420774 | 0.005891957 | 0.023595115 | 1 |
| HTN | monocyte | *GBA2* | -1.910125256 | 0.009623126 | 0.02668844 | 1 |
| HTN | monocyte | *NDST1* | -1.481276781 | 0.026983145 | 0.047401239 | 1 |
| HTN | monocyte | *ASNS* | 1.940040654 | 0.000728542 | 0.019858911 | 1 |
| HTN | monocyte | *AP3M2* | -2.84234859 | 0.011620465 | 0.028577038 | 1 |
| HTN | monocyte | *ST6GALNAC2* | -2.656618779 | 0.02256843 | 0.041986971 | 1 |
| HTN | monocyte | *EIF2B3* | -1.520418889 | 0.011432379 | 0.028446105 | 1 |
| HTN | monocyte | *WDR1* | -1.702099046 | 0.009652495 | 0.02668844 | 1 |
| HTN | monocyte | *SNX13* | -1.834328989 | 0.028519595 | 0.049191992 | 1 |
| HTN | monocyte | *VASH1* | 3.378147367 | 0.002379729 | 0.023595115 | 1 |
| HTN | monocyte | *TRIB2* | -1.636356301 | 0.024271721 | 0.044156876 | 1 |
| HTN | monocyte | *CPSF1* | 0.895867486 | 0.015853615 | 0.033684442 | 1 |
| HTN | monocyte | *CYBRD1* | -1.333387424 | 0.011149999 | 0.028118396 | 1 |
| HTN | monocyte | *EPN2* | -1.68768376 | 0.002238248 | 0.023595115 | 1 |
| HTN | monocyte | *AFF4* | -1.721526568 | 0.027505743 | 0.048058288 | 1 |
| HTN | monocyte | *TRHDE* | -0.6546363 | 0.002739831 | 0.023595115 | 1 |
| HTN | monocyte | *FCGR2B* | -2.700685856 | 0.012296476 | 0.029246324 | 1 |
| HTN | monocyte | *CRMP1* | -1.113494863 | 0.00563042 | 0.023595115 | 1 |
| HTN | monocyte | *NDE1* | 3.775298957 | 0.019455785 | 0.038071952 | 1 |
| HTN | monocyte | *AP1M1* | -3.079604077 | 0.016639739 | 0.034456332 | 1 |
| HTN | monocyte | *SCARB1* | -1.379237483 | 0.005753073 | 0.023595115 | 1 |
| HTN | monocyte | *MOV10L1* | -1.464168055 | 0.009850576 | 0.026779763 | 1 |
| HTN | monocyte | *ALPK1* | -1.257404182 | 0.00789185 | 0.025073967 | 1 |
| HTN | monocyte | *LLGL2* | -2.1868068 | 0.016323505 | 0.034182843 | 1 |
| HTN | monocyte | *PPP2R2C* | -3.769983476 | 0.008287476 | 0.025373581 | 1 |
| HTN | monocyte | *ZNF532* | -1.880327965 | 0.021599234 | 0.040841576 | 1 |
| HTN | monocyte | *TTC38* | -1.016207763 | 0.006711705 | 0.024005639 | 1 |
| HTN | monocyte | *ADD2* | 2.782553969 | 0.000615812 | 0.0181086 | 1 |
| HTN | monocyte | *FNDC3B* | -1.190789774 | 0.013868655 | 0.031408232 | 1 |
| HTN | monocyte | *MOCOS* | -1.584894654 | 0.00622214 | 0.023665784 | 1 |
| HTN | monocyte | *SEC31B* | -1.598241578 | 0.00691112 | 0.024148823 | 1 |
| HTN | monocyte | *MCM6* | -1.241677475 | 0.009863135 | 0.026779763 | 1 |
| HTN | monocyte | *MLH1* | -1.882310824 | 0.026069667 | 0.046367311 | 1 |
| HTN | monocyte | *FMO4* | -0.883547809 | 0.008383813 | 0.025487024 | 1 |
| HTN | monocyte | *RGS11* | -1.143097305 | 0.004050238 | 0.023595115 | 1 |
| HTN | monocyte | *SPAG5* | -5.599426316 | 0.001000403 | 0.021669448 | 1 |
| HTN | monocyte | *ANKRD13A* | -1.815621765 | 0.026800268 | 0.047199601 | 1 |
| HTN | monocyte | *PAG1* | 0.915972921 | 0.003011919 | 0.023595115 | 1 |
| HTN | monocyte | *GPATCH1* | -1.690351837 | 0.011738967 | 0.02867949 | 1 |
| HTN | monocyte | *ICAM3* | 3.879174293 | 0.004811592 | 0.023595115 | 1 |
| HTN | monocyte | *XAB2* | -2.101821266 | 0.008295513 | 0.025373581 | 1 |
| HTN | monocyte | *LRCH4* | 1.796238233 | 0.001475781 | 0.023595115 | 1 |
| HTN | monocyte | *SIRT6* | -1.775435427 | 0.025876166 | 0.046158513 | 1 |
| HTN | monocyte | *AMPH* | -1.215635862 | 0.009647509 | 0.02668844 | 1 |
| HTN | monocyte | *MCCC1* | -1.384142911 | 0.007426535 | 0.024522157 | 1 |
| HTN | monocyte | *FGF20* | 3.822111226 | 0.023730501 | 0.043449203 | 1 |
| HTN | monocyte | *PCM1* | -1.693407603 | 0.024515146 | 0.044413403 | 1 |

| | | | | | | |
|---|---|---|---|---|---|---|
| HTN | monocyte | *FKBP7* | -1.419015153 | 0.00253188 | 0.023595115 | 1 |
| HTN | monocyte | *SP140* | 1.509213082 | 0.010678181 | 0.027602843 | 1 |
| HTN | monocyte | *RAPGEF3* | -0.743822727 | 0.02593016 | 0.046203869 | 1 |
| HTN | monocyte | *DUSP13* | 15.09738703 | 0.000203629 | 0.010739192 | 0.988005656 |
| HTN | monocyte | *SCGN* | -8.747554659 | 0.006482574 | 0.023864166 | 1 |
| HTN | monocyte | *PGM1* | -1.356296341 | 0.005418146 | 0.023595115 | 1 |
| HTN | monocyte | *RABL2B* | -2.40630693 | 0.010933616 | 0.027890132 | 1 |
| HTN | monocyte | *SCTR* | -1.005653128 | 0.00513969 | 0.023595115 | 1 |
| HTN | monocyte | *RIF1* | -1.619899235 | 0.017196301 | 0.035103675 | 1 |
| HTN | monocyte | *KCNN2* | -3.522325595 | 0.002895003 | 0.023595115 | 1 |
| HTN | monocyte | *PSEN1* | 3.084900264 | 0.015621235 | 0.033404245 | 1 |
| HTN | monocyte | *MOK* | -1.26876613 | 0.017764434 | 0.03574991 | 1 |
| HTN | monocyte | *STK17B* | 1.870380799 | 0.00161585 | 0.023595115 | 1 |
| HTN | monocyte | *PCDHB4* | 6.845966537 | 0.004992229 | 0.023595115 | 1 |
| HTN | monocyte | *CCNT2* | -2.03083169 | 0.006607417 | 0.02395053 | 1 |
| HTN | monocyte | *LYRM2* | -2.612703038 | 0.002230016 | 0.023595115 | 1 |
| HTN | monocyte | *ULK2* | -2.123516283 | 0.014796903 | 0.032436518 | 1 |
| HTN | monocyte | *PIBF1* | -1.668198099 | 0.028683082 | 0.049403732 | 1 |
| HTN | monocyte | *OXCT1* | -1.498480594 | 0.003537573 | 0.023595115 | 1 |
| HTN | monocyte | *SLC27A5* | -1.339805294 | 0.009347313 | 0.026350386 | 1 |
| HTN | monocyte | *ZNF264* | -1.472507675 | 0.007040381 | 0.024238452 | 1 |
| HTN | monocyte | *PPIE* | -1.181593289 | 0.012449722 | 0.029382558 | 1 |
| HTN | monocyte | *NOA1* | 3.053428312 | 0.012952444 | 0.0302286 | 1 |
| HTN | monocyte | *GSTP1* | -2.265441957 | 0.026499763 | 0.046840382 | 1 |
| HTN | monocyte | *CAD* | 3.409729087 | 0.000331126 | 0.014588925 | 1 |
| HTN | monocyte | *HACE1* | -1.938361787 | 0.015362266 | 0.033143504 | 1 |
| HTN | monocyte | *SEH1L* | -2.343267464 | 0.012131693 | 0.029051118 | 1 |
| HTN | monocyte | *PILRA* | 0.658665077 | 0.00108464 | 0.022782137 | 1 |
| HTN | monocyte | *ZNF213* | -2.006944341 | 0.027668822 | 0.048273688 | 1 |
| HTN | monocyte | *RRN3* | -2.082305667 | 0.006623003 | 0.02395053 | 1 |
| HTN | monocyte | *USP40* | -1.316868643 | 0.009286667 | 0.026288744 | 1 |
| HTN | monocyte | *POMGNT1* | -1.794858339 | 0.011654223 | 0.028586902 | 1 |
| HTN | monocyte | *NME8* | 3.257258919 | 0.00336409 | 0.023595115 | 1 |
| HTN | monocyte | *ITPKC* | -1.878607382 | 0.003115334 | 0.023595115 | 1 |
| HTN | monocyte | *ALG9* | -3.974306976 | 0.02283186 | 0.042218058 | 1 |
| HTN | monocyte | *ACOX3* | -1.653288139 | 0.007909049 | 0.025073967 | 1 |
| HTN | monocyte | *TRIP6* | -1.463277021 | 0.012188205 | 0.029117267 | 1 |
| HTN | monocyte | *ADAMTS2* | -1.072725709 | 0.005478595 | 0.023595115 | 1 |
| HTN | monocyte | *UIMC1* | -1.883998793 | 0.027022551 | 0.047439938 | 1 |
| HTN | monocyte | *LPCAT2* | 2.352502753 | 0.003899236 | 0.023595115 | 1 |
| HTN | monocyte | *TXNDC16* | -2.15678531 | 0.00814174 | 0.025290474 | 1 |
| HTN | monocyte | *SF3B2* | -2.595218218 | 0.026334523 | 0.046599237 | 1 |
| HTN | monocyte | *PHACTR3* | -2.077024185 | 0.005247024 | 0.023595115 | 1 |
| HTN | monocyte | *AAMDC* | -2.072021615 | 0.000148918 | 0.009031904 | 0.722552285 |
| HTN | monocyte | *ASAP3* | -1.244890586 | 0.006392723 | 0.023797215 | 1 |
| HTN | monocyte | *ANKRD10* | -1.488638684 | 0.015099262 | 0.032779249 | 1 |
| HTN | monocyte | *DOCK3* | -1.019724707 | 0.00556212 | 0.023595115 | 1 |
| HTN | monocyte | *ARHGAP28* | -1.114641117 | 0.006911346 | 0.024148823 | 1 |
| HTN | monocyte | *PPP1R13B* | -2.602265673 | 0.002478138 | 0.023595115 | 1 |
| HTN | monocyte | *EBF4* | -1.955346998 | 0.00912576 | 0.026216909 | 1 |

| | | | | | | |
|---|---|---|---|---|---|---|
| HTN | monocyte | *CPXM1* | -1.217364296 | 0.00925228 | 0.026254852 | 1 |
| HTN | monocyte | *MAVS* | -2.116760916 | 0.002697443 | 0.023595115 | 1 |
| HTN | monocyte | *ESF1* | -1.724535125 | 0.015952501 | 0.03374074 | 1 |
| HTN | monocyte | *RBBP9* | -2.138520323 | 0.001183367 | 0.02330522 | 1 |
| HTN | monocyte | *TMEM230* | -1.928498092 | 0.014757811 | 0.032409041 | 1 |
| HTN | monocyte | *RPH3A* | -2.833324869 | 0.017898761 | 0.035878799 | 1 |
| HTN | monocyte | *KIF16B* | -1.94409486 | 0.021714419 | 0.041043381 | 1 |
| HTN | monocyte | *TBX5* | 1.701887038 | 0.010740727 | 0.027661364 | 1 |
| HTN | monocyte | *FXYD5* | 2.599706934 | 0.006217004 | 0.023665784 | 1 |
| HTN | monocyte | *ZNF302* | -1.430312935 | 0.009557398 | 0.026650066 | 1 |
| HTN | monocyte | *CDIP1* | -1.158498914 | 0.005093734 | 0.023595115 | 1 |
| HTN | monocyte | *KCNH4* | -2.117075063 | 0.011756718 | 0.028708402 | 1 |
| HTN | monocyte | *GANAB* | -2.847250276 | 0.021424343 | 0.040621692 | 1 |
| HTN | monocyte | *ANKRD24* | -1.616594896 | 0.004847569 | 0.023595115 | 1 |
| HTN | monocyte | *DHX32* | -1.823531034 | 0.010504859 | 0.027479049 | 1 |
| HTN | monocyte | *SPTLC1* | -2.732012546 | 0.016449487 | 0.034328134 | 1 |
| HTN | monocyte | *MAEA* | -1.492596861 | 0.007158823 | 0.024238452 | 1 |
| HTN | monocyte | *LYZ* | 0.704522588 | 0.00195214 | 0.023595115 | 1 |
| HTN | monocyte | *MUL1* | -3.227165702 | 0.000106667 | 0.007289407 | 0.517547922 |
| HTN | monocyte | *RAB11FIP3* | -1.210053678 | 0.010177159 | 0.027100648 | 1 |
| HTN | monocyte | *GNPTG* | -2.063631848 | 0.016645845 | 0.034456332 | 1 |
| HTN | monocyte | *PABPC4* | -5.200908045 | 1.85E-05 | 0.002082378 | 0.089542257 |
| HTN | monocyte | *CERS4* | -1.569177719 | 0.008681349 | 0.025839644 | 1 |
| HTN | monocyte | *USP48* | -1.786269231 | 0.018309479 | 0.036393934 | 1 |
| HTN | monocyte | *DTX2* | -2.271225116 | 0.008877031 | 0.025962238 | 1 |
| HTN | monocyte | *LAMB1* | -1.027667866 | 0.002905448 | 0.023595115 | 1 |
| HTN | monocyte | *ABCC6* | -1.260373762 | 0.001862563 | 0.023595115 | 1 |
| HTN | monocyte | *RAPGEF4* | -2.224927937 | 0.014369725 | 0.031982525 | 1 |
| HTN | monocyte | *RGS17* | -0.824719441 | 0.028286053 | 0.048876043 | 1 |
| HTN | monocyte | *HAUS4* | 1.761597606 | 0.003345249 | 0.023595115 | 1 |
| HTN | monocyte | *SCFD1* | -1.804279204 | 0.025542472 | 0.045780981 | 1 |
| HTN | monocyte | *SUPT16H* | -2.212345445 | 0.020098976 | 0.038821748 | 1 |
| HTN | monocyte | *TOX4* | -1.970040069 | 0.008758168 | 0.025848597 | 1 |
| HTN | monocyte | *CAPN3* | -2.814790517 | 0.006588681 | 0.023928355 | 1 |
| HTN | monocyte | *COMT* | -1.976251251 | 0.016920216 | 0.034713272 | 1 |
| HTN | monocyte | *SEC22C* | -2.299257601 | 0.019118011 | 0.037567848 | 1 |
| HTN | monocyte | *XYLB* | -1.76894902 | 0.008671282 | 0.025839644 | 1 |
| HTN | monocyte | *GABRP* | 5.357557359 | 0.026581945 | 0.046945924 | 1 |
| HTN | monocyte | *CDC6* | 1.731506395 | 0.018047409 | 0.036005768 | 1 |
| HTN | monocyte | *MSH2* | -1.752302279 | 0.009452597 | 0.026526317 | 1 |
| HTN | monocyte | *PSMD5* | -1.66753403 | 0.011110222 | 0.028084661 | 1 |
| HTN | monocyte | *CRAT* | -1.458390894 | 0.005115941 | 0.023595115 | 1 |
| HTN | monocyte | *NANS* | 4.45422364 | 0.006379919 | 0.023797215 | 1 |
| HTN | monocyte | *CWF19L1* | -1.550379165 | 0.009669884 | 0.02668844 | 1 |
| HTN | monocyte | *DSP* | -1.360784605 | 0.005327922 | 0.023595115 | 1 |
| HTN | monocyte | *IFT74* | -1.532679954 | 0.027722712 | 0.048332949 | 1 |
| HTN | monocyte | *ACOT7* | 2.783652045 | 0.016429907 | 0.034302026 | 1 |
| HTN | monocyte | *CDC7* | -1.363547112 | 0.013067355 | 0.030344279 | 1 |
| HTN | monocyte | *SCD* | -1.379690673 | 0.00023028 | 0.011401229 | 1 |
| HTN | monocyte | *ERMP1* | -1.612596303 | 0.010360034 | 0.027318959 | 1 |

| | | | | | | |
|---|---|---|---|---|---|---|
| HTN | monocyte | *RAB18* | -2.116493477 | 0.028126832 | 0.048791054 | 1 |
| HTN | monocyte | *PRTFDC1* | -1.095406706 | 0.014501276 | 0.03215962 | 1 |
| HTN | monocyte | *TSPAN15* | -0.999067033 | 0.005727689 | 0.023595115 | 1 |
| HTN | monocyte | *SETD1A* | 5.475753129 | 0.015856841 | 0.033684442 | 1 |
| HTN | monocyte | *CIRBP* | -1.678083935 | 0.0186848 | 0.036943215 | 1 |
| HTN | monocyte | *MTAP* | -3.836500307 | 0.001180062 | 0.02330522 | 1 |
| HTN | monocyte | *POLR2E* | -2.60187023 | 0.024624516 | 0.044531551 | 1 |
| HTN | monocyte | *MADCAM1* | -2.573265683 | 0.004355438 | 0.023595115 | 1 |
| HTN | monocyte | *CECR2* | -1.231649427 | 0.00426224 | 0.023595115 | 1 |
| HTN | monocyte | *SMARCB1* | -1.093097083 | 0.015573582 | 0.033395675 | 1 |
| HTN | monocyte | *BCL2L13* | -1.241377381 | 0.018665092 | 0.036926464 | 1 |
| HTN | monocyte | *DDTL* | -2.050917659 | 0.01776391 | 0.03574991 | 1 |
| HTN | monocyte | *DDT* | 1.740495543 | 0.005508455 | 0.023595115 | 1 |
| HTN | monocyte | *SPECC1L* | -3.022552424 | 0.003769649 | 0.023595115 | 1 |
| HTN | monocyte | *TOP3B* | -3.38765881 | 0.01494196 | 0.032627539 | 1 |
| HTN | monocyte | *CYTH4* | 0.712683339 | 0.00312625 | 0.023595115 | 1 |
| HTN | monocyte | *CARD10* | -1.376173696 | 0.007527586 | 0.024638032 | 1 |
| HTN | monocyte | *SH3BP1* | 0.653118215 | 0.003284072 | 0.023595115 | 1 |
| HTN | monocyte | *HPS4* | -1.952589241 | 0.01735365 | 0.035260232 | 1 |
| HTN | monocyte | *TRIOBP* | -3.029831077 | 0.012106827 | 0.029032068 | 1 |
| HTN | monocyte | *EIF3L* | 2.203352841 | 0.022364952 | 0.041811545 | 1 |
| HTN | monocyte | *TTC28* | -1.294055547 | 0.015973306 | 0.03374074 | 1 |
| HTN | monocyte | *CENPM* | -4.669184238 | 0.004917065 | 0.023595115 | 1 |
| HTN | monocyte | *KDELR3* | -1.526608742 | 0.004743178 | 0.023595115 | 1 |
| HTN | monocyte | *CYP2D6* | -3.430539637 | 0.002412868 | 0.023595115 | 1 |
| HTN | monocyte | *RTCB* | -1.603807513 | 0.009558913 | 0.026650066 | 1 |
| HTN | monocyte | *RAB36* | -2.747351772 | 0.00294748 | 0.023595115 | 1 |
| HTN | monocyte | *TIMP3* | -1.263195452 | 0.013763599 | 0.031249874 | 1 |
| HTN | monocyte | *CYB5R3* | -2.143154329 | 0.015956799 | 0.03374074 | 1 |
| HTN | monocyte | *RHBDD3* | -1.307646784 | 0.006143804 | 0.023595115 | 1 |
| HTN | monocyte | *CHKB* | 7.670070936 | 0.001470136 | 0.023595115 | 1 |
| HTN | monocyte | *THOC5* | -4.025620581 | 0.000193345 | 0.010423452 | 0.938110686 |
| HTN | monocyte | *TTLL12* | -1.718580615 | 0.002878672 | 0.023595115 | 1 |
| HTN | monocyte | *KIAA0930* | 1.332466856 | 0.002462461 | 0.023595115 | 1 |
| HTN | monocyte | *FAM118A* | 2.837655275 | 0.000398164 | 0.01501035 | 1 |
| HTN | monocyte | *L3MBTL2* | -1.343659831 | 0.004134328 | 0.023595115 | 1 |
| HTN | monocyte | *ZC3H7B* | -1.912105439 | 0.026941158 | 0.047361775 | 1 |
| HTN | monocyte | *ACO2* | -1.964406757 | 0.004289551 | 0.023595115 | 1 |
| HTN | monocyte | *DESI1* | -1.750153598 | 0.01899169 | 0.037415876 | 1 |
| HTN | monocyte | *MLC1* | 0.831857917 | 0.002600163 | 0.023595115 | 1 |
| HTN | monocyte | *HDAC10* | -1.421492687 | 0.005641009 | 0.023595115 | 1 |
| HTN | monocyte | *SDR39U1* | -1.968466093 | 0.011528277 | 0.028508147 | 1 |
| HTN | monocyte | *RBM23* | -1.667426171 | 0.005643962 | 0.023595115 | 1 |
| HTN | monocyte | *PYGL* | -1.120034614 | 0.001326383 | 0.023595115 | 1 |
| HTN | monocyte | *CNIH1* | -1.813778048 | 0.015155965 | 0.032858239 | 1 |
| HTN | monocyte | *ATP6V1D* | -1.844512456 | 0.014272374 | 0.031926951 | 1 |
| HTN | monocyte | *GSTZ1* | -1.400140315 | 0.006577247 | 0.023914328 | 1 |
| HTN | monocyte | *LGMN* | -0.886736443 | 0.022821954 | 0.042215829 | 1 |
| HTN | monocyte | *MTHFD1* | -1.11914742 | 0.009859695 | 0.026779763 | 1 |
| HTN | monocyte | *YY1* | -1.820468104 | 0.013241615 | 0.030507273 | 1 |

| | | | | | | |
|---|---|---|---|---|---|---|
| HTN | monocyte | *EFS* | -1.298255123 | 0.004815425 | 0.023595115 | 1 |
| HTN | monocyte | *CINP* | -2.989014687 | 0.021434524 | 0.040622233 | 1 |
| HTN | monocyte | *KIAA0391* | 1.34874193 | 0.002773743 | 0.023595115 | 1 |
| HTN | monocyte | *EMC9* | -1.922451301 | 0.004981043 | 0.023595115 | 1 |
| HTN | monocyte | *PLTP* | -1.222081031 | 0.00286192 | 0.023595115 | 1 |
| HTN | monocyte | *UQCC1* | -1.532007529 | 0.009077661 | 0.026214745 | 1 |
| HTN | monocyte | *IFT52* | -1.807217712 | 0.004993476 | 0.023595115 | 1 |
| HTN | monocyte | *PABPC1L* | -1.695369039 | 0.007133835 | 0.024238452 | 1 |
| HTN | monocyte | *DOK5* | -1.370106739 | 0.0054575 | 0.023595115 | 1 |
| HTN | monocyte | *NELFCD* | -2.316463032 | 0.009108984 | 0.026216909 | 1 |
| HTN | monocyte | *CTSZ* | -1.973187636 | 0.017002536 | 0.034837966 | 1 |
| HTN | monocyte | *MTG2* | -2.108556736 | 0.026601242 | 0.046945924 | 1 |
| HTN | monocyte | *BIRC7* | -6.412026256 | 0.00051514 | 0.016722594 | 1 |
| HTN | monocyte | *NKAIN4* | -3.03596172 | 0.006966821 | 0.024187912 | 1 |
| HTN | monocyte | *PTK6* | -3.940778634 | 0.005816205 | 0.023595115 | 1 |
| HTN | monocyte | *GMEB2* | -2.198431652 | 0.006702282 | 0.024005639 | 1 |
| HTN | monocyte | *CDC25B* | -1.446079551 | 0.007485668 | 0.024607358 | 1 |
| HTN | monocyte | *NDUFAF5* | -1.935497052 | 0.005012827 | 0.023595115 | 1 |
| HTN | monocyte | *TRIB3* | -3.614101791 | 0.008723137 | 0.025839644 | 1 |
| HTN | monocyte | *RASSF2* | 0.608897062 | 0.002342579 | 0.023595115 | 1 |
| HTN | monocyte | *ANGPT4* | -2.717506853 | 0.003469363 | 0.023595115 | 1 |
| HTN | monocyte | *RSPO4* | 2.839347594 | 0.00493297 | 0.023595115 | 1 |
| HTN | monocyte | *SIRPB1* | 0.515752988 | 0.007198042 | 0.024287135 | 1 |
| HTN | monocyte | *MROH8* | -1.410597129 | 0.00440181 | 0.023595115 | 1 |
| HTN | monocyte | *BPI* | -1.329798499 | 0.005789652 | 0.023595115 | 1 |
| HTN | monocyte | *ACTR5* | -1.575035922 | 0.013396116 | 0.030691157 | 1 |
| HTN | monocyte | *DNTTIP1* | -1.84978926 | 0.000182943 | 0.009973488 | 0.887640426 |
| HTN | monocyte | *MAP1LC3A* | -1.786242576 | 0.01008679 | 0.027007023 | 1 |
| HTN | monocyte | *MYOM1* | -1.061953669 | 0.006095574 | 0.023595115 | 1 |
| HTN | monocyte | *POLI* | -2.385219063 | 8.82E-06 | 0.001198503 | 0.04280798 |
| HTN | monocyte | *MIB1* | -1.630625598 | 0.015966378 | 0.03374074 | 1 |
| HTN | monocyte | *NALCN* | -1.077297207 | 0.012977085 | 0.030254871 | 1 |
| HTN | monocyte | *ACP5* | -1.348073382 | 0.013205204 | 0.030491729 | 1 |
| HTN | monocyte | *UGGT2* | -1.487639512 | 0.017038446 | 0.034882084 | 1 |
| HTN | monocyte | *SUPT20H* | -1.733216781 | 0.022745856 | 0.042171529 | 1 |
| HTN | monocyte | *MRPS31* | -1.885564592 | 0.015605292 | 0.033395675 | 1 |
| HTN | monocyte | *RGCC* | -1.253928386 | 0.004359592 | 0.023595115 | 1 |
| HTN | monocyte | *KATNAL1* | -1.460018339 | 0.009824324 | 0.026779763 | 1 |
| HTN | monocyte | *CLN5* | -2.358806574 | 0.028585763 | 0.049288601 | 1 |
| HTN | monocyte | *LONP2* | -1.462841149 | 0.021205425 | 0.040316897 | 1 |
| HTN | monocyte | *PLLP* | -1.035863185 | 0.004891998 | 0.023595115 | 1 |
| HTN | monocyte | *DHODH* | -1.333855541 | 0.020466412 | 0.039272886 | 1 |
| HTN | monocyte | *CCDC113* | -4.820551679 | 0.002156151 | 0.023595115 | 1 |
| HTN | monocyte | *NPRL3* | -1.084328986 | 0.012543907 | 0.029539829 | 1 |
| HTN | monocyte | *NAGPA* | -1.661492955 | 0.009999442 | 0.026909204 | 1 |
| HTN | monocyte | *ZNF500* | -1.595307219 | 0.001235198 | 0.023595115 | 1 |
| HTN | monocyte | *NME4* | -3.17287334 | 0.003468535 | 0.023595115 | 1 |
| HTN | monocyte | *LMF1* | -2.040877303 | 0.019459695 | 0.038071952 | 1 |
| HTN | monocyte | *MTHFSD* | 3.625560449 | 1.43E-05 | 0.001730185 | 0.069207403 |
| HTN | monocyte | *HAGHL* | 3.432219166 | 0.000475507 | 0.016247607 | 1 |

| | | | | | | |
|---|---|---|---|---|---|---|
| HTN | monocyte | *RHBDL1* | -4.517598897 | 0.012456703 | 0.029382558 | 1 |
| HTN | monocyte | *UBE2I* | -2.172659945 | 0.017098671 | 0.034961127 | 1 |
| HTN | monocyte | *CAPN15* | -2.37086942 | 0.006555499 | 0.02387934 | 1 |
| HTN | monocyte | *EARS2* | -1.67098807 | 0.020243582 | 0.038979314 | 1 |
| HTN | monocyte | *CPPED1* | 0.714928134 | 0.003194048 | 0.023595115 | 1 |
| HTN | monocyte | *PYCARD* | 8.715749037 | 7.01E-05 | 0.005633811 | 0.339982357 |
| HTN | monocyte | *CDIPT* | -2.010415386 | 0.022347875 | 0.041801279 | 1 |
| HTN | monocyte | *KAT8* | -1.489365103 | 0.006763081 | 0.02405092 | 1 |
| HTN | monocyte | *TMC5* | -1.239962441 | 0.003878737 | 0.023595115 | 1 |
| HTN | monocyte | *KNOP1* | -2.502099655 | 0.011917298 | 0.028877239 | 1 |
| HTN | monocyte | *IQCH* | -1.982321142 | 0.015879917 | 0.033690142 | 1 |
| HTN | monocyte | *LACTB* | -3.571975391 | 0.007753906 | 0.025006477 | 1 |
| HTN | monocyte | *AP3B2* | -1.368836928 | 0.003949684 | 0.023595115 | 1 |
| HTN | monocyte | *FAH* | -0.996120449 | 0.00496834 | 0.023595115 | 1 |
| HTN | monocyte | *RPAP1* | -2.538718709 | 0.001648351 | 0.023595115 | 1 |
| HTN | monocyte | *SLC30A4* | -1.154588359 | 0.006739441 | 0.024040665 | 1 |
| HTN | monocyte | *ZDHHC2* | -2.836036574 | 0.019318703 | 0.037857168 | 1 |
| HTN | monocyte | *ZFAND1* | -1.62773057 | 0.023091376 | 0.042551977 | 1 |
| HTN | monocyte | *EYA1* | -1.069510787 | 0.011829336 | 0.028782381 | 1 |
| HTN | monocyte | *IMPAD1* | -2.73437036 | 0.015020927 | 0.032711642 | 1 |
| HTN | monocyte | *SFRP1* | -1.28858484 | 0.005995643 | 0.023595115 | 1 |
| HTN | monocyte | *ARMC1* | -2.736199236 | 0.024430342 | 0.044329102 | 1 |
| HTN | monocyte | *TSTA3* | 3.500017801 | 0.000397314 | 0.01501035 | 1 |
| HTN | monocyte | *KLHDC4* | -1.847974586 | 0.007839959 | 0.025025974 | 1 |
| HTN | monocyte | *NFKBIB* | -1.989153946 | 0.018432948 | 0.036560986 | 1 |
| HTN | monocyte | *SARS2* | -2.068416805 | 0.00151104 | 0.023595115 | 1 |
| HTN | monocyte | *SNRNP70* | -1.708529358 | 0.014116838 | 0.031695926 | 1 |
| HTN | monocyte | *PPP1R37* | -2.735755289 | 0.013889387 | 0.031408232 | 1 |
| HTN | monocyte | *FCGRT* | -1.200018536 | 0.01514437 | 0.03284778 | 1 |
| HTN | monocyte | *ERCC2* | -1.584178558 | 0.005699345 | 0.023595115 | 1 |
| HTN | monocyte | *CD37* | 0.944629062 | 0.024639358 | 0.044541789 | 1 |
| HTN | monocyte | *AMH* | 3.398154995 | 0.003462874 | 0.023595115 | 1 |
| HTN | monocyte | *FCER2* | -4.174069432 | 0.005294413 | 0.023595115 | 1 |
| HTN | monocyte | *CLEC4M* | -4.088854405 | 0.005130887 | 0.023595115 | 1 |
| HTN | monocyte | *MED25* | -2.075909454 | 0.009073646 | 0.026214745 | 1 |
| HTN | monocyte | *TIMM44* | -1.904903737 | 0.027932254 | 0.048563258 | 1 |
| HTN | monocyte | *FAM32A* | -3.576602957 | 0.008113806 | 0.025252204 | 1 |
| HTN | monocyte | *OLFM2* | -1.044233142 | 0.008800701 | 0.02589483 | 1 |
| HTN | monocyte | *RASAL3* | 0.793005145 | 0.003104666 | 0.023595115 | 1 |
| HTN | monocyte | *EPHX3* | 0.523336728 | 0.003794056 | 0.023595115 | 1 |
| HTN | monocyte | *PDCD5* | -1.928023441 | 0.019094773 | 0.037539643 | 1 |
| HTN | monocyte | *ANKRD27* | -2.480454604 | 0.023551651 | 0.043186929 | 1 |
| HTN | monocyte | *CEACAM4* | 2.006245637 | 0.000900448 | 0.020511604 | 1 |
| HTN | monocyte | *MRPL4* | -1.592572678 | 0.023309858 | 0.042840694 | 1 |
| HTN | monocyte | *GRIN2D* | -2.847253006 | 0.003719017 | 0.023595115 | 1 |
| HTN | monocyte | *CCDC114* | 3.909042861 | 0.003668121 | 0.023595115 | 1 |
| HTN | monocyte | *LIG1* | -2.45583817 | 0.0074083 | 0.024485743 | 1 |
| HTN | monocyte | *CAPS* | -3.189970712 | 0.010390616 | 0.027326031 | 1 |
| HTN | monocyte | *PLEKHA4* | -1.172398829 | 0.004821603 | 0.023595115 | 1 |
| HTN | monocyte | *TNPO2* | -1.869973823 | 0.0038207 | 0.023595115 | 1 |

| | | | | | | |
|---|---|---|---|---|---|---|
| HTN | monocyte | *GCDH* | -3.705900425 | 0.017461547 | 0.03534561 | 1 |
| HTN | monocyte | *DNASE2* | 3.967166362 | 0.000669751 | 0.018676049 | 1 |
| HTN | monocyte | *TFPT* | 1.836833112 | 0.00298018 | 0.023595115 | 1 |
| HTN | monocyte | *RAB3A* | -5.135816473 | 0.005021798 | 0.023595115 | 1 |
| HTN | monocyte | *ETV2* | -13.16975643 | 0.00444489 | 0.023595115 | 1 |
| HTN | monocyte | *MAG* | 2.893392156 | 0.003871904 | 0.023595115 | 1 |
| HTN | monocyte | *ZNF14* | -1.74163053 | 0.007571235 | 0.024721152 | 1 |
| HTN | monocyte | *ZNF85* | -3.299939528 | 0.021223384 | 0.040335236 | 1 |
| HTN | monocyte | *ETHE1* | -1.636369895 | 0.007369728 | 0.024485743 | 1 |
| HTN | monocyte | *SMG9* | 5.098934271 | 0.006075979 | 0.023595115 | 1 |
| HTN | monocyte | *GTPBP10* | -1.447228981 | 0.006530008 | 0.023864166 | 1 |
| HTN | monocyte | *RASA4* | -8.486301989 | 0.004853611 | 0.023595115 | 1 |
| HTN | monocyte | *BET1* | -1.866501917 | 0.013153374 | 0.030434035 | 1 |
| HTN | monocyte | *DNAH11* | -2.716777056 | 0.022223064 | 0.041647859 | 1 |
| HTN | monocyte | *STEAP1B* | 3.49726984 | 0.001245595 | 0.023595115 | 1 |
| HTN | monocyte | *PTN* | -1.29122481 | 0.00555216 | 0.023595115 | 1 |
| HTN | monocyte | *ADAP1* | -4.143490781 | 0.005209843 | 0.023595115 | 1 |
| HTN | monocyte | *MET* | -1.227515749 | 0.005739149 | 0.023595115 | 1 |
| HTN | monocyte | *RNF32* | -2.369549712 | 0.0040916 | 0.023595115 | 1 |
| HTN | monocyte | *IQCE* | -1.680728191 | 0.00721801 | 0.024303806 | 1 |
| HTN | monocyte | *VIPR2* | -0.85839728 | 0.006155704 | 0.023595115 | 1 |
| HTN | monocyte | *CPVL* | 2.634394206 | 0.009200352 | 0.026254852 | 1 |
| HTN | monocyte | *CHN2* | 2.51235531 | 0.000844462 | 0.020511604 | 1 |
| HTN | monocyte | *ABHD11* | 1.402504049 | 0.001683144 | 0.023595115 | 1 |
| HTN | monocyte | *CRHR2* | 1.350161934 | 0.013515489 | 0.030874365 | 1 |
| HTN | monocyte | *EPHB6* | -0.950816596 | 0.015679359 | 0.033474034 | 1 |
| HTN | monocyte | *CHCHD2* | -1.384501211 | 0.011467254 | 0.02848908 | 1 |
| HTN | monocyte | *TAF6* | -3.382519336 | 0.004983138 | 0.023595115 | 1 |
| HTN | monocyte | *TFR2* | 4.41922138 | 0.00313234 | 0.023595115 | 1 |
| HTN | monocyte | *ZNF862* | -1.669886381 | 0.017919716 | 0.035878799 | 1 |
| HTN | monocyte | *MEST* | -0.824050372 | 0.000602897 | 0.017946357 | 1 |
| HTN | monocyte | *POU6F2* | -1.022656571 | 0.00293737 | 0.023595115 | 1 |
| HTN | monocyte | *TSPAN13* | -1.350034259 | 0.007406321 | 0.024485743 | 1 |
| HTN | monocyte | *RARRES2* | -1.653512401 | 0.010343998 | 0.027291507 | 1 |
| HTN | monocyte | *TMEM176B* | -1.449466101 | 0.019296262 | 0.03784376 | 1 |
| HTN | monocyte | *URGCP* | -1.819694727 | 0.00906618 | 0.026214745 | 1 |
| HTN | monocyte | *AEBP1* | -1.403444636 | 0.006511874 | 0.023864166 | 1 |
| HTN | monocyte | *GCK* | -7.735250387 | 0.005004351 | 0.023595115 | 1 |
| HTN | monocyte | *SPATA6L* | -0.953880894 | 0.00638396 | 0.023797215 | 1 |
| HTN | monocyte | *FKTN* | -1.438619274 | 0.008232171 | 0.025373581 | 1 |
| HTN | monocyte | *MEGF9* | 1.311307642 | 0.003162816 | 0.023595115 | 1 |
| HTN | monocyte | *CORO2A* | 5.1823113 | 0.000738857 | 0.019916297 | 1 |
| HTN | monocyte | *OGN* | -1.430657027 | 0.001377846 | 0.023595115 | 1 |
| HTN | monocyte | *ECM2* | -0.896606026 | 0.006859505 | 0.024118623 | 1 |
| HTN | monocyte | *PTGR1* | -1.01056382 | 0.00613853 | 0.023595115 | 1 |
| HTN | monocyte | *PLGRKT* | -2.958930717 | 0.001297458 | 0.023595115 | 1 |
| HTN | monocyte | *DOCK8* | 1.136192437 | 0.002542765 | 0.023595115 | 1 |
| HTN | monocyte | *KANK1* | -1.175807748 | 0.010528978 | 0.027495479 | 1 |
| HTN | monocyte | *FUBP3* | -2.001876835 | 0.010894941 | 0.02788959 | 1 |
| HTN | monocyte | *RGP1* | -1.848120409 | 0.007819351 | 0.025020002 | 1 |

| | | | | | | |
|---|---|---|---|---|---|---|
| HTN | monocyte | *PTGDS* | -1.700122505 | 0.006421353 | 0.023819881 | 1 |
| HTN | monocyte | *HPS1* | -2.012116087 | 0.007938873 | 0.025074621 | 1 |
| HTN | monocyte | *PHYH* | -1.509948464 | 0.011513527 | 0.028507713 | 1 |
| HTN | monocyte | *PKD2L1* | 0.796485436 | 0.005065278 | 0.023595115 | 1 |
| HTN | monocyte | *CUBN* | -2.012936201 | 0.019816393 | 0.038510682 | 1 |
| HTN | monocyte | *TRDMT1* | -2.137176157 | 0.021859993 | 0.041172245 | 1 |
| HTN | monocyte | *PLEKHA1* | -1.403213972 | 0.007756672 | 0.025006477 | 1 |
| HTN | monocyte | *BMPR1A* | -1.16187682 | 0.008057413 | 0.02515738 | 1 |
| HTN | monocyte | *LIPA* | -2.225388085 | 0.01183447 | 0.028782381 | 1 |
| HTN | monocyte | *ANKRD26* | -1.369036551 | 0.009005272 | 0.026132523 | 1 |
| HTN | monocyte | *LHPP* | -2.611471715 | 0.007828789 | 0.025020002 | 1 |
| HTN | monocyte | *PITRM1* | -2.270964627 | 0.00714004 | 0.024238452 | 1 |
| HTN | monocyte | *DNAJC12* | 1.641721329 | 0.003042475 | 0.023595115 | 1 |
| HTN | monocyte | *PBLD* | -1.207133539 | 0.006186927 | 0.023616295 | 1 |
| HTN | monocyte | *KRT23* | 3.655800416 | 0.000956659 | 0.021292238 | 1 |
| HTN | monocyte | *WNT3* | -2.217293931 | 0.004636303 | 0.023595115 | 1 |
| HTN | monocyte | *ASPA* | -0.783497396 | 0.008758263 | 0.025848597 | 1 |
| HTN | monocyte | *RAD51C* | -1.370089547 | 0.000924385 | 0.020668742 | 1 |
| HTN | monocyte | *PNPO* | -2.799268769 | 0.001010966 | 0.021745206 | 1 |
| HTN | monocyte | *TRIM16L* | 1.734231179 | 0.0036615 | 0.023595115 | 1 |
| HTN | monocyte | *CDK5RAP3* | -1.547469342 | 0.0046908 | 0.023595115 | 1 |
| HTN | monocyte | *C17orf75* | -2.291849483 | 0.008559961 | 0.025783919 | 1 |
| HTN | monocyte | *PSMD11* | -1.295860566 | 0.007861477 | 0.025067673 | 1 |
| HTN | monocyte | *PEX12* | -3.026758594 | 0.001366589 | 0.023595115 | 1 |
| HTN | monocyte | *DHX58* | 1.820495108 | 0.002266416 | 0.023595115 | 1 |
| HTN | monocyte | *HSD17B1* | -2.003306384 | 0.005267708 | 0.023595115 | 1 |
| HTN | monocyte | *CNTNAP1* | -1.511750392 | 0.017263478 | 0.035135234 | 1 |
| HTN | monocyte | *EZH1* | -1.247015091 | 0.008335934 | 0.025426186 | 1 |
| HTN | monocyte | *SGCA* | 3.075929125 | 0.00286525 | 0.023595115 | 1 |
| HTN | monocyte | *ABCC3* | 0.608981708 | 0.003690256 | 0.023595115 | 1 |
| HTN | monocyte | *EFTUD2* | 3.50285924 | 0.010628692 | 0.027588558 | 1 |
| HTN | monocyte | *HLF* | -1.388583277 | 0.01725531 | 0.035133346 | 1 |
| HTN | monocyte | *FAM20A* | -1.516073105 | 0.009776568 | 0.026725639 | 1 |
| HTN | monocyte | *MYH3* | -2.262493836 | 0.005412634 | 0.023595115 | 1 |
| HTN | monocyte | *SLAIN2* | -1.643769745 | 0.017654782 | 0.035588285 | 1 |
| HTN | monocyte | *OCIAD1* | -1.631750342 | 0.010314817 | 0.027258982 | 1 |
| HTN | monocyte | *UBE2D3* | -1.592770291 | 0.017129908 | 0.035010241 | 1 |
| HTN | monocyte | *KLHL2* | 3.157855304 | 0.011915656 | 0.028877239 | 1 |
| HTN | monocyte | *CPE* | -1.331283504 | 0.013909269 | 0.031433523 | 1 |
| HTN | monocyte | *WFS1* | -1.49727361 | 0.005924945 | 0.023595115 | 1 |
| HTN | monocyte | *GRPEL1* | -3.28731914 | 0.014536846 | 0.032192049 | 1 |
| HTN | monocyte | *GAR1* | -3.441391486 | 0.028071018 | 0.048747523 | 1 |
| HTN | monocyte | *AADAT* | -1.861814405 | 0.004369502 | 0.023595115 | 1 |
| HTN | monocyte | *TRIM2* | -1.863369745 | 0.007974503 | 0.025076015 | 1 |
| HTN | monocyte | *BST1* | 4.150440812 | 0.025924579 | 0.046203869 | 1 |
| HTN | monocyte | *FAM149A* | -0.93721434 | 0.005499298 | 0.023595115 | 1 |
| HTN | monocyte | *DDX25* | 2.077943861 | 0.027313222 | 0.047773523 | 1 |
| HTN | monocyte | *CTSC* | -1.815976247 | 0.007821699 | 0.025020002 | 1 |
| HTN | monocyte | *TECTA* | -2.168308592 | 0.005384487 | 0.023595115 | 1 |
| HTN | monocyte | *SC5D* | -3.180995214 | 0.017905776 | 0.035878799 | 1 |

| | | | | | | |
|---|---|---|---|---|---|---|
| HTN | monocyte | *B3GAT1* | 4.813262745 | 0.002358834 | 0.023595115 | 1 |
| HTN | monocyte | *P2RX3* | 3.463113179 | 0.003366254 | 0.023595115 | 1 |
| HTN | monocyte | *SNX15* | -5.057380079 | 0.000415628 | 0.015162607 | 1 |
| HTN | monocyte | *UNC93B1* | 1.880577382 | 0.001751326 | 0.023595115 | 1 |
| HTN | monocyte | *DCPS* | -2.496501196 | 0.007300887 | 0.024429382 | 1 |
| HTN | monocyte | *FOXRED1* | -2.255107453 | 0.007383668 | 0.024485743 | 1 |
| HTN | monocyte | *NRXN2* | -2.51591173 | 0.004164708 | 0.023595115 | 1 |
| HTN | monocyte | *ST3GAL4* | 3.087008663 | 0.001623262 | 0.023595115 | 1 |
| HTN | monocyte | *CCND1* | -8.471254299 | 2.10E-06 | 0.000407965 | 0.010199134 |
| HTN | monocyte | *CCDC86* | 1.523465637 | 0.009309195 | 0.026321802 | 1 |
| HTN | monocyte | *PANX1* | -2.055019547 | 0.010509122 | 0.027479049 | 1 |
| HTN | monocyte | *CEP164* | -1.881110637 | 0.017304859 | 0.035204686 | 1 |
| HTN | monocyte | *GALNT18* | -1.945654033 | 0.015834466 | 0.033678697 | 1 |
| HTN | monocyte | *KIAA1549L* | 15.34323979 | 0.024842498 | 0.044775557 | 1 |
| HTN | monocyte | *SLC22A18* | 1.698754446 | 0.001816907 | 0.023595115 | 1 |
| HTN | monocyte | *NDUFS8* | -1.906646089 | 0.006154294 | 0.023595115 | 1 |
| HTN | monocyte | *TCIRG1* | 3.062892465 | 0.007769916 | 0.025007212 | 1 |
| HTN | monocyte | *PRDM4* | 4.864152471 | 0.01601973 | 0.033779979 | 1 |
| HTN | monocyte | *ASIC1* | -3.217612143 | 0.005354406 | 0.023595115 | 1 |
| HTN | monocyte | *KCTD10* | -3.728109479 | 0.002096575 | 0.023595115 | 1 |
| HTN | monocyte | *IL23A* | -8.244824424 | 0.005330945 | 0.023595115 | 1 |
| HTN | monocyte | *ACSS3* | -0.613125655 | 0.009058917 | 0.026214745 | 1 |
| HTN | monocyte | *LTA4H* | 2.516886157 | 0.005133588 | 0.023595115 | 1 |
| HTN | monocyte | *AKAP3* | -1.554387296 | 0.003441098 | 0.023595115 | 1 |
| HTN | monocyte | *ACAD10* | -1.35474233 | 0.002155433 | 0.023595115 | 1 |
| HTN | monocyte | *LTBR* | 2.441842535 | 0.009667005 | 0.02668844 | 1 |
| HTN | monocyte | *DDX55* | -1.813567881 | 0.021563228 | 0.040821218 | 1 |
| HTN | monocyte | *ENDOU* | 1.391263749 | 0.003551864 | 0.023595115 | 1 |
| HTN | monocyte | *VDR* | 1.095797063 | 0.003284925 | 0.023595115 | 1 |
| HTN | monocyte | *STX2* | -1.104959958 | 0.012252849 | 0.029207568 | 1 |
| HTN | monocyte | *MDM1* | 2.842491576 | 0.003942127 | 0.023595115 | 1 |
| HTN | monocyte | *NUP107* | -1.796421108 | 0.01161561 | 0.028577038 | 1 |
| HTN | monocyte | *KRR1* | -1.948350421 | 0.011227733 | 0.028146849 | 1 |
| HTN | monocyte | *GNB3* | -2.316976162 | 0.002952781 | 0.023595115 | 1 |
| HTN | monocyte | *CDCA3* | 1.368860402 | 0.021511695 | 0.040739556 | 1 |
| HTN | monocyte | *CHPT1* | -1.293024149 | 0.007298781 | 0.024429382 | 1 |
| HTN | monocyte | *SPSB2* | -1.745024219 | 0.005984399 | 0.023595115 | 1 |
| HTN | monocyte | *ATN1* | -1.028852676 | 0.022680506 | 0.042098629 | 1 |
| HTN | monocyte | *C12orf57* | -1.278540579 | 0.006143112 | 0.023595115 | 1 |
| HTN | monocyte | *SUDS3* | -1.535365789 | 0.008137411 | 0.025290474 | 1 |
| HTN | monocyte | *GOLT1B* | 4.103657157 | 0.021267484 | 0.040403224 | 1 |
| HTN | monocyte | *C2CD5* | -1.715079167 | 0.017873847 | 0.035878799 | 1 |
| HTN | monocyte | *FRK* | -1.139262103 | 0.005517942 | 0.023595115 | 1 |
| HTN | monocyte | *RWDD1* | -1.890531915 | 0.007368791 | 0.024485743 | 1 |
| HTN | monocyte | *FAM184A* | -1.134450953 | 0.010279789 | 0.027210876 | 1 |
| HTN | monocyte | *GABRR2* | 1.560995682 | 0.002081856 | 0.023595115 | 1 |
| HTN | monocyte | *HDDC2* | -1.101128238 | 0.023635583 | 0.04329175 | 1 |
| HTN | monocyte | *UST* | -1.636030276 | 0.004595776 | 0.023595115 | 1 |
| HTN | monocyte | *MTRF1L* | -1.724629893 | 0.019063316 | 0.037504578 | 1 |
| HTN | monocyte | *SLC26A8* | 1.692590917 | 0.001597032 | 0.023595115 | 1 |

| | | | | | | |
|---|---|---|---|---|---|---|
| HTN | monocyte | *PHACTR1* | -1.902686028 | 0.013527675 | 0.03088766 | 1 |
| HTN | monocyte | *MDGA1* | -1.403031495 | 0.004320662 | 0.023595115 | 1 |
| HTN | monocyte | *CD83* | 1.213929625 | 0.003040757 | 0.023595115 | 1 |
| HTN | monocyte | *MDN1* | -1.482894649 | 0.009560091 | 0.026650066 | 1 |
| HTN | monocyte | *GLP1R* | -11.20961277 | 0.004361115 | 0.023595115 | 1 |
| HTN | monocyte | *BVES* | -1.957747551 | 0.000629293 | 0.018265624 | 1 |
| HTN | monocyte | *COL9A1* | 1.481313737 | 0.004210654 | 0.023595115 | 1 |
| HTN | monocyte | *WASF1* | -1.126151193 | 0.003483974 | 0.023595115 | 1 |
| HTN | monocyte | *ALDH5A1* | -1.03059753 | 0.006651933 | 0.023960787 | 1 |
| HTN | monocyte | *VNN2* | 0.88239009 | 0.002462837 | 0.023595115 | 1 |
| HTN | monocyte | *C6orf62* | -2.647822136 | 0.010241572 | 0.027124512 | 1 |
| HTN | monocyte | *PHACTR2* | -1.900294671 | 0.028189854 | 0.048791054 | 1 |
| HTN | monocyte | *CUTA* | -2.080091362 | 0.000658797 | 0.018584208 | 1 |
| HTN | monocyte | *PACRG* | -1.176245694 | 0.005430671 | 0.023595115 | 1 |
| HTN | monocyte | *GUCA1B* | -2.99935014 | 0.028031483 | 0.048696296 | 1 |
| HTN | monocyte | *IMPG1* | -4.324024833 | 0.001359093 | 0.023595115 | 1 |
| HTN | monocyte | *BTN2A1* | 5.080344608 | 0.0092864 | 0.026288744 | 1 |
| HTN | monocyte | *LY86* | 0.583083673 | 0.003484868 | 0.023595115 | 1 |
| HTN | monocyte | *PCDHB2* | -5.140785665 | 0.004447577 | 0.023595115 | 1 |
| HTN | monocyte | *NUDT12* | -2.264221088 | 0.012262122 | 0.029207568 | 1 |
| HTN | monocyte | *CEP72* | 2.437519185 | 0.002093328 | 0.023595115 | 1 |
| HTN | monocyte | *KIF20A* | -13.33547046 | 0.004396025 | 0.023595115 | 1 |
| HTN | monocyte | *PFDN1* | -1.250747816 | 0.008822393 | 0.025896099 | 1 |
| HTN | monocyte | *SPARC* | -1.36913223 | 0.009751796 | 0.02671918 | 1 |
| HTN | monocyte | *PCDHB15* | -2.181694548 | 0.014739016 | 0.032396802 | 1 |
| HTN | monocyte | *HAVCR1* | 1.12047817 | 0.003693103 | 0.023595115 | 1 |
| HTN | monocyte | *RNF130* | 2.985429081 | 0.019635956 | 0.038329817 | 1 |
| HTN | monocyte | *BTNL8* | -1.128883771 | 0.022797651 | 0.042203053 | 1 |
| HTN | monocyte | *MSH3* | -1.308450295 | 0.009164964 | 0.02623769 | 1 |
| HTN | monocyte | *RASGRF2* | -0.977530249 | 0.009094182 | 0.026216909 | 1 |
| HTN | monocyte | *DROSHA* | -1.867362566 | 0.011585583 | 0.028549135 | 1 |
| HTN | monocyte | *CDH6* | -1.76777328 | 0.01237018 | 0.029302872 | 1 |
| HTN | monocyte | *LMNB1* | 0.711841605 | 0.00387994 | 0.023595115 | 1 |
| HTN | monocyte | *SLC12A7* | -1.941126724 | 0.008282548 | 0.025373581 | 1 |
| HTN | monocyte | *SKP1* | -1.905126326 | 0.01597107 | 0.03374074 | 1 |
| HTN | monocyte | *SMAD5* | -1.960374195 | 0.01973046 | 0.038431229 | 1 |
| HTN | monocyte | *UNC5A* | -5.256064947 | 0.005413484 | 0.023595115 | 1 |
| HTN | monocyte | *ACTR8* | -4.588208938 | 0.009905931 | 0.026830579 | 1 |
| HTN | monocyte | *TBCCD1* | -2.354307445 | 0.011196075 | 0.028135587 | 1 |
| HTN | monocyte | *CEP70* | -1.7424945 | 0.022811246 | 0.042212115 | 1 |
| HTN | monocyte | *HYAL1* | -2.197911897 | 0.000310767 | 0.014091997 | 1 |
| HTN | monocyte | *CBLB* | -1.239198158 | 0.017431099 | 0.0353381 | 1 |
| HTN | monocyte | *IQCG* | -2.10098913 | 0.020920272 | 0.039914234 | 1 |
| HTN | monocyte | *SLC41A3* | -1.459408119 | 0.004540758 | 0.023595115 | 1 |
| HTN | monocyte | *PLXNA1* | -1.18893897 | 0.009898231 | 0.026830579 | 1 |
| HTN | monocyte | *KIAA1257* | -3.178783457 | 0.005469247 | 0.023595115 | 1 |
| HTN | monocyte | *NEK11* | -0.912099984 | 0.008874487 | 0.025962238 | 1 |
| HTN | monocyte | *PLSCR4* | -1.459395502 | 0.023017549 | 0.042480467 | 1 |
| HTN | monocyte | *HEMK1* | -1.11803487 | 0.005372669 | 0.023595115 | 1 |
| HTN | monocyte | *COMMD2* | -2.171980418 | 0.01403394 | 0.031656288 | 1 |

| | | | | | | |
|---|---|---|---|---|---|---|
| HTN | monocyte | *ABHD14B* | -2.77217786 | 0.010120745 | 0.027038216 | 1 |
| HTN | monocyte | *ARHGEF26* | -1.136268793 | 0.005872367 | 0.023595115 | 1 |
| HTN | monocyte | *VIPR1* | 1.115689713 | 0.002902565 | 0.023595115 | 1 |
| HTN | monocyte | *DNAH1* | 3.715386002 | 0.001189877 | 0.02330522 | 1 |
| HTN | monocyte | *NEK4* | -4.019521788 | 0.002106503 | 0.023595115 | 1 |
| HTN | monocyte | *ADAM23* | -2.906200792 | 0.004440222 | 0.023595115 | 1 |
| HTN | monocyte | *LMAN2L* | -1.359779098 | 0.009908771 | 0.026830579 | 1 |
| HTN | monocyte | *SLC35F5* | -2.065330601 | 0.004704633 | 0.023595115 | 1 |
| HTN | monocyte | *STEAP3* | -0.595769673 | 0.027921364 | 0.048563258 | 1 |
| HTN | monocyte | *OTOF* | -5.996451147 | 0.01974407 | 0.038442306 | 1 |
| HTN | monocyte | *CENPA* | -1.451332469 | 0.009144507 | 0.02623769 | 1 |
| HTN | monocyte | *NRBP1* | -2.236534594 | 0.022321403 | 0.041783738 | 1 |
| HTN | monocyte | *INO80B* | -1.492140642 | 0.02571008 | 0.045980578 | 1 |
| HTN | monocyte | *GRB14* | -1.177732788 | 0.00510901 | 0.023595115 | 1 |
| HTN | monocyte | *RTN4* | -1.371142313 | 0.013808881 | 0.031323371 | 1 |
| HTN | monocyte | *GALNT3* | 0.907845478 | 0.002260177 | 0.023595115 | 1 |
| HTN | monocyte | *ACADL* | -0.78132937 | 0.008272347 | 0.025373581 | 1 |
| HTN | monocyte | *IGFBP2* | -1.87975209 | 0.009761003 | 0.02671918 | 1 |
| HTN | monocyte | *WNT6* | -2.594307581 | 0.001110782 | 0.022966009 | 1 |
| HTN | monocyte | *IL1RL2* | -2.894127269 | 0.007825275 | 0.025020002 | 1 |
| HTN | monocyte | *IL18R1* | -1.270691713 | 0.002707954 | 0.023595115 | 1 |
| HTN | monocyte | *MLPH* | -1.358420827 | 0.015227748 | 0.032964034 | 1 |
| HTN | monocyte | *HDLBP* | -1.720713543 | 0.025268781 | 0.045397695 | 1 |
| HTN | monocyte | *PASK* | -1.704732718 | 0.004425408 | 0.023595115 | 1 |
| HTN | monocyte | *TPO* | -1.367905158 | 0.004670645 | 0.023595115 | 1 |
| HTN | monocyte | *STRN* | -1.848156584 | 0.02660785 | 0.046945924 | 1 |
| HTN | monocyte | *PRKD3* | -1.41714454 | 0.018378378 | 0.03647112 | 1 |
| HTN | monocyte | *DARS* | -1.925950843 | 0.021381609 | 0.040588249 | 1 |
| HTN | monocyte | *WIPF1* | 3.196141579 | 0.005948787 | 0.023595115 | 1 |
| HTN | monocyte | *PNO1* | -3.717364246 | 0.02474059 | 0.044674858 | 1 |
| HTN | monocyte | *AAK1* | -1.568145293 | 0.020807265 | 0.039793793 | 1 |
| HTN | monocyte | *NFE2L2* | -1.43576336 | 0.019138763 | 0.037580445 | 1 |
| HTN | monocyte | *MSH6* | -2.121161337 | 0.016371429 | 0.034238868 | 1 |
| HTN | monocyte | *MARK1* | -1.297702709 | 0.008226501 | 0.025373581 | 1 |
| HTN | monocyte | *MORN1* | -2.172294075 | 0.000382328 | 0.01501035 | 1 |
| HTN | monocyte | *RALGPS2* | -1.222936856 | 0.005857525 | 0.023595115 | 1 |
| HTN | monocyte | *WRAP73* | -2.15014981 | 0.002814533 | 0.023595115 | 1 |
| HTN | monocyte | *ICMT* | -3.711154834 | 0.020230039 | 0.038979314 | 1 |
| HTN | monocyte | *PARK7* | -2.156774769 | 0.01614376 | 0.03392357 | 1 |
| HTN | monocyte | *KIAA1324* | 10.70042858 | 0.003558156 | 0.023595115 | 1 |
| HTN | monocyte | *MECR* | -2.899292005 | 0.005154588 | 0.023595115 | 1 |
| HTN | monocyte | *KCNC4* | -1.580404065 | 0.00461951 | 0.023595115 | 1 |
| HTN | monocyte | *GON4L* | -1.720147139 | 0.027864996 | 0.048511289 | 1 |
| HTN | monocyte | *DOCK7* | -1.584973115 | 0.012929749 | 0.030204691 | 1 |
| HTN | monocyte | *DNAJC6* | -1.666551102 | 0.003488495 | 0.023595115 | 1 |
| HTN | monocyte | *LEPR* | -1.693155865 | 0.012272668 | 0.029218344 | 1 |
| HTN | monocyte | *PRG4* | -1.187575573 | 0.005330072 | 0.023595115 | 1 |
| HTN | monocyte | *NCF2* | 0.585857442 | 0.001891396 | 0.023595115 | 1 |
| HTN | monocyte | *TNNI3K* | 4.884116786 | 0.000395595 | 0.01501035 | 1 |
| HTN | monocyte | *TMEM9* | -3.355219135 | 0.002087399 | 0.023595115 | 1 |

| | | | | | | |
|---|---|---|---|---|---|---|
| HTN | monocyte | *WARS2* | -1.75806632 | 0.007142814 | 0.024238452 | 1 |
| HTN | monocyte | *OSCP1* | -1.05831812 | 0.006670593 | 0.023986397 | 1 |
| HTN | monocyte | *MRPS15* | -1.881780681 | 0.012109501 | 0.029032068 | 1 |
| HTN | monocyte | *EXOC8* | -1.865736579 | 0.007385767 | 0.024485743 | 1 |
| HTN | monocyte | *TBCE* | -1.809253122 | 0.011863033 | 0.028818926 | 1 |
| HTN | monocyte | *ACADM* | -1.237178947 | 0.01184878 | 0.028802745 | 1 |
| HTN | monocyte | *SLAMF1* | 0.744584484 | 0.003762311 | 0.023595115 | 1 |
| HTN | monocyte | *PADI2* | -2.790836338 | 0.008682111 | 0.025839644 | 1 |
| HTN | monocyte | *CTBS* | 1.803294283 | 0.028988454 | 0.049753087 | 1 |
| HTN | monocyte | *ZNHIT6* | -2.048238078 | 0.020330319 | 0.039066419 | 1 |
| HTN | monocyte | *GBP3* | -1.76555811 | 0.012252092 | 0.029207568 | 1 |
| HTN | monocyte | *KIF17* | 2.609799847 | 0.007094959 | 0.024238452 | 1 |
| HTN | monocyte | *EBNA1BP2* | -1.262662398 | 0.006744535 | 0.024040665 | 1 |
| HTN | monocyte | *PTBP2* | -1.316968446 | 0.011705084 | 0.028654424 | 1 |
| HTN | monocyte | *SERPINC1* | 5.069344097 | 0.001287685 | 0.023595115 | 1 |
| HTN | monocyte | *MARC2* | -1.418033607 | 0.008732234 | 0.025839644 | 1 |
| HTN | monocyte | *ESYT2* | -1.544225596 | 0.028207047 | 0.048791655 | 1 |
| HTN | monocyte | *RCN2* | -1.846436354 | 0.026106481 | 0.046375 | 1 |
| HTN | monocyte | *IFT46* | -2.901078654 | 0.006756028 | 0.024050072 | 1 |
| HTN | monocyte | *DDX59* | -3.793770609 | 0.002402383 | 0.023595115 | 1 |
| HTN | monocyte | *CASC1* | -1.385681706 | 0.001302913 | 0.023595115 | 1 |
| HTN | monocyte | *LRMP* | 1.359364713 | 0.001596742 | 0.023595115 | 1 |
| HTN | monocyte | *SGIP1* | -1.037138938 | 0.005220002 | 0.023595115 | 1 |
| HTN | monocyte | *AKAP7* | -1.645935037 | 0.020071089 | 0.038783324 | 1 |
| HTN | monocyte | *MED28* | -2.396765442 | 0.024530679 | 0.044413403 | 1 |
| HTN | monocyte | *SPP1* | -1.773805336 | 0.00911924 | 0.026216909 | 1 |
| HTN | monocyte | *MFSD1* | -1.956377104 | 0.017214958 | 0.035122418 | 1 |
| HTN | monocyte | *WDR35* | -1.512695608 | 0.012453911 | 0.029382558 | 1 |
| HTN | monocyte | *DNAH7* | -1.448272692 | 0.003586016 | 0.023595115 | 1 |
| HTN | monocyte | *C2orf40* | -1.658522576 | 0.009587276 | 0.026672858 | 1 |
| HTN | monocyte | *CPSF3* | -1.892788253 | 0.003902214 | 0.023595115 | 1 |
| HTN | monocyte | *SENP5* | -1.896130378 | 0.02176541 | 0.041091739 | 1 |
| HTN | monocyte | *CCDC92* | -1.675829497 | 0.00565265 | 0.023595115 | 1 |
| HTN | monocyte | *CTNNAL1* | -1.331119838 | 0.007797391 | 0.025020002 | 1 |
| HTN | monocyte | *TRIM32* | -1.691086551 | 0.004132741 | 0.023595115 | 1 |
| HTN | monocyte | *PPP6C* | -1.970239773 | 0.016964614 | 0.034774949 | 1 |
| HTN | monocyte | *HDHD3* | -1.297471976 | 0.001281615 | 0.023595115 | 1 |
| HTN | monocyte | *RBM18* | 3.779341697 | 0.005730222 | 0.023595115 | 1 |
| HTN | monocyte | *DCAF4* | -3.244440104 | 0.002291104 | 0.023595115 | 1 |
| HTN | monocyte | *IFI27L2* | -3.522200631 | 0.01489825 | 0.032576075 | 1 |
| HTN | monocyte | *NEK9* | -1.751080958 | 0.023052831 | 0.042513241 | 1 |
| HTN | monocyte | *TTLL5* | -1.333020084 | 0.008847994 | 0.025924194 | 1 |
| HTN | monocyte | *FLVCR2* | 1.246840238 | 0.004083923 | 0.023595115 | 1 |
| HTN | monocyte | *PPP4R4* | -1.653985059 | 0.007097405 | 0.024238452 | 1 |
| HTN | monocyte | *ESRRB* | -1.525153239 | 0.025732449 | 0.045987473 | 1 |
| HTN | monocyte | *COQ6* | -1.703134308 | 0.008633976 | 0.025828042 | 1 |
| HTN | monocyte | *DNMT3A* | -2.070806622 | 0.024376963 | 0.044273639 | 1 |
| HTN | monocyte | *CNRIP1* | -1.624289129 | 0.006403225 | 0.023797215 | 1 |
| HTN | monocyte | *CRIPT* | -1.915752043 | 0.009080957 | 0.026214745 | 1 |
| HTN | monocyte | *EPCAM* | -2.925781943 | 0.021999737 | 0.041373149 | 1 |

| | | | | | | |
|---|---|---|---|---|---|---|
| HTN | monocyte | *PYROXD2* | -1.501278336 | 0.014196863 | 0.031826319 | 1 |
| HTN | monocyte | *MXI1* | -1.777418744 | 0.01291984 | 0.03019608 | 1 |
| HTN | monocyte | *C10orf88* | -1.886639906 | 0.0041338 | 0.023595115 | 1 |
| HTN | monocyte | *HELLS* | -1.811402555 | 0.001112414 | 0.022966009 | 1 |
| HTN | monocyte | *KANSL1* | -1.544733984 | 0.017917595 | 0.035878799 | 1 |
| HTN | monocyte | *HOXB3* | -1.536962334 | 0.008337399 | 0.025426186 | 1 |
| HTN | monocyte | *PCDHB10* | 2.455190191 | 0.001211387 | 0.023439031 | 1 |
| HTN | monocyte | *PCDHB12* | -2.720276152 | 0.001266787 | 0.023595115 | 1 |
| HTN | monocyte | *MASTL* | -1.996650916 | 0.002806261 | 0.023595115 | 1 |
| HTN | monocyte | *MRC1* | -1.015140312 | 0.0070291 | 0.024238452 | 1 |
| HTN | monocyte | *IQSEC3* | 5.256561121 | 0.003864275 | 0.023595115 | 1 |
| HTN | monocyte | *CCDC77* | 1.647454867 | 0.007384104 | 0.024485743 | 1 |
| HTN | monocyte | *UFM1* | -2.623727784 | 0.012899752 | 0.030176778 | 1 |
| HTN | monocyte | *ALG5* | -1.975007814 | 0.004525574 | 0.023595115 | 1 |
| HTN | monocyte | *SIL1* | -1.598235672 | 0.01631523 | 0.034180266 | 1 |
| HTN | monocyte | *ARL1* | -1.656568863 | 0.003797058 | 0.023595115 | 1 |
| HTN | monocyte | *APAF1* | 2.996599742 | 0.016790441 | 0.034563946 | 1 |
| HTN | monocyte | *EPHX2* | -1.344854749 | 0.002239995 | 0.023595115 | 1 |
| HTN | monocyte | *AKAP1* | -1.166610593 | 0.004876378 | 0.023595115 | 1 |
| HTN | monocyte | *TBX2* | -2.860042599 | 2.71E-07 | 9.39E-05 | 0.001314352 |
| HTN | monocyte | *TRIM6* | -3.508289461 | 0.004077103 | 0.023595115 | 1 |
| HTN | monocyte | *ECHDC2* | -0.788121536 | 0.004824558 | 0.023595115 | 1 |
| HTN | monocyte | *PYROXD1* | 2.191864517 | 0.012374605 | 0.029302872 | 1 |
| HTN | monocyte | *ZNF549* | -1.489951192 | 0.007074766 | 0.024238452 | 1 |
| HTN | monocyte | *ZNF211* | 2.029069582 | 0.005736395 | 0.023595115 | 1 |
| HTN | monocyte | *PDZRN3* | -0.537077755 | 0.012677413 | 0.029744105 | 1 |
| HTN | monocyte | *LHX4* | -1.340909661 | 0.005176011 | 0.023595115 | 1 |
| HTN | monocyte | *SEC22A* | -1.863444606 | 0.022445387 | 0.04183827 | 1 |
| HTN | monocyte | *CD80* | 1.08741287 | 0.008958025 | 0.026057755 | 1 |
| HTN | monocyte | *MAPK8IP1* | -4.37725499 | 0.000138089 | 0.008589875 | 0.670010219 |
| HTN | monocyte | *CAT* | -1.077762606 | 0.005233505 | 0.023595115 | 1 |
| HTN | monocyte | *TNFSF10* | -1.781515439 | 0.017152982 | 0.03504264 | 1 |
| HTN | monocyte | *ZNF639* | -2.06991982 | 0.008697479 | 0.025839644 | 1 |
| HTN | monocyte | *CPXM2* | -1.925482281 | 0.007971288 | 0.025076015 | 1 |
| HTN | monocyte | *FLT3* | 1.563906673 | 0.001936493 | 0.023595115 | 1 |
| HTN | monocyte | *MTIF3* | -2.019119281 | 0.007818844 | 0.025020002 | 1 |
| HTN | monocyte | *FMOD* | -2.249341686 | 0.019469817 | 0.038076401 | 1 |
| HTN | monocyte | *CD244* | 0.327601698 | 0.003274508 | 0.023595115 | 1 |
| HTN | monocyte | *SERAC1* | -2.303951463 | 0.004758363 | 0.023595115 | 1 |
| HTN | monocyte | *LDB3* | -1.336275714 | 0.009227836 | 0.026254852 | 1 |
| HTN | monocyte | *FAM35A* | -1.955658612 | 0.00696043 | 0.024187912 | 1 |
| HTN | monocyte | *NAA60* | -1.987392517 | 0.006384036 | 0.023797215 | 1 |
| HTN | monocyte | *ODF2L* | -1.759149887 | 0.01349384 | 0.03083943 | 1 |
| HTN | monocyte | *RWDD3* | -1.991642495 | 0.007761697 | 0.025006477 | 1 |
| HTN | monocyte | *EEPD1* | -1.179471783 | 0.003736212 | 0.023595115 | 1 |
| HTN | monocyte | *SLC25A51* | -1.968183737 | 0.001961115 | 0.023595115 | 1 |
| HTN | monocyte | *TRIM24* | -2.098341999 | 0.011546078 | 0.028509706 | 1 |
| HTN | monocyte | *C7orf49* | -2.366647732 | 0.001406207 | 0.023595115 | 1 |
| HTN | monocyte | *RBM19* | -1.343923468 | 0.006778211 | 0.02405092 | 1 |
| HTN | monocyte | *DDX54* | 4.995485701 | 0.001471178 | 0.023595115 | 1 |

| | | | | | | |
|---|---|---|---|---|---|---|
| HTN | monocyte | *SSPN* | -1.578499741 | 0.004342873 | 0.023595115 | 1 |
| HTN | monocyte | *EBPL* | -1.421786918 | 0.022669445 | 0.042094201 | 1 |
| HTN | monocyte | *CENPK* | -2.705670331 | 0.028324078 | 0.048924325 | 1 |
| HTN | monocyte | *ITIH5* | -0.854455461 | 0.022660558 | 0.042094201 | 1 |
| HTN | monocyte | *TSFM* | -2.234684419 | 0.020927826 | 0.039914234 | 1 |
| HTN | monocyte | *ARHGAP9* | 0.867225259 | 0.003923616 | 0.023595115 | 1 |
| HTN | monocyte | *PDE1B* | -2.851579623 | 0.015266287 | 0.033008924 | 1 |
| HTN | monocyte | *CDK2* | -2.430892308 | 0.00032711 | 0.014560912 | 1 |
| HTN | monocyte | *DBH* | -34.7510174 | 0.004515924 | 0.023595115 | 1 |
| HTN | monocyte | *NDUFAF4* | 4.773298886 | 0.00226839 | 0.023595115 | 1 |
| HTN | monocyte | *USP45* | -1.437745584 | 0.009426937 | 0.026481384 | 1 |
| HTN | monocyte | *TTC21B* | -2.269796261 | 0.008190718 | 0.025373581 | 1 |
| HTN | monocyte | *ACVR1C* | -0.605167373 | 0.007772233 | 0.025007212 | 1 |
| HTN | monocyte | *SLC36A1* | 1.914909127 | 0.014664333 | 0.032341519 | 1 |
| HTN | monocyte | *PLA2G12A* | -1.922682204 | 0.012258382 | 0.029207568 | 1 |
| HTN | monocyte | *C4BPB* | -1.751491075 | 0.01238905 | 0.029321564 | 1 |
| HTN | monocyte | *RAB38* | -1.985512267 | 0.016547221 | 0.034411074 | 1 |
| HTN | monocyte | *DAW1* | -3.022410935 | 0.003966252 | 0.023595115 | 1 |
| HTN | monocyte | *OBSL1* | -1.50429643 | 0.003392921 | 0.023595115 | 1 |
| HTN | monocyte | *SLC12A4* | -3.117608996 | 0.012650266 | 0.029694772 | 1 |
| HTN | monocyte | *FAM210B* | -1.92359472 | 0.009219506 | 0.026254852 | 1 |
| HTN | monocyte | *WFDC3* | -1.893175734 | 2.12E-05 | 0.002330865 | 0.102868665 |
| HTN | monocyte | *MATN4* | -3.829122956 | 0.00518864 | 0.023595115 | 1 |
| HTN | monocyte | *TOX2* | -1.977172152 | 0.000139896 | 0.008592107 | 0.67877644 |
| HTN | monocyte | *PEPD* | -2.186910954 | 0.026925048 | 0.04735061 | 1 |
| HTN | monocyte | *CHST8* | -2.163952869 | 0.01008505 | 0.027007023 | 1 |
| HTN | monocyte | *STAMBP* | -1.878084848 | 0.012224156 | 0.029188782 | 1 |
| HTN | monocyte | *NAGK* | 2.672697855 | 0.003125253 | 0.023595115 | 1 |
| HTN | monocyte | *SNRNP27* | -1.520271565 | 0.006919731 | 0.024148823 | 1 |
| HTN | monocyte | *ATP8A1* | -1.494640838 | 0.006893397 | 0.024148823 | 1 |
| HTN | monocyte | *USP22* | -1.764945548 | 0.021441372 | 0.040622233 | 1 |
| HTN | monocyte | *HIF3A* | -1.990937166 | 0.026362419 | 0.046631593 | 1 |
| HTN | monocyte | *ZNF576* | 4.032816491 | 0.003485109 | 0.023595115 | 1 |
| HTN | monocyte | *BTN2A2* | 3.078356786 | 0.000103248 | 0.007156543 | 0.500957996 |
| HTN | monocyte | *MRS2* | -1.742496291 | 0.018895724 | 0.037253983 | 1 |
| HTN | monocyte | *SERPINB6* | -1.667950146 | 0.01820998 | 0.036255569 | 1 |
| HTN | monocyte | *PEX6* | -1.418784686 | 0.016031573 | 0.03378414 | 1 |
| HTN | monocyte | *ZNF391* | -2.225527743 | 6.40E-05 | 0.005444156 | 0.310316899 |
| HTN | monocyte | *RPS10* | 3.398128005 | 0.008213569 | 0.025373581 | 1 |
| HTN | monocyte | *TCP11* | 0.80336725 | 0.000870148 | 0.020511604 | 1 |
| HTN | monocyte | *GNMT* | 1.131484479 | 0.002438197 | 0.023595115 | 1 |
| HTN | monocyte | *TREM1* | 1.380493848 | 0.003845749 | 0.023595115 | 1 |
| HTN | monocyte | *MEA1* | 3.543108869 | 0.008587554 | 0.025783919 | 1 |
| HTN | monocyte | *KLHL31* | -0.865602724 | 0.007914454 | 0.025073967 | 1 |
| HTN | monocyte | *GLO1* | -1.367055789 | 0.01366242 | 0.031084791 | 1 |
| HTN | monocyte | *KCNK17* | -4.095876783 | 0.00602419 | 0.023595115 | 1 |
| HTN | monocyte | *RIOK1* | -1.463847867 | 0.008856562 | 0.025933639 | 1 |
| HTN | monocyte | *SH3TC1* | 1.365453102 | 0.002802957 | 0.023595115 | 1 |
| HTN | monocyte | *MT1G* | -6.645589073 | 0.003676023 | 0.023595115 | 1 |
| HTN | monocyte | *GOT2* | -1.408623051 | 0.009171295 | 0.02623769 | 1 |

| | | | | | | |
|---|---|---|---|---|---|---|
| HTN | monocyte | *ABCC4* | -1.477207582 | 0.009748561 | 0.02671918 | 1 |
| HTN | monocyte | *ATP5S* | -2.489327105 | 0.004900205 | 0.023595115 | 1 |
| HTN | monocyte | *HS3ST3B1* | 4.133043387 | 0.023204097 | 0.04267865 | 1 |
| HTN | monocyte | *MRPS7* | -1.885146972 | 0.011531853 | 0.028508147 | 1 |
| HTN | monocyte | *GGA3* | -1.874017255 | 0.008916225 | 0.026029796 | 1 |
| HTN | monocyte | *NUP85* | -2.639066024 | 0.011126134 | 0.028102031 | 1 |
| HTN | monocyte | *NT5C* | 2.437669118 | 0.004247607 | 0.023595115 | 1 |
| HTN | monocyte | *OPRL1* | 0.596876119 | 0.003520296 | 0.023595115 | 1 |
| HTN | monocyte | *PPDPF* | 2.907511712 | 0.003772668 | 0.023595115 | 1 |
| HTN | monocyte | *IL1B* | 0.358265126 | 0.00321612 | 0.023595115 | 1 |
| HTN | monocyte | *PAX8* | -1.761161005 | 0.009398908 | 0.026436812 | 1 |
| HTN | monocyte | *GTF2F1* | -1.842523267 | 0.008366838 | 0.025464826 | 1 |
| HTN | monocyte | *TNFSF9* | -23.97540074 | 0.015461578 | 0.033284629 | 1 |
| HTN | monocyte | *SH2D3A* | 1.700796604 | 0.007460684 | 0.024580667 | 1 |
| HTN | monocyte | *SNRPD2* | -2.025468137 | 0.009205603 | 0.026254852 | 1 |
| HTN | monocyte | *SDCBP2* | -3.301891579 | 0.003277532 | 0.023595115 | 1 |
| HTN | monocyte | *PSMF1* | -2.235317971 | 0.012444638 | 0.029382558 | 1 |
| HTN | monocyte | *DTD1* | -2.993326125 | 0.021589399 | 0.040841576 | 1 |
| HTN | monocyte | *RBCK1* | -2.03814464 | 0.007170893 | 0.024238452 | 1 |
| HTN | monocyte | *RRBP1* | -1.638516802 | 0.024663895 | 0.044569541 | 1 |
| HTN | monocyte | *FLRT3* | -1.174965484 | 0.005037292 | 0.023595115 | 1 |
| HTN | monocyte | *PCSK2* | -4.777385546 | 0.012131873 | 0.029051118 | 1 |
| HTN | monocyte | *LAMP5* | -25.99434743 | 0.007541889 | 0.024642599 | 1 |
| HTN | monocyte | *MGME1* | 2.864399789 | 0.015641418 | 0.033432671 | 1 |
| HTN | monocyte | *ITPA* | -2.166207875 | 0.028268557 | 0.048863213 | 1 |
| HTN | monocyte | *MRPS26* | 3.094139369 | 0.019849478 | 0.038539283 | 1 |
| HTN | monocyte | *UROD* | -1.571370803 | 0.000918237 | 0.020626325 | 1 |
| HTN | monocyte | *PRR12* | -1.751117198 | 0.002101235 | 0.023595115 | 1 |
| HTN | monocyte | *ZNF384* | -1.317841027 | 0.00591707 | 0.023595115 | 1 |
| HTN | monocyte | *EMG1* | -2.952821538 | 0.016400632 | 0.034255647 | 1 |
| HTN | monocyte | *KTN1* | -1.400202102 | 0.012945168 | 0.030226158 | 1 |
| HTN | monocyte | *ZBTB1* | -1.568449708 | 0.010950268 | 0.027890132 | 1 |
| HTN | monocyte | *WDR60* | -1.334761925 | 0.01595433 | 0.03374074 | 1 |
| HTN | monocyte | *AIF1L* | -3.259529055 | 0.011668819 | 0.028586902 | 1 |
| HTN | monocyte | *FGD3* | 0.722759288 | 0.002059048 | 0.023595115 | 1 |
| HTN | monocyte | *ABHD8* | -5.333069056 | 1.51E-05 | 0.001781674 | 0.073048635 |
| HTN | monocyte | *YEATS4* | 2.800048128 | 0.012371333 | 0.029302872 | 1 |
| HTN | monocyte | *LRRC61* | -3.17056122 | 0.019725709 | 0.038431229 | 1 |
| HTN | monocyte | *IDUA* | -1.716208743 | 0.004556448 | 0.023595115 | 1 |
| HTN | monocyte | *PLA2G5* | -1.086182566 | 0.024690222 | 0.044600504 | 1 |
| HTN | monocyte | *GFER* | -4.077205249 | 0.000247433 | 0.012005439 | 1 |
| HTN | monocyte | *PNKD* | -1.941080405 | 0.014204994 | 0.031826319 | 1 |
| HTN | monocyte | *STYXL1* | -2.470576852 | 0.00038472 | 0.01501035 | 1 |
| HTN | monocyte | *PEX1* | -1.039945954 | 0.005714796 | 0.023595115 | 1 |
| HTN | monocyte | *KDR* | -1.860359151 | 0.010933318 | 0.027890132 | 1 |
| HTN | monocyte | *DGCR6L* | 2.792424962 | 0.025822702 | 0.046080084 | 1 |
| HTN | monocyte | *MGAT3* | -2.432879742 | 0.006711847 | 0.024005639 | 1 |
| HTN | monocyte | *ATF4* | -1.680480543 | 0.007016076 | 0.024238452 | 1 |
| HTN | monocyte | *A4GALT* | -2.034205839 | 0.000882208 | 0.020511604 | 1 |
| HTN | monocyte | *CDC42EP1* | -2.534342581 | 0.01655306 | 0.034411074 | 1 |

| | | | | | | |
|---|---|---|---|---|---|---|
| HTN | monocyte | *CDHR3* | -1.050425533 | 0.006028215 | 0.023595115 | 1 |
| HTN | monocyte | *PRKRIP1* | -1.902362178 | 0.023810641 | 0.043579492 | 1 |
| HTN | monocyte | *OPN1SW* | -2.612177968 | 0.009126208 | 0.026216909 | 1 |
| HTN | monocyte | *HOXD1* | -4.253551269 | 0.019877523 | 0.038547458 | 1 |
| HTN | monocyte | *HOXD3* | -1.027354278 | 0.025977364 | 0.046241902 | 1 |
| HTN | monocyte | *MTX2* | -1.873916072 | 0.019990893 | 0.038673422 | 1 |
| HTN | monocyte | *GAD1* | -4.620121911 | 0.000451269 | 0.015752203 | 1 |
| HTN | monocyte | *PSMG2* | -1.258717425 | 0.01167161 | 0.028586902 | 1 |
| HTN | monocyte | *TWSG1* | -1.65394571 | 0.012006921 | 0.028978626 | 1 |
| HTN | monocyte | *EIF2AK4* | -2.036974649 | 0.001694044 | 0.023595115 | 1 |
| HTN | monocyte | *KNSTRN* | -1.34814438 | 0.000522828 | 0.016799742 | 1 |
| HTN | monocyte | *ARPP19* | -2.316166115 | 0.028162034 | 0.048791054 | 1 |
| HTN | monocyte | *CALML4* | -5.21014904 | 0.002756384 | 0.023595115 | 1 |
| HTN | monocyte | *ANAPC13* | -1.80865628 | 0.001802858 | 0.023595115 | 1 |
| HTN | monocyte | *BBOX1* | 16.29458846 | 0.008027973 | 0.025113943 | 1 |
| HTN | monocyte | *SERGEF* | 4.175333437 | 0.00056973 | 0.017385729 | 1 |
| HTN | monocyte | *DCTD* | -1.170526704 | 0.003936891 | 0.023595115 | 1 |
| HTN | monocyte | *USP6* | 1.034592138 | 0.01747115 | 0.035350299 | 1 |
| HTN | monocyte | *PLD2* | 4.686161263 | 0.006790162 | 0.02405092 | 1 |
| HTN | monocyte | *PUS7L* | -1.873455338 | 0.008565706 | 0.025783919 | 1 |
| HTN | monocyte | *AP1M2* | -2.352733902 | 0.005760196 | 0.023595115 | 1 |
| HTN | monocyte | *KLK8* | -9.253650942 | 0.005520695 | 0.023595115 | 1 |
| HTN | monocyte | *RIPK3* | 2.961227344 | 7.08E-05 | 0.005633811 | 0.343662471 |
| HTN | monocyte | *ADCY4* | 8.042848442 | 2.77E-05 | 0.002857219 | 0.134289312 |
| HTN | monocyte | *BCL2L2* | -2.916557332 | 0.014987151 | 0.032682093 | 1 |
| HTN | monocyte | *PARP2* | -1.959675398 | 0.011264646 | 0.028158713 | 1 |
| HTN | monocyte | *RNASE1* | -0.956467115 | 0.010138935 | 0.027044591 | 1 |
| HTN | monocyte | *EPB41L4A* | -1.398044106 | 0.011663103 | 0.028586902 | 1 |
| HTN | monocyte | *CDO1* | -0.947433253 | 0.015013141 | 0.032711642 | 1 |
| HTN | monocyte | *RHBDF2* | 2.765753446 | 0.000444667 | 0.015748338 | 1 |
| HTN | monocyte | *TMEM8A* | -2.79708204 | 0.018481657 | 0.036616169 | 1 |
| HTN | monocyte | *MAU2* | -1.999365509 | 0.007406558 | 0.024485743 | 1 |
| HTN | monocyte | *SHC2* | -1.331179815 | 0.006524072 | 0.023864166 | 1 |
| HTN | monocyte | *ERMARD* | -2.084984459 | 0.013146637 | 0.030432959 | 1 |
| HTN | monocyte | *PHF10* | -1.962091718 | 0.009929778 | 0.026846837 | 1 |
| HTN | monocyte | *XPO7* | -2.128641852 | 0.020132916 | 0.038871831 | 1 |
| HTN | monocyte | *DDA1* | -1.697342575 | 0.012595901 | 0.029610131 | 1 |
| HTN | monocyte | *MAP1S* | 5.80826544 | 0.01032527 | 0.027271752 | 1 |
| HTN | monocyte | *SSBP4* | -1.772188673 | 0.026287023 | 0.046532155 | 1 |
| HTN | monocyte | *KIAA1683* | -3.183258001 | 0.004680709 | 0.023595115 | 1 |
| HTN | monocyte | *CYP2E1* | -2.505927869 | 0.005350254 | 0.023595115 | 1 |
| HTN | monocyte | *PNPLA7* | -0.892837237 | 0.005278721 | 0.023595115 | 1 |
| HTN | monocyte | *PAK4* | -1.468428002 | 0.006017072 | 0.023595115 | 1 |
| HTN | monocyte | *ZNF337* | -1.460325566 | 0.025694668 | 0.045978047 | 1 |
| HTN | monocyte | *POMT1* | -1.547947795 | 0.007875747 | 0.025073967 | 1 |
| HTN | monocyte | *YIPF2* | 3.876182806 | 0.013891605 | 0.031408232 | 1 |
| HTN | monocyte | *NPAS1* | 2.239488974 | 0.002997005 | 0.023595115 | 1 |
| HTN | monocyte | *MAP3K10* | -2.190254487 | 0.005129401 | 0.023595115 | 1 |
| HTN | monocyte | *LRRC47* | -2.357488687 | 0.017641734 | 0.035576764 | 1 |
| HTN | monocyte | *THEMIS2* | 0.713443224 | 0.001597066 | 0.023595115 | 1 |

| | | | | | | |
|---|---|---|---|---|---|---|
| HTN | monocyte | *ANGPTL6* | -3.772293198 | 0.012685461 | 0.029748602 | 1 |
| HTN | monocyte | *HSD17B3* | -1.773177275 | 0.005082982 | 0.023595115 | 1 |
| HTN | monocyte | *TTLL9* | 9.627943423 | 0.009778931 | 0.026725639 | 1 |
| HTN | monocyte | *COX4I2* | -3.188470659 | 0.002196843 | 0.023595115 | 1 |
| HTN | monocyte | *IDO1* | 2.83194056 | 0.003112406 | 0.023595115 | 1 |
| HTN | monocyte | *HAUS8* | -7.013997862 | 4.60E-08 | 2.23E-05 | 0.000223271 |
| HTN | monocyte | *MRPS25* | -2.50851111 | 0.005059183 | 0.023595115 | 1 |
| HTN | monocyte | *SLC6A6* | 2.463020288 | 0.003557037 | 0.023595115 | 1 |
| HTN | monocyte | *KCNC3* | 1.301464334 | 0.012142871 | 0.02905188 | 1 |
| HTN | monocyte | *NAPSA* | 2.549778359 | 0.000486012 | 0.016310293 | 1 |
| HTN | monocyte | *PDLIM4* | 2.862814949 | 0.001126581 | 0.023030817 | 1 |
| HTN | monocyte | *KIF3A* | -1.508804213 | 0.010706472 | 0.027617119 | 1 |
| HTN | monocyte | *NDUFA2* | -1.217710434 | 0.0016914 | 0.023595115 | 1 |
| HTN | monocyte | *BARX1* | 4.287770939 | 0.003216588 | 0.023595115 | 1 |
| HTN | monocyte | *NINJ1* | -1.754455294 | 0.012982407 | 0.030254871 | 1 |
| HTN | monocyte | *MAP1B* | -0.938792258 | 0.010898323 | 0.02788959 | 1 |
| HTN | monocyte | *CKMT2* | -1.420915892 | 0.004689397 | 0.023595115 | 1 |
| HTN | monocyte | *CHD1L* | -1.940007589 | 0.010696899 | 0.027617119 | 1 |
| HTN | monocyte | *PRKAB2* | -1.995227794 | 0.00986153 | 0.026779763 | 1 |
| HTN | monocyte | *ZNF304* | -1.705046628 | 0.020785744 | 0.039783996 | 1 |
| HTN | monocyte | *RHPN2* | -0.9631771 | 0.017506578 | 0.035392466 | 1 |
| HTN | monocyte | *GCH1* | 0.569213556 | 0.001617517 | 0.023595115 | 1 |
| HTN | monocyte | *TRIM21* | -1.761591899 | 0.004285498 | 0.023595115 | 1 |
| HTN | monocyte | *SPATA6* | -1.337552898 | 0.011189528 | 0.028135587 | 1 |
| HTN | monocyte | *LRRC41* | -2.109587589 | 0.014472412 | 0.032137366 | 1 |
| HTN | monocyte | *CCT6B* | -2.967875253 | 0.003422733 | 0.023595115 | 1 |
| HTN | monocyte | *CNGA4* | -2.299234438 | 0.011014438 | 0.027980132 | 1 |
| HTN | monocyte | *TRIM22* | -1.989448222 | 0.015278353 | 0.033020297 | 1 |
| HTN | monocyte | *RRP8* | 3.74316625 | 0.0097185 | 0.02671918 | 1 |
| HTN | monocyte | *TIMM10B* | 4.522401431 | 0.006155998 | 0.023595115 | 1 |
| HTN | monocyte | *RPA1* | -2.101878456 | 0.006338175 | 0.02374425 | 1 |
| HTN | monocyte | *SERPINF1* | -1.533258076 | 0.008816624 | 0.02589483 | 1 |
| HTN | monocyte | *TMEM128* | -1.499474014 | 0.007452308 | 0.024580667 | 1 |
| HTN | monocyte | *SEC61G* | -2.202641953 | 0.00626916 | 0.023704653 | 1 |
| HTN | monocyte | *FIGNL1* | -2.172796575 | 0.028762695 | 0.049515188 | 1 |
| HTN | monocyte | *EIF5A* | 0.932091711 | 0.015306955 | 0.033067385 | 1 |
| HTN | monocyte | *RGS22* | -0.873653683 | 0.007142112 | 0.024238452 | 1 |
| HTN | monocyte | *PRMT7* | -2.080674519 | 0.003247303 | 0.023595115 | 1 |
| HTN | monocyte | *IGHMBP2* | -2.003363713 | 0.00342422 | 0.023595115 | 1 |
| HTN | monocyte | *OSER1* | -2.269247563 | 0.008423275 | 0.025487024 | 1 |
| HTN | monocyte | *BHMT2* | -0.865091938 | 0.001191198 | 0.02330522 | 1 |
| HTN | monocyte | *ZBED3* | -1.185037103 | 0.006546415 | 0.023864166 | 1 |
| HTN | monocyte | *RSG1* | -11.91741017 | 0.000133634 | 0.008531487 | 0.648392991 |
| HTN | monocyte | *CASP9* | -2.111488752 | 0.009738548 | 0.02671918 | 1 |
| HTN | monocyte | *ATP8A2* | -2.705447191 | 0.004847559 | 0.023595115 | 1 |
| HTN | monocyte | *WASF3* | -1.375467003 | 0.008047769 | 0.025143448 | 1 |
| HTN | monocyte | *MPRIP* | -1.77773333 | 0.016719305 | 0.034504301 | 1 |
| HTN | monocyte | *DSTYK* | -2.168540197 | 0.02337143 | 0.042937591 | 1 |
| HTN | monocyte | *COG6* | -1.844115366 | 0.008358347 | 0.025458066 | 1 |
| HTN | monocyte | *CNDP2* | -1.853690614 | 0.009190702 | 0.026254852 | 1 |

| | | | | | | |
|---|---|---|---|---|---|---|
| HTN | monocyte | *PDZD2* | -0.877926848 | 0.006904908 | 0.024148823 | 1 |
| HTN | monocyte | *FAM83F* | -7.134421885 | 0.007928334 | 0.025073967 | 1 |
| HTN | monocyte | *SFTPD* | 1.33619419 | 0.00571459 | 0.023595115 | 1 |
| HTN | monocyte | *IPO8* | -2.842057274 | 0.028917683 | 0.049698576 | 1 |
| HTN | monocyte | *LARS* | -1.964943005 | 0.003768681 | 0.023595115 | 1 |
| HTN | monocyte | *SPINK5* | -2.719173775 | 0.005744772 | 0.023595115 | 1 |
| HTN | monocyte | *IMPA1* | 4.221593012 | 0.000299432 | 0.013706097 | 1 |
| HTN | monocyte | *LRRCC1* | -1.578283559 | 0.004999786 | 0.023595115 | 1 |
| HTN | monocyte | *RRAS2* | -1.101217108 | 0.005079579 | 0.023595115 | 1 |
| HTN | monocyte | *CATSPERB* | -0.933747974 | 0.005797045 | 0.023595115 | 1 |
| HTN | monocyte | *COX16* | 9.401851246 | 0.00156791 | 0.023595115 | 1 |
| HTN | monocyte | *TTC9* | 3.931214851 | 0.00364597 | 0.023595115 | 1 |
| HTN | monocyte | *ELP3* | -1.818071894 | 0.009179504 | 0.026245699 | 1 |
| HTN | monocyte | *PEBP4* | -1.241359753 | 0.005644378 | 0.023595115 | 1 |
| HTN | monocyte | *CTIF* | -0.836108779 | 0.010904487 | 0.027890132 | 1 |
| HTN | monocyte | *THUMPD3* | -2.407837934 | 0.008018295 | 0.025099849 | 1 |
| HTN | monocyte | *CNTN6* | -1.765729375 | 0.006435296 | 0.02382428 | 1 |
| HTN | monocyte | *CHL1* | -0.851402722 | 0.010489353 | 0.027465914 | 1 |
| HTN | monocyte | *VAV3* | -1.97579069 | 0.002377693 | 0.023595115 | 1 |
| HTN | monocyte | *TRIM45* | -2.235055861 | 0.003852137 | 0.023595115 | 1 |
| HTN | monocyte | *PPHLN1* | -1.488999818 | 0.013021331 | 0.030301917 | 1 |
| HTN | monocyte | *YWHAQ* | -1.573347808 | 0.009740301 | 0.02671918 | 1 |
| HTN | monocyte | *KIDINS220* | -1.634985256 | 0.018359136 | 0.036447842 | 1 |
| HTN | monocyte | *LPIN1* | -1.949801209 | 0.009868552 | 0.026779763 | 1 |
| HTN | monocyte | *TMEM241* | -1.424405932 | 0.001117061 | 0.022966009 | 1 |
| HTN | monocyte | *SOX5* | -0.931633445 | 0.012408241 | 0.029325274 | 1 |
| HTN | monocyte | *KLRD1* | -1.818996944 | 0.013659981 | 0.031084791 | 1 |
| HTN | monocyte | *HOOK1* | -1.596403758 | 0.000984392 | 0.021612081 | 1 |
| HTN | monocyte | *ZCCHC11* | -1.154011936 | 0.016633333 | 0.034456332 | 1 |
| HTN | monocyte | *PRPF38A* | -1.633379822 | 0.009799044 | 0.026755746 | 1 |
| HTN | monocyte | *ELP2* | -1.873295507 | 0.020180241 | 0.038922443 | 1 |
| HTN | monocyte | *TPGS2* | -1.851368656 | 0.017624334 | 0.035556453 | 1 |
| HTN | monocyte | *TIMM10* | 2.497133149 | 0.00320297 | 0.023595115 | 1 |
| HTN | monocyte | *FADS2* | 3.959331752 | 0.015828414 | 0.033678697 | 1 |
| HTN | monocyte | *CLOCK* | -1.545562898 | 0.0125637 | 0.029548751 | 1 |
| HTN | monocyte | *CLDN10* | 8.230100893 | 0.000546402 | 0.017259229 | 1 |
| HTN | monocyte | *ARGLU1* | -1.446934898 | 0.024531723 | 0.044413403 | 1 |
| HTN | monocyte | *ERCC5* | -2.196674312 | 0.003467079 | 0.023595115 | 1 |
| HTN | monocyte | *CARS2* | -2.083351718 | 0.019698448 | 0.038430588 | 1 |
| HTN | monocyte | *KLB* | -0.77115575 | 0.004356473 | 0.023595115 | 1 |
| HTN | monocyte | *UBQLN1* | -1.475383921 | 0.01094615 | 0.027890132 | 1 |
| HTN | monocyte | *GOLM1* | -1.395755007 | 0.003884966 | 0.023595115 | 1 |
| HTN | monocyte | *ADAM19* | -1.029068566 | 0.024040418 | 0.043900681 | 1 |
| HTN | monocyte | *USP30* | -1.679034752 | 0.0004881 | 0.016310293 | 1 |
| HTN | monocyte | *DTX1* | -0.972289017 | 0.005171235 | 0.023595115 | 1 |
| HTN | monocyte | *DMTF1* | -1.651652907 | 0.014648926 | 0.032322232 | 1 |
| HTN | monocyte | *TMEM60* | -2.195524352 | 0.018022557 | 0.035970978 | 1 |
| HTN | monocyte | *POM121C* | -2.469671352 | 0.001369745 | 0.023595115 | 1 |
| HTN | monocyte | *TES* | 1.636153223 | 0.012554757 | 0.029542036 | 1 |
| HTN | monocyte | *MTO1* | -1.632974389 | 0.01456981 | 0.032235621 | 1 |

| | | | | | | |
|---|---|---|---|---|---|---|
| HTN | monocyte | *KHDC1* | -0.984473291 | 0.003676876 | 0.023595115 | 1 |
| HTN | monocyte | *PAN2* | -1.723279173 | 0.011772478 | 0.028732426 | 1 |
| HTN | monocyte | *MAP7* | -1.467369911 | 0.01435265 | 0.031973856 | 1 |
| HTN | monocyte | *NHSL1* | -2.293503078 | 0.022423513 | 0.04183827 | 1 |
| HTN | monocyte | *AHI1* | -1.061048655 | 0.020533442 | 0.039363201 | 1 |
| HTN | monocyte | *MICAL1* | 2.089007365 | 0.000361231 | 0.01501035 | 1 |
| HTN | monocyte | *CCT7* | -2.752866202 | 0.004068643 | 0.023595115 | 1 |
| HTN | monocyte | *SMYD5* | 1.463852543 | 0.006116349 | 0.023595115 | 1 |
| HTN | monocyte | *AGT* | 4.042407198 | 0.000125793 | 0.008137974 | 0.610348015 |
| HTN | monocyte | *CAPN9* | -4.993991554 | 0.000996399 | 0.021669448 | 1 |
| HTN | monocyte | *MRPL44* | -1.822176809 | 0.00706905 | 0.024238452 | 1 |
| HTN | monocyte | *SLC19A3* | -0.656877358 | 0.004034595 | 0.023595115 | 1 |
| HTN | monocyte | *TSGA10* | -0.816645989 | 0.002859812 | 0.023595115 | 1 |
| HTN | monocyte | *ANKRD36* | -1.541677598 | 0.009987869 | 0.026892975 | 1 |
| HTN | monocyte | *STAB2* | -1.032522325 | 0.005306395 | 0.023595115 | 1 |
| HTN | monocyte | *APPL2* | -1.711680733 | 0.013181616 | 0.030482557 | 1 |
| HTN | monocyte | *SLC41A2* | -1.561195752 | 0.005414656 | 0.023595115 | 1 |
| HTN | monocyte | *RNASEH2B* | -1.647371838 | 0.003705197 | 0.023595115 | 1 |
| HTN | monocyte | *TBC1D4* | -1.534089599 | 0.005283437 | 0.023595115 | 1 |
| HTN | monocyte | *SUCLA2* | -1.698398434 | 0.01150026 | 0.028507713 | 1 |
| HTN | monocyte | *RCBTB2* | 1.591966357 | 0.00323286 | 0.023595115 | 1 |
| HTN | monocyte | *C7orf25* | 2.511815889 | 0.003639981 | 0.023595115 | 1 |
| HTN | monocyte | *NUPL2* | -1.747868728 | 0.01147233 | 0.02848908 | 1 |
| HTN | monocyte | *AOAH* | 0.331431383 | 0.003121911 | 0.023595115 | 1 |
| HTN | monocyte | *TBRG4* | 2.375726921 | 0.002354261 | 0.023595115 | 1 |
| HTN | monocyte | *MTHFS* | 1.588347418 | 0.004850711 | 0.023595115 | 1 |
| HTN | monocyte | *RTP4* | 1.999836888 | 0.011296665 | 0.028195174 | 1 |
| HTN | monocyte | *NDUFB5* | -2.626171587 | 0.007479028 | 0.024602198 | 1 |
| HTN | monocyte | *SCN2A* | -1.206228211 | 0.014041896 | 0.031659517 | 1 |
| HTN | monocyte | *IMP4* | 2.536187011 | 0.011473137 | 0.02848908 | 1 |
| HTN | monocyte | *NIPSNAP3A* | -2.270911111 | 0.003411688 | 0.023595115 | 1 |
| HTN | monocyte | *TOR1B* | -1.863524235 | 0.012355021 | 0.029302872 | 1 |
| HTN | monocyte | *SLC2A8* | 3.499764948 | 0.001579098 | 0.023595115 | 1 |
| HTN | monocyte | *ANP32B* | -2.22544319 | 0.015839816 | 0.033678697 | 1 |
| HTN | monocyte | *DSCC1* | -1.031619176 | 0.003889486 | 0.023595115 | 1 |
| HTN | monocyte | *POLR1E* | 0.91924159 | 0.023590388 | 0.043241617 | 1 |
| HTN | monocyte | *IL11RA* | -2.266228906 | 0.000586495 | 0.017674986 | 1 |
| HTN | monocyte | *APTX* | -1.667675644 | 0.010130972 | 0.027038216 | 1 |
| HTN | monocyte | *DENND4C* | -1.505244814 | 0.008171613 | 0.025334612 | 1 |
| HTN | monocyte | *GMPR* | -1.079539579 | 0.006678817 | 0.023986397 | 1 |
| HTN | monocyte | *TMEM14B* | 3.464494065 | 0.012046556 | 0.028978626 | 1 |
| HTN | monocyte | *FRS3* | -2.702520157 | 0.01137395 | 0.028349555 | 1 |
| HTN | monocyte | *LRRC1* | -1.24441535 | 0.003133533 | 0.023595115 | 1 |
| HTN | monocyte | *TUBB2B* | -2.309283715 | 0.016805811 | 0.034580913 | 1 |
| HTN | monocyte | *NRM* | 1.259387713 | 0.003406665 | 0.023595115 | 1 |
| HTN | monocyte | *TLR2* | 0.782822891 | 0.004432658 | 0.023595115 | 1 |
| HTN | monocyte | *NUMA1* | -1.953328231 | 0.015906612 | 0.033717292 | 1 |
| HTN | monocyte | *SYTL2* | -1.211947403 | 0.001720128 | 0.023595115 | 1 |
| HTN | monocyte | *RAB30* | -2.000667693 | 0.009726505 | 0.02671918 | 1 |
| HTN | monocyte | *NARS2* | -1.802391997 | 0.014102542 | 0.031686282 | 1 |

| | | | | | | |
|---|---|---|---|---|---|---|
| HTN | monocyte | *GGH* | -1.856773926 | 0.001347847 | 0.023595115 | 1 |
| HTN | monocyte | *SULF1* | -0.798955693 | 0.004249118 | 0.023595115 | 1 |
| HTN | monocyte | *TGS1* | -1.908623771 | 0.0143162 | 0.031973856 | 1 |
| HTN | monocyte | *NEK1* | -1.199683602 | 0.008034361 | 0.025117732 | 1 |
| HTN | monocyte | *TRPC6* | -1.083190931 | 0.014712316 | 0.032390024 | 1 |
| HTN | monocyte | *PPP2R1B* | -0.831372454 | 0.0004249 | 0.01520058 | 1 |
| HTN | monocyte | *FDX1* | -1.697692437 | 0.025234148 | 0.045363499 | 1 |
| HTN | monocyte | *FXYD6* | -1.884619921 | 0.003041606 | 0.023595115 | 1 |
| HTN | monocyte | *FXYD2* | -2.273168444 | 0.014791083 | 0.032436518 | 1 |
| HTN | monocyte | *MAP2K5* | -1.472577077 | 0.027200103 | 0.047649798 | 1 |
| HTN | monocyte | *NDUFAF1* | -2.114065714 | 0.025189982 | 0.045300887 | 1 |
| HTN | monocyte | *PARP6* | -1.708128901 | 0.006866492 | 0.024120446 | 1 |
| HTN | monocyte | *ZNF280D* | -1.484462082 | 0.019848024 | 0.038539283 | 1 |
| HTN | monocyte | *RSL24D1* | 2.782876133 | 0.007515398 | 0.024638032 | 1 |
| HTN | monocyte | *SLC44A5* | -0.922148992 | 0.005535877 | 0.023595115 | 1 |
| HTN | monocyte | *RTCA* | -1.736316341 | 0.027100718 | 0.047538931 | 1 |
| HTN | monocyte | *KHK* | -2.817831101 | 0.000667141 | 0.018676049 | 1 |
| HTN | monocyte | *DYNC2LI1* | -0.82477471 | 0.024200924 | 0.044110776 | 1 |
| HTN | monocyte | *LHCGR* | -2.80457158 | 0.001308032 | 0.023595115 | 1 |
| HTN | monocyte | *THUMPD2* | -1.643382726 | 0.008258542 | 0.025373581 | 1 |
| HTN | monocyte | *ATRAID* | -2.855499106 | 0.009431128 | 0.026481384 | 1 |
| HTN | monocyte | *CENPO* | -2.462652389 | 0.015591646 | 0.033395675 | 1 |
| HTN | monocyte | *LRPPRC* | -1.825022003 | 0.016383742 | 0.034241688 | 1 |
| HTN | monocyte | *DTNB* | -1.3119353 | 0.026602669 | 0.046945924 | 1 |
| HTN | monocyte | *BTBD16* | -2.051695837 | 0.005211092 | 0.023595115 | 1 |
| HTN | monocyte | *TACC2* | -1.04481143 | 0.004186257 | 0.023595115 | 1 |
| HTN | monocyte | *CALHM2* | -2.268480628 | 0.004222964 | 0.023595115 | 1 |
| HTN | monocyte | *PLCE1* | -1.180003747 | 0.006111622 | 0.023595115 | 1 |
| HTN | monocyte | *RBP4* | -1.43917045 | 0.009258461 | 0.026254852 | 1 |
| HTN | monocyte | *ASCC1* | -1.732356137 | 0.003034424 | 0.023595115 | 1 |
| HTN | monocyte | *SMARCAL1* | -1.626854375 | 0.01018502 | 0.027100648 | 1 |
| HTN | monocyte | *BARD1* | -3.418785802 | 0.018993261 | 0.037415876 | 1 |
| HTN | monocyte | *STAT4* | 1.752520271 | 0.002736884 | 0.023595115 | 1 |
| HTN | monocyte | *SSB* | -2.086292217 | 0.017242704 | 0.035122418 | 1 |
| HTN | monocyte | *ITGAV* | -1.88138507 | 0.015110142 | 0.032788197 | 1 |
| HTN | monocyte | *SENP7* | -1.71746348 | 0.01066965 | 0.02759549 | 1 |
| HTN | monocyte | *COX17* | 4.525534368 | 0.012237441 | 0.029206131 | 1 |
| HTN | monocyte | *PCDH10* | 4.755896035 | 0.022042423 | 0.041421316 | 1 |
| HTN | monocyte | *AP1AR* | -1.963092494 | 0.009782509 | 0.026725639 | 1 |
| HTN | monocyte | *KIAA1109* | -1.701754135 | 0.028925964 | 0.049698576 | 1 |
| HTN | monocyte | *BMPR1B* | -0.93366119 | 0.005700222 | 0.023595115 | 1 |
| HTN | monocyte | *MMRN1* | -0.827701777 | 0.003270464 | 0.023595115 | 1 |
| HTN | monocyte | *PRDM5* | -1.464756417 | 0.008426418 | 0.025487024 | 1 |
| HTN | monocyte | *TRPC3* | -2.349253757 | 0.025370638 | 0.045544496 | 1 |
| HTN | monocyte | *NAAA* | 0.729150803 | 0.003735395 | 0.023595115 | 1 |
| HTN | monocyte | *ANXA3* | -0.921570423 | 0.009105119 | 0.026216909 | 1 |
| HTN | monocyte | *ENPEP* | -1.814928235 | 0.017235769 | 0.035122418 | 1 |
| HTN | monocyte | *FBN2* | 0.605316618 | 8.74E-05 | 0.006490259 | 0.424200137 |
| HTN | monocyte | *PARVG* | 0.665309434 | 0.005200443 | 0.023595115 | 1 |
| HTN | monocyte | *B4GALNT3* | 2.448663472 | 0.004120646 | 0.023595115 | 1 |

| | | | | | | |
|---|---|---|---|---|---|---|
| HTN | monocyte | *KIF21A* | -1.00979882 | 0.00651458 | 0.023864166 | 1 |
| HTN | monocyte | *ALG10* | 1.718164317 | 0.027939484 | 0.048563258 | 1 |
| HTN | monocyte | *ZCRB1* | -2.052202094 | 0.011867315 | 0.028818926 | 1 |
| HTN | monocyte | *CLSTN3* | -1.281022717 | 0.004505389 | 0.023595115 | 1 |
| HTN | monocyte | *VAMP1* | -1.29213812 | 0.015255457 | 0.033000212 | 1 |
| HTN | monocyte | *TAPBPL* | -1.303477637 | 0.020951285 | 0.039943276 | 1 |
| HTN | monocyte | *PEX5* | -1.671658466 | 0.008256548 | 0.025373581 | 1 |
| HTN | monocyte | *PPFIA2* | -1.575868997 | 0.008416977 | 0.025487024 | 1 |
| HTN | monocyte | *MARCH9* | -1.546965351 | 0.005347388 | 0.023595115 | 1 |
| HTN | monocyte | *LGR5* | -3.267588206 | 0.004644122 | 0.023595115 | 1 |
| HTN | monocyte | *POC1B* | 3.554653334 | 0.027820764 | 0.048455938 | 1 |
| HTN | monocyte | *AMDHD1* | -2.112629961 | 0.000893889 | 0.020511604 | 1 |
| HTN | monocyte | *RITA1* | -2.49660574 | 0.0059858 | 0.023595115 | 1 |
| HTN | monocyte | *SDSL* | -1.885924895 | 0.018829912 | 0.037139323 | 1 |
| HTN | monocyte | *GLTP* | -1.935827356 | 0.007280766 | 0.024429382 | 1 |
| HTN | monocyte | *FOXN4* | -3.682659188 | 0.003092425 | 0.023595115 | 1 |
| HTN | monocyte | *BRCA2* | -1.609998892 | 0.003863808 | 0.023595115 | 1 |
| HTN | monocyte | *CERS5* | -1.892938472 | 0.022911391 | 0.042348979 | 1 |
| HTN | monocyte | *LMBR1L* | -2.150397619 | 0.004898114 | 0.023595115 | 1 |
| HTN | monocyte | *ZNF740* | -1.426959332 | 0.004289603 | 0.023595115 | 1 |
| HTN | monocyte | *HNRNPA1L2* | 2.883882049 | 0.008815361 | 0.02589483 | 1 |
| HTN | monocyte | *TMX1* | -1.838364142 | 0.010114581 | 0.027038216 | 1 |
| HTN | monocyte | *RTN1* | 3.011689665 | 3.24E-05 | 0.003208353 | 0.157209317 |
| HTN | monocyte | *C14orf37* | -1.643942272 | 0.014909929 | 0.032586927 | 1 |
| HTN | monocyte | *EFCAB11* | -1.718871032 | 0.003170223 | 0.023595115 | 1 |
| HTN | monocyte | *SLC24A4* | 0.249261325 | 0.003491393 | 0.023595115 | 1 |
| HTN | monocyte | *C14orf79* | -1.339158492 | 0.005898991 | 0.023595115 | 1 |
| HTN | monocyte | *WARS* | 4.314588452 | 0.025989626 | 0.046241902 | 1 |
| HTN | monocyte | *SRP14* | -2.690003308 | 0.016666696 | 0.034480686 | 1 |
| HTN | monocyte | *HMG20A* | -1.78879317 | 0.015598081 | 0.033395675 | 1 |
| HTN | monocyte | *SCAPER* | -1.827815002 | 0.005474813 | 0.023595115 | 1 |
| HTN | monocyte | *NEIL1* | -1.995871616 | 0.008923098 | 0.026034197 | 1 |
| HTN | monocyte | *PIF1* | -1.75665047 | 0.014280489 | 0.031930384 | 1 |
| HTN | monocyte | *CYP11A1* | -2.242808525 | 0.004065363 | 0.023595115 | 1 |
| HTN | monocyte | *BBS4* | -1.786938273 | 0.010232511 | 0.027124512 | 1 |
| HTN | monocyte | *ULK3* | -1.879249499 | 0.020448205 | 0.039261848 | 1 |
| HTN | monocyte | *PCSK6* | -0.991772242 | 0.005444495 | 0.023595115 | 1 |
| HTN | monocyte | *CELF6* | 3.897736409 | 0.001762835 | 0.023595115 | 1 |
| HTN | monocyte | *HAPLN3* | -3.879343041 | 0.00021276 | 0.010880596 | 1 |
| HTN | monocyte | *UNC45A* | -3.033440824 | 0.009537614 | 0.026641624 | 1 |
| HTN | monocyte | *CRTC3* | -1.553647332 | 0.014703208 | 0.032390024 | 1 |
| HTN | monocyte | *ITGAX* | 0.237172389 | 0.003449915 | 0.023595115 | 1 |
| HTN | monocyte | *TGFB1I1* | -3.846826617 | 0.005842886 | 0.023595115 | 1 |
| HTN | monocyte | *DHX38* | -1.580401053 | 0.00728696 | 0.024429382 | 1 |
| HTN | monocyte | *GCSH* | -1.772245099 | 0.001705525 | 0.023595115 | 1 |
| HTN | monocyte | *CMTM3* | 2.352238701 | 0.004639354 | 0.023595115 | 1 |
| HTN | monocyte | *GFOD2* | -2.492401689 | 0.014587193 | 0.032244675 | 1 |
| HTN | monocyte | *PRPSAP2* | -1.105037625 | 0.007494089 | 0.024618363 | 1 |
| HTN | monocyte | *RASL10B* | -5.661149732 | 0.00171116 | 0.023595115 | 1 |
| HTN | monocyte | *UNC45B* | 2.032016043 | 0.003183206 | 0.023595115 | 1 |

| | | | | | | |
|---|---|---|---|---|---|---|
| HTN | monocyte | *C17orf80* | -3.486807324 | 0.020740577 | 0.039713212 | 1 |
| HTN | monocyte | *SGSM2* | -1.968893799 | 0.010390875 | 0.027326031 | 1 |
| HTN | monocyte | *LRRC46* | -4.695941573 | 4.97E-05 | 0.004548273 | 0.241058465 |
| HTN | monocyte | *SCRN2* | -1.166397741 | 2.16E-05 | 0.002330865 | 0.104888904 |
| HTN | monocyte | *RHBDL3* | -5.045604423 | 0.019998272 | 0.038673422 | 1 |
| HTN | monocyte | *IMPA2* | 2.223717397 | 0.002543062 | 0.023595115 | 1 |
| HTN | monocyte | *GNAL* | -1.877640288 | 0.01082465 | 0.027847932 | 1 |
| HTN | monocyte | *GALNT1* | -1.793965418 | 0.025084763 | 0.045128391 | 1 |
| HTN | monocyte | *C18orf8* | -5.54906937 | 0.002132603 | 0.023595115 | 1 |
| HTN | monocyte | *SLC14A1* | -1.368033939 | 0.005028893 | 0.023595115 | 1 |
| HTN | monocyte | *CCDC40* | -1.069598421 | 0.002584841 | 0.023595115 | 1 |
| HTN | monocyte | *CARD14* | -2.133312915 | 0.025743841 | 0.045990102 | 1 |
| HTN | monocyte | *ANAPC11* | -2.458010421 | 0.009479722 | 0.026571699 | 1 |
| HTN | monocyte | *TRIM65* | 1.899850782 | 0.011533696 | 0.028508147 | 1 |
| HTN | monocyte | *SECTM1* | 1.857245243 | 0.010195912 | 0.027105649 | 1 |
| HTN | monocyte | *RNF157* | -2.34193024 | 0.00199006 | 0.023595115 | 1 |
| HTN | monocyte | *WDR45B* | -2.308108716 | 0.011168724 | 0.028135587 | 1 |
| HTN | monocyte | *RNF165* | -3.263485042 | 0.003205721 | 0.023595115 | 1 |
| HTN | monocyte | *MAPK4* | 1.556278424 | 0.00329226 | 0.023595115 | 1 |
| HTN | monocyte | *FBXO15* | -2.14706765 | 0.00883443 | 0.025915752 | 1 |
| HTN | monocyte | *CBLN2* | -2.095661234 | 0.005290204 | 0.023595115 | 1 |
| HTN | monocyte | *ERBB2* | -1.633936247 | 0.010920738 | 0.027890132 | 1 |
| HTN | monocyte | *MIEN1* | -4.519734471 | 0.001610869 | 0.023595115 | 1 |
| HTN | monocyte | *IGFBP4* | -1.188953618 | 0.026779081 | 0.047187585 | 1 |
| HTN | monocyte | *TXNL4A* | -3.356841749 | 0.01064542 | 0.027588558 | 1 |
| HTN | monocyte | *CACNA1A* | -1.408338259 | 0.004184586 | 0.023595115 | 1 |
| HTN | monocyte | *SAMD1* | -3.02050691 | 0.004058321 | 0.023595115 | 1 |
| HTN | monocyte | *NFIC* | -1.250656083 | 0.013619243 | 0.031038312 | 1 |
| HTN | monocyte | *PRDM15* | -2.042660514 | 0.011283422 | 0.028176615 | 1 |
| HTN | monocyte | *DUS3L* | 2.434800596 | 0.000612344 | 0.0181086 | 1 |
| HTN | monocyte | *DPP9* | -1.858082277 | 0.009838782 | 0.026779763 | 1 |
| HTN | monocyte | *SIRT3* | -1.391596416 | 0.016638998 | 0.034456332 | 1 |
| HTN | monocyte | *HUNK* | -1.116888228 | 0.003632934 | 0.023595115 | 1 |
| HTN | monocyte | *SAE1* | -3.506571741 | 0.013923754 | 0.031451608 | 1 |
| HTN | monocyte | *EVI5L* | -3.328890547 | 0.025559171 | 0.045780981 | 1 |
| HTN | monocyte | *SLC47A1* | 2.329973614 | 0.014098165 | 0.031686282 | 1 |
| HTN | monocyte | *CTU1* | 4.311394435 | 0.00459078 | 0.023595115 | 1 |
| HTN | monocyte | *MMEL1* | -2.692340603 | 0.000337321 | 0.01461324 | 1 |
| HTN | monocyte | *ARHGEF19* | 0.805243977 | 0.01792461 | 0.035878799 | 1 |
| HTN | monocyte | *PEX14* | -1.523426054 | 0.010853416 | 0.027878663 | 1 |
| HTN | monocyte | *ZNF593* | 2.754123081 | 0.004938386 | 0.023595115 | 1 |
| HTN | monocyte | *C1orf216* | 1.743205823 | 0.021845428 | 0.041172245 | 1 |
| HTN | monocyte | *ITGB3BP* | -1.335056931 | 0.017937194 | 0.035886387 | 1 |
| HTN | monocyte | *PIGK* | -2.57623148 | 0.009300614 | 0.026312873 | 1 |
| HTN | monocyte | *RPS8* | -1.672800621 | 0.014807685 | 0.032436518 | 1 |
| HTN | monocyte | *SLC44A3* | -2.327360875 | 0.019317307 | 0.037857168 | 1 |
| HTN | monocyte | *GPR161* | -1.480111012 | 0.006450148 | 0.02383558 | 1 |
| HTN | monocyte | *ATP1B1* | -1.70154424 | 0.0099625 | 0.026884342 | 1 |
| HTN | monocyte | *NME7* | -2.738159258 | 0.002267691 | 0.023595115 | 1 |
| HTN | monocyte | *XCL1* | -4.506406204 | 0.013956012 | 0.031498674 | 1 |

| | | | | | | |
|---|---|---|---|---|---|---|
| HTN | monocyte | *MAEL* | -3.095972827 | 0.005152223 | 0.023595115 | 1 |
| HTN | monocyte | *ARNT* | -2.864378258 | 0.003672654 | 0.023595115 | 1 |
| HTN | monocyte | *POGZ* | -1.960308646 | 0.009650174 | 0.02668844 | 1 |
| HTN | monocyte | *GABPB2* | -1.417175085 | 0.012009082 | 0.028978626 | 1 |
| HTN | monocyte | *SMYD2* | -1.382387856 | 0.006748431 | 0.024040665 | 1 |
| HTN | monocyte | *RAB13* | -2.824792006 | 0.011164257 | 0.028135587 | 1 |
| HTN | monocyte | *KCNN3* | -1.445448623 | 0.00346084 | 0.023595115 | 1 |
| HTN | monocyte | *C1orf43* | -2.289398188 | 0.005653049 | 0.023595115 | 1 |
| HTN | monocyte | *RIT1* | -1.827228927 | 0.005001602 | 0.023595115 | 1 |
| HTN | monocyte | *FLG* | -1.590006441 | 0.005932518 | 0.023595115 | 1 |
| HTN | monocyte | *ACP1* | -1.292178127 | 0.005877917 | 0.023595115 | 1 |
| HTN | monocyte | *CNIH4* | 3.056733858 | 0.010569705 | 0.02752821 | 1 |
| HTN | monocyte | *MBOAT2* | 3.128184929 | 0.005343683 | 0.023595115 | 1 |
| HTN | monocyte | *PYCR2* | 3.075289356 | 0.004174158 | 0.023595115 | 1 |
| HTN | monocyte | *OSR1* | 3.155335022 | 0.01996546 | 0.038656189 | 1 |
| HTN | monocyte | *RHOB* | -1.885131223 | 0.009741902 | 0.02671918 | 1 |
| HTN | monocyte | *GALM* | -1.990081403 | 0.015090001 | 0.032773807 | 1 |
| HTN | monocyte | *WDPCP* | -1.018932557 | 0.00793251 | 0.025073967 | 1 |
| HTN | monocyte | *ABHD1* | -1.948992665 | 0.001510092 | 0.023595115 | 1 |
| HTN | monocyte | *ZNF514* | -4.111559455 | 0.002375758 | 0.023595115 | 1 |
| HTN | monocyte | *EXOC6B* | -1.395254039 | 0.018790305 | 0.03709492 | 1 |
| HTN | monocyte | *TEX261* | -1.452338005 | 0.005156404 | 0.023595115 | 1 |
| HTN | monocyte | *THNSL2* | -1.390722672 | 0.000602521 | 0.017946357 | 1 |
| HTN | monocyte | *RABL2A* | -3.725766638 | 0.000474744 | 0.016247607 | 1 |
| HTN | monocyte | *FBLN7* | -1.056102833 | 0.016494485 | 0.034375171 | 1 |
| HTN | monocyte | *FAHD2B* | -3.187157486 | 0.0170347 | 0.034882084 | 1 |
| HTN | monocyte | *LYG1* | -1.300583413 | 0.00435865 | 0.023595115 | 1 |
| HTN | monocyte | *THSD7B* | -1.825888913 | 0.005720092 | 0.023595115 | 1 |
| HTN | monocyte | *POLR2D* | -1.142017146 | 0.009968905 | 0.026886676 | 1 |
| HTN | monocyte | *GALNT13* | -0.878164606 | 0.00499427 | 0.023595115 | 1 |
| HTN | monocyte | *PHOSPHO2* | 2.135590044 | 0.016615177 | 0.034456332 | 1 |
| HTN | monocyte | *NBEAL1* | -1.511530464 | 0.012979243 | 0.030254871 | 1 |
| HTN | monocyte | *SUMF1* | -1.395181341 | 0.01691646 | 0.034713272 | 1 |
| HTN | monocyte | *ACKR3* | -1.27960027 | 0.012279595 | 0.029220497 | 1 |
| HTN | monocyte | *ESPNL* | -17.11251064 | 0.005067168 | 0.023595115 | 1 |
| HTN | monocyte | *ANKMY1* | -1.586029534 | 0.014972015 | 0.032663767 | 1 |
| HTN | monocyte | *TAMM41* | -1.694167361 | 0.027912994 | 0.048563258 | 1 |
| HTN | monocyte | *STK11IP* | 3.429337713 | 0.005435286 | 0.023595115 | 1 |
| HTN | monocyte | *EAF1* | -1.639129219 | 0.008955494 | 0.026057755 | 1 |
| HTN | monocyte | *CNTN4* | -1.254806959 | 0.007966898 | 0.025076015 | 1 |
| HTN | monocyte | *SLC25A38* | -1.605664618 | 0.003083918 | 0.023595115 | 1 |
| HTN | monocyte | *CTDSPL* | -2.578328932 | 0.008580608 | 0.025783919 | 1 |
| HTN | monocyte | *PTPRG* | -1.039509194 | 0.007051472 | 0.024238452 | 1 |
| HTN | monocyte | *SLC25A26* | -1.748742847 | 0.005218622 | 0.023595115 | 1 |
| HTN | monocyte | *ARL6IP5* | 2.865351063 | 0.011608569 | 0.028576751 | 1 |
| HTN | monocyte | *PLA1A* | 1.966661564 | 0.004949428 | 0.023595115 | 1 |
| HTN | monocyte | *ADPRH* | 2.798422276 | 0.017556607 | 0.035464053 | 1 |
| HTN | monocyte | *BOC* | -0.752028978 | 0.006134867 | 0.023595115 | 1 |
| HTN | monocyte | *MED12L* | -1.349565247 | 0.007293463 | 0.024429382 | 1 |
| HTN | monocyte | *EIF2A* | -1.561697565 | 0.001677975 | 0.023595115 | 1 |

| | | | | | | |
|---|---|---|---|---|---|---|
| HTN | monocyte | *NCEH1* | -2.844843095 | 0.005157403 | 0.023595115 | 1 |
| HTN | monocyte | *AMT* | -1.994643838 | 0.000284382 | 0.013267537 | 1 |
| HTN | monocyte | *CCDC39* | -1.323636432 | 0.00396257 | 0.023595115 | 1 |
| HTN | monocyte | *MUC4* | -1.688390053 | 0.003501478 | 0.023595115 | 1 |
| HTN | monocyte | *CENPC* | -1.720069236 | 0.010414928 | 0.027359627 | 1 |
| HTN | monocyte | *SCD5* | -1.076991626 | 0.00510441 | 0.023595115 | 1 |
| HTN | monocyte | *PYURF* | -1.357865034 | 0.004890009 | 0.023595115 | 1 |
| HTN | monocyte | *CISD2* | 4.524353114 | 0.000532771 | 0.017006613 | 1 |
| HTN | monocyte | *DDIT4L* | -3.290722903 | 0.015991251 | 0.033743802 | 1 |
| HTN | monocyte | *METTL14* | -1.750610965 | 0.011827414 | 0.028782381 | 1 |
| HTN | monocyte | *USP53* | -1.045019902 | 0.011634814 | 0.02857976 | 1 |
| HTN | monocyte | *CBR4* | -1.424323652 | 0.013360748 | 0.030679768 | 1 |
| HTN | monocyte | *MARCH6* | -1.583026723 | 0.017354415 | 0.035260232 | 1 |
| HTN | monocyte | *MYO10* | -1.619806017 | 0.008406303 | 0.025487024 | 1 |
| HTN | monocyte | *FAM105A* | 1.608922544 | 0.001134453 | 0.023030817 | 1 |
| HTN | monocyte | *GTF2H2* | -1.700427955 | 0.005467987 | 0.023595115 | 1 |
| HTN | monocyte | *SPATA9* | -0.875389121 | 0.00503977 | 0.023595115 | 1 |
| HTN | monocyte | *TSLP* | -1.95937863 | 0.000201931 | 0.010739192 | 0.979768227 |
| HTN | monocyte | *FEM1C* | -1.993949462 | 0.008506838 | 0.02565269 | 1 |
| HTN | monocyte | *YIPF5* | 3.903121271 | 0.01262217 | 0.029657515 | 1 |
| HTN | monocyte | *CXCL14* | -2.608720373 | 0.010239708 | 0.027124512 | 1 |
| HTN | monocyte | *BOD1* | 3.880425505 | 0.001105556 | 0.022966009 | 1 |
| HTN | monocyte | *FAM50B* | -3.335805972 | 0.004585867 | 0.023595115 | 1 |
| HTN | monocyte | *MYLK4* | -2.211708269 | 0.010121078 | 0.027038216 | 1 |
| HTN | monocyte | *GFOD1* | -1.509321155 | 0.021993241 | 0.041373149 | 1 |
| HTN | monocyte | *TRIM7* | -3.305693258 | 0.001625256 | 0.023595115 | 1 |
| HTN | monocyte | *RNF44* | -2.554035271 | 0.012070951 | 0.029008545 | 1 |
| HTN | monocyte | *MUT* | -1.213615368 | 0.005227408 | 0.023595115 | 1 |
| HTN | monocyte | *DOK3* | 0.418792946 | 0.014425342 | 0.03206219 | 1 |
| HTN | monocyte | *DAAM2* | -1.665363077 | 0.012026964 | 0.028978626 | 1 |
| HTN | monocyte | *MLIP* | -1.625969104 | 0.011375778 | 0.028349555 | 1 |
| HTN | monocyte | *ANO7* | 0.640850538 | 0.005765136 | 0.023595115 | 1 |
| HTN | monocyte | *RPL7L1* | -2.707183001 | 0.009949836 | 0.02686511 | 1 |
| HTN | monocyte | *RNF217* | -1.452215997 | 0.01254773 | 0.029539829 | 1 |
| HTN | monocyte | *ARHGAP18* | -1.284547867 | 0.014761744 | 0.032409041 | 1 |
| HTN | monocyte | *ABRACL* | 1.696892275 | 0.002096674 | 0.023595115 | 1 |
| HTN | monocyte | *SLC18B1* | -2.856082621 | 0.000163421 | 0.009328469 | 0.792919877 |
| HTN | monocyte | *PNLDC1* | 1.032034858 | 0.007623088 | 0.024857003 | 1 |
| HTN | monocyte | *WTAP* | -1.923946127 | 0.014342309 | 0.031973856 | 1 |
| HTN | monocyte | *SLC22A3* | -0.900463677 | 0.005142898 | 0.023595115 | 1 |
| HTN | monocyte | *GNA12* | -2.991636999 | 0.011071827 | 0.028052483 | 1 |
| HTN | monocyte | *SDK1* | -0.887841186 | 0.02430477 | 0.044183868 | 1 |
| HTN | monocyte | *CCZ1B* | -2.478169058 | 0.012891853 | 0.030176778 | 1 |
| HTN | monocyte | *POMZP3* | -1.285365313 | 0.018020178 | 0.035970978 | 1 |
| HTN | monocyte | *TMEM209* | -1.463513093 | 0.006923127 | 0.024148823 | 1 |
| HTN | monocyte | *EPHA1* | -3.381514826 | 0.005154427 | 0.023595115 | 1 |
| HTN | monocyte | *NCAPG2* | -1.088511926 | 0.010918123 | 0.027890132 | 1 |
| HTN | monocyte | *FBXO25* | -1.996298637 | 0.020878628 | 0.039867415 | 1 |
| HTN | monocyte | *CHMP7* | 5.068917687 | 0.003198197 | 0.023595115 | 1 |
| HTN | monocyte | *TACC1* | -1.403565356 | 0.027586718 | 0.048147754 | 1 |

| | | | | | | |
|---|---|---|---|---|---|---|
| HTN | monocyte | *ADHFE1* | -1.24984228 | 6.80E-05 | 0.005633811 | 0.329993111 |
| HTN | monocyte | *DPYS* | -0.77092375 | 0.005102301 | 0.023595115 | 1 |
| HTN | monocyte | *SLC39A4* | 0.625794494 | 0.005669239 | 0.023595115 | 1 |
| HTN | monocyte | *AK3* | -1.41561901 | 0.00812896 | 0.025283149 | 1 |
| HTN | monocyte | *C9orf72* | 1.87392422 | 0.004035975 | 0.023595115 | 1 |
| HTN | monocyte | *ALAD* | -1.45131685 | 0.02813856 | 0.048791054 | 1 |
| HTN | monocyte | *WDR31* | -0.901779765 | 0.021829134 | 0.041163994 | 1 |
| HTN | monocyte | *SURF1* | 2.766823541 | 0.003704388 | 0.023595115 | 1 |
| HTN | monocyte | *SURF6* | -2.076743145 | 0.010519913 | 0.027486602 | 1 |
| HTN | monocyte | *REXO4* | -1.812095632 | 0.008265509 | 0.025373581 | 1 |
| HTN | monocyte | *SH3GLB2* | 4.957558565 | 0.002652072 | 0.023595115 | 1 |
| HTN | monocyte | *INPP5E* | -3.24517602 | 0.016092534 | 0.03385992 | 1 |
| HTN | monocyte | *SLC39A12* | 5.152768774 | 0.000836971 | 0.020511604 | 1 |
| HTN | monocyte | *TMEM236* | -1.786734725 | 0.016583221 | 0.034444259 | 1 |
| HTN | monocyte | *RSU1* | -1.592229714 | 0.013654647 | 0.031084791 | 1 |
| HTN | monocyte | *ST8SIA6* | -1.211602214 | 0.005595414 | 0.023595115 | 1 |
| HTN | monocyte | *PARD3* | -1.106913421 | 0.009383088 | 0.026407624 | 1 |
| HTN | monocyte | *RGR* | -18.21689897 | 0.000558396 | 0.017299633 | 1 |
| HTN | monocyte | *FRA10AC1* | -1.511566531 | 0.015863345 | 0.033684442 | 1 |
| HTN | monocyte | *PDCD11* | -1.477591496 | 0.009233935 | 0.026254852 | 1 |
| HTN | monocyte | *GAS2* | -1.375528121 | 0.006296059 | 0.023717764 | 1 |
| HTN | monocyte | *LIN7C* | -2.38890032 | 0.014079087 | 0.031684476 | 1 |
| HTN | monocyte | *HSD17B12* | -3.041733889 | 1.09E-09 | 1.32E-06 | 5.27E-06 |
| HTN | monocyte | *EIF3M* | -3.031775901 | 0.009817438 | 0.026779763 | 1 |
| HTN | monocyte | *PTPRJ* | 0.919796328 | 0.003907814 | 0.023595115 | 1 |
| HTN | monocyte | *ENDOD1* | -2.239087505 | 0.014634135 | 0.032314157 | 1 |
| HTN | monocyte | *CCDC82* | -1.900779436 | 0.016576332 | 0.034444259 | 1 |
| HTN | monocyte | *CAPN5* | -2.46424541 | 0.013197466 | 0.030491729 | 1 |
| HTN | monocyte | *INTS4* | -1.756683989 | 0.022381596 | 0.04181575 | 1 |
| HTN | monocyte | *DRD2* | -10.56394644 | 0.005004784 | 0.023595115 | 1 |
| HTN | monocyte | *NPAT* | -2.306375501 | 0.027059373 | 0.047483572 | 1 |
| HTN | monocyte | *ST14* | -4.481216083 | 0.016592114 | 0.034447984 | 1 |
| HTN | monocyte | *FADS1* | 1.739616324 | 0.00391255 | 0.023595115 | 1 |
| HTN | monocyte | *TMC2* | 1.253800684 | 0.003947985 | 0.023595115 | 1 |
| HTN | monocyte | *INCENP* | -1.751727477 | 0.002567678 | 0.023595115 | 1 |
| HTN | monocyte | *PLCH2* | 7.103436042 | 0.005197362 | 0.023595115 | 1 |
| HTN | monocyte | *CCDC15* | 1.446268403 | 0.014368165 | 0.031982525 | 1 |
| HTN | monocyte | *CHEK1* | -6.081839171 | 0.021196494 | 0.040315716 | 1 |
| HTN | monocyte | *SCN2B* | 1.246566633 | 0.005638446 | 0.023595115 | 1 |
| HTN | monocyte | *SPATA25* | -5.191159359 | 0.00087276 | 0.020511604 | 1 |
| HTN | monocyte | *CABLES2* | -2.415691439 | 0.000735131 | 0.019916297 | 1 |
| HTN | monocyte | *TRPT1* | -1.877480641 | 0.000260431 | 0.012510987 | 1 |
| HTN | monocyte | *NUDT22* | -1.973377441 | 0.024799788 | 0.044715188 | 1 |
| HTN | monocyte | *TBX6* | 1.416838709 | 2.50E-05 | 0.002638137 | 0.121354319 |
| HTN | monocyte | *ALDOA* | -2.46703756 | 0.00869303 | 0.025839644 | 1 |
| HTN | monocyte | *ANXA8L1* | -0.808627687 | 0.006075075 | 0.023595115 | 1 |
| HTN | monocyte | *CWC15* | -1.693448378 | 0.020259171 | 0.038991471 | 1 |
| HTN | monocyte | *FCGR1A* | 1.399098664 | 0.014342787 | 0.031973856 | 1 |
| HTN | monocyte | *TMEM218* | -2.139182397 | 0.010291984 | 0.027228303 | 1 |
| HTN | monocyte | *FAM124A* | -2.150434889 | 0.01201929 | 0.028978626 | 1 |

| | | | | | | |
|---|---|---|---|---|---|---|
| HTN | monocyte | *LYPD6B* | -2.038392169 | 0.017371808 | 0.035260232 | 1 |
| HTN | monocyte | *CCDC102B* | -1.448731274 | 0.008285791 | 0.025373581 | 1 |
| HTN | monocyte | *RGS18* | 0.321008893 | 0.003701559 | 0.023595115 | 1 |
| HTN | monocyte | *PRSS23* | -0.882202127 | 0.008712404 | 0.025839644 | 1 |
| HTN | monocyte | *PPP1R1C* | 1.468286761 | 0.003313494 | 0.023595115 | 1 |
| HTN | monocyte | *FAM173B* | -1.791175715 | 0.015785179 | 0.033645753 | 1 |
| HTN | monocyte | *GPR158* | -1.912116829 | 0.005817303 | 0.023595115 | 1 |
| HTN | monocyte | *UBE3B* | -1.47619129 | 0.016080328 | 0.033848916 | 1 |
| HTN | monocyte | *DLG5* | -1.080924039 | 0.008317987 | 0.025414907 | 1 |
| HTN | monocyte | *EXT2* | -1.9382389 | 0.023007962 | 0.042480467 | 1 |
| HTN | monocyte | *TMEM18* | 4.600073953 | 0.010311035 | 0.027258982 | 1 |
| HTN | monocyte | *NDUFC2* | -2.120941165 | 0.007521765 | 0.024638032 | 1 |
| HTN | monocyte | *ADAMTS12* | -1.034168092 | 0.001784893 | 0.023595115 | 1 |
| HTN | monocyte | *NUBPL* | -1.543147285 | 0.013387496 | 0.030691157 | 1 |
| HTN | monocyte | *VIPAS39* | -1.730275392 | 0.007698545 | 0.024968811 | 1 |
| HTN | monocyte | *ANKRD50* | 4.294058221 | 0.003961353 | 0.023595115 | 1 |
| HTN | monocyte | *UPF2* | -1.854299417 | 0.0239774 | 0.043824751 | 1 |
| HTN | monocyte | *QDPR* | -1.673618855 | 0.023410754 | 0.042977291 | 1 |
| HTN | monocyte | *FAM160B1* | -1.833064647 | 0.016103727 | 0.033868784 | 1 |
| HTN | monocyte | *ZNF827* | -1.667326717 | 0.004831112 | 0.023595115 | 1 |
| HTN | monocyte | *PIGF* | -1.811939933 | 0.011909132 | 0.028877239 | 1 |
| HTN | monocyte | *MFSD6* | -2.066784845 | 0.013123892 | 0.030409324 | 1 |
| HTN | monocyte | *RNF144A* | -1.613829332 | 0.010511359 | 0.027479049 | 1 |
| HTN | monocyte | *BICD1* | -1.980570774 | 0.028220309 | 0.048797199 | 1 |
| HTN | monocyte | *GUF1* | -1.806379596 | 0.012402865 | 0.029325274 | 1 |
| HTN | monocyte | *GABRA2* | -4.311824365 | 0.001643883 | 0.023595115 | 1 |
| HTN | monocyte | *CACUL1* | -1.892028429 | 0.021867452 | 0.041172245 | 1 |
| HTN | monocyte | *TIAL1* | -1.852707726 | 0.019535898 | 0.038159491 | 1 |
| HTN | monocyte | *TMEM56* | -2.258960035 | 0.005236878 | 0.023595115 | 1 |
| HTN | monocyte | *MGAT5* | -2.023074589 | 0.027944892 | 0.048563258 | 1 |
| HTN | monocyte | *TMEM178A* | -2.342052484 | 0.005412448 | 0.023595115 | 1 |
| HTN | monocyte | *EPG5* | -2.011158558 | 0.01736085 | 0.035260232 | 1 |
| HTN | monocyte | *HAUS1* | -1.420047046 | 0.006733782 | 0.024040665 | 1 |
| HTN | monocyte | *KCNK13* | 0.638878868 | 0.002864169 | 0.023595115 | 1 |
| HTN | monocyte | *ATG10* | -1.161686482 | 0.020853672 | 0.039862752 | 1 |
| HTN | monocyte | *NMT2* | -1.983177896 | 0.006437269 | 0.02382428 | 1 |
| HTN | monocyte | *ZNF837* | -2.352642992 | 0.010864619 | 0.027878663 | 1 |
| HTN | monocyte | *CCDC50* | -1.546760946 | 0.010085479 | 0.027007023 | 1 |
| HTN | monocyte | *PLEKHH2* | -1.393425485 | 0.000806494 | 0.020419313 | 1 |
| HTN | monocyte | *SPARCL1* | -1.528185433 | 0.026102668 | 0.046375 | 1 |
| HTN | monocyte | *GPD1L* | -1.281266904 | 0.007898316 | 0.025073967 | 1 |
| HTN | monocyte | *CCNO* | -14.59135837 | 0.005409985 | 0.023595115 | 1 |
| HTN | monocyte | *CLEC4F* | -2.033031289 | 0.019977277 | 0.038663641 | 1 |
| HTN | monocyte | *SLC30A6* | -2.026047506 | 0.006061028 | 0.023595115 | 1 |
| HTN | monocyte | *RASGRP3* | -0.934144562 | 0.005875798 | 0.023595115 | 1 |
| HTN | monocyte | *GPR180* | -1.215039805 | 0.003253922 | 0.023595115 | 1 |
| HTN | monocyte | *TCTEX1D1* | -4.200715487 | 0.006100248 | 0.023595115 | 1 |
| HTN | monocyte | *IFIT5* | 1.33331043 | 0.007676146 | 0.024938402 | 1 |
| HTN | monocyte | *PRDM8* | -5.765392625 | 0.019124605 | 0.037567848 | 1 |
| HTN | monocyte | *GGPS1* | -1.697727237 | 0.023073537 | 0.04253526 | 1 |

| | | | | | | |
|---|---|---|---|---|---|---|
| HTN | monocyte | *SREK1IP1* | -2.231946489 | 0.027132552 | 0.047577573 | 1 |
| HTN | monocyte | *MR1* | -2.327893547 | 0.028398015 | 0.049017137 | 1 |
| HTN | monocyte | *CENPH* | -2.372034569 | 0.002765986 | 0.023595115 | 1 |
| HTN | monocyte | *CARHSP1* | -1.655091904 | 0.009342486 | 0.026350386 | 1 |
| HTN | monocyte | *CAST* | -2.42984181 | 0.004726672 | 0.023595115 | 1 |
| HTN | monocyte | *CLGN* | -5.06088118 | 0.011717849 | 0.028671206 | 1 |
| HTN | monocyte | *CETN3* | -1.815778185 | 0.004644367 | 0.023595115 | 1 |
| HTN | monocyte | *MERTK* | -0.852076805 | 0.006405433 | 0.023797215 | 1 |
| HTN | monocyte | *SLC25A27* | -0.891186732 | 0.024266518 | 0.044156876 | 1 |
| HTN | monocyte | *FRMD1* | -2.016550883 | 0.0055342 | 0.023595115 | 1 |
| HTN | monocyte | *INO80C* | -2.215046652 | 0.00589992 | 0.023595115 | 1 |
| HTN | monocyte | *LPCAT1* | 2.238298959 | 0.003067025 | 0.023595115 | 1 |
| HTN | monocyte | *ADPRHL1* | -2.4124086 | 0.006788496 | 0.02405092 | 1 |
| HTN | monocyte | *CFDP1* | -1.695011966 | 0.018720956 | 0.036999626 | 1 |
| HTN | monocyte | *ZDHHC7* | 3.221388403 | 0.010413892 | 0.027359627 | 1 |
| HTN | monocyte | *JAZF1* | -2.302111167 | 0.024580134 | 0.04448445 | 1 |
| HTN | monocyte | *FBXO36* | -1.791168925 | 0.010618289 | 0.027580267 | 1 |
| HTN | monocyte | *MCOLN2* | 0.844098732 | 0.003046815 | 0.023595115 | 1 |
| HTN | monocyte | *SEMA3D* | -1.620256144 | 0.013333559 | 0.03065466 | 1 |
| HTN | monocyte | *ANKRD29* | -2.045050861 | 0.012136565 | 0.029051118 | 1 |
| HTN | monocyte | *UBASH3B* | 0.464563252 | 0.00361952 | 0.023595115 | 1 |
| HTN | monocyte | *ROBO4* | -2.502655883 | 0.025271883 | 0.045397695 | 1 |
| HTN | monocyte | *ROBO3* | -1.408462835 | 0.021825203 | 0.041163994 | 1 |
| HTN | monocyte | *PRKCA* | -1.76546736 | 0.024254197 | 0.044156876 | 1 |
| HTN | monocyte | *LRRK1* | -2.21440855 | 0.010545465 | 0.027523721 | 1 |
| HTN | monocyte | *ABCA5* | -0.731850985 | 0.022216119 | 0.041647859 | 1 |
| HTN | monocyte | *ENPP3* | -1.233336928 | 0.00433258 | 0.023595115 | 1 |
| HTN | monocyte | *MIA3* | -2.062002469 | 0.022370728 | 0.041811545 | 1 |
| HTN | monocyte | *PGM5* | -1.686915309 | 0.015498348 | 0.03333244 | 1 |
| HTN | monocyte | *CCSAP* | -1.379265002 | 0.005296856 | 0.023595115 | 1 |
| HTN | monocyte | *GBP5* | 1.164019517 | 0.005873923 | 0.023595115 | 1 |
| HTN | monocyte | *PDE1C* | -1.54412152 | 0.009503503 | 0.026576944 | 1 |
| HTN | monocyte | *XPC* | -2.08259994 | 0.00582802 | 0.023595115 | 1 |
| HTN | monocyte | *FLCN* | -1.627770188 | 0.005813609 | 0.023595115 | 1 |
| HTN | monocyte | *OXNAD1* | -1.806278385 | 0.020271489 | 0.03899502 | 1 |
| HTN | monocyte | *APCDD1* | -0.747255579 | 0.007277534 | 0.024429382 | 1 |
| HTN | monocyte | *MPPE1* | -1.801066368 | 0.014638606 | 0.032314157 | 1 |
| HTN | monocyte | *RAB6B* | -1.91545461 | 0.002655609 | 0.023595115 | 1 |
| HTN | monocyte | *CYP2U1* | -2.230135246 | 0.024865694 | 0.044800723 | 1 |
| HTN | monocyte | *MARVELD1* | -1.742717417 | 0.005918243 | 0.023595115 | 1 |
| HTN | monocyte | *GOLGA7B* | 1.38351497 | 0.003415648 | 0.023595115 | 1 |
| HTN | monocyte | *MOV10* | 3.274528133 | 0.000796433 | 0.020419313 | 1 |
| HTN | monocyte | *CNOT8* | -2.756509352 | 0.02437171 | 0.044273639 | 1 |
| HTN | monocyte | *ZKSCAN2* | -2.253033116 | 0.002024825 | 0.023595115 | 1 |
| HTN | monocyte | *KDM8* | -2.1833168 | 0.006426637 | 0.023821268 | 1 |
| HTN | monocyte | *ALS2CR12* | 3.127337665 | 0.004499291 | 0.023595115 | 1 |
| HTN | monocyte | *DEPTOR* | -1.080137054 | 0.025732892 | 0.045987473 | 1 |
| HTN | monocyte | *RRAGA* | -1.366427126 | 0.004918165 | 0.023595115 | 1 |
| HTN | monocyte | *PSD3* | -1.419609662 | 0.007870452 | 0.025073822 | 1 |
| HTN | monocyte | *KCNMA1* | -1.221772636 | 0.002242318 | 0.023595115 | 1 |

| | | | | | | |
|---|---|---|---|---|---|---|
| HTN | monocyte | *NDUFAF6* | -1.805307707 | 0.000799489 | 0.020419313 | 1 |
| HTN | monocyte | *C8orf37* | -2.30508346 | 0.002080768 | 0.023595115 | 1 |
| HTN | monocyte | *PPEF2* | -4.230628922 | 0.016675043 | 0.034480686 | 1 |
| HTN | monocyte | *CCT8* | -1.812656276 | 0.005657239 | 0.023595115 | 1 |
| HTN | monocyte | *NAA11* | -2.433618832 | 0.001673522 | 0.023595115 | 1 |
| HTN | monocyte | *SFXN2* | 4.849362922 | 4.21E-09 | 3.41E-06 | 2.04E-05 |
| HTN | monocyte | *TDRD9* | -5.667311381 | 0.001932372 | 0.023595115 | 1 |
| HTN | monocyte | *PCDH1* | -1.533285495 | 0.002705713 | 0.023595115 | 1 |
| HTN | monocyte | *TYSND1* | -0.835059118 | 0.011278312 | 0.028176615 | 1 |
| HTN | monocyte | *NODAL* | -2.314899526 | 0.005512115 | 0.023595115 | 1 |
| HTN | monocyte | *ATAD2* | -1.993775498 | 0.020411291 | 0.039206485 | 1 |
| HTN | monocyte | *FBRS* | 2.738269917 | 0.008634189 | 0.025828042 | 1 |
| HTN | monocyte | *GHRL* | -1.246800202 | 0.009121934 | 0.026216909 | 1 |
| HTN | monocyte | *NMNAT2* | -1.798981151 | 0.004388866 | 0.023595115 | 1 |
| HTN | monocyte | *SLC6A1* | 1.850073524 | 0.0036807 | 0.023595115 | 1 |
| HTN | monocyte | *STEAP2* | -1.196882147 | 0.002690908 | 0.023595115 | 1 |
| HTN | monocyte | *CLDN12* | -3.223994383 | 0.017905108 | 0.035878799 | 1 |
| HTN | monocyte | *MMP14* | -2.299576166 | 0.016865819 | 0.034660293 | 1 |
| HTN | monocyte | *GATAD1* | -1.708072533 | 0.017240007 | 0.035122418 | 1 |
| HTN | monocyte | *TMED6* | -8.997792565 | 0.002848546 | 0.023595115 | 1 |
| HTN | monocyte | *CLEC18A* | -6.091630968 | 5.17E-06 | 0.000784532 | 0.025105026 |
| HTN | monocyte | *AASDH* | -1.69785956 | 0.015937409 | 0.03374074 | 1 |
| HTN | monocyte | *ETS2* | 2.331651465 | 0.001668812 | 0.023595115 | 1 |
| HTN | monocyte | *LCA5L* | -1.260912421 | 0.006628747 | 0.02395053 | 1 |
| HTN | monocyte | *DGKI* | -0.923294678 | 0.003927052 | 0.023595115 | 1 |
| HTN | monocyte | *SNX22* | 5.983259218 | 0.0167628 | 0.034551021 | 1 |
| HTN | monocyte | *SLC34A2* | -26.92226664 | 0.010467987 | 0.027439587 | 1 |
| HTN | monocyte | *AP3S2* | -1.089488642 | 0.00833045 | 0.025426186 | 1 |
| HTN | monocyte | *C12orf43* | -1.756661658 | 0.002426051 | 0.023595115 | 1 |
| HTN | monocyte | *LDLRAP1* | -1.712312789 | 0.007383579 | 0.024485743 | 1 |
| HTN | monocyte | *EXTL1* | -19.25174115 | 0.005082799 | 0.023595115 | 1 |
| HTN | monocyte | *LRRC43* | -5.711471594 | 0.000297481 | 0.013706097 | 1 |
| HTN | monocyte | *DZIP1L* | -1.552121143 | 0.010014434 | 0.026934608 | 1 |
| HTN | monocyte | *FANCC* | -1.465111523 | 0.014949653 | 0.032629652 | 1 |
| HTN | monocyte | *ESYT3* | -2.978780371 | 0.003751795 | 0.023595115 | 1 |
| HTN | monocyte | *CLSTN2* | -0.765187097 | 0.004319308 | 0.023595115 | 1 |
| HTN | monocyte | *COLEC12* | -0.928722203 | 0.006483798 | 0.023864166 | 1 |
| HTN | monocyte | *MITD1* | -1.825574582 | 0.02752993 | 0.048067063 | 1 |
| HTN | monocyte | *TSPAN33* | -2.825537417 | 0.000639322 | 0.018354964 | 1 |
| HTN | monocyte | *FAM86C1* | 12.98852183 | 0.01692961 | 0.034717864 | 1 |
| HTN | monocyte | *PPP1R9A* | -1.39905042 | 0.011509145 | 0.028507713 | 1 |
| HTN | monocyte | *DYNC1I1* | -0.854041827 | 0.005965542 | 0.023595115 | 1 |
| HTN | monocyte | *COPG2* | -0.906299829 | 0.011023738 | 0.027980937 | 1 |
| HTN | monocyte | *ELK4* | 4.250741397 | 0.015681467 | 0.033474034 | 1 |
| HTN | monocyte | *DUSP23* | -1.511092738 | 0.003505197 | 0.023595115 | 1 |
| HTN | monocyte | *F11R* | -1.710315539 | 0.000898054 | 0.020511604 | 1 |
| HTN | monocyte | *FGF17* | -9.094875587 | 0.003121348 | 0.023595115 | 1 |
| HTN | monocyte | *CDA* | 0.62807031 | 0.003909431 | 0.023595115 | 1 |
| HTN | monocyte | *SLC5A11* | -2.558819457 | 6.26E-08 | 2.76E-05 | 0.000303542 |
| HTN | monocyte | *CCAR2* | -2.371326438 | 0.025570169 | 0.045780981 | 1 |

| | | | | | | |
|---|---|---|---|---|---|---|
| HTN | monocyte | *IFNAR2* | 1.110852286 | 0.019422502 | 0.038029855 | 1 |
| HTN | monocyte | *SV2A* | 2.14414104 | 0.00115406 | 0.023234426 | 1 |
| HTN | monocyte | *CBR1* | -1.105884939 | 0.008707721 | 0.025839644 | 1 |
| HTN | monocyte | *HLCS* | -1.148758851 | 0.014335136 | 0.031973856 | 1 |
| HTN | monocyte | *ARHGAP27* | 4.660157152 | 0.007721388 | 0.02497075 | 1 |
| HTN | monocyte | *IRX6* | -1.187691904 | 0.002916678 | 0.023595115 | 1 |
| HTN | monocyte | *HK2* | -0.888904287 | 0.004755824 | 0.023595115 | 1 |
| HTN | monocyte | *ACE* | -1.452844685 | 0.005093037 | 0.023595115 | 1 |
| HTN | monocyte | *TEPP* | 1.33782759 | 0.005499337 | 0.023595115 | 1 |
| HTN | monocyte | *CTBP1* | -4.393123622 | 0.006539855 | 0.023864166 | 1 |
| HTN | monocyte | *LRRC36* | -2.099503135 | 0.000162037 | 0.009328469 | 0.78620532 |
| HTN | monocyte | *ZFYVE28* | -1.386433341 | 0.006161379 | 0.023595115 | 1 |
| HTN | monocyte | *C16orf86* | -1.369792651 | 0.024465626 | 0.044359946 | 1 |
| HTN | monocyte | *ZNF233* | -3.236750723 | 0.011236695 | 0.028146849 | 1 |
| HTN | monocyte | *ATAD3B* | 2.508249248 | 0.000269449 | 0.012692888 | 1 |
| HTN | monocyte | *SSU72* | -2.026237697 | 0.013818764 | 0.031331141 | 1 |
| HTN | monocyte | *CPAMD8* | -1.518181503 | 0.022580662 | 0.041993627 | 1 |
| HTN | monocyte | *ANKLE1* | -7.585845147 | 0.003372342 | 0.023595115 | 1 |
| HTN | monocyte | *PDE9A* | -1.898261417 | 0.021326277 | 0.04049906 | 1 |
| HTN | monocyte | *HSF2BP* | -2.643270865 | 0.000322779 | 0.014501148 | 1 |
| HTN | monocyte | *CSTB* | -2.089976139 | 0.018132606 | 0.036146016 | 1 |
| HTN | monocyte | *C21orf33* | -3.435567019 | 0.001990096 | 0.023595115 | 1 |
| HTN | monocyte | *C21orf2* | -2.204989439 | 0.006223731 | 0.023665784 | 1 |
| HTN | monocyte | *SPATC1L* | 0.810843171 | 0.009754824 | 0.02671918 | 1 |
| HTN | monocyte | *VAV2* | 3.384532762 | 0.009252021 | 0.026254852 | 1 |
| HTN | monocyte | *PCNT* | -2.104157399 | 0.001638405 | 0.023595115 | 1 |
| HTN | monocyte | *S100B* | -1.673372585 | 8.80E-07 | 0.000224758 | 0.004270407 |
| HTN | monocyte | *ZNF208* | -2.396491766 | 0.00051698 | 0.016722594 | 1 |
| HTN | monocyte | *ADAMTS13* | -1.252264123 | 0.015491852 | 0.03333244 | 1 |
| HTN | monocyte | *CACFD1* | 5.207782925 | 0.007530544 | 0.024638032 | 1 |
| HTN | monocyte | *RDH13* | -1.250370265 | 0.01691955 | 0.034713272 | 1 |
| HTN | monocyte | *TLCD1* | -6.493637245 | 0.013223161 | 0.030491729 | 1 |
| HTN | monocyte | *CCR5* | 3.679062803 | 0.000160344 | 0.009328469 | 0.777987098 |
| HTN | monocyte | *NBEAL2* | 1.186633733 | 0.002134545 | 0.023595115 | 1 |
| HTN | monocyte | *PTH1R* | -2.218112751 | 0.002364704 | 0.023595115 | 1 |
| HTN | monocyte | *FGFR4* | 3.50350069 | 0.002181942 | 0.023595115 | 1 |
| HTN | monocyte | *TONSL* | 2.363503077 | 0.001937024 | 0.023595115 | 1 |
| HTN | monocyte | *MUM1* | -1.47862599 | 0.009082231 | 0.026214745 | 1 |
| HTN | monocyte | *ZNF333* | -2.400088579 | 0.018146828 | 0.03615951 | 1 |
| HTN | monocyte | *SH2B2* | 0.469302817 | 0.005181434 | 0.023595115 | 1 |
| HTN | monocyte | *LRWD1* | 1.523922537 | 0.018588238 | 0.03679728 | 1 |
| HTN | monocyte | *ABCF3* | 3.655098754 | 3.52E-07 | 0.000107291 | 0.001706568 |
| HTN | monocyte | *FBXO27* | -1.181853923 | 0.002884362 | 0.023595115 | 1 |
| HTN | monocyte | *U2AF1L4* | -1.130740303 | 0.00407721 | 0.023595115 | 1 |
| HTN | monocyte | *BDH1* | -2.245165119 | 0.008967205 | 0.026068832 | 1 |
| HTN | monocyte | *PGAP3* | 3.086872311 | 0.003453176 | 0.023595115 | 1 |
| HTN | monocyte | *TMEM143* | -2.361790852 | 0.016500366 | 0.034375171 | 1 |
| HTN | monocyte | *ALDH16A1* | 2.254377156 | 0.000495338 | 0.01634951 | 1 |
| HTN | monocyte | *SIGLEC11* | 0.839212004 | 0.015398037 | 0.03319026 | 1 |
| HTN | monocyte | *PLCD3* | 4.629653086 | 0.007148635 | 0.024238452 | 1 |

| | | | | | | |
|---|---|---|---|---|---|---|
| HTN | monocyte | *AQP5* | -8.004669899 | 0.013060558 | 0.030344279 | 1 |
| HTN | monocyte | *ASGR2* | 0.781243486 | 0.003501262 | 0.023595115 | 1 |
| HTN | monocyte | *POLR3K* | 0.897944691 | 0.003548775 | 0.023595115 | 1 |
| HTN | monocyte | *WDR90* | -1.170834415 | 0.015610344 | 0.033395675 | 1 |
| HTN | monocyte | *CCDC78* | 1.06768418 | 0.001017805 | 0.021755015 | 1 |
| HTN | monocyte | *MEIOB* | -1.525266671 | 0.017784614 | 0.035760857 | 1 |
| HTN | monocyte | *ASRGL1* | 3.248414778 | 0.003914111 | 0.023595115 | 1 |
| HTN | monocyte | *RPS6KA4* | 1.495490842 | 0.003124859 | 0.023595115 | 1 |
| HTN | monocyte | *TPCN2* | -1.84136987 | 0.02388153 | 0.043692755 | 1 |
| HTN | monocyte | *PDZK1IP1* | -2.121217422 | 0.00482069 | 0.023595115 | 1 |
| HTN | monocyte | *COA7* | 6.379249751 | 0.009326954 | 0.026350386 | 1 |
| HTN | monocyte | *SLC1A7* | -1.529763343 | 0.006771849 | 0.02405092 | 1 |
| HTN | monocyte | *C1orf123* | -3.325753793 | 0.022418367 | 0.04183827 | 1 |
| HTN | monocyte | *USP24* | -1.745430748 | 0.009479638 | 0.026571699 | 1 |
| HTN | monocyte | *ZSWIM5* | -1.475946797 | 0.011229394 | 0.028146849 | 1 |
| HTN | monocyte | *AK4* | -1.383787736 | 0.011176172 | 0.028135587 | 1 |
| HTN | monocyte | *LAPTM5* | 2.576629525 | 0.003979236 | 0.023595115 | 1 |
| HTN | monocyte | *SYNC* | -1.274321323 | 0.002970344 | 0.023595115 | 1 |
| HTN | monocyte | *CAMK2N1* | 4.021824859 | 0.002228734 | 0.023595115 | 1 |
| HTN | monocyte | *OMA1* | -3.268233384 | 0.018348383 | 0.036441406 | 1 |
| HTN | monocyte | *NEXN* | -1.413502484 | 0.025816798 | 0.046080084 | 1 |
| HTN | monocyte | *LRRIQ3* | -1.02002339 | 0.004980514 | 0.023595115 | 1 |
| HTN | monocyte | *TYW3* | -1.017132012 | 0.005507849 | 0.023595115 | 1 |
| HTN | monocyte | *HFM1* | -1.405837435 | 0.002149043 | 0.023595115 | 1 |
| HTN | monocyte | *ZNF496* | -2.993673183 | 0.025518327 | 0.045772615 | 1 |
| HTN | monocyte | *FCRLB* | 2.348235284 | 0.003896904 | 0.023595115 | 1 |
| HTN | monocyte | *TDRD5* | -4.210531859 | 0.005025279 | 0.023595115 | 1 |
| HTN | monocyte | *ACP6* | -1.521968967 | 0.00543034 | 0.023595115 | 1 |
| HTN | monocyte | *PKDCC* | -1.180997853 | 0.020926175 | 0.039914234 | 1 |
| HTN | monocyte | *HAAO* | -1.978597188 | 0.009370746 | 0.026388194 | 1 |
| HTN | monocyte | *DUSP19* | 3.125327477 | 0.005303647 | 0.023595115 | 1 |
| HTN | monocyte | *NUP35* | -2.259474078 | 0.009608438 | 0.02668844 | 1 |
| HTN | monocyte | *C2orf48* | 5.069166554 | 0.002214365 | 0.023595115 | 1 |
| HTN | monocyte | *NOSTRIN* | -1.320987559 | 0.001738587 | 0.023595115 | 1 |
| HTN | monocyte | *RPRD2* | -1.794909155 | 0.026112222 | 0.046375 | 1 |
| HTN | monocyte | *CTSS* | 0.573440235 | 0.013957574 | 0.031498674 | 1 |
| HTN | monocyte | *C1QTNF7* | -1.135828207 | 0.008477798 | 0.025596935 | 1 |
| HTN | monocyte | *LYSMD1* | -4.668270881 | 1.54E-06 | 0.000339795 | 0.007475487 |
| HTN | monocyte | *RNF149* | 2.52567233 | 0.007985189 | 0.025077113 | 1 |
| HTN | monocyte | *S100A9* | -1.705146901 | 0.005762581 | 0.023595115 | 1 |
| HTN | monocyte | *TDRD10* | -1.641246152 | 0.008735639 | 0.025839644 | 1 |
| HTN | monocyte | *DCAF16* | -3.447386061 | 0.001298951 | 0.023595115 | 1 |
| HTN | monocyte | *GNPDA2* | -1.646346432 | 0.008625336 | 0.025828042 | 1 |
| HTN | monocyte | *DAPL1* | 1.372427114 | 0.001677757 | 0.023595115 | 1 |
| HTN | monocyte | *CLDN1* | -0.841561968 | 0.005746522 | 0.023595115 | 1 |
| HTN | monocyte | *DCST2* | -1.710963884 | 0.028659214 | 0.049380152 | 1 |
| HTN | monocyte | *SLC22A15* | -1.824029726 | 0.01880083 | 0.037097043 | 1 |
| HTN | monocyte | *LRRC58* | -2.258425828 | 0.011646712 | 0.028583634 | 1 |
| HTN | monocyte | *LMOD1* | -2.31876269 | 0.000559412 | 0.017299633 | 1 |
| HTN | monocyte | *CCT3* | 2.29760076 | 0.004890378 | 0.023595115 | 1 |

| | | | | | | |
|---|---|---|---|---|---|---|
| HTN | monocyte | *SERPINI1* | -1.577455934 | 0.004229719 | 0.023595115 | 1 |
| HTN | monocyte | *SUCLG1* | -2.093700077 | 0.019798042 | 0.038501043 | 1 |
| HTN | monocyte | *GTPBP8* | -2.499433019 | 0.012033261 | 0.028978626 | 1 |
| HTN | monocyte | *WDFY3* | -1.43702782 | 0.021780638 | 0.041104494 | 1 |
| HTN | monocyte | *C4orf36* | -0.985577484 | 0.005150868 | 0.023595115 | 1 |
| HTN | monocyte | *PPM1K* | -1.259206278 | 0.005466483 | 0.023595115 | 1 |
| HTN | monocyte | *TIPARP* | -2.476135812 | 0.026164201 | 0.046416345 | 1 |
| HTN | monocyte | *RPP14* | -1.32781354 | 0.003861846 | 0.023595115 | 1 |
| HTN | monocyte | *DNASE1L3* | 1.164951037 | 0.00226727 | 0.023595115 | 1 |
| HTN | monocyte | *IL17RC* | -3.266623527 | 0.002367745 | 0.023595115 | 1 |
| HTN | monocyte | *CRELD1* | -2.183712751 | 0.013456479 | 0.030775364 | 1 |
| HTN | monocyte | *TTC14* | -1.770111974 | 0.008104417 | 0.025239173 | 1 |
| HTN | monocyte | *CCDC158* | -1.672881049 | 0.015891748 | 0.033700508 | 1 |
| HTN | monocyte | *CPA3* | -1.746928443 | 0.00525597 | 0.023595115 | 1 |
| HTN | monocyte | *SLC4A1AP* | 3.171354604 | 0.006456176 | 0.023839702 | 1 |
| HTN | monocyte | *CDCP1* | -1.417669629 | 0.011090203 | 0.028056466 | 1 |
| HTN | monocyte | *CLEC3B* | -2.396363261 | 0.011233336 | 0.028146849 | 1 |
| HTN | monocyte | *TPRA1* | -3.749487604 | 0.0024429 | 0.023595115 | 1 |
| HTN | monocyte | *DNALI1* | -3.022181541 | 0.000211612 | 0.010880596 | 1 |
| HTN | monocyte | *RPN1* | -2.227258179 | 0.015394538 | 0.03319026 | 1 |
| HTN | monocyte | *RFT1* | -2.763414355 | 8.83E-05 | 0.006490259 | 0.428357121 |
| HTN | monocyte | *SFMBT1* | 4.531319371 | 0.021736026 | 0.041068224 | 1 |
| HTN | monocyte | *PIGX* | 1.372062502 | 0.020601116 | 0.039477336 | 1 |
| HTN | monocyte | *ABLIM2* | -1.450667886 | 0.012060303 | 0.028997319 | 1 |
| HTN | monocyte | *BDH2* | -2.118727337 | 0.001895494 | 0.023595115 | 1 |
| HTN | monocyte | *CDC25A* | -25.56008597 | 2.21E-09 | 2.14E-06 | 1.07E-05 |
| HTN | monocyte | *ZNF589* | -6.595313143 | 2.60E-06 | 0.000485219 | 0.012615689 |
| HTN | monocyte | *ATRIP* | -2.434208677 | 0.0015864 | 0.023595115 | 1 |
| HTN | monocyte | *RNF123* | 2.749733615 | 0.007429425 | 0.024522157 | 1 |
| HTN | monocyte | *MFSD8* | -2.018144757 | 0.01330497 | 0.030634456 | 1 |
| HTN | monocyte | *RAD54L2* | -2.165451494 | 0.027178407 | 0.047640762 | 1 |
| HTN | monocyte | *ETNPPL* | -9.381929411 | 0.005321628 | 0.023595115 | 1 |
| HTN | monocyte | *PRSS12* | -1.606993451 | 0.018546806 | 0.036730247 | 1 |
| HTN | monocyte | *CEP44* | -2.09127252 | 0.002775382 | 0.023595115 | 1 |
| HTN | monocyte | *HPGD* | -1.771371625 | 0.00506068 | 0.023595115 | 1 |
| HTN | monocyte | *TMEM144* | -2.728527161 | 0.007164894 | 0.024238452 | 1 |
| HTN | monocyte | *ITGA2* | -1.216050629 | 0.005443969 | 0.023595115 | 1 |
| HTN | monocyte | *MOCS2* | -1.90410205 | 0.007172243 | 0.024238452 | 1 |
| HTN | monocyte | *RNF180* | -1.662994259 | 0.011046931 | 0.028009441 | 1 |
| HTN | monocyte | *CCDC112* | 2.784359467 | 0.002331124 | 0.023595115 | 1 |
| HTN | monocyte | *CMBL* | -1.439939239 | 0.011596448 | 0.028561404 | 1 |
| HTN | monocyte | *C5orf63* | -1.183825873 | 0.00480126 | 0.023595115 | 1 |
| HTN | monocyte | *WDR41* | -1.669971145 | 0.016714656 | 0.034504301 | 1 |
| HTN | monocyte | *CASP3* | -1.948423375 | 0.007191752 | 0.024282798 | 1 |
| HTN | monocyte | *PRIMPOL* | -1.370270052 | 0.006526356 | 0.023864166 | 1 |
| HTN | monocyte | *ERAP1* | -2.444253725 | 0.000218009 | 0.01101852 | 1 |
| HTN | monocyte | *UTP15* | 2.121772368 | 0.003559398 | 0.023595115 | 1 |
| HTN | monocyte | *TLR3* | -1.931841973 | 0.005916666 | 0.023595115 | 1 |
| HTN | monocyte | *KLKB1* | -1.407768189 | 0.003390134 | 0.023595115 | 1 |
| HTN | monocyte | *CCDC127* | -1.763899482 | 0.001500797 | 0.023595115 | 1 |

| | | | | | | |
|---|---|---|---|---|---|---|
| HTN | monocyte | *RAET1E* | -1.343557421 | 0.005116071 | 0.023595115 | 1 |
| HTN | monocyte | *RPS14* | -1.954735976 | 0.022011676 | 0.041379563 | 1 |
| HTN | monocyte | *RP9* | -2.940513221 | 0.01832969 | 0.036419187 | 1 |
| HTN | monocyte | *CAMLG* | -2.721759016 | 0.005630482 | 0.023595115 | 1 |
| HTN | monocyte | *RELL2* | 1.433628185 | 0.00624201 | 0.023683186 | 1 |
| HTN | monocyte | *MIOS* | 3.376027103 | 0.025768911 | 0.046017945 | 1 |
| HTN | monocyte | *USP49* | -1.886148505 | 0.011559682 | 0.028514274 | 1 |
| HTN | monocyte | *CHMP4C* | -3.88030267 | 0.005404643 | 0.023595115 | 1 |
| HTN | monocyte | *CTSB* | -1.578435443 | 0.002402235 | 0.023595115 | 1 |
| HTN | monocyte | *TNFRSF11B* | -1.971341032 | 0.006501071 | 0.023864166 | 1 |
| HTN | monocyte | *DEFB1* | -7.408314973 | 0.00113135 | 0.023030817 | 1 |
| HTN | monocyte | *SUN1* | -1.567480855 | 0.011259648 | 0.028158713 | 1 |
| HTN | monocyte | *FASTK* | -1.846853299 | 0.00750862 | 0.024638032 | 1 |
| HTN | monocyte | *ALDH7A1* | -1.362880321 | 0.027476121 | 0.048023826 | 1 |
| HTN | monocyte | *YWHAZ* | -2.173653128 | 0.027266876 | 0.047742177 | 1 |
| HTN | monocyte | *SLC25A32* | -2.182871849 | 0.006397869 | 0.023797215 | 1 |
| HTN | monocyte | *INTS8* | -2.034483979 | 0.010994425 | 0.027958569 | 1 |
| HTN | monocyte | *RPP25L* | -4.329410804 | 5.27E-05 | 0.004735705 | 0.255728047 |
| HTN | monocyte | *C9orf24* | -1.231313042 | 0.00657989 | 0.023914328 | 1 |
| HTN | monocyte | *NUDT2* | -2.228317634 | 0.001834977 | 0.023595115 | 1 |
| HTN | monocyte | *TMEM65* | -1.76944201 | 0.013874694 | 0.031408232 | 1 |
| HTN | monocyte | *CCDC171* | -1.727545135 | 0.022430317 | 0.04183827 | 1 |
| HTN | monocyte | *ZMAT4* | -2.396857598 | 0.005589136 | 0.023595115 | 1 |
| HTN | monocyte | *C8orf34* | -0.773059662 | 0.006629341 | 0.02395053 | 1 |
| HTN | monocyte | *RASEF* | -1.457183879 | 0.013879565 | 0.031408232 | 1 |
| HTN | monocyte | *ANKS6* | -1.432760762 | 0.010663078 | 0.027593203 | 1 |
| HTN | monocyte | *AQP3* | -3.941173917 | 0.005059619 | 0.023595115 | 1 |
| HTN | monocyte | *TRMT10B* | -1.559240965 | 0.011791194 | 0.028738324 | 1 |
| HTN | monocyte | *PIGO* | 3.325759415 | 0.001452097 | 0.023595115 | 1 |
| HTN | monocyte | *MARCH8* | -2.136766339 | 0.015078283 | 0.032773807 | 1 |
| HTN | monocyte | *TSHR* | -0.885944909 | 0.005269523 | 0.023595115 | 1 |
| HTN | monocyte | *CFL2* | -2.382903578 | 0.010127475 | 0.027038216 | 1 |
| HTN | monocyte | *PGM2L1* | -1.507956767 | 0.002876293 | 0.023595115 | 1 |
| HTN | monocyte | *SLC16A9* | -1.696248399 | 0.005087949 | 0.023595115 | 1 |
| HTN | monocyte | *GJB2* | -7.363142704 | 0.005290914 | 0.023595115 | 1 |
| HTN | monocyte | *REEP3* | -2.423576773 | 0.004295407 | 0.023595115 | 1 |
| HTN | monocyte | *LRR1* | -2.597606209 | 0.007542098 | 0.024642599 | 1 |
| HTN | monocyte | *NOXRED1* | -1.531761304 | 0.005799275 | 0.023595115 | 1 |
| HTN | monocyte | *AKR1E2* | -1.54440455 | 0.005697743 | 0.023595115 | 1 |
| HTN | monocyte | *OXGR1* | -9.848990069 | 0.00303486 | 0.023595115 | 1 |
| HTN | monocyte | *VDAC2* | -3.044581026 | 0.007122004 | 0.024238452 | 1 |
| HTN | monocyte | *QSOX2* | -2.418847975 | 0.022348005 | 0.041801279 | 1 |
| HTN | monocyte | *CLEC1B* | 0.743609963 | 0.002294743 | 0.023595115 | 1 |
| HTN | monocyte | *RET* | -2.086196451 | 0.005148846 | 0.023595115 | 1 |
| HTN | monocyte | *CASP7* | -3.30576016 | 0.000568273 | 0.017385729 | 1 |
| HTN | monocyte | *PPP1R36* | -1.741803405 | 0.006278445 | 0.023704653 | 1 |
| HTN | monocyte | *VWA2* | -3.418754671 | 0.006626056 | 0.02395053 | 1 |
| HTN | monocyte | *TRUB1* | -2.857731152 | 0.002865018 | 0.023595115 | 1 |
| HTN | monocyte | *OTOGL* | 0.648891953 | 0.005559454 | 0.023595115 | 1 |
| HTN | monocyte | *PACSIN3* | -2.645267622 | 0.000479095 | 0.016255736 | 1 |

| | | | | | | |
|---|---|---|---|---|---|---|
| HTN | monocyte | *TTC7B* | -1.76288259 | 0.014106012 | 0.031686282 | 1 |
| HTN | monocyte | *SLC39A13* | -1.579333847 | 0.021406025 | 0.040616961 | 1 |
| HTN | monocyte | *PSMC3* | -2.343679803 | 0.007083896 | 0.024238452 | 1 |
| HTN | monocyte | *RAPSN* | 2.409558278 | 0.009637779 | 0.02668844 | 1 |
| HTN | monocyte | *IFI27L1* | -2.100545081 | 0.024441604 | 0.044332958 | 1 |
| HTN | monocyte | *C1QL3* | -1.445127114 | 0.014496787 | 0.03215962 | 1 |
| HTN | monocyte | *ARL5B* | -2.604039284 | 0.000514889 | 0.016722594 | 1 |
| HTN | monocyte | *LIPC* | -1.666459303 | 0.014523545 | 0.032177279 | 1 |
| HTN | monocyte | *CEP57* | -1.911750266 | 0.003122834 | 0.023595115 | 1 |
| HTN | monocyte | *JAM3* | -1.507937222 | 0.003333941 | 0.023595115 | 1 |
| HTN | monocyte | *ZFYVE19* | -2.555336917 | 0.003163924 | 0.023595115 | 1 |
| HTN | monocyte | *TRMT61A* | 2.40741589 | 0.004626658 | 0.023595115 | 1 |
| HTN | monocyte | *LARP6* | -1.630212777 | 0.009729067 | 0.02671918 | 1 |
| HTN | monocyte | *API5* | -1.55889359 | 0.008702091 | 0.025839644 | 1 |
| HTN | monocyte | *ASPG* | -7.660085154 | 0.006543211 | 0.023864166 | 1 |
| HTN | monocyte | *COX11* | -1.487020004 | 0.005893558 | 0.023595115 | 1 |
| HTN | monocyte | *ZNF202* | -1.533342936 | 0.005163027 | 0.023595115 | 1 |
| HTN | monocyte | *STXBP4* | -2.171882346 | 0.023985647 | 0.043824751 | 1 |
| HTN | monocyte | *APBB1* | -1.487853453 | 0.006962235 | 0.024187912 | 1 |
| HTN | monocyte | *NUDT13* | -3.008180222 | 0.006917574 | 0.024148823 | 1 |
| HTN | monocyte | *WDR88* | 2.731503201 | 0.004038434 | 0.023595115 | 1 |
| HTN | monocyte | *PLD4* | 0.455367567 | 0.004104041 | 0.023595115 | 1 |
| HTN | monocyte | *XRRA1* | -2.289394381 | 0.002347825 | 0.023595115 | 1 |
| HTN | monocyte | *TRIM66* | -2.553013093 | 6.58E-06 | 0.000958035 | 0.03191837 |
| HTN | monocyte | *CENPN* | 1.431017382 | 0.00316135 | 0.023595115 | 1 |
| HTN | monocyte | *C16orf46* | 7.257738621 | 0.015016571 | 0.032711642 | 1 |
| HTN | monocyte | *TMEM41B* | -2.286710049 | 0.024944738 | 0.044926454 | 1 |
| HTN | monocyte | *PRKCB* | 0.344967329 | 0.003151221 | 0.023595115 | 1 |
| HTN | monocyte | *NDST2* | -11.80109075 | 0.005310256 | 0.023595115 | 1 |
| HTN | monocyte | *CLEC4D* | 0.361498831 | 0.003514812 | 0.023595115 | 1 |
| HTN | monocyte | *ZSCAN21* | -2.0122402 | 0.001553797 | 0.023595115 | 1 |
| HTN | monocyte | *RIMKLB* | -4.425321533 | 2.80E-06 | 0.000502994 | 0.013580838 |
| HTN | monocyte | *TMED3* | -1.623122309 | 0.003306889 | 0.023595115 | 1 |
| HTN | monocyte | *SEC11C* | 2.740746364 | 0.003499804 | 0.023595115 | 1 |
| HTN | monocyte | *GALR1* | -2.968277114 | 0.003771636 | 0.023595115 | 1 |
| HTN | monocyte | *CHRFAM7A* | 2.48107103 | 0.006112603 | 0.023595115 | 1 |
| HTN | monocyte | *ACSM1* | -1.561970561 | 0.013665264 | 0.031084791 | 1 |
| HTN | monocyte | *SLFN5* | -1.616646871 | 0.011026288 | 0.027980937 | 1 |
| HTN | monocyte | *CATSPER2* | -2.769624415 | 0.026243044 | 0.046482276 | 1 |
| HTN | monocyte | *SAAL1* | -2.304884242 | 0.008658869 | 0.025839644 | 1 |
| HTN | monocyte | *FAM96A* | 3.63219238 | 0.008712805 | 0.025839644 | 1 |
| HTN | monocyte | *ANPEP* | 1.360496513 | 0.001073033 | 0.022636322 | 1 |
| HTN | monocyte | *STX3* | -2.780224686 | 0.016180914 | 0.033986925 | 1 |
| HTN | monocyte | *PIP4K2C* | -2.352698436 | 0.012036414 | 0.028978626 | 1 |
| HTN | monocyte | *MS4A7* | 0.662560479 | 0.019888247 | 0.038552846 | 1 |
| HTN | monocyte | *DIS3L* | -1.166865463 | 0.004849563 | 0.023595115 | 1 |
| HTN | monocyte | *EPB42* | -1.93149581 | 0.022297035 | 0.041754231 | 1 |
| HTN | monocyte | *SMAD3* | -2.001050115 | 0.024113313 | 0.044000674 | 1 |
| HTN | monocyte | *RCCD1* | -1.933527265 | 0.003827201 | 0.023595115 | 1 |
| HTN | monocyte | *SAMD14* | 2.211418522 | 0.003042064 | 0.023595115 | 1 |

| | | | | | | |
|---|---|---|---|---|---|---|
| HTN | monocyte | *URM1* | -2.096639641 | 0.005082688 | 0.023595115 | 1 |
| HTN | monocyte | *ISLR2* | 1.598966729 | 0.028196644 | 0.048791054 | 1 |
| HTN | monocyte | *SP2* | -2.137071276 | 0.010055758 | 0.027007023 | 1 |
| HTN | monocyte | *COQ7* | -2.155806258 | 0.010374574 | 0.027326031 | 1 |
| HTN | monocyte | *GPRC5B* | -2.813808865 | 0.008573695 | 0.025783919 | 1 |
| HTN | monocyte | *TBC1D2B* | -1.223886454 | 0.004784215 | 0.023595115 | 1 |
| HTN | monocyte | *LOXHD1* | -0.904027689 | 0.005821167 | 0.023595115 | 1 |
| HTN | monocyte | *KATNAL2* | -0.912280631 | 0.011798586 | 0.028738324 | 1 |
| HTN | monocyte | *ZNF91* | -1.678862702 | 0.008393879 | 0.025487024 | 1 |
| HTN | monocyte | *POP5* | 3.229679233 | 0.011201801 | 0.028135587 | 1 |
| HTN | monocyte | *MYO5B* | -0.96888315 | 0.004555815 | 0.023595115 | 1 |
| HTN | monocyte | *ART5* | 1.872519898 | 0.007808899 | 0.025020002 | 1 |
| HTN | monocyte | *RRM1* | -1.41900454 | 0.010942138 | 0.027890132 | 1 |
| HTN | monocyte | *ZNF23* | -4.071806197 | 0.000450006 | 0.015752203 | 1 |
| HTN | monocyte | *VKORC1* | 2.45860438 | 0.003307667 | 0.023595115 | 1 |
| HTN | monocyte | *SMG8* | -3.647667223 | 0.002174452 | 0.023595115 | 1 |
| HTN | monocyte | *RAB8A* | 1.713012495 | 0.001708841 | 0.023595115 | 1 |
| HTN | monocyte | *JSRP1* | 1.872494416 | 0.002185894 | 0.023595115 | 1 |
| HTN | monocyte | *SPATA33* | -1.876459102 | 0.005733605 | 0.023595115 | 1 |
| HTN | monocyte | *PROCA1* | 1.856689259 | 0.007917592 | 0.025073967 | 1 |
| HTN | monocyte | *TP53I13* | -1.061033389 | 0.006161082 | 0.023595115 | 1 |
| HTN | monocyte | *ZNF528* | -2.419119775 | 0.014633011 | 0.032314157 | 1 |
| HTN | monocyte | *SERTAD3* | -2.281090036 | 0.004818019 | 0.023595115 | 1 |
| HTN | monocyte | *TMC4* | 2.567845195 | 0.00469407 | 0.023595115 | 1 |
| HTN | monocyte | *TTYH1* | -1.667357575 | 0.00510633 | 0.023595115 | 1 |
| HTN | monocyte | *LENG8* | -1.622173287 | 0.016519347 | 0.034399945 | 1 |
| HTN | monocyte | *KIR3DL1* | 7.310658272 | 0.003484121 | 0.023595115 | 1 |
| HTN | monocyte | *DNAAF3* | -1.705297538 | 0.02666895 | 0.047036622 | 1 |
| HTN | monocyte | *CHAF1A* | -1.88828698 | 0.012629977 | 0.029661495 | 1 |
| HTN | monocyte | *FAM57A* | -2.03213581 | 0.008104302 | 0.025239173 | 1 |
| HTN | monocyte | *KLK5* | -2.230515772 | 0.002573879 | 0.023595115 | 1 |
| HTN | monocyte | *KLK6* | 1.969782339 | 0.00347403 | 0.023595115 | 1 |
| HTN | monocyte | *ZNF83* | -1.589410125 | 0.024381526 | 0.044273639 | 1 |
| HTN | monocyte | *SPRYD3* | -1.627038368 | 0.003226534 | 0.023595115 | 1 |
| HTN | monocyte | *ZNF558* | -1.095125906 | 0.009684644 | 0.026713982 | 1 |
| HTN | monocyte | *NDUFV1* | -1.424887517 | 0.007724876 | 0.02497075 | 1 |
| HTN | monocyte | *PRDX2* | -2.409527401 | 0.021165578 | 0.0402727 | 1 |
| HTN | monocyte | *ZNF232* | -1.112258527 | 0.027530578 | 0.048067063 | 1 |
| HTN | monocyte | *MIS12* | -1.476715663 | 0.00634224 | 0.02374425 | 1 |
| HTN | monocyte | *MGAT5B* | -3.57331513 | 0.004447582 | 0.023595115 | 1 |
| HTN | monocyte | *GSDMA* | -2.16214135 | 0.005373609 | 0.023595115 | 1 |
| HTN | monocyte | *TMEM99* | -1.295491904 | 0.02441276 | 0.044313772 | 1 |
| HTN | monocyte | *MLST8* | 1.12207919 | 0.003227231 | 0.023595115 | 1 |
| HTN | monocyte | *BSCL2* | 1.003855892 | 0.004278093 | 0.023595115 | 1 |
| HTN | monocyte | *ATG16L2* | 3.624726397 | 0.022110822 | 0.041485579 | 1 |
| HTN | monocyte | *LTBP3* | -1.914182914 | 0.010593668 | 0.027545809 | 1 |
| HTN | monocyte | *SCARA5* | -0.988425706 | 0.004292535 | 0.023595115 | 1 |
| HTN | monocyte | *KIAA1586* | -1.599512519 | 0.003371611 | 0.023595115 | 1 |
| HTN | monocyte | *FAM83B* | -13.08509978 | 0.005779205 | 0.023595115 | 1 |
| HTN | monocyte | *POLR2J3* | -1.078383506 | 0.008158072 | 0.025308801 | 1 |

| | | | | | | |
|---|---|---|---|---|---|---|
| HTN | monocyte | *COA6* | 3.199289487 | 0.003561244 | 0.023595115 | 1 |
| HTN | monocyte | *PXK* | -1.8761394 | 0.007737456 | 0.024994763 | 1 |
| HTN | monocyte | *PCMTD1* | -1.67781724 | 0.02266578 | 0.042094201 | 1 |
| HTN | monocyte | *ACOX2* | -2.56477333 | 0.005917579 | 0.023595115 | 1 |
| HTN | monocyte | *FAM107A* | -1.850389408 | 0.008006368 | 0.025094896 | 1 |
| HTN | monocyte | *SCN11A* | -1.086344977 | 0.025914168 | 0.046203869 | 1 |
| HTN | monocyte | *DTYMK* | -1.980807993 | 0.01070543 | 0.027617119 | 1 |
| HTN | monocyte | *ING5* | -1.778390804 | 0.019638896 | 0.038329817 | 1 |
| HTN | monocyte | *RFWD3* | -1.917554567 | 0.01603563 | 0.03378414 | 1 |
| HTN | monocyte | *KLHL30* | -10.57565491 | 0.007066073 | 0.024238452 | 1 |
| HTN | monocyte | *CCDC110* | -2.541772021 | 0.005032911 | 0.023595115 | 1 |
| HTN | monocyte | *SLC20A2* | -1.865885697 | 0.010853217 | 0.027878663 | 1 |
| HTN | monocyte | *TMUB2* | 2.623553354 | 0.017458494 | 0.03534561 | 1 |
| HTN | monocyte | *NBPF9* | -1.912276659 | 0.005647218 | 0.023595115 | 1 |
| HTN | monocyte | *NDUFS5* | -2.245037725 | 0.015990505 | 0.033743802 | 1 |
| HTN | monocyte | *ZNF30* | -1.176663596 | 0.021155378 | 0.040269084 | 1 |
| HTN | monocyte | *GSTM4* | -2.067587842 | 0.002923465 | 0.023595115 | 1 |
| HTN | monocyte | *TCTN2* | -1.225158588 | 0.000375898 | 0.01501035 | 1 |
| HTN | monocyte | *PPIP5K1* | -1.804743402 | 0.006106754 | 0.023595115 | 1 |
| HTN | monocyte | *ZNF507* | -2.288016437 | 0.015995619 | 0.033743802 | 1 |
| HTN | monocyte | *ATOH8* | 3.341165594 | 0.002040126 | 0.023595115 | 1 |
| HTN | monocyte | *MFF* | -1.79258351 | 0.017679039 | 0.035622383 | 1 |
| HTN | monocyte | *LGALS9* | 1.208828228 | 0.000644354 | 0.018390619 | 1 |
| HTN | monocyte | *CPLX1* | -1.041192785 | 0.023617129 | 0.043274287 | 1 |
| HTN | monocyte | *SIGLEC7* | 0.956837547 | 0.002661356 | 0.023595115 | 1 |
| HTN | monocyte | *MFSD7* | 2.483063272 | 0.003191071 | 0.023595115 | 1 |
| HTN | monocyte | *KLK7* | -2.019768284 | 0.005019586 | 0.023595115 | 1 |
| HTN | monocyte | *IRS1* | -0.857588579 | 0.000855078 | 0.020511604 | 1 |
| HTN | monocyte | *PCSK9* | 1.740891184 | 0.003292234 | 0.023595115 | 1 |
| HTN | monocyte | *XPO6* | -2.333152586 | 0.014250351 | 0.031892391 | 1 |
| HTN | monocyte | *NPIPB3* | -2.266405401 | 0.025699189 | 0.045978047 | 1 |
| HTN | monocyte | *SH3TC2* | -1.466308905 | 0.003216324 | 0.023595115 | 1 |
| HTN | monocyte | *PGM2* | -1.679251996 | 0.016777214 | 0.034563946 | 1 |
| HTN | monocyte | *SNUPN* | -2.132592798 | 0.002356051 | 0.023595115 | 1 |
| HTN | monocyte | *RNASE6* | 2.058744885 | 0.003977005 | 0.023595115 | 1 |
| HTN | monocyte | *CD52* | 0.468838191 | 0.00800015 | 0.025091616 | 1 |
| HTN | monocyte | *SLC38A11* | -1.950435456 | 0.01093071 | 0.027890132 | 1 |
| HTN | monocyte | *CCDC8* | -1.701726767 | 0.015795983 | 0.033645753 | 1 |
| HTN | monocyte | *HINT1* | -1.759641184 | 0.008796666 | 0.02589483 | 1 |
| HTN | monocyte | *DTWD2* | -2.275041448 | 0.01799108 | 0.035967335 | 1 |
| HTN | monocyte | *CLIC3* | -1.713755427 | 0.005281618 | 0.023595115 | 1 |
| HTN | monocyte | *INO80E* | -4.470617392 | 1.99E-08 | 1.21E-05 | 9.65E-05 |
| HTN | monocyte | *DFFB* | -4.481267195 | 0.025988065 | 0.046241902 | 1 |
| HTN | monocyte | *NFU1* | -1.051664335 | 0.020653815 | 0.039562697 | 1 |
| HTN | monocyte | *LUZP1* | -2.086143033 | 0.026156509 | 0.046416345 | 1 |
| HTN | monocyte | *LRRC45* | -2.05355981 | 0.00247872 | 0.023595115 | 1 |
| HTN | monocyte | *CHRNA5* | 1.652732609 | 0.007957797 | 0.025076015 | 1 |
| HTN | monocyte | *UGP2* | -1.159406602 | 0.006665591 | 0.023986397 | 1 |
| HTN | monocyte | *CSGALNACT2* | 3.092136654 | 0.007721691 | 0.02497075 | 1 |
| HTN | monocyte | *AVEN* | -2.087526059 | 0.014681016 | 0.032363602 | 1 |

| | | | | | | |
|---|---|---|---|---|---|---|
| HTN | monocyte | *CTNND2* | -1.390769958 | 0.001866857 | 0.023595115 | 1 |
| HTN | monocyte | *CALML6* | -22.851404 | 0.013692633 | 0.031132453 | 1 |
| HTN | monocyte | *TOR1AIP2* | -1.9934766 | 0.026824848 | 0.047208619 | 1 |
| HTN | monocyte | *ZFPM2* | -1.050830404 | 0.006258415 | 0.023704653 | 1 |
| HTN | monocyte | *MYO7B* | 5.611461428 | 8.19E-07 | 0.000220692 | 0.003972459 |
| HTN | monocyte | *NIPA1* | -1.845410099 | 0.005411144 | 0.023595115 | 1 |
| HTN | monocyte | *RNF150* | -0.886218967 | 0.004396537 | 0.023595115 | 1 |
| HTN | monocyte | *USP38* | -1.899726289 | 0.007521684 | 0.024638032 | 1 |
| HTN | monocyte | *ANKK1* | -4.444403884 | 0.00302782 | 0.023595115 | 1 |
| HTN | monocyte | *CNGB3* | -1.162955308 | 0.028191049 | 0.048791054 | 1 |
| HTN | monocyte | *ELP5* | -1.917117832 | 0.002042431 | 0.023595115 | 1 |
| HTN | monocyte | *ADORA2B* | 1.686922068 | 0.000361444 | 0.01501035 | 1 |
| HTN | monocyte | *MGMT* | -1.358444449 | 0.010787346 | 0.027766687 | 1 |
| HTN | monocyte | *NFXL1* | -1.836303382 | 0.017383033 | 0.035260232 | 1 |
| HTN | monocyte | *DNAJC18* | -4.168141721 | 2.99E-05 | 0.003023993 | 0.145151651 |
| HTN | monocyte | *SPATA24* | -2.691817742 | 0.003152913 | 0.023595115 | 1 |
| HTN | monocyte | *HSD17B13* | -3.063282733 | 0.00471059 | 0.023595115 | 1 |
| HTN | monocyte | *ELOVL6* | -1.3511722 | 0.007169904 | 0.024238452 | 1 |
| HTN | monocyte | *ARL6IP1* | -2.798353879 | 0.000984271 | 0.021612081 | 1 |
| HTN | monocyte | *SMAGP* | -1.909074574 | 0.001669854 | 0.023595115 | 1 |
| HTN | monocyte | *CDH2* | -2.157446204 | 0.004831582 | 0.023595115 | 1 |
| HTN | monocyte | *EMB* | -2.668882884 | 0.003711586 | 0.023595115 | 1 |
| HTN | monocyte | *SLC26A5* | -1.892302955 | 0.012042747 | 0.028978626 | 1 |
| HTN | monocyte | *GTSF1* | -5.524209444 | 0.001932584 | 0.023595115 | 1 |
| HTN | monocyte | *ACYP2* | -1.391139074 | 0.015812619 | 0.033665128 | 1 |
| HTN | monocyte | *TMEM133* | -3.048989643 | 0.018307596 | 0.036393934 | 1 |
| HTN | monocyte | *HOXB9* | -5.77097866 | 0.005615124 | 0.023595115 | 1 |
| HTN | monocyte | *TTLL6* | -17.22360657 | 7.20E-11 | 1.16E-07 | 3.49E-07 |
| HTN | monocyte | *TRIAP1* | 1.932254976 | 0.003548263 | 0.023595115 | 1 |
| HTN | monocyte | *TSEN34* | -1.522276314 | 0.005319398 | 0.023595115 | 1 |
| HTN | monocyte | *MSANTD4* | -2.079599213 | 0.009487982 | 0.026576944 | 1 |
| HTN | monocyte | *PAQR8* | -2.779352994 | 7.98E-05 | 0.006048139 | 0.387080892 |
| HTN | monocyte | *ZNF160* | -1.806487671 | 0.006394063 | 0.023797215 | 1 |
| HTN | monocyte | *ZNF415* | -2.675002223 | 0.025552069 | 0.045780981 | 1 |
| HTN | monocyte | *S1PR1* | -1.305337655 | 0.006324219 | 0.02374425 | 1 |
| HTN | monocyte | *PYGO1* | -1.411516836 | 0.009342211 | 0.026350386 | 1 |
| HTN | monocyte | *FPR1* | 1.913648553 | 0.005250768 | 0.023595115 | 1 |
| HTN | monocyte | *FEZ2* | -1.229533416 | 0.01672591 | 0.034504301 | 1 |
| HTN | monocyte | *SOX7* | -2.110129114 | 0.014315477 | 0.031973856 | 1 |
| HTN | monocyte | *TRMT61B* | -1.781145471 | 0.008755737 | 0.025848597 | 1 |
| HTN | monocyte | *ATP6V0E2* | -2.296065249 | 0.005805473 | 0.023595115 | 1 |
| HTN | monocyte | *MORN4* | -1.126580565 | 0.002425621 | 0.023595115 | 1 |
| HTN | monocyte | *SCAND1* | -2.761716265 | 0.00238708 | 0.023595115 | 1 |
| HTN | monocyte | *PDE7B* | -1.667460848 | 0.016333241 | 0.034184275 | 1 |
| HTN | monocyte | *MCC* | -1.469947251 | 0.011953033 | 0.028925743 | 1 |
| HTN | monocyte | *HOPX* | 1.652268579 | 0.023110123 | 0.042570355 | 1 |
| HTN | monocyte | *PPID* | -1.747838117 | 0.01307215 | 0.030344279 | 1 |
| HTN | monocyte | *NMUR1* | 5.198905022 | 0.002928927 | 0.023595115 | 1 |
| HTN | monocyte | *PIK3CD* | 1.96391352 | 0.002601064 | 0.023595115 | 1 |
| HTN | monocyte | *ANO5* | -1.096014894 | 0.006820706 | 0.02405092 | 1 |

| | | | | | | |
|---|---|---|---|---|---|---|
| HTN | monocyte | *VAT1L* | -1.51348076 | 0.00473946 | 0.023595115 | 1 |
| HTN | monocyte | *PAH* | -7.484758124 | 0.005044376 | 0.023595115 | 1 |
| HTN | monocyte | *PWWP2B* | 2.664656004 | 0.004785839 | 0.023595115 | 1 |
| HTN | monocyte | *EXOSC10* | -1.521773602 | 0.007682551 | 0.024938402 | 1 |
| HTN | monocyte | *FBXW10* | -3.318590426 | 0.005043234 | 0.023595115 | 1 |
| HTN | monocyte | *ATPAF2* | 4.685327615 | 0.001169991 | 0.02330522 | 1 |
| HTN | monocyte | *ANKRD20A4* | 0.89381939 | 0.003450156 | 0.023595115 | 1 |
| HTN | monocyte | *SERF1A* | 0.848229129 | 0.003685937 | 0.023595115 | 1 |
| HTN | monocyte | *NME6* | 3.426761625 | 1.18E-06 | 0.000272223 | 0.005716678 |
| HTN | monocyte | *CD8B* | 0.945223015 | 0.005136308 | 0.023595115 | 1 |
| HTN | monocyte | *TEFM* | -1.196067568 | 0.01633828 | 0.034184275 | 1 |
| HTN | monocyte | *MALT1* | -2.072019338 | 0.023120315 | 0.042571424 | 1 |
| HTN | monocyte | *MBOAT1* | 2.759550126 | 0.009987198 | 0.026892975 | 1 |
| HTN | monocyte | *CXCR6* | 1.97633299 | 0.003193099 | 0.023595115 | 1 |
| HTN | monocyte | *PAIP1* | -1.739146779 | 0.008260935 | 0.025373581 | 1 |
| HTN | monocyte | *MACROD2* | -0.898105822 | 0.022560305 | 0.041986971 | 1 |
| HTN | monocyte | *HINFP* | -2.194968193 | 0.018792109 | 0.03709492 | 1 |
| HTN | monocyte | *TP53RK* | -1.761900929 | 0.008273354 | 0.025373581 | 1 |
| HTN | monocyte | *CLEC12A* | 0.21762904 | 0.001901167 | 0.023595115 | 1 |
| HTN | monocyte | *SUCLG2* | -1.686980992 | 0.024225043 | 0.044121588 | 1 |
| HTN | monocyte | *ARNT2* | -0.896139272 | 0.018668717 | 0.036926464 | 1 |
| HTN | monocyte | *GTPBP2* | -2.767619081 | 0.00413823 | 0.023595115 | 1 |
| HTN | monocyte | *AFF1* | -1.225956545 | 0.011575331 | 0.028538368 | 1 |
| HTN | monocyte | *FIBP* | 2.294276139 | 0.000819089 | 0.020485662 | 1 |
| HTN | monocyte | *CTSW* | -0.514831906 | 0.020876796 | 0.039867415 | 1 |
| HTN | monocyte | *SNTG2* | -0.823566153 | 0.006097009 | 0.023595115 | 1 |
| HTN | monocyte | *SMPDL3A* | -1.175831961 | 0.01126141 | 0.028158713 | 1 |
| HTN | monocyte | *EFEMP2* | -1.67743915 | 0.027717988 | 0.048332949 | 1 |
| HTN | monocyte | *ZFAND4* | -1.747984663 | 0.001777847 | 0.023595115 | 1 |
| HTN | monocyte | *SLFN11* | -1.342321651 | 0.004252202 | 0.023595115 | 1 |
| HTN | monocyte | *FAM71D* | -1.942307628 | 0.004042096 | 0.023595115 | 1 |
| HTN | monocyte | *ZNF596* | -2.205728016 | 0.010085696 | 0.027007023 | 1 |
| HTN | monocyte | *CFL1* | 2.510353266 | 0.010655597 | 0.027588558 | 1 |
| HTN | monocyte | *RAB43* | 9.227832288 | 0.004673274 | 0.023595115 | 1 |
| HTN | monocyte | *CBWD1* | -2.745729914 | 0.013343496 | 0.03065466 | 1 |
| HTN | monocyte | *SNX32* | 1.228659303 | 0.000688128 | 0.018970451 | 1 |
| HTN | monocyte | *ZNF621* | -1.754575466 | 0.010448879 | 0.027419124 | 1 |
| HTN | monocyte | *RNASEH2C* | -2.843739528 | 0.004414398 | 0.023595115 | 1 |
| HTN | monocyte | *MYEOV* | -12.49703969 | 0.00088551 | 0.020511604 | 1 |
| HTN | monocyte | *MRGPRF* | -1.350982004 | 0.00224486 | 0.023595115 | 1 |
| HTN | monocyte | *LCLAT1* | -1.960431693 | 0.028919778 | 0.049698576 | 1 |
| HTN | monocyte | *SH3RF3* | -1.921004368 | 0.010879173 | 0.027884704 | 1 |
| HTN | monocyte | *GXYLT2* | -0.851806241 | 0.006691345 | 0.024005639 | 1 |
| HTN | monocyte | *DCAKD* | -2.59696344 | 0.002444636 | 0.023595115 | 1 |
| HTN | monocyte | *TRMT112* | -2.107881489 | 0.024787459 | 0.044714326 | 1 |
| HTN | monocyte | *NOC3L* | -3.821745418 | 0.001180439 | 0.02330522 | 1 |
| HTN | monocyte | *RHOD* | -1.639516783 | 0.013381263 | 0.030691157 | 1 |
| HTN | monocyte | *PARP15* | 2.124958093 | 0.001296927 | 0.023595115 | 1 |
| HTN | monocyte | *VANGL1* | -1.554521671 | 0.005300842 | 0.023595115 | 1 |
| HTN | monocyte | *ZNF483* | -1.335830934 | 0.007682548 | 0.024938402 | 1 |

| | | | | | | |
|---|---|---|---|---|---|---|
| HTN | monocyte | *MMRN2* | -1.125913863 | 0.007759851 | 0.025006477 | 1 |
| HTN | monocyte | *MAP3K11* | 5.768408003 | 0.000804623 | 0.020419313 | 1 |
| HTN | monocyte | *KCNK7* | -15.26691137 | 0.014055165 | 0.03166001 | 1 |
| HTN | monocyte | *NDNF* | -2.314229427 | 0.009197056 | 0.026254852 | 1 |
| HTN | monocyte | *ARV1* | -1.887623618 | 0.006842138 | 0.024108972 | 1 |
| HTN | monocyte | *CCDC36* | -1.35682155 | 0.000812227 | 0.020419313 | 1 |
| HTN | monocyte | *PPP1R14B* | -1.457553871 | 0.017435987 | 0.0353381 | 1 |
| HTN | monocyte | *PEAK1* | -1.65379022 | 0.017444474 | 0.035340537 | 1 |
| HTN | monocyte | *TNFRSF10D* | 1.630430684 | 0.002282175 | 0.023595115 | 1 |
| HTN | monocyte | *MST1* | -3.187467747 | 3.54E-07 | 0.000107291 | 0.001716663 |
| HTN | monocyte | *GMPPB* | 1.114826968 | 0.008290147 | 0.025373581 | 1 |
| HTN | monocyte | *SULT1B1* | 2.588951386 | 0.007310689 | 0.024429382 | 1 |
| HTN | monocyte | *NUDT4* | -1.973855512 | 0.009255089 | 0.026254852 | 1 |
| HTN | monocyte | *SCAI* | -1.567149853 | 0.016476999 | 0.034355995 | 1 |
| HTN | monocyte | *UQCRH* | -1.948641408 | 0.01663349 | 0.034456332 | 1 |
| HTN | monocyte | *C11orf80* | -1.580089812 | 0.000235658 | 0.011549646 | 1 |
| HTN | monocyte | *CD7* | -2.090669947 | 0.007083061 | 0.024238452 | 1 |
| HTN | monocyte | *TUBB8* | 0.566902705 | 0.002705804 | 0.023595115 | 1 |
| HTN | monocyte | *GOLIM4* | -2.189068239 | 0.024212268 | 0.044114879 | 1 |
| HTN | monocyte | *XXYLT1* | 3.05689734 | 0.010643153 | 0.027588558 | 1 |
| HTN | monocyte | *UBXN2A* | -1.287821772 | 0.011895537 | 0.02887301 | 1 |
| HTN | monocyte | *SLC25A30* | 2.231152188 | 0.0034405 | 0.023595115 | 1 |
| HTN | monocyte | *CTU2* | -4.513675829 | 0.000213037 | 0.010880596 | 1 |
| HTN | monocyte | *GLIS1* | -1.188157087 | 0.005037678 | 0.023595115 | 1 |
| HTN | monocyte | *RALGAPA1* | -1.510037186 | 0.004539564 | 0.023595115 | 1 |
| HTN | monocyte | *SNAPC5* | -1.509528966 | 0.019371908 | 0.037946103 | 1 |
| HTN | monocyte | *ANKRD36C* | -1.912940568 | 0.017780934 | 0.035760857 | 1 |
| HTN | monocyte | *ZNF497* | -7.985192429 | 0.01806654 | 0.036029122 | 1 |
| HTN | monocyte | *SH3PXD2B* | -1.552492437 | 0.012744058 | 0.029857157 | 1 |
| HTN | monocyte | *KIAA1551* | -1.890908142 | 0.026085877 | 0.046375 | 1 |
| HTN | monocyte | *FGFBP3* | 9.221764185 | 0.023430111 | 0.042996558 | 1 |
| HTN | monocyte | *C4orf32* | -2.04124633 | 0.016221313 | 0.034042305 | 1 |
| HTN | monocyte | *RSRC1* | -2.369060597 | 0.005239395 | 0.023595115 | 1 |
| HTN | monocyte | *KCTD13* | 4.030755313 | 0.000221539 | 0.011081506 | 1 |
| HTN | monocyte | *SLC22A1* | -4.050115887 | 0.00916605 | 0.02623769 | 1 |
| HTN | monocyte | *CHST2* | 2.091314195 | 0.016386892 | 0.034241688 | 1 |
| HTN | monocyte | *TVP23C* | 6.211851303 | 0.004860937 | 0.023595115 | 1 |
| HTN | monocyte | *SH3BP5L* | -2.771034038 | 0.008896053 | 0.02599588 | 1 |
| HTN | monocyte | *CADM2* | -1.241663506 | 0.001397799 | 0.023595115 | 1 |
| HTN | monocyte | *PSMD2* | -2.679398807 | 0.005806875 | 0.023595115 | 1 |
| HTN | monocyte | *FAM182B* | -1.234840816 | 0.024779848 | 0.044714326 | 1 |
| HTN | monocyte | *CCNE2* | -2.047478193 | 1.38E-05 | 0.001718243 | 0.06701147 |
| HTN | monocyte | *ZNF519* | -1.10444102 | 0.005520579 | 0.023595115 | 1 |
| HTN | monocyte | *BANF1* | 1.721444517 | 0.008814187 | 0.02589483 | 1 |
| HTN | monocyte | *CHRNA7* | 2.15164741 | 0.003645429 | 0.023595115 | 1 |
| HTN | monocyte | *TMEM9B* | 6.966515906 | 0.000351082 | 0.01501035 | 1 |
| HTN | monocyte | *SCUBE2* | -2.010371539 | 0.010187125 | 0.027100648 | 1 |
| HTN | monocyte | *EIF1AD* | -2.215711439 | 0.006339749 | 0.02374425 | 1 |
| HTN | monocyte | *CLTB* | 3.623379655 | 0.008808577 | 0.02589483 | 1 |
| HTN | monocyte | *PPP2R2D* | -3.575706879 | 0.004397568 | 0.023595115 | 1 |

| | | | | | | |
|---|---|---|---|---|---|---|
| HTN | monocyte | *LRRC25* | 0.479389362 | 0.002926591 | 0.023595115 | 1 |
| HTN | monocyte | *KCNE3* | 1.366972146 | 0.002850209 | 0.023595115 | 1 |
| HTN | monocyte | *C11orf68* | -4.22558376 | 0.022720035 | 0.04213976 | 1 |
| HTN | monocyte | *MRPL48* | -2.272877103 | 0.009654858 | 0.02668844 | 1 |
| HTN | monocyte | *RPS6KB2* | 4.290994543 | 0.017806367 | 0.035789765 | 1 |
| HTN | monocyte | *RMI2* | -1.968800229 | 0.028607643 | 0.049308804 | 1 |
| HTN | monocyte | *TOM1L2* | -1.279131563 | 0.00624295 | 0.023683186 | 1 |
| HTN | monocyte | *TEX26* | -4.923805853 | 0.021126444 | 0.040245585 | 1 |
| HTN | monocyte | *GPR156* | -2.301204718 | 0.003804043 | 0.023595115 | 1 |
| HTN | monocyte | *PRIMA1* | -1.257683904 | 0.004960371 | 0.023595115 | 1 |
| HTN | monocyte | *A2M* | -1.527822436 | 0.016898596 | 0.03471295 | 1 |
| HTN | monocyte | *DOK7* | 7.243542692 | 0.002403732 | 0.023595115 | 1 |
| HTN | monocyte | *ORAI3* | 8.301645068 | 1.69E-08 | 1.17E-05 | 8.20E-05 |
| HTN | monocyte | *JAKMIP2* | 0.584506043 | 0.005651133 | 0.023595115 | 1 |
| HTN | monocyte | *CCDC57* | -1.697294842 | 0.014447185 | 0.032096036 | 1 |
| HTN | monocyte | *CIDEA* | -0.726789146 | 0.004776578 | 0.023595115 | 1 |
| HTN | monocyte | *SLC35G1* | -2.385199728 | 0.011383361 | 0.028353218 | 1 |
| HTN | monocyte | *OR4N2* | -3.463945474 | 0.022533418 | 0.041970114 | 1 |
| HTN | monocyte | *CDC26* | -1.727020518 | 0.010437876 | 0.027405074 | 1 |
| HTN | monocyte | *VPS37D* | -2.441767618 | 0.004629745 | 0.023595115 | 1 |
| HTN | monocyte | *WDR25* | 3.718334366 | 0.027324034 | 0.047775212 | 1 |
| HTN | monocyte | *PLA2G16* | -0.768023966 | 0.016834016 | 0.034624267 | 1 |
| HTN | monocyte | *RMDN1* | -1.887380406 | 0.010240601 | 0.027124512 | 1 |
| HTN | monocyte | *RNF152* | -2.60008858 | 0.002659295 | 0.023595115 | 1 |
| HTN | monocyte | *LRRC37A* | 1.554806511 | 0.012031014 | 0.028978626 | 1 |
| HTN | monocyte | *BDNF* | -1.786850004 | 0.002766089 | 0.023595115 | 1 |
| HTN | monocyte | *BOK* | -0.981005152 | 0.005109349 | 0.023595115 | 1 |
| HTN | monocyte | *RUFY1* | -1.773269117 | 0.023506821 | 0.043121019 | 1 |
| HTN | monocyte | *VSIG10* | -0.939990464 | 0.005798235 | 0.023595115 | 1 |
| HTN | monocyte | *C8G* | -31.63124023 | 0.0049528 | 0.023595115 | 1 |
| HTN | monocyte | *MUC20* | -1.853317941 | 0.012394585 | 0.029321564 | 1 |
| HTN | monocyte | *SHMT1* | -1.136129258 | 0.008739253 | 0.025839644 | 1 |
| HTN | monocyte | *DPP7* | 2.823706247 | 0.005251219 | 0.023595115 | 1 |
| HTN | monocyte | *MTHFR* | -4.214882426 | 0.000365074 | 0.01501035 | 1 |
| HTN | monocyte | *C19orf18* | -1.480901189 | 0.009937712 | 0.026847314 | 1 |
| HTN | monocyte | *WDR73* | -1.157344195 | 0.00690463 | 0.024148823 | 1 |
| HTN | monocyte | *DSCAML1* | -0.899774785 | 0.00551442 | 0.023595115 | 1 |
| HTN | monocyte | *AP006621.1* | -1.278680749 | 0.006716878 | 0.024005639 | 1 |
| HTN | monocyte | *MIEF2* | -2.383267297 | 0.006187885 | 0.023616295 | 1 |
| HTN | monocyte | *NAP1L5* | 1.220517439 | 0.003054712 | 0.023595115 | 1 |
| HTN | monocyte | *RBM44* | 4.161091671 | 0.001631978 | 0.023595115 | 1 |
| HTN | monocyte | *SLC25A22* | -3.479645834 | 0.000724589 | 0.019858911 | 1 |
| HTN | monocyte | *ACAD9* | -1.58862122 | 0.021744841 | 0.041068887 | 1 |
| HTN | monocyte | *IL17RA* | 2.453904644 | 0.010087391 | 0.027007023 | 1 |
| HTN | monocyte | *MBOAT4* | 1.276076869 | 0.003374633 | 0.023595115 | 1 |
| HTN | monocyte | *AGTRAP* | 2.984871457 | 0.006526921 | 0.023864166 | 1 |
| HTN | monocyte | *THAP5* | -1.594830025 | 0.005183438 | 0.023595115 | 1 |
| HTN | monocyte | *NAALADL2* | -0.915926255 | 0.009931228 | 0.026846837 | 1 |
| HTN | monocyte | *SLC35G5* | 4.413682262 | 0.003332549 | 0.023595115 | 1 |
| HTN | monocyte | *FAM87B* | 9.034025943 | 0.003442181 | 0.023595115 | 1 |

| | | | | | | |
|---|---|---|---|---|---|---|
| HTN | monocyte | *PCDHB9* | 3.431795784 | 0.000547799 | 0.017259229 | 1 |
| HTN | monocyte | *CENPBD1* | 0.897884373 | 0.003395345 | 0.023595115 | 1 |
| HTN | monocyte | *ODF3B* | 1.018421548 | 0.000420392 | 0.01520058 | 1 |
| HTN | monocyte | *ADAMTSL1* | -0.99328698 | 0.006449097 | 0.02383558 | 1 |
| HTN | monocyte | *DALRD3* | -1.749382037 | 0.002669863 | 0.023595115 | 1 |
| HTN | monocyte | *ZNF114* | -2.962224603 | 0.021894022 | 0.041206282 | 1 |
| HTN | monocyte | *SH2B1* | -1.282171109 | 0.007065113 | 0.024238452 | 1 |
| HTN | monocyte | *VN1R1* | -4.552402885 | 0.003474723 | 0.023595115 | 1 |
| HTN | monocyte | *PLEC* | -2.515617704 | 0.001652118 | 0.023595115 | 1 |
| HTN | monocyte | *SH2D4B* | -4.483140629 | 0.00792213 | 0.025073967 | 1 |
| HTN | monocyte | *RNF212* | -1.11222104 | 0.006991199 | 0.0242295 | 1 |
| HTN | monocyte | *PRSS36* | -2.363308834 | 0.005890196 | 0.023595115 | 1 |
| HTN | monocyte | *GEN1* | -2.834427227 | 0.000401945 | 0.01501035 | 1 |
| HTN | monocyte | *TMPRSS9* | -2.157397105 | 0.005715457 | 0.023595115 | 1 |
| HTN | monocyte | *TMEM11* | 4.981189883 | 0.012498994 | 0.029453675 | 1 |
| HTN | monocyte | *ZFAND2A* | -3.555782127 | 0.001103447 | 0.022966009 | 1 |
| HTN | monocyte | *ZNF223* | -3.941491576 | 0.003725615 | 0.023595115 | 1 |
| HTN | monocyte | *GLDC* | -2.243906231 | 0.006258456 | 0.023704653 | 1 |
| HTN | monocyte | *P4HTM* | -2.115533006 | 0.003479444 | 0.023595115 | 1 |
| HTN | monocyte | *CA8* | -1.747985184 | 0.005864942 | 0.023595115 | 1 |
| HTN | monocyte | *NSUN3* | -1.910627032 | 0.015046589 | 0.032752825 | 1 |
| HTN | monocyte | *RPP25* | 2.226217574 | 0.011255586 | 0.028158713 | 1 |
| HTN | monocyte | *STX19* | -1.199973658 | 0.006086254 | 0.023595115 | 1 |
| HTN | monocyte | *MPI* | -8.589009677 | 6.20E-12 | 1.51E-08 | 3.01E-08 |
| HTN | monocyte | *EFCAB13* | -1.724516263 | 0.010206517 | 0.027105649 | 1 |
| HTN | monocyte | *EXOSC4* | 4.261882291 | 0.003783894 | 0.023595115 | 1 |
| HTN | monocyte | *ZNF852* | -4.338124396 | 0.002533453 | 0.023595115 | 1 |
| HTN | monocyte | *HYI* | -4.926633209 | 9.63E-07 | 0.000233663 | 0.004673254 |
| HTN | monocyte | *C17orf62* | 4.288031941 | 0.004936379 | 0.023595115 | 1 |
| HTN | monocyte | *CCDC89* | -4.392906507 | 0.00536416 | 0.023595115 | 1 |
| HTN | monocyte | *HTR1F* | -10.95474671 | 0.022707657 | 0.042132907 | 1 |
| HTN | monocyte | *ZNF664* | -1.873733959 | 0.00700847 | 0.024238452 | 1 |
| HTN | monocyte | *CDH4* | -2.823068316 | 0.001911252 | 0.023595115 | 1 |
| HTN | monocyte | *HLA-DQB1* | -1.92191039 | 0.000795783 | 0.020419313 | 1 |
| HTN | monocyte | *PACS2* | -1.998265896 | 0.015432755 | 0.033250323 | 1 |
| HTN | monocyte | *LINC00174* | -1.306408064 | 0.001748488 | 0.023595115 | 1 |
| HTN | monocyte | *GEMIN4* | -2.612649405 | 0.002853821 | 0.023595115 | 1 |
| HTN | monocyte | *ZFPM1* | -1.939417697 | 0.005674523 | 0.023595115 | 1 |
| HTN | monocyte | *FCER1A* | -2.417446667 | 0.002197282 | 0.023595115 | 1 |
| HTN | monocyte | *SERTAD2* | -1.648491744 | 0.015602718 | 0.033395675 | 1 |
| HTN | monocyte | *AKAP5* | -1.886498735 | 0.003433088 | 0.023595115 | 1 |
| HTN | monocyte | *GIPC3* | 24.95486574 | 0.005662994 | 0.023595115 | 1 |
| HTN | monocyte | *PDXDC1* | -1.54477143 | 0.019507034 | 0.038133814 | 1 |
| HTN | monocyte | *C1orf194* | 0.750636524 | 0.006287693 | 0.023704653 | 1 |
| HTN | monocyte | *SEPHS2* | 7.978287506 | 0.001555718 | 0.023595115 | 1 |
| HTN | monocyte | *SSC5D* | -1.093902504 | 0.025019275 | 0.045027271 | 1 |
| HTN | monocyte | *SEPT1* | -1.289279039 | 0.004971507 | 0.023595115 | 1 |
| HTN | monocyte | *EXOC3* | -2.157405727 | 0.020183196 | 0.038922443 | 1 |
| HTN | monocyte | *CSNK1A1L* | 2.221425515 | 0.003994236 | 0.023595115 | 1 |
| HTN | monocyte | *HCLS1* | 0.987970456 | 0.013337568 | 0.03065466 | 1 |

| | | | | | | |
|---|---|---|---|---|---|---|
| HTN | monocyte | *MTURN* | -1.398210951 | 0.010187877 | 0.027100648 | 1 |
| HTN | monocyte | *MCFD2* | -1.66891492 | 0.022091662 | 0.041465665 | 1 |
| HTN | monocyte | *GLIPR1L2* | -1.345277834 | 0.013403661 | 0.030691157 | 1 |
| HTN | monocyte | *DEFB119* | -19.95848759 | 0.009909398 | 0.026830579 | 1 |
| HTN | monocyte | *TSPYL5* | -2.319481408 | 0.001373409 | 0.023595115 | 1 |
| HTN | monocyte | *MB21D2* | -1.369013591 | 0.009000749 | 0.026132523 | 1 |
| HTN | monocyte | *SLC47A2* | -1.609543028 | 0.005549092 | 0.023595115 | 1 |
| HTN | monocyte | *OR51E1* | -2.558795572 | 0.002824345 | 0.023595115 | 1 |
| HTN | monocyte | *BHLHE22* | 1.704662758 | 0.013077827 | 0.030344279 | 1 |
| HTN | monocyte | *ZNF443* | 2.687411092 | 0.005198232 | 0.023595115 | 1 |
| HTN | monocyte | *CAPS2* | -1.558179034 | 0.020804896 | 0.039793793 | 1 |
| HTN | monocyte | *KCTD2* | 1.973999301 | 0.010575553 | 0.02752821 | 1 |
| HTN | monocyte | *D2HGDH* | -1.696178124 | 0.001338538 | 0.023595115 | 1 |
| HTN | monocyte | *LSMEM1* | -2.393455205 | 0.003871974 | 0.023595115 | 1 |
| HTN | monocyte | *NQO1* | -2.472487375 | 0.017240697 | 0.035122418 | 1 |
| HTN | monocyte | *RPH3AL* | -1.465992583 | 0.014580987 | 0.032244675 | 1 |
| HTN | monocyte | *ADIPOQ* | -0.633802811 | 0.00286429 | 0.023595115 | 1 |
| HTN | monocyte | *MUC16* | -1.661319349 | 0.008430395 | 0.025487024 | 1 |
| HTN | monocyte | *SLC25A41* | 2.986397475 | 0.003136886 | 0.023595115 | 1 |
| HTN | monocyte | *DDX60L* | 2.461925039 | 0.003226346 | 0.023595115 | 1 |
| HTN | monocyte | *OGFOD3* | -1.093859021 | 0.022436931 | 0.04183827 | 1 |
| HTN | monocyte | *TMEM45A* | -1.014392221 | 0.007016647 | 0.024238452 | 1 |
| HTN | monocyte | *SGSH* | 5.563977341 | 0.008465586 | 0.02557598 | 1 |
| HTN | monocyte | *ZBTB20* | -1.181532914 | 0.01557099 | 0.033395675 | 1 |
| HTN | monocyte | *AMIGO1* | 1.511831841 | 0.005513552 | 0.023595115 | 1 |
| HTN | monocyte | *TMEM252* | -52.69928275 | 0.005602765 | 0.023595115 | 1 |
| HTN | monocyte | *ODF3L2* | -4.863615041 | 0.01355679 | 0.030925032 | 1 |
| HTN | monocyte | *CLDN7* | -2.806783701 | 1.01E-05 | 0.001286005 | 0.04886818 |
| HTN | monocyte | *ZNF329* | -1.572963237 | 0.007306626 | 0.024429382 | 1 |
| HTN | monocyte | *ADO* | 2.606846407 | 1.90E-06 | 0.000384059 | 0.009217405 |
| HTN | monocyte | *MGAT4C* | -1.027927982 | 0.001794013 | 0.023595115 | 1 |
| HTN | monocyte | *WRB* | -1.581065144 | 0.011552477 | 0.028510997 | 1 |
| HTN | monocyte | *TDRKH* | 1.517177334 | 0.002713261 | 0.023595115 | 1 |
| HTN | monocyte | *ZNF708* | -1.979920248 | 0.001950269 | 0.023595115 | 1 |
| HTN | monocyte | *MRPL41* | -1.599153309 | 0.006204564 | 0.023648504 | 1 |
| HTN | monocyte | *ARL6IP4* | 2.306156099 | 0.001677275 | 0.023595115 | 1 |
| HTN | monocyte | *HHIPL1* | -1.26010875 | 0.006111074 | 0.023595115 | 1 |
| HTN | monocyte | *FAM153B* | -1.062656742 | 0.007031943 | 0.024238452 | 1 |
| HTN | monocyte | *BACE2* | -0.924865204 | 0.024605663 | 0.044514049 | 1 |
| HTN | monocyte | *SYNM* | -1.546825936 | 0.005025999 | 0.023595115 | 1 |
| HTN | monocyte | *FIGN* | -0.64176175 | 0.005405108 | 0.023595115 | 1 |
| HTN | monocyte | *CLN8* | 4.509060508 | 4.07E-06 | 0.000636752 | 0.0197393 |
| HTN | monocyte | *NXPH4* | 10.62030895 | 0.003211932 | 0.023595115 | 1 |
| HTN | monocyte | *NPLOC4* | -1.693675803 | 0.014828675 | 0.03246784 | 1 |
| HTN | monocyte | *KPNA2* | 0.951962728 | 0.00192296 | 0.023595115 | 1 |
| HTN | monocyte | *FES* | -2.546616831 | 3.41E-06 | 0.000570871 | 0.016555252 |
| HTN | monocyte | *CAV3* | -1.891048752 | 0.00085037 | 0.020511604 | 1 |
| HTN | monocyte | *MXRA7* | -1.970410152 | 0.001541835 | 0.023595115 | 1 |
| HTN | monocyte | *MFSD5* | -2.70392711 | 0.011090747 | 0.028056466 | 1 |
| HTN | monocyte | *ADI1* | -2.139137428 | 0.012102552 | 0.029032068 | 1 |

| | | | | | | |
|---|---|---|---|---|---|---|
| HTN | monocyte | *CLEC4G* | -6.786301154 | 0.002305194 | 0.023595115 | 1 |
| HTN | monocyte | *TRAK1* | -2.123293427 | 0.023292742 | 0.042825457 | 1 |
| HTN | monocyte | *SKA2* | -1.678911504 | 0.006486935 | 0.023864166 | 1 |
| HTN | monocyte | *BRICD5* | 1.176747232 | 0.003346893 | 0.023595115 | 1 |
| HTN | monocyte | *GRID1* | -3.683506493 | 0.002410373 | 0.023595115 | 1 |
| HTN | monocyte | *ACBD3* | -1.646290233 | 0.00973051 | 0.02671918 | 1 |
| HTN | monocyte | *PLCXD3* | -1.267727968 | 0.006651602 | 0.023960787 | 1 |
| HTN | monocyte | *GPIHBP1* | -1.560124352 | 0.01087107 | 0.027878663 | 1 |
| HTN | monocyte | *VMO1* | 1.443653222 | 0.003267166 | 0.023595115 | 1 |
| HTN | monocyte | *ALG12* | -2.706495042 | 0.01139776 | 0.028374517 | 1 |
| HTN | monocyte | *ZNF721* | -1.317820521 | 0.0266814 | 0.047041481 | 1 |
| HTN | monocyte | *ODF3L1* | -10.97246001 | 9.80E-05 | 0.006889102 | 0.475348044 |
| HTN | monocyte | *HMGN4* | -1.722437933 | 0.012694839 | 0.029756211 | 1 |
| HTN | monocyte | *ZNF320* | 2.536620383 | 6.71E-06 | 0.000958035 | 0.032573193 |
| HTN | monocyte | *C12orf60* | -2.194521386 | 0.00950084 | 0.026576944 | 1 |
| HTN | monocyte | *WBP2NL* | -2.725062008 | 0.003445651 | 0.023595115 | 1 |
| HTN | monocyte | *BEGAIN* | 4.499751565 | 0.007830843 | 0.025020002 | 1 |
| HTN | monocyte | *ARHGEF37* | -1.667517852 | 0.007165652 | 0.024238452 | 1 |
| HTN | monocyte | *CSMD1* | -2.074039273 | 0.009576002 | 0.026656777 | 1 |
| HTN | monocyte | *FANCF* | 6.405764904 | 0.002978399 | 0.023595115 | 1 |
| HTN | monocyte | *RUVBL2* | -1.958473077 | 0.006813834 | 0.02405092 | 1 |
| HTN | monocyte | *PTTG1IP* | -1.813141429 | 0.028494083 | 0.049165466 | 1 |
| HTN | monocyte | *PLGLB1* | 1.267983239 | 0.005761102 | 0.023595115 | 1 |
| HTN | monocyte | *ZNF623* | -2.364346771 | 0.029085416 | 0.049901852 | 1 |
| HTN | monocyte | *CCDC125* | -1.913504283 | 0.00631979 | 0.02374425 | 1 |
| HTN | monocyte | *FHL3* | -1.592894957 | 0.015182631 | 0.032901351 | 1 |
| HTN | monocyte | *C19orf71* | 4.618041859 | 0.011515894 | 0.028507713 | 1 |
| HTN | monocyte | *LRIT3* | -9.26520318 | 0.00509893 | 0.023595115 | 1 |
| HTN | monocyte | *GPR132* | 2.075994531 | 0.007173623 | 0.024238452 | 1 |
| HTN | monocyte | *AC131971.1* | -9.641724029 | 0.000406405 | 0.01501035 | 1 |
| HTN | monocyte | *SERHL2* | -1.582684265 | 0.005962466 | 0.023595115 | 1 |
| HTN | monocyte | *PCBP3* | -0.710056557 | 0.024070633 | 0.043939319 | 1 |
| HTN | monocyte | *SFXN4* | -2.003616763 | 0.013228584 | 0.030491729 | 1 |
| HTN | monocyte | *MRPL54* | -1.059959489 | 0.0242922 | 0.044177569 | 1 |
| HTN | monocyte | *HMCES* | -1.843953258 | 0.009152399 | 0.02623769 | 1 |
| HTN | monocyte | *NDUFB1* | -1.936379681 | 0.017495781 | 0.035385382 | 1 |
| HTN | monocyte | *TRMT12* | 2.90217673 | 0.002583821 | 0.023595115 | 1 |
| HTN | monocyte | *FIGLA* | 2.820617013 | 0.006971413 | 0.024187912 | 1 |
| HTN | monocyte | *ACSM2A* | 18.76397468 | 0.001501526 | 0.023595115 | 1 |
| HTN | monocyte | *KREMEN1* | -1.285907065 | 0.000575109 | 0.017440176 | 1 |
| HTN | monocyte | *ZNF703* | 1.09661533 | 0.01120696 | 0.028135587 | 1 |
| HTN | monocyte | *NPIPA5* | -1.037070583 | 0.01471677 | 0.032390024 | 1 |
| HTN | monocyte | *EMILIN3* | -6.754257478 | 0.000778242 | 0.020410967 | 1 |
| HTN | monocyte | *NUDT14* | -2.311245824 | 0.00145937 | 0.023595115 | 1 |
| HTN | monocyte | *TTC32* | -1.938657561 | 0.010583999 | 0.027535422 | 1 |
| HTN | monocyte | *PP2D1* | -2.129586156 | 0.007217363 | 0.024303806 | 1 |
| HTN | monocyte | *NIPSNAP1* | -1.663670478 | 0.017886127 | 0.035878799 | 1 |
| HTN | monocyte | *CNTN2* | -1.708565328 | 0.025372377 | 0.045544496 | 1 |
| HTN | monocyte | *CRELD2* | -1.30880332 | 0.00707561 | 0.024238452 | 1 |
| HTN | monocyte | *GOLGA6L4* | -3.030606471 | 0.000426067 | 0.01520058 | 1 |

| | | | | | | |
|---|---|---|---|---|---|---|
| HTN | monocyte | *PGP* | 3.869839784 | 0.010091453 | 0.027007023 | 1 |
| HTN | monocyte | *ALDH1A3* | -1.475760417 | 0.000176852 | 0.009750992 | 0.858087281 |
| HTN | monocyte | *CLECL1* | -2.10772114 | 0.002045745 | 0.023595115 | 1 |
| HTN | monocyte | *SS18L1* | -2.770865792 | 0.010202743 | 0.027105649 | 1 |
| HTN | monocyte | *COPB2* | -2.019380219 | 0.022606153 | 0.042024925 | 1 |
| HTN | monocyte | *WDR27* | -1.179544168 | 0.011195241 | 0.028135587 | 1 |
| HTN | monocyte | *TXNRD2* | -1.367704241 | 0.009157773 | 0.02623769 | 1 |
| HTN | monocyte | *TMEM255B* | -1.302110448 | 0.007372073 | 0.024485743 | 1 |
| HTN | monocyte | *HDDC3* | -3.339278629 | 0.007389039 | 0.024485743 | 1 |
| HTN | monocyte | *ZFP1* | -2.140870364 | 0.008491888 | 0.025623532 | 1 |
| HTN | monocyte | *DUSP8* | 2.827763382 | 0.003014591 | 0.023595115 | 1 |
| HTN | monocyte | *SNN* | -1.853129175 | 0.005454029 | 0.023595115 | 1 |
| HTN | monocyte | *KRBA2* | 2.715818217 | 0.006050208 | 0.023595115 | 1 |
| HTN | monocyte | *SEPT5* | -3.717855242 | 0.010964248 | 0.02789645 | 1 |
| HTN | monocyte | *RNLS* | -1.619726487 | 0.016717182 | 0.034504301 | 1 |
| HTN | monocyte | *TCTE3* | -1.515259718 | 0.005893652 | 0.023595115 | 1 |
| HTN | monocyte | *UBE2G2* | -1.457352782 | 0.015533017 | 0.033364983 | 1 |
| HTN | monocyte | *TMED9* | -2.146873771 | 0.019034785 | 0.037482457 | 1 |
| HTN | monocyte | *DRD1* | -29.3157482 | 0.006063907 | 0.023595115 | 1 |
| HTN | monocyte | *BTBD6* | -1.068135081 | 0.022083868 | 0.041465665 | 1 |
| HTN | monocyte | *NUTM2A* | -5.884220424 | 1.68E-05 | 0.001945544 | 0.081712865 |
| HTN | monocyte | *WT1* | -0.724860879 | 0.003636753 | 0.023595115 | 1 |
| HTN | monocyte | *ZFP90* | -2.094055682 | 0.015559835 | 0.033395675 | 1 |
| HTN | monocyte | *NOC4L* | 2.308113075 | 0.002401792 | 0.023595115 | 1 |
| HTN | monocyte | *SORCS2* | -1.743944384 | 0.0046541 | 0.023595115 | 1 |
| HTN | monocyte | *BRI3BP* | -3.991673816 | 0.018196365 | 0.036243335 | 1 |
| HTN | monocyte | *SEMA4B* | 3.461723519 | 0.005842173 | 0.023595115 | 1 |
| HTN | monocyte | *CIB1* | -2.535081186 | 0.016208341 | 0.034029802 | 1 |
| HTN | monocyte | *SGCZ* | -0.841486095 | 0.006813351 | 0.02405092 | 1 |
| HTN | monocyte | *C6orf120* | 2.964707995 | 0.010565969 | 0.02752821 | 1 |
| HTN | monocyte | *GP1BA* | 1.325894082 | 0.00286926 | 0.023595115 | 1 |
| HTN | monocyte | *CCDC137* | 3.671701631 | 0.005358521 | 0.023595115 | 1 |
| HTN | monocyte | *CDK10* | -2.713988808 | 9.11E-06 | 0.001198503 | 0.0442049 |
| HTN | monocyte | *TNFAIP8L1* | -2.244559785 | 0.003381609 | 0.023595115 | 1 |
| HTN | monocyte | *TARSL2* | -1.709444854 | 0.007204837 | 0.024293169 | 1 |
| HTN | monocyte | *SH3BGR* | -1.579604194 | 0.009844005 | 0.026779763 | 1 |
| HTN | monocyte | *SV2B* | -1.46905303 | 0.003443349 | 0.023595115 | 1 |
| HTN | monocyte | *AHNAK2* | 5.778042141 | 0.010891297 | 0.02788959 | 1 |
| HTN | monocyte | *SP1* | -1.502998258 | 0.016787186 | 0.034563946 | 1 |
| HTN | monocyte | *PSMD13* | -3.392545823 | 0.005270551 | 0.023595115 | 1 |
| HTN | monocyte | *PMEL* | -1.555227818 | 0.008930618 | 0.02604048 | 1 |
| HTN | monocyte | *MYBL1* | 1.641318989 | 0.01660402 | 0.034456332 | 1 |
| HTN | monocyte | *ADARB2* | 1.907083488 | 0.003865709 | 0.023595115 | 1 |
| HTN | monocyte | *DMWD* | -1.985808439 | 0.00735912 | 0.024485743 | 1 |
| HTN | monocyte | *PIGP* | -1.089818792 | 0.009974456 | 0.0268867 | 1 |
| HTN | monocyte | *ARL17A* | -1.293375419 | 0.002251374 | 0.023595115 | 1 |
| HTN | monocyte | *EVI2B* | 1.183063678 | 0.000743537 | 0.019931729 | 1 |
| HTN | monocyte | *NPIPB4* | -1.345471559 | 0.004217679 | 0.023595115 | 1 |
| HTN | monocyte | *ZNF829* | 4.714914867 | 0.000168238 | 0.009382678 | 0.816292996 |
| HTN | monocyte | *IFITM1* | -4.307844324 | 0.000653865 | 0.018552953 | 1 |

| | | | | | | |
|---|---|---|---|---|---|---|
| HTN | monocyte | *LINC00839* | -4.204266764 | 0.003071778 | 0.023595115 | 1 |
| HTN | monocyte | *RNPC3* | -1.061011967 | 0.014396029 | 0.032026379 | 1 |
| HTN | monocyte | *RASA3* | -2.005911349 | 0.006070376 | 0.023595115 | 1 |
| HTN | monocyte | *ZNF529* | -1.614313052 | 0.015964339 | 0.03374074 | 1 |
| HTN | monocyte | *KRT5* | 0.776624673 | 0.006242223 | 0.023683186 | 1 |
| HTN | monocyte | *WWOX* | -1.087039551 | 0.012042122 | 0.028978626 | 1 |
| HTN | monocyte | *FFAR4* | -1.044148158 | 0.011209004 | 0.028135587 | 1 |
| HTN | monocyte | *CYP4F12* | -1.470685041 | 0.008233892 | 0.025373581 | 1 |
| HTN | monocyte | *ZNF555* | 3.617417047 | 0.014510502 | 0.032163067 | 1 |
| HTN | monocyte | *ZNF197* | -1.959779427 | 0.01492999 | 0.03261608 | 1 |
| HTN | monocyte | *GNG2* | 2.519928148 | 0.005914567 | 0.023595115 | 1 |
| HTN | monocyte | *TMEM222* | -2.124429382 | 0.005966954 | 0.023595115 | 1 |
| HTN | monocyte | *SEPT10* | -2.436683071 | 0.022278351 | 0.041735352 | 1 |
| HTN | monocyte | *FAM86B1* | -3.460806153 | 0.021594563 | 0.040841576 | 1 |
| HTN | monocyte | *SMYD4* | -1.573651251 | 0.001316928 | 0.023595115 | 1 |
| HTN | monocyte | *MIR22HG* | -1.085482182 | 0.004281141 | 0.023595115 | 1 |
| HTN | monocyte | *CYP27C1* | -2.080681017 | 0.003075485 | 0.023595115 | 1 |
| HTN | monocyte | *ZNF732* | 2.509065831 | 0.006870251 | 0.024120446 | 1 |
| HTN | monocyte | *HYAL3* | -2.876159294 | 0.0130721 | 0.030344279 | 1 |
| HTN | monocyte | *VSIG10L* | -4.539600856 | 0.017943193 | 0.035886387 | 1 |
| HTN | monocyte | *TRABD2A* | 2.080935322 | 0.013855646 | 0.031400091 | 1 |
| HTN | monocyte | *MAPT* | -1.024758949 | 0.005268535 | 0.023595115 | 1 |
| HTN | monocyte | *TMEM17* | 1.38533296 | 0.006189315 | 0.023616295 | 1 |
| HTN | monocyte | *TNFRSF18* | -4.640439248 | 0.013005386 | 0.03029387 | 1 |
| HTN | monocyte | *C1QL4* | -15.48698689 | 0.005068426 | 0.023595115 | 1 |
| HTN | monocyte | *RTN4RL2* | -3.594736978 | 0.00081097 | 0.020419313 | 1 |
| HTN | monocyte | *TMEM232* | -0.809782196 | 0.00630113 | 0.02371845 | 1 |
| HTN | monocyte | *EFCAB6* | -1.296459513 | 0.009336441 | 0.026350386 | 1 |
| HTN | monocyte | *TMPRSS6* | -9.099505245 | 0.005144444 | 0.023595115 | 1 |
| HTN | monocyte | *CYP4A11* | -4.340011209 | 0.028189484 | 0.048791054 | 1 |
| HTN | monocyte | *TMEM216* | -2.053252071 | 0.015787291 | 0.033645753 | 1 |
| HTN | monocyte | *MRPS21* | -2.333904157 | 0.028911235 | 0.049698576 | 1 |
| HTN | monocyte | *ANGPTL5* | -2.020531595 | 0.019069207 | 0.037504578 | 1 |
| HTN | monocyte | *WDR86* | -0.80961501 | 0.008207123 | 0.025373581 | 1 |
| HTN | monocyte | *LUZP2* | 5.502193529 | 0.007817971 | 0.025020002 | 1 |
| HTN | monocyte | *GJA4* | -1.852639675 | 0.004115191 | 0.023595115 | 1 |
| HTN | monocyte | *IFT140* | -1.75861028 | 0.008939607 | 0.026051035 | 1 |
| HTN | monocyte | *EXD3* | -1.739785932 | 0.010061122 | 0.027007023 | 1 |
| HTN | monocyte | *C17orf97* | 0.59808484 | 0.005603783 | 0.023595115 | 1 |
| HTN | monocyte | *C2orf88* | -1.012113044 | 0.005119189 | 0.023595115 | 1 |
| HTN | monocyte | *KBTBD12* | -2.530443413 | 0.001890966 | 0.023595115 | 1 |
| HTN | monocyte | *FANCA* | 2.320873267 | 0.000374727 | 0.01501035 | 1 |
| HTN | monocyte | *DNAH17* | 4.578308008 | 0.01738141 | 0.035260232 | 1 |
| HTN | monocyte | *FANCM* | -2.082195144 | 0.003930275 | 0.023595115 | 1 |
| HTN | monocyte | *ZFP69* | -3.020173681 | 0.01150011 | 0.028507713 | 1 |
| HTN | monocyte | *TMEM220* | -1.149799608 | 0.019873682 | 0.038547458 | 1 |
| HTN | monocyte | *FAM122A* | -2.253444084 | 0.006575184 | 0.023914328 | 1 |
| HTN | monocyte | *GFRAL* | -3.558719924 | 0.004935088 | 0.023595115 | 1 |
| HTN | monocyte | *ZSCAN23* | -1.484355724 | 0.004282049 | 0.023595115 | 1 |
| HTN | monocyte | *MESP2* | -3.222244401 | 0.02649642 | 0.046840382 | 1 |

| | | | | | | |
|---|---|---|---|---|---|---|
| HTN | monocyte | *MAPK12* | 1.247030231 | 0.006351143 | 0.02375925 | 1 |
| HTN | monocyte | *LINC00265* | 5.111387653 | 0.020244821 | 0.038979314 | 1 |
| HTN | monocyte | *LAMTOR4* | -2.442497384 | 0.017376057 | 0.035260232 | 1 |
| HTN | monocyte | *PRKAR1B* | -1.381624633 | 0.009563896 | 0.026650066 | 1 |
| HTN | monocyte | *NCR3LG1* | -2.1704763 | 0.004370449 | 0.023595115 | 1 |
| HTN | monocyte | *ZNF793* | -2.970315957 | 0.001068543 | 0.022636322 | 1 |
| HTN | monocyte | *RUFY4* | 0.506062604 | 0.003500713 | 0.023595115 | 1 |
| HTN | monocyte | *ZNF669* | -2.292294437 | 0.008371063 | 0.025464826 | 1 |
| HTN | monocyte | *CENPP* | -1.616678702 | 0.000773124 | 0.020386954 | 1 |
| HTN | monocyte | *C3orf62* | -2.280103508 | 0.002971667 | 0.023595115 | 1 |
| HTN | monocyte | *ENO4* | -1.327290153 | 0.009657386 | 0.02668844 | 1 |
| HTN | monocyte | *ZNF559* | -1.838774787 | 5.40E-05 | 0.004764576 | 0.262051704 |
| HTN | monocyte | *H3F3C* | -49.77589439 | 0.001045427 | 0.022247423 | 1 |
| HTN | monocyte | *JAKMIP3* | -1.290853338 | 0.015071027 | 0.032773807 | 1 |
| HTN | monocyte | *PDCD1* | 5.704789373 | 0.006946997 | 0.024187912 | 1 |
| HTN | monocyte | *SELL* | 1.101034389 | 0.004621142 | 0.023595115 | 1 |
| HTN | monocyte | *C19orf54* | 6.837605284 | 0.006803842 | 0.02405092 | 1 |
| HTN | monocyte | *C22orf34* | -1.304556381 | 0.009283212 | 0.026288744 | 1 |
| HTN | monocyte | *COL25A1* | 2.749011489 | 0.006120693 | 0.023595115 | 1 |
| HTN | monocyte | *NBR1* | -1.917400544 | 0.008608593 | 0.025815139 | 1 |
| HTN | monocyte | *NDOR1* | 1.88782009 | 0.003277016 | 0.023595115 | 1 |
| HTN | monocyte | *FAM72B* | 1.154844344 | 0.00240022 | 0.023595115 | 1 |
| HTN | monocyte | *RHCE* | -2.402222366 | 0.00360628 | 0.023595115 | 1 |
| HTN | monocyte | *SLC4A5* | -3.81112471 | 0.006546399 | 0.023864166 | 1 |
| HTN | monocyte | *UROS* | -1.727497819 | 0.007832481 | 0.025020002 | 1 |
| HTN | monocyte | *ZBED6CL* | 2.609866182 | 0.005806937 | 0.023595115 | 1 |
| HTN | monocyte | *OSTN* | -1.416474736 | 0.00970191 | 0.02671918 | 1 |
| HTN | monocyte | *FSIP2* | -1.542857473 | 0.006032139 | 0.023595115 | 1 |
| HTN | monocyte | *NOXA1* | -1.430786371 | 0.005619279 | 0.023595115 | 1 |
| HTN | monocyte | *FZD9* | 3.748537956 | 0.002141225 | 0.023595115 | 1 |
| HTN | monocyte | *MTF1* | 3.178091348 | 0.002738973 | 0.023595115 | 1 |
| HTN | monocyte | *FAM26F* | 0.316947819 | 0.009866297 | 0.026779763 | 1 |
| HTN | monocyte | *ENTPD8* | -141.0259912 | 0.004257053 | 0.023595115 | 1 |
| HTN | monocyte | *FBF1* | -1.748839602 | 0.000151633 | 0.009083012 | 0.735723966 |
| HTN | monocyte | *ASTL* | 10.98142647 | 0.004139616 | 0.023595115 | 1 |
| HTN | monocyte | *MSL1* | -1.832650335 | 0.000893535 | 0.020511604 | 1 |
| HTN | monocyte | *LRRK2* | 1.418849567 | 0.002150123 | 0.023595115 | 1 |
| HTN | monocyte | *FAM196A* | -1.501507022 | 0.011795174 | 0.028738324 | 1 |
| HTN | monocyte | *UTS2B* | -3.75946543 | 0.016686037 | 0.034480686 | 1 |
| HTN | monocyte | *C9orf152* | -3.616109802 | 0.008207638 | 0.025373581 | 1 |
| HTN | monocyte | *NELFB* | -3.200688649 | 0.025503609 | 0.045763133 | 1 |
| HTN | monocyte | *SLC15A5* | -2.071134304 | 0.020057758 | 0.038773004 | 1 |
| HTN | monocyte | *ADAT2* | -1.150983658 | 0.005422938 | 0.023595115 | 1 |
| HTN | monocyte | *VSTM1* | 0.305717109 | 0.003140235 | 0.023595115 | 1 |
| HTN | monocyte | *PLAC9* | -1.506820745 | 0.00685979 | 0.024118623 | 1 |
| HTN | monocyte | *FAM47E* | -1.226598667 | 0.028120975 | 0.048791054 | 1 |
| HTN | monocyte | *TSPYL1* | -2.002929017 | 0.013568186 | 0.030936483 | 1 |
| HTN | monocyte | *FHIT* | -1.145656051 | 0.0158769 | 0.033690142 | 1 |
| HTN | monocyte | *SLC35E2B* | -1.518291166 | 0.019788656 | 0.038498219 | 1 |
| HTN | monocyte | *KIAA0408* | -1.378194693 | 0.014855625 | 0.032503268 | 1 |

| | | | | | | |
|---|---|---|---|---|---|---|
| HTN | monocyte | *BLOC1S2* | -1.658081231 | 0.007304077 | 0.024429382 | 1 |
| HTN | monocyte | *AKR1C3* | -1.651769456 | 0.007387829 | 0.024485743 | 1 |
| HTN | monocyte | *WDSUB1* | -2.136728899 | 0.022951656 | 0.042407249 | 1 |
| HTN | monocyte | *COLCA1* | -5.209907928 | 9.14E-06 | 0.001198503 | 0.044344608 |
| HTN | monocyte | *SEMA4A* | 1.255659455 | 0.004837636 | 0.023595115 | 1 |
| HTN | monocyte | *SIRPB2* | 0.584399439 | 0.00628105 | 0.023704653 | 1 |
| HTN | monocyte | *PPIA* | -1.924845077 | 0.0161253 | 0.03389946 | 1 |
| HTN | monocyte | *ZNF471* | -2.057766116 | 0.010646139 | 0.027588558 | 1 |
| HTN | monocyte | *ZNF493* | -1.574352614 | 0.006644972 | 0.023960787 | 1 |
| HTN | monocyte | *GTF2IRD2* | -2.432159415 | 0.019190324 | 0.037666445 | 1 |
| HTN | monocyte | *SUPT3H* | -1.619604753 | 0.017886516 | 0.035878799 | 1 |
| HTN | monocyte | *GIMAP5* | -2.232931679 | 0.007666183 | 0.024938402 | 1 |
| HTN | monocyte | *ZKSCAN7* | -1.131144083 | 0.019273729 | 0.037814853 | 1 |
| HTN | monocyte | *CPNE4* | -1.90646527 | 0.004367829 | 0.023595115 | 1 |
| HTN | monocyte | *ZNF565* | -1.701525203 | 0.005373764 | 0.023595115 | 1 |
| HTN | monocyte | *ZNF34* | 2.806346004 | 0.001012864 | 0.021745206 | 1 |
| HTN | monocyte | *ZNF781* | -1.43379006 | 0.002501531 | 0.023595115 | 1 |
| HTN | monocyte | *ZNF124* | -1.809353614 | 0.01417794 | 0.031818393 | 1 |
| HTN | monocyte | *PIK3R4* | -1.781069938 | 0.010698018 | 0.027617119 | 1 |
| HTN | monocyte | *RFX8* | -0.976319039 | 0.005658948 | 0.023595115 | 1 |
| HTN | monocyte | *C20orf96* | -1.352741506 | 0.004787777 | 0.023595115 | 1 |
| HTN | monocyte | *SULT1A1* | -2.339693615 | 0.002812235 | 0.023595115 | 1 |
| HTN | monocyte | *MYO18A* | -2.401063036 | 0.012104531 | 0.029032068 | 1 |
| HTN | monocyte | *SPTSSB* | -4.42597584 | 0.004867039 | 0.023595115 | 1 |
| HTN | monocyte | *CACNA1H* | -2.278999573 | 0.005445778 | 0.023595115 | 1 |
| HTN | monocyte | *SULF2* | 2.150207008 | 0.000405307 | 0.01501035 | 1 |
| HTN | monocyte | *XRCC2* | -2.715659163 | 0.005179592 | 0.023595115 | 1 |
| HTN | monocyte | *MYO6* | -1.51703693 | 0.004446061 | 0.023595115 | 1 |
| HTN | monocyte | *ADH1B* | -0.530041718 | 0.005271622 | 0.023595115 | 1 |
| HTN | monocyte | *ZKSCAN5* | -1.169198873 | 0.008587102 | 0.025783919 | 1 |
| HTN | monocyte | *ZFP62* | -1.420620058 | 0.016307508 | 0.034178845 | 1 |
| HTN | monocyte | *ZNF33B* | -1.310376846 | 0.011139736 | 0.028107124 | 1 |
| HTN | monocyte | *AMZ2* | -1.5297257 | 0.002623815 | 0.023595115 | 1 |
| HTN | monocyte | *ZNF431* | -1.593625999 | 0.018438733 | 0.036560986 | 1 |
| HTN | monocyte | *COL27A1* | -1.101202092 | 0.019858612 | 0.038541594 | 1 |
| HTN | monocyte | *ZNF239* | -1.955181722 | 0.00429546 | 0.023595115 | 1 |
| HTN | monocyte | *ANKRD36B* | -1.310418209 | 0.001668982 | 0.023595115 | 1 |
| HTN | monocyte | *FAM3C* | -2.664232372 | 0.013335866 | 0.03065466 | 1 |
| HTN | monocyte | *HIST1H3E* | 3.209183086 | 0.000166875 | 0.009382678 | 0.809675422 |
| HTN | monocyte | *WDR5B* | 5.954938952 | 0.013138429 | 0.030428476 | 1 |
| HTN | monocyte | *ZNF138* | -1.368372651 | 0.014738708 | 0.032396802 | 1 |
| HTN | monocyte | *ZNF429* | -1.084851061 | 0.00568008 | 0.023595115 | 1 |
| HTN | monocyte | *ZNF100* | -3.259510543 | 5.90E-05 | 0.005110226 | 0.286172659 |
| HTN | monocyte | *ZNF420* | -1.334263531 | 0.008299232 | 0.025373581 | 1 |
| HTN | monocyte | *ZMYM1* | -1.409566591 | 0.007724452 | 0.02497075 | 1 |
| HTN | monocyte | *IGF2R* | -2.117069869 | 0.009254449 | 0.026254852 | 1 |
| HTN | monocyte | *GAL3ST4* | -2.641471301 | 0.009351889 | 0.026350386 | 1 |
| HTN | monocyte | *ZNF772* | 0.598441972 | 0.024790094 | 0.044714326 | 1 |
| HTN | monocyte | *ADAM32* | -1.976112525 | 0.007151697 | 0.024238452 | 1 |
| HTN | monocyte | *PIWIL2* | -1.698926562 | 0.004498693 | 0.023595115 | 1 |

| | | | | | | |
|---|---|---|---|---|---|---|
| HTN | monocyte | *TBC1D9B* | -2.031189357 | 0.01709734 | 0.034961127 | 1 |
| HTN | monocyte | *FAM110D* | -1.833883898 | 0.015088934 | 0.032773807 | 1 |
| HTN | monocyte | *C6orf141* | -1.221036745 | 0.000849631 | 0.020511604 | 1 |
| HTN | monocyte | *CCL4L2* | 0.487657763 | 0.003075774 | 0.023595115 | 1 |
| HTN | monocyte | *RAMP2-AS1* | -1.009376076 | 0.005132264 | 0.023595115 | 1 |
| HTN | monocyte | *ZNF720* | -1.611282484 | 0.008798275 | 0.02589483 | 1 |
| HTN | monocyte | *ZNF98* | -1.160785942 | 0.024134158 | 0.044022155 | 1 |
| HTN | monocyte | *ZNF786* | 1.694603621 | 0.011505283 | 0.028507713 | 1 |
| HTN | monocyte | *ZNF517* | -1.733181284 | 0.002643064 | 0.023595115 | 1 |
| HTN | monocyte | *ZNF675* | -2.028459683 | 0.006643129 | 0.023960787 | 1 |
| HTN | monocyte | *SLC22A5* | -0.962355187 | 0.011964612 | 0.028939331 | 1 |
| HTN | monocyte | *SHPK* | -2.471036577 | 0.021089473 | 0.040190937 | 1 |
| HTN | monocyte | *GSTK1* | -3.012066558 | 0.000982158 | 0.021612081 | 1 |
| HTN | monocyte | *GYPE* | -2.000862231 | 0.017622015 | 0.035556453 | 1 |
| HTN | monocyte | *PCDHB11* | -2.393508853 | 0.000108785 | 0.0073309 | 0.527824789 |
| HTN | monocyte | *SLC2A10* | -1.120630577 | 0.006805425 | 0.02405092 | 1 |
| HTN | monocyte | *RPF2* | -2.177993936 | 0.012371879 | 0.029302872 | 1 |
| HTN | monocyte | *MYO5A* | -2.073505657 | 0.008157409 | 0.025308801 | 1 |
| HTN | monocyte | *PIGN* | -1.943221725 | 0.007684029 | 0.024938402 | 1 |
| HTN | monocyte | *HHLA3* | -1.654161568 | 0.003505977 | 0.023595115 | 1 |
| HTN | monocyte | *TOPORS* | -2.053731124 | 0.010610021 | 0.027573553 | 1 |
| HTN | monocyte | *BCO2* | -1.22694826 | 0.001287638 | 0.023595115 | 1 |
| HTN | monocyte | *ENTPD6* | -1.302449792 | 0.002853286 | 0.023595115 | 1 |
| HTN | monocyte | *ENPP1* | -0.908764614 | 0.013107713 | 0.030386346 | 1 |
| HTN | monocyte | *MFAP5* | -3.259351477 | 7.23E-05 | 0.005658641 | 0.35083572 |
| HTN | monocyte | *DNAH10* | -1.439775683 | 0.007916732 | 0.025073967 | 1 |
| HTN | monocyte | *NMB* | -1.508083875 | 0.003706076 | 0.023595115 | 1 |
| HTN | monocyte | *FAM114A1* | -1.625066321 | 0.015462398 | 0.033284629 | 1 |
| HTN | monocyte | *EME2* | -1.612504306 | 0.005070024 | 0.023595115 | 1 |
| HTN | monocyte | *AC122129.1* | 1.604833089 | 0.004391578 | 0.023595115 | 1 |
| HTN | monocyte | *OCLN* | -2.131243116 | 0.002671074 | 0.023595115 | 1 |
| HTN | monocyte | *SGTB* | -3.608398134 | 0.003588798 | 0.023595115 | 1 |
| HTN | monocyte | *ADH5* | -1.295681278 | 0.008103204 | 0.025239173 | 1 |
| HTN | monocyte | *ZNF677* | -1.701958335 | 0.005826919 | 0.023595115 | 1 |
| HTN | monocyte | *FCHSD1* | -2.457790173 | 0.005525529 | 0.023595115 | 1 |
| HTN | monocyte | *MPZL1* | -1.482874997 | 0.01423074 | 0.031863198 | 1 |
| HTN | monocyte | *DLGAP2* | -1.108591168 | 0.019950513 | 0.03864267 | 1 |
| HTN | monocyte | *ZNF273* | -2.166963178 | 0.00605141 | 0.023595115 | 1 |
| HTN | monocyte | *ZNF667* | -1.062066903 | 0.012321281 | 0.029290962 | 1 |
| HTN | monocyte | *ZNF649* | 2.14183294 | 0.006129887 | 0.023595115 | 1 |
| HTN | monocyte | *ADH4* | -1.26751498 | 0.000679935 | 0.018851689 | 1 |
| HTN | monocyte | *ZNF544* | -2.059066574 | 0.008735171 | 0.025839644 | 1 |
| HTN | monocyte | *MIER1* | -2.369949872 | 0.017197328 | 0.035103675 | 1 |
| HTN | monocyte | *SVIP* | -1.805457244 | 0.00859384 | 0.025786834 | 1 |
| HTN | monocyte | *ZNF334* | -1.380756235 | 0.006268503 | 0.023704653 | 1 |
| HTN | monocyte | *SULT1C2* | -1.177057315 | 0.020193446 | 0.038926738 | 1 |
| HTN | monocyte | *HELZ* | -1.482411378 | 0.019715673 | 0.038431229 | 1 |
| HTN | monocyte | *TMEM116* | -4.800381176 | 1.44E-13 | 6.98E-10 | 6.98E-10 |
| HTN | monocyte | *UCKL1* | -3.384150745 | 0.015587743 | 0.033395675 | 1 |
| HTN | monocyte | *ZNF442* | 6.192292176 | 0.000408361 | 0.01501035 | 1 |

| | | | | | | |
|---|---|---|---|---|---|---|
| HTN | monocyte | *GFPT1* | -1.427251151 | 0.010461446 | 0.027437263 | 1 |
| HTN | monocyte | *ZNF587* | -2.484285894 | 0.027203203 | 0.047649798 | 1 |
| HTN | monocyte | *FLVCR1-AS1* | -1.09601094 | 0.004332728 | 0.023595115 | 1 |
| HTN | monocyte | *HLA-DRB5* | -0.714143912 | 0.007019155 | 0.024238452 | 1 |
| HTN | monocyte | *CNGA1* | -1.518153226 | 0.014719704 | 0.032390024 | 1 |
| HTN | monocyte | *NCOA6* | -2.348196841 | 0.001627482 | 0.023595115 | 1 |
| HTN | monocyte | *FAM19A2* | -1.363032965 | 0.010071262 | 0.027007023 | 1 |
| HTN | monocyte | *TTC37* | -1.614152688 | 0.011931305 | 0.028887571 | 1 |
| HTN | monocyte | *TUSC1* | -1.933758384 | 0.004734408 | 0.023595115 | 1 |
| HTN | monocyte | *PAPSS2* | -1.198345753 | 0.007928989 | 0.025073967 | 1 |
| HTN | monocyte | *CEP290* | -1.689261202 | 0.007462625 | 0.024580667 | 1 |
| HTN | monocyte | *ANKRD13B* | 3.209788426 | 0.003514496 | 0.023595115 | 1 |
| HTN | monocyte | *ECI2* | -2.657261689 | 0.009500687 | 0.026576944 | 1 |
| HTN | monocyte | *UNC13B* | -1.778439711 | 0.009932002 | 0.026846837 | 1 |
| HTN | monocyte | *PPP1R14C* | 13.16382289 | 0.004814224 | 0.023595115 | 1 |
| HTN | monocyte | *GPATCH3* | 5.064312605 | 0.007360314 | 0.024485743 | 1 |
| HTN | monocyte | *COLGALT2* | 4.201601583 | 0.022413435 | 0.04183827 | 1 |
| HTN | monocyte | *RCSD1* | 1.787569508 | 0.003300715 | 0.023595115 | 1 |
| HTN | monocyte | *RYR3* | -0.778060703 | 0.011639243 | 0.02857976 | 1 |
| HTN | monocyte | *ITPRIPL1* | -1.833954081 | 0.000559778 | 0.017299633 | 1 |
| HTN | monocyte | *PRMT6* | -3.970056388 | 0.003235468 | 0.023595115 | 1 |
| HTN | monocyte | *SHISA4* | -6.876175945 | 4.72E-05 | 0.004404753 | 0.229047144 |
| HTN | monocyte | *MAP3K3* | -2.609089277 | 0.019525074 | 0.03815371 | 1 |
| HTN | monocyte | *CCDC167* | -2.064196335 | 0.004902618 | 0.023595115 | 1 |
| HTN | monocyte | *MFAP3L* | -1.517427701 | 0.016683286 | 0.034480686 | 1 |
| HTN | monocyte | *NAGA* | 3.613073617 | 0.00536838 | 0.023595115 | 1 |
| HTN | monocyte | *SMG5* | -3.56968736 | 0.005076514 | 0.023595115 | 1 |
| HTN | monocyte | *AL590822.1* | -1.750633809 | 0.001234636 | 0.023595115 | 1 |
| HTN | monocyte | *AP001007.1* | -2.861163061 | 0.018277202 | 0.036359566 | 1 |
| HTN | monocyte | *EFCAB2* | -1.346740141 | 0.004481186 | 0.023595115 | 1 |
| HTN | monocyte | *CHML* | 3.122588774 | 0.020859754 | 0.039862752 | 1 |
| HTN | monocyte | *TATDN3* | 1.843200367 | 0.01434686 | 0.031973856 | 1 |
| HTN | monocyte | *SAMD5* | -1.202439105 | 0.009243779 | 0.026254852 | 1 |
| HTN | monocyte | *GPR52* | -45.24841144 | 0.007593807 | 0.024778179 | 1 |
| HTN | monocyte | *FAM229B* | -1.28961446 | 0.003146814 | 0.023595115 | 1 |
| HTN | monocyte | *DDO* | -2.764661345 | 0.002035467 | 0.023595115 | 1 |
| HTN | monocyte | *RBM20* | -1.659883235 | 0.005969711 | 0.023595115 | 1 |
| HTN | monocyte | *C6orf163* | -1.922946518 | 0.002195733 | 0.023595115 | 1 |
| HTN | monocyte | *PCMTD2* | -1.260860947 | 0.007380872 | 0.024485743 | 1 |
| HTN | monocyte | *SAMD13* | -1.670515935 | 0.005247373 | 0.023595115 | 1 |
| HTN | monocyte | *EFCAB7* | -1.52184204 | 0.00960138 | 0.02668844 | 1 |
| HTN | monocyte | *IFIT1B* | -7.522063626 | 0.005712993 | 0.023595115 | 1 |
| HTN | monocyte | *LIPN* | 0.790115742 | 0.003416463 | 0.023595115 | 1 |
| HTN | monocyte | *INPP5B* | -1.854890771 | 0.001708712 | 0.023595115 | 1 |
| HTN | monocyte | *NEU4* | 3.067012976 | 0.005165605 | 0.023595115 | 1 |
| HTN | monocyte | *C2orf72* | 3.0766683 | 0.003278248 | 0.023595115 | 1 |
| HTN | monocyte | *RUFY2* | -1.660946304 | 0.015231938 | 0.032964034 | 1 |
| HTN | monocyte | *ASAH2B* | -1.702711459 | 0.015202947 | 0.032930668 | 1 |
| HTN | monocyte | *SYT15* | -3.318238077 | 0.010185555 | 0.027100648 | 1 |
| HTN | monocyte | *LINC00619* | -13.59996157 | 0.004911041 | 0.023595115 | 1 |

| | | | | | | |
|---|---|---|---|---|---|---|
| HTN | monocyte | *DAXX* | -2.496089731 | 0.023164126 | 0.042621289 | 1 |
| HTN | monocyte | *BMPR2* | -1.473619062 | 0.016850099 | 0.03464266 | 1 |
| HTN | monocyte | *TCEA3* | -0.925844742 | 0.005068856 | 0.023595115 | 1 |
| HTN | monocyte | *HSD17B8* | -4.001634996 | 1.62E-06 | 0.000342076 | 0.007867746 |
| HTN | monocyte | *OXLD1* | 1.990419197 | 0.001640511 | 0.023595115 | 1 |
| HTN | monocyte | *SMIM5* | -3.898525182 | 0.004266256 | 0.023595115 | 1 |
| HTN | monocyte | *STK19* | -1.504202311 | 0.011795231 | 0.028738324 | 1 |
| HTN | monocyte | *SKIV2L* | -1.477789891 | 0.000882251 | 0.020511604 | 1 |
| HTN | monocyte | *LAYN* | -1.888819461 | 0.000490788 | 0.016310293 | 1 |
| HTN | monocyte | *LY6G5C* | 1.67864127 | 0.007863355 | 0.025067673 | 1 |
| HTN | monocyte | *GPANK1* | -1.849994396 | 0.006817158 | 0.02405092 | 1 |
| HTN | monocyte | *LST1* | 0.271843732 | 0.003750491 | 0.023595115 | 1 |
| HTN | monocyte | *ZNF814* | -1.711714 | 0.025566371 | 0.045780981 | 1 |
| HTN | monocyte | *MICA* | -2.388363855 | 0.008221251 | 0.025373581 | 1 |
| HTN | monocyte | *HLA-C* | -3.786662912 | 4.26E-08 | 2.23E-05 | 0.00020682 |
| HTN | monocyte | *POU5F1* | -1.272163173 | 0.000135941 | 0.008566049 | 0.659585757 |
| HTN | monocyte | *CCHCR1* | -1.47324203 | 0.000458761 | 0.015899361 | 1 |
| HTN | monocyte | *PSORS1C2* | 2.980705076 | 0.008620408 | 0.025828042 | 1 |
| HTN | monocyte | *DHX16* | -1.654913498 | 0.015796584 | 0.033645753 | 1 |
| HTN | monocyte | *LILRB3* | 0.368932743 | 0.00329217 | 0.023595115 | 1 |
| HTN | monocyte | *ZNF468* | -2.132709004 | 0.01318691 | 0.030482557 | 1 |
| HTN | monocyte | *RNF39* | 2.7988656 | 0.011631035 | 0.02857976 | 1 |
| HTN | monocyte | *MALRD1* | -0.864856355 | 0.004371255 | 0.023595115 | 1 |
| HTN | monocyte | *FAM196B* | 1.159089277 | 0.00295459 | 0.023595115 | 1 |
| HTN | monocyte | *MROH6* | 0.69640022 | 0.004801854 | 0.023595115 | 1 |
| HTN | monocyte | *ZBTB48* | 3.691842664 | 0.00547454 | 0.023595115 | 1 |
| HTN | monocyte | *MZT1* | 4.834636011 | 0.007982459 | 0.025077113 | 1 |
| HTN | monocyte | *ZNF425* | -1.716188758 | 0.002229063 | 0.023595115 | 1 |
| HTN | monocyte | *C12orf73* | -2.865460532 | 0.011735225 | 0.02867949 | 1 |
| HTN | monocyte | *PCDHA7* | 5.125669839 | 0.004188582 | 0.023595115 | 1 |
| HTN | monocyte | *PCDHA4* | -3.689635205 | 0.004714347 | 0.023595115 | 1 |
| HTN | monocyte | *PKHD1L1* | -0.645465102 | 0.022773773 | 0.042191044 | 1 |
| HTN | monocyte | *SLC35B4* | -1.682802719 | 0.007395934 | 0.024485743 | 1 |
| HTN | monocyte | *TMEM231* | 4.036191627 | 0.004574363 | 0.023595115 | 1 |
| HTN | monocyte | *SCGB2B2* | -1.166138757 | 0.006538314 | 0.023864166 | 1 |
| HTN | monocyte | *CCDC144NL* | 1.549345121 | 0.003177045 | 0.023595115 | 1 |
| HTN | monocyte | *GOLGA6L10* | -4.085644642 | 0.002027438 | 0.023595115 | 1 |
| HTN | monocyte | *NT5M* | -1.522658886 | 0.004317206 | 0.023595115 | 1 |
| HTN | monocyte | *PRR13* | -3.464545109 | 0.00062819 | 0.018265624 | 1 |
| HTN | monocyte | *MT1A* | 2.181729035 | 0.003655253 | 0.023595115 | 1 |
| HTN | monocyte | *MT1M* | -4.174568353 | 0.026156271 | 0.046416345 | 1 |
| HTN | monocyte | *CFI* | -2.122143739 | 0.021413476 | 0.040616961 | 1 |
| HTN | monocyte | *SMN2* | -2.219265139 | 0.000391863 | 0.01501035 | 1 |
| HTN | monocyte | *KLRC2* | 1.975036312 | 0.002312366 | 0.023595115 | 1 |
| HTN | monocyte | *C1QTNF9B* | 2.542253052 | 0.003584748 | 0.023595115 | 1 |
| HTN | monocyte | *ZDHHC11B* | -2.16604925 | 0.005503656 | 0.023595115 | 1 |
| HTN | monocyte | *LNP1* | -1.396053302 | 0.010856908 | 0.027878663 | 1 |
| HTN | monocyte | *METTL6* | -1.815652549 | 0.019943263 | 0.03864267 | 1 |
| HTN | monocyte | *SLC48A1* | -1.965937857 | 0.009643959 | 0.02668844 | 1 |
| HTN | monocyte | *C17orf51* | -1.946168459 | 0.005099688 | 0.023595115 | 1 |

| | | | | | | |
|---|---|---|---|---|---|---|
| HTN | monocyte | *NUP62* | -2.201262159 | 0.000999595 | 0.021669448 | 1 |
| HTN | monocyte | *FGFR1OP* | -1.375468329 | 0.008573744 | 0.025783919 | 1 |
| HTN | monocyte | *ZNF254* | -2.030094878 | 0.026203889 | 0.046452783 | 1 |
| HTN | monocyte | *TCTEX1D2* | -3.009407115 | 0.013388648 | 0.030691157 | 1 |
| HTN | monocyte | *AC022498.1* | -7.148060539 | 0.007832931 | 0.025020002 | 1 |
| HTN | monocyte | *NOTCH2NL* | 1.5667524 | 0.001422372 | 0.023595115 | 1 |
| HTN | monocyte | *SUPT4H1* | 6.151378127 | 0.000160142 | 0.009328469 | 0.777007438 |
| HTN | monocyte | *GSTM2* | -0.941183992 | 0.003267254 | 0.023595115 | 1 |
| HTN | monocyte | *ARHGAP19* | 2.449947531 | 0.008696894 | 0.025839644 | 1 |
| HTN | monocyte | *LCAT* | -2.869748687 | 0.006974215 | 0.024187912 | 1 |
| HTN | monocyte | *ERV3-1* | -0.46762576 | 0.026202773 | 0.046452783 | 1 |
| HTN | monocyte | *GBP7* | 1.480972659 | 0.01455448 | 0.032216394 | 1 |
| HTN | monocyte | *TMEM110* | 1.79857723 | 0.000268434 | 0.012692888 | 1 |
| HTN | monocyte | *TMX2* | -1.399504673 | 0.011376048 | 0.028349555 | 1 |
| HTN | monocyte | *LBH* | -3.201911009 | 0.023128251 | 0.042571424 | 1 |
| HTN | monocyte | *NCKIPSD* | -2.480296765 | 0.004065485 | 0.023595115 | 1 |
| HTN | monocyte | *ATF6B* | 7.044826601 | 1.47E-07 | 5.95E-05 | 0.00071367 |
| HTN | monocyte | *SLC35F6* | -2.693621806 | 0.004648565 | 0.023595115 | 1 |
| HTN | monocyte | *EMP2* | -2.062729453 | 0.014742815 | 0.032396802 | 1 |
| HTN | monocyte | *PPM1N* | 1.626871558 | 0.000114338 | 0.00749688 | 0.554769101 |
| HTN | monocyte | *SLC23A3* | 2.672785376 | 0.009225527 | 0.026254852 | 1 |
| HTN | monocyte | *LTB4R* | -5.843360102 | 0.025980505 | 0.046241902 | 1 |
| HTN | monocyte | *IRF9* | -1.961501161 | 0.012359525 | 0.029302872 | 1 |
| HTN | monocyte | *ZNF726* | 1.738172112 | 0.006676237 | 0.023986397 | 1 |
| HTN | monocyte | *REPIN1* | -3.486339271 | 0.015729175 | 0.033561106 | 1 |
| HTN | monocyte | *ZNF891* | -1.482121962 | 0.008839897 | 0.025916121 | 1 |
| HTN | monocyte | *TSPAN4* | -1.573572334 | 0.013227317 | 0.030491729 | 1 |
| HTN | monocyte | *CPNE1* | -1.541142317 | 0.007041802 | 0.024238452 | 1 |
| HTN | monocyte | *PAXIP1-AS2* | -1.397578077 | 0.007652264 | 0.024918647 | 1 |
| HTN | monocyte | *LYRM4* | -1.601936316 | 0.009534121 | 0.026641624 | 1 |
| HTN | monocyte | *SH3D21* | 2.917866958 | 0.00090572 | 0.020535291 | 1 |
| HTN | monocyte | *FIS1* | -3.098557025 | 0.001154023 | 0.023234426 | 1 |
| HTN | monocyte | *SOGA3* | -1.388521401 | 0.013015686 | 0.030301917 | 1 |
| HTN | monocyte | *GNAT3* | -1.384360517 | 0.005377847 | 0.023595115 | 1 |
| HTN | monocyte | *PLIN5* | -0.706881603 | 0.005982783 | 0.023595115 | 1 |
| HTN | monocyte | *EML6* | -1.586045586 | 0.007063638 | 0.024238452 | 1 |
| HTN | monocyte | *ARHGEF33* | -2.090704052 | 0.004446078 | 0.023595115 | 1 |
| HTN | monocyte | *AC005082.1* | -6.871908568 | 0.000851062 | 0.020511604 | 1 |
| HTN | monocyte | *C5orf49* | 7.856646508 | 0.00381969 | 0.023595115 | 1 |
| HTN | monocyte | *NPEPL1* | -1.315449455 | 0.005567717 | 0.023595115 | 1 |
| HTN | monocyte | *TMEM242* | -1.949824409 | 0.020475868 | 0.039272886 | 1 |
| HTN | monocyte | *SLC35E2* | -2.430767394 | 0.007961608 | 0.025076015 | 1 |
| HTN | monocyte | *TSTD1* | -3.796076046 | 0.014805056 | 0.032436518 | 1 |
| HTN | monocyte | *AC009403.2* | 3.145338113 | 0.010370784 | 0.027326031 | 1 |
| HTN | monocyte | *PAM16* | -1.565270531 | 0.003573508 | 0.023595115 | 1 |
| HTN | monocyte | *NBPF1* | -1.352160978 | 0.014858254 | 0.032503268 | 1 |
| HTN | monocyte | *FAM228B* | -2.0125224 | 0.000794746 | 0.020419313 | 1 |
| HTN | monocyte | *RNU6ATAC35I* | -14.45191827 | 0.012473291 | 0.029407389 | 1 |
| HTN | monocyte | *FANCG* | 1.816234139 | 0.028368956 | 0.048984404 | 1 |
| HTN | monocyte | *AP4M1* | -3.746922291 | 3.75E-05 | 0.003568157 | 0.181975996 |

| | | | | | | |
|---|---|---|---|---|---|---|
| HTN | monocyte | *ZNF880* | -2.303778974 | 0.005206775 | 0.023595115 | 1 |
| HTN | monocyte | *APOL6* | -1.071676137 | 0.006604048 | 0.02395053 | 1 |
| HTN | monocyte | *TIAF1* | 8.427036292 | 7.96E-07 | 0.000220692 | 0.003862605 |
| HTN | monocyte | *RAB6C* | -32.49450691 | 0.017880451 | 0.035878799 | 1 |
| HTN | monocyte | *AC062017.1* | -1.307632313 | 0.005034472 | 0.023595115 | 1 |
| HTN | monocyte | *ADRA2B* | -6.99347252 | 0.00121253 | 0.023439031 | 1 |
| HTN | monocyte | *EXOSC6* | -5.098291925 | 7.68E-05 | 0.005912181 | 0.372467425 |
| HTN | monocyte | *LINC00571* | -1.980124155 | 0.002306962 | 0.023595115 | 1 |
| HTN | monocyte | *LINC01133* | -1.062812033 | 0.004649225 | 0.023595115 | 1 |
| HTN | monocyte | *LINC01117* | -1.295737841 | 0.005540315 | 0.023595115 | 1 |
| HTN | monocyte | *LINC00184* | 38.66180091 | 0.003273875 | 0.023595115 | 1 |
| HTN | monocyte | *AL049840.1* | -2.983198703 | 0.000769499 | 0.020386954 | 1 |
| HTN | monocyte | *PLEKHM1* | -3.871185341 | 0.001199235 | 0.023368216 | 1 |
| HTN | monocyte | *LINC00092* | -2.123990208 | 0.009164338 | 0.02623769 | 1 |
| HTN | monocyte | *AP000350.6* | 1.293005877 | 0.002199575 | 0.023595115 | 1 |
| HTN | monocyte | *CYP4A22-AS1* | -2.672921571 | 0.007703904 | 0.0249695 | 1 |
| HTN | monocyte | *MIAT* | -4.729511263 | 0.005656722 | 0.023595115 | 1 |
| HTN | monocyte | *ERCC6* | -1.680764973 | 0.011998294 | 0.028978626 | 1 |
| HTN | monocyte | *LINC00115* | -2.266069595 | 0.004168656 | 0.023595115 | 1 |
| HTN | monocyte | *HAR1A* | -18.02861955 | 0.002906698 | 0.023595115 | 1 |
| HTN | monocyte | *LINC00937* | 0.43865647 | 0.0061682 | 0.023602609 | 1 |
| HTN | monocyte | *FAM138B* | 19.22001373 | 0.003273189 | 0.023595115 | 1 |
| HTN | monocyte | *AC017074.2* | -22.77639293 | 0.004817358 | 0.023595115 | 1 |
| HTN | monocyte | *AC009299.3* | 3.932160863 | 0.004222936 | 0.023595115 | 1 |
| HTN | monocyte | *SLC2A1-AS1* | -2.481223392 | 0.006716788 | 0.024005639 | 1 |
| HTN | monocyte | *POLR2J2* | 4.352294535 | 0.002591296 | 0.023595115 | 1 |
| HTN | monocyte | *LINC00578* | -1.898628461 | 0.001931648 | 0.023595115 | 1 |
| HTN | monocyte | *C19orf24* | 1.12678144 | 0.002295989 | 0.023595115 | 1 |
| HTN | monocyte | *ARL17B* | -1.661555206 | 0.004536093 | 0.023595115 | 1 |
| HTN | monocyte | *CASC20* | 4.057133374 | 0.004385711 | 0.023595115 | 1 |
| HTN | monocyte | *PSMG3-AS1* | -1.972712809 | 0.004748405 | 0.023595115 | 1 |
| HTN | monocyte | *AC112229.4* | 1.200783021 | 0.002785746 | 0.023595115 | 1 |
| HTN | monocyte | *KLHL7-AS1* | 2.087080182 | 0.001166682 | 0.02330522 | 1 |
| HTN | monocyte | *LINC00332* | -2.177469324 | 0.016539188 | 0.034411074 | 1 |
| HTN | monocyte | *DIRC3* | -0.840695641 | 0.006071822 | 0.023595115 | 1 |
| HTN | monocyte | *AC107021.1* | -0.979612914 | 0.00697186 | 0.024187912 | 1 |
| HTN | monocyte | *TMA7* | 5.536226017 | 0.000113619 | 0.00749688 | 0.551279275 |
| HTN | monocyte | *AC009495.3* | 1.133022254 | 0.002148068 | 0.023595115 | 1 |
| HTN | monocyte | *LINC00665* | -2.270918704 | 0.007916353 | 0.025073967 | 1 |
| HTN | monocyte | *LINC00342* | 6.797643266 | 0.010390133 | 0.027326031 | 1 |
| HTN | monocyte | *LINC00702* | -2.478722404 | 0.005892257 | 0.023595115 | 1 |
| HTN | monocyte | *RPS28* | 5.249074947 | 0.027823216 | 0.048455938 | 1 |
| HTN | monocyte | *PINLYP* | -1.264678948 | 0.004795274 | 0.023595115 | 1 |
| HTN | monocyte | *ANKRD65* | 1.596294975 | 0.003191737 | 0.023595115 | 1 |
| HTN | monocyte | *LINC00957* | -3.255094264 | 0.016282344 | 0.034140853 | 1 |
| HTN | monocyte | *KIAA0040* | -1.64408806 | 0.005817896 | 0.023595115 | 1 |
| HTN | monocyte | *LINC00607* | 3.773887808 | 0.008014411 | 0.025099849 | 1 |
| HTN | monocyte | *LINC00941* | -4.859495537 | 0.017723016 | 0.035696171 | 1 |
| HTN | monocyte | *AC019117.2* | -12.68984392 | 0.000892933 | 0.020511604 | 1 |
| HTN | monocyte | *ZBED5* | -1.554607741 | 0.02678372 | 0.047187585 | 1 |

| | | | | | | |
|---|---|---|---|---|---|---|
| HTN | monocyte | *LINC00987* | -1.631147753 | 0.012161943 | 0.029071921 | 1 |
| HTN | monocyte | *LINC00959* | -3.323234063 | 0.02779721 | 0.048445425 | 1 |
| HTN | monocyte | *HLA-DQA2* | 1.906881404 | 1.79E-07 | 6.68E-05 | 0.000867836 |
| HTN | monocyte | *LINC01143* | -18.01958328 | 0.024183122 | 0.044094893 | 1 |
| HTN | monocyte | *LINC00211* | 2.093530239 | 0.008357027 | 0.025458066 | 1 |
| HTN | monocyte | *LRRC37A2* | -2.557819155 | 0.004441253 | 0.023595115 | 1 |
| HTN | monocyte | *LINC00707* | 3.222858765 | 0.003432144 | 0.023595115 | 1 |
| HTN | monocyte | *TXNDC5* | 1.844069929 | 0.003459122 | 0.023595115 | 1 |
| HTN | monocyte | *LINC01119* | -2.126452127 | 0.006785359 | 0.02405092 | 1 |
| HTN | monocyte | *C1orf226* | -4.037519406 | 0.003300536 | 0.023595115 | 1 |
| HTN | monocyte | *LILRA2* | 0.474322424 | 0.006039935 | 0.023595115 | 1 |
| HTN | monocyte | *ISY1* | 1.585247384 | 0.015512079 | 0.03334719 | 1 |
| HTN | monocyte | *PNMA2* | 1.107093441 | 0.012351768 | 0.029302872 | 1 |
| HTN | monocyte | *KRBOX1* | -20.97250658 | 0.004892276 | 0.023595115 | 1 |
| HTN | monocyte | *ARHGEF25* | -1.319613067 | 0.021136281 | 0.040248522 | 1 |
| HTN | monocyte | *LINC00886* | -0.838677216 | 0.012110657 | 0.029032068 | 1 |
| HTN | monocyte | *LINC00877* | 0.267652881 | 0.003242301 | 0.023595115 | 1 |
| HTN | monocyte | *INMT* | -1.160809809 | 0.003869658 | 0.023595115 | 1 |
| HTN | monocyte | *ATP5O* | -1.764439058 | 0.00239968 | 0.023595115 | 1 |
| HTN | monocyte | *C8orf58* | -2.440674077 | 0.006147482 | 0.023595115 | 1 |
| HTN | monocyte | *HOGA1* | -5.806907897 | 0.010652901 | 0.027588558 | 1 |
| HTN | monocyte | *LINC00969* | -1.423622119 | 0.01289422 | 0.030176778 | 1 |
| HTN | monocyte | *DECR2* | -1.909882723 | 0.005023209 | 0.023595115 | 1 |
| HTN | monocyte | *ZNF709* | 6.16580704 | 0.010556121 | 0.02752821 | 1 |
| HTN | monocyte | *ERVW-1* | -4.572268665 | 0.00469983 | 0.023595115 | 1 |
| HTN | monocyte | *MRPL33* | -1.669253651 | 0.017537667 | 0.03544055 | 1 |
| HTN | monocyte | *STON1* | -2.106619656 | 0.007173398 | 0.024238452 | 1 |
| HTN | monocyte | *VSIG8* | -2.909245529 | 0.001254953 | 0.023595115 | 1 |
| HTN | monocyte | *FGF14-IT1* | -4.028526704 | 0.00509566 | 0.023595115 | 1 |
| HTN | monocyte | *PALM2* | -1.173366758 | 0.005182109 | 0.023595115 | 1 |
| HTN | monocyte | *NBPF15* | -1.379463905 | 0.006191246 | 0.023616295 | 1 |
| HTN | monocyte | *WFDC6* | -14.3345423 | 9.31E-05 | 0.006642646 | 0.451699934 |
| HTN | monocyte | *UPK3B* | -1.52550283 | 0.005452913 | 0.023595115 | 1 |
| HTN | monocyte | *LEFTY1* | -32.7120523 | 0.000363602 | 0.01501035 | 1 |
| HTN | monocyte | *TTC4* | -1.578218505 | 0.008395378 | 0.025487024 | 1 |
| HTN | monocyte | *APOBEC3D* | -7.434585703 | 0.00598491 | 0.023595115 | 1 |
| HTN | monocyte | *TMEM199* | -1.775117942 | 0.002808742 | 0.023595115 | 1 |
| HTN | monocyte | *LILRA6* | 0.720962765 | 0.001729456 | 0.023595115 | 1 |
| HTN | monocyte | *CCDC13* | 0.990520161 | 0.006360529 | 0.023776031 | 1 |
| HTN | monocyte | *C4A* | -3.77109661 | 0.028969425 | 0.049738022 | 1 |
| HTN | monocyte | *RAB30-AS1* | -1.604129188 | 0.003668146 | 0.023595115 | 1 |
| HTN | monocyte | *LINC00926* | -2.095278945 | 0.004638603 | 0.023595115 | 1 |
| HTN | monocyte | *DPH6-AS1* | -0.99309483 | 0.006156666 | 0.023595115 | 1 |
| HTN | monocyte | *CDK11B* | 4.024474222 | 0.014415393 | 0.032054761 | 1 |
| HTN | monocyte | *ABHD14A* | -1.690003719 | 0.006511677 | 0.023864166 | 1 |
| HTN | monocyte | *SRP14-AS1* | -1.205093681 | 0.001889648 | 0.023595115 | 1 |
| HTN | monocyte | *GDNF-AS1* | -2.342113403 | 0.005317921 | 0.023595115 | 1 |
| HTN | monocyte | *CCDC153* | 2.496442372 | 0.008899249 | 0.02599588 | 1 |
| HTN | monocyte | *C5orf17* | -1.128254482 | 0.005904356 | 0.023595115 | 1 |
| HTN | monocyte | *LINC00939* | 2.547480663 | 0.003630675 | 0.023595115 | 1 |

| | | | | | | |
|---|---|---|---|---|---|---|
| HTN | monocyte | *LINC01060* | -1.103214187 | 0.011545916 | 0.028509706 | 1 |
| HTN | monocyte | *LINC00942* | 9.619480027 | 0.014197852 | 0.031826319 | 1 |
| HTN | monocyte | *PDCD6* | -2.392734377 | 0.023989806 | 0.043824751 | 1 |
| HTN | monocyte | *GOLGA8K* | -2.855153869 | 0.018268053 | 0.036356273 | 1 |
| HTN | monocyte | *TMEM158* | -5.591370129 | 0.006017535 | 0.023595115 | 1 |
| HTN | monocyte | *DNAH10OS* | -12.67036284 | 0.008957121 | 0.026057755 | 1 |
| HTN | monocyte | *PCDHA10* | 1.620023653 | 0.000391961 | 0.01501035 | 1 |
| HTN | monocyte | *SMIM20* | -1.210066249 | 0.005155251 | 0.023595115 | 1 |
| HTN | monocyte | *GLI4* | -3.128417794 | 0.022074009 | 0.041464612 | 1 |
| HTN | monocyte | *SNHG18* | -1.529449636 | 0.004355651 | 0.023595115 | 1 |
| HTN | monocyte | *ALG1L2* | -2.195157508 | 0.007244846 | 0.024377248 | 1 |
| HTN | monocyte | *LINC01094* | -1.511758807 | 0.00533203 | 0.023595115 | 1 |
| HTN | monocyte | *PCDHGB6* | 1.544041344 | 0.006718808 | 0.024005639 | 1 |
| HTN | monocyte | *SLC10A5* | 2.319251652 | 0.024529599 | 0.044413403 | 1 |
| HTN | monocyte | *UTP14C* | -1.594978989 | 0.009709041 | 0.02671918 | 1 |
| HTN | monocyte | *PCDHGB2* | -7.404102645 | 0.002682083 | 0.023595115 | 1 |
| HTN | monocyte | *LYN* | 0.399204599 | 0.002438157 | 0.023595115 | 1 |
| HTN | monocyte | *SIGLEC14* | -3.691076694 | 0.018003158 | 0.035970978 | 1 |
| HTN | monocyte | *HMBS* | 6.394098511 | 3.32E-05 | 0.003221349 | 0.161067463 |
| HTN | monocyte | *AC093668.2* | -1.720438146 | 0.008198647 | 0.025373581 | 1 |
| HTN | monocyte | *CLEC12B* | 0.494516706 | 0.002148415 | 0.023595115 | 1 |
| HTN | monocyte | *KIAA1147* | -1.44718466 | 0.013066056 | 0.030344279 | 1 |
| HTN | monocyte | *ZBED6* | -2.940878933 | 0.003532724 | 0.023595115 | 1 |
| HTN | monocyte | *CNPY2* | -1.755271241 | 0.009123206 | 0.026216909 | 1 |
| HTN | monocyte | *LINC00592* | -4.758236159 | 0.026249265 | 0.046482276 | 1 |
| HTN | monocyte | *RTEL1* | 1.62881599 | 0.005012317 | 0.023595115 | 1 |
| HTN | monocyte | *LINC00640* | 1.452788689 | 0.002499265 | 0.023595115 | 1 |
| HTN | monocyte | *HIF1A-AS2* | -1.773685586 | 0.010484707 | 0.027465914 | 1 |
| HTN | monocyte | *SLC25A21-AS1* | -1.523038945 | 0.007947984 | 0.025074621 | 1 |
| HTN | monocyte | *MC1R* | 6.499291977 | 0.002515834 | 0.023595115 | 1 |
| HTN | monocyte | *LINC00639* | -1.052428846 | 0.002912982 | 0.023595115 | 1 |
| HTN | monocyte | *LINC00924* | -1.165142498 | 0.005421602 | 0.023595115 | 1 |
| HTN | monocyte | *AC083843.1* | 8.706309388 | 0.00347401 | 0.023595115 | 1 |
| HTN | monocyte | *PWRN1* | -1.975320377 | 0.000914938 | 0.020626325 | 1 |
| HTN | monocyte | *C19orf84* | -15.80654516 | 0.013759475 | 0.031249874 | 1 |
| HTN | monocyte | *LINC00667* | -1.273561952 | 0.02681575 | 0.047208619 | 1 |
| HTN | monocyte | *LINC00672* | -2.902962836 | 0.005734147 | 0.023595115 | 1 |
| HTN | monocyte | *LINC00909* | -1.881682577 | 0.00876577 | 0.025855025 | 1 |
| HTN | monocyte | *DYNLL2* | -3.273383492 | 0.010570397 | 0.02752821 | 1 |
| HTN | monocyte | *LINC00663* | -1.164542189 | 0.013554461 | 0.030925032 | 1 |
| HTN | monocyte | *S1PR2* | -2.128092513 | 0.006909433 | 0.024148823 | 1 |
| HTN | monocyte | *NDUFA7* | -8.246892091 | 0.005506396 | 0.023595115 | 1 |
| HTN | monocyte | *ZNF728* | -2.62105754 | 0.013799739 | 0.031317275 | 1 |
| HTN | monocyte | *FBXO17* | -0.852442819 | 0.009627522 | 0.02668844 | 1 |
| HTN | monocyte | *SNHG8* | -1.547817462 | 0.017609408 | 0.035555909 | 1 |
| HTN | monocyte | *AP001172.2* | 1.88252929 | 0.006122796 | 0.023595115 | 1 |
| HTN | monocyte | *HCG17* | -1.394823866 | 0.007268487 | 0.024429382 | 1 |
| HTN | monocyte | *YTHDF3-AS1* | -3.700078443 | 0.004579192 | 0.023595115 | 1 |
| HTN | monocyte | *MILR1* | 0.924876616 | 0.002480617 | 0.023595115 | 1 |
| HTN | monocyte | *GTF2H5* | -1.739751355 | 0.009666577 | 0.02668844 | 1 |

| | | | | | | |
|---|---|---|---|---|---|---|
| HTN | monocyte | *FGF14-AS2* | -2.11486296 | 0.006335386 | 0.02374425 | 1 |
| HTN | monocyte | *CASC15* | -1.50918903 | 0.009763624 | 0.02671918 | 1 |
| HTN | monocyte | *GRIN2B* | -0.65679657 | 0.026514583 | 0.04684951 | 1 |
| HTN | monocyte | *PAXIP1-AS1* | -1.858850574 | 0.008640327 | 0.025830477 | 1 |
| HTN | nk_cell | *NFYA* | -2.883456359 | 7.04E-06 | 2.20E-05 | 0.034162655 |
| HTN | nk_cell | *ENPP4* | -1.711710004 | 0.000423912 | 0.000697938 | 1 |
| HTN | nk_cell | *SEMA3F* | -3.886767321 | 3.31E-08 | 1.91E-06 | 0.000160655 |
| HTN | nk_cell | *KRIT1* | -2.003094892 | 3.22E-05 | 7.44E-05 | 0.156145692 |
| HTN | nk_cell | *RAD52* | -2.527436461 | 3.69E-06 | 1.38E-05 | 0.017893504 |
| HTN | nk_cell | *HS3ST1* | -1.852147804 | 0.007251667 | 0.009655622 | 1 |
| HTN | nk_cell | *AOC1* | -2.482893453 | 5.28E-06 | 1.80E-05 | 0.025628875 |
| HTN | nk_cell | *SNX11* | -1.145449348 | 9.75E-05 | 0.000188689 | 0.47304216 |
| HTN | nk_cell | *TMEM176A* | -3.16920854 | 1.41E-05 | 3.82E-05 | 0.068345951 |
| HTN | nk_cell | *M6PR* | -2.554134387 | 0.003091234 | 0.004352486 | 1 |
| HTN | nk_cell | *CYP26B1* | -1.273960484 | 2.72E-06 | 1.11E-05 | 0.01318759 |
| HTN | nk_cell | *ALS2* | -2.155079118 | 1.76E-05 | 4.58E-05 | 0.08560381 |
| HTN | nk_cell | *CD38* | 0.858650907 | 2.95E-07 | 4.13E-06 | 0.001430997 |
| HTN | nk_cell | *RBM6* | -2.019133319 | 1.41E-09 | 2.27E-07 | 6.82E-06 |
| HTN | nk_cell | *RECQL* | -2.583089789 | 0.005630055 | 0.00760073 | 1 |
| HTN | nk_cell | *ARHGAP33* | -3.086427142 | 0.000103648 | 0.000199327 | 0.502901868 |
| HTN | nk_cell | *SLC22A16* | 0.988484275 | 0.001513783 | 0.002235883 | 1 |
| HTN | nk_cell | *CDC27* | -1.523721725 | 0.000776622 | 0.001215538 | 1 |
| HTN | nk_cell | *POLR2J* | 3.220231315 | 2.74E-07 | 4.07E-06 | 0.001330071 |
| HTN | nk_cell | *MEOX1* | -1.783026505 | 4.11E-07 | 4.24E-06 | 0.001992365 |
| HTN | nk_cell | *LIG3* | -2.066154905 | 8.47E-06 | 2.55E-05 | 0.041076662 |
| HTN | nk_cell | *CIAPIN1* | 4.016775801 | 0.000694157 | 0.001094232 | 1 |
| HTN | nk_cell | *PON1* | -6.023171306 | 1.54E-06 | 7.63E-06 | 0.00748019 |
| HTN | nk_cell | *GCFC2* | -2.096581819 | 0.000104402 | 0.0002003 | 0.506557785 |
| HTN | nk_cell | *RHBDD2* | -1.673767433 | 2.70E-05 | 6.42E-05 | 0.130913194 |
| HTN | nk_cell | *PDK2* | -1.713707081 | 8.60E-05 | 0.000169095 | 0.417157298 |
| HTN | nk_cell | *GGNBP2* | -2.230823051 | 1.76E-05 | 4.57E-05 | 0.085214083 |
| HTN | nk_cell | *ITGA2B* | -1.426493714 | 4.79E-07 | 4.32E-06 | 0.002322573 |
| HTN | nk_cell | *TMEM98* | -1.59799299 | 1.48E-05 | 3.97E-05 | 0.071603331 |
| HTN | nk_cell | *ABCC8* | -1.873756073 | 4.22E-05 | 9.29E-05 | 0.204752983 |
| HTN | nk_cell | *ZNF263* | -2.38806938 | 0.018255503 | 0.022988762 | 1 |
| HTN | nk_cell | *SPATA20* | -1.981322458 | 0.010261061 | 0.013397919 | 1 |
| HTN | nk_cell | *BAIAP2L1* | -1.834143968 | 1.49E-06 | 7.51E-06 | 0.007242876 |
| HTN | nk_cell | *AGK* | 2.968505022 | 2.33E-05 | 5.72E-05 | 0.113069556 |
| HTN | nk_cell | *FARP2* | -1.62487476 | 8.22E-06 | 2.50E-05 | 0.039864141 |
| HTN | nk_cell | *USH1C* | -12.85025738 | 1.28E-06 | 6.85E-06 | 0.006223393 |
| HTN | nk_cell | *IFRD1* | -1.380910756 | 7.62E-05 | 0.000152341 | 0.369579098 |
| HTN | nk_cell | *VPS41* | -1.961625581 | 3.14E-05 | 7.32E-05 | 0.152540751 |
| HTN | nk_cell | *ELAC2* | -2.266365879 | 3.27E-06 | 1.27E-05 | 0.015876363 |
| HTN | nk_cell | *SCIN* | -1.842297068 | 1.43E-07 | 3.91E-06 | 0.000694957 |
| HTN | nk_cell | *CCDC124* | -1.793015636 | 0.003855562 | 0.005332721 | 1 |
| HTN | nk_cell | *CEACAM21* | 0.361602908 | 1.98E-07 | 4.07E-06 | 0.000962244 |
| HTN | nk_cell | *CD79B* | 1.661725421 | 0.000869504 | 0.00134615 | 1 |
| HTN | nk_cell | *ST7L* | -2.267406328 | 2.08E-06 | 9.19E-06 | 0.010092594 |
| HTN | nk_cell | *PAX6* | 1.869992475 | 0.00020978 | 0.000372022 | 1 |
| HTN | nk_cell | *PIGQ* | -1.640147519 | 0.012442632 | 0.016032224 | 1 |

| | | | | | | |
|---|---|---|---|---|---|---|
| HTN | nk_cell | *FMO3* | -1.314960172 | 1.03E-06 | 6.04E-06 | 0.005000189 |
| HTN | nk_cell | *PSMB1* | 3.048124383 | 9.55E-05 | 0.000185198 | 0.463550362 |
| HTN | nk_cell | *NADK* | -2.165119863 | 3.90E-07 | 4.24E-06 | 0.00189141 |
| HTN | nk_cell | *DLEC1* | -1.928194738 | 3.67E-07 | 4.24E-06 | 0.00177945 |
| HTN | nk_cell | *CYB561* | -1.849121673 | 1.31E-06 | 6.91E-06 | 0.006348916 |
| HTN | nk_cell | *SS18L2* | -2.748592582 | 9.34E-06 | 2.75E-05 | 0.045319284 |
| HTN | nk_cell | *MGST1* | -0.844231693 | 0.000562276 | 0.000903067 | 1 |
| HTN | nk_cell | *MMP25* | 1.491694653 | 7.50E-05 | 0.000150417 | 0.363707962 |
| HTN | nk_cell | *IL32* | 0.250284855 | 2.83E-07 | 4.07E-06 | 0.001370771 |
| HTN | nk_cell | *PKD1* | -1.660075525 | 1.61E-06 | 7.81E-06 | 0.007814774 |
| HTN | nk_cell | *MAPK8IP2* | 6.156934246 | 9.49E-05 | 0.000184314 | 0.46060026 |
| HTN | nk_cell | *MED24* | -2.944064229 | 0.000165942 | 0.000302916 | 0.805151281 |
| HTN | nk_cell | *TRAF3IP3* | 0.285022131 | 1.53E-07 | 4.02E-06 | 0.000743101 |
| HTN | nk_cell | *POMT2* | -1.087630582 | 3.29E-06 | 1.27E-05 | 0.015970414 |
| HTN | nk_cell | *MLXIPL* | -1.160502855 | 1.62E-07 | 4.07E-06 | 0.000787389 |
| HTN | nk_cell | *DYRK4* | -1.595696555 | 5.83E-05 | 0.000121163 | 0.282920093 |
| HTN | nk_cell | *NCAPD2* | 1.21782097 | 0.003339553 | 0.0046696 | 1 |
| HTN | nk_cell | *FUZ* | 4.946002686 | 0.000123373 | 0.000232468 | 0.59860445 |
| HTN | nk_cell | *SLC6A13* | 1.539388352 | 0.027753666 | 0.034186542 | 1 |
| HTN | nk_cell | *LRRC23* | -2.412064418 | 1.48E-05 | 3.98E-05 | 0.071866349 |
| HTN | nk_cell | *CLCN6* | -3.322131169 | 3.77E-05 | 8.47E-05 | 0.183038857 |
| HTN | nk_cell | *SLC6A7* | -32.70950854 | 1.34E-06 | 7.05E-06 | 0.006524725 |
| HTN | nk_cell | *TSPAN9* | -1.756957883 | 3.71E-05 | 8.34E-05 | 0.179990455 |
| HTN | nk_cell | *UTP18* | -2.133381465 | 0.000321352 | 0.000543844 | 1 |
| HTN | nk_cell | *TTC19* | -2.093616272 | 0.000241564 | 0.00042252 | 1 |
| HTN | nk_cell | *LARS2* | -2.375376755 | 0.001295481 | 0.00193763 | 1 |
| HTN | nk_cell | *DCN* | -1.399535019 | 5.92E-06 | 1.95E-05 | 0.028700257 |
| HTN | nk_cell | *PPP5C* | 2.487941821 | 0.006736064 | 0.00900865 | 1 |
| HTN | nk_cell | *CEP68* | -1.211490195 | 0.002570773 | 0.003666488 | 1 |
| HTN | nk_cell | *ERCC1* | -2.204083478 | 0.000622358 | 0.000990709 | 1 |
| HTN | nk_cell | *CD22* | 0.543912396 | 0.005309048 | 0.007187361 | 1 |
| HTN | nk_cell | *MAP4K5* | -2.043698049 | 0.00039656 | 0.000659167 | 1 |
| HTN | nk_cell | *EHD3* | -3.817877085 | 1.73E-06 | 8.16E-06 | 0.008396573 |
| HTN | nk_cell | *MAN2B2* | -2.608586228 | 0.000401322 | 0.00066526 | 1 |
| HTN | nk_cell | *SLC25A39* | 2.750919036 | 1.94E-05 | 4.92E-05 | 0.093907323 |
| HTN | nk_cell | *RNF14* | -2.500920873 | 5.64E-05 | 0.000117965 | 0.273478802 |
| HTN | nk_cell | *DDX11* | -1.433384321 | 3.72E-07 | 4.24E-06 | 0.001807013 |
| HTN | nk_cell | *TACC3* | -2.992903767 | 0.000178091 | 0.000322063 | 0.864096241 |
| HTN | nk_cell | *ZC3H3* | -1.746789112 | 0.001803361 | 0.002621302 | 1 |
| HTN | nk_cell | *MDH1* | -2.169615553 | 0.000244189 | 0.000426036 | 1 |
| HTN | nk_cell | *MTMR11* | -0.978303788 | 0.001145278 | 0.001732198 | 1 |
| HTN | nk_cell | *COX15* | -2.144837425 | 8.24E-06 | 2.50E-05 | 0.039993051 |
| HTN | nk_cell | *CCDC88C* | 0.383646739 | 6.83E-07 | 4.83E-06 | 0.003314917 |
| HTN | nk_cell | *CHDH* | -3.865004299 | 1.65E-05 | 4.35E-05 | 0.080094631 |
| HTN | nk_cell | *CLCA4* | -5.263155267 | 5.67E-07 | 4.50E-06 | 0.002753288 |
| HTN | nk_cell | *RUFY3* | -1.818718096 | 0.000161173 | 0.000294877 | 0.782013218 |
| HTN | nk_cell | *SLC11A1* | -1.037907781 | 1.23E-07 | 3.73E-06 | 0.00059642 |
| HTN | nk_cell | *WWTR1* | -1.641152144 | 7.84E-06 | 2.41E-05 | 0.038060633 |
| HTN | nk_cell | *AGPS* | -1.664258833 | 0.004678331 | 0.006392357 | 1 |
| HTN | nk_cell | *TTC27* | -1.996839294 | 2.46E-05 | 5.96E-05 | 0.119203398 |

| | | | | | | |
|---|---|---|---|---|---|---|
| HTN | nk_cell | *VSIG2* | -1.55478801 | 1.41E-06 | 7.26E-06 | 0.006864829 |
| HTN | nk_cell | *HGF* | -2.935748467 | 0.001306366 | 0.001951595 | 1 |
| HTN | nk_cell | *SLC7A9* | -1.081598562 | 0.000490197 | 0.000797063 | 1 |
| HTN | nk_cell | *OSBPL5* | 1.829247724 | 6.11E-07 | 4.55E-06 | 0.002963388 |
| HTN | nk_cell | *CPS1* | -1.902482889 | 0.000112579 | 0.000214209 | 0.546233707 |
| HTN | nk_cell | *NLRP2* | 0.94066349 | 2.08E-05 | 5.21E-05 | 0.10104141 |
| HTN | nk_cell | *RNH1* | -1.390564581 | 0.017437751 | 0.022044806 | 1 |
| HTN | nk_cell | *GLRX2* | -3.11015 | 3.64E-06 | 1.37E-05 | 0.017676097 |
| HTN | nk_cell | *ABCC2* | -2.930592788 | 3.49E-06 | 1.33E-05 | 0.016928505 |
| HTN | nk_cell | *UBR2* | -1.632767448 | 0.000816019 | 0.001271051 | 1 |
| HTN | nk_cell | *RRAGD* | -1.616683006 | 1.25E-06 | 6.78E-06 | 0.006072769 |
| HTN | nk_cell | *HSF2* | -2.309675233 | 0.000229072 | 0.000403286 | 1 |
| HTN | nk_cell | *TOMM34* | 1.701478804 | 2.78E-07 | 4.07E-06 | 0.001350229 |
| HTN | nk_cell | *SEC63* | -2.009140598 | 0.000571187 | 0.000916468 | 1 |
| HTN | nk_cell | *FAS* | -2.681985189 | 0.00147544 | 0.002181907 | 1 |
| HTN | nk_cell | *RNASET2* | -2.656531941 | 1.06E-06 | 6.13E-06 | 0.005125956 |
| HTN | nk_cell | *AGPAT4* | -2.433533748 | 5.35E-05 | 0.000113013 | 0.259477207 |
| HTN | nk_cell | *BTN3A1* | 1.389161068 | 0.00010067 | 0.000194215 | 0.488450544 |
| HTN | nk_cell | *MIPEP* | -2.327568023 | 4.09E-06 | 1.49E-05 | 0.019867574 |
| HTN | nk_cell | *INSRR* | 4.582750183 | 0.00019588 | 0.00035031 | 0.950408278 |
| HTN | nk_cell | *B4GALT7* | -1.79751502 | 3.24E-07 | 4.19E-06 | 0.001571183 |
| HTN | nk_cell | *SH2D2A* | 0.320704563 | 2.92E-07 | 4.12E-06 | 0.001416799 |
| HTN | nk_cell | *VRK2* | -1.753596947 | 1.47E-05 | 3.96E-05 | 0.071443391 |
| HTN | nk_cell | *VEZT* | -2.018924566 | 9.36E-05 | 0.000182576 | 0.454248775 |
| HTN | nk_cell | *BRD9* | -2.358055794 | 0.000207578 | 0.000369062 | 1 |
| HTN | nk_cell | *ANK1* | 1.649481446 | 3.03E-07 | 4.15E-06 | 0.001470422 |
| HTN | nk_cell | *RABEP1* | -2.03079964 | 0.001175249 | 0.001773658 | 1 |
| HTN | nk_cell | *BAK1* | 1.706336807 | 6.81E-06 | 2.15E-05 | 0.03305219 |
| HTN | nk_cell | *IKZF2* | 2.050353561 | 2.57E-06 | 1.07E-05 | 0.012486576 |
| HTN | nk_cell | *GRN* | -2.106854651 | 0.000198369 | 0.000354377 | 0.962487352 |
| HTN | nk_cell | *IFT88* | -2.038318665 | 4.94E-05 | 0.000106101 | 0.239789303 |
| HTN | nk_cell | *CHPF2* | 1.700688077 | 1.24E-06 | 6.75E-06 | 0.005998228 |
| HTN | nk_cell | *UBA6* | -1.66624084 | 0.025858461 | 0.031990121 | 1 |
| HTN | nk_cell | *ATP6V0A1* | -1.783036448 | 5.18E-05 | 0.000110065 | 0.251498018 |
| HTN | nk_cell | *PIAS1* | -1.680299164 | 0.000429915 | 0.000706621 | 1 |
| HTN | nk_cell | *ASTE1* | -2.686028453 | 4.05E-05 | 9.00E-05 | 0.196479032 |
| HTN | nk_cell | *MYOC* | -1.11417284 | 3.08E-06 | 1.21E-05 | 0.014922399 |
| HTN | nk_cell | *SH3YL1* | -2.858514868 | 0.000287272 | 0.000491656 | 1 |
| HTN | nk_cell | *VCL* | -2.204767917 | 0.000146362 | 0.000271051 | 0.710150023 |
| HTN | nk_cell | *TBC1D23* | -2.22358041 | 7.87E-05 | 0.000156706 | 0.381891371 |
| HTN | nk_cell | *MYOM2* | 3.103005538 | 1.62E-07 | 4.07E-06 | 0.00078606 |
| HTN | nk_cell | *TUBG2* | -1.628074058 | 7.51E-07 | 5.09E-06 | 0.003643818 |
| HTN | nk_cell | *NSUN2* | -2.499364001 | 0.000788291 | 0.001232212 | 1 |
| HTN | nk_cell | *MRI1* | -1.74112595 | 0.001173825 | 0.001772059 | 1 |
| HTN | nk_cell | *AGA* | -3.12590547 | 0.00036397 | 0.00060938 | 1 |
| HTN | nk_cell | *PI4K2B* | -8.071183791 | 0.000145918 | 0.000270433 | 0.707993973 |
| HTN | nk_cell | *BOD1L1* | -1.764150108 | 2.72E-05 | 6.46E-05 | 0.132048629 |
| HTN | nk_cell | *PHLPP2* | -1.917813156 | 5.94E-05 | 0.000123064 | 0.288214913 |
| HTN | nk_cell | *SPDL1* | 1.355925425 | 9.90E-06 | 2.88E-05 | 0.048055457 |
| HTN | nk_cell | *PQLC2* | -3.134322671 | 1.09E-06 | 6.23E-06 | 0.005294879 |

| | | | | | | |
|---|---|---|---|---|---|---|
| HTN | nk_cell | *CTNS* | -1.282000663 | 0.036254344 | 0.044153131 | 1 |
| HTN | nk_cell | *INPP4A* | 1.492472634 | 2.93E-05 | 6.90E-05 | 0.142361212 |
| HTN | nk_cell | *PSMA4* | 2.913156546 | 6.81E-06 | 2.15E-05 | 0.033061799 |
| HTN | nk_cell | *TDP1* | -3.266769075 | 0.001024643 | 0.001564369 | 1 |
| HTN | nk_cell | *SPATA7* | -1.958214209 | 2.08E-06 | 9.19E-06 | 0.010084147 |
| HTN | nk_cell | *MED17* | 3.469916296 | 0.001503902 | 0.002222641 | 1 |
| HTN | nk_cell | *CAPG* | -1.091292905 | 9.16E-06 | 2.71E-05 | 0.044440045 |
| HTN | nk_cell | *AP2S1* | -3.195796278 | 0.003896657 | 0.005380359 | 1 |
| HTN | nk_cell | *TRIT1* | -1.74427773 | 0.00045065 | 0.000738202 | 1 |
| HTN | nk_cell | *CTNNA1* | -1.639827477 | 1.19E-05 | 3.34E-05 | 0.057948787 |
| HTN | nk_cell | *CNTLN* | -1.626809308 | 1.69E-05 | 4.43E-05 | 0.082023344 |
| HTN | nk_cell | *YTHDC2* | -2.118326675 | 0.000852548 | 0.001324109 | 1 |
| HTN | nk_cell | *ARAP2* | 1.329142648 | 2.14E-07 | 4.07E-06 | 0.001037045 |
| HTN | nk_cell | *DTNBP1* | -2.084584262 | 0.013712208 | 0.01756379 | 1 |
| HTN | nk_cell | *ANO2* | -1.290605836 | 1.12E-06 | 6.33E-06 | 0.00542274 |
| HTN | nk_cell | *SNX29* | -1.829091506 | 0.000153649 | 0.000282314 | 0.74550564 |
| HTN | nk_cell | *UTS2* | 3.39845307 | 3.66E-05 | 8.27E-05 | 0.177677595 |
| HTN | nk_cell | *EPN3* | -3.688287212 | 3.38E-05 | 7.72E-05 | 0.164136176 |
| HTN | nk_cell | *RCN1* | -3.057462483 | 4.69E-05 | 0.000101491 | 0.227644093 |
| HTN | nk_cell | *JKAMP* | -2.408645241 | 8.71E-05 | 0.000171002 | 0.422544764 |
| HTN | nk_cell | *ARHGEF5* | -1.279009506 | 6.77E-06 | 2.15E-05 | 0.032863113 |
| HTN | nk_cell | *MCUR1* | -1.819276684 | 2.51E-06 | 1.05E-05 | 0.012195203 |
| HTN | nk_cell | *MAPK9* | -2.443194389 | 0.012386644 | 0.015967056 | 1 |
| HTN | nk_cell | *FAM160A2* | -2.265651391 | 3.37E-05 | 7.69E-05 | 0.163272783 |
| HTN | nk_cell | *CYBA* | 1.321516238 | 5.77E-07 | 4.54E-06 | 0.002798893 |
| HTN | nk_cell | *THOC3* | 3.302090573 | 3.93E-07 | 4.24E-06 | 0.001905809 |
| HTN | nk_cell | *FSTL4* | -1.345790214 | 2.02E-05 | 5.08E-05 | 0.098190523 |
| HTN | nk_cell | *MRTO4* | 3.560803079 | 0.00472166 | 0.006447929 | 1 |
| HTN | nk_cell | *LY75* | 1.28818789 | 2.98E-07 | 4.13E-06 | 0.001445665 |
| HTN | nk_cell | *SDCCAG8* | -0.804407961 | 0.01029411 | 0.013437455 | 1 |
| HTN | nk_cell | *KIF1B* | -1.36781214 | 0.000356697 | 0.000598028 | 1 |
| HTN | nk_cell | *TBC1D22A* | -1.526511875 | 0.012096976 | 0.015631032 | 1 |
| HTN | nk_cell | *GALC* | -2.479975861 | 2.51E-05 | 6.06E-05 | 0.12175434 |
| HTN | nk_cell | *FAM114A2* | -2.240537055 | 0.010346946 | 0.013499161 | 1 |
| HTN | nk_cell | *NPFFR2* | -1.705648323 | 3.69E-07 | 4.24E-06 | 0.00179027 |
| HTN | nk_cell | *PCGF2* | -1.463436844 | 0.000759093 | 0.001190022 | 1 |
| HTN | nk_cell | *MSH4* | -1.016858849 | 0.0090792 | 0.011909239 | 1 |
| HTN | nk_cell | *GDI2* | -2.167415792 | 0.000806328 | 0.001257166 | 1 |
| HTN | nk_cell | *PITHD1* | -1.360636418 | 1.79E-06 | 8.32E-06 | 0.008673312 |
| HTN | nk_cell | *LAMC2* | -1.532515951 | 0.000124138 | 0.000233818 | 0.602315424 |
| HTN | nk_cell | *RASGRF1* | -3.509812639 | 2.03E-05 | 5.10E-05 | 0.098439405 |
| HTN | nk_cell | *CROCC* | -1.34293152 | 0.003675721 | 0.005106248 | 1 |
| HTN | nk_cell | *FLYWCH1* | -1.233645662 | 0.029315514 | 0.036028083 | 1 |
| HTN | nk_cell | *ALDH18A1* | -2.369751135 | 1.06E-05 | 3.05E-05 | 0.051449045 |
| HTN | nk_cell | *DNAJC25* | -1.321921418 | 4.34E-07 | 4.25E-06 | 0.002105896 |
| HTN | nk_cell | *OGFR* | -2.256588422 | 0.00153322 | 0.002261003 | 1 |
| HTN | nk_cell | *GNA15* | -1.230903662 | 1.63E-05 | 4.29E-05 | 0.078882262 |
| HTN | nk_cell | *PIGV* | -3.256786365 | 3.24E-09 | 4.04E-07 | 1.57E-05 |
| HTN | nk_cell | *RIMBP2* | -1.490072964 | 5.19E-07 | 4.37E-06 | 0.002517763 |
| HTN | nk_cell | *PRDM6* | -2.091442516 | 1.61E-06 | 7.81E-06 | 0.007809833 |

| | | | | | | |
|---|---|---|---|---|---|---|
| HTN | nk_cell | *SPAG4* | -4.20525883 | 1.54E-06 | 7.63E-06 | 0.007488482 |
| HTN | nk_cell | *NCKAP1* | -1.719140321 | 2.92E-06 | 1.16E-05 | 0.014166154 |
| HTN | nk_cell | *LTK* | -8.499956747 | 4.29E-07 | 4.24E-06 | 0.002083908 |
| HTN | nk_cell | *AHRR* | -1.955822318 | 3.51E-07 | 4.24E-06 | 0.001703978 |
| HTN | nk_cell | *ADCK1* | -2.053823076 | 1.54E-06 | 7.63E-06 | 0.007466461 |
| HTN | nk_cell | *RNF4* | 2.850283913 | 0.000960637 | 0.001477341 | 1 |
| HTN | nk_cell | *CASP8* | 1.661803003 | 4.48E-07 | 4.28E-06 | 0.002175042 |
| HTN | nk_cell | *TM7SF3* | -2.081586708 | 0.005024841 | 0.006823546 | 1 |
| HTN | nk_cell | *CDON* | -0.652362701 | 1.44E-05 | 3.89E-05 | 0.069777178 |
| HTN | nk_cell | *TMEM161A* | -3.502722567 | 0.001016433 | 0.001553791 | 1 |
| HTN | nk_cell | *LPAR2* | 2.319161618 | 1.51E-07 | 4.00E-06 | 0.000731455 |
| HTN | nk_cell | *CNN2* | 1.159237802 | 6.03E-07 | 4.54E-06 | 0.002927295 |
| HTN | nk_cell | *ABCA7* | -2.991930436 | 0.00151377 | 0.002235883 | 1 |
| HTN | nk_cell | *BTBD1* | 2.865347759 | 0.000308906 | 0.000525346 | 1 |
| HTN | nk_cell | *FAR2* | -2.103787107 | 0.00076774 | 0.001202801 | 1 |
| HTN | nk_cell | *BCAS1* | -2.650419043 | 1.56E-06 | 7.65E-06 | 0.007573063 |
| HTN | nk_cell | *CHI3L2* | -3.448319862 | 3.68E-05 | 8.30E-05 | 0.178513495 |
| HTN | nk_cell | *PMS1* | -2.084136999 | 0.003279787 | 0.004592649 | 1 |
| HTN | nk_cell | *ANKRD44* | 1.053235293 | 2.35E-07 | 4.07E-06 | 0.001139522 |
| HTN | nk_cell | *ADAT1* | -1.653504246 | 0.000234671 | 0.000411947 | 1 |
| HTN | nk_cell | *MAP2K4* | -1.853805642 | 1.06E-05 | 3.05E-05 | 0.051588881 |
| HTN | nk_cell | *TMEM206* | -1.707421774 | 0.000179803 | 0.000324675 | 0.872401911 |
| HTN | nk_cell | *SEC61A2* | -2.040292001 | 2.46E-06 | 1.04E-05 | 0.011951826 |
| HTN | nk_cell | *TIE1* | -3.827766913 | 4.74E-07 | 4.32E-06 | 0.002300613 |
| HTN | nk_cell | *ZMYND12* | -0.754716835 | 0.004522368 | 0.006189712 | 1 |
| HTN | nk_cell | *SLC9A3* | -1.40197398 | 2.94E-05 | 6.91E-05 | 0.142665049 |
| HTN | nk_cell | *NGEF* | -3.413121948 | 1.03E-07 | 3.38E-06 | 0.000500148 |
| HTN | nk_cell | *SPI1* | -1.896873204 | 4.68E-06 | 1.64E-05 | 0.02268878 |
| HTN | nk_cell | *ZNRD1* | -2.232719657 | 0.000323683 | 0.000547027 | 1 |
| HTN | nk_cell | *ATXN3* | -1.926516085 | 2.73E-06 | 1.11E-05 | 0.013225139 |
| HTN | nk_cell | *GOLGA5* | -2.431149737 | 3.68E-05 | 8.30E-05 | 0.178627725 |
| HTN | nk_cell | *FGFR2* | -1.689285214 | 2.62E-07 | 4.07E-06 | 0.001272031 |
| HTN | nk_cell | *FECH* | -1.190102703 | 1.77E-06 | 8.28E-06 | 0.008598577 |
| HTN | nk_cell | *MYO9A* | -1.53686856 | 8.69E-06 | 2.60E-05 | 0.042169693 |
| HTN | nk_cell | *EVI5* | -1.598822899 | 2.98E-05 | 6.99E-05 | 0.144621915 |
| HTN | nk_cell | *STOML1* | -5.307438234 | 7.05E-06 | 2.20E-05 | 0.034196972 |
| HTN | nk_cell | *METTL22* | -1.49746425 | 7.84E-07 | 5.21E-06 | 0.003804862 |
| HTN | nk_cell | *TP53BP1* | -2.182258782 | 1.12E-05 | 3.17E-05 | 0.054153242 |
| HTN | nk_cell | *DHX8* | -2.601064694 | 2.15E-05 | 5.32E-05 | 0.104083386 |
| HTN | nk_cell | *ROGDI* | 6.784014957 | 0.00036415 | 0.000609471 | 1 |
| HTN | nk_cell | *CBFB* | -2.283268442 | 0.001368089 | 0.002032445 | 1 |
| HTN | nk_cell | *IFI35* | -2.657752218 | 0.000320984 | 0.000543411 | 1 |
| HTN | nk_cell | *HEATR6* | -1.735699927 | 4.06E-05 | 9.01E-05 | 0.196919941 |
| HTN | nk_cell | *POLR1A* | -2.7895064 | 1.31E-08 | 1.01E-06 | 6.37E-05 |
| HTN | nk_cell | *IP6K2* | -2.53280123 | 2.46E-05 | 5.97E-05 | 0.119567136 |
| HTN | nk_cell | *STON1-GTF2A* | -0.730364994 | 0.000110316 | 0.000210234 | 0.535255025 |
| HTN | nk_cell | *CYFIP1* | -1.994220268 | 1.61E-05 | 4.26E-05 | 0.078191587 |
| HTN | nk_cell | *IFT80* | -3.546360022 | 4.78E-07 | 4.32E-06 | 0.002319935 |
| HTN | nk_cell | *NUCKS1* | -1.720396179 | 0.019848892 | 0.024917678 | 1 |
| HTN | nk_cell | *CLEC2D* | 1.410893354 | 6.09E-07 | 4.55E-06 | 0.002954003 |

| | | | | | | |
|---|---|---|---|---|---|---|
| HTN | nk_cell | *DRD4* | -4.815279685 | 5.55E-06 | 1.86E-05 | 0.026921264 |
| HTN | nk_cell | *PLA2G10* | 3.214494101 | 3.47E-07 | 4.24E-06 | 0.001685827 |
| HTN | nk_cell | *NEDD4* | -1.540362715 | 0.003272277 | 0.004583455 | 1 |
| HTN | nk_cell | *GNB5* | -2.342245781 | 3.30E-06 | 1.28E-05 | 0.016028114 |
| HTN | nk_cell | *LRP6* | -1.886685948 | 5.84E-06 | 1.93E-05 | 0.028316061 |
| HTN | nk_cell | *CLTCL1* | -2.455227994 | 1.83E-05 | 4.70E-05 | 0.088590619 |
| HTN | nk_cell | *ZXDC* | -2.090216648 | 0.000886103 | 0.0013701 | 1 |
| HTN | nk_cell | *WIPI1* | 2.082515318 | 0.001817773 | 0.002641459 | 1 |
| HTN | nk_cell | *GBA2* | -2.638193279 | 1.51E-05 | 4.04E-05 | 0.073036173 |
| HTN | nk_cell | *NDST1* | -1.323980918 | 5.26E-06 | 1.80E-05 | 0.025545531 |
| HTN | nk_cell | *ASNS* | -1.908105878 | 0.000136143 | 0.000254064 | 0.660567494 |
| HTN | nk_cell | *AP3M2* | -3.220761226 | 0.000310728 | 0.000528259 | 1 |
| HTN | nk_cell | *ST6GALNAC2* | -1.57684194 | 1.04E-06 | 6.09E-06 | 0.005069049 |
| HTN | nk_cell | *EIF2B3* | -2.090956481 | 0.000105448 | 0.000201908 | 0.511634931 |
| HTN | nk_cell | *MAP4K4* | -2.001161243 | 0.000527717 | 0.000852168 | 1 |
| HTN | nk_cell | *SNX13* | -2.035264382 | 0.0002407 | 0.000421616 | 1 |
| HTN | nk_cell | *RPS6KA2* | -1.791653128 | 0.000316487 | 0.000537247 | 1 |
| HTN | nk_cell | *ING3* | -2.224834816 | 3.88E-06 | 1.43E-05 | 0.018819254 |
| HTN | nk_cell | *VASH1* | -1.935815067 | 0.008592982 | 0.011314287 | 1 |
| HTN | nk_cell | *TRIB2* | 2.396549347 | 8.52E-05 | 0.000168051 | 0.413573733 |
| HTN | nk_cell | *CPSF1* | -2.476080868 | 5.34E-06 | 1.81E-05 | 0.025917175 |
| HTN | nk_cell | *CYBRD1* | -1.598345158 | 1.25E-05 | 3.47E-05 | 0.060836519 |
| HTN | nk_cell | *RDH11* | -2.576187417 | 0.001916401 | 0.002776464 | 1 |
| HTN | nk_cell | *EPN2* | -1.998137657 | 8.42E-07 | 5.42E-06 | 0.004083647 |
| HTN | nk_cell | *ALDH3A2* | -1.541073727 | 4.14E-05 | 9.14E-05 | 0.200713525 |
| HTN | nk_cell | *AFF4* | -1.817512415 | 5.84E-05 | 0.000121273 | 0.283415091 |
| HTN | nk_cell | *TRHDE* | -1.41159388 | 3.70E-07 | 4.24E-06 | 0.001795301 |
| HTN | nk_cell | *FCGR2B* | -2.622057282 | 3.30E-06 | 1.28E-05 | 0.015997348 |
| HTN | nk_cell | *CRMP1* | -1.680720154 | 1.46E-06 | 7.39E-06 | 0.00709243 |
| HTN | nk_cell | *NDE1* | -2.386564931 | 7.16E-05 | 0.000144511 | 0.347403949 |
| HTN | nk_cell | *PVR* | -3.660197458 | 5.79E-07 | 4.54E-06 | 0.002810675 |
| HTN | nk_cell | *SCARB1* | -1.978751196 | 4.83E-06 | 1.68E-05 | 0.023413702 |
| HTN | nk_cell | *MOV10L1* | -1.209568984 | 1.06E-06 | 6.14E-06 | 0.005143473 |
| HTN | nk_cell | *ALPK1* | -2.007116952 | 0.00046496 | 0.000759336 | 1 |
| HTN | nk_cell | *LLGL2* | 0.711242926 | 2.57E-07 | 4.07E-06 | 0.001246006 |
| HTN | nk_cell | *ADAM11* | -1.844059466 | 1.90E-06 | 8.69E-06 | 0.009240707 |
| HTN | nk_cell | *CD5L* | -2.270628519 | 0.005603048 | 0.007566376 | 1 |
| HTN | nk_cell | *PPP2R2C* | -1.764342213 | 4.35E-07 | 4.26E-06 | 0.002112554 |
| HTN | nk_cell | *ZNF532* | -1.983559033 | 2.13E-05 | 5.31E-05 | 0.103539867 |
| HTN | nk_cell | *ZZEF1* | -1.752073296 | 0.009024853 | 0.011841154 | 1 |
| HTN | nk_cell | *TACR2* | -10.48322541 | 0.004139359 | 0.005691179 | 1 |
| HTN | nk_cell | *TTC38* | 1.454978466 | 7.33E-07 | 5.01E-06 | 0.003555583 |
| HTN | nk_cell | *GRAMD4* | -2.431611401 | 0.002481825 | 0.003550063 | 1 |
| HTN | nk_cell | *VPS9D1* | 2.698112735 | 0.021143638 | 0.026481397 | 1 |
| HTN | nk_cell | *FNDC3B* | -1.543996374 | 8.78E-05 | 0.000172258 | 0.425993159 |
| HTN | nk_cell | *MOCOS* | -1.392490143 | 6.16E-07 | 4.56E-06 | 0.002991106 |
| HTN | nk_cell | *SEC31B* | -2.184399649 | 5.69E-05 | 0.000118814 | 0.276005437 |
| HTN | nk_cell | *KIFAP3* | -2.069848621 | 9.72E-05 | 0.000188275 | 0.471817932 |
| HTN | nk_cell | *MKRN2* | -2.194564728 | 0.000165184 | 0.000301759 | 0.801471158 |
| HTN | nk_cell | *MCM6* | -2.469228645 | 0.000198137 | 0.000354092 | 0.961359718 |

| | | | | | | |
|---|---|---|---|---|---|---|
| HTN | nk_cell | *MLH1* | -1.858088265 | 0.025009833 | 0.03103522 | 1 |
| HTN | nk_cell | *FMO4* | -1.998917769 | 8.90E-05 | 0.00017447 | 0.431813138 |
| HTN | nk_cell | *RGS11* | -1.759139983 | 9.04E-05 | 0.000177004 | 0.438439898 |
| HTN | nk_cell | *SPAG5* | 2.860203943 | 4.12E-06 | 1.49E-05 | 0.019994549 |
| HTN | nk_cell | *ANKRD13A* | -1.732849501 | 6.45E-06 | 2.07E-05 | 0.031306844 |
| HTN | nk_cell | *PAG1* | 2.769970616 | 0.000346885 | 0.000583797 | 1 |
| HTN | nk_cell | *GPATCH1* | -2.69060524 | 0.000239232 | 0.000419498 | 1 |
| HTN | nk_cell | *ICAM3* | 0.518132 | 2.66E-07 | 4.07E-06 | 0.001291759 |
| HTN | nk_cell | *NT5C2* | -1.301636949 | 0.004570641 | 0.006254019 | 1 |
| HTN | nk_cell | *XAB2* | -3.638908621 | 1.26E-05 | 3.48E-05 | 0.061045818 |
| HTN | nk_cell | *IL4R* | -2.216592116 | 1.96E-05 | 4.98E-05 | 0.095295589 |
| HTN | nk_cell | *DYNC1I2* | -1.977730262 | 2.44E-05 | 5.94E-05 | 0.118617581 |
| HTN | nk_cell | *LRCH4* | -2.389861852 | 0.000304672 | 0.00051869 | 1 |
| HTN | nk_cell | *SIRT6* | -2.49162498 | 0.011402856 | 0.014789269 | 1 |
| HTN | nk_cell | *GPR137B* | -1.569735525 | 0.002673166 | 0.003800234 | 1 |
| HTN | nk_cell | *NAALAD2* | -0.799955347 | 0.011782087 | 0.015252585 | 1 |
| HTN | nk_cell | *AMPH* | 2.948712954 | 1.68E-07 | 4.07E-06 | 0.000812715 |
| HTN | nk_cell | *MCCC1* | -1.537004535 | 6.51E-06 | 2.09E-05 | 0.03157989 |
| HTN | nk_cell | *ACER3* | -1.445832425 | 8.59E-06 | 2.57E-05 | 0.041668426 |
| HTN | nk_cell | *PIK3C3* | -2.166197673 | 0.000610565 | 0.000974156 | 1 |
| HTN | nk_cell | *RBFOX1* | -1.104881594 | 0.003250696 | 0.004559019 | 1 |
| HTN | nk_cell | *PCM1* | -1.769413896 | 6.66E-05 | 0.000135755 | 0.322961127 |
| HTN | nk_cell | *MYH7B* | -1.620422102 | 1.97E-07 | 4.07E-06 | 0.000953783 |
| HTN | nk_cell | *CLUL1* | -2.039853886 | 1.52E-06 | 7.55E-06 | 0.007354169 |
| HTN | nk_cell | *FKBP7* | -2.268512713 | 1.06E-06 | 6.15E-06 | 0.00516323 |
| HTN | nk_cell | *SP140* | 0.741093829 | 7.45E-07 | 5.07E-06 | 0.003616798 |
| HTN | nk_cell | *SAR1A* | -2.218340318 | 1.03E-05 | 2.98E-05 | 0.050098656 |
| HTN | nk_cell | *RAPGEF3* | -0.815898725 | 0.006851382 | 0.009157826 | 1 |
| HTN | nk_cell | *FDFT1* | 3.570000224 | 5.77E-05 | 0.000120096 | 0.280183131 |
| HTN | nk_cell | *SCGN* | 2.92405735 | 2.57E-07 | 4.07E-06 | 0.001245194 |
| HTN | nk_cell | *PGM1* | -1.945997538 | 1.99E-06 | 8.96E-06 | 0.009655168 |
| HTN | nk_cell | *DDX1* | -2.356843452 | 0.000872601 | 0.001350514 | 1 |
| HTN | nk_cell | *STX7* | -1.764878453 | 6.01E-06 | 1.97E-05 | 0.029179014 |
| HTN | nk_cell | *RABL2B* | -3.209965587 | 0.00010608 | 0.000202958 | 0.514701287 |
| HTN | nk_cell | *DCT* | -1.196934318 | 2.18E-05 | 5.38E-05 | 0.105593659 |
| HTN | nk_cell | *SCTR* | -1.576313361 | 9.51E-07 | 5.82E-06 | 0.004612531 |
| HTN | nk_cell | *RIF1* | -1.829443846 | 1.71E-05 | 4.46E-05 | 0.08290735 |
| HTN | nk_cell | *KCNN2* | 2.054940383 | 1.26E-06 | 6.79E-06 | 0.006093363 |
| HTN | nk_cell | *PSEN1* | -1.972803359 | 0.000128253 | 0.000240822 | 0.622283953 |
| HTN | nk_cell | *MOK* | -1.495343241 | 2.31E-05 | 5.68E-05 | 0.112263553 |
| HTN | nk_cell | *STK17B* | 1.454950661 | 2.29E-07 | 4.07E-06 | 0.0011109 |
| HTN | nk_cell | *PCDHB4* | 2.084396521 | 3.19E-07 | 4.18E-06 | 0.001545695 |
| HTN | nk_cell | *CCNT2* | -2.395039118 | 0.00046443 | 0.000758725 | 1 |
| HTN | nk_cell | *SEMA5B* | -1.536919865 | 7.75E-06 | 2.39E-05 | 0.037605738 |
| HTN | nk_cell | *DOPEY1* | -1.748106905 | 0.000247335 | 0.000431059 | 1 |
| HTN | nk_cell | *LYRM2* | -2.518991752 | 9.12E-07 | 5.69E-06 | 0.004424857 |
| HTN | nk_cell | *ZCCHC6* | -1.858012247 | 0.012230582 | 0.015791055 | 1 |
| HTN | nk_cell | *ULK2* | -1.546476539 | 1.29E-06 | 6.86E-06 | 0.006244676 |
| HTN | nk_cell | *PIBF1* | -1.799680672 | 0.001740711 | 0.002536315 | 1 |
| HTN | nk_cell | *OXCT1* | -2.475155254 | 1.14E-05 | 3.24E-05 | 0.055494831 |

| | | | | | | |
|---|---|---|---|---|---|---|
| HTN | nk_cell | *SLC27A5* | 3.31961545 | 7.50E-07 | 5.09E-06 | 0.003638358 |
| HTN | nk_cell | *ZNF586* | -1.777900375 | 0.011864344 | 0.015342696 | 1 |
| HTN | nk_cell | *ZNF264* | -2.334975337 | 6.94E-06 | 2.18E-05 | 0.033651237 |
| HTN | nk_cell | *RPS5* | 2.206733912 | 0.001076192 | 0.001637405 | 1 |
| HTN | nk_cell | *CHMP2B* | -3.146497451 | 0.001702041 | 0.0024837 | 1 |
| HTN | nk_cell | *NOA1* | -3.478902104 | 2.59E-06 | 1.07E-05 | 0.012550824 |
| HTN | nk_cell | *WBP11* | 3.282113718 | 3.96E-05 | 8.81E-05 | 0.192236424 |
| HTN | nk_cell | *NCOA1* | -1.443198666 | 0.017684688 | 0.022316283 | 1 |
| HTN | nk_cell | *CD59* | -1.507240918 | 0.004494943 | 0.006153911 | 1 |
| HTN | nk_cell | *HACE1* | -1.765834139 | 3.34E-06 | 1.28E-05 | 0.016221622 |
| HTN | nk_cell | *SEH1L* | -2.062561143 | 6.35E-06 | 2.06E-05 | 0.030826853 |
| HTN | nk_cell | *OVGP1* | -2.165988367 | 9.99E-06 | 2.90E-05 | 0.048469225 |
| HTN | nk_cell | *SLC25A24* | -2.092443706 | 4.27E-05 | 9.37E-05 | 0.207238711 |
| HTN | nk_cell | *ZNF213* | -2.046161876 | 0.000107534 | 0.000205504 | 0.521755503 |
| HTN | nk_cell | *CPNE3* | -2.391634514 | 2.51E-05 | 6.05E-05 | 0.121590771 |
| HTN | nk_cell | *RRN3* | -2.035807239 | 1.23E-06 | 6.74E-06 | 0.005951858 |
| HTN | nk_cell | *DDHD2* | -1.512950362 | 0.001333396 | 0.0019876 | 1 |
| HTN | nk_cell | *EPS15* | -1.575493158 | 0.000887219 | 0.001371388 | 1 |
| HTN | nk_cell | *USP40* | -1.657064525 | 6.02E-06 | 1.97E-05 | 0.029226141 |
| HTN | nk_cell | *POMGNT1* | -1.772158262 | 2.36E-06 | 1.01E-05 | 0.011469088 |
| HTN | nk_cell | *MAST2* | -1.916464852 | 0.001959986 | 0.002836222 | 1 |
| HTN | nk_cell | *EIF2AK1* | -2.219901383 | 1.42E-05 | 3.85E-05 | 0.068999795 |
| HTN | nk_cell | *NME8* | 4.433543668 | 2.45E-07 | 4.07E-06 | 0.001189584 |
| HTN | nk_cell | *ITPKC* | -1.506816206 | 2.64E-07 | 4.07E-06 | 0.001279881 |
| HTN | nk_cell | *TMED2* | -2.015564872 | 0.003645333 | 0.005070859 | 1 |
| HTN | nk_cell | *ALG9* | -2.961963159 | 2.03E-06 | 9.02E-06 | 0.009826725 |
| HTN | nk_cell | *ACOX3* | -2.545737056 | 0.000130522 | 0.000244514 | 0.633290703 |
| HTN | nk_cell | *HSD17B14* | -1.422164381 | 1.87E-06 | 8.58E-06 | 0.009069908 |
| HTN | nk_cell | *TRIP6* | -2.607583944 | 0.000140764 | 0.000261781 | 0.682985956 |
| HTN | nk_cell | *ACHE* | -2.349043181 | 0.00460524 | 0.006299584 | 1 |
| HTN | nk_cell | *SRRT* | -1.928129731 | 3.21E-06 | 1.25E-05 | 0.015555041 |
| HTN | nk_cell | *ADAMTS2* | -1.69824931 | 2.11E-06 | 9.29E-06 | 0.010249736 |
| HTN | nk_cell | *PSMC5* | 2.938823064 | 1.39E-05 | 3.78E-05 | 0.067441152 |
| HTN | nk_cell | *UIMC1* | -1.973497601 | 0.006173598 | 0.008295292 | 1 |
| HTN | nk_cell | *LPCAT2* | -2.721088955 | 0.000487693 | 0.000793789 | 1 |
| HTN | nk_cell | *OGFOD1* | -3.08092339 | 0.000241119 | 0.000422044 | 1 |
| HTN | nk_cell | *TXNDC16* | -2.312007303 | 0.001197507 | 0.001804442 | 1 |
| HTN | nk_cell | *SF3B2* | 1.842953011 | 0.010308322 | 0.013452388 | 1 |
| HTN | nk_cell | *AAMDC* | -2.352669989 | 1.28E-08 | 1.01E-06 | 6.19E-05 |
| HTN | nk_cell | *GP6* | 1.377957933 | 0.000120985 | 0.000228589 | 0.587017712 |
| HTN | nk_cell | *ASAP3* | -1.220327828 | 7.75E-07 | 5.19E-06 | 0.003760196 |
| HTN | nk_cell | *EDEM2* | -2.064406335 | 3.76E-06 | 1.41E-05 | 0.018253955 |
| HTN | nk_cell | *ANKRD10* | -1.747578528 | 1.95E-05 | 4.94E-05 | 0.094484175 |
| HTN | nk_cell | *DOCK3* | -1.857145248 | 2.52E-06 | 1.06E-05 | 0.012231191 |
| HTN | nk_cell | *ARHGAP28* | -1.500368591 | 2.95E-06 | 1.17E-05 | 0.014292242 |
| HTN | nk_cell | *PPP1R13B* | -2.561648683 | 0.000224646 | 0.000395926 | 1 |
| HTN | nk_cell | *NSFL1C* | -1.772762712 | 0.000170959 | 0.000310671 | 0.82949089 |
| HTN | nk_cell | *ZNF343* | -2.07555001 | 2.87E-05 | 6.77E-05 | 0.139390299 |
| HTN | nk_cell | *EBF4* | -2.696859496 | 5.30E-06 | 1.81E-05 | 0.025727467 |
| HTN | nk_cell | *CPXM1* | -1.645921802 | 5.25E-06 | 1.79E-05 | 0.025454829 |

| | | | | | | |
|---|---|---|---|---|---|---|
| HTN | nk_cell | *MAVS* | -2.602929615 | 6.23E-06 | 2.03E-05 | 0.03024817 |
| HTN | nk_cell | *XRN2* | -2.093686141 | 0.003863507 | 0.005339144 | 1 |
| HTN | nk_cell | *ESF1* | -1.848601686 | 1.13E-05 | 3.22E-05 | 0.055069072 |
| HTN | nk_cell | *RBBP9* | -3.134426411 | 3.59E-06 | 1.35E-05 | 0.017440735 |
| HTN | nk_cell | *TMEM230* | -2.22488289 | 5.48E-05 | 0.000115234 | 0.265728643 |
| HTN | nk_cell | *KIF16B* | -2.267118553 | 0.000299029 | 0.000509979 | 1 |
| HTN | nk_cell | *TBX5* | -6.042034455 | 8.45E-08 | 3.01E-06 | 0.00040979 |
| HTN | nk_cell | *FXYD5* | 1.988789063 | 1.25E-06 | 6.76E-06 | 0.006041195 |
| HTN | nk_cell | *ZNF302* | -1.54025623 | 0.012482519 | 0.016077829 | 1 |
| HTN | nk_cell | *GRAMD1A* | -2.318166618 | 1.31E-05 | 3.60E-05 | 0.063797159 |
| HTN | nk_cell | *CDIP1* | -2.830886059 | 6.09E-06 | 1.99E-05 | 0.029547488 |
| HTN | nk_cell | *GANAB* | -2.328288144 | 4.88E-06 | 1.69E-05 | 0.023655526 |
| HTN | nk_cell | *MLF2* | -1.348583514 | 3.36E-05 | 7.69E-05 | 0.163000469 |
| HTN | nk_cell | *ANKRD24* | -1.961928061 | 5.49E-07 | 4.45E-06 | 0.002665321 |
| HTN | nk_cell | *DHX32* | -1.651212888 | 2.31E-06 | 9.92E-06 | 0.011194572 |
| HTN | nk_cell | *SPTLC1* | -1.970300839 | 0.029000632 | 0.035659165 | 1 |
| HTN | nk_cell | *MRPS33* | -1.634034038 | 2.37E-06 | 1.01E-05 | 0.011503744 |
| HTN | nk_cell | *MUL1* | -2.993587241 | 1.44E-07 | 3.91E-06 | 0.000696755 |
| HTN | nk_cell | *SPG21* | -2.373426759 | 2.78E-05 | 6.58E-05 | 0.135023165 |
| HTN | nk_cell | *CHRD* | -1.922890654 | 4.92E-07 | 4.32E-06 | 0.002386446 |
| HTN | nk_cell | *RAB11FIP3* | -2.088554465 | 0.002015305 | 0.0029076 | 1 |
| HTN | nk_cell | *GNPTG* | -2.136609788 | 6.65E-06 | 2.12E-05 | 0.032275556 |
| HTN | nk_cell | *PABPC4* | -4.045335982 | 1.38E-08 | 1.03E-06 | 6.67E-05 |
| HTN | nk_cell | *USP48* | -1.9484393 | 0.000145659 | 0.00027011 | 0.706739286 |
| HTN | nk_cell | *PDPR* | -2.104007148 | 4.69E-06 | 1.64E-05 | 0.022748798 |
| HTN | nk_cell | *GLG1* | -1.908339527 | 0.002108547 | 0.00303761 | 1 |
| HTN | nk_cell | *DTX2* | -3.721862818 | 0.000207673 | 0.000369094 | 1 |
| HTN | nk_cell | *LAMB1* | -1.330027156 | 2.27E-06 | 9.81E-06 | 0.011038182 |
| HTN | nk_cell | *ABCC6* | -1.064379586 | 1.30E-07 | 3.82E-06 | 0.000630851 |
| HTN | nk_cell | *RAPGEF4* | -1.094779534 | 4.94E-07 | 4.32E-06 | 0.00239537 |
| HTN | nk_cell | *TF* | -2.945627948 | 3.10E-06 | 1.22E-05 | 0.015050447 |
| HTN | nk_cell | *RGS17* | -1.434347506 | 0.001639218 | 0.002402866 | 1 |
| HTN | nk_cell | *HAUS4* | 1.324157937 | 5.97E-05 | 0.000123665 | 0.289869798 |
| HTN | nk_cell | *OSGEP* | -1.998804571 | 0.01812576 | 0.022843165 | 1 |
| HTN | nk_cell | *SCFD1* | -1.99069726 | 0.000403936 | 0.000668678 | 1 |
| HTN | nk_cell | *SUPT16H* | -2.592006155 | 0.001105376 | 0.001677074 | 1 |
| HTN | nk_cell | *CAPN3* | 3.440589956 | 1.57E-06 | 7.69E-06 | 0.007639008 |
| HTN | nk_cell | *TBX15* | 2.859681551 | 8.21E-06 | 2.50E-05 | 0.039821541 |
| HTN | nk_cell | *UNC13D* | 1.577427806 | 2.87E-06 | 1.15E-05 | 0.01392576 |
| HTN | nk_cell | *MFSD11* | -2.09145426 | 0.000177431 | 0.00032099 | 0.860895468 |
| HTN | nk_cell | *COMT* | -2.334780303 | 5.39E-05 | 0.000113578 | 0.261342686 |
| HTN | nk_cell | *LRRFIP2* | -1.848034623 | 0.000189564 | 0.000340653 | 0.919762656 |
| HTN | nk_cell | *SEC22C* | -2.55662109 | 3.37E-05 | 7.70E-05 | 0.163688905 |
| HTN | nk_cell | *XYLB* | -1.890892221 | 2.00E-06 | 8.99E-06 | 0.009717955 |
| HTN | nk_cell | *GABRP* | 4.761406382 | 6.00E-06 | 1.97E-05 | 0.029128199 |
| HTN | nk_cell | *CDC6* | -3.561428639 | 1.73E-07 | 4.07E-06 | 0.000837939 |
| HTN | nk_cell | *FMO2* | -0.605579714 | 0.026946121 | 0.033250911 | 1 |
| HTN | nk_cell | *MSH2* | -2.338849039 | 1.10E-05 | 3.13E-05 | 0.053325929 |
| HTN | nk_cell | *PSMD5* | -2.429168832 | 7.72E-05 | 0.000154228 | 0.374620053 |
| HTN | nk_cell | *CRAT* | -2.263637107 | 9.50E-07 | 5.82E-06 | 0.004608861 |

| | | | | | | |
|---|---|---|---|---|---|---|
| HTN | nk_cell | *PDE6C* | -4.01175376 | 0.001216401 | 0.001829496 | 1 |
| HTN | nk_cell | *CWF19L1* | -1.361081146 | 0.001284282 | 0.001922658 | 1 |
| HTN | nk_cell | *CRTAC1* | -1.062268147 | 0.005866045 | 0.007903929 | 1 |
| HTN | nk_cell | *WAC* | -1.755470249 | 0.000189701 | 0.000340769 | 0.920430167 |
| HTN | nk_cell | *IFT74* | -1.470632972 | 8.66E-06 | 2.59E-05 | 0.041997427 |
| HTN | nk_cell | *IL12RB1* | 0.471975788 | 1.03E-06 | 6.04E-06 | 0.004983051 |
| HTN | nk_cell | *SCD* | -3.224239096 | 4.83E-09 | 4.89E-07 | 2.35E-05 |
| HTN | nk_cell | *ERMP1* | -2.331140142 | 0.000975806 | 0.001496381 | 1 |
| HTN | nk_cell | *RAB18* | -2.323552279 | 0.001329632 | 0.001983164 | 1 |
| HTN | nk_cell | *PRTFDC1* | -1.490517957 | 0.03391992 | 0.041455781 | 1 |
| HTN | nk_cell | *TSPAN15* | -1.979812427 | 4.49E-06 | 1.59E-05 | 0.021776197 |
| HTN | nk_cell | *CATSPERG* | -1.39341973 | 0.007544931 | 0.010018611 | 1 |
| HTN | nk_cell | *CIRBP* | -1.745499533 | 1.50E-05 | 4.02E-05 | 0.072633213 |
| HTN | nk_cell | *MTAP* | -3.166439088 | 2.37E-07 | 4.07E-06 | 0.001149042 |
| HTN | nk_cell | *POLR2E* | -1.882861829 | 2.10E-06 | 9.24E-06 | 0.010170257 |
| HTN | nk_cell | *MADCAM1* | -1.150121666 | 2.42E-07 | 4.07E-06 | 0.001175414 |
| HTN | nk_cell | *SERPIND1* | -2.406933746 | 0.000115649 | 0.000219363 | 0.561129634 |
| HTN | nk_cell | *SNAP29* | -1.399561052 | 0.010451608 | 0.013621059 | 1 |
| HTN | nk_cell | *CECR2* | -1.062582232 | 4.23E-07 | 4.24E-06 | 0.002050746 |
| HTN | nk_cell | *BCL2L13* | -1.513769963 | 2.76E-05 | 6.54E-05 | 0.133938308 |
| HTN | nk_cell | *DDTL* | 1.359912498 | 4.25E-05 | 9.35E-05 | 0.206322397 |
| HTN | nk_cell | *SUSD2* | -0.915419027 | 0.005911005 | 0.00795788 | 1 |
| HTN | nk_cell | *SPECC1L* | -2.563132995 | 9.61E-07 | 5.84E-06 | 0.004660582 |
| HTN | nk_cell | *PPIL2* | -1.92630396 | 0.015290558 | 0.019462169 | 1 |
| HTN | nk_cell | *PES1* | 2.048156583 | 0.004414472 | 0.00605057 | 1 |
| HTN | nk_cell | *SLC35E4* | -1.651097344 | 2.41E-07 | 4.07E-06 | 0.001167024 |
| HTN | nk_cell | *TOP3B* | -2.711889145 | 0.032020883 | 0.039203968 | 1 |
| HTN | nk_cell | *CRYBB3* | 3.002151044 | 0.005171797 | 0.007011333 | 1 |
| HTN | nk_cell | *CYTH4* | 1.161150792 | 3.10E-07 | 4.17E-06 | 0.001504324 |
| HTN | nk_cell | *CARD10* | -1.83415901 | 8.75E-06 | 2.61E-05 | 0.042444085 |
| HTN | nk_cell | *LRP5L* | -2.152513853 | 0.000146363 | 0.000271051 | 0.710154619 |
| HTN | nk_cell | *SH3BP1* | 0.686081849 | 2.65E-07 | 4.07E-06 | 0.001287666 |
| HTN | nk_cell | *HPS4* | -1.600673568 | 2.19E-06 | 9.56E-06 | 0.01064744 |
| HTN | nk_cell | *TRIOBP* | -2.157560258 | 1.03E-06 | 6.04E-06 | 0.004988106 |
| HTN | nk_cell | *ANKRD54* | -1.309795668 | 1.36E-06 | 7.10E-06 | 0.006598349 |
| HTN | nk_cell | *EIF3L* | -2.752913157 | 2.47E-06 | 1.04E-05 | 0.011982041 |
| HTN | nk_cell | *CCDC134* | -2.284814306 | 0.003606985 | 0.005021834 | 1 |
| HTN | nk_cell | *TTC28* | -1.446857165 | 2.01E-05 | 5.07E-05 | 0.097474212 |
| HTN | nk_cell | *CENPM* | 1.553104157 | 4.33E-05 | 9.49E-05 | 0.209953241 |
| HTN | nk_cell | *CYP2D6* | -2.591385958 | 1.83E-07 | 4.07E-06 | 0.000886028 |
| HTN | nk_cell | *CBY1* | -1.607965014 | 0.003131933 | 0.004404678 | 1 |
| HTN | nk_cell | *RTCB* | -2.288245565 | 8.32E-05 | 0.000164455 | 0.403476633 |
| HTN | nk_cell | *RAB36* | -2.130500193 | 3.64E-07 | 4.24E-06 | 0.001766675 |
| HTN | nk_cell | *TIMP3* | -1.481492937 | 1.80E-05 | 4.64E-05 | 0.08714504 |
| HTN | nk_cell | *PPP6R2* | -2.006542527 | 0.000679317 | 0.001072233 | 1 |
| HTN | nk_cell | *CYB5R3* | -2.145236391 | 1.40E-05 | 3.80E-05 | 0.067778582 |
| HTN | nk_cell | *RHBDD3* | -2.806201067 | 0.00010433 | 0.0002003 | 0.506209656 |
| HTN | nk_cell | *TOM1* | -2.065711936 | 0.000165123 | 0.000301759 | 0.801177192 |
| HTN | nk_cell | *CHKB* | -2.01261874 | 1.60E-05 | 4.24E-05 | 0.077600563 |
| HTN | nk_cell | *THOC5* | -3.16625238 | 9.95E-07 | 5.95E-06 | 0.004828561 |

| | | | | | | |
|---|---|---|---|---|---|---|
| HTN | nk_cell | *TSPO* | 2.605454639 | 0.007100606 | 0.009467475 | 1 |
| HTN | nk_cell | *TTLL12* | -3.554316351 | 9.59E-07 | 5.84E-06 | 0.004652415 |
| HTN | nk_cell | *MTMR3* | -1.58624764 | 0.000732956 | 0.001150906 | 1 |
| HTN | nk_cell | *APOL4* | -1.658220673 | 3.32E-07 | 4.24E-06 | 0.001613278 |
| HTN | nk_cell | *SAMM50* | -2.680993403 | 8.48E-06 | 2.55E-05 | 0.041139936 |
| HTN | nk_cell | *FOXRED2* | -1.217688828 | 7.57E-06 | 2.34E-05 | 0.036719238 |
| HTN | nk_cell | *IFT27* | -3.340121921 | 0.00270132 | 0.003836887 | 1 |
| HTN | nk_cell | *KIAA0930* | -2.469831979 | 0.005797402 | 0.007815781 | 1 |
| HTN | nk_cell | *SLC25A17* | -2.166556535 | 0.000149189 | 0.000275547 | 0.7238632 |
| HTN | nk_cell | *L3MBTL2* | 2.801665882 | 3.61E-05 | 8.18E-05 | 0.175351456 |
| HTN | nk_cell | *ZC3H7B* | -2.389992807 | 0.000390329 | 0.00065037 | 1 |
| HTN | nk_cell | *ACO2* | -1.796238989 | 6.17E-07 | 4.56E-06 | 0.002993658 |
| HTN | nk_cell | *DESI1* | -2.546050421 | 0.000154858 | 0.000283965 | 0.751370926 |
| HTN | nk_cell | *MLC1* | 0.503392791 | 2.16E-07 | 4.07E-06 | 0.001049014 |
| HTN | nk_cell | *HDAC10* | -2.327628732 | 1.64E-06 | 7.91E-06 | 0.007952745 |
| HTN | nk_cell | *KHNYN* | -2.171566241 | 0.000358685 | 0.000601153 | 1 |
| HTN | nk_cell | *SDR39U1* | -1.503052012 | 2.73E-06 | 1.11E-05 | 0.013251537 |
| HTN | nk_cell | *AP4S1* | -2.139561927 | 6.39E-07 | 4.62E-06 | 0.003098713 |
| HTN | nk_cell | *NIN* | 2.39181062 | 2.23E-05 | 5.51E-05 | 0.108367004 |
| HTN | nk_cell | *PYGL* | -1.21457 | 9.67E-07 | 5.86E-06 | 0.004691513 |
| HTN | nk_cell | *CNIH1* | -2.009287845 | 6.66E-06 | 2.12E-05 | 0.032310236 |
| HTN | nk_cell | *ATP6V1D* | -1.828554104 | 4.65E-06 | 1.63E-05 | 0.022561721 |
| HTN | nk_cell | *PLEK2* | -2.210927988 | 0.001254784 | 0.001881983 | 1 |
| HTN | nk_cell | *GSTZ1* | -1.129652243 | 1.04E-06 | 6.09E-06 | 0.005065273 |
| HTN | nk_cell | *TMED8* | -1.607042268 | 5.78E-06 | 1.92E-05 | 0.028037218 |
| HTN | nk_cell | *LGMN* | -1.423925429 | 0.00134675 | 0.002003812 | 1 |
| HTN | nk_cell | *MTHFD1* | -1.40878477 | 3.33E-06 | 1.28E-05 | 0.016151303 |
| HTN | nk_cell | *TELO2* | -1.773535263 | 3.84E-07 | 4.24E-06 | 0.001862762 |
| HTN | nk_cell | *GSKIP* | -2.964231689 | 3.32E-06 | 1.28E-05 | 0.016107396 |
| HTN | nk_cell | *PSMB5* | -1.471248622 | 0.01182986 | 0.015306261 | 1 |
| HTN | nk_cell | *YY1* | -2.3778394 | 0.001005881 | 0.001539116 | 1 |
| HTN | nk_cell | *CCNB1IP1* | -2.37304956 | 6.32E-06 | 2.05E-05 | 0.030652431 |
| HTN | nk_cell | *EFS* | -2.822261014 | 2.56E-06 | 1.07E-05 | 0.012442796 |
| HTN | nk_cell | *CINP* | -1.617317393 | 7.88E-07 | 5.21E-06 | 0.003823886 |
| HTN | nk_cell | *KIAA0391* | 4.893568958 | 2.33E-06 | 1.00E-05 | 0.011307336 |
| HTN | nk_cell | *EMC9* | -1.893315895 | 1.50E-06 | 7.51E-06 | 0.007257962 |
| HTN | nk_cell | *PLTP* | -1.103298832 | 0.001137685 | 0.001722884 | 1 |
| HTN | nk_cell | *TRPC4AP* | -1.702005204 | 0.000113291 | 0.000215396 | 0.549689509 |
| HTN | nk_cell | *PYGB* | -1.955023132 | 0.000503105 | 0.00081641 | 1 |
| HTN | nk_cell | *CD40* | -1.638552989 | 1.01E-06 | 5.99E-06 | 0.004911875 |
| HTN | nk_cell | *UQCC1* | -1.940307942 | 3.09E-05 | 7.19E-05 | 0.149788036 |
| HTN | nk_cell | *IFT52* | -2.107056483 | 0.003768132 | 0.005223708 | 1 |
| HTN | nk_cell | *NDRG3* | -2.307966843 | 0.005903782 | 0.00795036 | 1 |
| HTN | nk_cell | *PABPC1L* | -2.254035565 | 2.08E-06 | 9.19E-06 | 0.010099271 |
| HTN | nk_cell | *DOK5* | -1.637757766 | 7.80E-07 | 5.20E-06 | 0.003782739 |
| HTN | nk_cell | *CSTF1* | 3.576523748 | 0.024274484 | 0.03016128 | 1 |
| HTN | nk_cell | *NELFCD* | -2.823652442 | 0.001087691 | 0.001652827 | 1 |
| HTN | nk_cell | *CTSZ* | -2.191608435 | 1.83E-05 | 4.70E-05 | 0.088710196 |
| HTN | nk_cell | *MTG2* | -2.111742214 | 1.92E-05 | 4.88E-05 | 0.092956643 |
| HTN | nk_cell | *TCFL5* | -2.816043007 | 1.14E-05 | 3.23E-05 | 0.055381222 |

| | | | | | | |
|---|---|---|---|---|---|---|
| HTN | nk_cell | *DIDO1* | -1.888145786 | 0.003232429 | 0.004538062 | 1 |
| HTN | nk_cell | *BIRC7* | -3.857545369 | 0.000144681 | 0.000268552 | 0.701994176 |
| HTN | nk_cell | *NKAIN4* | -2.389827229 | 0.000952478 | 0.001465334 | 1 |
| HTN | nk_cell | *PTK6* | -3.12128624 | 5.06E-07 | 4.37E-06 | 0.002455951 |
| HTN | nk_cell | *GMEB2* | -2.132243828 | 9.99E-07 | 5.95E-06 | 0.004846052 |
| HTN | nk_cell | *CDC25B* | 2.513151963 | 6.48E-06 | 2.08E-05 | 0.03143683 |
| HTN | nk_cell | *RNF24* | -2.195205995 | 0.000138386 | 0.000257655 | 0.671449086 |
| HTN | nk_cell | *NDUFAF5* | -1.7177811 | 7.87E-05 | 0.000156703 | 0.381729688 |
| HTN | nk_cell | *TRIB3* | 6.426895079 | 6.39E-06 | 2.06E-05 | 0.03098145 |
| HTN | nk_cell | *RASSF2* | -1.714020367 | 0.011411272 | 0.01479623 | 1 |
| HTN | nk_cell | *ANGPT4* | -2.088843849 | 2.46E-07 | 4.07E-06 | 0.001194559 |
| HTN | nk_cell | *RSPO4* | -1.150092437 | 0.000115557 | 0.000219273 | 0.560680253 |
| HTN | nk_cell | *SIRPB1* | -3.96071309 | 0.000153918 | 0.000282669 | 0.74681226 |
| HTN | nk_cell | *SEC23B* | 2.398010829 | 0.00063017 | 0.001000468 | 1 |
| HTN | nk_cell | *CRNKL1* | -2.70724839 | 9.49E-05 | 0.000184234 | 0.460216557 |
| HTN | nk_cell | *MROH8* | -1.839395393 | 0.001735295 | 0.002529943 | 1 |
| HTN | nk_cell | *MANBAL* | -1.293250301 | 0.001070208 | 0.001628811 | 1 |
| HTN | nk_cell | *ACTR5* | 4.336009739 | 5.78E-06 | 1.92E-05 | 0.028031785 |
| HTN | nk_cell | *DNTTIP1* | -2.317935281 | 0.000625859 | 0.00099483 | 1 |
| HTN | nk_cell | *MAP1LC3A* | -2.207800517 | 5.86E-06 | 1.94E-05 | 0.028430317 |
| HTN | nk_cell | *RBFA* | 3.054542528 | 0.004090695 | 0.005630653 | 1 |
| HTN | nk_cell | *METTL4* | -2.666589644 | 2.26E-05 | 5.57E-05 | 0.109615666 |
| HTN | nk_cell | *MYOM1* | -1.600947695 | 2.02E-06 | 9.02E-06 | 0.009816524 |
| HTN | nk_cell | *LAMA1* | -1.377816741 | 9.14E-06 | 2.70E-05 | 0.044338074 |
| HTN | nk_cell | *RNF125* | 4.154554661 | 0.00137971 | 0.002047202 | 1 |
| HTN | nk_cell | *POLI* | -1.530639888 | 2.15E-07 | 4.07E-06 | 0.001042309 |
| HTN | nk_cell | *MIB1* | -1.991032814 | 8.53E-05 | 0.000168125 | 0.413922654 |
| HTN | nk_cell | *NALCN* | -1.691353457 | 5.75E-05 | 0.000119766 | 0.278935221 |
| HTN | nk_cell | *STK24* | -2.08141545 | 0.000131062 | 0.000245431 | 0.635912917 |
| HTN | nk_cell | *DNAJC3* | -2.173589466 | 0.000791011 | 0.001236065 | 1 |
| HTN | nk_cell | *UGGT2* | -1.650694776 | 2.09E-05 | 5.22E-05 | 0.101345957 |
| HTN | nk_cell | *ARHGEF7* | -1.869979547 | 2.56E-05 | 6.16E-05 | 0.124345357 |
| HTN | nk_cell | *PARP4* | -2.408101742 | 5.60E-05 | 0.000117384 | 0.271626561 |
| HTN | nk_cell | *SUPT20H* | -2.059712487 | 9.53E-05 | 0.000184805 | 0.462382413 |
| HTN | nk_cell | *MRPS31* | -2.096048353 | 1.54E-05 | 4.12E-05 | 0.074797563 |
| HTN | nk_cell | *RGCC* | -1.060739021 | 0.037221188 | 0.045296515 | 1 |
| HTN | nk_cell | *DGKH* | -1.780023503 | 0.001738891 | 0.002534424 | 1 |
| HTN | nk_cell | *KATNAL1* | -2.041557682 | 2.55E-05 | 6.14E-05 | 0.123805041 |
| HTN | nk_cell | *INTS6* | -2.015676807 | 0.000573462 | 0.00091951 | 1 |
| HTN | nk_cell | *CLN5* | -2.620374215 | 0.001283404 | 0.001921937 | 1 |
| HTN | nk_cell | *MGRN1* | -1.521888957 | 0.011267975 | 0.014622149 | 1 |
| HTN | nk_cell | *LONP2* | -1.696417883 | 0.000115925 | 0.000219715 | 0.562469333 |
| HTN | nk_cell | *PLLP* | -1.357967375 | 2.40E-06 | 1.02E-05 | 0.011641403 |
| HTN | nk_cell | *DHODH* | -1.693857156 | 3.19E-05 | 7.40E-05 | 0.154985707 |
| HTN | nk_cell | *CCDC113* | 2.155613299 | 0.000174613 | 0.000316482 | 0.847222512 |
| HTN | nk_cell | *PSMD7* | 2.53151358 | 0.010527592 | 0.013715479 | 1 |
| HTN | nk_cell | *COG4* | -2.417348953 | 0.000667281 | 0.001054609 | 1 |
| HTN | nk_cell | *MON1B* | -3.641288432 | 0.00038778 | 0.00064701 | 1 |
| HTN | nk_cell | *NPRL3* | -1.44346777 | 0.028099231 | 0.034585862 | 1 |
| HTN | nk_cell | *TAF1C* | 1.425807633 | 0.002291871 | 0.003293885 | 1 |

| | | | | | | |
|---|---|---|---|---|---|---|
| HTN | nk_cell | *NAGPA* | -1.957022066 | 2.32E-05 | 5.70E-05 | 0.112577999 |
| HTN | nk_cell | *ZNF500* | -2.214816776 | 1.12E-07 | 3.53E-06 | 0.000541195 |
| HTN | nk_cell | *NME4* | -2.677301843 | 2.83E-07 | 4.07E-06 | 0.001371719 |
| HTN | nk_cell | *NOMO3* | 1.412045619 | 0.033707257 | 0.041206251 | 1 |
| HTN | nk_cell | *LMF1* | -2.186737687 | 1.99E-05 | 5.02E-05 | 0.096400546 |
| HTN | nk_cell | *NARFL* | -1.922785351 | 0.000400658 | 0.000664841 | 1 |
| HTN | nk_cell | *SLC7A5* | 2.309280569 | 0.000287227 | 0.000491656 | 1 |
| HTN | nk_cell | *RHBDL1* | -3.786434318 | 1.60E-06 | 7.80E-06 | 0.007779022 |
| HTN | nk_cell | *NUBP1* | -2.84255136 | 5.12E-06 | 1.77E-05 | 0.024840051 |
| HTN | nk_cell | *CPPED1* | -1.8702073 | 3.82E-06 | 1.42E-05 | 0.01851571 |
| HTN | nk_cell | *HMOX2* | 2.444712201 | 1.21E-06 | 6.69E-06 | 0.005894545 |
| HTN | nk_cell | *TOX3* | -2.453278316 | 0.001680073 | 0.002454596 | 1 |
| HTN | nk_cell | *RRN3P2* | 2.474633627 | 7.83E-05 | 0.000156056 | 0.379683928 |
| HTN | nk_cell | *PYCARD* | 1.57161996 | 3.66E-08 | 1.98E-06 | 0.00017764 |
| HTN | nk_cell | *CDIPT* | 2.886617811 | 1.31E-06 | 6.92E-06 | 0.006365234 |
| HTN | nk_cell | *KAT8* | -1.999916537 | 2.07E-06 | 9.18E-06 | 0.010066483 |
| HTN | nk_cell | *TMC5* | -1.486336757 | 4.76E-07 | 4.32E-06 | 0.002311146 |
| HTN | nk_cell | *C16orf62* | -1.79005788 | 9.06E-06 | 2.69E-05 | 0.04394398 |
| HTN | nk_cell | *RNF40* | -2.965572835 | 3.95E-05 | 8.81E-05 | 0.191785195 |
| HTN | nk_cell | *KNOP1* | -2.72804107 | 1.78E-05 | 4.61E-05 | 0.08623837 |
| HTN | nk_cell | *IQCH* | -2.235584972 | 1.36E-05 | 3.70E-05 | 0.065779077 |
| HTN | nk_cell | *LACTB* | -2.404808101 | 5.63E-07 | 4.48E-06 | 0.002730653 |
| HTN | nk_cell | *HERC1* | -1.64544816 | 0.005006108 | 0.006801915 | 1 |
| HTN | nk_cell | *TRIP4* | -1.942153011 | 0.001180373 | 0.001780837 | 1 |
| HTN | nk_cell | *TTC23* | -1.833657731 | 3.77E-06 | 1.41E-05 | 0.018312571 |
| HTN | nk_cell | *FAH* | -1.685701394 | 1.13E-06 | 6.40E-06 | 0.005506276 |
| HTN | nk_cell | *SLC30A4* | -3.00737218 | 0.00035558 | 0.000596361 | 1 |
| HTN | nk_cell | *ZFAND1* | -2.248349626 | 0.000151053 | 0.000278316 | 0.732906881 |
| HTN | nk_cell | *EYA1* | -1.012001706 | 1.91E-06 | 8.72E-06 | 0.009274704 |
| HTN | nk_cell | *IMPAD1* | -1.660100245 | 7.21E-07 | 4.96E-06 | 0.00349684 |
| HTN | nk_cell | *SFRP1* | -3.056976892 | 0.000170352 | 0.000309801 | 0.826549866 |
| HTN | nk_cell | *RAB2A* | -1.770336898 | 0.000209518 | 0.000371694 | 1 |
| HTN | nk_cell | *ARMC1* | -2.32653341 | 6.44E-06 | 2.07E-05 | 0.031225532 |
| HTN | nk_cell | *SPAG1* | -2.21530542 | 0.001396624 | 0.0020704 | 1 |
| HTN | nk_cell | *CHRAC1* | -4.650482331 | 0.002946871 | 0.004164905 | 1 |
| HTN | nk_cell | *SNX16* | -1.799590461 | 0.027000932 | 0.033310074 | 1 |
| HTN | nk_cell | *TSTA3* | -1.301134426 | 1.12E-05 | 3.18E-05 | 0.0543137 |
| HTN | nk_cell | *R3HCC1* | -1.6886252 | 0.034105346 | 0.041671906 | 1 |
| HTN | nk_cell | *ERICH1* | -1.747073051 | 1.83E-05 | 4.70E-05 | 0.088632368 |
| HTN | nk_cell | *TUSC3* | -1.474469551 | 1.37E-06 | 7.14E-06 | 0.006664642 |
| HTN | nk_cell | *KLHDC4* | 2.242106215 | 2.70E-05 | 6.44E-05 | 0.131237155 |
| HTN | nk_cell | *MAN2B1* | -2.462986089 | 0.000285264 | 0.000488909 | 1 |
| HTN | nk_cell | *TULP2* | 8.600200743 | 1.98E-07 | 4.07E-06 | 0.000958466 |
| HTN | nk_cell | *NFKBIB* | 1.551112492 | 3.10E-07 | 4.17E-06 | 0.001505077 |
| HTN | nk_cell | *SARS2* | -1.562623071 | 1.17E-06 | 6.53E-06 | 0.005676149 |
| HTN | nk_cell | *SNRNP70* | -1.791413204 | 8.01E-06 | 2.45E-05 | 0.038880451 |
| HTN | nk_cell | *LIN7B* | -1.899548775 | 7.31E-08 | 2.73E-06 | 0.000354489 |
| HTN | nk_cell | *FCGRT* | -1.624069172 | 0.000208317 | 0.000370095 | 1 |
| HTN | nk_cell | *ARHGEF18* | -3.468623292 | 0.001330011 | 0.001983164 | 1 |
| HTN | nk_cell | *PEX11G* | -1.473460294 | 0.0007647 | 0.001198425 | 1 |

| | | | | | | |
|---|---|---|---|---|---|---|
| HTN | nk_cell | *ERCC2* | -3.977682686 | 3.32E-05 | 7.63E-05 | 0.161113909 |
| HTN | nk_cell | *CD37* | 1.013442061 | 1.18E-05 | 3.31E-05 | 0.05724164 |
| HTN | nk_cell | *AMH* | -24.67908706 | 1.55E-06 | 7.63E-06 | 0.007510395 |
| HTN | nk_cell | *DKKL1* | -1.438312526 | 0.008251805 | 0.010896191 | 1 |
| HTN | nk_cell | *FCER2* | -9.280779928 | 3.91E-06 | 1.44E-05 | 0.018965824 |
| HTN | nk_cell | *CLEC4M* | -3.207473352 | 4.29E-07 | 4.24E-06 | 0.002081801 |
| HTN | nk_cell | *TLE6* | -2.086766298 | 0.000138241 | 0.000257484 | 0.670744719 |
| HTN | nk_cell | *NOVA2* | -1.193306263 | 4.09E-05 | 9.05E-05 | 0.198361844 |
| HTN | nk_cell | *SGTA* | -2.833559463 | 0.021878157 | 0.027351925 | 1 |
| HTN | nk_cell | *KIR3DX1* | 0.235284055 | 2.84E-07 | 4.07E-06 | 0.001378406 |
| HTN | nk_cell | *MED25* | -1.887137369 | 1.28E-06 | 6.85E-06 | 0.006227134 |
| HTN | nk_cell | *TIMM44* | -2.147337576 | 0.012361503 | 0.015941707 | 1 |
| HTN | nk_cell | *IL27RA* | 0.608200131 | 2.50E-07 | 4.07E-06 | 0.001213943 |
| HTN | nk_cell | *FAM32A* | 4.194686272 | 6.35E-05 | 0.000130113 | 0.307978478 |
| HTN | nk_cell | *C19orf44* | -1.230666469 | 9.19E-05 | 0.000179708 | 0.446036028 |
| HTN | nk_cell | *OLFM2* | -1.702427272 | 3.96E-05 | 8.81E-05 | 0.192156115 |
| HTN | nk_cell | *RASAL3* | 0.303536122 | 2.70E-07 | 4.07E-06 | 0.001311599 |
| HTN | nk_cell | *PDCD5* | -1.337582035 | 1.56E-06 | 7.65E-06 | 0.007577659 |
| HTN | nk_cell | *ANKRD27* | -2.435169524 | 0.004952756 | 0.00673697 | 1 |
| HTN | nk_cell | *POLR2I* | -3.884806601 | 0.000134919 | 0.000251993 | 0.654626455 |
| HTN | nk_cell | *CCDC9* | -3.246060574 | 4.70E-05 | 0.00010161 | 0.228013644 |
| HTN | nk_cell | *TGFB1* | 2.661622273 | 0.000230592 | 0.000405521 | 1 |
| HTN | nk_cell | *ICAM5* | -3.250284046 | 3.70E-09 | 4.18E-07 | 1.80E-05 |
| HTN | nk_cell | *CD33* | -0.955233486 | 0.004110524 | 0.005656343 | 1 |
| HTN | nk_cell | *RABAC1* | 3.627497356 | 2.71E-07 | 4.07E-06 | 0.001316975 |
| HTN | nk_cell | *GRIN2D* | -4.965946367 | 1.62E-06 | 7.83E-06 | 0.007848051 |
| HTN | nk_cell | *CCDC114* | -4.400209705 | 0.001800779 | 0.002618334 | 1 |
| HTN | nk_cell | *LIG1* | 2.645370365 | 3.44E-06 | 1.31E-05 | 0.016707656 |
| HTN | nk_cell | *ZNF175* | -1.994468729 | 2.58E-06 | 1.07E-05 | 0.012514679 |
| HTN | nk_cell | *PLA2G4C* | -0.704682895 | 0.038136033 | 0.046386571 | 1 |
| HTN | nk_cell | *CAPS* | -1.628409053 | 5.26E-07 | 4.40E-06 | 0.002554347 |
| HTN | nk_cell | *MIER2* | -2.750991827 | 2.28E-06 | 9.81E-06 | 0.011047478 |
| HTN | nk_cell | *PLEKHA4* | -1.157374437 | 4.48E-07 | 4.28E-06 | 0.002174486 |
| HTN | nk_cell | *TNPO2* | -2.402810446 | 1.24E-06 | 6.75E-06 | 0.005999009 |
| HTN | nk_cell | *GCDH* | -3.185116476 | 2.50E-05 | 6.04E-05 | 0.121286239 |
| HTN | nk_cell | *TFPT* | -4.327559662 | 0.000348135 | 0.000585292 | 1 |
| HTN | nk_cell | *RAB3A* | -8.741129744 | 1.24E-06 | 6.76E-06 | 0.006028506 |
| HTN | nk_cell | *UPK1A* | -2.326763394 | 0.019843157 | 0.024916925 | 1 |
| HTN | nk_cell | *COPE* | 2.808780348 | 7.27E-05 | 0.000146316 | 0.352620945 |
| HTN | nk_cell | *ETV2* | -16.33819109 | 0.029817231 | 0.036635402 | 1 |
| HTN | nk_cell | *LSR* | -0.978989214 | 3.26E-05 | 7.52E-05 | 0.158069741 |
| HTN | nk_cell | *ZNF14* | -3.166412777 | 0.0001448 | 0.000268669 | 0.702569002 |
| HTN | nk_cell | *ZNF85* | 3.784121857 | 0.000650676 | 0.001029371 | 1 |
| HTN | nk_cell | *SMG9* | -2.760095663 | 0.008857927 | 0.011637873 | 1 |
| HTN | nk_cell | *GTPBP10* | -1.673713771 | 0.017740273 | 0.022380604 | 1 |
| HTN | nk_cell | *RASA4* | -6.616818539 | 4.06E-07 | 4.24E-06 | 0.001971827 |
| HTN | nk_cell | *BET1* | -1.828032754 | 4.02E-06 | 1.47E-05 | 0.019506008 |
| HTN | nk_cell | *SP4* | 2.632999053 | 8.22E-07 | 5.33E-06 | 0.003990665 |
| HTN | nk_cell | *DNAH11* | -3.125786391 | 0.000140623 | 0.000261619 | 0.682301523 |
| HTN | nk_cell | *STEAP1B* | -2.446080961 | 0.000388669 | 0.000648272 | 1 |

| | | | | | | |
|---|---|---|---|---|---|---|
| HTN | nk_cell | *PTN* | -2.309376216 | 5.16E-06 | 1.78E-05 | 0.025059929 |
| HTN | nk_cell | *ADAP1* | -10.70837049 | 1.05E-05 | 3.02E-05 | 0.050861544 |
| HTN | nk_cell | *MET* | -1.272160393 | 1.51E-06 | 7.55E-06 | 0.007320089 |
| HTN | nk_cell | *RNF32* | -2.916631561 | 9.02E-07 | 5.69E-06 | 0.004374651 |
| HTN | nk_cell | *IQCE* | -2.209273391 | 2.59E-06 | 1.08E-05 | 0.012590892 |
| HTN | nk_cell | *VIPR2* | -1.316783414 | 1.71E-06 | 8.10E-06 | 0.008275846 |
| HTN | nk_cell | *HIBADH* | -1.470484935 | 0.000152759 | 0.000281072 | 0.741186237 |
| HTN | nk_cell | *CPVL* | -2.042716332 | 0.000130446 | 0.000244466 | 0.632922668 |
| HTN | nk_cell | *GRB10* | -2.001953365 | 2.26E-05 | 5.58E-05 | 0.109836843 |
| HTN | nk_cell | *ABHD11* | -1.376486806 | 0.00030166 | 0.000513924 | 1 |
| HTN | nk_cell | *PLEKHA8* | -1.483862779 | 1.09E-06 | 6.22E-06 | 0.005267695 |
| HTN | nk_cell | *CRHR2* | 1.751073475 | 3.70E-05 | 8.33E-05 | 0.179753637 |
| HTN | nk_cell | *CHCHD2* | 2.185370702 | 0.010731152 | 0.013959128 | 1 |
| HTN | nk_cell | *CCL24* | 3.465579343 | 1.95E-05 | 4.94E-05 | 0.09439846 |
| HTN | nk_cell | *ZKSCAN1* | -1.911640015 | 0.000532103 | 0.00085887 | 1 |
| HTN | nk_cell | *EIF3B* | -2.681937909 | 0.00014277 | 0.000265106 | 0.692721855 |
| HTN | nk_cell | *TAF6* | -2.858157418 | 5.76E-09 | 5.45E-07 | 2.79E-05 |
| HTN | nk_cell | *TFR2* | 0.857598743 | 2.74E-07 | 4.07E-06 | 0.001328749 |
| HTN | nk_cell | *MOSPD3* | -1.443812519 | 2.86E-07 | 4.07E-06 | 0.001385927 |
| HTN | nk_cell | *RBM28* | -1.613622519 | 0.002012874 | 0.002904957 | 1 |
| HTN | nk_cell | *IMPDH1* | -2.804737259 | 2.90E-07 | 4.10E-06 | 0.001405315 |
| HTN | nk_cell | *C1GALT1* | -2.31109453 | 0.000370364 | 0.000619443 | 1 |
| HTN | nk_cell | *NRF1* | -1.778463019 | 0.008003009 | 0.010577663 | 1 |
| HTN | nk_cell | *ZNF862* | -2.139143114 | 0.000551251 | 0.000888004 | 1 |
| HTN | nk_cell | *MEST* | -1.477002481 | 5.84E-09 | 5.45E-07 | 2.83E-05 |
| HTN | nk_cell | *ACTR3C* | 4.011064906 | 0.0001124 | 0.000213953 | 0.545364943 |
| HTN | nk_cell | *POU6F2* | -1.222281677 | 2.00E-07 | 4.07E-06 | 0.000969247 |
| HTN | nk_cell | *TSPAN13* | -1.406704424 | 1.03E-06 | 6.04E-06 | 0.004977111 |
| HTN | nk_cell | *RARRES2* | -2.109251169 | 0.001672068 | 0.002443637 | 1 |
| HTN | nk_cell | *TMEM176B* | -1.105617629 | 0.011692401 | 0.01514052 | 1 |
| HTN | nk_cell | *COA1* | -1.887410923 | 0.002294685 | 0.003295002 | 1 |
| HTN | nk_cell | *BLVRA* | -2.432309407 | 0.000165487 | 0.000302199 | 0.80294229 |
| HTN | nk_cell | *URGCP* | -2.545941558 | 8.25E-05 | 0.00016338 | 0.40028028 |
| HTN | nk_cell | *AEBP1* | -2.880124224 | 7.14E-05 | 0.000144157 | 0.346368214 |
| HTN | nk_cell | *GCK* | -8.535992495 | 5.46E-07 | 4.44E-06 | 0.002648836 |
| HTN | nk_cell | *SPATA6L* | -1.677493723 | 2.74E-06 | 1.11E-05 | 0.013309187 |
| HTN | nk_cell | *FKTN* | -2.322536804 | 5.09E-05 | 0.000108607 | 0.246971616 |
| HTN | nk_cell | *NMRK1* | -1.927110273 | 6.88E-05 | 0.000139758 | 0.333673675 |
| HTN | nk_cell | *MEGF9* | -2.335134089 | 0.001446309 | 0.002140131 | 1 |
| HTN | nk_cell | *TRIM14* | 1.683262725 | 1.25E-06 | 6.78E-06 | 0.006082672 |
| HTN | nk_cell | *CORO2A* | 0.954015782 | 1.40E-07 | 3.89E-06 | 0.000677271 |
| HTN | nk_cell | *C5* | -1.473839497 | 1.93E-06 | 8.80E-06 | 0.009373964 |
| HTN | nk_cell | *OGN* | -2.039307112 | 2.53E-07 | 4.07E-06 | 0.001227108 |
| HTN | nk_cell | *ECM2* | -1.918148764 | 0.000148903 | 0.000275217 | 0.722476875 |
| HTN | nk_cell | *PTGR1* | -1.803982936 | 2.46E-05 | 5.97E-05 | 0.119518151 |
| HTN | nk_cell | *SUSD1* | 0.933968196 | 0.000639418 | 0.001013875 | 1 |
| HTN | nk_cell | *PLGRKT* | -2.373500476 | 0.000973986 | 0.001495141 | 1 |
| HTN | nk_cell | *DOCK8* | 0.724498158 | 0.015201629 | 0.019364217 | 1 |
| HTN | nk_cell | *KANK1* | -1.619737029 | 5.44E-05 | 0.000114429 | 0.263759826 |
| HTN | nk_cell | *FUBP3* | -2.03351479 | 5.92E-06 | 1.95E-05 | 0.028707404 |

| | | | | | | |
|---|---|---|---|---|---|---|
| HTN | nk_cell | *RGP1* | -3.107948573 | 0.009659747 | 0.012636585 | 1 |
| HTN | nk_cell | *HPS1* | -2.765361002 | 9.47E-05 | 0.000183937 | 0.459289942 |
| HTN | nk_cell | *PHYH* | -1.795792514 | 5.21E-05 | 0.000110424 | 0.252649189 |
| HTN | nk_cell | *DNMBP* | -1.598669367 | 8.24E-06 | 2.50E-05 | 0.039983664 |
| HTN | nk_cell | *PKD2L1* | 2.092752434 | 1.01E-05 | 2.93E-05 | 0.049108729 |
| HTN | nk_cell | *CUBN* | -2.540064131 | 0.001269641 | 0.001903089 | 1 |
| HTN | nk_cell | *TRDMT1* | -2.161054682 | 8.97E-06 | 2.67E-05 | 0.043515449 |
| HTN | nk_cell | *ATE1* | -1.510388536 | 0.000968503 | 0.001487552 | 1 |
| HTN | nk_cell | *PLEKHA1* | -1.998376997 | 0.00739101 | 0.009830368 | 1 |
| HTN | nk_cell | *CDH23* | -1.527622196 | 2.85E-06 | 1.14E-05 | 0.013837927 |
| HTN | nk_cell | *SPOCK2* | 0.932463768 | 4.26E-05 | 9.36E-05 | 0.206905043 |
| HTN | nk_cell | *BMPR1A* | -1.542402036 | 5.44E-06 | 1.83E-05 | 0.026407106 |
| HTN | nk_cell | *LIPA* | -2.014089288 | 5.86E-05 | 0.000121537 | 0.284154428 |
| HTN | nk_cell | *SFXN3* | -2.435697802 | 2.91E-06 | 1.16E-05 | 0.014142653 |
| HTN | nk_cell | *ANKRD26* | -1.860828347 | 1.86E-05 | 4.77E-05 | 0.090296221 |
| HTN | nk_cell | *LHPP* | -2.823126226 | 0.000274543 | 0.00047237 | 1 |
| HTN | nk_cell | *PITRM1* | -2.129628188 | 1.17E-06 | 6.53E-06 | 0.00566541 |
| HTN | nk_cell | *RPL28* | 1.576818461 | 0.000723119 | 0.001137129 | 1 |
| HTN | nk_cell | *DNAJC12* | -2.557515414 | 1.89E-06 | 8.64E-06 | 0.009163954 |
| HTN | nk_cell | *PBLD* | -2.758229676 | 2.38E-05 | 5.82E-05 | 0.115716594 |
| HTN | nk_cell | *TSPAN14* | -2.19897338 | 0.000232848 | 0.000409043 | 1 |
| HTN | nk_cell | *PSMB3* | 3.299161055 | 0.000249963 | 0.000435341 | 1 |
| HTN | nk_cell | *RPL19* | 2.001065163 | 0.000672521 | 0.001062199 | 1 |
| HTN | nk_cell | *RAPGEFL1* | -1.88901145 | 6.06E-07 | 4.55E-06 | 0.00294136 |
| HTN | nk_cell | *WNT3* | -1.747522662 | 4.77E-07 | 4.32E-06 | 0.002313238 |
| HTN | nk_cell | *ASPA* | -1.1652832 | 5.02E-06 | 1.74E-05 | 0.024351384 |
| HTN | nk_cell | *RAD51C* | -3.043015461 | 3.15E-05 | 7.32E-05 | 0.15288792 |
| HTN | nk_cell | *TRIM37* | -2.634377868 | 0.000862684 | 0.001336871 | 1 |
| HTN | nk_cell | *PNPO* | 3.654749588 | 0.005593482 | 0.007557665 | 1 |
| HTN | nk_cell | *TRIM16L* | -1.307209086 | 3.71E-07 | 4.24E-06 | 0.001797745 |
| HTN | nk_cell | *CDK5RAP3* | -3.794283406 | 2.86E-05 | 6.75E-05 | 0.138798293 |
| HTN | nk_cell | *PIGL* | -2.070695495 | 0.000782505 | 0.001223956 | 1 |
| HTN | nk_cell | *MED13* | -1.982185536 | 7.22E-05 | 0.000145543 | 0.350177425 |
| HTN | nk_cell | *CHRNE* | 1.034889478 | 0.00010407 | 0.000200059 | 0.504949584 |
| HTN | nk_cell | *RAI1* | 3.04262556 | 0.001069364 | 0.001628037 | 1 |
| HTN | nk_cell | *GOSR1* | -2.053031131 | 2.04E-05 | 5.12E-05 | 0.098940538 |
| HTN | nk_cell | *DRG2* | -2.035804357 | 3.08E-06 | 1.21E-05 | 0.014946533 |
| HTN | nk_cell | *ICAM2* | 1.919442865 | 5.70E-07 | 4.50E-06 | 0.002765879 |
| HTN | nk_cell | *DDX5* | -1.219971692 | 0.011027821 | 0.014321999 | 1 |
| HTN | nk_cell | *C17orf75* | -2.12480379 | 1.15E-05 | 3.24E-05 | 0.055624808 |
| HTN | nk_cell | *PSMD11* | -2.252160942 | 0.000340648 | 0.000573698 | 1 |
| HTN | nk_cell | *PEX12* | 1.686002 | 2.50E-06 | 1.05E-05 | 0.012124235 |
| HTN | nk_cell | *DHX58* | -1.691025461 | 3.32E-06 | 1.28E-05 | 0.016126722 |
| HTN | nk_cell | *RAB5C* | 2.498534561 | 0.000148339 | 0.000274291 | 0.719739211 |
| HTN | nk_cell | *NAGLU* | -1.91813505 | 0.015446372 | 0.019645032 | 1 |
| HTN | nk_cell | *CNTNAP1* | -1.473117843 | 3.77E-06 | 1.41E-05 | 0.01829639 |
| HTN | nk_cell | *EZH1* | -1.749827133 | 9.03E-06 | 2.68E-05 | 0.043809643 |
| HTN | nk_cell | *EFTUD2* | -2.420043079 | 1.44E-05 | 3.89E-05 | 0.069916305 |
| HTN | nk_cell | *HLF* | -1.725930274 | 6.40E-05 | 0.000131043 | 0.310572334 |
| HTN | nk_cell | *EFNB3* | -2.476302823 | 1.01E-06 | 5.98E-06 | 0.004887687 |

| | | | | | | |
|---|---|---|---|---|---|---|
| HTN | nk_cell | *FAM20A* | -1.818017849 | 5.43E-06 | 1.83E-05 | 0.026333613 |
| HTN | nk_cell | *DPH1* | -1.482763211 | 2.99E-07 | 4.13E-06 | 0.001451207 |
| HTN | nk_cell | *DHRS7B* | -2.318134721 | 0.001567598 | 0.002306941 | 1 |
| HTN | nk_cell | *MYH3* | 5.760220983 | 2.52E-05 | 6.08E-05 | 0.122421973 |
| HTN | nk_cell | *OCIAD1* | -2.17955352 | 0.000281815 | 0.000483421 | 1 |
| HTN | nk_cell | *UBE2D3* | -1.865800908 | 7.99E-05 | 0.00015873 | 0.387618811 |
| HTN | nk_cell | *NDUFC1* | 3.177217455 | 8.54E-05 | 0.00016818 | 0.414227994 |
| HTN | nk_cell | *KLHL2* | -2.727220492 | 0.00024236 | 0.000423454 | 1 |
| HTN | nk_cell | *CPE* | -1.515832248 | 2.41E-05 | 5.87E-05 | 0.116804151 |
| HTN | nk_cell | *WFS1* | -3.621673577 | 0.000186204 | 0.000334988 | 0.903463934 |
| HTN | nk_cell | *GRPEL1* | 3.413929969 | 0.028200584 | 0.034701809 | 1 |
| HTN | nk_cell | *AADAT* | -1.56295827 | 3.94E-07 | 4.24E-06 | 0.00191019 |
| HTN | nk_cell | *TRIM2* | -2.158116145 | 9.12E-06 | 2.70E-05 | 0.04425494 |
| HTN | nk_cell | *BST1* | -2.089414292 | 8.68E-06 | 2.59E-05 | 0.042103476 |
| HTN | nk_cell | *KLHL5* | -1.910157684 | 1.09E-05 | 3.12E-05 | 0.05300483 |
| HTN | nk_cell | *FAM149A* | -1.946188551 | 3.59E-06 | 1.35E-05 | 0.017409623 |
| HTN | nk_cell | *DDX25* | -1.698533682 | 1.08E-05 | 3.08E-05 | 0.052178614 |
| HTN | nk_cell | *HTATIP2* | -1.330808308 | 0.022306247 | 0.027865579 | 1 |
| HTN | nk_cell | *ELP4* | -1.886207878 | 0.000406809 | 0.000672517 | 1 |
| HTN | nk_cell | *TECTA* | -1.710961191 | 4.45E-07 | 4.28E-06 | 0.002161538 |
| HTN | nk_cell | *SC5D* | 2.944109662 | 0.003472836 | 0.004846189 | 1 |
| HTN | nk_cell | *B3GAT1* | 0.311985159 | 2.79E-07 | 4.07E-06 | 0.001355006 |
| HTN | nk_cell | *P2RX3* | 2.027378824 | 2.74E-07 | 4.07E-06 | 0.001330568 |
| HTN | nk_cell | *DNAJC4* | 2.640586405 | 6.09E-06 | 1.99E-05 | 0.029546365 |
| HTN | nk_cell | *SNX15* | -3.206002625 | 3.00E-06 | 1.19E-05 | 0.014575371 |
| HTN | nk_cell | *UNC93B1* | -2.362670148 | 0.000150949 | 0.000278269 | 0.732403766 |
| HTN | nk_cell | *PUS3* | -1.761542636 | 0.000137253 | 0.000255839 | 0.665950148 |
| HTN | nk_cell | *FOXRED1* | -1.361842938 | 4.25E-07 | 4.24E-06 | 0.002061865 |
| HTN | nk_cell | *NRXN2* | -6.542996932 | 4.06E-06 | 1.48E-05 | 0.019712388 |
| HTN | nk_cell | *CCND1* | -4.511681297 | 1.38E-10 | 3.05E-08 | 6.72E-07 |
| HTN | nk_cell | *CCDC86* | -5.452221925 | 8.56E-06 | 2.57E-05 | 0.04155707 |
| HTN | nk_cell | *PANX1* | -2.987259117 | 2.37E-05 | 5.80E-05 | 0.115121803 |
| HTN | nk_cell | *CEP164* | -2.041339803 | 1.87E-05 | 4.78E-05 | 0.090647373 |
| HTN | nk_cell | *KIAA1549L* | -3.475326755 | 3.81E-07 | 4.24E-06 | 0.001848524 |
| HTN | nk_cell | *ACCS* | -1.289383539 | 0.000737343 | 0.00115742 | 1 |
| HTN | nk_cell | *SLC22A18* | 1.254907997 | 0.00019402 | 0.000347632 | 0.941387018 |
| HTN | nk_cell | *SLC35F2* | -2.822606345 | 1.21E-05 | 3.37E-05 | 0.05856491 |
| HTN | nk_cell | *C11orf21* | 0.962974259 | 1.21E-06 | 6.69E-06 | 0.005877047 |
| HTN | nk_cell | *ELMOD1* | 1.797739974 | 2.41E-06 | 1.02E-05 | 0.011700391 |
| HTN | nk_cell | *RPS13* | 1.806400458 | 0.018414977 | 0.023171543 | 1 |
| HTN | nk_cell | *NDUFS8* | -2.568062642 | 0.016509681 | 0.020926064 | 1 |
| HTN | nk_cell | *TCIRG1* | 2.426107808 | 0.031575544 | 0.038697787 | 1 |
| HTN | nk_cell | *GTF2H1* | -2.22621367 | 1.69E-05 | 4.43E-05 | 0.082175542 |
| HTN | nk_cell | *PRDM4* | -2.032609161 | 0.000506469 | 0.000821319 | 1 |
| HTN | nk_cell | *CORO1C* | -1.549303861 | 2.36E-06 | 1.01E-05 | 0.011465655 |
| HTN | nk_cell | *ASIC1* | -2.109709583 | 3.98E-07 | 4.24E-06 | 0.001932719 |
| HTN | nk_cell | *KCTD10* | -2.045880765 | 9.98E-08 | 3.32E-06 | 0.000484004 |
| HTN | nk_cell | *IL23A* | 16.25823892 | 6.92E-07 | 4.87E-06 | 0.003356946 |
| HTN | nk_cell | *ACSS3* | -1.385633632 | 9.94E-05 | 0.000191847 | 0.482111559 |
| HTN | nk_cell | *LTA4H* | -1.442268982 | 0.002379957 | 0.003410382 | 1 |

| | | | | | | |
|---|---|---|---|---|---|---|
| HTN | nk_cell | *PARP11* | -2.389556884 | 5.08E-05 | 0.000108561 | 0.246649869 |
| HTN | nk_cell | *GPN3* | -2.200256345 | 1.72E-06 | 8.12E-06 | 0.008331143 |
| HTN | nk_cell | *VPS29* | -2.271025793 | 0.000160262 | 0.000293431 | 0.77759294 |
| HTN | nk_cell | *AKAP3* | -1.21173338 | 1.82E-06 | 8.42E-06 | 0.008831034 |
| HTN | nk_cell | *ACAD10* | -3.026129171 | 1.25E-05 | 3.46E-05 | 0.060483671 |
| HTN | nk_cell | *LTBR* | -2.04650773 | 7.83E-06 | 2.41E-05 | 0.037969025 |
| HTN | nk_cell | *CDK2AP1* | 2.94038226 | 6.78E-07 | 4.81E-06 | 0.003291335 |
| HTN | nk_cell | *ENDOU* | -2.950709866 | 4.53E-07 | 4.28E-06 | 0.002196564 |
| HTN | nk_cell | *VDR* | -1.805490534 | 4.70E-07 | 4.32E-06 | 0.002281864 |
| HTN | nk_cell | *STX2* | -1.569515741 | 0.000559664 | 0.000899765 | 1 |
| HTN | nk_cell | *CAND1* | -2.337545111 | 0.000836234 | 0.001300034 | 1 |
| HTN | nk_cell | *MDM1* | -2.928764116 | 4.21E-05 | 9.28E-05 | 0.20425455 |
| HTN | nk_cell | *NUP107* | -2.245844301 | 6.48E-05 | 0.000132438 | 0.314275777 |
| HTN | nk_cell | *KRR1* | -2.576070696 | 7.96E-05 | 0.000158225 | 0.386226858 |
| HTN | nk_cell | *GNB3* | -1.776051408 | 2.89E-05 | 6.81E-05 | 0.140398613 |
| HTN | nk_cell | *CDCA3* | -3.916475724 | 8.46E-07 | 5.44E-06 | 0.004106879 |
| HTN | nk_cell | *CHPT1* | -1.828905774 | 0.000285731 | 0.000489537 | 1 |
| HTN | nk_cell | *GNPTAB* | 2.366672357 | 5.05E-05 | 0.000107887 | 0.244795424 |
| HTN | nk_cell | *SPSB2* | -1.806498855 | 2.54E-06 | 1.06E-05 | 0.012339412 |
| HTN | nk_cell | *ATN1* | -1.568240116 | 0.000326768 | 0.000551663 | 1 |
| HTN | nk_cell | *C12orf57* | 3.468511336 | 6.45E-06 | 2.07E-05 | 0.031304167 |
| HTN | nk_cell | *NT5DC3* | -2.198373979 | 0.006924239 | 0.009250112 | 1 |
| HTN | nk_cell | *SUDS3* | -1.955262376 | 3.76E-06 | 1.40E-05 | 0.018219895 |
| HTN | nk_cell | *GOLT1B* | 3.363398806 | 0.000244369 | 0.000426196 | 1 |
| HTN | nk_cell | *C2CD5* | -2.070567105 | 0.000179385 | 0.000324162 | 0.870376169 |
| HTN | nk_cell | *FRK* | -1.862699941 | 1.29E-06 | 6.88E-06 | 0.006275819 |
| HTN | nk_cell | *RWDD1* | 1.793510011 | 0.039618546 | 0.04812949 | 1 |
| HTN | nk_cell | *TMEM14C* | -2.71537379 | 3.17E-05 | 7.36E-05 | 0.15380386 |
| HTN | nk_cell | *SMIM8* | -1.543565692 | 5.19E-05 | 0.000110187 | 0.251998083 |
| HTN | nk_cell | *FAM184A* | -1.580168271 | 6.23E-06 | 2.03E-05 | 0.030219201 |
| HTN | nk_cell | *RNGTT* | -2.135758336 | 0.000103239 | 0.000198697 | 0.500914644 |
| HTN | nk_cell | *HDDC2* | 2.619248935 | 0.001138055 | 0.001722884 | 1 |
| HTN | nk_cell | *UST* | -1.240217298 | 4.92E-07 | 4.32E-06 | 0.002388315 |
| HTN | nk_cell | *TULP1* | -3.7447211 | 9.43E-07 | 5.82E-06 | 0.004573342 |
| HTN | nk_cell | *RNF8* | -2.95240985 | 1.81E-05 | 4.66E-05 | 0.087724856 |
| HTN | nk_cell | *PHACTR1* | -1.88503362 | 0.001246599 | 0.001870284 | 1 |
| HTN | nk_cell | *MDGA1* | -2.514870039 | 9.75E-07 | 5.90E-06 | 0.004731575 |
| HTN | nk_cell | *CD83* | -2.479175842 | 2.22E-06 | 9.65E-06 | 0.010778305 |
| HTN | nk_cell | *MDN1* | -1.777135216 | 6.00E-06 | 1.97E-05 | 0.029090965 |
| HTN | nk_cell | *GLP1R* | -8.839948825 | 6.45E-06 | 2.07E-05 | 0.031286454 |
| HTN | nk_cell | *BVES* | -1.446544679 | 1.85E-07 | 4.07E-06 | 0.000898024 |
| HTN | nk_cell | *COL9A1* | -3.336951819 | 3.39E-07 | 4.24E-06 | 0.001645903 |
| HTN | nk_cell | *WASF1* | -1.682229371 | 6.94E-07 | 4.87E-06 | 0.003367453 |
| HTN | nk_cell | *ALDH5A1* | -1.465726492 | 0.03732236 | 0.045408248 | 1 |
| HTN | nk_cell | *VNN1* | -1.102660781 | 0.017516573 | 0.022121399 | 1 |
| HTN | nk_cell | *VNN2* | 2.124478595 | 1.86E-07 | 4.07E-06 | 0.000902666 |
| HTN | nk_cell | *C6orf62* | -2.503657286 | 7.49E-05 | 0.000150339 | 0.363369178 |
| HTN | nk_cell | *HBS1L* | -2.150143819 | 1.28E-05 | 3.52E-05 | 0.061888246 |
| HTN | nk_cell | *PEX7* | -1.420316142 | 0.010105047 | 0.013197763 | 1 |
| HTN | nk_cell | *FIG4* | -2.371146376 | 0.0010178 | 0.0015549 | 1 |

| | | | | | | |
|---|---|---|---|---|---|---|
| HTN | nk_cell | *PERP* | -1.933534607 | 0.000397209 | 0.00066002 | 1 |
| HTN | nk_cell | *PHACTR2* | -1.954400938 | 0.001078867 | 0.001640961 | 1 |
| HTN | nk_cell | *CUTA* | 1.34295843 | 7.76E-05 | 0.000154892 | 0.376541723 |
| HTN | nk_cell | *PACRG* | -1.237585682 | 5.97E-07 | 4.54E-06 | 0.002894543 |
| HTN | nk_cell | *QKI* | -1.60264938 | 5.24E-05 | 0.000111022 | 0.254241465 |
| HTN | nk_cell | *FAM120B* | -1.783612821 | 0.001008697 | 0.001542451 | 1 |
| HTN | nk_cell | *GUCA1B* | -1.654958155 | 4.18E-07 | 4.24E-06 | 0.002028617 |
| HTN | nk_cell | *PRPH2* | 0.706328359 | 0.00026297 | 0.00045447 | 1 |
| HTN | nk_cell | *TMEM30A* | -2.21806314 | 6.55E-06 | 2.10E-05 | 0.031765831 |
| HTN | nk_cell | *IMPG1* | -3.151708347 | 9.24E-08 | 3.18E-06 | 0.000448359 |
| HTN | nk_cell | *BTN2A1* | 2.99127373 | 0.005968755 | 0.00803117 | 1 |
| HTN | nk_cell | *LY86* | -3.697380312 | 0.004763692 | 0.006499841 | 1 |
| HTN | nk_cell | *PRSS16* | -3.202149223 | 0.001785444 | 0.002597593 | 1 |
| HTN | nk_cell | *PCDHB2* | -2.509775463 | 2.56E-07 | 4.07E-06 | 0.001244485 |
| HTN | nk_cell | *NUDT12* | -2.863278581 | 2.55E-05 | 6.14E-05 | 0.123852073 |
| HTN | nk_cell | *CEP72* | 4.510937892 | 1.04E-05 | 3.01E-05 | 0.050598912 |
| HTN | nk_cell | *NME5* | -0.831914509 | 0.003285257 | 0.004598981 | 1 |
| HTN | nk_cell | *KIF20A* | 4.391712932 | 1.96E-06 | 8.91E-06 | 0.009526822 |
| HTN | nk_cell | *PFDN1* | -1.874912892 | 2.87E-05 | 6.77E-05 | 0.139232822 |
| HTN | nk_cell | *SPARC* | -1.921580786 | 0.000104589 | 0.000200579 | 0.507465113 |
| HTN | nk_cell | *IK* | 3.157887895 | 9.56E-07 | 5.84E-06 | 0.004636098 |
| HTN | nk_cell | *HAVCR1* | 2.707684288 | 0.005475442 | 0.007406424 | 1 |
| HTN | nk_cell | *RNF130* | -1.150372732 | 0.020083409 | 0.025192529 | 1 |
| HTN | nk_cell | *THBS4* | -1.281703083 | 2.10E-06 | 9.26E-06 | 0.010204831 |
| HTN | nk_cell | *BTNL8* | -0.968100626 | 3.34E-05 | 7.65E-05 | 0.16204647 |
| HTN | nk_cell | *MSH3* | -1.774756164 | 0.0001941 | 0.000347646 | 0.941772227 |
| HTN | nk_cell | *RASGRF2* | -1.071145195 | 2.81E-06 | 1.13E-05 | 0.013655369 |
| HTN | nk_cell | *DROSHA* | -2.063501065 | 4.08E-05 | 9.03E-05 | 0.197830253 |
| HTN | nk_cell | *CDH6* | -0.912702926 | 6.22E-07 | 4.57E-06 | 0.0030173 |
| HTN | nk_cell | *LMNB1* | -2.381611762 | 0.034341928 | 0.041929299 | 1 |
| HTN | nk_cell | *SLC12A7* | -2.592699158 | 0.024575987 | 0.030520268 | 1 |
| HTN | nk_cell | *SKP1* | -2.19527855 | 2.20E-05 | 5.44E-05 | 0.106984107 |
| HTN | nk_cell | *PPWD1* | -2.324793816 | 0.001088111 | 0.001652948 | 1 |
| HTN | nk_cell | *TXNDC15* | -2.295855359 | 6.99E-06 | 2.19E-05 | 0.033930177 |
| HTN | nk_cell | *SMAD5* | -2.087705671 | 0.006932734 | 0.009258912 | 1 |
| HTN | nk_cell | *ERGIC1* | -2.30979242 | 0.001486968 | 0.002198285 | 1 |
| HTN | nk_cell | *ATP6V0E1* | -2.286224197 | 0.000292612 | 0.000500265 | 1 |
| HTN | nk_cell | *BNIP1* | 2.031308666 | 2.23E-08 | 1.38E-06 | 0.000108298 |
| HTN | nk_cell | *UNC5A* | -4.135103914 | 4.52E-07 | 4.28E-06 | 0.002194082 |
| HTN | nk_cell | *ACTR8* | -3.393525756 | 9.48E-07 | 5.82E-06 | 0.004601026 |
| HTN | nk_cell | *TBCCD1* | -3.351817758 | 0.000228757 | 0.000403024 | 1 |
| HTN | nk_cell | *CLDN16* | -1.139806924 | 6.63E-06 | 2.12E-05 | 0.032192026 |
| HTN | nk_cell | *CEP70* | -2.068492472 | 0.006126315 | 0.00823632 | 1 |
| HTN | nk_cell | *HYAL1* | -2.727544687 | 0.000573088 | 0.000919214 | 1 |
| HTN | nk_cell | *C3orf14* | -2.280965707 | 0.000260112 | 0.000450255 | 1 |
| HTN | nk_cell | *IQCG* | -2.316600075 | 3.45E-05 | 7.84E-05 | 0.16716297 |
| HTN | nk_cell | *SLC41A3* | -1.661389799 | 7.36E-07 | 5.02E-06 | 0.003571436 |
| HTN | nk_cell | *PLXNA1* | -2.229777569 | 2.15E-05 | 5.33E-05 | 0.104308133 |
| HTN | nk_cell | *SCAP* | -2.235098844 | 0.000804544 | 0.001255192 | 1 |
| HTN | nk_cell | *KIAA1257* | -2.888771937 | 5.31E-07 | 4.40E-06 | 0.002577132 |

| | | | | | | |
|---|---|---|---|---|---|---|
| HTN | nk_cell | *NEK11* | -1.407022718 | 9.21E-06 | 2.72E-05 | 0.044673695 |
| HTN | nk_cell | *PLSCR4* | -1.431634903 | 9.85E-06 | 2.87E-05 | 0.047809478 |
| HTN | nk_cell | *HEMK1* | -2.878869723 | 5.32E-05 | 0.000112412 | 0.257986475 |
| HTN | nk_cell | *AADAC* | -0.661442796 | 0.000552584 | 0.000889561 | 1 |
| HTN | nk_cell | *ABHD14B* | -3.193936284 | 0.000291901 | 0.000499225 | 1 |
| HTN | nk_cell | *ARHGEF26* | -2.056021021 | 4.20E-06 | 1.51E-05 | 0.020402133 |
| HTN | nk_cell | *VIPR1* | -1.806375302 | 8.54E-07 | 5.47E-06 | 0.004145162 |
| HTN | nk_cell | *DNAH1* | -1.642177944 | 0.000100097 | 0.000193186 | 0.485670487 |
| HTN | nk_cell | *NKTR* | -1.746977668 | 0.0004327 | 0.000710856 | 1 |
| HTN | nk_cell | *NEK4* | -2.244285299 | 1.35E-07 | 3.84E-06 | 0.000652609 |
| HTN | nk_cell | *ADAM23* | -3.188829939 | 2.82E-06 | 1.13E-05 | 0.013671701 |
| HTN | nk_cell | *LMAN2L* | -1.764576575 | 4.62E-06 | 1.63E-05 | 0.022435835 |
| HTN | nk_cell | *ACTR1B* | -2.336197276 | 0.0290768 | 0.035743763 | 1 |
| HTN | nk_cell | *SLC35F5* | -2.174845995 | 2.43E-06 | 1.02E-05 | 0.011768887 |
| HTN | nk_cell | *STEAP3* | -0.424670789 | 0.014987357 | 0.019111342 | 1 |
| HTN | nk_cell | *OTOF* | 3.961858915 | 0.00134427 | 0.002000736 | 1 |
| HTN | nk_cell | *CENPA* | -1.553481262 | 2.04E-06 | 9.07E-06 | 0.009893116 |
| HTN | nk_cell | *NRBP1* | -2.631184278 | 0.001173172 | 0.001771625 | 1 |
| HTN | nk_cell | *ITGB6* | -1.417255642 | 7.42E-07 | 5.05E-06 | 0.00359931 |
| HTN | nk_cell | *PPM1G* | 2.849555456 | 4.93E-07 | 4.32E-06 | 0.002391365 |
| HTN | nk_cell | *MOGS* | -1.950133142 | 1.81E-05 | 4.66E-05 | 0.08771666 |
| HTN | nk_cell | *GRB14* | -2.485635571 | 2.76E-06 | 1.11E-05 | 0.013389302 |
| HTN | nk_cell | *RTN4* | -1.58524632 | 3.43E-05 | 7.81E-05 | 0.166330429 |
| HTN | nk_cell | *GALNT3* | 0.71193693 | 1.82E-07 | 4.07E-06 | 0.000885368 |
| HTN | nk_cell | *POLE4* | 3.810587476 | 0.000656601 | 0.001038405 | 1 |
| HTN | nk_cell | *ACADL* | -1.799510531 | 0.000984394 | 0.001507188 | 1 |
| HTN | nk_cell | *MRPL19* | -2.006140726 | 4.21E-06 | 1.51E-05 | 0.020420161 |
| HTN | nk_cell | *IGFBP2* | -2.436410856 | 0.000416482 | 0.000687337 | 1 |
| HTN | nk_cell | *GNLY* | 0.099783976 | 2.75E-07 | 4.07E-06 | 0.001332097 |
| HTN | nk_cell | *CHMP3* | -2.647995063 | 0.000439572 | 0.000720542 | 1 |
| HTN | nk_cell | *WNT6* | -1.988568184 | 5.87E-08 | 2.37E-06 | 0.000284823 |
| HTN | nk_cell | *IL1RL2* | -2.45223923 | 1.31E-06 | 6.91E-06 | 0.006342506 |
| HTN | nk_cell | *IL18R1* | 1.283726029 | 2.26E-06 | 9.77E-06 | 0.010983763 |
| HTN | nk_cell | *MLPH* | -1.85282716 | 1.92E-05 | 4.89E-05 | 0.093176111 |
| HTN | nk_cell | *HDLBP* | -1.989646897 | 0.000612737 | 0.000976997 | 1 |
| HTN | nk_cell | *PASK* | -1.749615022 | 2.72E-05 | 6.46E-05 | 0.131998816 |
| HTN | nk_cell | *TPO* | -2.491875333 | 7.48E-06 | 2.32E-05 | 0.036303227 |
| HTN | nk_cell | *TAF1B* | -2.041752222 | 9.26E-05 | 0.000180792 | 0.449268794 |
| HTN | nk_cell | *NOL10* | -2.22699841 | 0.000103369 | 0.000198868 | 0.501544076 |
| HTN | nk_cell | *STRN* | -2.002341461 | 4.20E-05 | 9.27E-05 | 0.203948635 |
| HTN | nk_cell | *CEBPZ* | -1.971921381 | 0.006285144 | 0.008435828 | 1 |
| HTN | nk_cell | *PRKD3* | -1.935112834 | 0.00197255 | 0.002851852 | 1 |
| HTN | nk_cell | *SLC25A12* | -2.2408002 | 0.000323913 | 0.000547223 | 1 |
| HTN | nk_cell | *DARS* | -2.059538999 | 0.006515688 | 0.008728359 | 1 |
| HTN | nk_cell | *WIPF1* | 1.972505461 | 1.05E-06 | 6.12E-06 | 0.005113029 |
| HTN | nk_cell | *PNO1* | -3.640114771 | 1.28E-05 | 3.53E-05 | 0.06221308 |
| HTN | nk_cell | *ORC4* | -1.724389525 | 0.000395225 | 0.000657624 | 1 |
| HTN | nk_cell | *CD207* | -2.157750147 | 0.012751647 | 0.016398354 | 1 |
| HTN | nk_cell | *GRIN3B* | 1.033068671 | 3.98E-07 | 4.24E-06 | 0.001932404 |
| HTN | nk_cell | *NFE2L2* | -1.694150332 | 5.38E-05 | 0.000113578 | 0.261244244 |

| | | | | | | |
|---|---|---|---|---|---|---|
| HTN | nk_cell | *MSH6* | -2.237870529 | 9.24E-06 | 2.72E-05 | 0.044832887 |
| HTN | nk_cell | *ALMS1* | -2.089045307 | 3.00E-05 | 7.03E-05 | 0.145751426 |
| HTN | nk_cell | *DNAJC16* | -1.842009289 | 1.27E-06 | 6.83E-06 | 0.006179683 |
| HTN | nk_cell | *MARK1* | -1.880893659 | 9.74E-06 | 2.85E-05 | 0.047273162 |
| HTN | nk_cell | *MORN1* | -1.940701977 | 1.77E-08 | 1.21E-06 | 8.59E-05 |
| HTN | nk_cell | *GPX7* | 0.765494598 | 3.39E-07 | 4.24E-06 | 0.001645175 |
| HTN | nk_cell | *RALGPS2* | -1.042444922 | 6.65E-07 | 4.74E-06 | 0.003225547 |
| HTN | nk_cell | *CEP104* | -2.372009139 | 0.000391772 | 0.000652326 | 1 |
| HTN | nk_cell | *FAM20B* | -2.679360336 | 0.000207246 | 0.000368606 | 1 |
| HTN | nk_cell | *WRAP73* | 2.74362857 | 3.20E-05 | 7.41E-05 | 0.155302764 |
| HTN | nk_cell | *ICMT* | -3.844269288 | 1.35E-05 | 3.70E-05 | 0.065688658 |
| HTN | nk_cell | *PARK7* | -2.633312879 | 0.000427208 | 0.000702888 | 1 |
| HTN | nk_cell | *KIAA1324* | 1.209516864 | 1.06E-07 | 3.44E-06 | 0.000516133 |
| HTN | nk_cell | *MECR* | -1.975885542 | 3.65E-07 | 4.24E-06 | 0.001772263 |
| HTN | nk_cell | *KCNC4* | -1.986877492 | 1.57E-05 | 4.18E-05 | 0.076372643 |
| HTN | nk_cell | *EDEM3* | -2.351703509 | 0.001523658 | 0.002248415 | 1 |
| HTN | nk_cell | *CAPZA1* | -2.526063818 | 2.09E-05 | 5.22E-05 | 0.101383265 |
| HTN | nk_cell | *ASH1L* | -1.579510974 | 0.000274139 | 0.000471842 | 1 |
| HTN | nk_cell | *GON4L* | -1.835391259 | 4.90E-05 | 0.000105338 | 0.23774892 |
| HTN | nk_cell | *DOCK7* | -1.832323102 | 1.29E-05 | 3.55E-05 | 0.062650066 |
| HTN | nk_cell | *FBXO2* | 1.459478384 | 1.11E-07 | 3.53E-06 | 0.000537659 |
| HTN | nk_cell | *DNAJC6* | 2.098419621 | 0.01430513 | 0.018265392 | 1 |
| HTN | nk_cell | *LEPR* | -1.985752088 | 2.05E-05 | 5.15E-05 | 0.099602122 |
| HTN | nk_cell | *IVNS1ABP* | 1.534193487 | 0.011798787 | 0.01527013 | 1 |
| HTN | nk_cell | *PRG4* | -1.720796169 | 1.07E-06 | 6.15E-06 | 0.005176198 |
| HTN | nk_cell | *BCAS2* | -1.677426426 | 0.000278588 | 0.000478312 | 1 |
| HTN | nk_cell | *CFHR3* | -3.3935945 | 9.82E-05 | 0.000189744 | 0.476637828 |
| HTN | nk_cell | *CRYZ* | -0.790198367 | 0.027562178 | 0.033976546 | 1 |
| HTN | nk_cell | *TMEM9* | -2.453760543 | 4.20E-07 | 4.24E-06 | 0.002038238 |
| HTN | nk_cell | *WARS2* | -1.989506332 | 6.13E-06 | 2.00E-05 | 0.029758877 |
| HTN | nk_cell | *OSCP1* | -1.625955512 | 6.57E-06 | 2.10E-05 | 0.031890927 |
| HTN | nk_cell | *MRPS15* | -2.606622588 | 2.96E-05 | 6.96E-05 | 0.143825883 |
| HTN | nk_cell | *GNPAT* | -2.525006659 | 0.00438778 | 0.006015685 | 1 |
| HTN | nk_cell | *C1orf109* | -1.647659106 | 8.49E-07 | 5.45E-06 | 0.004119277 |
| HTN | nk_cell | *TBCE* | -2.461177964 | 3.86E-05 | 8.63E-05 | 0.187430479 |
| HTN | nk_cell | *ACADM* | -1.886421073 | 9.08E-05 | 0.000177752 | 0.440646652 |
| HTN | nk_cell | *PADI2* | 1.878643848 | 0.000134766 | 0.000251881 | 0.653884283 |
| HTN | nk_cell | *SDHB* | -2.53757597 | 0.000231363 | 0.000406729 | 1 |
| HTN | nk_cell | *KLHL12* | -2.789496341 | 0.000360859 | 0.000604588 | 1 |
| HTN | nk_cell | *ZNHIT6* | -2.298961809 | 0.000510657 | 0.000827281 | 1 |
| HTN | nk_cell | *KIF17* | -5.05529473 | 6.53E-06 | 2.09E-05 | 0.031679891 |
| HTN | nk_cell | *CD46* | -1.278048398 | 0.020435068 | 0.025620401 | 1 |
| HTN | nk_cell | *EBNA1BP2* | -1.571630927 | 1.24E-06 | 6.76E-06 | 0.006026821 |
| HTN | nk_cell | *ERI3* | -2.146122189 | 6.00E-05 | 0.000124028 | 0.290970319 |
| HTN | nk_cell | *PRRC2C* | -1.3823747 | 0.035143014 | 0.042864229 | 1 |
| HTN | nk_cell | *ABCD3* | -2.266329219 | 3.44E-05 | 7.84E-05 | 0.167078329 |
| HTN | nk_cell | *PTBP2* | -1.707049787 | 0.000171301 | 0.000311067 | 0.831153363 |
| HTN | nk_cell | *IRF6* | -1.846134066 | 5.69E-06 | 1.90E-05 | 0.027609115 |
| HTN | nk_cell | *SERPINC1* | -3.069884468 | 0.003899107 | 0.005382209 | 1 |
| HTN | nk_cell | *MTFR1L* | -1.966679622 | 1.84E-06 | 8.51E-06 | 0.008939923 |

| | | | | | | |
|---|---|---|---|---|---|---|
| HTN | nk_cell | *RPS6KA1* | 0.873688222 | 4.76E-07 | 4.32E-06 | 0.002308935 |
| HTN | nk_cell | *MARC2* | -1.612671787 | 2.38E-06 | 1.01E-05 | 0.011550956 |
| HTN | nk_cell | *SLC5A9* | -1.247621174 | 9.74E-07 | 5.90E-06 | 0.004725959 |
| HTN | nk_cell | *ESYT2* | -1.732285227 | 0.003953778 | 0.005449924 | 1 |
| HTN | nk_cell | *CD3EAP* | -1.79420228 | 8.28E-06 | 2.51E-05 | 0.040190374 |
| HTN | nk_cell | *RCN2* | -2.243049204 | 0.000121739 | 0.000229924 | 0.590675861 |
| HTN | nk_cell | *A4GNT* | -1.509551145 | 0.003776469 | 0.005232276 | 1 |
| HTN | nk_cell | *IFT46* | -1.868339962 | 3.92E-07 | 4.24E-06 | 0.001902742 |
| HTN | nk_cell | *APOA1* | -6.813218243 | 0.000457614 | 0.000748599 | 1 |
| HTN | nk_cell | *DDX59* | -2.941642064 | 1.66E-06 | 7.94E-06 | 0.008043682 |
| HTN | nk_cell | *CAMSAP2* | -1.487283811 | 4.33E-05 | 9.49E-05 | 0.210080377 |
| HTN | nk_cell | *KLF7* | -1.728585275 | 0.000353571 | 0.000593608 | 1 |
| HTN | nk_cell | *CASC1* | -1.829690164 | 9.44E-06 | 2.77E-05 | 0.045795303 |
| HTN | nk_cell | *LRMP* | 1.504862626 | 0.000610755 | 0.000974156 | 1 |
| HTN | nk_cell | *ELOVL4* | -1.560632595 | 0.01737231 | 0.021974129 | 1 |
| HTN | nk_cell | *CNR1* | -1.224428806 | 2.58E-06 | 1.07E-05 | 0.012531004 |
| HTN | nk_cell | *AKAP7* | -2.104099727 | 0.000299833 | 0.000511171 | 1 |
| HTN | nk_cell | *ALDH8A1* | -1.035227747 | 0.00052246 | 0.000844992 | 1 |
| HTN | nk_cell | *MED28* | -1.963026185 | 4.43E-06 | 1.58E-05 | 0.02151807 |
| HTN | nk_cell | *VAMP8* | 2.830911562 | 0.000453578 | 0.000742497 | 1 |
| HTN | nk_cell | *FOXO3* | -1.715574115 | 0.000183179 | 0.000330157 | 0.888783659 |
| HTN | nk_cell | *SPP1* | 2.005527864 | 6.01E-07 | 4.54E-06 | 0.002914333 |
| HTN | nk_cell | *MFSD1* | -2.110653914 | 1.62E-05 | 4.29E-05 | 0.078715473 |
| HTN | nk_cell | *UBN1* | -2.118515496 | 0.001790639 | 0.002604371 | 1 |
| HTN | nk_cell | *WDR35* | -2.170021279 | 5.14E-05 | 0.000109321 | 0.249200888 |
| HTN | nk_cell | *DNAH7* | -2.511828089 | 4.83E-06 | 1.68E-05 | 0.023422804 |
| HTN | nk_cell | *C2orf40* | -2.734953865 | 0.000436373 | 0.000716024 | 1 |
| HTN | nk_cell | *CPSF3* | -2.71794933 | 4.50E-05 | 9.80E-05 | 0.218423397 |
| HTN | nk_cell | *PIGZ* | -0.745850137 | 0.017398449 | 0.022000854 | 1 |
| HTN | nk_cell | *SENP5* | -1.805085618 | 8.29E-06 | 2.51E-05 | 0.040208826 |
| HTN | nk_cell | *CCDC92* | -2.296347245 | 1.59E-05 | 4.23E-05 | 0.077358333 |
| HTN | nk_cell | *FKBP15* | -1.141034766 | 0.015312444 | 0.019484914 | 1 |
| HTN | nk_cell | *CTNNAL1* | -1.363089173 | 0.004368201 | 0.005990534 | 1 |
| HTN | nk_cell | *RAB14* | -2.27657473 | 0.007625623 | 0.010114687 | 1 |
| HTN | nk_cell | *TRIM32* | 4.726856466 | 0.000136475 | 0.000254586 | 0.662177824 |
| HTN | nk_cell | *PPP6C* | -2.084932111 | 1.17E-05 | 3.29E-05 | 0.056637664 |
| HTN | nk_cell | *HDHD3* | -1.020046877 | 0.000830123 | 0.001291362 | 1 |
| HTN | nk_cell | *HSDL2* | -1.071284073 | 0.00018254 | 0.000329251 | 0.885685682 |
| HTN | nk_cell | *MAPKAP1* | -2.047233324 | 0.000414564 | 0.000684637 | 1 |
| HTN | nk_cell | *INVS* | -1.473815899 | 0.003228421 | 0.004533805 | 1 |
| HTN | nk_cell | *DENND1A* | -1.722393079 | 0.000154176 | 0.000283035 | 0.748062033 |
| HTN | nk_cell | *KDSR* | -1.99538929 | 6.14E-06 | 2.00E-05 | 0.029800383 |
| HTN | nk_cell | *DCAF4* | -2.498906546 | 1.87E-07 | 4.07E-06 | 0.000905094 |
| HTN | nk_cell | *IFI27L2* | 3.242387324 | 0.00033417 | 0.000563374 | 1 |
| HTN | nk_cell | *NEK9* | -1.977362336 | 2.75E-05 | 6.51E-05 | 0.133347794 |
| HTN | nk_cell | *ACOT2* | 0.663211793 | 0.018022856 | 0.02271938 | 1 |
| HTN | nk_cell | *TTLL5* | -2.149709797 | 0.000353825 | 0.000593829 | 1 |
| HTN | nk_cell | *FLVCR2* | -2.58588776 | 9.01E-06 | 2.68E-05 | 0.043715816 |
| HTN | nk_cell | *PPP4R4* | -1.533610725 | 4.61E-06 | 1.62E-05 | 0.022371238 |
| HTN | nk_cell | *ALDH6A1* | -1.396667513 | 0.000547446 | 0.00088246 | 1 |

| | | | | | | |
|---|---|---|---|---|---|---|
| HTN | nk_cell | *COQ6* | -2.633843007 | 0.016474587 | 0.020887038 | 1 |
| HTN | nk_cell | *DNMT3A* | -2.123527021 | 5.55E-05 | 0.000116561 | 0.269176881 |
| HTN | nk_cell | *CNRIP1* | -1.4535629 | 0.038835668 | 0.04720207 | 1 |
| HTN | nk_cell | *CRIPT* | -2.356527206 | 0.034701382 | 0.042336209 | 1 |
| HTN | nk_cell | *EPCAM* | -2.587436205 | 5.24E-06 | 1.79E-05 | 0.025427279 |
| HTN | nk_cell | *TECTB* | -1.804568002 | 0.005536846 | 0.00748531 | 1 |
| HTN | nk_cell | *ELOVL3* | -3.006499612 | 7.55E-05 | 0.000151265 | 0.366212609 |
| HTN | nk_cell | *PPP1R3C* | -1.755033967 | 9.79E-07 | 5.92E-06 | 0.004750273 |
| HTN | nk_cell | *PYROXD2* | -1.532834349 | 4.09E-06 | 1.49E-05 | 0.019839187 |
| HTN | nk_cell | *MXI1* | -1.853850958 | 5.37E-06 | 1.82E-05 | 0.026054939 |
| HTN | nk_cell | *C10orf88* | -2.152169621 | 0.013348054 | 0.017124473 | 1 |
| HTN | nk_cell | *HELLS* | 1.897751881 | 0.002738761 | 0.003886653 | 1 |
| HTN | nk_cell | *TCTN3* | -1.958613412 | 2.68E-06 | 1.10E-05 | 0.01298758 |
| HTN | nk_cell | *WDR11* | -1.96604418 | 6.52E-05 | 0.00013311 | 0.316137145 |
| HTN | nk_cell | *KANSL1* | -1.636058785 | 0.002587501 | 0.003688178 | 1 |
| HTN | nk_cell | *HOXB3* | -1.999537093 | 6.81E-06 | 2.15E-05 | 0.033039944 |
| HTN | nk_cell | *PCDHB10* | -3.095339022 | 0.000161005 | 0.00029468 | 0.781197151 |
| HTN | nk_cell | *GORAB* | -2.574725577 | 5.76E-05 | 0.000119966 | 0.27971568 |
| HTN | nk_cell | *SNX19* | -2.400217135 | 2.86E-07 | 4.07E-06 | 0.001387096 |
| HTN | nk_cell | *SLC10A7* | -2.217829591 | 5.52E-05 | 0.000116108 | 0.267976861 |
| HTN | nk_cell | *MASTL* | -2.475765774 | 1.74E-05 | 4.53E-05 | 0.084429954 |
| HTN | nk_cell | *MRC1* | -2.051283229 | 0.000347392 | 0.000584448 | 1 |
| HTN | nk_cell | *IQSEC3* | -1.324253553 | 5.14E-07 | 4.37E-06 | 0.0024963 |
| HTN | nk_cell | *CCDC77* | -3.603769944 | 0.000129906 | 0.000243548 | 0.630301665 |
| HTN | nk_cell | *DNAJC15* | -2.244281996 | 0.009128638 | 0.011969246 | 1 |
| HTN | nk_cell | *UFM1* | -2.583558873 | 1.39E-05 | 3.79E-05 | 0.067637894 |
| HTN | nk_cell | *ALG5* | -2.643345223 | 2.19E-05 | 5.40E-05 | 0.106020838 |
| HTN | nk_cell | *SIL1* | -1.602236845 | 6.70E-06 | 2.13E-05 | 0.032486997 |
| HTN | nk_cell | *ARL1* | -3.226490426 | 3.15E-05 | 7.32E-05 | 0.152784074 |
| HTN | nk_cell | *APAF1* | -2.737783879 | 0.000643262 | 0.001019637 | 1 |
| HTN | nk_cell | *EPHX2* | -2.253045144 | 1.09E-06 | 6.22E-06 | 0.005270672 |
| HTN | nk_cell | *AKAP1* | -1.498937006 | 8.07E-07 | 5.28E-06 | 0.00391609 |
| HTN | nk_cell | *SCPEP1* | -1.16453012 | 0.014131058 | 0.018062143 | 1 |
| HTN | nk_cell | *TBX2* | -1.791425271 | 4.82E-12 | 1.80E-09 | 2.34E-08 |
| HTN | nk_cell | *TEX14* | -1.128601251 | 0.004755404 | 0.006492184 | 1 |
| HTN | nk_cell | *TRIM6* | 7.783045436 | 0.000251601 | 0.000437865 | 1 |
| HTN | nk_cell | *ADCY7* | 1.148596867 | 9.08E-07 | 5.69E-06 | 0.004404609 |
| HTN | nk_cell | *ECHDC2* | -1.638552281 | 1.70E-05 | 4.44E-05 | 0.08240469 |
| HTN | nk_cell | *ZNF549* | -2.967796732 | 0.00321173 | 0.004512979 | 1 |
| HTN | nk_cell | *A1BG* | 2.69914084 | 5.45E-07 | 4.44E-06 | 0.00264193 |
| HTN | nk_cell | *ZSCAN18* | -2.444422751 | 1.19E-05 | 3.34E-05 | 0.057821402 |
| HTN | nk_cell | *PDZRN3* | -1.006580963 | 0.003044814 | 0.004292108 | 1 |
| HTN | nk_cell | *LHX4* | -2.264144702 | 1.27E-06 | 6.81E-06 | 0.006150423 |
| HTN | nk_cell | *SEC22A* | -2.091128341 | 6.72E-05 | 0.000137 | 0.326197135 |
| HTN | nk_cell | *POPDC2* | -1.902677891 | 1.15E-07 | 3.57E-06 | 0.000560299 |
| HTN | nk_cell | *CD80* | -1.718904044 | 5.91E-05 | 0.000122591 | 0.286985739 |
| HTN | nk_cell | *MAPK8IP1* | 1.562837963 | 8.23E-06 | 2.50E-05 | 0.039933129 |
| HTN | nk_cell | *CAT* | -1.36689248 | 4.02E-06 | 1.47E-05 | 0.019515948 |
| HTN | nk_cell | *PILRB* | 1.192371221 | 9.43E-05 | 0.000183485 | 0.457611456 |
| HTN | nk_cell | *ZCCHC17* | -2.451274009 | 2.40E-05 | 5.85E-05 | 0.116491436 |

| | | | | | | |
|---|---|---|---|---|---|---|
| HTN | nk_cell | *FABP3* | -2.430661299 | 1.63E-07 | 4.07E-06 | 0.000789082 |
| HTN | nk_cell | *TNFSF10* | -2.122619036 | 6.22E-05 | 0.000128028 | 0.301911926 |
| HTN | nk_cell | *ZNF639* | -3.025180352 | 1.93E-05 | 4.92E-05 | 0.093817673 |
| HTN | nk_cell | *CPXM2* | -1.084065345 | 4.59E-07 | 4.29E-06 | 0.002229409 |
| HTN | nk_cell | *FLT3* | 2.377542752 | 9.75E-08 | 3.29E-06 | 0.000473067 |
| HTN | nk_cell | *MTIF3* | -2.340583007 | 1.34E-05 | 3.65E-05 | 0.064850762 |
| HTN | nk_cell | *FMOD* | -1.851879453 | 2.38E-06 | 1.01E-05 | 0.01156202 |
| HTN | nk_cell | *CD244* | 0.254537343 | 2.62E-07 | 4.07E-06 | 0.001272189 |
| HTN | nk_cell | *RBBP6* | -2.135808749 | 8.66E-05 | 0.000170085 | 0.419940807 |
| HTN | nk_cell | *SERAC1* | 2.961539575 | 9.79E-05 | 0.000189279 | 0.474769322 |
| HTN | nk_cell | *LDB3* | -1.095487185 | 9.99E-07 | 5.95E-06 | 0.004845605 |
| HTN | nk_cell | *OPN4* | -6.606551952 | 1.58E-05 | 4.19E-05 | 0.076550124 |
| HTN | nk_cell | *FAM35A* | -2.36623419 | 3.49E-05 | 7.93E-05 | 0.169165032 |
| HTN | nk_cell | *ODF2L* | -2.085414108 | 3.53E-05 | 8.02E-05 | 0.171216885 |
| HTN | nk_cell | *SPATA1* | -2.02503124 | 5.64E-05 | 0.000117991 | 0.273740055 |
| HTN | nk_cell | *PMS2* | -2.535821489 | 0.000510336 | 0.000827038 | 1 |
| HTN | nk_cell | *EEPD1* | -1.45950752 | 1.75E-06 | 8.21E-06 | 0.008506986 |
| HTN | nk_cell | *KLHL7* | -2.059522653 | 1.52E-05 | 4.07E-05 | 0.0739242 |
| HTN | nk_cell | *NT5C3A* | -1.787824606 | 0.030999151 | 0.038020192 | 1 |
| HTN | nk_cell | *CCZ1* | -1.636238589 | 0.000479987 | 0.000781771 | 1 |
| HTN | nk_cell | *POLM* | -0.924871985 | 0.010606731 | 0.013812093 | 1 |
| HTN | nk_cell | *SMU1* | -1.947595331 | 7.81E-06 | 2.40E-05 | 0.03787249 |
| HTN | nk_cell | *GLIPR2* | 2.531049708 | 1.90E-06 | 8.69E-06 | 0.009234563 |
| HTN | nk_cell | *SLC25A51* | 0.640807079 | 0.012313932 | 0.015890213 | 1 |
| HTN | nk_cell | *TRIM24* | -2.500129839 | 9.20E-05 | 0.000179814 | 0.446477603 |
| HTN | nk_cell | *C7orf49* | -1.306486514 | 0.013300952 | 0.017073074 | 1 |
| HTN | nk_cell | *CISD1* | -1.67860017 | 0.000154472 | 0.000283365 | 0.749499417 |
| HTN | nk_cell | *RBM19* | -1.945498383 | 3.04E-06 | 1.20E-05 | 0.014762896 |
| HTN | nk_cell | *DDX54* | 3.875807655 | 0.002458368 | 0.003519623 | 1 |
| HTN | nk_cell | *SSPN* | -1.504577074 | 8.17E-07 | 5.32E-06 | 0.003965575 |
| HTN | nk_cell | *CCDC91* | -1.813002314 | 0.000620844 | 0.000988623 | 1 |
| HTN | nk_cell | *EBPL* | -1.846578643 | 0.000322823 | 0.000545952 | 1 |
| HTN | nk_cell | *CENPK* | 0.641923182 | 2.39E-07 | 4.07E-06 | 0.001159483 |
| HTN | nk_cell | *ITIH5* | -1.127597731 | 0.004864884 | 0.006630454 | 1 |
| HTN | nk_cell | *TSFM* | -2.236829586 | 1.72E-05 | 4.48E-05 | 0.083338595 |
| HTN | nk_cell | *ARHGAP9* | 0.684963974 | 3.62E-07 | 4.24E-06 | 0.001754939 |
| HTN | nk_cell | *PFDN5* | 2.517067372 | 0.000345692 | 0.000581991 | 1 |
| HTN | nk_cell | *PDE1B* | -3.21890354 | 1.51E-05 | 4.05E-05 | 0.073249972 |
| HTN | nk_cell | *CDK2* | -2.331698326 | 1.27E-08 | 1.01E-06 | 6.15E-05 |
| HTN | nk_cell | *SMUG1* | -1.337740745 | 0.015691014 | 0.019935271 | 1 |
| HTN | nk_cell | *SARDH* | -1.730919384 | 5.90E-05 | 0.000122337 | 0.286147221 |
| HTN | nk_cell | *DBH* | -26.90818607 | 3.91E-07 | 4.24E-06 | 0.001899456 |
| HTN | nk_cell | *NDUFAF4* | 3.23926172 | 1.79E-07 | 4.07E-06 | 0.0008695 |
| HTN | nk_cell | *USP45* | -1.943843596 | 1.69E-05 | 4.43E-05 | 0.081973003 |
| HTN | nk_cell | *TTC21B* | -2.322046278 | 8.68E-06 | 2.59E-05 | 0.042135627 |
| HTN | nk_cell | *ACVR1C* | -0.928209084 | 2.71E-05 | 6.45E-05 | 0.131724892 |
| HTN | nk_cell | *PLA2G12A* | -2.578733547 | 0.000221477 | 0.000390784 | 1 |
| HTN | nk_cell | *C4BPA* | -1.463570798 | 5.24E-06 | 1.79E-05 | 0.025414208 |
| HTN | nk_cell | *C4BPB* | -1.473513537 | 2.91E-06 | 1.16E-05 | 0.014132335 |
| HTN | nk_cell | *RAB38* | -2.120597849 | 6.61E-06 | 2.11E-05 | 0.032077812 |

| | | | | | | |
|---|---|---|---|---|---|---|
| HTN | nk_cell | *DAW1* | -2.363055047 | 3.06E-07 | 4.16E-06 | 0.001482504 |
| HTN | nk_cell | *OBSL1* | -2.975256584 | 4.66E-06 | 1.64E-05 | 0.02262776 |
| HTN | nk_cell | *SLC12A4* | -3.36320136 | 1.98E-05 | 5.00E-05 | 0.095829795 |
| HTN | nk_cell | *FAM210B* | -2.877843231 | 1.70E-05 | 4.44E-05 | 0.082274993 |
| HTN | nk_cell | *WFDC3* | -1.192411041 | 1.30E-08 | 1.01E-06 | 6.32E-05 |
| HTN | nk_cell | *SLC12A5* | 4.019609259 | 1.15E-05 | 3.25E-05 | 0.055916272 |
| HTN | nk_cell | *MATN4* | 3.469106124 | 4.07E-07 | 4.24E-06 | 0.001974928 |
| HTN | nk_cell | *VAPB* | -1.524328358 | 0.0094713 | 0.012400093 | 1 |
| HTN | nk_cell | *SRSF6* | -2.129606437 | 4.46E-05 | 9.74E-05 | 0.216554687 |
| HTN | nk_cell | *CSE1L* | -2.389253296 | 0.001424982 | 0.00210986 | 1 |
| HTN | nk_cell | *PTGIS* | -1.139181089 | 0.000161338 | 0.000295066 | 0.78281121 |
| HTN | nk_cell | *RNF114* | -2.192334177 | 3.93E-06 | 1.45E-05 | 0.019091755 |
| HTN | nk_cell | *NEURL2* | -1.363304164 | 5.38E-06 | 1.82E-05 | 0.026100685 |
| HTN | nk_cell | *MTRR* | -2.116811897 | 1.56E-05 | 4.15E-05 | 0.075534808 |
| HTN | nk_cell | *PEPD* | -2.046027173 | 7.93E-06 | 2.43E-05 | 0.03849956 |
| HTN | nk_cell | *CHST8* | -1.863712038 | 1.20E-06 | 6.67E-06 | 0.005833795 |
| HTN | nk_cell | *STAMBP* | -2.010594085 | 5.13E-06 | 1.77E-05 | 0.024882036 |
| HTN | nk_cell | *NAGK* | 2.066196829 | 0.026427697 | 0.032636087 | 1 |
| HTN | nk_cell | *SNRNP27* | -3.056971933 | 0.003256955 | 0.004564629 | 1 |
| HTN | nk_cell | *ATP8A1* | 1.079830394 | 6.14E-07 | 4.55E-06 | 0.002980804 |
| HTN | nk_cell | *USP22* | -2.069940581 | 0.003481642 | 0.004855685 | 1 |
| HTN | nk_cell | *HIF3A* | -1.332782966 | 1.42E-06 | 7.27E-06 | 0.006875873 |
| HTN | nk_cell | *ZNF576* | -1.415748523 | 2.00E-06 | 8.97E-06 | 0.0096819 |
| HTN | nk_cell | *BTN2A2* | 1.177496101 | 0.005688106 | 0.00767483 | 1 |
| HTN | nk_cell | *SERPINB6* | -1.775547918 | 1.08E-05 | 3.08E-05 | 0.052250156 |
| HTN | nk_cell | *PEX6* | -1.241959551 | 0.000154316 | 0.000283185 | 0.748741655 |
| HTN | nk_cell | *NQO2* | -2.089625449 | 8.00E-06 | 2.44E-05 | 0.03882467 |
| HTN | nk_cell | *ZNF391* | -1.578592433 | 3.70E-09 | 4.18E-07 | 1.80E-05 |
| HTN | nk_cell | *RPS10* | -2.315772542 | 6.85E-07 | 4.83E-06 | 0.003325585 |
| HTN | nk_cell | *MED20* | -1.881264788 | 0.034160582 | 0.041728888 | 1 |
| HTN | nk_cell | *TCP11* | 2.282955492 | 0.013656698 | 0.017497306 | 1 |
| HTN | nk_cell | *GNMT* | 1.72841196 | 2.52E-07 | 4.07E-06 | 0.001221611 |
| HTN | nk_cell | *TREM1* | -1.969183115 | 2.20E-06 | 9.58E-06 | 0.010679415 |
| HTN | nk_cell | *MEA1* | 1.844518184 | 1.52E-07 | 4.00E-06 | 0.000735528 |
| HTN | nk_cell | *KLHL31* | -1.259420729 | 0.001007042 | 0.001540407 | 1 |
| HTN | nk_cell | *GLO1* | -2.098143787 | 0.008539216 | 0.0112496 | 1 |
| HTN | nk_cell | *KCNK17* | -3.764052874 | 1.68E-05 | 4.42E-05 | 0.081482428 |
| HTN | nk_cell | *RREB1* | -1.572220372 | 8.85E-06 | 2.64E-05 | 0.042947863 |
| HTN | nk_cell | *RIOK1* | 1.885130199 | 0.034252948 | 0.041831187 | 1 |
| HTN | nk_cell | *ATXN1* | -1.538428169 | 0.003387946 | 0.004734537 | 1 |
| HTN | nk_cell | *RUNX2* | 2.543248885 | 0.007636217 | 0.010120438 | 1 |
| HTN | nk_cell | *CXCL6* | -2.974093708 | 3.83E-07 | 4.24E-06 | 0.00185599 |
| HTN | nk_cell | *SH3TC1* | -1.583245159 | 0.000115917 | 0.000219715 | 0.562428933 |
| HTN | nk_cell | *BBS2* | -1.05463005 | 0.000725471 | 0.001139892 | 1 |
| HTN | nk_cell | *MT1G* | -5.724969037 | 0.00045968 | 0.000751725 | 1 |
| HTN | nk_cell | *CLYBL* | -2.091093251 | 0.008680747 | 0.011423647 | 1 |
| HTN | nk_cell | *TMTC4* | -2.63648027 | 2.68E-06 | 1.10E-05 | 0.012991775 |
| HTN | nk_cell | *ATP5S* | -2.104313536 | 0.001596153 | 0.002346118 | 1 |
| HTN | nk_cell | *TEKT3* | -1.224909053 | 1.77E-05 | 4.59E-05 | 0.085764165 |
| HTN | nk_cell | *HS3ST3B1* | 0.445807097 | 3.27E-07 | 4.21E-06 | 0.00158428 |

| | | | | | | |
|---|---|---|---|---|---|---|
| HTN | nk_cell | *GGA3* | -2.747742263 | 0.000452297 | 0.00074065 | 1 |
| HTN | nk_cell | *NUP85* | -1.841098101 | 7.66E-07 | 5.16E-06 | 0.003715978 |
| HTN | nk_cell | *NT5C* | 3.259329729 | 0.000170014 | 0.000309302 | 0.824907198 |
| HTN | nk_cell | *DDX31* | -2.323249958 | 8.57E-06 | 2.57E-05 | 0.041581167 |
| HTN | nk_cell | *OPRL1* | -3.753985037 | 5.19E-07 | 4.37E-06 | 0.00251857 |
| HTN | nk_cell | *IL1B* | -4.588673881 | 1.02E-05 | 2.94E-05 | 0.049293624 |
| HTN | nk_cell | *PAX8* | -1.423487096 | 3.20E-06 | 1.25E-05 | 0.015515914 |
| HTN | nk_cell | *POLR1B* | 1.498422256 | 0.004670691 | 0.006383717 | 1 |
| HTN | nk_cell | *CCDC93* | -2.167560737 | 5.67E-05 | 0.000118474 | 0.274978844 |
| HTN | nk_cell | *PSD4* | 0.746241647 | 1.99E-07 | 4.07E-06 | 0.000963313 |
| HTN | nk_cell | *GTF2F1* | -3.045331082 | 0.000195209 | 0.000349374 | 0.94715277 |
| HTN | nk_cell | *TNFSF9* | -21.19743774 | 5.85E-06 | 1.93E-05 | 0.028360467 |
| HTN | nk_cell | *SH2D3A* | -3.234857253 | 1.08E-05 | 3.09E-05 | 0.052419667 |
| HTN | nk_cell | *GPR108* | -1.452930839 | 0.030232312 | 0.037117201 | 1 |
| HTN | nk_cell | *SNRPD2* | 3.564246473 | 4.81E-05 | 0.000103736 | 0.233302342 |
| HTN | nk_cell | *SDCBP2* | 2.268173859 | 2.05E-06 | 9.12E-06 | 0.009968543 |
| HTN | nk_cell | *PSMF1* | -2.307139929 | 9.79E-05 | 0.000189279 | 0.474901216 |
| HTN | nk_cell | *RRBP1* | -1.903944937 | 3.99E-05 | 8.86E-05 | 0.193395649 |
| HTN | nk_cell | *FLRT3* | -2.655970474 | 3.40E-06 | 1.30E-05 | 0.01649039 |
| HTN | nk_cell | *PCSK2* | -7.204852351 | 0.014267574 | 0.018222234 | 1 |
| HTN | nk_cell | *MKKS* | -1.853591254 | 0.019596262 | 0.024638782 | 1 |
| HTN | nk_cell | *LAMP5* | 17.73004354 | 2.56E-07 | 4.07E-06 | 0.001243656 |
| HTN | nk_cell | *MGME1* | -2.048374401 | 2.65E-05 | 6.31E-05 | 0.128481778 |
| HTN | nk_cell | *ITPA* | -2.424370838 | 8.54E-05 | 0.000168185 | 0.414407322 |
| HTN | nk_cell | *MCM8* | -1.048274218 | 0.025820241 | 0.031956346 | 1 |
| HTN | nk_cell | *NCLN* | -3.049983891 | 4.39E-05 | 9.61E-05 | 0.213096819 |
| HTN | nk_cell | *KLC1* | -2.187045669 | 0.004922334 | 0.006701224 | 1 |
| HTN | nk_cell | *XRCC3* | -2.064384021 | 1.17E-07 | 3.59E-06 | 0.000567693 |
| HTN | nk_cell | *TUBGCP3* | -1.850084883 | 0.026786787 | 0.033062704 | 1 |
| HTN | nk_cell | *MCF2L* | -1.489340249 | 0.000221059 | 0.000390312 | 1 |
| HTN | nk_cell | *PROZ* | -3.194813193 | 0.007722449 | 0.010226343 | 1 |
| HTN | nk_cell | *IGFLR1* | -1.475067797 | 0.041173921 | 0.049931483 | 1 |
| HTN | nk_cell | *PRDX5* | 4.078444091 | 1.97E-05 | 5.00E-05 | 0.095713447 |
| HTN | nk_cell | *IRF3* | 2.924830238 | 0.000174802 | 0.000316707 | 0.848140742 |
| HTN | nk_cell | *TSKS* | -1.463170566 | 1.46E-08 | 1.05E-06 | 7.07E-05 |
| HTN | nk_cell | *ZNF384* | -2.841059251 | 0.00048147 | 0.000783924 | 1 |
| HTN | nk_cell | *EMG1* | -2.281145837 | 0.023575283 | 0.029337592 | 1 |
| HTN | nk_cell | *ATG14* | -1.706133979 | 0.000295888 | 0.000505155 | 1 |
| HTN | nk_cell | *KTN1* | -1.888306808 | 0.000417886 | 0.000689185 | 1 |
| HTN | nk_cell | *L3HYPDH* | -0.956843102 | 8.48E-05 | 0.000167398 | 0.411479703 |
| HTN | nk_cell | *ZBTB1* | -2.070635548 | 6.88E-05 | 0.000139758 | 0.33382512 |
| HTN | nk_cell | *RHOT1* | -1.515520762 | 0.000209119 | 0.000371258 | 1 |
| HTN | nk_cell | *WDR60* | -1.732551423 | 0.000175891 | 0.000318322 | 0.853421539 |
| HTN | nk_cell | *AIF1L* | -1.476570301 | 5.17E-07 | 4.37E-06 | 0.002510284 |
| HTN | nk_cell | *FGD3* | 0.703388207 | 1.37E-07 | 3.87E-06 | 0.000664172 |
| HTN | nk_cell | *ABHD8* | -1.902911333 | 1.67E-09 | 2.53E-07 | 8.10E-06 |
| HTN | nk_cell | *ATP13A4* | 3.857274798 | 4.95E-08 | 2.19E-06 | 0.000240216 |
| HTN | nk_cell | *HELB* | 2.952317083 | 0.000541351 | 0.000872926 | 1 |
| HTN | nk_cell | *RAB3IP* | 3.085334725 | 1.34E-06 | 7.02E-06 | 0.006478224 |
| HTN | nk_cell | *YEATS4* | -1.112000065 | 0.001204061 | 0.001813454 | 1 |

| | | | | | | |
|---|---|---|---|---|---|---|
| HTN | nk_cell | *TAS2R4* | -1.219063535 | 5.64E-06 | 1.89E-05 | 0.027344411 |
| HTN | nk_cell | *LRRC61* | -2.53372339 | 1.11E-06 | 6.27E-06 | 0.005362646 |
| HTN | nk_cell | *IDUA* | -2.636638053 | 1.30E-06 | 6.91E-06 | 0.006318341 |
| HTN | nk_cell | *EMC1* | -2.994300527 | 0.000605095 | 0.000966847 | 1 |
| HTN | nk_cell | *PLA2G5* | -0.958538173 | 5.09E-05 | 0.00010857 | 0.246778524 |
| HTN | nk_cell | *UBR4* | -1.916804642 | 0.000204574 | 0.000364523 | 0.992594853 |
| HTN | nk_cell | *GFER* | -4.156967279 | 0.000802311 | 0.001252111 | 1 |
| HTN | nk_cell | *WDR24* | -1.418546147 | 2.58E-05 | 6.18E-05 | 0.125054845 |
| HTN | nk_cell | *PNKD* | -2.107195173 | 7.87E-06 | 2.42E-05 | 0.038184284 |
| HTN | nk_cell | *RNF6* | -1.590090215 | 1.29E-06 | 6.87E-06 | 0.006261047 |
| HTN | nk_cell | *HIP1* | -1.715950721 | 0.005156264 | 0.006992229 | 1 |
| HTN | nk_cell | *STYXL1* | -1.95049709 | 2.70E-07 | 4.07E-06 | 0.001307792 |
| HTN | nk_cell | *STEAP4* | -1.455616374 | 2.61E-06 | 1.08E-05 | 0.012646036 |
| HTN | nk_cell | *PEX1* | -2.581047704 | 9.42E-05 | 0.000183354 | 0.456919041 |
| HTN | nk_cell | *SRD5A3* | -2.032819277 | 0.001245583 | 0.001869338 | 1 |
| HTN | nk_cell | *KDR* | -2.275289554 | 0.019784877 | 0.024850175 | 1 |
| HTN | nk_cell | *DGCR6L* | -1.205327761 | 0.000449121 | 0.000735946 | 1 |
| HTN | nk_cell | *VPREB3* | -3.608425326 | 0.017112344 | 0.021667299 | 1 |
| HTN | nk_cell | *MGAT3* | -2.439733198 | 2.13E-06 | 9.35E-06 | 0.010348694 |
| HTN | nk_cell | *A4GALT* | -1.997171835 | 8.34E-08 | 3.01E-06 | 0.000404785 |
| HTN | nk_cell | *CDC42EP1* | -3.293170278 | 0.000101232 | 0.000195144 | 0.491177344 |
| HTN | nk_cell | *SPECC1* | -1.948216161 | 0.007190965 | 0.009580055 | 1 |
| HTN | nk_cell | *CDHR3* | -1.878363647 | 5.49E-06 | 1.85E-05 | 0.026634403 |
| HTN | nk_cell | *PRKRIP1* | -2.147123952 | 0.000488063 | 0.000793859 | 1 |
| HTN | nk_cell | *IRF5* | -1.277618474 | 3.21E-07 | 4.18E-06 | 0.001555814 |
| HTN | nk_cell | *OPN1SW* | -1.487273358 | 1.26E-07 | 3.79E-06 | 0.000613418 |
| HTN | nk_cell | *HOXD1* | -3.335443604 | 0.000626198 | 0.00099483 | 1 |
| HTN | nk_cell | *MTX2* | -1.815662165 | 7.92E-06 | 2.43E-05 | 0.038431394 |
| HTN | nk_cell | *GAD1* | -3.269309975 | 2.24E-08 | 1.38E-06 | 0.000108718 |
| HTN | nk_cell | *ORMDL1* | -1.691819164 | 0.001695464 | 0.002475867 | 1 |
| HTN | nk_cell | *HOXD9* | 0.907735544 | 0.003861054 | 0.005337275 | 1 |
| HTN | nk_cell | *HOXD10* | -2.488637443 | 3.85E-05 | 8.61E-05 | 0.187022428 |
| HTN | nk_cell | *PSMG2* | -2.01735778 | 0.000170821 | 0.000310537 | 0.828824521 |
| HTN | nk_cell | *TWSG1* | -2.649425752 | 0.000421357 | 0.000694438 | 1 |
| HTN | nk_cell | *EIF2AK4* | -2.756332242 | 1.37E-06 | 7.14E-06 | 0.00666372 |
| HTN | nk_cell | *TMOD2* | -2.166093868 | 2.97E-05 | 6.96E-05 | 0.143957059 |
| HTN | nk_cell | *IVD* | -1.445616755 | 0.011615952 | 0.015049559 | 1 |
| HTN | nk_cell | *KNSTRN* | -1.114801176 | 5.54E-06 | 1.86E-05 | 0.026871009 |
| HTN | nk_cell | *ARPP19* | 2.682034834 | 0.006256849 | 0.008400174 | 1 |
| HTN | nk_cell | *CALML4* | -3.979688204 | 2.12E-07 | 4.07E-06 | 0.00102745 |
| HTN | nk_cell | *ANAPC13* | -1.556369615 | 2.70E-06 | 1.10E-05 | 0.013124364 |
| HTN | nk_cell | *BBOX1* | 2.405725407 | 2.80E-07 | 4.07E-06 | 0.001359799 |
| HTN | nk_cell | *SERGEF* | 2.367693675 | 0.007689909 | 0.010186034 | 1 |
| HTN | nk_cell | *DCTD* | -2.109625961 | 0.019643306 | 0.024691534 | 1 |
| HTN | nk_cell | *RPAIN* | -1.986501271 | 1.65E-06 | 7.94E-06 | 0.008011197 |
| HTN | nk_cell | *USP6* | -1.349478765 | 0.000204335 | 0.00036423 | 0.991433613 |
| HTN | nk_cell | *PLD2* | -3.722233584 | 2.18E-05 | 5.39E-05 | 0.105775123 |
| HTN | nk_cell | *PUS7L* | -2.283120554 | 0.003649016 | 0.005074527 | 1 |
| HTN | nk_cell | *AP1M2* | -2.007792633 | 0.000493065 | 0.000800653 | 1 |
| HTN | nk_cell | *MTUS1* | -1.796891499 | 9.10E-06 | 2.70E-05 | 0.044170317 |

| | | | | | | |
|---|---|---|---|---|---|---|
| HTN | nk_cell | *KLK10* | -1.489876687 | 0.001242208 | 0.001864849 | 1 |
| HTN | nk_cell | *KLK8* | 3.178554036 | 2.72E-07 | 4.07E-06 | 0.001319763 |
| HTN | nk_cell | *RIPK3* | 2.791374637 | 6.89E-08 | 2.61E-06 | 0.000334133 |
| HTN | nk_cell | *BCL2L2* | -2.614933702 | 4.43E-05 | 9.67E-05 | 0.21496074 |
| HTN | nk_cell | *DTD2* | -1.389824422 | 1.44E-06 | 7.35E-06 | 0.006997958 |
| HTN | nk_cell | *PARP2* | 3.490749072 | 0.001226023 | 0.001841692 | 1 |
| HTN | nk_cell | *MIS18BP1* | -1.875214083 | 9.65E-06 | 2.83E-05 | 0.046842499 |
| HTN | nk_cell | *RNASE1* | -1.706914206 | 0.001271044 | 0.001904604 | 1 |
| HTN | nk_cell | *EPB41L4A* | -1.617732135 | 8.23E-06 | 2.50E-05 | 0.039924086 |
| HTN | nk_cell | *CDO1* | -1.298269569 | 0.000401127 | 0.000665164 | 1 |
| HTN | nk_cell | *REEP5* | -2.170991581 | 8.29E-05 | 0.000164064 | 0.402121574 |
| HTN | nk_cell | *RHBDF2* | 2.064595413 | 2.64E-07 | 4.07E-06 | 0.001278742 |
| HTN | nk_cell | *MAU2* | -2.41138025 | 1.79E-05 | 4.64E-05 | 0.087087983 |
| HTN | nk_cell | *SHC2* | -2.063730115 | 5.02E-06 | 1.74E-05 | 0.024362246 |
| HTN | nk_cell | *ERMARD* | -2.31959894 | 1.03E-05 | 2.97E-05 | 0.049863553 |
| HTN | nk_cell | *PHF10* | -2.238846731 | 4.39E-06 | 1.57E-05 | 0.021317529 |
| HTN | nk_cell | *GALNT8* | -1.793261857 | 1.24E-07 | 3.74E-06 | 0.000602404 |
| HTN | nk_cell | *PRKCSH* | 2.440327545 | 0.001374007 | 0.002040613 | 1 |
| HTN | nk_cell | *XPO7* | -2.383387271 | 0.000782023 | 0.001223597 | 1 |
| HTN | nk_cell | *FAM98C* | 2.86612462 | 0.000966065 | 0.001484277 | 1 |
| HTN | nk_cell | *ATP8B3* | -1.97021834 | 1.48E-09 | 2.32E-07 | 7.19E-06 |
| HTN | nk_cell | *MRPL34* | -0.669185001 | 0.038402472 | 0.046698946 | 1 |
| HTN | nk_cell | *TULP4* | -1.691236899 | 6.60E-05 | 0.000134638 | 0.320035695 |
| HTN | nk_cell | *QRSL1* | -1.272190084 | 0.000625615 | 0.00099483 | 1 |
| HTN | nk_cell | *RSPH3* | -0.76165151 | 0.017528902 | 0.022131208 | 1 |
| HTN | nk_cell | *STK33* | -1.339575697 | 1.83E-05 | 4.70E-05 | 0.088704734 |
| HTN | nk_cell | *NDUFA10* | -1.279054906 | 0.003063743 | 0.004316284 | 1 |
| HTN | nk_cell | *MAP1S* | -3.475698486 | 4.68E-06 | 1.64E-05 | 0.022721172 |
| HTN | nk_cell | *PGPEP1* | 3.539940159 | 3.62E-08 | 1.98E-06 | 0.000175852 |
| HTN | nk_cell | *KIAA1683* | -2.795933002 | 2.57E-06 | 1.07E-05 | 0.01246694 |
| HTN | nk_cell | *LSM4* | 1.195793926 | 0.015225988 | 0.019385068 | 1 |
| HTN | nk_cell | *TRPM4* | -2.148454983 | 5.81E-09 | 5.45E-07 | 2.82E-05 |
| HTN | nk_cell | *UBAC1* | 2.666332508 | 0.00234941 | 0.003371588 | 1 |
| HTN | nk_cell | *ATXN10* | -1.87681009 | 6.00E-05 | 0.000124087 | 0.291231904 |
| HTN | nk_cell | *CYP2E1* | -1.970750034 | 1.96E-06 | 8.91E-06 | 0.009530352 |
| HTN | nk_cell | *PNPLA7* | -1.728793671 | 2.38E-06 | 1.01E-05 | 0.011526223 |
| HTN | nk_cell | *ZNF337* | -1.762997087 | 7.92E-05 | 0.000157536 | 0.384247526 |
| HTN | nk_cell | *CEP85* | -1.44792566 | 0.00015065 | 0.000277824 | 0.73095369 |
| HTN | nk_cell | *POMT1* | -2.542923391 | 1.40E-05 | 3.80E-05 | 0.067714419 |
| HTN | nk_cell | *YIPF2* | 2.232897949 | 0.036950736 | 0.044989955 | 1 |
| HTN | nk_cell | *NPAS1* | 4.629865677 | 4.26E-05 | 9.36E-05 | 0.20685654 |
| HTN | nk_cell | *MAP3K10* | -2.179609346 | 9.91E-07 | 5.94E-06 | 0.004810292 |
| HTN | nk_cell | *LRRC47* | -1.79569707 | 1.97E-06 | 8.92E-06 | 0.009552254 |
| HTN | nk_cell | *THEMIS2* | 1.519449832 | 6.19E-08 | 2.44E-06 | 0.0003004 |
| HTN | nk_cell | *ZNF317* | -2.588229254 | 0.001709877 | 0.002494384 | 1 |
| HTN | nk_cell | *ANGPTL6* | 1.624390495 | 2.32E-05 | 5.70E-05 | 0.112717258 |
| HTN | nk_cell | *DNMT1* | 2.475492148 | 1.76E-06 | 8.25E-06 | 0.008554394 |
| HTN | nk_cell | *C12orf65* | -2.083189744 | 0.003608269 | 0.005022181 | 1 |
| HTN | nk_cell | *NOL11* | -2.868067783 | 0.000254207 | 0.00044145 | 1 |
| HTN | nk_cell | *PKDREJ* | -1.823394912 | 0.016354291 | 0.020739943 | 1 |

| | | | | | | |
|---|---|---|---|---|---|---|
| HTN | nk_cell | *HSD17B3* | -5.360331123 | 0.000561994 | 0.000902912 | 1 |
| HTN | nk_cell | *SLC35D2* | -2.517003187 | 0.000138995 | 0.000258689 | 0.674402852 |
| HTN | nk_cell | *ULBP2* | -10.55183284 | 5.07E-05 | 0.000108276 | 0.245803183 |
| HTN | nk_cell | *TTLL9* | -2.501033792 | 1.23E-06 | 6.75E-06 | 0.005988176 |
| HTN | nk_cell | *ATP6V1E1* | -2.582458659 | 0.002201038 | 0.003167092 | 1 |
| HTN | nk_cell | *IDO1* | 5.703771599 | 1.47E-06 | 7.41E-06 | 0.007125858 |
| HTN | nk_cell | *HAUS8* | -4.806928851 | 1.01E-06 | 5.98E-06 | 0.004887921 |
| HTN | nk_cell | *MRPS25* | -1.94806412 | 5.70E-07 | 4.50E-06 | 0.00276721 |
| HTN | nk_cell | *GALNT15* | -1.238432564 | 6.34E-07 | 4.62E-06 | 0.00307482 |
| HTN | nk_cell | *KCNC3* | -2.855294033 | 5.53E-08 | 2.29E-06 | 0.000268409 |
| HTN | nk_cell | *NAPSA* | 3.785739651 | 3.42E-05 | 7.79E-05 | 0.165806591 |
| HTN | nk_cell | *KIF3A* | -2.598969118 | 0.000867545 | 0.001343546 | 1 |
| HTN | nk_cell | *ACLY* | 2.528313747 | 0.005314488 | 0.007192719 | 1 |
| HTN | nk_cell | *NDUFA2* | -2.912073388 | 5.51E-07 | 4.45E-06 | 0.002672099 |
| HTN | nk_cell | *EXOC4* | -1.280668208 | 0.003910258 | 0.005392999 | 1 |
| HTN | nk_cell | *PPFIA1* | -1.654224592 | 6.79E-05 | 0.000138213 | 0.329500304 |
| HTN | nk_cell | *BARX1* | -4.095787446 | 3.72E-06 | 1.39E-05 | 0.01807274 |
| HTN | nk_cell | *NINJ1* | -1.28032738 | 2.06E-06 | 9.12E-06 | 0.009976335 |
| HTN | nk_cell | *NPHP4* | -1.344865548 | 5.36E-07 | 4.42E-06 | 0.002599072 |
| HTN | nk_cell | *MAP1B* | -1.533731779 | 6.80E-05 | 0.000138361 | 0.32999113 |
| HTN | nk_cell | *CKMT2* | -2.285555473 | 0.003417182 | 0.004772644 | 1 |
| HTN | nk_cell | *KHDRBS3* | -2.238624315 | 5.72E-06 | 1.90E-05 | 0.027748428 |
| HTN | nk_cell | *CHD1L* | -2.093926663 | 0.006847168 | 0.009154714 | 1 |
| HTN | nk_cell | *ZNF304* | -1.825963442 | 0.000212572 | 0.000376286 | 1 |
| HTN | nk_cell | *ZNF132* | -1.47774451 | 0.003904853 | 0.005387076 | 1 |
| HTN | nk_cell | *RHPN2* | -1.017770004 | 0.006485331 | 0.008692493 | 1 |
| HTN | nk_cell | *GCH1* | 3.732309192 | 1.69E-05 | 4.43E-05 | 0.082020546 |
| HTN | nk_cell | *MATN3* | -2.669932947 | 2.30E-07 | 4.07E-06 | 0.001115064 |
| HTN | nk_cell | *TRIM21* | 2.612597172 | 1.10E-06 | 6.25E-06 | 0.005338094 |
| HTN | nk_cell | *SPATA6* | -1.328000189 | 2.65E-06 | 1.09E-05 | 0.012855648 |
| HTN | nk_cell | *LRRC41* | -2.473185313 | 2.17E-05 | 5.37E-05 | 0.105234717 |
| HTN | nk_cell | *GAS2L2* | 4.172510979 | 1.89E-05 | 4.82E-05 | 0.091557334 |
| HTN | nk_cell | *CCT6B* | -2.487710801 | 2.75E-05 | 6.51E-05 | 0.133226246 |
| HTN | nk_cell | *CNGA4* | 2.808371659 | 7.54E-05 | 0.000151111 | 0.365688939 |
| HTN | nk_cell | *IMMT* | -2.001852082 | 0.020878141 | 0.026155626 | 1 |
| HTN | nk_cell | *CARD6* | -1.744613643 | 0.020032912 | 0.025135683 | 1 |
| HTN | nk_cell | *MYBBP1A* | -1.795402908 | 0.000582028 | 0.000932559 | 1 |
| HTN | nk_cell | *RPA1* | -2.220460718 | 0.002947709 | 0.004164905 | 1 |
| HTN | nk_cell | *SERPINF1* | -2.474565889 | 0.000527775 | 0.000852168 | 1 |
| HTN | nk_cell | *TMEM128* | -2.262407357 | 5.28E-05 | 0.000111766 | 0.256221875 |
| HTN | nk_cell | *FIGNL1* | -2.311526242 | 1.99E-05 | 5.02E-05 | 0.096413973 |
| HTN | nk_cell | *DDC* | -5.350337609 | 0.000473273 | 0.000772132 | 1 |
| HTN | nk_cell | *EIF5A* | 1.420926679 | 9.18E-05 | 0.000179509 | 0.445360827 |
| HTN | nk_cell | *RGS22* | -1.666241107 | 3.20E-05 | 7.41E-05 | 0.155197171 |
| HTN | nk_cell | *REEP2* | -3.550158972 | 4.66E-07 | 4.32E-06 | 0.002262119 |
| HTN | nk_cell | *PRMT7* | -2.084512882 | 8.18E-05 | 0.000162155 | 0.397117323 |
| HTN | nk_cell | *IGHMBP2* | -1.895764511 | 1.77E-05 | 4.60E-05 | 0.086089304 |
| HTN | nk_cell | *MUTYH* | 4.121085189 | 1.46E-07 | 3.95E-06 | 0.000707548 |
| HTN | nk_cell | *OSER1* | -2.982840556 | 0.002000306 | 0.002887678 | 1 |
| HTN | nk_cell | *DMGDH* | -1.790802655 | 0.010858441 | 0.014113355 | 1 |

| | | | | | | |
|---|---|---|---|---|---|---|
| HTN | nk_cell | *BHMT2* | -0.959237218 | 1.21E-05 | 3.37E-05 | 0.058505037 |
| HTN | nk_cell | *ZBED3* | -1.996653378 | 3.11E-06 | 1.22E-05 | 0.015069544 |
| HTN | nk_cell | *FBXO44* | -1.90452598 | 1.26E-06 | 6.79E-06 | 0.0061235 |
| HTN | nk_cell | *RSG1* | -8.620561714 | 2.27E-08 | 1.38E-06 | 0.000110152 |
| HTN | nk_cell | *CASP9* | -1.458939932 | 0.00275833 | 0.003912136 | 1 |
| HTN | nk_cell | *ATP8A2* | -4.46804167 | 0.000928425 | 0.001432343 | 1 |
| HTN | nk_cell | *ALOX5AP* | 1.067113401 | 3.15E-07 | 4.17E-06 | 0.001530196 |
| HTN | nk_cell | *WASF3* | -1.885552535 | 5.81E-06 | 1.93E-05 | 0.028213292 |
| HTN | nk_cell | *GPR12* | -6.068286942 | 0.001533307 | 0.002261003 | 1 |
| HTN | nk_cell | *PEMT* | -3.326112464 | 2.79E-05 | 6.59E-05 | 0.135303807 |
| HTN | nk_cell | *MPRIP* | -2.004640165 | 0.007289731 | 0.009703641 | 1 |
| HTN | nk_cell | *DSTYK* | -2.487954978 | 0.000492197 | 0.000799512 | 1 |
| HTN | nk_cell | *COG6* | -2.356171538 | 0.000168104 | 0.000305941 | 0.815638349 |
| HTN | nk_cell | *EPSTI1* | -2.150591183 | 0.000598092 | 0.000956475 | 1 |
| HTN | nk_cell | *BTBD2* | -2.456702305 | 4.49E-05 | 9.78E-05 | 0.217680146 |
| HTN | nk_cell | *CNDP2* | -2.853617635 | 4.24E-05 | 9.33E-05 | 0.205627166 |
| HTN | nk_cell | *PDZD2* | -1.539483326 | 1.56E-05 | 4.16E-05 | 0.075827035 |
| HTN | nk_cell | *GSTT2B* | -1.031818211 | 0.000137489 | 0.000256181 | 0.667095435 |
| HTN | nk_cell | *SLC2A11* | 3.088184379 | 3.15E-08 | 1.86E-06 | 0.000152908 |
| HTN | nk_cell | *GIMAP4* | 0.972676576 | 6.67E-07 | 4.75E-06 | 0.003234497 |
| HTN | nk_cell | *ADCK2* | -1.353540594 | 0.009234426 | 0.012103034 | 1 |
| HTN | nk_cell | *SFTPD* | -0.690435922 | 0.000263016 | 0.00045447 | 1 |
| HTN | nk_cell | *IPO8* | -2.785981988 | 0.000149821 | 0.0002764 | 0.726933153 |
| HTN | nk_cell | *LARS* | -1.72676738 | 6.06E-06 | 1.98E-05 | 0.029410718 |
| HTN | nk_cell | *SPINK5* | -1.889625581 | 4.82E-07 | 4.32E-06 | 0.002340749 |
| HTN | nk_cell | *LRRCC1* | 2.362340505 | 0.013962719 | 0.017861089 | 1 |
| HTN | nk_cell | *CATSPERB* | -1.887145943 | 0.005268502 | 0.007136452 | 1 |
| HTN | nk_cell | *COX16* | 1.102204274 | 2.28E-07 | 4.07E-06 | 0.001104848 |
| HTN | nk_cell | *TTC9* | 5.106389399 | 1.10E-05 | 3.13E-05 | 0.053344164 |
| HTN | nk_cell | *ELP3* | -2.45993898 | 0.001285786 | 0.001923723 | 1 |
| HTN | nk_cell | *PEBP4* | -1.720925508 | 9.54E-07 | 5.84E-06 | 0.004627486 |
| HTN | nk_cell | *CTIF* | -1.26279144 | 1.19E-05 | 3.34E-05 | 0.057828691 |
| HTN | nk_cell | *MRO* | -4.527510513 | 0.000919298 | 0.001419165 | 1 |
| HTN | nk_cell | *THUMPD3* | -3.274114999 | 0.000202279 | 0.000360831 | 0.981459233 |
| HTN | nk_cell | *CNTN6* | -2.273082997 | 1.46E-06 | 7.39E-06 | 0.007075168 |
| HTN | nk_cell | *CHL1* | -1.309925255 | 6.83E-06 | 2.16E-05 | 0.033162743 |
| HTN | nk_cell | *GSTM5* | -1.694946688 | 1.42E-07 | 3.91E-06 | 0.000688327 |
| HTN | nk_cell | *GSTM3* | -1.093695458 | 5.38E-05 | 0.00011349 | 0.260800602 |
| HTN | nk_cell | *VAV3* | 0.953835911 | 1.61E-05 | 4.26E-05 | 0.078070839 |
| HTN | nk_cell | *PSRC1* | -2.938185071 | 4.06E-05 | 9.01E-05 | 0.197199103 |
| HTN | nk_cell | *TRIM45* | -4.852474383 | 1.41E-05 | 3.84E-05 | 0.06863592 |
| HTN | nk_cell | *CEPT1* | -1.742304777 | 1.64E-05 | 4.33E-05 | 0.07951881 |
| HTN | nk_cell | *NAPG* | -2.242505843 | 1.56E-05 | 4.15E-05 | 0.075464158 |
| HTN | nk_cell | *PPHLN1* | -1.846123872 | 2.37E-05 | 5.80E-05 | 0.114980064 |
| HTN | nk_cell | *FKBP11* | 4.052921634 | 8.95E-06 | 2.67E-05 | 0.043440311 |
| HTN | nk_cell | *YWHAQ* | -2.120665242 | 1.85E-05 | 4.74E-05 | 0.089617898 |
| HTN | nk_cell | *KIDINS220* | -1.951209652 | 0.000153667 | 0.000282314 | 0.745590027 |
| HTN | nk_cell | *LPIN1* | -2.212853106 | 0.002104478 | 0.003032648 | 1 |
| HTN | nk_cell | *KIAA1468* | -1.847964568 | 0.013300581 | 0.017073074 | 1 |
| HTN | nk_cell | *IL15RA* | -2.828177645 | 1.79E-05 | 4.64E-05 | 0.087035396 |

| | | | | | | |
|---|---|---|---|---|---|---|
| HTN | nk_cell | *TMEM241* | -1.491221662 | 1.02E-07 | 3.38E-06 | 0.000496526 |
| HTN | nk_cell | *SOX5* | -1.260664649 | 1.99E-05 | 5.03E-05 | 0.096760255 |
| HTN | nk_cell | *KLRD1* | 0.426939599 | 2.74E-07 | 4.07E-06 | 0.00132815 |
| HTN | nk_cell | *ACP2* | 7.468617179 | 1.16E-11 | 3.50E-09 | 5.60E-08 |
| HTN | nk_cell | *HOOK1* | -1.394438379 | 4.03E-06 | 1.47E-05 | 0.01953328 |
| HTN | nk_cell | *ZCCHC11* | -1.564816379 | 0.000832376 | 0.001294451 | 1 |
| HTN | nk_cell | *PRPF38A* | 2.649995033 | 0.028850967 | 0.03548413 | 1 |
| HTN | nk_cell | *ELP2* | -2.207938541 | 0.000198935 | 0.000355126 | 0.965233493 |
| HTN | nk_cell | *DHX34* | -2.888495722 | 0.00021706 | 0.000383531 | 1 |
| HTN | nk_cell | *FADS2* | -1.696745311 | 7.33E-07 | 5.01E-06 | 0.003557648 |
| HTN | nk_cell | *CLOCK* | -2.102757783 | 0.000278155 | 0.000477738 | 1 |
| HTN | nk_cell | *GGACT* | -2.011724189 | 9.93E-06 | 2.88E-05 | 0.048203644 |
| HTN | nk_cell | *CLDN10* | -2.00809391 | 0.000124519 | 0.000234446 | 0.60416755 |
| HTN | nk_cell | *ARGLU1* | -1.659330539 | 0.001144239 | 0.001731166 | 1 |
| HTN | nk_cell | *ERCC5* | -3.741419405 | 2.00E-06 | 8.99E-06 | 0.009726272 |
| HTN | nk_cell | *CARS2* | -2.174831375 | 0.000277004 | 0.000476097 | 1 |
| HTN | nk_cell | *KLB* | -0.95896709 | 1.52E-06 | 7.55E-06 | 0.00735179 |
| HTN | nk_cell | *WDR36* | -2.457298428 | 4.06E-05 | 9.01E-05 | 0.196941855 |
| HTN | nk_cell | *UBQLN1* | -2.225183249 | 0.000410272 | 0.000677781 | 1 |
| HTN | nk_cell | *GOLM1* | -1.556763717 | 0.003757518 | 0.005210483 | 1 |
| HTN | nk_cell | *USP30* | -1.861406574 | 5.53E-08 | 2.29E-06 | 0.000268229 |
| HTN | nk_cell | *MSI1* | 4.931101703 | 4.13E-05 | 9.13E-05 | 0.200221713 |
| HTN | nk_cell | *P2RX4* | -1.657635937 | 0.00739553 | 0.009833683 | 1 |
| HTN | nk_cell | *DTX1* | -1.287318606 | 7.23E-07 | 4.96E-06 | 0.003509077 |
| HTN | nk_cell | *DMTF1* | -1.946304703 | 5.71E-05 | 0.00011907 | 0.27683726 |
| HTN | nk_cell | *TMEM60* | 5.269618052 | 5.42E-06 | 1.83E-05 | 0.026276491 |
| HTN | nk_cell | *TES* | 1.726288678 | 7.64E-06 | 2.36E-05 | 0.037054159 |
| HTN | nk_cell | *MTO1* | -1.941467996 | 6.99E-05 | 0.000141804 | 0.339302203 |
| HTN | nk_cell | *NT5E* | -1.451357354 | 0.000615244 | 0.000980028 | 1 |
| HTN | nk_cell | *AKIRIN2* | -2.530679833 | 7.53E-05 | 0.000151085 | 0.365475566 |
| HTN | nk_cell | *MAP3K7* | -1.872805451 | 9.80E-06 | 2.86E-05 | 0.047544615 |
| HTN | nk_cell | *PRR5L* | 1.688888538 | 5.34E-07 | 4.41E-06 | 0.002589937 |
| HTN | nk_cell | *PHF21A* | -1.834414077 | 0.000751401 | 0.001178726 | 1 |
| HTN | nk_cell | *CAPRIN1* | -1.835612938 | 0.021619475 | 0.027056408 | 1 |
| HTN | nk_cell | *B4GALNT1* | -3.774037102 | 0.000548093 | 0.00088321 | 1 |
| HTN | nk_cell | *FAIM2* | -2.248436573 | 4.64E-08 | 2.13E-06 | 0.000225367 |
| HTN | nk_cell | *KRT7* | -1.136423476 | 0.001327446 | 0.001980556 | 1 |
| HTN | nk_cell | *SLC26A10* | -2.439941047 | 1.64E-05 | 4.33E-05 | 0.07958273 |
| HTN | nk_cell | *MAP7* | -1.385065017 | 0.023000857 | 0.028681613 | 1 |
| HTN | nk_cell | *NHSL1* | -1.680884847 | 1.74E-06 | 8.18E-06 | 0.00843198 |
| HTN | nk_cell | *AHI1* | -1.262473106 | 0.000434386 | 0.000713246 | 1 |
| HTN | nk_cell | *SMPD2* | -1.490851707 | 7.23E-07 | 4.96E-06 | 0.003507981 |
| HTN | nk_cell | *CCT7* | 2.208879246 | 0.020431362 | 0.025620401 | 1 |
| HTN | nk_cell | *SMYD5* | -2.389855643 | 9.21E-06 | 2.72E-05 | 0.044691394 |
| HTN | nk_cell | *MDM2* | -1.954878026 | 0.00125959 | 0.001888606 | 1 |
| HTN | nk_cell | *MPHOSPH6* | 3.181734508 | 1.19E-05 | 3.34E-05 | 0.057698811 |
| HTN | nk_cell | *CCDC102A* | 3.457600202 | 5.13E-05 | 0.000109277 | 0.248822924 |
| HTN | nk_cell | *AGT* | 1.382232836 | 0.015399809 | 0.019590948 | 1 |
| HTN | nk_cell | *KCNK1* | -0.927820069 | 4.32E-05 | 9.49E-05 | 0.209818591 |
| HTN | nk_cell | *CAPN9* | 4.051655201 | 0.000241016 | 0.000422018 | 1 |

| | | | | | | |
|---|---|---|---|---|---|---|
| HTN | nk_cell | *NTPCR* | -2.734769724 | 8.49E-05 | 0.000167453 | 0.411935097 |
| HTN | nk_cell | *PIGC* | -2.627126629 | 7.04E-05 | 0.000142453 | 0.341745429 |
| HTN | nk_cell | *SP110* | 2.759038259 | 8.13E-05 | 0.000161362 | 0.394689519 |
| HTN | nk_cell | *MRPL44* | 2.694693601 | 5.59E-07 | 4.48E-06 | 0.002713259 |
| HTN | nk_cell | *USP37* | -1.79251675 | 0.001137756 | 0.001722884 | 1 |
| HTN | nk_cell | *SLC19A3* | -0.989271896 | 5.32E-07 | 4.40E-06 | 0.002578841 |
| HTN | nk_cell | *TMBIM1* | -2.012526087 | 3.80E-06 | 1.41E-05 | 0.018415752 |
| HTN | nk_cell | *TSGA10* | -1.179284343 | 3.51E-06 | 1.33E-05 | 0.017027168 |
| HTN | nk_cell | *GCC2* | -1.732592546 | 0.001086017 | 0.001650801 | 1 |
| HTN | nk_cell | *ANKRD36* | -2.302548417 | 0.000242344 | 0.000423454 | 1 |
| HTN | nk_cell | *EPC2* | -1.96004152 | 0.000126055 | 0.000236847 | 0.611620652 |
| HTN | nk_cell | *STAB2* | -2.536467747 | 7.66E-06 | 2.36E-05 | 0.037149021 |
| HTN | nk_cell | *APPL2* | -2.054732899 | 6.22E-05 | 0.000128028 | 0.302017574 |
| HTN | nk_cell | *DRAM1* | -2.216385003 | 8.40E-06 | 2.54E-05 | 0.04074159 |
| HTN | nk_cell | *SLC41A2* | -1.62854708 | 1.04E-06 | 6.09E-06 | 0.005057962 |
| HTN | nk_cell | *NEK3* | -1.582305072 | 0.000167966 | 0.000305915 | 0.814969079 |
| HTN | nk_cell | *CKAP2* | -0.680997574 | 0.025824574 | 0.031956346 | 1 |
| HTN | nk_cell | *TBC1D4* | -1.639576124 | 1.54E-06 | 7.63E-06 | 0.007487728 |
| HTN | nk_cell | *SUCLA2* | -1.784455523 | 3.86E-06 | 1.43E-05 | 0.018732524 |
| HTN | nk_cell | *RCBTB2* | 2.586872557 | 0.005035581 | 0.006834305 | 1 |
| HTN | nk_cell | *SCRN1* | -2.698053327 | 0.00039909 | 0.000662692 | 1 |
| HTN | nk_cell | *C7orf25* | -4.02858286 | 0.001085941 | 0.001650801 | 1 |
| HTN | nk_cell | *CHST12* | 1.737502879 | 4.06E-06 | 1.48E-05 | 0.019717799 |
| HTN | nk_cell | *NUPL2* | -2.446506102 | 0.000115213 | 0.000218706 | 0.559011849 |
| HTN | nk_cell | *ZDHHC4* | 3.970672464 | 0.00303171 | 0.004274879 | 1 |
| HTN | nk_cell | *AOAH* | 0.303561539 | 2.53E-07 | 4.07E-06 | 0.001225873 |
| HTN | nk_cell | *MTHFS* | -1.71333419 | 0.015169279 | 0.019328084 | 1 |
| HTN | nk_cell | *MYCBPAP* | 1.410808893 | 0.000142065 | 0.000263897 | 0.689298068 |
| HTN | nk_cell | *BRIP1* | 1.3190447 | 9.51E-07 | 5.82E-06 | 0.004612347 |
| HTN | nk_cell | *RTP4* | -2.961086578 | 4.41E-07 | 4.28E-06 | 0.002141831 |
| HTN | nk_cell | *NDUFB5* | -3.053024044 | 3.69E-05 | 8.30E-05 | 0.178803577 |
| HTN | nk_cell | *SCN2A* | -2.007513235 | 5.02E-05 | 0.000107373 | 0.243521692 |
| HTN | nk_cell | *RPS6KC1* | -2.136614672 | 0.000274709 | 0.000472488 | 1 |
| HTN | nk_cell | *CCDC115* | 1.926113688 | 9.34E-07 | 5.80E-06 | 0.004532572 |
| HTN | nk_cell | *IMP4* | -1.644623766 | 2.07E-07 | 4.07E-06 | 0.001006082 |
| HTN | nk_cell | *NIPSNAP3A* | -1.881003494 | 3.44E-07 | 4.24E-06 | 0.001668873 |
| HTN | nk_cell | *ODF2* | -3.067726786 | 6.39E-05 | 0.000130885 | 0.310067385 |
| HTN | nk_cell | *TOR1B* | -2.858024319 | 7.24E-05 | 0.000145978 | 0.351514342 |
| HTN | nk_cell | *SLC2A8* | -1.633596802 | 1.54E-06 | 7.63E-06 | 0.007456172 |
| HTN | nk_cell | *ZNF189* | -2.759345624 | 0.011876776 | 0.01535059 | 1 |
| HTN | nk_cell | *STX17* | -2.001566428 | 1.60E-05 | 4.25E-05 | 0.07781845 |
| HTN | nk_cell | *PRPF4* | -0.913314125 | 0.005498935 | 0.007436129 | 1 |
| HTN | nk_cell | *ANP32B* | -2.531421634 | 4.52E-05 | 9.82E-05 | 0.2192735 |
| HTN | nk_cell | *DSCC1* | 0.641169161 | 0.007732706 | 0.010237132 | 1 |
| HTN | nk_cell | *NOV* | -2.232528008 | 5.91E-06 | 1.95E-05 | 0.028691756 |
| HTN | nk_cell | *IL11RA* | -3.480919893 | 9.80E-07 | 5.92E-06 | 0.004756247 |
| HTN | nk_cell | *APTX* | -2.350991926 | 0.000157158 | 0.000287964 | 0.762528582 |
| HTN | nk_cell | *TMEM8B* | -1.823793459 | 0.000153107 | 0.000281605 | 0.742875222 |
| HTN | nk_cell | *GRHPR* | -1.875141328 | 5.48E-05 | 0.000115254 | 0.265892011 |
| HTN | nk_cell | *DENND4C* | -1.871053707 | 2.37E-05 | 5.80E-05 | 0.115066398 |

| | | | | | | |
|---|---|---|---|---|---|---|
| HTN | nk_cell | *GMPR* | -1.41671938 | 1.71E-06 | 8.12E-06 | 0.008316548 |
| HTN | nk_cell | *TMEM14B* | -1.587295007 | 2.76E-06 | 1.11E-05 | 0.013401267 |
| HTN | nk_cell | *FRS3* | -3.009236062 | 6.89E-06 | 2.17E-05 | 0.033419825 |
| HTN | nk_cell | *LRRC1* | -1.709010948 | 6.95E-07 | 4.87E-06 | 0.003372627 |
| HTN | nk_cell | *RIPK1* | -1.949431169 | 0.000417643 | 0.000689019 | 1 |
| HTN | nk_cell | *TUBB2B* | -1.495067838 | 0.017515544 | 0.022121399 | 1 |
| HTN | nk_cell | *TCF19* | 3.227196969 | 6.70E-08 | 2.56E-06 | 0.000324961 |
| HTN | nk_cell | *FLOT1* | -2.157608246 | 0.000860047 | 0.00133321 | 1 |
| HTN | nk_cell | *NRM* | 10.73914657 | 3.45E-06 | 1.31E-05 | 0.016719983 |
| HTN | nk_cell | *TLR2* | 3.726691783 | 0.000149797 | 0.0002764 | 0.726812626 |
| HTN | nk_cell | *NUMA1* | -2.026484471 | 1.23E-05 | 3.43E-05 | 0.059765403 |
| HTN | nk_cell | *SYTL2* | 1.403347321 | 0.000129019 | 0.000242073 | 0.625999938 |
| HTN | nk_cell | *RAB30* | -1.832041114 | 1.63E-06 | 7.89E-06 | 0.007927107 |
| HTN | nk_cell | *PRCP* | -1.995079662 | 5.72E-06 | 1.90E-05 | 0.027761105 |
| HTN | nk_cell | *NARS2* | -1.946256964 | 8.54E-06 | 2.57E-05 | 0.041456814 |
| HTN | nk_cell | *GGH* | -1.488316264 | 0.003748184 | 0.005199024 | 1 |
| HTN | nk_cell | *SULF1* | -1.325587054 | 3.66E-06 | 1.37E-05 | 0.017748268 |
| HTN | nk_cell | *NEK1* | -2.008319661 | 0.0002522 | 0.00043866 | 1 |
| HTN | nk_cell | *TRPC6* | -1.319499211 | 7.06E-06 | 2.21E-05 | 0.034268222 |
| HTN | nk_cell | *MMP27* | -4.104263538 | 0.003233323 | 0.004538062 | 1 |
| HTN | nk_cell | *POU2F3* | -2.374676859 | 5.33E-06 | 1.81E-05 | 0.025843828 |
| HTN | nk_cell | *PPP2R1B* | -1.28706667 | 3.65E-09 | 4.18E-07 | 1.77E-05 |
| HTN | nk_cell | *FXYD6* | -1.967908717 | 5.74E-06 | 1.91E-05 | 0.027846518 |
| HTN | nk_cell | *MAP2K5* | -1.685111696 | 5.28E-05 | 0.000111766 | 0.256278802 |
| HTN | nk_cell | *NUSAP1* | 3.447402912 | 1.36E-09 | 2.27E-07 | 6.58E-06 |
| HTN | nk_cell | *NDUFAF1* | -2.092583413 | 1.02E-05 | 2.95E-05 | 0.04962626 |
| HTN | nk_cell | *PARP6* | -2.665631967 | 4.86E-05 | 0.000104611 | 0.235584219 |
| HTN | nk_cell | *ZNF280D* | -1.71151904 | 6.35E-05 | 0.000130116 | 0.308113937 |
| HTN | nk_cell | *SPTBN5* | 0.810938754 | 7.73E-05 | 0.000154324 | 0.37500663 |
| HTN | nk_cell | *SLC44A5* | -1.405954287 | 1.38E-06 | 7.17E-06 | 0.006705699 |
| HTN | nk_cell | *DBT* | -2.55184388 | 7.84E-05 | 0.000156211 | 0.380216589 |
| HTN | nk_cell | *RTCA* | -1.886094686 | 0.026371565 | 0.032591654 | 1 |
| HTN | nk_cell | *KHK* | -2.051876379 | 4.19E-08 | 2.12E-06 | 0.000203388 |
| HTN | nk_cell | *DYNC2LI1* | -1.369410182 | 0.005658767 | 0.007637368 | 1 |
| HTN | nk_cell | *LHCGR* | -2.096503182 | 6.61E-08 | 2.55E-06 | 0.00032085 |
| HTN | nk_cell | *THUMPD2* | -2.073965424 | 8.21E-06 | 2.50E-05 | 0.039818769 |
| HTN | nk_cell | *ATRAID* | 2.327396616 | 0.001621464 | 0.002378278 | 1 |
| HTN | nk_cell | *CENPO* | -2.199082343 | 3.31E-06 | 1.28E-05 | 0.016082755 |
| HTN | nk_cell | *LRPPRC* | -2.13868852 | 7.03E-05 | 0.000142362 | 0.34119761 |
| HTN | nk_cell | *DTNB* | -1.53882716 | 0.025426693 | 0.03153638 | 1 |
| HTN | nk_cell | *CYP2C8* | -4.502898873 | 0.006708612 | 0.008976885 | 1 |
| HTN | nk_cell | *MYOF* | -1.445315574 | 9.19E-06 | 2.71E-05 | 0.044580183 |
| HTN | nk_cell | *BTBD16* | -2.266518772 | 1.02E-06 | 6.03E-06 | 0.004950195 |
| HTN | nk_cell | *TACC2* | -1.892805057 | 2.04E-06 | 9.07E-06 | 0.009894672 |
| HTN | nk_cell | *CALHM2* | -3.177694188 | 6.90E-06 | 2.17E-05 | 0.033474837 |
| HTN | nk_cell | *ARL3* | 1.753954562 | 0.039807536 | 0.048334876 | 1 |
| HTN | nk_cell | *PLCE1* | -1.79513012 | 3.88E-05 | 8.65E-05 | 0.188059189 |
| HTN | nk_cell | *RBP4* | -2.22684856 | 2.63E-05 | 6.27E-05 | 0.127384185 |
| HTN | nk_cell | *ASCC1* | -1.665759272 | 5.86E-07 | 4.54E-06 | 0.002844992 |
| HTN | nk_cell | *DNA2* | -0.692804802 | 0.005865689 | 0.007903929 | 1 |

| | | | | | | |
|---|---|---|---|---|---|---|
| HTN | nk_cell | *ATIC* | -1.522430115 | 5.16E-05 | 0.000109681 | 0.250510566 |
| HTN | nk_cell | *SMARCAL1* | -2.280240845 | 0.010408757 | 0.013568858 | 1 |
| HTN | nk_cell | *BARD1* | 2.362685327 | 8.37E-06 | 2.53E-05 | 0.040621352 |
| HTN | nk_cell | *STAT4* | 0.378170858 | 2.63E-07 | 4.07E-06 | 0.00127829 |
| HTN | nk_cell | *SSB* | -1.959461167 | 9.22E-06 | 2.72E-05 | 0.044716855 |
| HTN | nk_cell | *FASTKD1* | -2.294145192 | 2.63E-05 | 6.28E-05 | 0.127630644 |
| HTN | nk_cell | *ITGAV* | -2.004980615 | 1.52E-05 | 4.06E-05 | 0.07356224 |
| HTN | nk_cell | *SLC35A5* | -2.135348231 | 3.25E-05 | 7.52E-05 | 0.15789151 |
| HTN | nk_cell | *SENP7* | -1.929310243 | 1.16E-05 | 3.27E-05 | 0.056252093 |
| HTN | nk_cell | *COX17* | 3.656615366 | 1.95E-05 | 4.95E-05 | 0.094588674 |
| HTN | nk_cell | *SPPL2A* | -2.551965436 | 0.000782821 | 0.001224057 | 1 |
| HTN | nk_cell | *PPCDC* | -1.53541804 | 0.002671585 | 0.003800214 | 1 |
| HTN | nk_cell | *PCDH10* | 2.373603777 | 6.04E-07 | 4.54E-06 | 0.002931211 |
| HTN | nk_cell | *AP1AR* | -3.028526699 | 9.43E-05 | 0.000183461 | 0.457367606 |
| HTN | nk_cell | *PRKG2* | 1.955368933 | 4.50E-08 | 2.12E-06 | 0.000218395 |
| HTN | nk_cell | *KIAA1109* | -1.832105223 | 0.002111988 | 0.003041663 | 1 |
| HTN | nk_cell | *BMPR1B* | -1.179810537 | 9.82E-07 | 5.92E-06 | 0.00476571 |
| HTN | nk_cell | *MMRN1* | 1.19553623 | 0.006466448 | 0.008671976 | 1 |
| HTN | nk_cell | *PRDM5* | -1.573113278 | 1.88E-06 | 8.61E-06 | 0.009113546 |
| HTN | nk_cell | *TRPC3* | 0.400179542 | 2.34E-07 | 4.07E-06 | 0.00113678 |
| HTN | nk_cell | *NAAA* | -1.843425176 | 0.006942668 | 0.009269627 | 1 |
| HTN | nk_cell | *NUP54* | -1.654687925 | 0.007923081 | 0.010480586 | 1 |
| HTN | nk_cell | *CNOT6L* | 2.554239619 | 0.000121963 | 0.000230079 | 0.591763831 |
| HTN | nk_cell | *ANXA3* | -1.692206779 | 7.05E-05 | 0.000142611 | 0.342265438 |
| HTN | nk_cell | *PPA2* | -1.881049097 | 0.000254668 | 0.000441934 | 1 |
| HTN | nk_cell | *ENPEP* | -1.666660775 | 7.21E-07 | 4.96E-06 | 0.003497858 |
| HTN | nk_cell | *PAPSS1* | -1.682396416 | 7.44E-06 | 2.31E-05 | 0.036113562 |
| HTN | nk_cell | *PARVG* | 1.129201526 | 1.39E-06 | 7.20E-06 | 0.006746777 |
| HTN | nk_cell | *B4GALNT3* | 1.638316329 | 4.09E-07 | 4.24E-06 | 0.001986511 |
| HTN | nk_cell | *PDE6H* | -5.091976878 | 0.011670851 | 0.01511665 | 1 |
| HTN | nk_cell | *ERP27* | 0.525757472 | 8.87E-07 | 5.64E-06 | 0.004304396 |
| HTN | nk_cell | *KIF21A* | -1.892946304 | 0.01072703 | 0.013957509 | 1 |
| HTN | nk_cell | *ALG10* | -2.134912183 | 1.50E-07 | 4.00E-06 | 0.000728474 |
| HTN | nk_cell | *ZCRB1* | -2.840455568 | 0.001395796 | 0.002069805 | 1 |
| HTN | nk_cell | *C1RL* | -1.068480502 | 0.011252466 | 0.01460593 | 1 |
| HTN | nk_cell | *CLSTN3* | 2.240830103 | 1.89E-06 | 8.64E-06 | 0.009165574 |
| HTN | nk_cell | *KLRG1* | 0.700156773 | 1.75E-06 | 8.21E-06 | 0.008500552 |
| HTN | nk_cell | *VAMP1* | 2.41527735 | 6.09E-07 | 4.55E-06 | 0.00295279 |
| HTN | nk_cell | *PEX5* | -1.705488723 | 2.47E-06 | 1.04E-05 | 0.01197845 |
| HTN | nk_cell | *PPFIA2* | -1.455058269 | 1.15E-06 | 6.44E-06 | 0.005559279 |
| HTN | nk_cell | *MARCH9* | -3.37768678 | 4.58E-06 | 1.62E-05 | 0.022235341 |
| HTN | nk_cell | *LGR5* | -3.188948505 | 0.002523111 | 0.003604869 | 1 |
| HTN | nk_cell | *AMDHD1* | -1.62931956 | 3.29E-05 | 7.57E-05 | 0.159645994 |
| HTN | nk_cell | *RITA1* | -1.913611647 | 5.06E-07 | 4.37E-06 | 0.002453561 |
| HTN | nk_cell | *SDSL* | 2.97471548 | 4.65E-05 | 0.000100627 | 0.225404778 |
| HTN | nk_cell | *GLTP* | 2.532712894 | 7.96E-07 | 5.23E-06 | 0.003860805 |
| HTN | nk_cell | *FOXN4* | -1.679938274 | 1.85E-07 | 4.07E-06 | 0.000895351 |
| HTN | nk_cell | *MTMR6* | 3.192657491 | 2.15E-05 | 5.32E-05 | 0.104092443 |
| HTN | nk_cell | *CCDC65* | -2.188024617 | 0.000242647 | 0.000423803 | 1 |
| HTN | nk_cell | *BRCA2* | -4.745192268 | 0.000625285 | 0.000994715 | 1 |

| | | | | | | |
|---|---|---|---|---|---|---|
| HTN | nk_cell | *KANSL2* | -2.140266997 | 1.32E-05 | 3.62E-05 | 0.06422437 |
| HTN | nk_cell | *CERS5* | -2.253002817 | 0.000351998 | 0.000591376 | 1 |
| HTN | nk_cell | *LMBR1L* | -3.069967564 | 0.000258163 | 0.00044704 | 1 |
| HTN | nk_cell | *ZNF740* | 2.465650346 | 0.001062544 | 0.001618161 | 1 |
| HTN | nk_cell | *WDFY2* | -1.90945501 | 4.40E-05 | 9.62E-05 | 0.213364752 |
| HTN | nk_cell | *HNRNPA1L2* | -3.585240267 | 2.70E-07 | 4.07E-06 | 0.00131213 |
| HTN | nk_cell | *RB1* | -1.69074885 | 0.00134316 | 0.002000224 | 1 |
| HTN | nk_cell | *VPS33A* | -2.603384211 | 2.07E-06 | 9.18E-06 | 0.010051934 |
| HTN | nk_cell | *CBLN3* | -0.472656795 | 0.005591452 | 0.007557027 | 1 |
| HTN | nk_cell | *NOVA1* | -0.582802928 | 0.008804823 | 0.011571236 | 1 |
| HTN | nk_cell | *FITM1* | 1.089794527 | 5.72E-08 | 2.35E-06 | 0.000277604 |
| HTN | nk_cell | *RTN1* | -0.911983319 | 0.005900635 | 0.007948329 | 1 |
| HTN | nk_cell | *C14orf37* | -2.073568393 | 0.00018938 | 0.000340449 | 0.918872655 |
| HTN | nk_cell | *RAB15* | -3.64639079 | 0.004165902 | 0.005722807 | 1 |
| HTN | nk_cell | *EFCAB11* | -1.994549076 | 3.85E-07 | 4.24E-06 | 0.001868897 |
| HTN | nk_cell | *SLC24A4* | 2.326482064 | 3.90E-07 | 4.24E-06 | 0.001890507 |
| HTN | nk_cell | *C14orf79* | -3.132442429 | 6.47E-05 | 0.000132382 | 0.314009769 |
| HTN | nk_cell | *WARS* | -2.222497148 | 0.000135095 | 0.000252205 | 0.655481634 |
| HTN | nk_cell | *DUOXA1* | -1.888028247 | 0.000110754 | 0.000210985 | 0.537379349 |
| HTN | nk_cell | *SERF2* | 2.310411708 | 0.002547375 | 0.003636323 | 1 |
| HTN | nk_cell | *HMG20A* | -2.313079961 | 0.00184806 | 0.002681417 | 1 |
| HTN | nk_cell | *SCAPER* | -1.942686403 | 1.14E-05 | 3.22E-05 | 0.055174958 |
| HTN | nk_cell | *NEIL1* | -2.218308664 | 9.93E-06 | 2.88E-05 | 0.048194259 |
| HTN | nk_cell | *MAN2C1* | -0.851551146 | 0.000407169 | 0.000672884 | 1 |
| HTN | nk_cell | *PIF1* | 1.784728693 | 1.63E-07 | 4.07E-06 | 0.000791139 |
| HTN | nk_cell | *CYP11A1* | -1.757204775 | 5.81E-07 | 4.54E-06 | 0.002820147 |
| HTN | nk_cell | *BBS4* | -2.414127417 | 0.000281709 | 0.000483421 | 1 |
| HTN | nk_cell | *ULK3* | 5.589871864 | 4.42E-05 | 9.65E-05 | 0.214229449 |
| HTN | nk_cell | *PCSK6* | -1.956920595 | 2.39E-06 | 1.02E-05 | 0.01161673 |
| HTN | nk_cell | *CELF6* | 7.975651213 | 1.31E-07 | 3.84E-06 | 0.000637485 |
| HTN | nk_cell | *HAPLN3* | -1.562072118 | 2.18E-08 | 1.38E-06 | 0.000105932 |
| HTN | nk_cell | *MFGE8* | -2.334783644 | 0.002492871 | 0.003564813 | 1 |
| HTN | nk_cell | *UNC45A* | -2.108055334 | 8.47E-07 | 5.44E-06 | 0.004109614 |
| HTN | nk_cell | *CRTC3* | -1.775074877 | 3.34E-05 | 7.66E-05 | 0.162168585 |
| HTN | nk_cell | *ITGAX* | 0.317400116 | 2.66E-07 | 4.07E-06 | 0.001289508 |
| HTN | nk_cell | *TGFB1I1* | -5.114137592 | 9.51E-05 | 0.000184499 | 0.461248682 |
| HTN | nk_cell | *DHX38* | 2.396511201 | 0.039973558 | 0.048524319 | 1 |
| HTN | nk_cell | *NUDT7* | -2.396359133 | 2.56E-06 | 1.07E-05 | 0.012406212 |
| HTN | nk_cell | *GCSH* | -1.639155159 | 9.54E-08 | 3.26E-06 | 0.000463025 |
| HTN | nk_cell | *CMTM3* | -2.401316402 | 2.09E-06 | 9.22E-06 | 0.010136714 |
| HTN | nk_cell | *ZCCHC14* | -2.004502884 | 1.91E-05 | 4.87E-05 | 0.092590672 |
| HTN | nk_cell | *TLDC1* | -2.049007699 | 1.65E-06 | 7.94E-06 | 0.008026904 |
| HTN | nk_cell | *RPL3L* | 6.34351773 | 2.53E-05 | 6.10E-05 | 0.122816384 |
| HTN | nk_cell | *TIGD7* | -2.2719505 | 6.25E-07 | 4.59E-06 | 0.003032554 |
| HTN | nk_cell | *GALNS* | -2.585080757 | 1.59E-05 | 4.22E-05 | 0.07720253 |
| HTN | nk_cell | *GAS8* | -1.870552733 | 4.49E-06 | 1.59E-05 | 0.021773693 |
| HTN | nk_cell | *NCOR1* | -1.740958145 | 0.001055877 | 0.001609018 | 1 |
| HTN | nk_cell | *KSR1* | -2.364474047 | 6.26E-05 | 0.000128511 | 0.303519171 |
| HTN | nk_cell | *GFOD2* | -2.327594221 | 0.001975406 | 0.00285513 | 1 |
| HTN | nk_cell | *PRPSAP2* | -2.000607951 | 0.003504079 | 0.004885572 | 1 |

| | | | | | | |
|---|---|---|---|---|---|---|
| HTN | nk_cell | *RASL10B* | -4.22942265 | 1.35E-07 | 3.84E-06 | 0.000653107 |
| HTN | nk_cell | *UNC45B* | -5.55191147 | 4.38E-06 | 1.57E-05 | 0.021265393 |
| HTN | nk_cell | *C17orf80* | -1.808466447 | 5.96E-07 | 4.54E-06 | 0.002892594 |
| HTN | nk_cell | *VPS53* | -2.3426207 | 5.59E-07 | 4.48E-06 | 0.002714688 |
| HTN | nk_cell | *SGSM2* | -2.421841995 | 8.08E-06 | 2.47E-05 | 0.039206572 |
| HTN | nk_cell | *LRRC46* | -3.178685275 | 1.04E-08 | 8.67E-07 | 5.03E-05 |
| HTN | nk_cell | *SCRN2* | -1.247634326 | 6.81E-07 | 4.82E-06 | 0.003303878 |
| HTN | nk_cell | *SSH2* | -1.713889382 | 0.000395424 | 0.00065773 | 1 |
| HTN | nk_cell | *RHBDL3* | 2.665380067 | 3.05E-05 | 7.12E-05 | 0.147812535 |
| HTN | nk_cell | *G6PC3* | 4.141915534 | 0.00048798 | 0.000793859 | 1 |
| HTN | nk_cell | *CLTC* | -1.856603834 | 0.000335489 | 0.000565205 | 1 |
| HTN | nk_cell | *IMPA2* | -2.554857358 | 7.30E-05 | 0.000146759 | 0.353982696 |
| HTN | nk_cell | *GNAL* | -2.729533821 | 0.000304906 | 0.000518908 | 1 |
| HTN | nk_cell | *RPRD1A* | -1.845497773 | 0.000951401 | 0.00146453 | 1 |
| HTN | nk_cell | *GALNT1* | -2.056956721 | 9.05E-05 | 0.00017722 | 0.43914997 |
| HTN | nk_cell | *C18orf8* | -2.596470854 | 0.008418664 | 0.011099825 | 1 |
| HTN | nk_cell | *NPC1* | 1.837009492 | 0.018414012 | 0.023171543 | 1 |
| HTN | nk_cell | *SLC14A1* | -1.790390297 | 1.07E-05 | 3.07E-05 | 0.051939147 |
| HTN | nk_cell | *CCDC40* | -1.5258612 | 3.54E-07 | 4.24E-06 | 0.001719336 |
| HTN | nk_cell | *ARHGDIA* | 3.013192509 | 0.000255195 | 0.000442608 | 1 |
| HTN | nk_cell | *TMC6* | 1.130986952 | 3.78E-07 | 4.24E-06 | 0.001832589 |
| HTN | nk_cell | *CARD14* | -1.675072162 | 2.75E-06 | 1.11E-05 | 0.013354389 |
| HTN | nk_cell | *ANAPC11* | 2.941977205 | 0.01937367 | 0.024365227 | 1 |
| HTN | nk_cell | *TBCD* | -1.718541052 | 0.040097169 | 0.048662197 | 1 |
| HTN | nk_cell | *NARF* | -2.297077939 | 0.003790186 | 0.005249781 | 1 |
| HTN | nk_cell | *SECTM1* | -1.189479399 | 1.95E-06 | 8.85E-06 | 0.00945464 |
| HTN | nk_cell | *ZNF750* | 1.365035951 | 2.28E-07 | 4.07E-06 | 0.001105735 |
| HTN | nk_cell | *WDR45B* | -2.714981313 | 0.00017557 | 0.000317979 | 0.851866071 |
| HTN | nk_cell | *RNF165* | 2.053585621 | 0.000982443 | 0.001504676 | 1 |
| HTN | nk_cell | *MAPK4* | -5.627862996 | 2.02E-06 | 9.02E-06 | 0.009813134 |
| HTN | nk_cell | *FBXO15* | -3.024996262 | 4.77E-06 | 1.66E-05 | 0.02312395 |
| HTN | nk_cell | *CBLN2* | -1.638813805 | 4.33E-07 | 4.25E-06 | 0.002099553 |
| HTN | nk_cell | *NT5C3B* | -0.69293512 | 0.012568412 | 0.016171318 | 1 |
| HTN | nk_cell | *PIP4K2B* | -2.618975124 | 0.000229014 | 0.000403286 | 1 |
| HTN | nk_cell | *ERBB2* | -1.097074535 | 8.28E-07 | 5.35E-06 | 0.004019767 |
| HTN | nk_cell | *PNMT* | -4.408118728 | 1.24E-05 | 3.45E-05 | 0.060222744 |
| HTN | nk_cell | *IGFBP4* | -1.436848433 | 4.13E-05 | 9.13E-05 | 0.200351676 |
| HTN | nk_cell | *TXNL4A* | -1.66985315 | 2.64E-07 | 4.07E-06 | 0.00128086 |
| HTN | nk_cell | *CACNA1A* | -1.515770032 | 3.94E-07 | 4.24E-06 | 0.001913941 |
| HTN | nk_cell | *SAMD1* | -4.806986299 | 1.71E-05 | 4.47E-05 | 0.08315208 |
| HTN | nk_cell | *NFIC* | -1.527979986 | 1.46E-05 | 3.94E-05 | 0.070915897 |
| HTN | nk_cell | *PRDM15* | -1.976103159 | 2.02E-05 | 5.08E-05 | 0.098154607 |
| HTN | nk_cell | *MVB12A* | -2.271922503 | 0.00096207 | 0.001479076 | 1 |
| HTN | nk_cell | *DUS3L* | 1.893153552 | 3.67E-05 | 8.28E-05 | 0.178044539 |
| HTN | nk_cell | *DPP9* | 2.911428973 | 0.00384372 | 0.005317859 | 1 |
| HTN | nk_cell | *TMEM91* | -1.0273699 | 1.30E-05 | 3.57E-05 | 0.06299717 |
| HTN | nk_cell | *SIRT3* | -2.072065082 | 0.0084717 | 0.011163685 | 1 |
| HTN | nk_cell | *HUNK* | -1.053473862 | 2.98E-05 | 6.99E-05 | 0.144606038 |
| HTN | nk_cell | *COL6A1* | -0.878167715 | 0.00115517 | 0.001746071 | 1 |
| HTN | nk_cell | *IFNAR1* | -1.757690126 | 2.13E-05 | 5.31E-05 | 0.103541167 |

| | | | | | | |
|---|---|---|---|---|---|---|
| HTN | nk_cell | *DOPEY2* | -1.455572096 | 0.010653292 | 0.013869002 | 1 |
| HTN | nk_cell | *URB1* | -1.954149393 | 0.000133735 | 0.000250148 | 0.648882837 |
| HTN | nk_cell | *GEMIN7* | -3.397378025 | 2.16E-07 | 4.07E-06 | 0.001049288 |
| HTN | nk_cell | *SLC6A3* | -1.284247035 | 0.023245165 | 0.028963929 | 1 |
| HTN | nk_cell | *CAPN10* | 2.345738426 | 0.006484593 | 0.008692493 | 1 |
| HTN | nk_cell | *CACNG8* | 2.478213925 | 5.73E-06 | 1.91E-05 | 0.027794869 |
| HTN | nk_cell | *ZNF787* | -3.117836303 | 0.000252933 | 0.000439395 | 1 |
| HTN | nk_cell | *FBN3* | -1.62525538 | 0.000209404 | 0.000371627 | 1 |
| HTN | nk_cell | *EVI5L* | 4.021373878 | 2.18E-05 | 5.39E-05 | 0.105932689 |
| HTN | nk_cell | *SLC47A1* | 2.935589571 | 0.000467814 | 0.00076374 | 1 |
| HTN | nk_cell | *CTU1* | -1.304268992 | 4.17E-07 | 4.24E-06 | 0.002021594 |
| HTN | nk_cell | *RERE* | -1.376947307 | 5.68E-05 | 0.00011878 | 0.275806131 |
| HTN | nk_cell | *MMEL1* | -1.887500824 | 1.47E-06 | 7.43E-06 | 0.007156572 |
| HTN | nk_cell | *EPHA2* | -2.098392598 | 4.94E-07 | 4.32E-06 | 0.002396951 |
| HTN | nk_cell | *PEX14* | -1.803191091 | 1.88E-05 | 4.81E-05 | 0.091325881 |
| HTN | nk_cell | *ZNF593* | -2.834030499 | 5.11E-07 | 4.37E-06 | 0.002479263 |
| HTN | nk_cell | *HSPG2* | -1.134045388 | 0.000623209 | 0.000991738 | 1 |
| HTN | nk_cell | *ITGB3BP* | -1.425819864 | 3.59E-05 | 8.14E-05 | 0.174313033 |
| HTN | nk_cell | *PIGK* | -2.542308272 | 1.27E-05 | 3.51E-05 | 0.061524258 |
| HTN | nk_cell | *RPS8* | 1.748777198 | 0.021671554 | 0.02711459 | 1 |
| HTN | nk_cell | *CYP4B1* | -1.032447388 | 4.96E-05 | 0.000106349 | 0.240668255 |
| HTN | nk_cell | *SLC44A3* | -2.657878297 | 1.15E-05 | 3.24E-05 | 0.055588046 |
| HTN | nk_cell | *STRIP1* | -2.735376062 | 1.51E-05 | 4.05E-05 | 0.073437761 |
| HTN | nk_cell | *GPR161* | -2.462168356 | 1.20E-05 | 3.37E-05 | 0.058456671 |
| HTN | nk_cell | *ALDH9A1* | -2.710121435 | 1.57E-05 | 4.18E-05 | 0.076241566 |
| HTN | nk_cell | *ATP1B1* | -2.14723184 | 0.001603296 | 0.002355904 | 1 |
| HTN | nk_cell | *TIPRL* | 2.28718828 | 0.025478463 | 0.031592513 | 1 |
| HTN | nk_cell | *NME7* | -1.514417846 | 1.29E-07 | 3.82E-06 | 0.000627527 |
| HTN | nk_cell | *XCL1* | 1.660061723 | 4.91E-07 | 4.32E-06 | 0.002382433 |
| HTN | nk_cell | *XCL2* | 0.505002516 | 3.24E-07 | 4.19E-06 | 0.001572373 |
| HTN | nk_cell | *MAEL* | -2.921888068 | 4.73E-07 | 4.32E-06 | 0.002295434 |
| HTN | nk_cell | *RFWD2* | -1.692196945 | 0.004871422 | 0.006635637 | 1 |
| HTN | nk_cell | *RGS16* | 3.19568444 | 2.58E-05 | 6.18E-05 | 0.125013442 |
| HTN | nk_cell | *TARS2* | -2.527147139 | 9.63E-05 | 0.000186609 | 0.467455333 |
| HTN | nk_cell | *SNX27* | -1.653578629 | 0.000841799 | 0.001308267 | 1 |
| HTN | nk_cell | *SETDB1* | -2.282700731 | 4.88E-05 | 0.000105092 | 0.236987506 |
| HTN | nk_cell | *SEMA6C* | -1.183569319 | 1.20E-06 | 6.65E-06 | 0.005802412 |
| HTN | nk_cell | *ARNT* | -2.109325122 | 3.69E-07 | 4.24E-06 | 0.001788211 |
| HTN | nk_cell | *POGZ* | -1.949317028 | 8.36E-06 | 2.53E-05 | 0.040571906 |
| HTN | nk_cell | *GABPB2* | -1.644520833 | 0.002531101 | 0.003614156 | 1 |
| HTN | nk_cell | *DYRK3* | -1.771207222 | 2.98E-07 | 4.13E-06 | 0.001446412 |
| HTN | nk_cell | *EIF2D* | -2.535690839 | 2.27E-05 | 5.58E-05 | 0.109960266 |
| HTN | nk_cell | *TAF1A* | -2.244738619 | 2.01E-05 | 5.07E-05 | 0.097662216 |
| HTN | nk_cell | *RAB13* | 4.254648508 | 6.05E-05 | 0.000124984 | 0.293586747 |
| HTN | nk_cell | *S100A8* | -3.036967681 | 0.000239816 | 0.000420371 | 1 |
| HTN | nk_cell | *CREB3L4* | -5.248336617 | 1.49E-11 | 4.26E-09 | 7.24E-08 |
| HTN | nk_cell | *KCNN3* | -1.645848644 | 1.52E-06 | 7.57E-06 | 0.007378465 |
| HTN | nk_cell | *C1orf43* | -2.641022571 | 4.96E-06 | 1.72E-05 | 0.024072664 |
| HTN | nk_cell | *RIT1* | -2.710451868 | 1.80E-06 | 8.37E-06 | 0.008744394 |
| HTN | nk_cell | *FLG* | -1.246833193 | 2.17E-05 | 5.37E-05 | 0.105238719 |

| | | | | | | |
|---|---|---|---|---|---|---|
| HTN | nk_cell | *SCCPDH* | -2.185316461 | 4.19E-06 | 1.51E-05 | 0.020325163 |
| HTN | nk_cell | *CEP170* | -1.759161966 | 0.000265802 | 0.000458632 | 1 |
| HTN | nk_cell | *ACP1* | -2.592092305 | 1.92E-05 | 4.89E-05 | 0.093016526 |
| HTN | nk_cell | *CNIH4* | -2.145493144 | 3.58E-06 | 1.35E-05 | 0.017387863 |
| HTN | nk_cell | *MBOAT2* | -1.855749597 | 1.66E-06 | 7.94E-06 | 0.008046535 |
| HTN | nk_cell | *PYCR2* | 2.71455987 | 3.02E-06 | 1.20E-05 | 0.014629173 |
| HTN | nk_cell | *PLEKHA6* | -0.695499801 | 1.33E-05 | 3.65E-05 | 0.064685679 |
| HTN | nk_cell | *OSR1* | -0.961018109 | 5.11E-05 | 0.000109052 | 0.248093873 |
| HTN | nk_cell | *RHOB* | -2.284690472 | 5.37E-06 | 1.82E-05 | 0.026055716 |
| HTN | nk_cell | *GALM* | -2.276843686 | 1.65E-05 | 4.34E-05 | 0.079827546 |
| HTN | nk_cell | *WDPCP* | -1.416490921 | 4.17E-05 | 9.21E-05 | 0.20249348 |
| HTN | nk_cell | *VPS54* | -1.808599528 | 2.16E-05 | 5.36E-05 | 0.104821144 |
| HTN | nk_cell | *REG3G* | 8.060033187 | 0.00229289 | 0.003294375 | 1 |
| HTN | nk_cell | *ABHD1* | -4.525319738 | 9.89E-07 | 5.94E-06 | 0.004796544 |
| HTN | nk_cell | *CIAO1* | -2.030285672 | 0.000977316 | 0.001497769 | 1 |
| HTN | nk_cell | *ZNF514* | -3.141033631 | 3.94E-06 | 1.45E-05 | 0.019123777 |
| HTN | nk_cell | *EXOC6B* | -1.587246354 | 4.55E-05 | 9.89E-05 | 0.220969736 |
| HTN | nk_cell | *TEX261* | -4.359702538 | 0.00139979 | 0.00207446 | 1 |
| HTN | nk_cell | *RALB* | -2.866153404 | 0.000718363 | 0.001130187 | 1 |
| HTN | nk_cell | *RABL2A* | -2.898847365 | 5.48E-08 | 2.29E-06 | 0.000265751 |
| HTN | nk_cell | *FBLN7* | -1.302576197 | 4.94E-05 | 0.000106089 | 0.239654285 |
| HTN | nk_cell | *CNGA3* | -17.59091626 | 4.66E-05 | 0.000100993 | 0.226326209 |
| HTN | nk_cell | *FAHD2B* | -2.059129805 | 2.66E-07 | 4.07E-06 | 0.00128867 |
| HTN | nk_cell | *THSD7B* | -1.075676823 | 3.85E-07 | 4.24E-06 | 0.001870339 |
| HTN | nk_cell | *GPR17* | -0.448034787 | 0.007607183 | 0.010092987 | 1 |
| HTN | nk_cell | *POLR2D* | -2.284982404 | 0.007738911 | 0.010242552 | 1 |
| HTN | nk_cell | *GALNT13* | -2.055475079 | 4.17E-06 | 1.51E-05 | 0.020241869 |
| HTN | nk_cell | *PHOSPHO2* | -5.689105737 | 0.000354133 | 0.00059414 | 1 |
| HTN | nk_cell | *METTL21A* | -2.057293349 | 0.004135125 | 0.005686969 | 1 |
| HTN | nk_cell | *NBEAL1* | -1.835856581 | 7.39E-05 | 0.000148472 | 0.358559242 |
| HTN | nk_cell | *SUMF1* | -1.70737262 | 0.001300624 | 0.001944124 | 1 |
| HTN | nk_cell | *ACKR3* | -1.652270375 | 0.000128686 | 0.000241541 | 0.624382653 |
| HTN | nk_cell | *ESPNL* | -13.36343109 | 4.50E-07 | 4.28E-06 | 0.002183449 |
| HTN | nk_cell | *ANKMY1* | -1.671888029 | 0.025773207 | 0.031925352 | 1 |
| HTN | nk_cell | *TAMM41* | -2.102932597 | 0.001306426 | 0.001951595 | 1 |
| HTN | nk_cell | *STK11IP* | -2.963448541 | 1.48E-05 | 3.98E-05 | 0.071807334 |
| HTN | nk_cell | *EAF1* | 1.582902477 | 2.37E-07 | 4.07E-06 | 0.001152163 |
| HTN | nk_cell | *CNTN4* | -1.25372108 | 1.34E-06 | 7.03E-06 | 0.006502542 |
| HTN | nk_cell | *SLC25A38* | 1.495778635 | 1.22E-06 | 6.73E-06 | 0.005934533 |
| HTN | nk_cell | *CTDSPL* | -1.983479763 | 1.20E-06 | 6.66E-06 | 0.005812751 |
| HTN | nk_cell | *PTPRG* | -1.506782595 | 2.18E-05 | 5.39E-05 | 0.105849232 |
| HTN | nk_cell | *SLC25A26* | -2.367181062 | 0.000523796 | 0.000846798 | 1 |
| HTN | nk_cell | *LIMD1* | -1.864191932 | 0.000273659 | 0.000471182 | 1 |
| HTN | nk_cell | *PLA1A* | -3.538840533 | 5.59E-07 | 4.48E-06 | 0.002714445 |
| HTN | nk_cell | *ADPRH* | 2.474565831 | 5.36E-05 | 0.000113192 | 0.26000151 |
| HTN | nk_cell | *IGSF11* | -0.917207239 | 3.96E-06 | 1.45E-05 | 0.019217988 |
| HTN | nk_cell | *BOC* | -1.075508206 | 1.50E-06 | 7.52E-06 | 0.007269292 |
| HTN | nk_cell | *MED12L* | -2.332264125 | 7.42E-06 | 2.30E-05 | 0.035982133 |
| HTN | nk_cell | *EIF2A* | -1.935451589 | 9.41E-06 | 2.77E-05 | 0.045669091 |
| HTN | nk_cell | *NCEH1* | -2.03472751 | 4.52E-07 | 4.28E-06 | 0.002190923 |

| | | | | | | |
|---|---|---|---|---|---|---|
| HTN | nk_cell | *AMT* | -1.358589317 | 2.48E-05 | 6.01E-05 | 0.120526724 |
| HTN | nk_cell | *CCDC39* | -1.989670743 | 3.36E-06 | 1.29E-05 | 0.016309942 |
| HTN | nk_cell | *MUC4* | -2.310154289 | 4.94E-07 | 4.32E-06 | 0.002398338 |
| HTN | nk_cell | *DGKQ* | 1.69059827 | 3.99E-07 | 4.24E-06 | 0.00193699 |
| HTN | nk_cell | *CENPC* | -1.939956994 | 2.56E-05 | 6.14E-05 | 0.123993996 |
| HTN | nk_cell | *SCD5* | -2.357538281 | 3.09E-06 | 1.22E-05 | 0.015004868 |
| HTN | nk_cell | *TRMT10A* | -1.339732347 | 7.05E-07 | 4.90E-06 | 0.003418657 |
| HTN | nk_cell | *PYURF* | 3.388700919 | 0.004490251 | 0.006149674 | 1 |
| HTN | nk_cell | *DDIT4L* | -2.815542894 | 2.81E-06 | 1.13E-05 | 0.013649312 |
| HTN | nk_cell | *METTL14* | -2.42823611 | 0.00427478 | 0.005865733 | 1 |
| HTN | nk_cell | *USP53* | -1.592015634 | 5.42E-05 | 0.000114135 | 0.262967831 |
| HTN | nk_cell | *MARCH1* | -1.700453096 | 0.000438091 | 0.0007186 | 1 |
| HTN | nk_cell | *CBR4* | -1.897386553 | 0.000429552 | 0.000706264 | 1 |
| HTN | nk_cell | *MARCH6* | -1.936635903 | 0.000122995 | 0.000231846 | 0.596770433 |
| HTN | nk_cell | *NKD2* | 3.053723921 | 1.87E-05 | 4.79E-05 | 0.09092764 |
| HTN | nk_cell | *MYO10* | -1.035228074 | 6.01E-07 | 4.54E-06 | 0.002915373 |
| HTN | nk_cell | *FAM105A* | 1.650502977 | 0.000211627 | 0.000374886 | 1 |
| HTN | nk_cell | *BHMT* | -1.020748614 | 0.008703312 | 0.011450236 | 1 |
| HTN | nk_cell | *GTF2H2* | -3.496222451 | 4.08E-06 | 1.49E-05 | 0.019794556 |
| HTN | nk_cell | *SPATA9* | -1.141153895 | 6.52E-07 | 4.69E-06 | 0.003162748 |
| HTN | nk_cell | *TSLP* | -1.426547915 | 8.99E-06 | 2.67E-05 | 0.043595942 |
| HTN | nk_cell | *FEM1C* | 3.714772732 | 0.000666459 | 0.001053652 | 1 |
| HTN | nk_cell | *COMMD10* | -1.586089127 | 0.002856177 | 0.00404264 | 1 |
| HTN | nk_cell | *YIPF5* | -2.874172081 | 0.002253949 | 0.003240344 | 1 |
| HTN | nk_cell | *CXCL14* | -1.562138785 | 5.28E-07 | 4.40E-06 | 0.002563137 |
| HTN | nk_cell | *PCYOX1L* | -2.278337991 | 5.42E-07 | 4.44E-06 | 0.002627649 |
| HTN | nk_cell | *BOD1* | 4.160494202 | 1.85E-06 | 8.53E-06 | 0.008976626 |
| HTN | nk_cell | *TBC1D7* | 3.825233226 | 4.10E-06 | 1.49E-05 | 0.019906498 |
| HTN | nk_cell | *TRIM7* | -2.379884887 | 1.59E-07 | 4.07E-06 | 0.000769762 |
| HTN | nk_cell | *FAM193B* | -2.413599884 | 1.82E-05 | 4.70E-05 | 0.088416727 |
| HTN | nk_cell | *RNF44* | 3.120602682 | 1.33E-06 | 7.00E-06 | 0.006445601 |
| HTN | nk_cell | *MUT* | -2.529346329 | 1.96E-05 | 4.97E-05 | 0.094997447 |
| HTN | nk_cell | *DOK3* | -1.158205541 | 5.83E-08 | 2.37E-06 | 0.000282753 |
| HTN | nk_cell | *ABT1* | 2.056725741 | 2.99E-09 | 3.92E-07 | 1.45E-05 |
| HTN | nk_cell | *DAAM2* | -2.063258818 | 2.43E-05 | 5.90E-05 | 0.117880581 |
| HTN | nk_cell | *MLIP* | -1.2646308 | 1.16E-06 | 6.52E-06 | 0.005644147 |
| HTN | nk_cell | *ANO7* | -1.810381233 | 1.81E-06 | 8.41E-06 | 0.0088029 |
| HTN | nk_cell | *TCTE1* | 2.276094789 | 2.67E-06 | 1.09E-05 | 0.012959069 |
| HTN | nk_cell | *RPL7L1* | -3.055802681 | 1.84E-05 | 4.72E-05 | 0.089178165 |
| HTN | nk_cell | *TPBG* | -1.718191449 | 1.69E-06 | 8.05E-06 | 0.008210207 |
| HTN | nk_cell | *IRAK1BP1* | -1.329609527 | 0.003676036 | 0.005106248 | 1 |
| HTN | nk_cell | *PHIP* | -1.731874246 | 0.002026014 | 0.002922182 | 1 |
| HTN | nk_cell | *PNRC1* | -2.128531634 | 5.57E-06 | 1.87E-05 | 0.027021071 |
| HTN | nk_cell | *RNF217* | -1.289218641 | 2.02E-06 | 9.02E-06 | 0.009806333 |
| HTN | nk_cell | *ARHGAP18* | -1.616931889 | 0.000948175 | 0.001460049 | 1 |
| HTN | nk_cell | *ABRACL* | 1.063443492 | 2.22E-07 | 4.07E-06 | 0.001078606 |
| HTN | nk_cell | *SLC18B1* | -2.455876218 | 1.38E-08 | 1.03E-06 | 6.67E-05 |
| HTN | nk_cell | *PNLDC1* | -5.27823281 | 1.68E-05 | 4.42E-05 | 0.081596196 |
| HTN | nk_cell | *WTAP* | -2.09704963 | 7.26E-05 | 0.000146226 | 0.352257926 |
| HTN | nk_cell | *SLC22A3* | -2.056728611 | 3.79E-06 | 1.41E-05 | 0.018391316 |

| | | | | | | |
|---|---|---|---|---|---|---|
| HTN | nk_cell | *GNA12* | -2.656012327 | 1.92E-05 | 4.89E-05 | 0.093081117 |
| HTN | nk_cell | *SDK1* | -1.120887236 | 0.001216256 | 0.001829496 | 1 |
| HTN | nk_cell | *RBAK* | 3.52602379 | 1.17E-06 | 6.53E-06 | 0.005669077 |
| HTN | nk_cell | *POMZP3* | -1.657998237 | 0.013453774 | 0.017250981 | 1 |
| HTN | nk_cell | *PSPH* | -1.165157859 | 0.00042211 | 0.000695206 | 1 |
| HTN | nk_cell | *GIGYF1* | -2.028467109 | 4.99E-05 | 0.000106808 | 0.241919248 |
| HTN | nk_cell | *TMEM209* | -2.867438187 | 0.001978015 | 0.002858048 | 1 |
| HTN | nk_cell | *EPHA1* | 4.485349046 | 4.12E-07 | 4.24E-06 | 0.002001372 |
| HTN | nk_cell | *NCAPG2* | 3.95234948 | 3.19E-05 | 7.40E-05 | 0.154729213 |
| HTN | nk_cell | *FBXO25* | -2.358586227 | 5.69E-05 | 0.000118886 | 0.276292048 |
| HTN | nk_cell | *CSGALNACT1* | -0.965498952 | 0.002976189 | 0.004200253 | 1 |
| HTN | nk_cell | *CHMP7* | 3.871546619 | 9.87E-06 | 2.87E-05 | 0.047894773 |
| HTN | nk_cell | *DOCK5* | -1.901969986 | 0.00064364 | 0.001019903 | 1 |
| HTN | nk_cell | *TACC1* | -1.562930594 | 4.91E-05 | 0.000105486 | 0.238187195 |
| HTN | nk_cell | *ADHFE1* | -1.442317217 | 1.37E-07 | 3.87E-06 | 0.000665495 |
| HTN | nk_cell | *RPL7* | -2.152367084 | 3.96E-05 | 8.81E-05 | 0.192248806 |
| HTN | nk_cell | *ATP6V0D2* | -3.674962866 | 3.16E-06 | 1.23E-05 | 0.015341862 |
| HTN | nk_cell | *DPYS* | -0.94580375 | 6.22E-07 | 4.57E-06 | 0.003016189 |
| HTN | nk_cell | *LRP12* | -2.294983134 | 3.92E-06 | 1.45E-05 | 0.019007719 |
| HTN | nk_cell | *GSDMC* | -5.799028559 | 0.023100166 | 0.028798049 | 1 |
| HTN | nk_cell | *SLC39A4* | -1.486580798 | 0.000229255 | 0.000403462 | 1 |
| HTN | nk_cell | *AK3* | -2.351838409 | 8.59E-05 | 0.000169095 | 0.416990418 |
| HTN | nk_cell | *C9orf72* | -0.827283826 | 0.003594389 | 0.005008609 | 1 |
| HTN | nk_cell | *FBXO10* | -1.774996048 | 0.000202613 | 0.000361293 | 0.983078985 |
| HTN | nk_cell | *ZNF462* | -1.206404897 | 8.62E-06 | 2.58E-05 | 0.041826415 |
| HTN | nk_cell | *WDR31* | -1.360779803 | 0.000469124 | 0.00076562 | 1 |
| HTN | nk_cell | *POLE3* | 1.374024428 | 4.96E-08 | 2.19E-06 | 0.000240771 |
| HTN | nk_cell | *SURF1* | 1.974520064 | 0.001964955 | 0.002842564 | 1 |
| HTN | nk_cell | *SURF6* | -1.583481309 | 1.21E-06 | 6.69E-06 | 0.005859329 |
| HTN | nk_cell | *MED22* | 1.964831937 | 6.36E-06 | 2.06E-05 | 0.030848342 |
| HTN | nk_cell | *REXO4* | 3.349428717 | 2.35E-05 | 5.76E-05 | 0.114108413 |
| HTN | nk_cell | *SH3GLB2* | -2.907015085 | 0.00449058 | 0.006149674 | 1 |
| HTN | nk_cell | *INPP5E* | -2.323151063 | 1.55E-06 | 7.63E-06 | 0.007508721 |
| HTN | nk_cell | *SLC39A12* | -2.449412363 | 2.69E-06 | 1.10E-05 | 0.013049691 |
| HTN | nk_cell | *ST8SIA6* | 0.434777128 | 2.70E-07 | 4.07E-06 | 0.001308308 |
| HTN | nk_cell | *PARD3* | -1.320399547 | 5.39E-06 | 1.82E-05 | 0.02615718 |
| HTN | nk_cell | *CDHR1* | 0.757386752 | 3.50E-07 | 4.24E-06 | 0.001698148 |
| HTN | nk_cell | *RGR* | -13.08820917 | 2.04E-07 | 4.07E-06 | 0.000991903 |
| HTN | nk_cell | *FRA10AC1* | -1.958003349 | 0.000146529 | 0.000271254 | 0.710956571 |
| HTN | nk_cell | *ADD3* | -1.870926267 | 0.000818337 | 0.001274253 | 1 |
| HTN | nk_cell | *LRRC27* | -1.306324226 | 2.64E-06 | 1.09E-05 | 0.012809274 |
| HTN | nk_cell | *PAOX* | 4.566576016 | 6.18E-05 | 0.000127373 | 0.299834868 |
| HTN | nk_cell | *PDCD11* | 2.624817879 | 0.009810177 | 0.012826457 | 1 |
| HTN | nk_cell | *GAS2* | -2.855084457 | 2.66E-05 | 6.34E-05 | 0.129201699 |
| HTN | nk_cell | *LIN7C* | -2.670753942 | 0.013514285 | 0.017319416 | 1 |
| HTN | nk_cell | *ZNF215* | -0.635526155 | 0.007668583 | 0.010160559 | 1 |
| HTN | nk_cell | *HSD17B12* | -2.249178727 | 8.31E-14 | 1.01E-10 | 4.03E-10 |
| HTN | nk_cell | *APIP* | -0.666001092 | 0.04071267 | 0.049396818 | 1 |
| HTN | nk_cell | *EIF3M* | -2.588060218 | 0.001700424 | 0.002482087 | 1 |
| HTN | nk_cell | *PTPRJ* | 1.877407433 | 9.67E-06 | 2.83E-05 | 0.04694148 |

| | | | | | | |
|---|---|---|---|---|---|---|
| HTN | nk_cell | *CCDC82* | -1.370021946 | 0.004951967 | 0.00673697 | 1 |
| HTN | nk_cell | *CAPN5* | -1.583059137 | 9.34E-07 | 5.80E-06 | 0.004529473 |
| HTN | nk_cell | *INTS4* | -2.186172695 | 0.000322587 | 0.000545743 | 1 |
| HTN | nk_cell | *RPS3* | 2.039786623 | 6.36E-06 | 2.06E-05 | 0.030875351 |
| HTN | nk_cell | *DRD2* | -8.292429767 | 3.98E-07 | 4.24E-06 | 0.00193248 |
| HTN | nk_cell | *ATM* | 1.831532351 | 0.004113175 | 0.005658385 | 1 |
| HTN | nk_cell | *SLX4IP* | -2.129946536 | 5.26E-06 | 1.80E-05 | 0.025522575 |
| HTN | nk_cell | *ST14* | -3.022345359 | 1.16E-06 | 6.50E-06 | 0.005624526 |
| HTN | nk_cell | *FADS1* | 2.594088394 | 7.10E-06 | 2.22E-05 | 0.034449081 |
| HTN | nk_cell | *TMC2* | -3.661806382 | 5.17E-07 | 4.37E-06 | 0.002508873 |
| HTN | nk_cell | *ROM1* | -1.31961138 | 0.00010262 | 0.000197584 | 0.49791261 |
| HTN | nk_cell | *EML3* | -1.600689957 | 0.001159282 | 0.001751739 | 1 |
| HTN | nk_cell | *PLCH2* | 0.301391542 | 2.76E-07 | 4.07E-06 | 0.001338171 |
| HTN | nk_cell | *CPSF7* | -2.302928129 | 0.001630167 | 0.002390321 | 1 |
| HTN | nk_cell | *CCDC15* | -1.315520238 | 4.90E-07 | 4.32E-06 | 0.002376378 |
| HTN | nk_cell | *CHEK1* | -1.755916104 | 4.13E-07 | 4.24E-06 | 0.002002357 |
| HTN | nk_cell | *DUSP15* | -2.760048186 | 0.015223021 | 0.019385068 | 1 |
| HTN | nk_cell | *SPATA25* | -3.863507867 | 1.03E-06 | 6.04E-06 | 0.004985975 |
| HTN | nk_cell | *CABLES2* | -2.791109886 | 0.00012191 | 0.000230069 | 0.591507884 |
| HTN | nk_cell | *TRPT1* | -3.008155399 | 7.34E-09 | 6.67E-07 | 3.56E-05 |
| HTN | nk_cell | *NUDT22* | -2.238502525 | 0.004261779 | 0.005849549 | 1 |
| HTN | nk_cell | *ALDOA* | 2.075794207 | 0.008756625 | 0.011511011 | 1 |
| HTN | nk_cell | *ANXA8L1* | -0.697931074 | 0.015102846 | 0.019253549 | 1 |
| HTN | nk_cell | *CWC15* | 2.08139202 | 0.0032527 | 0.004559983 | 1 |
| HTN | nk_cell | *FCGR1A* | -2.061652725 | 0.008447707 | 0.011135092 | 1 |
| HTN | nk_cell | *TMEM218* | -2.881037567 | 4.49E-05 | 9.78E-05 | 0.21774342 |
| HTN | nk_cell | *FAM124A* | -2.256836835 | 3.15E-06 | 1.23E-05 | 0.0152922 |
| HTN | nk_cell | *LYPD6B* | -1.847045777 | 4.47E-06 | 1.59E-05 | 0.021673554 |
| HTN | nk_cell | *WDR17* | -2.547483782 | 8.09E-05 | 0.000160641 | 0.392607367 |
| HTN | nk_cell | *CCDC102B* | -1.507733832 | 5.17E-06 | 1.78E-05 | 0.025061618 |
| HTN | nk_cell | *RGS18* | 1.484795826 | 0.009753307 | 0.012755538 | 1 |
| HTN | nk_cell | *PRSS23* | -1.397995757 | 2.06E-05 | 5.16E-05 | 0.099875882 |
| HTN | nk_cell | *PPP1R1C* | -2.138083201 | 5.30E-07 | 4.40E-06 | 0.002569914 |
| HTN | nk_cell | *FAM173B* | -2.258322835 | 0.014244542 | 0.018197609 | 1 |
| HTN | nk_cell | *GPR158* | -2.528603615 | 1.06E-06 | 6.14E-06 | 0.005136851 |
| HTN | nk_cell | *UBE3B* | -1.29222558 | 0.032372054 | 0.039623917 | 1 |
| HTN | nk_cell | *ANK3* | 1.54757246 | 6.12E-07 | 4.55E-06 | 0.002969802 |
| HTN | nk_cell | *PLBD2* | -3.25260109 | 3.78E-06 | 1.41E-05 | 0.018350706 |
| HTN | nk_cell | *DLG5* | -1.656258745 | 9.35E-05 | 0.000182455 | 0.453765886 |
| HTN | nk_cell | *GXYLT1* | -2.470306912 | 0.001555049 | 0.002290558 | 1 |
| HTN | nk_cell | *EXT2* | -2.060704566 | 0.000552218 | 0.000889267 | 1 |
| HTN | nk_cell | *TMEM18* | -3.646366174 | 0.000241738 | 0.000422671 | 1 |
| HTN | nk_cell | *NDUFC2* | -2.918312638 | 0.023414018 | 0.029166833 | 1 |
| HTN | nk_cell | *ADAMTS12* | -1.683314882 | 5.19E-07 | 4.37E-06 | 0.002516617 |
| HTN | nk_cell | *NUBPL* | -2.055199859 | 0.000264759 | 0.000456994 | 1 |
| HTN | nk_cell | *VIPAS39* | -1.869905061 | 1.59E-06 | 7.75E-06 | 0.007711931 |
| HTN | nk_cell | *ANKRD50* | -2.296809955 | 5.55E-05 | 0.000116561 | 0.269255943 |
| HTN | nk_cell | *UPF2* | -2.034824961 | 0.000877236 | 0.001356821 | 1 |
| HTN | nk_cell | *C4orf33* | 3.393453405 | 0.000414884 | 0.000684933 | 1 |
| HTN | nk_cell | *PTPRO* | -1.963492602 | 5.74E-06 | 1.91E-05 | 0.027830366 |

| | | | | | | |
|---|---|---|---|---|---|---|
| HTN | nk_cell | *EPS8* | -1.707803387 | 7.18E-05 | 0.000144778 | 0.348191375 |
| HTN | nk_cell | *QDPR* | -1.43126264 | 3.79E-06 | 1.41E-05 | 0.018386844 |
| HTN | nk_cell | *FAM160B1* | -2.019691335 | 1.16E-05 | 3.27E-05 | 0.056212436 |
| HTN | nk_cell | *ZNF827* | -2.13537463 | 0.010529711 | 0.013715479 | 1 |
| HTN | nk_cell | *AKR1C2* | -1.29349705 | 2.69E-06 | 1.10E-05 | 0.013070381 |
| HTN | nk_cell | *DPYSL4* | 0.99749298 | 0.000668535 | 0.001056245 | 1 |
| HTN | nk_cell | *PIGF* | -2.280318234 | 2.12E-05 | 5.28E-05 | 0.102841317 |
| HTN | nk_cell | *INPP1* | -2.788737268 | 8.35E-07 | 5.39E-06 | 0.004053309 |
| HTN | nk_cell | *MFSD6* | 2.671575857 | 0.002039909 | 0.002941349 | 1 |
| HTN | nk_cell | *RNF144A* | -1.504683232 | 1.93E-06 | 8.80E-06 | 0.009380095 |
| HTN | nk_cell | *BICD1* | -2.124189856 | 6.78E-05 | 0.000137981 | 0.328808405 |
| HTN | nk_cell | *GUF1* | -2.029773573 | 6.76E-06 | 2.14E-05 | 0.032776347 |
| HTN | nk_cell | *GABRA2* | 2.463259644 | 3.42E-06 | 1.31E-05 | 0.016609103 |
| HTN | nk_cell | *PARP8* | 1.025926701 | 3.83E-05 | 8.57E-05 | 0.186038604 |
| HTN | nk_cell | *CACUL1* | -1.91553036 | 0.006988844 | 0.009328712 | 1 |
| HTN | nk_cell | *BEND6* | -1.45692623 | 4.15E-07 | 4.24E-06 | 0.002011553 |
| HTN | nk_cell | *TIAL1* | -2.211617346 | 0.001376779 | 0.002044105 | 1 |
| HTN | nk_cell | *TMEM56* | -4.036014458 | 1.60E-06 | 7.79E-06 | 0.007761943 |
| HTN | nk_cell | *PTPN14* | -1.644147214 | 2.28E-05 | 5.61E-05 | 0.110696613 |
| HTN | nk_cell | *MGAT5* | -2.097298184 | 0.000243743 | 0.00042541 | 1 |
| HTN | nk_cell | *TMEM178A* | 2.263604953 | 3.36E-07 | 4.24E-06 | 0.001630963 |
| HTN | nk_cell | *EPG5* | -2.313051636 | 0.001661785 | 0.002430806 | 1 |
| HTN | nk_cell | *ATG10* | -1.06230587 | 0.000218301 | 0.000385583 | 1 |
| HTN | nk_cell | *CWF19L2* | -1.404069608 | 0.006506247 | 0.00871812 | 1 |
| HTN | nk_cell | *NMT2* | -2.355110712 | 0.00013493 | 0.000251993 | 0.654678676 |
| HTN | nk_cell | *ZNF837* | -1.994628331 | 4.75E-06 | 1.66E-05 | 0.023043287 |
| HTN | nk_cell | *CCDC50* | -1.418359544 | 6.96E-06 | 2.19E-05 | 0.033785365 |
| HTN | nk_cell | *PLEKHH2* | -1.407928654 | 6.45E-08 | 2.51E-06 | 0.000312785 |
| HTN | nk_cell | *SPARCL1* | -1.605502666 | 2.89E-05 | 6.81E-05 | 0.140299944 |
| HTN | nk_cell | *CCNO* | -10.26787757 | 4.29E-07 | 4.24E-06 | 0.002080934 |
| HTN | nk_cell | *CLEC4F* | -2.365472442 | 2.14E-05 | 5.32E-05 | 0.103923404 |
| HTN | nk_cell | *SLC30A6* | -2.233847151 | 6.35E-06 | 2.06E-05 | 0.03083379 |
| HTN | nk_cell | *RASGRP3* | -2.238179617 | 0.006919424 | 0.009246226 | 1 |
| HTN | nk_cell | *GPR180* | -1.587605068 | 7.02E-06 | 2.20E-05 | 0.034049719 |
| HTN | nk_cell | *TCTEX1D1* | -3.504946585 | 2.02E-06 | 9.02E-06 | 0.009813543 |
| HTN | nk_cell | *IFIT5* | -1.079217437 | 0.004613797 | 0.006307733 | 1 |
| HTN | nk_cell | *PRDM8* | 1.588734871 | 4.95E-05 | 0.000106328 | 0.240407598 |
| HTN | nk_cell | *ZNF117* | -0.431765136 | 0.000295874 | 0.000505155 | 1 |
| HTN | nk_cell | *MED21* | -1.6621723 | 0.033233803 | 0.040647948 | 1 |
| HTN | nk_cell | *JAKMIP1* | 0.252333743 | 2.85E-07 | 4.07E-06 | 0.001382665 |
| HTN | nk_cell | *SREK1IP1* | -2.449695385 | 0.000105695 | 0.000202301 | 0.512834004 |
| HTN | nk_cell | *MR1* | 1.981472785 | 0.038412161 | 0.046699024 | 1 |
| HTN | nk_cell | *CENPH* | -2.986232787 | 3.08E-07 | 4.17E-06 | 0.001494543 |
| HTN | nk_cell | *CARHSP1* | -2.1129885 | 0.000462356 | 0.000755845 | 1 |
| HTN | nk_cell | *CAST* | -2.122421581 | 5.36E-06 | 1.82E-05 | 0.026021977 |
| HTN | nk_cell | *SCOC* | -2.556659941 | 0.001297877 | 0.001940616 | 1 |
| HTN | nk_cell | *CLGN* | 1.329509945 | 4.46E-07 | 4.28E-06 | 0.002163734 |
| HTN | nk_cell | *SYCP2L* | -1.166324641 | 5.90E-07 | 4.54E-06 | 0.002862496 |
| HTN | nk_cell | *MERTK* | -0.593200525 | 9.37E-05 | 0.000182612 | 0.454520928 |
| HTN | nk_cell | *TMEM87B* | -2.385528907 | 0.000149694 | 0.000276376 | 0.726317408 |

| | | | | | | |
|---|---|---|---|---|---|---|
| HTN | nk_cell | *PLA2R1* | -0.932743307 | 0.000976102 | 0.001496381 | 1 |
| HTN | nk_cell | *SLC25A27* | -1.155982546 | 0.001184951 | 0.001786632 | 1 |
| HTN | nk_cell | *FRMD1* | -1.611889951 | 4.77E-07 | 4.32E-06 | 0.002315083 |
| HTN | nk_cell | *INO80C* | -3.504601131 | 6.90E-06 | 2.17E-05 | 0.033487314 |
| HTN | nk_cell | *LPCAT1* | 0.91062544 | 2.84E-07 | 4.07E-06 | 0.001377479 |
| HTN | nk_cell | *NMRAL1* | 2.418843291 | 9.31E-05 | 0.000181712 | 0.451735279 |
| HTN | nk_cell | *ADPRHL1* | -2.445386215 | 1.05E-06 | 6.09E-06 | 0.005073843 |
| HTN | nk_cell | *CFDP1* | -1.993568341 | 9.80E-05 | 0.000189422 | 0.475449181 |
| HTN | nk_cell | *ZDHHC7* | -1.777244154 | 2.64E-06 | 1.09E-05 | 0.01278848 |
| HTN | nk_cell | *FBXO36* | -2.224033501 | 5.65E-06 | 1.89E-05 | 0.027401928 |
| HTN | nk_cell | *MCOLN2* | 0.444954522 | 2.62E-07 | 4.07E-06 | 0.001273282 |
| HTN | nk_cell | *GDPD1* | 3.266916431 | 0.000151089 | 0.000278316 | 0.733085281 |
| HTN | nk_cell | *SEMA3D* | -1.302392706 | 1.55E-06 | 7.63E-06 | 0.007534675 |
| HTN | nk_cell | *ANKRD29* | -2.144091058 | 5.19E-06 | 1.78E-05 | 0.02518174 |
| HTN | nk_cell | *DNAAF1* | -3.036932561 | 4.44E-08 | 2.12E-06 | 0.000215199 |
| HTN | nk_cell | *ANKH* | -2.109625845 | 7.59E-05 | 0.000151868 | 0.368279433 |
| HTN | nk_cell | *UBASH3B* | 1.241555087 | 1.37E-06 | 7.13E-06 | 0.006638059 |
| HTN | nk_cell | *ROBO4* | -2.389357251 | 1.05E-05 | 3.02E-05 | 0.050872671 |
| HTN | nk_cell | *ROBO3* | -0.990817159 | 1.87E-06 | 8.58E-06 | 0.009077017 |
| HTN | nk_cell | *CC2D1B* | -1.95226527 | 0.003773131 | 0.005229144 | 1 |
| HTN | nk_cell | *PRKCA* | -1.964071557 | 0.000116819 | 0.000221063 | 0.566804427 |
| HTN | nk_cell | *LRRK1* | -1.984135613 | 2.35E-06 | 1.01E-05 | 0.011417002 |
| HTN | nk_cell | *ENPP3* | -1.393327985 | 4.52E-07 | 4.28E-06 | 0.002194688 |
| HTN | nk_cell | *MIA3* | -2.247128018 | 8.79E-05 | 0.000172403 | 0.426525363 |
| HTN | nk_cell | *PGM5* | -2.010400137 | 6.19E-05 | 0.00012748 | 0.300215554 |
| HTN | nk_cell | *CCSAP* | -3.893836185 | 3.81E-05 | 8.53E-05 | 0.184983485 |
| HTN | nk_cell | *GBP5* | 0.359873193 | 3.34E-07 | 4.24E-06 | 0.001619501 |
| HTN | nk_cell | *ATP5G3* | 1.999565327 | 0.000141792 | 0.000263492 | 0.687976779 |
| HTN | nk_cell | *LY96* | -1.888558099 | 1.70E-05 | 4.44E-05 | 0.082344741 |
| HTN | nk_cell | *PDE1C* | -0.940835987 | 6.11E-07 | 4.55E-06 | 0.0029627 |
| HTN | nk_cell | *RABGEF1* | -1.183369527 | 6.78E-06 | 2.15E-05 | 0.032912019 |
| HTN | nk_cell | *MRPL39* | -0.979990316 | 0.027814384 | 0.034252637 | 1 |
| HTN | nk_cell | *XPC* | -2.735289479 | 4.51E-05 | 9.82E-05 | 0.219009473 |
| HTN | nk_cell | *FLCN* | -2.220917787 | 1.45E-05 | 3.91E-05 | 0.07025374 |
| HTN | nk_cell | *OXNAD1* | -2.148528419 | 0.00019191 | 0.000343978 | 0.931149519 |
| HTN | nk_cell | *APCDD1* | -0.862132614 | 1.51E-06 | 7.55E-06 | 0.007316184 |
| HTN | nk_cell | *ANKRD40* | -1.660067539 | 2.33E-06 | 1.00E-05 | 0.011319225 |
| HTN | nk_cell | *CYP2U1* | -1.478141012 | 1.57E-06 | 7.68E-06 | 0.007626699 |
| HTN | nk_cell | *PROM2* | -4.442323961 | 0.001149507 | 0.001738051 | 1 |
| HTN | nk_cell | *MARVELD1* | -1.373428283 | 5.19E-07 | 4.37E-06 | 0.002517032 |
| HTN | nk_cell | *GOLGA7B* | 1.838264965 | 2.63E-07 | 4.07E-06 | 0.001278035 |
| HTN | nk_cell | *TRMT44* | -2.868605271 | 0.002945386 | 0.004164048 | 1 |
| HTN | nk_cell | *MOV10* | -2.39081202 | 0.00016293 | 0.000297866 | 0.790535045 |
| HTN | nk_cell | *HEATR3* | -1.079845335 | 0.011079308 | 0.014385015 | 1 |
| HTN | nk_cell | *CNOT8* | -2.896312386 | 0.000249971 | 0.000435341 | 1 |
| HTN | nk_cell | *LRGUK* | -0.95717775 | 1.89E-07 | 4.07E-06 | 0.000916039 |
| HTN | nk_cell | *SETD9* | -0.731891832 | 3.66E-05 | 8.26E-05 | 0.177431642 |
| HTN | nk_cell | *NUP205* | 3.017843909 | 5.98E-05 | 0.000123788 | 0.290282609 |
| HTN | nk_cell | *PDIA4* | 2.851297363 | 1.74E-06 | 8.18E-06 | 0.008441691 |
| HTN | nk_cell | *KDM8* | 4.451554716 | 0.000267524 | 0.000461439 | 1 |

| | | | | | | |
|---|---|---|---|---|---|---|
| HTN | nk_cell | *ALS2CR12* | -3.090543591 | 0.000323684 | 0.000547027 | 1 |
| HTN | nk_cell | *FZD7* | -0.907891187 | 1.72E-06 | 8.14E-06 | 0.00836112 |
| HTN | nk_cell | *DEPTOR* | -1.173322236 | 1.51E-05 | 4.05E-05 | 0.073421575 |
| HTN | nk_cell | *RRAGA* | -3.586969617 | 0.007554368 | 0.010028397 | 1 |
| HTN | nk_cell | *TMBIM4* | -2.090677293 | 3.75E-05 | 8.43E-05 | 0.181980184 |
| HTN | nk_cell | *PSD3* | -1.57176912 | 1.74E-05 | 4.53E-05 | 0.084552138 |
| HTN | nk_cell | *UGT2B4* | -0.792587945 | 0.000296138 | 0.000505403 | 1 |
| HTN | nk_cell | *MMP16* | -1.613431022 | 2.20E-08 | 1.38E-06 | 0.000106827 |
| HTN | nk_cell | *KCNMA1* | -1.298542219 | 2.57E-06 | 1.07E-05 | 0.01245613 |
| HTN | nk_cell | *NDUFAF6* | -2.60009941 | 1.92E-07 | 4.07E-06 | 0.000931048 |
| HTN | nk_cell | *DRAM2* | -1.167335476 | 0.007632324 | 0.010120088 | 1 |
| HTN | nk_cell | *C8orf37* | -1.566121615 | 2.17E-07 | 4.07E-06 | 0.001050695 |
| HTN | nk_cell | *PPEF2* | -3.705889953 | 1.16E-05 | 3.26E-05 | 0.056097838 |
| HTN | nk_cell | *ART3* | -2.864746608 | 1.55E-08 | 1.11E-06 | 7.51E-05 |
| HTN | nk_cell | *N6AMT1* | -2.622132676 | 0.000533253 | 0.00086044 | 1 |
| HTN | nk_cell | *RWDD2B* | -2.019680471 | 0.000606916 | 0.000968988 | 1 |
| HTN | nk_cell | *NAA11* | -1.794966318 | 1.08E-07 | 3.46E-06 | 0.000521768 |
| HTN | nk_cell | *SFXN2* | 1.063009167 | 0.011867874 | 0.015343172 | 1 |
| HTN | nk_cell | *C14orf2* | -2.124451014 | 0.000626382 | 0.00099483 | 1 |
| HTN | nk_cell | *TDRD9* | 5.62871122 | 4.52E-05 | 9.82E-05 | 0.219368007 |
| HTN | nk_cell | *RPL30* | 2.108587951 | 0.001100953 | 0.00167141 | 1 |
| HTN | nk_cell | *HKDC1* | -5.26965007 | 7.72E-06 | 2.38E-05 | 0.037434179 |
| HTN | nk_cell | *TYSND1* | -1.281148331 | 5.01E-05 | 0.000107219 | 0.243065603 |
| HTN | nk_cell | *NODAL* | -1.990629315 | 4.98E-07 | 4.35E-06 | 0.002416666 |
| HTN | nk_cell | *MED19* | -1.376592269 | 0.000974063 | 0.001495141 | 1 |
| HTN | nk_cell | *FBRS* | -2.687378175 | 0.000405979 | 0.000671602 | 1 |
| HTN | nk_cell | *SASS6* | -1.266328665 | 4.86E-07 | 4.32E-06 | 0.002356507 |
| HTN | nk_cell | *ITGAD* | 0.319633143 | 1.76E-07 | 4.07E-06 | 0.000854674 |
| HTN | nk_cell | *MPV17L* | -1.684692284 | 1.45E-05 | 3.91E-05 | 0.070332133 |
| HTN | nk_cell | *RPUSD3* | -2.538992619 | 7.80E-06 | 2.40E-05 | 0.037848374 |
| HTN | nk_cell | *GHRL* | -2.47391272 | 0.00063197 | 0.00100272 | 1 |
| HTN | nk_cell | *NMNAT2* | -2.981217054 | 0.001520906 | 0.002245719 | 1 |
| HTN | nk_cell | *SLC6A1* | -3.242563336 | 3.60E-07 | 4.24E-06 | 0.001747384 |
| HTN | nk_cell | *FCHO2* | -1.594209921 | 0.000506833 | 0.000821636 | 1 |
| HTN | nk_cell | *TIMP4* | 1.26047131 | 0.025813109 | 0.031956346 | 1 |
| HTN | nk_cell | *NECAP2* | -2.735772881 | 6.49E-05 | 0.00013267 | 0.314958301 |
| HTN | nk_cell | *STEAP2* | -1.452628907 | 1.26E-06 | 6.79E-06 | 0.006112764 |
| HTN | nk_cell | *CLDN12* | -3.618981498 | 3.01E-05 | 7.04E-05 | 0.145975674 |
| HTN | nk_cell | *MMP14* | -1.386421995 | 9.59E-07 | 5.84E-06 | 0.004653656 |
| HTN | nk_cell | *GATAD1* | -2.044666668 | 4.88E-05 | 0.000104945 | 0.236546688 |
| HTN | nk_cell | *TMED6* | -6.843309979 | 9.87E-07 | 5.93E-06 | 0.004787425 |
| HTN | nk_cell | *CLEC18A* | -3.731304328 | 3.91E-07 | 4.24E-06 | 0.00189955 |
| HTN | nk_cell | *DHRS4* | -2.430529662 | 1.57E-06 | 7.67E-06 | 0.007607215 |
| HTN | nk_cell | *DHRS1* | -1.323768184 | 2.58E-05 | 6.18E-05 | 0.125115975 |
| HTN | nk_cell | *AASDH* | -2.097364563 | 3.98E-05 | 8.85E-05 | 0.193091985 |
| HTN | nk_cell | *ETS2* | 1.164928076 | 0.007041324 | 0.009391013 | 1 |
| HTN | nk_cell | *LCA5L* | -1.444281536 | 3.79E-05 | 8.50E-05 | 0.18408996 |
| HTN | nk_cell | *MX1* | -1.194548754 | 0.000348497 | 0.000585697 | 1 |
| HTN | nk_cell | *SLC38A10* | -2.509358301 | 0.000514512 | 0.000832692 | 1 |
| HTN | nk_cell | *C9orf43* | -1.416550686 | 0.000606887 | 0.000968988 | 1 |

| | | | | | | |
|---|---|---|---|---|---|---|
| HTN | nk_cell | *DGKI* | -1.01971299 | 6.35E-07 | 4.62E-06 | 0.003079923 |
| HTN | nk_cell | *SNX22* | 3.317382835 | 7.08E-07 | 4.91E-06 | 0.003433037 |
| HTN | nk_cell | *SLC34A2* | 8.936756548 | 3.40E-07 | 4.24E-06 | 0.001650909 |
| HTN | nk_cell | *SPPL3* | -1.861008159 | 0.000696664 | 0.001097829 | 1 |
| HTN | nk_cell | *C12orf43* | -1.699162241 | 2.13E-07 | 4.07E-06 | 0.001032252 |
| HTN | nk_cell | *PEX10* | -2.935120893 | 0.004147613 | 0.005700911 | 1 |
| HTN | nk_cell | *KRTCAP3* | -1.753613278 | 3.07E-05 | 7.17E-05 | 0.149035687 |
| HTN | nk_cell | *PAFAH2* | 1.729350906 | 0.001431889 | 0.00211944 | 1 |
| HTN | nk_cell | *EXTL1* | -15.11828652 | 4.16E-07 | 4.24E-06 | 0.002019384 |
| HTN | nk_cell | *WDR66* | -1.309020136 | 9.05E-06 | 2.69E-05 | 0.04392919 |
| HTN | nk_cell | *RHPN1* | -1.779541299 | 2.71E-05 | 6.45E-05 | 0.131667459 |
| HTN | nk_cell | *DZIP1L* | -1.804206951 | 3.79E-06 | 1.41E-05 | 0.018409553 |
| HTN | nk_cell | *FANCC* | -1.535216654 | 5.23E-06 | 1.79E-05 | 0.0253636 |
| HTN | nk_cell | *ESYT3* | -2.328209253 | 3.09E-07 | 4.17E-06 | 0.001501563 |
| HTN | nk_cell | *CLSTN2* | -1.887987481 | 3.94E-06 | 1.45E-05 | 0.019136551 |
| HTN | nk_cell | *COLEC12* | -1.162320013 | 4.98E-06 | 1.72E-05 | 0.024158741 |
| HTN | nk_cell | *TSPAN33* | -3.623661168 | 5.14E-08 | 2.19E-06 | 0.000249295 |
| HTN | nk_cell | *FAM86C1* | -7.447019855 | 0.006997266 | 0.009337385 | 1 |
| HTN | nk_cell | *PPP1R9A* | -1.99206466 | 0.000109685 | 0.000209277 | 0.532190653 |
| HTN | nk_cell | *DYNC1I1* | -1.74696806 | 1.45E-05 | 3.91E-05 | 0.070290655 |
| HTN | nk_cell | *COPG2* | -0.945002661 | 7.03E-05 | 0.000142362 | 0.341242398 |
| HTN | nk_cell | *ELK4* | 2.91747673 | 0.004999591 | 0.006794962 | 1 |
| HTN | nk_cell | *DUSP23* | -1.475737949 | 4.18E-07 | 4.24E-06 | 0.002026837 |
| HTN | nk_cell | *RNF166* | 0.853677054 | 1.57E-05 | 4.18E-05 | 0.076258055 |
| HTN | nk_cell | *SPATA2L* | 3.367004758 | 3.95E-08 | 2.04E-06 | 0.000191668 |
| HTN | nk_cell | *FGF17* | -7.158873745 | 3.63E-07 | 4.24E-06 | 0.001761192 |
| HTN | nk_cell | *CDA* | -1.151431132 | 0.000236259 | 0.000414586 | 1 |
| HTN | nk_cell | *SLC5A11* | -1.61762021 | 2.84E-12 | 1.25E-09 | 1.38E-08 |
| HTN | nk_cell | *CCAR2* | -2.758643354 | 0.001845003 | 0.00267782 | 1 |
| HTN | nk_cell | *CDC42SE2* | 1.59884043 | 6.42E-05 | 0.000131367 | 0.311471661 |
| HTN | nk_cell | *RAPGEF6* | 3.045218056 | 0.001381411 | 0.002049101 | 1 |
| HTN | nk_cell | *IFNAR2* | 1.197868131 | 0.007131396 | 0.009503304 | 1 |
| HTN | nk_cell | *SV2A* | -2.670675512 | 0.034699549 | 0.042336209 | 1 |
| HTN | nk_cell | *CLIC6* | -2.969593972 | 2.57E-05 | 6.17E-05 | 0.124778099 |
| HTN | nk_cell | *CBR1* | -1.541175163 | 0.007204427 | 0.009595355 | 1 |
| HTN | nk_cell | *CBR3* | -1.285183126 | 0.000511959 | 0.000829114 | 1 |
| HTN | nk_cell | *CHAF1B* | -1.972501945 | 0.002351399 | 0.003373444 | 1 |
| HTN | nk_cell | *SIM2* | 0.831301998 | 6.65E-06 | 2.12E-05 | 0.032267651 |
| HTN | nk_cell | *ARHGAP27* | -2.865631006 | 0.000859556 | 0.001332874 | 1 |
| HTN | nk_cell | *IRX6* | -0.899829047 | 3.92E-07 | 4.24E-06 | 0.001900938 |
| HTN | nk_cell | *HK2* | -1.761807607 | 2.73E-06 | 1.11E-05 | 0.013252972 |
| HTN | nk_cell | *AMFR* | 3.187621308 | 3.70E-05 | 8.33E-05 | 0.179648072 |
| HTN | nk_cell | *TMEM69* | -2.959021672 | 7.04E-05 | 0.000142424 | 0.341533643 |
| HTN | nk_cell | *ACE* | -1.760871849 | 6.53E-07 | 4.69E-06 | 0.003168687 |
| HTN | nk_cell | *TEPP* | 1.525693599 | 6.64E-07 | 4.74E-06 | 0.003223376 |
| HTN | nk_cell | *CHCHD6* | -1.988546073 | 0.000148953 | 0.000275217 | 0.722719834 |
| HTN | nk_cell | *CTBP1* | -3.643426665 | 4.89E-05 | 0.000105092 | 0.237086871 |
| HTN | nk_cell | *LRRC36* | -1.317078659 | 1.81E-08 | 1.22E-06 | 8.80E-05 |
| HTN | nk_cell | *ZFYVE28* | 1.420959873 | 9.57E-07 | 5.84E-06 | 0.004641407 |
| HTN | nk_cell | *C16orf86* | -1.710696812 | 2.61E-05 | 6.24E-05 | 0.126615182 |

| | | | | | | |
|---|---|---|---|---|---|---|
| HTN | nk_cell | *RGS12* | -1.828605621 | 2.42E-05 | 5.89E-05 | 0.117430981 |
| HTN | nk_cell | *LYPD5* | -2.397020546 | 3.83E-08 | 2.01E-06 | 0.000185779 |
| HTN | nk_cell | *ZNF233* | -3.410967803 | 4.63E-05 | 0.000100311 | 0.224560139 |
| HTN | nk_cell | *ATAD3B* | 2.277149075 | 0.014345814 | 0.018312521 | 1 |
| HTN | nk_cell | *SSU72* | 2.810503397 | 0.005043644 | 0.006843333 | 1 |
| HTN | nk_cell | *CPAMD8* | -1.162764653 | 2.22E-06 | 9.65E-06 | 0.010794655 |
| HTN | nk_cell | *ANKLE1* | -6.679877295 | 5.14E-05 | 0.000109321 | 0.249213558 |
| HTN | nk_cell | *CCDC58* | -2.324947173 | 2.00E-06 | 8.99E-06 | 0.009714676 |
| HTN | nk_cell | *CILP2* | 3.230850108 | 0.000134332 | 0.000251168 | 0.651780994 |
| HTN | nk_cell | *SLC37A1* | 3.773706145 | 8.60E-05 | 0.000169095 | 0.417102147 |
| HTN | nk_cell | *PDE9A* | -2.126411121 | 5.27E-05 | 0.00011162 | 0.25572175 |
| HTN | nk_cell | *HSF2BP* | -2.557272153 | 9.62E-09 | 8.19E-07 | 4.67E-05 |
| HTN | nk_cell | *CSTB* | -1.960310352 | 0.000296702 | 0.000506188 | 1 |
| HTN | nk_cell | *C21orf33* | 1.223531288 | 0.029861155 | 0.036680082 | 1 |
| HTN | nk_cell | *C21orf2* | -1.19496481 | 1.14E-07 | 3.55E-06 | 0.000553971 |
| HTN | nk_cell | *FTCD* | -1.138827485 | 0.0093335 | 0.012226281 | 1 |
| HTN | nk_cell | *LSS* | -0.700339391 | 0.020646907 | 0.025879306 | 1 |
| HTN | nk_cell | *VAV2* | -1.348838483 | 9.54E-06 | 2.80E-05 | 0.046306878 |
| HTN | nk_cell | *MCM3AP* | -1.827004851 | 1.09E-05 | 3.11E-05 | 0.052882262 |
| HTN | nk_cell | *PCNT* | -1.472481046 | 0.02160139 | 0.027040749 | 1 |
| HTN | nk_cell | *DIP2A* | 1.788581913 | 0.0134278 | 0.017222227 | 1 |
| HTN | nk_cell | *S100B* | -1.261978823 | 3.25E-09 | 4.04E-07 | 1.58E-05 |
| HTN | nk_cell | *ZNF208* | -2.064754631 | 4.58E-08 | 2.12E-06 | 0.000222054 |
| HTN | nk_cell | *ADAMTS13* | -1.23214564 | 4.19E-06 | 1.51E-05 | 0.020318074 |
| HTN | nk_cell | *CACFD1* | -6.445701274 | 4.33E-06 | 1.55E-05 | 0.021021583 |
| HTN | nk_cell | *GPSM1* | 2.856043284 | 0.002855151 | 0.004042368 | 1 |
| HTN | nk_cell | *RDH13* | 2.380192598 | 3.55E-05 | 8.07E-05 | 0.172458529 |
| HTN | nk_cell | *TAOK1* | -1.850231404 | 0.000167127 | 0.000304621 | 0.810901345 |
| HTN | nk_cell | *TLCD1* | -5.452384842 | 4.62E-05 | 0.000100243 | 0.22422487 |
| HTN | nk_cell | *S100A1* | -3.935986499 | 1.09E-05 | 3.12E-05 | 0.052952406 |
| HTN | nk_cell | *ZBTB7B* | -1.972525793 | 1.39E-06 | 7.20E-06 | 0.006759491 |
| HTN | nk_cell | *VPS11* | 2.278221523 | 0.001670542 | 0.002442143 | 1 |
| HTN | nk_cell | *PAQR6* | 1.175015122 | 0.017802713 | 0.022453539 | 1 |
| HTN | nk_cell | *CCR5* | 0.651078216 | 8.03E-08 | 2.97E-06 | 0.000389442 |
| HTN | nk_cell | *NBEAL2* | 1.921983097 | 2.69E-07 | 4.07E-06 | 0.00130465 |
| HTN | nk_cell | *PTH1R* | -2.657075628 | 8.56E-06 | 2.57E-05 | 0.041547758 |
| HTN | nk_cell | *FCRL3* | 0.263576589 | 1.72E-07 | 4.07E-06 | 0.000832723 |
| HTN | nk_cell | *FGFR4* | 5.064106871 | 3.46E-07 | 4.24E-06 | 0.001679302 |
| HTN | nk_cell | *LY6K* | -1.363422214 | 0.024004004 | 0.029839845 | 1 |
| HTN | nk_cell | *CPSF4* | 2.720547513 | 4.94E-08 | 2.19E-06 | 0.000239568 |
| HTN | nk_cell | *TONSL* | -3.383875779 | 0.001853983 | 0.002688442 | 1 |
| HTN | nk_cell | *MUM1* | -1.732296273 | 4.09E-06 | 1.49E-05 | 0.019820915 |
| HTN | nk_cell | *ZNF333* | 3.185737058 | 0.000320815 | 0.000543315 | 1 |
| HTN | nk_cell | *SH2B2* | -3.09517019 | 2.66E-06 | 1.09E-05 | 0.012907016 |
| HTN | nk_cell | *RPL8* | 2.058581992 | 0.005289589 | 0.007163016 | 1 |
| HTN | nk_cell | *LRWD1* | 1.441115847 | 2.00E-05 | 5.04E-05 | 0.096969436 |
| HTN | nk_cell | *DVL3* | -2.358722391 | 5.19E-06 | 1.78E-05 | 0.02518296 |
| HTN | nk_cell | *ABCF3* | 4.976074375 | 8.51E-11 | 2.17E-08 | 4.13E-07 |
| HTN | nk_cell | *PCYT1A* | 2.113405208 | 0.001117543 | 0.001693945 | 1 |
| HTN | nk_cell | *FBXO27* | -1.018583549 | 4.22E-07 | 4.24E-06 | 0.002048563 |

| | | | | | | |
|---|---|---|---|---|---|---|
| HTN | nk_cell | *U2AF1L4* | -0.843713281 | 5.16E-05 | 0.000109681 | 0.250467703 |
| HTN | nk_cell | *BDH1* | -3.182740458 | 0.000234249 | 0.000411356 | 1 |
| HTN | nk_cell | *LRRC56* | -2.069217919 | 0.000347547 | 0.000584506 | 1 |
| HTN | nk_cell | *PGAP3* | 4.540335662 | 0.001557079 | 0.002292852 | 1 |
| HTN | nk_cell | *ZNF577* | -1.720814618 | 2.62E-06 | 1.08E-05 | 0.012714774 |
| HTN | nk_cell | *TMEM143* | -2.306289003 | 4.53E-06 | 1.60E-05 | 0.021997982 |
| HTN | nk_cell | *CCL16* | -2.894203084 | 7.91E-06 | 2.43E-05 | 0.038386782 |
| HTN | nk_cell | *CCDC155* | -1.390432295 | 0.023782441 | 0.029580211 | 1 |
| HTN | nk_cell | *DCD* | -5.797758595 | 0.002959633 | 0.004178103 | 1 |
| HTN | nk_cell | *SIGLEC11* | -1.243558755 | 5.29E-05 | 0.000111872 | 0.256633687 |
| HTN | nk_cell | *ASB16* | -1.913857006 | 1.14E-06 | 6.43E-06 | 0.005539317 |
| HTN | nk_cell | *PLCD3* | -1.614767959 | 5.66E-07 | 4.50E-06 | 0.002748387 |
| HTN | nk_cell | *AQP5* | -5.299044518 | 0.002682287 | 0.003812085 | 1 |
| HTN | nk_cell | *IP6K3* | -13.40195193 | 1.71E-06 | 8.12E-06 | 0.008313736 |
| HTN | nk_cell | *LEMD2* | -2.735801666 | 2.42E-06 | 1.02E-05 | 0.011764841 |
| HTN | nk_cell | *ASGR2* | 8.293582346 | 2.17E-07 | 4.07E-06 | 0.001052463 |
| HTN | nk_cell | *POLR3K* | 5.423246435 | 0.000582369 | 0.000932559 | 1 |
| HTN | nk_cell | *SNRNP25* | -2.661428107 | 2.63E-05 | 6.27E-05 | 0.127403119 |
| HTN | nk_cell | *WDR90* | -1.594234659 | 2.35E-05 | 5.75E-05 | 0.113792497 |
| HTN | nk_cell | *JMJD8* | 2.157106664 | 9.99E-06 | 2.90E-05 | 0.048476449 |
| HTN | nk_cell | *CCDC78* | 4.94171814 | 3.83E-06 | 1.42E-05 | 0.018597963 |
| HTN | nk_cell | *MEIOB* | -1.334062634 | 2.83E-06 | 1.14E-05 | 0.013734366 |
| HTN | nk_cell | *HS3ST6* | -1.480563716 | 0.022542325 | 0.028153246 | 1 |
| HTN | nk_cell | *SHANK2* | -1.179165893 | 5.82E-07 | 4.54E-06 | 0.002824453 |
| HTN | nk_cell | *TTC9C* | -2.658044045 | 0.000600955 | 0.000960736 | 1 |
| HTN | nk_cell | *RPS6KA4* | 3.814421202 | 0.023548767 | 0.02932465 | 1 |
| HTN | nk_cell | *TPCN2* | -1.532767026 | 3.10E-06 | 1.22E-05 | 0.015030189 |
| HTN | nk_cell | *PDZK1IP1* | -3.11717873 | 7.80E-07 | 5.20E-06 | 0.003782196 |
| HTN | nk_cell | *CMPK1* | -1.778027138 | 3.40E-05 | 7.76E-05 | 0.165065126 |
| HTN | nk_cell | *COA7* | -5.182152991 | 9.38E-05 | 0.000182685 | 0.455067976 |
| HTN | nk_cell | *SLC1A7* | -1.329997122 | 2.63E-06 | 1.08E-05 | 0.012751869 |
| HTN | nk_cell | *C1orf123* | -3.577647768 | 0.00027016 | 0.000465323 | 1 |
| HTN | nk_cell | *USP24* | -1.767483254 | 0.017182421 | 0.021750355 | 1 |
| HTN | nk_cell | *ZSWIM5* | -1.987407631 | 3.04E-05 | 7.10E-05 | 0.147377795 |
| HTN | nk_cell | *AK4* | -1.773465193 | 2.42E-05 | 5.89E-05 | 0.117410051 |
| HTN | nk_cell | *RBP7* | -2.814816235 | 3.32E-05 | 7.63E-05 | 0.160987527 |
| HTN | nk_cell | *SLC25A34* | -2.078812144 | 2.31E-07 | 4.07E-06 | 0.001120237 |
| HTN | nk_cell | *PDPN* | -2.105969656 | 1.68E-06 | 8.02E-06 | 0.008145665 |
| HTN | nk_cell | *LAPTM5* | 3.075441008 | 6.03E-05 | 0.000124678 | 0.292744523 |
| HTN | nk_cell | *SDC3* | -2.51870581 | 6.38E-06 | 2.06E-05 | 0.030949384 |
| HTN | nk_cell | *SYNC* | -2.005154898 | 7.72E-07 | 5.19E-06 | 0.003747429 |
| HTN | nk_cell | *CAMK2N1* | -1.512655642 | 3.35E-05 | 7.68E-05 | 0.162729976 |
| HTN | nk_cell | *MEGF6* | -1.274906412 | 8.21E-07 | 5.33E-06 | 0.003985884 |
| HTN | nk_cell | *OMA1* | -2.037456219 | 9.11E-07 | 5.69E-06 | 0.004422141 |
| HTN | nk_cell | *TM2D1* | -2.178373872 | 0.000214717 | 0.000379806 | 1 |
| HTN | nk_cell | *NEXN* | -1.867745083 | 0.002669893 | 0.003798921 | 1 |
| HTN | nk_cell | *LRRIQ3* | -0.799479663 | 4.00E-07 | 4.24E-06 | 0.001940751 |
| HTN | nk_cell | *TYW3* | -2.093001237 | 3.82E-06 | 1.42E-05 | 0.018543719 |
| HTN | nk_cell | *WDR63* | -1.362997004 | 6.29E-05 | 0.000129116 | 0.305102257 |
| HTN | nk_cell | *HFM1* | -2.914012424 | 4.73E-06 | 1.65E-05 | 0.0229434 |

| | | | | | | |
|---|---|---|---|---|---|---|
| HTN | nk_cell | *ARPC5* | -2.097166908 | 0.006447354 | 0.00864876 | 1 |
| HTN | nk_cell | *ZNF496* | -2.886657497 | 4.09E-05 | 9.05E-05 | 0.198344514 |
| HTN | nk_cell | *TRIM58* | -1.763830568 | 3.66E-05 | 8.26E-05 | 0.17736483 |
| HTN | nk_cell | *FCRLB* | -3.396228383 | 5.13E-07 | 4.37E-06 | 0.002486779 |
| HTN | nk_cell | *FLVCR1* | -2.544595184 | 7.22E-06 | 2.25E-05 | 0.035032745 |
| HTN | nk_cell | *TDRD5* | -2.449951764 | 3.64E-07 | 4.24E-06 | 0.001765914 |
| HTN | nk_cell | *ACP6* | -2.301143931 | 1.80E-06 | 8.37E-06 | 0.008739323 |
| HTN | nk_cell | *CNST* | -1.992188159 | 2.57E-05 | 6.17E-05 | 0.124713692 |
| HTN | nk_cell | *PKDCC* | -1.191963569 | 9.44E-06 | 2.77E-05 | 0.04580972 |
| HTN | nk_cell | *HAAO* | -3.150303537 | 0.000180713 | 0.000326076 | 0.876819364 |
| HTN | nk_cell | *B3GALNT2* | -2.045014769 | 4.33E-06 | 1.55E-05 | 0.021019638 |
| HTN | nk_cell | *MRPL55* | 2.438182743 | 0.004060795 | 0.005591083 | 1 |
| HTN | nk_cell | *KIAA1841* | -2.395299421 | 0.000374656 | 0.000625759 | 1 |
| HTN | nk_cell | *KCNJ3* | -3.625995562 | 3.13E-06 | 1.23E-05 | 0.015202487 |
| HTN | nk_cell | *CLHC1* | -0.64478193 | 0.039678433 | 0.048190176 | 1 |
| HTN | nk_cell | *DUSP19* | -2.228125275 | 1.21E-06 | 6.69E-06 | 0.005887707 |
| HTN | nk_cell | *NUP35* | -1.74897882 | 9.47E-07 | 5.82E-06 | 0.004592908 |
| HTN | nk_cell | *C2orf48* | 0.812799192 | 2.54E-07 | 4.07E-06 | 0.001232461 |
| HTN | nk_cell | *CCDC74A* | -2.943924483 | 0.000206994 | 0.000368293 | 1 |
| HTN | nk_cell | *NOSTRIN* | -1.525937093 | 1.73E-07 | 4.07E-06 | 0.00083773 |
| HTN | nk_cell | *INHBB* | -1.646989289 | 1.42E-06 | 7.27E-06 | 0.00688797 |
| HTN | nk_cell | *RPRD2* | -1.950677004 | 0.00027643 | 0.00047528 | 1 |
| HTN | nk_cell | *PACRGL* | -2.29504654 | 0.001366282 | 0.002030383 | 1 |
| HTN | nk_cell | *C1QTNF7* | -1.624161072 | 7.58E-06 | 2.34E-05 | 0.036756391 |
| HTN | nk_cell | *LYSMD1* | -3.130760399 | 2.10E-09 | 2.99E-07 | 1.02E-05 |
| HTN | nk_cell | *ERCC3* | -2.283389855 | 0.000559939 | 0.000899909 | 1 |
| HTN | nk_cell | *RNF149* | -1.936942716 | 0.000974677 | 0.001495141 | 1 |
| HTN | nk_cell | *DHX57* | -2.444914642 | 6.88E-05 | 0.000139758 | 0.33388082 |
| HTN | nk_cell | *S100A9* | -1.972303311 | 3.28E-06 | 1.27E-05 | 0.015922264 |
| HTN | nk_cell | *TDRD10* | -1.943211439 | 3.33E-06 | 1.28E-05 | 0.016141305 |
| HTN | nk_cell | *GNPDA2* | -2.28634698 | 5.40E-05 | 0.000113783 | 0.262042481 |
| HTN | nk_cell | *NIPAL1* | -1.82540046 | 9.75E-06 | 2.85E-05 | 0.047311382 |
| HTN | nk_cell | *ANTXR2* | -1.54016085 | 3.77E-05 | 8.47E-05 | 0.18312198 |
| HTN | nk_cell | *GPR155* | -2.585244331 | 0.000522306 | 0.000844992 | 1 |
| HTN | nk_cell | *DAPL1* | -2.147813739 | 1.24E-05 | 3.46E-05 | 0.060366943 |
| HTN | nk_cell | *PBXIP1* | -2.941011132 | 0.003241969 | 0.004548882 | 1 |
| HTN | nk_cell | *CLDN1* | -1.79770384 | 1.29E-05 | 3.56E-05 | 0.062758854 |
| HTN | nk_cell | *DCST2* | -1.565338994 | 5.91E-06 | 1.95E-05 | 0.028692583 |
| HTN | nk_cell | *POGLUT1* | -1.841746769 | 1.49E-05 | 4.01E-05 | 0.072529042 |
| HTN | nk_cell | *SLC22A15* | -2.015337126 | 1.43E-05 | 3.88E-05 | 0.069514991 |
| HTN | nk_cell | *LRRC58* | -2.203288891 | 3.31E-06 | 1.28E-05 | 0.016074345 |
| HTN | nk_cell | *LMOD1* | -1.100398584 | 4.56E-08 | 2.12E-06 | 0.000221394 |
| HTN | nk_cell | *TMEM169* | 1.634325027 | 4.19E-07 | 4.24E-06 | 0.002034471 |
| HTN | nk_cell | *ARPC2* | 1.92203664 | 7.86E-06 | 2.41E-05 | 0.038133243 |
| HTN | nk_cell | *SERPINI1* | -2.133005167 | 7.12E-07 | 4.93E-06 | 0.003454856 |
| HTN | nk_cell | *SUCLG1* | -2.116401184 | 1.83E-05 | 4.70E-05 | 0.088595955 |
| HTN | nk_cell | *PRKCI* | -1.398771492 | 1.74E-05 | 4.53E-05 | 0.084335437 |
| HTN | nk_cell | *AIM2* | -2.204708277 | 5.74E-07 | 4.52E-06 | 0.002782622 |
| HTN | nk_cell | *EFHB* | -0.582426214 | 0.003126248 | 0.004397957 | 1 |
| HTN | nk_cell | *GTPBP8* | -3.375193353 | 9.57E-05 | 0.000185388 | 0.464211972 |

| | | | | | | |
|---|---|---|---|---|---|---|
| HTN | nk_cell | *WDFY3* | -1.667624061 | 6.76E-05 | 0.000137639 | 0.327856323 |
| HTN | nk_cell | *PTPN13* | -0.943934695 | 0.002703058 | 0.003838231 | 1 |
| HTN | nk_cell | *C4orf36* | -1.710395 | 1.35E-06 | 7.07E-06 | 0.006551136 |
| HTN | nk_cell | *TIPARP* | -2.682398922 | 5.64E-05 | 0.000117965 | 0.273560149 |
| HTN | nk_cell | *RPP14* | 2.799144283 | 0.035488823 | 0.043264263 | 1 |
| HTN | nk_cell | *DNASE1L3* | -1.718442525 | 3.70E-05 | 8.33E-05 | 0.179582753 |
| HTN | nk_cell | *IL17RC* | -3.070376899 | 5.53E-07 | 4.46E-06 | 0.002683592 |
| HTN | nk_cell | *CRELD1* | -2.485867585 | 1.20E-05 | 3.36E-05 | 0.058292956 |
| HTN | nk_cell | *TTC14* | -2.100240893 | 0.000126087 | 0.000236847 | 0.611775822 |
| HTN | nk_cell | *CCDC158* | -3.042875377 | 0.000371019 | 0.000620111 | 1 |
| HTN | nk_cell | *CPA3* | -3.792764411 | 3.25E-06 | 1.26E-05 | 0.015748785 |
| HTN | nk_cell | *SLC4A1AP* | -2.695800515 | 1.27E-05 | 3.51E-05 | 0.061699609 |
| HTN | nk_cell | *KIAA1143* | -1.494237681 | 8.56E-06 | 2.57E-05 | 0.041546449 |
| HTN | nk_cell | *CLEC3B* | -2.803415863 | 2.80E-06 | 1.13E-05 | 0.013570602 |
| HTN | nk_cell | *NMNAT3* | -1.55846725 | 9.75E-06 | 2.85E-05 | 0.047296176 |
| HTN | nk_cell | *TPRA1* | -3.255378027 | 6.38E-07 | 4.62E-06 | 0.003095912 |
| HTN | nk_cell | *MEAF6* | -3.084405425 | 4.17E-05 | 9.20E-05 | 0.202302291 |
| HTN | nk_cell | *SNIP1* | -2.973942956 | 1.43E-05 | 3.88E-05 | 0.069562619 |
| HTN | nk_cell | *DNALI1* | -1.862954994 | 1.74E-08 | 1.21E-06 | 8.46E-05 |
| HTN | nk_cell | *SENP2* | -1.658968884 | 0.000124646 | 0.000234593 | 0.604780944 |
| HTN | nk_cell | *IFT122* | -2.111648883 | 9.46E-05 | 0.000183848 | 0.458883474 |
| HTN | nk_cell | *RFT1* | -3.531776514 | 4.07E-07 | 4.24E-06 | 0.001975537 |
| HTN | nk_cell | *SFMBT1* | -2.141779214 | 3.94E-06 | 1.45E-05 | 0.019102144 |
| HTN | nk_cell | *UVSSA* | -1.838697063 | 0.004609251 | 0.006303294 | 1 |
| HTN | nk_cell | *LRPAP1* | -2.572864718 | 4.34E-05 | 9.51E-05 | 0.210623824 |
| HTN | nk_cell | *ABLIM2* | -2.247453806 | 0.004019032 | 0.005536724 | 1 |
| HTN | nk_cell | *ERMAP* | -1.306011985 | 6.38E-07 | 4.62E-06 | 0.003097225 |
| HTN | nk_cell | *ZNF691* | 2.617009924 | 0.001029599 | 0.001570455 | 1 |
| HTN | nk_cell | *METAP1* | 2.133001462 | 0.002866715 | 0.004056372 | 1 |
| HTN | nk_cell | *BDH2* | -1.744794601 | 1.66E-07 | 4.07E-06 | 0.000803386 |
| HTN | nk_cell | *CDC25A* | -7.829200451 | 6.78E-13 | 4.70E-10 | 3.29E-09 |
| HTN | nk_cell | *ZNF589* | -3.83495552 | 3.33E-12 | 1.34E-09 | 1.61E-08 |
| HTN | nk_cell | *ATRIP* | -1.000125111 | 0.037217956 | 0.045296515 | 1 |
| HTN | nk_cell | *HSPA4L* | -2.490788612 | 0.001647937 | 0.00241346 | 1 |
| HTN | nk_cell | *MFSD8* | -2.272718618 | 2.17E-05 | 5.37E-05 | 0.105254975 |
| HTN | nk_cell | *MST1R* | -0.794975615 | 0.016146919 | 0.020487671 | 1 |
| HTN | nk_cell | *RAD54L2* | -2.36592011 | 0.000316568 | 0.000537247 | 1 |
| HTN | nk_cell | *TEX264* | -2.282622829 | 0.001361632 | 0.002024093 | 1 |
| HTN | nk_cell | *ETNPPL* | 10.50282863 | 3.41E-07 | 4.24E-06 | 0.001655463 |
| HTN | nk_cell | *PRSS12* | -1.245187721 | 4.70E-06 | 1.64E-05 | 0.022782864 |
| HTN | nk_cell | *HMGB2* | -2.142783311 | 0.000101188 | 0.000195137 | 0.490964789 |
| HTN | nk_cell | *ANXA5* | -2.08963517 | 0.001285075 | 0.001923252 | 1 |
| HTN | nk_cell | *MAP9* | 2.968235593 | 1.46E-06 | 7.39E-06 | 0.007069318 |
| HTN | nk_cell | *CEP44* | -2.365439332 | 7.69E-07 | 5.17E-06 | 0.003729996 |
| HTN | nk_cell | *HPGD* | -2.689384911 | 8.92E-07 | 5.65E-06 | 0.004328451 |
| HTN | nk_cell | *C4orf45* | -1.426268654 | 5.98E-06 | 1.97E-05 | 0.029013898 |
| HTN | nk_cell | *TMEM144* | -2.807583093 | 5.17E-06 | 1.78E-05 | 0.025106595 |
| HTN | nk_cell | *IL15* | 2.456865925 | 0.005035024 | 0.006834305 | 1 |
| HTN | nk_cell | *ARFIP1* | -1.934284317 | 0.000205675 | 0.00036635 | 0.997936687 |
| HTN | nk_cell | *LSM6* | -1.879417713 | 3.06E-05 | 7.15E-05 | 0.148518496 |

| | | | | | | |
|---|---|---|---|---|---|---|
| HTN | nk_cell | *ITGA2* | -1.667251332 | 2.11E-05 | 5.27E-05 | 0.102591679 |
| HTN | nk_cell | *MOCS2* | -3.591332338 | 2.59E-05 | 6.20E-05 | 0.125771133 |
| HTN | nk_cell | *LMBRD2* | -2.818924409 | 1.22E-05 | 3.40E-05 | 0.059244082 |
| HTN | nk_cell | *RNF180* | -2.163357139 | 1.67E-05 | 4.40E-05 | 0.081081007 |
| HTN | nk_cell | *F2RL2* | 1.141176703 | 2.64E-07 | 4.07E-06 | 0.001280862 |
| HTN | nk_cell | *CMBL* | -1.98792997 | 7.37E-06 | 2.29E-05 | 0.035742331 |
| HTN | nk_cell | *C5orf63* | -2.731767936 | 0.00097937 | 0.001500444 | 1 |
| HTN | nk_cell | *WDR41* | -1.881293311 | 2.52E-05 | 6.07E-05 | 0.122129816 |
| HTN | nk_cell | *NDUFS4* | -1.84750845 | 6.44E-06 | 2.07E-05 | 0.031240391 |
| HTN | nk_cell | *SERINC5* | -1.961466604 | 1.79E-05 | 4.64E-05 | 0.086974096 |
| HTN | nk_cell | *PRIMPOL* | -2.31700732 | 7.56E-05 | 0.000151465 | 0.366848116 |
| HTN | nk_cell | *ERAP1* | -3.090589277 | 6.26E-06 | 2.03E-05 | 0.03039737 |
| HTN | nk_cell | *ERAP2* | 1.699837687 | 2.00E-07 | 4.07E-06 | 0.000972234 |
| HTN | nk_cell | *UTP15* | -2.542347768 | 9.30E-07 | 5.79E-06 | 0.004512655 |
| HTN | nk_cell | *TLR3* | -2.711952724 | 2.56E-06 | 1.07E-05 | 0.012396998 |
| HTN | nk_cell | *GRIK2* | -1.155573301 | 1.40E-07 | 3.89E-06 | 0.000679875 |
| HTN | nk_cell | *DCBLD1* | -1.970287215 | 7.25E-06 | 2.26E-05 | 0.035185768 |
| HTN | nk_cell | *PDSS2* | -1.848691428 | 0.002817347 | 0.003991173 | 1 |
| HTN | nk_cell | *RAET1E* | -1.59696496 | 6.13E-07 | 4.55E-06 | 0.0029734 |
| HTN | nk_cell | *DAGLB* | -1.608720641 | 5.61E-07 | 4.48E-06 | 0.002721215 |
| HTN | nk_cell | *STK17A* | 0.691060485 | 3.15E-07 | 4.17E-06 | 0.001528524 |
| HTN | nk_cell | *GALNT10* | 2.382544928 | 4.71E-05 | 0.000101853 | 0.228659064 |
| HTN | nk_cell | *SAP30L* | 3.104996811 | 0.012513437 | 0.0161091 | 1 |
| HTN | nk_cell | *RPS14* | 1.974398373 | 0.000252434 | 0.000438739 | 1 |
| HTN | nk_cell | *RP9* | 3.050288996 | 2.18E-06 | 9.50E-06 | 0.010573931 |
| HTN | nk_cell | *CAMLG* | -2.408809956 | 2.65E-06 | 1.09E-05 | 0.012863316 |
| HTN | nk_cell | *RELL2* | 0.871958574 | 4.74E-07 | 4.32E-06 | 0.002298979 |
| HTN | nk_cell | *MIOS* | -2.259900376 | 5.31E-06 | 1.81E-05 | 0.025787642 |
| HTN | nk_cell | *USP49* | -2.193805294 | 6.74E-06 | 2.14E-05 | 0.032686435 |
| HTN | nk_cell | *CHMP4C* | -9.321274502 | 0.027950756 | 0.034411841 | 1 |
| HTN | nk_cell | *SLC13A4* | -2.320481404 | 1.19E-05 | 3.34E-05 | 0.057748438 |
| HTN | nk_cell | *BRI3* | -2.288440428 | 0.000281988 | 0.000483465 | 1 |
| HTN | nk_cell | *CTSB* | -1.336150788 | 3.79E-05 | 8.50E-05 | 0.183985343 |
| HTN | nk_cell | *SOX17* | -1.933703744 | 0.000683529 | 0.00107818 | 1 |
| HTN | nk_cell | *RAD21* | -2.271726847 | 0.000857501 | 0.001330936 | 1 |
| HTN | nk_cell | *TNFRSF11B* | -1.570789437 | 6.01E-07 | 4.54E-06 | 0.002915915 |
| HTN | nk_cell | *DEFB1* | -5.606953662 | 1.12E-06 | 6.35E-06 | 0.005438458 |
| HTN | nk_cell | *SUN1* | -1.719613155 | 1.17E-05 | 3.29E-05 | 0.056804993 |
| HTN | nk_cell | *OXR1* | -1.578387907 | 1.99E-05 | 5.03E-05 | 0.096658714 |
| HTN | nk_cell | *GPER1* | 3.539538337 | 0.00666144 | 0.008918683 | 1 |
| HTN | nk_cell | *FASTK* | -2.904109337 | 6.40E-06 | 2.06E-05 | 0.031049446 |
| HTN | nk_cell | *ALDH7A1* | -1.845482963 | 0.00029053 | 0.000497056 | 1 |
| HTN | nk_cell | *YWHAZ* | -2.126882612 | 0.000102346 | 0.000197214 | 0.49658436 |
| HTN | nk_cell | *FZD6* | -1.397414401 | 0.000390073 | 0.000650167 | 1 |
| HTN | nk_cell | *SLC25A32* | -2.567990834 | 0.004245818 | 0.00583094 | 1 |
| HTN | nk_cell | *DCSTAMP* | -13.50854244 | 2.11E-05 | 5.27E-05 | 0.102448264 |
| HTN | nk_cell | *INTS8* | -2.518577214 | 5.07E-05 | 0.000108276 | 0.245894306 |
| HTN | nk_cell | *RPP25L* | -3.107131938 | 2.57E-09 | 3.56E-07 | 1.25E-05 |
| HTN | nk_cell | *C9orf24* | 4.261097076 | 1.94E-05 | 4.94E-05 | 0.094319048 |
| HTN | nk_cell | *NUDT2* | -1.494478613 | 0.000262435 | 0.000453789 | 1 |

| | | | | | | |
|---|---|---|---|---|---|---|
| HTN | nk_cell | *CCDC171* | -2.075461085 | 0.000646883 | 0.001024111 | 1 |
| HTN | nk_cell | *ZMAT4* | 1.68482224 | 2.71E-07 | 4.07E-06 | 0.001312735 |
| HTN | nk_cell | *C8orf34* | -1.355156617 | 8.32E-06 | 2.52E-05 | 0.040373952 |
| HTN | nk_cell | *RASEF* | -1.006584539 | 0.010384532 | 0.013544557 | 1 |
| HTN | nk_cell | *ANKS6* | -1.629797776 | 3.97E-06 | 1.45E-05 | 0.019238855 |
| HTN | nk_cell | *AQP7* | -0.694411237 | 0.001316542 | 0.001965495 | 1 |
| HTN | nk_cell | *NOL6* | -1.731960675 | 8.50E-07 | 5.45E-06 | 0.004122482 |
| HTN | nk_cell | *AQP3* | -3.088656253 | 4.19E-07 | 4.24E-06 | 0.00203473 |
| HTN | nk_cell | *PIGO* | 2.05534661 | 1.73E-07 | 4.07E-06 | 0.00083865 |
| HTN | nk_cell | *WRN* | -1.799729194 | 0.000391055 | 0.000651356 | 1 |
| HTN | nk_cell | *TSHR* | -1.909736412 | 7.97E-06 | 2.44E-05 | 0.038681099 |
| HTN | nk_cell | *CFL2* | -2.95698129 | 1.26E-05 | 3.50E-05 | 0.061319998 |
| HTN | nk_cell | *PGM2L1* | -2.436005473 | 0.001522131 | 0.002246845 | 1 |
| HTN | nk_cell | *SLC16A9* | -3.927273938 | 4.23E-06 | 1.52E-05 | 0.020544766 |
| HTN | nk_cell | *GJB2* | -16.04076273 | 3.55E-06 | 1.34E-05 | 0.017204457 |
| HTN | nk_cell | *CRYL1* | -2.22034921 | 0.000102615 | 0.000197584 | 0.497887944 |
| HTN | nk_cell | *REEP3* | -2.250426865 | 1.51E-06 | 7.55E-06 | 0.007325036 |
| HTN | nk_cell | *LRR1* | -2.511760426 | 1.03E-06 | 6.05E-06 | 0.005016941 |
| HTN | nk_cell | *C10orf25* | -0.959409146 | 0.001695653 | 0.002475867 | 1 |
| HTN | nk_cell | *TMEM63C* | 8.512490749 | 1.57E-06 | 7.67E-06 | 0.007596633 |
| HTN | nk_cell | *NOXRED1* | -2.679147213 | 1.40E-05 | 3.80E-05 | 0.067851643 |
| HTN | nk_cell | *AKR1E2* | -1.539600756 | 7.03E-07 | 4.90E-06 | 0.003413135 |
| HTN | nk_cell | *OXGR1* | 2.184630291 | 1.23E-05 | 3.43E-05 | 0.059756759 |
| HTN | nk_cell | *ATP5C1* | -2.165814976 | 0.00131388 | 0.001962125 | 1 |
| HTN | nk_cell | *PDZD8* | -1.749878946 | 0.019889101 | 0.024961697 | 1 |
| HTN | nk_cell | *CLEC1B* | -2.173937681 | 0.001913665 | 0.002773328 | 1 |
| HTN | nk_cell | *SNAPC4* | -1.384780618 | 1.68E-06 | 8.02E-06 | 0.00813851 |
| HTN | nk_cell | *TSC1* | -1.52114592 | 0.00141945 | 0.002102953 | 1 |
| HTN | nk_cell | *STOX1* | -0.867847413 | 0.006129743 | 0.008238646 | 1 |
| HTN | nk_cell | *RET* | -2.120137429 | 5.25E-07 | 4.40E-06 | 0.002546814 |
| HTN | nk_cell | *CASP7* | -2.697880297 | 2.15E-06 | 9.42E-06 | 0.010444106 |
| HTN | nk_cell | *PPP1R36* | -1.674991242 | 8.10E-07 | 5.29E-06 | 0.003929233 |
| HTN | nk_cell | *VWA2* | -2.732304239 | 7.76E-07 | 5.19E-06 | 0.003766344 |
| HTN | nk_cell | *METTL3* | -2.653508746 | 1.42E-05 | 3.84E-05 | 0.068773624 |
| HTN | nk_cell | *TRUB1* | -3.434725785 | 7.27E-06 | 2.26E-05 | 0.035259331 |
| HTN | nk_cell | *HSPA12A* | -0.992082181 | 0.002697 | 0.003831873 | 1 |
| HTN | nk_cell | *UBTD1* | -1.568923457 | 1.40E-06 | 7.20E-06 | 0.00677734 |
| HTN | nk_cell | *TTC7B* | -2.185796614 | 9.12E-05 | 0.000178389 | 0.442403979 |
| HTN | nk_cell | *PSMC3* | -2.91577703 | 4.17E-06 | 1.51E-05 | 0.020237632 |
| HTN | nk_cell | *RAPSN* | -1.69928046 | 0.000597483 | 0.000955815 | 1 |
| HTN | nk_cell | *TC2N* | 1.452139708 | 0.000194831 | 0.000348826 | 0.945317615 |
| HTN | nk_cell | *CPSF2* | -1.188123771 | 0.000114898 | 0.000218194 | 0.557484447 |
| HTN | nk_cell | *IFI27L1* | -1.900907943 | 4.41E-06 | 1.57E-05 | 0.021413407 |
| HTN | nk_cell | *C1QL3* | -1.802604069 | 0.000857757 | 0.001330936 | 1 |
| HTN | nk_cell | *LIPC* | -2.162609599 | 5.90E-05 | 0.000122337 | 0.28626879 |
| HTN | nk_cell | *CEP57* | -2.695816282 | 0.000232017 | 0.000407731 | 1 |
| HTN | nk_cell | *TCP11L2* | -1.576197968 | 2.04E-06 | 9.07E-06 | 0.009905806 |
| HTN | nk_cell | *JAM3* | -2.200321205 | 1.15E-06 | 6.47E-06 | 0.005588548 |
| HTN | nk_cell | *IKBIP* | 3.182721252 | 0.001338097 | 0.001993383 | 1 |
| HTN | nk_cell | *ZFYVE19* | -2.598775449 | 4.48E-06 | 1.59E-05 | 0.021727437 |

| | | | | | | |
|---|---|---|---|---|---|---|
| HTN | nk_cell | *TRMT61A* | -1.519751363 | 3.98E-07 | 4.24E-06 | 0.00193147 |
| HTN | nk_cell | *LARP6* | -2.059941059 | 5.33E-06 | 1.81E-05 | 0.025870688 |
| HTN | nk_cell | *API5* | -2.562060529 | 0.000260507 | 0.000450616 | 1 |
| HTN | nk_cell | *ASPG* | 6.657515147 | 0.000311026 | 0.00052858 | 1 |
| HTN | nk_cell | *COX11* | -2.648549505 | 5.38E-05 | 0.000113495 | 0.260925944 |
| HTN | nk_cell | *ZNF202* | -2.514643176 | 1.41E-06 | 7.22E-06 | 0.006818375 |
| HTN | nk_cell | *STXBP4* | -2.323132905 | 6.32E-05 | 0.000129642 | 0.306603849 |
| HTN | nk_cell | *WBP1L* | -1.880125487 | 0.001210606 | 0.001821917 | 1 |
| HTN | nk_cell | *APBB1* | -2.829593855 | 2.89E-05 | 6.81E-05 | 0.14030074 |
| HTN | nk_cell | *NUDT13* | -2.368756059 | 1.19E-05 | 3.34E-05 | 0.057927547 |
| HTN | nk_cell | *WDR88* | -2.41629136 | 1.98E-06 | 8.92E-06 | 0.009583983 |
| HTN | nk_cell | *CYB5R2* | -1.083867763 | 0.00289164 | 0.004089256 | 1 |
| HTN | nk_cell | *SERPINB8* | -1.188796878 | 0.001990929 | 0.002875852 | 1 |
| HTN | nk_cell | *XRRA1* | -2.069743829 | 5.87E-07 | 4.54E-06 | 0.002848887 |
| HTN | nk_cell | *TRIM66* | -1.662856502 | 1.09E-09 | 1.97E-07 | 5.31E-06 |
| HTN | nk_cell | *CENPN* | -3.478881614 | 1.34E-06 | 7.02E-06 | 0.00648677 |
| HTN | nk_cell | *C16orf46* | -3.484488327 | 0.009564449 | 0.012515294 | 1 |
| HTN | nk_cell | *TMEM41B* | -2.475102379 | 6.67E-05 | 0.000135909 | 0.323464233 |
| HTN | nk_cell | *TMX3* | -2.192118171 | 0.001102743 | 0.001673603 | 1 |
| HTN | nk_cell | *PRKCB* | 0.571186252 | 2.60E-07 | 4.07E-06 | 0.001261844 |
| HTN | nk_cell | *NDST2* | 4.347928186 | 2.81E-07 | 4.07E-06 | 0.001361223 |
| HTN | nk_cell | *CCDC68* | -1.955823836 | 0.003426152 | 0.004783796 | 1 |
| HTN | nk_cell | *CLEC4D* | -4.584954775 | 0.000121825 | 0.000229999 | 0.591096364 |
| HTN | nk_cell | *ZSCAN21* | -1.698094856 | 0.004255494 | 0.005842574 | 1 |
| HTN | nk_cell | *RIMKLB* | -2.360363889 | 1.65E-10 | 3.48E-08 | 8.00E-07 |
| HTN | nk_cell | *TMED3* | -1.41033012 | 0.000183754 | 0.000330949 | 0.891575298 |
| HTN | nk_cell | *SEC11C* | 4.026508294 | 5.60E-05 | 0.000117429 | 0.271848793 |
| HTN | nk_cell | *GALR1* | -2.117851577 | 0.000119993 | 0.000226804 | 0.582205652 |
| HTN | nk_cell | *CENPV* | -2.217957371 | 0.000382177 | 0.000638101 | 1 |
| HTN | nk_cell | *ZNF592* | -1.804712231 | 4.98E-05 | 0.000106796 | 0.241785921 |
| HTN | nk_cell | *ACSM1* | -1.856964999 | 7.53E-06 | 2.33E-05 | 0.036555503 |
| HTN | nk_cell | *SLFN5* | 2.480012582 | 0.000934351 | 0.001441027 | 1 |
| HTN | nk_cell | *SAAL1* | 4.196352901 | 7.85E-05 | 0.000156468 | 0.380999666 |
| HTN | nk_cell | *LDHC* | 1.533717954 | 4.07E-05 | 9.02E-05 | 0.197515551 |
| HTN | nk_cell | *FAM96A* | -1.651768028 | 1.98E-06 | 8.92E-06 | 0.009585893 |
| HTN | nk_cell | *FAM111A* | -2.327907724 | 0.000166872 | 0.000304269 | 0.809660842 |
| HTN | nk_cell | *TMEM170A* | -2.64354028 | 5.39E-07 | 4.43E-06 | 0.00261752 |
| HTN | nk_cell | *STAT6* | -2.25696945 | 0.005014744 | 0.00681174 | 1 |
| HTN | nk_cell | *STX3* | 2.477230915 | 0.013499137 | 0.017304573 | 1 |
| HTN | nk_cell | *PIP4K2C* | 3.577063414 | 8.38E-06 | 2.53E-05 | 0.040643943 |
| HTN | nk_cell | *MTMR10* | -2.047610684 | 3.82E-05 | 8.55E-05 | 0.185553522 |
| HTN | nk_cell | *NYAP1* | -0.705725824 | 0.031968086 | 0.03915909 | 1 |
| HTN | nk_cell | *MS4A7* | -1.312228125 | 0.00836453 | 0.011031448 | 1 |
| HTN | nk_cell | *MS4A14* | -1.089255242 | 1.65E-06 | 7.94E-06 | 0.00802479 |
| HTN | nk_cell | *CCNDBP1* | 2.372996639 | 0.015782626 | 0.020041168 | 1 |
| HTN | nk_cell | *SMAD3* | -2.042587872 | 0.002359439 | 0.003382978 | 1 |
| HTN | nk_cell | *MAP1A* | -1.67757454 | 3.33E-05 | 7.65E-05 | 0.161753802 |
| HTN | nk_cell | *RCCD1* | -2.230218715 | 4.77E-07 | 4.32E-06 | 0.002314251 |
| HTN | nk_cell | *AKTIP* | 1.98313116 | 1.75E-06 | 8.21E-06 | 0.008476179 |
| HTN | nk_cell | *MEI1* | 1.274332233 | 2.28E-07 | 4.07E-06 | 0.001106834 |

| | | | | | | |
|---|---|---|---|---|---|---|
| HTN | nk_cell | *SAMD14* | 2.040357173 | 5.57E-07 | 4.48E-06 | 0.002702513 |
| HTN | nk_cell | *ACSF2* | -1.905134921 | 0.025782626 | 0.031928867 | 1 |
| HTN | nk_cell | *URM1* | -2.380949884 | 0.011284593 | 0.014639798 | 1 |
| HTN | nk_cell | *ENDOG* | -1.732114254 | 0.001053555 | 0.001605984 | 1 |
| HTN | nk_cell | *ISLR2* | -2.595496997 | 1.81E-07 | 4.07E-06 | 0.000880447 |
| HTN | nk_cell | *SP2* | -2.82242554 | 1.45E-05 | 3.90E-05 | 0.070113622 |
| HTN | nk_cell | *COQ7* | -2.404286029 | 6.32E-06 | 2.05E-05 | 0.030679581 |
| HTN | nk_cell | *GPRC5B* | -1.985143792 | 1.08E-06 | 6.19E-06 | 0.00523795 |
| HTN | nk_cell | *TBC1D2B* | -1.364792945 | 1.02E-06 | 6.04E-06 | 0.004968812 |
| HTN | nk_cell | *LOXHD1* | -2.287266638 | 9.45E-05 | 0.000183788 | 0.458551407 |
| HTN | nk_cell | *KATNAL2* | -1.455417993 | 0.000799307 | 0.001247825 | 1 |
| HTN | nk_cell | *ZNF91* | -2.001955568 | 1.36E-05 | 3.71E-05 | 0.066066083 |
| HTN | nk_cell | *ENGASE* | -1.429513439 | 2.78E-05 | 6.58E-05 | 0.134888518 |
| HTN | nk_cell | *MYO5B* | -1.711671584 | 1.43E-06 | 7.30E-06 | 0.006940196 |
| HTN | nk_cell | *RRM1* | -0.962020381 | 0.026410567 | 0.032623236 | 1 |
| HTN | nk_cell | *ZNF23* | -2.880691665 | 2.00E-08 | 1.33E-06 | 9.72E-05 |
| HTN | nk_cell | *ZNF229* | -1.848367385 | 0.002186578 | 0.00314722 | 1 |
| HTN | nk_cell | *SMG8* | -2.799745173 | 5.87E-07 | 4.54E-06 | 0.002847239 |
| HTN | nk_cell | *RAB8A* | 2.011677993 | 2.29E-07 | 4.07E-06 | 0.001110226 |
| HTN | nk_cell | *JSRP1* | -1.958328914 | 0.0155938 | 0.019816951 | 1 |
| HTN | nk_cell | *RPL13* | 1.950768005 | 0.001538604 | 0.002267024 | 1 |
| HTN | nk_cell | *TP53I13* | -2.594619816 | 0.000152635 | 0.000280951 | 0.74058573 |
| HTN | nk_cell | *TUBA1C* | -1.425427647 | 0.002576844 | 0.003674067 | 1 |
| HTN | nk_cell | *ZNF528* | -1.818741081 | 1.42E-06 | 7.27E-06 | 0.006894555 |
| HTN | nk_cell | *SERTAD3* | -4.44190465 | 2.74E-06 | 1.11E-05 | 0.013277848 |
| HTN | nk_cell | *TMC4* | -2.491444066 | 1.28E-06 | 6.85E-06 | 0.006221194 |
| HTN | nk_cell | *TTYH1* | -2.614723662 | 9.47E-07 | 5.82E-06 | 0.004592941 |
| HTN | nk_cell | *LENG8* | -1.700846676 | 6.55E-06 | 2.10E-05 | 0.031800249 |
| HTN | nk_cell | *CDC42EP5* | -2.009427315 | 7.84E-06 | 2.41E-05 | 0.038049334 |
| HTN | nk_cell | *KIR3DL1* | 2.967910627 | 2.07E-07 | 4.07E-06 | 0.001006075 |
| HTN | nk_cell | *C19orf33* | -1.374501829 | 0.000453993 | 0.000742925 | 1 |
| HTN | nk_cell | *DNAAF3* | -1.518473653 | 6.60E-05 | 0.000134678 | 0.320264248 |
| HTN | nk_cell | *CHAF1A* | -1.080615021 | 0.009541829 | 0.012489062 | 1 |
| HTN | nk_cell | *FAM57A* | -2.203923005 | 4.06E-05 | 9.01E-05 | 0.197045671 |
| HTN | nk_cell | *KIFC2* | -1.406469114 | 4.82E-07 | 4.32E-06 | 0.00233936 |
| HTN | nk_cell | *SRR* | 1.192930294 | 0.003373313 | 0.004715446 | 1 |
| HTN | nk_cell | *CYB5D2* | -2.316407896 | 0.000238631 | 0.000418597 | 1 |
| HTN | nk_cell | *C19orf48* | -0.910077215 | 0.006099748 | 0.008202876 | 1 |
| HTN | nk_cell | *KLK5* | -1.68102099 | 2.14E-07 | 4.07E-06 | 0.001040383 |
| HTN | nk_cell | *KLK6* | -8.375733738 | 4.59E-07 | 4.29E-06 | 0.002228805 |
| HTN | nk_cell | *ZNF83* | -1.896534969 | 0.000224483 | 0.000395782 | 1 |
| HTN | nk_cell | *SPRYD3* | -4.324716308 | 4.55E-06 | 1.61E-05 | 0.022090226 |
| HTN | nk_cell | *NDUFV1* | -2.528355306 | 0.001334526 | 0.001988673 | 1 |
| HTN | nk_cell | *MIS12* | -3.103320497 | 0.000859383 | 0.001332874 | 1 |
| HTN | nk_cell | *CD300C* | -0.889924196 | 0.011863233 | 0.015342696 | 1 |
| HTN | nk_cell | *ATP5H* | 3.344601807 | 0.001954415 | 0.002829005 | 1 |
| HTN | nk_cell | *MGAT5B* | -2.790512517 | 4.17E-07 | 4.24E-06 | 0.002023889 |
| HTN | nk_cell | *GSDMA* | -1.713146801 | 6.75E-06 | 2.14E-05 | 0.032763204 |
| HTN | nk_cell | *TMEM99* | -1.736612632 | 0.000974527 | 0.001495141 | 1 |
| HTN | nk_cell | *ZNF598* | -2.570073943 | 1.06E-08 | 8.70E-07 | 5.13E-05 |

| | | | | | | |
|---|---|---|---|---|---|---|
| HTN | nk_cell | *MLST8* | -1.543510314 | 5.21E-07 | 4.37E-06 | 0.002526021 |
| HTN | nk_cell | *ECI1* | -2.751551795 | 4.21E-05 | 9.28E-05 | 0.204179675 |
| HTN | nk_cell | *ABCA3* | -2.46602271 | 5.83E-05 | 0.000121163 | 0.283036513 |
| HTN | nk_cell | *ATG16L2* | -2.482679857 | 0.005736676 | 0.007738214 | 1 |
| HTN | nk_cell | *RPSA* | 2.461566382 | 5.14E-05 | 0.000109321 | 0.249251004 |
| HTN | nk_cell | *ULK4* | -1.568017932 | 0.000222781 | 0.000392924 | 1 |
| HTN | nk_cell | *NAALADL1* | 1.85086787 | 9.96E-07 | 5.95E-06 | 0.004830974 |
| HTN | nk_cell | *SCARA5* | -0.905510122 | 4.86E-07 | 4.32E-06 | 0.00235618 |
| HTN | nk_cell | *KIAA1586* | -2.291453038 | 2.68E-06 | 1.10E-05 | 0.013012139 |
| HTN | nk_cell | *FAM83B* | -10.41818609 | 6.13E-07 | 4.55E-06 | 0.002975469 |
| HTN | nk_cell | *COA6* | 2.897580593 | 7.90E-07 | 5.22E-06 | 0.003833876 |
| HTN | nk_cell | *PXK* | -2.090521892 | 4.39E-06 | 1.57E-05 | 0.02128569 |
| HTN | nk_cell | *PCMTD1* | -1.923095861 | 0.000796544 | 0.001243911 | 1 |
| HTN | nk_cell | *ACOX2* | -1.495934285 | 3.69E-07 | 4.24E-06 | 0.001789809 |
| HTN | nk_cell | *SCN11A* | -1.120831195 | 8.41E-05 | 0.000166069 | 0.408031875 |
| HTN | nk_cell | *SEPT2* | -1.073983458 | 0.005954053 | 0.00801361 | 1 |
| HTN | nk_cell | *DTYMK* | -3.484731695 | 0.002603485 | 0.003707693 | 1 |
| HTN | nk_cell | *ING5* | -1.861253406 | 1.05E-05 | 3.03E-05 | 0.051074349 |
| HTN | nk_cell | *RFWD3* | -2.646461758 | 0.000534165 | 0.000861625 | 1 |
| HTN | nk_cell | *KLHL30* | -8.48129274 | 7.39E-07 | 5.04E-06 | 0.003586176 |
| HTN | nk_cell | *CCDC110* | -2.091854473 | 4.25E-07 | 4.24E-06 | 0.002060043 |
| HTN | nk_cell | *FNTA* | -1.849580398 | 6.77E-06 | 2.14E-05 | 0.032824179 |
| HTN | nk_cell | *SERINC2* | -6.209215586 | 4.12E-09 | 4.55E-07 | 2.00E-05 |
| HTN | nk_cell | *SLC20A2* | -2.228815835 | 0.000723244 | 0.001137129 | 1 |
| HTN | nk_cell | *TMUB2* | -2.098255732 | 9.64E-07 | 5.85E-06 | 0.004677821 |
| HTN | nk_cell | *ZSWIM1* | 0.76521329 | 0.003251073 | 0.004559019 | 1 |
| HTN | nk_cell | *NBPF9* | -1.388275839 | 4.45E-07 | 4.28E-06 | 0.002161065 |
| HTN | nk_cell | *ADAM9* | -1.842676973 | 6.24E-05 | 0.00012822 | 0.302598837 |
| HTN | nk_cell | *ZNF30* | -1.419387214 | 4.62E-05 | 0.000100243 | 0.224243251 |
| HTN | nk_cell | *SLC16A4* | -1.076598867 | 0.025638729 | 0.031774997 | 1 |
| HTN | nk_cell | *NPNT* | -1.283930775 | 0.000702404 | 0.001106515 | 1 |
| HTN | nk_cell | *TCTN2* | -1.005202362 | 5.08E-08 | 2.19E-06 | 0.000246415 |
| HTN | nk_cell | *PPIP5K1* | -2.1762568 | 0.008252989 | 0.010896191 | 1 |
| HTN | nk_cell | *ADAL* | -1.264490815 | 8.93E-05 | 0.000174972 | 0.433231408 |
| HTN | nk_cell | *SNTB2* | -2.006266885 | 8.48E-05 | 0.000167398 | 0.411632039 |
| HTN | nk_cell | *ZNF507* | -2.4731626 | 0.002373062 | 0.003401505 | 1 |
| HTN | nk_cell | *STX18* | -2.277239195 | 7.80E-05 | 0.000155569 | 0.378343555 |
| HTN | nk_cell | *ZBTB49* | -3.053275478 | 9.49E-06 | 2.79E-05 | 0.046069319 |
| HTN | nk_cell | *GFM1* | -1.442208771 | 0.000396072 | 0.000658581 | 1 |
| HTN | nk_cell | *CEP120* | -2.62652203 | 2.01E-05 | 5.07E-05 | 0.097672541 |
| HTN | nk_cell | *MFF* | -2.186793999 | 0.000191759 | 0.000343961 | 0.930413325 |
| HTN | nk_cell | *CPLX1* | -1.226630214 | 7.92E-05 | 0.000157536 | 0.384387766 |
| HTN | nk_cell | *MFSD7* | -3.912190526 | 1.78E-06 | 8.29E-06 | 0.008633875 |
| HTN | nk_cell | *KLK7* | 2.247275912 | 5.21E-07 | 4.37E-06 | 0.002527279 |
| HTN | nk_cell | *IRS1* | -1.14542524 | 3.78E-05 | 8.48E-05 | 0.183523302 |
| HTN | nk_cell | *UPF3A* | -1.135071765 | 0.020699962 | 0.025939105 | 1 |
| HTN | nk_cell | *HSPBAP1* | -2.025301382 | 0.001353848 | 0.002013138 | 1 |
| HTN | nk_cell | *UBE2V2* | -2.385130703 | 4.87E-05 | 0.000104785 | 0.236080027 |
| HTN | nk_cell | *CPT1C* | 5.437392735 | 2.76E-12 | 1.25E-09 | 1.34E-08 |
| HTN | nk_cell | *PCSK9* | 8.874619804 | 8.03E-07 | 5.27E-06 | 0.003898548 |

| | | | | | | |
|---|---|---|---|---|---|---|
| HTN | nk_cell | *XPO6* | -2.531921377 | 4.84E-05 | 0.000104285 | 0.234745765 |
| HTN | nk_cell | *RGS14* | 0.653281414 | 1.06E-05 | 3.04E-05 | 0.051216455 |
| HTN | nk_cell | *RAB24* | -2.615901587 | 3.85E-08 | 2.01E-06 | 0.000186939 |
| HTN | nk_cell | *THBS3* | -1.878621013 | 4.78E-06 | 1.67E-05 | 0.023201879 |
| HTN | nk_cell | *SH3TC2* | -1.566501605 | 0.010771924 | 0.01400841 | 1 |
| HTN | nk_cell | *RNASE6* | -1.647650096 | 0.010856542 | 0.014113355 | 1 |
| HTN | nk_cell | *CD52* | 0.347598471 | 7.89E-07 | 5.21E-06 | 0.003826255 |
| HTN | nk_cell | *SLC38A11* | -1.189512028 | 6.38E-07 | 4.62E-06 | 0.003096674 |
| HTN | nk_cell | *CCDC8* | -1.556687445 | 3.22E-06 | 1.25E-05 | 0.01562075 |
| HTN | nk_cell | *ZEB2* | -1.744316853 | 8.17E-05 | 0.000161924 | 0.396390051 |
| HTN | nk_cell | *DTWD2* | -2.28183707 | 3.32E-05 | 7.63E-05 | 0.161158383 |
| HTN | nk_cell | *CLIC3* | 1.460072536 | 2.98E-07 | 4.13E-06 | 0.00144814 |
| HTN | nk_cell | *INO80E* | -3.658585099 | 8.42E-09 | 7.43E-07 | 4.08E-05 |
| HTN | nk_cell | *NFU1* | -1.766570141 | 0.00307016 | 0.004324069 | 1 |
| HTN | nk_cell | *C15orf40* | -2.11976389 | 1.88E-05 | 4.80E-05 | 0.091121553 |
| HTN | nk_cell | *PROKR1* | 3.143400456 | 0.004774081 | 0.006508525 | 1 |
| HTN | nk_cell | *RGPD8* | -0.843942542 | 0.000210065 | 0.000372255 | 1 |
| HTN | nk_cell | *LRRC45* | 1.253923902 | 0.000119813 | 0.000226553 | 0.581334012 |
| HTN | nk_cell | *CHRNA5* | 4.547369001 | 0.007633867 | 0.010120088 | 1 |
| HTN | nk_cell | *UGP2* | -1.70565817 | 1.02E-05 | 2.94E-05 | 0.049362671 |
| HTN | nk_cell | *AVEN* | -2.016980949 | 4.94E-06 | 1.72E-05 | 0.023981087 |
| HTN | nk_cell | *CTNND2* | -1.623751403 | 1.87E-07 | 4.07E-06 | 0.000904936 |
| HTN | nk_cell | *CALML6* | -19.3768611 | 1.69E-06 | 8.03E-06 | 0.00817734 |
| HTN | nk_cell | *MUC3A* | -2.761456605 | 8.23E-07 | 5.33E-06 | 0.003991707 |
| HTN | nk_cell | *TPST1* | -1.137471534 | 8.10E-06 | 2.47E-05 | 0.039322353 |
| HTN | nk_cell | *TOR1AIP2* | -2.063317474 | 2.92E-05 | 6.87E-05 | 0.141691303 |
| HTN | nk_cell | *OTUD3* | -1.866437733 | 0.001231314 | 0.001849067 | 1 |
| HTN | nk_cell | *ZFPM2* | -1.121385744 | 2.61E-06 | 1.08E-05 | 0.012661253 |
| HTN | nk_cell | *MAP3K2* | -1.820255946 | 0.000306248 | 0.000521008 | 1 |
| HTN | nk_cell | *MYO7B* | -1.096381971 | 0.001025967 | 0.001565899 | 1 |
| HTN | nk_cell | *UBE2E3* | -2.19681775 | 1.56E-05 | 4.15E-05 | 0.075455142 |
| HTN | nk_cell | *NIPA1* | 2.652857964 | 0.008276205 | 0.01092387 | 1 |
| HTN | nk_cell | *HNRNPA3* | -1.79835568 | 0.003312344 | 0.004635562 | 1 |
| HTN | nk_cell | *RNF150* | -1.598484672 | 5.61E-06 | 1.88E-05 | 0.027217207 |
| HTN | nk_cell | *HOXD4* | 3.671175431 | 0.000254512 | 0.000441822 | 1 |
| HTN | nk_cell | *ANKK1* | -3.783688681 | 4.83E-06 | 1.68E-05 | 0.023420254 |
| HTN | nk_cell | *PDCD6IP* | -1.886379791 | 3.09E-05 | 7.19E-05 | 0.149714215 |
| HTN | nk_cell | *CRTAP* | -2.224477511 | 0.000523926 | 0.000846798 | 1 |
| HTN | nk_cell | *CNGB3* | -1.657989352 | 0.002786049 | 0.003949141 | 1 |
| HTN | nk_cell | *ELP5* | -3.027734518 | 3.03E-06 | 1.20E-05 | 0.014700577 |
| HTN | nk_cell | *NFRKB* | -2.926501438 | 0.000479622 | 0.000781439 | 1 |
| HTN | nk_cell | *SEMA3E* | -1.210824013 | 2.23E-06 | 9.67E-06 | 0.010843013 |
| HTN | nk_cell | *MGMT* | -1.923265806 | 0.001143683 | 0.001730864 | 1 |
| HTN | nk_cell | *NFXL1* | -2.275598403 | 3.68E-05 | 8.30E-05 | 0.178426898 |
| HTN | nk_cell | *DNAJC18* | -2.798338951 | 2.05E-09 | 2.99E-07 | 9.94E-06 |
| HTN | nk_cell | *SPATA24* | -2.171387362 | 2.24E-06 | 9.70E-06 | 0.010887965 |
| HTN | nk_cell | *HSD17B13* | -7.980998244 | 2.49E-05 | 6.02E-05 | 0.120854943 |
| HTN | nk_cell | *ELOVL6* | 2.731030707 | 2.12E-05 | 5.28E-05 | 0.102928353 |
| HTN | nk_cell | *ARL6IP1* | -2.189587933 | 0.015895803 | 0.020179601 | 1 |
| HTN | nk_cell | *SMAGP* | -1.083150552 | 0.003690014 | 0.005124198 | 1 |

| | | | | | | |
|---|---|---|---|---|---|---|
| HTN | nk_cell | *CDH2* | -1.70878146 | 3.52E-07 | 4.24E-06 | 0.00170634 |
| HTN | nk_cell | *EMB* | 1.201390926 | 0.000252465 | 0.000438739 | 1 |
| HTN | nk_cell | *STAT2* | -2.47073518 | 0.000180488 | 0.000325792 | 0.87572897 |
| HTN | nk_cell | *SLC26A5* | -2.670231303 | 0.00089704 | 0.001385686 | 1 |
| HTN | nk_cell | *GTSF1* | -4.157187139 | 2.48E-07 | 4.07E-06 | 0.001205438 |
| HTN | nk_cell | *ARMC10* | -2.499656934 | 0.000269256 | 0.000464261 | 1 |
| HTN | nk_cell | *ACYP2* | -1.667375293 | 8.72E-05 | 0.000171212 | 0.423236477 |
| HTN | nk_cell | *TRABD* | 2.728831954 | 2.42E-06 | 1.02E-05 | 0.011745679 |
| HTN | nk_cell | *TMEM133* | -2.789385592 | 3.16E-05 | 7.33E-05 | 0.153125724 |
| HTN | nk_cell | *HOXB9* | -4.586347988 | 7.96E-07 | 5.23E-06 | 0.003859782 |
| HTN | nk_cell | *TTLL6* | -5.282107513 | 1.50E-12 | 9.08E-10 | 7.26E-09 |
| HTN | nk_cell | *KCNS3* | -0.781524719 | 6.57E-05 | 0.000134075 | 0.318561532 |
| HTN | nk_cell | *FOXN2* | -2.097727681 | 0.004473038 | 0.006129111 | 1 |
| HTN | nk_cell | *GPR27* | -1.94173821 | 0.000277547 | 0.000476862 | 1 |
| HTN | nk_cell | *TRIAP1* | -2.486751705 | 1.08E-06 | 6.18E-06 | 0.005226132 |
| HTN | nk_cell | *RPS9* | 1.308607951 | 0.032999996 | 0.04038234 | 1 |
| HTN | nk_cell | *TSEN34* | -3.93852003 | 9.23E-05 | 0.000180194 | 0.447601052 |
| HTN | nk_cell | *MSANTD4* | 3.877621391 | 2.47E-05 | 5.98E-05 | 0.119851668 |
| HTN | nk_cell | *NDUFA3* | 2.313291683 | 1.13E-06 | 6.38E-06 | 0.005478159 |
| HTN | nk_cell | *PAQR8* | -3.974469749 | 4.41E-08 | 2.12E-06 | 0.000213871 |
| HTN | nk_cell | *NUDT6* | -1.735827022 | 5.13E-05 | 0.000109258 | 0.24867108 |
| HTN | nk_cell | *ZNF160* | -1.806484502 | 3.94E-06 | 1.45E-05 | 0.019100474 |
| HTN | nk_cell | *ZNF415* | -3.050582871 | 7.01E-05 | 0.000142011 | 0.340116346 |
| HTN | nk_cell | *S1PR1* | -2.564216347 | 3.21E-05 | 7.43E-05 | 0.155673433 |
| HTN | nk_cell | *PYGO1* | -2.418476986 | 3.69E-05 | 8.31E-05 | 0.179078454 |
| HTN | nk_cell | *TSNARE1* | 2.635863899 | 0.003219842 | 0.004523067 | 1 |
| HTN | nk_cell | *FPR1* | -2.75727235 | 3.01E-06 | 1.19E-05 | 0.014582665 |
| HTN | nk_cell | *FEZ2* | -1.701859428 | 0.001652834 | 0.002419901 | 1 |
| HTN | nk_cell | *SOX7* | -3.7090506 | 0.013880827 | 0.017765701 | 1 |
| HTN | nk_cell | *TRMT61B* | -2.234157634 | 0.000371217 | 0.000620228 | 1 |
| HTN | nk_cell | *MFN1* | -1.629422275 | 5.95E-05 | 0.000123186 | 0.288623835 |
| HTN | nk_cell | *ATP6V0E2* | 2.039910476 | 2.50E-05 | 6.04E-05 | 0.121322469 |
| HTN | nk_cell | *MORN4* | -1.345280595 | 1.20E-05 | 3.36E-05 | 0.058205309 |
| HTN | nk_cell | *SCAND1* | 2.657306157 | 0.000243325 | 0.000424833 | 1 |
| HTN | nk_cell | *SOSTDC1* | -2.761526124 | 3.82E-05 | 8.54E-05 | 0.185227895 |
| HTN | nk_cell | *GAA* | -0.698895734 | 0.028311498 | 0.034829459 | 1 |
| HTN | nk_cell | *CANT1* | -1.255081008 | 0.012574327 | 0.016174611 | 1 |
| HTN | nk_cell | *PGAM1* | -1.713486368 | 2.63E-05 | 6.28E-05 | 0.127687677 |
| HTN | nk_cell | *LURAP1* | -1.015987148 | 0.00238517 | 0.003416842 | 1 |
| HTN | nk_cell | *PDE7B* | -1.640913276 | 7.94E-06 | 2.43E-05 | 0.038506352 |
| HTN | nk_cell | *MRPL36* | 2.401887956 | 7.09E-08 | 2.67E-06 | 0.000344094 |
| HTN | nk_cell | *MCC* | -1.932468192 | 4.50E-05 | 9.80E-05 | 0.218547334 |
| HTN | nk_cell | *ZNF561* | -1.374260996 | 0.014953362 | 0.019073005 | 1 |
| HTN | nk_cell | *HOPX* | 0.34045254 | 4.55E-07 | 4.29E-06 | 0.002206181 |
| HTN | nk_cell | *PPID* | -2.786144608 | 0.001848586 | 0.002681417 | 1 |
| HTN | nk_cell | *ETFDH* | -0.711227603 | 0.022875513 | 0.028539982 | 1 |
| HTN | nk_cell | *TBCA* | -2.068401721 | 0.000316747 | 0.000537362 | 1 |
| HTN | nk_cell | *ZNF584* | -0.876542299 | 0.000122312 | 0.000230649 | 0.593458928 |
| HTN | nk_cell | *NMUR1* | 0.442826141 | 2.72E-07 | 4.07E-06 | 0.001317977 |
| HTN | nk_cell | *PIK3CD* | 1.205164497 | 3.13E-07 | 4.17E-06 | 0.001519641 |

| | | | | | | |
|---|---|---|---|---|---|---|
| HTN | nk_cell | *ANO5* | -1.375143095 | 1.36E-06 | 7.11E-06 | 0.006615085 |
| HTN | nk_cell | *VAT1L* | -3.160766646 | 2.61E-06 | 1.08E-05 | 0.012653026 |
| HTN | nk_cell | *CAMTA1* | -1.627886896 | 0.015451829 | 0.019646822 | 1 |
| HTN | nk_cell | *LGALS4* | -1.904079017 | 0.001130344 | 0.001712814 | 1 |
| HTN | nk_cell | *PAH* | -5.869711657 | 6.51E-07 | 4.69E-06 | 0.003158532 |
| HTN | nk_cell | *RHNO1* | 0.925494065 | 1.77E-07 | 4.07E-06 | 0.000860177 |
| HTN | nk_cell | *METTL18* | 0.755813964 | 0.000205899 | 0.000366614 | 0.999022732 |
| HTN | nk_cell | *PWWP2B* | -1.639795432 | 1.71E-06 | 8.12E-06 | 0.008320898 |
| HTN | nk_cell | *EXOSC10* | -2.494941226 | 3.61E-05 | 8.18E-05 | 0.175234758 |
| HTN | nk_cell | *C3AR1* | -1.247388522 | 0.017217699 | 0.021789326 | 1 |
| HTN | nk_cell | *PRNP* | -2.128557363 | 0.000421637 | 0.000694663 | 1 |
| HTN | nk_cell | *TVP23B* | -1.901240303 | 0.007998308 | 0.01057433 | 1 |
| HTN | nk_cell | *FBXW10* | -2.604515759 | 4.15E-07 | 4.24E-06 | 0.002015245 |
| HTN | nk_cell | *SRGAP2C* | -1.305691444 | 1.21E-06 | 6.69E-06 | 0.005884682 |
| HTN | nk_cell | *ATPAF2* | -1.998538555 | 0.000742472 | 0.001165095 | 1 |
| HTN | nk_cell | *ZNF554* | 3.599896413 | 1.46E-06 | 7.39E-06 | 0.007085728 |
| HTN | nk_cell | *SERF1A* | -8.244372972 | 3.01E-07 | 4.15E-06 | 0.001461957 |
| HTN | nk_cell | *CD8B* | 0.540980458 | 4.10E-07 | 4.24E-06 | 0.001989309 |
| HTN | nk_cell | *MTBP* | -2.274854877 | 1.08E-05 | 3.09E-05 | 0.052418438 |
| HTN | nk_cell | *TEFM* | -1.739331389 | 5.73E-05 | 0.000119407 | 0.277980125 |
| HTN | nk_cell | *MALT1* | -2.220110767 | 0.004773141 | 0.006508525 | 1 |
| HTN | nk_cell | *MBOAT1* | -2.100325077 | 6.33E-06 | 2.05E-05 | 0.030719357 |
| HTN | nk_cell | *CXCR6* | 1.399516724 | 2.62E-07 | 4.07E-06 | 0.001269612 |
| HTN | nk_cell | *PAIP1* | -2.793876873 | 0.000118585 | 0.000224317 | 0.575372198 |
| HTN | nk_cell | *C1QTNF4* | -0.938601211 | 0.004866483 | 0.006630771 | 1 |
| HTN | nk_cell | *MACROD2* | -1.224403133 | 0.000116204 | 0.000220072 | 0.563823952 |
| HTN | nk_cell | *BSG* | -2.489899437 | 0.000153446 | 0.000282122 | 0.744519152 |
| HTN | nk_cell | *HINFP* | -1.645274322 | 1.75E-06 | 8.21E-06 | 0.00850051 |
| HTN | nk_cell | *TP53RK* | 1.375005893 | 1.91E-07 | 4.07E-06 | 0.000927738 |
| HTN | nk_cell | *SUCLG2* | -1.648315197 | 0.016729923 | 0.021199683 | 1 |
| HTN | nk_cell | *ARNT2* | -1.144851602 | 5.15E-05 | 0.000109495 | 0.24986713 |
| HTN | nk_cell | *GTPBP2* | -2.700093803 | 5.59E-05 | 0.000117314 | 0.27134717 |
| HTN | nk_cell | *FGGY* | -1.671058929 | 0.006676573 | 0.008936478 | 1 |
| HTN | nk_cell | *MANEA* | -2.822054093 | 2.06E-05 | 5.17E-05 | 0.100105249 |
| HTN | nk_cell | *AFF1* | -1.635273338 | 4.45E-05 | 9.72E-05 | 0.216083699 |
| HTN | nk_cell | *FIBP* | 1.631240074 | 3.57E-07 | 4.24E-06 | 0.001733661 |
| HTN | nk_cell | *CTSW* | 0.166802142 | 1.79E-07 | 4.07E-06 | 0.000870276 |
| HTN | nk_cell | *SNTG2* | -1.351253064 | 1.61E-05 | 4.26E-05 | 0.07822465 |
| HTN | nk_cell | *RAD9A* | -1.015933599 | 0.008119708 | 0.010726061 | 1 |
| HTN | nk_cell | *EFEMP2* | -1.316813481 | 4.16E-07 | 4.24E-06 | 0.002017816 |
| HTN | nk_cell | *ZFAND4* | -2.708672207 | 1.09E-06 | 6.24E-06 | 0.005309355 |
| HTN | nk_cell | *ZNF738* | 1.355543385 | 0.018165561 | 0.022881439 | 1 |
| HTN | nk_cell | *SLFN11* | -3.09634816 | 0.000295104 | 0.000504171 | 1 |
| HTN | nk_cell | *FAM71D* | -2.176638897 | 0.031684163 | 0.0388211 | 1 |
| HTN | nk_cell | *FUT10* | -0.835981875 | 0.023799239 | 0.029593518 | 1 |
| HTN | nk_cell | *CFL1* | 1.702312078 | 6.69E-07 | 4.76E-06 | 0.0032465 |
| HTN | nk_cell | *FAM192A* | -2.271020088 | 0.000806314 | 0.001257166 | 1 |
| HTN | nk_cell | *CBWD1* | -2.17006655 | 1.46E-06 | 7.39E-06 | 0.007092844 |
| HTN | nk_cell | *DCP2* | -2.514643895 | 2.51E-05 | 6.06E-05 | 0.121803415 |
| HTN | nk_cell | *SNX32* | -1.974287297 | 0.001669143 | 0.002440832 | 1 |

| | | | | | | |
|---|---|---|---|---|---|---|
| HTN | nk_cell | *RNASEH2C* | -2.020289893 | 0.000263864 | 0.000455612 | 1 |
| HTN | nk_cell | *MYEOV* | -6.258472055 | 5.76E-09 | 5.45E-07 | 2.80E-05 |
| HTN | nk_cell | *ANKRD13D* | 2.539535459 | 0.00274672 | 0.003896808 | 1 |
| HTN | nk_cell | *MRGPRF* | -1.211125049 | 1.97E-07 | 4.07E-06 | 0.00095591 |
| HTN | nk_cell | *LCLAT1* | -2.119884118 | 0.003604675 | 0.005021093 | 1 |
| HTN | nk_cell | *SH3RF3* | -2.051768062 | 1.09E-05 | 3.12E-05 | 0.05302101 |
| HTN | nk_cell | *GXYLT2* | -1.518091305 | 5.04E-06 | 1.74E-05 | 0.024460561 |
| HTN | nk_cell | *DCAKD* | -3.214051184 | 2.02E-05 | 5.08E-05 | 0.098111538 |
| HTN | nk_cell | *TRMT112* | -1.660103663 | 0.026743162 | 0.033017257 | 1 |
| HTN | nk_cell | *NOC3L* | -2.901445167 | 1.63E-07 | 4.07E-06 | 0.000791227 |
| HTN | nk_cell | *RHOD* | -1.453753457 | 2.02E-06 | 9.02E-06 | 0.009815252 |
| HTN | nk_cell | *PARP15* | 0.393757214 | 8.79E-08 | 3.09E-06 | 0.000426674 |
| HTN | nk_cell | *AHSA2* | -1.312117346 | 0.001204235 | 0.001813454 | 1 |
| HTN | nk_cell | *VANGL1* | -2.086251427 | 1.41E-06 | 7.22E-06 | 0.006818633 |
| HTN | nk_cell | *IQCB1* | -1.760222968 | 3.63E-05 | 8.22E-05 | 0.176363643 |
| HTN | nk_cell | *ZNF483* | -1.714084676 | 6.97E-06 | 2.19E-05 | 0.033820478 |
| HTN | nk_cell | *SLC2A14* | -1.33286922 | 4.68E-06 | 1.64E-05 | 0.022720933 |
| HTN | nk_cell | *MMRN2* | -2.249533935 | 0.000557943 | 0.000897296 | 1 |
| HTN | nk_cell | *MAP3K11* | -1.74079623 | 0.00631432 | 0.008472644 | 1 |
| HTN | nk_cell | *KCNK7* | 3.599351001 | 3.46E-07 | 4.24E-06 | 0.001680965 |
| HTN | nk_cell | *NDNF* | -1.841460002 | 1.28E-06 | 6.85E-06 | 0.00622866 |
| HTN | nk_cell | *ARV1* | -1.853529912 | 2.43E-06 | 1.02E-05 | 0.011778172 |
| HTN | nk_cell | *CCDC36* | -1.680987374 | 1.94E-07 | 4.07E-06 | 0.000942039 |
| HTN | nk_cell | *EHBP1L1* | -1.555574812 | 0.011539873 | 0.014954985 | 1 |
| HTN | nk_cell | *PPP1R14B* | 7.326223963 | 4.72E-06 | 1.65E-05 | 0.02289383 |
| HTN | nk_cell | *PEAK1* | -1.732389144 | 2.06E-05 | 5.15E-05 | 0.09976037 |
| HTN | nk_cell | *MST1* | -1.887278053 | 1.82E-12 | 9.84E-10 | 8.85E-09 |
| HTN | nk_cell | *GMPPB* | -3.92565662 | 3.83E-08 | 2.01E-06 | 0.000185824 |
| HTN | nk_cell | *NUDT18* | -7.701441109 | 5.00E-06 | 1.73E-05 | 0.024254822 |
| HTN | nk_cell | *SULT1B1* | 5.483225075 | 0.002783047 | 0.003946038 | 1 |
| HTN | nk_cell | *NUDT4* | -1.706215435 | 1.25E-06 | 6.76E-06 | 0.006051539 |
| HTN | nk_cell | *SCAI* | -1.814635677 | 0.00025146 | 0.000437776 | 1 |
| HTN | nk_cell | *HSPB7* | 2.394443629 | 8.40E-06 | 2.54E-05 | 0.040772903 |
| HTN | nk_cell | *UQCRH* | -2.399881466 | 0.000269759 | 0.000464963 | 1 |
| HTN | nk_cell | *C11orf80* | -4.320905259 | 2.57E-08 | 1.54E-06 | 0.000124833 |
| HTN | nk_cell | *CD7* | 0.197987594 | 2.72E-07 | 4.07E-06 | 0.00131794 |
| HTN | nk_cell | *RNF213* | 1.902369182 | 0.001538359 | 0.002267024 | 1 |
| HTN | nk_cell | *GPR160* | 4.415924417 | 4.80E-07 | 4.32E-06 | 0.002329827 |
| HTN | nk_cell | *GOLIM4* | -2.041144262 | 8.40E-06 | 2.54E-05 | 0.040777619 |
| HTN | nk_cell | *HOXB2* | -2.227603389 | 1.52E-05 | 4.06E-05 | 0.073635261 |
| HTN | nk_cell | *XXYLT1* | -1.869954571 | 2.41E-06 | 1.02E-05 | 0.011675146 |
| HTN | nk_cell | *UBXN2A* | -1.981613004 | 0.00037095 | 0.000620111 | 1 |
| HTN | nk_cell | *NRROS* | 1.30422172 | 0.000104369 | 0.0002003 | 0.506396228 |
| HTN | nk_cell | *CEP19* | -1.571170334 | 0.002600326 | 0.003704281 | 1 |
| HTN | nk_cell | *SLC25A30* | -3.706551801 | 0.000494789 | 0.000803183 | 1 |
| HTN | nk_cell | *CTSF* | -2.06854347 | 0.001660018 | 0.002428953 | 1 |
| HTN | nk_cell | *CTU2* | 1.094234804 | 0.003124853 | 0.00439727 | 1 |
| HTN | nk_cell | *SNX31* | -1.868805864 | 5.47E-06 | 1.84E-05 | 0.026516346 |
| HTN | nk_cell | *GLIS1* | -1.358806293 | 5.48E-07 | 4.45E-06 | 0.00266066 |
| HTN | nk_cell | *RALGAPA1* | -1.981267523 | 3.52E-05 | 7.99E-05 | 0.170669698 |

| | | | | | | |
|---|---|---|---|---|---|---|
| HTN | nk_cell | *SNAPC5* | 2.688675203 | 0.02620129 | 0.032397721 | 1 |
| HTN | nk_cell | *ANKRD36C* | -2.313322639 | 0.00011382 | 0.000216317 | 0.552256919 |
| HTN | nk_cell | *TMEM81* | 3.677399472 | 3.28E-05 | 7.55E-05 | 0.159040599 |
| HTN | nk_cell | *MSL2* | -2.151215388 | 0.010390945 | 0.013549278 | 1 |
| HTN | nk_cell | *ZNF497* | 7.85149767 | 1.12E-07 | 3.53E-06 | 0.00054322 |
| HTN | nk_cell | *TRAM1L1* | -3.050026384 | 3.37E-05 | 7.69E-05 | 0.163393665 |
| HTN | nk_cell | *ANGEL2* | -2.37061011 | 0.000865473 | 0.001340765 | 1 |
| HTN | nk_cell | *SH3PXD2B* | -1.225258417 | 1.35E-06 | 7.08E-06 | 0.006574115 |
| HTN | nk_cell | *LARP7* | -2.518307809 | 0.001017752 | 0.0015549 | 1 |
| HTN | nk_cell | *FGFBP3* | 2.871232052 | 3.70E-07 | 4.24E-06 | 0.001796273 |
| HTN | nk_cell | *C4orf32* | -1.612075572 | 2.72E-06 | 1.11E-05 | 0.013174483 |
| HTN | nk_cell | *HRAS* | 2.465761838 | 0.000110142 | 0.000210067 | 0.534409483 |
| HTN | nk_cell | *PCP2* | 1.994812494 | 0.001210175 | 0.001821833 | 1 |
| HTN | nk_cell | *CEP135* | -2.19184024 | 0.014221825 | 0.018173373 | 1 |
| HTN | nk_cell | *NLRP6* | 7.557960415 | 0.001588057 | 0.002334925 | 1 |
| HTN | nk_cell | *RSRC1* | -2.133252587 | 2.66E-06 | 1.09E-05 | 0.012885415 |
| HTN | nk_cell | *PTDSS2* | -1.826978959 | 2.99E-05 | 7.00E-05 | 0.144948782 |
| HTN | nk_cell | *SEZ6L2* | 3.831880527 | 4.37E-08 | 2.12E-06 | 0.000212183 |
| HTN | nk_cell | *DHX36* | -2.024193238 | 0.003891273 | 0.005374454 | 1 |
| HTN | nk_cell | *SLC22A1* | -3.481340581 | 1.05E-06 | 6.11E-06 | 0.005093285 |
| HTN | nk_cell | *CHST2* | 0.538247826 | 4.81E-07 | 4.32E-06 | 0.002335791 |
| HTN | nk_cell | *TVP23C* | 4.992747246 | 1.70E-06 | 8.06E-06 | 0.008232694 |
| HTN | nk_cell | *CADM2* | -1.993382024 | 6.96E-07 | 4.87E-06 | 0.003376262 |
| HTN | nk_cell | *PSMD2* | -2.042162266 | 4.91E-07 | 4.32E-06 | 0.002382239 |
| HTN | nk_cell | *ATG13* | -2.680256993 | 0.000490888 | 0.00079792 | 1 |
| HTN | nk_cell | *CCNE2* | -1.903557821 | 5.48E-11 | 1.48E-08 | 2.66E-07 |
| HTN | nk_cell | *ZNF519* | -1.697789658 | 3.52E-06 | 1.33E-05 | 0.017098757 |
| HTN | nk_cell | *BANF1* | 3.49693681 | 0.024388127 | 0.030294724 | 1 |
| HTN | nk_cell | *CHRNA7* | -2.385682535 | 1.08E-05 | 3.10E-05 | 0.052558935 |
| HTN | nk_cell | *TMEM9B* | -2.012934675 | 0.000255754 | 0.000443344 | 1 |
| HTN | nk_cell | *SCUBE2* | -1.854168791 | 1.65E-06 | 7.94E-06 | 0.008016401 |
| HTN | nk_cell | *EIF1AD* | -2.232153321 | 3.03E-07 | 4.15E-06 | 0.001472333 |
| HTN | nk_cell | *EIF3F* | 2.559688096 | 0.000964152 | 0.001481807 | 1 |
| HTN | nk_cell | *ZNF25* | -2.553350095 | 0.000281863 | 0.000483421 | 1 |
| HTN | nk_cell | *CLTB* | -2.8603528 | 0.002672522 | 0.003800234 | 1 |
| HTN | nk_cell | *PPP2R2D* | -2.354276232 | 0.002952244 | 0.004170099 | 1 |
| HTN | nk_cell | *MCTP1* | -1.740829263 | 2.02E-05 | 5.08E-05 | 0.098136774 |
| HTN | nk_cell | *LRRC25* | -1.367398244 | 0.007559909 | 0.010033008 | 1 |
| HTN | nk_cell | *UBQLNL* | -1.549500428 | 0.012973212 | 0.016678861 | 1 |
| HTN | nk_cell | *KCNE3* | -2.032650731 | 0.025545616 | 0.031667688 | 1 |
| HTN | nk_cell | *UCP2* | 1.895919101 | 0.004692672 | 0.006410148 | 1 |
| HTN | nk_cell | *C11orf68* | 4.173819615 | 1.10E-06 | 6.24E-06 | 0.005317987 |
| HTN | nk_cell | *MRPL48* | -2.891212183 | 6.93E-05 | 0.000140531 | 0.336008582 |
| HTN | nk_cell | *RAB6A* | -2.06308868 | 0.000463369 | 0.000757247 | 1 |
| HTN | nk_cell | *TMEM70* | -2.556068155 | 4.27E-06 | 1.53E-05 | 0.020734467 |
| HTN | nk_cell | *RPS6KB2* | 1.882656774 | 7.16E-07 | 4.95E-06 | 0.003474954 |
| HTN | nk_cell | *TOM1L2* | -1.801468022 | 5.77E-06 | 1.91E-05 | 0.027988201 |
| HTN | nk_cell | *TEX26* | -4.433188974 | 9.51E-06 | 2.79E-05 | 0.046139493 |
| HTN | nk_cell | *GPR156* | -4.006167041 | 2.74E-05 | 6.49E-05 | 0.13281795 |
| HTN | nk_cell | *TTLL11* | -2.473110408 | 0.000125132 | 0.000235418 | 0.607141897 |

| | | | | | | |
|---|---|---|---|---|---|---|
| HTN | nk_cell | *PRIMA1* | -1.170634297 | 4.58E-07 | 4.29E-06 | 0.002220892 |
| HTN | nk_cell | *CTDNEP1* | 4.161294985 | 2.68E-09 | 3.61E-07 | 1.30E-05 |
| HTN | nk_cell | *HOXD8* | -1.644919066 | 8.64E-05 | 0.000169785 | 0.419029193 |
| HTN | nk_cell | *ZDHHC21* | -1.938681312 | 0.001043007 | 0.001590405 | 1 |
| HTN | nk_cell | *DOK7* | 5.173022485 | 1.97E-07 | 4.07E-06 | 0.000956698 |
| HTN | nk_cell | *ORAI3* | 2.1303245 | 1.06E-10 | 2.46E-08 | 5.16E-07 |
| HTN | nk_cell | *B3GALT6* | 7.295700825 | 0.026379712 | 0.03259342 | 1 |
| HTN | nk_cell | *ZNF613* | -1.941721906 | 3.94E-06 | 1.45E-05 | 0.019114597 |
| HTN | nk_cell | *JAKMIP2* | 0.246674772 | 4.20E-07 | 4.24E-06 | 0.00203929 |
| HTN | nk_cell | *TCP11L1* | -1.639742771 | 0.002335302 | 0.003352333 | 1 |
| HTN | nk_cell | *CCDC57* | -1.770279288 | 3.61E-05 | 8.17E-05 | 0.175011634 |
| HTN | nk_cell | *MYPOP* | -1.965159895 | 3.34E-07 | 4.24E-06 | 0.001618553 |
| HTN | nk_cell | *CIDEA* | -1.097100421 | 8.89E-07 | 5.64E-06 | 0.004312642 |
| HTN | nk_cell | *SLC35G1* | -2.560685509 | 3.91E-06 | 1.44E-05 | 0.018968446 |
| HTN | nk_cell | *OR4N2* | -3.081837087 | 4.56E-06 | 1.61E-05 | 0.022106836 |
| HTN | nk_cell | *TAC4* | -3.090907757 | 4.25E-05 | 9.34E-05 | 0.206018002 |
| HTN | nk_cell | *CDC26* | -1.666229493 | 4.16E-06 | 1.51E-05 | 0.020207053 |
| HTN | nk_cell | *SPRYD4* | -1.795835226 | 2.59E-05 | 6.20E-05 | 0.125632158 |
| HTN | nk_cell | *VPS37D* | -1.841135636 | 2.51E-05 | 6.06E-05 | 0.121865393 |
| HTN | nk_cell | *ZNF575* | -0.623962343 | 0.027119576 | 0.033447937 | 1 |
| HTN | nk_cell | *WDR25* | -2.56407711 | 2.38E-05 | 5.81E-05 | 0.115444497 |
| HTN | nk_cell | *PLA2G16* | -1.080323548 | 0.000131824 | 0.000246764 | 0.639611234 |
| HTN | nk_cell | *B3GNT5* | -1.901907498 | 1.52E-06 | 7.55E-06 | 0.007353559 |
| HTN | nk_cell | *RMDN1* | -1.918404629 | 3.37E-05 | 7.69E-05 | 0.163276781 |
| HTN | nk_cell | *RNF152* | -1.891504437 | 2.30E-07 | 4.07E-06 | 0.001117946 |
| HTN | nk_cell | *LRRC37A* | -1.242345407 | 0.007110472 | 0.009478025 | 1 |
| HTN | nk_cell | *BDNF* | -2.413587607 | 2.80E-07 | 4.07E-06 | 0.001360256 |
| HTN | nk_cell | *BOK* | -1.705287684 | 1.23E-06 | 6.75E-06 | 0.00597239 |
| HTN | nk_cell | *RUFY1* | -1.828897172 | 2.14E-05 | 5.31E-05 | 0.103695178 |
| HTN | nk_cell | *VSIG10* | -1.572460607 | 7.01E-06 | 2.20E-05 | 0.034022025 |
| HTN | nk_cell | *ANKLE2* | -2.508117877 | 1.30E-05 | 3.58E-05 | 0.063259718 |
| HTN | nk_cell | *C8G* | -22.55953177 | 4.12E-07 | 4.24E-06 | 0.002000331 |
| HTN | nk_cell | *MUC20* | 3.560479261 | 1.97E-07 | 4.07E-06 | 0.000957764 |
| HTN | nk_cell | *LY6H* | -1.811987791 | 0.001936514 | 0.002803929 | 1 |
| HTN | nk_cell | *SHMT1* | -2.189661032 | 0.003859851 | 0.005337132 | 1 |
| HTN | nk_cell | *DPP7* | -2.465007097 | 0.007377963 | 0.009815705 | 1 |
| HTN | nk_cell | *MTHFR* | -3.16127916 | 3.62E-08 | 1.98E-06 | 0.000175458 |
| HTN | nk_cell | *C19orf18* | -1.980321847 | 1.37E-05 | 3.74E-05 | 0.066562747 |
| HTN | nk_cell | *TMEM80* | -2.655513587 | 3.88E-06 | 1.43E-05 | 0.018816911 |
| HTN | nk_cell | *WDR73* | -2.089187527 | 2.64E-05 | 6.30E-05 | 0.128167917 |
| HTN | nk_cell | *DSCAML1* | -1.120791763 | 8.03E-07 | 5.27E-06 | 0.00389437 |
| HTN | nk_cell | *TALDO1* | -2.893291886 | 0.000432784 | 0.000710856 | 1 |
| HTN | nk_cell | *AP006621.1* | -1.302396644 | 7.75E-07 | 5.19E-06 | 0.003762282 |
| HTN | nk_cell | *MAN1B1* | -3.162105019 | 7.11E-05 | 0.00014374 | 0.345119929 |
| HTN | nk_cell | *FBXO39* | -1.359369291 | 0.007908457 | 0.010464094 | 1 |
| HTN | nk_cell | *MIEF2* | -2.480543073 | 8.10E-07 | 5.29E-06 | 0.003931447 |
| HTN | nk_cell | *NAP1L5* | -2.064155719 | 6.60E-07 | 4.73E-06 | 0.003204088 |
| HTN | nk_cell | *RBM44* | 4.452987845 | 0.000474464 | 0.000773814 | 1 |
| HTN | nk_cell | *SLC25A22* | 1.923058222 | 1.49E-05 | 4.01E-05 | 0.072387272 |
| HTN | nk_cell | *GBA* | -1.757033963 | 5.11E-07 | 4.37E-06 | 0.002477168 |

| | | | | | | |
|---|---|---|---|---|---|---|
| HTN | nk_cell | *ACAD9* | -1.386793451 | 7.58E-05 | 0.000151779 | 0.367912329 |
| HTN | nk_cell | *IL17RA* | -2.203127819 | 0.000156356 | 0.000286603 | 0.758637574 |
| HTN | nk_cell | *MBOAT4* | -2.847187218 | 1.43E-05 | 3.88E-05 | 0.069591347 |
| HTN | nk_cell | *AGTRAP* | 1.914636395 | 7.73E-07 | 5.19E-06 | 0.003748804 |
| HTN | nk_cell | *THAP5* | -3.260712644 | 0.007528563 | 0.009999613 | 1 |
| HTN | nk_cell | *NAALADL2* | -1.382696931 | 6.12E-05 | 0.000126386 | 0.297133167 |
| HTN | nk_cell | *CD151* | -1.844242111 | 0.003904045 | 0.005387076 | 1 |
| HTN | nk_cell | *FAM87B* | -9.237784724 | 5.00E-07 | 4.35E-06 | 0.002426828 |
| HTN | nk_cell | *PCDHB9* | 3.608394492 | 0.000626365 | 0.00099483 | 1 |
| HTN | nk_cell | *CENPBD1* | 5.012797867 | 6.35E-07 | 4.62E-06 | 0.003082902 |
| HTN | nk_cell | *RPS27* | 1.651209506 | 7.83E-07 | 5.20E-06 | 0.003798052 |
| HTN | nk_cell | *RIC8A* | 1.905091403 | 1.77E-06 | 8.28E-06 | 0.008590202 |
| HTN | nk_cell | *ASB8* | 2.960643931 | 0.012363694 | 0.015941707 | 1 |
| HTN | nk_cell | *ODF3B* | -0.935188463 | 0.000111299 | 0.00021194 | 0.540022335 |
| HTN | nk_cell | *DPY19L2* | -0.990582551 | 1.43E-06 | 7.30E-06 | 0.006940049 |
| HTN | nk_cell | *ADAMTSL1* | -1.19308904 | 1.55E-06 | 7.63E-06 | 0.007521443 |
| HTN | nk_cell | *GRAMD1C* | -1.58146538 | 6.19E-05 | 0.000127533 | 0.300467823 |
| HTN | nk_cell | *DALRD3* | -2.595401551 | 1.03E-06 | 6.04E-06 | 0.004991526 |
| HTN | nk_cell | *ZNF114* | -3.74098259 | 0.008096987 | 0.01069896 | 1 |
| HTN | nk_cell | *SPINK6* | -4.322872621 | 2.42E-05 | 5.89E-05 | 0.117323561 |
| HTN | nk_cell | *SH2B1* | -2.265376954 | 2.20E-05 | 5.43E-05 | 0.106741222 |
| HTN | nk_cell | *VN1R1* | -5.195507208 | 1.12E-05 | 3.19E-05 | 0.054501329 |
| HTN | nk_cell | *KDELC2* | -3.768114823 | 0.001192322 | 0.001797187 | 1 |
| HTN | nk_cell | *PLEC* | -2.309825802 | 6.01E-06 | 1.97E-05 | 0.029173289 |
| HTN | nk_cell | *SH2D4B* | -3.597926289 | 9.48E-07 | 5.82E-06 | 0.004601153 |
| HTN | nk_cell | *PRSS36* | -4.828865633 | 0.000404545 | 0.000669458 | 1 |
| HTN | nk_cell | *WDR6* | -0.94760033 | 0.031478462 | 0.038588554 | 1 |
| HTN | nk_cell | *TMPRSS9* | -1.310588791 | 3.57E-07 | 4.24E-06 | 0.001730745 |
| HTN | nk_cell | *TMEM11* | -1.498264047 | 8.96E-07 | 5.67E-06 | 0.004348335 |
| HTN | nk_cell | *ZFAND2A* | -2.11848741 | 8.88E-08 | 3.10E-06 | 0.000430904 |
| HTN | nk_cell | *ZNF223* | -3.144231216 | 7.36E-06 | 2.29E-05 | 0.035703787 |
| HTN | nk_cell | *FAM220A* | -2.979432547 | 6.68E-06 | 2.13E-05 | 0.03241373 |
| HTN | nk_cell | *NT5DC1* | -2.086958014 | 0.001352103 | 0.00201116 | 1 |
| HTN | nk_cell | *GLDC* | -1.782856019 | 6.45E-07 | 4.66E-06 | 0.003131853 |
| HTN | nk_cell | *P4HTM* | -2.207523131 | 4.87E-07 | 4.32E-06 | 0.002365161 |
| HTN | nk_cell | *CA8* | -2.735168442 | 4.55E-06 | 1.61E-05 | 0.022084741 |
| HTN | nk_cell | *ZNF713* | -0.962579552 | 0.002293951 | 0.003294923 | 1 |
| HTN | nk_cell | *PARP10* | -2.087820627 | 8.28E-07 | 5.35E-06 | 0.004017097 |
| HTN | nk_cell | *NSUN3* | -2.279877356 | 3.23E-05 | 7.46E-05 | 0.156721048 |
| HTN | nk_cell | *RPP25* | 2.161767514 | 5.72E-12 | 1.88E-09 | 2.78E-08 |
| HTN | nk_cell | *STX19* | -1.755773848 | 4.25E-06 | 1.52E-05 | 0.020610375 |
| HTN | nk_cell | *MPI* | -6.579051105 | 3.03E-17 | 7.36E-14 | 1.47E-13 |
| HTN | nk_cell | *TRIM73* | -4.048908099 | 0.018140861 | 0.022856261 | 1 |
| HTN | nk_cell | *EFCAB13* | -1.79007269 | 1.10E-05 | 3.13E-05 | 0.05334001 |
| HTN | nk_cell | *EXOSC4* | -4.572033166 | 0.001874538 | 0.002717436 | 1 |
| HTN | nk_cell | *ZNF852* | -3.08101498 | 0.001838901 | 0.002669832 | 1 |
| HTN | nk_cell | *HYI* | -2.59253449 | 5.82E-12 | 1.88E-09 | 2.82E-08 |
| HTN | nk_cell | *C17orf62* | 3.994795861 | 7.33E-05 | 0.000147235 | 0.355424627 |
| HTN | nk_cell | *GAK* | -2.218360509 | 0.005767971 | 0.007778264 | 1 |
| HTN | nk_cell | *CTC1* | 4.477662774 | 3.39E-07 | 4.24E-06 | 0.001642529 |

| | | | | | | |
|---|---|---|---|---|---|---|
| HTN | nk_cell | *C3orf38* | -2.110187214 | 8.96E-06 | 2.67E-05 | 0.043489941 |
| HTN | nk_cell | *CCDC89* | -3.465861364 | 4.45E-07 | 4.28E-06 | 0.002157025 |
| HTN | nk_cell | *HTR1F* | -4.245561362 | 2.27E-08 | 1.38E-06 | 0.000110202 |
| HTN | nk_cell | *ZNF664* | -2.757653978 | 4.18E-06 | 1.51E-05 | 0.020298325 |
| HTN | nk_cell | *CDH4* | -2.250213615 | 1.49E-07 | 3.99E-06 | 0.000721404 |
| HTN | nk_cell | *NSUN7* | -1.084369106 | 0.000874731 | 0.001353378 | 1 |
| HTN | nk_cell | *PACS2* | -2.13700923 | 1.00E-05 | 2.91E-05 | 0.048729872 |
| HTN | nk_cell | *LINC00174* | -1.671769317 | 0.000419857 | 0.0006922 | 1 |
| HTN | nk_cell | *GEMIN4* | -3.837669298 | 4.19E-06 | 1.51E-05 | 0.020334431 |
| HTN | nk_cell | *LBX2* | 1.249622542 | 8.67E-05 | 0.000170227 | 0.420460641 |
| HTN | nk_cell | *ZFPM1* | 1.303484301 | 2.23E-06 | 9.67E-06 | 0.010831695 |
| HTN | nk_cell | *FCER1A* | -1.85190136 | 4.46E-07 | 4.28E-06 | 0.00216273 |
| HTN | nk_cell | *RPRML* | 1.59104579 | 1.32E-07 | 3.84E-06 | 0.000642677 |
| HTN | nk_cell | *SERTAD2* | -2.059535709 | 0.007404663 | 0.009840434 | 1 |
| HTN | nk_cell | *AKAP5* | 0.968568149 | 4.33E-07 | 4.25E-06 | 0.002101903 |
| HTN | nk_cell | *GIPC3* | 6.311449051 | 2.86E-07 | 4.07E-06 | 0.001389893 |
| HTN | nk_cell | *PDXDC1* | -1.689691546 | 1.80E-05 | 4.65E-05 | 0.08732739 |
| HTN | nk_cell | *C1orf194* | -3.603825664 | 0.003660813 | 0.005088016 | 1 |
| HTN | nk_cell | *SEPHS2* | -2.005221975 | 4.79E-05 | 0.000103479 | 0.232621816 |
| HTN | nk_cell | *ZADH2* | -2.423588939 | 6.21E-05 | 0.000127899 | 0.301458926 |
| HTN | nk_cell | *SEPT1* | 3.237227287 | 4.51E-06 | 1.60E-05 | 0.021881984 |
| HTN | nk_cell | *TDRD6* | -0.789183195 | 0.003723628 | 0.005168675 | 1 |
| HTN | nk_cell | *CSNK1A1L* | 7.834606288 | 3.16E-06 | 1.23E-05 | 0.015343451 |
| HTN | nk_cell | *LYNX1* | -1.369679069 | 1.14E-05 | 3.22E-05 | 0.055074461 |
| HTN | nk_cell | *FAHD1* | -0.662166599 | 0.008755351 | 0.011511011 | 1 |
| HTN | nk_cell | *HCLS1* | 1.087438307 | 1.30E-05 | 3.57E-05 | 0.063020146 |
| HTN | nk_cell | *MTURN* | -1.609421848 | 4.12E-06 | 1.49E-05 | 0.019970651 |
| HTN | nk_cell | *CCDC66* | -1.956146676 | 9.22E-06 | 2.72E-05 | 0.044715498 |
| HTN | nk_cell | *DEFB124* | -11.14227036 | 9.88E-06 | 2.87E-05 | 0.047915618 |
| HTN | nk_cell | *GLIPR1L2* | -1.26712246 | 2.78E-06 | 1.12E-05 | 0.013505212 |
| HTN | nk_cell | *DEFB119* | -16.54013418 | 1.45E-06 | 7.37E-06 | 0.007042007 |
| HTN | nk_cell | *TSPYL5* | -2.324362101 | 4.28E-07 | 4.24E-06 | 0.002074388 |
| HTN | nk_cell | *MB21D2* | -1.766666095 | 4.61E-06 | 1.62E-05 | 0.022362524 |
| HTN | nk_cell | *SLC47A2* | -2.472483972 | 2.42E-05 | 5.89E-05 | 0.11740787 |
| HTN | nk_cell | *CHST13* | -8.690179077 | 1.65E-06 | 7.94E-06 | 0.007993878 |
| HTN | nk_cell | *SLC36A4* | -2.257409986 | 9.83E-06 | 2.86E-05 | 0.04769153 |
| HTN | nk_cell | *OR51E1* | -1.976893603 | 1.39E-06 | 7.20E-06 | 0.006768517 |
| HTN | nk_cell | *PSMG4* | -2.011104109 | 9.42E-06 | 2.77E-05 | 0.045704583 |
| HTN | nk_cell | *ZNF443* | -3.577430912 | 2.54E-07 | 4.07E-06 | 0.001230736 |
| HTN | nk_cell | *CAPS2* | -2.050517694 | 0.000188289 | 0.000338613 | 0.913578806 |
| HTN | nk_cell | *KCTD2* | -1.600025097 | 5.44E-07 | 4.44E-06 | 0.002638775 |
| HTN | nk_cell | *D2HGDH* | -2.305807532 | 9.07E-07 | 5.69E-06 | 0.004399534 |
| HTN | nk_cell | *CMTR2* | -2.397002612 | 0.002592854 | 0.003694722 | 1 |
| HTN | nk_cell | *ST20* | 0.747969453 | 0.000630345 | 0.001000468 | 1 |
| HTN | nk_cell | *MRPL14* | -2.653630777 | 0.004150157 | 0.005702793 | 1 |
| HTN | nk_cell | *NQO1* | -2.460536193 | 0.000569972 | 0.000914822 | 1 |
| HTN | nk_cell | *FKRP* | -1.201180609 | 0.001422359 | 0.002106619 | 1 |
| HTN | nk_cell | *RPH3AL* | -1.937843595 | 0.00255197 | 0.003641812 | 1 |
| HTN | nk_cell | *OR52N4* | -2.879933665 | 0.030277465 | 0.037163233 | 1 |
| HTN | nk_cell | *ADIPOQ* | -0.776985886 | 3.15E-05 | 7.32E-05 | 0.152941503 |

| | | | | | | |
|---|---|---|---|---|---|---|
| HTN | nk_cell | *F2R* | 0.575196626 | 8.34E-08 | 3.01E-06 | 0.000404838 |
| HTN | nk_cell | *MUC16* | -2.744371361 | 5.19E-05 | 0.000110099 | 0.251685953 |
| HTN | nk_cell | *SLC25A41* | 15.55864953 | 2.18E-06 | 9.50E-06 | 0.010576333 |
| HTN | nk_cell | *TMEM136* | -1.925792831 | 3.36E-05 | 7.68E-05 | 0.162890677 |
| HTN | nk_cell | *DDX60L* | -2.273539267 | 0.001094611 | 0.001662301 | 1 |
| HTN | nk_cell | *TMEM45A* | -1.940202465 | 9.59E-06 | 2.81E-05 | 0.046533576 |
| HTN | nk_cell | *SGSH* | -3.72688908 | 7.33E-06 | 2.28E-05 | 0.035543644 |
| HTN | nk_cell | *ZFP41* | 2.641473202 | 4.55E-09 | 4.80E-07 | 2.21E-05 |
| HTN | nk_cell | *ZBTB20* | -1.410345679 | 0.000171305 | 0.000311067 | 0.831171874 |
| HTN | nk_cell | *AMIGO1* | -1.918931424 | 6.29E-05 | 0.000129188 | 0.305400027 |
| HTN | nk_cell | *TMEM252* | -41.20956605 | 5.14E-07 | 4.37E-06 | 0.002495164 |
| HTN | nk_cell | *ODF3L2* | 3.105743722 | 1.49E-06 | 7.48E-06 | 0.007207382 |
| HTN | nk_cell | *RELL1* | -1.998702268 | 0.000175648 | 0.000318002 | 0.852245844 |
| HTN | nk_cell | *RFX7* | -1.981257222 | 0.003258518 | 0.004565501 | 1 |
| HTN | nk_cell | *CLDN7* | -1.798711714 | 2.76E-10 | 5.57E-08 | 1.34E-06 |
| HTN | nk_cell | *ZNF329* | -1.70828344 | 1.37E-06 | 7.14E-06 | 0.006661003 |
| HTN | nk_cell | *ADO* | 5.763389835 | 1.32E-13 | 1.28E-10 | 6.42E-10 |
| HTN | nk_cell | *MGAT4C* | -1.207348092 | 0.005204468 | 0.007051683 | 1 |
| HTN | nk_cell | *IDH2* | -3.263350617 | 0.002209731 | 0.003178658 | 1 |
| HTN | nk_cell | *PTCHD3* | -1.755996348 | 0.008893017 | 0.011680811 | 1 |
| HTN | nk_cell | *WRB* | -1.522010172 | 8.13E-05 | 0.000161362 | 0.394692239 |
| HTN | nk_cell | *FAM181B* | -3.184556312 | 0.000952528 | 0.001465334 | 1 |
| HTN | nk_cell | *TDRKH* | -1.839975775 | 7.22E-05 | 0.000145634 | 0.350540864 |
| HTN | nk_cell | *ZNF708* | -2.293350141 | 5.82E-06 | 1.93E-05 | 0.028223919 |
| HTN | nk_cell | *ERCC6L2* | -1.957596934 | 0.000401644 | 0.000665521 | 1 |
| HTN | nk_cell | *MRPL41* | -2.866382809 | 6.15E-06 | 2.00E-05 | 0.029838442 |
| HTN | nk_cell | *HHIPL1* | -2.562687469 | 1.17E-05 | 3.29E-05 | 0.056674275 |
| HTN | nk_cell | *FAM153B* | -2.013560352 | 8.51E-06 | 2.56E-05 | 0.041276548 |
| HTN | nk_cell | *BACE2* | -1.124142226 | 0.000704037 | 0.001108726 | 1 |
| HTN | nk_cell | *SYNM* | -3.467953519 | 0.011424172 | 0.014808999 | 1 |
| HTN | nk_cell | *FIGN* | -1.79914471 | 0.000166141 | 0.000303166 | 0.806117361 |
| HTN | nk_cell | *IZUMO1* | 1.567214971 | 0.002495797 | 0.003567945 | 1 |
| HTN | nk_cell | *FBXL6* | 5.179317459 | 0.000136649 | 0.000254812 | 0.663020649 |
| HTN | nk_cell | *NXPH4* | 16.05503422 | 3.85E-07 | 4.24E-06 | 0.001869907 |
| HTN | nk_cell | *NPLOC4* | -1.859015761 | 0.000686804 | 0.001082994 | 1 |
| HTN | nk_cell | *CAPN12* | 1.023971069 | 1.04E-07 | 3.38E-06 | 0.000503301 |
| HTN | nk_cell | *KPNA2* | -2.482668542 | 0.000129645 | 0.000243154 | 0.629039781 |
| HTN | nk_cell | *ORAI1* | 2.168356927 | 4.42E-08 | 2.12E-06 | 0.000214224 |
| HTN | nk_cell | *FES* | -4.19260684 | 6.53E-15 | 1.06E-11 | 3.17E-11 |
| HTN | nk_cell | *GLRX5* | -3.205368146 | 0.010999316 | 0.014289252 | 1 |
| HTN | nk_cell | *CAV3* | -1.54545519 | 8.79E-08 | 3.09E-06 | 0.000426355 |
| HTN | nk_cell | *MXRA7* | -1.120624572 | 0.035686258 | 0.043483105 | 1 |
| HTN | nk_cell | *MFSD5* | 1.643392768 | 1.28E-05 | 3.53E-05 | 0.062099129 |
| HTN | nk_cell | *ADI1* | -2.820857495 | 0.002478504 | 0.003546359 | 1 |
| HTN | nk_cell | *CLEC4G* | -5.284163127 | 3.02E-07 | 4.15E-06 | 0.001464728 |
| HTN | nk_cell | *TRAK1* | -1.97758657 | 0.000993472 | 0.001520608 | 1 |
| HTN | nk_cell | *SKA2* | -2.184465263 | 0.025688719 | 0.031828821 | 1 |
| HTN | nk_cell | *TTC3* | -0.792359716 | 0.019660077 | 0.024706215 | 1 |
| HTN | nk_cell | *PPP1R27* | 2.841726024 | 2.40E-07 | 4.07E-06 | 0.001163083 |
| HTN | nk_cell | *BRICD5* | -2.421820812 | 4.78E-07 | 4.32E-06 | 0.002317427 |

| | | | | | | |
|---|---|---|---|---|---|---|
| HTN | nk_cell | *GRID1* | -1.351904415 | 9.74E-08 | 3.29E-06 | 0.000472367 |
| HTN | nk_cell | *ACBD3* | -2.26925535 | 0.001318704 | 0.001968118 | 1 |
| HTN | nk_cell | *PLCXD3* | -2.563372052 | 0.000182681 | 0.000329383 | 0.886369679 |
| HTN | nk_cell | *GPIHBP1* | -1.212675472 | 0.000151479 | 0.000278929 | 0.734978338 |
| HTN | nk_cell | *VMO1* | 2.036063653 | 0.002464635 | 0.003527554 | 1 |
| HTN | nk_cell | *ALG12* | 2.671983926 | 1.18E-06 | 6.58E-06 | 0.005727891 |
| HTN | nk_cell | *COL18A1* | -1.573127883 | 0.000605176 | 0.000966847 | 1 |
| HTN | nk_cell | *ZNF721* | -1.496931363 | 0.017922742 | 0.02259905 | 1 |
| HTN | nk_cell | *CEP63* | -1.572700998 | 5.98E-07 | 4.54E-06 | 0.00290127 |
| HTN | nk_cell | *HMGN4* | -2.021704063 | 3.64E-05 | 8.23E-05 | 0.176606963 |
| HTN | nk_cell | *ZNF320* | 0.842688965 | 0.008718286 | 0.011466827 | 1 |
| HTN | nk_cell | *C12orf60* | -2.719715195 | 6.37E-06 | 2.06E-05 | 0.030905513 |
| HTN | nk_cell | *ABAT* | -2.820946551 | 0.001972109 | 0.002851852 | 1 |
| HTN | nk_cell | *LYSMD4* | -1.970124259 | 3.59E-07 | 4.24E-06 | 0.001740859 |
| HTN | nk_cell | *WBP2NL* | -2.523540111 | 7.58E-07 | 5.11E-06 | 0.003676281 |
| HTN | nk_cell | *BEGAIN* | -2.102510831 | 9.92E-07 | 5.94E-06 | 0.004814458 |
| HTN | nk_cell | *ARHGEF37* | -3.23134832 | 0.000184064 | 0.000331383 | 0.893077031 |
| HTN | nk_cell | *CSMD1* | -1.972880866 | 1.61E-06 | 7.81E-06 | 0.007812759 |
| HTN | nk_cell | *RABIF* | 3.627673804 | 1.01E-06 | 5.98E-06 | 0.004890565 |
| HTN | nk_cell | *FANCF* | 4.455266464 | 6.20E-07 | 4.57E-06 | 0.003010072 |
| HTN | nk_cell | *SMDT1* | 1.404146163 | 0.003815746 | 0.005283675 | 1 |
| HTN | nk_cell | *C2CD4C* | -1.202771395 | 0.001029213 | 0.001570359 | 1 |
| HTN | nk_cell | *RUVBL2* | 3.034843188 | 1.87E-06 | 8.58E-06 | 0.00905302 |
| HTN | nk_cell | *GDPGP1* | -1.704909107 | 0.001774764 | 0.00258283 | 1 |
| HTN | nk_cell | *PTTG1IP* | -1.766150445 | 0.00283606 | 0.00401651 | 1 |
| HTN | nk_cell | *PLGLB1* | -1.413750462 | 0.016094865 | 0.020426965 | 1 |
| HTN | nk_cell | *ZNF623* | -1.548957975 | 1.30E-06 | 6.89E-06 | 0.00629483 |
| HTN | nk_cell | *CCDC125* | -1.730685467 | 9.41E-07 | 5.82E-06 | 0.004565714 |
| HTN | nk_cell | *C10orf107* | -0.92133665 | 0.000255239 | 0.000442608 | 1 |
| HTN | nk_cell | *FHL3* | -1.582952022 | 0.00165351 | 0.00242016 | 1 |
| HTN | nk_cell | *C19orf71* | -1.52403715 | 3.99E-07 | 4.24E-06 | 0.001935987 |
| HTN | nk_cell | *LRIT3* | -7.265810775 | 4.22E-07 | 4.24E-06 | 0.00204787 |
| HTN | nk_cell | *NPIPA1* | -0.621117068 | 0.003652278 | 0.005077608 | 1 |
| HTN | nk_cell | *SF3A3* | -1.994595363 | 0.002499401 | 0.003572046 | 1 |
| HTN | nk_cell | *GPR132* | 2.35636097 | 0.000646929 | 0.001024111 | 1 |
| HTN | nk_cell | *PSMG1* | -2.261211504 | 0.001561408 | 0.002298529 | 1 |
| HTN | nk_cell | *AC131971.1* | -7.038414158 | 3.44E-08 | 1.96E-06 | 0.000166772 |
| HTN | nk_cell | *SERHL2* | -0.920706389 | 4.09E-07 | 4.24E-06 | 0.001985147 |
| HTN | nk_cell | *PCBP3* | -0.934783374 | 0.000300642 | 0.000512369 | 1 |
| HTN | nk_cell | *MRPL54* | 2.080758397 | 2.27E-05 | 5.59E-05 | 0.11013902 |
| HTN | nk_cell | *HMCES* | -2.194689545 | 0.009975289 | 0.013035309 | 1 |
| HTN | nk_cell | *NDUFB1* | -2.259626977 | 4.57E-05 | 9.92E-05 | 0.221635579 |
| HTN | nk_cell | *FAM19A1* | 0.983872306 | 4.59E-07 | 4.29E-06 | 0.002228311 |
| HTN | nk_cell | *TRMT12* | 3.465982408 | 0.030359502 | 0.037254502 | 1 |
| HTN | nk_cell | *ALYREF* | 3.028735212 | 4.99E-05 | 0.000106825 | 0.242066581 |
| HTN | nk_cell | *MRGPRX2* | 3.260568242 | 1.90E-07 | 4.07E-06 | 0.000922778 |
| HTN | nk_cell | *UPP1* | 1.485850033 | 2.05E-07 | 4.07E-06 | 0.000992479 |
| HTN | nk_cell | *FIGLA* | -5.493387387 | 9.92E-08 | 3.32E-06 | 0.000481485 |
| HTN | nk_cell | *ACSM2A* | -9.211093192 | 8.81E-07 | 5.61E-06 | 0.00427617 |
| HTN | nk_cell | *KREMEN1* | -1.615314236 | 6.47E-08 | 2.51E-06 | 0.000314148 |

| | | | | | | |
|---|---|---|---|---|---|---|
| HTN | nk_cell | *ZNF703* | 2.177685887 | 0.009319696 | 0.012211495 | 1 |
| HTN | nk_cell | *EMILIN3* | -5.077364439 | 5.50E-07 | 4.45E-06 | 0.002667471 |
| HTN | nk_cell | *FAM162B* | -1.609534687 | 0.003724173 | 0.005168675 | 1 |
| HTN | nk_cell | *BTBD9* | -1.766441949 | 0.000317675 | 0.00053856 | 1 |
| HTN | nk_cell | *NUDT14* | 3.227950733 | 0.003323628 | 0.004648672 | 1 |
| HTN | nk_cell | *FAM3B* | -0.465269139 | 0.014021538 | 0.017931604 | 1 |
| HTN | nk_cell | *NPIPA7* | -2.151617876 | 0.02625458 | 0.032455343 | 1 |
| HTN | nk_cell | *PP2D1* | -2.85422174 | 2.85E-06 | 1.14E-05 | 0.013851662 |
| HTN | nk_cell | *DENND5A* | -2.119102875 | 1.28E-05 | 3.52E-05 | 0.061960745 |
| HTN | nk_cell | *CNTN2* | -1.559487198 | 5.61E-06 | 1.88E-05 | 0.027234316 |
| HTN | nk_cell | *CRELD2* | 3.156140509 | 3.35E-06 | 1.28E-05 | 0.016246214 |
| HTN | nk_cell | *GOLGA6L4* | -2.138427813 | 5.99E-08 | 2.40E-06 | 0.000290838 |
| HTN | nk_cell | *PGP* | -1.902589187 | 0.033493664 | 0.040955458 | 1 |
| HTN | nk_cell | *OAF* | -0.946202126 | 0.000474936 | 0.000774324 | 1 |
| HTN | nk_cell | *ALDH1A3* | -1.069283512 | 8.69E-09 | 7.53E-07 | 4.22E-05 |
| HTN | nk_cell | *CLECL1* | -1.612722061 | 1.47E-07 | 3.97E-06 | 0.000714146 |
| HTN | nk_cell | *MROH7* | -1.525172181 | 0.000513954 | 0.000832068 | 1 |
| HTN | nk_cell | *PKP3* | 11.62716857 | 2.74E-06 | 1.11E-05 | 0.013286671 |
| HTN | nk_cell | *A3GALT2* | -2.629575031 | 8.87E-10 | 1.65E-07 | 4.30E-06 |
| HTN | nk_cell | *SS18L1* | -1.792250063 | 8.23E-07 | 5.33E-06 | 0.003994278 |
| HTN | nk_cell | *TOP1MT* | -1.49362594 | 6.10E-08 | 2.43E-06 | 0.000296064 |
| HTN | nk_cell | *WDR27* | -1.793817128 | 0.000213183 | 0.00037723 | 1 |
| HTN | nk_cell | *TXNRD2* | -1.864856555 | 4.10E-06 | 1.49E-05 | 0.019879544 |
| HTN | nk_cell | *TMEM255B* | 3.736265223 | 0.009991801 | 0.01305337 | 1 |
| HTN | nk_cell | *HDDC3* | -2.808314161 | 2.01E-06 | 9.00E-06 | 0.009748077 |
| HTN | nk_cell | *ZFP1* | 2.718291482 | 3.80E-06 | 1.41E-05 | 0.018426599 |
| HTN | nk_cell | *DUSP8* | -2.183203444 | 1.14E-05 | 3.22E-05 | 0.055143328 |
| HTN | nk_cell | *SNN* | 4.793146378 | 1.87E-06 | 8.58E-06 | 0.009057937 |
| HTN | nk_cell | *KRBA2* | -2.560708665 | 8.59E-06 | 2.57E-05 | 0.04167638 |
| HTN | nk_cell | *SEPT5* | -3.834167516 | 0.000825741 | 0.00128537 | 1 |
| HTN | nk_cell | *RNLS* | -1.599825983 | 5.39E-06 | 1.82E-05 | 0.026135245 |
| HTN | nk_cell | *NDUFA12* | 2.71671522 | 0.004938066 | 0.006720757 | 1 |
| HTN | nk_cell | *TCTE3* | -2.314656972 | 2.13E-06 | 9.35E-06 | 0.010326341 |
| HTN | nk_cell | *UBE2G2* | -1.784308196 | 0.00672884 | 0.00900147 | 1 |
| HTN | nk_cell | *TMED9* | 3.240517779 | 0.000491137 | 0.000798057 | 1 |
| HTN | nk_cell | *DRD1* | -23.31495396 | 5.06E-07 | 4.37E-06 | 0.002457105 |
| HTN | nk_cell | *WT1* | -1.103418833 | 7.56E-07 | 5.11E-06 | 0.00366921 |
| HTN | nk_cell | *NOC4L* | 3.479082937 | 1.86E-06 | 8.58E-06 | 0.009040871 |
| HTN | nk_cell | *SORCS2* | -1.650365667 | 4.51E-07 | 4.28E-06 | 0.002187109 |
| HTN | nk_cell | *BRI3BP* | -3.12357266 | 0.02507652 | 0.031110016 | 1 |
| HTN | nk_cell | *MAFF* | -1.329233018 | 1.68E-07 | 4.07E-06 | 0.00081321 |
| HTN | nk_cell | *SEMA4B* | -2.972596179 | 0.000125271 | 0.000235586 | 0.607813147 |
| HTN | nk_cell | *SGCZ* | -1.184323507 | 2.12E-05 | 5.28E-05 | 0.102956181 |
| HTN | nk_cell | *C6orf120* | -2.647605341 | 0.023552795 | 0.02932465 | 1 |
| HTN | nk_cell | *SIGIRR* | 1.183946366 | 1.75E-07 | 4.07E-06 | 0.000848275 |
| HTN | nk_cell | *IFITM2* | 1.942626033 | 1.07E-06 | 6.16E-06 | 0.005190769 |
| HTN | nk_cell | *GP1BA* | -1.635068691 | 1.33E-05 | 3.65E-05 | 0.064710842 |
| HTN | nk_cell | *CDK10* | -1.50870221 | 0.001644156 | 0.002408649 | 1 |
| HTN | nk_cell | *ATP6V0A2* | 2.833601341 | 4.69E-05 | 0.000101423 | 0.227391184 |
| HTN | nk_cell | *C14orf80* | 2.216325985 | 3.37E-05 | 7.69E-05 | 0.163368323 |

| | | | | | | |
|---|---|---|---|---|---|---|
| HTN | nk_cell | *TNFAIP8L1* | -4.638519771 | 4.39E-05 | 9.61E-05 | 0.212876839 |
| HTN | nk_cell | *TARSL2* | -2.129323814 | 2.35E-05 | 5.75E-05 | 0.113814398 |
| HTN | nk_cell | *SH3BGR* | -1.077319518 | 0.002000112 | 0.002887678 | 1 |
| HTN | nk_cell | *FAM174B* | -1.952952397 | 4.65E-06 | 1.63E-05 | 0.022556118 |
| HTN | nk_cell | *SV2B* | -1.62313634 | 5.44E-07 | 4.44E-06 | 0.002641249 |
| HTN | nk_cell | *AHNAK2* | -3.290047834 | 0.003833317 | 0.005306166 | 1 |
| HTN | nk_cell | *SP1* | -1.912185648 | 2.58E-05 | 6.18E-05 | 0.125169651 |
| HTN | nk_cell | *PCGF3* | -2.053383529 | 0.000613036 | 0.000977152 | 1 |
| HTN | nk_cell | *PSMD13* | -3.174905249 | 1.74E-05 | 4.53E-05 | 0.084280432 |
| HTN | nk_cell | *PMEL* | -1.29213386 | 2.78E-06 | 1.12E-05 | 0.013478103 |
| HTN | nk_cell | *C6orf201* | -2.903549103 | 2.23E-06 | 9.65E-06 | 0.01080154 |
| HTN | nk_cell | *MYBL1* | 0.156096699 | 3.47E-07 | 4.24E-06 | 0.001682757 |
| HTN | nk_cell | *ADARB2* | 2.421258323 | 5.27E-07 | 4.40E-06 | 0.002555582 |
| HTN | nk_cell | *DMWD* | -1.947874878 | 1.73E-06 | 8.16E-06 | 0.008396135 |
| HTN | nk_cell | *SLC52A2* | 2.91495794 | 1.13E-07 | 3.54E-06 | 0.000549254 |
| HTN | nk_cell | *PIGP* | -0.859488603 | 0.000935581 | 0.001442465 | 1 |
| HTN | nk_cell | *EVI2B* | 1.229042255 | 3.52E-06 | 1.33E-05 | 0.017059259 |
| HTN | nk_cell | *ZNF829* | -1.330698787 | 0.031060453 | 0.038085752 | 1 |
| HTN | nk_cell | *IFITM1* | 0.832031254 | 1.06E-06 | 6.15E-06 | 0.005163741 |
| HTN | nk_cell | *LINC00839* | -3.210267396 | 2.10E-07 | 4.07E-06 | 0.001019961 |
| HTN | nk_cell | *RTN4RL1* | -1.251486918 | 4.06E-06 | 1.48E-05 | 0.019697744 |
| HTN | nk_cell | *RNPC3* | -1.257129383 | 1.07E-05 | 3.08E-05 | 0.05206255 |
| HTN | nk_cell | *RASA3* | 1.652305891 | 0.000216776 | 0.000383168 | 1 |
| HTN | nk_cell | *ZNF529* | -2.113488713 | 3.33E-05 | 7.64E-05 | 0.161438812 |
| HTN | nk_cell | *WWOX* | -1.422080484 | 4.26E-05 | 9.35E-05 | 0.206530351 |
| HTN | nk_cell | *CYP4Z1* | -0.579981164 | 0.003167649 | 0.004453617 | 1 |
| HTN | nk_cell | *POLR1D* | -2.099546591 | 0.000158412 | 0.000290152 | 0.768612811 |
| HTN | nk_cell | *FFAR4* | -1.115782185 | 2.13E-06 | 9.35E-06 | 0.010350451 |
| HTN | nk_cell | *CYP4F12* | -2.067357818 | 4.39E-06 | 1.57E-05 | 0.02129171 |
| HTN | nk_cell | *ZNF749* | -0.724328542 | 0.012346249 | 0.015927679 | 1 |
| HTN | nk_cell | *GPAT2* | -3.537148275 | 0.000104844 | 0.000200909 | 0.508701363 |
| HTN | nk_cell | *TOR3A* | -2.944983505 | 2.81E-05 | 6.63E-05 | 0.136136965 |
| HTN | nk_cell | *ZNF555* | -2.04005887 | 2.46E-06 | 1.04E-05 | 0.011920125 |
| HTN | nk_cell | *GOLGA8R* | 3.472330154 | 0.000634209 | 0.001005944 | 1 |
| HTN | nk_cell | *CCDC30* | -1.071043968 | 0.002956465 | 0.004174845 | 1 |
| HTN | nk_cell | *GLDN* | -0.272368642 | 0.023593674 | 0.029352951 | 1 |
| HTN | nk_cell | *ZNF197* | -2.194776786 | 1.69E-05 | 4.43E-05 | 0.082041435 |
| HTN | nk_cell | *FAM228A* | 1.233551109 | 2.60E-06 | 1.08E-05 | 0.012601112 |
| HTN | nk_cell | *RPS23* | 1.643955774 | 0.01212614 | 0.015664545 | 1 |
| HTN | nk_cell | *GNG2* | 0.861530708 | 3.51E-07 | 4.24E-06 | 0.001702387 |
| HTN | nk_cell | *BTN3A2* | 0.653551403 | 0.002615019 | 0.003723026 | 1 |
| HTN | nk_cell | *TMEM222* | -3.554248314 | 0.001106582 | 0.00167838 | 1 |
| HTN | nk_cell | *SEPT10* | -2.085884311 | 4.85E-06 | 1.69E-05 | 0.02354543 |
| HTN | nk_cell | *FAM86B1* | -3.453887599 | 0.000252238 | 0.00043866 | 1 |
| HTN | nk_cell | *SMYD4* | -2.21446109 | 1.92E-07 | 4.07E-06 | 0.000929729 |
| HTN | nk_cell | *CEACAM19* | -1.2192951 | 4.76E-05 | 0.000102843 | 0.230984881 |
| HTN | nk_cell | *MIR22HG* | -1.978063503 | 8.72E-07 | 5.57E-06 | 0.004230747 |
| HTN | nk_cell | *CYP27C1* | -1.643531539 | 3.50E-07 | 4.24E-06 | 0.001698559 |
| HTN | nk_cell | *BCR* | -2.388930763 | 0.000773256 | 0.001210661 | 1 |
| HTN | nk_cell | *FOXI2* | -3.244804914 | 0.000107538 | 0.000205504 | 0.521774239 |

| | | | | | | |
|---|---|---|---|---|---|---|
| HTN | nk_cell | *ZNF732* | -1.893466555 | 2.55E-07 | 4.07E-06 | 0.001235355 |
| HTN | nk_cell | *HYAL3* | -1.762456841 | 0.00560065 | 0.007565244 | 1 |
| HTN | nk_cell | *VSIG10L* | -3.524187551 | 1.82E-06 | 8.42E-06 | 0.008829877 |
| HTN | nk_cell | *TRABD2A* | -0.860410261 | 0.001460011 | 0.002159748 | 1 |
| HTN | nk_cell | *QRFPR* | -3.953327634 | 6.17E-05 | 0.000127183 | 0.299133711 |
| HTN | nk_cell | *MAPT* | -1.58626563 | 1.07E-06 | 6.17E-06 | 0.005204187 |
| HTN | nk_cell | *TMEM17* | -1.747454481 | 0.000116565 | 0.000220668 | 0.565571773 |
| HTN | nk_cell | *TNFRSF18* | 0.670730846 | 2.93E-07 | 4.12E-06 | 0.001421401 |
| HTN | nk_cell | *C1QL4* | -12.17565847 | 4.09E-07 | 4.24E-06 | 0.001985927 |
| HTN | nk_cell | *RTN4RL2* | -2.578953461 | 5.14E-08 | 2.19E-06 | 0.000249613 |
| HTN | nk_cell | *TMEM232* | -1.406683627 | 1.01E-05 | 2.92E-05 | 0.048887967 |
| HTN | nk_cell | *EFCAB6* | -2.276110105 | 8.03E-05 | 0.00015948 | 0.389608998 |
| HTN | nk_cell | *TMPRSS6* | -10.50955853 | 5.97E-07 | 4.54E-06 | 0.00289851 |
| HTN | nk_cell | *CYP4A11* | 2.577711532 | 3.16E-07 | 4.17E-06 | 0.001532908 |
| HTN | nk_cell | *TMEM216* | 2.042926119 | 6.11E-06 | 2.00E-05 | 0.029654585 |
| HTN | nk_cell | *C3orf70* | -1.100683134 | 3.50E-06 | 1.33E-05 | 0.016997462 |
| HTN | nk_cell | *ENTPD5* | -1.716295157 | 0.002403417 | 0.003441966 | 1 |
| HTN | nk_cell | *MRPS21* | -1.750334372 | 0.00599037 | 0.00805802 | 1 |
| HTN | nk_cell | *ANGPTL5* | 1.903722736 | 0.008333735 | 0.010993823 | 1 |
| HTN | nk_cell | *WDR86* | -1.195790557 | 1.24E-05 | 3.45E-05 | 0.060228872 |
| HTN | nk_cell | *EPOR* | -1.135269161 | 3.52E-06 | 1.33E-05 | 0.017101142 |
| HTN | nk_cell | *LUZP2* | -1.275950542 | 4.10E-07 | 4.24E-06 | 0.001987527 |
| HTN | nk_cell | *GJA4* | -5.012981233 | 6.84E-06 | 2.16E-05 | 0.033181971 |
| HTN | nk_cell | *IFT140* | -1.686345922 | 1.86E-06 | 8.55E-06 | 0.009006972 |
| HTN | nk_cell | *SBK2* | 6.032194935 | 2.67E-07 | 4.07E-06 | 0.001297738 |
| HTN | nk_cell | *ZNF385C* | 1.684868761 | 4.45E-08 | 2.12E-06 | 0.00021592 |
| HTN | nk_cell | *EXD3* | -2.560648128 | 8.15E-05 | 0.000161553 | 0.395321069 |
| HTN | nk_cell | *SAMD11* | -1.433769248 | 1.15E-05 | 3.25E-05 | 0.055830646 |
| HTN | nk_cell | *TRPV2* | 1.347393169 | 2.20E-07 | 4.07E-06 | 0.001066281 |
| HTN | nk_cell | *C2orf88* | -1.554780805 | 1.39E-06 | 7.20E-06 | 0.00676312 |
| HTN | nk_cell | *KBTBD12* | -2.192164574 | 5.94E-07 | 4.54E-06 | 0.002880321 |
| HTN | nk_cell | *FANCA* | 1.120976916 | 3.29E-08 | 1.91E-06 | 0.000159831 |
| HTN | nk_cell | *DNAH17* | -1.09320789 | 6.88E-06 | 2.17E-05 | 0.033366259 |
| HTN | nk_cell | *FANCM* | -2.919243223 | 6.92E-05 | 0.000140412 | 0.335584506 |
| HTN | nk_cell | *ZFP69* | 2.042858968 | 4.11E-07 | 4.24E-06 | 0.001994359 |
| HTN | nk_cell | *TMEM220* | -1.486832589 | 4.49E-05 | 9.78E-05 | 0.217887992 |
| HTN | nk_cell | *P2RX2* | -1.430666675 | 0.022917514 | 0.028585033 | 1 |
| HTN | nk_cell | *FAM122A* | -3.406167227 | 3.20E-06 | 1.25E-05 | 0.015525211 |
| HTN | nk_cell | *GFRAL* | -2.52737576 | 4.47E-07 | 4.28E-06 | 0.00216715 |
| HTN | nk_cell | *LDLRAD2* | -1.314644381 | 3.66E-06 | 1.38E-05 | 0.017782039 |
| HTN | nk_cell | *CPSF4L* | -5.015378945 | 6.55E-07 | 4.70E-06 | 0.003176295 |
| HTN | nk_cell | *KLHL17* | -1.1832167 | 0.021870874 | 0.027349866 | 1 |
| HTN | nk_cell | *ZSCAN23* | -1.177320075 | 3.09E-05 | 7.19E-05 | 0.149742062 |
| HTN | nk_cell | *OR7D2* | 5.651076166 | 0.000167806 | 0.000305744 | 0.814195407 |
| HTN | nk_cell | *TMEM221* | -2.957178538 | 0.000209898 | 0.000372095 | 1 |
| HTN | nk_cell | *MESP2* | 2.187823449 | 1.07E-05 | 3.08E-05 | 0.052034216 |
| HTN | nk_cell | *MAPK12* | -5.390144313 | 9.38E-05 | 0.000182684 | 0.454882288 |
| HTN | nk_cell | *FAM166A* | -3.875272253 | 0.009383425 | 0.012288362 | 1 |
| HTN | nk_cell | *LINC00265* | 3.681067801 | 3.00E-06 | 1.19E-05 | 0.014560504 |
| HTN | nk_cell | *PRKAR1B* | -2.131205785 | 4.42E-05 | 9.67E-05 | 0.214691698 |

| | | | | | | |
|---|---|---|---|---|---|---|
| HTN | nk_cell | *NCR3LG1* | -2.649897612 | 4.57E-07 | 4.29E-06 | 0.002216726 |
| HTN | nk_cell | *ZNF793* | -2.003760676 | 1.74E-05 | 4.53E-05 | 0.084525528 |
| HTN | nk_cell | *AGAP4* | 1.913458664 | 0.007402749 | 0.009840434 | 1 |
| HTN | nk_cell | *RUFY4* | -4.430282603 | 1.23E-06 | 6.74E-06 | 0.005962237 |
| HTN | nk_cell | *CENPP* | -1.660253172 | 2.41E-07 | 4.07E-06 | 0.001170532 |
| HTN | nk_cell | *C3orf62* | -2.633251753 | 5.41E-06 | 1.83E-05 | 0.026270309 |
| HTN | nk_cell | *ENO4* | -1.524286391 | 3.28E-06 | 1.27E-05 | 0.015914912 |
| HTN | nk_cell | *ZNF559* | -1.521798477 | 3.27E-05 | 7.53E-05 | 0.158426357 |
| HTN | nk_cell | *ZP3* | -1.230775022 | 0.001586869 | 0.002333885 | 1 |
| HTN | nk_cell | *H3F3C* | -36.01094903 | 9.00E-08 | 3.12E-06 | 0.000436499 |
| HTN | nk_cell | *JAKMIP3* | -1.212983233 | 0.000842257 | 0.001308559 | 1 |
| HTN | nk_cell | *PDCD1* | 1.109137804 | 3.34E-07 | 4.24E-06 | 0.001622112 |
| HTN | nk_cell | *SELL* | 0.609036763 | 3.14E-07 | 4.17E-06 | 0.001522344 |
| HTN | nk_cell | *C19orf54* | -3.013872602 | 2.15E-06 | 9.40E-06 | 0.010415694 |
| HTN | nk_cell | *C22orf34* | 2.160779503 | 5.93E-07 | 4.54E-06 | 0.002876984 |
| HTN | nk_cell | *COL25A1* | -1.034065075 | 8.90E-07 | 5.65E-06 | 0.004318692 |
| HTN | nk_cell | *SRSF10* | -1.422004323 | 0.013153357 | 0.016901506 | 1 |
| HTN | nk_cell | *NBR1* | -2.271522807 | 0.004990819 | 0.006784941 | 1 |
| HTN | nk_cell | *NDOR1* | -1.384639208 | 5.38E-07 | 4.43E-06 | 0.002611483 |
| HTN | nk_cell | *FAM72B* | -3.046542099 | 0.000104164 | 0.00020016 | 0.505405128 |
| HTN | nk_cell | *GOLGA8M* | -2.852413223 | 4.13E-06 | 1.49E-05 | 0.020027598 |
| HTN | nk_cell | *RHCE* | -1.852092363 | 2.80E-07 | 4.07E-06 | 0.00135738 |
| HTN | nk_cell | *IDO2* | -2.789891615 | 1.94E-06 | 8.83E-06 | 0.009425709 |
| HTN | nk_cell | *SLC4A5* | -3.031990942 | 6.83E-07 | 4.83E-06 | 0.003313344 |
| HTN | nk_cell | *UROS* | 2.547679036 | 2.87E-06 | 1.15E-05 | 0.013923834 |
| HTN | nk_cell | *ZBED6CL* | -3.658703388 | 7.19E-06 | 2.24E-05 | 0.034896918 |
| HTN | nk_cell | *OSTN* | -1.186823342 | 1.13E-06 | 6.38E-06 | 0.005486147 |
| HTN | nk_cell | *FSIP2* | -1.62955732 | 1.27E-06 | 6.83E-06 | 0.006182366 |
| HTN | nk_cell | *NOXA1* | -3.06404758 | 0.013285094 | 0.017061746 | 1 |
| HTN | nk_cell | *MTF1* | 2.613065407 | 8.74E-07 | 5.57E-06 | 0.004240606 |
| HTN | nk_cell | *ZDHHC11* | -2.542445535 | 0.00137799 | 0.002045276 | 1 |
| HTN | nk_cell | *ENTPD8* | -109.6301145 | 3.41E-07 | 4.24E-06 | 0.00165291 |
| HTN | nk_cell | *RPL14* | 2.519471504 | 0.000185207 | 0.000333318 | 0.898625407 |
| HTN | nk_cell | *FBF1* | -3.955594318 | 2.41E-06 | 1.02E-05 | 0.011688679 |
| HTN | nk_cell | *ASTL* | -3.403812615 | 4.12E-07 | 4.24E-06 | 0.002000892 |
| HTN | nk_cell | *MSL1* | -2.770105774 | 7.48E-06 | 2.32E-05 | 0.036277856 |
| HTN | nk_cell | *LRRK2* | -1.569843383 | 0.000329234 | 0.000555246 | 1 |
| HTN | nk_cell | *FAM196A* | -2.099683461 | 3.56E-05 | 8.08E-05 | 0.172762318 |
| HTN | nk_cell | *UTS2B* | -1.741625627 | 3.20E-07 | 4.18E-06 | 0.001552088 |
| HTN | nk_cell | *C9orf152* | -4.267214742 | 3.05E-06 | 1.21E-05 | 0.014820298 |
| HTN | nk_cell | *NELFB* | 3.518073008 | 3.26E-05 | 7.52E-05 | 0.158013113 |
| HTN | nk_cell | *SLC15A5* | -1.361900944 | 1.30E-06 | 6.91E-06 | 0.006315541 |
| HTN | nk_cell | *LIPI* | -1.349153943 | 1.27E-05 | 3.51E-05 | 0.061757281 |
| HTN | nk_cell | *ADAT2* | -2.633511288 | 1.61E-05 | 4.26E-05 | 0.078003455 |
| HTN | nk_cell | *ALKBH2* | 2.039175235 | 0.007988385 | 0.010564089 | 1 |
| HTN | nk_cell | *LITAF* | 2.575286058 | 0.001618421 | 0.002374532 | 1 |
| HTN | nk_cell | *SF3B3* | -2.141153362 | 0.004962721 | 0.006748633 | 1 |
| HTN | nk_cell | *PLAC9* | -2.053112065 | 5.97E-06 | 1.97E-05 | 0.028973534 |
| HTN | nk_cell | *ZNF573* | -2.379371312 | 0.000110283 | 0.000210234 | 0.535091464 |
| HTN | nk_cell | *FAM47E* | -1.113630179 | 4.91E-06 | 1.71E-05 | 0.023830031 |

| | | | | | | |
|---|---|---|---|---|---|---|
| HTN | nk_cell | *ZNF600* | 1.905893349 | 5.86E-07 | 4.54E-06 | 0.002844919 |
| HTN | nk_cell | *TSPYL1* | -3.08341907 | 8.32E-05 | 0.000164455 | 0.403544504 |
| HTN | nk_cell | *PNRC2* | -2.499767061 | 5.26E-06 | 1.80E-05 | 0.025499996 |
| HTN | nk_cell | *FHIT* | -1.388116797 | 1.98E-05 | 5.01E-05 | 0.096018787 |
| HTN | nk_cell | *ZKSCAN3* | -1.543969647 | 0.000327198 | 0.000552197 | 1 |
| HTN | nk_cell | *KIAA0408* | -1.278647993 | 1.98E-05 | 5.01E-05 | 0.096115228 |
| HTN | nk_cell | *HLA-DRB1* | -0.906316216 | 0.013942471 | 0.017839892 | 1 |
| HTN | nk_cell | *AKR1C3* | -1.431138924 | 8.13E-07 | 5.30E-06 | 0.003946259 |
| HTN | nk_cell | *WDSUB1* | -2.295299534 | 0.000190943 | 0.00034275 | 0.926454374 |
| HTN | nk_cell | *ZNF79* | 0.831791385 | 0.012249819 | 0.015811684 | 1 |
| HTN | nk_cell | *COLCA1* | -4.287355235 | 3.43E-06 | 1.31E-05 | 0.016660875 |
| HTN | nk_cell | *MPHOSPH8* | -1.681039967 | 0.000400817 | 0.000664877 | 1 |
| HTN | nk_cell | *SIRPB2* | -1.211145062 | 0.000608153 | 0.000970645 | 1 |
| HTN | nk_cell | *SRGAP3* | -1.448630418 | 5.15E-05 | 0.000109487 | 0.249738742 |
| HTN | nk_cell | *XPNPEP3* | -1.241872059 | 3.35E-06 | 1.28E-05 | 0.016250585 |
| HTN | nk_cell | *PPIA* | -2.426801471 | 0.000941403 | 0.001450981 | 1 |
| HTN | nk_cell | *ZNF471* | -1.856038415 | 1.67E-06 | 7.98E-06 | 0.008086702 |
| HTN | nk_cell | *ZNF493* | -2.313399192 | 7.53E-06 | 2.33E-05 | 0.036525008 |
| HTN | nk_cell | *GTF2IRD2* | -1.739195824 | 1.54E-06 | 7.63E-06 | 0.007483936 |
| HTN | nk_cell | *SUPT3H* | -2.010780497 | 0.00026998 | 0.000465179 | 1 |
| HTN | nk_cell | *GIMAP5* | 1.402771298 | 1.26E-06 | 6.79E-06 | 0.006099872 |
| HTN | nk_cell | *STK31* | -2.17332703 | 3.12E-06 | 1.22E-05 | 0.01514611 |
| HTN | nk_cell | *ZKSCAN7* | 4.260960148 | 3.54E-07 | 4.24E-06 | 0.001715241 |
| HTN | nk_cell | *CD55* | -1.978837191 | 1.78E-05 | 4.61E-05 | 0.086420847 |
| HTN | nk_cell | *CPNE4* | -1.024861973 | 2.98E-07 | 4.13E-06 | 0.00144729 |
| HTN | nk_cell | *ZNF565* | -2.0488147 | 0.000505232 | 0.000819588 | 1 |
| HTN | nk_cell | *ZNF34* | -2.993046831 | 0.000303644 | 0.000517122 | 1 |
| HTN | nk_cell | *ZNF781* | -1.157356162 | 1.63E-06 | 7.89E-06 | 0.007928197 |
| HTN | nk_cell | *EVL* | 1.923983137 | 3.12E-07 | 4.17E-06 | 0.00151526 |
| HTN | nk_cell | *ZNF124* | -2.415435657 | 0.003097308 | 0.004359773 | 1 |
| HTN | nk_cell | *PIK3R4* | -2.394026365 | 2.11E-05 | 5.26E-05 | 0.102202444 |
| HTN | nk_cell | *ZNF605* | -1.331096335 | 0.001343514 | 0.002000224 | 1 |
| HTN | nk_cell | *RFX8* | -1.362118088 | 2.48E-05 | 6.01E-05 | 0.120484438 |
| HTN | nk_cell | *C20orf96* | -3.321181509 | 5.70E-06 | 1.90E-05 | 0.027666572 |
| HTN | nk_cell | *SULT1A1* | -1.718755601 | 2.22E-07 | 4.07E-06 | 0.001078061 |
| HTN | nk_cell | *TPK1* | -2.052512593 | 0.008601089 | 0.01132189 | 1 |
| HTN | nk_cell | *SPTSSB* | -3.561621568 | 7.82E-07 | 5.20E-06 | 0.003792699 |
| HTN | nk_cell | *CACNA1H* | -1.800608926 | 4.72E-07 | 4.32E-06 | 0.002291944 |
| HTN | nk_cell | *XRCC2* | 4.26640378 | 5.00E-07 | 4.35E-06 | 0.00242775 |
| HTN | nk_cell | *MYO6* | -2.132504305 | 4.17E-06 | 1.51E-05 | 0.020255094 |
| HTN | nk_cell | *ZNF846* | -2.204790519 | 4.08E-05 | 9.03E-05 | 0.197776749 |
| HTN | nk_cell | *ADH1B* | -1.318505617 | 1.25E-05 | 3.46E-05 | 0.060625573 |
| HTN | nk_cell | *ZKSCAN5* | -1.884558798 | 6.09E-06 | 1.99E-05 | 0.029541588 |
| HTN | nk_cell | *ZNF502* | -2.680887425 | 2.42E-05 | 5.89E-05 | 0.117596009 |
| HTN | nk_cell | *TRAPPC4* | -1.660352924 | 0.002566594 | 0.003661604 | 1 |
| HTN | nk_cell | *ZFP62* | -2.448678258 | 0.00127622 | 0.001911769 | 1 |
| HTN | nk_cell | *ZNF33B* | -1.765218723 | 0.000241348 | 0.000422295 | 1 |
| HTN | nk_cell | *AMZ2* | 1.538925327 | 3.33E-06 | 1.28E-05 | 0.01614396 |
| HTN | nk_cell | *ZNF431* | -1.776481931 | 0.016892884 | 0.021400594 | 1 |
| HTN | nk_cell | *HLA-DQA1* | -1.472064495 | 0.000725929 | 0.001140241 | 1 |

| | | | | | | |
|---|---|---|---|---|---|---|
| HTN | nk_cell | *GM2A* | -0.952659411 | 0.003885137 | 0.005367507 | 1 |
| HTN | nk_cell | *SPRR2B* | -9.754200712 | 0.021561736 | 0.026998075 | 1 |
| HTN | nk_cell | *ANKRD36B* | -3.122266135 | 1.18E-05 | 3.31E-05 | 0.057073587 |
| HTN | nk_cell | *FAM3C* | 2.058229958 | 0.004366242 | 0.005989541 | 1 |
| HTN | nk_cell | *SLC39A10* | 2.541744549 | 0.000852811 | 0.001324109 | 1 |
| HTN | nk_cell | *AP2A1* | -2.880029012 | 0.000146829 | 0.000271706 | 0.712412817 |
| HTN | nk_cell | *HIST1H3E* | 2.880114689 | 4.99E-13 | 4.03E-10 | 2.42E-09 |
| HTN | nk_cell | *FUT11* | 2.066731842 | 7.80E-06 | 2.40E-05 | 0.037842712 |
| HTN | nk_cell | *WDR5B* | -1.859731192 | 0.000578086 | 0.000926619 | 1 |
| HTN | nk_cell | *NPIPB9* | -4.787901934 | 0.000208388 | 0.000370095 | 1 |
| HTN | nk_cell | *ZNF138* | -2.031189134 | 0.000245669 | 0.000428309 | 1 |
| HTN | nk_cell | *ZNF470* | 3.383585094 | 0.023218119 | 0.028937661 | 1 |
| HTN | nk_cell | *ZNF100* | -2.768845747 | 1.40E-08 | 1.03E-06 | 6.77E-05 |
| HTN | nk_cell | *ANXA6* | 2.241885288 | 2.47E-06 | 1.04E-05 | 0.011989553 |
| HTN | nk_cell | *ZNF420* | -2.382792489 | 5.82E-05 | 0.000120896 | 0.282170236 |
| HTN | nk_cell | *ZNF763* | -1.724164981 | 1.63E-06 | 7.88E-06 | 0.007904988 |
| HTN | nk_cell | *ZMYM1* | -2.553597684 | 0.000206306 | 0.000367204 | 1 |
| HTN | nk_cell | *GAL3ST4* | -3.496446094 | 3.53E-05 | 8.02E-05 | 0.17148506 |
| HTN | nk_cell | *SLC25A29* | -1.641015295 | 7.91E-05 | 0.000157408 | 0.383759821 |
| HTN | nk_cell | *ZNF772* | -1.96365878 | 1.45E-06 | 7.36E-06 | 0.007017073 |
| HTN | nk_cell | *ZNF257* | -3.28801725 | 3.05E-05 | 7.13E-05 | 0.148114332 |
| HTN | nk_cell | *ADAM32* | -1.581103836 | 6.99E-07 | 4.88E-06 | 0.003391206 |
| HTN | nk_cell | *LRRC8B* | -1.942540748 | 9.12E-06 | 2.70E-05 | 0.044232464 |
| HTN | nk_cell | *ABCB8* | -2.429007365 | 1.64E-05 | 4.33E-05 | 0.079554554 |
| HTN | nk_cell | *SND1* | -1.763291717 | 0.000793734 | 0.001239921 | 1 |
| HTN | nk_cell | *SULT1A2* | 3.35154618 | 0.001107499 | 0.001679246 | 1 |
| HTN | nk_cell | *NEK5* | -1.065125668 | 0.017443527 | 0.022046365 | 1 |
| HTN | nk_cell | *PIWIL2* | -2.513009303 | 6.17E-05 | 0.000127183 | 0.299262103 |
| HTN | nk_cell | *TBC1D9B* | -2.119833253 | 0.000108287 | 0.000206773 | 0.525409948 |
| HTN | nk_cell | *FAM110D* | 2.647788933 | 8.73E-07 | 5.57E-06 | 0.004237767 |
| HTN | nk_cell | *CCL4L2* | 0.347032184 | 2.40E-07 | 4.07E-06 | 0.001163496 |
| HTN | nk_cell | *ZNF165* | 2.002653386 | 2.60E-07 | 4.07E-06 | 0.001260695 |
| HTN | nk_cell | *RAMP2-AS1* | -1.416214609 | 7.95E-07 | 5.23E-06 | 0.003857458 |
| HTN | nk_cell | *ZNF786* | 1.902582166 | 5.61E-05 | 0.000117464 | 0.272163175 |
| HTN | nk_cell | *ZNF517* | -1.730503646 | 1.67E-07 | 4.07E-06 | 0.000812187 |
| HTN | nk_cell | *ZNF675* | -2.952218927 | 0.0026386 | 0.003755497 | 1 |
| HTN | nk_cell | *SLC22A5* | -1.856324356 | 0.000263464 | 0.000455083 | 1 |
| HTN | nk_cell | *VEPH1* | -1.154789277 | 5.46E-07 | 4.44E-06 | 0.00264992 |
| HTN | nk_cell | *SHPK* | -1.690030366 | 4.85E-07 | 4.32E-06 | 0.002351092 |
| HTN | nk_cell | *MAP3K5* | -1.580210713 | 0.017372786 | 0.021974129 | 1 |
| HTN | nk_cell | *GSTK1* | -2.643821304 | 0.000212219 | 0.000375798 | 1 |
| HTN | nk_cell | *GYPE* | -2.645740337 | 0.000114357 | 0.000217251 | 0.554859505 |
| HTN | nk_cell | *PCDHB11* | -1.860524208 | 9.00E-07 | 5.69E-06 | 0.004367804 |
| HTN | nk_cell | *SLC2A10* | -1.32949469 | 1.33E-06 | 7.01E-06 | 0.006467469 |
| HTN | nk_cell | *ZNF665* | -3.16283675 | 0.001754878 | 0.002555423 | 1 |
| HTN | nk_cell | *RPF2* | -1.962759323 | 2.33E-06 | 9.99E-06 | 0.011287107 |
| HTN | nk_cell | *ATG7* | -1.393227503 | 0.001060093 | 0.001614936 | 1 |
| HTN | nk_cell | *RAB40C* | 3.083050175 | 0.003413124 | 0.00476835 | 1 |
| HTN | nk_cell | *PIGN* | -1.765440562 | 2.28E-06 | 9.83E-06 | 0.011077974 |
| HTN | nk_cell | *HHLA3* | -1.465838515 | 1.10E-06 | 6.25E-06 | 0.005334174 |

| | | | | | | |
|---|---|---|---|---|---|---|
| HTN | nk_cell | *TOPORS* | -2.829261023 | 0.003605418 | 0.005021093 | 1 |
| HTN | nk_cell | *BCO2* | -1.311170665 | 3.33E-05 | 7.65E-05 | 0.161810697 |
| HTN | nk_cell | *ENTPD6* | -2.08510151 | 3.79E-07 | 4.24E-06 | 0.001838507 |
| HTN | nk_cell | *ENPP1* | -1.594003132 | 4.63E-05 | 0.000100311 | 0.224596104 |
| HTN | nk_cell | *MFAP5* | -1.686658058 | 3.42E-09 | 4.14E-07 | 1.66E-05 |
| HTN | nk_cell | *DNAH10* | -2.441150213 | 0.001838948 | 0.002669832 | 1 |
| HTN | nk_cell | *NMB* | 2.289074661 | 1.83E-05 | 4.70E-05 | 0.088866465 |
| HTN | nk_cell | *FAM114A1* | -1.769899313 | 8.34E-06 | 2.53E-05 | 0.04048022 |
| HTN | nk_cell | *PSAP* | -1.608473204 | 0.00068111 | 0.001074714 | 1 |
| HTN | nk_cell | *TXNRD3* | -1.438622927 | 1.46E-06 | 7.39E-06 | 0.007098261 |
| HTN | nk_cell | *EME2* | -1.252038183 | 4.10E-07 | 4.24E-06 | 0.001989542 |
| HTN | nk_cell | *ZNF780A* | -0.856764863 | 0.00064788 | 0.001025282 | 1 |
| HTN | nk_cell | *AC122129.1* | -4.042543977 | 2.04E-07 | 4.07E-06 | 0.000989017 |
| HTN | nk_cell | *CCDC180* | -1.613382707 | 1.76E-05 | 4.57E-05 | 0.085352935 |
| HTN | nk_cell | *OCLN* | -1.599912856 | 3.22E-07 | 4.19E-06 | 0.001562106 |
| HTN | nk_cell | *ADH5* | -2.255048454 | 0.000327858 | 0.000553118 | 1 |
| HTN | nk_cell | *ZNF677* | -2.364691564 | 1.74E-06 | 8.17E-06 | 0.008419219 |
| HTN | nk_cell | *ZNF311* | -0.906957786 | 0.022182735 | 0.027725562 | 1 |
| HTN | nk_cell | *FCHSD1* | 2.325897046 | 0.000948189 | 0.001460049 | 1 |
| HTN | nk_cell | *ZNF71* | -1.707827552 | 0.030380624 | 0.037270995 | 1 |
| HTN | nk_cell | *RPL12* | 1.367361385 | 0.022235394 | 0.027784222 | 1 |
| HTN | nk_cell | *MPZL1* | -1.555192924 | 5.69E-06 | 1.90E-05 | 0.027619142 |
| HTN | nk_cell | *VPS13A* | -1.986057613 | 0.001223193 | 0.00183801 | 1 |
| HTN | nk_cell | *NOL8* | -2.17637983 | 0.000829884 | 0.001291362 | 1 |
| HTN | nk_cell | *DLGAP2* | -0.90555122 | 2.35E-06 | 1.01E-05 | 0.011390323 |
| HTN | nk_cell | *MRPL42* | -1.489725234 | 0.000179785 | 0.000324675 | 0.872316901 |
| HTN | nk_cell | *ZNF560* | -1.506004785 | 0.040826732 | 0.049522825 | 1 |
| HTN | nk_cell | *AGAP9* | -2.045382318 | 1.73E-08 | 1.21E-06 | 8.38E-05 |
| HTN | nk_cell | *ZNF273* | -2.797499692 | 2.87E-05 | 6.77E-05 | 0.139172346 |
| HTN | nk_cell | *MAK16* | -2.532414651 | 1.10E-05 | 3.14E-05 | 0.053472677 |
| HTN | nk_cell | *ZNF667* | -1.160466495 | 3.03E-06 | 1.20E-05 | 0.014705632 |
| HTN | nk_cell | *SFI1* | 1.619396664 | 2.49E-06 | 1.05E-05 | 0.01208058 |
| HTN | nk_cell | *ADH4* | -1.225543704 | 3.60E-08 | 1.98E-06 | 0.00017481 |
| HTN | nk_cell | *HIBCH* | -1.228244791 | 0.000438635 | 0.000719249 | 1 |
| HTN | nk_cell | *ZNF544* | -2.128225583 | 1.69E-06 | 8.03E-06 | 0.008182395 |
| HTN | nk_cell | *SOWAHC* | -4.97997038 | 2.40E-05 | 5.85E-05 | 0.116368791 |
| HTN | nk_cell | *AC135178.1* | 11.14581078 | 9.04E-07 | 5.69E-06 | 0.004384359 |
| HTN | nk_cell | *MIER1* | -2.355087008 | 6.26E-05 | 0.000128511 | 0.303542959 |
| HTN | nk_cell | *MAN1A2* | -1.843259199 | 0.008591337 | 0.011314287 | 1 |
| HTN | nk_cell | *SVIP* | -2.853511268 | 0.00071352 | 0.001123297 | 1 |
| HTN | nk_cell | *DDRGK1* | -2.466500685 | 1.35E-05 | 3.69E-05 | 0.065619107 |
| HTN | nk_cell | *ZNF334* | -1.668348102 | 1.68E-06 | 8.02E-06 | 0.008161197 |
| HTN | nk_cell | *HSD17B11* | -2.157228563 | 0.000401754 | 0.000665521 | 1 |
| HTN | nk_cell | *SULT1C2* | 1.177151181 | 0.00033537 | 0.000565201 | 1 |
| HTN | nk_cell | *DDX42* | -1.917629383 | 1.38E-05 | 3.76E-05 | 0.066989857 |
| HTN | nk_cell | *HELZ* | -1.760561128 | 0.001604315 | 0.002356687 | 1 |
| HTN | nk_cell | *TMEM116* | -2.8963137 | 1.04E-17 | 5.04E-14 | 5.04E-14 |
| HTN | nk_cell | *UCKL1* | -2.246624262 | 9.16E-07 | 5.71E-06 | 0.004443015 |
| HTN | nk_cell | *GFPT1* | -2.038376434 | 6.35E-05 | 0.000130113 | 0.307907536 |
| HTN | nk_cell | *ZNF568* | -3.084370901 | 3.21E-05 | 7.44E-05 | 0.155957462 |

| | | | | | | |
|---|---|---|---|---|---|---|
| HTN | nk_cell | *ZNF587* | -2.625110833 | 0.000191417 | 0.000343474 | 0.928754956 |
| HTN | nk_cell | *SH3BGRL2* | -1.228520392 | 3.80E-05 | 8.51E-05 | 0.184333426 |
| HTN | nk_cell | *HLA-DRB5* | -0.368210586 | 0.013764797 | 0.017621846 | 1 |
| HTN | nk_cell | *CNGA1* | -1.46238079 | 0.000171472 | 0.000311254 | 0.831981326 |
| HTN | nk_cell | *ZNF28* | 3.304226444 | 2.16E-06 | 9.44E-06 | 0.010489722 |
| HTN | nk_cell | *C20orf203* | -2.374304946 | 1.51E-06 | 7.55E-06 | 0.007328438 |
| HTN | nk_cell | *ZNF534* | -1.341527549 | 0.001172027 | 0.001770447 | 1 |
| HTN | nk_cell | *NCOA6* | -2.116631523 | 0.0017322 | 0.002526189 | 1 |
| HTN | nk_cell | *FAM19A2* | -1.281696162 | 1.77E-06 | 8.28E-06 | 0.008609076 |
| HTN | nk_cell | *TTC37* | -1.891552385 | 0.00517638 | 0.007015585 | 1 |
| HTN | nk_cell | *TUSC1* | -1.884818393 | 4.40E-07 | 4.28E-06 | 0.002134464 |
| HTN | nk_cell | *PAPSS2* | -1.552148763 | 3.56E-06 | 1.34E-05 | 0.017261543 |
| HTN | nk_cell | *CEP290* | -2.184819785 | 5.61E-05 | 0.000117432 | 0.271972253 |
| HTN | nk_cell | *ANKRD13B* | -2.179560134 | 6.03E-07 | 4.54E-06 | 0.002926636 |
| HTN | nk_cell | *ECI2* | -2.794090863 | 0.00013195 | 0.000246904 | 0.640220985 |
| HTN | nk_cell | *UNC13B* | -1.938665546 | 4.96E-06 | 1.72E-05 | 0.024065285 |
| HTN | nk_cell | *PPP1R14C* | -9.483932685 | 2.33E-06 | 9.99E-06 | 0.01129069 |
| HTN | nk_cell | *GPATCH3* | -5.378839333 | 5.72E-05 | 0.000119212 | 0.277406261 |
| HTN | nk_cell | *RPL10A* | 2.025252177 | 0.01694021 | 0.021454947 | 1 |
| HTN | nk_cell | *COLGALT2* | 1.775909846 | 2.87E-07 | 4.07E-06 | 0.001391926 |
| HTN | nk_cell | *RCSD1* | 1.538594434 | 6.58E-07 | 4.71E-06 | 0.003191861 |
| HTN | nk_cell | *GRIN3A* | -1.538327895 | 3.32E-05 | 7.63E-05 | 0.160903804 |
| HTN | nk_cell | *TMEM184B* | -1.964381764 | 0.002986896 | 0.004214138 | 1 |
| HTN | nk_cell | *SCAMP5* | -0.899287536 | 0.000397434 | 0.000660167 | 1 |
| HTN | nk_cell | *SFT2D1* | -2.553885644 | 0.000104818 | 0.000200909 | 0.508577269 |
| HTN | nk_cell | *HMGN2* | -1.917662972 | 3.11E-06 | 1.22E-05 | 0.015074131 |
| HTN | nk_cell | *DENND4B* | -2.961956064 | 0.000645375 | 0.001022319 | 1 |
| HTN | nk_cell | *RYR3* | -1.424636653 | 0.000946993 | 0.001459133 | 1 |
| HTN | nk_cell | *TYW1* | -1.865539372 | 0.000105221 | 0.000201552 | 0.510531033 |
| HTN | nk_cell | *ITPRIPL1* | -1.09603152 | 0.005351849 | 0.007241263 | 1 |
| HTN | nk_cell | *PRMT6* | 1.626309556 | 3.88E-07 | 4.24E-06 | 0.001881357 |
| HTN | nk_cell | *SHISA4* | -4.863039348 | 1.17E-09 | 2.03E-07 | 5.67E-06 |
| HTN | nk_cell | *MAP3K3* | -2.011466722 | 0.008971147 | 0.011777057 | 1 |
| HTN | nk_cell | *C1orf174* | -2.012195369 | 2.09E-05 | 5.23E-05 | 0.101517379 |
| HTN | nk_cell | *DZIP3* | -2.298558949 | 0.00025616 | 0.000443888 | 1 |
| HTN | nk_cell | *KIAA0753* | -2.330290538 | 1.18E-06 | 6.57E-06 | 0.00571369 |
| HTN | nk_cell | *TBKBP1* | -1.297527236 | 0.034635177 | 0.042276699 | 1 |
| HTN | nk_cell | *CCDC167* | 4.491393943 | 0.035868859 | 0.043694628 | 1 |
| HTN | nk_cell | *ZFP2* | -3.063029094 | 9.47E-06 | 2.78E-05 | 0.045953146 |
| HTN | nk_cell | *L3MBTL3* | 1.725378715 | 0.017504761 | 0.022117995 | 1 |
| HTN | nk_cell | *MFAP3L* | -2.812193497 | 0.005090971 | 0.006905617 | 1 |
| HTN | nk_cell | *NAGA* | -2.308341164 | 0.000814824 | 0.001269597 | 1 |
| HTN | nk_cell | *SMG5* | -2.806424383 | 3.38E-06 | 1.29E-05 | 0.016407137 |
| HTN | nk_cell | *TGM2* | 1.200483305 | 0.035228484 | 0.042957679 | 1 |
| HTN | nk_cell | *PJA2* | -2.094335337 | 0.000109496 | 0.000208999 | 0.531275376 |
| HTN | nk_cell | *AL590822.1* | -1.813936956 | 4.30E-08 | 2.12E-06 | 0.000208765 |
| HTN | nk_cell | *AP001007.1* | -2.416394414 | 3.13E-05 | 7.28E-05 | 0.151661098 |
| HTN | nk_cell | *EFCAB2* | -1.682573527 | 2.89E-06 | 1.15E-05 | 0.014003746 |
| HTN | nk_cell | *CHML* | -2.363198107 | 1.69E-05 | 4.43E-05 | 0.081867039 |
| HTN | nk_cell | *TATDN3* | -1.606732117 | 0.000756526 | 0.001186382 | 1 |

| | | | | | | |
|---|---|---|---|---|---|---|
| HTN | nk_cell | *SAMD5* | -2.295052834 | 0.000319125 | 0.00054083 | 1 |
| HTN | nk_cell | *ECT2L* | -1.709996128 | 9.11E-07 | 5.69E-06 | 0.004420942 |
| HTN | nk_cell | *GPR52* | 12.48472029 | 2.22E-06 | 9.65E-06 | 0.010779884 |
| HTN | nk_cell | *FAM229B* | -2.538145833 | 1.01E-06 | 5.99E-06 | 0.004914292 |
| HTN | nk_cell | *DDO* | -2.676417785 | 1.00E-06 | 5.96E-06 | 0.004858077 |
| HTN | nk_cell | *RBM20* | -1.311350429 | 5.94E-07 | 4.54E-06 | 0.002884498 |
| HTN | nk_cell | *PCMTD2* | -2.364479268 | 0.000362937 | 0.00060786 | 1 |
| HTN | nk_cell | *SAMD13* | -1.969668931 | 5.89E-07 | 4.54E-06 | 0.002857118 |
| HTN | nk_cell | *EFCAB7* | -1.909003818 | 5.39E-06 | 1.82E-05 | 0.026132696 |
| HTN | nk_cell | *IFIT1B* | 3.378118605 | 2.68E-07 | 4.07E-06 | 0.001302333 |
| HTN | nk_cell | *LIPN* | -3.76877256 | 6.34E-07 | 4.62E-06 | 0.003074703 |
| HTN | nk_cell | *INPP5B* | -1.274200455 | 0.00102229 | 0.001561269 | 1 |
| HTN | nk_cell | *NEU4* | -3.032214457 | 3.26E-05 | 7.52E-05 | 0.158202513 |
| HTN | nk_cell | *C2orf72* | -8.986960582 | 1.27E-05 | 3.50E-05 | 0.061408076 |
| HTN | nk_cell | *RUFY2* | -1.826119279 | 1.28E-05 | 3.52E-05 | 0.061969692 |
| HTN | nk_cell | *PHACTR4* | -1.61246377 | 0.000313741 | 0.000533007 | 1 |
| HTN | nk_cell | *ASAH2B* | -2.597234477 | 3.17E-05 | 7.36E-05 | 0.153848943 |
| HTN | nk_cell | *AGAP6* | -1.439164789 | 0.000399823 | 0.000663681 | 1 |
| HTN | nk_cell | *SYT15* | -2.741239555 | 5.65E-06 | 1.89E-05 | 0.027401125 |
| HTN | nk_cell | *LINC00619* | -10.57613958 | 4.53E-07 | 4.28E-06 | 0.002196216 |
| HTN | nk_cell | *BMPR2* | -1.651511294 | 2.03E-05 | 5.11E-05 | 0.098687412 |
| HTN | nk_cell | *TCEA3* | -2.279277268 | 1.77E-05 | 4.60E-05 | 0.085969003 |
| HTN | nk_cell | *HSD17B8* | -2.492260773 | 1.06E-10 | 2.46E-08 | 5.13E-07 |
| HTN | nk_cell | *TAP2* | 1.185709618 | 8.38E-05 | 0.000165622 | 0.406768348 |
| HTN | nk_cell | *COL15A1* | -1.079229477 | 7.42E-05 | 0.000148995 | 0.359972582 |
| HTN | nk_cell | *PRRT1* | 1.72280984 | 0.000394176 | 0.000656104 | 1 |
| HTN | nk_cell | *SMIM5* | -3.098348254 | 4.76E-07 | 4.32E-06 | 0.002309593 |
| HTN | nk_cell | *STK19* | -1.102334981 | 1.84E-06 | 8.48E-06 | 0.008907672 |
| HTN | nk_cell | *SKIV2L* | -1.126458179 | 1.22E-07 | 3.72E-06 | 0.000591431 |
| HTN | nk_cell | *LAYN* | -4.184441677 | 1.12E-06 | 6.36E-06 | 0.005453668 |
| HTN | nk_cell | *BAG6* | 2.037405667 | 0.0276479 | 0.034064909 | 1 |
| HTN | nk_cell | *LST1* | 1.772610933 | 0.003179337 | 0.004468755 | 1 |
| HTN | nk_cell | *ZNF814* | -2.052390391 | 0.000527447 | 0.000852168 | 1 |
| HTN | nk_cell | *MICB* | -0.427200652 | 0.024009636 | 0.029839845 | 1 |
| HTN | nk_cell | *MICA* | -2.099459175 | 4.78E-05 | 0.000103123 | 0.231716329 |
| HTN | nk_cell | *POU5F1* | -0.756957979 | 3.14E-05 | 7.31E-05 | 0.152320887 |
| HTN | nk_cell | *DHX16* | -2.352305055 | 0.000674695 | 0.001065285 | 1 |
| HTN | nk_cell | *HLA-E* | 1.444162713 | 0.006194599 | 0.008321206 | 1 |
| HTN | nk_cell | *ZNF468* | -1.886139476 | 1.55E-06 | 7.63E-06 | 0.007530441 |
| HTN | nk_cell | *HLA-G* | 1.533174352 | 4.16E-05 | 9.20E-05 | 0.20203686 |
| HTN | nk_cell | *ZFP57* | -0.723933093 | 0.015726754 | 0.019975448 | 1 |
| HTN | nk_cell | *FAM153C* | -0.770923819 | 0.000317317 | 0.000538141 | 1 |
| HTN | nk_cell | *TRIM27* | -1.628165975 | 0.033199931 | 0.040616759 | 1 |
| HTN | nk_cell | *MALRD1* | -2.13923606 | 3.35E-06 | 1.28E-05 | 0.016273067 |
| HTN | nk_cell | *FAM196B* | 1.98268732 | 9.87E-07 | 5.93E-06 | 0.004788525 |
| HTN | nk_cell | *MROH6* | -1.906916358 | 1.80E-05 | 4.65E-05 | 0.087448267 |
| HTN | nk_cell | *DCTN1* | -3.35909977 | 0.001767227 | 0.002572633 | 1 |
| HTN | nk_cell | *FAM216A* | -2.801319718 | 0.000724768 | 0.001139156 | 1 |
| HTN | nk_cell | *ZBTB48* | -2.00989666 | 0.013738354 | 0.017592635 | 1 |
| HTN | nk_cell | *MZT1* | 4.051351504 | 9.80E-06 | 2.86E-05 | 0.047546681 |

| | | | | | | |
|---|---|---|---|---|---|---|
| HTN | nk_cell | *ZNF155* | -2.212644913 | 5.83E-07 | 4.54E-06 | 0.002828831 |
| HTN | nk_cell | *ZNF783* | -3.156417575 | 5.71E-05 | 0.0001191 | 0.277025539 |
| HTN | nk_cell | *ZNF425* | -2.374640311 | 7.06E-07 | 4.91E-06 | 0.003425163 |
| HTN | nk_cell | *C12orf73* | -2.104725694 | 1.30E-06 | 6.91E-06 | 0.006329264 |
| HTN | nk_cell | *PCDHGA1* | -1.73985346 | 0.000389659 | 0.000649699 | 1 |
| HTN | nk_cell | *PCDHA7* | -20.16664554 | 0.001608578 | 0.002361519 | 1 |
| HTN | nk_cell | *PCDHA4* | -2.901907157 | 6.07E-07 | 4.55E-06 | 0.002947063 |
| HTN | nk_cell | *TRIM13* | -1.899138632 | 0.00038403 | 0.000640975 | 1 |
| HTN | nk_cell | *PKHD1L1* | -0.816522611 | 0.004759459 | 0.006495891 | 1 |
| HTN | nk_cell | *SLFN12L* | 0.355514655 | 6.73E-07 | 4.78E-06 | 0.003264175 |
| HTN | nk_cell | *SYCE1L* | 0.836871903 | 0.000191854 | 0.000343978 | 0.930876256 |
| HTN | nk_cell | *TMEM231* | -1.601482453 | 1.21E-06 | 6.69E-06 | 0.005881573 |
| HTN | nk_cell | *LINC00654* | -2.554145012 | 0.000403404 | 0.000668026 | 1 |
| HTN | nk_cell | *SCGB2B2* | -1.597509751 | 1.98E-06 | 8.92E-06 | 0.009612167 |
| HTN | nk_cell | *CCDC144NL* | 5.323332696 | 0.002711232 | 0.003848712 | 1 |
| HTN | nk_cell | *TMEM170B* | -0.806480575 | 0.001744876 | 0.002541621 | 1 |
| HTN | nk_cell | *GOLGA6L10* | -3.438381302 | 1.30E-07 | 3.82E-06 | 0.000628552 |
| HTN | nk_cell | *NT5M* | -1.419867834 | 3.63E-07 | 4.24E-06 | 0.001760602 |
| HTN | nk_cell | *IPO7* | -2.089628534 | 1.57E-05 | 4.18E-05 | 0.076300432 |
| HTN | nk_cell | *PRR13* | 1.922940068 | 0.024975776 | 0.031000886 | 1 |
| HTN | nk_cell | *TECPR1* | -3.025640426 | 5.77E-05 | 0.000119966 | 0.279761248 |
| HTN | nk_cell | *MT1M* | 3.277707727 | 9.10E-07 | 5.69E-06 | 0.004415957 |
| HTN | nk_cell | *CFI* | -1.537263957 | 1.45E-06 | 7.37E-06 | 0.007038102 |
| HTN | nk_cell | *ATP6AP1L* | -1.049403883 | 0.000555063 | 0.00089296 | 1 |
| HTN | nk_cell | *RGL3* | -1.18355161 | 3.50E-06 | 1.33E-05 | 0.016979719 |
| HTN | nk_cell | *SERF1B* | -1.706957731 | 0.000568881 | 0.000913372 | 1 |
| HTN | nk_cell | *LIN52* | -1.452203238 | 0.014845432 | 0.01894032 | 1 |
| HTN | nk_cell | *KLRC2* | 0.130970989 | 2.78E-07 | 4.07E-06 | 0.001350108 |
| HTN | nk_cell | *LINC00487* | 4.116899042 | 7.00E-05 | 0.000141836 | 0.339555267 |
| HTN | nk_cell | *RFPL3S* | 1.143218026 | 0.000771449 | 0.001208221 | 1 |
| HTN | nk_cell | *C1QTNF9B* | -9.58024541 | 4.15E-07 | 4.24E-06 | 0.002012128 |
| HTN | nk_cell | *ZDHHC11B* | -2.041090599 | 5.94E-07 | 4.54E-06 | 0.002884211 |
| HTN | nk_cell | *HBA1* | 0.902994873 | 1.74E-07 | 4.07E-06 | 0.000845413 |
| HTN | nk_cell | *COL6A6* | -1.672622179 | 2.76E-05 | 6.53E-05 | 0.133700297 |
| HTN | nk_cell | *HLA-A* | 1.504057664 | 5.22E-06 | 1.79E-05 | 0.025348238 |
| HTN | nk_cell | *LNP1* | -1.722956102 | 0.03014392 | 0.037018046 | 1 |
| HTN | nk_cell | *PRSS50* | -3.297184391 | 0.007321422 | 0.009743154 | 1 |
| HTN | nk_cell | *METTL6* | -2.120115476 | 0.009918193 | 0.01296419 | 1 |
| HTN | nk_cell | *SACM1L* | 2.49136244 | 0.003834175 | 0.005306166 | 1 |
| HTN | nk_cell | *SLC48A1* | -1.962856706 | 1.98E-06 | 8.92E-06 | 0.009604792 |
| HTN | nk_cell | *TAS2R50* | -20.42060948 | 3.31E-05 | 7.61E-05 | 0.160563429 |
| HTN | nk_cell | *TAS2R14* | 2.107914872 | 6.43E-07 | 4.65E-06 | 0.00312097 |
| HTN | nk_cell | *AC084018.1* | -1.566387213 | 6.90E-07 | 4.86E-06 | 0.003345812 |
| HTN | nk_cell | *OR2A42* | -1.806270993 | 0.000221487 | 0.000390784 | 1 |
| HTN | nk_cell | *ZNF580* | -1.504435594 | 7.28E-05 | 0.00014651 | 0.353235726 |
| HTN | nk_cell | *AL021068.1* | -2.113503901 | 0.002525989 | 0.003607918 | 1 |
| HTN | nk_cell | *FGFR1OP* | 4.315220045 | 5.76E-05 | 0.000119966 | 0.279541349 |
| HTN | nk_cell | *FAM106A* | -3.197376378 | 0.024712086 | 0.030681433 | 1 |
| HTN | nk_cell | *ZNF254* | -2.137403174 | 0.000112889 | 0.000214714 | 0.547736634 |
| HTN | nk_cell | *TCTEX1D2* | -2.600687653 | 3.15E-06 | 1.23E-05 | 0.015302224 |

| | | | | | | |
|---|---|---|---|---|---|---|
| HTN | nk_cell | *AC022498.1* | 4.046912007 | 2.67E-07 | 4.07E-06 | 0.001296876 |
| HTN | nk_cell | *LINGO4* | -2.162289295 | 0.01832216 | 0.023066715 | 1 |
| HTN | nk_cell | *ASIC3* | -2.11284765 | 0.004018162 | 0.005536724 | 1 |
| HTN | nk_cell | *GIMAP1* | 2.249306508 | 4.47E-06 | 1.59E-05 | 0.021674509 |
| HTN | nk_cell | *DNLZ* | 0.794647599 | 2.72E-05 | 6.46E-05 | 0.132026681 |
| HTN | nk_cell | *NOTCH2NL* | 1.355455931 | 1.85E-07 | 4.07E-06 | 0.000895322 |
| HTN | nk_cell | *SUPT4H1* | 2.250076743 | 3.67E-08 | 1.98E-06 | 0.000178088 |
| HTN | nk_cell | *MXD3* | 1.026764109 | 0.03896142 | 0.047343053 | 1 |
| HTN | nk_cell | *GSTM2* | -1.656163818 | 4.04E-06 | 1.48E-05 | 0.019594257 |
| HTN | nk_cell | *ARHGAP19* | -2.258496427 | 2.82E-07 | 4.07E-06 | 0.001369096 |
| HTN | nk_cell | *LCAT* | -3.808825239 | 0.004873609 | 0.006636753 | 1 |
| HTN | nk_cell | *GPC2* | -5.587227088 | 3.87E-05 | 8.64E-05 | 0.187921754 |
| HTN | nk_cell | *ERV3-1* | -0.611163565 | 0.00025638 | 0.000444112 | 1 |
| HTN | nk_cell | *GBP7* | -0.928832077 | 0.027252303 | 0.033603094 | 1 |
| HTN | nk_cell | *SRA1* | -2.576359415 | 0.000320025 | 0.000542166 | 1 |
| HTN | nk_cell | *TMEM110* | -1.488334505 | 0.022651823 | 0.028275443 | 1 |
| HTN | nk_cell | *TMX2* | 2.746300401 | 0.018737472 | 0.023571225 | 1 |
| HTN | nk_cell | *LBH* | 1.117720761 | 7.21E-07 | 4.96E-06 | 0.003500423 |
| HTN | nk_cell | *PPP1CB* | -1.924684338 | 0.001604848 | 0.002356756 | 1 |
| HTN | nk_cell | *NCKIPSD* | -2.358985216 | 3.56E-05 | 8.08E-05 | 0.172968972 |
| HTN | nk_cell | *SLC35F6* | -3.629870546 | 0.003570655 | 0.004976965 | 1 |
| HTN | nk_cell | *EMP2* | -2.174794975 | 1.87E-05 | 4.78E-05 | 0.09070299 |
| HTN | nk_cell | *PPM1N* | 2.523405281 | 7.42E-09 | 6.67E-07 | 3.60E-05 |
| HTN | nk_cell | *SLC23A3* | -1.082310231 | 0.000891931 | 0.001378232 | 1 |
| HTN | nk_cell | *LTB4R* | -5.200579851 | 0.0001716 | 0.000311369 | 0.832600983 |
| HTN | nk_cell | *IRF9* | 3.578999213 | 0.001219388 | 0.00183286 | 1 |
| HTN | nk_cell | *GALT* | 4.431133885 | 1.87E-05 | 4.78E-05 | 0.090595154 |
| HTN | nk_cell | *ZNF726* | -4.82534846 | 3.77E-05 | 8.46E-05 | 0.182725308 |
| HTN | nk_cell | *ZNF90* | -1.815335809 | 3.06E-06 | 1.21E-05 | 0.014839654 |
| HTN | nk_cell | *REPIN1* | -2.44509146 | 1.24E-06 | 6.76E-06 | 0.006025628 |
| HTN | nk_cell | *ZNF891* | -2.342769175 | 1.15E-05 | 3.24E-05 | 0.055645607 |
| HTN | nk_cell | *TSPAN4* | -2.097728432 | 0.000108241 | 0.000206766 | 0.525185956 |
| HTN | nk_cell | *CPNE1* | -0.930545098 | 0.015540663 | 0.019754597 | 1 |
| HTN | nk_cell | *PAXIP1-AS2* | -1.819267726 | 2.25E-06 | 9.72E-06 | 0.010911141 |
| HTN | nk_cell | *LYRM4* | -2.349187874 | 5.22E-05 | 0.000110754 | 0.253514932 |
| HTN | nk_cell | *PRCD* | -2.430103612 | 2.30E-05 | 5.65E-05 | 0.111434888 |
| HTN | nk_cell | *SH3D21* | -1.415343708 | 4.41E-06 | 1.57E-05 | 0.021396523 |
| HTN | nk_cell | *FIS1* | -1.981380119 | 8.44E-08 | 3.01E-06 | 0.000409602 |
| HTN | nk_cell | *ANG* | -3.357208874 | 1.41E-05 | 3.84E-05 | 0.068589336 |
| HTN | nk_cell | *COLCA2* | -1.550222035 | 1.09E-06 | 6.22E-06 | 0.005278343 |
| HTN | nk_cell | *MBLAC1* | -0.899982433 | 0.004768127 | 0.006504063 | 1 |
| HTN | nk_cell | *LCNL1* | -2.782755438 | 3.65E-06 | 1.37E-05 | 0.017723828 |
| HTN | nk_cell | *GNAT3* | -2.240329777 | 1.20E-06 | 6.67E-06 | 0.005831849 |
| HTN | nk_cell | *PLIN5* | -0.717560337 | 1.31E-06 | 6.91E-06 | 0.006334888 |
| HTN | nk_cell | *STARD10* | -0.634192665 | 0.035656945 | 0.043458301 | 1 |
| HTN | nk_cell | *NUTM2D* | -13.22513757 | 7.14E-05 | 0.000144157 | 0.346410306 |
| HTN | nk_cell | *CPEB1* | -0.854298669 | 5.12E-07 | 4.37E-06 | 0.002483044 |
| HTN | nk_cell | *EML6* | -3.413628934 | 0.000107201 | 0.000205021 | 0.52013948 |
| HTN | nk_cell | *ARHGEF33* | -2.742760391 | 0.000178164 | 0.000322076 | 0.86445175 |
| HTN | nk_cell | *IFRD2* | -1.65696528 | 0.000435857 | 0.000715418 | 1 |

| | | | | | | |
|---|---|---|---|---|---|---|
| HTN | nk_cell | *EVPLL* | -2.360315037 | 3.22E-05 | 7.45E-05 | 0.156363179 |
| HTN | nk_cell | *AC005082.1* | -4.933870457 | 4.99E-08 | 2.19E-06 | 0.000242282 |
| HTN | nk_cell | *SMTNL1* | -1.073148634 | 8.37E-05 | 0.000165494 | 0.406286758 |
| HTN | nk_cell | *C5orf49* | -10.49571378 | 5.69E-07 | 4.50E-06 | 0.002762269 |
| HTN | nk_cell | *VPS16* | -2.598593112 | 8.66E-06 | 2.59E-05 | 0.042028904 |
| HTN | nk_cell | *NPEPL1* | -2.910968265 | 6.12E-06 | 2.00E-05 | 0.029705826 |
| HTN | nk_cell | *SLC35E2* | -2.616188377 | 1.66E-05 | 4.36E-05 | 0.080306502 |
| HTN | nk_cell | *LINC01139* | -0.723702212 | 0.000911509 | 0.001407588 | 1 |
| HTN | nk_cell | *TSTD1* | 3.326269955 | 0.000428085 | 0.000704091 | 1 |
| HTN | nk_cell | *AC009403.2* | -2.248898282 | 7.95E-06 | 2.43E-05 | 0.03857303 |
| HTN | nk_cell | *AC016773.1* | 2.967068981 | 4.25E-10 | 8.25E-08 | 2.06E-06 |
| HTN | nk_cell | *AP000350.4* | -0.610669378 | 0.008900645 | 0.011687666 | 1 |
| HTN | nk_cell | *RNASEK* | -2.178130825 | 0.000145688 | 0.00027011 | 0.706877778 |
| HTN | nk_cell | *FAM228B* | -1.85466345 | 7.31E-05 | 0.000146989 | 0.354683898 |
| HTN | nk_cell | *RNU6ATAC35I* | -12.25762292 | 2.80E-06 | 1.13E-05 | 0.013585004 |
| HTN | nk_cell | *PPP3R1* | -2.011352307 | 1.32E-05 | 3.62E-05 | 0.064032766 |
| HTN | nk_cell | *FANCG* | -2.064603636 | 0.000215333 | 0.000380756 | 1 |
| HTN | nk_cell | *AP4M1* | -2.276386748 | 4.83E-09 | 4.89E-07 | 2.34E-05 |
| HTN | nk_cell | *ZNF880* | -1.923006546 | 1.57E-05 | 4.18E-05 | 0.076406001 |
| HTN | nk_cell | *FXYD7* | -1.709294673 | 5.21E-07 | 4.37E-06 | 0.002526763 |
| HTN | nk_cell | *XKR9* | -0.631834064 | 0.013127917 | 0.016873285 | 1 |
| HTN | nk_cell | *SLC12A8* | -1.010693723 | 0.003934463 | 0.005424841 | 1 |
| HTN | nk_cell | *APOL6* | -2.149918912 | 0.002816884 | 0.003991173 | 1 |
| HTN | nk_cell | *OR2A1* | -1.823994208 | 0.000582323 | 0.000932559 | 1 |
| HTN | nk_cell | *UBA52* | 2.17666354 | 0.001533586 | 0.002261003 | 1 |
| HTN | nk_cell | *TIAF1* | 5.52930628 | 1.40E-07 | 3.89E-06 | 0.000680785 |
| HTN | nk_cell | *RAB6C* | -28.32480788 | 3.06E-06 | 1.21E-05 | 0.014867036 |
| HTN | nk_cell | *AC062017.1* | -1.304126501 | 4.89E-07 | 4.32E-06 | 0.002370857 |
| HTN | nk_cell | *ADRA2B* | -6.254820693 | 4.26E-07 | 4.24E-06 | 0.002067889 |
| HTN | nk_cell | *EXOSC6* | -3.851569523 | 4.25E-09 | 4.58E-07 | 2.06E-05 |
| HTN | nk_cell | *ZNF844* | -1.901155735 | 2.61E-05 | 6.23E-05 | 0.126465005 |
| HTN | nk_cell | *LINC00571* | -2.901182481 | 6.95E-06 | 2.18E-05 | 0.033705287 |
| HTN | nk_cell | *CERS1* | -8.321935341 | 0.000613377 | 0.000977374 | 1 |
| HTN | nk_cell | *C5orf66* | -2.766905246 | 4.83E-05 | 0.000104162 | 0.23436466 |
| HTN | nk_cell | *LINC01133* | -0.978315565 | 4.13E-07 | 4.24E-06 | 0.002004508 |
| HTN | nk_cell | *LINC01117* | -3.627626043 | 7.70E-05 | 0.000153973 | 0.373846282 |
| HTN | nk_cell | *LINC00240* | -0.979444775 | 2.99E-05 | 7.01E-05 | 0.145202852 |
| HTN | nk_cell | *LINC00184* | -25.9012979 | 4.00E-07 | 4.24E-06 | 0.001943068 |
| HTN | nk_cell | *AL049840.1* | -3.634137985 | 1.23E-05 | 3.42E-05 | 0.059522084 |
| HTN | nk_cell | *PLEKHM1* | -3.09560352 | 2.15E-07 | 4.07E-06 | 0.001044091 |
| HTN | nk_cell | *LINC00092* | 0.749314374 | 0.016279797 | 0.020650869 | 1 |
| HTN | nk_cell | *AP000350.6* | -0.954431369 | 0.000324165 | 0.000547458 | 1 |
| HTN | nk_cell | *SFTA1P* | 1.950199917 | 1.42E-06 | 7.27E-06 | 0.006899319 |
| HTN | nk_cell | *CYP4A22-AS1* | -2.898672417 | 1.56E-06 | 7.65E-06 | 0.007566913 |
| HTN | nk_cell | *FAM225B* | -2.799093054 | 0.023562474 | 0.029329174 | 1 |
| HTN | nk_cell | *MIAT* | 0.898138342 | 5.10E-07 | 4.37E-06 | 0.002472744 |
| HTN | nk_cell | *ERCC6* | -2.086494416 | 0.01403216 | 0.017940458 | 1 |
| HTN | nk_cell | *LINC00694* | -3.164840043 | 4.22E-05 | 9.29E-05 | 0.204642919 |
| HTN | nk_cell | *LINC00115* | 2.961411964 | 1.55E-06 | 7.63E-06 | 0.007501058 |
| HTN | nk_cell | *HAR1A* | 16.68938601 | 2.84E-05 | 6.71E-05 | 0.137932679 |

| | | | | | | |
|---|---|---|---|---|---|---|
| HTN | nk_cell | *LINC00937* | -1.339388958 | 3.15E-07 | 4.17E-06 | 0.001530592 |
| HTN | nk_cell | *FTCDNL1* | -0.872651826 | 0.02270949 | 0.028340135 | 1 |
| HTN | nk_cell | *FAM138B* | -14.81928148 | 4.10E-07 | 4.24E-06 | 0.001991337 |
| HTN | nk_cell | *AC017074.2* | -18.93064971 | 5.07E-06 | 1.75E-05 | 0.024587302 |
| HTN | nk_cell | *AC009299.3* | -2.413214746 | 1.80E-06 | 8.37E-06 | 0.008756667 |
| HTN | nk_cell | *SCAMP4* | -2.620141865 | 0.013229123 | 0.016994362 | 1 |
| HTN | nk_cell | *SLC2A1-AS1* | -1.99408844 | 6.98E-07 | 4.88E-06 | 0.003388566 |
| HTN | nk_cell | *LINC01068* | -1.835017953 | 0.013329742 | 0.017105503 | 1 |
| HTN | nk_cell | *AC005884.1* | -2.752033867 | 0.003634841 | 0.005057714 | 1 |
| HTN | nk_cell | *HCG20* | 2.291820184 | 0.001618077 | 0.002374532 | 1 |
| HTN | nk_cell | *C14orf144* | -2.667372024 | 1.53E-05 | 4.10E-05 | 0.074388163 |
| HTN | nk_cell | *POLR2J2* | 2.918147349 | 2.06E-07 | 4.07E-06 | 0.001000062 |
| HTN | nk_cell | *LINC00578* | -1.171851008 | 1.34E-07 | 3.84E-06 | 0.000651833 |
| HTN | nk_cell | *C19orf24* | -2.280788891 | 5.72E-06 | 1.90E-05 | 0.027761422 |
| HTN | nk_cell | *ARL17B* | -2.090169584 | 3.48E-06 | 1.32E-05 | 0.016874128 |
| HTN | nk_cell | *DHFR* | -0.730549358 | 0.000593967 | 0.000950504 | 1 |
| HTN | nk_cell | *PATL2* | 1.421967438 | 3.08E-07 | 4.17E-06 | 0.001494248 |
| HTN | nk_cell | *CASC20* | -3.192747192 | 7.87E-07 | 5.21E-06 | 0.003820099 |
| HTN | nk_cell | *PSMG3-AS1* | -2.200416343 | 7.12E-07 | 4.93E-06 | 0.003454243 |
| HTN | nk_cell | *AC112229.4* | -2.210731401 | 0.01436843 | 0.018336565 | 1 |
| HTN | nk_cell | *KLHL7-AS1* | -1.777475375 | 2.38E-05 | 5.81E-05 | 0.115357157 |
| HTN | nk_cell | *LINC00332* | -2.009907614 | 0.000189769 | 0.000340769 | 0.920758429 |
| HTN | nk_cell | *AC092171.4* | -2.339030747 | 9.08E-07 | 5.69E-06 | 0.00440743 |
| HTN | nk_cell | *RPS18* | 1.928963029 | 0.003465654 | 0.004837558 | 1 |
| HTN | nk_cell | *DIRC3* | -1.162629661 | 1.79E-06 | 8.32E-06 | 0.008667462 |
| HTN | nk_cell | *TAPBP* | 2.452878061 | 1.25E-06 | 6.76E-06 | 0.006045307 |
| HTN | nk_cell | *AC107021.1* | -1.626053205 | 6.59E-06 | 2.11E-05 | 0.031957536 |
| HTN | nk_cell | *TMA7* | 2.774407721 | 2.22E-08 | 1.38E-06 | 0.000107622 |
| HTN | nk_cell | *MTRNR2L4* | -3.41187483 | 0.000240394 | 0.000421233 | 1 |
| HTN | nk_cell | *LINC00865* | -1.481011374 | 3.95E-05 | 8.80E-05 | 0.19163349 |
| HTN | nk_cell | *AC009495.3* | 0.553431411 | 1.93E-07 | 4.07E-06 | 0.000934464 |
| HTN | nk_cell | *GOLGA8N* | -0.980943845 | 0.017661169 | 0.022292402 | 1 |
| HTN | nk_cell | *LINC00665* | -2.462178967 | 8.32E-05 | 0.000164455 | 0.403572707 |
| HTN | nk_cell | *LYRM9* | -1.860007504 | 3.45E-06 | 1.31E-05 | 0.01675975 |
| HTN | nk_cell | *LINC00342* | 4.28671358 | 0.006639528 | 0.0088918 | 1 |
| HTN | nk_cell | *LINC00702* | -1.968460644 | 5.07E-07 | 4.37E-06 | 0.00246234 |
| HTN | nk_cell | *GPX1* | -1.593657704 | 0.027600398 | 0.03401502 | 1 |
| HTN | nk_cell | *RPS28* | -2.56668875 | 0.010710706 | 0.013940007 | 1 |
| HTN | nk_cell | *TRIM26* | -2.096662729 | 5.56E-05 | 0.00011671 | 0.269716085 |
| HTN | nk_cell | *ZNF736* | -0.967620821 | 0.015148865 | 0.019307143 | 1 |
| HTN | nk_cell | *PINLYP* | -1.896839561 | 1.98E-06 | 8.92E-06 | 0.009593002 |
| HTN | nk_cell | *ANKRD65* | -8.303194524 | 8.88E-06 | 2.65E-05 | 0.043107705 |
| HTN | nk_cell | *ZSCAN31* | -0.794928387 | 0.007016694 | 0.009360737 | 1 |
| HTN | nk_cell | *AC009495.2* | -27.08207204 | 0.000174214 | 0.000315878 | 0.845288657 |
| HTN | nk_cell | *LINC00957* | -3.957857369 | 1.87E-05 | 4.79E-05 | 0.090821791 |
| HTN | nk_cell | *LINC00484* | 0.933449249 | 1.79E-05 | 4.64E-05 | 0.087019454 |
| HTN | nk_cell | *KIAA0040* | 2.124058627 | 3.19E-06 | 1.24E-05 | 0.015460919 |
| HTN | nk_cell | *LINC00607* | -1.247523148 | 7.03E-06 | 2.20E-05 | 0.03411512 |
| HTN | nk_cell | *AP001468.1* | -1.326837908 | 0.001926022 | 0.00278957 | 1 |
| HTN | nk_cell | *LINC00941* | 1.553282024 | 9.45E-07 | 5.82E-06 | 0.004583071 |

| | | | | | | |
|---|---|---|---|---|---|---|
| HTN | nk_cell | *AC019117.2* | -9.228764264 | 4.59E-08 | 2.12E-06 | 0.000222701 |
| HTN | nk_cell | *ZBED5* | -1.84321013 | 5.59E-05 | 0.000117297 | 0.271189969 |
| HTN | nk_cell | *LINC00987* | 1.404537986 | 2.40E-07 | 4.07E-06 | 0.001165992 |
| HTN | nk_cell | *GS1-124K5.4* | 0.780423184 | 0.001832947 | 0.002662712 | 1 |
| HTN | nk_cell | *LINC00959* | -3.139902559 | 1.22E-05 | 3.40E-05 | 0.059167474 |
| HTN | nk_cell | *IRGM* | 1.355867398 | 7.57E-05 | 0.000151516 | 0.367123511 |
| HTN | nk_cell | *LINC01143* | -6.522210987 | 5.62E-07 | 4.48E-06 | 0.00272645 |
| HTN | nk_cell | *LINC00211* | 3.430337958 | 4.02E-06 | 1.47E-05 | 0.019488852 |
| HTN | nk_cell | *LRRC37A2* | -2.385405567 | 1.07E-06 | 6.16E-06 | 0.005193019 |
| HTN | nk_cell | *TXNDC5* | -5.257210231 | 4.57E-07 | 4.29E-06 | 0.00221674 |
| HTN | nk_cell | *LINC01119* | -2.212060375 | 9.21E-07 | 5.74E-06 | 0.004467838 |
| HTN | nk_cell | *NME1* | 1.560795244 | 6.75E-06 | 2.14E-05 | 0.032755442 |
| HTN | nk_cell | *TNFSF12* | 2.4929068 | 0.007524558 | 0.00999703 | 1 |
| HTN | nk_cell | *CDRT4* | -2.728685301 | 0.000104285 | 0.0002003 | 0.505991675 |
| HTN | nk_cell | *APOBEC3G* | 0.31255439 | 1.40E-06 | 7.22E-06 | 0.006797876 |
| HTN | nk_cell | *C1orf226* | 3.907808511 | 2.66E-06 | 1.09E-05 | 0.012916014 |
| HTN | nk_cell | *LILRA2* | -1.127877563 | 0.003481547 | 0.004855685 | 1 |
| HTN | nk_cell | *PSMB9* | 1.284107864 | 2.42E-06 | 1.02E-05 | 0.011730775 |
| HTN | nk_cell | *L1TD1* | -2.430056012 | 2.81E-05 | 6.63E-05 | 0.13619326 |
| HTN | nk_cell | *C1QTNF9* | 1.022137059 | 3.19E-07 | 4.18E-06 | 0.001546093 |
| HTN | nk_cell | *ISY1* | -1.042423675 | 4.65E-07 | 4.32E-06 | 0.002254546 |
| HTN | nk_cell | *PNMA2* | 2.288968114 | 0.009008287 | 0.011822615 | 1 |
| HTN | nk_cell | *KRBOX1* | -16.45135613 | 3.90E-07 | 4.24E-06 | 0.001890601 |
| HTN | nk_cell | *ARHGEF25* | -1.175820049 | 5.19E-06 | 1.78E-05 | 0.025164625 |
| HTN | nk_cell | *MIF* | 0.983286672 | 0.026033597 | 0.032198575 | 1 |
| HTN | nk_cell | *NSUN6* | -1.60219018 | 0.012161102 | 0.015705527 | 1 |
| HTN | nk_cell | *LINC00877* | -1.565758541 | 1.40E-06 | 7.20E-06 | 0.006775435 |
| HTN | nk_cell | *TDGF1* | -2.840700505 | 6.19E-06 | 2.01E-05 | 0.030034151 |
| HTN | nk_cell | *CD302* | -1.649530496 | 1.98E-06 | 8.92E-06 | 0.009588187 |
| HTN | nk_cell | *INMT* | -2.290663058 | 0.000168026 | 0.000305915 | 0.815263647 |
| HTN | nk_cell | *ATP5O* | -3.460412019 | 4.52E-06 | 1.60E-05 | 0.021935754 |
| HTN | nk_cell | *C8orf58* | -3.510672815 | 2.16E-06 | 9.43E-06 | 0.010462401 |
| HTN | nk_cell | *HOGA1* | -2.078957903 | 3.99E-07 | 4.24E-06 | 0.001935664 |
| HTN | nk_cell | *LINC00969* | -1.269731221 | 5.72E-06 | 1.90E-05 | 0.027738465 |
| HTN | nk_cell | *MRPL20* | -2.263856045 | 0.000314579 | 0.000534245 | 1 |
| HTN | nk_cell | *DECR2* | -1.497740561 | 4.07E-07 | 4.24E-06 | 0.001977075 |
| HTN | nk_cell | *ZNF709* | 11.55839297 | 2.37E-05 | 5.79E-05 | 0.114764077 |
| HTN | nk_cell | *ERVW-1* | 4.790788861 | 3.08E-06 | 1.21E-05 | 0.014957267 |
| HTN | nk_cell | *MRPL33* | -2.151179556 | 0.000195948 | 0.00035031 | 0.95074122 |
| HTN | nk_cell | *STON1* | -2.961095357 | 0.00091963 | 0.001419225 | 1 |
| HTN | nk_cell | *VSIG8* | -3.287078368 | 8.29E-08 | 3.01E-06 | 0.00040211 |
| HTN | nk_cell | *FGF14-IT1* | -3.159659941 | 4.30E-07 | 4.24E-06 | 0.002086711 |
| HTN | nk_cell | *PALM2* | -0.961369994 | 4.29E-07 | 4.24E-06 | 0.002083614 |
| HTN | nk_cell | *NBPF15* | -2.261959764 | 3.60E-06 | 1.36E-05 | 0.017477068 |
| HTN | nk_cell | *WFDC6* | -8.640723611 | 1.70E-05 | 4.44E-05 | 0.082519944 |
| HTN | nk_cell | *UPK3B* | -2.693103955 | 1.29E-05 | 3.56E-05 | 0.062746784 |
| HTN | nk_cell | *NME1-NME2* | -3.978103865 | 0.022635663 | 0.028262542 | 1 |
| HTN | nk_cell | *LEFTY1* | 7.668900153 | 4.41E-06 | 1.57E-05 | 0.021386794 |
| HTN | nk_cell | *TTC4* | 1.961577051 | 0.012568433 | 0.016171318 | 1 |
| HTN | nk_cell | *APOBEC3D* | 1.061114494 | 2.76E-07 | 4.07E-06 | 0.001338569 |

| | | | | | | |
|---|---|---|---|---|---|---|
| HTN | nk_cell | *TMEM199* | 1.099394804 | 3.01E-05 | 7.04E-05 | 0.145866633 |
| HTN | nk_cell | *SPRR2F* | -3.195741624 | 0.00058868 | 0.000942353 | 1 |
| HTN | nk_cell | *P2RY11* | 0.860115898 | 7.24E-07 | 4.96E-06 | 0.003514214 |
| HTN | nk_cell | *DBNDD2* | -7.274693292 | 5.39E-05 | 0.000113656 | 0.261636311 |
| HTN | nk_cell | *LILRA6* | -0.920531846 | 0.00158108 | 0.002326077 | 1 |
| HTN | nk_cell | *SCARF2* | -2.345133307 | 6.02E-06 | 1.97E-05 | 0.029194212 |
| HTN | nk_cell | *APOBEC3C* | 0.874795533 | 2.44E-07 | 4.07E-06 | 0.001184262 |
| HTN | nk_cell | *C4A* | -2.392467694 | 0.012443808 | 0.016032224 | 1 |
| HTN | nk_cell | *CRYBB2* | -1.858630208 | 0.032017459 | 0.039203968 | 1 |
| HTN | nk_cell | *NEAT1* | -1.119421453 | 4.96E-05 | 0.000106337 | 0.240533857 |
| HTN | nk_cell | *RAB30-AS1* | -2.033671673 | 3.32E-06 | 1.28E-05 | 0.016085982 |
| HTN | nk_cell | *DNAJC3-AS1* | -2.669830291 | 6.61E-06 | 2.11E-05 | 0.032076916 |
| HTN | nk_cell | *LINC00926* | -2.463236184 | 6.01E-07 | 4.54E-06 | 0.002916178 |
| HTN | nk_cell | *DPH6-AS1* | -1.560736541 | 5.28E-06 | 1.80E-05 | 0.025622151 |
| HTN | nk_cell | *CDK11B* | -2.984476197 | 6.99E-05 | 0.000141804 | 0.339337206 |
| HTN | nk_cell | *ABHD14A* | -3.170564711 | 0.002221729 | 0.00319497 | 1 |
| HTN | nk_cell | *SRP14-AS1* | -0.75247673 | 0.012497156 | 0.016092411 | 1 |
| HTN | nk_cell | *GDNF-AS1* | -1.845849511 | 5.37E-07 | 4.42E-06 | 0.002605128 |
| HTN | nk_cell | *CCDC153* | -1.679969374 | 4.11E-07 | 4.24E-06 | 0.00199327 |
| HTN | nk_cell | *C5orf17* | -2.462159424 | 1.86E-05 | 4.76E-05 | 0.090051203 |
| HTN | nk_cell | *TMEM150C* | -1.442716957 | 0.00118452 | 0.001786538 | 1 |
| HTN | nk_cell | *LINC00939* | -2.764049075 | 1.69E-05 | 4.43E-05 | 0.082148777 |
| HTN | nk_cell | *LINC01060* | -1.772667003 | 4.39E-05 | 9.61E-05 | 0.213009326 |
| HTN | nk_cell | *LINC00942* | -6.341682671 | 1.84E-07 | 4.07E-06 | 0.000893801 |
| HTN | nk_cell | *PDCD6* | -2.653062551 | 0.002114119 | 0.00304383 | 1 |
| HTN | nk_cell | *GOLGA8K* | -2.387588819 | 0.000173225 | 0.000314201 | 0.840487649 |
| HTN | nk_cell | *TMEM158* | -4.462013184 | 5.85E-07 | 4.54E-06 | 0.002840328 |
| HTN | nk_cell | *DNAH10OS* | -7.967533678 | 6.02E-07 | 4.54E-06 | 0.002923195 |
| HTN | nk_cell | *PCDHA10* | -1.093759459 | 0.00071736 | 0.001128976 | 1 |
| HTN | nk_cell | *ZNF718* | 2.838889848 | 0.000148087 | 0.00027393 | 0.718519067 |
| HTN | nk_cell | *SMIM20* | -3.584581103 | 0.002044731 | 0.002947426 | 1 |
| HTN | nk_cell | *LINC00989* | -1.235956293 | 4.94E-07 | 4.32E-06 | 0.002396832 |
| HTN | nk_cell | *GLI4* | -1.432894258 | 6.34E-07 | 4.62E-06 | 0.003074234 |
| HTN | nk_cell | *SNHG18* | -2.442804732 | 5.57E-06 | 1.87E-05 | 0.027031149 |
| HTN | nk_cell | *ALG1L2* | -3.338586337 | 6.37E-06 | 2.06E-05 | 0.03091668 |
| HTN | nk_cell | *ZNF550* | 4.01202445 | 0.000195339 | 0.000349478 | 0.94778444 |
| HTN | nk_cell | *LINC01094* | -1.663749246 | 5.65E-07 | 4.50E-06 | 0.002742519 |
| HTN | nk_cell | *TMEM200B* | 4.396449694 | 1.31E-05 | 3.59E-05 | 0.06345803 |
| HTN | nk_cell | *SLC10A5* | -4.052848508 | 0.000183511 | 0.000330633 | 0.890394206 |
| HTN | nk_cell | *PCDHGA6* | -0.987444463 | 3.27E-05 | 7.53E-05 | 0.158605929 |
| HTN | nk_cell | *UTP14C* | 4.098085989 | 1.22E-05 | 3.41E-05 | 0.059343452 |
| HTN | nk_cell | *PCDHGB2* | -5.634808143 | 1.69E-07 | 4.07E-06 | 0.000819424 |
| HTN | nk_cell | *LYN* | 1.539843644 | 3.70E-06 | 1.39E-05 | 0.017937313 |
| HTN | nk_cell | *SIGLEC14* | -3.131132009 | 0.000116152 | 0.000220059 | 0.563570427 |
| HTN | nk_cell | *MEX3A* | -2.060878532 | 0.000365483 | 0.000611491 | 1 |
| HTN | nk_cell | *BRK1* | 2.048923107 | 0.000260245 | 0.000450325 | 1 |
| HTN | nk_cell | *SNHG9* | -2.01574938 | 2.45E-05 | 5.95E-05 | 0.118906961 |
| HTN | nk_cell | *PCDHA3* | -3.445518095 | 0.000353213 | 0.000593212 | 1 |
| HTN | nk_cell | *ZNF432* | -1.92933935 | 1.26E-06 | 6.79E-06 | 0.006127642 |
| HTN | nk_cell | *HMBS* | -1.288847497 | 3.91E-05 | 8.72E-05 | 0.189806169 |

| | | | | | | |
|---|---|---|---|---|---|---|
| HTN | nk_cell | *TAS2R31* | 1.393780939 | 6.08E-05 | 0.000125603 | 0.295167181 |
| HTN | nk_cell | *AC093668.2* | -1.749208366 | 1.23E-06 | 6.75E-06 | 0.005988134 |
| HTN | nk_cell | *KIAA1147* | -1.930317017 | 2.59E-05 | 6.20E-05 | 0.125676743 |
| HTN | nk_cell | *ZBED6* | -2.252077807 | 2.75E-07 | 4.07E-06 | 0.001334872 |
| HTN | nk_cell | *CNPY2* | -2.820134292 | 0.000286016 | 0.000489852 | 1 |
| HTN | nk_cell | *LINC00592* | -4.262521503 | 9.52E-05 | 0.000184678 | 0.461879175 |
| HTN | nk_cell | *CHURC1* | -1.760855103 | 0.010999661 | 0.014289252 | 1 |
| HTN | nk_cell | *RTEL1* | 2.307151897 | 0.000476033 | 0.000775852 | 1 |
| HTN | nk_cell | *PDF* | 6.600154591 | 0.002872853 | 0.004063872 | 1 |
| HTN | nk_cell | *LINC00640* | -1.11257707 | 0.003010955 | 0.004246847 | 1 |
| HTN | nk_cell | *HIF1A-AS2* | -2.396312778 | 0.003053758 | 0.004303467 | 1 |
| HTN | nk_cell | *LINC00638* | -1.934373214 | 0.003313319 | 0.00463559 | 1 |
| HTN | nk_cell | *SLC25A21-AS1* | 3.413646182 | 0.001216774 | 0.001829496 | 1 |
| HTN | nk_cell | *RNASE4* | -4.715448012 | 5.29E-07 | 4.40E-06 | 0.002565366 |
| HTN | nk_cell | *MC1R* | 4.962640341 | 0.000125854 | 0.000236592 | 0.610644645 |
| HTN | nk_cell | *LINC00639* | 1.848743595 | 1.01E-05 | 2.93E-05 | 0.049055843 |
| HTN | nk_cell | *SMIM6* | 6.341651452 | 0.002356623 | 0.003379939 | 1 |
| HTN | nk_cell | *LINC00924* | -1.204704807 | 0.000293217 | 0.000501124 | 1 |
| HTN | nk_cell | *ITGB3* | -1.284347348 | 0.008308157 | 0.010963061 | 1 |
| HTN | nk_cell | *MRPL46* | 3.499428441 | 2.70E-06 | 1.10E-05 | 0.013082157 |
| HTN | nk_cell | *UBE2Q2L* | -3.74253703 | 2.02E-05 | 5.08E-05 | 0.09812078 |
| HTN | nk_cell | *IQCH-AS1* | -2.049118349 | 0.00912988 | 0.011969246 | 1 |
| HTN | nk_cell | *AC083843.1* | -19.45497645 | 4.06E-07 | 4.24E-06 | 0.001968991 |
| HTN | nk_cell | *PWRN1* | -1.423594522 | 5.01E-08 | 2.19E-06 | 0.000243132 |
| HTN | nk_cell | *LINC01003* | 2.922087368 | 6.81E-05 | 0.000138543 | 0.33056375 |
| HTN | nk_cell | *C15orf65* | -2.829810955 | 0.000406174 | 0.000671696 | 1 |
| HTN | nk_cell | *GOLGA8H* | -0.558045234 | 0.019782016 | 0.024850175 | 1 |
| HTN | nk_cell | *LINC00662* | -1.974339695 | 7.68E-05 | 0.000153515 | 0.372581311 |
| HTN | nk_cell | *C19orf84* | 0.404761966 | 2.80E-07 | 4.07E-06 | 0.001359207 |
| HTN | nk_cell | *LINC00675* | -1.470099471 | 0.006222545 | 0.008356432 | 1 |
| HTN | nk_cell | *LINC00667* | -1.644535444 | 0.000229734 | 0.000404158 | 1 |
| HTN | nk_cell | *LINC00909* | -2.488763679 | 0.000553568 | 0.000890849 | 1 |
| HTN | nk_cell | *DYNLL2* | 4.545750815 | 8.44E-06 | 2.55E-05 | 0.040958875 |
| HTN | nk_cell | *STRADA* | -2.079803616 | 0.004025068 | 0.005543466 | 1 |
| HTN | nk_cell | *MYO15B* | -1.779836819 | 0.000140876 | 0.000261889 | 0.683530248 |
| HTN | nk_cell | *ILF3-AS1* | -1.655341192 | 0.021715709 | 0.02716283 | 1 |
| HTN | nk_cell | *ZNF582-AS1* | 5.525011888 | 0.0037462 | 0.005197759 | 1 |
| HTN | nk_cell | *ZNF224* | -2.677542079 | 9.82E-05 | 0.000189717 | 0.476379407 |
| HTN | nk_cell | *LIN37* | -4.283034547 | 0.000198627 | 0.000354708 | 0.963740464 |
| HTN | nk_cell | *NDUFA7* | 1.406675948 | 2.75E-07 | 4.07E-06 | 0.001334661 |
| HTN | nk_cell | *LINC00664* | 2.665930198 | 3.32E-07 | 4.24E-06 | 0.001609626 |
| HTN | nk_cell | *SCGB1B2P* | -3.023630004 | 0.000426422 | 0.000701831 | 1 |
| HTN | nk_cell | *ZNF728* | -1.558878371 | 7.52E-07 | 5.09E-06 | 0.00364678 |
| HTN | nk_cell | *TRABD2B* | -2.875429871 | 3.45E-05 | 7.84E-05 | 0.167235768 |
| HTN | nk_cell | *AC093157.1* | -1.689918497 | 1.60E-06 | 7.80E-06 | 0.007780743 |
| HTN | nk_cell | *FBXO17* | -1.474736676 | 2.10E-05 | 5.24E-05 | 0.101878238 |
| HTN | nk_cell | *ZNF587B* | -1.329093719 | 4.28E-07 | 4.24E-06 | 0.002075587 |
| HTN | nk_cell | *SNHG8* | -1.944260689 | 0.000806788 | 0.00125748 | 1 |
| HTN | nk_cell | *HCG17* | -2.036639234 | 4.51E-05 | 9.81E-05 | 0.218698673 |
| HTN | nk_cell | *GTF2H5* | -1.726821487 | 1.25E-05 | 3.46E-05 | 0.060494484 |

| | | | | | | |
|---|---|---|---|---|---|---|
| HTN | nk_cell | *FGF14-AS2* | -1.684186678 | 5.93E-07 | 4.54E-06 | 0.002876792 |
| HTN | nk_cell | *CASC15* | -1.528975154 | 2.12E-06 | 9.30E-06 | 0.010262309 |
| HTN | nk_cell | *AP000347.2* | -2.389800881 | 0.001643349 | 0.002408194 | 1 |
| HTN | nk_cell | *GRIN2B* | -0.960076736 | 0.000201174 | 0.000358991 | 0.97609637 |
| HTN | nk_cell | *PAXIP1-AS1* | -3.147204205 | 0.000166656 | 0.000303991 | 0.808616409 |
| HTN | pericyte | *SEMA3F* | -7.975821262 | 1.68E-05 | 0.001015923 | 0.081273863 |
| HTN | pericyte | *RAD52* | -7.446021986 | 0.00073776 | 0.013318553 | 1 |
| HTN | pericyte | *HS3ST1* | -2.481984419 | 0.012095844 | 0.048303732 | 1 |
| HTN | pericyte | *M6PR* | -7.262377614 | 8.94E-05 | 0.00344405 | 0.433950311 |
| HTN | pericyte | *RBM6* | -3.962519043 | 5.17E-07 | 6.78E-05 | 0.002509552 |
| HTN | pericyte | *RECQL* | -7.104837563 | 0.004071591 | 0.027025118 | 1 |
| HTN | pericyte | *POLR2J* | 7.111298887 | 0.011624084 | 0.047000047 | 1 |
| HTN | pericyte | *MEOX1* | -3.704016204 | 0.001756909 | 0.022027195 | 1 |
| HTN | pericyte | *PON1* | -9.501288234 | 0.005028213 | 0.030382178 | 1 |
| HTN | pericyte | *GGNBP2* | -6.921155508 | 0.005482078 | 0.031735351 | 1 |
| HTN | pericyte | *ABCC8* | 1.463672662 | 0.008204811 | 0.038500008 | 1 |
| HTN | pericyte | *SCIN* | -4.556423175 | 0.000264157 | 0.006890799 | 1 |
| HTN | pericyte | *CD79B* | -7.977027188 | 4.88E-06 | 0.000372658 | 0.023660196 |
| HTN | pericyte | *ST7L* | -4.380820665 | 0.000293336 | 0.00733642 | 1 |
| HTN | pericyte | *NADK* | -7.001182592 | 0.00016603 | 0.005098592 | 0.805577558 |
| HTN | pericyte | *DLEC1* | -4.507824107 | 0.002401245 | 0.023442335 | 1 |
| HTN | pericyte | *MMP25* | -4.050359104 | 0.004115624 | 0.027168715 | 1 |
| HTN | pericyte | *IL32* | -4.399668731 | 0.002961294 | 0.023712861 | 1 |
| HTN | pericyte | *MLXIPL* | -1.985371489 | 0.000215225 | 0.00601954 | 1 |
| HTN | pericyte | *CLCN6* | -8.485333872 | 6.08E-05 | 0.002644087 | 0.294759619 |
| HTN | pericyte | *TTC19* | -5.591406887 | 0.008940949 | 0.040430088 | 1 |
| HTN | pericyte | *CEP68* | 5.815683225 | 0.004061441 | 0.027025118 | 1 |
| HTN | pericyte | *EHD3* | -5.603203786 | 0.004013547 | 0.026897416 | 1 |
| HTN | pericyte | *CLCA4* | -7.470902439 | 0.011510552 | 0.046618697 | 1 |
| HTN | pericyte | *HGF* | 1.573606845 | 0.006393302 | 0.034659555 | 1 |
| HTN | pericyte | *GLRX2* | -7.827388994 | 0.002884805 | 0.02367523 | 1 |
| HTN | pericyte | *GCLM* | -5.970880831 | 0.012719427 | 0.049371727 | 1 |
| HTN | pericyte | *RNASET2* | -4.696616988 | 4.52E-05 | 0.00212984 | 0.219373547 |
| HTN | pericyte | *MIPEP* | -7.380224008 | 0.001724154 | 0.021785405 | 1 |
| HTN | pericyte | *INSRR* | -8.750348968 | 0.000287622 | 0.007305639 | 1 |
| HTN | pericyte | *B4GALT7* | -7.447376596 | 0.006232544 | 0.034131269 | 1 |
| HTN | pericyte | *GRN* | -4.019308529 | 0.008160713 | 0.038500008 | 1 |
| HTN | pericyte | *CENPQ* | -1.759346226 | 0.00469381 | 0.029272961 | 1 |
| HTN | pericyte | *UBA6* | -4.484742991 | 0.012643761 | 0.049235575 | 1 |
| HTN | pericyte | *SH3YL1* | -7.819951368 | 0.000639141 | 0.012305994 | 1 |
| HTN | pericyte | *ADSS* | -6.618800706 | 0.01162007 | 0.047000047 | 1 |
| HTN | pericyte | *DNAH5* | -3.849895322 | 0.00421445 | 0.027381347 | 1 |
| HTN | pericyte | *INPP4A* | -5.715133634 | 0.004557519 | 0.028606836 | 1 |
| HTN | pericyte | *SPATA7* | -7.308616061 | 0.00220015 | 0.023308138 | 1 |
| HTN | pericyte | *MED17* | -8.989412372 | 0.004024634 | 0.026899796 | 1 |
| HTN | pericyte | *ANO2* | -5.687572338 | 0.006001223 | 0.033344492 | 1 |
| HTN | pericyte | *RCN1* | -6.926049978 | 0.009045936 | 0.040535121 | 1 |
| HTN | pericyte | *THOC3* | -10.79947108 | 0.00305585 | 0.023886115 | 1 |
| HTN | pericyte | *LY75* | -8.569339363 | 0.002949336 | 0.023712861 | 1 |
| HTN | pericyte | *SDCCAG8* | 4.375089313 | 0.00196154 | 0.022752995 | 1 |

| | | | | | | |
|---|---|---|---|---|---|---|
| HTN | pericyte | *GALC* | -5.966704141 | 0.006511096 | 0.03502421 | 1 |
| HTN | pericyte | *ITIH4* | 1.537721494 | 0.001801738 | 0.0223011 | 1 |
| HTN | pericyte | *NPFFR2* | -6.181772414 | 0.001924539 | 0.022609835 | 1 |
| HTN | pericyte | *IL17RB* | -5.578308269 | 0.00084089 | 0.014416964 | 1 |
| HTN | pericyte | *DNAJC25* | -8.493200063 | 0.002488044 | 0.023519329 | 1 |
| HTN | pericyte | *PIGV* | -5.172421086 | 1.57E-07 | 2.83E-05 | 0.000764158 |
| HTN | pericyte | *RIMBP2* | -6.604934125 | 0.002353721 | 0.023376742 | 1 |
| HTN | pericyte | *SPAG4* | -5.707380522 | 0.013028308 | 0.049971029 | 1 |
| HTN | pericyte | *LTK* | -24.74353753 | 0.002936158 | 0.023704224 | 1 |
| HTN | pericyte | *ADCK1* | -7.341838191 | 0.003959089 | 0.02677323 | 1 |
| HTN | pericyte | *SLC12A2* | 0.910716044 | 0.00492552 | 0.030023393 | 1 |
| HTN | pericyte | *BCAS1* | -3.916651029 | 0.005421909 | 0.031467827 | 1 |
| HTN | pericyte | *UHRF1BP1* | 3.70447051 | 0.001950093 | 0.022752995 | 1 |
| HTN | pericyte | *MCM10* | 0.998841315 | 0.001360058 | 0.01890831 | 1 |
| HTN | pericyte | *TIE1* | -5.876146767 | 6.76E-07 | 8.41E-05 | 0.003278289 |
| HTN | pericyte | *SLC9A3* | -1.654001768 | 0.007597146 | 0.037121203 | 1 |
| HTN | pericyte | *NGEF* | -9.177302007 | 0.000631245 | 0.012271001 | 1 |
| HTN | pericyte | *ATXN3* | -4.319938342 | 0.001510572 | 0.020137909 | 1 |
| HTN | pericyte | *FGFR2* | -2.646473863 | 0.006889618 | 0.035790607 | 1 |
| HTN | pericyte | *MTFR1* | -6.215949343 | 0.011308647 | 0.046108871 | 1 |
| HTN | pericyte | *METTL22* | -7.5003221 | 0.007666175 | 0.037305125 | 1 |
| HTN | pericyte | *ROGDI* | -8.907957127 | 0.002543106 | 0.023519329 | 1 |
| HTN | pericyte | *POLR1A* | -6.188765924 | 2.67E-06 | 0.000227351 | 0.012958988 |
| HTN | pericyte | *IP6K2* | -7.306227492 | 0.005338089 | 0.031271844 | 1 |
| HTN | pericyte | *IFT80* | -16.57623317 | 2.38E-05 | 0.001315306 | 0.115716223 |
| HTN | pericyte | *DRD4* | -5.187085895 | 0.004172874 | 0.027381347 | 1 |
| HTN | pericyte | *PIGB* | -5.924390888 | 0.010543377 | 0.044037653 | 1 |
| HTN | pericyte | *ASNS* | 5.720317624 | 0.001324836 | 0.01847157 | 1 |
| HTN | pericyte | *AP3M2* | -8.703555268 | 0.00253211 | 0.023519329 | 1 |
| HTN | pericyte | *ST6GALNAC2* | -3.42859103 | 0.009411534 | 0.041400512 | 1 |
| HTN | pericyte | *EPN2* | -5.55728425 | 0.001892154 | 0.0224925 | 1 |
| HTN | pericyte | *TRHDE* | -2.531689286 | 0.000302422 | 0.007383636 | 1 |
| HTN | pericyte | *CRMP1* | -7.327363233 | 0.009652273 | 0.041803673 | 1 |
| HTN | pericyte | *AP1M1* | -9.234373053 | 0.001843216 | 0.022363374 | 1 |
| HTN | pericyte | *PVR* | -6.104094826 | 0.000110414 | 0.00399797 | 0.535727994 |
| HTN | pericyte | *SCARB1* | -5.63895901 | 0.006309722 | 0.034321491 | 1 |
| HTN | pericyte | *ALPK1* | -4.196212217 | 0.011231012 | 0.045830839 | 1 |
| HTN | pericyte | *ADAM11* | -2.140452006 | 0.00691373 | 0.0358138 | 1 |
| HTN | pericyte | *PPP2R2C* | -16.36752142 | 0.008967291 | 0.04051145 | 1 |
| HTN | pericyte | *SPAG5* | -11.72708528 | 0.000337088 | 0.008056899 | 1 |
| HTN | pericyte | *XAB2* | -6.35231268 | 0.008378532 | 0.038902044 | 1 |
| HTN | pericyte | *SIRT6* | 5.528458549 | 0.008190069 | 0.038500008 | 1 |
| HTN | pericyte | *MYH7B* | -5.214608383 | 0.005704417 | 0.032409638 | 1 |
| HTN | pericyte | *SAR1A* | -5.581668726 | 0.003675257 | 0.026071244 | 1 |
| HTN | pericyte | *DUSP13* | 15.14859086 | 0.004362174 | 0.027927924 | 1 |
| HTN | pericyte | *FDFT1* | 5.87088643 | 0.008432557 | 0.039003588 | 1 |
| HTN | pericyte | *SCGN* | 1.898266501 | 0.004251642 | 0.027507529 | 1 |
| HTN | pericyte | *PGM1* | -6.501754173 | 0.002317028 | 0.023376742 | 1 |
| HTN | pericyte | *RABL2B* | -7.308373745 | 0.00250863 | 0.023519329 | 1 |
| HTN | pericyte | *SCTR* | -3.627830798 | 0.00242824 | 0.023519329 | 1 |

| | | | | | | |
|---|---|---|---|---|---|---|
| HTN | pericyte | *KCNN2* | -6.980789129 | 0.001792823 | 0.02224751 | 1 |
| HTN | pericyte | *CCNT2* | -6.064744732 | 0.010173322 | 0.043034836 | 1 |
| HTN | pericyte | *LYRM2* | -6.635051358 | 0.000848005 | 0.014473383 | 1 |
| HTN | pericyte | *ULK2* | -5.815963709 | 0.011354422 | 0.046217831 | 1 |
| HTN | pericyte | *PLOD1* | 6.397194229 | 0.000689142 | 0.012910095 | 1 |
| HTN | pericyte | *OXCT1* | -6.3310232 | 0.000455201 | 0.00985997 | 1 |
| HTN | pericyte | *NOA1* | -8.038550988 | 0.001566347 | 0.020431711 | 1 |
| HTN | pericyte | *WBP11* | -6.182108572 | 0.003710218 | 0.026106374 | 1 |
| HTN | pericyte | *CPNE3* | -6.081138126 | 0.009810026 | 0.042148419 | 1 |
| HTN | pericyte | *MTIF2* | -5.837066475 | 0.002898679 | 0.02367523 | 1 |
| HTN | pericyte | *SNX10* | -5.492022696 | 7.90E-09 | 2.56E-06 | 3.83E-05 |
| HTN | pericyte | *ITPKC* | -5.536137063 | 0.000909537 | 0.015217497 | 1 |
| HTN | pericyte | *ALG9* | -6.231645506 | 0.002622731 | 0.023519329 | 1 |
| HTN | pericyte | *LPCAT2* | -6.792202286 | 0.009820242 | 0.042148419 | 1 |
| HTN | pericyte | *SF3B2* | -4.717426632 | 0.009396889 | 0.041380441 | 1 |
| HTN | pericyte | *AAMDC* | -6.538765777 | 7.67E-07 | 9.08E-05 | 0.00372298 |
| HTN | pericyte | *ASAP3* | -5.61541184 | 0.003191534 | 0.024319694 | 1 |
| HTN | pericyte | *DOCK3* | -3.459076239 | 0.002468952 | 0.023519329 | 1 |
| HTN | pericyte | *ARHGAP28* | -4.503556465 | 0.004811759 | 0.029495473 | 1 |
| HTN | pericyte | *PPP1R13B* | -6.256392638 | 0.00037337 | 0.008585735 | 1 |
| HTN | pericyte | *EBF4* | -6.654172564 | 0.012844907 | 0.049620613 | 1 |
| HTN | pericyte | *MAVS* | -5.155355539 | 0.011147891 | 0.045606719 | 1 |
| HTN | pericyte | *SNX5* | -7.181384203 | 0.012812787 | 0.049575472 | 1 |
| HTN | pericyte | *ANKRD24* | -9.903721521 | 0.002102675 | 0.02292625 | 1 |
| HTN | pericyte | *SPTLC1* | -6.573187683 | 0.003972944 | 0.02677323 | 1 |
| HTN | pericyte | *YPEL3* | 5.015195146 | 0.00725641 | 0.036435078 | 1 |
| HTN | pericyte | *MUL1* | -4.700963106 | 2.09E-05 | 0.001192687 | 0.101378432 |
| HTN | pericyte | *PABPC4* | -10.1452515 | 6.29E-08 | 1.57E-05 | 0.000305007 |
| HTN | pericyte | *CERS4* | -6.507158897 | 0.007053104 | 0.035947121 | 1 |
| HTN | pericyte | *DTX2* | -5.769436982 | 0.002625483 | 0.023519329 | 1 |
| HTN | pericyte | *LAMB1* | -2.349786543 | 0.012920094 | 0.049713159 | 1 |
| HTN | pericyte | *ABCC6* | -2.937364459 | 0.00041545 | 0.009162558 | 1 |
| HTN | pericyte | *RAPGEF4* | -4.793975373 | 0.001442636 | 0.019606915 | 1 |
| HTN | pericyte | *TF* | -6.246315356 | 0.002464975 | 0.023519329 | 1 |
| HTN | pericyte | *TOX4* | -6.952158387 | 0.011458446 | 0.046485268 | 1 |
| HTN | pericyte | *TBX15* | 0.957877375 | 0.001857584 | 0.02236476 | 1 |
| HTN | pericyte | *XYLB* | -7.542180863 | 0.011136526 | 0.045598669 | 1 |
| HTN | pericyte | *MSH2* | -7.077257769 | 0.012838094 | 0.049620613 | 1 |
| HTN | pericyte | *NANS* | -6.024724855 | 0.007630901 | 0.037248624 | 1 |
| HTN | pericyte | *DSP* | -6.537027255 | 0.003005503 | 0.02386694 | 1 |
| HTN | pericyte | *SCD* | -7.917683703 | 8.18E-08 | 1.68E-05 | 0.000396939 |
| HTN | pericyte | *MAST3* | -3.210862439 | 0.002857528 | 0.02367523 | 1 |
| HTN | pericyte | *SETD1A* | 9.851198242 | 0.00058025 | 0.011730712 | 1 |
| HTN | pericyte | *MARCH2* | 3.458803878 | 0.008137699 | 0.038500008 | 1 |
| HTN | pericyte | *MTAP* | -9.689843298 | 6.92E-05 | 0.002844458 | 0.335646002 |
| HTN | pericyte | *MADCAM1* | -6.167180546 | 0.000730896 | 0.013312983 | 1 |
| HTN | pericyte | *MED15* | -5.816156543 | 0.006987878 | 0.0358138 | 1 |
| HTN | pericyte | *SPECC1L* | -3.841589523 | 0.008335938 | 0.038778495 | 1 |
| HTN | pericyte | *PES1* | -4.939683787 | 0.007409273 | 0.03668346 | 1 |
| HTN | pericyte | *LGALS2* | -3.601445013 | 0.001411629 | 0.01934809 | 1 |

| | | | | | | |
|---|---|---|---|---|---|---|
| HTN | pericyte | *SH3BP1* | -11.16124228 | 0.007156708 | 0.036208913 | 1 |
| HTN | pericyte | *TRIOBP* | -7.127589616 | 0.002455276 | 0.023519329 | 1 |
| HTN | pericyte | *EIF3L* | -7.099585284 | 0.007832378 | 0.037701091 | 1 |
| HTN | pericyte | *CYP2D6* | -6.925537751 | 0.001311494 | 0.01847157 | 1 |
| HTN | pericyte | *RAB36* | -4.096866272 | 0.012470841 | 0.049034456 | 1 |
| HTN | pericyte | *THOC5* | -5.116927662 | 0.000525509 | 0.010850088 | 1 |
| HTN | pericyte | *TSPO* | -6.93236756 | 0.012060293 | 0.048201436 | 1 |
| HTN | pericyte | *TTLL12* | -6.15789886 | 0.00066832 | 0.012617472 | 1 |
| HTN | pericyte | *APOL4* | -3.184487983 | 0.004513606 | 0.028441579 | 1 |
| HTN | pericyte | *SAMM50* | -6.370404917 | 0.003602475 | 0.025973602 | 1 |
| HTN | pericyte | *IFT27* | -8.273267664 | 0.00627144 | 0.034227583 | 1 |
| HTN | pericyte | *FAM118A* | 2.346843306 | 0.010457466 | 0.043816601 | 1 |
| HTN | pericyte | *L3MBTL2* | -6.102911137 | 0.009535078 | 0.041604495 | 1 |
| HTN | pericyte | *PYGL* | -1.939868679 | 0.001831267 | 0.022363374 | 1 |
| HTN | pericyte | *EFS* | -10.18173423 | 0.005095587 | 0.030407778 | 1 |
| HTN | pericyte | *CD40* | -6.784006038 | 0.006674469 | 0.035407327 | 1 |
| HTN | pericyte | *PABPC1L* | -7.788509639 | 0.012597975 | 0.049215278 | 1 |
| HTN | pericyte | *DOK5* | -5.126173447 | 0.002301685 | 0.023376742 | 1 |
| HTN | pericyte | *CSTF1* | -6.04426034 | 0.002927271 | 0.023704224 | 1 |
| HTN | pericyte | *BIRC7* | -5.933530309 | 0.00060514 | 0.012082883 | 1 |
| HTN | pericyte | *PTK6* | -8.87699201 | 0.003658827 | 0.026071244 | 1 |
| HTN | pericyte | *RNF24* | -4.180635229 | 0.005508164 | 0.031765065 | 1 |
| HTN | pericyte | *ANGPT4* | 2.056398749 | 0.008490559 | 0.039211167 | 1 |
| HTN | pericyte | *SIRPB1* | -4.918740945 | 0.008294149 | 0.038658223 | 1 |
| HTN | pericyte | *CRNKL1* | -4.675733203 | 0.006547562 | 0.035132512 | 1 |
| HTN | pericyte | *DNTTIP1* | -5.297887775 | 1.37E-05 | 0.000860982 | 0.06629562 |
| HTN | pericyte | *LAMA1* | 6.165939288 | 0.006763531 | 0.035515857 | 1 |
| HTN | pericyte | *POLI* | -2.341216477 | 2.58E-05 | 0.001392841 | 0.125355652 |
| HTN | pericyte | *PARP4* | -6.689695367 | 0.004188533 | 0.027381347 | 1 |
| HTN | pericyte | *CCDC113* | -6.061109207 | 0.005099699 | 0.030407778 | 1 |
| HTN | pericyte | *ZNF500* | -5.760911508 | 0.000140973 | 0.004590617 | 0.684001939 |
| HTN | pericyte | *NME4* | -11.04607615 | 0.000150957 | 0.004756134 | 0.732444577 |
| HTN | pericyte | *SLC7A5* | -5.32445166 | 0.002374912 | 0.023376742 | 1 |
| HTN | pericyte | *UBE2I* | -6.705157153 | 0.00980312 | 0.042148419 | 1 |
| HTN | pericyte | *CAPN15* | -7.112911034 | 0.011920681 | 0.047800945 | 1 |
| HTN | pericyte | *TOX3* | 3.418865289 | 0.000732598 | 0.013312983 | 1 |
| HTN | pericyte | *RRN3P2* | -5.706775291 | 0.004948435 | 0.030125227 | 1 |
| HTN | pericyte | *TMC5* | -2.960728087 | 0.002316139 | 0.023376742 | 1 |
| HTN | pericyte | *LACTB* | -9.146224983 | 0.000600347 | 0.012036706 | 1 |
| HTN | pericyte | *TTC23* | -6.374814725 | 0.010956046 | 0.045142399 | 1 |
| HTN | pericyte | *FAH* | -4.643735256 | 0.003037727 | 0.023886115 | 1 |
| HTN | pericyte | *RPAP1* | -7.40902978 | 0.000118781 | 0.004036836 | 0.576323881 |
| HTN | pericyte | *SFRP1* | -6.163736181 | 0.004493395 | 0.028387958 | 1 |
| HTN | pericyte | *SPAG1* | -3.923282749 | 0.008612228 | 0.039585488 | 1 |
| HTN | pericyte | *TSTA3* | 4.682478736 | 0.001523849 | 0.020256757 | 1 |
| HTN | pericyte | *R3HCC1* | -3.697591289 | 0.008277765 | 0.038618959 | 1 |
| HTN | pericyte | *KLHDC4* | -4.563116707 | 0.009042118 | 0.040535121 | 1 |
| HTN | pericyte | *MAN2B1* | -3.869642961 | 0.002913049 | 0.02367523 | 1 |
| HTN | pericyte | *LIN7B* | -14.56525774 | 0.001005639 | 0.01584344 | 1 |
| HTN | pericyte | *PPP1R37* | -3.21032137 | 0.007815638 | 0.037701091 | 1 |

| | | | | | | |
|---|---|---|---|---|---|---|
| HTN | pericyte | *ARHGEF18* | 3.986177658 | 0.00820386 | 0.038500008 | 1 |
| HTN | pericyte | *ERCC2* | -7.446770061 | 0.006194899 | 0.034040371 | 1 |
| HTN | pericyte | *CD37* | -9.616793318 | 7.50E-06 | 0.000519976 | 0.036398328 |
| HTN | pericyte | *AMH* | -31.27033905 | 0.002762178 | 0.023519329 | 1 |
| HTN | pericyte | *FCER2* | -11.72068257 | 0.0026179 | 0.023519329 | 1 |
| HTN | pericyte | *CLEC4M* | -29.96064392 | 0.004306126 | 0.027673274 | 1 |
| HTN | pericyte | *IL27RA* | -5.998099767 | 0.00994459 | 0.042399955 | 1 |
| HTN | pericyte | *FAM32A* | -12.26377418 | 0.005564665 | 0.03188882 | 1 |
| HTN | pericyte | *OLFM2* | 1.695621318 | 0.009600248 | 0.041738713 | 1 |
| HTN | pericyte | *RASAL3* | -6.932775754 | 0.005327939 | 0.031271844 | 1 |
| HTN | pericyte | *POLR2I* | -6.956425926 | 0.001541104 | 0.020319118 | 1 |
| HTN | pericyte | *ICAM5* | -5.383575741 | 7.36E-05 | 0.002950525 | 0.357013486 |
| HTN | pericyte | *GRIN2D* | -8.495184997 | 0.000928786 | 0.01543311 | 1 |
| HTN | pericyte | *CAPS* | -7.221004748 | 0.002270026 | 0.023376742 | 1 |
| HTN | pericyte | *PLEKHA4* | -7.139482272 | 0.001923895 | 0.022609835 | 1 |
| HTN | pericyte | *TNPO2* | -8.538922998 | 0.000289294 | 0.007305639 | 1 |
| HTN | pericyte | *GCDH* | -6.441896003 | 0.00408803 | 0.027097163 | 1 |
| HTN | pericyte | *RAB3A* | -11.85695618 | 0.002564909 | 0.023519329 | 1 |
| HTN | pericyte | *UPK1A* | 5.958940212 | 0.006936306 | 0.0358138 | 1 |
| HTN | pericyte | *ZNF85* | -9.320305941 | 0.005011557 | 0.030349051 | 1 |
| HTN | pericyte | *RASA4* | -19.65329676 | 0.002375254 | 0.023376742 | 1 |
| HTN | pericyte | *ADAP1* | 7.213568598 | 0.007760267 | 0.03755768 | 1 |
| HTN | pericyte | *VIPR2* | -3.638870524 | 0.004752136 | 0.02937244 | 1 |
| HTN | pericyte | *EIF3B* | -7.131472296 | 0.009117635 | 0.040731344 | 1 |
| HTN | pericyte | *TAF6* | -3.933473203 | 0.000805055 | 0.014077604 | 1 |
| HTN | pericyte | *TFR2* | -4.608086133 | 0.005016475 | 0.030349051 | 1 |
| HTN | pericyte | *IMPDH1* | -6.724595442 | 0.002847953 | 0.02367523 | 1 |
| HTN | pericyte | *MEST* | -4.895727632 | 2.42E-06 | 0.000213395 | 0.011736709 |
| HTN | pericyte | *POU6F2* | -3.477837244 | 0.000379818 | 0.008692815 | 1 |
| HTN | pericyte | *AEBP1* | -7.667517126 | 0.012911024 | 0.049713159 | 1 |
| HTN | pericyte | *GCK* | -17.9819751 | 0.002450672 | 0.023519329 | 1 |
| HTN | pericyte | *FKTN* | -6.291158577 | 0.009984917 | 0.042497208 | 1 |
| HTN | pericyte | *OGN* | -5.782180048 | 0.000109465 | 0.003993428 | 0.531125961 |
| HTN | pericyte | *ECM2* | -4.617884362 | 0.009830743 | 0.042148419 | 1 |
| HTN | pericyte | *PTGR1* | -3.491878203 | 0.009675606 | 0.04184139 | 1 |
| HTN | pericyte | *HPS1* | -5.04199299 | 0.003790359 | 0.026406882 | 1 |
| HTN | pericyte | *CDH23* | -4.488538935 | 0.004051244 | 0.027000873 | 1 |
| HTN | pericyte | *LIPA* | -3.184111164 | 0.012784239 | 0.049504493 | 1 |
| HTN | pericyte | *PITRM1* | -8.394011355 | 0.00861682 | 0.039585488 | 1 |
| HTN | pericyte | *DNAJC12* | 3.387749218 | 0.006621215 | 0.035303446 | 1 |
| HTN | pericyte | *RAD51C* | -6.354899575 | 6.27E-05 | 0.002646543 | 0.30435239 |
| HTN | pericyte | *PNPO* | -19.14894484 | 2.39E-05 | 0.001315306 | 0.115746917 |
| HTN | pericyte | *RAI1* | -6.135469057 | 0.00399155 | 0.026810347 | 1 |
| HTN | pericyte | *ICAM2* | -6.684901285 | 0.004677481 | 0.029208669 | 1 |
| HTN | pericyte | *PEX12* | -6.888129836 | 0.001113242 | 0.0167473 | 1 |
| HTN | pericyte | *SGCA* | 3.263880482 | 0.000344381 | 0.008125648 | 1 |
| HTN | pericyte | *EFTUD2* | -6.988091295 | 0.008939944 | 0.040430088 | 1 |
| HTN | pericyte | *FAM20A* | -5.584668032 | 0.005067039 | 0.030407778 | 1 |
| HTN | pericyte | *DPH1* | -6.368035299 | 0.000707469 | 0.013051853 | 1 |
| HTN | pericyte | *DHRS7B* | -6.188211465 | 0.008081413 | 0.038424723 | 1 |

| | | | | | | |
|---|---|---|---|---|---|---|
| HTN | pericyte | *KLHL2* | -6.713618664 | 0.005869771 | 0.032886984 | 1 |
| HTN | pericyte | *GRPEL1* | -10.88581033 | 0.012559507 | 0.049132942 | 1 |
| HTN | pericyte | *GAR1* | -8.233358543 | 0.005397401 | 0.031400705 | 1 |
| HTN | pericyte | *TRIM2* | -6.118668705 | 0.009575394 | 0.041705396 | 1 |
| HTN | pericyte | *FAM149A* | -3.97722816 | 0.004512611 | 0.028441579 | 1 |
| HTN | pericyte | *SC5D* | -4.60192918 | 0.001749688 | 0.021993482 | 1 |
| HTN | pericyte | *P2RX3* | -39.32945359 | 0.003699255 | 0.02608835 | 1 |
| HTN | pericyte | *SNX15* | -4.226943088 | 0.006613989 | 0.035303446 | 1 |
| HTN | pericyte | *CCND1* | -8.531726648 | 6.59E-05 | 0.002731877 | 0.319629561 |
| HTN | pericyte | *CCDC86* | -7.811158967 | 0.001172038 | 0.017180444 | 1 |
| HTN | pericyte | *GALNT18* | -4.582173248 | 0.010276187 | 0.043254826 | 1 |
| HTN | pericyte | *KIAA1549L* | -10.49112479 | 0.001964466 | 0.022752995 | 1 |
| HTN | pericyte | *C11orf21* | -9.938552 | 0.00020002 | 0.005727878 | 0.970497944 |
| HTN | pericyte | *ASIC1* | -11.95913816 | 0.001956348 | 0.022752995 | 1 |
| HTN | pericyte | *KCTD10* | -8.018229138 | 0.0001264 | 0.004200631 | 0.613292098 |
| HTN | pericyte | *IL23A* | -18.93057395 | 0.002936072 | 0.023704224 | 1 |
| HTN | pericyte | *ACAD10* | -7.466271705 | 2.80E-05 | 0.001492474 | 0.135815141 |
| HTN | pericyte | *ENDOU* | -8.275695983 | 0.002910142 | 0.02367523 | 1 |
| HTN | pericyte | *VDR* | -5.851620235 | 0.002729253 | 0.023519329 | 1 |
| HTN | pericyte | *NUP107* | -5.98123564 | 0.009981389 | 0.042497208 | 1 |
| HTN | pericyte | *BTN3A3* | -3.441785374 | 0.009814228 | 0.042148419 | 1 |
| HTN | pericyte | *FRK* | -7.513718526 | 0.007012792 | 0.035854655 | 1 |
| HTN | pericyte | *UST* | -3.046146625 | 0.006492345 | 0.034988583 | 1 |
| HTN | pericyte | *TULP1* | -4.644009377 | 0.001978169 | 0.022833649 | 1 |
| HTN | pericyte | *MRPL18* | -4.259116542 | 0.011803084 | 0.047412156 | 1 |
| HTN | pericyte | *PHACTR1* | -4.022746581 | 0.007296277 | 0.036435078 | 1 |
| HTN | pericyte | *COL9A1* | 1.254907048 | 0.005487626 | 0.031735351 | 1 |
| HTN | pericyte | *C6orf62* | -7.530351044 | 0.000744495 | 0.013329474 | 1 |
| HTN | pericyte | *CUTA* | -5.94052313 | 0.000168944 | 0.005155462 | 0.81971839 |
| HTN | pericyte | *IMPG1* | -14.16278803 | 0.000146325 | 0.004640332 | 0.709970782 |
| HTN | pericyte | *LY86* | -6.932030809 | 0.002099511 | 0.02292625 | 1 |
| HTN | pericyte | *NUDT12* | -7.841943915 | 0.007724187 | 0.03751022 | 1 |
| HTN | pericyte | *ARSB* | -6.979462336 | 0.010120427 | 0.042923349 | 1 |
| HTN | pericyte | *THBS4* | 0.78288076 | 0.00410735 | 0.027151036 | 1 |
| HTN | pericyte | *CDH6* | 0.940075932 | 0.007386488 | 0.03664544 | 1 |
| HTN | pericyte | *SLC12A7* | -8.441895747 | 0.010166897 | 0.043034836 | 1 |
| HTN | pericyte | *ATP6V0E1* | -6.042033153 | 0.006487666 | 0.034988583 | 1 |
| HTN | pericyte | *UNC5A* | -12.14719553 | 0.002952951 | 0.023712861 | 1 |
| HTN | pericyte | *ACTR8* | -8.278158407 | 0.000304354 | 0.007383636 | 1 |
| HTN | pericyte | *TBCCD1* | -5.950386533 | 0.002697096 | 0.023519329 | 1 |
| HTN | pericyte | *PCCB* | 6.671338158 | 4.77E-05 | 0.002206448 | 0.231677063 |
| HTN | pericyte | *HYAL1* | -4.866520902 | 0.000401708 | 0.009003382 | 1 |
| HTN | pericyte | *KIAA1257* | 5.184675355 | 0.008323895 | 0.038759634 | 1 |
| HTN | pericyte | *ABHD14B* | -6.038180975 | 0.007877253 | 0.037842013 | 1 |
| HTN | pericyte | *ARHGEF26* | -5.42101119 | 0.002950007 | 0.023712861 | 1 |
| HTN | pericyte | *VIPR1* | 0.520343021 | 0.004257501 | 0.027507529 | 1 |
| HTN | pericyte | *NEK4* | -6.830813685 | 0.003319903 | 0.024819987 | 1 |
| HTN | pericyte | *ADAM23* | -4.233802306 | 0.000863527 | 0.014548028 | 1 |
| HTN | pericyte | *SLC35F5* | -5.9923976 | 0.002646489 | 0.023519329 | 1 |
| HTN | pericyte | *GRB14* | -5.82409032 | 0.002723384 | 0.023519329 | 1 |

| | | | | | | |
|---|---|---|---|---|---|---|
| HTN | pericyte | *MRPL19* | -6.906429027 | 0.006630456 | 0.035313909 | 1 |
| HTN | pericyte | *GNLY* | -2.513578371 | 0.007020326 | 0.035855393 | 1 |
| HTN | pericyte | *WNT6* | -5.262758403 | 0.00811534 | 0.038500008 | 1 |
| HTN | pericyte | *TPO* | -4.236528481 | 0.001995356 | 0.022833649 | 1 |
| HTN | pericyte | *MSH6* | -5.725674575 | 0.006496741 | 0.034988583 | 1 |
| HTN | pericyte | *DNAJC16* | -6.732727813 | 0.004601952 | 0.028811186 | 1 |
| HTN | pericyte | *FAM20B* | -6.522753049 | 0.005660712 | 0.032274704 | 1 |
| HTN | pericyte | *WRAP73* | -8.650986517 | 0.000989025 | 0.015820885 | 1 |
| HTN | pericyte | *MECR* | -8.321135884 | 0.002255896 | 0.023376742 | 1 |
| HTN | pericyte | *CAPZA1* | -8.477181166 | 0.000386783 | 0.008810654 | 1 |
| HTN | pericyte | *DNAJC6* | -4.88127795 | 0.012566773 | 0.049132942 | 1 |
| HTN | pericyte | *LEPR* | -5.008713842 | 0.012718313 | 0.049371727 | 1 |
| HTN | pericyte | *PRG4* | -5.106777131 | 0.002099064 | 0.02292625 | 1 |
| HTN | pericyte | *TMEM9* | -4.746442412 | 0.000742478 | 0.013329474 | 1 |
| HTN | pericyte | *C1orf109* | -4.129074947 | 0.003412653 | 0.025396001 | 1 |
| HTN | pericyte | *GBP3* | -1.816561204 | 0.008351612 | 0.038814198 | 1 |
| HTN | pericyte | *EBNA1BP2* | -4.824747764 | 0.008003458 | 0.038243432 | 1 |
| HTN | pericyte | *CCDC181* | -4.370735296 | 0.002272 | 0.023376742 | 1 |
| HTN | pericyte | *IRF6* | -2.403802803 | 0.001114876 | 0.0167473 | 1 |
| HTN | pericyte | *RPS6KA1* | -6.470297533 | 0.001479951 | 0.019919162 | 1 |
| HTN | pericyte | *DDX59* | -6.033022997 | 0.002080849 | 0.02292625 | 1 |
| HTN | pericyte | *CASC1* | -2.345028186 | 0.001367408 | 0.018956181 | 1 |
| HTN | pericyte | *CNR1* | 0.535615212 | 0.004199822 | 0.027381347 | 1 |
| HTN | pericyte | *SGIP1* | -7.857344806 | 0.008790404 | 0.040047923 | 1 |
| HTN | pericyte | *DNAH7* | -4.967849307 | 0.009650157 | 0.041803673 | 1 |
| HTN | pericyte | *CPSF3* | -4.84774264 | 0.010830883 | 0.044839118 | 1 |
| HTN | pericyte | *TRIM32* | -8.620654072 | 0.001206139 | 0.017476015 | 1 |
| HTN | pericyte | *DCAF4* | -7.838579266 | 0.00050727 | 0.010608936 | 1 |
| HTN | pericyte | *FLVCR2* | -5.113606692 | 0.009791507 | 0.042148419 | 1 |
| HTN | pericyte | *CNRIP1* | -3.434820651 | 0.012226976 | 0.048587457 | 1 |
| HTN | pericyte | *HELLS* | -7.294710414 | 6.37E-05 | 0.002662602 | 0.308861858 |
| HTN | pericyte | *PCDHB12* | -2.837736475 | 0.002656496 | 0.023519329 | 1 |
| HTN | pericyte | *SNX19* | -6.434234299 | 0.000412983 | 0.009149744 | 1 |
| HTN | pericyte | *IQSEC3* | 5.254855312 | 0.011387596 | 0.046275221 | 1 |
| HTN | pericyte | *CCDC77* | -8.709887426 | 0.00098083 | 0.015786801 | 1 |
| HTN | pericyte | *ALG5* | -6.247897504 | 0.001562218 | 0.020431711 | 1 |
| HTN | pericyte | *ZFP30* | 4.580958817 | 0.012467679 | 0.049034456 | 1 |
| HTN | pericyte | *ARL1* | -7.642230375 | 0.002928887 | 0.023704224 | 1 |
| HTN | pericyte | *EPHX2* | -4.596706345 | 0.009873658 | 0.042185921 | 1 |
| HTN | pericyte | *TEX14* | 4.142874416 | 1.63E-06 | 0.000152261 | 0.007913835 |
| HTN | pericyte | *TRIM6* | -14.12519909 | 0.00153319 | 0.020269862 | 1 |
| HTN | pericyte | *ECHDC2* | -4.071386441 | 0.004215554 | 0.027381347 | 1 |
| HTN | pericyte | *LHX4* | -7.663277287 | 0.00274975 | 0.023519329 | 1 |
| HTN | pericyte | *POPDC2* | -3.829165891 | 0.001082841 | 0.01652184 | 1 |
| HTN | pericyte | *MAPK8IP1* | -16.3530927 | 2.90E-07 | 4.26E-05 | 0.001407227 |
| HTN | pericyte | *FABP3* | -19.84586484 | 0.000266625 | 0.006917999 | 1 |
| HTN | pericyte | *TNFSF10* | -5.344985586 | 0.007658505 | 0.037305125 | 1 |
| HTN | pericyte | *CPXM2* | -3.244258365 | 0.012164833 | 0.048459583 | 1 |
| HTN | pericyte | *SERAC1* | -7.532454297 | 0.003097647 | 0.024009239 | 1 |
| HTN | pericyte | *OPN4* | 3.883119796 | 0.004900962 | 0.029948951 | 1 |

| | | | | | | |
|---|---|---|---|---|---|---|
| HTN | pericyte | *FAM35A* | -5.884352363 | 0.007271583 | 0.036435078 | 1 |
| HTN | pericyte | *EEPD1* | -4.195968779 | 0.006006407 | 0.033344492 | 1 |
| HTN | pericyte | *SLC25A51* | -1.659312078 | 0.012105852 | 0.04830394 | 1 |
| HTN | pericyte | *C7orf49* | -2.559012286 | 0.010969268 | 0.045142399 | 1 |
| HTN | pericyte | *SSPN* | -3.746271222 | 0.009505899 | 0.041552345 | 1 |
| HTN | pericyte | *ARHGAP9* | -12.3957117 | 0.006987716 | 0.0358138 | 1 |
| HTN | pericyte | *PDE1B* | 2.111802085 | 0.007342297 | 0.036548752 | 1 |
| HTN | pericyte | *CDK2* | -5.376135324 | 0.002022777 | 0.02292625 | 1 |
| HTN | pericyte | *DBH* | -82.15143092 | 0.001848251 | 0.022363374 | 1 |
| HTN | pericyte | *TTC21B* | -7.65590595 | 0.002766279 | 0.023519329 | 1 |
| HTN | pericyte | *DAW1* | -5.866335379 | 0.003288681 | 0.024662563 | 1 |
| HTN | pericyte | *OBSL1* | -7.566768965 | 0.00034596 | 0.008125648 | 1 |
| HTN | pericyte | *FAM210B* | -4.75057705 | 0.005288357 | 0.031177529 | 1 |
| HTN | pericyte | *WFDC3* | -1.755006528 | 3.23E-05 | 0.001670145 | 0.156529018 |
| HTN | pericyte | *MATN4* | -8.32373203 | 0.003570245 | 0.025973602 | 1 |
| HTN | pericyte | *TOX2* | -2.237762016 | 0.000686256 | 0.012905864 | 1 |
| HTN | pericyte | *RNF114* | -7.204081233 | 0.005672468 | 0.032303771 | 1 |
| HTN | pericyte | *PEPD* | -5.905744614 | 0.009023278 | 0.040535121 | 1 |
| HTN | pericyte | *SNRNP27* | -7.151663908 | 0.006406193 | 0.034690678 | 1 |
| HTN | pericyte | *ATP8A1* | -6.483861321 | 0.002985464 | 0.023785667 | 1 |
| HTN | pericyte | *BTN2A2* | 3.281256804 | 2.21E-05 | 0.001244079 | 0.10699079 |
| HTN | pericyte | *SNRPC* | -8.642316197 | 2.51E-07 | 3.80E-05 | 0.00121559 |
| HTN | pericyte | *ZNF391* | -2.028660143 | 0.000104901 | 0.003895403 | 0.508981051 |
| HTN | pericyte | *TCP11* | 4.318674658 | 0.0058212 | 0.032788699 | 1 |
| HTN | pericyte | *TMTC4* | -8.894730619 | 0.001578175 | 0.020474082 | 1 |
| HTN | pericyte | *ABCC4* | -4.660040855 | 0.012010569 | 0.048081916 | 1 |
| HTN | pericyte | *GRK4* | 1.229012521 | 0.008128956 | 0.038500008 | 1 |
| HTN | pericyte | *NUP85* | -6.268074088 | 0.005566742 | 0.03188882 | 1 |
| HTN | pericyte | *NT5C* | -7.457892412 | 0.007443038 | 0.036738169 | 1 |
| HTN | pericyte | *DDX31* | -4.450877707 | 0.006948213 | 0.0358138 | 1 |
| HTN | pericyte | *OPRL1* | 4.369910695 | 0.005543745 | 0.031832248 | 1 |
| HTN | pericyte | *DTD1* | -7.010369733 | 0.001206609 | 0.017476015 | 1 |
| HTN | pericyte | *RBCK1* | -6.362572129 | 0.008646743 | 0.039585488 | 1 |
| HTN | pericyte | *FLRT3* | -4.339313168 | 0.002245692 | 0.023376742 | 1 |
| HTN | pericyte | *PCSK2* | 5.446360275 | 0.003603397 | 0.025973602 | 1 |
| HTN | pericyte | *LAMP5* | -49.09163527 | 0.010415362 | 0.043689185 | 1 |
| HTN | pericyte | *UROD* | -7.33105945 | 0.00071168 | 0.013079812 | 1 |
| HTN | pericyte | *PRR12* | -6.62496805 | 0.000858174 | 0.014508229 | 1 |
| HTN | pericyte | *TSKS* | -6.249982922 | 5.24E-06 | 0.000382721 | 0.025431123 |
| HTN | pericyte | *TRAP1* | 3.570784129 | 3.33E-05 | 0.001700049 | 0.161504671 |
| HTN | pericyte | *EMG1* | -5.721004575 | 0.009477528 | 0.041552345 | 1 |
| HTN | pericyte | *ABHD8* | -8.415659848 | 1.78E-07 | 3.00E-05 | 0.000864411 |
| HTN | pericyte | *LRRC61* | -4.025299044 | 0.001828449 | 0.022363374 | 1 |
| HTN | pericyte | *EMC1* | -7.039350368 | 0.007375627 | 0.03664544 | 1 |
| HTN | pericyte | *PNKD* | -6.030513848 | 0.008660021 | 0.039602659 | 1 |
| HTN | pericyte | *STYXL1* | -3.057013507 | 0.001856821 | 0.02236476 | 1 |
| HTN | pericyte | *STEAP4* | 0.59384123 | 0.004181456 | 0.027381347 | 1 |
| HTN | pericyte | *KDR* | -5.196838333 | 0.003483025 | 0.025722434 | 1 |
| HTN | pericyte | *MGAT3* | 1.786086563 | 0.011678379 | 0.047141009 | 1 |
| HTN | pericyte | *ATF4* | -5.998436481 | 0.002622463 | 0.023519329 | 1 |

| | | | | | | |
|---|---|---|---|---|---|---|
| HTN | pericyte | *EIF2AK4* | -8.144865109 | 0.001220747 | 0.017523865 | 1 |
| HTN | pericyte | *CALML4* | -10.03096716 | 0.001864572 | 0.022392469 | 1 |
| HTN | pericyte | *BBOX1* | -9.966169444 | 0.002853227 | 0.02367523 | 1 |
| HTN | pericyte | *DCTD* | -4.557004052 | 0.007526567 | 0.036999899 | 1 |
| HTN | pericyte | *RPAIN* | -4.903442047 | 0.005093435 | 0.030407778 | 1 |
| HTN | pericyte | *KRI1* | -2.674484545 | 0.010048029 | 0.042653577 | 1 |
| HTN | pericyte | *KLK8* | -21.21397051 | 0.003117902 | 0.024031844 | 1 |
| HTN | pericyte | *PRKCSH* | -9.173260722 | 0.001457604 | 0.019699985 | 1 |
| HTN | pericyte | *ATP8B3* | -3.283369296 | 5.32E-05 | 0.002412194 | 0.258104778 |
| HTN | pericyte | *PGPEP1* | 5.312644317 | 0.000344459 | 0.008125648 | 1 |
| HTN | pericyte | *KIAA1683* | -4.55068135 | 0.001847465 | 0.022363374 | 1 |
| HTN | pericyte | *PNPLA7* | -5.86017243 | 0.003962071 | 0.02677323 | 1 |
| HTN | pericyte | *PAK4* | -7.1894992 | 0.010003263 | 0.042537978 | 1 |
| HTN | pericyte | *NOL11* | -5.659699649 | 0.013024109 | 0.049971029 | 1 |
| HTN | pericyte | *HSD17B3* | -6.043034289 | 0.002457684 | 0.023519329 | 1 |
| HTN | pericyte | *TTLL9* | -5.767416453 | 0.007561652 | 0.037022338 | 1 |
| HTN | pericyte | *COX4I2* | 1.038424646 | 0.006867226 | 0.035715938 | 1 |
| HTN | pericyte | *CHMP1A* | -8.44279217 | 0.008056181 | 0.038397436 | 1 |
| HTN | pericyte | *IDO1* | -5.174362014 | 0.003082538 | 0.023955563 | 1 |
| HTN | pericyte | *HAUS8* | -9.135969201 | 4.82E-07 | 6.50E-05 | 0.002339926 |
| HTN | pericyte | *MRPS25* | -8.141815323 | 0.00085313 | 0.014473383 | 1 |
| HTN | pericyte | *KCNC3* | -6.735418221 | 0.000348338 | 0.008125648 | 1 |
| HTN | pericyte | *NDUFA2* | -5.357765693 | 0.007183998 | 0.036271341 | 1 |
| HTN | pericyte | *C1orf159* | -5.624001601 | 0.003995029 | 0.026810347 | 1 |
| HTN | pericyte | *CKMT2* | -3.865064308 | 0.009833473 | 0.042148419 | 1 |
| HTN | pericyte | *KHDRBS3* | 1.655975221 | 0.007803013 | 0.037701091 | 1 |
| HTN | pericyte | *TRIM21* | -11.16776612 | 0.00221108 | 0.023372896 | 1 |
| HTN | pericyte | *CCT6B* | -3.61164419 | 0.002076925 | 0.02292625 | 1 |
| HTN | pericyte | *IMMT* | -6.384651746 | 0.00711366 | 0.03616544 | 1 |
| HTN | pericyte | *REEP2* | -5.637535475 | 0.00461837 | 0.028876717 | 1 |
| HTN | pericyte | *PRMT7* | -3.469787879 | 0.003110969 | 0.024031844 | 1 |
| HTN | pericyte | *RSG1* | -12.506248 | 0.000401888 | 0.009003382 | 1 |
| HTN | pericyte | *CASP9* | -2.340146056 | 0.004209454 | 0.027381347 | 1 |
| HTN | pericyte | *ALOX5AP* | -7.445431941 | 0.00568972 | 0.032364035 | 1 |
| HTN | pericyte | *DSTYK* | -6.402771463 | 0.011742381 | 0.047281354 | 1 |
| HTN | pericyte | *CCNA1* | 13.34961012 | 0.005132087 | 0.030478442 | 1 |
| HTN | pericyte | *GIMAP4* | -3.658629939 | 0.010763851 | 0.044714216 | 1 |
| HTN | pericyte | *IPO8* | -8.060842239 | 0.002138699 | 0.023008801 | 1 |
| HTN | pericyte | *SPINK5* | -5.213198782 | 0.003829721 | 0.026468005 | 1 |
| HTN | pericyte | *COX16* | 3.878771471 | 0.003908175 | 0.026670131 | 1 |
| HTN | pericyte | *TTC9* | -6.278667152 | 0.002688064 | 0.023519329 | 1 |
| HTN | pericyte | *ELP3* | -6.956501726 | 0.004994728 | 0.030330941 | 1 |
| HTN | pericyte | *PEBP4* | -6.405107354 | 0.004734748 | 0.029362627 | 1 |
| HTN | pericyte | *THUMPD3* | -8.779122342 | 0.005857516 | 0.032856262 | 1 |
| HTN | pericyte | *GSTM5* | -2.961995597 | 0.008421607 | 0.038990114 | 1 |
| HTN | pericyte | *VAV3* | -3.04114857 | 0.001183524 | 0.017296562 | 1 |
| HTN | pericyte | *TRIM45* | -7.189553932 | 0.003195563 | 0.024319694 | 1 |
| HTN | pericyte | *LPIN1* | -4.984352488 | 0.003985469 | 0.026810347 | 1 |
| HTN | pericyte | *IL15RA* | -6.332796639 | 0.004428218 | 0.028196477 | 1 |
| HTN | pericyte | *TMEM241* | -4.906348597 | 0.000105173 | 0.003895403 | 0.510297783 |

| | | | | | | |
|---|---|---|---|---|---|---|
| HTN | pericyte | *ACP2* | 15.75782449 | 6.48E-08 | 1.57E-05 | 0.000314636 |
| HTN | pericyte | *DHX34* | -6.877308733 | 0.005727116 | 0.032448526 | 1 |
| HTN | pericyte | *ERCC5* | -5.445540311 | 0.002561538 | 0.023519329 | 1 |
| HTN | pericyte | *KLB* | -1.675632347 | 0.012906264 | 0.049713159 | 1 |
| HTN | pericyte | *USP30* | -3.771438296 | 7.25E-05 | 0.002932802 | 0.351936229 |
| HTN | pericyte | *DTX1* | -6.77920727 | 0.004716441 | 0.029338682 | 1 |
| HTN | pericyte | *POM121C* | -8.089815638 | 5.55E-05 | 0.002449055 | 0.269396089 |
| HTN | pericyte | *TES* | -8.469244798 | 0.000226874 | 0.006219165 | 1 |
| HTN | pericyte | *B4GALNT1* | 2.231246054 | 0.001791275 | 0.02224751 | 1 |
| HTN | pericyte | *FAIM2* | -5.676673922 | 0.000300523 | 0.007383636 | 1 |
| HTN | pericyte | *CCT7* | -9.091428123 | 0.000616149 | 0.01210347 | 1 |
| HTN | pericyte | *MPHOSPH6* | -6.208333281 | 0.006675809 | 0.035407327 | 1 |
| HTN | pericyte | *AGT* | 3.505667516 | 0.000424775 | 0.009313697 | 1 |
| HTN | pericyte | *CAPN9* | -12.95434136 | 0.00079517 | 0.014042101 | 1 |
| HTN | pericyte | *NTPCR* | -6.401748087 | 0.012887449 | 0.049705803 | 1 |
| HTN | pericyte | *SLC19A3* | -2.907126054 | 0.000953026 | 0.015701809 | 1 |
| HTN | pericyte | *TSGA10* | -1.724775652 | 0.00929135 | 0.041245774 | 1 |
| HTN | pericyte | *STAB2* | -3.342916568 | 0.002731741 | 0.023519329 | 1 |
| HTN | pericyte | *DRAM1* | -6.804223819 | 0.01235836 | 0.04882961 | 1 |
| HTN | pericyte | *TBC1D4* | -5.585180151 | 0.00103147 | 0.01604068 | 1 |
| HTN | pericyte | *RCBTB2* | -6.419782545 | 0.009865119 | 0.042185921 | 1 |
| HTN | pericyte | *SCRN1* | -5.578231779 | 0.012297328 | 0.048787109 | 1 |
| HTN | pericyte | *CHST12* | -8.328065148 | 0.005119694 | 0.030458325 | 1 |
| HTN | pericyte | *KDELR2* | -7.690905635 | 0.007140662 | 0.03616544 | 1 |
| HTN | pericyte | *NMT1* | 4.677150338 | 7.86E-05 | 0.003072014 | 0.381248968 |
| HTN | pericyte | *RTP4* | -5.156115311 | 0.002511206 | 0.023519329 | 1 |
| HTN | pericyte | *NDUFB5* | -8.864653908 | 0.001317442 | 0.01847157 | 1 |
| HTN | pericyte | *IMP4* | -11.54237079 | 0.003562832 | 0.025973602 | 1 |
| HTN | pericyte | *ODF2* | -7.219732395 | 0.002176076 | 0.023205098 | 1 |
| HTN | pericyte | *ZNF189* | -7.578234154 | 0.008902625 | 0.040407425 | 1 |
| HTN | pericyte | *DENND4C* | -5.731860138 | 0.005537441 | 0.031832248 | 1 |
| HTN | pericyte | *GMPR* | -3.379270659 | 0.012749478 | 0.049448814 | 1 |
| HTN | pericyte | *CAPN11* | 2.246072358 | 0.003610217 | 0.025973602 | 1 |
| HTN | pericyte | *LRRC1* | -3.871207645 | 0.002547558 | 0.023519329 | 1 |
| HTN | pericyte | *TCF19* | 4.76638849 | 0.00061591 | 0.01210347 | 1 |
| HTN | pericyte | *TLR2* | -9.112822714 | 0.001035861 | 0.016057502 | 1 |
| HTN | pericyte | *RAB30* | -7.663470205 | 0.00577425 | 0.032585753 | 1 |
| HTN | pericyte | *SULF1* | -3.081892223 | 0.004601279 | 0.028811186 | 1 |
| HTN | pericyte | *TRPC6* | 0.91034639 | 0.002674389 | 0.023519329 | 1 |
| HTN | pericyte | *PPP2R1B* | -4.335894161 | 1.31E-06 | 0.00013534 | 0.006334814 |
| HTN | pericyte | *NUSAP1* | 10.55145387 | 5.51E-06 | 0.000392835 | 0.026712809 |
| HTN | pericyte | *SLC44A5* | -5.174725773 | 0.008236955 | 0.038529454 | 1 |
| HTN | pericyte | *DBT* | -5.683996393 | 0.006751987 | 0.035511559 | 1 |
| HTN | pericyte | *KHK* | -4.900262861 | 0.001500091 | 0.02010619 | 1 |
| HTN | pericyte | *ADCY3* | 3.12962135 | 0.001728806 | 0.021787441 | 1 |
| HTN | pericyte | *LHCGR* | -5.276775422 | 0.000475336 | 0.010108697 | 1 |
| HTN | pericyte | *CENPO* | 2.272300301 | 0.008513202 | 0.039227023 | 1 |
| HTN | pericyte | *BTBD16* | -4.802253679 | 0.012966685 | 0.049813424 | 1 |
| HTN | pericyte | *TACC2* | -5.530547634 | 0.000790916 | 0.014042101 | 1 |
| HTN | pericyte | *ARL3* | 4.797080745 | 0.006755393 | 0.035511559 | 1 |

| | | | | | | |
|---|---|---|---|---|---|---|
| HTN | pericyte | *SMARCAL1* | -6.180611424 | 0.01233567 | 0.04882961 | 1 |
| HTN | pericyte | *STAT4* | 1.628601125 | 0.006172688 | 0.033956784 | 1 |
| HTN | pericyte | *COX17* | -8.507490687 | 0.009044607 | 0.040535121 | 1 |
| HTN | pericyte | *SPPL2A* | -7.838850683 | 0.000347016 | 0.008125648 | 1 |
| HTN | pericyte | *AP1AR* | -7.475954195 | 0.012561857 | 0.049132942 | 1 |
| HTN | pericyte | *BMPR1B* | -3.312186507 | 0.012408355 | 0.048907668 | 1 |
| HTN | pericyte | *MMRN1* | -1.894834227 | 0.00178848 | 0.02224751 | 1 |
| HTN | pericyte | *TRPC3* | 2.135643366 | 0.003049251 | 0.023886115 | 1 |
| HTN | pericyte | *FBN2* | 2.087501463 | 1.22E-05 | 0.000790252 | 0.058999458 |
| HTN | pericyte | *PARVG* | -8.171841963 | 0.000258566 | 0.006818272 | 1 |
| HTN | pericyte | *PDE6H* | 3.400755425 | 0.00969976 | 0.041908493 | 1 |
| HTN | pericyte | *KIF21A* | -5.097490567 | 0.005071591 | 0.030407778 | 1 |
| HTN | pericyte | *ALG10* | -4.225759553 | 0.000627914 | 0.012271001 | 1 |
| HTN | pericyte | *CLSTN3* | -6.016718866 | 0.004740796 | 0.029362627 | 1 |
| HTN | pericyte | *PPFIA2* | 2.220700888 | 0.012209757 | 0.048561619 | 1 |
| HTN | pericyte | *AMDHD1* | -1.94357839 | 0.008786565 | 0.040047923 | 1 |
| HTN | pericyte | *FOXN4* | -4.213701403 | 0.00129024 | 0.018308461 | 1 |
| HTN | pericyte | *HNRNPA1L2* | -8.732815871 | 0.00205069 | 0.02292625 | 1 |
| HTN | pericyte | *FITM1* | 11.77741101 | 0.000103347 | 0.003887147 | 0.501441945 |
| HTN | pericyte | *RTN1* | 3.464823696 | 0.005892372 | 0.032937547 | 1 |
| HTN | pericyte | *SLC24A4* | -6.356095394 | 0.003495936 | 0.025739424 | 1 |
| HTN | pericyte | *SCAPER* | -5.11692384 | 0.002546695 | 0.023519329 | 1 |
| HTN | pericyte | *CYP11A1* | -3.234797848 | 0.009435456 | 0.041468144 | 1 |
| HTN | pericyte | *PCSK6* | -6.349946513 | 0.006834847 | 0.035658794 | 1 |
| HTN | pericyte | *HAPLN3* | -6.000557228 | 0.000632265 | 0.012271001 | 1 |
| HTN | pericyte | *ITGAX* | -9.542397835 | 0.012342183 | 0.04882961 | 1 |
| HTN | pericyte | *NUDT7* | -3.667710468 | 0.003027009 | 0.023886115 | 1 |
| HTN | pericyte | *GCSH* | -6.168482542 | 0.00097781 | 0.015786801 | 1 |
| HTN | pericyte | *CMTM3* | -6.345060613 | 0.006151412 | 0.033916647 | 1 |
| HTN | pericyte | *TIGD7* | -7.268689088 | 0.00072622 | 0.013296678 | 1 |
| HTN | pericyte | *DEF8* | 6.586769387 | 0.000982608 | 0.015786801 | 1 |
| HTN | pericyte | *GALNS* | -5.229034857 | 0.006485502 | 0.034988583 | 1 |
| HTN | pericyte | *C17orf80* | -5.949506273 | 0.002085688 | 0.02292625 | 1 |
| HTN | pericyte | *VPS53* | -6.277410215 | 0.000226251 | 0.006219165 | 1 |
| HTN | pericyte | *LRRC46* | -5.414179993 | 0.000126199 | 0.004200631 | 0.612319811 |
| HTN | pericyte | *SCRN2* | -1.816037765 | 7.91E-05 | 0.003072014 | 0.384001778 |
| HTN | pericyte | *RHBDL3* | -6.839448891 | 0.003265253 | 0.024639201 | 1 |
| HTN | pericyte | *IMPA2* | 2.421155578 | 0.006365263 | 0.034546149 | 1 |
| HTN | pericyte | *C18orf8* | -11.63966275 | 6.89E-08 | 1.59E-05 | 0.000334246 |
| HTN | pericyte | *NPC1* | -9.044763969 | 0.00051972 | 0.010776417 | 1 |
| HTN | pericyte | *CCDC40* | -3.769695739 | 0.002012358 | 0.022920094 | 1 |
| HTN | pericyte | *ANAPC11* | -8.210310719 | 0.002687692 | 0.023519329 | 1 |
| HTN | pericyte | *TRIM65* | -6.937768076 | 0.00050994 | 0.010619013 | 1 |
| HTN | pericyte | *RNF157* | -3.117708885 | 0.005500219 | 0.031765065 | 1 |
| HTN | pericyte | *RNF165* | -5.773828568 | 0.000809597 | 0.014077604 | 1 |
| HTN | pericyte | *MAPK4* | 6.569451096 | 0.009398408 | 0.041380441 | 1 |
| HTN | pericyte | *CBLN2* | -4.703165258 | 0.00257678 | 0.023519329 | 1 |
| HTN | pericyte | *MIEN1* | -7.503593567 | 0.00011777 | 0.004036836 | 0.571417831 |
| HTN | pericyte | *PFKL* | -7.459823155 | 0.005011937 | 0.030349051 | 1 |
| HTN | pericyte | *GEMIN7* | -5.867615532 | 0.000638812 | 0.012305994 | 1 |

| | | | | | | |
|---|---|---|---|---|---|---|
| HTN | pericyte | *SLC47A1* | -4.191051894 | 0.003523288 | 0.025862317 | 1 |
| HTN | pericyte | *MMEL1* | -2.379322857 | 0.001837547 | 0.022363374 | 1 |
| HTN | pericyte | *ZNF593* | -12.6675722 | 0.008880167 | 0.040343232 | 1 |
| HTN | pericyte | *EVA1B* | 3.604536309 | 0.001993701 | 0.022833649 | 1 |
| HTN | pericyte | *ALDH9A1* | -7.963304938 | 0.012159753 | 0.048459583 | 1 |
| HTN | pericyte | *NME7* | -7.606277347 | 0.000244347 | 0.006586507 | 1 |
| HTN | pericyte | *ARNT* | -7.508130913 | 0.000261044 | 0.006846408 | 1 |
| HTN | pericyte | *POGZ* | -5.425995137 | 0.012489888 | 0.049069583 | 1 |
| HTN | pericyte | *DYRK3* | -10.1760518 | 0.00116725 | 0.017162107 | 1 |
| HTN | pericyte | *SMYD2* | -7.097327243 | 0.007139698 | 0.03616544 | 1 |
| HTN | pericyte | *RAB13* | -5.388441859 | 0.002296064 | 0.023376742 | 1 |
| HTN | pericyte | *S100A8* | -7.0288782 | 0.005655074 | 0.032274704 | 1 |
| HTN | pericyte | *CREB3L4* | -10.51090944 | 4.97E-09 | 1.85E-06 | 2.41E-05 |
| HTN | pericyte | *C1orf43* | -9.375890943 | 0.001060189 | 0.016330279 | 1 |
| HTN | pericyte | *RIT1* | -9.290232841 | 0.001695416 | 0.02153444 | 1 |
| HTN | pericyte | *SCCPDH* | -7.269865094 | 0.009303971 | 0.041264048 | 1 |
| HTN | pericyte | *GALM* | -6.254369999 | 0.007094912 | 0.036122258 | 1 |
| HTN | pericyte | *ABHD1* | -7.758275101 | 0.000231753 | 0.006317212 | 1 |
| HTN | pericyte | *ZNF514* | -5.592526163 | 0.002019975 | 0.02292625 | 1 |
| HTN | pericyte | *TEX261* | -8.519755945 | 0.002904064 | 0.02367523 | 1 |
| HTN | pericyte | *THNSL2* | -2.582515187 | 0.003155459 | 0.024148714 | 1 |
| HTN | pericyte | *RABL2A* | -5.267592337 | 0.003590369 | 0.025973602 | 1 |
| HTN | pericyte | *THSD7B* | 0.572312007 | 0.005928675 | 0.033102337 | 1 |
| HTN | pericyte | *GALNT13* | -3.140393248 | 0.002165413 | 0.023142253 | 1 |
| HTN | pericyte | *PHOSPHO2* | -11.14960077 | 0.002738059 | 0.023519329 | 1 |
| HTN | pericyte | *ESPNL* | -39.75453814 | 0.002476428 | 0.023519329 | 1 |
| HTN | pericyte | *SLC25A38* | -7.51646769 | 0.00127115 | 0.018140061 | 1 |
| HTN | pericyte | *CTDSPL* | -6.542032299 | 0.003539187 | 0.025868726 | 1 |
| HTN | pericyte | *SLC25A26* | -6.177410573 | 0.002242319 | 0.023376742 | 1 |
| HTN | pericyte | *PLA1A* | -7.976185185 | 0.006816248 | 0.035623078 | 1 |
| HTN | pericyte | *EIF2A* | -3.375452405 | 0.005825198 | 0.032788699 | 1 |
| HTN | pericyte | *NCEH1* | -4.465821809 | 0.003852117 | 0.026473757 | 1 |
| HTN | pericyte | *AMT* | -1.964170978 | 0.000964113 | 0.01574363 | 1 |
| HTN | pericyte | *CCDC39* | -5.057571817 | 0.000658646 | 0.012581577 | 1 |
| HTN | pericyte | *SCD5* | -8.582511035 | 0.011226799 | 0.045830839 | 1 |
| HTN | pericyte | *PYURF* | -8.273445779 | 0.006165542 | 0.033955972 | 1 |
| HTN | pericyte | *DDIT4L* | 1.495930886 | 0.003385531 | 0.025271685 | 1 |
| HTN | pericyte | *GTF2H2* | -7.586100746 | 0.002731922 | 0.023519329 | 1 |
| HTN | pericyte | *SPATA9* | -3.691948991 | 0.002256157 | 0.023376742 | 1 |
| HTN | pericyte | *TSLP* | -1.689932884 | 0.005287867 | 0.031177529 | 1 |
| HTN | pericyte | *FAM50B* | -7.129186768 | 0.002851654 | 0.02367523 | 1 |
| HTN | pericyte | *HIGD2A* | -5.955175707 | 0.000961375 | 0.01574363 | 1 |
| HTN | pericyte | *DOK3* | -2.16038677 | 0.006260973 | 0.034209733 | 1 |
| HTN | pericyte | *IRAK1BP1* | 7.112091512 | 0.001896008 | 0.0224925 | 1 |
| HTN | pericyte | *RNF217* | -4.958340692 | 0.010810095 | 0.044791271 | 1 |
| HTN | pericyte | *SLC18B1* | -4.852300885 | 0.000706363 | 0.013051853 | 1 |
| HTN | pericyte | *SLC22A3* | -4.196687473 | 0.002522853 | 0.023519329 | 1 |
| HTN | pericyte | *GNA12* | -5.938394227 | 0.01260947 | 0.049215835 | 1 |
| HTN | pericyte | *RBAK* | -9.396983247 | 0.003286704 | 0.024662563 | 1 |
| HTN | pericyte | *EPHA1* | 2.79316308 | 0.005386895 | 0.031400705 | 1 |

| | | | | | | |
|---|---|---|---|---|---|---|
| HTN | pericyte | *ADHFE1* | -2.579346005 | 1.22E-05 | 0.000790252 | 0.05926891 |
| HTN | pericyte | *DPYS* | -3.009458222 | 0.002366069 | 0.023376742 | 1 |
| HTN | pericyte | *ST8SIA6* | -3.650676791 | 0.003491071 | 0.025739424 | 1 |
| HTN | pericyte | *RGR* | -19.72166147 | 0.002707308 | 0.023519329 | 1 |
| HTN | pericyte | *PAOX* | -8.022918479 | 0.003426549 | 0.025460358 | 1 |
| HTN | pericyte | *LIN7C* | -7.278161744 | 0.004775954 | 0.029398444 | 1 |
| HTN | pericyte | *HSD17B12* | -4.101082563 | 3.28E-10 | 1.49E-07 | 1.59E-06 |
| HTN | pericyte | *EIF3M* | -7.690579577 | 0.000156386 | 0.004876146 | 0.758785928 |
| HTN | pericyte | *TNKS1BP1* | 5.741988055 | 1.37E-06 | 0.000135446 | 0.006633751 |
| HTN | pericyte | *PTPRJ* | -6.079946419 | 0.010273282 | 0.043254826 | 1 |
| HTN | pericyte | *DRD2* | 11.045073 | 0.006206655 | 0.03406639 | 1 |
| HTN | pericyte | *SLX4IP* | -4.601742014 | 0.008229186 | 0.038529454 | 1 |
| HTN | pericyte | *CHEK1* | -11.63679672 | 0.002096569 | 0.02292625 | 1 |
| HTN | pericyte | *DUSP15* | 4.558907545 | 0.000692589 | 0.012924777 | 1 |
| HTN | pericyte | *SPATA25* | -4.758967643 | 0.004403764 | 0.028077611 | 1 |
| HTN | pericyte | *CABLES2* | -3.687618565 | 0.000812393 | 0.014077604 | 1 |
| HTN | pericyte | *TRPT1* | -3.733274427 | 0.008072401 | 0.038424723 | 1 |
| HTN | pericyte | *TBX6* | 1.921481307 | 0.0004771 | 0.010108697 | 1 |
| HTN | pericyte | *ALDOA* | -6.462361193 | 0.001983311 | 0.022833649 | 1 |
| HTN | pericyte | *PPP1R1C* | -9.253199532 | 0.002289031 | 0.023376742 | 1 |
| HTN | pericyte | *GPR158* | -5.572074047 | 0.00904772 | 0.040535121 | 1 |
| HTN | pericyte | *NDUFC2* | -8.771246913 | 0.003971751 | 0.02677323 | 1 |
| HTN | pericyte | *VIPAS39* | -6.843933815 | 0.006991503 | 0.0358138 | 1 |
| HTN | pericyte | *INPP1* | -7.393583794 | 0.003041703 | 0.023886115 | 1 |
| HTN | pericyte | *GABRA2* | -7.918442983 | 0.001290497 | 0.018308461 | 1 |
| HTN | pericyte | *TMEM56* | -9.764292182 | 0.00267562 | 0.023519329 | 1 |
| HTN | pericyte | *TMEM178A* | -5.512592991 | 0.002685061 | 0.023519329 | 1 |
| HTN | pericyte | *NMT2* | -5.299454631 | 0.010675074 | 0.04442149 | 1 |
| HTN | pericyte | *PLEKHH2* | -3.69832491 | 0.001132447 | 0.01687464 | 1 |
| HTN | pericyte | *CCNO* | -28.89324013 | 0.003056534 | 0.023886115 | 1 |
| HTN | pericyte | *CLEC4F* | 5.990491324 | 0.007639479 | 0.037253018 | 1 |
| HTN | pericyte | *SLC30A6* | -5.426443683 | 0.009354906 | 0.041319857 | 1 |
| HTN | pericyte | *RASGRP3* | -3.680767756 | 0.006717289 | 0.03548317 | 1 |
| HTN | pericyte | *PRDM8* | -10.15140127 | 3.69E-05 | 0.001816267 | 0.179255825 |
| HTN | pericyte | *JAKMIP1* | -11.69819691 | 0.009029582 | 0.040535121 | 1 |
| HTN | pericyte | *CAST* | -6.039373832 | 0.000665897 | 0.012617472 | 1 |
| HTN | pericyte | *CETN3* | -4.046256648 | 0.008196907 | 0.038500008 | 1 |
| HTN | pericyte | *FRMD1* | -3.610902537 | 0.007875366 | 0.037842013 | 1 |
| HTN | pericyte | *LPCAT1* | -8.118091466 | 0.00659776 | 0.035255874 | 1 |
| HTN | pericyte | *JAZF1* | -5.16023388 | 0.001566487 | 0.020431711 | 1 |
| HTN | pericyte | *FBXO36* | -5.265893243 | 0.006945859 | 0.0358138 | 1 |
| HTN | pericyte | *GDPD1* | -8.608936754 | 0.002984905 | 0.023785667 | 1 |
| HTN | pericyte | *SEMA3D* | -5.436385125 | 0.008158701 | 0.038500008 | 1 |
| HTN | pericyte | *DNAAF1* | -7.139805829 | 0.000954665 | 0.015701809 | 1 |
| HTN | pericyte | *ROBO4* | -7.140142456 | 0.010711972 | 0.044536836 | 1 |
| HTN | pericyte | *NEIL2* | 4.816911738 | 0.000220223 | 0.006105841 | 1 |
| HTN | pericyte | *CCSAP* | -5.760904093 | 0.002662789 | 0.023519329 | 1 |
| HTN | pericyte | *PDE1C* | 0.646245506 | 0.005048111 | 0.030407778 | 1 |
| HTN | pericyte | *XPC* | -7.436330161 | 0.004788633 | 0.029410692 | 1 |
| HTN | pericyte | *GOLGA7B* | -15.98279308 | 0.003829377 | 0.026468005 | 1 |

| | | | | | | |
|---|---|---|---|---|---|---|
| HTN | pericyte | *MOV10* | 8.056316421 | 0.009127409 | 0.040731344 | 1 |
| HTN | pericyte | *CNOT8* | -7.055785395 | 0.002446663 | 0.023519329 | 1 |
| HTN | pericyte | *LRGUK* | -2.323969905 | 0.005455305 | 0.031623824 | 1 |
| HTN | pericyte | *ZKSCAN2* | -3.871967644 | 0.00402652 | 0.026899796 | 1 |
| HTN | pericyte | *KDM8* | -5.577167713 | 0.004030534 | 0.026899796 | 1 |
| HTN | pericyte | *MMP16* | -2.600494551 | 0.000994512 | 0.015820885 | 1 |
| HTN | pericyte | *NDUFAF6* | -10.37781875 | 1.63E-06 | 0.000152261 | 0.007917557 |
| HTN | pericyte | *ART3* | -3.382371288 | 0.003276882 | 0.024662563 | 1 |
| HTN | pericyte | *NAA11* | -12.48927938 | 0.000247777 | 0.00664206 | 1 |
| HTN | pericyte | *SFXN2* | 4.192803243 | 3.37E-07 | 4.67E-05 | 0.001633568 |
| HTN | pericyte | *TDRD9* | -8.871582986 | 0.000404521 | 0.009003382 | 1 |
| HTN | pericyte | *NODAL* | -7.810721521 | 0.002155234 | 0.023084315 | 1 |
| HTN | pericyte | *NMNAT2* | -8.102212597 | 0.001951121 | 0.022752995 | 1 |
| HTN | pericyte | *SLC6A1* | 2.835013915 | 0.005942874 | 0.033143477 | 1 |
| HTN | pericyte | *NECAP2* | -6.701618286 | 0.005101204 | 0.030407778 | 1 |
| HTN | pericyte | *STEAP2* | -2.527030113 | 0.008543219 | 0.039327986 | 1 |
| HTN | pericyte | *CLDN12* | -8.628668207 | 0.003840971 | 0.026468005 | 1 |
| HTN | pericyte | *TMED6* | -10.66290455 | 0.010959589 | 0.045142399 | 1 |
| HTN | pericyte | *CLEC18A* | -5.040314494 | 1.46E-05 | 0.000899584 | 0.071067125 |
| HTN | pericyte | *SLC38A10* | -6.463859561 | 0.012960373 | 0.049813424 | 1 |
| HTN | pericyte | *EXTL1* | -44.52347215 | 0.00256366 | 0.023519329 | 1 |
| HTN | pericyte | *LRRC43* | -8.405396187 | 3.89E-05 | 0.001848882 | 0.188585993 |
| HTN | pericyte | *CLSTN2* | -5.604351373 | 0.011876342 | 0.047662539 | 1 |
| HTN | pericyte | *TSPAN33* | -5.819981562 | 0.000112487 | 0.004035815 | 0.545787723 |
| HTN | pericyte | *FAM86C1* | -9.139496422 | 0.005375042 | 0.031400705 | 1 |
| HTN | pericyte | *RNF166* | -3.301472839 | 0.002720132 | 0.023519329 | 1 |
| HTN | pericyte | *F11R* | -2.764772674 | 0.007240991 | 0.036435078 | 1 |
| HTN | pericyte | *SLC5A11* | -3.575178506 | 1.30E-08 | 3.72E-06 | 6.32E-05 |
| HTN | pericyte | *CDC42SE2* | -7.601140189 | 0.000193998 | 0.005636402 | 0.941279214 |
| HTN | pericyte | *SV2A* | 6.60386178 | 0.000706221 | 0.013051853 | 1 |
| HTN | pericyte | *IRX6* | -2.117765204 | 0.004151591 | 0.027368912 | 1 |
| HTN | pericyte | *HK2* | -3.130584032 | 0.003130281 | 0.024031844 | 1 |
| HTN | pericyte | *TEPP* | -15.35776072 | 0.000160808 | 0.004969677 | 0.780239263 |
| HTN | pericyte | *CTBP1* | -9.152252163 | 1.44E-05 | 0.0008935 | 0.069692992 |
| HTN | pericyte | *LRRC36* | -1.774520327 | 0.008181016 | 0.038500008 | 1 |
| HTN | pericyte | *LYPD5* | -8.797051812 | 0.000287638 | 0.007305639 | 1 |
| HTN | pericyte | *ANKLE1* | -8.108909136 | 0.009346708 | 0.041319857 | 1 |
| HTN | pericyte | *CCDC58* | -9.103842621 | 0.008185903 | 0.038500008 | 1 |
| HTN | pericyte | *S100B* | -1.852560961 | 5.13E-06 | 0.000382708 | 0.024875994 |
| HTN | pericyte | *ZNF208* | -8.180558543 | 3.71E-05 | 0.001816267 | 0.179810403 |
| HTN | pericyte | *CACFD1* | -6.809050333 | 0.002333196 | 0.023376742 | 1 |
| HTN | pericyte | *CCR5* | 4.902773938 | 0.006285014 | 0.034227583 | 1 |
| HTN | pericyte | *LY6K* | 3.757121647 | 0.000197942 | 0.005716762 | 0.960415985 |
| HTN | pericyte | *ZNF333* | -7.420427131 | 0.011804181 | 0.047412156 | 1 |
| HTN | pericyte | *ABCF3* | 7.436885618 | 2.07E-07 | 3.34E-05 | 0.001002195 |
| HTN | pericyte | *FBXO27* | -1.771358166 | 0.009141189 | 0.040731344 | 1 |
| HTN | pericyte | *U2AF1L4* | -1.006100645 | 0.009099401 | 0.040691516 | 1 |
| HTN | pericyte | *TMEM143* | -5.610719834 | 0.008922802 | 0.040423377 | 1 |
| HTN | pericyte | *PLCD3* | -5.801092665 | 0.002190452 | 0.023256174 | 1 |
| HTN | pericyte | *LEMD2* | -8.2227007 | 0.009083714 | 0.040658836 | 1 |

| | | | | | | |
|---|---|---|---|---|---|---|
| HTN | pericyte | *ASGR2* | 31.37130649 | 0.007457448 | 0.036771889 | 1 |
| HTN | pericyte | *TTC9C* | -5.114266688 | 0.007513377 | 0.036972522 | 1 |
| HTN | pericyte | *RPS6KA4* | -6.323895309 | 0.002808341 | 0.02367523 | 1 |
| HTN | pericyte | *PDZK1IP1* | -6.936856668 | 0.002101129 | 0.02292625 | 1 |
| HTN | pericyte | *COA7* | -7.399170096 | 0.002389056 | 0.023420475 | 1 |
| HTN | pericyte | *C1orf123* | -6.965192748 | 0.001829123 | 0.022363374 | 1 |
| HTN | pericyte | *SYNC* | -6.363016073 | 0.001202123 | 0.017476015 | 1 |
| HTN | pericyte | *CAMK2N1* | 2.065547317 | 0.001717596 | 0.021759201 | 1 |
| HTN | pericyte | *LRRIQ3* | -2.899752428 | 0.002353463 | 0.023376742 | 1 |
| HTN | pericyte | *HFM1* | -6.490363561 | 0.000115038 | 0.004035815 | 0.55816363 |
| HTN | pericyte | *ZNF496* | -5.418836474 | 0.005405797 | 0.031411889 | 1 |
| HTN | pericyte | *FCRLB* | -9.092911981 | 0.003792211 | 0.026406882 | 1 |
| HTN | pericyte | *MRPL55* | 7.810089382 | 5.73E-06 | 0.000403156 | 0.027817757 |
| HTN | pericyte | *H3F3A* | -3.92361742 | 0.005622802 | 0.032171977 | 1 |
| HTN | pericyte | *C1QTNF7* | -4.389779743 | 0.012285793 | 0.048781235 | 1 |
| HTN | pericyte | *LYSMD1* | -4.731037054 | 9.23E-07 | 0.00010416 | 0.004478866 |
| HTN | pericyte | *VPS72* | -5.89871456 | 0.002907442 | 0.02367523 | 1 |
| HTN | pericyte | *DCAF16* | -13.75419097 | 1.26E-05 | 0.00080655 | 0.06129782 |
| HTN | pericyte | *NIPAL1* | 1.145303767 | 0.003772041 | 0.026371674 | 1 |
| HTN | pericyte | *DAPL1* | 2.437198035 | 0.001382031 | 0.01910432 | 1 |
| HTN | pericyte | *CLDN1* | -3.312894089 | 0.007228934 | 0.036432899 | 1 |
| HTN | pericyte | *LINC01116* | -6.670177944 | 0.000108385 | 0.003983979 | 0.525885186 |
| HTN | pericyte | *SERPINI1* | -9.531420378 | 0.00206539 | 0.02292625 | 1 |
| HTN | pericyte | *AIM2* | -5.382025375 | 0.002759238 | 0.023519329 | 1 |
| HTN | pericyte | *GTPBP8* | -7.152682695 | 0.003171605 | 0.024234058 | 1 |
| HTN | pericyte | *C4orf36* | -6.687548032 | 0.003754981 | 0.026290283 | 1 |
| HTN | pericyte | *PPM1K* | -5.235178577 | 0.012525412 | 0.049129589 | 1 |
| HTN | pericyte | *DCLK3* | 1.716569513 | 0.006867883 | 0.035715938 | 1 |
| HTN | pericyte | *RPP14* | -10.62287872 | 0.002134014 | 0.023008801 | 1 |
| HTN | pericyte | *IL17RC* | -7.200388426 | 0.00021587 | 0.00601954 | 1 |
| HTN | pericyte | *TTC14* | -4.971703282 | 0.011944984 | 0.047858845 | 1 |
| HTN | pericyte | *CCDC158* | 7.908106618 | 0.00368264 | 0.02608492 | 1 |
| HTN | pericyte | *CDCP1* | -2.512034291 | 0.01096747 | 0.045142399 | 1 |
| HTN | pericyte | *TPRA1* | -8.675966717 | 0.001846881 | 0.022363374 | 1 |
| HTN | pericyte | *SNIP1* | -8.617292975 | 0.007821929 | 0.037701091 | 1 |
| HTN | pericyte | *DNALI1* | -4.941713762 | 0.000146057 | 0.004640332 | 0.70866815 |
| HTN | pericyte | *RFT1* | -6.371395655 | 1.72E-05 | 0.001019217 | 0.083375515 |
| HTN | pericyte | *METAP1* | -7.828471859 | 0.00104065 | 0.016080368 | 1 |
| HTN | pericyte | *CDC25A* | -25.10024638 | 1.71E-11 | 1.18E-08 | 8.28E-08 |
| HTN | pericyte | *ZNF589* | -9.416702472 | 8.98E-12 | 7.26E-09 | 4.36E-08 |
| HTN | pericyte | *RNF123* | 6.583396365 | 6.12E-07 | 7.82E-05 | 0.002970546 |
| HTN | pericyte | *MFSD8* | -5.688227433 | 0.011370083 | 0.046242786 | 1 |
| HTN | pericyte | *ETNPPL* | -21.59856313 | 0.002844627 | 0.02367523 | 1 |
| HTN | pericyte | *HPGD* | 2.658968377 | 0.007887379 | 0.037853177 | 1 |
| HTN | pericyte | *TMEM144* | -4.319515218 | 0.000472201 | 0.010093035 | 1 |
| HTN | pericyte | *LMBRD2* | -6.718081276 | 0.001211486 | 0.017480633 | 1 |
| HTN | pericyte | *F2RL2* | -8.243495351 | 0.003675336 | 0.026071244 | 1 |
| HTN | pericyte | *C5orf63* | -4.400502251 | 0.010278816 | 0.043254826 | 1 |
| HTN | pericyte | *ERAP1* | -9.622509651 | 1.11E-06 | 0.000119441 | 0.005374828 |
| HTN | pericyte | *ERAP2* | 1.610489615 | 0.012434026 | 0.048969069 | 1 |

| | | | | | | |
|---|---|---|---|---|---|---|
| HTN | pericyte | *UTP15* | -6.454897736 | 0.002600757 | 0.023519329 | 1 |
| HTN | pericyte | *TLR3* | -7.69692782 | 0.005065457 | 0.030407778 | 1 |
| HTN | pericyte | *CCDC127* | -9.237020589 | 0.00782536 | 0.037701091 | 1 |
| HTN | pericyte | *GRIK2* | -2.049938279 | 0.000830167 | 0.01433441 | 1 |
| HTN | pericyte | *DCBLD1* | 0.904343056 | 0.009501931 | 0.041552345 | 1 |
| HTN | pericyte | *RAET1E* | -8.348876182 | 0.004205554 | 0.027381347 | 1 |
| HTN | pericyte | *DAGLB* | -2.260646466 | 0.005751944 | 0.032527312 | 1 |
| HTN | pericyte | *RP9* | -8.444990594 | 0.00564447 | 0.032257911 | 1 |
| HTN | pericyte | *MIOS* | -5.725912885 | 0.002710102 | 0.023519329 | 1 |
| HTN | pericyte | *CHMP4C* | -9.770146464 | 0.003070822 | 0.023915939 | 1 |
| HTN | pericyte | *DEFB1* | -6.84253917 | 0.005343025 | 0.031271844 | 1 |
| HTN | pericyte | *GPER1* | 2.566453619 | 0.001821833 | 0.022363374 | 1 |
| HTN | pericyte | *SLC25A32* | -7.933277834 | 0.001531231 | 0.020269862 | 1 |
| HTN | pericyte | *DCSTAMP* | 79.43681653 | 0.000233839 | 0.006338473 | 1 |
| HTN | pericyte | *INTS8* | -6.974458315 | 0.010987655 | 0.045179747 | 1 |
| HTN | pericyte | *RPP25L* | -7.840803695 | 2.55E-05 | 0.001392558 | 0.123937649 |
| HTN | pericyte | *NUDT2* | -2.476945659 | 0.000655674 | 0.012574434 | 1 |
| HTN | pericyte | *ZMAT4* | -5.508356758 | 0.003202862 | 0.024319694 | 1 |
| HTN | pericyte | *NOL6* | -3.022635352 | 0.005384709 | 0.031400705 | 1 |
| HTN | pericyte | *SLC16A9* | -13.52635869 | 0.009498763 | 0.041552345 | 1 |
| HTN | pericyte | *LRR1* | -9.867503438 | 0.003146354 | 0.024117075 | 1 |
| HTN | pericyte | *OXGR1* | -8.682239211 | 0.007763882 | 0.03755768 | 1 |
| HTN | pericyte | *VDAC2* | -10.79646802 | 0.000992804 | 0.015820885 | 1 |
| HTN | pericyte | *CASP7* | -3.855797163 | 0.004363019 | 0.027927924 | 1 |
| HTN | pericyte | *PPP1R36* | -4.248711552 | 0.011439873 | 0.046448757 | 1 |
| HTN | pericyte | *TRUB1* | -5.803926667 | 0.000170766 | 0.005178474 | 0.828555855 |
| HTN | pericyte | *ARL5B* | -14.73831384 | 9.16E-07 | 0.00010416 | 0.004444528 |
| HTN | pericyte | *CEP57* | -8.195227669 | 0.00058622 | 0.011802233 | 1 |
| HTN | pericyte | *ZFYVE19* | -3.556356582 | 0.00208783 | 0.02292625 | 1 |
| HTN | pericyte | *TRMT61A* | -7.762115209 | 0.004157486 | 0.027370584 | 1 |
| HTN | pericyte | *ZNF202* | -4.670845578 | 0.003222506 | 0.024430623 | 1 |
| HTN | pericyte | *NUDT13* | -3.744779492 | 0.001107858 | 0.016745563 | 1 |
| HTN | pericyte | *WDR88* | -4.697802838 | 0.002294202 | 0.023376742 | 1 |
| HTN | pericyte | *CYB5R2* | 4.292142487 | 7.68E-05 | 0.003028107 | 0.372457182 |
| HTN | pericyte | *XRRA1* | -5.821785136 | 0.00084076 | 0.014416964 | 1 |
| HTN | pericyte | *TRIM66* | -2.69184284 | 1.00E-06 | 0.000110813 | 0.004875788 |
| HTN | pericyte | *CENPN* | -5.703756511 | 0.004775559 | 0.029398444 | 1 |
| HTN | pericyte | *NDST2* | -27.94370304 | 0.002464992 | 0.023519329 | 1 |
| HTN | pericyte | *RIMKLB* | -8.695024141 | 7.16E-09 | 2.48E-06 | 3.47E-05 |
| HTN | pericyte | *TMED3* | -2.02745979 | 0.006232409 | 0.034131269 | 1 |
| HTN | pericyte | *CENPV* | 7.479506183 | 0.008242698 | 0.038529454 | 1 |
| HTN | pericyte | *FAM96A* | -4.32499875 | 0.008035389 | 0.038335998 | 1 |
| HTN | pericyte | *ANPEP* | 1.43610456 | 0.012317836 | 0.048828547 | 1 |
| HTN | pericyte | *STAT6* | -6.518093911 | 0.007291709 | 0.036435078 | 1 |
| HTN | pericyte | *NYAP1* | 2.836743195 | 0.000258035 | 0.006818272 | 1 |
| HTN | pericyte | *MS4A7* | -2.585819243 | 0.005207711 | 0.030797343 | 1 |
| HTN | pericyte | *MS4A14* | -1.290434972 | 0.009398443 | 0.041380441 | 1 |
| HTN | pericyte | *SMAD3* | -5.49771453 | 0.00887459 | 0.040343232 | 1 |
| HTN | pericyte | *MAP1A* | 2.892248918 | 0.005731325 | 0.032448526 | 1 |
| HTN | pericyte | *URM1* | -5.698322127 | 0.008805448 | 0.040078833 | 1 |

| | | | | | | |
|---|---|---|---|---|---|---|
| HTN | pericyte | *ISLR2* | -4.797606264 | 0.002362343 | 0.023376742 | 1 |
| HTN | pericyte | *SP2* | -5.540701828 | 0.002626783 | 0.023519329 | 1 |
| HTN | pericyte | *GPRC5B* | -5.091840781 | 0.006712588 | 0.03548317 | 1 |
| HTN | pericyte | *TBC1D2B* | -4.924296272 | 0.003618746 | 0.025973602 | 1 |
| HTN | pericyte | *LOXHD1* | -2.303876525 | 0.004766871 | 0.029398444 | 1 |
| HTN | pericyte | *POP5* | -6.552623986 | 0.010794449 | 0.04476467 | 1 |
| HTN | pericyte | *MYO5B* | -4.24245402 | 0.003023368 | 0.023886115 | 1 |
| HTN | pericyte | *ZNF23* | -8.510868648 | 5.44E-05 | 0.00242362 | 0.26417453 |
| HTN | pericyte | *SMG8* | -4.185489369 | 0.007282875 | 0.036435078 | 1 |
| HTN | pericyte | *SPATA33* | -10.69996494 | 0.00430194 | 0.027673274 | 1 |
| HTN | pericyte | *TMC4* | 2.625343908 | 0.011336291 | 0.046182773 | 1 |
| HTN | pericyte | *CHAF1A* | -2.273430385 | 0.000966942 | 0.01574363 | 1 |
| HTN | pericyte | *SRR* | 2.318464654 | 0.000997955 | 0.015823785 | 1 |
| HTN | pericyte | *KLK5* | -4.70268755 | 0.001165997 | 0.017162107 | 1 |
| HTN | pericyte | *KLK6* | -24.24476067 | 0.003241384 | 0.02450575 | 1 |
| HTN | pericyte | *MGAT5B* | -6.779834107 | 0.004257658 | 0.027507529 | 1 |
| HTN | pericyte | *ZNF598* | -7.593039379 | 7.94E-06 | 0.00054245 | 0.038513919 |
| HTN | pericyte | *ECI1* | -4.970475055 | 0.001987189 | 0.022833649 | 1 |
| HTN | pericyte | *PXK* | -6.507331695 | 0.007730878 | 0.03751022 | 1 |
| HTN | pericyte | *FAM107A* | -3.539389535 | 0.012622673 | 0.049215835 | 1 |
| HTN | pericyte | *CCDC110* | -12.56246944 | 0.00223447 | 0.023376742 | 1 |
| HTN | pericyte | *SERINC2* | -11.48411963 | 1.79E-05 | 0.001044724 | 0.086712131 |
| HTN | pericyte | *SLC20A2* | -6.334629604 | 0.003514718 | 0.025838503 | 1 |
| HTN | pericyte | *NBPF9* | -7.724017315 | 0.002222498 | 0.023376742 | 1 |
| HTN | pericyte | *GSTM4* | -10.70869553 | 0.000464256 | 0.009967133 | 1 |
| HTN | pericyte | *TCTN2* | -1.449327117 | 0.006812682 | 0.035623078 | 1 |
| HTN | pericyte | *BTNL3* | 2.022605441 | 0.007503095 | 0.036959406 | 1 |
| HTN | pericyte | *MFSD7* | -12.88292069 | 0.003310292 | 0.024786325 | 1 |
| HTN | pericyte | *KLK7* | -9.782532886 | 0.008212594 | 0.038500008 | 1 |
| HTN | pericyte | *IRS1* | -1.588466628 | 0.002114628 | 0.022953412 | 1 |
| HTN | pericyte | *C8orf46* | 1.002607909 | 0.010602974 | 0.044195688 | 1 |
| HTN | pericyte | *CPT1C* | 9.00187875 | 8.33E-08 | 1.68E-05 | 0.00040418 |
| HTN | pericyte | *RGS14* | -2.607612987 | 0.001599056 | 0.020634628 | 1 |
| HTN | pericyte | *RAB24* | -26.70021877 | 7.19E-07 | 8.72E-05 | 0.00348896 |
| HTN | pericyte | *SH3TC2* | -2.461442329 | 0.010182551 | 0.043036357 | 1 |
| HTN | pericyte | *SNUPN* | -5.543074528 | 0.007334999 | 0.036548752 | 1 |
| HTN | pericyte | *SLC38A11* | 0.809502749 | 0.006723774 | 0.03548317 | 1 |
| HTN | pericyte | *INO80E* | -7.253298246 | 7.29E-12 | 7.26E-09 | 3.54E-08 |
| HTN | pericyte | *DFFB* | -12.29130923 | 0.003443179 | 0.025505811 | 1 |
| HTN | pericyte | *RGPD8* | -1.466537937 | 0.000138017 | 0.004524716 | 0.669657934 |
| HTN | pericyte | *CHRNA5* | -11.07595866 | 0.000143538 | 0.00461223 | 0.696446698 |
| HTN | pericyte | *CTNND2* | -3.713537884 | 0.002503326 | 0.023519329 | 1 |
| HTN | pericyte | *UBE2E3* | -6.182572424 | 0.009597823 | 0.041738713 | 1 |
| HTN | pericyte | *NIPA1* | -6.147356529 | 0.001005725 | 0.01584344 | 1 |
| HTN | pericyte | *RNF150* | -4.074006763 | 0.002798885 | 0.02367523 | 1 |
| HTN | pericyte | *DNAJC18* | -6.986729922 | 3.01E-06 | 0.00024727 | 0.014588938 |
| HTN | pericyte | *HSD17B13* | -6.988042119 | 0.002360616 | 0.023376742 | 1 |
| HTN | pericyte | *ELOVL6* | -6.453580824 | 0.006640485 | 0.035328546 | 1 |
| HTN | pericyte | *ARL6IP1* | -6.168556838 | 0.000316647 | 0.007643629 | 1 |
| HTN | pericyte | *CDH2* | -5.605076897 | 0.011099271 | 0.04552296 | 1 |

| | | | | | | |
|---|---|---|---|---|---|---|
| HTN | pericyte | *EMB* | -4.320846578 | 0.000323529 | 0.007771104 | 1 |
| HTN | pericyte | *STAT2* | -6.484104766 | 0.007962242 | 0.038174701 | 1 |
| HTN | pericyte | *GTSF1* | -8.078100289 | 0.003652777 | 0.026071244 | 1 |
| HTN | pericyte | *TTLL6* | -17.37270426 | 8.46E-12 | 7.26E-09 | 4.11E-08 |
| HTN | pericyte | *MSANTD4* | -7.678865546 | 0.007431436 | 0.036736748 | 1 |
| HTN | pericyte | *PAQR8* | -5.994009975 | 9.18E-05 | 0.00350721 | 0.445415718 |
| HTN | pericyte | *ZNF160* | -3.933888642 | 0.005850149 | 0.032856262 | 1 |
| HTN | pericyte | *FPR1* | -4.291373719 | 0.001634453 | 0.020924443 | 1 |
| HTN | pericyte | *TRMT61B* | -3.837213325 | 0.012713542 | 0.049371727 | 1 |
| HTN | pericyte | *MRPL36* | 3.18914029 | 0.000502173 | 0.010547799 | 1 |
| HTN | pericyte | *NMUR1* | -7.513241396 | 0.004067413 | 0.027025118 | 1 |
| HTN | pericyte | *RGS19* | -4.324422001 | 3.38E-10 | 1.49E-07 | 1.64E-06 |
| HTN | pericyte | *ANO5* | -3.00167409 | 0.006989709 | 0.0358138 | 1 |
| HTN | pericyte | *VAT1L* | -4.66073795 | 0.002389665 | 0.023420475 | 1 |
| HTN | pericyte | *PAH* | -12.45285238 | 0.00927419 | 0.0412073 | 1 |
| HTN | pericyte | *FBXW10* | -11.83822365 | 0.00287648 | 0.02367523 | 1 |
| HTN | pericyte | *ZNF554* | -4.790696671 | 0.00675334 | 0.035511559 | 1 |
| HTN | pericyte | *ANKRD20A4* | -13.287122 | 0.002757558 | 0.023519329 | 1 |
| HTN | pericyte | *SERF1A* | -24.76048215 | 0.001691476 | 0.02153444 | 1 |
| HTN | pericyte | *NME6* | 9.79424871 | 2.14E-07 | 3.35E-05 | 0.00103742 |
| HTN | pericyte | *MALT1* | -5.995376085 | 0.008085664 | 0.038424723 | 1 |
| HTN | pericyte | *MANEA* | -6.600284172 | 0.006847101 | 0.035684356 | 1 |
| HTN | pericyte | *CTSW* | -1.008589265 | 1.05E-05 | 0.000697776 | 0.050937655 |
| HTN | pericyte | *EFEMP2* | 5.457192599 | 0.00440318 | 0.028077611 | 1 |
| HTN | pericyte | *ZNF738* | 3.003340263 | 0.003712572 | 0.026106374 | 1 |
| HTN | pericyte | *SLFN11* | -8.132555532 | 0.007382301 | 0.03664544 | 1 |
| HTN | pericyte | *FAM71D* | -3.423101802 | 0.003107176 | 0.024031844 | 1 |
| HTN | pericyte | *ZNF596* | -6.810540009 | 0.003693527 | 0.026085867 | 1 |
| HTN | pericyte | *RAB43* | 17.30853941 | 0.00871299 | 0.039807371 | 1 |
| HTN | pericyte | *CBWD1* | -8.750265326 | 0.005161728 | 0.030617001 | 1 |
| HTN | pericyte | *MYEOV* | -12.49021564 | 7.65E-05 | 0.003028107 | 0.371119673 |
| HTN | pericyte | *SH3RF3* | -5.655991296 | 0.002394179 | 0.023420475 | 1 |
| HTN | pericyte | *GXYLT2* | -3.888142855 | 0.00699742 | 0.0358138 | 1 |
| HTN | pericyte | *DCAKD* | -11.10556178 | 3.63E-05 | 0.001816267 | 0.176065706 |
| HTN | pericyte | *NOC3L* | -10.58722205 | 2.59E-06 | 0.000224121 | 0.01255075 |
| HTN | pericyte | *ARV1* | -3.426220241 | 0.010517965 | 0.04401265 | 1 |
| HTN | pericyte | *PPP1R14B* | 8.547182667 | 0.000183188 | 0.005361456 | 0.88882804 |
| HTN | pericyte | *MST1* | -3.176096195 | 1.79E-07 | 3.00E-05 | 0.000869761 |
| HTN | pericyte | *GMPPB* | -6.076425055 | 0.000156776 | 0.004876146 | 0.760678849 |
| HTN | pericyte | *SULT1B1* | -7.486206362 | 0.001438377 | 0.019603944 | 1 |
| HTN | pericyte | *C11orf80* | -11.88122239 | 1.57E-09 | 6.36E-07 | 7.63E-06 |
| HTN | pericyte | *CTU2* | -4.567090136 | 6.14E-05 | 0.002644087 | 0.298138033 |
| HTN | pericyte | *GLIS1* | -5.744330941 | 0.002314293 | 0.023376742 | 1 |
| HTN | pericyte | *RALGAPA1* | -4.491350826 | 0.012377892 | 0.04886699 | 1 |
| HTN | pericyte | *FGFBP3* | -13.80363071 | 0.002856409 | 0.02367523 | 1 |
| HTN | pericyte | *RSRC1* | -7.729479155 | 0.000932913 | 0.015448786 | 1 |
| HTN | pericyte | *SEZ6L2* | 5.245645042 | 3.91E-06 | 0.000316302 | 0.018978148 |
| HTN | pericyte | *TVP23C* | -9.239235153 | 0.002756941 | 0.023519329 | 1 |
| HTN | pericyte | *CADM2* | -3.645250957 | 0.003278564 | 0.024662563 | 1 |
| HTN | pericyte | *CCNE2* | -9.641740531 | 7.94E-11 | 4.82E-08 | 3.85E-07 |

| | | | | | | |
|---|---|---|---|---|---|---|
| HTN | pericyte | *BANF1* | -7.659902639 | 0.000894191 | 0.015012514 | 1 |
| HTN | pericyte | *TMEM9B* | 8.971321125 | 0.001400319 | 0.019257661 | 1 |
| HTN | pericyte | *EIF1AD* | -7.594293415 | 0.010781021 | 0.044747232 | 1 |
| HTN | pericyte | *PPP2R2D* | -8.008403524 | 0.000119807 | 0.004036836 | 0.581304368 |
| HTN | pericyte | *MCTP1* | -4.87014253 | 0.012705749 | 0.049371727 | 1 |
| HTN | pericyte | *MRPL48* | -5.985291351 | 0.002186557 | 0.023256174 | 1 |
| HTN | pericyte | *TOM1L2* | -5.394725425 | 0.010396423 | 0.04367398 | 1 |
| HTN | pericyte | *PRIMA1* | -4.118821865 | 0.002239174 | 0.023376742 | 1 |
| HTN | pericyte | *ORAI3* | 8.65601931 | 3.32E-07 | 4.67E-05 | 0.001612944 |
| HTN | pericyte | *B3GALT6* | -9.947227455 | 0.002627565 | 0.023519329 | 1 |
| HTN | pericyte | *CIDEA* | -3.85069145 | 0.007553734 | 0.037022338 | 1 |
| HTN | pericyte | *BOK* | -7.719670555 | 0.009645588 | 0.041803673 | 1 |
| HTN | pericyte | *C8G* | -68.16482672 | 0.002143573 | 0.023010218 | 1 |
| HTN | pericyte | *SHMT1* | -3.321468446 | 0.006714722 | 0.03548317 | 1 |
| HTN | pericyte | *MTHFR* | -11.70795584 | 8.87E-08 | 1.72E-05 | 0.000430214 |
| HTN | pericyte | *TMEM80* | -6.016466029 | 0.002695816 | 0.023519329 | 1 |
| HTN | pericyte | *MIEF2* | -9.193526484 | 0.004546723 | 0.028589746 | 1 |
| HTN | pericyte | *NAP1L5* | 5.591719322 | 0.0093273 | 0.041319857 | 1 |
| HTN | pericyte | *SLC25A22* | -6.716720048 | 0.000171932 | 0.00518146 | 0.834215106 |
| HTN | pericyte | *THAP5* | -9.344110325 | 0.009141886 | 0.040731344 | 1 |
| HTN | pericyte | *SLC35G5* | -22.56270136 | 0.00276692 | 0.023519329 | 1 |
| HTN | pericyte | *FAM87B* | -21.31334348 | 0.002313907 | 0.023376742 | 1 |
| HTN | pericyte | *PCDHB9* | 4.704837346 | 0.009344672 | 0.041319857 | 1 |
| HTN | pericyte | *CENPBD1* | 4.593349644 | 0.007435179 | 0.036736748 | 1 |
| HTN | pericyte | *RIC8A* | -6.632452803 | 0.002506092 | 0.023519329 | 1 |
| HTN | pericyte | *ODF3B* | 3.460262224 | 0.007291954 | 0.036435078 | 1 |
| HTN | pericyte | *DPY19L2* | 0.728007656 | 0.005101387 | 0.030407778 | 1 |
| HTN | pericyte | *SH2B1* | -6.409115539 | 0.009239859 | 0.041092389 | 1 |
| HTN | pericyte | *KDELC2* | -10.38077244 | 0.003436313 | 0.025493869 | 1 |
| HTN | pericyte | *SH2D4B* | 13.93499547 | 0.012057616 | 0.048201436 | 1 |
| HTN | pericyte | *GEN1* | -2.921758928 | 0.000299163 | 0.007383636 | 1 |
| HTN | pericyte | *TMEM11* | -6.103144926 | 0.009877 | 0.042185921 | 1 |
| HTN | pericyte | *ZFAND2A* | -8.384045495 | 0.000101658 | 0.003853469 | 0.493244073 |
| HTN | pericyte | *P4HTM* | -9.731361508 | 0.008389423 | 0.038915375 | 1 |
| HTN | pericyte | *CA8* | -3.502388365 | 0.006025623 | 0.033412941 | 1 |
| HTN | pericyte | *ERN1* | -6.405014884 | 0.00481947 | 0.029495473 | 1 |
| HTN | pericyte | *PARP10* | -5.979748786 | 0.001134579 | 0.01687464 | 1 |
| HTN | pericyte | *RPP25* | 4.435394782 | 4.17E-06 | 0.000326518 | 0.020244093 |
| HTN | pericyte | *MPI* | -14.26177715 | 1.13E-17 | 5.49E-14 | 5.49E-14 |
| HTN | pericyte | *TRIM73* | -10.89618952 | 0.000213457 | 0.00601954 | 1 |
| HTN | pericyte | *ZNF852* | -4.922236568 | 0.000202936 | 0.005758152 | 0.98464403 |
| HTN | pericyte | *HYI* | -3.10961779 | 0.000490116 | 0.010339327 | 1 |
| HTN | pericyte | *CCDC89* | -9.969088691 | 0.003037657 | 0.023886115 | 1 |
| HTN | pericyte | *SPTY2D1* | -7.587174566 | 0.00712622 | 0.03616544 | 1 |
| HTN | pericyte | *ZNF664* | -7.557284468 | 0.003124954 | 0.024031844 | 1 |
| HTN | pericyte | *CDH4* | -3.582013289 | 0.000460341 | 0.009927007 | 1 |
| HTN | pericyte | *HLA-DQB1* | -1.760250471 | 0.001873731 | 0.022392469 | 1 |
| HTN | pericyte | *PACS2* | -6.432144411 | 0.008499956 | 0.039211167 | 1 |
| HTN | pericyte | *GEMIN4* | -6.03191829 | 0.001401062 | 0.019257661 | 1 |
| HTN | pericyte | *ZFPM1* | -3.047707855 | 0.006330302 | 0.034394877 | 1 |

| | | | | | | |
|---|---|---|---|---|---|---|
| HTN | pericyte | *FCER1A* | -3.023694217 | 0.005518951 | 0.031765065 | 1 |
| HTN | pericyte | *AKAP5* | -6.987160595 | 0.000576017 | 0.011693872 | 1 |
| HTN | pericyte | *GIPC3* | -35.81606758 | 0.003967419 | 0.02677323 | 1 |
| HTN | pericyte | *SEPHS2* | 8.585643309 | 0.0029938 | 0.023812983 | 1 |
| HTN | pericyte | *SEPT1* | -7.384798955 | 0.002093993 | 0.02292625 | 1 |
| HTN | pericyte | *MTURN* | -4.963139546 | 0.008648107 | 0.039585488 | 1 |
| HTN | pericyte | *DEFB124* | 2.02711895 | 0.004476304 | 0.028316853 | 1 |
| HTN | pericyte | *TSPYL5* | -3.623042956 | 0.001557485 | 0.020431711 | 1 |
| HTN | pericyte | *SLC36A4* | -6.14389714 | 0.008403999 | 0.038945752 | 1 |
| HTN | pericyte | *D2HGDH* | -4.5477073 | 0.002848679 | 0.02367523 | 1 |
| HTN | pericyte | *MUC16* | -4.029519607 | 0.003740932 | 0.02622977 | 1 |
| HTN | pericyte | *SLC25A41* | -12.84074895 | 0.004450085 | 0.028219466 | 1 |
| HTN | pericyte | *SGSH* | -11.39236036 | 0.00725533 | 0.036435078 | 1 |
| HTN | pericyte | *TMEM252* | -118.295531 | 0.003735444 | 0.026229195 | 1 |
| HTN | pericyte | *ZNF329* | -6.433001492 | 0.003573993 | 0.025973602 | 1 |
| HTN | pericyte | *ADO* | 8.823246079 | 1.04E-10 | 5.62E-08 | 5.06E-07 |
| HTN | pericyte | *MGAT4C* | -2.082276374 | 0.00363839 | 0.026037563 | 1 |
| HTN | pericyte | *ZNF708* | -4.486475219 | 0.003540155 | 0.025868726 | 1 |
| HTN | pericyte | *FAM153B* | -3.206411236 | 0.007127581 | 0.03616544 | 1 |
| HTN | pericyte | *KCNA4* | -11.5154214 | 0.008990881 | 0.040525766 | 1 |
| HTN | pericyte | *CLN8* | 4.408007617 | 1.37E-06 | 0.000135446 | 0.006636866 |
| HTN | pericyte | *FES* | -13.07846114 | 4.20E-12 | 6.79E-09 | 2.04E-08 |
| HTN | pericyte | *CAV3* | -4.112504086 | 0.001026025 | 0.016007307 | 1 |
| HTN | pericyte | *MFSD5* | -3.010674736 | 0.00585701 | 0.032856262 | 1 |
| HTN | pericyte | *ADI1* | -6.607006753 | 0.005993865 | 0.033344492 | 1 |
| HTN | pericyte | *BRICD5* | -6.150485142 | 0.002847224 | 0.02367523 | 1 |
| HTN | pericyte | *PLCXD3* | -4.508507348 | 0.00267927 | 0.023519329 | 1 |
| HTN | pericyte | *GPIHBP1* | -1.514542388 | 0.008207653 | 0.038500008 | 1 |
| HTN | pericyte | *ALG12* | -9.467866159 | 0.00655856 | 0.035132512 | 1 |
| HTN | pericyte | *COL18A1* | 3.863176735 | 0.002335142 | 0.023376742 | 1 |
| HTN | pericyte | *CEP63* | -3.875326783 | 0.005317987 | 0.031271844 | 1 |
| HTN | pericyte | *ODF3L1* | -11.0150929 | 0.000426142 | 0.009313697 | 1 |
| HTN | pericyte | *ZNF320* | 2.164662246 | 0.000119574 | 0.004036836 | 0.580174001 |
| HTN | pericyte | *C12orf60* | -8.290525144 | 0.005189737 | 0.030745548 | 1 |
| HTN | pericyte | *LYSMD4* | -5.572572907 | 0.005882692 | 0.032921365 | 1 |
| HTN | pericyte | *WBP2NL* | -7.308279218 | 0.000181617 | 0.005361456 | 0.881208031 |
| HTN | pericyte | *ARHGEF37* | -6.096690416 | 0.003129453 | 0.024031844 | 1 |
| HTN | pericyte | *CSMD1* | -8.182939223 | 0.012356223 | 0.04882961 | 1 |
| HTN | pericyte | *FANCF* | -5.997440898 | 0.005330893 | 0.031271844 | 1 |
| HTN | pericyte | *CCDC125* | -5.65069566 | 0.004302535 | 0.027673274 | 1 |
| HTN | pericyte | *LRIT3* | -21.5270012 | 0.002545192 | 0.023519329 | 1 |
| HTN | pericyte | *NPIPA1* | 1.733310351 | 0.007973352 | 0.038190232 | 1 |
| HTN | pericyte | *AC131971.1* | -12.30232735 | 0.001672249 | 0.021351977 | 1 |
| HTN | pericyte | *ACSM2A* | -23.67949122 | 0.01024005 | 0.043228567 | 1 |
| HTN | pericyte | *KREMEN1* | -2.699979834 | 0.005075849 | 0.030407778 | 1 |
| HTN | pericyte | *FAM162B* | 0.866278601 | 0.002804651 | 0.02367523 | 1 |
| HTN | pericyte | *NUDT14* | -10.94978749 | 0.000273353 | 0.007054822 | 1 |
| HTN | pericyte | *TTC32* | -5.164858149 | 0.006820659 | 0.035623078 | 1 |
| HTN | pericyte | *PP2D1* | -9.682168566 | 0.004919024 | 0.030021513 | 1 |
| HTN | pericyte | *GOLGA6L4* | -3.83095927 | 0.011767263 | 0.047342256 | 1 |

| | | | | | | |
|---|---|---|---|---|---|---|
| HTN | pericyte | *ALDH1A3* | -3.388231853 | 4.60E-05 | 0.002145685 | 0.223151273 |
| HTN | pericyte | *A3GALT2* | -3.914846869 | 4.14E-06 | 0.000326518 | 0.020087348 |
| HTN | pericyte | *SS18L1* | -6.58105327 | 0.012541412 | 0.049132942 | 1 |
| HTN | pericyte | *TOP1MT* | -2.585658715 | 0.002128739 | 0.023003652 | 1 |
| HTN | pericyte | *ZFP1* | -3.968507082 | 0.009726001 | 0.041984481 | 1 |
| HTN | pericyte | *RNLS* | -4.588204182 | 0.012628548 | 0.049215835 | 1 |
| HTN | pericyte | *DRD1* | -67.58692243 | 0.003630382 | 0.02601863 | 1 |
| HTN | pericyte | *NUTM2A* | -5.21007033 | 5.38E-05 | 0.002415956 | 0.260923276 |
| HTN | pericyte | *WT1* | -2.501065163 | 0.004191185 | 0.027381347 | 1 |
| HTN | pericyte | *BRI3BP* | -9.73753409 | 0.001510758 | 0.020137909 | 1 |
| HTN | pericyte | *MAFF* | -2.752217937 | 0.007558279 | 0.037022338 | 1 |
| HTN | pericyte | *C6orf120* | -4.922152015 | 0.003841763 | 0.026468005 | 1 |
| HTN | pericyte | *CDK10* | -3.538448271 | 3.24E-05 | 0.001670145 | 0.156993601 |
| HTN | pericyte | *C14orf80* | -5.174512197 | 0.004701765 | 0.029284932 | 1 |
| HTN | pericyte | *TNFAIP8L1* | -10.57301494 | 0.006547329 | 0.035132512 | 1 |
| HTN | pericyte | *FAM174B* | -6.200617232 | 0.010245848 | 0.043228567 | 1 |
| HTN | pericyte | *SV2B* | -3.144155056 | 0.010044323 | 0.042653577 | 1 |
| HTN | pericyte | *MYBL1* | -4.733410992 | 0.000119224 | 0.004036836 | 0.578472838 |
| HTN | pericyte | *SLC52A2* | 6.031949314 | 0.00951883 | 0.041570982 | 1 |
| HTN | pericyte | *ARL17A* | -3.733649542 | 0.006994497 | 0.0358138 | 1 |
| HTN | pericyte | *IFITM1* | -6.821709402 | 0.000575999 | 0.011693872 | 1 |
| HTN | pericyte | *FFAR3* | 4.13604396 | 0.008008105 | 0.038243432 | 1 |
| HTN | pericyte | *LINC00839* | -10.33370084 | 0.001165072 | 0.017162107 | 1 |
| HTN | pericyte | *RASA3* | -4.22299802 | 0.00200547 | 0.022895385 | 1 |
| HTN | pericyte | *TMEM222* | -9.852771199 | 0.002344155 | 0.023376742 | 1 |
| HTN | pericyte | *SMYD4* | -5.085006831 | 0.006587256 | 0.035238549 | 1 |
| HTN | pericyte | *MIR22HG* | -8.651138441 | 0.00361618 | 0.025973602 | 1 |
| HTN | pericyte | *FOXI2* | -4.075374643 | 0.006130555 | 0.033916647 | 1 |
| HTN | pericyte | *ZNF732* | -7.928674033 | 0.00204613 | 0.02292625 | 1 |
| HTN | pericyte | *C1QL4* | -35.79563123 | 0.002543211 | 0.023519329 | 1 |
| HTN | pericyte | *TMPRSS6* | -21.11234566 | 0.002588145 | 0.023519329 | 1 |
| HTN | pericyte | *CYP4A11* | 4.785924568 | 0.01057411 | 0.044114859 | 1 |
| HTN | pericyte | *MRPS21* | -3.234889893 | 0.011202253 | 0.045790505 | 1 |
| HTN | pericyte | *EPOR* | 2.631928087 | 0.010959935 | 0.045142399 | 1 |
| HTN | pericyte | *LUZP2* | -12.92693786 | 0.006497262 | 0.034988583 | 1 |
| HTN | pericyte | *GJA4* | 1.581605733 | 0.006935522 | 0.0358138 | 1 |
| HTN | pericyte | *SBK2* | 3.714820213 | 0.004441373 | 0.028207424 | 1 |
| HTN | pericyte | *DHRS4L2* | -2.220278466 | 0.010942986 | 0.045142399 | 1 |
| HTN | pericyte | *C2orf88* | -6.041408028 | 0.002334879 | 0.023376742 | 1 |
| HTN | pericyte | *KBTBD12* | -3.523201382 | 0.012210465 | 0.048561619 | 1 |
| HTN | pericyte | *FANCM* | -6.097985419 | 0.001296448 | 0.018339262 | 1 |
| HTN | pericyte | *ZFP69* | -7.329889342 | 0.002887287 | 0.02367523 | 1 |
| HTN | pericyte | *GFRAL* | 3.272686377 | 0.009829392 | 0.042148419 | 1 |
| HTN | pericyte | *KLHL17* | 4.994226239 | 0.000806508 | 0.014077604 | 1 |
| HTN | pericyte | *MAPK12* | -7.800844719 | 0.003396063 | 0.025311361 | 1 |
| HTN | pericyte | *AGRN* | -3.588195177 | 0.003031727 | 0.023886115 | 1 |
| HTN | pericyte | *ZNF626* | -9.012549064 | 0.007231014 | 0.036432899 | 1 |
| HTN | pericyte | *NCR3LG1* | -9.965047055 | 0.001137264 | 0.01687464 | 1 |
| HTN | pericyte | *ZNF793* | -2.45057061 | 0.007403572 | 0.03668346 | 1 |
| HTN | pericyte | *RUFY4* | -6.384796074 | 0.002842713 | 0.02367523 | 1 |

| | | | | | | |
|---|---|---|---|---|---|---|
| HTN | pericyte | *CENPP* | -3.218247481 | 0.001214133 | 0.017480633 | 1 |
| HTN | pericyte | *C3orf62* | -5.349238977 | 0.003883044 | 0.026573384 | 1 |
| HTN | pericyte | *ZNF559* | -1.989677982 | 0.002691468 | 0.023519329 | 1 |
| HTN | pericyte | *H3F3C* | -95.60399701 | 0.00039608 | 0.008980278 | 1 |
| HTN | pericyte | *C19orf54* | -6.771398166 | 0.007179491 | 0.036271341 | 1 |
| HTN | pericyte | *COL25A1* | 0.438174872 | 0.00478058 | 0.029398444 | 1 |
| HTN | pericyte | *NDOR1* | -7.484364896 | 0.002767835 | 0.023519329 | 1 |
| HTN | pericyte | *FAM72B* | 5.180944239 | 0.004224783 | 0.027404608 | 1 |
| HTN | pericyte | *SLC4A5* | 8.089862569 | 0.009505998 | 0.041552345 | 1 |
| HTN | pericyte | *FSIP2* | -4.794400954 | 0.006728054 | 0.03548317 | 1 |
| HTN | pericyte | *NOXA1* | -7.370083588 | 0.002984097 | 0.023785667 | 1 |
| HTN | pericyte | *MTF1* | -4.086655935 | 0.009943931 | 0.042399955 | 1 |
| HTN | pericyte | *FAM26F* | -3.04981984 | 0.002042781 | 0.02292625 | 1 |
| HTN | pericyte | *ENTPD8* | -322.3599934 | 0.001964861 | 0.022752995 | 1 |
| HTN | pericyte | *FBF1* | -9.211605286 | 1.23E-08 | 3.72E-06 | 5.99E-05 |
| HTN | pericyte | *ASTL* | -10.0352 | 0.002606798 | 0.023519329 | 1 |
| HTN | pericyte | *MSL1* | -5.829962146 | 0.000297961 | 0.007383636 | 1 |
| HTN | pericyte | *UTS2B* | -3.610130958 | 0.001324618 | 0.01847157 | 1 |
| HTN | pericyte | *ADAT2* | -5.765599268 | 0.004744497 | 0.029362627 | 1 |
| HTN | pericyte | *LITAF* | -5.486807405 | 0.006963243 | 0.0358138 | 1 |
| HTN | pericyte | *HLA-DRB1* | -2.561133096 | 0.000113972 | 0.004035815 | 0.552993501 |
| HTN | pericyte | *COLCA1* | -6.900515269 | 2.27E-06 | 0.000207624 | 0.011004069 |
| HTN | pericyte | *SEMA4A* | -6.154199363 | 0.003242517 | 0.02450575 | 1 |
| HTN | pericyte | *HRCT1* | 4.267930355 | 0.000553646 | 0.011338333 | 1 |
| HTN | pericyte | *ZNF493* | -7.479330335 | 0.010878131 | 0.044996327 | 1 |
| HTN | pericyte | *C20orf96* | -10.10543574 | 0.005775711 | 0.032585753 | 1 |
| HTN | pericyte | *SULT1A1* | -5.825865875 | 0.000142331 | 0.004603939 | 0.690590866 |
| HTN | pericyte | *TPK1* | -5.817909409 | 0.006141215 | 0.033916647 | 1 |
| HTN | pericyte | *CACNA1H* | -7.635709391 | 0.00729903 | 0.036435078 | 1 |
| HTN | pericyte | *XRCC2* | -6.288430874 | 0.002642951 | 0.023519329 | 1 |
| HTN | pericyte | *ZNF846* | -5.21046394 | 0.010611702 | 0.044195688 | 1 |
| HTN | pericyte | *ADH1B* | -3.054799507 | 0.002961664 | 0.023712861 | 1 |
| HTN | pericyte | *AMZ2* | -5.982970477 | 0.003935183 | 0.02677323 | 1 |
| HTN | pericyte | *HLA-DQA1* | -2.081017973 | 0.011657658 | 0.047096551 | 1 |
| HTN | pericyte | *ZNF239* | -5.446842994 | 0.008131217 | 0.038500008 | 1 |
| HTN | pericyte | *PCDHB16* | 2.636301798 | 0.002744944 | 0.023519329 | 1 |
| HTN | pericyte | *HIST1H3E* | 4.626558773 | 7.77E-08 | 1.68E-05 | 0.00037692 |
| HTN | pericyte | *ZNF429* | -5.235203621 | 0.002439259 | 0.023519329 | 1 |
| HTN | pericyte | *ZNF100* | -7.418196681 | 1.56E-06 | 0.000151617 | 0.007580855 |
| HTN | pericyte | *GMFB* | -7.051902797 | 0.008003455 | 0.038243432 | 1 |
| HTN | pericyte | *ZMYM1* | -6.226904432 | 0.006248268 | 0.034178803 | 1 |
| HTN | pericyte | *ZNF257* | -4.731270049 | 0.002565823 | 0.023519329 | 1 |
| HTN | pericyte | *ADAM32* | -5.387548001 | 0.001871389 | 0.022392469 | 1 |
| HTN | pericyte | *PIWIL2* | -3.185863808 | 0.01147988 | 0.046533317 | 1 |
| HTN | pericyte | *C6orf141* | -1.321157827 | 0.001019771 | 0.01596106 | 1 |
| HTN | pericyte | *ZNF720* | -5.676862407 | 0.009637219 | 0.041803673 | 1 |
| HTN | pericyte | *ZNF517* | -12.72344065 | 0.006146526 | 0.033916647 | 1 |
| HTN | pericyte | *MAP3K5* | -4.344143758 | 0.010316709 | 0.043376664 | 1 |
| HTN | pericyte | *GSTK1* | -5.999000303 | 0.000608955 | 0.01210347 | 1 |
| HTN | pericyte | *PCDHB11* | -2.487242978 | 0.000290599 | 0.007305639 | 1 |

| | | | | | | |
|---|---|---|---|---|---|---|
| HTN | pericyte | *ZNF665* | -5.785835605 | 0.003085785 | 0.023955563 | 1 |
| HTN | pericyte | *RPF2* | -6.403987243 | 0.011078387 | 0.045475746 | 1 |
| HTN | pericyte | *BCO2* | -1.751287882 | 0.003894809 | 0.026616358 | 1 |
| HTN | pericyte | *ENTPD6* | -6.179264763 | 0.000979625 | 0.015786801 | 1 |
| HTN | pericyte | *ENPP1* | 4.111029058 | 0.006059038 | 0.033559878 | 1 |
| HTN | pericyte | *MFAP5* | -5.817977962 | 2.99E-06 | 0.00024727 | 0.014514007 |
| HTN | pericyte | *FAM114A1* | -4.69177911 | 0.008643008 | 0.039585488 | 1 |
| HTN | pericyte | *RPS26* | -3.765962622 | 4.92E-06 | 0.000372658 | 0.023850098 |
| HTN | pericyte | *EME2* | -12.74827365 | 0.007592306 | 0.037121203 | 1 |
| HTN | pericyte | *AC122129.1* | -16.15315675 | 0.000303438 | 0.007383636 | 1 |
| HTN | pericyte | *OCLN* | -3.207662236 | 0.003943201 | 0.02677323 | 1 |
| HTN | pericyte | *SGTB* | -7.262739451 | 0.003829588 | 0.026468005 | 1 |
| HTN | pericyte | *AGAP9* | -8.624149195 | 0.000760018 | 0.013557386 | 1 |
| HTN | pericyte | *ZNF273* | -5.502712337 | 0.002640352 | 0.023519329 | 1 |
| HTN | pericyte | *SFI1* | -7.682317801 | 0.000289757 | 0.007305639 | 1 |
| HTN | pericyte | *ADH4* | -2.75152784 | 0.005066296 | 0.030407778 | 1 |
| HTN | pericyte | *ZNF607* | 3.936429699 | 0.003795801 | 0.026406882 | 1 |
| HTN | pericyte | *RPS6KL1* | 8.172673102 | 0.00081213 | 0.014077604 | 1 |
| HTN | pericyte | *STYX* | -7.419027764 | 0.002484253 | 0.023519329 | 1 |
| HTN | pericyte | *TMEM116* | -5.570093565 | 6.46E-16 | 1.57E-12 | 3.13E-12 |
| HTN | pericyte | *UCKL1* | -10.73925909 | 0.001879953 | 0.022411632 | 1 |
| HTN | pericyte | *ZNF568* | -7.532505876 | 0.004105996 | 0.027151036 | 1 |
| HTN | pericyte | *TMA16* | -6.61019724 | 0.0082527 | 0.038539076 | 1 |
| HTN | pericyte | *HLA-DRB5* | -1.081631128 | 3.63E-05 | 0.001816267 | 0.176248647 |
| HTN | pericyte | *NCOA6* | -5.364962514 | 0.001588764 | 0.020556487 | 1 |
| HTN | pericyte | *TUSC1* | -16.2354636 | 0.004275551 | 0.027586403 | 1 |
| HTN | pericyte | *ANKRD13B* | -12.23797732 | 0.003197891 | 0.024319694 | 1 |
| HTN | pericyte | *PPP1R14C* | 0.677171771 | 0.004372118 | 0.027949299 | 1 |
| HTN | pericyte | *GRIN3A* | 3.329432591 | 0.004441565 | 0.028207424 | 1 |
| HTN | pericyte | *PNP* | -1.45019892 | 0.001016084 | 0.015954822 | 1 |
| HTN | pericyte | *DENND4B* | -8.121028296 | 1.72E-05 | 0.001019217 | 0.08357581 |
| HTN | pericyte | *PRMT6* | -17.24583791 | 0.000437071 | 0.009509733 | 1 |
| HTN | pericyte | *SHISA4* | -18.17182773 | 4.97E-08 | 1.34E-05 | 0.000241339 |
| HTN | pericyte | *MAP3K3* | -6.683273656 | 0.00289353 | 0.02367523 | 1 |
| HTN | pericyte | *KIAA0753* | -7.100860671 | 0.001577556 | 0.020474082 | 1 |
| HTN | pericyte | *CCDC167* | -9.144410842 | 0.002904838 | 0.02367523 | 1 |
| HTN | pericyte | *SMG5* | -5.732471671 | 0.003057147 | 0.023886115 | 1 |
| HTN | pericyte | *TGM2* | 2.857923349 | 0.005727794 | 0.032448526 | 1 |
| HTN | pericyte | *AL590822.1* | -5.666735508 | 3.79E-05 | 0.00183849 | 0.183848975 |
| HTN | pericyte | *GPR52* | -49.97161719 | 0.012407852 | 0.048907668 | 1 |
| HTN | pericyte | *DDO* | -4.003789171 | 0.003659332 | 0.026071244 | 1 |
| HTN | pericyte | *RBM20* | 1.85198046 | 0.005517218 | 0.031765065 | 1 |
| HTN | pericyte | *IFIT1B* | -17.00562796 | 0.003529914 | 0.025868726 | 1 |
| HTN | pericyte | *LINC00619* | -28.82181234 | 0.003480318 | 0.025722434 | 1 |
| HTN | pericyte | *HSD17B8* | -4.960467038 | 2.84E-05 | 0.001498369 | 0.137849948 |
| HTN | pericyte | *AGER* | 2.525115116 | 0.00756006 | 0.037022338 | 1 |
| HTN | pericyte | *PRRT1* | 3.533882531 | 3.85E-05 | 0.001847413 | 0.18658876 |
| HTN | pericyte | *LAYN* | -7.180329788 | 6.22E-05 | 0.002646543 | 0.301859401 |
| HTN | pericyte | *MICA* | -2.928602841 | 0.004820674 | 0.029495473 | 1 |
| HTN | pericyte | *HLA-C* | -4.088313394 | 1.12E-07 | 2.09E-05 | 0.000543422 |

| | | | | | | |
|---|---|---|---|---|---|---|
| HTN | pericyte | *POU5F1* | -1.021429139 | 0.000795873 | 0.014042101 | 1 |
| HTN | pericyte | *PSORS1C2* | 3.027621867 | 0.004742843 | 0.029362627 | 1 |
| HTN | pericyte | *HLA-E* | -3.878765921 | 0.011035189 | 0.045336779 | 1 |
| HTN | pericyte | *ZNF468* | 10.86294864 | 0.001094858 | 0.016652832 | 1 |
| HTN | pericyte | *MALRD1* | -3.53103797 | 0.001243433 | 0.017796863 | 1 |
| HTN | pericyte | *DCTN1* | -8.409956512 | 0.008501679 | 0.039211167 | 1 |
| HTN | pericyte | *ZNF155* | -5.926566472 | 0.009220183 | 0.041042502 | 1 |
| HTN | pericyte | *ZNF425* | -4.78329765 | 0.003798847 | 0.026406882 | 1 |
| HTN | pericyte | *PCDHA7* | 7.677260805 | 0.005955918 | 0.033178087 | 1 |
| HTN | pericyte | *SCGB2B2* | -6.0288855 | 0.006285403 | 0.034227583 | 1 |
| HTN | pericyte | *GOLGA6L10* | -8.579602783 | 0.000738395 | 0.013318553 | 1 |
| HTN | pericyte | *PRR13* | -7.537015304 | 4.93E-05 | 0.002254702 | 0.238998465 |
| HTN | pericyte | *MT1A* | -5.238519742 | 0.006898614 | 0.035799012 | 1 |
| HTN | pericyte | *CFI* | -4.941542366 | 0.009359135 | 0.041319857 | 1 |
| HTN | pericyte | *KLRC2* | -6.307107708 | 0.008995517 | 0.040525766 | 1 |
| HTN | pericyte | *C1QTNF9B* | -29.49716271 | 0.001902517 | 0.022514662 | 1 |
| HTN | pericyte | *ZDHHC11B* | -8.446094369 | 0.002113953 | 0.022953412 | 1 |
| HTN | pericyte | *HBA1* | 1.76101753 | 0.007753023 | 0.03755768 | 1 |
| HTN | pericyte | *SACM1L* | -6.370178039 | 0.008632029 | 0.039585488 | 1 |
| HTN | pericyte | *TAS2R50* | 21.55054674 | 0.001630208 | 0.020924443 | 1 |
| HTN | pericyte | *AC022498.1* | -13.97535521 | 0.009496381 | 0.041552345 | 1 |
| HTN | pericyte | *MXD3* | -5.250445521 | 7.13E-05 | 0.002905098 | 0.345706617 |
| HTN | pericyte | *GSTM2* | -2.842760286 | 0.005397084 | 0.031400705 | 1 |
| HTN | pericyte | *GPC2* | 3.502301855 | 0.003878675 | 0.026573384 | 1 |
| HTN | pericyte | *SRA1* | -5.401093788 | 0.010418052 | 0.043689185 | 1 |
| HTN | pericyte | *ATF6B* | 9.100929151 | 2.39E-06 | 0.000213395 | 0.011575184 |
| HTN | pericyte | *SLC35F6* | -8.123704001 | 0.001482032 | 0.019919162 | 1 |
| HTN | pericyte | *PPM1N* | 2.965055374 | 0.007038234 | 0.035909054 | 1 |
| HTN | pericyte | *LTB4R* | -9.738412596 | 1.31E-06 | 0.00013534 | 0.006360959 |
| HTN | pericyte | *ZNF726* | -5.964840856 | 0.002077839 | 0.02292625 | 1 |
| HTN | pericyte | *REPIN1* | -5.359289539 | 0.002360208 | 0.023376742 | 1 |
| HTN | pericyte | *PRCD* | 4.021744041 | 0.001920847 | 0.022609835 | 1 |
| HTN | pericyte | *SH3D21* | 2.697721819 | 0.002126292 | 0.023003652 | 1 |
| HTN | pericyte | *FIS1* | -3.249734343 | 0.005211174 | 0.030797343 | 1 |
| HTN | pericyte | *GNAT3* | -3.822282023 | 0.002876207 | 0.02367523 | 1 |
| HTN | pericyte | *CPEB1* | -2.256480008 | 0.004315413 | 0.027696272 | 1 |
| HTN | pericyte | *EML6* | -5.669257685 | 0.01277253 | 0.049498656 | 1 |
| HTN | pericyte | *AC005082.1* | -15.19258472 | 0.000115618 | 0.004035815 | 0.560978295 |
| HTN | pericyte | *SLC35E2* | -3.905506832 | 0.004163832 | 0.027375221 | 1 |
| HTN | pericyte | *TSTD1* | -9.958496421 | 0.001106566 | 0.016745563 | 1 |
| HTN | pericyte | *PAM16* | -4.18722327 | 0.001118374 | 0.016747989 | 1 |
| HTN | pericyte | *AC016773.1* | 4.927087072 | 0.00018343 | 0.005361456 | 0.890001751 |
| HTN | pericyte | *AP4M1* | -4.040493581 | 0.001426448 | 0.019496125 | 1 |
| HTN | pericyte | *APOL6* | -4.946418875 | 0.003864226 | 0.026519414 | 1 |
| HTN | pericyte | *TIAF1* | 7.138406749 | 0.00018029 | 0.005361456 | 0.874768329 |
| HTN | pericyte | *EXOSC6* | -11.0586964 | 5.28E-06 | 0.000382721 | 0.02564233 |
| HTN | pericyte | *LINC00571* | -4.019366804 | 0.001074134 | 0.016440696 | 1 |
| HTN | pericyte | *C5orf66* | -6.342305317 | 0.002075916 | 0.02292625 | 1 |
| HTN | pericyte | *LINC01133* | -3.466811225 | 0.001768412 | 0.022114271 | 1 |
| HTN | pericyte | *LINC01117* | -4.640394717 | 0.003606076 | 0.025973602 | 1 |

| | | | | | | |
|---|---|---|---|---|---|---|
| HTN | pericyte | *LINC00184* | -121.5649612 | 0.002338201 | 0.023376742 | 1 |
| HTN | pericyte | *AL049840.1* | -6.591774741 | 0.007344401 | 0.036548752 | 1 |
| HTN | pericyte | *PLEKHM1* | -9.194579303 | 8.75E-06 | 0.000589944 | 0.042475943 |
| HTN | pericyte | *LINC00115* | -9.246818182 | 0.001322039 | 0.01847157 | 1 |
| HTN | pericyte | *HAR1A* | -34.71082532 | 0.002074168 | 0.02292625 | 1 |
| HTN | pericyte | *FAM138B* | -43.87441082 | 0.002520004 | 0.023519329 | 1 |
| HTN | pericyte | *AC009299.3* | -4.108972053 | 0.003069747 | 0.023915939 | 1 |
| HTN | pericyte | *C14orf144* | 1.443951026 | 0.003825322 | 0.026468005 | 1 |
| HTN | pericyte | *LINC00578* | -6.036733042 | 0.000133362 | 0.00440186 | 0.647073438 |
| HTN | pericyte | *CASC20* | -5.014112194 | 0.002301457 | 0.023376742 | 1 |
| HTN | pericyte | *PSMG3-AS1* | -5.103037476 | 0.002365078 | 0.023376742 | 1 |
| HTN | pericyte | *AC092171.4* | -10.16452445 | 0.003688423 | 0.026085867 | 1 |
| HTN | pericyte | *DIRC3* | -3.351072416 | 0.002776829 | 0.0235545 | 1 |
| HTN | pericyte | *AC009495.3* | 9.818213238 | 0.00954802 | 0.041623532 | 1 |
| HTN | pericyte | *LINC00702* | -10.71468084 | 0.009658268 | 0.041803673 | 1 |
| HTN | pericyte | *AC019117.2* | -26.17351413 | 0.000179815 | 0.005361456 | 0.872460576 |
| HTN | pericyte | *HLA-DQA2* | 1.447553628 | 0.000114522 | 0.004035815 | 0.555659361 |
| HTN | pericyte | *LINC01143* | 18.11458328 | 0.012507755 | 0.049100024 | 1 |
| HTN | pericyte | *LINC00211* | -16.34071205 | 6.16E-05 | 0.002644087 | 0.298781837 |
| HTN | pericyte | *LRRC37A2* | -7.58803697 | 0.000365289 | 0.008439918 | 1 |
| HTN | pericyte | *TXNDC5* | -15.54543339 | 0.002749964 | 0.023519329 | 1 |
| HTN | pericyte | *TNFSF12* | 4.923125118 | 0.00285562 | 0.02367523 | 1 |
| HTN | pericyte | *C1orf226* | -9.110071061 | 0.002588138 | 0.023519329 | 1 |
| HTN | pericyte | *KRBOX1* | -48.85203828 | 0.002301855 | 0.023376742 | 1 |
| HTN | pericyte | *ATP5O* | -7.591666935 | 0.00055383 | 0.011338333 | 1 |
| HTN | pericyte | *HOGA1* | -7.063635253 | 0.002312744 | 0.023376742 | 1 |
| HTN | pericyte | *ZNF709* | -33.80361466 | 1.97E-05 | 0.00114008 | 0.095766748 |
| HTN | pericyte | *ERVW-1* | -8.967023541 | 0.003954431 | 0.02677323 | 1 |
| HTN | pericyte | *VSIG8* | -10.04109481 | 0.000661233 | 0.012581577 | 1 |
| HTN | pericyte | *FGF14-IT1* | -9.251608801 | 0.002671442 | 0.023519329 | 1 |
| HTN | pericyte | *PALM2* | -4.007647034 | 0.00767323 | 0.037305125 | 1 |
| HTN | pericyte | *WFDC6* | -12.84269758 | 0.000200688 | 0.005727878 | 0.973739313 |
| HTN | pericyte | *UPK3B* | -8.323342019 | 0.008988587 | 0.040525766 | 1 |
| HTN | pericyte | *PLA2G4B* | 8.892346996 | 0.002580387 | 0.023519329 | 1 |
| HTN | pericyte | *LEFTY1* | -38.70073649 | 0.001066979 | 0.016382854 | 1 |
| HTN | pericyte | *JMJD7* | 2.954604507 | 0.001457171 | 0.019699985 | 1 |
| HTN | pericyte | *APOBEC3D* | -17.00003687 | 0.00366658 | 0.026071244 | 1 |
| HTN | pericyte | *TMEM199* | -2.360652461 | 0.006677186 | 0.035407327 | 1 |
| HTN | pericyte | *APOBEC3C* | -4.016518228 | 0.011708352 | 0.047222714 | 1 |
| HTN | pericyte | *RAB30-AS1* | -5.318978826 | 0.003602576 | 0.025973602 | 1 |
| HTN | pericyte | *DNAJC3-AS1* | -5.600166783 | 0.003590366 | 0.025973602 | 1 |
| HTN | pericyte | *BCKDHA* | 3.648884879 | 0.003845825 | 0.026468005 | 1 |
| HTN | pericyte | *ABHD14A* | -7.592298652 | 0.003925268 | 0.026749156 | 1 |
| HTN | pericyte | *GDNF-AS1* | -5.666045302 | 0.008913139 | 0.040417335 | 1 |
| HTN | pericyte | *LINC00942* | -19.43246852 | 0.000614671 | 0.01210347 | 1 |
| HTN | pericyte | *PDCD6* | -6.994095982 | 0.010546528 | 0.044037653 | 1 |
| HTN | pericyte | *TMEM158* | -10.74755009 | 0.006560193 | 0.035132512 | 1 |
| HTN | pericyte | *DNAH10OS* | -17.99903203 | 0.006957513 | 0.0358138 | 1 |
| HTN | pericyte | *PCDHA10* | 1.964701559 | 0.000404053 | 0.009003382 | 1 |
| HTN | pericyte | *LINC00989* | 0.474701559 | 0.004455093 | 0.028219466 | 1 |

| | | | | | | |
|---|---|---|---|---|---|---|
| HTN | pericyte | *GLI4* | -6.224978463 | 0.004548904 | 0.028589746 | 1 |
| HTN | pericyte | *ALG1L2* | -7.952022709 | 0.012864888 | 0.049658263 | 1 |
| HTN | pericyte | *ZNF550* | -7.662402434 | 0.00161313 | 0.020761032 | 1 |
| HTN | pericyte | *LINC01094* | -7.030365108 | 0.002898291 | 0.02367523 | 1 |
| HTN | pericyte | *PCDHGB2* | 2.725818086 | 0.00678128 | 0.035570562 | 1 |
| HTN | pericyte | *TAS2R31* | -3.923146599 | 0.008748578 | 0.039932362 | 1 |
| HTN | pericyte | *ZBED6* | -21.29215228 | 0.000852175 | 0.014473383 | 1 |
| HTN | pericyte | *CNPY2* | -6.659609084 | 0.005122422 | 0.030458325 | 1 |
| HTN | pericyte | *RNASE4* | -10.54088682 | 0.006803823 | 0.035623078 | 1 |
| HTN | pericyte | *MC1R* | 5.767834392 | 0.010522398 | 0.04401265 | 1 |
| HTN | pericyte | *LINC00639* | -6.248923574 | 0.000251489 | 0.006704523 | 1 |
| HTN | pericyte | *AC083843.1* | -57.8315399 | 0.002414075 | 0.023519329 | 1 |
| HTN | pericyte | *LINC00672* | -13.50160044 | 0.004986408 | 0.03031836 | 1 |
| HTN | pericyte | *ZNF224* | -7.020226646 | 0.01112592 | 0.04559372 | 1 |
| HTN | pericyte | *LIN37* | 8.968421191 | 0.000925168 | 0.015425822 | 1 |
| HTN | pericyte | *NDUFA7* | 23.33775819 | 0.011724892 | 0.047250146 | 1 |
| HTN | pericyte | *AC026740.1* | 5.719865645 | 0.000357104 | 0.008290289 | 1 |
| HTN | pericyte | *ZNF587B* | -12.04267004 | 0.005332271 | 0.031271844 | 1 |
| HTN | pericyte | *CASC15* | 1.77556049 | 0.010160956 | 0.043034836 | 1 |
| HTN | pericyte | *PAXIP1-AS1* | -7.594049501 | 0.009860847 | 0.042185921 | 1 |
| HTN | SMC | *SEMA3F* | -6.766530032 | 1.43E-06 | 9.47E-05 | 0.00691583 |
| HTN | SMC | *KRIT1* | -6.918903562 | 0.000387377 | 0.001866487 | 1 |
| HTN | SMC | *RAD52* | -8.430269416 | 7.44E-06 | 0.000269234 | 0.036077321 |
| HTN | SMC | *HS3ST1* | -2.888653086 | 0.002026745 | 0.005734311 | 1 |
| HTN | SMC | *TMEM176A* | -7.490833515 | 0.000371207 | 0.001822973 | 1 |
| HTN | SMC | *M6PR* | -7.334205805 | 1.54E-07 | 1.74E-05 | 0.000747525 |
| HTN | SMC | *CYP26B1* | 5.534256563 | 0.020144564 | 0.035855255 | 1 |
| HTN | SMC | *DBNDD1* | 2.536849472 | 0.000485965 | 0.002179209 | 1 |
| HTN | SMC | *ALS2* | -7.53694351 | 0.000253042 | 0.001446125 | 1 |
| HTN | SMC | *CD38* | -5.554698707 | 0.000458895 | 0.002102512 | 1 |
| HTN | SMC | *RBM6* | -3.884145992 | 3.12E-08 | 4.58E-06 | 0.000151211 |
| HTN | SMC | *RECQL* | -7.398729733 | 3.26E-05 | 0.000629438 | 0.157988931 |
| HTN | SMC | *POLR2J* | -5.324479746 | 0.000956057 | 0.00333007 | 1 |
| HTN | SMC | *MEOX1* | -3.98759396 | 0.000224148 | 0.001362865 | 1 |
| HTN | SMC | *LIG3* | -3.086491315 | 0.022755547 | 0.03987357 | 1 |
| HTN | SMC | *PON1* | -9.121104578 | 0.000811449 | 0.002971188 | 1 |
| HTN | SMC | *GCFC2* | -7.149805724 | 0.000257002 | 0.001461871 | 1 |
| HTN | SMC | *PDK2* | -2.457521581 | 0.017478473 | 0.031857833 | 1 |
| HTN | SMC | *GGNBP2* | -6.853904112 | 0.001299292 | 0.004163913 | 1 |
| HTN | SMC | *ITGA2B* | -4.264573275 | 0.000540002 | 0.002294301 | 1 |
| HTN | SMC | *ABCC8* | 1.299232079 | 0.00474732 | 0.010991265 | 1 |
| HTN | SMC | *SYNRG* | -6.915000713 | 0.000167463 | 0.001169111 | 0.812531825 |
| HTN | SMC | *ZNF263* | -8.813756567 | 0.000841174 | 0.003043532 | 1 |
| HTN | SMC | *SPATA20* | -3.061944771 | 0.028119729 | 0.048007363 | 1 |
| HTN | SMC | *BAIAP2L1* | -4.891148728 | 0.000115921 | 0.000993383 | 0.562448619 |
| HTN | SMC | *USH1C* | 5.952041896 | 0.004992293 | 0.011414989 | 1 |
| HTN | SMC | *VPS41* | -6.689725269 | 0.000456699 | 0.002098395 | 1 |
| HTN | SMC | *ELAC2* | -3.392134999 | 0.015560639 | 0.028947709 | 1 |
| HTN | SMC | *SCIN* | -5.139358389 | 2.76E-05 | 0.000569929 | 0.134137893 |
| HTN | SMC | *CD79B* | -7.009397466 | 0.000242598 | 0.001416471 | 1 |

| | | | | | | |
|---|---|---|---|---|---|---|
| HTN | SMC | *ST7L* | -4.68762514 | 1.45E-05 | 0.000407783 | 0.070138699 |
| HTN | SMC | *PAX6* | 1.81324553 | 0.026715178 | 0.045867672 | 1 |
| HTN | SMC | *TSR3* | -10.42608701 | 0.001614295 | 0.004823006 | 1 |
| HTN | SMC | *FMO3* | -2.350221387 | 0.001555833 | 0.004709234 | 1 |
| HTN | SMC | *PSMB1* | -7.897505057 | 0.000198751 | 0.00127558 | 0.964338484 |
| HTN | SMC | *NADK* | -6.834160187 | 4.29E-06 | 0.000182555 | 0.0208113 |
| HTN | SMC | *DLEC1* | -5.14718064 | 0.001653219 | 0.004906066 | 1 |
| HTN | SMC | *SS18L2* | -6.344648339 | 0.000250539 | 0.001439693 | 1 |
| HTN | SMC | *IL32* | -7.64679843 | 0.000152307 | 0.001131693 | 0.738995254 |
| HTN | SMC | *MED24* | -8.163893611 | 0.000149022 | 0.00112132 | 0.723055067 |
| HTN | SMC | *MLXIPL* | -1.960713685 | 6.57E-05 | 0.000834038 | 0.31860242 |
| HTN | SMC | *ZNF200* | -2.430457599 | 0.019870494 | 0.035497657 | 1 |
| HTN | SMC | *LRRC23* | -3.270888274 | 0.029304542 | 0.049784887 | 1 |
| HTN | SMC | *CLCN6* | -7.898420422 | 4.59E-05 | 0.000758453 | 0.222596304 |
| HTN | SMC | *SLC6A7* | -57.50293731 | 0.003330568 | 0.008351377 | 1 |
| HTN | SMC | *TSPAN9* | -3.587461137 | 0.006188491 | 0.013667073 | 1 |
| HTN | SMC | *UTP18* | -3.734132713 | 0.019033956 | 0.034242771 | 1 |
| HTN | SMC | *TTC19* | -6.756373057 | 0.000149154 | 0.00112132 | 0.723694101 |
| HTN | SMC | *DPF1* | 1.966942078 | 0.007510906 | 0.015983734 | 1 |
| HTN | SMC | *LARS2* | -6.658369667 | 0.000272679 | 0.001509827 | 1 |
| HTN | SMC | *DCN* | -4.476470463 | 6.82E-05 | 0.000836037 | 0.331034724 |
| HTN | SMC | *ZBTB32* | -8.5121272 | 0.001510156 | 0.004640453 | 1 |
| HTN | SMC | *ALOX5* | -2.367694729 | 0.000106445 | 0.000950786 | 0.516471468 |
| HTN | SMC | *MAP4K5* | -6.750987006 | 0.000207858 | 0.001311479 | 1 |
| HTN | SMC | *EHD3* | 1.558924476 | 0.004346933 | 0.010278421 | 1 |
| HTN | SMC | *MAN2B2* | -6.484558432 | 0.002704852 | 0.007155911 | 1 |
| HTN | SMC | *RNF14* | -6.280790227 | 0.007111244 | 0.015307789 | 1 |
| HTN | SMC | *DDX11* | -3.35125462 | 0.007144804 | 0.015366397 | 1 |
| HTN | SMC | *TACC3* | -4.937342061 | 0.002957871 | 0.007615417 | 1 |
| HTN | SMC | *MDH1* | -4.672765778 | 0.002773914 | 0.007290916 | 1 |
| HTN | SMC | *COX15* | -5.666363932 | 0.000125736 | 0.001028322 | 0.610069695 |
| HTN | SMC | *CCDC88C* | -3.600051435 | 0.000496717 | 0.002202348 | 1 |
| HTN | SMC | *CHDH* | -4.41963202 | 0.004671765 | 0.010892553 | 1 |
| HTN | SMC | *CLCA4* | -7.545648925 | 0.001919562 | 0.005494816 | 1 |
| HTN | SMC | *RUFY3* | -5.402520913 | 0.002791275 | 0.00731673 | 1 |
| HTN | SMC | *SLC11A1* | -1.548866073 | 0.001801018 | 0.005229527 | 1 |
| HTN | SMC | *WWTR1* | -6.582315411 | 0.00061336 | 0.00245255 | 1 |
| HTN | SMC | *AGPS* | -3.639046215 | 0.014077992 | 0.026626859 | 1 |
| HTN | SMC | *TTC27* | -6.571439131 | 0.001213537 | 0.003943792 | 1 |
| HTN | SMC | *ZDHHC6* | -9.122304459 | 7.99E-05 | 0.000867401 | 0.387728162 |
| HTN | SMC | *GLRX2* | -9.697687405 | 0.015064302 | 0.028188196 | 1 |
| HTN | SMC | *GCLM* | -7.122580212 | 0.000260941 | 0.001475067 | 1 |
| HTN | SMC | *RRAGD* | -4.450628632 | 0.002129629 | 0.005955597 | 1 |
| HTN | SMC | *HSF2* | -4.033576069 | 0.00578059 | 0.012889441 | 1 |
| HTN | SMC | *TOMM34* | -6.711502815 | 0.00229345 | 0.00629922 | 1 |
| HTN | SMC | *RNASET2* | -4.812718867 | 2.89E-06 | 0.00014601 | 0.014016971 |
| HTN | SMC | *AGPAT4* | -8.362157199 | 0.000143226 | 0.001090942 | 0.694930138 |
| HTN | SMC | *MIPEP* | -8.083833817 | 9.40E-06 | 0.000312306 | 0.04559669 |
| HTN | SMC | *INSRR* | -4.217854629 | 0.000151522 | 0.001127586 | 0.735186119 |
| HTN | SMC | *B4GALT7* | -8.551980682 | 0.001575953 | 0.004740559 | 1 |

| | | | | | | |
|---|---|---|---|---|---|---|
| HTN | SMC | *SH2D2A* | -7.718883427 | 0.00080906 | 0.00297081 | 1 |
| HTN | SMC | *VRK2* | -4.711376558 | 0.018410091 | 0.033256054 | 1 |
| HTN | SMC | *VEZT* | -5.666026298 | 0.006562307 | 0.014323128 | 1 |
| HTN | SMC | *BRD9* | -6.351068727 | 0.005414587 | 0.012219337 | 1 |
| HTN | SMC | *RABEP1* | -4.824017613 | 0.01308601 | 0.025086258 | 1 |
| HTN | SMC | *IKZF2* | -7.331552469 | 0.002261618 | 0.006244323 | 1 |
| HTN | SMC | *GRN* | -4.49919243 | 3.44E-05 | 0.000641172 | 0.166704814 |
| HTN | SMC | *CENPQ* | -1.870240112 | 0.000699453 | 0.002675893 | 1 |
| HTN | SMC | *IFT88* | -5.8036371 | 0.001537643 | 0.004685283 | 1 |
| HTN | SMC | *CHPF2* | -3.692278714 | 0.007204764 | 0.015413367 | 1 |
| HTN | SMC | *UBA6* | -4.864516513 | 0.002007483 | 0.005696085 | 1 |
| HTN | SMC | *PIAS1* | -4.074718919 | 0.015852054 | 0.02936776 | 1 |
| HTN | SMC | *MYOC* | -2.842331664 | 0.013839476 | 0.026271181 | 1 |
| HTN | SMC | *SH3YL1* | -8.692393875 | 1.83E-06 | 0.000106855 | 0.008868967 |
| HTN | SMC | *ADSS* | -7.95625912 | 0.002473756 | 0.006645302 | 1 |
| HTN | SMC | *TBC1D23* | -7.184286575 | 8.44E-05 | 0.000876014 | 0.409339005 |
| HTN | SMC | *MYOM2* | 3.320219199 | 0.012239967 | 0.023723247 | 1 |
| HTN | SMC | *TUBG2* | -4.658369636 | 0.000920927 | 0.003251201 | 1 |
| HTN | SMC | *AGA* | -7.133307575 | 0.016089075 | 0.029693493 | 1 |
| HTN | SMC | *BOD1L1* | -4.174153874 | 0.0072521 | 0.015500965 | 1 |
| HTN | SMC | *DNAH5* | -4.22242027 | 0.00061342 | 0.00245255 | 1 |
| HTN | SMC | *PHLPP2* | -3.344368829 | 0.003730795 | 0.009123901 | 1 |
| HTN | SMC | *CTNS* | -3.26460482 | 0.000737499 | 0.002775793 | 1 |
| HTN | SMC | *INPP4A* | -6.237559048 | 0.000272488 | 0.001509827 | 1 |
| HTN | SMC | *PSMA4* | -9.573197295 | 0.000399179 | 0.001913851 | 1 |
| HTN | SMC | *LSG1* | -1.748213028 | 0.020586242 | 0.036574313 | 1 |
| HTN | SMC | *TDP1* | -9.837972167 | 4.78E-05 | 0.000762524 | 0.231807397 |
| HTN | SMC | *SPATA7* | -6.335617649 | 0.004015531 | 0.009645225 | 1 |
| HTN | SMC | *MED17* | -10.76202587 | 0.000546598 | 0.002310188 | 1 |
| HTN | SMC | *AP2S1* | -10.85892823 | 0.000807986 | 0.00297081 | 1 |
| HTN | SMC | *CUL7* | -5.989608953 | 0.004796152 | 0.011075153 | 1 |
| HTN | SMC | *CTNNA1* | -5.73006322 | 0.002629887 | 0.007011106 | 1 |
| HTN | SMC | *CNTLN* | -3.969308587 | 0.026889973 | 0.046151451 | 1 |
| HTN | SMC | *YTHDC2* | -6.794406964 | 0.001140663 | 0.003777813 | 1 |
| HTN | SMC | *ARAP2* | -4.047663024 | 0.001947545 | 0.005553892 | 1 |
| HTN | SMC | *DTNBP1* | -6.944691939 | 0.002952625 | 0.00761344 | 1 |
| HTN | SMC | *ANO2* | -7.7696103 | 0.000139143 | 0.001073329 | 0.675123848 |
| HTN | SMC | *SNX29* | -5.755652017 | 0.000270681 | 0.00150483 | 1 |
| HTN | SMC | *MRPS10* | -1.928383298 | 0.003936178 | 0.009490549 | 1 |
| HTN | SMC | *EPN3* | -3.839429329 | 0.025227802 | 0.043731795 | 1 |
| HTN | SMC | *RCN1* | -8.299467059 | 0.000214538 | 0.00132773 | 1 |
| HTN | SMC | *JKAMP* | -6.131543906 | 7.09E-05 | 0.000837934 | 0.34390551 |
| HTN | SMC | *MCUR1* | -5.950341879 | 0.015310205 | 0.028564551 | 1 |
| HTN | SMC | *MAPK9* | -9.555459804 | 0.000190753 | 0.001250601 | 0.925534977 |
| HTN | SMC | *FAM160A2* | -4.616178426 | 0.008167921 | 0.01709696 | 1 |
| HTN | SMC | *CYBA* | -6.293063168 | 7.78E-05 | 0.000851198 | 0.377659158 |
| HTN | SMC | *THOC3* | -3.988553553 | 0.000621502 | 0.002459648 | 1 |
| HTN | SMC | *LY75* | -14.61025284 | 0.000166498 | 0.001165725 | 0.807847355 |
| HTN | SMC | *TBC1D22A* | -5.368992546 | 0.001146889 | 0.003790672 | 1 |
| HTN | SMC | *GALC* | -7.573939953 | 0.000164774 | 0.001162037 | 0.799485631 |

| | | | | | | |
|---|---|---|---|---|---|---|
| HTN | SMC | *ITIH4* | 1.760617968 | 4.93E-05 | 0.000773529 | 0.239020581 |
| HTN | SMC | *NPFFR2* | -4.965576049 | 0.000312638 | 0.001641688 | 1 |
| HTN | SMC | *GDI2* | -4.963505139 | 0.017520429 | 0.03191033 | 1 |
| HTN | SMC | *ATP11B* | -4.193711353 | 0.017574608 | 0.031996997 | 1 |
| HTN | SMC | *RASGRF1* | -3.944949547 | 0.002973821 | 0.007642467 | 1 |
| HTN | SMC | *DNAJC25* | -10.4861817 | 5.39E-05 | 0.000806822 | 0.261732338 |
| HTN | SMC | *OGFR* | -5.135610433 | 0.00717296 | 0.015386031 | 1 |
| HTN | SMC | *PIGV* | -2.962110503 | 0.000751939 | 0.002817287 | 1 |
| HTN | SMC | *RIMBP2* | -7.801595177 | 8.44E-05 | 0.000876014 | 0.409582488 |
| HTN | SMC | *SPAG4* | -5.768839878 | 0.002700142 | 0.007153341 | 1 |
| HTN | SMC | *LTK* | -41.67635483 | 0.00019575 | 0.001271662 | 0.949777888 |
| HTN | SMC | *AHRR* | 3.565331384 | 0.028908762 | 0.049215899 | 1 |
| HTN | SMC | *RNF4* | -8.745925227 | 0.000232932 | 0.001383335 | 1 |
| HTN | SMC | *CASP8* | -6.158941319 | 0.003377485 | 0.008442844 | 1 |
| HTN | SMC | *TM7SF3* | -6.750700718 | 0.001565957 | 0.00472808 | 1 |
| HTN | SMC | *TMEM161A* | -10.76508971 | 0.000921352 | 0.003251201 | 1 |
| HTN | SMC | *LPAR2* | -4.512836649 | 0.013810881 | 0.026227159 | 1 |
| HTN | SMC | *SUGP2* | -7.111047236 | 0.000270389 | 0.00150483 | 1 |
| HTN | SMC | *SLC12A2* | -7.736806211 | 0.000719096 | 0.002726438 | 1 |
| HTN | SMC | *ABCA7* | -5.092410239 | 6.25E-05 | 0.000827638 | 0.303438888 |
| HTN | SMC | *BTBD1* | -6.540764881 | 0.001524756 | 0.004658765 | 1 |
| HTN | SMC | *FAR2* | -4.972888047 | 0.000807028 | 0.00297081 | 1 |
| HTN | SMC | *BCAS1* | -5.586548772 | 0.007046992 | 0.015209966 | 1 |
| HTN | SMC | *SBNO2* | -7.324944998 | 0.000814668 | 0.002978725 | 1 |
| HTN | SMC | *PMS1* | -5.819549287 | 0.000489414 | 0.002184577 | 1 |
| HTN | SMC | *MCM10* | 1.114590424 | 0.02358395 | 0.041220938 | 1 |
| HTN | SMC | *ADAT1* | -2.166613154 | 0.001637037 | 0.004869958 | 1 |
| HTN | SMC | *TIE1* | -4.119081273 | 2.90E-07 | 2.93E-05 | 0.001406882 |
| HTN | SMC | *SLC9A3* | -1.554535917 | 0.005964318 | 0.013220132 | 1 |
| HTN | SMC | *NGEF* | -7.226948451 | 0.000130959 | 0.001043376 | 0.63541483 |
| HTN | SMC | *SPI1* | -3.343302809 | 0.000929835 | 0.003271618 | 1 |
| HTN | SMC | *ATXN3* | -5.014453055 | 1.54E-05 | 0.000422289 | 0.074745141 |
| HTN | SMC | *GOLGA5* | -7.1177208 | 0.00022771 | 0.001377231 | 1 |
| HTN | SMC | *FGFR2* | -2.373069269 | 8.91E-05 | 0.000888243 | 0.432310961 |
| HTN | SMC | *MTFR1* | -7.487001956 | 0.000196486 | 0.001271662 | 0.953349009 |
| HTN | SMC | *MYO9A* | -5.236157362 | 0.000617665 | 0.002454805 | 1 |
| HTN | SMC | *TRAM1* | -7.297933203 | 0.000369148 | 0.001814699 | 1 |
| HTN | SMC | *EVI5* | -4.51599576 | 0.003315583 | 0.008334 | 1 |
| HTN | SMC | *STOML1* | -9.512402799 | 0.000134438 | 0.001055488 | 0.652291502 |
| HTN | SMC | *METTL22* | -8.164359849 | 6.81E-05 | 0.000836037 | 0.33045752 |
| HTN | SMC | *TP53BP1* | -5.187536532 | 0.018182306 | 0.032955005 | 1 |
| HTN | SMC | *ROGDI* | -19.9290035 | 0.000847396 | 0.003059199 | 1 |
| HTN | SMC | *CBFB* | -7.658838059 | 6.24E-05 | 0.000827638 | 0.302662335 |
| HTN | SMC | *FGFR3* | 2.650982423 | 0.014651163 | 0.027553273 | 1 |
| HTN | SMC | *IFI35* | -4.638658916 | 0.016792892 | 0.030793314 | 1 |
| HTN | SMC | *POLR1A* | -6.200797722 | 5.77E-08 | 7.57E-06 | 0.000280038 |
| HTN | SMC | *IP6K2* | -7.994334921 | 5.96E-06 | 0.000227562 | 0.028900396 |
| HTN | SMC | *SRBD1* | -6.060249947 | 0.001471081 | 0.004566657 | 1 |
| HTN | SMC | *CYFIP1* | -6.299975874 | 0.002804284 | 0.00733179 | 1 |
| HTN | SMC | *IFT80* | -8.664860711 | 0.004959822 | 0.011362161 | 1 |

| | | | | | | |
|---|---|---|---|---|---|---|
| HTN | SMC | *NUCKS1* | -6.936885336 | 0.000403245 | 0.001929517 | 1 |
| HTN | SMC | *CLEC2D* | -5.130155188 | 0.012801048 | 0.024611155 | 1 |
| HTN | SMC | *DRD4* | -3.921961782 | 0.000803758 | 0.002965655 | 1 |
| HTN | SMC | *PLA2G10* | -5.448329168 | 0.000509267 | 0.002232127 | 1 |
| HTN | SMC | *PIGB* | -6.848722143 | 0.002453528 | 0.006624663 | 1 |
| HTN | SMC | *GNB5* | -3.932115215 | 0.002063782 | 0.005808647 | 1 |
| HTN | SMC | *LRP6* | -7.560018703 | 6.85E-05 | 0.000836037 | 0.332386849 |
| HTN | SMC | *DAPP1* | -1.83707442 | 0.001948212 | 0.005553892 | 1 |
| HTN | SMC | *DGCR2* | -6.954711195 | 0.000988285 | 0.003408073 | 1 |
| HTN | SMC | *ZXDC* | -6.410413379 | 0.000372119 | 0.001823761 | 1 |
| HTN | SMC | *WIPI1* | -3.862375942 | 0.006409698 | 0.014046908 | 1 |
| HTN | SMC | *GBA2* | -6.880203108 | 0.022819494 | 0.039971186 | 1 |
| HTN | SMC | *AP3M2* | -10.0465041 | 1.97E-05 | 0.000481979 | 0.095431799 |
| HTN | SMC | *ST6GALNAC2* | -3.956624291 | 0.001188606 | 0.003888818 | 1 |
| HTN | SMC | *EIF2B3* | -6.775028888 | 8.57E-05 | 0.000876014 | 0.415763975 |
| HTN | SMC | *MAP4K4* | -6.598611913 | 0.000503857 | 0.002218434 | 1 |
| HTN | SMC | *SNX13* | -6.738740887 | 0.000515286 | 0.002236542 | 1 |
| HTN | SMC | *RPS6KA2* | -4.425333106 | 0.002262473 | 0.006244323 | 1 |
| HTN | SMC | *ING3* | -9.33911079 | 0.000581895 | 0.002388625 | 1 |
| HTN | SMC | *CPSF1* | -3.441921687 | 0.003754487 | 0.009172595 | 1 |
| HTN | SMC | *CYBRD1* | -5.045492049 | 0.001827795 | 0.005291446 | 1 |
| HTN | SMC | *RDH11* | -6.780262429 | 0.005421418 | 0.012229065 | 1 |
| HTN | SMC | *EPN2* | -4.688593073 | 0.001720586 | 0.005050384 | 1 |
| HTN | SMC | *AFF4* | -5.004441947 | 0.007601089 | 0.016105015 | 1 |
| HTN | SMC | *TRHDE* | -2.639018959 | 1.58E-05 | 0.000427306 | 0.076475518 |
| HTN | SMC | *NFATC3* | -5.005358007 | 0.003703672 | 0.009071286 | 1 |
| HTN | SMC | *TRNT1* | -6.066477592 | 0.005274152 | 0.011952446 | 1 |
| HTN | SMC | *CRMP1* | -8.962758116 | 7.74E-05 | 0.000851198 | 0.375748436 |
| HTN | SMC | *NDE1* | 1.418149652 | 0.01020041 | 0.020459855 | 1 |
| HTN | SMC | *AP1M1* | -11.14706327 | 4.07E-05 | 0.000707069 | 0.197272169 |
| HTN | SMC | *PVR* | -7.484015195 | 6.27E-05 | 0.000827638 | 0.304105992 |
| HTN | SMC | *SCARB1* | -5.591342485 | 3.91E-05 | 0.000689568 | 0.189567379 |
| HTN | SMC | *MOV10L1* | -3.964873736 | 0.005240696 | 0.011887732 | 1 |
| HTN | SMC | *ALPK1* | -5.291981796 | 0.000379354 | 0.00184247 | 1 |
| HTN | SMC | *LLGL2* | -8.071233363 | 0.001762478 | 0.005154636 | 1 |
| HTN | SMC | *ADAM11* | -1.847138858 | 0.012545494 | 0.024193456 | 1 |
| HTN | SMC | *PPP2R2C* | -15.56426266 | 5.13E-05 | 0.000794475 | 0.248670763 |
| HTN | SMC | *ZNF532* | -6.956984717 | 0.001233016 | 0.003988396 | 1 |
| HTN | SMC | *ZZEF1* | -5.035883543 | 0.003465508 | 0.008605243 | 1 |
| HTN | SMC | *TTC38* | -5.070722936 | 0.002359902 | 0.006435458 | 1 |
| HTN | SMC | *GRAMD4* | -5.473540833 | 0.000720823 | 0.002728108 | 1 |
| HTN | SMC | *VPS9D1* | -6.532322016 | 0.001227564 | 0.003976061 | 1 |
| HTN | SMC | *FNDC3B* | -5.231632836 | 0.000224452 | 0.001363007 | 1 |
| HTN | SMC | *MOCOS* | -4.642747034 | 0.003141853 | 0.007992092 | 1 |
| HTN | SMC | *SEC31B* | -3.429996817 | 0.006899132 | 0.014953353 | 1 |
| HTN | SMC | *MLH1* | -5.243513036 | 0.002638189 | 0.007029376 | 1 |
| HTN | SMC | *FMO4* | -4.363673299 | 0.018346924 | 0.033179008 | 1 |
| HTN | SMC | *RGS11* | -2.439234144 | 0.009465163 | 0.019183363 | 1 |
| HTN | SMC | *SPAG5* | -8.562384566 | 9.02E-05 | 0.000888243 | 0.437584227 |
| HTN | SMC | *PAG1* | -7.075099453 | 0.001500127 | 0.004625724 | 1 |

| | | | | | | |
|---|---|---|---|---|---|---|
| HTN | SMC | *XAB2* | -3.837387296 | 0.000688212 | 0.002653042 | 1 |
| HTN | SMC | *DYNC1I2* | -6.374653787 | 0.003584785 | 0.008827153 | 1 |
| HTN | SMC | *APBB1IP* | -2.582349334 | 0.006204657 | 0.013693924 | 1 |
| HTN | SMC | *LRCH4* | 3.730906214 | 0.013249101 | 0.02531888 | 1 |
| HTN | SMC | *SIRT6* | -3.448721897 | 0.019315444 | 0.034710568 | 1 |
| HTN | SMC | *AMPH* | 3.471574184 | 0.0137115 | 0.026058832 | 1 |
| HTN | SMC | *MCCC1* | -3.348066184 | 0.000490381 | 0.002186885 | 1 |
| HTN | SMC | *ACER3* | -3.588200532 | 0.002483822 | 0.006665655 | 1 |
| HTN | SMC | *PIK3C3* | -6.92705373 | 0.000565192 | 0.002345861 | 1 |
| HTN | SMC | *SYNJ2* | -2.787391659 | 0.001295492 | 0.004154478 | 1 |
| HTN | SMC | *FGF20* | -10.64705453 | 0.00620826 | 0.013693924 | 1 |
| HTN | SMC | *PCM1* | -5.781279509 | 0.000510038 | 0.00223349 | 1 |
| HTN | SMC | *MYH7B* | -5.833455001 | 0.000318353 | 0.001657346 | 1 |
| HTN | SMC | *XRCC5* | -5.772906617 | 0.003205527 | 0.008120239 | 1 |
| HTN | SMC | *SP140* | -4.229580393 | 0.000468396 | 0.002133951 | 1 |
| HTN | SMC | *SAR1A* | -7.857350774 | 0.000249733 | 0.001438619 | 1 |
| HTN | SMC | *FDFT1* | 4.888094153 | 0.007003711 | 0.015130011 | 1 |
| HTN | SMC | *SCGN* | -14.63264357 | 0.000948004 | 0.003313916 | 1 |
| HTN | SMC | *PGM1* | -7.276329833 | 2.75E-05 | 0.000569929 | 0.133270917 |
| HTN | SMC | *DDX1* | -5.613970734 | 0.002694142 | 0.007145664 | 1 |
| HTN | SMC | *DNM2* | -5.343351128 | 0.006807119 | 0.014810825 | 1 |
| HTN | SMC | *STX7* | -6.540261593 | 0.000356274 | 0.001787632 | 1 |
| HTN | SMC | *RABL2B* | -7.145492806 | 4.25E-05 | 0.000731096 | 0.206169113 |
| HTN | SMC | *SCTR* | -3.999149772 | 0.000157391 | 0.001140149 | 0.763662796 |
| HTN | SMC | *RIF1* | -4.99329275 | 0.015905309 | 0.029432708 | 1 |
| HTN | SMC | *KCNN2* | -10.00967971 | 0.000810557 | 0.00297081 | 1 |
| HTN | SMC | *PSEN1* | -5.701702551 | 0.003661395 | 0.008981338 | 1 |
| HTN | SMC | *PCDHB4* | -4.506457542 | 0.000489329 | 0.002184577 | 1 |
| HTN | SMC | *ME2* | -7.013353664 | 0.006636593 | 0.014465745 | 1 |
| HTN | SMC | *CCNT2* | -6.692572304 | 0.000660416 | 0.002573074 | 1 |
| HTN | SMC | *SEMA5B* | 0.641389804 | 0.007083152 | 0.015254227 | 1 |
| HTN | SMC | *LYRM2* | -7.401869983 | 1.61E-05 | 0.000427306 | 0.078257711 |
| HTN | SMC | *ZCCHC6* | -6.65765401 | 0.001622275 | 0.004834937 | 1 |
| HTN | SMC | *ULK2* | -6.864126328 | 0.000409092 | 0.001942185 | 1 |
| HTN | SMC | *PIBF1* | -5.725240752 | 0.001643281 | 0.004882547 | 1 |
| HTN | SMC | *OXCT1* | -7.283283549 | 0.000252038 | 0.001442083 | 1 |
| HTN | SMC | *ZNF586* | -4.980500478 | 0.001300831 | 0.004166094 | 1 |
| HTN | SMC | *RPS5* | -5.613902001 | 0.002134222 | 0.005965004 | 1 |
| HTN | SMC | *CHMP2B* | -10.08390207 | 0.000210553 | 0.001314937 | 1 |
| HTN | SMC | *NOA1* | -10.07918997 | 6.44E-05 | 0.000829756 | 0.312534602 |
| HTN | SMC | *NCOA1* | -5.688707202 | 0.000462465 | 0.002113438 | 1 |
| HTN | SMC | *CD82* | 1.879478725 | 0.015239871 | 0.028472797 | 1 |
| HTN | SMC | *HACE1* | -7.981525657 | 0.000137828 | 0.001071047 | 0.668742804 |
| HTN | SMC | *SEH1L* | -4.397352989 | 0.00303612 | 0.00777375 | 1 |
| HTN | SMC | *OVGP1* | -2.242366008 | 0.021449122 | 0.037857818 | 1 |
| HTN | SMC | *SLC25A24* | -5.818297055 | 0.009068924 | 0.018597811 | 1 |
| HTN | SMC | *ZNF213* | -4.897247847 | 0.001053347 | 0.003569021 | 1 |
| HTN | SMC | *CPNE3* | -7.434728176 | 4.46E-05 | 0.000752126 | 0.216612309 |
| HTN | SMC | *RRN3* | -8.930914792 | 0.000196878 | 0.001271662 | 0.955250774 |
| HTN | SMC | *MTIF2* | -7.420878146 | 5.42E-05 | 0.000806822 | 0.263023931 |

| | | | | | | |
|---|---|---|---|---|---|---|
| HTN | SMC | *EPS15* | -3.562730985 | 0.007200919 | 0.015413367 | 1 |
| HTN | SMC | *USP40* | -6.942295831 | 0.000204105 | 0.001294533 | 0.99031801 |
| HTN | SMC | *EIF2AK1* | -7.911397848 | 0.000290091 | 0.001567395 | 1 |
| HTN | SMC | *NME8* | -48.69380066 | 0.002278959 | 0.006279107 | 1 |
| HTN | SMC | *SNX10* | -5.680680264 | 2.72E-10 | 1.20E-07 | 1.32E-06 |
| HTN | SMC | *ITPKC* | -6.234357065 | 0.000690739 | 0.002657784 | 1 |
| HTN | SMC | *TMED2* | -3.868996793 | 0.005363979 | 0.012122043 | 1 |
| HTN | SMC | *SRRT* | -7.602684956 | 0.005454036 | 0.012291213 | 1 |
| HTN | SMC | *ADAMTS2* | -6.55045737 | 2.74E-05 | 0.000569929 | 0.132860577 |
| HTN | SMC | *UIMC1* | -5.802640228 | 0.007162577 | 0.015380334 | 1 |
| HTN | SMC | *LPCAT2* | -6.776382646 | 7.13E-05 | 0.000837934 | 0.346066787 |
| HTN | SMC | *OGFOD1* | -5.340937246 | 0.010757385 | 0.021359516 | 1 |
| HTN | SMC | *TXNDC16* | -3.80914767 | 0.021239395 | 0.03758335 | 1 |
| HTN | SMC | *AAMDC* | -5.905370162 | 1.01E-06 | 6.98E-05 | 0.004887783 |
| HTN | SMC | *ASAP3* | -4.817644407 | 7.37E-05 | 0.000847247 | 0.357447613 |
| HTN | SMC | *EDEM2* | -5.612515291 | 0.002244746 | 0.006209524 | 1 |
| HTN | SMC | *ARHGAP28* | -5.189050689 | 8.31E-05 | 0.000876014 | 0.4032752 |
| HTN | SMC | *PPP1R13B* | -6.162605979 | 1.64E-06 | 0.000100629 | 0.007949691 |
| HTN | SMC | *MAVS* | -7.017015939 | 2.25E-05 | 0.000515555 | 0.109347895 |
| HTN | SMC | *XRN2* | -6.856431823 | 0.000641987 | 0.002518127 | 1 |
| HTN | SMC | *SNX5* | -7.964730885 | 0.004612773 | 0.010780914 | 1 |
| HTN | SMC | *ESF1* | -5.873385766 | 0.001179659 | 0.003869982 | 1 |
| HTN | SMC | *TMEM230* | -6.468059288 | 0.001129718 | 0.003749244 | 1 |
| HTN | SMC | *RPH3A* | 11.97501687 | 0.00253012 | 0.006767444 | 1 |
| HTN | SMC | *KIF16B* | -5.534230603 | 0.015868431 | 0.02938688 | 1 |
| HTN | SMC | *TBX5* | -11.13628681 | 0.000285202 | 0.00155483 | 1 |
| HTN | SMC | *FXYD5* | -4.511321754 | 0.000220082 | 0.001344061 | 1 |
| HTN | SMC | *ZNF302* | -3.683479288 | 0.007518713 | 0.015987365 | 1 |
| HTN | SMC | *GRAMD1A* | -8.398971947 | 7.65E-05 | 0.000851198 | 0.370944653 |
| HTN | SMC | *CDIP1* | -10.15501415 | 5.40E-05 | 0.000806822 | 0.261895174 |
| HTN | SMC | *GANAB* | -8.405971657 | 6.12E-05 | 0.000827638 | 0.297139132 |
| HTN | SMC | *ANKRD24* | -3.203078163 | 0.000418481 | 0.001971331 | 1 |
| HTN | SMC | *DHX32* | -5.967258156 | 0.000878419 | 0.003138504 | 1 |
| HTN | SMC | *SPTLC1* | -7.334165974 | 0.000329221 | 0.001697537 | 1 |
| HTN | SMC | *MRPS33* | -4.723283042 | 0.01601546 | 0.029602671 | 1 |
| HTN | SMC | *MAEA* | -5.053284127 | 0.004695207 | 0.010920971 | 1 |
| HTN | SMC | *LYZ* | -2.973760924 | 0.018206588 | 0.03297438 | 1 |
| HTN | SMC | *MUL1* | -2.950650257 | 0.006709889 | 0.014606439 | 1 |
| HTN | SMC | *SPG21* | -5.718849482 | 0.000218295 | 0.001341927 | 1 |
| HTN | SMC | *RAB11FIP3* | -5.339018335 | 0.001872988 | 0.005399725 | 1 |
| HTN | SMC | *GNPTG* | -7.795681406 | 0.000532711 | 0.002275276 | 1 |
| HTN | SMC | *PABPC4* | -9.980389785 | 2.78E-09 | 6.74E-07 | 1.35E-05 |
| HTN | SMC | *USP48* | -5.214425956 | 0.000527138 | 0.002261427 | 1 |
| HTN | SMC | *PDPR* | -7.004330116 | 0.000162619 | 0.001160337 | 0.789029195 |
| HTN | SMC | *GLG1* | -5.931959626 | 0.002443344 | 0.006604516 | 1 |
| HTN | SMC | *DTX2* | -4.436867949 | 0.000197223 | 0.001271662 | 0.956926131 |
| HTN | SMC | *LAMB1* | -2.727833766 | 0.000255068 | 0.001454597 | 1 |
| HTN | SMC | *ABCC6* | -3.91402435 | 2.81E-05 | 0.000569929 | 0.136385228 |
| HTN | SMC | *RAPGEF4* | -5.785843885 | 2.78E-05 | 0.000569929 | 0.13507518 |
| HTN | SMC | *TF* | -5.878847784 | 0.000131374 | 0.001043376 | 0.637424371 |

| | | | | | | |
|---|---|---|---|---|---|---|
| HTN | SMC | *OSGEP* | -4.782222849 | 0.000565741 | 0.002346132 | 1 |
| HTN | SMC | *SCFD1* | -6.16240457 | 0.001550567 | 0.004700863 | 1 |
| HTN | SMC | *SUPT16H* | -8.634585363 | 0.000155349 | 0.001140149 | 0.753751119 |
| HTN | SMC | *TOX4* | -8.32654401 | 0.00032868 | 0.001696546 | 1 |
| HTN | SMC | *CAPN3* | -6.576894125 | 0.002562063 | 0.006845336 | 1 |
| HTN | SMC | *MFSD11* | -4.630795622 | 0.000156034 | 0.001140149 | 0.757075581 |
| HTN | SMC | *SEC22C* | -9.070047117 | 0.000357702 | 0.001791816 | 1 |
| HTN | SMC | *XYLB* | -9.312982023 | 8.98E-05 | 0.000888243 | 0.435482658 |
| HTN | SMC | *CDC6* | -5.99044959 | 0.000313485 | 0.001643575 | 1 |
| HTN | SMC | *MSH2* | -8.946789522 | 0.000739144 | 0.002775793 | 1 |
| HTN | SMC | *PSMD5* | -7.025292198 | 0.006234697 | 0.013737852 | 1 |
| HTN | SMC | *NANS* | -8.212459508 | 0.000257689 | 0.001464062 | 1 |
| HTN | SMC | *PDE6C* | -7.765550145 | 0.013396362 | 0.025570081 | 1 |
| HTN | SMC | *CWF19L1* | -2.037939551 | 0.007547173 | 0.016032747 | 1 |
| HTN | SMC | *WAC* | -5.384352855 | 0.002063913 | 0.005808647 | 1 |
| HTN | SMC | *DSP* | -8.426401245 | 9.05E-05 | 0.000888775 | 0.439054884 |
| HTN | SMC | *IL12RB1* | -2.26521598 | 0.004568279 | 0.010697533 | 1 |
| HTN | SMC | *ACOT7* | -5.20695303 | 0.0003383 | 0.001729645 | 1 |
| HTN | SMC | *SCD* | -7.28770608 | 1.96E-10 | 9.52E-08 | 9.52E-07 |
| HTN | SMC | *ERMP1* | -6.415667623 | 0.000524475 | 0.002257599 | 1 |
| HTN | SMC | *RAB18* | -7.128876015 | 0.002461023 | 0.006628022 | 1 |
| HTN | SMC | *TSPAN15* | -6.491387956 | 7.23E-05 | 0.000845677 | 0.350929706 |
| HTN | SMC | *CIRBP* | -4.76262844 | 0.005625697 | 0.01259632 | 1 |
| HTN | SMC | *MTAP* | -6.86797205 | 6.85E-07 | 5.28E-05 | 0.003325446 |
| HTN | SMC | *MADCAM1* | -5.033480243 | 3.92E-05 | 0.000689568 | 0.190017802 |
| HTN | SMC | *MED15* | -7.166641526 | 0.00092708 | 0.003264289 | 1 |
| HTN | SMC | *CECR2* | -4.988028945 | 7.50E-05 | 0.000847247 | 0.364139729 |
| HTN | SMC | *BCL2L13* | -3.968937824 | 0.003093662 | 0.007887782 | 1 |
| HTN | SMC | *DDT* | -6.332359744 | 0.000248373 | 0.001436361 | 1 |
| HTN | SMC | *CABIN1* | -5.962593913 | 0.004789642 | 0.011066354 | 1 |
| HTN | SMC | *SUSD2* | 0.585209252 | 0.021654879 | 0.038179315 | 1 |
| HTN | SMC | *SPECC1L* | -4.133108844 | 0.00188099 | 0.005413145 | 1 |
| HTN | SMC | *PPIL2* | -3.968589192 | 0.023769687 | 0.041500727 | 1 |
| HTN | SMC | *PES1* | -5.335312463 | 9.09E-05 | 0.000890467 | 0.441185274 |
| HTN | SMC | *SLC35E4* | -3.96066336 | 0.001140098 | 0.003777813 | 1 |
| HTN | SMC | *TOP3B* | -7.703371347 | 0.000156657 | 0.001140149 | 0.76010121 |
| HTN | SMC | *CYTH4* | -6.613634001 | 0.000111261 | 0.000970929 | 0.539836488 |
| HTN | SMC | *CARD10* | -2.384516344 | 0.003435789 | 0.008544567 | 1 |
| HTN | SMC | *LGALS2* | -3.358599822 | 0.004742527 | 0.010988893 | 1 |
| HTN | SMC | *SH3BP1* | -16.3722566 | 0.000220224 | 0.001344061 | 1 |
| HTN | SMC | *HPS4* | -6.844004441 | 0.005459268 | 0.012291586 | 1 |
| HTN | SMC | *TRIOBP* | -8.156645255 | 3.03E-06 | 0.000150762 | 0.014719497 |
| HTN | SMC | *ANKRD54* | -2.629251692 | 0.02543911 | 0.044050878 | 1 |
| HTN | SMC | *EIF3L* | -8.031126649 | 0.000497026 | 0.002202348 | 1 |
| HTN | SMC | *TTC28* | -4.959757029 | 0.00170238 | 0.005012103 | 1 |
| HTN | SMC | *CENPM* | -3.963420725 | 0.008274442 | 0.017268138 | 1 |
| HTN | SMC | *KDELR3* | -5.895386257 | 0.021713384 | 0.038268558 | 1 |
| HTN | SMC | *CYP2D6* | -5.471279846 | 0.00026766 | 0.001494463 | 1 |
| HTN | SMC | *RAB36* | -3.870515836 | 0.019691039 | 0.035242897 | 1 |
| HTN | SMC | *PPP6R2* | -6.623491917 | 0.000525627 | 0.002257599 | 1 |

| | | | | | | |
|---|---|---|---|---|---|---|
| HTN | SMC | *RHBDD3* | -6.228039115 | 0.003767303 | 0.009199272 | 1 |
| HTN | SMC | *TOM1* | -3.489685958 | 0.004835817 | 0.011135921 | 1 |
| HTN | SMC | *CHKB* | -2.524267621 | 0.009598892 | 0.01940576 | 1 |
| HTN | SMC | *THOC5* | -5.310458979 | 1.33E-06 | 8.96E-05 | 0.006451118 |
| HTN | SMC | *TSPO* | -8.436423813 | 0.001521364 | 0.004651328 | 1 |
| HTN | SMC | *TTLL12* | -9.337698428 | 5.82E-05 | 0.000817716 | 0.282190483 |
| HTN | SMC | *APOL4* | -3.402246496 | 0.000808622 | 0.00297081 | 1 |
| HTN | SMC | *SAMM50* | -8.931029 | 0.00114155 | 0.003778173 | 1 |
| HTN | SMC | *IFT27* | -8.853763218 | 0.006896871 | 0.014953353 | 1 |
| HTN | SMC | *KIAA0930* | -4.518235814 | 0.001501399 | 0.004625724 | 1 |
| HTN | SMC | *FAM118A* | 1.917907162 | 0.019517396 | 0.035008653 | 1 |
| HTN | SMC | *L3MBTL2* | -7.468766233 | 0.000371724 | 0.001823663 | 1 |
| HTN | SMC | *ACO2* | -8.793929093 | 2.16E-05 | 0.000506636 | 0.104873689 |
| HTN | SMC | *POLR3H* | -10.05005083 | 0.000700234 | 0.002675893 | 1 |
| HTN | SMC | *DESI1* | -5.487861744 | 0.016545651 | 0.030431956 | 1 |
| HTN | SMC | *HDAC10* | -9.171695855 | 0.000209795 | 0.001313543 | 1 |
| HTN | SMC | *RBM23* | -5.330891201 | 0.00109048 | 0.003666672 | 1 |
| HTN | SMC | *AP4S1* | -8.362171509 | 0.000150103 | 0.00112219 | 0.728301373 |
| HTN | SMC | *NIN* | -7.311222907 | 0.000142404 | 0.001090656 | 0.690942826 |
| HTN | SMC | *PYGL* | -2.086145894 | 0.000149608 | 0.00112132 | 0.725899801 |
| HTN | SMC | *ATP6V1D* | -7.819255279 | 0.000114474 | 0.00098569 | 0.555427979 |
| HTN | SMC | *GSTZ1* | -3.283498275 | 0.00805327 | 0.016900721 | 1 |
| HTN | SMC | *MTHFD1* | -4.787464894 | 0.000687811 | 0.002653042 | 1 |
| HTN | SMC | *TELO2* | -9.486543483 | 0.002980478 | 0.007655521 | 1 |
| HTN | SMC | *GSKIP* | -10.97753456 | 5.86E-05 | 0.000817716 | 0.284242439 |
| HTN | SMC | *YY1* | -5.863351078 | 0.014768889 | 0.027742412 | 1 |
| HTN | SMC | *KIAA0391* | -6.243565947 | 0.025873384 | 0.044659431 | 1 |
| HTN | SMC | *PLTP* | -1.351303238 | 0.021261547 | 0.037608832 | 1 |
| HTN | SMC | *TRPC4AP* | -4.31893516 | 0.010727985 | 0.021341608 | 1 |
| HTN | SMC | *ABHD12* | -4.882898045 | 0.004293459 | 0.010161886 | 1 |
| HTN | SMC | *UQCC1* | -5.049718168 | 0.002288485 | 0.006291065 | 1 |
| HTN | SMC | *PABPC1L* | -5.353086102 | 0.000236691 | 0.001395412 | 1 |
| HTN | SMC | *DOK5* | -6.134841037 | 9.49E-05 | 0.000903 | 0.460463792 |
| HTN | SMC | *CSTF1* | -10.41924239 | 0.000103018 | 0.000942633 | 0.499843795 |
| HTN | SMC | *CTSZ* | -7.812139346 | 0.000107039 | 0.000950786 | 0.519351226 |
| HTN | SMC | *MTG2* | -6.337398244 | 0.000365507 | 0.001809686 | 1 |
| HTN | SMC | *TCFL5* | -4.679886859 | 0.000789791 | 0.002925242 | 1 |
| HTN | SMC | *DIDO1* | -4.818338759 | 0.015662618 | 0.029105714 | 1 |
| HTN | SMC | *PTK6* | 7.03416507 | 0.015781392 | 0.029270379 | 1 |
| HTN | SMC | *GMEB2* | -11.5438593 | 0.000628768 | 0.002476284 | 1 |
| HTN | SMC | *C20orf27* | -4.133523663 | 0.008158392 | 0.017084384 | 1 |
| HTN | SMC | *CDC25B* | -4.294588659 | 0.015767864 | 0.029256472 | 1 |
| HTN | SMC | *TRIB3* | -9.239742738 | 0.000197486 | 0.001271662 | 0.958203284 |
| HTN | SMC | *ANGPT4* | 0.419139769 | 0.002805288 | 0.00733179 | 1 |
| HTN | SMC | *SIRPB1* | -4.793783067 | 0.004485365 | 0.010538978 | 1 |
| HTN | SMC | *CRNKL1* | -5.385754294 | 0.00020022 | 0.001281046 | 0.971466795 |
| HTN | SMC | *MROH8* | -2.8652706 | 0.00514696 | 0.011715907 | 1 |
| HTN | SMC | *TTI1* | -8.815407164 | 0.000219876 | 0.001344061 | 1 |
| HTN | SMC | *DNTTIP1* | -5.349541729 | 5.97E-07 | 4.99E-05 | 0.002894416 |
| HTN | SMC | *MAP1LC3A* | -8.498436036 | 0.000602109 | 0.002429934 | 1 |

| | | | | | | |
|---|---|---|---|---|---|---|
| HTN | SMC | *RBFA* | -4.570760831 | 0.006525886 | 0.01426931 | 1 |
| HTN | SMC | *METTL4* | -8.386109757 | 0.003023036 | 0.007751946 | 1 |
| HTN | SMC | *LPIN2* | -6.934249465 | 0.001032773 | 0.003513798 | 1 |
| HTN | SMC | *MYOM1* | 0.911247375 | 0.007597902 | 0.016105015 | 1 |
| HTN | SMC | *CEP192* | -4.100323612 | 0.002412682 | 0.006543507 | 1 |
| HTN | SMC | *POLI* | -2.231045807 | 1.04E-05 | 0.000334062 | 0.050443298 |
| HTN | SMC | *MIB1* | -6.587035406 | 0.000943173 | 0.003305786 | 1 |
| HTN | SMC | *NALCN* | -1.943086091 | 0.006107599 | 0.013513027 | 1 |
| HTN | SMC | *STK24* | -6.235492233 | 0.003349982 | 0.008387055 | 1 |
| HTN | SMC | *DNAJC3* | -4.744722053 | 0.027106924 | 0.04645807 | 1 |
| HTN | SMC | *PARP4* | -7.960650671 | 2.45E-05 | 0.000537075 | 0.118798144 |
| HTN | SMC | *SUPT20H* | -6.027483277 | 0.001482413 | 0.004587346 | 1 |
| HTN | SMC | *MRPS31* | -7.013780825 | 0.003007984 | 0.007718001 | 1 |
| HTN | SMC | *RGCC* | -1.996912869 | 0.014382855 | 0.027143373 | 1 |
| HTN | SMC | *KATNAL1* | -5.889749596 | 0.008582933 | 0.017796748 | 1 |
| HTN | SMC | *INTS6* | -6.146859253 | 0.000713669 | 0.002711607 | 1 |
| HTN | SMC | *LONP2* | -3.43655505 | 0.018817238 | 0.033890586 | 1 |
| HTN | SMC | *PLLP* | -2.787628897 | 0.003091975 | 0.007887625 | 1 |
| HTN | SMC | *CCDC113* | -5.418889411 | 5.68E-05 | 0.000817716 | 0.275553223 |
| HTN | SMC | *PSMD7* | -7.079557862 | 0.003572667 | 0.008808222 | 1 |
| HTN | SMC | *COG4* | -5.695036636 | 0.000164046 | 0.001162037 | 0.795952423 |
| HTN | SMC | *NPRL3* | -3.398825164 | 0.014622173 | 0.027509416 | 1 |
| HTN | SMC | *ZNF500* | -6.291187067 | 4.87E-06 | 0.000196769 | 0.023612294 |
| HTN | SMC | *NME4* | -14.83650729 | 4.54E-06 | 0.000189428 | 0.022018893 |
| HTN | SMC | *NOMO3* | -2.765736675 | 0.016992414 | 0.031071961 | 1 |
| HTN | SMC | *LMF1* | -7.345280511 | 7.79E-05 | 0.000851198 | 0.377931807 |
| HTN | SMC | *MTHFSD* | 2.034025394 | 0.008453819 | 0.017589165 | 1 |
| HTN | SMC | *CLCN7* | -2.247437955 | 0.01026623 | 0.020583367 | 1 |
| HTN | SMC | *HAGHL* | 2.754600323 | 0.001515926 | 0.004642097 | 1 |
| HTN | SMC | *SLC7A5* | -4.819361268 | 0.000428924 | 0.002004689 | 1 |
| HTN | SMC | *RHBDL1* | -7.016972305 | 0.004012014 | 0.009641551 | 1 |
| HTN | SMC | *NUBP1* | -7.080581237 | 2.00E-05 | 0.000486513 | 0.09681613 |
| HTN | SMC | *UBE2I* | -6.815917239 | 0.008100067 | 0.01696957 | 1 |
| HTN | SMC | *EARS2* | -2.459887782 | 0.005799438 | 0.012925528 | 1 |
| HTN | SMC | *CPPED1* | -7.065440566 | 0.000481416 | 0.002162804 | 1 |
| HTN | SMC | *HMOX2* | -5.551891917 | 0.014955116 | 0.028016302 | 1 |
| HTN | SMC | *TOX3* | 4.827394706 | 0.017889053 | 0.032484163 | 1 |
| HTN | SMC | *RRN3P2* | -6.769799765 | 0.000903814 | 0.003205631 | 1 |
| HTN | SMC | *RBL2* | -4.772397763 | 0.005811302 | 0.012946023 | 1 |
| HTN | SMC | *CDIPT* | -5.158857991 | 0.004424208 | 0.010430641 | 1 |
| HTN | SMC | *KAT8* | -10.00209645 | 0.000401742 | 0.001924235 | 1 |
| HTN | SMC | *TMC5* | -4.482594499 | 0.000312453 | 0.001641688 | 1 |
| HTN | SMC | *C16orf62* | -6.896911864 | 0.000939 | 0.00329908 | 1 |
| HTN | SMC | *RNF40* | -7.270537841 | 0.025197741 | 0.043695296 | 1 |
| HTN | SMC | *KNOP1* | -9.77750249 | 0.000161521 | 0.001157608 | 0.783700952 |
| HTN | SMC | *IQCH* | -6.229469618 | 0.000109913 | 0.000967877 | 0.533300291 |
| HTN | SMC | *LACTB* | -7.528308725 | 0.001411641 | 0.004428458 | 1 |
| HTN | SMC | *HERC1* | -5.573608028 | 0.001118119 | 0.003720928 | 1 |
| HTN | SMC | *AP3B2* | -1.420744149 | 0.008916544 | 0.018323624 | 1 |
| HTN | SMC | *TTC23* | -7.626977385 | 7.44E-05 | 0.000847247 | 0.360986211 |

| | | | | | | |
|---|---|---|---|---|---|---|
| HTN | SMC | *FAH* | -6.180618092 | 9.14E-05 | 0.000890467 | 0.443290995 |
| HTN | SMC | *RPAP1* | -8.295555698 | 0.000178878 | 0.001210482 | 0.867915872 |
| HTN | SMC | *SLC30A4* | -7.871987977 | 0.012652165 | 0.024379787 | 1 |
| HTN | SMC | *EYA1* | -1.772434508 | 0.003937444 | 0.009490549 | 1 |
| HTN | SMC | *IMPAD1* | -9.125215965 | 1.46E-05 | 0.000410113 | 0.07094962 |
| HTN | SMC | *SFRP1* | -8.434766266 | 0.000106524 | 0.000950786 | 0.516853673 |
| HTN | SMC | *NIPAL2* | -6.193981808 | 0.002372205 | 0.006452615 | 1 |
| HTN | SMC | *RAB2A* | -4.968616933 | 0.003114702 | 0.007936206 | 1 |
| HTN | SMC | *ARMC1* | -4.601968069 | 0.007696501 | 0.016243333 | 1 |
| HTN | SMC | *SPAG1* | -4.943554447 | 0.004444701 | 0.010473866 | 1 |
| HTN | SMC | *GSDMD* | -7.620127561 | 0.001885583 | 0.005416726 | 1 |
| HTN | SMC | *R3HCC1* | -3.224318485 | 0.000107294 | 0.000950786 | 0.520591837 |
| HTN | SMC | *ERICH1* | -6.005268533 | 0.00024995 | 0.001438619 | 1 |
| HTN | SMC | *TUSC3* | -8.424041944 | 0.000513069 | 0.002236542 | 1 |
| HTN | SMC | *KLHDC4* | -5.552142418 | 0.000771679 | 0.002871309 | 1 |
| HTN | SMC | *MAN2B1* | -3.230921468 | 0.013579739 | 0.025879377 | 1 |
| HTN | SMC | *TULP2* | -10.13840888 | 0.003855698 | 0.009342086 | 1 |
| HTN | SMC | *SARS2* | -1.991310428 | 0.003385605 | 0.00845382 | 1 |
| HTN | SMC | *SNRNP70* | -4.924480394 | 0.020006876 | 0.035675619 | 1 |
| HTN | SMC | *FCGRT* | -3.050642283 | 0.026452008 | 0.045512461 | 1 |
| HTN | SMC | *ERCC2* | -11.47043711 | 0.001379432 | 0.004351759 | 1 |
| HTN | SMC | *CD37* | -10.1907515 | 1.15E-08 | 2.54E-06 | 5.60E-05 |
| HTN | SMC | *AMH* | 22.44653315 | 0.015706382 | 0.02915769 | 1 |
| HTN | SMC | *FCER2* | -15.83873512 | 0.000177615 | 0.001205299 | 0.861788659 |
| HTN | SMC | *CLEC4M* | -20.70855233 | 9.49E-05 | 0.000903 | 0.460321475 |
| HTN | SMC | *SGTA* | -6.448679784 | 0.000305317 | 0.001619413 | 1 |
| HTN | SMC | *MED25* | -4.970448498 | 0.000124575 | 0.001022735 | 0.604436386 |
| HTN | SMC | *IL27RA* | -5.493456682 | 0.000573907 | 0.002363597 | 1 |
| HTN | SMC | *FAM32A* | -12.33621529 | 1.62E-05 | 0.000427306 | 0.078833348 |
| HTN | SMC | *RASAL3* | -3.422986132 | 0.00070923 | 0.002703126 | 1 |
| HTN | SMC | *ANKRD27* | -8.896281941 | 0.000178257 | 0.001207968 | 0.864905024 |
| HTN | SMC | *POLR2I* | -7.915603586 | 0.000187389 | 0.001238705 | 0.909209477 |
| HTN | SMC | *CCDC9* | -4.813468973 | 0.00109927 | 0.003686012 | 1 |
| HTN | SMC | *TGFB1* | -7.282630885 | 0.002188901 | 0.006086275 | 1 |
| HTN | SMC | *ICAM5* | -5.025719561 | 1.33E-05 | 0.000389012 | 0.064323786 |
| HTN | SMC | *CD33* | -1.392386081 | 0.001185563 | 0.0038841 | 1 |
| HTN | SMC | *GRIN2D* | -9.855496457 | 5.05E-05 | 0.000788116 | 0.245104023 |
| HTN | SMC | *CARD8* | -6.201233587 | 0.000208511 | 0.001312184 | 1 |
| HTN | SMC | *LIG1* | -7.075029301 | 0.000121655 | 0.001013054 | 0.590268553 |
| HTN | SMC | *ZNF175* | -5.412804816 | 0.02910304 | 0.049498057 | 1 |
| HTN | SMC | *PLA2G4C* | -1.55258628 | 0.003438854 | 0.008547806 | 1 |
| HTN | SMC | *MIER2* | -7.501303243 | 0.000265922 | 0.001489899 | 1 |
| HTN | SMC | *TNPO2* | -7.641725324 | 0.001534072 | 0.004678389 | 1 |
| HTN | SMC | *GCDH* | -6.340369224 | 2.26E-05 | 0.000515555 | 0.109459226 |
| HTN | SMC | *TFPT* | -8.770495314 | 0.000138507 | 0.001071831 | 0.672038267 |
| HTN | SMC | *RAB3A* | -8.734925179 | 0.000558674 | 0.002332876 | 1 |
| HTN | SMC | *UPK1A* | 4.797319788 | 0.022750184 | 0.03987357 | 1 |
| HTN | SMC | *COPE* | -4.676894104 | 0.003893775 | 0.009408664 | 1 |
| HTN | SMC | *ETV2* | -20.00208479 | 0.000874314 | 0.003131136 | 1 |
| HTN | SMC | *ZNF85* | -11.7067781 | 0.000620144 | 0.002458746 | 1 |

| | | | | | | |
|---|---|---|---|---|---|---|
| HTN | SMC | *ETHE1* | -8.564460368 | 0.012242995 | 0.023723247 | 1 |
| HTN | SMC | *SMG9* | -6.586584308 | 0.014987188 | 0.028065548 | 1 |
| HTN | SMC | *GTPBP10* | -4.308187306 | 0.000349562 | 0.001764904 | 1 |
| HTN | SMC | *RASA4* | -35.66735883 | 0.000118334 | 0.001000276 | 0.574158558 |
| HTN | SMC | *BET1* | -8.010009961 | 0.000176697 | 0.001202931 | 0.857332726 |
| HTN | SMC | *DNAH11* | -9.840516055 | 9.03E-05 | 0.000888243 | 0.437903717 |
| HTN | SMC | *PTN* | -6.667766546 | 0.011077057 | 0.021919201 | 1 |
| HTN | SMC | *ADAP1* | -8.234982756 | 0.000480149 | 0.002161116 | 1 |
| HTN | SMC | *MET* | -2.823696669 | 0.000441885 | 0.002053663 | 1 |
| HTN | SMC | *RNF32* | -12.69795799 | 6.23E-05 | 0.000827638 | 0.302183355 |
| HTN | SMC | *CPVL* | -6.351990976 | 0.000121403 | 0.001013054 | 0.589048811 |
| HTN | SMC | *CHN2* | -2.782968989 | 0.027068796 | 0.046409116 | 1 |
| HTN | SMC | *GRB10* | -6.652417737 | 0.001913971 | 0.005485286 | 1 |
| HTN | SMC | *PLEKHA8* | -6.450219606 | 0.024026807 | 0.041874305 | 1 |
| HTN | SMC | *ZKSCAN1* | -4.916129148 | 0.013606746 | 0.025920664 | 1 |
| HTN | SMC | *EIF3B* | -8.768423992 | 7.78E-05 | 0.000851198 | 0.377345232 |
| HTN | SMC | *TAF6* | -3.460736884 | 0.00101685 | 0.003479625 | 1 |
| HTN | SMC | *MOSPD3* | -4.283921681 | 0.018936855 | 0.034093366 | 1 |
| HTN | SMC | *IMPDH1* | -7.181917981 | 0.000571346 | 0.002363597 | 1 |
| HTN | SMC | *C1GALT1* | -7.371388483 | 0.000521136 | 0.002253612 | 1 |
| HTN | SMC | *ZNF862* | -6.191083696 | 0.001880308 | 0.005413145 | 1 |
| HTN | SMC | *MEST* | -4.1698217 | 3.85E-08 | 5.34E-06 | 0.000186769 |
| HTN | SMC | *ACTR3C* | -3.202693071 | 0.009977014 | 0.020086504 | 1 |
| HTN | SMC | *POU6F2* | -11.72298197 | 3.80E-07 | 3.54E-05 | 0.001842577 |
| HTN | SMC | *TSPAN13* | -4.573277685 | 0.001075181 | 0.003630326 | 1 |
| HTN | SMC | *RARRES2* | -6.937432784 | 0.000261146 | 0.001475067 | 1 |
| HTN | SMC | *COA1* | -6.1833554 | 0.000491432 | 0.002189557 | 1 |
| HTN | SMC | *BLVRA* | -5.866579266 | 0.000608903 | 0.002441651 | 1 |
| HTN | SMC | *URGCP* | -6.960404964 | 4.73E-05 | 0.00076177 | 0.229671331 |
| HTN | SMC | *AEBP1* | 1.388707247 | 0.011349074 | 0.02236625 | 1 |
| HTN | SMC | *GCK* | 13.2874938 | 0.021191919 | 0.03751302 | 1 |
| HTN | SMC | *FKTN* | -7.314836724 | 0.002364427 | 0.006437822 | 1 |
| HTN | SMC | *NMRK1* | -4.09037727 | 0.002743806 | 0.007239233 | 1 |
| HTN | SMC | *MEGF9* | -7.812265039 | 0.00022969 | 0.001377575 | 1 |
| HTN | SMC | *TRIM14* | -2.661259693 | 0.011630108 | 0.022818149 | 1 |
| HTN | SMC | *C5* | -6.132397597 | 0.002688511 | 0.007139933 | 1 |
| HTN | SMC | *OGN* | -6.470190312 | 4.18E-06 | 0.000181808 | 0.020291994 |
| HTN | SMC | *ECM2* | -6.236869845 | 0.000856774 | 0.003088461 | 1 |
| HTN | SMC | *PTGR1* | -4.431318915 | 0.002175922 | 0.006061047 | 1 |
| HTN | SMC | *PLGRKT* | -3.229102677 | 0.014028807 | 0.026557852 | 1 |
| HTN | SMC | *KANK1* | -4.196332051 | 0.001804062 | 0.005235233 | 1 |
| HTN | SMC | *NCS1* | 0.908372204 | 0.00206174 | 0.005808647 | 1 |
| HTN | SMC | *FUBP3* | -7.223694818 | 0.00071961 | 0.002726438 | 1 |
| HTN | SMC | *RGP1* | -9.391977412 | 0.000980131 | 0.003389571 | 1 |
| HTN | SMC | *PTGDS* | -8.593414339 | 4.55E-05 | 0.00075533 | 0.220556442 |
| HTN | SMC | *HPS1* | -5.669373564 | 0.010151606 | 0.020383368 | 1 |
| HTN | SMC | *CUBN* | -5.620551817 | 0.012392843 | 0.023937132 | 1 |
| HTN | SMC | *PLEKHA1* | -5.88352205 | 0.000592849 | 0.002411151 | 1 |
| HTN | SMC | *CDH23* | -4.258989844 | 9.12E-05 | 0.000890467 | 0.442599139 |
| HTN | SMC | *SPOCK2* | -1.536055381 | 0.020765685 | 0.036852635 | 1 |

| | | | | | | |
|---|---|---|---|---|---|---|
| HTN | SMC | *LIPA* | -3.678480147 | 0.001171972 | 0.003852579 | 1 |
| HTN | SMC | *SFXN3* | -10.8883096 | 0.001463473 | 0.004552981 | 1 |
| HTN | SMC | *TNKS2* | -5.567262284 | 0.014081723 | 0.026626859 | 1 |
| HTN | SMC | *ANKRD26* | -5.887104846 | 0.000168813 | 0.001173469 | 0.819081639 |
| HTN | SMC | *ACBD5* | -7.515241385 | 0.002960155 | 0.007615417 | 1 |
| HTN | SMC | *LHPP* | -6.080269577 | 0.000208949 | 0.001313238 | 1 |
| HTN | SMC | *PITRM1* | -9.331972337 | 0.000733005 | 0.00276129 | 1 |
| HTN | SMC | *RPL28* | -5.175182364 | 0.00118711 | 0.003886544 | 1 |
| HTN | SMC | *DNAJC12* | -4.26321226 | 0.005657166 | 0.012654942 | 1 |
| HTN | SMC | *TSPAN14* | -5.486561283 | 7.27E-05 | 0.000847247 | 0.35292358 |
| HTN | SMC | *TADA2A* | -1.732233638 | 0.026550102 | 0.045632694 | 1 |
| HTN | SMC | *PSMB3* | -7.64619927 | 0.000478436 | 0.002159009 | 1 |
| HTN | SMC | *RPL19* | -5.709703108 | 0.002874609 | 0.007460204 | 1 |
| HTN | SMC | *RAPGEFL1* | -6.412229342 | 0.000333214 | 0.001710851 | 1 |
| HTN | SMC | *WNT3* | -4.418146933 | 0.010424767 | 0.02083236 | 1 |
| HTN | SMC | *TRIM37* | -5.511986143 | 0.025711253 | 0.044442822 | 1 |
| HTN | SMC | *PNPO* | -8.76076796 | 2.46E-06 | 0.00013247 | 0.011922271 |
| HTN | SMC | *TRIM16L* | -8.085640389 | 7.09E-05 | 0.000837934 | 0.344049801 |
| HTN | SMC | *PIGL* | -5.1209283 | 0.016483547 | 0.030340733 | 1 |
| HTN | SMC | *MED13* | -5.532138348 | 0.002695088 | 0.007145664 | 1 |
| HTN | SMC | *ENO3* | 2.970098333 | 0.004850008 | 0.011158007 | 1 |
| HTN | SMC | *RAI1* | -7.055649041 | 0.013183285 | 0.025242818 | 1 |
| HTN | SMC | *GOSR1* | -7.362790601 | 0.000877284 | 0.003136759 | 1 |
| HTN | SMC | *ALDH3A1* | -7.366449401 | 0.007217457 | 0.015433717 | 1 |
| HTN | SMC | *ICAM2* | -8.58432335 | 8.45E-05 | 0.000876014 | 0.40980976 |
| HTN | SMC | *PSMD11* | -5.333912619 | 0.002616356 | 0.006978868 | 1 |
| HTN | SMC | *PEX12* | -7.38085734 | 0.002772827 | 0.007290916 | 1 |
| HTN | SMC | *RAB5C* | -4.671455435 | 0.004048079 | 0.00969461 | 1 |
| HTN | SMC | *HSD17B1* | -4.055126717 | 0.001087639 | 0.003659655 | 1 |
| HTN | SMC | *EZH1* | -5.112939316 | 0.000606266 | 0.002438018 | 1 |
| HTN | SMC | *SGCA* | 1.222215895 | 0.000122352 | 0.001013054 | 0.59364966 |
| HTN | SMC | *ABCC3* | -1.980977191 | 0.023551585 | 0.041179204 | 1 |
| HTN | SMC | *EFTUD2* | -9.104617288 | 0.000157515 | 0.001140149 | 0.764265107 |
| HTN | SMC | *EFNB3* | -3.800453176 | 0.01116113 | 0.022058576 | 1 |
| HTN | SMC | *FAM20A* | -6.377465226 | 5.61E-05 | 0.000817716 | 0.272330471 |
| HTN | SMC | *DPH1* | -7.780637481 | 2.23E-05 | 0.000515555 | 0.108123417 |
| HTN | SMC | *DHRS7B* | -7.925362367 | 0.000352927 | 0.001775279 | 1 |
| HTN | SMC | *TMEM97* | -2.847626482 | 0.003712466 | 0.009088237 | 1 |
| HTN | SMC | *SLAIN2* | -5.442307499 | 0.013884528 | 0.026346395 | 1 |
| HTN | SMC | *OCIAD1* | -7.416242919 | 0.000189498 | 0.001247553 | 0.919446438 |
| HTN | SMC | *UBE2D3* | -5.829521908 | 0.001922591 | 0.005500242 | 1 |
| HTN | SMC | *NDUFC1* | -8.479164612 | 0.002749382 | 0.007246062 | 1 |
| HTN | SMC | *KLHL2* | -8.162763873 | 6.73E-05 | 0.000836037 | 0.326338346 |
| HTN | SMC | *WFS1* | -10.82103737 | 7.45E-05 | 0.000847247 | 0.361233608 |
| HTN | SMC | *GRPEL1* | -8.833022476 | 5.74E-05 | 0.000817716 | 0.278330647 |
| HTN | SMC | *AADAT* | 2.145010886 | 0.015708615 | 0.02915769 | 1 |
| HTN | SMC | *TRIM2* | -7.043261946 | 3.51E-05 | 0.000647573 | 0.170311737 |
| HTN | SMC | *STIM2* | -7.098674402 | 0.000314176 | 0.001644425 | 1 |
| HTN | SMC | *BST1* | -5.834252843 | 0.003230185 | 0.008154453 | 1 |
| HTN | SMC | *UFSP2* | -8.648623493 | 0.000632818 | 0.00248618 | 1 |

| | | | | | | |
|---|---|---|---|---|---|---|
| HTN | SMC | *KLHL5* | -7.292732023 | 0.000113431 | 0.000982803 | 0.550369487 |
| HTN | SMC | *FAM149A* | -4.228167857 | 0.000236392 | 0.001395343 | 1 |
| HTN | SMC | *DDX25* | -2.056133727 | 0.010710576 | 0.021315717 | 1 |
| HTN | SMC | *CTSC* | -2.873755773 | 0.00284318 | 0.007412739 | 1 |
| HTN | SMC | *ELP4* | -5.239741733 | 0.001957806 | 0.005577964 | 1 |
| HTN | SMC | *TECTA* | -11.95777501 | 0.000504902 | 0.002221021 | 1 |
| HTN | SMC | *SC5D* | -8.449562114 | 0.000809518 | 0.00297081 | 1 |
| HTN | SMC | *B3GAT1* | -15.50534914 | 0.002295346 | 0.00629922 | 1 |
| HTN | SMC | *SNX15* | -4.36916455 | 0.001419404 | 0.00444606 | 1 |
| HTN | SMC | *DCPS* | -6.834723591 | 6.29E-05 | 0.000827638 | 0.305164427 |
| HTN | SMC | *FOXRED1* | -4.351352581 | 0.002970674 | 0.007638425 | 1 |
| HTN | SMC | *NRXN2* | -20.63701205 | 8.45E-06 | 0.000291476 | 0.041003343 |
| HTN | SMC | *CEP164* | -4.915067975 | 0.026567848 | 0.045647026 | 1 |
| HTN | SMC | *GALNT18* | -4.835002974 | 0.006590024 | 0.014377156 | 1 |
| HTN | SMC | *KIAA1549L* | -7.662773348 | 0.000446996 | 0.002067517 | 1 |
| HTN | SMC | *SLC35F2* | -7.826156562 | 0.001055457 | 0.003573675 | 1 |
| HTN | SMC | *C11orf21* | -8.898663431 | 3.46E-05 | 0.000642066 | 0.168011127 |
| HTN | SMC | *TCIRG1* | -8.316470945 | 0.001336435 | 0.004254846 | 1 |
| HTN | SMC | *GTF2H1* | -5.682560252 | 0.005216146 | 0.011844081 | 1 |
| HTN | SMC | *CORO1C* | -5.412621525 | 0.019404297 | 0.03484443 | 1 |
| HTN | SMC | *ASIC1* | 4.206915271 | 0.016241299 | 0.029928897 | 1 |
| HTN | SMC | *IL23A* | -14.05428856 | 0.000610556 | 0.002446259 | 1 |
| HTN | SMC | *ACSS3* | -2.710611924 | 0.00988878 | 0.0199254 | 1 |
| HTN | SMC | *PARP11* | -6.959352174 | 6.18E-05 | 0.000827638 | 0.299764947 |
| HTN | SMC | *VPS29* | -6.231508003 | 0.004618479 | 0.010789052 | 1 |
| HTN | SMC | *AKAP3* | -1.664498894 | 0.003224906 | 0.008153471 | 1 |
| HTN | SMC | *ACAD10* | -8.505462974 | 1.76E-07 | 1.94E-05 | 0.000854551 |
| HTN | SMC | *DDX55* | -5.48430774 | 0.008911026 | 0.018323624 | 1 |
| HTN | SMC | *ENDOU* | -7.73098126 | 0.000479236 | 0.002159009 | 1 |
| HTN | SMC | *VDR* | -19.76539937 | 0.001007018 | 0.003458271 | 1 |
| HTN | SMC | *CAND1* | -7.780074525 | 0.000501657 | 0.002210755 | 1 |
| HTN | SMC | *MDM1* | -6.408280796 | 9.72E-05 | 0.000914024 | 0.471636458 |
| HTN | SMC | *NUP107* | -6.490011672 | 0.002690849 | 0.007142232 | 1 |
| HTN | SMC | *KRR1* | -5.770286617 | 0.019534618 | 0.035026596 | 1 |
| HTN | SMC | *GNB3* | -1.918841543 | 0.008827795 | 0.018216582 | 1 |
| HTN | SMC | *CDCA3* | -4.814344102 | 0.002427742 | 0.006569661 | 1 |
| HTN | SMC | *CHPT1* | -3.931139689 | 0.000574781 | 0.002363597 | 1 |
| HTN | SMC | *GNPTAB* | -7.034843819 | 0.000304984 | 0.001619413 | 1 |
| HTN | SMC | *C12orf57* | -9.960957502 | 0.001596178 | 0.00478656 | 1 |
| HTN | SMC | *NT5DC3* | 2.344422439 | 0.019883523 | 0.03550786 | 1 |
| HTN | SMC | *SUDS3* | -6.928618131 | 0.009160045 | 0.01874506 | 1 |
| HTN | SMC | *GOLT1B* | -7.266366985 | 0.001342925 | 0.004262545 | 1 |
| HTN | SMC | *C2CD5* | -6.325537757 | 0.002336301 | 0.00637914 | 1 |
| HTN | SMC | *BTN3A3* | -4.406873153 | 1.31E-05 | 0.000389012 | 0.063691696 |
| HTN | SMC | *FRK* | 0.909612342 | 0.004682846 | 0.010902671 | 1 |
| HTN | SMC | *RWDD1* | -5.660489494 | 0.000665085 | 0.002581595 | 1 |
| HTN | SMC | *TMEM14C* | -9.740712988 | 0.000372547 | 0.001824013 | 1 |
| HTN | SMC | *FAM184A* | -4.765667233 | 0.003206591 | 0.008120239 | 1 |
| HTN | SMC | *RNGTT* | -6.061429789 | 0.005842849 | 0.012998397 | 1 |
| HTN | SMC | *UST* | -3.327936167 | 9.59E-05 | 0.000908214 | 0.465433592 |

| | | | | | | |
|---|---|---|---|---|---|---|
| HTN | SMC | *RNF8* | -9.696743711 | 3.88E-05 | 0.000689568 | 0.188406457 |
| HTN | SMC | *MDGA1* | -9.338976899 | 0.008853154 | 0.018232386 | 1 |
| HTN | SMC | *MDN1* | -5.961680305 | 0.000179722 | 0.001212811 | 0.8720111 |
| HTN | SMC | *GLP1R* | -10.10114058 | 0.008217815 | 0.017179163 | 1 |
| HTN | SMC | *WASF1* | -6.27322735 | 0.000190993 | 0.001250601 | 0.926695652 |
| HTN | SMC | *ACOT13* | -6.725073503 | 0.012925847 | 0.024808628 | 1 |
| HTN | SMC | *C6orf62* | -8.332434753 | 2.31E-06 | 0.000127244 | 0.011197464 |
| HTN | SMC | *HBS1L* | -6.545251147 | 0.000453547 | 0.002087867 | 1 |
| HTN | SMC | *FIG4* | -6.737910046 | 0.000366729 | 0.001809686 | 1 |
| HTN | SMC | *PHACTR2* | -5.226425869 | 0.003472346 | 0.008617813 | 1 |
| HTN | SMC | *CUTA* | -4.427625047 | 0.003211139 | 0.008127514 | 1 |
| HTN | SMC | *PACRG* | -4.200335504 | 0.00015398 | 0.001136201 | 0.747110453 |
| HTN | SMC | *QKI* | -5.548900924 | 0.000171984 | 0.001186677 | 0.83446852 |
| HTN | SMC | *FAM120B* | -4.078118661 | 0.015635124 | 0.029065755 | 1 |
| HTN | SMC | *GUCA1B* | -3.124386979 | 0.000446021 | 0.002064973 | 1 |
| HTN | SMC | *DNPH1* | -2.879871341 | 0.008534546 | 0.017719135 | 1 |
| HTN | SMC | *EXOC2* | -6.655414561 | 0.000385225 | 0.001859815 | 1 |
| HTN | SMC | *TMEM30A* | -8.45936398 | 0.001789294 | 0.005198595 | 1 |
| HTN | SMC | *IMPG1* | -17.64347125 | 1.62E-05 | 0.000427306 | 0.078775239 |
| HTN | SMC | *LY86* | -5.207526998 | 0.00012858 | 0.001039835 | 0.623871002 |
| HTN | SMC | *NUDT12* | -8.363221015 | 5.84E-05 | 0.000817716 | 0.283408844 |
| HTN | SMC | *DAP* | -4.169727439 | 0.002458529 | 0.006627102 | 1 |
| HTN | SMC | *KIF20A* | -14.41569675 | 0.004157694 | 0.009898494 | 1 |
| HTN | SMC | *PFDN1* | -5.419338322 | 0.000771125 | 0.002871309 | 1 |
| HTN | SMC | *SPARC* | -6.160050506 | 0.000646125 | 0.002530266 | 1 |
| HTN | SMC | *PCDHB15* | -2.269822626 | 0.010758998 | 0.021359516 | 1 |
| HTN | SMC | *ARSB* | -7.425348852 | 0.006903444 | 0.014953353 | 1 |
| HTN | SMC | *BTNL8* | -1.091011951 | 0.012322974 | 0.023849648 | 1 |
| HTN | SMC | *MSH3* | -3.802020205 | 0.007167134 | 0.015380334 | 1 |
| HTN | SMC | *RASGRF2* | -2.016234802 | 0.00078734 | 0.002918392 | 1 |
| HTN | SMC | *DROSHA* | -5.349845399 | 0.000487817 | 0.002184577 | 1 |
| HTN | SMC | *CDH6* | 0.539354705 | 0.004035963 | 0.009675143 | 1 |
| HTN | SMC | *LMNB1* | -4.061565242 | 0.000602475 | 0.002429934 | 1 |
| HTN | SMC | *SLC12A7* | -10.60162306 | 0.000103154 | 0.000942633 | 0.500502876 |
| HTN | SMC | *SKP1* | -6.609342329 | 0.007642274 | 0.016156999 | 1 |
| HTN | SMC | *PPWD1* | -6.638695356 | 0.005024245 | 0.01147723 | 1 |
| HTN | SMC | *TXNDC15* | -7.649955865 | 0.009258044 | 0.018881896 | 1 |
| HTN | SMC | *SMAD5* | -7.691960673 | 0.000314712 | 0.001645453 | 1 |
| HTN | SMC | *ERGIC1* | -7.746862076 | 0.000137588 | 0.001071047 | 0.66757668 |
| HTN | SMC | *ATP6V0E1* | -6.442219425 | 0.002813725 | 0.007340933 | 1 |
| HTN | SMC | *BNIP1* | 2.180830155 | 0.012232484 | 0.023721828 | 1 |
| HTN | SMC | *UNC5A* | -17.69092346 | 0.000249417 | 0.001438619 | 1 |
| HTN | SMC | *ACTR8* | -7.957877511 | 0.003828915 | 0.009310417 | 1 |
| HTN | SMC | *TBCCD1* | -10.39839067 | 7.42E-05 | 0.000847247 | 0.35979235 |
| HTN | SMC | *PCCB* | 4.792016997 | 0.001083914 | 0.003652186 | 1 |
| HTN | SMC | *CEP70* | -5.06799829 | 0.014528086 | 0.027364237 | 1 |
| HTN | SMC | *HYAL1* | -4.766515829 | 0.000164814 | 0.001162037 | 0.799678499 |
| HTN | SMC | *CBLB* | -4.753690291 | 0.001620718 | 0.004833266 | 1 |
| HTN | SMC | *IQCG* | -8.089453094 | 0.000406814 | 0.001935158 | 1 |
| HTN | SMC | *NCBP2* | -5.74185031 | 0.002081769 | 0.005848722 | 1 |

| | | | | | | |
|---|---|---|---|---|---|---|
| HTN | SMC | *KIAA1257* | -7.190959635 | 0.000337841 | 0.001729121 | 1 |
| HTN | SMC | *NEK11* | -3.885632884 | 0.003551411 | 0.008764724 | 1 |
| HTN | SMC | *PLSCR4* | -5.219680333 | 0.000286691 | 0.001557696 | 1 |
| HTN | SMC | *HEMK1* | -6.594649784 | 0.003884177 | 0.009390148 | 1 |
| HTN | SMC | *COMMD2* | -8.207395759 | 0.00120431 | 0.003924321 | 1 |
| HTN | SMC | *ABHD14B* | -7.157761083 | 0.002250106 | 0.006220806 | 1 |
| HTN | SMC | *ARHGEF26* | -8.416659239 | 6.94E-05 | 0.000836037 | 0.336922984 |
| HTN | SMC | *DNAH1* | -2.385572772 | 0.016668524 | 0.030611536 | 1 |
| HTN | SMC | *NKTR* | -4.887178522 | 0.003781816 | 0.009225426 | 1 |
| HTN | SMC | *NEK4* | -8.830028632 | 2.25E-06 | 0.00012532 | 0.010902836 |
| HTN | SMC | *ADAM23* | -7.642499364 | 0.001997513 | 0.005677757 | 1 |
| HTN | SMC | *LMAN2L* | -5.416885326 | 0.013210687 | 0.025265373 | 1 |
| HTN | SMC | *SLC35F5* | -5.937434455 | 0.000600241 | 0.002428998 | 1 |
| HTN | SMC | *OTOF* | -5.543522515 | 0.022059531 | 0.038822213 | 1 |
| HTN | SMC | *CENPA* | -3.139314591 | 0.017912641 | 0.032514828 | 1 |
| HTN | SMC | *ITGB6* | -3.833727752 | 0.001782228 | 0.005190499 | 1 |
| HTN | SMC | *MOGS* | -3.697981368 | 0.000739099 | 0.002775793 | 1 |
| HTN | SMC | *GRB14* | -6.885973708 | 8.38E-05 | 0.000876014 | 0.406470901 |
| HTN | SMC | *RTN4* | -4.962588656 | 0.000806011 | 0.00297081 | 1 |
| HTN | SMC | *GALNT3* | -3.541680844 | 0.011563883 | 0.022715773 | 1 |
| HTN | SMC | *POLE4* | -7.743581122 | 0.015508149 | 0.02887396 | 1 |
| HTN | SMC | *ACADL* | -2.782326184 | 0.004099742 | 0.009774913 | 1 |
| HTN | SMC | *MRPL19* | -5.939713645 | 0.009420225 | 0.019122238 | 1 |
| HTN | SMC | *IGFBP2* | -4.240100463 | 0.011739014 | 0.022968203 | 1 |
| HTN | SMC | *GNLY* | -1.96389303 | 0.00079355 | 0.002935648 | 1 |
| HTN | SMC | *CHMP3* | -5.021762428 | 3.15E-05 | 0.000616069 | 0.15278522 |
| HTN | SMC | *IL1RL2* | -4.308780486 | 0.002136532 | 0.005968021 | 1 |
| HTN | SMC | *IL18R1* | -6.445825863 | 3.11E-05 | 0.00061356 | 0.150935859 |
| HTN | SMC | *MLPH* | 3.848758409 | 0.023195203 | 0.040599973 | 1 |
| HTN | SMC | *HDLBP* | -5.482607407 | 0.003189305 | 0.008093361 | 1 |
| HTN | SMC | *PASK* | -2.032336194 | 0.002142429 | 0.005974847 | 1 |
| HTN | SMC | *TPO* | -6.951852518 | 0.000177018 | 0.001202931 | 0.858892745 |
| HTN | SMC | *TAF1B* | -5.012624331 | 0.001252997 | 0.004045459 | 1 |
| HTN | SMC | *NOL10* | -5.603976856 | 0.013733287 | 0.026090018 | 1 |
| HTN | SMC | *STRN* | -6.898646943 | 0.000525709 | 0.002257599 | 1 |
| HTN | SMC | *CEBPZ* | -5.277649771 | 0.013632621 | 0.025949579 | 1 |
| HTN | SMC | *PRKD3* | -4.930232442 | 0.01525746 | 0.028494687 | 1 |
| HTN | SMC | *SLC25A12* | -5.096772797 | 0.012027286 | 0.023389336 | 1 |
| HTN | SMC | *DARS* | -7.036237088 | 0.001558561 | 0.00471455 | 1 |
| HTN | SMC | *WIPF1* | -5.360559203 | 0.00545908 | 0.012291586 | 1 |
| HTN | SMC | *PNO1* | -6.668069801 | 0.000107385 | 0.000950786 | 0.521030923 |
| HTN | SMC | *AAK1* | -5.303364057 | 0.004461484 | 0.010498118 | 1 |
| HTN | SMC | *NFE2L2* | -4.822192197 | 0.001884564 | 0.005416726 | 1 |
| HTN | SMC | *MSH6* | -5.868727708 | 0.021433713 | 0.037853195 | 1 |
| HTN | SMC | *ALMS1* | -5.485803644 | 0.020155847 | 0.035862181 | 1 |
| HTN | SMC | *DNAJC16* | -8.148492633 | 6.34E-05 | 0.000827638 | 0.307675401 |
| HTN | SMC | *MORN1* | -6.06058837 | 1.90E-06 | 0.000109935 | 0.009234526 |
| HTN | SMC | *RALGPS2* | -4.436845119 | 0.004742479 | 0.010988893 | 1 |
| HTN | SMC | *WRAP73* | -9.744798557 | 2.04E-05 | 0.000491528 | 0.098797226 |
| HTN | SMC | *ICMT* | -11.19982396 | 0.007628771 | 0.016142519 | 1 |

| | | | | | | |
|---|---|---|---|---|---|---|
| HTN | SMC | *KIAA1324* | -3.343185441 | 0.018720655 | 0.033754225 | 1 |
| HTN | SMC | *MECR* | -9.755725935 | 9.57E-06 | 0.000313646 | 0.046419539 |
| HTN | SMC | *EDEM3* | -8.226429505 | 0.000134038 | 0.001055488 | 0.650352542 |
| HTN | SMC | *CAPZA1* | -8.785662395 | 7.60E-06 | 0.000273327 | 0.036899206 |
| HTN | SMC | *ASH1L* | -3.474778333 | 0.025057066 | 0.043480965 | 1 |
| HTN | SMC | *GON4L* | -4.105035465 | 0.017931068 | 0.032536103 | 1 |
| HTN | SMC | *DOCK7* | -6.460476516 | 0.001682604 | 0.004971981 | 1 |
| HTN | SMC | *FBXO6* | -3.243024833 | 0.016508595 | 0.030375314 | 1 |
| HTN | SMC | *DNAJC6* | 0.809221014 | 0.007519202 | 0.015987365 | 1 |
| HTN | SMC | *LEPR* | -5.790729099 | 0.002959255 | 0.007615417 | 1 |
| HTN | SMC | *IVNS1ABP* | -2.879533123 | 0.002038981 | 0.005761874 | 1 |
| HTN | SMC | *PRG4* | -8.327313352 | 4.17E-05 | 0.000719923 | 0.20229833 |
| HTN | SMC | *TMEM9* | -4.761916354 | 0.001251789 | 0.004045459 | 1 |
| HTN | SMC | *OSCP1* | -3.179165079 | 0.001473946 | 0.004572624 | 1 |
| HTN | SMC | *EXOC8* | -6.105939372 | 0.00155411 | 0.004706954 | 1 |
| HTN | SMC | *ACADM* | -5.278988714 | 0.005361006 | 0.012120969 | 1 |
| HTN | SMC | *PADI2* | 1.183311807 | 0.011689482 | 0.022906852 | 1 |
| HTN | SMC | *SDHB* | -8.121715226 | 0.000753745 | 0.002819716 | 1 |
| HTN | SMC | *KLHL12* | -5.433786049 | 0.009725232 | 0.019644807 | 1 |
| HTN | SMC | *ZNHIT6* | -7.2769549 | 0.000340084 | 0.001734441 | 1 |
| HTN | SMC | *GBP3* | -1.934160946 | 0.001888707 | 0.005419283 | 1 |
| HTN | SMC | *KIF17* | -8.740925273 | 0.000501522 | 0.002210755 | 1 |
| HTN | SMC | *EBNA1BP2* | -6.572356145 | 0.022327649 | 0.039265587 | 1 |
| HTN | SMC | *ERI3* | -5.919002561 | 0.009462109 | 0.019183363 | 1 |
| HTN | SMC | *PRDX1* | -5.04754833 | 0.007067629 | 0.015240088 | 1 |
| HTN | SMC | *PRRC2C* | -4.564134013 | 0.010116992 | 0.020326147 | 1 |
| HTN | SMC | *ABCD3* | -7.977897938 | 0.000216537 | 0.00133839 | 1 |
| HTN | SMC | *PTBP2* | -3.671158734 | 0.012003913 | 0.023371985 | 1 |
| HTN | SMC | *DARS2* | -5.426871954 | 0.0004252 | 0.001997165 | 1 |
| HTN | SMC | *IRF6* | -2.283887636 | 0.000597106 | 0.002424389 | 1 |
| HTN | SMC | *MTFR1L* | -7.46453329 | 0.000810666 | 0.00297081 | 1 |
| HTN | SMC | *RPS6KA1* | -9.622159278 | 9.39E-05 | 0.000903 | 0.455833568 |
| HTN | SMC | *MARC2* | -6.125107304 | 0.000105191 | 0.000950786 | 0.510386786 |
| HTN | SMC | *SLC5A9* | -5.56428989 | 0.019936722 | 0.035589762 | 1 |
| HTN | SMC | *IFT46* | -3.850585657 | 0.012027143 | 0.023389336 | 1 |
| HTN | SMC | *DDX59* | -5.84154372 | 0.00066352 | 0.002577581 | 1 |
| HTN | SMC | *CASC1* | -2.626035299 | 0.000562171 | 0.002341332 | 1 |
| HTN | SMC | *CNR1* | 0.785636318 | 0.002907916 | 0.007532946 | 1 |
| HTN | SMC | *AKAP7* | -5.886521394 | 0.001585038 | 0.004759038 | 1 |
| HTN | SMC | *ALDH8A1* | -1.180809931 | 0.016554574 | 0.030436829 | 1 |
| HTN | SMC | *MED28* | -6.282572585 | 0.000578236 | 0.002375614 | 1 |
| HTN | SMC | *VAMP8* | -7.520485194 | 0.00034948 | 0.001764904 | 1 |
| HTN | SMC | *FOXO3* | -5.119494828 | 0.003275407 | 0.008251441 | 1 |
| HTN | SMC | *MFSD1* | -6.545341593 | 4.55E-05 | 0.00075533 | 0.220530031 |
| HTN | SMC | *UBN1* | -5.45345446 | 0.015391805 | 0.028679354 | 1 |
| HTN | SMC | *DNAH7* | -3.680536045 | 0.000114234 | 0.00098569 | 0.554263579 |
| HTN | SMC | *C2orf40* | 1.153937064 | 0.006557922 | 0.014319999 | 1 |
| HTN | SMC | *CPSF3* | -5.871896679 | 0.000175629 | 0.001200263 | 0.852150623 |
| HTN | SMC | *SENP5* | -6.744781151 | 0.001517388 | 0.004642097 | 1 |
| HTN | SMC | *CCDC92* | -6.110562284 | 0.001416391 | 0.00443949 | 1 |

| | | | | | | |
|---|---|---|---|---|---|---|
| HTN | SMC | *RAB14* | -7.206564936 | 0.004022021 | 0.009651259 | 1 |
| HTN | SMC | *HDHD3* | -1.497500446 | 0.002852489 | 0.00742504 | 1 |
| HTN | SMC | *MAPKAP1* | -6.808638498 | 0.000342304 | 0.001737301 | 1 |
| HTN | SMC | *INVS* | -3.191714379 | 0.014939465 | 0.027997793 | 1 |
| HTN | SMC | *DENND1A* | -5.401197167 | 0.001604244 | 0.004802914 | 1 |
| HTN | SMC | *KDSR* | -8.122505989 | 0.000140027 | 0.001078427 | 0.679409261 |
| HTN | SMC | *DCAF4* | -8.902345482 | 5.27E-05 | 0.000806078 | 0.255526818 |
| HTN | SMC | *IFI27L2* | -6.754868053 | 0.008866277 | 0.018251666 | 1 |
| HTN | SMC | *NEK9* | -6.14725466 | 0.001787455 | 0.005198595 | 1 |
| HTN | SMC | *ACOT2* | 1.190877108 | 0.00707037 | 0.015240088 | 1 |
| HTN | SMC | *TTLL5* | -6.716780954 | 8.50E-05 | 0.000876014 | 0.412196328 |
| HTN | SMC | *FLVCR2* | -6.134275777 | 0.004822387 | 0.011118112 | 1 |
| HTN | SMC | *COQ6* | -9.348793791 | 0.001586019 | 0.004759038 | 1 |
| HTN | SMC | *DNMT3A* | -5.551561775 | 0.000629749 | 0.002478136 | 1 |
| HTN | SMC | *CNRIP1* | -3.980391741 | 0.000536221 | 0.002284237 | 1 |
| HTN | SMC | *CRIPT* | -6.618861823 | 6.75E-05 | 0.000836037 | 0.327563113 |
| HTN | SMC | *SLC17A5* | -5.989057698 | 0.001859904 | 0.005368385 | 1 |
| HTN | SMC | *PPP1R3C* | 1.150247978 | 0.003620421 | 0.008896162 | 1 |
| HTN | SMC | *MXI1* | -7.281313011 | 0.000694649 | 0.002668595 | 1 |
| HTN | SMC | *C10orf88* | -4.463910369 | 0.000234507 | 0.001389292 | 1 |
| HTN | SMC | *HELLS* | -7.712537607 | 4.71E-05 | 0.00076177 | 0.228392864 |
| HTN | SMC | *WDR11* | -5.97876014 | 0.00068885 | 0.002653042 | 1 |
| HTN | SMC | *KANSL1* | -4.863599977 | 0.002856236 | 0.007430809 | 1 |
| HTN | SMC | *PCDHB10* | 7.373229826 | 0.004740609 | 0.010988893 | 1 |
| HTN | SMC | *PCDHB12* | -2.792269648 | 0.000692374 | 0.002661962 | 1 |
| HTN | SMC | *SNX19* | -7.025897367 | 8.43E-06 | 0.000291476 | 0.040889777 |
| HTN | SMC | *SLC10A7* | -6.058986514 | 0.007685731 | 0.016234726 | 1 |
| HTN | SMC | *MASTL* | -4.170438785 | 0.00025073 | 0.001439693 | 1 |
| HTN | SMC | *MRC1* | -6.171781826 | 0.000235001 | 0.001389748 | 1 |
| HTN | SMC | *IQSEC3* | -4.034300088 | 0.000585159 | 0.002395942 | 1 |
| HTN | SMC | *CCDC77* | -7.83823757 | 7.98E-06 | 0.000282729 | 0.038733808 |
| HTN | SMC | *DNAJC15* | -7.681922226 | 0.000175636 | 0.001200263 | 0.852187056 |
| HTN | SMC | *UFM1* | -9.121998188 | 1.40E-05 | 0.000397041 | 0.067894045 |
| HTN | SMC | *ALG5* | -6.149910213 | 0.001705304 | 0.005014626 | 1 |
| HTN | SMC | *SIL1* | -6.564906847 | 0.000329965 | 0.001697761 | 1 |
| HTN | SMC | *ARL1* | -7.645588076 | 0.001744316 | 0.005110762 | 1 |
| HTN | SMC | *APAF1* | -7.984363572 | 0.000111199 | 0.000970929 | 0.539538853 |
| HTN | SMC | *ADRA1A* | 1.015129111 | 0.008561893 | 0.017760712 | 1 |
| HTN | SMC | *EPHX2* | -5.594754488 | 2.43E-05 | 0.000537075 | 0.11789742 |
| HTN | SMC | *TBX2* | 0.316733566 | 0.028384575 | 0.048391411 | 1 |
| HTN | SMC | *TEX14* | 4.919837289 | 6.49E-07 | 5.10E-05 | 0.003147246 |
| HTN | SMC | *ADCY7* | -3.651123615 | 0.002772971 | 0.007290916 | 1 |
| HTN | SMC | *ECHDC2* | -5.026796855 | 6.00E-05 | 0.000822062 | 0.291009843 |
| HTN | SMC | *PYROXD1* | -3.585083957 | 0.007203633 | 0.015413367 | 1 |
| HTN | SMC | *ZNF549* | -9.575239893 | 0.001499853 | 0.004625724 | 1 |
| HTN | SMC | *A1BG* | 2.285499388 | 0.006421988 | 0.014058978 | 1 |
| HTN | SMC | *ZNF211* | -2.73727116 | 0.008538266 | 0.017719275 | 1 |
| HTN | SMC | *SEC22A* | -5.243750758 | 0.016893797 | 0.030931586 | 1 |
| HTN | SMC | *MAPK8IP1* | -14.76033126 | 2.55E-08 | 3.99E-06 | 0.000123641 |
| HTN | SMC | *CAT* | -3.056362391 | 0.000535165 | 0.002281742 | 1 |

| | | | | | | |
|---|---|---|---|---|---|---|
| HTN | SMC | *FABP3* | -20.88949555 | 2.78E-05 | 0.000569929 | 0.134841865 |
| HTN | SMC | *TNFSF10* | -6.40608825 | 8.46E-05 | 0.000876014 | 0.410637839 |
| HTN | SMC | *ZNF639* | -8.562039082 | 0.00893704 | 0.01835062 | 1 |
| HTN | SMC | *CPXM2* | -3.67856454 | 0.000475223 | 0.002152032 | 1 |
| HTN | SMC | *FLT3* | -2.92641044 | 0.005269734 | 0.011948014 | 1 |
| HTN | SMC | *MTIF3* | -8.048576452 | 2.26E-05 | 0.000515555 | 0.109813175 |
| HTN | SMC | *CD244* | -5.32896589 | 0.00133924 | 0.004255399 | 1 |
| HTN | SMC | *RBBP6* | -7.047113805 | 0.000289431 | 0.001565572 | 1 |
| HTN | SMC | *LDB3* | 0.347670772 | 0.00228708 | 0.006290766 | 1 |
| HTN | SMC | *OPN4* | 1.557613889 | 0.002284724 | 0.006288724 | 1 |
| HTN | SMC | *FAM35A* | -7.024660882 | 2.54E-05 | 0.000550673 | 0.123027282 |
| HTN | SMC | *NAA60* | -6.088410481 | 0.000144972 | 0.001097623 | 0.703403103 |
| HTN | SMC | *ODF2L* | -6.955988626 | 4.42E-05 | 0.000749874 | 0.214463966 |
| HTN | SMC | *SPATA1* | -6.02396041 | 0.006964187 | 0.01507182 | 1 |
| HTN | SMC | *EEPD1* | -4.423578613 | 1.23E-05 | 0.000374297 | 0.059513205 |
| HTN | SMC | *KLHL7* | -7.381668252 | 0.001411721 | 0.004428458 | 1 |
| HTN | SMC | *NT5C3A* | -7.158705555 | 0.000130189 | 0.00104255 | 0.631679069 |
| HTN | SMC | *SMU1* | -6.245465068 | 0.012376901 | 0.023924626 | 1 |
| HTN | SMC | *SLC25A51* | -1.899506645 | 0.000724922 | 0.002739346 | 1 |
| HTN | SMC | *TRIM24* | -6.695882312 | 0.002332932 | 0.006373528 | 1 |
| HTN | SMC | *C7orf49* | -3.46903578 | 3.30E-05 | 0.000635347 | 0.160107515 |
| HTN | SMC | *RBM19* | -7.446961316 | 0.006710172 | 0.014606439 | 1 |
| HTN | SMC | *DDX54* | -5.268329997 | 0.008607576 | 0.017832605 | 1 |
| HTN | SMC | *SSPN* | -4.328435853 | 0.000782511 | 0.002904931 | 1 |
| HTN | SMC | *CCDC91* | -5.703451206 | 0.000573518 | 0.002363597 | 1 |
| HTN | SMC | *CENPK* | -3.687956421 | 0.004082275 | 0.009747637 | 1 |
| HTN | SMC | *ARHGAP9* | -3.1173012 | 0.000288773 | 0.001565506 | 1 |
| HTN | SMC | *PFDN5* | -7.795168735 | 0.000980824 | 0.003389571 | 1 |
| HTN | SMC | *CDK2* | -6.298062503 | 5.79E-06 | 0.000222793 | 0.028071973 |
| HTN | SMC | *SARDH* | 2.15776281 | 0.004956986 | 0.011361028 | 1 |
| HTN | SMC | *DBH* | 21.28992159 | 0.004607629 | 0.010779274 | 1 |
| HTN | SMC | *USP45* | -7.01155914 | 0.000696953 | 0.002675327 | 1 |
| HTN | SMC | *TTC21B* | -7.658528769 | 0.000405009 | 0.001929517 | 1 |
| HTN | SMC | *ACVR1C* | -1.578011784 | 0.004118293 | 0.009814322 | 1 |
| HTN | SMC | *PLA2G12A* | -6.321126787 | 7.60E-05 | 0.000849747 | 0.368790103 |
| HTN | SMC | *C4BPB* | -2.130371452 | 0.011671658 | 0.022881166 | 1 |
| HTN | SMC | *RAB38* | -3.481389898 | 0.001819439 | 0.005273547 | 1 |
| HTN | SMC | *DNPEP* | -6.749971811 | 0.009439861 | 0.01914808 | 1 |
| HTN | SMC | *OBSL1* | -8.743393885 | 4.36E-06 | 0.000183865 | 0.021144522 |
| HTN | SMC | *WFDC3* | -1.659259685 | 1.19E-05 | 0.000369253 | 0.05797266 |
| HTN | SMC | *MATN4* | -12.01675392 | 0.000450602 | 0.002080231 | 1 |
| HTN | SMC | *VAPB* | -2.746011427 | 0.014235893 | 0.026907889 | 1 |
| HTN | SMC | *TOX2* | -2.300918645 | 7.04E-05 | 0.000837934 | 0.341509728 |
| HTN | SMC | *SRSF6* | -6.155828796 | 0.003132987 | 0.007975474 | 1 |
| HTN | SMC | *CSE1L* | -6.055728266 | 0.00507043 | 0.011566396 | 1 |
| HTN | SMC | *RNF114* | -7.829797152 | 1.64E-05 | 0.000427306 | 0.079375729 |
| HTN | SMC | *PEPD* | -7.557650481 | 0.000134335 | 0.001055488 | 0.651795718 |
| HTN | SMC | *STAMBP* | -5.656107664 | 0.028413746 | 0.04842413 | 1 |
| HTN | SMC | *NAGK* | -3.728051749 | 0.003748065 | 0.009161516 | 1 |
| HTN | SMC | *ATP8A1* | -7.222583192 | 2.36E-05 | 0.000530042 | 0.114489054 |

| | | | | | | |
|---|---|---|---|---|---|---|
| HTN | SMC | *USP22* | -5.704763801 | 0.009741707 | 0.019669898 | 1 |
| HTN | SMC | *HIF3A* | -4.520989878 | 0.002738076 | 0.007228044 | 1 |
| HTN | SMC | *BTN2A2* | 1.695218997 | 0.008801065 | 0.018171389 | 1 |
| HTN | SMC | *MRS2* | -2.976321371 | 0.011494053 | 0.022606057 | 1 |
| HTN | SMC | *SNRPC* | -8.764395004 | 7.60E-08 | 9.71E-06 | 0.000368904 |
| HTN | SMC | *SERPINB6* | -5.223498358 | 0.008655011 | 0.017900303 | 1 |
| HTN | SMC | *NQO2* | -7.672850865 | 6.92E-05 | 0.000836037 | 0.335794274 |
| HTN | SMC | *ZNF391* | -1.937830234 | 4.74E-05 | 0.00076177 | 0.22986843 |
| HTN | SMC | *MED20* | -3.216365055 | 0.025367367 | 0.04394233 | 1 |
| HTN | SMC | *TCP11* | 3.757094885 | 0.008448706 | 0.017586068 | 1 |
| HTN | SMC | *TREM1* | -2.959491623 | 0.008239412 | 0.017216894 | 1 |
| HTN | SMC | *KLHL31* | -1.512905832 | 0.010331297 | 0.020688176 | 1 |
| HTN | SMC | *KCNK17* | 1.36234792 | 0.010505434 | 0.020967654 | 1 |
| HTN | SMC | *RREB1* | -5.339985545 | 0.000169529 | 0.001176761 | 0.82255563 |
| HTN | SMC | *RUNX2* | -5.773023199 | 0.023609223 | 0.041250252 | 1 |
| HTN | SMC | *SH3TC1* | -2.071275982 | 0.019827518 | 0.03543393 | 1 |
| HTN | SMC | *MT1G* | -7.045331029 | 0.003550997 | 0.008764724 | 1 |
| HTN | SMC | *GOT2* | 3.6432905 | 0.022058607 | 0.038822213 | 1 |
| HTN | SMC | *TMTC4* | -10.7023693 | 0.000242431 | 0.001416471 | 1 |
| HTN | SMC | *ABCC4* | -4.467815143 | 0.000202523 | 0.001287864 | 0.982640136 |
| HTN | SMC | *ATP5S* | -4.133090745 | 0.000294497 | 0.001578896 | 1 |
| HTN | SMC | *GRK4* | 1.195424591 | 0.004989435 | 0.011413832 | 1 |
| HTN | SMC | *HS3ST3B1* | -3.43347188 | 0.000818674 | 0.002986621 | 1 |
| HTN | SMC | *MRPS7* | -9.07716323 | 0.002586132 | 0.006905839 | 1 |
| HTN | SMC | *GGA3* | -7.758395877 | 0.00020589 | 0.001304152 | 0.998980547 |
| HTN | SMC | *NUP85* | -5.938114398 | 0.005379776 | 0.012152082 | 1 |
| HTN | SMC | *NT5C* | -8.678931653 | 0.003214818 | 0.008132585 | 1 |
| HTN | SMC | *OPRL1* | -7.48131743 | 0.000590082 | 0.002407971 | 1 |
| HTN | SMC | *IL1B* | -6.818662877 | 0.029101786 | 0.049498057 | 1 |
| HTN | SMC | *PAX8* | -2.019252775 | 0.016996081 | 0.031071961 | 1 |
| HTN | SMC | *CCDC93* | -5.811801138 | 0.002059789 | 0.005807146 | 1 |
| HTN | SMC | *SH2D3A* | -4.847026163 | 0.015884193 | 0.029404848 | 1 |
| HTN | SMC | *SNRPD2* | -9.680272642 | 0.003157878 | 0.008022003 | 1 |
| HTN | SMC | *SDCBP2* | -5.165893029 | 0.000561638 | 0.002341122 | 1 |
| HTN | SMC | *PSMF1* | -5.254520146 | 0.002206941 | 0.006115694 | 1 |
| HTN | SMC | *RBCK1* | -5.764408536 | 0.010951359 | 0.021723628 | 1 |
| HTN | SMC | *RRBP1* | -5.419886168 | 0.006014999 | 0.013320299 | 1 |
| HTN | SMC | *FLRT3* | -2.274348504 | 0.000471204 | 0.00214272 | 1 |
| HTN | SMC | *PCSK2* | -7.271342499 | 0.003940265 | 0.009492635 | 1 |
| HTN | SMC | *MKKS* | -4.622979797 | 0.00843834 | 0.017572029 | 1 |
| HTN | SMC | *LAMP5* | -41.06248939 | 0.001575807 | 0.004740559 | 1 |
| HTN | SMC | *MGME1* | -4.087785941 | 0.013336232 | 0.025466205 | 1 |
| HTN | SMC | *ITPA* | -5.790318278 | 0.011770019 | 0.022999651 | 1 |
| HTN | SMC | *XRCC3* | -5.635875515 | 0.000738381 | 0.002775793 | 1 |
| HTN | SMC | *TUBGCP3* | -7.351257993 | 0.000758621 | 0.002831406 | 1 |
| HTN | SMC | *PRDX5* | -4.952625239 | 0.020276931 | 0.036064394 | 1 |
| HTN | SMC | *IRF3* | -8.68262864 | 0.000305714 | 0.001619413 | 1 |
| HTN | SMC | *TSKS* | -7.061816362 | 3.01E-07 | 2.98E-05 | 0.001458106 |
| HTN | SMC | *TRAP1* | 2.223437324 | 0.003256033 | 0.00821116 | 1 |
| HTN | SMC | *EMG1* | -6.903946636 | 0.00029516 | 0.001580703 | 1 |

| | | | | | | |
|---|---|---|---|---|---|---|
| HTN | SMC | *ZBTB1* | -6.42562016 | 0.000837675 | 0.003035398 | 1 |
| HTN | SMC | *WDR60* | -4.000691749 | 0.003913705 | 0.009452115 | 1 |
| HTN | SMC | *AIF1L* | -6.737187668 | 0.000115994 | 0.000993383 | 0.56280147 |
| HTN | SMC | *CANX* | -6.960279373 | 0.000321611 | 0.001668936 | 1 |
| HTN | SMC | *ABHD8* | -3.600782283 | 0.005006203 | 0.011441402 | 1 |
| HTN | SMC | *HELB* | -8.678937627 | 0.000128801 | 0.001039835 | 0.624940821 |
| HTN | SMC | *RAB3IP* | -11.01895214 | 0.000221663 | 0.001351139 | 1 |
| HTN | SMC | *LRRC61* | -3.584360545 | 9.33E-05 | 0.000903 | 0.452542071 |
| HTN | SMC | *IDUA* | -7.887094031 | 0.000840559 | 0.003043532 | 1 |
| HTN | SMC | *EMC1* | -7.334836766 | 5.56E-05 | 0.000817658 | 0.269690603 |
| HTN | SMC | *GFER* | -8.089391177 | 5.82E-05 | 0.000817716 | 0.282291618 |
| HTN | SMC | *WFIKKN1* | 4.693139796 | 0.003529645 | 0.008725771 | 1 |
| HTN | SMC | *PNKD* | -7.419127633 | 0.00116156 | 0.003823535 | 1 |
| HTN | SMC | *RNF6* | -7.680131337 | 0.013248458 | 0.02531888 | 1 |
| HTN | SMC | *PTPN12* | -5.321243884 | 0.004063881 | 0.009719522 | 1 |
| HTN | SMC | *STYXL1* | -3.404345118 | 2.73E-05 | 0.000569929 | 0.132377466 |
| HTN | SMC | *STEAP4* | 0.919197696 | 0.005714868 | 0.012766362 | 1 |
| HTN | SMC | *PEX1* | -8.411503972 | 0.001105959 | 0.003693812 | 1 |
| HTN | SMC | *KDR* | -3.812965298 | 0.000165253 | 0.001162037 | 0.801805237 |
| HTN | SMC | *ATF4* | -7.251276415 | 0.023038942 | 0.040341013 | 1 |
| HTN | SMC | *CDHR3* | -7.324611839 | 6.06E-05 | 0.000827638 | 0.294066372 |
| HTN | SMC | *PRKRIP1* | -6.089927161 | 0.003414834 | 0.008501167 | 1 |
| HTN | SMC | *IRF5* | -1.802370143 | 0.001986503 | 0.005650925 | 1 |
| HTN | SMC | *OPN1SW* | -2.93738214 | 0.00101764 | 0.003479625 | 1 |
| HTN | SMC | *MTX2* | -6.940081129 | 0.001434962 | 0.004483218 | 1 |
| HTN | SMC | *GAD1* | 1.136436862 | 0.009571527 | 0.019369068 | 1 |
| HTN | SMC | *ORMDL1* | -3.481033732 | 0.02350653 | 0.04113007 | 1 |
| HTN | SMC | *EIF2AK4* | -8.70541878 | 2.06E-05 | 0.00049543 | 0.100076924 |
| HTN | SMC | *KNSTRN* | -1.054639507 | 0.020035725 | 0.035700822 | 1 |
| HTN | SMC | *ARPP19* | -7.809145967 | 0.000119384 | 0.001005578 | 0.579251999 |
| HTN | SMC | *VPS13C* | -4.878772139 | 0.005426838 | 0.012235604 | 1 |
| HTN | SMC | *CALML4* | -8.075217675 | 0.000355569 | 0.001785945 | 1 |
| HTN | SMC | *ANAPC13* | -2.078696806 | 0.003694873 | 0.009054306 | 1 |
| HTN | SMC | *SUMF2* | -8.447708044 | 2.57E-05 | 0.000552295 | 0.124818568 |
| HTN | SMC | *BBOX1* | -7.368787494 | 0.00060649 | 0.002438018 | 1 |
| HTN | SMC | *RPAIN* | -5.707106416 | 8.19E-05 | 0.000876014 | 0.397310276 |
| HTN | SMC | *PLD2* | -13.1551678 | 0.000900658 | 0.00320171 | 1 |
| HTN | SMC | *PHF20L1* | -5.340898278 | 0.006276118 | 0.013816571 | 1 |
| HTN | SMC | *PUS7L* | -6.365260936 | 0.000150734 | 0.001123782 | 0.731358977 |
| HTN | SMC | *AP1M2* | -2.420091902 | 0.00653558 | 0.014277638 | 1 |
| HTN | SMC | *MTUS1* | -4.757068843 | 0.009270427 | 0.018883816 | 1 |
| HTN | SMC | *KLK8* | -15.74319652 | 0.000654469 | 0.002554694 | 1 |
| HTN | SMC | *RIPK3* | 2.23604139 | 0.0257829 | 0.04452267 | 1 |
| HTN | SMC | *BCL2L2* | -5.392302726 | 0.000835988 | 0.003033818 | 1 |
| HTN | SMC | *PARP2* | -4.841543721 | 0.000777625 | 0.002891214 | 1 |
| HTN | SMC | *MIS18BP1* | -5.561628536 | 0.000127772 | 0.001039637 | 0.619949171 |
| HTN | SMC | *RNASE1* | -4.367255676 | 0.008588273 | 0.017800213 | 1 |
| HTN | SMC | *EPB41L4A* | -6.517671478 | 0.000245003 | 0.001421998 | 1 |
| HTN | SMC | *CDO1* | -1.431139401 | 0.019093838 | 0.034337769 | 1 |
| HTN | SMC | *REEP5* | -6.062714081 | 0.007083218 | 0.015254227 | 1 |

| | | | | | | |
|---|---|---|---|---|---|---|
| HTN | SMC | *MAU2* | -7.826069984 | 0.000479147 | 0.002159009 | 1 |
| HTN | SMC | *ERMARD* | -7.407140832 | 0.000953616 | 0.003323955 | 1 |
| HTN | SMC | *PHF10* | -9.091133274 | 0.000105311 | 0.000950786 | 0.510968582 |
| HTN | SMC | *GALNT8* | -7.552951619 | 3.85E-05 | 0.000689173 | 0.186765878 |
| HTN | SMC | *PRKCSH* | -8.355831778 | 1.58E-06 | 9.82E-05 | 0.00765784 |
| HTN | SMC | *XPO7* | -7.867585949 | 5.70E-05 | 0.000817716 | 0.276425113 |
| HTN | SMC | *ATP8B3* | -3.023852411 | 5.66E-05 | 0.000817716 | 0.274719757 |
| HTN | SMC | *DDA1* | -6.019707667 | 0.011786244 | 0.023022084 | 1 |
| HTN | SMC | *KIAA1683* | -5.780646211 | 0.001100109 | 0.003686278 | 1 |
| HTN | SMC | *LSM4* | -2.735312785 | 0.003850991 | 0.009337834 | 1 |
| HTN | SMC | *TRPM4* | -7.855628831 | 4.79E-08 | 6.46E-06 | 0.000232624 |
| HTN | SMC | *ATXN10* | -5.191882121 | 0.008409871 | 0.017520263 | 1 |
| HTN | SMC | *PNPLA7* | -6.050354538 | 5.41E-05 | 0.000806822 | 0.26232553 |
| HTN | SMC | *PAK4* | -6.355552856 | 7.03E-05 | 0.000837934 | 0.341269467 |
| HTN | SMC | *NPAS1* | -3.953176898 | 0.000960106 | 0.003338651 | 1 |
| HTN | SMC | *MAP3K10* | -6.936801276 | 0.0007261 | 0.002741663 | 1 |
| HTN | SMC | *DNMT1* | -8.875379617 | 0.000130704 | 0.001043376 | 0.634176739 |
| HTN | SMC | *NOL11* | -7.000154366 | 0.004934948 | 0.011315863 | 1 |
| HTN | SMC | *HSD17B3* | -13.94092578 | 0.008646109 | 0.01789715 | 1 |
| HTN | SMC | *SLC35D2* | -8.132692216 | 0.000944316 | 0.003305786 | 1 |
| HTN | SMC | *TTLL9* | -4.980112004 | 0.00100178 | 0.003447261 | 1 |
| HTN | SMC | *ATP6V1E1* | -7.614800118 | 0.000549637 | 0.002313255 | 1 |
| HTN | SMC | *CHMP1A* | -6.653011748 | 6.79E-05 | 0.000836037 | 0.329546089 |
| HTN | SMC | *IDO1* | -19.85110912 | 0.000121293 | 0.001013054 | 0.588512277 |
| HTN | SMC | *HAUS8* | -7.503279967 | 1.50E-06 | 9.66E-05 | 0.007281895 |
| HTN | SMC | *MRPS25* | -9.127406133 | 2.41E-06 | 0.000131513 | 0.011704636 |
| HTN | SMC | *GALNT15* | -9.606891864 | 0.000228819 | 0.001377231 | 1 |
| HTN | SMC | *SLC6A6* | -7.057532218 | 0.000100951 | 0.000932982 | 0.489815293 |
| HTN | SMC | *KCNC3* | -7.0781492 | 0.00166821 | 0.004941487 | 1 |
| HTN | SMC | *PDLIM4* | -6.372976335 | 0.012260535 | 0.023747752 | 1 |
| HTN | SMC | *NDUFA2* | -6.118202883 | 0.00047547 | 0.002152032 | 1 |
| HTN | SMC | *C1orf159* | -7.83772178 | 8.84E-05 | 0.000888243 | 0.428911047 |
| HTN | SMC | *PPFIA1* | -4.61763153 | 0.008200199 | 0.017149727 | 1 |
| HTN | SMC | *BARX1* | -6.126616603 | 0.011416916 | 0.022472567 | 1 |
| HTN | SMC | *NPHP4* | -4.509465733 | 0.000310981 | 0.001640089 | 1 |
| HTN | SMC | *TRIM21* | -12.37442655 | 0.003830089 | 0.009310417 | 1 |
| HTN | SMC | *SPATA6* | -5.729735031 | 0.003546483 | 0.008761475 | 1 |
| HTN | SMC | *LRRC41* | -8.147627853 | 0.002362232 | 0.006435458 | 1 |
| HTN | SMC | *CCT6B* | -3.692517203 | 0.000731269 | 0.002756891 | 1 |
| HTN | SMC | *CNGA4* | -2.97175839 | 0.004183988 | 0.009936716 | 1 |
| HTN | SMC | *TRIM22* | -5.777107614 | 7.23E-05 | 0.000845677 | 0.350956125 |
| HTN | SMC | *IMMT* | -6.337506489 | 0.001768191 | 0.005162012 | 1 |
| HTN | SMC | *CARD6* | -2.813441714 | 0.0061508 | 0.013596211 | 1 |
| HTN | SMC | *RPA1* | -4.728488081 | 0.004480859 | 0.010537499 | 1 |
| HTN | SMC | *SERPINF1* | -6.461409851 | 5.35E-05 | 0.000806822 | 0.259809634 |
| HTN | SMC | *FIGNL1* | -5.105945466 | 0.011951571 | 0.023307485 | 1 |
| HTN | SMC | *EIF5A* | -3.531405353 | 0.006255967 | 0.013778461 | 1 |
| HTN | SMC | *RGS22* | -1.919301098 | 0.001507009 | 0.004634646 | 1 |
| HTN | SMC | *PRMT7* | -3.897251651 | 0.000313675 | 0.001643575 | 1 |
| HTN | SMC | *CTNNBL1* | -6.468691923 | 0.005923944 | 0.013154681 | 1 |

| | | | | | | |
|---|---|---|---|---|---|---|
| HTN | SMC | *OSER1* | -7.270671163 | 0.003934448 | 0.009490549 | 1 |
| HTN | SMC | *SERINC3* | -4.635245919 | 0.003203265 | 0.008120239 | 1 |
| HTN | SMC | *DMGDH* | -3.669380007 | 0.000319876 | 0.001663491 | 1 |
| HTN | SMC | *BHMT2* | -1.315662022 | 0.001050635 | 0.00356232 | 1 |
| HTN | SMC | *AP3B1* | -6.311741249 | 0.000605924 | 0.002438018 | 1 |
| HTN | SMC | *ZBED3* | -6.284161926 | 0.000457935 | 0.002102081 | 1 |
| HTN | SMC | *FBXO44* | -7.69574652 | 0.010615334 | 0.021169585 | 1 |
| HTN | SMC | *RSG1* | -11.78790079 | 0.000114577 | 0.00098569 | 0.555929202 |
| HTN | SMC | *CASP9* | -2.361649208 | 0.003523697 | 0.008722948 | 1 |
| HTN | SMC | *ATP8A2* | -7.086609801 | 0.018191226 | 0.03295886 | 1 |
| HTN | SMC | *ALOX5AP* | -5.381237085 | 0.000161336 | 0.001157608 | 0.78280081 |
| HTN | SMC | *WASF3* | -5.397753239 | 5.98E-05 | 0.000821986 | 0.29016115 |
| HTN | SMC | *PEMT* | -6.820025347 | 8.37E-05 | 0.000876014 | 0.406047628 |
| HTN | SMC | *SCO1* | -4.86669417 | 0.007162416 | 0.015380334 | 1 |
| HTN | SMC | *DSTYK* | -7.475207944 | 7.12E-05 | 0.000837934 | 0.345249441 |
| HTN | SMC | *COG6* | -7.044935174 | 0.000156642 | 0.001140149 | 0.760029132 |
| HTN | SMC | *EPSTI1* | -5.271381848 | 0.000300334 | 0.001599583 | 1 |
| HTN | SMC | *CSNK1G2* | -9.920868495 | 0.001887829 | 0.005419283 | 1 |
| HTN | SMC | *CNDP2* | -5.137797959 | 0.000195712 | 0.001271662 | 0.949592763 |
| HTN | SMC | *PDZD2* | -3.891489833 | 6.41E-05 | 0.000829756 | 0.310951716 |
| HTN | SMC | *FAM83F* | -10.96592827 | 0.002120987 | 0.00593485 | 1 |
| HTN | SMC | *GIMAP4* | -3.879880058 | 0.000546297 | 0.002310188 | 1 |
| HTN | SMC | *IPO8* | -5.574347395 | 0.017754011 | 0.032263094 | 1 |
| HTN | SMC | *LARS* | -3.133381484 | 0.002451739 | 0.006623519 | 1 |
| HTN | SMC | *SPINK5* | -12.05421566 | 0.000976996 | 0.003382734 | 1 |
| HTN | SMC | *LRRCC1* | -5.843504443 | 0.00272357 | 0.007197593 | 1 |
| HTN | SMC | *RRAS2* | -5.916905661 | 7.36E-05 | 0.000847247 | 0.357004658 |
| HTN | SMC | *CATSPERB* | -4.302944189 | 0.001512561 | 0.004641266 | 1 |
| HTN | SMC | *COX16* | -5.681323006 | 0.001274723 | 0.004098711 | 1 |
| HTN | SMC | *TTC9* | -5.149172661 | 0.00026349 | 0.001484848 | 1 |
| HTN | SMC | *ELP3* | -8.008502268 | 0.000699417 | 0.002675893 | 1 |
| HTN | SMC | *CTIF* | -3.469581771 | 0.005729113 | 0.012792295 | 1 |
| HTN | SMC | *MRO* | -4.591545804 | 0.001256728 | 0.004054285 | 1 |
| HTN | SMC | *THUMPD3* | -10.09011223 | 0.000436335 | 0.002031762 | 1 |
| HTN | SMC | *CNTN6* | -4.077058382 | 0.000926856 | 0.003264289 | 1 |
| HTN | SMC | *CHL1* | -2.418585675 | 0.002949721 | 0.00761279 | 1 |
| HTN | SMC | *GSTM5* | -3.295957164 | 0.000493344 | 0.002196058 | 1 |
| HTN | SMC | *VAV3* | -2.556100754 | 6.20E-05 | 0.000827638 | 0.300915451 |
| HTN | SMC | *TRIM45* | -3.702671801 | 0.000451497 | 0.002082381 | 1 |
| HTN | SMC | *CEPT1* | -5.705494353 | 0.000173046 | 0.001187282 | 0.839619227 |
| HTN | SMC | *PPHLN1* | -6.308847209 | 0.000874421 | 0.003131136 | 1 |
| HTN | SMC | *FKBP11* | -5.83460156 | 0.000272653 | 0.001509827 | 1 |
| HTN | SMC | *KIDINS220* | -5.339459379 | 0.007187362 | 0.015406042 | 1 |
| HTN | SMC | *LPIN1* | -5.93390621 | 0.000126528 | 0.001031791 | 0.613915609 |
| HTN | SMC | *NARS* | -6.177594316 | 0.003726802 | 0.009118731 | 1 |
| HTN | SMC | *KIAA1468* | -7.072965106 | 0.000281399 | 0.001541024 | 1 |
| HTN | SMC | *IL15RA* | -7.969099591 | 0.000219946 | 0.001344061 | 1 |
| HTN | SMC | *TMEM241* | -5.069415109 | 5.42E-07 | 4.61E-05 | 0.002629776 |
| HTN | SMC | *KLRD1* | -8.190315577 | 0.000480828 | 0.002162165 | 1 |
| HTN | SMC | *HOOK1* | -1.742273386 | 0.00477841 | 0.011045662 | 1 |

| | | | | | | |
|---|---|---|---|---|---|---|
| HTN | SMC | *PRPF38A* | -7.08500022 | 0.012986862 | 0.024915877 | 1 |
| HTN | SMC | *ELP2* | -6.494617843 | 0.003575454 | 0.008810615 | 1 |
| HTN | SMC | *TIMM10* | -6.32998274 | 0.001647391 | 0.004891762 | 1 |
| HTN | SMC | *DHX34* | -8.961094185 | 9.49E-05 | 0.000903 | 0.460566079 |
| HTN | SMC | *TMEM258* | -4.103569183 | 0.027731375 | 0.047427786 | 1 |
| HTN | SMC | *CLOCK* | -5.558128326 | 0.008830462 | 0.018216582 | 1 |
| HTN | SMC | *CLDN10* | 6.202671699 | 0.001715899 | 0.005039674 | 1 |
| HTN | SMC | *ARGLU1* | -4.656544347 | 0.006417721 | 0.014058142 | 1 |
| HTN | SMC | *ERCC5* | -5.921973131 | 0.000350926 | 0.001769952 | 1 |
| HTN | SMC | *CARS2* | -5.16029385 | 0.001027601 | 0.00350135 | 1 |
| HTN | SMC | *KLB* | -1.816518492 | 0.002457667 | 0.006627102 | 1 |
| HTN | SMC | *WDR36* | -8.524545564 | 0.000265769 | 0.001489899 | 1 |
| HTN | SMC | *OSTF1* | -7.359669673 | 0.000104557 | 0.000950786 | 0.507310303 |
| HTN | SMC | *UBQLN1* | -7.180237075 | 0.000365848 | 0.001809686 | 1 |
| HTN | SMC | *USP30* | -3.909589591 | 9.02E-06 | 0.000301812 | 0.043762697 |
| HTN | SMC | *DTX1* | -4.281565648 | 0.000138185 | 0.001071047 | 0.670475667 |
| HTN | SMC | *DMTF1* | -6.444822319 | 0.000606159 | 0.002438018 | 1 |
| HTN | SMC | *TMEM60* | -6.688397342 | 0.002396954 | 0.006509174 | 1 |
| HTN | SMC | *POM121C* | -8.651152257 | 4.18E-06 | 0.000181808 | 0.020283433 |
| HTN | SMC | *SRPK2* | -5.500423685 | 0.001766256 | 0.005159465 | 1 |
| HTN | SMC | *TES* | -9.430943419 | 3.25E-05 | 0.000629438 | 0.157622563 |
| HTN | SMC | *AKIRIN2* | -7.346125445 | 0.000409835 | 0.001943812 | 1 |
| HTN | SMC | *MAP3K7* | -6.7244036 | 0.003347316 | 0.008384707 | 1 |
| HTN | SMC | *PRR5L* | -2.836025432 | 0.014370899 | 0.027131362 | 1 |
| HTN | SMC | *PHF21A* | -6.113834054 | 0.000420761 | 0.001980147 | 1 |
| HTN | SMC | *CAPRIN1* | -6.099669103 | 0.004678238 | 0.010897172 | 1 |
| HTN | SMC | *ATP5G2* | -6.660506112 | 0.002794993 | 0.007318569 | 1 |
| HTN | SMC | *B4GALNT1* | 1.946764436 | 0.001113329 | 0.003710076 | 1 |
| HTN | SMC | *FAIM2* | -6.63156878 | 0.000317562 | 0.001656784 | 1 |
| HTN | SMC | *PAN2* | -5.245046595 | 0.001542743 | 0.004693033 | 1 |
| HTN | SMC | *SLC26A10* | 3.092107438 | 0.003030999 | 0.007764735 | 1 |
| HTN | SMC | *NHSL1* | -5.743042226 | 9.95E-06 | 0.000321952 | 0.048292819 |
| HTN | SMC | *SMPD2* | -2.029440462 | 0.009561691 | 0.019362824 | 1 |
| HTN | SMC | *MICAL1* | 1.478848069 | 0.000660769 | 0.002573074 | 1 |
| HTN | SMC | *CCT7* | -10.2711255 | 5.12E-06 | 0.000203448 | 0.024820632 |
| HTN | SMC | *USP15* | -5.842163788 | 0.001040569 | 0.003530658 | 1 |
| HTN | SMC | *MDM2* | -4.812045118 | 0.009407671 | 0.019109588 | 1 |
| HTN | SMC | *MPHOSPH6* | -5.142107889 | 0.00016233 | 0.001159977 | 0.787624711 |
| HTN | SMC | *AGT* | 3.101663484 | 0.0006163 | 0.002454805 | 1 |
| HTN | SMC | *CAPN9* | -9.608706377 | 3.67E-05 | 0.000672209 | 0.178135511 |
| HTN | SMC | *NTPCR* | -6.130109033 | 0.000133489 | 0.001054634 | 0.647689956 |
| HTN | SMC | *PIGC* | -8.234529205 | 0.0013458 | 0.004265069 | 1 |
| HTN | SMC | *SP110* | -6.146379392 | 0.004534361 | 0.010633504 | 1 |
| HTN | SMC | *SLC19A3* | -2.925420676 | 4.91E-05 | 0.000773529 | 0.238336188 |
| HTN | SMC | *TMBIM1* | -8.585113413 | 8.91E-05 | 0.000888243 | 0.432373189 |
| HTN | SMC | *TSGA10* | -1.842049048 | 0.003770025 | 0.009201289 | 1 |
| HTN | SMC | *ANKRD36* | -6.425948599 | 0.005471429 | 0.012313254 | 1 |
| HTN | SMC | *EPC2* | -6.246866879 | 0.00105649 | 0.003574678 | 1 |
| HTN | SMC | *STAB2* | -2.191668886 | 0.000471142 | 0.00214272 | 1 |
| HTN | SMC | *APPL2* | -5.82243459 | 0.001879962 | 0.005413145 | 1 |

| | | | | | | |
|---|---|---|---|---|---|---|
| HTN | SMC | *DRAM1* | -7.690070569 | 6.56E-05 | 0.000834038 | 0.318430205 |
| HTN | SMC | *SLC41A2* | -5.672095248 | 6.30E-05 | 0.000827638 | 0.305628826 |
| HTN | SMC | *RNASEH2B* | -6.968612825 | 1.06E-05 | 0.000336521 | 0.05164146 |
| HTN | SMC | *TBC1D4* | -6.436956088 | 1.12E-05 | 0.000349468 | 0.054516936 |
| HTN | SMC | *SUCLA2* | -7.897106564 | 0.000135943 | 0.001065582 | 0.659595196 |
| HTN | SMC | *RCBTB2* | -6.607907327 | 7.11E-05 | 0.000837934 | 0.344791792 |
| HTN | SMC | *SETDB2* | -6.2335676 | 0.015274564 | 0.028515653 | 1 |
| HTN | SMC | *SCRN1* | -7.068252613 | 0.002654961 | 0.007066303 | 1 |
| HTN | SMC | *CHST12* | -9.931196545 | 4.22E-06 | 0.000181808 | 0.020495212 |
| HTN | SMC | *NUPL2* | -7.991337457 | 0.000106477 | 0.000950786 | 0.516628546 |
| HTN | SMC | *ZDHHC4* | -5.250004887 | 0.000233676 | 0.001386058 | 1 |
| HTN | SMC | *AOAH* | -2.143928039 | 0.008845476 | 0.018224921 | 1 |
| HTN | SMC | *TBRG4* | -6.173984742 | 0.000798273 | 0.002949903 | 1 |
| HTN | SMC | *NMT1* | 3.116147403 | 0.002889393 | 0.00748896 | 1 |
| HTN | SMC | *BRIP1* | -3.754956402 | 0.007691863 | 0.016240608 | 1 |
| HTN | SMC | *RTP4* | -5.992885893 | 0.000951734 | 0.003321745 | 1 |
| HTN | SMC | *NDUFB5* | -10.11664131 | 6.33E-05 | 0.000827638 | 0.306968946 |
| HTN | SMC | *RPS6KC1* | -6.526272154 | 0.000613643 | 0.00245255 | 1 |
| HTN | SMC | *IMP4* | -12.54537023 | 0.000255321 | 0.001454597 | 1 |
| HTN | SMC | *NIPSNAP3A* | -7.278226787 | 0.000601825 | 0.002429934 | 1 |
| HTN | SMC | *ODF2* | -10.38569292 | 8.39E-05 | 0.000876014 | 0.407019222 |
| HTN | SMC | *TOR1B* | -7.235980022 | 0.00802742 | 0.016853761 | 1 |
| HTN | SMC | *SLC2A8* | -5.015317123 | 0.005505448 | 0.012378329 | 1 |
| HTN | SMC | *ZNF189* | -10.20078279 | 0.000723344 | 0.002735515 | 1 |
| HTN | SMC | *STX17* | -7.250449231 | 0.000702432 | 0.002679402 | 1 |
| HTN | SMC | *ANP32B* | -7.865358343 | 0.000919061 | 0.003251201 | 1 |
| HTN | SMC | *DERL1* | -6.065771257 | 0.003683986 | 0.009032188 | 1 |
| HTN | SMC | *NOV* | 1.398586923 | 0.003353888 | 0.008392504 | 1 |
| HTN | SMC | *IL11RA* | -6.324334563 | 1.70E-05 | 0.000440884 | 0.082445252 |
| HTN | SMC | *APTX* | -7.311238014 | 8.99E-05 | 0.000888243 | 0.436058517 |
| HTN | SMC | *DENND4C* | -6.85165697 | 2.45E-05 | 0.000537075 | 0.118966693 |
| HTN | SMC | *TMEM14B* | -4.883660493 | 0.004823501 | 0.011118112 | 1 |
| HTN | SMC | *LRRC1* | -3.693125129 | 0.007818445 | 0.016472882 | 1 |
| HTN | SMC | *TUBB2B* | -2.22324437 | 0.016600331 | 0.030509396 | 1 |
| HTN | SMC | *NRM* | -6.673698596 | 0.000616331 | 0.002454805 | 1 |
| HTN | SMC | *TLR2* | -3.803144778 | 0.00016392 | 0.001162037 | 0.795337709 |
| HTN | SMC | *MYO7A* | -2.510216742 | 0.003075732 | 0.007864857 | 1 |
| HTN | SMC | *NUMA1* | -6.990607148 | 0.001482473 | 0.004587346 | 1 |
| HTN | SMC | *SYTL2* | 0.545679939 | 0.009262157 | 0.018882347 | 1 |
| HTN | SMC | *RAB30* | -7.901146719 | 0.004727917 | 0.010976007 | 1 |
| HTN | SMC | *PRCP* | -4.816574727 | 0.002268234 | 0.006253108 | 1 |
| HTN | SMC | *NARS2* | -6.539788875 | 0.005511898 | 0.012385087 | 1 |
| HTN | SMC | *GGH* | -2.404924556 | 0.00155113 | 0.004700863 | 1 |
| HTN | SMC | *SULF1* | -3.432226808 | 0.000105736 | 0.000950786 | 0.513032367 |
| HTN | SMC | *TGS1* | -7.727164653 | 0.000150779 | 0.001123782 | 0.731581961 |
| HTN | SMC | *NEK1* | -5.994525794 | 0.001673533 | 0.004948192 | 1 |
| HTN | SMC | *TRPC6* | 1.164097375 | 0.002723574 | 0.007197593 | 1 |
| HTN | SMC | *SLC37A4* | 3.862944469 | 0.008533687 | 0.017719135 | 1 |
| HTN | SMC | *POU2F3* | -2.772931247 | 0.01064643 | 0.02121416 | 1 |
| HTN | SMC | *PPP2R1B* | -4.367537573 | 3.66E-08 | 5.22E-06 | 0.00017754 |

| | | | | | | |
|---|---|---|---|---|---|---|
| HTN | SMC | *FXYD6* | -2.885975415 | 0.003003932 | 0.007711682 | 1 |
| HTN | SMC | *MAP2K5* | -4.791631668 | 0.012073724 | 0.023451444 | 1 |
| HTN | SMC | *NUSAP1* | 9.112833582 | 0.00020981 | 0.001313543 | 1 |
| HTN | SMC | *NDUFAF1* | -3.880551539 | 0.02259031 | 0.039655639 | 1 |
| HTN | SMC | *ADAM10* | -4.759599923 | 0.008715954 | 0.018018665 | 1 |
| HTN | SMC | *ZNF280D* | -5.848842885 | 0.000458378 | 0.002102129 | 1 |
| HTN | SMC | *SLC44A5* | -6.559610771 | 0.000182243 | 0.001221551 | 0.884242002 |
| HTN | SMC | *DBT* | -6.277776761 | 0.017330075 | 0.031599219 | 1 |
| HTN | SMC | *KHK* | -4.351211442 | 0.004751598 | 0.010994161 | 1 |
| HTN | SMC | *ADCY3* | 1.79967948 | 0.004285595 | 0.010148222 | 1 |
| HTN | SMC | *LHCGR* | -4.287347132 | 9.47E-05 | 0.000903 | 0.459293353 |
| HTN | SMC | *THUMPD2* | -7.88738333 | 0.000357846 | 0.001791816 | 1 |
| HTN | SMC | *ACTR2* | -6.914401296 | 0.000297289 | 0.001588595 | 1 |
| HTN | SMC | *ATRAID* | -8.069657412 | 8.58E-05 | 0.000876014 | 0.41650218 |
| HTN | SMC | *CENPO* | 2.252702228 | 0.028271986 | 0.048233361 | 1 |
| HTN | SMC | *LRPPRC* | -5.14697388 | 0.017584371 | 0.032002764 | 1 |
| HTN | SMC | *ARL3* | 3.499897352 | 0.015093304 | 0.028231576 | 1 |
| HTN | SMC | *RBP4* | -7.039045795 | 0.000410678 | 0.001944008 | 1 |
| HTN | SMC | *ASCC1* | -4.713301485 | 0.002072032 | 0.005828115 | 1 |
| HTN | SMC | *RPS24* | -4.543474448 | 0.014787304 | 0.027766253 | 1 |
| HTN | SMC | *SMARCAL1* | -8.127251737 | 0.001016724 | 0.003479625 | 1 |
| HTN | SMC | *SSB* | -3.962125037 | 0.008467248 | 0.017602008 | 1 |
| HTN | SMC | *FASTKD1* | -5.019185834 | 7.04E-05 | 0.000837934 | 0.341575269 |
| HTN | SMC | *ITGAV* | -6.70676858 | 0.000635867 | 0.002496136 | 1 |
| HTN | SMC | *SLC35A5* | -5.024554773 | 0.002045462 | 0.005776219 | 1 |
| HTN | SMC | *SENP7* | -5.513830961 | 0.004705683 | 0.010934854 | 1 |
| HTN | SMC | *COX17* | -9.888079331 | 0.004382666 | 0.01034275 | 1 |
| HTN | SMC | *SPPL2A* | -9.154814505 | 4.39E-07 | 3.88E-05 | 0.00213192 |
| HTN | SMC | *PCDH10* | 1.10780783 | 0.002886404 | 0.007485213 | 1 |
| HTN | SMC | *AP1AR* | -9.391667832 | 0.001445029 | 0.004505963 | 1 |
| HTN | SMC | *KIAA1109* | -5.34947257 | 0.004911105 | 0.011277179 | 1 |
| HTN | SMC | *BMPR1B* | -3.955825065 | 0.016785507 | 0.030791409 | 1 |
| HTN | SMC | *MMRN1* | -1.318619892 | 0.000365061 | 0.001809686 | 1 |
| HTN | SMC | *PRDM5* | -7.900801396 | 0.001154385 | 0.003807666 | 1 |
| HTN | SMC | *TRPC3* | 1.994411521 | 0.002322349 | 0.00635177 | 1 |
| HTN | SMC | *CNOT6L* | -4.269336059 | 0.027595998 | 0.047212899 | 1 |
| HTN | SMC | *ANXA3* | -3.09969462 | 0.006336621 | 0.013911895 | 1 |
| HTN | SMC | *PPA2* | -5.126934547 | 0.005587707 | 0.012528445 | 1 |
| HTN | SMC | *FBN2* | 1.110908847 | 0.007581861 | 0.016078316 | 1 |
| HTN | SMC | *PARVG* | -5.894097973 | 1.29E-05 | 0.000389012 | 0.062424583 |
| HTN | SMC | *B4GALNT3* | -4.095987561 | 0.00068896 | 0.002653042 | 1 |
| HTN | SMC | *ERP27* | -4.571546515 | 0.006118094 | 0.013530078 | 1 |
| HTN | SMC | *KIF21A* | -7.146907158 | 0.000182558 | 0.001221754 | 0.88577176 |
| HTN | SMC | *ALG10* | -4.731230481 | 0.00865113 | 0.017899908 | 1 |
| HTN | SMC | *ZCRB1* | -7.408441607 | 0.00022657 | 0.001374147 | 1 |
| HTN | SMC | *CLSTN3* | -5.606704445 | 0.000170583 | 0.001182386 | 0.827670218 |
| HTN | SMC | *PEX5* | -2.897213189 | 0.014333283 | 0.02707088 | 1 |
| HTN | SMC | *PPFIA2* | 0.81198609 | 0.005940805 | 0.013180058 | 1 |
| HTN | SMC | *MARCH9* | -11.88304315 | 5.81E-05 | 0.000817716 | 0.281923436 |
| HTN | SMC | *LGR5* | 1.321305558 | 0.008897508 | 0.018308189 | 1 |

| | | | | | | |
|---|---|---|---|---|---|---|
| HTN | SMC | *POC1B* | -8.041967706 | 0.000908625 | 0.003220341 | 1 |
| HTN | SMC | *AMDHD1* | -1.916597704 | 0.004261346 | 0.010105596 | 1 |
| HTN | SMC | *RITA1* | -15.29446635 | 5.15E-05 | 0.00079539 | 0.249752546 |
| HTN | SMC | *SDSL* | -2.520748766 | 0.011234628 | 0.022185761 | 1 |
| HTN | SMC | *GLTP* | -12.09318895 | 0.000121729 | 0.001013054 | 0.590627991 |
| HTN | SMC | *FOXN4* | -3.426984831 | 0.000239816 | 0.001405298 | 1 |
| HTN | SMC | *MTMR6* | -8.453558328 | 0.000623552 | 0.002462978 | 1 |
| HTN | SMC | *CCDC65* | -4.528944061 | 0.029392185 | 0.049898839 | 1 |
| HTN | SMC | *ACVRL1* | -11.84335662 | 0.000294404 | 0.001578896 | 1 |
| HTN | SMC | *BRCA2* | -12.75646862 | 0.000710054 | 0.002703126 | 1 |
| HTN | SMC | *CERS5* | -5.135082094 | 0.02652825 | 0.045611293 | 1 |
| HTN | SMC | *ESYT1* | -5.236653349 | 0.0263881 | 0.045434727 | 1 |
| HTN | SMC | *ZNF740* | -4.787705978 | 0.000239368 | 0.001404371 | 1 |
| HTN | SMC | *WDFY2* | -4.758522727 | 0.01096307 | 0.021729091 | 1 |
| HTN | SMC | *HNRNPA1L2* | -8.302099287 | 0.000312185 | 0.001641688 | 1 |
| HTN | SMC | *TMX1* | -8.48607272 | 7.49E-05 | 0.000847247 | 0.363209494 |
| HTN | SMC | *C14orf37* | -6.97082171 | 0.000211186 | 0.001315636 | 1 |
| HTN | SMC | *EFCAB11* | -3.500676696 | 0.000450337 | 0.002080231 | 1 |
| HTN | SMC | *SLC24A4* | -3.140689625 | 0.000551019 | 0.002316761 | 1 |
| HTN | SMC | *C14orf79* | -5.8810248 | 0.010003733 | 0.020131942 | 1 |
| HTN | SMC | *WARS* | -5.454312604 | 0.001759728 | 0.005149699 | 1 |
| HTN | SMC | *SERF2* | -7.087338627 | 0.000901388 | 0.00320171 | 1 |
| HTN | SMC | *SRP14* | -5.551727058 | 0.010188214 | 0.020443843 | 1 |
| HTN | SMC | *HMG20A* | -7.230106019 | 0.002202521 | 0.006110139 | 1 |
| HTN | SMC | *SCAPER* | -4.857006685 | 0.000952297 | 0.003321745 | 1 |
| HTN | SMC | *NEIL1* | -4.440488318 | 0.000360257 | 0.001798319 | 1 |
| HTN | SMC | *CYP11A1* | -3.114179928 | 0.001986908 | 0.005650925 | 1 |
| HTN | SMC | *BBS4* | -6.166804714 | 0.000249597 | 0.001438619 | 1 |
| HTN | SMC | *PCSK6* | -6.345900303 | 0.000117376 | 0.000995642 | 0.569506977 |
| HTN | SMC | *POLG* | -8.792624682 | 0.000141953 | 0.0010898 | 0.688753788 |
| HTN | SMC | *MFGE8* | 1.652289629 | 0.013553757 | 0.025840011 | 1 |
| HTN | SMC | *UNC45A* | -11.06514014 | 2.61E-05 | 0.000556439 | 0.126868075 |
| HTN | SMC | *IQGAP1* | -4.134681476 | 0.004357345 | 0.010298022 | 1 |
| HTN | SMC | *ITGAX* | -3.820886292 | 0.000548951 | 0.002313255 | 1 |
| HTN | SMC | *DHX38* | -9.260799593 | 0.002183085 | 0.006073582 | 1 |
| HTN | SMC | *NUDT7* | -6.054437289 | 0.000142738 | 0.001090656 | 0.692566592 |
| HTN | SMC | *GCSH* | -6.685303747 | 0.000189263 | 0.001247553 | 0.918303272 |
| HTN | SMC | *CMTM3* | -6.925914116 | 0.000137876 | 0.001071047 | 0.668975228 |
| HTN | SMC | *ZCCHC14* | -6.160248304 | 0.003455089 | 0.008583765 | 1 |
| HTN | SMC | *TLDC1* | -6.21079242 | 8.22E-05 | 0.000876014 | 0.398652729 |
| HTN | SMC | *TIGD7* | -5.758979646 | 0.017655321 | 0.032107803 | 1 |
| HTN | SMC | *DEF8* | 3.952902943 | 0.024665232 | 0.042894518 | 1 |
| HTN | SMC | *GALNS* | -5.802023236 | 0.000172474 | 0.001187009 | 0.836841589 |
| HTN | SMC | *NCOR1* | -5.482609564 | 0.001618334 | 0.004831758 | 1 |
| HTN | SMC | *KSR1* | -7.895914595 | 0.000157846 | 0.001140149 | 0.765871212 |
| HTN | SMC | *GFOD2* | -4.355923858 | 0.002789636 | 0.00731673 | 1 |
| HTN | SMC | *PRPSAP2* | -3.847877847 | 0.010113588 | 0.020326147 | 1 |
| HTN | SMC | *RASL10B* | -8.566659352 | 0.000193096 | 0.001260968 | 0.936899443 |
| HTN | SMC | *UNC45B* | 3.86272699 | 0.003248847 | 0.008197298 | 1 |
| HTN | SMC | *C17orf80* | -6.133509366 | 6.46E-05 | 0.000829756 | 0.313211127 |

| | | | | | | |
|---|---|---|---|---|---|---|
| HTN | SMC | *VPS53* | -6.927335778 | 1.72E-06 | 0.000103291 | 0.008366541 |
| HTN | SMC | *SGSM2* | -7.174074803 | 0.0005646 | 0.002345411 | 1 |
| HTN | SMC | *LRRC46* | -4.674406272 | 0.000807083 | 0.00297081 | 1 |
| HTN | SMC | *SCRN2* | -1.812831286 | 1.11E-05 | 0.000347754 | 0.053901846 |
| HTN | SMC | *SSH2* | -5.560226897 | 0.000120807 | 0.001013054 | 0.586155341 |
| HTN | SMC | *RHBDL3* | -5.466652229 | 0.000149529 | 0.00112132 | 0.725512805 |
| HTN | SMC | *G6PC3* | -6.618305251 | 0.01280267 | 0.024611155 | 1 |
| HTN | SMC | *CLTC* | -4.427163179 | 0.001507311 | 0.004634646 | 1 |
| HTN | SMC | *IMPA2* | -6.246432554 | 0.012613944 | 0.024315794 | 1 |
| HTN | SMC | *GALNT1* | -6.594483944 | 0.00071982 | 0.002726438 | 1 |
| HTN | SMC | *C18orf8* | -6.971466647 | 0.000109217 | 0.000963489 | 0.529918979 |
| HTN | SMC | *NPC1* | -9.452076409 | 7.72E-07 | 5.77E-05 | 0.003744422 |
| HTN | SMC | *CCDC40* | -3.97701294 | 0.004062331 | 0.009719522 | 1 |
| HTN | SMC | *CARD14* | -3.255661614 | 0.001779492 | 0.005185642 | 1 |
| HTN | SMC | *TBCD* | -5.829277704 | 0.003605591 | 0.008866868 | 1 |
| HTN | SMC | *NARF* | -5.762554081 | 0.000238801 | 0.001402741 | 1 |
| HTN | SMC | *TRIM65* | -7.893102169 | 1.23E-05 | 0.000374297 | 0.05944563 |
| HTN | SMC | *SECTM1* | -2.482242322 | 0.015456687 | 0.028789192 | 1 |
| HTN | SMC | *RNF157* | -3.508475709 | 0.000477119 | 0.002157486 | 1 |
| HTN | SMC | *ZNF750* | -8.735536259 | 0.018704693 | 0.033737982 | 1 |
| HTN | SMC | *WDR45B* | -7.829701907 | 0.000325638 | 0.00168443 | 1 |
| HTN | SMC | *RNF165* | -4.400135817 | 2.10E-05 | 0.000499016 | 0.101799334 |
| HTN | SMC | *MAPK4* | 1.146448333 | 0.003199115 | 0.008114012 | 1 |
| HTN | SMC | *CBLN2* | -8.990717661 | 0.000117047 | 0.000995642 | 0.567913839 |
| HTN | SMC | *PNMT* | -4.918607311 | 0.025973604 | 0.044784621 | 1 |
| HTN | SMC | *TXNL4A* | -7.008283322 | 0.000949413 | 0.003316451 | 1 |
| HTN | SMC | *CACNA1A* | 0.687117326 | 0.004376935 | 0.010334253 | 1 |
| HTN | SMC | *SAMD1* | -6.698001334 | 8.18E-05 | 0.000876014 | 0.397033083 |
| HTN | SMC | *NFIC* | -5.330371142 | 0.000570117 | 0.00236226 | 1 |
| HTN | SMC | *PRDM15* | -2.952115543 | 0.007449532 | 0.015874013 | 1 |
| HTN | SMC | *PFKL* | -7.288802791 | 4.50E-05 | 0.000754445 | 0.218417905 |
| HTN | SMC | *MVB12A* | -3.590776028 | 0.002198722 | 0.006106581 | 1 |
| HTN | SMC | *DPP9* | -7.208822638 | 0.012685985 | 0.024426069 | 1 |
| HTN | SMC | *HUNK* | -1.203972961 | 0.009863424 | 0.019882565 | 1 |
| HTN | SMC | *IFNAR1* | -3.154531166 | 0.016352694 | 0.03012273 | 1 |
| HTN | SMC | *SAE1* | -7.689284238 | 0.001441581 | 0.004500999 | 1 |
| HTN | SMC | *GEMIN7* | -4.373645624 | 0.015986013 | 0.029564609 | 1 |
| HTN | SMC | *CAPN10* | -2.533654169 | 0.02468415 | 0.042912038 | 1 |
| HTN | SMC | *EVI5L* | -12.2454856 | 0.000131016 | 0.001043376 | 0.635689393 |
| HTN | SMC | *CTU1* | -14.67731835 | 0.000103355 | 0.000942633 | 0.501480789 |
| HTN | SMC | *RERE* | -3.753759818 | 0.016760539 | 0.030757238 | 1 |
| HTN | SMC | *MMEL1* | -1.869995201 | 0.008464449 | 0.017602008 | 1 |
| HTN | SMC | *PEX14* | -4.279016124 | 0.00098824 | 0.003408073 | 1 |
| HTN | SMC | *C1orf216* | -2.65190079 | 0.009097555 | 0.018632898 | 1 |
| HTN | SMC | *PLK4* | -3.957844519 | 0.013336691 | 0.025466205 | 1 |
| HTN | SMC | *RPS8* | -6.21300784 | 0.002091574 | 0.005869471 | 1 |
| HTN | SMC | *STRIP1* | -9.828080238 | 0.000318182 | 0.001657346 | 1 |
| HTN | SMC | *ALDH9A1* | -9.084547383 | 5.89E-05 | 0.000817716 | 0.286003388 |
| HTN | SMC | *ATP1B1* | -4.490404106 | 0.00284818 | 0.007421787 | 1 |
| HTN | SMC | *TIPRL* | -5.721840768 | 0.000960585 | 0.003338651 | 1 |

| | | | | | | |
|---|---|---|---|---|---|---|
| HTN | SMC | *NME7* | -8.438994992 | 4.24E-07 | 3.81E-05 | 0.002054869 |
| HTN | SMC | *MAEL* | -28.99519816 | 0.002000546 | 0.005683051 | 1 |
| HTN | SMC | *RFWD2* | -6.062500363 | 0.000238245 | 0.001401168 | 1 |
| HTN | SMC | *TARS2* | -3.995560756 | 0.002930449 | 0.007575141 | 1 |
| HTN | SMC | *SETDB1* | -7.204954574 | 0.000246277 | 0.001427639 | 1 |
| HTN | SMC | *SEMA6C* | -4.634968853 | 0.004874116 | 0.011202848 | 1 |
| HTN | SMC | *ARNT* | -8.28142435 | 5.32E-07 | 4.61E-05 | 0.002581605 |
| HTN | SMC | *EIF2D* | -7.316466367 | 0.004978947 | 0.011395213 | 1 |
| HTN | SMC | *SMYD2* | -4.528434647 | 0.00013139 | 0.001043376 | 0.637502764 |
| HTN | SMC | *S100A8* | -7.703635827 | 6.98E-05 | 0.000837934 | 0.338669597 |
| HTN | SMC | *CREB3L4* | -10.22092012 | 5.03E-10 | 2.03E-07 | 2.44E-06 |
| HTN | SMC | *KCNN3* | -3.347904705 | 0.000598103 | 0.002424389 | 1 |
| HTN | SMC | *C1orf43* | -10.31264506 | 2.74E-06 | 0.000141251 | 0.013287297 |
| HTN | SMC | *RIT1* | -8.532178203 | 0.009087104 | 0.018619353 | 1 |
| HTN | SMC | *FLG* | -1.353103834 | 0.013162075 | 0.025212154 | 1 |
| HTN | SMC | *SCCPDH* | -9.339794789 | 0.00026478 | 0.00148866 | 1 |
| HTN | SMC | *CEP170* | -4.579538083 | 0.01062678 | 0.021183704 | 1 |
| HTN | SMC | *ACP1* | -8.814591271 | 0.001345743 | 0.004265069 | 1 |
| HTN | SMC | *CNIH4* | -8.624239352 | 0.000865816 | 0.003112541 | 1 |
| HTN | SMC | *PYCR2* | -3.939643919 | 0.009908171 | 0.019956183 | 1 |
| HTN | SMC | *GALM* | -8.544192134 | 0.000299424 | 0.001597571 | 1 |
| HTN | SMC | *WDPCP* | -3.518039297 | 0.000891588 | 0.003180872 | 1 |
| HTN | SMC | *VPS54* | -6.292366169 | 0.001332395 | 0.004244769 | 1 |
| HTN | SMC | *ABHD1* | -3.270469444 | 7.51E-05 | 0.000847247 | 0.364312754 |
| HTN | SMC | *ZNF514* | -6.935993187 | 6.00E-06 | 0.000227565 | 0.029128309 |
| HTN | SMC | *EXOC6B* | -5.32792347 | 0.001150094 | 0.003798678 | 1 |
| HTN | SMC | *TEX261* | -11.99957677 | 5.56E-05 | 0.000817658 | 0.269703051 |
| HTN | SMC | *THNSL2* | -2.727576313 | 0.000153485 | 0.001136201 | 0.744709752 |
| HTN | SMC | *RALB* | -9.184979924 | 0.000110568 | 0.00097012 | 0.536476318 |
| HTN | SMC | *RABL2A* | -6.670447994 | 2.61E-05 | 0.000556439 | 0.126730006 |
| HTN | SMC | *LIPT1* | -2.093719772 | 0.011549768 | 0.022697236 | 1 |
| HTN | SMC | *FAHD2B* | -3.876901543 | 0.001629231 | 0.00485269 | 1 |
| HTN | SMC | *POLR2D* | -3.715287224 | 0.009764075 | 0.019698665 | 1 |
| HTN | SMC | *GALNT13* | -3.548475607 | 0.000138826 | 0.001072583 | 0.673582281 |
| HTN | SMC | *METTL21A* | -5.360609805 | 0.006423898 | 0.014058978 | 1 |
| HTN | SMC | *SUMF1* | -5.669152007 | 0.000651995 | 0.002547086 | 1 |
| HTN | SMC | *ESPNL* | -29.25783714 | 0.000542947 | 0.002300768 | 1 |
| HTN | SMC | *ANKMY1* | -4.453253534 | 0.005765561 | 0.01286184 | 1 |
| HTN | SMC | *STK11IP* | -5.551978683 | 0.002285041 | 0.006288724 | 1 |
| HTN | SMC | *CTDSPL* | -6.607358872 | 0.000347892 | 0.001761973 | 1 |
| HTN | SMC | *SLC25A26* | -6.860443302 | 0.000176859 | 0.001202931 | 0.858120778 |
| HTN | SMC | *ARL6IP5* | -7.509207656 | 0.000864648 | 0.003112219 | 1 |
| HTN | SMC | *PLA1A* | -8.483358411 | 0.001512824 | 0.004641266 | 1 |
| HTN | SMC | *BOC* | -3.53967632 | 0.000130052 | 0.00104255 | 0.631014201 |
| HTN | SMC | *EIF2A* | -4.075094682 | 5.47E-05 | 0.000811912 | 0.265495223 |
| HTN | SMC | *NCEH1* | -4.138582892 | 0.002875223 | 0.007460204 | 1 |
| HTN | SMC | *AMT* | -1.986414412 | 4.65E-05 | 0.000760128 | 0.225758086 |
| HTN | SMC | *CCDC39* | -6.405199848 | 1.63E-05 | 0.000427306 | 0.079172476 |
| HTN | SMC | *MUC4* | -2.838860399 | 0.000309923 | 0.001638066 | 1 |
| HTN | SMC | *CENPC* | -6.307688933 | 5.21E-05 | 0.000801884 | 0.252593458 |

| | | | | | | |
|---|---|---|---|---|---|---|
| HTN | SMC | *PYURF* | -8.424310548 | 0.009243902 | 0.018868915 | 1 |
| HTN | SMC | *DDIT4L* | 2.952569032 | 0.002873274 | 0.007460204 | 1 |
| HTN | SMC | *METTL14* | -6.840706391 | 0.008358828 | 0.017436386 | 1 |
| HTN | SMC | *USP53* | -4.06868773 | 0.00385676 | 0.009342086 | 1 |
| HTN | SMC | *MARCH1* | -3.507396839 | 0.005216571 | 0.011844081 | 1 |
| HTN | SMC | *MARCH6* | -5.57701709 | 0.005176722 | 0.01177013 | 1 |
| HTN | SMC | *GTF2H2* | -13.26792549 | 0.005530769 | 0.012412252 | 1 |
| HTN | SMC | *SPATA9* | -3.250741955 | 0.000167259 | 0.001169111 | 0.811539664 |
| HTN | SMC | *TSLP* | -1.908406189 | 0.000163609 | 0.001162037 | 0.793831635 |
| HTN | SMC | *FEM1C* | -10.89355499 | 0.000984864 | 0.003401109 | 1 |
| HTN | SMC | *COMMD10* | -4.339215205 | 0.001575685 | 0.004740559 | 1 |
| HTN | SMC | *YIPF5* | -8.088045086 | 0.000291761 | 0.001572918 | 1 |
| HTN | SMC | *CXCL14* | -4.556332074 | 0.002059147 | 0.005807146 | 1 |
| HTN | SMC | *FBXO38* | -6.399465484 | 0.000326744 | 0.001688351 | 1 |
| HTN | SMC | *BOD1* | -4.874781017 | 0.002850171 | 0.00742299 | 1 |
| HTN | SMC | *FAM50B* | -13.36090372 | 0.001513818 | 0.004641266 | 1 |
| HTN | SMC | *MYLK4* | -3.824445186 | 0.001253159 | 0.004045459 | 1 |
| HTN | SMC | *FAM193B* | -8.359092744 | 8.11E-06 | 0.000285237 | 0.039362725 |
| HTN | SMC | *MUT* | -5.897954773 | 0.010579819 | 0.021107434 | 1 |
| HTN | SMC | *DOK3* | -2.342409403 | 0.000163999 | 0.001162037 | 0.795722241 |
| HTN | SMC | *MLIP* | -4.263009754 | 0.021587873 | 0.038088858 | 1 |
| HTN | SMC | *ANO7* | -2.634627245 | 0.014017723 | 0.026547225 | 1 |
| HTN | SMC | *RPL7L1* | -10.6898089 | 0.000100414 | 0.000932454 | 0.487210635 |
| HTN | SMC | *TPBG* | -6.969084739 | 0.00039714 | 0.001907844 | 1 |
| HTN | SMC | *PHIP* | -4.806288523 | 0.009228597 | 0.018848761 | 1 |
| HTN | SMC | *RNF217* | -6.442018369 | 0.000184313 | 0.001226806 | 0.894286536 |
| HTN | SMC | *ARHGAP18* | -3.935274654 | 0.000538273 | 0.002288957 | 1 |
| HTN | SMC | *SLC18B1* | -5.997973718 | 2.68E-07 | 2.76E-05 | 0.001298178 |
| HTN | SMC | *WTAP* | -4.218323135 | 0.01181076 | 0.02305141 | 1 |
| HTN | SMC | *SLC22A3* | 0.845442762 | 0.005958369 | 0.013212983 | 1 |
| HTN | SMC | *GNA12* | -7.568157958 | 0.000110541 | 0.00097012 | 0.536347076 |
| HTN | SMC | *CCZ1B* | -3.514088291 | 0.007610882 | 0.016118724 | 1 |
| HTN | SMC | *RBAK* | -5.920698046 | 0.000182937 | 0.001222605 | 0.887611129 |
| HTN | SMC | *EPHA1* | -26.77320043 | 7.35E-05 | 0.000847247 | 0.356596207 |
| HTN | SMC | *FBXO25* | -6.729097658 | 0.008845773 | 0.018224921 | 1 |
| HTN | SMC | *BIN3* | -6.989987839 | 0.00385852 | 0.009342086 | 1 |
| HTN | SMC | *DOCK5* | -4.873291625 | 0.00010051 | 0.000932454 | 0.487673362 |
| HTN | SMC | *TACC1* | -3.910425204 | 0.024241863 | 0.04221878 | 1 |
| HTN | SMC | *ADHFE1* | -2.562926417 | 2.18E-06 | 0.000122783 | 0.010559373 |
| HTN | SMC | *ATP6V0D2* | -5.756113084 | 0.01042027 | 0.020831953 | 1 |
| HTN | SMC | *DPYS* | -3.350582934 | 0.000133108 | 0.00105357 | 0.645838702 |
| HTN | SMC | *LRP12* | -10.28735467 | 0.000280309 | 0.001540273 | 1 |
| HTN | SMC | *ZNF462* | -3.762407503 | 0.026253377 | 0.045234868 | 1 |
| HTN | SMC | *ALAD* | -1.80389715 | 0.003591815 | 0.008837469 | 1 |
| HTN | SMC | *SURF6* | -6.795877638 | 0.003308239 | 0.008325507 | 1 |
| HTN | SMC | *MED22* | -4.159304625 | 0.000920232 | 0.003251201 | 1 |
| HTN | SMC | *SEC16A* | -5.285358122 | 0.024512369 | 0.042674566 | 1 |
| HTN | SMC | *SLC39A12* | -4.172182148 | 0.006225238 | 0.013723242 | 1 |
| HTN | SMC | *ST8SIA6* | -9.770982432 | 0.000230185 | 0.001378826 | 1 |
| HTN | SMC | *RGR* | -19.48719972 | 0.00054139 | 0.002296175 | 1 |

| | | | | | | |
|---|---|---|---|---|---|---|
| HTN | SMC | *ADD3* | -6.006794069 | 0.000112045 | 0.000976019 | 0.543642703 |
| HTN | SMC | *PAOX* | -13.52153873 | 0.000498783 | 0.002206163 | 1 |
| HTN | SMC | *PDCD11* | -6.094226973 | 0.002080652 | 0.005848722 | 1 |
| HTN | SMC | *RGS10* | -4.271248666 | 0.000404822 | 0.001929517 | 1 |
| HTN | SMC | *GAS2* | -7.047992478 | 0.000119583 | 0.001005578 | 0.580218275 |
| HTN | SMC | *LIN7C* | -8.140201175 | 5.63E-05 | 0.000817716 | 0.27293509 |
| HTN | SMC | *HSD17B12* | -3.884057376 | 5.32E-11 | 3.69E-08 | 2.58E-07 |
| HTN | SMC | *EIF3M* | -9.052286928 | 3.54E-06 | 0.000162031 | 0.017175283 |
| HTN | SMC | *TNKS1BP1* | 3.625841503 | 0.000432649 | 0.002016536 | 1 |
| HTN | SMC | *PTPRJ* | -6.559201526 | 1.90E-05 | 0.000473276 | 0.092098275 |
| HTN | SMC | *INTS4* | -5.84898728 | 0.000523303 | 0.002257599 | 1 |
| HTN | SMC | *DRD2* | -18.18326072 | 0.00051373 | 0.002236542 | 1 |
| HTN | SMC | *NPAT* | -8.503753325 | 0.000336077 | 0.001721906 | 1 |
| HTN | SMC | *ATM* | -5.724791832 | 0.00066337 | 0.002577581 | 1 |
| HTN | SMC | *SLX4IP* | -5.566724321 | 0.000235157 | 0.001389748 | 1 |
| HTN | SMC | *ST14* | -4.364536376 | 0.000498797 | 0.002206163 | 1 |
| HTN | SMC | *FADS1* | -5.937701917 | 0.005830542 | 0.012976968 | 1 |
| HTN | SMC | *TMC2* | -4.049244693 | 0.000362655 | 0.001802871 | 1 |
| HTN | SMC | *INCENP* | -9.674254542 | 4.23E-06 | 0.000181808 | 0.020544345 |
| HTN | SMC | *PLCH2* | -6.623132121 | 0.027868606 | 0.047628911 | 1 |
| HTN | SMC | *CPSF7* | -6.490870054 | 0.003318485 | 0.008334 | 1 |
| HTN | SMC | *CCDC15* | -3.214280466 | 0.001215992 | 0.003949124 | 1 |
| HTN | SMC | *DUSP15* | 7.227424927 | 0.000523008 | 0.002257599 | 1 |
| HTN | SMC | *SPATA25* | -4.791366072 | 0.001103405 | 0.003692221 | 1 |
| HTN | SMC | *CABLES2* | -3.729473534 | 0.000196794 | 0.001271662 | 0.954846126 |
| HTN | SMC | *TRPT1* | -5.261778291 | 1.68E-06 | 0.000102152 | 0.008172184 |
| HTN | SMC | *ALDOA* | -5.093324072 | 0.014575837 | 0.027432878 | 1 |
| HTN | SMC | *TMEM218* | -7.085710903 | 0.00068373 | 0.0026455 | 1 |
| HTN | SMC | *SAP18* | -9.714323893 | 0.000511924 | 0.002236542 | 1 |
| HTN | SMC | *FAM124A* | -7.566360894 | 0.000995628 | 0.003430956 | 1 |
| HTN | SMC | *LYPD6B* | -2.606694714 | 0.010153875 | 0.020383368 | 1 |
| HTN | SMC | *RGS18* | -2.020345237 | 0.019650464 | 0.035208291 | 1 |
| HTN | SMC | *PRSS23* | -4.379388179 | 0.00026634 | 0.001490521 | 1 |
| HTN | SMC | *PPP1R1C* | -10.89833105 | 8.53E-05 | 0.000876014 | 0.413768748 |
| HTN | SMC | *FAM173B* | -6.880466088 | 0.001938397 | 0.005538928 | 1 |
| HTN | SMC | *GPR158* | -7.902944557 | 0.004499289 | 0.010561467 | 1 |
| HTN | SMC | *PLBD2* | -9.075645602 | 3.16E-05 | 0.000616361 | 0.153474002 |
| HTN | SMC | *EXT2* | -3.818410793 | 0.016839868 | 0.030855634 | 1 |
| HTN | SMC | *NDUFC2* | -8.967172145 | 0.011102568 | 0.021960725 | 1 |
| HTN | SMC | *ADAMTS12* | -4.7042631 | 3.22E-06 | 0.000153937 | 0.015619857 |
| HTN | SMC | *VIPAS39* | -9.016167859 | 0.00131712 | 0.0042127 | 1 |
| HTN | SMC | *UPF2* | -6.876700576 | 0.000219849 | 0.001344061 | 1 |
| HTN | SMC | *PTPRO* | -3.546926088 | 0.001173552 | 0.003855163 | 1 |
| HTN | SMC | *FAM160B1* | -7.24853072 | 0.00041632 | 0.001964964 | 1 |
| HTN | SMC | *ZNF827* | -8.142137829 | 2.46E-05 | 0.000537075 | 0.119230737 |
| HTN | SMC | *AKR1C2* | -2.870198225 | 0.002917959 | 0.007546874 | 1 |
| HTN | SMC | *PIGF* | -8.348877126 | 0.000472546 | 0.002146812 | 1 |
| HTN | SMC | *INPP1* | -8.451335437 | 8.98E-05 | 0.000888243 | 0.435622867 |
| HTN | SMC | *MFSD6* | -7.469922065 | 0.003658719 | 0.008981338 | 1 |
| HTN | SMC | *RNF144A* | -7.632751017 | 0.000106792 | 0.000950786 | 0.51815607 |

| | | | | | | |
|---|---|---|---|---|---|---|
| HTN | SMC | *BICD1* | -7.236309287 | 0.000153783 | 0.001136201 | 0.746154836 |
| HTN | SMC | *GABRA2* | -6.392335008 | 0.000218205 | 0.001341927 | 1 |
| HTN | SMC | *PARP8* | -3.583343271 | 0.00034192 | 0.001737169 | 1 |
| HTN | SMC | *CACUL1* | -6.46015898 | 0.000462809 | 0.002113438 | 1 |
| HTN | SMC | *TIAL1* | -6.670247322 | 0.002813639 | 0.007340933 | 1 |
| HTN | SMC | *PTPN14* | -5.34345172 | 0.003861921 | 0.009345656 | 1 |
| HTN | SMC | *MGAT5* | -6.602490832 | 0.000143027 | 0.001090942 | 0.693964862 |
| HTN | SMC | *TMEM178A* | -3.632713841 | 0.000591738 | 0.002410673 | 1 |
| HTN | SMC | *EPG5* | -7.773458322 | 0.000157774 | 0.001140149 | 0.765520246 |
| HTN | SMC | *KCNK13* | -3.083266857 | 0.001895568 | 0.005435755 | 1 |
| HTN | SMC | *NMT2* | -6.42519133 | 0.00026089 | 0.001475067 | 1 |
| HTN | SMC | *CCDC50* | -2.625811893 | 0.008519727 | 0.017703519 | 1 |
| HTN | SMC | *PLEKHH2* | -3.800442922 | 0.000168328 | 0.001171775 | 0.816727493 |
| HTN | SMC | *MBNL1* | -3.855267788 | 0.01134486 | 0.02236625 | 1 |
| HTN | SMC | *GPD1L* | -4.798584717 | 0.002294938 | 0.00629922 | 1 |
| HTN | SMC | *CLEC4F* | -2.964463908 | 0.013662885 | 0.025996781 | 1 |
| HTN | SMC | *SLC30A6* | -4.693283738 | 0.009225888 | 0.018848761 | 1 |
| HTN | SMC | *RASGRP3* | -2.730699933 | 0.000445606 | 0.002064973 | 1 |
| HTN | SMC | *GPR180* | -2.735865058 | 0.001857665 | 0.005365113 | 1 |
| HTN | SMC | *TCTEX1D1* | -4.289332979 | 0.006458599 | 0.014128548 | 1 |
| HTN | SMC | *PRDM8* | -5.764625606 | 1.80E-06 | 0.00010626 | 0.008713356 |
| HTN | SMC | *MED21* | -3.522538805 | 0.022043456 | 0.038822087 | 1 |
| HTN | SMC | *JAKMIP1* | -9.292994118 | 9.92E-05 | 0.000925638 | 0.481331673 |
| HTN | SMC | *SREK1IP1* | -7.497961862 | 0.002499576 | 0.006704226 | 1 |
| HTN | SMC | *CENPH* | -5.159577929 | 8.78E-05 | 0.000888243 | 0.426225952 |
| HTN | SMC | *CARHSP1* | -4.38162691 | 0.011311997 | 0.022311305 | 1 |
| HTN | SMC | *CAST* | -6.83482748 | 3.05E-06 | 0.000150762 | 0.014774673 |
| HTN | SMC | *SCOC* | -5.190590431 | 0.016165703 | 0.029812236 | 1 |
| HTN | SMC | *CLGN* | -5.698476124 | 0.011141165 | 0.022028089 | 1 |
| HTN | SMC | *CETN3* | -4.634982633 | 0.000361324 | 0.00179994 | 1 |
| HTN | SMC | *SYCP2L* | -3.203806956 | 0.005341075 | 0.012081537 | 1 |
| HTN | SMC | *MERTK* | -0.621025136 | 0.012306191 | 0.023826672 | 1 |
| HTN | SMC | *TMEM87B* | -7.987938011 | 0.000440643 | 0.002049857 | 1 |
| HTN | SMC | *CFDP1* | -6.366363931 | 0.001562514 | 0.004723562 | 1 |
| HTN | SMC | *ZDHHC7* | -7.907105879 | 7.57E-05 | 0.000849747 | 0.36738589 |
| HTN | SMC | *JAZF1* | -5.846666949 | 2.40E-05 | 0.000537036 | 0.116536769 |
| HTN | SMC | *FBXO36* | -8.752824236 | 0.000429281 | 0.002004689 | 1 |
| HTN | SMC | *MCOLN2* | -3.125527523 | 0.025527863 | 0.044188795 | 1 |
| HTN | SMC | *GDPD1* | -10.92880595 | 9.51E-05 | 0.000903 | 0.461432835 |
| HTN | SMC | *SEMA3D* | -4.514543337 | 7.54E-05 | 0.000848694 | 0.365787296 |
| HTN | SMC | *DNAAF1* | -5.865828674 | 8.64E-05 | 0.000878791 | 0.419183179 |
| HTN | SMC | *ANKH* | -7.136450178 | 0.000229294 | 0.001377231 | 1 |
| HTN | SMC | *UBASH3B* | -2.674069573 | 0.012763126 | 0.024554594 | 1 |
| HTN | SMC | *ROBO4* | -8.10417149 | 7.48E-05 | 0.000847247 | 0.363082298 |
| HTN | SMC | *ROBO3* | -3.645679862 | 0.020385369 | 0.036243975 | 1 |
| HTN | SMC | *LRRK1* | -8.633754714 | 3.90E-05 | 0.000689568 | 0.189098595 |
| HTN | SMC | *ENPP3* | -5.7145963 | 9.90E-05 | 0.000925598 | 0.480385577 |
| HTN | SMC | *MIA3* | -5.63091696 | 0.009311893 | 0.018951891 | 1 |
| HTN | SMC | *NEIL2* | 7.10489274 | 2.08E-08 | 3.74E-06 | 0.000100891 |
| HTN | SMC | *PGM5* | -6.258495203 | 0.000584936 | 0.002395942 | 1 |

| | | | | | | |
|---|---|---|---|---|---|---|
| HTN | SMC | *CCSAP* | -12.97592433 | 9.66E-05 | 0.000910242 | 0.468774882 |
| HTN | SMC | *LY96* | -3.453386243 | 0.003295007 | 0.008296509 | 1 |
| HTN | SMC | *PDE1C* | 1.196094672 | 0.009759486 | 0.019697597 | 1 |
| HTN | SMC | *XPC* | -8.675708014 | 7.76E-05 | 0.000851198 | 0.376526238 |
| HTN | SMC | *FLCN* | -5.310500857 | 0.00118506 | 0.0038841 | 1 |
| HTN | SMC | *OXNAD1* | -7.078360167 | 0.000230743 | 0.001378826 | 1 |
| HTN | SMC | *APCDD1* | -2.200214221 | 0.004738146 | 0.010988893 | 1 |
| HTN | SMC | *MPPE1* | -2.216619113 | 0.012698373 | 0.024439708 | 1 |
| HTN | SMC | *RAB6B* | -4.924580571 | 0.008791044 | 0.01816616 | 1 |
| HTN | SMC | *CYP2U1* | -3.833793604 | 0.003824299 | 0.009310336 | 1 |
| HTN | SMC | *PROM2* | -8.271560008 | 1.72E-05 | 0.000441573 | 0.083358869 |
| HTN | SMC | *MARCKS* | -4.749869126 | 0.000974395 | 0.003376975 | 1 |
| HTN | SMC | *GOLGA7B* | -29.88502772 | 0.000280677 | 0.001540546 | 1 |
| HTN | SMC | *TRMT44* | -9.469210156 | 3.97E-05 | 0.000693618 | 0.192602441 |
| HTN | SMC | *CNOT8* | -9.134154476 | 0.000368379 | 0.001812751 | 1 |
| HTN | SMC | *LRGUK* | -2.295353552 | 0.009195491 | 0.018809663 | 1 |
| HTN | SMC | *NUP205* | -8.346002298 | 0.000145007 | 0.001097623 | 0.703576356 |
| HTN | SMC | *ZKSCAN2* | -3.36400855 | 0.000404163 | 0.001929517 | 1 |
| HTN | SMC | *PDIA4* | -6.738271254 | 0.000105823 | 0.000950786 | 0.513454468 |
| HTN | SMC | *KDM8* | -9.041932401 | 0.007620956 | 0.016133018 | 1 |
| HTN | SMC | *ALS2CR12* | -6.496129296 | 0.008389506 | 0.017485346 | 1 |
| HTN | SMC | *DEPTOR* | -2.405782151 | 0.02533508 | 0.043902074 | 1 |
| HTN | SMC | *RRAGA* | -11.05755975 | 0.001193668 | 0.003900118 | 1 |
| HTN | SMC | *TMBIM4* | -4.76846502 | 0.00036701 | 0.001809686 | 1 |
| HTN | SMC | *PSD3* | -3.290740904 | 0.001925212 | 0.005504497 | 1 |
| HTN | SMC | *MMP16* | -2.301269569 | 0.001338396 | 0.004255399 | 1 |
| HTN | SMC | *NDUFAF6* | -9.485668208 | 8.16E-10 | 3.05E-07 | 3.96E-06 |
| HTN | SMC | *C8orf37* | -2.937815746 | 0.000285967 | 0.001556812 | 1 |
| HTN | SMC | *WHAMM* | -2.601316374 | 0.020675835 | 0.036706604 | 1 |
| HTN | SMC | *N6AMT1* | -4.665928985 | 0.000383005 | 0.001850936 | 1 |
| HTN | SMC | *CCT8* | -1.906226157 | 0.015690619 | 0.029146587 | 1 |
| HTN | SMC | *NAA11* | 0.609214036 | 0.004316246 | 0.010210836 | 1 |
| HTN | SMC | *SFXN2* | 2.901929635 | 5.92E-05 | 0.000817716 | 0.287018167 |
| HTN | SMC | *SUPV3L1* | -7.604989052 | 0.004676801 | 0.010897172 | 1 |
| HTN | SMC | *NODAL* | -11.91447447 | 5.81E-05 | 0.000817716 | 0.282124261 |
| HTN | SMC | *ZDHHC5* | -9.221748728 | 0.001514246 | 0.004641266 | 1 |
| HTN | SMC | *ATAD2* | -3.746490935 | 0.011808288 | 0.02305141 | 1 |
| HTN | SMC | *FBRS* | -4.478084659 | 0.028354665 | 0.048357411 | 1 |
| HTN | SMC | *SASS6* | -6.476819582 | 0.000616067 | 0.002454805 | 1 |
| HTN | SMC | *MPV17L* | 4.751128286 | 0.001328587 | 0.004237371 | 1 |
| HTN | SMC | *GHRL* | -4.795770176 | 0.010982825 | 0.021759357 | 1 |
| HTN | SMC | *SLC6A1* | 0.670654109 | 0.002664906 | 0.007088884 | 1 |
| HTN | SMC | *NECAP2* | -9.419865956 | 0.00019649 | 0.001271662 | 0.953370055 |
| HTN | SMC | *STEAP2* | -2.718627785 | 0.000340668 | 0.001734441 | 1 |
| HTN | SMC | *CLDN12* | -8.555970493 | 0.028058917 | 0.047920403 | 1 |
| HTN | SMC | *MMP14* | -7.612517559 | 0.000293798 | 0.001578634 | 1 |
| HTN | SMC | *GATAD1* | -6.72339882 | 0.001494089 | 0.004620344 | 1 |
| HTN | SMC | *TMED6* | -10.38766709 | 0.002093426 | 0.005871274 | 1 |
| HTN | SMC | *DHRS4* | -7.840576893 | 7.47E-05 | 0.000847247 | 0.362324871 |
| HTN | SMC | *AASDH* | -6.032325261 | 0.003228116 | 0.008153471 | 1 |

| | | | | | | |
|---|---|---|---|---|---|---|
| HTN | SMC | *SLC38A10* | -7.197721467 | 6.46E-05 | 0.000829756 | 0.313647782 |
| HTN | SMC | *DGKI* | -2.255105446 | 0.000573163 | 0.002363597 | 1 |
| HTN | SMC | *SLC34A2* | -35.97136519 | 0.005224408 | 0.011856327 | 1 |
| HTN | SMC | *SPPL3* | -5.488748044 | 0.002197917 | 0.006106581 | 1 |
| HTN | SMC | *PEX10* | -6.340749151 | 0.003621171 | 0.008896162 | 1 |
| HTN | SMC | *KRTCAP3* | -2.190653239 | 0.004611937 | 0.010780914 | 1 |
| HTN | SMC | *PAFAH2* | -3.218153873 | 0.018512385 | 0.033405799 | 1 |
| HTN | SMC | *EXTL1* | -32.85474676 | 0.000555923 | 0.002326902 | 1 |
| HTN | SMC | *WDR66* | -2.018858827 | 0.02705252 | 0.046397607 | 1 |
| HTN | SMC | *LRRC43* | -6.406377147 | 1.33E-05 | 0.000389012 | 0.06446547 |
| HTN | SMC | *EYA3* | -7.108376068 | 0.000156674 | 0.001140149 | 0.760183725 |
| HTN | SMC | *DZIP1L* | -4.943371589 | 0.001108858 | 0.003697718 | 1 |
| HTN | SMC | *ESYT3* | -4.514855427 | 0.000616666 | 0.002454805 | 1 |
| HTN | SMC | *COLEC12* | -2.804980516 | 0.004367475 | 0.010316938 | 1 |
| HTN | SMC | *MITD1* | -5.791852715 | 0.00250967 | 0.006721936 | 1 |
| HTN | SMC | *TSPAN33* | -7.715819368 | 1.90E-05 | 0.000473276 | 0.092288809 |
| HTN | SMC | *FAM86C1* | -21.13262701 | 0.007295797 | 0.015587498 | 1 |
| HTN | SMC | *DYNC1I1* | 0.967394925 | 0.006604012 | 0.014401198 | 1 |
| HTN | SMC | *ELK4* | -7.089518173 | 0.009230143 | 0.018848761 | 1 |
| HTN | SMC | *DUSP23* | -5.255243829 | 0.00087664 | 0.003136759 | 1 |
| HTN | SMC | *RNF166* | -3.387509536 | 8.86E-05 | 0.000888243 | 0.429697243 |
| HTN | SMC | *F11R* | -3.044251973 | 0.001017547 | 0.003479625 | 1 |
| HTN | SMC | *SPATA2L* | -3.237400117 | 0.017164864 | 0.031345096 | 1 |
| HTN | SMC | *FGF17* | -11.54655144 | 0.001197958 | 0.003911503 | 1 |
| HTN | SMC | *SLC5A11* | -2.781229949 | 2.65E-06 | 0.000139919 | 0.012875451 |
| HTN | SMC | *CCAR2* | -6.596506187 | 0.006530427 | 0.014272806 | 1 |
| HTN | SMC | *CDC42SE2* | -8.098140824 | 8.13E-07 | 5.98E-05 | 0.003946192 |
| HTN | SMC | *IFNAR2* | -2.929884184 | 0.000506961 | 0.002226042 | 1 |
| HTN | SMC | *SV2A* | 4.320085274 | 0.000460863 | 0.002109537 | 1 |
| HTN | SMC | *CHAF1B* | -2.697983641 | 0.001374377 | 0.004340371 | 1 |
| HTN | SMC | *ARHGAP27* | -5.794641893 | 0.000901267 | 0.00320171 | 1 |
| HTN | SMC | *IRX6* | -2.123456693 | 0.000701906 | 0.002679402 | 1 |
| HTN | SMC | *HK2* | -2.796949967 | 0.000241708 | 0.001414677 | 1 |
| HTN | SMC | *TMEM69* | -7.882039594 | 0.000958752 | 0.003337063 | 1 |
| HTN | SMC | *ACE* | -6.97641975 | 0.000123327 | 0.00101896 | 0.598380438 |
| HTN | SMC | *TEPP* | -9.905859108 | 5.29E-05 | 0.000806822 | 0.256612743 |
| HTN | SMC | *CTBP1* | -10.94562552 | 1.05E-07 | 1.25E-05 | 0.000510806 |
| HTN | SMC | *LRRC36* | -1.992456235 | 0.000902932 | 0.003204847 | 1 |
| HTN | SMC | *RGS12* | -4.138296865 | 0.023921681 | 0.041721063 | 1 |
| HTN | SMC | *LYPD5* | -8.861580517 | 2.25E-05 | 0.000515555 | 0.1091184 |
| HTN | SMC | *ZNF233* | -5.42449918 | 0.000454413 | 0.002089869 | 1 |
| HTN | SMC | *SSU72* | -6.222674265 | 0.022355381 | 0.039285878 | 1 |
| HTN | SMC | *ANKLE1* | -8.336561936 | 0.001641581 | 0.004880484 | 1 |
| HTN | SMC | *CCDC58* | -9.298129996 | 5.59E-05 | 0.000817716 | 0.270988789 |
| HTN | SMC | *PDE9A* | -3.9596893 | 0.003928766 | 0.009483768 | 1 |
| HTN | SMC | *HSF2BP* | -3.967127873 | 1.58E-05 | 0.000427306 | 0.076713683 |
| HTN | SMC | *CSTB* | -2.624131947 | 0.024850536 | 0.043185816 | 1 |
| HTN | SMC | *C21orf2* | -1.912558245 | 0.016624711 | 0.030542634 | 1 |
| HTN | SMC | *MCM3AP* | -3.313024552 | 0.011300206 | 0.022297112 | 1 |
| HTN | SMC | *PCNT* | -3.767273424 | 0.000529002 | 0.002261977 | 1 |

| | | | | | | |
|---|---|---|---|---|---|---|
| HTN | SMC | *S100B* | -1.797335327 | 9.44E-07 | 6.64E-05 | 0.004580014 |
| HTN | SMC | *ZNF208* | -7.817075112 | 1.98E-08 | 3.69E-06 | 9.59E-05 |
| HTN | SMC | *TAOK1* | -5.920921678 | 0.000835639 | 0.003033818 | 1 |
| HTN | SMC | *TLCD1* | -5.900414272 | 0.016086861 | 0.029693493 | 1 |
| HTN | SMC | *S100A1* | -5.144050795 | 0.027467459 | 0.04704275 | 1 |
| HTN | SMC | *ZBTB7B* | -9.381628936 | 0.006003029 | 0.013299861 | 1 |
| HTN | SMC | *VPS11* | -4.527913173 | 0.000415458 | 0.001962807 | 1 |
| HTN | SMC | *PAQR6* | -3.143682061 | 0.002783826 | 0.00730905 | 1 |
| HTN | SMC | *PTH1R* | -4.885931379 | 0.001656783 | 0.004910637 | 1 |
| HTN | SMC | *LY6K* | 4.006510984 | 0.000310856 | 0.001640089 | 1 |
| HTN | SMC | *IER2* | -6.67121485 | 0.001823859 | 0.005283202 | 1 |
| HTN | SMC | *MUM1* | -6.376187398 | 0.000822599 | 0.002993156 | 1 |
| HTN | SMC | *ZNF333* | -6.332872946 | 0.027944224 | 0.047741329 | 1 |
| HTN | SMC | *RPL8* | -5.519457451 | 0.009330527 | 0.018981853 | 1 |
| HTN | SMC | *DVL3* | -4.649006818 | 0.011376638 | 0.022411468 | 1 |
| HTN | SMC | *ABCF3* | 5.950719234 | 4.22E-06 | 0.000181808 | 0.020455214 |
| HTN | SMC | *U2AF1L4* | -1.086869504 | 0.001323822 | 0.004225779 | 1 |
| HTN | SMC | *BDH1* | -5.737086095 | 0.007858778 | 0.016535469 | 1 |
| HTN | SMC | *ZNF577* | -4.78084628 | 0.000618001 | 0.002454805 | 1 |
| HTN | SMC | *TMEM143* | -6.58551312 | 0.000293144 | 0.001576868 | 1 |
| HTN | SMC | *PLCD3* | -3.2496518 | 0.000361013 | 0.00179994 | 1 |
| HTN | SMC | *AQP5* | -6.608323886 | 0.017022149 | 0.031096185 | 1 |
| HTN | SMC | *IP6K3* | 1.104798471 | 0.00232657 | 0.006359729 | 1 |
| HTN | SMC | *LEMD2* | -9.58984738 | 7.90E-06 | 0.0002818 | 0.038324758 |
| HTN | SMC | *ASGR2* | -14.41404551 | 0.000968703 | 0.00336205 | 1 |
| HTN | SMC | *POLR3K* | -4.128042969 | 0.000427954 | 0.002004278 | 1 |
| HTN | SMC | *SHANK2* | -7.681156961 | 8.93E-05 | 0.000888243 | 0.433182023 |
| HTN | SMC | *RPS6KA4* | -9.288345143 | 8.48E-05 | 0.000876014 | 0.411412077 |
| HTN | SMC | *PDZK1IP1* | -3.612023831 | 0.000514364 | 0.002236542 | 1 |
| HTN | SMC | *COA7* | -3.665821691 | 0.000442312 | 0.002053681 | 1 |
| HTN | SMC | *SLC1A7* | -1.795461858 | 0.014688423 | 0.027601948 | 1 |
| HTN | SMC | *USP24* | -7.403188575 | 0.000130211 | 0.00104255 | 0.631785053 |
| HTN | SMC | *PDPN* | -5.376316747 | 0.000105298 | 0.000950786 | 0.510904808 |
| HTN | SMC | *LAPTM5* | -6.690379679 | 7.67E-05 | 0.000851198 | 0.37220183 |
| HTN | SMC | *SDC3* | -6.445102887 | 0.000382434 | 0.001850936 | 1 |
| HTN | SMC | *SYNC* | -7.178036413 | 3.97E-05 | 0.000693618 | 0.192825777 |
| HTN | SMC | *MEGF6* | -3.990525329 | 0.000888196 | 0.003171101 | 1 |
| HTN | SMC | *TM2D1* | -6.983542667 | 0.001118071 | 0.003720928 | 1 |
| HTN | SMC | *NEXN* | 1.847597605 | 0.021438701 | 0.037853195 | 1 |
| HTN | SMC | *LRRIQ3* | -2.778870532 | 0.000278803 | 0.001535474 | 1 |
| HTN | SMC | *WDR63* | -1.930478253 | 0.004492634 | 0.010550949 | 1 |
| HTN | SMC | *HFM1* | -7.185676003 | 0.000251234 | 0.001440884 | 1 |
| HTN | SMC | *ZNF496* | -6.926092646 | 0.001267909 | 0.004079507 | 1 |
| HTN | SMC | *FCRLB* | -7.100720239 | 0.000697569 | 0.002675579 | 1 |
| HTN | SMC | *FLVCR1* | -7.807049833 | 0.005504677 | 0.012378329 | 1 |
| HTN | SMC | *TDRD5* | -7.946450048 | 0.002593151 | 0.006920776 | 1 |
| HTN | SMC | *CNST* | -6.685339813 | 0.001502502 | 0.004625724 | 1 |
| HTN | SMC | *PM20D1* | -2.965414956 | 0.005936635 | 0.013176831 | 1 |
| HTN | SMC | *HAAO* | -9.254910481 | 0.000197926 | 0.001271967 | 0.960335191 |
| HTN | SMC | *MRPL55* | 3.108161982 | 0.019011177 | 0.034214477 | 1 |

| | | | | | | |
|---|---|---|---|---|---|---|
| HTN | SMC | *KCNJ3* | 1.392841529 | 0.002646155 | 0.00704673 | 1 |
| HTN | SMC | *NUP35* | -5.949635707 | 0.029427181 | 0.049940777 | 1 |
| HTN | SMC | *NOSTRIN* | -5.036053583 | 3.40E-05 | 0.000641172 | 0.164746966 |
| HTN | SMC | *INHBB* | -3.300370654 | 0.003881448 | 0.009388227 | 1 |
| HTN | SMC | *RPRD2* | -5.631074561 | 0.00209866 | 0.005882552 | 1 |
| HTN | SMC | *CTSS* | -1.298051118 | 0.010738118 | 0.021348248 | 1 |
| HTN | SMC | *C1QTNF7* | -5.602975086 | 0.004923336 | 0.011294576 | 1 |
| HTN | SMC | *LYSMD1* | -2.206780107 | 0.00740481 | 0.015799533 | 1 |
| HTN | SMC | *VPS72* | -10.59105577 | 0.00154847 | 0.004700863 | 1 |
| HTN | SMC | *RNF149* | -4.921200509 | 0.01158151 | 0.022741193 | 1 |
| HTN | SMC | *DHX57* | -7.922805061 | 0.000214473 | 0.00132773 | 1 |
| HTN | SMC | *S100A9* | -5.37622998 | 0.001343246 | 0.004262545 | 1 |
| HTN | SMC | *TDRD10* | -8.759175845 | 0.000119492 | 0.001005578 | 0.579773194 |
| HTN | SMC | *DCAF16* | -8.765383387 | 2.35E-08 | 3.94E-06 | 0.000114181 |
| HTN | SMC | *NIPAL1* | 2.556290551 | 0.020426873 | 0.036304464 | 1 |
| HTN | SMC | *ANTXR2* | -3.810140685 | 0.007125986 | 0.015332721 | 1 |
| HTN | SMC | *GPR155* | -8.210843805 | 3.84E-05 | 0.000689173 | 0.186546686 |
| HTN | SMC | *CLDN1* | -4.789518812 | 0.000917221 | 0.003248435 | 1 |
| HTN | SMC | *LINC01116* | -4.797041781 | 3.41E-05 | 0.000641172 | 0.165384935 |
| HTN | SMC | *POGLUT1* | -2.480831393 | 0.005159548 | 0.01173658 | 1 |
| HTN | SMC | *SLC22A15* | -4.086475914 | 0.004918296 | 0.011288351 | 1 |
| HTN | SMC | *LRRC58* | -8.860933631 | 0.000322841 | 0.00167353 | 1 |
| HTN | SMC | *LMOD1* | 0.509693084 | 0.012525433 | 0.024169481 | 1 |
| HTN | SMC | *ARPC2* | -7.047772165 | 0.000321349 | 0.001668936 | 1 |
| HTN | SMC | *GLB1L* | -3.43911777 | 0.008188441 | 0.017132521 | 1 |
| HTN | SMC | *SERPINI1* | 1.364592964 | 0.020622895 | 0.03662602 | 1 |
| HTN | SMC | *SUCLG1* | -5.888987576 | 0.000591422 | 0.002410673 | 1 |
| HTN | SMC | *PRKCI* | -2.534573344 | 0.014569465 | 0.027431527 | 1 |
| HTN | SMC | *WDFY3* | -4.076728543 | 0.021050882 | 0.037304193 | 1 |
| HTN | SMC | *C4orf36* | -8.456751183 | 5.31E-05 | 0.000806822 | 0.257750719 |
| HTN | SMC | *TIPARP* | -7.4126601 | 0.000200922 | 0.001281046 | 0.974875838 |
| HTN | SMC | *RPP14* | -9.964774593 | 0.003570426 | 0.008807171 | 1 |
| HTN | SMC | *IL17RC* | -9.54415845 | 1.40E-05 | 0.000397041 | 0.067718516 |
| HTN | SMC | *CRELD1* | -6.871400414 | 0.013963941 | 0.026455697 | 1 |
| HTN | SMC | *TTC14* | -6.218709602 | 0.000121722 | 0.001013054 | 0.590594138 |
| HTN | SMC | *CCDC158* | -3.045135251 | 0.007843577 | 0.016510645 | 1 |
| HTN | SMC | *CPA3* | -3.047470233 | 0.000515806 | 0.002236542 | 1 |
| HTN | SMC | *SLC4A1AP* | -6.643951913 | 0.027551448 | 0.047153308 | 1 |
| HTN | SMC | *CDCP1* | -3.009387603 | 0.001764029 | 0.005156067 | 1 |
| HTN | SMC | *FYCO1* | -4.800475755 | 0.003846062 | 0.009337834 | 1 |
| HTN | SMC | *TPRA1* | -8.895079214 | 0.000248358 | 0.001436361 | 1 |
| HTN | SMC | *MEAF6* | -9.01227921 | 0.001276935 | 0.004103105 | 1 |
| HTN | SMC | *SNIP1* | -11.21515677 | 0.000136674 | 0.001069585 | 0.663142651 |
| HTN | SMC | *DNALI1* | -5.010579823 | 2.30E-05 | 0.000519068 | 0.111599584 |
| HTN | SMC | *RFT1* | -5.622508724 | 2.44E-05 | 0.000537075 | 0.118357093 |
| HTN | SMC | *SFMBT1* | -6.300602704 | 0.00015791 | 0.001140149 | 0.766180385 |
| HTN | SMC | *LRPAP1* | -7.868570545 | 0.000488444 | 0.002184577 | 1 |
| HTN | SMC | *PIGX* | -3.065676276 | 0.004648785 | 0.0108494 | 1 |
| HTN | SMC | *ERMAP* | -3.648501988 | 0.000260995 | 0.001475067 | 1 |
| HTN | SMC | *METAP1* | -8.537286903 | 3.69E-06 | 0.000167415 | 0.017913408 |

| | | | | | | |
|---|---|---|---|---|---|---|
| HTN | SMC | *BDH2* | -7.118529812 | 6.45E-05 | 0.000829756 | 0.31295524 |
| HTN | SMC | *CDC25A* | -7.763270065 | 0.005910022 | 0.013129775 | 1 |
| HTN | SMC | *ZNF589* | -9.643787822 | 3.75E-13 | 4.55E-10 | 1.82E-09 |
| HTN | SMC | *ATRIP* | -1.664127277 | 0.024955797 | 0.043337697 | 1 |
| HTN | SMC | *RNF123* | 4.864876374 | 3.62E-05 | 0.000666191 | 0.175874481 |
| HTN | SMC | *MFSD8* | -7.202674101 | 0.00070037 | 0.002675893 | 1 |
| HTN | SMC | *RAD54L2* | -6.320806998 | 0.002474868 | 0.006645302 | 1 |
| HTN | SMC | *TEX264* | -4.200610283 | 0.001789066 | 0.005198595 | 1 |
| HTN | SMC | *ETNPPL* | -28.19349149 | 0.000283999 | 0.001551763 | 1 |
| HTN | SMC | *PRSS12* | -1.742276337 | 0.006893803 | 0.014953353 | 1 |
| HTN | SMC | *ANXA5* | -4.462368136 | 0.026061442 | 0.044920112 | 1 |
| HTN | SMC | *CEP44* | -8.061222101 | 1.31E-05 | 0.000389012 | 0.063498095 |
| HTN | SMC | *HPGD* | -3.082194938 | 0.000488977 | 0.002184577 | 1 |
| HTN | SMC | *TMEM144* | -4.965127578 | 1.64E-05 | 0.000427306 | 0.07947899 |
| HTN | SMC | *IL15* | -5.245217209 | 0.002176081 | 0.006061047 | 1 |
| HTN | SMC | *ARFIP1* | -5.314701312 | 0.005581615 | 0.012520571 | 1 |
| HTN | SMC | *LSM6* | -3.842726928 | 0.007745157 | 0.016331813 | 1 |
| HTN | SMC | *MOCS2* | -11.08932733 | 0.000207711 | 0.001311479 | 1 |
| HTN | SMC | *LMBRD2* | -8.005400309 | 2.15E-05 | 0.000505599 | 0.104153441 |
| HTN | SMC | *RNF180* | 1.460289306 | 0.016845955 | 0.030855634 | 1 |
| HTN | SMC | *PGGT1B* | -6.637703295 | 0.000688812 | 0.002653042 | 1 |
| HTN | SMC | *F2RL2* | -6.846511667 | 0.000563878 | 0.00234515 | 1 |
| HTN | SMC | *CMBL* | -5.863001044 | 0.007553855 | 0.016032941 | 1 |
| HTN | SMC | *C5orf63* | -4.76154882 | 0.018303355 | 0.033112557 | 1 |
| HTN | SMC | *WDR41* | -5.057655914 | 0.009635239 | 0.019471128 | 1 |
| HTN | SMC | *NDUFS4* | -6.786467896 | 0.001574401 | 0.004740559 | 1 |
| HTN | SMC | *SERINC5* | -6.796306377 | 0.000117903 | 0.000998368 | 0.572064827 |
| HTN | SMC | *CASP3* | -6.40380162 | 0.001443062 | 0.004502726 | 1 |
| HTN | SMC | *PRIMPOL* | -7.35894893 | 0.000181632 | 0.001220605 | 0.881276462 |
| HTN | SMC | *ERAP1* | -10.31830391 | 1.25E-10 | 7.07E-08 | 6.09E-07 |
| HTN | SMC | *CCDC127* | -8.945648344 | 8.41E-07 | 6.00E-05 | 0.004079914 |
| HTN | SMC | *GRIK2* | -2.094316239 | 0.000730002 | 0.002754253 | 1 |
| HTN | SMC | *PDSS2* | -6.079060361 | 0.001352846 | 0.004284602 | 1 |
| HTN | SMC | *RAET1E* | -5.18634168 | 0.000179424 | 0.001212485 | 0.870564455 |
| HTN | SMC | *DAGLB* | -2.407195658 | 0.001105349 | 0.003693812 | 1 |
| HTN | SMC | *STK17A* | -4.965490801 | 0.01252808 | 0.024169481 | 1 |
| HTN | SMC | *GALNT10* | -5.012019568 | 0.00334383 | 0.0083803 | 1 |
| HTN | SMC | *SAP30L* | -4.606054015 | 0.003329335 | 0.008351377 | 1 |
| HTN | SMC | *RPS14* | -5.122198968 | 0.006895706 | 0.014953353 | 1 |
| HTN | SMC | *MIOS* | -8.695259138 | 0.000366375 | 0.001809686 | 1 |
| HTN | SMC | *USP49* | -8.738636687 | 0.000185041 | 0.001228209 | 0.897820711 |
| HTN | SMC | *CHMP4C* | 0.957531235 | 0.002747206 | 0.007244264 | 1 |
| HTN | SMC | *SLC13A4* | -8.981658777 | 0.000190573 | 0.001250601 | 0.924661271 |
| HTN | SMC | *BRI3* | -6.870906542 | 0.002777801 | 0.007297179 | 1 |
| HTN | SMC | *CTSB* | -2.028024085 | 0.001604601 | 0.004802914 | 1 |
| HTN | SMC | *RAD21* | -6.771329479 | 0.002456678 | 0.006627102 | 1 |
| HTN | SMC | *TNFRSF11B* | -10.91511376 | 0.000824561 | 0.002996831 | 1 |
| HTN | SMC | *DEFB1* | -7.043604466 | 0.001086077 | 0.003656937 | 1 |
| HTN | SMC | *OXR1* | -5.202022757 | 0.003843167 | 0.009337529 | 1 |
| HTN | SMC | *INTS1* | -10.8729509 | 4.31E-05 | 0.000739239 | 0.209234883 |

| | | | | | | |
|---|---|---|---|---|---|---|
| HTN | SMC | *SLC4A2* | -7.926825516 | 0.000661815 | 0.002575083 | 1 |
| HTN | SMC | *FASTK* | -8.990841016 | 0.00011286 | 0.0009796 | 0.547596595 |
| HTN | SMC | *ALDH7A1* | -4.231795261 | 0.024224653 | 0.042203955 | 1 |
| HTN | SMC | *YWHAZ* | -4.59423623 | 0.00754162 | 0.016028008 | 1 |
| HTN | SMC | *SLC25A32* | -8.482182887 | 8.79E-06 | 0.000298214 | 0.04264456 |
| HTN | SMC | *INTS8* | -8.67208784 | 8.92E-05 | 0.000888243 | 0.432720169 |
| HTN | SMC | *C9orf24* | -6.529730424 | 0.000608271 | 0.002441134 | 1 |
| HTN | SMC | *NUDT2* | -2.569714517 | 8.21E-05 | 0.000876014 | 0.398412585 |
| HTN | SMC | *TMEM65* | -7.513723196 | 0.001040541 | 0.003530658 | 1 |
| HTN | SMC | *CCDC171* | -5.3397205 | 0.00272571 | 0.007199317 | 1 |
| HTN | SMC | *ZMAT4* | -17.77640603 | 0.00010657 | 0.000950786 | 0.517077136 |
| HTN | SMC | *C8orf34* | -2.443788248 | 0.002105323 | 0.00589782 | 1 |
| HTN | SMC | *ANKS6* | -4.245009971 | 0.025016742 | 0.04342799 | 1 |
| HTN | SMC | *TRMT10B* | -6.788264851 | 0.000159964 | 0.001151189 | 0.776147104 |
| HTN | SMC | *VCP* | -4.426249084 | 0.014917199 | 0.027966866 | 1 |
| HTN | SMC | *PIGO* | -5.45907207 | 0.01500085 | 0.028080295 | 1 |
| HTN | SMC | *MARCH8* | -8.00594917 | 0.001392049 | 0.004385857 | 1 |
| HTN | SMC | *TSHR* | -5.41136807 | 0.000231589 | 0.001378826 | 1 |
| HTN | SMC | *PGM2L1* | -4.32384983 | 0.000499906 | 0.002207048 | 1 |
| HTN | SMC | *SLC16A9* | 1.280480618 | 0.004833373 | 0.011135577 | 1 |
| HTN | SMC | *GJB2* | 9.057400314 | 0.007997128 | 0.016797431 | 1 |
| HTN | SMC | *CRYL1* | -5.610576367 | 0.000622987 | 0.002462978 | 1 |
| HTN | SMC | *REEP3* | -9.364306131 | 2.49E-06 | 0.000132689 | 0.012074688 |
| HTN | SMC | *MICU2* | -5.406226455 | 0.007061763 | 0.01523507 | 1 |
| HTN | SMC | *LRR1* | -5.115273349 | 0.000217346 | 0.001341684 | 1 |
| HTN | SMC | *NOXRED1* | -4.29231979 | 0.021119246 | 0.037407554 | 1 |
| HTN | SMC | *AKR1E2* | -7.75239614 | 0.002467938 | 0.006637715 | 1 |
| HTN | SMC | *OXGR1* | -8.725442514 | 0.002947121 | 0.007610129 | 1 |
| HTN | SMC | *ATP5C1* | -5.380736018 | 0.017219694 | 0.031433391 | 1 |
| HTN | SMC | *VDAC2* | -11.93835425 | 3.51E-06 | 0.000162031 | 0.017021039 |
| HTN | SMC | *PDZD8* | -5.056002532 | 0.000564054 | 0.00234515 | 1 |
| HTN | SMC | *QSOX2* | -6.436897374 | 0.00314281 | 0.007992092 | 1 |
| HTN | SMC | *RET* | -9.544748814 | 9.62E-05 | 0.000908214 | 0.466822172 |
| HTN | SMC | *CASP7* | -4.88587773 | 3.78E-05 | 0.000687368 | 0.183527285 |
| HTN | SMC | *PPP1R36* | -5.964837363 | 0.011875923 | 0.023169272 | 1 |
| HTN | SMC | *METTL3* | -7.780772923 | 0.005298507 | 0.011996432 | 1 |
| HTN | SMC | *TRUB1* | -9.413190563 | 3.16E-06 | 0.000153181 | 0.015318096 |
| HTN | SMC | *UBTD1* | -7.089697835 | 5.74E-05 | 0.000817716 | 0.278344215 |
| HTN | SMC | *OTOGL* | 1.4691913 | 0.021391363 | 0.037797121 | 1 |
| HTN | SMC | *PACSIN3* | -1.879330993 | 0.022413289 | 0.039373382 | 1 |
| HTN | SMC | *TTC7B* | -5.783473969 | 0.008093768 | 0.016966257 | 1 |
| HTN | SMC | *SLC39A13* | 5.310715442 | 0.0151531 | 0.028321588 | 1 |
| HTN | SMC | *TC2N* | -2.20337062 | 0.01613716 | 0.029770912 | 1 |
| HTN | SMC | *IFI27L1* | -5.534370467 | 0.000764123 | 0.002849751 | 1 |
| HTN | SMC | *PTER* | -4.078403541 | 0.002201448 | 0.006110139 | 1 |
| HTN | SMC | *ARL5B* | -11.89789632 | 1.50E-09 | 4.57E-07 | 7.27E-06 |
| HTN | SMC | *LIPC* | -2.39432786 | 0.00942318 | 0.019122238 | 1 |
| HTN | SMC | *CEP57* | -8.963518045 | 6.25E-06 | 0.000233172 | 0.03031237 |
| HTN | SMC | *TCP11L2* | -7.765399314 | 0.000116086 | 0.000993383 | 0.563248113 |
| HTN | SMC | *ZFYVE19* | -3.724279199 | 0.005186356 | 0.011786509 | 1 |

| | | | | | | |
|---|---|---|---|---|---|---|
| HTN | SMC | *TRMT61A* | -10.44469533 | 0.000793811 | 0.002935648 | 1 |
| HTN | SMC | *ASPG* | -9.497014065 | 0.00365986 | 0.008981338 | 1 |
| HTN | SMC | *COX11* | -5.760176014 | 0.017235779 | 0.031450922 | 1 |
| HTN | SMC | *ZNF202* | 2.617544493 | 0.025898551 | 0.044686973 | 1 |
| HTN | SMC | *STXBP4* | -6.9482526 | 0.001694575 | 0.005001264 | 1 |
| HTN | SMC | *WBP1L* | -4.493193969 | 0.014817501 | 0.027812192 | 1 |
| HTN | SMC | *NUDT13* | -3.841836269 | 0.000312475 | 0.001641688 | 1 |
| HTN | SMC | *WDR88* | -6.602747596 | 8.47E-05 | 0.000876014 | 0.411206421 |
| HTN | SMC | *CYB5R2* | 3.324460014 | 5.38E-05 | 0.000806822 | 0.260870279 |
| HTN | SMC | *XRRA1* | -6.316294961 | 2.28E-05 | 0.000517778 | 0.110804471 |
| HTN | SMC | *TRIM66* | -2.573687627 | 3.20E-07 | 3.10E-05 | 0.001552252 |
| HTN | SMC | *CENPN* | -4.383644591 | 0.00087204 | 0.003127226 | 1 |
| HTN | SMC | *C16orf46* | 5.799830069 | 0.025065181 | 0.043480965 | 1 |
| HTN | SMC | *TMEM41B* | -7.85524541 | 0.001670008 | 0.004943796 | 1 |
| HTN | SMC | *TMX3* | -6.254936959 | 0.002800448 | 0.007328897 | 1 |
| HTN | SMC | *PRKCB* | -4.10671692 | 0.001885305 | 0.005416726 | 1 |
| HTN | SMC | *NDST2* | -20.62151806 | 0.000515738 | 0.002236542 | 1 |
| HTN | SMC | *CCDC68* | -2.959245354 | 0.011704189 | 0.022926413 | 1 |
| HTN | SMC | *ZSCAN21* | -2.366450909 | 0.022641901 | 0.039727027 | 1 |
| HTN | SMC | *RIMKLB* | -6.174179131 | 6.10E-07 | 5.02E-05 | 0.002961386 |
| HTN | SMC | *TMED3* | -2.089296653 | 0.002420669 | 0.006554176 | 1 |
| HTN | SMC | *SEC11C* | -11.98315151 | 0.000362622 | 0.001802871 | 1 |
| HTN | SMC | *GALR1* | -2.420264294 | 0.006998846 | 0.015126236 | 1 |
| HTN | SMC | *ACSM1* | -2.379947756 | 0.00563851 | 0.012619027 | 1 |
| HTN | SMC | *SLFN5* | -6.259155315 | 9.40E-05 | 0.000903 | 0.45631994 |
| HTN | SMC | *CATSPER2* | -9.179583063 | 0.000555008 | 0.002326902 | 1 |
| HTN | SMC | *SAAL1* | -12.54511852 | 0.000123566 | 0.00101896 | 0.599543681 |
| HTN | SMC | *FAM96A* | -6.653649232 | 0.000970515 | 0.003365931 | 1 |
| HTN | SMC | *FAM111A* | -4.518038441 | 0.00034062 | 0.001734441 | 1 |
| HTN | SMC | *TMEM170A* | -9.242751377 | 4.75E-05 | 0.00076177 | 0.230586443 |
| HTN | SMC | *STAT6* | -7.674127807 | 4.34E-05 | 0.000739239 | 0.210394509 |
| HTN | SMC | *STX3* | -6.706924417 | 0.000132866 | 0.001053374 | 0.644665021 |
| HTN | SMC | *MTMR10* | -5.293112333 | 0.002806087 | 0.00733179 | 1 |
| HTN | SMC | *MS4A7* | -2.879536353 | 2.90E-05 | 0.000580436 | 0.140465611 |
| HTN | SMC | *MS4A14* | -1.400625981 | 0.001199448 | 0.003913733 | 1 |
| HTN | SMC | *CCNDBP1* | -6.002948675 | 0.001469938 | 0.004566029 | 1 |
| HTN | SMC | *EPB42* | -7.396029756 | 0.000166151 | 0.001164975 | 0.806162498 |
| HTN | SMC | *SMAD3* | -6.358015844 | 0.001020573 | 0.003487197 | 1 |
| HTN | SMC | *MAP1A* | 2.074965906 | 0.003086714 | 0.007879215 | 1 |
| HTN | SMC | *RCCD1* | -8.111012951 | 0.001465187 | 0.004554189 | 1 |
| HTN | SMC | *AKTIP* | -5.992774738 | 0.000464961 | 0.002120291 | 1 |
| HTN | SMC | *CNPY4* | 4.844658254 | 0.001739443 | 0.005100181 | 1 |
| HTN | SMC | *MEI1* | -3.541723024 | 0.014071813 | 0.026626859 | 1 |
| HTN | SMC | *ENDOG* | -1.927486001 | 0.0245804 | 0.042777655 | 1 |
| HTN | SMC | *ISLR2* | -4.490546133 | 0.000306923 | 0.00162398 | 1 |
| HTN | SMC | *SP2* | -9.072013844 | 5.78E-05 | 0.000817716 | 0.280490947 |
| HTN | SMC | *COQ7* | -7.776454512 | 0.000128592 | 0.001039835 | 0.623926662 |
| HTN | SMC | *TBC1D2B* | -5.371056527 | 2.01E-05 | 0.000488158 | 0.097631625 |
| HTN | SMC | *LOXHD1* | -3.057943917 | 0.00057257 | 0.002363597 | 1 |
| HTN | SMC | *ZNF91* | -7.257152418 | 0.000107052 | 0.000950786 | 0.519414213 |

| | | | | | | |
|---|---|---|---|---|---|---|
| HTN | SMC | *POP5* | -6.555609352 | 8.48E-05 | 0.000876014 | 0.411491955 |
| HTN | SMC | *ZNF23* | -12.35044446 | 2.95E-05 | 0.000586475 | 0.143099948 |
| HTN | SMC | *VKORC1* | -13.37982544 | 0.000269986 | 0.00150483 | 1 |
| HTN | SMC | *RAB8A* | -2.424348005 | 0.019685258 | 0.035242897 | 1 |
| HTN | SMC | *SPATA33* | -7.446470639 | 3.34E-05 | 0.000639497 | 0.161989929 |
| HTN | SMC | *RPL13* | -5.415927598 | 0.003828437 | 0.009310417 | 1 |
| HTN | SMC | *TP53I13* | -5.583599636 | 0.000386725 | 0.001865198 | 1 |
| HTN | SMC | *ZNF528* | -10.00029722 | 0.000423155 | 0.001989486 | 1 |
| HTN | SMC | *LENG8* | -6.629718863 | 0.001476251 | 0.004576851 | 1 |
| HTN | SMC | *CDC42EP5* | -2.757804499 | 0.001539667 | 0.004686615 | 1 |
| HTN | SMC | *CHAF1A* | -2.260492306 | 8.68E-05 | 0.000880996 | 0.42111619 |
| HTN | SMC | *SRR* | 2.048993934 | 0.00146386 | 0.004552981 | 1 |
| HTN | SMC | *KLK5* | -8.230013162 | 0.000427466 | 0.002004278 | 1 |
| HTN | SMC | *KLK6* | 7.416436938 | 0.004168032 | 0.009913378 | 1 |
| HTN | SMC | *ZNF83* | -4.84048305 | 0.007441228 | 0.015863286 | 1 |
| HTN | SMC | *NDUFV1* | -6.474379655 | 0.012381438 | 0.023924626 | 1 |
| HTN | SMC | *NUDT8* | -2.201727021 | 0.003407406 | 0.008491905 | 1 |
| HTN | SMC | *PRDX2* | -9.245566025 | 0.000378564 | 0.001841989 | 1 |
| HTN | SMC | *GSDMA* | -2.05358518 | 0.007822242 | 0.016472882 | 1 |
| HTN | SMC | *ZNF598* | -8.874840069 | 2.41E-07 | 2.54E-05 | 0.001168224 |
| HTN | SMC | *MLST8* | -4.259712579 | 0.000656576 | 0.002560858 | 1 |
| HTN | SMC | *ECI1* | -5.377952928 | 0.000211228 | 0.001315636 | 1 |
| HTN | SMC | *ABCA3* | -3.957536187 | 0.004773076 | 0.011038592 | 1 |
| HTN | SMC | *LTBP3* | -5.552604712 | 0.006365088 | 0.013961758 | 1 |
| HTN | SMC | *SCARA5* | -2.988273893 | 0.001033265 | 0.003513798 | 1 |
| HTN | SMC | *FAM83B* | -20.35433189 | 0.001095262 | 0.003675112 | 1 |
| HTN | SMC | *COA6* | -12.33260027 | 6.94E-05 | 0.000836037 | 0.336830516 |
| HTN | SMC | *PXK* | -7.764907282 | 3.00E-05 | 0.000594747 | 0.145712903 |
| HTN | SMC | *PCMTD1* | -6.226087333 | 0.000106197 | 0.000950786 | 0.515266611 |
| HTN | SMC | *ACOX2* | -6.698800191 | 8.26E-05 | 0.000876014 | 0.400669159 |
| HTN | SMC | *FAM107A* | -4.505113409 | 0.002953111 | 0.00761344 | 1 |
| HTN | SMC | *DTYMK* | -6.083670709 | 0.001530967 | 0.004671856 | 1 |
| HTN | SMC | *ING5* | -5.495680839 | 0.00052578 | 0.002257599 | 1 |
| HTN | SMC | *KLHL30* | 6.141690672 | 0.022225203 | 0.039099595 | 1 |
| HTN | SMC | *CCDC110* | -8.993633152 | 0.000181598 | 0.001220605 | 0.881111491 |
| HTN | SMC | *FNTA* | -4.02763637 | 0.014854774 | 0.02787137 | 1 |
| HTN | SMC | *SERINC2* | -11.62272155 | 3.32E-06 | 0.000156411 | 0.016110302 |
| HTN | SMC | *TMUB2* | -8.784303068 | 1.93E-05 | 0.000474842 | 0.09354396 |
| HTN | SMC | *NDUFS5* | -6.739746401 | 0.001575345 | 0.004740559 | 1 |
| HTN | SMC | *NPNT* | 1.029915225 | 0.003949116 | 0.00950452 | 1 |
| HTN | SMC | *GSTM4* | -7.564695929 | 0.016408427 | 0.030213924 | 1 |
| HTN | SMC | *TCTN2* | -1.24183224 | 0.011437512 | 0.022503976 | 1 |
| HTN | SMC | *PPIP5K1* | -3.814499343 | 0.017503941 | 0.031892272 | 1 |
| HTN | SMC | *ADAL* | -1.429822273 | 0.021376276 | 0.037784222 | 1 |
| HTN | SMC | *SNTB2* | -6.313543122 | 0.000185634 | 0.001228781 | 0.900696648 |
| HTN | SMC | *STX18* | -7.597831861 | 0.00084374 | 0.003048271 | 1 |
| HTN | SMC | *ATOH8* | 2.406227459 | 0.009270662 | 0.018883816 | 1 |
| HTN | SMC | *TMEM129* | 4.541118124 | 0.009253805 | 0.018881187 | 1 |
| HTN | SMC | *CEP120* | -7.768874319 | 0.00214267 | 0.005974847 | 1 |
| HTN | SMC | *MFF* | -5.799071913 | 0.009207449 | 0.018826187 | 1 |

| | | | | | | |
|---|---|---|---|---|---|---|
| HTN | SMC | *MFSD7* | -6.785832201 | 0.000229009 | 0.001377231 | 1 |
| HTN | SMC | *MAP2K1* | -7.796269158 | 0.000329779 | 0.001697761 | 1 |
| HTN | SMC | *KLK7* | -12.35711771 | 0.000515123 | 0.002236542 | 1 |
| HTN | SMC | *IRS1* | -1.646880601 | 0.000574824 | 0.002363597 | 1 |
| HTN | SMC | *C8orf46* | 0.981933475 | 0.006903211 | 0.014953353 | 1 |
| HTN | SMC | *HSPBAP1* | -4.099987604 | 0.001527585 | 0.004664469 | 1 |
| HTN | SMC | *UBE2V2* | -7.074458038 | 0.000537897 | 0.002288957 | 1 |
| HTN | SMC | *CPT1C* | 3.926225822 | 0.004857525 | 0.011170005 | 1 |
| HTN | SMC | *PCSK9* | -11.22287096 | 0.010362179 | 0.020732904 | 1 |
| HTN | SMC | *XPO6* | -8.131961533 | 0.000528005 | 0.002261977 | 1 |
| HTN | SMC | *RAB24* | -9.006432958 | 3.24E-06 | 0.000153937 | 0.01570161 |
| HTN | SMC | *THBS3* | -5.619427819 | 0.000893413 | 0.003185039 | 1 |
| HTN | SMC | *NPIPB3* | -2.907766949 | 0.021319659 | 0.037697881 | 1 |
| HTN | SMC | *SH3TC2* | -2.416052751 | 0.014251824 | 0.026927512 | 1 |
| HTN | SMC | *SNUPN* | -6.66681291 | 3.41E-05 | 0.000641172 | 0.165679814 |
| HTN | SMC | *CD52* | -1.435111634 | 0.028627932 | 0.048772024 | 1 |
| HTN | SMC | *SLC38A11* | 0.586813678 | 0.003487541 | 0.008651099 | 1 |
| HTN | SMC | *ZEB2* | -4.846884385 | 0.000171267 | 0.001183741 | 0.830986362 |
| HTN | SMC | *HINT1* | -8.074417731 | 0.001746917 | 0.005115294 | 1 |
| HTN | SMC | *DTWD2* | -3.767946264 | 0.025751207 | 0.044496031 | 1 |
| HTN | SMC | *CLIC3* | -4.025209333 | 0.000410451 | 0.001944008 | 1 |
| HTN | SMC | *INO80E* | -4.855025712 | 6.32E-07 | 5.10E-05 | 0.003068499 |
| HTN | SMC | *DFFB* | -10.20399687 | 3.36E-06 | 0.000156644 | 0.016291002 |
| HTN | SMC | *C15orf40* | -4.063800859 | 0.00268647 | 0.007138418 | 1 |
| HTN | SMC | *RGPD8* | -1.183209657 | 0.000818581 | 0.002986621 | 1 |
| HTN | SMC | *LUZP1* | -7.099306118 | 0.002005194 | 0.00569292 | 1 |
| HTN | SMC | *LRRC45* | -2.659548215 | 0.002030514 | 0.00574129 | 1 |
| HTN | SMC | *CHRNA5* | -8.251825419 | 3.35E-05 | 0.000639497 | 0.162432203 |
| HTN | SMC | *UGP2* | -5.84668404 | 5.91E-05 | 0.000817716 | 0.286989625 |
| HTN | SMC | *HNRNPF* | -6.179791816 | 0.00020025 | 0.001281046 | 0.971615219 |
| HTN | SMC | *AVEN* | -5.503092915 | 4.96E-05 | 0.000776428 | 0.240692761 |
| HTN | SMC | *CTNND2* | -3.933228034 | 0.000428587 | 0.002004689 | 1 |
| HTN | SMC | *CALML6* | -35.99446135 | 0.004212002 | 0.009998352 | 1 |
| HTN | SMC | *MUC3A* | -2.140655139 | 0.027760908 | 0.047461566 | 1 |
| HTN | SMC | *TPST1* | -2.753494417 | 0.008262338 | 0.017257367 | 1 |
| HTN | SMC | *TOR1AIP2* | -6.048800067 | 0.000102629 | 0.000942633 | 0.49795438 |
| HTN | SMC | *ZFPM2* | -3.786459627 | 0.000157578 | 0.001140149 | 0.764568297 |
| HTN | SMC | *MAP3K2* | -4.020122105 | 0.011739724 | 0.022968203 | 1 |
| HTN | SMC | *UBE2E3* | -7.578619909 | 7.58E-05 | 0.000849747 | 0.368016357 |
| HTN | SMC | *NIPA1* | -9.015534345 | 1.80E-05 | 0.000456819 | 0.087252472 |
| HTN | SMC | *RNF150* | -3.675809407 | 0.012462107 | 0.024061339 | 1 |
| HTN | SMC | *ANKK1* | -4.72034124 | 0.002859913 | 0.007436387 | 1 |
| HTN | SMC | *PDCD6IP* | -6.802833939 | 0.000265635 | 0.001489899 | 1 |
| HTN | SMC | *ELP5* | -6.133181455 | 0.000820409 | 0.002989783 | 1 |
| HTN | SMC | *STX8* | -5.808576483 | 0.001120422 | 0.003726037 | 1 |
| HTN | SMC | *NFRKB* | -7.969694402 | 0.006306031 | 0.013863553 | 1 |
| HTN | SMC | *ADORA2B* | 1.521919434 | 0.009578214 | 0.019372028 | 1 |
| HTN | SMC | *MGMT* | -6.097176763 | 0.001035634 | 0.003518835 | 1 |
| HTN | SMC | *NFXL1* | -5.79044296 | 0.010740122 | 0.021348248 | 1 |
| HTN | SMC | *DNAJC18* | -4.777987351 | 0.000341174 | 0.001735198 | 1 |

| | | | | | | |
|---|---|---|---|---|---|---|
| HTN | SMC | *HSD17B13* | -5.161740121 | 0.000529131 | 0.002261977 | 1 |
| HTN | SMC | *ELOVL6* | -7.918817095 | 7.39E-05 | 0.000847247 | 0.358745653 |
| HTN | SMC | *ARL6IP1* | -4.626892817 | 0.004552797 | 0.010671581 | 1 |
| HTN | SMC | *SERPINB9* | -5.620781392 | 0.00071088 | 0.002703127 | 1 |
| HTN | SMC | *SMAGP* | -1.807732847 | 0.0013292 | 0.004237371 | 1 |
| HTN | SMC | *CDH2* | -3.451500224 | 0.000687413 | 0.002653042 | 1 |
| HTN | SMC | *EMB* | -4.312334174 | 4.06E-06 | 0.000181808 | 0.019686256 |
| HTN | SMC | *STAT2* | -8.025371086 | 9.41E-05 | 0.000903 | 0.456491043 |
| HTN | SMC | *SLC26A5* | -6.881448298 | 0.000138003 | 0.001071047 | 0.669589063 |
| HTN | SMC | *ACYP2* | -4.634262922 | 0.003024409 | 0.007751946 | 1 |
| HTN | SMC | *TRABD* | -5.250200291 | 0.001708645 | 0.005021407 | 1 |
| HTN | SMC | *TMEM133* | -3.020250561 | 0.019423615 | 0.034866216 | 1 |
| HTN | SMC | *HOXB9* | -7.837949015 | 0.002792962 | 0.007317198 | 1 |
| HTN | SMC | *TTLL6* | -17.35021767 | 2.35E-09 | 6.33E-07 | 1.14E-05 |
| HTN | SMC | *FOXN2* | -7.371353424 | 0.000547459 | 0.00231181 | 1 |
| HTN | SMC | *TRIAP1* | -7.360338726 | 0.000857991 | 0.003090552 | 1 |
| HTN | SMC | *TSEN34* | -8.678270606 | 0.000618255 | 0.002454805 | 1 |
| HTN | SMC | *NDUFA3* | -10.97850102 | 0.000208159 | 0.001311674 | 1 |
| HTN | SMC | *PAQR8* | -4.135607608 | 1.58E-06 | 9.82E-05 | 0.007661419 |
| HTN | SMC | *NUDT6* | -2.99163031 | 0.023651557 | 0.041309343 | 1 |
| HTN | SMC | *ZNF160* | -4.624886434 | 0.000209773 | 0.001313543 | 1 |
| HTN | SMC | *ZNF415* | -4.868508574 | 0.00237627 | 0.006459196 | 1 |
| HTN | SMC | *S1PR1* | -4.637919132 | 0.018802645 | 0.033876878 | 1 |
| HTN | SMC | *FPR1* | -6.967217591 | 0.011026519 | 0.021828098 | 1 |
| HTN | SMC | *SOX7* | -3.274571382 | 0.004839355 | 0.011138781 | 1 |
| HTN | SMC | *TRMT61B* | -4.028511178 | 0.015514779 | 0.028875224 | 1 |
| HTN | SMC | *GIMAP8* | -3.167898136 | 0.015330255 | 0.028575643 | 1 |
| HTN | SMC | *MORN4* | -1.613437393 | 0.013534122 | 0.025812719 | 1 |
| HTN | SMC | *SCAND1* | -10.26965508 | 8.53E-06 | 0.000291476 | 0.041389561 |
| HTN | SMC | *PDE7B* | -6.099717839 | 7.87E-05 | 0.00085619 | 0.381860932 |
| HTN | SMC | *MCC* | -5.99018899 | 6.55E-05 | 0.000834038 | 0.317819323 |
| HTN | SMC | *PPID* | -7.098215635 | 0.001064629 | 0.003599708 | 1 |
| HTN | SMC | *TBCA* | -5.819675926 | 0.005669348 | 0.012676348 | 1 |
| HTN | SMC | *PIK3CD* | -5.084896952 | 0.000597866 | 0.002424389 | 1 |
| HTN | SMC | *RGS19* | -3.942202891 | 1.99E-11 | 1.61E-08 | 9.64E-08 |
| HTN | SMC | *ANO5* | -3.503849111 | 0.000786937 | 0.002918392 | 1 |
| HTN | SMC | *PAH* | -10.8781137 | 0.001496221 | 0.00462399 | 1 |
| HTN | SMC | *PWWP2B* | -3.309783412 | 0.002361347 | 0.006435458 | 1 |
| HTN | SMC | *EXOSC10* | -5.664866939 | 0.02013184 | 0.035845756 | 1 |
| HTN | SMC | *TRAPPC12* | -7.258746197 | 0.001082748 | 0.003650794 | 1 |
| HTN | SMC | *C3AR1* | -2.420008179 | 0.002258301 | 0.006239908 | 1 |
| HTN | SMC | *FBXW10* | -9.658544235 | 0.000291114 | 0.001571176 | 1 |
| HTN | SMC | *ZNF554* | -6.28471361 | 0.000473646 | 0.002149795 | 1 |
| HTN | SMC | *ANKRD20A4* | -7.673280773 | 0.000442739 | 0.002053699 | 1 |
| HTN | SMC | *SERF1A* | -18.34373389 | 0.000397665 | 0.001908479 | 1 |
| HTN | SMC | *EIF2AK3* | -6.70005008 | 0.002764169 | 0.007277129 | 1 |
| HTN | SMC | *NME6* | 9.201170924 | 9.62E-08 | 1.17E-05 | 0.000466917 |
| HTN | SMC | *MTBP* | -4.936827237 | 0.022676951 | 0.039764571 | 1 |
| HTN | SMC | *MALT1* | -7.839290148 | 0.000149756 | 0.00112132 | 0.726615319 |
| HTN | SMC | *MBOAT1* | -7.354297549 | 3.47E-05 | 0.000642066 | 0.168221307 |

| | | | | | | |
|---|---|---|---|---|---|---|
| HTN | SMC | *CXCR6* | -4.208788302 | 0.001819079 | 0.005273547 | 1 |
| HTN | SMC | *PAIP1* | -7.550847839 | 0.005065154 | 0.011559796 | 1 |
| HTN | SMC | *BSG* | -5.819344136 | 0.016962243 | 0.031033485 | 1 |
| HTN | SMC | *TP53RK* | -8.214162786 | 0.004798016 | 0.011075153 | 1 |
| HTN | SMC | *SUCLG2* | -5.853993411 | 0.002702406 | 0.007153341 | 1 |
| HTN | SMC | *GTPBP2* | -4.635682727 | 0.00456827 | 0.010697533 | 1 |
| HTN | SMC | *MANEA* | -8.184209971 | 0.000155657 | 0.001140149 | 0.755247526 |
| HTN | SMC | *AFF1* | -5.578211797 | 0.00086602 | 0.003112541 | 1 |
| HTN | SMC | *CTSW* | -0.983935867 | 7.73E-07 | 5.77E-05 | 0.003750436 |
| HTN | SMC | *SNTG2* | -1.875892451 | 0.001368039 | 0.004324251 | 1 |
| HTN | SMC | *EFEMP2* | -4.204803324 | 0.027546508 | 0.047153308 | 1 |
| HTN | SMC | *ZFAND4* | -6.515994197 | 2.68E-06 | 0.000139919 | 0.013012489 |
| HTN | SMC | *ZNF738* | 2.076218461 | 0.026516348 | 0.04560699 | 1 |
| HTN | SMC | *SLFN11* | -9.849564584 | 2.78E-05 | 0.000569929 | 0.134820157 |
| HTN | SMC | *FAM71D* | -3.341717196 | 0.000553282 | 0.002322255 | 1 |
| HTN | SMC | *ZNF596* | -8.729120022 | 6.81E-05 | 0.000836037 | 0.330439097 |
| HTN | SMC | *CFL1* | -10.31672673 | 3.13E-05 | 0.000615364 | 0.151994835 |
| HTN | SMC | *FAM192A* | -4.809374814 | 0.008920115 | 0.018323624 | 1 |
| HTN | SMC | *DCP2* | -7.297557012 | 0.001227128 | 0.003976061 | 1 |
| HTN | SMC | *NADSYN1* | -4.323485061 | 0.007731538 | 0.016310183 | 1 |
| HTN | SMC | *RNASEH2C* | -3.365174252 | 0.007167032 | 0.015380334 | 1 |
| HTN | SMC | *MYEOV* | -11.26871158 | 0.000621288 | 0.002459648 | 1 |
| HTN | SMC | *ANKRD13D* | -6.123154708 | 0.006165185 | 0.013621802 | 1 |
| HTN | SMC | *MRGPRF* | 1.01809332 | 0.022563324 | 0.039622602 | 1 |
| HTN | SMC | *LCLAT1* | 1.718557132 | 0.025910852 | 0.044692304 | 1 |
| HTN | SMC | *GXYLT2* | -3.634416206 | 0.000211962 | 0.001318513 | 1 |
| HTN | SMC | *DCAKD* | -8.383564818 | 9.29E-08 | 1.16E-05 | 0.000450836 |
| HTN | SMC | *NOC3L* | -11.99595205 | 1.33E-08 | 2.68E-06 | 6.44E-05 |
| HTN | SMC | *PARP14* | -1.8371658 | 0.017317763 | 0.031588642 | 1 |
| HTN | SMC | *ZNF483* | -3.524247658 | 0.012197169 | 0.023662801 | 1 |
| HTN | SMC | *MMRN2* | -3.873495123 | 0.007573857 | 0.016068367 | 1 |
| HTN | SMC | *KCNK7* | -14.48147466 | 0.016908584 | 0.030946983 | 1 |
| HTN | SMC | *NDNF* | -4.70642121 | 0.004518451 | 0.010601318 | 1 |
| HTN | SMC | *ARV1* | -3.570897663 | 0.019981049 | 0.035642666 | 1 |
| HTN | SMC | *CCDC36* | -3.365938873 | 1.33E-05 | 0.000389012 | 0.064575989 |
| HTN | SMC | *PPP1R14B* | 6.328010158 | 0.001942245 | 0.005546659 | 1 |
| HTN | SMC | *ZNF417* | 3.012394712 | 0.027388959 | 0.046924869 | 1 |
| HTN | SMC | *PEAK1* | -4.75134491 | 0.005672584 | 0.012677743 | 1 |
| HTN | SMC | *MST1* | -3.114334256 | 8.18E-09 | 1.89E-06 | 3.97E-05 |
| HTN | SMC | *GMPPB* | -6.885251762 | 1.38E-05 | 0.000396487 | 0.066921007 |
| HTN | SMC | *NUDT18* | -7.534092661 | 0.000411593 | 0.001946443 | 1 |
| HTN | SMC | *SULT1B1* | -5.862823394 | 0.000251839 | 0.001442083 | 1 |
| HTN | SMC | *SCAI* | -4.156445015 | 0.002362063 | 0.006435458 | 1 |
| HTN | SMC | *UQCRH* | -7.474106881 | 0.000822933 | 0.002993156 | 1 |
| HTN | SMC | *C11orf80* | -10.39880949 | 9.90E-10 | 3.43E-07 | 4.81E-06 |
| HTN | SMC | *RNF213* | -5.243030121 | 0.000771423 | 0.002871309 | 1 |
| HTN | SMC | *XXYLT1* | -7.226840475 | 3.43E-05 | 0.000641172 | 0.16637037 |
| HTN | SMC | *UBXN2A* | -5.606566839 | 0.000513273 | 0.002236542 | 1 |
| HTN | SMC | *LEMD3* | -6.56923623 | 0.007642106 | 0.016156999 | 1 |
| HTN | SMC | *SNX31* | -3.282379736 | 2.71E-08 | 4.11E-06 | 0.000131384 |

| | | | | | | |
|---|---|---|---|---|---|---|
| HTN | SMC | *GLIS1* | -5.315178217 | 0.000117175 | 0.000995642 | 0.568530926 |
| HTN | SMC | *RALGAPA1* | -4.758718067 | 0.001538264 | 0.004685283 | 1 |
| HTN | SMC | *MSL2* | -7.831667765 | 0.000977451 | 0.003382734 | 1 |
| HTN | SMC | *ZNF497* | -11.4716811 | 0.010009326 | 0.020134846 | 1 |
| HTN | SMC | *ANGEL2* | -5.711543014 | 0.019372199 | 0.03479967 | 1 |
| HTN | SMC | *SH3PXD2B* | -6.464007596 | 0.000625346 | 0.002464809 | 1 |
| HTN | SMC | *KIAA1551* | -4.310851722 | 0.000317023 | 0.001655756 | 1 |
| HTN | SMC | *LARP7* | -8.074486224 | 0.000284807 | 0.001554424 | 1 |
| HTN | SMC | *FGFBP3* | -20.35538123 | 0.003813696 | 0.009289183 | 1 |
| HTN | SMC | *C4orf32* | -4.287281169 | 0.020016587 | 0.035679823 | 1 |
| HTN | SMC | *HRAS* | -3.33918322 | 0.005283077 | 0.011967081 | 1 |
| HTN | SMC | *CEP135* | -8.108428793 | 0.000378876 | 0.001841989 | 1 |
| HTN | SMC | *RSRC1* | -8.907766285 | 6.51E-07 | 5.10E-05 | 0.003159411 |
| HTN | SMC | *DHX36* | -6.052054181 | 0.005073663 | 0.011568332 | 1 |
| HTN | SMC | *FBXW8* | -5.305178962 | 0.016208139 | 0.029879138 | 1 |
| HTN | SMC | *SLC22A1* | -4.137838538 | 0.000506364 | 0.002225431 | 1 |
| HTN | SMC | *CHST2* | -9.521456417 | 0.00067252 | 0.002606282 | 1 |
| HTN | SMC | *TVP23C* | -4.942867745 | 0.000378493 | 0.001841989 | 1 |
| HTN | SMC | *PACS1* | -4.812909891 | 0.00157336 | 0.004740559 | 1 |
| HTN | SMC | *SH3BP5L* | -13.97580742 | 0.001499425 | 0.004625724 | 1 |
| HTN | SMC | *CADM2* | -4.252600434 | 1.92E-05 | 0.000474842 | 0.093092068 |
| HTN | SMC | *CCNE2* | -8.643020436 | 9.55E-13 | 9.26E-10 | 4.63E-09 |
| HTN | SMC | *ZNF519* | -4.13700723 | 0.000218492 | 0.001341927 | 1 |
| HTN | SMC | *BANF1* | -9.730483165 | 0.000100878 | 0.000932982 | 0.489458154 |
| HTN | SMC | *CHRNA7* | -3.414230302 | 0.028137952 | 0.048021577 | 1 |
| HTN | SMC | *SCUBE2* | -8.385369518 | 0.002024256 | 0.005733618 | 1 |
| HTN | SMC | *ZNF25* | -5.211727682 | 0.001319904 | 0.004216046 | 1 |
| HTN | SMC | *CLTB* | -7.077527152 | 0.018109727 | 0.032847999 | 1 |
| HTN | SMC | *PPP2R2D* | -8.672454729 | 1.36E-07 | 1.58E-05 | 0.000661863 |
| HTN | SMC | *MCTP1* | -6.364060771 | 0.000217796 | 0.001341927 | 1 |
| HTN | SMC | *KCNE3* | -3.708600086 | 0.002701808 | 0.007153341 | 1 |
| HTN | SMC | *UCP2* | -2.563376496 | 0.00398003 | 0.009569428 | 1 |
| HTN | SMC | *C11orf68* | 2.843839819 | 0.015988775 | 0.029564609 | 1 |
| HTN | SMC | *MRPL48* | -7.637933468 | 0.003322417 | 0.00833956 | 1 |
| HTN | SMC | *RAB6A* | -6.78081795 | 0.00024501 | 0.001421998 | 1 |
| HTN | SMC | *TMEM70* | -6.021470731 | 0.000607952 | 0.002441134 | 1 |
| HTN | SMC | *RPS6KB2* | -5.880520184 | 0.013626679 | 0.025948448 | 1 |
| HTN | SMC | *RMI2* | -2.231957192 | 0.020049278 | 0.035711857 | 1 |
| HTN | SMC | *TOM1L2* | -5.001644144 | 0.007920261 | 0.016650393 | 1 |
| HTN | SMC | *TEX26* | -5.146933079 | 0.026951337 | 0.046240412 | 1 |
| HTN | SMC | *GPR156* | -3.944998766 | 0.000556308 | 0.002326902 | 1 |
| HTN | SMC | *PRIMA1* | -2.145504626 | 0.000546538 | 0.002310188 | 1 |
| HTN | SMC | *HOXD8* | -2.0406079 | 0.003970169 | 0.009550451 | 1 |
| HTN | SMC | *ZDHHC21* | -5.555095501 | 0.001411915 | 0.004428458 | 1 |
| HTN | SMC | *DOK7* | -12.74750295 | 0.004820354 | 0.011118112 | 1 |
| HTN | SMC | *B3GALT6* | -28.45120053 | 0.000517338 | 0.002239184 | 1 |
| HTN | SMC | *JAKMIP2* | -4.223329785 | 0.012837689 | 0.0246687 | 1 |
| HTN | SMC | *CCDC57* | -2.941980591 | 0.018119655 | 0.032853725 | 1 |
| HTN | SMC | *MYPOP* | -11.72224392 | 2.87E-05 | 0.000577552 | 0.139189981 |
| HTN | SMC | *CIDEA* | -4.758855241 | 0.000182276 | 0.001221551 | 0.884402781 |

| | | | | | | |
|---|---|---|---|---|---|---|
| HTN | SMC | *SLC35G1* | -3.577503849 | 0.000286207 | 0.001556812 | 1 |
| HTN | SMC | *OR4N2* | -4.764527682 | 0.014467099 | 0.027259947 | 1 |
| HTN | SMC | *DNAJC30* | -5.64547165 | 0.002106555 | 0.005897867 | 1 |
| HTN | SMC | *VPS37D* | -2.047193672 | 0.017008064 | 0.031082158 | 1 |
| HTN | SMC | *WDR25* | -5.32684654 | 8.33E-05 | 0.000876014 | 0.404081593 |
| HTN | SMC | *PLA2G16* | -1.736308712 | 0.026450801 | 0.045512461 | 1 |
| HTN | SMC | *B3GNT5* | -4.431119682 | 0.000173074 | 0.001187282 | 0.839756835 |
| HTN | SMC | *RMDN1* | -4.469042631 | 0.001258525 | 0.004057386 | 1 |
| HTN | SMC | *BDNF* | -7.187308112 | 0.008993797 | 0.018459349 | 1 |
| HTN | SMC | *BOK* | -8.315887558 | 5.37E-05 | 0.000806822 | 0.260441741 |
| HTN | SMC | *RUFY1* | -4.139430205 | 0.007498119 | 0.015970532 | 1 |
| HTN | SMC | *VSIG10* | -3.675241543 | 0.002814125 | 0.007340933 | 1 |
| HTN | SMC | *ANKLE2* | -8.086262236 | 1.60E-05 | 0.000427306 | 0.077430342 |
| HTN | SMC | *C8G* | 26.95093917 | 0.006644721 | 0.014476959 | 1 |
| HTN | SMC | *MUC20* | -4.308967248 | 0.024883205 | 0.043227107 | 1 |
| HTN | SMC | *SHMT1* | -4.688545473 | 0.000302965 | 0.001611829 | 1 |
| HTN | SMC | *DPP7* | -8.606420058 | 0.000166025 | 0.001164975 | 0.805554613 |
| HTN | SMC | *MTHFR* | -5.461497081 | 0.004449161 | 0.010479287 | 1 |
| HTN | SMC | *C19orf18* | -3.330748039 | 0.015132993 | 0.028294906 | 1 |
| HTN | SMC | *TMEM80* | -6.659465012 | 4.81E-05 | 0.000765887 | 0.233595562 |
| HTN | SMC | *WDR73* | -6.577010123 | 0.002473563 | 0.006645302 | 1 |
| HTN | SMC | *DSCAML1* | -6.82332335 | 5.56E-05 | 0.000817658 | 0.269827022 |
| HTN | SMC | *TALDO1* | -8.795795508 | 7.76E-05 | 0.000851198 | 0.376300154 |
| HTN | SMC | *MIEF2* | -12.82783499 | 0.000418316 | 0.001971331 | 1 |
| HTN | SMC | *SLC25A22* | -8.369829498 | 7.12E-05 | 0.000837934 | 0.345672456 |
| HTN | SMC | *C18orf32* | -2.948653889 | 0.011605917 | 0.022779898 | 1 |
| HTN | SMC | *GBA* | -5.548866388 | 0.000210574 | 0.001314937 | 1 |
| HTN | SMC | *IL17RA* | -5.467562802 | 0.00150209 | 0.004625724 | 1 |
| HTN | SMC | *THAP5* | -11.14655598 | 4.76E-05 | 0.00076177 | 0.23081618 |
| HTN | SMC | *NAALADL2* | -3.660496466 | 0.002510336 | 0.006721936 | 1 |
| HTN | SMC | *SLC35G5* | -16.6642975 | 0.000586095 | 0.00239775 | 1 |
| HTN | SMC | *FAM87B* | -27.88866064 | 0.000213683 | 0.00132582 | 1 |
| HTN | SMC | *CENPBD1* | -5.08778035 | 0.000525285 | 0.002257599 | 1 |
| HTN | SMC | *RPS27* | -5.999891972 | 0.010306926 | 0.020647895 | 1 |
| HTN | SMC | *RIC8A* | -7.795245025 | 0.000164453 | 0.001162037 | 0.79792702 |
| HTN | SMC | *DPY19L2* | 0.957482232 | 0.025662859 | 0.04437498 | 1 |
| HTN | SMC | *ADAMTSL1* | -3.427164513 | 0.000184577 | 0.001226806 | 0.895568642 |
| HTN | SMC | *GRAMD1C* | -2.307436125 | 0.012378272 | 0.023924626 | 1 |
| HTN | SMC | *DALRD3* | -8.703919912 | 0.000167924 | 0.00117064 | 0.814765312 |
| HTN | SMC | *ZNF114* | -4.200037983 | 0.029300198 | 0.049784887 | 1 |
| HTN | SMC | *SH2B1* | -8.102677714 | 0.000801386 | 0.002959152 | 1 |
| HTN | SMC | *VN1R1* | -6.063574334 | 0.001012195 | 0.003473248 | 1 |
| HTN | SMC | *KDELC2* | -8.407402854 | 1.49E-05 | 0.00041313 | 0.072297681 |
| HTN | SMC | *PLEC* | -2.923289159 | 0.017677988 | 0.03213698 | 1 |
| HTN | SMC | *SH2D4B* | -10.86745019 | 0.001945374 | 0.005552326 | 1 |
| HTN | SMC | *RNF212* | -3.860085376 | 0.008385894 | 0.017485328 | 1 |
| HTN | SMC | *PRSS36* | -4.808690267 | 0.000896332 | 0.003190757 | 1 |
| HTN | SMC | *GEN1* | -2.853105365 | 0.000102527 | 0.000942633 | 0.497461679 |
| HTN | SMC | *TMPRSS9* | -3.960961937 | 0.018353941 | 0.03317933 | 1 |
| HTN | SMC | *TMEM11* | -9.182592066 | 0.001603681 | 0.004802914 | 1 |

| | | | | | | |
|---|---|---|---|---|---|---|
| HTN | SMC | *ZFAND2A* | -8.502148722 | 0.002266084 | 0.006250733 | 1 |
| HTN | SMC | *FAM220A* | -4.686151381 | 0.001770845 | 0.00516665 | 1 |
| HTN | SMC | *NT5DC1* | -5.591077225 | 0.008094988 | 0.016966257 | 1 |
| HTN | SMC | *GLDC* | -5.852372167 | 0.00109311 | 0.003670429 | 1 |
| HTN | SMC | *P4HTM* | -12.12649495 | 3.84E-05 | 0.000689173 | 0.186344069 |
| HTN | SMC | *CA8* | -5.305782803 | 0.001582576 | 0.004754587 | 1 |
| HTN | SMC | *ERN1* | -7.340002365 | 4.91E-05 | 0.000773529 | 0.238154184 |
| HTN | SMC | *PARP10* | -7.61106762 | 2.82E-05 | 0.000569929 | 0.136782955 |
| HTN | SMC | *RPP25* | 2.674190473 | 0.001151787 | 0.003801682 | 1 |
| HTN | SMC | *MPI* | -10.40563833 | 9.07E-18 | 2.20E-14 | 4.40E-14 |
| HTN | SMC | *TRIM73* | -9.810040736 | 0.000264774 | 0.00148866 | 1 |
| HTN | SMC | *EFCAB13* | -4.159077865 | 0.00102324 | 0.00348894 | 1 |
| HTN | SMC | *ZNF852* | -6.273768599 | 0.000600852 | 0.002429444 | 1 |
| HTN | SMC | *HYI* | -5.319498481 | 1.51E-09 | 4.57E-07 | 7.32E-06 |
| HTN | SMC | *C17orf62* | -8.409371601 | 0.000176091 | 0.001201676 | 0.854391919 |
| HTN | SMC | *C3orf38* | -4.146844627 | 0.000515315 | 0.002236542 | 1 |
| HTN | SMC | *CCDC89* | -8.122958133 | 0.000592261 | 0.002410779 | 1 |
| HTN | SMC | *HTR1F* | 1.468544788 | 0.009842963 | 0.019849566 | 1 |
| HTN | SMC | *SPTY2D1* | -10.18874877 | 0.000390104 | 0.001877762 | 1 |
| HTN | SMC | *ZNF664* | -10.30190731 | 0.004904482 | 0.011267305 | 1 |
| HTN | SMC | *HLA-DQB1* | -1.949741175 | 0.000123905 | 0.00101896 | 0.601186505 |
| HTN | SMC | *PACS2* | -8.352390248 | 0.000496985 | 0.002202348 | 1 |
| HTN | SMC | *LINC00174* | -2.698651946 | 0.002534943 | 0.006776609 | 1 |
| HTN | SMC | *GEMIN4* | -6.60331401 | 0.000157631 | 0.001140149 | 0.76482537 |
| HTN | SMC | *LBX2* | -2.269213969 | 0.012087921 | 0.023460238 | 1 |
| HTN | SMC | *FCER1A* | -2.879578631 | 0.00110169 | 0.003689028 | 1 |
| HTN | SMC | *SERTAD2* | -6.252510845 | 0.000741825 | 0.002783709 | 1 |
| HTN | SMC | *AKAP5* | -3.735319784 | 0.00020216 | 0.001287247 | 0.980882013 |
| HTN | SMC | *GIPC3* | 2.866839762 | 0.003165592 | 0.00803739 | 1 |
| HTN | SMC | *PDXDC1* | -4.986958072 | 0.013196719 | 0.025248613 | 1 |
| HTN | SMC | *C1orf194* | -4.772202512 | 0.011724933 | 0.022957778 | 1 |
| HTN | SMC | *ZADH2* | -6.732546455 | 0.000207768 | 0.001311479 | 1 |
| HTN | SMC | *EXOC3* | -5.940780107 | 0.021124629 | 0.037407554 | 1 |
| HTN | SMC | *HCLS1* | -2.959488996 | 0.00027718 | 0.001528269 | 1 |
| HTN | SMC | *MTURN* | -6.964477797 | 0.000587128 | 0.002397932 | 1 |
| HTN | SMC | *CCDC66* | -6.585711471 | 0.000946628 | 0.003311492 | 1 |
| HTN | SMC | *DEFB124* | 4.736528628 | 0.004068498 | 0.009719522 | 1 |
| HTN | SMC | *MB21D2* | -6.328025957 | 0.000281863 | 0.001541823 | 1 |
| HTN | SMC | *SLC47A2* | -3.75660422 | 0.00092001 | 0.003251201 | 1 |
| HTN | SMC | *CHST13* | 4.825359775 | 0.003850271 | 0.009337834 | 1 |
| HTN | SMC | *SLC36A4* | -6.616786172 | 0.005736965 | 0.012803934 | 1 |
| HTN | SMC | *ZDHHC20* | -7.389519423 | 0.000404006 | 0.001929517 | 1 |
| HTN | SMC | *PSMG4* | -5.561673265 | 0.000944181 | 0.003305786 | 1 |
| HTN | SMC | *ZNF443* | -19.39661165 | 0.003512489 | 0.008699642 | 1 |
| HTN | SMC | *CAPS2* | -5.431115349 | 0.011978297 | 0.023331472 | 1 |
| HTN | SMC | *KCTD2* | -6.721661388 | 0.000143689 | 0.001091044 | 0.697177081 |
| HTN | SMC | *D2HGDH* | -5.369555201 | 1.51E-05 | 0.000416075 | 0.07322924 |
| HTN | SMC | *CMTR2* | -5.951980751 | 0.004482563 | 0.010537499 | 1 |
| HTN | SMC | *LSMEM1* | -2.699568634 | 0.00123275 | 0.003988396 | 1 |
| HTN | SMC | *NQO1* | -4.481417021 | 0.001482322 | 0.004587346 | 1 |

| | | | | | | |
|---|---|---|---|---|---|---|
| HTN | SMC | *RPH3AL* | -3.749712957 | 0.012060912 | 0.023435941 | 1 |
| HTN | SMC | *ADIPOQ* | -1.050320029 | 0.007988822 | 0.016787251 | 1 |
| HTN | SMC | *MUC16* | -8.064315672 | 0.000276089 | 0.001523987 | 1 |
| HTN | SMC | *SLC25A41* | -9.76592275 | 0.000843546 | 0.003048271 | 1 |
| HTN | SMC | *TMEM136* | 2.482934484 | 0.0038481 | 0.009337834 | 1 |
| HTN | SMC | *DDX60L* | -6.739206688 | 0.000373296 | 0.00182584 | 1 |
| HTN | SMC | *TMEM45A* | -4.399336815 | 0.003226532 | 0.008153471 | 1 |
| HTN | SMC | *SGSH* | -13.34956879 | 6.72E-05 | 0.000836037 | 0.325854874 |
| HTN | SMC | *ZBTB20* | -4.350151654 | 0.001656002 | 0.004910637 | 1 |
| HTN | SMC | *C3orf58* | -8.770944658 | 6.28E-05 | 0.000827638 | 0.304941765 |
| HTN | SMC | *TMEM252* | -88.2669993 | 0.000756428 | 0.00282757 | 1 |
| HTN | SMC | *ODF3L2* | -5.446655778 | 0.014450755 | 0.027247073 | 1 |
| HTN | SMC | *RFX7* | -6.448273912 | 0.001739654 | 0.005100181 | 1 |
| HTN | SMC | *CLDN7* | -4.646399821 | 1.57E-08 | 3.05E-06 | 7.62E-05 |
| HTN | SMC | *ZNF329* | -9.688621602 | 0.000431438 | 0.002012825 | 1 |
| HTN | SMC | *MGAT4C* | -1.515766315 | 0.000229349 | 0.001377231 | 1 |
| HTN | SMC | *IDH2* | -10.7636572 | 9.52E-06 | 0.000313646 | 0.0462062 |
| HTN | SMC | *FAM181B* | 3.277306634 | 0.001158728 | 0.003816803 | 1 |
| HTN | SMC | *ZNF708* | -5.043080572 | 0.0001855 | 0.001228781 | 0.900046026 |
| HTN | SMC | *MRPL41* | -11.59244424 | 0.000932188 | 0.003277519 | 1 |
| HTN | SMC | *HHIPL1* | -5.778486986 | 0.004693358 | 0.010920971 | 1 |
| HTN | SMC | *KCNA4* | -8.165717236 | 0.000103247 | 0.000942633 | 0.500953532 |
| HTN | SMC | *FIGN* | -4.862186566 | 0.007915922 | 0.016648484 | 1 |
| HTN | SMC | *C8orf33* | -7.477968383 | 0.007649871 | 0.016166016 | 1 |
| HTN | SMC | *FBXL6* | -4.623857635 | 0.001290903 | 0.0041425 | 1 |
| HTN | SMC | *KBTBD3* | -7.709622401 | 0.026323411 | 0.045339435 | 1 |
| HTN | SMC | *NXPH4* | -22.0015202 | 0.000586735 | 0.002397932 | 1 |
| HTN | SMC | *NPLOC4* | -3.714439257 | 0.004092073 | 0.009766227 | 1 |
| HTN | SMC | *FES* | -16.15863167 | 6.23E-22 | 3.03E-18 | 3.03E-18 |
| HTN | SMC | *GLRX5* | -10.48087412 | 6.63E-05 | 0.000836037 | 0.321781027 |
| HTN | SMC | *MXRA7* | -2.577122389 | 0.002307931 | 0.006323026 | 1 |
| HTN | SMC | *ADI1* | -8.983976328 | 0.000752515 | 0.002817287 | 1 |
| HTN | SMC | *CLEC4G* | -8.443854128 | 0.000718277 | 0.002726438 | 1 |
| HTN | SMC | *TRAK1* | -4.378565983 | 0.004977775 | 0.011395213 | 1 |
| HTN | SMC | *BRICD5* | -16.19899235 | 8.80E-05 | 0.000888243 | 0.426907365 |
| HTN | SMC | *PLCXD3* | -7.221835478 | 6.26E-05 | 0.000827638 | 0.30369601 |
| HTN | SMC | *GPIHBP1* | -1.549562597 | 0.000852475 | 0.003075247 | 1 |
| HTN | SMC | *COL18A1* | 2.344485888 | 0.009403622 | 0.019109588 | 1 |
| HTN | SMC | *CEP63* | -3.96295119 | 0.002088544 | 0.005864362 | 1 |
| HTN | SMC | *ODF3L1* | -10.51211929 | 0.000172181 | 0.001186677 | 0.835420363 |
| HTN | SMC | *ZNF320* | 1.183526995 | 0.016023683 | 0.029606592 | 1 |
| HTN | SMC | *ABAT* | -5.827601799 | 4.46E-05 | 0.000752126 | 0.21631497 |
| HTN | SMC | *LYSMD4* | -6.088741738 | 4.60E-05 | 0.000758453 | 0.222985127 |
| HTN | SMC | *WBP2NL* | -9.350794761 | 3.14E-06 | 0.000153181 | 0.015223108 |
| HTN | SMC | *CSMD1* | -10.82702571 | 0.000494179 | 0.00219776 | 1 |
| HTN | SMC | *FANCF* | -8.450372145 | 0.000380516 | 0.001846261 | 1 |
| HTN | SMC | *RUVBL2* | -6.873556614 | 0.000243594 | 0.001420122 | 1 |
| HTN | SMC | *ZNF623* | -4.37339448 | 0.002010302 | 0.005700751 | 1 |
| HTN | SMC | *CCDC125* | -6.712257636 | 8.95E-05 | 0.000888243 | 0.434298872 |
| HTN | SMC | *C19orf71* | -4.365506092 | 0.000499493 | 0.002207048 | 1 |

| | | | | | | |
|---|---|---|---|---|---|---|
| HTN | SMC | *LRIT3* | -15.93209832 | 0.000535048 | 0.002281742 | 1 |
| HTN | SMC | *NPIPA1* | 1.503450724 | 0.010927344 | 0.021684856 | 1 |
| HTN | SMC | *SF3A3* | -3.384323798 | 0.013668134 | 0.025996781 | 1 |
| HTN | SMC | *GPR132* | -3.15571688 | 0.00239734 | 0.006509174 | 1 |
| HTN | SMC | *SERHL2* | -4.728196198 | 0.0001426 | 0.001090656 | 0.69189317 |
| HTN | SMC | *HMCES* | -5.913607296 | 0.001550844 | 0.004700863 | 1 |
| HTN | SMC | *RP1L1* | -4.632390307 | 0.007325701 | 0.015644498 | 1 |
| HTN | SMC | *NDUFB1* | -5.547101113 | 0.008274613 | 0.017268138 | 1 |
| HTN | SMC | *FAM19A1* | -4.852231916 | 0.000170909 | 0.001182956 | 0.829252066 |
| HTN | SMC | *UPP1* | -3.610166114 | 0.00940907 | 0.019109588 | 1 |
| HTN | SMC | *FIGLA* | -12.35042154 | 7.87E-05 | 0.00085619 | 0.3817959 |
| HTN | SMC | *ACSM2A* | -18.75462951 | 0.001692464 | 0.004998073 | 1 |
| HTN | SMC | *KREMEN1* | -3.207830224 | 0.000165235 | 0.001162037 | 0.801722232 |
| HTN | SMC | *NPIPA5* | -0.851136008 | 0.028849181 | 0.049131705 | 1 |
| HTN | SMC | *BTBD9* | -5.737451992 | 0.000529054 | 0.002261977 | 1 |
| HTN | SMC | *NUDT14* | -9.484633861 | 0.001705095 | 0.005014626 | 1 |
| HTN | SMC | *TTC32* | -9.696473791 | 0.00112704 | 0.003742915 | 1 |
| HTN | SMC | *PP2D1* | -7.009607743 | 9.47E-05 | 0.000903 | 0.459672668 |
| HTN | SMC | *DENND5A* | -5.654116665 | 0.009297512 | 0.018930561 | 1 |
| HTN | SMC | *NIPSNAP1* | -3.292007261 | 0.025540954 | 0.044195689 | 1 |
| HTN | SMC | *CRELD2* | -5.821062644 | 0.00250807 | 0.006721936 | 1 |
| HTN | SMC | *ALDH1A3* | -3.420362523 | 9.92E-06 | 0.000321952 | 0.048144093 |
| HTN | SMC | *CLECL1* | -3.54047254 | 0.000678329 | 0.002626697 | 1 |
| HTN | SMC | *A3GALT2* | -3.639978411 | 1.49E-06 | 9.66E-05 | 0.007253692 |
| HTN | SMC | *SS18L1* | -6.900062399 | 4.51E-05 | 0.000754445 | 0.218789148 |
| HTN | SMC | *TOP1MT* | -2.632749458 | 0.000195427 | 0.001271662 | 0.948213496 |
| HTN | SMC | *COPB2* | -7.023014432 | 0.000107639 | 0.000951299 | 0.522263234 |
| HTN | SMC | *WDR27* | -4.057500299 | 0.002466481 | 0.006637474 | 1 |
| HTN | SMC | *TMEM255B* | -3.353685169 | 0.001168219 | 0.003842846 | 1 |
| HTN | SMC | *HDDC3* | -7.929825325 | 0.001580532 | 0.004751388 | 1 |
| HTN | SMC | *TCTE3* | -11.40461713 | 0.000187803 | 0.001239754 | 0.911219036 |
| HTN | SMC | *TMED9* | -5.891162304 | 0.019693714 | 0.035242897 | 1 |
| HTN | SMC | *DRD1* | 1.489041915 | 0.002301016 | 0.006307643 | 1 |
| HTN | SMC | *NUTM2A* | -4.951099255 | 2.26E-05 | 0.000515555 | 0.109574583 |
| HTN | SMC | *WT1* | -2.91008726 | 0.00038096 | 0.001846573 | 1 |
| HTN | SMC | *SORCS2* | -6.490412068 | 0.000130033 | 0.00104255 | 0.630919152 |
| HTN | SMC | *BRI3BP* | -7.942062817 | 2.54E-05 | 0.000550673 | 0.123350744 |
| HTN | SMC | *UBOX5* | -4.52065913 | 0.000263173 | 0.001484785 | 1 |
| HTN | SMC | *MAFF* | -2.741886092 | 0.004140295 | 0.009861911 | 1 |
| HTN | SMC | *C6orf120* | -6.87961757 | 0.001702379 | 0.005012103 | 1 |
| HTN | SMC | *CDK10* | -3.482186927 | 6.60E-06 | 0.000244574 | 0.032039187 |
| HTN | SMC | *ATP6V0A2* | -5.668748658 | 0.000223457 | 0.001360366 | 1 |
| HTN | SMC | *TNFAIP8L1* | -6.946531496 | 2.65E-05 | 0.000561014 | 0.128472093 |
| HTN | SMC | *SP140L* | -5.678220858 | 0.004066975 | 0.009719522 | 1 |
| HTN | SMC | *TARSL2* | -5.367833112 | 0.001217107 | 0.003950101 | 1 |
| HTN | SMC | *SH3BGR* | -1.48937679 | 0.005900721 | 0.013115115 | 1 |
| HTN | SMC | *FAM174B* | -7.882204533 | 0.000358631 | 0.001793894 | 1 |
| HTN | SMC | *SV2B* | -3.473546103 | 0.005143659 | 0.011715907 | 1 |
| HTN | SMC | *AHNAK2* | -6.773286998 | 0.006356003 | 0.013948135 | 1 |
| HTN | SMC | *SP1* | -6.010774273 | 0.005625768 | 0.01259632 | 1 |

| | | | | | | |
|---|---|---|---|---|---|---|
| HTN | SMC | *PCGF3* | -4.447124543 | 0.010020878 | 0.02014973 | 1 |
| HTN | SMC | *PSMD13* | -6.322631431 | 0.001461208 | 0.004550565 | 1 |
| HTN | SMC | *C6orf201* | -4.511034459 | 0.001630983 | 0.004854926 | 1 |
| HTN | SMC | *MYBL1* | -11.5247765 | 1.03E-06 | 7.02E-05 | 0.004986033 |
| HTN | SMC | *ADARB2* | -4.191452847 | 0.001550804 | 0.004700863 | 1 |
| HTN | SMC | *SLC52A2* | -2.899730804 | 0.01035146 | 0.020720001 | 1 |
| HTN | SMC | *ARL17A* | -3.408474105 | 0.012084341 | 0.023460238 | 1 |
| HTN | SMC | *ZNF829* | 3.523473113 | 0.00540467 | 0.012202634 | 1 |
| HTN | SMC | *IFITM1* | -6.019253192 | 6.88E-05 | 0.000836037 | 0.333730566 |
| HTN | SMC | *LINC00839* | -6.970715635 | 0.000295734 | 0.00158203 | 1 |
| HTN | SMC | *RNPC3* | -3.746924538 | 0.003368296 | 0.008424213 | 1 |
| HTN | SMC | *RASA3* | -4.863749383 | 0.000153883 | 0.001136201 | 0.746638984 |
| HTN | SMC | *ZNF529* | -6.184504664 | 0.000617073 | 0.002454805 | 1 |
| HTN | SMC | *WWOX* | -4.74425654 | 0.00136041 | 0.004305747 | 1 |
| HTN | SMC | *POLR1D* | -5.456663844 | 0.00349127 | 0.008655924 | 1 |
| HTN | SMC | *FFAR4* | -3.000442893 | 0.004273273 | 0.010128931 | 1 |
| HTN | SMC | *CYP4F12* | -5.505051446 | 0.001564337 | 0.00472613 | 1 |
| HTN | SMC | *ZNF555* | -5.561385383 | 0.013125942 | 0.025152872 | 1 |
| HTN | SMC | *GLDN* | 0.808244793 | 0.003316935 | 0.008334 | 1 |
| HTN | SMC | *ZNF197* | -5.100396742 | 0.009114789 | 0.018660319 | 1 |
| HTN | SMC | *RPS23* | -4.029833093 | 0.018513643 | 0.033405799 | 1 |
| HTN | SMC | *GNG2* | -6.121640721 | 7.44E-05 | 0.000847247 | 0.361071914 |
| HTN | SMC | *TMEM222* | -8.178831117 | 1.89E-05 | 0.000473276 | 0.091768835 |
| HTN | SMC | *SEPT10* | -6.609750397 | 0.002207044 | 0.006115694 | 1 |
| HTN | SMC | *SMYD4* | -4.658872116 | 0.003585803 | 0.008827153 | 1 |
| HTN | SMC | *MIR22HG* | -8.547519759 | 0.003411426 | 0.008497042 | 1 |
| HTN | SMC | *CYP27C1* | -2.616627816 | 0.001192334 | 0.003898386 | 1 |
| HTN | SMC | *BCR* | -7.082122358 | 0.000162038 | 0.001159599 | 0.786208047 |
| HTN | SMC | *FOXI2* | -4.006033778 | 0.002917088 | 0.007546874 | 1 |
| HTN | SMC | *ZNF732* | -11.19826727 | 0.000926347 | 0.003264289 | 1 |
| HTN | SMC | *QRFPR* | 9.134637718 | 0.001391074 | 0.004385636 | 1 |
| HTN | SMC | *C1QL4* | -26.40822376 | 0.000555504 | 0.002326902 | 1 |
| HTN | SMC | *EFCAB6* | -6.848161056 | 0.000815572 | 0.002979786 | 1 |
| HTN | SMC | *TMPRSS6* | 6.652386759 | 0.004748082 | 0.010991265 | 1 |
| HTN | SMC | *MRPS21* | -3.08061588 | 0.010303358 | 0.020647895 | 1 |
| HTN | SMC | *WDR86* | -1.981944685 | 0.006209116 | 0.013693924 | 1 |
| HTN | SMC | *LUZP2* | -3.194663769 | 0.000463022 | 0.002113438 | 1 |
| HTN | SMC | *IFT140* | -6.150356842 | 0.017803963 | 0.032341755 | 1 |
| HTN | SMC | *SAMD11* | 5.292818635 | 0.005310051 | 0.012016963 | 1 |
| HTN | SMC | *C2orf88* | -4.942120007 | 3.37E-05 | 0.000641172 | 0.163623727 |
| HTN | SMC | *FANCM* | -6.969682419 | 0.000112852 | 0.0009796 | 0.54755956 |
| HTN | SMC | *ZFP69* | -8.924018627 | 8.20E-05 | 0.000876014 | 0.398007452 |
| HTN | SMC | *MAPK12* | -6.874473724 | 0.00015858 | 0.001143283 | 0.769429363 |
| HTN | SMC | *AGRN* | -13.61814502 | 0.000125855 | 0.001028322 | 0.610648077 |
| HTN | SMC | *ZNF626* | -8.175487512 | 6.88E-05 | 0.000836037 | 0.33400736 |
| HTN | SMC | *PRKAR1B* | -4.285901151 | 0.003498338 | 0.008669018 | 1 |
| HTN | SMC | *ZNF793* | -2.359045574 | 0.004180329 | 0.009932887 | 1 |
| HTN | SMC | *RUFY4* | -6.519740136 | 0.000377644 | 0.001841989 | 1 |
| HTN | SMC | *CENPP* | -3.703836894 | 8.51E-06 | 0.000291476 | 0.041272987 |
| HTN | SMC | *C3orf62* | -4.969276387 | 4.64E-05 | 0.000760128 | 0.225252793 |

| | | | | | | |
|---|---|---|---|---|---|---|
| HTN | SMC | *ENO4* | -4.24607781 | 0.006313606 | 0.013873919 | 1 |
| HTN | SMC | *ZNF559* | -2.325661451 | 8.56E-05 | 0.000876014 | 0.415356412 |
| HTN | SMC | *H3F3C* | -75.81539141 | 9.02E-05 | 0.000888243 | 0.437837175 |
| HTN | SMC | *JAKMIP3* | -1.434166661 | 0.010460025 | 0.02088991 | 1 |
| HTN | SMC | *PDCD1* | -10.12467038 | 0.019719614 | 0.035267073 | 1 |
| HTN | SMC | *C19orf54* | -6.089051253 | 0.000494762 | 0.002198339 | 1 |
| HTN | SMC | *C22orf34* | -6.216288733 | 0.00040523 | 0.001929517 | 1 |
| HTN | SMC | *COL25A1* | 1.270555051 | 0.014661912 | 0.027562804 | 1 |
| HTN | SMC | *NBR1* | -6.663379344 | 0.002407953 | 0.006534333 | 1 |
| HTN | SMC | *GOLGA8M* | -3.400604627 | 0.000513373 | 0.002236542 | 1 |
| HTN | SMC | *RHCE* | -9.13445267 | 0.003086165 | 0.007879215 | 1 |
| HTN | SMC | *IDO2* | -4.899872217 | 0.005148045 | 0.011715907 | 1 |
| HTN | SMC | *SLC4A5* | 2.720953382 | 0.004172765 | 0.009919772 | 1 |
| HTN | SMC | *UROS* | -9.124490295 | 0.000339431 | 0.001733601 | 1 |
| HTN | SMC | *ZBED6CL* | -4.81972218 | 0.016087813 | 0.029693493 | 1 |
| HTN | SMC | *OSTN* | -2.028597182 | 0.00326927 | 0.00824026 | 1 |
| HTN | SMC | *FSIP2* | -3.240207237 | 0.000125891 | 0.001028322 | 0.610823546 |
| HTN | SMC | *NOXA1* | -11.16723177 | 0.001179623 | 0.003869982 | 1 |
| HTN | SMC | *FZD9* | 39.93459992 | 0.004283324 | 0.010147797 | 1 |
| HTN | SMC | *FAM26F* | -3.255344344 | 0.000184566 | 0.001226806 | 0.895512033 |
| HTN | SMC | *ENTPD8* | 156.1271583 | 0.011246563 | 0.022200295 | 1 |
| HTN | SMC | *FBF1* | -8.883261449 | 1.31E-10 | 7.07E-08 | 6.37E-07 |
| HTN | SMC | *ASTL* | 4.204416539 | 0.006290091 | 0.013841053 | 1 |
| HTN | SMC | *MSL1* | -6.33356265 | 5.24E-06 | 0.000206614 | 0.025413527 |
| HTN | SMC | *UTS2B* | -5.560109836 | 0.000349157 | 0.001764904 | 1 |
| HTN | SMC | *NELFB* | -10.86570345 | 0.00094155 | 0.003305619 | 1 |
| HTN | SMC | *SLC15A5* | -2.430067999 | 0.003407613 | 0.008491905 | 1 |
| HTN | SMC | *ADAT2* | -7.47327569 | 0.000111105 | 0.000970929 | 0.539082774 |
| HTN | SMC | *LITAF* | -7.736759004 | 0.00047442 | 0.002151296 | 1 |
| HTN | SMC | *SF3B3* | -6.133063107 | 0.008799154 | 0.018171389 | 1 |
| HTN | SMC | *ZNF573* | -4.964298547 | 0.004099637 | 0.009774913 | 1 |
| HTN | SMC | *FHIT* | -4.923181767 | 0.000651994 | 0.002547086 | 1 |
| HTN | SMC | *RRP7A* | -6.334059988 | 0.000332447 | 0.001708722 | 1 |
| HTN | SMC | *KIAA0408* | -1.431180146 | 0.029339452 | 0.049826749 | 1 |
| HTN | SMC | *BLOC1S2* | -2.314520534 | 0.002309893 | 0.00632483 | 1 |
| HTN | SMC | *HLA-DRB1* | -2.704122005 | 4.61E-06 | 0.000189428 | 0.022352446 |
| HTN | SMC | *AKR1C3* | -3.642145605 | 0.0001801 | 0.001213674 | 0.873845047 |
| HTN | SMC | *WDSUB1* | -5.384913585 | 0.00697421 | 0.015086431 | 1 |
| HTN | SMC | *COLCA1* | -7.07175818 | 4.17E-07 | 3.81E-05 | 0.002021404 |
| HTN | SMC | *SEMA4A* | -6.800034237 | 0.000366707 | 0.001809686 | 1 |
| HTN | SMC | *SIRPB2* | -1.465463552 | 0.007011679 | 0.015140484 | 1 |
| HTN | SMC | *SRGAP3* | -2.592576813 | 0.024664307 | 0.042894518 | 1 |
| HTN | SMC | *XPNPEP3* | -1.995944583 | 0.009572759 | 0.019369068 | 1 |
| HTN | SMC | *PPIA* | -4.809762767 | 0.010418295 | 0.020831953 | 1 |
| HTN | SMC | *ZNF471* | -9.084708867 | 6.79E-05 | 0.000836037 | 0.329316817 |
| HTN | SMC | *ZNF493* | -9.2549089 | 6.44E-05 | 0.000829756 | 0.312360706 |
| HTN | SMC | *GTF2IRD2* | -8.344356336 | 0.000553132 | 0.002322255 | 1 |
| HTN | SMC | *GIMAP5* | -3.75066483 | 0.01369661 | 0.026040733 | 1 |
| HTN | SMC | *STK31* | -5.239556727 | 0.006887569 | 0.014953353 | 1 |
| HTN | SMC | *CD55* | -6.342093646 | 6.81E-05 | 0.000836037 | 0.330586266 |

| | | | | | | |
|---|---|---|---|---|---|---|
| HTN | SMC | *CPNE4* | -3.904832192 | 0.001262915 | 0.004068834 | 1 |
| HTN | SMC | *ZNF565* | -3.297801045 | 0.025556547 | 0.044206904 | 1 |
| HTN | SMC | *ZNF124* | -7.578320344 | 0.000558698 | 0.002332876 | 1 |
| HTN | SMC | *PIK3R4* | -8.055729172 | 0.002228146 | 0.006167121 | 1 |
| HTN | SMC | *RFX8* | -1.862071259 | 0.003057172 | 0.007823523 | 1 |
| HTN | SMC | *C20orf96* | -7.403759354 | 6.94E-05 | 0.000836037 | 0.336661173 |
| HTN | SMC | *SULT1A1* | -5.202546571 | 7.10E-06 | 0.000259096 | 0.034459772 |
| HTN | SMC | *TPK1* | -7.257859411 | 0.000231031 | 0.001378826 | 1 |
| HTN | SMC | *CACNA1H* | 0.518928302 | 0.0027562 | 0.007260087 | 1 |
| HTN | SMC | *XRCC2* | -7.637763293 | 0.000288342 | 0.001564914 | 1 |
| HTN | SMC | *MYO6* | -7.205083237 | 0.000228898 | 0.001377231 | 1 |
| HTN | SMC | *ADH1B* | -2.868114995 | 7.79E-05 | 0.000851198 | 0.37773652 |
| HTN | SMC | *ZNF502* | -4.544859991 | 0.004065745 | 0.009719522 | 1 |
| HTN | SMC | *TRAPPC4* | -2.255522581 | 0.029105102 | 0.049498057 | 1 |
| HTN | SMC | *TOMM7* | -7.842868042 | 0.000710323 | 0.002703126 | 1 |
| HTN | SMC | *AMZ2* | -6.328442862 | 0.001432348 | 0.004477932 | 1 |
| HTN | SMC | *HLA-DQA1* | -2.734703606 | 6.14E-05 | 0.000827638 | 0.29773096 |
| HTN | SMC | *COL27A1* | 1.185230478 | 0.023524008 | 0.041145814 | 1 |
| HTN | SMC | *ZNF239* | -3.160445953 | 0.000560295 | 0.002337532 | 1 |
| HTN | SMC | *ANKRD36B* | -6.468993887 | 0.00035308 | 0.001775279 | 1 |
| HTN | SMC | *SLC39A10* | -7.823341965 | 0.000122297 | 0.001013054 | 0.593386413 |
| HTN | SMC | *AP2A1* | -7.553960866 | 1.38E-05 | 0.000396487 | 0.06700622 |
| HTN | SMC | *PCDHB16* | 2.067547235 | 0.011975869 | 0.023331472 | 1 |
| HTN | SMC | *HIST1H3E* | 3.228663736 | 2.15E-05 | 0.000505599 | 0.104115524 |
| HTN | SMC | *FUT11* | -6.099556725 | 0.000583801 | 0.002394424 | 1 |
| HTN | SMC | *ZNF429* | -6.891217349 | 3.82E-05 | 0.000689173 | 0.185350724 |
| HTN | SMC | *ZNF470* | -6.51225575 | 0.0043989 | 0.010376016 | 1 |
| HTN | SMC | *ZNF100* | -7.394112215 | 2.73E-09 | 6.74E-07 | 1.32E-05 |
| HTN | SMC | *GMFB* | -9.662987939 | 0.000352496 | 0.001775279 | 1 |
| HTN | SMC | *ZMYM1* | -8.540869214 | 0.000305726 | 0.001619413 | 1 |
| HTN | SMC | *IGF2R* | -6.402100176 | 0.001864052 | 0.005377159 | 1 |
| HTN | SMC | *ZNF772* | -2.498346362 | 0.000323322 | 0.001674236 | 1 |
| HTN | SMC | *ZNF257* | -5.480770584 | 0.000942224 | 0.003305619 | 1 |
| HTN | SMC | *ADAM32* | -6.041725971 | 6.87E-05 | 0.000836037 | 0.333242092 |
| HTN | SMC | *SND1* | -4.818520719 | 0.000836745 | 0.003034294 | 1 |
| HTN | SMC | *PIWIL2* | -3.174012596 | 0.001607163 | 0.004807618 | 1 |
| HTN | SMC | *ENTPD4* | -6.778635388 | 0.004032552 | 0.009671745 | 1 |
| HTN | SMC | *TBC1D9B* | -3.517836336 | 0.022344111 | 0.039280299 | 1 |
| HTN | SMC | *C6orf141* | -1.265727064 | 0.000347843 | 0.001761973 | 1 |
| HTN | SMC | *RAMP2-AS1* | -2.881006909 | 0.000289306 | 0.001565572 | 1 |
| HTN | SMC | *ZNF720* | -7.021220353 | 0.000148261 | 0.00111876 | 0.719362652 |
| HTN | SMC | *ZNF786* | -3.973896993 | 0.001984485 | 0.005650658 | 1 |
| HTN | SMC | *ZNF517* | -6.950144474 | 1.30E-05 | 0.000389012 | 0.063044097 |
| HTN | SMC | *ZNF675* | -8.220238499 | 0.001207758 | 0.003930277 | 1 |
| HTN | SMC | *VEPH1* | -6.178703284 | 0.000203709 | 0.001293711 | 0.988395512 |
| HTN | SMC | *SHPK* | -2.548356124 | 0.012024371 | 0.023389336 | 1 |
| HTN | SMC | *MAP3K5* | -4.676852891 | 0.010695928 | 0.02130404 | 1 |
| HTN | SMC | *GSTK1* | -4.801907982 | 0.001284602 | 0.004125008 | 1 |
| HTN | SMC | *GYPE* | -3.152892488 | 0.019142606 | 0.034412718 | 1 |
| HTN | SMC | *PCDHB11* | -2.125329995 | 0.001033427 | 0.003513798 | 1 |

| | | | | | | |
|---|---|---|---|---|---|---|
| HTN | SMC | *ZNF665* | -10.3246018 | 0.001106164 | 0.003693812 | 1 |
| HTN | SMC | *RPF2* | -7.999816768 | 0.001007118 | 0.003458271 | 1 |
| HTN | SMC | *MYO5A* | -4.163225784 | 0.000299627 | 0.001597571 | 1 |
| HTN | SMC | *RAB40C* | -9.87360789 | 0.000154085 | 0.001136201 | 0.747620378 |
| HTN | SMC | *TOPORS* | -10.01453555 | 0.000173247 | 0.001187282 | 0.840595338 |
| HTN | SMC | *BCO2* | -1.707706251 | 0.003426505 | 0.008525846 | 1 |
| HTN | SMC | *ENTPD6* | -7.547570171 | 0.000137422 | 0.001071047 | 0.666771339 |
| HTN | SMC | *MFAP5* | -5.79358224 | 1.20E-08 | 2.54E-06 | 5.84E-05 |
| HTN | SMC | *NMB* | -4.719915392 | 0.000811994 | 0.002971188 | 1 |
| HTN | SMC | *FAM114A1* | -6.772623367 | 0.000363406 | 0.001804758 | 1 |
| HTN | SMC | *RPS26* | -3.435731446 | 2.17E-08 | 3.76E-06 | 0.0001053 |
| HTN | SMC | *PSAP* | -4.49557479 | 0.002046439 | 0.005776219 | 1 |
| HTN | SMC | *TXNRD3* | -3.632152248 | 0.000599408 | 0.002427651 | 1 |
| HTN | SMC | *EME2* | -16.49341531 | 0.000189824 | 0.001248001 | 0.921024654 |
| HTN | SMC | *ZNF780A* | -1.026238584 | 0.011022367 | 0.021828098 | 1 |
| HTN | SMC | *ZNF181* | -3.728877939 | 0.009074315 | 0.018601004 | 1 |
| HTN | SMC | *SGTB* | -7.8560739 | 0.000382696 | 0.001850936 | 1 |
| HTN | SMC | *ADH5* | -6.609274399 | 0.000231323 | 0.001378826 | 1 |
| HTN | SMC | *ZNF677* | -11.52331621 | 9.39E-05 | 0.000903 | 0.455436251 |
| HTN | SMC | *FCHSD1* | -4.202566945 | 0.001844903 | 0.005337787 | 1 |
| HTN | SMC | *RPL12* | -3.869802955 | 0.002866048 | 0.007448347 | 1 |
| HTN | SMC | *MPZL1* | -5.578843868 | 0.000129888 | 0.00104255 | 0.630215473 |
| HTN | SMC | *VPS13A* | -5.42297787 | 0.005045271 | 0.011519838 | 1 |
| HTN | SMC | *NOL8* | -5.113086027 | 0.018736814 | 0.033770809 | 1 |
| HTN | SMC | *DLGAP2* | -3.038924249 | 0.007188639 | 0.015406042 | 1 |
| HTN | SMC | *AGAP9* | -5.42862251 | 4.77E-06 | 0.000194457 | 0.023140337 |
| HTN | SMC | *ZNF273* | -5.477589145 | 6.50E-05 | 0.000832115 | 0.315371731 |
| HTN | SMC | *SFI1* | -10.62861917 | 5.07E-06 | 0.00020334 | 0.024604175 |
| HTN | SMC | *ADH4* | -3.143389043 | 6.79E-05 | 0.000836037 | 0.329467727 |
| HTN | SMC | *ZNF544* | -9.372181645 | 7.78E-05 | 0.000851198 | 0.377410972 |
| HTN | SMC | *SOWAHC* | -9.925627648 | 0.002228118 | 0.006167121 | 1 |
| HTN | SMC | *MIER1* | -7.007227784 | 0.000427695 | 0.002004278 | 1 |
| HTN | SMC | *MAN1A2* | -5.753738439 | 0.005519041 | 0.012391665 | 1 |
| HTN | SMC | *SVIP* | -7.967621408 | 0.004697671 | 0.010921465 | 1 |
| HTN | SMC | *DDRGK1* | -7.026037743 | 0.011536539 | 0.022680424 | 1 |
| HTN | SMC | *ZNF334* | -4.234698234 | 0.00048436 | 0.002174021 | 1 |
| HTN | SMC | *HSD17B11* | -5.680039768 | 0.001516443 | 0.004642097 | 1 |
| HTN | SMC | *SULT1C2* | -1.28798873 | 0.021003201 | 0.037233296 | 1 |
| HTN | SMC | *DDX42* | -5.149153344 | 0.021598869 | 0.038094407 | 1 |
| HTN | SMC | *HELZ* | -5.873323443 | 0.000228881 | 0.001377231 | 1 |
| HTN | SMC | *TMEM116* | -5.205995385 | 8.01E-16 | 1.29E-12 | 3.88E-12 |
| HTN | SMC | *UCKL1* | -11.67703972 | 9.99E-05 | 0.000930246 | 0.484658009 |
| HTN | SMC | *GFPT1* | -6.515668621 | 0.001429207 | 0.004473878 | 1 |
| HTN | SMC | *ZNF568* | -9.224659564 | 2.77E-06 | 0.000141251 | 0.013418863 |
| HTN | SMC | *HLA-DRB5* | -0.99589472 | 2.82E-05 | 0.000569929 | 0.13670561 |
| HTN | SMC | *NCOA6* | -6.213306641 | 1.72E-05 | 0.000441573 | 0.08345723 |
| HTN | SMC | *FAM19A2* | -4.625900183 | 0.000192398 | 0.001258104 | 0.93351323 |
| HTN | SMC | *TTC37* | -6.054355864 | 0.001021531 | 0.003488014 | 1 |
| HTN | SMC | *TUSC1* | -19.42832806 | 3.69E-05 | 0.000673182 | 0.179066498 |
| HTN | SMC | *PAPSS2* | -5.037825626 | 6.13E-05 | 0.000827638 | 0.297543615 |

| | | | | | | |
|---|---|---|---|---|---|---|
| HTN | SMC | *CEP290* | -6.275705309 | 0.000541241 | 0.002296175 | 1 |
| HTN | SMC | *ANKRD13B* | -8.319934703 | 0.000114081 | 0.00098569 | 0.553519926 |
| HTN | SMC | *ECI2* | -6.841104603 | 2.81E-05 | 0.000569929 | 0.136232817 |
| HTN | SMC | *PPP1R14C* | -7.831239068 | 0.000573736 | 0.002363597 | 1 |
| HTN | SMC | *COLGALT2* | -3.528798797 | 0.001318327 | 0.004213784 | 1 |
| HTN | SMC | *PNP* | -1.497222267 | 0.000123781 | 0.00101896 | 0.600583428 |
| HTN | SMC | *SFT2D1* | -8.399090663 | 0.000549708 | 0.002313255 | 1 |
| HTN | SMC | *HMGN2* | -5.40592079 | 0.000868542 | 0.003119295 | 1 |
| HTN | SMC | *DENND4B* | -6.694016057 | 2.44E-08 | 3.94E-06 | 0.000118307 |
| HTN | SMC | *TYW1* | -4.606883573 | 0.011395669 | 0.022439849 | 1 |
| HTN | SMC | *ITPRIPL1* | -1.531697017 | 0.012686252 | 0.024426069 | 1 |
| HTN | SMC | *PRMT6* | -6.833343377 | 0.000244252 | 0.001420997 | 1 |
| HTN | SMC | *MAP3K3* | -7.6997791 | 4.58E-06 | 0.000189428 | 0.022246065 |
| HTN | SMC | *C1orf174* | -3.586399281 | 0.025784976 | 0.04452267 | 1 |
| HTN | SMC | *DZIP3* | -6.220029982 | 0.006891429 | 0.014953353 | 1 |
| HTN | SMC | *KIAA0753* | -7.866836153 | 1.07E-05 | 0.000336521 | 0.051807851 |
| HTN | SMC | *CCDC167* | -15.20668802 | 0.001430804 | 0.004475991 | 1 |
| HTN | SMC | *ZFP2* | -6.93685676 | 4.72E-05 | 0.00076177 | 0.229232109 |
| HTN | SMC | *MFAP3L* | 1.993864986 | 0.001772733 | 0.005169052 | 1 |
| HTN | SMC | *SMG5* | -6.968382742 | 1.07E-05 | 0.000336521 | 0.051824203 |
| HTN | SMC | *PJA2* | -4.53811361 | 0.019552413 | 0.035045551 | 1 |
| HTN | SMC | *AL590822.1* | -7.127528679 | 8.33E-07 | 6.00E-05 | 0.004039318 |
| HTN | SMC | *EFCAB2* | -4.715228501 | 0.000391531 | 0.001882765 | 1 |
| HTN | SMC | *TATDN3* | -2.367702618 | 0.010462177 | 0.02088991 | 1 |
| HTN | SMC | *SAMD5* | -4.31059925 | 0.002960156 | 0.007615417 | 1 |
| HTN | SMC | *GPR52* | -37.16583674 | 0.000255424 | 0.001454597 | 1 |
| HTN | SMC | *FAM229B* | -6.458734474 | 0.003387104 | 0.00845382 | 1 |
| HTN | SMC | *RBM20* | 1.018945051 | 0.003809366 | 0.009283296 | 1 |
| HTN | SMC | *C6orf163* | -2.139806312 | 0.001374928 | 0.004340371 | 1 |
| HTN | SMC | *PCMTD2* | -6.493887243 | 5.91E-05 | 0.000817716 | 0.286831603 |
| HTN | SMC | *SAMD13* | -9.461867358 | 0.000243809 | 0.001420122 | 1 |
| HTN | SMC | *EFCAB7* | -5.1526963 | 0.02079607 | 0.036893065 | 1 |
| HTN | SMC | *IFIT1B* | -25.34972245 | 0.000334258 | 0.001714395 | 1 |
| HTN | SMC | *LIPN* | -5.694273082 | 0.000667012 | 0.002587006 | 1 |
| HTN | SMC | *INPP5B* | -2.07490004 | 0.002026864 | 0.005734311 | 1 |
| HTN | SMC | *RUFY2* | -5.240550822 | 0.013193802 | 0.025248613 | 1 |
| HTN | SMC | *SYT15* | -3.728784624 | 0.011659785 | 0.022867129 | 1 |
| HTN | SMC | *LINC00619* | -22.29827546 | 0.000648342 | 0.002536898 | 1 |
| HTN | SMC | *BMPR2* | -5.570288333 | 0.002419341 | 0.006554176 | 1 |
| HTN | SMC | *HSD17B8* | -4.417247598 | 3.60E-07 | 3.42E-05 | 0.001745848 |
| HTN | SMC | *PRRT1* | 1.853519322 | 0.008614467 | 0.017839264 | 1 |
| HTN | SMC | *STK19* | -2.907552691 | 0.016716638 | 0.030688282 | 1 |
| HTN | SMC | *LAYN* | -7.395252613 | 2.03E-06 | 0.000115675 | 0.009832367 |
| HTN | SMC | *LST1* | -2.903371912 | 0.001206918 | 0.003930178 | 1 |
| HTN | SMC | *ZNF814* | -5.417536595 | 0.001695933 | 0.005002229 | 1 |
| HTN | SMC | *MICB* | -0.811944483 | 0.004456337 | 0.010491095 | 1 |
| HTN | SMC | *MICA* | -2.595774147 | 0.012899495 | 0.024767847 | 1 |
| HTN | SMC | *HLA-C* | -4.122329304 | 1.94E-09 | 5.53E-07 | 9.40E-06 |
| HTN | SMC | *POU5F1* | -0.616944011 | 0.020828236 | 0.036936624 | 1 |
| HTN | SMC | *PSORS1C2* | 2.964000031 | 0.002300119 | 0.006307643 | 1 |

| | | | | | | |
|---|---|---|---|---|---|---|
| HTN | SMC | *DHX16* | -4.338364776 | 0.006964349 | 0.01507182 | 1 |
| HTN | SMC | *LILRB3* | -5.901848877 | 0.007822191 | 0.016472882 | 1 |
| HTN | SMC | *HLA-E* | -4.60627045 | 5.96E-05 | 0.000821755 | 0.289257798 |
| HTN | SMC | *ZNF468* | -6.476118212 | 0.012350128 | 0.023892671 | 1 |
| HTN | SMC | *TRIM27* | -3.595780135 | 0.023851492 | 0.041628575 | 1 |
| HTN | SMC | *MALRD1* | -5.958877206 | 2.09E-05 | 0.000499016 | 0.101340585 |
| HTN | SMC | *FAM196B* | -4.621786933 | 0.00027412 | 0.001514842 | 1 |
| HTN | SMC | *ZBTB48* | -3.925213041 | 0.003115926 | 0.007936206 | 1 |
| HTN | SMC | *MZT1* | -9.122447816 | 0.018396355 | 0.033243617 | 1 |
| HTN | SMC | *ZNF155* | -4.746124498 | 0.018237036 | 0.033017201 | 1 |
| HTN | SMC | *ZNF783* | -8.934461378 | 1.49E-05 | 0.00041313 | 0.072228734 |
| HTN | SMC | *C12orf73* | -6.205904552 | 5.11E-05 | 0.000794475 | 0.247924327 |
| HTN | SMC | *TRIM13* | -5.219833268 | 0.000779415 | 0.002895652 | 1 |
| HTN | SMC | *SLFN12L* | -5.424442303 | 4.90E-05 | 0.000773529 | 0.237756251 |
| HTN | SMC | *SLC35B4* | -9.500311571 | 0.001409792 | 0.004428458 | 1 |
| HTN | SMC | *LINC00654* | -3.080003517 | 0.006297001 | 0.013849976 | 1 |
| HTN | SMC | *SCGB2B2* | -6.642609109 | 6.11E-05 | 0.000827638 | 0.296379243 |
| HTN | SMC | *CCDC144NL* | -15.50069652 | 8.26E-05 | 0.000876014 | 0.400890109 |
| HTN | SMC | *GOLGA6L10* | -7.267375314 | 0.000140865 | 0.001083168 | 0.683478755 |
| HTN | SMC | *IPO7* | -5.669895699 | 0.014439879 | 0.027240395 | 1 |
| HTN | SMC | *PRR13* | -5.699982466 | 0.005885192 | 0.013086596 | 1 |
| HTN | SMC | *MT1A* | -4.240820828 | 0.001209399 | 0.003932978 | 1 |
| HTN | SMC | *CFI* | -6.213205586 | 0.000272902 | 0.001509827 | 1 |
| HTN | SMC | *NAP1L4* | -5.275242109 | 0.000747084 | 0.002801276 | 1 |
| HTN | SMC | *SMN2* | -2.216185696 | 0.006330966 | 0.013905771 | 1 |
| HTN | SMC | *KLRC2* | -4.155666605 | 0.001363115 | 0.004311496 | 1 |
| HTN | SMC | *C1QTNF9B* | -21.32577264 | 0.000452786 | 0.002086341 | 1 |
| HTN | SMC | *COL6A6* | -3.451933079 | 0.001393042 | 0.004386138 | 1 |
| HTN | SMC | *HLA-A* | -3.89595306 | 0.002932557 | 0.007576554 | 1 |
| HTN | SMC | *METTL6* | -5.818889729 | 0.001447379 | 0.004510392 | 1 |
| HTN | SMC | *SACM1L* | -7.895023137 | 0.001133425 | 0.003758972 | 1 |
| HTN | SMC | *SLC48A1* | -8.425422802 | 6.85E-05 | 0.000836037 | 0.332354987 |
| HTN | SMC | *AC084018.1* | -1.854568078 | 0.006849152 | 0.014895602 | 1 |
| HTN | SMC | *C17orf51* | -19.42749797 | 9.89E-05 | 0.000925598 | 0.479824671 |
| HTN | SMC | *NUP62* | -4.031705621 | 0.000869933 | 0.00312198 | 1 |
| HTN | SMC | *ZNF254* | -5.654891935 | 0.004639384 | 0.010832672 | 1 |
| HTN | SMC | *TCTEX1D2* | -3.651696403 | 0.009534556 | 0.019315936 | 1 |
| HTN | SMC | *AC022498.1* | -11.83261042 | 0.001550779 | 0.004700863 | 1 |
| HTN | SMC | *GIMAP1* | -3.681678643 | 0.007389959 | 0.015774782 | 1 |
| HTN | SMC | *MXD3* | -4.999153403 | 4.34E-05 | 0.000739239 | 0.210683057 |
| HTN | SMC | *GSTM2* | -3.183918234 | 0.000700409 | 0.002675893 | 1 |
| HTN | SMC | *ARHGAP19* | -8.309645351 | 0.000281077 | 0.001541 | 1 |
| HTN | SMC | *SRA1* | -6.023892525 | 0.004167613 | 0.009913378 | 1 |
| HTN | SMC | *TMX2* | -3.741870973 | 0.008077189 | 0.01694359 | 1 |
| HTN | SMC | *LBH* | 0.48322659 | 0.004021968 | 0.009651259 | 1 |
| HTN | SMC | *NCKIPSD* | -3.889007914 | 0.001619216 | 0.004831758 | 1 |
| HTN | SMC | *SLC35F6* | -9.208588198 | 3.92E-05 | 0.000689568 | 0.190320699 |
| HTN | SMC | *EMP2* | -6.879397601 | 0.001022728 | 0.00348894 | 1 |
| HTN | SMC | *LTB4R* | -9.886441774 | 0.000117043 | 0.000995642 | 0.567892704 |
| HTN | SMC | *IRF9* | -8.522391487 | 0.002138409 | 0.005969827 | 1 |

| | | | | | | |
|---|---|---|---|---|---|---|
| HTN | SMC | *ZNF726* | -5.358435251 | 0.000213428 | 0.00132582 | 1 |
| HTN | SMC | *REPIN1* | -12.84782959 | 0.003087054 | 0.007879215 | 1 |
| HTN | SMC | *ZNF891* | -7.841536608 | 0.006045207 | 0.013381088 | 1 |
| HTN | SMC | *SMIM7* | -6.65438909 | 0.001302307 | 0.004168069 | 1 |
| HTN | SMC | *TSPAN4* | -5.673227021 | 0.00099942 | 0.003441578 | 1 |
| HTN | SMC | *PAXIP1-AS2* | -7.04858382 | 0.00023228 | 0.001381153 | 1 |
| HTN | SMC | *FIS1* | -3.930875218 | 9.62E-05 | 0.000908214 | 0.466717524 |
| HTN | SMC | *ANG* | -8.321240816 | 6.62E-05 | 0.000836037 | 0.320962792 |
| HTN | SMC | *GNAT3* | -2.359089901 | 0.000624374 | 0.002462978 | 1 |
| HTN | SMC | *PLIN5* | -1.529409749 | 0.009063585 | 0.01859472 | 1 |
| HTN | SMC | *C5orf49* | -20.8572886 | 0.00106674 | 0.003604332 | 1 |
| HTN | SMC | *NPEPL1* | -4.884604906 | 0.000197616 | 0.001271662 | 0.958833414 |
| HTN | SMC | *TMEM242* | -5.640643622 | 0.001673002 | 0.004948192 | 1 |
| HTN | SMC | *SLC35E2* | -5.044184696 | 0.001411959 | 0.004428458 | 1 |
| HTN | SMC | *TSTD1* | -10.40729567 | 8.87E-06 | 0.000298909 | 0.04304293 |
| HTN | SMC | *AC009403.2* | -3.057080116 | 0.018435854 | 0.033290199 | 1 |
| HTN | SMC | *AC016773.1* | 4.441329962 | 7.51E-05 | 0.000847247 | 0.364316319 |
| HTN | SMC | *RNASEK* | -3.759558818 | 0.006854174 | 0.014899843 | 1 |
| HTN | SMC | *FAM228B* | -2.21147428 | 0.011761196 | 0.022991669 | 1 |
| HTN | SMC | *RNU6ATAC35I* | -18.55896863 | 0.007550459 | 0.016032747 | 1 |
| HTN | SMC | *PPP3R1* | -3.245820695 | 0.011750287 | 0.022979603 | 1 |
| HTN | SMC | *AP4M1* | -3.463880685 | 0.001701645 | 0.005012103 | 1 |
| HTN | SMC | *ZNF880* | -2.52486432 | 0.013032798 | 0.024994124 | 1 |
| HTN | SMC | *FXYD7* | -4.01097276 | 0.002179343 | 0.00606665 | 1 |
| HTN | SMC | *APOL6* | -4.153737346 | 8.47E-05 | 0.000876014 | 0.410922038 |
| HTN | SMC | *TIAF1* | 4.027288571 | 0.014454651 | 0.027247073 | 1 |
| HTN | SMC | *RAB6C* | -46.39015615 | 0.008917757 | 0.018323624 | 1 |
| HTN | SMC | *AC062017.1* | -5.435449285 | 0.000133677 | 0.001054634 | 0.648599739 |
| HTN | SMC | *ADRA2B* | 1.628075195 | 0.008844599 | 0.018224921 | 1 |
| HTN | SMC | *EXOSC6* | -11.03539038 | 2.28E-07 | 2.46E-05 | 0.00110593 |
| HTN | SMC | *LINC00571* | -5.217072376 | 0.000574178 | 0.002363597 | 1 |
| HTN | SMC | *CERS1* | 4.158640722 | 0.017594299 | 0.032008827 | 1 |
| HTN | SMC | *OSER1-AS1* | 3.0767433 | 0.01390083 | 0.026367017 | 1 |
| HTN | SMC | *C5orf66* | -8.464992556 | 0.000183871 | 0.001226806 | 0.892141522 |
| HTN | SMC | *LINC01133* | -4.463247863 | 8.59E-05 | 0.000876014 | 0.416567316 |
| HTN | SMC | *LINC01117* | -4.5763841 | 0.000279479 | 0.00153745 | 1 |
| HTN | SMC | *LINC00184* | -56.36927088 | 0.000549002 | 0.002313255 | 1 |
| HTN | SMC | *AL049840.1* | -8.590347904 | 4.63E-05 | 0.000760128 | 0.224651211 |
| HTN | SMC | *PLEKHM1* | -7.29499082 | 0.000644426 | 0.002525652 | 1 |
| HTN | SMC | *CYP4A22-AS1* | -8.821815846 | 8.13E-05 | 0.000876014 | 0.394588024 |
| HTN | SMC | *MIAT* | -10.01767947 | 5.72E-05 | 0.000817716 | 0.277637379 |
| HTN | SMC | *ERCC6* | -5.333273639 | 0.000143457 | 0.001090989 | 0.696051159 |
| HTN | SMC | *LINC00694* | 3.213411966 | 0.001919243 | 0.005494816 | 1 |
| HTN | SMC | *LINC00115* | -14.12664268 | 0.000145592 | 0.00110033 | 0.706411636 |
| HTN | SMC | *HAR1A* | -41.02000781 | 0.001201651 | 0.003918286 | 1 |
| HTN | SMC | *LINC00937* | -2.314770713 | 0.001155692 | 0.003809387 | 1 |
| HTN | SMC | *FAM138B* | 9.951486223 | 0.0037971 | 0.009258054 | 1 |
| HTN | SMC | *AC017074.2* | -21.14205229 | 0.004726125 | 0.010976007 | 1 |
| HTN | SMC | *AC009299.3* | -7.859452111 | 0.0001221 | 0.001013054 | 0.592430305 |
| HTN | SMC | *SCAMP4* | -9.467533901 | 9.20E-05 | 0.00089488 | 0.446544996 |

| | | | | | | |
|---|---|---|---|---|---|---|
| HTN | SMC | *SLC2A1-AS1* | -3.889523812 | 0.001266054 | 0.004076242 | 1 |
| HTN | SMC | *AC005884.1* | -5.49643689 | 0.001855221 | 0.00536444 | 1 |
| HTN | SMC | *AC108463.2* | -5.418005865 | 0.01583647 | 0.029350097 | 1 |
| HTN | SMC | *POLR2J2* | 8.03444668 | 0.001144387 | 0.00378498 | 1 |
| HTN | SMC | *LINC00578* | -6.769895597 | 5.48E-06 | 0.000214301 | 0.026573267 |
| HTN | SMC | *ARL17B* | -6.971060372 | 1.80E-05 | 0.000456819 | 0.087113149 |
| HTN | SMC | *PATL2* | -3.678475668 | 0.010701265 | 0.021305925 | 1 |
| HTN | SMC | *CASC20* | -3.441695039 | 0.000508744 | 0.002231851 | 1 |
| HTN | SMC | *LINC00332* | -2.123773492 | 0.024010996 | 0.041861787 | 1 |
| HTN | SMC | *RPS18* | -5.114907735 | 0.006381154 | 0.013990674 | 1 |
| HTN | SMC | *DIRC3* | -5.366966667 | 0.000200741 | 0.001281046 | 0.97399384 |
| HTN | SMC | *AC107021.1* | -3.512707782 | 0.000408705 | 0.001942185 | 1 |
| HTN | SMC | *GOLGA8N* | 1.854075272 | 0.015318418 | 0.028564551 | 1 |
| HTN | SMC | *LYRM9* | -2.954865048 | 0.002020679 | 0.005726832 | 1 |
| HTN | SMC | *LINC00342* | -7.317350322 | 0.003318147 | 0.008334 | 1 |
| HTN | SMC | *LINC00702* | 0.293379697 | 0.002416678 | 0.006550683 | 1 |
| HTN | SMC | *RPS28* | -6.395077232 | 0.005513559 | 0.012385087 | 1 |
| HTN | SMC | *TRIM26* | -4.097703205 | 0.007505161 | 0.015978518 | 1 |
| HTN | SMC | *PINLYP* | -4.068866751 | 0.014889661 | 0.027926029 | 1 |
| HTN | SMC | *ANKRD65* | -6.051018344 | 0.000757691 | 0.002830112 | 1 |
| HTN | SMC | *ZSCAN31* | -1.134205742 | 0.016801734 | 0.030797889 | 1 |
| HTN | SMC | *LINC00957* | 2.20720191 | 0.003076566 | 0.007864857 | 1 |
| HTN | SMC | *LINC00607* | -2.467414405 | 0.010960774 | 0.021729091 | 1 |
| HTN | SMC | *AC019117.2* | -20.27143474 | 4.17E-05 | 0.000719923 | 0.202235839 |
| HTN | SMC | *ZBED5* | -4.449456479 | 0.004247394 | 0.010077435 | 1 |
| HTN | SMC | *CDKN2AIPNL* | -3.733849042 | 0.00237252 | 0.006452615 | 1 |
| HTN | SMC | *LINC00959* | -3.128417431 | 0.000359342 | 0.0017956 | 1 |
| HTN | SMC | *HLA-DQA2* | 0.985742717 | 0.002910959 | 0.007536806 | 1 |
| HTN | SMC | *LINC01143* | -11.0430938 | 0.000624096 | 0.002462978 | 1 |
| HTN | SMC | *LINC00211* | -12.18214977 | 1.88E-05 | 0.000473276 | 0.091031106 |
| HTN | SMC | *LRRC37A2* | -9.699849495 | 6.17E-06 | 0.000231963 | 0.02992321 |
| HTN | SMC | *TXNDC5* | -31.56176441 | 0.000127973 | 0.001039637 | 0.620927217 |
| HTN | SMC | *LINC01119* | -3.491863138 | 0.001078045 | 0.003637465 | 1 |
| HTN | SMC | *TNFSF12* | 7.765986696 | 1.51E-06 | 9.66E-05 | 0.007344276 |
| HTN | SMC | *C1orf226* | -10.81085712 | 0.000478263 | 0.002159009 | 1 |
| HTN | SMC | *L1TD1* | -2.722115353 | 0.011976321 | 0.023331472 | 1 |
| HTN | SMC | *C1QTNF9* | -9.358278462 | 0.000375363 | 0.001834098 | 1 |
| HTN | SMC | *PNMA2* | -7.750893052 | 5.61E-06 | 0.000217703 | 0.027212871 |
| HTN | SMC | *KRBOX1* | -35.81552048 | 0.000516804 | 0.002238867 | 1 |
| HTN | SMC | *ARHGEF25* | 1.346510539 | 0.006990245 | 0.01511438 | 1 |
| HTN | SMC | *NSUN6* | -4.056837965 | 0.015317633 | 0.028564551 | 1 |
| HTN | SMC | *TDGF1* | -3.454998882 | 0.001219945 | 0.003956666 | 1 |
| HTN | SMC | *CD302* | -6.435011144 | 0.003530232 | 0.008725771 | 1 |
| HTN | SMC | *INMT* | -3.347665092 | 0.021948891 | 0.038669578 | 1 |
| HTN | SMC | *PLEKHO2* | -3.882928302 | 0.000128133 | 0.001039637 | 0.62170292 |
| HTN | SMC | *C8orf58* | -9.538300178 | 0.002461603 | 0.006628022 | 1 |
| HTN | SMC | *PI4KA* | -6.835338851 | 0.000260611 | 0.001475067 | 1 |
| HTN | SMC | *LINC00969* | -2.871452213 | 0.002311394 | 0.006325373 | 1 |
| HTN | SMC | *LINC00960* | -1.026977736 | 0.007150753 | 0.015372376 | 1 |
| HTN | SMC | *DECR2* | -7.115844652 | 0.000164625 | 0.001162037 | 0.798761751 |

| | | | | | | |
|---|---|---|---|---|---|---|
| HTN | SMC | *ZNF709* | -22.46423866 | 1.38E-05 | 0.000396487 | 0.066808481 |
| HTN | SMC | *ERVW-1* | -7.339414019 | 0.000620261 | 0.002458746 | 1 |
| HTN | SMC | *C7orf73* | -5.301701395 | 0.005820496 | 0.012960555 | 1 |
| HTN | SMC | *FGF14-IT1* | -9.816672936 | 0.000378092 | 0.001841989 | 1 |
| HTN | SMC | *WFDC6* | -11.60312136 | 0.000231604 | 0.001378826 | 1 |
| HTN | SMC | *UPK3B* | -10.16463773 | 9.37E-05 | 0.000903 | 0.454821011 |
| HTN | SMC | *IL10RB* | -6.768195638 | 0.001857564 | 0.005365113 | 1 |
| HTN | SMC | *PLA2G4B* | 7.068617215 | 0.007417684 | 0.015820045 | 1 |
| HTN | SMC | *LEFTY1* | -35.66544879 | 0.000267026 | 0.001492638 | 1 |
| HTN | SMC | *TTC4* | -2.702252648 | 0.022647353 | 0.039727027 | 1 |
| HTN | SMC | *JMJD7* | 2.037423565 | 0.013487102 | 0.025733157 | 1 |
| HTN | SMC | *APOBEC3D* | 11.35494581 | 0.009369747 | 0.019053651 | 1 |
| HTN | SMC | *TMEM199* | -2.345144646 | 0.001091429 | 0.003667321 | 1 |
| HTN | SMC | *DBNDD2* | 2.170517887 | 0.003529478 | 0.008725771 | 1 |
| HTN | SMC | *SCARF2* | -5.417618537 | 0.015565658 | 0.028947709 | 1 |
| HTN | SMC | *APOBEC3C* | -4.904409626 | 0.001784642 | 0.00519441 | 1 |
| HTN | SMC | *CCDC13* | -2.496063556 | 0.003381437 | 0.00844837 | 1 |
| HTN | SMC | *NEAT1* | -3.587347099 | 0.00267048 | 0.007099819 | 1 |
| HTN | SMC | *RAB30-AS1* | -5.939263464 | 0.000196955 | 0.001271662 | 0.955627701 |
| HTN | SMC | *UBAP1L* | -8.096922343 | 0.003944551 | 0.009498244 | 1 |
| HTN | SMC | *DNAJC3-AS1* | -5.855237877 | 0.000117028 | 0.000995642 | 0.567817979 |
| HTN | SMC | *LINC00926* | -20.42590694 | 2.91E-05 | 0.000580779 | 0.141129192 |
| HTN | SMC | *DPH6-AS1* | -5.50478603 | 0.000596406 | 0.002423585 | 1 |
| HTN | SMC | *CDK11B* | -8.794119981 | 0.004661249 | 0.01087326 | 1 |
| HTN | SMC | *ABHD14A* | -10.44574984 | 0.000292206 | 0.001573568 | 1 |
| HTN | SMC | *SRP14-AS1* | -1.350522504 | 0.002442269 | 0.006604516 | 1 |
| HTN | SMC | *GDNF-AS1* | -6.674273884 | 0.001609777 | 0.00481247 | 1 |
| HTN | SMC | *CCDC153* | -5.214970188 | 0.00089414 | 0.003185291 | 1 |
| HTN | SMC | *C5orf17* | -8.475988165 | 9.14E-05 | 0.000890467 | 0.443452724 |
| HTN | SMC | *LINC01060* | -2.653747032 | 0.012877711 | 0.024735809 | 1 |
| HTN | SMC | *LINC00942* | -19.26910183 | 8.19E-05 | 0.000876014 | 0.397295858 |
| HTN | SMC | *PDCD6* | -7.416393658 | 0.007198532 | 0.015413367 | 1 |
| HTN | SMC | *TMEM158* | -9.909228812 | 0.001126526 | 0.003742915 | 1 |
| HTN | SMC | *DNAH10OS* | -15.05972421 | 0.001338837 | 0.004255399 | 1 |
| HTN | SMC | *SMIM20* | -9.822329184 | 5.75E-05 | 0.000817716 | 0.279188365 |
| HTN | SMC | *LINC00989* | 0.414821836 | 0.002790205 | 0.00731673 | 1 |
| HTN | SMC | *GLI4* | -10.56652558 | 0.000237061 | 0.001395897 | 1 |
| HTN | SMC | *ALG1L2* | -8.074146475 | 0.000227762 | 0.001377231 | 1 |
| HTN | SMC | *ZNF550* | -5.324827689 | 9.75E-05 | 0.00091456 | 0.472827717 |
| HTN | SMC | *LINC01094* | -3.798853906 | 0.000368019 | 0.001812751 | 1 |
| HTN | SMC | *PCDHGB2* | -12.4005218 | 0.000200644 | 0.001281046 | 0.973526932 |
| HTN | SMC | *LYN* | -1.65596033 | 0.019763693 | 0.035332881 | 1 |
| HTN | SMC | *ARMS2* | -4.532736 | 0.004044888 | 0.009691751 | 1 |
| HTN | SMC | *BRK1* | -3.23870049 | 0.029218123 | 0.049672857 | 1 |
| HTN | SMC | *SNHG9* | -3.900642399 | 0.013943305 | 0.026437247 | 1 |
| HTN | SMC | *ZNF432* | -4.208829378 | 0.013956371 | 0.026451684 | 1 |
| HTN | SMC | *ZBED6* | -14.16248646 | 2.55E-05 | 0.000550877 | 0.123947436 |
| HTN | SMC | *CNPY2* | -9.416926984 | 0.000270758 | 0.00150483 | 1 |
| HTN | SMC | *HIF1A-AS2* | -7.257230839 | 0.003149417 | 0.008004699 | 1 |
| HTN | SMC | *SLC25A21-AS1* | -3.683214212 | 0.001402876 | 0.004414237 | 1 |

| | | | | | | |
|---|---|---|---|---|---|---|
| HTN | SMC | *RNASE4* | -8.960048408 | 0.001108694 | 0.003697718 | 1 |
| HTN | SMC | *LINC00639* | -3.156732527 | 5.24E-05 | 0.000804267 | 0.254148492 |
| HTN | SMC | *UBE2Q2L* | 6.046644352 | 0.023866577 | 0.041639925 | 1 |
| HTN | SMC | *IQCH-AS1* | -3.979932395 | 0.0158315 | 0.029350097 | 1 |
| HTN | SMC | *AC083843.1* | -42.55977168 | 0.000525155 | 0.002257599 | 1 |
| HTN | SMC | *PWRN1* | -5.319644753 | 7.08E-06 | 0.000259096 | 0.03435707 |
| HTN | SMC | *LINC01003* | -5.112680521 | 0.000155799 | 0.001140149 | 0.755938733 |
| HTN | SMC | *GOLGA8H* | 1.103308778 | 0.019698833 | 0.035242897 | 1 |
| HTN | SMC | *C19orf84* | -26.34846248 | 0.003395669 | 0.008470841 | 1 |
| HTN | SMC | *LINC00672* | 5.450419434 | 0.01827275 | 0.03306952 | 1 |
| HTN | SMC | *LINC00909* | -6.766067652 | 8.59E-05 | 0.000876014 | 0.416982634 |
| HTN | SMC | *STRADA* | -3.846812556 | 0.019949815 | 0.035600038 | 1 |
| HTN | SMC | *ILF3-AS1* | -3.385223985 | 0.026606412 | 0.045697102 | 1 |
| HTN | SMC | *ZNF582-AS1* | -12.57655044 | 6.35E-05 | 0.000827638 | 0.30788123 |
| HTN | SMC | *ZNF285* | -2.356594897 | 0.005613671 | 0.012580846 | 1 |
| HTN | SMC | *ZNF224* | -9.030990162 | 0.000160151 | 0.001151189 | 0.777052908 |
| HTN | SMC | *LIN37* | 3.132798093 | 0.001004692 | 0.003454831 | 1 |
| HTN | SMC | *NDUFA7* | -14.28100118 | 0.000631917 | 0.002484654 | 1 |
| HTN | SMC | *LINC00664* | 3.861071536 | 0.002517025 | 0.00673613 | 1 |
| HTN | SMC | *ZNF728* | -7.197068404 | 0.001015649 | 0.003479625 | 1 |
| HTN | SMC | *TRABD2B* | 1.51263636 | 0.012046036 | 0.023416413 | 1 |
| HTN | SMC | *AC093157.1* | -8.158922694 | 0.000963539 | 0.003346523 | 1 |
| HTN | SMC | *ZNF587B* | -12.54046685 | 0.011212678 | 0.02215143 | 1 |
| HTN | SMC | *SNHG8* | -3.284616857 | 0.010748794 | 0.021356736 | 1 |
| HTN | SMC | *HCG17* | -4.084772312 | 0.019499571 | 0.034989615 | 1 |
| HTN | SMC | *YTHDF3-AS1* | -4.894922815 | 0.004592336 | 0.010748681 | 1 |
| HTN | SMC | *GTF2H5* | -2.944426987 | 0.005111849 | 0.011649926 | 1 |
| HTN | SMC | *FGF14-AS2* | -5.584528855 | 0.000820773 | 0.002989783 | 1 |
| HTN | SMC | *PAXIP1-AS1* | -9.05898871 | 0.000102373 | 0.000942633 | 0.49671184 |
| HTN | t_cell | *SEMA3F* | -3.151927726 | 0.000408337 | 0.00899518 | 1 |
| HTN | t_cell | *RBM6* | -1.974954375 | 0.000403578 | 0.008988159 | 1 |
| HTN | t_cell | *MEOX1* | -1.897579958 | 0.008802702 | 0.047950379 | 1 |
| HTN | t_cell | *SYNRG* | 3.182110174 | 0.001167744 | 0.014129416 | 1 |
| HTN | t_cell | *CEACAM21* | 0.780366304 | 0.001774103 | 0.017389789 | 1 |
| HTN | t_cell | *CD79B* | -4.701704935 | 0.001842936 | 0.017883853 | 1 |
| HTN | t_cell | *TSR3* | 11.3152075 | 0.000243177 | 0.007332694 | 1 |
| HTN | t_cell | *PIGQ* | 3.555450287 | 0.008565589 | 0.047435111 | 1 |
| HTN | t_cell | *SS18L2* | 5.323380734 | 0.001713968 | 0.017171831 | 1 |
| HTN | t_cell | *IL32* | 0.907501935 | 0.002327126 | 0.02031219 | 1 |
| HTN | t_cell | *RPS20* | 4.801363456 | 0.001753202 | 0.017349326 | 1 |
| HTN | t_cell | *TRAF3IP3* | 0.581107406 | 0.000935725 | 0.012421218 | 1 |
| HTN | t_cell | *PPP5C* | 4.108806661 | 0.005456686 | 0.034788754 | 1 |
| HTN | t_cell | *CEP68* | 7.078010276 | 5.21E-07 | 8.42E-05 | 0.002525607 |
| HTN | t_cell | *MAN2B2* | 6.026458115 | 0.00447991 | 0.030400729 | 1 |
| HTN | t_cell | *TACC3* | 4.742781826 | 7.43E-05 | 0.003498558 | 0.360351499 |
| HTN | t_cell | *NUDCD3* | 3.103336458 | 0.004776741 | 0.031584994 | 1 |
| HTN | t_cell | *GRAMD1B* | 1.35366 | 0.007956499 | 0.045417571 | 1 |
| HTN | t_cell | *RNH1* | 3.882805823 | 0.003276904 | 0.024536323 | 1 |
| HTN | t_cell | *GCLM* | 6.329925347 | 0.000299305 | 0.00820468 | 1 |
| HTN | t_cell | *UBR2* | 3.738347622 | 0.005901419 | 0.036662846 | 1 |

| | | | | | | |
|---|---|---|---|---|---|---|
| HTN | t_cell | *INSRR* | -3.481748397 | 0.003546333 | 0.025934257 | 1 |
| HTN | t_cell | *SH2D2A* | 1.082375903 | 0.003535457 | 0.025912443 | 1 |
| HTN | t_cell | *SNX1* | 5.908347166 | 0.000393383 | 0.008960997 | 1 |
| HTN | t_cell | *IKZF2* | 4.133859396 | 0.000936967 | 0.012421218 | 1 |
| HTN | t_cell | *CENPQ* | -1.170550897 | 0.006784003 | 0.040940276 | 1 |
| HTN | t_cell | *ADSS* | 5.949847419 | 0.00498579 | 0.032690613 | 1 |
| HTN | t_cell | *AGA* | 8.741484651 | 0.000351923 | 0.008756578 | 1 |
| HTN | t_cell | *PNKP* | 2.713252018 | 0.002114388 | 0.019263747 | 1 |
| HTN | t_cell | *TDP1* | 4.861067075 | 0.00097435 | 0.012573256 | 1 |
| HTN | t_cell | *ARAP2* | 1.792679853 | 0.000292482 | 0.008144892 | 1 |
| HTN | t_cell | *CYBA* | 4.65345387 | 0.000535905 | 0.009873328 | 1 |
| HTN | t_cell | *THOC3* | 2.226041756 | 0.001852724 | 0.017907204 | 1 |
| HTN | t_cell | *FSTL4* | 2.125758918 | 0.007227542 | 0.043081123 | 1 |
| HTN | t_cell | *ANAPC4* | 7.984837911 | 3.49E-06 | 0.000385172 | 0.016947548 |
| HTN | t_cell | *LY75* | 4.546618122 | 0.001602387 | 0.016684077 | 1 |
| HTN | t_cell | *SDCCAG8* | 5.057627794 | 3.44E-07 | 6.43E-05 | 0.001670939 |
| HTN | t_cell | *TBC1D22A* | 4.119854361 | 0.00451448 | 0.030518825 | 1 |
| HTN | t_cell | *ITIH4* | 1.104217621 | 0.001746766 | 0.017349326 | 1 |
| HTN | t_cell | *NPFFR2* | -2.195824971 | 0.008793864 | 0.047950379 | 1 |
| HTN | t_cell | *ATP11B* | 4.005443469 | 0.002915994 | 0.022856872 | 1 |
| HTN | t_cell | *DNAJC25* | 9.993968063 | 0.001686659 | 0.017059213 | 1 |
| HTN | t_cell | *PIGV* | -1.816173592 | 0.008668186 | 0.047684853 | 1 |
| HTN | t_cell | *LTK* | 1.005343481 | 0.003569273 | 0.02600317 | 1 |
| HTN | t_cell | *EIF4B* | 3.315685756 | 0.002103528 | 0.019263747 | 1 |
| HTN | t_cell | *HAGH* | 4.621485938 | 8.60E-05 | 0.003935741 | 0.417188495 |
| HTN | t_cell | *RNF4* | 5.876942952 | 0.005283383 | 0.034043789 | 1 |
| HTN | t_cell | *CASP8* | 2.15558341 | 0.000547388 | 0.009873328 | 1 |
| HTN | t_cell | *LPAR2* | 2.927055603 | 0.000618267 | 0.010192716 | 1 |
| HTN | t_cell | *SLC12A2* | 6.836348361 | 0.000433435 | 0.00917827 | 1 |
| HTN | t_cell | *CNN2* | 1.953458915 | 0.008167386 | 0.04591907 | 1 |
| HTN | t_cell | *BTBD1* | 5.917327871 | 0.001545523 | 0.016399464 | 1 |
| HTN | t_cell | *FAR2* | 4.139268435 | 0.000387236 | 0.008960997 | 1 |
| HTN | t_cell | *CHI3L2* | 5.637574741 | 0.000391611 | 0.008960997 | 1 |
| HTN | t_cell | *UHRF1BP1* | 3.981052776 | 4.06E-06 | 0.000419179 | 0.019701431 |
| HTN | t_cell | *ANKRD44* | 1.357758828 | 0.00051783 | 0.009873328 | 1 |
| HTN | t_cell | *TIE1* | -3.04127244 | 5.87E-05 | 0.002874853 | 0.284610459 |
| HTN | t_cell | *NGEF* | -6.188450861 | 0.007263482 | 0.043189233 | 1 |
| HTN | t_cell | *MTFR1* | 2.285838511 | 0.009134398 | 0.048972486 | 1 |
| HTN | t_cell | *HEATR6* | 5.880589515 | 0.000158267 | 0.005730697 | 0.767913399 |
| HTN | t_cell | *POLR1A* | -3.283295315 | 0.000332453 | 0.00850409 | 1 |
| HTN | t_cell | *SRBD1* | 4.884266765 | 0.005110131 | 0.033191908 | 1 |
| HTN | t_cell | *CLEC2D* | 1.620867157 | 0.000913387 | 0.012349099 | 1 |
| HTN | t_cell | *PLA2G10* | 8.781857218 | 0.001070701 | 0.013458659 | 1 |
| HTN | t_cell | *WDR1* | 6.09100614 | 0.00782776 | 0.045177826 | 1 |
| HTN | t_cell | *NFATC3* | 3.081454071 | 0.005479821 | 0.034832622 | 1 |
| HTN | t_cell | *NDE1* | 5.616975213 | 0.001479187 | 0.016079706 | 1 |
| HTN | t_cell | *PVR* | -2.717529938 | 0.001415518 | 0.015644861 | 1 |
| HTN | t_cell | *LLGL2* | 2.427105472 | 0.000757904 | 0.011326355 | 1 |
| HTN | t_cell | *SDHA* | 4.405900226 | 0.000969108 | 0.012573256 | 1 |
| HTN | t_cell | *ZZEF1* | 4.893051147 | 0.003576532 | 0.026016989 | 1 |

| | | | | | | |
|---|---|---|---|---|---|---|
| HTN | t_cell | *VPS9D1* | 7.495273742 | 0.000260385 | 0.00761076 | 1 |
| HTN | t_cell | *MCM6* | 3.96501432 | 0.002106878 | 0.019263747 | 1 |
| HTN | t_cell | *SPAG5* | -5.767664566 | 0.001546905 | 0.016399464 | 1 |
| HTN | t_cell | *ANKRD13A* | 7.481670527 | 0.001392856 | 0.015560606 | 1 |
| HTN | t_cell | *PAG1* | 1.822525453 | 0.000846168 | 0.011988677 | 1 |
| HTN | t_cell | *ICAM3* | 1.854054206 | 0.001634293 | 0.016871469 | 1 |
| HTN | t_cell | *XAB2* | 10.17647091 | 0.000267146 | 0.007681101 | 1 |
| HTN | t_cell | *APBB1IP* | 2.258065833 | 0.00206408 | 0.019003639 | 1 |
| HTN | t_cell | *LRCH4* | 4.096883914 | 0.001144921 | 0.01390596 | 1 |
| HTN | t_cell | *AMPH* | 7.490962369 | 0.0011985 | 0.01435833 | 1 |
| HTN | t_cell | *XRCC5* | 4.428553171 | 0.006014198 | 0.037125812 | 1 |
| HTN | t_cell | *SP140* | 0.846438851 | 0.005945862 | 0.036797606 | 1 |
| HTN | t_cell | *FDFT1* | 8.015274493 | 2.94E-07 | 5.70E-05 | 0.001425825 |
| HTN | t_cell | *DNM2* | 5.729153011 | 0.000312959 | 0.008435989 | 1 |
| HTN | t_cell | *RABL2B* | 6.449711672 | 0.008730551 | 0.0478111 | 1 |
| HTN | t_cell | *KCNN2* | -3.448221138 | 0.004643256 | 0.030989101 | 1 |
| HTN | t_cell | *STK17B* | 1.876552473 | 0.000319631 | 0.008451766 | 1 |
| HTN | t_cell | *ZNF506* | 5.694422037 | 0.00083325 | 0.011988677 | 1 |
| HTN | t_cell | *ME2* | 5.600332968 | 0.007441436 | 0.043658822 | 1 |
| HTN | t_cell | *CAD* | 6.036959626 | 0.002963057 | 0.023100903 | 1 |
| HTN | t_cell | *CD82* | 1.797961328 | 0.002689587 | 0.0217861 | 1 |
| HTN | t_cell | *DDHD2* | 3.851458713 | 0.005851052 | 0.036396547 | 1 |
| HTN | t_cell | *SNX10* | -3.173789168 | 5.57E-05 | 0.002758081 | 0.270291945 |
| HTN | t_cell | *SRRT* | 6.615084263 | 0.002125227 | 0.019263747 | 1 |
| HTN | t_cell | *AAMDC* | -3.026156008 | 0.001295823 | 0.015139453 | 1 |
| HTN | t_cell | *TBX5* | -6.954340614 | 0.000922935 | 0.012349099 | 1 |
| HTN | t_cell | *FXYD5* | 3.283984032 | 0.00235792 | 0.020429695 | 1 |
| HTN | t_cell | *GRAMD1A* | 5.949836479 | 0.003251367 | 0.024428605 | 1 |
| HTN | t_cell | *YPEL3* | 5.921417374 | 5.41E-06 | 0.000514821 | 0.026255853 |
| HTN | t_cell | *LYZ* | 4.247794581 | 0.000900502 | 0.01227313 | 1 |
| HTN | t_cell | *MUL1* | -2.466934123 | 0.005621785 | 0.035378603 | 1 |
| HTN | t_cell | *PABPC4* | -4.80299724 | 0.000388928 | 0.008960997 | 1 |
| HTN | t_cell | *CERS4* | 5.816501592 | 0.001290426 | 0.015123544 | 1 |
| HTN | t_cell | *GLG1* | 4.560332832 | 0.002428521 | 0.02070858 | 1 |
| HTN | t_cell | *PPP2R3C* | 4.53835764 | 0.000884546 | 0.012123781 | 1 |
| HTN | t_cell | *UNC13D* | 1.603238051 | 0.002679401 | 0.02173989 | 1 |
| HTN | t_cell | *NUP50* | 3.263151839 | 0.00748298 | 0.043826994 | 1 |
| HTN | t_cell | *NANS* | 7.508922438 | 0.000498765 | 0.009758406 | 1 |
| HTN | t_cell | *ITPR3* | 4.340356511 | 3.20E-05 | 0.001933659 | 0.155029291 |
| HTN | t_cell | *SCD* | -3.247914434 | 0.002367068 | 0.020435969 | 1 |
| HTN | t_cell | *SETD1A* | 10.10547271 | 2.77E-07 | 5.59E-05 | 0.001342367 |
| HTN | t_cell | *MARCH2* | 4.027691947 | 2.99E-05 | 0.001883968 | 0.145065515 |
| HTN | t_cell | *SERPIND1* | 4.745685711 | 0.000497553 | 0.009758406 | 1 |
| HTN | t_cell | *CYTH4* | 1.967003762 | 0.000996018 | 0.012717571 | 1 |
| HTN | t_cell | *SH3BP1* | 1.737318922 | 0.001720014 | 0.017171831 | 1 |
| HTN | t_cell | *RAB36* | -2.361683797 | 0.006740995 | 0.040731391 | 1 |
| HTN | t_cell | *FAM118A* | 2.053738076 | 0.001657023 | 0.017059213 | 1 |
| HTN | t_cell | *NIN* | 3.695570121 | 0.000763338 | 0.011326355 | 1 |
| HTN | t_cell | *GSKIP* | 8.109843423 | 0.000610154 | 0.010181068 | 1 |
| HTN | t_cell | *PSMB5* | 3.325834682 | 0.003994624 | 0.028130503 | 1 |

| | | | | | | |
|---|---|---|---|---|---|---|
| HTN | t_cell | *BIRC7* | -3.76362207 | 0.002181106 | 0.019346851 | 1 |
| HTN | t_cell | *PTK6* | 5.389957525 | 0.001322642 | 0.015279665 | 1 |
| HTN | t_cell | *GMEB2* | 6.519167547 | 0.008502901 | 0.047203746 | 1 |
| HTN | t_cell | *RNF24* | 4.413561537 | 0.007377296 | 0.043658822 | 1 |
| HTN | t_cell | *TRIB3* | 12.95887366 | 0.000597648 | 0.010162236 | 1 |
| HTN | t_cell | *RASSF2* | 1.866683479 | 0.000619714 | 0.010192716 | 1 |
| HTN | t_cell | *DNTTIP1* | -2.850495197 | 0.001684653 | 0.017059213 | 1 |
| HTN | t_cell | *LPIN2* | 4.6837958 | 0.00078861 | 0.011576575 | 1 |
| HTN | t_cell | *RNF125* | 4.542690117 | 0.000560512 | 0.009998542 | 1 |
| HTN | t_cell | *POLI* | -1.367074483 | 0.000612754 | 0.010181783 | 1 |
| HTN | t_cell | *STK24* | 4.702686614 | 0.006428994 | 0.039286496 | 1 |
| HTN | t_cell | *MGRN1* | 4.920696249 | 0.00166726 | 0.017059213 | 1 |
| HTN | t_cell | *PSMD7* | 6.044021103 | 0.00453974 | 0.030518825 | 1 |
| HTN | t_cell | *MTHFSD* | 2.24049377 | 0.000233059 | 0.007202575 | 1 |
| HTN | t_cell | *HAGHL* | 2.462187232 | 0.000432394 | 0.00917827 | 1 |
| HTN | t_cell | *HMOX2* | 3.973837102 | 0.000476792 | 0.00953704 | 1 |
| HTN | t_cell | *RBL2* | 3.101110164 | 0.000361355 | 0.008766464 | 1 |
| HTN | t_cell | *PYCARD* | 7.530859057 | 7.95E-07 | 0.000110202 | 0.003857058 |
| HTN | t_cell | *HERC1* | 3.903708653 | 0.008354624 | 0.04680905 | 1 |
| HTN | t_cell | *CHRAC1* | 10.93187013 | 0.003325457 | 0.024785123 | 1 |
| HTN | t_cell | *SNX16* | 6.35339281 | 0.00154013 | 0.016399464 | 1 |
| HTN | t_cell | *TNFRSF10A* | 1.213707737 | 0.004202793 | 0.029110503 | 1 |
| HTN | t_cell | *ERICH1* | 4.116581961 | 0.006189086 | 0.038060136 | 1 |
| HTN | t_cell | *IL27RA* | 1.456679112 | 0.001539941 | 0.016399464 | 1 |
| HTN | t_cell | *RASAL3* | 0.676974203 | 0.002594566 | 0.021409585 | 1 |
| HTN | t_cell | *TGFB1* | 4.910173611 | 0.000544409 | 0.009873328 | 1 |
| HTN | t_cell | *ICAM5* | -3.535080992 | 9.54E-05 | 0.004151182 | 0.462967094 |
| HTN | t_cell | *RABAC1* | 7.929664484 | 6.51E-06 | 0.000585228 | 0.031602298 |
| HTN | t_cell | *CARD8* | 3.891259655 | 0.002475537 | 0.020889229 | 1 |
| HTN | t_cell | *LIG1* | 4.190299078 | 0.004211783 | 0.029110503 | 1 |
| HTN | t_cell | *PLA2G4C* | -1.129411269 | 0.006679845 | 0.040513257 | 1 |
| HTN | t_cell | *DNASE2* | 6.201987337 | 0.002757326 | 0.022130428 | 1 |
| HTN | t_cell | *SP4* | 3.816450374 | 0.000105485 | 0.004481533 | 0.511810802 |
| HTN | t_cell | *ADAP1* | 9.887098534 | 0.000968068 | 0.012573256 | 1 |
| HTN | t_cell | *CHN2* | 5.041551713 | 6.85E-05 | 0.003290268 | 0.332317103 |
| HTN | t_cell | *TAF6* | -2.51703626 | 0.002789384 | 0.022223465 | 1 |
| HTN | t_cell | *TFR2* | 3.079269142 | 0.001582905 | 0.016552277 | 1 |
| HTN | t_cell | *NRF1* | 4.397794532 | 0.002739846 | 0.022082613 | 1 |
| HTN | t_cell | *MEST* | -2.505483275 | 0.001113764 | 0.013750594 | 1 |
| HTN | t_cell | *ACTR3C* | 6.489522247 | 0.000196154 | 0.006609299 | 0.951739123 |
| HTN | t_cell | *COA1* | 4.489834303 | 0.001912649 | 0.018232171 | 1 |
| HTN | t_cell | *TRIM14* | 2.453679282 | 0.002404932 | 0.020689239 | 1 |
| HTN | t_cell | *CORO2A* | 1.803658076 | 0.00057761 | 0.010154224 | 1 |
| HTN | t_cell | *CUBN* | 4.492890536 | 0.000941222 | 0.012443616 | 1 |
| HTN | t_cell | *ATE1* | 3.732817854 | 0.005719399 | 0.035807125 | 1 |
| HTN | t_cell | *SPOCK2* | 0.85197021 | 0.00384886 | 0.027382209 | 1 |
| HTN | t_cell | *SFXN3* | 7.738542303 | 0.001357324 | 0.015388209 | 1 |
| HTN | t_cell | *ACBD5* | 5.227502701 | 0.007407284 | 0.043658822 | 1 |
| HTN | t_cell | *TFAM* | 7.333143709 | 0.003249247 | 0.024428605 | 1 |
| HTN | t_cell | *PNPO* | -6.274839709 | 0.00047369 | 0.00953704 | 1 |

| | | | | | | |
|---|---|---|---|---|---|---|
| HTN | t_cell | *DRG2* | 6.227835057 | 0.000642712 | 0.010434098 | 1 |
| HTN | t_cell | *ICAM2* | 7.591847765 | 0.001630169 | 0.016864772 | 1 |
| HTN | t_cell | *DDX5* | 3.728229057 | 0.000871485 | 0.012012667 | 1 |
| HTN | t_cell | *SGCA* | 1.971387421 | 0.004430303 | 0.030106203 | 1 |
| HTN | t_cell | *SLAIN2* | 4.835120099 | 0.008772599 | 0.047950379 | 1 |
| HTN | t_cell | *GRPEL1* | 8.618331864 | 0.007735541 | 0.044735213 | 1 |
| HTN | t_cell | *GAR1* | 9.17937556 | 0.001850328 | 0.017907204 | 1 |
| HTN | t_cell | *STIM2* | 4.487148769 | 0.000907978 | 0.012340357 | 1 |
| HTN | t_cell | *B3GAT1* | 2.179035991 | 0.002616387 | 0.021512816 | 1 |
| HTN | t_cell | *DCPS* | 5.257865287 | 0.003493286 | 0.025673285 | 1 |
| HTN | t_cell | *CCND1* | -6.833432402 | 2.27E-06 | 0.00026277 | 0.011036334 |
| HTN | t_cell | *CCDC86* | -3.427380354 | 0.002342465 | 0.020368529 | 1 |
| HTN | t_cell | *KIAA1549L* | -5.094343371 | 0.009072473 | 0.048748217 | 1 |
| HTN | t_cell | *SLC35F2* | 8.660473229 | 0.000149986 | 0.005471682 | 0.727733726 |
| HTN | t_cell | *RPS13* | 3.964961449 | 0.002141086 | 0.019277303 | 1 |
| HTN | t_cell | *PRDM4* | 6.826988417 | 0.000375006 | 0.008900183 | 1 |
| HTN | t_cell | *KCTD10* | -3.384038088 | 0.007922188 | 0.045417571 | 1 |
| HTN | t_cell | *PRR4* | 3.628844646 | 0.000686561 | 0.010815564 | 1 |
| HTN | t_cell | *PARP11* | 5.458793294 | 0.000859864 | 0.011988677 | 1 |
| HTN | t_cell | *GPN3* | 8.614121805 | 0.003377731 | 0.025097626 | 1 |
| HTN | t_cell | *KRR1* | 4.697267643 | 0.001883411 | 0.018059902 | 1 |
| HTN | t_cell | *GNPTAB* | 4.744353582 | 0.004534579 | 0.030518825 | 1 |
| HTN | t_cell | *RNGTT* | 4.18506892 | 0.00054686 | 0.009873328 | 1 |
| HTN | t_cell | *TULP1* | -3.304914932 | 0.001708109 | 0.017171831 | 1 |
| HTN | t_cell | *CD83* | 5.163935838 | 0.00061897 | 0.010192716 | 1 |
| HTN | t_cell | *BVES* | -1.405754008 | 0.004541342 | 0.030518825 | 1 |
| HTN | t_cell | *VNN2* | 3.255884637 | 0.00182701 | 0.017764831 | 1 |
| HTN | t_cell | *PHACTR2* | 3.025792905 | 0.002382789 | 0.020535157 | 1 |
| HTN | t_cell | *EXOC2* | 5.208508123 | 0.000602582 | 0.010162236 | 1 |
| HTN | t_cell | *IMPG1* | -6.445037305 | 0.001821902 | 0.017750742 | 1 |
| HTN | t_cell | *BTN2A1* | 6.71944247 | 2.27E-05 | 0.001527754 | 0.109998304 |
| HTN | t_cell | *CEP72* | 5.838185209 | 0.000459701 | 0.009475372 | 1 |
| HTN | t_cell | *IK* | 4.608760371 | 5.34E-05 | 0.002673106 | 0.259291288 |
| HTN | t_cell | *RNF130* | 3.543548104 | 0.002608305 | 0.021486412 | 1 |
| HTN | t_cell | *ARSB* | 5.805814338 | 0.008984565 | 0.048598786 | 1 |
| HTN | t_cell | *LMNB1* | 1.966133856 | 0.005484774 | 0.034832622 | 1 |
| HTN | t_cell | *SKP1* | 5.752392792 | 0.002458474 | 0.020854049 | 1 |
| HTN | t_cell | *TXNDC15* | 6.90182939 | 0.002463855 | 0.020863218 | 1 |
| HTN | t_cell | *PCCB* | 4.660691812 | 1.86E-05 | 0.001346963 | 0.090394074 |
| HTN | t_cell | *CBLB* | 3.241698145 | 0.000333013 | 0.00850409 | 1 |
| HTN | t_cell | *VIPR1* | 10.48363286 | 0.000515171 | 0.009873328 | 1 |
| HTN | t_cell | *ADAM23* | 1.522165314 | 0.005376078 | 0.034412574 | 1 |
| HTN | t_cell | *ACTR1B* | 7.765876319 | 9.58E-05 | 0.004151182 | 0.464932435 |
| HTN | t_cell | *PPM1G* | 8.15003488 | 4.14E-05 | 0.002256082 | 0.200791272 |
| HTN | t_cell | *GALNT3* | 1.738882655 | 0.000836017 | 0.011988677 | 1 |
| HTN | t_cell | *GNLY* | 2.02229096 | 0.002002017 | 0.018733594 | 1 |
| HTN | t_cell | *TPO* | -1.180790778 | 0.009017241 | 0.048667021 | 1 |
| HTN | t_cell | *CEBPZ* | 6.845301369 | 0.000226191 | 0.007126499 | 1 |
| HTN | t_cell | *WIPF1* | 3.007393365 | 0.000137615 | 0.005282043 | 0.667706717 |
| HTN | t_cell | *AAK1* | 3.566334704 | 0.000522559 | 0.009873328 | 1 |

| | | | | | | |
|---|---|---|---|---|---|---|
| HTN | t_cell | *MORN1* | -3.118858848 | 0.002674153 | 0.021733653 | 1 |
| HTN | t_cell | *KIAA1324* | 3.343715638 | 0.000249378 | 0.007423215 | 1 |
| HTN | t_cell | *RAP1A* | 4.322912929 | 0.000269035 | 0.007681101 | 1 |
| HTN | t_cell | *FBXO2* | 2.216289469 | 0.002417972 | 0.02069412 | 1 |
| HTN | t_cell | *EXOC8* | 8.4826498 | 0.001465044 | 0.016009899 | 1 |
| HTN | t_cell | *SLAMF1* | 0.349308484 | 0.001768861 | 0.017373513 | 1 |
| HTN | t_cell | *KIF17* | -2.860019608 | 0.008384731 | 0.046815552 | 1 |
| HTN | t_cell | *PRRC2C* | 4.742761164 | 0.001129392 | 0.013865559 | 1 |
| HTN | t_cell | *IRF6* | -1.727567306 | 0.001042869 | 0.013185341 | 1 |
| HTN | t_cell | *RPS6KA1* | 1.921580414 | 0.00424153 | 0.029191351 | 1 |
| HTN | t_cell | *ESYT2* | 3.947709021 | 0.009069023 | 0.048748217 | 1 |
| HTN | t_cell | *APOA1* | 7.241170442 | 0.002245067 | 0.01983569 | 1 |
| HTN | t_cell | *LRMP* | 1.497559941 | 0.009047922 | 0.048724217 | 1 |
| HTN | t_cell | *ELOVL4* | 3.496677007 | 0.000246644 | 0.007387128 | 1 |
| HTN | t_cell | *FOXO3* | 4.103423084 | 0.004076788 | 0.028528962 | 1 |
| HTN | t_cell | *UBN1* | 6.032281988 | 0.002141481 | 0.019277303 | 1 |
| HTN | t_cell | *FKBP15* | 2.645029945 | 0.00809619 | 0.045627875 | 1 |
| HTN | t_cell | *ACOT2* | 1.224275531 | 0.000415148 | 0.009032732 | 1 |
| HTN | t_cell | *PCDHB12* | -1.946979565 | 0.002546336 | 0.021228213 | 1 |
| HTN | t_cell | *MRC1* | 4.557490295 | 0.004060678 | 0.028512897 | 1 |
| HTN | t_cell | *IQSEC3* | 10.95548505 | 0.002558549 | 0.021277196 | 1 |
| HTN | t_cell | *ZFP30* | 6.115189377 | 1.84E-06 | 0.000228563 | 0.008913957 |
| HTN | t_cell | *TBX2* | -3.945744543 | 2.90E-09 | 1.17E-06 | 1.41E-05 |
| HTN | t_cell | *TEX14* | 6.977841626 | 1.17E-07 | 2.57E-05 | 0.000566121 |
| HTN | t_cell | *ZNF549* | 6.690900389 | 0.000610612 | 0.010181068 | 1 |
| HTN | t_cell | *LHX4* | 7.904890327 | 0.000792132 | 0.011576575 | 1 |
| HTN | t_cell | *MAPK8IP1* | -9.123390358 | 1.51E-05 | 0.001126083 | 0.073195422 |
| HTN | t_cell | *ZCCHC17* | 5.78492883 | 0.00784001 | 0.045177826 | 1 |
| HTN | t_cell | *FABP3* | -3.565982928 | 0.004404535 | 0.030057389 | 1 |
| HTN | t_cell | *CD244* | 1.549272876 | 0.001401481 | 0.015560606 | 1 |
| HTN | t_cell | *OPN4* | 6.058440841 | 0.008254088 | 0.046352817 | 1 |
| HTN | t_cell | *PMS2* | 5.420827224 | 0.004113659 | 0.028595237 | 1 |
| HTN | t_cell | *GLIPR2* | 2.850966014 | 0.000845034 | 0.011988677 | 1 |
| HTN | t_cell | *CCDC91* | 4.232274643 | 0.000734436 | 0.011097787 | 1 |
| HTN | t_cell | *CENPK* | 1.260741094 | 0.001739884 | 0.017334528 | 1 |
| HTN | t_cell | *ARHGAP9* | 2.044643617 | 0.002886651 | 0.022663476 | 1 |
| HTN | t_cell | *CDK2* | -3.752790011 | 0.000985343 | 0.012647836 | 1 |
| HTN | t_cell | *DBH* | -35.42649684 | 0.008017541 | 0.045452802 | 1 |
| HTN | t_cell | *NDUFAF4* | 4.454440383 | 0.000322677 | 0.008451766 | 1 |
| HTN | t_cell | *WFDC3* | -1.255675652 | 2.63E-05 | 0.001724545 | 0.127616316 |
| HTN | t_cell | *CSE1L* | 5.573286329 | 0.001190408 | 0.014332155 | 1 |
| HTN | t_cell | *ATP8A1* | 2.677144017 | 0.001331918 | 0.015291977 | 1 |
| HTN | t_cell | *BTN2A2* | 2.736081837 | 6.14E-07 | 9.31E-05 | 0.002980221 |
| HTN | t_cell | *ZNF391* | -1.540761265 | 3.25E-05 | 0.001933659 | 0.157590444 |
| HTN | t_cell | *TCP11* | 2.981582938 | 0.007951188 | 0.045417571 | 1 |
| HTN | t_cell | *ATXN1* | 3.696493555 | 0.000326468 | 0.008470705 | 1 |
| HTN | t_cell | *RUNX2* | 1.851781945 | 0.001312498 | 0.01519867 | 1 |
| HTN | t_cell | *GOT2* | 8.586039175 | 3.20E-05 | 0.001933659 | 0.155326457 |
| HTN | t_cell | *CLYBL* | 5.119647443 | 0.00046088 | 0.009475372 | 1 |
| HTN | t_cell | *GRK4* | 0.998255553 | 0.002726967 | 0.022015383 | 1 |

| | | | | | | |
|---|---|---|---|---|---|---|
| HTN | t_cell | *HS3ST3B1* | 0.644202017 | 0.003629142 | 0.026231565 | 1 |
| HTN | t_cell | *IL1B* | 13.86273993 | 6.36E-05 | 0.003085354 | 0.308535423 |
| HTN | t_cell | *PSD4* | 1.469877783 | 0.001109296 | 0.013730365 | 1 |
| HTN | t_cell | *SH2D3A* | 1.164053751 | 0.004967174 | 0.032612621 | 1 |
| HTN | t_cell | *GPR108* | 5.936157697 | 1.97E-05 | 0.001363933 | 0.095475284 |
| HTN | t_cell | *SDCBP2* | -3.377704533 | 0.008096213 | 0.045627875 | 1 |
| HTN | t_cell | *UROD* | -3.794793226 | 0.000917643 | 0.012349099 | 1 |
| HTN | t_cell | *PRDX5* | 7.69428811 | 0.000289994 | 0.008133232 | 1 |
| HTN | t_cell | *IRF3* | 5.481891266 | 0.009029828 | 0.048680809 | 1 |
| HTN | t_cell | *TSKS* | -3.740614329 | 9.09E-05 | 0.004083084 | 0.440973101 |
| HTN | t_cell | *TRAP1* | 2.749419932 | 4.99E-06 | 0.000484073 | 0.024203637 |
| HTN | t_cell | *ZNF484* | 7.464340742 | 0.000734481 | 0.011097787 | 1 |
| HTN | t_cell | *FGD3* | 1.226824443 | 0.000671013 | 0.010605073 | 1 |
| HTN | t_cell | *ABHD8* | -4.571942683 | 3.81E-05 | 0.002151383 | 0.185018929 |
| HTN | t_cell | *ATP13A4* | 2.264614329 | 0.000593794 | 0.010162236 | 1 |
| HTN | t_cell | *HELB* | 1.400989583 | 0.00171131 | 0.017171831 | 1 |
| HTN | t_cell | *RAB3IP* | 3.076984125 | 0.000840675 | 0.011988677 | 1 |
| HTN | t_cell | *YEATS4* | 2.267536484 | 0.002932928 | 0.02295253 | 1 |
| HTN | t_cell | *GFER* | -4.887434654 | 0.000858527 | 0.011988677 | 1 |
| HTN | t_cell | *SMARCA4* | 5.538010984 | 0.001619915 | 0.016794498 | 1 |
| HTN | t_cell | *GAD1* | -4.676330589 | 0.000401316 | 0.008988159 | 1 |
| HTN | t_cell | *ORMDL1* | 3.974198114 | 0.004232512 | 0.029170668 | 1 |
| HTN | t_cell | *VPS13C* | 4.457649932 | 0.001395369 | 0.015560606 | 1 |
| HTN | t_cell | *BBOX1* | 29.10730928 | 0.000707788 | 0.010867676 | 1 |
| HTN | t_cell | *SERGEF* | 5.418480928 | 8.23E-06 | 0.000700159 | 0.03990909 |
| HTN | t_cell | *RIPK3* | 2.854191583 | 0.000173251 | 0.006047578 | 0.840613409 |
| HTN | t_cell | *REEP5* | 4.81438267 | 0.007937848 | 0.045417571 | 1 |
| HTN | t_cell | *RHBDF2* | 2.259003294 | 0.001542815 | 0.016399464 | 1 |
| HTN | t_cell | *TMEM8A* | 8.462378717 | 0.005927919 | 0.036756978 | 1 |
| HTN | t_cell | *CDC16* | 4.739947431 | 0.000124001 | 0.004991087 | 0.601654454 |
| HTN | t_cell | *ATP8B3* | -2.135862974 | 0.000307987 | 0.00839523 | 1 |
| HTN | t_cell | *DDA1* | 6.646731891 | 0.008004413 | 0.045452802 | 1 |
| HTN | t_cell | *LSM7* | 3.686525389 | 0.002966171 | 0.023100903 | 1 |
| HTN | t_cell | *ACTN4* | 5.040748562 | 0.002137872 | 0.019277303 | 1 |
| HTN | t_cell | *SSBP4* | 3.570254405 | 0.000342056 | 0.008644045 | 1 |
| HTN | t_cell | *PGPEP1* | 4.149628189 | 0.000145467 | 0.005421382 | 0.705805329 |
| HTN | t_cell | *TRPM4* | -3.625402649 | 0.000115415 | 0.004745727 | 0.559995775 |
| HTN | t_cell | *ZNF337* | 3.780974837 | 0.007918164 | 0.045417571 | 1 |
| HTN | t_cell | *THEMIS2* | 2.976631588 | 0.0034146 | 0.02521711 | 1 |
| HTN | t_cell | *DNMT1* | 5.566779446 | 0.000383076 | 0.008960997 | 1 |
| HTN | t_cell | *C12orf65* | 3.563907516 | 0.002116248 | 0.019263747 | 1 |
| HTN | t_cell | *SLC35D2* | 5.469183306 | 0.009371383 | 0.04991213 | 1 |
| HTN | t_cell | *ULBP2* | 16.76833862 | 0.007243455 | 0.043122996 | 1 |
| HTN | t_cell | *HAUS8* | -6.582337273 | 5.76E-08 | 1.33E-05 | 0.000279619 |
| HTN | t_cell | *SLC6A6* | 4.489302303 | 0.000474695 | 0.00953704 | 1 |
| HTN | t_cell | *NAPSA* | 3.227345621 | 0.007712827 | 0.044694225 | 1 |
| HTN | t_cell | *KIF3A* | 6.955171726 | 0.00128023 | 0.015053256 | 1 |
| HTN | t_cell | *EXOC4* | 4.249951677 | 0.0008306 | 0.011988677 | 1 |
| HTN | t_cell | *PRKAB2* | 6.483593328 | 0.001517879 | 0.016329815 | 1 |
| HTN | t_cell | *GCH1* | 4.167557384 | 6.23E-06 | 0.000569926 | 0.030206067 |

| | | | | | | |
|---|---|---|---|---|---|---|
| HTN | t_cell | *MATN3* | -2.77356761 | 0.004622001 | 0.030889737 | 1 |
| HTN | t_cell | *TRIM21* | 3.370211821 | 0.002542693 | 0.021228213 | 1 |
| HTN | t_cell | *TRIM22* | 4.238758671 | 0.000540322 | 0.009873328 | 1 |
| HTN | t_cell | *REEP2* | -3.576914491 | 0.005331231 | 0.034306544 | 1 |
| HTN | t_cell | *AP3B1* | 4.226458778 | 0.006656231 | 0.040420568 | 1 |
| HTN | t_cell | *RSG1* | -8.444102909 | 0.000475158 | 0.00953704 | 1 |
| HTN | t_cell | *ALOX5AP* | 2.137267365 | 0.001281326 | 0.015053256 | 1 |
| HTN | t_cell | *EPSTI1* | 5.088885802 | 0.004920332 | 0.03239274 | 1 |
| HTN | t_cell | *CSNK1G2* | 6.162069043 | 0.000527846 | 0.009873328 | 1 |
| HTN | t_cell | *GIMAP4* | 1.747630209 | 0.005707485 | 0.035778702 | 1 |
| HTN | t_cell | *IMPA1* | 3.270829098 | 0.000198453 | 0.006640647 | 0.962893861 |
| HTN | t_cell | *CATSPERB* | 2.993972986 | 0.001359445 | 0.015388209 | 1 |
| HTN | t_cell | *TTC9* | 1.612267753 | 0.001866475 | 0.017980217 | 1 |
| HTN | t_cell | *GSTM5* | -1.993025361 | 0.006521831 | 0.039703796 | 1 |
| HTN | t_cell | *FKBP11* | 7.975996046 | 0.000314711 | 0.00843633 | 1 |
| HTN | t_cell | *KLRD1* | 3.583061162 | 0.000405586 | 0.008988159 | 1 |
| HTN | t_cell | *ACP2* | 13.55617665 | 2.25E-11 | 2.18E-08 | 1.09E-07 |
| HTN | t_cell | *ZCCHC11* | 3.526260163 | 0.003555196 | 0.025939564 | 1 |
| HTN | t_cell | *PRPF38A* | 7.148928957 | 0.001818384 | 0.017750742 | 1 |
| HTN | t_cell | *TIMM10* | 5.116470551 | 0.00262794 | 0.021538456 | 1 |
| HTN | t_cell | *OSTF1* | 4.607927085 | 0.000499146 | 0.009758406 | 1 |
| HTN | t_cell | *UBQLN1* | 5.250797624 | 0.002795367 | 0.022234628 | 1 |
| HTN | t_cell | *ADAM19* | 1.118248603 | 0.00066044 | 0.01058442 | 1 |
| HTN | t_cell | *USP30* | -2.035920922 | 0.006877681 | 0.041339446 | 1 |
| HTN | t_cell | *TMEM60* | 7.799429469 | 3.41E-05 | 0.001971842 | 0.165634711 |
| HTN | t_cell | *PRR5L* | 3.172362345 | 0.004719521 | 0.031325742 | 1 |
| HTN | t_cell | *FAIM2* | -3.485079765 | 0.001186868 | 0.014325086 | 1 |
| HTN | t_cell | *MICAL1* | 2.789671673 | 0.000124469 | 0.004991087 | 0.603921586 |
| HTN | t_cell | *USP15* | 4.383687127 | 0.001104774 | 0.013709375 | 1 |
| HTN | t_cell | *AGT* | 2.414818971 | 0.000670335 | 0.010605073 | 1 |
| HTN | t_cell | *CAPN9* | -4.999983077 | 0.001043522 | 0.013185341 | 1 |
| HTN | t_cell | *SP110* | 4.386278852 | 0.000295486 | 0.008146014 | 1 |
| HTN | t_cell | *MRPL44* | 7.216101719 | 0.000376038 | 0.008900183 | 1 |
| HTN | t_cell | *USP37* | 4.503123627 | 0.004650819 | 0.030996942 | 1 |
| HTN | t_cell | *GCC2* | 5.671237869 | 9.47E-05 | 0.004151182 | 0.459599924 |
| HTN | t_cell | *MRPS9* | 3.535009132 | 0.001433638 | 0.015773269 | 1 |
| HTN | t_cell | *ANKRD36* | 5.132588134 | 0.003584025 | 0.02603247 | 1 |
| HTN | t_cell | *SETDB2* | 4.899520309 | 0.002055531 | 0.018960903 | 1 |
| HTN | t_cell | *AOAH* | 1.066611712 | 0.001681998 | 0.017059213 | 1 |
| HTN | t_cell | *NMT1* | 4.823258485 | 1.08E-08 | 3.74E-06 | 5.24E-05 |
| HTN | t_cell | *TOR1B* | 6.982573489 | 0.002858214 | 0.022586406 | 1 |
| HTN | t_cell | *ANP32B* | 4.896464799 | 0.009084708 | 0.048759961 | 1 |
| HTN | t_cell | *RIPK1* | 4.066693589 | 0.004210489 | 0.029110503 | 1 |
| HTN | t_cell | *SLC37A4* | 3.315657414 | 0.002256404 | 0.01983569 | 1 |
| HTN | t_cell | *PPP2R1B* | -1.994937379 | 0.005350962 | 0.034387907 | 1 |
| HTN | t_cell | *FDX1* | 4.796815103 | 0.002528017 | 0.021221349 | 1 |
| HTN | t_cell | *NUSAP1* | 5.657176646 | 1.02E-06 | 0.000133674 | 0.004945927 |
| HTN | t_cell | *ADAM10* | 4.928686721 | 0.001989936 | 0.018675376 | 1 |
| HTN | t_cell | *RTCA* | 5.544431929 | 0.002170854 | 0.019346851 | 1 |
| HTN | t_cell | *KHK* | -2.862608179 | 0.00130424 | 0.015139453 | 1 |

| | | | | | | |
|---|---|---|---|---|---|---|
| HTN | t_cell | *ADCY3* | 6.388991559 | 0.000696672 | 0.010834138 | 1 |
| HTN | t_cell | *LHCGR* | -3.435464168 | 0.002565367 | 0.021277196 | 1 |
| HTN | t_cell | *ACTR2* | 4.864322389 | 0.001040345 | 0.013185341 | 1 |
| HTN | t_cell | *DTNB* | 5.659498367 | 0.000238005 | 0.007308865 | 1 |
| HTN | t_cell | *STAT4* | 0.734904242 | 0.001898582 | 0.018133701 | 1 |
| HTN | t_cell | *PRKG2* | 2.223884056 | 0.000330862 | 0.00850409 | 1 |
| HTN | t_cell | *TRPC3* | 2.583758632 | 0.0015788 | 0.016552277 | 1 |
| HTN | t_cell | *G3BP2* | 5.372843623 | 7.52E-05 | 0.003506989 | 0.364726843 |
| HTN | t_cell | *CNOT6L* | 4.342701899 | 0.000217441 | 0.007008867 | 1 |
| HTN | t_cell | *ENPEP* | -1.486586477 | 0.002594263 | 0.021409585 | 1 |
| HTN | t_cell | *FBN2* | 1.198891897 | 0.000435174 | 0.00917827 | 1 |
| HTN | t_cell | *B4GALNT3* | 1.398211809 | 0.002949619 | 0.023045978 | 1 |
| HTN | t_cell | *KIF21A* | 4.879076886 | 0.002113069 | 0.019263747 | 1 |
| HTN | t_cell | *ALG10* | -2.743597308 | 0.007090869 | 0.042367165 | 1 |
| HTN | t_cell | *KLRG1* | 0.809087336 | 0.007719242 | 0.044694225 | 1 |
| HTN | t_cell | *VAMP1* | 3.123648299 | 0.003065204 | 0.023569524 | 1 |
| HTN | t_cell | *MARCH9* | 7.14913177 | 0.000509154 | 0.009873328 | 1 |
| HTN | t_cell | *POC1B* | 5.799100169 | 0.000525836 | 0.009873328 | 1 |
| HTN | t_cell | *RITA1* | 12.75697442 | 0.002663222 | 0.021717565 | 1 |
| HTN | t_cell | *GLTP* | 8.765373988 | 0.002169327 | 0.019346851 | 1 |
| HTN | t_cell | *FOXN4* | -2.481820546 | 0.004056267 | 0.028512897 | 1 |
| HTN | t_cell | *MTMR6* | 6.469596261 | 0.002437655 | 0.020713662 | 1 |
| HTN | t_cell | *CCDC65* | 5.237628199 | 0.00256443 | 0.021277196 | 1 |
| HTN | t_cell | *ESYT1* | 6.184193068 | 0.002256657 | 0.01983569 | 1 |
| HTN | t_cell | *HNRNPA1L2* | -4.490586028 | 0.004689181 | 0.031166992 | 1 |
| HTN | t_cell | *FITM1* | 8.178209672 | 0.000605294 | 0.010162236 | 1 |
| HTN | t_cell | *RTN1* | 2.389723874 | 0.004421465 | 0.030106203 | 1 |
| HTN | t_cell | *RAB15* | 8.570906177 | 0.003763634 | 0.027053555 | 1 |
| HTN | t_cell | *ULK3* | 12.77172588 | 7.82E-07 | 0.000110202 | 0.003794406 |
| HTN | t_cell | *ITGAX* | 4.639011497 | 0.000373093 | 0.008900183 | 1 |
| HTN | t_cell | *GCSH* | -3.112113819 | 0.005031591 | 0.032901994 | 1 |
| HTN | t_cell | *CMTM3* | 4.692150635 | 0.005108362 | 0.033191908 | 1 |
| HTN | t_cell | *TLDC1* | 4.473471237 | 0.000573212 | 0.010113538 | 1 |
| HTN | t_cell | *DEF8* | 3.904406084 | 0.001879122 | 0.018054458 | 1 |
| HTN | t_cell | *TCF25* | 5.37314394 | 0.000232836 | 0.007202575 | 1 |
| HTN | t_cell | *MYO19* | 6.869212389 | 0.000178042 | 0.006126663 | 0.863859514 |
| HTN | t_cell | *RASL10B* | -5.709261804 | 0.003497535 | 0.025673285 | 1 |
| HTN | t_cell | *UNC45B* | 3.52001739 | 0.00264924 | 0.02163992 | 1 |
| HTN | t_cell | *LRRC46* | -3.634841878 | 0.000106219 | 0.004481533 | 0.515376273 |
| HTN | t_cell | *SCRN2* | -1.062331951 | 0.001481712 | 0.016079706 | 1 |
| HTN | t_cell | *SSH2* | 3.931108047 | 0.001678829 | 0.017059213 | 1 |
| HTN | t_cell | *RHBDL3* | -2.660869465 | 0.004564704 | 0.030633394 | 1 |
| HTN | t_cell | *TMC6* | 1.453813779 | 0.001717978 | 0.017171831 | 1 |
| HTN | t_cell | *TBCD* | 4.965555954 | 0.00135227 | 0.015388209 | 1 |
| HTN | t_cell | *MMEL1* | -1.727878235 | 0.001257358 | 0.01484355 | 1 |
| HTN | t_cell | *RPS8* | 4.358867627 | 0.003226767 | 0.024348789 | 1 |
| HTN | t_cell | *TIPRL* | 5.895847977 | 0.000927369 | 0.012361522 | 1 |
| HTN | t_cell | *XCL1* | 0.882825535 | 0.003901445 | 0.027594479 | 1 |
| HTN | t_cell | *SNX27* | 5.412254237 | 0.000962879 | 0.012573256 | 1 |
| HTN | t_cell | *SMYD2* | 5.621283636 | 0.000469571 | 0.00953704 | 1 |

| | | | | | | |
|---|---|---|---|---|---|---|
| HTN | t_cell | *CREB3L4* | -4.36065077 | 9.73E-07 | 0.000131079 | 0.004718833 |
| HTN | t_cell | *GALM* | 4.934544283 | 0.000483958 | 0.009584338 | 1 |
| HTN | t_cell | *ABHD1* | -3.988535629 | 0.003713385 | 0.026771685 | 1 |
| HTN | t_cell | *CIAO1* | 5.816529531 | 0.002417339 | 0.02069412 | 1 |
| HTN | t_cell | *THNSL2* | -1.571846418 | 0.008080584 | 0.045627875 | 1 |
| HTN | t_cell | *PHOSPHO2* | -3.682032325 | 0.00148638 | 0.016079706 | 1 |
| HTN | t_cell | *METTL21A* | 7.138311713 | 3.97E-05 | 0.002190959 | 0.192804364 |
| HTN | t_cell | *ESPNL* | 29.47201579 | 0.00140106 | 0.015560606 | 1 |
| HTN | t_cell | *ANKMY1* | 3.465827653 | 0.006863572 | 0.041317682 | 1 |
| HTN | t_cell | *TAMM41* | 5.049389181 | 0.00508189 | 0.033132618 | 1 |
| HTN | t_cell | *EAF1* | 6.540935258 | 0.000142627 | 0.005377706 | 0.6920255 |
| HTN | t_cell | *ARL6IP5* | 4.967248448 | 0.006171912 | 0.038002687 | 1 |
| HTN | t_cell | *MED12L* | 7.477508058 | 0.001251144 | 0.014806224 | 1 |
| HTN | t_cell | *CISD2* | 11.29587512 | 5.90E-06 | 0.000550815 | 0.028642364 |
| HTN | t_cell | *METTL14* | 5.607863205 | 0.008038098 | 0.045508581 | 1 |
| HTN | t_cell | *FAM105A* | 1.663308852 | 0.003983687 | 0.028094259 | 1 |
| HTN | t_cell | *TSLP* | -1.382991588 | 0.000591411 | 0.010162236 | 1 |
| HTN | t_cell | *MYLK4* | 7.516621941 | 0.009262681 | 0.049496178 | 1 |
| HTN | t_cell | *RNF44* | 3.91344116 | 0.000779891 | 0.011501608 | 1 |
| HTN | t_cell | *ABT1* | 2.711915454 | 0.000645808 | 0.010434098 | 1 |
| HTN | t_cell | *IRAK1BP1* | 6.189848501 | 3.20E-05 | 0.001933659 | 0.155194089 |
| HTN | t_cell | *ABRACL* | 1.941494141 | 0.007839001 | 0.045177826 | 1 |
| HTN | t_cell | *SLC18B1* | -2.891469397 | 0.003183696 | 0.024125993 | 1 |
| HTN | t_cell | *GIGYF1* | 5.427905377 | 0.003275722 | 0.024536323 | 1 |
| HTN | t_cell | *EPHA1* | 13.08165034 | 0.000591448 | 0.010162236 | 1 |
| HTN | t_cell | *NCAPG2* | 5.2273269 | 0.004232302 | 0.029170668 | 1 |
| HTN | t_cell | *ADHFE1* | -1.469548992 | 0.000646762 | 0.010434098 | 1 |
| HTN | t_cell | *POLE3* | 2.807742708 | 0.000834229 | 0.011988677 | 1 |
| HTN | t_cell | *SEC16A* | 5.179323765 | 0.007099032 | 0.042367165 | 1 |
| HTN | t_cell | *ST8SIA6* | 7.391936971 | 0.000401803 | 0.008988159 | 1 |
| HTN | t_cell | *CDHR1* | 1.439424863 | 0.005850978 | 0.036396547 | 1 |
| HTN | t_cell | *RGR* | -13.07057046 | 0.002572738 | 0.021301916 | 1 |
| HTN | t_cell | *PAOX* | 8.751525534 | 0.000647293 | 0.010434098 | 1 |
| HTN | t_cell | *RGS10* | 5.301457889 | 0.000706771 | 0.010867676 | 1 |
| HTN | t_cell | *HSD17B12* | -2.766531201 | 1.44E-09 | 6.96E-07 | 6.96E-06 |
| HTN | t_cell | *TNKS1BP1* | 3.598027324 | 3.08E-06 | 0.000348061 | 0.014966627 |
| HTN | t_cell | *PTPRJ* | 3.322948957 | 0.001762823 | 0.017349326 | 1 |
| HTN | t_cell | *CAPN5* | 10.89475359 | 0.007617354 | 0.04440186 | 1 |
| HTN | t_cell | *RPS3* | 4.149999554 | 0.003141042 | 0.023945159 | 1 |
| HTN | t_cell | *TTC12* | 4.348897948 | 1.92E-05 | 0.001346963 | 0.092940416 |
| HTN | t_cell | *NPAT* | 6.026443051 | 0.001762071 | 0.017349326 | 1 |
| HTN | t_cell | *ATM* | 3.306763307 | 0.002172376 | 0.019346851 | 1 |
| HTN | t_cell | *PLCH2* | 1.150749285 | 0.003059366 | 0.023561972 | 1 |
| HTN | t_cell | *CHEK1* | 5.286518789 | 0.001104276 | 0.013709375 | 1 |
| HTN | t_cell | *SPATA25* | -3.396149122 | 0.003306921 | 0.02468489 | 1 |
| HTN | t_cell | *TRPT1* | -3.156420606 | 0.001686641 | 0.017059213 | 1 |
| HTN | t_cell | *ANK3* | 0.769034784 | 0.001687639 | 0.017059213 | 1 |
| HTN | t_cell | *GXYLT1* | 5.138934001 | 0.007941303 | 0.045417571 | 1 |
| HTN | t_cell | *MFSD6* | 4.277810351 | 0.000850913 | 0.011988677 | 1 |
| HTN | t_cell | *BICD1* | 4.900521705 | 0.005632863 | 0.035402398 | 1 |

| | | | | | | |
|---|---|---|---|---|---|---|
| HTN | t_cell | *MGAT5* | 3.305984642 | 0.004424284 | 0.030106203 | 1 |
| HTN | t_cell | *EPG5* | 4.760103541 | 0.008407435 | 0.046888362 | 1 |
| HTN | t_cell | *CWF19L2* | 6.532507922 | 1.13E-05 | 0.000914033 | 0.054842004 |
| HTN | t_cell | *PLEKHH2* | -1.992345089 | 0.005119 | 0.033205064 | 1 |
| HTN | t_cell | *MBNL1* | 2.945565809 | 0.00054074 | 0.009873328 | 1 |
| HTN | t_cell | *CCNO* | 9.069699551 | 0.002759462 | 0.022130428 | 1 |
| HTN | t_cell | *GGPS1* | 5.187928974 | 0.001143236 | 0.01390596 | 1 |
| HTN | t_cell | *MED21* | 7.554434962 | 0.000111813 | 0.004676879 | 0.542517964 |
| HTN | t_cell | *JAKMIP1* | 0.635276373 | 0.002837667 | 0.022497324 | 1 |
| HTN | t_cell | *MR1* | 4.08691293 | 0.008452503 | 0.047031589 | 1 |
| HTN | t_cell | *TMEM87B* | 5.609700991 | 0.002639784 | 0.021599046 | 1 |
| HTN | t_cell | *LPCAT1* | 5.115978764 | 0.000403591 | 0.008988159 | 1 |
| HTN | t_cell | *MCOLN2* | 1.121133703 | 0.002128064 | 0.019263747 | 1 |
| HTN | t_cell | *DNAAF1* | -4.499398003 | 0.006314335 | 0.038634495 | 1 |
| HTN | t_cell | *ANKH* | 4.19705975 | 0.00582691 | 0.036339548 | 1 |
| HTN | t_cell | *UBASH3B* | 1.47608521 | 0.00660401 | 0.040153703 | 1 |
| HTN | t_cell | *PRKCA* | 4.498882394 | 0.0021276 | 0.019263747 | 1 |
| HTN | t_cell | *ABCA5* | 4.381861691 | 0.000368899 | 0.008900183 | 1 |
| HTN | t_cell | *NEIL2* | 5.731400531 | 1.92E-08 | 5.47E-06 | 9.30E-05 |
| HTN | t_cell | *CCSAP* | 4.261038065 | 0.000669178 | 0.010605073 | 1 |
| HTN | t_cell | *GBP5* | 0.605526182 | 0.003633061 | 0.026231565 | 1 |
| HTN | t_cell | *OXNAD1* | 2.744388379 | 0.000517837 | 0.009873328 | 1 |
| HTN | t_cell | *PROM2* | -5.834346734 | 0.000115109 | 0.004745727 | 0.558507193 |
| HTN | t_cell | *GOLGA7B* | 3.368246421 | 0.00242256 | 0.02069412 | 1 |
| HTN | t_cell | *NUP205* | 5.762857559 | 0.008573906 | 0.047435111 | 1 |
| HTN | t_cell | *RRAGA* | 7.122222962 | 0.006884228 | 0.041339446 | 1 |
| HTN | t_cell | *MMP16* | -1.628802086 | 0.002318201 | 0.020303087 | 1 |
| HTN | t_cell | *ART3* | -2.539112236 | 0.002020182 | 0.018773924 | 1 |
| HTN | t_cell | *NAA11* | -3.92294851 | 0.002331799 | 0.02031219 | 1 |
| HTN | t_cell | *SFXN2* | 3.303352569 | 1.83E-08 | 5.47E-06 | 8.89E-05 |
| HTN | t_cell | *TDRD9* | -8.629548576 | 0.007437206 | 0.043658822 | 1 |
| HTN | t_cell | *RPL30* | 4.590600987 | 0.000604588 | 0.010162236 | 1 |
| HTN | t_cell | *HKDC1* | 1.754955654 | 0.001867689 | 0.017980217 | 1 |
| HTN | t_cell | *MED19* | 4.437525864 | 0.000436954 | 0.00917827 | 1 |
| HTN | t_cell | *ITGAD* | 1.09518743 | 0.001146411 | 0.01390596 | 1 |
| HTN | t_cell | *VPS8* | 5.742672282 | 7.89E-05 | 0.003645321 | 0.382758738 |
| HTN | t_cell | *NTAN1* | 5.592476796 | 0.000852763 | 0.011988677 | 1 |
| HTN | t_cell | *CLEC18A* | -3.318784 | 5.30E-05 | 0.002673106 | 0.257140419 |
| HTN | t_cell | *SLC34A2* | 9.873026739 | 0.00386314 | 0.027403447 | 1 |
| HTN | t_cell | *RHPN1* | 3.136416744 | 0.00092 | 0.012349099 | 1 |
| HTN | t_cell | *LRRC43* | -5.577912934 | 0.000323996 | 0.008451766 | 1 |
| HTN | t_cell | *ELK4* | 6.774922758 | 0.003121955 | 0.023930059 | 1 |
| HTN | t_cell | *SPATA2L* | 6.43723452 | 1.44E-05 | 0.001106229 | 0.069692422 |
| HTN | t_cell | *SLC5A11* | -2.162984924 | 4.45E-06 | 0.000450129 | 0.021606208 |
| HTN | t_cell | *CDC42SE2* | 1.3250849 | 0.006090199 | 0.037547199 | 1 |
| HTN | t_cell | *RAPGEF6* | 2.79532226 | 0.008805397 | 0.047950379 | 1 |
| HTN | t_cell | *ARHGAP27* | 6.772451155 | 0.001083429 | 0.013542169 | 1 |
| HTN | t_cell | *THEM4* | 2.881026118 | 0.006816189 | 0.041083418 | 1 |
| HTN | t_cell | *LRRC36* | -1.381860208 | 0.003143301 | 0.023945159 | 1 |
| HTN | t_cell | *ZFYVE28* | 1.739807725 | 0.002195922 | 0.019442721 | 1 |

| | | | | | | |
|---|---|---|---|---|---|---|
| HTN | t_cell | *LYPD5* | -3.361987387 | 0.000731881 | 0.011097787 | 1 |
| HTN | t_cell | *CCDC107* | 5.026637654 | 0.000269123 | 0.007681101 | 1 |
| HTN | t_cell | *ATAD3B* | 4.433162443 | 0.000632888 | 0.010363951 | 1 |
| HTN | t_cell | *SLC37A1* | 6.590541284 | 0.000319552 | 0.008451766 | 1 |
| HTN | t_cell | *HSF2BP* | -3.600611651 | 0.003615242 | 0.026206993 | 1 |
| HTN | t_cell | *S100B* | -1.104055957 | 0.000163559 | 0.005805807 | 0.793587083 |
| HTN | t_cell | *PRMT2* | 3.690206946 | 0.001953593 | 0.018477261 | 1 |
| HTN | t_cell | *ZNF208* | -3.631759303 | 0.003165836 | 0.024038552 | 1 |
| HTN | t_cell | *FDPS* | 5.254151631 | 0.002974754 | 0.023125811 | 1 |
| HTN | t_cell | *CCR5* | 1.463620406 | 0.000360793 | 0.008766464 | 1 |
| HTN | t_cell | *NBEAL2* | 3.075549352 | 0.000603074 | 0.010162236 | 1 |
| HTN | t_cell | *FCRL3* | 1.230155506 | 0.001222479 | 0.014502371 | 1 |
| HTN | t_cell | *CPSF4* | 4.551496027 | 0.007881857 | 0.045311336 | 1 |
| HTN | t_cell | *RPL8* | 4.244930402 | 0.005463526 | 0.034788754 | 1 |
| HTN | t_cell | *ABCF3* | 5.345935403 | 1.38E-07 | 2.92E-05 | 0.000670545 |
| HTN | t_cell | *BDH1* | 3.541270831 | 0.008660676 | 0.047684853 | 1 |
| HTN | t_cell | *ALDH16A1* | 2.354575589 | 0.00235064 | 0.020403049 | 1 |
| HTN | t_cell | *POLR3K* | 6.027750961 | 0.003187296 | 0.024125993 | 1 |
| HTN | t_cell | *CCDC78* | 4.764675725 | 0.000568517 | 0.010067315 | 1 |
| HTN | t_cell | *PDZK1IP1* | 11.65477438 | 0.005373728 | 0.034412574 | 1 |
| HTN | t_cell | *ZSWIM5* | 4.163357123 | 0.002023653 | 0.018773924 | 1 |
| HTN | t_cell | *SLC25A34* | -2.702863382 | 0.00645804 | 0.039414353 | 1 |
| HTN | t_cell | *LAPTM5* | 5.720935868 | 0.000471816 | 0.00953704 | 1 |
| HTN | t_cell | *HFM1* | -2.32966336 | 0.005735477 | 0.035861512 | 1 |
| HTN | t_cell | *FCRLB* | 6.559192037 | 0.00518756 | 0.033560056 | 1 |
| HTN | t_cell | *MRPL55* | 7.754461508 | 7.89E-10 | 4.89E-07 | 3.83E-06 |
| HTN | t_cell | *NOSTRIN* | -1.998981449 | 0.004778109 | 0.031584994 | 1 |
| HTN | t_cell | *LYSMD1* | -3.168854114 | 1.91E-06 | 0.000231175 | 0.009247012 |
| HTN | t_cell | *RNF149* | 4.505801395 | 0.001760896 | 0.017349326 | 1 |
| HTN | t_cell | *GPR155* | 5.166722351 | 0.008096785 | 0.045627875 | 1 |
| HTN | t_cell | *PBXIP1* | 2.932807006 | 0.000432286 | 0.00917827 | 1 |
| HTN | t_cell | *DCST2* | 4.715379584 | 0.003618855 | 0.026206993 | 1 |
| HTN | t_cell | *LINC01116* | -3.979011039 | 0.000857461 | 0.011988677 | 1 |
| HTN | t_cell | *LMOD1* | -2.699513502 | 0.00085066 | 0.011988677 | 1 |
| HTN | t_cell | *ARPC2* | 4.033866805 | 0.001947536 | 0.018455945 | 1 |
| HTN | t_cell | *GTPBP8* | 7.207861714 | 0.000851418 | 0.011988677 | 1 |
| HTN | t_cell | *TIPARP* | 5.804746409 | 0.006980804 | 0.041867566 | 1 |
| HTN | t_cell | *IL17RC* | -3.043659043 | 0.008725529 | 0.0478111 | 1 |
| HTN | t_cell | *CCDC158* | 8.133242416 | 0.000414558 | 0.009032732 | 1 |
| HTN | t_cell | *DNALI1* | -2.89681377 | 0.000387377 | 0.008960997 | 1 |
| HTN | t_cell | *RFT1* | -2.755710037 | 0.00899706 | 0.048612176 | 1 |
| HTN | t_cell | *SFMBT1* | 4.828098782 | 0.000254958 | 0.007497321 | 1 |
| HTN | t_cell | *FAM208A* | 6.80628852 | 4.96E-05 | 0.00253469 | 0.240795578 |
| HTN | t_cell | *CDC25A* | -13.421788 | 1.08E-09 | 5.83E-07 | 5.25E-06 |
| HTN | t_cell | *ZNF589* | -4.975595052 | 3.64E-06 | 0.000390538 | 0.017684802 |
| HTN | t_cell | *ATRIP* | -1.474057359 | 0.008889381 | 0.048245277 | 1 |
| HTN | t_cell | *RNF123* | 6.559525665 | 5.75E-13 | 9.30E-10 | 2.79E-09 |
| HTN | t_cell | *HMGB2* | 4.975913268 | 0.004667483 | 0.031065331 | 1 |
| HTN | t_cell | *MAP9* | 7.176315154 | 0.008514522 | 0.047214241 | 1 |
| HTN | t_cell | *MOCS2* | 7.312457227 | 0.00060196 | 0.010162236 | 1 |

| | | | | | | |
|---|---|---|---|---|---|---|
| HTN | t_cell | *F2RL2* | 1.85281194 | 0.002253782 | 0.01983569 | 1 |
| HTN | t_cell | *SERINC5* | 3.507684126 | 0.008858043 | 0.048129032 | 1 |
| HTN | t_cell | *ERAP2* | 1.865325558 | 2.84E-05 | 0.001823249 | 0.137975251 |
| HTN | t_cell | *GRIK2* | -1.250126423 | 0.003861853 | 0.027403447 | 1 |
| HTN | t_cell | *STK17A* | 1.399911403 | 0.001413935 | 0.015644861 | 1 |
| HTN | t_cell | *GALNT10* | 4.253702358 | 0.001496704 | 0.01613779 | 1 |
| HTN | t_cell | *SAP30L* | 8.121558945 | 0.000135611 | 0.00526388 | 0.657985059 |
| HTN | t_cell | *RPS14* | 3.582202539 | 0.008166918 | 0.04591907 | 1 |
| HTN | t_cell | *RPP25L* | -4.655714126 | 0.000175358 | 0.006077421 | 0.850838919 |
| HTN | t_cell | *TMEM65* | 5.0963308 | 0.000754729 | 0.011326355 | 1 |
| HTN | t_cell | *AQP3* | 5.4277107 | 0.001587209 | 0.016561583 | 1 |
| HTN | t_cell | *GJB2* | 9.395116977 | 0.001522415 | 0.016342387 | 1 |
| HTN | t_cell | *MICU2* | 5.336041171 | 0.000693907 | 0.010834138 | 1 |
| HTN | t_cell | *OXGR1* | -6.391743711 | 0.00558562 | 0.035278491 | 1 |
| HTN | t_cell | *OTOGL* | -2.688266572 | 0.004281196 | 0.029380991 | 1 |
| HTN | t_cell | *PACSIN3* | -1.752956259 | 0.004271476 | 0.029355807 | 1 |
| HTN | t_cell | *TC2N* | 1.146884238 | 0.002020201 | 0.018773924 | 1 |
| HTN | t_cell | *ARL5B* | -5.565268681 | 0.000277205 | 0.007819763 | 1 |
| HTN | t_cell | *TRIM66* | -1.637090174 | 4.62E-05 | 0.002432491 | 0.224024276 |
| HTN | t_cell | *RPL27A* | 5.439454139 | 0.002162909 | 0.019346851 | 1 |
| HTN | t_cell | *PRKCB* | 1.25518646 | 0.001074622 | 0.013473042 | 1 |
| HTN | t_cell | *NDST2* | 38.70548143 | 0.000736498 | 0.011097787 | 1 |
| HTN | t_cell | *RIMKLB* | -4.46330136 | 2.12E-06 | 0.000250592 | 0.010274273 |
| HTN | t_cell | *SLFN5* | 3.355792502 | 0.000634397 | 0.010363951 | 1 |
| HTN | t_cell | *STX3* | 3.317209058 | 0.003085981 | 0.023691739 | 1 |
| HTN | t_cell | *PIP4K2C* | 5.686733227 | 0.001131684 | 0.013865559 | 1 |
| HTN | t_cell | *SMAD3* | 3.586202814 | 0.002815875 | 0.022361086 | 1 |
| HTN | t_cell | *AKTIP* | 2.190104957 | 0.003252448 | 0.024428605 | 1 |
| HTN | t_cell | *CNPY4* | 6.641896689 | 1.20E-05 | 0.000952301 | 0.058090348 |
| HTN | t_cell | *MEI1* | 1.630742468 | 0.001582136 | 0.016552277 | 1 |
| HTN | t_cell | *RAB8A* | 3.706612701 | 0.001398616 | 0.015560606 | 1 |
| HTN | t_cell | *RPL13* | 3.868334145 | 0.003203634 | 0.024211886 | 1 |
| HTN | t_cell | *TP53I13* | 5.228221964 | 0.003147968 | 0.023945159 | 1 |
| HTN | t_cell | *SRR* | 1.985334268 | 8.80E-05 | 0.003992533 | 0.427201047 |
| HTN | t_cell | *ATP5H* | 6.962254558 | 0.002421002 | 0.02069412 | 1 |
| HTN | t_cell | *ATG16L2* | 6.162509849 | 0.001085718 | 0.013542169 | 1 |
| HTN | t_cell | *RPSA* | 3.28534397 | 0.000589222 | 0.010162236 | 1 |
| HTN | t_cell | *PDHB* | 7.931816606 | 0.000211926 | 0.006901095 | 1 |
| HTN | t_cell | *DTYMK* | 7.468723933 | 0.000340546 | 0.008644045 | 1 |
| HTN | t_cell | *MLKL* | 2.147672412 | 0.007003273 | 0.041950468 | 1 |
| HTN | t_cell | *SERINC2* | -7.708849317 | 9.50E-05 | 0.004151182 | 0.460999285 |
| HTN | t_cell | *TCTN2* | -0.993840327 | 0.008018878 | 0.045452802 | 1 |
| HTN | t_cell | *TMEM129* | 4.76902017 | 0.005367514 | 0.034412574 | 1 |
| HTN | t_cell | *MAP2K1* | 5.130117394 | 0.007418198 | 0.043658822 | 1 |
| HTN | t_cell | *UPF3A* | 3.065681429 | 0.006725887 | 0.04069078 | 1 |
| HTN | t_cell | *C8orf46* | 0.871153178 | 0.002436955 | 0.020713662 | 1 |
| HTN | t_cell | *CPT1C* | 4.031854696 | 0.000219569 | 0.007008867 | 1 |
| HTN | t_cell | *XPO6* | 3.772206553 | 0.003385078 | 0.025113762 | 1 |
| HTN | t_cell | *HINT1* | 5.426848853 | 0.004310632 | 0.029541221 | 1 |
| HTN | t_cell | *CLIC3* | 10.62249543 | 0.000551591 | 0.009875722 | 1 |

| | | | | | | |
|---|---|---|---|---|---|---|
| HTN | t_cell | *INO80E* | -4.615200671 | 3.12E-08 | 8.40E-06 | 0.000151246 |
| HTN | t_cell | *RGPD8* | -1.012345938 | 0.000219273 | 0.007008867 | 1 |
| HTN | t_cell | *LUZP1* | 5.097068544 | 0.00156027 | 0.016493316 | 1 |
| HTN | t_cell | *CSGALNACT2* | 4.516102552 | 0.000543009 | 0.009873328 | 1 |
| HTN | t_cell | *MUC3A* | -2.411669645 | 0.001703588 | 0.017171831 | 1 |
| HTN | t_cell | *STX8* | 4.349921604 | 0.00472983 | 0.031351275 | 1 |
| HTN | t_cell | *DNAJC18* | -3.844717851 | 4.70E-05 | 0.002432491 | 0.228055715 |
| HTN | t_cell | *HSD17B13* | -3.059410059 | 0.0091854 | 0.04919157 | 1 |
| HTN | t_cell | *ELOVL6* | 4.604524681 | 0.000527535 | 0.009873328 | 1 |
| HTN | t_cell | *SERPINB9* | 4.303371741 | 0.000923411 | 0.012349099 | 1 |
| HTN | t_cell | *GTSF1* | 1.385329673 | 0.008449595 | 0.047031589 | 1 |
| HTN | t_cell | *TRABD* | 4.973490856 | 0.000968574 | 0.012573256 | 1 |
| HTN | t_cell | *TTLL6* | -13.32057949 | 1.13E-10 | 9.14E-08 | 5.48E-07 |
| HTN | t_cell | *NDUFA3* | 6.961373243 | 0.002317662 | 0.020303087 | 1 |
| HTN | t_cell | *TSNARE1* | 4.254523132 | 0.002155615 | 0.019346851 | 1 |
| HTN | t_cell | *MFN1* | 4.047126275 | 0.005931722 | 0.036756978 | 1 |
| HTN | t_cell | *MRPL36* | 2.373663826 | 0.000387802 | 0.008960997 | 1 |
| HTN | t_cell | *PPID* | 7.979701112 | 0.000663162 | 0.01058442 | 1 |
| HTN | t_cell | *NMUR1* | 9.519736089 | 0.000243315 | 0.007332694 | 1 |
| HTN | t_cell | *PIK3CD* | 2.133467917 | 0.000542283 | 0.009873328 | 1 |
| HTN | t_cell | *RGS19* | -2.003739393 | 2.86E-05 | 0.001823249 | 0.138566887 |
| HTN | t_cell | *SERF1A* | -10.68456658 | 0.007594603 | 0.044342975 | 1 |
| HTN | t_cell | *NME6* | 9.250498613 | 1.16E-11 | 1.40E-08 | 5.60E-08 |
| HTN | t_cell | *CD8B* | 0.729793349 | 0.005798238 | 0.036207271 | 1 |
| HTN | t_cell | *MALT1* | 5.433872556 | 0.002755919 | 0.022130428 | 1 |
| HTN | t_cell | *MBOAT1* | 5.032307072 | 0.005049123 | 0.032972202 | 1 |
| HTN | t_cell | *CXCR6* | 0.703175259 | 0.00300273 | 0.023199435 | 1 |
| HTN | t_cell | *TP53RK* | 7.277374194 | 0.000159658 | 0.005738216 | 0.77465921 |
| HTN | t_cell | *SUCLG2* | 4.493074054 | 0.003762494 | 0.027053555 | 1 |
| HTN | t_cell | *FGGY* | 5.113799192 | 0.002053326 | 0.018960903 | 1 |
| HTN | t_cell | *FIBP* | 5.998361503 | 0.000208747 | 0.006843529 | 1 |
| HTN | t_cell | *CTSW* | -0.51223559 | 0.003148601 | 0.023945159 | 1 |
| HTN | t_cell | *ZNF738* | 2.762108463 | 0.000163931 | 0.005805807 | 0.795395542 |
| HTN | t_cell | *MYEOV* | -7.83859835 | 0.000103464 | 0.004442553 | 0.50200854 |
| HTN | t_cell | *ANKRD13D* | 4.501248521 | 0.004080606 | 0.028528962 | 1 |
| HTN | t_cell | *LCLAT1* | 3.888556146 | 0.00236555 | 0.020435969 | 1 |
| HTN | t_cell | *PARP15* | 1.029995804 | 0.000423866 | 0.009140439 | 1 |
| HTN | t_cell | *AHSA2* | 3.369001702 | 0.001987948 | 0.018675376 | 1 |
| HTN | t_cell | *CCDC36* | -2.027095345 | 0.004832522 | 0.031857874 | 1 |
| HTN | t_cell | *MST1* | -1.9819359 | 4.86E-06 | 0.000481276 | 0.023582545 |
| HTN | t_cell | *SULT1B1* | -3.753663803 | 0.005390432 | 0.034458994 | 1 |
| HTN | t_cell | *CD7* | 1.374428246 | 0.002176833 | 0.019346851 | 1 |
| HTN | t_cell | *RNF213* | 3.409640668 | 0.001670082 | 0.017059213 | 1 |
| HTN | t_cell | *SNX31* | -2.423036218 | 4.52E-07 | 8.12E-05 | 0.002192116 |
| HTN | t_cell | *SNAPC5* | 6.108765587 | 0.00040569 | 0.008988159 | 1 |
| HTN | t_cell | *ANKRD36C* | 5.274889607 | 0.003774224 | 0.027058063 | 1 |
| HTN | t_cell | *MSL2* | 5.516191352 | 0.003897025 | 0.027594479 | 1 |
| HTN | t_cell | *SEZ6L2* | 4.15348462 | 1.90E-05 | 0.001346963 | 0.092032663 |
| HTN | t_cell | *KCTD13* | 4.541494752 | 3.62E-05 | 0.002066288 | 0.175634515 |
| HTN | t_cell | *DHX36* | 4.671934822 | 0.007039857 | 0.042117614 | 1 |

| | | | | | | |
|---|---|---|---|---|---|---|
| HTN | t_cell | *PACS1* | 2.806408387 | 0.001358762 | 0.015388209 | 1 |
| HTN | t_cell | *CADM2* | -1.705224847 | 0.002850965 | 0.022565878 | 1 |
| HTN | t_cell | *ARHGAP1* | 3.986726987 | 0.004517633 | 0.030518825 | 1 |
| HTN | t_cell | *CCNE2* | -5.895287465 | 1.51E-08 | 4.89E-06 | 7.33E-05 |
| HTN | t_cell | *TMEM9B* | 5.761515698 | 0.004524124 | 0.030518825 | 1 |
| HTN | t_cell | *CTDNEP1* | 6.281974541 | 4.31E-05 | 0.002309755 | 0.209000705 |
| HTN | t_cell | *A2M* | 3.019256637 | 0.000458618 | 0.009475372 | 1 |
| HTN | t_cell | *ORAI3* | 6.02833081 | 6.48E-07 | 9.53E-05 | 0.003144372 |
| HTN | t_cell | *JAKMIP2* | 0.537187426 | 0.005971074 | 0.036906562 | 1 |
| HTN | t_cell | *CRLF3* | 5.348112423 | 1.47E-05 | 0.001112753 | 0.071216192 |
| HTN | t_cell | *C8G* | -29.29077838 | 0.008966436 | 0.048570505 | 1 |
| HTN | t_cell | *MTHFR* | -5.418617494 | 0.002327788 | 0.02031219 | 1 |
| HTN | t_cell | *NIM1K* | 4.432887794 | 0.001445809 | 0.015871193 | 1 |
| HTN | t_cell | *SLC25A22* | -4.059948116 | 0.000837951 | 0.011988677 | 1 |
| HTN | t_cell | *IL17RA* | 3.516220501 | 0.007865659 | 0.045271857 | 1 |
| HTN | t_cell | *MBOAT4* | 1.990572237 | 0.003778758 | 0.027058063 | 1 |
| HTN | t_cell | *SLC35G5* | 20.05601153 | 0.000992014 | 0.012699875 | 1 |
| HTN | t_cell | *FAM87B* | 6.435061772 | 0.002498111 | 0.021006646 | 1 |
| HTN | t_cell | *RPS27* | 5.541469758 | 0.000729163 | 0.011097787 | 1 |
| HTN | t_cell | *ASB8* | 6.931221731 | 0.001475981 | 0.016079706 | 1 |
| HTN | t_cell | *WDR6* | 2.432612615 | 0.002493919 | 0.021006646 | 1 |
| HTN | t_cell | *GEN1* | -1.791092444 | 0.001946074 | 0.018455945 | 1 |
| HTN | t_cell | *FAM220A* | 6.811091708 | 0.002774592 | 0.022208839 | 1 |
| HTN | t_cell | *ERN1* | 2.638282965 | 0.000873964 | 0.012012667 | 1 |
| HTN | t_cell | *NSUN3* | 5.328129207 | 0.005518421 | 0.035000496 | 1 |
| HTN | t_cell | *RPP25* | 2.254898124 | 0.000483429 | 0.009584338 | 1 |
| HTN | t_cell | *MPI* | -8.876048284 | 3.79E-13 | 9.19E-10 | 1.84E-09 |
| HTN | t_cell | *TRIM73* | -7.737858004 | 0.000230392 | 0.007202575 | 1 |
| HTN | t_cell | *HYI* | -3.3950747 | 1.22E-06 | 0.000156235 | 0.005936928 |
| HTN | t_cell | *C17orf62* | 7.120236932 | 0.000221086 | 0.007011157 | 1 |
| HTN | t_cell | *GAK* | 6.266422873 | 0.000959572 | 0.012573256 | 1 |
| HTN | t_cell | *CTC1* | 6.021607042 | 4.81E-07 | 8.34E-05 | 0.002335616 |
| HTN | t_cell | *HTR1F* | -9.190564315 | 0.000535757 | 0.009873328 | 1 |
| HTN | t_cell | *CDH4* | -2.395647682 | 0.003780991 | 0.027058063 | 1 |
| HTN | t_cell | *GEMIN4* | -2.943472592 | 0.008640169 | 0.047684853 | 1 |
| HTN | t_cell | *ZADH2* | 5.556753747 | 0.007488165 | 0.043826994 | 1 |
| HTN | t_cell | *SEPT1* | 2.681270228 | 0.001201896 | 0.014363546 | 1 |
| HTN | t_cell | *ZDHHC20* | 4.973902356 | 0.000814692 | 0.011870534 | 1 |
| HTN | t_cell | *ZNF443* | -4.672367752 | 0.005591616 | 0.035278491 | 1 |
| HTN | t_cell | *F2R* | 1.415508603 | 0.000546435 | 0.009873328 | 1 |
| HTN | t_cell | *DDX60L* | 5.565900502 | 0.00524016 | 0.033855201 | 1 |
| HTN | t_cell | *RFX7* | 4.658473717 | 0.004110277 | 0.028595237 | 1 |
| HTN | t_cell | *CLDN7* | -4.89965572 | 5.13E-07 | 8.42E-05 | 0.002489819 |
| HTN | t_cell | *ADO* | 6.517368146 | 8.06E-10 | 4.89E-07 | 3.91E-06 |
| HTN | t_cell | *IDH2* | 3.930624425 | 0.002469492 | 0.020874517 | 1 |
| HTN | t_cell | *MRPL41* | 7.895966715 | 0.005598606 | 0.035278491 | 1 |
| HTN | t_cell | *KBTBD3* | 7.534389976 | 0.009238965 | 0.049423877 | 1 |
| HTN | t_cell | *CLN8* | 2.853729922 | 2.47E-05 | 0.001644547 | 0.120051946 |
| HTN | t_cell | *CAPN12* | 2.372881018 | 0.000348701 | 0.00873041 | 1 |
| HTN | t_cell | *KPNA2* | 6.327270159 | 0.000134596 | 0.00526388 | 0.653059814 |

| | | | | | | |
|---|---|---|---|---|---|---|
| HTN | t_cell | *ORAI1* | 3.57149868 | 0.000146373 | 0.005421382 | 0.710200987 |
| HTN | t_cell | *FES* | -8.554810356 | 2.26E-13 | 9.19E-10 | 1.10E-09 |
| HTN | t_cell | *GLRX5* | 7.840720124 | 0.008493917 | 0.047203746 | 1 |
| HTN | t_cell | *BRICD5* | 11.23778195 | 0.000477638 | 0.00953704 | 1 |
| HTN | t_cell | *ZNF721* | 4.948598597 | 0.001548012 | 0.016399464 | 1 |
| HTN | t_cell | *ODF3L1* | -7.907392567 | 0.000200667 | 0.006668727 | 0.973634137 |
| HTN | t_cell | *ZNF320* | 1.503362612 | 0.00019136 | 0.006538585 | 0.928479123 |
| HTN | t_cell | *RABIF* | 4.929725477 | 0.000500792 | 0.009758406 | 1 |
| HTN | t_cell | *FANCF* | 10.40036793 | 0.000168871 | 0.005937397 | 0.819360761 |
| HTN | t_cell | *GPR132* | 1.898173266 | 0.00567368 | 0.035612804 | 1 |
| HTN | t_cell | *FAM19A1* | 1.77955811 | 0.002035171 | 0.018844753 | 1 |
| HTN | t_cell | *UPP1* | 4.251259661 | 0.003408043 | 0.02521711 | 1 |
| HTN | t_cell | *FIGLA* | -7.677695244 | 0.001805212 | 0.017659053 | 1 |
| HTN | t_cell | *KREMEN1* | -1.764216742 | 0.007289577 | 0.043291344 | 1 |
| HTN | t_cell | *EMILIN3* | -4.529952601 | 0.002671209 | 0.021733653 | 1 |
| HTN | t_cell | *BTBD9* | 3.691619008 | 0.006291817 | 0.038545321 | 1 |
| HTN | t_cell | *GOLGA6L4* | -2.747814397 | 0.002866552 | 0.022615461 | 1 |
| HTN | t_cell | *ACOT1* | 1.981159943 | 0.004402098 | 0.030057389 | 1 |
| HTN | t_cell | *ALDH1A3* | -2.003936237 | 0.000360715 | 0.008766464 | 1 |
| HTN | t_cell | *A3GALT2* | -1.996602603 | 0.001134507 | 0.013865559 | 1 |
| HTN | t_cell | *NDUFA12* | 4.88679918 | 0.008605995 | 0.047558415 | 1 |
| HTN | t_cell | *TCTE3* | 6.784373762 | 0.001304265 | 0.015139453 | 1 |
| HTN | t_cell | *NUTM2A* | -3.253544374 | 0.000370696 | 0.008900183 | 1 |
| HTN | t_cell | *NOC4L* | 3.898287889 | 0.003916014 | 0.027657204 | 1 |
| HTN | t_cell | *TMEM106A* | 2.202418071 | 0.006718059 | 0.04069078 | 1 |
| HTN | t_cell | *SIGIRR* | 2.549555317 | 0.000601196 | 0.010162236 | 1 |
| HTN | t_cell | *CCDC137* | 2.770013517 | 0.00837086 | 0.046815552 | 1 |
| HTN | t_cell | *CDK10* | -1.951660456 | 0.001336439 | 0.0152934 | 1 |
| HTN | t_cell | *SP140L* | 4.387914536 | 0.001384214 | 0.015560606 | 1 |
| HTN | t_cell | *MYBL1* | 0.495223115 | 0.005463172 | 0.034788754 | 1 |
| HTN | t_cell | *ADARB2* | 3.81276838 | 0.005163328 | 0.033447885 | 1 |
| HTN | t_cell | *C11orf87* | 6.463011292 | 0.00481182 | 0.031764557 | 1 |
| HTN | t_cell | *SLC52A2* | 6.214308337 | 0.000773157 | 0.011437062 | 1 |
| HTN | t_cell | *EVI2B* | 1.136765691 | 0.002620378 | 0.021512816 | 1 |
| HTN | t_cell | *IFITM1* | -3.953061595 | 0.00088997 | 0.012163754 | 1 |
| HTN | t_cell | *CCDC30* | 2.741559244 | 0.003491527 | 0.025673285 | 1 |
| HTN | t_cell | *GNG2* | 2.640038555 | 0.000549429 | 0.009873437 | 1 |
| HTN | t_cell | *FOXI2* | -2.973234474 | 0.005009819 | 0.032803838 | 1 |
| HTN | t_cell | *TRABD2A* | 1.437676193 | 0.000360956 | 0.008766464 | 1 |
| HTN | t_cell | *TNFRSF18* | 1.13353634 | 0.003808228 | 0.027172825 | 1 |
| HTN | t_cell | *RTN4RL2* | -5.59812645 | 0.001423072 | 0.015692598 | 1 |
| HTN | t_cell | *RHD* | 2.851556244 | 0.00743486 | 0.043658822 | 1 |
| HTN | t_cell | *TRPV2* | 2.052658898 | 0.001363753 | 0.015388209 | 1 |
| HTN | t_cell | *KBTBD12* | -2.104058744 | 0.008652022 | 0.047684853 | 1 |
| HTN | t_cell | *MAPK12* | -2.850958775 | 0.003437663 | 0.025348849 | 1 |
| HTN | t_cell | *ZNF793* | -1.941814891 | 0.001362807 | 0.015388209 | 1 |
| HTN | t_cell | *ZNF559* | -1.517667522 | 0.000982327 | 0.012642577 | 1 |
| HTN | t_cell | *H3F3C* | -61.31072013 | 0.001984023 | 0.018675376 | 1 |
| HTN | t_cell | *PDCD1* | 1.048407103 | 0.004080249 | 0.028528962 | 1 |
| HTN | t_cell | *SELL* | 1.070695035 | 0.006496857 | 0.039601445 | 1 |

| | | | | | | |
|---|---|---|---|---|---|---|
| HTN | t_cell | *C22orf34* | 3.70524176 | 0.000193319 | 0.006559323 | 0.937983199 |
| HTN | t_cell | *NDOR1* | 8.892898388 | 0.002534559 | 0.021228213 | 1 |
| HTN | t_cell | *RHCE* | -2.664334415 | 0.008786136 | 0.047950379 | 1 |
| HTN | t_cell | *FAM26F* | -2.238048232 | 0.000420556 | 0.009109535 | 1 |
| HTN | t_cell | *ENTPD8* | -141.3014299 | 0.007979898 | 0.045444207 | 1 |
| HTN | t_cell | *RPL14* | 5.200854565 | 0.003137394 | 0.023945159 | 1 |
| HTN | t_cell | *FBF1* | -5.08203971 | 1.24E-05 | 0.000971582 | 0.060238053 |
| HTN | t_cell | *LITAF* | 5.59685083 | 0.000973972 | 0.012573256 | 1 |
| HTN | t_cell | *ZNF600* | 2.344584913 | 0.001330466 | 0.015291977 | 1 |
| HTN | t_cell | *COLCA1* | -3.753166377 | 1.83E-05 | 0.001346963 | 0.088999647 |
| HTN | t_cell | *MPHOSPH8* | 4.357633443 | 0.002178937 | 0.019346851 | 1 |
| HTN | t_cell | *SUPT3H* | 4.571180168 | 0.001194836 | 0.014349858 | 1 |
| HTN | t_cell | *EVL* | 2.379508586 | 0.000135108 | 0.00526388 | 0.655543767 |
| HTN | t_cell | *TPK1* | 3.968198275 | 0.008704043 | 0.047773775 | 1 |
| HTN | t_cell | *ZFP62* | 8.071366189 | 0.000121606 | 0.004958257 | 0.590032558 |
| HTN | t_cell | *TOMM7* | 5.484869091 | 0.001575711 | 0.016552277 | 1 |
| HTN | t_cell | *SLC39A10* | 4.397213007 | 0.000761636 | 0.011326355 | 1 |
| HTN | t_cell | *HIST1H3E* | 3.40376263 | 3.37E-08 | 8.60E-06 | 0.000163366 |
| HTN | t_cell | *WDR5B* | 4.460282734 | 0.008679084 | 0.047690731 | 1 |
| HTN | t_cell | *ZNF138* | 4.505579365 | 0.00383849 | 0.027348539 | 1 |
| HTN | t_cell | *ZNF100* | -3.307932162 | 0.002983579 | 0.023125811 | 1 |
| HTN | t_cell | *ANXA6* | 4.006292938 | 0.000545196 | 0.009873328 | 1 |
| HTN | t_cell | *GAL3ST4* | 4.103267673 | 0.000695041 | 0.010834138 | 1 |
| HTN | t_cell | *ZNF772* | -1.587337352 | 0.008969327 | 0.048570505 | 1 |
| HTN | t_cell | *SND1* | 5.105618251 | 0.000349072 | 0.00873041 | 1 |
| HTN | t_cell | *SULT1A2* | 3.620077817 | 0.003294083 | 0.024626948 | 1 |
| HTN | t_cell | *ENTPD4* | 4.945005647 | 0.001568677 | 0.016546136 | 1 |
| HTN | t_cell | *C6orf141* | -0.895209933 | 0.002003861 | 0.018733594 | 1 |
| HTN | t_cell | *CCL4L2* | 6.156771823 | 0.000149325 | 0.005471682 | 0.724523407 |
| HTN | t_cell | *MAP3K5* | 3.98975275 | 0.001390019 | 0.015560606 | 1 |
| HTN | t_cell | *PCDHB11* | -1.675034373 | 0.000602806 | 0.010162236 | 1 |
| HTN | t_cell | *ATG7* | 5.030460697 | 0.00025191 | 0.00745286 | 1 |
| HTN | t_cell | *MFAP5* | -2.983916324 | 0.0003126 | 0.008435989 | 1 |
| HTN | t_cell | *RPS26* | -2.413213431 | 4.33E-05 | 0.002309755 | 0.210187711 |
| HTN | t_cell | *EME2* | 10.36192274 | 0.001488003 | 0.016079706 | 1 |
| HTN | t_cell | *ATAD3A* | 4.79313165 | 0.006270671 | 0.038464341 | 1 |
| HTN | t_cell | *VPS13A* | 4.397071129 | 0.002873164 | 0.022630828 | 1 |
| HTN | t_cell | *NOL8* | 7.052026848 | 0.000390995 | 0.008960997 | 1 |
| HTN | t_cell | *AGAP9* | -5.701263293 | 0.000511057 | 0.009873328 | 1 |
| HTN | t_cell | *SFI1* | 2.138728646 | 0.004499434 | 0.030490578 | 1 |
| HTN | t_cell | *MAN1A2* | 4.770460267 | 0.001121165 | 0.013806839 | 1 |
| HTN | t_cell | *HSD17B11* | 6.622651323 | 0.001014526 | 0.012919892 | 1 |
| HTN | t_cell | *STYX* | 5.27357748 | 0.001966286 | 0.018561124 | 1 |
| HTN | t_cell | *TMEM116* | -2.322108728 | 5.68E-07 | 8.89E-05 | 0.002756306 |
| HTN | t_cell | *HLA-DRB5* | -0.673476052 | 0.000274116 | 0.007777831 | 1 |
| HTN | t_cell | *RPL10A* | 4.931154207 | 0.001449993 | 0.015881185 | 1 |
| HTN | t_cell | *COLGALT2* | 2.711739839 | 0.000442946 | 0.009263679 | 1 |
| HTN | t_cell | *RCSD1* | 1.604822606 | 0.000967623 | 0.012573256 | 1 |
| HTN | t_cell | *PRMT6* | -4.274072287 | 0.005087345 | 0.033132618 | 1 |
| HTN | t_cell | *SHISA4* | -7.960614018 | 7.47E-06 | 0.000658863 | 0.03623745 |

| | | | | | | |
|---|---|---|---|---|---|---|
| HTN | t_cell | *CCDC167* | 4.072346606 | 0.001750079 | 0.017349326 | 1 |
| HTN | t_cell | *L3MBTL3* | 2.585127874 | 0.008010708 | 0.045452802 | 1 |
| HTN | t_cell | *TGM2* | 2.460206103 | 0.000870601 | 0.012012667 | 1 |
| HTN | t_cell | *AL590822.1* | -4.015325955 | 0.000696474 | 0.010834138 | 1 |
| HTN | t_cell | *ECT2L* | 8.991639746 | 0.007663528 | 0.044537374 | 1 |
| HTN | t_cell | *GPR52* | 8.518869018 | 0.007979628 | 0.045444207 | 1 |
| HTN | t_cell | *LIPN* | 23.23736349 | 0.000358674 | 0.008766464 | 1 |
| HTN | t_cell | *ASAH2B* | 6.825012653 | 0.000872121 | 0.012012667 | 1 |
| HTN | t_cell | *HSD17B8* | -3.575138497 | 2.15E-05 | 0.001466791 | 0.104142188 |
| HTN | t_cell | *PRRT1* | 1.525917897 | 0.003050236 | 0.023529009 | 1 |
| HTN | t_cell | *LAYN* | -3.897644258 | 0.002778394 | 0.022208839 | 1 |
| HTN | t_cell | *C6orf48* | 3.77096738 | 0.001935904 | 0.018417656 | 1 |
| HTN | t_cell | *HLA-C* | -1.681680244 | 0.001056571 | 0.013315543 | 1 |
| HTN | t_cell | *POU5F1* | -0.768897053 | 0.00043697 | 0.00917827 | 1 |
| HTN | t_cell | *PSORS1C2* | 2.029162119 | 0.007664614 | 0.044537374 | 1 |
| HTN | t_cell | *FAM196B* | 2.346301116 | 0.00079045 | 0.011576575 | 1 |
| HTN | t_cell | *SLFN12L* | 0.59724807 | 0.007409911 | 0.043658822 | 1 |
| HTN | t_cell | *TMEM170B* | 1.740025702 | 0.008306711 | 0.046594404 | 1 |
| HTN | t_cell | *GOLGA6L10* | -4.078673825 | 0.002995506 | 0.023180537 | 1 |
| HTN | t_cell | *NAP1L4* | 4.522758744 | 0.000762064 | 0.011326355 | 1 |
| HTN | t_cell | *SMN2* | -1.748183094 | 0.004104892 | 0.028595237 | 1 |
| HTN | t_cell | *KLRC2* | 0.774457814 | 0.002543075 | 0.021228213 | 1 |
| HTN | t_cell | *C1QTNF9B* | 67.85054117 | 0.002019011 | 0.018773924 | 1 |
| HTN | t_cell | *HLA-A* | 2.450547867 | 0.007389079 | 0.043658822 | 1 |
| HTN | t_cell | *SACM1L* | 4.983604999 | 0.000724055 | 0.011082381 | 1 |
| HTN | t_cell | *TAS2R14* | 1.845457939 | 0.005270433 | 0.034005508 | 1 |
| HTN | t_cell | *FGFR1OP* | 6.314490089 | 0.002104453 | 0.019263747 | 1 |
| HTN | t_cell | *SUPT4H1* | 13.89151692 | 5.66E-08 | 1.33E-05 | 0.000274753 |
| HTN | t_cell | *MXD3* | -3.314715659 | 0.000262154 | 0.007616594 | 1 |
| HTN | t_cell | *TMEM110* | 6.102649587 | 7.75E-06 | 0.000671697 | 0.037615046 |
| HTN | t_cell | *LBH* | 1.35667544 | 0.004098424 | 0.028595237 | 1 |
| HTN | t_cell | *ATF6B* | 7.712633787 | 6.57E-09 | 2.45E-06 | 3.19E-05 |
| HTN | t_cell | *PPM1N* | 3.095361033 | 3.95E-05 | 0.002190959 | 0.19150935 |
| HTN | t_cell | *GALT* | 7.863271691 | 4.71E-05 | 0.002432491 | 0.228654171 |
| HTN | t_cell | *ANG* | 7.225647978 | 0.000599312 | 0.010162236 | 1 |
| HTN | t_cell | *ARHGEF33* | -1.635374707 | 0.007564599 | 0.044221007 | 1 |
| HTN | t_cell | *AC005082.1* | -7.102003769 | 0.000863459 | 0.012004301 | 1 |
| HTN | t_cell | *UBXN2B* | 5.053371719 | 7.40E-05 | 0.003498558 | 0.3589389 |
| HTN | t_cell | *GOLGA8B* | 2.735375882 | 0.000126886 | 0.005046304 | 0.615649082 |
| HTN | t_cell | *VPS16* | 8.182011262 | 0.004588095 | 0.030747839 | 1 |
| HTN | t_cell | *TMEM242* | 4.590590426 | 0.007704858 | 0.044694225 | 1 |
| HTN | t_cell | *TSTD1* | 3.49782721 | 0.003549123 | 0.025934257 | 1 |
| HTN | t_cell | *PAM16* | -2.331280931 | 0.009295679 | 0.049617858 | 1 |
| HTN | t_cell | *AC016773.1* | 4.107350416 | 9.35E-06 | 0.000781847 | 0.045347124 |
| HTN | t_cell | *AP4M1* | -3.055129361 | 0.000323556 | 0.008451766 | 1 |
| HTN | t_cell | *UBA52* | 3.735800652 | 0.000565779 | 0.010055538 | 1 |
| HTN | t_cell | *TIAF1* | 6.028171694 | 1.01E-05 | 0.000833906 | 0.049200434 |
| HTN | t_cell | *ADRA2B* | -5.462970881 | 0.005546079 | 0.035084189 | 1 |
| HTN | t_cell | *MIAT* | 1.046370826 | 0.004140062 | 0.028737598 | 1 |
| HTN | t_cell | *HAR1A* | 6.839855855 | 0.007440487 | 0.043658822 | 1 |

| | | | | | | |
|---|---|---|---|---|---|---|
| HTN | t_cell | *AC108463.2* | -4.991161739 | 0.000409715 | 0.00899518 | 1 |
| HTN | t_cell | *LINC00578* | -2.844650942 | 0.002983668 | 0.023125811 | 1 |
| HTN | t_cell | *PATL2* | 2.544211727 | 0.00070461 | 0.010867676 | 1 |
| HTN | t_cell | *RPS18* | 3.971450935 | 0.001333163 | 0.015291977 | 1 |
| HTN | t_cell | *TAPBP* | 4.406400537 | 0.000243024 | 0.007332694 | 1 |
| HTN | t_cell | *TMA7* | 7.711732172 | 3.70E-06 | 0.000390538 | 0.017964747 |
| HTN | t_cell | *AC009495.3* | 1.623718709 | 0.001216717 | 0.014469385 | 1 |
| HTN | t_cell | *LINC00342* | 7.741969681 | 0.000203188 | 0.006706577 | 0.985866773 |
| HTN | t_cell | *GPX1* | 5.585640743 | 3.35E-05 | 0.001960732 | 0.162740746 |
| HTN | t_cell | *KIAA0040* | 3.174421902 | 0.003414137 | 0.02521711 | 1 |
| HTN | t_cell | *AC019117.2* | -12.84780392 | 0.000923892 | 0.012349099 | 1 |
| HTN | t_cell | *LINC00987* | 1.817796838 | 0.001889698 | 0.018084451 | 1 |
| HTN | t_cell | *HLA-DQA2* | 0.942256601 | 0.000293767 | 0.008144892 | 1 |
| HTN | t_cell | *LINC00211* | -9.446406416 | 0.000445923 | 0.009285916 | 1 |
| HTN | t_cell | *TNFSF12* | 7.88895523 | 2.33E-09 | 1.03E-06 | 1.13E-05 |
| HTN | t_cell | *CDRT4* | 5.370305623 | 0.007411251 | 0.043658822 | 1 |
| HTN | t_cell | *C1orf226* | -3.921115669 | 0.00623188 | 0.038274785 | 1 |
| HTN | t_cell | *PSMB9* | 1.381055028 | 0.004338764 | 0.029692078 | 1 |
| HTN | t_cell | *C1QTNF9* | -6.020504286 | 0.00762299 | 0.04440186 | 1 |
| HTN | t_cell | *PNMA2* | -2.883872629 | 0.000662508 | 0.01058442 | 1 |
| HTN | t_cell | *KRBOX1* | -21.06775779 | 0.009313675 | 0.049659285 | 1 |
| HTN | t_cell | *NSUN6* | 5.314812506 | 0.001617971 | 0.016794498 | 1 |
| HTN | t_cell | *TDGF1* | -2.510915771 | 0.002785338 | 0.022223465 | 1 |
| HTN | t_cell | *MRPL20* | 6.81710687 | 0.000142977 | 0.005377706 | 0.693724063 |
| HTN | t_cell | *LINC00960* | -0.870175084 | 0.00379227 | 0.027098811 | 1 |
| HTN | t_cell | *ZNF709* | -53.35695184 | 3.27E-05 | 0.001933659 | 0.158560053 |
| HTN | t_cell | *ERVW-1* | -4.437676639 | 0.008378822 | 0.046815552 | 1 |
| HTN | t_cell | *WFDC6* | -9.420339955 | 0.000138256 | 0.005282043 | 0.670819407 |
| HTN | t_cell | *PLA2G4B* | 5.819452175 | 0.004959521 | 0.0326065 | 1 |
| HTN | t_cell | *LEFTY1* | -24.24607537 | 0.001684856 | 0.017059213 | 1 |
| HTN | t_cell | *APOBEC3D* | 2.74508891 | 0.002724793 | 0.022015383 | 1 |
| HTN | t_cell | *APOBEC3C* | 4.401252376 | 0.001299217 | 0.015139453 | 1 |
| HTN | t_cell | *CDK11B* | 7.043733626 | 0.00287801 | 0.022632257 | 1 |
| HTN | t_cell | *LINC00942* | -9.436806602 | 0.003332137 | 0.024796821 | 1 |
| HTN | t_cell | *ZNF718* | 5.910422897 | 0.000969918 | 0.012573256 | 1 |
| HTN | t_cell | *SNHG9* | 4.165800158 | 0.004601011 | 0.030791866 | 1 |
| HTN | t_cell | *ZBED6* | -4.41991712 | 0.008857825 | 0.048129032 | 1 |
| HTN | t_cell | *LINC00639* | -1.680994823 | 0.005527844 | 0.03501449 | 1 |
| HTN | t_cell | *PWRN1* | -3.482050514 | 0.0007043 | 0.010867676 | 1 |
| HTN | t_cell | *LINC00672* | 4.317974648 | 0.001209827 | 0.014422798 | 1 |
| Obesity | adipocyte | *SEMA3F* | -1.097127395 | 1.35E-06 | 8.19E-05 | 0.006555416 |
| Obesity | adipocyte | *RBM6* | -0.854998803 | 7.02E-24 | 1.14E-20 | 3.41E-20 |
| Obesity | adipocyte | *ARHGAP33* | 0.48238336 | 0.002157539 | 0.014682162 | 1 |
| Obesity | adipocyte | *CDC27* | 0.705255807 | 0.000829715 | 0.007550452 | 1 |
| Obesity | adipocyte | *POLR2J* | 1.923104895 | 0.002471893 | 0.016273573 | 1 |
| Obesity | adipocyte | *MEOX1* | 1.066142576 | 0.007608129 | 0.036226343 | 1 |
| Obesity | adipocyte | *RHBDD2* | 0.680851559 | 0.004496782 | 0.024844992 | 1 |
| Obesity | adipocyte | *ZNF263* | 1.770147907 | 0.000377177 | 0.004441901 | 1 |
| Obesity | adipocyte | *BAIAP2L1* | 1.669375379 | 0.000133354 | 0.002246638 | 0.647031608 |
| Obesity | adipocyte | *AGK* | 0.725768657 | 0.008018714 | 0.03732363 | 1 |

| | | | | | | |
|---|---|---|---|---|---|---|
| Obesity | adipocyte | *TSR3* | 2.029438159 | 0.004424419 | 0.024590243 | 1 |
| Obesity | adipocyte | *FMO3* | -0.376671159 | 0.002131664 | 0.01454688 | 1 |
| Obesity | adipocyte | *CYB561* | 0.808285823 | 9.38E-05 | 0.001887835 | 0.455224682 |
| Obesity | adipocyte | *SS18L2* | 1.451312452 | 0.001201258 | 0.009633887 | 1 |
| Obesity | adipocyte | *IL32* | 2.259538596 | 1.38E-06 | 8.28E-05 | 0.006709141 |
| Obesity | adipocyte | *PKD1* | 1.760211132 | 0.000125189 | 0.002177124 | 0.607417494 |
| Obesity | adipocyte | *SLC6A13* | -0.350020132 | 0.006751182 | 0.033289367 | 1 |
| Obesity | adipocyte | *CLCN6* | 1.125342886 | 2.75E-05 | 0.000873517 | 0.133648083 |
| Obesity | adipocyte | *DCN* | 0.694300805 | 0.00052498 | 0.005455908 | 1 |
| Obesity | adipocyte | *ZBTB32* | -1.085377771 | 0.010110733 | 0.044396156 | 1 |
| Obesity | adipocyte | *MAP4K5* | 1.101986006 | 0.006330182 | 0.031894125 | 1 |
| Obesity | adipocyte | *EHD3* | 1.188164301 | 0.005815684 | 0.029860001 | 1 |
| Obesity | adipocyte | *MTMR11* | 0.247085258 | 0.006591806 | 0.032735516 | 1 |
| Obesity | adipocyte | *NUDCD3* | 0.598208102 | 0.000997176 | 0.008475282 | 1 |
| Obesity | adipocyte | *CHDH* | 2.643893937 | 0.000232522 | 0.003294385 | 1 |
| Obesity | adipocyte | *CLCA4* | -0.953990535 | 0.0002501 | 0.003413789 | 1 |
| Obesity | adipocyte | *RUFY3* | 0.999222105 | 0.005341913 | 0.028138025 | 1 |
| Obesity | adipocyte | *TTC27* | 1.290278769 | 0.001191852 | 0.009606802 | 1 |
| Obesity | adipocyte | *HGF* | 1.35919887 | 8.67E-05 | 0.001790003 | 0.420650605 |
| Obesity | adipocyte | *OSBPL5* | 1.322653453 | 0.000681549 | 0.006627001 | 1 |
| Obesity | adipocyte | *RNH1* | 0.622575185 | 0.006292975 | 0.031787934 | 1 |
| Obesity | adipocyte | *FAS* | 1.369794662 | 3.08E-05 | 0.000922495 | 0.149311996 |
| Obesity | adipocyte | *AGPAT4* | 1.305629621 | 0.000107882 | 0.002021023 | 0.523444993 |
| Obesity | adipocyte | *SH2D2A* | 2.490170275 | 2.22E-05 | 0.000765564 | 0.107525024 |
| Obesity | adipocyte | *SNX1* | 1.178372876 | 0.001859836 | 0.013189377 | 1 |
| Obesity | adipocyte | *ANK1* | 3.810882995 | 0.000470608 | 0.005071018 | 1 |
| Obesity | adipocyte | *RABEP1* | 1.369874526 | 0.000112974 | 0.00205413 | 0.54815102 |
| Obesity | adipocyte | *CENPQ* | -0.168414603 | 0.010455477 | 0.045511831 | 1 |
| Obesity | adipocyte | *ATP6V0A1* | 1.212840441 | 0.000144714 | 0.00232726 | 0.70215333 |
| Obesity | adipocyte | *PIAS1* | 1.630441797 | 7.93E-08 | 6.87E-06 | 0.000384687 |
| Obesity | adipocyte | *MYOC* | 0.864373774 | 0.001586897 | 0.011900502 | 1 |
| Obesity | adipocyte | *SH3YL1* | -1.007503186 | 0.000990376 | 0.008445179 | 1 |
| Obesity | adipocyte | *ADSS* | 1.638270696 | 0.001382432 | 0.01072231 | 1 |
| Obesity | adipocyte | *NSUN2* | 1.230466021 | 0.001185646 | 0.009592089 | 1 |
| Obesity | adipocyte | *PI4K2B* | 3.679435957 | 7.28E-05 | 0.001597861 | 0.353127232 |
| Obesity | adipocyte | *BOD1L1* | 0.601615984 | 0.009142003 | 0.041182155 | 1 |
| Obesity | adipocyte | *DNAH5* | 1.913279958 | 0.000266731 | 0.003540206 | 1 |
| Obesity | adipocyte | *LSG1* | 0.318909234 | 0.000403158 | 0.004646369 | 1 |
| Obesity | adipocyte | *TDP1* | 1.916305931 | 0.001744507 | 0.012717987 | 1 |
| Obesity | adipocyte | *ARAP2* | 0.822038738 | 0.000867109 | 0.007776737 | 1 |
| Obesity | adipocyte | *DTNBP1* | 1.123234757 | 0.00925312 | 0.04137893 | 1 |
| Obesity | adipocyte | *ANO2* | 1.380271854 | 5.64E-05 | 0.001367377 | 0.273475314 |
| Obesity | adipocyte | *UTS2* | -0.544501045 | 0.005373865 | 0.028138025 | 1 |
| Obesity | adipocyte | *JKAMP* | 0.740648455 | 0.006417553 | 0.032101 | 1 |
| Obesity | adipocyte | *MAPK9* | 1.50646301 | 0.008674575 | 0.03959458 | 1 |
| Obesity | adipocyte | *CYBA* | 1.438701137 | 0.00029981 | 0.003856703 | 1 |
| Obesity | adipocyte | *ANAPC4* | 1.490713988 | 5.42E-06 | 0.000252858 | 0.02629721 |
| Obesity | adipocyte | *LY75* | 2.537852514 | 8.66E-06 | 0.000385342 | 0.042002224 |
| Obesity | adipocyte | *OPN3* | 0.346682539 | 0.005989982 | 0.030528772 | 1 |
| Obesity | adipocyte | *SDCCAG8* | 0.664249626 | 0.000163778 | 0.00256627 | 0.794648913 |

| | | | | | | |
|---|---|---|---|---|---|---|
| Obesity | adipocyte | *TBC1D22A* | 1.008786038 | 0.002723149 | 0.017500291 | 1 |
| Obesity | adipocyte | *ITIH4* | 0.338560131 | 3.93E-11 | 7.34E-09 | 1.91E-07 |
| Obesity | adipocyte | *NPFFR2* | 2.141722616 | 0.000261617 | 0.003528633 | 1 |
| Obesity | adipocyte | *ATP11B* | 0.998959285 | 0.000913476 | 0.008014798 | 1 |
| Obesity | adipocyte | *CROCC* | 0.296521318 | 0.008243718 | 0.038130144 | 1 |
| Obesity | adipocyte | *RIMBP2* | 1.765204143 | 0.000133105 | 0.002246638 | 0.645825466 |
| Obesity | adipocyte | *PRDM6* | 1.980055078 | 3.38E-05 | 0.000993083 | 0.163858679 |
| Obesity | adipocyte | *LTK* | 10.95505828 | 2.77E-05 | 0.0008739 | 0.134580614 |
| Obesity | adipocyte | *TM7SF3* | 1.088177781 | 0.009187089 | 0.041207992 | 1 |
| Obesity | adipocyte | *LPAR2* | 1.40023617 | 8.42E-05 | 0.001745534 | 0.408455058 |
| Obesity | adipocyte | *SLC12A2* | 1.39678788 | 0.007805573 | 0.036723823 | 1 |
| Obesity | adipocyte | *ABCA7* | -0.516114359 | 0.006932596 | 0.034011076 | 1 |
| Obesity | adipocyte | *BTBD1* | 1.569980878 | 0.000145695 | 0.002333037 | 0.706910332 |
| Obesity | adipocyte | *UHRF1BP1* | 1.352476865 | 6.69E-23 | 8.12E-20 | 3.25E-19 |
| Obesity | adipocyte | *ANKRD44* | 0.639326253 | 0.00674579 | 0.033289367 | 1 |
| Obesity | adipocyte | *TRAM1* | 1.036666433 | 0.002355395 | 0.015785051 | 1 |
| Obesity | adipocyte | *EVI5* | 0.672553447 | 0.00682107 | 0.03356575 | 1 |
| Obesity | adipocyte | *TP53BP1* | 1.260728867 | 0.002048176 | 0.014130834 | 1 |
| Obesity | adipocyte | *ROGDI* | 3.298124093 | 0.009959351 | 0.044009811 | 1 |
| Obesity | adipocyte | *SRBD1* | 1.230641666 | 0.00382354 | 0.022164653 | 1 |
| Obesity | adipocyte | *CYFIP1* | 1.257150907 | 0.001706887 | 0.012529222 | 1 |
| Obesity | adipocyte | *LRP6* | 1.190451306 | 0.008383045 | 0.03855406 | 1 |
| Obesity | adipocyte | *WIPI1* | -0.541826307 | 0.004657327 | 0.025447466 | 1 |
| Obesity | adipocyte | *MAP4K4* | 0.93821134 | 0.008596466 | 0.039275005 | 1 |
| Obesity | adipocyte | *WDR1* | 1.382136123 | 0.001021355 | 0.008588589 | 1 |
| Obesity | adipocyte | *SNX13* | 1.140567685 | 0.007251857 | 0.03522123 | 1 |
| Obesity | adipocyte | *RPS6KA2* | 0.695585079 | 0.010114506 | 0.044396156 | 1 |
| Obesity | adipocyte | *CYBRD1* | 0.899323466 | 0.00149963 | 0.011369073 | 1 |
| Obesity | adipocyte | *AFF4* | 1.449268602 | 6.03E-05 | 0.001441243 | 0.292605975 |
| Obesity | adipocyte | *TRHDE* | -0.554965789 | 9.66E-05 | 0.001890159 | 0.46875931 |
| Obesity | adipocyte | *FCGR2B* | 1.302109439 | 0.002962887 | 0.018467765 | 1 |
| Obesity | adipocyte | *NDE1* | 1.275292845 | 0.000448324 | 0.004942243 | 1 |
| Obesity | adipocyte | *PVR* | 0.611450111 | 0.009272777 | 0.041428649 | 1 |
| Obesity | adipocyte | *ALPK1* | 0.796525196 | 0.003814269 | 0.022163873 | 1 |
| Obesity | adipocyte | *SDHA* | 0.596830565 | 0.011211743 | 0.048098478 | 1 |
| Obesity | adipocyte | *SMARCE1* | 0.859247943 | 0.005666545 | 0.029217935 | 1 |
| Obesity | adipocyte | *CD5L* | 0.51396913 | 0.003700971 | 0.021713558 | 1 |
| Obesity | adipocyte | *PPP2R2C* | 2.837635829 | 0.000321972 | 0.004015963 | 1 |
| Obesity | adipocyte | *ZZEF1* | 1.165075652 | 0.001520834 | 0.011511837 | 1 |
| Obesity | adipocyte | *TIPIN* | 0.296667998 | 0.00164008 | 0.012112129 | 1 |
| Obesity | adipocyte | *ADD2* | 1.304894286 | 4.94E-05 | 0.001269024 | 0.23984561 |
| Obesity | adipocyte | *KIFAP3* | 1.170045242 | 0.001452914 | 0.011095722 | 1 |
| Obesity | adipocyte | *FMO4* | 1.043474579 | 0.00121838 | 0.009739015 | 1 |
| Obesity | adipocyte | *SPAG5* | -1.383658239 | 0.00582587 | 0.029880679 | 1 |
| Obesity | adipocyte | *PAG1* | 1.089823544 | 0.001056048 | 0.008770334 | 1 |
| Obesity | adipocyte | *NT5C2* | 0.786885929 | 1.29E-05 | 0.000525917 | 0.062584064 |
| Obesity | adipocyte | *IL4R* | 1.340643037 | 2.07E-05 | 0.000722591 | 0.100440094 |
| Obesity | adipocyte | *DYNC1I2* | 1.248221241 | 0.005489746 | 0.028515578 | 1 |
| Obesity | adipocyte | *AMPH* | 1.089854118 | 0.000246391 | 0.003377088 | 1 |
| Obesity | adipocyte | *XRCC5* | 0.992815817 | 0.010521596 | 0.045703477 | 1 |

| | | | | | | |
|---|---|---|---|---|---|---|
| Obesity | adipocyte | *FDFT1* | 1.124693702 | 1.69E-05 | 0.000641362 | 0.082094336 |
| Obesity | adipocyte | *SCGN* | 9.45691856 | 0.000113189 | 0.00205413 | 0.549194358 |
| Obesity | adipocyte | *PSEN1* | 1.135257224 | 0.002545251 | 0.016621211 | 1 |
| Obesity | adipocyte | *ME2* | 1.616511905 | 0.000932729 | 0.008081427 | 1 |
| Obesity | adipocyte | *DOPEY1* | 0.703184607 | 0.000470085 | 0.005071018 | 1 |
| Obesity | adipocyte | *ZCCHC6* | 1.311816354 | 0.001690707 | 0.012429258 | 1 |
| Obesity | adipocyte | *PIBF1* | 1.009835037 | 0.008364956 | 0.03850737 | 1 |
| Obesity | adipocyte | *ZNF586* | 0.888647972 | 0.003129435 | 0.019166741 | 1 |
| Obesity | adipocyte | *ZNF264* | 1.595847954 | 0.007891291 | 0.036922413 | 1 |
| Obesity | adipocyte | *PPIE* | 0.467543731 | 0.005412092 | 0.028235385 | 1 |
| Obesity | adipocyte | *GSTP1* | 1.326568977 | 2.16E-07 | 1.66E-05 | 0.001046921 |
| Obesity | adipocyte | *NCOA1* | 0.917491934 | 0.010647286 | 0.046084416 | 1 |
| Obesity | adipocyte | *CD59* | 0.476004444 | 0.00707821 | 0.034550779 | 1 |
| Obesity | adipocyte | *MAST2* | 1.270332397 | 1.24E-05 | 0.000516209 | 0.06039647 |
| Obesity | adipocyte | *NME8* | 20.3719139 | 1.12E-06 | 6.89E-05 | 0.005442124 |
| Obesity | adipocyte | *LPCAT2* | 1.351079681 | 6.67E-05 | 0.001534004 | 0.323674869 |
| Obesity | adipocyte | *OGFOD1* | -0.925134397 | 0.001777976 | 0.012799963 | 1 |
| Obesity | adipocyte | *DOCK3* | 1.578465292 | 0.00232301 | 0.015632791 | 1 |
| Obesity | adipocyte | *ARHGAP28* | 0.959932865 | 0.003437961 | 0.020593813 | 1 |
| Obesity | adipocyte | *NSFL1C* | 0.855936599 | 0.001018267 | 0.008577486 | 1 |
| Obesity | adipocyte | *EBF4* | 1.140787599 | 0.002790492 | 0.017791679 | 1 |
| Obesity | adipocyte | *XRN2* | 1.112058695 | 0.004151794 | 0.023441543 | 1 |
| Obesity | adipocyte | *FXYD5* | 1.05660577 | 0.000678322 | 0.006608873 | 1 |
| Obesity | adipocyte | *GRAMD1A* | 1.590825912 | 0.003380817 | 0.020352013 | 1 |
| Obesity | adipocyte | *CDIP1* | 2.140554238 | 0.000902891 | 0.007965144 | 1 |
| Obesity | adipocyte | *ANKRD24* | -0.810285595 | 0.00367402 | 0.021581529 | 1 |
| Obesity | adipocyte | *YPEL3* | 1.345674214 | 2.11E-10 | 3.21E-08 | 1.03E-06 |
| Obesity | adipocyte | *LYZ* | 0.937160532 | 1.60E-05 | 0.000621651 | 0.077706333 |
| Obesity | adipocyte | *PABPC4* | -1.125926297 | 1.94E-07 | 1.57E-05 | 0.000942092 |
| Obesity | adipocyte | *GLG1* | 1.121728197 | 0.005485818 | 0.028515578 | 1 |
| Obesity | adipocyte | *DTX2* | 2.261200776 | 0.001843998 | 0.013099674 | 1 |
| Obesity | adipocyte | *TF* | -0.815489472 | 0.007018213 | 0.034326987 | 1 |
| Obesity | adipocyte | *RGS17* | -1.137765526 | 3.12E-10 | 4.59E-08 | 1.51E-06 |
| Obesity | adipocyte | *PPP2R3C* | 1.053284978 | 3.04E-05 | 0.000922495 | 0.1474629 |
| Obesity | adipocyte | *TBX15* | -0.725794587 | 1.00E-06 | 6.33E-05 | 0.004854333 |
| Obesity | adipocyte | *COMT* | 1.490015406 | 0.001368528 | 0.010675396 | 1 |
| Obesity | adipocyte | *LRRFIP2* | 1.342055525 | 0.00061036 | 0.00605617 | 1 |
| Obesity | adipocyte | *MSH2* | 1.457318437 | 0.011780029 | 0.049874956 | 1 |
| Obesity | adipocyte | *NANS* | 1.735395069 | 0.000817077 | 0.007502305 | 1 |
| Obesity | adipocyte | *ITPR3* | 0.85955107 | 1.36E-08 | 1.47E-06 | 6.62E-05 |
| Obesity | adipocyte | *DSP* | 1.861699644 | 8.22E-05 | 0.001726722 | 0.398872716 |
| Obesity | adipocyte | *IFT74* | 0.868871674 | 0.001808571 | 0.01288574 | 1 |
| Obesity | adipocyte | *SETD1A* | 1.811416794 | 8.07E-08 | 6.87E-06 | 0.000391406 |
| Obesity | adipocyte | *CECR2* | -0.416919302 | 0.004594658 | 0.025190149 | 1 |
| Obesity | adipocyte | *SMARCB1* | 0.421239758 | 0.001626495 | 0.012066902 | 1 |
| Obesity | adipocyte | *BCL2L13* | 0.652818818 | 0.005381716 | 0.028138025 | 1 |
| Obesity | adipocyte | *CABIN1* | 1.172341212 | 0.001391933 | 0.010744369 | 1 |
| Obesity | adipocyte | *CYTH4* | 1.633384792 | 3.68E-05 | 0.001049088 | 0.178344908 |
| Obesity | adipocyte | *SH3BP1* | 2.805698921 | 1.10E-05 | 0.000471541 | 0.05328409 |
| Obesity | adipocyte | *ANKRD54* | -0.594746454 | 0.000141267 | 0.002315631 | 0.685426683 |

| | | | | | | |
|---|---|---|---|---|---|---|
| Obesity | adipocyte | *TTC28* | 0.888670857 | 0.006135441 | 0.031106749 | 1 |
| Obesity | adipocyte | *CENPM* | 0.775424299 | 1.22E-05 | 0.00051024 | 0.059187897 |
| Obesity | adipocyte | *KDELR3* | -0.856118702 | 0.010918542 | 0.047132354 | 1 |
| Obesity | adipocyte | *DESI1* | 1.020066831 | 0.010622914 | 0.04601998 | 1 |
| Obesity | adipocyte | *NIN* | 1.180981758 | 0.002018074 | 0.013988138 | 1 |
| Obesity | adipocyte | *GSKIP* | 2.496616534 | 0.000819047 | 0.007502305 | 1 |
| Obesity | adipocyte | *PSMB5* | 0.485255154 | 0.006966709 | 0.03414391 | 1 |
| Obesity | adipocyte | *ABHD12* | 1.239447032 | 1.56E-06 | 9.02E-05 | 0.007573047 |
| Obesity | adipocyte | *CD40* | 1.685958934 | 0.004683609 | 0.025533564 | 1 |
| Obesity | adipocyte | *NDRG3* | 1.911791818 | 1.92E-05 | 0.000691702 | 0.093351928 |
| Obesity | adipocyte | *PABPC1L* | 1.581315705 | 0.007653261 | 0.036298752 | 1 |
| Obesity | adipocyte | *DOK5* | 1.554382811 | 0.000841226 | 0.007600794 | 1 |
| Obesity | adipocyte | *CSTF1* | 1.95183969 | 0.009995895 | 0.044050936 | 1 |
| Obesity | adipocyte | *NELFCD* | 1.225939688 | 0.008715186 | 0.039705243 | 1 |
| Obesity | adipocyte | *CTSZ* | 1.514876889 | 0.000421153 | 0.00477438 | 1 |
| Obesity | adipocyte | *DIDO1* | 1.00215416 | 0.007116461 | 0.034667739 | 1 |
| Obesity | adipocyte | *PTK6* | 3.342529285 | 0.010458716 | 0.045511831 | 1 |
| Obesity | adipocyte | *RNF24* | 1.023791886 | 0.001563048 | 0.011776255 | 1 |
| Obesity | adipocyte | *RASSF2* | 0.567721312 | 0.00088249 | 0.007856593 | 1 |
| Obesity | adipocyte | *ANGPT4* | 2.022958197 | 0.001875361 | 0.013264212 | 1 |
| Obesity | adipocyte | *MANBAL* | 0.573768641 | 0.001484919 | 0.01129283 | 1 |
| Obesity | adipocyte | *METTL4* | 1.643716958 | 0.002444822 | 0.016181881 | 1 |
| Obesity | adipocyte | *PARP4* | 1.006915923 | 0.001768038 | 0.012799963 | 1 |
| Obesity | adipocyte | *DGKH* | 0.608638665 | 0.001171735 | 0.009514295 | 1 |
| Obesity | adipocyte | *CLN5* | 1.353959067 | 0.009182528 | 0.041207992 | 1 |
| Obesity | adipocyte | *ELMO3* | 0.30723694 | 0.001805842 | 0.012885216 | 1 |
| Obesity | adipocyte | *CCDC113* | -0.734172012 | 0.000730984 | 0.006968045 | 1 |
| Obesity | adipocyte | *COG4* | -0.668364696 | 0.002949929 | 0.018452052 | 1 |
| Obesity | adipocyte | *NME4* | -1.293426946 | 0.00363175 | 0.021410997 | 1 |
| Obesity | adipocyte | *CLCN7* | 0.342239734 | 0.003532431 | 0.020967771 | 1 |
| Obesity | adipocyte | *RHBDL1* | -2.267249156 | 0.000110752 | 0.002048785 | 0.537367853 |
| Obesity | adipocyte | *NUBP1* | 1.508920742 | 0.000186469 | 0.002845111 | 0.904745343 |
| Obesity | adipocyte | *CLUAP1* | 0.371728879 | 0.00058416 | 0.005880382 | 1 |
| Obesity | adipocyte | *PYCARD* | 2.484067916 | 3.55E-13 | 1.01E-10 | 1.72E-09 |
| Obesity | adipocyte | *TRIP4* | 1.069597126 | 0.005414655 | 0.028235385 | 1 |
| Obesity | adipocyte | *FAH* | -0.617177338 | 0.00525902 | 0.027948264 | 1 |
| Obesity | adipocyte | *RPAP1* | -0.849909063 | 0.003188578 | 0.019411516 | 1 |
| Obesity | adipocyte | *SLC30A4* | 2.12077135 | 0.000643202 | 0.00630468 | 1 |
| Obesity | adipocyte | *NIPAL2* | 1.247922931 | 0.002166393 | 0.014701174 | 1 |
| Obesity | adipocyte | *TUSC3* | 1.25781304 | 8.27E-05 | 0.001728601 | 0.401035359 |
| Obesity | adipocyte | *AMH* | 15.30899514 | 0.000104484 | 0.001988073 | 0.506958539 |
| Obesity | adipocyte | *FCER2* | 5.603463348 | 0.000164491 | 0.00256627 | 0.798110062 |
| Obesity | adipocyte | *CLEC4M* | 4.560541722 | 0.000265966 | 0.003540206 | 1 |
| Obesity | adipocyte | *NOVA2* | 0.286952748 | 0.00245973 | 0.016215502 | 1 |
| Obesity | adipocyte | *IL27RA* | 1.601253763 | 0.000135069 | 0.002259841 | 0.655353895 |
| Obesity | adipocyte | *RASAL3* | 2.937139817 | 1.54E-06 | 9.02E-05 | 0.007494396 |
| Obesity | adipocyte | *TGFB1* | 1.455695208 | 0.002829013 | 0.017896181 | 1 |
| Obesity | adipocyte | *RABAC1* | 0.920456877 | 0.002458179 | 0.016215502 | 1 |
| Obesity | adipocyte | *PLEKHA4* | 1.464622272 | 0.000371614 | 0.004393015 | 1 |
| Obesity | adipocyte | *GCDH* | -0.774868707 | 0.001385591 | 0.01072231 | 1 |

| | | | | | | |
|---|---|---|---|---|---|---|
| Obesity | adipocyte | *RAB3A* | 9.190380007 | 0.000775157 | 0.007260737 | 1 |
| Obesity | adipocyte | *DNAH11* | 1.85990011 | 0.000315765 | 0.003969148 | 1 |
| Obesity | adipocyte | *PTN* | 1.244662444 | 0.002361427 | 0.01580365 | 1 |
| Obesity | adipocyte | *VIPR2* | 1.137932604 | 1.26E-05 | 0.000517643 | 0.061081895 |
| Obesity | adipocyte | *CPVL* | 1.058241458 | 0.000279239 | 0.003661585 | 1 |
| Obesity | adipocyte | *GRB10* | 1.42204256 | 0.001029204 | 0.008609822 | 1 |
| Obesity | adipocyte | *PLEKHA8* | 1.575313087 | 0.004764505 | 0.025945428 | 1 |
| Obesity | adipocyte | *TAF6* | -0.321257682 | 0.008821731 | 0.040040261 | 1 |
| Obesity | adipocyte | *MEST* | -0.501328755 | 4.69E-06 | 0.000223112 | 0.022757386 |
| Obesity | adipocyte | *POU6F2* | -0.631464792 | 0.003721027 | 0.021778556 | 1 |
| Obesity | adipocyte | *TSPAN13* | 1.689960903 | 0.001752086 | 0.01274531 | 1 |
| Obesity | adipocyte | *COA1* | 1.055793943 | 0.006639098 | 0.032870308 | 1 |
| Obesity | adipocyte | *AEBP1* | 1.472812693 | 0.000232888 | 0.003294385 | 1 |
| Obesity | adipocyte | *GCK* | 12.5646298 | 0.001417333 | 0.010915713 | 1 |
| Obesity | adipocyte | *TBL2* | 0.686920072 | 0.00119194 | 0.009606802 | 1 |
| Obesity | adipocyte | *SPATA6L* | 0.792971893 | 0.010543746 | 0.045755156 | 1 |
| Obesity | adipocyte | *TRIM14* | 0.584149824 | 0.000361971 | 0.004347234 | 1 |
| Obesity | adipocyte | *OGN* | 0.698816719 | 0.000190393 | 0.0028689 | 0.923785757 |
| Obesity | adipocyte | *RGP1* | 1.893427395 | 0.000238522 | 0.003319488 | 1 |
| Obesity | adipocyte | *PTGDS* | 1.859746249 | 0.000257934 | 0.003495803 | 1 |
| Obesity | adipocyte | *DNMBP* | 0.938993467 | 0.000429549 | 0.004824471 | 1 |
| Obesity | adipocyte | *PKD2L1* | 0.514818504 | 0.006598191 | 0.032735516 | 1 |
| Obesity | adipocyte | *LIPA* | 0.417278598 | 0.004050144 | 0.023146404 | 1 |
| Obesity | adipocyte | *SFXN3* | 2.072921429 | 0.000350961 | 0.004267829 | 1 |
| Obesity | adipocyte | *TNKS2* | 1.276011914 | 0.006296003 | 0.031787934 | 1 |
| Obesity | adipocyte | *TFAM* | 1.790657068 | 0.007860185 | 0.036905011 | 1 |
| Obesity | adipocyte | *RPL28* | 0.646486052 | 0.00537865 | 0.028138025 | 1 |
| Obesity | adipocyte | *DNAJC12* | 0.909764773 | 0.00880528 | 0.040003012 | 1 |
| Obesity | adipocyte | *RAD51C* | -0.657867528 | 0.000529109 | 0.005473856 | 1 |
| Obesity | adipocyte | *TRIM37* | 1.264844508 | 0.006479121 | 0.032342281 | 1 |
| Obesity | adipocyte | *PNPO* | -1.312148227 | 0.007721682 | 0.036452195 | 1 |
| Obesity | adipocyte | *TRIM16L* | -1.311622195 | 0.007723177 | 0.036452195 | 1 |
| Obesity | adipocyte | *CHRNE* | 0.202033481 | 0.000525882 | 0.005455908 | 1 |
| Obesity | adipocyte | *RAI1* | 1.718881965 | 0.002274821 | 0.015393908 | 1 |
| Obesity | adipocyte | *ICAM2* | 1.932221422 | 4.79E-05 | 0.001259605 | 0.232194308 |
| Obesity | adipocyte | *FAM20A* | 1.310899618 | 0.000283252 | 0.003694455 | 1 |
| Obesity | adipocyte | *GAR1* | 2.166172684 | 0.001943784 | 0.013628961 | 1 |
| Obesity | adipocyte | *STIM2* | 1.422756202 | 0.000768944 | 0.007216475 | 1 |
| Obesity | adipocyte | *BST1* | 1.147012926 | 0.000187876 | 0.002857598 | 0.911573694 |
| Obesity | adipocyte | *UFSP2* | 1.53571546 | 0.004606408 | 0.02522606 | 1 |
| Obesity | adipocyte | *SC5D* | -0.922210552 | 0.005097999 | 0.027334773 | 1 |
| Obesity | adipocyte | *B3GAT1* | 19.82290472 | 1.25E-11 | 2.68E-09 | 6.08E-08 |
| Obesity | adipocyte | *P2RX3* | 16.9709471 | 1.03E-05 | 0.00044484 | 0.049822132 |
| Obesity | adipocyte | *NRXN2* | 3.157803845 | 0.000817368 | 0.007502305 | 1 |
| Obesity | adipocyte | *SLC22A18* | 0.192875972 | 0.003889576 | 0.022386222 | 1 |
| Obesity | adipocyte | *SLC35F2* | 1.682456625 | 0.000330413 | 0.004110678 | 1 |
| Obesity | adipocyte | *PRDM4* | 1.14274226 | 0.000830985 | 0.007550452 | 1 |
| Obesity | adipocyte | *ASIC1* | 3.417044998 | 0.000230336 | 0.003287029 | 1 |
| Obesity | adipocyte | *IL23A* | 15.62412248 | 0.000117575 | 0.002088842 | 0.57047197 |
| Obesity | adipocyte | *DDX55* | 1.288922707 | 0.000334392 | 0.004149535 | 1 |

| | | | | | | |
|---|---|---|---|---|---|---|
| Obesity | adipocyte | *VDR* | 2.185329931 | 0.000102308 | 0.001975563 | 0.496400533 |
| Obesity | adipocyte | *CAND1* | 1.371480494 | 0.00414161 | 0.023441543 | 1 |
| Obesity | adipocyte | *C2CD5* | 1.2802046 | 0.000932697 | 0.008081427 | 1 |
| Obesity | adipocyte | *BTN3A3* | 0.973078424 | 1.25E-12 | 3.19E-10 | 6.06E-09 |
| Obesity | adipocyte | *FRK* | 1.924942511 | 0.002114669 | 0.014476445 | 1 |
| Obesity | adipocyte | *RNGTT* | 1.220407283 | 0.005775668 | 0.029685952 | 1 |
| Obesity | adipocyte | *GABRR2* | 0.621623344 | 0.003200916 | 0.019462212 | 1 |
| Obesity | adipocyte | *TULP1* | -0.553399414 | 0.0004945 | 0.005243227 | 1 |
| Obesity | adipocyte | *SLC26A8* | -0.285011271 | 0.009025656 | 0.040737195 | 1 |
| Obesity | adipocyte | *COL9A1* | 1.453881115 | 0.006601625 | 0.032735516 | 1 |
| Obesity | adipocyte | *FIG4* | 1.446467719 | 0.00152881 | 0.011554185 | 1 |
| Obesity | adipocyte | *PACRG* | 1.817109015 | 0.000588691 | 0.005901508 | 1 |
| Obesity | adipocyte | *TMEM30A* | 1.475461387 | 0.00773236 | 0.03646007 | 1 |
| Obesity | adipocyte | *IMPG1* | -1.795575374 | 0.001382677 | 0.01072231 | 1 |
| Obesity | adipocyte | *LY86* | 1.677732432 | 0.003965964 | 0.022745695 | 1 |
| Obesity | adipocyte | *PRSS16* | 0.773742622 | 0.000465607 | 0.005051097 | 1 |
| Obesity | adipocyte | *DAP* | 0.623930877 | 0.004410498 | 0.024540982 | 1 |
| Obesity | adipocyte | *NME5* | 0.293624679 | 0.003292757 | 0.019896503 | 1 |
| Obesity | adipocyte | *PFDN1* | 0.761428741 | 0.010552354 | 0.045755156 | 1 |
| Obesity | adipocyte | *TMCO6* | -0.718457819 | 0.000147903 | 0.002360611 | 0.717625676 |
| Obesity | adipocyte | *IK* | -0.543826649 | 0.007404754 | 0.035642727 | 1 |
| Obesity | adipocyte | *ARSB* | 1.577290939 | 0.003072672 | 0.018990597 | 1 |
| Obesity | adipocyte | *SLC12A7* | 1.919745559 | 9.55E-05 | 0.001887835 | 0.46353145 |
| Obesity | adipocyte | *TXNDC15* | 1.776578158 | 0.000615471 | 0.006069645 | 1 |
| Obesity | adipocyte | *SMAD5* | 1.292836481 | 0.006468642 | 0.032323225 | 1 |
| Obesity | adipocyte | *ERGIC1* | 1.42121481 | 0.002150124 | 0.014652247 | 1 |
| Obesity | adipocyte | *ATP6V0E1* | 1.474385663 | 0.000508455 | 0.005363095 | 1 |
| Obesity | adipocyte | *BNIP1* | -0.27259844 | 0.007396291 | 0.035637343 | 1 |
| Obesity | adipocyte | *UNC5A* | 8.744281321 | 0.000434896 | 0.00485162 | 1 |
| Obesity | adipocyte | *PCCB* | 0.750802856 | 2.46E-05 | 0.000812964 | 0.119505753 |
| Obesity | adipocyte | *CBLB* | 0.970864328 | 0.000826028 | 0.007533628 | 1 |
| Obesity | adipocyte | *PLXNA1* | 1.428343089 | 0.000221438 | 0.003183834 | 1 |
| Obesity | adipocyte | *SCAP* | 1.351794026 | 0.0003941 | 0.004585544 | 1 |
| Obesity | adipocyte | *KIAA1257* | 2.717064036 | 9.52E-05 | 0.001887835 | 0.461888476 |
| Obesity | adipocyte | *VIPR1* | 1.912437462 | 0.000109563 | 0.002044622 | 0.531601767 |
| Obesity | adipocyte | *NKTR* | 1.052915059 | 0.000744977 | 0.007073634 | 1 |
| Obesity | adipocyte | *ACTR1B* | 2.077785743 | 0.000198555 | 0.002958252 | 0.963388604 |
| Obesity | adipocyte | *POLE4* | 1.874278446 | 0.000901176 | 0.007964495 | 1 |
| Obesity | adipocyte | *GGCX* | 0.760028712 | 0.003556895 | 0.02104641 | 1 |
| Obesity | adipocyte | *CHMP3* | -0.68494616 | 0.000319556 | 0.003996103 | 1 |
| Obesity | adipocyte | *IL1RL2* | 0.938393745 | 0.002496017 | 0.016387923 | 1 |
| Obesity | adipocyte | *NOL10* | 1.204961801 | 0.010419926 | 0.045424511 | 1 |
| Obesity | adipocyte | *STRN* | 1.190774139 | 0.007881837 | 0.036913779 | 1 |
| Obesity | adipocyte | *WIPF1* | 1.20954893 | 0.001017087 | 0.008577486 | 1 |
| Obesity | adipocyte | *CAPZA1* | 1.08173741 | 0.002951111 | 0.018452052 | 1 |
| Obesity | adipocyte | *LEPR* | 0.896451963 | 0.004336304 | 0.02423934 | 1 |
| Obesity | adipocyte | *PRG4* | 1.557597888 | 0.000834473 | 0.007553846 | 1 |
| Obesity | adipocyte | *WARS2* | 0.734296696 | 0.001939617 | 0.013619425 | 1 |
| Obesity | adipocyte | *EXOC8* | 1.387107849 | 0.007710529 | 0.036452195 | 1 |
| Obesity | adipocyte | *GNPAT* | 1.162217965 | 0.003810735 | 0.022163873 | 1 |

| | | | | | | |
|---|---|---|---|---|---|---|
| Obesity | adipocyte | *MFAP2* | -0.222176816 | 0.009206402 | 0.041207992 | 1 |
| Obesity | adipocyte | *EBNA1BP2* | 1.334667893 | 0.000800189 | 0.007423552 | 1 |
| Obesity | adipocyte | *NSUN4* | -0.933224651 | 0.000308615 | 0.003930183 | 1 |
| Obesity | adipocyte | *PRRC2C* | 1.409277561 | 4.25E-05 | 0.00117194 | 0.206261468 |
| Obesity | adipocyte | *SERPINC1* | 1.531166083 | 5.27E-08 | 4.83E-06 | 0.00025573 |
| Obesity | adipocyte | *SLC5A9* | 1.049768887 | 0.002422299 | 0.016056528 | 1 |
| Obesity | adipocyte | *KLF7* | 1.141199811 | 4.31E-05 | 0.001176187 | 0.209361199 |
| Obesity | adipocyte | *SGIP1* | 1.703276398 | 8.92E-05 | 0.001818245 | 0.432742282 |
| Obesity | adipocyte | *MFSD1* | 1.366788844 | 0.00100923 | 0.008557091 | 1 |
| Obesity | adipocyte | *WDR35* | 1.261246022 | 0.002038649 | 0.014090491 | 1 |
| Obesity | adipocyte | *C2orf40* | 1.249025563 | 0.000767378 | 0.007215734 | 1 |
| Obesity | adipocyte | *CCDC92* | 1.351379399 | 4.85E-06 | 0.000228688 | 0.023554862 |
| Obesity | adipocyte | *CTNNAL1* | 0.325377499 | 0.003080297 | 0.018990597 | 1 |
| Obesity | adipocyte | *RAB14* | 1.522445343 | 0.001818881 | 0.01294019 | 1 |
| Obesity | adipocyte | *TRIM32* | 2.337562222 | 0.001309302 | 0.010379574 | 1 |
| Obesity | adipocyte | *MAPKAP1* | 1.062846393 | 0.009978423 | 0.044013915 | 1 |
| Obesity | adipocyte | *INVS* | 1.114982144 | 5.76E-08 | 5.08E-06 | 0.000279375 |
| Obesity | adipocyte | *DENND1A* | 1.213349114 | 0.000371182 | 0.004393015 | 1 |
| Obesity | adipocyte | *CNRIP1* | -0.537744968 | 0.001332815 | 0.010498081 | 1 |
| Obesity | adipocyte | *PPP1R3C* | 2.388931656 | 2.72E-05 | 0.000869641 | 0.132185391 |
| Obesity | adipocyte | *MXI1* | 1.177631828 | 0.004248689 | 0.023859538 | 1 |
| Obesity | adipocyte | *GORAB* | 1.335755449 | 0.009978378 | 0.044013915 | 1 |
| Obesity | adipocyte | *SNX19* | -0.939842928 | 1.50E-05 | 0.000603979 | 0.072559689 |
| Obesity | adipocyte | *SLC10A7* | 1.200181561 | 0.009719614 | 0.043068096 | 1 |
| Obesity | adipocyte | *MASTL* | -0.423446285 | 0.010121315 | 0.044396156 | 1 |
| Obesity | adipocyte | *MRC1* | 0.985963997 | 0.001091313 | 0.009003769 | 1 |
| Obesity | adipocyte | *DNAJC15* | 1.454827893 | 0.001440489 | 0.011058946 | 1 |
| Obesity | adipocyte | *APAF1* | 1.047561434 | 0.007603488 | 0.036226343 | 1 |
| Obesity | adipocyte | *ADRA1A* | 0.129500041 | 0.003822115 | 0.022164653 | 1 |
| Obesity | adipocyte | *TEX14* | 0.78256363 | 6.86E-05 | 0.001537561 | 0.333057165 |
| Obesity | adipocyte | *A1BG* | 1.353589163 | 7.58E-05 | 0.001649115 | 0.367752656 |
| Obesity | adipocyte | *CPXM2* | 0.650038586 | 1.21E-05 | 0.00051024 | 0.058807418 |
| Obesity | adipocyte | *CD244* | 1.629220155 | 2.55E-05 | 0.000830644 | 0.123729084 |
| Obesity | adipocyte | *OPN4* | 2.462346882 | 2.75E-06 | 0.000148173 | 0.013351749 |
| Obesity | adipocyte | *ODF2L* | 1.213300306 | 0.006108842 | 0.031004289 | 1 |
| Obesity | adipocyte | *NT5C3A* | 1.061441367 | 0.011527658 | 0.0491261 | 1 |
| Obesity | adipocyte | *SMU1* | 1.276099332 | 0.008028189 | 0.03732363 | 1 |
| Obesity | adipocyte | *SLC25A51* | 0.179227909 | 0.008692679 | 0.039639922 | 1 |
| Obesity | adipocyte | *DDX54* | 1.17787959 | 0.00466909 | 0.02548304 | 1 |
| Obesity | adipocyte | *CCDC91* | 1.10295983 | 0.002955557 | 0.018456066 | 1 |
| Obesity | adipocyte | *ARHGAP9* | 1.441513344 | 0.00450721 | 0.024851116 | 1 |
| Obesity | adipocyte | *PFDN5* | 1.286364102 | 0.005299706 | 0.02801222 | 1 |
| Obesity | adipocyte | *SMUG1* | 0.322367892 | 0.008892395 | 0.040210533 | 1 |
| Obesity | adipocyte | *SLC12A4* | 1.698458649 | 0.004497317 | 0.024844992 | 1 |
| Obesity | adipocyte | *MATN4* | 2.459010065 | 0.007272024 | 0.03528386 | 1 |
| Obesity | adipocyte | *CSE1L* | 1.213541231 | 0.001286744 | 0.010234892 | 1 |
| Obesity | adipocyte | *PTGIS* | 0.553632032 | 4.00E-05 | 0.001121108 | 0.19395161 |
| Obesity | adipocyte | *NEURL2* | 0.281970496 | 0.005329845 | 0.028138025 | 1 |
| Obesity | adipocyte | *CHST8* | 0.682077846 | 0.008302728 | 0.038366511 | 1 |
| Obesity | adipocyte | *NAGK* | 0.73822432 | 0.002608617 | 0.016966499 | 1 |

| | | | | | | |
|---|---|---|---|---|---|---|
| Obesity | adipocyte | *ATP8A1* | 1.384017683 | 3.10E-05 | 0.000923023 | 0.150452692 |
| Obesity | adipocyte | *BTN2A2* | 0.392909722 | 8.90E-07 | 5.84E-05 | 0.004318992 |
| Obesity | adipocyte | *SNRPC* | -1.868236473 | 2.50E-20 | 1.74E-17 | 1.21E-16 |
| Obesity | adipocyte | *TCP11* | 0.433079675 | 0.002647178 | 0.017125477 | 1 |
| Obesity | adipocyte | *TREM1* | 0.748727501 | 0.000124015 | 0.002164461 | 0.601720139 |
| Obesity | adipocyte | *KLHL31* | -0.214634903 | 0.008084578 | 0.037501314 | 1 |
| Obesity | adipocyte | *GLO1* | 1.333152441 | 0.000608412 | 0.006049207 | 1 |
| Obesity | adipocyte | *ATXN1* | 0.897836379 | 0.004528735 | 0.024884965 | 1 |
| Obesity | adipocyte | *CXCL6* | 1.763892462 | 0.011263631 | 0.048235777 | 1 |
| Obesity | adipocyte | *GOT2* | 1.978571345 | 9.73E-06 | 0.000425481 | 0.04722839 |
| Obesity | adipocyte | *ABCC4* | 0.842662376 | 0.001023361 | 0.008590564 | 1 |
| Obesity | adipocyte | *GRK4* | 0.170212398 | 0.000425815 | 0.004793632 | 1 |
| Obesity | adipocyte | *GGA3* | 1.315406615 | 0.000794114 | 0.007395475 | 1 |
| Obesity | adipocyte | *PPDPF* | 0.744130434 | 0.003435864 | 0.020593813 | 1 |
| Obesity | adipocyte | *PSD4* | 1.145412477 | 0.004025244 | 0.023031231 | 1 |
| Obesity | adipocyte | *RRBP1* | 1.036172403 | 0.002130769 | 0.01454688 | 1 |
| Obesity | adipocyte | *FLRT3* | 1.909010917 | 0.000750803 | 0.007087344 | 1 |
| Obesity | adipocyte | *KLC1* | 1.397011433 | 0.004776823 | 0.025983347 | 1 |
| Obesity | adipocyte | *TUBGCP3* | 1.300363464 | 0.004088666 | 0.023284282 | 1 |
| Obesity | adipocyte | *COX6B1* | 1.621247735 | 0.000940196 | 0.008131608 | 1 |
| Obesity | adipocyte | *IRF3* | 1.419247604 | 0.007381077 | 0.03559939 | 1 |
| Obesity | adipocyte | *KTN1* | 1.007588266 | 0.011458596 | 0.048898073 | 1 |
| Obesity | adipocyte | *AIF1L* | 1.981645123 | 7.03E-05 | 0.001564812 | 0.341129029 |
| Obesity | adipocyte | *CANX* | 1.713539058 | 1.93E-05 | 0.000691702 | 0.093404071 |
| Obesity | adipocyte | *ZNF484* | 1.492607661 | 0.001221968 | 0.009751622 | 1 |
| Obesity | adipocyte | *HELB* | 1.513674359 | 0.001141393 | 0.009292008 | 1 |
| Obesity | adipocyte | *YEATS4* | 0.360074645 | 0.001782832 | 0.012801304 | 1 |
| Obesity | adipocyte | *HIP1* | 1.22976796 | 0.000445378 | 0.004937388 | 1 |
| Obesity | adipocyte | *STYXL1* | 0.292437781 | 0.005357433 | 0.028138025 | 1 |
| Obesity | adipocyte | *KDR* | 1.331818224 | 6.73E-05 | 0.00153723 | 0.326759949 |
| Obesity | adipocyte | *RIBC2* | 0.436083145 | 0.000749325 | 0.007087344 | 1 |
| Obesity | adipocyte | *SPECC1* | 1.323026724 | 0.001055879 | 0.008770334 | 1 |
| Obesity | adipocyte | *TWSG1* | 1.463754384 | 0.004951388 | 0.02675505 | 1 |
| Obesity | adipocyte | *EIF2AK4* | 1.394356541 | 4.81E-05 | 0.001259605 | 0.233549336 |
| Obesity | adipocyte | *TMOD2* | 1.06125392 | 0.00045103 | 0.004942243 | 1 |
| Obesity | adipocyte | *ARPP19* | 1.51546049 | 0.006025434 | 0.030659999 | 1 |
| Obesity | adipocyte | *VPS13C* | 0.992844516 | 0.00307944 | 0.018990597 | 1 |
| Obesity | adipocyte | *SERGEF* | 0.841416363 | 0.001765508 | 0.012799963 | 1 |
| Obesity | adipocyte | *RPAIN* | -0.88074459 | 5.40E-05 | 0.001351276 | 0.262147618 |
| Obesity | adipocyte | *PLD2* | 2.187904394 | 0.000118404 | 0.002089082 | 0.574497465 |
| Obesity | adipocyte | *AP1M2* | -0.373596003 | 0.001446478 | 0.011069895 | 1 |
| Obesity | adipocyte | *MTUS1* | 0.872306939 | 0.002687534 | 0.017340312 | 1 |
| Obesity | adipocyte | *KLK8* | 8.332712931 | 6.38E-05 | 0.001482155 | 0.309383073 |
| Obesity | adipocyte | *RNASE1* | 0.891814355 | 0.001319919 | 0.010422457 | 1 |
| Obesity | adipocyte | *SHC2* | 1.011905219 | 1.93E-05 | 0.000691702 | 0.093879341 |
| Obesity | adipocyte | *PRKCSH* | 0.919855949 | 0.007492641 | 0.035907734 | 1 |
| Obesity | adipocyte | *XPO7* | 1.268023713 | 0.006396284 | 0.032055192 | 1 |
| Obesity | adipocyte | *ATP8B3* | -0.533718545 | 4.88E-10 | 6.76E-08 | 2.37E-06 |
| Obesity | adipocyte | *ACTN4* | 1.011888078 | 0.00639853 | 0.032055192 | 1 |
| Obesity | adipocyte | *STK33* | 0.852873128 | 5.50E-08 | 4.94E-06 | 0.000266806 |

| | | | | | | |
|---|---|---|---|---|---|---|
| Obesity | adipocyte | *MAP1S* | 2.705867048 | 0.003221628 | 0.01956363 | 1 |
| Obesity | adipocyte | *SSBP4* | 0.92281552 | 0.001270936 | 0.010125751 | 1 |
| Obesity | adipocyte | *PGPEP1* | 0.534720913 | 0.001196813 | 0.009621291 | 1 |
| Obesity | adipocyte | *ATXN10* | 1.079681691 | 0.007279894 | 0.035286758 | 1 |
| Obesity | adipocyte | *CYP2E1* | -0.35407242 | 0.000200818 | 0.002970685 | 0.974370226 |
| Obesity | adipocyte | *NPAS1* | -1.544864999 | 7.18E-05 | 0.001586418 | 0.348423288 |
| Obesity | adipocyte | *THEMIS2* | 0.400961794 | 0.00777802 | 0.036639758 | 1 |
| Obesity | adipocyte | *DNMT1* | 1.710667148 | 0.001012681 | 0.008560151 | 1 |
| Obesity | adipocyte | *C12orf65* | 0.804296775 | 0.000629942 | 0.006187201 | 1 |
| Obesity | adipocyte | *NOL11* | 1.367057602 | 0.00054592 | 0.00560916 | 1 |
| Obesity | adipocyte | *PKDREJ* | -0.454459917 | 0.000741594 | 0.007055317 | 1 |
| Obesity | adipocyte | *TTLL9* | 2.389830863 | 0.002982083 | 0.018550084 | 1 |
| Obesity | adipocyte | *IDO1* | 2.707052081 | 2.25E-05 | 0.000767693 | 0.109012411 |
| Obesity | adipocyte | *GALNT15* | 1.413444413 | 5.59E-05 | 0.001363471 | 0.271141001 |
| Obesity | adipocyte | *SLC6A6* | 1.406545705 | 6.79E-05 | 0.00153723 | 0.329465825 |
| Obesity | adipocyte | *PDLIM4* | 2.213711142 | 0.001905221 | 0.013416736 | 1 |
| Obesity | adipocyte | *KIF3A* | 1.88449276 | 4.20E-05 | 0.001164011 | 0.203701938 |
| Obesity | adipocyte | *NDUFA2* | 0.88510228 | 6.16E-05 | 0.001450846 | 0.29887429 |
| Obesity | adipocyte | *EXOC4* | 1.168271318 | 4.37E-05 | 0.001183438 | 0.211835374 |
| Obesity | adipocyte | *PRKAB2* | 1.81139699 | 0.000232816 | 0.003294385 | 1 |
| Obesity | adipocyte | *TRIM21* | 1.35005275 | 0.008578455 | 0.039229656 | 1 |
| Obesity | adipocyte | *SPATA6* | 1.013655678 | 0.002700306 | 0.017399578 | 1 |
| Obesity | adipocyte | *TRIM22* | 1.316061575 | 0.00011625 | 0.002081342 | 0.564043791 |
| Obesity | adipocyte | *CARD6* | 0.57194972 | 0.002386761 | 0.015929253 | 1 |
| Obesity | adipocyte | *MYBBP1A* | 0.62380073 | 0.000340854 | 0.004197519 | 1 |
| Obesity | adipocyte | *RGS22* | -0.405939386 | 0.002115375 | 0.014476445 | 1 |
| Obesity | adipocyte | *IGHMBP2* | 0.526588537 | 1.15E-05 | 0.000489704 | 0.055826231 |
| Obesity | adipocyte | *MUTYH* | 0.879788313 | 0.000549144 | 0.005621194 | 1 |
| Obesity | adipocyte | *DMGDH* | -0.485607726 | 0.009685572 | 0.042956485 | 1 |
| Obesity | adipocyte | *AP3B1* | 1.124286545 | 0.00446345 | 0.024764727 | 1 |
| Obesity | adipocyte | *ZBED3* | -1.313984357 | 0.000262033 | 0.003528633 | 1 |
| Obesity | adipocyte | *ALOX5AP* | 1.967050176 | 2.22E-05 | 0.000765564 | 0.107944555 |
| Obesity | adipocyte | *DSTYK* | 1.528952422 | 0.000911834 | 0.008014798 | 1 |
| Obesity | adipocyte | *EPSTI1* | 1.046597926 | 0.000139168 | 0.002296752 | 0.675245179 |
| Obesity | adipocyte | *GIMAP4* | 0.633023832 | 0.001205374 | 0.009650952 | 1 |
| Obesity | adipocyte | *LARS* | -0.381227798 | 0.003105836 | 0.019075337 | 1 |
| Obesity | adipocyte | *COX16* | 3.794815879 | 0.004208766 | 0.023662728 | 1 |
| Obesity | adipocyte | *TTC9* | 1.811944125 | 5.59E-05 | 0.001363471 | 0.271330739 |
| Obesity | adipocyte | *GSTM5* | 0.463656909 | 0.000111053 | 0.002048785 | 0.53883058 |
| Obesity | adipocyte | *NAPG* | 1.277942422 | 0.00415439 | 0.023441543 | 1 |
| Obesity | adipocyte | *FKBP11* | 2.034070006 | 0.000484808 | 0.005169861 | 1 |
| Obesity | adipocyte | *YWHAQ* | 1.480991549 | 0.000729441 | 0.006968045 | 1 |
| Obesity | adipocyte | *NARS* | 0.87557875 | 0.010279106 | 0.044940644 | 1 |
| Obesity | adipocyte | *KIAA1468* | 1.174448099 | 0.008749028 | 0.03978471 | 1 |
| Obesity | adipocyte | *IL15RA* | 1.244728669 | 0.000267048 | 0.003540206 | 1 |
| Obesity | adipocyte | *KLRD1* | 1.659785491 | 0.001639813 | 0.012112129 | 1 |
| Obesity | adipocyte | *ACP2* | 1.236880771 | 0.000200821 | 0.002970685 | 0.974384818 |
| Obesity | adipocyte | *PRPF38A* | 1.538093894 | 0.002395112 | 0.015963028 | 1 |
| Obesity | adipocyte | *ARGLU1* | 1.089327055 | 0.00105271 | 0.008770334 | 1 |
| Obesity | adipocyte | *OSTF1* | 1.537156873 | 0.000276934 | 0.003649079 | 1 |

| | | | | | | |
|---|---|---|---|---|---|---|
| Obesity | adipocyte | *DTX1* | -0.593756674 | 0.009494281 | 0.042262617 | 1 |
| Obesity | adipocyte | *POM121C* | -1.879155798 | 1.93E-12 | 4.69E-10 | 9.38E-09 |
| Obesity | adipocyte | *MAP3K7* | 1.339086815 | 0.003754175 | 0.021919682 | 1 |
| Obesity | adipocyte | *PHF21A* | 1.057979389 | 0.00523608 | 0.027914074 | 1 |
| Obesity | adipocyte | *NHSL1* | 0.743975064 | 0.000851093 | 0.007661419 | 1 |
| Obesity | adipocyte | *MICAL1* | 0.527722942 | 0.000114922 | 0.002067903 | 0.557599966 |
| Obesity | adipocyte | *MPHOSPH6* | 1.565968228 | 0.003845164 | 0.022263406 | 1 |
| Obesity | adipocyte | *NTPCR* | 1.595445685 | 0.004522486 | 0.024878801 | 1 |
| Obesity | adipocyte | *PIGC* | 1.602223672 | 0.001092997 | 0.009003769 | 1 |
| Obesity | adipocyte | *SP110* | 1.398590201 | 1.52E-05 | 0.000603979 | 0.073685409 |
| Obesity | adipocyte | *USP37* | 1.249007301 | 2.71E-05 | 0.000869641 | 0.131644882 |
| Obesity | adipocyte | *SLC19A3* | -0.381643982 | 0.002058154 | 0.014144705 | 1 |
| Obesity | adipocyte | *MRPS9* | 0.550823323 | 0.002002464 | 0.013939675 | 1 |
| Obesity | adipocyte | *ANKRD36* | 1.436199073 | 0.000713847 | 0.006858588 | 1 |
| Obesity | adipocyte | *DRAM1* | 1.39305749 | 9.33E-05 | 0.001887835 | 0.452856342 |
| Obesity | adipocyte | *SLC41A2* | 0.662872161 | 0.005364401 | 0.028138025 | 1 |
| Obesity | adipocyte | *RCBTB2* | 1.614247967 | 0.00195238 | 0.013669474 | 1 |
| Obesity | adipocyte | *SCRN1* | 0.9525914 | 0.008323867 | 0.038415274 | 1 |
| Obesity | adipocyte | *KDELR2* | 1.953804192 | 0.000803699 | 0.007441881 | 1 |
| Obesity | adipocyte | *STX17* | 1.548925246 | 0.001655638 | 0.012208445 | 1 |
| Obesity | adipocyte | *DSCC1* | 0.155481613 | 0.008355957 | 0.038502472 | 1 |
| Obesity | adipocyte | *DENND4C* | 0.893747764 | 0.009387483 | 0.041902547 | 1 |
| Obesity | adipocyte | *TCF19* | 0.607774451 | 0.000117773 | 0.002088842 | 0.571434876 |
| Obesity | adipocyte | *FLOT1* | 0.696704564 | 0.000765621 | 0.007213195 | 1 |
| Obesity | adipocyte | *TLR2* | 1.199209443 | 0.005541972 | 0.028697598 | 1 |
| Obesity | adipocyte | *TRPC6* | 0.633616497 | 0.003465586 | 0.020682688 | 1 |
| Obesity | adipocyte | *SLC37A4* | 0.999377768 | 8.73E-09 | 9.85E-07 | 4.24E-05 |
| Obesity | adipocyte | *FDX1* | 0.727267235 | 0.011141468 | 0.047848224 | 1 |
| Obesity | adipocyte | *MAP2K5* | 1.406776491 | 0.000466383 | 0.005051097 | 1 |
| Obesity | adipocyte | *NUSAP1* | 1.09743921 | 0.000457764 | 0.004991173 | 1 |
| Obesity | adipocyte | *NDUFAF1* | 0.848726529 | 0.000164446 | 0.00256627 | 0.797892685 |
| Obesity | adipocyte | *ADAM10* | 1.626380681 | 7.57E-07 | 5.10E-05 | 0.003671012 |
| Obesity | adipocyte | *SLC44A5* | 1.350330387 | 4.21E-06 | 0.000206534 | 0.020446891 |
| Obesity | adipocyte | *ADCY3* | 1.58979442 | 1.48E-06 | 8.78E-05 | 0.007201593 |
| Obesity | adipocyte | *LHCGR* | -0.463348293 | 0.005299921 | 0.02801222 | 1 |
| Obesity | adipocyte | *ACTR2* | 1.327948769 | 0.001385081 | 0.01072231 | 1 |
| Obesity | adipocyte | *ATRAID* | 1.13628237 | 0.000397098 | 0.00459838 | 1 |
| Obesity | adipocyte | *CENPO* | 2.955146545 | 4.23E-21 | 4.10E-18 | 2.05E-17 |
| Obesity | adipocyte | *DTNB* | 0.777816613 | 0.010218677 | 0.044748213 | 1 |
| Obesity | adipocyte | *CYP2C8* | 0.971003104 | 0.011280654 | 0.04824573 | 1 |
| Obesity | adipocyte | *CALHM2* | 0.708748913 | 0.00271256 | 0.017455359 | 1 |
| Obesity | adipocyte | *RPS24* | 1.035027956 | 0.000512946 | 0.005392915 | 1 |
| Obesity | adipocyte | *STAT4* | 2.300466742 | 5.79E-06 | 0.000265225 | 0.028113831 |
| Obesity | adipocyte | *COX17* | 2.207874451 | 0.000367961 | 0.004386597 | 1 |
| Obesity | adipocyte | *KIAA1109* | 1.140424867 | 0.002412947 | 0.016037832 | 1 |
| Obesity | adipocyte | *PRDM5* | 1.406273081 | 0.00547543 | 0.028505137 | 1 |
| Obesity | adipocyte | *G3BP2* | 0.637910607 | 0.00413725 | 0.023441543 | 1 |
| Obesity | adipocyte | *ANXA3* | 0.689101292 | 0.000171624 | 0.00264355 | 0.832718407 |
| Obesity | adipocyte | *CLSTN3* | 1.127935628 | 0.001027113 | 0.008607174 | 1 |
| Obesity | adipocyte | *KLRG1* | 0.239356266 | 0.000985561 | 0.008433762 | 1 |

| | | | | | | |
|---|---|---|---|---|---|---|
| Obesity | adipocyte | *MARCH9* | 2.65144391 | 0.001786167 | 0.012801304 | 1 |
| Obesity | adipocyte | *GLTP* | 2.370780194 | 0.000277517 | 0.003649079 | 1 |
| Obesity | adipocyte | *ACVRL1* | 1.801706288 | 0.000129188 | 0.002213269 | 0.626818091 |
| Obesity | adipocyte | *VPS33A* | 1.028651624 | 2.30E-06 | 0.000125233 | 0.011145733 |
| Obesity | adipocyte | *DIAPH3* | 0.409967661 | 4.29E-05 | 0.001176187 | 0.20838877 |
| Obesity | adipocyte | *NOVA1* | 0.211048658 | 0.002585513 | 0.016838807 | 1 |
| Obesity | adipocyte | *FITM1* | 1.044195925 | 0.002526343 | 0.016542264 | 1 |
| Obesity | adipocyte | *TMX1* | 1.615720099 | 0.004363986 | 0.024338001 | 1 |
| Obesity | adipocyte | *SLC24A4* | 2.243463091 | 3.08E-05 | 0.000922495 | 0.149444248 |
| Obesity | adipocyte | *WARS* | 1.301368763 | 0.000364996 | 0.004367521 | 1 |
| Obesity | adipocyte | *SERF2* | 1.278265829 | 0.004058451 | 0.023166591 | 1 |
| Obesity | adipocyte | *CYP11A1* | 0.538687913 | 0.00177195 | 0.012799963 | 1 |
| Obesity | adipocyte | *ABHD2* | 1.189934796 | 0.003959502 | 0.022735509 | 1 |
| Obesity | adipocyte | *ITGAX* | 2.538940456 | 3.93E-06 | 0.000197072 | 0.019070942 |
| Obesity | adipocyte | *TIGD7* | 0.982810922 | 0.00614645 | 0.031130038 | 1 |
| Obesity | adipocyte | *KSR1* | 1.458996374 | 0.002808233 | 0.017834483 | 1 |
| Obesity | adipocyte | *MYO19* | 2.362508819 | 4.94E-11 | 8.88E-09 | 2.40E-07 |
| Obesity | adipocyte | *GALNT1* | 1.160426127 | 0.004517422 | 0.024878801 | 1 |
| Obesity | adipocyte | *C18orf8* | -1.797690018 | 2.83E-11 | 5.72E-09 | 1.37E-07 |
| Obesity | adipocyte | *TMC6* | 0.59833481 | 0.011016384 | 0.047428125 | 1 |
| Obesity | adipocyte | *TRIM65* | 1.288829836 | 0.000203876 | 0.002997588 | 0.989204076 |
| Obesity | adipocyte | *CBX8* | 0.391894636 | 0.005209763 | 0.027808326 | 1 |
| Obesity | adipocyte | *RNF165* | 0.54768726 | 0.008402914 | 0.038608842 | 1 |
| Obesity | adipocyte | *CBLN2* | 2.049610531 | 0.000198761 | 0.002958252 | 0.964389994 |
| Obesity | adipocyte | *IGFBP4* | 0.813994261 | 0.000287229 | 0.003736282 | 1 |
| Obesity | adipocyte | *DPP9* | 1.584059491 | 0.006928901 | 0.034011076 | 1 |
| Obesity | adipocyte | *COL6A1* | 0.32638019 | 5.44E-05 | 0.001352594 | 0.26375574 |
| Obesity | adipocyte | *SAE1* | -2.21869079 | 2.04E-07 | 1.59E-05 | 0.000987633 |
| Obesity | adipocyte | *RPS8* | 1.464122785 | 9.61E-05 | 0.001887835 | 0.466295229 |
| Obesity | adipocyte | *STRIP1* | 2.135225467 | 0.000615206 | 0.006069645 | 1 |
| Obesity | adipocyte | *GPR161* | 0.612747392 | 0.006310964 | 0.031830349 | 1 |
| Obesity | adipocyte | *TIPRL* | 1.577095965 | 1.78E-05 | 0.000659132 | 0.086486115 |
| Obesity | adipocyte | *MAEL* | 5.115214148 | 0.004117406 | 0.023395947 | 1 |
| Obesity | adipocyte | *SNX27* | 0.892369886 | 0.005042707 | 0.02715562 | 1 |
| Obesity | adipocyte | *SEMA6C* | 2.99534 | 6.66E-11 | 1.15E-08 | 3.23E-07 |
| Obesity | adipocyte | *DYRK3* | 1.244755969 | 0.001862062 | 0.013189377 | 1 |
| Obesity | adipocyte | *SMYD2* | 1.822582055 | 0.000372121 | 0.004393015 | 1 |
| Obesity | adipocyte | *RAB13* | 2.421638787 | 0.000778922 | 0.007281947 | 1 |
| Obesity | adipocyte | *S100A8* | 1.843131946 | 0.000234936 | 0.003304092 | 1 |
| Obesity | adipocyte | *RIT1* | 2.201164389 | 8.78E-05 | 0.001796487 | 0.425767308 |
| Obesity | adipocyte | *CEP170* | 1.546809378 | 0.000245613 | 0.003375956 | 1 |
| Obesity | adipocyte | *MBOAT2* | 1.536176196 | 0.00010351 | 0.001985097 | 0.502229626 |
| Obesity | adipocyte | *PLEKHA6* | 0.432122952 | 0.006078743 | 0.030883836 | 1 |
| Obesity | adipocyte | *ABHD1* | -0.466771801 | 0.005007954 | 0.026998438 | 1 |
| Obesity | adipocyte | *RALB* | 1.662449806 | 0.000140172 | 0.002305474 | 0.680114775 |
| Obesity | adipocyte | *FAHD2B* | -0.573696483 | 0.000391854 | 0.004570374 | 1 |
| Obesity | adipocyte | *METTL21A* | 1.479460791 | 0.000707092 | 0.006819773 | 1 |
| Obesity | adipocyte | *NBEAL1* | 1.187136279 | 0.00012865 | 0.002213269 | 0.62420764 |
| Obesity | adipocyte | *ESPNL* | 20.14385283 | 0.001888564 | 0.013333973 | 1 |
| Obesity | adipocyte | *TAMM41* | 0.968489934 | 0.0116775 | 0.049596312 | 1 |

| | | | | | | |
|---|---|---|---|---|---|---|
| Obesity | adipocyte | *CNTN4* | 0.980346458 | 0.000389246 | 0.004550901 | 1 |
| Obesity | adipocyte | *CTDSPL* | 0.781174199 | 0.005339224 | 0.028138025 | 1 |
| Obesity | adipocyte | *LIMD1* | 1.289367412 | 3.19E-07 | 2.38E-05 | 0.001550038 |
| Obesity | adipocyte | *BOC* | 1.093838374 | 0.001789672 | 0.012807504 | 1 |
| Obesity | adipocyte | *MED12L* | 1.713501764 | 0.000181716 | 0.002781345 | 0.881686364 |
| Obesity | adipocyte | *AMT* | -0.16969077 | 0.007453905 | 0.03582934 | 1 |
| Obesity | adipocyte | *CCDC39* | 1.165286893 | 8.22E-06 | 0.000369129 | 0.039865881 |
| Obesity | adipocyte | *SCD5* | 1.344125074 | 0.000112632 | 0.00205413 | 0.54649248 |
| Obesity | adipocyte | *TRMT10A* | -0.911806478 | 0.002054205 | 0.014137591 | 1 |
| Obesity | adipocyte | *CISD2* | 1.898972965 | 0.001172619 | 0.009514295 | 1 |
| Obesity | adipocyte | *DDIT4L* | 1.152673646 | 0.007496812 | 0.035907734 | 1 |
| Obesity | adipocyte | *USP53* | 0.662841572 | 0.007931468 | 0.037038962 | 1 |
| Obesity | adipocyte | *MYO10* | 1.203097586 | 0.000137171 | 0.002287132 | 0.665555518 |
| Obesity | adipocyte | *BHMT* | -0.224251886 | 0.007621897 | 0.036236563 | 1 |
| Obesity | adipocyte | *SPATA9* | -0.428700791 | 0.002761562 | 0.017676912 | 1 |
| Obesity | adipocyte | *FEM1C* | 1.81628699 | 0.011731884 | 0.049757954 | 1 |
| Obesity | adipocyte | *YIPF5* | 1.836800687 | 8.10E-05 | 0.001717094 | 0.393214516 |
| Obesity | adipocyte | *GFOD1* | 1.359226468 | 0.000318738 | 0.003996103 | 1 |
| Obesity | adipocyte | *ABT1* | 0.349965409 | 0.00240752 | 0.016023713 | 1 |
| Obesity | adipocyte | *MLIP* | -0.656938886 | 0.001345064 | 0.010560278 | 1 |
| Obesity | adipocyte | *TPBG* | 0.726862002 | 0.007927479 | 0.037038962 | 1 |
| Obesity | adipocyte | *IRAK1BP1* | 1.033762903 | 0.000407934 | 0.004668155 | 1 |
| Obesity | adipocyte | *PHIP* | 1.103572464 | 0.00429375 | 0.024029152 | 1 |
| Obesity | adipocyte | *PNRC1* | 1.677501937 | 0.000593897 | 0.005929192 | 1 |
| Obesity | adipocyte | *ARHGAP18* | 0.660961859 | 0.000626264 | 0.006163556 | 1 |
| Obesity | adipocyte | *SLC22A3* | -0.474753735 | 0.003857117 | 0.022279441 | 1 |
| Obesity | adipocyte | *RBAK* | 2.558134106 | 0.011050847 | 0.047534317 | 1 |
| Obesity | adipocyte | *EPHA1* | 3.025557811 | 0.000166201 | 0.002584641 | 0.806408029 |
| Obesity | adipocyte | *CSGALNACT1* | 0.311774981 | 0.007230775 | 0.035154029 | 1 |
| Obesity | adipocyte | *DOCK5* | 0.933909547 | 0.001010555 | 0.008557091 | 1 |
| Obesity | adipocyte | *ADHFE1* | -0.264540818 | 3.53E-05 | 0.001025016 | 0.171177648 |
| Obesity | adipocyte | *DPYS* | 1.099764507 | 0.000188642 | 0.002859079 | 0.91528872 |
| Obesity | adipocyte | *ZNF462* | 1.177666764 | 0.000193222 | 0.002902521 | 0.937514421 |
| Obesity | adipocyte | *RSU1* | 1.230787557 | 0.002289318 | 0.015470431 | 1 |
| Obesity | adipocyte | *ST8SIA6* | 2.065980718 | 4.53E-06 | 0.000219794 | 0.021979357 |
| Obesity | adipocyte | *RGR* | -2.243937111 | 0.002250693 | 0.015251901 | 1 |
| Obesity | adipocyte | *ADD3* | 1.16547275 | 0.001623039 | 0.012059701 | 1 |
| Obesity | adipocyte | *MTG1* | 0.442456003 | 0.000220177 | 0.003183029 | 1 |
| Obesity | adipocyte | *PDCD11* | 1.317593657 | 0.000143469 | 0.002320923 | 0.696109954 |
| Obesity | adipocyte | *RGS10* | 1.088234552 | 0.000880593 | 0.007856593 | 1 |
| Obesity | adipocyte | *HSD17B12* | -0.42881654 | 3.22E-10 | 4.59E-08 | 1.56E-06 |
| Obesity | adipocyte | *TNKS1BP1* | 0.660130802 | 5.18E-07 | 3.55E-05 | 0.002513333 |
| Obesity | adipocyte | *PTPRJ* | 1.075093714 | 0.001122327 | 0.009198528 | 1 |
| Obesity | adipocyte | *ENDOD1* | 1.593268351 | 5.36E-05 | 0.00134842 | 0.260245033 |
| Obesity | adipocyte | *CAPN5* | 1.625375591 | 0.004900153 | 0.026564853 | 1 |
| Obesity | adipocyte | *INTS4* | 1.227569347 | 0.001454427 | 0.011095722 | 1 |
| Obesity | adipocyte | *TTC12* | 0.736795759 | 4.10E-06 | 0.000203185 | 0.019912083 |
| Obesity | adipocyte | *DRD2* | -6.17151478 | 0.002532101 | 0.01655762 | 1 |
| Obesity | adipocyte | *ATM* | 0.809943185 | 0.007305337 | 0.035374747 | 1 |
| Obesity | adipocyte | *ST14* | 1.419605691 | 0.001361975 | 0.01064139 | 1 |

| | | | | | | |
|---|---|---|---|---|---|---|
| Obesity | adipocyte | *TMC2* | -1.514567572 | 0.005069039 | 0.027267157 | 1 |
| Obesity | adipocyte | *CHEK1* | 3.751639836 | 0.000482688 | 0.005169861 | 1 |
| Obesity | adipocyte | *TRPT1* | -0.399100728 | 0.008032198 | 0.03732363 | 1 |
| Obesity | adipocyte | *NUDT22* | -0.914595957 | 9.61E-05 | 0.001887835 | 0.466064592 |
| Obesity | adipocyte | *TBX6* | 0.371512729 | 8.01E-11 | 1.30E-08 | 3.89E-07 |
| Obesity | adipocyte | *FAM124A* | 1.373056848 | 0.003530632 | 0.020967771 | 1 |
| Obesity | adipocyte | *LYPD6B* | -0.446226917 | 0.007877873 | 0.036913779 | 1 |
| Obesity | adipocyte | *RGS18* | 0.519808169 | 1.79E-05 | 0.000659132 | 0.087005444 |
| Obesity | adipocyte | *PPP1R1C* | 2.667964558 | 0.000129548 | 0.002213269 | 0.628568435 |
| Obesity | adipocyte | *TMEM18* | 2.164363614 | 0.003457531 | 0.020660027 | 1 |
| Obesity | adipocyte | *ANKRD50* | 1.441917335 | 0.000410016 | 0.004680932 | 1 |
| Obesity | adipocyte | *PTPRO* | 0.513013924 | 0.004360974 | 0.024338001 | 1 |
| Obesity | adipocyte | *AKR1C2* | -0.386564709 | 0.01004953 | 0.044206998 | 1 |
| Obesity | adipocyte | *DPYSL4* | 0.180796952 | 0.001638224 | 0.012112129 | 1 |
| Obesity | adipocyte | *INPP1* | 0.839578569 | 0.002658348 | 0.017174837 | 1 |
| Obesity | adipocyte | *MFSD6* | 1.391942039 | 0.002899976 | 0.018249911 | 1 |
| Obesity | adipocyte | *BICD1* | 1.285578231 | 0.000169909 | 0.002625466 | 0.82439618 |
| Obesity | adipocyte | *TIAL1* | 1.525926647 | 0.00079999 | 0.007423552 | 1 |
| Obesity | adipocyte | *POC5* | -0.523581448 | 0.000101203 | 0.00196414 | 0.491034943 |
| Obesity | adipocyte | *CWF19L2* | 1.108571123 | 0.000352853 | 0.004279881 | 1 |
| Obesity | adipocyte | *SPARCL1* | 0.913438893 | 0.000424679 | 0.004791954 | 1 |
| Obesity | adipocyte | *CLEC4F* | 0.853240656 | 0.0029093 | 0.018261222 | 1 |
| Obesity | adipocyte | *JAKMIP1* | 2.535650706 | 0.007811003 | 0.036723823 | 1 |
| Obesity | adipocyte | *TMEM87B* | 1.422152233 | 0.000434905 | 0.00485162 | 1 |
| Obesity | adipocyte | *PLA2R1* | 0.481892794 | 4.85E-05 | 0.001259605 | 0.235257365 |
| Obesity | adipocyte | *LPCAT1* | 1.909378128 | 3.64E-05 | 0.001045408 | 0.17667398 |
| Obesity | adipocyte | *ZDHHC7* | 1.418391407 | 0.007364077 | 0.035552739 | 1 |
| Obesity | adipocyte | *JAZF1* | 1.176492912 | 3.21E-09 | 3.89E-07 | 1.56E-05 |
| Obesity | adipocyte | *GDPD1* | 1.278012002 | 0.002422378 | 0.016056528 | 1 |
| Obesity | adipocyte | *SEMA3D* | 1.104734661 | 0.000153252 | 0.002422085 | 0.743579998 |
| Obesity | adipocyte | *ANKRD29* | 1.492349198 | 6.43E-05 | 0.001485895 | 0.312037915 |
| Obesity | adipocyte | *ANKH* | 1.139788098 | 0.002162413 | 0.014694718 | 1 |
| Obesity | adipocyte | *ROBO4* | 1.382289608 | 7.19E-05 | 0.001586418 | 0.349011869 |
| Obesity | adipocyte | *ROBO3* | 0.662463041 | 0.003882041 | 0.022370149 | 1 |
| Obesity | adipocyte | *PRKCA* | 1.128757339 | 0.003706972 | 0.021722499 | 1 |
| Obesity | adipocyte | *LRRK1* | 1.01881365 | 0.000311832 | 0.003950409 | 1 |
| Obesity | adipocyte | *NEIL2* | 0.510587308 | 0.000653586 | 0.006380683 | 1 |
| Obesity | adipocyte | *CCSAP* | 2.471347996 | 0.002851008 | 0.01801184 | 1 |
| Obesity | adipocyte | *PDE1C* | 1.219616311 | 0.000115073 | 0.002067903 | 0.558333687 |
| Obesity | adipocyte | *MARVELD1* | 1.684485919 | 0.000578591 | 0.00584859 | 1 |
| Obesity | adipocyte | *NUP205* | 1.357457792 | 0.008149958 | 0.037732441 | 1 |
| Obesity | adipocyte | *PDIA4* | 2.005987721 | 0.000349663 | 0.004262727 | 1 |
| Obesity | adipocyte | *DEPTOR* | 0.448426286 | 0.00509851 | 0.027334773 | 1 |
| Obesity | adipocyte | *MMP16* | -0.23475562 | 0.004643985 | 0.025403173 | 1 |
| Obesity | adipocyte | *RWDD2B* | -0.480400547 | 0.00244796 | 0.016181881 | 1 |
| Obesity | adipocyte | *RPL30* | 1.144756003 | 0.000985376 | 0.008433762 | 1 |
| Obesity | adipocyte | *NODAL* | -0.988024567 | 0.002950225 | 0.018452052 | 1 |
| Obesity | adipocyte | *ZDHHC5* | 1.59730846 | 0.010473858 | 0.045536883 | 1 |
| Obesity | adipocyte | *ITGAD* | 0.496503563 | 0.000597827 | 0.005956174 | 1 |
| Obesity | adipocyte | *VPS8* | 0.754890903 | 0.010778645 | 0.046569888 | 1 |

| | | | | | | |
|---|---|---|---|---|---|---|
| Obesity | adipocyte | *NTAN1* | 1.060813931 | 8.00E-05 | 0.001713183 | 0.388010436 |
| Obesity | adipocyte | *SLC6A1* | 3.140826817 | 5.36E-05 | 0.00134842 | 0.259936355 |
| Obesity | adipocyte | *FCHO2* | 0.916100544 | 0.000353716 | 0.004279881 | 1 |
| Obesity | adipocyte | *NECAP2* | 1.736189961 | 0.001547852 | 0.011679906 | 1 |
| Obesity | adipocyte | *STEAP2* | -0.331907129 | 0.003121163 | 0.019145237 | 1 |
| Obesity | adipocyte | *CLEC18A* | -1.083048579 | 2.03E-19 | 1.23E-16 | 9.87E-16 |
| Obesity | adipocyte | *AASDH* | 0.983577333 | 0.007064548 | 0.034518818 | 1 |
| Obesity | adipocyte | *ETS2* | 0.303172426 | 0.000130152 | 0.002215777 | 0.631496545 |
| Obesity | adipocyte | *DGKI* | -0.343910308 | 0.000699935 | 0.006765111 | 1 |
| Obesity | adipocyte | *EXTL1* | 13.2867395 | 0.000203135 | 0.002995785 | 0.985613331 |
| Obesity | adipocyte | *DYNC1I1* | 0.744301621 | 0.00049493 | 0.005243227 | 1 |
| Obesity | adipocyte | *ELK4* | 1.706942452 | 0.001134961 | 0.009279468 | 1 |
| Obesity | adipocyte | *SLC5A11* | -0.355220942 | 1.99E-07 | 1.58E-05 | 0.000966559 |
| Obesity | adipocyte | *CDC42SE2* | 0.793562276 | 0.007605619 | 0.036226343 | 1 |
| Obesity | adipocyte | *HK2* | -0.366754054 | 0.005987871 | 0.030528772 | 1 |
| Obesity | adipocyte | *ACE* | 1.616819976 | 6.33E-05 | 0.001482155 | 0.307112708 |
| Obesity | adipocyte | *CILP2* | -0.869815128 | 8.17E-05 | 0.001724478 | 0.396629889 |
| Obesity | adipocyte | *SLC37A1* | 1.184264528 | 0.00083276 | 0.007552433 | 1 |
| Obesity | adipocyte | *PDE9A* | 0.622628847 | 0.002623165 | 0.017038281 | 1 |
| Obesity | adipocyte | *VAV2* | 0.600699914 | 0.000874885 | 0.007831997 | 1 |
| Obesity | adipocyte | *ZNF208* | 0.63219256 | 0.005417796 | 0.028235385 | 1 |
| Obesity | adipocyte | *GPSM1* | 0.764197966 | 4.82E-05 | 0.001259605 | 0.233952445 |
| Obesity | adipocyte | *PAQR6* | -0.344831055 | 0.005573054 | 0.028797078 | 1 |
| Obesity | adipocyte | *NBEAL2* | 0.990247855 | 9.53E-05 | 0.001887835 | 0.462503021 |
| Obesity | adipocyte | *IER2* | 1.041384384 | 0.006366128 | 0.03200876 | 1 |
| Obesity | adipocyte | *TONSL* | -0.890759976 | 0.003512762 | 0.020912787 | 1 |
| Obesity | adipocyte | *RPL8* | 1.283252811 | 0.000822735 | 0.00751772 | 1 |
| Obesity | adipocyte | *LRWD1* | -0.381390559 | 0.003088438 | 0.019016626 | 1 |
| Obesity | adipocyte | *ABCF3* | 0.489415152 | 0.001122091 | 0.009198528 | 1 |
| Obesity | adipocyte | *PGAP3* | 2.541242964 | 0.003596132 | 0.021239138 | 1 |
| Obesity | adipocyte | *ASGR2* | 15.7534003 | 1.54E-07 | 1.27E-05 | 0.00074694 |
| Obesity | adipocyte | *HS3ST6* | 0.442846128 | 0.004117918 | 0.023395947 | 1 |
| Obesity | adipocyte | *SHANK2* | 1.2767602 | 0.000238768 | 0.003319488 | 1 |
| Obesity | adipocyte | *ASRGL1* | 1.823785081 | 6.81E-05 | 0.00153723 | 0.330504477 |
| Obesity | adipocyte | *RPS6KA4* | 2.850085588 | 0.001197704 | 0.009621291 | 1 |
| Obesity | adipocyte | *PDZK1IP1* | 2.308467139 | 0.000781654 | 0.007293435 | 1 |
| Obesity | adipocyte | *PDPN* | 1.286902486 | 0.000517078 | 0.005418712 | 1 |
| Obesity | adipocyte | *LAPTM5* | 1.609893167 | 0.000107282 | 0.002021023 | 0.5205304 |
| Obesity | adipocyte | *CAMK2N1* | 0.707168604 | 2.04E-05 | 0.000718391 | 0.099137891 |
| Obesity | adipocyte | *MEGF6* | 0.56663359 | 0.007458292 | 0.03582934 | 1 |
| Obesity | adipocyte | *LRRIQ3* | -0.409213814 | 0.00172822 | 0.012628498 | 1 |
| Obesity | adipocyte | *ZNF496* | 0.993308773 | 0.000499837 | 0.005283683 | 1 |
| Obesity | adipocyte | *TDRD5* | -0.952508674 | 0.005287116 | 0.02801222 | 1 |
| Obesity | adipocyte | *PM20D1* | 0.465120509 | 0.003448242 | 0.020629926 | 1 |
| Obesity | adipocyte | *CLHC1* | 0.20826426 | 0.001677786 | 0.012352987 | 1 |
| Obesity | adipocyte | *RPRD2* | 0.899969298 | 0.009440442 | 0.042061549 | 1 |
| Obesity | adipocyte | *TDRD10* | 1.185796589 | 0.00011346 | 0.00205413 | 0.550506918 |
| Obesity | adipocyte | *ANTXR2* | 1.08441413 | 1.75E-06 | 9.99E-05 | 0.00849305 |
| Obesity | adipocyte | *PBXIP1* | 2.036295719 | 0.000924081 | 0.008049621 | 1 |
| Obesity | adipocyte | *CLDN1* | 0.75042639 | 0.000445783 | 0.004937388 | 1 |

| | | | | | | |
|---|---|---|---|---|---|---|
| Obesity | adipocyte | *LINC01116* | 1.341612508 | 0.000155787 | 0.002454145 | 0.755876767 |
| Obesity | adipocyte | *ARPC2* | 1.080039345 | 0.003410039 | 0.020502489 | 1 |
| Obesity | adipocyte | *AIM2* | 1.667293312 | 0.000813607 | 0.007502305 | 1 |
| Obesity | adipocyte | *WDFY3* | 0.904643477 | 0.007477418 | 0.035885691 | 1 |
| Obesity | adipocyte | *CCDC158* | -0.551702617 | 0.000696582 | 0.006756163 | 1 |
| Obesity | adipocyte | *CPA3* | 2.558113479 | 0.000918095 | 0.008015193 | 1 |
| Obesity | adipocyte | *CLEC3B* | 2.570960938 | 2.25E-06 | 0.000123926 | 0.010905521 |
| Obesity | adipocyte | *DNALI1* | -0.668274478 | 4.43E-05 | 0.001186962 | 0.21484013 |
| Obesity | adipocyte | *RPN1* | 1.785726304 | 0.000647914 | 0.006338067 | 1 |
| Obesity | adipocyte | *RFT1* | -0.7854817 | 8.56E-07 | 5.69E-05 | 0.004152839 |
| Obesity | adipocyte | *RNF123* | 1.347402966 | 1.58E-20 | 1.28E-17 | 7.67E-17 |
| Obesity | adipocyte | *MST1R* | 1.22287337 | 3.96E-25 | 9.61E-22 | 1.92E-21 |
| Obesity | adipocyte | *TEX264* | -0.600270832 | 0.003102171 | 0.019075337 | 1 |
| Obesity | adipocyte | *ETNPPL* | 15.10948605 | 0.000244951 | 0.003375956 | 1 |
| Obesity | adipocyte | *HPGD* | 2.014096686 | 0.000471358 | 0.005071018 | 1 |
| Obesity | adipocyte | *TMEM144* | -0.847930499 | 0.00989683 | 0.043783747 | 1 |
| Obesity | adipocyte | *IL15* | 1.09364731 | 1.53E-05 | 0.000604927 | 0.074406082 |
| Obesity | adipocyte | *ARFIP1* | 1.387946744 | 0.000144854 | 0.00232726 | 0.702832418 |
| Obesity | adipocyte | *F2RL2* | 2.801230708 | 3.75E-05 | 0.001063557 | 0.181868191 |
| Obesity | adipocyte | *ERAP1* | -0.797704524 | 0.001311352 | 0.010379574 | 1 |
| Obesity | adipocyte | *DCBLD1* | 0.490669127 | 0.008513263 | 0.038968256 | 1 |
| Obesity | adipocyte | *RPS14* | 1.130091455 | 0.001442906 | 0.01106 | 1 |
| Obesity | adipocyte | *CHMP4C* | 3.472134496 | 6.76E-05 | 0.00153723 | 0.327856139 |
| Obesity | adipocyte | *BRI3* | 1.345488719 | 0.005371801 | 0.028138025 | 1 |
| Obesity | adipocyte | *RAD21* | 1.304551125 | 0.003598222 | 0.021239138 | 1 |
| Obesity | adipocyte | *DCSTAMP* | 6.811126011 | 0.000557981 | 0.005699632 | 1 |
| Obesity | adipocyte | *RPP25L* | -0.771945508 | 8.72E-05 | 0.00179209 | 0.422933336 |
| Obesity | adipocyte | *TMEM65* | 1.447689332 | 0.00037085 | 0.004393015 | 1 |
| Obesity | adipocyte | *ZMAT4* | 4.197136331 | 5.79E-05 | 0.001398486 | 0.28109572 |
| Obesity | adipocyte | *C8orf34* | -0.460316575 | 0.000546812 | 0.00560916 | 1 |
| Obesity | adipocyte | *ANKS6* | 1.065407838 | 8.36E-05 | 0.001740156 | 0.405456302 |
| Obesity | adipocyte | *AQP3* | 5.837425621 | 0.000313561 | 0.003961975 | 1 |
| Obesity | adipocyte | *WRN* | 1.094745161 | 0.00734443 | 0.035528588 | 1 |
| Obesity | adipocyte | *PGM2L1* | 0.473143812 | 0.006492033 | 0.032373429 | 1 |
| Obesity | adipocyte | *SLC16A9* | 2.405023265 | 0.000142769 | 0.002320923 | 0.692717455 |
| Obesity | adipocyte | *GJB2* | 5.99667967 | 6.88E-05 | 0.001537561 | 0.333650706 |
| Obesity | adipocyte | *TSC1* | 0.391942895 | 0.006997985 | 0.034262589 | 1 |
| Obesity | adipocyte | *RET* | 1.854208793 | 0.000220424 | 0.003183029 | 1 |
| Obesity | adipocyte | *VWA2* | 1.146656567 | 0.006605126 | 0.032735516 | 1 |
| Obesity | adipocyte | *SLC39A13* | 2.914285316 | 3.58E-14 | 1.33E-11 | 1.73E-10 |
| Obesity | adipocyte | *RAPSN* | -1.124888613 | 9.65E-18 | 5.20E-15 | 4.68E-14 |
| Obesity | adipocyte | *TCP11L2* | 1.464892334 | 0.003663244 | 0.021570459 | 1 |
| Obesity | adipocyte | *IKBIP* | 1.219873114 | 0.001616015 | 0.012042262 | 1 |
| Obesity | adipocyte | *RPL27A* | 1.485180008 | 0.000614398 | 0.006069645 | 1 |
| Obesity | adipocyte | *PRKCB* | 0.835590884 | 7.94E-05 | 0.001711405 | 0.385066027 |
| Obesity | adipocyte | *CCDC68* | 0.704613878 | 5.77E-06 | 0.000265225 | 0.028011869 |
| Obesity | adipocyte | *ZSCAN21* | -0.386017781 | 0.001392871 | 0.010744369 | 1 |
| Obesity | adipocyte | *SEC11C* | 2.186150536 | 9.52E-05 | 0.001887835 | 0.462089595 |
| Obesity | adipocyte | *STAT6* | 1.14759059 | 0.005846809 | 0.029956405 | 1 |
| Obesity | adipocyte | *PIP4K2C* | 2.581128397 | 1.23E-08 | 1.36E-06 | 5.98E-05 |

| | | | | | | |
|---|---|---|---|---|---|---|
| Obesity | adipocyte | *CCNDBP1* | 1.052571586 | 0.003418281 | 0.020526611 | 1 |
| Obesity | adipocyte | *ISLR2* | 0.592247995 | 0.00274457 | 0.017591354 | 1 |
| Obesity | adipocyte | *GPRC5B* | 0.768512981 | 0.000451239 | 0.004942243 | 1 |
| Obesity | adipocyte | *LOXHD1* | 0.931600395 | 1.69E-05 | 0.000641362 | 0.08195999 |
| Obesity | adipocyte | *VKORC1* | 2.332050843 | 0.000137798 | 0.002289715 | 0.668596767 |
| Obesity | adipocyte | *SMG8* | -0.546149967 | 0.002729298 | 0.017516607 | 1 |
| Obesity | adipocyte | *SERTAD3* | 3.011594993 | 0.00286487 | 0.018075875 | 1 |
| Obesity | adipocyte | *FAM57A* | 0.398655221 | 0.005662455 | 0.029217935 | 1 |
| Obesity | adipocyte | *SRR* | 0.311615352 | 2.28E-05 | 0.000774645 | 0.110774277 |
| Obesity | adipocyte | *KLK6* | 8.653046184 | 8.03E-05 | 0.001713183 | 0.389832102 |
| Obesity | adipocyte | *ZNF83* | 0.937517441 | 0.003361859 | 0.020263032 | 1 |
| Obesity | adipocyte | *NUDT8* | -0.311129695 | 0.001600505 | 0.011984025 | 1 |
| Obesity | adipocyte | *ZNF598* | 0.757588886 | 0.005364743 | 0.028138025 | 1 |
| Obesity | adipocyte | *ATG16L2* | 1.696509536 | 0.00036546 | 0.004367521 | 1 |
| Obesity | adipocyte | *KIAA1586* | -0.66758455 | 0.002504407 | 0.016420787 | 1 |
| Obesity | adipocyte | *NT5DC2* | -0.507311331 | 4.90E-05 | 0.001263461 | 0.237530663 |
| Obesity | adipocyte | *COA6* | 2.444798608 | 0.007641043 | 0.036276264 | 1 |
| Obesity | adipocyte | *PDHB* | 1.192101964 | 0.002965043 | 0.018467765 | 1 |
| Obesity | adipocyte | *FAM107A* | 1.00036127 | 0.004500979 | 0.024844992 | 1 |
| Obesity | adipocyte | *KLHL30* | -1.189482576 | 0.000590641 | 0.005908842 | 1 |
| Obesity | adipocyte | *CCDC110* | 3.360300987 | 0.000272431 | 0.003601736 | 1 |
| Obesity | adipocyte | *FNTA* | 1.065611726 | 3.78E-06 | 0.000193007 | 0.018335638 |
| Obesity | adipocyte | *ADAM9* | 0.92814722 | 0.001777435 | 0.012799963 | 1 |
| Obesity | adipocyte | *NDUFS5* | 1.140213582 | 0.00097487 | 0.008371803 | 1 |
| Obesity | adipocyte | *DNAJC21* | 1.591662035 | 0.007158371 | 0.034836927 | 1 |
| Obesity | adipocyte | *NPNT* | 0.448845287 | 7.81E-05 | 0.00169174 | 0.378949735 |
| Obesity | adipocyte | *CPLX1* | 0.376901045 | 0.00047316 | 0.005079142 | 1 |
| Obesity | adipocyte | *MFSD7* | 3.836223591 | 0.000523723 | 0.005455908 | 1 |
| Obesity | adipocyte | *MAP2K1* | 1.315497826 | 0.007720833 | 0.036452195 | 1 |
| Obesity | adipocyte | *KLK7* | 2.067201723 | 3.56E-05 | 0.001028334 | 0.17276018 |
| Obesity | adipocyte | *C8orf46* | 0.122412653 | 0.002898288 | 0.018249911 | 1 |
| Obesity | adipocyte | *THBS3* | 1.241998932 | 0.002798479 | 0.01779583 | 1 |
| Obesity | adipocyte | *GPRIN1* | 1.09555154 | 0.000381559 | 0.00448262 | 1 |
| Obesity | adipocyte | *PGM2* | 1.05238948 | 0.000412579 | 0.004688129 | 1 |
| Obesity | adipocyte | *CLIC3* | 2.255811475 | 5.53E-05 | 0.001363471 | 0.268413757 |
| Obesity | adipocyte | *INO80E* | -0.971827544 | 3.94E-15 | 1.74E-12 | 1.91E-11 |
| Obesity | adipocyte | *RGPD8* | -0.116516041 | 0.003539284 | 0.020967771 | 1 |
| Obesity | adipocyte | *CSGALNACT2* | 1.577586198 | 0.000544186 | 0.005605925 | 1 |
| Obesity | adipocyte | *CALML6* | -7.352413391 | 2.46E-05 | 0.000812964 | 0.119365559 |
| Obesity | adipocyte | *OTUD3* | 0.491188869 | 0.00524108 | 0.027914074 | 1 |
| Obesity | adipocyte | *MYO7B* | 0.51036666 | 0.001783658 | 0.012801304 | 1 |
| Obesity | adipocyte | *CNGB3* | 0.924663723 | 3.14E-05 | 0.000929298 | 0.152404913 |
| Obesity | adipocyte | *ADORA2B* | -0.343348568 | 3.53E-07 | 2.60E-05 | 0.001713204 |
| Obesity | adipocyte | *HSD17B13* | 2.510899901 | 0.000465107 | 0.005051097 | 1 |
| Obesity | adipocyte | *SERPINB9* | 0.792754556 | 0.00954965 | 0.042415284 | 1 |
| Obesity | adipocyte | *TTLL6* | -2.435469259 | 7.11E-13 | 1.92E-10 | 3.45E-09 |
| Obesity | adipocyte | *TSNARE1* | 1.060558748 | 1.78E-06 | 0.000100207 | 0.008653014 |
| Obesity | adipocyte | *FPR1* | 0.825221074 | 0.007538839 | 0.036057403 | 1 |
| Obesity | adipocyte | *SOX7* | 1.040182589 | 0.011336305 | 0.048418797 | 1 |
| Obesity | adipocyte | *TRMT61B* | 0.930764057 | 3.04E-05 | 0.000922495 | 0.147288174 |

| | | | | | | |
|---|---|---|---|---|---|---|
| Obesity | adipocyte | *GIMAP8* | 0.943682552 | 0.000222494 | 0.00318448 | 1 |
| Obesity | adipocyte | *PDE7B* | 0.801250773 | 0.002316808 | 0.01561271 | 1 |
| Obesity | adipocyte | *MCC* | 1.171263286 | 0.00495178 | 0.02675505 | 1 |
| Obesity | adipocyte | *PIK3CD* | 1.456261796 | 3.59E-07 | 2.60E-05 | 0.001744084 |
| Obesity | adipocyte | *RGS19* | 0.259266635 | 0.000399621 | 0.004616569 | 1 |
| Obesity | adipocyte | *CAMTA1* | 1.392121231 | 2.86E-05 | 0.000883891 | 0.138727585 |
| Obesity | adipocyte | *FBXW10* | -1.489959898 | 0.000446726 | 0.004937388 | 1 |
| Obesity | adipocyte | *ZNF554* | -1.224879973 | 0.007108867 | 0.03466555 | 1 |
| Obesity | adipocyte | *SERF1A* | 5.54212801 | 0.008107676 | 0.037572536 | 1 |
| Obesity | adipocyte | *EIF2AK3* | 0.922829983 | 0.007985046 | 0.037253311 | 1 |
| Obesity | adipocyte | *MALT1* | 1.472082382 | 0.001360171 | 0.01064139 | 1 |
| Obesity | adipocyte | *MBOAT1* | 1.244933056 | 0.00552582 | 0.02864453 | 1 |
| Obesity | adipocyte | *C1QTNF4* | -0.537743914 | 2.58E-15 | 1.25E-12 | 1.25E-11 |
| Obesity | adipocyte | *SUCLG2* | 1.033603396 | 0.011558211 | 0.049174752 | 1 |
| Obesity | adipocyte | *ARNT2* | 0.362103261 | 0.00161821 | 0.012042262 | 1 |
| Obesity | adipocyte | *FIBP* | 0.849950374 | 0.002995708 | 0.018610981 | 1 |
| Obesity | adipocyte | *CTSW* | -0.129808886 | 3.88E-08 | 3.69E-06 | 0.000188283 |
| Obesity | adipocyte | *EFEMP2* | 1.840440996 | 2.24E-07 | 1.70E-05 | 0.001086124 |
| Obesity | adipocyte | *SLFN11* | 1.105155887 | 0.000294999 | 0.003806745 | 1 |
| Obesity | adipocyte | *FUT10* | -0.333822159 | 0.000336268 | 0.004157514 | 1 |
| Obesity | adipocyte | *SNX32* | 0.328460459 | 1.94E-05 | 0.000691702 | 0.094071511 |
| Obesity | adipocyte | *NADSYN1* | 0.720842827 | 0.004124242 | 0.02340447 | 1 |
| Obesity | adipocyte | *MRGPRF* | 0.935400231 | 2.78E-06 | 0.000148173 | 0.01348376 |
| Obesity | adipocyte | *GXYLT2* | 1.05109922 | 0.010384683 | 0.045311583 | 1 |
| Obesity | adipocyte | *NOC3L* | 0.93323522 | 0.008873413 | 0.040162126 | 1 |
| Obesity | adipocyte | *MMRN2* | 0.771718955 | 0.000896343 | 0.007949496 | 1 |
| Obesity | adipocyte | *NDNF* | 0.901191874 | 0.000897841 | 0.007949496 | 1 |
| Obesity | adipocyte | *MST1* | -0.181992367 | 0.003976626 | 0.022779915 | 1 |
| Obesity | adipocyte | *CD7* | 1.60764274 | 0.000345328 | 0.004240847 | 1 |
| Obesity | adipocyte | *GOLIM4* | 1.276395407 | 0.002009631 | 0.013969524 | 1 |
| Obesity | adipocyte | *HOXB2* | 0.93627978 | 0.000134124 | 0.002251799 | 0.650769856 |
| Obesity | adipocyte | *SLC25A30* | 1.689016268 | 0.000572683 | 0.005813088 | 1 |
| Obesity | adipocyte | *GLIS1* | -0.467722502 | 0.001745709 | 0.012717987 | 1 |
| Obesity | adipocyte | *ANKRD36C* | 1.249509834 | 0.000958314 | 0.008246507 | 1 |
| Obesity | adipocyte | *SH3PXD2B* | 0.885906244 | 0.001970962 | 0.01375987 | 1 |
| Obesity | adipocyte | *SRP72* | 0.578712634 | 0.000534398 | 0.005516809 | 1 |
| Obesity | adipocyte | *CEP135* | 1.559155239 | 0.000111846 | 0.00205413 | 0.54267511 |
| Obesity | adipocyte | *SEZ6L2* | 0.772639502 | 2.71E-08 | 2.68E-06 | 0.000131253 |
| Obesity | adipocyte | *KCTD13* | 1.106877558 | 8.14E-09 | 9.40E-07 | 3.95E-05 |
| Obesity | adipocyte | *FBXW8* | 0.961193676 | 0.010758537 | 0.046524441 | 1 |
| Obesity | adipocyte | *CADM2* | 0.90354149 | 7.24E-11 | 1.21E-08 | 3.51E-07 |
| Obesity | adipocyte | *BANF1* | -1.234931879 | 0.00030046 | 0.003856703 | 1 |
| Obesity | adipocyte | *EIF1AD* | 1.30143451 | 0.0059372 | 0.030387442 | 1 |
| Obesity | adipocyte | *CLTB* | 1.714316295 | 0.004804602 | 0.026105186 | 1 |
| Obesity | adipocyte | *MCTP1* | 1.156336431 | 0.00010755 | 0.002021023 | 0.521831795 |
| Obesity | adipocyte | *RPS6KB2* | 1.154033654 | 0.000574662 | 0.005820998 | 1 |
| Obesity | adipocyte | *A2M* | 1.202005901 | 0.000243719 | 0.003369018 | 1 |
| Obesity | adipocyte | *DOK7* | 2.950396014 | 0.006701713 | 0.033112741 | 1 |
| Obesity | adipocyte | *ORAI3* | 1.35586414 | 7.20E-14 | 2.33E-11 | 3.49E-10 |
| Obesity | adipocyte | *B3GALT6* | 5.120961578 | 0.000143503 | 0.002320923 | 0.696277024 |

| | | | | | | |
|---|---|---|---|---|---|---|
| Obesity | adipocyte | *MYPOP* | 1.246558313 | 0.006688011 | 0.033078725 | 1 |
| Obesity | adipocyte | *CIDEA* | -0.39062331 | 0.006885972 | 0.033850798 | 1 |
| Obesity | adipocyte | *DNAJC30* | -0.969353716 | 0.002343466 | 0.015742859 | 1 |
| Obesity | adipocyte | *SPRYD4* | -0.434335196 | 0.000388433 | 0.004550901 | 1 |
| Obesity | adipocyte | *RMDN1* | 0.707615725 | 0.000347901 | 0.004251932 | 1 |
| Obesity | adipocyte | *BOK* | -0.560610092 | 0.005123663 | 0.027439309 | 1 |
| Obesity | adipocyte | *RUFY1* | 1.078369374 | 1.80E-06 | 0.000100207 | 0.008717972 |
| Obesity | adipocyte | *ANKLE2* | 1.363444325 | 0.001345033 | 0.010560278 | 1 |
| Obesity | adipocyte | *C8G* | -17.30586368 | 0.003179699 | 0.019381783 | 1 |
| Obesity | adipocyte | *LY6H* | 0.711232523 | 0.0009974 | 0.008475282 | 1 |
| Obesity | adipocyte | *TMEM80* | -0.655229501 | 0.011285842 | 0.04824573 | 1 |
| Obesity | adipocyte | *DSCAML1* | 1.099262591 | 0.000152208 | 0.002413444 | 0.738513948 |
| Obesity | adipocyte | *MAN1B1* | 1.852259423 | 0.005954022 | 0.030441428 | 1 |
| Obesity | adipocyte | *CD151* | 1.296023786 | 2.42E-05 | 0.000808395 | 0.117217266 |
| Obesity | adipocyte | *SLC35G5* | 17.42178235 | 0.000238298 | 0.003319488 | 1 |
| Obesity | adipocyte | *FAM87B* | -4.052314298 | 0.002909278 | 0.018261222 | 1 |
| Obesity | adipocyte | *RPS27* | 1.492372835 | 0.000861163 | 0.007737707 | 1 |
| Obesity | adipocyte | *ADAMTSL1* | 0.651443118 | 0.006291424 | 0.031787934 | 1 |
| Obesity | adipocyte | *SPINK6* | -0.991647746 | 0.001433222 | 0.011020593 | 1 |
| Obesity | adipocyte | *VN1R1* | -0.779103405 | 0.011563972 | 0.049174752 | 1 |
| Obesity | adipocyte | *ERN1* | 1.227909782 | 0.004154931 | 0.023441543 | 1 |
| Obesity | adipocyte | *NSUN3* | 1.609390726 | 0.000336748 | 0.004157514 | 1 |
| Obesity | adipocyte | *RPP25* | -0.353251717 | 0.000395805 | 0.004594366 | 1 |
| Obesity | adipocyte | *STX19* | 1.128719308 | 2.27E-08 | 2.33E-06 | 0.00010994 |
| Obesity | adipocyte | *MPI* | 0.718159111 | 0.000102605 | 0.001975563 | 0.497841884 |
| Obesity | adipocyte | *TRIM73* | -2.313448996 | 2.72E-14 | 1.10E-11 | 1.32E-10 |
| Obesity | adipocyte | *ZNF852* | -0.958979804 | 1.78E-05 | 0.000659132 | 0.086275781 |
| Obesity | adipocyte | *C17orf62* | 2.414735804 | 0.00034612 | 0.004240847 | 1 |
| Obesity | adipocyte | *GAK* | 1.210708983 | 0.010996592 | 0.04742708 | 1 |
| Obesity | adipocyte | *TUFM* | 1.348953534 | 1.27E-11 | 2.68E-09 | 6.17E-08 |
| Obesity | adipocyte | *C3orf38* | 1.053614441 | 1.01E-09 | 1.32E-07 | 4.88E-06 |
| Obesity | adipocyte | *CCDC89* | 3.199264902 | 0.000314439 | 0.003962754 | 1 |
| Obesity | adipocyte | *PACS2* | 1.357116388 | 0.010129131 | 0.044396156 | 1 |
| Obesity | adipocyte | *RPRML* | 1.952257438 | 7.59E-06 | 0.00034411 | 0.036819734 |
| Obesity | adipocyte | *SERTAD2* | 1.263219881 | 0.000918476 | 0.008015193 | 1 |
| Obesity | adipocyte | *GIPC3* | 13.45153203 | 2.01E-05 | 0.000710692 | 0.097364739 |
| Obesity | adipocyte | *MCFD2* | 0.890580649 | 0.007361825 | 0.035552739 | 1 |
| Obesity | adipocyte | *MB21D2* | 1.086367575 | 0.009170466 | 0.041207992 | 1 |
| Obesity | adipocyte | *SLC36A4* | 1.087143843 | 0.003672421 | 0.021581529 | 1 |
| Obesity | adipocyte | *ZDHHC20* | 1.34640906 | 0.003258392 | 0.019762148 | 1 |
| Obesity | adipocyte | *CMTR2* | 1.050013768 | 0.002345855 | 0.015742859 | 1 |
| Obesity | adipocyte | *MRPL14* | 0.862370152 | 0.006028368 | 0.030659999 | 1 |
| Obesity | adipocyte | *NQO1* | -0.58227887 | 0.004467595 | 0.024764727 | 1 |
| Obesity | adipocyte | *MUC16* | 1.386538957 | 3.47E-05 | 0.001013996 | 0.168323329 |
| Obesity | adipocyte | *SLC25A41* | 6.711563638 | 1.58E-05 | 0.00061726 | 0.076540216 |
| Obesity | adipocyte | *ZNF322* | -0.628563701 | 0.006388305 | 0.032055192 | 1 |
| Obesity | adipocyte | *DDX60L* | 1.268543202 | 0.000110118 | 0.002047091 | 0.534290687 |
| Obesity | adipocyte | *SGSH* | 2.54722709 | 0.007864753 | 0.036905011 | 1 |
| Obesity | adipocyte | *ZFP41* | 0.607903953 | 0.003265328 | 0.019779491 | 1 |
| Obesity | adipocyte | *AMIGO1* | 0.360445656 | 0.009203402 | 0.041207992 | 1 |

| | | | | | | |
|---|---|---|---|---|---|---|
| Obesity | adipocyte | *TMEM252* | 98.33476938 | 0.000126657 | 0.002194777 | 0.614537507 |
| Obesity | adipocyte | *RFX7* | 1.17994699 | 0.005762797 | 0.02965121 | 1 |
| Obesity | adipocyte | *TDRKH* | -0.405849767 | 0.010322302 | 0.045079938 | 1 |
| Obesity | adipocyte | *HHIPL1* | 0.829120582 | 0.00307806 | 0.018990597 | 1 |
| Obesity | adipocyte | *FAM153B* | 1.025668538 | 0.000356937 | 0.004308102 | 1 |
| Obesity | adipocyte | *KCNA4* | 2.516012299 | 0.000221792 | 0.003183834 | 1 |
| Obesity | adipocyte | *CLN8* | 0.318654932 | 0.001381804 | 0.01072231 | 1 |
| Obesity | adipocyte | *NXPH4* | -2.544655771 | 0.00095858 | 0.008246507 | 1 |
| Obesity | adipocyte | *ORAI1* | 0.648805596 | 0.004993127 | 0.026948443 | 1 |
| Obesity | adipocyte | *FES* | 0.578640071 | 0.00846582 | 0.038839669 | 1 |
| Obesity | adipocyte | *CAV3* | -0.420143992 | 0.003472143 | 0.020696361 | 1 |
| Obesity | adipocyte | *PPP1R27* | -2.217338426 | 0.000174714 | 0.00268264 | 0.847714315 |
| Obesity | adipocyte | *PLCXD3* | 1.574491179 | 0.000563387 | 0.005730724 | 1 |
| Obesity | adipocyte | *COL18A1* | 1.145523406 | 3.96E-07 | 2.83E-05 | 0.001921528 |
| Obesity | adipocyte | *C12orf60* | 2.012313134 | 0.001610041 | 0.012019112 | 1 |
| Obesity | adipocyte | *LYSMD4* | 0.770990684 | 0.001498973 | 0.011369073 | 1 |
| Obesity | adipocyte | *CSMD1* | 1.979782604 | 0.003894058 | 0.022386222 | 1 |
| Obesity | adipocyte | *FANCF* | 2.857505487 | 0.002063828 | 0.014163642 | 1 |
| Obesity | adipocyte | *PLGLB1* | 0.367848171 | 0.002050311 | 0.014130834 | 1 |
| Obesity | adipocyte | *LRIT3* | -6.992094266 | 0.004265527 | 0.023926403 | 1 |
| Obesity | adipocyte | *PSMG1* | 0.834992571 | 4.85E-05 | 0.001259605 | 0.2355461 |
| Obesity | adipocyte | *UPP1* | 1.084032891 | 9.21E-06 | 0.000406032 | 0.044663533 |
| Obesity | adipocyte | *ACSM2A* | -6.595205639 | 0.000513505 | 0.005392915 | 1 |
| Obesity | adipocyte | *NPIPA5* | 0.126995645 | 0.005267743 | 0.02796399 | 1 |
| Obesity | adipocyte | *EMILIN3* | 0.85166881 | 0.000311583 | 0.003950409 | 1 |
| Obesity | adipocyte | *DENND5A* | 1.160107594 | 0.005161594 | 0.027581558 | 1 |
| Obesity | adipocyte | *CRELD2* | 1.04994198 | 0.001584463 | 0.011900502 | 1 |
| Obesity | adipocyte | *PGP* | -0.792349894 | 0.011014169 | 0.047428125 | 1 |
| Obesity | adipocyte | *ACOT1* | 0.29172247 | 0.002815582 | 0.017845477 | 1 |
| Obesity | adipocyte | *TMEM255B* | 1.2057576 | 0.00203722 | 0.014090491 | 1 |
| Obesity | adipocyte | *SNN* | 3.092034927 | 0.000169092 | 0.002621197 | 0.820434538 |
| Obesity | adipocyte | *DRD1* | 28.82946033 | 0.000360701 | 0.004342737 | 1 |
| Obesity | adipocyte | *NUTM2A* | -0.604428314 | 4.68E-06 | 0.000223112 | 0.022703291 |
| Obesity | adipocyte | *ZFP90* | 1.470064215 | 0.007625212 | 0.036236563 | 1 |
| Obesity | adipocyte | *MAFF* | -0.346121677 | 0.002581249 | 0.016833629 | 1 |
| Obesity | adipocyte | *SIGIRR* | 0.786790864 | 0.009899169 | 0.043783747 | 1 |
| Obesity | adipocyte | *IFITM2* | 1.508736758 | 0.00052625 | 0.005455908 | 1 |
| Obesity | adipocyte | *GP1BA* | 0.657669252 | 2.37E-05 | 0.000796984 | 0.114765658 |
| Obesity | adipocyte | *PCGF3* | 0.811420995 | 0.002643644 | 0.017125445 | 1 |
| Obesity | adipocyte | *LINC00839* | 1.882023335 | 0.008740745 | 0.039784327 | 1 |
| Obesity | adipocyte | *C15orf41* | 1.161391349 | 3.94E-05 | 0.001112769 | 0.191396347 |
| Obesity | adipocyte | *POLR1D* | 1.082450465 | 0.003163317 | 0.019330498 | 1 |
| Obesity | adipocyte | *FFAR4* | -0.622977296 | 0.00179606 | 0.012834292 | 1 |
| Obesity | adipocyte | *RPS23* | 0.745634659 | 0.004909873 | 0.026587837 | 1 |
| Obesity | adipocyte | *BTN3A2* | -0.268073528 | 5.14E-10 | 6.93E-08 | 2.49E-06 |
| Obesity | adipocyte | *MAPT* | -0.586509115 | 0.006507557 | 0.03241752 | 1 |
| Obesity | adipocyte | *C1QL4* | 10.70320924 | 0.000236781 | 0.003319488 | 1 |
| Obesity | adipocyte | *TMPRSS6* | 12.37032432 | 0.001610145 | 0.012019112 | 1 |
| Obesity | adipocyte | *CYP4A11* | 0.959254187 | 0.00801633 | 0.03732363 | 1 |
| Obesity | adipocyte | *ANGPTL5* | -0.41733819 | 0.003765991 | 0.021962246 | 1 |

| | | | | | | |
|---|---|---|---|---|---|---|
| Obesity | adipocyte | *GJA4* | 2.609290078 | 2.84E-05 | 0.000883891 | 0.137583822 |
| Obesity | adipocyte | *SBK2* | 7.269880258 | 0.00010603 | 0.002009597 | 0.514456922 |
| Obesity | adipocyte | *GFRAL* | 1.520347842 | 0.000302508 | 0.003872746 | 1 |
| Obesity | adipocyte | *ZSCAN23* | 0.218480548 | 0.00042346 | 0.004789346 | 1 |
| Obesity | adipocyte | *AGRN* | 2.185166672 | 8.05E-05 | 0.001713183 | 0.390605675 |
| Obesity | adipocyte | *NCR3LG1* | 1.877774116 | 0.00011796 | 0.002088842 | 0.572342744 |
| Obesity | adipocyte | *RUFY4* | -0.860089806 | 0.001778066 | 0.012799963 | 1 |
| Obesity | adipocyte | *CENPP* | 0.514072884 | 3.39E-06 | 0.000175235 | 0.016472059 |
| Obesity | adipocyte | *PLSCR1* | 0.958241436 | 0.000434966 | 0.00485162 | 1 |
| Obesity | adipocyte | *PDCD1* | 1.693394684 | 0.004812533 | 0.026119029 | 1 |
| Obesity | adipocyte | *C22orf34* | 1.496721977 | 1.66E-05 | 0.000637628 | 0.080341133 |
| Obesity | adipocyte | *GOLGA8M* | -0.562178078 | 0.001484604 | 0.01129283 | 1 |
| Obesity | adipocyte | *FZD9* | -5.05230149 | 0.001091591 | 0.009003769 | 1 |
| Obesity | adipocyte | *SKOR1* | 0.987970351 | 0.000234924 | 0.003304092 | 1 |
| Obesity | adipocyte | *MTF1* | 1.385102642 | 0.001759882 | 0.012782857 | 1 |
| Obesity | adipocyte | *ENTPD8* | 54.02351564 | 0.004402215 | 0.024523016 | 1 |
| Obesity | adipocyte | *RPL14* | 1.434509235 | 0.000812896 | 0.007502305 | 1 |
| Obesity | adipocyte | *FBF1* | 0.725162508 | 0.000264734 | 0.003540206 | 1 |
| Obesity | adipocyte | *ASTL* | -2.446114701 | 0.003855534 | 0.022279441 | 1 |
| Obesity | adipocyte | *FAM196A* | 1.437038386 | 0.001321066 | 0.010422457 | 1 |
| Obesity | adipocyte | *UTS2B* | -0.473459175 | 0.005141583 | 0.027504918 | 1 |
| Obesity | adipocyte | *PLAC9* | 0.772627256 | 0.008498019 | 0.038935211 | 1 |
| Obesity | adipocyte | *TSPYL1* | 1.464700321 | 0.005568872 | 0.028797078 | 1 |
| Obesity | adipocyte | *ZKSCAN3* | -0.901263649 | 2.87E-08 | 2.79E-06 | 0.000139347 |
| Obesity | adipocyte | *KIAA0408* | -0.221158246 | 0.005289774 | 0.02801222 | 1 |
| Obesity | adipocyte | *SEMA4A* | 1.200005889 | 0.000250476 | 0.003413789 | 1 |
| Obesity | adipocyte | *PPIA* | 0.783517758 | 0.003734884 | 0.021833321 | 1 |
| Obesity | adipocyte | *CD55* | 1.313726901 | 0.000212203 | 0.00309192 | 1 |
| Obesity | adipocyte | *CPNE4* | 0.549357744 | 0.001141269 | 0.009292008 | 1 |
| Obesity | adipocyte | *ZNF605* | 0.599154412 | 0.000455996 | 0.004983088 | 1 |
| Obesity | adipocyte | *SULT1A1* | -0.906647071 | 0.004160868 | 0.023447771 | 1 |
| Obesity | adipocyte | *TPK1* | 1.221684826 | 0.000262538 | 0.003528633 | 1 |
| Obesity | adipocyte | *MYO18A* | 0.696874756 | 0.011763869 | 0.049850037 | 1 |
| Obesity | adipocyte | *CACNA1H* | 2.129320302 | 1.28E-07 | 1.07E-05 | 0.000622197 |
| Obesity | adipocyte | *SULF2* | 0.402947864 | 0.006597455 | 0.032735516 | 1 |
| Obesity | adipocyte | *ZFP62* | 1.401796141 | 0.000411776 | 0.004688129 | 1 |
| Obesity | adipocyte | *TOMM7* | 1.444721971 | 0.00281732 | 0.017845477 | 1 |
| Obesity | adipocyte | *ZNF33B* | 0.417302127 | 0.011532281 | 0.0491261 | 1 |
| Obesity | adipocyte | *ZNF239* | 0.822129756 | 0.008469161 | 0.038839669 | 1 |
| Obesity | adipocyte | *FAM3C* | 0.83265613 | 0.001186161 | 0.009592089 | 1 |
| Obesity | adipocyte | *SLC39A10* | 1.417644485 | 0.00095544 | 0.008246507 | 1 |
| Obesity | adipocyte | *ZNF138* | 0.84385011 | 0.006365539 | 0.03200876 | 1 |
| Obesity | adipocyte | *GMFB* | 1.607772527 | 0.00906565 | 0.040879677 | 1 |
| Obesity | adipocyte | *SLC25A29* | 0.440255065 | 0.009554803 | 0.042415284 | 1 |
| Obesity | adipocyte | *SND1* | 1.177505055 | 0.000893467 | 0.007939747 | 1 |
| Obesity | adipocyte | *SULT1A2* | 2.330306454 | 2.50E-30 | 1.21E-26 | 1.21E-26 |
| Obesity | adipocyte | *ZNF165* | -0.370744676 | 0.000138694 | 0.002296738 | 0.672944371 |
| Obesity | adipocyte | *MRPL21* | -0.253905236 | 0.000404971 | 0.004656203 | 1 |
| Obesity | adipocyte | *SLC2A10* | 0.775657933 | 0.009417973 | 0.042000003 | 1 |
| Obesity | adipocyte | *DNAH10* | 1.545952461 | 2.39E-13 | 7.24E-11 | 1.16E-09 |

| | | | | | | |
|---|---|---|---|---|---|---|
| Obesity | adipocyte | *FAM114A1* | 1.506097607 | 0.001044462 | 0.008722425 | 1 |
| Obesity | adipocyte | *RPS26* | -0.308811314 | 0.000303591 | 0.003876376 | 1 |
| Obesity | adipocyte | *PSAP* | 0.813599895 | 0.003292847 | 0.019896503 | 1 |
| Obesity | adipocyte | *ZNF780A* | -0.187935361 | 5.85E-05 | 0.001404312 | 0.283671031 |
| Obesity | adipocyte | *SGTB* | 0.704691678 | 0.009149704 | 0.041182155 | 1 |
| Obesity | adipocyte | *MPZL1* | 1.184188451 | 0.002365966 | 0.015812215 | 1 |
| Obesity | adipocyte | *DLGAP2* | 0.771504873 | 0.000293188 | 0.00379346 | 1 |
| Obesity | adipocyte | *ADH4* | -0.350737127 | 0.000443709 | 0.004937388 | 1 |
| Obesity | adipocyte | *SOWAHC* | 2.245538276 | 3.27E-06 | 0.000170739 | 0.015878738 |
| Obesity | adipocyte | *MIER1* | 1.024263294 | 0.003301403 | 0.019923393 | 1 |
| Obesity | adipocyte | *HSD17B11* | 1.620264761 | 0.000131595 | 0.002232518 | 0.638500073 |
| Obesity | adipocyte | *DDX42* | 1.01912677 | 0.011234336 | 0.048152824 | 1 |
| Obesity | adipocyte | *SH3BGRL2* | 0.355302973 | 0.002016493 | 0.013988138 | 1 |
| Obesity | adipocyte | *PAPSS2* | 1.259467864 | 0.000486721 | 0.005178884 | 1 |
| Obesity | adipocyte | *ANKRD13B* | 2.915236226 | 0.000241861 | 0.00335289 | 1 |
| Obesity | adipocyte | *PPP1R14C* | 7.797344447 | 0.000521036 | 0.005448424 | 1 |
| Obesity | adipocyte | *RPL10A* | 1.36340664 | 0.001073146 | 0.008885503 | 1 |
| Obesity | adipocyte | *COLGALT2* | 0.791822554 | 0.003012639 | 0.018668356 | 1 |
| Obesity | adipocyte | *RCSD1* | 1.34743103 | 6.06E-05 | 0.001441243 | 0.294013636 |
| Obesity | adipocyte | *SHISA4* | -4.333790074 | 2.97E-09 | 3.70E-07 | 1.44E-05 |
| Obesity | adipocyte | *C6orf163* | -0.263692199 | 0.004184045 | 0.02355103 | 1 |
| Obesity | adipocyte | *PCMTD2* | 1.364394099 | 0.004471125 | 0.024764727 | 1 |
| Obesity | adipocyte | *SAMD13* | 1.65006513 | 0.000749811 | 0.007087344 | 1 |
| Obesity | adipocyte | *IFIT1B* | 7.279172467 | 0.000927349 | 0.008063615 | 1 |
| Obesity | adipocyte | *LIPN* | 4.335923298 | 5.57E-05 | 0.001363471 | 0.27030304 |
| Obesity | adipocyte | *NOTCH4* | -0.080339105 | 1.43E-08 | 1.51E-06 | 6.92E-05 |
| Obesity | adipocyte | *SKIV2L* | -0.34222765 | 1.06E-06 | 6.57E-05 | 0.005121501 |
| Obesity | adipocyte | *GPANK1* | -1.180114382 | 2.97E-11 | 5.76E-09 | 1.44E-07 |
| Obesity | adipocyte | *LST1* | 0.439121944 | 0.0038731 | 0.022345163 | 1 |
| Obesity | adipocyte | *ZNF814* | 0.902627859 | 0.006401789 | 0.032055192 | 1 |
| Obesity | adipocyte | *HLA-C* | 0.27089984 | 0.000708402 | 0.006819773 | 1 |
| Obesity | adipocyte | *POU5F1* | 0.124820621 | 7.38E-05 | 0.001612358 | 0.357943403 |
| Obesity | adipocyte | *CCHCR1* | 0.257203937 | 2.86E-06 | 0.000150768 | 0.013870619 |
| Obesity | adipocyte | *ABCF1* | 0.687554893 | 0.000290779 | 0.003772356 | 1 |
| Obesity | adipocyte | *HLA-E* | 0.665802975 | 9.59E-05 | 0.001887835 | 0.465381289 |
| Obesity | adipocyte | *RNF39* | -0.765382719 | 1.77E-05 | 0.000659132 | 0.085801836 |
| Obesity | adipocyte | *ZFP57* | 0.309446059 | 3.35E-09 | 3.96E-07 | 1.62E-05 |
| Obesity | adipocyte | *FAM196B* | 0.88076515 | 0.001111625 | 0.009141703 | 1 |
| Obesity | adipocyte | *DCTN1* | 2.149368258 | 0.001586128 | 0.011900502 | 1 |
| Obesity | adipocyte | *TRIM13* | 0.907048839 | 0.002479322 | 0.016300364 | 1 |
| Obesity | adipocyte | *SPIRE2* | -0.254800195 | 0.01168355 | 0.049596312 | 1 |
| Obesity | adipocyte | *SLC35B4* | 1.907268364 | 0.001310823 | 0.010379574 | 1 |
| Obesity | adipocyte | *TECPR1* | 1.511896884 | 0.004289656 | 0.024029152 | 1 |
| Obesity | adipocyte | *C15orf59* | 0.456996863 | 0.004540263 | 0.024920085 | 1 |
| Obesity | adipocyte | *CFI* | 0.720905191 | 0.002293223 | 0.015475266 | 1 |
| Obesity | adipocyte | *SMN2* | -0.277533401 | 0.002641927 | 0.017125445 | 1 |
| Obesity | adipocyte | *C17orf107* | 0.541491335 | 0.000149225 | 0.002373907 | 0.724041785 |
| Obesity | adipocyte | *KLRC2* | 4.888846857 | 2.02E-09 | 2.58E-07 | 9.81E-06 |
| Obesity | adipocyte | *C1QTNF9B* | 15.76982123 | 0.000559757 | 0.005705759 | 1 |
| Obesity | adipocyte | *SLC48A1* | 1.678190135 | 0.000127831 | 0.002207242 | 0.620234867 |

| | | | | | | |
|---|---|---|---|---|---|---|
| Obesity | adipocyte | *TAS2R50* | -4.117824135 | 0.005495067 | 0.028515578 | 1 |
| Obesity | adipocyte | *AC084018.1* | -0.343323673 | 4.79E-05 | 0.001259605 | 0.23223487 |
| Obesity | adipocyte | *ASIC3* | 0.4668842 | 0.002795209 | 0.01779583 | 1 |
| Obesity | adipocyte | *GIMAP1* | 0.537705153 | 0.007551991 | 0.036065219 | 1 |
| Obesity | adipocyte | *SUPT4H1* | 3.013208034 | 5.19E-07 | 3.55E-05 | 0.00251848 |
| Obesity | adipocyte | *SRA1* | 0.899320767 | 0.004085552 | 0.023284282 | 1 |
| Obesity | adipocyte | *TMEM110* | 1.330102052 | 1.36E-10 | 2.12E-08 | 6.58E-07 |
| Obesity | adipocyte | *EMP2* | 1.029622065 | 0.005718386 | 0.029453937 | 1 |
| Obesity | adipocyte | *SLC23A3* | 0.339720338 | 0.001136027 | 0.009279468 | 1 |
| Obesity | adipocyte | *IRF9* | 1.866653403 | 0.000104429 | 0.001988073 | 0.506690473 |
| Obesity | adipocyte | *GALT* | 1.233693044 | 0.000205399 | 0.00301086 | 0.996594758 |
| Obesity | adipocyte | *ZNF891* | 1.45063843 | 0.008329115 | 0.038415274 | 1 |
| Obesity | adipocyte | *TSPAN4* | 1.290109776 | 1.52E-05 | 0.000603979 | 0.073546837 |
| Obesity | adipocyte | *PAXIP1-AS2* | 1.615484496 | 0.002786594 | 0.017791679 | 1 |
| Obesity | adipocyte | *LYRM4* | 1.300555968 | 0.010281145 | 0.044940644 | 1 |
| Obesity | adipocyte | *GNAT3* | 2.5316573 | 0.001923265 | 0.013524177 | 1 |
| Obesity | adipocyte | *EVPLL* | -0.791122694 | 0.001719838 | 0.012586203 | 1 |
| Obesity | adipocyte | *AC005082.1* | -1.938113045 | 0.001358501 | 0.01064139 | 1 |
| Obesity | adipocyte | *PAM16* | 1.321823228 | 0.009020893 | 0.040737195 | 1 |
| Obesity | adipocyte | *NBPF1* | 0.405853615 | 0.010046289 | 0.044206998 | 1 |
| Obesity | adipocyte | *RNU6ATAC35I* | -3.772181021 | 3.06E-05 | 0.000922495 | 0.148658245 |
| Obesity | adipocyte | *AC062017.1* | -0.86627554 | 0.008038581 | 0.03732363 | 1 |
| Obesity | adipocyte | *EXOSC6* | -1.90850556 | 2.36E-12 | 5.45E-10 | 1.15E-08 |
| Obesity | adipocyte | *C4B* | 0.787491311 | 5.15E-05 | 0.001315524 | 0.249949491 |
| Obesity | adipocyte | *LINC01117* | 1.403398455 | 6.14E-05 | 0.001450846 | 0.298041948 |
| Obesity | adipocyte | *LINC00240* | 0.255165237 | 0.000697617 | 0.006756163 | 1 |
| Obesity | adipocyte | *LINC00184* | -6.761758621 | 0.000819502 | 0.007502305 | 1 |
| Obesity | adipocyte | *AL049840.1* | -1.223449771 | 0.000266798 | 0.003540206 | 1 |
| Obesity | adipocyte | *CYP4A22-AS1* | 2.806145841 | 0.000881102 | 0.007856593 | 1 |
| Obesity | adipocyte | *SCAMP4* | 1.666199521 | 0.008850011 | 0.040103421 | 1 |
| Obesity | adipocyte | *SLC2A1-AS1* | -0.628858906 | 0.005088836 | 0.027334773 | 1 |
| Obesity | adipocyte | *AC005884.1* | -0.646873132 | 0.009199256 | 0.041207992 | 1 |
| Obesity | adipocyte | *HCG20* | -0.699670371 | 4.31E-07 | 3.03E-05 | 0.002092271 |
| Obesity | adipocyte | *LINC00578* | 1.016074042 | 0.003132569 | 0.019166741 | 1 |
| Obesity | adipocyte | *CASC20* | 1.764528754 | 0.00021038 | 0.003074586 | 1 |
| Obesity | adipocyte | *LINC00332* | -0.362046664 | 0.000987329 | 0.008434014 | 1 |
| Obesity | adipocyte | *RPS18* | 1.41003344 | 2.86E-05 | 0.000883891 | 0.138770946 |
| Obesity | adipocyte | *TAPBP* | 1.095040823 | 1.00E-06 | 6.33E-05 | 0.004871442 |
| Obesity | adipocyte | *TMA7* | 1.122276809 | 0.000143303 | 0.002320923 | 0.695307656 |
| Obesity | adipocyte | *LINC00702* | 2.78712845 | 5.30E-05 | 0.001345953 | 0.257077037 |
| Obesity | adipocyte | *ZSCAN31* | 0.317251353 | 2.31E-08 | 2.33E-06 | 0.00011193 |
| Obesity | adipocyte | *KIAA0040* | 1.019852035 | 0.000585553 | 0.005882199 | 1 |
| Obesity | adipocyte | *LINC00607* | 0.695181745 | 0.000580492 | 0.005855609 | 1 |
| Obesity | adipocyte | *LINC01143* | 4.528699988 | 4.40E-05 | 0.001185399 | 0.213371793 |
| Obesity | adipocyte | *LINC00211* | 1.927845425 | 0.009535748 | 0.042408296 | 1 |
| Obesity | adipocyte | *TXNDC5* | 9.575324052 | 6.38E-05 | 0.001482155 | 0.30977041 |
| Obesity | adipocyte | *TNFSF12* | 0.658093942 | 0.000730597 | 0.006968045 | 1 |
| Obesity | adipocyte | *PSMB9* | 0.413951301 | 4.56E-08 | 4.25E-06 | 0.000221207 |
| Obesity | adipocyte | *C1QTNF9* | 2.656295047 | 0.011143548 | 0.047848224 | 1 |
| Obesity | adipocyte | *ARHGEF25* | 0.434697472 | 0.00040643 | 0.004661939 | 1 |

| | | | | | | |
|---|---|---|---|---|---|---|
| Obesity | adipocyte | *ATP5O* | 0.812010517 | 0.006805587 | 0.03352356 | 1 |
| Obesity | adipocyte | *HOGA1* | -1.49328994 | 0.002786856 | 0.017791679 | 1 |
| Obesity | adipocyte | *PI4KA* | 1.264783516 | 0.000483938 | 0.005169861 | 1 |
| Obesity | adipocyte | *FGF14-IT1* | 3.249697217 | 0.000279977 | 0.003661585 | 1 |
| Obesity | adipocyte | *PALM2* | -0.514217896 | 0.000907196 | 0.007988594 | 1 |
| Obesity | adipocyte | *NBPF15* | 1.796443215 | 0.001987943 | 0.013858477 | 1 |
| Obesity | adipocyte | *UPK3B* | 1.417396369 | 0.000450395 | 0.004942243 | 1 |
| Obesity | adipocyte | *APOBEC3D* | 5.338605002 | 2.55E-05 | 0.000830644 | 0.123765947 |
| Obesity | adipocyte | *C4A* | -0.791667214 | 9.48E-07 | 6.13E-05 | 0.004597589 |
| Obesity | adipocyte | *SRP14-AS1* | 0.147706289 | 0.007663204 | 0.036310417 | 1 |
| Obesity | adipocyte | *LINC00942* | -4.727573597 | 9.84E-05 | 0.001916605 | 0.477234578 |
| Obesity | adipocyte | *TMEM158* | 2.135111136 | 0.005257509 | 0.027948264 | 1 |
| Obesity | adipocyte | *DNAH10OS* | 5.320762234 | 3.94E-06 | 0.000197072 | 0.019115952 |
| Obesity | adipocyte | *ZNF718* | 1.203138134 | 0.005985058 | 0.030528772 | 1 |
| Obesity | adipocyte | *SMIM20* | 2.22122502 | 0.00189072 | 0.013333973 | 1 |
| Obesity | adipocyte | *GLI4* | -1.191493137 | 0.003005507 | 0.018647978 | 1 |
| Obesity | adipocyte | *LINC01094* | 1.663227409 | 0.000119699 | 0.002104278 | 0.580780618 |
| Obesity | adipocyte | *PCDHGB2* | 3.632142053 | 0.000123453 | 0.002162438 | 0.598995402 |
| Obesity | adipocyte | *DPP3* | 0.252074755 | 0.00072808 | 0.006968045 | 1 |
| Obesity | adipocyte | *BRK1* | -0.584875594 | 0.001709641 | 0.012530482 | 1 |
| Obesity | adipocyte | *SNHG9* | 0.694852455 | 0.007542923 | 0.036057403 | 1 |
| Obesity | adipocyte | *PCDHA3* | -0.666039486 | 0.003790939 | 0.022081195 | 1 |
| Obesity | adipocyte | *HMBS* | 0.928708839 | 2.59E-05 | 0.000837689 | 0.125653279 |
| Obesity | adipocyte | *AC093668.2* | -0.708891171 | 0.003170364 | 0.019349191 | 1 |
| Obesity | adipocyte | *PDF* | -1.629974098 | 0.00091827 | 0.008015193 | 1 |
| Obesity | adipocyte | *LINC00639* | 0.831472133 | 0.001963803 | 0.013729644 | 1 |
| Obesity | adipocyte | *AC083843.1* | -5.103847772 | 0.00084398 | 0.007611507 | 1 |
| Obesity | adipocyte | *C19orf84* | 43.67652772 | 4.80E-14 | 1.66E-11 | 2.33E-10 |
| Obesity | adipocyte | *LINC00672* | 3.323214746 | 0.000189152 | 0.002859079 | 0.917764442 |
| Obesity | adipocyte | *MYO15B* | 0.62995876 | 0.008852177 | 0.040103421 | 1 |
| Obesity | adipocyte | *LIN37* | -1.359326919 | 0.003535396 | 0.020967771 | 1 |
| Obesity | adipocyte | *LINC00664* | -7.413023263 | 0.000251254 | 0.003414801 | 1 |
| Obesity | adipocyte | *TRABD2B* | 1.350242356 | 4.11E-05 | 0.00114725 | 0.199621508 |
| Obesity | adipocyte | *AP001172.2* | 0.232771383 | 0.001057429 | 0.008770334 | 1 |
| Obesity | adipocyte | *GTF2H5* | 0.511926163 | 0.000215139 | 0.003125308 | 1 |
| Obesity | adipocyte | *CASC15* | 1.294944303 | 0.000194723 | 0.002916037 | 0.944796081 |
| Obesity | ASPC | *SEMA3F* | -0.701126534 | 1.05E-09 | 1.49E-07 | 5.08E-06 |
| Obesity | ASPC | *RBM6* | -0.426830672 | 5.91E-25 | 1.43E-21 | 2.87E-21 |
| Obesity | ASPC | *GGNBP2* | -0.763165262 | 0.002968553 | 0.048165781 | 1 |
| Obesity | ASPC | *FMO3* | -0.185056543 | 0.001395992 | 0.028700641 | 1 |
| Obesity | ASPC | *IL32* | 1.027381432 | 0.001502803 | 0.030255601 | 1 |
| Obesity | ASPC | *PKD1* | 0.736468484 | 0.002978099 | 0.048165781 | 1 |
| Obesity | ASPC | *CLCN6* | 0.471585356 | 0.000570104 | 0.014871735 | 1 |
| Obesity | ASPC | *NUDCD3* | 0.286403281 | 0.00156848 | 0.031317953 | 1 |
| Obesity | ASPC | *RABEP1* | 0.689009809 | 0.00062512 | 0.015767112 | 1 |
| Obesity | ASPC | *PIAS1* | 0.83137778 | 3.17E-06 | 0.000202357 | 0.01537916 |
| Obesity | ASPC | *SH3YL1* | -0.600041164 | 0.000177905 | 0.006078828 | 0.863193588 |
| Obesity | ASPC | *LSG1* | 0.135321634 | 0.001999218 | 0.037165537 | 1 |
| Obesity | ASPC | *ANAPC4* | 0.631599704 | 0.000416662 | 0.011543555 | 1 |
| Obesity | ASPC | *SDCCAG8* | 0.301180079 | 0.000737969 | 0.018083968 | 1 |

| | | | | | | |
|---|---|---|---|---|---|---|
| Obesity | ASPC | *ITIH4* | 0.161800327 | 5.55E-11 | 1.17E-08 | 2.69E-07 |
| Obesity | ASPC | *ATP11B* | 0.54350706 | 0.000882896 | 0.02059524 | 1 |
| Obesity | ASPC | *UHRF1BP1* | 0.675354948 | 1.02E-23 | 1.65E-20 | 4.94E-20 |
| Obesity | ASPC | *AFF4* | 0.720836264 | 0.000877816 | 0.02059524 | 1 |
| Obesity | ASPC | *TIPIN* | 0.157542702 | 0.000597924 | 0.015234691 | 1 |
| Obesity | ASPC | *NT5C2* | 0.35262622 | 0.000106536 | 0.004038369 | 0.516911276 |
| Obesity | ASPC | *FDFT1* | 0.862036572 | 2.01E-09 | 2.71E-07 | 9.76E-06 |
| Obesity | ASPC | *DOPEY1* | 0.305629447 | 0.002553977 | 0.044338413 | 1 |
| Obesity | ASPC | *GSTP1* | 0.57293367 | 1.17E-05 | 0.000602663 | 0.056650332 |
| Obesity | ASPC | *MAST2* | 0.509142946 | 0.000958405 | 0.021831845 | 1 |
| Obesity | ASPC | *OGFOD1* | -0.57249965 | 0.000125927 | 0.004593959 | 0.610996542 |
| Obesity | ASPC | *NSFL1C* | 0.398224525 | 0.002347191 | 0.041716382 | 1 |
| Obesity | ASPC | *YPEL3* | 0.63395779 | 1.73E-09 | 2.40E-07 | 8.39E-06 |
| Obesity | ASPC | *LYZ* | 0.368938699 | 0.001289338 | 0.026849222 | 1 |
| Obesity | ASPC | *PABPC4* | -0.6398154 | 2.25E-09 | 2.95E-07 | 1.09E-05 |
| Obesity | ASPC | *RGS17* | -0.634197257 | 1.33E-08 | 1.54E-06 | 6.45E-05 |
| Obesity | ASPC | *PPP2R3C* | 0.509451367 | 8.44E-05 | 0.003330409 | 0.409640338 |
| Obesity | ASPC | *TBX15* | -0.312564649 | 2.19E-05 | 0.001031664 | 0.106261386 |
| Obesity | ASPC | *ITPR3* | 0.307814164 | 6.42E-05 | 0.002638927 | 0.311393406 |
| Obesity | ASPC | *SETD1A* | 0.967944009 | 1.39E-08 | 1.57E-06 | 6.76E-05 |
| Obesity | ASPC | *BCL2L13* | 0.551729025 | 2.72E-05 | 0.001242795 | 0.131736306 |
| Obesity | ASPC | *ANKRD54* | -0.27525017 | 0.000313073 | 0.009369493 | 1 |
| Obesity | ASPC | *CENPM* | 0.357906085 | 2.83E-05 | 0.001284181 | 0.137407347 |
| Obesity | ASPC | *KDELR3* | -0.498421968 | 0.002655405 | 0.045207107 | 1 |
| Obesity | ASPC | *DESI1* | 0.639233469 | 0.002320363 | 0.04169779 | 1 |
| Obesity | ASPC | *ABHD12* | 0.597646006 | 4.68E-06 | 0.000283997 | 0.022719722 |
| Obesity | ASPC | *ELMO3* | 0.14629046 | 0.002027168 | 0.037541288 | 1 |
| Obesity | ASPC | *CLUAP1* | 0.172731257 | 0.000973204 | 0.021962719 | 1 |
| Obesity | ASPC | *RRN3P2* | -0.440043828 | 0.002339649 | 0.041716382 | 1 |
| Obesity | ASPC | *PYCARD* | 1.27274566 | 3.08E-10 | 4.67E-08 | 1.49E-06 |
| Obesity | ASPC | *KNOP1* | -1.291205383 | 2.92E-05 | 0.001312458 | 0.141745469 |
| Obesity | ASPC | *LIN7B* | -1.146427803 | 0.000320317 | 0.009419258 | 1 |
| Obesity | ASPC | *RASAL3* | 1.517755012 | 0.00195869 | 0.036693291 | 1 |
| Obesity | ASPC | *MEST* | -0.214284499 | 0.000496481 | 0.013163536 | 1 |
| Obesity | ASPC | *RGP1* | 0.839474846 | 0.001843825 | 0.035386989 | 1 |
| Obesity | ASPC | *RAD51C* | -0.34786162 | 0.000195621 | 0.006591356 | 0.949155242 |
| Obesity | ASPC | *PNPO* | -0.988285111 | 7.61E-05 | 0.00302797 | 0.369412376 |
| Obesity | ASPC | *CHRNE* | 0.098196683 | 0.000468608 | 0.012492772 | 1 |
| Obesity | ASPC | *B3GAT1* | 4.030426613 | 0.000245171 | 0.007930469 | 1 |
| Obesity | ASPC | *DDX55* | 0.704649035 | 0.000197177 | 0.006597963 | 0.956704564 |
| Obesity | ASPC | *BTN3A3* | 0.447600638 | 7.83E-11 | 1.41E-08 | 3.80E-07 |
| Obesity | ASPC | *GABRR2* | 0.343837659 | 0.000820171 | 0.019700338 | 1 |
| Obesity | ASPC | *TULP1* | -0.27077007 | 0.000401135 | 0.011250336 | 1 |
| Obesity | ASPC | *CUTA* | -0.297522138 | 0.001272103 | 0.026677225 | 1 |
| Obesity | ASPC | *PRSS16* | 0.351235976 | 0.001058518 | 0.023239502 | 1 |
| Obesity | ASPC | *NME5* | 0.19235165 | 0.000104283 | 0.004002663 | 0.505982919 |
| Obesity | ASPC | *TMCO6* | -0.30944441 | 0.000992492 | 0.02201747 | 1 |
| Obesity | ASPC | *IK* | -0.319175435 | 0.00141325 | 0.02892395 | 1 |
| Obesity | ASPC | *PCCB* | 0.441535346 | 8.20E-07 | 6.22E-05 | 0.003977789 |
| Obesity | ASPC | *SCAP* | 0.779071902 | 0.000151014 | 0.005250637 | 0.732718744 |

| | | | | | | |
|---|---|---|---|---|---|---|
| Obesity | ASPC | *CHMP3* | -0.413010112 | 1.14E-05 | 0.000593519 | 0.055197306 |
| Obesity | ASPC | *NSUN4* | -0.469007125 | 0.000322666 | 0.009431182 | 1 |
| Obesity | ASPC | *PRRC2C* | 0.617637262 | 0.003021318 | 0.048541173 | 1 |
| Obesity | ASPC | *SERPINC1* | 0.905624205 | 7.58E-11 | 1.41E-08 | 3.68E-07 |
| Obesity | ASPC | *CCDC92* | 0.693598918 | 7.16E-06 | 0.000413363 | 0.034722522 |
| Obesity | ASPC | *INVS* | 0.511714325 | 1.13E-06 | 8.20E-05 | 0.005497324 |
| Obesity | ASPC | *SNX19* | -0.483132159 | 8.25E-06 | 0.000465319 | 0.040017468 |
| Obesity | ASPC | *ADRA1A* | 0.069601763 | 0.001372802 | 0.028343977 | 1 |
| Obesity | ASPC | *TEX14* | 0.405349897 | 2.33E-05 | 0.001086502 | 0.112996205 |
| Obesity | ASPC | *OPN4* | 1.17689431 | 3.14E-06 | 0.000202357 | 0.015255137 |
| Obesity | ASPC | *SLC25A51* | 0.103277504 | 0.001852493 | 0.035386989 | 1 |
| Obesity | ASPC | *PEPD* | 1.038431503 | 0.000745369 | 0.018173509 | 1 |
| Obesity | ASPC | *BTN2A2* | 0.168482199 | 1.37E-05 | 0.000701766 | 0.06666777 |
| Obesity | ASPC | *SNRPC* | -0.96044023 | 3.04E-22 | 2.95E-19 | 1.48E-18 |
| Obesity | ASPC | *TCP11* | 0.207370647 | 0.002811914 | 0.046628802 | 1 |
| Obesity | ASPC | *TREM1* | 0.292782596 | 0.00310106 | 0.049332269 | 1 |
| Obesity | ASPC | *GRK4* | 0.088954629 | 0.000144068 | 0.005065356 | 0.699019069 |
| Obesity | ASPC | *CANX* | 0.773000017 | 0.002576872 | 0.0444946 | 1 |
| Obesity | ASPC | *YEATS4* | 0.167509353 | 0.002643993 | 0.045171314 | 1 |
| Obesity | ASPC | *RIBC2* | 0.21322647 | 0.000627175 | 0.015767112 | 1 |
| Obesity | ASPC | *EIF2AK4* | 0.615602185 | 0.001172154 | 0.025203632 | 1 |
| Obesity | ASPC | *RPAIN* | -0.422373161 | 0.000161188 | 0.005546686 | 0.78208269 |
| Obesity | ASPC | *AP1M2* | -0.186330578 | 0.000978553 | 0.021981192 | 1 |
| Obesity | ASPC | *ATP8B3* | -0.269309742 | 8.23E-11 | 1.43E-08 | 3.99E-07 |
| Obesity | ASPC | *TULP4* | 0.479634685 | 0.00029662 | 0.009051566 | 1 |
| Obesity | ASPC | *STK33* | 0.365500387 | 8.86E-06 | 0.000490296 | 0.043010442 |
| Obesity | ASPC | *PGPEP1* | 0.268771499 | 0.000863929 | 0.020447726 | 1 |
| Obesity | ASPC | *CYP2E1* | -0.145103203 | 0.001949505 | 0.036662783 | 1 |
| Obesity | ASPC | *NPAS1* | -1.068930025 | 6.97E-08 | 6.99E-06 | 0.000338207 |
| Obesity | ASPC | *C12orf65* | 0.398439369 | 0.000640539 | 0.01602008 | 1 |
| Obesity | ASPC | *PKDREJ* | -0.20317694 | 0.001839836 | 0.035386989 | 1 |
| Obesity | ASPC | *NDUFA2* | 0.420321391 | 0.000109572 | 0.004089562 | 0.531643038 |
| Obesity | ASPC | *EXOC4* | 0.574131498 | 0.000599717 | 0.015234691 | 1 |
| Obesity | ASPC | *MYBBP1A* | 0.27837656 | 0.001009613 | 0.022266557 | 1 |
| Obesity | ASPC | *IGHMBP2* | 0.248334529 | 2.00E-05 | 0.000951621 | 0.097065381 |
| Obesity | ASPC | *MUTYH* | 0.39031694 | 0.001807391 | 0.035249452 | 1 |
| Obesity | ASPC | *LARS* | -0.21084247 | 0.000844298 | 0.020081046 | 1 |
| Obesity | ASPC | *ACP2* | 0.560710098 | 0.000644863 | 0.016045518 | 1 |
| Obesity | ASPC | *TMEM60* | -0.980069567 | 0.003018531 | 0.048541173 | 1 |
| Obesity | ASPC | *POM121C* | -1.027861446 | 2.40E-14 | 9.70E-12 | 1.16E-10 |
| Obesity | ASPC | *MICAL1* | 0.203428179 | 0.002558688 | 0.044338413 | 1 |
| Obesity | ASPC | *USP37* | 0.535840798 | 0.000577271 | 0.014978174 | 1 |
| Obesity | ASPC | *SCN2A* | 0.755509939 | 3.69E-07 | 3.26E-05 | 0.001790411 |
| Obesity | ASPC | *TCF19* | 0.243183247 | 0.002851552 | 0.046900777 | 1 |
| Obesity | ASPC | *SLC37A4* | 0.468184664 | 2.52E-08 | 2.65E-06 | 0.000122089 |
| Obesity | ASPC | *MAP2K5* | 0.834674461 | 0.000993781 | 0.02201747 | 1 |
| Obesity | ASPC | *NUSAP1* | 0.525926736 | 0.00056662 | 0.014860754 | 1 |
| Obesity | ASPC | *NDUFAF1* | 0.497624057 | 1.08E-05 | 0.000568929 | 0.0523415 |
| Obesity | ASPC | *ADAM10* | 0.78059808 | 0.000314762 | 0.009369493 | 1 |
| Obesity | ASPC | *RSL24D1* | -0.545372961 | 0.002188711 | 0.039625462 | 1 |

| | | | | | | |
|---|---|---|---|---|---|---|
| Obesity | ASPC | *SLC44A5* | 0.755903439 | 0.001787417 | 0.035249452 | 1 |
| Obesity | ASPC | *CENPO* | 1.326843326 | 1.07E-16 | 6.47E-14 | 5.18E-13 |
| Obesity | ASPC | *STAT4* | 1.054258014 | 0.001238701 | 0.026245305 | 1 |
| Obesity | ASPC | *KLRG1* | 0.113785942 | 0.00126252 | 0.026633692 | 1 |
| Obesity | ASPC | *VPS33A* | 0.490698836 | 3.63E-06 | 0.000224762 | 0.017629654 |
| Obesity | ASPC | *DIAPH3* | 0.178922239 | 0.000237831 | 0.007744674 | 1 |
| Obesity | ASPC | *CYP11A1* | 0.245754729 | 0.002963995 | 0.048165781 | 1 |
| Obesity | ASPC | *MYO19* | 1.20702525 | 2.60E-10 | 4.08E-08 | 1.26E-06 |
| Obesity | ASPC | *C18orf8* | -0.993091283 | 2.40E-13 | 8.74E-11 | 1.16E-09 |
| Obesity | ASPC | *NPC1* | -0.982045205 | 6.30E-05 | 0.002613767 | 0.305810727 |
| Obesity | ASPC | *CBX8* | 0.209597356 | 0.001967336 | 0.036713509 | 1 |
| Obesity | ASPC | *COL6A1* | 0.123497911 | 0.001804963 | 0.035249452 | 1 |
| Obesity | ASPC | *SAE1* | -1.535407627 | 6.32E-11 | 1.28E-08 | 3.06E-07 |
| Obesity | ASPC | *SEMA6C* | 1.364948791 | 3.79E-07 | 3.28E-05 | 0.001836595 |
| Obesity | ASPC | *FAHD2B* | -0.245242845 | 0.002110877 | 0.03879536 | 1 |
| Obesity | ASPC | *NBEAL1* | 0.599612124 | 0.000584059 | 0.014993932 | 1 |
| Obesity | ASPC | *LIMD1* | 0.513494135 | 0.000221425 | 0.007308519 | 1 |
| Obesity | ASPC | *ABT1* | 0.178544298 | 0.001421208 | 0.02892395 | 1 |
| Obesity | ASPC | *SDK1* | -0.157442996 | 0.002825406 | 0.046628802 | 1 |
| Obesity | ASPC | *ADHFE1* | -0.112447628 | 0.000404279 | 0.011273355 | 1 |
| Obesity | ASPC | *ST8SIA6* | 1.004057795 | 0.001745773 | 0.034715125 | 1 |
| Obesity | ASPC | *RGR* | -1.436979795 | 3.45E-05 | 0.001508608 | 0.167455528 |
| Obesity | ASPC | *MTG1* | 0.21109473 | 0.000272464 | 0.008438865 | 1 |
| Obesity | ASPC | *PDCD11* | 0.551678006 | 0.002175194 | 0.039625462 | 1 |
| Obesity | ASPC | *HSD17B12* | -0.214435024 | 7.81E-11 | 1.41E-08 | 3.79E-07 |
| Obesity | ASPC | *TNKS1BP1* | 0.334982294 | 1.99E-07 | 1.86E-05 | 0.000965413 |
| Obesity | ASPC | *TTC12* | 0.327303318 | 3.01E-05 | 0.001338445 | 0.14589051 |
| Obesity | ASPC | *NUDT22* | -0.386053571 | 0.002458647 | 0.043222307 | 1 |
| Obesity | ASPC | *TBX6* | 0.184791476 | 2.00E-11 | 4.63E-09 | 9.72E-08 |
| Obesity | ASPC | *RGS18* | 0.221547243 | 0.000185753 | 0.006302607 | 0.901272845 |
| Obesity | ASPC | *DPYSL4* | 0.085752068 | 0.001936443 | 0.036558844 | 1 |
| Obesity | ASPC | *POC5* | -0.292022161 | 8.99E-06 | 0.000490296 | 0.04363636 |
| Obesity | ASPC | *CWF19L2* | 0.565286126 | 0.000903997 | 0.020745624 | 1 |
| Obesity | ASPC | *PLA2R1* | 0.181528816 | 0.001909014 | 0.03632367 | 1 |
| Obesity | ASPC | *JAZF1* | 0.518932188 | 2.09E-06 | 0.000145053 | 0.010153733 |
| Obesity | ASPC | *NEIL2* | 0.300253592 | 4.51E-05 | 0.001894967 | 0.218764334 |
| Obesity | ASPC | *ALS2CR12* | 0.566500759 | 0.002702339 | 0.045685539 | 1 |
| Obesity | ASPC | *MMP16* | -0.143656051 | 0.000359099 | 0.010309749 | 1 |
| Obesity | ASPC | *RWDD2B* | -0.231310104 | 0.002682159 | 0.04550292 | 1 |
| Obesity | ASPC | *ITGAD* | 0.217902514 | 0.001811024 | 0.035249452 | 1 |
| Obesity | ASPC | *STEAP2* | -0.179073042 | 0.001072747 | 0.023445802 | 1 |
| Obesity | ASPC | *TMED6* | 0.947672129 | 0.000108476 | 0.004080042 | 0.526325424 |
| Obesity | ASPC | *CLEC18A* | -0.506469773 | 3.00E-18 | 2.08E-15 | 1.46E-14 |
| Obesity | ASPC | *ETS2* | 0.128430513 | 0.000892931 | 0.020630965 | 1 |
| Obesity | ASPC | *LRRC43* | 0.471970575 | 0.002368372 | 0.041939199 | 1 |
| Obesity | ASPC | *SLC5A11* | -0.167478719 | 3.67E-07 | 3.26E-05 | 0.001779997 |
| Obesity | ASPC | *CILP2* | -0.453924003 | 1.52E-05 | 0.000743607 | 0.07361709 |
| Obesity | ASPC | *NBEAL2* | 0.40934099 | 0.001155443 | 0.025027723 | 1 |
| Obesity | ASPC | *ABCF3* | 0.240118751 | 0.000968291 | 0.021953959 | 1 |
| Obesity | ASPC | *CAMK2N1* | 0.236759921 | 0.002753681 | 0.046071931 | 1 |

| | | | | | | |
|---|---|---|---|---|---|---|
| Obesity | ASPC | *CLHC1* | 0.096623729 | 0.002553358 | 0.044338413 | 1 |
| Obesity | ASPC | *ANTXR2* | 0.436378623 | 0.000131503 | 0.004726315 | 0.638052499 |
| Obesity | ASPC | *DCST2* | 0.36732175 | 0.00263209 | 0.045126849 | 1 |
| Obesity | ASPC | *LMOD1* | -0.577108252 | 7.06E-08 | 6.99E-06 | 0.000342324 |
| Obesity | ASPC | *DNALI1* | -0.353272454 | 8.22E-06 | 0.000465319 | 0.03986383 |
| Obesity | ASPC | *RFT1* | -0.393024748 | 4.36E-07 | 3.71E-05 | 0.002114542 |
| Obesity | ASPC | *RNF123* | 0.639150659 | 1.31E-19 | 1.06E-16 | 6.38E-16 |
| Obesity | ASPC | *MST1R* | 0.572394224 | 1.00E-22 | 1.22E-19 | 4.87E-19 |
| Obesity | ASPC | *TEX264* | -0.320297905 | 0.00127558 | 0.026677225 | 1 |
| Obesity | ASPC | *ERAP1* | -0.472313428 | 0.000270671 | 0.008438865 | 1 |
| Obesity | ASPC | *GALNT10* | -0.532828489 | 0.000769462 | 0.018667147 | 1 |
| Obesity | ASPC | *RPP25L* | -0.34297695 | 0.000441231 | 0.011964297 | 1 |
| Obesity | ASPC | *ANKS6* | 0.512475687 | 0.000141672 | 0.005017475 | 0.687394021 |
| Obesity | ASPC | *SLC39A13* | 1.145737339 | 3.80E-09 | 4.85E-07 | 1.84E-05 |
| Obesity | ASPC | *RAPSN* | -0.51185821 | 1.37E-15 | 6.67E-13 | 6.67E-12 |
| Obesity | ASPC | *CCDC68* | 0.291489019 | 0.000305331 | 0.009259162 | 1 |
| Obesity | ASPC | *ZSCAN21* | -0.180794872 | 0.002221801 | 0.040075013 | 1 |
| Obesity | ASPC | *PIP4K2C* | 1.16397523 | 2.71E-06 | 0.000180222 | 0.013156214 |
| Obesity | ASPC | *SMG8* | -0.274135648 | 0.001816233 | 0.035249452 | 1 |
| Obesity | ASPC | *SRR* | 0.153006562 | 1.66E-05 | 0.000805336 | 0.080533562 |
| Obesity | ASPC | *KIAA1586* | -0.33086161 | 0.002731976 | 0.045967859 | 1 |
| Obesity | ASPC | *NT5DC2* | -0.280950929 | 3.66E-06 | 0.000224762 | 0.017756166 |
| Obesity | ASPC | *FNTA* | 0.470308361 | 4.02E-05 | 0.001712541 | 0.195229727 |
| Obesity | ASPC | *NDUFS5* | 0.550790311 | 0.00218289 | 0.039625462 | 1 |
| Obesity | ASPC | *C8orf46* | 0.07262049 | 0.000295757 | 0.009051566 | 1 |
| Obesity | ASPC | *GPRIN1* | 0.507513946 | 0.000669772 | 0.016580284 | 1 |
| Obesity | ASPC | *INO80E* | -0.46190139 | 2.15E-14 | 9.47E-12 | 1.04E-10 |
| Obesity | ASPC | *MYO7B* | 0.235969985 | 0.002777717 | 0.04631438 | 1 |
| Obesity | ASPC | *CNGB3* | 0.39815838 | 0.000332881 | 0.009671499 | 1 |
| Obesity | ASPC | *ADORA2B* | -0.161653341 | 6.62E-07 | 5.17E-05 | 0.00321399 |
| Obesity | ASPC | *TTLL6* | -1.133178685 | 2.64E-12 | 7.12E-10 | 1.28E-08 |
| Obesity | ASPC | *TSNARE1* | 0.493812199 | 6.82E-06 | 0.000403632 | 0.033097855 |
| Obesity | ASPC | *TRMT61B* | 0.485817576 | 1.46E-05 | 0.000732444 | 0.071047079 |
| Obesity | ASPC | *PIK3CD* | 0.695454951 | 0.000268566 | 0.008438865 | 1 |
| Obesity | ASPC | *RGS19* | 0.113788358 | 0.001351984 | 0.028033453 | 1 |
| Obesity | ASPC | *CAMTA1* | 0.626390871 | 0.000906445 | 0.020745624 | 1 |
| Obesity | ASPC | *C1QTNF4* | -0.262087417 | 1.11E-15 | 6.00E-13 | 5.40E-12 |
| Obesity | ASPC | *CTSW* | -0.06798368 | 5.15E-09 | 6.25E-07 | 2.50E-05 |
| Obesity | ASPC | *EFEMP2* | 0.928102002 | 4.96E-07 | 4.15E-05 | 0.002408619 |
| Obesity | ASPC | *FUT10* | -0.162643098 | 0.000318618 | 0.009419258 | 1 |
| Obesity | ASPC | *SNX32* | 0.143823229 | 7.19E-05 | 0.002883635 | 0.348919781 |
| Obesity | ASPC | *MRGPRF* | 0.39023336 | 6.67E-05 | 0.002697858 | 0.323742929 |
| Obesity | ASPC | *MST1* | -0.093806153 | 0.002174515 | 0.039625462 | 1 |
| Obesity | ASPC | *HOXB2* | 0.424613253 | 0.000458074 | 0.012307221 | 1 |
| Obesity | ASPC | *SRP72* | 0.244467036 | 0.002944148 | 0.04809766 | 1 |
| Obesity | ASPC | *SEZ6L2* | 0.337517794 | 9.09E-07 | 6.78E-05 | 0.004410004 |
| Obesity | ASPC | *KCTD13* | 0.5022388 | 1.12E-07 | 1.09E-05 | 0.000544493 |
| Obesity | ASPC | *CADM2* | 0.420071532 | 8.41E-10 | 1.24E-07 | 4.08E-06 |
| Obesity | ASPC | *BANF1* | -0.841392645 | 1.66E-06 | 0.000117524 | 0.008045982 |
| Obesity | ASPC | *RPS6KB2* | 0.523681856 | 0.001561634 | 0.031310111 | 1 |

| | | | | | | |
|---|---|---|---|---|---|---|
| Obesity | ASPC | *ORAI3* | 0.609522882 | 6.74E-12 | 1.72E-09 | 3.27E-08 |
| Obesity | ASPC | *CCDC57* | -0.236504442 | 0.002922232 | 0.047900919 | 1 |
| Obesity | ASPC | *MYPOP* | 0.709523204 | 0.00307988 | 0.049234279 | 1 |
| Obesity | ASPC | *SPRYD4* | -0.181684276 | 0.002606023 | 0.044838378 | 1 |
| Obesity | ASPC | *RMDN1* | 0.317309523 | 0.001424737 | 0.02892395 | 1 |
| Obesity | ASPC | *RUFY1* | 0.514208061 | 7.05E-06 | 0.000412332 | 0.034223557 |
| Obesity | ASPC | *AP006621.1* | 0.56774549 | 1.67E-06 | 0.000117524 | 0.008109173 |
| Obesity | ASPC | *CD151* | 0.540693963 | 0.001085269 | 0.023613124 | 1 |
| Obesity | ASPC | *RPP25* | -0.184470932 | 0.000123835 | 0.004551857 | 0.600845074 |
| Obesity | ASPC | *STX19* | 0.497286788 | 5.91E-07 | 4.70E-05 | 0.002865338 |
| Obesity | ASPC | *MPI* | 0.373827224 | 3.40E-05 | 0.001500864 | 0.165095042 |
| Obesity | ASPC | *TRIM73* | -1.073953327 | 2.88E-13 | 8.75E-11 | 1.40E-09 |
| Obesity | ASPC | *ZNF852* | -0.456049317 | 3.70E-05 | 0.001588547 | 0.17950579 |
| Obesity | ASPC | *TUFM* | 0.658344183 | 1.57E-11 | 3.80E-09 | 7.61E-08 |
| Obesity | ASPC | *C3orf38* | 0.476860538 | 2.06E-08 | 2.23E-06 | 0.000100193 |
| Obesity | ASPC | *RPRML* | 0.913245798 | 1.39E-05 | 0.000703411 | 0.067527454 |
| Obesity | ASPC | *CMTR2* | 0.593961253 | 0.000988877 | 0.02201747 | 1 |
| Obesity | ASPC | *ZFP41* | 0.359187589 | 0.00036131 | 0.010312222 | 1 |
| Obesity | ASPC | *PPP1R27* | -1.035834448 | 0.000259203 | 0.008241987 | 1 |
| Obesity | ASPC | *GRID1* | -0.410927287 | 3.53E-05 | 0.001530929 | 0.171464083 |
| Obesity | ASPC | *COL18A1* | 0.453453017 | 0.000223147 | 0.007315614 | 1 |
| Obesity | ASPC | *LYSMD4* | 0.374483046 | 0.001848607 | 0.035386989 | 1 |
| Obesity | ASPC | *PLGLB1* | 0.191547899 | 0.000889071 | 0.020630965 | 1 |
| Obesity | ASPC | *PSMG1* | 0.389263038 | 9.61E-05 | 0.003730306 | 0.466288248 |
| Obesity | ASPC | *UPP1* | 0.439970655 | 0.000441387 | 0.011964297 | 1 |
| Obesity | ASPC | *EMILIN3* | 0.395497439 | 0.000514547 | 0.013568389 | 1 |
| Obesity | ASPC | *ACOT1* | 0.189088309 | 9.22E-05 | 0.00360749 | 0.447328794 |
| Obesity | ASPC | *OAF* | -0.115299175 | 0.003084753 | 0.049234279 | 1 |
| Obesity | ASPC | *NUTM2A* | -0.275920784 | 1.51E-05 | 0.000743607 | 0.073110196 |
| Obesity | ASPC | *MAFF* | -0.198349995 | 0.000385678 | 0.010879717 | 1 |
| Obesity | ASPC | *GP1BA* | 0.397246864 | 2.62E-07 | 2.40E-05 | 0.001271755 |
| Obesity | ASPC | *ATP6V0A2* | -0.54591697 | 0.002737987 | 0.045967859 | 1 |
| Obesity | ASPC | *BTN3A2* | -0.132874063 | 1.51E-10 | 2.44E-08 | 7.32E-07 |
| Obesity | ASPC | *ZSCAN23* | 0.099704016 | 0.000882232 | 0.02059524 | 1 |
| Obesity | ASPC | *CENPP* | 0.245473685 | 5.96E-06 | 0.00035689 | 0.028908059 |
| Obesity | ASPC | *OSTN* | 0.216761775 | 0.002346562 | 0.041716382 | 1 |
| Obesity | ASPC | *SKOR1* | 0.421234982 | 0.001230475 | 0.026185377 | 1 |
| Obesity | ASPC | *FBF1* | 0.357652512 | 0.000259898 | 0.008241987 | 1 |
| Obesity | ASPC | *ZKSCAN3* | -0.440713153 | 3.73E-08 | 3.85E-06 | 0.00018106 |
| Obesity | ASPC | *CPNE4* | 0.37339581 | 9.88E-06 | 0.00052675 | 0.047934265 |
| Obesity | ASPC | *ZNF605* | 0.324937693 | 0.000114726 | 0.004249231 | 0.556649201 |
| Obesity | ASPC | *CACNA1H* | 1.121762446 | 4.53E-05 | 0.001894967 | 0.219816139 |
| Obesity | ASPC | *ZFP62* | 0.629750626 | 0.002053804 | 0.037889957 | 1 |
| Obesity | ASPC | *SULT1A2* | 1.095409109 | 1.69E-28 | 8.19E-25 | 8.19E-25 |
| Obesity | ASPC | *ZNF165* | -0.168244683 | 0.000368637 | 0.010459815 | 1 |
| Obesity | ASPC | *MRPL21* | -0.123721777 | 0.000354541 | 0.010239493 | 1 |
| Obesity | ASPC | *MFAP5* | -0.29222173 | 1.91E-05 | 0.000917818 | 0.092699665 |
| Obesity | ASPC | *DNAH10* | 0.749140279 | 2.52E-13 | 8.74E-11 | 1.22E-09 |
| Obesity | ASPC | *RPS26* | -0.1446709 | 0.000459111 | 0.012307221 | 1 |
| Obesity | ASPC | *ZNF780A* | -0.089939904 | 6.48E-05 | 0.002642681 | 0.31447906 |

| | | | | | | |
|---|---|---|---|---|---|---|
| Obesity | ASPC | *TMEM184B* | -0.390469597 | 0.002819225 | 0.046628802 | 1 |
| Obesity | ASPC | *SHISA4* | -2.621594715 | 8.96E-13 | 2.56E-10 | 4.35E-09 |
| Obesity | ASPC | *MAP3K3* | -0.623080623 | 0.000421107 | 0.011543555 | 1 |
| Obesity | ASPC | *NOTCH4* | -0.038811656 | 1.66E-08 | 1.83E-06 | 8.07E-05 |
| Obesity | ASPC | *SKIV2L* | -0.165064438 | 1.02E-06 | 7.47E-05 | 0.00493246 |
| Obesity | ASPC | *GPANK1* | -0.594688163 | 2.14E-11 | 4.71E-09 | 1.04E-07 |
| Obesity | ASPC | *POU5F1* | 0.058870313 | 0.000104769 | 0.004002663 | 0.508338165 |
| Obesity | ASPC | *CCHCR1* | 0.123121067 | 3.45E-06 | 0.000217653 | 0.016759313 |
| Obesity | ASPC | *ABCF1* | 0.321440358 | 0.000581285 | 0.014993932 | 1 |
| Obesity | ASPC | *RNF39* | -0.379184885 | 8.92E-06 | 0.000490296 | 0.043275573 |
| Obesity | ASPC | *ZFP57* | 0.148113985 | 5.28E-09 | 6.25E-07 | 2.56E-05 |
| Obesity | ASPC | *C17orf107* | 0.251802083 | 0.000254085 | 0.008164371 | 1 |
| Obesity | ASPC | *KLRC2* | 3.313577112 | 2.23E-06 | 0.000152707 | 0.010842196 |
| Obesity | ASPC | *AC084018.1* | -0.154147133 | 0.000151502 | 0.005250637 | 0.735089214 |
| Obesity | ASPC | *AC022498.1* | 1.957668989 | 0.002416296 | 0.042632249 | 1 |
| Obesity | ASPC | *SUPT4H1* | 1.299881427 | 0.000133227 | 0.004753071 | 0.646417673 |
| Obesity | ASPC | *SRA1* | 0.553730595 | 0.000777315 | 0.018763831 | 1 |
| Obesity | ASPC | *TMEM110* | 0.646799233 | 9.47E-11 | 1.58E-08 | 4.60E-07 |
| Obesity | ASPC | *PPP1CB* | -0.430051842 | 0.00068426 | 0.01685293 | 1 |
| Obesity | ASPC | *GALT* | 0.715261559 | 2.55E-05 | 0.001177602 | 0.123648245 |
| Obesity | ASPC | *EXOSC6* | -0.983203408 | 2.79E-13 | 8.75E-11 | 1.35E-09 |
| Obesity | ASPC | *C4B* | 0.362397748 | 0.000129603 | 0.004692802 | 0.628835433 |
| Obesity | ASPC | *LINC00240* | 0.131201465 | 0.000309144 | 0.009316551 | 1 |
| Obesity | ASPC | *AL049840.1* | -0.778413406 | 9.21E-06 | 0.00049647 | 0.044682257 |
| Obesity | ASPC | *HCG20* | -0.335035287 | 5.77E-07 | 4.66E-05 | 0.002798948 |
| Obesity | ASPC | *TAPBP* | 0.439135451 | 0.000273063 | 0.008438865 | 1 |
| Obesity | ASPC | *TMA7* | 0.477409988 | 0.001500914 | 0.030255601 | 1 |
| Obesity | ASPC | *ZSCAN31* | 0.144885145 | 1.78E-07 | 1.69E-05 | 0.000862886 |
| Obesity | ASPC | *LINC01143* | 3.89795414 | 0.002495057 | 0.043704026 | 1 |
| Obesity | ASPC | *PSMB9* | 0.192467078 | 5.46E-07 | 4.49E-05 | 0.00264787 |
| Obesity | ASPC | *ISY1* | -0.201719164 | 0.001220138 | 0.026079775 | 1 |
| Obesity | ASPC | *C4A* | -0.380203167 | 6.72E-07 | 5.17E-05 | 0.003259675 |
| Obesity | ASPC | *DNAH10OS* | 3.343058269 | 4.24E-09 | 5.28E-07 | 2.06E-05 |
| Obesity | ASPC | *DPP3* | 0.120158258 | 0.000832091 | 0.019888208 | 1 |
| Obesity | ASPC | *BRK1* | -0.322569236 | 0.000420136 | 0.011543555 | 1 |
| Obesity | ASPC | *PCDHA3* | -0.413321296 | 0.000219407 | 0.007291538 | 1 |
| Obesity | ASPC | *HMBS* | 0.505061695 | 2.64E-06 | 0.000178175 | 0.012828618 |
| Obesity | ASPC | *C19orf84* | 11.57582226 | 3.08E-06 | 0.000201699 | 0.014925696 |
| Obesity | ASPC | *NDUFA7* | 7.536807637 | 0.001921508 | 0.036418574 | 1 |
| Obesity | ASPC | *AP001172.2* | 0.11104497 | 0.001173953 | 0.025203632 | 1 |
| Obesity | ASPC | *GTF2H5* | 0.213226405 | 0.001759548 | 0.034846235 | 1 |
| Obesity | b_cell | *RBM6* | -1.53093848 | 0.000108741 | 0.020292726 | 0.527610871 |
| Obesity | b_cell | *ITIH4* | 5.863199263 | 7.53E-09 | 5.22E-06 | 3.66E-05 |
| Obesity | b_cell | *BTN3A3* | 8.095178409 | 1.68E-05 | 0.004064518 | 0.081290361 |
| Obesity | b_cell | *TULP1* | -10.81127692 | 0.000102685 | 0.019929103 | 0.498227586 |
| Obesity | b_cell | *SNRPC* | -3.524311705 | 0.000209846 | 0.037710118 | 1 |
| Obesity | b_cell | *ATP8B3* | -9.74579889 | 2.72E-10 | 2.64E-07 | 1.32E-06 |
| Obesity | b_cell | *SLC37A4* | 9.816132571 | 5.43E-05 | 0.011444772 | 0.263229751 |
| Obesity | b_cell | *HSD17B12* | -3.284190059 | 4.14E-06 | 0.001256225 | 0.0200996 |
| Obesity | b_cell | *TBX6* | 5.221232095 | 7.70E-10 | 6.23E-07 | 3.74E-06 |

| | | | | | | |
|---|---|---|---|---|---|---|
| Obesity | b_cell | *CLEC18A* | -22.66979777 | 2.78E-17 | 1.35E-13 | 1.35E-13 |
| Obesity | b_cell | *SLC5A11* | -3.699579267 | 7.08E-06 | 0.002019604 | 0.034333274 |
| Obesity | b_cell | *CILP2* | -16.67289215 | 1.28E-05 | 0.00327129 | 0.062154512 |
| Obesity | b_cell | *RNF123* | 8.958576158 | 1.02E-08 | 6.18E-06 | 4.95E-05 |
| Obesity | b_cell | *RAPSN* | -11.96932732 | 4.40E-11 | 5.34E-08 | 2.13E-07 |
| Obesity | b_cell | *INO80E* | -6.333658224 | 1.15E-06 | 0.000464892 | 0.005578699 |
| Obesity | b_cell | *TTLL6* | -12.13300102 | 9.66E-06 | 0.002604676 | 0.046884176 |
| Obesity | b_cell | *CTSW* | -4.079213027 | 2.92E-14 | 7.07E-11 | 1.41E-10 |
| Obesity | b_cell | *ORAI3* | 10.80469229 | 1.52E-06 | 0.000567361 | 0.007375694 |
| Obesity | b_cell | *RPP25* | -6.546821961 | 6.14E-05 | 0.012422618 | 0.29814283 |
| Obesity | b_cell | *TRIM73* | -46.2400483 | 5.52E-14 | 8.92E-11 | 2.68E-10 |
| Obesity | b_cell | *BTN3A2* | -4.542652522 | 1.71E-08 | 9.23E-06 | 8.30E-05 |
| Obesity | b_cell | *ZNF780A* | -3.416185116 | 2.71E-05 | 0.006267449 | 0.131616436 |
| Obesity | b_cell | *NOTCH4* | -1.199501692 | 1.02E-06 | 0.000448882 | 0.004937701 |
| Obesity | b_cell | *RNF39* | -11.32480594 | 3.51E-05 | 0.007740011 | 0.170280242 |
| Obesity | b_cell | *ZFP57* | 6.264604383 | 3.61E-08 | 1.75E-05 | 0.000175323 |
| Obesity | b_cell | *HCG20* | -10.68542246 | 4.12E-06 | 0.001256225 | 0.020010699 |
| Obesity | b_cell | *C4A* | -16.2434207 | 1.72E-06 | 0.000596191 | 0.008346679 |
| Obesity | b_cell | *HMBS* | 8.287431137 | 0.000281837 | 0.048838332 | 1 |
| Obesity | dendritic_cell | *RBM6* | -4.394403682 | 8.38E-14 | 6.77E-11 | 4.06E-10 |
| Obesity | dendritic_cell | *LSG1* | 2.324571755 | 0.001207665 | 0.041001804 | 1 |
| Obesity | dendritic_cell | *UHRF1BP1* | 7.004921142 | 2.03E-15 | 2.46E-12 | 9.84E-12 |
| Obesity | dendritic_cell | *TIPIN* | 2.174403476 | 0.00028102 | 0.01704384 | 1 |
| Obesity | dendritic_cell | *FDFT1* | 2.68827301 | 0.000828739 | 0.032691403 | 1 |
| Obesity | dendritic_cell | *GSTP1* | 3.627078536 | 0.001063193 | 0.038211961 | 1 |
| Obesity | dendritic_cell | *SNX10* | -1.680139426 | 0.001574285 | 0.048040446 | 1 |
| Obesity | dendritic_cell | *YPEL3* | 2.977429968 | 7.45E-05 | 0.006570828 | 0.361395522 |
| Obesity | dendritic_cell | *PABPC4* | -9.342215816 | 5.53E-10 | 2.07E-07 | 2.69E-06 |
| Obesity | dendritic_cell | *ITPR3* | 2.632318444 | 0.000222173 | 0.014971979 | 1 |
| Obesity | dendritic_cell | *IL12RB1* | -2.392393201 | 0.000511784 | 0.023373013 | 1 |
| Obesity | dendritic_cell | *SETD1A* | 2.516187676 | 0.001266593 | 0.041900586 | 1 |
| Obesity | dendritic_cell | *CENPM* | 6.544653458 | 7.19E-06 | 0.000830854 | 0.034895849 |
| Obesity | dendritic_cell | *KDELR3* | -7.461134868 | 0.000135012 | 0.010398057 | 0.65507757 |
| Obesity | dendritic_cell | *CYP2D6* | -4.533933138 | 0.001380605 | 0.044957681 | 1 |
| Obesity | dendritic_cell | *ELMO3* | 2.633257681 | 0.000498666 | 0.023373013 | 1 |
| Obesity | dendritic_cell | *RHBDL1* | -20.90950252 | 0.001505516 | 0.046945771 | 1 |
| Obesity | dendritic_cell | *CLUAP1* | 1.85761837 | 0.00052718 | 0.023373013 | 1 |
| Obesity | dendritic_cell | *PYCARD* | 7.990409769 | 1.98E-05 | 0.002186929 | 0.096224864 |
| Obesity | dendritic_cell | *PKD2L1* | 4.428121174 | 0.001452504 | 0.046805898 | 1 |
| Obesity | dendritic_cell | *RAD51C* | -4.644066048 | 0.000921397 | 0.034389382 | 1 |
| Obesity | dendritic_cell | *PNPO* | -4.834397854 | 0.000278179 | 0.01704384 | 1 |
| Obesity | dendritic_cell | *CHRNE* | 1.66781279 | 0.000912009 | 0.034389382 | 1 |
| Obesity | dendritic_cell | *BTN3A3* | 5.303141575 | 2.01E-06 | 0.000304168 | 0.009733376 |
| Obesity | dendritic_cell | *GABRR2* | 3.256230527 | 0.000594049 | 0.025379444 | 1 |
| Obesity | dendritic_cell | *CUTA* | -5.455935634 | 6.79E-05 | 0.006219585 | 0.329638004 |
| Obesity | dendritic_cell | *PRSS16* | 6.311303608 | 0.000190112 | 0.013264927 | 0.92242239 |
| Obesity | dendritic_cell | *NME5* | 1.96980261 | 0.000155068 | 0.011406758 | 0.752391786 |
| Obesity | dendritic_cell | *PCCB* | 1.580627845 | 0.000808133 | 0.032139832 | 1 |
| Obesity | dendritic_cell | *CHMP3* | -6.967685104 | 3.65E-07 | 8.42E-05 | 0.001769069 |
| Obesity | dendritic_cell | *INVS* | 2.970199012 | 0.000917888 | 0.034389382 | 1 |

| | | | | | | |
|---|---|---|---|---|---|---|
| Obesity | dendritic_cell | *SNX19* | -7.087956992 | 2.67E-06 | 0.000368347 | 0.012978851 |
| Obesity | dendritic_cell | *ADRA1A* | 0.929643116 | 0.000251419 | 0.016051138 | 1 |
| Obesity | dendritic_cell | *OPN4* | 16.08197046 | 1.67E-06 | 0.000266226 | 0.008103224 |
| Obesity | dendritic_cell | *SLC25A51* | 1.734745396 | 0.000305134 | 0.017837447 | 1 |
| Obesity | dendritic_cell | *SNRPC* | -5.710836079 | 3.88E-07 | 8.55E-05 | 0.001881032 |
| Obesity | dendritic_cell | *KCNK17* | 4.743345107 | 0.001481617 | 0.046805898 | 1 |
| Obesity | dendritic_cell | *GRK4* | 1.629907528 | 6.08E-05 | 0.005677101 | 0.295209277 |
| Obesity | dendritic_cell | *YEATS4* | 2.921511586 | 0.00103282 | 0.037678531 | 1 |
| Obesity | dendritic_cell | *RIBC2* | 3.924276362 | 0.000448585 | 0.022302354 | 1 |
| Obesity | dendritic_cell | *ATP8B3* | -5.015654126 | 1.55E-12 | 9.40E-10 | 7.52E-09 |
| Obesity | dendritic_cell | *CYP2E1* | -2.634201571 | 0.00163733 | 0.049343627 | 1 |
| Obesity | dendritic_cell | *MYBBP1A* | 2.753036866 | 0.000914021 | 0.034389382 | 1 |
| Obesity | dendritic_cell | *IGHMBP2* | 3.75964861 | 7.81E-05 | 0.00676856 | 0.379039354 |
| Obesity | dendritic_cell | *GSTM5* | 2.681525956 | 0.000360871 | 0.020133421 | 1 |
| Obesity | dendritic_cell | *DSCC1* | 1.21779129 | 0.001007397 | 0.037029465 | 1 |
| Obesity | dendritic_cell | *DERL1* | -4.788083667 | 0.000901096 | 0.034389382 | 1 |
| Obesity | dendritic_cell | *TCF19* | 3.557529106 | 0.00026488 | 0.016690888 | 1 |
| Obesity | dendritic_cell | *CENPO* | 6.023437811 | 4.91E-07 | 9.92E-05 | 0.002380938 |
| Obesity | dendritic_cell | *KLRG1* | 1.931253849 | 0.000372834 | 0.020154422 | 1 |
| Obesity | dendritic_cell | *VPS33A* | 6.731037459 | 2.61E-05 | 0.002582888 | 0.126561532 |
| Obesity | dendritic_cell | *DIAPH3* | 1.523071335 | 0.000482808 | 0.023373013 | 1 |
| Obesity | dendritic_cell | *FITM1* | 9.073814148 | 0.000762608 | 0.030996178 | 1 |
| Obesity | dendritic_cell | *DUOXA1* | -2.711200228 | 0.001218857 | 0.0410687 | 1 |
| Obesity | dendritic_cell | *CYP11A1* | 4.543741079 | 0.000394963 | 0.020606052 | 1 |
| Obesity | dendritic_cell | *MYO19* | 4.95463773 | 0.000534708 | 0.023373013 | 1 |
| Obesity | dendritic_cell | *COL6A1* | 1.564107559 | 0.000499907 | 0.023373013 | 1 |
| Obesity | dendritic_cell | *MMEL1* | -2.143681476 | 0.000361007 | 0.020133421 | 1 |
| Obesity | dendritic_cell | *PHOSPHO2* | -6.197257944 | 0.000787661 | 0.031584536 | 1 |
| Obesity | dendritic_cell | *PCYOX1L* | -8.194708289 | 0.000837084 | 0.032754271 | 1 |
| Obesity | dendritic_cell | *ABT1* | 2.48412233 | 0.000486667 | 0.023373013 | 1 |
| Obesity | dendritic_cell | *MTG1* | 2.710006662 | 0.000155162 | 0.011406758 | 0.752846049 |
| Obesity | dendritic_cell | *HSD17B12* | -1.539998992 | 0.000391397 | 0.020606052 | 1 |
| Obesity | dendritic_cell | *TNKS1BP1* | 2.511811907 | 2.39E-05 | 0.002465062 | 0.115857911 |
| Obesity | dendritic_cell | *TTC12* | 3.941121475 | 0.000155102 | 0.011406758 | 0.752555091 |
| Obesity | dendritic_cell | *TBX6* | 3.330723718 | 3.37E-12 | 1.48E-09 | 1.63E-08 |
| Obesity | dendritic_cell | *NEIL2* | 2.203557279 | 0.000334027 | 0.019294024 | 1 |
| Obesity | dendritic_cell | *RWDD2B* | -4.647485742 | 0.000296235 | 0.017744744 | 1 |
| Obesity | dendritic_cell | *ITGAD* | 3.099577047 | 0.000507098 | 0.023373013 | 1 |
| Obesity | dendritic_cell | *CLEC18A* | -9.431120537 | 3.10E-17 | 7.53E-14 | 1.51E-13 |
| Obesity | dendritic_cell | *CILP2* | -5.750772933 | 0.001360885 | 0.044614968 | 1 |
| Obesity | dendritic_cell | *GPSM1* | 3.061764127 | 0.001204739 | 0.041001804 | 1 |
| Obesity | dendritic_cell | *FGFR4* | -6.771036938 | 0.001164508 | 0.040358527 | 1 |
| Obesity | dendritic_cell | *TONSL* | -4.187227179 | 0.0007666 | 0.030996178 | 1 |
| Obesity | dendritic_cell | *ABCF3* | 3.655848569 | 0.000404439 | 0.020845646 | 1 |
| Obesity | dendritic_cell | *HS3ST6* | 3.819426254 | 0.001208421 | 0.041001804 | 1 |
| Obesity | dendritic_cell | *CLHC1* | 1.668038355 | 0.001509386 | 0.046945771 | 1 |
| Obesity | dendritic_cell | *RNF123* | 6.284320699 | 2.54E-13 | 1.76E-10 | 1.23E-09 |
| Obesity | dendritic_cell | *MST1R* | 6.259508098 | 9.68E-17 | 1.57E-13 | 4.70E-13 |
| Obesity | dendritic_cell | *RPP25L* | -5.18050935 | 2.59E-05 | 0.002582888 | 0.125507649 |
| Obesity | dendritic_cell | *SLC39A13* | 4.257658025 | 0.00017159 | 0.012243445 | 0.83255428 |

| | | | | | | |
|---|---|---|---|---|---|---|
| Obesity | dendritic_cell | *RAPSN* | -6.345345011 | 4.37E-14 | 4.24E-11 | 2.12E-10 |
| Obesity | dendritic_cell | *SPDYE6* | 2.404941733 | 0.000596302 | 0.025379444 | 1 |
| Obesity | dendritic_cell | *MS4A7* | -1.974866288 | 0.001269453 | 0.041900586 | 1 |
| Obesity | dendritic_cell | *ISLR2* | 3.747560458 | 0.001130924 | 0.04005287 | 1 |
| Obesity | dendritic_cell | *SMG8* | -2.561195874 | 0.0004296 | 0.021712721 | 1 |
| Obesity | dendritic_cell | *SRR* | 2.734340671 | 7.19E-06 | 0.000830854 | 0.034881101 |
| Obesity | dendritic_cell | *NT5DC2* | -5.353653333 | 5.59E-07 | 0.000108563 | 0.002714075 |
| Obesity | dendritic_cell | *FNTA* | 3.597265136 | 0.000762399 | 0.030996178 | 1 |
| Obesity | dendritic_cell | *NPNT* | 1.71905299 | 0.001481396 | 0.046805898 | 1 |
| Obesity | dendritic_cell | *CPLX1* | 1.961208576 | 0.001158285 | 0.040358527 | 1 |
| Obesity | dendritic_cell | *IRS1* | 1.067829367 | 0.000998192 | 0.036971204 | 1 |
| Obesity | dendritic_cell | *C8orf46* | 1.370049987 | 8.65E-05 | 0.007367186 | 0.419929574 |
| Obesity | dendritic_cell | *GPRIN1* | 9.444072079 | 0.000274347 | 0.01704384 | 1 |
| Obesity | dendritic_cell | *INO80E* | -2.793339793 | 0.000103745 | 0.008389523 | 0.503371397 |
| Obesity | dendritic_cell | *MYO7B* | 2.73491244 | 0.000863409 | 0.033514091 | 1 |
| Obesity | dendritic_cell | *ADORA2B* | -1.673693091 | 1.63E-06 | 0.000266226 | 0.007905593 |
| Obesity | dendritic_cell | *C1QTNF4* | -3.916955472 | 2.75E-12 | 1.48E-09 | 1.33E-08 |
| Obesity | dendritic_cell | *CTSW* | -1.201644006 | 8.50E-09 | 2.58E-06 | 4.12E-05 |
| Obesity | dendritic_cell | *EFEMP2* | 3.850663655 | 0.001247545 | 0.041745424 | 1 |
| Obesity | dendritic_cell | *SEZ6L2* | 2.510776537 | 3.83E-05 | 0.003720574 | 0.18602868 |
| Obesity | dendritic_cell | *KCTD13* | 4.643250847 | 1.27E-05 | 0.001436732 | 0.061779494 |
| Obesity | dendritic_cell | *CADM2* | 4.520789741 | 4.54E-07 | 9.57E-05 | 0.002202152 |
| Obesity | dendritic_cell | *ORAI3* | 4.168681013 | 0.000510159 | 0.023373013 | 1 |
| Obesity | dendritic_cell | *RUFY1* | 3.48519895 | 0.001560923 | 0.047934152 | 1 |
| Obesity | dendritic_cell | *LY6H* | 4.465959613 | 0.000761875 | 0.030996178 | 1 |
| Obesity | dendritic_cell | *STX19* | 2.560099161 | 0.000228279 | 0.015172712 | 1 |
| Obesity | dendritic_cell | *MPI* | 3.614189142 | 0.000191374 | 0.013264927 | 0.928544871 |
| Obesity | dendritic_cell | *TRIM73* | -18.24456112 | 3.06E-12 | 1.48E-09 | 1.48E-08 |
| Obesity | dendritic_cell | *ZNF852* | -7.960060742 | 4.06E-06 | 0.000522686 | 0.01971038 |
| Obesity | dendritic_cell | *TUFM* | 3.991018174 | 2.09E-06 | 0.000307975 | 0.010163182 |
| Obesity | dendritic_cell | *C3orf38* | 5.566514061 | 2.18E-06 | 0.000311268 | 0.010583096 |
| Obesity | dendritic_cell | *HLA-DQB1* | -0.6027835 | 0.001612331 | 0.048893949 | 1 |
| Obesity | dendritic_cell | *RPRML* | 17.18291655 | 2.73E-06 | 0.000368347 | 0.013260483 |
| Obesity | dendritic_cell | *ZNF322* | -5.555996823 | 0.000357228 | 0.020133421 | 1 |
| Obesity | dendritic_cell | *CLDN7* | -4.461879562 | 0.000677708 | 0.028593371 | 1 |
| Obesity | dendritic_cell | *CAV3* | -3.411429382 | 0.000559613 | 0.024243215 | 1 |
| Obesity | dendritic_cell | *PLGLB1* | 3.564488604 | 0.000496858 | 0.023373013 | 1 |
| Obesity | dendritic_cell | *PSMG1* | 4.698026516 | 0.000299891 | 0.017744744 | 1 |
| Obesity | dendritic_cell | *EMILIN3* | 7.330300572 | 7.31E-05 | 0.006569432 | 0.354749334 |
| Obesity | dendritic_cell | *PGP* | -4.661200367 | 0.000389852 | 0.020606052 | 1 |
| Obesity | dendritic_cell | *TOP1MT* | -2.299668779 | 0.001147827 | 0.040356931 | 1 |
| Obesity | dendritic_cell | *NUTM2A* | -5.221426167 | 1.70E-06 | 0.000266226 | 0.008253001 |
| Obesity | dendritic_cell | *GP1BA* | 6.629456778 | 9.03E-07 | 0.000168468 | 0.004380171 |
| Obesity | dendritic_cell | *BTN3A2* | -2.397130658 | 7.66E-11 | 3.10E-08 | 3.72E-07 |
| Obesity | dendritic_cell | *ZSCAN23* | 1.816336069 | 0.000372979 | 0.020154422 | 1 |
| Obesity | dendritic_cell | *CENPP* | 3.080561051 | 9.90E-05 | 0.008141885 | 0.480371223 |
| Obesity | dendritic_cell | *OSTN* | 1.982770529 | 0.001485595 | 0.046805898 | 1 |
| Obesity | dendritic_cell | *ZKSCAN3* | -7.505313076 | 1.81E-09 | 6.27E-07 | 8.77E-06 |
| Obesity | dendritic_cell | *CPNE4* | 5.088948326 | 0.000241475 | 0.015621799 | 1 |
| Obesity | dendritic_cell | *ZNF605* | 3.106602517 | 0.000408148 | 0.020845646 | 1 |

| | | | | | | |
|---|---|---|---|---|---|---|
| Obesity | dendritic_cell | *SULT1A2* | 15.48220237 | 6.32E-24 | 3.07E-20 | 3.07E-20 |
| Obesity | dendritic_cell | *DNAH10* | 6.872253914 | 4.27E-09 | 1.38E-06 | 2.07E-05 |
| Obesity | dendritic_cell | *RPS26* | -2.790847147 | 0.000128818 | 0.01008106 | 0.625025715 |
| Obesity | dendritic_cell | *HLA-DRB5* | -0.650144049 | 0.000755565 | 0.030996178 | 1 |
| Obesity | dendritic_cell | *SHISA4* | -9.308737425 | 5.86E-06 | 0.000710482 | 0.028419293 |
| Obesity | dendritic_cell | *C6orf163* | -2.230395092 | 0.000526274 | 0.023373013 | 1 |
| Obesity | dendritic_cell | *NOTCH4* | -0.658389404 | 5.20E-08 | 1.40E-05 | 0.000252525 |
| Obesity | dendritic_cell | *SKIV2L* | -2.062086239 | 0.000237173 | 0.015550864 | 1 |
| Obesity | dendritic_cell | *GPANK1* | -7.852365217 | 1.34E-08 | 3.81E-06 | 6.48E-05 |
| Obesity | dendritic_cell | *POU5F1* | 1.172418537 | 2.24E-05 | 0.002367197 | 0.108891068 |
| Obesity | dendritic_cell | *CCHCR1* | 2.152787051 | 4.79E-06 | 0.000596139 | 0.023249422 |
| Obesity | dendritic_cell | *RNF39* | -5.031645643 | 0.000529544 | 0.023373013 | 1 |
| Obesity | dendritic_cell | *ZFP57* | 2.074865872 | 1.65E-06 | 0.000266226 | 0.00801838 |
| Obesity | dendritic_cell | *TMEM110* | 9.145902966 | 8.72E-08 | 2.23E-05 | 0.000423168 |
| Obesity | dendritic_cell | *PPP1CB* | -5.630359421 | 0.000162186 | 0.011745191 | 0.786927775 |
| Obesity | dendritic_cell | *SLC23A3* | 2.405834526 | 0.000532872 | 0.023373013 | 1 |
| Obesity | dendritic_cell | *SLC35E2* | -2.668795443 | 0.001126083 | 0.04005287 | 1 |
| Obesity | dendritic_cell | *ADRA2B* | -4.633674298 | 0.001534874 | 0.04743445 | 1 |
| Obesity | dendritic_cell | *LINC00240* | 2.008462091 | 9.78E-05 | 0.008141885 | 0.474421898 |
| Obesity | dendritic_cell | *HCG20* | -1.233381295 | 0.000373845 | 0.020154422 | 1 |
| Obesity | dendritic_cell | *ZSCAN31* | 1.967522372 | 1.83E-07 | 4.45E-05 | 0.000890123 |
| Obesity | dendritic_cell | *PSMB9* | 3.21898948 | 1.69E-06 | 0.000266226 | 0.008205947 |
| Obesity | dendritic_cell | *ARHGEF25* | 2.010976584 | 0.001481524 | 0.046805898 | 1 |
| Obesity | dendritic_cell | *C4A* | -6.625900177 | 4.09E-06 | 0.000522686 | 0.019862067 |
| Obesity | dendritic_cell | *DNAH10OS* | 17.22580965 | 4.11E-05 | 0.003907976 | 0.199306771 |
| Obesity | dendritic_cell | *PCDHA3* | -7.576569997 | 2.14E-05 | 0.002307041 | 0.103816863 |
| Obesity | dendritic_cell | *HMBS* | 3.060243052 | 0.000197635 | 0.013506016 | 0.958927128 |
| Obesity | dendritic_cell | *AC093668.2* | -6.225305182 | 0.001046361 | 0.037887627 | 1 |
| Obesity | dendritic_cell | *STRADA* | -4.366558232 | 0.00045046 | 0.022302354 | 1 |
| Obesity | dendritic_cell | *AP001172.2* | 2.180343306 | 0.000122332 | 0.009730396 | 0.593554153 |
| Obesity | endometrium | *NFYA* | 4.237796444 | 1.17E-06 | 2.73E-06 | 0.00565879 |
| Obesity | endometrium | *ENPP4* | 6.865488865 | 3.71E-07 | 1.10E-06 | 0.00179879 |
| Obesity | endometrium | *SEMA3F* | 2.871157049 | 0.004615082 | 0.005787639 | 1 |
| Obesity | endometrium | *KRIT1* | 3.776666345 | 0.035364404 | 0.04078633 | 1 |
| Obesity | endometrium | *RAD52* | 9.019572103 | 1.75E-08 | 1.11E-07 | 8.50E-05 |
| Obesity | endometrium | *HS3ST1* | 8.844163969 | 2.55E-06 | 4.92E-06 | 0.012368954 |
| Obesity | endometrium | *AOC1* | 5.047509842 | 0.009448161 | 0.011446311 | 1 |
| Obesity | endometrium | *SNX11* | 2.583446373 | 7.55E-05 | 0.000114816 | 0.366377781 |
| Obesity | endometrium | *TMEM176A* | 8.049136826 | 2.02E-07 | 6.78E-07 | 0.000979736 |
| Obesity | endometrium | *M6PR* | 7.31862534 | 1.64E-08 | 1.06E-07 | 7.94E-05 |
| Obesity | endometrium | *CYP26B1* | 5.978585367 | 1.27E-07 | 4.73E-07 | 0.000617355 |
| Obesity | endometrium | *DBNDD1* | 2.814552828 | 0.017965332 | 0.021255252 | 1 |
| Obesity | endometrium | *ALS2* | 12.5955785 | 4.87E-09 | 5.40E-08 | 2.36E-05 |
| Obesity | endometrium | *CD38* | 7.893403846 | 5.92E-08 | 2.63E-07 | 0.00028704 |
| Obesity | endometrium | *RBM6* | -2.012680644 | 0.022491117 | 0.026397411 | 1 |
| Obesity | endometrium | *RECQL* | 13.04339402 | 8.62E-09 | 7.23E-08 | 4.18E-05 |
| Obesity | endometrium | *ARHGAP33* | 5.558341323 | 4.68E-08 | 2.24E-07 | 0.000226871 |
| Obesity | endometrium | *CDC27* | 5.119244156 | 2.33E-09 | 4.01E-08 | 1.13E-05 |
| Obesity | endometrium | *POLR2J* | 4.192636339 | 9.93E-07 | 2.40E-06 | 0.004818625 |
| Obesity | endometrium | *MEOX1* | -1.173581314 | 0.000317032 | 0.000445737 | 1 |

| | | | | | | |
|---|---|---|---|---|---|---|
| Obesity | endometrium | *LIG3* | -2.319637382 | 0.007296021 | 0.008928195 | 1 |
| Obesity | endometrium | *CIAPIN1* | 3.60532667 | 1.87E-06 | 3.77E-06 | 0.009082933 |
| Obesity | endometrium | *PON1* | 13.33905615 | 2.20E-06 | 4.33E-06 | 0.010688279 |
| Obesity | endometrium | *GCFC2* | 5.954377179 | 1.18E-08 | 8.63E-08 | 5.75E-05 |
| Obesity | endometrium | *ZNF195* | 10.00266204 | 5.73E-08 | 2.59E-07 | 0.000278012 |
| Obesity | endometrium | *PDK2* | 5.963613908 | 4.37E-05 | 6.80E-05 | 0.21223057 |
| Obesity | endometrium | *GGNBP2* | 9.234323054 | 7.43E-07 | 1.93E-06 | 0.003605567 |
| Obesity | endometrium | *ITGA2B* | 5.287570159 | 3.02E-07 | 9.37E-07 | 0.001464711 |
| Obesity | endometrium | *ASB4* | -2.212833205 | 0.005523905 | 0.006865263 | 1 |
| Obesity | endometrium | *TMEM98* | 3.632041445 | 1.97E-06 | 3.94E-06 | 0.009567057 |
| Obesity | endometrium | *ABCC8* | 6.413999823 | 4.49E-05 | 6.98E-05 | 0.2180231 |
| Obesity | endometrium | *SYNRG* | 4.713798806 | 5.28E-08 | 2.44E-07 | 0.000255968 |
| Obesity | endometrium | *ZNF263* | 5.098481103 | 4.91E-08 | 2.31E-07 | 0.000238017 |
| Obesity | endometrium | *SPATA20* | 4.006476632 | 1.38E-06 | 3.12E-06 | 0.006681831 |
| Obesity | endometrium | *BAIAP2L1* | 4.761997733 | 2.60E-07 | 8.32E-07 | 0.001263284 |
| Obesity | endometrium | *AGK* | 10.02881238 | 4.34E-06 | 7.83E-06 | 0.021046268 |
| Obesity | endometrium | *FARP2* | 4.133592844 | 1.38E-07 | 5.02E-07 | 0.000668872 |
| Obesity | endometrium | *USH1C* | -8.927164404 | 0.000311782 | 0.000439119 | 1 |
| Obesity | endometrium | *IFRD1* | 4.276504883 | 6.92E-07 | 1.83E-06 | 0.003358352 |
| Obesity | endometrium | *VPS41* | 8.464706509 | 2.90E-09 | 4.29E-08 | 1.41E-05 |
| Obesity | endometrium | *ELAC2* | 5.659800315 | 0.000209906 | 0.000301677 | 1 |
| Obesity | endometrium | *SCIN* | 5.04056158 | 0.032260201 | 0.037339336 | 1 |
| Obesity | endometrium | *CCDC124* | 4.222844315 | 2.74E-06 | 5.24E-06 | 0.013291991 |
| Obesity | endometrium | *CEACAM21* | 4.435452839 | 3.82E-05 | 5.98E-05 | 0.18524419 |
| Obesity | endometrium | *CD79B* | -2.165516618 | 4.06E-05 | 6.33E-05 | 0.197092723 |
| Obesity | endometrium | *ST7L* | 3.63555439 | 1.93E-06 | 3.87E-06 | 0.009356339 |
| Obesity | endometrium | *PAX6* | 2.738141562 | 0.025150052 | 0.029354836 | 1 |
| Obesity | endometrium | *TSR3* | 4.891776036 | 1.33E-06 | 3.03E-06 | 0.006434057 |
| Obesity | endometrium | *PIGQ* | 11.2304598 | 8.53E-07 | 2.14E-06 | 0.004136434 |
| Obesity | endometrium | *FMO3* | 2.551079675 | 0.002593289 | 0.003333149 | 1 |
| Obesity | endometrium | *PSMB1* | 9.045326256 | 5.09E-09 | 5.56E-08 | 2.47E-05 |
| Obesity | endometrium | *NADK* | 3.035714255 | 3.22E-06 | 6.03E-06 | 0.015647678 |
| Obesity | endometrium | *DLEC1* | -2.336704464 | 0.006056225 | 0.007496123 | 1 |
| Obesity | endometrium | *CYB561* | 7.624643098 | 6.07E-09 | 6.00E-08 | 2.94E-05 |
| Obesity | endometrium | *SS18L2* | 6.926740929 | 3.11E-08 | 1.65E-07 | 0.000150919 |
| Obesity | endometrium | *MGST1* | 2.472459056 | 3.07E-06 | 5.77E-06 | 0.014894053 |
| Obesity | endometrium | *MMP25* | -3.658869093 | 0.00107967 | 0.00143877 | 1 |
| Obesity | endometrium | *IL32* | 4.166073176 | 7.77E-07 | 1.99E-06 | 0.003767713 |
| Obesity | endometrium | *PKD1* | 6.558591961 | 5.99E-09 | 5.97E-08 | 2.91E-05 |
| Obesity | endometrium | *MAPK8IP2* | 20.48167843 | 7.02E-07 | 1.84E-06 | 0.003406568 |
| Obesity | endometrium | *MED24* | 3.052992514 | 1.69E-06 | 3.49E-06 | 0.008189081 |
| Obesity | endometrium | *RPS20* | 10.17103527 | 3.99E-09 | 4.83E-08 | 1.94E-05 |
| Obesity | endometrium | *TRAF3IP3* | 3.690772196 | 1.18E-06 | 2.76E-06 | 0.005737363 |
| Obesity | endometrium | *POMT2* | 3.412979878 | 3.24E-06 | 6.06E-06 | 0.015735984 |
| Obesity | endometrium | *MLXIPL* | 2.47804886 | 8.67E-08 | 3.52E-07 | 0.000420686 |
| Obesity | endometrium | *DYRK4* | 4.074756742 | 4.17E-05 | 6.49E-05 | 0.20215536 |
| Obesity | endometrium | *NCAPD2* | 4.120626371 | 0.003198893 | 0.004076971 | 1 |
| Obesity | endometrium | *FUZ* | 5.373212089 | 8.07E-07 | 2.05E-06 | 0.003913465 |
| Obesity | endometrium | *ZNF200* | 4.097416521 | 7.86E-06 | 1.36E-05 | 0.038121487 |
| Obesity | endometrium | *LRRC23* | 9.344177876 | 6.46E-09 | 6.17E-08 | 3.13E-05 |

| | | | | | | |
|---|---|---|---|---|---|---|
| Obesity | endometrium | *CLCN6* | 4.336619992 | 2.44E-08 | 1.39E-07 | 0.000118438 |
| Obesity | endometrium | *SLC6A7* | 113.4731761 | 1.47E-05 | 2.42E-05 | 0.071412151 |
| Obesity | endometrium | *TSPAN9* | 5.060496605 | 4.88E-08 | 2.30E-07 | 0.000237014 |
| Obesity | endometrium | *UTP18* | 5.076020198 | 9.34E-08 | 3.72E-07 | 0.000453118 |
| Obesity | endometrium | *TTC19* | 7.169913531 | 1.06E-09 | 3.68E-08 | 5.16E-06 |
| Obesity | endometrium | *LARS2* | 4.655550981 | 0.023705456 | 0.027735441 | 1 |
| Obesity | endometrium | *DCN* | 5.603190917 | 0.000120389 | 0.000178632 | 0.584127903 |
| Obesity | endometrium | *PPP5C* | 7.17980642 | 3.96E-08 | 1.98E-07 | 0.000192123 |
| Obesity | endometrium | *CEP68* | 8.139083648 | 5.65E-05 | 8.68E-05 | 0.273974704 |
| Obesity | endometrium | *ZBTB32* | 3.838265251 | 5.89E-06 | 1.04E-05 | 0.028593403 |
| Obesity | endometrium | *ERCC1* | 4.744518934 | 1.51E-06 | 3.29E-06 | 0.007322816 |
| Obesity | endometrium | *CD22* | 1.826367957 | 0.018155607 | 0.02146467 | 1 |
| Obesity | endometrium | *ALOX5* | 2.316943768 | 0.000118119 | 0.000175587 | 0.573114443 |
| Obesity | endometrium | *MAP4K5* | 9.207461438 | 1.47E-09 | 3.98E-08 | 7.11E-06 |
| Obesity | endometrium | *EHD3* | -2.128499926 | 0.000443397 | 0.000612924 | 1 |
| Obesity | endometrium | *MAN2B2* | 5.655093801 | 8.61E-08 | 3.50E-07 | 0.000417802 |
| Obesity | endometrium | *SLC25A39* | 3.542617609 | 1.08E-05 | 1.83E-05 | 0.052628433 |
| Obesity | endometrium | *RNF14* | 8.473358984 | 4.52E-09 | 5.16E-08 | 2.19E-05 |
| Obesity | endometrium | *DDX11* | 3.79879669 | 0.000373758 | 0.000521113 | 1 |
| Obesity | endometrium | *TACC3* | 4.03500237 | 1.95E-06 | 3.90E-06 | 0.009444398 |
| Obesity | endometrium | *ZC3H3* | 5.631070964 | 9.68E-07 | 2.36E-06 | 0.004695153 |
| Obesity | endometrium | *MDH1* | 4.055726417 | 4.84E-08 | 2.29E-07 | 0.000235037 |
| Obesity | endometrium | *MTMR11* | 3.08659962 | 0.024401398 | 0.028515313 | 1 |
| Obesity | endometrium | *COX15* | 9.639494399 | 0.000167689 | 0.000243675 | 0.813629312 |
| Obesity | endometrium | *CCDC88C* | 2.86758475 | 7.10E-06 | 1.24E-05 | 0.034468329 |
| Obesity | endometrium | *NUDCD3* | 9.88442856 | 1.63E-11 | 2.93E-09 | 7.91E-08 |
| Obesity | endometrium | *CHDH* | 8.585896592 | 1.11E-06 | 2.63E-06 | 0.005375775 |
| Obesity | endometrium | *CLCA4* | 8.309949612 | 0.023639019 | 0.02766438 | 1 |
| Obesity | endometrium | *RUFY3* | 8.595399151 | 1.36E-09 | 3.98E-08 | 6.58E-06 |
| Obesity | endometrium | *SLC11A1* | 2.03809848 | 0.000938567 | 0.001258687 | 1 |
| Obesity | endometrium | *WWTR1* | 7.865613482 | 1.90E-09 | 4.00E-08 | 9.22E-06 |
| Obesity | endometrium | *AGPS* | 8.329377863 | 2.97E-09 | 4.29E-08 | 1.44E-05 |
| Obesity | endometrium | *TTC27* | 4.985491672 | 1.69E-08 | 1.09E-07 | 8.21E-05 |
| Obesity | endometrium | *MARCO* | 1.69701799 | 0.000118777 | 0.000176403 | 0.576307796 |
| Obesity | endometrium | *HGF* | 4.792908148 | 2.23E-07 | 7.34E-07 | 0.001080874 |
| Obesity | endometrium | *SLC7A9* | 3.347984206 | 0.000668655 | 0.000907247 | 1 |
| Obesity | endometrium | *OSBPL5* | 6.638467436 | 1.13E-08 | 8.42E-08 | 5.47E-05 |
| Obesity | endometrium | *CPS1* | 6.199445603 | 0.000439196 | 0.000607463 | 1 |
| Obesity | endometrium | *SLC45A4* | 4.882740577 | 3.29E-07 | 1.00E-06 | 0.001598598 |
| Obesity | endometrium | *ZDHHC6* | 3.443619335 | 6.64E-07 | 1.77E-06 | 0.003222745 |
| Obesity | endometrium | *GRAMD1B* | 2.771273343 | 9.72E-06 | 1.65E-05 | 0.047180959 |
| Obesity | endometrium | *RNH1* | 9.305485884 | 1.45E-10 | 1.40E-08 | 7.04E-07 |
| Obesity | endometrium | *GLRX2* | 4.294966449 | 1.79E-06 | 3.64E-06 | 0.00866782 |
| Obesity | endometrium | *ABCC2* | 5.372993726 | 4.51E-06 | 8.11E-06 | 0.021889814 |
| Obesity | endometrium | *GCLM* | 3.13500587 | 1.87E-06 | 3.76E-06 | 0.009057582 |
| Obesity | endometrium | *UBR2* | 3.394266966 | 1.82E-07 | 6.27E-07 | 0.000884902 |
| Obesity | endometrium | *RRAGD* | 4.346164394 | 1.70E-06 | 3.50E-06 | 0.008244894 |
| Obesity | endometrium | *HSF2* | 3.049313614 | 2.92E-06 | 5.52E-06 | 0.014149527 |
| Obesity | endometrium | *TYMP* | 2.973292933 | 8.79E-07 | 2.20E-06 | 0.004265292 |
| Obesity | endometrium | *TOMM34* | 4.622001088 | 4.37E-06 | 7.88E-06 | 0.021199771 |

| | | | | | | |
|---|---|---|---|---|---|---|
| Obesity | endometrium | *SEC63* | 5.850876057 | 1.52E-08 | 1.02E-07 | 7.38E-05 |
| Obesity | endometrium | *FAS* | 4.599027162 | 1.22E-07 | 4.58E-07 | 0.000591062 |
| Obesity | endometrium | *RNASET2* | 5.18174522 | 0.000377226 | 0.000525496 | 1 |
| Obesity | endometrium | *AGPAT4* | 3.317740724 | 5.56E-07 | 1.52E-06 | 0.002696723 |
| Obesity | endometrium | *BTN3A1* | 3.102084981 | 0.000443154 | 0.000612762 | 1 |
| Obesity | endometrium | *MIPEP* | 6.861216231 | 0.002589581 | 0.003329266 | 1 |
| Obesity | endometrium | *INSRR* | -4.320806858 | 0.001982179 | 0.002577045 | 1 |
| Obesity | endometrium | *B4GALT7* | 5.59763664 | 8.92E-07 | 2.22E-06 | 0.004329242 |
| Obesity | endometrium | *SH2D2A* | 13.66709276 | 1.98E-07 | 6.69E-07 | 0.000962007 |
| Obesity | endometrium | *VRK2* | 9.937116198 | 4.58E-09 | 5.18E-08 | 2.22E-05 |
| Obesity | endometrium | *VEZT* | 8.605128674 | 1.69E-09 | 3.98E-08 | 8.19E-06 |
| Obesity | endometrium | *BRD9* | 11.79985032 | 2.43E-08 | 1.39E-07 | 0.000118049 |
| Obesity | endometrium | *SNX1* | 3.155829316 | 1.86E-07 | 6.38E-07 | 0.000903978 |
| Obesity | endometrium | *ANK1* | 5.48639725 | 1.44E-06 | 3.19E-06 | 0.006971221 |
| Obesity | endometrium | *RABEP1* | 4.335412915 | 1.47E-08 | 1.00E-07 | 7.14E-05 |
| Obesity | endometrium | *BAK1* | 5.673513649 | 0.000234562 | 0.000335622 | 1 |
| Obesity | endometrium | *IKZF2* | 10.32355582 | 2.23E-09 | 4.00E-08 | 1.08E-05 |
| Obesity | endometrium | *GRN* | 2.223819844 | 2.06E-06 | 4.10E-06 | 0.010005667 |
| Obesity | endometrium | *IFT88* | 6.047375964 | 1.59E-08 | 1.04E-07 | 7.73E-05 |
| Obesity | endometrium | *CHPF2* | 8.602660407 | 0.001240684 | 0.001645653 | 1 |
| Obesity | endometrium | *UBA6* | 7.046517626 | 9.15E-06 | 1.56E-05 | 0.044400845 |
| Obesity | endometrium | *ATP6V0A1* | 5.860580256 | 2.86E-09 | 4.29E-08 | 1.39E-05 |
| Obesity | endometrium | *PIAS1* | 5.336719495 | 5.04E-10 | 2.47E-08 | 2.45E-06 |
| Obesity | endometrium | *MYOC* | 3.978941509 | 7.22E-07 | 1.88E-06 | 0.003503427 |
| Obesity | endometrium | *SH3YL1* | 3.522586159 | 7.76E-06 | 1.35E-05 | 0.037633451 |
| Obesity | endometrium | *VCL* | 5.051506764 | 0.008462449 | 0.010303589 | 1 |
| Obesity | endometrium | *ADSS* | 4.125433795 | 2.50E-07 | 8.03E-07 | 0.001211792 |
| Obesity | endometrium | *TBC1D23* | 10.75248042 | 1.76E-09 | 3.98E-08 | 8.54E-06 |
| Obesity | endometrium | *MYOM2* | 5.981109025 | 5.04E-05 | 7.78E-05 | 0.244339794 |
| Obesity | endometrium | *TUBG2* | -2.779979159 | 0.036209165 | 0.041720938 | 1 |
| Obesity | endometrium | *FLT4* | 1.51461021 | 0.028364352 | 0.032955899 | 1 |
| Obesity | endometrium | *NSUN2* | 10.17072726 | 3.65E-10 | 2.30E-08 | 1.77E-06 |
| Obesity | endometrium | *MFAP3* | 6.140251622 | 1.37E-06 | 3.11E-06 | 0.006669878 |
| Obesity | endometrium | *AGA* | 7.304708128 | 2.42E-07 | 7.85E-07 | 0.001173527 |
| Obesity | endometrium | *PI4K2B* | 8.137923879 | 1.50E-06 | 3.28E-06 | 0.00727569 |
| Obesity | endometrium | *BOD1L1* | 7.523364633 | 2.17E-09 | 4.00E-08 | 1.05E-05 |
| Obesity | endometrium | *DNAH5* | 5.464366836 | 9.82E-07 | 2.38E-06 | 0.004764406 |
| Obesity | endometrium | *PNKP* | 5.336268457 | 1.54E-08 | 1.03E-07 | 7.48E-05 |
| Obesity | endometrium | *PHLPP2* | 2.988266409 | 9.67E-07 | 2.36E-06 | 0.004692971 |
| Obesity | endometrium | *SPDL1* | 5.515528799 | 0.011681772 | 0.014029693 | 1 |
| Obesity | endometrium | *CTNS* | 5.24313712 | 9.51E-05 | 0.000143094 | 0.461636884 |
| Obesity | endometrium | *INPP4A* | 2.850136036 | 4.27E-07 | 1.24E-06 | 0.002073731 |
| Obesity | endometrium | *PSMA4* | 5.063664062 | 3.14E-07 | 9.64E-07 | 0.001521228 |
| Obesity | endometrium | *LSG1* | 3.178660096 | 2.31E-08 | 1.34E-07 | 0.000111928 |
| Obesity | endometrium | *TDP1* | 7.030196187 | 2.87E-08 | 1.56E-07 | 0.00013927 |
| Obesity | endometrium | *SPATA7* | 4.810214571 | 0.022606707 | 0.026520247 | 1 |
| Obesity | endometrium | *MED17* | 3.547703528 | 1.36E-06 | 3.09E-06 | 0.006595773 |
| Obesity | endometrium | *CAPG* | 3.797396034 | 4.72E-06 | 8.47E-06 | 0.022896211 |
| Obesity | endometrium | *AP2S1* | 5.476524566 | 2.33E-07 | 7.62E-07 | 0.001132916 |
| Obesity | endometrium | *TRIT1* | 3.433458258 | 1.44E-06 | 3.20E-06 | 0.006988287 |

| | | | | | | |
|---|---|---|---|---|---|---|
| Obesity | endometrium | *CUL7* | 7.826107992 | 4.59E-07 | 1.31E-06 | 0.002226356 |
| Obesity | endometrium | *CTNNA1* | 6.593736204 | 3.59E-09 | 4.68E-08 | 1.74E-05 |
| Obesity | endometrium | *CNTLN* | 4.496064054 | 6.32E-08 | 2.77E-07 | 0.00030671 |
| Obesity | endometrium | *YTHDC2* | 5.074256529 | 2.81E-08 | 1.53E-07 | 0.000136397 |
| Obesity | endometrium | *POLR2B* | 4.023561806 | 1.79E-07 | 6.19E-07 | 0.000870476 |
| Obesity | endometrium | *ARAP2* | 3.589987982 | 2.59E-07 | 8.29E-07 | 0.0012579 |
| Obesity | endometrium | *DTNBP1* | 11.292701 | 4.95E-07 | 1.39E-06 | 0.002404022 |
| Obesity | endometrium | *ANO2* | 2.414786816 | 1.48E-06 | 3.26E-06 | 0.007200979 |
| Obesity | endometrium | *SNX29* | 2.469708714 | 5.14E-07 | 1.43E-06 | 0.002493888 |
| Obesity | endometrium | *MRPS10* | 2.108636273 | 0.000596988 | 0.000813191 | 1 |
| Obesity | endometrium | *UTS2* | 4.50993406 | 0.004133512 | 0.005212007 | 1 |
| Obesity | endometrium | *EPN3* | 8.026581679 | 0.004076833 | 0.005143213 | 1 |
| Obesity | endometrium | *RCN1* | 2.642732082 | 7.37E-06 | 1.28E-05 | 0.035743482 |
| Obesity | endometrium | *JKAMP* | 12.46820687 | 4.29E-10 | 2.40E-08 | 2.08E-06 |
| Obesity | endometrium | *MCUR1* | 5.818674479 | 1.98E-07 | 6.69E-07 | 0.000962216 |
| Obesity | endometrium | *LAMC3* | -0.710844136 | 0.000176919 | 0.000256165 | 0.858408714 |
| Obesity | endometrium | *MAPK9* | 14.1569397 | 4.20E-08 | 2.07E-07 | 0.000203669 |
| Obesity | endometrium | *FAM160A2* | 5.863859836 | 1.70E-07 | 5.94E-07 | 0.000824383 |
| Obesity | endometrium | *CYBA* | 4.472023982 | 6.81E-08 | 2.94E-07 | 0.000330582 |
| Obesity | endometrium | *THOC3* | 36.94141499 | 5.02E-09 | 5.53E-08 | 2.44E-05 |
| Obesity | endometrium | *ALX4* | 7.205262674 | 6.30E-06 | 1.11E-05 | 0.030563923 |
| Obesity | endometrium | *FSTL4* | 3.165716296 | 0.006722402 | 0.008280552 | 1 |
| Obesity | endometrium | *AKR7A2* | 11.3309394 | 8.97E-09 | 7.32E-08 | 4.35E-05 |
| Obesity | endometrium | *MRTO4* | 4.3455481 | 8.81E-07 | 2.20E-06 | 0.004273868 |
| Obesity | endometrium | *ANAPC4* | 4.654045767 | 1.74E-08 | 1.11E-07 | 8.43E-05 |
| Obesity | endometrium | *LY75* | 7.500996039 | 6.97E-07 | 1.84E-06 | 0.003382014 |
| Obesity | endometrium | *OPN3* | 8.135239376 | 4.11E-06 | 7.45E-06 | 0.019921268 |
| Obesity | endometrium | *SDCCAG8* | 6.696690952 | 3.29E-11 | 5.31E-09 | 1.59E-07 |
| Obesity | endometrium | *KIF1B* | 9.467645567 | 6.34E-10 | 2.72E-08 | 3.08E-06 |
| Obesity | endometrium | *TBC1D22A* | 5.194607725 | 9.36E-09 | 7.47E-08 | 4.54E-05 |
| Obesity | endometrium | *FAM114A2* | 6.773742019 | 1.95E-07 | 6.62E-07 | 0.000947253 |
| Obesity | endometrium | *GINM1* | 5.873285454 | 5.02E-08 | 2.35E-07 | 0.000243732 |
| Obesity | endometrium | *ITIH4* | 5.663064217 | 2.20E-13 | 8.91E-11 | 1.07E-09 |
| Obesity | endometrium | *NPFFR2* | 6.514560809 | 1.64E-06 | 3.44E-06 | 0.007981143 |
| Obesity | endometrium | *PCGF2* | 3.442362344 | 1.82E-05 | 2.96E-05 | 0.088178516 |
| Obesity | endometrium | *IL17RB* | 8.601707865 | 6.21E-05 | 9.50E-05 | 0.301475975 |
| Obesity | endometrium | *MSH4* | 3.064382013 | 1.24E-06 | 2.86E-06 | 0.006019382 |
| Obesity | endometrium | *GDI2* | 4.934592952 | 1.32E-07 | 4.87E-07 | 0.000641828 |
| Obesity | endometrium | *PITHD1* | 7.010952288 | 5.68E-08 | 2.57E-07 | 0.000275399 |
| Obesity | endometrium | *ATP11B* | 5.403994349 | 2.03E-09 | 4.00E-08 | 9.85E-06 |
| Obesity | endometrium | *RASGRF1* | 9.209865338 | 7.96E-07 | 2.03E-06 | 0.003862234 |
| Obesity | endometrium | *CROCC* | 7.448770685 | 2.42E-08 | 1.38E-07 | 0.00011758 |
| Obesity | endometrium | *FLYWCH1* | 6.449736933 | 3.56E-05 | 5.60E-05 | 0.172930988 |
| Obesity | endometrium | *DNAJC25* | 4.527241313 | 1.58E-06 | 3.36E-06 | 0.00768286 |
| Obesity | endometrium | *OGFR* | 6.355441661 | 0.003763216 | 0.00476615 | 1 |
| Obesity | endometrium | *GNA15* | 3.04993341 | 0.001033647 | 0.001380092 | 1 |
| Obesity | endometrium | *PIGV* | -2.608668434 | 0.007009003 | 0.008603006 | 1 |
| Obesity | endometrium | *RIMBP2* | 4.284268589 | 1.62E-06 | 3.41E-06 | 0.007880213 |
| Obesity | endometrium | *PRDM6* | -2.260212524 | 0.007366395 | 0.009009768 | 1 |
| Obesity | endometrium | *SPAG4* | 7.167212726 | 0.015877673 | 0.018867544 | 1 |

| | | | | | | |
|---|---|---|---|---|---|---|
| Obesity | endometrium | *NCKAP1* | 5.033662712 | 4.48E-08 | 2.16E-07 | 0.000217491 |
| Obesity | endometrium | *LTK* | 32.79508579 | 1.31E-06 | 3.00E-06 | 0.006358744 |
| Obesity | endometrium | *EIF4B* | 9.417635215 | 4.40E-08 | 2.13E-07 | 0.000213545 |
| Obesity | endometrium | *SLC6A16* | 3.232107976 | 2.27E-06 | 4.44E-06 | 0.011011936 |
| Obesity | endometrium | *AHRR* | 11.85013599 | 2.21E-06 | 4.33E-06 | 0.010704548 |
| Obesity | endometrium | *ADCK1* | 3.372495851 | 1.67E-06 | 3.47E-06 | 0.008114375 |
| Obesity | endometrium | *HAGH* | 6.421917225 | 3.75E-08 | 1.90E-07 | 0.000181788 |
| Obesity | endometrium | *RNF4* | 6.592063651 | 2.13E-08 | 1.27E-07 | 0.000103318 |
| Obesity | endometrium | *CASP8* | 11.15069517 | 1.90E-06 | 3.83E-06 | 0.009227465 |
| Obesity | endometrium | *TM7SF3* | 3.516614395 | 2.49E-07 | 8.02E-07 | 0.001207996 |
| Obesity | endometrium | *CDON* | 1.629304778 | 3.46E-06 | 6.42E-06 | 0.016795441 |
| Obesity | endometrium | *TMEM161A* | 4.345693347 | 1.21E-06 | 2.81E-06 | 0.00585852 |
| Obesity | endometrium | *LPAR2* | 6.990429862 | 1.52E-07 | 5.43E-07 | 0.00073621 |
| Obesity | endometrium | *SUGP2* | 3.904174882 | 3.66E-07 | 1.09E-06 | 0.001776468 |
| Obesity | endometrium | *CNN2* | 9.660547836 | 5.46E-08 | 2.51E-07 | 0.000264792 |
| Obesity | endometrium | *ABCA7* | 3.798659546 | 2.96E-05 | 4.69E-05 | 0.143624128 |
| Obesity | endometrium | *DDX20* | 4.375804417 | 4.32E-07 | 1.25E-06 | 0.002098086 |
| Obesity | endometrium | *BTBD1* | 6.655968938 | 6.31E-09 | 6.12E-08 | 3.06E-05 |
| Obesity | endometrium | *FAR2* | 9.940479166 | 6.74E-09 | 6.29E-08 | 3.27E-05 |
| Obesity | endometrium | *BCAS1* | 5.785237764 | 6.05E-06 | 1.07E-05 | 0.029330373 |
| Obesity | endometrium | *CHI3L2* | -6.973853138 | 0.012812236 | 0.015345586 | 1 |
| Obesity | endometrium | *SBNO2* | 6.325177327 | 1.81E-08 | 1.14E-07 | 8.79E-05 |
| Obesity | endometrium | *PMS1* | 7.530310474 | 2.89E-09 | 4.29E-08 | 1.40E-05 |
| Obesity | endometrium | *UHRF1BP1* | 7.1826231 | 3.54E-18 | 3.43E-15 | 1.72E-14 |
| Obesity | endometrium | *WDR18* | 3.193455507 | 0.00032611 | 0.000457572 | 1 |
| Obesity | endometrium | *ANKRD44* | 2.245908339 | 7.61E-07 | 1.96E-06 | 0.003692789 |
| Obesity | endometrium | *MAP2K4* | 4.1807774 | 8.85E-08 | 3.56E-07 | 0.000429224 |
| Obesity | endometrium | *TMEM206* | 3.129670054 | 2.08E-06 | 4.13E-06 | 0.010101675 |
| Obesity | endometrium | *TIE1* | 5.848537984 | 2.05E-05 | 3.31E-05 | 0.099300279 |
| Obesity | endometrium | *ZMYND12* | -1.847913386 | 0.015498076 | 0.018448642 | 1 |
| Obesity | endometrium | *NGEF* | 10.80347463 | 1.77E-05 | 2.88E-05 | 0.085762814 |
| Obesity | endometrium | *SPI1* | 6.615849678 | 3.87E-06 | 7.07E-06 | 0.018767288 |
| Obesity | endometrium | *ZNRD1* | -2.560661119 | 0.00027949 | 0.000395937 | 1 |
| Obesity | endometrium | *ATXN3* | 5.081495088 | 5.24E-09 | 5.66E-08 | 2.54E-05 |
| Obesity | endometrium | *GOLGA5* | 12.30283487 | 2.56E-06 | 4.93E-06 | 0.012411918 |
| Obesity | endometrium | *FGFR2* | 3.55115951 | 6.78E-07 | 1.80E-06 | 0.00329166 |
| Obesity | endometrium | *MTFR1* | 7.793352285 | 1.70E-07 | 5.94E-07 | 0.000826055 |
| Obesity | endometrium | *FECH* | 4.334281467 | 0.000238691 | 0.000341027 | 1 |
| Obesity | endometrium | *MYO9A* | 5.769753143 | 8.84E-09 | 7.32E-08 | 4.29E-05 |
| Obesity | endometrium | *TRAM1* | 4.323328726 | 1.04E-07 | 4.04E-07 | 0.000502559 |
| Obesity | endometrium | *EVI5* | 5.377814544 | 7.17E-09 | 6.50E-08 | 3.48E-05 |
| Obesity | endometrium | *METTL22* | 10.23516819 | 3.87E-08 | 1.95E-07 | 0.000187871 |
| Obesity | endometrium | *TP53BP1* | 4.926400051 | 3.31E-08 | 1.73E-07 | 0.000160543 |
| Obesity | endometrium | *RRP15* | 5.068803342 | 2.27E-06 | 4.44E-06 | 0.011031156 |
| Obesity | endometrium | *DHX8* | 13.49574245 | 1.73E-09 | 3.98E-08 | 8.38E-06 |
| Obesity | endometrium | *ROGDI* | 20.08394647 | 0.000469652 | 0.000647557 | 1 |
| Obesity | endometrium | *CBFB* | 7.587153512 | 5.76E-09 | 5.88E-08 | 2.79E-05 |
| Obesity | endometrium | *FGFR3* | -1.807962619 | 0.000138952 | 0.000204549 | 0.674194498 |
| Obesity | endometrium | *IFI35* | 8.908578421 | 8.40E-08 | 3.44E-07 | 0.000407632 |
| Obesity | endometrium | *HEATR6* | 11.47652771 | 4.97E-08 | 2.33E-07 | 0.000241302 |

| | | | | | | |
|---|---|---|---|---|---|---|
| Obesity | endometrium | *POLR1A* | 2.857244998 | 7.01E-06 | 1.22E-05 | 0.034020677 |
| Obesity | endometrium | *IP6K2* | 5.433675559 | 8.14E-08 | 3.37E-07 | 0.000394779 |
| Obesity | endometrium | *SRBD1* | 5.572793077 | 1.03E-08 | 7.97E-08 | 4.99E-05 |
| Obesity | endometrium | *CYFIP1* | 5.430172039 | 1.52E-08 | 1.02E-07 | 7.39E-05 |
| Obesity | endometrium | *IFT80* | 7.189033973 | 7.03E-08 | 3.02E-07 | 0.000341214 |
| Obesity | endometrium | *NUCKS1* | 4.598610616 | 5.31E-08 | 2.45E-07 | 0.000257419 |
| Obesity | endometrium | *CLEC2D* | 4.803145333 | 7.24E-08 | 3.08E-07 | 0.00035108 |
| Obesity | endometrium | *DRD4* | -2.790247358 | 0.00103176 | 0.001378331 | 1 |
| Obesity | endometrium | *PLA2G10* | 9.388182297 | 1.50E-06 | 3.28E-06 | 0.007291652 |
| Obesity | endometrium | *NEDD4* | 8.565127989 | 1.19E-06 | 2.79E-06 | 0.005794424 |
| Obesity | endometrium | *PIGB* | 5.266199008 | 8.21E-08 | 3.38E-07 | 0.000398214 |
| Obesity | endometrium | *GNB5* | 6.710453634 | 0.000151834 | 0.000222098 | 0.736700586 |
| Obesity | endometrium | *LRP6* | 4.660224588 | 7.53E-08 | 3.17E-07 | 0.000365253 |
| Obesity | endometrium | *DAPP1* | 1.538010373 | 0.036982493 | 0.042579822 | 1 |
| Obesity | endometrium | *CLTCL1* | 2.842514478 | 7.71E-07 | 1.99E-06 | 0.003742097 |
| Obesity | endometrium | *DGCR2* | 10.56653159 | 2.41E-09 | 4.02E-08 | 1.17E-05 |
| Obesity | endometrium | *ZXDC* | 6.203736264 | 1.11E-08 | 8.34E-08 | 5.39E-05 |
| Obesity | endometrium | *WIPI1* | 4.558748837 | 0.030181135 | 0.035024843 | 1 |
| Obesity | endometrium | *GBA2* | 14.21244155 | 5.62E-07 | 1.53E-06 | 0.00272647 |
| Obesity | endometrium | *NDST1* | 12.75249503 | 7.03E-09 | 6.44E-08 | 3.41E-05 |
| Obesity | endometrium | *ASNS* | -2.640239059 | 0.001015915 | 0.001357537 | 1 |
| Obesity | endometrium | *AP3M2* | 13.59722316 | 4.03E-07 | 1.18E-06 | 0.001955669 |
| Obesity | endometrium | *ST6GALNAC2* | 4.227706885 | 1.66E-06 | 3.45E-06 | 0.008031687 |
| Obesity | endometrium | *EIF2B3* | 7.54250905 | 4.13E-09 | 4.89E-08 | 2.00E-05 |
| Obesity | endometrium | *MAP4K4* | 9.076700669 | 5.34E-09 | 5.71E-08 | 2.59E-05 |
| Obesity | endometrium | *WDR1* | 4.047566961 | 4.85E-07 | 1.37E-06 | 0.002354102 |
| Obesity | endometrium | *SNX13* | 5.068884559 | 2.27E-08 | 1.33E-07 | 0.000110054 |
| Obesity | endometrium | *RPS6KA2* | 3.966402788 | 1.78E-07 | 6.16E-07 | 0.000863718 |
| Obesity | endometrium | *ING3* | 3.797176489 | 7.33E-07 | 1.91E-06 | 0.003557365 |
| Obesity | endometrium | *VASH1* | 4.640416332 | 1.76E-06 | 3.60E-06 | 0.008544832 |
| Obesity | endometrium | *TRIB2* | 3.806409383 | 3.11E-06 | 5.82E-06 | 0.015069847 |
| Obesity | endometrium | *CPSF1* | 2.875943396 | 0.000277074 | 0.000392859 | 1 |
| Obesity | endometrium | *CYBRD1* | 7.940820755 | 3.22E-10 | 2.19E-08 | 1.56E-06 |
| Obesity | endometrium | *RDH11* | 3.700282112 | 1.39E-06 | 3.14E-06 | 0.006750743 |
| Obesity | endometrium | *EPN2* | 4.97564178 | 6.13E-08 | 2.70E-07 | 0.000297188 |
| Obesity | endometrium | *ALDH3A2* | 5.472201428 | 1.90E-08 | 1.18E-07 | 9.24E-05 |
| Obesity | endometrium | *AFF4* | 5.832261103 | 1.38E-09 | 3.98E-08 | 6.70E-06 |
| Obesity | endometrium | *TRHDE* | 3.023176179 | 1.81E-06 | 3.67E-06 | 0.008774304 |
| Obesity | endometrium | *FCGR2B* | 3.366942145 | 1.72E-06 | 3.53E-06 | 0.008363897 |
| Obesity | endometrium | *NFATC3* | 4.13387302 | 5.53E-08 | 2.53E-07 | 0.000268414 |
| Obesity | endometrium | *TRNT1* | 2.487498986 | 1.40E-06 | 3.14E-06 | 0.006786346 |
| Obesity | endometrium | *NDE1* | 8.002898978 | 5.23E-09 | 5.66E-08 | 2.54E-05 |
| Obesity | endometrium | *AP1M1* | 9.8506673 | 6.52E-09 | 6.17E-08 | 3.16E-05 |
| Obesity | endometrium | *PVR* | 14.31238458 | 6.33E-09 | 6.12E-08 | 3.07E-05 |
| Obesity | endometrium | *SCARB1* | 3.44966165 | 7.36E-07 | 1.92E-06 | 0.003573486 |
| Obesity | endometrium | *MOV10L1* | -1.881465592 | 0.001485261 | 0.001954567 | 1 |
| Obesity | endometrium | *ALPK1* | 4.245919511 | 2.71E-08 | 1.49E-07 | 0.000131258 |
| Obesity | endometrium | *LLGL2* | 4.219474357 | 7.59E-07 | 1.96E-06 | 0.003681339 |
| Obesity | endometrium | *SDHA* | 5.10970534 | 1.37E-08 | 9.51E-08 | 6.64E-05 |
| Obesity | endometrium | *SMARCE1* | 12.63177573 | 4.40E-07 | 1.27E-06 | 0.00213631 |

| | | | | | | |
|---|---|---|---|---|---|---|
| Obesity | endometrium | *CD5L* | 11.00664138 | 7.86E-06 | 1.36E-05 | 0.038126829 |
| Obesity | endometrium | *ZNF532* | 3.86921905 | 1.44E-07 | 5.21E-07 | 0.000700757 |
| Obesity | endometrium | *ZZEF1* | 8.750177339 | 4.14E-10 | 2.39E-08 | 2.01E-06 |
| Obesity | endometrium | *TACR2* | 42.54457961 | 0.000212515 | 0.000305247 | 1 |
| Obesity | endometrium | *TIPIN* | 7.468265008 | 1.92E-09 | 4.00E-08 | 9.30E-06 |
| Obesity | endometrium | *TTC38* | 6.510416871 | 1.15E-08 | 8.49E-08 | 5.58E-05 |
| Obesity | endometrium | *GRAMD4* | 7.189812088 | 2.05E-08 | 1.24E-07 | 9.97E-05 |
| Obesity | endometrium | *ADD2* | 5.709950552 | 2.27E-07 | 7.48E-07 | 0.001103326 |
| Obesity | endometrium | *VPS9D1* | 3.173890608 | 8.72E-07 | 2.18E-06 | 0.00423174 |
| Obesity | endometrium | *FNDC3B* | 5.032256795 | 4.44E-09 | 5.09E-08 | 2.15E-05 |
| Obesity | endometrium | *CACNG4* | 6.785330249 | 4.78E-06 | 8.57E-06 | 0.023177076 |
| Obesity | endometrium | *MOCOS* | 5.168431725 | 3.09E-07 | 9.54E-07 | 0.001500925 |
| Obesity | endometrium | *SEC31B* | 7.477025556 | 2.15E-09 | 4.00E-08 | 1.04E-05 |
| Obesity | endometrium | *KIFAP3* | 5.653527571 | 1.98E-08 | 1.21E-07 | 9.59E-05 |
| Obesity | endometrium | *MKRN2* | 11.55524168 | 2.87E-09 | 4.29E-08 | 1.39E-05 |
| Obesity | endometrium | *FMO4* | 5.255799131 | 2.60E-08 | 1.45E-07 | 0.000125999 |
| Obesity | endometrium | *RGS11* | 3.115500241 | 1.14E-06 | 2.68E-06 | 0.005524711 |
| Obesity | endometrium | *SPAG5* | 11.24647495 | 0.000326766 | 0.000458228 | 1 |
| Obesity | endometrium | *ANKRD13A* | 8.763131517 | 0.001034659 | 0.001381064 | 1 |
| Obesity | endometrium | *PAG1* | 2.824671586 | 1.15E-06 | 2.70E-06 | 0.00557118 |
| Obesity | endometrium | *GPATCH1* | 14.13540878 | 1.93E-09 | 4.00E-08 | 9.39E-06 |
| Obesity | endometrium | *ICAM3* | -1.242329914 | 0.000322216 | 0.000452502 | 1 |
| Obesity | endometrium | *NT5C2* | 8.425589794 | 2.75E-12 | 6.07E-10 | 1.34E-08 |
| Obesity | endometrium | *XAB2* | 5.05200041 | 1.15E-06 | 2.70E-06 | 0.005592158 |
| Obesity | endometrium | *IL4R* | 9.08822224 | 1.82E-09 | 3.98E-08 | 8.85E-06 |
| Obesity | endometrium | *DYNC1I2* | 8.331237085 | 2.23E-09 | 4.00E-08 | 1.08E-05 |
| Obesity | endometrium | *APBB1IP* | 2.774577456 | 4.60E-07 | 1.31E-06 | 0.002231696 |
| Obesity | endometrium | *LRCH4* | 6.458128508 | 1.97E-06 | 3.94E-06 | 0.009568311 |
| Obesity | endometrium | *SIRT6* | -2.168721303 | 0.001680754 | 0.002198119 | 1 |
| Obesity | endometrium | *GPR137B* | 4.640197287 | 3.17E-07 | 9.72E-07 | 0.001537095 |
| Obesity | endometrium | *AMPH* | 15.28982386 | 7.74E-06 | 1.34E-05 | 0.037560983 |
| Obesity | endometrium | *MCCC1* | 8.106085538 | 2.93E-08 | 1.58E-07 | 0.000142217 |
| Obesity | endometrium | *ACER3* | 4.880903046 | 2.09E-09 | 4.00E-08 | 1.02E-05 |
| Obesity | endometrium | *PIK3C3* | 6.174330496 | 6.48E-08 | 2.83E-07 | 0.000314627 |
| Obesity | endometrium | *SYNJ2* | 3.51339945 | 1.49E-05 | 2.45E-05 | 0.072402451 |
| Obesity | endometrium | *RBFOX1* | 2.773698311 | 4.74E-07 | 1.34E-06 | 0.002300786 |
| Obesity | endometrium | *FGF20* | 17.98802284 | 0.005049839 | 0.006305152 | 1 |
| Obesity | endometrium | *PCM1* | 8.197677287 | 3.74E-09 | 4.72E-08 | 1.82E-05 |
| Obesity | endometrium | *MYH7B* | -2.850722737 | 0.011344413 | 0.013644792 | 1 |
| Obesity | endometrium | *CLUL1* | 7.15080028 | 5.47E-06 | 9.72E-06 | 0.026518341 |
| Obesity | endometrium | *FKBP7* | -1.180195727 | 0.000435736 | 0.000603193 | 1 |
| Obesity | endometrium | *XRCC5* | 6.093893772 | 8.92E-09 | 7.32E-08 | 4.33E-05 |
| Obesity | endometrium | *SP140* | 3.39043026 | 2.28E-06 | 4.45E-06 | 0.011050808 |
| Obesity | endometrium | *SAR1A* | 7.553244464 | 6.32E-09 | 6.12E-08 | 3.07E-05 |
| Obesity | endometrium | *RAPGEF3* | 2.957147836 | 1.10E-05 | 1.86E-05 | 0.053537133 |
| Obesity | endometrium | *FDFT1* | 5.344137416 | 4.96E-10 | 2.46E-08 | 2.41E-06 |
| Obesity | endometrium | *SCGN* | 27.3580935 | 1.08E-06 | 2.58E-06 | 0.005257894 |
| Obesity | endometrium | *PGM1* | 5.459554915 | 1.11E-07 | 4.27E-07 | 0.000539885 |
| Obesity | endometrium | *DDX1* | 3.142604225 | 1.87E-06 | 3.78E-06 | 0.009095111 |
| Obesity | endometrium | *DNM2* | 3.576756367 | 5.50E-07 | 1.51E-06 | 0.00266993 |

| | | | | | | |
|---|---|---|---|---|---|---|
| Obesity | endometrium | *STX7* | 6.238471104 | 5.82E-09 | 5.92E-08 | 2.82E-05 |
| Obesity | endometrium | *RABL2B* | 3.822124065 | 1.01E-06 | 2.43E-06 | 0.004893158 |
| Obesity | endometrium | *DCT* | 4.992198158 | 3.54E-07 | 1.06E-06 | 0.001718845 |
| Obesity | endometrium | *SCTR* | 3.030581991 | 1.65E-06 | 3.44E-06 | 0.008000367 |
| Obesity | endometrium | *RIF1* | 9.567187069 | 1.44E-09 | 3.98E-08 | 6.98E-06 |
| Obesity | endometrium | *KCNN2* | -5.893945537 | 0.002454738 | 0.00316261 | 1 |
| Obesity | endometrium | *PSEN1* | 3.91580047 | 8.37E-08 | 3.44E-07 | 0.000406317 |
| Obesity | endometrium | *MOK* | 7.446354156 | 1.46E-09 | 3.98E-08 | 7.11E-06 |
| Obesity | endometrium | *STK17B* | 4.885257055 | 1.34E-07 | 4.89E-07 | 0.00064943 |
| Obesity | endometrium | *ZNF506* | 8.485871955 | 9.34E-10 | 3.49E-08 | 4.53E-06 |
| Obesity | endometrium | *PCDHB4* | 6.869782837 | 1.55E-06 | 3.32E-06 | 0.007519245 |
| Obesity | endometrium | *ME2* | 3.31961922 | 5.60E-07 | 1.53E-06 | 0.002715826 |
| Obesity | endometrium | *CCNT2* | 4.453592257 | 4.25E-07 | 1.23E-06 | 0.002064366 |
| Obesity | endometrium | *SEMA5B* | -2.026133862 | 0.000859952 | 0.001158059 | 1 |
| Obesity | endometrium | *DOPEY1* | 10.6830219 | 5.31E-10 | 2.53E-08 | 2.58E-06 |
| Obesity | endometrium | *LYRM2* | 4.843167527 | 5.36E-08 | 2.47E-07 | 0.000259889 |
| Obesity | endometrium | *ZCCHC6* | 3.845049511 | 1.26E-07 | 4.70E-07 | 0.000611787 |
| Obesity | endometrium | *ULK2* | 9.825065233 | 1.42E-09 | 3.98E-08 | 6.88E-06 |
| Obesity | endometrium | *PLOD1* | 13.03115156 | 5.92E-10 | 2.63E-08 | 2.87E-06 |
| Obesity | endometrium | *PIBF1* | 4.435265134 | 2.56E-08 | 1.44E-07 | 0.000124248 |
| Obesity | endometrium | *OXCT1* | 4.055524067 | 3.86E-07 | 1.14E-06 | 0.001875149 |
| Obesity | endometrium | *SLC27A5* | 4.361756558 | 1.32E-06 | 3.01E-06 | 0.006388977 |
| Obesity | endometrium | *ZNF586* | 8.826458476 | 2.05E-10 | 1.77E-08 | 9.93E-07 |
| Obesity | endometrium | *ZNF264* | 3.578589476 | 6.76E-07 | 1.79E-06 | 0.003278211 |
| Obesity | endometrium | *RPS5* | 10.65605646 | 1.81E-08 | 1.14E-07 | 8.78E-05 |
| Obesity | endometrium | *CHMP2B* | 6.179365179 | 7.91E-08 | 3.29E-07 | 0.000383675 |
| Obesity | endometrium | *PPIE* | 9.248877837 | 5.85E-10 | 2.63E-08 | 2.84E-06 |
| Obesity | endometrium | *NOA1* | -1.5746311 | 0.000563546 | 0.000770666 | 1 |
| Obesity | endometrium | *GSTP1* | 14.46544888 | 2.62E-10 | 1.96E-08 | 1.27E-06 |
| Obesity | endometrium | *WBP11* | 3.000751708 | 1.54E-06 | 3.32E-06 | 0.007467497 |
| Obesity | endometrium | *NCOA1* | 6.977547984 | 1.38E-09 | 3.98E-08 | 6.70E-06 |
| Obesity | endometrium | *CAD* | 3.455619214 | 2.28E-06 | 4.45E-06 | 0.011046286 |
| Obesity | endometrium | *CD59* | 3.341184487 | 1.83E-07 | 6.29E-07 | 0.000889301 |
| Obesity | endometrium | *CD82* | 4.884443571 | 7.75E-08 | 3.24E-07 | 0.000376027 |
| Obesity | endometrium | *HACE1* | 6.366496415 | 1.50E-08 | 1.01E-07 | 7.29E-05 |
| Obesity | endometrium | *SEH1L* | 8.896987769 | 3.68E-07 | 1.10E-06 | 0.001786706 |
| Obesity | endometrium | *OVGP1* | 2.892806158 | 0.041950458 | 0.048141822 | 1 |
| Obesity | endometrium | *SLC25A24* | 3.31863601 | 2.37E-07 | 7.73E-07 | 0.001150137 |
| Obesity | endometrium | *PILRA* | 1.961811144 | 4.32E-06 | 7.80E-06 | 0.02094803 |
| Obesity | endometrium | *CPNE3* | 6.575700847 | 1.13E-08 | 8.43E-08 | 5.50E-05 |
| Obesity | endometrium | *RRN3* | 4.889128781 | 5.41E-07 | 1.49E-06 | 0.00262685 |
| Obesity | endometrium | *MTIF2* | 13.52890414 | 3.30E-09 | 4.49E-08 | 1.60E-05 |
| Obesity | endometrium | *DDHD2* | 3.876598344 | 6.81E-07 | 1.80E-06 | 0.003303378 |
| Obesity | endometrium | *EPS15* | 4.979220727 | 2.27E-06 | 4.44E-06 | 0.011004326 |
| Obesity | endometrium | *USP40* | 5.050532719 | 2.62E-08 | 1.46E-07 | 0.000127199 |
| Obesity | endometrium | *POMGNT1* | 8.242254467 | 1.51E-07 | 5.43E-07 | 0.00073421 |
| Obesity | endometrium | *MAST2* | 7.544992868 | 2.27E-10 | 1.93E-08 | 1.10E-06 |
| Obesity | endometrium | *EIF2AK1* | 5.341402837 | 0.018751023 | 0.022141631 | 1 |
| Obesity | endometrium | *NME8* | -5.036528449 | 0.000139915 | 0.000205905 | 0.678868827 |
| Obesity | endometrium | *SNX10* | 2.170658214 | 8.27E-05 | 0.000124933 | 0.401160901 |

| | | | | | | |
|---|---|---|---|---|---|---|
| Obesity | endometrium | *ITPKC* | 7.19568031 | 0.001447195 | 0.001907577 | 1 |
| Obesity | endometrium | *TMED2* | 9.268871548 | 6.17E-06 | 1.09E-05 | 0.029916595 |
| Obesity | endometrium | *ALG9* | 6.836826194 | 3.09E-07 | 9.53E-07 | 0.001497498 |
| Obesity | endometrium | *ACOX3* | 9.461834408 | 1.21E-08 | 8.73E-08 | 5.88E-05 |
| Obesity | endometrium | *HSD17B14* | 4.730885359 | 2.40E-06 | 4.66E-06 | 0.011655343 |
| Obesity | endometrium | *TRIP6* | 6.086251805 | 1.11E-06 | 2.63E-06 | 0.005371147 |
| Obesity | endometrium | *ACHE* | -1.551628779 | 7.65E-05 | 0.000116153 | 0.37099209 |
| Obesity | endometrium | *SRRT* | 4.254863247 | 5.39E-07 | 1.49E-06 | 0.002614473 |
| Obesity | endometrium | *ADAMTS2* | 9.849819898 | 1.55E-05 | 2.54E-05 | 0.075146102 |
| Obesity | endometrium | *PSMC5* | 4.179557341 | 2.77E-06 | 5.29E-06 | 0.013453732 |
| Obesity | endometrium | *UIMC1* | 4.952284786 | 3.72E-08 | 1.89E-07 | 0.0001803 |
| Obesity | endometrium | *LPCAT2* | 2.713316609 | 1.52E-06 | 3.31E-06 | 0.007390155 |
| Obesity | endometrium | *OGFOD1* | 4.616622743 | 1.90E-05 | 3.08E-05 | 0.091967247 |
| Obesity | endometrium | *TXNDC16* | 9.118578022 | 5.61E-07 | 1.53E-06 | 0.002723386 |
| Obesity | endometrium | *SF3B2* | 7.593181462 | 4.30E-07 | 1.24E-06 | 0.00208563 |
| Obesity | endometrium | *PHACTR3* | 3.33376615 | 0.007335296 | 0.008973994 | 1 |
| Obesity | endometrium | *AAMDC* | 4.675052933 | 7.11E-06 | 1.24E-05 | 0.034512437 |
| Obesity | endometrium | *ASAP3* | 3.053618103 | 1.00E-06 | 2.42E-06 | 0.004864653 |
| Obesity | endometrium | *EDEM2* | 6.37321391 | 5.44E-07 | 1.50E-06 | 0.002638786 |
| Obesity | endometrium | *ANKRD10* | 5.256321897 | 3.47E-08 | 1.79E-07 | 0.000168492 |
| Obesity | endometrium | *DOCK3* | 10.59645417 | 2.23E-09 | 4.00E-08 | 1.08E-05 |
| Obesity | endometrium | *ARHGAP28* | 4.109136463 | 5.88E-08 | 2.62E-07 | 0.000285095 |
| Obesity | endometrium | *PPP1R13B* | 3.168335469 | 8.10E-06 | 1.40E-05 | 0.039287099 |
| Obesity | endometrium | *NSFL1C* | 3.054633863 | 2.73E-07 | 8.61E-07 | 0.001322692 |
| Obesity | endometrium | *ZNF343* | 10.4623896 | 4.21E-07 | 1.22E-06 | 0.002041171 |
| Obesity | endometrium | *EBF4* | 16.20147807 | 1.50E-08 | 1.01E-07 | 7.27E-05 |
| Obesity | endometrium | *CPXM1* | -2.18948129 | 0.003767103 | 0.004769829 | 1 |
| Obesity | endometrium | *MAVS* | 7.229466733 | 5.86E-09 | 5.92E-08 | 2.84E-05 |
| Obesity | endometrium | *XRN2* | 6.793039566 | 4.82E-09 | 5.35E-08 | 2.34E-05 |
| Obesity | endometrium | *SNX5* | 12.32394308 | 1.72E-09 | 3.98E-08 | 8.33E-06 |
| Obesity | endometrium | *ESF1* | 6.646555644 | 2.09E-08 | 1.25E-07 | 0.000101592 |
| Obesity | endometrium | *RBBP9* | -2.96148075 | 0.003675094 | 0.004660626 | 1 |
| Obesity | endometrium | *TMEM230* | 5.667282495 | 3.55E-08 | 1.82E-07 | 0.000172233 |
| Obesity | endometrium | *RPH3A* | 10.66747212 | 3.75E-07 | 1.11E-06 | 0.001817181 |
| Obesity | endometrium | *KIF16B* | 2.620852188 | 9.16E-07 | 2.26E-06 | 0.004444034 |
| Obesity | endometrium | *TBX5* | -1.450803454 | 0.000124916 | 0.00018501 | 0.606091212 |
| Obesity | endometrium | *FXYD5* | 3.42884277 | 1.02E-06 | 2.45E-06 | 0.004941237 |
| Obesity | endometrium | *ZNF302* | 7.050840071 | 9.99E-06 | 1.69E-05 | 0.048455214 |
| Obesity | endometrium | *GRAMD1A* | 6.156006663 | 3.00E-08 | 1.61E-07 | 0.000145778 |
| Obesity | endometrium | *CDIP1* | -2.340902176 | 0.014337441 | 0.017100605 | 1 |
| Obesity | endometrium | *GANAB* | 11.12983423 | 5.10E-08 | 2.38E-07 | 0.000247443 |
| Obesity | endometrium | *MLF2* | 11.28849379 | 8.14E-08 | 3.37E-07 | 0.00039519 |
| Obesity | endometrium | *DHX32* | 6.849332952 | 3.01E-08 | 1.61E-07 | 0.000145815 |
| Obesity | endometrium | *SPTLC1* | 5.161390931 | 1.07E-07 | 4.13E-07 | 0.000517234 |
| Obesity | endometrium | *YPEL3* | 12.93712038 | 4.54E-10 | 2.45E-08 | 2.20E-06 |
| Obesity | endometrium | *MRPS33* | 7.002442711 | 6.96E-07 | 1.83E-06 | 0.003377429 |
| Obesity | endometrium | *MAEA* | 3.533128094 | 1.28E-06 | 2.94E-06 | 0.006201208 |
| Obesity | endometrium | *STRN4* | 3.12405281 | 1.12E-05 | 1.88E-05 | 0.054197221 |
| Obesity | endometrium | *LYZ* | 2.531485718 | 3.23E-07 | 9.86E-07 | 0.001569516 |
| Obesity | endometrium | *MUL1* | 3.706501933 | 6.89E-06 | 1.20E-05 | 0.033451931 |

| | | | | | | |
|---|---|---|---|---|---|---|
| Obesity | endometrium | *SPG21* | 7.053173654 | 1.98E-07 | 6.69E-07 | 0.00096095 |
| Obesity | endometrium | *CHRD* | -1.239391588 | 0.000125503 | 0.000185823 | 0.60894202 |
| Obesity | endometrium | *GNPTG* | 5.291491373 | 1.58E-07 | 5.61E-07 | 0.000767363 |
| Obesity | endometrium | *PABPC4* | 2.526307401 | 0.000502359 | 0.000691206 | 1 |
| Obesity | endometrium | *USP48* | 4.761105976 | 1.26E-07 | 4.70E-07 | 0.000611367 |
| Obesity | endometrium | *GLG1* | 5.477883525 | 1.16E-08 | 8.55E-08 | 5.65E-05 |
| Obesity | endometrium | *DTX2* | 3.703086723 | 1.58E-06 | 3.36E-06 | 0.007660416 |
| Obesity | endometrium | *LAMB1* | 3.791331894 | 1.45E-08 | 9.90E-08 | 7.03E-05 |
| Obesity | endometrium | *ABCC6* | 8.284524858 | 4.10E-07 | 1.20E-06 | 0.001986945 |
| Obesity | endometrium | *TF* | 6.244710839 | 2.87E-07 | 8.98E-07 | 0.001390902 |
| Obesity | endometrium | *ALKBH5* | 5.10308522 | 0.024715478 | 0.028875391 | 1 |
| Obesity | endometrium | *SPAG7* | 3.039262309 | 3.79E-07 | 1.12E-06 | 0.001837352 |
| Obesity | endometrium | *RGS17* | 3.577200692 | 0.000765903 | 0.001035142 | 1 |
| Obesity | endometrium | *PPP2R3C* | 7.30104821 | 7.59E-11 | 9.21E-09 | 3.68E-07 |
| Obesity | endometrium | *HAUS4* | 2.412186142 | 0.004671469 | 0.005853813 | 1 |
| Obesity | endometrium | *OSGEP* | 4.467709208 | 2.84E-07 | 8.93E-07 | 0.001379561 |
| Obesity | endometrium | *SCFD1* | 7.911199806 | 1.98E-09 | 4.00E-08 | 9.62E-06 |
| Obesity | endometrium | *SUPT16H* | 7.269504229 | 1.19E-08 | 8.63E-08 | 5.75E-05 |
| Obesity | endometrium | *TOX4* | 3.005093295 | 7.82E-07 | 2.00E-06 | 0.003792278 |
| Obesity | endometrium | *CAPN3* | -3.509104253 | 0.000383724 | 0.000533934 | 1 |
| Obesity | endometrium | *UNC13D* | 2.412294404 | 0.001304924 | 0.001727555 | 1 |
| Obesity | endometrium | *MFSD11* | 6.167408315 | 7.24E-08 | 3.08E-07 | 0.000351083 |
| Obesity | endometrium | *NUP50* | 3.646877008 | 1.13E-06 | 2.67E-06 | 0.005490671 |
| Obesity | endometrium | *COMT* | 8.010864393 | 4.74E-09 | 5.28E-08 | 2.30E-05 |
| Obesity | endometrium | *LRRFIP2* | 5.285286458 | 8.99E-09 | 7.32E-08 | 4.36E-05 |
| Obesity | endometrium | *SEC22C* | 9.552353095 | 0.00021 | 0.000301723 | 1 |
| Obesity | endometrium | *XYLB* | 9.197691475 | 5.95E-09 | 5.97E-08 | 2.89E-05 |
| Obesity | endometrium | *GABRP* | 7.524405059 | 0.000143919 | 0.000211157 | 0.698295884 |
| Obesity | endometrium | *CDC6* | 6.577442492 | 0.007244885 | 0.008874573 | 1 |
| Obesity | endometrium | *FMO2* | 3.740744683 | 1.33E-07 | 4.89E-07 | 0.000647732 |
| Obesity | endometrium | *MSH2* | 3.354061947 | 7.20E-07 | 1.88E-06 | 0.003493821 |
| Obesity | endometrium | *CRAT* | 4.361619458 | 1.63E-06 | 3.42E-06 | 0.007929139 |
| Obesity | endometrium | *NANS* | 8.240507035 | 2.40E-08 | 1.38E-07 | 0.000116563 |
| Obesity | endometrium | *PDE6C* | -2.887045813 | 0.00294457 | 0.003765697 | 1 |
| Obesity | endometrium | *CRTAC1* | -2.69752566 | 0.002568042 | 0.003302449 | 1 |
| Obesity | endometrium | *WAC* | 6.317662311 | 5.42E-09 | 5.76E-08 | 2.63E-05 |
| Obesity | endometrium | *ITPR3* | 5.271734201 | 2.49E-10 | 1.94E-08 | 1.21E-06 |
| Obesity | endometrium | *DSP* | 4.123084408 | 1.53E-06 | 3.31E-06 | 0.00741238 |
| Obesity | endometrium | *IFT74* | 5.383048006 | 9.41E-09 | 7.48E-08 | 4.57E-05 |
| Obesity | endometrium | *ACOT7* | 4.852533611 | 2.17E-06 | 4.27E-06 | 0.010520951 |
| Obesity | endometrium | *SCD* | 3.888989577 | 4.00E-07 | 1.17E-06 | 0.001938632 |
| Obesity | endometrium | *RAB18* | 6.888091091 | 9.00E-09 | 7.32E-08 | 4.37E-05 |
| Obesity | endometrium | *TSPAN15* | 8.074937237 | 4.18E-09 | 4.89E-08 | 2.03E-05 |
| Obesity | endometrium | *MAST3* | 2.641414299 | 3.45E-05 | 5.43E-05 | 0.167412525 |
| Obesity | endometrium | *OCEL1* | 2.937035404 | 0.00028523 | 0.000403598 | 1 |
| Obesity | endometrium | *CATSPERG* | 3.25263676 | 0.000111757 | 0.000166435 | 0.542245468 |
| Obesity | endometrium | *SETD1A* | 4.219794013 | 3.85E-08 | 1.95E-07 | 0.000187027 |
| Obesity | endometrium | *CIRBP* | 9.733839975 | 5.78E-08 | 2.59E-07 | 0.00028041 |
| Obesity | endometrium | *MARCH2* | 2.784289499 | 1.58E-06 | 3.36E-06 | 0.007661352 |
| Obesity | endometrium | *MTAP* | -2.484218011 | 0.025115493 | 0.029321552 | 1 |

| | | | | | | |
|---|---|---|---|---|---|---|
| Obesity | endometrium | *POLR2E* | 8.354889323 | 3.74E-08 | 1.90E-07 | 0.000181489 |
| Obesity | endometrium | *MADCAM1* | 13.65230655 | 2.61E-06 | 5.02E-06 | 0.012665514 |
| Obesity | endometrium | *MED15* | 9.617909341 | 9.36E-06 | 1.60E-05 | 0.045415134 |
| Obesity | endometrium | *SNAP29* | 4.840119547 | 4.30E-05 | 6.68E-05 | 0.20844542 |
| Obesity | endometrium | *LZTR1* | 5.448754892 | 1.38E-06 | 3.13E-06 | 0.006713283 |
| Obesity | endometrium | *MMP11* | -0.714123689 | 0.000102471 | 0.000153643 | 0.497188318 |
| Obesity | endometrium | *CECR2* | 7.28105076 | 9.90E-08 | 3.90E-07 | 0.00048056 |
| Obesity | endometrium | *SMARCB1* | 3.39895559 | 5.43E-08 | 2.49E-07 | 0.000263451 |
| Obesity | endometrium | *BCL2L13* | 10.72713398 | 1.63E-12 | 4.40E-10 | 7.93E-09 |
| Obesity | endometrium | *DDTL* | 3.664675567 | 0.006638011 | 0.008182833 | 1 |
| Obesity | endometrium | *DDT* | -2.557606336 | 0.019253504 | 0.022701822 | 1 |
| Obesity | endometrium | *OSM* | -2.283297645 | 0.000133043 | 0.000196208 | 0.645524262 |
| Obesity | endometrium | *CABIN1* | 8.451817322 | 8.42E-10 | 3.24E-08 | 4.09E-06 |
| Obesity | endometrium | *SPECC1L* | -2.215194082 | 0.001226969 | 0.001627906 | 1 |
| Obesity | endometrium | *PPIL2* | 8.632700734 | 7.64E-05 | 0.000116084 | 0.370655915 |
| Obesity | endometrium | *PES1* | 3.402256441 | 2.25E-06 | 4.41E-06 | 0.010925413 |
| Obesity | endometrium | *SLC35E4* | -1.917950693 | 0.028409777 | 0.033000775 | 1 |
| Obesity | endometrium | *TOP3B* | 7.673261762 | 0.012766188 | 0.015294208 | 1 |
| Obesity | endometrium | *CRYBB3* | -4.287140904 | 0.02667541 | 0.031090341 | 1 |
| Obesity | endometrium | *CYTH4* | 3.229392988 | 1.45E-06 | 3.21E-06 | 0.007042081 |
| Obesity | endometrium | *CARD10* | 3.172944699 | 0.000345109 | 0.000482834 | 1 |
| Obesity | endometrium | *LRP5L* | 3.461656769 | 2.28E-06 | 4.46E-06 | 0.011086217 |
| Obesity | endometrium | *LGALS2* | 5.516213284 | 2.96E-05 | 4.69E-05 | 0.143455283 |
| Obesity | endometrium | *SH3BP1* | 5.752299559 | 1.30E-06 | 2.98E-06 | 0.006294701 |
| Obesity | endometrium | *HPS4* | 8.180846858 | 4.06E-09 | 4.85E-08 | 1.97E-05 |
| Obesity | endometrium | *TRIOBP* | 5.378365424 | 3.62E-07 | 1.08E-06 | 0.001754463 |
| Obesity | endometrium | *ANKRD54* | -4.27385507 | 0.000502703 | 0.000691206 | 1 |
| Obesity | endometrium | *EIF3L* | 3.258658265 | 1.60E-06 | 3.39E-06 | 0.007781553 |
| Obesity | endometrium | *PICK1* | 3.149498909 | 2.63E-05 | 4.19E-05 | 0.12745475 |
| Obesity | endometrium | *TTC28* | 3.914555965 | 2.51E-08 | 1.42E-07 | 0.000121769 |
| Obesity | endometrium | *CENPM* | 14.4007447 | 1.37E-08 | 9.50E-08 | 6.63E-05 |
| Obesity | endometrium | *KDELR3* | -2.576590436 | 0.000509774 | 0.000700289 | 1 |
| Obesity | endometrium | *CYP2D6* | -9.626688332 | 0.004904074 | 0.00613104 | 1 |
| Obesity | endometrium | *CBY1* | 3.220726877 | 4.75E-05 | 7.36E-05 | 0.230561852 |
| Obesity | endometrium | *RTCB* | 7.989287976 | 1.10E-08 | 8.33E-08 | 5.36E-05 |
| Obesity | endometrium | *RAB36* | 5.115537234 | 0.001146004 | 0.001524654 | 1 |
| Obesity | endometrium | *TIMP3* | 5.144956047 | 6.46E-09 | 6.17E-08 | 3.13E-05 |
| Obesity | endometrium | *PPP6R2* | 5.280688842 | 1.43E-08 | 9.78E-08 | 6.93E-05 |
| Obesity | endometrium | *CYB5R3* | 4.602405981 | 0.022941143 | 0.026886576 | 1 |
| Obesity | endometrium | *RHBDD3* | 7.389751015 | 8.99E-08 | 3.61E-07 | 0.000436195 |
| Obesity | endometrium | *TOM1* | 2.560138659 | 3.77E-06 | 6.92E-06 | 0.018297271 |
| Obesity | endometrium | *CHKB* | -2.983177027 | 0.032415061 | 0.037491746 | 1 |
| Obesity | endometrium | *THOC5* | 3.947612781 | 3.53E-05 | 5.55E-05 | 0.171501949 |
| Obesity | endometrium | *ARSA* | 3.14472945 | 6.22E-06 | 1.10E-05 | 0.030203284 |
| Obesity | endometrium | *TSPO* | 7.094552605 | 1.40E-08 | 9.71E-08 | 6.82E-05 |
| Obesity | endometrium | *TTLL12* | -2.048791145 | 0.001682704 | 0.002199483 | 1 |
| Obesity | endometrium | *MTMR3* | 7.375317354 | 1.04E-07 | 4.06E-07 | 0.000506727 |
| Obesity | endometrium | *APOL4* | 4.66947086 | 1.07E-06 | 2.56E-06 | 0.0052013 |
| Obesity | endometrium | *SAMM50* | 9.769306251 | 2.67E-08 | 1.48E-07 | 0.000129441 |
| Obesity | endometrium | *FOXRED2* | 3.991606705 | 4.76E-06 | 8.55E-06 | 0.023114578 |

| | | | | | | |
|---|---|---|---|---|---|---|
| Obesity | endometrium | *IFT27* | 3.998490743 | 1.54E-06 | 3.32E-06 | 0.007496302 |
| Obesity | endometrium | *KIAA0930* | 4.922436527 | 8.06E-07 | 2.05E-06 | 0.003911496 |
| Obesity | endometrium | *SLC25A17* | 5.84451399 | 9.02E-08 | 3.62E-07 | 0.000437817 |
| Obesity | endometrium | *FAM118A* | -1.827041497 | 0.022916818 | 0.026864556 | 1 |
| Obesity | endometrium | *L3MBTL2* | 10.62711875 | 0.000229111 | 0.000328113 | 1 |
| Obesity | endometrium | *ZC3H7B* | 10.02770153 | 3.40E-09 | 4.54E-08 | 1.65E-05 |
| Obesity | endometrium | *ACO2* | 10.56574135 | 1.84E-06 | 3.72E-06 | 0.008942219 |
| Obesity | endometrium | *POLR3H* | -1.917614565 | 0.00481505 | 0.006027508 | 1 |
| Obesity | endometrium | *DESI1* | 4.860513741 | 1.00E-07 | 3.93E-07 | 0.000487136 |
| Obesity | endometrium | *MLC1* | 7.524566268 | 0.000288485 | 0.000407846 | 1 |
| Obesity | endometrium | *HDAC10* | 6.21413987 | 2.99E-07 | 9.30E-07 | 0.001451431 |
| Obesity | endometrium | *KHNYN* | 4.562553597 | 0.021636411 | 0.02540655 | 1 |
| Obesity | endometrium | *SDR39U1* | -1.762261698 | 0.022546111 | 0.026455558 | 1 |
| Obesity | endometrium | *RBM23* | 6.681505495 | 8.16E-08 | 3.37E-07 | 0.000395989 |
| Obesity | endometrium | *AP4S1* | 5.404469402 | 3.73E-07 | 1.11E-06 | 0.001807713 |
| Obesity | endometrium | *NIN* | 10.05655973 | 2.11E-09 | 4.00E-08 | 1.02E-05 |
| Obesity | endometrium | *PYGL* | 2.48466912 | 0.000205668 | 0.000295937 | 0.997900947 |
| Obesity | endometrium | *CNIH1* | 3.784332791 | 8.88E-07 | 2.22E-06 | 0.004310782 |
| Obesity | endometrium | *ATP6V1D* | 4.066988738 | 1.67E-07 | 5.86E-07 | 0.000811127 |
| Obesity | endometrium | *PLEK2* | -3.079821052 | 0.002385263 | 0.003078823 | 1 |
| Obesity | endometrium | *GSTZ1* | -2.051061026 | 0.005768674 | 0.007156637 | 1 |
| Obesity | endometrium | *TMED8* | 8.580438563 | 8.41E-10 | 3.24E-08 | 4.08E-06 |
| Obesity | endometrium | *LGMN* | 2.040440275 | 1.58E-06 | 3.36E-06 | 0.007678067 |
| Obesity | endometrium | *MTHFD1* | 12.99070295 | 1.20E-08 | 8.67E-08 | 5.82E-05 |
| Obesity | endometrium | *TELO2* | 13.79502192 | 2.25E-08 | 1.32E-07 | 0.000109382 |
| Obesity | endometrium | *GSKIP* | -2.236149848 | 0.003194624 | 0.0040726 | 1 |
| Obesity | endometrium | *PSMB5* | 11.80636347 | 3.27E-09 | 4.49E-08 | 1.59E-05 |
| Obesity | endometrium | *YY1* | 4.076284457 | 1.65E-07 | 5.83E-07 | 0.000801571 |
| Obesity | endometrium | *CCNB1IP1* | 5.217626305 | 1.44E-07 | 5.21E-07 | 0.000699208 |
| Obesity | endometrium | *CINP* | 18.01701956 | 4.71E-09 | 5.26E-08 | 2.28E-05 |
| Obesity | endometrium | *KIAA0391* | -4.111327217 | 0.004897675 | 0.006124619 | 1 |
| Obesity | endometrium | *EMC9* | 4.535363835 | 5.56E-05 | 8.55E-05 | 0.269706315 |
| Obesity | endometrium | *PLTP* | 2.828343774 | 5.20E-07 | 1.44E-06 | 0.002521661 |
| Obesity | endometrium | *TRPC4AP* | 8.809170976 | 6.24E-10 | 2.70E-08 | 3.03E-06 |
| Obesity | endometrium | *PYGB* | 6.923099751 | 1.01E-07 | 3.97E-07 | 0.000492262 |
| Obesity | endometrium | *ABHD12* | 4.878852832 | 3.21E-09 | 4.46E-08 | 1.56E-05 |
| Obesity | endometrium | *CD40* | 3.434817454 | 1.64E-06 | 3.43E-06 | 0.007970077 |
| Obesity | endometrium | *UQCC1* | 4.14500686 | 1.11E-07 | 4.27E-07 | 0.000539337 |
| Obesity | endometrium | *NDRG3* | 3.670847906 | 3.02E-07 | 9.37E-07 | 0.001465439 |
| Obesity | endometrium | *PABPC1L* | 10.8637431 | 2.04E-08 | 1.24E-07 | 9.92E-05 |
| Obesity | endometrium | *DOK5* | 5.242727337 | 4.36E-07 | 1.26E-06 | 0.002114252 |
| Obesity | endometrium | *CSTF1* | 4.304396415 | 1.25E-06 | 2.89E-06 | 0.006087914 |
| Obesity | endometrium | *NELFCD* | 3.989122234 | 2.47E-07 | 7.96E-07 | 0.001196595 |
| Obesity | endometrium | *CTSZ* | 2.553054368 | 1.34E-06 | 3.05E-06 | 0.006481977 |
| Obesity | endometrium | *MTG2* | 4.434542821 | 0.028027284 | 0.032587679 | 1 |
| Obesity | endometrium | *TCFL5* | -2.701722175 | 0.008676861 | 0.010548768 | 1 |
| Obesity | endometrium | *DIDO1* | 3.342702913 | 1.84E-07 | 6.30E-07 | 0.000890627 |
| Obesity | endometrium | *GID8* | 11.45373356 | 1.41E-05 | 2.33E-05 | 0.068384369 |
| Obesity | endometrium | *NKAIN4* | 5.748316036 | 0.001288326 | 0.001707445 | 1 |
| Obesity | endometrium | *PTK6* | -1.666456076 | 0.000181304 | 0.000261968 | 0.879689234 |

| | | | | | | |
|---|---|---|---|---|---|---|
| Obesity | endometrium | *GMEB2* | 7.492531182 | 1.02E-07 | 3.98E-07 | 0.000494411 |
| Obesity | endometrium | *C20orf27* | 4.172384349 | 8.82E-07 | 2.20E-06 | 0.004279221 |
| Obesity | endometrium | *CDC25B* | 7.254600246 | 3.23E-07 | 9.86E-07 | 0.001568956 |
| Obesity | endometrium | *RNF24* | 4.436072909 | 8.55E-08 | 3.49E-07 | 0.000414911 |
| Obesity | endometrium | *NDUFAF5* | 3.638373535 | 5.52E-06 | 9.82E-06 | 0.026795953 |
| Obesity | endometrium | *TRIB3* | 6.463204529 | 1.35E-06 | 3.07E-06 | 0.006549646 |
| Obesity | endometrium | *RASSF2* | 4.410643985 | 2.44E-07 | 7.91E-07 | 0.001186039 |
| Obesity | endometrium | *ANGPT4* | 7.962681055 | 1.04E-06 | 2.50E-06 | 0.005058841 |
| Obesity | endometrium | *RSPO4* | 4.243381159 | 0.000100287 | 0.000150462 | 0.486593613 |
| Obesity | endometrium | *SIRPB1* | -2.416056304 | 0.00022632 | 0.000324212 | 1 |
| Obesity | endometrium | *CRNKL1* | 3.714123403 | 2.51E-06 | 4.85E-06 | 0.012167463 |
| Obesity | endometrium | *MANBAL* | 9.001561079 | 3.31E-10 | 2.20E-08 | 1.60E-06 |
| Obesity | endometrium | *TTI1* | 2.869792872 | 2.04E-06 | 4.06E-06 | 0.009889565 |
| Obesity | endometrium | *BPI* | -0.848517476 | 0.000185094 | 0.000267126 | 0.898077168 |
| Obesity | endometrium | *ACTR5* | 5.228200713 | 1.35E-06 | 3.08E-06 | 0.006569551 |
| Obesity | endometrium | *DNTTIP1* | 2.749199229 | 7.84E-06 | 1.36E-05 | 0.038024686 |
| Obesity | endometrium | *MAP1LC3A* | 5.765159668 | 1.20E-07 | 4.52E-07 | 0.000580734 |
| Obesity | endometrium | *RBFA* | 3.494118076 | 3.38E-06 | 6.28E-06 | 0.016395776 |
| Obesity | endometrium | *METTL4* | 2.818924849 | 1.53E-06 | 3.31E-06 | 0.007435317 |
| Obesity | endometrium | *LPIN2* | 5.557726791 | 7.82E-08 | 3.26E-07 | 0.000379481 |
| Obesity | endometrium | *MYOM1* | 3.299732793 | 4.98E-07 | 1.40E-06 | 0.002415508 |
| Obesity | endometrium | *CEP192* | 3.36538888 | 5.36E-06 | 9.55E-06 | 0.026008411 |
| Obesity | endometrium | *LAMA1* | -1.60836031 | 0.000159366 | 0.000232275 | 0.773244166 |
| Obesity | endometrium | *RNF125* | 3.838097705 | 1.52E-06 | 3.31E-06 | 0.007370348 |
| Obesity | endometrium | *POLI* | 3.186003831 | 5.96E-07 | 1.61E-06 | 0.002893147 |
| Obesity | endometrium | *MIB1* | 5.09353042 | 2.23E-08 | 1.31E-07 | 0.000108129 |
| Obesity | endometrium | *NALCN* | 3.485857016 | 3.98E-06 | 7.25E-06 | 0.019306479 |
| Obesity | endometrium | *STK24* | 10.80750283 | 5.47E-08 | 2.51E-07 | 0.000265472 |
| Obesity | endometrium | *ACP5* | 4.094864408 | 1.17E-05 | 1.97E-05 | 0.056982886 |
| Obesity | endometrium | *DNAJC3* | 8.749235852 | 3.63E-09 | 4.70E-08 | 1.76E-05 |
| Obesity | endometrium | *UGGT2* | 7.178754508 | 5.94E-08 | 2.64E-07 | 0.000288095 |
| Obesity | endometrium | *ARHGEF7* | 5.604087368 | 1.28E-08 | 9.12E-08 | 6.23E-05 |
| Obesity | endometrium | *PARP4* | 7.853997748 | 1.68E-09 | 3.98E-08 | 8.16E-06 |
| Obesity | endometrium | *SUPT20H* | 9.265088035 | 1.59E-05 | 2.60E-05 | 0.077016346 |
| Obesity | endometrium | *MRPS31* | 10.23716809 | 1.38E-09 | 3.98E-08 | 6.70E-06 |
| Obesity | endometrium | *RGCC* | 3.780982067 | 0.000110934 | 0.000165351 | 0.538252065 |
| Obesity | endometrium | *DGKH* | 7.435794661 | 1.59E-09 | 3.98E-08 | 7.73E-06 |
| Obesity | endometrium | *KATNAL1* | 7.992982205 | 2.99E-09 | 4.29E-08 | 1.45E-05 |
| Obesity | endometrium | *INTS6* | 9.319113495 | 2.71E-06 | 5.19E-06 | 0.013144112 |
| Obesity | endometrium | *CLN5* | 4.891510077 | 9.71E-08 | 3.84E-07 | 0.000471163 |
| Obesity | endometrium | *MGRN1* | 3.943224603 | 1.61E-07 | 5.70E-07 | 0.000780969 |
| Obesity | endometrium | *LONP2* | 5.475452345 | 1.61E-08 | 1.05E-07 | 7.84E-05 |
| Obesity | endometrium | *PLLP* | 3.818900889 | 1.58E-05 | 2.59E-05 | 0.076645307 |
| Obesity | endometrium | *DHODH* | 4.541961554 | 1.30E-07 | 4.80E-07 | 0.000628357 |
| Obesity | endometrium | *CCDC113* | 4.654376789 | 0.000257685 | 0.000366224 | 1 |
| Obesity | endometrium | *PSMD7* | 3.936774394 | 3.14E-07 | 9.64E-07 | 0.001521669 |
| Obesity | endometrium | *COG4* | 6.368521736 | 6.08E-05 | 9.30E-05 | 0.294802452 |
| Obesity | endometrium | *MON1B* | 5.616071156 | 2.69E-07 | 8.52E-07 | 0.001305577 |
| Obesity | endometrium | *NPRL3* | 6.115438997 | 0.001384772 | 0.001830767 | 1 |
| Obesity | endometrium | *TAF1C* | 2.452619147 | 0.0005655 | 0.000772686 | 1 |

| | | | | | | |
|---|---|---|---|---|---|---|
| Obesity | endometrium | *ZNF500* | 12.21964407 | 3.83E-08 | 1.94E-07 | 0.000186056 |
| Obesity | endometrium | *NOMO3* | 4.556644261 | 9.31E-05 | 0.000140159 | 0.451732782 |
| Obesity | endometrium | *LMF1* | 10.12311757 | 1.62E-05 | 2.65E-05 | 0.078490735 |
| Obesity | endometrium | *NARFL* | 6.918583764 | 1.13E-07 | 4.32E-07 | 0.000549228 |
| Obesity | endometrium | *MTHFSD* | 2.738132591 | 6.86E-06 | 1.20E-05 | 0.033267337 |
| Obesity | endometrium | *CLCN7* | 3.448151979 | 6.94E-08 | 2.99E-07 | 0.000336838 |
| Obesity | endometrium | *HAGHL* | 4.546331334 | 0.000428167 | 0.000593054 | 1 |
| Obesity | endometrium | *SLC7A5* | 7.871572161 | 6.72E-05 | 0.000102426 | 0.32581569 |
| Obesity | endometrium | *RHBDL1* | -2.43101736 | 0.000147828 | 0.000216499 | 0.717261445 |
| Obesity | endometrium | *NUBP1* | 15.18397396 | 6.08E-11 | 8.04E-09 | 2.95E-07 |
| Obesity | endometrium | *CAPN15* | -2.168063933 | 0.004290165 | 0.005398309 | 1 |
| Obesity | endometrium | *CLUAP1* | 8.110553093 | 1.73E-10 | 1.58E-08 | 8.39E-07 |
| Obesity | endometrium | *CPPED1* | 5.100308 | 2.21E-07 | 7.30E-07 | 0.001073377 |
| Obesity | endometrium | *HMOX2* | 10.8789752 | 9.36E-08 | 3.72E-07 | 0.000453962 |
| Obesity | endometrium | *TOX3* | -5.365547851 | 0.005986752 | 0.007415808 | 1 |
| Obesity | endometrium | *RRN3P2* | 2.993843266 | 1.98E-05 | 3.21E-05 | 0.096016567 |
| Obesity | endometrium | *RBL2* | 5.499157916 | 2.03E-08 | 1.23E-07 | 9.84E-05 |
| Obesity | endometrium | *PYCARD* | 5.267122504 | 2.10E-08 | 1.25E-07 | 0.000101991 |
| Obesity | endometrium | *CDIPT* | 4.188591927 | 3.48E-06 | 6.45E-06 | 0.016906193 |
| Obesity | endometrium | *TMC5* | 3.168871495 | 2.53E-05 | 4.03E-05 | 0.122571416 |
| Obesity | endometrium | *C16orf62* | 6.886275573 | 5.30E-09 | 5.69E-08 | 2.57E-05 |
| Obesity | endometrium | *RNF40* | 6.613807512 | 7.45E-08 | 3.15E-07 | 0.000361451 |
| Obesity | endometrium | *KNOP1* | 2.610619037 | 6.66E-06 | 1.17E-05 | 0.032325604 |
| Obesity | endometrium | *IQCH* | 3.210965352 | 7.04E-07 | 1.85E-06 | 0.003413643 |
| Obesity | endometrium | *LACTB* | 4.050288655 | 6.07E-07 | 1.63E-06 | 0.002946272 |
| Obesity | endometrium | *HERC1* | 7.66612975 | 2.24E-09 | 4.00E-08 | 1.09E-05 |
| Obesity | endometrium | *TRIP4* | 4.307683326 | 4.88E-08 | 2.30E-07 | 0.000236642 |
| Obesity | endometrium | *AP3B2* | 2.181118432 | 0.004721266 | 0.00591316 | 1 |
| Obesity | endometrium | *CTSH* | 2.322924259 | 8.73E-06 | 1.50E-05 | 0.04237754 |
| Obesity | endometrium | *TTC23* | 8.829080951 | 2.98E-09 | 4.29E-08 | 1.44E-05 |
| Obesity | endometrium | *FAH* | 4.289904282 | 5.46E-07 | 1.50E-06 | 0.002651426 |
| Obesity | endometrium | *SLC30A4* | 3.324914824 | 1.14E-06 | 2.68E-06 | 0.005531011 |
| Obesity | endometrium | *ZDHHC2* | 3.04068048 | 1.21E-05 | 2.02E-05 | 0.058583287 |
| Obesity | endometrium | *EYA1* | -1.640686816 | 0.00552382 | 0.006865263 | 1 |
| Obesity | endometrium | *IMPAD1* | 10.82360065 | 7.35E-09 | 6.60E-08 | 3.57E-05 |
| Obesity | endometrium | *SFRP1* | 9.76979069 | 8.33E-09 | 7.12E-08 | 4.04E-05 |
| Obesity | endometrium | *NIPAL2* | 9.785739153 | 2.50E-05 | 3.99E-05 | 0.121124106 |
| Obesity | endometrium | *RAB2A* | 6.071631777 | 4.17E-08 | 2.06E-07 | 0.000202465 |
| Obesity | endometrium | *ARMC1* | 3.508934405 | 7.76E-07 | 1.99E-06 | 0.00376592 |
| Obesity | endometrium | *SPAG1* | 4.387361326 | 1.06E-07 | 4.13E-07 | 0.000516732 |
| Obesity | endometrium | *CHRAC1* | -2.004655898 | 0.00109006 | 0.001451818 | 1 |
| Obesity | endometrium | *SNX16* | 4.932586129 | 1.86E-07 | 6.37E-07 | 0.000901875 |
| Obesity | endometrium | *SQLE* | 3.584615254 | 1.54E-06 | 3.32E-06 | 0.007462205 |
| Obesity | endometrium | *R3HCC1* | 9.124149039 | 5.96E-06 | 1.06E-05 | 0.028922298 |
| Obesity | endometrium | *TNFRSF10A* | 4.166184383 | 1.06E-06 | 2.52E-06 | 0.005119548 |
| Obesity | endometrium | *ERICH1* | 7.717727431 | 3.17E-08 | 1.68E-07 | 0.000153581 |
| Obesity | endometrium | *TUSC3* | 2.538096735 | 1.54E-06 | 3.32E-06 | 0.007488419 |
| Obesity | endometrium | *KLHDC4* | 7.090050536 | 0.001284242 | 0.001702497 | 1 |
| Obesity | endometrium | *MAN2B1* | 4.559009851 | 0.000244407 | 0.000348783 | 1 |
| Obesity | endometrium | *TULP2* | 21.00461491 | 4.21E-06 | 7.62E-06 | 0.020407292 |

| | | | | | | |
|---|---|---|---|---|---|---|
| Obesity | endometrium | *NFKBIB* | 4.724977603 | 2.08E-06 | 4.13E-06 | 0.010114029 |
| Obesity | endometrium | *SARS2* | 2.669550658 | 0.009658749 | 0.011681019 | 1 |
| Obesity | endometrium | *SNRNP70* | 10.58110935 | 1.31E-08 | 9.23E-08 | 6.36E-05 |
| Obesity | endometrium | *LIN7B* | 13.09220799 | 3.89E-06 | 7.11E-06 | 0.018885099 |
| Obesity | endometrium | *PPP1R37* | 6.394479359 | 6.59E-05 | 0.000100572 | 0.319716912 |
| Obesity | endometrium | *FCGRT* | 2.701623113 | 5.31E-07 | 1.47E-06 | 0.002578398 |
| Obesity | endometrium | *PEX11G* | -2.231112464 | 0.001036496 | 0.001383135 | 1 |
| Obesity | endometrium | *ERCC2* | 4.631058028 | 1.96E-06 | 3.93E-06 | 0.009525188 |
| Obesity | endometrium | *CD37* | 3.411364525 | 5.13E-06 | 9.17E-06 | 0.024887308 |
| Obesity | endometrium | *AMH* | 41.05257931 | 1.59E-06 | 3.37E-06 | 0.007701751 |
| Obesity | endometrium | *FCER2* | 12.64942718 | 1.60E-06 | 3.39E-06 | 0.007762139 |
| Obesity | endometrium | *CLEC4M* | 58.85681395 | 9.23E-07 | 2.27E-06 | 0.004478645 |
| Obesity | endometrium | *TLE6* | 4.740642487 | 0.036788624 | 0.04236848 | 1 |
| Obesity | endometrium | *NOVA2* | 4.002226535 | 1.33E-07 | 4.88E-07 | 0.000645815 |
| Obesity | endometrium | *SGTA* | 7.878938749 | 2.07E-07 | 6.90E-07 | 0.001004787 |
| Obesity | endometrium | *KIR3DX1* | 21.87730834 | 3.28E-05 | 5.16E-05 | 0.158907429 |
| Obesity | endometrium | *MED25* | 14.90104769 | 3.04E-06 | 5.71E-06 | 0.014740242 |
| Obesity | endometrium | *TIMM44* | 10.33439521 | 3.14E-09 | 4.44E-08 | 1.52E-05 |
| Obesity | endometrium | *IL27RA* | 5.300742691 | 8.51E-07 | 2.14E-06 | 0.004130727 |
| Obesity | endometrium | *FAM32A* | 5.90650215 | 6.50E-07 | 1.74E-06 | 0.003156024 |
| Obesity | endometrium | *OLFM2* | 4.631517587 | 0.020496748 | 0.024109144 | 1 |
| Obesity | endometrium | *RASAL3* | 5.369811779 | 1.10E-06 | 2.61E-06 | 0.005321696 |
| Obesity | endometrium | *AURKC* | 4.757901875 | 0.014845946 | 0.017689717 | 1 |
| Obesity | endometrium | *ANKRD27* | 8.693399095 | 1.50E-09 | 3.98E-08 | 7.28E-06 |
| Obesity | endometrium | *TGFB1* | 4.383965 | 1.12E-07 | 4.29E-07 | 0.000543377 |
| Obesity | endometrium | *CEACAM4* | 6.584206133 | 0.008815257 | 0.010711652 | 1 |
| Obesity | endometrium | *MRPL4* | 5.840823325 | 2.45E-07 | 7.92E-07 | 0.001189449 |
| Obesity | endometrium | *CD33* | 1.790334856 | 0.005690746 | 0.007067188 | 1 |
| Obesity | endometrium | *RABAC1* | 6.238101757 | 3.34E-08 | 1.74E-07 | 0.000162024 |
| Obesity | endometrium | *GRIN2D* | -2.366648063 | 0.000581183 | 0.000793221 | 1 |
| Obesity | endometrium | *CCDC114* | -7.541625745 | 0.002240442 | 0.002901154 | 1 |
| Obesity | endometrium | *CARD8* | 4.06425713 | 1.62E-07 | 5.74E-07 | 0.000787855 |
| Obesity | endometrium | *LIG1* | -2.875837713 | 0.035974653 | 0.041460574 | 1 |
| Obesity | endometrium | *ZNF175* | 3.715099111 | 2.74E-06 | 5.24E-06 | 0.013288842 |
| Obesity | endometrium | *PLA2G4C* | 3.130398229 | 9.95E-05 | 0.000149307 | 0.482560538 |
| Obesity | endometrium | *CAPS* | -1.564589459 | 0.000640104 | 0.000869968 | 1 |
| Obesity | endometrium | *MIER2* | 3.547442403 | 1.31E-06 | 2.99E-06 | 0.006345783 |
| Obesity | endometrium | *PLEKHA4* | 7.022683914 | 9.95E-08 | 3.90E-07 | 0.000482555 |
| Obesity | endometrium | *TNPO2* | 12.09091179 | 2.93E-07 | 9.15E-07 | 0.001422901 |
| Obesity | endometrium | *GCDH* | 2.891761028 | 1.19E-05 | 1.99E-05 | 0.057553788 |
| Obesity | endometrium | *TFPT* | 4.443726341 | 1.68E-06 | 3.48E-06 | 0.008165988 |
| Obesity | endometrium | *RAB3A* | 15.4264133 | 1.57E-06 | 3.35E-06 | 0.00761933 |
| Obesity | endometrium | *COPE* | 7.380395251 | 0.003683507 | 0.004667636 | 1 |
| Obesity | endometrium | *ETV2* | 38.7535977 | 1.41E-06 | 3.16E-06 | 0.006829661 |
| Obesity | endometrium | *LSR* | 4.077362007 | 1.62E-05 | 2.65E-05 | 0.078447786 |
| Obesity | endometrium | *ZNF14* | 12.50232128 | 2.12E-09 | 4.00E-08 | 1.03E-05 |
| Obesity | endometrium | *ZNF85* | 10.85101931 | 9.48E-09 | 7.53E-08 | 4.60E-05 |
| Obesity | endometrium | *ETHE1* | 5.877717419 | 3.53E-07 | 1.06E-06 | 0.001713619 |
| Obesity | endometrium | *SMG9* | 3.52237041 | 2.28E-06 | 4.45E-06 | 0.011043761 |
| Obesity | endometrium | *GTPBP10* | 8.878150191 | 9.26E-07 | 2.28E-06 | 0.004492239 |

| | | | | | | |
|---|---|---|---|---|---|---|
| Obesity | endometrium | *RASA4* | -2.183812455 | 0.000145758 | 0.000213661 | 0.707216418 |
| Obesity | endometrium | *BET1* | 5.330076073 | 4.79E-08 | 2.27E-07 | 0.000232565 |
| Obesity | endometrium | *SP4* | 6.25930527 | 0.013200933 | 0.015787756 | 1 |
| Obesity | endometrium | *DNAH11* | 3.990854228 | 6.41E-07 | 1.71E-06 | 0.003108045 |
| Obesity | endometrium | *STEAP1B* | 7.405966715 | 4.66E-07 | 1.32E-06 | 0.002259081 |
| Obesity | endometrium | *PTN* | 3.770812832 | 5.16E-07 | 1.44E-06 | 0.002501681 |
| Obesity | endometrium | *ADAP1* | 12.53124166 | 1.54E-06 | 3.32E-06 | 0.007489995 |
| Obesity | endometrium | *MET* | 1.99587023 | 1.45E-05 | 2.39E-05 | 0.070430584 |
| Obesity | endometrium | *RNF32* | 3.912176215 | 2.88E-06 | 5.46E-06 | 0.013953038 |
| Obesity | endometrium | *IQCE* | 3.42222649 | 1.24E-06 | 2.86E-06 | 0.005993242 |
| Obesity | endometrium | *VIPR2* | 9.094491006 | 5.60E-10 | 2.59E-08 | 2.72E-06 |
| Obesity | endometrium | *HIBADH* | 5.711347951 | 1.26E-07 | 4.70E-07 | 0.000610599 |
| Obesity | endometrium | *CPVL* | 2.071665028 | 1.65E-06 | 3.44E-06 | 0.008016342 |
| Obesity | endometrium | *CHN2* | 2.88458273 | 1.45E-06 | 3.21E-06 | 0.007021613 |
| Obesity | endometrium | *GRB10* | 2.656589823 | 5.97E-07 | 1.61E-06 | 0.002896015 |
| Obesity | endometrium | *ABHD11* | 4.199420933 | 0.00046041 | 0.000635717 | 1 |
| Obesity | endometrium | *PLEKHA8* | 12.01390988 | 1.27E-09 | 3.98E-08 | 6.14E-06 |
| Obesity | endometrium | *EPHB6* | 3.456974869 | 1.55E-06 | 3.32E-06 | 0.007524132 |
| Obesity | endometrium | *CHCHD2* | 3.889599627 | 5.31E-07 | 1.47E-06 | 0.002577467 |
| Obesity | endometrium | *CCL24* | -1.437905171 | 5.81E-05 | 8.92E-05 | 0.281721579 |
| Obesity | endometrium | *ZKSCAN1* | 3.876495018 | 1.40E-07 | 5.07E-07 | 0.000677167 |
| Obesity | endometrium | *EIF3B* | 11.45636348 | 8.52E-06 | 1.47E-05 | 0.04133975 |
| Obesity | endometrium | *TFR2* | -4.803925043 | 0.005949558 | 0.007371618 | 1 |
| Obesity | endometrium | *MOSPD3* | -2.08978542 | 0.003894569 | 0.004923514 | 1 |
| Obesity | endometrium | *RBM28* | 8.233694102 | 2.41E-05 | 3.86E-05 | 0.116919737 |
| Obesity | endometrium | *C1GALT1* | 8.520559273 | 2.43E-09 | 4.02E-08 | 1.18E-05 |
| Obesity | endometrium | *NRF1* | 4.979533361 | 2.06E-08 | 1.24E-07 | 9.99E-05 |
| Obesity | endometrium | *ZNF862* | 4.27655686 | 5.78E-08 | 2.59E-07 | 0.000280631 |
| Obesity | endometrium | *MEST* | 3.15248192 | 0.005338024 | 0.006646162 | 1 |
| Obesity | endometrium | *ACTR3C* | 3.720014175 | 2.90E-06 | 5.49E-06 | 0.014052782 |
| Obesity | endometrium | *POU6F2* | -1.878586289 | 0.006255756 | 0.007727324 | 1 |
| Obesity | endometrium | *TSPAN13* | 8.062589542 | 6.10E-08 | 2.69E-07 | 0.00029606 |
| Obesity | endometrium | *RARRES2* | 5.945169785 | 2.22E-08 | 1.31E-07 | 0.000107704 |
| Obesity | endometrium | *TMEM176B* | 3.16778406 | 6.73E-05 | 0.000102657 | 0.326654565 |
| Obesity | endometrium | *COA1* | 5.482614372 | 8.07E-09 | 7.03E-08 | 3.92E-05 |
| Obesity | endometrium | *BLVRA* | 10.68243869 | 2.13E-09 | 4.00E-08 | 1.03E-05 |
| Obesity | endometrium | *URGCP* | 7.720813559 | 5.47E-09 | 5.77E-08 | 2.65E-05 |
| Obesity | endometrium | *GCK* | 118.4747038 | 3.81E-08 | 1.93E-07 | 0.000185038 |
| Obesity | endometrium | *BCL7B* | 3.62643967 | 8.16E-06 | 1.41E-05 | 0.039607041 |
| Obesity | endometrium | *SPATA6L* | 4.728131146 | 2.10E-07 | 7.00E-07 | 0.001020861 |
| Obesity | endometrium | *SLC1A1* | 2.396005247 | 1.16E-06 | 2.72E-06 | 0.005629973 |
| Obesity | endometrium | *FKTN* | 6.85570335 | 1.01E-08 | 7.86E-08 | 4.88E-05 |
| Obesity | endometrium | *NMRK1* | 9.659457799 | 2.02E-07 | 6.79E-07 | 0.000981147 |
| Obesity | endometrium | *MEGF9* | 5.680171702 | 2.90E-08 | 1.57E-07 | 0.000140688 |
| Obesity | endometrium | *TRIM14* | 3.00236938 | 2.90E-07 | 9.08E-07 | 0.001408711 |
| Obesity | endometrium | *CORO2A* | 4.823118539 | 2.91E-06 | 5.51E-06 | 0.014120617 |
| Obesity | endometrium | *C5* | 13.6210122 | 2.63E-07 | 8.36E-07 | 0.001275077 |
| Obesity | endometrium | *OGN* | 4.073227691 | 2.05E-08 | 1.24E-07 | 9.93E-05 |
| Obesity | endometrium | *ECM2* | 9.604506876 | 6.99E-09 | 6.42E-08 | 3.39E-05 |
| Obesity | endometrium | *PTGR1* | 7.344188894 | 1.18E-06 | 2.75E-06 | 0.005712891 |

| | | | | | | |
|---|---|---|---|---|---|---|
| Obesity | endometrium | *SUSD1* | 2.512263477 | 2.88E-06 | 5.46E-06 | 0.013960366 |
| Obesity | endometrium | *PLGRKT* | -2.225437377 | 0.002990813 | 0.00382282 | 1 |
| Obesity | endometrium | *DOCK8* | 1.60381275 | 1.27E-05 | 2.11E-05 | 0.061467007 |
| Obesity | endometrium | *KANK1* | 5.232256744 | 8.14E-08 | 3.37E-07 | 0.000394913 |
| Obesity | endometrium | *NCS1* | 3.814535066 | 0.000956692 | 0.001280869 | 1 |
| Obesity | endometrium | *FUBP3* | 10.09395035 | 2.69E-09 | 4.24E-08 | 1.31E-05 |
| Obesity | endometrium | *RGP1* | 3.964927772 | 5.81E-07 | 1.57E-06 | 0.002817102 |
| Obesity | endometrium | *HPS1* | 6.160617169 | 1.75E-06 | 3.59E-06 | 0.008508371 |
| Obesity | endometrium | *PHYH* | 7.835861244 | 6.02E-08 | 2.67E-07 | 0.000292111 |
| Obesity | endometrium | *DNMBP* | 4.062503062 | 5.98E-08 | 2.65E-07 | 0.000290232 |
| Obesity | endometrium | *PKD2L1* | 8.981223529 | 0.001899193 | 0.002472467 | 1 |
| Obesity | endometrium | *CUBN* | 6.020805084 | 3.61E-07 | 1.08E-06 | 0.001753326 |
| Obesity | endometrium | *TRDMT1* | 13.79171218 | 7.40E-10 | 2.94E-08 | 3.59E-06 |
| Obesity | endometrium | *ATE1* | 8.172357039 | 1.97E-09 | 4.00E-08 | 9.58E-06 |
| Obesity | endometrium | *PLEKHA1* | 10.68097854 | 3.06E-07 | 9.47E-07 | 0.00148507 |
| Obesity | endometrium | *CDH23* | 4.462670443 | 9.11E-08 | 3.64E-07 | 0.000441808 |
| Obesity | endometrium | *SPOCK2* | 2.957482196 | 0.000190751 | 0.000275126 | 0.925524827 |
| Obesity | endometrium | *BMPR1A* | 9.334091565 | 2.20E-07 | 7.27E-07 | 0.001067999 |
| Obesity | endometrium | *LIPA* | 2.652933842 | 2.30E-07 | 7.53E-07 | 0.001115087 |
| Obesity | endometrium | *SFXN3* | 3.43278196 | 1.57E-06 | 3.35E-06 | 0.007628858 |
| Obesity | endometrium | *TNKS2* | 4.702285254 | 5.73E-08 | 2.59E-07 | 0.000278161 |
| Obesity | endometrium | *ANKRD26* | 6.2841331 | 8.30E-09 | 7.12E-08 | 4.03E-05 |
| Obesity | endometrium | *ACBD5* | 11.69830869 | 8.83E-06 | 1.51E-05 | 0.042864431 |
| Obesity | endometrium | *LHPP* | 11.38339781 | 2.83E-06 | 5.39E-06 | 0.013745125 |
| Obesity | endometrium | *PITRM1* | 12.38862211 | 5.35E-08 | 2.47E-07 | 0.000259426 |
| Obesity | endometrium | *TFAM* | 12.1964301 | 2.19E-09 | 4.00E-08 | 1.06E-05 |
| Obesity | endometrium | *RPL28* | 8.875348625 | 1.67E-09 | 3.98E-08 | 8.08E-06 |
| Obesity | endometrium | *PBLD* | 3.782883706 | 1.02E-06 | 2.44E-06 | 0.004925203 |
| Obesity | endometrium | *TSPAN14* | 6.614304439 | 5.22E-06 | 9.31E-06 | 0.025303388 |
| Obesity | endometrium | *TADA2A* | 2.393268906 | 9.47E-05 | 0.000142421 | 0.459306424 |
| Obesity | endometrium | *PSMB3* | 10.86568502 | 4.11E-08 | 2.04E-07 | 0.000199454 |
| Obesity | endometrium | *RPL19* | 9.669645187 | 8.07E-08 | 3.35E-07 | 0.000391514 |
| Obesity | endometrium | *RAPGEFL1* | 5.501086853 | 1.20E-06 | 2.81E-06 | 0.005839748 |
| Obesity | endometrium | *WNT3* | 5.654310369 | 2.98E-05 | 4.72E-05 | 0.14464841 |
| Obesity | endometrium | *ASPA* | 3.558523681 | 0.034949899 | 0.040341624 | 1 |
| Obesity | endometrium | *RAD51C* | 2.692679485 | 8.97E-05 | 0.000135227 | 0.435429471 |
| Obesity | endometrium | *TRIM37* | 3.99523733 | 2.16E-07 | 7.15E-07 | 0.001047285 |
| Obesity | endometrium | *PNPO* | 11.67641925 | 0.002815157 | 0.003607803 | 1 |
| Obesity | endometrium | *TRIM16L* | 4.920140593 | 2.27E-06 | 4.44E-06 | 0.01100174 |
| Obesity | endometrium | *CDK5RAP3* | -2.834659204 | 0.001812465 | 0.002364 | 1 |
| Obesity | endometrium | *PIGL* | 7.519539476 | 4.97E-09 | 5.50E-08 | 2.41E-05 |
| Obesity | endometrium | *MED13* | 5.350767611 | 5.79E-08 | 2.59E-07 | 0.000281066 |
| Obesity | endometrium | *ENO3* | 6.667337402 | 8.92E-05 | 0.000134404 | 0.432647245 |
| Obesity | endometrium | *CHRNE* | 3.663438537 | 3.59E-05 | 5.64E-05 | 0.174405117 |
| Obesity | endometrium | *RAI1* | 4.14292476 | 3.25E-07 | 9.90E-07 | 0.001578652 |
| Obesity | endometrium | *GOSR1* | 11.59683612 | 6.27E-09 | 6.12E-08 | 3.04E-05 |
| Obesity | endometrium | *DRG2* | 3.758210895 | 9.58E-07 | 2.34E-06 | 0.004646823 |
| Obesity | endometrium | *FTSJ3* | -1.767376286 | 0.040791739 | 0.046867515 | 1 |
| Obesity | endometrium | *ALDH3A1* | -8.343926932 | 0.012637329 | 0.015147312 | 1 |
| Obesity | endometrium | *ICAM2* | 9.657278137 | 6.02E-09 | 5.98E-08 | 2.92E-05 |

| | | | | | | |
|---|---|---|---|---|---|---|
| Obesity | endometrium | *DDX5* | 4.200138638 | 1.87E-07 | 6.41E-07 | 0.00090956 |
| Obesity | endometrium | *C17orf75* | 3.463944586 | 1.39E-06 | 3.13E-06 | 0.006726049 |
| Obesity | endometrium | *PSMD11* | 5.769089899 | 4.81E-08 | 2.28E-07 | 0.000233391 |
| Obesity | endometrium | *DHX58* | -2.48837185 | 0.005708475 | 0.00708739 | 1 |
| Obesity | endometrium | *RAB5C* | 3.568325528 | 1.18E-05 | 1.98E-05 | 0.057263621 |
| Obesity | endometrium | *NAGLU* | 3.943807536 | 6.88E-07 | 1.82E-06 | 0.003338575 |
| Obesity | endometrium | *HSD17B1* | 6.341814471 | 6.03E-07 | 1.62E-06 | 0.002926054 |
| Obesity | endometrium | *EZH1* | 2.815840751 | 4.73E-07 | 1.34E-06 | 0.00229411 |
| Obesity | endometrium | *ABCC3* | 5.747151437 | 3.32E-08 | 1.73E-07 | 0.000160962 |
| Obesity | endometrium | *EFTUD2* | 3.095162341 | 1.02E-06 | 2.45E-06 | 0.004932152 |
| Obesity | endometrium | *HLF* | 2.624853651 | 1.45E-05 | 2.39E-05 | 0.070225895 |
| Obesity | endometrium | *EFNB3* | -3.16326321 | 0.006168597 | 0.007625486 | 1 |
| Obesity | endometrium | *DPH1* | -2.097358421 | 0.002900846 | 0.003713695 | 1 |
| Obesity | endometrium | *DHRS7B* | 3.537033015 | 2.99E-07 | 9.30E-07 | 0.001449908 |
| Obesity | endometrium | *RCVRN* | -4.511606017 | 0.000229885 | 0.000329124 | 1 |
| Obesity | endometrium | *MYH3* | 5.640878578 | 1.41E-06 | 3.15E-06 | 0.006819298 |
| Obesity | endometrium | *SLAIN2* | 11.61503608 | 3.53E-09 | 4.65E-08 | 1.71E-05 |
| Obesity | endometrium | *OCIAD1* | 9.424642223 | 5.53E-09 | 5.80E-08 | 2.68E-05 |
| Obesity | endometrium | *UBE2D3* | 4.923454236 | 1.87E-08 | 1.17E-07 | 9.07E-05 |
| Obesity | endometrium | *KLHL2* | 7.965841295 | 2.40E-08 | 1.38E-07 | 0.000116543 |
| Obesity | endometrium | *CPE* | 3.981604089 | 8.88E-07 | 2.22E-06 | 0.004308669 |
| Obesity | endometrium | *WFS1* | 18.60917867 | 3.98E-08 | 1.99E-07 | 0.000192922 |
| Obesity | endometrium | *GAR1* | -2.047411501 | 0.002278437 | 0.002947994 | 1 |
| Obesity | endometrium | *AADAT* | -1.273092874 | 0.000324829 | 0.000456039 | 1 |
| Obesity | endometrium | *TRIM2* | 3.825520665 | 2.46E-07 | 7.95E-07 | 0.001194176 |
| Obesity | endometrium | *STIM2* | 4.847193003 | 2.77E-08 | 1.52E-07 | 0.00013436 |
| Obesity | endometrium | *BST1* | 4.78471321 | 1.36E-07 | 4.95E-07 | 0.00065746 |
| Obesity | endometrium | *UFSP2* | 6.11323557 | 3.09E-08 | 1.64E-07 | 0.000149732 |
| Obesity | endometrium | *KLHL5* | 3.253795137 | 3.47E-07 | 1.05E-06 | 0.001682844 |
| Obesity | endometrium | *FAM149A* | -1.701432365 | 0.00424042 | 0.005339869 | 1 |
| Obesity | endometrium | *DDX25* | 3.238809194 | 0.000753658 | 0.001019729 | 1 |
| Obesity | endometrium | *CTSC* | 2.280571628 | 3.10E-06 | 5.82E-06 | 0.015061249 |
| Obesity | endometrium | *ELP4* | 8.032876641 | 3.08E-08 | 1.64E-07 | 0.000149519 |
| Obesity | endometrium | *TECTA* | 8.176613489 | 9.07E-07 | 2.24E-06 | 0.004402407 |
| Obesity | endometrium | *SC5D* | 7.542839289 | 8.62E-07 | 2.16E-06 | 0.00418436 |
| Obesity | endometrium | *B3GAT1* | -7.461687524 | 0.000564572 | 0.000771635 | 1 |
| Obesity | endometrium | *P2RX3* | -2.577006894 | 0.000129398 | 0.000191123 | 0.627838064 |
| Obesity | endometrium | *DNAJC4* | 3.869132589 | 1.30E-05 | 2.16E-05 | 0.06311387 |
| Obesity | endometrium | *UNC93B1* | 3.561961493 | 3.11E-06 | 5.82E-06 | 0.01506572 |
| Obesity | endometrium | *PUS3* | 5.52990045 | 1.03E-05 | 1.74E-05 | 0.049979374 |
| Obesity | endometrium | *DCPS* | 18.43419668 | 1.38E-08 | 9.59E-08 | 6.71E-05 |
| Obesity | endometrium | *NRXN2* | -1.967793511 | 0.000384631 | 0.000535042 | 1 |
| Obesity | endometrium | *ST3GAL4* | 4.147113562 | 9.63E-06 | 1.64E-05 | 0.046709342 |
| Obesity | endometrium | *CCND1* | 6.476181177 | 5.82E-07 | 1.58E-06 | 0.00282232 |
| Obesity | endometrium | *CCDC86* | 7.084955734 | 0.000132778 | 0.000195877 | 0.644238366 |
| Obesity | endometrium | *PANX1* | -1.071200124 | 0.00030314 | 0.00042732 | 1 |
| Obesity | endometrium | *CEP164* | 7.570827773 | 6.45E-09 | 6.17E-08 | 3.13E-05 |
| Obesity | endometrium | *GALNT18* | 2.296109792 | 1.94E-06 | 3.89E-06 | 0.009409191 |
| Obesity | endometrium | *KIAA1549L* | 13.31802842 | 1.44E-06 | 3.20E-06 | 0.006990402 |
| Obesity | endometrium | *ACCS* | 3.066803755 | 1.47E-06 | 3.25E-06 | 0.00715613 |

| | | | | | | |
|---|---|---|---|---|---|---|
| Obesity | endometrium | *SLC22A18* | 3.99531109 | 1.66E-07 | 5.84E-07 | 0.000805922 |
| Obesity | endometrium | *C11orf21* | 11.11723976 | 0.00046068 | 0.000635909 | 1 |
| Obesity | endometrium | *ELMOD1* | -6.716748943 | 0.006449961 | 0.007959108 | 1 |
| Obesity | endometrium | *RPS13* | 7.977414006 | 4.86E-05 | 7.52E-05 | 0.23570904 |
| Obesity | endometrium | *NDUFS8* | 9.20000267 | 2.36E-08 | 1.36E-07 | 0.000114286 |
| Obesity | endometrium | *GTF2H1* | 11.35600248 | 7.23E-10 | 2.94E-08 | 3.51E-06 |
| Obesity | endometrium | *PTPN5* | 3.042048987 | 0.015951554 | 0.018941982 | 1 |
| Obesity | endometrium | *PRDM4* | 14.89484575 | 6.85E-09 | 6.33E-08 | 3.32E-05 |
| Obesity | endometrium | *COQ5* | 2.067188919 | 3.10E-05 | 4.89E-05 | 0.1502004 |
| Obesity | endometrium | *CORO1C* | 4.654858772 | 7.11E-08 | 3.04E-07 | 0.000344892 |
| Obesity | endometrium | *ASIC1* | 8.080950963 | 1.69E-06 | 3.49E-06 | 0.008200003 |
| Obesity | endometrium | *CAPRIN2* | 7.16054163 | 5.63E-07 | 1.53E-06 | 0.002732371 |
| Obesity | endometrium | *KCTD10* | 3.857671439 | 1.40E-05 | 2.31E-05 | 0.067721613 |
| Obesity | endometrium | *IL23A* | 24.95164304 | 1.50E-06 | 3.28E-06 | 0.007282978 |
| Obesity | endometrium | *LTA4H* | 2.311844222 | 2.58E-06 | 4.97E-06 | 0.012518487 |
| Obesity | endometrium | *PRR4* | 8.43668432 | 2.99E-07 | 9.30E-07 | 0.001451024 |
| Obesity | endometrium | *PARP11* | 9.933473688 | 1.93E-09 | 4.00E-08 | 9.38E-06 |
| Obesity | endometrium | *GPN3* | 13.78574938 | 6.35E-09 | 6.12E-08 | 3.08E-05 |
| Obesity | endometrium | *VPS29* | 6.369200122 | 7.42E-09 | 6.64E-08 | 3.60E-05 |
| Obesity | endometrium | *AKAP3* | 2.999516864 | 0.000491146 | 0.000676423 | 1 |
| Obesity | endometrium | *ACAD10* | 6.06976583 | 1.53E-08 | 1.02E-07 | 7.40E-05 |
| Obesity | endometrium | *LTBR* | 5.410339801 | 6.22E-08 | 2.73E-07 | 0.000301612 |
| Obesity | endometrium | *DDX55* | 13.52161019 | 9.94E-12 | 2.01E-09 | 4.82E-08 |
| Obesity | endometrium | *ENDOU* | -4.079551631 | 0.000502734 | 0.000691206 | 1 |
| Obesity | endometrium | *VDR* | 11.20563124 | 1.27E-07 | 4.72E-07 | 0.000615026 |
| Obesity | endometrium | *STX2* | 4.921767321 | 8.80E-08 | 3.55E-07 | 0.000426873 |
| Obesity | endometrium | *CAND1* | 5.359163269 | 3.04E-08 | 1.63E-07 | 0.000147443 |
| Obesity | endometrium | *NUP107* | 8.663701187 | 5.55E-10 | 2.59E-08 | 2.69E-06 |
| Obesity | endometrium | *KRR1* | 11.26953217 | 6.81E-06 | 1.19E-05 | 0.033029555 |
| Obesity | endometrium | *CDCA3* | -1.228982899 | 0.000156344 | 0.000228282 | 0.758579698 |
| Obesity | endometrium | *CHPT1* | 4.060506475 | 1.71E-07 | 5.98E-07 | 0.000831727 |
| Obesity | endometrium | *GNPTAB* | 8.323647684 | 3.88E-09 | 4.80E-08 | 1.88E-05 |
| Obesity | endometrium | *SPSB2* | 5.368064292 | 7.40E-07 | 1.92E-06 | 0.003588185 |
| Obesity | endometrium | *C12orf57* | 14.8945417 | 4.37E-09 | 5.04E-08 | 2.12E-05 |
| Obesity | endometrium | *NT5DC3* | 11.5394938 | 1.35E-06 | 3.07E-06 | 0.006557179 |
| Obesity | endometrium | *GOLT1B* | 11.13209135 | 3.69E-08 | 1.88E-07 | 0.000179107 |
| Obesity | endometrium | *C2CD5* | 3.851808454 | 1.12E-07 | 4.29E-07 | 0.000545394 |
| Obesity | endometrium | *BTN3A3* | 8.044574688 | 5.88E-14 | 3.08E-11 | 2.85E-10 |
| Obesity | endometrium | *RWDD1* | 7.483078072 | 3.01E-07 | 9.36E-07 | 0.001461414 |
| Obesity | endometrium | *RSPH4A* | -2.345214014 | 0.000757544 | 0.001024701 | 1 |
| Obesity | endometrium | *SMIM8* | 7.642822988 | 3.41E-09 | 4.54E-08 | 1.65E-05 |
| Obesity | endometrium | *FAM184A* | 5.469551814 | 2.33E-08 | 1.34E-07 | 0.000112933 |
| Obesity | endometrium | *RNGTT* | 8.42772141 | 2.41E-09 | 4.02E-08 | 1.17E-05 |
| Obesity | endometrium | *GABRR2* | 8.281870872 | 8.45E-09 | 7.13E-08 | 4.10E-05 |
| Obesity | endometrium | *HDDC2* | 6.301781199 | 3.25E-07 | 9.90E-07 | 0.001577935 |
| Obesity | endometrium | *UST* | -1.313882418 | 0.007851268 | 0.009578665 | 1 |
| Obesity | endometrium | *MTRF1L* | 4.415092576 | 0.015304875 | 0.018227603 | 1 |
| Obesity | endometrium | *TULP1* | -6.51384162 | 0.007443523 | 0.009097223 | 1 |
| Obesity | endometrium | *SLC26A8* | 2.313860819 | 0.016606582 | 0.019700522 | 1 |
| Obesity | endometrium | *MRPL18* | 4.663517928 | 1.92E-07 | 6.54E-07 | 0.000931434 |

| | | | | | | |
|---|---|---|---|---|---|---|
| Obesity | endometrium | *RNF8* | 3.583370645 | 7.17E-07 | 1.88E-06 | 0.003480841 |
| Obesity | endometrium | *PHACTR1* | 2.405392989 | 2.40E-06 | 4.66E-06 | 0.011665101 |
| Obesity | endometrium | *MDGA1* | 4.051671008 | 2.83E-06 | 5.39E-06 | 0.013741907 |
| Obesity | endometrium | *CD83* | 4.087526727 | 2.23E-06 | 4.37E-06 | 0.010805765 |
| Obesity | endometrium | *MDN1* | 8.289165792 | 4.03E-08 | 2.01E-07 | 0.000195438 |
| Obesity | endometrium | *COL9A1* | 12.73020377 | 8.91E-07 | 2.22E-06 | 0.004322685 |
| Obesity | endometrium | *WASF1* | 10.08757502 | 4.41E-09 | 5.07E-08 | 2.14E-05 |
| Obesity | endometrium | *VNN1* | 2.883482855 | 0.009597368 | 0.011609681 | 1 |
| Obesity | endometrium | *VNN2* | 8.397532585 | 2.70E-06 | 5.18E-06 | 0.013114855 |
| Obesity | endometrium | *ACOT13* | 5.168834025 | 8.94E-08 | 3.60E-07 | 0.000433676 |
| Obesity | endometrium | *C6orf62* | 3.761555554 | 8.22E-07 | 2.08E-06 | 0.003986086 |
| Obesity | endometrium | *HBS1L* | 9.729237016 | 6.06E-09 | 6.00E-08 | 2.94E-05 |
| Obesity | endometrium | *PEX7* | 3.04625503 | 3.33E-06 | 6.20E-06 | 0.016144685 |
| Obesity | endometrium | *FIG4* | 12.99023551 | 3.66E-09 | 4.71E-08 | 1.78E-05 |
| Obesity | endometrium | *PERP* | 3.692224017 | 1.94E-05 | 3.15E-05 | 0.094111868 |
| Obesity | endometrium | *PHACTR2* | 5.703066891 | 2.13E-08 | 1.27E-07 | 0.000103191 |
| Obesity | endometrium | *PACRG* | 3.145853628 | 1.63E-06 | 3.42E-06 | 0.007909872 |
| Obesity | endometrium | *QKI* | 3.966820795 | 3.00E-08 | 1.61E-07 | 0.000145446 |
| Obesity | endometrium | *FAM120B* | 5.953864809 | 1.22E-08 | 8.80E-08 | 5.94E-05 |
| Obesity | endometrium | *GUCA1B* | -3.337263334 | 0.003104702 | 0.003962128 | 1 |
| Obesity | endometrium | *PRPH2* | -1.963680982 | 0.001411425 | 0.001863973 | 1 |
| Obesity | endometrium | *DNPH1* | 3.235950845 | 1.95E-05 | 3.17E-05 | 0.094781327 |
| Obesity | endometrium | *EXOC2* | 4.045931477 | 1.44E-07 | 5.20E-07 | 0.000697636 |
| Obesity | endometrium | *TMEM30A* | 9.830320268 | 0.000142122 | 0.000208709 | 0.689575165 |
| Obesity | endometrium | *IMPG1* | -10.28123924 | 0.002809991 | 0.003602134 | 1 |
| Obesity | endometrium | *BTN2A1* | 6.538137341 | 1.24E-06 | 2.86E-06 | 0.005998782 |
| Obesity | endometrium | *LY86* | 3.886080701 | 1.90E-06 | 3.82E-06 | 0.009200156 |
| Obesity | endometrium | *PRSS16* | 13.87558192 | 3.16E-07 | 9.70E-07 | 0.001533703 |
| Obesity | endometrium | *PCDHB2* | 7.370916561 | 0.000110118 | 0.000164246 | 0.534290615 |
| Obesity | endometrium | *NUDT12* | 3.541926748 | 9.62E-07 | 2.34E-06 | 0.004667905 |
| Obesity | endometrium | *CEP72* | 10.29636947 | 2.49E-07 | 8.02E-07 | 0.001209042 |
| Obesity | endometrium | *KIF20A* | -7.121805587 | 0.000105013 | 0.000157261 | 0.509525136 |
| Obesity | endometrium | *PFDN1* | 6.505076397 | 1.81E-09 | 3.98E-08 | 8.77E-06 |
| Obesity | endometrium | *TMCO6* | 3.006069711 | 0.000284721 | 0.000402994 | 1 |
| Obesity | endometrium | *SPARC* | 5.513386973 | 1.70E-08 | 1.09E-07 | 8.26E-05 |
| Obesity | endometrium | *IK* | 5.482931377 | 0.003869629 | 0.004893261 | 1 |
| Obesity | endometrium | *PCDHB15* | 2.940655111 | 0.014317468 | 0.017080982 | 1 |
| Obesity | endometrium | *HAVCR1* | 5.954596442 | 0.007146535 | 0.008760735 | 1 |
| Obesity | endometrium | *RNF130* | 4.137980823 | 9.49E-08 | 3.76E-07 | 0.000460426 |
| Obesity | endometrium | *ARSB* | 2.954031115 | 1.21E-06 | 2.81E-06 | 0.00586398 |
| Obesity | endometrium | *THBS4* | 4.283508817 | 3.17E-07 | 9.72E-07 | 0.001538135 |
| Obesity | endometrium | *BTNL8* | 1.227505664 | 0.03302578 | 0.03818901 | 1 |
| Obesity | endometrium | *MSH3* | 6.345306872 | 0.00024957 | 0.000355628 | 1 |
| Obesity | endometrium | *RASGRF2* | 1.957502992 | 1.08E-06 | 2.58E-06 | 0.005240401 |
| Obesity | endometrium | *DROSHA* | 6.850513079 | 0.000388739 | 0.000540293 | 1 |
| Obesity | endometrium | *CDH6* | 3.236258072 | 2.39E-06 | 4.64E-06 | 0.011587969 |
| Obesity | endometrium | *LMNB1* | 9.817449877 | 5.33E-07 | 1.47E-06 | 0.002588443 |
| Obesity | endometrium | *SLC12A7* | 3.17479756 | 1.43E-06 | 3.19E-06 | 0.006943771 |
| Obesity | endometrium | *SKP1* | 7.085647155 | 9.03E-09 | 7.32E-08 | 4.38E-05 |
| Obesity | endometrium | *PPWD1* | 5.19769815 | 4.62E-08 | 2.22E-07 | 0.000224117 |

| | | | | | | |
|---|---|---|---|---|---|---|
| Obesity | endometrium | *TXNDC15* | 11.6660025 | 1.79E-09 | 3.98E-08 | 8.67E-06 |
| Obesity | endometrium | *SMAD5* | 5.837601132 | 1.78E-08 | 1.13E-07 | 8.62E-05 |
| Obesity | endometrium | *ERGIC1* | 4.40984631 | 0.027158739 | 0.031615691 | 1 |
| Obesity | endometrium | *ATP6V0E1* | 11.95579171 | 3.94E-10 | 2.34E-08 | 1.91E-06 |
| Obesity | endometrium | *BNIP1* | 2.110787552 | 0.007797863 | 0.009518297 | 1 |
| Obesity | endometrium | *UNC5A* | 15.95688403 | 1.63E-06 | 3.42E-06 | 0.007899548 |
| Obesity | endometrium | *ACTR8* | -2.939394454 | 0.004310723 | 0.005421366 | 1 |
| Obesity | endometrium | *TBCCD1* | 10.22581339 | 7.15E-09 | 6.50E-08 | 3.47E-05 |
| Obesity | endometrium | *CLDN16* | 4.651796813 | 3.48E-06 | 6.44E-06 | 0.016866151 |
| Obesity | endometrium | *NPHP3* | 3.80090585 | 4.45E-05 | 6.91E-05 | 0.215904319 |
| Obesity | endometrium | *PCCB* | 3.541226463 | 1.03E-08 | 7.97E-08 | 4.98E-05 |
| Obesity | endometrium | *CEP70* | 10.57490481 | 3.26E-08 | 1.71E-07 | 0.000158278 |
| Obesity | endometrium | *HYAL1* | -3.915757649 | 0.033128895 | 0.038299118 | 1 |
| Obesity | endometrium | *CYB561D2* | 3.899633216 | 7.18E-07 | 1.88E-06 | 0.003483596 |
| Obesity | endometrium | *C3orf14* | 3.639354211 | 0.001595945 | 0.002092852 | 1 |
| Obesity | endometrium | *CBLB* | 9.145963052 | 1.77E-08 | 1.12E-07 | 8.60E-05 |
| Obesity | endometrium | *IQCG* | 5.144625443 | 5.62E-08 | 2.55E-07 | 0.000272647 |
| Obesity | endometrium | *NCBP2* | 12.98708237 | 1.21E-06 | 2.81E-06 | 0.005859614 |
| Obesity | endometrium | *SLC41A3* | 10.63363888 | 2.28E-05 | 3.67E-05 | 0.110868037 |
| Obesity | endometrium | *PLXNA1* | 4.433502425 | 4.65E-07 | 1.32E-06 | 0.002255222 |
| Obesity | endometrium | *SCAP* | 12.42020513 | 2.13E-11 | 3.69E-09 | 1.03E-07 |
| Obesity | endometrium | *KIAA1257* | 8.39228887 | 1.54E-06 | 3.32E-06 | 0.007494989 |
| Obesity | endometrium | *PLSCR4* | 5.883813508 | 8.93E-09 | 7.32E-08 | 4.33E-05 |
| Obesity | endometrium | *HEMK1* | 4.04809599 | 3.70E-07 | 1.10E-06 | 0.001796529 |
| Obesity | endometrium | *COMMD2* | 11.4008572 | 2.46E-09 | 4.04E-08 | 1.19E-05 |
| Obesity | endometrium | *AADAC* | 1.622493711 | 0.004661872 | 0.005843297 | 1 |
| Obesity | endometrium | *ABHD14B* | 6.508421491 | 3.44E-07 | 1.04E-06 | 0.001669298 |
| Obesity | endometrium | *ARHGEF26* | 5.783888757 | 5.36E-08 | 2.47E-07 | 0.000260297 |
| Obesity | endometrium | *VIPR1* | 5.512820637 | 8.45E-07 | 2.12E-06 | 0.004100715 |
| Obesity | endometrium | *NKTR* | 5.228881522 | 4.16E-09 | 4.89E-08 | 2.02E-05 |
| Obesity | endometrium | *NEK4* | 2.728897947 | 1.26E-05 | 2.11E-05 | 0.061310794 |
| Obesity | endometrium | *ADAM23* | 3.758907032 | 4.08E-06 | 7.42E-06 | 0.019811009 |
| Obesity | endometrium | *LMAN2L* | 6.065597326 | 8.62E-08 | 3.50E-07 | 0.000418139 |
| Obesity | endometrium | *ACTR1B* | 4.328041209 | 2.85E-07 | 8.95E-07 | 0.001383932 |
| Obesity | endometrium | *SLC35F5* | 6.523538471 | 1.87E-08 | 1.17E-07 | 9.05E-05 |
| Obesity | endometrium | *STEAP3* | 1.359863081 | 0.001431857 | 0.001888899 | 1 |
| Obesity | endometrium | *CENPA* | 4.740867754 | 9.09E-07 | 2.25E-06 | 0.004412425 |
| Obesity | endometrium | *NRBP1* | 3.72141395 | 5.80E-07 | 1.57E-06 | 0.002815622 |
| Obesity | endometrium | *ITGB6* | -2.090931223 | 0.000639117 | 0.00086887 | 1 |
| Obesity | endometrium | *PPM1G* | 3.82132547 | 5.97E-07 | 1.61E-06 | 0.002899029 |
| Obesity | endometrium | *INO80B* | 6.637061214 | 0.000160438 | 0.000233627 | 0.778443806 |
| Obesity | endometrium | *MOGS* | 5.944508844 | 0.000596576 | 0.000812871 | 1 |
| Obesity | endometrium | *GRB14* | 3.531530692 | 1.54E-06 | 3.32E-06 | 0.007487835 |
| Obesity | endometrium | *RTN4* | 5.190845472 | 1.42E-08 | 9.78E-08 | 6.91E-05 |
| Obesity | endometrium | *GALNT3* | 5.50347805 | 2.39E-07 | 7.77E-07 | 0.001160793 |
| Obesity | endometrium | *POLE4* | 11.45507102 | 2.69E-09 | 4.24E-08 | 1.31E-05 |
| Obesity | endometrium | *ACADL* | 10.26153636 | 2.61E-07 | 8.34E-07 | 0.00126866 |
| Obesity | endometrium | *MRPL19* | 9.028145199 | 3.82E-09 | 4.77E-08 | 1.85E-05 |
| Obesity | endometrium | *DNAH6* | 2.061495672 | 0.003224835 | 0.004107876 | 1 |
| Obesity | endometrium | *IGFBP2* | 6.821237603 | 1.94E-07 | 6.60E-07 | 0.000943418 |

| | | | | | | |
|---|---|---|---|---|---|---|
| Obesity | endometrium | *GGCX* | 8.779838395 | 0.00025498 | 0.000362486 | 1 |
| Obesity | endometrium | *GNLY* | 2.723789506 | 1.81E-06 | 3.68E-06 | 0.008791806 |
| Obesity | endometrium | *CHMP3* | 2.698713289 | 0.000298746 | 0.000421371 | 1 |
| Obesity | endometrium | *WNT6* | -1.885102715 | 0.000118292 | 0.000175789 | 0.573951639 |
| Obesity | endometrium | *IL1RL2* | -3.722426087 | 0.010539683 | 0.012705228 | 1 |
| Obesity | endometrium | *IL18R1* | 5.282586172 | 1.30E-06 | 2.98E-06 | 0.006313087 |
| Obesity | endometrium | *MLPH* | 4.421052259 | 4.66E-06 | 8.37E-06 | 0.022616644 |
| Obesity | endometrium | *HDLBP* | 6.50361396 | 5.74E-09 | 5.87E-08 | 2.78E-05 |
| Obesity | endometrium | *PASK* | -1.914914454 | 0.011554284 | 0.013886892 | 1 |
| Obesity | endometrium | *TPO* | 3.006244185 | 2.53E-06 | 4.89E-06 | 0.012293535 |
| Obesity | endometrium | *TAF1B* | 10.46549466 | 1.04E-08 | 8.01E-08 | 5.07E-05 |
| Obesity | endometrium | *NOL10* | 5.211911651 | 2.27E-08 | 1.33E-07 | 0.000109937 |
| Obesity | endometrium | *STRN* | 6.439905795 | 9.81E-09 | 7.75E-08 | 4.76E-05 |
| Obesity | endometrium | *CEBPZ* | 8.217297297 | 1.46E-08 | 9.96E-08 | 7.08E-05 |
| Obesity | endometrium | *PRKD3* | 3.845984571 | 2.69E-07 | 8.52E-07 | 0.001307205 |
| Obesity | endometrium | *SLC25A12* | 9.066759704 | 1.76E-09 | 3.98E-08 | 8.52E-06 |
| Obesity | endometrium | *DARS* | 5.561276225 | 2.31E-08 | 1.34E-07 | 0.000112197 |
| Obesity | endometrium | *WIPF1* | 3.199247584 | 2.75E-07 | 8.68E-07 | 0.00133647 |
| Obesity | endometrium | *PNO1* | -1.34384322 | 0.000577835 | 0.000789095 | 1 |
| Obesity | endometrium | *ORC4* | 7.127588495 | 2.77E-09 | 4.28E-08 | 1.34E-05 |
| Obesity | endometrium | *AAK1* | 5.782210179 | 6.03E-09 | 5.98E-08 | 2.93E-05 |
| Obesity | endometrium | *CD207* | 8.851063851 | 0.002286966 | 0.002956664 | 1 |
| Obesity | endometrium | *GRIN3B* | 2.833876462 | 0.015793179 | 0.018772294 | 1 |
| Obesity | endometrium | *NFE2L2* | 7.767687128 | 1.87E-09 | 4.00E-08 | 9.09E-06 |
| Obesity | endometrium | *MSH6* | 6.248121479 | 9.20E-08 | 3.68E-07 | 0.000446607 |
| Obesity | endometrium | *ALMS1* | 3.598473139 | 2.61E-07 | 8.33E-07 | 0.001266466 |
| Obesity | endometrium | *DNAJC16* | 3.765607443 | 8.90E-07 | 2.22E-06 | 0.004316297 |
| Obesity | endometrium | *MARK1* | 5.269861319 | 1.07E-07 | 4.13E-07 | 0.000516752 |
| Obesity | endometrium | *MORN1* | 3.42340575 | 2.17E-05 | 3.50E-05 | 0.105262609 |
| Obesity | endometrium | *RALGPS2* | 6.12417936 | 5.76E-08 | 2.59E-07 | 0.000279699 |
| Obesity | endometrium | *CEP104* | 7.392447602 | 1.14E-07 | 4.34E-07 | 0.000554056 |
| Obesity | endometrium | *FAM20B* | 7.409498357 | 0.002444744 | 0.003150571 | 1 |
| Obesity | endometrium | *WRAP73* | 7.343254946 | 8.96E-07 | 2.22E-06 | 0.004344973 |
| Obesity | endometrium | *ICMT* | 4.471688458 | 1.53E-06 | 3.31E-06 | 0.007414849 |
| Obesity | endometrium | *PARK7* | 9.0461948 | 1.08E-08 | 8.19E-08 | 5.22E-05 |
| Obesity | endometrium | *KIAA1324* | -5.607871905 | 0.0067137 | 0.008271934 | 1 |
| Obesity | endometrium | *MECR* | 6.464613448 | 5.98E-09 | 5.97E-08 | 2.90E-05 |
| Obesity | endometrium | *EDEM3* | 8.537919982 | 1.58E-08 | 1.04E-07 | 7.67E-05 |
| Obesity | endometrium | *RAP1A* | 5.39037887 | 1.97E-08 | 1.21E-07 | 9.56E-05 |
| Obesity | endometrium | *CAPZA1* | 4.275469739 | 3.34E-08 | 1.74E-07 | 0.000162265 |
| Obesity | endometrium | *ASH1L* | 7.195271226 | 3.25E-10 | 2.19E-08 | 1.58E-06 |
| Obesity | endometrium | *GON4L* | 6.259430769 | 1.07E-07 | 4.14E-07 | 0.000519858 |
| Obesity | endometrium | *DOCK7* | 5.64495186 | 1.43E-08 | 9.78E-08 | 6.93E-05 |
| Obesity | endometrium | *FBXO2* | 4.571255412 | 0.001096056 | 0.001459403 | 1 |
| Obesity | endometrium | *FBXO6* | 4.607863972 | 2.56E-05 | 4.09E-05 | 0.124145797 |
| Obesity | endometrium | *DNAJC6* | 3.212349807 | 2.10E-05 | 3.40E-05 | 0.102087538 |
| Obesity | endometrium | *LEPR* | 4.472092833 | 5.64E-08 | 2.56E-07 | 0.000273753 |
| Obesity | endometrium | *IVNS1ABP* | 4.848583061 | 1.73E-07 | 6.01E-07 | 0.000839424 |
| Obesity | endometrium | *MFN2* | 3.25148938 | 9.55E-07 | 2.33E-06 | 0.004633177 |
| Obesity | endometrium | *PRG4* | 3.018933087 | 1.70E-06 | 3.50E-06 | 0.008236412 |

| | | | | | | |
|---|---|---|---|---|---|---|
| Obesity | endometrium | *NCF2* | 2.887813852 | 3.24E-05 | 5.10E-05 | 0.157020587 |
| Obesity | endometrium | *BCAS2* | 4.659139541 | 1.00E-06 | 2.42E-06 | 0.004867584 |
| Obesity | endometrium | *TNNI3K* | 7.166005563 | 0.008973462 | 0.010895705 | 1 |
| Obesity | endometrium | *CFHR3* | 7.923408831 | 0.006901754 | 0.008484244 | 1 |
| Obesity | endometrium | *CRYZ* | 2.943531896 | 1.53E-06 | 3.31E-06 | 0.007402443 |
| Obesity | endometrium | *TMEM9* | 3.335627584 | 1.72E-05 | 2.80E-05 | 0.083319332 |
| Obesity | endometrium | *WARS2* | 5.992070298 | 4.65E-09 | 5.22E-08 | 2.26E-05 |
| Obesity | endometrium | *OSCP1* | 3.842498795 | 1.13E-06 | 2.67E-06 | 0.005494441 |
| Obesity | endometrium | *MRPS15* | 3.233529901 | 1.73E-06 | 3.55E-06 | 0.008400996 |
| Obesity | endometrium | *EXOC8* | 5.983479375 | 8.98E-07 | 2.23E-06 | 0.004358372 |
| Obesity | endometrium | *GNPAT* | 12.44491116 | 5.55E-10 | 2.59E-08 | 2.69E-06 |
| Obesity | endometrium | *C1orf109* | 6.458562953 | 1.15E-07 | 4.37E-07 | 0.000558869 |
| Obesity | endometrium | *TBCE* | 13.79297455 | 6.31E-09 | 6.12E-08 | 3.06E-05 |
| Obesity | endometrium | *ACADM* | 8.897113885 | 1.40E-09 | 3.98E-08 | 6.79E-06 |
| Obesity | endometrium | *SLAMF1* | 4.489009355 | 0.000246055 | 0.000350928 | 1 |
| Obesity | endometrium | *SDHB* | 6.221095342 | 4.33E-08 | 2.10E-07 | 0.000210011 |
| Obesity | endometrium | *MFAP2* | -3.330870768 | 0.018353617 | 0.021682919 | 1 |
| Obesity | endometrium | *CTBS* | 3.090257215 | 2.51E-05 | 4.01E-05 | 0.121741436 |
| Obesity | endometrium | *KLHL12* | 2.942268866 | 6.18E-06 | 1.09E-05 | 0.029971161 |
| Obesity | endometrium | *ZNHIT6* | 8.551789678 | 5.26E-09 | 5.66E-08 | 2.55E-05 |
| Obesity | endometrium | *GBP3* | 2.292165103 | 0.004530138 | 0.005685523 | 1 |
| Obesity | endometrium | *KIF17* | 6.544244345 | 3.81E-05 | 5.97E-05 | 0.184830141 |
| Obesity | endometrium | *CD46* | 4.97886926 | 9.49E-08 | 3.76E-07 | 0.000460365 |
| Obesity | endometrium | *EBNA1BP2* | 5.169115054 | 2.15E-07 | 7.12E-07 | 0.001041446 |
| Obesity | endometrium | *ERI3* | 3.082987132 | 4.99E-07 | 1.40E-06 | 0.002419097 |
| Obesity | endometrium | *PRDX1* | 2.716594521 | 2.50E-06 | 4.84E-06 | 0.012140132 |
| Obesity | endometrium | *CCDC181* | 5.114602895 | 1.34E-05 | 2.22E-05 | 0.064803189 |
| Obesity | endometrium | *NSUN4* | 6.074502861 | 0.000105858 | 0.00015838 | 0.513624803 |
| Obesity | endometrium | *DR1* | 8.630657112 | 1.30E-05 | 2.16E-05 | 0.062899813 |
| Obesity | endometrium | *PRRC2C* | 5.460292387 | 2.42E-09 | 4.02E-08 | 1.17E-05 |
| Obesity | endometrium | *ABCD3* | 6.372794322 | 1.71E-08 | 1.09E-07 | 8.28E-05 |
| Obesity | endometrium | *VAMP4* | 4.007420988 | 2.05E-07 | 6.86E-07 | 0.000996388 |
| Obesity | endometrium | *PTBP2* | 5.733174505 | 1.93E-09 | 4.00E-08 | 9.38E-06 |
| Obesity | endometrium | *DARS2* | -2.113579181 | 0.004437416 | 0.005577808 | 1 |
| Obesity | endometrium | *SERPINC1* | -1.896574501 | 0.006355084 | 0.007846023 | 1 |
| Obesity | endometrium | *MTFR1L* | 12.46347372 | 0.000128392 | 0.00018981 | 0.622957329 |
| Obesity | endometrium | *RPS6KA1* | 4.207437344 | 4.25E-06 | 7.69E-06 | 0.020605415 |
| Obesity | endometrium | *RPA2* | 2.977363788 | 1.54E-06 | 3.32E-06 | 0.007494205 |
| Obesity | endometrium | *SLC5A9* | 3.63710654 | 8.39E-07 | 2.11E-06 | 0.004069506 |
| Obesity | endometrium | *ESYT2* | 5.370074097 | 5.61E-08 | 2.55E-07 | 0.000272207 |
| Obesity | endometrium | *CD3EAP* | 3.977210537 | 1.03E-06 | 2.47E-06 | 0.005001074 |
| Obesity | endometrium | *RCN2* | 4.405948108 | 2.15E-07 | 7.14E-07 | 0.001044773 |
| Obesity | endometrium | *IFT46* | 6.258549095 | 1.97E-08 | 1.21E-07 | 9.56E-05 |
| Obesity | endometrium | *APOA1* | 22.99955208 | 0.000392604 | 0.00054504 | 1 |
| Obesity | endometrium | *DDX59* | 3.470485394 | 1.41E-06 | 3.16E-06 | 0.00684677 |
| Obesity | endometrium | *CAMSAP2* | 7.389881046 | 4.27E-09 | 4.96E-08 | 2.07E-05 |
| Obesity | endometrium | *KLF7* | 5.440642046 | 9.37E-09 | 7.47E-08 | 4.55E-05 |
| Obesity | endometrium | *CASC1* | 2.884249463 | 9.11E-05 | 0.000137279 | 0.442176032 |
| Obesity | endometrium | *LRMP* | 2.890434974 | 2.73E-06 | 5.22E-06 | 0.013240147 |
| Obesity | endometrium | *ELOVL4* | 9.941924972 | 4.44E-06 | 8.00E-06 | 0.021562201 |

| | | | | | | |
|---|---|---|---|---|---|---|
| Obesity | endometrium | *CNR1* | 7.482490003 | 0.000168058 | 0.000244063 | 0.815415721 |
| Obesity | endometrium | *AKAP7* | 6.491180239 | 1.96E-08 | 1.21E-07 | 9.51E-05 |
| Obesity | endometrium | *MED28* | 10.87226916 | 1.93E-06 | 3.87E-06 | 0.009353185 |
| Obesity | endometrium | *TMEM5* | 12.67585774 | 2.67E-09 | 4.24E-08 | 1.30E-05 |
| Obesity | endometrium | *VAMP8* | 13.58840027 | 3.39E-09 | 4.54E-08 | 1.64E-05 |
| Obesity | endometrium | *FOXO3* | 6.216571977 | 6.29E-09 | 6.12E-08 | 3.05E-05 |
| Obesity | endometrium | *SPP1* | -1.621367149 | 0.000946965 | 0.001269598 | 1 |
| Obesity | endometrium | *MFSD1* | 6.16413807 | 1.53E-08 | 1.02E-07 | 7.42E-05 |
| Obesity | endometrium | *UBN1* | 10.0121493 | 1.25E-09 | 3.98E-08 | 6.07E-06 |
| Obesity | endometrium | *DNAH7* | 2.931528335 | 1.26E-05 | 2.10E-05 | 0.061176717 |
| Obesity | endometrium | *C2orf40* | 7.114524224 | 1.61E-08 | 1.05E-07 | 7.83E-05 |
| Obesity | endometrium | *CPSF3* | 7.479919308 | 2.68E-05 | 4.27E-05 | 0.12988455 |
| Obesity | endometrium | *PIGZ* | 3.742405386 | 0.000580185 | 0.000792082 | 1 |
| Obesity | endometrium | *SENP5* | 5.667035861 | 1.59E-08 | 1.04E-07 | 7.71E-05 |
| Obesity | endometrium | *CCDC92* | 2.644600191 | 1.76E-07 | 6.10E-07 | 0.000852888 |
| Obesity | endometrium | *FKBP15* | 6.634121749 | 6.97E-09 | 6.42E-08 | 3.38E-05 |
| Obesity | endometrium | *CTNNAL1* | 3.290791781 | 3.14E-08 | 1.66E-07 | 0.000152194 |
| Obesity | endometrium | *RAB14* | 8.498239187 | 3.55E-09 | 4.67E-08 | 1.72E-05 |
| Obesity | endometrium | *TRIM32* | -2.43743837 | 0.002848678 | 0.003648834 | 1 |
| Obesity | endometrium | *PPP6C* | 8.551897165 | 3.34E-09 | 4.54E-08 | 1.62E-05 |
| Obesity | endometrium | *HDHD3* | 1.509562543 | 0.018839229 | 0.022238496 | 1 |
| Obesity | endometrium | *RBM18* | 3.417233239 | 1.49E-06 | 3.26E-06 | 0.007205811 |
| Obesity | endometrium | *HSDL2* | 4.037645132 | 9.49E-07 | 2.32E-06 | 0.00460227 |
| Obesity | endometrium | *MAPKAP1* | 7.084497872 | 2.44E-09 | 4.02E-08 | 1.18E-05 |
| Obesity | endometrium | *INVS* | 4.612626143 | 2.49E-10 | 1.94E-08 | 1.21E-06 |
| Obesity | endometrium | *DENND1A* | 6.392594265 | 1.98E-09 | 4.00E-08 | 9.60E-06 |
| Obesity | endometrium | *KDSR* | 4.504645471 | 1.45E-07 | 5.24E-07 | 0.000704977 |
| Obesity | endometrium | *DCAF4* | 13.6544104 | 1.72E-09 | 3.98E-08 | 8.36E-06 |
| Obesity | endometrium | *IFI27L2* | 4.12832796 | 1.79E-06 | 3.65E-06 | 0.008707467 |
| Obesity | endometrium | *NEK9* | 9.496070876 | 2.22E-05 | 3.58E-05 | 0.107906152 |
| Obesity | endometrium | *ACOT2* | 2.013826806 | 0.000851046 | 0.001146384 | 1 |
| Obesity | endometrium | *TTLL5* | 9.381302975 | 3.28E-09 | 4.49E-08 | 1.59E-05 |
| Obesity | endometrium | *FLVCR2* | 3.479870353 | 2.79E-06 | 5.32E-06 | 0.013541946 |
| Obesity | endometrium | *PPP4R4* | 3.920750563 | 2.19E-05 | 3.53E-05 | 0.106098235 |
| Obesity | endometrium | *ALDH6A1* | 4.865397444 | 9.74E-06 | 1.66E-05 | 0.047281008 |
| Obesity | endometrium | *ESRRB* | 5.274346986 | 5.02E-06 | 8.97E-06 | 0.024335008 |
| Obesity | endometrium | *EIF2B2* | 2.447645514 | 0.000110961 | 0.000165351 | 0.538382329 |
| Obesity | endometrium | *COQ6* | 4.82723566 | 3.53E-07 | 1.06E-06 | 0.001712796 |
| Obesity | endometrium | *DNMT3A* | 3.546289042 | 3.40E-07 | 1.03E-06 | 0.001649681 |
| Obesity | endometrium | *CNRIP1* | 2.75426094 | 3.85E-05 | 6.02E-05 | 0.186561775 |
| Obesity | endometrium | *CRIPT* | 9.364308051 | 6.80E-09 | 6.31E-08 | 3.30E-05 |
| Obesity | endometrium | *EPCAM* | 12.77954747 | 2.95E-06 | 5.56E-06 | 0.014307036 |
| Obesity | endometrium | *SLC17A5* | 9.561579067 | 1.99E-08 | 1.21E-07 | 9.64E-05 |
| Obesity | endometrium | *TECTB* | 5.25928436 | 0.01239755 | 0.014867255 | 1 |
| Obesity | endometrium | *ELOVL3* | 7.40881214 | 0.002933006 | 0.003752886 | 1 |
| Obesity | endometrium | *PYROXD2* | 9.212352698 | 8.59E-05 | 0.000129665 | 0.416873748 |
| Obesity | endometrium | *MXI1* | 5.86921257 | 1.88E-08 | 1.17E-07 | 9.13E-05 |
| Obesity | endometrium | *HELLS* | 9.143635188 | 0.000112634 | 0.00016769 | 0.546501797 |
| Obesity | endometrium | *TCTN3* | 9.127717371 | 3.20E-07 | 9.81E-07 | 0.001554818 |
| Obesity | endometrium | *WDR11* | 9.526256376 | 1.11E-09 | 3.83E-08 | 5.40E-06 |

| | | | | | | |
|---|---|---|---|---|---|---|
| Obesity | endometrium | *KANSL1* | 5.185225011 | 8.98E-09 | 7.32E-08 | 4.36E-05 |
| Obesity | endometrium | *HOXB3* | 4.361919551 | 1.15E-07 | 4.38E-07 | 0.000560271 |
| Obesity | endometrium | *CAAP1* | 7.210177353 | 3.01E-06 | 5.66E-06 | 0.014582053 |
| Obesity | endometrium | *PCDHB10* | -1.889889704 | 0.000116844 | 0.000173797 | 0.566927118 |
| Obesity | endometrium | *PCDHB12* | 3.043081756 | 0.021972102 | 0.025794493 | 1 |
| Obesity | endometrium | *GORAB* | -2.26629302 | 0.013046896 | 0.015615081 | 1 |
| Obesity | endometrium | *ACAT2* | 5.384351648 | 3.14E-07 | 9.65E-07 | 0.001525308 |
| Obesity | endometrium | *SNX19* | 3.405519555 | 9.80E-05 | 0.000147231 | 0.475557332 |
| Obesity | endometrium | *SLC10A7* | 5.163559709 | 3.89E-08 | 1.96E-07 | 0.000188515 |
| Obesity | endometrium | *MASTL* | 4.916147047 | 0.002935705 | 0.003755349 | 1 |
| Obesity | endometrium | *MRC1* | 2.102590906 | 1.65E-06 | 3.44E-06 | 0.00801719 |
| Obesity | endometrium | *IQSEC3* | 25.10383749 | 1.69E-07 | 5.91E-07 | 0.000819195 |
| Obesity | endometrium | *DNAJC15* | 7.167223991 | 6.62E-09 | 6.23E-08 | 3.21E-05 |
| Obesity | endometrium | *UFM1* | 4.537286079 | 9.44E-08 | 3.75E-07 | 0.000458113 |
| Obesity | endometrium | *ALG5* | 9.460705316 | 1.42E-08 | 9.78E-08 | 6.91E-05 |
| Obesity | endometrium | *SIL1* | 9.012777969 | 3.68E-09 | 4.71E-08 | 1.78E-05 |
| Obesity | endometrium | *ZFP30* | 2.778575314 | 6.64E-07 | 1.77E-06 | 0.003220252 |
| Obesity | endometrium | *ARL1* | 12.26236891 | 1.59E-08 | 1.04E-07 | 7.70E-05 |
| Obesity | endometrium | *APAF1* | 11.39267224 | 2.15E-09 | 4.00E-08 | 1.05E-05 |
| Obesity | endometrium | *ADRA1A* | 3.369769285 | 8.33E-09 | 7.12E-08 | 4.04E-05 |
| Obesity | endometrium | *EPHX2* | 8.509326721 | 1.69E-08 | 1.09E-07 | 8.21E-05 |
| Obesity | endometrium | *AKAP1* | -1.664220632 | 0.019408369 | 0.022873307 | 1 |
| Obesity | endometrium | *SCPEP1* | 2.861039998 | 2.32E-07 | 7.58E-07 | 0.001124166 |
| Obesity | endometrium | *TEX14* | -1.033051145 | 0.001581199 | 0.002074628 | 1 |
| Obesity | endometrium | *TRIM6* | 9.039496061 | 2.05E-06 | 4.08E-06 | 0.009961461 |
| Obesity | endometrium | *ADCY7* | 2.684774058 | 2.85E-06 | 5.41E-06 | 0.013810763 |
| Obesity | endometrium | *ECHDC2* | 5.834155529 | 2.05E-07 | 6.86E-07 | 0.000994025 |
| Obesity | endometrium | *PYROXD1* | 6.422504703 | 2.88E-09 | 4.29E-08 | 1.40E-05 |
| Obesity | endometrium | *ZNF549* | 5.801360317 | 1.32E-07 | 4.87E-07 | 0.000640803 |
| Obesity | endometrium | *A1BG* | -2.687821187 | 0.005466177 | 0.006798742 | 1 |
| Obesity | endometrium | *ZSCAN18* | 5.302280448 | 8.03E-08 | 3.34E-07 | 0.00038961 |
| Obesity | endometrium | *ZNF211* | 4.578614335 | 6.80E-07 | 1.80E-06 | 0.003299438 |
| Obesity | endometrium | *PDZRN3* | 3.363345345 | 8.80E-07 | 2.20E-06 | 0.004268383 |
| Obesity | endometrium | *LHX4* | 5.646277557 | 3.95E-07 | 1.16E-06 | 0.001914115 |
| Obesity | endometrium | *SEC22A* | 5.001757287 | 5.94E-08 | 2.64E-07 | 0.000288445 |
| Obesity | endometrium | *POPDC2* | 4.976938171 | 0.00015957 | 0.000232503 | 0.774236061 |
| Obesity | endometrium | *CD80* | 3.520847805 | 0.004861325 | 0.006082298 | 1 |
| Obesity | endometrium | *MAPK8IP1* | 9.84466956 | 7.33E-07 | 1.91E-06 | 0.003556269 |
| Obesity | endometrium | *CAT* | 4.185365137 | 4.68E-08 | 2.24E-07 | 0.000227279 |
| Obesity | endometrium | *PILRB* | 2.812765567 | 8.99E-07 | 2.23E-06 | 0.004363531 |
| Obesity | endometrium | *ZCCHC17* | 7.95043731 | 8.38E-09 | 7.13E-08 | 4.07E-05 |
| Obesity | endometrium | *FABP3* | 6.01114367 | 3.45E-06 | 6.41E-06 | 0.016752328 |
| Obesity | endometrium | *TNFSF10* | 5.348065877 | 4.21E-08 | 2.07E-07 | 0.000204278 |
| Obesity | endometrium | *ZNF639* | 3.313257999 | 1.83E-06 | 3.71E-06 | 0.008891131 |
| Obesity | endometrium | *CPXM2* | 2.845989219 | 1.28E-07 | 4.77E-07 | 0.000622918 |
| Obesity | endometrium | *FLT3* | 6.424802423 | 1.81E-06 | 3.67E-06 | 0.008771379 |
| Obesity | endometrium | *MTIF3* | 7.866434526 | 2.38E-09 | 4.02E-08 | 1.15E-05 |
| Obesity | endometrium | *FMOD* | 4.827126812 | 2.63E-07 | 8.36E-07 | 0.001274395 |
| Obesity | endometrium | *CD244* | 5.285044286 | 6.78E-07 | 1.80E-06 | 0.003290117 |
| Obesity | endometrium | *SERAC1* | 3.523399848 | 2.16E-06 | 4.26E-06 | 0.010472039 |

| | | | | | | |
|---|---|---|---|---|---|---|
| Obesity | endometrium | *LDB3* | 4.388669788 | 1.15E-06 | 2.70E-06 | 0.005565898 |
| Obesity | endometrium | *OPN4* | 35.54951085 | 3.94E-11 | 5.98E-09 | 1.91E-07 |
| Obesity | endometrium | *FAM35A* | 7.696247657 | 4.07E-09 | 4.85E-08 | 1.97E-05 |
| Obesity | endometrium | *ODF2L* | 10.69936628 | 2.08E-09 | 4.00E-08 | 1.01E-05 |
| Obesity | endometrium | *SPATA1* | 6.012011239 | 1.81E-08 | 1.14E-07 | 8.81E-05 |
| Obesity | endometrium | *RWDD3* | 3.262259178 | 0.00867485 | 0.010548768 | 1 |
| Obesity | endometrium | *EEPD1* | 3.679381549 | 3.91E-08 | 1.97E-07 | 0.000189949 |
| Obesity | endometrium | *KLHL7* | 5.936220707 | 2.03E-08 | 1.23E-07 | 9.85E-05 |
| Obesity | endometrium | *NT5C3A* | 5.89657572 | 2.46E-08 | 1.39E-07 | 0.00011919 |
| Obesity | endometrium | *CCZ1* | 6.33093206 | 1.16E-08 | 8.54E-08 | 5.63E-05 |
| Obesity | endometrium | *SMU1* | 13.82010105 | 2.01E-09 | 4.00E-08 | 9.74E-06 |
| Obesity | endometrium | *GLIPR2* | -2.354450929 | 0.006315339 | 0.007798937 | 1 |
| Obesity | endometrium | *SLC25A51* | 4.257193061 | 6.32E-08 | 2.77E-07 | 0.000306632 |
| Obesity | endometrium | *TRIM24* | 9.575088277 | 1.83E-09 | 3.98E-08 | 8.88E-06 |
| Obesity | endometrium | *C7orf49* | 5.940079948 | 3.20E-09 | 4.46E-08 | 1.55E-05 |
| Obesity | endometrium | *RBM19* | 13.65913767 | 9.76E-08 | 3.85E-07 | 0.000473621 |
| Obesity | endometrium | *DDX54* | 10.91205009 | 3.43E-09 | 4.56E-08 | 1.66E-05 |
| Obesity | endometrium | *CCDC91* | 10.03596983 | 6.53E-09 | 6.17E-08 | 3.17E-05 |
| Obesity | endometrium | *EBPL* | 6.676642059 | 0.001635377 | 0.002142238 | 1 |
| Obesity | endometrium | *ATP7B* | 2.226171814 | 9.85E-07 | 2.39E-06 | 0.004777671 |
| Obesity | endometrium | *CENPK* | 8.394323513 | 7.73E-07 | 1.99E-06 | 0.003752607 |
| Obesity | endometrium | *ITIH5* | 3.266684412 | 1.42E-08 | 9.76E-08 | 6.88E-05 |
| Obesity | endometrium | *TSFM* | 4.482175087 | 2.77E-06 | 5.29E-06 | 0.013456823 |
| Obesity | endometrium | *ARHGAP9* | 4.750592716 | 2.16E-06 | 4.26E-06 | 0.010477866 |
| Obesity | endometrium | *PFDN5* | 11.64157413 | 3.56E-09 | 4.67E-08 | 1.73E-05 |
| Obesity | endometrium | *PDE1B* | 3.681709547 | 1.70E-06 | 3.51E-06 | 0.008271662 |
| Obesity | endometrium | *CDK2* | 4.62422403 | 7.08E-05 | 0.000107884 | 0.343717961 |
| Obesity | endometrium | *SMUG1* | 8.164589495 | 3.00E-09 | 4.29E-08 | 1.45E-05 |
| Obesity | endometrium | *SARDH* | 5.720156226 | 4.54E-05 | 7.05E-05 | 0.220419947 |
| Obesity | endometrium | *DBH* | 102.8828242 | 1.82E-06 | 3.70E-06 | 0.008854883 |
| Obesity | endometrium | *NDUFAF4* | 9.101370949 | 7.59E-08 | 3.18E-07 | 0.000368063 |
| Obesity | endometrium | *USP45* | 4.362833793 | 7.04E-08 | 3.02E-07 | 0.000341742 |
| Obesity | endometrium | *TTC21B* | 6.813671189 | 2.41E-08 | 1.38E-07 | 0.000117009 |
| Obesity | endometrium | *ACVR1C* | 1.862635107 | 2.16E-06 | 4.26E-06 | 0.010490504 |
| Obesity | endometrium | *SLC36A1* | 3.99882598 | 7.80E-07 | 2.00E-06 | 0.003784373 |
| Obesity | endometrium | *PLA2G12A* | 12.80970734 | 2.88E-09 | 4.29E-08 | 1.40E-05 |
| Obesity | endometrium | *C4BPA* | 5.492256344 | 1.20E-05 | 2.02E-05 | 0.058428972 |
| Obesity | endometrium | *C4BPB* | 5.328011596 | 5.49E-06 | 9.77E-06 | 0.026657306 |
| Obesity | endometrium | *RAB38* | 9.393670664 | 9.25E-08 | 3.69E-07 | 0.000448862 |
| Obesity | endometrium | *DAW1* | 8.439879196 | 3.78E-06 | 6.94E-06 | 0.018353766 |
| Obesity | endometrium | *DNPEP* | 4.675478247 | 1.97E-07 | 6.68E-07 | 0.000958133 |
| Obesity | endometrium | *OBSL1* | 3.4692684 | 8.44E-07 | 2.12E-06 | 0.004095365 |
| Obesity | endometrium | *SLC12A4* | 6.279693538 | 1.30E-07 | 4.80E-07 | 0.00062918 |
| Obesity | endometrium | *FAM210B* | 13.44558693 | 6.94E-09 | 6.40E-08 | 3.37E-05 |
| Obesity | endometrium | *SLC12A5* | -7.445279256 | 0.006679852 | 0.008232319 | 1 |
| Obesity | endometrium | *MATN4* | 29.81842199 | 1.29E-08 | 9.12E-08 | 6.24E-05 |
| Obesity | endometrium | *VAPB* | 2.780574343 | 9.04E-07 | 2.24E-06 | 0.00438438 |
| Obesity | endometrium | *TOX2* | -1.275781843 | 0.001517857 | 0.001995298 | 1 |
| Obesity | endometrium | *SRSF6* | 5.140380788 | 9.78E-08 | 3.86E-07 | 0.000474685 |
| Obesity | endometrium | *CSE1L* | 4.669576915 | 8.29E-08 | 3.41E-07 | 0.000402353 |

| | | | | | | |
|---|---|---|---|---|---|---|
| Obesity | endometrium | *PTGIS* | 2.704409736 | 9.92E-08 | 3.90E-07 | 0.000481358 |
| Obesity | endometrium | *RNF114* | 9.61085047 | 1.24E-09 | 3.98E-08 | 6.01E-06 |
| Obesity | endometrium | *NEURL2* | 6.889066285 | 1.43E-06 | 3.19E-06 | 0.006948213 |
| Obesity | endometrium | *PEPD* | 9.9951299 | 3.42E-10 | 2.24E-08 | 1.66E-06 |
| Obesity | endometrium | *STAMBP* | 12.95297282 | 4.06E-09 | 4.85E-08 | 1.97E-05 |
| Obesity | endometrium | *NAGK* | 3.996890759 | 1.55E-07 | 5.52E-07 | 0.000750766 |
| Obesity | endometrium | *SNRNP27* | 3.75473107 | 9.23E-07 | 2.27E-06 | 0.004477174 |
| Obesity | endometrium | *ATP8A1* | 10.87158288 | 5.96E-10 | 2.63E-08 | 2.89E-06 |
| Obesity | endometrium | *USP22* | 6.024211196 | 2.30E-08 | 1.34E-07 | 0.000111511 |
| Obesity | endometrium | *HIF3A* | 3.603026888 | 1.64E-06 | 3.42E-06 | 0.007934432 |
| Obesity | endometrium | *BTN2A2* | 5.676231762 | 1.25E-11 | 2.43E-09 | 6.06E-08 |
| Obesity | endometrium | *MRS2* | 3.012581212 | 6.51E-07 | 1.74E-06 | 0.003158163 |
| Obesity | endometrium | *SNRPC* | -7.531516072 | 0.001080999 | 0.001440145 | 1 |
| Obesity | endometrium | *SERPINB6* | 6.431197591 | 9.32E-09 | 7.47E-08 | 4.52E-05 |
| Obesity | endometrium | *PEX6* | 3.652384877 | 7.19E-05 | 0.000109373 | 0.348682517 |
| Obesity | endometrium | *NQO2* | 12.41553832 | 2.29E-07 | 7.52E-07 | 0.001112283 |
| Obesity | endometrium | *ZNF391* | 2.316513826 | 6.37E-05 | 9.74E-05 | 0.309166087 |
| Obesity | endometrium | *RPS10* | -1.450803368 | 0.00025227 | 0.000359054 | 1 |
| Obesity | endometrium | *MED20* | 5.517066027 | 7.39E-08 | 3.13E-07 | 0.000358564 |
| Obesity | endometrium | *TCP11* | 7.082657225 | 0.004516325 | 0.00567112 | 1 |
| Obesity | endometrium | *TREM1* | 8.604371354 | 3.89E-08 | 1.96E-07 | 0.000188768 |
| Obesity | endometrium | *MEA1* | 5.215331201 | 0.001215366 | 0.001612953 | 1 |
| Obesity | endometrium | *KLHL31* | 3.152788348 | 0.000168616 | 0.000244728 | 0.818124684 |
| Obesity | endometrium | *GLO1* | 13.38753558 | 8.15E-11 | 9.64E-09 | 3.95E-07 |
| Obesity | endometrium | *KCNK17* | -2.319553581 | 0.002465582 | 0.003175738 | 1 |
| Obesity | endometrium | *RREB1* | 7.042321273 | 1.34E-09 | 3.98E-08 | 6.49E-06 |
| Obesity | endometrium | *RIOK1* | 3.334889912 | 1.30E-06 | 2.98E-06 | 0.006300633 |
| Obesity | endometrium | *ATXN1* | 8.595981992 | 1.77E-09 | 3.98E-08 | 8.59E-06 |
| Obesity | endometrium | *RUNX2* | 5.619928086 | 1.29E-07 | 4.77E-07 | 0.000623984 |
| Obesity | endometrium | *CXCL6* | 8.181718383 | 2.89E-06 | 5.48E-06 | 0.014029161 |
| Obesity | endometrium | *SH3TC1* | 3.060970096 | 1.65E-06 | 3.44E-06 | 0.007992877 |
| Obesity | endometrium | *BBS2* | 4.808994183 | 3.33E-08 | 1.73E-07 | 0.000161423 |
| Obesity | endometrium | *MT1G* | 12.88553842 | 0.000533069 | 0.000730636 | 1 |
| Obesity | endometrium | *GOT2* | 4.070495745 | 8.32E-07 | 2.10E-06 | 0.004034464 |
| Obesity | endometrium | *CLYBL* | 7.841936688 | 4.53E-09 | 5.16E-08 | 2.20E-05 |
| Obesity | endometrium | *TMTC4* | 5.703928023 | 0.032338783 | 0.037421363 | 1 |
| Obesity | endometrium | *ABCC4* | 9.040846836 | 5.67E-09 | 5.87E-08 | 2.75E-05 |
| Obesity | endometrium | *ATP5S* | 5.993526709 | 1.71E-05 | 2.80E-05 | 0.08312366 |
| Obesity | endometrium | *GRK4* | 3.125730229 | 7.75E-10 | 3.06E-08 | 3.76E-06 |
| Obesity | endometrium | *TEKT3* | 4.360886087 | 6.65E-06 | 1.17E-05 | 0.032265071 |
| Obesity | endometrium | *HS3ST3B1* | 5.87879512 | 1.07E-05 | 1.81E-05 | 0.051998254 |
| Obesity | endometrium | *GGA3* | 11.39281933 | 4.85E-10 | 2.45E-08 | 2.35E-06 |
| Obesity | endometrium | *NUP85* | 5.892489122 | 1.98E-07 | 6.69E-07 | 0.000960424 |
| Obesity | endometrium | *NT5C* | 13.02353622 | 2.27E-09 | 4.01E-08 | 1.10E-05 |
| Obesity | endometrium | *OPRL1* | 12.46979643 | 1.58E-06 | 3.36E-06 | 0.007670587 |
| Obesity | endometrium | *IL1B* | 17.09953189 | 2.79E-05 | 4.44E-05 | 0.135408374 |
| Obesity | endometrium | *PAX8* | 3.607044963 | 2.33E-05 | 3.75E-05 | 0.113264202 |
| Obesity | endometrium | *POLR1B* | 3.320453153 | 1.53E-06 | 3.31E-06 | 0.007420374 |
| Obesity | endometrium | *CCDC93* | 5.874513027 | 1.88E-08 | 1.17E-07 | 9.11E-05 |
| Obesity | endometrium | *PSD4* | 14.64042714 | 1.58E-08 | 1.04E-07 | 7.67E-05 |

| | | | | | | |
|---|---|---|---|---|---|---|
| Obesity | endometrium | *GTF2F1* | 8.128176484 | 4.26E-08 | 2.08E-07 | 0.000206872 |
| Obesity | endometrium | *TNFSF9* | 52.67754962 | 0.001150333 | 0.001529574 | 1 |
| Obesity | endometrium | *SH2D3A* | 4.72447771 | 2.39E-05 | 3.83E-05 | 0.115902839 |
| Obesity | endometrium | *SNRPD2* | 7.546012143 | 3.35E-08 | 1.74E-07 | 0.000162499 |
| Obesity | endometrium | *SDCBP2* | -4.596014969 | 0.003919691 | 0.004952693 | 1 |
| Obesity | endometrium | *PSMF1* | 3.604435773 | 1.29E-06 | 2.97E-06 | 0.006273458 |
| Obesity | endometrium | *DTD1* | 10.30690001 | 5.94E-06 | 1.05E-05 | 0.028820669 |
| Obesity | endometrium | *RBCK1* | 5.184205392 | 1.38E-07 | 5.03E-07 | 0.000671208 |
| Obesity | endometrium | *RRBP1* | 5.570080268 | 2.44E-08 | 1.39E-07 | 0.000118226 |
| Obesity | endometrium | *FLRT3* | 3.843214639 | 1.23E-06 | 2.85E-06 | 0.005976265 |
| Obesity | endometrium | *PCSK2* | 15.56200479 | 2.67E-06 | 5.12E-06 | 0.012947846 |
| Obesity | endometrium | *MKKS* | 10.1330095 | 2.28E-07 | 7.49E-07 | 0.001105941 |
| Obesity | endometrium | *LAMP5* | 73.26446148 | 1.60E-05 | 2.62E-05 | 0.077489296 |
| Obesity | endometrium | *MGME1* | 5.418866348 | 1.06E-07 | 4.11E-07 | 0.000514356 |
| Obesity | endometrium | *ITPA* | 6.008248403 | 6.08E-08 | 2.69E-07 | 0.000294815 |
| Obesity | endometrium | *NCLN* | 3.419983402 | 1.86E-06 | 3.75E-06 | 0.009013124 |
| Obesity | endometrium | *UROD* | 10.66769456 | 4.38E-07 | 1.26E-06 | 0.002124894 |
| Obesity | endometrium | *KLC1* | 3.179997668 | 4.86E-07 | 1.37E-06 | 0.002355661 |
| Obesity | endometrium | *XRCC3* | 6.879671462 | 5.49E-06 | 9.77E-06 | 0.026657152 |
| Obesity | endometrium | *TUBGCP3* | 2.911165812 | 6.21E-07 | 1.67E-06 | 0.003011515 |
| Obesity | endometrium | *MCF2L* | 2.923708808 | 2.76E-06 | 5.28E-06 | 0.013409359 |
| Obesity | endometrium | *PROZ* | 7.203736831 | 0.038688651 | 0.044493324 | 1 |
| Obesity | endometrium | *IGFLR1* | 5.550308922 | 0.006991238 | 0.008583372 | 1 |
| Obesity | endometrium | *COX6B1* | 8.001389237 | 0.00088037 | 0.001183913 | 1 |
| Obesity | endometrium | *PRDX5* | 14.45453947 | 4.85E-07 | 1.37E-06 | 0.002352402 |
| Obesity | endometrium | *IRF3* | 13.12763997 | 8.63E-10 | 3.30E-08 | 4.19E-06 |
| Obesity | endometrium | *PRR12* | 8.265844474 | 1.41E-08 | 9.75E-08 | 6.85E-05 |
| Obesity | endometrium | *TSKS* | -1.894450774 | 0.002963895 | 0.003789412 | 1 |
| Obesity | endometrium | *TRAP1* | 4.509131183 | 1.17E-08 | 8.59E-08 | 5.70E-05 |
| Obesity | endometrium | *ZNF384* | 12.48239464 | 1.02E-05 | 1.73E-05 | 0.049673453 |
| Obesity | endometrium | *EMG1* | 4.300674569 | 7.92E-07 | 2.02E-06 | 0.003842039 |
| Obesity | endometrium | *ATG14* | 4.837826725 | 3.64E-08 | 1.86E-07 | 0.000176624 |
| Obesity | endometrium | *KTN1* | 8.006270988 | 1.59E-09 | 3.98E-08 | 7.71E-06 |
| Obesity | endometrium | *L3HYPDH* | 2.923232039 | 0.000947388 | 0.001269814 | 1 |
| Obesity | endometrium | *ZBTB1* | 6.562093557 | 0.000801857 | 0.001081928 | 1 |
| Obesity | endometrium | *RHOT1* | 7.929387443 | 8.89E-10 | 3.37E-08 | 4.31E-06 |
| Obesity | endometrium | *WDR60* | 7.981049621 | 7.45E-09 | 6.64E-08 | 3.61E-05 |
| Obesity | endometrium | *AIF1L* | 4.31397168 | 1.54E-06 | 3.32E-06 | 0.007495918 |
| Obesity | endometrium | *CANX* | 5.930365075 | 5.65E-09 | 5.87E-08 | 2.74E-05 |
| Obesity | endometrium | *ZNF484* | 9.354658575 | 0.001352011 | 0.001788429 | 1 |
| Obesity | endometrium | *FGD3* | 2.969542176 | 8.04E-06 | 1.39E-05 | 0.038990843 |
| Obesity | endometrium | *ABHD8* | 3.002410849 | 6.63E-06 | 1.16E-05 | 0.032181373 |
| Obesity | endometrium | *ATP13A4* | 7.50804415 | 1.84E-06 | 3.72E-06 | 0.00894725 |
| Obesity | endometrium | *HELB* | 3.834541467 | 3.47E-07 | 1.05E-06 | 0.00168525 |
| Obesity | endometrium | *RAB3IP* | 5.358673513 | 3.76E-07 | 1.12E-06 | 0.001824508 |
| Obesity | endometrium | *TAS2R4* | 5.038662854 | 1.82E-06 | 3.69E-06 | 0.008833988 |
| Obesity | endometrium | *LRRC61* | 3.338863691 | 2.98E-05 | 4.72E-05 | 0.144730256 |
| Obesity | endometrium | *IDUA* | 8.237488068 | 1.02E-06 | 2.44E-06 | 0.004924799 |
| Obesity | endometrium | *EMC1* | 5.175802472 | 1.55E-07 | 5.52E-07 | 0.000751061 |
| Obesity | endometrium | *PLA2G5* | 1.926263788 | 0.012159088 | 0.014588501 | 1 |

| | | | | | | |
|---|---|---|---|---|---|---|
| Obesity | endometrium | *UBR4* | 6.247933549 | 2.69E-07 | 8.52E-07 | 0.001305806 |
| Obesity | endometrium | *GFER* | 10.79348441 | 0.00103928 | 0.001386469 | 1 |
| Obesity | endometrium | *WFIKKN1* | 5.977096186 | 0.037955056 | 0.043660012 | 1 |
| Obesity | endometrium | *WDR24* | 5.168900162 | 4.26E-05 | 6.63E-05 | 0.206819433 |
| Obesity | endometrium | *SMARCA4* | 7.724974992 | 1.74E-08 | 1.11E-07 | 8.46E-05 |
| Obesity | endometrium | *PNKD* | 9.215927967 | 3.87E-09 | 4.80E-08 | 1.88E-05 |
| Obesity | endometrium | *RNF6* | 16.85790331 | 5.99E-09 | 5.97E-08 | 2.90E-05 |
| Obesity | endometrium | *HIP1* | 6.950380453 | 2.09E-09 | 4.00E-08 | 1.01E-05 |
| Obesity | endometrium | *PTPN12* | 6.326144945 | 1.49E-08 | 1.01E-07 | 7.24E-05 |
| Obesity | endometrium | *STYXL1* | 5.970721215 | 2.02E-05 | 3.27E-05 | 0.09809775 |
| Obesity | endometrium | *STEAP4* | 3.072811771 | 9.25E-06 | 1.58E-05 | 0.044896193 |
| Obesity | endometrium | *PEX1* | 5.36251837 | 7.10E-08 | 3.04E-07 | 0.000344731 |
| Obesity | endometrium | *KDR* | 3.506689591 | 5.86E-07 | 1.59E-06 | 0.002842546 |
| Obesity | endometrium | *VPREB3* | -10.9126419 | 0.008132635 | 0.009911968 | 1 |
| Obesity | endometrium | *MGAT3* | -2.24669184 | 0.004960758 | 0.00619552 | 1 |
| Obesity | endometrium | *ATF4* | 5.323778639 | 1.86E-07 | 6.37E-07 | 0.000902306 |
| Obesity | endometrium | *A4GALT* | 3.948283481 | 8.00E-06 | 1.38E-05 | 0.038820938 |
| Obesity | endometrium | *CDC42EP1* | 16.34212897 | 3.51E-08 | 1.81E-07 | 0.000170368 |
| Obesity | endometrium | *SPECC1* | 6.89894168 | 5.22E-09 | 5.66E-08 | 2.53E-05 |
| Obesity | endometrium | *CDHR3* | 8.972528379 | 2.56E-09 | 4.16E-08 | 1.24E-05 |
| Obesity | endometrium | *PRKRIP1* | 5.352553494 | 7.08E-08 | 3.03E-07 | 0.000343565 |
| Obesity | endometrium | *IRF5* | 2.760372894 | 0.000480802 | 0.000662554 | 1 |
| Obesity | endometrium | *OPN1SW* | -2.175224386 | 0.000206955 | 0.000297701 | 1 |
| Obesity | endometrium | *HOXD1* | -3.111821785 | 0.000104568 | 0.000156642 | 0.507363651 |
| Obesity | endometrium | *HOXD3* | -1.105711288 | 0.037939807 | 0.04365282 | 1 |
| Obesity | endometrium | *MTX2* | 10.87686448 | 1.53E-09 | 3.98E-08 | 7.40E-06 |
| Obesity | endometrium | *GAD1* | 8.631447966 | 0.000144794 | 0.000212312 | 0.702541711 |
| Obesity | endometrium | *ORMDL1* | 5.435484877 | 3.42E-08 | 1.77E-07 | 0.000166077 |
| Obesity | endometrium | *HOXD9* | 2.095399852 | 0.02917504 | 0.033873485 | 1 |
| Obesity | endometrium | *HOXD10* | -2.172277935 | 0.000271323 | 0.000384817 | 1 |
| Obesity | endometrium | *PSMG2* | 8.588508742 | 2.04E-07 | 6.84E-07 | 0.000990782 |
| Obesity | endometrium | *TWSG1* | 3.801671352 | 3.90E-07 | 1.15E-06 | 0.001890646 |
| Obesity | endometrium | *WDFY4* | 1.8838824 | 0.000773595 | 0.001045248 | 1 |
| Obesity | endometrium | *EIF2AK4* | 4.879731165 | 1.17E-08 | 8.55E-08 | 5.66E-05 |
| Obesity | endometrium | *TMOD2* | 11.67882452 | 1.81E-06 | 3.68E-06 | 0.00879597 |
| Obesity | endometrium | *IVD* | 5.210095654 | 5.32E-08 | 2.46E-07 | 0.000257906 |
| Obesity | endometrium | *KNSTRN* | -1.681761717 | 0.006765808 | 0.008323454 | 1 |
| Obesity | endometrium | *ARPP19* | 10.45316287 | 2.12E-09 | 4.00E-08 | 1.03E-05 |
| Obesity | endometrium | *VPS13C* | 3.945889089 | 4.42E-08 | 2.14E-07 | 0.000214539 |
| Obesity | endometrium | *CALML4* | -2.822063978 | 0.000208188 | 0.000299297 | 1 |
| Obesity | endometrium | *ANAPC13* | -2.082679002 | 0.004017976 | 0.005071597 | 1 |
| Obesity | endometrium | *SUMF2* | 7.137372708 | 1.13E-08 | 8.42E-08 | 5.47E-05 |
| Obesity | endometrium | *SERGEF* | 4.862101958 | 2.78E-08 | 1.52E-07 | 0.000134881 |
| Obesity | endometrium | *DCTD* | 9.291591357 | 5.57E-08 | 2.54E-07 | 0.000270058 |
| Obesity | endometrium | *RPAIN* | 5.911895632 | 0.000608842 | 0.000828872 | 1 |
| Obesity | endometrium | *USP6* | -2.078362959 | 0.043176561 | 0.04951375 | 1 |
| Obesity | endometrium | *PHF20L1* | 10.94105068 | 5.11E-09 | 5.57E-08 | 2.48E-05 |
| Obesity | endometrium | *PUS7L* | 5.584783384 | 8.78E-08 | 3.55E-07 | 0.000425813 |
| Obesity | endometrium | *KRI1* | 4.686439126 | 0.002841196 | 0.003640212 | 1 |
| Obesity | endometrium | *AP1M2* | -6.102664169 | 1.25E-05 | 2.08E-05 | 0.060630605 |

| | | | | | | |
|---|---|---|---|---|---|---|
| Obesity | endometrium | *MTUS1* | 7.943095611 | 1.22E-09 | 3.98E-08 | 5.93E-06 |
| Obesity | endometrium | *KLK10* | 3.574508342 | 0.024913409 | 0.029092626 | 1 |
| Obesity | endometrium | *KLK8* | -17.2787769 | 0.006979032 | 0.008570555 | 1 |
| Obesity | endometrium | *RIPK3* | 4.695677867 | 0.00032507 | 0.000456245 | 1 |
| Obesity | endometrium | *ADCY4* | 4.04592443 | 1.52E-06 | 3.31E-06 | 0.007387167 |
| Obesity | endometrium | *BCL2L2* | 2.976739202 | 1.55E-06 | 3.32E-06 | 0.007497888 |
| Obesity | endometrium | *DTD2* | -2.216187993 | 0.012978545 | 0.015537108 | 1 |
| Obesity | endometrium | *PARP2* | 3.709879924 | 1.77E-06 | 3.60E-06 | 0.008567251 |
| Obesity | endometrium | *MIS18BP1* | 5.092224003 | 1.42E-08 | 9.76E-08 | 6.88E-05 |
| Obesity | endometrium | *RNASE1* | 2.245266979 | 6.86E-07 | 1.81E-06 | 0.003328935 |
| Obesity | endometrium | *EPB41L4A* | 6.417771386 | 4.99E-09 | 5.51E-08 | 2.42E-05 |
| Obesity | endometrium | *CDO1* | 4.960602139 | 0.000922627 | 0.001238679 | 1 |
| Obesity | endometrium | *REEP5* | 10.60911822 | 1.82E-09 | 3.98E-08 | 8.84E-06 |
| Obesity | endometrium | *RHBDF2* | 2.945772485 | 1.63E-05 | 2.66E-05 | 0.078994246 |
| Obesity | endometrium | *TMEM8A* | 6.349900815 | 2.97E-07 | 9.26E-07 | 0.001442548 |
| Obesity | endometrium | *MAU2* | 7.618796185 | 1.32E-08 | 9.23E-08 | 6.39E-05 |
| Obesity | endometrium | *SHC2* | 14.89083749 | 6.68E-11 | 8.31E-09 | 3.24E-07 |
| Obesity | endometrium | *ERMARD* | 5.2071365 | 2.24E-07 | 7.37E-07 | 0.001086638 |
| Obesity | endometrium | *PHF10* | 3.156431512 | 1.43E-06 | 3.18E-06 | 0.006926733 |
| Obesity | endometrium | *PRKCSH* | 11.26860843 | 7.97E-05 | 0.000120728 | 0.386813957 |
| Obesity | endometrium | *CDC16* | 5.421228488 | 1.84E-06 | 3.72E-06 | 0.008934586 |
| Obesity | endometrium | *XPO7* | 9.212425378 | 2.04E-09 | 4.00E-08 | 9.88E-06 |
| Obesity | endometrium | *FAM98C* | 8.755763358 | 0.000154536 | 0.00022571 | 0.749809534 |
| Obesity | endometrium | *ATP8B3* | -2.567018966 | 0.015565665 | 0.018524554 | 1 |
| Obesity | endometrium | *PLVAP* | 3.652617795 | 4.24E-06 | 7.67E-06 | 0.020548955 |
| Obesity | endometrium | *TULP4* | 8.385712581 | 4.76E-11 | 7.00E-09 | 2.31E-07 |
| Obesity | endometrium | *QRSL1* | 2.514544126 | 1.37E-05 | 2.27E-05 | 0.06649581 |
| Obesity | endometrium | *RSPH3* | 3.641704591 | 9.94E-06 | 1.69E-05 | 0.048206399 |
| Obesity | endometrium | *ACTN4* | 4.154181621 | 1.07E-07 | 4.14E-07 | 0.000518761 |
| Obesity | endometrium | *STK33* | 4.153818523 | 3.75E-09 | 4.72E-08 | 1.82E-05 |
| Obesity | endometrium | *NDUFA10* | 4.539956093 | 0.002049124 | 0.002661229 | 1 |
| Obesity | endometrium | *MAP1S* | 4.520967873 | 1.58E-06 | 3.36E-06 | 0.007687526 |
| Obesity | endometrium | *SSBP4* | 6.097204656 | 0.007197373 | 0.008820827 | 1 |
| Obesity | endometrium | *PGPEP1* | 8.622221282 | 1.02E-10 | 1.16E-08 | 4.97E-07 |
| Obesity | endometrium | *KIAA1683* | -1.793953701 | 0.000841017 | 0.001133504 | 1 |
| Obesity | endometrium | *TRPM4* | 6.410880617 | 0.00012013 | 0.000178303 | 0.582872255 |
| Obesity | endometrium | *UBAC1* | 10.91504753 | 2.40E-09 | 4.02E-08 | 1.16E-05 |
| Obesity | endometrium | *ATXN10* | 8.844189979 | 1.64E-09 | 3.98E-08 | 7.96E-06 |
| Obesity | endometrium | *CYP2E1* | -5.335314373 | 7.98E-05 | 0.000120744 | 0.386985821 |
| Obesity | endometrium | *PNPLA7* | 6.430413498 | 2.11E-08 | 1.26E-07 | 0.000102255 |
| Obesity | endometrium | *PAK4* | 5.735316341 | 1.11E-07 | 4.26E-07 | 0.000537631 |
| Obesity | endometrium | *ZNF337* | 9.073232588 | 4.38E-09 | 5.04E-08 | 2.12E-05 |
| Obesity | endometrium | *CEP85* | 3.602252403 | 8.51E-06 | 1.46E-05 | 0.04130092 |
| Obesity | endometrium | *POMT1* | -2.697720409 | 0.01654799 | 0.019635816 | 1 |
| Obesity | endometrium | *CHMP2A* | 8.636961123 | 5.78E-08 | 2.59E-07 | 0.000280648 |
| Obesity | endometrium | *NPAS1* | -5.074627856 | 0.000141088 | 0.000207506 | 0.684560813 |
| Obesity | endometrium | *MAP3K10* | -1.818074263 | 0.00192645 | 0.002506606 | 1 |
| Obesity | endometrium | *LRRC47* | 3.576937707 | 4.42E-07 | 1.27E-06 | 0.002146716 |
| Obesity | endometrium | *THEMIS2* | 4.171052394 | 2.89E-07 | 9.05E-07 | 0.001403173 |
| Obesity | endometrium | *ZNF317* | -2.894413769 | 0.011115343 | 0.013379222 | 1 |

| | | | | | | |
|---|---|---|---|---|---|---|
| Obesity | endometrium | *ANGPTL6* | 6.752187819 | 0.003837761 | 0.004855493 | 1 |
| Obesity | endometrium | *DNMT1* | 6.058688509 | 7.16E-08 | 3.05E-07 | 0.000347289 |
| Obesity | endometrium | *C12orf65* | 6.077393939 | 0.003689752 | 0.004674328 | 1 |
| Obesity | endometrium | *PKDREJ* | -4.371459093 | 0.036989685 | 0.042579822 | 1 |
| Obesity | endometrium | *HSD17B3* | 13.71300745 | 1.59E-08 | 1.04E-07 | 7.70E-05 |
| Obesity | endometrium | *SLC35D2* | 3.293145384 | 6.41E-07 | 1.71E-06 | 0.003110994 |
| Obesity | endometrium | *ULBP2* | 36.96510399 | 7.86E-05 | 0.000119204 | 0.381453527 |
| Obesity | endometrium | *TTLL9* | 6.811918256 | 1.06E-06 | 2.53E-06 | 0.005123317 |
| Obesity | endometrium | *COX4I2* | 6.014785744 | 1.15E-05 | 1.93E-05 | 0.055756654 |
| Obesity | endometrium | *ATP6V1E1* | 5.419556627 | 9.73E-07 | 2.36E-06 | 0.00471859 |
| Obesity | endometrium | *CHMP1A* | 9.552451818 | 1.40E-08 | 9.71E-08 | 6.80E-05 |
| Obesity | endometrium | *IDO1* | 6.823496459 | 1.39E-06 | 3.14E-06 | 0.006749707 |
| Obesity | endometrium | *HAUS8* | 4.321802792 | 1.56E-06 | 3.34E-06 | 0.007589293 |
| Obesity | endometrium | *MRPS25* | 5.169873463 | 1.60E-07 | 5.66E-07 | 0.000775318 |
| Obesity | endometrium | *GALNT15* | 2.777891082 | 1.40E-06 | 3.14E-06 | 0.006789222 |
| Obesity | endometrium | *SLC6A6* | 3.084227666 | 1.41E-06 | 3.16E-06 | 0.006852912 |
| Obesity | endometrium | *KCNC3* | -4.560142964 | 0.00046677 | 0.000644132 | 1 |
| Obesity | endometrium | *NAPSA* | 7.949705732 | 0.000795942 | 0.001074544 | 1 |
| Obesity | endometrium | *PDLIM4* | 7.023195504 | 1.00E-06 | 2.42E-06 | 0.004865981 |
| Obesity | endometrium | *KIF3A* | 15.59183477 | 1.54E-09 | 3.98E-08 | 7.46E-06 |
| Obesity | endometrium | *ACLY* | 2.744890472 | 3.65E-06 | 6.72E-06 | 0.01770096 |
| Obesity | endometrium | *EXOC4* | 5.841983341 | 2.78E-10 | 2.05E-08 | 1.35E-06 |
| Obesity | endometrium | *C1orf159* | 10.17615273 | 8.87E-09 | 7.32E-08 | 4.31E-05 |
| Obesity | endometrium | *PPFIA1* | 5.966823367 | 7.97E-09 | 6.98E-08 | 3.87E-05 |
| Obesity | endometrium | *BARX1* | 14.64275597 | 1.40E-05 | 2.32E-05 | 0.068166016 |
| Obesity | endometrium | *NINJ1* | 3.512708934 | 9.34E-07 | 2.29E-06 | 0.004529911 |
| Obesity | endometrium | *CKMT2* | 6.045049933 | 4.39E-06 | 7.92E-06 | 0.021317396 |
| Obesity | endometrium | *KHDRBS3* | 4.917756015 | 1.57E-07 | 5.57E-07 | 0.000759858 |
| Obesity | endometrium | *CHD1L* | 8.63619612 | 5.12E-06 | 9.15E-06 | 0.024821024 |
| Obesity | endometrium | *PRKAB2* | 4.887764351 | 1.70E-07 | 5.94E-07 | 0.000825601 |
| Obesity | endometrium | *ZNF304* | 4.471786389 | 2.63E-05 | 4.19E-05 | 0.127606193 |
| Obesity | endometrium | *ZNF132* | 3.771418147 | 0.000392432 | 0.000544956 | 1 |
| Obesity | endometrium | *RHPN2* | 2.722024085 | 2.19E-05 | 3.53E-05 | 0.106318102 |
| Obesity | endometrium | *GCH1* | 3.929109792 | 2.78E-06 | 5.30E-06 | 0.013495505 |
| Obesity | endometrium | *MATN3* | 6.375913106 | 0.000552247 | 0.000756585 | 1 |
| Obesity | endometrium | *TRIM21* | 7.044544345 | 7.81E-07 | 2.00E-06 | 0.003789994 |
| Obesity | endometrium | *SPATA6* | 6.473071037 | 0.000516402 | 0.000708993 | 1 |
| Obesity | endometrium | *CCT6B* | 4.8570492 | 8.48E-06 | 1.46E-05 | 0.041136232 |
| Obesity | endometrium | *CNGA4* | 5.242224566 | 0.000250886 | 0.000357399 | 1 |
| Obesity | endometrium | *TRIM22* | 6.328077782 | 0.005213455 | 0.006497736 | 1 |
| Obesity | endometrium | *TIMM10B* | 9.219836029 | 4.45E-07 | 1.28E-06 | 0.002160981 |
| Obesity | endometrium | *IMMT* | 6.482502793 | 8.05E-09 | 7.03E-08 | 3.91E-05 |
| Obesity | endometrium | *CARD6* | 6.616246099 | 1.13E-08 | 8.43E-08 | 5.51E-05 |
| Obesity | endometrium | *MYBBP1A* | 5.935845676 | 6.70E-09 | 6.26E-08 | 3.25E-05 |
| Obesity | endometrium | *RPA1* | 7.408611446 | 1.04E-08 | 8.01E-08 | 5.05E-05 |
| Obesity | endometrium | *SEC61G* | 10.78162457 | 2.70E-07 | 8.52E-07 | 0.00130798 |
| Obesity | endometrium | *FIGNL1* | -2.063356658 | 0.000694116 | 0.000941266 | 1 |
| Obesity | endometrium | *EIF5A* | 3.918427175 | 0.000142819 | 0.000209669 | 0.692956914 |
| Obesity | endometrium | *RGS22* | -2.23117017 | 0.027277768 | 0.031739024 | 1 |
| Obesity | endometrium | *REEP2* | -0.945813292 | 0.000167603 | 0.000243622 | 0.81321126 |

| | | | | | | |
|---|---|---|---|---|---|---|
| Obesity | endometrium | *PRMT7* | 3.935462871 | 4.70E-06 | 8.43E-06 | 0.022794454 |
| Obesity | endometrium | *IGHMBP2* | 9.773898948 | 1.08E-12 | 3.34E-10 | 5.26E-09 |
| Obesity | endometrium | *MUTYH* | 4.276736542 | 1.08E-07 | 4.17E-07 | 0.000526032 |
| Obesity | endometrium | *CTNNBL1* | 6.937339626 | 2.90E-08 | 1.57E-07 | 0.000140833 |
| Obesity | endometrium | *OSER1* | 5.934587514 | 6.49E-08 | 2.83E-07 | 0.000314961 |
| Obesity | endometrium | *SERINC3* | 3.359044122 | 8.15E-07 | 2.06E-06 | 0.003952911 |
| Obesity | endometrium | *DMGDH* | 2.294763983 | 1.37E-05 | 2.27E-05 | 0.066478433 |
| Obesity | endometrium | *AP3B1* | 9.567785708 | 2.66E-09 | 4.24E-08 | 1.29E-05 |
| Obesity | endometrium | *ZBED3* | 3.536823529 | 5.38E-06 | 9.58E-06 | 0.026104134 |
| Obesity | endometrium | *FBXO44* | 4.115222678 | 9.46E-06 | 1.61E-05 | 0.04588637 |
| Obesity | endometrium | *RSG1* | 14.82900301 | 0.002602917 | 0.003344638 | 1 |
| Obesity | endometrium | *CASP9* | 2.527030101 | 0.000353249 | 0.000493511 | 1 |
| Obesity | endometrium | *ATP8A2* | -1.753983074 | 0.000897569 | 0.001206038 | 1 |
| Obesity | endometrium | *ALOX5AP* | 3.689957722 | 9.88E-07 | 2.39E-06 | 0.004795125 |
| Obesity | endometrium | *WASF3* | 10.62840307 | 3.10E-06 | 5.82E-06 | 0.015047464 |
| Obesity | endometrium | *GPR12* | 16.78082585 | 0.003400754 | 0.004324019 | 1 |
| Obesity | endometrium | *SCO1* | 7.663322671 | 1.61E-07 | 5.70E-07 | 0.000781872 |
| Obesity | endometrium | *MPRIP* | 6.839240748 | 6.40E-09 | 6.16E-08 | 3.11E-05 |
| Obesity | endometrium | *DSTYK* | 10.62851903 | 1.59E-09 | 3.98E-08 | 7.71E-06 |
| Obesity | endometrium | *CCNA1* | 22.35481921 | 0.006785064 | 0.008345027 | 1 |
| Obesity | endometrium | *COG6* | 9.662775847 | 4.21E-09 | 4.91E-08 | 2.04E-05 |
| Obesity | endometrium | *EPSTI1* | 3.695210423 | 1.98E-07 | 6.69E-07 | 0.000959306 |
| Obesity | endometrium | *BTBD2* | 12.31512426 | 6.05E-10 | 2.64E-08 | 2.93E-06 |
| Obesity | endometrium | *CSNK1G2* | -1.951136379 | 0.006100102 | 0.007546582 | 1 |
| Obesity | endometrium | *CNDP2* | 4.996754624 | 3.80E-07 | 1.13E-06 | 0.00184498 |
| Obesity | endometrium | *PDZD2* | 8.712151544 | 3.76E-09 | 4.72E-08 | 1.82E-05 |
| Obesity | endometrium | *GSTT2B* | -1.798007192 | 0.020400064 | 0.02400124 | 1 |
| Obesity | endometrium | *FAM83F* | 20.33742617 | 1.12E-05 | 1.89E-05 | 0.054453882 |
| Obesity | endometrium | *GIMAP4* | 3.322534164 | 4.36E-07 | 1.26E-06 | 0.002116656 |
| Obesity | endometrium | *ADCK2* | 3.738648838 | 1.22E-05 | 2.03E-05 | 0.059026019 |
| Obesity | endometrium | *IPO8* | 12.47867213 | 3.13E-08 | 1.66E-07 | 0.000152028 |
| Obesity | endometrium | *LARS* | 2.983577622 | 0.026919008 | 0.03135919 | 1 |
| Obesity | endometrium | *SPINK5* | 7.197258536 | 1.78E-06 | 3.62E-06 | 0.008626471 |
| Obesity | endometrium | *IMPA1* | 2.74991962 | 1.48E-05 | 2.43E-05 | 0.071739087 |
| Obesity | endometrium | *LRRCC1* | 12.94025048 | 1.69E-08 | 1.09E-07 | 8.19E-05 |
| Obesity | endometrium | *RRAS2* | 8.768426743 | 1.19E-09 | 3.96E-08 | 5.75E-06 |
| Obesity | endometrium | *CATSPERB* | 2.582774605 | 1.40E-06 | 3.14E-06 | 0.006777263 |
| Obesity | endometrium | *COX16* | 23.41830888 | 0.009497972 | 0.011498044 | 1 |
| Obesity | endometrium | *TTC9* | 4.932700177 | 1.44E-06 | 3.20E-06 | 0.007007827 |
| Obesity | endometrium | *ELP3* | 7.053390218 | 1.87E-08 | 1.17E-07 | 9.08E-05 |
| Obesity | endometrium | *PEBP4* | 3.777954415 | 1.68E-06 | 3.48E-06 | 0.008151461 |
| Obesity | endometrium | *CTIF* | 3.415504673 | 4.37E-07 | 1.26E-06 | 0.002119334 |
| Obesity | endometrium | *MRO* | 9.824752638 | 4.20E-07 | 1.22E-06 | 0.00203625 |
| Obesity | endometrium | *THUMPD3* | -2.092607199 | 0.006039239 | 0.007477006 | 1 |
| Obesity | endometrium | *CNTN6* | 5.385831221 | 2.17E-06 | 4.27E-06 | 0.010512167 |
| Obesity | endometrium | *CHL1* | 4.999188975 | 1.29E-07 | 4.78E-07 | 0.000625319 |
| Obesity | endometrium | *GSTM3* | -1.720412997 | 0.001961527 | 0.002551563 | 1 |
| Obesity | endometrium | *VAV3* | 2.198181876 | 4.65E-06 | 8.35E-06 | 0.022564492 |
| Obesity | endometrium | *PSRC1* | -4.117310656 | 0.004645117 | 0.005823801 | 1 |
| Obesity | endometrium | *TRIM45* | -3.917859731 | 0.005944247 | 0.007366919 | 1 |

| | | | | | | |
|---|---|---|---|---|---|---|
| Obesity | endometrium | *CEPT1* | 6.706751593 | 6.34E-09 | 6.12E-08 | 3.08E-05 |
| Obesity | endometrium | *NAPG* | 9.706746394 | 1.13E-09 | 3.86E-08 | 5.48E-06 |
| Obesity | endometrium | *PPHLN1* | 5.421509841 | 1.64E-08 | 1.06E-07 | 7.95E-05 |
| Obesity | endometrium | *FKBP11* | 3.829521139 | 1.29E-06 | 2.96E-06 | 0.00624984 |
| Obesity | endometrium | *YWHAQ* | 4.735711151 | 5.23E-08 | 2.43E-07 | 0.000253991 |
| Obesity | endometrium | *KIDINS220* | 9.275920676 | 1.76E-09 | 3.98E-08 | 8.54E-06 |
| Obesity | endometrium | *LPIN1* | 6.61661143 | 1.89E-08 | 1.17E-07 | 9.18E-05 |
| Obesity | endometrium | *NARS* | 10.26284768 | 3.95E-10 | 2.34E-08 | 1.92E-06 |
| Obesity | endometrium | *KIAA1468* | 8.813491514 | 1.79E-09 | 3.98E-08 | 8.66E-06 |
| Obesity | endometrium | *IL15RA* | 6.657794562 | 1.60E-08 | 1.04E-07 | 7.75E-05 |
| Obesity | endometrium | *TMEM241* | 5.48371298 | 1.38E-06 | 3.12E-06 | 0.006699072 |
| Obesity | endometrium | *SOX5* | 3.915986565 | 6.99E-08 | 3.00E-07 | 0.000339175 |
| Obesity | endometrium | *KLRD1* | 7.719325521 | 5.86E-09 | 5.92E-08 | 2.84E-05 |
| Obesity | endometrium | *ACP2* | 18.7118506 | 1.36E-11 | 2.54E-09 | 6.61E-08 |
| Obesity | endometrium | *HOOK1* | 1.85911577 | 0.018842221 | 0.022238496 | 1 |
| Obesity | endometrium | *ZCCHC11* | 4.478727076 | 1.57E-08 | 1.04E-07 | 7.60E-05 |
| Obesity | endometrium | *PRPF38A* | 13.32430914 | 4.92E-10 | 2.46E-08 | 2.39E-06 |
| Obesity | endometrium | *ELP2* | 6.282307578 | 1.13E-08 | 8.42E-08 | 5.46E-05 |
| Obesity | endometrium | *TPGS2* | 8.35528864 | 1.04E-08 | 8.01E-08 | 5.06E-05 |
| Obesity | endometrium | *TIMM10* | -2.065727785 | 0.000955162 | 0.001279174 | 1 |
| Obesity | endometrium | *DHX34* | 11.53044683 | 1.72E-09 | 3.98E-08 | 8.33E-06 |
| Obesity | endometrium | *FADS2* | -2.135494402 | 0.005313704 | 0.006620979 | 1 |
| Obesity | endometrium | *CLOCK* | 6.211084743 | 8.45E-09 | 7.13E-08 | 4.10E-05 |
| Obesity | endometrium | *GGACT* | 7.901930273 | 4.64E-06 | 8.33E-06 | 0.022489893 |
| Obesity | endometrium | *CLDN10* | 7.08610245 | 1.23E-05 | 2.05E-05 | 0.059619224 |
| Obesity | endometrium | *ARGLU1* | 4.839184448 | 8.94E-09 | 7.32E-08 | 4.34E-05 |
| Obesity | endometrium | *ERCC5* | -3.727243013 | 0.003806429 | 0.004817108 | 1 |
| Obesity | endometrium | *CARS2* | 4.962432053 | 0.009217338 | 0.011169461 | 1 |
| Obesity | endometrium | *KLB* | 3.693425169 | 2.59E-08 | 1.45E-07 | 0.000125471 |
| Obesity | endometrium | *WDR36* | 9.829262973 | 2.47E-09 | 4.04E-08 | 1.20E-05 |
| Obesity | endometrium | *OSTF1* | 4.463037511 | 1.09E-07 | 4.21E-07 | 0.000530781 |
| Obesity | endometrium | *UBQLN1* | 7.113496419 | 7.18E-09 | 6.50E-08 | 3.48E-05 |
| Obesity | endometrium | *GOLM1* | 3.111906165 | 4.44E-05 | 6.90E-05 | 0.215355444 |
| Obesity | endometrium | *ADAM19* | 2.584801249 | 3.80E-05 | 5.95E-05 | 0.184165995 |
| Obesity | endometrium | *USP30* | 4.592746561 | 0.00301824 | 0.003856861 | 1 |
| Obesity | endometrium | *MSI1* | 8.111936179 | 8.13E-06 | 1.40E-05 | 0.039459259 |
| Obesity | endometrium | *P2RX4* | 2.710371903 | 2.21E-05 | 3.56E-05 | 0.107150137 |
| Obesity | endometrium | *DTX1* | 2.941675913 | 1.53E-06 | 3.31E-06 | 0.007430395 |
| Obesity | endometrium | *DMTF1* | 6.035213695 | 7.84E-09 | 6.88E-08 | 3.81E-05 |
| Obesity | endometrium | *TMEM60* | 4.06039129 | 7.80E-06 | 1.35E-05 | 0.037850314 |
| Obesity | endometrium | *POM121C* | 3.009098706 | 0.000562603 | 0.000769875 | 1 |
| Obesity | endometrium | *SRPK2* | 7.669504826 | 1.45E-09 | 3.98E-08 | 7.05E-06 |
| Obesity | endometrium | *TES* | 3.552265133 | 4.31E-06 | 7.79E-06 | 0.020910823 |
| Obesity | endometrium | *NT5E* | 3.104392518 | 3.48E-06 | 6.44E-06 | 0.016875608 |
| Obesity | endometrium | *AKIRIN2* | 5.51531061 | 4.96E-08 | 2.33E-07 | 0.000240779 |
| Obesity | endometrium | *MAP3K7* | 12.06888386 | 3.03E-09 | 4.33E-08 | 1.47E-05 |
| Obesity | endometrium | *PRR5L* | 9.229032181 | 4.62E-07 | 1.32E-06 | 0.002240848 |
| Obesity | endometrium | *PHF21A* | 6.823384869 | 2.54E-09 | 4.15E-08 | 1.23E-05 |
| Obesity | endometrium | *CAPRIN1* | 10.70381062 | 6.19E-08 | 2.72E-07 | 0.000300524 |
| Obesity | endometrium | *ATP5G2* | 11.48782484 | 9.47E-10 | 3.50E-08 | 4.59E-06 |

| | | | | | | |
|---|---|---|---|---|---|---|
| Obesity | endometrium | *B4GALNT1* | 12.14608574 | 0.001446876 | 0.001907577 | 1 |
| Obesity | endometrium | *FAIM2* | 7.349579863 | 0.016473591 | 0.019552315 | 1 |
| Obesity | endometrium | *KRT7* | 4.774611176 | 0.000171334 | 0.000248524 | 0.831313363 |
| Obesity | endometrium | *SLC26A10* | 13.69792278 | 3.49E-05 | 5.49E-05 | 0.169260821 |
| Obesity | endometrium | *MAP7* | 5.835393155 | 2.46E-05 | 3.94E-05 | 0.119492071 |
| Obesity | endometrium | *NHSL1* | 2.409759566 | 7.14E-07 | 1.87E-06 | 0.00346271 |
| Obesity | endometrium | *AHI1* | 4.458865956 | 0.000106776 | 0.000159605 | 0.518077899 |
| Obesity | endometrium | *SMPD2* | 2.112024952 | 0.017529018 | 0.020759286 | 1 |
| Obesity | endometrium | *MICAL1* | -1.288178614 | 0.009484356 | 0.011484426 | 1 |
| Obesity | endometrium | *CCT7* | 5.087946536 | 3.46E-07 | 1.04E-06 | 0.001676556 |
| Obesity | endometrium | *SMYD5* | 11.15385406 | 9.52E-06 | 1.62E-05 | 0.046176078 |
| Obesity | endometrium | *USP15* | 4.916179611 | 1.77E-08 | 1.12E-07 | 8.60E-05 |
| Obesity | endometrium | *MDM2* | 9.694769087 | 3.68E-09 | 4.71E-08 | 1.79E-05 |
| Obesity | endometrium | *KLHL36* | 2.8882972 | 9.36E-07 | 2.29E-06 | 0.004539193 |
| Obesity | endometrium | *MPHOSPH6* | 3.642903327 | 8.34E-07 | 2.10E-06 | 0.004046678 |
| Obesity | endometrium | *CCDC102A* | 9.973486329 | 6.16E-06 | 1.09E-05 | 0.029902399 |
| Obesity | endometrium | *AGT* | 2.783094879 | 0.024352877 | 0.028472328 | 1 |
| Obesity | endometrium | *ZNF670* | 10.48054124 | 8.12E-07 | 2.06E-06 | 0.003941449 |
| Obesity | endometrium | *CAPN9* | -1.715496003 | 0.000160049 | 0.000233131 | 0.776559639 |
| Obesity | endometrium | *NTPCR* | 15.40642864 | 7.50E-09 | 6.64E-08 | 3.64E-05 |
| Obesity | endometrium | *PIGC* | 9.711669779 | 1.59E-09 | 3.98E-08 | 7.71E-06 |
| Obesity | endometrium | *SP110* | 7.263172962 | 2.77E-09 | 4.28E-08 | 1.34E-05 |
| Obesity | endometrium | *MRPL44* | 4.343732962 | 1.44E-06 | 3.19E-06 | 0.006965215 |
| Obesity | endometrium | *USP37* | 7.608823632 | 2.46E-10 | 1.94E-08 | 1.19E-06 |
| Obesity | endometrium | *SLC19A3* | 3.850976751 | 1.46E-07 | 5.25E-07 | 0.000706342 |
| Obesity | endometrium | *TMBIM1* | 9.951865948 | 2.78E-09 | 4.28E-08 | 1.35E-05 |
| Obesity | endometrium | *TSGA10* | 2.865833642 | 5.99E-05 | 9.17E-05 | 0.290750005 |
| Obesity | endometrium | *GCC2* | 6.613392392 | 1.03E-08 | 7.97E-08 | 4.99E-05 |
| Obesity | endometrium | *MRPS9* | 7.574956663 | 2.44E-10 | 1.94E-08 | 1.18E-06 |
| Obesity | endometrium | *ANKRD36* | 8.144624647 | 2.92E-09 | 4.29E-08 | 1.42E-05 |
| Obesity | endometrium | *EPC2* | 8.340935711 | 1.78E-09 | 3.98E-08 | 8.65E-06 |
| Obesity | endometrium | *STAB2* | 7.34546385 | 0.008188676 | 0.009977764 | 1 |
| Obesity | endometrium | *APPL2* | 8.008816017 | 1.90E-08 | 1.18E-07 | 9.22E-05 |
| Obesity | endometrium | *DRAM1* | 4.681624228 | 1.31E-07 | 4.84E-07 | 0.000635932 |
| Obesity | endometrium | *SLC41A2* | 3.527939376 | 1.86E-07 | 6.37E-07 | 0.000902001 |
| Obesity | endometrium | *NEK3* | 7.063055737 | 5.98E-08 | 2.65E-07 | 0.000290288 |
| Obesity | endometrium | *RNASEH2B* | 4.206917152 | 3.11E-07 | 9.59E-07 | 0.001509146 |
| Obesity | endometrium | *CKAP2* | -1.912724538 | 0.010554216 | 0.012716428 | 1 |
| Obesity | endometrium | *TBC1D4* | 5.114214762 | 3.16E-09 | 4.44E-08 | 1.53E-05 |
| Obesity | endometrium | *SUCLA2* | 9.236802137 | 2.29E-09 | 4.01E-08 | 1.11E-05 |
| Obesity | endometrium | *RCBTB1* | 2.865189257 | 1.06E-05 | 1.80E-05 | 0.051624448 |
| Obesity | endometrium | *RCBTB2* | 3.466657627 | 4.83E-07 | 1.36E-06 | 0.002344139 |
| Obesity | endometrium | *SETDB2* | 5.713392389 | 6.65E-08 | 2.88E-07 | 0.000322525 |
| Obesity | endometrium | *SCRN1* | 4.915074813 | 1.94E-07 | 6.59E-07 | 0.000941061 |
| Obesity | endometrium | *CHST12* | 6.003645739 | 7.89E-08 | 3.29E-07 | 0.000383038 |
| Obesity | endometrium | *KDELR2* | 5.068610194 | 2.07E-07 | 6.89E-07 | 0.001003492 |
| Obesity | endometrium | *NUPL2* | 7.592106338 | 7.06E-09 | 6.45E-08 | 3.42E-05 |
| Obesity | endometrium | *ZDHHC4* | -2.45640421 | 0.010047551 | 0.01213308 | 1 |
| Obesity | endometrium | *AOAH* | 2.173750701 | 2.40E-06 | 4.66E-06 | 0.011647112 |
| Obesity | endometrium | *TBRG4* | 4.156160501 | 2.93E-06 | 5.53E-06 | 0.014211745 |

| | | | | | | |
|---|---|---|---|---|---|---|
| Obesity | endometrium | *MTHFS* | 4.460956168 | 1.03E-05 | 1.74E-05 | 0.05003385 |
| Obesity | endometrium | *NMT1* | 8.068943419 | 1.54E-09 | 3.98E-08 | 7.49E-06 |
| Obesity | endometrium | *MYCBPAP* | 2.744342036 | 0.00200717 | 0.002608139 | 1 |
| Obesity | endometrium | *RTP4* | 6.822435101 | 5.30E-05 | 8.18E-05 | 0.257131787 |
| Obesity | endometrium | *NDUFB5* | 4.684931614 | 1.72E-07 | 6.00E-07 | 0.000836484 |
| Obesity | endometrium | *SCN2A* | 6.226896795 | 2.59E-09 | 4.19E-08 | 1.26E-05 |
| Obesity | endometrium | *RPS6KC1* | 9.141470282 | 3.65E-09 | 4.70E-08 | 1.77E-05 |
| Obesity | endometrium | *SMPD4* | 5.253700569 | 1.82E-07 | 6.27E-07 | 0.000884903 |
| Obesity | endometrium | *IMP4* | 12.0429596 | 4.77E-08 | 2.26E-07 | 0.000231237 |
| Obesity | endometrium | *ODF2* | 9.92031424 | 2.39E-08 | 1.38E-07 | 0.000116189 |
| Obesity | endometrium | *TOR1B* | 12.00519798 | 1.56E-08 | 1.03E-07 | 7.57E-05 |
| Obesity | endometrium | *SLC2A8* | 5.829555041 | 7.45E-07 | 1.93E-06 | 0.003614558 |
| Obesity | endometrium | *ZNF189* | 14.70266281 | 9.87E-10 | 3.56E-08 | 4.79E-06 |
| Obesity | endometrium | *STX17* | 10.81863764 | 3.14E-10 | 2.18E-08 | 1.52E-06 |
| Obesity | endometrium | *ANP32B* | 12.62790152 | 4.85E-08 | 2.29E-07 | 0.000235084 |
| Obesity | endometrium | *DSCC1* | 3.150764723 | 9.53E-08 | 3.78E-07 | 0.000462578 |
| Obesity | endometrium | *DERL1* | 11.13143594 | 9.02E-06 | 1.54E-05 | 0.043747957 |
| Obesity | endometrium | *NOV* | 6.048439564 | 2.35E-06 | 4.57E-06 | 0.011401586 |
| Obesity | endometrium | *IL11RA* | 3.205646627 | 0.000119752 | 0.000177795 | 0.581034547 |
| Obesity | endometrium | *APTX* | 7.774038022 | 1.03E-08 | 7.97E-08 | 5.00E-05 |
| Obesity | endometrium | *TMEM8B* | 5.217319293 | 2.62E-07 | 8.36E-07 | 0.001273248 |
| Obesity | endometrium | *GRHPR* | -1.643292081 | 0.019018451 | 0.022430122 | 1 |
| Obesity | endometrium | *DENND4C* | 3.795643206 | 5.35E-08 | 2.47E-07 | 0.000259463 |
| Obesity | endometrium | *GMPR* | 8.728490332 | 2.19E-07 | 7.25E-07 | 0.001064256 |
| Obesity | endometrium | *FRS3* | 4.137304239 | 8.51E-07 | 2.14E-06 | 0.004127925 |
| Obesity | endometrium | *IRF4* | 3.370390475 | 2.86E-05 | 4.54E-05 | 0.138711725 |
| Obesity | endometrium | *LRRC1* | 6.775072249 | 8.52E-08 | 3.48E-07 | 0.000413151 |
| Obesity | endometrium | *RIPK1* | 5.731923491 | 3.61E-08 | 1.85E-07 | 0.000175089 |
| Obesity | endometrium | *TCF19* | 11.05432099 | 1.35E-06 | 3.06E-06 | 0.006527557 |
| Obesity | endometrium | *FLOT1* | 4.35470928 | 7.57E-08 | 3.18E-07 | 0.000367524 |
| Obesity | endometrium | *TPMT* | 3.422700902 | 2.98E-06 | 5.61E-06 | 0.014453299 |
| Obesity | endometrium | *NRM* | 11.90596449 | 1.40E-06 | 3.15E-06 | 0.006816302 |
| Obesity | endometrium | *TLR2* | 5.116214692 | 1.03E-06 | 2.48E-06 | 0.005014934 |
| Obesity | endometrium | *MYO7A* | 3.81700226 | 4.62E-05 | 7.17E-05 | 0.224344483 |
| Obesity | endometrium | *NUMA1* | 5.188303259 | 2.29E-08 | 1.33E-07 | 0.000110959 |
| Obesity | endometrium | *SYTL2* | 2.444505467 | 1.92E-05 | 3.12E-05 | 0.093321286 |
| Obesity | endometrium | *RAB30* | 3.358067129 | 8.93E-07 | 2.22E-06 | 0.004331061 |
| Obesity | endometrium | *PRCP* | 9.777703253 | 3.01E-08 | 1.61E-07 | 0.000146079 |
| Obesity | endometrium | *NARS2* | 10.42628225 | 3.68E-06 | 6.78E-06 | 0.017860164 |
| Obesity | endometrium | *GGH* | 3.052938674 | 0.001424986 | 0.001880346 | 1 |
| Obesity | endometrium | *SULF1* | 3.243256834 | 5.16E-07 | 1.44E-06 | 0.002502688 |
| Obesity | endometrium | *NEK1* | 5.335230683 | 1.84E-08 | 1.15E-07 | 8.92E-05 |
| Obesity | endometrium | *TRPC6* | 11.24038735 | 3.48E-06 | 6.44E-06 | 0.016876437 |
| Obesity | endometrium | *MMP27* | 18.11194262 | 3.44E-05 | 5.41E-05 | 0.166839962 |
| Obesity | endometrium | *POU2F3* | -2.865924733 | 0.007416918 | 0.009066991 | 1 |
| Obesity | endometrium | *PPP2R1B* | 2.891869087 | 3.82E-07 | 1.13E-06 | 0.001853235 |
| Obesity | endometrium | *FDX1* | 6.308204476 | 2.18E-08 | 1.29E-07 | 0.000105942 |
| Obesity | endometrium | *FXYD6* | 4.047937238 | 0.017307058 | 0.020506434 | 1 |
| Obesity | endometrium | *FXYD2* | 7.393257479 | 3.80E-06 | 6.96E-06 | 0.018419111 |
| Obesity | endometrium | *MAP2K5* | 5.311543965 | 3.90E-09 | 4.80E-08 | 1.89E-05 |

| | | | | | | |
|---|---|---|---|---|---|---|
| Obesity | endometrium | *NUSAP1* | 19.16856162 | 3.85E-11 | 5.98E-09 | 1.87E-07 |
| Obesity | endometrium | *PARP6* | 10.6611137 | 5.44E-09 | 5.76E-08 | 2.64E-05 |
| Obesity | endometrium | *ADAM10* | 3.425669615 | 5.89E-08 | 2.62E-07 | 0.000285578 |
| Obesity | endometrium | *ZNF280D* | 6.839959713 | 1.90E-09 | 4.00E-08 | 9.21E-06 |
| Obesity | endometrium | *SPTBN5* | 1.315266166 | 0.021146449 | 0.024861297 | 1 |
| Obesity | endometrium | *SLC44A5* | 13.97947124 | 2.63E-10 | 1.96E-08 | 1.28E-06 |
| Obesity | endometrium | *DBT* | 12.72957827 | 6.60E-09 | 6.22E-08 | 3.20E-05 |
| Obesity | endometrium | *RTCA* | 6.593961578 | 1.98E-08 | 1.21E-07 | 9.58E-05 |
| Obesity | endometrium | *KHK* | 5.534898896 | 7.83E-05 | 0.00011876 | 0.379675212 |
| Obesity | endometrium | *ADCY3* | 3.610054347 | 1.52E-07 | 5.43E-07 | 0.000735263 |
| Obesity | endometrium | *DYNC2LI1* | 7.357927573 | 8.82E-09 | 7.32E-08 | 4.28E-05 |
| Obesity | endometrium | *LHCGR* | 8.39765855 | 0.000167976 | 0.000244018 | 0.815020957 |
| Obesity | endometrium | *THUMPD2* | 9.408460114 | 2.32E-09 | 4.01E-08 | 1.13E-05 |
| Obesity | endometrium | *ACTR2* | 4.649603604 | 4.13E-08 | 2.05E-07 | 0.000200479 |
| Obesity | endometrium | *ATRAID* | 8.658427105 | 6.86E-10 | 2.89E-08 | 3.33E-06 |
| Obesity | endometrium | *CENPO* | -1.049185481 | 0.004603351 | 0.005774421 | 1 |
| Obesity | endometrium | *LRPPRC* | 10.82966123 | 6.54E-09 | 6.17E-08 | 3.17E-05 |
| Obesity | endometrium | *DTNB* | 10.2992075 | 3.40E-09 | 4.54E-08 | 1.65E-05 |
| Obesity | endometrium | *CYP2C8* | 22.66112462 | 1.00E-05 | 1.69E-05 | 0.048527711 |
| Obesity | endometrium | *MYOF* | 4.405125035 | 4.91E-06 | 8.79E-06 | 0.023818383 |
| Obesity | endometrium | *BTBD16* | 5.948108147 | 3.37E-06 | 6.27E-06 | 0.016342046 |
| Obesity | endometrium | *TACC2* | 6.396429367 | 8.26E-09 | 7.12E-08 | 4.01E-05 |
| Obesity | endometrium | *CALHM2* | 13.90922564 | 4.16E-07 | 1.21E-06 | 0.002017976 |
| Obesity | endometrium | *ARL3* | 3.945192846 | 1.63E-07 | 5.77E-07 | 0.000792443 |
| Obesity | endometrium | *PLCE1* | 2.965014006 | 8.66E-07 | 2.17E-06 | 0.004200987 |
| Obesity | endometrium | *ASCC1* | 4.279654515 | 0.027133386 | 0.031593758 | 1 |
| Obesity | endometrium | *RPS24* | 8.019805801 | 5.69E-10 | 2.61E-08 | 2.76E-06 |
| Obesity | endometrium | *ATIC* | 3.302421119 | 4.00E-06 | 7.28E-06 | 0.01941848 |
| Obesity | endometrium | *SMARCAL1* | 3.604883778 | 4.21E-07 | 1.22E-06 | 0.002044522 |
| Obesity | endometrium | *BARD1* | 4.333584135 | 5.06E-07 | 1.42E-06 | 0.002456705 |
| Obesity | endometrium | *STAT4* | 2.867045688 | 9.24E-07 | 2.27E-06 | 0.004480894 |
| Obesity | endometrium | *SSB* | 5.87887557 | 9.70E-09 | 7.68E-08 | 4.71E-05 |
| Obesity | endometrium | *FASTKD1* | 9.597389464 | 2.95E-09 | 4.29E-08 | 1.43E-05 |
| Obesity | endometrium | *ITGAV* | 11.06617985 | 1.13E-08 | 8.43E-08 | 5.49E-05 |
| Obesity | endometrium | *SLC35A5* | 4.548290227 | 4.60E-07 | 1.31E-06 | 0.002232748 |
| Obesity | endometrium | *SENP7* | 4.571714014 | 7.28E-08 | 3.09E-07 | 0.000353064 |
| Obesity | endometrium | *COX17* | 4.991507916 | 3.44E-07 | 1.04E-06 | 0.001670189 |
| Obesity | endometrium | *SPPL2A* | 7.386505823 | 1.25E-08 | 8.93E-08 | 6.06E-05 |
| Obesity | endometrium | *PPCDC* | 6.436731294 | 2.41E-05 | 3.86E-05 | 0.11689732 |
| Obesity | endometrium | *SEMA7A* | 7.023047056 | 0.001249598 | 0.001657024 | 1 |
| Obesity | endometrium | *PCDH10* | 10.86411014 | 0.006203849 | 0.007665158 | 1 |
| Obesity | endometrium | *AP1AR* | 13.60456393 | 4.98E-06 | 8.90E-06 | 0.024141327 |
| Obesity | endometrium | *KIAA1109* | 6.889577543 | 2.69E-09 | 4.24E-08 | 1.31E-05 |
| Obesity | endometrium | *BMPR1B* | 4.406565972 | 0.035641264 | 0.041095868 | 1 |
| Obesity | endometrium | *MMRN1* | 1.851257168 | 0.00014364 | 0.000210811 | 0.696941329 |
| Obesity | endometrium | *PRDM5* | 12.62410163 | 5.72E-09 | 5.87E-08 | 2.78E-05 |
| Obesity | endometrium | *TRPC3* | -6.709366827 | 0.02699202 | 0.031436697 | 1 |
| Obesity | endometrium | *NAAA* | 4.140048493 | 7.01E-07 | 1.84E-06 | 0.003402554 |
| Obesity | endometrium | *NUP54* | 6.83697735 | 2.53E-07 | 8.12E-07 | 0.001229946 |
| Obesity | endometrium | *CNOT6L* | 3.773421833 | 5.25E-07 | 1.46E-06 | 0.002545703 |

| | | | | | | |
|---|---|---|---|---|---|---|
| Obesity | endometrium | *ANXA3* | 2.647669034 | 5.16E-07 | 1.44E-06 | 0.002501765 |
| Obesity | endometrium | *PPA2* | 10.09774887 | 2.32E-09 | 4.01E-08 | 1.13E-05 |
| Obesity | endometrium | *ENPEP* | -1.454691032 | 0.001032616 | 0.001379095 | 1 |
| Obesity | endometrium | *PAPSS1* | 7.318847443 | 2.06E-08 | 1.24E-07 | 9.99E-05 |
| Obesity | endometrium | *FBN2* | 1.562456704 | 0.018071855 | 0.02137607 | 1 |
| Obesity | endometrium | *PARVG* | 3.711784805 | 6.40E-06 | 1.13E-05 | 0.031046601 |
| Obesity | endometrium | *PDE6H* | 15.40151783 | 0.007623053 | 0.009311947 | 1 |
| Obesity | endometrium | *ERP27* | 7.809032783 | 3.77E-06 | 6.93E-06 | 0.018312955 |
| Obesity | endometrium | *KIF21A* | 10.94457187 | 7.53E-09 | 6.66E-08 | 3.65E-05 |
| Obesity | endometrium | *ALG10* | -3.090043311 | 0.037540669 | 0.043203825 | 1 |
| Obesity | endometrium | *C1RL* | 5.361238259 | 1.09E-06 | 2.60E-06 | 0.005296712 |
| Obesity | endometrium | *CLSTN3* | -2.31899471 | 0.022813413 | 0.026756268 | 1 |
| Obesity | endometrium | *KLRG1* | 3.017180622 | 9.05E-09 | 7.33E-08 | 4.39E-05 |
| Obesity | endometrium | *VAMP1* | -2.990509004 | 0.009588646 | 0.011602023 | 1 |
| Obesity | endometrium | *PEX5* | -1.98529553 | 0.003021997 | 0.003860645 | 1 |
| Obesity | endometrium | *MARCH9* | 11.66717114 | 0.003422757 | 0.004349716 | 1 |
| Obesity | endometrium | *LGR5* | 6.502831232 | 0.000784049 | 0.001059078 | 1 |
| Obesity | endometrium | *POC1B* | 10.70243096 | 1.99E-09 | 4.00E-08 | 9.65E-06 |
| Obesity | endometrium | *AMDHD1* | -1.346013117 | 5.44E-05 | 8.38E-05 | 0.263831359 |
| Obesity | endometrium | *RITA1* | 18.86216032 | 0.000400072 | 0.000555089 | 1 |
| Obesity | endometrium | *SDSL* | 5.80265529 | 2.40E-05 | 3.85E-05 | 0.11654413 |
| Obesity | endometrium | *GLTP* | 3.74959559 | 1.53E-06 | 3.31E-06 | 0.007424321 |
| Obesity | endometrium | *FOXN4* | 5.802852065 | 3.84E-06 | 7.04E-06 | 0.018640228 |
| Obesity | endometrium | *MTMR6* | 3.255034458 | 1.50E-06 | 3.27E-06 | 0.007255512 |
| Obesity | endometrium | *CCDC65* | 11.80106901 | 1.15E-08 | 8.48E-08 | 5.57E-05 |
| Obesity | endometrium | *ACVRL1* | 11.49889956 | 6.66E-09 | 6.25E-08 | 3.23E-05 |
| Obesity | endometrium | *BRCA2* | 9.80631018 | 1.55E-07 | 5.54E-07 | 0.000754079 |
| Obesity | endometrium | *KANSL2* | 7.795014419 | 1.67E-07 | 5.85E-07 | 0.000808459 |
| Obesity | endometrium | *CERS5* | 5.362575679 | 2.75E-08 | 1.51E-07 | 0.000133446 |
| Obesity | endometrium | *LMBR1L* | 12.28133231 | 2.18E-07 | 7.22E-07 | 0.001057809 |
| Obesity | endometrium | *ESYT1* | 8.547150886 | 1.84E-09 | 3.98E-08 | 8.91E-06 |
| Obesity | endometrium | *ZNF740* | 12.39508894 | 1.20E-08 | 8.67E-08 | 5.81E-05 |
| Obesity | endometrium | *WDFY2* | 3.424541521 | 3.76E-07 | 1.12E-06 | 0.00182438 |
| Obesity | endometrium | *HNRNPA1L2* | 23.89954533 | 0.000150585 | 0.000220404 | 0.730640316 |
| Obesity | endometrium | *RB1* | 6.703074884 | 8.42E-09 | 7.13E-08 | 4.09E-05 |
| Obesity | endometrium | *DIAPH3* | 8.210346974 | 9.69E-10 | 3.54E-08 | 4.70E-06 |
| Obesity | endometrium | *GRTP1* | 3.124607071 | 0.010885672 | 0.013109277 | 1 |
| Obesity | endometrium | *NOVA1* | 4.182624186 | 5.24E-10 | 2.52E-08 | 2.54E-06 |
| Obesity | endometrium | *FITM1* | 22.15217027 | 1.28E-06 | 2.95E-06 | 0.006219592 |
| Obesity | endometrium | *TMX1* | 10.08122448 | 2.78E-09 | 4.28E-08 | 1.35E-05 |
| Obesity | endometrium | *RTN1* | 2.025811664 | 1.72E-06 | 3.53E-06 | 0.008346518 |
| Obesity | endometrium | *C14orf37* | 7.346154439 | 3.10E-09 | 4.39E-08 | 1.50E-05 |
| Obesity | endometrium | *RAB15* | 8.51968609 | 4.24E-08 | 2.07E-07 | 0.000205773 |
| Obesity | endometrium | *EFCAB11* | 6.213857517 | 1.24E-05 | 2.06E-05 | 0.059954722 |
| Obesity | endometrium | *PTGR2* | 2.420966235 | 5.52E-05 | 8.50E-05 | 0.267931247 |
| Obesity | endometrium | *SLC24A4* | 4.695721334 | 1.45E-06 | 3.21E-06 | 0.007023554 |
| Obesity | endometrium | *C14orf79* | -2.207908349 | 0.003368573 | 0.004284224 | 1 |
| Obesity | endometrium | *WARS* | 13.29494677 | 2.57E-07 | 8.23E-07 | 0.001249135 |
| Obesity | endometrium | *SERF2* | 10.79555517 | 2.23E-09 | 4.00E-08 | 1.08E-05 |
| Obesity | endometrium | *ZSCAN29* | 5.741619782 | 1.54E-08 | 1.03E-07 | 7.49E-05 |

| | | | | | | |
|---|---|---|---|---|---|---|
| Obesity | endometrium | *SRP14* | 5.576612849 | 0.008600627 | 0.010466578 | 1 |
| Obesity | endometrium | *UBE2Q2* | 5.18324398 | 1.91E-07 | 6.53E-07 | 0.000927731 |
| Obesity | endometrium | *HMG20A* | 4.817944922 | 6.72E-08 | 2.91E-07 | 0.000326225 |
| Obesity | endometrium | *SCAPER* | 6.770467987 | 2.45E-05 | 3.93E-05 | 0.118996662 |
| Obesity | endometrium | *NEIL1* | 4.546516452 | 0.039935858 | 0.045916773 | 1 |
| Obesity | endometrium | *PIF1* | 10.12139767 | 0.000660209 | 0.000896539 | 1 |
| Obesity | endometrium | *CYP11A1* | 7.623897589 | 8.92E-09 | 7.32E-08 | 4.33E-05 |
| Obesity | endometrium | *BBS4* | 6.623289074 | 1.32E-08 | 9.23E-08 | 6.40E-05 |
| Obesity | endometrium | *ULK3* | 5.950555603 | 6.21E-07 | 1.67E-06 | 0.003014335 |
| Obesity | endometrium | *CELF6* | 25.22218843 | 0.000352563 | 0.000492695 | 1 |
| Obesity | endometrium | *HAPLN3* | -1.435736497 | 0.000391003 | 0.000543283 | 1 |
| Obesity | endometrium | *POLG* | 8.939990474 | 4.62E-09 | 5.20E-08 | 2.24E-05 |
| Obesity | endometrium | *ABHD2* | 7.236870288 | 0.002307545 | 0.002982474 | 1 |
| Obesity | endometrium | *MFGE8* | 8.070891409 | 4.76E-08 | 2.26E-07 | 0.000231138 |
| Obesity | endometrium | *UNC45A* | 3.034293255 | 1.34E-06 | 3.05E-06 | 0.006486133 |
| Obesity | endometrium | *IQGAP1* | 4.423445315 | 4.07E-08 | 2.02E-07 | 0.000197261 |
| Obesity | endometrium | *CRTC3* | 4.226375288 | 1.10E-07 | 4.24E-07 | 0.000535633 |
| Obesity | endometrium | *ITGAX* | 4.810409963 | 1.23E-06 | 2.85E-06 | 0.005980693 |
| Obesity | endometrium | *TGFB1I1* | -1.652931706 | 0.000478219 | 0.000659182 | 1 |
| Obesity | endometrium | *DHX38* | 3.640954553 | 7.69E-07 | 1.98E-06 | 0.003731413 |
| Obesity | endometrium | *NUDT7* | 8.563668485 | 4.06E-08 | 2.02E-07 | 0.000197056 |
| Obesity | endometrium | *GCSH* | 4.602339754 | 2.94E-06 | 5.56E-06 | 0.01427755 |
| Obesity | endometrium | *CMTM3* | 9.04911822 | 0.0044666 | 0.005610133 | 1 |
| Obesity | endometrium | *ZCCHC14* | 5.034577959 | 8.70E-08 | 3.53E-07 | 0.0004221 |
| Obesity | endometrium | *TLDC1* | 10.28175681 | 6.46E-09 | 6.17E-08 | 3.14E-05 |
| Obesity | endometrium | *RPL3L* | 16.01019288 | 2.42E-05 | 3.88E-05 | 0.117404204 |
| Obesity | endometrium | *TIGD7* | 3.9719304 | 2.05E-07 | 6.86E-07 | 0.000995852 |
| Obesity | endometrium | *DEF8* | 3.793457195 | 3.89E-06 | 7.11E-06 | 0.018878756 |
| Obesity | endometrium | *TCF25* | 4.090304868 | 1.32E-07 | 4.87E-07 | 0.000641461 |
| Obesity | endometrium | *GALNS* | 12.63522682 | 5.26E-07 | 1.46E-06 | 0.002553586 |
| Obesity | endometrium | *GAS8* | 11.30903143 | 4.15E-08 | 2.05E-07 | 0.000201166 |
| Obesity | endometrium | *NCOR1* | 6.553927104 | 2.92E-09 | 4.29E-08 | 1.42E-05 |
| Obesity | endometrium | *KSR1* | 5.577304206 | 3.54E-08 | 1.82E-07 | 0.00017179 |
| Obesity | endometrium | *GFOD2* | 3.054589614 | 1.12E-06 | 2.66E-06 | 0.005455195 |
| Obesity | endometrium | *PRPSAP2* | 6.432117775 | 3.25E-08 | 1.71E-07 | 0.000157839 |
| Obesity | endometrium | *MYO19* | 6.86983269 | 1.27E-10 | 1.34E-08 | 6.18E-07 |
| Obesity | endometrium | *RASL10B* | 14.04427795 | 6.48E-06 | 1.14E-05 | 0.031452249 |
| Obesity | endometrium | *UNC45B* | 16.87193099 | 0.000213998 | 0.000307194 | 1 |
| Obesity | endometrium | *C17orf80* | 4.128245684 | 4.31E-07 | 1.25E-06 | 0.002093581 |
| Obesity | endometrium | *VPS53* | 8.672722305 | 4.72E-07 | 1.34E-06 | 0.002290739 |
| Obesity | endometrium | *SGSM2* | 12.94562558 | 6.81E-09 | 6.31E-08 | 3.31E-05 |
| Obesity | endometrium | *LRRC46* | 4.57449337 | 0.013992837 | 0.016701905 | 1 |
| Obesity | endometrium | *SCRN2* | 2.371386423 | 2.06E-07 | 6.89E-07 | 0.001001711 |
| Obesity | endometrium | *SSH2* | 6.455720361 | 2.34E-09 | 4.01E-08 | 1.13E-05 |
| Obesity | endometrium | *RHBDL3* | -1.857025235 | 0.000750517 | 0.001016045 | 1 |
| Obesity | endometrium | *CLTC* | 6.280655502 | 6.09E-09 | 6.01E-08 | 2.96E-05 |
| Obesity | endometrium | *IMPA2* | 3.078000354 | 1.18E-06 | 2.76E-06 | 0.005722717 |
| Obesity | endometrium | *GNAL* | 6.328548986 | 3.36E-08 | 1.74E-07 | 0.000163181 |
| Obesity | endometrium | *RPRD1A* | 9.939689061 | 3.65E-08 | 1.86E-07 | 0.000176907 |
| Obesity | endometrium | *GALNT1* | 4.564925457 | 6.33E-08 | 2.77E-07 | 0.000306916 |

| | | | | | | |
|---|---|---|---|---|---|---|
| Obesity | endometrium | *C18orf8* | -4.624081813 | 0.027444391 | 0.031925242 | 1 |
| Obesity | endometrium | *NPC1* | 7.119558547 | 4.72E-07 | 1.34E-06 | 0.002288073 |
| Obesity | endometrium | *SLC14A1* | 3.540252216 | 2.25E-05 | 3.63E-05 | 0.109401654 |
| Obesity | endometrium | *CCDC40* | 2.763324627 | 8.37E-06 | 1.44E-05 | 0.04060571 |
| Obesity | endometrium | *TMC6* | 3.440912888 | 7.98E-07 | 2.03E-06 | 0.003870543 |
| Obesity | endometrium | *CARD14* | -2.177520981 | 0.006168575 | 0.007625486 | 1 |
| Obesity | endometrium | *TBCD* | 6.736289917 | 2.28E-08 | 1.33E-07 | 0.000110724 |
| Obesity | endometrium | *FN3KRP* | 2.82698866 | 2.96E-06 | 5.59E-06 | 0.014368047 |
| Obesity | endometrium | *NARF* | 8.96474317 | 0.0001467 | 0.000214911 | 0.71178673 |
| Obesity | endometrium | *TRIM65* | 12.11090393 | 3.09E-05 | 4.89E-05 | 0.149982203 |
| Obesity | endometrium | *CBX8* | 9.182853025 | 3.50E-06 | 6.47E-06 | 0.016983993 |
| Obesity | endometrium | *SECTM1* | -1.846352411 | 0.005086879 | 0.006346888 | 1 |
| Obesity | endometrium | *RNF157* | 3.58118802 | 1.38E-07 | 5.02E-07 | 0.000668461 |
| Obesity | endometrium | *ZNF750* | 10.28729768 | 1.64E-05 | 2.68E-05 | 0.079500418 |
| Obesity | endometrium | *WDR45B* | 3.363822883 | 8.05E-07 | 2.05E-06 | 0.003907005 |
| Obesity | endometrium | *RNF165* | 4.097891298 | 4.57E-07 | 1.30E-06 | 0.002216538 |
| Obesity | endometrium | *MAPK4* | 8.751165769 | 1.57E-06 | 3.35E-06 | 0.007616205 |
| Obesity | endometrium | *FBXO15* | -3.998613886 | 0.031673779 | 0.036678085 | 1 |
| Obesity | endometrium | *CBLN2* | -3.86651484 | 0.009785427 | 0.011822434 | 1 |
| Obesity | endometrium | *NT5C3B* | -1.220825687 | 0.008966582 | 0.010890076 | 1 |
| Obesity | endometrium | *PIP4K2B* | 10.96199511 | 6.87E-08 | 2.96E-07 | 0.0003331 |
| Obesity | endometrium | *ERBB2* | -1.603895997 | 0.001572412 | 0.002063658 | 1 |
| Obesity | endometrium | *MIEN1* | 8.295690431 | 6.54E-08 | 2.85E-07 | 0.000317262 |
| Obesity | endometrium | *PNMT* | 12.26751554 | 0.000306189 | 0.000431493 | 1 |
| Obesity | endometrium | *IGFBP4* | 8.202946603 | 1.00E-09 | 3.57E-08 | 4.86E-06 |
| Obesity | endometrium | *TXNL4A* | 12.15845757 | 4.18E-07 | 1.22E-06 | 0.00202913 |
| Obesity | endometrium | *CACNA1A* | 5.507514453 | 1.11E-06 | 2.63E-06 | 0.005395194 |
| Obesity | endometrium | *SAMD1* | -3.367392212 | 0.004678707 | 0.00586137 | 1 |
| Obesity | endometrium | *NFIC* | 8.494299008 | 4.48E-10 | 2.44E-08 | 2.17E-06 |
| Obesity | endometrium | *PRDM15* | 5.208457446 | 0.002172318 | 0.002816698 | 1 |
| Obesity | endometrium | *PFKL* | 3.234838505 | 1.80E-06 | 3.67E-06 | 0.008755264 |
| Obesity | endometrium | *MVB12A* | 10.01977245 | 2.33E-05 | 3.75E-05 | 0.11309986 |
| Obesity | endometrium | *DUS3L* | 3.184893161 | 0.000104439 | 0.000156498 | 0.50673921 |
| Obesity | endometrium | *DPP9* | 4.714971033 | 2.01E-07 | 6.77E-07 | 0.000977073 |
| Obesity | endometrium | *SIRT3* | 10.20835422 | 2.46E-10 | 1.94E-08 | 1.19E-06 |
| Obesity | endometrium | *HUNK* | 2.512547297 | 6.45E-05 | 9.85E-05 | 0.312744502 |
| Obesity | endometrium | *COL6A1* | 5.614608461 | 1.33E-07 | 4.88E-07 | 0.000645278 |
| Obesity | endometrium | *IFNAR1* | 4.365468588 | 8.23E-07 | 2.08E-06 | 0.003992093 |
| Obesity | endometrium | *TRPM2* | 2.178749212 | 1.11E-05 | 1.87E-05 | 0.053801824 |
| Obesity | endometrium | *TMEM50B* | 4.917034647 | 3.81E-07 | 1.13E-06 | 0.001847369 |
| Obesity | endometrium | *DOPEY2* | 2.46365601 | 3.30E-06 | 6.15E-06 | 0.015988359 |
| Obesity | endometrium | *URB1* | 6.145561853 | 9.94E-07 | 2.40E-06 | 0.004823386 |
| Obesity | endometrium | *SAE1* | 4.032182464 | 1.30E-05 | 2.16E-05 | 0.063190213 |
| Obesity | endometrium | *GEMIN7* | -1.993671077 | 0.000281588 | 0.000398794 | 1 |
| Obesity | endometrium | *SLC6A3* | 5.365418442 | 0.000467083 | 0.000644381 | 1 |
| Obesity | endometrium | *CACNG8* | 8.041708275 | 0.001469093 | 0.00193434 | 1 |
| Obesity | endometrium | *ZNF787* | 4.019979433 | 5.38E-07 | 1.49E-06 | 0.002611382 |
| Obesity | endometrium | *FBN3* | 3.908008873 | 0.003619381 | 0.00459598 | 1 |
| Obesity | endometrium | *EVI5L* | 4.28380399 | 9.72E-07 | 2.36E-06 | 0.004717055 |
| Obesity | endometrium | *CTU1* | 5.01996735 | 1.49E-06 | 3.26E-06 | 0.007228939 |

| | | | | | | |
|---|---|---|---|---|---|---|
| Obesity | endometrium | *RERE* | 4.882465751 | 5.25E-09 | 5.66E-08 | 2.54E-05 |
| Obesity | endometrium | *EPHA2* | 3.468212137 | 1.54E-05 | 2.52E-05 | 0.074498559 |
| Obesity | endometrium | *ARHGEF19* | -1.553869636 | 0.018226843 | 0.021538394 | 1 |
| Obesity | endometrium | *PEX14* | 3.337873978 | 2.49E-07 | 8.02E-07 | 0.00120681 |
| Obesity | endometrium | *CNKSR1* | 2.255306293 | 0.01744912 | 0.02066971 | 1 |
| Obesity | endometrium | *ZNF593* | -1.490947532 | 0.000292965 | 0.000413818 | 1 |
| Obesity | endometrium | *C1orf216* | 7.961995591 | 6.30E-07 | 1.69E-06 | 0.003055044 |
| Obesity | endometrium | *EVA1B* | -2.184874017 | 0.01070527 | 0.012895226 | 1 |
| Obesity | endometrium | *PLK4* | -1.91454448 | 0.00034388 | 0.000481393 | 1 |
| Obesity | endometrium | *NBPF3* | 2.986438271 | 8.83E-06 | 1.51E-05 | 0.042839924 |
| Obesity | endometrium | *HSPG2* | 3.503552694 | 7.95E-07 | 2.03E-06 | 0.003857984 |
| Obesity | endometrium | *ITGB3BP* | 4.611502008 | 4.12E-07 | 1.20E-06 | 0.001999771 |
| Obesity | endometrium | *PIGK* | 7.733844718 | 1.25E-07 | 4.68E-07 | 0.000608094 |
| Obesity | endometrium | *RPS8* | 6.614439741 | 2.33E-09 | 4.01E-08 | 1.13E-05 |
| Obesity | endometrium | *CYP4B1* | 4.778381769 | 0.009162497 | 0.011111331 | 1 |
| Obesity | endometrium | *SLC44A3* | 3.949917262 | 1.97E-06 | 3.94E-06 | 0.009558256 |
| Obesity | endometrium | *STRIP1* | 8.981721636 | 8.30E-09 | 7.12E-08 | 4.03E-05 |
| Obesity | endometrium | *GPR161* | 3.712371086 | 1.01E-06 | 2.42E-06 | 0.004880142 |
| Obesity | endometrium | *ATP1B1* | 3.935629925 | 7.69E-07 | 1.98E-06 | 0.003730187 |
| Obesity | endometrium | *TIPRL* | 4.059823141 | 1.21E-07 | 4.56E-07 | 0.000588721 |
| Obesity | endometrium | *NME7* | 7.133907489 | 1.58E-07 | 5.61E-07 | 0.000766743 |
| Obesity | endometrium | *XCL1* | -4.562628927 | 0.001553213 | 0.002040117 | 1 |
| Obesity | endometrium | *MAEL* | 9.344743134 | 1.70E-06 | 3.51E-06 | 0.008259314 |
| Obesity | endometrium | *RFWD2* | 7.090570729 | 2.26E-09 | 4.00E-08 | 1.09E-05 |
| Obesity | endometrium | *RGS16* | 4.947850342 | 4.29E-06 | 7.77E-06 | 0.020838321 |
| Obesity | endometrium | *TARS2* | -2.419705495 | 0.01376508 | 0.016450287 | 1 |
| Obesity | endometrium | *SNX27* | 4.498318011 | 5.22E-08 | 2.43E-07 | 0.000253501 |
| Obesity | endometrium | *SETDB1* | 7.297137855 | 4.55E-09 | 5.17E-08 | 2.21E-05 |
| Obesity | endometrium | *SEMA6C* | 5.243162853 | 9.72E-08 | 3.84E-07 | 0.000471588 |
| Obesity | endometrium | *ARNT* | 6.694725129 | 2.42E-08 | 1.38E-07 | 0.000117548 |
| Obesity | endometrium | *POGZ* | 5.014828203 | 2.00E-08 | 1.22E-07 | 9.72E-05 |
| Obesity | endometrium | *GABPB2* | 3.315756544 | 1.64E-06 | 3.42E-06 | 0.007949339 |
| Obesity | endometrium | *DYRK3* | 4.454007052 | 1.61E-06 | 3.39E-06 | 0.007817698 |
| Obesity | endometrium | *TAF1A* | -2.307693452 | 0.015576779 | 0.018528691 | 1 |
| Obesity | endometrium | *SMYD2* | 4.036588224 | 4.28E-07 | 1.24E-06 | 0.002076621 |
| Obesity | endometrium | *ADAM15* | 6.148531444 | 1.10E-06 | 2.62E-06 | 0.005359833 |
| Obesity | endometrium | *RAB13* | 3.900170879 | 1.57E-06 | 3.35E-06 | 0.007628936 |
| Obesity | endometrium | *S100A8* | 3.032559148 | 1.55E-06 | 3.33E-06 | 0.007532169 |
| Obesity | endometrium | *CREB3L4* | 4.554448581 | 3.62E-05 | 5.68E-05 | 0.175459789 |
| Obesity | endometrium | *KCNN3* | 2.763380311 | 2.20E-05 | 3.55E-05 | 0.106982363 |
| Obesity | endometrium | *C1orf43* | 11.43227533 | 2.98E-09 | 4.29E-08 | 1.45E-05 |
| Obesity | endometrium | *RIT1* | 3.656829416 | 7.91E-07 | 2.02E-06 | 0.003837754 |
| Obesity | endometrium | *FLG* | -1.566063409 | 8.71E-05 | 0.000131451 | 0.422745864 |
| Obesity | endometrium | *SCCPDH* | 4.832425497 | 1.81E-07 | 6.24E-07 | 0.00087956 |
| Obesity | endometrium | *CEP170* | 3.674994862 | 9.91E-08 | 3.90E-07 | 0.000480792 |
| Obesity | endometrium | *ACP1* | 13.529149 | 2.05E-07 | 6.85E-07 | 0.000992627 |
| Obesity | endometrium | *CNIH4* | 9.946615226 | 0.000174931 | 0.000253438 | 0.848764226 |
| Obesity | endometrium | *MBOAT2* | 3.047492621 | 1.48E-06 | 3.25E-06 | 0.00717253 |
| Obesity | endometrium | *PLEKHA6* | 5.127149275 | 7.47E-08 | 3.15E-07 | 0.00036253 |
| Obesity | endometrium | *OSR1* | 5.684385597 | 3.13E-06 | 5.87E-06 | 0.015202155 |

| | | | | | | |
|---|---|---|---|---|---|---|
| Obesity | endometrium | *RHOB* | 5.377973738 | 1.33E-07 | 4.88E-07 | 0.000644409 |
| Obesity | endometrium | *GALM* | 4.55644433 | 1.42E-07 | 5.14E-07 | 0.000688291 |
| Obesity | endometrium | *WDPCP* | 5.665958401 | 2.84E-09 | 4.29E-08 | 1.38E-05 |
| Obesity | endometrium | *VPS54* | 5.752458221 | 8.39E-09 | 7.13E-08 | 4.07E-05 |
| Obesity | endometrium | *ABHD1* | 13.29305512 | 2.36E-05 | 3.79E-05 | 0.114678568 |
| Obesity | endometrium | *CIAO1* | 9.889937381 | 2.03E-08 | 1.23E-07 | 9.83E-05 |
| Obesity | endometrium | *ZNF514* | 6.656963453 | 0.000122896 | 0.000182241 | 0.596291178 |
| Obesity | endometrium | *EXOC6B* | 7.837487169 | 1.75E-09 | 3.98E-08 | 8.47E-06 |
| Obesity | endometrium | *TEX261* | 4.383027268 | 1.59E-06 | 3.37E-06 | 0.00771521 |
| Obesity | endometrium | *THNSL2* | 2.914245946 | 2.68E-06 | 5.15E-06 | 0.013018129 |
| Obesity | endometrium | *RALB* | 10.4273187 | 2.67E-09 | 4.24E-08 | 1.30E-05 |
| Obesity | endometrium | *RABL2A* | 5.154044641 | 0.023327462 | 0.027312946 | 1 |
| Obesity | endometrium | *LIPT1* | -2.151061153 | 0.00672993 | 0.008285618 | 1 |
| Obesity | endometrium | *CNGA3* | 60.9212026 | 0.000135456 | 0.000199585 | 0.657232015 |
| Obesity | endometrium | *FAHD2B* | -2.913790211 | 5.88E-05 | 9.02E-05 | 0.28525884 |
| Obesity | endometrium | *LYG1* | 4.497768075 | 1.05E-06 | 2.51E-06 | 0.005083394 |
| Obesity | endometrium | *GPR17* | 2.003101996 | 0.000341533 | 0.000478383 | 1 |
| Obesity | endometrium | *GALNT13* | -0.966917312 | 0.000745811 | 0.001009956 | 1 |
| Obesity | endometrium | *PHOSPHO2* | 5.093446426 | 2.47E-05 | 3.95E-05 | 0.119655383 |
| Obesity | endometrium | *METTL21A* | 8.884704362 | 9.53E-10 | 3.50E-08 | 4.62E-06 |
| Obesity | endometrium | *NBEAL1* | 5.610529873 | 6.46E-10 | 2.75E-08 | 3.14E-06 |
| Obesity | endometrium | *SUMF1* | 4.301671309 | 2.28E-08 | 1.33E-07 | 0.000110608 |
| Obesity | endometrium | *ACKR3* | 3.733277407 | 7.22E-07 | 1.88E-06 | 0.003502483 |
| Obesity | endometrium | *ESPNL* | 51.05051735 | 2.03E-06 | 4.05E-06 | 0.009855158 |
| Obesity | endometrium | *ANKMY1* | 8.368808217 | 5.80E-08 | 2.59E-07 | 0.000281247 |
| Obesity | endometrium | *TAMM41* | 5.27218091 | 2.28E-08 | 1.33E-07 | 0.00011069 |
| Obesity | endometrium | *STK11IP* | -1.508715805 | 0.000493281 | 0.000678978 | 1 |
| Obesity | endometrium | *EAF1* | 13.8029503 | 8.19E-09 | 7.09E-08 | 3.97E-05 |
| Obesity | endometrium | *CNTN4* | 3.717397651 | 0.034953872 | 0.040341624 | 1 |
| Obesity | endometrium | *SLC25A38* | 4.496238717 | 2.19E-06 | 4.30E-06 | 0.010602242 |
| Obesity | endometrium | *CTDSPL* | 7.102839938 | 4.06E-09 | 4.85E-08 | 1.97E-05 |
| Obesity | endometrium | *PTPRG* | 4.07988885 | 3.17E-08 | 1.68E-07 | 0.000154044 |
| Obesity | endometrium | *ARL6IP5* | 3.785373772 | 1.91E-07 | 6.53E-07 | 0.000928584 |
| Obesity | endometrium | *LIMD1* | 5.712600692 | 1.26E-09 | 3.98E-08 | 6.12E-06 |
| Obesity | endometrium | *ZNF660* | -1.274594047 | 0.000300475 | 0.000423687 | 1 |
| Obesity | endometrium | *PLA1A* | 12.9484549 | 1.94E-06 | 3.88E-06 | 0.009396819 |
| Obesity | endometrium | *ADPRH* | 5.57260467 | 1.73E-05 | 2.82E-05 | 0.083926942 |
| Obesity | endometrium | *IGSF11* | -1.443569545 | 0.006171058 | 0.007626585 | 1 |
| Obesity | endometrium | *BOC* | 6.185415466 | 5.73E-09 | 5.87E-08 | 2.78E-05 |
| Obesity | endometrium | *MED12L* | 3.141297764 | 1.35E-06 | 3.06E-06 | 0.006526137 |
| Obesity | endometrium | *EIF2A* | 2.449021882 | 1.79E-05 | 2.92E-05 | 0.086932684 |
| Obesity | endometrium | *NCEH1* | 2.357022542 | 1.38E-05 | 2.29E-05 | 0.067054475 |
| Obesity | endometrium | *CCDC39* | 7.291461479 | 8.80E-11 | 1.02E-08 | 4.27E-07 |
| Obesity | endometrium | *MUC4* | 4.759727176 | 2.61E-06 | 5.02E-06 | 0.012665614 |
| Obesity | endometrium | *DGKQ* | -2.70582228 | 0.008583811 | 0.010448733 | 1 |
| Obesity | endometrium | *CENPC* | 5.778512769 | 5.61E-08 | 2.55E-07 | 0.000272114 |
| Obesity | endometrium | *SCD5* | 4.977140902 | 2.91E-07 | 9.08E-07 | 0.001410607 |
| Obesity | endometrium | *PYURF* | 18.02544736 | 2.55E-08 | 1.43E-07 | 0.000123855 |
| Obesity | endometrium | *CISD2* | 5.936082241 | 4.31E-08 | 2.09E-07 | 0.000208943 |
| Obesity | endometrium | *DDIT4L* | -4.491313701 | 0.003108709 | 0.003966199 | 1 |

| | | | | | | |
|---|---|---|---|---|---|---|
| Obesity | endometrium | *METTL14* | 11.09705779 | 1.16E-05 | 1.95E-05 | 0.056458024 |
| Obesity | endometrium | *USP53* | 7.725362714 | 4.14E-10 | 2.39E-08 | 2.01E-06 |
| Obesity | endometrium | *MARCH1* | 3.002806037 | 2.07E-06 | 4.10E-06 | 0.01002375 |
| Obesity | endometrium | *CBR4* | 9.648985939 | 5.49E-09 | 5.77E-08 | 2.66E-05 |
| Obesity | endometrium | *CYP4V2* | 3.141067734 | 0.00151494 | 0.001992002 | 1 |
| Obesity | endometrium | *MARCH6* | 5.885118574 | 7.45E-09 | 6.64E-08 | 3.61E-05 |
| Obesity | endometrium | *NKD2* | 7.862440222 | 0.002257595 | 0.002922586 | 1 |
| Obesity | endometrium | *MYO10* | 5.11843393 | 5.77E-08 | 2.59E-07 | 0.000279998 |
| Obesity | endometrium | *FAM105A* | 2.517912344 | 4.00E-05 | 6.24E-05 | 0.19406271 |
| Obesity | endometrium | *GTF2H2* | 17.21120746 | 2.84E-09 | 4.29E-08 | 1.38E-05 |
| Obesity | endometrium | *SPATA9* | 2.629287352 | 1.68E-06 | 3.48E-06 | 0.008153107 |
| Obesity | endometrium | *FEM1C* | 7.088079577 | 5.80E-08 | 2.59E-07 | 0.000281268 |
| Obesity | endometrium | *COMMD10* | 4.183233405 | 9.95E-08 | 3.90E-07 | 0.000482874 |
| Obesity | endometrium | *YIPF5* | 8.047147633 | 5.06E-09 | 5.55E-08 | 2.45E-05 |
| Obesity | endometrium | *CXCL14* | 7.356462045 | 2.56E-08 | 1.44E-07 | 0.000124405 |
| Obesity | endometrium | *FBXO38* | 5.907746291 | 2.73E-08 | 1.50E-07 | 0.000132324 |
| Obesity | endometrium | *BOD1* | 3.802478528 | 1.38E-06 | 3.13E-06 | 0.006714318 |
| Obesity | endometrium | *FAM50B* | 9.167671164 | 8.14E-06 | 1.40E-05 | 0.039494572 |
| Obesity | endometrium | *MYLK4* | -2.353143163 | 0.000987741 | 0.00132098 | 1 |
| Obesity | endometrium | *TBC1D7* | 3.585316499 | 2.98E-06 | 5.61E-06 | 0.014442199 |
| Obesity | endometrium | *GFOD1* | 10.68173193 | 8.82E-07 | 2.20E-06 | 0.004280178 |
| Obesity | endometrium | *TRIM7* | 6.886253475 | 3.94E-05 | 6.15E-05 | 0.191286914 |
| Obesity | endometrium | *HIGD2A* | 11.30664689 | 0.00017151 | 0.000248705 | 0.832166186 |
| Obesity | endometrium | *FAM193B* | 3.321662938 | 4.65E-07 | 1.32E-06 | 0.002257719 |
| Obesity | endometrium | *MUT* | 2.859310992 | 2.66E-06 | 5.11E-06 | 0.012901064 |
| Obesity | endometrium | *DOK3* | 2.686184767 | 0.000516705 | 0.000709209 | 1 |
| Obesity | endometrium | *ABT1* | 7.79691078 | 9.06E-10 | 3.41E-08 | 4.40E-06 |
| Obesity | endometrium | *DAAM2* | 2.6089558 | 1.08E-06 | 2.58E-06 | 0.005240809 |
| Obesity | endometrium | *MLIP* | 9.31097254 | 0.000386495 | 0.000537482 | 1 |
| Obesity | endometrium | *ANO7* | 5.991214793 | 9.09E-06 | 1.55E-05 | 0.044119656 |
| Obesity | endometrium | *RPL7L1* | 7.08156223 | 2.55E-08 | 1.43E-07 | 0.000123701 |
| Obesity | endometrium | *TPBG* | 2.81455578 | 2.09E-06 | 4.15E-06 | 0.010151368 |
| Obesity | endometrium | *IRAK1BP1* | 3.963842896 | 4.24E-08 | 2.07E-07 | 0.000205579 |
| Obesity | endometrium | *PHIP* | 6.712827754 | 1.97E-09 | 4.00E-08 | 9.58E-06 |
| Obesity | endometrium | *PNRC1* | 3.708785109 | 4.55E-07 | 1.30E-06 | 0.002207425 |
| Obesity | endometrium | *RNF217* | 6.042882177 | 1.01E-08 | 7.86E-08 | 4.88E-05 |
| Obesity | endometrium | *ARHGAP18* | 3.166671577 | 1.02E-07 | 4.00E-07 | 0.000496689 |
| Obesity | endometrium | *ABRACL* | 5.345410181 | 4.28E-06 | 7.75E-06 | 0.020766141 |
| Obesity | endometrium | *SLC18B1* | 4.047729618 | 1.10E-05 | 1.85E-05 | 0.053350084 |
| Obesity | endometrium | *PNLDC1* | -4.532798313 | 0.022822864 | 0.026760884 | 1 |
| Obesity | endometrium | *WTAP* | 8.656005856 | 3.84E-09 | 4.78E-08 | 1.86E-05 |
| Obesity | endometrium | *SLC22A3* | 2.710890989 | 1.65E-06 | 3.44E-06 | 0.008001537 |
| Obesity | endometrium | *GNA12* | 5.62330616 | 0.006752492 | 0.008309178 | 1 |
| Obesity | endometrium | *SDK1* | -1.917284086 | 0.00508719 | 0.006346888 | 1 |
| Obesity | endometrium | *CCZ1B* | 5.017718574 | 0.002200046 | 0.002851128 | 1 |
| Obesity | endometrium | *RBAK* | 9.96734939 | 5.52E-08 | 2.52E-07 | 0.000267688 |
| Obesity | endometrium | *POMZP3* | 3.528020671 | 6.25E-07 | 1.68E-06 | 0.003034234 |
| Obesity | endometrium | *PSPH* | 4.249407647 | 0.004143671 | 0.005223458 | 1 |
| Obesity | endometrium | *GIGYF1* | 11.6822436 | 2.46E-08 | 1.39E-07 | 0.000119361 |
| Obesity | endometrium | *TRIM4* | 1.99522129 | 0.00034727 | 0.000485578 | 1 |

| | | | | | | |
|---|---|---|---|---|---|---|
| Obesity | endometrium | *TMEM209* | 5.722187671 | 8.71E-08 | 3.53E-07 | 0.000422639 |
| Obesity | endometrium | *EPHA1* | 10.14295664 | 1.76E-06 | 3.59E-06 | 0.008526247 |
| Obesity | endometrium | *NCAPG2* | 3.594225314 | 1.62E-06 | 3.41E-06 | 0.007874295 |
| Obesity | endometrium | *FBXO25* | 10.27997954 | 1.63E-09 | 3.98E-08 | 7.90E-06 |
| Obesity | endometrium | *CSGALNACT1* | 4.684297355 | 1.58E-08 | 1.04E-07 | 7.67E-05 |
| Obesity | endometrium | *BIN3* | 4.827658658 | 3.26E-07 | 9.91E-07 | 0.001580137 |
| Obesity | endometrium | *CHMP7* | 5.838476575 | 2.06E-06 | 4.10E-06 | 0.010009722 |
| Obesity | endometrium | *DOCK5* | 3.769513117 | 1.08E-07 | 4.16E-07 | 0.000524199 |
| Obesity | endometrium | *TACC1* | 4.805364266 | 3.05E-08 | 1.63E-07 | 0.000147854 |
| Obesity | endometrium | *ADHFE1* | 1.264112203 | 0.035915514 | 0.041402251 | 1 |
| Obesity | endometrium | *RPL7* | -2.129311486 | 0.00568095 | 0.007056828 | 1 |
| Obesity | endometrium | *ATP6V0D2* | 14.3122084 | 2.77E-06 | 5.29E-06 | 0.013458749 |
| Obesity | endometrium | *DPYS* | 2.323837582 | 1.63E-06 | 3.42E-06 | 0.00792243 |
| Obesity | endometrium | *LRP12* | 15.47467571 | 8.04E-08 | 3.34E-07 | 0.000390326 |
| Obesity | endometrium | *MAL2* | 4.784870997 | 0.000179333 | 0.000259275 | 0.870125968 |
| Obesity | endometrium | *SLC39A4* | -1.407611637 | 0.000437016 | 0.000604792 | 1 |
| Obesity | endometrium | *AK3* | 4.235068391 | 1.33E-07 | 4.88E-07 | 0.000646446 |
| Obesity | endometrium | *HAUS6* | 8.250748051 | 1.20E-08 | 8.67E-08 | 5.82E-05 |
| Obesity | endometrium | *C9orf72* | 2.554538214 | 2.39E-06 | 4.64E-06 | 0.011598339 |
| Obesity | endometrium | *FBXO10* | 7.183312199 | 2.04E-05 | 3.30E-05 | 0.098913157 |
| Obesity | endometrium | *ZNF462* | 3.224700586 | 1.81E-07 | 6.24E-07 | 0.000879436 |
| Obesity | endometrium | *ALAD* | 5.223414703 | 1.20E-07 | 4.52E-07 | 0.000580447 |
| Obesity | endometrium | *WDR31* | 5.654167757 | 0.001634942 | 0.002142238 | 1 |
| Obesity | endometrium | *POLE3* | 3.752834606 | 8.11E-05 | 0.000122666 | 0.393444123 |
| Obesity | endometrium | *MED22* | 3.882271643 | 2.04E-05 | 3.31E-05 | 0.099214468 |
| Obesity | endometrium | *REXO4* | 4.029461821 | 6.03E-07 | 1.62E-06 | 0.002924234 |
| Obesity | endometrium | *SH3GLB2* | 12.58795226 | 3.96E-09 | 4.82E-08 | 1.92E-05 |
| Obesity | endometrium | *INPP5E* | 18.87782575 | 1.38E-07 | 5.02E-07 | 0.000668606 |
| Obesity | endometrium | *SEC16A* | 10.55544064 | 5.35E-05 | 8.25E-05 | 0.259623334 |
| Obesity | endometrium | *SLC39A12* | 30.30605003 | 2.28E-06 | 4.45E-06 | 0.011066365 |
| Obesity | endometrium | *TMEM236* | 5.496193927 | 1.28E-05 | 2.13E-05 | 0.061938811 |
| Obesity | endometrium | *RSU1* | 3.977231755 | 9.88E-08 | 3.89E-07 | 0.000479181 |
| Obesity | endometrium | *ST8SIA6* | 19.44605962 | 1.04E-08 | 8.01E-08 | 5.04E-05 |
| Obesity | endometrium | *PARD3* | 6.805566785 | 1.19E-09 | 3.96E-08 | 5.78E-06 |
| Obesity | endometrium | *FRA10AC1* | 5.744575493 | 4.93E-08 | 2.32E-07 | 0.000239213 |
| Obesity | endometrium | *ADD3* | 8.168892405 | 1.38E-09 | 3.98E-08 | 6.70E-06 |
| Obesity | endometrium | *LRRC27* | 3.352076796 | 1.91E-05 | 3.11E-05 | 0.092848252 |
| Obesity | endometrium | *MTG1* | 8.373822827 | 6.13E-11 | 8.04E-09 | 2.98E-07 |
| Obesity | endometrium | *PAOX* | 17.95803346 | 1.42E-09 | 3.98E-08 | 6.89E-06 |
| Obesity | endometrium | *PDCD11* | 6.095523022 | 1.35E-08 | 9.41E-08 | 6.54E-05 |
| Obesity | endometrium | *RGS10* | 3.375543082 | 1.39E-06 | 3.13E-06 | 0.006735643 |
| Obesity | endometrium | *LIN7C* | 3.32416352 | 1.27E-06 | 2.92E-06 | 0.00615067 |
| Obesity | endometrium | *ZNF215* | 3.037634982 | 0.000158362 | 0.00023102 | 0.768372039 |
| Obesity | endometrium | *APIP* | 2.445370149 | 1.75E-05 | 2.85E-05 | 0.08479733 |
| Obesity | endometrium | *EIF3M* | 11.19819965 | 1.91E-08 | 1.18E-07 | 9.28E-05 |
| Obesity | endometrium | *TNKS1BP1* | 11.21586886 | 1.86E-15 | 1.50E-12 | 9.02E-12 |
| Obesity | endometrium | *PTPRJ* | 2.245873355 | 1.34E-06 | 3.06E-06 | 0.006513646 |
| Obesity | endometrium | *ENDOD1* | 3.480627644 | 1.12E-06 | 2.64E-06 | 0.005415135 |
| Obesity | endometrium | *CCDC82* | 3.119087427 | 6.35E-05 | 9.71E-05 | 0.308301419 |
| Obesity | endometrium | *CAPN5* | -2.356274167 | 0.000785233 | 0.001060381 | 1 |

| | | | | | | |
|---|---|---|---|---|---|---|
| Obesity | endometrium | *INTS4* | 6.000938729 | 0.002745722 | 0.003522539 | 1 |
| Obesity | endometrium | *RPS3* | 10.15164067 | 9.47E-08 | 3.76E-07 | 0.000459466 |
| Obesity | endometrium | *TTC12* | 10.1464546 | 4.91E-13 | 1.83E-10 | 2.38E-09 |
| Obesity | endometrium | *DRD2* | 31.79767057 | 1.82E-06 | 3.70E-06 | 0.008852912 |
| Obesity | endometrium | *NPAT* | 6.757257663 | 1.91E-08 | 1.18E-07 | 9.29E-05 |
| Obesity | endometrium | *ATM* | 5.138494697 | 1.24E-08 | 8.90E-08 | 6.02E-05 |
| Obesity | endometrium | *SLX4IP* | 4.729500646 | 1.73E-08 | 1.10E-07 | 8.37E-05 |
| Obesity | endometrium | *ST14* | 6.444962295 | 1.22E-06 | 2.84E-06 | 0.005939113 |
| Obesity | endometrium | *FADS1* | 6.512809312 | 3.73E-06 | 6.86E-06 | 0.018112287 |
| Obesity | endometrium | *TMC2* | 6.718316501 | 3.46E-06 | 6.42E-06 | 0.016789508 |
| Obesity | endometrium | *EML3* | 3.335168588 | 2.69E-06 | 5.16E-06 | 0.01304634 |
| Obesity | endometrium | *INCENP* | 4.419899116 | 8.33E-06 | 1.44E-05 | 0.040435895 |
| Obesity | endometrium | *PLCH2* | 13.64432541 | 0.001304304 | 0.001727205 | 1 |
| Obesity | endometrium | *CPSF7* | 11.56863805 | 2.43E-07 | 7.87E-07 | 0.001177429 |
| Obesity | endometrium | *CHEK1* | 6.742239941 | 1.61E-06 | 3.39E-06 | 0.007818659 |
| Obesity | endometrium | *DUSP15* | -1.191404851 | 0.000139995 | 0.00020596 | 0.679257563 |
| Obesity | endometrium | *SPATA25* | 8.616134227 | 0.000378507 | 0.000527128 | 1 |
| Obesity | endometrium | *CABLES2* | 4.128887142 | 0.002150896 | 0.00279116 | 1 |
| Obesity | endometrium | *TRPT1* | 3.103930905 | 0.000451046 | 0.000623319 | 1 |
| Obesity | endometrium | *NUDT22* | -3.415999737 | 0.018645172 | 0.022022 | 1 |
| Obesity | endometrium | *TBX6* | 6.529211883 | 2.24E-15 | 1.55E-12 | 1.09E-11 |
| Obesity | endometrium | *ALDOA* | 10.58370477 | 3.35E-06 | 6.24E-06 | 0.016265567 |
| Obesity | endometrium | *ANXA8L1* | 1.970023338 | 4.37E-05 | 6.80E-05 | 0.212215085 |
| Obesity | endometrium | *CWC15* | 4.714623157 | 1.73E-07 | 6.01E-07 | 0.000838748 |
| Obesity | endometrium | *FCGR1A* | -2.931645662 | 0.000131603 | 0.000194262 | 0.638539739 |
| Obesity | endometrium | *TMEM218* | 4.977940709 | 2.26E-07 | 7.44E-07 | 0.001097017 |
| Obesity | endometrium | *SAP18* | 3.98562236 | 5.79E-07 | 1.57E-06 | 0.002807115 |
| Obesity | endometrium | *FAM124A* | 18.24576934 | 8.38E-07 | 2.11E-06 | 0.004063802 |
| Obesity | endometrium | *LYPD6B* | -5.109295798 | 0.021240184 | 0.024959403 | 1 |
| Obesity | endometrium | *WDR17* | 3.055878157 | 5.61E-06 | 9.97E-06 | 0.027237685 |
| Obesity | endometrium | *CCDC102B* | 2.42735658 | 1.45E-06 | 3.21E-06 | 0.007023005 |
| Obesity | endometrium | *RGS18* | 5.508837247 | 2.79E-09 | 4.28E-08 | 1.35E-05 |
| Obesity | endometrium | *PRSS23* | 3.875920181 | 6.59E-08 | 2.86E-07 | 0.000319726 |
| Obesity | endometrium | *PPP1R1C* | 6.012082106 | 1.61E-06 | 3.39E-06 | 0.00780402 |
| Obesity | endometrium | *FAM173B* | 2.84396714 | 1.70E-06 | 3.51E-06 | 0.008255099 |
| Obesity | endometrium | *GPR158* | 5.846006976 | 1.53E-06 | 3.31E-06 | 0.007406955 |
| Obesity | endometrium | *UBE3B* | 5.588336497 | 4.10E-09 | 4.87E-08 | 1.99E-05 |
| Obesity | endometrium | *ANK3* | 3.550118935 | 4.11E-07 | 1.20E-06 | 0.001995105 |
| Obesity | endometrium | *PLBD2* | 3.691691938 | 6.84E-07 | 1.81E-06 | 0.003320028 |
| Obesity | endometrium | *DLG5* | 5.113448284 | 3.26E-07 | 9.92E-07 | 0.001582961 |
| Obesity | endometrium | *GXYLT1* | 3.840536389 | 3.96E-07 | 1.16E-06 | 0.001919445 |
| Obesity | endometrium | *EXT2* | 3.83372561 | 2.01E-07 | 6.77E-07 | 0.00097664 |
| Obesity | endometrium | *TMEM18* | 13.75900899 | 2.36E-09 | 4.01E-08 | 1.15E-05 |
| Obesity | endometrium | *NDUFC2* | 4.210280249 | 1.01E-06 | 2.42E-06 | 0.004880512 |
| Obesity | endometrium | *ADAMTS12* | 6.191137454 | 2.41E-05 | 3.86E-05 | 0.117011898 |
| Obesity | endometrium | *NUBPL* | 5.027800487 | 2.51E-08 | 1.42E-07 | 0.000121673 |
| Obesity | endometrium | *VIPAS39* | 4.404702511 | 5.07E-07 | 1.42E-06 | 0.002462048 |
| Obesity | endometrium | *ANKRD50* | 9.67966092 | 1.47E-09 | 3.98E-08 | 7.11E-06 |
| Obesity | endometrium | *UPF2* | 5.017131014 | 2.79E-08 | 1.52E-07 | 0.000135131 |
| Obesity | endometrium | *C4orf33* | 4.687416427 | 5.45E-07 | 1.50E-06 | 0.002646397 |

| | | | | | | |
|---|---|---|---|---|---|---|
| Obesity | endometrium | *PTPRO* | 3.362796484 | 6.18E-07 | 1.66E-06 | 0.002997195 |
| Obesity | endometrium | *EPS8* | 5.358165814 | 7.50E-09 | 6.64E-08 | 3.64E-05 |
| Obesity | endometrium | *QDPR* | 6.907801937 | 9.08E-08 | 3.64E-07 | 0.000440668 |
| Obesity | endometrium | *FAM160B1* | 10.70807947 | 2.64E-06 | 5.08E-06 | 0.012809825 |
| Obesity | endometrium | *ZNF827* | 2.985890782 | 7.41E-07 | 1.93E-06 | 0.00359443 |
| Obesity | endometrium | *AKR1C2* | 5.426849305 | 3.91E-06 | 7.14E-06 | 0.018967966 |
| Obesity | endometrium | *DPYSL4* | 3.157544287 | 0.00034041 | 0.000476948 | 1 |
| Obesity | endometrium | *PIGF* | 10.91707202 | 1.59E-09 | 3.98E-08 | 7.73E-06 |
| Obesity | endometrium | *INPP1* | 8.507864592 | 0.000957395 | 0.001281457 | 1 |
| Obesity | endometrium | *MFSD6* | 6.229497532 | 3.17E-08 | 1.68E-07 | 0.000153615 |
| Obesity | endometrium | *RNF144A* | 8.296553617 | 0.000109516 | 0.000163449 | 0.531371682 |
| Obesity | endometrium | *CENPU* | -1.205550261 | 0.000343808 | 0.000481393 | 1 |
| Obesity | endometrium | *BICD1* | 3.939542999 | 1.29E-07 | 4.78E-07 | 0.000625296 |
| Obesity | endometrium | *CCDC122* | 3.269842796 | 0.005337387 | 0.006646162 | 1 |
| Obesity | endometrium | *GABRA2* | 12.79390287 | 7.45E-07 | 1.93E-06 | 0.003616396 |
| Obesity | endometrium | *PARP8* | 4.409562805 | 2.27E-06 | 4.44E-06 | 0.010994356 |
| Obesity | endometrium | *CACUL1* | 5.208977076 | 3.28E-08 | 1.72E-07 | 0.00015896 |
| Obesity | endometrium | *BEND6* | -1.350381164 | 7.85E-05 | 0.00011913 | 0.38109718 |
| Obesity | endometrium | *TIAL1* | 6.461790543 | 5.87E-09 | 5.92E-08 | 2.85E-05 |
| Obesity | endometrium | *TMEM56* | 22.17116911 | 0.000531058 | 0.000728085 | 1 |
| Obesity | endometrium | *PTPN14* | 5.218753769 | 1.49E-08 | 1.01E-07 | 7.25E-05 |
| Obesity | endometrium | *MGAT5* | 3.342410127 | 3.67E-07 | 1.09E-06 | 0.00178109 |
| Obesity | endometrium | *TMEM178A* | 6.446906181 | 1.49E-06 | 3.26E-06 | 0.007230844 |
| Obesity | endometrium | *HAUS1* | 3.211825393 | 7.87E-06 | 1.36E-05 | 0.038208237 |
| Obesity | endometrium | *SPC25* | 3.038506652 | 8.33E-05 | 0.000125845 | 0.404213829 |
| Obesity | endometrium | *KCNK13* | 5.509808516 | 3.19E-06 | 5.96E-06 | 0.015456043 |
| Obesity | endometrium | *ATG10* | 2.814746536 | 0.002409627 | 0.003106966 | 1 |
| Obesity | endometrium | *POC5* | 2.641989222 | 0.01411362 | 0.016841929 | 1 |
| Obesity | endometrium | *CWF19L2* | 7.299726231 | 1.96E-10 | 1.73E-08 | 9.50E-07 |
| Obesity | endometrium | *ZNF773* | 3.658633926 | 2.36E-05 | 3.79E-05 | 0.11443301 |
| Obesity | endometrium | *NMT2* | 4.417421651 | 9.92E-07 | 2.40E-06 | 0.004811599 |
| Obesity | endometrium | *ZNF837* | 5.011918984 | 0.000862368 | 0.001160991 | 1 |
| Obesity | endometrium | *CCDC50* | 6.646349195 | 4.50E-08 | 2.16E-07 | 0.000218437 |
| Obesity | endometrium | *SPARCL1* | 4.654030881 | 1.84E-08 | 1.15E-07 | 8.91E-05 |
| Obesity | endometrium | *MBNL1* | 4.542019848 | 7.26E-09 | 6.56E-08 | 3.52E-05 |
| Obesity | endometrium | *GPD1L* | 9.202979882 | 5.22E-10 | 2.52E-08 | 2.53E-06 |
| Obesity | endometrium | *CCNO* | 37.02979462 | 6.12E-06 | 1.08E-05 | 0.029676522 |
| Obesity | endometrium | *CLEC4F* | 7.526214143 | 4.76E-07 | 1.34E-06 | 0.002308781 |
| Obesity | endometrium | *SLC30A6* | 9.758005223 | 7.04E-07 | 1.85E-06 | 0.003418063 |
| Obesity | endometrium | *RASGRP3* | 3.186460088 | 2.03E-06 | 4.04E-06 | 0.009841549 |
| Obesity | endometrium | *GPR180* | 3.803090372 | 1.42E-05 | 2.34E-05 | 0.068658728 |
| Obesity | endometrium | *TCTEX1D1* | 7.729877584 | 0.00166209 | 0.002175469 | 1 |
| Obesity | endometrium | *IFIT5* | 3.073167719 | 0.001535068 | 0.002017376 | 1 |
| Obesity | endometrium | *PRDM8* | 8.548700603 | 0.003356836 | 0.004270416 | 1 |
| Obesity | endometrium | *GGPS1* | 7.826588596 | 7.59E-09 | 6.70E-08 | 3.68E-05 |
| Obesity | endometrium | *ZNF117* | 2.469940969 | 3.35E-07 | 1.02E-06 | 0.001625756 |
| Obesity | endometrium | *MED21* | 6.991345631 | 1.83E-08 | 1.15E-07 | 8.90E-05 |
| Obesity | endometrium | *JAKMIP1* | 4.882750834 | 1.37E-06 | 3.10E-06 | 0.006627181 |
| Obesity | endometrium | *SREK1IP1* | 6.180262152 | 2.45E-08 | 1.39E-07 | 0.000119109 |
| Obesity | endometrium | *MR1* | 3.519991498 | 5.56E-07 | 1.52E-06 | 0.00269975 |

| | | | | | | |
|---|---|---|---|---|---|---|
| Obesity | endometrium | *CENPH* | -3.359342808 | 0.001419312 | 0.001873877 | 1 |
| Obesity | endometrium | *CARHSP1* | 2.239319566 | 2.05E-06 | 4.07E-06 | 0.009934833 |
| Obesity | endometrium | *CAST* | 5.279289194 | 8.16E-08 | 3.37E-07 | 0.000395712 |
| Obesity | endometrium | *SCOC* | 9.639535673 | 0.000819926 | 0.001105692 | 1 |
| Obesity | endometrium | *CLGN* | 9.589028115 | 0.003676258 | 0.004660884 | 1 |
| Obesity | endometrium | *CETN3* | 11.08749851 | 1.05E-09 | 3.67E-08 | 5.10E-06 |
| Obesity | endometrium | *MERTK* | 1.307534669 | 7.09E-05 | 0.000107887 | 0.343835142 |
| Obesity | endometrium | *TMEM87B* | 6.972735643 | 1.06E-08 | 8.07E-08 | 5.12E-05 |
| Obesity | endometrium | *PLA2R1* | 3.220763616 | 0.007946377 | 0.009689826 | 1 |
| Obesity | endometrium | *SLC25A27* | 3.802705993 | 2.63E-07 | 8.37E-07 | 0.001276657 |
| Obesity | endometrium | *LPCAT1* | 3.609467607 | 1.44E-06 | 3.19E-06 | 0.00697011 |
| Obesity | endometrium | *NMRAL1* | 3.194169417 | 2.86E-05 | 4.54E-05 | 0.138861392 |
| Obesity | endometrium | *ADPRHL1* | 7.059126525 | 5.87E-06 | 1.04E-05 | 0.028485135 |
| Obesity | endometrium | *CFDP1* | 4.054757417 | 7.59E-08 | 3.18E-07 | 0.000368107 |
| Obesity | endometrium | *ZDHHC7* | 12.22929708 | 2.01E-09 | 4.00E-08 | 9.76E-06 |
| Obesity | endometrium | *FBXO36* | 5.926109338 | 1.14E-07 | 4.33E-07 | 0.000551035 |
| Obesity | endometrium | *MCOLN2* | 6.000087622 | 1.05E-06 | 2.51E-06 | 0.005097105 |
| Obesity | endometrium | *GDPD1* | 3.335750223 | 1.84E-06 | 3.72E-06 | 0.008938824 |
| Obesity | endometrium | *SEMA3D* | 7.924614495 | 2.70E-09 | 4.24E-08 | 1.31E-05 |
| Obesity | endometrium | *PPP2R5E* | 6.064521693 | 8.41E-09 | 7.13E-08 | 4.08E-05 |
| Obesity | endometrium | *ANKRD29* | 2.682198096 | 1.49E-06 | 3.27E-06 | 0.007239041 |
| Obesity | endometrium | *DNAAF1* | -5.652772594 | 0.001806019 | 0.00235686 | 1 |
| Obesity | endometrium | *ANKH* | 3.71405276 | 1.38E-07 | 5.02E-07 | 0.000669613 |
| Obesity | endometrium | *UBASH3B* | 3.744271956 | 3.24E-06 | 6.05E-06 | 0.015704844 |
| Obesity | endometrium | *ROBO4* | 4.784695451 | 1.67E-07 | 5.87E-07 | 0.000812428 |
| Obesity | endometrium | *ROBO3* | 3.118829121 | 5.29E-07 | 1.47E-06 | 0.002564799 |
| Obesity | endometrium | *CC2D1B* | 6.613033375 | 8.31E-09 | 7.12E-08 | 4.03E-05 |
| Obesity | endometrium | *PRKCA* | 5.842043395 | 1.01E-08 | 7.86E-08 | 4.88E-05 |
| Obesity | endometrium | *LRRK1* | 2.331749147 | 1.57E-06 | 3.35E-06 | 0.007634158 |
| Obesity | endometrium | *ABCA5* | 3.768348186 | 1.55E-07 | 5.52E-07 | 0.000750921 |
| Obesity | endometrium | *MIA3* | 7.474732168 | 2.85E-09 | 4.29E-08 | 1.38E-05 |
| Obesity | endometrium | *NEIL2* | 7.947578682 | 1.32E-10 | 1.34E-08 | 6.40E-07 |
| Obesity | endometrium | *PGM5* | 9.070046423 | 6.55E-06 | 1.15E-05 | 0.031765378 |
| Obesity | endometrium | *CCSAP* | 4.108294306 | 1.58E-06 | 3.35E-06 | 0.007645391 |
| Obesity | endometrium | *GBP5* | 3.127324856 | 0.000179025 | 0.000258906 | 0.868627975 |
| Obesity | endometrium | *ATP5G3* | 5.451034483 | 2.72E-08 | 1.50E-07 | 0.000131756 |
| Obesity | endometrium | *LY96* | 2.751696561 | 8.71E-06 | 1.50E-05 | 0.042251423 |
| Obesity | endometrium | *PDE1C* | 2.583575066 | 1.53E-06 | 3.31E-06 | 0.007419226 |
| Obesity | endometrium | *XPC* | 10.89650217 | 1.01E-08 | 7.88E-08 | 4.90E-05 |
| Obesity | endometrium | *FLCN* | 3.228155914 | 1.16E-06 | 2.72E-06 | 0.005623922 |
| Obesity | endometrium | *OXNAD1* | 4.606644677 | 6.62E-08 | 2.88E-07 | 0.000321363 |
| Obesity | endometrium | *APCDD1* | 5.700963182 | 0.00021563 | 0.000309446 | 1 |
| Obesity | endometrium | *MPPE1* | 4.245610331 | 3.56E-06 | 6.56E-06 | 0.017251945 |
| Obesity | endometrium | *RAB6B* | 8.084698627 | 0.006625601 | 0.00816961 | 1 |
| Obesity | endometrium | *ANKRD40* | 7.135938927 | 7.58E-08 | 3.18E-07 | 0.000367741 |
| Obesity | endometrium | *CYP2U1* | 5.79055179 | 0.010033644 | 0.012119303 | 1 |
| Obesity | endometrium | *PROM2* | 9.916673202 | 0.001460403 | 0.001924464 | 1 |
| Obesity | endometrium | *AK9* | 3.519373955 | 1.29E-07 | 4.77E-07 | 0.000623792 |
| Obesity | endometrium | *MARCKS* | 3.508342048 | 8.00E-07 | 2.04E-06 | 0.003882756 |
| Obesity | endometrium | *MARVELD1* | 11.58290701 | 0.000927099 | 0.001244338 | 1 |

| | | | | | | |
|---|---|---|---|---|---|---|
| Obesity | endometrium | *GOLGA7B* | 21.63801371 | 1.14E-06 | 2.68E-06 | 0.005520286 |
| Obesity | endometrium | *TRMT44* | 9.48908314 | 3.76E-09 | 4.72E-08 | 1.82E-05 |
| Obesity | endometrium | *MOV10* | 13.64061611 | 3.51E-06 | 6.49E-06 | 0.01704029 |
| Obesity | endometrium | *HEATR3* | 2.807200231 | 1.02E-05 | 1.73E-05 | 0.049712162 |
| Obesity | endometrium | *CNOT8* | 13.25352774 | 5.56E-08 | 2.54E-07 | 0.000269882 |
| Obesity | endometrium | *LRGUK* | 3.130298788 | 0.026736624 | 0.031154203 | 1 |
| Obesity | endometrium | *SETD9* | 2.311563888 | 0.000392291 | 0.000544918 | 1 |
| Obesity | endometrium | *NUP205* | 3.689751951 | 5.50E-07 | 1.51E-06 | 0.002666764 |
| Obesity | endometrium | *ZKSCAN2* | -1.541492857 | 0.001349539 | 0.001786132 | 1 |
| Obesity | endometrium | *KDM8* | 4.741466185 | 1.20E-06 | 2.79E-06 | 0.005800874 |
| Obesity | endometrium | *ALS2CR12* | 9.67961005 | 1.27E-09 | 3.98E-08 | 6.18E-06 |
| Obesity | endometrium | *FZD7* | 5.055943169 | 2.38E-06 | 4.62E-06 | 0.011551964 |
| Obesity | endometrium | *DEPTOR* | 4.659394776 | 4.29E-08 | 2.09E-07 | 0.00020833 |
| Obesity | endometrium | *RRAGA* | 4.081280912 | 1.71E-06 | 3.51E-06 | 0.008289117 |
| Obesity | endometrium | *TMBIM4* | 8.766793133 | 1.75E-09 | 3.98E-08 | 8.50E-06 |
| Obesity | endometrium | *PSD3* | 5.958669768 | 7.58E-06 | 1.32E-05 | 0.036786954 |
| Obesity | endometrium | *UGT2B4* | 2.757951969 | 5.80E-05 | 8.91E-05 | 0.281392299 |
| Obesity | endometrium | *MMP16* | 1.607622476 | 0.041475124 | 0.04761886 | 1 |
| Obesity | endometrium | *KCNMA1* | 1.882265426 | 1.00E-05 | 1.70E-05 | 0.048712754 |
| Obesity | endometrium | *NDUFAF6* | 6.577277615 | 3.58E-09 | 4.68E-08 | 1.74E-05 |
| Obesity | endometrium | *DRAM2* | 4.764562637 | 3.18E-07 | 9.73E-07 | 0.001541188 |
| Obesity | endometrium | *C8orf37* | 3.522573302 | 1.68E-05 | 2.75E-05 | 0.081583602 |
| Obesity | endometrium | *PPEF2* | 9.106944152 | 0.001433719 | 0.001890841 | 1 |
| Obesity | endometrium | *ART3* | 6.459460632 | 4.10E-05 | 6.39E-05 | 0.199004931 |
| Obesity | endometrium | *WHAMM* | 3.251621975 | 2.79E-06 | 5.32E-06 | 0.013548264 |
| Obesity | endometrium | *N6AMT1* | 4.082549824 | 3.34E-06 | 6.22E-06 | 0.016196469 |
| Obesity | endometrium | *RWDD2B* | 2.997185511 | 0.000554874 | 0.000759878 | 1 |
| Obesity | endometrium | *CCT8* | 2.072081646 | 0.000157542 | 0.000229962 | 0.764395252 |
| Obesity | endometrium | *NAA11* | -5.233978809 | 0.007223101 | 0.008850123 | 1 |
| Obesity | endometrium | *SFXN2* | 3.008822415 | 0.000381829 | 0.000531603 | 1 |
| Obesity | endometrium | *TDRD9* | 9.022640587 | 8.20E-06 | 1.41E-05 | 0.039796479 |
| Obesity | endometrium | *RPL30* | 6.062232556 | 3.95E-09 | 4.82E-08 | 1.91E-05 |
| Obesity | endometrium | *SUPV3L1* | 13.94056644 | 2.09E-08 | 1.25E-07 | 0.00010149 |
| Obesity | endometrium | *HKDC1* | -3.534939387 | 0.000146555 | 0.000214764 | 0.711084909 |
| Obesity | endometrium | *ZDHHC5* | 8.043639433 | 1.29E-08 | 9.14E-08 | 6.27E-05 |
| Obesity | endometrium | *MED19* | -3.085851064 | 0.009014116 | 0.01094233 | 1 |
| Obesity | endometrium | *WDYHV1* | 2.768689649 | 0.013970664 | 0.016679542 | 1 |
| Obesity | endometrium | *ATAD2* | 9.669123785 | 1.39E-06 | 3.14E-06 | 0.006753415 |
| Obesity | endometrium | *FBRS* | 3.369655737 | 1.20E-06 | 2.79E-06 | 0.005808928 |
| Obesity | endometrium | *SASS6* | -3.048419275 | 0.009096708 | 0.011037066 | 1 |
| Obesity | endometrium | *ITGAD* | 7.880073127 | 2.53E-08 | 1.43E-07 | 0.000122944 |
| Obesity | endometrium | *VPS8* | 5.082494778 | 1.70E-08 | 1.09E-07 | 8.24E-05 |
| Obesity | endometrium | *MPV17L* | 6.193454644 | 3.69E-05 | 5.79E-05 | 0.17919628 |
| Obesity | endometrium | *RPUSD3* | 5.154698747 | 5.85E-07 | 1.59E-06 | 0.002840531 |
| Obesity | endometrium | *GHRL* | 13.65266743 | 5.76E-07 | 1.56E-06 | 0.002794525 |
| Obesity | endometrium | *NTAN1* | 7.312693497 | 0.001567965 | 0.002058936 | 1 |
| Obesity | endometrium | *NMNAT2* | 3.621205252 | 8.32E-07 | 2.10E-06 | 0.004038349 |
| Obesity | endometrium | *SLC6A1* | 12.57551798 | 9.93E-07 | 2.40E-06 | 0.004819557 |
| Obesity | endometrium | *FCHO2* | 4.170787436 | 1.85E-08 | 1.16E-07 | 8.98E-05 |
| Obesity | endometrium | *TIMP4* | 2.496701651 | 5.83E-05 | 8.95E-05 | 0.282968838 |

| | | | | | | |
|---|---|---|---|---|---|---|
| Obesity | endometrium | *NECAP2* | 3.784809319 | 5.00E-07 | 1.40E-06 | 0.002425506 |
| Obesity | endometrium | *STEAP2* | 1.744990094 | 0.000241554 | 0.000344813 | 1 |
| Obesity | endometrium | *CLDN12* | 5.504127243 | 3.97E-07 | 1.16E-06 | 0.001925104 |
| Obesity | endometrium | *MMP14* | 10.37487511 | 1.14E-08 | 8.46E-08 | 5.54E-05 |
| Obesity | endometrium | *GATAD1* | 3.333964679 | 4.19E-07 | 1.22E-06 | 0.002034635 |
| Obesity | endometrium | *TMED6* | 30.20335476 | 1.36E-09 | 3.98E-08 | 6.61E-06 |
| Obesity | endometrium | *CLEC18A* | -2.365042167 | 9.25E-09 | 7.43E-08 | 4.49E-05 |
| Obesity | endometrium | *DHRS4* | 4.031963344 | 1.51E-06 | 3.29E-06 | 0.007310609 |
| Obesity | endometrium | *AASDH* | 9.653710891 | 1.56E-10 | 1.46E-08 | 7.57E-07 |
| Obesity | endometrium | *ETS2* | 3.222603088 | 1.20E-08 | 8.67E-08 | 5.82E-05 |
| Obesity | endometrium | *LCA5L* | -1.762721551 | 0.001180939 | 0.00156941 | 1 |
| Obesity | endometrium | *MX1* | 3.607805369 | 1.43E-06 | 3.18E-06 | 0.006917992 |
| Obesity | endometrium | *SLC38A10* | 11.51400413 | 1.93E-06 | 3.88E-06 | 0.009372519 |
| Obesity | endometrium | *DGKI* | 2.30722421 | 3.51E-05 | 5.51E-05 | 0.170129342 |
| Obesity | endometrium | *SNX22* | -4.70804398 | 0.000661117 | 0.000897521 | 1 |
| Obesity | endometrium | *SLC34A2* | -51.71210701 | 0.003325411 | 0.004231549 | 1 |
| Obesity | endometrium | *AP3S2* | 14.25512827 | 2.67E-08 | 1.48E-07 | 0.000129664 |
| Obesity | endometrium | *SPPL3* | 7.387705578 | 1.15E-09 | 3.89E-08 | 5.60E-06 |
| Obesity | endometrium | *C12orf43* | -1.773030221 | 0.000309461 | 0.000435977 | 1 |
| Obesity | endometrium | *LDLRAP1* | 9.973914245 | 6.48E-06 | 1.14E-05 | 0.031423602 |
| Obesity | endometrium | *KRTCAP3* | 4.761511966 | 0.002283706 | 0.002954023 | 1 |
| Obesity | endometrium | *PAFAH2* | 6.071956451 | 7.08E-06 | 1.23E-05 | 0.03433927 |
| Obesity | endometrium | *EXTL1* | 57.96402204 | 1.71E-06 | 3.52E-06 | 0.008312321 |
| Obesity | endometrium | *WDR66* | 7.344254975 | 1.88E-05 | 3.05E-05 | 0.091087745 |
| Obesity | endometrium | *RHPN1* | 6.267228178 | 7.75E-05 | 0.000117673 | 0.376083811 |
| Obesity | endometrium | *EYA3* | 5.15970857 | 3.50E-08 | 1.80E-07 | 0.000169617 |
| Obesity | endometrium | *DZIP1L* | 10.47036822 | 5.05E-07 | 1.41E-06 | 0.002450053 |
| Obesity | endometrium | *ESYT3* | -3.590760203 | 0.009706227 | 0.011735513 | 1 |
| Obesity | endometrium | *CLSTN2* | 2.245954012 | 1.92E-06 | 3.85E-06 | 0.009303188 |
| Obesity | endometrium | *COLEC12* | 1.83753053 | 1.52E-06 | 3.31E-06 | 0.007381891 |
| Obesity | endometrium | *MITD1* | 10.74939834 | 5.94E-10 | 2.63E-08 | 2.88E-06 |
| Obesity | endometrium | *TSPAN33* | 6.210675272 | 1.37E-05 | 2.27E-05 | 0.066329515 |
| Obesity | endometrium | *FAM86C1* | -1.759321002 | 0.000259425 | 0.000368589 | 1 |
| Obesity | endometrium | *PPP1R9A* | 5.701115763 | 3.71E-08 | 1.89E-07 | 0.000180237 |
| Obesity | endometrium | *DYNC1I1* | 5.315945142 | 2.16E-08 | 1.28E-07 | 0.000104843 |
| Obesity | endometrium | *COPG2* | 1.858434807 | 0.001613247 | 0.002114391 | 1 |
| Obesity | endometrium | *ELK4* | 14.67620556 | 2.08E-09 | 4.00E-08 | 1.01E-05 |
| Obesity | endometrium | *DUSP23* | 9.663864845 | 1.80E-05 | 2.93E-05 | 0.087364661 |
| Obesity | endometrium | *RNF166* | 3.402456382 | 1.32E-05 | 2.20E-05 | 0.064269614 |
| Obesity | endometrium | *F11R* | 3.504209168 | 9.00E-06 | 1.54E-05 | 0.043671385 |
| Obesity | endometrium | *FGF17* | -2.07029083 | 0.000193065 | 0.000278216 | 0.936752572 |
| Obesity | endometrium | *CDA* | 2.508313928 | 0.012872408 | 0.01541385 | 1 |
| Obesity | endometrium | *SLC5A11* | -3.036100323 | 0.002311319 | 0.002986556 | 1 |
| Obesity | endometrium | *CCAR2* | -2.399264876 | 0.028169422 | 0.032745097 | 1 |
| Obesity | endometrium | *CDC42SE2* | 2.330650422 | 8.69E-07 | 2.18E-06 | 0.00421858 |
| Obesity | endometrium | *RAPGEF6* | 4.308996189 | 0.00014169 | 0.000208222 | 0.687480984 |
| Obesity | endometrium | *IFNAR2* | 2.183455193 | 3.37E-06 | 6.27E-06 | 0.016363186 |
| Obesity | endometrium | *SV2A* | -2.959231632 | 0.000237236 | 0.000339048 | 1 |
| Obesity | endometrium | *CLIC6* | -2.849983936 | 0.000937675 | 0.001257837 | 1 |
| Obesity | endometrium | *CBR1* | -2.697194896 | 0.004288758 | 0.005397939 | 1 |

| | | | | | | |
|---|---|---|---|---|---|---|
| Obesity | endometrium | *CBR3* | 5.429172095 | 8.22E-07 | 2.08E-06 | 0.003987322 |
| Obesity | endometrium | *CHAF1B* | 4.338841821 | 0.000738839 | 0.001000795 | 1 |
| Obesity | endometrium | *HLCS* | 4.844697059 | 4.78E-08 | 2.27E-07 | 0.000232043 |
| Obesity | endometrium | *ARHGAP27* | 10.84245493 | 0.000413939 | 0.000574001 | 1 |
| Obesity | endometrium | *IRX6* | 3.020193324 | 1.08E-05 | 1.82E-05 | 0.05244931 |
| Obesity | endometrium | *HK2* | 5.018994131 | 1.72E-07 | 6.00E-07 | 0.000835652 |
| Obesity | endometrium | *THEM4* | 2.891547998 | 1.99E-06 | 3.97E-06 | 0.009644127 |
| Obesity | endometrium | *AMFR* | 3.264805714 | 6.33E-07 | 1.70E-06 | 0.003073573 |
| Obesity | endometrium | *TMEM69* | 4.353673612 | 9.13E-07 | 2.25E-06 | 0.00442874 |
| Obesity | endometrium | *ACE* | 8.644307448 | 7.38E-08 | 3.13E-07 | 0.0003581 |
| Obesity | endometrium | *TEPP* | 13.79993885 | 1.55E-05 | 2.54E-05 | 0.075171584 |
| Obesity | endometrium | *CHCHD6* | 10.94354853 | 2.65E-08 | 1.47E-07 | 0.000128507 |
| Obesity | endometrium | *CTBP1* | 3.444242948 | 1.51E-06 | 3.29E-06 | 0.007334195 |
| Obesity | endometrium | *LRRC36* | 3.513804835 | 1.72E-06 | 3.53E-06 | 0.008337064 |
| Obesity | endometrium | *ZFYVE28* | 4.001860207 | 5.17E-07 | 1.44E-06 | 0.00251068 |
| Obesity | endometrium | *C16orf86* | 4.600686744 | 1.28E-05 | 2.14E-05 | 0.062289346 |
| Obesity | endometrium | *RGS12* | 5.527775359 | 8.25E-08 | 3.39E-07 | 0.00040012 |
| Obesity | endometrium | *LYPD5* | 7.120736757 | 2.83E-05 | 4.50E-05 | 0.137520519 |
| Obesity | endometrium | *CCDC107* | 10.45724058 | 5.22E-08 | 2.43E-07 | 0.000253172 |
| Obesity | endometrium | *ZNF233* | 8.341204979 | 3.21E-07 | 9.82E-07 | 0.001559116 |
| Obesity | endometrium | *SSU72* | 10.3779696 | 6.28E-09 | 6.12E-08 | 3.05E-05 |
| Obesity | endometrium | *CPAMD8* | 7.102531637 | 3.40E-06 | 6.32E-06 | 0.016502889 |
| Obesity | endometrium | *ANKLE1* | 9.574571365 | 0.02135711 | 0.025090726 | 1 |
| Obesity | endometrium | *CCDC58* | 17.52783364 | 2.22E-07 | 7.31E-07 | 0.001075621 |
| Obesity | endometrium | *SLC37A1* | 3.878076172 | 9.55E-07 | 2.33E-06 | 0.004633665 |
| Obesity | endometrium | *PDE9A* | 3.027110667 | 8.11E-07 | 2.06E-06 | 0.003935888 |
| Obesity | endometrium | *NDUFV3* | 2.52283193 | 8.35E-05 | 0.000126082 | 0.405100688 |
| Obesity | endometrium | *HSF2BP* | 5.40999495 | 0.042248987 | 0.048472945 | 1 |
| Obesity | endometrium | *CSTB* | 3.305891243 | 8.92E-07 | 2.22E-06 | 0.004329864 |
| Obesity | endometrium | *C21orf2* | -1.477218033 | 8.06E-05 | 0.000121969 | 0.391033373 |
| Obesity | endometrium | *LSS* | 4.127104844 | 3.94E-06 | 7.18E-06 | 0.019093627 |
| Obesity | endometrium | *PCNT* | 5.763275867 | 2.79E-05 | 4.44E-05 | 0.135280459 |
| Obesity | endometrium | *DIP2A* | 6.833264359 | 5.83E-09 | 5.92E-08 | 2.83E-05 |
| Obesity | endometrium | *PRMT2* | 6.969259783 | 1.71E-09 | 3.98E-08 | 8.32E-06 |
| Obesity | endometrium | *ZNF208* | 3.358231943 | 2.43E-07 | 7.87E-07 | 0.001179068 |
| Obesity | endometrium | *ADAMTS13* | -1.658535006 | 0.000759198 | 0.001026653 | 1 |
| Obesity | endometrium | *CACFD1* | 8.735026998 | 2.92E-06 | 5.52E-06 | 0.014178611 |
| Obesity | endometrium | *GPSM1* | 6.092323737 | 2.61E-08 | 1.46E-07 | 0.000126582 |
| Obesity | endometrium | *RDH13* | 7.096336402 | 0.00031461 | 0.00044246 | 1 |
| Obesity | endometrium | *TAOK1* | 3.797028875 | 7.14E-08 | 3.04E-07 | 0.000346378 |
| Obesity | endometrium | *S100A1* | 16.62112533 | 4.02E-07 | 1.18E-06 | 0.001951434 |
| Obesity | endometrium | *ZBTB7B* | 4.915026562 | 9.11E-07 | 2.25E-06 | 0.004421706 |
| Obesity | endometrium | *FDPS* | 5.12894619 | 1.13E-07 | 4.31E-07 | 0.000547798 |
| Obesity | endometrium | *CCR5* | 7.563866403 | 4.43E-06 | 7.98E-06 | 0.021486308 |
| Obesity | endometrium | *NBEAL2* | 6.693220468 | 9.88E-09 | 7.80E-08 | 4.79E-05 |
| Obesity | endometrium | *FCRL3* | 4.710905522 | 0.000816751 | 0.001101717 | 1 |
| Obesity | endometrium | *FGFR4* | 9.903761124 | 0.010061718 | 0.012147165 | 1 |
| Obesity | endometrium | *LY6K* | 8.231673896 | 0.000312754 | 0.000440283 | 1 |
| Obesity | endometrium | *IER2* | 5.526288512 | 7.11E-08 | 3.04E-07 | 0.000344754 |
| Obesity | endometrium | *VPS28* | 8.622989821 | 2.21E-05 | 3.57E-05 | 0.107469522 |

| | | | | | | |
|---|---|---|---|---|---|---|
| Obesity | endometrium | *TONSL* | 5.831961371 | 0.000349119 | 0.000488022 | 1 |
| Obesity | endometrium | *MUM1* | 5.3649343 | 2.99E-08 | 1.61E-07 | 0.000145297 |
| Obesity | endometrium | *ZNF333* | 3.31512461 | 9.79E-07 | 2.38E-06 | 0.004748166 |
| Obesity | endometrium | *SH2B2* | 6.476135426 | 3.17E-05 | 5.00E-05 | 0.153757027 |
| Obesity | endometrium | *RPL8* | 8.578330684 | 2.78E-05 | 4.42E-05 | 0.134729812 |
| Obesity | endometrium | *ABCF3* | 5.488051597 | 1.05E-08 | 8.03E-08 | 5.09E-05 |
| Obesity | endometrium | *PCYT1A* | 4.587541482 | 5.08E-05 | 7.85E-05 | 0.246624675 |
| Obesity | endometrium | *DMKN* | 1.140720583 | 0.003663277 | 0.004646855 | 1 |
| Obesity | endometrium | *LRRC56* | 7.696930943 | 0.000142102 | 0.000208709 | 0.689479404 |
| Obesity | endometrium | *ZNF577* | 6.769815409 | 4.17E-09 | 4.89E-08 | 2.02E-05 |
| Obesity | endometrium | *TMEM143* | 12.68345307 | 3.25E-09 | 4.48E-08 | 1.58E-05 |
| Obesity | endometrium | *CCL16* | -3.583022271 | 0.001078294 | 0.00143733 | 1 |
| Obesity | endometrium | *ALDH16A1* | 6.031126864 | 0.000164654 | 0.000239478 | 0.798899838 |
| Obesity | endometrium | *DCD* | -10.81256403 | 0.00445373 | 0.005595416 | 1 |
| Obesity | endometrium | *ASB16* | 3.822267694 | 2.34E-06 | 4.56E-06 | 0.011354265 |
| Obesity | endometrium | *DBF4B* | 1.919943273 | 0.019639962 | 0.023135007 | 1 |
| Obesity | endometrium | *PLCD3* | 20.50341508 | 2.36E-09 | 4.01E-08 | 1.14E-05 |
| Obesity | endometrium | *IP6K3* | 46.03269078 | 1.85E-05 | 3.01E-05 | 0.089692571 |
| Obesity | endometrium | *CXCL16* | 2.09159331 | 0.002206627 | 0.002858893 | 1 |
| Obesity | endometrium | *ASGR2* | 27.24271388 | 7.08E-07 | 1.85E-06 | 0.003433035 |
| Obesity | endometrium | *SNRNP25* | 8.02697169 | 2.04E-07 | 6.83E-07 | 0.000988328 |
| Obesity | endometrium | *WDR90* | 3.384759405 | 5.98E-05 | 9.15E-05 | 0.289933565 |
| Obesity | endometrium | *JMJD8* | 4.842333686 | 2.57E-07 | 8.23E-07 | 0.001247028 |
| Obesity | endometrium | *CCDC78* | 6.691758266 | 0.027231046 | 0.031692261 | 1 |
| Obesity | endometrium | *MEIOB* | 5.48939073 | 7.95E-07 | 2.03E-06 | 0.003857314 |
| Obesity | endometrium | *HS3ST6* | 9.517422552 | 1.85E-06 | 3.74E-06 | 0.008998731 |
| Obesity | endometrium | *SHANK2* | 3.134727316 | 1.37E-06 | 3.10E-06 | 0.006637024 |
| Obesity | endometrium | *ASRGL1* | 4.320595117 | 1.56E-06 | 3.33E-06 | 0.007553415 |
| Obesity | endometrium | *RPS6KA4* | 6.591848271 | 3.05E-07 | 9.44E-07 | 0.001479867 |
| Obesity | endometrium | *TPCN2* | 6.353923256 | 4.83E-08 | 2.29E-07 | 0.000234485 |
| Obesity | endometrium | *PDZK1IP1* | 6.321872125 | 1.79E-06 | 3.64E-06 | 0.008672554 |
| Obesity | endometrium | *CMPK1* | 7.241135968 | 2.64E-07 | 8.38E-07 | 0.001279915 |
| Obesity | endometrium | *COA7* | -2.661327472 | 0.002191499 | 0.00284081 | 1 |
| Obesity | endometrium | *SLC1A7* | 3.330780587 | 0.000368773 | 0.000514311 | 1 |
| Obesity | endometrium | *C1orf123* | 3.563363311 | 1.68E-06 | 3.48E-06 | 0.008158732 |
| Obesity | endometrium | *USP24* | 4.09061075 | 7.46E-08 | 3.15E-07 | 0.000362178 |
| Obesity | endometrium | *ZSWIM5* | 5.390367913 | 2.30E-08 | 1.34E-07 | 0.000111783 |
| Obesity | endometrium | *AK4* | 3.985042347 | 6.61E-07 | 1.76E-06 | 0.003209581 |
| Obesity | endometrium | *RBP7* | 10.20404867 | 9.00E-09 | 7.32E-08 | 4.37E-05 |
| Obesity | endometrium | *SLC25A34* | -1.326493821 | 0.000154527 | 0.00022571 | 0.749767198 |
| Obesity | endometrium | *PDPN* | 3.297162956 | 1.83E-06 | 3.70E-06 | 0.008863719 |
| Obesity | endometrium | *LAPTM5* | 2.761021727 | 1.52E-06 | 3.30E-06 | 0.007356774 |
| Obesity | endometrium | *SDC3* | 3.338883713 | 1.62E-06 | 3.40E-06 | 0.007846507 |
| Obesity | endometrium | *SYNC* | 3.026442118 | 2.21E-06 | 4.33E-06 | 0.010702842 |
| Obesity | endometrium | *CAMK2N1* | 7.945222238 | 4.03E-09 | 4.85E-08 | 1.95E-05 |
| Obesity | endometrium | *MEGF6* | 11.51192141 | 8.23E-08 | 3.39E-07 | 0.000399187 |
| Obesity | endometrium | *TM2D1* | 5.166493003 | 2.99E-08 | 1.61E-07 | 0.000145133 |
| Obesity | endometrium | *NEXN* | 2.874326991 | 6.01E-07 | 1.62E-06 | 0.002918048 |
| Obesity | endometrium | *LRRIQ3* | 3.055536081 | 1.72E-06 | 3.53E-06 | 0.008340052 |
| Obesity | endometrium | *TYW3* | 5.628073596 | 3.12E-07 | 9.60E-07 | 0.001512651 |

| | | | | | | |
|---|---|---|---|---|---|---|
| Obesity | endometrium | *WDR63* | 2.373608728 | 9.40E-06 | 1.60E-05 | 0.045595277 |
| Obesity | endometrium | *HFM1* | 10.3769132 | 9.37E-06 | 1.60E-05 | 0.045453325 |
| Obesity | endometrium | *ARPC5* | 6.631473385 | 1.25E-07 | 4.67E-07 | 0.000604912 |
| Obesity | endometrium | *ZNF496* | 14.58569429 | 6.63E-08 | 2.88E-07 | 0.000321451 |
| Obesity | endometrium | *FCRLB* | -2.981632218 | 0.000314304 | 0.000442159 | 1 |
| Obesity | endometrium | *KLHDC9* | 3.058162667 | 0.01588109 | 0.018867544 | 1 |
| Obesity | endometrium | *TDRD5* | -5.608245135 | 0.001204216 | 0.001598592 | 1 |
| Obesity | endometrium | *BROX* | 3.420120055 | 1.22E-05 | 2.04E-05 | 0.059113296 |
| Obesity | endometrium | *CNST* | 5.30803964 | 2.20E-08 | 1.30E-07 | 0.000106742 |
| Obesity | endometrium | *PM20D1* | 5.04252842 | 9.68E-08 | 3.83E-07 | 0.000469547 |
| Obesity | endometrium | *PKDCC* | 4.741954019 | 3.38E-07 | 1.02E-06 | 0.001639945 |
| Obesity | endometrium | *B3GALNT2* | 5.174249001 | 1.18E-07 | 4.48E-07 | 0.000574373 |
| Obesity | endometrium | *MRPL55* | 10.14042927 | 1.36E-08 | 9.47E-08 | 6.60E-05 |
| Obesity | endometrium | *KCNJ3* | -1.788450679 | 0.000127735 | 0.000188954 | 0.619769524 |
| Obesity | endometrium | *CLHC1* | 3.504662211 | 4.79E-09 | 5.33E-08 | 2.32E-05 |
| Obesity | endometrium | *DUSP19* | -4.064888707 | 0.013893363 | 0.016595421 | 1 |
| Obesity | endometrium | *C2orf48* | 36.03155121 | 7.74E-07 | 1.99E-06 | 0.003755961 |
| Obesity | endometrium | *CCDC74A* | 8.254951465 | 0.001635875 | 0.002142312 | 1 |
| Obesity | endometrium | *H3F3A* | 8.915965791 | 1.23E-06 | 2.84E-06 | 0.005959424 |
| Obesity | endometrium | *NOSTRIN* | 5.414736643 | 2.33E-07 | 7.61E-07 | 0.00112998 |
| Obesity | endometrium | *INHBB* | 6.471767761 | 2.13E-06 | 4.22E-06 | 0.010352472 |
| Obesity | endometrium | *RPRD2* | 7.203803669 | 4.79E-10 | 2.45E-08 | 2.32E-06 |
| Obesity | endometrium | *CTSS* | 1.494258464 | 0.00023488 | 0.000335978 | 1 |
| Obesity | endometrium | *PACRGL* | 5.762247362 | 3.54E-08 | 1.82E-07 | 0.000171594 |
| Obesity | endometrium | *C1QTNF7* | 5.517147198 | 1.50E-08 | 1.01E-07 | 7.26E-05 |
| Obesity | endometrium | *LYSMD1* | 3.43635794 | 0.024359323 | 0.028473003 | 1 |
| Obesity | endometrium | *VPS72* | 7.435431842 | 4.48E-08 | 2.16E-07 | 0.000217408 |
| Obesity | endometrium | *ERCC3* | 3.353235445 | 6.10E-07 | 1.64E-06 | 0.002958353 |
| Obesity | endometrium | *RNF149* | 3.509417221 | 2.61E-07 | 8.32E-07 | 0.00126406 |
| Obesity | endometrium | *DHX57* | 5.198944499 | 8.50E-08 | 3.48E-07 | 0.000412431 |
| Obesity | endometrium | *S100A9* | 8.281191391 | 1.56E-06 | 3.34E-06 | 0.007568116 |
| Obesity | endometrium | *TDRD10* | 2.658017668 | 1.60E-06 | 3.39E-06 | 0.007770312 |
| Obesity | endometrium | *DCAF16* | 6.661282612 | 2.72E-07 | 8.58E-07 | 0.001317898 |
| Obesity | endometrium | *GNPDA2* | 8.05442351 | 3.99E-09 | 4.83E-08 | 1.94E-05 |
| Obesity | endometrium | *NIPAL1* | 4.026501681 | 3.73E-06 | 6.86E-06 | 0.018118409 |
| Obesity | endometrium | *ANTXR2* | 9.022364331 | 1.74E-12 | 4.43E-10 | 8.42E-09 |
| Obesity | endometrium | *CGGBP1* | 3.391797226 | 8.72E-05 | 0.00013149 | 0.42300325 |
| Obesity | endometrium | *GPR155* | 4.112865068 | 2.53E-07 | 8.12E-07 | 0.00122873 |
| Obesity | endometrium | *DAPL1* | 8.987550789 | 2.92E-06 | 5.52E-06 | 0.014161432 |
| Obesity | endometrium | *PBXIP1* | 3.366655122 | 1.03E-06 | 2.48E-06 | 0.005005678 |
| Obesity | endometrium | *CLDN1* | 2.352008697 | 9.55E-07 | 2.33E-06 | 0.004634264 |
| Obesity | endometrium | *DCST2* | 7.4621922 | 3.29E-08 | 1.72E-07 | 0.000159423 |
| Obesity | endometrium | *LINC01116* | 15.82402356 | 7.36E-09 | 6.60E-08 | 3.57E-05 |
| Obesity | endometrium | *POGLUT1* | 3.940760783 | 9.82E-06 | 1.67E-05 | 0.04765077 |
| Obesity | endometrium | *SLC22A15* | 10.5925764 | 9.11E-09 | 7.37E-08 | 4.42E-05 |
| Obesity | endometrium | *LRRC58* | 10.58359002 | 2.72E-09 | 4.26E-08 | 1.32E-05 |
| Obesity | endometrium | *LMOD1* | 2.053487887 | 7.56E-05 | 0.00011486 | 0.366631866 |
| Obesity | endometrium | *TMEM169* | 4.963809529 | 4.30E-05 | 6.69E-05 | 0.208810468 |
| Obesity | endometrium | *ARPC2* | 8.438871559 | 2.23E-09 | 4.00E-08 | 1.08E-05 |
| Obesity | endometrium | *GLB1L* | 3.237473563 | 9.19E-05 | 0.000138462 | 0.44612419 |

| | | | | | | |
|---|---|---|---|---|---|---|
| Obesity | endometrium | *SERPINI1* | 12.33667124 | 9.89E-10 | 3.56E-08 | 4.80E-06 |
| Obesity | endometrium | *SUCLG1* | 10.86743837 | 1.46E-09 | 3.98E-08 | 7.08E-06 |
| Obesity | endometrium | *SPTA1* | -2.155814837 | 0.000106586 | 0.00015937 | 0.517155417 |
| Obesity | endometrium | *PRKCI* | 4.265134527 | 6.32E-06 | 1.11E-05 | 0.03067034 |
| Obesity | endometrium | *EFHB* | 2.223034332 | 2.51E-05 | 4.01E-05 | 0.12191918 |
| Obesity | endometrium | *GTPBP8* | 7.038143171 | 6.98E-08 | 3.00E-07 | 0.000338889 |
| Obesity | endometrium | *WDFY3* | 4.953608013 | 1.30E-08 | 9.16E-08 | 6.30E-05 |
| Obesity | endometrium | *PTPN13* | 3.533050192 | 1.09E-06 | 2.59E-06 | 0.005282661 |
| Obesity | endometrium | *C4orf36* | 9.717033082 | 0.000387272 | 0.000538408 | 1 |
| Obesity | endometrium | *PPM1K* | 5.949173834 | 1.88E-08 | 1.17E-07 | 9.14E-05 |
| Obesity | endometrium | *TIPARP* | -2.026985854 | 0.042489523 | 0.048737391 | 1 |
| Obesity | endometrium | *DCLK3* | 14.35954195 | 3.10E-05 | 4.89E-05 | 0.150339371 |
| Obesity | endometrium | *RPL9* | -1.196133919 | 0.021198026 | 0.024915897 | 1 |
| Obesity | endometrium | *RPP14* | 5.036174769 | 5.51E-07 | 1.51E-06 | 0.002674008 |
| Obesity | endometrium | *DNASE1L3* | -3.47775029 | 0.00363957 | 0.00461799 | 1 |
| Obesity | endometrium | *IL17RC* | 3.52988065 | 4.05E-06 | 7.36E-06 | 0.019632581 |
| Obesity | endometrium | *CRELD1* | 3.501486822 | 1.02E-06 | 2.45E-06 | 0.004955048 |
| Obesity | endometrium | *TTC14* | 6.396059709 | 2.37E-07 | 7.73E-07 | 0.001149886 |
| Obesity | endometrium | *RCHY1* | 3.472394862 | 1.02E-06 | 2.46E-06 | 0.004972586 |
| Obesity | endometrium | *CCDC158* | 6.137048439 | 6.54E-06 | 1.15E-05 | 0.031721743 |
| Obesity | endometrium | *SLC4A1AP* | 13.50633414 | 3.65E-06 | 6.72E-06 | 0.017697258 |
| Obesity | endometrium | *KIAA1143* | 5.018678437 | 1.08E-05 | 1.82E-05 | 0.052336863 |
| Obesity | endometrium | *CDCP1* | 3.195524848 | 1.22E-05 | 2.04E-05 | 0.059309645 |
| Obesity | endometrium | *CLEC3B* | 7.826270136 | 4.06E-07 | 1.19E-06 | 0.001968724 |
| Obesity | endometrium | *FYCO1* | 3.399476328 | 1.07E-05 | 1.81E-05 | 0.052022834 |
| Obesity | endometrium | *NMNAT3* | 4.973842056 | 1.68E-07 | 5.88E-07 | 0.000814623 |
| Obesity | endometrium | *SNIP1* | 4.611802296 | 4.72E-07 | 1.34E-06 | 0.002289289 |
| Obesity | endometrium | *RPN1* | 2.821597701 | 1.55E-06 | 3.32E-06 | 0.007506525 |
| Obesity | endometrium | *SENP2* | 7.818095731 | 3.74E-06 | 6.87E-06 | 0.01813218 |
| Obesity | endometrium | *IFT122* | 5.462334756 | 2.74E-07 | 8.65E-07 | 0.001331555 |
| Obesity | endometrium | *RFT1* | 2.225026985 | 0.001496864 | 0.001969302 | 1 |
| Obesity | endometrium | *SFMBT1* | 8.452792546 | 9.38E-09 | 7.47E-08 | 4.55E-05 |
| Obesity | endometrium | *UVSSA* | 6.594435809 | 3.94E-07 | 1.16E-06 | 0.00191084 |
| Obesity | endometrium | *FAM208A* | 5.604698477 | 1.95E-08 | 1.20E-07 | 9.48E-05 |
| Obesity | endometrium | *LRPAP1* | 5.588074268 | 7.30E-08 | 3.09E-07 | 0.000354031 |
| Obesity | endometrium | *PIGX* | 3.15120637 | 5.40E-05 | 8.33E-05 | 0.262206656 |
| Obesity | endometrium | *ABLIM2* | 4.474794131 | 3.86E-07 | 1.14E-06 | 0.001874403 |
| Obesity | endometrium | *METAP1* | 2.798256219 | 2.02E-06 | 4.03E-06 | 0.009813363 |
| Obesity | endometrium | *BDH2* | 9.89495674 | 3.08E-07 | 9.53E-07 | 0.001495431 |
| Obesity | endometrium | *CDC25A* | 18.02189885 | 4.85E-06 | 8.68E-06 | 0.023513115 |
| Obesity | endometrium | *ATRIP* | -2.082209923 | 5.91E-05 | 9.07E-05 | 0.286988988 |
| Obesity | endometrium | *RNF123* | 14.5296818 | 7.53E-24 | 3.66E-20 | 3.66E-20 |
| Obesity | endometrium | *MFSD8* | 8.593882571 | 1.33E-08 | 9.30E-08 | 6.46E-05 |
| Obesity | endometrium | *MST1R* | 9.31770433 | 3.85E-20 | 6.22E-17 | 1.87E-16 |
| Obesity | endometrium | *RAD54L2* | 8.371242652 | 2.90E-08 | 1.57E-07 | 0.000140648 |
| Obesity | endometrium | *TEX264* | 4.582660274 | 0.000240901 | 0.000343982 | 1 |
| Obesity | endometrium | *ETNPPL* | 28.53160457 | 1.59E-06 | 3.38E-06 | 0.007730766 |
| Obesity | endometrium | *PRSS12* | 3.549190871 | 0.023184792 | 0.027153868 | 1 |
| Obesity | endometrium | *HMGB2* | 9.916012579 | 7.06E-08 | 3.02E-07 | 0.000342427 |
| Obesity | endometrium | *ANXA5* | 3.444112906 | 1.10E-06 | 2.60E-06 | 0.005315598 |

| | | | | | | |
|---|---|---|---|---|---|---|
| Obesity | endometrium | *MAP9* | 5.557525002 | 2.65E-07 | 8.40E-07 | 0.001283565 |
| Obesity | endometrium | *HPGD* | -2.66746226 | 0.001809034 | 0.002360159 | 1 |
| Obesity | endometrium | *C4orf45* | 10.79200049 | 1.57E-08 | 1.04E-07 | 7.64E-05 |
| Obesity | endometrium | *TMEM144* | 2.938376532 | 4.02E-06 | 7.31E-06 | 0.019482466 |
| Obesity | endometrium | *IL15* | 3.974188492 | 9.99E-08 | 3.92E-07 | 0.000484824 |
| Obesity | endometrium | *ARFIP1* | 6.613436055 | 1.36E-09 | 3.98E-08 | 6.60E-06 |
| Obesity | endometrium | *LSM6* | 5.109608197 | 1.40E-06 | 3.15E-06 | 0.006814403 |
| Obesity | endometrium | *ITGA2* | 3.478221099 | 1.43E-06 | 3.19E-06 | 0.006938585 |
| Obesity | endometrium | *MOCS2* | 4.289361742 | 1.47E-06 | 3.23E-06 | 0.007113946 |
| Obesity | endometrium | *NDUFAF2* | 5.601469878 | 1.10E-08 | 8.30E-08 | 5.32E-05 |
| Obesity | endometrium | *LMBRD2* | 7.978458717 | 1.92E-07 | 6.53E-07 | 0.000929539 |
| Obesity | endometrium | *RNF180* | 3.620851277 | 7.20E-07 | 1.88E-06 | 0.003493149 |
| Obesity | endometrium | *PGGT1B* | 4.870643524 | 1.41E-07 | 5.13E-07 | 0.000686176 |
| Obesity | endometrium | *F2RL2* | 9.735015103 | 1.34E-06 | 3.06E-06 | 0.006508965 |
| Obesity | endometrium | *CCDC112* | 3.213218081 | 1.67E-06 | 3.47E-06 | 0.008088273 |
| Obesity | endometrium | *C5orf63* | 6.074175318 | 1.32E-07 | 4.86E-07 | 0.00063886 |
| Obesity | endometrium | *F2RL1* | 10.92127152 | 0.006026018 | 0.007462542 | 1 |
| Obesity | endometrium | *WDR41* | 11.04076009 | 2.56E-09 | 4.16E-08 | 1.24E-05 |
| Obesity | endometrium | *NDUFS4* | 6.782408188 | 0.000652165 | 0.000886112 | 1 |
| Obesity | endometrium | *SERINC5* | 3.643100402 | 1.79E-07 | 6.17E-07 | 0.000866372 |
| Obesity | endometrium | *CASP3* | 5.100674069 | 6.60E-07 | 1.76E-06 | 0.003202269 |
| Obesity | endometrium | *PRIMPOL* | 6.525002389 | 1.29E-08 | 9.12E-08 | 6.24E-05 |
| Obesity | endometrium | *ERAP1* | 3.304534893 | 5.34E-06 | 9.53E-06 | 0.025928343 |
| Obesity | endometrium | *ERAP2* | 3.765063632 | 5.18E-07 | 1.44E-06 | 0.002515246 |
| Obesity | endometrium | *UTP15* | -2.383069338 | 0.001129555 | 0.001503182 | 1 |
| Obesity | endometrium | *TLR3* | 13.27165735 | 3.96E-08 | 1.98E-07 | 0.000192013 |
| Obesity | endometrium | *KLKB1* | 2.083858382 | 0.007124857 | 0.008738576 | 1 |
| Obesity | endometrium | *NSA2* | 3.666445377 | 8.29E-07 | 2.09E-06 | 0.004020245 |
| Obesity | endometrium | *CCDC127* | 7.967044569 | 5.66E-07 | 1.54E-06 | 0.002747638 |
| Obesity | endometrium | *GRIK2* | 2.125409488 | 5.49E-05 | 8.46E-05 | 0.266489956 |
| Obesity | endometrium | *PDSS2* | 4.174778638 | 4.95E-08 | 2.32E-07 | 0.000240235 |
| Obesity | endometrium | *RAET1E* | 4.034854478 | 1.77E-06 | 3.60E-06 | 0.008564974 |
| Obesity | endometrium | *DAGLB* | 4.676232414 | 1.53E-05 | 2.51E-05 | 0.074218355 |
| Obesity | endometrium | *STK17A* | 7.877222609 | 8.89E-09 | 7.32E-08 | 4.31E-05 |
| Obesity | endometrium | *GALNT10* | 3.319378885 | 3.26E-06 | 6.09E-06 | 0.015826391 |
| Obesity | endometrium | *SAP30L* | 3.456490771 | 1.45E-06 | 3.21E-06 | 0.007050368 |
| Obesity | endometrium | *RPS14* | 9.671641055 | 2.74E-09 | 4.27E-08 | 1.33E-05 |
| Obesity | endometrium | *RP9* | 5.496817033 | 2.68E-07 | 8.49E-07 | 0.00129815 |
| Obesity | endometrium | *CAMLG* | 11.43892126 | 1.24E-06 | 2.86E-06 | 0.006014763 |
| Obesity | endometrium | *RELL2* | 5.529283332 | 0.013475762 | 0.016108499 | 1 |
| Obesity | endometrium | *MIOS* | 4.69704829 | 1.50E-07 | 5.39E-07 | 0.000727996 |
| Obesity | endometrium | *CHMP4C* | 11.71879645 | 1.53E-06 | 3.31E-06 | 0.007435908 |
| Obesity | endometrium | *SLC13A4* | 3.622405889 | 4.93E-07 | 1.39E-06 | 0.002390562 |
| Obesity | endometrium | *BRI3* | 10.04819562 | 6.33E-06 | 1.11E-05 | 0.030708453 |
| Obesity | endometrium | *CTSB* | 1.420078812 | 0.000288427 | 0.000407846 | 1 |
| Obesity | endometrium | *SOX17* | 5.906187474 | 4.00E-06 | 7.28E-06 | 0.019411009 |
| Obesity | endometrium | *TNFRSF11B* | 6.125742574 | 1.47E-06 | 3.24E-06 | 0.007146549 |
| Obesity | endometrium | *DEFB1* | 9.768247899 | 0.005873949 | 0.007283517 | 1 |
| Obesity | endometrium | *SUN1* | 9.16623359 | 1.52E-08 | 1.02E-07 | 7.40E-05 |
| Obesity | endometrium | *OXR1* | 7.833843161 | 1.05E-05 | 1.78E-05 | 0.051153058 |

| | | | | | | |
|---|---|---|---|---|---|---|
| Obesity | endometrium | *GPER1* | 10.21003493 | 2.30E-07 | 7.53E-07 | 0.001115047 |
| Obesity | endometrium | *INTS1* | 17.04638163 | 1.14E-07 | 4.34E-07 | 0.000554207 |
| Obesity | endometrium | *FASTK* | 8.499922467 | 0.010289005 | 0.012415382 | 1 |
| Obesity | endometrium | *ALDH7A1* | 8.686257798 | 2.07E-09 | 4.00E-08 | 1.00E-05 |
| Obesity | endometrium | *YWHAZ* | 6.712858155 | 1.22E-06 | 2.82E-06 | 0.005899272 |
| Obesity | endometrium | *SLC25A32* | 6.748720728 | 1.04E-07 | 4.06E-07 | 0.00050576 |
| Obesity | endometrium | *DCSTAMP* | -5.318326492 | 0.000178626 | 0.000258482 | 0.866691309 |
| Obesity | endometrium | *RPP25L* | -4.657149244 | 0.000130378 | 0.000192511 | 0.632591725 |
| Obesity | endometrium | *C9orf24* | 3.939695922 | 7.57E-07 | 1.96E-06 | 0.003673775 |
| Obesity | endometrium | *TMEM65* | 8.029587773 | 3.64E-09 | 4.70E-08 | 1.76E-05 |
| Obesity | endometrium | *CCDC171* | 10.16685992 | 5.80E-08 | 2.60E-07 | 0.000281626 |
| Obesity | endometrium | *METTL2B* | 4.498850212 | 2.97E-06 | 5.59E-06 | 0.014394796 |
| Obesity | endometrium | *ZMAT4* | 7.319510576 | 1.45E-06 | 3.21E-06 | 0.007041733 |
| Obesity | endometrium | *C8orf34* | 2.301730537 | 2.48E-05 | 3.97E-05 | 0.120488637 |
| Obesity | endometrium | *ANKS6* | 8.591304497 | 1.06E-10 | 1.17E-08 | 5.15E-07 |
| Obesity | endometrium | *AQP7* | 2.530473492 | 1.14E-06 | 2.68E-06 | 0.005528926 |
| Obesity | endometrium | *NOL6* | -2.177658394 | 0.033681222 | 0.038928368 | 1 |
| Obesity | endometrium | *AQP3* | 11.84638264 | 1.69E-06 | 3.50E-06 | 0.008214973 |
| Obesity | endometrium | *TRMT10B* | 4.955691772 | 6.78E-08 | 2.93E-07 | 0.000328841 |
| Obesity | endometrium | *VCP* | 6.456692813 | 1.83E-08 | 1.15E-07 | 8.86E-05 |
| Obesity | endometrium | *PIGO* | 4.953630266 | 3.50E-06 | 6.47E-06 | 0.016980079 |
| Obesity | endometrium | *WRN* | 5.324779424 | 1.09E-08 | 8.28E-08 | 5.29E-05 |
| Obesity | endometrium | *MARCH8* | 8.997405404 | 3.45E-09 | 4.57E-08 | 1.67E-05 |
| Obesity | endometrium | *TSHR* | 6.810532195 | 3.60E-08 | 1.84E-07 | 0.000174572 |
| Obesity | endometrium | *CFL2* | 16.96329268 | 7.27E-09 | 6.56E-08 | 3.53E-05 |
| Obesity | endometrium | *PGM2L1* | 3.950848182 | 4.82E-07 | 1.36E-06 | 0.002336706 |
| Obesity | endometrium | *SLC16A9* | 5.110324594 | 1.61E-06 | 3.39E-06 | 0.007793567 |
| Obesity | endometrium | *GJB2* | 22.23583972 | 1.46E-06 | 3.23E-06 | 0.007106974 |
| Obesity | endometrium | *CRYL1* | 7.202034239 | 6.11E-08 | 2.69E-07 | 0.000296363 |
| Obesity | endometrium | *REEP3* | 9.560291613 | 2.12E-07 | 7.05E-07 | 0.001028188 |
| Obesity | endometrium | *MICU2* | 9.061234396 | 2.08E-09 | 4.00E-08 | 1.01E-05 |
| Obesity | endometrium | *LRR1* | 24.05237845 | 2.75E-08 | 1.51E-07 | 0.000133405 |
| Obesity | endometrium | *TMEM63C* | -3.608207349 | 0.000296255 | 0.0004181 | 1 |
| Obesity | endometrium | *NOXRED1* | 4.63419864 | 1.67E-06 | 3.48E-06 | 0.008125011 |
| Obesity | endometrium | *AKR1E2* | 13.9314083 | 3.42E-06 | 6.35E-06 | 0.016594948 |
| Obesity | endometrium | *ATP5C1* | 8.5738177 | 4.65E-09 | 5.22E-08 | 2.26E-05 |
| Obesity | endometrium | *VDAC2* | 9.929685371 | 1.76E-07 | 6.10E-07 | 0.000853381 |
| Obesity | endometrium | *PDZD8* | 9.560726913 | 8.38E-09 | 7.13E-08 | 4.07E-05 |
| Obesity | endometrium | *QSOX2* | 4.316934438 | 1.67E-06 | 3.47E-06 | 0.008098781 |
| Obesity | endometrium | *CLEC1B* | -2.891123004 | 0.000192213 | 0.000277071 | 0.932619809 |
| Obesity | endometrium | *SNAPC4* | -2.498718298 | 0.040962823 | 0.04705294 | 1 |
| Obesity | endometrium | *TSC1* | 4.337874657 | 1.18E-08 | 8.63E-08 | 5.75E-05 |
| Obesity | endometrium | *STOX1* | 3.102065626 | 5.64E-07 | 1.54E-06 | 0.002737264 |
| Obesity | endometrium | *RET* | 5.606691522 | 1.46E-06 | 3.23E-06 | 0.007086455 |
| Obesity | endometrium | *CASP7* | 4.104314302 | 0.000424935 | 0.000588913 | 1 |
| Obesity | endometrium | *PPP1R36* | -3.792202952 | 0.03453508 | 0.039886743 | 1 |
| Obesity | endometrium | *VWA2* | 11.0834014 | 4.71E-07 | 1.34E-06 | 0.002283646 |
| Obesity | endometrium | *METTL3* | 14.47232049 | 1.92E-07 | 6.53E-07 | 0.000930077 |
| Obesity | endometrium | *TRUB1* | 3.085603745 | 2.20E-06 | 4.33E-06 | 0.010683526 |
| Obesity | endometrium | *HSPA12A* | 3.550041094 | 3.16E-05 | 4.99E-05 | 0.153312121 |

| | | | | | | |
|---|---|---|---|---|---|---|
| Obesity | endometrium | *UBTD1* | 12.37069855 | 1.83E-09 | 3.98E-08 | 8.89E-06 |
| Obesity | endometrium | *OTOGL* | 7.648704494 | 3.51E-06 | 6.49E-06 | 0.017042842 |
| Obesity | endometrium | *TTC7B* | 7.19107552 | 1.27E-08 | 9.04E-08 | 6.15E-05 |
| Obesity | endometrium | *RAPSN* | -5.828756304 | 0.000331129 | 0.000464201 | 1 |
| Obesity | endometrium | *TC2N* | 2.106653616 | 2.26E-06 | 4.44E-06 | 0.010985609 |
| Obesity | endometrium | *CPSF2* | 3.639921489 | 0.000223667 | 0.000320506 | 1 |
| Obesity | endometrium | *PTER* | 4.314979976 | 0.030020808 | 0.034847119 | 1 |
| Obesity | endometrium | *C1QL3* | -1.781927275 | 0.030441716 | 0.03528727 | 1 |
| Obesity | endometrium | *ARL5B* | 4.435239888 | 4.60E-07 | 1.31E-06 | 0.002231868 |
| Obesity | endometrium | *LIPC* | 4.787643146 | 1.07E-05 | 1.80E-05 | 0.051753932 |
| Obesity | endometrium | *CEP57* | 3.561260251 | 3.82E-07 | 1.13E-06 | 0.001852662 |
| Obesity | endometrium | *TCP11L2* | 7.230914392 | 8.08E-09 | 7.03E-08 | 3.92E-05 |
| Obesity | endometrium | *JAM3* | 5.401527266 | 9.95E-07 | 2.40E-06 | 0.004827068 |
| Obesity | endometrium | *IKBIP* | 6.797739383 | 1.29E-08 | 9.12E-08 | 6.25E-05 |
| Obesity | endometrium | *ZFYVE19* | 4.592485313 | 1.92E-07 | 6.55E-07 | 0.000933317 |
| Obesity | endometrium | *TRMT61A* | 7.204938387 | 3.62E-07 | 1.08E-06 | 0.001756938 |
| Obesity | endometrium | *LARP6* | 6.003040326 | 1.25E-07 | 4.68E-07 | 0.000607391 |
| Obesity | endometrium | *API5* | 8.632754237 | 4.05E-09 | 4.85E-08 | 1.96E-05 |
| Obesity | endometrium | *ASPG* | 24.39064564 | 2.49E-07 | 8.02E-07 | 0.001210297 |
| Obesity | endometrium | *GABRB3* | 2.937262588 | 0.001708641 | 0.002232783 | 1 |
| Obesity | endometrium | *COX11* | 11.78943574 | 9.35E-07 | 2.29E-06 | 0.00453429 |
| Obesity | endometrium | *STXBP4* | 9.001576541 | 6.51E-09 | 6.17E-08 | 3.16E-05 |
| Obesity | endometrium | *WBP1L* | 10.06050085 | 6.91E-10 | 2.89E-08 | 3.35E-06 |
| Obesity | endometrium | *APBB1* | 9.628118681 | 3.97E-07 | 1.16E-06 | 0.001924867 |
| Obesity | endometrium | *CYB5R2* | 4.85363958 | 0.000178983 | 0.000258906 | 0.868424885 |
| Obesity | endometrium | *XRRA1* | 7.671489364 | 8.08E-06 | 1.40E-05 | 0.039212344 |
| Obesity | endometrium | *RPL27A* | 11.57174688 | 2.97E-07 | 9.24E-07 | 0.001438911 |
| Obesity | endometrium | *CDYL2* | 2.89947707 | 0.000988499 | 0.00132163 | 1 |
| Obesity | endometrium | *CENPN* | 8.044394513 | 1.71E-06 | 3.52E-06 | 0.008303507 |
| Obesity | endometrium | *C16orf46* | 13.55980015 | 0.001399894 | 0.001850255 | 1 |
| Obesity | endometrium | *TMEM41B* | 5.095882757 | 7.78E-08 | 3.25E-07 | 0.000377447 |
| Obesity | endometrium | *TMX3* | 5.40070479 | 6.59E-08 | 2.86E-07 | 0.000319733 |
| Obesity | endometrium | *PRKCB* | 2.419446792 | 7.14E-07 | 1.87E-06 | 0.003465881 |
| Obesity | endometrium | *NDST2* | -3.837074438 | 0.000161577 | 0.000235215 | 0.783970755 |
| Obesity | endometrium | *CCDC68* | 7.781485012 | 4.21E-10 | 2.40E-08 | 2.04E-06 |
| Obesity | endometrium | *CLEC4D* | 9.690993365 | 0.013924585 | 0.01662862 | 1 |
| Obesity | endometrium | *ZSCAN21* | 2.273548233 | 0.00075994 | 0.001027369 | 1 |
| Obesity | endometrium | *RIMKLB* | 4.981259708 | 4.07E-05 | 6.34E-05 | 0.197628224 |
| Obesity | endometrium | *TMED3* | -1.344255527 | 0.007016964 | 0.008610599 | 1 |
| Obesity | endometrium | *SEC11C* | 3.698473615 | 1.38E-06 | 3.13E-06 | 0.006719757 |
| Obesity | endometrium | *FAM96B* | 6.972364388 | 0.003575439 | 0.004542558 | 1 |
| Obesity | endometrium | *CHRFAM7A* | 5.376743202 | 0.008243056 | 0.010041503 | 1 |
| Obesity | endometrium | *ZNF592* | 5.327274863 | 2.43E-07 | 7.87E-07 | 0.00117791 |
| Obesity | endometrium | *ACSM1* | 5.03900155 | 4.84E-06 | 8.68E-06 | 0.023506819 |
| Obesity | endometrium | *SLFN5* | 7.250003535 | 1.10E-08 | 8.30E-08 | 5.32E-05 |
| Obesity | endometrium | *CATSPER2* | 3.399268622 | 1.22E-06 | 2.84E-06 | 0.005939913 |
| Obesity | endometrium | *ZNF667-AS1* | 4.88124747 | 9.70E-07 | 2.36E-06 | 0.00470453 |
| Obesity | endometrium | *SAAL1* | 8.564349022 | 0.010522841 | 0.012688078 | 1 |
| Obesity | endometrium | *YPEL4* | 8.395830163 | 8.13E-06 | 1.40E-05 | 0.039425886 |
| Obesity | endometrium | *LDHC* | 3.301392824 | 0.00056265 | 0.000769875 | 1 |

| | | | | | | |
|---|---|---|---|---|---|---|
| Obesity | endometrium | *FAM96A* | 10.35096441 | 0.000159243 | 0.000232165 | 0.772645638 |
| Obesity | endometrium | *FAM111A* | 5.790577025 | 3.85E-06 | 7.04E-06 | 0.018660829 |
| Obesity | endometrium | *TMEM170A* | 8.316246912 | 0.005746862 | 0.007132266 | 1 |
| Obesity | endometrium | *ANPEP* | 2.167460788 | 0.002526566 | 0.003249973 | 1 |
| Obesity | endometrium | *DCTN5* | 4.020125757 | 8.73E-08 | 3.54E-07 | 0.000423764 |
| Obesity | endometrium | *STAT6* | 5.396514374 | 3.21E-08 | 1.70E-07 | 0.000155957 |
| Obesity | endometrium | *STX3* | 9.667488024 | 3.21E-07 | 9.82E-07 | 0.001559731 |
| Obesity | endometrium | *PIP4K2C* | 5.013184431 | 8.21E-08 | 3.38E-07 | 0.000398393 |
| Obesity | endometrium | *NYAP1* | 3.221592353 | 0.002760893 | 0.003541067 | 1 |
| Obesity | endometrium | *MS4A7* | 1.903468489 | 0.000116822 | 0.000173797 | 0.566819475 |
| Obesity | endometrium | *DIS3L* | 4.984212244 | 3.07E-07 | 9.50E-07 | 0.001490227 |
| Obesity | endometrium | *CCNDBP1* | 3.427376846 | 3.36E-07 | 1.02E-06 | 0.001631005 |
| Obesity | endometrium | *EPB42* | 6.082935946 | 1.32E-08 | 9.23E-08 | 6.39E-05 |
| Obesity | endometrium | *SMAD3* | 7.404145983 | 1.11E-08 | 8.35E-08 | 5.40E-05 |
| Obesity | endometrium | *RCCD1* | 4.873394584 | 1.15E-06 | 2.70E-06 | 0.005579747 |
| Obesity | endometrium | *CNPY4* | 5.228134904 | 2.43E-07 | 7.87E-07 | 0.00117928 |
| Obesity | endometrium | *MEI1* | 5.604332459 | 1.43E-07 | 5.18E-07 | 0.00069485 |
| Obesity | endometrium | *SAMD14* | 10.56356435 | 5.89E-07 | 1.59E-06 | 0.002859975 |
| Obesity | endometrium | *PIP5KL1* | 6.166593201 | 0.000221803 | 0.000318 | 1 |
| Obesity | endometrium | *ACSF2* | 3.552450719 | 1.41E-06 | 3.16E-06 | 0.006852672 |
| Obesity | endometrium | *URM1* | 6.25263301 | 3.16E-06 | 5.91E-06 | 0.015313047 |
| Obesity | endometrium | *ENDOG* | 5.348537325 | 3.83E-07 | 1.13E-06 | 0.001860621 |
| Obesity | endometrium | *ISLR2* | 9.886396752 | 4.62E-08 | 2.22E-07 | 0.000224362 |
| Obesity | endometrium | *SP2* | 13.39291356 | 1.73E-09 | 3.98E-08 | 8.39E-06 |
| Obesity | endometrium | *GPRC5B* | 3.276672083 | 6.52E-07 | 1.74E-06 | 0.003165455 |
| Obesity | endometrium | *FBXO22* | 2.623094916 | 1.40E-06 | 3.14E-06 | 0.006787857 |
| Obesity | endometrium | *TBC1D2B* | 7.127378733 | 5.31E-05 | 8.20E-05 | 0.257880591 |
| Obesity | endometrium | *LOXHD1* | 2.8795957 | 5.70E-07 | 1.55E-06 | 0.002765672 |
| Obesity | endometrium | *KATNAL2* | 2.957051513 | 1.90E-06 | 3.83E-06 | 0.009236739 |
| Obesity | endometrium | *ZNF91* | 5.448116894 | 1.11E-08 | 8.34E-08 | 5.38E-05 |
| Obesity | endometrium | *POP5* | 3.037913534 | 3.59E-06 | 6.63E-06 | 0.017442814 |
| Obesity | endometrium | *MYO5B* | 2.690708176 | 2.78E-06 | 5.30E-06 | 0.013490198 |
| Obesity | endometrium | *ZNF23* | 9.023512526 | 7.00E-06 | 1.22E-05 | 0.033955011 |
| Obesity | endometrium | *RAB8A* | 7.660291857 | 3.84E-07 | 1.13E-06 | 0.001863531 |
| Obesity | endometrium | *SPATA33* | 8.898423873 | 2.09E-08 | 1.25E-07 | 0.000101608 |
| Obesity | endometrium | *RPL13* | 6.220023342 | 8.58E-09 | 7.20E-08 | 4.16E-05 |
| Obesity | endometrium | *TP53I13* | 3.290493956 | 6.89E-07 | 1.82E-06 | 0.00334322 |
| Obesity | endometrium | *TUBA1C* | 5.81556072 | 8.06E-06 | 1.39E-05 | 0.039094667 |
| Obesity | endometrium | *TMC4* | 8.608708439 | 4.65E-08 | 2.23E-07 | 0.000225753 |
| Obesity | endometrium | *TTYH1* | 5.488060011 | 1.20E-06 | 2.81E-06 | 0.005843472 |
| Obesity | endometrium | *LENG8* | 7.809994667 | 4.57E-09 | 5.18E-08 | 2.22E-05 |
| Obesity | endometrium | *CDC42EP5* | -2.201150458 | 0.000659839 | 0.000896288 | 1 |
| Obesity | endometrium | *C19orf33* | 5.779614448 | 2.08E-05 | 3.36E-05 | 0.100773297 |
| Obesity | endometrium | *DNAAF3* | 3.298010127 | 0.006904658 | 0.008485664 | 1 |
| Obesity | endometrium | *FAM57A* | 5.81715394 | 1.05E-07 | 4.08E-07 | 0.000510065 |
| Obesity | endometrium | *SERPINF2* | 5.351453992 | 2.01E-05 | 3.25E-05 | 0.097382297 |
| Obesity | endometrium | *SRR* | 5.967388138 | 3.23E-11 | 5.31E-09 | 1.57E-07 |
| Obesity | endometrium | *TRPV3* | 5.146094663 | 2.98E-06 | 5.61E-06 | 0.014453681 |
| Obesity | endometrium | *CYB5D2* | 6.011021888 | 2.10E-08 | 1.25E-07 | 0.000102025 |
| Obesity | endometrium | *C19orf48* | 3.616244492 | 1.28E-06 | 2.95E-06 | 0.006221241 |

| | | | | | | |
|---|---|---|---|---|---|---|
| Obesity | endometrium | *KLK5* | -2.799214333 | 0.000552313 | 0.000756585 | 1 |
| Obesity | endometrium | *KLK6* | 32.29133835 | 1.52E-06 | 3.30E-06 | 0.007354085 |
| Obesity | endometrium | *ZNF83* | 6.127953909 | 2.63E-09 | 4.24E-08 | 1.28E-05 |
| Obesity | endometrium | *SPRYD3* | 4.39844206 | 5.97E-06 | 1.06E-05 | 0.028990246 |
| Obesity | endometrium | *SOAT2* | -0.641352282 | 6.91E-05 | 0.000105403 | 0.335496432 |
| Obesity | endometrium | *ZNF558* | 8.824414463 | 1.51E-07 | 5.43E-07 | 0.000734602 |
| Obesity | endometrium | *NDUFV1* | 12.02160546 | 1.72E-09 | 3.98E-08 | 8.36E-06 |
| Obesity | endometrium | *NUDT8* | -3.571610728 | 5.94E-05 | 9.10E-05 | 0.288122666 |
| Obesity | endometrium | *PRDX2* | 6.534321585 | 6.81E-08 | 2.94E-07 | 0.000330507 |
| Obesity | endometrium | *MIS12* | 5.113415497 | 1.74E-07 | 6.05E-07 | 0.00084575 |
| Obesity | endometrium | *MGAT5B* | -1.292211409 | 0.000188621 | 0.000272134 | 0.915187342 |
| Obesity | endometrium | *GSDMA* | 5.091302045 | 3.65E-05 | 5.73E-05 | 0.177139967 |
| Obesity | endometrium | *TMEM99* | 4.039074164 | 6.41E-06 | 1.13E-05 | 0.031114939 |
| Obesity | endometrium | *MLST8* | 8.958816689 | 4.16E-07 | 1.21E-06 | 0.002018738 |
| Obesity | endometrium | *ECI1* | 9.92731579 | 2.19E-08 | 1.29E-07 | 0.0001061 |
| Obesity | endometrium | *ABCA3* | 6.659591022 | 0.000865922 | 0.001165451 | 1 |
| Obesity | endometrium | *ATG16L2* | 3.30857256 | 6.97E-07 | 1.84E-06 | 0.003381625 |
| Obesity | endometrium | *RPSA* | 5.771162117 | 6.20E-08 | 2.73E-07 | 0.000301018 |
| Obesity | endometrium | *ULK4* | 4.663055074 | 0.004059693 | 0.005122921 | 1 |
| Obesity | endometrium | *LTBP3* | 2.728498174 | 1.55E-06 | 3.32E-06 | 0.007510649 |
| Obesity | endometrium | *NAALADL1* | -2.628377125 | 0.028262187 | 0.032845061 | 1 |
| Obesity | endometrium | *KIAA1586* | 4.177827371 | 2.90E-05 | 4.60E-05 | 0.140525259 |
| Obesity | endometrium | *FAM83B* | -8.60305949 | 0.000162903 | 0.000237004 | 0.790407741 |
| Obesity | endometrium | *POLR2J3* | 3.051054461 | 7.88E-05 | 0.000119414 | 0.382242757 |
| Obesity | endometrium | *NT5DC2* | -6.970300031 | 0.000268449 | 0.000380853 | 1 |
| Obesity | endometrium | *COA6* | 4.296113275 | 1.50E-06 | 3.27E-06 | 0.007266079 |
| Obesity | endometrium | *PDHB* | 11.34238537 | 4.45E-10 | 2.44E-08 | 2.16E-06 |
| Obesity | endometrium | *PXK* | 3.585138414 | 7.03E-07 | 1.84E-06 | 0.003408774 |
| Obesity | endometrium | *PCMTD1* | 4.168750782 | 4.92E-08 | 2.31E-07 | 0.000238627 |
| Obesity | endometrium | *ACOX2* | 3.26929283 | 2.82E-06 | 5.36E-06 | 0.013659791 |
| Obesity | endometrium | *FAM107A* | 13.00548899 | 0.001753386 | 0.002289405 | 1 |
| Obesity | endometrium | *SCN11A* | 3.199972013 | 8.26E-05 | 0.000124917 | 0.400983529 |
| Obesity | endometrium | *SEPT2* | 4.339181216 | 1.98E-08 | 1.21E-07 | 9.62E-05 |
| Obesity | endometrium | *ING5* | 5.809635136 | 8.18E-09 | 7.09E-08 | 3.97E-05 |
| Obesity | endometrium | *MLKL* | 5.325114685 | 0.000367678 | 0.000512931 | 1 |
| Obesity | endometrium | *KLHL30* | 14.89708491 | 5.23E-06 | 9.34E-06 | 0.02538894 |
| Obesity | endometrium | *CCDC110* | 11.42190572 | 3.57E-07 | 1.07E-06 | 0.001731715 |
| Obesity | endometrium | *FNTA* | 4.737023953 | 7.35E-09 | 6.60E-08 | 3.56E-05 |
| Obesity | endometrium | *SERINC2* | 8.385909019 | 0.00320306 | 0.00408121 | 1 |
| Obesity | endometrium | *SLC20A2* | 6.276429336 | 7.69E-09 | 6.76E-08 | 3.73E-05 |
| Obesity | endometrium | *TMUB2* | 2.86364066 | 2.82E-06 | 5.37E-06 | 0.013696047 |
| Obesity | endometrium | *ZSWIM1* | 2.663206481 | 0.000991245 | 0.001324936 | 1 |
| Obesity | endometrium | *NBPF9* | 8.730261872 | 1.12E-07 | 4.29E-07 | 0.000545082 |
| Obesity | endometrium | *ADAM9* | 3.649120772 | 2.47E-07 | 7.98E-07 | 0.001200339 |
| Obesity | endometrium | *NDUFS5* | 7.73102775 | 0.000879621 | 0.001183233 | 1 |
| Obesity | endometrium | *SLC16A4* | 5.665875046 | 8.81E-09 | 7.32E-08 | 4.28E-05 |
| Obesity | endometrium | *DNAJC21* | 7.94158783 | 1.10E-08 | 8.31E-08 | 5.33E-05 |
| Obesity | endometrium | *NPNT* | 3.056181812 | 3.00E-08 | 1.61E-07 | 0.000145387 |
| Obesity | endometrium | *GSTM4* | 4.622431075 | 6.77E-07 | 1.80E-06 | 0.003285848 |
| Obesity | endometrium | *PPIP5K1* | 3.380922014 | 1.64E-05 | 2.68E-05 | 0.079388836 |

| | | | | | | |
|---|---|---|---|---|---|---|
| Obesity | endometrium | *ADAL* | 4.431996403 | 8.89E-07 | 2.22E-06 | 0.004312282 |
| Obesity | endometrium | *SNTB2* | 6.640033305 | 6.69E-09 | 6.26E-08 | 3.25E-05 |
| Obesity | endometrium | *ZNF507* | 6.349797447 | 2.13E-07 | 7.06E-07 | 0.001031192 |
| Obesity | endometrium | *STX18* | 4.687605253 | 0.019864728 | 0.023394092 | 1 |
| Obesity | endometrium | *ZBTB49* | 8.16616424 | 3.90E-08 | 1.96E-07 | 0.000189422 |
| Obesity | endometrium | *GFM1* | 8.032911369 | 7.07E-09 | 6.45E-08 | 3.43E-05 |
| Obesity | endometrium | *ATOH8* | 3.360287522 | 2.47E-06 | 4.78E-06 | 0.011992744 |
| Obesity | endometrium | *BTNL3* | 5.47272692 | 5.09E-05 | 7.87E-05 | 0.247050061 |
| Obesity | endometrium | *TMEM129* | -2.15394884 | 0.006386014 | 0.007882203 | 1 |
| Obesity | endometrium | *CEP120* | 4.797466606 | 1.20E-07 | 4.53E-07 | 0.000583696 |
| Obesity | endometrium | *MFF* | 3.596249478 | 2.06E-07 | 6.87E-07 | 0.000997762 |
| Obesity | endometrium | *LGALS9* | 4.313725324 | 3.03E-06 | 5.70E-06 | 0.014693951 |
| Obesity | endometrium | *CPLX1* | 4.265622675 | 5.01E-08 | 2.34E-07 | 0.000243114 |
| Obesity | endometrium | *SIGLEC7* | 2.859458148 | 0.034361853 | 0.039696121 | 1 |
| Obesity | endometrium | *MFSD7* | 10.73568275 | 0.040269859 | 0.046289826 | 1 |
| Obesity | endometrium | *MAP2K1* | 4.327254004 | 1.11E-07 | 4.27E-07 | 0.000540606 |
| Obesity | endometrium | *KLK7* | 6.324887152 | 5.60E-07 | 1.53E-06 | 0.002719426 |
| Obesity | endometrium | *IRS1* | 4.360212844 | 7.12E-08 | 3.04E-07 | 0.000345547 |
| Obesity | endometrium | *UPF3A* | 4.265113486 | 7.01E-07 | 1.84E-06 | 0.003403604 |
| Obesity | endometrium | *C8orf46* | 2.986417628 | 2.37E-08 | 1.37E-07 | 0.000115203 |
| Obesity | endometrium | *HSPBAP1* | 2.367043139 | 8.58E-06 | 1.47E-05 | 0.041633009 |
| Obesity | endometrium | *UBE2V2* | 8.70746065 | 9.99E-09 | 7.86E-08 | 4.84E-05 |
| Obesity | endometrium | *CPT1C* | 4.972064895 | 2.41E-05 | 3.86E-05 | 0.11695278 |
| Obesity | endometrium | *PCSK9* | 23.67138523 | 9.93E-05 | 0.000149076 | 0.481666138 |
| Obesity | endometrium | *XPO6* | 9.778786858 | 2.67E-08 | 1.48E-07 | 0.000129428 |
| Obesity | endometrium | *RGS14* | -1.231629937 | 0.000521719 | 0.000715686 | 1 |
| Obesity | endometrium | *RAB24* | 8.841013141 | 3.78E-06 | 6.93E-06 | 0.018342151 |
| Obesity | endometrium | *THBS3* | 15.09946351 | 2.52E-10 | 1.94E-08 | 1.22E-06 |
| Obesity | endometrium | *NPIPB3* | 5.515602 | 0.001423018 | 0.00187826 | 1 |
| Obesity | endometrium | *SH3TC2* | 3.448446463 | 4.62E-05 | 7.16E-05 | 0.223932446 |
| Obesity | endometrium | *GPRIN1* | 21.19719508 | 3.18E-06 | 5.95E-06 | 0.01541191 |
| Obesity | endometrium | *PGM2* | 3.662590655 | 7.86E-07 | 2.01E-06 | 0.003811916 |
| Obesity | endometrium | *P2RY12* | 4.925366259 | 1.09E-06 | 2.60E-06 | 0.005291579 |
| Obesity | endometrium | *SNUPN* | 11.3749839 | 1.91E-10 | 1.72E-08 | 9.27E-07 |
| Obesity | endometrium | *RNASE6* | -1.624271884 | 0.00473278 | 0.005926052 | 1 |
| Obesity | endometrium | *CD52* | 2.770041099 | 6.61E-06 | 1.16E-05 | 0.032055602 |
| Obesity | endometrium | *SLC38A11* | -1.559750731 | 0.001841626 | 0.002399454 | 1 |
| Obesity | endometrium | *CCDC8* | 5.241706953 | 1.68E-05 | 2.74E-05 | 0.081437287 |
| Obesity | endometrium | *ZEB2* | 3.636499493 | 4.82E-07 | 1.36E-06 | 0.002340069 |
| Obesity | endometrium | *HINT1* | 10.20860998 | 3.76E-09 | 4.72E-08 | 1.83E-05 |
| Obesity | endometrium | *DTWD2* | 5.63970513 | 2.86E-07 | 8.95E-07 | 0.001385273 |
| Obesity | endometrium | *INO80E* | -6.270260074 | 0.000141341 | 0.000207813 | 0.685784401 |
| Obesity | endometrium | *DFFB* | -2.422591452 | 0.031338212 | 0.036298163 | 1 |
| Obesity | endometrium | *NFU1* | 11.19747433 | 3.26E-09 | 4.49E-08 | 1.58E-05 |
| Obesity | endometrium | *C15orf40* | 7.808927187 | 0.000127249 | 0.000188293 | 0.617411321 |
| Obesity | endometrium | *LUZP1* | 4.607606216 | 8.41E-08 | 3.45E-07 | 0.000408263 |
| Obesity | endometrium | *LRRC45* | 3.117689678 | 0.005554459 | 0.006901468 | 1 |
| Obesity | endometrium | *CHRNA5* | 10.9899819 | 3.87E-06 | 7.08E-06 | 0.018780807 |
| Obesity | endometrium | *UGP2* | 8.683054053 | 2.93E-10 | 2.11E-08 | 1.42E-06 |
| Obesity | endometrium | *HNRNPF* | 10.24923267 | 1.17E-08 | 8.55E-08 | 5.66E-05 |

| | | | | | | |
|---|---|---|---|---|---|---|
| Obesity | endometrium | *CSGALNACT2* | 8.083696501 | 4.49E-09 | 5.14E-08 | 2.18E-05 |
| Obesity | endometrium | *AVEN* | 5.353617757 | 0.014657756 | 0.017474061 | 1 |
| Obesity | endometrium | *CTNND2* | 2.921250249 | 0.000161848 | 0.000235538 | 0.78528447 |
| Obesity | endometrium | *CALML6* | 65.916601 | 5.12E-05 | 7.91E-05 | 0.248485068 |
| Obesity | endometrium | *MUC3A* | -1.990025062 | 0.041166277 | 0.04727545 | 1 |
| Obesity | endometrium | *TPST1* | 5.620228594 | 2.03E-07 | 6.80E-07 | 0.000983644 |
| Obesity | endometrium | *TOR1AIP2* | 6.072430384 | 3.63E-08 | 1.85E-07 | 0.000176072 |
| Obesity | endometrium | *OTUD3* | 4.99532025 | 0.007113185 | 0.008726466 | 1 |
| Obesity | endometrium | *ZFPM2* | 3.936616322 | 1.57E-08 | 1.04E-07 | 7.62E-05 |
| Obesity | endometrium | *MAP3K2* | 4.428663832 | 2.19E-08 | 1.29E-07 | 0.000106238 |
| Obesity | endometrium | *MYO7B* | 6.962109574 | 2.69E-08 | 1.48E-07 | 0.000130486 |
| Obesity | endometrium | *UBE2E3* | 9.748174144 | 2.14E-07 | 7.10E-07 | 0.001037919 |
| Obesity | endometrium | *NIPA1* | 3.512830736 | 9.25E-07 | 2.27E-06 | 0.004485731 |
| Obesity | endometrium | *HNRNPA3* | 7.120573805 | 6.51E-09 | 6.17E-08 | 3.16E-05 |
| Obesity | endometrium | *RNF150* | 6.376143352 | 0.000109351 | 0.000163303 | 0.530572861 |
| Obesity | endometrium | *USP38* | 8.985628312 | 9.80E-07 | 2.38E-06 | 0.004753082 |
| Obesity | endometrium | *ANKK1* | 5.840179356 | 0.016743719 | 0.019858354 | 1 |
| Obesity | endometrium | *PDCD6IP* | 5.150398885 | 1.97E-08 | 1.21E-07 | 9.57E-05 |
| Obesity | endometrium | *CRTAP* | 8.405879709 | 5.84E-08 | 2.61E-07 | 0.000283338 |
| Obesity | endometrium | *CNGB3* | 3.081067399 | 1.32E-07 | 4.86E-07 | 0.000639259 |
| Obesity | endometrium | *ELP5* | 7.727902854 | 0.002514869 | 0.003235785 | 1 |
| Obesity | endometrium | *STX8* | 3.483470765 | 1.46E-07 | 5.27E-07 | 0.00071022 |
| Obesity | endometrium | *NFRKB* | 7.388224288 | 4.09E-08 | 2.03E-07 | 0.000198469 |
| Obesity | endometrium | *ADORA2B* | -4.153882314 | 5.01E-05 | 7.74E-05 | 0.242887581 |
| Obesity | endometrium | *MGMT* | 6.101090139 | 5.87E-09 | 5.92E-08 | 2.85E-05 |
| Obesity | endometrium | *NFXL1* | 11.94512831 | 2.30E-09 | 4.01E-08 | 1.12E-05 |
| Obesity | endometrium | *DNAJC18* | 6.771383281 | 0.000874744 | 0.001176998 | 1 |
| Obesity | endometrium | *HSD17B13* | 9.129491205 | 1.43E-06 | 3.18E-06 | 0.00692891 |
| Obesity | endometrium | *ELOVL6* | 8.184238938 | 5.91E-09 | 5.95E-08 | 2.87E-05 |
| Obesity | endometrium | *ARL6IP1* | 7.727560849 | 0.000826944 | 0.001114847 | 1 |
| Obesity | endometrium | *SERPINB9* | 2.821260943 | 1.26E-06 | 2.90E-06 | 0.006116498 |
| Obesity | endometrium | *SMAGP* | -2.622816802 | 0.003849982 | 0.004869685 | 1 |
| Obesity | endometrium | *CDH2* | -1.123190046 | 0.000278654 | 0.000394868 | 1 |
| Obesity | endometrium | *EMB* | 2.474650303 | 9.28E-07 | 2.28E-06 | 0.004501723 |
| Obesity | endometrium | *STAT2* | 8.384279715 | 5.51E-09 | 5.78E-08 | 2.67E-05 |
| Obesity | endometrium | *SLC26A5* | 14.35496316 | 8.69E-09 | 7.26E-08 | 4.22E-05 |
| Obesity | endometrium | *GTSF1* | 10.84903754 | 0.00032048 | 0.000450193 | 1 |
| Obesity | endometrium | *ARMC10* | 10.30112333 | 1.11E-08 | 8.34E-08 | 5.38E-05 |
| Obesity | endometrium | *ACYP2* | 6.813690696 | 4.75E-10 | 2.45E-08 | 2.30E-06 |
| Obesity | endometrium | *TRABD* | 3.572060821 | 1.30E-06 | 2.98E-06 | 0.0062979 |
| Obesity | endometrium | *TMEM133* | 5.106278961 | 0.013099892 | 0.015674643 | 1 |
| Obesity | endometrium | *HOXB9* | 18.21752344 | 5.74E-06 | 1.02E-05 | 0.027848448 |
| Obesity | endometrium | *KCNS3* | -1.782094981 | 0.015353106 | 0.018280558 | 1 |
| Obesity | endometrium | *FOXN2* | 3.226489438 | 3.63E-07 | 1.09E-06 | 0.00176267 |
| Obesity | endometrium | *GPR27* | 5.056177268 | 0.001479805 | 0.001947915 | 1 |
| Obesity | endometrium | *TRIAP1* | 6.563254731 | 1.14E-06 | 2.68E-06 | 0.005513269 |
| Obesity | endometrium | *RPS9* | 5.88354449 | 0.000176505 | 0.000255643 | 0.856402548 |
| Obesity | endometrium | *TSEN34* | 5.592069951 | 6.16E-07 | 1.65E-06 | 0.002988816 |
| Obesity | endometrium | *MSANTD4* | 3.578971344 | 1.46E-06 | 3.22E-06 | 0.007075297 |
| Obesity | endometrium | *NDUFA3* | 15.61913061 | 1.29E-09 | 3.98E-08 | 6.26E-06 |

| | | | | | | |
|---|---|---|---|---|---|---|
| Obesity | endometrium | *NUDT6* | 7.428763347 | 1.54E-05 | 2.52E-05 | 0.074628477 |
| Obesity | endometrium | *ZNF160* | 2.830953587 | 4.86E-07 | 1.37E-06 | 0.002359812 |
| Obesity | endometrium | *ZNF415* | 11.30513552 | 1.47E-06 | 3.24E-06 | 0.007138077 |
| Obesity | endometrium | *PYGO1* | 4.375445475 | 3.21E-07 | 9.82E-07 | 0.001556952 |
| Obesity | endometrium | *TSNARE1* | 3.551758289 | 3.40E-08 | 1.76E-07 | 0.000164824 |
| Obesity | endometrium | *FPR1* | 3.61853744 | 2.13E-06 | 4.21E-06 | 0.010346652 |
| Obesity | endometrium | *FEZ2* | 5.603344244 | 2.41E-08 | 1.38E-07 | 0.000117046 |
| Obesity | endometrium | *SOX7* | -4.366012617 | 0.017063232 | 0.020232356 | 1 |
| Obesity | endometrium | *TRMT61B* | 11.30028708 | 5.93E-13 | 2.05E-10 | 2.88E-09 |
| Obesity | endometrium | *MFN1* | 10.9051893 | 3.92E-08 | 1.97E-07 | 0.000190237 |
| Obesity | endometrium | *GIMAP8* | 5.288577813 | 5.19E-08 | 2.42E-07 | 0.000252013 |
| Obesity | endometrium | *ATP6V0E2* | -2.958082082 | 0.000799465 | 0.001078999 | 1 |
| Obesity | endometrium | *MORN4* | 2.634866295 | 0.004824382 | 0.006037633 | 1 |
| Obesity | endometrium | *SCAND1* | 5.700650904 | 2.09E-06 | 4.15E-06 | 0.010162074 |
| Obesity | endometrium | *SOSTDC1* | 9.785455061 | 2.12E-05 | 3.43E-05 | 0.103079486 |
| Obesity | endometrium | *GAA* | 2.13242782 | 0.013124848 | 0.015700632 | 1 |
| Obesity | endometrium | *PGAM1* | 4.340310483 | 3.92E-05 | 6.12E-05 | 0.190022441 |
| Obesity | endometrium | *LURAP1* | -2.787068159 | 0.000216316 | 0.000310247 | 1 |
| Obesity | endometrium | *PDE7B* | 5.819425555 | 1.36E-08 | 9.46E-08 | 6.58E-05 |
| Obesity | endometrium | *MCC* | 3.006889057 | 3.24E-07 | 9.87E-07 | 0.00157181 |
| Obesity | endometrium | *ZNF561* | 3.367382779 | 0.032048594 | 0.037103263 | 1 |
| Obesity | endometrium | *HOPX* | 5.607036939 | 2.68E-05 | 4.27E-05 | 0.129871546 |
| Obesity | endometrium | *PPID* | 3.578747379 | 8.11E-07 | 2.06E-06 | 0.003934673 |
| Obesity | endometrium | *ETFDH* | 3.073011448 | 0.000636713 | 0.000865844 | 1 |
| Obesity | endometrium | *TBCA* | 10.16448826 | 2.19E-09 | 4.00E-08 | 1.06E-05 |
| Obesity | endometrium | *NMUR1* | 5.926679985 | 1.33E-06 | 3.03E-06 | 0.006428938 |
| Obesity | endometrium | *PIK3CD* | 4.557315541 | 2.88E-08 | 1.56E-07 | 0.000139509 |
| Obesity | endometrium | *RGS19* | 3.864065604 | 1.95E-08 | 1.20E-07 | 9.45E-05 |
| Obesity | endometrium | *ANO5* | 3.362175854 | 1.37E-06 | 3.10E-06 | 0.006648795 |
| Obesity | endometrium | *VAT1L* | 4.406560163 | 2.59E-06 | 4.99E-06 | 0.012584742 |
| Obesity | endometrium | *CAMTA1* | 6.506769465 | 1.04E-09 | 3.67E-08 | 5.03E-06 |
| Obesity | endometrium | *LGALS4* | 7.256234887 | 6.36E-07 | 1.70E-06 | 0.003086147 |
| Obesity | endometrium | *LRRC34* | 2.381587855 | 0.00159595 | 0.002092852 | 1 |
| Obesity | endometrium | *PAH* | 17.40546499 | 0.000149105 | 0.000218303 | 0.723457627 |
| Obesity | endometrium | *SPATA5L1* | 4.917540587 | 2.16E-07 | 7.15E-07 | 0.001047217 |
| Obesity | endometrium | *NXNL1* | -1.351520199 | 0.027773472 | 0.032300308 | 1 |
| Obesity | endometrium | *RHNO1* | 6.766939901 | 0.00056137 | 0.000768557 | 1 |
| Obesity | endometrium | *METTL18* | 2.12883242 | 0.001181878 | 0.001570228 | 1 |
| Obesity | endometrium | *PWWP2B* | -2.422526745 | 0.005336774 | 0.006646162 | 1 |
| Obesity | endometrium | *EXOSC10* | 8.854338067 | 1.79E-08 | 1.13E-07 | 8.67E-05 |
| Obesity | endometrium | *TRAPPC12* | 4.175815454 | 1.33E-07 | 4.88E-07 | 0.000646232 |
| Obesity | endometrium | *C3AR1* | 2.382732602 | 5.85E-05 | 8.98E-05 | 0.283816707 |
| Obesity | endometrium | *PRNP* | 5.672200291 | 1.99E-07 | 6.70E-07 | 0.000965839 |
| Obesity | endometrium | *FBXW10* | -1.467738733 | 0.000177213 | 0.000256515 | 0.859838119 |
| Obesity | endometrium | *SRGAP2C* | 2.593942338 | 1.43E-06 | 3.18E-06 | 0.006914545 |
| Obesity | endometrium | *ATPAF2* | 8.860675894 | 2.74E-05 | 4.36E-05 | 0.132723024 |
| Obesity | endometrium | *ZNF554* | -2.779801271 | 0.002872501 | 0.003678378 | 1 |
| Obesity | endometrium | *ANKRD20A4* | 21.18086426 | 6.79E-08 | 2.93E-07 | 0.000329224 |
| Obesity | endometrium | *EIF2AK3* | 5.555487581 | 5.23E-08 | 2.43E-07 | 0.000253793 |
| Obesity | endometrium | *NME6* | 4.875577565 | 2.12E-06 | 4.20E-06 | 0.01029982 |

| | | | | | | |
|---|---|---|---|---|---|---|
| Obesity | endometrium | *CD8B* | 3.366144303 | 0.00879001 | 0.01068365 | 1 |
| Obesity | endometrium | *SLFN12* | 2.825130852 | 1.35E-05 | 2.23E-05 | 0.065287254 |
| Obesity | endometrium | *TEFM* | 8.554473982 | 7.96E-05 | 0.000120543 | 0.385978917 |
| Obesity | endometrium | *MALT1* | 10.01514299 | 1.56E-09 | 3.98E-08 | 7.55E-06 |
| Obesity | endometrium | *MBOAT1* | 11.80828396 | 3.65E-09 | 4.70E-08 | 1.77E-05 |
| Obesity | endometrium | *CXCR6* | 18.4230943 | 5.95E-05 | 9.12E-05 | 0.288844856 |
| Obesity | endometrium | *PAIP1* | 7.911380083 | 2.25E-08 | 1.32E-07 | 0.000108933 |
| Obesity | endometrium | *C1QTNF4* | -5.598630643 | 9.23E-09 | 7.43E-08 | 4.48E-05 |
| Obesity | endometrium | *MACROD2* | 3.920348119 | 7.02E-09 | 6.44E-08 | 3.41E-05 |
| Obesity | endometrium | *BSG* | 5.014530991 | 0.018165572 | 0.021471219 | 1 |
| Obesity | endometrium | *HINFP* | 6.478481765 | 0.017160814 | 0.020338121 | 1 |
| Obesity | endometrium | *TP53RK* | 25.14949818 | 2.62E-08 | 1.46E-07 | 0.000127341 |
| Obesity | endometrium | *SUCLG2* | 9.933472369 | 5.29E-08 | 2.45E-07 | 0.00025678 |
| Obesity | endometrium | *ARNT2* | 5.797070501 | 1.64E-08 | 1.06E-07 | 7.94E-05 |
| Obesity | endometrium | *FGGY* | 7.045638321 | 0.000425108 | 0.000588985 | 1 |
| Obesity | endometrium | *MANEA* | 3.034268462 | 3.34E-06 | 6.22E-06 | 0.016189878 |
| Obesity | endometrium | *AFF1* | 5.904967768 | 3.86E-09 | 4.80E-08 | 1.87E-05 |
| Obesity | endometrium | *FIBP* | 5.387907245 | 4.35E-07 | 1.26E-06 | 0.002110567 |
| Obesity | endometrium | *CTSW* | -1.397032036 | 0.000128841 | 0.000190417 | 0.62513836 |
| Obesity | endometrium | *SMPDL3A* | 3.2043692 | 2.37E-06 | 4.60E-06 | 0.01147639 |
| Obesity | endometrium | *RAD9A* | 7.166238056 | 5.69E-07 | 1.55E-06 | 0.002761288 |
| Obesity | endometrium | *EFEMP2* | 3.820747532 | 1.19E-07 | 4.49E-07 | 0.00057649 |
| Obesity | endometrium | *ZFAND4* | 7.86327615 | 0.000229974 | 0.000329154 | 1 |
| Obesity | endometrium | *SLFN11* | 2.913801201 | 1.41E-06 | 3.16E-06 | 0.006842897 |
| Obesity | endometrium | *FAM71D* | 4.868547123 | 6.25E-06 | 1.10E-05 | 0.030310928 |
| Obesity | endometrium | *FUT10* | -2.932804169 | 0.000948057 | 0.00127036 | 1 |
| Obesity | endometrium | *ZNF596* | 4.80150784 | 4.49E-07 | 1.29E-06 | 0.002180715 |
| Obesity | endometrium | *CFL1* | 12.49460243 | 1.52E-08 | 1.02E-07 | 7.37E-05 |
| Obesity | endometrium | *FAM192A* | 8.945309677 | 3.86E-05 | 6.03E-05 | 0.187153318 |
| Obesity | endometrium | *RAB43* | -10.17435666 | 9.53E-05 | 0.000143227 | 0.462336252 |
| Obesity | endometrium | *CBWD1* | 9.821623839 | 1.29E-08 | 9.12E-08 | 6.25E-05 |
| Obesity | endometrium | *SNX32* | 4.302288643 | 1.00E-08 | 7.86E-08 | 4.87E-05 |
| Obesity | endometrium | *ZNF621* | 11.3605291 | 3.52E-09 | 4.65E-08 | 1.71E-05 |
| Obesity | endometrium | *ANKRD13D* | 2.546924545 | 2.58E-06 | 4.97E-06 | 0.012507822 |
| Obesity | endometrium | *MRGPRF* | 4.353066501 | 1.74E-08 | 1.11E-07 | 8.46E-05 |
| Obesity | endometrium | *SH3RF3* | 3.12604129 | 8.04E-07 | 2.05E-06 | 0.003902433 |
| Obesity | endometrium | *GXYLT2* | 10.22126052 | 3.52E-10 | 2.28E-08 | 1.71E-06 |
| Obesity | endometrium | *DCAKD* | 4.507001422 | 3.48E-07 | 1.05E-06 | 0.001686976 |
| Obesity | endometrium | *TRMT112* | -2.752869351 | 0.004450283 | 0.005592533 | 1 |
| Obesity | endometrium | *NOC3L* | -1.800024661 | 0.015606537 | 0.018559539 | 1 |
| Obesity | endometrium | *RHOD* | -2.910357668 | 0.010547088 | 0.012710997 | 1 |
| Obesity | endometrium | *PARP14* | 3.013027079 | 7.50E-07 | 1.94E-06 | 0.003641419 |
| Obesity | endometrium | *PARP15* | 2.835058163 | 9.03E-06 | 1.55E-05 | 0.04382906 |
| Obesity | endometrium | *AHSA2* | 5.028278417 | 1.07E-07 | 4.14E-07 | 0.000520119 |
| Obesity | endometrium | *IQCB1* | 5.175115131 | 9.59E-07 | 2.34E-06 | 0.004652059 |
| Obesity | endometrium | *ZNF483* | 8.273798791 | 1.16E-07 | 4.40E-07 | 0.000564167 |
| Obesity | endometrium | *SLC2A14* | 5.632270206 | 0.000344096 | 0.000481555 | 1 |
| Obesity | endometrium | *MMRN2* | 4.526255689 | 0.023800238 | 0.027839623 | 1 |
| Obesity | endometrium | *MZT2A* | 4.208470412 | 9.44E-07 | 2.31E-06 | 0.004580618 |
| Obesity | endometrium | *MAP3K11* | 8.651812734 | 3.81E-07 | 1.13E-06 | 0.001848482 |

| | | | | | | |
|---|---|---|---|---|---|---|
| Obesity | endometrium | *KCNK7* | 24.3879402 | 0.016173559 | 0.019200907 | 1 |
| Obesity | endometrium | *CCDC36* | 2.355298701 | 1.29E-05 | 2.15E-05 | 0.062828766 |
| Obesity | endometrium | *EHBP1L1* | 2.989127959 | 4.35E-06 | 7.85E-06 | 0.021113316 |
| Obesity | endometrium | *PPP1R14B* | 4.866214048 | 6.13E-06 | 1.08E-05 | 0.029719638 |
| Obesity | endometrium | *ZNF417* | 5.235776226 | 5.62E-05 | 8.64E-05 | 0.272545558 |
| Obesity | endometrium | *PEAK1* | 4.056191668 | 5.65E-08 | 2.56E-07 | 0.000274005 |
| Obesity | endometrium | *TNFRSF10D* | 8.951030837 | 2.54E-07 | 8.12E-07 | 0.001230013 |
| Obesity | endometrium | *GMPPB* | 6.713394431 | 1.90E-06 | 3.83E-06 | 0.009238692 |
| Obesity | endometrium | *NUDT18* | 10.30149797 | 1.80E-06 | 3.66E-06 | 0.008735373 |
| Obesity | endometrium | *SULT1B1* | 17.21181726 | 1.94E-05 | 3.15E-05 | 0.094172284 |
| Obesity | endometrium | *SCAI* | 5.55574207 | 1.04E-07 | 4.06E-07 | 0.000505814 |
| Obesity | endometrium | *HSPB7* | 3.637189412 | 0.000108235 | 0.000161687 | 0.525158397 |
| Obesity | endometrium | *UQCRH* | 12.22626817 | 1.93E-09 | 4.00E-08 | 9.36E-06 |
| Obesity | endometrium | *C11orf80* | 2.3608496 | 6.50E-06 | 1.14E-05 | 0.031545558 |
| Obesity | endometrium | *CD7* | 6.752408481 | 5.53E-07 | 1.52E-06 | 0.002685195 |
| Obesity | endometrium | *RNF213* | 5.555884328 | 2.50E-08 | 1.41E-07 | 0.000121154 |
| Obesity | endometrium | *TUBB8* | 6.757412293 | 2.92E-06 | 5.52E-06 | 0.014176528 |
| Obesity | endometrium | *GPR160* | 4.783418289 | 7.25E-06 | 1.26E-05 | 0.035191257 |
| Obesity | endometrium | *GOLIM4* | 2.710783969 | 1.10E-06 | 2.62E-06 | 0.005351536 |
| Obesity | endometrium | *UBXN2A* | 6.386754466 | 3.21E-09 | 4.46E-08 | 1.56E-05 |
| Obesity | endometrium | *NRROS* | 3.009009838 | 0.000616077 | 0.000838487 | 1 |
| Obesity | endometrium | *SLC25A30* | 4.268047971 | 8.31E-07 | 2.10E-06 | 0.004031595 |
| Obesity | endometrium | *CTSF* | 7.844696042 | 7.97E-08 | 3.32E-07 | 0.00038687 |
| Obesity | endometrium | *LEMD3* | 6.366229501 | 2.15E-08 | 1.27E-07 | 0.000104105 |
| Obesity | endometrium | *CTU2* | 6.492910084 | 4.00E-06 | 7.28E-06 | 0.019389367 |
| Obesity | endometrium | *GLIS1* | 3.566335981 | 1.70E-06 | 3.50E-06 | 0.008228602 |
| Obesity | endometrium | *RALGAPA1* | 4.656354241 | 3.22E-08 | 1.70E-07 | 0.000156397 |
| Obesity | endometrium | *SNAPC5* | 11.59792231 | 1.94E-05 | 3.15E-05 | 0.094236474 |
| Obesity | endometrium | *ANKRD36C* | 9.338833262 | 3.84E-05 | 6.01E-05 | 0.186462487 |
| Obesity | endometrium | *MSL2* | 7.769124932 | 5.43E-09 | 5.76E-08 | 2.64E-05 |
| Obesity | endometrium | *ZNF497* | 25.58359036 | 1.35E-05 | 2.24E-05 | 0.065494401 |
| Obesity | endometrium | *TRAM1L1* | 7.770970676 | 0.015709155 | 0.018676996 | 1 |
| Obesity | endometrium | *ANGEL2* | 4.110558136 | 1.43E-07 | 5.18E-07 | 0.000694366 |
| Obesity | endometrium | *ZNF266* | 2.824713061 | 6.10E-07 | 1.64E-06 | 0.002960292 |
| Obesity | endometrium | *SH3PXD2B* | 3.807748129 | 2.68E-07 | 8.50E-07 | 0.001301094 |
| Obesity | endometrium | *KIAA1551* | 4.946532768 | 4.22E-08 | 2.07E-07 | 0.000204792 |
| Obesity | endometrium | *LARP7* | 5.715568441 | 4.47E-08 | 2.16E-07 | 0.000217064 |
| Obesity | endometrium | *FGFBP3* | 6.942922571 | 1.48E-06 | 3.25E-06 | 0.007170717 |
| Obesity | endometrium | *C4orf32* | 10.42163714 | 1.04E-09 | 3.67E-08 | 5.06E-06 |
| Obesity | endometrium | *HRAS* | 5.447181746 | 3.69E-07 | 1.10E-06 | 0.001791715 |
| Obesity | endometrium | *SRP72* | 10.88298856 | 1.50E-09 | 3.98E-08 | 7.26E-06 |
| Obesity | endometrium | *NLRP6* | -12.51326395 | 1.77E-05 | 2.89E-05 | 0.086070077 |
| Obesity | endometrium | *RSRC1* | 7.1034942 | 3.07E-08 | 1.64E-07 | 0.000148902 |
| Obesity | endometrium | *PTDSS2* | 10.76070051 | 3.24E-09 | 4.48E-08 | 1.57E-05 |
| Obesity | endometrium | *SEZ6L2* | 5.811781649 | 2.96E-10 | 2.11E-08 | 1.44E-06 |
| Obesity | endometrium | *KCTD13* | 3.660539385 | 8.86E-09 | 7.32E-08 | 4.30E-05 |
| Obesity | endometrium | *DHX36* | 9.708365568 | 1.67E-09 | 3.98E-08 | 8.13E-06 |
| Obesity | endometrium | *FBXW8* | 12.42676914 | 1.80E-09 | 3.98E-08 | 8.74E-06 |
| Obesity | endometrium | *SLC22A1* | 6.291319446 | 2.06E-06 | 4.09E-06 | 0.00998475 |
| Obesity | endometrium | *CHST2* | 12.45845973 | 1.14E-05 | 1.92E-05 | 0.055464297 |

| | | | | | | |
|---|---|---|---|---|---|---|
| Obesity | endometrium | *TVP23C* | 6.077255991 | 1.52E-06 | 3.31E-06 | 0.007394456 |
| Obesity | endometrium | *PACS1* | 5.27450371 | 4.67E-07 | 1.33E-06 | 0.002268025 |
| Obesity | endometrium | *SH3BP5L* | 4.454599402 | 1.42E-06 | 3.17E-06 | 0.006866969 |
| Obesity | endometrium | *CADM2* | 2.817649649 | 3.22E-09 | 4.46E-08 | 1.56E-05 |
| Obesity | endometrium | *PSMD2* | 14.48139378 | 1.14E-08 | 8.43E-08 | 5.51E-05 |
| Obesity | endometrium | *FAM182B* | 4.093708732 | 0.0043071 | 0.005418213 | 1 |
| Obesity | endometrium | *ARHGAP1* | 5.506795374 | 8.61E-08 | 3.50E-07 | 0.000417532 |
| Obesity | endometrium | *ATG13* | 8.438794858 | 9.21E-09 | 7.43E-08 | 4.47E-05 |
| Obesity | endometrium | *CCNE2* | -1.805358002 | 0.00492837 | 0.006156655 | 1 |
| Obesity | endometrium | *ZNF519* | 4.041375654 | 1.36E-06 | 3.09E-06 | 0.006610889 |
| Obesity | endometrium | *BANF1* | 4.704412374 | 0.00032034 | 0.000450128 | 1 |
| Obesity | endometrium | *CHRNA7* | -2.499389552 | 0.000519212 | 0.000712449 | 1 |
| Obesity | endometrium | *TMEM9B* | 7.698846688 | 2.23E-08 | 1.31E-07 | 0.000108251 |
| Obesity | endometrium | *SCUBE2* | 3.706420868 | 3.55E-06 | 6.56E-06 | 0.017235986 |
| Obesity | endometrium | *EIF1AD* | 10.12021862 | 1.04E-08 | 8.01E-08 | 5.06E-05 |
| Obesity | endometrium | *EIF3F* | 5.573849825 | 1.82E-06 | 3.69E-06 | 0.008823548 |
| Obesity | endometrium | *ZNF25* | 7.522674056 | 0.000103992 | 0.000155875 | 0.504567413 |
| Obesity | endometrium | *PPP2R2D* | 2.604978525 | 5.32E-07 | 1.47E-06 | 0.002580025 |
| Obesity | endometrium | *MCTP1* | 1.935205141 | 1.52E-06 | 3.30E-06 | 0.007366625 |
| Obesity | endometrium | *LRRC25* | 3.41826107 | 0.000963397 | 0.001289135 | 1 |
| Obesity | endometrium | *UBQLNL* | -1.347616311 | 0.000109677 | 0.000163638 | 0.532152154 |
| Obesity | endometrium | *KCNE3* | 5.069603111 | 2.23E-05 | 3.59E-05 | 0.108113866 |
| Obesity | endometrium | *UCP2* | 2.817673026 | 9.11E-06 | 1.56E-05 | 0.044190481 |
| Obesity | endometrium | *C11orf68* | 5.883358937 | 1.42E-06 | 3.17E-06 | 0.00686807 |
| Obesity | endometrium | *PAAF1* | 2.260186624 | 0.00675212 | 0.008309178 | 1 |
| Obesity | endometrium | *MRPL48* | 6.067289949 | 1.79E-07 | 6.17E-07 | 0.000867109 |
| Obesity | endometrium | *RAB6A* | 5.767132173 | 1.52E-08 | 1.02E-07 | 7.37E-05 |
| Obesity | endometrium | *TMEM70* | 4.160671875 | 7.50E-07 | 1.94E-06 | 0.003637623 |
| Obesity | endometrium | *RPS6KB2* | -1.522727359 | 0.002167006 | 0.002810562 | 1 |
| Obesity | endometrium | *RMI2* | -2.388745263 | 0.00687851 | 0.008457813 | 1 |
| Obesity | endometrium | *TOM1L2* | 10.59227905 | 1.53E-09 | 3.98E-08 | 7.44E-06 |
| Obesity | endometrium | *TEX26* | 11.7656157 | 0.000845764 | 0.001139586 | 1 |
| Obesity | endometrium | *GPR156* | 9.0785683 | 0.006540834 | 0.008069191 | 1 |
| Obesity | endometrium | *TTLL11* | 12.68321354 | 2.96E-08 | 1.60E-07 | 0.000143413 |
| Obesity | endometrium | *CTDNEP1* | 8.099336557 | 1.49E-08 | 1.01E-07 | 7.21E-05 |
| Obesity | endometrium | *HOXD8* | 3.169071031 | 0.003141951 | 0.004007556 | 1 |
| Obesity | endometrium | *ZDHHC21* | 5.301742654 | 2.68E-08 | 1.48E-07 | 0.000129884 |
| Obesity | endometrium | *A2M* | 3.761573384 | 7.62E-08 | 3.19E-07 | 0.00036964 |
| Obesity | endometrium | *DOK7* | 28.41764465 | 1.64E-06 | 3.42E-06 | 0.007944852 |
| Obesity | endometrium | *ORAI3* | 20.12193097 | 5.33E-20 | 6.46E-17 | 2.59E-16 |
| Obesity | endometrium | *B3GALT6* | 8.585627741 | 1.53E-06 | 3.31E-06 | 0.007414911 |
| Obesity | endometrium | *ZNF613* | 11.42654292 | 2.88E-07 | 9.01E-07 | 0.001397225 |
| Obesity | endometrium | *JAKMIP2* | 3.26192703 | 5.97E-05 | 9.15E-05 | 0.289899426 |
| Obesity | endometrium | *TCP11L1* | 10.21673579 | 2.08E-08 | 1.25E-07 | 0.000100712 |
| Obesity | endometrium | *CCDC57* | 2.838437803 | 3.99E-05 | 6.23E-05 | 0.193663127 |
| Obesity | endometrium | *CIDEA* | 3.175547844 | 5.39E-07 | 1.49E-06 | 0.002613877 |
| Obesity | endometrium | *SMIM19* | 7.354877418 | 3.95E-09 | 4.82E-08 | 1.91E-05 |
| Obesity | endometrium | *ZNF404* | 2.977520962 | 7.22E-05 | 0.000109922 | 0.350542377 |
| Obesity | endometrium | *ZNF135* | 3.127828337 | 3.85E-05 | 6.02E-05 | 0.186849269 |
| Obesity | endometrium | *OR4N2* | 12.30656826 | 2.78E-06 | 5.30E-06 | 0.013469015 |

| | | | | | | |
|---|---|---|---|---|---|---|
| Obesity | endometrium | *TAC4* | 11.24856678 | 3.75E-09 | 4.72E-08 | 1.82E-05 |
| Obesity | endometrium | *CDC26* | -1.816950892 | 0.000951264 | 0.001274305 | 1 |
| Obesity | endometrium | *CRLF3* | 3.844337695 | 1.12E-07 | 4.29E-07 | 0.000544996 |
| Obesity | endometrium | *DNAJC30* | 5.241605358 | 9.43E-06 | 1.61E-05 | 0.045730152 |
| Obesity | endometrium | *SPRYD4* | 2.582286614 | 0.004173531 | 0.005259734 | 1 |
| Obesity | endometrium | *VPS37D* | 5.212970242 | 0.000216195 | 0.000310166 | 1 |
| Obesity | endometrium | *ZNF575* | 2.031478585 | 0.007262409 | 0.008893793 | 1 |
| Obesity | endometrium | *WDR25* | -1.939707442 | 0.035015199 | 0.040402793 | 1 |
| Obesity | endometrium | *PLA2G16* | 3.90218474 | 3.72E-06 | 6.85E-06 | 0.018051539 |
| Obesity | endometrium | *RMDN1* | 10.88892744 | 1.22E-10 | 1.31E-08 | 5.90E-07 |
| Obesity | endometrium | *RNF152* | 5.29980775 | 4.49E-07 | 1.28E-06 | 0.002177885 |
| Obesity | endometrium | *LRRC37A* | -1.807866696 | 0.004554848 | 0.005715057 | 1 |
| Obesity | endometrium | *BDNF* | 5.011160305 | 1.67E-06 | 3.47E-06 | 0.008102686 |
| Obesity | endometrium | *BOK* | 2.951643181 | 1.64E-06 | 3.42E-06 | 0.007938791 |
| Obesity | endometrium | *PFN4* | 2.933742349 | 0.00056415 | 0.000771276 | 1 |
| Obesity | endometrium | *RUFY1* | 6.493705255 | 6.49E-11 | 8.29E-09 | 3.15E-07 |
| Obesity | endometrium | *VSIG10* | 4.883510011 | 8.39E-08 | 3.44E-07 | 0.000407276 |
| Obesity | endometrium | *C8G* | 86.29600346 | 1.81E-06 | 3.68E-06 | 0.008803611 |
| Obesity | endometrium | *MUC20* | 5.795766482 | 7.40E-06 | 1.28E-05 | 0.035888334 |
| Obesity | endometrium | *LY6H* | 11.25093106 | 8.17E-08 | 3.37E-07 | 0.000396485 |
| Obesity | endometrium | *SHMT1* | 5.457447301 | 4.89E-08 | 2.30E-07 | 0.000237304 |
| Obesity | endometrium | *DPP7* | 8.838066968 | 5.58E-09 | 5.83E-08 | 2.71E-05 |
| Obesity | endometrium | *MTHFR* | 9.307071865 | 0.000244691 | 0.000349085 | 1 |
| Obesity | endometrium | *C19orf18* | 3.764612792 | 3.98E-06 | 7.26E-06 | 0.019323174 |
| Obesity | endometrium | *TMEM80* | 6.025110479 | 6.91E-06 | 1.21E-05 | 0.033511421 |
| Obesity | endometrium | *WDR73* | 7.926673023 | 0.000331217 | 0.000464201 | 1 |
| Obesity | endometrium | *DSCAML1* | 3.518548714 | 3.30E-07 | 1.00E-06 | 0.001602159 |
| Obesity | endometrium | *TALDO1* | 3.801169129 | 5.41E-07 | 1.49E-06 | 0.00262283 |
| Obesity | endometrium | *AP006621.1* | 16.62515198 | 6.35E-14 | 3.08E-11 | 3.08E-10 |
| Obesity | endometrium | *MAN1B1* | 6.276875618 | 9.19E-08 | 3.67E-07 | 0.000445749 |
| Obesity | endometrium | *SAMD9L* | 4.297804272 | 7.24E-08 | 3.08E-07 | 0.000351462 |
| Obesity | endometrium | *MIEF2* | 4.957847329 | 2.25E-06 | 4.41E-06 | 0.010915296 |
| Obesity | endometrium | *NAP1L5* | -1.718561019 | 0.000409269 | 0.000567688 | 1 |
| Obesity | endometrium | *NIM1K* | 7.804305845 | 7.56E-07 | 1.95E-06 | 0.003667311 |
| Obesity | endometrium | *RBM44* | 9.136985509 | 9.78E-06 | 1.66E-05 | 0.047472776 |
| Obesity | endometrium | *SLC25A22* | -1.675197786 | 0.000164841 | 0.00023968 | 0.799810749 |
| Obesity | endometrium | *GBA* | 18.70600645 | 1.68E-07 | 5.90E-07 | 0.000817489 |
| Obesity | endometrium | *ACAD9* | 3.170638236 | 1.29E-05 | 2.15E-05 | 0.062709738 |
| Obesity | endometrium | *IL17RA* | 3.559969594 | 6.83E-07 | 1.81E-06 | 0.003312658 |
| Obesity | endometrium | *MBOAT4* | 10.2684869 | 9.40E-06 | 1.60E-05 | 0.045612852 |
| Obesity | endometrium | *AGTRAP* | 3.465982327 | 6.30E-06 | 1.11E-05 | 0.030578098 |
| Obesity | endometrium | *THAP5* | 3.538761032 | 1.65E-06 | 3.45E-06 | 0.008028098 |
| Obesity | endometrium | *NAALADL2* | 5.873731894 | 4.26E-08 | 2.08E-07 | 0.00020655 |
| Obesity | endometrium | *CD151* | 4.297844862 | 3.25E-08 | 1.71E-07 | 0.000157879 |
| Obesity | endometrium | *SLC35G5* | 29.35830531 | 1.62E-06 | 3.41E-06 | 0.007869955 |
| Obesity | endometrium | *FAM87B* | 27.39332621 | 1.54E-06 | 3.32E-06 | 0.007457349 |
| Obesity | endometrium | *PCDHB9* | 5.882118695 | 0.000753555 | 0.001019729 | 1 |
| Obesity | endometrium | *CENPBD1* | 7.928519679 | 1.48E-06 | 3.25E-06 | 0.007186417 |
| Obesity | endometrium | *RPS27* | 9.608240059 | 1.65E-05 | 2.70E-05 | 0.080032037 |
| Obesity | endometrium | *RIC8A* | 4.344972602 | 3.30E-06 | 6.16E-06 | 0.016021204 |

| | | | | | | |
|---|---|---|---|---|---|---|
| Obesity | endometrium | *ODF3B* | 5.167635315 | 1.16E-06 | 2.71E-06 | 0.005616271 |
| Obesity | endometrium | *DPY19L2* | 4.585106461 | 1.44E-05 | 2.38E-05 | 0.070033566 |
| Obesity | endometrium | *ADAMTSL1* | 2.093224623 | 1.40E-06 | 3.14E-06 | 0.006789168 |
| Obesity | endometrium | *IMPDH2* | 4.226555901 | 1.36E-06 | 3.08E-06 | 0.006578411 |
| Obesity | endometrium | *GRAMD1C* | 3.290913896 | 3.43E-05 | 5.40E-05 | 0.166359617 |
| Obesity | endometrium | *DALRD3* | 5.20367473 | 9.06E-07 | 2.24E-06 | 0.004398059 |
| Obesity | endometrium | *ZNF114* | 8.581676914 | 0.000101847 | 0.000152754 | 0.4941599 |
| Obesity | endometrium | *SPINK6* | 9.410799266 | 0.009717351 | 0.011743111 | 1 |
| Obesity | endometrium | *ZNF454* | 3.400114256 | 2.85E-05 | 4.53E-05 | 0.138234172 |
| Obesity | endometrium | *SH2B1* | 9.95410066 | 2.03E-09 | 4.00E-08 | 9.86E-06 |
| Obesity | endometrium | *VN1R1* | 7.874974314 | 0.00269498 | 0.00345958 | 1 |
| Obesity | endometrium | *KDELC2* | 4.386770789 | 9.44E-07 | 2.31E-06 | 0.00458235 |
| Obesity | endometrium | *PLEC* | 3.686722105 | 5.31E-06 | 9.47E-06 | 0.025780742 |
| Obesity | endometrium | *SH2D4B* | 13.89341179 | 2.31E-06 | 4.49E-06 | 0.011190871 |
| Obesity | endometrium | *RNF212* | 10.39279093 | 3.07E-05 | 4.85E-05 | 0.148795523 |
| Obesity | endometrium | *PRSS36* | 6.637137931 | 9.44E-06 | 1.61E-05 | 0.045783071 |
| Obesity | endometrium | *WDR6* | 4.708869419 | 0.002513678 | 0.00323511 | 1 |
| Obesity | endometrium | *GEN1* | 3.022067593 | 0.011225614 | 0.0135086 | 1 |
| Obesity | endometrium | *TMPRSS9* | 8.49215422 | 3.49E-07 | 1.05E-06 | 0.001695677 |
| Obesity | endometrium | *ZFAND2A* | 5.115661554 | 2.54E-06 | 4.89E-06 | 0.012302908 |
| Obesity | endometrium | *FAM220A* | 5.499208999 | 8.62E-06 | 1.48E-05 | 0.041811876 |
| Obesity | endometrium | *NT5DC1* | 9.002148333 | 1.56E-09 | 3.98E-08 | 7.55E-06 |
| Obesity | endometrium | *GLDC* | 6.933958037 | 1.36E-06 | 3.08E-06 | 0.006584246 |
| Obesity | endometrium | *P4HTM* | 12.13435009 | 4.00E-05 | 6.24E-05 | 0.19414577 |
| Obesity | endometrium | *ERN1* | 9.212653358 | 5.70E-09 | 5.87E-08 | 2.77E-05 |
| Obesity | endometrium | *GPR35* | 2.953800636 | 0.006099605 | 0.007546582 | 1 |
| Obesity | endometrium | *PARP10* | 9.695981099 | 1.14E-07 | 4.33E-07 | 0.000551082 |
| Obesity | endometrium | *NSUN3* | 12.3159651 | 1.34E-07 | 4.89E-07 | 0.000648574 |
| Obesity | endometrium | *STX19* | 6.08136324 | 3.94E-10 | 2.34E-08 | 1.91E-06 |
| Obesity | endometrium | *MPI* | 4.755483346 | 1.06E-08 | 8.12E-08 | 5.17E-05 |
| Obesity | endometrium | *TRIM73* | -29.69680897 | 1.47E-10 | 1.40E-08 | 7.12E-07 |
| Obesity | endometrium | *EFCAB13* | 2.757498253 | 4.18E-06 | 7.58E-06 | 0.020281357 |
| Obesity | endometrium | *EXOSC4* | 10.25525114 | 0.023186001 | 0.027153868 | 1 |
| Obesity | endometrium | *ZNF852* | -5.357423489 | 0.007286119 | 0.008920578 | 1 |
| Obesity | endometrium | *HYI* | -2.247088423 | 0.000606577 | 0.000826021 | 1 |
| Obesity | endometrium | *C17orf62* | 3.485209185 | 1.24E-06 | 2.87E-06 | 0.006032336 |
| Obesity | endometrium | *GAK* | 3.44098229 | 4.64E-07 | 1.32E-06 | 0.002251738 |
| Obesity | endometrium | *TUFM* | 7.033517462 | 2.29E-12 | 5.55E-10 | 1.11E-08 |
| Obesity | endometrium | *C3orf38* | 7.467577479 | 1.39E-12 | 3.98E-10 | 6.76E-09 |
| Obesity | endometrium | *CCDC89* | 13.33031179 | 1.66E-06 | 3.45E-06 | 0.00805466 |
| Obesity | endometrium | *HTR1F* | 11.62500095 | 8.25E-05 | 0.000124798 | 0.40047718 |
| Obesity | endometrium | *SPTY2D1* | 11.64168274 | 2.99E-09 | 4.29E-08 | 1.45E-05 |
| Obesity | endometrium | *ZNF664* | 9.678156702 | 0.003234682 | 0.004119338 | 1 |
| Obesity | endometrium | *CDH4* | 5.926726448 | 4.47E-06 | 8.04E-06 | 0.021688704 |
| Obesity | endometrium | *HLA-DQB1* | 2.01330748 | 0.000581501 | 0.00079337 | 1 |
| Obesity | endometrium | *LINC00174* | 3.287264784 | 0.001078121 | 0.00143733 | 1 |
| Obesity | endometrium | *GEMIN4* | 5.633073613 | 4.44E-07 | 1.27E-06 | 0.002152013 |
| Obesity | endometrium | *LBX2* | 5.189564721 | 0.000458043 | 0.000632628 | 1 |
| Obesity | endometrium | *ZFPM1* | 5.100282018 | 1.99E-07 | 6.70E-07 | 0.000965423 |
| Obesity | endometrium | *FCER1A* | 4.171866499 | 0.001405387 | 0.001857009 | 1 |

| | | | | | | |
|---|---|---|---|---|---|---|
| Obesity | endometrium | *RPRML* | 34.59800063 | 7.51E-08 | 3.16E-07 | 0.000364516 |
| Obesity | endometrium | *SERTAD2* | 2.896822663 | 5.55E-07 | 1.52E-06 | 0.002691408 |
| Obesity | endometrium | *AKAP5* | 8.447725273 | 2.19E-07 | 7.24E-07 | 0.001062591 |
| Obesity | endometrium | *GIPC3* | 21.26016293 | 1.39E-06 | 3.14E-06 | 0.006759355 |
| Obesity | endometrium | *PDXDC1* | 6.016706964 | 7.48E-09 | 6.64E-08 | 3.63E-05 |
| Obesity | endometrium | *C1orf194* | -1.216950698 | 8.50E-05 | 0.000128373 | 0.412591027 |
| Obesity | endometrium | *SEPHS2* | 8.545611005 | 0.011614987 | 0.013956394 | 1 |
| Obesity | endometrium | *SEPT1* | 4.065268438 | 1.36E-06 | 3.08E-06 | 0.006583106 |
| Obesity | endometrium | *EXOC3* | 9.704192164 | 0.000251014 | 0.000357475 | 1 |
| Obesity | endometrium | *TDRD6* | 1.982782102 | 0.004885483 | 0.006110947 | 1 |
| Obesity | endometrium | *CSNK1A1L* | -19.04491837 | 0.011284128 | 0.013575648 | 1 |
| Obesity | endometrium | *LYNX1* | 5.182928665 | 4.38E-06 | 7.90E-06 | 0.021252535 |
| Obesity | endometrium | *HCLS1* | 2.590841939 | 8.76E-06 | 1.50E-05 | 0.042493851 |
| Obesity | endometrium | *MTURN* | 10.8783346 | 3.47E-08 | 1.79E-07 | 0.000168415 |
| Obesity | endometrium | *CCDC66* | 7.343410758 | 4.18E-09 | 4.89E-08 | 2.03E-05 |
| Obesity | endometrium | *DEFB124* | 27.60748729 | 1.76E-06 | 3.59E-06 | 0.008515444 |
| Obesity | endometrium | *DEFB119* | 63.72386081 | 4.09E-06 | 7.42E-06 | 0.01982966 |
| Obesity | endometrium | *TSPYL5* | -1.644653316 | 0.000506772 | 0.00069656 | 1 |
| Obesity | endometrium | *MB21D2* | 11.97321535 | 1.36E-10 | 1.35E-08 | 6.60E-07 |
| Obesity | endometrium | *SLC47A2* | 4.044791487 | 2.90E-06 | 5.49E-06 | 0.014066913 |
| Obesity | endometrium | *CHST13* | 33.61900382 | 1.21E-06 | 2.82E-06 | 0.005878957 |
| Obesity | endometrium | *SLC36A4* | 12.13309592 | 2.89E-09 | 4.29E-08 | 1.40E-05 |
| Obesity | endometrium | *ZDHHC20* | 6.073416419 | 1.32E-08 | 9.23E-08 | 6.39E-05 |
| Obesity | endometrium | *OR51E1* | 4.16887812 | 0.002402359 | 0.003099241 | 1 |
| Obesity | endometrium | *PSMG4* | 8.859746015 | 4.40E-08 | 2.13E-07 | 0.000213376 |
| Obesity | endometrium | *BHLHE22* | 5.180115458 | 0.002373085 | 0.003064735 | 1 |
| Obesity | endometrium | *ZNF443* | 12.15191704 | 1.17E-05 | 1.97E-05 | 0.056894425 |
| Obesity | endometrium | *CAPS2* | 3.461079899 | 1.63E-07 | 5.76E-07 | 0.000790642 |
| Obesity | endometrium | *KCTD2* | -2.176110167 | 0.024086887 | 0.028168131 | 1 |
| Obesity | endometrium | *D2HGDH* | 6.704789809 | 3.25E-06 | 6.06E-06 | 0.015756079 |
| Obesity | endometrium | *CMTR2* | 6.824231413 | 0.002687581 | 0.003451599 | 1 |
| Obesity | endometrium | *ST20* | -1.315679134 | 0.000313251 | 0.000440804 | 1 |
| Obesity | endometrium | *MRPL14* | 11.08648433 | 0.000182324 | 0.000263363 | 0.884634912 |
| Obesity | endometrium | *LSMEM1* | 3.912821188 | 6.66E-06 | 1.17E-05 | 0.032331949 |
| Obesity | endometrium | *NQO1* | 2.745920374 | 3.78E-05 | 5.92E-05 | 0.183183555 |
| Obesity | endometrium | *FKRP* | 3.458366827 | 6.55E-08 | 2.85E-07 | 0.000317829 |
| Obesity | endometrium | *RPH3AL* | 2.662315539 | 9.12E-07 | 2.25E-06 | 0.004423303 |
| Obesity | endometrium | *ADIPOQ* | 2.523923701 | 1.21E-05 | 2.02E-05 | 0.058469663 |
| Obesity | endometrium | *F2R* | -1.923323826 | 0.001681708 | 0.002198773 | 1 |
| Obesity | endometrium | *MUC16* | 3.090773974 | 1.09E-06 | 2.60E-06 | 0.005299542 |
| Obesity | endometrium | *PENK* | 8.398794966 | 0.02148129 | 0.025230506 | 1 |
| Obesity | endometrium | *SLC25A41* | 18.52039766 | 6.75E-07 | 1.79E-06 | 0.003274677 |
| Obesity | endometrium | *TMEM136* | 4.414289058 | 1.93E-06 | 3.88E-06 | 0.009387173 |
| Obesity | endometrium | *ZNF322* | 6.462432766 | 0.00051015 | 0.000700609 | 1 |
| Obesity | endometrium | *DDX60L* | 8.294999036 | 3.38E-09 | 4.54E-08 | 1.64E-05 |
| Obesity | endometrium | *OGFOD3* | 5.434077576 | 0.007397897 | 0.009046018 | 1 |
| Obesity | endometrium | *TMEM45A* | -1.39985404 | 0.00112397 | 0.00149616 | 1 |
| Obesity | endometrium | *SGSH* | 6.608890726 | 3.56E-07 | 1.07E-06 | 0.001725981 |
| Obesity | endometrium | *ZFP41* | 10.34854355 | 4.09E-09 | 4.87E-08 | 1.99E-05 |
| Obesity | endometrium | *ZBTB20* | 4.388342023 | 7.14E-09 | 6.50E-08 | 3.46E-05 |

| | | | | | | |
|---|---|---|---|---|---|---|
| Obesity | endometrium | *C3orf58* | 6.366204931 | 5.61E-08 | 2.55E-07 | 0.000272171 |
| Obesity | endometrium | *TMEM252* | -28.17276156 | 0.000247856 | 0.000353394 | 1 |
| Obesity | endometrium | *ODF3L2* | 8.245979586 | 0.011415276 | 0.013726622 | 1 |
| Obesity | endometrium | *RELL1* | 8.610440809 | 4.71E-10 | 2.45E-08 | 2.28E-06 |
| Obesity | endometrium | *RFX7* | 4.363277089 | 5.70E-08 | 2.58E-07 | 0.000276658 |
| Obesity | endometrium | *CLDN7* | -5.030104809 | 0.000353646 | 0.000493923 | 1 |
| Obesity | endometrium | *ADO* | 5.103274045 | 5.42E-06 | 9.64E-06 | 0.026276114 |
| Obesity | endometrium | *MGAT4C* | 2.697329463 | 2.46E-06 | 4.77E-06 | 0.011957248 |
| Obesity | endometrium | *IDH2* | 3.137951682 | 1.86E-06 | 3.75E-06 | 0.009033428 |
| Obesity | endometrium | *FAM181B* | -2.394871909 | 0.000592411 | 0.000807637 | 1 |
| Obesity | endometrium | *TDRKH* | 3.351856257 | 0.001463834 | 0.001928461 | 1 |
| Obesity | endometrium | *ZNF708* | 6.615856006 | 4.19E-06 | 7.60E-06 | 0.020332905 |
| Obesity | endometrium | *ERCC6L2* | 6.513905707 | 9.32E-09 | 7.47E-08 | 4.52E-05 |
| Obesity | endometrium | *MRPL41* | 6.351874821 | 1.41E-07 | 5.12E-07 | 0.000684872 |
| Obesity | endometrium | *ARL6IP4* | -2.376378044 | 0.000153006 | 0.000223744 | 0.742383281 |
| Obesity | endometrium | *HHIPL1* | 3.92229825 | 1.12E-06 | 2.64E-06 | 0.005426627 |
| Obesity | endometrium | *FAM153B* | 2.990457991 | 1.47E-06 | 3.23E-06 | 0.007112052 |
| Obesity | endometrium | *BACE2* | 4.357847283 | 0.000266226 | 0.00037792 | 1 |
| Obesity | endometrium | *SYNM* | 7.982475114 | 1.20E-07 | 4.53E-07 | 0.000583518 |
| Obesity | endometrium | *KCNA4* | 4.606647363 | 1.59E-06 | 3.38E-06 | 0.007736104 |
| Obesity | endometrium | *FIGN* | 4.866630673 | 1.50E-08 | 1.01E-07 | 7.27E-05 |
| Obesity | endometrium | *C8orf33* | 8.017117664 | 6.10E-08 | 2.69E-07 | 0.000295937 |
| Obesity | endometrium | *FBXL6* | -2.73313681 | 0.004238204 | 0.005338465 | 1 |
| Obesity | endometrium | *KBTBD3* | 3.760916849 | 1.74E-06 | 3.56E-06 | 0.008425922 |
| Obesity | endometrium | *CLN8* | 3.540034139 | 8.74E-08 | 3.54E-07 | 0.000423854 |
| Obesity | endometrium | *NXPH4* | 38.86569994 | 1.63E-06 | 3.41E-06 | 0.007894531 |
| Obesity | endometrium | *NPLOC4* | 3.398526559 | 2.37E-06 | 4.61E-06 | 0.011503415 |
| Obesity | endometrium | *CAPN12* | 3.534352551 | 0.000141704 | 0.000208222 | 0.68754824 |
| Obesity | endometrium | *ORAI1* | 5.78444744 | 1.47E-07 | 5.27E-07 | 0.000711056 |
| Obesity | endometrium | *FES* | 10.59799792 | 3.07E-09 | 4.37E-08 | 1.49E-05 |
| Obesity | endometrium | *GLRX5* | 11.35015362 | 6.12E-09 | 6.02E-08 | 2.97E-05 |
| Obesity | endometrium | *CAV3* | 2.848079888 | 0.000221853 | 0.000318 | 1 |
| Obesity | endometrium | *MXRA7* | 5.040754312 | 5.37E-07 | 1.48E-06 | 0.002604726 |
| Obesity | endometrium | *MFSD5* | 6.075126364 | 0.000124742 | 0.000184808 | 0.60524635 |
| Obesity | endometrium | *ADI1* | 5.63523453 | 1.21E-07 | 4.56E-07 | 0.000588818 |
| Obesity | endometrium | *CLEC4G* | -4.98546766 | 7.85E-05 | 0.000119081 | 0.380821522 |
| Obesity | endometrium | *TRAK1* | 5.264112672 | 3.21E-07 | 9.82E-07 | 0.001559586 |
| Obesity | endometrium | *SKA2* | 11.45113376 | 1.64E-09 | 3.98E-08 | 7.98E-06 |
| Obesity | endometrium | *TTC3* | 3.059393972 | 2.69E-07 | 8.52E-07 | 0.001306195 |
| Obesity | endometrium | *BRICD5* | 8.075804522 | 1.55E-06 | 3.33E-06 | 0.007543483 |
| Obesity | endometrium | *GRID1* | -3.467697816 | 0.00537554 | 0.006691154 | 1 |
| Obesity | endometrium | *ACBD3* | 10.74802312 | 4.72E-08 | 2.25E-07 | 0.000228864 |
| Obesity | endometrium | *PLCXD3* | 2.767271459 | 1.14E-06 | 2.68E-06 | 0.00551524 |
| Obesity | endometrium | *GPIHBP1* | 2.781793564 | 4.77E-05 | 7.39E-05 | 0.231444645 |
| Obesity | endometrium | *VMO1* | -2.448097208 | 0.019956184 | 0.023490394 | 1 |
| Obesity | endometrium | *ALG12* | 3.781292172 | 1.42E-06 | 3.17E-06 | 0.006891601 |
| Obesity | endometrium | *COL18A1* | 11.60906089 | 2.75E-12 | 6.07E-10 | 1.34E-08 |
| Obesity | endometrium | *ZNF721* | 7.911665457 | 1.66E-09 | 3.98E-08 | 8.05E-06 |
| Obesity | endometrium | *C11orf54* | 2.91582673 | 0.000277994 | 0.000394049 | 1 |
| Obesity | endometrium | *CEP63* | 5.769971946 | 0.000722332 | 0.000978709 | 1 |

| | | | | | | |
|---|---|---|---|---|---|---|
| Obesity | endometrium | *ZNF320* | -1.173541041 | 0.025333966 | 0.029555279 | 1 |
| Obesity | endometrium | *C12orf60* | 3.558247389 | 1.28E-06 | 2.94E-06 | 0.006189568 |
| Obesity | endometrium | *ABAT* | 5.873818063 | 8.53E-08 | 3.48E-07 | 0.000413983 |
| Obesity | endometrium | *WBP2NL* | 5.270139619 | 2.84E-07 | 8.93E-07 | 0.001377386 |
| Obesity | endometrium | *BEGAIN* | 5.678612056 | 1.52E-05 | 2.50E-05 | 0.073797181 |
| Obesity | endometrium | *ARHGEF37* | 9.028765645 | 1.19E-08 | 8.66E-08 | 5.78E-05 |
| Obesity | endometrium | *RABIF* | 4.277621544 | 2.92E-06 | 5.52E-06 | 0.014150137 |
| Obesity | endometrium | *FANCF* | -2.68567254 | 0.001757999 | 0.00229481 | 1 |
| Obesity | endometrium | *C2CD4C* | 2.901620964 | 0.005067696 | 0.00632582 | 1 |
| Obesity | endometrium | *RUVBL2* | -2.587819218 | 0.01203081 | 0.014441734 | 1 |
| Obesity | endometrium | *PTTG1IP* | 2.980121728 | 4.06E-06 | 7.37E-06 | 0.01967508 |
| Obesity | endometrium | *PLGLB1* | 8.319914315 | 1.21E-06 | 2.82E-06 | 0.00588463 |
| Obesity | endometrium | *ZNF623* | 8.475269052 | 0.000204927 | 0.000294959 | 0.994306574 |
| Obesity | endometrium | *CCDC125* | 5.856029435 | 4.12E-08 | 2.04E-07 | 0.000199876 |
| Obesity | endometrium | *C10orf107* | 3.318914012 | 9.34E-07 | 2.29E-06 | 0.004533786 |
| Obesity | endometrium | *FHL3* | 4.438075344 | 5.02E-05 | 7.76E-05 | 0.243488788 |
| Obesity | endometrium | *C19orf71* | -3.261222872 | 0.005101331 | 0.006362894 | 1 |
| Obesity | endometrium | *LRIT3* | 27.66296778 | 2.10E-06 | 4.16E-06 | 0.010189082 |
| Obesity | endometrium | *NPIPA1* | 1.903562522 | 0.009710873 | 0.011738205 | 1 |
| Obesity | endometrium | *GPR132* | 3.481478963 | 8.88E-06 | 1.52E-05 | 0.043077211 |
| Obesity | endometrium | *AC131971.1* | 13.76480811 | 0.002063017 | 0.002678555 | 1 |
| Obesity | endometrium | *SERHL2* | 7.136492627 | 8.28E-08 | 3.40E-07 | 0.000401729 |
| Obesity | endometrium | *PCBP3* | 2.96111024 | 6.51E-06 | 1.14E-05 | 0.031573456 |
| Obesity | endometrium | *SFXN4* | 3.828898214 | 6.71E-07 | 1.78E-06 | 0.003254312 |
| Obesity | endometrium | *MRPL54* | 4.383980068 | 0.00729111 | 0.008924436 | 1 |
| Obesity | endometrium | *RP1L1* | 6.626646946 | 0.012349335 | 0.014813096 | 1 |
| Obesity | endometrium | *NDUFB1* | 9.397836135 | 5.41E-09 | 5.76E-08 | 2.63E-05 |
| Obesity | endometrium | *TRMT12* | 5.400210709 | 8.55E-06 | 1.47E-05 | 0.041489801 |
| Obesity | endometrium | *ALYREF* | -3.439055987 | 0.011804281 | 0.014173316 | 1 |
| Obesity | endometrium | *MRGPRX2* | 22.31316309 | 0.003907088 | 0.004938055 | 1 |
| Obesity | endometrium | *UPP1* | 5.564408836 | 0.019569742 | 0.02305789 | 1 |
| Obesity | endometrium | *FIGLA* | 19.17233279 | 2.18E-06 | 4.29E-06 | 0.010574301 |
| Obesity | endometrium | *ACSM2A* | 34.99465764 | 4.30E-06 | 7.78E-06 | 0.020884591 |
| Obesity | endometrium | *KREMEN1* | -1.384329987 | 0.019912469 | 0.023444625 | 1 |
| Obesity | endometrium | *ZNF703* | 6.367737403 | 4.33E-07 | 1.25E-06 | 0.002102173 |
| Obesity | endometrium | *NPIPA5* | 2.824519847 | 1.38E-05 | 2.29E-05 | 0.067026125 |
| Obesity | endometrium | *EMILIN3* | 14.27006968 | 7.96E-05 | 0.00012059 | 0.386250565 |
| Obesity | endometrium | *FAM162B* | 5.363969468 | 7.08E-06 | 1.23E-05 | 0.034329669 |
| Obesity | endometrium | *BTBD9* | 7.625543221 | 2.69E-09 | 4.24E-08 | 1.31E-05 |
| Obesity | endometrium | *NUDT14* | 9.236917721 | 9.58E-06 | 1.63E-05 | 0.046496509 |
| Obesity | endometrium | *NPIPA7* | -2.941092752 | 0.002022666 | 0.002627571 | 1 |
| Obesity | endometrium | *TTC32* | -1.456126043 | 0.001071518 | 0.001429083 | 1 |
| Obesity | endometrium | *PP2D1* | 12.27354947 | 7.68E-09 | 6.76E-08 | 3.73E-05 |
| Obesity | endometrium | *DENND5A* | 2.716195923 | 6.98E-07 | 1.84E-06 | 0.003384605 |
| Obesity | endometrium | *NIPSNAP1* | -2.282406338 | 0.003633622 | 0.004611649 | 1 |
| Obesity | endometrium | *CNTN2* | 5.719445744 | 1.11E-05 | 1.87E-05 | 0.053996249 |
| Obesity | endometrium | *GOLGA6L4* | -1.694561807 | 0.000168261 | 0.000244286 | 0.816403388 |
| Obesity | endometrium | *PGP* | 4.768935217 | 4.34E-06 | 7.83E-06 | 0.02103509 |
| Obesity | endometrium | *ACOT1* | 7.83782024 | 1.25E-08 | 8.93E-08 | 6.05E-05 |
| Obesity | endometrium | *OAF* | 1.826858509 | 0.00215219 | 0.002792093 | 1 |

| | | | | | | |
|---|---|---|---|---|---|---|
| Obesity | endometrium | *ALDH1A3* | 2.355279127 | 6.59E-05 | 0.000100549 | 0.319543593 |
| Obesity | endometrium | *CLECL1* | 5.392437134 | 3.69E-06 | 6.79E-06 | 0.017898697 |
| Obesity | endometrium | *MROH7* | 5.470494645 | 0.000184875 | 0.000266889 | 0.897012561 |
| Obesity | endometrium | *PKP3* | -1.70163374 | 7.35E-05 | 0.000111782 | 0.356585267 |
| Obesity | endometrium | *A3GALT2* | 4.664978967 | 0.00023711 | 0.000338975 | 1 |
| Obesity | endometrium | *SS18L1* | 7.595300058 | 1.55E-08 | 1.03E-07 | 7.51E-05 |
| Obesity | endometrium | *COPB2* | 4.298521856 | 1.24E-07 | 4.66E-07 | 0.000603337 |
| Obesity | endometrium | *WDR27* | 8.611500059 | 4.59E-10 | 2.45E-08 | 2.23E-06 |
| Obesity | endometrium | *TMEM255B* | 4.066145385 | 1.71E-06 | 3.52E-06 | 0.00830969 |
| Obesity | endometrium | *HDDC3* | 8.94815321 | 0.002481904 | 0.003195913 | 1 |
| Obesity | endometrium | *ZFP1* | 10.32511794 | 0.001605566 | 0.002104892 | 1 |
| Obesity | endometrium | *DUSP8* | -1.244543483 | 0.000249556 | 0.000355628 | 1 |
| Obesity | endometrium | *FAM19A3* | -13.8053389 | 0.000197476 | 0.000284319 | 0.958155241 |
| Obesity | endometrium | *SNN* | 5.621179688 | 1.60E-06 | 3.39E-06 | 0.007768518 |
| Obesity | endometrium | *KRBA2* | 5.585654832 | 0.007690186 | 0.009391589 | 1 |
| Obesity | endometrium | *SEPT5* | 8.827433228 | 2.94E-05 | 4.66E-05 | 0.142730362 |
| Obesity | endometrium | *RNLS* | 8.615396645 | 5.66E-06 | 1.00E-05 | 0.027441429 |
| Obesity | endometrium | *NDUFA12* | 5.198504119 | 9.10E-08 | 3.64E-07 | 0.000441562 |
| Obesity | endometrium | *TCTE3* | 9.668941094 | 2.06E-08 | 1.24E-07 | 9.97E-05 |
| Obesity | endometrium | *UBE2G2* | 5.586935937 | 2.64E-08 | 1.47E-07 | 0.000128 |
| Obesity | endometrium | *TMED9* | 9.811724169 | 2.33E-08 | 1.34E-07 | 0.000113095 |
| Obesity | endometrium | *DRD1* | 88.9881715 | 2.20E-06 | 4.33E-06 | 0.010684566 |
| Obesity | endometrium | *BTBD6* | 3.455381529 | 1.99E-05 | 3.23E-05 | 0.096785131 |
| Obesity | endometrium | *NUTM2A* | -7.279879419 | 0.000364853 | 0.000509137 | 1 |
| Obesity | endometrium | *WT1* | 1.88329008 | 1.44E-05 | 2.38E-05 | 0.069803084 |
| Obesity | endometrium | *ZFP90* | 4.491030248 | 1.70E-07 | 5.94E-07 | 0.00082595 |
| Obesity | endometrium | *FAM227A* | 2.855982611 | 0.000133254 | 0.00019646 | 0.646549515 |
| Obesity | endometrium | *NOC4L* | 6.358549767 | 1.60E-05 | 2.63E-05 | 0.077865835 |
| Obesity | endometrium | *NDUFA6* | 3.713677186 | 3.82E-07 | 1.13E-06 | 0.001852225 |
| Obesity | endometrium | *SORCS2* | 9.874042095 | 7.98E-07 | 2.03E-06 | 0.003869942 |
| Obesity | endometrium | *TMEM106A* | 2.676899927 | 2.83E-05 | 4.50E-05 | 0.137401404 |
| Obesity | endometrium | *BRI3BP* | 8.824089725 | 0.008614351 | 0.01048065 | 1 |
| Obesity | endometrium | *UBOX5* | 3.312964664 | 2.39E-06 | 4.64E-06 | 0.011595556 |
| Obesity | endometrium | *MAFF* | -2.517728378 | 3.97E-05 | 6.19E-05 | 0.192509712 |
| Obesity | endometrium | *SEMA4B* | 8.344363251 | 3.28E-08 | 1.72E-07 | 0.000158984 |
| Obesity | endometrium | *CIB1* | -1.782189723 | 0.040611681 | 0.04667169 | 1 |
| Obesity | endometrium | *SGCZ* | 2.482595178 | 3.89E-06 | 7.11E-06 | 0.018893055 |
| Obesity | endometrium | *C6orf120* | 4.199787431 | 4.12E-06 | 7.48E-06 | 0.019997425 |
| Obesity | endometrium | *SIGIRR* | 5.641568449 | 1.97E-07 | 6.66E-07 | 0.0009539 |
| Obesity | endometrium | *IFITM2* | 4.978071887 | 1.23E-07 | 4.61E-07 | 0.000596171 |
| Obesity | endometrium | *GP1BA* | 13.51547702 | 8.72E-09 | 7.27E-08 | 4.23E-05 |
| Obesity | endometrium | *CCDC137* | 7.094354073 | 1.19E-07 | 4.48E-07 | 0.000575208 |
| Obesity | endometrium | *CDK10* | 2.771355422 | 1.45E-05 | 2.39E-05 | 0.070459725 |
| Obesity | endometrium | *ATP6V0A2* | 8.449787439 | 3.40E-06 | 6.31E-06 | 0.016485259 |
| Obesity | endometrium | *C14orf80* | 7.331423623 | 2.22E-06 | 4.35E-06 | 0.010753639 |
| Obesity | endometrium | *TNFAIP8L1* | 14.09999789 | 0.00015359 | 0.000224532 | 0.745220255 |
| Obesity | endometrium | *SP140L* | 2.863275571 | 1.23E-06 | 2.85E-06 | 0.005987412 |
| Obesity | endometrium | *TARSL2* | 7.332661265 | 2.28E-09 | 4.01E-08 | 1.11E-05 |
| Obesity | endometrium | *SH3BGR* | -2.293509172 | 0.002695221 | 0.00345958 | 1 |
| Obesity | endometrium | *FAM174B* | 6.207556057 | 2.88E-08 | 1.56E-07 | 0.000139558 |

| | | | | | | |
|---|---|---|---|---|---|---|
| Obesity | endometrium | *SV2B* | 3.691287184 | 1.88E-05 | 3.06E-05 | 0.091208817 |
| Obesity | endometrium | *AHNAK2* | 5.288645557 | 1.56E-06 | 3.34E-06 | 0.007587786 |
| Obesity | endometrium | *SP1* | 2.904078093 | 5.10E-07 | 1.42E-06 | 0.002472506 |
| Obesity | endometrium | *PCGF3* | 3.422215516 | 1.47E-07 | 5.30E-07 | 0.000714988 |
| Obesity | endometrium | *PSMD13* | 7.821276262 | 0.001301595 | 0.001724089 | 1 |
| Obesity | endometrium | *PMEL* | 3.476107748 | 0.020554248 | 0.024170919 | 1 |
| Obesity | endometrium | *C6orf201* | 4.597353167 | 2.51E-06 | 4.85E-06 | 0.012163856 |
| Obesity | endometrium | *MYBL1* | 3.812659357 | 2.03E-05 | 3.28E-05 | 0.098480529 |
| Obesity | endometrium | *ADARB2* | 8.764935063 | 4.42E-07 | 1.27E-06 | 0.002146594 |
| Obesity | endometrium | *DMWD* | 7.661522898 | 0.004914731 | 0.006141198 | 1 |
| Obesity | endometrium | *SLC52A2* | 4.715708069 | 2.24E-07 | 7.37E-07 | 0.001086562 |
| Obesity | endometrium | *PIGP* | -1.288919338 | 0.030235307 | 0.035070932 | 1 |
| Obesity | endometrium | *ARL17A* | 4.961071497 | 5.58E-07 | 1.53E-06 | 0.002708954 |
| Obesity | endometrium | *EVI2B* | 2.881192624 | 3.87E-05 | 6.04E-05 | 0.187632611 |
| Obesity | endometrium | *NPIPB4* | 3.789538091 | 1.46E-06 | 3.23E-06 | 0.00709768 |
| Obesity | endometrium | *ZNF829* | 5.037571658 | 3.58E-06 | 6.60E-06 | 0.017355887 |
| Obesity | endometrium | *IFITM1* | 9.868316167 | 3.38E-06 | 6.28E-06 | 0.016381179 |
| Obesity | endometrium | *LINC00839* | 11.88327323 | 1.69E-06 | 3.50E-06 | 0.008217184 |
| Obesity | endometrium | *RTN4RL1* | 9.017398913 | 5.41E-06 | 9.63E-06 | 0.026253606 |
| Obesity | endometrium | *RNPC3* | 4.606029436 | 9.54E-09 | 7.56E-08 | 4.63E-05 |
| Obesity | endometrium | *RASA3* | 8.714439203 | 1.53E-07 | 5.45E-07 | 0.00074023 |
| Obesity | endometrium | *ZNF529* | 13.06237645 | 2.13E-08 | 1.27E-07 | 0.000103513 |
| Obesity | endometrium | *C15orf41* | 3.63120481 | 9.67E-08 | 3.83E-07 | 0.000468964 |
| Obesity | endometrium | *WWOX* | 6.576690316 | 1.58E-09 | 3.98E-08 | 7.64E-06 |
| Obesity | endometrium | *CYP4Z1* | 1.429796136 | 0.030509198 | 0.03535482 | 1 |
| Obesity | endometrium | *FFAR4* | 3.376942841 | 3.04E-06 | 5.72E-06 | 0.014763477 |
| Obesity | endometrium | *CYP4F12* | 3.950987456 | 5.02E-07 | 1.41E-06 | 0.002433377 |
| Obesity | endometrium | *ZNF749* | -1.441227069 | 0.000526518 | 0.000722065 | 1 |
| Obesity | endometrium | *NBPF12* | 3.718015721 | 0.001710315 | 0.002234369 | 1 |
| Obesity | endometrium | *TOR3A* | 3.797720163 | 2.84E-06 | 5.39E-06 | 0.013771236 |
| Obesity | endometrium | *ZNF555* | 8.31715267 | 4.24E-08 | 2.07E-07 | 0.000205593 |
| Obesity | endometrium | *GOLGA8R* | -6.360183622 | 5.58E-05 | 8.58E-05 | 0.270576262 |
| Obesity | endometrium | *CCDC30* | 6.407864582 | 4.29E-08 | 2.09E-07 | 0.000208373 |
| Obesity | endometrium | *GLDN* | 1.672782892 | 9.00E-06 | 1.54E-05 | 0.043682503 |
| Obesity | endometrium | *ZNF197* | 7.069431334 | 3.61E-08 | 1.85E-07 | 0.000175173 |
| Obesity | endometrium | *RPS23* | 9.211297795 | 2.08E-09 | 4.00E-08 | 1.01E-05 |
| Obesity | endometrium | *GNG2* | 2.631963939 | 7.64E-07 | 1.97E-06 | 0.003705046 |
| Obesity | endometrium | *BTN3A2* | -1.649268379 | 0.00587188 | 0.007282812 | 1 |
| Obesity | endometrium | *TMEM222* | 5.762371842 | 2.84E-07 | 8.92E-07 | 0.001375576 |
| Obesity | endometrium | *SEPT10* | 4.56000194 | 2.94E-07 | 9.17E-07 | 0.001426656 |
| Obesity | endometrium | *FAM86B1* | -5.499394087 | 0.014837551 | 0.017684058 | 1 |
| Obesity | endometrium | *SMYD4* | 4.423434063 | 1.07E-07 | 4.14E-07 | 0.000520302 |
| Obesity | endometrium | *CEACAM19* | -1.73912961 | 0.00031279 | 0.000440283 | 1 |
| Obesity | endometrium | *CYP27C1* | 5.083835149 | 0.00057658 | 0.000787604 | 1 |
| Obesity | endometrium | *BCR* | 6.499145225 | 5.61E-09 | 5.86E-08 | 2.72E-05 |
| Obesity | endometrium | *FOXI2* | -5.342071979 | 0.012440833 | 0.014915473 | 1 |
| Obesity | endometrium | *ZNF732* | 7.033034736 | 1.74E-06 | 3.56E-06 | 0.00842889 |
| Obesity | endometrium | *VSIG10L* | -1.429742039 | 0.000253924 | 0.000361302 | 1 |
| Obesity | endometrium | *TRABD2A* | 5.418754753 | 5.18E-07 | 1.44E-06 | 0.002514676 |
| Obesity | endometrium | *QRFPR* | 16.07698547 | 1.46E-05 | 2.40E-05 | 0.070813081 |

| | | | | | | |
|---|---|---|---|---|---|---|
| Obesity | endometrium | *MAPT* | 4.426431487 | 3.04E-07 | 9.41E-07 | 0.001474377 |
| Obesity | endometrium | *TMEM17* | -3.181930645 | 0.00574756 | 0.007132266 | 1 |
| Obesity | endometrium | *TNFRSF18* | 13.51314479 | 1.50E-05 | 2.47E-05 | 0.072782731 |
| Obesity | endometrium | *C1QL4* | 46.78628347 | 1.72E-06 | 3.53E-06 | 0.008332891 |
| Obesity | endometrium | *RTN4RL2* | 7.563611163 | 4.70E-05 | 7.28E-05 | 0.227962469 |
| Obesity | endometrium | *TMEM232* | 5.894604064 | 9.03E-07 | 2.24E-06 | 0.00438171 |
| Obesity | endometrium | *EFCAB6* | 6.691009076 | 2.08E-08 | 1.25E-07 | 0.000100779 |
| Obesity | endometrium | *RHD* | 7.295838672 | 2.10E-06 | 4.16E-06 | 0.010207073 |
| Obesity | endometrium | *TMPRSS6* | 27.21221012 | 2.02E-06 | 4.03E-06 | 0.009815688 |
| Obesity | endometrium | *CYP4A11* | 15.53023375 | 3.83E-07 | 1.13E-06 | 0.00185807 |
| Obesity | endometrium | *TMEM216* | -1.036044633 | 0.000467416 | 0.000644657 | 1 |
| Obesity | endometrium | *ENTPD5* | 7.29318756 | 6.00E-06 | 1.06E-05 | 0.029119269 |
| Obesity | endometrium | *ANGPTL5* | 3.507548467 | 0.014355341 | 0.017117747 | 1 |
| Obesity | endometrium | *WDR86* | -1.404962583 | 0.000932839 | 0.001251697 | 1 |
| Obesity | endometrium | *EPOR* | -1.971093522 | 0.005930254 | 0.007351454 | 1 |
| Obesity | endometrium | *LUZP2* | 10.35554694 | 5.49E-08 | 2.51E-07 | 0.000266278 |
| Obesity | endometrium | *GJA4* | 5.498771872 | 1.11E-06 | 2.63E-06 | 0.005399207 |
| Obesity | endometrium | *SBK2* | 36.26463752 | 1.44E-06 | 3.20E-06 | 0.006982383 |
| Obesity | endometrium | *ZNF385C* | 5.741071516 | 7.00E-05 | 0.000106617 | 0.339467658 |
| Obesity | endometrium | *EXD3* | 3.503428535 | 5.09E-07 | 1.42E-06 | 0.002470787 |
| Obesity | endometrium | *SAMD11* | 8.022611974 | 7.90E-07 | 2.02E-06 | 0.003832198 |
| Obesity | endometrium | *TRPV2* | 3.511433156 | 6.83E-06 | 1.19E-05 | 0.03314711 |
| Obesity | endometrium | *C2orf88* | 6.024805427 | 3.36E-08 | 1.74E-07 | 0.000163072 |
| Obesity | endometrium | *FANCA* | 2.437291624 | 0.001981721 | 0.002577045 | 1 |
| Obesity | endometrium | *DNAH17* | 3.935223241 | 9.52E-06 | 1.62E-05 | 0.046192561 |
| Obesity | endometrium | *FANCM* | 4.303240899 | 1.43E-06 | 3.19E-06 | 0.006958486 |
| Obesity | endometrium | *ZFP69* | 4.912271072 | 9.58E-07 | 2.34E-06 | 0.004647736 |
| Obesity | endometrium | *TMEM220* | 8.31006913 | 2.79E-08 | 1.52E-07 | 0.000135435 |
| Obesity | endometrium | *P2RX2* | -3.557309854 | 0.000502672 | 0.000691206 | 1 |
| Obesity | endometrium | *FAM122A* | -2.131190881 | 0.001411093 | 0.001863973 | 1 |
| Obesity | endometrium | *GFRAL* | 10.05613929 | 4.76E-07 | 1.34E-06 | 0.002307856 |
| Obesity | endometrium | *LDLRAD2* | 5.846919135 | 5.68E-05 | 8.73E-05 | 0.275708559 |
| Obesity | endometrium | *CPSF4L* | 18.49780806 | 1.68E-06 | 3.48E-06 | 0.008145167 |
| Obesity | endometrium | *KLHL17* | -1.753266155 | 0.001639676 | 0.002146711 | 1 |
| Obesity | endometrium | *ZSCAN23* | 4.516354473 | 4.12E-07 | 1.20E-06 | 0.001999104 |
| Obesity | endometrium | *TMEM221* | 7.542893164 | 0.00199917 | 0.002598439 | 1 |
| Obesity | endometrium | *MAPK12* | 11.42259817 | 1.00E-05 | 1.70E-05 | 0.048591173 |
| Obesity | endometrium | *AGRN* | 19.54444528 | 1.46E-05 | 2.41E-05 | 0.071069567 |
| Obesity | endometrium | *FAM166A* | -9.336428786 | 0.010389304 | 0.012533293 | 1 |
| Obesity | endometrium | *ZNF626* | 13.11222513 | 2.85E-09 | 4.29E-08 | 1.38E-05 |
| Obesity | endometrium | *LINC00265* | 4.731391633 | 1.07E-06 | 2.55E-06 | 0.005176965 |
| Obesity | endometrium | *LAMTOR4* | 4.849842935 | 1.04E-07 | 4.05E-07 | 0.000504388 |
| Obesity | endometrium | *PRKAR1B* | 10.33033733 | 6.91E-06 | 1.21E-05 | 0.033519116 |
| Obesity | endometrium | *NCR3LG1* | -1.680679312 | 0.000398364 | 0.000552878 | 1 |
| Obesity | endometrium | *ZNF793* | 2.720810813 | 3.94E-05 | 6.15E-05 | 0.191129917 |
| Obesity | endometrium | *AGAP4* | 5.096975175 | 0.017809563 | 0.021076097 | 1 |
| Obesity | endometrium | *RUFY4* | 7.906962399 | 1.57E-06 | 3.35E-06 | 0.007612407 |
| Obesity | endometrium | *CENPP* | 8.338393577 | 9.03E-14 | 3.98E-11 | 4.38E-10 |
| Obesity | endometrium | *PLSCR1* | 3.089393399 | 4.36E-07 | 1.26E-06 | 0.002113099 |
| Obesity | endometrium | *ENO4* | 8.232691444 | 4.69E-08 | 2.24E-07 | 0.000227748 |

| | | | | | | |
|---|---|---|---|---|---|---|
| Obesity | endometrium | *ZNF559* | 2.771848736 | 0.000169689 | 0.000246212 | 0.82333326 |
| Obesity | endometrium | *ZP3* | 5.31824045 | 5.25E-06 | 9.36E-06 | 0.025454147 |
| Obesity | endometrium | *H3F3C* | 110.7849175 | 2.43E-05 | 3.89E-05 | 0.117757554 |
| Obesity | endometrium | *JAKMIP3* | -2.533985817 | 2.31E-05 | 3.71E-05 | 0.111869393 |
| Obesity | endometrium | *PDCD1* | 30.7797039 | 2.27E-06 | 4.44E-06 | 0.011025813 |
| Obesity | endometrium | *SELL* | 4.576573141 | 0.000254621 | 0.000362081 | 1 |
| Obesity | endometrium | *C19orf54* | 5.857875333 | 6.32E-07 | 1.69E-06 | 0.003066969 |
| Obesity | endometrium | *C22orf34* | 2.767147012 | 1.22E-06 | 2.83E-06 | 0.005919294 |
| Obesity | endometrium | *SRSF10* | 6.35356191 | 4.72E-08 | 2.25E-07 | 0.000228971 |
| Obesity | endometrium | *NBR1* | 7.093081484 | 8.57E-09 | 7.20E-08 | 4.16E-05 |
| Obesity | endometrium | *NDOR1* | -2.217544674 | 0.003683222 | 0.004667636 | 1 |
| Obesity | endometrium | *FAM72B* | -2.769782208 | 0.000282169 | 0.000399499 | 1 |
| Obesity | endometrium | *GOLGA8M* | 4.315066215 | 2.84E-06 | 5.40E-06 | 0.01380151 |
| Obesity | endometrium | *RHCE* | 10.81946668 | 4.43E-07 | 1.27E-06 | 0.002147076 |
| Obesity | endometrium | *IDO2* | 10.57042151 | 4.89E-06 | 8.76E-06 | 0.023739609 |
| Obesity | endometrium | *SLC4A5* | 12.07719169 | 8.91E-07 | 2.22E-06 | 0.004322796 |
| Obesity | endometrium | *UROS* | 5.209614043 | 1.57E-07 | 5.59E-07 | 0.000763162 |
| Obesity | endometrium | *ZBED6CL* | 12.22312531 | 3.20E-05 | 5.04E-05 | 0.155077941 |
| Obesity | endometrium | *OSTN* | 5.236484526 | 2.55E-08 | 1.43E-07 | 0.000123663 |
| Obesity | endometrium | *FSIP2* | 4.182489081 | 1.97E-07 | 6.68E-07 | 0.000957199 |
| Obesity | endometrium | *NOXA1* | -1.750762092 | 0.001872753 | 0.002438701 | 1 |
| Obesity | endometrium | *FZD9* | 34.93601201 | 2.98E-05 | 4.72E-05 | 0.144603065 |
| Obesity | endometrium | *SKOR1* | 17.12834529 | 1.63E-08 | 1.06E-07 | 7.90E-05 |
| Obesity | endometrium | *MTF1* | 3.983854694 | 3.32E-07 | 1.01E-06 | 0.00161179 |
| Obesity | endometrium | *ZDHHC11* | 6.86720322 | 0.00019125 | 0.000275764 | 0.927945467 |
| Obesity | endometrium | *FAM26F* | 4.127863281 | 1.47E-05 | 2.42E-05 | 0.071376745 |
| Obesity | endometrium | *ENTPD8* | 400.8752631 | 4.30E-06 | 7.78E-06 | 0.020881738 |
| Obesity | endometrium | *RPL14* | 11.172821 | 2.33E-07 | 7.61E-07 | 0.001130488 |
| Obesity | endometrium | *FBF1* | 5.744179117 | 0.002384932 | 0.003078823 | 1 |
| Obesity | endometrium | *MSL1* | 2.977453078 | 2.20E-06 | 4.32E-06 | 0.010654876 |
| Obesity | endometrium | *FAM196A* | 3.961982042 | 1.58E-07 | 5.60E-07 | 0.000764586 |
| Obesity | endometrium | *UTS2B* | 4.411696251 | 1.36E-05 | 2.26E-05 | 0.066159732 |
| Obesity | endometrium | *C9orf152* | 11.30361445 | 1.70E-06 | 3.50E-06 | 0.008231043 |
| Obesity | endometrium | *NELFB* | 3.400548389 | 1.80E-06 | 3.66E-06 | 0.008732108 |
| Obesity | endometrium | *SLC15A5* | 5.209295954 | 2.42E-06 | 4.68E-06 | 0.011719438 |
| Obesity | endometrium | *ADAT2* | 8.770789972 | 7.40E-08 | 3.13E-07 | 0.000358931 |
| Obesity | endometrium | *ALKBH2* | 6.526390984 | 3.40E-07 | 1.03E-06 | 0.001651875 |
| Obesity | endometrium | *LITAF* | 5.699881858 | 2.66E-08 | 1.48E-07 | 0.000129177 |
| Obesity | endometrium | *VSTM1* | -0.960559117 | 0.000194476 | 0.000280165 | 0.943595274 |
| Obesity | endometrium | *SF3B3* | 12.72838162 | 4.14E-08 | 2.05E-07 | 0.000200643 |
| Obesity | endometrium | *PLAC9* | 6.285958831 | 1.55E-08 | 1.03E-07 | 7.51E-05 |
| Obesity | endometrium | *ZNF573* | 6.109670224 | 3.84E-07 | 1.13E-06 | 0.001861333 |
| Obesity | endometrium | *FAM47E* | 4.704888535 | 1.46E-06 | 3.23E-06 | 0.007100867 |
| Obesity | endometrium | *ZNF600* | 2.266257051 | 0.000153664 | 0.000224572 | 0.745578373 |
| Obesity | endometrium | *FHIT* | 6.118349499 | 1.63E-09 | 3.98E-08 | 7.93E-06 |
| Obesity | endometrium | *ZKSCAN3* | -6.189902129 | 0.000123326 | 0.000182822 | 0.598377854 |
| Obesity | endometrium | *SLC35E2B* | 5.566354785 | 6.15E-09 | 6.04E-08 | 2.99E-05 |
| Obesity | endometrium | *KIAA0408* | 2.298898955 | 0.013331243 | 0.015939673 | 1 |
| Obesity | endometrium | *HLA-DRB1* | 2.15515996 | 1.31E-06 | 3.00E-06 | 0.006353865 |
| Obesity | endometrium | *AKR1C3* | 5.030071711 | 1.88E-07 | 6.43E-07 | 0.00091304 |

| | | | | | | |
|---|---|---|---|---|---|---|
| Obesity | endometrium | *WDSUB1* | 10.40826627 | 6.03E-07 | 1.62E-06 | 0.002924321 |
| Obesity | endometrium | *ZNF79* | -1.295274039 | 0.009570789 | 0.011583305 | 1 |
| Obesity | endometrium | *COLCA1* | -2.386781783 | 0.00167551 | 0.002191851 | 1 |
| Obesity | endometrium | *SEMA4A* | -2.779452452 | 0.025237111 | 0.029449365 | 1 |
| Obesity | endometrium | *HRCT1* | 3.426382968 | 0.002264927 | 0.002931296 | 1 |
| Obesity | endometrium | *MPHOSPH8* | 5.377827387 | 5.08E-09 | 5.56E-08 | 2.46E-05 |
| Obesity | endometrium | *SIRPB2* | 2.414786352 | 0.000150702 | 0.000220509 | 0.731208006 |
| Obesity | endometrium | *SRGAP3* | 5.351379401 | 0.002211717 | 0.002864723 | 1 |
| Obesity | endometrium | *XPNPEP3* | 3.439638043 | 2.20E-05 | 3.55E-05 | 0.106729967 |
| Obesity | endometrium | *ZNF107* | 5.948008644 | 6.71E-06 | 1.17E-05 | 0.032580014 |
| Obesity | endometrium | *PPIA* | 10.58874182 | 7.36E-10 | 2.94E-08 | 3.57E-06 |
| Obesity | endometrium | *ZNF471* | 3.973414084 | 5.81E-07 | 1.57E-06 | 0.002818782 |
| Obesity | endometrium | *ZNF493* | 12.06713818 | 8.31E-09 | 7.12E-08 | 4.03E-05 |
| Obesity | endometrium | *GTF2IRD2* | 8.517793865 | 9.18E-09 | 7.41E-08 | 4.45E-05 |
| Obesity | endometrium | *SUPT3H* | 9.505144104 | 1.91E-09 | 4.00E-08 | 9.26E-06 |
| Obesity | endometrium | *GIMAP5* | 7.116500036 | 4.51E-06 | 8.11E-06 | 0.02187935 |
| Obesity | endometrium | *STK31* | 5.82513566 | 4.41E-07 | 1.27E-06 | 0.002141657 |
| Obesity | endometrium | *ZKSCAN7* | 8.978128006 | 2.20E-09 | 4.00E-08 | 1.07E-05 |
| Obesity | endometrium | *CD55* | 3.316171632 | 3.09E-07 | 9.54E-07 | 0.00150021 |
| Obesity | endometrium | *CPNE4* | 4.285411484 | 1.08E-08 | 8.23E-08 | 5.25E-05 |
| Obesity | endometrium | *ZNF565* | 4.192290156 | 4.34E-08 | 2.10E-07 | 0.000210494 |
| Obesity | endometrium | *LONP1* | 4.324562171 | 0.011460179 | 0.013777202 | 1 |
| Obesity | endometrium | *ZNF34* | 4.457957275 | 1.58E-06 | 3.36E-06 | 0.007673715 |
| Obesity | endometrium | *EVL* | 6.82048901 | 0.000354851 | 0.000495464 | 1 |
| Obesity | endometrium | *PIK3R4* | 12.6398736 | 9.04E-07 | 2.24E-06 | 0.004385067 |
| Obesity | endometrium | *ZNF605* | 13.09910925 | 5.00E-11 | 7.13E-09 | 2.42E-07 |
| Obesity | endometrium | *RFX8* | 2.637406256 | 1.45E-05 | 2.39E-05 | 0.070493695 |
| Obesity | endometrium | *C20orf96* | 5.16081054 | 5.32E-07 | 1.47E-06 | 0.002583183 |
| Obesity | endometrium | *TPK1* | 6.65592642 | 4.62E-09 | 5.20E-08 | 2.24E-05 |
| Obesity | endometrium | *MYO18A* | 5.981765817 | 9.28E-08 | 3.70E-07 | 0.000450183 |
| Obesity | endometrium | *SPTSSB* | -1.988795955 | 9.64E-05 | 0.000144917 | 0.467935951 |
| Obesity | endometrium | *CACNA1H* | 7.59701712 | 1.30E-07 | 4.80E-07 | 0.000630677 |
| Obesity | endometrium | *SULF2* | 8.315177831 | 4.65E-08 | 2.23E-07 | 0.000225683 |
| Obesity | endometrium | *XRCC2* | -1.532875248 | 0.000252241 | 0.000359054 | 1 |
| Obesity | endometrium | *MYO6* | 9.919739886 | 2.39E-07 | 7.77E-07 | 0.0011587 |
| Obesity | endometrium | *ZNF846* | 7.887026728 | 4.22E-08 | 2.07E-07 | 0.00020491 |
| Obesity | endometrium | *ADH1B* | 6.305069132 | 1.83E-09 | 3.98E-08 | 8.89E-06 |
| Obesity | endometrium | *ZNF502* | 4.295686256 | 1.13E-06 | 2.67E-06 | 0.00550191 |
| Obesity | endometrium | *TRAPPC4* | 4.358489441 | 1.38E-06 | 3.13E-06 | 0.006712031 |
| Obesity | endometrium | *ZNF33B* | 3.843288587 | 1.15E-07 | 4.35E-07 | 0.00055638 |
| Obesity | endometrium | *AMZ2* | 4.384287469 | 7.76E-07 | 1.99E-06 | 0.003763056 |
| Obesity | endometrium | *ZNF431* | 10.72173379 | 5.98E-09 | 5.97E-08 | 2.90E-05 |
| Obesity | endometrium | *HLA-DQA1* | 2.596596355 | 1.23E-05 | 2.06E-05 | 0.059776753 |
| Obesity | endometrium | *ZNF239* | 6.26036894 | 1.64E-07 | 5.77E-07 | 0.000793305 |
| Obesity | endometrium | *SPRR2B* | -7.358250494 | 9.37E-05 | 0.00014095 | 0.454421677 |
| Obesity | endometrium | *FAM3C* | 6.903542399 | 4.36E-09 | 5.04E-08 | 2.12E-05 |
| Obesity | endometrium | *SLC39A10* | 10.19986388 | 1.47E-09 | 3.98E-08 | 7.14E-06 |
| Obesity | endometrium | *AP2A1* | 2.677675839 | 1.09E-06 | 2.60E-06 | 0.005292706 |
| Obesity | endometrium | *PCDHB16* | 3.470566861 | 0.005189554 | 0.006469608 | 1 |
| Obesity | endometrium | *HIST1H3E* | 4.734265576 | 0.000266467 | 0.000378151 | 1 |

| | | | | | | |
|---|---|---|---|---|---|---|
| Obesity | endometrium | *FUT11* | 3.571052598 | 1.08E-06 | 2.58E-06 | 0.005249293 |
| Obesity | endometrium | *WDR5B* | 5.433107133 | 2.05E-06 | 4.07E-06 | 0.009933578 |
| Obesity | endometrium | *NPIPB9* | 10.48466469 | 0.006557804 | 0.008088069 | 1 |
| Obesity | endometrium | *ZNF138* | 8.481328195 | 3.85E-10 | 2.34E-08 | 1.87E-06 |
| Obesity | endometrium | *ZNF429* | 5.376238569 | 5.10E-08 | 2.38E-07 | 0.000247686 |
| Obesity | endometrium | *ZNF470* | 19.32274308 | 9.29E-07 | 2.28E-06 | 0.004505879 |
| Obesity | endometrium | *ZNF100* | 2.870354285 | 8.82E-06 | 1.51E-05 | 0.042815614 |
| Obesity | endometrium | *ANXA6* | 6.984323058 | 1.14E-07 | 4.34E-07 | 0.000554853 |
| Obesity | endometrium | *GMFB* | 2.949355645 | 1.61E-06 | 3.39E-06 | 0.007820132 |
| Obesity | endometrium | *ZNF420* | 12.14350748 | 8.24E-07 | 2.08E-06 | 0.003996509 |
| Obesity | endometrium | *ZNF763* | -2.172690682 | 0.002284855 | 0.002954722 | 1 |
| Obesity | endometrium | *ZMYM1* | 13.09124279 | 6.79E-09 | 6.31E-08 | 3.29E-05 |
| Obesity | endometrium | *IGF2R* | 7.047153469 | 1.15E-08 | 8.48E-08 | 5.57E-05 |
| Obesity | endometrium | *SLC25A29* | 3.116089947 | 2.39E-07 | 7.77E-07 | 0.001160384 |
| Obesity | endometrium | *ZNF257* | 9.029968652 | 9.52E-05 | 0.000143094 | 0.461764868 |
| Obesity | endometrium | *ADAM32* | -2.289025846 | 0.017594128 | 0.02083131 | 1 |
| Obesity | endometrium | *LRRC8B* | 5.540366686 | 3.39E-08 | 1.76E-07 | 0.00016455 |
| Obesity | endometrium | *ABCB8* | 10.03623126 | 0.000117455 | 0.000174652 | 0.569890909 |
| Obesity | endometrium | *SND1* | 5.517314207 | 5.63E-09 | 5.87E-08 | 2.73E-05 |
| Obesity | endometrium | *SULT1A2* | 14.75137114 | 4.28E-12 | 9.02E-10 | 2.08E-08 |
| Obesity | endometrium | *NEK5* | 3.209294115 | 0.000121533 | 0.000180275 | 0.589679156 |
| Obesity | endometrium | *ENTPD4* | 5.452552779 | 1.24E-07 | 4.65E-07 | 0.00060221 |
| Obesity | endometrium | *TBC1D9B* | 7.0479693 | 1.07E-07 | 4.14E-07 | 0.000521336 |
| Obesity | endometrium | *FAM110D* | 10.11560444 | 0.000914531 | 0.00122815 | 1 |
| Obesity | endometrium | *CCL4L2* | 10.77950521 | 1.50E-06 | 3.27E-06 | 0.007256611 |
| Obesity | endometrium | *ZNF165* | -4.611153348 | 1.21E-06 | 2.81E-06 | 0.0058714 |
| Obesity | endometrium | *RAMP2-AS1* | -1.957468937 | 0.007813523 | 0.009535013 | 1 |
| Obesity | endometrium | *ZNF720* | 3.146497754 | 9.71E-07 | 2.36E-06 | 0.004711379 |
| Obesity | endometrium | *ZNF98* | 2.639242234 | 3.02E-05 | 4.77E-05 | 0.146358205 |
| Obesity | endometrium | *ZNF517* | 5.004883955 | 7.68E-07 | 1.98E-06 | 0.003727946 |
| Obesity | endometrium | *ZNF675* | 4.158665543 | 1.21E-06 | 2.81E-06 | 0.005869236 |
| Obesity | endometrium | *VEPH1* | 3.241088278 | 3.04E-06 | 5.71E-06 | 0.014737458 |
| Obesity | endometrium | *SHPK* | 4.817194734 | 0.009063534 | 0.010999566 | 1 |
| Obesity | endometrium | *MAP3K5* | 8.506287726 | 7.38E-10 | 2.94E-08 | 3.58E-06 |
| Obesity | endometrium | *GSTK1* | 4.272869394 | 2.38E-07 | 7.75E-07 | 0.001154444 |
| Obesity | endometrium | *GYPE* | 6.839089785 | 2.34E-06 | 4.55E-06 | 0.011330074 |
| Obesity | endometrium | *PCDHB11* | -1.25600777 | 0.000110384 | 0.000164593 | 0.535584945 |
| Obesity | endometrium | *SLC2A10* | 4.286821395 | 3.27E-07 | 9.94E-07 | 0.001587901 |
| Obesity | endometrium | *ZNF665* | 3.230415366 | 1.54E-06 | 3.32E-06 | 0.007482581 |
| Obesity | endometrium | *RPF2* | 9.250250447 | 3.87E-09 | 4.80E-08 | 1.88E-05 |
| Obesity | endometrium | *MYO5A* | 2.44783893 | 1.05E-06 | 2.51E-06 | 0.005082632 |
| Obesity | endometrium | *ATG7* | 4.748076467 | 7.46E-08 | 3.15E-07 | 0.000361743 |
| Obesity | endometrium | *RAB40C* | 4.808883 | 2.28E-07 | 7.50E-07 | 0.001107936 |
| Obesity | endometrium | *PIGN* | 5.438491734 | 0.001165087 | 0.001548768 | 1 |
| Obesity | endometrium | *HHLA3* | -2.477259779 | 0.023439786 | 0.027437839 | 1 |
| Obesity | endometrium | *TOPORS* | 14.49681653 | 3.14E-07 | 9.64E-07 | 0.001522527 |
| Obesity | endometrium | *BCO2* | 1.908032385 | 0.000105347 | 0.000157711 | 0.511141669 |
| Obesity | endometrium | *ENTPD6* | 3.084290782 | 8.51E-06 | 1.46E-05 | 0.041293185 |
| Obesity | endometrium | *ENPP1* | -0.9669544 | 0.00048548 | 0.00066881 | 1 |
| Obesity | endometrium | *MFAP5* | 1.984528227 | 0.000113552 | 0.000169005 | 0.550956251 |

| | | | | | | |
|---|---|---|---|---|---|---|
| Obesity | endometrium | *PDCD1LG2* | 2.185770591 | 3.95E-06 | 7.20E-06 | 0.01915642 |
| Obesity | endometrium | *DNAH10* | 23.85108812 | 1.36E-20 | 3.29E-17 | 6.58E-17 |
| Obesity | endometrium | *FAM114A1* | 9.143362944 | 7.22E-10 | 2.94E-08 | 3.50E-06 |
| Obesity | endometrium | *PSAP* | 2.238875867 | 6.60E-07 | 1.76E-06 | 0.003201128 |
| Obesity | endometrium | *TXNRD3* | 3.557898108 | 4.37E-07 | 1.26E-06 | 0.002120882 |
| Obesity | endometrium | *ATAD3A* | 7.113987727 | 0.018903855 | 0.022305813 | 1 |
| Obesity | endometrium | *AC122129.1* | 11.17671795 | 9.60E-07 | 2.34E-06 | 0.004655874 |
| Obesity | endometrium | *OCLN* | -2.326597105 | 0.000508136 | 0.000698237 | 1 |
| Obesity | endometrium | *ZNF181* | 5.180715423 | 5.73E-06 | 1.02E-05 | 0.027787075 |
| Obesity | endometrium | *SGTB* | 3.9197608 | 8.14E-07 | 2.06E-06 | 0.003947529 |
| Obesity | endometrium | *ADH5* | 10.1396884 | 2.82E-09 | 4.29E-08 | 1.37E-05 |
| Obesity | endometrium | *HRNR* | 1.448010062 | 0.003945993 | 0.004982034 | 1 |
| Obesity | endometrium | *ZNF677* | 3.549399344 | 1.14E-06 | 2.68E-06 | 0.005525908 |
| Obesity | endometrium | *ZNF311* | 4.269981196 | 1.12E-05 | 1.89E-05 | 0.054458816 |
| Obesity | endometrium | *FCHSD1* | 8.906600684 | 0.000254563 | 0.000362081 | 1 |
| Obesity | endometrium | *RPL12* | 3.968048956 | 0.019320366 | 0.022775125 | 1 |
| Obesity | endometrium | *MPZL1* | 8.680285936 | 1.39E-09 | 3.98E-08 | 6.76E-06 |
| Obesity | endometrium | *VPS13A* | 3.916501009 | 1.55E-07 | 5.52E-07 | 0.000750645 |
| Obesity | endometrium | *NOL8* | 3.938666322 | 1.49E-06 | 3.26E-06 | 0.007231708 |
| Obesity | endometrium | *DLGAP2* | 7.666945202 | 4.31E-10 | 2.40E-08 | 2.09E-06 |
| Obesity | endometrium | *ZNF560* | -2.215459383 | 0.006147398 | 0.007603154 | 1 |
| Obesity | endometrium | *AGAP9* | 5.957214641 | 2.78E-05 | 4.43E-05 | 0.134954736 |
| Obesity | endometrium | *ZNF273* | 3.835907664 | 2.52E-06 | 4.88E-06 | 0.012243813 |
| Obesity | endometrium | *MAK16* | 3.931863411 | 1.17E-06 | 2.74E-06 | 0.005673139 |
| Obesity | endometrium | *SFI1* | 4.652641329 | 3.11E-07 | 9.59E-07 | 0.001510419 |
| Obesity | endometrium | *ZNF649* | 3.593749694 | 2.91E-05 | 4.61E-05 | 0.141098743 |
| Obesity | endometrium | *ADH4* | -2.928702308 | 0.000428779 | 0.000593732 | 1 |
| Obesity | endometrium | *HIBCH* | 3.056628632 | 5.26E-06 | 9.39E-06 | 0.025527342 |
| Obesity | endometrium | *SOWAHC* | 7.730467568 | 1.67E-07 | 5.85E-07 | 0.000808394 |
| Obesity | endometrium | *AC135178.1* | 23.37207139 | 0.043491824 | 0.0498635 | 1 |
| Obesity | endometrium | *MIER1* | 10.25369955 | 3.53E-07 | 1.06E-06 | 0.001713259 |
| Obesity | endometrium | *MAN1A2* | 9.861544913 | 1.83E-09 | 3.98E-08 | 8.86E-06 |
| Obesity | endometrium | *DDRGK1* | 13.06974287 | 2.18E-09 | 4.00E-08 | 1.06E-05 |
| Obesity | endometrium | *ZNF607* | 5.585927604 | 0.00045725 | 0.000631713 | 1 |
| Obesity | endometrium | *ZNF334* | 2.918611131 | 3.50E-06 | 6.47E-06 | 0.016965913 |
| Obesity | endometrium | *HSD17B11* | 4.500278737 | 8.52E-08 | 3.48E-07 | 0.000413598 |
| Obesity | endometrium | *SULT1C2* | 3.231898566 | 0.000106406 | 0.00015915 | 0.516284022 |
| Obesity | endometrium | *RPS6KL1* | 13.48815867 | 2.61E-05 | 4.16E-05 | 0.126593294 |
| Obesity | endometrium | *DDX42* | 5.80272708 | 1.03E-08 | 7.98E-08 | 5.01E-05 |
| Obesity | endometrium | *HELZ* | 7.116049951 | 2.15E-09 | 4.00E-08 | 1.04E-05 |
| Obesity | endometrium | *TMEM116* | 2.506359375 | 0.018958375 | 0.022364706 | 1 |
| Obesity | endometrium | *UCKL1* | 7.068233254 | 0.008930101 | 0.010848485 | 1 |
| Obesity | endometrium | *GFPT1* | 10.98307055 | 8.01E-09 | 7.01E-08 | 3.89E-05 |
| Obesity | endometrium | *ZNF568* | 2.73810225 | 1.55E-06 | 3.33E-06 | 0.007542787 |
| Obesity | endometrium | *ZNF587* | 8.910901987 | 6.55E-08 | 2.85E-07 | 0.000317856 |
| Obesity | endometrium | *FLVCR1-AS1* | 2.358474843 | 0.000118673 | 0.000176302 | 0.575800747 |
| Obesity | endometrium | *SH3BGRL2* | 7.935544209 | 8.46E-09 | 7.13E-08 | 4.11E-05 |
| Obesity | endometrium | *TMA16* | 8.384418646 | 5.47E-09 | 5.77E-08 | 2.65E-05 |
| Obesity | endometrium | *CNGA1* | 4.618984423 | 3.75E-05 | 5.87E-05 | 0.181755036 |
| Obesity | endometrium | *ZNF28* | 7.635371907 | 9.26E-08 | 3.69E-07 | 0.000449102 |

| | | | | | | |
|---|---|---|---|---|---|---|
| Obesity | endometrium | *C20orf203* | 8.797847603 | 0.000596586 | 0.000812871 | 1 |
| Obesity | endometrium | *NCOA6* | 4.993416815 | 2.71E-06 | 5.20E-06 | 0.013161013 |
| Obesity | endometrium | *FAM19A2* | 8.070635744 | 8.73E-05 | 0.000131582 | 0.423432103 |
| Obesity | endometrium | *TTC37* | 6.054996167 | 3.22E-08 | 1.70E-07 | 0.000156137 |
| Obesity | endometrium | *TUSC1* | -1.257070915 | 0.00028681 | 0.000405714 | 1 |
| Obesity | endometrium | *PAPSS2* | 9.746712939 | 1.68E-09 | 3.98E-08 | 8.16E-06 |
| Obesity | endometrium | *CEP290* | 8.509729185 | 4.28E-09 | 4.96E-08 | 2.07E-05 |
| Obesity | endometrium | *ANKRD13B* | 5.423732617 | 1.62E-06 | 3.40E-06 | 0.007837092 |
| Obesity | endometrium | *ECI2* | 3.654405231 | 5.17E-07 | 1.44E-06 | 0.002509505 |
| Obesity | endometrium | *UNC13B* | 4.769713812 | 1.73E-07 | 6.01E-07 | 0.000838555 |
| Obesity | endometrium | *PPP1R14C* | -1.634513781 | 0.000173338 | 0.00025128 | 0.841034387 |
| Obesity | endometrium | *GPATCH3* | 5.349251313 | 1.70E-06 | 3.50E-06 | 0.008230717 |
| Obesity | endometrium | *RPL10A* | 8.228448214 | 2.41E-09 | 4.02E-08 | 1.17E-05 |
| Obesity | endometrium | *COLGALT2* | 13.16299976 | 4.65E-08 | 2.23E-07 | 0.000225583 |
| Obesity | endometrium | *RCSD1* | 2.399682642 | 1.48E-06 | 3.26E-06 | 0.007194333 |
| Obesity | endometrium | *GRIN3A* | -1.054033108 | 0.000132614 | 0.000195695 | 0.643444801 |
| Obesity | endometrium | *TMEM184B* | 4.774096703 | 7.85E-06 | 1.36E-05 | 0.038082268 |
| Obesity | endometrium | *SCAMP5* | 1.851297798 | 0.007562777 | 0.009240643 | 1 |
| Obesity | endometrium | *SFT2D1* | 7.711969017 | 6.80E-09 | 6.31E-08 | 3.30E-05 |
| Obesity | endometrium | *HMGN2* | 4.755102912 | 9.09E-07 | 2.25E-06 | 0.004410807 |
| Obesity | endometrium | *DENND4B* | 2.767237826 | 3.01E-06 | 5.66E-06 | 0.014602222 |
| Obesity | endometrium | *RYR3* | 2.666838596 | 7.43E-07 | 1.93E-06 | 0.003606505 |
| Obesity | endometrium | *TYW1* | 4.892814163 | 8.82E-08 | 3.56E-07 | 0.000428026 |
| Obesity | endometrium | *PRMT6* | 10.69291631 | 5.86E-06 | 1.04E-05 | 0.028412018 |
| Obesity | endometrium | *SHISA4* | -16.45362017 | 2.56E-06 | 4.94E-06 | 0.012424219 |
| Obesity | endometrium | *MAP3K3* | 2.122208318 | 7.42E-06 | 1.29E-05 | 0.036024273 |
| Obesity | endometrium | *C1orf174* | 4.078364108 | 1.64E-06 | 3.42E-06 | 0.007941671 |
| Obesity | endometrium | *DZIP3* | 4.138403601 | 1.83E-07 | 6.29E-07 | 0.000889261 |
| Obesity | endometrium | *KIAA0753* | 11.84235 | 1.23E-07 | 4.60E-07 | 0.000594734 |
| Obesity | endometrium | *TBKBP1* | 4.588932046 | 3.86E-06 | 7.06E-06 | 0.018710909 |
| Obesity | endometrium | *ZFP2* | 8.804294555 | 4.22E-08 | 2.07E-07 | 0.000204626 |
| Obesity | endometrium | *L3MBTL3* | 9.847099402 | 3.14E-08 | 1.66E-07 | 0.000152322 |
| Obesity | endometrium | *MFAP3L* | 5.348950774 | 1.43E-06 | 3.18E-06 | 0.006919156 |
| Obesity | endometrium | *NAGA* | 4.384657046 | 3.82E-06 | 7.00E-06 | 0.018543262 |
| Obesity | endometrium | *SMG5* | 7.417459335 | 2.81E-05 | 4.47E-05 | 0.136484496 |
| Obesity | endometrium | *TGM2* | -1.527189763 | 0.003253258 | 0.004141907 | 1 |
| Obesity | endometrium | *PJA2* | 9.943050774 | 1.50E-07 | 5.38E-07 | 0.00072578 |
| Obesity | endometrium | *AL590822.1* | 4.552009157 | 1.76E-06 | 3.60E-06 | 0.008561038 |
| Obesity | endometrium | *AP001007.1* | 9.018222757 | 4.09E-06 | 7.42E-06 | 0.019836608 |
| Obesity | endometrium | *EFCAB2* | 3.240866847 | 2.31E-07 | 7.55E-07 | 0.001118874 |
| Obesity | endometrium | *CHML* | 8.947532967 | 1.30E-07 | 4.80E-07 | 0.000630653 |
| Obesity | endometrium | *TATDN3* | 3.31571438 | 3.65E-07 | 1.09E-06 | 0.00176994 |
| Obesity | endometrium | *CR1* | 1.385885267 | 0.002653583 | 0.003408839 | 1 |
| Obesity | endometrium | *SAMD5* | 2.920113514 | 2.01E-06 | 4.02E-06 | 0.009768685 |
| Obesity | endometrium | *ECT2L* | 5.603348316 | 8.10E-07 | 2.06E-06 | 0.003930504 |
| Obesity | endometrium | *GPR52* | 109.3485301 | 3.92E-05 | 6.13E-05 | 0.190294057 |
| Obesity | endometrium | *FAM229B* | 3.365509715 | 1.08E-05 | 1.82E-05 | 0.052415517 |
| Obesity | endometrium | *DDO* | 7.044755017 | 3.08E-06 | 5.78E-06 | 0.014926054 |
| Obesity | endometrium | *RBM20* | 4.765055718 | 6.52E-06 | 1.15E-05 | 0.031652067 |
| Obesity | endometrium | *PCMTD2* | 4.923336331 | 5.99E-08 | 2.65E-07 | 0.000290479 |

| | | | | | | |
|---|---|---|---|---|---|---|
| Obesity | endometrium | *SAMD13* | -2.035024042 | 0.001291266 | 0.001710875 | 1 |
| Obesity | endometrium | *EFCAB7* | 10.22803201 | 2.16E-09 | 4.00E-08 | 1.05E-05 |
| Obesity | endometrium | *IFIT1B* | 22.55990701 | 2.15E-06 | 4.25E-06 | 0.010451723 |
| Obesity | endometrium | *LIPN* | 10.19282394 | 1.51E-06 | 3.29E-06 | 0.007310939 |
| Obesity | endometrium | *INPP5B* | 2.317642333 | 0.003161463 | 0.004031384 | 1 |
| Obesity | endometrium | *NEU4* | -3.009587622 | 0.000289037 | 0.000408508 | 1 |
| Obesity | endometrium | *C2orf72* | -4.544825316 | 0.00013734 | 0.000202238 | 0.666375646 |
| Obesity | endometrium | *RUFY2* | 10.56484981 | 3.75E-09 | 4.72E-08 | 1.82E-05 |
| Obesity | endometrium | *PHACTR4* | 4.809215535 | 1.36E-07 | 4.96E-07 | 0.00065956 |
| Obesity | endometrium | *ASAH2B* | -2.248541302 | 0.001196911 | 0.001589765 | 1 |
| Obesity | endometrium | *AGAP6* | 3.409166866 | 2.37E-06 | 4.60E-06 | 0.011480234 |
| Obesity | endometrium | *SYT15* | 8.972635118 | 2.15E-05 | 3.47E-05 | 0.10419246 |
| Obesity | endometrium | *LINC00619* | 38.35121756 | 3.85E-06 | 7.05E-06 | 0.018676873 |
| Obesity | endometrium | *DAXX* | 4.023116351 | 2.95E-06 | 5.56E-06 | 0.014293568 |
| Obesity | endometrium | *BMPR2* | 5.319651842 | 7.49E-09 | 6.64E-08 | 3.63E-05 |
| Obesity | endometrium | *TCEA3* | 3.089629921 | 3.55E-06 | 6.55E-06 | 0.017217393 |
| Obesity | endometrium | *HSD17B8* | 4.048879378 | 0.001147542 | 0.001526281 | 1 |
| Obesity | endometrium | *OXLD1* | 3.475875881 | 1.45E-05 | 2.39E-05 | 0.070323305 |
| Obesity | endometrium | *TAP2* | 2.727874014 | 0.001571338 | 0.002062806 | 1 |
| Obesity | endometrium | *COL15A1* | 6.607565619 | 8.98E-08 | 3.61E-07 | 0.000435907 |
| Obesity | endometrium | *NOTCH4* | -1.286851413 | 8.66E-08 | 3.52E-07 | 0.000420319 |
| Obesity | endometrium | *AGER* | 4.658840231 | 9.91E-06 | 1.68E-05 | 0.048076752 |
| Obesity | endometrium | *SMIM5* | 10.38113194 | 6.87E-06 | 1.20E-05 | 0.033331714 |
| Obesity | endometrium | *STK19* | 3.62499 | 1.22E-06 | 2.84E-06 | 0.005940885 |
| Obesity | endometrium | *C6orf48* | 4.808661918 | 0.028423543 | 0.033008863 | 1 |
| Obesity | endometrium | *GPANK1* | 2.40091035 | 0.001511182 | 0.0019876 | 1 |
| Obesity | endometrium | *BAG6* | 4.381117354 | 7.21E-07 | 1.88E-06 | 0.00349878 |
| Obesity | endometrium | *LST1* | 3.23513985 | 7.47E-07 | 1.94E-06 | 0.003626122 |
| Obesity | endometrium | *ZNF814* | 8.586232871 | 3.10E-10 | 2.18E-08 | 1.50E-06 |
| Obesity | endometrium | *MICA* | 3.420860734 | 0.00018026 | 0.000260537 | 0.874622015 |
| Obesity | endometrium | *HLA-C* | 4.834266087 | 8.64E-09 | 7.23E-08 | 4.19E-05 |
| Obesity | endometrium | *POU5F1* | 2.167914123 | 6.18E-06 | 1.09E-05 | 0.029981462 |
| Obesity | endometrium | *CCHCR1* | 3.296598824 | 3.41E-09 | 4.54E-08 | 1.65E-05 |
| Obesity | endometrium | *PSORS1C1* | -0.673834251 | 0.03639435 | 0.041924356 | 1 |
| Obesity | endometrium | *DHX16* | 11.10087302 | 6.07E-08 | 2.68E-07 | 0.000294347 |
| Obesity | endometrium | *ABCF1* | 7.920524982 | 3.77E-10 | 2.34E-08 | 1.83E-06 |
| Obesity | endometrium | *LILRB3* | 5.445257212 | 2.02E-06 | 4.03E-06 | 0.009801996 |
| Obesity | endometrium | *HLA-E* | 7.104383442 | 4.65E-10 | 2.45E-08 | 2.26E-06 |
| Obesity | endometrium | *ZNF468* | -3.906016728 | 0.00146841 | 0.001933965 | 1 |
| Obesity | endometrium | *RNF39* | -11.88892895 | 2.73E-07 | 8.61E-07 | 0.001324674 |
| Obesity | endometrium | *HCG9* | 4.346639572 | 0.00363203 | 0.004610834 | 1 |
| Obesity | endometrium | *HLA-G* | 4.213720641 | 4.28E-06 | 7.74E-06 | 0.02074639 |
| Obesity | endometrium | *ZFP57* | 4.338521271 | 1.51E-07 | 5.42E-07 | 0.000732752 |
| Obesity | endometrium | *MALRD1* | 5.169681298 | 5.79E-08 | 2.59E-07 | 0.000281098 |
| Obesity | endometrium | *FAM196B* | 2.722735412 | 1.69E-06 | 3.49E-06 | 0.008195173 |
| Obesity | endometrium | *MROH6* | 3.967614686 | 0.019980716 | 0.023513566 | 1 |
| Obesity | endometrium | *DCTN1* | 17.8687034 | 2.42E-08 | 1.38E-07 | 0.000117208 |
| Obesity | endometrium | *ZBTB48* | 8.621693555 | 4.00E-09 | 4.83E-08 | 1.94E-05 |
| Obesity | endometrium | *MZT1* | 5.398984351 | 6.73E-07 | 1.79E-06 | 0.003266757 |
| Obesity | endometrium | *ZNF155* | 4.366822498 | 2.54E-07 | 8.14E-07 | 0.001233344 |

| | | | | | | |
|---|---|---|---|---|---|---|
| Obesity | endometrium | *ZNF425* | 6.588771185 | 2.77E-06 | 5.29E-06 | 0.013433926 |
| Obesity | endometrium | *C12orf73* | 10.93447373 | 5.82E-08 | 2.60E-07 | 0.000282348 |
| Obesity | endometrium | *PCDHGA1* | -2.735115416 | 6.27E-05 | 9.59E-05 | 0.304326496 |
| Obesity | endometrium | *PCDHA4* | 7.621899946 | 0.000663019 | 0.000899851 | 1 |
| Obesity | endometrium | *TRIM13* | 8.503945334 | 7.23E-10 | 2.94E-08 | 3.51E-06 |
| Obesity | endometrium | *SPIRE2* | -2.706547606 | 0.000293397 | 0.000414308 | 1 |
| Obesity | endometrium | *PKHD1L1* | 2.210747849 | 8.44E-07 | 2.12E-06 | 0.004095653 |
| Obesity | endometrium | *SLFN12L* | 2.289066478 | 2.57E-05 | 4.11E-05 | 0.124886483 |
| Obesity | endometrium | *SLC35B4* | 5.449120679 | 1.03E-07 | 4.04E-07 | 0.000501768 |
| Obesity | endometrium | *SYCE1L* | 2.121727118 | 0.013797543 | 0.016485023 | 1 |
| Obesity | endometrium | *TMEM231* | 6.078003496 | 8.90E-07 | 2.22E-06 | 0.004316019 |
| Obesity | endometrium | *SCGB2B2* | 5.067928394 | 7.60E-08 | 3.18E-07 | 0.000368764 |
| Obesity | endometrium | *CCDC144NL* | 5.097092977 | 1.57E-06 | 3.35E-06 | 0.007610969 |
| Obesity | endometrium | *TMEM170B* | 2.245291182 | 1.40E-05 | 2.31E-05 | 0.067912701 |
| Obesity | endometrium | *GOLGA6L10* | 11.42329384 | 7.11E-07 | 1.86E-06 | 0.003450165 |
| Obesity | endometrium | *NT5M* | 6.392507749 | 5.43E-07 | 1.49E-06 | 0.002633027 |
| Obesity | endometrium | *IPO7* | 11.6979952 | 5.19E-07 | 1.44E-06 | 0.002520122 |
| Obesity | endometrium | *PRR13* | 6.260953373 | 0.000134038 | 0.000197555 | 0.650351848 |
| Obesity | endometrium | *TECPR1* | 4.314790004 | 2.81E-07 | 8.86E-07 | 0.001365829 |
| Obesity | endometrium | *MT1A* | -1.648413963 | 0.000237115 | 0.000338975 | 1 |
| Obesity | endometrium | *C15orf59* | 9.218399133 | 0.000128188 | 0.000189567 | 0.621969603 |
| Obesity | endometrium | *MT1M* | 4.762416481 | 2.12E-06 | 4.20E-06 | 0.010294647 |
| Obesity | endometrium | *CFI* | 2.616445765 | 1.13E-06 | 2.67E-06 | 0.005486375 |
| Obesity | endometrium | *ATP6AP1L* | 3.757485107 | 4.56E-05 | 7.08E-05 | 0.221415048 |
| Obesity | endometrium | *RGL3* | 2.870215235 | 2.73E-06 | 5.23E-06 | 0.013258018 |
| Obesity | endometrium | *NAP1L4* | 4.326292659 | 1.66E-07 | 5.85E-07 | 0.000807101 |
| Obesity | endometrium | *LIN52* | 3.143443592 | 3.49E-07 | 1.05E-06 | 0.001692206 |
| Obesity | endometrium | *C17orf107* | 9.454390313 | 1.27E-05 | 2.11E-05 | 0.061518591 |
| Obesity | endometrium | *KLRC2* | 8.049739701 | 5.98E-07 | 1.61E-06 | 0.002903578 |
| Obesity | endometrium | *LINC00487* | 8.656291816 | 0.023154487 | 0.027130058 | 1 |
| Obesity | endometrium | *RFPL3S* | 3.411105658 | 0.008389941 | 0.01021787 | 1 |
| Obesity | endometrium | *C1QTNF9B* | -2.310150941 | 0.000129035 | 0.000190646 | 0.626080045 |
| Obesity | endometrium | *ZDHHC11B* | -2.301691405 | 0.004187588 | 0.005276078 | 1 |
| Obesity | endometrium | *HCG27* | 3.168874166 | 0.000136354 | 0.000200846 | 0.661588364 |
| Obesity | endometrium | *COL6A6* | 9.175861637 | 7.06E-07 | 1.85E-06 | 0.003423381 |
| Obesity | endometrium | *HLA-A* | 5.270713357 | 4.00E-07 | 1.17E-06 | 0.001942522 |
| Obesity | endometrium | *LNP1* | 4.256160882 | 7.41E-06 | 1.29E-05 | 0.035930877 |
| Obesity | endometrium | *PRSS50* | 12.11792336 | 0.003030241 | 0.003870158 | 1 |
| Obesity | endometrium | *METTL6* | 8.973801075 | 5.71E-09 | 5.87E-08 | 2.77E-05 |
| Obesity | endometrium | *SACM1L* | 4.122293222 | 1.94E-07 | 6.60E-07 | 0.000943481 |
| Obesity | endometrium | *SLC48A1* | 12.73253148 | 1.45E-09 | 3.98E-08 | 7.03E-06 |
| Obesity | endometrium | *TAS2R50* | 73.76363212 | 6.65E-05 | 0.00010144 | 0.322577719 |
| Obesity | endometrium | *AC084018.1* | -4.612092694 | 7.74E-07 | 1.99E-06 | 0.003756187 |
| Obesity | endometrium | *C17orf51* | 5.86471936 | 1.61E-06 | 3.39E-06 | 0.007805721 |
| Obesity | endometrium | *OR2A42* | -2.330537225 | 3.02E-05 | 4.77E-05 | 0.146297331 |
| Obesity | endometrium | *NUP62* | 3.954020648 | 0.000144435 | 0.00021185 | 0.700798458 |
| Obesity | endometrium | *AL021068.1* | 8.064507825 | 0.00032645 | 0.000457917 | 1 |
| Obesity | endometrium | *FGFR1OP* | 18.31111078 | 1.77E-07 | 6.12E-07 | 0.000857518 |
| Obesity | endometrium | *ZNF254* | 8.340473325 | 5.09E-08 | 2.38E-07 | 0.000247035 |
| Obesity | endometrium | *TCTEX1D2* | 10.60717992 | 2.07E-07 | 6.89E-07 | 0.001002082 |

| | | | | | | |
|---|---|---|---|---|---|---|
| Obesity | endometrium | *AC022498.1* | 24.80213231 | 1.26E-07 | 4.71E-07 | 0.000612882 |
| Obesity | endometrium | *LINGO4* | 5.455701855 | 0.035070316 | 0.04045677 | 1 |
| Obesity | endometrium | *GIMAP1* | 10.48892899 | 1.21E-08 | 8.70E-08 | 5.86E-05 |
| Obesity | endometrium | *NOTCH2NL* | 4.171257058 | 0.009458323 | 0.011455762 | 1 |
| Obesity | endometrium | *SUPT4H1* | 6.197224652 | 8.80E-08 | 3.55E-07 | 0.00042695 |
| Obesity | endometrium | *MXD3* | 5.328460519 | 7.76E-07 | 1.99E-06 | 0.003763966 |
| Obesity | endometrium | *GSTM2* | 6.412162626 | 0.000108167 | 0.000161634 | 0.524824754 |
| Obesity | endometrium | *ARHGAP19* | 5.675541446 | 3.22E-05 | 5.08E-05 | 0.156366521 |
| Obesity | endometrium | *LCAT* | 4.006223434 | 1.84E-06 | 3.72E-06 | 0.008940055 |
| Obesity | endometrium | *GPC2* | -5.189923548 | 0.000124205 | 0.000184069 | 0.602643289 |
| Obesity | endometrium | *ERV3-1* | 2.515194981 | 1.00E-08 | 7.86E-08 | 4.87E-05 |
| Obesity | endometrium | *GBP7* | -0.977074686 | 0.000240602 | 0.000343657 | 1 |
| Obesity | endometrium | *SRA1* | 6.301541895 | 2.94E-09 | 4.29E-08 | 1.43E-05 |
| Obesity | endometrium | *TMEM110* | 17.20623555 | 4.41E-15 | 2.68E-12 | 2.14E-11 |
| Obesity | endometrium | *TMX2* | 5.588628406 | 3.18E-06 | 5.96E-06 | 0.015442841 |
| Obesity | endometrium | *LBH* | 3.270974677 | 6.26E-06 | 1.10E-05 | 0.030365139 |
| Obesity | endometrium | *PPP1CB* | 4.037272121 | 6.86E-06 | 1.20E-05 | 0.033294505 |
| Obesity | endometrium | *NCKIPSD* | -2.430870099 | 0.009158112 | 0.011108789 | 1 |
| Obesity | endometrium | *ATF6B* | 9.337103398 | 2.51E-07 | 8.07E-07 | 0.001219299 |
| Obesity | endometrium | *SLC35F6* | 3.962230615 | 1.92E-06 | 3.86E-06 | 0.009332938 |
| Obesity | endometrium | *EMP2* | 6.876993297 | 5.18E-09 | 5.64E-08 | 2.51E-05 |
| Obesity | endometrium | *PPM1N* | 4.289265258 | 3.19E-05 | 5.03E-05 | 0.154769668 |
| Obesity | endometrium | *SLC23A3* | -1.176111892 | 0.005167157 | 0.006443343 | 1 |
| Obesity | endometrium | *LTB4R* | 4.587781034 | 2.66E-06 | 5.12E-06 | 0.012929539 |
| Obesity | endometrium | *IRF9* | 7.12414573 | 2.76E-08 | 1.51E-07 | 0.000133843 |
| Obesity | endometrium | *GALT* | 3.660834817 | 1.66E-07 | 5.84E-07 | 0.000803694 |
| Obesity | endometrium | *ZNF90* | 3.82421205 | 3.77E-06 | 6.92E-06 | 0.018293708 |
| Obesity | endometrium | *REPIN1* | 22.06690162 | 1.23E-08 | 8.86E-08 | 5.99E-05 |
| Obesity | endometrium | *ZNF891* | 13.79078434 | 2.30E-06 | 4.48E-06 | 0.011141901 |
| Obesity | endometrium | *SMIM7* | 6.095371673 | 5.68E-08 | 2.57E-07 | 0.000275555 |
| Obesity | endometrium | *TSPAN4* | 5.433354844 | 5.69E-09 | 5.87E-08 | 2.76E-05 |
| Obesity | endometrium | *PAXIP1-AS2* | 10.56794455 | 1.38E-09 | 3.98E-08 | 6.71E-06 |
| Obesity | endometrium | *LYRM4* | 5.058802075 | 5.43E-08 | 2.49E-07 | 0.000263415 |
| Obesity | endometrium | *PRCD* | 11.59754632 | 0.001546208 | 0.002031466 | 1 |
| Obesity | endometrium | *SH3D21* | 6.746976701 | 1.43E-06 | 3.19E-06 | 0.006939948 |
| Obesity | endometrium | *FIS1* | 5.863436592 | 7.75E-05 | 0.000117619 | 0.3757939 |
| Obesity | endometrium | *ANG* | -2.621611022 | 0.026197276 | 0.030540409 | 1 |
| Obesity | endometrium | *COLCA2* | 3.204941413 | 0.003418763 | 0.004345779 | 1 |
| Obesity | endometrium | *SOGA3* | 3.037909759 | 0.000885759 | 0.00119083 | 1 |
| Obesity | endometrium | *LCNL1* | 7.665928657 | 0.000297727 | 0.000420055 | 1 |
| Obesity | endometrium | *GNAT3* | 4.203480837 | 1.55E-06 | 3.32E-06 | 0.007520983 |
| Obesity | endometrium | *PLIN5* | 2.700101412 | 2.05E-05 | 3.32E-05 | 0.099604371 |
| Obesity | endometrium | *SEC14L6* | 1.133925286 | 0.01521055 | 0.018119713 | 1 |
| Obesity | endometrium | *CPEB1* | 6.208329895 | 0.00308026 | 0.003931971 | 1 |
| Obesity | endometrium | *EML6* | 14.5482124 | 6.53E-09 | 6.17E-08 | 3.17E-05 |
| Obesity | endometrium | *ARHGEF33* | 4.068726607 | 0.000109452 | 0.000163403 | 0.531059369 |
| Obesity | endometrium | *IFRD2* | 5.815996577 | 3.38E-05 | 5.32E-05 | 0.16390534 |
| Obesity | endometrium | *EVPLL* | 8.66305227 | 0.012091425 | 0.014510906 | 1 |
| Obesity | endometrium | *AC005082.1* | 14.57614304 | 4.72E-05 | 7.32E-05 | 0.229090691 |
| Obesity | endometrium | *SMTNL1* | 1.795255601 | 0.024889149 | 0.029071293 | 1 |

| | | | | | | |
|---|---|---|---|---|---|---|
| Obesity | endometrium | *ACSM4* | 3.1215698 | 0.007861351 | 0.009588556 | 1 |
| Obesity | endometrium | *UBXN2B* | 9.408259964 | 9.06E-06 | 1.55E-05 | 0.043935135 |
| Obesity | endometrium | *C5orf49* | 42.35635654 | 4.00E-07 | 1.17E-06 | 0.001940326 |
| Obesity | endometrium | *GOLGA8B* | 3.141291697 | 2.38E-07 | 7.75E-07 | 0.001155194 |
| Obesity | endometrium | *VPS16* | 14.59654457 | 1.60E-05 | 2.63E-05 | 0.077817331 |
| Obesity | endometrium | *MYL5* | -4.47134763 | 0.011620137 | 0.013959125 | 1 |
| Obesity | endometrium | *NPEPL1* | 3.704037477 | 1.96E-06 | 3.92E-06 | 0.009491483 |
| Obesity | endometrium | *TMEM242* | 12.0934064 | 1.03E-08 | 7.97E-08 | 4.97E-05 |
| Obesity | endometrium | *SLC35E2* | 3.128671427 | 3.42E-05 | 5.38E-05 | 0.165770819 |
| Obesity | endometrium | *LINC01139* | -1.259806308 | 0.003037894 | 0.003878911 | 1 |
| Obesity | endometrium | *TSTD1* | 9.830045919 | 0.002368431 | 0.003059538 | 1 |
| Obesity | endometrium | *AC009403.2* | -4.471351276 | 0.002902004 | 0.003714197 | 1 |
| Obesity | endometrium | *PAM16* | 18.82516064 | 3.16E-09 | 4.44E-08 | 1.53E-05 |
| Obesity | endometrium | *AC016773.1* | 5.255700785 | 1.07E-05 | 1.81E-05 | 0.051994716 |
| Obesity | endometrium | *AP000350.4* | -1.155754946 | 0.010922164 | 0.01314996 | 1 |
| Obesity | endometrium | *RNASEK* | 9.845142966 | 4.40E-06 | 7.93E-06 | 0.021350355 |
| Obesity | endometrium | *NBPF1* | 8.444655458 | 1.66E-05 | 2.71E-05 | 0.080461185 |
| Obesity | endometrium | *FAM228B* | 2.893222437 | 1.34E-05 | 2.23E-05 | 0.065209021 |
| Obesity | endometrium | *RNU6ATAC35I* | 28.13427493 | 0.004326925 | 0.005440332 | 1 |
| Obesity | endometrium | *FANCG* | -2.737465115 | 0.00026241 | 0.000372721 | 1 |
| Obesity | endometrium | *AP4M1* | 4.083281013 | 0.001723059 | 0.002250413 | 1 |
| Obesity | endometrium | *ZNF880* | 2.568104141 | 8.11E-05 | 0.000122666 | 0.393512306 |
| Obesity | endometrium | *TIGD1* | 6.168912712 | 1.15E-05 | 1.94E-05 | 0.055994349 |
| Obesity | endometrium | *FXYD7* | 5.336419053 | 1.51E-05 | 2.48E-05 | 0.073148765 |
| Obesity | endometrium | *SLC12A8* | 3.115753311 | 5.23E-05 | 8.08E-05 | 0.253951392 |
| Obesity | endometrium | *APOL6* | 11.27334265 | 6.80E-08 | 2.94E-07 | 0.000329856 |
| Obesity | endometrium | *OR2A1* | 5.332750461 | 0.000627215 | 0.000853407 | 1 |
| Obesity | endometrium | *UBA52* | 8.269242925 | 3.91E-09 | 4.80E-08 | 1.90E-05 |
| Obesity | endometrium | *TIAF1* | 6.71996739 | 3.50E-05 | 5.50E-05 | 0.169661727 |
| Obesity | endometrium | *RAB6C* | 107.0303391 | 7.11E-06 | 1.24E-05 | 0.034517486 |
| Obesity | endometrium | *AC062017.1* | 3.925564482 | 1.68E-06 | 3.48E-06 | 0.0081544 |
| Obesity | endometrium | *ADRA2B* | 7.240287849 | 0.041804569 | 0.047985751 | 1 |
| Obesity | endometrium | *EXOSC6* | -7.197117129 | 0.008011233 | 0.009766458 | 1 |
| Obesity | endometrium | *ZNF844* | 6.646911403 | 0.010104954 | 0.012196328 | 1 |
| Obesity | endometrium | *LINC00571* | -2.431868761 | 0.000899096 | 0.001207755 | 1 |
| Obesity | endometrium | *OSER1-AS1* | 8.121927614 | 3.17E-09 | 4.45E-08 | 1.54E-05 |
| Obesity | endometrium | *C5orf66* | 3.601914214 | 5.55E-07 | 1.52E-06 | 0.002690495 |
| Obesity | endometrium | *LINC01133* | 2.689861638 | 2.83E-06 | 5.38E-06 | 0.013726684 |
| Obesity | endometrium | *LINC01117* | 3.953489177 | 1.33E-06 | 3.04E-06 | 0.006457614 |
| Obesity | endometrium | *RASAL2-AS1* | -5.275690461 | 0.005414393 | 0.006737788 | 1 |
| Obesity | endometrium | *LINC00240* | 5.329451527 | 1.16E-09 | 3.89E-08 | 5.61E-06 |
| Obesity | endometrium | *LINC00184* | 98.96341105 | 1.80E-06 | 3.66E-06 | 0.008744412 |
| Obesity | endometrium | *AL049840.1* | 3.214994419 | 1.13E-05 | 1.91E-05 | 0.055062242 |
| Obesity | endometrium | *PLEKHM1* | 7.629361779 | 2.78E-08 | 1.52E-07 | 0.000134986 |
| Obesity | endometrium | *AP000350.6* | 3.904664938 | 1.22E-05 | 2.04E-05 | 0.059346694 |
| Obesity | endometrium | *SFTA1P* | -9.009407477 | 0.001374867 | 0.001818167 | 1 |
| Obesity | endometrium | *CYP4A22-AS1* | 4.68385626 | 1.60E-06 | 3.39E-06 | 0.007779172 |
| Obesity | endometrium | *FAM225B* | -5.846964304 | 0.001921032 | 0.002500228 | 1 |
| Obesity | endometrium | *MIAT* | 5.242898301 | 1.47E-06 | 3.25E-06 | 0.007155181 |
| Obesity | endometrium | *ERCC6* | 5.550880394 | 4.02E-08 | 2.00E-07 | 0.000194854 |

| | | | | | | |
|---|---|---|---|---|---|---|
| Obesity | endometrium | *LINC00694* | 13.16838723 | 2.13E-06 | 4.21E-06 | 0.010337699 |
| Obesity | endometrium | *LINC00115* | -1.330410024 | 0.000317681 | 0.00044652 | 1 |
| Obesity | endometrium | *HAR1A* | -1.60373847 | 0.000105429 | 0.000157785 | 0.511540266 |
| Obesity | endometrium | *FTCDNL1* | 2.826160269 | 0.000213893 | 0.000307135 | 1 |
| Obesity | endometrium | *FAM138B* | 56.81064586 | 1.76E-06 | 3.59E-06 | 0.008526443 |
| Obesity | endometrium | *ZNF717* | 3.517407744 | 6.66E-06 | 1.17E-05 | 0.03231717 |
| Obesity | endometrium | *AC017074.2* | 38.93731368 | 0.001816051 | 0.002368041 | 1 |
| Obesity | endometrium | *AC009299.3* | 14.69963142 | 3.38E-09 | 4.54E-08 | 1.64E-05 |
| Obesity | endometrium | *SCAMP4* | 6.738428295 | 4.20E-08 | 2.07E-07 | 0.000203776 |
| Obesity | endometrium | *SLC2A1-AS1* | -3.292441118 | 0.000893569 | 0.001200996 | 1 |
| Obesity | endometrium | *LINC01068* | 6.487438492 | 0.003784536 | 0.004790652 | 1 |
| Obesity | endometrium | *AC005884.1* | 8.55107219 | 0.000346686 | 0.000484901 | 1 |
| Obesity | endometrium | *AC108463.2* | 6.642404797 | 0.006934154 | 0.008519755 | 1 |
| Obesity | endometrium | *HCG20* | -7.737468303 | 0.000492493 | 0.000678086 | 1 |
| Obesity | endometrium | *C14orf144* | -3.63381055 | 0.00035966 | 0.000502033 | 1 |
| Obesity | endometrium | *LINC00578* | 5.809730478 | 4.35E-07 | 1.26E-06 | 0.002111054 |
| Obesity | endometrium | *C19orf24* | -1.394440999 | 0.000437216 | 0.000604897 | 1 |
| Obesity | endometrium | *ZNF883* | 4.432683474 | 3.03E-07 | 9.39E-07 | 0.001469133 |
| Obesity | endometrium | *ARL17B* | 8.902906752 | 2.02E-05 | 3.28E-05 | 0.09822159 |
| Obesity | endometrium | *PATL2* | 3.722877298 | 4.18E-07 | 1.22E-06 | 0.002028853 |
| Obesity | endometrium | *CASC20* | 6.070612158 | 1.52E-06 | 3.31E-06 | 0.007393278 |
| Obesity | endometrium | *PSMG3-AS1* | 10.63750132 | 1.70E-08 | 1.09E-07 | 8.26E-05 |
| Obesity | endometrium | *AC112229.4* | 5.635369027 | 5.62E-05 | 8.64E-05 | 0.272814881 |
| Obesity | endometrium | *KLHL7-AS1* | 7.60271913 | 1.99E-06 | 3.97E-06 | 0.009636627 |
| Obesity | endometrium | *AC092171.4* | 4.637244443 | 1.61E-06 | 3.39E-06 | 0.007818898 |
| Obesity | endometrium | *RPS18* | 7.822373541 | 3.61E-10 | 2.30E-08 | 1.75E-06 |
| Obesity | endometrium | *DIRC3* | 2.262648297 | 5.63E-07 | 1.53E-06 | 0.002729949 |
| Obesity | endometrium | *TAPBP* | 5.835833942 | 8.38E-10 | 3.24E-08 | 4.07E-06 |
| Obesity | endometrium | *AC107021.1* | 6.995401643 | 1.39E-07 | 5.07E-07 | 0.000676685 |
| Obesity | endometrium | *TMA7* | 5.835830401 | 8.31E-09 | 7.12E-08 | 4.03E-05 |
| Obesity | endometrium | *LINC00865* | -1.045317083 | 0.00019696 | 0.00028366 | 0.955650853 |
| Obesity | endometrium | *AC009495.3* | 18.54711388 | 0.000711112 | 0.000963776 | 1 |
| Obesity | endometrium | *HLA-DQB2* | 1.420162503 | 0.000679682 | 0.00092195 | 1 |
| Obesity | endometrium | *LYRM9* | -2.550596674 | 0.002416209 | 0.003114625 | 1 |
| Obesity | endometrium | *LINC01166* | 4.163343492 | 0.030443634 | 0.03528727 | 1 |
| Obesity | endometrium | *LINC00342* | 8.412495738 | 0.03064695 | 0.035505969 | 1 |
| Obesity | endometrium | *LINC00702* | 7.810752959 | 7.85E-07 | 2.01E-06 | 0.003809782 |
| Obesity | endometrium | *GPX1* | 4.668234829 | 2.86E-07 | 8.96E-07 | 0.001386412 |
| Obesity | endometrium | *RPS28* | 14.15648864 | 9.06E-06 | 1.55E-05 | 0.043962611 |
| Obesity | endometrium | *TRIM26* | 8.421511821 | 0.000111653 | 0.000166331 | 0.541741261 |
| Obesity | endometrium | *ZNF736* | 3.03883166 | 0.032411089 | 0.037491746 | 1 |
| Obesity | endometrium | *PINLYP* | 4.223300536 | 4.59E-06 | 8.25E-06 | 0.02227387 |
| Obesity | endometrium | *ANKRD65* | 16.14958332 | 3.50E-07 | 1.05E-06 | 0.001699941 |
| Obesity | endometrium | *ZSCAN31* | 5.284017452 | 1.10E-12 | 3.34E-10 | 5.34E-09 |
| Obesity | endometrium | *AC009495.2* | 66.82546289 | 0.004912295 | 0.006139736 | 1 |
| Obesity | endometrium | *LINC00957* | 10.3884667 | 8.12E-06 | 1.40E-05 | 0.039395428 |
| Obesity | endometrium | *LINC00484* | -2.15004825 | 0.002505362 | 0.003225262 | 1 |
| Obesity | endometrium | *KIAA0040* | 3.223519472 | 9.69E-07 | 2.36E-06 | 0.004702272 |
| Obesity | endometrium | *LINC00607* | 12.29340272 | 3.24E-08 | 1.70E-07 | 0.000157031 |
| Obesity | endometrium | *LINC00941* | 11.66167883 | 0.000374866 | 0.000522508 | 1 |

| | | | | | | |
|---|---|---|---|---|---|---|
| Obesity | endometrium | *AC019117.2* | 24.00836664 | 0.000295729 | 0.000417479 | 1 |
| Obesity | endometrium | *ZBTB22* | 6.649281055 | 8.63E-07 | 2.16E-06 | 0.004186097 |
| Obesity | endometrium | *ZBED5* | 7.412743498 | 6.40E-08 | 2.80E-07 | 0.000310296 |
| Obesity | endometrium | *CDKN2AIPNL* | 9.141197037 | 7.17E-06 | 1.25E-05 | 0.034811 |
| Obesity | endometrium | *LINC00987* | 5.811232617 | 3.73E-06 | 6.86E-06 | 0.018114572 |
| Obesity | endometrium | *GS1-124K5.4* | 3.402950323 | 5.15E-06 | 9.20E-06 | 0.024986347 |
| Obesity | endometrium | *LINC00959* | -1.836013344 | 0.002791664 | 0.003579586 | 1 |
| Obesity | endometrium | *HLA-DQA2* | 1.909306401 | 3.63E-05 | 5.70E-05 | 0.176339813 |
| Obesity | endometrium | *IRGM* | 4.034826868 | 2.88E-05 | 4.58E-05 | 0.139880926 |
| Obesity | endometrium | *LINC01143* | -12.74420031 | 0.01590885 | 0.018895897 | 1 |
| Obesity | endometrium | *LINC00211* | -1.460127432 | 0.000183043 | 0.000264323 | 0.888125556 |
| Obesity | endometrium | *LRRC37A2* | 7.007215905 | 1.79E-07 | 6.17E-07 | 0.000867633 |
| Obesity | endometrium | *LINC00707* | 17.93858249 | 0.00240003 | 0.003097061 | 1 |
| Obesity | endometrium | *TXNDC5* | 20.25600419 | 1.48E-06 | 3.26E-06 | 0.007203126 |
| Obesity | endometrium | *LINC01119* | -3.601921651 | 0.006962284 | 0.008552152 | 1 |
| Obesity | endometrium | *NME1* | 5.722960735 | 5.19E-05 | 8.01E-05 | 0.251891535 |
| Obesity | endometrium | *TNFSF12* | 5.091700527 | 0.025465914 | 0.029702071 | 1 |
| Obesity | endometrium | *CDRT4* | 10.25933255 | 4.06E-05 | 6.33E-05 | 0.197175037 |
| Obesity | endometrium | *APOBEC3G* | 2.059759976 | 0.000563126 | 0.000770309 | 1 |
| Obesity | endometrium | *C1orf226* | 13.9198081 | 4.81E-05 | 7.44E-05 | 0.233349507 |
| Obesity | endometrium | *LILRA2* | 2.708864857 | 0.010513225 | 0.012679634 | 1 |
| Obesity | endometrium | *PSMB9* | 5.680471671 | 6.13E-11 | 8.04E-09 | 2.98E-07 |
| Obesity | endometrium | *PPIL3* | 3.5475475 | 0.001351151 | 0.001787778 | 1 |
| Obesity | endometrium | *C1QTNF9* | 15.40377023 | 3.48E-06 | 6.44E-06 | 0.016871049 |
| Obesity | endometrium | *ISY1* | 2.632725477 | 0.009191613 | 0.011143855 | 1 |
| Obesity | endometrium | *PNMA2* | 5.913531264 | 9.94E-06 | 1.69E-05 | 0.048240108 |
| Obesity | endometrium | *KRBOX1* | 62.94994904 | 1.77E-06 | 3.61E-06 | 0.00858563 |
| Obesity | endometrium | *ARHGEF25* | 5.330326343 | 4.21E-08 | 2.07E-07 | 0.000204159 |
| Obesity | endometrium | *LINC00886* | 2.185820557 | 5.89E-05 | 9.03E-05 | 0.285567385 |
| Obesity | endometrium | *NSUN6* | 8.928109922 | 2.05E-09 | 4.00E-08 | 9.95E-06 |
| Obesity | endometrium | *HLA-DOB* | -1.726292176 | 0.003921352 | 0.004953501 | 1 |
| Obesity | endometrium | *LINC00877* | 4.470487185 | 0.000173425 | 0.000251332 | 0.841459022 |
| Obesity | endometrium | *TDGF1* | 5.279146088 | 0.00361611 | 0.004593028 | 1 |
| Obesity | endometrium | *CD302* | 4.096435809 | 7.52E-07 | 1.94E-06 | 0.003647397 |
| Obesity | endometrium | *INMT* | 5.897132787 | 0.004525076 | 0.005680638 | 1 |
| Obesity | endometrium | *PLEKHO2* | 4.339940185 | 4.86E-07 | 1.37E-06 | 0.002359503 |
| Obesity | endometrium | *C8orf58* | 7.017217831 | 7.08E-06 | 1.23E-05 | 0.034332227 |
| Obesity | endometrium | *HOGA1* | -2.079194241 | 0.000376704 | 0.000524919 | 1 |
| Obesity | endometrium | *PI4KA* | 4.692435417 | 2.31E-08 | 1.34E-07 | 0.000112023 |
| Obesity | endometrium | *LINC00969* | 4.991384815 | 4.28E-08 | 2.08E-07 | 0.000207445 |
| Obesity | endometrium | *BGLAP* | -5.876446749 | 6.56E-05 | 0.000100122 | 0.318086617 |
| Obesity | endometrium | *MRPL20* | 9.260617347 | 2.94E-09 | 4.29E-08 | 1.43E-05 |
| Obesity | endometrium | *ZNF709* | -6.869880886 | 0.000264357 | 0.000375376 | 1 |
| Obesity | endometrium | *ERVW-1* | 13.06590454 | 3.20E-06 | 5.99E-06 | 0.015537482 |
| Obesity | endometrium | *MRPL33* | 4.606764667 | 1.66E-07 | 5.84E-07 | 0.000805015 |
| Obesity | endometrium | *STON1* | 9.301453551 | 3.94E-08 | 1.97E-07 | 0.000191065 |
| Obesity | endometrium | *VSIG8* | 7.337286572 | 2.90E-06 | 5.50E-06 | 0.014086318 |
| Obesity | endometrium | *C7orf73* | 5.217142522 | 7.46E-08 | 3.15E-07 | 0.000361886 |
| Obesity | endometrium | *FGF14-IT1* | 12.1656359 | 1.65E-06 | 3.44E-06 | 0.007996618 |
| Obesity | endometrium | *PALM2* | 3.319084084 | 6.54E-06 | 1.15E-05 | 0.031745769 |

| | | | | | | |
|---|---|---|---|---|---|---|
| Obesity | endometrium | *NBPF15* | 12.57522798 | 1.98E-09 | 4.00E-08 | 9.60E-06 |
| Obesity | endometrium | *UPK3B* | 2.788290726 | 1.61E-06 | 3.39E-06 | 0.007806382 |
| Obesity | endometrium | *IL10RB* | 2.985923863 | 1.24E-06 | 2.86E-06 | 0.006014222 |
| Obesity | endometrium | *LEFTY1* | -38.90015066 | 0.00070797 | 0.000959785 | 1 |
| Obesity | endometrium | *TTC4* | 4.921565216 | 1.71E-06 | 3.51E-06 | 0.00827957 |
| Obesity | endometrium | *APOBEC3D* | -13.44915849 | 0.007137365 | 0.008751704 | 1 |
| Obesity | endometrium | *TMEM199* | 4.691558749 | 2.17E-06 | 4.28E-06 | 0.010546239 |
| Obesity | endometrium | *SPRR2F* | 8.417927625 | 0.003269148 | 0.004161046 | 1 |
| Obesity | endometrium | *P2RY11* | 2.804109775 | 0.002113686 | 0.002743607 | 1 |
| Obesity | endometrium | *DBNDD2* | 16.24716889 | 0.005422424 | 0.006746051 | 1 |
| Obesity | endometrium | *LILRA6* | 3.140639963 | 0.000581619 | 0.00079337 | 1 |
| Obesity | endometrium | *SCARF2* | -1.127169133 | 0.000421098 | 0.000583762 | 1 |
| Obesity | endometrium | *APOBEC3C* | 5.088327736 | 5.79E-06 | 1.03E-05 | 0.028097924 |
| Obesity | endometrium | *CCDC13* | -1.848027461 | 0.002724989 | 0.003496865 | 1 |
| Obesity | endometrium | *C4A* | -11.88745262 | 3.77E-08 | 1.91E-07 | 0.000182902 |
| Obesity | endometrium | *CRYBB2* | 5.39979588 | 0.030189148 | 0.035025764 | 1 |
| Obesity | endometrium | *NEAT1* | 4.040791608 | 4.18E-09 | 4.89E-08 | 2.03E-05 |
| Obesity | endometrium | *RAB30-AS1* | 6.321645324 | 1.78E-07 | 6.14E-07 | 0.00086151 |
| Obesity | endometrium | *UBAP1L* | 4.759834014 | 2.85E-07 | 8.94E-07 | 0.001380776 |
| Obesity | endometrium | *LINC00926* | 9.477231365 | 2.92E-07 | 9.11E-07 | 0.001415857 |
| Obesity | endometrium | *DPH6-AS1* | 7.396232636 | 2.35E-09 | 4.01E-08 | 1.14E-05 |
| Obesity | endometrium | *BCKDHA* | 5.658782717 | 0.001822648 | 0.002376004 | 1 |
| Obesity | endometrium | *ABHD14A* | 4.964660941 | 1.57E-07 | 5.57E-07 | 0.000760369 |
| Obesity | endometrium | *GDNF-AS1* | -0.733462567 | 0.00015898 | 0.000231851 | 0.771369926 |
| Obesity | endometrium | *CCDC153* | -2.263137396 | 0.001666507 | 0.002180662 | 1 |
| Obesity | endometrium | *TMEM150C* | 3.579056508 | 2.66E-06 | 5.12E-06 | 0.012920574 |
| Obesity | endometrium | *LINC00939* | 9.69410498 | 2.99E-05 | 4.73E-05 | 0.145019782 |
| Obesity | endometrium | *LINC01060* | 8.315835958 | 1.34E-05 | 2.23E-05 | 0.065131462 |
| Obesity | endometrium | *LINC00942* | -3.146973253 | 0.000127001 | 0.000187983 | 0.616209814 |
| Obesity | endometrium | *PDCD6* | 14.02324559 | 1.19E-08 | 8.63E-08 | 5.75E-05 |
| Obesity | endometrium | *TMEM158* | 17.43681115 | 8.98E-07 | 2.23E-06 | 0.004355164 |
| Obesity | endometrium | *DNAH10OS* | 40.31914557 | 1.32E-10 | 1.34E-08 | 6.41E-07 |
| Obesity | endometrium | *PCDHA10* | 4.018798739 | 0.000584514 | 0.000797094 | 1 |
| Obesity | endometrium | *ZNF718* | 3.19761264 | 4.54E-07 | 1.30E-06 | 0.002203781 |
| Obesity | endometrium | *LINC00989* | 4.558040499 | 3.94E-06 | 7.19E-06 | 0.019120817 |
| Obesity | endometrium | *ATP6V1E2* | 2.871655552 | 0.000631941 | 0.000859596 | 1 |
| Obesity | endometrium | *GLI4* | -2.489272949 | 0.003940123 | 0.004975918 | 1 |
| Obesity | endometrium | *SNHG18* | 3.63334288 | 1.04E-05 | 1.76E-05 | 0.0506098 |
| Obesity | endometrium | *ALG1L2* | 4.493984082 | 1.11E-06 | 2.63E-06 | 0.005395248 |
| Obesity | endometrium | *ZNF550* | 3.797271508 | 1.56E-06 | 3.34E-06 | 0.007574394 |
| Obesity | endometrium | *LINC01094* | 4.581591301 | 1.62E-06 | 3.40E-06 | 0.007846583 |
| Obesity | endometrium | *TMEM200B* | -2.551726914 | 0.030336784 | 0.035180229 | 1 |
| Obesity | endometrium | *PCDHGB6* | 8.172822789 | 2.08E-06 | 4.13E-06 | 0.010105726 |
| Obesity | endometrium | *SLC10A5* | 6.760682545 | 0.034799666 | 0.040182766 | 1 |
| Obesity | endometrium | *PCDHGA6* | -1.498090259 | 0.015574338 | 0.018528691 | 1 |
| Obesity | endometrium | *UTP14C* | 4.109767691 | 3.11E-06 | 5.83E-06 | 0.015097732 |
| Obesity | endometrium | *PCDHGB2* | 21.7277809 | 5.04E-07 | 1.41E-06 | 0.002442985 |
| Obesity | endometrium | *LYN* | 2.167668524 | 2.53E-06 | 4.89E-06 | 0.012286072 |
| Obesity | endometrium | *SIGLEC14* | 5.066705734 | 0.034161826 | 0.039474441 | 1 |
| Obesity | endometrium | *ARMS2* | 6.220713633 | 0.009210226 | 0.011163631 | 1 |

| | | | | | | |
|---|---|---|---|---|---|---|
| Obesity | endometrium | *FPGT* | 3.067487933 | 5.48E-05 | 8.43E-05 | 0.265698584 |
| Obesity | endometrium | *MEX3A* | -2.065304224 | 0.000208173 | 0.000299297 | 1 |
| Obesity | endometrium | *DPP3* | 5.164589318 | 4.46E-08 | 2.16E-07 | 0.000216582 |
| Obesity | endometrium | *SNHG9* | -1.686161297 | 0.018084031 | 0.021385259 | 1 |
| Obesity | endometrium | *ZNF432* | 6.553886164 | 2.42E-07 | 7.86E-07 | 0.001174854 |
| Obesity | endometrium | *HMBS* | -1.229444641 | 0.00240494 | 0.003101746 | 1 |
| Obesity | endometrium | *TAS2R31* | 5.694792182 | 0.000977992 | 0.001308304 | 1 |
| Obesity | endometrium | *AC093668.2* | -5.485639938 | 0.012671507 | 0.015184527 | 1 |
| Obesity | endometrium | *KIAA1147* | 5.76071885 | 1.70E-08 | 1.09E-07 | 8.27E-05 |
| Obesity | endometrium | *GATC* | 3.475002065 | 0.017661565 | 0.020906053 | 1 |
| Obesity | endometrium | *ZBED6* | 8.438841541 | 1.99E-06 | 3.98E-06 | 0.009662697 |
| Obesity | endometrium | *CNPY2* | 3.243238776 | 8.96E-07 | 2.22E-06 | 0.004345205 |
| Obesity | endometrium | *PDF* | -7.488411715 | 9.21E-06 | 1.57E-05 | 0.044695771 |
| Obesity | endometrium | *LINC00640* | 3.066343835 | 0.007704314 | 0.009406475 | 1 |
| Obesity | endometrium | *HIF1A-AS2* | 6.520082224 | 1.66E-08 | 1.07E-07 | 8.06E-05 |
| Obesity | endometrium | *LINC00638* | 11.68847443 | 7.50E-07 | 1.94E-06 | 0.003636977 |
| Obesity | endometrium | *SLC25A21-AS1* | 4.634399564 | 5.49E-07 | 1.51E-06 | 0.002662302 |
| Obesity | endometrium | *RNASE4* | 17.05483731 | 3.35E-06 | 6.24E-06 | 0.016256592 |
| Obesity | endometrium | *MC1R* | 9.758376159 | 1.87E-05 | 3.04E-05 | 0.090563909 |
| Obesity | endometrium | *LINC00639* | 2.948909433 | 1.62E-06 | 3.41E-06 | 0.00788253 |
| Obesity | endometrium | *LINC00924* | 2.229200365 | 0.001202998 | 0.001597413 | 1 |
| Obesity | endometrium | *MRPL46* | -2.70651299 | 0.005469519 | 0.006801155 | 1 |
| Obesity | endometrium | *IQCH-AS1* | 4.273982262 | 6.91E-07 | 1.82E-06 | 0.003354057 |
| Obesity | endometrium | *PWRN1* | 4.751455622 | 4.88E-06 | 8.74E-06 | 0.023676411 |
| Obesity | endometrium | *LINC01003* | 4.466936404 | 2.34E-06 | 4.56E-06 | 0.01136541 |
| Obesity | endometrium | *C15orf65* | 8.176111812 | 0.006727016 | 0.008284133 | 1 |
| Obesity | endometrium | *GOLGA8H* | -0.931884111 | 0.004280716 | 0.005389215 | 1 |
| Obesity | endometrium | *LINC00662* | 6.305460897 | 8.90E-08 | 3.58E-07 | 0.000431912 |
| Obesity | endometrium | *C19orf84* | 56.49230112 | 2.49E-07 | 8.02E-07 | 0.001209523 |
| Obesity | endometrium | *LINC00675* | -3.362548565 | 9.95E-05 | 0.000149355 | 0.482864059 |
| Obesity | endometrium | *LINC00667* | 7.039534064 | 4.78E-05 | 7.40E-05 | 0.231797771 |
| Obesity | endometrium | *LINC00672* | 20.27280595 | 3.99E-08 | 1.99E-07 | 0.000193792 |
| Obesity | endometrium | *LINC00909* | 10.18446213 | 1.47E-08 | 9.99E-08 | 7.11E-05 |
| Obesity | endometrium | *DYNLL2* | 4.592592551 | 2.23E-06 | 4.38E-06 | 0.010843289 |
| Obesity | endometrium | *STRADA* | 5.449676103 | 8.93E-06 | 1.53E-05 | 0.043322347 |
| Obesity | endometrium | *MYO15B* | 8.674412996 | 3.69E-09 | 4.71E-08 | 1.79E-05 |
| Obesity | endometrium | *LINC00663* | 2.119922681 | 0.004082991 | 0.005149641 | 1 |
| Obesity | endometrium | *ILF3-AS1* | 9.317639943 | 1.30E-08 | 9.20E-08 | 6.33E-05 |
| Obesity | endometrium | *S1PR2* | 5.620510271 | 1.29E-05 | 2.15E-05 | 0.062554199 |
| Obesity | endometrium | *ZNF224* | 12.54304955 | 1.56E-09 | 3.98E-08 | 7.57E-06 |
| Obesity | endometrium | *LIN37* | -1.774007429 | 7.08E-05 | 0.000107821 | 0.343409314 |
| Obesity | endometrium | *NDUFA7* | 25.20482976 | 1.21E-06 | 2.82E-06 | 0.005878066 |
| Obesity | endometrium | *AC004466.1* | 6.728166576 | 0.001847082 | 0.002405917 | 1 |
| Obesity | endometrium | *LINC00664* | -100.8992425 | 0.017114398 | 0.020288067 | 1 |
| Obesity | endometrium | *SCGB1B2P* | 9.202589601 | 0.001840719 | 0.002398918 | 1 |
| Obesity | endometrium | *AC026740.1* | 13.64848161 | 0.025687095 | 0.029952845 | 1 |
| Obesity | endometrium | *ZNF728* | 17.06677766 | 2.26E-09 | 4.00E-08 | 1.10E-05 |
| Obesity | endometrium | *TRABD2B* | 3.118784706 | 1.31E-06 | 2.99E-06 | 0.006335373 |
| Obesity | endometrium | *AC093157.1* | 3.004235007 | 1.59E-06 | 3.38E-06 | 0.00773569 |
| Obesity | endometrium | *FBXO17* | 3.764387908 | 1.95E-07 | 6.62E-07 | 0.000946344 |

| | | | | | | |
|---|---|---|---|---|---|---|
| Obesity | endometrium | *ZNF587B* | 4.85437181 | 1.66E-06 | 3.45E-06 | 0.00805052 |
| Obesity | endometrium | *SNHG8* | 2.692738029 | 1.19E-05 | 1.99E-05 | 0.05753489 |
| Obesity | endometrium | *AP001172.2* | 3.869454898 | 0.000383479 | 0.000533746 | 1 |
| Obesity | endometrium | *HCG17* | 4.473993694 | 5.48E-08 | 2.51E-07 | 0.000266073 |
| Obesity | endometrium | *YTHDF3-AS1* | 7.951365211 | 0.000292075 | 0.000412681 | 1 |
| Obesity | endometrium | *MILR1* | 2.590545085 | 0.00015805 | 0.000230634 | 0.766859075 |
| Obesity | endometrium | *GTF2H5* | 5.275286786 | 1.84E-09 | 3.98E-08 | 8.94E-06 |
| Obesity | endometrium | *FGF14-AS2* | 6.385426214 | 2.61E-06 | 5.03E-06 | 0.012678347 |
| Obesity | endometrium | *CASC15* | 2.563006943 | 1.11E-06 | 2.62E-06 | 0.005361708 |
| Obesity | endometrium | *GRIN2B* | 7.512443288 | 5.40E-09 | 5.76E-08 | 2.62E-05 |
| Obesity | endometrium | *AP000350.5* | 2.36443309 | 0.008617167 | 0.010481448 | 1 |
| Obesity | endometrium | *PAXIP1-AS1* | 6.031931147 | 8.96E-08 | 3.60E-07 | 0.000434742 |
| Obesity | endothelial | *NFYA* | -1.617412022 | 2.50E-06 | 3.13E-05 | 0.012150236 |
| Obesity | endothelial | *ENPP4* | 0.481756953 | 0.00219089 | 0.004265729 | 1 |
| Obesity | endothelial | *SEMA3F* | 0.18940648 | 0.003656793 | 0.006650209 | 1 |
| Obesity | endothelial | *KRIT1* | -0.995987396 | 0.000150082 | 0.000468896 | 0.728195924 |
| Obesity | endothelial | *RAD52* | -1.137944673 | 1.07E-05 | 6.93E-05 | 0.051878342 |
| Obesity | endothelial | *AOC1* | -1.346380916 | 0.000145847 | 0.000458322 | 0.707648712 |
| Obesity | endothelial | *SNX11* | -0.444660507 | 0.008304691 | 0.013622163 | 1 |
| Obesity | endothelial | *TMEM176A* | -1.941557171 | 2.17E-06 | 2.92E-05 | 0.010525189 |
| Obesity | endothelial | *M6PR* | -0.722797503 | 0.01216962 | 0.019214772 | 1 |
| Obesity | endothelial | *DBNDD1* | -0.442237199 | 0.008131841 | 0.01336575 | 1 |
| Obesity | endothelial | *ALS2* | -0.791624683 | 0.012981191 | 0.020357059 | 1 |
| Obesity | endothelial | *CD38* | -0.980649843 | 3.19E-06 | 3.43E-05 | 0.015500769 |
| Obesity | endothelial | *RBM6* | -1.458044602 | 2.72E-25 | 1.32E-21 | 1.32E-21 |
| Obesity | endothelial | *RECQL* | -1.053091293 | 0.002944315 | 0.00549666 | 1 |
| Obesity | endothelial | *POLR2J* | -1.504710568 | 0.003055981 | 0.00567673 | 1 |
| Obesity | endothelial | *MEOX1* | 0.150358792 | 0.0001015 | 0.00034391 | 0.492478533 |
| Obesity | endothelial | *GCFC2* | -1.010811974 | 0.000105124 | 0.000353064 | 0.510063304 |
| Obesity | endothelial | *PDK2* | -0.754576983 | 0.000349574 | 0.000915345 | 1 |
| Obesity | endothelial | *GGNBP2* | -1.252966983 | 3.90E-07 | 1.52E-05 | 0.00189014 |
| Obesity | endothelial | *ITGA2B* | -1.273685173 | 0.000903922 | 0.002005409 | 1 |
| Obesity | endothelial | *TMEM98* | -0.963256392 | 8.01E-05 | 0.000293171 | 0.388451822 |
| Obesity | endothelial | *ABCC8* | -0.522911637 | 0.028782563 | 0.041737297 | 1 |
| Obesity | endothelial | *SYNRG* | -0.988118104 | 8.72E-05 | 0.000310779 | 0.422969857 |
| Obesity | endothelial | *ZNF263* | -1.048757612 | 0.002929376 | 0.005477199 | 1 |
| Obesity | endothelial | *MAP3K9* | -0.339442908 | 0.018149994 | 0.027468425 | 1 |
| Obesity | endothelial | *BAIAP2L1* | 0.422313415 | 0.003993012 | 0.007186237 | 1 |
| Obesity | endothelial | *FARP2* | -1.033585775 | 0.000134826 | 0.00042953 | 0.654174515 |
| Obesity | endothelial | *USH1C* | 3.814219518 | 0.033204335 | 0.047364155 | 1 |
| Obesity | endothelial | *IFRD1* | -0.66262554 | 0.000289906 | 0.000783675 | 1 |
| Obesity | endothelial | *VPS41* | -0.76206901 | 0.006142357 | 0.010435125 | 1 |
| Obesity | endothelial | *ELAC2* | -1.127637083 | 2.18E-05 | 0.000114765 | 0.10584962 |
| Obesity | endothelial | *SCIN* | -1.327878969 | 6.03E-06 | 4.69E-05 | 0.029247173 |
| Obesity | endothelial | *NOS2* | -1.744052371 | 0.004987088 | 0.008700954 | 1 |
| Obesity | endothelial | *CD79B* | 0.39780582 | 0.015171471 | 0.023391158 | 1 |
| Obesity | endothelial | *ST7L* | -0.531925855 | 0.011464545 | 0.018220102 | 1 |
| Obesity | endothelial | *TSR3* | -2.057954881 | 0.000313012 | 0.00083447 | 1 |
| Obesity | endothelial | *PIGQ* | -0.838649204 | 0.00046512 | 0.001144983 | 1 |
| Obesity | endothelial | *FMO3* | -1.011268436 | 6.49E-08 | 4.43E-06 | 0.000314792 |

| | | | | | | |
|---|---|---|---|---|---|---|
| Obesity | endothelial | *PSMB1* | -0.83420071 | 0.018072621 | 0.027376945 | 1 |
| Obesity | endothelial | *NADK* | -1.450649089 | 0.000285045 | 0.000773944 | 1 |
| Obesity | endothelial | *DLEC1* | -0.906978865 | 1.20E-05 | 7.48E-05 | 0.058448397 |
| Obesity | endothelial | *CYB561* | 0.542706681 | 0.000295197 | 0.000794836 | 1 |
| Obesity | endothelial | *MGST1* | -0.516130917 | 5.00E-06 | 4.29E-05 | 0.024274382 |
| Obesity | endothelial | *MMP25* | -0.844027813 | 0.005120085 | 0.008894612 | 1 |
| Obesity | endothelial | *IL32* | 0.699316094 | 0.026692948 | 0.03899855 | 1 |
| Obesity | endothelial | *PKD1* | -1.084961472 | 0.000368274 | 0.000955477 | 1 |
| Obesity | endothelial | *MAPK8IP2* | -1.723902801 | 0.026922398 | 0.039262841 | 1 |
| Obesity | endothelial | *MED24* | -1.490018841 | 1.47E-05 | 8.70E-05 | 0.071321059 |
| Obesity | endothelial | *TRAF3IP3* | -0.618578028 | 0.010423097 | 0.016773753 | 1 |
| Obesity | endothelial | *POMT2* | -1.135791117 | 0.000121336 | 0.000394038 | 0.588723293 |
| Obesity | endothelial | *DYRK4* | -0.722922805 | 0.001772413 | 0.003543365 | 1 |
| Obesity | endothelial | *FUZ* | -1.629037453 | 7.43E-05 | 0.000278055 | 0.360359844 |
| Obesity | endothelial | *SLC6A13* | -0.892712634 | 8.49E-06 | 5.94E-05 | 0.041195625 |
| Obesity | endothelial | *ZNF200* | -0.472759353 | 0.028884861 | 0.041873123 | 1 |
| Obesity | endothelial | *LRRC23* | -0.632669771 | 0.007911997 | 0.013053047 | 1 |
| Obesity | endothelial | *CLCN6* | -0.621568287 | 0.03343295 | 0.047668726 | 1 |
| Obesity | endothelial | *SLC6A7* | 5.462085545 | 0.000188796 | 0.000562412 | 0.916036462 |
| Obesity | endothelial | *TSPAN9* | 0.316140387 | 0.00595096 | 0.010163344 | 1 |
| Obesity | endothelial | *UTP18* | -0.831029137 | 0.002664685 | 0.005042532 | 1 |
| Obesity | endothelial | *DPF1* | -1.369947627 | 0.003999218 | 0.007194737 | 1 |
| Obesity | endothelial | *LARS2* | -0.80045041 | 0.011416925 | 0.018156316 | 1 |
| Obesity | endothelial | *DCN* | -0.668134261 | 0.000166874 | 0.000508106 | 0.809674786 |
| Obesity | endothelial | *ZBTB32* | -2.084749569 | 1.20E-06 | 2.57E-05 | 0.005831181 |
| Obesity | endothelial | *ERCC1* | -1.082079382 | 4.98E-05 | 0.000207207 | 0.241810635 |
| Obesity | endothelial | *CD22* | 0.2431008 | 0.018827374 | 0.028413816 | 1 |
| Obesity | endothelial | *ALOX5* | -0.864885395 | 3.66E-07 | 1.46E-05 | 0.001774042 |
| Obesity | endothelial | *MAP4K5* | -0.813189847 | 0.002022983 | 0.003975502 | 1 |
| Obesity | endothelial | *MAN2B2* | -1.305754118 | 6.34E-05 | 0.000247826 | 0.307552047 |
| Obesity | endothelial | *DDX11* | -0.774598804 | 0.005444119 | 0.009373621 | 1 |
| Obesity | endothelial | *ZC3H3* | -0.56812092 | 0.018115366 | 0.027433132 | 1 |
| Obesity | endothelial | *MDH1* | -0.705518223 | 0.004173747 | 0.007453448 | 1 |
| Obesity | endothelial | *COX15* | -1.27461925 | 8.36E-05 | 0.000301906 | 0.405761121 |
| Obesity | endothelial | *CCDC88C* | 0.331633724 | 0.003578493 | 0.006532298 | 1 |
| Obesity | endothelial | *CHDH* | -3.151865856 | 1.82E-05 | 0.000101436 | 0.088451768 |
| Obesity | endothelial | *CLCA4* | -2.37874846 | 2.47E-06 | 3.13E-05 | 0.011972097 |
| Obesity | endothelial | *RUFY3* | -0.86036582 | 0.000296836 | 0.000797039 | 1 |
| Obesity | endothelial | *SLC11A1* | -0.303109994 | 0.00893087 | 0.014555788 | 1 |
| Obesity | endothelial | *AGPS* | -0.751125471 | 0.002979844 | 0.005548058 | 1 |
| Obesity | endothelial | *TTC27* | -0.915380518 | 0.000394907 | 0.001007938 | 1 |
| Obesity | endothelial | *VSIG2* | 0.461165527 | 0.007271683 | 0.012124469 | 1 |
| Obesity | endothelial | *MARCO* | -0.427629458 | 0.00239639 | 0.004606689 | 1 |
| Obesity | endothelial | *HGF* | -1.348986873 | 0.000289225 | 0.000782666 | 1 |
| Obesity | endothelial | *CPS1* | -0.765407882 | 0.001297337 | 0.002712055 | 1 |
| Obesity | endothelial | *ZDHHC6* | -0.838602504 | 0.01833493 | 0.02773101 | 1 |
| Obesity | endothelial | *GRAMD1B* | -0.424442434 | 0.012541446 | 0.019747312 | 1 |
| Obesity | endothelial | *RNH1* | -0.668698658 | 0.005658253 | 0.009714737 | 1 |
| Obesity | endothelial | *ABCC2* | -1.904365908 | 7.59E-07 | 2.24E-05 | 0.003682143 |
| Obesity | endothelial | *GCLM* | -1.350837258 | 0.000766519 | 0.001739546 | 1 |

| | | | | | | |
|---|---|---|---|---|---|---|
| Obesity | endothelial | *UBR2* | -0.815390765 | 0.000324757 | 0.000858704 | 1 |
| Obesity | endothelial | *RRAGD* | -0.803223325 | 7.31E-06 | 5.39E-05 | 0.035483254 |
| Obesity | endothelial | *HSF2* | -0.93542964 | 0.00040123 | 0.001021389 | 1 |
| Obesity | endothelial | *TYMP* | -0.522531718 | 0.006265795 | 0.010622514 | 1 |
| Obesity | endothelial | *TOMM34* | -1.722486926 | 0.001389809 | 0.002875631 | 1 |
| Obesity | endothelial | *SEC63* | -1.009279604 | 4.76E-05 | 0.000199366 | 0.231065352 |
| Obesity | endothelial | *FAS* | -0.983765195 | 4.28E-06 | 3.91E-05 | 0.020746407 |
| Obesity | endothelial | *RNASET2* | -0.752798351 | 7.03E-05 | 0.000267321 | 0.341101897 |
| Obesity | endothelial | *MIPEP* | -1.233038157 | 4.15E-06 | 3.88E-05 | 0.020115809 |
| Obesity | endothelial | *INSRR* | -1.264960299 | 7.74E-07 | 2.24E-05 | 0.003755061 |
| Obesity | endothelial | *VRK2* | -0.661529962 | 0.015479949 | 0.023793552 | 1 |
| Obesity | endothelial | *VEZT* | -0.642094281 | 0.017624077 | 0.026739218 | 1 |
| Obesity | endothelial | *BRD9* | -0.863970183 | 0.00419864 | 0.007492387 | 1 |
| Obesity | endothelial | *SNX1* | -0.901492808 | 0.00019637 | 0.000580615 | 0.952788924 |
| Obesity | endothelial | *ANK1* | 0.492618305 | 0.000402662 | 0.001023961 | 1 |
| Obesity | endothelial | *RABEP1* | -0.732884447 | 0.003645781 | 0.006633529 | 1 |
| Obesity | endothelial | *IKZF2* | -1.175873177 | 1.83E-05 | 0.000101625 | 0.08902332 |
| Obesity | endothelial | *GRN* | -0.775100725 | 0.000372472 | 0.000962318 | 1 |
| Obesity | endothelial | *CENPQ* | -0.415124079 | 0.001792163 | 0.003579899 | 1 |
| Obesity | endothelial | *IFT88* | -0.861289429 | 0.001124198 | 0.002407152 | 1 |
| Obesity | endothelial | *CHPF2* | -0.722467823 | 0.030101993 | 0.043468712 | 1 |
| Obesity | endothelial | *UBA6* | -0.918401689 | 0.000402121 | 0.00102312 | 1 |
| Obesity | endothelial | *ATP6V0A1* | -0.798295497 | 0.001751644 | 0.003506178 | 1 |
| Obesity | endothelial | *PIAS1* | -0.525283581 | 0.019814134 | 0.029727327 | 1 |
| Obesity | endothelial | *ASTE1* | -1.553463198 | 3.00E-06 | 3.39E-05 | 0.014564636 |
| Obesity | endothelial | *MYOC* | -1.064835557 | 6.42E-05 | 0.000249776 | 0.311406351 |
| Obesity | endothelial | *SH3YL1* | -1.528394339 | 3.72E-08 | 3.36E-06 | 0.000180455 |
| Obesity | endothelial | *VCL* | -1.082427802 | 2.74E-05 | 0.000133636 | 0.133101591 |
| Obesity | endothelial | *TBC1D23* | -1.105897493 | 3.23E-05 | 0.000151401 | 0.156831379 |
| Obesity | endothelial | *MYOM2* | -1.132476122 | 4.54E-05 | 0.000191924 | 0.220424217 |
| Obesity | endothelial | *TUBG2* | -1.216977956 | 0.000109849 | 0.000365813 | 0.532989261 |
| Obesity | endothelial | *NSUN2* | -0.974493783 | 0.002626448 | 0.004985288 | 1 |
| Obesity | endothelial | *MRI1* | -0.882336716 | 0.000211371 | 0.000613014 | 1 |
| Obesity | endothelial | *AGA* | -1.851133055 | 2.82E-06 | 3.32E-05 | 0.013665568 |
| Obesity | endothelial | *PI4K2B* | 1.140229686 | 0.0046765 | 0.008233083 | 1 |
| Obesity | endothelial | *BOD1L1* | -0.653925373 | 0.010883691 | 0.017405296 | 1 |
| Obesity | endothelial | *DNAH5* | -1.01642893 | 8.34E-06 | 5.85E-05 | 0.04047154 |
| Obesity | endothelial | *PHLPP2* | -0.69319282 | 0.000254763 | 0.000708372 | 1 |
| Obesity | endothelial | *PQLC2* | -1.269583453 | 2.10E-06 | 2.89E-05 | 0.010187591 |
| Obesity | endothelial | *CTNS* | -0.606572838 | 0.00312569 | 0.005790701 | 1 |
| Obesity | endothelial | *INPP4A* | -0.571550876 | 0.030345098 | 0.043715682 | 1 |
| Obesity | endothelial | *PSMA4* | -1.519718114 | 3.64E-05 | 0.000164507 | 0.176680752 |
| Obesity | endothelial | *TDP1* | -1.453528796 | 0.000114233 | 0.000376952 | 0.554256161 |
| Obesity | endothelial | *SPATA7* | -1.272867536 | 6.39E-06 | 4.90E-05 | 0.031015243 |
| Obesity | endothelial | *MED17* | -1.525091406 | 0.000274204 | 0.000750811 | 1 |
| Obesity | endothelial | *CAPG* | -0.739141515 | 0.000184598 | 0.000552882 | 0.895668273 |
| Obesity | endothelial | *AP2S1* | -1.460177581 | 1.95E-06 | 2.81E-05 | 0.009451756 |
| Obesity | endothelial | *TRIT1* | -0.626009125 | 0.009411444 | 0.015262141 | 1 |
| Obesity | endothelial | *CUL7* | -1.289666319 | 4.43E-06 | 4.02E-05 | 0.021509223 |
| Obesity | endothelial | *CNTLN* | -0.589684295 | 0.019199095 | 0.028881317 | 1 |

| | | | | | | |
|---|---|---|---|---|---|---|
| Obesity | endothelial | *YTHDC2* | -0.933591132 | 0.000594481 | 0.001405664 | 1 |
| Obesity | endothelial | *POLR2B* | -0.898458069 | 0.002971514 | 0.005543171 | 1 |
| Obesity | endothelial | *ARAP2* | -0.73024156 | 0.009280747 | 0.015055227 | 1 |
| Obesity | endothelial | *DTNBP1* | -0.796643645 | 0.010044876 | 0.016213486 | 1 |
| Obesity | endothelial | *ANO2* | 0.107136246 | 0.000116797 | 0.000383164 | 0.566699428 |
| Obesity | endothelial | *RRM2B* | -1.425504522 | 6.79E-07 | 2.20E-05 | 0.00329553 |
| Obesity | endothelial | *SNX29* | -0.868648477 | 3.69E-05 | 0.000165914 | 0.179021636 |
| Obesity | endothelial | *MRPS10* | -0.372817457 | 0.014572677 | 0.022568347 | 1 |
| Obesity | endothelial | *UTS2* | -1.57457964 | 2.38E-05 | 0.000121919 | 0.115508527 |
| Obesity | endothelial | *EPN3* | -0.93454074 | 0.00487016 | 0.008530692 | 1 |
| Obesity | endothelial | *JKAMP* | -0.684024265 | 0.014766349 | 0.02282457 | 1 |
| Obesity | endothelial | *ARHGEF5* | -0.723386332 | 0.000205577 | 0.000600518 | 0.997460782 |
| Obesity | endothelial | *LAMC3* | -0.639726974 | 0.002182286 | 0.004250684 | 1 |
| Obesity | endothelial | *MAPK9* | -1.096543713 | 0.002527087 | 0.004819744 | 1 |
| Obesity | endothelial | *FAM160A2* | -0.805532661 | 0.010866777 | 0.017389711 | 1 |
| Obesity | endothelial | *CYBA* | -1.06455175 | 5.95E-05 | 0.000236199 | 0.288871823 |
| Obesity | endothelial | *THOC3* | -2.678528544 | 0.004130865 | 0.007395925 | 1 |
| Obesity | endothelial | *ALX4* | -0.720991498 | 0.001682843 | 0.003392253 | 1 |
| Obesity | endothelial | *FSTL4* | -1.054914191 | 2.68E-05 | 0.000131605 | 0.130025434 |
| Obesity | endothelial | *AKR7A2* | -1.494499702 | 9.80E-05 | 0.000335019 | 0.475391965 |
| Obesity | endothelial | *MRTO4* | -1.309803657 | 0.000781041 | 0.001764252 | 1 |
| Obesity | endothelial | *ANAPC4* | -0.724590317 | 0.014161449 | 0.021980599 | 1 |
| Obesity | endothelial | *KIF1B* | -0.524137238 | 0.026754418 | 0.039064832 | 1 |
| Obesity | endothelial | *TBC1D22A* | -0.687098122 | 0.003382563 | 0.006214387 | 1 |
| Obesity | endothelial | *PLEKHH1* | -0.846066952 | 0.00013813 | 0.000438331 | 0.67020741 |
| Obesity | endothelial | *GALC* | -1.413239582 | 6.99E-07 | 2.20E-05 | 0.00339273 |
| Obesity | endothelial | *ITIH4* | 0.669100693 | 3.80E-10 | 7.68E-08 | 1.84E-06 |
| Obesity | endothelial | *PCGF2* | 0.351653736 | 0.014243675 | 0.022101154 | 1 |
| Obesity | endothelial | *IL17RB* | -1.866103952 | 2.79E-05 | 0.00013475 | 0.135220448 |
| Obesity | endothelial | *GDI2* | -0.863787459 | 0.003986454 | 0.007177096 | 1 |
| Obesity | endothelial | *PITHD1* | -1.160433535 | 0.001325321 | 0.002761038 | 1 |
| Obesity | endothelial | *ATP11B* | -0.659741559 | 0.006517823 | 0.010999818 | 1 |
| Obesity | endothelial | *LAMC2* | -0.473819287 | 0.011070129 | 0.017674322 | 1 |
| Obesity | endothelial | *RASGRF1* | 1.039165281 | 0.012099725 | 0.019123083 | 1 |
| Obesity | endothelial | *ALDH18A1* | -0.969341664 | 0.000618769 | 0.0014534 | 1 |
| Obesity | endothelial | *DNAJC25* | -1.901733494 | 0.00126857 | 0.002666504 | 1 |
| Obesity | endothelial | *OGFR* | -0.912531316 | 0.002995825 | 0.005575658 | 1 |
| Obesity | endothelial | *GNA15* | -0.39898477 | 0.029746411 | 0.04301925 | 1 |
| Obesity | endothelial | *PIGV* | -0.633783925 | 0.001467901 | 0.003018589 | 1 |
| Obesity | endothelial | *PRDM6* | -1.185230915 | 3.44E-06 | 3.56E-05 | 0.016673898 |
| Obesity | endothelial | *SPAG4* | -1.582788403 | 0.001313045 | 0.002742189 | 1 |
| Obesity | endothelial | *SLC6A16* | -1.4952126 | 3.78E-05 | 0.000168422 | 0.183411734 |
| Obesity | endothelial | *AHRR* | -2.27006895 | 1.88E-07 | 9.10E-06 | 0.000909898 |
| Obesity | endothelial | *ADCK1* | -1.559276067 | 3.07E-06 | 3.39E-05 | 0.01491921 |
| Obesity | endothelial | *HAGH* | -0.627322289 | 0.01544361 | 0.023750363 | 1 |
| Obesity | endothelial | *RNF4* | -1.262871014 | 5.60E-05 | 0.000224837 | 0.271760139 |
| Obesity | endothelial | *CASP8* | -1.340775923 | 5.93E-06 | 4.67E-05 | 0.028755395 |
| Obesity | endothelial | *TM7SF3* | -0.638921202 | 0.030449177 | 0.043852599 | 1 |
| Obesity | endothelial | *CDON* | -0.433577434 | 3.42E-05 | 0.000156959 | 0.166159283 |
| Obesity | endothelial | *TMEM161A* | -1.527060383 | 6.27E-07 | 2.12E-05 | 0.003044224 |

| | | | | | | |
|---|---|---|---|---|---|---|
| Obesity | endothelial | *LPAR2* | -0.999121377 | 0.000245047 | 0.00068647 | 1 |
| Obesity | endothelial | *SUGP2* | -1.203402636 | 1.75E-05 | 9.89E-05 | 0.085124312 |
| Obesity | endothelial | *CNN2* | -0.838481673 | 0.000915144 | 0.002026599 | 1 |
| Obesity | endothelial | *ABCA7* | -1.436242521 | 6.14E-06 | 4.75E-05 | 0.029767848 |
| Obesity | endothelial | *DDX20* | -0.906321518 | 0.006483378 | 0.010950555 | 1 |
| Obesity | endothelial | *BCAS1* | -1.207524241 | 1.49E-05 | 8.80E-05 | 0.072325637 |
| Obesity | endothelial | *CHI3L2* | -1.309788598 | 2.38E-05 | 0.000121919 | 0.115579211 |
| Obesity | endothelial | *SBNO2* | -1.136949382 | 2.04E-05 | 0.000109347 | 0.098849952 |
| Obesity | endothelial | *PMS1* | -0.975769298 | 0.000709059 | 0.001631439 | 1 |
| Obesity | endothelial | *UHRF1BP1* | 0.510357344 | 0.006741935 | 0.011322903 | 1 |
| Obesity | endothelial | *WDR18* | -0.367697892 | 0.025501289 | 0.037370055 | 1 |
| Obesity | endothelial | *ANKRD44* | -0.753692528 | 0.001047636 | 0.002264201 | 1 |
| Obesity | endothelial | *ADAT1* | -0.528353179 | 0.00674479 | 0.011323779 | 1 |
| Obesity | endothelial | *MAP2K4* | -0.926757022 | 0.000533018 | 0.001281567 | 1 |
| Obesity | endothelial | *SEC61A2* | -1.365506146 | 2.32E-06 | 3.03E-05 | 0.011264221 |
| Obesity | endothelial | *TIE1* | 0.161198723 | 0.000346042 | 0.000907566 | 1 |
| Obesity | endothelial | *ZMYND12* | -0.319528043 | 0.00915233 | 0.01487177 | 1 |
| Obesity | endothelial | *SLC9A3* | -0.262055305 | 0.034310782 | 0.048781862 | 1 |
| Obesity | endothelial | *SPI1* | -0.784763354 | 2.49E-05 | 0.000125515 | 0.120995999 |
| Obesity | endothelial | *ZNRD1* | -1.023316168 | 8.83E-06 | 6.04E-05 | 0.042822379 |
| Obesity | endothelial | *ATXN3* | -0.522680306 | 0.007740886 | 0.012809951 | 1 |
| Obesity | endothelial | *GOLGA5* | -1.15691121 | 0.000851243 | 0.001901581 | 1 |
| Obesity | endothelial | *MTFR1* | -1.251034139 | 3.33E-06 | 3.51E-05 | 0.016151615 |
| Obesity | endothelial | *MYO9A* | -0.75883087 | 0.000441227 | 0.001101897 | 1 |
| Obesity | endothelial | *TRAM1* | -0.866737962 | 0.004153402 | 0.007428052 | 1 |
| Obesity | endothelial | *EVI5* | -0.777817929 | 0.000174448 | 0.000527694 | 0.846420522 |
| Obesity | endothelial | *STOML1* | -3.143760317 | 6.27E-05 | 0.000245665 | 0.30437924 |
| Obesity | endothelial | *METTL22* | -1.634311992 | 1.51E-06 | 2.57E-05 | 0.007325601 |
| Obesity | endothelial | *TP53BP1* | -1.008156424 | 0.00022884 | 0.000651985 | 1 |
| Obesity | endothelial | *RRP15* | -0.698433724 | 0.001213056 | 0.002569073 | 1 |
| Obesity | endothelial | *DHX8* | -1.195660711 | 0.001294357 | 0.002706992 | 1 |
| Obesity | endothelial | *ROGDI* | -2.494558566 | 1.81E-06 | 2.68E-05 | 0.008759521 |
| Obesity | endothelial | *CBFB* | -1.012136546 | 0.000413877 | 0.001047539 | 1 |
| Obesity | endothelial | *HEATR6* | -1.186218856 | 0.00016537 | 0.000504707 | 0.802375314 |
| Obesity | endothelial | *POLR1A* | -0.898355256 | 5.00E-05 | 0.000207902 | 0.242829072 |
| Obesity | endothelial | *IP6K2* | -0.64957607 | 0.032425248 | 0.046368201 | 1 |
| Obesity | endothelial | *SRBD1* | -0.875414649 | 0.001170073 | 0.00249109 | 1 |
| Obesity | endothelial | *CYFIP1* | -0.773917213 | 0.005290763 | 0.009151794 | 1 |
| Obesity | endothelial | *IFT80* | -1.681523312 | 4.11E-05 | 0.000179567 | 0.199499339 |
| Obesity | endothelial | *NUCKS1* | -0.924940309 | 0.00039327 | 0.001004817 | 1 |
| Obesity | endothelial | *CLEC2D* | -0.986785024 | 0.000409091 | 0.00103759 | 1 |
| Obesity | endothelial | *DRD4* | -1.971046779 | 3.79E-06 | 3.77E-05 | 0.018394678 |
| Obesity | endothelial | *PLA2G10* | -3.638645573 | 0.003365042 | 0.006186882 | 1 |
| Obesity | endothelial | *PIGB* | -1.041412921 | 0.001291674 | 0.002702544 | 1 |
| Obesity | endothelial | *GNB5* | -0.584233908 | 0.021343435 | 0.031834721 | 1 |
| Obesity | endothelial | *DAPP1* | -0.498298942 | 0.000611669 | 0.001438593 | 1 |
| Obesity | endothelial | *CLTCL1* | -1.018167047 | 0.000319071 | 0.0008469 | 1 |
| Obesity | endothelial | *DGCR2* | -1.007484002 | 0.000377151 | 0.000971304 | 1 |
| Obesity | endothelial | *ZXDC* | -0.907054099 | 0.001351023 | 0.002806149 | 1 |
| Obesity | endothelial | *WIPI1* | -1.412877696 | 3.74E-08 | 3.36E-06 | 0.000181503 |

| | | | | | | |
|---|---|---|---|---|---|---|
| Obesity | endothelial | *GBA2* | -1.250250309 | 0.001602 | 0.0032509 | 1 |
| Obesity | endothelial | *NDST1* | 0.448330412 | 0.031374407 | 0.045024733 | 1 |
| Obesity | endothelial | *ASNS* | -0.958370709 | 0.00730337 | 0.01216894 | 1 |
| Obesity | endothelial | *EIF2B3* | -1.003354267 | 4.03E-06 | 3.85E-05 | 0.019549266 |
| Obesity | endothelial | *SNX13* | -0.876958079 | 0.000819338 | 0.001836226 | 1 |
| Obesity | endothelial | *TRIB2* | 0.390685693 | 0.032201439 | 0.046129726 | 1 |
| Obesity | endothelial | *CPSF1* | -0.742890913 | 0.004084031 | 0.007325589 | 1 |
| Obesity | endothelial | *CYBRD1* | -0.766485787 | 0.000626433 | 0.001468476 | 1 |
| Obesity | endothelial | *RDH11* | -1.541046752 | 1.83E-05 | 0.000101521 | 0.088725438 |
| Obesity | endothelial | *EPN2* | -0.549325585 | 0.021660611 | 0.032278036 | 1 |
| Obesity | endothelial | *ALDH3A2* | -0.806596216 | 0.00017581 | 0.000530492 | 0.853030726 |
| Obesity | endothelial | *AFF4* | -0.67734124 | 0.005673294 | 0.009733671 | 1 |
| Obesity | endothelial | *TRHDE* | -0.806189561 | 4.99E-08 | 4.01E-06 | 0.000242156 |
| Obesity | endothelial | *FCGR2B* | -1.385521657 | 2.43E-06 | 3.10E-05 | 0.011771891 |
| Obesity | endothelial | *NFATC3* | -0.787614763 | 0.001260264 | 0.0026563 | 1 |
| Obesity | endothelial | *CRMP1* | -0.920653326 | 1.30E-06 | 2.57E-05 | 0.006305659 |
| Obesity | endothelial | *AP1M1* | -1.316826389 | 0.000493856 | 0.001204116 | 1 |
| Obesity | endothelial | *PVR* | 0.651340175 | 0.025601085 | 0.03750497 | 1 |
| Obesity | endothelial | *MOV10L1* | -1.175956373 | 3.55E-05 | 0.000160904 | 0.172328416 |
| Obesity | endothelial | *ALPK1* | -0.594367114 | 0.021824308 | 0.032479916 | 1 |
| Obesity | endothelial | *SDHA* | -0.609070093 | 0.029074526 | 0.042097762 | 1 |
| Obesity | endothelial | *ADAM11* | -0.524039705 | 0.00132996 | 0.002768325 | 1 |
| Obesity | endothelial | *PPP2R2C* | -1.168431117 | 2.72E-06 | 3.28E-05 | 0.013215426 |
| Obesity | endothelial | *ZNF532* | -0.831952454 | 0.003736217 | 0.006771806 | 1 |
| Obesity | endothelial | *ZZEF1* | -0.837958001 | 0.001695204 | 0.0034115 | 1 |
| Obesity | endothelial | *TACR2* | 4.515091854 | 0.000188823 | 0.000562412 | 0.916169271 |
| Obesity | endothelial | *TTC38* | -0.9207667 | 0.000434887 | 0.001089912 | 1 |
| Obesity | endothelial | *GRAMD4* | -1.188592045 | 0.000114282 | 0.000376952 | 0.554497015 |
| Obesity | endothelial | *ADD2* | 0.296582854 | 7.98E-05 | 0.000292909 | 0.387116759 |
| Obesity | endothelial | *VPS9D1* | -1.165512743 | 0.002014466 | 0.003963581 | 1 |
| Obesity | endothelial | *FNDC3B* | -0.735760556 | 0.00016703 | 0.000508106 | 0.810428365 |
| Obesity | endothelial | *MOCOS* | -0.963929526 | 0.004881533 | 0.008541362 | 1 |
| Obesity | endothelial | *KIFAP3* | -0.935948476 | 0.000678992 | 0.001576302 | 1 |
| Obesity | endothelial | *MCM6* | -1.087127719 | 0.000314696 | 0.000837119 | 1 |
| Obesity | endothelial | *MLH1* | -1.147533551 | 3.39E-05 | 0.000156224 | 0.1643475 |
| Obesity | endothelial | *FMO4* | -0.965580347 | 0.000780367 | 0.001764081 | 1 |
| Obesity | endothelial | *RGS11* | -0.479633651 | 0.005214223 | 0.009038732 | 1 |
| Obesity | endothelial | *SPAG5* | -3.590111344 | 1.72E-07 | 8.98E-06 | 0.000835577 |
| Obesity | endothelial | *ANKRD13A* | -1.17627887 | 0.001613812 | 0.003270767 | 1 |
| Obesity | endothelial | *PAG1* | -1.051343745 | 2.07E-06 | 2.88E-05 | 0.010027014 |
| Obesity | endothelial | *GPATCH1* | -1.250718692 | 0.000506574 | 0.001227721 | 1 |
| Obesity | endothelial | *ICAM3* | -1.121257679 | 0.020199347 | 0.030258485 | 1 |
| Obesity | endothelial | *XAB2* | -1.693339483 | 0.006852395 | 0.01146872 | 1 |
| Obesity | endothelial | *IL4R* | -1.207334358 | 5.99E-05 | 0.000237485 | 0.290682125 |
| Obesity | endothelial | *APBB1IP* | -0.654308559 | 3.29E-05 | 0.000153294 | 0.159425565 |
| Obesity | endothelial | *LRCH4* | -0.960906413 | 0.000420287 | 0.001058791 | 1 |
| Obesity | endothelial | *SIRT6* | -1.338846376 | 5.37E-05 | 0.000218877 | 0.260463957 |
| Obesity | endothelial | *GPR137B* | -0.790352036 | 0.001944725 | 0.003840378 | 1 |
| Obesity | endothelial | *NAALAD2* | -0.429667606 | 0.000930993 | 0.002054196 | 1 |
| Obesity | endothelial | *AMPH* | -0.977673259 | 1.02E-05 | 6.72E-05 | 0.049692234 |

| | | | | | | |
|---|---|---|---|---|---|---|
| Obesity | endothelial | *MCCC1* | -0.623277125 | 0.000217554 | 0.000626825 | 1 |
| Obesity | endothelial | *PIK3C3* | -1.029940721 | 0.000230794 | 0.000656781 | 1 |
| Obesity | endothelial | *SYNJ2* | 0.264526779 | 0.002530968 | 0.004825249 | 1 |
| Obesity | endothelial | *RBFOX1* | -0.477417486 | 0.002402216 | 0.004614232 | 1 |
| Obesity | endothelial | *PCM1* | -0.798927021 | 0.000356452 | 0.000931203 | 1 |
| Obesity | endothelial | *MYH7B* | -0.936870279 | 0.010469132 | 0.016836668 | 1 |
| Obesity | endothelial | *FKBP7* | -1.018472569 | 0.000392038 | 0.001002196 | 1 |
| Obesity | endothelial | *XRCC5* | -0.683167901 | 0.00949254 | 0.015373099 | 1 |
| Obesity | endothelial | *SAR1A* | -1.136001916 | 2.19E-05 | 0.000114765 | 0.106042868 |
| Obesity | endothelial | *RAPGEF3* | 0.216472879 | 0.000361847 | 0.000940879 | 1 |
| Obesity | endothelial | *SCGN* | -6.13849533 | 5.95E-06 | 4.67E-05 | 0.02888145 |
| Obesity | endothelial | *PGM1* | -1.199525241 | 1.97E-06 | 2.82E-05 | 0.009556696 |
| Obesity | endothelial | *DDX1* | -1.150236887 | 4.17E-05 | 0.000180741 | 0.202249446 |
| Obesity | endothelial | *DNM2* | -1.127031553 | 4.13E-05 | 0.000179866 | 0.200190314 |
| Obesity | endothelial | *STX7* | -0.842171317 | 0.000798814 | 0.001798535 | 1 |
| Obesity | endothelial | *RABL2B* | -1.228673906 | 8.95E-07 | 2.40E-05 | 0.004340915 |
| Obesity | endothelial | *SCTR* | -0.930184563 | 1.35E-06 | 2.57E-05 | 0.006567695 |
| Obesity | endothelial | *RIF1* | -0.995313125 | 0.0001293 | 0.000415472 | 0.627363113 |
| Obesity | endothelial | *KCNN2* | 0.326827497 | 0.000152059 | 0.00047355 | 0.737790461 |
| Obesity | endothelial | *PSEN1* | -0.910840172 | 0.000552662 | 0.001320293 | 1 |
| Obesity | endothelial | *MOK* | -0.704851564 | 0.001114136 | 0.002391941 | 1 |
| Obesity | endothelial | *STK17B* | -1.030771583 | 2.51E-05 | 0.00012572 | 0.121948452 |
| Obesity | endothelial | *PCDHB4* | 1.026285115 | 0.0304877 | 0.043882029 | 1 |
| Obesity | endothelial | *ME2* | -1.130469889 | 0.000513823 | 0.001242187 | 1 |
| Obesity | endothelial | *CCNT2* | -1.104932636 | 5.10E-05 | 0.00021082 | 0.247291566 |
| Obesity | endothelial | *LYRM2* | -0.776469752 | 0.006098589 | 0.010378939 | 1 |
| Obesity | endothelial | *ZCCHC6* | -1.03263047 | 1.98E-05 | 0.000107191 | 0.095936019 |
| Obesity | endothelial | *ULK2* | -0.913898732 | 1.05E-05 | 6.85E-05 | 0.050968521 |
| Obesity | endothelial | *PLOD1* | -1.054925149 | 0.0002984 | 0.000800795 | 1 |
| Obesity | endothelial | *PIBF1* | -0.6594095 | 0.006073542 | 0.010350835 | 1 |
| Obesity | endothelial | *OXCT1* | -1.056380454 | 0.00020472 | 0.000598375 | 0.993302261 |
| Obesity | endothelial | *ZNF586* | -0.700852161 | 0.008969732 | 0.01460441 | 1 |
| Obesity | endothelial | *ZNF264* | -1.297984807 | 0.000373346 | 0.000964064 | 1 |
| Obesity | endothelial | *CHMP2B* | -1.204474233 | 0.001984815 | 0.003913174 | 1 |
| Obesity | endothelial | *NOA1* | -1.003763337 | 0.032635992 | 0.046628337 | 1 |
| Obesity | endothelial | *WBP11* | -1.00363601 | 0.011500868 | 0.018265863 | 1 |
| Obesity | endothelial | *NCOA1* | -0.728227155 | 0.000921764 | 0.002038468 | 1 |
| Obesity | endothelial | *CAD* | -1.504684208 | 7.22E-06 | 5.37E-05 | 0.035044248 |
| Obesity | endothelial | *CD59* | 0.36148571 | 0.015158899 | 0.023379204 | 1 |
| Obesity | endothelial | *CD82* | 0.655926212 | 0.001065437 | 0.0022996 | 1 |
| Obesity | endothelial | *HACE1* | -0.792090612 | 0.013004285 | 0.020380101 | 1 |
| Obesity | endothelial | *SEH1L* | -0.890322656 | 0.001888304 | 0.003747261 | 1 |
| Obesity | endothelial | *SLC25A24* | -0.960738155 | 0.000664988 | 0.001547492 | 1 |
| Obesity | endothelial | *ZNF213* | -1.164096994 | 1.00E-06 | 2.45E-05 | 0.004873458 |
| Obesity | endothelial | *CPNE3* | -0.986014788 | 0.000606548 | 0.001428627 | 1 |
| Obesity | endothelial | *RRN3* | -1.313186939 | 0.000453986 | 0.001123275 | 1 |
| Obesity | endothelial | *MTIF2* | -1.437052615 | 4.80E-06 | 4.19E-05 | 0.023284396 |
| Obesity | endothelial | *DDHD2* | -1.092235448 | 3.34E-06 | 3.51E-05 | 0.016188408 |
| Obesity | endothelial | *EPS15* | -0.961919514 | 3.63E-06 | 3.68E-05 | 0.01758911 |
| Obesity | endothelial | *USP40* | -0.9970593 | 1.04E-05 | 6.78E-05 | 0.050256585 |

| | | | | | | |
|---|---|---|---|---|---|---|
| Obesity | endothelial | *POMGNT1* | -0.938902467 | 0.0123389 | 0.019450403 | 1 |
| Obesity | endothelial | *EIF2AK1* | -1.234288457 | 0.000110148 | 0.000366555 | 0.534437379 |
| Obesity | endothelial | *NME8* | -8.093447692 | 4.44E-05 | 0.000188161 | 0.215444226 |
| Obesity | endothelial | *EPDR1* | -0.472971179 | 0.001192734 | 0.002532667 | 1 |
| Obesity | endothelial | *SNX10* | -0.858844434 | 6.07E-07 | 2.09E-05 | 0.00294324 |
| Obesity | endothelial | *ITPKC* | -0.670254326 | 0.028896126 | 0.041876942 | 1 |
| Obesity | endothelial | *TMED2* | -1.117588908 | 0.000295679 | 0.000795693 | 1 |
| Obesity | endothelial | *ALG9* | -2.182753066 | 1.02E-05 | 6.71E-05 | 0.049525413 |
| Obesity | endothelial | *ACOX3* | -1.129454352 | 0.000236296 | 0.000667351 | 1 |
| Obesity | endothelial | *HSD17B14* | -1.163265182 | 8.67E-06 | 6.00E-05 | 0.042059471 |
| Obesity | endothelial | *TRIP6* | -1.489468503 | 9.73E-08 | 6.18E-06 | 0.000471952 |
| Obesity | endothelial | *ACHE* | -1.270447572 | 4.33E-05 | 0.000185308 | 0.210183708 |
| Obesity | endothelial | *SRRT* | -1.328518879 | 0.000390957 | 0.00100099 | 1 |
| Obesity | endothelial | *ADAMTS2* | -0.76963616 | 1.33E-06 | 2.57E-05 | 0.006449067 |
| Obesity | endothelial | *PSMC5* | -1.651523304 | 2.56E-05 | 0.00012728 | 0.12397045 |
| Obesity | endothelial | *LPCAT2* | 0.35301171 | 0.002254811 | 0.004369147 | 1 |
| Obesity | endothelial | *OGFOD1* | -1.770864106 | 2.04E-07 | 9.62E-06 | 0.000990482 |
| Obesity | endothelial | *TXNDC16* | -0.999251974 | 9.96E-05 | 0.000339025 | 0.483110134 |
| Obesity | endothelial | *SF3B2* | -1.084988284 | 0.000117346 | 0.000384703 | 0.569360799 |
| Obesity | endothelial | *AAMDC* | -0.920557982 | 7.21E-06 | 5.37E-05 | 0.034996303 |
| Obesity | endothelial | *GP6* | -0.679711286 | 3.12E-06 | 3.39E-05 | 0.015126725 |
| Obesity | endothelial | *ASAP3* | -1.280030989 | 1.77E-05 | 9.92E-05 | 0.085900817 |
| Obesity | endothelial | *EDEM2* | -1.287264923 | 1.17E-05 | 7.35E-05 | 0.056806632 |
| Obesity | endothelial | *ANKRD10* | -0.884117229 | 0.000520535 | 0.001255912 | 1 |
| Obesity | endothelial | *DOCK3* | -0.948721064 | 1.39E-06 | 2.57E-05 | 0.006720119 |
| Obesity | endothelial | *ARHGAP28* | -0.633124346 | 0.018528161 | 0.027988367 | 1 |
| Obesity | endothelial | *ZNF343* | -0.838104287 | 0.007432474 | 0.012337449 | 1 |
| Obesity | endothelial | *CPXM1* | -1.083542112 | 2.20E-05 | 0.000115 | 0.106545985 |
| Obesity | endothelial | *MAVS* | -0.85693431 | 0.00075089 | 0.001712085 | 1 |
| Obesity | endothelial | *SNX5* | -1.356645307 | 2.16E-05 | 0.000114183 | 0.104819595 |
| Obesity | endothelial | *ESF1* | -1.045201516 | 6.54E-05 | 0.000253253 | 0.317325427 |
| Obesity | endothelial | *TMEM230* | -1.061684909 | 8.29E-05 | 0.000300233 | 0.402011717 |
| Obesity | endothelial | *RPH3A* | -1.751840744 | 5.12E-05 | 0.000211454 | 0.248458857 |
| Obesity | endothelial | *KIF16B* | -0.955546354 | 0.001137508 | 0.002433506 | 1 |
| Obesity | endothelial | *TBX5* | -1.73527175 | 0.000226023 | 0.000646236 | 1 |
| Obesity | endothelial | *FXYD5* | 0.540162486 | 0.01555681 | 0.023871488 | 1 |
| Obesity | endothelial | *ZNF302* | -0.945415379 | 2.20E-05 | 0.000115 | 0.106703918 |
| Obesity | endothelial | *KCNH4* | 0.317480863 | 0.008717105 | 0.01423129 | 1 |
| Obesity | endothelial | *GANAB* | -1.284802225 | 2.18E-05 | 0.000114765 | 0.106006621 |
| Obesity | endothelial | *MLF2* | -1.012141911 | 0.000248795 | 0.000694165 | 1 |
| Obesity | endothelial | *ANKRD24* | -1.021220145 | 2.51E-06 | 3.13E-05 | 0.012201341 |
| Obesity | endothelial | *DHX32* | -0.906373401 | 0.001733645 | 0.003478761 | 1 |
| Obesity | endothelial | *SPTLC1* | -1.028613991 | 0.000592588 | 0.001401871 | 1 |
| Obesity | endothelial | *MRPS33* | -1.271748584 | 0.000572017 | 0.001360502 | 1 |
| Obesity | endothelial | *MAEA* | -0.940454073 | 0.002841198 | 0.005330817 | 1 |
| Obesity | endothelial | *STRN4* | -1.399671624 | 9.94E-06 | 6.63E-05 | 0.048245441 |
| Obesity | endothelial | *SPG21* | -1.230355379 | 3.30E-05 | 0.000153785 | 0.160241605 |
| Obesity | endothelial | *RAB11FIP3* | -1.024622915 | 0.00038756 | 0.000994417 | 1 |
| Obesity | endothelial | *GNPTG* | -1.383592128 | 4.46E-05 | 0.000189019 | 0.216616257 |
| Obesity | endothelial | *PABPC4* | -2.079786587 | 1.15E-13 | 4.64E-11 | 5.56E-10 |

| | | | | | | |
|---|---|---|---|---|---|---|
| Obesity | endothelial | *CERS4* | -0.954800661 | 1.50E-06 | 2.57E-05 | 0.007254827 |
| Obesity | endothelial | *USP48* | -0.943256181 | 7.99E-05 | 0.000293092 | 0.387760739 |
| Obesity | endothelial | *PDPR* | -1.069354523 | 0.000809345 | 0.001817187 | 1 |
| Obesity | endothelial | *GLG1* | -0.810945518 | 0.001500002 | 0.003074782 | 1 |
| Obesity | endothelial | *DTX2* | -1.69630761 | 3.82E-06 | 3.77E-05 | 0.018523038 |
| Obesity | endothelial | *LAMB1* | -0.344574646 | 0.01872472 | 0.028267686 | 1 |
| Obesity | endothelial | *ABCC6* | -0.926657169 | 3.69E-05 | 0.0001659 | 0.178840519 |
| Obesity | endothelial | *TF* | -1.767270498 | 2.28E-06 | 3.01E-05 | 0.011068398 |
| Obesity | endothelial | *ALKBH5* | -1.04766421 | 0.001689579 | 0.003401593 | 1 |
| Obesity | endothelial | *RGS17* | -1.380486081 | 6.44E-12 | 2.08E-09 | 3.12E-08 |
| Obesity | endothelial | *OSGEP* | -0.84369549 | 0.010236529 | 0.016500876 | 1 |
| Obesity | endothelial | *SCFD1* | -0.859275504 | 0.000592199 | 0.001401634 | 1 |
| Obesity | endothelial | *SUPT16H* | -1.201991169 | 0.00023806 | 0.000671159 | 1 |
| Obesity | endothelial | *TOX4* | -1.085417065 | 0.000453322 | 0.001122234 | 1 |
| Obesity | endothelial | *CAPN3* | -2.632104594 | 5.49E-06 | 4.47E-05 | 0.026633199 |
| Obesity | endothelial | *TBX15* | -1.481502196 | 6.28E-11 | 1.45E-08 | 3.05E-07 |
| Obesity | endothelial | *UNC13D* | -0.595917316 | 0.0024321 | 0.004656886 | 1 |
| Obesity | endothelial | *MFSD11* | -0.832005131 | 0.000146779 | 0.000460357 | 0.712172459 |
| Obesity | endothelial | *COMT* | -1.151904807 | 0.000117705 | 0.000385622 | 0.571106834 |
| Obesity | endothelial | *XYLB* | -1.317451747 | 3.45E-06 | 3.56E-05 | 0.016749289 |
| Obesity | endothelial | *GABRP* | 0.201721733 | 0.000102322 | 0.000345488 | 0.496465661 |
| Obesity | endothelial | *CDC6* | -1.439978666 | 2.23E-06 | 2.99E-05 | 0.010841751 |
| Obesity | endothelial | *PSMD5* | -1.223801705 | 8.66E-05 | 0.000309343 | 0.420153489 |
| Obesity | endothelial | *CRAT* | -2.021924368 | 0.000259579 | 0.000719701 | 1 |
| Obesity | endothelial | *NANS* | -1.357565068 | 0.001859731 | 0.003701154 | 1 |
| Obesity | endothelial | *PDE6C* | -2.066848188 | 4.71E-06 | 4.14E-05 | 0.022831809 |
| Obesity | endothelial | *CWF19L1* | -0.490579438 | 0.003611108 | 0.006586879 | 1 |
| Obesity | endothelial | *WAC* | -0.748587147 | 0.002052155 | 0.004023054 | 1 |
| Obesity | endothelial | *DSP* | -1.924419086 | 0.00016504 | 0.000504529 | 0.80077449 |
| Obesity | endothelial | *IFT74* | -0.849266208 | 0.000293252 | 0.000791797 | 1 |
| Obesity | endothelial | *IL12RB1* | -0.899262965 | 4.55E-05 | 0.000191971 | 0.220766574 |
| Obesity | endothelial | *ACOT7* | -1.448067524 | 6.26E-05 | 0.000245162 | 0.30351099 |
| Obesity | endothelial | *SCD* | -0.712125068 | 0.00193896 | 0.003835237 | 1 |
| Obesity | endothelial | *ERMP1* | -1.249267661 | 5.48E-06 | 4.47E-05 | 0.026579537 |
| Obesity | endothelial | *RAB18* | -1.105421881 | 0.0001409 | 0.00044407 | 0.683647481 |
| Obesity | endothelial | *PRTFDC1* | -0.725131387 | 1.18E-05 | 7.37E-05 | 0.057158628 |
| Obesity | endothelial | *MAST3* | -0.7848396 | 1.85E-05 | 0.000102234 | 0.089761644 |
| Obesity | endothelial | *OCEL1* | -1.007410084 | 1.07E-05 | 6.93E-05 | 0.052070991 |
| Obesity | endothelial | *CATSPERG* | -0.636288007 | 0.001475453 | 0.003029579 | 1 |
| Obesity | endothelial | *SETD1A* | -0.674494188 | 0.026274025 | 0.038421209 | 1 |
| Obesity | endothelial | *CIRBP* | -0.764499825 | 0.0010329 | 0.002235338 | 1 |
| Obesity | endothelial | *MTAP* | -1.431997547 | 1.60E-05 | 9.27E-05 | 0.07768864 |
| Obesity | endothelial | *POLR2E* | -1.295388914 | 0.000203292 | 0.000595276 | 0.986372447 |
| Obesity | endothelial | *MADCAM1* | -1.130885596 | 3.70E-06 | 3.73E-05 | 0.017976423 |
| Obesity | endothelial | *MED15* | -1.19589827 | 1.09E-05 | 6.99E-05 | 0.052920026 |
| Obesity | endothelial | *SERPIND1* | -1.771376829 | 1.10E-07 | 6.51E-06 | 0.000533434 |
| Obesity | endothelial | *SNAP29* | -0.95347996 | 3.19E-05 | 0.00015022 | 0.154656542 |
| Obesity | endothelial | *LZTR1* | -1.12590131 | 0.001715475 | 0.003446577 | 1 |
| Obesity | endothelial | *MMP11* | -1.138599714 | 0.002117032 | 0.0041402 | 1 |
| Obesity | endothelial | *CECR2* | -1.082411819 | 0.000626493 | 0.001468476 | 1 |

| | | | | | | |
|---|---|---|---|---|---|---|
| Obesity | endothelial | *BCL2L13* | -0.572624874 | 0.009672987 | 0.015639232 | 1 |
| Obesity | endothelial | *CABIN1* | -0.858989804 | 0.004034103 | 0.007249433 | 1 |
| Obesity | endothelial | *PPIL2* | -1.080586187 | 6.58E-05 | 0.000254136 | 0.319449567 |
| Obesity | endothelial | *PES1* | 0.34620444 | 0.000780602 | 0.001764081 | 1 |
| Obesity | endothelial | *TOP3B* | -0.999480663 | 0.026904599 | 0.039248681 | 1 |
| Obesity | endothelial | *CRYBB3* | 0.545872489 | 8.98E-05 | 0.000316495 | 0.435496847 |
| Obesity | endothelial | *CYTH4* | -1.189287909 | 1.91E-06 | 2.78E-05 | 0.009267458 |
| Obesity | endothelial | *CARD10* | 0.231562753 | 0.002934436 | 0.005482435 | 1 |
| Obesity | endothelial | *LRP5L* | -0.702018128 | 0.01576853 | 0.024165795 | 1 |
| Obesity | endothelial | *LGALS2* | -0.740998152 | 0.000221109 | 0.000634429 | 1 |
| Obesity | endothelial | *SH3BP1* | -1.385044385 | 2.88E-06 | 3.33E-05 | 0.013964852 |
| Obesity | endothelial | *HPS4* | -1.173823554 | 9.76E-05 | 0.000334445 | 0.473573437 |
| Obesity | endothelial | *TRIOBP* | -1.188939602 | 3.52E-05 | 0.000159794 | 0.170659565 |
| Obesity | endothelial | *ANKRD54* | -1.441721637 | 3.10E-09 | 4.53E-07 | 1.51E-05 |
| Obesity | endothelial | *EIF3L* | -1.254132258 | 0.000176021 | 0.000530799 | 0.854056304 |
| Obesity | endothelial | *CCDC134* | -0.899147047 | 0.006124099 | 0.010411397 | 1 |
| Obesity | endothelial | *PICK1* | -0.956334229 | 0.000178404 | 0.000536649 | 0.865614431 |
| Obesity | endothelial | *CENPM* | 0.879447849 | 0.013000613 | 0.020380101 | 1 |
| Obesity | endothelial | *SLC5A4* | 0.087552718 | 0.033956904 | 0.048330565 | 1 |
| Obesity | endothelial | *KDELR3* | -2.309438995 | 2.58E-08 | 2.45E-06 | 0.000124971 |
| Obesity | endothelial | *CYP2D6* | -2.759783224 | 5.46E-08 | 4.08E-06 | 0.000264996 |
| Obesity | endothelial | *CBY1* | -0.853082191 | 8.63E-05 | 0.000308787 | 0.418714546 |
| Obesity | endothelial | *RTCB* | -1.016562231 | 0.000788508 | 0.001779461 | 1 |
| Obesity | endothelial | *RAB36* | -1.25825391 | 3.88E-06 | 3.78E-05 | 0.018809264 |
| Obesity | endothelial | *PPP6R2* | -0.909882828 | 0.000185815 | 0.000555842 | 0.901575561 |
| Obesity | endothelial | *RHBDD3* | -1.324253057 | 0.00016572 | 0.000505072 | 0.804073905 |
| Obesity | endothelial | *TOM1* | -0.793941636 | 2.27E-05 | 0.000117989 | 0.11008342 |
| Obesity | endothelial | *CHKB* | -1.25791756 | 0.000299679 | 0.000801326 | 1 |
| Obesity | endothelial | *THOC5* | -1.022077431 | 2.54E-05 | 0.000126791 | 0.123368104 |
| Obesity | endothelial | *TSPO* | -0.777512708 | 0.027391718 | 0.039899315 | 1 |
| Obesity | endothelial | *TTLL12* | -1.207387304 | 0.01576529 | 0.024165795 | 1 |
| Obesity | endothelial | *MTMR3* | -0.852002103 | 7.42E-05 | 0.000278055 | 0.360158191 |
| Obesity | endothelial | *SAMM50* | -1.238324126 | 0.002073311 | 0.004061245 | 1 |
| Obesity | endothelial | *FOXRED2* | -0.561775121 | 0.000695144 | 0.001607644 | 1 |
| Obesity | endothelial | *IFT27* | -1.506985348 | 0.000769065 | 0.001744507 | 1 |
| Obesity | endothelial | *KIAA0930* | -1.122998044 | 9.44E-07 | 2.44E-05 | 0.004577956 |
| Obesity | endothelial | *SLC25A17* | -0.864001347 | 0.002550452 | 0.004856669 | 1 |
| Obesity | endothelial | *L3MBTL2* | -1.095253039 | 0.002431923 | 0.004656886 | 1 |
| Obesity | endothelial | *ZC3H7B* | -1.197288888 | 0.000140433 | 0.000443609 | 0.681383078 |
| Obesity | endothelial | *ACO2* | -1.160176217 | 4.66E-06 | 4.13E-05 | 0.022633711 |
| Obesity | endothelial | *POLR3H* | -1.379449343 | 7.09E-06 | 5.30E-05 | 0.034402314 |
| Obesity | endothelial | *DESI1* | -0.872955698 | 0.030342552 | 0.043715682 | 1 |
| Obesity | endothelial | *HDAC10* | -1.94021248 | 3.32E-05 | 0.000154241 | 0.161072798 |
| Obesity | endothelial | *SDR39U1* | -0.407750001 | 0.013683605 | 0.021313917 | 1 |
| Obesity | endothelial | *RBM23* | -1.00260574 | 9.31E-05 | 0.000323462 | 0.451876902 |
| Obesity | endothelial | *AP4S1* | -1.229327843 | 0.00067689 | 0.001572926 | 1 |
| Obesity | endothelial | *NIN* | -0.75291484 | 0.013438998 | 0.020980058 | 1 |
| Obesity | endothelial | *PYGL* | -0.461207174 | 5.91E-05 | 0.000234802 | 0.286693112 |
| Obesity | endothelial | *CNIH1* | -1.330982866 | 0.000418459 | 0.001055776 | 1 |
| Obesity | endothelial | *ATP6V1D* | -1.140389003 | 6.60E-05 | 0.00025464 | 0.320337034 |

| | | | | | | |
|---|---|---|---|---|---|---|
| Obesity | endothelial | *GSTZ1* | -0.903148015 | 0.000854961 | 0.001908129 | 1 |
| Obesity | endothelial | *TMED8* | -0.794484933 | 0.006706091 | 0.011270507 | 1 |
| Obesity | endothelial | *LGMN* | -0.791080424 | 2.19E-05 | 0.000115 | 0.106478169 |
| Obesity | endothelial | *MTHFD1* | -1.059598642 | 8.43E-06 | 5.90E-05 | 0.040918145 |
| Obesity | endothelial | *TELO2* | -1.552063849 | 0.00174667 | 0.003499109 | 1 |
| Obesity | endothelial | *YY1* | -1.047539637 | 0.000585931 | 0.001391549 | 1 |
| Obesity | endothelial | *CCNB1IP1* | -1.426330914 | 1.21E-05 | 7.49E-05 | 0.058543877 |
| Obesity | endothelial | *EFS* | -0.871809748 | 3.11E-06 | 3.39E-05 | 0.015081022 |
| Obesity | endothelial | *KIAA0391* | -1.894145612 | 0.000471698 | 0.001157066 | 1 |
| Obesity | endothelial | *TRPC4AP* | -0.703296442 | 0.004818604 | 0.008452592 | 1 |
| Obesity | endothelial | *PYGB* | -0.510261028 | 0.023052474 | 0.034100794 | 1 |
| Obesity | endothelial | *UQCC1* | -0.899298059 | 0.000206545 | 0.000601532 | 1 |
| Obesity | endothelial | *IFT52* | -0.77580502 | 0.003417431 | 0.006271322 | 1 |
| Obesity | endothelial | *NDRG3* | -0.975930117 | 0.006171274 | 0.01047325 | 1 |
| Obesity | endothelial | *PABPC1L* | -1.306192491 | 1.11E-06 | 2.57E-05 | 0.005409843 |
| Obesity | endothelial | *DOK5* | -1.735730797 | 0.000103573 | 0.000348742 | 0.502537062 |
| Obesity | endothelial | *CSTF1* | -1.609221749 | 0.00322135 | 0.005947486 | 1 |
| Obesity | endothelial | *NELFCD* | -1.09757396 | 0.00117191 | 0.002493906 | 1 |
| Obesity | endothelial | *CTSZ* | -1.126888333 | 0.000266993 | 0.000734382 | 1 |
| Obesity | endothelial | *MTG2* | -1.070808422 | 4.96E-05 | 0.000206397 | 0.240659237 |
| Obesity | endothelial | *TCFL5* | -1.440032094 | 0.000163955 | 0.000502217 | 0.795511882 |
| Obesity | endothelial | *DIDO1* | -0.872309093 | 0.001233244 | 0.002608413 | 1 |
| Obesity | endothelial | *GID8* | -1.407830823 | 9.80E-06 | 6.56E-05 | 0.047544238 |
| Obesity | endothelial | *PTK6* | -3.676789471 | 6.63E-07 | 2.19E-05 | 0.00321868 |
| Obesity | endothelial | *GMEB2* | -1.769022492 | 2.74E-06 | 3.28E-05 | 0.013304437 |
| Obesity | endothelial | *C20orf27* | -0.732500359 | 0.015905318 | 0.024352351 | 1 |
| Obesity | endothelial | *RNF24* | -0.964683209 | 5.39E-06 | 4.47E-05 | 0.026134535 |
| Obesity | endothelial | *NDUFAF5* | -0.540617711 | 0.001324951 | 0.002761038 | 1 |
| Obesity | endothelial | *TRIB3* | -2.646966185 | 0.00121242 | 0.002568849 | 1 |
| Obesity | endothelial | *ANGPT4* | -2.379184149 | 1.85E-05 | 0.000102164 | 0.089597934 |
| Obesity | endothelial | *RSPO4* | -0.59156416 | 0.004178855 | 0.007459826 | 1 |
| Obesity | endothelial | *SEC23B* | -0.861478762 | 0.000205849 | 0.000600901 | 0.998777199 |
| Obesity | endothelial | *CRNKL1* | -0.881753223 | 0.00355725 | 0.006498411 | 1 |
| Obesity | endothelial | *MROH8* | -0.698218431 | 0.0001402 | 0.000443158 | 0.680248151 |
| Obesity | endothelial | *MANBAL* | -0.542592756 | 0.019005421 | 0.028646879 | 1 |
| Obesity | endothelial | *TTI1* | -1.380852458 | 7.05E-05 | 0.000267967 | 0.342193532 |
| Obesity | endothelial | *BPI* | -0.298835015 | 0.004241835 | 0.007560895 | 1 |
| Obesity | endothelial | *DNTTIP1* | -0.675527316 | 0.004114503 | 0.007371373 | 1 |
| Obesity | endothelial | *MAP1LC3A* | -1.336547347 | 2.90E-06 | 3.33E-05 | 0.01407169 |
| Obesity | endothelial | *RBFA* | -1.364619831 | 0.00018699 | 0.000558667 | 0.90727566 |
| Obesity | endothelial | *LPIN2* | -1.159791069 | 1.79E-06 | 2.68E-05 | 0.008665363 |
| Obesity | endothelial | *MYOM1* | -1.014589122 | 1.82E-06 | 2.68E-05 | 0.008810999 |
| Obesity | endothelial | *CEP192* | -0.891650777 | 0.000156621 | 0.000483407 | 0.759925379 |
| Obesity | endothelial | *LAMA1* | -1.737083863 | 4.86E-07 | 1.78E-05 | 0.002358754 |
| Obesity | endothelial | *RNF125* | 0.578257029 | 0.02274249 | 0.03366277 | 1 |
| Obesity | endothelial | *MIB1* | -0.834248937 | 0.001623564 | 0.003286543 | 1 |
| Obesity | endothelial | *NALCN* | -0.784434972 | 0.014160708 | 0.021980599 | 1 |
| Obesity | endothelial | *STK24* | -0.996183109 | 0.000134124 | 0.000427707 | 0.650770701 |
| Obesity | endothelial | *ACP5* | -0.675988001 | 7.81E-05 | 0.000288234 | 0.378738854 |
| Obesity | endothelial | *DNAJC3* | -0.968067364 | 0.000761955 | 0.001732429 | 1 |

| | | | | | | |
|---|---|---|---|---|---|---|
| Obesity | endothelial | *UGGT2* | -0.965098829 | 9.21E-05 | 0.000322282 | 0.446879565 |
| Obesity | endothelial | *SUPT20H* | -1.196452797 | 1.93E-05 | 0.000105366 | 0.093670282 |
| Obesity | endothelial | *MRPS31* | -0.860451853 | 0.005598774 | 0.009616018 | 1 |
| Obesity | endothelial | *DGKH* | 0.373719361 | 0.016715982 | 0.025513037 | 1 |
| Obesity | endothelial | *KATNAL1* | -1.04682622 | 0.000121411 | 0.000394038 | 0.589087052 |
| Obesity | endothelial | *INTS6* | -1.005981187 | 0.000151276 | 0.000471718 | 0.733992766 |
| Obesity | endothelial | *CLN5* | -0.894051577 | 0.010987456 | 0.017553671 | 1 |
| Obesity | endothelial | *MGRN1* | -0.997197966 | 0.000102922 | 0.000347272 | 0.499377211 |
| Obesity | endothelial | *LONP2* | -0.666913928 | 0.003420888 | 0.006275293 | 1 |
| Obesity | endothelial | *DHODH* | -0.829304957 | 0.000717886 | 0.001649235 | 1 |
| Obesity | endothelial | *PSMD7* | -1.158667436 | 0.000494776 | 0.001205668 | 1 |
| Obesity | endothelial | *COG4* | -1.235334138 | 2.38E-05 | 0.000121825 | 0.115246605 |
| Obesity | endothelial | *MON1B* | -1.737328982 | 8.19E-06 | 5.79E-05 | 0.039758607 |
| Obesity | endothelial | *NPRL3* | -1.00110489 | 4.11E-06 | 3.87E-05 | 0.019952624 |
| Obesity | endothelial | *NAGPA* | -0.902105076 | 0.00060066 | 0.00141883 | 1 |
| Obesity | endothelial | *ZNF500* | -0.589943394 | 0.027211542 | 0.039648769 | 1 |
| Obesity | endothelial | *NME4* | -1.96472284 | 2.63E-07 | 1.17E-05 | 0.001274777 |
| Obesity | endothelial | *LMF1* | -0.894075733 | 0.005179839 | 0.008985549 | 1 |
| Obesity | endothelial | *NARFL* | -1.000334387 | 0.00423471 | 0.0075512 | 1 |
| Obesity | endothelial | *RHBDL1* | -3.648689435 | 1.46E-07 | 7.95E-06 | 0.000707208 |
| Obesity | endothelial | *NUBP1* | -1.11561686 | 0.000258679 | 0.000717651 | 1 |
| Obesity | endothelial | *UBE2I* | -1.196304917 | 0.000183181 | 0.000549657 | 0.888794754 |
| Obesity | endothelial | *CAPN15* | -1.510058545 | 0.000272415 | 0.000746757 | 1 |
| Obesity | endothelial | *EARS2* | -0.764842246 | 0.000139322 | 0.000441535 | 0.675989579 |
| Obesity | endothelial | *CPPED1* | -1.329658163 | 1.04E-05 | 6.78E-05 | 0.050268654 |
| Obesity | endothelial | *HMOX2* | -0.976143121 | 0.007804977 | 0.012898415 | 1 |
| Obesity | endothelial | *RRN3P2* | -1.221612235 | 0.000874233 | 0.001946664 | 1 |
| Obesity | endothelial | *RBL2* | -0.758245196 | 0.003582803 | 0.006537705 | 1 |
| Obesity | endothelial | *CDIPT* | -1.438164574 | 0.000194638 | 0.000576899 | 0.944383704 |
| Obesity | endothelial | *KAT8* | -1.497666364 | 6.22E-05 | 0.000244456 | 0.301903379 |
| Obesity | endothelial | *TMC5* | -1.147050033 | 7.11E-08 | 4.72E-06 | 0.000344813 |
| Obesity | endothelial | *C16orf62* | -1.024871155 | 6.57E-05 | 0.000253909 | 0.318899574 |
| Obesity | endothelial | *RNF40* | -1.562610775 | 2.42E-05 | 0.000123058 | 0.117274419 |
| Obesity | endothelial | *KNOP1* | -0.965714496 | 0.0059681 | 0.010189028 | 1 |
| Obesity | endothelial | *IQCH* | -1.187737028 | 0.00044035 | 0.001100762 | 1 |
| Obesity | endothelial | *LACTB* | -1.317413731 | 1.99E-05 | 0.00010749 | 0.096741091 |
| Obesity | endothelial | *HERC1* | -0.724513904 | 0.001930755 | 0.003820564 | 1 |
| Obesity | endothelial | *TRIP4* | -0.823281075 | 0.005431261 | 0.009354803 | 1 |
| Obesity | endothelial | *AP3B2* | -0.342346906 | 0.006826991 | 0.011438038 | 1 |
| Obesity | endothelial | *TTC23* | -1.044370253 | 0.000868793 | 0.001935437 | 1 |
| Obesity | endothelial | *FAH* | -1.01423055 | 9.52E-07 | 2.44E-05 | 0.00461707 |
| Obesity | endothelial | *RPAP1* | -1.806866165 | 5.36E-08 | 4.06E-06 | 0.000260032 |
| Obesity | endothelial | *ZFAND1* | -0.758186124 | 0.034639003 | 0.049185966 | 1 |
| Obesity | endothelial | *EYA1* | -0.73082864 | 0.003494868 | 0.006401322 | 1 |
| Obesity | endothelial | *IMPAD1* | -1.419183032 | 3.14E-05 | 0.00014818 | 0.152180609 |
| Obesity | endothelial | *SFRP1* | -1.316801632 | 2.27E-06 | 3.01E-05 | 0.011037479 |
| Obesity | endothelial | *NIPAL2* | -0.84926443 | 0.007322642 | 0.012188494 | 1 |
| Obesity | endothelial | *RAB2A* | -0.919029046 | 7.90E-05 | 0.000291097 | 0.383374758 |
| Obesity | endothelial | *ARMC1* | -0.971821569 | 0.000228797 | 0.000651985 | 1 |
| Obesity | endothelial | *CHRAC1* | -2.310290394 | 8.74E-05 | 0.000311501 | 0.424264849 |

| | | | | | | |
|---|---|---|---|---|---|---|
| Obesity | endothelial | *SNX16* | -1.314365467 | 0.000111145 | 0.000368608 | 0.539273572 |
| Obesity | endothelial | *SQLE* | -0.79299197 | 0.004156597 | 0.00742829 | 1 |
| Obesity | endothelial | *TNFRSF10A* | 0.29294823 | 5.60E-05 | 0.000224837 | 0.271747133 |
| Obesity | endothelial | *ERICH1* | -0.82057516 | 0.001427073 | 0.002941443 | 1 |
| Obesity | endothelial | *TUSC3* | 0.283043932 | 0.000961732 | 0.002107644 | 1 |
| Obesity | endothelial | *KLHDC4* | -0.762282889 | 0.012285908 | 0.01939207 | 1 |
| Obesity | endothelial | *MAN2B1* | -1.057073765 | 1.09E-07 | 6.51E-06 | 0.000530982 |
| Obesity | endothelial | *TULP2* | -4.204830002 | 3.13E-06 | 3.39E-05 | 0.015178482 |
| Obesity | endothelial | *SNRNP70* | -1.051584298 | 1.91E-05 | 0.000104627 | 0.092770716 |
| Obesity | endothelial | *LIN7B* | -1.605136386 | 0.001973641 | 0.003892727 | 1 |
| Obesity | endothelial | *PPP1R37* | -1.049418671 | 0.000518174 | 0.001251458 | 1 |
| Obesity | endothelial | *FCGRT* | -0.572825805 | 0.014678508 | 0.022710497 | 1 |
| Obesity | endothelial | *ARHGEF18* | -1.760121702 | 0.000346918 | 0.000909371 | 1 |
| Obesity | endothelial | *PEX11G* | -0.907197166 | 0.003000621 | 0.005582443 | 1 |
| Obesity | endothelial | *ERCC2* | -1.801414657 | 1.66E-06 | 2.63E-05 | 0.008038337 |
| Obesity | endothelial | *CD37* | -1.257726677 | 3.19E-05 | 0.00015022 | 0.154726745 |
| Obesity | endothelial | *AMH* | -3.585941136 | 9.63E-06 | 6.48E-05 | 0.046711588 |
| Obesity | endothelial | *FCER2* | -1.675609852 | 4.99E-06 | 4.29E-05 | 0.024224708 |
| Obesity | endothelial | *CLEC4M* | -1.254079147 | 7.39E-06 | 5.42E-05 | 0.035842828 |
| Obesity | endothelial | *NOVA2* | 0.239546922 | 1.98E-05 | 0.000107241 | 0.096302293 |
| Obesity | endothelial | *KIR3DX1* | -2.033558763 | 0.003882009 | 0.007020316 | 1 |
| Obesity | endothelial | *MED25* | -1.537342641 | 4.21E-06 | 3.91E-05 | 0.020418214 |
| Obesity | endothelial | *TIMM44* | -1.18159107 | 0.000132271 | 0.000422882 | 0.641777538 |
| Obesity | endothelial | *IL27RA* | 0.523146347 | 0.000748891 | 0.001708332 | 1 |
| Obesity | endothelial | *C19orf44* | -0.379201732 | 0.019566365 | 0.029373763 | 1 |
| Obesity | endothelial | *OLFM2* | -0.924816413 | 5.84E-06 | 4.64E-05 | 0.028358088 |
| Obesity | endothelial | *RASAL3* | -2.001471026 | 0.003643989 | 0.006633529 | 1 |
| Obesity | endothelial | *ANKRD27* | -1.044659822 | 0.001118607 | 0.002398358 | 1 |
| Obesity | endothelial | *CCDC9* | -2.026519862 | 7.00E-05 | 0.00026663 | 0.339420442 |
| Obesity | endothelial | *TGFB1* | -0.931434473 | 0.002810586 | 0.005281551 | 1 |
| Obesity | endothelial | *MRPL4* | -1.146028689 | 0.000795103 | 0.001791011 | 1 |
| Obesity | endothelial | *CD33* | -0.360567125 | 0.001867537 | 0.003712123 | 1 |
| Obesity | endothelial | *GRIN2D* | -1.475736628 | 7.71E-07 | 2.24E-05 | 0.003742905 |
| Obesity | endothelial | *LIG1* | -1.792967166 | 6.70E-06 | 5.06E-05 | 0.032504068 |
| Obesity | endothelial | *ZNF175* | -1.301958936 | 0.001331212 | 0.002769744 | 1 |
| Obesity | endothelial | *CAPS* | -1.901773902 | 1.54E-06 | 2.57E-05 | 0.007496023 |
| Obesity | endothelial | *PLEKHA4* | -0.927331693 | 1.33E-06 | 2.57E-05 | 0.006470238 |
| Obesity | endothelial | *TNPO2* | -1.31905253 | 1.26E-06 | 2.57E-05 | 0.006094259 |
| Obesity | endothelial | *GCDH* | -1.527034652 | 5.75E-08 | 4.16E-06 | 0.00027887 |
| Obesity | endothelial | *DNASE2* | -1.407373804 | 0.000281533 | 0.000766371 | 1 |
| Obesity | endothelial | *TFPT* | -2.096742904 | 8.00E-05 | 0.000293171 | 0.388267414 |
| Obesity | endothelial | *UPK1A* | -1.361374761 | 0.001909524 | 0.003784726 | 1 |
| Obesity | endothelial | *COPE* | -1.103099421 | 0.001143323 | 0.002441639 | 1 |
| Obesity | endothelial | *ETV2* | -6.399515849 | 0.005588118 | 0.009601115 | 1 |
| Obesity | endothelial | *MAG* | 1.66968976 | 3.67E-05 | 0.000165528 | 0.178107783 |
| Obesity | endothelial | *LSR* | 0.307896582 | 0.013993853 | 0.021755263 | 1 |
| Obesity | endothelial | *ZNF14* | -1.529210337 | 6.75E-05 | 0.000258762 | 0.327502908 |
| Obesity | endothelial | *ETHE1* | -1.699113306 | 0.000181272 | 0.000544264 | 0.879530659 |
| Obesity | endothelial | *SMG9* | -1.238860563 | 0.004958755 | 0.008663983 | 1 |
| Obesity | endothelial | *GTPBP10* | -0.752565946 | 0.002315775 | 0.004464101 | 1 |

| | | | | | | |
|---|---|---|---|---|---|---|
| Obesity | endothelial | *RASA4* | 1.391722627 | 0.000162646 | 0.000499177 | 0.789157247 |
| Obesity | endothelial | *BET1* | -1.130224995 | 0.000173876 | 0.00052662 | 0.843645833 |
| Obesity | endothelial | *DNAH11* | -0.990151175 | 1.48E-06 | 2.57E-05 | 0.007194491 |
| Obesity | endothelial | *STEAP1B* | 0.472629746 | 0.000566407 | 0.001349806 | 1 |
| Obesity | endothelial | *PTN* | -1.250531999 | 4.19E-06 | 3.91E-05 | 0.020347213 |
| Obesity | endothelial | *ADAP1* | -2.316851398 | 2.97E-06 | 3.38E-05 | 0.014430981 |
| Obesity | endothelial | *MET* | 0.21364886 | 0.007145932 | 0.011931198 | 1 |
| Obesity | endothelial | *IQCE* | -1.511545015 | 5.46E-06 | 4.47E-05 | 0.02649757 |
| Obesity | endothelial | *VIPR2* | -0.794283383 | 5.63E-06 | 4.54E-05 | 0.027335525 |
| Obesity | endothelial | *HIBADH* | -0.987956803 | 2.00E-06 | 2.84E-05 | 0.009684421 |
| Obesity | endothelial | *CPVL* | -0.973959728 | 4.24E-06 | 3.91E-05 | 0.020558211 |
| Obesity | endothelial | *CHN2* | -0.925799375 | 7.63E-05 | 0.000283876 | 0.370174625 |
| Obesity | endothelial | *GRB10* | 0.331838768 | 0.012543532 | 0.019747312 | 1 |
| Obesity | endothelial | *ABHD11* | -0.534360551 | 0.014920011 | 0.023040067 | 1 |
| Obesity | endothelial | *PLEKHA8* | -1.253991164 | 0.000610111 | 0.001435624 | 1 |
| Obesity | endothelial | *EPHB6* | -0.446423687 | 0.007000794 | 0.011700948 | 1 |
| Obesity | endothelial | *CHCHD2* | -0.872924836 | 0.004802119 | 0.00842672 | 1 |
| Obesity | endothelial | *CCL24* | -1.391061915 | 0.000188649 | 0.000562412 | 0.915326625 |
| Obesity | endothelial | *ZKSCAN1* | -1.001260076 | 0.000222728 | 0.000638319 | 1 |
| Obesity | endothelial | *EIF3B* | -1.325195979 | 3.86E-05 | 0.000171098 | 0.18735209 |
| Obesity | endothelial | *TAF6* | -0.77679688 | 0.001263682 | 0.002660038 | 1 |
| Obesity | endothelial | *TFR2* | -1.878656724 | 3.99E-07 | 1.55E-05 | 0.001933575 |
| Obesity | endothelial | *RBM28* | -0.99973048 | 2.88E-05 | 0.000138356 | 0.139877991 |
| Obesity | endothelial | *C1GALT1* | -1.119333571 | 9.92E-06 | 6.63E-05 | 0.048133695 |
| Obesity | endothelial | *NRF1* | -0.874109103 | 0.000495637 | 0.001206639 | 1 |
| Obesity | endothelial | *ZNF862* | -0.957385762 | 0.000289146 | 0.000782666 | 1 |
| Obesity | endothelial | *MEST* | -0.786614448 | 1.71E-09 | 2.77E-07 | 8.30E-06 |
| Obesity | endothelial | *ACTR3C* | -0.767577445 | 2.26E-06 | 3.00E-05 | 0.010942624 |
| Obesity | endothelial | *POU6F2* | -0.659843387 | 1.40E-06 | 2.57E-05 | 0.006777689 |
| Obesity | endothelial | *TSPAN13* | 0.255803861 | 0.000156019 | 0.000481862 | 0.757005624 |
| Obesity | endothelial | *RARRES2* | -1.024659338 | 2.58E-05 | 0.000127898 | 0.12495587 |
| Obesity | endothelial | *TMEM176B* | -0.413686956 | 0.013020745 | 0.020399308 | 1 |
| Obesity | endothelial | *COA1* | -0.785216312 | 0.002003945 | 0.003944479 | 1 |
| Obesity | endothelial | *BLVRA* | -1.041492655 | 8.89E-05 | 0.000315162 | 0.431320649 |
| Obesity | endothelial | *URGCP* | -1.144801409 | 4.14E-05 | 0.000179918 | 0.200967979 |
| Obesity | endothelial | *AEBP1* | -1.250230428 | 1.92E-06 | 2.79E-05 | 0.009326373 |
| Obesity | endothelial | *GCK* | -9.447132892 | 1.21E-06 | 2.57E-05 | 0.005882951 |
| Obesity | endothelial | *SPATA6L* | 0.437056543 | 0.030927034 | 0.044474799 | 1 |
| Obesity | endothelial | *SLC1A1* | 0.274469504 | 0.006655544 | 0.011201075 | 1 |
| Obesity | endothelial | *FKTN* | -1.110643029 | 0.00020763 | 0.000603606 | 1 |
| Obesity | endothelial | *MEGF9* | -1.105908031 | 5.00E-06 | 4.29E-05 | 0.024269765 |
| Obesity | endothelial | *CORO2A* | 0.284511624 | 0.000184319 | 0.000552387 | 0.894315044 |
| Obesity | endothelial | *C5* | -1.019838781 | 6.28E-06 | 4.84E-05 | 0.030464569 |
| Obesity | endothelial | *OGN* | -0.569499581 | 0.010592307 | 0.017012206 | 1 |
| Obesity | endothelial | *ECM2* | -0.842814769 | 1.22E-06 | 2.57E-05 | 0.005896322 |
| Obesity | endothelial | *PTGR1* | -0.971986142 | 5.35E-07 | 1.89E-05 | 0.002594334 |
| Obesity | endothelial | *SUSD1* | -0.372611131 | 0.015879008 | 0.024319743 | 1 |
| Obesity | endothelial | *PLGRKT* | -1.021511799 | 7.70E-05 | 0.000285083 | 0.373458343 |
| Obesity | endothelial | *DOCK8* | -0.235897919 | 0.023842678 | 0.035183903 | 1 |
| Obesity | endothelial | *KANK1* | -0.863954489 | 1.73E-06 | 2.68E-05 | 0.008414784 |

| | | | | | | |
|---|---|---|---|---|---|---|
| Obesity | endothelial | *NCS1* | -0.803350868 | 5.13E-06 | 4.36E-05 | 0.024907189 |
| Obesity | endothelial | *FUBP3* | -1.070902438 | 5.25E-05 | 0.000215908 | 0.25477141 |
| Obesity | endothelial | *RGP1* | -1.286348954 | 0.003124104 | 0.005789974 | 1 |
| Obesity | endothelial | *PTGDS* | -1.556120593 | 8.65E-06 | 6.00E-05 | 0.041984462 |
| Obesity | endothelial | *HPS1* | -1.568982853 | 1.82E-07 | 9.10E-06 | 0.00088541 |
| Obesity | endothelial | *DNMBP* | -0.776542294 | 0.003883541 | 0.00702047 | 1 |
| Obesity | endothelial | *ATE1* | -0.718740781 | 0.002299999 | 0.004440746 | 1 |
| Obesity | endothelial | *PLEKHA1* | -0.890855244 | 0.001600283 | 0.003250134 | 1 |
| Obesity | endothelial | *BMPR1A* | -0.771957966 | 1.21E-06 | 2.57E-05 | 0.005875811 |
| Obesity | endothelial | *TNKS2* | -0.99972729 | 0.000451832 | 0.001119657 | 1 |
| Obesity | endothelial | *ANKRD26* | -0.944054444 | 7.07E-05 | 0.000268352 | 0.342954088 |
| Obesity | endothelial | *ACBD5* | -1.306846751 | 7.18E-05 | 0.000271285 | 0.348601115 |
| Obesity | endothelial | *LHPP* | -0.983173324 | 0.000102099 | 0.000345455 | 0.495382137 |
| Obesity | endothelial | *PITRM1* | -1.578973536 | 8.00E-06 | 5.68E-05 | 0.038803161 |
| Obesity | endothelial | *TFAM* | -1.271396287 | 0.00234877 | 0.004522313 | 1 |
| Obesity | endothelial | *PBLD* | -1.11445168 | 1.52E-05 | 8.94E-05 | 0.073901996 |
| Obesity | endothelial | *TADA2A* | -0.51022681 | 0.003743723 | 0.006782877 | 1 |
| Obesity | endothelial | *PSMB3* | -1.413696528 | 0.000242279 | 0.000680682 | 1 |
| Obesity | endothelial | *RAPGEFL1* | -1.500719351 | 4.56E-06 | 4.07E-05 | 0.022129042 |
| Obesity | endothelial | *WNT3* | -0.905580489 | 9.94E-06 | 6.63E-05 | 0.048213757 |
| Obesity | endothelial | *ASPA* | -0.675164842 | 4.38E-05 | 0.000185966 | 0.212372625 |
| Obesity | endothelial | *RAD51C* | -1.537330494 | 1.19E-08 | 1.31E-06 | 5.76E-05 |
| Obesity | endothelial | *TRIM37* | -1.178160388 | 5.58E-05 | 0.000224728 | 0.270572837 |
| Obesity | endothelial | *TRIM16L* | -2.247264771 | 7.87E-07 | 2.25E-05 | 0.003819012 |
| Obesity | endothelial | *CDK5RAP3* | -1.827587277 | 2.49E-07 | 1.13E-05 | 0.0012101 |
| Obesity | endothelial | *PIGL* | -1.041870852 | 8.03E-05 | 0.000293596 | 0.389723249 |
| Obesity | endothelial | *MED13* | -0.984405006 | 3.44E-05 | 0.000157204 | 0.166950971 |
| Obesity | endothelial | *CHRNE* | 0.432430635 | 0.000385838 | 0.000990521 | 1 |
| Obesity | endothelial | *RAI1* | -1.204015711 | 0.000881173 | 0.00195852 | 1 |
| Obesity | endothelial | *GOSR1* | -1.063859565 | 0.000154375 | 0.00047739 | 0.749025545 |
| Obesity | endothelial | *DRG2* | -1.374797479 | 0.000686148 | 0.001591392 | 1 |
| Obesity | endothelial | *FTSJ3* | -0.466601007 | 3.99E-05 | 0.000175354 | 0.193590875 |
| Obesity | endothelial | *ALDH3A1* | -1.478026662 | 0.005988349 | 0.010220004 | 1 |
| Obesity | endothelial | *ICAM2* | 0.254241409 | 0.000299754 | 0.000801326 | 1 |
| Obesity | endothelial | *DDX5* | -0.561689027 | 0.005058174 | 0.00880282 | 1 |
| Obesity | endothelial | *PSMD11* | -0.912381864 | 0.002142096 | 0.004182474 | 1 |
| Obesity | endothelial | *NAGLU* | -1.237725054 | 4.34E-06 | 3.95E-05 | 0.021044614 |
| Obesity | endothelial | *HSD17B1* | -1.038759908 | 0.001270052 | 0.002666504 | 1 |
| Obesity | endothelial | *CNTNAP1* | -0.916594047 | 0.026889531 | 0.039238497 | 1 |
| Obesity | endothelial | *EZH1* | -0.741934089 | 0.003188338 | 0.005895509 | 1 |
| Obesity | endothelial | *EFTUD2* | -1.315428747 | 9.21E-05 | 0.000322282 | 0.447005118 |
| Obesity | endothelial | *HLF* | -0.65685485 | 0.022080897 | 0.032783511 | 1 |
| Obesity | endothelial | *EFNB3* | -0.901642422 | 0.001741096 | 0.003490826 | 1 |
| Obesity | endothelial | *FAM20A* | -1.017343843 | 0.000553083 | 0.001320648 | 1 |
| Obesity | endothelial | *DPH1* | -1.701443661 | 0.000357828 | 0.000933432 | 1 |
| Obesity | endothelial | *DHRS7B* | -0.994311649 | 0.00096857 | 0.002119757 | 1 |
| Obesity | endothelial | *RCVRN* | -0.611727209 | 0.029420758 | 0.042569046 | 1 |
| Obesity | endothelial | *MYH3* | -2.460337535 | 4.98E-06 | 4.29E-05 | 0.024139597 |
| Obesity | endothelial | *GNRHR* | -0.556587256 | 0.013398631 | 0.020930508 | 1 |
| Obesity | endothelial | *SLAIN2* | -0.938186489 | 0.001363443 | 0.002830734 | 1 |

| | | | | | | |
|---|---|---|---|---|---|---|
| Obesity | endothelial | *OCIAD1* | -0.93192111 | 0.000549674 | 0.001314932 | 1 |
| Obesity | endothelial | *UBE2D3* | -0.861019121 | 0.000281624 | 0.000766371 | 1 |
| Obesity | endothelial | *NDUFC1* | -1.208562356 | 0.001533877 | 0.003130993 | 1 |
| Obesity | endothelial | *CPE* | -0.668445892 | 0.000988066 | 0.002152536 | 1 |
| Obesity | endothelial | *GRPEL1* | -1.973711208 | 8.52E-05 | 0.000306609 | 0.413309134 |
| Obesity | endothelial | *GAR1* | -1.358219491 | 0.009426025 | 0.01528068 | 1 |
| Obesity | endothelial | *AADAT* | -1.791296085 | 2.12E-06 | 2.89E-05 | 0.010304713 |
| Obesity | endothelial | *TRIM2* | -1.060022103 | 0.000119584 | 0.000390341 | 0.580223075 |
| Obesity | endothelial | *BST1* | -0.945027278 | 2.06E-05 | 0.000110016 | 0.099784624 |
| Obesity | endothelial | *UFSP2* | -1.11205069 | 0.000978864 | 0.002137466 | 1 |
| Obesity | endothelial | *FAM149A* | -1.33785438 | 6.57E-05 | 0.000253909 | 0.318850851 |
| Obesity | endothelial | *DDX25* | -0.887593608 | 9.14E-07 | 2.41E-05 | 0.004435319 |
| Obesity | endothelial | *CTSC* | -0.629741557 | 0.000318596 | 0.000846101 | 1 |
| Obesity | endothelial | *ELP4* | -0.815906611 | 0.001120623 | 0.002401617 | 1 |
| Obesity | endothelial | *TECTA* | -1.779978048 | 2.08E-06 | 2.88E-05 | 0.010076906 |
| Obesity | endothelial | *SC5D* | -1.722532465 | 4.93E-07 | 1.78E-05 | 0.002390703 |
| Obesity | endothelial | *B3GAT1* | -2.877040583 | 3.55E-05 | 0.000160904 | 0.172178579 |
| Obesity | endothelial | *UNC93B1* | -1.335090906 | 2.47E-05 | 0.000124619 | 0.119802067 |
| Obesity | endothelial | *DCPS* | -1.499231022 | 0.00042944 | 0.001078079 | 1 |
| Obesity | endothelial | *FOXRED1* | -0.622967326 | 0.002088643 | 0.004087976 | 1 |
| Obesity | endothelial | *NRXN2* | -0.912084341 | 5.40E-06 | 4.47E-05 | 0.026209233 |
| Obesity | endothelial | *ST3GAL4* | -1.200411205 | 0.00017318 | 0.000524839 | 0.840267013 |
| Obesity | endothelial | *CCND1* | 0.566976872 | 0.024545729 | 0.036100599 | 1 |
| Obesity | endothelial | *CCDC86* | -2.229403212 | 1.71E-05 | 9.72E-05 | 0.083120399 |
| Obesity | endothelial | *PANX1* | -1.839136093 | 2.90E-05 | 0.000139136 | 0.140805771 |
| Obesity | endothelial | *CEP164* | -1.099167643 | 3.85E-05 | 0.000171065 | 0.18697417 |
| Obesity | endothelial | *GALNT18* | 0.273498229 | 0.0035421 | 0.006482937 | 1 |
| Obesity | endothelial | *KIAA1549L* | 1.023723061 | 0.000293627 | 0.000792367 | 1 |
| Obesity | endothelial | *ELMOD1* | -1.110039141 | 0.004485108 | 0.007939345 | 1 |
| Obesity | endothelial | *NDUFS8* | -1.253915792 | 0.001234472 | 0.002609873 | 1 |
| Obesity | endothelial | *TCIRG1* | -1.257237867 | 0.00180598 | 0.003603048 | 1 |
| Obesity | endothelial | *GTF2H1* | -0.952240606 | 0.000152228 | 0.000473771 | 0.738608635 |
| Obesity | endothelial | *CORO1C* | -1.114544906 | 3.72E-05 | 0.000166444 | 0.180424804 |
| Obesity | endothelial | *ASIC1* | -1.422418377 | 3.01E-06 | 3.39E-05 | 0.014596694 |
| Obesity | endothelial | *IL23A* | 2.052354265 | 0.000327242 | 0.000863861 | 1 |
| Obesity | endothelial | *ACSS3* | -0.669845903 | 5.19E-06 | 4.38E-05 | 0.025177533 |
| Obesity | endothelial | *LTA4H* | -0.688075478 | 0.00203903 | 0.004000555 | 1 |
| Obesity | endothelial | *PRR4* | -0.538480564 | 0.031996444 | 0.045863145 | 1 |
| Obesity | endothelial | *PARP11* | -1.061303408 | 0.001003603 | 0.002182646 | 1 |
| Obesity | endothelial | *GPN3* | -1.88010182 | 5.93E-06 | 4.67E-05 | 0.02879255 |
| Obesity | endothelial | *VPS29* | -0.853298449 | 0.002645753 | 0.005010613 | 1 |
| Obesity | endothelial | *ACAD10* | -0.967144509 | 3.48E-05 | 0.000158603 | 0.168912202 |
| Obesity | endothelial | *DDX55* | -0.879641638 | 0.003197907 | 0.005908699 | 1 |
| Obesity | endothelial | *ENDOU* | -3.115106745 | 3.58E-07 | 1.45E-05 | 0.001737788 |
| Obesity | endothelial | *VDR* | -2.036849896 | 1.34E-06 | 2.57E-05 | 0.006508569 |
| Obesity | endothelial | *STX2* | -0.609725656 | 0.007404398 | 0.012307687 | 1 |
| Obesity | endothelial | *CAND1* | -1.087348534 | 0.000109148 | 0.000363977 | 0.529585857 |
| Obesity | endothelial | *MDM1* | -1.321705606 | 0.001235488 | 0.002610882 | 1 |
| Obesity | endothelial | *NUP107* | -0.891806114 | 0.001432541 | 0.002950453 | 1 |
| Obesity | endothelial | *KRR1* | -1.127153096 | 0.000794077 | 0.001789532 | 1 |

| | | | | | | |
|---|---|---|---|---|---|---|
| Obesity | endothelial | *CHPT1* | -0.769669468 | 3.37E-05 | 0.000155599 | 0.163379032 |
| Obesity | endothelial | *GNPTAB* | -1.073776736 | 1.72E-05 | 9.77E-05 | 0.083602705 |
| Obesity | endothelial | *ATN1* | -0.857388848 | 0.002489928 | 0.00475448 | 1 |
| Obesity | endothelial | *NT5DC3* | -1.073901905 | 7.63E-07 | 2.24E-05 | 0.003703389 |
| Obesity | endothelial | *SUDS3* | -1.250466268 | 0.000100678 | 0.000341603 | 0.488491709 |
| Obesity | endothelial | *GOLT1B* | -1.362177442 | 0.000376009 | 0.000968878 | 1 |
| Obesity | endothelial | *C2CD5* | -0.866627268 | 0.001302299 | 0.002721255 | 1 |
| Obesity | endothelial | *BTN3A3* | 0.969282112 | 1.39E-07 | 7.65E-06 | 0.000673611 |
| Obesity | endothelial | *FRK* | -1.046780347 | 1.25E-06 | 2.57E-05 | 0.006057872 |
| Obesity | endothelial | *TMEM14C* | -0.876437599 | 0.02177508 | 0.032438651 | 1 |
| Obesity | endothelial | *SMIM8* | -0.629967763 | 0.008228428 | 0.013506201 | 1 |
| Obesity | endothelial | *FAM184A* | 0.462843259 | 0.011793518 | 0.018669543 | 1 |
| Obesity | endothelial | *RNGTT* | -0.923884558 | 0.001048748 | 0.002265595 | 1 |
| Obesity | endothelial | *GABRR2* | 0.899886107 | 0.027499385 | 0.040044122 | 1 |
| Obesity | endothelial | *UST* | -0.664919764 | 2.87E-05 | 0.000138082 | 0.139325028 |
| Obesity | endothelial | *TULP1* | -0.787307717 | 0.013418921 | 0.020955457 | 1 |
| Obesity | endothelial | *MRPL18* | -0.825030238 | 0.026046497 | 0.038134461 | 1 |
| Obesity | endothelial | *RNF8* | -1.232961888 | 0.000276951 | 0.000756199 | 1 |
| Obesity | endothelial | *MDGA1* | -1.000689341 | 9.69E-07 | 2.45E-05 | 0.004702632 |
| Obesity | endothelial | *CD83* | -1.150675579 | 9.96E-07 | 2.45E-05 | 0.00483265 |
| Obesity | endothelial | *MDN1* | -1.08723051 | 9.91E-05 | 0.000337757 | 0.480628121 |
| Obesity | endothelial | *GLP1R* | -3.447606352 | 2.54E-06 | 3.15E-05 | 0.012308901 |
| Obesity | endothelial | *COL9A1* | -2.504190497 | 0.001017353 | 0.002205393 | 1 |
| Obesity | endothelial | *WASF1* | -0.832299342 | 1.30E-05 | 7.93E-05 | 0.062968673 |
| Obesity | endothelial | *VNN1* | -0.447145701 | 0.022479509 | 0.033306604 | 1 |
| Obesity | endothelial | *VNN2* | -0.876433905 | 0.010658606 | 0.017096051 | 1 |
| Obesity | endothelial | *ACOT13* | -1.294690324 | 3.02E-05 | 0.000143886 | 0.146763623 |
| Obesity | endothelial | *C6orf62* | -1.206507857 | 2.01E-06 | 2.84E-05 | 0.009741653 |
| Obesity | endothelial | *HBS1L* | -0.860289148 | 0.002261316 | 0.004379335 | 1 |
| Obesity | endothelial | *PEX7* | -0.551096162 | 0.006560751 | 0.011064569 | 1 |
| Obesity | endothelial | *FIG4* | -1.140676248 | 0.000109451 | 0.000364736 | 0.531055847 |
| Obesity | endothelial | *PERP* | 0.377821027 | 0.005459959 | 0.009390898 | 1 |
| Obesity | endothelial | *CUTA* | -1.336943302 | 6.81E-06 | 5.13E-05 | 0.033040118 |
| Obesity | endothelial | *PACRG* | -1.519326485 | 7.72E-06 | 5.56E-05 | 0.037461887 |
| Obesity | endothelial | *QKI* | -0.686567823 | 0.00158819 | 0.003230984 | 1 |
| Obesity | endothelial | *GUCA1B* | -1.176977629 | 1.52E-05 | 8.93E-05 | 0.073779432 |
| Obesity | endothelial | *PRPH2* | -0.321704529 | 0.000248675 | 0.000694165 | 1 |
| Obesity | endothelial | *DNPH1* | -0.773797757 | 0.000512674 | 0.001241114 | 1 |
| Obesity | endothelial | *IMPG1* | 0.907808122 | 0.017099881 | 0.026041627 | 1 |
| Obesity | endothelial | *BTN2A1* | -1.299781421 | 0.000447721 | 0.00111345 | 1 |
| Obesity | endothelial | *LY86* | -1.19402639 | 1.06E-06 | 2.56E-05 | 0.005142888 |
| Obesity | endothelial | *NUDT12* | -1.302023337 | 2.06E-06 | 2.88E-05 | 0.009972643 |
| Obesity | endothelial | *CEP72* | -1.836626728 | 3.88E-06 | 3.78E-05 | 0.018832914 |
| Obesity | endothelial | *KIF20A* | -3.682189489 | 1.23E-05 | 7.60E-05 | 0.059669495 |
| Obesity | endothelial | *PFDN1* | -0.720406486 | 0.005527449 | 0.009503609 | 1 |
| Obesity | endothelial | *TMCO6* | -1.704569399 | 1.08E-07 | 6.51E-06 | 0.000525566 |
| Obesity | endothelial | *SPARC* | -0.938009562 | 3.27E-05 | 0.000152532 | 0.158481019 |
| Obesity | endothelial | *IK* | -1.425861095 | 1.90E-06 | 2.77E-05 | 0.009196792 |
| Obesity | endothelial | *PCDHB15* | -0.764032632 | 0.000640656 | 0.001495892 | 1 |
| Obesity | endothelial | *HAVCR1* | -1.016980454 | 0.000129648 | 0.00041604 | 0.629053122 |

| | | | | | | |
|---|---|---|---|---|---|---|
| Obesity | endothelial | *RNF130* | -0.805049253 | 7.67E-05 | 0.000284485 | 0.372106374 |
| Obesity | endothelial | *ARSB* | -1.328437868 | 3.50E-05 | 0.000159243 | 0.169912399 |
| Obesity | endothelial | *THBS4* | -0.918807551 | 0.000193148 | 0.000573534 | 0.937154588 |
| Obesity | endothelial | *MSH3* | -0.885300669 | 6.43E-05 | 0.000249962 | 0.311952121 |
| Obesity | endothelial | *RASGRF2* | 0.276610891 | 0.003004963 | 0.00558838 | 1 |
| Obesity | endothelial | *DROSHA* | -0.681909318 | 0.009579245 | 0.015503168 | 1 |
| Obesity | endothelial | *CDH6* | -0.932753787 | 1.82E-05 | 0.000101436 | 0.088400915 |
| Obesity | endothelial | *LMNB1* | -1.216993344 | 5.28E-05 | 0.000216407 | 0.256009077 |
| Obesity | endothelial | *SKP1* | -0.980836912 | 0.002205257 | 0.004286822 | 1 |
| Obesity | endothelial | *PPWD1* | -1.060769149 | 0.000394409 | 0.001007195 | 1 |
| Obesity | endothelial | *TXNDC15* | -1.111214982 | 0.003815863 | 0.006905844 | 1 |
| Obesity | endothelial | *SMAD5* | -0.806591961 | 0.006835242 | 0.01144791 | 1 |
| Obesity | endothelial | *ERGIC1* | -1.053062817 | 0.000406942 | 0.001033219 | 1 |
| Obesity | endothelial | *ATP6V0E1* | -0.7286822 | 0.010566825 | 0.0169769 | 1 |
| Obesity | endothelial | *BNIP1* | -0.698658832 | 0.000502691 | 0.001220504 | 1 |
| Obesity | endothelial | *UNC5A* | -5.911553247 | 1.38E-06 | 2.57E-05 | 0.006707475 |
| Obesity | endothelial | *ACTR8* | -1.646610117 | 0.000282994 | 0.000769236 | 1 |
| Obesity | endothelial | *TBCCD1* | -1.628699937 | 3.20E-05 | 0.000150484 | 0.155149205 |
| Obesity | endothelial | *CLDN16* | -0.767379419 | 2.34E-05 | 0.000120712 | 0.113590105 |
| Obesity | endothelial | *NPHP3* | -0.53458283 | 0.003084016 | 0.005722236 | 1 |
| Obesity | endothelial | *CEP70* | -1.141041152 | 0.000168749 | 0.000513015 | 0.818772077 |
| Obesity | endothelial | *HYAL1* | 0.292959643 | 0.020262929 | 0.030344361 | 1 |
| Obesity | endothelial | *CYB561D2* | -1.027801598 | 0.00043639 | 0.001092551 | 1 |
| Obesity | endothelial | *CBLB* | -0.699105833 | 0.001319135 | 0.002751695 | 1 |
| Obesity | endothelial | *IQCG* | -1.186409935 | 4.22E-05 | 0.000182338 | 0.204947568 |
| Obesity | endothelial | *NCBP2* | -1.26323809 | 0.001066001 | 0.002299794 | 1 |
| Obesity | endothelial | *PLXNA1* | -1.146675018 | 2.53E-05 | 0.000126203 | 0.122669717 |
| Obesity | endothelial | *SCAP* | -0.880511931 | 0.004357856 | 0.007742336 | 1 |
| Obesity | endothelial | *KIAA1257* | 1.034033102 | 0.001650874 | 0.003334737 | 1 |
| Obesity | endothelial | *NEK11* | -0.902340519 | 9.58E-06 | 6.46E-05 | 0.046477786 |
| Obesity | endothelial | *PLSCR4* | -0.545858492 | 0.019439385 | 0.029219299 | 1 |
| Obesity | endothelial | *HEMK1* | -0.976419158 | 2.87E-05 | 0.000138082 | 0.139058422 |
| Obesity | endothelial | *COMMD2* | -1.033211348 | 0.002494251 | 0.004760859 | 1 |
| Obesity | endothelial | *ABHD14B* | -1.473434193 | 7.63E-06 | 5.51E-05 | 0.037021322 |
| Obesity | endothelial | *ARHGEF26* | -1.303062502 | 1.20E-05 | 7.47E-05 | 0.058257856 |
| Obesity | endothelial | *VIPR1* | -1.697160073 | 3.53E-06 | 3.62E-05 | 0.017108605 |
| Obesity | endothelial | *NEK4* | -1.32510337 | 7.90E-05 | 0.000291097 | 0.383353365 |
| Obesity | endothelial | *LMAN2L* | -1.207749111 | 1.13E-05 | 7.17E-05 | 0.05493114 |
| Obesity | endothelial | *SLC35F5* | -0.807679309 | 0.001571316 | 0.003202028 | 1 |
| Obesity | endothelial | *OTOF* | -1.789969563 | 0.005359789 | 0.009251404 | 1 |
| Obesity | endothelial | *NRBP1* | -1.181903867 | 0.00080536 | 0.001809078 | 1 |
| Obesity | endothelial | *ITGB6* | -1.231493373 | 1.45E-06 | 2.57E-05 | 0.007041133 |
| Obesity | endothelial | *PPM1G* | -1.282493248 | 0.000236238 | 0.000667351 | 1 |
| Obesity | endothelial | *MOGS* | -0.573427343 | 0.007345469 | 0.012222296 | 1 |
| Obesity | endothelial | *GRB14* | 0.204212432 | 0.000166932 | 0.000508106 | 0.809952777 |
| Obesity | endothelial | *RTN4* | -0.801155322 | 5.36E-05 | 0.000218701 | 0.260035428 |
| Obesity | endothelial | *ACADL* | -0.638903323 | 1.60E-05 | 9.27E-05 | 0.077757482 |
| Obesity | endothelial | *MRPL19* | -1.040261224 | 0.00049283 | 0.001202296 | 1 |
| Obesity | endothelial | *IGFBP2* | 0.418085875 | 0.028219263 | 0.041030825 | 1 |
| Obesity | endothelial | *GNLY* | -0.978194232 | 0.000148745 | 0.00046562 | 0.721710913 |

| | | | | | | |
|---|---|---|---|---|---|---|
| Obesity | endothelial | *CHMP3* | -1.370526474 | 2.87E-06 | 3.33E-05 | 0.01392668 |
| Obesity | endothelial | *WNT6* | -1.104976414 | 5.25E-07 | 1.89E-05 | 0.002547287 |
| Obesity | endothelial | *IL1RL2* | -1.27136619 | 0.001338539 | 0.002782602 | 1 |
| Obesity | endothelial | *MLPH* | 0.178200464 | 0.000122154 | 0.000396184 | 0.5926912 |
| Obesity | endothelial | *HDLBP* | -0.900248433 | 0.000453334 | 0.001122234 | 1 |
| Obesity | endothelial | *TPO* | 0.105417296 | 9.88E-05 | 0.000337268 | 0.479257386 |
| Obesity | endothelial | *TAF1B* | -1.097706185 | 4.14E-05 | 0.000179918 | 0.200689626 |
| Obesity | endothelial | *NOL10* | -0.870287047 | 0.003103099 | 0.005755442 | 1 |
| Obesity | endothelial | *STRN* | -0.762288767 | 0.008450144 | 0.013825689 | 1 |
| Obesity | endothelial | *CEBPZ* | -1.172555372 | 9.72E-05 | 0.000333385 | 0.471610182 |
| Obesity | endothelial | *SLC25A12* | -0.881314787 | 0.001261588 | 0.002657936 | 1 |
| Obesity | endothelial | *DARS* | -0.998123653 | 0.000480642 | 0.001177816 | 1 |
| Obesity | endothelial | *WIPF1* | -0.848024207 | 0.001750495 | 0.003505324 | 1 |
| Obesity | endothelial | *PNO1* | -1.550914878 | 0.004478539 | 0.007930609 | 1 |
| Obesity | endothelial | *ORC4* | -0.82085443 | 0.001594785 | 0.003241682 | 1 |
| Obesity | endothelial | *AAK1* | -0.677833364 | 0.005221113 | 0.009047443 | 1 |
| Obesity | endothelial | *CD207* | 0.349615932 | 4.36E-05 | 0.000185573 | 0.211552675 |
| Obesity | endothelial | *NFE2L2* | -0.764057325 | 0.001417044 | 0.002926989 | 1 |
| Obesity | endothelial | *MSH6* | -1.306570487 | 8.90E-05 | 0.000315162 | 0.431771861 |
| Obesity | endothelial | *ALMS1* | -0.742347246 | 0.015562497 | 0.023872663 | 1 |
| Obesity | endothelial | *DNAJC16* | -1.260978659 | 4.12E-05 | 0.000179818 | 0.199957343 |
| Obesity | endothelial | *MARK1* | -1.099737236 | 3.10E-06 | 3.39E-05 | 0.015053135 |
| Obesity | endothelial | *MORN1* | -1.116688472 | 1.59E-06 | 2.58E-05 | 0.007705513 |
| Obesity | endothelial | *RALGPS2* | -0.871581712 | 3.97E-06 | 3.81E-05 | 0.019252508 |
| Obesity | endothelial | *CEP104* | -1.195271893 | 0.000157944 | 0.000486876 | 0.766343162 |
| Obesity | endothelial | *FAM20B* | -1.110154441 | 0.001230291 | 0.002603303 | 1 |
| Obesity | endothelial | *ICMT* | -2.074124464 | 5.82E-05 | 0.000232269 | 0.282438741 |
| Obesity | endothelial | *PARK7* | -1.102013286 | 0.001110522 | 0.002386295 | 1 |
| Obesity | endothelial | *KIAA1324* | -2.460721423 | 0.000130145 | 0.000417334 | 0.631465358 |
| Obesity | endothelial | *MECR* | -0.971597589 | 9.24E-05 | 0.000322309 | 0.448332449 |
| Obesity | endothelial | *EDEM3* | -1.093249356 | 0.000213086 | 0.000616147 | 1 |
| Obesity | endothelial | *RAP1A* | -0.797000855 | 0.00185827 | 0.003699764 | 1 |
| Obesity | endothelial | *CAPZA1* | -0.850647319 | 0.000790146 | 0.001782328 | 1 |
| Obesity | endothelial | *ASH1L* | -0.627292193 | 0.002623126 | 0.004985276 | 1 |
| Obesity | endothelial | *GON4L* | -0.774931623 | 0.003078582 | 0.005716526 | 1 |
| Obesity | endothelial | *DOCK7* | -0.932813689 | 3.13E-06 | 3.39E-05 | 0.015203908 |
| Obesity | endothelial | *FBXO2* | -0.678214082 | 0.001161498 | 0.002476094 | 1 |
| Obesity | endothelial | *DNAJC6* | -1.426854584 | 5.56E-05 | 0.000224606 | 0.26997627 |
| Obesity | endothelial | *MFN2* | -0.9918208 | 7.89E-05 | 0.000291039 | 0.382716935 |
| Obesity | endothelial | *PRG4* | -1.16100033 | 1.36E-06 | 2.57E-05 | 0.006608612 |
| Obesity | endothelial | *NCF2* | -0.675292884 | 0.000456733 | 0.001128343 | 1 |
| Obesity | endothelial | *BCAS2* | -0.975627499 | 0.000860463 | 0.001918642 | 1 |
| Obesity | endothelial | *TNNI3K* | -1.272599986 | 0.000469078 | 0.001151805 | 1 |
| Obesity | endothelial | *CFHR3* | 0.766067232 | 0.005821639 | 0.009963524 | 1 |
| Obesity | endothelial | *TMEM9* | -1.113787519 | 6.72E-05 | 0.00025823 | 0.326143861 |
| Obesity | endothelial | *WARS2* | -0.779145926 | 0.001121587 | 0.002402623 | 1 |
| Obesity | endothelial | *OSCP1* | -0.688713225 | 6.87E-06 | 5.15E-05 | 0.033325728 |
| Obesity | endothelial | *MRPS15* | -1.480017935 | 3.75E-05 | 0.00016765 | 0.182093145 |
| Obesity | endothelial | *EXOC8* | -1.972601781 | 0.000264756 | 0.000729469 | 1 |
| Obesity | endothelial | *C1orf109* | -1.299808626 | 2.34E-05 | 0.000120712 | 0.113487571 |

| | | | | | | |
|---|---|---|---|---|---|---|
| Obesity | endothelial | *TBCE* | -1.286709755 | 5.68E-05 | 0.000227387 | 0.275593045 |
| Obesity | endothelial | *ACADM* | -0.878593982 | 0.001180817 | 0.00250956 | 1 |
| Obesity | endothelial | *SLAMF1* | -0.943858398 | 0.000142536 | 0.000448497 | 0.691583012 |
| Obesity | endothelial | *PADI2* | -1.177403197 | 2.15E-05 | 0.000113882 | 0.104429464 |
| Obesity | endothelial | *SDHB* | -1.282636472 | 1.45E-05 | 8.61E-05 | 0.070406929 |
| Obesity | endothelial | *MFAP2* | -0.506600627 | 0.004441247 | 0.007876071 | 1 |
| Obesity | endothelial | *CTBS* | -0.54135324 | 0.01184039 | 0.01873763 | 1 |
| Obesity | endothelial | *KLHL12* | -1.41705185 | 2.36E-05 | 0.000121326 | 0.114531536 |
| Obesity | endothelial | *ZNHIT6* | -0.89586911 | 0.002619894 | 0.004983035 | 1 |
| Obesity | endothelial | *KIF17* | 0.459515903 | 0.000970194 | 0.002121397 | 1 |
| Obesity | endothelial | *CD46* | -0.663801603 | 0.004517236 | 0.007984564 | 1 |
| Obesity | endothelial | *EBNA1BP2* | -1.397845108 | 2.51E-05 | 0.000125716 | 0.121751637 |
| Obesity | endothelial | *ERI3* | -1.049892914 | 0.000243719 | 0.000683145 | 1 |
| Obesity | endothelial | *NSUN4* | -1.646964909 | 1.00E-06 | 2.45E-05 | 0.004866335 |
| Obesity | endothelial | *DR1* | -0.808529975 | 0.01027513 | 0.0165576 | 1 |
| Obesity | endothelial | *PRRC2C* | -0.593755803 | 0.015489678 | 0.023798581 | 1 |
| Obesity | endothelial | *ABCD3* | -1.002213099 | 0.001553187 | 0.003167745 | 1 |
| Obesity | endothelial | *PTBP2* | -0.501810351 | 0.022133696 | 0.032851849 | 1 |
| Obesity | endothelial | *DARS2* | -1.441596772 | 9.41E-05 | 0.000325372 | 0.456497505 |
| Obesity | endothelial | *IRF6* | 0.289538975 | 1.44E-05 | 8.58E-05 | 0.070107343 |
| Obesity | endothelial | *SERPINC1* | 1.398840672 | 0.006093626 | 0.010377773 | 1 |
| Obesity | endothelial | *MTFR1L* | -1.495432782 | 0.000132303 | 0.000422882 | 0.641935127 |
| Obesity | endothelial | *RPS6KA1* | -1.582274673 | 6.63E-07 | 2.19E-05 | 0.003216431 |
| Obesity | endothelial | *RPA2* | -1.236782817 | 2.52E-05 | 0.000125732 | 0.12208606 |
| Obesity | endothelial | *MARC2* | -0.851169049 | 0.008881792 | 0.014485531 | 1 |
| Obesity | endothelial | *SLC5A9* | -1.07508075 | 0.006974297 | 0.011660679 | 1 |
| Obesity | endothelial | *ESYT2* | -0.803084861 | 0.001326992 | 0.002763333 | 1 |
| Obesity | endothelial | *CD3EAP* | -1.394162686 | 7.30E-05 | 0.000274567 | 0.354191028 |
| Obesity | endothelial | *RCN2* | -1.108925428 | 0.000666361 | 0.001549945 | 1 |
| Obesity | endothelial | *DDX59* | -0.654759593 | 0.033649108 | 0.047934666 | 1 |
| Obesity | endothelial | *CASC1* | -0.647852816 | 0.000290547 | 0.000784931 | 1 |
| Obesity | endothelial | *LRMP* | 0.311501858 | 0.031328904 | 0.044972734 | 1 |
| Obesity | endothelial | *SGIP1* | 0.424035914 | 0.002972936 | 0.005543691 | 1 |
| Obesity | endothelial | *AKAP7* | -1.060052341 | 1.33E-05 | 8.07E-05 | 0.0645272 |
| Obesity | endothelial | *MED28* | -1.141071591 | 0.00014619 | 0.000459102 | 0.709311884 |
| Obesity | endothelial | *TMEM5* | -1.108144699 | 0.000541214 | 0.001298294 | 1 |
| Obesity | endothelial | *VAMP8* | -1.181547069 | 2.44E-05 | 0.000123783 | 0.118336759 |
| Obesity | endothelial | *FOXO3* | -0.887707334 | 2.78E-05 | 0.00013462 | 0.134763988 |
| Obesity | endothelial | *MFSD1* | -1.047490416 | 4.73E-06 | 4.16E-05 | 0.022951143 |
| Obesity | endothelial | *UBN1* | -0.697063673 | 0.026777634 | 0.039086968 | 1 |
| Obesity | endothelial | *WDR35* | -1.109388151 | 2.37E-05 | 0.000121786 | 0.115087643 |
| Obesity | endothelial | *DNAH7* | -1.150748593 | 6.58E-08 | 4.43E-06 | 0.000319092 |
| Obesity | endothelial | *CPSF3* | -1.295062123 | 2.54E-07 | 1.14E-05 | 0.001232969 |
| Obesity | endothelial | *SENP5* | -0.869498496 | 0.001761521 | 0.003524494 | 1 |
| Obesity | endothelial | *CCDC92* | -0.626296684 | 0.015997113 | 0.02447745 | 1 |
| Obesity | endothelial | *FKBP15* | -0.606779301 | 0.0018848 | 0.003741838 | 1 |
| Obesity | endothelial | *CTNNAL1* | 0.438698808 | 0.00650976 | 0.010990034 | 1 |
| Obesity | endothelial | *PPP6C* | -1.162357931 | 6.83E-05 | 0.000261536 | 0.331627455 |
| Obesity | endothelial | *HDHD3* | -0.433755986 | 0.000102218 | 0.000345488 | 0.495960942 |
| Obesity | endothelial | *HSDL2* | -0.666765031 | 3.20E-05 | 0.000150578 | 0.155396462 |

| | | | | | | |
|---|---|---|---|---|---|---|
| Obesity | endothelial | *MAPKAP1* | -0.797045286 | 0.002278787 | 0.004405049 | 1 |
| Obesity | endothelial | *DENND1A* | -0.771430085 | 0.000730163 | 0.001672608 | 1 |
| Obesity | endothelial | *KDSR* | -1.225780975 | 1.57E-05 | 9.10E-05 | 0.076013286 |
| Obesity | endothelial | *NEK9* | -1.152680698 | 6.42E-05 | 0.000249776 | 0.311470528 |
| Obesity | endothelial | *TTLL5* | -0.929098775 | 0.000731065 | 0.001672608 | 1 |
| Obesity | endothelial | *FLVCR2* | -1.26278647 | 5.49E-06 | 4.47E-05 | 0.026615503 |
| Obesity | endothelial | *PPP4R4* | -0.56937565 | 0.010488812 | 0.016857143 | 1 |
| Obesity | endothelial | *ALDH6A1* | -0.637497359 | 0.005333327 | 0.009215563 | 1 |
| Obesity | endothelial | *EIF2B2* | -0.888168896 | 8.50E-05 | 0.000306216 | 0.412428037 |
| Obesity | endothelial | *COQ6* | -1.534636192 | 7.91E-05 | 0.000291276 | 0.383902319 |
| Obesity | endothelial | *DNMT3A* | -0.792521912 | 0.002975424 | 0.00554407 | 1 |
| Obesity | endothelial | *CNRIP1* | -0.776463338 | 0.00161326 | 0.003270767 | 1 |
| Obesity | endothelial | *CRIPT* | -1.32190717 | 0.0007355 | 0.001680946 | 1 |
| Obesity | endothelial | *SLC17A5* | -1.189830856 | 2.03E-05 | 0.000108995 | 0.09831388 |
| Obesity | endothelial | *TECTB* | -0.571120004 | 0.019128724 | 0.028796949 | 1 |
| Obesity | endothelial | *PPP1R3C* | -2.03749503 | 0.000126274 | 0.000407402 | 0.612682951 |
| Obesity | endothelial | *PYROXD2* | -0.878269398 | 0.000371751 | 0.000961993 | 1 |
| Obesity | endothelial | *MXI1* | -1.082737014 | 0.000116637 | 0.000382898 | 0.565923638 |
| Obesity | endothelial | *C10orf88* | -0.891069364 | 0.003544015 | 0.006483998 | 1 |
| Obesity | endothelial | *HELLS* | -1.352974424 | 5.34E-05 | 0.00021837 | 0.259292329 |
| Obesity | endothelial | *TCTN3* | -1.217127459 | 0.000100108 | 0.000339903 | 0.485722049 |
| Obesity | endothelial | *WDR11* | -0.795836985 | 0.004471626 | 0.00792126 | 1 |
| Obesity | endothelial | *KANSL1* | -0.705997522 | 0.000818642 | 0.001835513 | 1 |
| Obesity | endothelial | *CAAP1* | -1.053032504 | 5.27E-05 | 0.000216407 | 0.255881801 |
| Obesity | endothelial | *GORAB* | -1.168151531 | 0.007979956 | 0.013151748 | 1 |
| Obesity | endothelial | *ACAT2* | -0.918388896 | 0.000310391 | 0.000828848 | 1 |
| Obesity | endothelial | *SNX19* | -1.551733263 | 4.46E-08 | 3.73E-06 | 0.000216184 |
| Obesity | endothelial | *SLC10A7* | -0.749662898 | 0.01120467 | 0.017853879 | 1 |
| Obesity | endothelial | *MASTL* | -1.209025549 | 1.09E-05 | 6.98E-05 | 0.052695353 |
| Obesity | endothelial | *MRC1* | -0.994781237 | 6.12E-06 | 4.74E-05 | 0.029692342 |
| Obesity | endothelial | *CCDC77* | -1.18703762 | 1.08E-05 | 6.98E-05 | 0.052532076 |
| Obesity | endothelial | *UFM1* | -1.06989351 | 0.000331995 | 0.000874507 | 1 |
| Obesity | endothelial | *ALG5* | -0.834892848 | 0.006240667 | 0.010587313 | 1 |
| Obesity | endothelial | *SIL1* | -0.992179305 | 8.97E-06 | 6.13E-05 | 0.043504958 |
| Obesity | endothelial | *ARL1* | -1.313169208 | 0.000227115 | 0.000648428 | 1 |
| Obesity | endothelial | *APAF1* | -0.981971444 | 0.004243264 | 0.007560895 | 1 |
| Obesity | endothelial | *EPHX2* | -0.791151729 | 0.002031581 | 0.003990782 | 1 |
| Obesity | endothelial | *AKAP1* | -1.144926067 | 5.55E-05 | 0.000224116 | 0.269162806 |
| Obesity | endothelial | *SCPEP1* | -0.460162579 | 0.006621776 | 0.01115198 | 1 |
| Obesity | endothelial | *TBX2* | -0.724908367 | 0.000423243 | 0.001064579 | 1 |
| Obesity | endothelial | *TRIM6* | -3.749032133 | 0.000222611 | 0.000638319 | 1 |
| Obesity | endothelial | *ADCY7* | -0.870243214 | 5.06E-05 | 0.00020935 | 0.245358234 |
| Obesity | endothelial | *ECHDC2* | -0.83742675 | 3.10E-07 | 1.36E-05 | 0.001506199 |
| Obesity | endothelial | *A1BG* | -0.967370447 | 0.000103349 | 0.000348472 | 0.501451093 |
| Obesity | endothelial | *PDZRN3* | -0.596010095 | 4.55E-06 | 4.07E-05 | 0.022093176 |
| Obesity | endothelial | *LHX4* | -1.882626496 | 5.88E-06 | 4.66E-05 | 0.028508693 |
| Obesity | endothelial | *SEC22A* | -1.034351521 | 0.000200276 | 0.000588277 | 0.97173717 |
| Obesity | endothelial | *MAPK8IP1* | -1.546518752 | 0.009210343 | 0.014951015 | 1 |
| Obesity | endothelial | *CAT* | -0.513232299 | 0.005125385 | 0.008900633 | 1 |
| Obesity | endothelial | *ZCCHC17* | -1.264488184 | 0.000126621 | 0.000407946 | 0.614366628 |

| | | | | | | |
|---|---|---|---|---|---|---|
| Obesity | endothelial | *FABP3* | -3.047519449 | 4.45E-06 | 4.03E-05 | 0.021578266 |
| Obesity | endothelial | *ZNF639* | -1.615704424 | 3.94E-06 | 3.80E-05 | 0.019133559 |
| Obesity | endothelial | *CPXM2* | 0.276479249 | 6.64E-05 | 0.000256029 | 0.322340486 |
| Obesity | endothelial | *FLT3* | -1.757197198 | 4.72E-05 | 0.00019756 | 0.228773928 |
| Obesity | endothelial | *MTIF3* | -0.781449481 | 0.00795907 | 0.013121782 | 1 |
| Obesity | endothelial | *FMOD* | -1.132177996 | 0.000126196 | 0.000407402 | 0.612303176 |
| Obesity | endothelial | *CD244* | -1.292724618 | 3.80E-05 | 0.00016908 | 0.184579614 |
| Obesity | endothelial | *RBBP6* | -0.984865157 | 0.00043899 | 0.001097928 | 1 |
| Obesity | endothelial | *SERAC1* | -1.520177634 | 0.000103477 | 0.000348659 | 0.502069436 |
| Obesity | endothelial | *LDB3* | -1.111609178 | 3.02E-05 | 0.000143705 | 0.146291749 |
| Obesity | endothelial | *FAM35A* | -0.885906341 | 0.001017699 | 0.002205393 | 1 |
| Obesity | endothelial | *NAA60* | -1.773137022 | 0.001610198 | 0.003266171 | 1 |
| Obesity | endothelial | *SPATA1* | -0.827242791 | 0.006112371 | 0.010398746 | 1 |
| Obesity | endothelial | *RWDD3* | -0.915157735 | 0.000463361 | 0.001141812 | 1 |
| Obesity | endothelial | *PMS2* | -1.345058999 | 0.000278091 | 0.000758032 | 1 |
| Obesity | endothelial | *KLHL7* | -1.131833781 | 6.31E-05 | 0.000246953 | 0.306222251 |
| Obesity | endothelial | *NT5C3A* | -0.776530387 | 0.009597182 | 0.015521842 | 1 |
| Obesity | endothelial | *POLM* | -0.418124485 | 0.009221661 | 0.014964381 | 1 |
| Obesity | endothelial | *SMU1* | -1.161558656 | 0.000101614 | 0.000344056 | 0.493031942 |
| Obesity | endothelial | *GLIPR2* | -1.258789151 | 3.93E-05 | 0.000173137 | 0.190450197 |
| Obesity | endothelial | *CISD1* | -0.610648877 | 0.013134923 | 0.020558274 | 1 |
| Obesity | endothelial | *RBM19* | -1.321482708 | 2.46E-05 | 0.000124585 | 0.119383946 |
| Obesity | endothelial | *DDX54* | -1.315292197 | 0.000105671 | 0.000354331 | 0.512717325 |
| Obesity | endothelial | *SSPN* | -0.748833628 | 0.000200289 | 0.000588277 | 0.971799927 |
| Obesity | endothelial | *CCDC91* | -0.811183453 | 0.00021916 | 0.000630329 | 1 |
| Obesity | endothelial | *TSFM* | -0.958260683 | 0.004637958 | 0.008177097 | 1 |
| Obesity | endothelial | *ARHGAP9* | -1.579513867 | 1.24E-06 | 2.57E-05 | 0.006019931 |
| Obesity | endothelial | *PFDN5* | -0.666231222 | 0.031593548 | 0.045325811 | 1 |
| Obesity | endothelial | *PDE1B* | -1.187040988 | 1.22E-06 | 2.57E-05 | 0.005933582 |
| Obesity | endothelial | *CDK2* | -1.068284693 | 0.001334118 | 0.0027746 | 1 |
| Obesity | endothelial | *SARDH* | -0.755330441 | 0.00032272 | 0.000855182 | 1 |
| Obesity | endothelial | *DBH* | -9.20498227 | 7.91E-06 | 5.64E-05 | 0.038356027 |
| Obesity | endothelial | *USP45* | -0.989882532 | 0.000174933 | 0.000528503 | 0.848775725 |
| Obesity | endothelial | *TTC21B* | -0.972529504 | 0.00081171 | 0.001820812 | 1 |
| Obesity | endothelial | *ACVR1C* | -0.406926101 | 0.002678825 | 0.005061393 | 1 |
| Obesity | endothelial | *PLA2G12A* | -1.146100682 | 0.00035659 | 0.000931203 | 1 |
| Obesity | endothelial | *C4BPA* | -1.141585651 | 0.000130223 | 0.000417334 | 0.631843025 |
| Obesity | endothelial | *RAB38* | -0.963015386 | 3.25E-05 | 0.000151801 | 0.157569298 |
| Obesity | endothelial | *DAW1* | -1.207835702 | 1.73E-05 | 9.78E-05 | 0.083775801 |
| Obesity | endothelial | *DNPEP* | -1.275324389 | 0.000356407 | 0.000931203 | 1 |
| Obesity | endothelial | *OBSL1* | -1.066585748 | 4.09E-06 | 3.86E-05 | 0.019845192 |
| Obesity | endothelial | *FAM210B* | -1.475180003 | 0.000589963 | 0.001397638 | 1 |
| Obesity | endothelial | *WFDC3* | -0.207539305 | 0.017303959 | 0.026319376 | 1 |
| Obesity | endothelial | *MATN4* | -1.848270664 | 1.32E-05 | 8.04E-05 | 0.063955232 |
| Obesity | endothelial | *TOX2* | 0.227990263 | 0.001928544 | 0.003817746 | 1 |
| Obesity | endothelial | *SRSF6* | -1.18547092 | 2.51E-05 | 0.000125716 | 0.121563777 |
| Obesity | endothelial | *CSE1L* | -0.924713092 | 0.005352311 | 0.009241784 | 1 |
| Obesity | endothelial | *PTGIS* | 0.419072758 | 0.002363153 | 0.0045464 | 1 |
| Obesity | endothelial | *RNF114* | -1.115206218 | 0.000198987 | 0.000585853 | 0.965485885 |
| Obesity | endothelial | *PEPD* | -1.07006159 | 3.39E-05 | 0.000156224 | 0.164324841 |

| | | | | | | |
|---|---|---|---|---|---|---|
| Obesity | endothelial | *CHST8* | -0.779698348 | 0.013940315 | 0.021678977 | 1 |
| Obesity | endothelial | *STAMBP* | -1.250864015 | 2.57E-05 | 0.000127888 | 0.124690533 |
| Obesity | endothelial | *NAGK* | -0.723276335 | 0.016663324 | 0.025440669 | 1 |
| Obesity | endothelial | *SNRNP27* | -1.268504624 | 0.002775272 | 0.005219631 | 1 |
| Obesity | endothelial | *ATP8A1* | 0.333420528 | 0.002906243 | 0.005440235 | 1 |
| Obesity | endothelial | *USP22* | -1.080680519 | 9.34E-05 | 0.000323722 | 0.453211235 |
| Obesity | endothelial | *HIF3A* | 0.244938344 | 0.001927508 | 0.003817253 | 1 |
| Obesity | endothelial | *ZNF576* | -1.169396891 | 0.0015143 | 0.003098855 | 1 |
| Obesity | endothelial | *BTN2A2* | 0.632850709 | 1.91E-05 | 0.000104627 | 0.092804251 |
| Obesity | endothelial | *SNRPC* | -3.004786858 | 7.33E-23 | 1.78E-19 | 3.56E-19 |
| Obesity | endothelial | *NQO2* | -1.211489043 | 9.81E-06 | 6.56E-05 | 0.047583038 |
| Obesity | endothelial | *TCP11* | 0.728898116 | 0.031129524 | 0.04473947 | 1 |
| Obesity | endothelial | *GNMT* | -0.826817284 | 0.030645154 | 0.044095577 | 1 |
| Obesity | endothelial | *MEA1* | 0.506542303 | 0.006430232 | 0.010867115 | 1 |
| Obesity | endothelial | *KLHL31* | -0.588140086 | 5.99E-06 | 4.68E-05 | 0.029058234 |
| Obesity | endothelial | *RREB1* | -0.767283189 | 0.000414748 | 0.001048649 | 1 |
| Obesity | endothelial | *RIOK1* | -0.683123725 | 0.032159309 | 0.046082979 | 1 |
| Obesity | endothelial | *ATXN1* | -0.777932254 | 9.50E-05 | 0.000327789 | 0.460871567 |
| Obesity | endothelial | *RUNX2* | -1.343219783 | 1.23E-05 | 7.58E-05 | 0.059449104 |
| Obesity | endothelial | *CXCL6* | -1.33554786 | 2.82E-06 | 3.32E-05 | 0.013673305 |
| Obesity | endothelial | *SH3TC1* | -0.658926816 | 0.001993358 | 0.003926826 | 1 |
| Obesity | endothelial | *MT1G* | -1.423335266 | 0.000542708 | 0.001300356 | 1 |
| Obesity | endothelial | *CLYBL* | -1.00580717 | 0.00091333 | 0.002023506 | 1 |
| Obesity | endothelial | *ABCC4* | 0.254669134 | 0.000759747 | 0.001729031 | 1 |
| Obesity | endothelial | *ATP5S* | -0.842057915 | 0.000385744 | 0.000990521 | 1 |
| Obesity | endothelial | *GRK4* | 0.261500223 | 0.008001535 | 0.013182834 | 1 |
| Obesity | endothelial | *HS3ST3B1* | -1.781116971 | 2.73E-05 | 0.000133426 | 0.13249165 |
| Obesity | endothelial | *MRPS7* | -1.468380513 | 9.96E-06 | 6.63E-05 | 0.048320326 |
| Obesity | endothelial | *GGA3* | -1.112380356 | 0.000295873 | 0.000795775 | 1 |
| Obesity | endothelial | *NUP85* | -1.093273945 | 0.000764229 | 0.001736787 | 1 |
| Obesity | endothelial | *NT5C* | -1.136606289 | 1.28E-06 | 2.57E-05 | 0.00618969 |
| Obesity | endothelial | *DDX31* | -1.344390709 | 1.20E-06 | 2.57E-05 | 0.005819553 |
| Obesity | endothelial | *OPRL1* | -3.95414285 | 1.21E-06 | 2.57E-05 | 0.005890249 |
| Obesity | endothelial | *IL1B* | -4.251545599 | 2.40E-06 | 3.08E-05 | 0.011658457 |
| Obesity | endothelial | *PAX8* | -0.479553583 | 0.008281366 | 0.013588498 | 1 |
| Obesity | endothelial | *CCDC93* | -0.733067382 | 0.008674918 | 0.014167183 | 1 |
| Obesity | endothelial | *PSD4* | -1.218830947 | 6.82E-06 | 5.13E-05 | 0.03307503 |
| Obesity | endothelial | *GTF2F1* | -1.062698586 | 0.007310965 | 0.012173233 | 1 |
| Obesity | endothelial | *TNFSF9* | -7.425463213 | 0.000206204 | 0.000600901 | 1 |
| Obesity | endothelial | *SH2D3A* | -1.324299301 | 2.67E-06 | 3.25E-05 | 0.012947098 |
| Obesity | endothelial | *PSMF1* | -0.750439606 | 0.007416617 | 0.012323776 | 1 |
| Obesity | endothelial | *DTD1* | -0.842263417 | 0.007519816 | 0.012478163 | 1 |
| Obesity | endothelial | *FLRT3* | -0.981415392 | 1.58E-06 | 2.58E-05 | 0.007648845 |
| Obesity | endothelial | *MKKS* | -1.219615999 | 0.000154861 | 0.000478589 | 0.751385336 |
| Obesity | endothelial | *LAMP5* | -7.573556736 | 4.35E-06 | 3.95E-05 | 0.021099423 |
| Obesity | endothelial | *MGME1* | -0.767035624 | 0.011146617 | 0.017778891 | 1 |
| Obesity | endothelial | *ITPA* | -1.199222311 | 3.05E-05 | 0.000144836 | 0.148167009 |
| Obesity | endothelial | *MRPS26* | -0.838687947 | 0.001388695 | 0.002874551 | 1 |
| Obesity | endothelial | *NCLN* | -0.926811879 | 0.017097461 | 0.026041627 | 1 |
| Obesity | endothelial | *TUBGCP3* | -0.874004169 | 0.004048489 | 0.0072699 | 1 |

| | | | | | | |
|---|---|---|---|---|---|---|
| Obesity | endothelial | *MCF2L* | 0.17690433 | 0.000174773 | 0.000528349 | 0.847999665 |
| Obesity | endothelial | *COX6B1* | -0.997955025 | 0.003264677 | 0.006018318 | 1 |
| Obesity | endothelial | *IRF3* | -1.162886141 | 0.000441486 | 0.001101897 | 1 |
| Obesity | endothelial | *PRR12* | -0.969003287 | 0.001250182 | 0.002638487 | 1 |
| Obesity | endothelial | *TSKS* | -0.81218476 | 0.00225206 | 0.004367215 | 1 |
| Obesity | endothelial | *ZNF384* | -1.275059327 | 0.000286683 | 0.000777957 | 1 |
| Obesity | endothelial | *EMG1* | -1.265538404 | 1.52E-05 | 8.93E-05 | 0.073703853 |
| Obesity | endothelial | *ATG14* | -0.821504272 | 0.00021683 | 0.000625108 | 1 |
| Obesity | endothelial | *L3HYPDH* | -0.618539862 | 0.000164576 | 0.000503482 | 0.798522029 |
| Obesity | endothelial | *ZBTB1* | -1.010965774 | 2.77E-05 | 0.000134428 | 0.134428209 |
| Obesity | endothelial | *RHOT1* | -0.704666853 | 0.001801656 | 0.003595901 | 1 |
| Obesity | endothelial | *WDR60* | -0.683151935 | 0.005095545 | 0.0088615 | 1 |
| Obesity | endothelial | *AIF1L* | 0.220013799 | 0.000140879 | 0.00044407 | 0.683546 |
| Obesity | endothelial | *CANX* | -0.631231371 | 0.033496414 | 0.04773116 | 1 |
| Obesity | endothelial | *FGD3* | -0.999095066 | 1.93E-05 | 0.000105189 | 0.093407434 |
| Obesity | endothelial | *ABHD8* | -1.069440514 | 1.40E-05 | 8.40E-05 | 0.068048863 |
| Obesity | endothelial | *ATP13A4* | -1.478171205 | 3.53E-05 | 0.000160431 | 0.171500358 |
| Obesity | endothelial | *HELB* | -1.093143375 | 0.002222904 | 0.004317666 | 1 |
| Obesity | endothelial | *RAB3IP* | -1.771088822 | 4.32E-05 | 0.000185209 | 0.209817062 |
| Obesity | endothelial | *TAS2R4* | -0.930906725 | 0.002728584 | 0.005142799 | 1 |
| Obesity | endothelial | *LRRC61* | -0.78836447 | 2.51E-06 | 3.13E-05 | 0.012186509 |
| Obesity | endothelial | *IDUA* | -1.513082896 | 8.95E-07 | 2.40E-05 | 0.004341863 |
| Obesity | endothelial | *EMC1* | -1.229154129 | 0.000600926 | 0.00141883 | 1 |
| Obesity | endothelial | *UBR4* | -0.735273361 | 0.003549702 | 0.006488806 | 1 |
| Obesity | endothelial | *WFIKKN1* | -0.904587219 | 0.011442146 | 0.018190463 | 1 |
| Obesity | endothelial | *WDR24* | -1.227868702 | 0.000709765 | 0.001631439 | 1 |
| Obesity | endothelial | *SMARCA4* | -0.976886149 | 0.001737393 | 0.00348484 | 1 |
| Obesity | endothelial | *PNKD* | -1.135545528 | 0.000179368 | 0.000539216 | 0.870294031 |
| Obesity | endothelial | *STEAP4* | -1.253897252 | 6.04E-06 | 4.70E-05 | 0.029311466 |
| Obesity | endothelial | *PEX1* | -1.245667398 | 0.000120154 | 0.000391265 | 0.582985072 |
| Obesity | endothelial | *KDR* | 0.367782042 | 0.002845869 | 0.005337517 | 1 |
| Obesity | endothelial | *VPREB3* | -1.671388228 | 0.002624249 | 0.004985288 | 1 |
| Obesity | endothelial | *MGAT3* | 0.79984696 | 0.021815689 | 0.032479916 | 1 |
| Obesity | endothelial | *ATF4* | -1.401563549 | 0.000572711 | 0.001361486 | 1 |
| Obesity | endothelial | *A4GALT* | 0.376259928 | 0.014356566 | 0.022269201 | 1 |
| Obesity | endothelial | *CDC42EP1* | -1.067936123 | 0.022153723 | 0.032871519 | 1 |
| Obesity | endothelial | *RIBC2* | 0.54964715 | 0.035163764 | 0.049829025 | 1 |
| Obesity | endothelial | *SPECC1* | -0.96717226 | 0.000349079 | 0.000914541 | 1 |
| Obesity | endothelial | *CDHR3* | -1.112024808 | 4.92E-06 | 4.26E-05 | 0.023848529 |
| Obesity | endothelial | *PRKRIP1* | -1.116033365 | 1.77E-05 | 9.92E-05 | 0.085826057 |
| Obesity | endothelial | *OPN1SW* | -0.77333688 | 0.001491088 | 0.003059094 | 1 |
| Obesity | endothelial | *HOXD1* | 0.621767654 | 0.000692003 | 0.001601144 | 1 |
| Obesity | endothelial | *HOXD3* | 0.176521792 | 3.40E-05 | 0.000156557 | 0.164854613 |
| Obesity | endothelial | *MTX2* | -0.785863279 | 0.006540859 | 0.011034857 | 1 |
| Obesity | endothelial | *PDE11A* | -0.452660982 | 0.0126334 | 0.019858273 | 1 |
| Obesity | endothelial | *GAD1* | -1.412235626 | 1.71E-06 | 2.66E-05 | 0.008283935 |
| Obesity | endothelial | *ORMDL1* | -0.826792773 | 0.000935589 | 0.002063398 | 1 |
| Obesity | endothelial | *HOXD9* | 0.297291295 | 0.000342254 | 0.000898602 | 1 |
| Obesity | endothelial | *HOXD10* | 1.059604453 | 0.003276986 | 0.006037704 | 1 |
| Obesity | endothelial | *PSMG2* | -1.06813222 | 0.000251151 | 0.000699131 | 1 |

| | | | | | | |
|---|---|---|---|---|---|---|
| Obesity | endothelial | *TWSG1* | -1.247502316 | 1.69E-05 | 9.64E-05 | 0.082092156 |
| Obesity | endothelial | *WDFY4* | -0.566526789 | 3.30E-05 | 0.000153785 | 0.160244198 |
| Obesity | endothelial | *TMOD2* | -1.001667812 | 0.002448712 | 0.004684997 | 1 |
| Obesity | endothelial | *IVD* | -0.626485086 | 0.010954356 | 0.017512532 | 1 |
| Obesity | endothelial | *KNSTRN* | -0.244972993 | 0.020015237 | 0.030010484 | 1 |
| Obesity | endothelial | *ARPP19* | -1.081486067 | 0.0021353 | 0.004172563 | 1 |
| Obesity | endothelial | *VPS13C* | -0.605187702 | 0.011766879 | 0.018636609 | 1 |
| Obesity | endothelial | *CALML4* | 1.222445166 | 0.003646254 | 0.006633529 | 1 |
| Obesity | endothelial | *ANAPC13* | -0.398479862 | 0.018029439 | 0.027320062 | 1 |
| Obesity | endothelial | *SUMF2* | -1.125837459 | 0.000526003 | 0.001267843 | 1 |
| Obesity | endothelial | *BBOX1* | -3.161614736 | 2.24E-06 | 2.99E-05 | 0.010892645 |
| Obesity | endothelial | *DCTD* | -0.893333063 | 0.00545462 | 0.00938837 | 1 |
| Obesity | endothelial | *RPAIN* | -1.55356842 | 1.08E-08 | 1.27E-06 | 5.24E-05 |
| Obesity | endothelial | *PHF20L1* | -0.913209827 | 0.000744336 | 0.001699539 | 1 |
| Obesity | endothelial | *PUS7L* | -1.000923777 | 0.000706868 | 0.001629321 | 1 |
| Obesity | endothelial | *AP1M2* | -0.910941764 | 0.000162654 | 0.000499177 | 0.789198316 |
| Obesity | endothelial | *KLK8* | -6.903926504 | 2.62E-06 | 3.21E-05 | 0.01269168 |
| Obesity | endothelial | *RIPK3* | 0.441148468 | 0.024380168 | 0.035878852 | 1 |
| Obesity | endothelial | *ADCY4* | 0.19170751 | 0.000107792 | 0.000360198 | 0.523007331 |
| Obesity | endothelial | *BCL2L2* | -0.941651357 | 0.000170964 | 0.000518774 | 0.829518895 |
| Obesity | endothelial | *DTD2* | -0.421552685 | 0.024613549 | 0.036167456 | 1 |
| Obesity | endothelial | *PARP2* | -1.462222234 | 7.01E-05 | 0.000266945 | 0.340087855 |
| Obesity | endothelial | *MIS18BP1* | -0.648439662 | 0.009589644 | 0.015514823 | 1 |
| Obesity | endothelial | *EPB41L4A* | 0.29110875 | 0.013302418 | 0.0207936 | 1 |
| Obesity | endothelial | *REEP5* | -0.989755722 | 0.000478737 | 0.001173741 | 1 |
| Obesity | endothelial | *TMEM8A* | -1.908150856 | 5.35E-05 | 0.00021837 | 0.259423053 |
| Obesity | endothelial | *MAU2* | -1.12509487 | 8.36E-05 | 0.000301906 | 0.405742586 |
| Obesity | endothelial | *SHC2* | 0.821474915 | 0.001986204 | 0.003914322 | 1 |
| Obesity | endothelial | *SYT5* | -2.387233336 | 0.013171437 | 0.020602132 | 1 |
| Obesity | endothelial | *ERMARD* | -1.333597713 | 1.01E-05 | 6.68E-05 | 0.049234186 |
| Obesity | endothelial | *PHF10* | -0.886288985 | 0.018434479 | 0.027855526 | 1 |
| Obesity | endothelial | *PRKCSH* | -0.980138952 | 0.011310966 | 0.018005514 | 1 |
| Obesity | endothelial | *CDC16* | -1.01015625 | 6.57E-05 | 0.000253909 | 0.318909498 |
| Obesity | endothelial | *XPO7* | -0.914799429 | 0.002732637 | 0.005145674 | 1 |
| Obesity | endothelial | *FAM98C* | -0.920056286 | 0.002362797 | 0.0045464 | 1 |
| Obesity | endothelial | *ATP8B3* | -1.297588761 | 8.97E-15 | 4.83E-12 | 4.35E-11 |
| Obesity | endothelial | *PLVAP* | 0.169454789 | 6.44E-05 | 0.000250194 | 0.312492022 |
| Obesity | endothelial | *DDA1* | -1.278736552 | 0.007360611 | 0.012243293 | 1 |
| Obesity | endothelial | *QRSL1* | -0.463160818 | 0.02866399 | 0.041577781 | 1 |
| Obesity | endothelial | *STK33* | 0.659703554 | 0.002303372 | 0.004445489 | 1 |
| Obesity | endothelial | *NDUFA10* | -0.469008874 | 0.015229077 | 0.023472516 | 1 |
| Obesity | endothelial | *MAP1S* | -2.088436461 | 3.93E-06 | 3.80E-05 | 0.019066074 |
| Obesity | endothelial | *KIAA1683* | -1.246100497 | 8.61E-05 | 0.000308494 | 0.417701103 |
| Obesity | endothelial | *UBAC1* | -0.794553409 | 0.008899175 | 0.014509004 | 1 |
| Obesity | endothelial | *ATXN10* | -0.926432978 | 0.000101167 | 0.000343021 | 0.49086309 |
| Obesity | endothelial | *CYP2E1* | -0.459400693 | 0.019143695 | 0.028810548 | 1 |
| Obesity | endothelial | *PNPLA7* | -1.104438003 | 2.36E-06 | 3.05E-05 | 0.011455345 |
| Obesity | endothelial | *PAK4* | -1.116528587 | 0.003714159 | 0.006741371 | 1 |
| Obesity | endothelial | *ZNF337* | -0.701382967 | 0.013636222 | 0.021260587 | 1 |
| Obesity | endothelial | *CEP85* | -0.718832685 | 0.000371993 | 0.000962106 | 1 |

| | | | | | | |
|---|---|---|---|---|---|---|
| Obesity | endothelial | *POMT1* | -1.332923696 | 0.00100887 | 0.002190173 | 1 |
| Obesity | endothelial | *CHMP2A* | -0.74125585 | 0.029913883 | 0.043235675 | 1 |
| Obesity | endothelial | *YIPF2* | -0.999226489 | 0.002637211 | 0.004996387 | 1 |
| Obesity | endothelial | *NPAS1* | -3.680125839 | 4.95E-11 | 1.20E-08 | 2.40E-07 |
| Obesity | endothelial | *MAP3K10* | -1.129735423 | 0.002625711 | 0.004985288 | 1 |
| Obesity | endothelial | *THEMIS2* | -0.519386858 | 0.023262682 | 0.034390778 | 1 |
| Obesity | endothelial | *ZNF317* | -1.63046522 | 1.05E-05 | 6.86E-05 | 0.051080627 |
| Obesity | endothelial | *ANGPTL6* | 0.746524104 | 0.000464362 | 0.001143697 | 1 |
| Obesity | endothelial | *PKDREJ* | -1.052421657 | 0.000165514 | 0.000504759 | 0.803072152 |
| Obesity | endothelial | *HSD17B3* | 0.814538391 | 0.024599805 | 0.03615821 | 1 |
| Obesity | endothelial | *SLC35D2* | -1.0047404 | 0.002826717 | 0.005305698 | 1 |
| Obesity | endothelial | *ULBP2* | 1.252237376 | 8.30E-05 | 0.000300462 | 0.402891095 |
| Obesity | endothelial | *TTLL9* | 1.006649907 | 0.004970258 | 0.00867471 | 1 |
| Obesity | endothelial | *ATP6V1E1* | -1.543435756 | 1.80E-06 | 2.68E-05 | 0.008748136 |
| Obesity | endothelial | *CHMP1A* | -1.602725262 | 4.35E-05 | 0.000185308 | 0.211065493 |
| Obesity | endothelial | *IDO1* | 0.204668999 | 8.19E-05 | 0.000298 | 0.397449522 |
| Obesity | endothelial | *HAUS8* | -0.730542268 | 0.028408159 | 0.041243682 | 1 |
| Obesity | endothelial | *MRPS25* | -1.200444828 | 5.00E-06 | 4.29E-05 | 0.024272452 |
| Obesity | endothelial | *GALNT15* | 0.227877887 | 0.000324203 | 0.000858175 | 1 |
| Obesity | endothelial | *KCNC3* | -1.343706208 | 3.74E-06 | 3.75E-05 | 0.018155758 |
| Obesity | endothelial | *PDLIM4* | -2.476335042 | 0.000735 | 0.001680595 | 1 |
| Obesity | endothelial | *KIF3A* | -1.134313645 | 0.001172889 | 0.002494526 | 1 |
| Obesity | endothelial | *ACLY* | -1.233340867 | 0.000211725 | 0.000613674 | 1 |
| Obesity | endothelial | *EXOC4* | -0.470118135 | 0.02183625 | 0.032479916 | 1 |
| Obesity | endothelial | *C1orf159* | -1.471810614 | 2.97E-06 | 3.38E-05 | 0.014396277 |
| Obesity | endothelial | *PPFIA1* | -0.944216184 | 6.08E-05 | 0.000239986 | 0.295182835 |
| Obesity | endothelial | *BARX1* | -1.774867267 | 7.98E-05 | 0.000292909 | 0.387225381 |
| Obesity | endothelial | *NINJ1* | -0.584474233 | 0.003241881 | 0.005980839 | 1 |
| Obesity | endothelial | *NPHP4* | -1.103410657 | 0.000235114 | 0.000665931 | 1 |
| Obesity | endothelial | *MAP1B* | -0.905172846 | 5.65E-06 | 4.54E-05 | 0.027434488 |
| Obesity | endothelial | *CKMT2* | -1.081389803 | 0.000245565 | 0.000687525 | 1 |
| Obesity | endothelial | *KHDRBS3* | -1.143357423 | 1.76E-06 | 2.68E-05 | 0.008529187 |
| Obesity | endothelial | *CHD1L* | -1.191344077 | 4.23E-06 | 3.91E-05 | 0.020516482 |
| Obesity | endothelial | *ZNF132* | -0.498484588 | 0.031222564 | 0.04484662 | 1 |
| Obesity | endothelial | *RHPN2* | -0.365256109 | 0.004072727 | 0.007310718 | 1 |
| Obesity | endothelial | *GCH1* | 0.392408092 | 0.002896455 | 0.005426101 | 1 |
| Obesity | endothelial | *MATN3* | -0.964483383 | 0.000421581 | 0.0010615 | 1 |
| Obesity | endothelial | *TRIM21* | -1.509163378 | 0.018426695 | 0.027852437 | 1 |
| Obesity | endothelial | *SPATA6* | -0.960913365 | 8.30E-05 | 0.000300462 | 0.402919246 |
| Obesity | endothelial | *LRRC41* | -1.55149284 | 4.60E-06 | 4.08E-05 | 0.02232083 |
| Obesity | endothelial | *CCT6B* | -0.649898433 | 0.006286374 | 0.010646243 | 1 |
| Obesity | endothelial | *CNGA4* | -1.118636103 | 8.72E-06 | 6.02E-05 | 0.042332679 |
| Obesity | endothelial | *IMMT* | -0.856004648 | 0.003130234 | 0.005794695 | 1 |
| Obesity | endothelial | *RPA1* | -0.729025232 | 0.008321154 | 0.01363534 | 1 |
| Obesity | endothelial | *SERPINF1* | -1.123578405 | 3.20E-06 | 3.43E-05 | 0.015511496 |
| Obesity | endothelial | *TMEM128* | -0.788557018 | 0.002630323 | 0.004987232 | 1 |
| Obesity | endothelial | *FIGNL1* | -1.887358635 | 6.38E-08 | 4.43E-06 | 0.000309772 |
| Obesity | endothelial | *RGS22* | -0.773232276 | 4.31E-07 | 1.64E-05 | 0.002090344 |
| Obesity | endothelial | *REEP2* | 0.583497197 | 8.83E-05 | 0.000314256 | 0.428644623 |
| Obesity | endothelial | *PRMT7* | -0.781623628 | 0.000275739 | 0.000753313 | 1 |

| | | | | | | |
|---|---|---|---|---|---|---|
| Obesity | endothelial | *CTNNBL1* | -0.962723411 | 0.002886889 | 0.005410269 | 1 |
| Obesity | endothelial | *OSER1* | -1.046643708 | 0.005994172 | 0.010226344 | 1 |
| Obesity | endothelial | *DMGDH* | -1.178941501 | 1.37E-06 | 2.57E-05 | 0.006634828 |
| Obesity | endothelial | *BHMT2* | -0.3172414 | 0.000235607 | 0.000666569 | 1 |
| Obesity | endothelial | *AP3B1* | -0.772257666 | 0.002019304 | 0.003970872 | 1 |
| Obesity | endothelial | *ZBED3* | -1.909022271 | 9.78E-07 | 2.45E-05 | 0.004744675 |
| Obesity | endothelial | *FBXO44* | -1.747924366 | 4.13E-08 | 3.58E-06 | 0.000200304 |
| Obesity | endothelial | *CASP9* | -0.665269629 | 0.000113115 | 0.000373865 | 0.548833465 |
| Obesity | endothelial | *ATP8A2* | -1.355070388 | 0.006348673 | 0.010740503 | 1 |
| Obesity | endothelial | *ALOX5AP* | -1.549272094 | 4.84E-05 | 0.000201711 | 0.2347919 |
| Obesity | endothelial | *WASF3* | -1.011394587 | 2.16E-06 | 2.92E-05 | 0.010503373 |
| Obesity | endothelial | *GPR12* | 1.404553904 | 8.16E-05 | 0.000297261 | 0.395951729 |
| Obesity | endothelial | *PEMT* | -1.085652599 | 0.021035007 | 0.031422985 | 1 |
| Obesity | endothelial | *SCO1* | -1.220811566 | 0.000235208 | 0.000665931 | 1 |
| Obesity | endothelial | *MPRIP* | -0.82459004 | 0.004289475 | 0.007632025 | 1 |
| Obesity | endothelial | *DSTYK* | -1.040962489 | 0.001201154 | 0.002548316 | 1 |
| Obesity | endothelial | *COG6* | -0.80662523 | 0.004434374 | 0.007866758 | 1 |
| Obesity | endothelial | *EPSTI1* | -0.77230859 | 0.009696737 | 0.015672407 | 1 |
| Obesity | endothelial | *BTBD2* | -1.218899261 | 2.76E-05 | 0.000134115 | 0.133980948 |
| Obesity | endothelial | *CSNK1G2* | -1.525427782 | 9.30E-05 | 0.000323239 | 0.451011312 |
| Obesity | endothelial | *PDZD2* | -0.735675806 | 0.001014572 | 0.002201568 | 1 |
| Obesity | endothelial | *FAM83F* | 0.707161393 | 0.000127589 | 0.000410518 | 0.619060445 |
| Obesity | endothelial | *GIMAP4* | 0.340068225 | 0.00019852 | 0.000585373 | 0.96321851 |
| Obesity | endothelial | *ADCK2* | -0.577172989 | 0.035062025 | 0.049713895 | 1 |
| Obesity | endothelial | *IPO8* | -0.905131735 | 0.006120531 | 0.010408979 | 1 |
| Obesity | endothelial | *LARS* | -1.038509809 | 1.66E-07 | 8.77E-06 | 0.000806719 |
| Obesity | endothelial | *SPINK5* | 0.475846619 | 0.00023623 | 0.000667351 | 1 |
| Obesity | endothelial | *IMPA1* | -0.520798813 | 0.023701001 | 0.035006775 | 1 |
| Obesity | endothelial | *RRAS2* | -0.823251928 | 1.78E-05 | 9.97E-05 | 0.086416328 |
| Obesity | endothelial | *CATSPERB* | -0.889938028 | 4.32E-05 | 0.000185209 | 0.20968509 |
| Obesity | endothelial | *COX16* | 1.618781253 | 0.013489642 | 0.021052345 | 1 |
| Obesity | endothelial | *TTC9* | -2.078119623 | 0.001143042 | 0.002441639 | 1 |
| Obesity | endothelial | *ELP3* | -1.101960479 | 0.000344507 | 0.000904029 | 1 |
| Obesity | endothelial | *PEBP4* | -1.721508668 | 2.88E-06 | 3.33E-05 | 0.013975116 |
| Obesity | endothelial | *CTIF* | -0.875969779 | 3.07E-06 | 3.39E-05 | 0.014878357 |
| Obesity | endothelial | *MRO* | -0.791955317 | 0.000314548 | 0.000837119 | 1 |
| Obesity | endothelial | *THUMPD3* | -1.603413618 | 2.74E-06 | 3.28E-05 | 0.013281577 |
| Obesity | endothelial | *CNTN6* | -1.600288363 | 4.69E-06 | 4.14E-05 | 0.022745734 |
| Obesity | endothelial | *CHL1* | -0.8571857 | 5.88E-06 | 4.66E-05 | 0.028552436 |
| Obesity | endothelial | *GSTM3* | -0.417390739 | 0.002248659 | 0.004362452 | 1 |
| Obesity | endothelial | *VAV3* | -0.438715922 | 0.032828229 | 0.046861596 | 1 |
| Obesity | endothelial | *PSRC1* | -0.826471516 | 0.019082523 | 0.028736313 | 1 |
| Obesity | endothelial | *TRIM45* | -1.750252466 | 5.44E-06 | 4.47E-05 | 0.026379226 |
| Obesity | endothelial | *CEPT1* | -1.001163136 | 1.83E-05 | 0.000101521 | 0.088729037 |
| Obesity | endothelial | *NAPG* | -1.041381733 | 0.001072612 | 0.002312002 | 1 |
| Obesity | endothelial | *PPHLN1* | -0.931342164 | 0.000203886 | 0.000596655 | 0.989253503 |
| Obesity | endothelial | *FKBP11* | -1.5437285 | 3.87E-06 | 3.78E-05 | 0.018798792 |
| Obesity | endothelial | *YWHAQ* | -0.863911371 | 0.004613215 | 0.00813643 | 1 |
| Obesity | endothelial | *KIDINS220* | -0.880404735 | 0.000414616 | 0.001048649 | 1 |
| Obesity | endothelial | *LPIN1* | -0.902101551 | 1.08E-05 | 6.98E-05 | 0.052583727 |

| | | | | | | |
|---|---|---|---|---|---|---|
| Obesity | endothelial | *NARS* | -0.790570124 | 0.01700516 | 0.025913642 | 1 |
| Obesity | endothelial | *KIAA1468* | -0.852552706 | 0.002268753 | 0.004387402 | 1 |
| Obesity | endothelial | *IL15RA* | 0.517694869 | 0.011155753 | 0.017787615 | 1 |
| Obesity | endothelial | *TMEM241* | -0.810737238 | 1.44E-05 | 8.58E-05 | 0.070062595 |
| Obesity | endothelial | *SOX5* | -0.779644853 | 6.05E-05 | 0.00023913 | 0.293651229 |
| Obesity | endothelial | *KLRD1* | -0.812769417 | 0.015440948 | 0.023750363 | 1 |
| Obesity | endothelial | *ACP2* | -0.932712921 | 0.0014779 | 0.003033321 | 1 |
| Obesity | endothelial | *HOOK1* | -0.451844429 | 0.000875523 | 0.001946856 | 1 |
| Obesity | endothelial | *ZCCHC11* | -0.69519751 | 0.001426868 | 0.002941443 | 1 |
| Obesity | endothelial | *PRPF38A* | -1.322712447 | 0.000531438 | 0.001279635 | 1 |
| Obesity | endothelial | *ELP2* | -1.070490142 | 5.25E-05 | 0.000215908 | 0.254672823 |
| Obesity | endothelial | *TPGS2* | -1.018514581 | 0.004500792 | 0.007961299 | 1 |
| Obesity | endothelial | *DHX34* | -1.386552651 | 2.69E-05 | 0.000131801 | 0.130351539 |
| Obesity | endothelial | *FADS2* | -1.433048658 | 1.41E-06 | 2.57E-05 | 0.006856286 |
| Obesity | endothelial | *TMEM258* | -0.771706896 | 0.014815528 | 0.022893294 | 1 |
| Obesity | endothelial | *CLOCK* | -0.792342186 | 0.003922389 | 0.007074881 | 1 |
| Obesity | endothelial | *CLDN10* | 0.524732317 | 0.000399385 | 0.001018831 | 1 |
| Obesity | endothelial | *CARS2* | -0.923038745 | 0.00013374 | 0.000426912 | 0.648905668 |
| Obesity | endothelial | *WDR36* | -1.26982578 | 3.72E-05 | 0.000166444 | 0.180303926 |
| Obesity | endothelial | *UBQLN1* | -1.067205043 | 0.000139635 | 0.000442237 | 0.677506861 |
| Obesity | endothelial | *GOLM1* | -0.746322473 | 0.000333829 | 0.000878383 | 1 |
| Obesity | endothelial | *ADAM19* | -0.819274889 | 1.09E-05 | 6.98E-05 | 0.052800064 |
| Obesity | endothelial | *USP30* | -0.505718322 | 0.003900113 | 0.007039936 | 1 |
| Obesity | endothelial | *MSI1* | -1.212934027 | 0.004574182 | 0.008073456 | 1 |
| Obesity | endothelial | *P2RX4* | -1.093835904 | 5.48E-06 | 4.47E-05 | 0.026574791 |
| Obesity | endothelial | *DTX1* | -0.802798064 | 1.79E-06 | 2.68E-05 | 0.008698586 |
| Obesity | endothelial | *DMTF1* | -0.900280147 | 0.000191799 | 0.000569878 | 0.930611129 |
| Obesity | endothelial | *TMEM60* | -2.350209351 | 1.92E-08 | 1.90E-06 | 9.31E-05 |
| Obesity | endothelial | *POM121C* | -2.118875783 | 2.55E-14 | 1.13E-11 | 1.24E-10 |
| Obesity | endothelial | *SRPK2* | -0.740881945 | 0.001432684 | 0.002950453 | 1 |
| Obesity | endothelial | *MTO1* | -0.835736787 | 0.000311013 | 0.000830053 | 1 |
| Obesity | endothelial | *KHDC1* | -0.476659339 | 0.000357698 | 0.000933432 | 1 |
| Obesity | endothelial | *AKIRIN2* | -1.20508225 | 9.16E-05 | 0.000320944 | 0.44450708 |
| Obesity | endothelial | *MAP3K7* | -0.824320107 | 0.007780022 | 0.012861556 | 1 |
| Obesity | endothelial | *PRR5L* | -0.584334443 | 0.033867101 | 0.048216894 | 1 |
| Obesity | endothelial | *PHF21A* | -0.782702768 | 0.000958656 | 0.002101853 | 1 |
| Obesity | endothelial | *CAPRIN1* | -1.076584392 | 0.000317848 | 0.000844579 | 1 |
| Obesity | endothelial | *ATP5G2* | -0.886285301 | 0.004933979 | 0.008629831 | 1 |
| Obesity | endothelial | *B4GALNT1* | -1.539339835 | 0.031610948 | 0.045337369 | 1 |
| Obesity | endothelial | *FAIM2* | -1.184584118 | 4.01E-05 | 0.00017598 | 0.194590893 |
| Obesity | endothelial | *PAN2* | -1.313793054 | 8.52E-06 | 5.95E-05 | 0.041334212 |
| Obesity | endothelial | *SLC26A10* | -1.04609818 | 0.014388024 | 0.022310863 | 1 |
| Obesity | endothelial | *MAP7* | -0.80812734 | 1.24E-05 | 7.64E-05 | 0.060071083 |
| Obesity | endothelial | *NHSL1* | -0.843292955 | 4.21E-05 | 0.000181903 | 0.204276988 |
| Obesity | endothelial | *AHI1* | -0.721067065 | 2.17E-05 | 0.000114536 | 0.10537299 |
| Obesity | endothelial | *SMPD2* | -0.606880485 | 0.000927202 | 0.002047695 | 1 |
| Obesity | endothelial | *MICAL1* | 0.584438859 | 0.025815364 | 0.03780747 | 1 |
| Obesity | endothelial | *CCT7* | -0.956406917 | 0.012563641 | 0.019772554 | 1 |
| Obesity | endothelial | *USP15* | -0.725506594 | 0.002911946 | 0.005448809 | 1 |
| Obesity | endothelial | *MDM2* | -1.045050622 | 2.13E-05 | 0.000112748 | 0.103277001 |

| | | | | | | |
|---|---|---|---|---|---|---|
| Obesity | endothelial | *KLHL36* | -1.067597163 | 0.000238359 | 0.000671614 | 1 |
| Obesity | endothelial | *MPHOSPH6* | -1.293591957 | 3.62E-06 | 3.68E-05 | 0.017544237 |
| Obesity | endothelial | *CCDC102A* | -1.192710278 | 0.003216756 | 0.005941264 | 1 |
| Obesity | endothelial | *ZNF670* | -0.764126487 | 0.029426335 | 0.042569046 | 1 |
| Obesity | endothelial | *KCNK1* | -0.442975279 | 0.025083457 | 0.03681335 | 1 |
| Obesity | endothelial | *CAPN9* | -1.347736829 | 7.41E-06 | 5.43E-05 | 0.035971505 |
| Obesity | endothelial | *NTPCR* | -0.863131801 | 0.023797216 | 0.03513819 | 1 |
| Obesity | endothelial | *MRPL44* | -1.562451209 | 2.61E-06 | 3.21E-05 | 0.01268153 |
| Obesity | endothelial | *USP37* | -0.601643329 | 0.034314199 | 0.048781862 | 1 |
| Obesity | endothelial | *SLC19A3* | -0.794161295 | 7.89E-07 | 2.25E-05 | 0.003828041 |
| Obesity | endothelial | *TMBIM1* | -1.128963858 | 0.001281717 | 0.002686346 | 1 |
| Obesity | endothelial | *TSGA10* | -0.503922413 | 2.43E-05 | 0.000123522 | 0.117963063 |
| Obesity | endothelial | *GCC2* | -0.829169244 | 0.001421753 | 0.002934097 | 1 |
| Obesity | endothelial | *ANKRD36* | -0.648695571 | 0.030486069 | 0.043882029 | 1 |
| Obesity | endothelial | *EPC2* | -0.906814569 | 0.000539359 | 0.001294888 | 1 |
| Obesity | endothelial | *STAB2* | -1.370854077 | 0.000631997 | 0.001478519 | 1 |
| Obesity | endothelial | *APPL2* | -1.025489582 | 3.04E-05 | 0.000144396 | 0.147572911 |
| Obesity | endothelial | *DRAM1* | -1.108707194 | 0.001151464 | 0.002457467 | 1 |
| Obesity | endothelial | *SLC41A2* | -0.843424889 | 0.00010001 | 0.00033981 | 0.485248029 |
| Obesity | endothelial | *RNASEH2B* | -1.060939183 | 1.27E-05 | 7.78E-05 | 0.061608271 |
| Obesity | endothelial | *CKAP2* | -0.294488249 | 0.014047729 | 0.021825034 | 1 |
| Obesity | endothelial | *TBC1D4* | -0.73801974 | 0.000195491 | 0.000578368 | 0.948523081 |
| Obesity | endothelial | *SUCLA2* | -1.111015096 | 0.000206668 | 0.000601532 | 1 |
| Obesity | endothelial | *RCBTB2* | -0.781987951 | 0.021204303 | 0.031636924 | 1 |
| Obesity | endothelial | *SETDB2* | -1.034379511 | 0.002671057 | 0.005050651 | 1 |
| Obesity | endothelial | *SCRN1* | -1.10753076 | 3.06E-06 | 3.39E-05 | 0.014849188 |
| Obesity | endothelial | *C7orf25* | -1.904774776 | 0.000319303 | 0.000847053 | 1 |
| Obesity | endothelial | *CHST12* | -0.745593517 | 0.033485516 | 0.047729648 | 1 |
| Obesity | endothelial | *NUPL2* | -1.17955816 | 9.60E-05 | 0.000330442 | 0.465593472 |
| Obesity | endothelial | *ZDHHC4* | -1.42981184 | 9.16E-06 | 6.21E-05 | 0.044426771 |
| Obesity | endothelial | *AOAH* | -0.65086148 | 0.000804956 | 0.001809007 | 1 |
| Obesity | endothelial | *TBRG4* | -1.987180875 | 3.48E-06 | 3.58E-05 | 0.016879564 |
| Obesity | endothelial | *MTHFS* | -0.711371379 | 0.004801685 | 0.00842672 | 1 |
| Obesity | endothelial | *MYCBPAP* | -0.371441453 | 0.02291599 | 0.033909235 | 1 |
| Obesity | endothelial | *BRIP1* | -1.017512122 | 4.60E-06 | 4.08E-05 | 0.022297535 |
| Obesity | endothelial | *RTP4* | 0.699234483 | 0.011963796 | 0.018914414 | 1 |
| Obesity | endothelial | *NDUFB5* | -1.213314902 | 0.000373955 | 0.00096472 | 1 |
| Obesity | endothelial | *RPS6KC1* | -0.886352596 | 0.000336322 | 0.000883984 | 1 |
| Obesity | endothelial | *SMPD4* | -0.695105348 | 0.019487111 | 0.029272899 | 1 |
| Obesity | endothelial | *IMP4* | -1.694258659 | 0.001124907 | 0.002407609 | 1 |
| Obesity | endothelial | *NIPSNAP3A* | -0.801633459 | 0.020980322 | 0.031350947 | 1 |
| Obesity | endothelial | *ODF2* | -0.855300376 | 0.026738579 | 0.039053457 | 1 |
| Obesity | endothelial | *TOR1B* | -1.512837945 | 0.000267907 | 0.000736062 | 1 |
| Obesity | endothelial | *SLC2A8* | -1.634278134 | 0.000233764 | 0.00066329 | 1 |
| Obesity | endothelial | *ZNF189* | -1.192545346 | 0.004143365 | 0.007413766 | 1 |
| Obesity | endothelial | *STX17* | -0.717812302 | 0.022481271 | 0.033306604 | 1 |
| Obesity | endothelial | *ANP32B* | -0.85394695 | 0.013031271 | 0.020409208 | 1 |
| Obesity | endothelial | *DERL1* | -1.563763733 | 1.39E-05 | 8.37E-05 | 0.067573181 |
| Obesity | endothelial | *NOV* | 0.312226666 | 9.64E-05 | 0.000331493 | 0.467737125 |
| Obesity | endothelial | *POLR1E* | -0.352578042 | 0.021480982 | 0.032030032 | 1 |

| | | | | | | |
|---|---|---|---|---|---|---|
| Obesity | endothelial | *IL11RA* | -1.052406695 | 0.001116885 | 0.002396783 | 1 |
| Obesity | endothelial | *APTX* | -1.161423194 | 7.61E-06 | 5.50E-05 | 0.036929136 |
| Obesity | endothelial | *TMEM8B* | -1.084856737 | 0.000139773 | 0.000442385 | 0.678176174 |
| Obesity | endothelial | *GRHPR* | -0.599760474 | 0.023810144 | 0.035146584 | 1 |
| Obesity | endothelial | *DENND4C* | -0.778738316 | 0.000606332 | 0.001428627 | 1 |
| Obesity | endothelial | *GMPR* | -1.019317428 | 0.006429575 | 0.010867115 | 1 |
| Obesity | endothelial | *TMEM14B* | -1.160862806 | 0.000470115 | 0.001153768 | 1 |
| Obesity | endothelial | *CAPN11* | 0.30119905 | 0.001520861 | 0.003109658 | 1 |
| Obesity | endothelial | *LRRC1* | 0.322775981 | 0.006967826 | 0.011653874 | 1 |
| Obesity | endothelial | *RIPK1* | -0.809289969 | 0.003717083 | 0.006742163 | 1 |
| Obesity | endothelial | *TUBB2B* | -0.576080657 | 0.011606624 | 0.018403706 | 1 |
| Obesity | endothelial | *TCF19* | 0.702480204 | 0.024559855 | 0.036110429 | 1 |
| Obesity | endothelial | *VARS2* | -0.294744115 | 0.002148998 | 0.004194262 | 1 |
| Obesity | endothelial | *TLR2* | -1.37800207 | 3.47E-05 | 0.000158296 | 0.168426564 |
| Obesity | endothelial | *MYO7A* | -0.96559976 | 0.000153776 | 0.000476451 | 0.746121768 |
| Obesity | endothelial | *NUMA1* | -0.960575077 | 0.000492741 | 0.001202296 | 1 |
| Obesity | endothelial | *SYTL2* | -1.09869171 | 4.73E-07 | 1.75E-05 | 0.002293456 |
| Obesity | endothelial | *NARS2* | -1.125603325 | 5.50E-05 | 0.00022274 | 0.266643237 |
| Obesity | endothelial | *GGH* | -0.502933097 | 0.006757513 | 0.011341215 | 1 |
| Obesity | endothelial | *TGS1* | -1.193763314 | 1.10E-05 | 7.01E-05 | 0.053288927 |
| Obesity | endothelial | *NEK1* | -0.878425563 | 0.000481115 | 0.001178378 | 1 |
| Obesity | endothelial | *TRPC6* | -0.753596586 | 0.013913198 | 0.021643744 | 1 |
| Obesity | endothelial | *SLC37A4* | 1.236627111 | 0.000118797 | 0.000388675 | 0.576405151 |
| Obesity | endothelial | *PPP2R1B* | -0.496161246 | 0.000954141 | 0.002096149 | 1 |
| Obesity | endothelial | *FDX1* | -0.940193571 | 3.42E-05 | 0.000156798 | 0.165735249 |
| Obesity | endothelial | *FXYD2* | -1.059932276 | 0.000550472 | 0.00131571 | 1 |
| Obesity | endothelial | *MAP2K5* | -0.646160776 | 0.01021834 | 0.016480072 | 1 |
| Obesity | endothelial | *NUSAP1* | -0.775571545 | 0.003503554 | 0.006414808 | 1 |
| Obesity | endothelial | *PARP6* | -1.307427022 | 5.17E-06 | 4.37E-05 | 0.025084393 |
| Obesity | endothelial | *ZNF280D* | -0.699349551 | 0.00278214 | 0.005230121 | 1 |
| Obesity | endothelial | *RSL24D1* | -1.595993873 | 2.17E-05 | 0.000114427 | 0.105158637 |
| Obesity | endothelial | *DBT* | -1.335702077 | 1.54E-05 | 8.95E-05 | 0.074542643 |
| Obesity | endothelial | *RTCA* | -0.89940893 | 0.007175393 | 0.011976266 | 1 |
| Obesity | endothelial | *ADCY3* | -0.899414164 | 0.000150372 | 0.000469501 | 0.729605038 |
| Obesity | endothelial | *DYNC2LI1* | -0.846397381 | 0.000152907 | 0.000474669 | 0.741907158 |
| Obesity | endothelial | *LHCGR* | -1.083253208 | 1.65E-05 | 9.48E-05 | 0.080036737 |
| Obesity | endothelial | *THUMPD2* | -1.113565122 | 6.38E-05 | 0.000248783 | 0.309734607 |
| Obesity | endothelial | *ACTR2* | -0.724933626 | 0.008762664 | 0.014300857 | 1 |
| Obesity | endothelial | *LRPPRC* | -0.961267799 | 0.000502911 | 0.001220504 | 1 |
| Obesity | endothelial | *CYP2C8* | 2.359307158 | 0.002372656 | 0.004562873 | 1 |
| Obesity | endothelial | *BTBD16* | -1.247017559 | 7.15E-05 | 0.000270444 | 0.347028493 |
| Obesity | endothelial | *TACC2* | -0.931874407 | 1.01E-05 | 6.67E-05 | 0.048921008 |
| Obesity | endothelial | *PLCE1* | -0.782442776 | 3.87E-05 | 0.00017154 | 0.188008302 |
| Obesity | endothelial | *RBP4* | -1.16306802 | 9.30E-05 | 0.000323239 | 0.451241291 |
| Obesity | endothelial | *ASCC1* | -0.870802236 | 0.000970019 | 0.002121397 | 1 |
| Obesity | endothelial | *ATIC* | -0.75093209 | 0.000102266 | 0.000345488 | 0.496196415 |
| Obesity | endothelial | *SMARCAL1* | -1.208483888 | 0.000170224 | 0.000516851 | 0.825927834 |
| Obesity | endothelial | *STAT4* | -1.18084659 | 3.92E-05 | 0.000173055 | 0.190187602 |
| Obesity | endothelial | *SSB* | -0.67661328 | 0.005230688 | 0.009057565 | 1 |
| Obesity | endothelial | *FASTKD1* | -1.067317862 | 0.000239273 | 0.000673407 | 1 |

| | | | | | | |
|---|---|---|---|---|---|---|
| Obesity | endothelial | *SLC35A5* | -1.118157395 | 8.96E-05 | 0.000316163 | 0.434506547 |
| Obesity | endothelial | *SPPL2A* | -0.889237052 | 0.000774697 | 0.001753185 | 1 |
| Obesity | endothelial | *PPCDC* | -0.876692628 | 0.000500929 | 0.00121769 | 1 |
| Obesity | endothelial | *SEMA7A* | 0.579475348 | 9.59E-05 | 0.000330442 | 0.46543071 |
| Obesity | endothelial | *PCDH10* | -1.756175496 | 8.28E-05 | 0.000300181 | 0.401620753 |
| Obesity | endothelial | *AP1AR* | -1.247017281 | 0.001940955 | 0.003837617 | 1 |
| Obesity | endothelial | *BMPR1B* | -0.824438745 | 8.81E-06 | 6.04E-05 | 0.04275886 |
| Obesity | endothelial | *NAAA* | -0.786537001 | 0.002674085 | 0.005054405 | 1 |
| Obesity | endothelial | *NUP54* | -0.924881204 | 0.000235244 | 0.000665931 | 1 |
| Obesity | endothelial | *ANXA3* | 0.339709994 | 0.000527988 | 0.001271995 | 1 |
| Obesity | endothelial | *PPA2* | -0.626544562 | 0.021151325 | 0.031567588 | 1 |
| Obesity | endothelial | *ENPEP* | -0.487241284 | 0.000391751 | 0.00100199 | 1 |
| Obesity | endothelial | *PAPSS1* | -1.089727668 | 0.000239089 | 0.000673278 | 1 |
| Obesity | endothelial | *PARVG* | -1.765698505 | 6.35E-06 | 4.87E-05 | 0.030800424 |
| Obesity | endothelial | *B4GALNT3* | -1.117163476 | 2.73E-05 | 0.000133426 | 0.132439423 |
| Obesity | endothelial | *KIF21A* | -0.85985678 | 1.77E-06 | 2.68E-05 | 0.008576367 |
| Obesity | endothelial | *ALG10* | -1.095116357 | 0.000510771 | 0.001237275 | 1 |
| Obesity | endothelial | *ZCRB1* | -1.269041735 | 0.000372214 | 0.000962165 | 1 |
| Obesity | endothelial | *C1RL* | -0.455723673 | 0.0042104 | 0.007510611 | 1 |
| Obesity | endothelial | *VAMP1* | -0.967312525 | 0.002979851 | 0.005548058 | 1 |
| Obesity | endothelial | *PEX5* | -0.801413717 | 0.001089293 | 0.002343791 | 1 |
| Obesity | endothelial | *PPFIA2* | -1.078843312 | 1.66E-06 | 2.63E-05 | 0.008068212 |
| Obesity | endothelial | *MARCH9* | -1.631063978 | 0.004778622 | 0.008391558 | 1 |
| Obesity | endothelial | *LGR5* | -1.236761345 | 7.20E-05 | 0.000271706 | 0.349414083 |
| Obesity | endothelial | *POC1B* | -0.728141688 | 0.031139814 | 0.044741006 | 1 |
| Obesity | endothelial | *AMDHD1* | -0.392462222 | 0.015337294 | 0.023609311 | 1 |
| Obesity | endothelial | *SDSL* | -0.893219494 | 9.15E-05 | 0.000320851 | 0.444057301 |
| Obesity | endothelial | *FOXN4* | -0.903372213 | 8.76E-05 | 0.000311971 | 0.425216388 |
| Obesity | endothelial | *MTMR6* | -1.206158267 | 0.001563333 | 0.003187097 | 1 |
| Obesity | endothelial | *CCDC65* | -1.167300406 | 1.01E-05 | 6.67E-05 | 0.048784711 |
| Obesity | endothelial | *ACVRL1* | 0.480542636 | 0.002634519 | 0.004993237 | 1 |
| Obesity | endothelial | *BRCA2* | -1.980171128 | 1.55E-06 | 2.57E-05 | 0.007498679 |
| Obesity | endothelial | *KANSL2* | -1.095644121 | 0.000619328 | 0.001453789 | 1 |
| Obesity | endothelial | *CERS5* | -1.003487595 | 0.000652032 | 0.00152099 | 1 |
| Obesity | endothelial | *LMBR1L* | -0.945801324 | 0.00721182 | 0.012032927 | 1 |
| Obesity | endothelial | *ZNF740* | -1.049533058 | 4.32E-05 | 0.000185209 | 0.209841857 |
| Obesity | endothelial | *WDFY2* | -1.011548787 | 3.39E-06 | 3.55E-05 | 0.016458569 |
| Obesity | endothelial | *GRTP1* | -0.589395774 | 7.45E-05 | 0.000278428 | 0.361400066 |
| Obesity | endothelial | *CBLN3* | -0.227116873 | 0.001210618 | 0.00256615 | 1 |
| Obesity | endothelial | *FITM1* | 2.484756596 | 7.28E-06 | 5.39E-05 | 0.035325366 |
| Obesity | endothelial | *TMX1* | -1.092060666 | 0.002558562 | 0.004868292 | 1 |
| Obesity | endothelial | *RTN1* | -0.598148521 | 7.68E-05 | 0.000284782 | 0.372779644 |
| Obesity | endothelial | *RAB15* | -1.669316146 | 2.51E-06 | 3.13E-05 | 0.012172142 |
| Obesity | endothelial | *SLC24A4* | -1.516858599 | 1.72E-06 | 2.67E-05 | 0.008347944 |
| Obesity | endothelial | *C14orf79* | -1.347048937 | 4.64E-05 | 0.000195288 | 0.225362505 |
| Obesity | endothelial | *WARS* | 0.410324137 | 0.024091682 | 0.035518943 | 1 |
| Obesity | endothelial | *DUOXA1* | -0.922249562 | 0.000156719 | 0.000483407 | 0.760399159 |
| Obesity | endothelial | *DISP2* | -0.215783318 | 0.015714472 | 0.024098173 | 1 |
| Obesity | endothelial | *HMG20A* | -1.141708805 | 7.14E-05 | 0.00027032 | 0.346495945 |
| Obesity | endothelial | *SCAPER* | -0.611349925 | 0.005565192 | 0.009565112 | 1 |

| | | | | | | |
|---|---|---|---|---|---|---|
| Obesity | endothelial | *PIF1* | -0.939879296 | 7.27E-06 | 5.39E-05 | 0.035290176 |
| Obesity | endothelial | *BBS4* | -1.068296673 | 3.64E-05 | 0.000164463 | 0.176468933 |
| Obesity | endothelial | *ULK3* | -1.608844884 | 0.002096276 | 0.004101263 | 1 |
| Obesity | endothelial | *PCSK6* | -1.207562196 | 1.43E-06 | 2.57E-05 | 0.006956867 |
| Obesity | endothelial | *HAPLN3* | 0.382266284 | 0.000731163 | 0.001672608 | 1 |
| Obesity | endothelial | *POLG* | -1.128091778 | 0.000948014 | 0.002087007 | 1 |
| Obesity | endothelial | *ABHD2* | -1.072380861 | 0.001405799 | 0.002906237 | 1 |
| Obesity | endothelial | *MFGE8* | -1.224538426 | 0.000108545 | 0.000362213 | 0.526658097 |
| Obesity | endothelial | *UNC45A* | -1.226312055 | 0.001084702 | 0.002334948 | 1 |
| Obesity | endothelial | *IQGAP1* | -0.647518713 | 0.003294644 | 0.006066646 | 1 |
| Obesity | endothelial | *CRTC3* | -0.883388334 | 3.86E-05 | 0.000171098 | 0.187296696 |
| Obesity | endothelial | *ITGAX* | -2.108377963 | 1.01E-05 | 6.67E-05 | 0.04905626 |
| Obesity | endothelial | *DHX38* | -1.033870335 | 0.009446813 | 0.015309263 | 1 |
| Obesity | endothelial | *NUDT7* | -1.696707247 | 0.000233407 | 0.000662664 | 1 |
| Obesity | endothelial | *GCSH* | -1.023576488 | 3.32E-05 | 0.000154241 | 0.16121184 |
| Obesity | endothelial | *CMTM3* | -1.548682517 | 1.20E-05 | 7.47E-05 | 0.058279425 |
| Obesity | endothelial | *TLDC1* | -1.600617005 | 1.94E-05 | 0.000105741 | 0.094109342 |
| Obesity | endothelial | *TIGD7* | -1.009845504 | 0.003715257 | 0.006741371 | 1 |
| Obesity | endothelial | *DEF8* | -1.181831542 | 9.24E-05 | 0.000322309 | 0.44817871 |
| Obesity | endothelial | *TCF25* | -0.880285786 | 0.000274946 | 0.000751993 | 1 |
| Obesity | endothelial | *GALNS* | -1.215359394 | 1.40E-06 | 2.57E-05 | 0.006781769 |
| Obesity | endothelial | *GAS8* | -0.793290766 | 0.011298723 | 0.017997834 | 1 |
| Obesity | endothelial | *NCOR1* | -0.748321018 | 0.001166741 | 0.002485086 | 1 |
| Obesity | endothelial | *GFOD2* | 0.431666771 | 0.004962667 | 0.008665318 | 1 |
| Obesity | endothelial | *PRPSAP2* | -0.871007861 | 0.000704168 | 0.001624641 | 1 |
| Obesity | endothelial | *RASL10B* | -1.66199091 | 4.66E-05 | 0.000195558 | 0.226065304 |
| Obesity | endothelial | *UNC45B* | -2.790698533 | 8.22E-06 | 5.79E-05 | 0.039895359 |
| Obesity | endothelial | *VPS53* | -0.658369356 | 0.004744964 | 0.008335469 | 1 |
| Obesity | endothelial | *SGSM2* | -1.286681274 | 5.11E-05 | 0.000211135 | 0.247872803 |
| Obesity | endothelial | *SSH2* | -0.749104359 | 0.00062607 | 0.001468476 | 1 |
| Obesity | endothelial | *RHBDL3* | 0.478772228 | 0.000269097 | 0.000738495 | 1 |
| Obesity | endothelial | *G6PC3* | -1.270744973 | 0.010883499 | 0.017405296 | 1 |
| Obesity | endothelial | *CLTC* | -0.707414502 | 0.001686766 | 0.003398749 | 1 |
| Obesity | endothelial | *IMPA2* | -1.059692878 | 7.30E-06 | 5.39E-05 | 0.035442465 |
| Obesity | endothelial | *RPRD1A* | -0.930528306 | 0.00080132 | 0.001803341 | 1 |
| Obesity | endothelial | *GALNT1* | -0.882703514 | 0.002019809 | 0.003970872 | 1 |
| Obesity | endothelial | *ADCYAP1* | -0.688218664 | 0.003869941 | 0.007001102 | 1 |
| Obesity | endothelial | *C18orf8* | -2.213745628 | 1.86E-14 | 9.01E-12 | 9.01E-11 |
| Obesity | endothelial | *NPC1* | -1.446671622 | 9.81E-08 | 6.18E-06 | 0.00047611 |
| Obesity | endothelial | *CCDC40* | -0.830996458 | 2.82E-06 | 3.32E-05 | 0.013689657 |
| Obesity | endothelial | *ARHGDIA* | -0.711319188 | 0.02182469 | 0.032479916 | 1 |
| Obesity | endothelial | *CARD14* | -1.159530654 | 3.23E-05 | 0.000151367 | 0.156513598 |
| Obesity | endothelial | *ANAPC11* | -1.767161253 | 1.11E-05 | 7.07E-05 | 0.053902637 |
| Obesity | endothelial | *TBCD* | 0.348559951 | 0.027884265 | 0.040568052 | 1 |
| Obesity | endothelial | *NARF* | -1.106862888 | 0.000177069 | 0.000532965 | 0.85913916 |
| Obesity | endothelial | *CBX8* | 1.029337814 | 0.000283406 | 0.000769924 | 1 |
| Obesity | endothelial | *SECTM1* | -0.555463576 | 0.02502082 | 0.036732532 | 1 |
| Obesity | endothelial | *RNF157* | -0.394811581 | 0.017269654 | 0.026275434 | 1 |
| Obesity | endothelial | *WDR45B* | -1.073695821 | 0.001140868 | 0.002439616 | 1 |
| Obesity | endothelial | *MAPK4* | -4.194916227 | 0.000110859 | 0.000367911 | 0.537886312 |

| | | | | | | |
|---|---|---|---|---|---|---|
| Obesity | endothelial | *FBXO15* | -1.408018317 | 2.43E-05 | 0.000123389 | 0.117713562 |
| Obesity | endothelial | *CBLN2* | -1.035189331 | 3.13E-06 | 3.39E-05 | 0.015204966 |
| Obesity | endothelial | *PIP4K2B* | -0.861455441 | 0.016777663 | 0.025591078 | 1 |
| Obesity | endothelial | *ERBB2* | -1.370477074 | 0.000268872 | 0.000738295 | 1 |
| Obesity | endothelial | *MIEN1* | -1.210048157 | 0.008405241 | 0.01376383 | 1 |
| Obesity | endothelial | *PNMT* | 0.505686783 | 6.19E-05 | 0.000243558 | 0.300550251 |
| Obesity | endothelial | *IGFBP4* | 0.37564451 | 0.032386477 | 0.04632641 | 1 |
| Obesity | endothelial | *CACNA1A* | -2.027711753 | 8.60E-06 | 5.99E-05 | 0.041725364 |
| Obesity | endothelial | *SAMD1* | -2.182025648 | 3.38E-06 | 3.55E-05 | 0.016393958 |
| Obesity | endothelial | *NFIC* | -0.828127115 | 1.99E-05 | 0.000107281 | 0.096446028 |
| Obesity | endothelial | *PRDM15* | -0.92723693 | 1.43E-05 | 8.51E-05 | 0.069179705 |
| Obesity | endothelial | *PFKL* | -1.225349568 | 0.001681529 | 0.003391014 | 1 |
| Obesity | endothelial | *DPP9* | -0.893707781 | 0.017560823 | 0.026659923 | 1 |
| Obesity | endothelial | *SIRT3* | -0.999400182 | 0.000513124 | 0.001241114 | 1 |
| Obesity | endothelial | *DOPEY2* | -0.821875208 | 0.000197927 | 0.000584506 | 0.960342971 |
| Obesity | endothelial | *URB1* | -0.89501827 | 0.000721439 | 0.001655829 | 1 |
| Obesity | endothelial | *SAE1* | -1.815307489 | 1.23E-07 | 7.11E-06 | 0.000597499 |
| Obesity | endothelial | *GEMIN7* | -1.29361197 | 0.000245935 | 0.000688163 | 1 |
| Obesity | endothelial | *CAPN10* | -0.885510511 | 0.005062845 | 0.008807789 | 1 |
| Obesity | endothelial | *CACNG8* | 0.488873227 | 5.27E-05 | 0.000216407 | 0.255626831 |
| Obesity | endothelial | *EVI5L* | -1.724091773 | 0.000298816 | 0.000801024 | 1 |
| Obesity | endothelial | *SLC47A1* | -1.174679223 | 0.000603056 | 0.001422473 | 1 |
| Obesity | endothelial | *CTU1* | -1.810518355 | 0.005300515 | 0.009162129 | 1 |
| Obesity | endothelial | *RERE* | -0.632538834 | 0.002252914 | 0.004367215 | 1 |
| Obesity | endothelial | *EPHA2* | -1.301620749 | 0.001631226 | 0.003299169 | 1 |
| Obesity | endothelial | *PEX14* | -0.788802198 | 0.000266825 | 0.000734382 | 1 |
| Obesity | endothelial | *ZNF593* | -1.916256668 | 0.021075994 | 0.031474522 | 1 |
| Obesity | endothelial | *EVA1B* | 0.533024055 | 0.007681487 | 0.012724676 | 1 |
| Obesity | endothelial | *PLK4* | -1.207099144 | 4.02E-05 | 0.000176312 | 0.195176843 |
| Obesity | endothelial | *ITGB3BP* | -0.535568816 | 0.006347944 | 0.010740503 | 1 |
| Obesity | endothelial | *PIGK* | -0.981771489 | 0.000972908 | 0.002126374 | 1 |
| Obesity | endothelial | *SLC44A3* | -1.233730613 | 1.33E-06 | 2.57E-05 | 0.006470021 |
| Obesity | endothelial | *STRIP1* | -1.398691164 | 0.000661344 | 0.00153975 | 1 |
| Obesity | endothelial | *ALDH9A1* | -1.439824673 | 8.59E-05 | 0.000308067 | 0.41681474 |
| Obesity | endothelial | *ATP1B1* | -0.944365076 | 7.34E-06 | 5.40E-05 | 0.035605731 |
| Obesity | endothelial | *NME7* | -1.080940536 | 3.25E-05 | 0.000151801 | 0.157519743 |
| Obesity | endothelial | *XCL1* | 1.345143773 | 0.008607165 | 0.014061267 | 1 |
| Obesity | endothelial | *XCL2* | -0.923189649 | 0.000430917 | 0.001081081 | 1 |
| Obesity | endothelial | *MAEL* | -1.378231199 | 4.24E-06 | 3.91E-05 | 0.020550584 |
| Obesity | endothelial | *RFWD2* | -0.676854907 | 0.004171314 | 0.007451847 | 1 |
| Obesity | endothelial | *RGS16* | -0.82889418 | 1.53E-05 | 8.95E-05 | 0.074300381 |
| Obesity | endothelial | *TARS2* | -1.403672557 | 4.11E-05 | 0.000179453 | 0.199192643 |
| Obesity | endothelial | *SNX27* | -0.963709384 | 0.000366827 | 0.000952299 | 1 |
| Obesity | endothelial | *SETDB1* | -1.031295801 | 0.000558851 | 0.001333765 | 1 |
| Obesity | endothelial | *SEMA6C* | 0.768312345 | 0.000149848 | 0.000468772 | 0.727064702 |
| Obesity | endothelial | *ARNT* | -0.981074399 | 2.71E-05 | 0.000132593 | 0.131399854 |
| Obesity | endothelial | *POGZ* | -0.808748614 | 0.000631622 | 0.001478356 | 1 |
| Obesity | endothelial | *GABPB2* | -0.79444167 | 3.22E-06 | 3.45E-05 | 0.01561319 |
| Obesity | endothelial | *DYRK3* | 0.414770415 | 0.000446635 | 0.00111189 | 1 |
| Obesity | endothelial | *EIF2D* | -1.383960145 | 2.75E-05 | 0.000133648 | 0.133247303 |

| | | | | | | |
|---|---|---|---|---|---|---|
| Obesity | endothelial | *TAF1A* | -0.932256953 | 2.70E-05 | 0.000132377 | 0.131053507 |
| Obesity | endothelial | *SMYD2* | -1.408605284 | 2.08E-05 | 0.000110614 | 0.1007418 |
| Obesity | endothelial | *ADAM15* | 0.307295019 | 0.001118277 | 0.002398358 | 1 |
| Obesity | endothelial | *S100A8* | -1.410679039 | 4.80E-06 | 4.19E-05 | 0.023274714 |
| Obesity | endothelial | *KCNN3* | 0.253425431 | 0.016860969 | 0.025701986 | 1 |
| Obesity | endothelial | *C1orf43* | -1.185655033 | 0.00013994 | 0.000442626 | 0.678988906 |
| Obesity | endothelial | *FLG* | -0.406388183 | 0.000929895 | 0.002052706 | 1 |
| Obesity | endothelial | *SCCPDH* | -1.331614816 | 5.16E-06 | 4.37E-05 | 0.025035192 |
| Obesity | endothelial | *CEP170* | -0.935221496 | 0.000313681 | 0.000835794 | 1 |
| Obesity | endothelial | *ACP1* | -1.397838689 | 3.66E-05 | 0.000165008 | 0.177383172 |
| Obesity | endothelial | *CNIH4* | -1.413202466 | 7.14E-05 | 0.00027032 | 0.346549883 |
| Obesity | endothelial | *PYCR2* | -1.196741367 | 0.001512483 | 0.003096441 | 1 |
| Obesity | endothelial | *PLEKHA6* | -0.577707502 | 0.018589366 | 0.028072083 | 1 |
| Obesity | endothelial | *OSR1* | -0.58588073 | 0.016597743 | 0.025356502 | 1 |
| Obesity | endothelial | *GALM* | -1.231077412 | 8.35E-06 | 5.85E-05 | 0.040516047 |
| Obesity | endothelial | *WDPCP* | -0.584563466 | 0.000374197 | 0.00096472 | 1 |
| Obesity | endothelial | *VPS54* | -0.817006918 | 0.001468234 | 0.003018589 | 1 |
| Obesity | endothelial | *REG3G* | 1.911825082 | 0.020582821 | 0.030804394 | 1 |
| Obesity | endothelial | *ABHD1* | -1.088279249 | 4.86E-06 | 4.22E-05 | 0.023572166 |
| Obesity | endothelial | *CIAO1* | -1.227258623 | 0.000207172 | 0.000602636 | 1 |
| Obesity | endothelial | *ZNF514* | -1.301819583 | 2.95E-06 | 3.38E-05 | 0.014309771 |
| Obesity | endothelial | *EXOC6B* | -0.632942672 | 0.003703905 | 0.006725803 | 1 |
| Obesity | endothelial | *TEX261* | -2.142693757 | 2.09E-05 | 0.000111292 | 0.101498578 |
| Obesity | endothelial | *THNSL2* | -0.406671797 | 0.005661251 | 0.009716444 | 1 |
| Obesity | endothelial | *RALB* | 0.428776103 | 0.015295661 | 0.02356477 | 1 |
| Obesity | endothelial | *RABL2A* | -1.046813093 | 0.000111581 | 0.000369802 | 0.541389427 |
| Obesity | endothelial | *FBLN7* | -0.937867553 | 1.77E-07 | 9.04E-06 | 0.000861064 |
| Obesity | endothelial | *CNGA3* | -7.0064992 | 2.12E-05 | 0.000112406 | 0.102851701 |
| Obesity | endothelial | *FAHD2B* | -1.54950269 | 1.50E-07 | 8.10E-06 | 0.000729258 |
| Obesity | endothelial | *LYG1* | -0.606387242 | 0.009074172 | 0.014749709 | 1 |
| Obesity | endothelial | *THSD7B* | -0.811607126 | 0.010441241 | 0.016797382 | 1 |
| Obesity | endothelial | *GPR17* | 0.200831214 | 0.005906827 | 0.010095077 | 1 |
| Obesity | endothelial | *POLR2D* | -1.261116983 | 0.00043692 | 0.001093315 | 1 |
| Obesity | endothelial | *GALNT13* | -0.912606162 | 1.26E-06 | 2.57E-05 | 0.006137412 |
| Obesity | endothelial | *PHOSPHO2* | -2.069095452 | 0.000499016 | 0.001214255 | 1 |
| Obesity | endothelial | *NBEAL1* | -0.629242792 | 0.004115632 | 0.007371373 | 1 |
| Obesity | endothelial | *SUMF1* | -0.834684703 | 4.85E-05 | 0.000201981 | 0.23530789 |
| Obesity | endothelial | *ACKR3* | -0.683803962 | 0.000765737 | 0.001738584 | 1 |
| Obesity | endothelial | *ESPNL* | -12.4124651 | 1.15E-06 | 2.57E-05 | 0.005574811 |
| Obesity | endothelial | *ANKMY1* | -0.916978881 | 0.001017617 | 0.002205393 | 1 |
| Obesity | endothelial | *TAMM41* | -0.944053819 | 0.001158404 | 0.002470583 | 1 |
| Obesity | endothelial | *STK11IP* | -1.687992829 | 9.11E-05 | 0.000319598 | 0.441941245 |
| Obesity | endothelial | *EAF1* | -1.748892484 | 0.000200433 | 0.000588277 | 0.97249996 |
| Obesity | endothelial | *CNTN4* | -0.814650094 | 1.17E-06 | 2.57E-05 | 0.00568034 |
| Obesity | endothelial | *SLC25A38* | -1.605762096 | 3.31E-05 | 0.000154078 | 0.160703784 |
| Obesity | endothelial | *CTDSPL* | -0.871699511 | 0.000607036 | 0.001429082 | 1 |
| Obesity | endothelial | *SLC25A26* | -1.182451693 | 2.17E-06 | 2.92E-05 | 0.010507023 |
| Obesity | endothelial | *ARL6IP5* | -0.919313146 | 0.002280669 | 0.004406932 | 1 |
| Obesity | endothelial | *LIMD1* | -0.685981867 | 0.002822529 | 0.00530143 | 1 |
| Obesity | endothelial | *PLA1A* | 0.344146318 | 6.02E-05 | 0.000238229 | 0.292068657 |

| | | | | | | |
|---|---|---|---|---|---|---|
| Obesity | endothelial | *ADPRH* | 0.520264552 | 0.000847885 | 0.001894951 | 1 |
| Obesity | endothelial | *IGSF11* | -0.370367671 | 0.028338372 | 0.041179329 | 1 |
| Obesity | endothelial | *BOC* | -0.857987693 | 3.04E-06 | 3.39E-05 | 0.014739266 |
| Obesity | endothelial | *MED12L* | -0.989345966 | 2.18E-06 | 2.92E-05 | 0.010572814 |
| Obesity | endothelial | *EIF2A* | -0.873066008 | 9.84E-07 | 2.45E-05 | 0.004772388 |
| Obesity | endothelial | *NCEH1* | -1.042088871 | 2.58E-05 | 0.000128134 | 0.1253147 |
| Obesity | endothelial | *AMT* | -0.556352414 | 4.05E-06 | 3.85E-05 | 0.019640316 |
| Obesity | endothelial | *CCDC39* | -0.638899054 | 0.000227191 | 0.000648428 | 1 |
| Obesity | endothelial | *MUC4* | -1.298770066 | 5.36E-06 | 4.47E-05 | 0.025985648 |
| Obesity | endothelial | *DGKQ* | -0.732249956 | 0.008811293 | 0.014375384 | 1 |
| Obesity | endothelial | *CENPC* | -0.997027835 | 2.48E-05 | 0.000125141 | 0.120511184 |
| Obesity | endothelial | *TRMT10A* | -1.932498254 | 1.20E-05 | 7.47E-05 | 0.058184498 |
| Obesity | endothelial | *PYURF* | -1.336411696 | 0.002667609 | 0.005046098 | 1 |
| Obesity | endothelial | *CISD2* | -1.342626197 | 6.67E-05 | 0.000256506 | 0.323710955 |
| Obesity | endothelial | *DDIT4L* | 1.384096148 | 0.000709583 | 0.001631439 | 1 |
| Obesity | endothelial | *METTL14* | -1.279865734 | 5.71E-05 | 0.000228213 | 0.276822282 |
| Obesity | endothelial | *USP53* | -0.63834756 | 0.005849767 | 0.010008134 | 1 |
| Obesity | endothelial | *MARCH1* | -0.850062143 | 3.68E-07 | 1.46E-05 | 0.001783564 |
| Obesity | endothelial | *CBR4* | -0.819328283 | 0.000802201 | 0.001804487 | 1 |
| Obesity | endothelial | *CYP4V2* | -0.464677296 | 0.00627624 | 0.010632791 | 1 |
| Obesity | endothelial | *MARCH6* | -0.874832087 | 0.000513072 | 0.001241114 | 1 |
| Obesity | endothelial | *MYO10* | 0.350368698 | 0.005293074 | 0.009152528 | 1 |
| Obesity | endothelial | *FAM105A* | -0.492506169 | 0.003701996 | 0.006724854 | 1 |
| Obesity | endothelial | *GTF2H2* | -1.950336374 | 1.77E-06 | 2.68E-05 | 0.00859756 |
| Obesity | endothelial | *SPATA9* | -0.814729833 | 1.29E-06 | 2.57E-05 | 0.006252151 |
| Obesity | endothelial | *COMMD10* | -0.581746254 | 0.008955762 | 0.014586559 | 1 |
| Obesity | endothelial | *FBXO38* | -0.786350512 | 0.012949932 | 0.020321175 | 1 |
| Obesity | endothelial | *PCYOX1L* | -3.139485442 | 1.70E-06 | 2.65E-05 | 0.00822612 |
| Obesity | endothelial | *MYLK4* | -2.218697845 | 1.11E-05 | 7.07E-05 | 0.053954621 |
| Obesity | endothelial | *TBC1D7* | -0.948976378 | 0.012476054 | 0.01966022 | 1 |
| Obesity | endothelial | *GFOD1* | 0.241078738 | 0.000567237 | 0.001351121 | 1 |
| Obesity | endothelial | *FAM193B* | -1.104137107 | 5.41E-05 | 0.000219845 | 0.26227488 |
| Obesity | endothelial | *MUT* | -1.347863533 | 1.32E-05 | 8.06E-05 | 0.064152463 |
| Obesity | endothelial | *DOK3* | -0.426781305 | 0.004697035 | 0.008266236 | 1 |
| Obesity | endothelial | *DAAM2* | -0.779632208 | 2.10E-06 | 2.89E-05 | 0.010212924 |
| Obesity | endothelial | *RPL7L1* | -1.35652589 | 0.000141557 | 0.000445707 | 0.686834206 |
| Obesity | endothelial | *TPBG* | -1.049805775 | 0.002035226 | 0.003994707 | 1 |
| Obesity | endothelial | *IRAK1BP1* | -0.74155769 | 0.005194864 | 0.009008391 | 1 |
| Obesity | endothelial | *PHIP* | -0.778538561 | 0.000987945 | 0.002152536 | 1 |
| Obesity | endothelial | *RNF217* | -0.94168299 | 0.000137424 | 0.000436375 | 0.666781029 |
| Obesity | endothelial | *ARHGAP18* | -0.727049073 | 0.000223099 | 0.000638629 | 1 |
| Obesity | endothelial | *SLC18B1* | -0.95994624 | 1.38E-05 | 8.29E-05 | 0.066715139 |
| Obesity | endothelial | *WTAP* | -0.726234635 | 0.004103678 | 0.007355392 | 1 |
| Obesity | endothelial | *SLC22A3* | -0.834117682 | 1.37E-06 | 2.57E-05 | 0.006633551 |
| Obesity | endothelial | *GNA12* | -1.181711626 | 2.49E-06 | 3.13E-05 | 0.012084237 |
| Obesity | endothelial | *SDK1* | -0.792504471 | 5.12E-08 | 4.01E-06 | 0.000248341 |
| Obesity | endothelial | *CCZ1B* | -1.161771524 | 5.07E-06 | 4.33E-05 | 0.024617476 |
| Obesity | endothelial | *RBAK* | -2.255382884 | 8.05E-05 | 0.000294064 | 0.390810667 |
| Obesity | endothelial | *POMZP3* | -0.625625344 | 0.021831844 | 0.032479916 | 1 |
| Obesity | endothelial | *PSPH* | -0.588324608 | 0.000324525 | 0.000858557 | 1 |

| | | | | | | |
|---|---|---|---|---|---|---|
| Obesity | endothelial | *GIGYF1* | -0.95802947 | 0.007810274 | 0.012902774 | 1 |
| Obesity | endothelial | *TRIM4* | -0.552097183 | 0.000119898 | 0.000390696 | 0.581746426 |
| Obesity | endothelial | *TMEM209* | -1.365394373 | 0.00018842 | 0.000562246 | 0.914211492 |
| Obesity | endothelial | *NCAPG2* | -1.228360466 | 4.34E-05 | 0.000185308 | 0.210453557 |
| Obesity | endothelial | *CHMP7* | -1.459026767 | 5.92E-07 | 2.05E-05 | 0.002872256 |
| Obesity | endothelial | *DOCK5* | -0.913473024 | 5.34E-06 | 4.47E-05 | 0.025894661 |
| Obesity | endothelial | *ADHFE1* | -0.62855111 | 3.27E-09 | 4.53E-07 | 1.58E-05 |
| Obesity | endothelial | *RPL7* | -0.847658917 | 0.000840654 | 0.001880523 | 1 |
| Obesity | endothelial | *ATP6V0D2* | -1.593796217 | 8.99E-05 | 0.00031667 | 0.436370919 |
| Obesity | endothelial | *DPYS* | -0.853573948 | 1.36E-06 | 2.57E-05 | 0.006599755 |
| Obesity | endothelial | *LRP12* | -1.010434036 | 0.012115425 | 0.01913543 | 1 |
| Obesity | endothelial | *MAL2* | -0.536675744 | 7.11E-05 | 0.000269701 | 0.345216725 |
| Obesity | endothelial | *GSDMC* | -2.345577481 | 0.012314245 | 0.019424161 | 1 |
| Obesity | endothelial | *AK3* | -0.871069144 | 0.005263363 | 0.009107645 | 1 |
| Obesity | endothelial | *HAUS6* | -1.433643539 | 1.52E-05 | 8.93E-05 | 0.073701844 |
| Obesity | endothelial | *C9orf72* | -0.375936718 | 0.008064951 | 0.013264793 | 1 |
| Obesity | endothelial | *ALAD* | -0.66358795 | 0.007308654 | 0.012173233 | 1 |
| Obesity | endothelial | *MED22* | -1.40428349 | 3.63E-06 | 3.68E-05 | 0.017606291 |
| Obesity | endothelial | *REXO4* | -1.416234781 | 0.000389215 | 0.000997964 | 1 |
| Obesity | endothelial | *SH3GLB2* | -1.61269199 | 5.65E-05 | 0.000226484 | 0.274271682 |
| Obesity | endothelial | *INPP5E* | -1.66585056 | 0.001659523 | 0.00334942 | 1 |
| Obesity | endothelial | *SEC16A* | -1.411284872 | 6.38E-05 | 0.00024874 | 0.309432388 |
| Obesity | endothelial | *SLC39A12* | -2.210896228 | 0.000229277 | 0.000652847 | 1 |
| Obesity | endothelial | *TMEM236* | -0.733360007 | 0.003966808 | 0.00714703 | 1 |
| Obesity | endothelial | *ST8SIA6* | 0.185809828 | 0.000116153 | 0.000382085 | 0.563575631 |
| Obesity | endothelial | *PARD3* | -0.766951981 | 1.33E-05 | 8.06E-05 | 0.064350658 |
| Obesity | endothelial | *CDHR1* | -1.623470246 | 0.001142564 | 0.002441639 | 1 |
| Obesity | endothelial | *RGR* | -5.686821555 | 3.22E-09 | 4.53E-07 | 1.56E-05 |
| Obesity | endothelial | *FRA10AC1* | -0.840658125 | 0.002060828 | 0.004038424 | 1 |
| Obesity | endothelial | *ADD3* | -0.906783037 | 3.43E-05 | 0.000156959 | 0.166249841 |
| Obesity | endothelial | *LRRC27* | -0.769217353 | 6.48E-05 | 0.000251195 | 0.314244676 |
| Obesity | endothelial | *PAOX* | -1.99931496 | 2.66E-05 | 0.00013085 | 0.128887514 |
| Obesity | endothelial | *PDCD11* | -0.84208662 | 0.013143646 | 0.020565292 | 1 |
| Obesity | endothelial | *RGS10* | -1.119744456 | 3.03E-06 | 3.39E-05 | 0.014697356 |
| Obesity | endothelial | *GAS2* | -0.903569472 | 0.030228367 | 0.043599243 | 1 |
| Obesity | endothelial | *LIN7C* | -1.493779238 | 5.52E-05 | 0.000223384 | 0.267837588 |
| Obesity | endothelial | *HSD17B12* | -0.689304404 | 5.68E-08 | 4.16E-06 | 0.000275551 |
| Obesity | endothelial | *EIF3M* | -1.03956514 | 0.000220553 | 0.000633584 | 1 |
| Obesity | endothelial | *PTPRJ* | -0.972735778 | 0.000189958 | 0.000564753 | 0.921676295 |
| Obesity | endothelial | *ENDOD1* | 0.52777051 | 0.00659264 | 0.011106767 | 1 |
| Obesity | endothelial | *CCDC82* | -0.496177014 | 0.00764307 | 0.012665361 | 1 |
| Obesity | endothelial | *CAPN5* | 0.847775052 | 0.006582617 | 0.011093733 | 1 |
| Obesity | endothelial | *INTS4* | -0.902365384 | 0.001191792 | 0.002531776 | 1 |
| Obesity | endothelial | *DRD2* | -4.065402429 | 5.09E-06 | 4.33E-05 | 0.024681163 |
| Obesity | endothelial | *NPAT* | -1.215751693 | 0.000195409 | 0.000578368 | 0.948124899 |
| Obesity | endothelial | *ATM* | -0.84117389 | 0.000151419 | 0.000471858 | 0.734683449 |
| Obesity | endothelial | *ST14* | -2.229858101 | 3.94E-05 | 0.000173543 | 0.191070648 |
| Obesity | endothelial | *FADS1* | -1.609782987 | 1.84E-07 | 9.10E-06 | 0.000895181 |
| Obesity | endothelial | *TMC2* | -2.011454334 | 6.22E-07 | 2.12E-05 | 0.003019646 |
| Obesity | endothelial | *EML3* | -0.824204036 | 0.004377119 | 0.007773711 | 1 |

| | | | | | | |
|---|---|---|---|---|---|---|
| Obesity | endothelial | *INCENP* | -2.527813843 | 1.65E-06 | 2.63E-05 | 0.008019229 |
| Obesity | endothelial | *PLCH2* | -2.012324846 | 5.64E-06 | 4.54E-05 | 0.027372915 |
| Obesity | endothelial | *CPSF7* | -1.178951965 | 2.92E-05 | 0.00013966 | 0.141475426 |
| Obesity | endothelial | *CCDC15* | -0.928573319 | 1.08E-06 | 2.57E-05 | 0.00521683 |
| Obesity | endothelial | *CHEK1* | -1.450411953 | 2.08E-06 | 2.88E-05 | 0.01011516 |
| Obesity | endothelial | *CABLES2* | -1.038855309 | 1.40E-05 | 8.38E-05 | 0.067796658 |
| Obesity | endothelial | *TRPT1* | -1.225724936 | 3.37E-07 | 1.40E-05 | 0.001633002 |
| Obesity | endothelial | *NUDT22* | -1.494363557 | 5.74E-09 | 7.44E-07 | 2.79E-05 |
| Obesity | endothelial | *TBX6* | 0.713475672 | 1.82E-09 | 2.86E-07 | 8.85E-06 |
| Obesity | endothelial | *ALDOA* | -1.072838984 | 0.00063838 | 0.001492014 | 1 |
| Obesity | endothelial | *ANXA8L1* | -0.268688898 | 0.013794534 | 0.021479806 | 1 |
| Obesity | endothelial | *FCGR1A* | -0.658567388 | 0.022699148 | 0.033617015 | 1 |
| Obesity | endothelial | *TMEM218* | -1.285196585 | 0.000197871 | 0.000584506 | 0.960070707 |
| Obesity | endothelial | *SAP18* | -1.214599825 | 0.001847473 | 0.003681288 | 1 |
| Obesity | endothelial | *FAM124A* | 0.287904969 | 0.000234638 | 0.000665379 | 1 |
| Obesity | endothelial | *LYPD6B* | 0.37900601 | 0.008937884 | 0.014562329 | 1 |
| Obesity | endothelial | *WDR17* | -1.304487333 | 4.27E-06 | 3.91E-05 | 0.020720528 |
| Obesity | endothelial | *CCDC102B* | -0.814369342 | 5.73E-06 | 4.57E-05 | 0.027817088 |
| Obesity | endothelial | *PRSS23* | -0.736333811 | 0.000443118 | 0.001104834 | 1 |
| Obesity | endothelial | *PPP1R1C* | -2.755391879 | 3.10E-06 | 3.39E-05 | 0.015035521 |
| Obesity | endothelial | *FAM173B* | -1.342788619 | 3.81E-05 | 0.00016908 | 0.184635169 |
| Obesity | endothelial | *GPR158* | -0.921637548 | 1.12E-05 | 7.11E-05 | 0.054362521 |
| Obesity | endothelial | *ANK3* | -1.028050611 | 1.69E-05 | 9.64E-05 | 0.08212419 |
| Obesity | endothelial | *PLBD2* | -1.132643364 | 0.000269658 | 0.000739616 | 1 |
| Obesity | endothelial | *DLG5* | -0.790800829 | 1.53E-05 | 8.94E-05 | 0.073997564 |
| Obesity | endothelial | *TMEM18* | -1.760081176 | 5.42E-05 | 0.000220004 | 0.26285078 |
| Obesity | endothelial | *ADAMTS12* | -0.603415316 | 7.52E-06 | 5.48E-05 | 0.03647409 |
| Obesity | endothelial | *NUBPL* | -0.994657304 | 3.72E-05 | 0.000166525 | 0.180679894 |
| Obesity | endothelial | *VIPAS39* | -1.470363866 | 7.24E-05 | 0.00027292 | 0.351248454 |
| Obesity | endothelial | *UPF2* | -0.85422262 | 0.000958361 | 0.002101853 | 1 |
| Obesity | endothelial | *PTPRO* | -0.66050125 | 0.00035966 | 0.000936699 | 1 |
| Obesity | endothelial | *EPS8* | -0.759552557 | 9.51E-05 | 0.000328008 | 0.461507747 |
| Obesity | endothelial | *QDPR* | -1.048239449 | 9.35E-05 | 0.000323766 | 0.453595988 |
| Obesity | endothelial | *FAM160B1* | -0.963556911 | 0.001278532 | 0.002681988 | 1 |
| Obesity | endothelial | *ZNF827* | -1.037944398 | 4.06E-06 | 3.85E-05 | 0.019685959 |
| Obesity | endothelial | *AKR1C2* | -1.002599959 | 0.000165126 | 0.000504529 | 0.801191726 |
| Obesity | endothelial | *DPYSL4* | 0.341687778 | 0.004417469 | 0.007839634 | 1 |
| Obesity | endothelial | *PIGF* | -1.157560356 | 0.000232621 | 0.000661204 | 1 |
| Obesity | endothelial | *INPP1* | 0.452989084 | 0.002628686 | 0.004986076 | 1 |
| Obesity | endothelial | *CCDC122* | -0.603419488 | 0.000231063 | 0.000657162 | 1 |
| Obesity | endothelial | *GUF1* | -1.489146562 | 3.44E-05 | 0.000157104 | 0.166687742 |
| Obesity | endothelial | *GABRA2* | 0.462801114 | 6.19E-05 | 0.000243457 | 0.300182981 |
| Obesity | endothelial | *PARP8* | -0.737855685 | 4.32E-06 | 3.94E-05 | 0.020984018 |
| Obesity | endothelial | *CACUL1* | -0.990445867 | 7.80E-05 | 0.000288234 | 0.378653383 |
| Obesity | endothelial | *BEND6* | -1.093218122 | 1.14E-07 | 6.66E-06 | 0.00055241 |
| Obesity | endothelial | *TIAL1* | -0.912656891 | 0.001722003 | 0.003458262 | 1 |
| Obesity | endothelial | *TMEM56* | -1.74678639 | 1.80E-06 | 2.68E-05 | 0.008754708 |
| Obesity | endothelial | *MGAT5* | -0.893709512 | 0.000435314 | 0.001090421 | 1 |
| Obesity | endothelial | *TMEM178A* | 0.739798279 | 0.001026614 | 0.002222726 | 1 |
| Obesity | endothelial | *EPG5* | -1.057038442 | 0.000212211 | 0.000614348 | 1 |

| | | | | | | |
|---|---|---|---|---|---|---|
| Obesity | endothelial | *SPC25* | 0.131825139 | 4.80E-05 | 0.000200318 | 0.232769251 |
| Obesity | endothelial | *ATG10* | -0.29787023 | 0.026660794 | 0.038963305 | 1 |
| Obesity | endothelial | *POC5* | -1.296708902 | 1.44E-08 | 1.52E-06 | 6.99E-05 |
| Obesity | endothelial | *NMT2* | -0.785710922 | 0.003891639 | 0.007027255 | 1 |
| Obesity | endothelial | *PLEKHH2* | -0.567256266 | 5.19E-05 | 0.000213999 | 0.251735812 |
| Obesity | endothelial | *SPARCL1* | 0.238850803 | 0.002532194 | 0.004825689 | 1 |
| Obesity | endothelial | *MBNL1* | -0.447511991 | 0.023300145 | 0.03443567 | 1 |
| Obesity | endothelial | *CLEC4F* | 0.196482195 | 5.19E-05 | 0.000213999 | 0.251877065 |
| Obesity | endothelial | *SLC30A6* | -0.980564192 | 0.000181258 | 0.000544264 | 0.879464454 |
| Obesity | endothelial | *RASGRP3* | 0.336394782 | 0.006770353 | 0.011354909 | 1 |
| Obesity | endothelial | *GPR180* | -0.718127902 | 1.42E-05 | 8.47E-05 | 0.068756111 |
| Obesity | endothelial | *TCTEX1D1* | 0.779933831 | 0.001283159 | 0.002688206 | 1 |
| Obesity | endothelial | *PRDM8* | -1.776617668 | 1.38E-06 | 2.57E-05 | 0.006699758 |
| Obesity | endothelial | *GGPS1* | -0.868118662 | 0.006695859 | 0.01125721 | 1 |
| Obesity | endothelial | *MED21* | -0.867849134 | 0.011867661 | 0.018774663 | 1 |
| Obesity | endothelial | *JAKMIP1* | -1.587956837 | 2.68E-06 | 3.26E-05 | 0.013007451 |
| Obesity | endothelial | *SREK1IP1* | -1.215026301 | 2.46E-05 | 0.000124585 | 0.119477016 |
| Obesity | endothelial | *MR1* | -1.090024817 | 2.88E-05 | 0.000138277 | 0.139659991 |
| Obesity | endothelial | *CENPH* | -2.242088285 | 1.90E-05 | 0.000104178 | 0.092154596 |
| Obesity | endothelial | *CAST* | -0.737269671 | 0.000904853 | 0.002006558 | 1 |
| Obesity | endothelial | *CLGN* | -1.971583323 | 7.95E-06 | 5.67E-05 | 0.038589398 |
| Obesity | endothelial | *CETN3* | -0.669439146 | 0.003637791 | 0.006628075 | 1 |
| Obesity | endothelial | *SYCP2L* | -0.860505496 | 0.00137275 | 0.002845187 | 1 |
| Obesity | endothelial | *NMRAL1* | -0.979782768 | 0.000466106 | 0.001145666 | 1 |
| Obesity | endothelial | *ADPRHL1* | -1.520102402 | 2.47E-06 | 3.13E-05 | 0.011990719 |
| Obesity | endothelial | *CFDP1* | -0.93428279 | 0.000243672 | 0.000683145 | 1 |
| Obesity | endothelial | *ZDHHC7* | -1.209432281 | 5.44E-06 | 4.47E-05 | 0.026386974 |
| Obesity | endothelial | *JAZF1* | -0.424298891 | 0.030233561 | 0.043599243 | 1 |
| Obesity | endothelial | *FBXO36* | -1.18937091 | 1.51E-06 | 2.57E-05 | 0.007303756 |
| Obesity | endothelial | *MCOLN2* | -0.863027569 | 0.000261899 | 0.000724064 | 1 |
| Obesity | endothelial | *GDPD1* | -1.54917026 | 0.000116319 | 0.000382371 | 0.564379053 |
| Obesity | endothelial | *SEMA3D* | -0.889975195 | 1.97E-06 | 2.82E-05 | 0.009534246 |
| Obesity | endothelial | *PPP2R5E* | -0.946237477 | 0.000278067 | 0.000758032 | 1 |
| Obesity | endothelial | *ANKRD29* | 0.219736193 | 0.000326648 | 0.000862763 | 1 |
| Obesity | endothelial | *DNAAF1* | -1.436446913 | 1.90E-05 | 0.000104178 | 0.092197967 |
| Obesity | endothelial | *UBASH3B* | -0.631211372 | 0.010784585 | 0.017275274 | 1 |
| Obesity | endothelial | *ROBO4* | 0.311142644 | 0.000907882 | 0.002012355 | 1 |
| Obesity | endothelial | *ROBO3* | -0.864024448 | 0.000705118 | 0.001626061 | 1 |
| Obesity | endothelial | *PRKCA* | -0.930177626 | 4.14E-06 | 3.88E-05 | 0.020083763 |
| Obesity | endothelial | *LRRK1* | -1.080080911 | 0.000104307 | 0.000350726 | 0.506096937 |
| Obesity | endothelial | *ABCA5* | -0.81981827 | 5.78E-05 | 0.000230842 | 0.28024174 |
| Obesity | endothelial | *ENPP3* | -1.43730891 | 1.49E-06 | 2.57E-05 | 0.00724946 |
| Obesity | endothelial | *MIA3* | -0.959449025 | 0.000599463 | 0.001416754 | 1 |
| Obesity | endothelial | *CCSAP* | -1.711618778 | 1.99E-06 | 2.84E-05 | 0.009651646 |
| Obesity | endothelial | *GBP5* | -0.475516745 | 0.01210426 | 0.019124021 | 1 |
| Obesity | endothelial | *LY96* | -0.998934283 | 0.00027484 | 0.000751993 | 1 |
| Obesity | endothelial | *PDE1C* | -1.1287375 | 2.69E-06 | 3.26E-05 | 0.013055462 |
| Obesity | endothelial | *RABGEF1* | -0.397915376 | 0.002397737 | 0.004607453 | 1 |
| Obesity | endothelial | *XPC* | -1.120376459 | 0.000263667 | 0.000728124 | 1 |
| Obesity | endothelial | *FLCN* | -0.839083199 | 4.51E-06 | 4.05E-05 | 0.021881154 |

| | | | | | | |
|---|---|---|---|---|---|---|
| Obesity | endothelial | *OXNAD1* | -1.053590662 | 7.35E-05 | 0.000276054 | 0.35666177 |
| Obesity | endothelial | *APCDD1* | -0.729854475 | 7.22E-06 | 5.37E-05 | 0.035014207 |
| Obesity | endothelial | *ANKRD40* | -1.473627681 | 7.98E-05 | 0.000292909 | 0.387165943 |
| Obesity | endothelial | *CYP2U1* | -1.020595009 | 0.000134165 | 0.000427707 | 0.650970584 |
| Obesity | endothelial | *MARCKS* | -0.991750951 | 0.000688122 | 0.001594444 | 1 |
| Obesity | endothelial | *MARVELD1* | -1.812643357 | 9.33E-05 | 0.000323665 | 0.452807752 |
| Obesity | endothelial | *GOLGA7B* | -6.145637031 | 3.03E-06 | 3.39E-05 | 0.014714916 |
| Obesity | endothelial | *TRMT44* | -1.280069603 | 0.0002997 | 0.000801326 | 1 |
| Obesity | endothelial | *CNOT8* | -0.960973123 | 0.007396698 | 0.012299102 | 1 |
| Obesity | endothelial | *LRGUK* | -0.708676823 | 3.13E-05 | 0.000148107 | 0.151957372 |
| Obesity | endothelial | *SETD9* | -0.520156033 | 0.001173229 | 0.002494526 | 1 |
| Obesity | endothelial | *NUP205* | -0.959319261 | 0.007751272 | 0.012822766 | 1 |
| Obesity | endothelial | *PDIA4* | -1.431622881 | 0.00123881 | 0.002616764 | 1 |
| Obesity | endothelial | *KDM8* | -1.645598003 | 0.001703051 | 0.003425871 | 1 |
| Obesity | endothelial | *ALS2CR12* | -1.106069821 | 0.000247224 | 0.000690725 | 1 |
| Obesity | endothelial | *FZD7* | -0.874760258 | 2.92E-05 | 0.000139749 | 0.141704984 |
| Obesity | endothelial | *DEPTOR* | -0.570130567 | 0.011546852 | 0.018326897 | 1 |
| Obesity | endothelial | *PSD3* | -0.593667186 | 0.004247984 | 0.007563749 | 1 |
| Obesity | endothelial | *KCNMA1* | -0.463934355 | 0.000465597 | 0.001145508 | 1 |
| Obesity | endothelial | *NDUFAF6* | -0.704885305 | 0.007077026 | 0.011820218 | 1 |
| Obesity | endothelial | *DRAM2* | -0.414545251 | 0.022354223 | 0.033148745 | 1 |
| Obesity | endothelial | *C8orf37* | -0.615215537 | 0.005429977 | 0.009354803 | 1 |
| Obesity | endothelial | *WHAMM* | -0.807160738 | 0.000382096 | 0.000982474 | 1 |
| Obesity | endothelial | *RWDD2B* | -1.22595892 | 2.74E-05 | 0.000133636 | 0.132881115 |
| Obesity | endothelial | *CCT8* | -0.579666209 | 0.000737745 | 0.001685281 | 1 |
| Obesity | endothelial | *NAA11* | -2.580784263 | 7.03E-05 | 0.000267321 | 0.340920662 |
| Obesity | endothelial | *SFXN2* | -0.49819687 | 0.001469316 | 0.003019534 | 1 |
| Obesity | endothelial | *C14orf2* | -1.138465201 | 0.000149956 | 0.000468807 | 0.727588112 |
| Obesity | endothelial | *TDRD9* | -4.07857199 | 0.000131907 | 0.000422394 | 0.640014998 |
| Obesity | endothelial | *PCDH1* | 0.148954796 | 0.005261915 | 0.009107645 | 1 |
| Obesity | endothelial | *SUPV3L1* | -1.321179068 | 2.36E-06 | 3.05E-05 | 0.011444885 |
| Obesity | endothelial | *TYSND1* | -0.546175665 | 0.001472064 | 0.003023902 | 1 |
| Obesity | endothelial | *NODAL* | -2.607110916 | 2.42E-06 | 3.09E-05 | 0.011727885 |
| Obesity | endothelial | *ZDHHC5* | -1.252786872 | 0.000944408 | 0.002080958 | 1 |
| Obesity | endothelial | *MED19* | -1.146116129 | 0.000219099 | 0.000630329 | 1 |
| Obesity | endothelial | *ATAD2* | -0.782727386 | 8.50E-05 | 0.000306216 | 0.412473356 |
| Obesity | endothelial | *FBRS* | -1.094952682 | 0.000723549 | 0.001659886 | 1 |
| Obesity | endothelial | *SASS6* | -1.735255164 | 1.39E-06 | 2.57E-05 | 0.006766825 |
| Obesity | endothelial | *VPS8* | -0.825236391 | 0.000587419 | 0.001394402 | 1 |
| Obesity | endothelial | *MPV17L* | -1.478866351 | 0.00151132 | 0.003095367 | 1 |
| Obesity | endothelial | *RPUSD3* | -1.599221672 | 1.15E-05 | 7.27E-05 | 0.055875389 |
| Obesity | endothelial | *GHRL* | -1.033216915 | 0.002716118 | 0.005121884 | 1 |
| Obesity | endothelial | *NMNAT2* | 0.413727676 | 0.010279101 | 0.016558498 | 1 |
| Obesity | endothelial | *SLC6A1* | -4.31727912 | 2.06E-05 | 0.00011009 | 0.099962057 |
| Obesity | endothelial | *NECAP2* | -1.254364831 | 0.000486784 | 0.001189862 | 1 |
| Obesity | endothelial | *STEAP2* | -0.978689075 | 1.37E-08 | 1.47E-06 | 6.63E-05 |
| Obesity | endothelial | *CLDN12* | -1.5850454 | 1.82E-06 | 2.68E-05 | 0.008826776 |
| Obesity | endothelial | *MMP14* | -1.446553076 | 1.41E-06 | 2.57E-05 | 0.006835891 |
| Obesity | endothelial | *GATAD1* | -1.058279692 | 0.000146364 | 0.000459353 | 0.710159777 |
| Obesity | endothelial | *TMED6* | 3.231605297 | 0.001746273 | 0.003499109 | 1 |

| | | | | | | |
|---|---|---|---|---|---|---|
| Obesity | endothelial | *CLEC18A* | -2.357005937 | 2.09E-18 | 1.69E-15 | 1.02E-14 |
| Obesity | endothelial | *DHRS4* | -1.925129173 | 8.55E-06 | 5.96E-05 | 0.041497087 |
| Obesity | endothelial | *DHRS1* | -0.634182765 | 0.001081231 | 0.00232851 | 1 |
| Obesity | endothelial | *AASDH* | -0.923549537 | 0.00024647 | 0.000689265 | 1 |
| Obesity | endothelial | *ETS2* | 0.43059689 | 1.64E-05 | 9.46E-05 | 0.079670978 |
| Obesity | endothelial | *MX1* | 0.348994511 | 0.002238269 | 0.004344033 | 1 |
| Obesity | endothelial | *SLC38A10* | -1.21547976 | 2.63E-05 | 0.000129952 | 0.12781621 |
| Obesity | endothelial | *DGKI* | -0.832181147 | 1.11E-06 | 2.57E-05 | 0.005384123 |
| Obesity | endothelial | *SNX22* | 1.086111769 | 0.02431497 | 0.035804624 | 1 |
| Obesity | endothelial | *SLC34A2* | 6.067727018 | 0.008202994 | 0.01347357 | 1 |
| Obesity | endothelial | *AP3S2* | -1.431274503 | 1.74E-05 | 9.81E-05 | 0.084198699 |
| Obesity | endothelial | *SPPL3* | -0.800028178 | 0.00080313 | 0.00180574 | 1 |
| Obesity | endothelial | *C12orf43* | -1.285184423 | 0.003277682 | 0.006037704 | 1 |
| Obesity | endothelial | *PEX10* | -0.986581309 | 0.008314304 | 0.013633322 | 1 |
| Obesity | endothelial | *EXTL1* | -24.17630607 | 1.44E-06 | 2.57E-05 | 0.006968692 |
| Obesity | endothelial | *WDR66* | -0.724224596 | 0.000465805 | 0.001145508 | 1 |
| Obesity | endothelial | *RHPN1* | 0.615338251 | 0.009888497 | 0.015971699 | 1 |
| Obesity | endothelial | *LRRC43* | 1.290306088 | 0.033209575 | 0.047364155 | 1 |
| Obesity | endothelial | *DZIP1L* | -0.944078904 | 8.27E-07 | 2.32E-05 | 0.00401368 |
| Obesity | endothelial | *FANCC* | -0.853134845 | 0.002420248 | 0.004641519 | 1 |
| Obesity | endothelial | *ESYT3* | -1.124383939 | 0.000949502 | 0.002089335 | 1 |
| Obesity | endothelial | *CLSTN2* | -0.694136345 | 1.08E-06 | 2.57E-05 | 0.005247304 |
| Obesity | endothelial | *COLEC12* | -0.585612115 | 0.00012949 | 0.000415808 | 0.628286064 |
| Obesity | endothelial | *MITD1* | -0.729465698 | 0.021882623 | 0.032522442 | 1 |
| Obesity | endothelial | *TSPAN33* | -1.435932958 | 8.04E-05 | 0.000293596 | 0.389894993 |
| Obesity | endothelial | *FAM86C1* | -2.226694209 | 1.47E-06 | 2.57E-05 | 0.007118276 |
| Obesity | endothelial | *PPP1R9A* | -0.708859171 | 0.008020436 | 0.013205006 | 1 |
| Obesity | endothelial | *DYNC1I1* | -0.663296845 | 0.016450454 | 0.025147323 | 1 |
| Obesity | endothelial | *COPG2* | -0.499526346 | 3.84E-06 | 3.77E-05 | 0.01861551 |
| Obesity | endothelial | *DUSP23* | 0.450669339 | 0.017920613 | 0.02717213 | 1 |
| Obesity | endothelial | *SPATA2L* | -1.976413462 | 1.76E-07 | 9.04E-06 | 0.000852314 |
| Obesity | endothelial | *FGF17* | -2.114358585 | 0.004450194 | 0.007889054 | 1 |
| Obesity | endothelial | *CDA* | 0.242561997 | 0.00127945 | 0.002682754 | 1 |
| Obesity | endothelial | *SLC5A11* | -0.868883517 | 3.13E-11 | 7.98E-09 | 1.52E-07 |
| Obesity | endothelial | *CCAR2* | -1.230146833 | 0.002003777 | 0.003944479 | 1 |
| Obesity | endothelial | *CDC42SE2* | -0.818874504 | 0.001193746 | 0.002533708 | 1 |
| Obesity | endothelial | *RAPGEF6* | -1.806592112 | 1.85E-06 | 2.71E-05 | 0.008958479 |
| Obesity | endothelial | *CLIC6* | -0.829707718 | 0.025241219 | 0.037011301 | 1 |
| Obesity | endothelial | *CBR1* | -0.67831274 | 0.005402972 | 0.00932029 | 1 |
| Obesity | endothelial | *CHAF1B* | -0.735513613 | 0.000174182 | 0.000527218 | 0.845130323 |
| Obesity | endothelial | *HLCS* | -0.754859766 | 0.00018389 | 0.000551444 | 0.892235934 |
| Obesity | endothelial | *IRX6* | -0.577972259 | 0.000263365 | 0.000727705 | 1 |
| Obesity | endothelial | *HK2* | -0.840719747 | 2.63E-06 | 3.23E-05 | 0.012777175 |
| Obesity | endothelial | *THEM4* | -0.849393345 | 8.28E-05 | 0.000300181 | 0.401642565 |
| Obesity | endothelial | *AMFR* | -1.02532726 | 0.002198084 | 0.004276304 | 1 |
| Obesity | endothelial | *TMEM69* | -1.633860806 | 0.000298675 | 0.000801024 | 1 |
| Obesity | endothelial | *ACE* | 0.237560905 | 0.000152457 | 0.000473929 | 0.739720216 |
| Obesity | endothelial | *TEPP* | -4.19257753 | 0.000603979 | 0.001423959 | 1 |
| Obesity | endothelial | *CHCHD6* | -1.035953515 | 0.000514412 | 0.001242992 | 1 |
| Obesity | endothelial | *CTBP1* | -1.390647462 | 3.34E-05 | 0.000154638 | 0.162060972 |

| | | | | | | |
|---|---|---|---|---|---|---|
| Obesity | endothelial | *C16orf86* | -1.140646593 | 0.00036006 | 0.000937237 | 1 |
| Obesity | endothelial | *LYPD5* | 0.702677465 | 0.009480846 | 0.015359287 | 1 |
| Obesity | endothelial | *CCDC107* | -1.144904689 | 0.002966369 | 0.005535701 | 1 |
| Obesity | endothelial | *ATAD3B* | -0.910534194 | 0.001623628 | 0.003286543 | 1 |
| Obesity | endothelial | *SSU72* | -1.27860441 | 0.00035571 | 0.000930407 | 1 |
| Obesity | endothelial | *CPAMD8* | 0.265832225 | 0.000390313 | 0.000999894 | 1 |
| Obesity | endothelial | *ANKLE1* | -1.938554362 | 0.000590223 | 0.001397638 | 1 |
| Obesity | endothelial | *CCDC58* | -1.595103672 | 2.99E-06 | 3.39E-05 | 0.014489912 |
| Obesity | endothelial | *CILP2* | -2.178307028 | 1.78E-06 | 2.68E-05 | 0.008632591 |
| Obesity | endothelial | *PDE9A* | 0.308693655 | 0.000354362 | 0.000927381 | 1 |
| Obesity | endothelial | *HSF2BP* | -0.758644035 | 0.002141579 | 0.004182474 | 1 |
| Obesity | endothelial | *C21orf33* | -0.567676888 | 0.032525096 | 0.046483584 | 1 |
| Obesity | endothelial | *C21orf2* | -0.71366706 | 2.11E-05 | 0.000112132 | 0.102376768 |
| Obesity | endothelial | *MCM3AP* | -1.121578205 | 1.54E-06 | 2.57E-05 | 0.007494104 |
| Obesity | endothelial | *PCNT* | -0.732865141 | 0.000618072 | 0.001452949 | 1 |
| Obesity | endothelial | *S100B* | -0.23398037 | 0.006795428 | 0.011393026 | 1 |
| Obesity | endothelial | *ADAMTS13* | -0.9122087 | 0.000111704 | 0.000369958 | 0.541987922 |
| Obesity | endothelial | *CACFD1* | -3.173449792 | 0.000136607 | 0.000434633 | 0.662814976 |
| Obesity | endothelial | *RDH13* | -0.886126168 | 0.00063793 | 0.001491679 | 1 |
| Obesity | endothelial | *TAOK1* | -0.851482838 | 0.000531687 | 0.001279635 | 1 |
| Obesity | endothelial | *S100A1* | 1.533734918 | 5.39E-05 | 0.000219557 | 0.26171243 |
| Obesity | endothelial | *ZBTB7B* | -1.265497163 | 1.10E-06 | 2.57E-05 | 0.005341852 |
| Obesity | endothelial | *VPS11* | -1.262668849 | 1.76E-06 | 2.68E-05 | 0.008518611 |
| Obesity | endothelial | *PAQR6* | -0.923917611 | 0.000122675 | 0.000397343 | 0.595220028 |
| Obesity | endothelial | *NBEAL2* | 0.769233801 | 0.010631192 | 0.017063362 | 1 |
| Obesity | endothelial | *PTH1R* | -0.648195285 | 0.000403971 | 0.001026213 | 1 |
| Obesity | endothelial | *FGFR4* | -2.171595117 | 6.30E-07 | 2.12E-05 | 0.003054769 |
| Obesity | endothelial | *IER2* | -1.094105163 | 0.001796402 | 0.00358689 | 1 |
| Obesity | endothelial | *TONSL* | -2.637640056 | 3.27E-07 | 1.37E-05 | 0.001586476 |
| Obesity | endothelial | *MUM1* | -0.988791874 | 9.12E-06 | 6.21E-05 | 0.044267494 |
| Obesity | endothelial | *ZNF333* | -1.329951559 | 0.000240788 | 0.000676885 | 1 |
| Obesity | endothelial | *SH2B2* | -0.956782816 | 0.000325889 | 0.000861229 | 1 |
| Obesity | endothelial | *DVL3* | -0.990288966 | 2.20E-05 | 0.000115 | 0.10672045 |
| Obesity | endothelial | *PCYT1A* | -1.138587874 | 2.23E-05 | 0.000116275 | 0.108019206 |
| Obesity | endothelial | *BDH1* | -1.11423026 | 3.90E-06 | 3.78E-05 | 0.018924557 |
| Obesity | endothelial | *PGAP3* | -2.027244316 | 7.64E-05 | 0.000283942 | 0.37082804 |
| Obesity | endothelial | *ZNF577* | -0.75820014 | 0.003925374 | 0.007077635 | 1 |
| Obesity | endothelial | *TMEM143* | -1.300521723 | 2.06E-06 | 2.88E-05 | 0.010000492 |
| Obesity | endothelial | *CCL16* | 0.270887885 | 8.98E-05 | 0.000316583 | 0.435934109 |
| Obesity | endothelial | *DCD* | -1.976134835 | 0.006659142 | 0.011203244 | 1 |
| Obesity | endothelial | *SIGLEC11* | -0.414850629 | 0.00835456 | 0.013685458 | 1 |
| Obesity | endothelial | *ASB16* | -1.699797133 | 1.58E-06 | 2.58E-05 | 0.007677136 |
| Obesity | endothelial | *IP6K3* | -4.804383564 | 2.35E-05 | 0.00012075 | 0.11386758 |
| Obesity | endothelial | *LEMD2* | -1.612983894 | 1.12E-05 | 7.11E-05 | 0.054337609 |
| Obesity | endothelial | *ASGR2* | -3.473916635 | 1.14E-05 | 7.22E-05 | 0.055468237 |
| Obesity | endothelial | *SNRNP25* | -1.203233743 | 0.000222861 | 0.000638326 | 1 |
| Obesity | endothelial | *WDR90* | -1.201362764 | 8.28E-08 | 5.43E-06 | 0.000401629 |
| Obesity | endothelial | *MEIOB* | -0.783832687 | 0.004997498 | 0.008712849 | 1 |
| Obesity | endothelial | *SHANK2* | 0.381449037 | 0.001284188 | 0.002689201 | 1 |
| Obesity | endothelial | *ASRGL1* | 0.337549193 | 0.000299118 | 0.000801326 | 1 |

| | | | | | | |
|---|---|---|---|---|---|---|
| Obesity | endothelial | *TTC9C* | -0.836591455 | 0.005103329 | 0.008871857 | 1 |
| Obesity | endothelial | *RPS6KA4* | -1.538234305 | 0.008451565 | 0.013825689 | 1 |
| Obesity | endothelial | *TPCN2* | -1.001170448 | 4.24E-06 | 3.91E-05 | 0.020565213 |
| Obesity | endothelial | *CMPK1* | -1.039436141 | 0.000874931 | 0.001946856 | 1 |
| Obesity | endothelial | *COA7* | -2.200913296 | 2.48E-06 | 3.13E-05 | 0.012042692 |
| Obesity | endothelial | *SLC1A7* | -0.50566145 | 0.002312047 | 0.004458685 | 1 |
| Obesity | endothelial | *USP24* | -0.831966902 | 0.001422299 | 0.002934097 | 1 |
| Obesity | endothelial | *AK4* | -0.591525205 | 0.022458605 | 0.033293356 | 1 |
| Obesity | endothelial | *RBP7* | 0.28104345 | 0.000639288 | 0.001493416 | 1 |
| Obesity | endothelial | *PDPN* | -1.035505669 | 1.17E-06 | 2.57E-05 | 0.00567828 |
| Obesity | endothelial | *LAPTM5* | -1.253081079 | 4.00E-06 | 3.84E-05 | 0.019405994 |
| Obesity | endothelial | *SYNC* | -1.109162982 | 6.92E-07 | 2.20E-05 | 0.003356654 |
| Obesity | endothelial | *MEGF6* | -0.771067164 | 0.019202624 | 0.028881317 | 1 |
| Obesity | endothelial | *OMA1* | -1.014964039 | 0.008012778 | 0.013196876 | 1 |
| Obesity | endothelial | *TM2D1* | -0.980169435 | 0.000399624 | 0.001018905 | 1 |
| Obesity | endothelial | *NEXN* | -0.865525784 | 5.71E-06 | 4.56E-05 | 0.027701169 |
| Obesity | endothelial | *LRRIQ3* | -1.250281459 | 1.35E-06 | 2.57E-05 | 0.006533789 |
| Obesity | endothelial | *TYW3* | -0.84477759 | 0.025182843 | 0.036948036 | 1 |
| Obesity | endothelial | *HFM1* | -1.062280337 | 5.38E-05 | 0.000219234 | 0.261108286 |
| Obesity | endothelial | *ARPC5* | -1.176054439 | 1.06E-05 | 6.89E-05 | 0.051494587 |
| Obesity | endothelial | *FCRLB* | -1.871324906 | 6.44E-06 | 4.91E-05 | 0.031250562 |
| Obesity | endothelial | *FLVCR1* | -1.802989559 | 6.70E-06 | 5.06E-05 | 0.032506958 |
| Obesity | endothelial | *TDRD5* | -0.879437327 | 4.17E-06 | 3.89E-05 | 0.020222573 |
| Obesity | endothelial | *ACP6* | -1.561407062 | 3.80E-06 | 3.77E-05 | 0.018415552 |
| Obesity | endothelial | *PM20D1* | 0.649937012 | 0.021507948 | 0.032060388 | 1 |
| Obesity | endothelial | *PKDCC* | -0.727225847 | 7.50E-05 | 0.000280056 | 0.363792872 |
| Obesity | endothelial | *HAAO* | -1.163398817 | 4.56E-06 | 4.07E-05 | 0.022135468 |
| Obesity | endothelial | *B3GALNT2* | -0.595830173 | 0.003433771 | 0.006296545 | 1 |
| Obesity | endothelial | *MRPL55* | -0.716596773 | 0.011109251 | 0.017727662 | 1 |
| Obesity | endothelial | *DUSP19* | -1.667745791 | 0.00176793 | 0.003535859 | 1 |
| Obesity | endothelial | *NUP35* | -0.966892499 | 0.011768873 | 0.018636609 | 1 |
| Obesity | endothelial | *C2orf48* | -7.648873197 | 9.02E-05 | 0.000317134 | 0.437454849 |
| Obesity | endothelial | *CCDC74A* | -0.929341267 | 0.007425833 | 0.012334866 | 1 |
| Obesity | endothelial | *NOSTRIN* | 0.140362088 | 0.000112881 | 0.000373505 | 0.547700927 |
| Obesity | endothelial | *INHBB* | 0.467889111 | 0.000247277 | 0.000690725 | 1 |
| Obesity | endothelial | *RPRD2* | -0.764639311 | 0.001285771 | 0.002691356 | 1 |
| Obesity | endothelial | *CTSS* | -0.353254911 | 0.002900299 | 0.005431205 | 1 |
| Obesity | endothelial | *PACRGL* | -0.842980792 | 0.006412669 | 0.010844988 | 1 |
| Obesity | endothelial | *C1QTNF7* | -0.825179294 | 3.26E-06 | 3.48E-05 | 0.015803586 |
| Obesity | endothelial | *LYSMD1* | 0.437754732 | 0.034432218 | 0.048920973 | 1 |
| Obesity | endothelial | *VPS72* | -1.634511451 | 1.83E-05 | 0.000101625 | 0.08899027 |
| Obesity | endothelial | *ERCC3* | -1.156395675 | 0.000112929 | 0.000373505 | 0.547932033 |
| Obesity | endothelial | *RNF149* | -1.008675223 | 3.44E-06 | 3.56E-05 | 0.01668704 |
| Obesity | endothelial | *DHX57* | -1.218914704 | 2.32E-05 | 0.000120065 | 0.11245081 |
| Obesity | endothelial | *S100A9* | -1.095724076 | 8.17E-07 | 2.32E-05 | 0.003964951 |
| Obesity | endothelial | *DCAF16* | -1.543399491 | 4.32E-07 | 1.64E-05 | 0.002096411 |
| Obesity | endothelial | *GNPDA2* | -0.629399121 | 0.033982618 | 0.048352981 | 1 |
| Obesity | endothelial | *NIPAL1* | -1.331990528 | 1.70E-05 | 9.68E-05 | 0.082547064 |
| Obesity | endothelial | *ANTXR2* | -0.55182573 | 0.001642285 | 0.003320153 | 1 |
| Obesity | endothelial | *CGGBP1* | -0.60323023 | 0.000175074 | 0.0005286 | 0.849459891 |

| | | | | | | |
|---|---|---|---|---|---|---|
| Obesity | endothelial | *GPR155* | -1.242524435 | 3.57E-05 | 0.000161584 | 0.173218213 |
| Obesity | endothelial | *DAPL1* | -1.459120278 | 0.000153144 | 0.000475099 | 0.743055242 |
| Obesity | endothelial | *PBXIP1* | -1.438610105 | 5.02E-05 | 0.000208293 | 0.243494514 |
| Obesity | endothelial | *CLDN1* | -0.797646441 | 2.78E-05 | 0.00013462 | 0.134889433 |
| Obesity | endothelial | *LINC01116* | 0.33474258 | 6.24E-05 | 0.000244714 | 0.302710629 |
| Obesity | endothelial | *SLC22A15* | -0.928556449 | 0.000443008 | 0.001104834 | 1 |
| Obesity | endothelial | *LRRC58* | -1.268056924 | 0.000212123 | 0.000614348 | 1 |
| Obesity | endothelial | *LMOD1* | -1.398304446 | 2.36E-11 | 6.37E-09 | 1.15E-07 |
| Obesity | endothelial | *TMEM169* | -0.592977553 | 0.027138005 | 0.039565384 | 1 |
| Obesity | endothelial | *ARPC2* | -0.943035043 | 0.000403839 | 0.001026213 | 1 |
| Obesity | endothelial | *CCT3* | -0.702760418 | 0.028532466 | 0.041399379 | 1 |
| Obesity | endothelial | *GLB1L* | -1.262192221 | 2.48E-07 | 1.13E-05 | 0.001201467 |
| Obesity | endothelial | *SUCLG1* | -0.883102907 | 0.002305279 | 0.004447402 | 1 |
| Obesity | endothelial | *PRKCI* | -0.833263518 | 1.95E-05 | 0.000106014 | 0.094498849 |
| Obesity | endothelial | *AIM2* | 0.222436229 | 0.000110631 | 0.000367657 | 0.536779334 |
| Obesity | endothelial | *GTPBP8* | -1.627967262 | 0.000110824 | 0.000367911 | 0.537719762 |
| Obesity | endothelial | *WDFY3* | -0.663565489 | 0.004330245 | 0.007698918 | 1 |
| Obesity | endothelial | *PTPN13* | -0.590890175 | 4.66E-05 | 0.000195558 | 0.225974254 |
| Obesity | endothelial | *PPM1K* | -0.970778141 | 0.000922631 | 0.002039457 | 1 |
| Obesity | endothelial | *TIPARP* | -1.380159505 | 2.18E-05 | 0.000114765 | 0.105733345 |
| Obesity | endothelial | *RPP14* | -1.549723253 | 3.75E-06 | 3.75E-05 | 0.018216845 |
| Obesity | endothelial | *DNASE1L3* | 0.241987587 | 0.000220444 | 0.000633584 | 1 |
| Obesity | endothelial | *IL17RC* | -1.531326798 | 8.72E-06 | 6.02E-05 | 0.04229991 |
| Obesity | endothelial | *CRELD1* | -1.438518525 | 5.62E-06 | 4.54E-05 | 0.027252898 |
| Obesity | endothelial | *TTC14* | -1.042510503 | 9.15E-06 | 6.21E-05 | 0.044384766 |
| Obesity | endothelial | *CCDC158* | -1.056692481 | 1.80E-06 | 2.68E-05 | 0.008734822 |
| Obesity | endothelial | *CPA3* | -2.335680101 | 4.03E-06 | 3.85E-05 | 0.019556485 |
| Obesity | endothelial | *SLC4A1AP* | -1.474434223 | 2.46E-05 | 0.000124585 | 0.11931287 |
| Obesity | endothelial | *CDCP1* | -1.01254234 | 3.57E-07 | 1.45E-05 | 0.001733623 |
| Obesity | endothelial | *CLEC3B* | 1.35297949 | 0.004407913 | 0.007825538 | 1 |
| Obesity | endothelial | *FYCO1* | -1.298711821 | 4.26E-06 | 3.91E-05 | 0.020648042 |
| Obesity | endothelial | *NMNAT3* | -0.877947753 | 0.001576522 | 0.003209938 | 1 |
| Obesity | endothelial | *TPRA1* | -1.56570636 | 8.91E-05 | 0.000315285 | 0.432261904 |
| Obesity | endothelial | *MEAF6* | -0.936516522 | 0.012634478 | 0.019858273 | 1 |
| Obesity | endothelial | *SNIP1* | -1.393038708 | 0.001526325 | 0.003119515 | 1 |
| Obesity | endothelial | *DNALI1* | -1.862454544 | 1.88E-10 | 3.96E-08 | 9.12E-07 |
| Obesity | endothelial | *RPN1* | -0.989734503 | 0.00805604 | 0.013254631 | 1 |
| Obesity | endothelial | *SENP2* | -0.999223356 | 2.32E-05 | 0.000120195 | 0.112742662 |
| Obesity | endothelial | *IFT122* | -1.208025705 | 7.86E-06 | 5.62E-05 | 0.038134315 |
| Obesity | endothelial | *RFT1* | -1.788616843 | 1.43E-11 | 4.34E-09 | 6.95E-08 |
| Obesity | endothelial | *SFMBT1* | -1.210855994 | 3.06E-06 | 3.39E-05 | 0.01485141 |
| Obesity | endothelial | *UVSSA* | -0.95423852 | 7.73E-05 | 0.000285691 | 0.374826837 |
| Obesity | endothelial | *FAM208A* | -0.995647239 | 0.000198703 | 0.000585373 | 0.964108633 |
| Obesity | endothelial | *LRPAP1* | -1.070027868 | 0.001103925 | 0.002373169 | 1 |
| Obesity | endothelial | *PIGX* | -0.897720005 | 9.32E-05 | 0.000323517 | 0.452276754 |
| Obesity | endothelial | *ERMAP* | -1.124056514 | 5.86E-05 | 0.000233306 | 0.284166475 |
| Obesity | endothelial | *METAP1* | -0.780114237 | 0.021111796 | 0.031518287 | 1 |
| Obesity | endothelial | *BDH2* | -1.036837916 | 0.00010744 | 0.000359537 | 0.521300421 |
| Obesity | endothelial | *CDC25A* | 1.51304159 | 0.000198413 | 0.000585373 | 0.962699433 |
| Obesity | endothelial | *ZNF589* | -0.765128416 | 0.002044078 | 0.004008839 | 1 |

| | | | | | | |
|---|---|---|---|---|---|---|
| Obesity | endothelial | *ATRIP* | -0.453299179 | 0.007617222 | 0.012631155 | 1 |
| Obesity | endothelial | *RNF123* | 1.319469384 | 1.87E-07 | 9.10E-06 | 0.00090828 |
| Obesity | endothelial | *HSPA4L* | -1.035995409 | 0.002198044 | 0.004276304 | 1 |
| Obesity | endothelial | *MFSD8* | -1.083195517 | 0.000521924 | 0.001258637 | 1 |
| Obesity | endothelial | *MST1R* | 2.165966853 | 8.18E-20 | 9.92E-17 | 3.97E-16 |
| Obesity | endothelial | *RAD54L2* | -1.190548629 | 2.06E-05 | 0.000110176 | 0.100150308 |
| Obesity | endothelial | *TEX264* | -1.48156143 | 1.47E-06 | 2.57E-05 | 0.007132312 |
| Obesity | endothelial | *ETNPPL* | -2.480636602 | 1.00E-05 | 6.64E-05 | 0.048507873 |
| Obesity | endothelial | *ANXA5* | -1.144062218 | 4.78E-05 | 0.000199922 | 0.232013759 |
| Obesity | endothelial | *MAP9* | -1.0171653 | 2.14E-06 | 2.91E-05 | 0.01040473 |
| Obesity | endothelial | *CEP44* | -1.269785017 | 8.67E-06 | 6.00E-05 | 0.042060063 |
| Obesity | endothelial | *HPGD* | -2.651462479 | 1.39E-05 | 8.35E-05 | 0.067341042 |
| Obesity | endothelial | *TMEM144* | -1.376418791 | 6.46E-05 | 0.000250799 | 0.313499371 |
| Obesity | endothelial | *IL15* | 0.427130887 | 0.007720902 | 0.012781241 | 1 |
| Obesity | endothelial | *LSM6* | -0.703141583 | 0.01472285 | 0.022770745 | 1 |
| Obesity | endothelial | *ITGA2* | 0.381429415 | 0.000520411 | 0.001255912 | 1 |
| Obesity | endothelial | *MOCS2* | -1.425703333 | 0.006097661 | 0.010378939 | 1 |
| Obesity | endothelial | *NDUFAF2* | -0.810739421 | 0.000811609 | 0.001820812 | 1 |
| Obesity | endothelial | *LMBRD2* | -1.383232137 | 4.43E-05 | 0.000187818 | 0.214864045 |
| Obesity | endothelial | *PGGT1B* | -0.936148641 | 0.00312307 | 0.005789974 | 1 |
| Obesity | endothelial | *F2RL2* | -2.337931279 | 2.78E-06 | 3.30E-05 | 0.01348089 |
| Obesity | endothelial | *CMBL* | -0.864532197 | 4.11E-06 | 3.87E-05 | 0.019948779 |
| Obesity | endothelial | *C5orf63* | -0.992616938 | 4.01E-05 | 0.00017598 | 0.19463339 |
| Obesity | endothelial | *F2RL1* | -1.47952849 | 0.001575552 | 0.003209312 | 1 |
| Obesity | endothelial | *WDR41* | -0.949723734 | 0.00025014 | 0.000696715 | 1 |
| Obesity | endothelial | *NDUFS4* | -0.969648282 | 0.000289921 | 0.000783675 | 1 |
| Obesity | endothelial | *SERINC5* | -0.973909771 | 3.22E-05 | 0.000151156 | 0.156144409 |
| Obesity | endothelial | *PRIMPOL* | -1.046398829 | 9.64E-05 | 0.000331493 | 0.467609446 |
| Obesity | endothelial | *ERAP1* | -1.239545057 | 1.36E-07 | 7.62E-06 | 0.00066028 |
| Obesity | endothelial | *UTP15* | -2.005028681 | 7.02E-07 | 2.20E-05 | 0.003406103 |
| Obesity | endothelial | *CCDC127* | -1.813159836 | 5.48E-06 | 4.47E-05 | 0.026598179 |
| Obesity | endothelial | *GRIK2* | -0.356362785 | 0.009002659 | 0.014648189 | 1 |
| Obesity | endothelial | *PDSS2* | -0.867327293 | 0.000407734 | 0.001034689 | 1 |
| Obesity | endothelial | *RAET1E* | -1.575954251 | 1.24E-06 | 2.57E-05 | 0.006028684 |
| Obesity | endothelial | *STK17A* | -0.761640989 | 0.013043999 | 0.02042255 | 1 |
| Obesity | endothelial | *RP9* | -1.725092474 | 4.35E-05 | 0.000185308 | 0.210877971 |
| Obesity | endothelial | *MIOS* | -1.245527817 | 0.000120487 | 0.000392089 | 0.584604391 |
| Obesity | endothelial | *USP49* | -1.341387634 | 1.17E-05 | 7.35E-05 | 0.056977972 |
| Obesity | endothelial | *CHMP4C* | 0.792688744 | 0.000216678 | 0.000625044 | 1 |
| Obesity | endothelial | *SLC13A4* | -1.304505126 | 2.27E-05 | 0.000117989 | 0.110037106 |
| Obesity | endothelial | *CTSB* | -0.671470563 | 4.88E-07 | 1.78E-05 | 0.002369424 |
| Obesity | endothelial | *SOX17* | 0.163977688 | 7.32E-05 | 0.00027495 | 0.354961063 |
| Obesity | endothelial | *RAD21* | -0.834469176 | 0.006167034 | 0.010469716 | 1 |
| Obesity | endothelial | *OXR1* | -0.883573033 | 5.60E-05 | 0.000224837 | 0.271828368 |
| Obesity | endothelial | *INTS1* | -1.772681438 | 1.79E-05 | 9.98E-05 | 0.086613881 |
| Obesity | endothelial | *SLC4A2* | -1.221981048 | 1.57E-05 | 9.10E-05 | 0.076051234 |
| Obesity | endothelial | *FASTK* | -1.590918059 | 3.26E-06 | 3.48E-05 | 0.015828652 |
| Obesity | endothelial | *ALDH7A1* | -0.943804435 | 0.000952425 | 0.002093869 | 1 |
| Obesity | endothelial | *YWHAZ* | -1.161719328 | 1.55E-06 | 2.57E-05 | 0.007499942 |
| Obesity | endothelial | *FZD6* | 0.379261797 | 0.001004788 | 0.002183265 | 1 |

| | | | | | | |
|---|---|---|---|---|---|---|
| Obesity | endothelial | *SLC25A32* | -1.184173284 | 0.000336019 | 0.000883666 | 1 |
| Obesity | endothelial | *DCSTAMP* | -4.820521619 | 7.16E-05 | 0.000270444 | 0.347250486 |
| Obesity | endothelial | *INTS8* | -0.942315417 | 0.003127403 | 0.005791664 | 1 |
| Obesity | endothelial | *RPP25L* | -1.818278467 | 2.36E-09 | 3.58E-07 | 1.14E-05 |
| Obesity | endothelial | *C9orf24* | -1.400401237 | 4.56E-05 | 0.000192356 | 0.221473878 |
| Obesity | endothelial | *CCDC171* | -0.657846133 | 0.020533462 | 0.030740006 | 1 |
| Obesity | endothelial | *ZMAT4* | -3.467168624 | 4.69E-05 | 0.000196659 | 0.227534697 |
| Obesity | endothelial | *C8orf34* | -0.680544595 | 6.53E-08 | 4.43E-06 | 0.000316643 |
| Obesity | endothelial | *RASEF* | -0.437659491 | 0.003941222 | 0.00710357 | 1 |
| Obesity | endothelial | *NOL6* | -0.691242439 | 0.007430781 | 0.012337449 | 1 |
| Obesity | endothelial | *AQP3* | -5.647234446 | 3.44E-06 | 3.56E-05 | 0.016707135 |
| Obesity | endothelial | *TRMT10B* | -1.099959832 | 7.71E-05 | 0.000285256 | 0.373970666 |
| Obesity | endothelial | *VCP* | -0.681245898 | 0.017131229 | 0.026073 | 1 |
| Obesity | endothelial | *PIGO* | -1.972587489 | 2.61E-05 | 0.000128957 | 0.12639714 |
| Obesity | endothelial | *WRN* | -0.759777424 | 0.004342965 | 0.007718706 | 1 |
| Obesity | endothelial | *TSHR* | -0.830994364 | 1.52E-06 | 2.57E-05 | 0.007385132 |
| Obesity | endothelial | *CFL2* | -1.520096958 | 0.000147527 | 0.000462404 | 0.71580126 |
| Obesity | endothelial | *PGM2L1* | 0.526010467 | 0.004155094 | 0.00742829 | 1 |
| Obesity | endothelial | *SLC16A9* | -1.483602846 | 1.46E-06 | 2.57E-05 | 0.007107605 |
| Obesity | endothelial | *GJB2* | 1.504243432 | 0.000212619 | 0.000615164 | 1 |
| Obesity | endothelial | *CRYL1* | -1.039231676 | 0.000707257 | 0.001629446 | 1 |
| Obesity | endothelial | *REEP3* | -0.874946898 | 0.001957286 | 0.003862038 | 1 |
| Obesity | endothelial | *MICU2* | -1.003461265 | 3.88E-05 | 0.000171733 | 0.188390656 |
| Obesity | endothelial | *LRR1* | -1.83682767 | 1.45E-06 | 2.57E-05 | 0.007045353 |
| Obesity | endothelial | *NOXRED1* | -1.117760451 | 0.002176189 | 0.004242213 | 1 |
| Obesity | endothelial | *ATP5C1* | -0.952374435 | 0.001862871 | 0.003705884 | 1 |
| Obesity | endothelial | *VDAC2* | -1.746293581 | 1.79E-07 | 9.04E-06 | 0.000867502 |
| Obesity | endothelial | *PDZD8* | -0.710423763 | 0.009179068 | 0.014910224 | 1 |
| Obesity | endothelial | *QSOX2* | -1.832041837 | 0.000137422 | 0.000436375 | 0.666771943 |
| Obesity | endothelial | *CLEC1B* | -0.668368938 | 0.017393745 | 0.026422808 | 1 |
| Obesity | endothelial | *SNAPC4* | -0.727370818 | 0.01021358 | 0.016480072 | 1 |
| Obesity | endothelial | *STOX1* | -0.340210516 | 0.012575389 | 0.019784626 | 1 |
| Obesity | endothelial | *RET* | -1.170893428 | 3.07E-06 | 3.39E-05 | 0.014886794 |
| Obesity | endothelial | *PPP1R36* | -1.798811221 | 6.15E-05 | 0.000242589 | 0.298626523 |
| Obesity | endothelial | *VWA2* | 0.712365057 | 9.72E-05 | 0.000333385 | 0.471740181 |
| Obesity | endothelial | *METTL3* | -1.378292949 | 8.22E-05 | 0.000298616 | 0.398651742 |
| Obesity | endothelial | *TRUB1* | -1.258095473 | 0.000409819 | 0.001038892 | 1 |
| Obesity | endothelial | *HSPA12A* | -0.703404528 | 1.71E-06 | 2.66E-05 | 0.008293735 |
| Obesity | endothelial | *UBTD1* | -1.055272923 | 1.58E-06 | 2.58E-05 | 0.007679416 |
| Obesity | endothelial | *OTOGL* | -0.851325096 | 0.001549571 | 0.003161698 | 1 |
| Obesity | endothelial | *PACSIN3* | -0.670904217 | 0.000423745 | 0.00106529 | 1 |
| Obesity | endothelial | *TTC7B* | -0.852103012 | 0.003256532 | 0.006005585 | 1 |
| Obesity | endothelial | *RAPSN* | -2.375614379 | 2.14E-20 | 3.47E-17 | 1.04E-16 |
| Obesity | endothelial | *TC2N* | -0.585545697 | 0.000314887 | 0.000837169 | 1 |
| Obesity | endothelial | *IFI27L1* | -1.460992176 | 0.000161271 | 0.000495872 | 0.782486649 |
| Obesity | endothelial | *PTER* | -0.942947185 | 0.000282471 | 0.000768245 | 1 |
| Obesity | endothelial | *LIPC* | -0.883292339 | 0.001036008 | 0.002241066 | 1 |
| Obesity | endothelial | *CEP57* | -0.733515006 | 0.010834247 | 0.017349099 | 1 |
| Obesity | endothelial | *TRMT61A* | -1.152397716 | 3.92E-06 | 3.79E-05 | 0.018999785 |
| Obesity | endothelial | *LARP6* | -1.225853282 | 7.29E-05 | 0.000274419 | 0.353726704 |

| | | | | | | |
|---|---|---|---|---|---|---|
| Obesity | endothelial | *API5* | -1.086280276 | 0.000981931 | 0.002142235 | 1 |
| Obesity | endothelial | *ASPG* | -1.612989446 | 0.005863485 | 0.010024535 | 1 |
| Obesity | endothelial | *COX11* | -1.268296349 | 0.000105148 | 0.000353064 | 0.510177761 |
| Obesity | endothelial | *STXBP4* | -1.174420999 | 8.66E-06 | 6.00E-05 | 0.042034042 |
| Obesity | endothelial | *WBP1L* | -0.9148849 | 0.000966749 | 0.002117681 | 1 |
| Obesity | endothelial | *APBB1* | -1.533826543 | 0.000263889 | 0.000728321 | 1 |
| Obesity | endothelial | *NUDT13* | -0.776654383 | 0.000382058 | 0.000982474 | 1 |
| Obesity | endothelial | *WDR88* | -1.22589245 | 3.90E-06 | 3.78E-05 | 0.018903534 |
| Obesity | endothelial | *CYB5R2* | -0.564296856 | 0.000419425 | 0.001057169 | 1 |
| Obesity | endothelial | *SERPINB8* | -0.368504761 | 0.024183068 | 0.035632021 | 1 |
| Obesity | endothelial | *XRRA1* | -0.771491248 | 0.0008634 | 0.001924307 | 1 |
| Obesity | endothelial | *CDYL2* | -0.525043618 | 0.003154791 | 0.005837928 | 1 |
| Obesity | endothelial | *CENPN* | -2.123222143 | 3.80E-06 | 3.77E-05 | 0.018452436 |
| Obesity | endothelial | *C16orf46* | 1.317486858 | 0.028372491 | 0.041206446 | 1 |
| Obesity | endothelial | *TMEM41B* | -1.224496616 | 1.07E-05 | 6.92E-05 | 0.051761989 |
| Obesity | endothelial | *TMX3* | -1.095197812 | 0.000153978 | 0.000476467 | 0.747100674 |
| Obesity | endothelial | *PRKCB* | -0.780635976 | 7.57E-05 | 0.000281749 | 0.367118615 |
| Obesity | endothelial | *NDST2* | -4.318714916 | 6.18E-06 | 4.77E-05 | 0.030007219 |
| Obesity | endothelial | *CCDC68* | 0.198027163 | 2.34E-05 | 0.00012075 | 0.113768136 |
| Obesity | endothelial | *CLEC4D* | -1.656492244 | 0.012643039 | 0.019865293 | 1 |
| Obesity | endothelial | *ZSCAN21* | -0.993528174 | 7.79E-06 | 5.60E-05 | 0.037808198 |
| Obesity | endothelial | *TMED3* | -0.419324406 | 9.94E-05 | 0.000338625 | 0.482202527 |
| Obesity | endothelial | *CENPV* | -1.338963188 | 5.71E-07 | 1.99E-05 | 0.002769265 |
| Obesity | endothelial | *ZNF592* | -1.010458756 | 9.11E-05 | 0.000319598 | 0.442003991 |
| Obesity | endothelial | *ACSM1* | -1.148129169 | 0.000159448 | 0.000490889 | 0.773640474 |
| Obesity | endothelial | *CATSPER2* | -0.87079479 | 0.031236176 | 0.044852893 | 1 |
| Obesity | endothelial | *SAAL1* | -1.848222688 | 1.77E-05 | 9.92E-05 | 0.085812446 |
| Obesity | endothelial | *FAM96A* | -1.282117661 | 5.50E-06 | 4.47E-05 | 0.026682853 |
| Obesity | endothelial | *FAM111A* | -0.721798838 | 0.006150254 | 0.010444883 | 1 |
| Obesity | endothelial | *ANPEP* | -0.277258154 | 0.028424673 | 0.041255314 | 1 |
| Obesity | endothelial | *STAT6* | -1.098655364 | 5.03E-05 | 0.000208522 | 0.243970802 |
| Obesity | endothelial | *PIP4K2C* | -1.079116536 | 0.000532955 | 0.001281567 | 1 |
| Obesity | endothelial | *MTMR10* | -0.739459937 | 0.007697362 | 0.012746622 | 1 |
| Obesity | endothelial | *MS4A7* | -0.754420583 | 6.98E-07 | 2.20E-05 | 0.003385608 |
| Obesity | endothelial | *MS4A14* | -0.371310979 | 0.000206144 | 0.000600901 | 1 |
| Obesity | endothelial | *DIS3L* | -0.942193054 | 8.88E-05 | 0.000315162 | 0.430871977 |
| Obesity | endothelial | *CCNDBP1* | -0.944094853 | 0.003640354 | 0.006630255 | 1 |
| Obesity | endothelial | *EPB42* | -0.881527176 | 0.002426323 | 0.004649494 | 1 |
| Obesity | endothelial | *SMAD3* | -0.962397574 | 0.000215741 | 0.000623452 | 1 |
| Obesity | endothelial | *MAP1A* | -0.966792497 | 2.05E-05 | 0.000109754 | 0.099437005 |
| Obesity | endothelial | *RCCD1* | -1.423673541 | 2.04E-05 | 0.000109395 | 0.099002092 |
| Obesity | endothelial | *MEI1* | -0.792976131 | 0.011540012 | 0.018322035 | 1 |
| Obesity | endothelial | *PIP5KL1* | 0.656974829 | 0.005341774 | 0.009226873 | 1 |
| Obesity | endothelial | *ACSF2* | -1.144772703 | 1.49E-05 | 8.79E-05 | 0.072129108 |
| Obesity | endothelial | *URM1* | -1.222315546 | 0.000411348 | 0.00104168 | 1 |
| Obesity | endothelial | *ISLR2* | 0.999090338 | 1.71E-05 | 9.70E-05 | 0.082871892 |
| Obesity | endothelial | *SP2* | -1.435664163 | 0.000417688 | 0.001054435 | 1 |
| Obesity | endothelial | *COQ7* | -0.911714409 | 0.01324473 | 0.020710097 | 1 |
| Obesity | endothelial | *GPRC5B* | 0.442098956 | 0.000980496 | 0.002140066 | 1 |
| Obesity | endothelial | *FBXO22* | -1.2005221 | 6.05E-05 | 0.00023913 | 0.293649693 |

| | | | | | | |
|---|---|---|---|---|---|---|
| Obesity | endothelial | *TBC1D2B* | -0.938988023 | 3.17E-07 | 1.36E-05 | 0.001536916 |
| Obesity | endothelial | *LOXHD1* | -0.915843301 | 0.000542205 | 0.001299792 | 1 |
| Obesity | endothelial | *KATNAL2* | -0.703839634 | 0.000751384 | 0.001712406 | 1 |
| Obesity | endothelial | *ZNF91* | -0.866044985 | 0.000492862 | 0.001202296 | 1 |
| Obesity | endothelial | *POP5* | -1.570186249 | 1.97E-06 | 2.82E-05 | 0.00953895 |
| Obesity | endothelial | *MYO5B* | -0.831640592 | 1.53E-06 | 2.57E-05 | 0.007406846 |
| Obesity | endothelial | *ZNF23* | -1.747540444 | 0.004567405 | 0.008064429 | 1 |
| Obesity | endothelial | *ZNF229* | 0.724542915 | 0.000311856 | 0.000831843 | 1 |
| Obesity | endothelial | *SMG8* | -0.87834781 | 0.014664433 | 0.022695957 | 1 |
| Obesity | endothelial | *RAB8A* | -0.793536747 | 0.013813362 | 0.021495328 | 1 |
| Obesity | endothelial | *SPATA33* | -1.677763122 | 0.000127351 | 0.000410023 | 0.61790499 |
| Obesity | endothelial | *ZNF641* | -1.276997966 | 6.60E-06 | 5.01E-05 | 0.032036121 |
| Obesity | endothelial | *TP53I13* | -1.133103461 | 2.51E-05 | 0.000125716 | 0.121819239 |
| Obesity | endothelial | *ZNF528* | -1.00969028 | 0.011553992 | 0.018332234 | 1 |
| Obesity | endothelial | *TMC4* | -1.410594855 | 2.79E-05 | 0.00013475 | 0.135289001 |
| Obesity | endothelial | *TTYH1* | -1.301899597 | 1.67E-06 | 2.63E-05 | 0.008094887 |
| Obesity | endothelial | *LENG8* | -1.102974725 | 3.76E-05 | 0.00016765 | 0.182235334 |
| Obesity | endothelial | *CHAF1A* | -0.397115239 | 0.008481167 | 0.013869383 | 1 |
| Obesity | endothelial | *KIFC2* | -1.133805073 | 0.006269282 | 0.010624715 | 1 |
| Obesity | endothelial | *SERPINF2* | -0.51504981 | 0.006811832 | 0.011416583 | 1 |
| Obesity | endothelial | *SRR* | 0.465905811 | 0.001731702 | 0.003476301 | 1 |
| Obesity | endothelial | *CYB5D2* | -1.157035129 | 0.000256485 | 0.000712344 | 1 |
| Obesity | endothelial | *KLK5* | -1.751283474 | 1.58E-06 | 2.58E-05 | 0.007653979 |
| Obesity | endothelial | *KLK6* | -15.4228821 | 9.17E-06 | 6.21E-05 | 0.044483867 |
| Obesity | endothelial | *ZNF83* | -0.664210474 | 0.011204631 | 0.017853879 | 1 |
| Obesity | endothelial | *SPRYD3* | -2.167618997 | 4.57E-05 | 0.000192356 | 0.221594412 |
| Obesity | endothelial | *SOAT2* | -0.855640836 | 0.000314199 | 0.000836715 | 1 |
| Obesity | endothelial | *ZNF558* | -0.866892252 | 0.000698884 | 0.001613986 | 1 |
| Obesity | endothelial | *NDUFV1* | -1.036061347 | 0.001865864 | 0.003710316 | 1 |
| Obesity | endothelial | *NUDT8* | -0.619994437 | 0.00217999 | 0.004247916 | 1 |
| Obesity | endothelial | *PRDX2* | -1.400986574 | 0.00029487 | 0.000794836 | 1 |
| Obesity | endothelial | *ZNF232* | -0.376567494 | 0.004503053 | 0.007962395 | 1 |
| Obesity | endothelial | *MIS12* | -1.465377941 | 5.33E-05 | 0.000218313 | 0.258700764 |
| Obesity | endothelial | *ATP5H* | -1.143348157 | 0.003617955 | 0.006594409 | 1 |
| Obesity | endothelial | *MGAT5B* | 0.702332252 | 0.000210552 | 0.000611734 | 1 |
| Obesity | endothelial | *MLST8* | -1.85523331 | 4.82E-06 | 4.20E-05 | 0.023396683 |
| Obesity | endothelial | *ECI1* | -0.561545026 | 0.03516349 | 0.049829025 | 1 |
| Obesity | endothelial | *LTBP3* | -1.056327171 | 0.000448535 | 0.001114613 | 1 |
| Obesity | endothelial | *NAALADL1* | -0.67812301 | 0.013611583 | 0.021228994 | 1 |
| Obesity | endothelial | *SCARA5* | -0.748504968 | 0.000105594 | 0.000354315 | 0.512339761 |
| Obesity | endothelial | *KIAA1586* | -1.54033937 | 3.71E-07 | 1.46E-05 | 0.001800342 |
| Obesity | endothelial | *FAM83B* | -6.475229862 | 1.21E-06 | 2.57E-05 | 0.005868432 |
| Obesity | endothelial | *POLR2J3* | -0.53954247 | 0.000832875 | 0.001863982 | 1 |
| Obesity | endothelial | *NT5DC2* | -1.116611139 | 1.33E-05 | 8.06E-05 | 0.06435566 |
| Obesity | endothelial | *COA6* | -1.770025485 | 0.001615243 | 0.0032723 | 1 |
| Obesity | endothelial | *PXK* | -1.234598402 | 5.27E-06 | 4.44E-05 | 0.025579503 |
| Obesity | endothelial | *PCMTD1* | -0.556424342 | 0.027564675 | 0.040127153 | 1 |
| Obesity | endothelial | *ACOX2* | -1.295072177 | 4.74E-06 | 4.16E-05 | 0.023004037 |
| Obesity | endothelial | *FAM107A* | 0.273926422 | 9.47E-05 | 0.000327428 | 0.459708214 |
| Obesity | endothelial | *SCN11A* | 0.280969989 | 0.003019671 | 0.005611431 | 1 |

| | | | | | | |
|---|---|---|---|---|---|---|
| Obesity | endothelial | *DTYMK* | -1.852101907 | 0.000261797 | 0.000724064 | 1 |
| Obesity | endothelial | *ING5* | -0.917599101 | 0.000123559 | 0.000399673 | 0.59950878 |
| Obesity | endothelial | *RFWD3* | -1.212924814 | 0.000173117 | 0.000524839 | 0.839965425 |
| Obesity | endothelial | *KLHL30* | -2.212406406 | 2.84E-06 | 3.32E-05 | 0.013772237 |
| Obesity | endothelial | *CCDC110* | -2.039511878 | 1.49E-06 | 2.57E-05 | 0.007219199 |
| Obesity | endothelial | *SLC20A2* | -0.99025851 | 2.47E-05 | 0.000124619 | 0.119883867 |
| Obesity | endothelial | *TMUB2* | -1.43540293 | 1.09E-05 | 7.00E-05 | 0.053126474 |
| Obesity | endothelial | *NBPF9* | -1.31226364 | 1.37E-06 | 2.57E-05 | 0.006658052 |
| Obesity | endothelial | *ADAM9* | -0.945422865 | 0.001208541 | 0.002562868 | 1 |
| Obesity | endothelial | *NDUFS5* | -0.825276723 | 0.01041676 | 0.016769118 | 1 |
| Obesity | endothelial | *SLC16A4* | -0.418790397 | 0.032332998 | 0.046263553 | 1 |
| Obesity | endothelial | *DNAJC21* | -0.92508281 | 0.012703388 | 0.019953654 | 1 |
| Obesity | endothelial | *GSTM4* | -1.290813746 | 4.28E-05 | 0.000184076 | 0.207464905 |
| Obesity | endothelial | *TCTN2* | -0.383615206 | 0.00036108 | 0.000939388 | 1 |
| Obesity | endothelial | *PPIP5K1* | -1.25226056 | 2.60E-05 | 0.00012875 | 0.12604622 |
| Obesity | endothelial | *SNTB2* | -0.919304178 | 0.00077878 | 0.001761603 | 1 |
| Obesity | endothelial | *ZNF507* | -1.314812594 | 4.31E-05 | 0.0001852 | 0.209090308 |
| Obesity | endothelial | *STX18* | -1.017125026 | 0.000695588 | 0.001607905 | 1 |
| Obesity | endothelial | *ZBTB49* | -1.762774184 | 5.33E-05 | 0.000218313 | 0.258695429 |
| Obesity | endothelial | *ATOH8* | 0.417113682 | 0.010702577 | 0.017155237 | 1 |
| Obesity | endothelial | *BTNL3* | 0.48716702 | 1.05E-05 | 6.83E-05 | 0.050706611 |
| Obesity | endothelial | *TMEM129* | -1.156299341 | 0.000193624 | 0.000574595 | 0.93946359 |
| Obesity | endothelial | *CEP120* | -1.046799707 | 0.001651794 | 0.003335209 | 1 |
| Obesity | endothelial | *MFF* | -0.981293684 | 0.000451294 | 0.001119657 | 1 |
| Obesity | endothelial | *LGALS9* | 0.41363737 | 0.030239248 | 0.043599243 | 1 |
| Obesity | endothelial | *CPLX1* | 0.322118942 | 1.64E-05 | 9.46E-05 | 0.079570978 |
| Obesity | endothelial | *SIGLEC7* | -0.724414757 | 0.000537411 | 0.001291442 | 1 |
| Obesity | endothelial | *MFSD7* | -2.051448074 | 0.012624409 | 0.01985531 | 1 |
| Obesity | endothelial | *MAP2K1* | -0.988908534 | 0.001113061 | 0.00239069 | 1 |
| Obesity | endothelial | *KLK7* | -1.606969822 | 3.68E-05 | 0.0001659 | 0.178716932 |
| Obesity | endothelial | *HSPBAP1* | -1.093752067 | 2.64E-05 | 0.000129952 | 0.127872402 |
| Obesity | endothelial | *AFAP1L2* | 0.212593711 | 0.00011661 | 0.000382898 | 0.565794029 |
| Obesity | endothelial | *UBE2V2* | -1.217531238 | 6.16E-05 | 0.000242716 | 0.29902666 |
| Obesity | endothelial | *PCSK9* | -3.219270834 | 1.06E-05 | 6.87E-05 | 0.051218405 |
| Obesity | endothelial | *XPO6* | -1.253884952 | 2.94E-05 | 0.000140456 | 0.142703582 |
| Obesity | endothelial | *RAB24* | -3.072323406 | 6.43E-06 | 4.91E-05 | 0.031203153 |
| Obesity | endothelial | *THBS3* | -1.164263761 | 8.66E-05 | 0.000309343 | 0.420396822 |
| Obesity | endothelial | *NPIPB3* | -1.809807681 | 1.29E-05 | 7.92E-05 | 0.062827307 |
| Obesity | endothelial | *SH3TC2* | 0.323991225 | 0.002261874 | 0.004379335 | 1 |
| Obesity | endothelial | *P2RY12* | -0.82508854 | 2.23E-05 | 0.000116387 | 0.108240123 |
| Obesity | endothelial | *SNUPN* | -0.764944515 | 0.00338896 | 0.006223783 | 1 |
| Obesity | endothelial | *SLC38A11* | -0.801888758 | 0.000541313 | 0.001298294 | 1 |
| Obesity | endothelial | *CCDC8* | -1.053749 | 7.28E-05 | 0.000274289 | 0.353283859 |
| Obesity | endothelial | *ZEB2* | -0.754273608 | 9.40E-08 | 6.08E-06 | 0.000455965 |
| Obesity | endothelial | *DTWD2* | -0.935449939 | 7.56E-06 | 5.48E-05 | 0.036700834 |
| Obesity | endothelial | *INO80E* | -1.871966035 | 9.09E-17 | 6.13E-14 | 4.41E-13 |
| Obesity | endothelial | *DFFB* | -1.54175272 | 0.000567545 | 0.001351191 | 1 |
| Obesity | endothelial | *NFU1* | -0.998332701 | 0.0002873 | 0.000779195 | 1 |
| Obesity | endothelial | *C15orf40* | -0.744296295 | 0.004001195 | 0.007195626 | 1 |
| Obesity | endothelial | *PROKR1* | 0.776398606 | 0.007848499 | 0.01296151 | 1 |

| | | | | | | |
|---|---|---|---|---|---|---|
| Obesity | endothelial | *RGPD8* | -0.237394393 | 0.00446333 | 0.007909451 | 1 |
| Obesity | endothelial | *LRRC45* | -0.69640765 | 0.002655025 | 0.005026212 | 1 |
| Obesity | endothelial | *UGP2* | -0.743209776 | 2.94E-05 | 0.000140456 | 0.142655466 |
| Obesity | endothelial | *HNRNPF* | -0.863089373 | 0.004095406 | 0.007343277 | 1 |
| Obesity | endothelial | *CSGALNACT2* | -1.077929227 | 0.00104398 | 0.002257304 | 1 |
| Obesity | endothelial | *AVEN* | -0.738618078 | 0.024105773 | 0.035528922 | 1 |
| Obesity | endothelial | *CTNND2* | -1.22950895 | 3.16E-07 | 1.36E-05 | 0.001534425 |
| Obesity | endothelial | *CALML6* | -8.164261101 | 9.66E-07 | 2.45E-05 | 0.0046886 |
| Obesity | endothelial | *MUC3A* | 0.551870378 | 0.01540533 | 0.023706521 | 1 |
| Obesity | endothelial | *TOR1AIP2* | -1.133922461 | 1.40E-05 | 8.38E-05 | 0.067734564 |
| Obesity | endothelial | *ZFPM2* | -0.550253072 | 0.000415453 | 0.001049885 | 1 |
| Obesity | endothelial | *MAP3K2* | -0.6558816 | 0.005748884 | 0.00985291 | 1 |
| Obesity | endothelial | *MYO7B* | 0.389972942 | 1.17E-05 | 7.32E-05 | 0.05653387 |
| Obesity | endothelial | *UBE2E3* | -0.76419219 | 0.00573113 | 0.009825951 | 1 |
| Obesity | endothelial | *NIPA1* | -1.143057822 | 0.00308007 | 0.005717101 | 1 |
| Obesity | endothelial | *HNRNPA3* | -0.86563353 | 0.001381441 | 0.002860756 | 1 |
| Obesity | endothelial | *RNF150* | -0.75806537 | 7.54E-07 | 2.24E-05 | 0.003657263 |
| Obesity | endothelial | *HOXD4* | 1.041091272 | 0.000416755 | 0.001052625 | 1 |
| Obesity | endothelial | *CHRNB1* | -0.308995381 | 0.011479446 | 0.01823781 | 1 |
| Obesity | endothelial | *USP38* | -1.738608851 | 1.39E-06 | 2.57E-05 | 0.006723934 |
| Obesity | endothelial | *ANKK1* | -1.273868262 | 7.64E-05 | 0.000283942 | 0.370814368 |
| Obesity | endothelial | *PDCD6IP* | -0.796127769 | 0.002457065 | 0.004699125 | 1 |
| Obesity | endothelial | *CRTAP* | -1.259505597 | 1.97E-05 | 0.000106783 | 0.095420915 |
| Obesity | endothelial | *CNGB3* | -0.576150038 | 0.020070939 | 0.030075415 | 1 |
| Obesity | endothelial | *ELP5* | -0.88228096 | 0.010626427 | 0.017061357 | 1 |
| Obesity | endothelial | *STX8* | -0.789272174 | 0.001496026 | 0.003067928 | 1 |
| Obesity | endothelial | *NFRKB* | -1.395416219 | 1.66E-06 | 2.63E-05 | 0.008031754 |
| Obesity | endothelial | *SEMA3E* | 0.232683992 | 0.000110523 | 0.000367553 | 0.536259231 |
| Obesity | endothelial | *ADORA2B* | -0.803074328 | 3.70E-09 | 4.98E-07 | 1.79E-05 |
| Obesity | endothelial | *MGMT* | -0.728726337 | 0.004042527 | 0.007261881 | 1 |
| Obesity | endothelial | *METTL7B* | -1.372264976 | 0.001413625 | 0.00292117 | 1 |
| Obesity | endothelial | *DNAJC18* | 0.311780073 | 0.001094063 | 0.002353011 | 1 |
| Obesity | endothelial | *HSD17B13* | 0.680809542 | 0.000255625 | 0.000710361 | 1 |
| Obesity | endothelial | *ELOVL6* | -1.144835796 | 2.96E-06 | 3.38E-05 | 0.014378371 |
| Obesity | endothelial | *ARL6IP1* | 0.410028768 | 0.016796722 | 0.025612098 | 1 |
| Obesity | endothelial | *SERPINB9* | -1.149303062 | 4.18E-05 | 0.000180904 | 0.202792975 |
| Obesity | endothelial | *CDH2* | 0.899659594 | 0.008483992 | 0.013869383 | 1 |
| Obesity | endothelial | *EMB* | -0.614581126 | 9.87E-05 | 0.000337151 | 0.478754795 |
| Obesity | endothelial | *STAT2* | -1.159157537 | 2.39E-05 | 0.000122187 | 0.116077686 |
| Obesity | endothelial | *SLC26A5* | -0.9016389 | 0.011356578 | 0.018072192 | 1 |
| Obesity | endothelial | *ARMC10* | -0.872891378 | 0.002329677 | 0.004487333 | 1 |
| Obesity | endothelial | *ACYP2* | -0.719509946 | 0.000228742 | 0.000651985 | 1 |
| Obesity | endothelial | *TRABD* | -1.254577554 | 0.000457737 | 0.001130248 | 1 |
| Obesity | endothelial | *TMEM133* | -0.881687708 | 0.001899871 | 0.003768672 | 1 |
| Obesity | endothelial | *HOXB9* | -1.447563614 | 4.49E-05 | 0.000190137 | 0.218087279 |
| Obesity | endothelial | *TTLL6* | -5.547058492 | 1.01E-16 | 6.13E-14 | 4.90E-13 |
| Obesity | endothelial | *KCNS3* | -0.469381974 | 0.008142716 | 0.013379092 | 1 |
| Obesity | endothelial | *FOXN2* | -1.065308233 | 9.23E-05 | 0.000322309 | 0.447741914 |
| Obesity | endothelial | *TSEN34* | -1.48749661 | 0.00540354 | 0.00932029 | 1 |
| Obesity | endothelial | *NDUFA3* | -0.966939093 | 0.027904896 | 0.040585899 | 1 |

| | | | | | | |
|---|---|---|---|---|---|---|
| Obesity | endothelial | *PAQR8* | -0.726600802 | 0.021858666 | 0.032503294 | 1 |
| Obesity | endothelial | *NUDT6* | -1.06468432 | 0.000189476 | 0.000563666 | 0.919339724 |
| Obesity | endothelial | *ZNF160* | -0.664647469 | 0.002683638 | 0.005068513 | 1 |
| Obesity | endothelial | *ZNF415* | -1.565823654 | 1.00E-04 | 0.00033981 | 0.485152395 |
| Obesity | endothelial | *S1PR1* | 0.218501035 | 0.000118409 | 0.000387667 | 0.574522457 |
| Obesity | endothelial | *PYGO1* | -0.847936928 | 5.17E-06 | 4.37E-05 | 0.025099045 |
| Obesity | endothelial | *FPR1* | -1.210565783 | 2.49E-06 | 3.13E-05 | 0.012105135 |
| Obesity | endothelial | *FEZ2* | -0.728977028 | 0.003009329 | 0.005594354 | 1 |
| Obesity | endothelial | *SOX7* | 0.223162317 | 8.07E-05 | 0.000294229 | 0.39132411 |
| Obesity | endothelial | *MFN1* | -0.772664101 | 0.003413175 | 0.006265882 | 1 |
| Obesity | endothelial | *GIMAP8* | 0.238378919 | 0.000237426 | 0.000669762 | 1 |
| Obesity | endothelial | *ATP6V0E2* | -1.167958044 | 2.31E-05 | 0.000119755 | 0.111970893 |
| Obesity | endothelial | *MORN4* | -0.356101735 | 0.01834184 | 0.027732816 | 1 |
| Obesity | endothelial | *SOSTDC1* | -0.744745137 | 0.012497587 | 0.019687757 | 1 |
| Obesity | endothelial | *PGAM1* | -0.805853748 | 0.000459377 | 0.001133145 | 1 |
| Obesity | endothelial | *HOPX* | -1.040019077 | 0.003728888 | 0.006761048 | 1 |
| Obesity | endothelial | *PPID* | -1.332697271 | 0.00069615 | 0.001608438 | 1 |
| Obesity | endothelial | *ETFDH* | -0.398962575 | 0.000275691 | 0.000753313 | 1 |
| Obesity | endothelial | *TBCA* | -0.936150969 | 0.000450564 | 0.001118802 | 1 |
| Obesity | endothelial | *NMUR1* | -1.569197604 | 2.74E-06 | 3.28E-05 | 0.013301238 |
| Obesity | endothelial | *PIK3CD* | -0.840895499 | 8.03E-05 | 0.000293596 | 0.389775457 |
| Obesity | endothelial | *ANO5* | -1.223072381 | 3.04E-05 | 0.000144335 | 0.147365896 |
| Obesity | endothelial | *VAT1L* | 0.29335094 | 0.000264629 | 0.000729469 | 1 |
| Obesity | endothelial | *LRRC34* | -0.494724573 | 0.000140933 | 0.00044407 | 0.683808288 |
| Obesity | endothelial | *RHNO1* | -0.926312722 | 0.034766758 | 0.049320098 | 1 |
| Obesity | endothelial | *PWWP2B* | 0.69959892 | 0.029051389 | 0.042089382 | 1 |
| Obesity | endothelial | *EXOSC10* | -1.247264705 | 0.000186532 | 0.000557642 | 0.905053469 |
| Obesity | endothelial | *TRAPPC12* | -1.097959739 | 4.43E-05 | 0.000187818 | 0.214741852 |
| Obesity | endothelial | *C3AR1* | -0.663117527 | 9.48E-05 | 0.000327502 | 0.460139799 |
| Obesity | endothelial | *PRNP* | -0.945626918 | 0.000226457 | 0.000647097 | 1 |
| Obesity | endothelial | *TVP23B* | -0.998851277 | 0.000549876 | 0.001314932 | 1 |
| Obesity | endothelial | *FBXW10* | -1.84680826 | 1.30E-06 | 2.57E-05 | 0.006323554 |
| Obesity | endothelial | *SRGAP2C* | -0.650857725 | 0.001227436 | 0.002598395 | 1 |
| Obesity | endothelial | *ATPAF2* | -1.493279187 | 1.53E-05 | 8.95E-05 | 0.074405115 |
| Obesity | endothelial | *ZNF554* | -2.230756618 | 6.97E-07 | 2.20E-05 | 0.003380495 |
| Obesity | endothelial | *ANKRD20A4* | -3.968192365 | 0.003787688 | 0.006859971 | 1 |
| Obesity | endothelial | *SERF1A* | 3.627687906 | 0.006296598 | 0.010659838 | 1 |
| Obesity | endothelial | *EIF2AK3* | -1.059503659 | 0.000947383 | 0.002086564 | 1 |
| Obesity | endothelial | *MTBP* | -1.173293602 | 0.00054717 | 0.001309753 | 1 |
| Obesity | endothelial | *TEFM* | -1.05749264 | 0.000147847 | 0.000463109 | 0.717355299 |
| Obesity | endothelial | *MBOAT1* | -1.21031644 | 1.87E-05 | 0.000103149 | 0.090667639 |
| Obesity | endothelial | *CXCR6* | -1.41780414 | 4.76E-06 | 4.17E-05 | 0.023081913 |
| Obesity | endothelial | *PAIP1* | -1.203427544 | 0.000857472 | 0.001912853 | 1 |
| Obesity | endothelial | *C1QTNF4* | -1.186714587 | 4.17E-19 | 4.05E-16 | 2.03E-15 |
| Obesity | endothelial | *MACROD2* | -0.548063568 | 0.000379532 | 0.000976918 | 1 |
| Obesity | endothelial | *BSG* | -1.045349825 | 0.000941562 | 0.002075629 | 1 |
| Obesity | endothelial | *HINFP* | -1.309454253 | 0.00091957 | 0.002034543 | 1 |
| Obesity | endothelial | *SUCLG2* | -0.887633103 | 0.000341757 | 0.000897782 | 1 |
| Obesity | endothelial | *MANEA* | -1.563272672 | 1.16E-05 | 7.28E-05 | 0.056132122 |
| Obesity | endothelial | *FIBP* | 0.992729687 | 0.006848743 | 0.011466563 | 1 |

| | | | | | | |
|---|---|---|---|---|---|---|
| Obesity | endothelial | *CTSW* | -0.353509904 | 8.85E-13 | 3.07E-10 | 4.29E-09 |
| Obesity | endothelial | *SNTG2* | 0.220020711 | 0.008408367 | 0.013764304 | 1 |
| Obesity | endothelial | *RAD9A* | -0.401333622 | 0.034623 | 0.049177633 | 1 |
| Obesity | endothelial | *ZFAND4* | -1.432720931 | 2.12E-06 | 2.89E-05 | 0.01030234 |
| Obesity | endothelial | *FAM71D* | -1.051926996 | 0.000784052 | 0.001770227 | 1 |
| Obesity | endothelial | *FUT10* | -0.791061004 | 3.03E-06 | 3.39E-05 | 0.014700172 |
| Obesity | endothelial | *ZNF596* | -1.535096236 | 6.45E-06 | 4.91E-05 | 0.031290142 |
| Obesity | endothelial | *FAM192A* | -1.042477108 | 5.80E-05 | 0.000231733 | 0.281556083 |
| Obesity | endothelial | *CBWD1* | -1.703139337 | 8.05E-06 | 5.71E-05 | 0.039047727 |
| Obesity | endothelial | *DCP2* | -0.784831341 | 0.009182856 | 0.014911385 | 1 |
| Obesity | endothelial | *SNX32* | 0.349484735 | 0.01673261 | 0.025530385 | 1 |
| Obesity | endothelial | *ZNF621* | -1.037670147 | 0.006138826 | 0.01043278 | 1 |
| Obesity | endothelial | *MYEOV* | -1.778222658 | 0.006257629 | 0.010612378 | 1 |
| Obesity | endothelial | *ANKRD13D* | -0.938958296 | 0.004075674 | 0.007313302 | 1 |
| Obesity | endothelial | *MRGPRF* | -0.500491045 | 0.011612293 | 0.01840668 | 1 |
| Obesity | endothelial | *GXYLT2* | -0.784797163 | 4.98E-06 | 4.29E-05 | 0.024166224 |
| Obesity | endothelial | *DCAKD* | -1.1511723 | 5.40E-07 | 1.90E-05 | 0.002621897 |
| Obesity | endothelial | *NOC3L* | -1.176789052 | 8.58E-05 | 0.00030779 | 0.416131782 |
| Obesity | endothelial | *PARP15* | -0.324971391 | 0.023939041 | 0.035304628 | 1 |
| Obesity | endothelial | *AHSA2* | -0.74231355 | 0.000677449 | 0.001573471 | 1 |
| Obesity | endothelial | *IQCB1* | -1.177334003 | 3.76E-06 | 3.75E-05 | 0.018253396 |
| Obesity | endothelial | *ZNF483* | -0.822905122 | 3.96E-05 | 0.000174064 | 0.19199304 |
| Obesity | endothelial | *SLC2A14* | -0.483562746 | 0.004715813 | 0.008296274 | 1 |
| Obesity | endothelial | *MMRN2* | 0.242386183 | 9.90E-05 | 0.000337757 | 0.480306336 |
| Obesity | endothelial | *MZT2A* | -0.621162088 | 0.023860591 | 0.035199632 | 1 |
| Obesity | endothelial | *NDNF* | 0.283237464 | 7.41E-05 | 0.000277913 | 0.359481526 |
| Obesity | endothelial | *ARV1* | -0.668245598 | 0.027189092 | 0.039627959 | 1 |
| Obesity | endothelial | *CCDC36* | -0.700070943 | 0.000765344 | 0.001738506 | 1 |
| Obesity | endothelial | *PPP1R14B* | -2.445826359 | 1.66E-06 | 2.63E-05 | 0.008067996 |
| Obesity | endothelial | *ZNF417* | -1.401709627 | 8.19E-05 | 0.000298 | 0.39753231 |
| Obesity | endothelial | *PEAK1* | -0.554078599 | 0.016490287 | 0.025200274 | 1 |
| Obesity | endothelial | *TNFRSF10D* | 0.40711979 | 0.000562308 | 0.001340697 | 1 |
| Obesity | endothelial | *MST1* | -0.606629055 | 9.08E-07 | 2.41E-05 | 0.004406721 |
| Obesity | endothelial | *GMPPB* | -1.58309029 | 7.41E-05 | 0.000277913 | 0.359619952 |
| Obesity | endothelial | *SULT1B1* | 0.562174391 | 0.000446419 | 0.00111189 | 1 |
| Obesity | endothelial | *NUDT4* | -1.299962447 | 3.17E-05 | 0.00014947 | 0.153654978 |
| Obesity | endothelial | *SCAI* | -0.87594182 | 0.000103953 | 0.000349779 | 0.504380818 |
| Obesity | endothelial | *HSPB7* | -0.626171031 | 0.001807927 | 0.003605452 | 1 |
| Obesity | endothelial | *UQCRH* | -1.115606461 | 6.37E-05 | 0.000248717 | 0.309155316 |
| Obesity | endothelial | *C11orf80* | -0.947956131 | 0.000746365 | 0.001703369 | 1 |
| Obesity | endothelial | *CD7* | -1.503065733 | 9.30E-05 | 0.000323239 | 0.451037861 |
| Obesity | endothelial | *RNF213* | -0.764948173 | 0.001262506 | 0.002658715 | 1 |
| Obesity | endothelial | *GPR160* | 0.253864019 | 0.000176897 | 0.000532778 | 0.858305353 |
| Obesity | endothelial | *XXYLT1* | -1.265025008 | 8.66E-05 | 0.000309343 | 0.419984996 |
| Obesity | endothelial | *UBXN2A* | -0.921909777 | 0.000165392 | 0.000504707 | 0.802483695 |
| Obesity | endothelial | *CTSF* | -0.744711556 | 0.005459654 | 0.009390898 | 1 |
| Obesity | endothelial | *LEMD3* | -0.991354882 | 0.001849569 | 0.003683954 | 1 |
| Obesity | endothelial | *CTU2* | 0.561101402 | 0.030246301 | 0.043599243 | 1 |
| Obesity | endothelial | *GLIS1* | -0.985004366 | 1.44E-06 | 2.57E-05 | 0.006986431 |
| Obesity | endothelial | *RALGAPA1* | -0.525266952 | 0.018998746 | 0.028645716 | 1 |

| | | | | | | |
|---|---|---|---|---|---|---|
| Obesity | endothelial | *SNAPC5* | -0.982859974 | 0.015916817 | 0.02436227 | 1 |
| Obesity | endothelial | *ANKRD36C* | -0.690065376 | 0.025317352 | 0.03711172 | 1 |
| Obesity | endothelial | *MSL2* | -1.155013676 | 9.71E-05 | 0.000333322 | 0.470984494 |
| Obesity | endothelial | *ZNF497* | -2.82798701 | 5.67E-06 | 4.54E-05 | 0.02752412 |
| Obesity | endothelial | *ANGEL2* | -0.95471488 | 0.002497156 | 0.00476453 | 1 |
| Obesity | endothelial | *KIAA1551* | -0.479021458 | 0.034766268 | 0.049320098 | 1 |
| Obesity | endothelial | *LARP7* | -1.207834454 | 2.40E-05 | 0.000122203 | 0.116337267 |
| Obesity | endothelial | *C4orf32* | 0.298321972 | 0.000915611 | 0.002026708 | 1 |
| Obesity | endothelial | *PCP2* | -0.82395123 | 0.004874323 | 0.008534286 | 1 |
| Obesity | endothelial | *NLRP6* | -4.302166332 | 0.000686572 | 0.001591614 | 1 |
| Obesity | endothelial | *RSRC1* | -0.969206355 | 7.44E-05 | 0.000278226 | 0.360859185 |
| Obesity | endothelial | *PTDSS2* | -0.767588707 | 0.003232462 | 0.005965731 | 1 |
| Obesity | endothelial | *DHX36* | -0.97808756 | 0.000287671 | 0.000779766 | 1 |
| Obesity | endothelial | *FBXW8* | -0.79519416 | 0.010321593 | 0.01662143 | 1 |
| Obesity | endothelial | *SLC22A1* | -1.839379111 | 1.43E-06 | 2.57E-05 | 0.006915416 |
| Obesity | endothelial | *CHST2* | 1.299525227 | 0.003910065 | 0.007055275 | 1 |
| Obesity | endothelial | *PACS1* | -1.017987865 | 1.01E-05 | 6.68E-05 | 0.049225336 |
| Obesity | endothelial | *SH3BP5L* | -2.041337004 | 7.96E-06 | 5.67E-05 | 0.038639403 |
| Obesity | endothelial | *CADM2* | 0.149900841 | 9.65E-06 | 6.48E-05 | 0.046805384 |
| Obesity | endothelial | *PSMD2* | -1.088102251 | 0.006567951 | 0.011072863 | 1 |
| Obesity | endothelial | *ARHGAP1* | -0.951017173 | 8.62E-05 | 0.000308712 | 0.418304717 |
| Obesity | endothelial | *ATG13* | -1.292023174 | 5.62E-06 | 4.54E-05 | 0.027268 |
| Obesity | endothelial | *CCNE2* | -0.755492199 | 0.002733647 | 0.005145674 | 1 |
| Obesity | endothelial | *ZNF519* | -1.133685597 | 6.40E-07 | 2.14E-05 | 0.003107473 |
| Obesity | endothelial | *BANF1* | -1.039225985 | 0.023361176 | 0.034515355 | 1 |
| Obesity | endothelial | *CHRNA7* | 0.577933773 | 0.002135155 | 0.004172563 | 1 |
| Obesity | endothelial | *SCUBE2* | 0.385368696 | 0.001368691 | 0.0028392 | 1 |
| Obesity | endothelial | *EIF1AD* | -1.397176659 | 0.000629946 | 0.001475143 | 1 |
| Obesity | endothelial | *EIF3F* | -0.756336492 | 0.021949016 | 0.032607662 | 1 |
| Obesity | endothelial | *ZNF25* | -1.388525703 | 6.05E-06 | 4.70E-05 | 0.029366248 |
| Obesity | endothelial | *CLTB* | -1.401596256 | 6.48E-06 | 4.92E-05 | 0.031436691 |
| Obesity | endothelial | *PPP2R2D* | -0.740802176 | 0.004858406 | 0.008513177 | 1 |
| Obesity | endothelial | *MCTP1* | 0.232843853 | 0.001372595 | 0.002845187 | 1 |
| Obesity | endothelial | *LRRC25* | -0.762191063 | 0.000730944 | 0.001672608 | 1 |
| Obesity | endothelial | *UCP2* | -0.881018134 | 1.95E-05 | 0.000106014 | 0.094564215 |
| Obesity | endothelial | *PAAF1* | -0.275668996 | 0.018922639 | 0.028548708 | 1 |
| Obesity | endothelial | *MRPL48* | -1.465564811 | 1.35E-05 | 8.20E-05 | 0.065697603 |
| Obesity | endothelial | *RAB6A* | -0.649122726 | 0.014119296 | 0.021929201 | 1 |
| Obesity | endothelial | *TMEM70* | -1.263935086 | 0.002874527 | 0.005389182 | 1 |
| Obesity | endothelial | *TOM1L2* | -0.873831259 | 0.000163882 | 0.000502217 | 0.79515755 |
| Obesity | endothelial | *TEX26* | -1.525751391 | 5.50E-05 | 0.00022274 | 0.266842048 |
| Obesity | endothelial | *GPR156* | -0.965158682 | 2.07E-06 | 2.88E-05 | 0.010023171 |
| Obesity | endothelial | *TTLL11* | -1.029634526 | 0.001315304 | 0.002744885 | 1 |
| Obesity | endothelial | *PRIMA1* | -1.089498715 | 1.14E-06 | 2.57E-05 | 0.005520734 |
| Obesity | endothelial | *HOXD8* | 0.264465588 | 9.24E-05 | 0.000322309 | 0.448088404 |
| Obesity | endothelial | *ZDHHC21* | -0.696349986 | 0.006624221 | 0.011152227 | 1 |
| Obesity | endothelial | *A2M* | 0.279941846 | 0.009045572 | 0.014713078 | 1 |
| Obesity | endothelial | *DOK7* | 2.885079551 | 0.000578639 | 0.001374904 | 1 |
| Obesity | endothelial | *JAKMIP2* | -1.320890197 | 4.13E-08 | 3.58E-06 | 0.000200587 |
| Obesity | endothelial | *TCP11L1* | -0.688555862 | 0.004247029 | 0.007563749 | 1 |

| | | | | | | |
|---|---|---|---|---|---|---|
| Obesity | endothelial | *CCDC57* | -1.143208081 | 1.10E-08 | 1.27E-06 | 5.34E-05 |
| Obesity | endothelial | *MYPOP* | -1.135822857 | 0.0107294 | 0.017192552 | 1 |
| Obesity | endothelial | *CIDEA* | -0.951879305 | 5.78E-06 | 4.60E-05 | 0.028054393 |
| Obesity | endothelial | *SLC35G1* | -1.651439838 | 0.000159409 | 0.000490889 | 0.773454439 |
| Obesity | endothelial | *ZNF135* | -0.698255417 | 0.000448648 | 0.001114613 | 1 |
| Obesity | endothelial | *OR4N2* | -1.359343673 | 4.21E-05 | 0.000181903 | 0.204109231 |
| Obesity | endothelial | *TAC4* | -1.567937717 | 1.01E-05 | 6.67E-05 | 0.048938085 |
| Obesity | endothelial | *CDC26* | -0.73138937 | 0.000688792 | 0.001595236 | 1 |
| Obesity | endothelial | *CRLF3* | -0.84083385 | 0.000108309 | 0.000361677 | 0.525516464 |
| Obesity | endothelial | *DNAJC30* | -2.05809792 | 2.47E-07 | 1.13E-05 | 0.001200134 |
| Obesity | endothelial | *SPRYD4* | -1.13617481 | 1.78E-06 | 2.68E-05 | 0.008644168 |
| Obesity | endothelial | *WDR25* | -1.284549976 | 0.000264513 | 0.000729469 | 1 |
| Obesity | endothelial | *PLA2G16* | -0.7098277 | 2.39E-05 | 0.000122192 | 0.116204457 |
| Obesity | endothelial | *B3GNT5* | -1.42611903 | 3.36E-05 | 0.000155536 | 0.163156925 |
| Obesity | endothelial | *RNF152* | 0.33708736 | 0.034220159 | 0.048676696 | 1 |
| Obesity | endothelial | *LRRC37A* | -0.461476701 | 0.007608556 | 0.012621098 | 1 |
| Obesity | endothelial | *BDNF* | -1.172137337 | 1.54E-05 | 8.95E-05 | 0.074534815 |
| Obesity | endothelial | *BOK* | -1.406832398 | 5.44E-06 | 4.47E-05 | 0.026374362 |
| Obesity | endothelial | *PFN4* | -0.545623283 | 0.006011796 | 0.010252807 | 1 |
| Obesity | endothelial | *RUFY1* | -0.482346045 | 0.030012565 | 0.043352475 | 1 |
| Obesity | endothelial | *VSIG10* | -0.629624862 | 0.010982077 | 0.017551066 | 1 |
| Obesity | endothelial | *ANKLE2* | -0.984052699 | 0.002466377 | 0.004715076 | 1 |
| Obesity | endothelial | *C8G* | -18.70917013 | 1.62E-06 | 2.61E-05 | 0.007843285 |
| Obesity | endothelial | *MUC20* | -1.433405719 | 2.65E-06 | 3.24E-05 | 0.012861148 |
| Obesity | endothelial | *LY6H* | 1.38339458 | 0.000560753 | 0.001337646 | 1 |
| Obesity | endothelial | *SHMT1* | -1.13064301 | 6.46E-06 | 4.91E-05 | 0.031331175 |
| Obesity | endothelial | *DPP7* | -0.866650921 | 0.015522807 | 0.023834386 | 1 |
| Obesity | endothelial | *MTHFR* | -0.9723682 | 0.005044412 | 0.008782018 | 1 |
| Obesity | endothelial | *TMEM80* | -1.376462593 | 6.58E-08 | 4.43E-06 | 0.000319102 |
| Obesity | endothelial | *WDR73* | -1.069084282 | 0.000239939 | 0.00067489 | 1 |
| Obesity | endothelial | *DSCAML1* | -1.016286251 | 2.15E-06 | 2.91E-05 | 0.010423526 |
| Obesity | endothelial | *TALDO1* | -1.392690338 | 4.82E-05 | 0.000200919 | 0.233668308 |
| Obesity | endothelial | *MAN1B1* | -1.561279881 | 8.75E-06 | 6.02E-05 | 0.042431877 |
| Obesity | endothelial | *NAP1L5* | -1.854348616 | 0.013649485 | 0.021274431 | 1 |
| Obesity | endothelial | *RBM44* | 0.476654784 | 9.78E-05 | 0.000334569 | 0.474418069 |
| Obesity | endothelial | *C18orf32* | -0.838882047 | 0.000828255 | 0.001854497 | 1 |
| Obesity | endothelial | *ACAD9* | -0.349458496 | 0.03291826 | 0.046976293 | 1 |
| Obesity | endothelial | *IL17RA* | -1.111944001 | 1.67E-05 | 9.56E-05 | 0.080988148 |
| Obesity | endothelial | *MBOAT4* | 0.997696506 | 0.026540106 | 0.038798613 | 1 |
| Obesity | endothelial | *AGTRAP* | -0.844056995 | 0.015481501 | 0.023793552 | 1 |
| Obesity | endothelial | *NAALADL2* | -0.768995429 | 1.98E-05 | 0.000107241 | 0.096164109 |
| Obesity | endothelial | *FAM87B* | -12.89697625 | 2.74E-06 | 3.28E-05 | 0.013300836 |
| Obesity | endothelial | *PCDHB9* | -1.253277845 | 0.000152496 | 0.000473929 | 0.739909275 |
| Obesity | endothelial | *ASB8* | -1.40932756 | 1.76E-05 | 9.89E-05 | 0.085288405 |
| Obesity | endothelial | *ADAMTSL1* | -0.86683699 | 3.11E-05 | 0.000147451 | 0.150989981 |
| Obesity | endothelial | *GRAMD1C* | 0.269029225 | 0.0322441 | 0.046163581 | 1 |
| Obesity | endothelial | *DALRD3* | -1.458353616 | 9.15E-06 | 6.21E-05 | 0.044388695 |
| Obesity | endothelial | *ZNF114* | 0.544390222 | 0.000602266 | 0.001421302 | 1 |
| Obesity | endothelial | *SPINK6* | -2.127921134 | 3.17E-06 | 3.42E-05 | 0.015389521 |
| Obesity | endothelial | *SH2B1* | -1.079532947 | 2.36E-06 | 3.05E-05 | 0.011472739 |

| | | | | | | |
|---|---|---|---|---|---|---|
| Obesity | endothelial | *VN1R1* | -1.901865258 | 0.002411153 | 0.004627735 | 1 |
| Obesity | endothelial | *KDELC2* | -1.70203879 | 8.56E-05 | 0.000307442 | 0.415353983 |
| Obesity | endothelial | *SH2D4B* | 0.984579681 | 0.000272864 | 0.000747564 | 1 |
| Obesity | endothelial | *RNF212* | -0.783027732 | 6.87E-06 | 5.15E-05 | 0.033324213 |
| Obesity | endothelial | *PRSS36* | -1.366347131 | 1.49E-06 | 2.57E-05 | 0.007209864 |
| Obesity | endothelial | *TMPRSS9* | -0.848458653 | 1.51E-05 | 8.92E-05 | 0.073434482 |
| Obesity | endothelial | *TMEM11* | -1.68678291 | 0.000258692 | 0.000717651 | 1 |
| Obesity | endothelial | *FAM220A* | -2.224374718 | 0.000260985 | 0.000722775 | 1 |
| Obesity | endothelial | *NT5DC1* | -0.875899599 | 0.001267927 | 0.002666504 | 1 |
| Obesity | endothelial | *GLDC* | -1.94495202 | 0.000160196 | 0.000492878 | 0.777268997 |
| Obesity | endothelial | *LINC00314* | 0.186316414 | 0.000400721 | 0.001020628 | 1 |
| Obesity | endothelial | *P4HTM* | -1.511254999 | 9.40E-07 | 2.44E-05 | 0.004561373 |
| Obesity | endothelial | *CA8* | 0.20092632 | 0.000107718 | 0.000360197 | 0.522645969 |
| Obesity | endothelial | *ERN1* | -1.255941535 | 7.09E-05 | 0.000268979 | 0.344024393 |
| Obesity | endothelial | *PARP10* | -1.616020725 | 3.40E-06 | 3.56E-05 | 0.016509121 |
| Obesity | endothelial | *NSUN3* | -0.812586725 | 0.011310748 | 0.018005514 | 1 |
| Obesity | endothelial | *RPP25* | -0.812996214 | 9.02E-05 | 0.000317134 | 0.437645293 |
| Obesity | endothelial | *TRIM73* | -3.770637013 | 1.01E-09 | 1.69E-07 | 4.90E-06 |
| Obesity | endothelial | *EFCAB13* | -0.732269803 | 0.00282335 | 0.00530143 | 1 |
| Obesity | endothelial | *EXOSC4* | -2.547335131 | 0.000296631 | 0.000796929 | 1 |
| Obesity | endothelial | *ZNF852* | -2.073880948 | 6.28E-10 | 1.13E-07 | 3.05E-06 |
| Obesity | endothelial | *HYI* | -0.688962575 | 0.000429499 | 0.001078079 | 1 |
| Obesity | endothelial | *GAK* | -0.950942871 | 0.003800425 | 0.006880471 | 1 |
| Obesity | endothelial | *TUFM* | 0.706761148 | 0.020973142 | 0.031349872 | 1 |
| Obesity | endothelial | *CCDC89* | -2.33760334 | 3.89E-06 | 3.78E-05 | 0.018896642 |
| Obesity | endothelial | *HTR1F* | -2.391015735 | 2.76E-06 | 3.28E-05 | 0.013369335 |
| Obesity | endothelial | *SPTY2D1* | -1.494363163 | 7.85E-06 | 5.62E-05 | 0.038095568 |
| Obesity | endothelial | *ZNF664* | -1.523825613 | 1.84E-06 | 2.71E-05 | 0.008935637 |
| Obesity | endothelial | *CDH4* | -0.887823961 | 0.000123159 | 0.000398643 | 0.597566499 |
| Obesity | endothelial | *NSUN7* | -0.558814116 | 0.001322616 | 0.002757771 | 1 |
| Obesity | endothelial | *PACS2* | -0.785937245 | 0.017325656 | 0.026344118 | 1 |
| Obesity | endothelial | *LINC00174* | -0.719401198 | 8.38E-05 | 0.000302203 | 0.406463474 |
| Obesity | endothelial | *GEMIN4* | -1.096462324 | 0.001649331 | 0.003333009 | 1 |
| Obesity | endothelial | *RPRML* | 4.305057799 | 7.98E-09 | 9.68E-07 | 3.87E-05 |
| Obesity | endothelial | *AKAP5* | -1.498133931 | 3.56E-06 | 3.64E-05 | 0.01728352 |
| Obesity | endothelial | *GIPC3* | 1.132164633 | 0.000140945 | 0.00044407 | 0.683867375 |
| Obesity | endothelial | *PDXDC1* | -0.966143548 | 2.87E-05 | 0.000138082 | 0.139217246 |
| Obesity | endothelial | *SEPHS2* | -1.46099528 | 0.000954327 | 0.002096149 | 1 |
| Obesity | endothelial | *SSC5D* | -0.469069908 | 0.000200005 | 0.000588277 | 0.970424606 |
| Obesity | endothelial | *SEPT1* | -1.473327627 | 8.97E-07 | 2.40E-05 | 0.004352134 |
| Obesity | endothelial | *EXOC3* | -1.295596701 | 1.65E-05 | 9.48E-05 | 0.080033369 |
| Obesity | endothelial | *CSNK1A1L* | -2.990903813 | 0.034774125 | 0.049320098 | 1 |
| Obesity | endothelial | *HCLS1* | -0.568435299 | 0.009764186 | 0.015776167 | 1 |
| Obesity | endothelial | *MTURN* | -1.136125382 | 5.49E-05 | 0.000222687 | 0.266333693 |
| Obesity | endothelial | *CCDC66* | -0.936178998 | 0.000266893 | 0.000734382 | 1 |
| Obesity | endothelial | *DEFB124* | -2.775428645 | 3.77E-05 | 0.000168162 | 0.182960348 |
| Obesity | endothelial | *GLIPR1L2* | -0.873425106 | 0.010856091 | 0.017378341 | 1 |
| Obesity | endothelial | *DEFB119* | -6.965614121 | 4.37E-05 | 0.000185733 | 0.211921595 |
| Obesity | endothelial | *MB21D2* | -0.984202161 | 0.001004371 | 0.002183265 | 1 |
| Obesity | endothelial | *SLC47A2* | -0.84681986 | 1.88E-05 | 0.000103504 | 0.091083775 |

| | | | | | | |
|---|---|---|---|---|---|---|
| Obesity | endothelial | *CHST13* | -2.669218735 | 0.000216165 | 0.000623935 | 1 |
| Obesity | endothelial | *SLC36A4* | -0.837326913 | 0.01288304 | 0.020223594 | 1 |
| Obesity | endothelial | *OR51E1* | 0.222168129 | 0.000119629 | 0.000390341 | 0.580440223 |
| Obesity | endothelial | *PSMG4* | -1.028073156 | 0.000331192 | 0.000873339 | 1 |
| Obesity | endothelial | *BHLHE22* | -0.533914315 | 0.023120334 | 0.034190753 | 1 |
| Obesity | endothelial | *ZNF443* | 1.158773116 | 0.002485146 | 0.004747217 | 1 |
| Obesity | endothelial | *CAPS2* | -0.95546538 | 0.00049499 | 0.001205668 | 1 |
| Obesity | endothelial | *KCTD2* | -1.177302356 | 0.000418655 | 0.001055776 | 1 |
| Obesity | endothelial | *D2HGDH* | -0.900170609 | 1.63E-05 | 9.43E-05 | 0.079196307 |
| Obesity | endothelial | *CMTR2* | -0.880937304 | 0.005105165 | 0.00887187 | 1 |
| Obesity | endothelial | *MRPL14* | -1.025086708 | 0.004935651 | 0.008629831 | 1 |
| Obesity | endothelial | *LSMEM1* | -0.424604073 | 0.017335039 | 0.026350128 | 1 |
| Obesity | endothelial | *NQO1* | -1.475216263 | 3.25E-07 | 1.37E-05 | 0.001575817 |
| Obesity | endothelial | *F2R* | 0.303564345 | 0.000451778 | 0.001119657 | 1 |
| Obesity | endothelial | *MUC16* | -1.078890608 | 4.46E-06 | 4.03E-05 | 0.021655267 |
| Obesity | endothelial | *PENK* | -1.466805565 | 0.000771582 | 0.001748064 | 1 |
| Obesity | endothelial | *SLC25A41* | -2.338150815 | 1.38E-05 | 8.29E-05 | 0.066715937 |
| Obesity | endothelial | *ZNF322* | -1.610124819 | 2.49E-07 | 1.13E-05 | 0.001208255 |
| Obesity | endothelial | *TMEM45A* | -0.762001384 | 2.88E-06 | 3.33E-05 | 0.013988445 |
| Obesity | endothelial | *SGSH* | -1.958170703 | 2.30E-06 | 3.02E-05 | 0.011171304 |
| Obesity | endothelial | *TMEM252* | 3.24227319 | 6.66E-05 | 0.000256283 | 0.322916735 |
| Obesity | endothelial | *RFX7* | -0.820119482 | 0.002403899 | 0.004615639 | 1 |
| Obesity | endothelial | *CLDN7* | -1.048810575 | 1.89E-08 | 1.90E-06 | 9.18E-05 |
| Obesity | endothelial | *ZNF329* | -0.895864898 | 0.018267004 | 0.027636889 | 1 |
| Obesity | endothelial | *MGAT4C* | -0.365076689 | 0.004647908 | 0.00818869 | 1 |
| Obesity | endothelial | *IDH2* | -1.54242592 | 7.37E-06 | 5.42E-05 | 0.035771798 |
| Obesity | endothelial | *FAM181B* | 1.526697298 | 9.36E-05 | 0.000323876 | 0.4540745 |
| Obesity | endothelial | *TDRKH* | -1.200569062 | 7.95E-05 | 0.000292438 | 0.385725169 |
| Obesity | endothelial | *ZNF708* | -0.882629901 | 0.000153305 | 0.000475295 | 0.743836001 |
| Obesity | endothelial | *ERCC6L2* | -0.868353433 | 0.001829563 | 0.003647099 | 1 |
| Obesity | endothelial | *MRPL41* | -1.741673179 | 2.03E-05 | 0.000109213 | 0.098619297 |
| Obesity | endothelial | *HHIPL1* | -1.202567112 | 0.000628197 | 0.001471759 | 1 |
| Obesity | endothelial | *FAM153B* | -0.852177676 | 2.61E-06 | 3.21E-05 | 0.012674698 |
| Obesity | endothelial | *SYNM* | -1.876398291 | 7.66E-05 | 0.000284313 | 0.37159769 |
| Obesity | endothelial | *KCNA4* | -0.76331813 | 3.74E-06 | 3.75E-05 | 0.018140329 |
| Obesity | endothelial | *FIGN* | -0.682218101 | 1.55E-06 | 2.57E-05 | 0.007516541 |
| Obesity | endothelial | *IZUMO1* | 0.455097495 | 4.23E-05 | 0.000182569 | 0.205390056 |
| Obesity | endothelial | *FBXL6* | -1.389941976 | 0.015802007 | 0.024209453 | 1 |
| Obesity | endothelial | *NXPH4* | -5.605860929 | 4.68E-06 | 4.14E-05 | 0.022718816 |
| Obesity | endothelial | *NPLOC4* | -0.934467287 | 3.71E-05 | 0.000166374 | 0.179943944 |
| Obesity | endothelial | *CAPN12* | 0.318639218 | 0.03471456 | 0.049278831 | 1 |
| Obesity | endothelial | *ORAI1* | 0.531185621 | 8.23E-05 | 0.00029873 | 0.399103815 |
| Obesity | endothelial | *GLRX5* | -1.273936936 | 0.00777189 | 0.012852492 | 1 |
| Obesity | endothelial | *CAV3* | -1.171303693 | 2.02E-07 | 9.59E-06 | 0.000978347 |
| Obesity | endothelial | *ADI1* | -1.403746266 | 1.09E-05 | 6.98E-05 | 0.052732426 |
| Obesity | endothelial | *CLEC4G* | -2.709289886 | 8.54E-05 | 0.00030695 | 0.414382707 |
| Obesity | endothelial | *TRAK1* | -0.784403152 | 0.001151748 | 0.002457467 | 1 |
| Obesity | endothelial | *SKA2* | -1.253329662 | 1.68E-05 | 9.60E-05 | 0.081391457 |
| Obesity | endothelial | *TTC3* | -0.340737938 | 0.014604493 | 0.022610402 | 1 |
| Obesity | endothelial | *PPP1R27* | -5.015048149 | 5.82E-09 | 7.44E-07 | 2.83E-05 |

| | | | | | | |
|---|---|---|---|---|---|---|
| Obesity | endothelial | *BRICD5* | -2.371175169 | 0.026137313 | 0.03825588 | 1 |
| Obesity | endothelial | *GRID1* | -1.368234657 | 4.81E-10 | 8.97E-08 | 2.33E-06 |
| Obesity | endothelial | *ACBD3* | -0.950927521 | 0.001067552 | 0.002302117 | 1 |
| Obesity | endothelial | *PLCXD3* | 0.276528635 | 0.000853786 | 0.001906383 | 1 |
| Obesity | endothelial | *GPIHBP1* | 0.173190768 | 0.000136968 | 0.000435497 | 0.664567815 |
| Obesity | endothelial | *ALG12* | -1.594870271 | 3.65E-06 | 3.69E-05 | 0.017691813 |
| Obesity | endothelial | *ZNF721* | -0.709978489 | 0.005004569 | 0.008722043 | 1 |
| Obesity | endothelial | *CEP63* | -0.884855287 | 2.68E-05 | 0.000131605 | 0.129963281 |
| Obesity | endothelial | *ODF3L1* | -2.032435557 | 0.00012083 | 0.000392859 | 0.58626799 |
| Obesity | endothelial | *HMGN4* | -0.996649138 | 6.07E-05 | 0.000239576 | 0.294438724 |
| Obesity | endothelial | *C12orf60* | -1.28717366 | 1.50E-06 | 2.57E-05 | 0.007257013 |
| Obesity | endothelial | *ABAT* | -1.20342814 | 2.98E-06 | 3.38E-05 | 0.014434755 |
| Obesity | endothelial | *WBP2NL* | -1.305300252 | 7.45E-06 | 5.45E-05 | 0.036130844 |
| Obesity | endothelial | *BEGAIN* | -2.083387146 | 1.39E-06 | 2.57E-05 | 0.00676755 |
| Obesity | endothelial | *CSMD1* | 0.441632309 | 0.002412712 | 0.004628896 | 1 |
| Obesity | endothelial | *C2CD4C* | 0.252262923 | 0.000131976 | 0.000422394 | 0.640348726 |
| Obesity | endothelial | *RUVBL2* | -1.703393218 | 6.99E-05 | 0.000266568 | 0.339074624 |
| Obesity | endothelial | *ZNF623* | -0.896723004 | 0.001528099 | 0.003120847 | 1 |
| Obesity | endothelial | *CCDC125* | -1.015719834 | 4.14E-05 | 0.000179918 | 0.200682775 |
| Obesity | endothelial | *FHL3* | -0.503934444 | 0.014036216 | 0.021814132 | 1 |
| Obesity | endothelial | *LRIT3* | 3.959389353 | 0.024761687 | 0.036374117 | 1 |
| Obesity | endothelial | *GPR132* | -0.713803992 | 3.15E-06 | 3.41E-05 | 0.015292087 |
| Obesity | endothelial | *SERHL2* | -1.727195093 | 7.54E-06 | 5.48E-05 | 0.03659183 |
| Obesity | endothelial | *PCBP3* | -0.330553914 | 0.00449301 | 0.007950432 | 1 |
| Obesity | endothelial | *SFXN4* | -1.085777646 | 0.001021071 | 0.002211713 | 1 |
| Obesity | endothelial | *MRPL54* | -0.755806572 | 0.000997301 | 0.002170886 | 1 |
| Obesity | endothelial | *HMCES* | -1.299698541 | 1.16E-06 | 2.57E-05 | 0.00564005 |
| Obesity | endothelial | *NDUFB1* | -0.983755623 | 0.001584478 | 0.003224784 | 1 |
| Obesity | endothelial | *FAM19A1* | -1.159221397 | 5.29E-07 | 1.89E-05 | 0.002565627 |
| Obesity | endothelial | *TRMT12* | -1.637135458 | 7.32E-07 | 2.23E-05 | 0.003552369 |
| Obesity | endothelial | *MRGPRX2* | -2.537876975 | 0.005699416 | 0.009775032 | 1 |
| Obesity | endothelial | *UPP1* | 0.629344716 | 0.000122625 | 0.000397343 | 0.594977865 |
| Obesity | endothelial | *FIGLA* | 0.457763176 | 6.74E-05 | 0.00025864 | 0.326920479 |
| Obesity | endothelial | *ACSM2A* | -3.259417864 | 5.41E-06 | 4.47E-05 | 0.02625336 |
| Obesity | endothelial | *KREMEN1* | -0.380044575 | 0.024527429 | 0.036084623 | 1 |
| Obesity | endothelial | *FAM162B* | -0.861554375 | 1.89E-05 | 0.000104093 | 0.091748156 |
| Obesity | endothelial | *BTBD9* | -0.826947407 | 0.000138569 | 0.000439434 | 0.672334485 |
| Obesity | endothelial | *FAM3B* | -0.195594164 | 0.009969086 | 0.016096507 | 1 |
| Obesity | endothelial | *TTC32* | -1.654639823 | 1.26E-05 | 7.75E-05 | 0.061113973 |
| Obesity | endothelial | *PP2D1* | -1.908667328 | 0.000126284 | 0.000407402 | 0.6127322 |
| Obesity | endothelial | *DENND5A* | -0.770387707 | 0.007931436 | 0.013080668 | 1 |
| Obesity | endothelial | *NIPSNAP1* | -1.126279248 | 0.000975922 | 0.002132001 | 1 |
| Obesity | endothelial | *CNTN2* | -0.849155655 | 0.00523058 | 0.009057565 | 1 |
| Obesity | endothelial | *GOLGA6L4* | -1.005682701 | 9.28E-05 | 0.000323239 | 0.45041551 |
| Obesity | endothelial | *PGP* | -1.020803174 | 0.002686381 | 0.005069747 | 1 |
| Obesity | endothelial | *OAF* | -0.67865918 | 7.38E-07 | 2.24E-05 | 0.003581873 |
| Obesity | endothelial | *ALDH1A3* | -0.534418133 | 0.002298815 | 0.004440228 | 1 |
| Obesity | endothelial | *MROH7* | -0.452432614 | 0.028332218 | 0.041179329 | 1 |
| Obesity | endothelial | *PKP3* | -8.54542038 | 3.46E-06 | 3.57E-05 | 0.016807904 |
| Obesity | endothelial | *A3GALT2* | 0.553766793 | 0.001926866 | 0.003817253 | 1 |

| | | | | | | |
|---|---|---|---|---|---|---|
| Obesity | endothelial | *SS18L1* | -1.186862398 | 0.000389354 | 0.000997964 | 1 |
| Obesity | endothelial | *TOP1MT* | -0.768090908 | 1.15E-05 | 7.27E-05 | 0.056008597 |
| Obesity | endothelial | *COPB2* | -1.121544221 | 0.000162432 | 0.000499126 | 0.788120099 |
| Obesity | endothelial | *WDR27* | -0.668962339 | 0.00580757 | 0.009949976 | 1 |
| Obesity | endothelial | *TXNRD2* | -1.211365106 | 6.36E-05 | 0.000248482 | 0.308614447 |
| Obesity | endothelial | *TMEM255B* | 0.34875364 | 0.000368445 | 0.000955477 | 1 |
| Obesity | endothelial | *HDDC3* | -1.316504621 | 0.000295168 | 0.000794836 | 1 |
| Obesity | endothelial | *DUSP8* | 0.851819997 | 0.015099363 | 0.023309612 | 1 |
| Obesity | endothelial | *SNN* | -1.820274741 | 0.014398617 | 0.022320157 | 1 |
| Obesity | endothelial | *KRBA2* | -1.803425928 | 3.45E-05 | 0.000157436 | 0.167354323 |
| Obesity | endothelial | *SEPT5* | -1.236635834 | 0.000770789 | 0.001747602 | 1 |
| Obesity | endothelial | *RNLS* | -1.055085187 | 4.61E-07 | 1.72E-05 | 0.002236522 |
| Obesity | endothelial | *NDUFA12* | -0.749512438 | 0.032512063 | 0.046478647 | 1 |
| Obesity | endothelial | *TCTE3* | -1.511629169 | 1.20E-06 | 2.57E-05 | 0.005838137 |
| Obesity | endothelial | *UBE2G2* | -0.869719804 | 0.000726652 | 0.001666218 | 1 |
| Obesity | endothelial | *DRD1* | -7.978599128 | 7.81E-06 | 5.60E-05 | 0.037885779 |
| Obesity | endothelial | *BTBD6* | -0.940297098 | 0.003194685 | 0.005904995 | 1 |
| Obesity | endothelial | *NUTM2A* | -1.283914803 | 3.29E-06 | 3.49E-05 | 0.015961517 |
| Obesity | endothelial | *WT1* | -0.850670376 | 7.18E-07 | 2.22E-05 | 0.003481624 |
| Obesity | endothelial | *ZFP90* | -1.036369229 | 0.002627243 | 0.004985288 | 1 |
| Obesity | endothelial | *FAM227A* | -0.499388165 | 0.000729286 | 0.001671466 | 1 |
| Obesity | endothelial | *SORCS2* | -1.320050242 | 4.27E-07 | 1.64E-05 | 0.00207146 |
| Obesity | endothelial | *TMEM106A* | -0.711986665 | 0.000752247 | 0.001713568 | 1 |
| Obesity | endothelial | *BRI3BP* | -1.500915249 | 2.09E-06 | 2.88E-05 | 0.010135383 |
| Obesity | endothelial | *IFNE* | -1.878775428 | 0.015500952 | 0.023808363 | 1 |
| Obesity | endothelial | *UBOX5* | -1.636604371 | 2.34E-06 | 3.04E-05 | 0.011333297 |
| Obesity | endothelial | *MAFF* | -1.140224141 | 1.03E-07 | 6.42E-06 | 0.000500569 |
| Obesity | endothelial | *SEMA4B* | -0.994996011 | 1.60E-06 | 2.59E-05 | 0.007763761 |
| Obesity | endothelial | *CIB1* | -0.500365976 | 0.029943732 | 0.043265928 | 1 |
| Obesity | endothelial | *SGCZ* | -0.49041997 | 0.000771713 | 0.001748064 | 1 |
| Obesity | endothelial | *C6orf120* | -1.327370361 | 0.002150283 | 0.004195083 | 1 |
| Obesity | endothelial | *SIGIRR* | 0.612879476 | 0.004993929 | 0.008709756 | 1 |
| Obesity | endothelial | *ATP6V0A2* | -1.507637468 | 3.02E-05 | 0.000143738 | 0.146468819 |
| Obesity | endothelial | *TARSL2* | -0.791694452 | 0.001257681 | 0.002652007 | 1 |
| Obesity | endothelial | *SH3BGR* | -0.471259919 | 0.000120886 | 0.000392859 | 0.58653889 |
| Obesity | endothelial | *SV2B* | 0.208703293 | 0.000260892 | 0.000722775 | 1 |
| Obesity | endothelial | *SP1* | -1.061945912 | 5.64E-05 | 0.000225969 | 0.273422283 |
| Obesity | endothelial | *PSMD13* | -1.266065558 | 0.000249285 | 0.000695132 | 1 |
| Obesity | endothelial | *MYBL1* | -1.793181082 | 2.40E-08 | 2.33E-06 | 0.000116358 |
| Obesity | endothelial | *ADARB2* | 0.347292333 | 5.57E-05 | 0.000224676 | 0.27028552 |
| Obesity | endothelial | *DMWD* | -0.924512076 | 0.013552599 | 0.021143798 | 1 |
| Obesity | endothelial | *SLC52A2* | -0.870146853 | 0.025209225 | 0.036975562 | 1 |
| Obesity | endothelial | *ARL17A* | -0.770301558 | 0.000828029 | 0.001854497 | 1 |
| Obesity | endothelial | *GNB1L* | -0.783928271 | 0.00011929 | 0.000390022 | 0.578793067 |
| Obesity | endothelial | *ZNF829* | -0.680699181 | 0.026245941 | 0.038391711 | 1 |
| Obesity | endothelial | *IFITM1* | 0.813045006 | 0.000150519 | 0.000469657 | 0.730316359 |
| Obesity | endothelial | *LINC00839* | 1.561103077 | 0.003447125 | 0.006318643 | 1 |
| Obesity | endothelial | *RTN4RL1* | -0.668836976 | 0.021884949 | 0.032522442 | 1 |
| Obesity | endothelial | *RNPC3* | -0.740528643 | 8.92E-05 | 0.000315285 | 0.432570475 |
| Obesity | endothelial | *RASA3* | -0.762249307 | 1.98E-05 | 0.000107241 | 0.096218199 |

| | | | | | | |
|---|---|---|---|---|---|---|
| Obesity | endothelial | *ZNF529* | -1.082436955 | 0.000954927 | 0.002096519 | 1 |
| Obesity | endothelial | *WWOX* | -0.700697614 | 0.000163924 | 0.000502217 | 0.795357738 |
| Obesity | endothelial | *CYP4Z1* | -0.285968698 | 0.01380976 | 0.021495328 | 1 |
| Obesity | endothelial | *POLR1D* | -0.774683958 | 0.011625447 | 0.018421511 | 1 |
| Obesity | endothelial | *FFAR4* | -1.278217247 | 7.10E-07 | 2.21E-05 | 0.003443636 |
| Obesity | endothelial | *TOR3A* | -1.779031242 | 6.00E-05 | 0.000237844 | 0.29135917 |
| Obesity | endothelial | *ZNF555* | -1.055799879 | 0.00821597 | 0.013490316 | 1 |
| Obesity | endothelial | *GOLGA8R* | -1.380316086 | 0.003617583 | 0.006594409 | 1 |
| Obesity | endothelial | *CCDC30* | -0.583393583 | 0.004564889 | 0.008062919 | 1 |
| Obesity | endothelial | *ZNF197* | -1.083022475 | 0.000204249 | 0.000597356 | 0.991013974 |
| Obesity | endothelial | *GNG2* | -0.923275543 | 2.84E-06 | 3.32E-05 | 0.01376616 |
| Obesity | endothelial | *BTN3A2* | -0.513814668 | 4.67E-08 | 3.84E-06 | 0.000226777 |
| Obesity | endothelial | *TMEM222* | -1.412823681 | 0.001348213 | 0.002801512 | 1 |
| Obesity | endothelial | *FAM86B1* | -1.238431541 | 0.00065986 | 0.001537033 | 1 |
| Obesity | endothelial | *MIR22HG* | -1.012232516 | 2.84E-06 | 3.32E-05 | 0.013794258 |
| Obesity | endothelial | *CYP27C1* | -0.633684471 | 0.001943878 | 0.003840268 | 1 |
| Obesity | endothelial | *ZNF732* | 0.946617739 | 0.014726834 | 0.022770745 | 1 |
| Obesity | endothelial | *VSIG10L* | 1.113380189 | 0.000772193 | 0.001748335 | 1 |
| Obesity | endothelial | *TRABD2A* | 0.509336077 | 0.00098424 | 0.002146306 | 1 |
| Obesity | endothelial | *QRFPR* | -2.021394837 | 0.000485071 | 0.001186272 | 1 |
| Obesity | endothelial | *MAPT* | -1.134431358 | 1.46E-06 | 2.57E-05 | 0.007073299 |
| Obesity | endothelial | *TMEM17* | -0.717270048 | 0.003888983 | 0.00702584 | 1 |
| Obesity | endothelial | *TNFRSF18* | 1.378188987 | 0.001278431 | 0.002681988 | 1 |
| Obesity | endothelial | *C1QL4* | -4.090501398 | 9.31E-06 | 6.29E-05 | 0.045179876 |
| Obesity | endothelial | *TMEM232* | -0.703682281 | 0.000892931 | 0.001983745 | 1 |
| Obesity | endothelial | *EFCAB6* | -0.955637927 | 1.41E-06 | 2.57E-05 | 0.006840328 |
| Obesity | endothelial | *TMPRSS6* | -8.559838881 | 8.21E-07 | 2.32E-05 | 0.003985555 |
| Obesity | endothelial | *C3orf70* | -0.641049322 | 0.011939452 | 0.018882079 | 1 |
| Obesity | endothelial | *ENTPD5* | -1.105142525 | 1.74E-05 | 9.82E-05 | 0.084332597 |
| Obesity | endothelial | *ANGPTL5* | -0.827549678 | 9.22E-07 | 2.42E-05 | 0.004471645 |
| Obesity | endothelial | *WDR86* | -0.671176466 | 3.42E-06 | 3.56E-05 | 0.016614134 |
| Obesity | endothelial | *EPOR* | -0.655551352 | 0.000126497 | 0.000407817 | 0.613765318 |
| Obesity | endothelial | *LUZP2* | 0.695492506 | 0.005414809 | 0.009333091 | 1 |
| Obesity | endothelial | *GJA4* | 0.399531241 | 0.000211022 | 0.000612735 | 1 |
| Obesity | endothelial | *SBK2* | -3.133517603 | 1.07E-05 | 6.93E-05 | 0.051993641 |
| Obesity | endothelial | *SAMD11* | -0.993547016 | 0.000458419 | 0.001131358 | 1 |
| Obesity | endothelial | *C2orf88* | -1.11342958 | 2.43E-06 | 3.10E-05 | 0.011806042 |
| Obesity | endothelial | *KBTBD12* | -0.615802952 | 0.000288899 | 0.000782656 | 1 |
| Obesity | endothelial | *FANCA* | -0.72423429 | 0.000384575 | 0.000988325 | 1 |
| Obesity | endothelial | *DNAH17* | -1.204019387 | 0.000653055 | 0.001522644 | 1 |
| Obesity | endothelial | *FANCM* | -1.300341629 | 7.25E-06 | 5.39E-05 | 0.035200351 |
| Obesity | endothelial | *ZFP69* | -1.850283824 | 2.61E-06 | 3.21E-05 | 0.01268411 |
| Obesity | endothelial | *TMEM220* | -0.82488425 | 8.89E-05 | 0.000315162 | 0.431505823 |
| Obesity | endothelial | *FAM122A* | -2.275770956 | 6.23E-05 | 0.000244467 | 0.302160802 |
| Obesity | endothelial | *GFRAL* | -1.978341051 | 0.006031415 | 0.010282652 | 1 |
| Obesity | endothelial | *LDLRAD2* | 0.430592415 | 0.011567998 | 0.018348456 | 1 |
| Obesity | endothelial | *KLHL17* | -0.546132219 | 0.022338965 | 0.033136245 | 1 |
| Obesity | endothelial | *MESP2* | -0.915526028 | 4.31E-05 | 0.000185209 | 0.209301669 |
| Obesity | endothelial | *MAPK12* | 0.487223193 | 0.000296405 | 0.000796764 | 1 |
| Obesity | endothelial | *AGRN* | 0.572887225 | 0.001508606 | 0.003091113 | 1 |

| | | | | | | |
|---|---|---|---|---|---|---|
| Obesity | endothelial | *FAM166A* | -1.688794533 | 0.003161549 | 0.005848203 | 1 |
| Obesity | endothelial | *ZNF626* | -1.443866783 | 0.002266945 | 0.004387402 | 1 |
| Obesity | endothelial | *LINC00265* | -1.259143214 | 1.67E-05 | 9.56E-05 | 0.080916413 |
| Obesity | endothelial | *NCR3LG1* | -3.050789525 | 1.09E-05 | 7.00E-05 | 0.05303316 |
| Obesity | endothelial | *RUFY4* | -1.162649772 | 4.49E-06 | 4.04E-05 | 0.021802306 |
| Obesity | endothelial | *ZNF669* | 0.646674792 | 0.002208874 | 0.004292133 | 1 |
| Obesity | endothelial | *PLSCR1* | 0.353693061 | 0.032257708 | 0.046169439 | 1 |
| Obesity | endothelial | *C3orf62* | -1.012558195 | 9.97E-05 | 0.000339076 | 0.483522759 |
| Obesity | endothelial | *ENO4* | -0.77834752 | 5.57E-06 | 4.51E-05 | 0.02702075 |
| Obesity | endothelial | *JAKMIP3* | -0.61115973 | 2.29E-06 | 3.01E-05 | 0.011093236 |
| Obesity | endothelial | *PDCD1* | -1.992891524 | 0.010640785 | 0.017073112 | 1 |
| Obesity | endothelial | *C19orf54* | -1.451273904 | 1.43E-05 | 8.54E-05 | 0.069486847 |
| Obesity | endothelial | *C22orf34* | 0.320908595 | 0.00089985 | 0.001998203 | 1 |
| Obesity | endothelial | *COL25A1* | -1.057547909 | 0.000200406 | 0.000588277 | 0.972370842 |
| Obesity | endothelial | *SRSF10* | -0.852539826 | 0.00045182 | 0.001119657 | 1 |
| Obesity | endothelial | *NBR1* | -1.02772666 | 0.000370925 | 0.000960366 | 1 |
| Obesity | endothelial | *NDOR1* | -1.924120309 | 2.61E-05 | 0.000128957 | 0.126506746 |
| Obesity | endothelial | *FAM72B* | -1.193902272 | 0.026145551 | 0.038256397 | 1 |
| Obesity | endothelial | *RHCE* | -1.903313419 | 0.000793057 | 0.001788064 | 1 |
| Obesity | endothelial | *IDO2* | 1.165681187 | 0.032690216 | 0.04669206 | 1 |
| Obesity | endothelial | *SLC4A5* | -3.152557767 | 3.40E-05 | 0.000156653 | 0.165167479 |
| Obesity | endothelial | *UROS* | -1.412384917 | 4.17E-05 | 0.000180822 | 0.202520493 |
| Obesity | endothelial | *ZBED6CL* | -1.357797569 | 0.001952113 | 0.003853399 | 1 |
| Obesity | endothelial | *FSIP2* | -0.946345076 | 0.000502795 | 0.001220504 | 1 |
| Obesity | endothelial | *NOXA1* | -1.461406604 | 1.30E-06 | 2.57E-05 | 0.006325429 |
| Obesity | endothelial | *FZD9* | -5.732450749 | 7.70E-07 | 2.24E-05 | 0.003737995 |
| Obesity | endothelial | *MTF1* | -1.077829412 | 0.005137268 | 0.008914887 | 1 |
| Obesity | endothelial | *ENTPD8* | -37.50812293 | 5.50E-06 | 4.47E-05 | 0.026707005 |
| Obesity | endothelial | *FBF1* | -0.52207523 | 0.019052952 | 0.028709603 | 1 |
| Obesity | endothelial | *ASTL* | -3.89381008 | 1.27E-06 | 2.57E-05 | 0.006179924 |
| Obesity | endothelial | *MSL1* | -0.877355925 | 0.000466395 | 0.001145798 | 1 |
| Obesity | endothelial | *LRRK2* | -0.734558726 | 0.002444668 | 0.004679104 | 1 |
| Obesity | endothelial | *UTS2B* | -1.004557989 | 3.66E-06 | 3.70E-05 | 0.017743607 |
| Obesity | endothelial | *C9orf152* | -1.834379155 | 0.001687881 | 0.003399583 | 1 |
| Obesity | endothelial | *NELFB* | -1.505058622 | 0.000570309 | 0.001357107 | 1 |
| Obesity | endothelial | *SLC15A5* | -0.931297235 | 0.001629813 | 0.003297686 | 1 |
| Obesity | endothelial | *ADAT2* | -1.537183063 | 8.62E-07 | 2.40E-05 | 0.004180408 |
| Obesity | endothelial | *LITAF* | -1.146478022 | 4.35E-05 | 0.000185308 | 0.210871908 |
| Obesity | endothelial | *SF3B3* | -1.059615995 | 0.000441404 | 0.001101897 | 1 |
| Obesity | endothelial | *ZNF573* | -0.924323408 | 0.003299356 | 0.006073018 | 1 |
| Obesity | endothelial | *ZNF600* | -0.841697194 | 7.56E-05 | 0.000281669 | 0.366732397 |
| Obesity | endothelial | *TSPYL1* | -1.342749727 | 0.004845233 | 0.008493161 | 1 |
| Obesity | endothelial | *PNRC2* | -1.522226184 | 4.15E-05 | 0.000179935 | 0.201167644 |
| Obesity | endothelial | *FHIT* | -0.764258923 | 1.65E-05 | 9.48E-05 | 0.080087632 |
| Obesity | endothelial | *ZKSCAN3* | -1.768928325 | 1.45E-10 | 3.21E-08 | 7.06E-07 |
| Obesity | endothelial | *RRP7A* | -1.372754688 | 0.000499972 | 0.001215972 | 1 |
| Obesity | endothelial | *KIAA0408* | -0.576839613 | 2.62E-05 | 0.000129626 | 0.127293013 |
| Obesity | endothelial | *BLOC1S2* | -0.890131216 | 3.11E-06 | 3.39E-05 | 0.015104368 |
| Obesity | endothelial | *COLCA1* | -0.783447947 | 0.001879248 | 0.003732342 | 1 |
| Obesity | endothelial | *SEMA4A* | -1.037860237 | 0.001465982 | 0.003016516 | 1 |

| | | | | | | |
|---|---|---|---|---|---|---|
| Obesity | endothelial | *HRCT1* | 0.303803657 | 0.029184325 | 0.042244136 | 1 |
| Obesity | endothelial | *MPHOSPH8* | -0.748077611 | 0.001267784 | 0.002666504 | 1 |
| Obesity | endothelial | *SIRPB2* | -0.460810085 | 2.57E-05 | 0.000127898 | 0.124868667 |
| Obesity | endothelial | *SRGAP3* | -0.595744019 | 0.000484221 | 0.001185321 | 1 |
| Obesity | endothelial | *XPNPEP3* | -0.54265462 | 0.00040065 | 0.001020628 | 1 |
| Obesity | endothelial | *PPIA* | -0.81617219 | 0.010475716 | 0.016841674 | 1 |
| Obesity | endothelial | *ZNF471* | -1.423455714 | 9.08E-05 | 0.00031897 | 0.440497119 |
| Obesity | endothelial | *ZNF493* | -1.291577538 | 4.14E-05 | 0.000179918 | 0.200838989 |
| Obesity | endothelial | *GTF2IRD2* | -1.248151275 | 7.67E-06 | 5.53E-05 | 0.037235957 |
| Obesity | endothelial | *SUPT3H* | -0.709518366 | 0.008114172 | 0.013341228 | 1 |
| Obesity | endothelial | *GIMAP5* | 0.268278665 | 0.000195141 | 0.000578035 | 0.946822145 |
| Obesity | endothelial | *STK31* | -1.358826226 | 0.001374883 | 0.002848391 | 1 |
| Obesity | endothelial | *ZKSCAN7* | -0.753221425 | 0.000261835 | 0.000724064 | 1 |
| Obesity | endothelial | *CD55* | -0.791618746 | 0.006721821 | 0.011293032 | 1 |
| Obesity | endothelial | *CPNE4* | 0.823392305 | 1.69E-05 | 9.64E-05 | 0.082097797 |
| Obesity | endothelial | *ZNF565* | -0.640986125 | 0.006693551 | 0.01125721 | 1 |
| Obesity | endothelial | *ZNF34* | -1.196693205 | 0.020873942 | 0.031211207 | 1 |
| Obesity | endothelial | *EVL* | -0.750349413 | 0.003354872 | 0.006170522 | 1 |
| Obesity | endothelial | *ZNF124* | -1.31111215 | 1.54E-05 | 8.95E-05 | 0.074607596 |
| Obesity | endothelial | *PIK3R4* | -1.263746405 | 0.000133663 | 0.000426912 | 0.648531445 |
| Obesity | endothelial | *RFX8* | -0.527875941 | 0.001313448 | 0.002742189 | 1 |
| Obesity | endothelial | *C20orf96* | -1.763841242 | 7.31E-06 | 5.39E-05 | 0.035456371 |
| Obesity | endothelial | *SULT1A1* | -1.139561723 | 3.03E-08 | 2.83E-06 | 0.000146982 |
| Obesity | endothelial | *SPTSSB* | -2.218072252 | 4.78E-05 | 0.000199922 | 0.232109422 |
| Obesity | endothelial | *CACNA1H* | -1.462102734 | 0.000444363 | 0.00110737 | 1 |
| Obesity | endothelial | *XRCC2* | -1.420418235 | 3.42E-06 | 3.56E-05 | 0.016602771 |
| Obesity | endothelial | *ZNF846* | -1.153888661 | 1.59E-06 | 2.58E-05 | 0.007709734 |
| Obesity | endothelial | *ADH1B* | -0.67204914 | 2.83E-06 | 3.32E-05 | 0.013750928 |
| Obesity | endothelial | *AMZ2* | -1.002624488 | 0.002975409 | 0.00554407 | 1 |
| Obesity | endothelial | *ZNF431* | -0.787022904 | 0.006484119 | 0.010950555 | 1 |
| Obesity | endothelial | *COL27A1* | -0.509406333 | 1.33E-05 | 8.07E-05 | 0.064459749 |
| Obesity | endothelial | *ZNF239* | -0.773601698 | 0.000277639 | 0.000757653 | 1 |
| Obesity | endothelial | *SPRR2B* | -3.822462477 | 0.015136284 | 0.023359176 | 1 |
| Obesity | endothelial | *FAM3C* | -0.809874935 | 0.004963091 | 0.008665318 | 1 |
| Obesity | endothelial | *SLC39A10* | -0.869114576 | 0.00494392 | 0.008641174 | 1 |
| Obesity | endothelial | *AP2A1* | -0.76926445 | 0.007854204 | 0.012966519 | 1 |
| Obesity | endothelial | *HIST1H3E* | 0.445260894 | 0.015151193 | 0.023374749 | 1 |
| Obesity | endothelial | *WDR5B* | -1.305392712 | 0.002233056 | 0.004335649 | 1 |
| Obesity | endothelial | *ZNF138* | -0.831610512 | 0.002734037 | 0.005145674 | 1 |
| Obesity | endothelial | *ZNF429* | -1.179394415 | 2.24E-05 | 0.000116731 | 0.108676723 |
| Obesity | endothelial | *ZNF100* | -1.016342407 | 5.42E-05 | 0.000220004 | 0.262905261 |
| Obesity | endothelial | *ANXA6* | -1.111389521 | 6.30E-06 | 4.84E-05 | 0.030569702 |
| Obesity | endothelial | *GMFB* | -1.10032226 | 1.41E-06 | 2.57E-05 | 0.006837005 |
| Obesity | endothelial | *ZNF420* | -0.794575012 | 0.019562003 | 0.029373763 | 1 |
| Obesity | endothelial | *ZMYM1* | -1.089062251 | 0.001007084 | 0.002187276 | 1 |
| Obesity | endothelial | *IGF2R* | -0.888639244 | 0.001433268 | 0.002950453 | 1 |
| Obesity | endothelial | *GAL3ST4* | -1.563635846 | 4.55E-06 | 4.07E-05 | 0.022085635 |
| Obesity | endothelial | *ADAM32* | -1.146169153 | 0.010220226 | 0.016480072 | 1 |
| Obesity | endothelial | *ABCB8* | -1.126419527 | 8.19E-06 | 5.79E-05 | 0.039725834 |
| Obesity | endothelial | *SND1* | -0.852115099 | 0.00019862 | 0.000585373 | 0.963705889 |

| | | | | | | |
|---|---|---|---|---|---|---|
| Obesity | endothelial | *SULT1A2* | 0.607842776 | 0.015323691 | 0.023595859 | 1 |
| Obesity | endothelial | *NEK5* | -0.730248893 | 3.23E-05 | 0.000151401 | 0.156851441 |
| Obesity | endothelial | *TBC1D9B* | -0.794382617 | 0.003676835 | 0.006684153 | 1 |
| Obesity | endothelial | *FAM110D* | 0.192632917 | 5.54E-05 | 0.00022383 | 0.268595712 |
| Obesity | endothelial | *C6orf141* | 0.134275968 | 0.001660688 | 0.003350377 | 1 |
| Obesity | endothelial | *CCL4L2* | -2.695144722 | 0.022056645 | 0.032757527 | 1 |
| Obesity | endothelial | *ZNF165* | -0.809925758 | 9.30E-06 | 6.29E-05 | 0.045141994 |
| Obesity | endothelial | *RAMP2-AS1* | -1.441114594 | 0.000128282 | 0.000412476 | 0.622426531 |
| Obesity | endothelial | *ZNF720* | -1.217935181 | 8.36E-05 | 0.000301906 | 0.405660094 |
| Obesity | endothelial | *MRPL21* | -0.598037677 | 3.79E-05 | 0.00016889 | 0.18408971 |
| Obesity | endothelial | *ZNF98* | -1.069814353 | 1.53E-06 | 2.57E-05 | 0.007411176 |
| Obesity | endothelial | *ZNF517* | -1.257392733 | 0.031902656 | 0.045742224 | 1 |
| Obesity | endothelial | *ZNF675* | -1.360618184 | 0.000391154 | 0.00100099 | 1 |
| Obesity | endothelial | *SLC22A5* | -0.969268761 | 2.39E-05 | 0.000122088 | 0.115861231 |
| Obesity | endothelial | *VEPH1* | -1.243461779 | 7.03E-07 | 2.20E-05 | 0.003412779 |
| Obesity | endothelial | *MAP3K5* | -0.862547284 | 5.58E-05 | 0.00022482 | 0.270907631 |
| Obesity | endothelial | *SLC2A10* | -1.008305289 | 2.85E-05 | 0.000137546 | 0.138371695 |
| Obesity | endothelial | *ZNF665* | -1.539637975 | 9.68E-05 | 0.000332792 | 0.469902837 |
| Obesity | endothelial | *RPF2* | -1.22880694 | 0.000200538 | 0.000588277 | 0.973010879 |
| Obesity | endothelial | *MYO5A* | -0.68233012 | 0.000153917 | 0.000476467 | 0.746802936 |
| Obesity | endothelial | *ATG7* | -0.927322421 | 5.46E-06 | 4.47E-05 | 0.026496848 |
| Obesity | endothelial | *RAB40C* | -1.352362256 | 3.55E-06 | 3.63E-05 | 0.017216682 |
| Obesity | endothelial | *PIGN* | -1.023146801 | 5.69E-06 | 4.54E-05 | 0.027586109 |
| Obesity | endothelial | *HHLA3* | -0.523911301 | 0.015545989 | 0.02386243 | 1 |
| Obesity | endothelial | *TOPORS* | -1.38364369 | 0.000169444 | 0.000514803 | 0.822141007 |
| Obesity | endothelial | *BCO2* | -0.410259168 | 0.000952236 | 0.002093869 | 1 |
| Obesity | endothelial | *ENTPD6* | -1.127789783 | 0.001877027 | 0.003729457 | 1 |
| Obesity | endothelial | *ENPP1* | -0.790125164 | 5.95E-06 | 4.67E-05 | 0.02886356 |
| Obesity | endothelial | *CCDC154* | -0.71909911 | 3.95E-05 | 0.000174004 | 0.191752192 |
| Obesity | endothelial | *MFAP5* | -1.208524903 | 1.18E-08 | 1.31E-06 | 5.72E-05 |
| Obesity | endothelial | *PDCD1LG2* | -0.236519533 | 0.034412569 | 0.048907377 | 1 |
| Obesity | endothelial | *NMB* | -0.959395562 | 0.006079775 | 0.010357819 | 1 |
| Obesity | endothelial | *FAM114A1* | -1.030199313 | 0.000113881 | 0.000376141 | 0.552550533 |
| Obesity | endothelial | *RPS26* | -0.697953532 | 5.83E-05 | 0.000232526 | 0.282983875 |
| Obesity | endothelial | *PSAP* | -0.766579371 | 0.000503943 | 0.001221954 | 1 |
| Obesity | endothelial | *TXNRD3* | -1.010319439 | 8.96E-05 | 0.000316163 | 0.434528212 |
| Obesity | endothelial | *EME2* | -1.702345933 | 1.13E-06 | 2.57E-05 | 0.005481905 |
| Obesity | endothelial | *ZNF780A* | -0.470727585 | 9.45E-07 | 2.44E-05 | 0.004582813 |
| Obesity | endothelial | *ATAD3A* | -0.936401979 | 7.55E-05 | 0.000281483 | 0.366209843 |
| Obesity | endothelial | *AC122129.1* | -2.833442263 | 7.56E-06 | 5.48E-05 | 0.03667632 |
| Obesity | endothelial | *CCDC180* | -0.770193455 | 0.003975047 | 0.007159217 | 1 |
| Obesity | endothelial | *OCLN* | 0.332466299 | 0.000845654 | 0.001890837 | 1 |
| Obesity | endothelial | *SGTB* | -1.162206143 | 0.000588804 | 0.001396323 | 1 |
| Obesity | endothelial | *ADH5* | -1.06905051 | 6.02E-06 | 4.69E-05 | 0.029193299 |
| Obesity | endothelial | *ZNF311* | -0.505048983 | 0.003889408 | 0.00702584 | 1 |
| Obesity | endothelial | *MPZL1* | -0.981729144 | 3.43E-05 | 0.000156959 | 0.166376315 |
| Obesity | endothelial | *VPS13A* | -0.721427664 | 0.007283158 | 0.012139431 | 1 |
| Obesity | endothelial | *NOL8* | -1.507989152 | 1.44E-06 | 2.57E-05 | 0.00700383 |
| Obesity | endothelial | *DLGAP2* | -0.630063738 | 6.81E-05 | 0.000260778 | 0.33040512 |
| Obesity | endothelial | *MRPL42* | -0.999851063 | 1.26E-05 | 7.75E-05 | 0.061282487 |

| | | | | | | |
|---|---|---|---|---|---|---|
| Obesity | endothelial | *ZNF560* | 0.988662416 | 0.000902549 | 0.00200328 | 1 |
| Obesity | endothelial | *AGAP9* | -1.555969206 | 1.36E-05 | 8.20E-05 | 0.065761242 |
| Obesity | endothelial | *ZNF273* | -1.198275152 | 0.000537657 | 0.001291442 | 1 |
| Obesity | endothelial | *ZNF667* | -1.312257329 | 4.06E-06 | 3.85E-05 | 0.019683109 |
| Obesity | endothelial | *SFI1* | -0.958225383 | 0.008050792 | 0.013250489 | 1 |
| Obesity | endothelial | *ZNF649* | -0.660827986 | 0.017427926 | 0.026466447 | 1 |
| Obesity | endothelial | *ADH4* | -0.72352415 | 5.29E-08 | 4.06E-06 | 0.000256709 |
| Obesity | endothelial | *HIBCH* | -0.868378371 | 5.49E-06 | 4.47E-05 | 0.026654974 |
| Obesity | endothelial | *ZNF544* | -1.67984284 | 1.37E-05 | 8.28E-05 | 0.066505782 |
| Obesity | endothelial | *SOWAHC* | -1.509866241 | 0.019924711 | 0.029883987 | 1 |
| Obesity | endothelial | *AC135178.1* | 2.281778749 | 0.000370907 | 0.000960366 | 1 |
| Obesity | endothelial | *MAN1A2* | -0.893533115 | 0.000718538 | 0.00164995 | 1 |
| Obesity | endothelial | *SVIP* | 0.419182737 | 0.028374018 | 0.041206446 | 1 |
| Obesity | endothelial | *DDRGK1* | -1.334603517 | 0.000456537 | 0.001128343 | 1 |
| Obesity | endothelial | *ZNF334* | -1.058660211 | 4.48E-06 | 4.04E-05 | 0.021737656 |
| Obesity | endothelial | *SULT1C2* | -0.321706209 | 0.02784107 | 0.040517358 | 1 |
| Obesity | endothelial | *RPS6KL1* | -1.57844102 | 0.000143443 | 0.00045106 | 0.695986127 |
| Obesity | endothelial | *DDX42* | -1.008004764 | 0.000267612 | 0.000735666 | 1 |
| Obesity | endothelial | *STYX* | -1.164305829 | 0.002267999 | 0.004387402 | 1 |
| Obesity | endothelial | *HELZ* | -0.876097735 | 3.33E-05 | 0.000154241 | 0.161336521 |
| Obesity | endothelial | *UCKL1* | -1.331320787 | 0.00067528 | 0.001569936 | 1 |
| Obesity | endothelial | *GFPT1* | -0.950345825 | 6.19E-06 | 4.77E-05 | 0.030018148 |
| Obesity | endothelial | *ZNF568* | -0.939001459 | 0.004608563 | 0.00813118 | 1 |
| Obesity | endothelial | *ZNF587* | -1.166982898 | 2.74E-05 | 0.000133636 | 0.133020978 |
| Obesity | endothelial | *SH3BGRL2* | 0.343682751 | 3.91E-05 | 0.000172796 | 0.189729911 |
| Obesity | endothelial | *TMA16* | -1.249566972 | 9.27E-05 | 0.000323015 | 0.449637091 |
| Obesity | endothelial | *CNGA1* | -0.392829157 | 0.016323221 | 0.02496069 | 1 |
| Obesity | endothelial | *ZNF28* | -1.611831808 | 0.000211248 | 0.000613014 | 1 |
| Obesity | endothelial | *NCOA6* | -0.978233915 | 4.26E-05 | 0.000183696 | 0.206841441 |
| Obesity | endothelial | *FAM19A2* | -0.955980258 | 1.78E-06 | 2.68E-05 | 0.00861828 |
| Obesity | endothelial | *TTC37* | -0.894955242 | 0.000484438 | 0.001185321 | 1 |
| Obesity | endothelial | *TUSC1* | -1.8121351 | 8.99E-07 | 2.40E-05 | 0.004360906 |
| Obesity | endothelial | *PAPSS2* | -0.92213766 | 0.000655426 | 0.001527438 | 1 |
| Obesity | endothelial | *CEP290* | -0.798468389 | 0.003368402 | 0.006190714 | 1 |
| Obesity | endothelial | *ANKRD13B* | -2.499096307 | 3.29E-06 | 3.49E-05 | 0.015962663 |
| Obesity | endothelial | *PPP1R14C* | -6.258189473 | 2.81E-06 | 3.32E-05 | 0.01362022 |
| Obesity | endothelial | *GPATCH3* | -1.563321884 | 1.55E-06 | 2.57E-05 | 0.007512126 |
| Obesity | endothelial | *COLGALT2* | -0.892760772 | 4.27E-06 | 3.91E-05 | 0.02073168 |
| Obesity | endothelial | *RCSD1* | -1.151810672 | 5.20E-05 | 0.000214288 | 0.252431722 |
| Obesity | endothelial | *GRIN3A* | -1.031566103 | 2.76E-05 | 0.000134044 | 0.13377566 |
| Obesity | endothelial | *SCAMP5* | -0.651459619 | 5.98E-06 | 4.68E-05 | 0.028991147 |
| Obesity | endothelial | *SFT2D1* | -1.228397903 | 5.86E-05 | 0.000233441 | 0.284565078 |
| Obesity | endothelial | *HMGN2* | -1.135201808 | 6.85E-07 | 2.20E-05 | 0.003323155 |
| Obesity | endothelial | *DENND4B* | -1.049035356 | 8.54E-05 | 0.00030695 | 0.414307264 |
| Obesity | endothelial | *RYR3* | 0.148050645 | 9.78E-05 | 0.000334569 | 0.474418184 |
| Obesity | endothelial | *TYW1* | -1.026362309 | 1.69E-05 | 9.64E-05 | 0.082079857 |
| Obesity | endothelial | *PRMT6* | 1.19567546 | 0.004646082 | 0.008188446 | 1 |
| Obesity | endothelial | *MAP3K3* | -0.854943494 | 0.001365911 | 0.002834645 | 1 |
| Obesity | endothelial | *C1orf174* | -1.036352691 | 0.005406773 | 0.009322553 | 1 |
| Obesity | endothelial | *DZIP3* | -1.084029185 | 4.91E-06 | 4.26E-05 | 0.023799286 |

| | | | | | | |
|---|---|---|---|---|---|---|
| Obesity | endothelial | *KIAA0753* | -0.972085141 | 0.002034736 | 0.003994707 | 1 |
| Obesity | endothelial | *TBKBP1* | 0.462877694 | 0.012319698 | 0.019426446 | 1 |
| Obesity | endothelial | *NAGA* | -1.194489427 | 0.000119673 | 0.000390341 | 0.580654173 |
| Obesity | endothelial | *SMG5* | -1.295800309 | 3.76E-06 | 3.75E-05 | 0.018234419 |
| Obesity | endothelial | *TGM2* | 0.329772661 | 0.005029718 | 0.008762727 | 1 |
| Obesity | endothelial | *PJA2* | -0.977534184 | 0.000115437 | 0.000379986 | 0.560099576 |
| Obesity | endothelial | *AL590822.1* | -0.728221175 | 7.56E-06 | 5.48E-05 | 0.036662576 |
| Obesity | endothelial | *EFCAB2* | -0.66085765 | 0.003550469 | 0.006488806 | 1 |
| Obesity | endothelial | *TATDN3* | -0.441747239 | 0.033829606 | 0.048177649 | 1 |
| Obesity | endothelial | *SAMD5* | 0.344492149 | 0.001600174 | 0.003250134 | 1 |
| Obesity | endothelial | *ECT2L* | -2.132344199 | 1.26E-05 | 7.75E-05 | 0.061280186 |
| Obesity | endothelial | *C6orf163* | -0.680770784 | 0.000300306 | 0.00080236 | 1 |
| Obesity | endothelial | *PCMTD2* | -1.085678775 | 0.000232891 | 0.000661584 | 1 |
| Obesity | endothelial | *EFCAB7* | -1.111631454 | 0.000152571 | 0.000473929 | 0.740276703 |
| Obesity | endothelial | *IFIT1B* | 2.359841009 | 0.000957967 | 0.002101853 | 1 |
| Obesity | endothelial | *LIPN* | -1.626605858 | 4.07E-06 | 3.86E-05 | 0.019752285 |
| Obesity | endothelial | *INPP5B* | -0.593693986 | 1.24E-05 | 7.65E-05 | 0.060228541 |
| Obesity | endothelial | *NEU4* | -1.191796249 | 0.000299447 | 0.000801326 | 1 |
| Obesity | endothelial | *C2orf72* | -4.083632796 | 0.000114944 | 0.000378621 | 0.557708375 |
| Obesity | endothelial | *RUFY2* | -0.971791843 | 0.000189282 | 0.000563434 | 0.918397234 |
| Obesity | endothelial | *AGAP6* | -0.569587297 | 0.032813262 | 0.046854016 | 1 |
| Obesity | endothelial | *SYT15* | 0.426876188 | 5.59E-05 | 0.000224837 | 0.271200995 |
| Obesity | endothelial | *LINC00619* | 0.626713327 | 6.66E-05 | 0.00025634 | 0.323244948 |
| Obesity | endothelial | *TCEA3* | -0.989675764 | 3.54E-07 | 1.45E-05 | 0.001718674 |
| Obesity | endothelial | *OXLD1* | -0.677287704 | 0.008497198 | 0.013886294 | 1 |
| Obesity | endothelial | *SMIM5* | -1.273134269 | 0.003494224 | 0.006401322 | 1 |
| Obesity | endothelial | *SKIV2L* | -0.79849461 | 4.43E-10 | 8.59E-08 | 2.15E-06 |
| Obesity | endothelial | *GPANK1* | -1.727406548 | 7.39E-13 | 2.76E-10 | 3.59E-09 |
| Obesity | endothelial | *BAG6* | -0.763564367 | 0.01288358 | 0.020223594 | 1 |
| Obesity | endothelial | *LST1* | -0.650319284 | 0.003319185 | 0.006107199 | 1 |
| Obesity | endothelial | *ZNF814* | -0.805053306 | 0.002546429 | 0.004850912 | 1 |
| Obesity | endothelial | *MICA* | -0.760173718 | 0.000546396 | 0.001308545 | 1 |
| Obesity | endothelial | *HLA-C* | 0.279902196 | 0.010990943 | 0.017553671 | 1 |
| Obesity | endothelial | *POU5F1* | 0.242950615 | 0.000218116 | 0.000628071 | 1 |
| Obesity | endothelial | *CCHCR1* | 0.38899546 | 0.001519086 | 0.003107338 | 1 |
| Obesity | endothelial | *DHX16* | -0.887705366 | 0.008318412 | 0.01363534 | 1 |
| Obesity | endothelial | *LILRB3* | -1.307934805 | 0.017385121 | 0.026417979 | 1 |
| Obesity | endothelial | *HLA-E* | 0.333854978 | 0.004833193 | 0.008475118 | 1 |
| Obesity | endothelial | *ZNF468* | -3.36488212 | 1.89E-05 | 0.000104126 | 0.091943097 |
| Obesity | endothelial | *RNF39* | -1.60356137 | 1.08E-07 | 6.51E-06 | 0.000525126 |
| Obesity | endothelial | *HCG9* | -1.02285336 | 0.000293997 | 0.000792926 | 1 |
| Obesity | endothelial | *HLA-G* | 0.402642574 | 0.019077248 | 0.028736313 | 1 |
| Obesity | endothelial | *ZFP57* | 0.610851823 | 1.91E-07 | 9.17E-06 | 0.000926479 |
| Obesity | endothelial | *FAM153C* | -0.385101907 | 0.000358672 | 0.000934628 | 1 |
| Obesity | endothelial | *TRIM27* | -1.405016794 | 1.63E-07 | 8.68E-06 | 0.000789473 |
| Obesity | endothelial | *MALRD1* | -0.773599964 | 2.38E-06 | 3.06E-05 | 0.011544791 |
| Obesity | endothelial | *FAM196B* | -1.090032387 | 3.82E-06 | 3.77E-05 | 0.018535253 |
| Obesity | endothelial | *FAM216A* | -1.527729479 | 4.16E-06 | 3.88E-05 | 0.020160745 |
| Obesity | endothelial | *ZBTB48* | -0.989985512 | 0.000644719 | 0.001504655 | 1 |
| Obesity | endothelial | *ZNF783* | -1.625498256 | 4.09E-06 | 3.86E-05 | 0.019844153 |

| | | | | | | |
|---|---|---|---|---|---|---|
| Obesity | endothelial | *ZNF425* | -1.069130603 | 4.54E-05 | 0.000191924 | 0.220520185 |
| Obesity | endothelial | *C12orf73* | -1.154641604 | 0.002472928 | 0.004725737 | 1 |
| Obesity | endothelial | *PCDHA7* | -8.964277386 | 0.000164386 | 0.000503218 | 0.79760131 |
| Obesity | endothelial | *PCDHA4* | -2.549531763 | 8.88E-07 | 2.40E-05 | 0.004306334 |
| Obesity | endothelial | *SPIRE2* | -0.777658719 | 4.28E-05 | 0.000184076 | 0.207637605 |
| Obesity | endothelial | *PKHD1L1* | -0.311880557 | 0.009069206 | 0.014746577 | 1 |
| Obesity | endothelial | *SLFN12L* | -1.102982483 | 1.79E-05 | 1.00E-04 | 0.08685905 |
| Obesity | endothelial | *SLC35B4* | -1.081145536 | 0.007642246 | 0.012665361 | 1 |
| Obesity | endothelial | *TMEM231* | 0.803876567 | 0.024359245 | 0.035858937 | 1 |
| Obesity | endothelial | *SCGB2B2* | -1.205212668 | 8.21E-06 | 5.79E-05 | 0.039830586 |
| Obesity | endothelial | *CCDC144NL* | -1.263235166 | 1.94E-06 | 2.80E-05 | 0.009390122 |
| Obesity | endothelial | *TMEM170B* | -0.5902008 | 8.87E-05 | 0.000315059 | 0.430370899 |
| Obesity | endothelial | *NT5M* | -1.315671131 | 9.88E-07 | 2.45E-05 | 0.004795079 |
| Obesity | endothelial | *IPO7* | -1.178163062 | 4.03E-05 | 0.000176662 | 0.195741637 |
| Obesity | endothelial | *PRR13* | -1.904322496 | 1.12E-06 | 2.57E-05 | 0.005453493 |
| Obesity | endothelial | *TECPR1* | 0.451961183 | 0.020776444 | 0.031075002 | 1 |
| Obesity | endothelial | *MT1A* | -2.120113903 | 6.42E-06 | 4.91E-05 | 0.031150342 |
| Obesity | endothelial | *MT1M* | -2.35044887 | 6.68E-06 | 5.06E-05 | 0.032424782 |
| Obesity | endothelial | *ATP6AP1L* | -0.520198953 | 0.002173529 | 0.004238731 | 1 |
| Obesity | endothelial | *RGL3* | -0.623539679 | 0.001269026 | 0.002666504 | 1 |
| Obesity | endothelial | *NAP1L4* | -1.009099934 | 7.53E-05 | 0.00028103 | 0.365339484 |
| Obesity | endothelial | *SMN2* | -0.660804289 | 0.00048988 | 0.001196828 | 1 |
| Obesity | endothelial | *SERF1B* | -0.962181849 | 8.15E-05 | 0.000297092 | 0.395429675 |
| Obesity | endothelial | *C17orf107* | 0.714344282 | 0.013669631 | 0.021298988 | 1 |
| Obesity | endothelial | *KLRC2* | -2.058230581 | 2.29E-05 | 0.000118729 | 0.110892829 |
| Obesity | endothelial | *LINC00487* | 1.186014989 | 0.001420273 | 0.002932411 | 1 |
| Obesity | endothelial | *HCG27* | -0.528208833 | 0.000125829 | 0.000406744 | 0.610523263 |
| Obesity | endothelial | *COL6A6* | -0.814836193 | 1.76E-05 | 9.89E-05 | 0.085381389 |
| Obesity | endothelial | *LNP1* | -0.86273218 | 5.90E-05 | 0.000234625 | 0.286242048 |
| Obesity | endothelial | *PRSS50* | -1.60651154 | 0.000503093 | 0.001220504 | 1 |
| Obesity | endothelial | *METTL6* | -1.115027443 | 0.000201491 | 0.000590359 | 0.977635198 |
| Obesity | endothelial | *SACM1L* | -0.928509137 | 0.004005769 | 0.007201182 | 1 |
| Obesity | endothelial | *SLC48A1* | 0.400855501 | 0.002552836 | 0.004859303 | 1 |
| Obesity | endothelial | *TAS2R50* | -7.936237725 | 2.32E-05 | 0.000120065 | 0.112501268 |
| Obesity | endothelial | *AC084018.1* | -0.776918696 | 8.33E-06 | 5.85E-05 | 0.040414261 |
| Obesity | endothelial | *C17orf51* | -1.805297639 | 1.43E-06 | 2.57E-05 | 0.006914834 |
| Obesity | endothelial | *OR2A42* | -0.703186662 | 0.001791497 | 0.003579899 | 1 |
| Obesity | endothelial | *ZNF580* | -0.593407978 | 0.016204335 | 0.024786707 | 1 |
| Obesity | endothelial | *NUP62* | -1.010680315 | 1.97E-05 | 0.000106783 | 0.095464217 |
| Obesity | endothelial | *ZNF254* | -1.148795905 | 7.85E-06 | 5.62E-05 | 0.038107022 |
| Obesity | endothelial | *AC022498.1* | -3.848661535 | 0.002775484 | 0.005219631 | 1 |
| Obesity | endothelial | *GIMAP1* | 0.336150471 | 0.000279735 | 0.000762085 | 1 |
| Obesity | endothelial | *ARHGEF35* | -0.730475167 | 0.019618313 | 0.029442639 | 1 |
| Obesity | endothelial | *GSTM2* | -0.482201779 | 0.000709804 | 0.001631439 | 1 |
| Obesity | endothelial | *LCAT* | -1.873611797 | 0.000107446 | 0.000359537 | 0.521329117 |
| Obesity | endothelial | *GPC2* | -2.71938528 | 2.81E-05 | 0.000135622 | 0.136300095 |
| Obesity | endothelial | *CCL14* | 0.152562906 | 0.001269769 | 0.002666504 | 1 |
| Obesity | endothelial | *SRA1* | -0.770105884 | 0.011110845 | 0.017727662 | 1 |
| Obesity | endothelial | *TMEM110* | 2.594566419 | 7.36E-10 | 1.27E-07 | 3.57E-06 |
| Obesity | endothelial | *TMX2* | -1.064927899 | 0.010670816 | 0.017109981 | 1 |

| | | | | | | |
|---|---|---|---|---|---|---|
| Obesity | endothelial | *LBH* | -1.374092508 | 9.58E-07 | 2.45E-05 | 0.004650107 |
| Obesity | endothelial | *PPP1CB* | -1.272137902 | 7.28E-07 | 2.23E-05 | 0.00353362 |
| Obesity | endothelial | *NCKIPSD* | -0.933062605 | 4.64E-05 | 0.000195251 | 0.225123932 |
| Obesity | endothelial | *ATF6B* | -0.784850823 | 0.005947731 | 0.010161406 | 1 |
| Obesity | endothelial | *SLC35F6* | -1.266918685 | 0.007020509 | 0.011729859 | 1 |
| Obesity | endothelial | *ATP6V1G2* | -0.332448127 | 0.008973638 | 0.014605867 | 1 |
| Obesity | endothelial | *SLC23A3* | 0.734818229 | 0.000242883 | 0.000681588 | 1 |
| Obesity | endothelial | *LTB4R* | -1.596119331 | 0.000332737 | 0.000875986 | 1 |
| Obesity | endothelial | *IRF9* | 0.514371339 | 0.020739929 | 0.031029952 | 1 |
| Obesity | endothelial | *ZNF726* | -2.203652497 | 5.37E-06 | 4.47E-05 | 0.026034618 |
| Obesity | endothelial | *ZNF90* | -1.132349427 | 0.003550655 | 0.006488806 | 1 |
| Obesity | endothelial | *REPIN1* | -2.020490576 | 0.000176783 | 0.000532765 | 0.857751614 |
| Obesity | endothelial | *ZNF891* | -0.923301803 | 0.020051762 | 0.030055961 | 1 |
| Obesity | endothelial | *SMIM7* | -1.299060569 | 2.47E-05 | 0.000124602 | 0.119617882 |
| Obesity | endothelial | *TSPAN4* | -0.781739642 | 0.002923279 | 0.005467907 | 1 |
| Obesity | endothelial | *CPNE1* | -0.470759268 | 0.001077105 | 0.002320653 | 1 |
| Obesity | endothelial | *ARL16* | -0.363074414 | 0.002078219 | 0.004069217 | 1 |
| Obesity | endothelial | *PAXIP1-AS2* | -1.095441293 | 0.00487573 | 0.008534286 | 1 |
| Obesity | endothelial | *LYRM4* | -0.758352376 | 0.018128391 | 0.027444291 | 1 |
| Obesity | endothelial | *ANG* | -1.482285964 | 0.002930982 | 0.005478091 | 1 |
| Obesity | endothelial | *COLCA2* | -0.54902062 | 0.000454419 | 0.001123773 | 1 |
| Obesity | endothelial | *MBLAC1* | -0.383838352 | 0.014456351 | 0.022395343 | 1 |
| Obesity | endothelial | *SOGA3* | -0.473723872 | 0.001528263 | 0.003120847 | 1 |
| Obesity | endothelial | *LCNL1* | -1.166319391 | 3.41E-05 | 0.000156653 | 0.165379069 |
| Obesity | endothelial | *GNAT3* | -1.988718001 | 8.22E-06 | 5.79E-05 | 0.03990524 |
| Obesity | endothelial | *PLIN5* | -0.496895862 | 3.12E-05 | 0.000147518 | 0.151206189 |
| Obesity | endothelial | *STARD10* | -0.422115912 | 0.004143886 | 0.007413766 | 1 |
| Obesity | endothelial | *NUTM2D* | -4.221873873 | 0.000761116 | 0.001731334 | 1 |
| Obesity | endothelial | *CPEB1* | -0.786683823 | 6.75E-05 | 0.000258762 | 0.327593258 |
| Obesity | endothelial | *ARHGEF33* | -1.219179439 | 5.67E-06 | 4.54E-05 | 0.027519091 |
| Obesity | endothelial | *IFRD2* | -1.100075861 | 1.53E-05 | 8.95E-05 | 0.074351734 |
| Obesity | endothelial | *EVPLL* | -1.157024433 | 2.74E-06 | 3.28E-05 | 0.013316881 |
| Obesity | endothelial | *AC005082.1* | -4.558307357 | 1.63E-06 | 2.61E-05 | 0.007891206 |
| Obesity | endothelial | *ACSM4* | -0.562143718 | 0.001250956 | 0.002638974 | 1 |
| Obesity | endothelial | *UBXN2B* | -0.937921829 | 8.68E-05 | 0.00030984 | 0.421381991 |
| Obesity | endothelial | *C5orf49* | -4.556698527 | 6.51E-05 | 0.000252147 | 0.315687775 |
| Obesity | endothelial | *VPS16* | -1.629267403 | 1.23E-05 | 7.58E-05 | 0.059458981 |
| Obesity | endothelial | *NPEPL1* | -1.794828058 | 3.94E-06 | 3.80E-05 | 0.019127435 |
| Obesity | endothelial | *TMEM242* | -0.931884441 | 0.00504033 | 0.008778063 | 1 |
| Obesity | endothelial | *SLC35E2* | -1.12784066 | 4.26E-08 | 3.62E-06 | 0.000206607 |
| Obesity | endothelial | *TSTD1* | -1.678549686 | 1.50E-06 | 2.57E-05 | 0.007270297 |
| Obesity | endothelial | *AC009403.2* | -0.814207932 | 3.50E-05 | 0.000159243 | 0.169877805 |
| Obesity | endothelial | *PAM16* | -1.820574195 | 0.000410435 | 0.001039911 | 1 |
| Obesity | endothelial | *RNASEK* | -1.009206505 | 0.000691668 | 0.001601132 | 1 |
| Obesity | endothelial | *NBPF1* | -0.534205389 | 0.019462298 | 0.029244679 | 1 |
| Obesity | endothelial | *FAM228B* | -0.547092607 | 0.002621253 | 0.004983668 | 1 |
| Obesity | endothelial | *RNU6ATAC35I* | -5.102426674 | 2.93E-07 | 1.29E-05 | 0.001421206 |
| Obesity | endothelial | *PPP3R1* | -0.725903855 | 0.014439339 | 0.022376134 | 1 |
| Obesity | endothelial | *FANCG* | -0.890002159 | 0.000249672 | 0.000695812 | 1 |
| Obesity | endothelial | *AP4M1* | -0.795618301 | 0.001000413 | 0.002176684 | 1 |

| | | | | | | |
|---|---|---|---|---|---|---|
| Obesity | endothelial | *ZNF880* | -0.624091387 | 0.004282955 | 0.007623221 | 1 |
| Obesity | endothelial | *TIGD1* | -0.885244593 | 0.000924544 | 0.002042754 | 1 |
| Obesity | endothelial | *FXYD7* | -1.168923162 | 0.000447349 | 0.001113095 | 1 |
| Obesity | endothelial | *SLC12A8* | -0.883757988 | 1.24E-05 | 7.65E-05 | 0.060255891 |
| Obesity | endothelial | *APOL6* | -1.050527589 | 5.98E-06 | 4.68E-05 | 0.029038146 |
| Obesity | endothelial | *OR2A1* | -0.661812886 | 0.000364711 | 0.00094731 | 1 |
| Obesity | endothelial | *RAB6C* | -9.974745167 | 1.80E-05 | 0.000100539 | 0.087468514 |
| Obesity | endothelial | *AC062017.1* | -1.580659734 | 1.31E-06 | 2.57E-05 | 0.006337577 |
| Obesity | endothelial | *EXOSC6* | -2.625370447 | 1.56E-11 | 4.45E-09 | 7.56E-08 |
| Obesity | endothelial | *ZNF844* | -1.194841578 | 0.000681745 | 0.001581935 | 1 |
| Obesity | endothelial | *LINC00571* | -1.171096856 | 2.07E-06 | 2.88E-05 | 0.01004789 |
| Obesity | endothelial | *C5orf66* | -1.183643776 | 0.00032076 | 0.000850453 | 1 |
| Obesity | endothelial | *LINC01133* | -1.18741918 | 2.28E-06 | 3.01E-05 | 0.011085566 |
| Obesity | endothelial | *C4B* | 1.410017014 | 0.001164608 | 0.002481632 | 1 |
| Obesity | endothelial | *LINC01117* | 0.371484807 | 0.000431181 | 0.001081183 | 1 |
| Obesity | endothelial | *LINC00184* | -8.856708645 | 8.79E-06 | 6.04E-05 | 0.042660004 |
| Obesity | endothelial | *AL049840.1* | -1.675776241 | 1.29E-07 | 7.38E-06 | 0.000626916 |
| Obesity | endothelial | *PLEKHM1* | -1.212939561 | 0.000184835 | 0.00055325 | 0.89681782 |
| Obesity | endothelial | *SFTA1P* | -1.515217119 | 5.32E-06 | 4.47E-05 | 0.025800136 |
| Obesity | endothelial | *CYP4A22-AS1* | -1.783829304 | 1.46E-06 | 2.57E-05 | 0.007096578 |
| Obesity | endothelial | *MIAT* | -1.486156505 | 1.40E-05 | 8.40E-05 | 0.068147297 |
| Obesity | endothelial | *ERCC6* | -1.142358357 | 1.59E-05 | 9.24E-05 | 0.077297654 |
| Obesity | endothelial | *LINC00694* | 0.78697946 | 4.09E-05 | 0.00017898 | 0.198488555 |
| Obesity | endothelial | *LINC00115* | -2.306847756 | 5.03E-06 | 4.30E-05 | 0.024424363 |
| Obesity | endothelial | *HAR1A* | -5.668972216 | 9.69E-06 | 6.50E-05 | 0.046998996 |
| Obesity | endothelial | *LINC00937* | -0.536457951 | 0.017583651 | 0.026686229 | 1 |
| Obesity | endothelial | *FTCDNL1* | -0.363980071 | 0.012298961 | 0.01940636 | 1 |
| Obesity | endothelial | *FAM138B* | -12.31280572 | 2.01E-06 | 2.84E-05 | 0.009749926 |
| Obesity | endothelial | *AC009299.3* | -1.561287668 | 1.52E-06 | 2.57E-05 | 0.007364379 |
| Obesity | endothelial | *SCAMP4* | -1.435819857 | 7.46E-06 | 5.45E-05 | 0.036178018 |
| Obesity | endothelial | *SLC2A1-AS1* | -1.621717684 | 9.61E-06 | 6.48E-05 | 0.046641823 |
| Obesity | endothelial | *AC005884.1* | -1.550085306 | 2.12E-05 | 0.000112277 | 0.102621001 |
| Obesity | endothelial | *AC108463.2* | -1.802485021 | 1.11E-05 | 7.07E-05 | 0.053778302 |
| Obesity | endothelial | *HCG20* | -1.386379359 | 1.37E-07 | 7.62E-06 | 0.000663031 |
| Obesity | endothelial | *C14orf144* | 0.679217869 | 0.001239658 | 0.002617415 | 1 |
| Obesity | endothelial | *POLR2J2* | 4.438550742 | 0.000253587 | 0.000705507 | 1 |
| Obesity | endothelial | *LINC00578* | -0.874006054 | 5.90E-06 | 4.66E-05 | 0.028625489 |
| Obesity | endothelial | *ZNF883* | -0.780863578 | 0.012973398 | 0.020351415 | 1 |
| Obesity | endothelial | *ARL17B* | -0.948922658 | 0.000587814 | 0.001394657 | 1 |
| Obesity | endothelial | *DHFR* | -0.288111195 | 0.002423168 | 0.004645283 | 1 |
| Obesity | endothelial | *PATL2* | -0.917605294 | 0.000136138 | 0.000433426 | 0.660541512 |
| Obesity | endothelial | *CASC20* | -1.342420277 | 3.10E-06 | 3.39E-05 | 0.015059704 |
| Obesity | endothelial | *PSMG3-AS1* | -1.150196016 | 6.85E-05 | 0.000261716 | 0.332379188 |
| Obesity | endothelial | *KLHL7-AS1* | -0.776251371 | 0.000331584 | 0.000873899 | 1 |
| Obesity | endothelial | *LINC00332* | -0.898994068 | 2.08E-05 | 0.000110614 | 0.100769532 |
| Obesity | endothelial | *AC092171.4* | -2.183882291 | 4.59E-06 | 4.08E-05 | 0.022255203 |
| Obesity | endothelial | *DIRC3* | -0.789559111 | 1.43E-05 | 8.54E-05 | 0.069617103 |
| Obesity | endothelial | *AC107021.1* | -1.075035211 | 1.04E-06 | 2.53E-05 | 0.005055625 |
| Obesity | endothelial | *MTRNR2L4* | -1.018461894 | 0.004742877 | 0.008334821 | 1 |
| Obesity | endothelial | *LINC00865* | -0.565348679 | 0.02270465 | 0.033617015 | 1 |

| | | | | | | |
|---|---|---|---|---|---|---|
| Obesity | endothelial | *LYRM9* | -1.388179764 | 7.48E-06 | 5.46E-05 | 0.036282603 |
| Obesity | endothelial | *LINC01166* | -0.673904924 | 0.02423366 | 0.035695725 | 1 |
| Obesity | endothelial | *LINC00342* | -1.903911285 | 0.000589289 | 0.00139679 | 1 |
| Obesity | endothelial | *LINC00702* | -1.795664242 | 1.20E-05 | 7.46E-05 | 0.057983195 |
| Obesity | endothelial | *RPS28* | -0.937754596 | 0.030733158 | 0.044209096 | 1 |
| Obesity | endothelial | *TRIM26* | -1.053775177 | 0.00070097 | 0.001618033 | 1 |
| Obesity | endothelial | *PINLYP* | -1.098500442 | 2.08E-06 | 2.88E-05 | 0.010115629 |
| Obesity | endothelial | *ZSCAN31* | 0.487661613 | 4.46E-07 | 1.68E-05 | 0.002163369 |
| Obesity | endothelial | *AC009495.2* | -9.652044245 | 0.000370693 | 0.000960366 | 1 |
| Obesity | endothelial | *LINC00484* | -0.427509262 | 0.001593977 | 0.003241399 | 1 |
| Obesity | endothelial | *KIAA0040* | -1.061078565 | 0.002684842 | 0.005068814 | 1 |
| Obesity | endothelial | *LINC00607* | -0.664376149 | 0.002940742 | 0.005492101 | 1 |
| Obesity | endothelial | *AC019117.2* | -5.768801894 | 5.13E-08 | 4.01E-06 | 0.000248843 |
| Obesity | endothelial | *ZBED5* | -1.15874718 | 2.02E-05 | 0.000108875 | 0.098096343 |
| Obesity | endothelial | *LINC00987* | 0.325706586 | 0.000200879 | 0.000588922 | 0.974665701 |
| Obesity | endothelial | *GS1-124K5.4* | 0.232536554 | 0.029060732 | 0.04209035 | 1 |
| Obesity | endothelial | *LINC00959* | -1.562147217 | 5.34E-05 | 0.00021837 | 0.259166199 |
| Obesity | endothelial | *LINC01143* | 0.293223057 | 5.92E-05 | 0.000235193 | 0.287405547 |
| Obesity | endothelial | *LINC00211* | 0.724781266 | 4.35E-05 | 0.000185308 | 0.211022534 |
| Obesity | endothelial | *LRRC37A2* | -1.519814306 | 5.55E-06 | 4.50E-05 | 0.026918368 |
| Obesity | endothelial | *LINC00707* | -1.645891167 | 0.030940974 | 0.044481661 | 1 |
| Obesity | endothelial | *TXNDC5* | 2.173641035 | 0.000875356 | 0.001946856 | 1 |
| Obesity | endothelial | *LINC01119* | -1.586160062 | 2.51E-05 | 0.000125716 | 0.121590188 |
| Obesity | endothelial | *TNFSF12* | 0.780673628 | 0.007884317 | 0.013011805 | 1 |
| Obesity | endothelial | *CDRT4* | -1.2221807 | 0.030119875 | 0.043481592 | 1 |
| Obesity | endothelial | *PSMB9* | 0.541273608 | 8.85E-05 | 0.000314743 | 0.429623776 |
| Obesity | endothelial | *PPIL3* | -0.401860925 | 0.013395584 | 0.020930508 | 1 |
| Obesity | endothelial | *L1TD1* | -0.898911147 | 0.000422418 | 0.001063056 | 1 |
| Obesity | endothelial | *C1QTNF9* | 0.436730562 | 8.96E-05 | 0.000316163 | 0.434724416 |
| Obesity | endothelial | *ISY1* | -1.164723935 | 7.15E-09 | 8.89E-07 | 3.47E-05 |
| Obesity | endothelial | *PNMA2* | -1.212407093 | 1.89E-05 | 0.000104093 | 0.091809703 |
| Obesity | endothelial | *KRBOX1* | -5.534510366 | 8.98E-06 | 6.13E-05 | 0.043550209 |
| Obesity | endothelial | *LINC00886* | -0.465579518 | 0.003565885 | 0.006511732 | 1 |
| Obesity | endothelial | *NSUN6* | -0.938743226 | 0.000425501 | 0.001069151 | 1 |
| Obesity | endothelial | *LINC00877* | -0.828256788 | 0.004541225 | 0.008024044 | 1 |
| Obesity | endothelial | *CD302* | -1.242289549 | 3.70E-06 | 3.73E-05 | 0.017946583 |
| Obesity | endothelial | *INMT* | -0.701524269 | 0.015298645 | 0.02356477 | 1 |
| Obesity | endothelial | *C8orf58* | -1.665427148 | 0.032219362 | 0.046141778 | 1 |
| Obesity | endothelial | *HOGA1* | -1.404888346 | 2.90E-06 | 3.33E-05 | 0.014055711 |
| Obesity | endothelial | *PI4KA* | -0.81405291 | 0.002317542 | 0.004465732 | 1 |
| Obesity | endothelial | *LINC00969* | -0.531092217 | 0.001052129 | 0.002271887 | 1 |
| Obesity | endothelial | *AMACR* | -0.250338602 | 0.005820973 | 0.009963524 | 1 |
| Obesity | endothelial | *ARFGAP3* | -0.976923638 | 0.005132178 | 0.008909241 | 1 |
| Obesity | endothelial | *MRPL20* | -0.937661484 | 0.006867885 | 0.011490683 | 1 |
| Obesity | endothelial | *DECR2* | -2.31336959 | 1.33E-06 | 2.57E-05 | 0.00647086 |
| Obesity | endothelial | *ZNF709* | 3.958603181 | 0.009542508 | 0.015448864 | 1 |
| Obesity | endothelial | *MRPL33* | -0.954746571 | 0.001601152 | 0.00325054 | 1 |
| Obesity | endothelial | *STON1* | 0.476044916 | 0.030187889 | 0.043566816 | 1 |
| Obesity | endothelial | *VSIG8* | -1.644505717 | 0.000119709 | 0.000390341 | 0.580827339 |
| Obesity | endothelial | *C7orf73* | -0.980287194 | 0.000967316 | 0.002117969 | 1 |

| | | | | | | |
|---|---|---|---|---|---|---|
| Obesity | endothelial | *FGF14-IT1* | -1.884236707 | 3.83E-06 | 3.77E-05 | 0.018574549 |
| Obesity | endothelial | *PALM2* | -0.880808799 | 7.75E-07 | 2.24E-05 | 0.003759312 |
| Obesity | endothelial | *NBPF15* | -1.075541758 | 0.007260042 | 0.012109221 | 1 |
| Obesity | endothelial | *UPK3B* | -1.239170102 | 3.31E-06 | 3.50E-05 | 0.016064613 |
| Obesity | endothelial | *NME1-NME2* | -1.402203962 | 0.029827806 | 0.04312411 | 1 |
| Obesity | endothelial | *APOBEC3D* | -5.354115565 | 4.31E-06 | 3.94E-05 | 0.020930556 |
| Obesity | endothelial | *SCARF2* | -1.412605313 | 3.69E-05 | 0.000165962 | 0.179239312 |
| Obesity | endothelial | *APOBEC3C* | -1.670113605 | 9.73E-07 | 2.45E-05 | 0.004722437 |
| Obesity | endothelial | *CCDC13* | -0.598829071 | 0.001943371 | 0.003840268 | 1 |
| Obesity | endothelial | *C4A* | -1.666986886 | 9.01E-07 | 2.40E-05 | 0.00437239 |
| Obesity | endothelial | *NEAT1* | -0.577274244 | 0.000358662 | 0.000934628 | 1 |
| Obesity | endothelial | *RAB30-AS1* | -1.011941896 | 1.17E-05 | 7.35E-05 | 0.056941037 |
| Obesity | endothelial | *UBAP1L* | -1.424251285 | 1.05E-05 | 6.84E-05 | 0.050843706 |
| Obesity | endothelial | *DNAJC3-AS1* | -1.596364559 | 8.64E-07 | 2.40E-05 | 0.004193044 |
| Obesity | endothelial | *LINC00926* | -2.327375171 | 8.73E-07 | 2.40E-05 | 0.004233816 |
| Obesity | endothelial | *DPH6-AS1* | -0.847445425 | 2.68E-05 | 0.000131605 | 0.130024871 |
| Obesity | endothelial | *CDK11B* | -1.494647903 | 3.00E-05 | 0.000142947 | 0.145377399 |
| Obesity | endothelial | *ABHD14A* | -1.402952587 | 6.84E-05 | 0.000261639 | 0.33201997 |
| Obesity | endothelial | *GDNF-AS1* | -1.032299512 | 1.14E-05 | 7.22E-05 | 0.055364869 |
| Obesity | endothelial | *C5orf17* | -1.337890848 | 1.01E-05 | 6.67E-05 | 0.048885419 |
| Obesity | endothelial | *TMEM150C* | 0.300135977 | 0.000225903 | 0.000646236 | 1 |
| Obesity | endothelial | *LINC00939* | -1.076104418 | 0.001709513 | 0.003436023 | 1 |
| Obesity | endothelial | *LINC01060* | 0.258607858 | 0.000756324 | 0.001722048 | 1 |
| Obesity | endothelial | *PDCD6* | -0.907074888 | 0.011392968 | 0.018124157 | 1 |
| Obesity | endothelial | *TMEM158* | -3.299014212 | 0.004660967 | 0.008208717 | 1 |
| Obesity | endothelial | *DNAH10OS* | -1.830210932 | 0.002729329 | 0.005142799 | 1 |
| Obesity | endothelial | *PCDHA10* | -0.690251037 | 0.000187157 | 0.000558822 | 0.90808587 |
| Obesity | endothelial | *ZNF718* | -0.86126384 | 0.006761253 | 0.011343568 | 1 |
| Obesity | endothelial | *LINC00989* | -1.380284924 | 0.005860952 | 0.010023736 | 1 |
| Obesity | endothelial | *ATP6V1E2* | -0.335498193 | 0.018947582 | 0.028577454 | 1 |
| Obesity | endothelial | *GLI4* | -1.410893465 | 3.19E-07 | 1.36E-05 | 0.001549646 |
| Obesity | endothelial | *SNHG18* | -1.099283902 | 7.56E-07 | 2.24E-05 | 0.003667317 |
| Obesity | endothelial | *ALG1L2* | -1.469209837 | 1.75E-05 | 9.88E-05 | 0.084955691 |
| Obesity | endothelial | *LINC01094* | -1.787485566 | 1.49E-06 | 2.57E-05 | 0.00722451 |
| Obesity | endothelial | *TMEM200B* | -0.797555976 | 0.000374017 | 0.00096472 | 1 |
| Obesity | endothelial | *PCDHGB6* | 0.631902448 | 0.017787405 | 0.026978583 | 1 |
| Obesity | endothelial | *PCDHGB2* | -4.606516479 | 0.000194244 | 0.000576083 | 0.942472122 |
| Obesity | endothelial | *LYN* | -0.460837832 | 0.016634141 | 0.025404108 | 1 |
| Obesity | endothelial | *ARMS2* | -1.252378463 | 2.34E-05 | 0.000120712 | 0.113560223 |
| Obesity | endothelial | *FPGT* | -0.484306671 | 0.018021489 | 0.027316546 | 1 |
| Obesity | endothelial | *BRK1* | -1.281017279 | 7.04E-06 | 5.28E-05 | 0.0341823 |
| Obesity | endothelial | *SNHG9* | -0.812826619 | 0.00429393 | 0.00763715 | 1 |
| Obesity | endothelial | *PCDHA3* | -2.0396652 | 1.61E-08 | 1.66E-06 | 7.79E-05 |
| Obesity | endothelial | *AC093668.2* | -1.527156941 | 0.000362627 | 0.000942402 | 1 |
| Obesity | endothelial | *GATC* | -0.709906587 | 0.002201193 | 0.004280637 | 1 |
| Obesity | endothelial | *ZBED6* | -2.526612909 | 1.38E-06 | 2.57E-05 | 0.006676854 |
| Obesity | endothelial | *CNPY2* | -1.247387345 | 3.29E-06 | 3.49E-05 | 0.015952667 |
| Obesity | endothelial | *LINC00592* | -1.296071198 | 0.001403647 | 0.002903025 | 1 |
| Obesity | endothelial | *RTEL1* | -0.749135306 | 0.003682458 | 0.006691868 | 1 |
| Obesity | endothelial | *PDF* | 1.063634764 | 0.019364158 | 0.029115244 | 1 |

| | | | | | | |
|---|---|---|---|---|---|---|
| Obesity | endothelial | *HIF1A-AS2* | -1.139056508 | 0.000206037 | 0.000600901 | 0.999691789 |
| Obesity | endothelial | *RNASE4* | 1.802374075 | 0.000988427 | 0.002152536 | 1 |
| Obesity | endothelial | *LINC00639* | 0.128456058 | 0.000114566 | 0.000377633 | 0.555875312 |
| Obesity | endothelial | *LINC00924* | -0.342109274 | 0.017120588 | 0.02606498 | 1 |
| Obesity | endothelial | *MRPL46* | -1.512524951 | 0.001909453 | 0.003784726 | 1 |
| Obesity | endothelial | *UBE2Q2L* | -1.192302371 | 0.000709505 | 0.001631439 | 1 |
| Obesity | endothelial | *IQCH-AS1* | -0.811479144 | 0.005813457 | 0.009956546 | 1 |
| Obesity | endothelial | *AC083843.1* | -6.577897744 | 8.75E-06 | 6.02E-05 | 0.042440945 |
| Obesity | endothelial | *PWRN1* | -0.855176756 | 5.47E-06 | 4.47E-05 | 0.026520205 |
| Obesity | endothelial | *LINC00662* | -1.020621627 | 0.000236936 | 0.000668768 | 1 |
| Obesity | endothelial | *C19orf84* | -4.29317816 | 5.04E-05 | 0.000208821 | 0.244529713 |
| Obesity | endothelial | *LINC00675* | -0.842500181 | 0.000323031 | 0.000855537 | 1 |
| Obesity | endothelial | *LINC00667* | -0.87823422 | 0.00010632 | 0.000356261 | 0.51586539 |
| Obesity | endothelial | *LINC00672* | -4.244157468 | 2.51E-05 | 0.000125716 | 0.121684166 |
| Obesity | endothelial | *LINC00909* | -1.155750857 | 3.71E-05 | 0.000166374 | 0.180016919 |
| Obesity | endothelial | *DYNLL2* | -2.3279037 | 6.86E-05 | 0.000261806 | 0.332755921 |
| Obesity | endothelial | *STRADA* | -1.487236268 | 1.07E-06 | 2.57E-05 | 0.005210376 |
| Obesity | endothelial | *LINC00663* | -0.495908042 | 0.000462331 | 0.001139852 | 1 |
| Obesity | endothelial | *ILF3-AS1* | -0.999938693 | 1.47E-05 | 8.69E-05 | 0.07113909 |
| Obesity | endothelial | *ZNF224* | -0.802037803 | 0.024983349 | 0.036688622 | 1 |
| Obesity | endothelial | *LIN37* | -2.66441423 | 0.004719901 | 0.008300456 | 1 |
| Obesity | endothelial | *NDUFA7* | -4.01336772 | 5.04E-06 | 4.30E-05 | 0.024440766 |
| Obesity | endothelial | *AC004466.1* | 0.795307357 | 0.000328976 | 0.000867968 | 1 |
| Obesity | endothelial | *LINC00664* | 3.133528145 | 0.000242836 | 0.000681588 | 1 |
| Obesity | endothelial | *SCGB1B2P* | -1.360337506 | 0.004739382 | 0.008331696 | 1 |
| Obesity | endothelial | *ZNF728* | -1.438257566 | 0.001709446 | 0.003436023 | 1 |
| Obesity | endothelial | *TRABD2B* | 0.322848804 | 0.000618863 | 0.0014534 | 1 |
| Obesity | endothelial | *FBXO17* | -0.648848909 | 3.33E-05 | 0.000154548 | 0.161811316 |
| Obesity | endothelial | *ZNF587B* | -1.221980591 | 1.68E-06 | 2.63E-05 | 0.008131719 |
| Obesity | endothelial | *SNHG8* | -1.098507584 | 3.41E-05 | 0.000156653 | 0.165425711 |
| Obesity | endothelial | *HCG17* | -0.88209256 | 0.000215998 | 0.000623823 | 1 |
| Obesity | endothelial | *FGF14-AS2* | -2.284768872 | 8.83E-06 | 6.04E-05 | 0.042843133 |
| Obesity | endothelial | *CASC15* | -0.913542515 | 0.002689581 | 0.005073813 | 1 |
| Obesity | endothelial | *DOC2B* | 0.167854218 | 0.000220918 | 0.000634258 | 1 |
| Obesity | endothelial | *GRIN2B* | -0.36963726 | 0.014884289 | 0.022992222 | 1 |
| Obesity | endothelial | *PAXIP1-AS1* | -1.176224093 | 2.31E-06 | 3.02E-05 | 0.011202163 |
| Obesity | LEC | *SEMA3F* | -4.610203941 | 1.41E-08 | 2.45E-06 | 6.85E-05 |
| Obesity | LEC | *RBM6* | -2.697512425 | 2.63E-20 | 4.25E-17 | 1.27E-16 |
| Obesity | LEC | *CLCN6* | 2.719849555 | 0.000766839 | 0.026961605 | 1 |
| Obesity | LEC | *PIAS1* | 2.321930021 | 0.001145404 | 0.03680464 | 1 |
| Obesity | LEC | *LSG1* | 1.004680498 | 0.001740584 | 0.048536273 | 1 |
| Obesity | LEC | *ANAPC4* | 3.025127596 | 0.001302256 | 0.039990786 | 1 |
| Obesity | LEC | *SDCCAG8* | 1.633107856 | 0.001497762 | 0.04446132 | 1 |
| Obesity | LEC | *ITIH4* | 1.164906433 | 2.83E-10 | 7.22E-08 | 1.37E-06 |
| Obesity | LEC | *UHRF1BP1* | 4.282782524 | 5.38E-21 | 1.31E-17 | 2.61E-17 |
| Obesity | LEC | *TIPIN* | 1.174194515 | 0.00044265 | 0.017604427 | 1 |
| Obesity | LEC | *NT5C2* | 2.109460822 | 0.000160869 | 0.007728076 | 0.780535643 |
| Obesity | LEC | *FDFT1* | 2.552458383 | 7.02E-05 | 0.004258306 | 0.340664478 |
| Obesity | LEC | *GSTP1* | 3.143180505 | 4.59E-05 | 0.003006519 | 0.222482438 |
| Obesity | LEC | *MAST2* | 2.634558206 | 0.001450136 | 0.043765079 | 1 |

| | | | | | | |
|---|---|---|---|---|---|---|
| Obesity | LEC | *YPEL3* | 3.277503402 | 1.77E-07 | 2.46E-05 | 0.000860794 |
| Obesity | LEC | *PABPC4* | -3.120671051 | 5.02E-06 | 0.00048727 | 0.024363477 |
| Obesity | LEC | *RGS17* | -3.142212372 | 1.14E-05 | 0.000945968 | 0.055113715 |
| Obesity | LEC | *PPP2R3C* | 2.848439017 | 0.00015836 | 0.007683618 | 0.76836175 |
| Obesity | LEC | *ITPR3* | 2.575811062 | 3.84E-06 | 0.000413864 | 0.018623899 |
| Obesity | LEC | *SETD1A* | 2.995426703 | 0.000119217 | 0.006287413 | 0.578441987 |
| Obesity | LEC | *CENPM* | 2.538220004 | 4.52E-05 | 0.003003905 | 0.219285078 |
| Obesity | LEC | *ABHD12* | 2.653592673 | 0.00017859 | 0.00833193 | 0.866520682 |
| Obesity | LEC | *CLUAP1* | 1.288527329 | 0.000708433 | 0.025461596 | 1 |
| Obesity | LEC | *PYCARD* | 3.623321644 | 4.71E-05 | 0.003045399 | 0.228404911 |
| Obesity | LEC | *CHRNE* | 0.751558821 | 0.000403722 | 0.016188917 | 1 |
| Obesity | LEC | *DDX55* | 3.345355228 | 0.00088554 | 0.030046427 | 1 |
| Obesity | LEC | *BTN3A3* | 3.190697064 | 8.28E-11 | 2.36E-08 | 4.02E-07 |
| Obesity | LEC | *GABRR2* | 2.57824989 | 0.000599399 | 0.02272095 | 1 |
| Obesity | LEC | *TULP1* | -1.928922025 | 0.000507846 | 0.02003307 | 1 |
| Obesity | LEC | *PRSS16* | 2.453822923 | 0.001716213 | 0.048133338 | 1 |
| Obesity | LEC | *NME5* | 1.409810587 | 9.70E-05 | 0.005472268 | 0.470615022 |
| Obesity | LEC | *PCCB* | 2.381114188 | 1.06E-05 | 0.000909185 | 0.051368109 |
| Obesity | LEC | *SCAP* | 3.282752514 | 0.001452221 | 0.043765079 | 1 |
| Obesity | LEC | *CHMP3* | -2.293031812 | 0.000296027 | 0.012323798 | 1 |
| Obesity | LEC | *SERPINC1* | 5.288130851 | 8.93E-09 | 1.67E-06 | 4.33E-05 |
| Obesity | LEC | *CCDC92* | 3.143677308 | 0.000107765 | 0.005875004 | 0.522875318 |
| Obesity | LEC | *INVS* | 2.799394398 | 5.45E-06 | 0.000518399 | 0.026438329 |
| Obesity | LEC | *ADRA1A* | 0.485557075 | 0.001502815 | 0.04446132 | 1 |
| Obesity | LEC | *TEX14* | 2.208496739 | 0.000250245 | 0.010745049 | 1 |
| Obesity | LEC | *OPN4* | 8.477421941 | 4.83E-06 | 0.00047838 | 0.023440601 |
| Obesity | LEC | *SLC25A51* | 0.788195756 | 0.001641075 | 0.047025285 | 1 |
| Obesity | LEC | *BTN2A2* | 1.157267871 | 2.50E-05 | 0.00180744 | 0.121098448 |
| Obesity | LEC | *SNRPC* | -3.835732684 | 4.50E-12 | 1.56E-09 | 2.18E-08 |
| Obesity | LEC | *GRK4* | 0.691661342 | 7.13E-05 | 0.004272442 | 0.346067773 |
| Obesity | LEC | *RIBC2* | 1.575278964 | 0.000656758 | 0.024565234 | 1 |
| Obesity | LEC | *AP1M2* | -1.377834022 | 0.001125143 | 0.036803719 | 1 |
| Obesity | LEC | *ATP8B3* | -1.572164867 | 1.31E-08 | 2.35E-06 | 6.35E-05 |
| Obesity | LEC | *PGPEP1* | 1.947134699 | 0.000576978 | 0.022218238 | 1 |
| Obesity | LEC | *PKDREJ* | -1.57489181 | 0.001650187 | 0.047025285 | 1 |
| Obesity | LEC | *NDUFA2* | 2.475044603 | 0.000213919 | 0.009610495 | 1 |
| Obesity | LEC | *IGHMBP2* | 1.835340279 | 1.52E-05 | 0.001191237 | 0.073856691 |
| Obesity | LEC | *MUTYH* | 2.775467955 | 0.000945352 | 0.031633422 | 1 |
| Obesity | LEC | *GSTM5* | 1.304212324 | 0.001615557 | 0.046658833 | 1 |
| Obesity | LEC | *POM121C* | -4.163652944 | 1.15E-07 | 1.68E-05 | 0.000555795 |
| Obesity | LEC | *USP37* | 2.750163884 | 0.001231404 | 0.038301571 | 1 |
| Obesity | LEC | *TCF19* | 1.970987493 | 0.001190577 | 0.038004468 | 1 |
| Obesity | LEC | *SLC37A4* | 3.485191046 | 1.88E-08 | 3.05E-06 | 9.14E-05 |
| Obesity | LEC | *NUSAP1* | 3.75342012 | 0.000372055 | 0.015169848 | 1 |
| Obesity | LEC | *NDUFAF1* | 3.432013749 | 9.09E-06 | 0.000801996 | 0.044109798 |
| Obesity | LEC | *CENPO* | 6.537953464 | 1.77E-12 | 7.16E-10 | 8.59E-09 |
| Obesity | LEC | *KLRG1* | 0.823403051 | 0.001542643 | 0.04486468 | 1 |
| Obesity | LEC | *VPS33A* | 2.538389748 | 7.54E-05 | 0.004460512 | 0.365761958 |
| Obesity | LEC | *DIAPH3* | 1.131101652 | 0.000677656 | 0.025099138 | 1 |
| Obesity | LEC | *MYO19* | 3.945340727 | 5.57E-06 | 0.000519949 | 0.027037342 |

| | | | | | | |
|---|---|---|---|---|---|---|
| Obesity | LEC | *C18orf8* | -2.759673977 | 2.62E-05 | 0.001868726 | 0.127073385 |
| Obesity | LEC | *COL6A1* | 0.880622783 | 0.001130205 | 0.036803719 | 1 |
| Obesity | LEC | *SAE1* | -3.443432069 | 0.001516438 | 0.044592474 | 1 |
| Obesity | LEC | *SEMA6C* | 4.720370194 | 0.000126722 | 0.006611367 | 0.614857099 |
| Obesity | LEC | *LIMD1* | 2.472697476 | 0.000777474 | 0.027040407 | 1 |
| Obesity | LEC | *CCDC39* | 2.182769858 | 0.001657321 | 0.047025285 | 1 |
| Obesity | LEC | *ABT1* | 1.327277444 | 0.001144471 | 0.03680464 | 1 |
| Obesity | LEC | *RGR* | -8.382010403 | 0.000658178 | 0.024565234 | 1 |
| Obesity | LEC | *MTG1* | 1.609579463 | 0.000172775 | 0.008138864 | 0.838302968 |
| Obesity | LEC | *HSD17B12* | -1.503437524 | 6.59E-10 | 1.52E-07 | 3.20E-06 |
| Obesity | LEC | *TNKS1BP1* | 1.825959962 | 4.20E-06 | 0.00044336 | 0.02039456 |
| Obesity | LEC | *TTC12* | 2.196202003 | 3.54E-05 | 0.002416287 | 0.171556361 |
| Obesity | LEC | *TBX6* | 1.398523076 | 1.82E-11 | 5.90E-09 | 8.84E-08 |
| Obesity | LEC | *RGS18* | 1.545145079 | 0.00026869 | 0.01143581 | 1 |
| Obesity | LEC | *POC5* | -1.733762293 | 0.000184525 | 0.008526805 | 0.895314564 |
| Obesity | LEC | *PLA2R1* | 1.357727298 | 0.000685924 | 0.025196283 | 1 |
| Obesity | LEC | *JAZF1* | 1.964516069 | 0.000205285 | 0.009394408 | 0.996043103 |
| Obesity | LEC | *NEIL2* | 2.075751122 | 8.01E-05 | 0.004682137 | 0.388617341 |
| Obesity | LEC | *TMED6* | 6.911705709 | 0.000140766 | 0.007041179 | 0.68299432 |
| Obesity | LEC | *CLEC18A* | -3.722448665 | 8.75E-18 | 7.07E-15 | 4.24E-14 |
| Obesity | LEC | *ETS2* | 0.995487903 | 0.000536061 | 0.020807726 | 1 |
| Obesity | LEC | *SLC5A11* | -1.147346992 | 1.48E-06 | 0.000171051 | 0.007184127 |
| Obesity | LEC | *CILP2* | -2.933587712 | 0.000130458 | 0.006733858 | 0.632982657 |
| Obesity | LEC | *ABCF3* | 1.674780762 | 0.001039762 | 0.034319213 | 1 |
| Obesity | LEC | *ANTXR2* | 2.583113895 | 8.35E-05 | 0.00476603 | 0.405112585 |
| Obesity | LEC | *DNALI1* | -2.349495143 | 4.31E-05 | 0.002903313 | 0.20903854 |
| Obesity | LEC | *RFT1* | -2.342976906 | 1.88E-05 | 0.001385082 | 0.091415432 |
| Obesity | LEC | *RNF123* | 4.505873645 | 5.87E-19 | 5.70E-16 | 2.85E-15 |
| Obesity | LEC | *MST1R* | 4.304803021 | 7.13E-23 | 3.46E-19 | 3.46E-19 |
| Obesity | LEC | *ANKS6* | 3.531817526 | 5.76E-05 | 0.003579978 | 0.279238256 |
| Obesity | LEC | *SLC39A13* | 3.60124064 | 2.98E-05 | 0.002065969 | 0.144617819 |
| Obesity | LEC | *RAPSN* | -3.303952874 | 1.41E-13 | 8.55E-11 | 6.84E-10 |
| Obesity | LEC | *SRR* | 1.167052313 | 1.17E-05 | 0.000945968 | 0.056758058 |
| Obesity | LEC | *NT5DC2* | -1.979632473 | 1.16E-05 | 0.000945968 | 0.056464234 |
| Obesity | LEC | *FNTA* | 2.460928816 | 0.000242029 | 0.010579491 | 1 |
| Obesity | LEC | *C8orf46* | 0.543192586 | 0.000322965 | 0.013279867 | 1 |
| Obesity | LEC | *GPRIN1* | 3.780828221 | 0.000785799 | 0.027040407 | 1 |
| Obesity | LEC | *INO80E* | -3.15709014 | 7.29E-13 | 3.21E-10 | 3.54E-09 |
| Obesity | LEC | *ADORA2B* | -1.216464734 | 7.46E-07 | 9.28E-05 | 0.003619399 |
| Obesity | LEC | *TTLL6* | -7.433116288 | 5.09E-11 | 1.54E-08 | 2.47E-07 |
| Obesity | LEC | *TSNARE1* | 3.061088537 | 1.42E-05 | 0.001133093 | 0.069118654 |
| Obesity | LEC | *TRMT61B* | 2.767902104 | 5.18E-05 | 0.003262462 | 0.251209584 |
| Obesity | LEC | *RGS19* | 0.853229771 | 0.001329749 | 0.04057825 | 1 |
| Obesity | LEC | *C1QTNF4* | -1.910881607 | 5.88E-15 | 4.07E-12 | 2.85E-11 |
| Obesity | LEC | *CTSW* | -0.472778308 | 2.20E-08 | 3.44E-06 | 0.000106744 |
| Obesity | LEC | *EFEMP2* | 3.369520096 | 0.000100751 | 0.005618907 | 0.488844943 |
| Obesity | LEC | *FUT10* | -1.029161073 | 0.001703861 | 0.048064719 | 1 |
| Obesity | LEC | *SNX32* | 0.993385231 | 0.000134925 | 0.00681934 | 0.65465664 |
| Obesity | LEC | *MRGPRF* | 2.130202147 | 0.000297173 | 0.012323798 | 1 |
| Obesity | LEC | *SEZ6L2* | 1.593065182 | 8.14E-05 | 0.004701395 | 0.394917147 |

| | | | | | | |
|---|---|---|---|---|---|---|
| Obesity | LEC | *KCTD13* | 2.630563004 | 4.51E-06 | 0.000456376 | 0.021906026 |
| Obesity | LEC | *CADM2* | 2.528468109 | 1.77E-08 | 2.97E-06 | 8.61E-05 |
| Obesity | LEC | *BANF1* | -4.808346866 | 6.87E-05 | 0.004218286 | 0.333244599 |
| Obesity | LEC | *ORAI3* | 4.529752594 | 2.82E-12 | 1.05E-09 | 1.37E-08 |
| Obesity | LEC | *RMDN1* | 1.990901883 | 0.001241015 | 0.038352901 | 1 |
| Obesity | LEC | *RUFY1* | 2.91995371 | 2.79E-05 | 0.001964411 | 0.135544341 |
| Obesity | LEC | *AP006621.1* | 2.57828757 | 0.00017012 | 0.008092386 | 0.825423386 |
| Obesity | LEC | *RPP25* | -1.252725781 | 0.00039834 | 0.01610623 | 1 |
| Obesity | LEC | *STX19* | 2.796935632 | 7.69E-06 | 0.000691142 | 0.037321686 |
| Obesity | LEC | *MPI* | 1.881885273 | 0.000531204 | 0.020785515 | 1 |
| Obesity | LEC | *TRIM73* | -7.978409453 | 5.13E-13 | 2.77E-10 | 2.49E-09 |
| Obesity | LEC | *TUFM* | 4.062491312 | 2.71E-10 | 7.22E-08 | 1.31E-06 |
| Obesity | LEC | *C3orf38* | 2.487777565 | 1.35E-06 | 0.000163209 | 0.006528355 |
| Obesity | LEC | *RPRML* | 6.692238368 | 1.69E-05 | 0.001283227 | 0.082126525 |
| Obesity | LEC | *PPP1R27* | -7.977845889 | 0.000207173 | 0.009394408 | 1 |
| Obesity | LEC | *COL18A1* | 2.74165625 | 0.000228098 | 0.010061181 | 1 |
| Obesity | LEC | *PLGLB1* | 1.469086737 | 0.000741896 | 0.026468219 | 1 |
| Obesity | LEC | *PSMG1* | 2.437162907 | 0.000274611 | 0.011586204 | 1 |
| Obesity | LEC | *UPP1* | 2.83836279 | 0.000782588 | 0.027040407 | 1 |
| Obesity | LEC | *EMILIN3* | 2.757894663 | 0.00070551 | 0.025461596 | 1 |
| Obesity | LEC | *ACOT1* | 1.37212304 | 0.000109617 | 0.005909549 | 0.531859435 |
| Obesity | LEC | *NUTM2A* | -2.070132015 | 1.77E-05 | 0.001319925 | 0.085795139 |
| Obesity | LEC | *GP1BA* | 2.072684823 | 1.60E-05 | 0.001233713 | 0.077723934 |
| Obesity | LEC | *BTN3A2* | -0.957114232 | 4.10E-10 | 9.96E-08 | 1.99E-06 |
| Obesity | LEC | *CENPP* | 1.219914412 | 0.000113202 | 0.006035806 | 0.549258306 |
| Obesity | LEC | *FBF1* | 1.895452151 | 0.00123146 | 0.038301571 | 1 |
| Obesity | LEC | *ZKSCAN3* | -2.969191187 | 3.62E-07 | 4.75E-05 | 0.001758159 |
| Obesity | LEC | *ZNF605* | 2.178414466 | 0.000145235 | 0.007122967 | 0.704678067 |
| Obesity | LEC | *SULT1A2* | 5.185440145 | 5.70E-19 | 5.70E-16 | 2.76E-15 |
| Obesity | LEC | *ZNF165* | -1.311824363 | 0.000228052 | 0.010061181 | 1 |
| Obesity | LEC | *MRPL21* | -0.855938191 | 0.00087351 | 0.029846966 | 1 |
| Obesity | LEC | *DNAH10* | 5.345478895 | 5.78E-13 | 2.80E-10 | 2.80E-09 |
| Obesity | LEC | *RPS26* | -1.053896753 | 0.000690665 | 0.025196283 | 1 |
| Obesity | LEC | *ZNF780A* | -0.659401189 | 0.000107452 | 0.005875004 | 0.521357223 |
| Obesity | LEC | *NOTCH4* | -0.302904557 | 5.78E-09 | 1.12E-06 | 2.80E-05 |
| Obesity | LEC | *SKIV2L* | -1.135220535 | 4.42E-06 | 0.000456376 | 0.021453418 |
| Obesity | LEC | *GPANK1* | -3.765317941 | 2.93E-09 | 6.18E-07 | 1.42E-05 |
| Obesity | LEC | *HLA-C* | 0.887199601 | 0.0017936 | 0.049728847 | 1 |
| Obesity | LEC | *POU5F1* | 0.467177997 | 5.14E-05 | 0.003262462 | 0.249372887 |
| Obesity | LEC | *CCHCR1* | 0.892079951 | 3.04E-06 | 0.000335225 | 0.014749915 |
| Obesity | LEC | *RNF39* | -2.83075771 | 6.87E-06 | 0.000628941 | 0.033333879 |
| Obesity | LEC | *ZFP57* | 1.122038723 | 4.34E-09 | 8.77E-07 | 2.10E-05 |
| Obesity | LEC | *C17orf107* | 1.902744697 | 0.000249473 | 0.010745049 | 1 |
| Obesity | LEC | *AC084018.1* | -1.157213966 | 0.000134738 | 0.00681934 | 0.65374817 |
| Obesity | LEC | *SUPT4H1* | 5.711295548 | 0.000757108 | 0.026813773 | 1 |
| Obesity | LEC | *SRA1* | 2.894639339 | 0.001544188 | 0.04486468 | 1 |
| Obesity | LEC | *TMEM110* | 3.46489932 | 5.75E-08 | 8.72E-06 | 0.000278916 |
| Obesity | LEC | *GALT* | 2.829770472 | 0.001017358 | 0.033809718 | 1 |
| Obesity | LEC | *EXOSC6* | -5.358106306 | 2.75E-09 | 6.07E-07 | 1.34E-05 |
| Obesity | LEC | *C4B* | 2.601711002 | 0.000145337 | 0.007122967 | 0.705173736 |

| | | | | | | |
|---|---|---|---|---|---|---|
| Obesity | LEC | *LINC00240* | 0.751985632 | 0.001200854 | 0.038081987 | 1 |
| Obesity | LEC | *HCG20* | -2.56144575 | 4.85E-07 | 6.20E-05 | 0.002355596 |
| Obesity | LEC | *TAPBP* | 2.131597278 | 0.001218199 | 0.038301571 | 1 |
| Obesity | LEC | *ZSCAN31* | 0.995263554 | 3.06E-07 | 4.13E-05 | 0.001486145 |
| Obesity | LEC | *PSMB9* | 1.260394443 | 1.81E-06 | 0.000204423 | 0.008790191 |
| Obesity | LEC | *C4A* | -2.990345662 | 1.41E-06 | 0.000166744 | 0.006836488 |
| Obesity | LEC | *DNAH10OS* | 20.60873175 | 1.32E-07 | 1.88E-05 | 0.000639799 |
| Obesity | LEC | *DPP3* | 0.92279116 | 0.000594091 | 0.022697064 | 1 |
| Obesity | LEC | *PCDHA3* | -2.457763305 | 0.001489357 | 0.04446132 | 1 |
| Obesity | LEC | *HMBS* | 3.14575828 | 1.07E-05 | 0.000909185 | 0.051823557 |
| Obesity | LEC | *AP001172.2* | 0.805989056 | 0.000922896 | 0.031096477 | 1 |
| Obesity | macrophage | *SEMA3F* | -1.566490942 | 1.56E-07 | 2.04E-05 | 0.000756199 |
| Obesity | macrophage | *RBM6* | -1.333216088 | 2.04E-23 | 4.95E-20 | 9.90E-20 |
| Obesity | macrophage | *CLCN6* | 1.235892962 | 0.001524335 | 0.045098013 | 1 |
| Obesity | macrophage | *PIAS1* | 1.226599533 | 0.00166842 | 0.048648017 | 1 |
| Obesity | macrophage | *ITIH4* | 0.536747617 | 9.89E-11 | 2.20E-08 | 4.80E-07 |
| Obesity | macrophage | *UHRF1BP1* | 2.051208 | 2.37E-21 | 3.84E-18 | 1.15E-17 |
| Obesity | macrophage | *TIPIN* | 0.478222043 | 0.00130321 | 0.040020082 | 1 |
| Obesity | macrophage | *NT5C2* | 0.963724771 | 0.000385893 | 0.016569507 | 1 |
| Obesity | macrophage | *FDFT1* | 1.372785008 | 7.36E-05 | 0.00415289 | 0.357148516 |
| Obesity | macrophage | *GSTP1* | 1.562208129 | 6.85E-05 | 0.00400387 | 0.332321182 |
| Obesity | macrophage | *OGFOD1* | -1.387131844 | 0.000756402 | 0.026214747 | 1 |
| Obesity | macrophage | *YPEL3* | 1.671897675 | 1.40E-07 | 1.88E-05 | 0.000678442 |
| Obesity | macrophage | *PABPC4* | -1.267829003 | 6.76E-06 | 0.000537349 | 0.032778303 |
| Obesity | macrophage | *RGS17* | -1.122446201 | 1.66E-05 | 0.001223055 | 0.08072165 |
| Obesity | macrophage | *PPP2R3C* | 1.255287803 | 0.000660687 | 0.02447065 | 1 |
| Obesity | macrophage | *TBX15* | -1.053338003 | 1.60E-05 | 0.001197747 | 0.077853539 |
| Obesity | macrophage | *ITPR3* | 0.792219974 | 0.000365102 | 0.015816726 | 1 |
| Obesity | macrophage | *SETD1A* | 2.596606241 | 4.76E-07 | 5.43E-05 | 0.002311081 |
| Obesity | macrophage | *ANKRD54* | -0.857298608 | 0.00042831 | 0.018070973 | 1 |
| Obesity | macrophage | *CENPM* | 1.152026975 | 3.21E-05 | 0.002104345 | 0.155721498 |
| Obesity | macrophage | *ABHD12* | 1.061689013 | 0.000605012 | 0.022933723 | 1 |
| Obesity | macrophage | *RAD51C* | -0.966889568 | 0.000545746 | 0.021528141 | 1 |
| Obesity | macrophage | *CHRNE* | 0.326620167 | 0.00054008 | 0.021479264 | 1 |
| Obesity | macrophage | *B3GAT1* | 24.54743346 | 0.000103107 | 0.005322087 | 0.500276194 |
| Obesity | macrophage | *BTN3A3* | 1.453178865 | 1.45E-10 | 2.71E-08 | 7.05E-07 |
| Obesity | macrophage | *TULP1* | -0.903119615 | 0.000338529 | 0.014932216 | 1 |
| Obesity | macrophage | *NME5* | 0.515289659 | 0.000812861 | 0.027388895 | 1 |
| Obesity | macrophage | *TMCO6* | -1.027901464 | 0.000793238 | 0.027104169 | 1 |
| Obesity | macrophage | *PCCB* | 1.318662509 | 4.03E-06 | 0.000349518 | 0.019573018 |
| Obesity | macrophage | *CHMP3* | -1.22054878 | 3.89E-05 | 0.002451176 | 0.188740579 |
| Obesity | macrophage | *NSUN4* | -1.34186197 | 0.000811472 | 0.027388895 | 1 |
| Obesity | macrophage | *SERPINC1* | 2.848345758 | 7.26E-10 | 1.26E-07 | 3.52E-06 |
| Obesity | macrophage | *INVS* | 1.068198226 | 0.000148485 | 0.007232633 | 0.720448241 |
| Obesity | macrophage | *SNX19* | -1.322442144 | 6.04E-05 | 0.003661376 | 0.292910049 |
| Obesity | macrophage | *TEX14* | 1.145978144 | 0.000145373 | 0.007197463 | 0.705351332 |
| Obesity | macrophage | *OPN4* | 2.851926817 | 0.000141364 | 0.007071091 | 0.685895801 |
| Obesity | macrophage | *BTN2A2* | 0.528445466 | 2.74E-05 | 0.001819154 | 0.132798216 |
| Obesity | macrophage | *SNRPC* | -2.723493214 | 8.22E-19 | 9.58E-16 | 3.99E-15 |
| Obesity | macrophage | *GRK4* | 0.291071811 | 0.000188634 | 0.008973038 | 0.915249848 |

| | | | | | | |
|---|---|---|---|---|---|---|
| Obesity | macrophage | *RIBC2* | 0.688443334 | 0.000671419 | 0.024679743 | 1 |
| Obesity | macrophage | *RPAIN* | -1.082944053 | 0.000653301 | 0.024383214 | 1 |
| Obesity | macrophage | *AP1M2* | -0.630033484 | 0.00071314 | 0.025540736 | 1 |
| Obesity | macrophage | *ATP8B3* | -0.897516175 | 1.04E-10 | 2.20E-08 | 5.06E-07 |
| Obesity | macrophage | *PGPEP1* | 0.854509111 | 0.001093326 | 0.035131235 | 1 |
| Obesity | macrophage | *CYP2E1* | -0.488359461 | 0.001650296 | 0.048528699 | 1 |
| Obesity | macrophage | *NPAS1* | -2.779648004 | 4.81E-07 | 5.43E-05 | 0.002335699 |
| Obesity | macrophage | *NDUFA2* | 1.273377903 | 0.000158643 | 0.007621164 | 0.769737604 |
| Obesity | macrophage | *MYBBP1A* | 0.881986025 | 0.001391466 | 0.042044504 | 1 |
| Obesity | macrophage | *IGHMBP2* | 0.77051989 | 5.58E-05 | 0.003473905 | 0.270964552 |
| Obesity | macrophage | *LARS* | -0.639965185 | 0.001220473 | 0.037959854 | 1 |
| Obesity | macrophage | *POM121C* | -2.630166779 | 1.81E-11 | 5.87E-09 | 8.80E-08 |
| Obesity | macrophage | *SLC37A4* | 1.316943178 | 3.21E-07 | 4.01E-05 | 0.001558529 |
| Obesity | macrophage | *NUSAP1* | 1.410346628 | 0.001412307 | 0.04229947 | 1 |
| Obesity | macrophage | *NDUFAF1* | 1.424976466 | 6.29E-05 | 0.003768198 | 0.305224033 |
| Obesity | macrophage | *CENPO* | 2.421557817 | 1.12E-09 | 1.87E-07 | 5.42E-06 |
| Obesity | macrophage | *VPS33A* | 1.512132461 | 1.16E-05 | 0.000896935 | 0.056506914 |
| Obesity | macrophage | *DIAPH3* | 0.567922124 | 0.000439887 | 0.018370321 | 1 |
| Obesity | macrophage | *MYO19* | 2.398786833 | 1.32E-06 | 0.000121101 | 0.006418367 |
| Obesity | macrophage | *C18orf8* | -2.71092441 | 3.33E-11 | 9.50E-09 | 1.61E-07 |
| Obesity | macrophage | *SAE1* | -2.829857356 | 9.78E-07 | 9.69E-05 | 0.004746705 |
| Obesity | macrophage | *ADHFE1* | -0.35485595 | 0.000448228 | 0.018430512 | 1 |
| Obesity | macrophage | *RGR* | -4.055639968 | 0.000260744 | 0.011935191 | 1 |
| Obesity | macrophage | *MTG1* | 0.651012257 | 0.00057897 | 0.022654553 | 1 |
| Obesity | macrophage | *HSD17B12* | -0.713560396 | 8.64E-11 | 2.10E-08 | 4.19E-07 |
| Obesity | macrophage | *TNKS1BP1* | 0.706439179 | 6.57E-05 | 0.003887946 | 0.318811581 |
| Obesity | macrophage | *TTC12* | 0.903135611 | 0.000196288 | 0.009234361 | 0.952389636 |
| Obesity | macrophage | *TBX6* | 0.614856225 | 2.97E-11 | 9.01E-09 | 1.44E-07 |
| Obesity | macrophage | *POC5* | -0.912547153 | 2.05E-05 | 0.001423424 | 0.099639704 |
| Obesity | macrophage | *JAZF1* | 1.194859649 | 8.89E-05 | 0.004792355 | 0.431311959 |
| Obesity | macrophage | *NEIL2* | 0.963736981 | 7.18E-05 | 0.004144975 | 0.348177927 |
| Obesity | macrophage | *MMP16* | -0.449525764 | 0.000682701 | 0.024806352 | 1 |
| Obesity | macrophage | *TMED6* | 2.861000141 | 0.000426182 | 0.018070973 | 1 |
| Obesity | macrophage | *CLEC18A* | -1.701457842 | 3.00E-18 | 2.08E-15 | 1.46E-14 |
| Obesity | macrophage | *SLC5A11* | -0.560645272 | 3.78E-07 | 4.58E-05 | 0.001831778 |
| Obesity | macrophage | *CILP2* | -1.446111435 | 3.79E-05 | 0.002430969 | 0.183740506 |
| Obesity | macrophage | *ABCF3* | 0.79050037 | 0.001036191 | 0.033742269 | 1 |
| Obesity | macrophage | *CAMK2N1* | 0.85151361 | 0.001149906 | 0.036229491 | 1 |
| Obesity | macrophage | *ANTXR2* | 1.217582554 | 0.000246841 | 0.011406426 | 1 |
| Obesity | macrophage | *LMOD1* | -1.016483509 | 8.31E-05 | 0.004581761 | 0.403194945 |
| Obesity | macrophage | *DNALI1* | -1.033642648 | 2.33E-05 | 0.001566889 | 0.112816041 |
| Obesity | macrophage | *RFT1* | -1.24631118 | 8.18E-07 | 8.45E-05 | 0.003970094 |
| Obesity | macrophage | *RNF123* | 2.04666103 | 9.87E-19 | 9.58E-16 | 4.79E-15 |
| Obesity | macrophage | *MST1R* | 1.554438211 | 2.78E-18 | 2.08E-15 | 1.35E-14 |
| Obesity | macrophage | *RPP25L* | -1.115628305 | 0.000461996 | 0.018654914 | 1 |
| Obesity | macrophage | *SLC39A13* | 2.62363016 | 7.32E-07 | 7.72E-05 | 0.003552826 |
| Obesity | macrophage | *RAPSN* | -1.485389814 | 1.50E-13 | 7.28E-11 | 7.28E-10 |
| Obesity | macrophage | *PIP4K2C* | 2.260829555 | 0.000600188 | 0.022933723 | 1 |
| Obesity | macrophage | *SRR* | 0.509632384 | 1.73E-05 | 0.001237314 | 0.084137335 |
| Obesity | macrophage | *NT5DC2* | -0.928448409 | 5.20E-06 | 0.000427768 | 0.02523833 |

| | | | | | | |
|---|---|---|---|---|---|---|
| Obesity | macrophage | *FNTA* | 1.390044433 | 0.000102541 | 0.005322087 | 0.497530663 |
| Obesity | macrophage | *C8orf46* | 0.238988165 | 0.000332816 | 0.014814882 | 1 |
| Obesity | macrophage | *GPRIN1* | 1.64882691 | 0.000821902 | 0.02750253 | 1 |
| Obesity | macrophage | *INO80E* | -1.498749507 | 3.57E-14 | 1.93E-11 | 1.73E-10 |
| Obesity | macrophage | *ADORA2B* | -0.514726079 | 1.68E-06 | 0.000150709 | 0.008138284 |
| Obesity | macrophage | *TTLL6* | -3.555339809 | 1.01E-11 | 3.76E-09 | 4.89E-08 |
| Obesity | macrophage | *TSNARE1* | 1.476216662 | 1.85E-05 | 0.001301163 | 0.089780225 |
| Obesity | macrophage | *TRMT61B* | 1.067705693 | 0.00062363 | 0.02345622 | 1 |
| Obesity | macrophage | *C1QTNF4* | -0.871612191 | 2.12E-15 | 1.28E-12 | 1.03E-11 |
| Obesity | macrophage | *CTSW* | -0.217418729 | 9.61E-09 | 1.50E-06 | 4.66E-05 |
| Obesity | macrophage | *FUT10* | -0.520944791 | 0.000442978 | 0.018370321 | 1 |
| Obesity | macrophage | *SNX32* | 0.494546841 | 7.34E-05 | 0.00415289 | 0.356204456 |
| Obesity | macrophage | *MRGPRF* | 0.893543679 | 0.001328797 | 0.040549192 | 1 |
| Obesity | macrophage | *MST1* | -0.319494585 | 0.001674406 | 0.048648017 | 1 |
| Obesity | macrophage | *SEZ6L2* | 0.781184594 | 9.40E-05 | 0.004959174 | 0.456243986 |
| Obesity | macrophage | *KCTD13* | 1.528657687 | 4.57E-07 | 5.41E-05 | 0.00221869 |
| Obesity | macrophage | *CADM2* | 1.049608869 | 3.22E-07 | 4.01E-05 | 0.001563171 |
| Obesity | macrophage | *BANF1* | -2.533096761 | 4.99E-06 | 0.000424723 | 0.024232843 |
| Obesity | macrophage | *ORAI3* | 1.853748862 | 3.80E-11 | 1.03E-08 | 1.85E-07 |
| Obesity | macrophage | *RUFY1* | 1.306662223 | 0.000108369 | 0.005534779 | 0.525804032 |
| Obesity | macrophage | *RPP25* | -0.514426805 | 0.000729586 | 0.025625676 | 1 |
| Obesity | macrophage | *STX19* | 1.416500555 | 5.08E-06 | 0.000424723 | 0.024633951 |
| Obesity | macrophage | *MPI* | 1.070619537 | 0.000197934 | 0.009234361 | 0.960373539 |
| Obesity | macrophage | *TRIM73* | -3.631051301 | 1.74E-13 | 7.67E-11 | 8.44E-10 |
| Obesity | macrophage | *ZNF852* | -1.342493808 | 8.59E-05 | 0.004685184 | 0.41698137 |
| Obesity | macrophage | *TUFM* | 2.019308649 | 9.96E-11 | 2.20E-08 | 4.83E-07 |
| Obesity | macrophage | *C3orf38* | 1.432081566 | 8.91E-08 | 1.24E-05 | 0.000432347 |
| Obesity | macrophage | *RPRML* | 2.977296501 | 2.10E-05 | 0.00143613 | 0.101965251 |
| Obesity | macrophage | *ZFP41* | 1.157793438 | 0.000603232 | 0.022933723 | 1 |
| Obesity | macrophage | *PPP1R27* | -3.02094339 | 0.000715899 | 0.025540736 | 1 |
| Obesity | macrophage | *PLGLB1* | 0.634384962 | 0.001052279 | 0.03403772 | 1 |
| Obesity | macrophage | *PSMG1* | 1.068764367 | 0.000734124 | 0.025625676 | 1 |
| Obesity | macrophage | *EMILIN3* | 1.199887548 | 0.001276462 | 0.039448371 | 1 |
| Obesity | macrophage | *ACOT1* | 0.609607046 | 0.000149065 | 0.007232633 | 0.723263285 |
| Obesity | macrophage | *NUTM2A* | -0.942272106 | 1.28E-05 | 0.000967176 | 0.06189926 |
| Obesity | macrophage | *MAFF* | -0.597725821 | 0.000589394 | 0.022877926 | 1 |
| Obesity | macrophage | *GP1BA* | 1.233216594 | 1.16E-06 | 0.000108184 | 0.005625585 |
| Obesity | macrophage | *BTN3A2* | -0.446315372 | 1.38E-10 | 2.71E-08 | 6.71E-07 |
| Obesity | macrophage | *CENPP* | 0.750692122 | 1.70E-05 | 0.001229531 | 0.082378545 |
| Obesity | macrophage | *SKOR1* | 1.408604466 | 0.001195937 | 0.037436681 | 1 |
| Obesity | macrophage | *ZKSCAN3* | -1.404835349 | 6.97E-08 | 9.94E-06 | 0.000338114 |
| Obesity | macrophage | *CPNE4* | 1.151155845 | 3.81E-05 | 0.002430969 | 0.184753682 |
| Obesity | macrophage | *ZNF605* | 0.87347955 | 0.000775712 | 0.026693283 | 1 |
| Obesity | macrophage | *SULT1A2* | 3.497260428 | 3.35E-27 | 1.63E-23 | 1.63E-23 |
| Obesity | macrophage | *ZNF165* | -0.573079684 | 0.000304613 | 0.01368503 | 1 |
| Obesity | macrophage | *MRPL21* | -0.406489209 | 0.000456011 | 0.01859299 | 1 |
| Obesity | macrophage | *MFAP5* | -0.654005561 | 0.001034996 | 0.033742269 | 1 |
| Obesity | macrophage | *DNAH10* | 1.894319915 | 5.57E-10 | 1.00E-07 | 2.70E-06 |
| Obesity | macrophage | *RPS26* | -0.483992399 | 0.000465219 | 0.018654914 | 1 |
| Obesity | macrophage | *ZNF780A* | -0.304364646 | 5.73E-05 | 0.00351829 | 0.277944899 |

| | | | | | | |
|---|---|---|---|---|---|---|
| Obesity | macrophage | *SHISA4* | -8.402107327 | 1.92E-12 | 7.75E-10 | 9.29E-09 |
| Obesity | macrophage | *NOTCH4* | -0.130734639 | 1.39E-08 | 2.11E-06 | 6.74E-05 |
| Obesity | macrophage | *SKIV2L* | -0.561143681 | 6.41E-07 | 7.07E-05 | 0.003109938 |
| Obesity | macrophage | *GPANK1* | -1.858270495 | 6.59E-11 | 1.68E-08 | 3.20E-07 |
| Obesity | macrophage | *POU5F1* | 0.200869288 | 8.31E-05 | 0.004581761 | 0.403037605 |
| Obesity | macrophage | *CCHCR1* | 0.414158052 | 3.15E-06 | 0.00027806 | 0.015293273 |
| Obesity | macrophage | *RNF39* | -1.266500676 | 5.64E-06 | 0.000455969 | 0.027358158 |
| Obesity | macrophage | *ZFP57* | 0.496876354 | 5.41E-09 | 8.75E-07 | 2.62E-05 |
| Obesity | macrophage | *C17orf107* | 0.83603974 | 0.000303987 | 0.01368503 | 1 |
| Obesity | macrophage | *AC084018.1* | -0.533148106 | 9.08E-05 | 0.004843081 | 0.440720336 |
| Obesity | macrophage | *TMEM110* | 2.147301501 | 1.40E-10 | 2.71E-08 | 6.79E-07 |
| Obesity | macrophage | *GALT* | 1.601984222 | 0.000843909 | 0.028045527 | 1 |
| Obesity | macrophage | *EXOSC6* | -2.857435762 | 1.45E-11 | 5.03E-09 | 7.05E-08 |
| Obesity | macrophage | *C4B* | 1.210383537 | 0.000139707 | 0.007061008 | 0.677856796 |
| Obesity | macrophage | *LINC00240* | 0.360314039 | 0.001395129 | 0.042044504 | 1 |
| Obesity | macrophage | *AL049840.1* | -1.256658761 | 0.001463577 | 0.04356611 | 1 |
| Obesity | macrophage | *HCG20* | -1.117994997 | 6.56E-07 | 7.08E-05 | 0.003184555 |
| Obesity | macrophage | *TAPBP* | 1.155273306 | 0.001114906 | 0.035356366 | 1 |
| Obesity | macrophage | *ZSCAN31* | 0.440739208 | 8.79E-07 | 8.88E-05 | 0.004262557 |
| Obesity | macrophage | *PSMB9* | 0.618505158 | 1.12E-06 | 0.00010659 | 0.005436074 |
| Obesity | macrophage | *ISY1* | -0.702893946 | 0.000730305 | 0.025625676 | 1 |
| Obesity | macrophage | *C4A* | -1.238395984 | 1.00E-06 | 9.70E-05 | 0.004851284 |
| Obesity | macrophage | *DNAH10OS* | 10.24896337 | 3.66E-08 | 5.38E-06 | 0.000177678 |
| Obesity | macrophage | *DPP3* | 0.385439748 | 0.000966065 | 0.031886728 | 1 |
| Obesity | macrophage | *BRK1* | -0.989194243 | 0.000685089 | 0.024806352 | 1 |
| Obesity | macrophage | *PCDHA3* | -1.320459523 | 0.000342188 | 0.014957626 | 1 |
| Obesity | macrophage | *HMBS* | 1.593084664 | 7.91E-06 | 0.000619317 | 0.038397658 |
| Obesity | macrophage | *AP001172.2* | 0.373067163 | 0.00110698 | 0.035335972 | 1 |
| Obesity | mast_cell | *NFYA* | 2.322696741 | 0.000927309 | 0.001685764 | 1 |
| Obesity | mast_cell | *ENPP4* | 3.786777048 | 0.000845294 | 0.001572223 | 1 |
| Obesity | mast_cell | *SEMA3F* | -7.61351263 | 1.53E-08 | 2.48E-06 | 7.43E-05 |
| Obesity | mast_cell | *KRIT1* | 2.982054922 | 0.000219043 | 0.000976811 | 1 |
| Obesity | mast_cell | *RAD52* | 1.893714414 | 0.00064152 | 0.001292095 | 1 |
| Obesity | mast_cell | *HS3ST1* | 3.18037793 | 0.000792336 | 0.001499967 | 1 |
| Obesity | mast_cell | *SNX11* | 1.282397587 | 0.013370516 | 0.017590495 | 1 |
| Obesity | mast_cell | *M6PR* | 4.624507081 | 0.000416699 | 0.001080612 | 1 |
| Obesity | mast_cell | *CYP26B1* | 7.487273055 | 5.37E-06 | 0.000239147 | 0.026067019 |
| Obesity | mast_cell | *CD38* | 2.021485882 | 0.000599125 | 0.001235425 | 1 |
| Obesity | mast_cell | *RBM6* | -2.410165814 | 3.38E-05 | 0.000631284 | 0.164133885 |
| Obesity | mast_cell | *ARHGAP33* | 3.578769644 | 1.34E-05 | 0.000402498 | 0.064812534 |
| Obesity | mast_cell | *CDC27* | 3.061645007 | 2.23E-05 | 0.000536645 | 0.108205213 |
| Obesity | mast_cell | *POLR2J* | 4.776707333 | 0.000125811 | 0.000976811 | 0.610433263 |
| Obesity | mast_cell | *MEOX1* | 2.739621404 | 0.00090738 | 0.001656362 | 1 |
| Obesity | mast_cell | *LIG3* | 3.298331095 | 0.006230092 | 0.00869632 | 1 |
| Obesity | mast_cell | *CIAPIN1* | 5.881374755 | 0.000774484 | 0.001479447 | 1 |
| Obesity | mast_cell | *PON1* | -1.959148654 | 0.002053656 | 0.003252069 | 1 |
| Obesity | mast_cell | *GCFC2* | 3.140926204 | 0.000204628 | 0.000976811 | 0.992856573 |
| Obesity | mast_cell | *RHBDD2* | 2.661780736 | 7.52E-05 | 0.000934364 | 0.364635348 |
| Obesity | mast_cell | *ZNF195* | 4.743640805 | 0.000840964 | 0.00156689 | 1 |
| Obesity | mast_cell | *PDK2* | 2.315798909 | 0.024764334 | 0.031104465 | 1 |

| | | | | | | |
|---|---|---|---|---|---|---|
| Obesity | mast_cell | *GGNBP2* | 2.524892716 | 0.001964505 | 0.003133392 | 1 |
| Obesity | mast_cell | *TMEM98* | 1.559209814 | 0.001194293 | 0.002075005 | 1 |
| Obesity | mast_cell | *ABCC8* | 3.341544793 | 0.00390455 | 0.005706289 | 1 |
| Obesity | mast_cell | *SYNRG* | 3.116384599 | 0.000320111 | 0.000999138 | 1 |
| Obesity | mast_cell | *ZNF263* | 8.297517987 | 1.76E-05 | 0.000483458 | 0.085443883 |
| Obesity | mast_cell | *BAIAP2L1* | 3.179362473 | 0.000286806 | 0.000993978 | 1 |
| Obesity | mast_cell | *AGK* | 5.534021216 | 1.06E-05 | 0.000371097 | 0.051211327 |
| Obesity | mast_cell | *FARP2* | 2.647309424 | 0.000309545 | 0.000998908 | 1 |
| Obesity | mast_cell | *USH1C* | -3.005998562 | 0.002010546 | 0.003193182 | 1 |
| Obesity | mast_cell | *IFRD1* | 1.657832037 | 0.001232814 | 0.00212793 | 1 |
| Obesity | mast_cell | *VPS41* | 4.470088812 | 0.000245883 | 0.000976811 | 1 |
| Obesity | mast_cell | *SCIN* | -3.381403291 | 0.004721967 | 0.006766386 | 1 |
| Obesity | mast_cell | *CCDC124* | 3.468177776 | 0.013271753 | 0.017489013 | 1 |
| Obesity | mast_cell | *CEACAM21* | 2.27378425 | 0.005612 | 0.007920135 | 1 |
| Obesity | mast_cell | *CD79B* | -4.688204207 | 5.65E-05 | 0.000818974 | 0.274356395 |
| Obesity | mast_cell | *ST7L* | 2.09144921 | 0.007525597 | 0.010343965 | 1 |
| Obesity | mast_cell | *PAX6* | -2.257804888 | 0.010323195 | 0.013867353 | 1 |
| Obesity | mast_cell | *TSR3* | 13.01990762 | 0.000520547 | 0.001152325 | 1 |
| Obesity | mast_cell | *PSMB1* | 4.412700402 | 0.000215642 | 0.000976811 | 1 |
| Obesity | mast_cell | *NADK* | 4.345862383 | 0.001158842 | 0.00202374 | 1 |
| Obesity | mast_cell | *DLEC1* | 2.123939251 | 0.001329196 | 0.002271666 | 1 |
| Obesity | mast_cell | *CYB561* | 4.654274518 | 1.82E-05 | 0.00049351 | 0.088151232 |
| Obesity | mast_cell | *SS18L2* | 3.564774225 | 0.000109981 | 0.000976811 | 0.533629584 |
| Obesity | mast_cell | *MGST1* | 1.206559453 | 0.005298756 | 0.007521814 | 1 |
| Obesity | mast_cell | *IL32* | 3.218303674 | 0.000157677 | 0.000976811 | 0.765046723 |
| Obesity | mast_cell | *PKD1* | 8.138671797 | 5.66E-06 | 0.000243327 | 0.027445033 |
| Obesity | mast_cell | *MAPK8IP2* | 12.35496999 | 0.000139968 | 0.000976811 | 0.679125323 |
| Obesity | mast_cell | *MED24* | 2.743337324 | 0.000380117 | 0.001048903 | 1 |
| Obesity | mast_cell | *RPS20* | 2.226431621 | 0.000362627 | 0.001037687 | 1 |
| Obesity | mast_cell | *TRAF3IP3* | 2.193186043 | 0.000390911 | 0.001054382 | 1 |
| Obesity | mast_cell | *POMT2* | 2.037783576 | 0.000965094 | 0.001741405 | 1 |
| Obesity | mast_cell | *MLXIPL* | 1.867515476 | 3.03E-05 | 0.000621513 | 0.147157649 |
| Obesity | mast_cell | *DYRK4* | 2.18313285 | 0.005943628 | 0.008337231 | 1 |
| Obesity | mast_cell | *NCAPD2* | 2.384175168 | 0.000676998 | 0.001338003 | 1 |
| Obesity | mast_cell | *FUZ* | 5.127020956 | 0.000149396 | 0.000976811 | 0.724870735 |
| Obesity | mast_cell | *LRRC23* | 2.168697697 | 0.000134034 | 0.000976811 | 0.650334266 |
| Obesity | mast_cell | *CLCN6* | 2.229187094 | 4.94E-05 | 0.000765648 | 0.239621118 |
| Obesity | mast_cell | *SLC6A7* | 69.75405807 | 0.004855349 | 0.00694521 | 1 |
| Obesity | mast_cell | *TSPAN9* | 1.701805368 | 0.000355295 | 0.00103227 | 1 |
| Obesity | mast_cell | *UTP18* | 2.681314998 | 0.000149704 | 0.000976811 | 0.72636228 |
| Obesity | mast_cell | *TTC19* | 3.378064815 | 8.17E-05 | 0.000957283 | 0.396499033 |
| Obesity | mast_cell | *LARS2* | 2.327169422 | 0.000268488 | 0.000986064 | 1 |
| Obesity | mast_cell | *DCN* | 1.905872544 | 0.000132074 | 0.000976811 | 0.640823986 |
| Obesity | mast_cell | *PPP5C* | 3.815390903 | 0.000324942 | 0.001004947 | 1 |
| Obesity | mast_cell | *CEP68* | 4.971878556 | 0.00030183 | 0.000998908 | 1 |
| Obesity | mast_cell | *ZBTB32* | -3.757532909 | 0.004556643 | 0.006557152 | 1 |
| Obesity | mast_cell | *ERCC1* | 1.818215539 | 0.001462141 | 0.002459884 | 1 |
| Obesity | mast_cell | *CD22* | -0.190662441 | 0.002919699 | 0.004406339 | 1 |
| Obesity | mast_cell | *ALOX5* | -1.731371146 | 0.000654916 | 0.001311454 | 1 |
| Obesity | mast_cell | *MAP4K5* | 3.366788395 | 0.000175976 | 0.000976811 | 0.853837432 |

| | | | | | | |
|---|---|---|---|---|---|---|
| Obesity | mast_cell | *EHD3* | 4.828243576 | 0.000591378 | 0.001222568 | 1 |
| Obesity | mast_cell | *MAN2B2* | 5.704145893 | 0.000132509 | 0.000976811 | 0.642935445 |
| Obesity | mast_cell | *SLC25A39* | 2.734659235 | 0.015954463 | 0.0207592 | 1 |
| Obesity | mast_cell | *RNF14* | 2.381756386 | 0.000234861 | 0.000976811 | 1 |
| Obesity | mast_cell | *DDX11* | 1.478880763 | 0.027821521 | 0.034719655 | 1 |
| Obesity | mast_cell | *TACC3* | 3.238262471 | 0.000522333 | 0.001153408 | 1 |
| Obesity | mast_cell | *ZC3H3* | 2.060123909 | 0.001056669 | 0.001870475 | 1 |
| Obesity | mast_cell | *MDH1* | 2.956760091 | 4.60E-05 | 0.000754069 | 0.223375743 |
| Obesity | mast_cell | *MTMR11* | 2.947829063 | 0.000364958 | 0.001037687 | 1 |
| Obesity | mast_cell | *COX15* | 2.742717353 | 0.000294373 | 0.000998908 | 1 |
| Obesity | mast_cell | *CCDC88C* | 1.753463537 | 0.001116933 | 0.001960694 | 1 |
| Obesity | mast_cell | *NUDCD3* | 3.298934186 | 5.59E-06 | 0.000243327 | 0.0271358 |
| Obesity | mast_cell | *CHDH* | 5.349576274 | 0.000357783 | 0.001034916 | 1 |
| Obesity | mast_cell | *RUFY3* | 4.423486086 | 0.00016066 | 0.000976811 | 0.779524665 |
| Obesity | mast_cell | *WWTR1* | 2.101394278 | 0.000278251 | 0.000992498 | 1 |
| Obesity | mast_cell | *AGPS* | 4.891543541 | 0.000297821 | 0.000998908 | 1 |
| Obesity | mast_cell | *TTC27* | 1.961090317 | 0.000199012 | 0.000976811 | 0.965608384 |
| Obesity | mast_cell | *VSIG2* | -6.015732804 | 0.021456462 | 0.027203229 | 1 |
| Obesity | mast_cell | *MARCO* | 0.812301048 | 0.022273985 | 0.028169041 | 1 |
| Obesity | mast_cell | *HGF* | 6.943583634 | 9.79E-05 | 0.000976811 | 0.474808157 |
| Obesity | mast_cell | *OSBPL5* | 2.856934673 | 0.00017204 | 0.000976811 | 0.834737507 |
| Obesity | mast_cell | *SLC45A4* | 2.303527693 | 0.000496001 | 0.001138685 | 1 |
| Obesity | mast_cell | *ZDHHC6* | 4.858711026 | 0.000193522 | 0.000976811 | 0.938971073 |
| Obesity | mast_cell | *GRAMD1B* | 1.565874738 | 0.001993419 | 0.003171171 | 1 |
| Obesity | mast_cell | *RNH1* | 3.152931246 | 3.23E-05 | 0.000621513 | 0.156879948 |
| Obesity | mast_cell | *GLRX2* | 8.135854012 | 0.034398969 | 0.04240442 | 1 |
| Obesity | mast_cell | *ABCC2* | 6.244293328 | 0.002830406 | 0.004282236 | 1 |
| Obesity | mast_cell | *GCLM* | 4.257162546 | 0.000311202 | 0.000998908 | 1 |
| Obesity | mast_cell | *UBR2* | 2.506467275 | 0.000393737 | 0.001055175 | 1 |
| Obesity | mast_cell | *RRAGD* | 2.661650395 | 0.000537691 | 0.001169861 | 1 |
| Obesity | mast_cell | *HSF2* | 2.233054016 | 0.000895004 | 0.001639939 | 1 |
| Obesity | mast_cell | *TYMP* | 2.257441429 | 0.000587805 | 0.001217775 | 1 |
| Obesity | mast_cell | *TOMM34* | 2.747084281 | 0.001291263 | 0.002215421 | 1 |
| Obesity | mast_cell | *SEC63* | 4.628003227 | 0.000224347 | 0.000976811 | 1 |
| Obesity | mast_cell | *FAS* | 1.841836715 | 0.000343746 | 0.001027777 | 1 |
| Obesity | mast_cell | *RNASET2* | 1.405096165 | 0.002621815 | 0.004012948 | 1 |
| Obesity | mast_cell | *AGPAT4* | 6.758686945 | 0.000288233 | 0.000994669 | 1 |
| Obesity | mast_cell | *MIPEP* | 2.420531123 | 0.000571047 | 0.001195306 | 1 |
| Obesity | mast_cell | *INSRR* | 3.713353804 | 0.002232276 | 0.003494999 | 1 |
| Obesity | mast_cell | *VRK2* | 2.500362924 | 0.000300177 | 0.000998908 | 1 |
| Obesity | mast_cell | *VEZT* | 3.637262531 | 0.000177623 | 0.000976811 | 0.861825018 |
| Obesity | mast_cell | *BRD9* | 3.692998055 | 0.000190271 | 0.000976811 | 0.923194484 |
| Obesity | mast_cell | *SNX1* | 2.810694488 | 9.43E-05 | 0.000976811 | 0.457655377 |
| Obesity | mast_cell | *ANK1* | 3.41917895 | 0.00043586 | 0.001098433 | 1 |
| Obesity | mast_cell | *RABEP1* | 3.535663747 | 2.37E-05 | 0.000545024 | 0.11512579 |
| Obesity | mast_cell | *BAK1* | 2.874177646 | 0.009731213 | 0.01310823 | 1 |
| Obesity | mast_cell | *IKZF2* | 2.83079343 | 0.000272039 | 0.000987746 | 1 |
| Obesity | mast_cell | *GRN* | 1.776967477 | 0.000640361 | 0.001290831 | 1 |
| Obesity | mast_cell | *CENPQ* | -1.389021905 | 0.034280073 | 0.042268593 | 1 |
| Obesity | mast_cell | *IFT88* | 2.823200861 | 0.000287008 | 0.000993978 | 1 |

| | | | | | | |
|---|---|---|---|---|---|---|
| Obesity | mast_cell | *CHPF2* | 2.846409571 | 0.000283711 | 0.000993978 | 1 |
| Obesity | mast_cell | *UBA6* | 2.081140834 | 0.006466659 | 0.008994022 | 1 |
| Obesity | mast_cell | *ATP6V0A1* | 2.702718731 | 6.47E-05 | 0.000874503 | 0.313946565 |
| Obesity | mast_cell | *PIAS1* | 3.493835815 | 7.09E-06 | 0.000279503 | 0.034378912 |
| Obesity | mast_cell | *ASTE1* | 2.161053029 | 0.001561819 | 0.002595186 | 1 |
| Obesity | mast_cell | *MYOC* | 2.495375586 | 0.000233439 | 0.000976811 | 1 |
| Obesity | mast_cell | *SH3YL1* | 2.375032877 | 0.025542192 | 0.032030916 | 1 |
| Obesity | mast_cell | *VCL* | 2.8489139 | 0.00042831 | 0.001092079 | 1 |
| Obesity | mast_cell | *ADSS* | 5.581781622 | 0.000164161 | 0.000976811 | 0.796507292 |
| Obesity | mast_cell | *TBC1D23* | 2.206381927 | 0.000316904 | 0.000998908 | 1 |
| Obesity | mast_cell | *MYOM2* | -1.387745945 | 0.001894908 | 0.003036358 | 1 |
| Obesity | mast_cell | *TUBG2* | 1.666761178 | 0.003690837 | 0.005429939 | 1 |
| Obesity | mast_cell | *NSUN2* | 6.070674061 | 1.76E-05 | 0.000483458 | 0.085186899 |
| Obesity | mast_cell | *MFAP3* | 2.315400775 | 0.002263893 | 0.003542214 | 1 |
| Obesity | mast_cell | *MRI1* | 2.652189221 | 0.001291199 | 0.002215421 | 1 |
| Obesity | mast_cell | *AGA* | 2.360565168 | 0.000808809 | 0.001522242 | 1 |
| Obesity | mast_cell | *BOD1L1* | 5.252027237 | 0.000147142 | 0.000976811 | 0.713932145 |
| Obesity | mast_cell | *DNAH5* | 3.428443915 | 0.00030667 | 0.000998908 | 1 |
| Obesity | mast_cell | *PNKP* | 3.239607296 | 6.38E-05 | 0.000870117 | 0.309761565 |
| Obesity | mast_cell | *PHLPP2* | 2.074867368 | 0.000713584 | 0.001391373 | 1 |
| Obesity | mast_cell | *CTNS* | 1.711569642 | 0.00413391 | 0.006016116 | 1 |
| Obesity | mast_cell | *INPP4A* | 2.359349189 | 0.000192586 | 0.000976811 | 0.934426551 |
| Obesity | mast_cell | *PSMA4* | 2.974053258 | 0.000531544 | 0.001161155 | 1 |
| Obesity | mast_cell | *LSG1* | 3.430931777 | 4.08E-06 | 0.000195814 | 0.019777192 |
| Obesity | mast_cell | *TDP1* | 2.038095359 | 0.000366715 | 0.001037687 | 1 |
| Obesity | mast_cell | *SPATA7* | 1.503311436 | 0.001016078 | 0.00181317 | 1 |
| Obesity | mast_cell | *MED17* | 3.405406056 | 0.000262382 | 0.000983135 | 1 |
| Obesity | mast_cell | *AP2S1* | 2.591581274 | 0.000437665 | 0.001098433 | 1 |
| Obesity | mast_cell | *TRIT1* | 2.946761032 | 0.019101781 | 0.024499561 | 1 |
| Obesity | mast_cell | *CUL7* | 2.197320354 | 0.001033827 | 0.001838083 | 1 |
| Obesity | mast_cell | *CTNNA1* | 2.489845091 | 0.000227598 | 0.000976811 | 1 |
| Obesity | mast_cell | *CNTLN* | 3.471015409 | 0.000380908 | 0.001048903 | 1 |
| Obesity | mast_cell | *YTHDC2* | 3.35289856 | 0.000183804 | 0.000976811 | 0.891819367 |
| Obesity | mast_cell | *POLR2B* | 5.129946481 | 0.000281412 | 0.000993978 | 1 |
| Obesity | mast_cell | *ARAP2* | 1.356371498 | 0.000522933 | 0.00115383 | 1 |
| Obesity | mast_cell | *DTNBP1* | 5.31365252 | 0.000306045 | 0.000998908 | 1 |
| Obesity | mast_cell | *ANO2* | 1.499790396 | 0.000452726 | 0.001107978 | 1 |
| Obesity | mast_cell | *RRM2B* | 4.236993221 | 0.000819835 | 0.001535845 | 1 |
| Obesity | mast_cell | *SNX29* | 3.641848953 | 0.000167815 | 0.000976811 | 0.814239643 |
| Obesity | mast_cell | *EPN3* | -2.260546485 | 0.006879356 | 0.009517717 | 1 |
| Obesity | mast_cell | *RCN1* | 2.720327035 | 0.004857246 | 0.006945876 | 1 |
| Obesity | mast_cell | *JKAMP* | 2.567022857 | 6.57E-05 | 0.000883548 | 0.318960898 |
| Obesity | mast_cell | *MCUR1* | 3.014223997 | 0.000433645 | 0.001096396 | 1 |
| Obesity | mast_cell | *LAMC3* | -1.654871723 | 0.014576618 | 0.019068685 | 1 |
| Obesity | mast_cell | *MAPK9* | 6.948127703 | 0.000260308 | 0.00098093 | 1 |
| Obesity | mast_cell | *FAM160A2* | 1.984596276 | 0.000641336 | 0.001292095 | 1 |
| Obesity | mast_cell | *CYBA* | 2.598748357 | 0.000232676 | 0.000976811 | 1 |
| Obesity | mast_cell | *THOC3* | -4.410885705 | 0.011120213 | 0.014851438 | 1 |
| Obesity | mast_cell | *ALX4* | 3.988583324 | 0.001249465 | 0.002153606 | 1 |
| Obesity | mast_cell | *AKR7A2* | 2.286992332 | 0.000458373 | 0.001112606 | 1 |

| | | | | | | |
|---|---|---|---|---|---|---|
| Obesity | mast_cell | *ANAPC4* | 2.582964534 | 5.24E-05 | 0.000791601 | 0.254388815 |
| Obesity | mast_cell | *LY75* | 12.09048549 | 3.51E-05 | 0.000647264 | 0.170353251 |
| Obesity | mast_cell | *OPN3* | 2.75863278 | 2.52E-05 | 0.000550066 | 0.122078366 |
| Obesity | mast_cell | *SDCCAG8* | 2.896023544 | 6.61E-06 | 0.000269699 | 0.032094163 |
| Obesity | mast_cell | *KIF1B* | 3.456136106 | 4.64E-05 | 0.000755431 | 0.225118475 |
| Obesity | mast_cell | *TBC1D22A* | 2.195619543 | 0.000227239 | 0.000976811 | 1 |
| Obesity | mast_cell | *PLEKHH1* | 2.398524521 | 0.002673998 | 0.004083802 | 1 |
| Obesity | mast_cell | *FAM114A2* | 1.939562796 | 0.000743042 | 0.001435778 | 1 |
| Obesity | mast_cell | *GINM1* | 2.634196076 | 0.000367971 | 0.001037687 | 1 |
| Obesity | mast_cell | *ITIH4* | 3.516786315 | 7.20E-12 | 3.18E-09 | 3.49E-08 |
| Obesity | mast_cell | *PCGF2* | 1.892406061 | 0.00386674 | 0.005661262 | 1 |
| Obesity | mast_cell | *IL17RB* | 3.437370289 | 0.008027751 | 0.010987489 | 1 |
| Obesity | mast_cell | *MSH4* | 2.042333404 | 0.009267611 | 0.012542942 | 1 |
| Obesity | mast_cell | *GDI2* | 4.360952155 | 0.000729238 | 0.001413873 | 1 |
| Obesity | mast_cell | *PITHD1* | 6.314462416 | 0.000942636 | 0.001709783 | 1 |
| Obesity | mast_cell | *ATP11B* | 3.510913241 | 1.83E-05 | 0.00049351 | 0.088710026 |
| Obesity | mast_cell | *LAMC2* | 1.865360518 | 0.006659922 | 0.009251057 | 1 |
| Obesity | mast_cell | *RASGRF1* | 5.399219434 | 0.000241078 | 0.000976811 | 1 |
| Obesity | mast_cell | *CROCC* | 3.468050779 | 6.26E-05 | 0.000864985 | 0.303609828 |
| Obesity | mast_cell | *FLYWCH1* | 2.282101876 | 0.000192055 | 0.000976811 | 0.931849376 |
| Obesity | mast_cell | *ALDH18A1* | 3.149698287 | 0.000110086 | 0.000976811 | 0.534137133 |
| Obesity | mast_cell | *DNAJC25* | 2.802833142 | 0.00049968 | 0.001141453 | 1 |
| Obesity | mast_cell | *OGFR* | 3.162953997 | 0.000157962 | 0.000976811 | 0.766433651 |
| Obesity | mast_cell | *PIGV* | 1.439678867 | 0.032982949 | 0.040741668 | 1 |
| Obesity | mast_cell | *RIMBP2* | -3.179152818 | 0.016370291 | 0.021243287 | 1 |
| Obesity | mast_cell | *PRDM6* | 9.406315177 | 8.16E-05 | 0.000957283 | 0.395938839 |
| Obesity | mast_cell | *NCKAP1* | 2.27160088 | 0.000436953 | 0.001098433 | 1 |
| Obesity | mast_cell | *LTK* | 20.57693055 | 0.000390845 | 0.001054382 | 1 |
| Obesity | mast_cell | *EIF4B* | 3.798768997 | 7.34E-05 | 0.000924875 | 0.356076877 |
| Obesity | mast_cell | *SLC6A16* | 1.97224787 | 0.000711226 | 0.00138868 | 1 |
| Obesity | mast_cell | *AHRR* | 4.045648668 | 0.002102735 | 0.003322739 | 1 |
| Obesity | mast_cell | *ADCK1* | 2.086088623 | 0.000523323 | 0.001154019 | 1 |
| Obesity | mast_cell | *HAGH* | 4.086169257 | 0.001194453 | 0.002075005 | 1 |
| Obesity | mast_cell | *RNF4* | 5.48627325 | 0.000166068 | 0.000976811 | 0.805763105 |
| Obesity | mast_cell | *CASP8* | 1.528679714 | 0.000880388 | 0.001617433 | 1 |
| Obesity | mast_cell | *TM7SF3* | 4.380248341 | 0.000143886 | 0.000976811 | 0.698136797 |
| Obesity | mast_cell | *CDON* | 1.074937851 | 0.001549124 | 0.002580277 | 1 |
| Obesity | mast_cell | *TMEM161A* | 2.154986268 | 0.000732662 | 0.001419113 | 1 |
| Obesity | mast_cell | *LPAR2* | 4.570030622 | 4.45E-05 | 0.00074422 | 0.216055515 |
| Obesity | mast_cell | *SUGP2* | 2.290208399 | 0.000603612 | 0.001238886 | 1 |
| Obesity | mast_cell | *SLC12A2* | 2.594500088 | 0.000329524 | 0.001011931 | 1 |
| Obesity | mast_cell | *CNN2* | 3.547153461 | 0.000227831 | 0.000976811 | 1 |
| Obesity | mast_cell | *ABCA7* | 2.385970062 | 0.010463523 | 0.014043988 | 1 |
| Obesity | mast_cell | *DDX20* | 4.092306218 | 0.000103757 | 0.000976811 | 0.503427075 |
| Obesity | mast_cell | *BTBD1* | 5.222126044 | 5.13E-05 | 0.000782037 | 0.248687802 |
| Obesity | mast_cell | *FAR2* | 4.034660207 | 0.000342423 | 0.001027482 | 1 |
| Obesity | mast_cell | *BCAS1* | 3.299450709 | 0.001857201 | 0.002985798 | 1 |
| Obesity | mast_cell | *CHI3L2* | -2.593184396 | 0.002713563 | 0.004130261 | 1 |
| Obesity | mast_cell | *SBNO2* | 2.247578642 | 0.000261897 | 0.000983135 | 1 |
| Obesity | mast_cell | *PMS1* | 6.776530004 | 0.000300783 | 0.000998908 | 1 |

| | | | | | | |
|---|---|---|---|---|---|---|
| Obesity | mast_cell | *UHRF1BP1* | 5.783971061 | 5.14E-16 | 4.99E-13 | 2.50E-12 |
| Obesity | mast_cell | *WDR18* | 1.505135964 | 0.024513442 | 0.030805288 | 1 |
| Obesity | mast_cell | *ANKRD44* | 1.878408562 | 0.000207715 | 0.000976811 | 1 |
| Obesity | mast_cell | *ADAT1* | -1.349630589 | 0.005344272 | 0.007577559 | 1 |
| Obesity | mast_cell | *MAP2K4* | 2.267917581 | 0.000322228 | 0.001000927 | 1 |
| Obesity | mast_cell | *TMEM206* | 2.042766728 | 0.000680307 | 0.001342901 | 1 |
| Obesity | mast_cell | *SEC61A2* | 2.517860712 | 0.002647669 | 0.004048322 | 1 |
| Obesity | mast_cell | *TIE1* | 1.585946819 | 0.004200382 | 0.006108347 | 1 |
| Obesity | mast_cell | *NGEF* | -6.939304444 | 0.036977103 | 0.04543249 | 1 |
| Obesity | mast_cell | *SPI1* | 1.766783537 | 0.001675331 | 0.002736938 | 1 |
| Obesity | mast_cell | *ATXN3* | 2.830113607 | 7.58E-05 | 0.000935479 | 0.368015678 |
| Obesity | mast_cell | *GOLGA5* | 7.468925798 | 0.000672433 | 0.001332235 | 1 |
| Obesity | mast_cell | *FGFR2* | 2.145868026 | 0.000183114 | 0.000976811 | 0.888470792 |
| Obesity | mast_cell | *MTFR1* | 3.299674492 | 0.003620065 | 0.005340394 | 1 |
| Obesity | mast_cell | *FECH* | 1.561219805 | 0.001807145 | 0.002922462 | 1 |
| Obesity | mast_cell | *MYO9A* | 2.609102093 | 0.000314892 | 0.000998908 | 1 |
| Obesity | mast_cell | *TRAM1* | 4.762502607 | 0.000139273 | 0.000976811 | 0.675753218 |
| Obesity | mast_cell | *EVI5* | 1.964959431 | 0.000202366 | 0.000976811 | 0.981880025 |
| Obesity | mast_cell | *STOML1* | 4.012264935 | 0.000503971 | 0.001144783 | 1 |
| Obesity | mast_cell | *METTL22* | 2.483563397 | 0.00091085 | 0.00165895 | 1 |
| Obesity | mast_cell | *TP53BP1* | 4.403343228 | 7.02E-05 | 0.000898663 | 0.340593095 |
| Obesity | mast_cell | *RRP15* | 2.144609011 | 0.003125413 | 0.004681848 | 1 |
| Obesity | mast_cell | *DHX8* | 3.324645195 | 0.000259657 | 0.00098093 | 1 |
| Obesity | mast_cell | *ROGDI* | -4.910308898 | 0.027362127 | 0.034199135 | 1 |
| Obesity | mast_cell | *CBFB* | 4.300146916 | 0.000219173 | 0.000976811 | 1 |
| Obesity | mast_cell | *IFI35* | 3.428072694 | 0.000548534 | 0.001175269 | 1 |
| Obesity | mast_cell | *HEATR6* | 2.838942169 | 0.000565361 | 0.001190078 | 1 |
| Obesity | mast_cell | *POLR1A* | 1.847335034 | 0.003806333 | 0.005579556 | 1 |
| Obesity | mast_cell | *IP6K2* | 4.184836141 | 0.000553317 | 0.00118075 | 1 |
| Obesity | mast_cell | *STON1-GTF2A* | 1.045580341 | 0.006005195 | 0.008409006 | 1 |
| Obesity | mast_cell | *SRBD1* | 5.259636549 | 8.19E-05 | 0.000957283 | 0.397272244 |
| Obesity | mast_cell | *CYFIP1* | 2.667785121 | 0.000191736 | 0.000976811 | 0.930302555 |
| Obesity | mast_cell | *NUCKS1* | 4.44514248 | 0.00031614 | 0.000998908 | 1 |
| Obesity | mast_cell | *CLEC2D* | 1.445509607 | 0.000506103 | 0.001144962 | 1 |
| Obesity | mast_cell | *DRD4* | 11.29453447 | 0.000134824 | 0.000976811 | 0.654165752 |
| Obesity | mast_cell | *PLA2G10* | 18.708297 | 0.000139421 | 0.000976811 | 0.676471567 |
| Obesity | mast_cell | *NEDD4* | 3.534209455 | 0.012142258 | 0.016118806 | 1 |
| Obesity | mast_cell | *PIGB* | 1.993581022 | 0.000416012 | 0.001080612 | 1 |
| Obesity | mast_cell | *GNB5* | 2.679868883 | 0.016831735 | 0.021818749 | 1 |
| Obesity | mast_cell | *LRP6* | 2.297958781 | 0.000320637 | 0.000999138 | 1 |
| Obesity | mast_cell | *CLTCL1* | 2.019511974 | 0.000194949 | 0.000976811 | 0.945890918 |
| Obesity | mast_cell | *DGCR2* | 3.96820767 | 0.000171783 | 0.000976811 | 0.833490857 |
| Obesity | mast_cell | *ZXDC* | 3.935696127 | 0.000182347 | 0.000976811 | 0.884746728 |
| Obesity | mast_cell | *GBA2* | 3.455714315 | 0.000528442 | 0.001158084 | 1 |
| Obesity | mast_cell | *NDST1* | 4.331742833 | 0.000100462 | 0.000976811 | 0.487443819 |
| Obesity | mast_cell | *ASNS* | 4.49924044 | 0.002357695 | 0.003672403 | 1 |
| Obesity | mast_cell | *AP3M2* | 3.248241679 | 0.000592791 | 0.001224967 | 1 |
| Obesity | mast_cell | *ST6GALNAC2* | 4.849320586 | 0.000443749 | 0.001101314 | 1 |
| Obesity | mast_cell | *EIF2B3* | 2.735857449 | 0.000279884 | 0.000993978 | 1 |
| Obesity | mast_cell | *MAP4K4* | 3.671796508 | 0.000305734 | 0.000998908 | 1 |

| | | | | | | |
|---|---|---|---|---|---|---|
| Obesity | mast_cell | *WDR1* | 3.673150407 | 0.000257717 | 0.00098093 | 1 |
| Obesity | mast_cell | *SNX13* | 2.085368437 | 0.000294343 | 0.000998908 | 1 |
| Obesity | mast_cell | *RPS6KA2* | 2.539298454 | 0.000419757 | 0.00108267 | 1 |
| Obesity | mast_cell | *ING3* | 7.278014948 | 0.004503262 | 0.006490424 | 1 |
| Obesity | mast_cell | *TRIB2* | 2.811134069 | 0.00174825 | 0.002836012 | 1 |
| Obesity | mast_cell | *CYBRD1* | 2.746197137 | 6.36E-05 | 0.000869436 | 0.308649648 |
| Obesity | mast_cell | *RDH11* | 3.921356395 | 0.000834022 | 0.001557611 | 1 |
| Obesity | mast_cell | *EPN2* | 2.692261635 | 0.000545002 | 0.00117408 | 1 |
| Obesity | mast_cell | *ALDH3A2* | 2.064206265 | 0.000201449 | 0.000976811 | 0.977431167 |
| Obesity | mast_cell | *AFF4* | 3.300828463 | 3.27E-05 | 0.000621513 | 0.158441329 |
| Obesity | mast_cell | *TRHDE* | 1.799177327 | 0.003809612 | 0.005582675 | 1 |
| Obesity | mast_cell | *FCGR2B* | 2.073510773 | 0.000554265 | 0.001181069 | 1 |
| Obesity | mast_cell | *NFATC3* | 2.694997031 | 0.00017231 | 0.000976811 | 0.836049064 |
| Obesity | mast_cell | *TRNT1* | 2.484974332 | 0.000272348 | 0.000987746 | 1 |
| Obesity | mast_cell | *CRMP1* | 2.272539888 | 0.000399391 | 0.001064165 | 1 |
| Obesity | mast_cell | *NDE1* | 1.836664478 | 0.000327055 | 0.001007536 | 1 |
| Obesity | mast_cell | *AP1M1* | 2.281557652 | 0.000459009 | 0.001112606 | 1 |
| Obesity | mast_cell | *PVR* | 4.605926393 | 4.46E-05 | 0.00074422 | 0.216567972 |
| Obesity | mast_cell | *SCARB1* | 1.564760201 | 0.000800127 | 0.001511766 | 1 |
| Obesity | mast_cell | *MOV10L1* | 4.880797366 | 0.000239807 | 0.000976811 | 1 |
| Obesity | mast_cell | *ALPK1* | 3.467363266 | 4.91E-05 | 0.000765648 | 0.238283627 |
| Obesity | mast_cell | *LLGL2* | 2.862855761 | 0.000401909 | 0.001064698 | 1 |
| Obesity | mast_cell | *SDHA* | 3.498467538 | 3.00E-05 | 0.000621513 | 0.145785665 |
| Obesity | mast_cell | *SMARCE1* | 2.89459164 | 0.000111135 | 0.000976811 | 0.539229282 |
| Obesity | mast_cell | *ADAM11* | -1.4642512 | 0.001056672 | 0.001870475 | 1 |
| Obesity | mast_cell | *CD5L* | 6.419093609 | 0.000140677 | 0.000976811 | 0.682563286 |
| Obesity | mast_cell | *PPP2R2C* | 3.817092284 | 0.000554582 | 0.001181226 | 1 |
| Obesity | mast_cell | *ZNF532* | 2.388746841 | 0.000283816 | 0.000993978 | 1 |
| Obesity | mast_cell | *ZZEF1* | 2.860684186 | 9.63E-05 | 0.000976811 | 0.467266995 |
| Obesity | mast_cell | *TACR2* | 22.02955256 | 0.006177353 | 0.008635124 | 1 |
| Obesity | mast_cell | *TIPIN* | 3.150105246 | 9.76E-07 | 7.40E-05 | 0.004734455 |
| Obesity | mast_cell | *TTC38* | 2.483665431 | 0.000155907 | 0.000976811 | 0.756462644 |
| Obesity | mast_cell | *GRAMD4* | 5.435405439 | 0.00021952 | 0.000976811 | 1 |
| Obesity | mast_cell | *ADD2* | 3.346908703 | 9.94E-05 | 0.000976811 | 0.482413218 |
| Obesity | mast_cell | *VPS9D1* | 2.513298513 | 0.000192816 | 0.000976811 | 0.935542928 |
| Obesity | mast_cell | *FNDC3B* | 2.917587519 | 0.00013087 | 0.000976811 | 0.634981596 |
| Obesity | mast_cell | *CACNG4* | 3.696895477 | 0.001859126 | 0.002987902 | 1 |
| Obesity | mast_cell | *MOCOS* | 3.284226408 | 0.000101807 | 0.000976811 | 0.493968007 |
| Obesity | mast_cell | *SEC31B* | 4.508993026 | 1.46E-05 | 0.000430748 | 0.071073477 |
| Obesity | mast_cell | *KIFAP3* | 3.004383305 | 0.000258221 | 0.00098093 | 1 |
| Obesity | mast_cell | *MKRN2* | 2.11466454 | 0.000179007 | 0.000976811 | 0.86854108 |
| Obesity | mast_cell | *MLH1* | 4.822898776 | 0.012816725 | 0.016935389 | 1 |
| Obesity | mast_cell | *FMO4* | 6.355152607 | 1.25E-05 | 0.000388294 | 0.060573857 |
| Obesity | mast_cell | *RGS11* | 1.893592006 | 0.000318268 | 0.000998908 | 1 |
| Obesity | mast_cell | *SPAG5* | 4.934912578 | 0.039688321 | 0.048493512 | 1 |
| Obesity | mast_cell | *ANKRD13A* | 3.008256317 | 0.000401947 | 0.001064698 | 1 |
| Obesity | mast_cell | *GPATCH1* | 2.058937287 | 0.000379605 | 0.001048885 | 1 |
| Obesity | mast_cell | *ICAM3* | 4.688368018 | 0.001438359 | 0.002424078 | 1 |
| Obesity | mast_cell | *NT5C2* | 3.967268865 | 8.43E-07 | 6.49E-05 | 0.004088618 |
| Obesity | mast_cell | *XAB2* | 3.149908997 | 0.000376519 | 0.001048052 | 1 |

| | | | | | | |
|---|---|---|---|---|---|---|
| Obesity | mast_cell | *IL4R* | 3.327649851 | 0.000152343 | 0.000976811 | 0.739165971 |
| Obesity | mast_cell | *DYNC1I2* | 3.49673798 | 0.000184212 | 0.000976811 | 0.893796514 |
| Obesity | mast_cell | *APBB1IP* | 1.852369278 | 0.000458164 | 0.001112606 | 1 |
| Obesity | mast_cell | *LRCH4* | 5.407818257 | 0.001873878 | 0.003008623 | 1 |
| Obesity | mast_cell | *GPR137B* | 2.095883366 | 0.000568087 | 0.001192771 | 1 |
| Obesity | mast_cell | *NAALAD2* | 1.537699174 | 0.020010798 | 0.025550629 | 1 |
| Obesity | mast_cell | *AMPH* | 2.531102927 | 0.00030776 | 0.000998908 | 1 |
| Obesity | mast_cell | *MCCC1* | 2.173508652 | 0.000151824 | 0.000976811 | 0.736651173 |
| Obesity | mast_cell | *ACER3* | 4.871097244 | 0.000204459 | 0.000976811 | 0.99203664 |
| Obesity | mast_cell | *PIK3C3* | 2.319318556 | 0.000953219 | 0.001725753 | 1 |
| Obesity | mast_cell | *RBFOX1* | 1.553888563 | 0.000376425 | 0.001048052 | 1 |
| Obesity | mast_cell | *PCM1* | 2.565961484 | 0.000269288 | 0.000986064 | 1 |
| Obesity | mast_cell | *MYH7B* | 2.655050665 | 0.002333251 | 0.003636663 | 1 |
| Obesity | mast_cell | *FKBP7* | 2.158036596 | 0.001543005 | 0.002571893 | 1 |
| Obesity | mast_cell | *XRCC5* | 3.091989764 | 0.000219437 | 0.000976811 | 1 |
| Obesity | mast_cell | *SP140* | 2.2056046 | 0.00058129 | 0.001209443 | 1 |
| Obesity | mast_cell | *SAR1A* | 1.971392802 | 0.000333147 | 0.001016573 | 1 |
| Obesity | mast_cell | *RAPGEF3* | 1.460421782 | 0.024230118 | 0.030465025 | 1 |
| Obesity | mast_cell | *FDFT1* | 3.164171516 | 2.35E-06 | 0.000137224 | 0.011413785 |
| Obesity | mast_cell | *SCGN* | 17.23059493 | 0.00032317 | 0.001002436 | 1 |
| Obesity | mast_cell | *PGM1* | 3.068835871 | 0.00048262 | 0.001131243 | 1 |
| Obesity | mast_cell | *DDX1* | 2.835810215 | 0.001900581 | 0.003043439 | 1 |
| Obesity | mast_cell | *DNM2* | 2.57420231 | 0.000893888 | 0.001638513 | 1 |
| Obesity | mast_cell | *STX7* | 2.764990206 | 0.000185055 | 0.000976811 | 0.89788474 |
| Obesity | mast_cell | *RABL2B* | 3.544719878 | 0.000398027 | 0.001061493 | 1 |
| Obesity | mast_cell | *DCT* | 3.052408262 | 0.000127907 | 0.000976811 | 0.620603257 |
| Obesity | mast_cell | *SCTR* | 1.873011004 | 0.00052202 | 0.001153388 | 1 |
| Obesity | mast_cell | *RIF1* | 4.720828717 | 0.000140628 | 0.000976811 | 0.682325806 |
| Obesity | mast_cell | *KCNN2* | 4.832801295 | 0.004097651 | 0.005972305 | 1 |
| Obesity | mast_cell | *PSEN1* | 2.806121176 | 0.000130339 | 0.000976811 | 0.632406153 |
| Obesity | mast_cell | *MOK* | 2.868588124 | 4.79E-05 | 0.000760536 | 0.232601198 |
| Obesity | mast_cell | *STK17B* | 2.956709746 | 0.000493236 | 0.001138685 | 1 |
| Obesity | mast_cell | *ZNF506* | 6.21231314 | 1.57E-05 | 0.000455532 | 0.076073821 |
| Obesity | mast_cell | *PCDHB4* | 6.076362052 | 0.000332865 | 0.001016573 | 1 |
| Obesity | mast_cell | *ME2* | 2.951721061 | 0.000249322 | 0.000976811 | 1 |
| Obesity | mast_cell | *CCNT2* | 3.262096268 | 0.003072533 | 0.00461403 | 1 |
| Obesity | mast_cell | *SEMA5B* | -1.910053003 | 0.013133962 | 0.01732155 | 1 |
| Obesity | mast_cell | *DOPEY1* | 2.94062093 | 1.34E-05 | 0.000402498 | 0.065204748 |
| Obesity | mast_cell | *LYRM2* | 4.466371494 | 8.90E-05 | 0.000976811 | 0.431976822 |
| Obesity | mast_cell | *ZCCHC6* | 2.550699666 | 0.0002448 | 0.000976811 | 1 |
| Obesity | mast_cell | *ULK2* | 2.368781656 | 0.000211215 | 0.000976811 | 1 |
| Obesity | mast_cell | *PLOD1* | 3.991951489 | 6.90E-05 | 0.000898663 | 0.334640926 |
| Obesity | mast_cell | *PIBF1* | 2.48390946 | 0.000210776 | 0.000976811 | 1 |
| Obesity | mast_cell | *OXCT1* | 1.444879094 | 0.000857357 | 0.001585931 | 1 |
| Obesity | mast_cell | *SLC27A5* | 2.705630603 | 0.000394015 | 0.001055175 | 1 |
| Obesity | mast_cell | *ZNF586* | 4.197554457 | 2.42E-05 | 0.000545024 | 0.117411025 |
| Obesity | mast_cell | *ZNF264* | 3.044119885 | 0.000280902 | 0.000993978 | 1 |
| Obesity | mast_cell | *RPS5* | 3.029928896 | 0.000223084 | 0.000976811 | 1 |
| Obesity | mast_cell | *CHMP2B* | 3.670074117 | 0.00025834 | 0.00098093 | 1 |
| Obesity | mast_cell | *PPIE* | 2.944639548 | 2.40E-05 | 0.000545024 | 0.116375896 |

| | | | | | | |
|---|---|---|---|---|---|---|
| Obesity | mast_cell | *NOA1* | 3.885878526 | 0.001884786 | 0.003024133 | 1 |
| Obesity | mast_cell | *GSTP1* | 5.759149103 | 2.98E-07 | 3.21E-05 | 0.001445553 |
| Obesity | mast_cell | *WBP11* | 1.863008262 | 0.000477871 | 0.001129371 | 1 |
| Obesity | mast_cell | *NCOA1* | 3.403692463 | 0.000140823 | 0.000976811 | 0.683273204 |
| Obesity | mast_cell | *CAD* | 6.072055804 | 0.001650493 | 0.002706385 | 1 |
| Obesity | mast_cell | *CD59* | 2.572904985 | 0.000121529 | 0.000976811 | 0.589657429 |
| Obesity | mast_cell | *CD82* | -0.806512309 | 0.031074591 | 0.038521696 | 1 |
| Obesity | mast_cell | *HACE1* | 2.887977823 | 0.000227452 | 0.000976811 | 1 |
| Obesity | mast_cell | *SEH1L* | 3.823069941 | 0.003212463 | 0.004794484 | 1 |
| Obesity | mast_cell | *SLC25A24* | 3.67807042 | 7.83E-05 | 0.000944925 | 0.380101416 |
| Obesity | mast_cell | *PILRA* | 1.060138307 | 0.000926731 | 0.001685344 | 1 |
| Obesity | mast_cell | *ZNF213* | 2.303076555 | 0.003061822 | 0.004602219 | 1 |
| Obesity | mast_cell | *CPNE3* | 2.707021172 | 0.000225578 | 0.000976811 | 1 |
| Obesity | mast_cell | *RRN3* | 1.865583862 | 0.000871597 | 0.001605539 | 1 |
| Obesity | mast_cell | *MTIF2* | 6.394976682 | 0.000306858 | 0.000998908 | 1 |
| Obesity | mast_cell | *DDHD2* | 3.650538538 | 0.007353561 | 0.010118968 | 1 |
| Obesity | mast_cell | *EPS15* | 1.895027168 | 0.015556409 | 0.020268447 | 1 |
| Obesity | mast_cell | *USP40* | 2.295630237 | 0.000278756 | 0.000992498 | 1 |
| Obesity | mast_cell | *POMGNT1* | 2.655916686 | 0.00084541 | 0.001572223 | 1 |
| Obesity | mast_cell | *MAST2* | 3.817157299 | 1.11E-05 | 0.000373166 | 0.053757209 |
| Obesity | mast_cell | *EIF2AK1* | 6.890497605 | 0.000219422 | 0.000976811 | 1 |
| Obesity | mast_cell | *NME8* | 70.40559465 | 4.80E-05 | 0.000760536 | 0.232723991 |
| Obesity | mast_cell | *SNX10* | 1.103602141 | 0.015806193 | 0.020577314 | 1 |
| Obesity | mast_cell | *ITPKC* | 2.035861948 | 0.000146034 | 0.000976811 | 0.708557443 |
| Obesity | mast_cell | *TMED2* | 2.86600414 | 0.003428538 | 0.005076371 | 1 |
| Obesity | mast_cell | *ALG9* | 2.845537582 | 0.000475312 | 0.001127398 | 1 |
| Obesity | mast_cell | *ACOX3* | 4.007769701 | 0.000461116 | 0.001114555 | 1 |
| Obesity | mast_cell | *HSD17B14* | 2.879044413 | 0.000667646 | 0.001326001 | 1 |
| Obesity | mast_cell | *TRIP6* | 2.108537225 | 0.001838402 | 0.002965401 | 1 |
| Obesity | mast_cell | *ACHE* | -3.024751887 | 0.000721725 | 0.001403532 | 1 |
| Obesity | mast_cell | *SRRT* | 2.68546844 | 0.000503562 | 0.001144783 | 1 |
| Obesity | mast_cell | *ADAMTS2* | 1.994057038 | 0.000380484 | 0.001048903 | 1 |
| Obesity | mast_cell | *PSMC5* | 2.045425095 | 0.001267023 | 0.002177681 | 1 |
| Obesity | mast_cell | *UIMC1* | 3.227882077 | 0.000207784 | 0.000976811 | 1 |
| Obesity | mast_cell | *LPCAT2* | 2.015464273 | 0.000395868 | 0.001059433 | 1 |
| Obesity | mast_cell | *OGFOD1* | 1.87508069 | 0.012842152 | 0.016964367 | 1 |
| Obesity | mast_cell | *TXNDC16* | 2.363703078 | 0.000850089 | 0.001577296 | 1 |
| Obesity | mast_cell | *SF3B2* | 1.961567701 | 0.001305704 | 0.002237033 | 1 |
| Obesity | mast_cell | *AAMDC* | 1.604890143 | 0.0078522 | 0.010768478 | 1 |
| Obesity | mast_cell | *GP6* | -2.249239728 | 0.005514143 | 0.007795636 | 1 |
| Obesity | mast_cell | *ASAP3* | 1.542780766 | 0.000617309 | 0.001259538 | 1 |
| Obesity | mast_cell | *ANKRD10* | 3.129660429 | 0.00044641 | 0.001103405 | 1 |
| Obesity | mast_cell | *DOCK3* | 2.854032587 | 0.000303394 | 0.000998908 | 1 |
| Obesity | mast_cell | *ARHGAP28* | 1.871895062 | 0.000248807 | 0.000976811 | 1 |
| Obesity | mast_cell | *PPP1R13B* | 1.576594973 | 0.00630923 | 0.008796662 | 1 |
| Obesity | mast_cell | *NSFL1C* | 3.524329287 | 1.88E-05 | 0.000496117 | 0.091285614 |
| Obesity | mast_cell | *ZNF343* | 2.783531462 | 0.000387456 | 0.00105395 | 1 |
| Obesity | mast_cell | *EBF4* | 7.332656495 | 0.000816965 | 0.001532014 | 1 |
| Obesity | mast_cell | *MAVS* | 2.898864051 | 0.000156134 | 0.000976811 | 0.757561778 |
| Obesity | mast_cell | *XRN2* | 3.793159112 | 0.000118934 | 0.000976811 | 0.577069963 |

| | | | | | | |
|---|---|---|---|---|---|---|
| Obesity | mast_cell | *SNX5* | 7.779591147 | 0.001015964 | 0.00181317 | 1 |
| Obesity | mast_cell | *ESF1* | 4.383101968 | 0.000723104 | 0.00140565 | 1 |
| Obesity | mast_cell | *TMEM230* | 3.325705347 | 0.000225084 | 0.000976811 | 1 |
| Obesity | mast_cell | *RPH3A* | 6.862379464 | 0.00011745 | 0.000976811 | 0.569865997 |
| Obesity | mast_cell | *KIF16B* | 2.633192685 | 0.000257507 | 0.00098093 | 1 |
| Obesity | mast_cell | *TBX5* | 6.640043437 | 0.02751166 | 0.034377175 | 1 |
| Obesity | mast_cell | *ZNF302* | 2.716789497 | 0.018642446 | 0.023961099 | 1 |
| Obesity | mast_cell | *GRAMD1A* | 3.634602983 | 0.000195122 | 0.000976811 | 0.946730922 |
| Obesity | mast_cell | *GANAB* | 4.268136489 | 0.000962131 | 0.001739291 | 1 |
| Obesity | mast_cell | *MLF2* | 5.13787098 | 0.001201549 | 0.00208305 | 1 |
| Obesity | mast_cell | *ANKRD24* | 3.003012575 | 0.000503486 | 0.001144783 | 1 |
| Obesity | mast_cell | *DHX32* | 2.556607967 | 0.000446202 | 0.001103405 | 1 |
| Obesity | mast_cell | *SPTLC1* | 3.864166665 | 0.000356325 | 0.001033017 | 1 |
| Obesity | mast_cell | *YPEL3* | 3.746769712 | 1.42E-07 | 1.81E-05 | 0.000688274 |
| Obesity | mast_cell | *MRPS33* | 4.561925374 | 0.021287949 | 0.027006751 | 1 |
| Obesity | mast_cell | *STRN4* | 2.595346736 | 0.007642585 | 0.010489908 | 1 |
| Obesity | mast_cell | *LYZ* | 2.005284754 | 5.97E-05 | 0.000847545 | 0.289562255 |
| Obesity | mast_cell | *MUL1* | 2.346455986 | 0.002227338 | 0.00348975 | 1 |
| Obesity | mast_cell | *SPG21* | 2.586272429 | 0.001311323 | 0.002244282 | 1 |
| Obesity | mast_cell | *CHRD* | -2.309626645 | 0.003712916 | 0.005457571 | 1 |
| Obesity | mast_cell | *RAB11FIP3* | 4.288094253 | 0.000217893 | 0.000976811 | 1 |
| Obesity | mast_cell | *GNPTG* | 3.884853373 | 0.000309898 | 0.000998908 | 1 |
| Obesity | mast_cell | *CERS4* | 3.600597217 | 0.000218475 | 0.000976811 | 1 |
| Obesity | mast_cell | *USP48* | 3.574161406 | 0.002589448 | 0.003970924 | 1 |
| Obesity | mast_cell | *PDPR* | 2.732828503 | 0.000612714 | 0.001252797 | 1 |
| Obesity | mast_cell | *GLG1* | 3.027296928 | 0.000184795 | 0.000976811 | 0.896627199 |
| Obesity | mast_cell | *DTX2* | 5.699110467 | 0.000199025 | 0.000976811 | 0.96566751 |
| Obesity | mast_cell | *LAMB1* | 1.626332757 | 8.36E-05 | 0.000964174 | 0.405676402 |
| Obesity | mast_cell | *ABCC6* | 2.008233512 | 0.000747784 | 0.001440352 | 1 |
| Obesity | mast_cell | *RAPGEF4* | 1.392477096 | 0.000793183 | 0.001500984 | 1 |
| Obesity | mast_cell | *TF* | 9.868398816 | 0.000248135 | 0.000976811 | 1 |
| Obesity | mast_cell | *ALKBH5* | 6.083015174 | 0.00010823 | 0.000976811 | 0.525130852 |
| Obesity | mast_cell | *SPAG7* | 2.914084505 | 7.86E-05 | 0.000944925 | 0.381250371 |
| Obesity | mast_cell | *RGS17* | -7.259566467 | 3.00E-06 | 0.000158357 | 0.014568889 |
| Obesity | mast_cell | *PPP2R3C* | 3.992418808 | 3.06E-06 | 0.000159489 | 0.01483249 |
| Obesity | mast_cell | *SCFD1* | 2.857348718 | 0.00021062 | 0.000976811 | 1 |
| Obesity | mast_cell | *SUPT16H* | 2.66057616 | 0.00029358 | 0.000998908 | 1 |
| Obesity | mast_cell | *TOX4* | 2.522217916 | 0.000308279 | 0.000998908 | 1 |
| Obesity | mast_cell | *CAPN3* | -5.567201306 | 0.001852172 | 0.0029787 | 1 |
| Obesity | mast_cell | *UNC13D* | -1.54943506 | 0.000428552 | 0.001092087 | 1 |
| Obesity | mast_cell | *MFSD11* | 3.434443941 | 0.00158664 | 0.002622956 | 1 |
| Obesity | mast_cell | *NUP50* | 3.933418403 | 0.001201661 | 0.00208305 | 1 |
| Obesity | mast_cell | *COMT* | 5.044849065 | 0.000175447 | 0.000976811 | 0.851267651 |
| Obesity | mast_cell | *LRRFIP2* | 2.817313903 | 0.000117766 | 0.000976811 | 0.571398326 |
| Obesity | mast_cell | *SEC22C* | 4.463820225 | 0.000147623 | 0.000976811 | 0.716267901 |
| Obesity | mast_cell | *XYLB* | 2.868529964 | 0.000318901 | 0.000998908 | 1 |
| Obesity | mast_cell | *GABRP* | 3.55485112 | 0.02253211 | 0.02847026 | 1 |
| Obesity | mast_cell | *FMO2* | 1.783302125 | 0.000412129 | 0.001075765 | 1 |
| Obesity | mast_cell | *MSH2* | 4.200615031 | 0.000192722 | 0.000976811 | 0.935088782 |
| Obesity | mast_cell | *PSMD5* | 3.547606424 | 0.000225233 | 0.000976811 | 1 |

| | | | | | | |
|---|---|---|---|---|---|---|
| Obesity | mast_cell | *NANS* | 2.142163703 | 0.000460497 | 0.001114279 | 1 |
| Obesity | mast_cell | *CWF19L1* | 1.359326859 | 0.013537911 | 0.017791426 | 1 |
| Obesity | mast_cell | *WAC* | 4.202611011 | 0.000280882 | 0.000993978 | 1 |
| Obesity | mast_cell | *ITPR3* | 7.662567883 | 1.00E-08 | 1.87E-06 | 4.85E-05 |
| Obesity | mast_cell | *DSP* | 3.333502281 | 0.000366293 | 0.001037687 | 1 |
| Obesity | mast_cell | *IFT74* | 4.387444115 | 3.20E-05 | 0.000621513 | 0.155436925 |
| Obesity | mast_cell | *ACOT7* | -2.754851275 | 0.016602151 | 0.021538405 | 1 |
| Obesity | mast_cell | *CDC7* | 1.591700625 | 0.025064063 | 0.03146464 | 1 |
| Obesity | mast_cell | *SCD* | 3.501993356 | 0.007575127 | 0.010408192 | 1 |
| Obesity | mast_cell | *ERMP1* | 2.978822399 | 0.000854551 | 0.00158376 | 1 |
| Obesity | mast_cell | *RAB18* | 3.84227877 | 0.000184561 | 0.000976811 | 0.895489037 |
| Obesity | mast_cell | *TSPAN15* | 1.996204641 | 0.000323194 | 0.001002436 | 1 |
| Obesity | mast_cell | *MAST3* | 1.554302948 | 0.008588588 | 0.011685875 | 1 |
| Obesity | mast_cell | *SETD1A* | 5.471731471 | 4.08E-07 | 4.04E-05 | 0.001981064 |
| Obesity | mast_cell | *CIRBP* | 2.088888603 | 0.000163576 | 0.000976811 | 0.793669636 |
| Obesity | mast_cell | *MARCH2* | 3.424106708 | 0.001127459 | 0.00197774 | 1 |
| Obesity | mast_cell | *MTAP* | 2.281705569 | 0.00134385 | 0.002290257 | 1 |
| Obesity | mast_cell | *POLR2E* | 4.812962083 | 0.000496919 | 0.001138685 | 1 |
| Obesity | mast_cell | *MADCAM1* | 2.153499612 | 0.000642966 | 0.001293933 | 1 |
| Obesity | mast_cell | *MED15* | 3.46299819 | 0.000773478 | 0.001479273 | 1 |
| Obesity | mast_cell | *SERPIND1* | 2.106995999 | 0.006185087 | 0.008640956 | 1 |
| Obesity | mast_cell | *SNAP29* | 1.748932027 | 0.011843354 | 0.015743549 | 1 |
| Obesity | mast_cell | *MMP11* | 5.3696935 | 0.012066217 | 0.016022246 | 1 |
| Obesity | mast_cell | *CECR2* | 1.662982236 | 0.000804968 | 0.001517957 | 1 |
| Obesity | mast_cell | *SMARCB1* | 5.058869924 | 3.20E-05 | 0.000621513 | 0.155021474 |
| Obesity | mast_cell | *BCL2L13* | 3.920336838 | 2.53E-06 | 0.000141479 | 0.012265797 |
| Obesity | mast_cell | *DDT* | 2.548178955 | 0.001550528 | 0.002580972 | 1 |
| Obesity | mast_cell | *OSM* | -2.687437935 | 0.006416097 | 0.008935391 | 1 |
| Obesity | mast_cell | *CABIN1* | 4.224297323 | 4.98E-05 | 0.000765648 | 0.241515681 |
| Obesity | mast_cell | *PPIL2* | 1.929901771 | 0.000876562 | 0.001612848 | 1 |
| Obesity | mast_cell | *PES1* | 2.062572116 | 0.000662658 | 0.001319333 | 1 |
| Obesity | mast_cell | *SLC35E4* | 3.114192737 | 0.001550604 | 0.002580972 | 1 |
| Obesity | mast_cell | *TOP3B* | 2.522456691 | 0.000387093 | 0.00105395 | 1 |
| Obesity | mast_cell | *CRYBB3* | 4.641603367 | 0.008957441 | 0.012160465 | 1 |
| Obesity | mast_cell | *CYTH4* | 2.012498723 | 0.000445184 | 0.001103405 | 1 |
| Obesity | mast_cell | *LRP5L* | 2.197269395 | 0.0008492 | 0.001576854 | 1 |
| Obesity | mast_cell | *LGALS2* | 1.928752792 | 0.002871245 | 0.004342669 | 1 |
| Obesity | mast_cell | *SH3BP1* | 3.61132398 | 0.000387175 | 0.00105395 | 1 |
| Obesity | mast_cell | *HPS4* | 2.18825419 | 0.000296573 | 0.000998908 | 1 |
| Obesity | mast_cell | *TRIOBP* | 1.89025053 | 0.001598069 | 0.002640051 | 1 |
| Obesity | mast_cell | *EIF3L* | 2.629048341 | 0.000509688 | 0.001145411 | 1 |
| Obesity | mast_cell | *CCDC134* | 2.607690385 | 0.000181608 | 0.000976811 | 0.881163549 |
| Obesity | mast_cell | *PICK1* | 1.715706302 | 0.005759979 | 0.008112458 | 1 |
| Obesity | mast_cell | *TTC28* | 2.645716672 | 0.000195692 | 0.000976811 | 0.949498762 |
| Obesity | mast_cell | *CENPM* | 8.67340667 | 4.56E-07 | 4.43E-05 | 0.002212958 |
| Obesity | mast_cell | *KDELR3* | -6.359394452 | 0.002707094 | 0.004125257 | 1 |
| Obesity | mast_cell | *CYP2D6* | 3.691873842 | 0.023670586 | 0.029831087 | 1 |
| Obesity | mast_cell | *RTCB* | 3.592418508 | 0.000269367 | 0.000986064 | 1 |
| Obesity | mast_cell | *TIMP3* | 2.61293381 | 0.000191001 | 0.000976811 | 0.926735842 |
| Obesity | mast_cell | *PPP6R2* | 3.62158125 | 0.000166978 | 0.000976811 | 0.81017794 |

| | | | | | | |
|---|---|---|---|---|---|---|
| Obesity | mast_cell | *CYB5R3* | 2.131642326 | 0.000505151 | 0.001144783 | 1 |
| Obesity | mast_cell | *RHBDD3* | 2.137132974 | 0.000532019 | 0.001161204 | 1 |
| Obesity | mast_cell | *TOM1* | 1.833070721 | 0.001356276 | 0.002307381 | 1 |
| Obesity | mast_cell | *CHKB* | 4.557747636 | 0.000174021 | 0.000976811 | 0.84435018 |
| Obesity | mast_cell | *ARSA* | 1.821639091 | 0.001998885 | 0.003178823 | 1 |
| Obesity | mast_cell | *TSPO* | 3.223309221 | 0.000218211 | 0.000976811 | 1 |
| Obesity | mast_cell | *TTLL12* | 3.306538553 | 0.000365774 | 0.001037687 | 1 |
| Obesity | mast_cell | *MTMR3* | 2.849301662 | 0.001261266 | 0.002169324 | 1 |
| Obesity | mast_cell | *APOL4* | 3.475088716 | 0.000404773 | 0.001068171 | 1 |
| Obesity | mast_cell | *FOXRED2* | 2.265581795 | 0.001227424 | 0.002121129 | 1 |
| Obesity | mast_cell | *IFT27* | 2.08785896 | 0.000761224 | 0.001459865 | 1 |
| Obesity | mast_cell | *SLC25A17* | 2.529773554 | 0.000507282 | 0.001144962 | 1 |
| Obesity | mast_cell | *FAM118A* | 1.60602648 | 0.018769274 | 0.024098576 | 1 |
| Obesity | mast_cell | *L3MBTL2* | 8.406385173 | 0.000301709 | 0.000998908 | 1 |
| Obesity | mast_cell | *ZC3H7B* | 4.626027759 | 0.000192875 | 0.000976811 | 0.935831513 |
| Obesity | mast_cell | *ACO2* | 3.418761283 | 0.000534855 | 0.001165296 | 1 |
| Obesity | mast_cell | *POLR3H* | 3.259237985 | 0.000215096 | 0.000976811 | 1 |
| Obesity | mast_cell | *DESI1* | 9.720650161 | 7.43E-06 | 0.000290571 | 0.036030746 |
| Obesity | mast_cell | *MLC1* | 3.792535151 | 0.008669135 | 0.011788857 | 1 |
| Obesity | mast_cell | *HDAC10* | 6.562268888 | 0.000137516 | 0.000976811 | 0.667228273 |
| Obesity | mast_cell | *KHNYN* | 3.357674358 | 8.24E-05 | 0.000958567 | 0.399946659 |
| Obesity | mast_cell | *SDR39U1* | 1.404513978 | 0.011376438 | 0.015160252 | 1 |
| Obesity | mast_cell | *RBM23* | 3.406142567 | 0.001519474 | 0.002542237 | 1 |
| Obesity | mast_cell | *AP4S1* | 5.220976171 | 0.001918315 | 0.003068798 | 1 |
| Obesity | mast_cell | *NIN* | 3.746401933 | 0.000202326 | 0.000976811 | 0.981683706 |
| Obesity | mast_cell | *PYGL* | 0.936974435 | 0.017922336 | 0.023090593 | 1 |
| Obesity | mast_cell | *CNIH1* | 4.291619322 | 0.000429379 | 0.001092129 | 1 |
| Obesity | mast_cell | *ATP6V1D* | 5.690843984 | 0.000223615 | 0.000976811 | 1 |
| Obesity | mast_cell | *PLEK2* | 4.523689496 | 0.005934394 | 0.008326685 | 1 |
| Obesity | mast_cell | *GSTZ1* | 3.784549494 | 0.007592721 | 0.010427365 | 1 |
| Obesity | mast_cell | *TMED8* | 5.108774359 | 1.83E-05 | 0.00049351 | 0.088831738 |
| Obesity | mast_cell | *LGMN* | 1.255284313 | 0.000508499 | 0.001144962 | 1 |
| Obesity | mast_cell | *MTHFD1* | 2.914933134 | 0.000191471 | 0.000976811 | 0.929015004 |
| Obesity | mast_cell | *TELO2* | 2.570461953 | 0.00063335 | 0.00128149 | 1 |
| Obesity | mast_cell | *PSMB5* | 4.364761941 | 8.37E-06 | 0.00032238 | 0.040619875 |
| Obesity | mast_cell | *YY1* | 5.789602344 | 0.000212058 | 0.000976811 | 1 |
| Obesity | mast_cell | *CCNB1IP1* | 3.561785147 | 0.00025345 | 0.000977535 | 1 |
| Obesity | mast_cell | *EFS* | 2.818001098 | 0.000353741 | 0.001031837 | 1 |
| Obesity | mast_cell | *CINP* | 2.147685619 | 0.000347674 | 0.001029861 | 1 |
| Obesity | mast_cell | *KIAA0391* | 4.926338767 | 0.00054967 | 0.001175407 | 1 |
| Obesity | mast_cell | *EMC9* | -2.127091055 | 0.016332154 | 0.021205141 | 1 |
| Obesity | mast_cell | *PLTP* | 1.676872934 | 0.000221907 | 0.000976811 | 1 |
| Obesity | mast_cell | *TRPC4AP* | 4.106615433 | 7.44E-05 | 0.000932859 | 0.361016299 |
| Obesity | mast_cell | *PYGB* | 2.701393453 | 0.000210919 | 0.000976811 | 1 |
| Obesity | mast_cell | *ABHD12* | 5.208847327 | 5.27E-07 | 4.92E-05 | 0.002557004 |
| Obesity | mast_cell | *CD40* | 3.349313372 | 0.000342033 | 0.001027099 | 1 |
| Obesity | mast_cell | *UQCC1* | 3.049912274 | 0.000244247 | 0.000976811 | 1 |
| Obesity | mast_cell | *IFT52* | 1.64621353 | 0.000541174 | 0.001171675 | 1 |
| Obesity | mast_cell | *NDRG3* | 2.219434601 | 0.000200298 | 0.000976811 | 0.971845812 |
| Obesity | mast_cell | *PABPC1L* | 2.342682646 | 0.00060117 | 0.001237692 | 1 |

| | | | | | | |
|---|---|---|---|---|---|---|
| Obesity | mast_cell | *DOK5* | 8.659797172 | 0.008076828 | 0.011045444 | 1 |
| Obesity | mast_cell | *CSTF1* | 2.681947158 | 0.000402465 | 0.001064756 | 1 |
| Obesity | mast_cell | *NELFCD* | 6.030470848 | 6.63E-05 | 0.000885378 | 0.32153414 |
| Obesity | mast_cell | *CTSZ* | 1.924985206 | 0.00038738 | 0.00105395 | 1 |
| Obesity | mast_cell | *MTG2* | 2.072477989 | 0.000314388 | 0.000998908 | 1 |
| Obesity | mast_cell | *TCFL5* | -2.852111221 | 0.020526954 | 0.026140887 | 1 |
| Obesity | mast_cell | *DIDO1* | 5.282081133 | 7.29E-05 | 0.000923552 | 0.353720565 |
| Obesity | mast_cell | *GID8* | 4.369910879 | 0.001352432 | 0.002302804 | 1 |
| Obesity | mast_cell | *BIRC7* | -1.044891542 | 0.000969132 | 0.001746565 | 1 |
| Obesity | mast_cell | *PTK6* | 7.478184136 | 0.000802788 | 0.001514435 | 1 |
| Obesity | mast_cell | *GMEB2* | 5.117466285 | 0.000544327 | 0.00117408 | 1 |
| Obesity | mast_cell | *CDC25B* | 4.142023778 | 0.001692761 | 0.002757758 | 1 |
| Obesity | mast_cell | *RNF24* | 1.963143884 | 0.000313534 | 0.000998908 | 1 |
| Obesity | mast_cell | *NDUFAF5* | 1.840865231 | 0.003322216 | 0.004943083 | 1 |
| Obesity | mast_cell | *TRIB3* | 4.037330044 | 0.000410996 | 0.001075307 | 1 |
| Obesity | mast_cell | *RASSF2* | 3.303457311 | 4.91E-05 | 0.000765648 | 0.238306828 |
| Obesity | mast_cell | *ANGPT4* | 5.013394545 | 0.000307663 | 0.000998908 | 1 |
| Obesity | mast_cell | *SEC23B* | 2.853552788 | 0.004036156 | 0.005891525 | 1 |
| Obesity | mast_cell | *CRNKL1* | 3.480446999 | 0.021290359 | 0.027006751 | 1 |
| Obesity | mast_cell | *MROH8* | 1.801452178 | 0.002086386 | 0.003299591 | 1 |
| Obesity | mast_cell | *MANBAL* | 5.25502747 | 0.001307531 | 0.002239372 | 1 |
| Obesity | mast_cell | *TTI1* | 2.388480296 | 0.00052713 | 0.001157301 | 1 |
| Obesity | mast_cell | *ACTR5* | 2.45707762 | 0.000790009 | 0.001498485 | 1 |
| Obesity | mast_cell | *MAP1LC3A* | 5.018082867 | 0.000331258 | 0.001014048 | 1 |
| Obesity | mast_cell | *RBFA* | 5.225104057 | 0.005728019 | 0.008074476 | 1 |
| Obesity | mast_cell | *METTL4* | 2.357830814 | 0.000353119 | 0.001031837 | 1 |
| Obesity | mast_cell | *LPIN2* | 3.522880702 | 0.000521182 | 0.001152731 | 1 |
| Obesity | mast_cell | *MYOM1* | 1.554906328 | 0.00048989 | 0.001135803 | 1 |
| Obesity | mast_cell | *CEP192* | 1.662565888 | 0.007691063 | 0.010553461 | 1 |
| Obesity | mast_cell | *LAMA1* | 2.395966009 | 0.017499054 | 0.022581226 | 1 |
| Obesity | mast_cell | *RNF125* | 2.386408614 | 0.000466429 | 0.001116682 | 1 |
| Obesity | mast_cell | *POLI* | 1.642760827 | 0.000247573 | 0.000976811 | 1 |
| Obesity | mast_cell | *MIB1* | 4.350908517 | 0.000178091 | 0.000976811 | 0.864095511 |
| Obesity | mast_cell | *NALCN* | 2.081032407 | 0.001036058 | 0.001840701 | 1 |
| Obesity | mast_cell | *STK24* | 2.919205591 | 0.000206449 | 0.000976811 | 1 |
| Obesity | mast_cell | *ACP5* | 2.220337263 | 0.003265996 | 0.004868391 | 1 |
| Obesity | mast_cell | *DNAJC3* | 2.900106426 | 0.000269771 | 0.000986064 | 1 |
| Obesity | mast_cell | *UGGT2* | 2.37776093 | 0.001205158 | 0.002086876 | 1 |
| Obesity | mast_cell | *ARHGEF7* | 2.285554193 | 0.000265112 | 0.000985696 | 1 |
| Obesity | mast_cell | *PARP4* | 3.968904576 | 0.000104254 | 0.000976811 | 0.505839226 |
| Obesity | mast_cell | *SUPT20H* | 4.293567375 | 0.003924144 | 0.00573147 | 1 |
| Obesity | mast_cell | *MRPS31* | 3.757834883 | 0.000172491 | 0.000976811 | 0.836926789 |
| Obesity | mast_cell | *RGCC* | 1.284352264 | 0.00584899 | 0.0082235 | 1 |
| Obesity | mast_cell | *DGKH* | 2.723170293 | 7.52E-05 | 0.000934364 | 0.364783193 |
| Obesity | mast_cell | *KATNAL1* | 2.860677425 | 0.000234095 | 0.000976811 | 1 |
| Obesity | mast_cell | *INTS6* | 2.750109163 | 0.000565091 | 0.001190078 | 1 |
| Obesity | mast_cell | *CLN5* | 3.567662925 | 0.000159843 | 0.000976811 | 0.775557487 |
| Obesity | mast_cell | *MGRN1* | 4.062035054 | 0.000181755 | 0.000976811 | 0.881876475 |
| Obesity | mast_cell | *ELMO3* | 3.516529484 | 9.43E-06 | 0.000349581 | 0.045757005 |
| Obesity | mast_cell | *LONP2* | 3.401204371 | 0.00028028 | 0.000993978 | 1 |

| | | | | | | |
|---|---|---|---|---|---|---|
| Obesity | mast_cell | *PLLP* | 1.937391595 | 0.009757299 | 0.013139721 | 1 |
| Obesity | mast_cell | *DHODH* | 4.731425551 | 5.41E-05 | 0.000802079 | 0.262613095 |
| Obesity | mast_cell | *PSMD7* | 3.055110155 | 0.000252256 | 0.000976811 | 1 |
| Obesity | mast_cell | *MON1B* | 2.280632148 | 0.000482555 | 0.001131243 | 1 |
| Obesity | mast_cell | *NPRL3* | 2.082061032 | 0.001100437 | 0.001935939 | 1 |
| Obesity | mast_cell | *NME4* | 2.988007694 | 0.001518584 | 0.002542237 | 1 |
| Obesity | mast_cell | *NOMO3* | 2.263020804 | 0.013122883 | 0.017311644 | 1 |
| Obesity | mast_cell | *LMF1* | 2.747579998 | 0.000248272 | 0.000976811 | 1 |
| Obesity | mast_cell | *NARFL* | 2.315405016 | 0.000393028 | 0.001054923 | 1 |
| Obesity | mast_cell | *MTHFSD* | 1.794417338 | 0.003098939 | 0.004649367 | 1 |
| Obesity | mast_cell | *CLCN7* | 4.12725996 | 0.000211498 | 0.000976811 | 1 |
| Obesity | mast_cell | *SLC7A5* | 4.00131017 | 0.031164648 | 0.038613604 | 1 |
| Obesity | mast_cell | *RHBDL1* | 7.556976476 | 0.006609337 | 0.00918342 | 1 |
| Obesity | mast_cell | *NUBP1* | 4.86239236 | 2.84E-05 | 0.000600011 | 0.138002419 |
| Obesity | mast_cell | *UBE2I* | 2.440260788 | 0.000320828 | 0.000999138 | 1 |
| Obesity | mast_cell | *CAPN15* | 6.583832174 | 0.001049178 | 0.001859925 | 1 |
| Obesity | mast_cell | *CLUAP1* | 3.588732612 | 1.25E-06 | 8.82E-05 | 0.006087555 |
| Obesity | mast_cell | *EARS2* | 2.073304097 | 0.002279877 | 0.003562629 | 1 |
| Obesity | mast_cell | *CPPED1* | 4.433149099 | 0.000971484 | 0.001748383 | 1 |
| Obesity | mast_cell | *HMOX2* | 2.02454948 | 0.000963212 | 0.001739949 | 1 |
| Obesity | mast_cell | *TOX3* | -1.966688761 | 0.002726287 | 0.004146703 | 1 |
| Obesity | mast_cell | *RBL2* | 2.889173271 | 0.000249698 | 0.000976811 | 1 |
| Obesity | mast_cell | *PYCARD* | 4.663583134 | 1.58E-06 | 0.000104951 | 0.00766139 |
| Obesity | mast_cell | *CDIPT* | 3.749086408 | 0.005189703 | 0.007384293 | 1 |
| Obesity | mast_cell | *KAT8* | 2.988004561 | 0.000396807 | 0.001060771 | 1 |
| Obesity | mast_cell | *TMC5* | 1.744841286 | 0.005875279 | 0.008253287 | 1 |
| Obesity | mast_cell | *C16orf62* | 2.802670318 | 0.000229679 | 0.000976811 | 1 |
| Obesity | mast_cell | *RNF40* | 2.058945603 | 0.000467351 | 0.001116682 | 1 |
| Obesity | mast_cell | *LACTB* | 5.973539734 | 0.001201425 | 0.00208305 | 1 |
| Obesity | mast_cell | *HERC1* | 3.225374288 | 0.000218223 | 0.000976811 | 1 |
| Obesity | mast_cell | *TRIP4* | 3.07657561 | 6.71E-05 | 0.000885378 | 0.325393538 |
| Obesity | mast_cell | *AP3B2* | -1.202549525 | 0.033587169 | 0.041456359 | 1 |
| Obesity | mast_cell | *TTC23* | 2.13763474 | 0.000354681 | 0.001031837 | 1 |
| Obesity | mast_cell | *FAH* | 1.979560153 | 0.000632398 | 0.001280633 | 1 |
| Obesity | mast_cell | *RPAP1* | 1.734999248 | 0.004721374 | 0.006766386 | 1 |
| Obesity | mast_cell | *SLC30A4* | 4.431171353 | 0.000121964 | 0.000976811 | 0.591767886 |
| Obesity | mast_cell | *ZDHHC2* | 1.989395877 | 0.008788429 | 0.011944386 | 1 |
| Obesity | mast_cell | *ZFAND1* | 4.805325899 | 0.000286065 | 0.000993978 | 1 |
| Obesity | mast_cell | *EYA1* | 2.269284793 | 0.000258909 | 0.00098093 | 1 |
| Obesity | mast_cell | *IMPAD1* | 4.373251616 | 0.000425583 | 0.001090111 | 1 |
| Obesity | mast_cell | *SFRP1* | 1.929903875 | 0.000531125 | 0.001160818 | 1 |
| Obesity | mast_cell | *NIPAL2* | 1.896172769 | 0.000353099 | 0.001031837 | 1 |
| Obesity | mast_cell | *RAB2A* | 2.425134523 | 0.00098293 | 0.001763748 | 1 |
| Obesity | mast_cell | *ARMC1* | 2.640833075 | 0.00058231 | 0.001209489 | 1 |
| Obesity | mast_cell | *SPAG1* | 3.143355899 | 0.000131468 | 0.000976811 | 0.637882966 |
| Obesity | mast_cell | *CHRAC1* | 2.980088115 | 0.000495297 | 0.001138685 | 1 |
| Obesity | mast_cell | *SNX16* | 3.826771946 | 0.000227557 | 0.000976811 | 1 |
| Obesity | mast_cell | *GSDMD* | 2.928780444 | 0.001201267 | 0.00208305 | 1 |
| Obesity | mast_cell | *TSTA3* | 2.976318723 | 0.001653358 | 0.002709251 | 1 |
| Obesity | mast_cell | *SQLE* | 2.693467438 | 0.000458171 | 0.001112606 | 1 |

| | | | | | | |
|---|---|---|---|---|---|---|
| Obesity | mast_cell | *R3HCC1* | 3.914728472 | 0.007275043 | 0.010022291 | 1 |
| Obesity | mast_cell | *TNFRSF10A* | 2.483398393 | 0.000199695 | 0.000976811 | 0.968918359 |
| Obesity | mast_cell | *ERICH1* | 2.329518862 | 0.00087834 | 0.001614282 | 1 |
| Obesity | mast_cell | *TUSC3* | 2.821656926 | 0.000259627 | 0.00098093 | 1 |
| Obesity | mast_cell | *KLHDC4* | 4.714264259 | 0.019407336 | 0.024865169 | 1 |
| Obesity | mast_cell | *TULP2* | 12.33597531 | 0.001297148 | 0.002224192 | 1 |
| Obesity | mast_cell | *NFKBIB* | 5.161079641 | 0.00141458 | 0.002392312 | 1 |
| Obesity | mast_cell | *SARS2* | -1.341400057 | 0.032972623 | 0.040739284 | 1 |
| Obesity | mast_cell | *SNRNP70* | 3.566518094 | 0.000262055 | 0.000983135 | 1 |
| Obesity | mast_cell | *LIN7B* | 2.680945391 | 0.003671777 | 0.005406817 | 1 |
| Obesity | mast_cell | *PPP1R37* | -2.856997517 | 0.016331734 | 0.021205141 | 1 |
| Obesity | mast_cell | *FCGRT* | 2.28616449 | 0.000276036 | 0.000990625 | 1 |
| Obesity | mast_cell | *PEX11G* | 2.338485846 | 0.005335438 | 0.007569458 | 1 |
| Obesity | mast_cell | *ERCC2* | 2.839718423 | 0.000618979 | 0.001262311 | 1 |
| Obesity | mast_cell | *CD37* | -2.831020356 | 0.027817985 | 0.034719655 | 1 |
| Obesity | mast_cell | *AMH* | 25.40160277 | 0.000508531 | 0.001144962 | 1 |
| Obesity | mast_cell | *FCER2* | 7.806438573 | 0.000509848 | 0.001145411 | 1 |
| Obesity | mast_cell | *CLEC4M* | 24.86125334 | 0.000242216 | 0.000976811 | 1 |
| Obesity | mast_cell | *TLE6* | -2.259590924 | 0.003277427 | 0.004880931 | 1 |
| Obesity | mast_cell | *NOVA2* | 2.478743189 | 3.37E-05 | 0.000630711 | 0.163354145 |
| Obesity | mast_cell | *SGTA* | 6.130977476 | 0.001452561 | 0.002444616 | 1 |
| Obesity | mast_cell | *KIR3DX1* | 11.88474865 | 0.00316128 | 0.004728277 | 1 |
| Obesity | mast_cell | *MED25* | 3.87614279 | 0.000269545 | 0.000986064 | 1 |
| Obesity | mast_cell | *TIMM44* | 2.933266297 | 0.00025821 | 0.00098093 | 1 |
| Obesity | mast_cell | *IL27RA* | 2.934369888 | 0.000392031 | 0.001054889 | 1 |
| Obesity | mast_cell | *FAM32A* | 10.07607734 | 0.00096762 | 0.001744664 | 1 |
| Obesity | mast_cell | *C19orf44* | 1.708373847 | 0.00097445 | 0.00175136 | 1 |
| Obesity | mast_cell | *OLFM2* | 1.657279523 | 0.004093313 | 0.005967775 | 1 |
| Obesity | mast_cell | *PDCD5* | 2.14348193 | 0.000626816 | 0.001273048 | 1 |
| Obesity | mast_cell | *ANKRD27* | 7.031003961 | 6.29E-05 | 0.000865434 | 0.305289959 |
| Obesity | mast_cell | *POLR2I* | 2.549932228 | 0.00066018 | 0.001316561 | 1 |
| Obesity | mast_cell | *TGFB1* | 4.347927811 | 0.000158774 | 0.000976811 | 0.770372961 |
| Obesity | mast_cell | *MRPL4* | 3.077754445 | 0.000415893 | 0.001080612 | 1 |
| Obesity | mast_cell | *ICAM5* | -2.954271401 | 0.001487778 | 0.002496955 | 1 |
| Obesity | mast_cell | *RABAC1* | 2.288473272 | 0.000248612 | 0.000976811 | 1 |
| Obesity | mast_cell | *GRIN2D* | 3.603450696 | 0.001223282 | 0.002115239 | 1 |
| Obesity | mast_cell | *CARD8* | 4.836234698 | 0.000389363 | 0.001054382 | 1 |
| Obesity | mast_cell | *ZNF175* | 2.357607987 | 0.001102387 | 0.001938666 | 1 |
| Obesity | mast_cell | *PLA2G4C* | 1.514497819 | 0.00388724 | 0.005684415 | 1 |
| Obesity | mast_cell | *CAPS* | 3.959785879 | 0.000394059 | 0.001055175 | 1 |
| Obesity | mast_cell | *MIER2* | 4.401957534 | 0.00025182 | 0.000976811 | 1 |
| Obesity | mast_cell | *PLEKHA4* | 2.151360362 | 0.000582218 | 0.001209489 | 1 |
| Obesity | mast_cell | *TNPO2* | 2.007168144 | 0.000749565 | 0.001443209 | 1 |
| Obesity | mast_cell | *GCDH* | 2.237816357 | 0.005332079 | 0.007566904 | 1 |
| Obesity | mast_cell | *TFPT* | 4.946529291 | 0.000306047 | 0.000998908 | 1 |
| Obesity | mast_cell | *RAB3A* | 9.535882417 | 0.000495063 | 0.001138685 | 1 |
| Obesity | mast_cell | *UPK1A* | -1.514813856 | 0.001072967 | 0.001895171 | 1 |
| Obesity | mast_cell | *COPE* | 4.759705171 | 0.009792447 | 0.013176082 | 1 |
| Obesity | mast_cell | *ETV2* | 23.4746686 | 0.000384639 | 0.001052047 | 1 |
| Obesity | mast_cell | *MAG* | -2.379622448 | 0.010042664 | 0.013501525 | 1 |

| | | | | | | |
|---|---|---|---|---|---|---|
| Obesity | mast_cell | *LSR* | -1.255445445 | 0.015372211 | 0.020039218 | 1 |
| Obesity | mast_cell | *ZNF14* | 7.119232971 | 0.000191307 | 0.000976811 | 0.92821956 |
| Obesity | mast_cell | *ZNF85* | 2.209049748 | 0.000519328 | 0.001151636 | 1 |
| Obesity | mast_cell | *ETHE1* | 3.436208603 | 0.000516519 | 0.001150279 | 1 |
| Obesity | mast_cell | *GTPBP10* | 3.254879909 | 0.000552295 | 0.001179461 | 1 |
| Obesity | mast_cell | *RASA4* | 15.58906234 | 0.000545916 | 0.00117408 | 1 |
| Obesity | mast_cell | *BET1* | 5.726769009 | 0.000201573 | 0.000976811 | 0.978030549 |
| Obesity | mast_cell | *SP4* | 1.936331939 | 0.001275824 | 0.002191255 | 1 |
| Obesity | mast_cell | *DNAH11* | 4.208201531 | 0.00022558 | 0.000976811 | 1 |
| Obesity | mast_cell | *STEAP1B* | 3.876525839 | 0.000467518 | 0.001116682 | 1 |
| Obesity | mast_cell | *PTN* | 2.208923199 | 0.000426935 | 0.001091691 | 1 |
| Obesity | mast_cell | *ADAP1* | 7.758474839 | 0.00048628 | 0.001132608 | 1 |
| Obesity | mast_cell | *RNF32* | 3.778021139 | 0.000791425 | 0.001499309 | 1 |
| Obesity | mast_cell | *IQCE* | 3.149037829 | 0.000315743 | 0.000998908 | 1 |
| Obesity | mast_cell | *VIPR2* | 2.434341563 | 0.000160996 | 0.000976811 | 0.781153373 |
| Obesity | mast_cell | *HIBADH* | 2.058801512 | 0.001370204 | 0.002327811 | 1 |
| Obesity | mast_cell | *CPVL* | 1.742489318 | 0.00038916 | 0.001054382 | 1 |
| Obesity | mast_cell | *CHN2* | 1.961556491 | 0.002025687 | 0.003214073 | 1 |
| Obesity | mast_cell | *GRB10* | 1.749319849 | 0.000305336 | 0.000998908 | 1 |
| Obesity | mast_cell | *ABHD11* | 1.96260594 | 0.029375203 | 0.036536397 | 1 |
| Obesity | mast_cell | *PLEKHA8* | 1.792736599 | 0.00045612 | 0.00111155 | 1 |
| Obesity | mast_cell | *EPHB6* | 2.252652023 | 0.001347039 | 0.002294885 | 1 |
| Obesity | mast_cell | *CHCHD2* | 3.595468021 | 0.000475678 | 0.001127398 | 1 |
| Obesity | mast_cell | *ZKSCAN1* | 2.597611985 | 0.000237495 | 0.000976811 | 1 |
| Obesity | mast_cell | *EIF3B* | 2.348511619 | 0.000425194 | 0.001090111 | 1 |
| Obesity | mast_cell | *TFR2* | 3.641267753 | 0.001891956 | 0.00303263 | 1 |
| Obesity | mast_cell | *MOSPD3* | 2.760848601 | 0.000433377 | 0.001096322 | 1 |
| Obesity | mast_cell | *RBM28* | 2.735947378 | 0.001333767 | 0.002277873 | 1 |
| Obesity | mast_cell | *IMPDH1* | 3.146621178 | 0.000695879 | 0.001365894 | 1 |
| Obesity | mast_cell | *C1GALT1* | 2.718475183 | 0.000232159 | 0.000976811 | 1 |
| Obesity | mast_cell | *NRF1* | 3.249337281 | 0.000134446 | 0.000976811 | 0.652330495 |
| Obesity | mast_cell | *ZNF862* | 3.958941134 | 8.26E-05 | 0.000958567 | 0.400680822 |
| Obesity | mast_cell | *MEST* | 0.868680271 | 0.028919603 | 0.035997412 | 1 |
| Obesity | mast_cell | *ACTR3C* | 4.36366972 | 0.001919217 | 0.00306923 | 1 |
| Obesity | mast_cell | *POU6F2* | 2.307551948 | 0.00060874 | 0.001246806 | 1 |
| Obesity | mast_cell | *TSPAN13* | 2.67434901 | 0.000357027 | 0.001033588 | 1 |
| Obesity | mast_cell | *RARRES2* | 3.609705816 | 0.00040007 | 0.001064254 | 1 |
| Obesity | mast_cell | *COA1* | 2.845047071 | 0.000133769 | 0.000976811 | 0.649045186 |
| Obesity | mast_cell | *BLVRA* | 6.273291744 | 0.000301865 | 0.000998908 | 1 |
| Obesity | mast_cell | *URGCP* | 2.868970668 | 0.000205049 | 0.000976811 | 0.994895326 |
| Obesity | mast_cell | *AEBP1* | 3.573501457 | 0.000273401 | 0.000987746 | 1 |
| Obesity | mast_cell | *GCK* | 14.31269718 | 0.000556705 | 0.001182832 | 1 |
| Obesity | mast_cell | *TBL2* | 5.823694789 | 0.002336123 | 0.00363997 | 1 |
| Obesity | mast_cell | *SPATA6L* | 1.886425643 | 0.00041257 | 0.001076232 | 1 |
| Obesity | mast_cell | *SLC1A1* | 1.391998024 | 0.000292459 | 0.000998908 | 1 |
| Obesity | mast_cell | *FKTN* | 2.889432002 | 0.000239819 | 0.000976811 | 1 |
| Obesity | mast_cell | *NMRK1* | 1.792808401 | 0.000706651 | 0.001381414 | 1 |
| Obesity | mast_cell | *MEGF9* | 3.296066431 | 0.000213766 | 0.000976811 | 1 |
| Obesity | mast_cell | *TRIM14* | 1.947223829 | 7.77E-05 | 0.00094053 | 0.377152347 |
| Obesity | mast_cell | *CORO2A* | 2.902257021 | 0.000860909 | 0.001591288 | 1 |

| | | | | | | |
|---|---|---|---|---|---|---|
| Obesity | mast_cell | *C5* | 2.796942024 | 0.000230277 | 0.000976811 | 1 |
| Obesity | mast_cell | *OGN* | 2.534698337 | 3.82E-05 | 0.00068216 | 0.18554752 |
| Obesity | mast_cell | *ECM2* | 2.806436662 | 0.000313958 | 0.000998908 | 1 |
| Obesity | mast_cell | *PTGR1* | 1.548555429 | 0.001580485 | 0.002616232 | 1 |
| Obesity | mast_cell | *SUSD1* | 2.000250356 | 0.002088486 | 0.003301835 | 1 |
| Obesity | mast_cell | *DOCK8* | 1.163479237 | 0.012590124 | 0.016676845 | 1 |
| Obesity | mast_cell | *KANK1* | 1.631119548 | 0.00097341 | 0.001751199 | 1 |
| Obesity | mast_cell | *FUBP3* | 2.587030151 | 0.000289735 | 0.000996309 | 1 |
| Obesity | mast_cell | *PTGDS* | 3.966130822 | 0.000238183 | 0.000976811 | 1 |
| Obesity | mast_cell | *ATRNL1* | 1.153521299 | 0.006111489 | 0.008547981 | 1 |
| Obesity | mast_cell | *HPS1* | 2.199784835 | 0.003577934 | 0.005287887 | 1 |
| Obesity | mast_cell | *PHYH* | 3.785951737 | 0.00029071 | 0.000997541 | 1 |
| Obesity | mast_cell | *DNMBP* | 1.692145138 | 0.000193073 | 0.000976811 | 0.936790246 |
| Obesity | mast_cell | *CUBN* | 4.827628461 | 0.004526527 | 0.006518607 | 1 |
| Obesity | mast_cell | *TRDMT1* | 6.033418499 | 9.45E-05 | 0.000976811 | 0.458524584 |
| Obesity | mast_cell | *ATE1* | 3.765119075 | 0.0001488 | 0.000976811 | 0.721975302 |
| Obesity | mast_cell | *PLEKHA1* | 2.372434939 | 0.000294243 | 0.000998908 | 1 |
| Obesity | mast_cell | *CDH23* | 4.203775796 | 0.000270497 | 0.000986064 | 1 |
| Obesity | mast_cell | *SPOCK2* | 1.286694619 | 0.012353454 | 0.016376765 | 1 |
| Obesity | mast_cell | *BMPR1A* | 2.241957377 | 0.000333415 | 0.001016573 | 1 |
| Obesity | mast_cell | *LIPA* | 2.016459707 | 7.67E-05 | 0.000936854 | 0.371931044 |
| Obesity | mast_cell | *SFXN3* | 3.520068553 | 0.000294096 | 0.000998908 | 1 |
| Obesity | mast_cell | *TNKS2* | 1.949390054 | 0.000345695 | 0.00102869 | 1 |
| Obesity | mast_cell | *ANKRD26* | 4.739554392 | 0.00029514 | 0.000998908 | 1 |
| Obesity | mast_cell | *ACBD5* | 3.650961212 | 0.000288803 | 0.00099522 | 1 |
| Obesity | mast_cell | *LHPP* | 5.037664591 | 0.000417664 | 0.001082536 | 1 |
| Obesity | mast_cell | *PITRM1* | 5.70356421 | 0.021227077 | 0.026940565 | 1 |
| Obesity | mast_cell | *TFAM* | 3.816465778 | 0.000243011 | 0.000976811 | 1 |
| Obesity | mast_cell | *RPL28* | 2.381771837 | 0.000146447 | 0.000976811 | 0.710560064 |
| Obesity | mast_cell | *DNAJC12* | 6.838370672 | 5.61E-05 | 0.000817301 | 0.272307133 |
| Obesity | mast_cell | *PBLD* | 2.364834914 | 0.000326842 | 0.001007521 | 1 |
| Obesity | mast_cell | *TSPAN14* | 2.393600754 | 0.018188139 | 0.023420608 | 1 |
| Obesity | mast_cell | *RPL19* | 2.402487288 | 0.00048158 | 0.001130443 | 1 |
| Obesity | mast_cell | *RAPGEFL1* | 3.268203892 | 0.00043754 | 0.001098433 | 1 |
| Obesity | mast_cell | *WNT3* | 3.038671281 | 0.005868157 | 0.00824567 | 1 |
| Obesity | mast_cell | *ASPA* | 2.092402666 | 0.000285007 | 0.000993978 | 1 |
| Obesity | mast_cell | *TRIM37* | 4.063367224 | 0.000123268 | 0.000976811 | 0.598098296 |
| Obesity | mast_cell | *PNPO* | 2.873410694 | 0.022367997 | 0.028277625 | 1 |
| Obesity | mast_cell | *TRIM16L* | 3.034295163 | 0.00070582 | 0.001381207 | 1 |
| Obesity | mast_cell | *CDK5RAP3* | 2.549883037 | 0.002049157 | 0.003247872 | 1 |
| Obesity | mast_cell | *PIGL* | 2.308399009 | 0.000324176 | 0.001004343 | 1 |
| Obesity | mast_cell | *MED13* | 1.802737316 | 0.000812805 | 0.001527984 | 1 |
| Obesity | mast_cell | *ENO3* | 3.34728869 | 0.009844843 | 0.013242911 | 1 |
| Obesity | mast_cell | *RAI1* | 5.293943698 | 0.000160973 | 0.000976811 | 0.781041699 |
| Obesity | mast_cell | *GOSR1* | 3.609873709 | 0.000191804 | 0.000976811 | 0.930631882 |
| Obesity | mast_cell | *DRG2* | 2.357978878 | 0.000304702 | 0.000998908 | 1 |
| Obesity | mast_cell | *ICAM2* | 2.88884887 | 0.000289356 | 0.000995715 | 1 |
| Obesity | mast_cell | *DDX5* | 2.31314798 | 0.002690025 | 0.004101823 | 1 |
| Obesity | mast_cell | *C17orf75* | 2.155345187 | 0.000400036 | 0.001064254 | 1 |
| Obesity | mast_cell | *PSMD11* | 4.038170815 | 0.000259688 | 0.00098093 | 1 |

| | | | | | | |
|---|---|---|---|---|---|---|
| Obesity | mast_cell | *DHX58* | 3.085534515 | 0.000437562 | 0.001098433 | 1 |
| Obesity | mast_cell | *RAB5C* | 1.970704283 | 0.003292422 | 0.004901759 | 1 |
| Obesity | mast_cell | *NAGLU* | 3.23455196 | 0.000342879 | 0.001027777 | 1 |
| Obesity | mast_cell | *HSD17B1* | 3.292858733 | 0.000359193 | 0.001035264 | 1 |
| Obesity | mast_cell | *CNTNAP1* | 4.306136944 | 7.75E-05 | 0.00094053 | 0.376230626 |
| Obesity | mast_cell | *EZH1* | 3.301853393 | 0.000217048 | 0.000976811 | 1 |
| Obesity | mast_cell | *ABCC3* | 2.178162362 | 6.97E-05 | 0.000898663 | 0.338043393 |
| Obesity | mast_cell | *EFTUD2* | 4.852338045 | 0.000174468 | 0.000976811 | 0.84651639 |
| Obesity | mast_cell | *EFNB3* | 3.190154534 | 0.003374504 | 0.005010127 | 1 |
| Obesity | mast_cell | *FAM20A* | 2.016988116 | 0.000360062 | 0.001035657 | 1 |
| Obesity | mast_cell | *DPH1* | 10.2058149 | 9.19E-05 | 0.000976811 | 0.445791337 |
| Obesity | mast_cell | *DHRS7B* | 2.978318156 | 0.000237007 | 0.000976811 | 1 |
| Obesity | mast_cell | *MYH3* | 3.514417824 | 0.000435594 | 0.001098433 | 1 |
| Obesity | mast_cell | *TMEM97* | -1.231275158 | 0.008602815 | 0.011701951 | 1 |
| Obesity | mast_cell | *GNRHR* | 1.870822152 | 0.018748905 | 0.024078795 | 1 |
| Obesity | mast_cell | *SLAIN2* | 4.197883133 | 0.000140643 | 0.000976811 | 0.682400093 |
| Obesity | mast_cell | *OCIAD1* | 4.009302427 | 0.000390938 | 0.001054382 | 1 |
| Obesity | mast_cell | *UBE2D3* | 2.723001235 | 0.000226944 | 0.000976811 | 1 |
| Obesity | mast_cell | *NDUFC1* | 2.413209098 | 0.000538392 | 0.001170375 | 1 |
| Obesity | mast_cell | *KLHL2* | 4.157953002 | 0.000478984 | 0.001129371 | 1 |
| Obesity | mast_cell | *CPE* | 1.73056622 | 0.00206161 | 0.003262536 | 1 |
| Obesity | mast_cell | *WFS1* | 2.602065066 | 0.000425668 | 0.001090111 | 1 |
| Obesity | mast_cell | *GRPEL1* | 10.90446472 | 0.003683239 | 0.005422049 | 1 |
| Obesity | mast_cell | *GAR1* | 3.48692613 | 0.000369699 | 0.00103942 | 1 |
| Obesity | mast_cell | *AADAT* | 3.095057765 | 0.000795534 | 0.001504846 | 1 |
| Obesity | mast_cell | *TRIM2* | 3.286803894 | 0.000290168 | 0.000996679 | 1 |
| Obesity | mast_cell | *STIM2* | 2.680375086 | 0.000173379 | 0.000976811 | 0.841233971 |
| Obesity | mast_cell | *BST1* | 5.740757966 | 5.74E-05 | 0.000828263 | 0.278296297 |
| Obesity | mast_cell | *UFSP2* | 5.69663599 | 0.00015897 | 0.000976811 | 0.771322508 |
| Obesity | mast_cell | *KLHL5* | 4.914513438 | 0.000177249 | 0.000976811 | 0.860011721 |
| Obesity | mast_cell | *FAM149A* | 2.503997717 | 0.000505534 | 0.001144783 | 1 |
| Obesity | mast_cell | *HTATIP2* | 2.180833518 | 0.000375835 | 0.001047881 | 1 |
| Obesity | mast_cell | *CTSC* | 1.545757795 | 0.00156477 | 0.002598381 | 1 |
| Obesity | mast_cell | *ELP4* | 2.170128983 | 0.000746666 | 0.001439342 | 1 |
| Obesity | mast_cell | *SC5D* | 3.761153873 | 0.00467108 | 0.006703366 | 1 |
| Obesity | mast_cell | *B3GAT1* | 21.81646359 | 6.46E-05 | 0.000874503 | 0.313448223 |
| Obesity | mast_cell | *P2RX3* | 28.08593885 | 0.000349551 | 0.001031643 | 1 |
| Obesity | mast_cell | *DNAJC4* | 2.142704886 | 0.003137254 | 0.004696684 | 1 |
| Obesity | mast_cell | *UNC93B1* | 3.202308926 | 0.001064858 | 0.001882904 | 1 |
| Obesity | mast_cell | *FOXRED1* | 2.411035548 | 5.54E-05 | 0.000811438 | 0.268585829 |
| Obesity | mast_cell | *NRXN2* | -5.5896342 | 0.013612179 | 0.017879343 | 1 |
| Obesity | mast_cell | *CCND1* | 4.341653713 | 0.000671965 | 0.001331852 | 1 |
| Obesity | mast_cell | *CCDC86* | 3.534829239 | 0.019671013 | 0.025167305 | 1 |
| Obesity | mast_cell | *PANX1* | 4.458616877 | 0.000846993 | 0.001573358 | 1 |
| Obesity | mast_cell | *CEP164* | 2.732286789 | 0.000293122 | 0.000998908 | 1 |
| Obesity | mast_cell | *GALNT18* | 1.334041437 | 0.000713746 | 0.001391373 | 1 |
| Obesity | mast_cell | *KIAA1549L* | -4.016464466 | 0.003447129 | 0.005100784 | 1 |
| Obesity | mast_cell | *ACCS* | 2.110882969 | 0.000816612 | 0.001532014 | 1 |
| Obesity | mast_cell | *SLC22A18* | 2.394193151 | 0.000286073 | 0.000993978 | 1 |
| Obesity | mast_cell | *C11orf21* | -3.546547475 | 0.003182918 | 0.004753314 | 1 |

| | | | | | | |
|---|---|---|---|---|---|---|
| Obesity | mast_cell | *ELMOD1* | -3.07469635 | 0.002081668 | 0.003293202 | 1 |
| Obesity | mast_cell | *RPS13* | 3.029145883 | 0.001138302 | 0.001993877 | 1 |
| Obesity | mast_cell | *NDUFS8* | 1.886501256 | 0.000669166 | 0.001326846 | 1 |
| Obesity | mast_cell | *TCIRG1* | 2.46424622 | 0.000827834 | 0.001547843 | 1 |
| Obesity | mast_cell | *GTF2H1* | 2.72820858 | 0.000128713 | 0.000976811 | 0.624513371 |
| Obesity | mast_cell | *PRDM4* | 6.350308002 | 2.63E-05 | 0.000562671 | 0.127695293 |
| Obesity | mast_cell | *COQ5* | 1.085849269 | 0.004040939 | 0.005893188 | 1 |
| Obesity | mast_cell | *CORO1C* | 3.906024618 | 0.000186326 | 0.000976811 | 0.904052792 |
| Obesity | mast_cell | *ASIC1* | 4.976489009 | 0.000546146 | 0.00117408 | 1 |
| Obesity | mast_cell | *CAPRIN2* | 2.789433993 | 0.000960859 | 0.001737747 | 1 |
| Obesity | mast_cell | *KCTD10* | 1.634576981 | 0.004875275 | 0.006969604 | 1 |
| Obesity | mast_cell | *IL23A* | -4.124814734 | 0.00227921 | 0.003562629 | 1 |
| Obesity | mast_cell | *ACSS3* | 1.318486822 | 0.000522504 | 0.001153408 | 1 |
| Obesity | mast_cell | *LTA4H* | 1.51881179 | 0.00070626 | 0.001381207 | 1 |
| Obesity | mast_cell | *PRR4* | 3.741697694 | 0.005141205 | 0.007317432 | 1 |
| Obesity | mast_cell | *PARP11* | 3.174969956 | 0.000233604 | 0.000976811 | 1 |
| Obesity | mast_cell | *GPN3* | 2.228772526 | 0.000643279 | 0.001294025 | 1 |
| Obesity | mast_cell | *VPS29* | 3.436319246 | 0.000156542 | 0.000976811 | 0.759541655 |
| Obesity | mast_cell | *ACAD10* | 1.782432779 | 0.000312327 | 0.000998908 | 1 |
| Obesity | mast_cell | *LTBR* | 2.818169914 | 0.000171204 | 0.000976811 | 0.830679628 |
| Obesity | mast_cell | *CDK2AP1* | -1.378399505 | 0.037334718 | 0.045813872 | 1 |
| Obesity | mast_cell | *DDX55* | 3.475666234 | 2.40E-05 | 0.000545024 | 0.116578142 |
| Obesity | mast_cell | *ENDOU* | 6.242976289 | 0.00154303 | 0.002571893 | 1 |
| Obesity | mast_cell | *VDR* | 5.607130208 | 0.000348355 | 0.001029993 | 1 |
| Obesity | mast_cell | *STX2* | 2.298140154 | 0.000391912 | 0.001054889 | 1 |
| Obesity | mast_cell | *CAND1* | 2.685039773 | 0.000231532 | 0.000976811 | 1 |
| Obesity | mast_cell | *MDM1* | 7.692762401 | 0.000396196 | 0.001059725 | 1 |
| Obesity | mast_cell | *NUP107* | 2.576250652 | 0.000130097 | 0.000976811 | 0.631231528 |
| Obesity | mast_cell | *KRR1* | 3.961816176 | 0.000414522 | 0.001080439 | 1 |
| Obesity | mast_cell | *MRPL51* | 1.869496346 | 0.000615782 | 0.001256951 | 1 |
| Obesity | mast_cell | *CHPT1* | 2.745434182 | 0.000824008 | 0.001543067 | 1 |
| Obesity | mast_cell | *SPSB2* | 3.238553356 | 0.000202254 | 0.000976811 | 0.981334649 |
| Obesity | mast_cell | *ATN1* | 3.069768594 | 0.016051799 | 0.020875347 | 1 |
| Obesity | mast_cell | *NT5DC3* | 2.514312139 | 0.001075655 | 0.001897157 | 1 |
| Obesity | mast_cell | *SUDS3* | 4.682503103 | 0.000186739 | 0.000976811 | 0.906057753 |
| Obesity | mast_cell | *GOLT1B* | 6.02023463 | 0.003394011 | 0.005034467 | 1 |
| Obesity | mast_cell | *C2CD5* | 1.88043303 | 0.000338336 | 0.00102217 | 1 |
| Obesity | mast_cell | *BTN3A3* | 3.06855847 | 3.82E-08 | 5.45E-06 | 0.00018545 |
| Obesity | mast_cell | *FRK* | 2.134329477 | 0.000548944 | 0.001175269 | 1 |
| Obesity | mast_cell | *RWDD1* | 2.404040151 | 0.001262361 | 0.002170437 | 1 |
| Obesity | mast_cell | *TMEM14C* | 7.885708849 | 0.000401582 | 0.001064698 | 1 |
| Obesity | mast_cell | *SMIM8* | 3.157461187 | 0.00010713 | 0.000976811 | 0.519796221 |
| Obesity | mast_cell | *FAM184A* | 1.745810608 | 0.00024031 | 0.000976811 | 1 |
| Obesity | mast_cell | *RNGTT* | 3.302461734 | 0.000209098 | 0.000976811 | 1 |
| Obesity | mast_cell | *GABRR2* | 5.768783998 | 2.32E-06 | 0.000137034 | 0.011236772 |
| Obesity | mast_cell | *HDDC2* | 2.383759231 | 0.000973824 | 0.001751295 | 1 |
| Obesity | mast_cell | *UST* | 1.182431 | 0.001048029 | 0.001858565 | 1 |
| Obesity | mast_cell | *MTRF1L* | 2.324763447 | 0.000101943 | 0.000976811 | 0.494628303 |
| Obesity | mast_cell | *TULP1* | -3.641427417 | 0.017531188 | 0.022616678 | 1 |
| Obesity | mast_cell | *MRPL18* | 5.495416505 | 8.60E-05 | 0.000971162 | 0.417341101 |

| | | | | | | |
|---|---|---|---|---|---|---|
| Obesity | mast_cell | *RNF8* | 3.641031007 | 0.000318796 | 0.000998908 | 1 |
| Obesity | mast_cell | *PHACTR1* | 2.117260227 | 0.001686112 | 0.002748996 | 1 |
| Obesity | mast_cell | *MDGA1* | 2.449342451 | 0.00089365 | 0.001638513 | 1 |
| Obesity | mast_cell | *CD83* | 2.501953573 | 0.00068973 | 0.001356535 | 1 |
| Obesity | mast_cell | *MDN1* | 2.5010599 | 0.000909179 | 0.001658397 | 1 |
| Obesity | mast_cell | *COL9A1* | 7.856536666 | 0.000275559 | 0.000990086 | 1 |
| Obesity | mast_cell | *WASF1* | 1.68220592 | 0.00036716 | 0.001037687 | 1 |
| Obesity | mast_cell | *VNN2* | 4.877788156 | 0.000551217 | 0.001177677 | 1 |
| Obesity | mast_cell | *ACOT13* | 6.523559225 | 0.000210683 | 0.000976811 | 1 |
| Obesity | mast_cell | *C6orf62* | 2.172333611 | 0.001849659 | 0.002976632 | 1 |
| Obesity | mast_cell | *HBS1L* | 3.937638919 | 0.000290253 | 0.000996679 | 1 |
| Obesity | mast_cell | *PEX7* | 2.372315659 | 0.003049653 | 0.004586768 | 1 |
| Obesity | mast_cell | *FIG4* | 3.009135112 | 0.000185557 | 0.000976811 | 0.900320709 |
| Obesity | mast_cell | *PERP* | 2.348576931 | 0.009315983 | 0.012601379 | 1 |
| Obesity | mast_cell | *PHACTR2* | 3.726597876 | 0.000520287 | 0.001152325 | 1 |
| Obesity | mast_cell | *CUTA* | 1.361482541 | 0.023275699 | 0.029348672 | 1 |
| Obesity | mast_cell | *PACRG* | 8.061410389 | 0.000335461 | 0.001017877 | 1 |
| Obesity | mast_cell | *QKI* | 2.338617099 | 0.000212472 | 0.000976811 | 1 |
| Obesity | mast_cell | *FAM120B* | 2.727477376 | 0.000296564 | 0.000998908 | 1 |
| Obesity | mast_cell | *GUCA1B* | -3.323290284 | 0.005877574 | 0.008254121 | 1 |
| Obesity | mast_cell | *DNPH1* | 1.751928536 | 0.004746956 | 0.006798179 | 1 |
| Obesity | mast_cell | *EXOC2* | 4.282577818 | 0.000213825 | 0.000976811 | 1 |
| Obesity | mast_cell | *TMEM30A* | 4.270268426 | 0.000215761 | 0.000976811 | 1 |
| Obesity | mast_cell | *IMPG1* | 4.897498541 | 0.012632293 | 0.016719009 | 1 |
| Obesity | mast_cell | *BTN2A1* | 3.245963186 | 0.008076924 | 0.011045444 | 1 |
| Obesity | mast_cell | *LY86* | 2.395955327 | 0.000582111 | 0.001209489 | 1 |
| Obesity | mast_cell | *PCDHB2* | 3.638270801 | 0.020092994 | 0.025648831 | 1 |
| Obesity | mast_cell | *NUDT12* | 4.402393273 | 0.000215361 | 0.000976811 | 1 |
| Obesity | mast_cell | *CEP72* | 2.657396193 | 0.000982898 | 0.001763748 | 1 |
| Obesity | mast_cell | *DAP* | 4.257546578 | 6.80E-05 | 0.000889685 | 0.33007313 |
| Obesity | mast_cell | *NME5* | 2.853070055 | 6.66E-07 | 5.77E-05 | 0.00323056 |
| Obesity | mast_cell | *PFDN1* | 3.116811951 | 5.85E-05 | 0.000839117 | 0.283621447 |
| Obesity | mast_cell | *SPARC* | 1.887181553 | 0.00041631 | 0.001080612 | 1 |
| Obesity | mast_cell | *IK* | 1.33512557 | 0.012273199 | 0.016288173 | 1 |
| Obesity | mast_cell | *RNF130* | 2.487445043 | 0.000537313 | 0.001169601 | 1 |
| Obesity | mast_cell | *ARSB* | 2.012058204 | 0.000438 | 0.001098433 | 1 |
| Obesity | mast_cell | *THBS4* | 1.830403105 | 0.00038752 | 0.00105395 | 1 |
| Obesity | mast_cell | *MSH3* | 1.777763803 | 0.000992507 | 0.001776335 | 1 |
| Obesity | mast_cell | *RASGRF2* | 2.057991347 | 0.000231588 | 0.000976811 | 1 |
| Obesity | mast_cell | *DROSHA* | 2.807561795 | 0.000152157 | 0.000976811 | 0.738266865 |
| Obesity | mast_cell | *CDH6* | 1.663798503 | 0.001010767 | 0.001805685 | 1 |
| Obesity | mast_cell | *LMNB1* | 4.784929257 | 0.02978805 | 0.037021419 | 1 |
| Obesity | mast_cell | *SLC12A7* | 3.595735509 | 0.000227476 | 0.000976811 | 1 |
| Obesity | mast_cell | *SKP1* | 2.898367076 | 0.000235917 | 0.000976811 | 1 |
| Obesity | mast_cell | *PPWD1* | 2.290811283 | 0.000319998 | 0.000999138 | 1 |
| Obesity | mast_cell | *TXNDC15* | 2.248536656 | 0.000383518 | 0.001052047 | 1 |
| Obesity | mast_cell | *SMAD5* | 4.084877413 | 0.000160749 | 0.000976811 | 0.779954223 |
| Obesity | mast_cell | *ERGIC1* | 4.953374748 | 0.000172975 | 0.000976811 | 0.839274599 |
| Obesity | mast_cell | *ATP6V0E1* | 2.360601661 | 0.000123867 | 0.000976811 | 0.601004379 |
| Obesity | mast_cell | *BNIP1* | -3.068249812 | 0.00156961 | 0.002601895 | 1 |

| | | | | | | |
|---|---|---|---|---|---|---|
| Obesity | mast_cell | *UNC5A* | 9.928335615 | 0.00050286 | 0.001144783 | 1 |
| Obesity | mast_cell | *ACTR8* | 4.146895225 | 0.008120537 | 0.01109258 | 1 |
| Obesity | mast_cell | *TBCCD1* | 8.881371105 | 0.000313763 | 0.000998908 | 1 |
| Obesity | mast_cell | *CLDN16* | 2.71198553 | 0.001111127 | 0.001952621 | 1 |
| Obesity | mast_cell | *NPHP3* | 1.629303942 | 0.017096372 | 0.022096856 | 1 |
| Obesity | mast_cell | *PCCB* | 6.175193802 | 5.90E-09 | 1.36E-06 | 2.86E-05 |
| Obesity | mast_cell | *CEP70* | 2.11111437 | 0.000668476 | 0.001326427 | 1 |
| Obesity | mast_cell | *CBLB* | 2.186286052 | 0.000128999 | 0.000976811 | 0.625901737 |
| Obesity | mast_cell | *IQCG* | 3.429307355 | 0.000176731 | 0.000976811 | 0.857500329 |
| Obesity | mast_cell | *NCBP2* | 3.876290776 | 0.00103036 | 0.001833936 | 1 |
| Obesity | mast_cell | *PLXNA1* | 7.658628384 | 3.44E-05 | 0.000640166 | 0.167083452 |
| Obesity | mast_cell | *SCAP* | 4.03700186 | 1.88E-05 | 0.000496117 | 0.091117011 |
| Obesity | mast_cell | *KIAA1257* | 6.898061445 | 0.000365291 | 0.001037687 | 1 |
| Obesity | mast_cell | *NEK11* | 2.191604402 | 0.00041217 | 0.001075765 | 1 |
| Obesity | mast_cell | *PLSCR4* | 1.682160803 | 0.000435971 | 0.001098433 | 1 |
| Obesity | mast_cell | *HEMK1* | 4.378115545 | 7.09E-05 | 0.000904727 | 0.343796383 |
| Obesity | mast_cell | *COMMD2* | 7.016866216 | 0.001869501 | 0.003003051 | 1 |
| Obesity | mast_cell | *ABHD14B* | 2.093236452 | 0.000796035 | 0.001505207 | 1 |
| Obesity | mast_cell | *ARHGEF26* | 2.330197116 | 0.000368067 | 0.001037687 | 1 |
| Obesity | mast_cell | *VIPR1* | 3.112120591 | 0.000380329 | 0.001048903 | 1 |
| Obesity | mast_cell | *DNAH1* | 5.515122722 | 4.12E-05 | 0.0007135 | 0.199779864 |
| Obesity | mast_cell | *NKTR* | 2.85276328 | 8.68E-05 | 0.000971162 | 0.421335239 |
| Obesity | mast_cell | *ADAM23* | 2.242844075 | 0.001184916 | 0.002062867 | 1 |
| Obesity | mast_cell | *ACTR1B* | 2.308962319 | 0.000257939 | 0.00098093 | 1 |
| Obesity | mast_cell | *SLC35F5* | 3.250959377 | 0.000358326 | 0.001035264 | 1 |
| Obesity | mast_cell | *CENPA* | -1.570901925 | 0.020485944 | 0.026102364 | 1 |
| Obesity | mast_cell | *NRBP1* | 3.220456837 | 0.000351179 | 0.001031837 | 1 |
| Obesity | mast_cell | *PPM1G* | 3.178429757 | 0.000340299 | 0.001025052 | 1 |
| Obesity | mast_cell | *INO80B* | 2.633529371 | 0.032352923 | 0.040014373 | 1 |
| Obesity | mast_cell | *MOGS* | 2.858741572 | 0.000516582 | 0.001150279 | 1 |
| Obesity | mast_cell | *RTN4* | 2.631414352 | 0.00038915 | 0.001054382 | 1 |
| Obesity | mast_cell | *GALNT3* | -1.022643343 | 0.005950128 | 0.008343937 | 1 |
| Obesity | mast_cell | *ACADL* | 1.753611902 | 0.000272868 | 0.000987746 | 1 |
| Obesity | mast_cell | *MRPL19* | 1.817312506 | 0.000454375 | 0.001110332 | 1 |
| Obesity | mast_cell | *DNAH6* | 1.113212168 | 0.00123868 | 0.002136536 | 1 |
| Obesity | mast_cell | *IGFBP2* | 2.928326025 | 0.00066263 | 0.001319333 | 1 |
| Obesity | mast_cell | *GGCX* | 2.662592832 | 6.50E-05 | 0.00087635 | 0.315486004 |
| Obesity | mast_cell | *IL1RL2* | 5.861405894 | 0.000141031 | 0.000976811 | 0.684281709 |
| Obesity | mast_cell | *IL18R1* | -0.23272464 | 0.001669156 | 0.002729608 | 1 |
| Obesity | mast_cell | *MLPH* | -0.3989404 | 0.001643581 | 0.002696873 | 1 |
| Obesity | mast_cell | *HDLBP* | 3.630529105 | 0.000145742 | 0.000976811 | 0.707140232 |
| Obesity | mast_cell | *PASK* | 1.868203478 | 0.034917681 | 0.042989238 | 1 |
| Obesity | mast_cell | *TPO* | 1.823130995 | 0.000763265 | 0.001462564 | 1 |
| Obesity | mast_cell | *TAF1B* | 3.52255612 | 0.000330977 | 0.001014048 | 1 |
| Obesity | mast_cell | *NOL10* | 2.651289095 | 0.000168291 | 0.000976811 | 0.816546595 |
| Obesity | mast_cell | *STRN* | 5.148520459 | 0.000220052 | 0.000976811 | 1 |
| Obesity | mast_cell | *CEBPZ* | 4.449927056 | 0.000656615 | 0.001312378 | 1 |
| Obesity | mast_cell | *PRKD3* | 4.301636559 | 0.000780665 | 0.001486572 | 1 |
| Obesity | mast_cell | *SLC25A12* | 2.028130508 | 0.000300257 | 0.000998908 | 1 |
| Obesity | mast_cell | *DARS* | 2.282904486 | 0.00029463 | 0.000998908 | 1 |

| | | | | | | |
|---|---|---|---|---|---|---|
| Obesity | mast_cell | *WIPF1* | 3.318423246 | 0.000191451 | 0.000976811 | 0.92891982 |
| Obesity | mast_cell | *PNO1* | -3.45533897 | 0.033469898 | 0.041322124 | 1 |
| Obesity | mast_cell | *ORC4* | 2.582746048 | 0.000149353 | 0.000976811 | 0.724662527 |
| Obesity | mast_cell | *AAK1* | 2.51549639 | 0.000210694 | 0.000976811 | 1 |
| Obesity | mast_cell | *CD207* | 4.409396029 | 0.021549304 | 0.027313799 | 1 |
| Obesity | mast_cell | *NFE2L2* | 2.388679106 | 0.000197039 | 0.000976811 | 0.956030846 |
| Obesity | mast_cell | *MSH6* | 3.434052288 | 0.000608388 | 0.001246806 | 1 |
| Obesity | mast_cell | *ALMS1* | 6.352541259 | 0.000313621 | 0.000998908 | 1 |
| Obesity | mast_cell | *DNAJC16* | 2.566241591 | 0.000957302 | 0.001732498 | 1 |
| Obesity | mast_cell | *MARK1* | 1.479489715 | 0.000704073 | 0.00137971 | 1 |
| Obesity | mast_cell | *MORN1* | 1.736968967 | 0.006743169 | 0.009347958 | 1 |
| Obesity | mast_cell | *RALGPS2* | 2.968381161 | 0.00096491 | 0.001741405 | 1 |
| Obesity | mast_cell | *CEP104* | 2.359464248 | 0.001075332 | 0.001897157 | 1 |
| Obesity | mast_cell | *FAM20B* | 2.567784827 | 0.000423069 | 0.001086605 | 1 |
| Obesity | mast_cell | *WRAP73* | 2.257776212 | 0.001394303 | 0.002360495 | 1 |
| Obesity | mast_cell | *ICMT* | 4.739853207 | 0.00028411 | 0.000993978 | 1 |
| Obesity | mast_cell | *PARK7* | 4.231990635 | 0.000427109 | 0.001091691 | 1 |
| Obesity | mast_cell | *KIAA1324* | 4.648427793 | 0.002105424 | 0.003325364 | 1 |
| Obesity | mast_cell | *MECR* | 5.759269424 | 1.23E-05 | 0.000386737 | 0.059557501 |
| Obesity | mast_cell | *KCNC4* | 1.812875228 | 0.002274109 | 0.003555907 | 1 |
| Obesity | mast_cell | *EDEM3* | 3.567117282 | 0.00068902 | 0.001355939 | 1 |
| Obesity | mast_cell | *RAP1A* | 3.25897056 | 0.00025851 | 0.00098093 | 1 |
| Obesity | mast_cell | *CAPZA1* | 2.5104283 | 0.000124626 | 0.000976811 | 0.604687201 |
| Obesity | mast_cell | *ASH1L* | 3.280210451 | 4.58E-05 | 0.000754069 | 0.222328804 |
| Obesity | mast_cell | *GON4L* | 2.623449077 | 0.001438991 | 0.002424301 | 1 |
| Obesity | mast_cell | *DOCK7* | 2.756816533 | 0.000236288 | 0.000976811 | 1 |
| Obesity | mast_cell | *FBXO6* | 2.779974783 | 0.009035833 | 0.012256601 | 1 |
| Obesity | mast_cell | *DNAJC6* | 1.8883698 | 0.005533769 | 0.007818826 | 1 |
| Obesity | mast_cell | *LEPR* | 2.419079413 | 0.000218154 | 0.000976811 | 1 |
| Obesity | mast_cell | *IVNS1ABP* | 1.44295911 | 0.000580528 | 0.001208894 | 1 |
| Obesity | mast_cell | *MFN2* | 2.627867101 | 0.000213457 | 0.000976811 | 1 |
| Obesity | mast_cell | *PRG4* | 2.669897388 | 0.000392214 | 0.001054889 | 1 |
| Obesity | mast_cell | *NCF2* | 1.598833097 | 0.005381436 | 0.00762357 | 1 |
| Obesity | mast_cell | *BCAS2* | 3.314073339 | 0.001551693 | 0.002581897 | 1 |
| Obesity | mast_cell | *TMEM9* | 2.442025015 | 0.015258072 | 0.019895771 | 1 |
| Obesity | mast_cell | *WARS2* | 3.068037354 | 4.89E-05 | 0.000765648 | 0.237445761 |
| Obesity | mast_cell | *OSCP1* | 1.631462719 | 0.00088852 | 0.001631139 | 1 |
| Obesity | mast_cell | *MRPS15* | 7.67991938 | 0.004122036 | 0.006000635 | 1 |
| Obesity | mast_cell | *EXOC8* | 3.741988299 | 0.0002928 | 0.000998908 | 1 |
| Obesity | mast_cell | *GNPAT* | 2.660244568 | 0.000167251 | 0.000976811 | 0.811502438 |
| Obesity | mast_cell | *C1orf109* | 5.244264834 | 0.000141125 | 0.000976811 | 0.684738275 |
| Obesity | mast_cell | *TBCE* | 1.97512516 | 0.000409641 | 0.001074715 | 1 |
| Obesity | mast_cell | *ACADM* | 2.853805488 | 0.000188602 | 0.000976811 | 0.915099006 |
| Obesity | mast_cell | *SLAMF1* | 1.961272972 | 0.033809926 | 0.041699481 | 1 |
| Obesity | mast_cell | *PADI2* | -0.765949856 | 0.000698614 | 0.00137012 | 1 |
| Obesity | mast_cell | *SDHB* | 4.673413912 | 0.000352144 | 0.001031837 | 1 |
| Obesity | mast_cell | *MFAP2* | -2.258418125 | 0.015229424 | 0.019869095 | 1 |
| Obesity | mast_cell | *CTBS* | 1.338665146 | 0.007950427 | 0.010890873 | 1 |
| Obesity | mast_cell | *KLHL12* | 2.527036246 | 0.002569878 | 0.003952038 | 1 |
| Obesity | mast_cell | *ZNHIT6* | 3.012450501 | 0.000312541 | 0.000998908 | 1 |

| | | | | | | |
|---|---|---|---|---|---|---|
| Obesity | mast_cell | *KIF17* | 3.530351064 | 0.008000099 | 0.010955823 | 1 |
| Obesity | mast_cell | *CD46* | 2.527437411 | 0.000983502 | 0.001764123 | 1 |
| Obesity | mast_cell | *EBNA1BP2* | 4.740232548 | 0.000162678 | 0.000976811 | 0.789312178 |
| Obesity | mast_cell | *ERI3* | 2.760397348 | 0.000237059 | 0.000976811 | 1 |
| Obesity | mast_cell | *PRDX1* | 2.500142761 | 0.000846562 | 0.00157316 | 1 |
| Obesity | mast_cell | *DR1* | 2.320820464 | 0.002491846 | 0.003844336 | 1 |
| Obesity | mast_cell | *PRRC2C* | 4.08422787 | 2.39E-05 | 0.000545024 | 0.115771556 |
| Obesity | mast_cell | *ABCD3* | 2.582326702 | 0.000282575 | 0.000993978 | 1 |
| Obesity | mast_cell | *VAMP4* | 2.931779944 | 0.000154437 | 0.000976811 | 0.749330467 |
| Obesity | mast_cell | *PTBP2* | 3.426691945 | 5.41E-05 | 0.000802079 | 0.262268904 |
| Obesity | mast_cell | *DARS2* | 3.975414632 | 0.001641026 | 0.002693592 | 1 |
| Obesity | mast_cell | *SERPINC1* | 12.2383798 | 3.58E-06 | 0.000177485 | 0.017393505 |
| Obesity | mast_cell | *MTFR1L* | 6.439458431 | 0.000154345 | 0.000976811 | 0.748882152 |
| Obesity | mast_cell | *RPS6KA1* | 2.506042239 | 0.001273975 | 0.002188855 | 1 |
| Obesity | mast_cell | *RPA2* | 3.914166843 | 0.000257379 | 0.00098093 | 1 |
| Obesity | mast_cell | *MARC2* | 2.928193011 | 0.000385519 | 0.001052047 | 1 |
| Obesity | mast_cell | *SLC5A9* | 3.83199515 | 0.000133456 | 0.000976811 | 0.647526754 |
| Obesity | mast_cell | *ESYT2* | 2.221287082 | 0.000782475 | 0.001487241 | 1 |
| Obesity | mast_cell | *CD3EAP* | 2.482992901 | 0.000331031 | 0.001014048 | 1 |
| Obesity | mast_cell | *RCN2* | 3.491511599 | 0.000328274 | 0.001010651 | 1 |
| Obesity | mast_cell | *IFT46* | 4.629343315 | 0.006678904 | 0.009272115 | 1 |
| Obesity | mast_cell | *APOA1* | 11.13624167 | 0.017446835 | 0.02251983 | 1 |
| Obesity | mast_cell | *DDX59* | 3.17737347 | 0.000805505 | 0.001518379 | 1 |
| Obesity | mast_cell | *CAMSAP2* | 2.164102091 | 0.000271342 | 0.000987746 | 1 |
| Obesity | mast_cell | *KLF7* | 2.307581642 | 0.000164487 | 0.000976811 | 0.798091022 |
| Obesity | mast_cell | *LRMP* | 2.20207261 | 0.002713782 | 0.004130261 | 1 |
| Obesity | mast_cell | *ELOVL4* | 5.601516997 | 0.000305743 | 0.000998908 | 1 |
| Obesity | mast_cell | *CNR1* | 3.128561101 | 0.00670831 | 0.009304952 | 1 |
| Obesity | mast_cell | *SGIP1* | 4.015441845 | 0.000207896 | 0.000976811 | 1 |
| Obesity | mast_cell | *AKAP7* | 2.714475894 | 0.000337014 | 0.001020083 | 1 |
| Obesity | mast_cell | *MED28* | 2.210783072 | 0.000660761 | 0.001317116 | 1 |
| Obesity | mast_cell | *TMEM5* | 4.954117904 | 0.000139747 | 0.000976811 | 0.678050273 |
| Obesity | mast_cell | *VAMP8* | 2.874412112 | 0.000236109 | 0.000976811 | 1 |
| Obesity | mast_cell | *FOXO3* | 1.993131275 | 0.000360999 | 0.001035816 | 1 |
| Obesity | mast_cell | *SPP1* | 3.639951732 | 0.000199443 | 0.000976811 | 0.967699554 |
| Obesity | mast_cell | *MFSD1* | 1.77720677 | 0.000389547 | 0.001054382 | 1 |
| Obesity | mast_cell | *UBN1* | 1.915481875 | 0.000340625 | 0.001025052 | 1 |
| Obesity | mast_cell | *WDR35* | 1.621796327 | 0.000397749 | 0.001061493 | 1 |
| Obesity | mast_cell | *DNAH7* | 1.650450153 | 0.003619005 | 0.005340394 | 1 |
| Obesity | mast_cell | *C2orf40* | 2.852334538 | 0.000195413 | 0.000976811 | 0.948143914 |
| Obesity | mast_cell | *CPSF3* | 2.350280128 | 0.012846109 | 0.016964975 | 1 |
| Obesity | mast_cell | *PIGZ* | 2.080221668 | 0.001523972 | 0.002547128 | 1 |
| Obesity | mast_cell | *SENP5* | 2.809918233 | 0.000223034 | 0.000976811 | 1 |
| Obesity | mast_cell | *CCDC92* | 3.617195032 | 4.71E-06 | 0.00021961 | 0.022839417 |
| Obesity | mast_cell | *FKBP15* | 2.074402212 | 0.000139242 | 0.000976811 | 0.675604581 |
| Obesity | mast_cell | *CTNNAL1* | 2.409481192 | 1.30E-05 | 0.000398143 | 0.063304671 |
| Obesity | mast_cell | *RAB14* | 4.726685947 | 0.000130384 | 0.000976811 | 0.632623051 |
| Obesity | mast_cell | *TRIM32* | 3.189663997 | 0.000306982 | 0.000998908 | 1 |
| Obesity | mast_cell | *PPP6C* | 2.293460113 | 0.000325044 | 0.001004947 | 1 |
| Obesity | mast_cell | *RBM18* | 3.222365404 | 0.000292386 | 0.000998908 | 1 |

| | | | | | | |
|---|---|---|---|---|---|---|
| Obesity | mast_cell | *HSDL2* | 1.791310072 | 0.002326204 | 0.003626844 | 1 |
| Obesity | mast_cell | *MAPKAP1* | 2.528336104 | 0.000196744 | 0.000976811 | 0.954600866 |
| Obesity | mast_cell | *INVS* | 4.238272482 | 1.99E-07 | 2.48E-05 | 0.000966425 |
| Obesity | mast_cell | *DENND1A* | 2.04032391 | 0.000204046 | 0.000976811 | 0.990033434 |
| Obesity | mast_cell | *KDSR* | 2.847037342 | 0.000309072 | 0.000998908 | 1 |
| Obesity | mast_cell | *DCAF4* | 3.33995197 | 0.00013875 | 0.000976811 | 0.673217099 |
| Obesity | mast_cell | *IFI27L2* | 2.532942572 | 0.000541607 | 0.001171675 | 1 |
| Obesity | mast_cell | *NEK9* | 3.471029974 | 0.000560734 | 0.001185482 | 1 |
| Obesity | mast_cell | *ACOT2* | 0.967320327 | 0.018414664 | 0.023699721 | 1 |
| Obesity | mast_cell | *TTLL5* | 2.086031074 | 0.000390419 | 0.001054382 | 1 |
| Obesity | mast_cell | *FLVCR2* | 2.102177743 | 0.000846496 | 0.00157316 | 1 |
| Obesity | mast_cell | *PPP4R4* | 2.107271744 | 0.004220747 | 0.006133293 | 1 |
| Obesity | mast_cell | *ALDH6A1* | 1.502009164 | 0.002281279 | 0.003563673 | 1 |
| Obesity | mast_cell | *ESRRB* | 2.966246676 | 0.0010601 | 0.001875859 | 1 |
| Obesity | mast_cell | *EIF2B2* | 1.255185971 | 0.023740058 | 0.029903106 | 1 |
| Obesity | mast_cell | *COQ6* | 2.956732653 | 0.000392517 | 0.001054923 | 1 |
| Obesity | mast_cell | *DNMT3A* | 4.000898025 | 0.000541905 | 0.001171675 | 1 |
| Obesity | mast_cell | *CRIPT* | 2.830642831 | 0.000325474 | 0.001005082 | 1 |
| Obesity | mast_cell | *EPCAM* | -1.685038657 | 0.003443543 | 0.005097032 | 1 |
| Obesity | mast_cell | *SLC17A5* | 1.963862163 | 0.000613875 | 0.001253586 | 1 |
| Obesity | mast_cell | *PPP1R3C* | 3.144183139 | 0.000302532 | 0.000998908 | 1 |
| Obesity | mast_cell | *PYROXD2* | 1.984931026 | 0.000200987 | 0.000976811 | 0.975189177 |
| Obesity | mast_cell | *MXI1* | 3.373967798 | 0.000168603 | 0.000976811 | 0.818059556 |
| Obesity | mast_cell | *C10orf88* | 3.121152949 | 0.000909977 | 0.00165895 | 1 |
| Obesity | mast_cell | *HELLS* | 1.880569752 | 0.002574082 | 0.003952357 | 1 |
| Obesity | mast_cell | *TCTN3* | 3.0176126 | 0.001979521 | 0.003151127 | 1 |
| Obesity | mast_cell | *WDR11* | 2.826368474 | 0.000172036 | 0.000976811 | 0.834719043 |
| Obesity | mast_cell | *KANSL1* | 2.51727806 | 0.000286491 | 0.000993978 | 1 |
| Obesity | mast_cell | *HOXB3* | 3.438126457 | 0.000206185 | 0.000976811 | 1 |
| Obesity | mast_cell | *CAAP1* | 2.530774588 | 0.005834011 | 0.008204818 | 1 |
| Obesity | mast_cell | *GORAB* | 3.922960679 | 0.000279985 | 0.000993978 | 1 |
| Obesity | mast_cell | *ACAT2* | 2.348898884 | 0.00036048 | 0.001035657 | 1 |
| Obesity | mast_cell | *SLC10A7* | 4.761020927 | 0.000203137 | 0.000976811 | 0.985621281 |
| Obesity | mast_cell | *MASTL* | 1.379802315 | 0.015842022 | 0.020618426 | 1 |
| Obesity | mast_cell | *MRC1* | 1.300394257 | 0.000518367 | 0.001151636 | 1 |
| Obesity | mast_cell | *IQSEC3* | 3.276021655 | 0.000312993 | 0.000998908 | 1 |
| Obesity | mast_cell | *CCDC77* | 4.290109592 | 0.000172543 | 0.000976811 | 0.837177955 |
| Obesity | mast_cell | *DNAJC15* | 2.674187212 | 0.000197536 | 0.000976811 | 0.958443167 |
| Obesity | mast_cell | *UFM1* | 5.108458427 | 8.95E-05 | 0.000976811 | 0.434053322 |
| Obesity | mast_cell | *ALG5* | 1.934104055 | 0.000493734 | 0.001138685 | 1 |
| Obesity | mast_cell | *SIL1* | 2.703066604 | 0.000318601 | 0.000998908 | 1 |
| Obesity | mast_cell | *ZFP30* | 1.945066249 | 0.000175739 | 0.000976811 | 0.85268707 |
| Obesity | mast_cell | *ARL1* | 3.081202639 | 0.000434549 | 0.001097571 | 1 |
| Obesity | mast_cell | *APAF1* | 5.439514352 | 0.000166125 | 0.000976811 | 0.806036904 |
| Obesity | mast_cell | *ADRA1A* | 1.721075702 | 0.000115554 | 0.000976811 | 0.560667935 |
| Obesity | mast_cell | *EPHX2* | 3.538314044 | 0.0002608 | 0.00098093 | 1 |
| Obesity | mast_cell | *AKAP1* | 2.387413299 | 0.000260335 | 0.00098093 | 1 |
| Obesity | mast_cell | *SCPEP1* | 2.768957624 | 8.85E-05 | 0.000976811 | 0.429182574 |
| Obesity | mast_cell | *TEX14* | 6.780447952 | 0.000242315 | 0.000976811 | 1 |
| Obesity | mast_cell | *TRIM6* | 5.53240322 | 0.00063579 | 0.001283751 | 1 |

| | | | | | | |
|---|---|---|---|---|---|---|
| Obesity | mast_cell | *ADCY7* | 2.999598755 | 0.001638211 | 0.002692168 | 1 |
| Obesity | mast_cell | *ECHDC2* | 1.406616382 | 0.0015775 | 0.002612297 | 1 |
| Obesity | mast_cell | *PYROXD1* | 2.899203324 | 3.06E-05 | 0.000621513 | 0.148378035 |
| Obesity | mast_cell | *ZNF549* | 5.304436541 | 0.000230884 | 0.000976811 | 1 |
| Obesity | mast_cell | *A1BG* | -1.922995872 | 0.015252664 | 0.019894067 | 1 |
| Obesity | mast_cell | *ZSCAN18* | 3.605219839 | 0.000211824 | 0.000976811 | 1 |
| Obesity | mast_cell | *PDZRN3* | 1.589166008 | 0.002577682 | 0.003956631 | 1 |
| Obesity | mast_cell | *LHX4* | 2.532628854 | 0.00050221 | 0.001144783 | 1 |
| Obesity | mast_cell | *SEC22A* | 2.97375953 | 0.000364549 | 0.001037687 | 1 |
| Obesity | mast_cell | *POPDC2* | 2.409209675 | 0.017874561 | 0.023035158 | 1 |
| Obesity | mast_cell | *MAPK8IP1* | 6.082373106 | 0.00021927 | 0.000976811 | 1 |
| Obesity | mast_cell | *CAT* | 2.178193909 | 0.000252076 | 0.000976811 | 1 |
| Obesity | mast_cell | *PILRB* | 1.948358691 | 0.001720289 | 0.002797199 | 1 |
| Obesity | mast_cell | *ZCCHC17* | 3.827100401 | 0.000208996 | 0.000976811 | 1 |
| Obesity | mast_cell | *FABP3* | -3.980469838 | 0.004596457 | 0.006606046 | 1 |
| Obesity | mast_cell | *TNFSF10* | 2.30228005 | 0.000397024 | 0.001060771 | 1 |
| Obesity | mast_cell | *ZNF639* | 8.54008836 | 0.001662084 | 0.002721711 | 1 |
| Obesity | mast_cell | *CPXM2* | 2.15118106 | 3.07E-05 | 0.000621513 | 0.149004598 |
| Obesity | mast_cell | *FLT3* | 3.958700766 | 0.000495868 | 0.001138685 | 1 |
| Obesity | mast_cell | *MTIF3* | 4.378818446 | 9.95E-05 | 0.000976811 | 0.482581653 |
| Obesity | mast_cell | *FMOD* | 2.152422357 | 0.000317169 | 0.000998908 | 1 |
| Obesity | mast_cell | *CD244* | 3.400688083 | 0.000181831 | 0.000976811 | 0.882245953 |
| Obesity | mast_cell | *RBBP6* | 2.79534068 | 0.000313864 | 0.000998908 | 1 |
| Obesity | mast_cell | *SERAC1* | 2.161666692 | 0.000634145 | 0.001281728 | 1 |
| Obesity | mast_cell | *LDB3* | 2.702737675 | 0.000378408 | 0.001048564 | 1 |
| Obesity | mast_cell | *OPN4* | 23.12557098 | 1.00E-08 | 1.87E-06 | 4.87E-05 |
| Obesity | mast_cell | *FAM35A* | 3.502543003 | 0.000173918 | 0.000976811 | 0.843849687 |
| Obesity | mast_cell | *NAA60* | 2.664525085 | 0.000493535 | 0.001138685 | 1 |
| Obesity | mast_cell | *ODF2L* | 3.896084793 | 0.000155794 | 0.000976811 | 0.755913852 |
| Obesity | mast_cell | *SPATA1* | 2.810690137 | 0.000249617 | 0.000976811 | 1 |
| Obesity | mast_cell | *LRRC39* | 1.391414311 | 0.003641025 | 0.00536642 | 1 |
| Obesity | mast_cell | *PMS2* | 2.232321355 | 0.000493008 | 0.001138685 | 1 |
| Obesity | mast_cell | *EEPD1* | 2.588374833 | 0.000119505 | 0.000976811 | 0.579838092 |
| Obesity | mast_cell | *KLHL7* | 2.539298395 | 0.000263795 | 0.000984805 | 1 |
| Obesity | mast_cell | *NT5C3A* | 3.819170951 | 0.000246527 | 0.000976811 | 1 |
| Obesity | mast_cell | *CCZ1* | 4.64168153 | 3.34E-05 | 0.000630711 | 0.162278391 |
| Obesity | mast_cell | *SMU1* | 3.563740249 | 0.000120267 | 0.000976811 | 0.583535911 |
| Obesity | mast_cell | *SLC25A51* | 2.542769783 | 9.48E-06 | 0.000349581 | 0.045977051 |
| Obesity | mast_cell | *TRIM24* | 5.714700085 | 0.000161453 | 0.000976811 | 0.783368277 |
| Obesity | mast_cell | *C7orf49* | 3.25027438 | 2.11E-05 | 0.000524672 | 0.102336941 |
| Obesity | mast_cell | *CISD1* | 1.67843513 | 0.001561692 | 0.002595186 | 1 |
| Obesity | mast_cell | *RBM19* | 3.177656903 | 0.000266695 | 0.000985747 | 1 |
| Obesity | mast_cell | *DDX54* | 2.493169844 | 0.000240015 | 0.000976811 | 1 |
| Obesity | mast_cell | *SSPN* | 3.454985626 | 0.000305375 | 0.000998908 | 1 |
| Obesity | mast_cell | *CCDC91* | 2.169041619 | 0.000130191 | 0.000976811 | 0.631685598 |
| Obesity | mast_cell | *EBPL* | 2.493395649 | 0.000305243 | 0.000998908 | 1 |
| Obesity | mast_cell | *ATP7B* | 1.346790379 | 0.001580954 | 0.002616232 | 1 |
| Obesity | mast_cell | *CENPK* | 3.297719217 | 0.000691378 | 0.001358674 | 1 |
| Obesity | mast_cell | *ITIH5* | 2.623550699 | 6.72E-06 | 0.000271528 | 0.032617456 |
| Obesity | mast_cell | *TSFM* | 2.134913095 | 0.001472947 | 0.00247549 | 1 |

| | | | | | | |
|---|---|---|---|---|---|---|
| Obesity | mast_cell | *ARHGAP9* | 2.919975712 | 0.000652707 | 0.001308725 | 1 |
| Obesity | mast_cell | *PFDN5* | 3.250470584 | 0.00024887 | 0.000976811 | 1 |
| Obesity | mast_cell | *PDE1B* | 2.278530244 | 0.000530066 | 0.00115966 | 1 |
| Obesity | mast_cell | *CDK2* | -2.214979566 | 0.0024396 | 0.003779355 | 1 |
| Obesity | mast_cell | *SMUG1* | 2.129668002 | 3.15E-05 | 0.000621513 | 0.152831188 |
| Obesity | mast_cell | *SARDH* | 2.929549629 | 0.006758325 | 0.009366293 | 1 |
| Obesity | mast_cell | *DBH* | 63.19775311 | 0.000582021 | 0.001209489 | 1 |
| Obesity | mast_cell | *NDUFAF4* | 3.389935183 | 0.000310222 | 0.000998908 | 1 |
| Obesity | mast_cell | *USP45* | 3.376075276 | 0.000209303 | 0.000976811 | 1 |
| Obesity | mast_cell | *TTC21B* | 2.172545982 | 0.000564496 | 0.001190078 | 1 |
| Obesity | mast_cell | *ACVR1C* | 1.594627218 | 0.000730797 | 0.001416066 | 1 |
| Obesity | mast_cell | *PLA2G12A* | 2.255404661 | 0.000329943 | 0.001012576 | 1 |
| Obesity | mast_cell | *C4BPA* | 3.030936357 | 0.003157374 | 0.004723891 | 1 |
| Obesity | mast_cell | *C4BPB* | 3.035121461 | 0.00098461 | 0.001765457 | 1 |
| Obesity | mast_cell | *RAB38* | -1.945274353 | 0.01491963 | 0.019501628 | 1 |
| Obesity | mast_cell | *DAW1* | 4.987765954 | 0.000987107 | 0.001768628 | 1 |
| Obesity | mast_cell | *DNPEP* | 3.908460551 | 0.000135696 | 0.000976811 | 0.658399009 |
| Obesity | mast_cell | *OBSL1* | 6.677365609 | 0.000365701 | 0.001037687 | 1 |
| Obesity | mast_cell | *SLC12A4* | 3.085211283 | 0.00035866 | 0.001035264 | 1 |
| Obesity | mast_cell | *FAM210B* | 6.424577484 | 0.034480332 | 0.042493922 | 1 |
| Obesity | mast_cell | *SLC12A5* | -2.964935826 | 0.001217069 | 0.002105998 | 1 |
| Obesity | mast_cell | *MATN4* | 7.387857678 | 0.00028656 | 0.000993978 | 1 |
| Obesity | mast_cell | *VAPB* | 1.853758741 | 0.000415842 | 0.001080612 | 1 |
| Obesity | mast_cell | *TOX2* | 1.149504344 | 0.040330399 | 0.049204718 | 1 |
| Obesity | mast_cell | *SRSF6* | 1.99379922 | 0.000521965 | 0.001153388 | 1 |
| Obesity | mast_cell | *CSE1L* | 2.695735277 | 0.000144298 | 0.000976811 | 0.700135857 |
| Obesity | mast_cell | *PTGIS* | 3.888241059 | 6.32E-06 | 0.000259823 | 0.03065912 |
| Obesity | mast_cell | *RNF114* | 2.640530526 | 0.000207547 | 0.000976811 | 1 |
| Obesity | mast_cell | *NEURL2* | 2.890490689 | 1.68E-05 | 0.000474735 | 0.081654476 |
| Obesity | mast_cell | *MTRR* | 2.510100403 | 0.000139353 | 0.000976811 | 0.676139325 |
| Obesity | mast_cell | *PEPD* | 5.47276533 | 3.26E-05 | 0.000621513 | 0.158416239 |
| Obesity | mast_cell | *CHST8* | 5.084935349 | 2.05E-05 | 0.000522259 | 0.099420998 |
| Obesity | mast_cell | *STAMBP* | 3.907552312 | 0.00020539 | 0.000976811 | 0.996553571 |
| Obesity | mast_cell | *NAGK* | 3.090173315 | 6.13E-05 | 0.000858537 | 0.297624906 |
| Obesity | mast_cell | *SNRNP27* | 3.060264948 | 0.000317846 | 0.000998908 | 1 |
| Obesity | mast_cell | *ATP8A1* | 2.107998421 | 0.000243036 | 0.000976811 | 1 |
| Obesity | mast_cell | *USP22* | 2.712988919 | 0.000335981 | 0.001017877 | 1 |
| Obesity | mast_cell | *HIF3A* | 1.385653032 | 0.001334297 | 0.002277976 | 1 |
| Obesity | mast_cell | *ZNF576* | 2.419005311 | 0.00089552 | 0.001640265 | 1 |
| Obesity | mast_cell | *BTN2A2* | 3.429991439 | 2.17E-05 | 0.000529276 | 0.105325959 |
| Obesity | mast_cell | *MRS2* | 2.302325889 | 0.000179841 | 0.000976811 | 0.872589494 |
| Obesity | mast_cell | *SERPINB6* | 2.284627329 | 0.000272582 | 0.000987746 | 1 |
| Obesity | mast_cell | *PEX6* | 1.283037684 | 0.001883704 | 0.003023397 | 1 |
| Obesity | mast_cell | *NQO2* | 2.084302873 | 0.000377153 | 0.001048135 | 1 |
| Obesity | mast_cell | *ZNF391* | 1.256477166 | 0.003586923 | 0.005297946 | 1 |
| Obesity | mast_cell | *RPS10* | 4.873265378 | 0.001031418 | 0.001835145 | 1 |
| Obesity | mast_cell | *MED20* | 3.560611346 | 2.16E-05 | 0.000529276 | 0.105024381 |
| Obesity | mast_cell | *TREM1* | 7.712772443 | 5.27E-05 | 0.000791601 | 0.255557318 |
| Obesity | mast_cell | *KLHL31* | 1.214830492 | 0.017218608 | 0.022248917 | 1 |
| Obesity | mast_cell | *GLO1* | 3.361658903 | 5.96E-05 | 0.000847545 | 0.289001588 |

| | | | | | | |
|---|---|---|---|---|---|---|
| Obesity | mast_cell | *KCNK17* | 7.300844565 | 4.31E-05 | 0.000736534 | 0.209175664 |
| Obesity | mast_cell | *RREB1* | 2.289037866 | 0.000158651 | 0.000976811 | 0.769776262 |
| Obesity | mast_cell | *RIOK1* | 2.391722551 | 0.000391449 | 0.001054889 | 1 |
| Obesity | mast_cell | *ATXN1* | 3.473111634 | 0.000113941 | 0.000976811 | 0.552839599 |
| Obesity | mast_cell | *RUNX2* | 1.684139815 | 0.000652744 | 0.001308725 | 1 |
| Obesity | mast_cell | *CXCL6* | 4.944253333 | 0.000895959 | 0.001640451 | 1 |
| Obesity | mast_cell | *SH3TC1* | 1.856947216 | 0.000467446 | 0.001116682 | 1 |
| Obesity | mast_cell | *BBS2* | 2.814819175 | 6.73E-05 | 0.000885378 | 0.326360872 |
| Obesity | mast_cell | *MT1G* | -3.91044818 | 0.008106618 | 0.011076686 | 1 |
| Obesity | mast_cell | *GOT2* | 5.128277968 | 7.71E-05 | 0.000938986 | 0.37409901 |
| Obesity | mast_cell | *CLYBL* | 3.444562686 | 0.00020577 | 0.000976811 | 0.998397783 |
| Obesity | mast_cell | *TMTC4* | 1.96769034 | 0.00050562 | 0.001144783 | 1 |
| Obesity | mast_cell | *ABCC4* | 5.960623047 | 0.001342491 | 0.002289548 | 1 |
| Obesity | mast_cell | *ATP5S* | 2.332348517 | 0.010934314 | 0.014623289 | 1 |
| Obesity | mast_cell | *GRK4* | 2.135642903 | 2.85E-07 | 3.21E-05 | 0.001384138 |
| Obesity | mast_cell | *TEKT3* | 2.046822053 | 0.015077131 | 0.01968099 | 1 |
| Obesity | mast_cell | *HS3ST3B1* | 3.374089414 | 0.002521011 | 0.003883158 | 1 |
| Obesity | mast_cell | *MRPS7* | 2.214429165 | 0.000381642 | 0.001049541 | 1 |
| Obesity | mast_cell | *GGA3* | 2.756360218 | 0.000152857 | 0.000976811 | 0.741662483 |
| Obesity | mast_cell | *NUP85* | 1.794675624 | 0.000854401 | 0.00158376 | 1 |
| Obesity | mast_cell | *NT5C* | 7.006979695 | 0.000299459 | 0.000998908 | 1 |
| Obesity | mast_cell | *DDX31* | 2.3053901 | 0.011589934 | 0.015427808 | 1 |
| Obesity | mast_cell | *OPRL1* | 7.748751872 | 0.000484924 | 0.001131723 | 1 |
| Obesity | mast_cell | *PPDPF* | 2.940218271 | 6.35E-05 | 0.000869436 | 0.30811849 |
| Obesity | mast_cell | *IL1B* | 8.800245488 | 0.007924423 | 0.010861385 | 1 |
| Obesity | mast_cell | *PAX8* | 1.570207008 | 0.010934129 | 0.014623289 | 1 |
| Obesity | mast_cell | *POLR1B* | 2.403108601 | 0.000531757 | 0.001161155 | 1 |
| Obesity | mast_cell | *CCDC93* | 2.362071201 | 0.000253374 | 0.000977535 | 1 |
| Obesity | mast_cell | *PSD4* | 3.096193868 | 0.000441071 | 0.001100333 | 1 |
| Obesity | mast_cell | *GTF2F1* | 6.474664486 | 0.00928936 | 0.012568872 | 1 |
| Obesity | mast_cell | *GPR108* | 3.609316278 | 0.000150676 | 0.000976811 | 0.731078578 |
| Obesity | mast_cell | *SNRPD2* | 6.602245882 | 0.000160806 | 0.000976811 | 0.780228938 |
| Obesity | mast_cell | *SDCBP2* | -2.574632073 | 0.003883118 | 0.0056801 | 1 |
| Obesity | mast_cell | *PSMF1* | 3.471933482 | 0.021226523 | 0.026940565 | 1 |
| Obesity | mast_cell | *DTD1* | 2.344873444 | 0.000802617 | 0.001514435 | 1 |
| Obesity | mast_cell | *RBCK1* | 4.674076951 | 0.000305617 | 0.000998908 | 1 |
| Obesity | mast_cell | *RRBP1* | 2.92921699 | 0.000207529 | 0.000976811 | 1 |
| Obesity | mast_cell | *PCSK2* | -2.417803404 | 0.002396374 | 0.003725393 | 1 |
| Obesity | mast_cell | *MKKS* | 3.090916279 | 0.001433201 | 0.002417063 | 1 |
| Obesity | mast_cell | *LAMP5* | 41.13513655 | 0.003685883 | 0.005424296 | 1 |
| Obesity | mast_cell | *MGME1* | 3.235263287 | 0.000252599 | 0.000977361 | 1 |
| Obesity | mast_cell | *ITPA* | 3.270508049 | 0.000250334 | 0.000976811 | 1 |
| Obesity | mast_cell | *MCM8* | 2.043220119 | 0.027078932 | 0.033880087 | 1 |
| Obesity | mast_cell | *NCLN* | 5.485816307 | 0.00085595 | 0.001584536 | 1 |
| Obesity | mast_cell | *UROD* | 4.712676791 | 0.001920544 | 0.00307034 | 1 |
| Obesity | mast_cell | *KLC1* | 4.227906473 | 0.000142681 | 0.000976811 | 0.692288944 |
| Obesity | mast_cell | *TUBGCP3* | 3.47089926 | 0.000169666 | 0.000976811 | 0.823217044 |
| Obesity | mast_cell | *MCF2L* | 1.725043104 | 0.000733536 | 0.001419672 | 1 |
| Obesity | mast_cell | *PROZ* | -2.33092733 | 0.004576335 | 0.006581024 | 1 |
| Obesity | mast_cell | *COX6B1* | 3.905552478 | 7.97E-05 | 0.000949918 | 0.386596946 |

| | | | | | | |
|---|---|---|---|---|---|---|
| Obesity | mast_cell | *PRDX5* | 4.732802647 | 0.00023966 | 0.000976811 | 1 |
| Obesity | mast_cell | *IRF3* | 5.810792013 | 9.06E-05 | 0.000976811 | 0.439457564 |
| Obesity | mast_cell | *PRR12* | 5.78140397 | 0.00048995 | 0.001135803 | 1 |
| Obesity | mast_cell | *TRAP1* | 2.496786399 | 3.89E-05 | 0.000685908 | 0.188624695 |
| Obesity | mast_cell | *ZNF384* | 2.340669646 | 0.000451279 | 0.001106421 | 1 |
| Obesity | mast_cell | *EMG1* | 2.609649795 | 0.000589006 | 0.001219421 | 1 |
| Obesity | mast_cell | *ATG14* | 2.90890504 | 0.000163947 | 0.000976811 | 0.795471256 |
| Obesity | mast_cell | *KTN1* | 2.903416233 | 0.000192405 | 0.000976811 | 0.933548383 |
| Obesity | mast_cell | *ZBTB1* | 2.846937093 | 0.00030325 | 0.000998908 | 1 |
| Obesity | mast_cell | *RHOT1* | 2.11762044 | 0.000120735 | 0.000976811 | 0.585808079 |
| Obesity | mast_cell | *WDR60* | 2.875170077 | 0.000329389 | 0.001011931 | 1 |
| Obesity | mast_cell | *AIF1L* | 4.751332591 | 0.000260236 | 0.00098093 | 1 |
| Obesity | mast_cell | *CANX* | 2.91955706 | 8.42E-05 | 0.000964176 | 0.408779111 |
| Obesity | mast_cell | *ZNF484* | 6.051447942 | 6.04E-05 | 0.000854146 | 0.292972247 |
| Obesity | mast_cell | *FGD3* | 1.731022896 | 0.001960136 | 0.003127451 | 1 |
| Obesity | mast_cell | *ABHD8* | 2.775030483 | 0.008842344 | 0.012010933 | 1 |
| Obesity | mast_cell | *HELB* | 2.498107614 | 0.000226754 | 0.000976811 | 1 |
| Obesity | mast_cell | *RAB3IP* | 2.464329079 | 0.000401909 | 0.001064698 | 1 |
| Obesity | mast_cell | *YEATS4* | 3.029514325 | 6.24E-06 | 0.000258807 | 0.030280443 |
| Obesity | mast_cell | *TAS2R4* | 3.009165082 | 0.000540436 | 0.001171639 | 1 |
| Obesity | mast_cell | *LRRC61* | 1.805055024 | 0.006420345 | 0.008938742 | 1 |
| Obesity | mast_cell | *IDUA* | 3.174967598 | 0.00205895 | 0.003259389 | 1 |
| Obesity | mast_cell | *EMC1* | 2.499225536 | 0.000344817 | 0.00102869 | 1 |
| Obesity | mast_cell | *UBR4* | 2.472537825 | 0.001737778 | 0.002821751 | 1 |
| Obesity | mast_cell | *GFER* | 4.313612511 | 0.000196559 | 0.000976811 | 0.953705301 |
| Obesity | mast_cell | *WDR24* | 2.649849106 | 0.009620502 | 0.012980722 | 1 |
| Obesity | mast_cell | *SMARCA4* | 4.175276775 | 0.000419947 | 0.00108267 | 1 |
| Obesity | mast_cell | *PNKD* | 1.8682237 | 0.000424815 | 0.001090008 | 1 |
| Obesity | mast_cell | *RNF6* | 2.017414539 | 0.000384812 | 0.001052047 | 1 |
| Obesity | mast_cell | *HIP1* | 2.431866931 | 0.000198057 | 0.000976811 | 0.96097316 |
| Obesity | mast_cell | *PTPN12* | 2.568291851 | 0.000445465 | 0.001103405 | 1 |
| Obesity | mast_cell | *STYXL1* | 3.99219795 | 2.13E-05 | 0.000524672 | 0.103360365 |
| Obesity | mast_cell | *STEAP4* | 1.502054472 | 0.002988264 | 0.004501415 | 1 |
| Obesity | mast_cell | *PEX1* | 3.788432888 | 0.000216019 | 0.000976811 | 1 |
| Obesity | mast_cell | *SRD5A3* | 5.686418524 | 0.000328827 | 0.001011712 | 1 |
| Obesity | mast_cell | *KDR* | 1.69742047 | 0.00046892 | 0.001118034 | 1 |
| Obesity | mast_cell | *DGCR6L* | -1.587056877 | 0.004910201 | 0.007013334 | 1 |
| Obesity | mast_cell | *MGAT3* | 3.623200116 | 0.000145667 | 0.000976811 | 0.706777457 |
| Obesity | mast_cell | *ATF4* | 5.99016932 | 0.00017209 | 0.000976811 | 0.834982289 |
| Obesity | mast_cell | *A4GALT* | 1.995759937 | 0.002173128 | 0.003424495 | 1 |
| Obesity | mast_cell | *CDC42EP1* | 3.909804534 | 0.000637307 | 0.001285743 | 1 |
| Obesity | mast_cell | *RIBC2* | 5.226886471 | 4.58E-05 | 0.000754069 | 0.222193486 |
| Obesity | mast_cell | *SPECC1* | 4.417584472 | 0.000171371 | 0.000976811 | 0.831494111 |
| Obesity | mast_cell | *CDHR3* | 1.443103416 | 0.000512372 | 0.001145635 | 1 |
| Obesity | mast_cell | *PRKRIP1* | 2.838934593 | 0.000656739 | 0.001312378 | 1 |
| Obesity | mast_cell | *OPN1SW* | -2.480633107 | 0.032141255 | 0.039762716 | 1 |
| Obesity | mast_cell | *HOXD3* | 1.691136194 | 0.000840588 | 0.001566859 | 1 |
| Obesity | mast_cell | *MTX2* | 2.600085912 | 0.000218089 | 0.000976811 | 1 |
| Obesity | mast_cell | *GAD1* | 4.208281278 | 0.023107529 | 0.029151776 | 1 |
| Obesity | mast_cell | *ORMDL1* | 3.270976832 | 0.000370609 | 0.00103942 | 1 |

| | | | | | | |
|---|---|---|---|---|---|---|
| Obesity | mast_cell | *HOXD10* | 6.865269906 | 0.000335755 | 0.001017877 | 1 |
| Obesity | mast_cell | *PSMG2* | 2.88430952 | 0.001421115 | 0.002400853 | 1 |
| Obesity | mast_cell | *EIF2AK4* | 2.297671779 | 8.24E-05 | 0.000958567 | 0.399645158 |
| Obesity | mast_cell | *TMOD2* | 2.253908012 | 0.000358887 | 0.001035264 | 1 |
| Obesity | mast_cell | *IVD* | 2.719891216 | 0.000147797 | 0.000976811 | 0.717111156 |
| Obesity | mast_cell | *ARPP19* | 4.236963974 | 0.000187141 | 0.000976811 | 0.90800871 |
| Obesity | mast_cell | *VPS13C* | 4.427188652 | 0.000155593 | 0.000976811 | 0.754935112 |
| Obesity | mast_cell | *CALML4* | -4.806390508 | 0.002578773 | 0.003957055 | 1 |
| Obesity | mast_cell | *SUMF2* | 2.705415 | 0.000265873 | 0.000985747 | 1 |
| Obesity | mast_cell | *BBOX1* | 8.160117784 | 0.000465771 | 0.001116682 | 1 |
| Obesity | mast_cell | *SERGEF* | 2.058860184 | 0.00013642 | 0.000976811 | 0.661910898 |
| Obesity | mast_cell | *DCTD* | 4.389461762 | 0.000811113 | 0.001525988 | 1 |
| Obesity | mast_cell | *RPAIN* | 1.600720366 | 0.029921404 | 0.037168113 | 1 |
| Obesity | mast_cell | *PLD2* | 5.898099206 | 0.000188652 | 0.000976811 | 0.91534035 |
| Obesity | mast_cell | *PHF20L1* | 2.886521172 | 0.000185877 | 0.000976811 | 0.901874716 |
| Obesity | mast_cell | *PUS7L* | 3.662288897 | 0.000473553 | 0.001125761 | 1 |
| Obesity | mast_cell | *KRI1* | 1.895293277 | 0.000782149 | 0.001487241 | 1 |
| Obesity | mast_cell | *AP1M2* | -2.856355638 | 0.013881448 | 0.018208377 | 1 |
| Obesity | mast_cell | *MTUS1* | 1.727071297 | 0.000257927 | 0.00098093 | 1 |
| Obesity | mast_cell | *KLK10* | 2.476277063 | 0.007495349 | 0.010305308 | 1 |
| Obesity | mast_cell | *KLK8* | 17.62021217 | 0.00044598 | 0.001103405 | 1 |
| Obesity | mast_cell | *ADCY4* | 2.449451421 | 0.000508375 | 0.001144962 | 1 |
| Obesity | mast_cell | *BCL2L2* | 3.133425283 | 0.000344281 | 0.001027971 | 1 |
| Obesity | mast_cell | *MIS18BP1* | 2.838887247 | 0.000111383 | 0.000976811 | 0.540429715 |
| Obesity | mast_cell | *RNASE1* | 1.943672424 | 0.000220969 | 0.000976811 | 1 |
| Obesity | mast_cell | *EPB41L4A* | 2.660465294 | 0.000216316 | 0.000976811 | 1 |
| Obesity | mast_cell | *CDO1* | 1.856623374 | 0.000642277 | 0.001293082 | 1 |
| Obesity | mast_cell | *REEP5* | 5.07975374 | 0.000189452 | 0.000976811 | 0.91921971 |
| Obesity | mast_cell | *RHBDF2* | 1.638672604 | 0.003062869 | 0.004602367 | 1 |
| Obesity | mast_cell | *AANAT* | -1.956239828 | 0.004381769 | 0.006336913 | 1 |
| Obesity | mast_cell | *TMEM8A* | 3.279306746 | 0.000449137 | 0.001104518 | 1 |
| Obesity | mast_cell | *MAU2* | 2.047095363 | 0.00052919 | 0.0011592 | 1 |
| Obesity | mast_cell | *SHC2* | 2.938941758 | 4.43E-05 | 0.00074422 | 0.214768856 |
| Obesity | mast_cell | *ERMARD* | 2.637264887 | 0.000565096 | 0.001190078 | 1 |
| Obesity | mast_cell | *PHF10* | 4.382804333 | 0.000345047 | 0.00102869 | 1 |
| Obesity | mast_cell | *GALNT8* | 4.779132346 | 9.45E-05 | 0.000976811 | 0.458572299 |
| Obesity | mast_cell | *PRKCSH* | 6.408784031 | 9.65E-05 | 0.000976811 | 0.468225154 |
| Obesity | mast_cell | *CDC16* | 2.41300525 | 0.010633912 | 0.014245097 | 1 |
| Obesity | mast_cell | *XPO7* | 2.899641274 | 0.00023256 | 0.000976811 | 1 |
| Obesity | mast_cell | *FAM98C* | 3.337518438 | 0.002624824 | 0.004016288 | 1 |
| Obesity | mast_cell | *ATP8B3* | -4.275659551 | 9.75E-12 | 3.94E-09 | 4.73E-08 |
| Obesity | mast_cell | *PLVAP* | 1.851377256 | 0.000540797 | 0.001171639 | 1 |
| Obesity | mast_cell | *DDA1* | 5.05107104 | 0.000161532 | 0.000976811 | 0.783754719 |
| Obesity | mast_cell | *MRPL34* | 1.844023719 | 0.000374116 | 0.001044426 | 1 |
| Obesity | mast_cell | *LSM7* | 2.300235449 | 0.0003169 | 0.000998908 | 1 |
| Obesity | mast_cell | *TULP4* | 3.095632011 | 1.49E-05 | 0.00043435 | 0.072102051 |
| Obesity | mast_cell | *QRSL1* | 1.841419592 | 0.016052333 | 0.020875347 | 1 |
| Obesity | mast_cell | *RSPH3* | 1.630475354 | 0.00173027 | 0.002811544 | 1 |
| Obesity | mast_cell | *ACTN4* | 3.61190577 | 0.000210666 | 0.000976811 | 1 |
| Obesity | mast_cell | *STK33* | 2.802744719 | 2.40E-06 | 0.000137224 | 0.011664028 |

| | | | | | | |
|---|---|---|---|---|---|---|
| Obesity | mast_cell | *NDUFA10* | 2.375980082 | 0.001392355 | 0.00235966 | 1 |
| Obesity | mast_cell | *MAP1S* | 7.832123155 | 0.000182001 | 0.000976811 | 0.883068064 |
| Obesity | mast_cell | *SSBP4* | 4.708246823 | 0.000102064 | 0.000976811 | 0.495212317 |
| Obesity | mast_cell | *KIAA1683* | 3.924272943 | 0.020299744 | 0.025878707 | 1 |
| Obesity | mast_cell | *LSM4* | 2.559513671 | 0.002551148 | 0.003924595 | 1 |
| Obesity | mast_cell | *TRPM4* | 1.631392636 | 0.007282552 | 0.010029788 | 1 |
| Obesity | mast_cell | *UBAC1* | 2.535781141 | 0.000173221 | 0.000976811 | 0.840468081 |
| Obesity | mast_cell | *ATXN10* | 2.817908521 | 0.000211891 | 0.000976811 | 1 |
| Obesity | mast_cell | *CYP2E1* | -2.758914845 | 0.004979292 | 0.007107834 | 1 |
| Obesity | mast_cell | *PNPLA7* | 2.940994874 | 0.000275658 | 0.000990086 | 1 |
| Obesity | mast_cell | *PAK4* | 7.671153434 | 0.001203715 | 0.002085867 | 1 |
| Obesity | mast_cell | *ZNF337* | 3.124877837 | 0.000217959 | 0.000976811 | 1 |
| Obesity | mast_cell | *CEP85* | 2.69192748 | 0.017059148 | 0.022072264 | 1 |
| Obesity | mast_cell | *POMT1* | 2.686186098 | 0.001606917 | 0.002651058 | 1 |
| Obesity | mast_cell | *CHMP2A* | 2.635256642 | 0.000506952 | 0.001144962 | 1 |
| Obesity | mast_cell | *YIPF2* | 5.391797702 | 0.00013317 | 0.000976811 | 0.646138615 |
| Obesity | mast_cell | *MAP3K10* | 2.189923739 | 0.000974582 | 0.00175136 | 1 |
| Obesity | mast_cell | *LRRC47* | 5.932744346 | 2.60E-05 | 0.00056111 | 0.126249765 |
| Obesity | mast_cell | *THEMIS2* | 2.596645807 | 0.000103684 | 0.000976811 | 0.503072804 |
| Obesity | mast_cell | *ZNF317* | 1.939286276 | 0.00257306 | 0.003952038 | 1 |
| Obesity | mast_cell | *DNMT1* | 3.952767446 | 0.000247794 | 0.000976811 | 1 |
| Obesity | mast_cell | *C12orf65* | 2.813981301 | 2.02E-05 | 0.000522259 | 0.097992552 |
| Obesity | mast_cell | *NOL11* | 5.51521649 | 1.87E-05 | 0.000496117 | 0.090898417 |
| Obesity | mast_cell | *PKDREJ* | -4.217525612 | 0.002154535 | 0.003397401 | 1 |
| Obesity | mast_cell | *HSD17B3* | 3.331194823 | 0.000442855 | 0.00110084 | 1 |
| Obesity | mast_cell | *SLC35D2* | 3.560168219 | 0.000233002 | 0.000976811 | 1 |
| Obesity | mast_cell | *ULBP2* | 18.85085541 | 0.008451479 | 0.011509002 | 1 |
| Obesity | mast_cell | *TTLL9* | 4.264174587 | 0.000332041 | 0.001015802 | 1 |
| Obesity | mast_cell | *COX4I2* | 3.361607803 | 0.003324149 | 0.004944442 | 1 |
| Obesity | mast_cell | *ATP6V1E1* | 3.303452604 | 0.005130061 | 0.007303713 | 1 |
| Obesity | mast_cell | *CHMP1A* | 7.626227708 | 0.013246799 | 0.017460872 | 1 |
| Obesity | mast_cell | *IDO1* | 4.254395904 | 0.000411902 | 0.001075765 | 1 |
| Obesity | mast_cell | *MRPS25* | 2.641383832 | 0.000733194 | 0.001419576 | 1 |
| Obesity | mast_cell | *GALNT15* | 1.725184707 | 0.000442763 | 0.00110084 | 1 |
| Obesity | mast_cell | *SLC6A6* | 1.917501846 | 0.000441907 | 0.00110084 | 1 |
| Obesity | mast_cell | *KCNC3* | -5.170823119 | 0.009225803 | 0.012489843 | 1 |
| Obesity | mast_cell | *NAPSA* | 3.833138805 | 0.020006614 | 0.025550629 | 1 |
| Obesity | mast_cell | *PDLIM4* | 4.402012998 | 0.000313311 | 0.000998908 | 1 |
| Obesity | mast_cell | *ACLY* | 4.356980388 | 0.004592649 | 0.006602529 | 1 |
| Obesity | mast_cell | *NDUFA2* | 5.304536979 | 5.20E-07 | 4.92E-05 | 0.002525266 |
| Obesity | mast_cell | *EXOC4* | 2.915663026 | 2.44E-05 | 0.000545024 | 0.118572411 |
| Obesity | mast_cell | *C1orf159* | -2.647432846 | 0.025015951 | 0.031412369 | 1 |
| Obesity | mast_cell | *PPFIA1* | 3.579721259 | 0.000145532 | 0.000976811 | 0.706120383 |
| Obesity | mast_cell | *BARX1* | -3.958035663 | 0.004686519 | 0.006721546 | 1 |
| Obesity | mast_cell | *NINJ1* | 2.100196178 | 0.000310538 | 0.000998908 | 1 |
| Obesity | mast_cell | *NPHP4* | 1.823724715 | 0.00141984 | 0.002399535 | 1 |
| Obesity | mast_cell | *MAP1B* | 1.988377265 | 0.000724508 | 0.001406873 | 1 |
| Obesity | mast_cell | *CKMT2* | 2.012738772 | 0.00310952 | 0.004662359 | 1 |
| Obesity | mast_cell | *KHDRBS3* | 2.735573471 | 0.000318422 | 0.000998908 | 1 |
| Obesity | mast_cell | *CHD1L* | 2.076828609 | 0.002221395 | 0.003484711 | 1 |

| | | | | | | |
|---|---|---|---|---|---|---|
| Obesity | mast_cell | *PRKAB2* | 3.629090409 | 0.000123328 | 0.000976811 | 0.598389581 |
| Obesity | mast_cell | *ZNF304* | 2.414199183 | 0.004231252 | 0.006144878 | 1 |
| Obesity | mast_cell | *ZNF132* | 1.759953059 | 0.031686798 | 0.039230504 | 1 |
| Obesity | mast_cell | *RHPN2* | 1.436567281 | 0.004459282 | 0.0064375 | 1 |
| Obesity | mast_cell | *GCH1* | 2.019285199 | 0.001180318 | 0.002058556 | 1 |
| Obesity | mast_cell | *TRIM21* | 3.116500119 | 0.000725493 | 0.001408036 | 1 |
| Obesity | mast_cell | *SPATA6* | 2.42594339 | 0.00033381 | 0.001016573 | 1 |
| Obesity | mast_cell | *LRRC41* | 2.680554573 | 0.00104573 | 0.001856525 | 1 |
| Obesity | mast_cell | *CCT6B* | 2.480415987 | 0.028687855 | 0.035745627 | 1 |
| Obesity | mast_cell | *CNGA4* | 2.508405243 | 0.028737986 | 0.035798898 | 1 |
| Obesity | mast_cell | *TRIM22* | 2.102218753 | 0.000352953 | 0.001031837 | 1 |
| Obesity | mast_cell | *RRP8* | 5.224555221 | 0.000481495 | 0.001130443 | 1 |
| Obesity | mast_cell | *TIMM10B* | 3.337263323 | 0.000569547 | 0.001193196 | 1 |
| Obesity | mast_cell | *IMMT* | 3.311892661 | 0.0001109 | 0.000976811 | 0.53808806 |
| Obesity | mast_cell | *CARD6* | 6.120698587 | 0.000137975 | 0.000976811 | 0.669455727 |
| Obesity | mast_cell | *MYBBP1A* | 3.676265551 | 5.25E-06 | 0.000235708 | 0.025456457 |
| Obesity | mast_cell | *RPA1* | 1.968927745 | 0.000274677 | 0.000990086 | 1 |
| Obesity | mast_cell | *SERPINF1* | 4.921540082 | 0.000200781 | 0.000976811 | 0.974187566 |
| Obesity | mast_cell | *TMEM128* | 2.633248673 | 0.000439089 | 0.00109874 | 1 |
| Obesity | mast_cell | *SEC61G* | 3.084369965 | 0.000577947 | 0.00120507 | 1 |
| Obesity | mast_cell | *FIGNL1* | 2.495744896 | 0.008031517 | 0.010989543 | 1 |
| Obesity | mast_cell | *RGS22* | 1.775972015 | 0.005461931 | 0.007728577 | 1 |
| Obesity | mast_cell | *PRMT7* | 2.175327592 | 0.008868699 | 0.01204336 | 1 |
| Obesity | mast_cell | *IGHMBP2* | 5.601610714 | 7.78E-07 | 6.34E-05 | 0.003774552 |
| Obesity | mast_cell | *OSER1* | 7.321370663 | 8.57E-05 | 0.000971162 | 0.416017588 |
| Obesity | mast_cell | *SERINC3* | 2.610634706 | 0.000764291 | 0.001463434 | 1 |
| Obesity | mast_cell | *DMGDH* | 2.189807975 | 0.029949336 | 0.037193288 | 1 |
| Obesity | mast_cell | *AP3B1* | 2.463624858 | 0.000226457 | 0.000976811 | 1 |
| Obesity | mast_cell | *ZBED3* | 4.357143278 | 0.02733699 | 0.03417652 | 1 |
| Obesity | mast_cell | *FBXO44* | 2.376113256 | 0.002618161 | 0.00400862 | 1 |
| Obesity | mast_cell | *CASP9* | 1.114126079 | 0.037596926 | 0.046089006 | 1 |
| Obesity | mast_cell | *ATP8A2* | 6.790416436 | 7.32E-05 | 0.000924875 | 0.355298478 |
| Obesity | mast_cell | *ALOX5AP* | 4.901551639 | 9.61E-05 | 0.000976811 | 0.46635674 |
| Obesity | mast_cell | *WASF3* | 3.303660533 | 0.000226608 | 0.000976811 | 1 |
| Obesity | mast_cell | *PEMT* | 2.207774091 | 0.000496675 | 0.001138685 | 1 |
| Obesity | mast_cell | *SCO1* | 3.869893552 | 0.001520357 | 0.002542837 | 1 |
| Obesity | mast_cell | *MPRIP* | 2.644761657 | 0.000244547 | 0.000976811 | 1 |
| Obesity | mast_cell | *DSTYK* | 5.166261166 | 0.033158909 | 0.040948594 | 1 |
| Obesity | mast_cell | *COG6* | 4.030330104 | 0.0002411 | 0.000976811 | 1 |
| Obesity | mast_cell | *EPSTI1* | 3.046037265 | 0.000193599 | 0.000976811 | 0.939342233 |
| Obesity | mast_cell | *BTBD2* | 1.794767753 | 0.00036073 | 0.001035657 | 1 |
| Obesity | mast_cell | *CSNK1G2* | 3.715386229 | 0.000255961 | 0.00098093 | 1 |
| Obesity | mast_cell | *CNDP2* | 2.847311633 | 0.000464362 | 0.001116682 | 1 |
| Obesity | mast_cell | *PDZD2* | 1.229682188 | 0.000439069 | 0.00109874 | 1 |
| Obesity | mast_cell | *GSTT2B* | -0.945360357 | 0.022475149 | 0.028405684 | 1 |
| Obesity | mast_cell | *FAM83F* | -5.729433149 | 0.001968736 | 0.003137847 | 1 |
| Obesity | mast_cell | *GIMAP4* | 2.537325021 | 9.14E-05 | 0.000976811 | 0.443575673 |
| Obesity | mast_cell | *ADCK2* | 2.038432253 | 0.003472982 | 0.005135906 | 1 |
| Obesity | mast_cell | *IPO8* | 3.507253218 | 0.000307609 | 0.000998908 | 1 |
| Obesity | mast_cell | *SPINK5* | 4.428450194 | 0.000520411 | 0.001152325 | 1 |

| | | | | | | |
|---|---|---|---|---|---|---|
| Obesity | mast_cell | *IMPA1* | 1.880173958 | 0.011877763 | 0.015784964 | 1 |
| Obesity | mast_cell | *LRRCC1* | 2.495560255 | 0.000291761 | 0.000998325 | 1 |
| Obesity | mast_cell | *RRAS2* | 1.935533518 | 0.000228012 | 0.000976811 | 1 |
| Obesity | mast_cell | *CATSPERB* | 3.977723625 | 0.00054735 | 0.001175269 | 1 |
| Obesity | mast_cell | *COX16* | -2.194301067 | 0.00272941 | 0.004150141 | 1 |
| Obesity | mast_cell | *TTC9* | 3.068825242 | 0.000447807 | 0.001104518 | 1 |
| Obesity | mast_cell | *ELP3* | 2.339673257 | 0.000467698 | 0.001116682 | 1 |
| Obesity | mast_cell | *PEBP4* | 4.172528795 | 0.000313659 | 0.000998908 | 1 |
| Obesity | mast_cell | *CTIF* | 2.003378643 | 0.001047895 | 0.001858565 | 1 |
| Obesity | mast_cell | *MRO* | 6.055267944 | 0.000115039 | 0.000976811 | 0.558169893 |
| Obesity | mast_cell | *THUMPD3* | 5.222738097 | 0.000994861 | 0.001779235 | 1 |
| Obesity | mast_cell | *CNTN6* | 3.280246925 | 0.000656523 | 0.001312378 | 1 |
| Obesity | mast_cell | *CHL1* | 1.627300078 | 0.000517632 | 0.001151558 | 1 |
| Obesity | mast_cell | *GSTM5* | 3.434455299 | 4.24E-06 | 0.000201709 | 0.020574325 |
| Obesity | mast_cell | *GSTM3* | 0.988181083 | 0.040168567 | 0.049030914 | 1 |
| Obesity | mast_cell | *VAV3* | 2.181209275 | 0.005108316 | 0.007279163 | 1 |
| Obesity | mast_cell | *PSRC1* | -1.011062508 | 0.00267917 | 0.004089127 | 1 |
| Obesity | mast_cell | *TRIM45* | 7.292732929 | 0.039443289 | 0.048206256 | 1 |
| Obesity | mast_cell | *CEPT1* | 1.944799067 | 0.000354012 | 0.001031837 | 1 |
| Obesity | mast_cell | *NAPG* | 2.466326153 | 0.000222722 | 0.000976811 | 1 |
| Obesity | mast_cell | *PPHLN1* | 2.953587012 | 0.000217611 | 0.000976811 | 1 |
| Obesity | mast_cell | *FKBP11* | 10.77621741 | 0.000841251 | 0.00156689 | 1 |
| Obesity | mast_cell | *YWHAQ* | 3.401635777 | 0.000135311 | 0.000976811 | 0.656530979 |
| Obesity | mast_cell | *KIDINS220* | 3.133653361 | 0.000191287 | 0.000976811 | 0.928126157 |
| Obesity | mast_cell | *LPIN1* | 3.21964774 | 0.000302017 | 0.000998908 | 1 |
| Obesity | mast_cell | *NARS* | 3.442228196 | 6.74E-05 | 0.000885378 | 0.326796596 |
| Obesity | mast_cell | *KIAA1468* | 2.316212702 | 0.000280256 | 0.000993978 | 1 |
| Obesity | mast_cell | *IL15RA* | 2.896638029 | 0.000118651 | 0.000976811 | 0.57569703 |
| Obesity | mast_cell | *TMEM241* | 1.859578611 | 0.003114603 | 0.004668537 | 1 |
| Obesity | mast_cell | *SOX5* | 2.434880236 | 0.000367128 | 0.001037687 | 1 |
| Obesity | mast_cell | *KLRD1* | 2.617030758 | 0.000242714 | 0.000976811 | 1 |
| Obesity | mast_cell | *ACP2* | 5.546014782 | 8.08E-06 | 0.000313499 | 0.039187365 |
| Obesity | mast_cell | *ZCCHC11* | 2.892222965 | 0.000106502 | 0.000976811 | 0.516745882 |
| Obesity | mast_cell | *PRPF38A* | 7.126687913 | 5.30E-05 | 0.000791601 | 0.257270477 |
| Obesity | mast_cell | *ELP2* | 2.466165967 | 0.000275681 | 0.000990086 | 1 |
| Obesity | mast_cell | *TPGS2* | 6.199684173 | 0.000303538 | 0.000998908 | 1 |
| Obesity | mast_cell | *TIMM10* | 3.507926983 | 0.000755073 | 0.001450502 | 1 |
| Obesity | mast_cell | *DHX34* | 3.272836706 | 0.00024737 | 0.000976811 | 1 |
| Obesity | mast_cell | *FADS2* | 8.02299724 | 0.001847244 | 0.002975701 | 1 |
| Obesity | mast_cell | *TMEM258* | 2.97364011 | 0.001065537 | 0.001883419 | 1 |
| Obesity | mast_cell | *CLOCK* | 2.716395554 | 0.00022879 | 0.000976811 | 1 |
| Obesity | mast_cell | *GGACT* | 4.47703357 | 0.001074347 | 0.001896919 | 1 |
| Obesity | mast_cell | *ARGLU1* | 3.00199618 | 0.000145955 | 0.000976811 | 0.708171497 |
| Obesity | mast_cell | *CARS2* | 3.19729164 | 0.000461947 | 0.001114555 | 1 |
| Obesity | mast_cell | *KLB* | 1.868927592 | 3.73E-05 | 0.000675733 | 0.181096435 |
| Obesity | mast_cell | *WDR36* | 2.306588269 | 0.00034643 | 0.00102869 | 1 |
| Obesity | mast_cell | *OSTF1* | 2.335552383 | 0.000278671 | 0.000992498 | 1 |
| Obesity | mast_cell | *UBQLN1* | 2.335189944 | 0.000318392 | 0.000998908 | 1 |
| Obesity | mast_cell | *GOLM1* | 1.405785664 | 0.016292522 | 0.021165011 | 1 |
| Obesity | mast_cell | *ADAM19* | 1.497309442 | 0.013454873 | 0.017687089 | 1 |

| | | | | | | |
|---|---|---|---|---|---|---|
| Obesity | mast_cell | *USP30* | 2.185543597 | 0.000367991 | 0.001037687 | 1 |
| Obesity | mast_cell | *MSI1* | 4.60481044 | 0.001588246 | 0.002624717 | 1 |
| Obesity | mast_cell | *P2RX4* | 1.863609488 | 0.010543167 | 0.014135244 | 1 |
| Obesity | mast_cell | *DTX1* | 7.94120188 | 0.000319743 | 0.000999138 | 1 |
| Obesity | mast_cell | *DMTF1* | 3.934959645 | 0.000211621 | 0.000976811 | 1 |
| Obesity | mast_cell | *SRPK2* | 2.739662308 | 0.000179116 | 0.000976811 | 0.869071689 |
| Obesity | mast_cell | *TES* | 2.269932912 | 0.002963627 | 0.004467076 | 1 |
| Obesity | mast_cell | *MTO1* | 2.742677522 | 0.000202037 | 0.000976811 | 0.980283648 |
| Obesity | mast_cell | *KHDC1* | 1.579956913 | 0.003205963 | 0.004786256 | 1 |
| Obesity | mast_cell | *NT5E* | 2.747453252 | 0.002230796 | 0.00349381 | 1 |
| Obesity | mast_cell | *AKIRIN2* | 4.688023049 | 9.88E-05 | 0.000976811 | 0.47932682 |
| Obesity | mast_cell | *MAP3K7* | 3.155122593 | 0.000189919 | 0.000976811 | 0.921488803 |
| Obesity | mast_cell | *PRR5L* | 2.92152292 | 0.000657586 | 0.001312467 | 1 |
| Obesity | mast_cell | *PHF21A* | 2.700647203 | 0.000201075 | 0.000976811 | 0.975615955 |
| Obesity | mast_cell | *CAPRIN1* | 2.76879596 | 0.000661001 | 0.001317116 | 1 |
| Obesity | mast_cell | *ATP5G2* | 3.107466548 | 0.000176747 | 0.000976811 | 0.857574306 |
| Obesity | mast_cell | *FAIM2* | -2.18956417 | 0.002523912 | 0.003886392 | 1 |
| Obesity | mast_cell | *PAN2* | 4.271757293 | 0.001713807 | 0.002788528 | 1 |
| Obesity | mast_cell | *KRT7* | 2.363088914 | 0.006387593 | 0.008900804 | 1 |
| Obesity | mast_cell | *SLC26A10* | 5.673431926 | 0.027004094 | 0.033803887 | 1 |
| Obesity | mast_cell | *MAP7* | 1.412197899 | 0.002572433 | 0.003952038 | 1 |
| Obesity | mast_cell | *NHSL1* | 2.914448192 | 0.000185713 | 0.000976811 | 0.901078992 |
| Obesity | mast_cell | *AHI1* | 1.509103491 | 0.020463297 | 0.026080357 | 1 |
| Obesity | mast_cell | *MICAL1* | 3.314289479 | 5.57E-06 | 0.000243327 | 0.027023193 |
| Obesity | mast_cell | *CCT7* | 3.265498618 | 0.000525987 | 0.001156886 | 1 |
| Obesity | mast_cell | *SMYD5* | 2.973005818 | 0.000208258 | 0.000976811 | 1 |
| Obesity | mast_cell | *USP15* | 2.315316486 | 0.000251832 | 0.000976811 | 1 |
| Obesity | mast_cell | *MDM2* | 2.493722633 | 0.000301946 | 0.000998908 | 1 |
| Obesity | mast_cell | *KLHL36* | 3.833434087 | 9.36E-05 | 0.000976811 | 0.453948142 |
| Obesity | mast_cell | *CCDC102A* | 2.440932513 | 0.001886966 | 0.003026631 | 1 |
| Obesity | mast_cell | *ZNF670* | 6.535897718 | 0.005559471 | 0.007850569 | 1 |
| Obesity | mast_cell | *KCNK1* | -1.512229897 | 0.013245391 | 0.017460872 | 1 |
| Obesity | mast_cell | *CAPN9* | 5.986332963 | 0.005267422 | 0.007479523 | 1 |
| Obesity | mast_cell | *NTPCR* | 2.762262574 | 0.000227383 | 0.000976811 | 1 |
| Obesity | mast_cell | *PIGC* | 2.347541326 | 0.000271952 | 0.000987746 | 1 |
| Obesity | mast_cell | *SP110* | 3.574428851 | 7.55E-05 | 0.000934364 | 0.366270768 |
| Obesity | mast_cell | *MRPL44* | 2.703945011 | 0.000442092 | 0.00110084 | 1 |
| Obesity | mast_cell | *USP37* | 6.470421364 | 2.19E-06 | 0.00013132 | 0.010636898 |
| Obesity | mast_cell | *SLC19A3* | 1.58068447 | 0.000604791 | 0.001240781 | 1 |
| Obesity | mast_cell | *TMBIM1* | 3.130175045 | 0.00026442 | 0.00098538 | 1 |
| Obesity | mast_cell | *TSGA10* | 1.004560525 | 0.007130419 | 0.009837018 | 1 |
| Obesity | mast_cell | *GCC2* | 3.141893247 | 0.000347513 | 0.001029861 | 1 |
| Obesity | mast_cell | *MRPS9* | 4.170923787 | 5.18E-06 | 0.000234841 | 0.025128007 |
| Obesity | mast_cell | *ANKRD36* | 3.06437496 | 0.000212063 | 0.000976811 | 1 |
| Obesity | mast_cell | *EPC2* | 3.745173947 | 0.000159016 | 0.000976811 | 0.771545422 |
| Obesity | mast_cell | *STAB2* | 1.944873337 | 0.000483377 | 0.001131376 | 1 |
| Obesity | mast_cell | *APPL2* | 2.803233569 | 0.000649616 | 0.001304609 | 1 |
| Obesity | mast_cell | *DRAM1* | 2.332910337 | 0.000320588 | 0.000999138 | 1 |
| Obesity | mast_cell | *SLC41A2* | 2.821531394 | 0.000152702 | 0.000976811 | 0.740911925 |
| Obesity | mast_cell | *NEK3* | 2.737293787 | 0.00018435 | 0.000976811 | 0.894465956 |

| | | | | | | |
|---|---|---|---|---|---|---|
| Obesity | mast_cell | *RNASEH2B* | 2.171288477 | 0.000695896 | 0.001365894 | 1 |
| Obesity | mast_cell | *TBC1D4* | 3.141871691 | 3.79E-05 | 0.000679348 | 0.1841032 |
| Obesity | mast_cell | *SUCLA2* | 2.582058027 | 0.000286387 | 0.000993978 | 1 |
| Obesity | mast_cell | *RCBTB1* | 1.694719515 | 0.002726295 | 0.004146703 | 1 |
| Obesity | mast_cell | *RCBTB2* | 2.469741645 | 0.000302047 | 0.000998908 | 1 |
| Obesity | mast_cell | *SETDB2* | 3.143116709 | 0.000321362 | 0.001000159 | 1 |
| Obesity | mast_cell | *SCRN1* | 2.713762662 | 0.000356535 | 0.001033017 | 1 |
| Obesity | mast_cell | *CHST12* | 3.165995113 | 0.000375806 | 0.001047881 | 1 |
| Obesity | mast_cell | *KDELR2* | 5.192096878 | 0.000176053 | 0.000976811 | 0.854210603 |
| Obesity | mast_cell | *NUPL2* | 2.171375739 | 0.000350902 | 0.001031837 | 1 |
| Obesity | mast_cell | *AOAH* | 1.348622132 | 0.000602005 | 0.001238217 | 1 |
| Obesity | mast_cell | *TBRG4* | 2.523634283 | 0.000863326 | 0.001593934 | 1 |
| Obesity | mast_cell | *MTHFS* | 2.479002993 | 0.002408402 | 0.003739381 | 1 |
| Obesity | mast_cell | *NMT1* | 3.553411827 | 1.90E-05 | 0.000499059 | 0.092402751 |
| Obesity | mast_cell | *BRIP1* | 2.007979778 | 0.00411538 | 0.005995123 | 1 |
| Obesity | mast_cell | *RTP4* | 3.603585394 | 0.008103083 | 0.011074974 | 1 |
| Obesity | mast_cell | *NDUFB5* | 6.85352284 | 0.000188774 | 0.000976811 | 0.915932479 |
| Obesity | mast_cell | *SCN2A* | 12.13156588 | 1.22E-08 | 2.04E-06 | 5.91E-05 |
| Obesity | mast_cell | *RPS6KC1* | 2.488577884 | 0.000283043 | 0.000993978 | 1 |
| Obesity | mast_cell | *SMPD4* | 2.486940558 | 0.000272085 | 0.000987746 | 1 |
| Obesity | mast_cell | *CCDC115* | 2.323934692 | 0.000741303 | 0.001434132 | 1 |
| Obesity | mast_cell | *IMP4* | 3.736298784 | 0.000626626 | 0.001273048 | 1 |
| Obesity | mast_cell | *NIPSNAP3A* | 3.576853984 | 0.003366884 | 0.005000343 | 1 |
| Obesity | mast_cell | *ODF2* | 4.225563681 | 0.000688731 | 0.001355939 | 1 |
| Obesity | mast_cell | *TOR1B* | 2.526146776 | 0.000631591 | 0.001279531 | 1 |
| Obesity | mast_cell | *SLC2A8* | 3.192940521 | 0.000377717 | 0.001048135 | 1 |
| Obesity | mast_cell | *ZNF189* | 2.321832139 | 0.000376711 | 0.001048052 | 1 |
| Obesity | mast_cell | *STX17* | 2.813473225 | 0.00013595 | 0.000976811 | 0.659629253 |
| Obesity | mast_cell | *ANP32B* | 4.025601327 | 0.000456802 | 0.001112606 | 1 |
| Obesity | mast_cell | *DSCC1* | 1.908847581 | 1.76E-05 | 0.000483458 | 0.08557212 |
| Obesity | mast_cell | *DERL1* | 2.564177347 | 0.002391103 | 0.003719665 | 1 |
| Obesity | mast_cell | *NOV* | 3.618795875 | 0.000597687 | 0.001232983 | 1 |
| Obesity | mast_cell | *POLR1E* | 1.58961591 | 0.008337692 | 0.011366811 | 1 |
| Obesity | mast_cell | *APTX* | 3.956012451 | 0.00038121 | 0.00104914 | 1 |
| Obesity | mast_cell | *TMEM8B* | 2.198651182 | 0.000378024 | 0.001048135 | 1 |
| Obesity | mast_cell | *DENND4C* | 3.251571467 | 0.000156762 | 0.000976811 | 0.760610185 |
| Obesity | mast_cell | *GMPR* | -2.781248497 | 0.0173014 | 0.022343995 | 1 |
| Obesity | mast_cell | *FRS3* | 2.862454586 | 0.00027157 | 0.000987746 | 1 |
| Obesity | mast_cell | *IRF4* | 1.907119302 | 0.000663112 | 0.001319697 | 1 |
| Obesity | mast_cell | *LRRC1* | 1.744452324 | 0.000554907 | 0.0011814 | 1 |
| Obesity | mast_cell | *RIPK1* | 3.156173343 | 0.000302875 | 0.000998908 | 1 |
| Obesity | mast_cell | *TCF19* | 3.51797998 | 4.04E-05 | 0.000702302 | 0.195942131 |
| Obesity | mast_cell | *FLOT1* | 2.720481611 | 9.40E-05 | 0.000976811 | 0.456268111 |
| Obesity | mast_cell | *TPMT* | 2.529079176 | 0.005897357 | 0.008279506 | 1 |
| Obesity | mast_cell | *NRM* | 7.418818307 | 0.000431556 | 0.001094569 | 1 |
| Obesity | mast_cell | *TLR2* | 3.212977409 | 0.000301856 | 0.000998908 | 1 |
| Obesity | mast_cell | *MYO7A* | 2.023069304 | 0.00809362 | 0.011065158 | 1 |
| Obesity | mast_cell | *NUMA1* | 3.858884229 | 0.000115212 | 0.000976811 | 0.559007329 |
| Obesity | mast_cell | *RAB30* | 1.832933806 | 0.000540008 | 0.001171639 | 1 |
| Obesity | mast_cell | *PRCP* | 2.848222468 | 0.000299587 | 0.000998908 | 1 |

| | | | | | | |
|---|---|---|---|---|---|---|
| Obesity | mast_cell | *NARS2* | 2.890127527 | 0.000246772 | 0.000976811 | 1 |
| Obesity | mast_cell | *SULF1* | 2.16864779 | 0.003026059 | 0.004554107 | 1 |
| Obesity | mast_cell | *TGS1* | 5.137459309 | 0.000182302 | 0.000976811 | 0.884530966 |
| Obesity | mast_cell | *NEK1* | 2.879651533 | 0.000246013 | 0.000976811 | 1 |
| Obesity | mast_cell | *TRPC6* | 2.168108728 | 0.000178807 | 0.000976811 | 0.867572101 |
| Obesity | mast_cell | *MMP27* | 9.772681815 | 0.001335591 | 0.002278582 | 1 |
| Obesity | mast_cell | *SLC37A4* | 9.818509038 | 6.46E-11 | 2.24E-08 | 3.13E-07 |
| Obesity | mast_cell | *POU2F3* | 2.949903167 | 0.017078641 | 0.022086629 | 1 |
| Obesity | mast_cell | *PPP2R1B* | 1.750931465 | 0.000467203 | 0.001116682 | 1 |
| Obesity | mast_cell | *FDX1* | 2.217266977 | 0.000216716 | 0.000976811 | 1 |
| Obesity | mast_cell | *FXYD6* | 1.710641563 | 0.004679313 | 0.006713195 | 1 |
| Obesity | mast_cell | *FXYD2* | 4.32084106 | 0.000815108 | 0.001531712 | 1 |
| Obesity | mast_cell | *MAP2K5* | 2.799754559 | 6.75E-05 | 0.000885378 | 0.327589754 |
| Obesity | mast_cell | *NUSAP1* | 10.78211787 | 5.83E-07 | 5.25E-05 | 0.002828143 |
| Obesity | mast_cell | *NDUFAF1* | 2.602244827 | 1.14E-05 | 0.000375035 | 0.055505171 |
| Obesity | mast_cell | *PARP6* | 2.427218506 | 0.000478619 | 0.001129371 | 1 |
| Obesity | mast_cell | *ADAM10* | 4.921527064 | 1.17E-05 | 0.000377668 | 0.056583195 |
| Obesity | mast_cell | *ZNF280D* | 3.114862865 | 0.000153652 | 0.000976811 | 0.745521658 |
| Obesity | mast_cell | *RSL24D1* | 1.902567038 | 0.004241918 | 0.00615762 | 1 |
| Obesity | mast_cell | *SLC44A5* | 2.825960384 | 0.000113056 | 0.000976811 | 0.54854924 |
| Obesity | mast_cell | *DBT* | 1.925056456 | 0.0005297 | 0.00115966 | 1 |
| Obesity | mast_cell | *RTCA* | 6.70179747 | 0.000220603 | 0.000976811 | 1 |
| Obesity | mast_cell | *KHK* | -2.24274397 | 0.006695676 | 0.00929274 | 1 |
| Obesity | mast_cell | *ADCY3* | 3.468899906 | 3.54E-05 | 0.000647299 | 0.171534275 |
| Obesity | mast_cell | *DYNC2LI1* | 2.889087576 | 0.000274451 | 0.000990086 | 1 |
| Obesity | mast_cell | *LHCGR* | 3.548745004 | 0.016100936 | 0.020932942 | 1 |
| Obesity | mast_cell | *THUMPD2* | 2.044954458 | 0.000363169 | 0.001037687 | 1 |
| Obesity | mast_cell | *ACTR2* | 3.520923071 | 0.000165464 | 0.000976811 | 0.802830219 |
| Obesity | mast_cell | *ATRAID* | 6.368328129 | 9.01E-06 | 0.000338797 | 0.043704848 |
| Obesity | mast_cell | *CENPO* | 4.613183381 | 3.93E-08 | 5.45E-06 | 0.000190715 |
| Obesity | mast_cell | *LRPPRC* | 3.452632888 | 0.000225289 | 0.000976811 | 1 |
| Obesity | mast_cell | *DTNB* | 2.410306078 | 0.000259677 | 0.00098093 | 1 |
| Obesity | mast_cell | *CYP2C8* | 12.74308739 | 0.000338241 | 0.00102217 | 1 |
| Obesity | mast_cell | *MYOF* | 1.529008034 | 0.002801892 | 0.00424439 | 1 |
| Obesity | mast_cell | *BTBD16* | 3.5578483 | 0.000827273 | 0.00154739 | 1 |
| Obesity | mast_cell | *TACC2* | 2.984920938 | 0.000161763 | 0.000976811 | 0.7848763 |
| Obesity | mast_cell | *CALHM2* | 2.602826607 | 0.000223701 | 0.000976811 | 1 |
| Obesity | mast_cell | *ARL3* | 3.212095709 | 8.63E-05 | 0.000971162 | 0.418657339 |
| Obesity | mast_cell | *PLCE1* | 1.613156028 | 0.000790637 | 0.001498503 | 1 |
| Obesity | mast_cell | *RBP4* | 3.227635058 | 0.000170684 | 0.000976811 | 0.828157232 |
| Obesity | mast_cell | *ASCC1* | 3.094460497 | 0.009623334 | 0.012980933 | 1 |
| Obesity | mast_cell | *RPS24* | 4.554327973 | 4.72E-05 | 0.000758703 | 0.228864152 |
| Obesity | mast_cell | *SMARCAL1* | 3.580771094 | 0.000196044 | 0.000976811 | 0.951207797 |
| Obesity | mast_cell | *BARD1* | 2.745220926 | 0.000370353 | 0.00103942 | 1 |
| Obesity | mast_cell | *STAT4* | 2.22469978 | 0.000201091 | 0.000976811 | 0.975693951 |
| Obesity | mast_cell | *SSB* | 4.924386037 | 0.000137605 | 0.000976811 | 0.667661203 |
| Obesity | mast_cell | *FASTKD1* | 2.498428872 | 0.000311047 | 0.000998908 | 1 |
| Obesity | mast_cell | *ITGAV* | 5.593248576 | 0.00062874 | 0.001275241 | 1 |
| Obesity | mast_cell | *SLC35A5* | 4.357486156 | 0.009396988 | 0.012700352 | 1 |
| Obesity | mast_cell | *SENP7* | 3.284154569 | 0.000397756 | 0.001061493 | 1 |

| | | | | | | |
|---|---|---|---|---|---|---|
| Obesity | mast_cell | *COX17* | 2.402331324 | 0.000419747 | 0.00108267 | 1 |
| Obesity | mast_cell | *SPPL2A* | 3.860390123 | 0.000458968 | 0.001112606 | 1 |
| Obesity | mast_cell | *PPCDC* | 1.751020408 | 0.005963583 | 0.008360389 | 1 |
| Obesity | mast_cell | *SEMA7A* | -0.654482242 | 0.002580677 | 0.003958724 | 1 |
| Obesity | mast_cell | *AP1AR* | 7.821128584 | 0.000724603 | 0.001406873 | 1 |
| Obesity | mast_cell | *PRKG2* | -0.360517275 | 0.002297032 | 0.003585972 | 1 |
| Obesity | mast_cell | *KIAA1109* | 2.911489029 | 0.000190111 | 0.000976811 | 0.922417176 |
| Obesity | mast_cell | *BMPR1B* | 1.724359056 | 0.000590209 | 0.00122067 | 1 |
| Obesity | mast_cell | *MMRN1* | 0.922391685 | 0.023961022 | 0.030157945 | 1 |
| Obesity | mast_cell | *PRDM5* | 2.300267413 | 0.000333672 | 0.001016573 | 1 |
| Obesity | mast_cell | *TRPC3* | 2.982998855 | 0.038841943 | 0.04751919 | 1 |
| Obesity | mast_cell | *NAAA* | 3.251376059 | 0.001506528 | 0.002524059 | 1 |
| Obesity | mast_cell | *NUP54* | 2.255680297 | 0.001378887 | 0.002339286 | 1 |
| Obesity | mast_cell | *G3BP2* | 6.188659476 | 8.85E-05 | 0.000976811 | 0.429172235 |
| Obesity | mast_cell | *CNOT6L* | 3.001353269 | 0.001056621 | 0.001870475 | 1 |
| Obesity | mast_cell | *ANXA3* | 3.990979873 | 3.25E-05 | 0.000621513 | 0.157561088 |
| Obesity | mast_cell | *PPA2* | 5.175931145 | 0.000203885 | 0.000976811 | 0.989250916 |
| Obesity | mast_cell | *ENPEP* | 1.019357075 | 0.029835634 | 0.037071062 | 1 |
| Obesity | mast_cell | *PAPSS1* | 1.804433186 | 0.000553593 | 0.00118075 | 1 |
| Obesity | mast_cell | *PARVG* | 2.188429251 | 0.001721781 | 0.002798688 | 1 |
| Obesity | mast_cell | *B4GALNT3* | 4.738621111 | 0.000206261 | 0.000976811 | 1 |
| Obesity | mast_cell | *PDE6H* | 7.941283506 | 0.040331237 | 0.049204718 | 1 |
| Obesity | mast_cell | *KIF21A* | 1.803149251 | 0.000370288 | 0.00103942 | 1 |
| Obesity | mast_cell | *ALG10* | 2.833886734 | 0.001636185 | 0.002690938 | 1 |
| Obesity | mast_cell | *ZCRB1* | 3.371798543 | 0.000251701 | 0.000976811 | 1 |
| Obesity | mast_cell | *C1RL* | 1.590360525 | 0.000465237 | 0.001116682 | 1 |
| Obesity | mast_cell | *CLSTN3* | 3.980509617 | 0.000145602 | 0.000976811 | 0.706462515 |
| Obesity | mast_cell | *KLRG1* | 2.770281325 | 2.25E-05 | 0.000536798 | 0.109282954 |
| Obesity | mast_cell | *TAPBPL* | 1.640609037 | 0.002631789 | 0.004025675 | 1 |
| Obesity | mast_cell | *PEX5* | -1.784752368 | 0.015163904 | 0.019788936 | 1 |
| Obesity | mast_cell | *PPFIA2* | 1.862673862 | 0.000449906 | 0.001105848 | 1 |
| Obesity | mast_cell | *MARCH9* | 12.27028483 | 0.000674971 | 0.001336113 | 1 |
| Obesity | mast_cell | *LGR5* | -1.603480393 | 0.001391918 | 0.00235966 | 1 |
| Obesity | mast_cell | *POC1B* | 2.912125169 | 0.000269487 | 0.000986064 | 1 |
| Obesity | mast_cell | *AMDHD1* | -1.935772544 | 0.00148391 | 0.002491326 | 1 |
| Obesity | mast_cell | *RITA1* | 3.479705385 | 0.0005137 | 0.001146491 | 1 |
| Obesity | mast_cell | *GLTP* | 8.49330155 | 0.020703514 | 0.026344991 | 1 |
| Obesity | mast_cell | *FOXN4* | -1.992861235 | 0.008827475 | 0.011994094 | 1 |
| Obesity | mast_cell | *MTMR6* | 4.863663269 | 0.000429919 | 0.001092129 | 1 |
| Obesity | mast_cell | *CCDC65* | 2.828839099 | 0.000360548 | 0.001035657 | 1 |
| Obesity | mast_cell | *ACVRL1* | 9.348541014 | 0.000282518 | 0.000993978 | 1 |
| Obesity | mast_cell | *BRCA2* | 2.809726999 | 0.000750393 | 0.001444231 | 1 |
| Obesity | mast_cell | *KANSL2* | 3.788905479 | 0.001482771 | 0.002490276 | 1 |
| Obesity | mast_cell | *CERS5* | 6.020125067 | 0.000142904 | 0.000976811 | 0.693368663 |
| Obesity | mast_cell | *LMBR1L* | 4.112430545 | 0.000712747 | 0.00139109 | 1 |
| Obesity | mast_cell | *ESYT1* | 5.63250981 | 3.26E-05 | 0.000621513 | 0.158257304 |
| Obesity | mast_cell | *ZNF740* | 5.005518756 | 0.001969238 | 0.003137847 | 1 |
| Obesity | mast_cell | *WDFY2* | 2.828759649 | 0.000666143 | 0.001324099 | 1 |
| Obesity | mast_cell | *HNRNPA1L2* | -4.122410498 | 0.019726392 | 0.025227321 | 1 |
| Obesity | mast_cell | *RB1* | 2.707999266 | 0.000420964 | 0.001083564 | 1 |

| | | | | | | |
|---|---|---|---|---|---|---|
| Obesity | mast_cell | *VPS33A* | 5.988161862 | 8.84E-08 | 1.19E-05 | 0.000428698 |
| Obesity | mast_cell | *DIAPH3* | 2.840758377 | 1.21E-06 | 8.61E-05 | 0.005856639 |
| Obesity | mast_cell | *NOVA1* | 1.825625359 | 3.87E-06 | 0.000187911 | 0.018791139 |
| Obesity | mast_cell | *FITM1* | 12.5517316 | 8.37E-05 | 0.000964174 | 0.405909357 |
| Obesity | mast_cell | *TMX1* | 4.606616456 | 0.000180834 | 0.000976811 | 0.877408262 |
| Obesity | mast_cell | *RTN1* | 2.104616502 | 0.000408146 | 0.001073348 | 1 |
| Obesity | mast_cell | *C14orf37* | 3.675543677 | 0.000171673 | 0.000976811 | 0.832957092 |
| Obesity | mast_cell | *RAB15* | 2.376654097 | 0.000484075 | 0.001131723 | 1 |
| Obesity | mast_cell | *EFCAB11* | 2.43497979 | 0.003647062 | 0.005372054 | 1 |
| Obesity | mast_cell | *PTGR2* | 1.240964644 | 0.010338808 | 0.013884278 | 1 |
| Obesity | mast_cell | *SLC24A4* | 2.928587757 | 0.000440133 | 0.001099087 | 1 |
| Obesity | mast_cell | *C14orf79* | 4.285296445 | 0.000569438 | 0.001193196 | 1 |
| Obesity | mast_cell | *WARS* | 6.892118251 | 0.005638392 | 0.007955068 | 1 |
| Obesity | mast_cell | *SERF2* | 4.684725157 | 0.000174515 | 0.000976811 | 0.846744722 |
| Obesity | mast_cell | *ZSCAN29* | 3.012039614 | 2.07E-05 | 0.000522259 | 0.10027371 |
| Obesity | mast_cell | *SRP14* | 3.973624535 | 0.000248018 | 0.000976811 | 1 |
| Obesity | mast_cell | *UBE2Q2* | 3.174199746 | 0.000936533 | 0.001700621 | 1 |
| Obesity | mast_cell | *HMG20A* | 3.841075918 | 0.000155134 | 0.000976811 | 0.752710758 |
| Obesity | mast_cell | *SCAPER* | 2.13279341 | 0.001204912 | 0.002086876 | 1 |
| Obesity | mast_cell | *NEIL1* | 3.011521563 | 0.004759884 | 0.006814682 | 1 |
| Obesity | mast_cell | *MAN2C1* | 0.908402049 | 0.030911566 | 0.038329394 | 1 |
| Obesity | mast_cell | *PIF1* | 2.985757982 | 0.003720094 | 0.005466353 | 1 |
| Obesity | mast_cell | *CYP11A1* | 6.773611526 | 6.44E-05 | 0.000874503 | 0.312522814 |
| Obesity | mast_cell | *BBS4* | 2.831705091 | 0.000218606 | 0.000976811 | 1 |
| Obesity | mast_cell | *ULK3* | 2.377378987 | 0.000801504 | 0.001513647 | 1 |
| Obesity | mast_cell | *PCSK6* | 2.273243747 | 0.000475598 | 0.001127398 | 1 |
| Obesity | mast_cell | *CELF6* | 11.75750376 | 0.027564165 | 0.034433916 | 1 |
| Obesity | mast_cell | *HAPLN3* | 2.539235648 | 0.003095645 | 0.004645861 | 1 |
| Obesity | mast_cell | *POLG* | 4.043180254 | 0.000112053 | 0.000976811 | 0.543679191 |
| Obesity | mast_cell | *ABHD2* | 5.479050333 | 0.00034107 | 0.001025322 | 1 |
| Obesity | mast_cell | *MFGE8* | 5.305313386 | 0.002809396 | 0.004254429 | 1 |
| Obesity | mast_cell | *IQGAP1* | 1.95733744 | 0.000372271 | 0.00104107 | 1 |
| Obesity | mast_cell | *CRTC3* | 3.229190315 | 0.000557087 | 0.001182832 | 1 |
| Obesity | mast_cell | *ITGAX* | -1.546137267 | 0.0042165 | 0.006128957 | 1 |
| Obesity | mast_cell | *TGFB1I1* | 2.745892325 | 0.001183803 | 0.00206167 | 1 |
| Obesity | mast_cell | *DHX38* | 2.856161937 | 0.000313432 | 0.000998908 | 1 |
| Obesity | mast_cell | *NUDT7* | 2.231636587 | 0.000558262 | 0.001182832 | 1 |
| Obesity | mast_cell | *GCSH* | 2.710234365 | 0.000960918 | 0.001737747 | 1 |
| Obesity | mast_cell | *CMTM3* | 7.467723719 | 0.001765519 | 0.002861155 | 1 |
| Obesity | mast_cell | *ZCCHC14* | 5.672790508 | 0.000428273 | 0.001092079 | 1 |
| Obesity | mast_cell | *TLDC1* | 3.482981224 | 0.000291724 | 0.000998325 | 1 |
| Obesity | mast_cell | *RPL3L* | 8.601754199 | 0.001159104 | 0.00202374 | 1 |
| Obesity | mast_cell | *TIGD7* | 2.223970053 | 0.000123265 | 0.000976811 | 0.598083687 |
| Obesity | mast_cell | *DEF8* | 2.751919431 | 0.002124082 | 0.00335156 | 1 |
| Obesity | mast_cell | *TCF25* | 2.928475492 | 0.000659894 | 0.001316532 | 1 |
| Obesity | mast_cell | *GALNS* | 1.926286751 | 0.00080589 | 0.001518516 | 1 |
| Obesity | mast_cell | *GAS8* | 4.277390728 | 0.000247188 | 0.000976811 | 1 |
| Obesity | mast_cell | *NCOR1* | 2.183676352 | 0.000252991 | 0.000977535 | 1 |
| Obesity | mast_cell | *KSR1* | 5.651747867 | 0.00014478 | 0.000976811 | 0.702473643 |
| Obesity | mast_cell | *GFOD2* | 4.384065534 | 0.000379085 | 0.001048885 | 1 |

| | | | | | | |
|---|---|---|---|---|---|---|
| Obesity | mast_cell | *PRPSAP2* | 3.891622226 | 0.027709649 | 0.034597843 | 1 |
| Obesity | mast_cell | *MYO19* | 10.53957215 | 2.86E-09 | 7.30E-07 | 1.39E-05 |
| Obesity | mast_cell | *C17orf80* | 5.573490125 | 8.68E-05 | 0.000971162 | 0.421251153 |
| Obesity | mast_cell | *VPS53* | 3.583162768 | 0.000321936 | 0.001000663 | 1 |
| Obesity | mast_cell | *SGSM2* | 2.67037721 | 0.000371899 | 0.00104107 | 1 |
| Obesity | mast_cell | *SCRN2* | 1.596289618 | 0.000264062 | 0.000984805 | 1 |
| Obesity | mast_cell | *SSH2* | 2.625534891 | 0.000191447 | 0.000976811 | 0.928902029 |
| Obesity | mast_cell | *RHBDL3* | -1.698238154 | 0.00630008 | 0.00878643 | 1 |
| Obesity | mast_cell | *G6PC3* | 3.231476851 | 0.000429113 | 0.001092129 | 1 |
| Obesity | mast_cell | *CLTC* | 2.480400368 | 0.00019064 | 0.000976811 | 0.924984699 |
| Obesity | mast_cell | *IMPA2* | 4.298172627 | 0.000130595 | 0.000976811 | 0.633645372 |
| Obesity | mast_cell | *GNAL* | 5.61360301 | 0.000177196 | 0.000976811 | 0.859754707 |
| Obesity | mast_cell | *RPRD1A* | 2.779528172 | 0.000498952 | 0.001141403 | 1 |
| Obesity | mast_cell | *GALNT1* | 3.842592317 | 0.000180091 | 0.000976811 | 0.873802732 |
| Obesity | mast_cell | *ADCYAP1* | -2.545561594 | 0.008027052 | 0.010987489 | 1 |
| Obesity | mast_cell | *C18orf8* | 1.059238913 | 0.040895325 | 0.049830266 | 1 |
| Obesity | mast_cell | *NPC1* | 2.554193065 | 0.004200215 | 0.006108347 | 1 |
| Obesity | mast_cell | *SLC14A1* | 1.929438309 | 0.004290045 | 0.006217234 | 1 |
| Obesity | mast_cell | *CCDC40* | 1.591740069 | 0.00221673 | 0.003478517 | 1 |
| Obesity | mast_cell | *ARHGDIA* | 3.179696 | 0.004002367 | 0.005843961 | 1 |
| Obesity | mast_cell | *ANAPC11* | 4.164350804 | 0.00050638 | 0.001144962 | 1 |
| Obesity | mast_cell | *TBCD* | 3.841813743 | 0.00031758 | 0.000998908 | 1 |
| Obesity | mast_cell | *FN3KRP* | 1.762112886 | 0.000689148 | 0.001355939 | 1 |
| Obesity | mast_cell | *NARF* | 5.819149982 | 8.15E-05 | 0.000957283 | 0.395673922 |
| Obesity | mast_cell | *TRIM65* | 3.295405882 | 5.92E-05 | 0.000847286 | 0.28722988 |
| Obesity | mast_cell | *CBX8* | 5.29697898 | 8.57E-05 | 0.000971162 | 0.415897889 |
| Obesity | mast_cell | *SECTM1* | 2.704076804 | 0.00055779 | 0.001182832 | 1 |
| Obesity | mast_cell | *RNF157* | 2.303364719 | 4.22E-05 | 0.000728972 | 0.204841108 |
| Obesity | mast_cell | *ZNF750* | 7.758902236 | 0.012319975 | 0.016341311 | 1 |
| Obesity | mast_cell | *WDR45B* | 4.392739715 | 0.000313526 | 0.000998908 | 1 |
| Obesity | mast_cell | *RNF165* | 3.425830659 | 0.00024967 | 0.000976811 | 1 |
| Obesity | mast_cell | *MAPK4* | 5.442374117 | 0.000479028 | 0.001129371 | 1 |
| Obesity | mast_cell | *FBXO15* | -3.69609966 | 0.037539179 | 0.046029845 | 1 |
| Obesity | mast_cell | *CBLN2* | 3.624122629 | 0.00054842 | 0.001175269 | 1 |
| Obesity | mast_cell | *PIP4K2B* | 3.866381968 | 0.001105527 | 0.001943484 | 1 |
| Obesity | mast_cell | *ERBB2* | 2.803449856 | 0.000504041 | 0.001144783 | 1 |
| Obesity | mast_cell | *MIEN1* | 3.102254269 | 0.000224324 | 0.000976811 | 1 |
| Obesity | mast_cell | *PNMT* | 6.159677041 | 0.013007132 | 0.017168281 | 1 |
| Obesity | mast_cell | *IGFBP4* | 2.720081058 | 9.57E-05 | 0.000976811 | 0.464238872 |
| Obesity | mast_cell | *TXNL4A* | 4.018056101 | 0.000232997 | 0.000976811 | 1 |
| Obesity | mast_cell | *CACNA1A* | 2.222300254 | 0.001097458 | 0.0019321 | 1 |
| Obesity | mast_cell | *SAMD1* | 7.799690816 | 0.006917342 | 0.009564817 | 1 |
| Obesity | mast_cell | *NFIC* | 2.237753338 | 0.000152646 | 0.000976811 | 0.740637355 |
| Obesity | mast_cell | *MVB12A* | 5.70557283 | 0.001519419 | 0.002542237 | 1 |
| Obesity | mast_cell | *DUS3L* | 1.568275393 | 0.014108646 | 0.018481411 | 1 |
| Obesity | mast_cell | *DPP9* | 2.131395279 | 0.000410455 | 0.001075307 | 1 |
| Obesity | mast_cell | *TMEM91* | 1.987265281 | 0.002467225 | 0.003812413 | 1 |
| Obesity | mast_cell | *SIRT3* | 3.209708987 | 5.97E-05 | 0.000847545 | 0.28986051 |
| Obesity | mast_cell | *COL6A1* | 2.892671992 | 5.95E-07 | 5.25E-05 | 0.002887757 |
| Obesity | mast_cell | *IFNAR1* | 2.066665188 | 0.002016509 | 0.003201604 | 1 |

| | | | | | | |
|---|---|---|---|---|---|---|
| Obesity | mast_cell | *TMEM50B* | 2.018295917 | 0.000634326 | 0.001281728 | 1 |
| Obesity | mast_cell | *DOPEY2* | 2.345708486 | 0.00184535 | 0.002973643 | 1 |
| Obesity | mast_cell | *URB1* | 2.686135003 | 0.002745401 | 0.004170534 | 1 |
| Obesity | mast_cell | *SAE1* | 1.797166663 | 0.01164411 | 0.015495672 | 1 |
| Obesity | mast_cell | *GEMIN7* | 2.819657743 | 0.012291171 | 0.016307564 | 1 |
| Obesity | mast_cell | *SLC6A3* | 2.597671493 | 0.017079355 | 0.022086629 | 1 |
| Obesity | mast_cell | *CAPN10* | 2.380739053 | 0.001847854 | 0.002975701 | 1 |
| Obesity | mast_cell | *CACNG8* | 3.812497339 | 0.031138861 | 0.038591508 | 1 |
| Obesity | mast_cell | *ZNF787* | 7.530809582 | 4.37E-05 | 0.00074212 | 0.212246238 |
| Obesity | mast_cell | *SLC47A1* | 3.422474989 | 0.000898783 | 0.001644379 | 1 |
| Obesity | mast_cell | *RERE* | 2.609882161 | 0.000126889 | 0.000976811 | 0.615664418 |
| Obesity | mast_cell | *EPHA2* | 2.265359402 | 0.005246929 | 0.007456971 | 1 |
| Obesity | mast_cell | *ARHGEF19* | -1.111985214 | 0.001153765 | 0.002016596 | 1 |
| Obesity | mast_cell | *PEX14* | 3.116162922 | 0.000232943 | 0.000976811 | 1 |
| Obesity | mast_cell | *ZNF593* | 4.507036887 | 0.000483359 | 0.001131376 | 1 |
| Obesity | mast_cell | *C1orf216* | 2.860864907 | 0.000168251 | 0.000976811 | 0.81635208 |
| Obesity | mast_cell | *PLK4* | 3.256963222 | 0.004881906 | 0.006977027 | 1 |
| Obesity | mast_cell | *HSPG2* | 1.831812218 | 0.00261766 | 0.00400862 | 1 |
| Obesity | mast_cell | *ITGB3BP* | 2.23341043 | 0.001352636 | 0.002302804 | 1 |
| Obesity | mast_cell | *PIGK* | 3.025890469 | 0.001185745 | 0.002063075 | 1 |
| Obesity | mast_cell | *RPS8* | 4.694381805 | 6.15E-05 | 0.000858537 | 0.298220887 |
| Obesity | mast_cell | *CYP4B1* | 1.956924941 | 0.000862389 | 0.00159281 | 1 |
| Obesity | mast_cell | *SLC44A3* | 2.430781242 | 0.000602763 | 0.001238217 | 1 |
| Obesity | mast_cell | *STRIP1* | 9.875910751 | 0.000517242 | 0.00115122 | 1 |
| Obesity | mast_cell | *GPR161* | 2.305564229 | 0.000315426 | 0.000998908 | 1 |
| Obesity | mast_cell | *ALDH9A1* | 3.789889116 | 0.000246361 | 0.000976811 | 1 |
| Obesity | mast_cell | *ATP1B1* | 1.599932993 | 0.000826715 | 0.001546942 | 1 |
| Obesity | mast_cell | *TIPRL* | 8.814547794 | 0.000180884 | 0.000976811 | 0.877651266 |
| Obesity | mast_cell | *NME7* | 2.954893759 | 0.002878728 | 0.00435263 | 1 |
| Obesity | mast_cell | *XCL1* | 7.032091267 | 0.002739308 | 0.004162581 | 1 |
| Obesity | mast_cell | *MAEL* | 5.762762372 | 0.000547732 | 0.001175269 | 1 |
| Obesity | mast_cell | *RFWD2* | 3.804714684 | 0.00019319 | 0.000976811 | 0.937358628 |
| Obesity | mast_cell | *TARS2* | 3.980877341 | 0.000665941 | 0.001324099 | 1 |
| Obesity | mast_cell | *SNX27* | 3.079670632 | 9.33E-05 | 0.000976811 | 0.452728948 |
| Obesity | mast_cell | *SETDB1* | 3.221791784 | 0.000136437 | 0.000976811 | 0.661991738 |
| Obesity | mast_cell | *SEMA6C* | 19.96249315 | 1.21E-08 | 2.04E-06 | 5.86E-05 |
| Obesity | mast_cell | *ARNT* | 2.615471689 | 0.000562649 | 0.001188051 | 1 |
| Obesity | mast_cell | *POGZ* | 2.548019982 | 0.000262771 | 0.000983135 | 1 |
| Obesity | mast_cell | *GABPB2* | 1.831748093 | 0.001976632 | 0.003148594 | 1 |
| Obesity | mast_cell | *DYRK3* | 2.91199723 | 0.000457775 | 0.001112606 | 1 |
| Obesity | mast_cell | *EIF2D* | 2.573296266 | 0.000450771 | 0.001106294 | 1 |
| Obesity | mast_cell | *TAF1A* | 4.225078116 | 0.000574923 | 0.001201079 | 1 |
| Obesity | mast_cell | *SMYD2* | 5.561547218 | 0.000155107 | 0.000976811 | 0.752577024 |
| Obesity | mast_cell | *ADAM15* | 1.780456055 | 0.000381789 | 0.001049541 | 1 |
| Obesity | mast_cell | *RAB13* | 2.418480185 | 0.000487985 | 0.001134501 | 1 |
| Obesity | mast_cell | *S100A8* | 2.093896872 | 0.000439947 | 0.001099087 | 1 |
| Obesity | mast_cell | *CREB3L4* | 2.428048295 | 0.007413083 | 0.01019509 | 1 |
| Obesity | mast_cell | *KCNN3* | 1.334057837 | 0.005255012 | 0.00746578 | 1 |
| Obesity | mast_cell | *C1orf43* | 3.35722197 | 0.000272948 | 0.000987746 | 1 |
| Obesity | mast_cell | *RIT1* | 5.368365157 | 4.95E-05 | 0.000765648 | 0.2401733 |

| | | | | | | |
|---|---|---|---|---|---|---|
| Obesity | mast_cell | *SCCPDH* | 3.411497535 | 0.000283723 | 0.000993978 | 1 |
| Obesity | mast_cell | *CEP170* | 3.74776205 | 6.30E-05 | 0.000865434 | 0.30549831 |
| Obesity | mast_cell | *ACP1* | 3.193846186 | 0.000428324 | 0.001092079 | 1 |
| Obesity | mast_cell | *CNIH4* | 1.749687701 | 0.001126892 | 0.001977461 | 1 |
| Obesity | mast_cell | *MBOAT2* | 3.324297748 | 0.000267458 | 0.000985747 | 1 |
| Obesity | mast_cell | *PYCR2* | 2.466579833 | 0.001693191 | 0.002757758 | 1 |
| Obesity | mast_cell | *PLEKHA6* | 1.996348039 | 0.000177042 | 0.000976811 | 0.859008974 |
| Obesity | mast_cell | *RHOB* | 2.063851075 | 0.000467892 | 0.001116682 | 1 |
| Obesity | mast_cell | *GALM* | 1.591644206 | 0.000491283 | 0.001138351 | 1 |
| Obesity | mast_cell | *WDPCP* | 1.754520474 | 0.000174586 | 0.000976811 | 0.847092344 |
| Obesity | mast_cell | *VPS54* | 3.626250747 | 0.000149801 | 0.000976811 | 0.726835812 |
| Obesity | mast_cell | *ABHD1* | 2.926032527 | 0.001167712 | 0.002038036 | 1 |
| Obesity | mast_cell | *CIAO1* | 4.850616527 | 0.035608415 | 0.043795191 | 1 |
| Obesity | mast_cell | *ZNF514* | 2.070437984 | 0.014581397 | 0.019069795 | 1 |
| Obesity | mast_cell | *EXOC6B* | 2.732824322 | 0.000187863 | 0.000976811 | 0.911511885 |
| Obesity | mast_cell | *TEX261* | -2.289006015 | 0.006467466 | 0.008994022 | 1 |
| Obesity | mast_cell | *THNSL2* | 1.702636961 | 0.000677329 | 0.001338111 | 1 |
| Obesity | mast_cell | *RALB* | 6.676463591 | 0.000173147 | 0.000976811 | 0.840107728 |
| Obesity | mast_cell | *CNGA3* | 30.37698723 | 0.014031343 | 0.018390081 | 1 |
| Obesity | mast_cell | *LYG1* | 2.681735596 | 0.000486937 | 0.001132608 | 1 |
| Obesity | mast_cell | *THSD7B* | 1.835705319 | 0.000202056 | 0.000976811 | 0.980376702 |
| Obesity | mast_cell | *GPR17* | 0.928524081 | 0.02839942 | 0.035413514 | 1 |
| Obesity | mast_cell | *POLR2D* | 5.326276453 | 0.002170057 | 0.003420766 | 1 |
| Obesity | mast_cell | *GALNT13* | 1.629984349 | 0.000530811 | 0.001160656 | 1 |
| Obesity | mast_cell | *PHOSPHO2* | 2.809672004 | 0.005901198 | 0.008282503 | 1 |
| Obesity | mast_cell | *METTL21A* | 2.774410615 | 0.000131224 | 0.000976811 | 0.636700866 |
| Obesity | mast_cell | *NBEAL1* | 3.160569123 | 1.91E-05 | 0.000499059 | 0.092825025 |
| Obesity | mast_cell | *SUMF1* | 2.10002627 | 0.000244521 | 0.000976811 | 1 |
| Obesity | mast_cell | *ACKR3* | 3.00564419 | 0.011981887 | 0.015914623 | 1 |
| Obesity | mast_cell | *ESPNL* | 31.27307906 | 0.00063401 | 0.001281728 | 1 |
| Obesity | mast_cell | *TAMM41* | 4.31159293 | 7.22E-05 | 0.000918732 | 0.350454452 |
| Obesity | mast_cell | *STK11IP* | 5.400679546 | 0.011233289 | 0.01498541 | 1 |
| Obesity | mast_cell | *EAF1* | 2.890588843 | 0.000524963 | 0.001155681 | 1 |
| Obesity | mast_cell | *CNTN4* | 3.396892232 | 0.000257581 | 0.00098093 | 1 |
| Obesity | mast_cell | *SLC25A38* | 3.666611855 | 0.000599656 | 0.001235995 | 1 |
| Obesity | mast_cell | *CTDSPL* | 1.909740761 | 0.00031457 | 0.000998908 | 1 |
| Obesity | mast_cell | *PTPRG* | 2.613338769 | 0.000455772 | 0.00111155 | 1 |
| Obesity | mast_cell | *SLC25A26* | 1.678495358 | 0.001497655 | 0.002511679 | 1 |
| Obesity | mast_cell | *ARL6IP5* | 2.697454569 | 0.000226264 | 0.000976811 | 1 |
| Obesity | mast_cell | *LIMD1* | 3.375641132 | 8.81E-06 | 0.000334041 | 0.042757197 |
| Obesity | mast_cell | *PLA1A* | 7.62038707 | 0.000564311 | 0.001190078 | 1 |
| Obesity | mast_cell | *ADPRH* | 3.061776818 | 0.002597667 | 0.003982268 | 1 |
| Obesity | mast_cell | *IGSF11* | 1.972649985 | 0.00068195 | 0.001345596 | 1 |
| Obesity | mast_cell | *BOC* | 1.589352794 | 0.000385222 | 0.001052047 | 1 |
| Obesity | mast_cell | *MED12L* | 6.7671478 | 0.02188257 | 0.027714495 | 1 |
| Obesity | mast_cell | *NCEH1* | 1.352479845 | 0.003735417 | 0.005485545 | 1 |
| Obesity | mast_cell | *AMT* | -1.656894428 | 0.009190846 | 0.012449466 | 1 |
| Obesity | mast_cell | *CCDC39* | 3.196511473 | 1.13E-05 | 0.000373166 | 0.054704513 |
| Obesity | mast_cell | *MUC4* | 3.502679003 | 0.000761033 | 0.001459865 | 1 |
| Obesity | mast_cell | *SLC26A1* | 1.990846409 | 0.009641354 | 0.012998013 | 1 |

| | | | | | | |
|---|---|---|---|---|---|---|
| Obesity | mast_cell | *CENPC* | 2.107045623 | 0.000837829 | 0.001562916 | 1 |
| Obesity | mast_cell | *SCD5* | -2.545976927 | 0.022865732 | 0.028854235 | 1 |
| Obesity | mast_cell | *TRMT10A* | 2.937521636 | 0.002689518 | 0.004101823 | 1 |
| Obesity | mast_cell | *PYURF* | 9.64476344 | 0.001627616 | 0.002677922 | 1 |
| Obesity | mast_cell | *CISD2* | 4.341501897 | 7.37E-05 | 0.000926712 | 0.357710755 |
| Obesity | mast_cell | *DDIT4L* | 7.099959982 | 0.000264054 | 0.000984805 | 1 |
| Obesity | mast_cell | *METTL14* | 3.912899731 | 0.000287621 | 0.000994669 | 1 |
| Obesity | mast_cell | *USP53* | 2.446504154 | 8.39E-05 | 0.000964176 | 0.407214895 |
| Obesity | mast_cell | *MARCH1* | 1.09378452 | 0.001739551 | 0.002822844 | 1 |
| Obesity | mast_cell | *CBR4* | 2.484321985 | 0.000231452 | 0.000976811 | 1 |
| Obesity | mast_cell | *MARCH6* | 3.859926129 | 0.000150817 | 0.000976811 | 0.731765523 |
| Obesity | mast_cell | *NKD2* | 3.845055549 | 0.029355382 | 0.036521106 | 1 |
| Obesity | mast_cell | *MYO10* | 4.70110481 | 0.000159346 | 0.000976811 | 0.773147973 |
| Obesity | mast_cell | *FAM105A* | 1.353768757 | 0.006243196 | 0.008712105 | 1 |
| Obesity | mast_cell | *BHMT* | -2.073423607 | 0.000162133 | 0.000976811 | 0.786670936 |
| Obesity | mast_cell | *GTF2H2* | 2.917077069 | 0.000495693 | 0.001138685 | 1 |
| Obesity | mast_cell | *SPATA9* | 4.273937397 | 0.000224264 | 0.000976811 | 1 |
| Obesity | mast_cell | *TSLP* | -1.667324663 | 0.004120867 | 0.006000635 | 1 |
| Obesity | mast_cell | *FEM1C* | 3.243758549 | 0.000309442 | 0.000998908 | 1 |
| Obesity | mast_cell | *COMMD10* | 2.299378515 | 0.000744467 | 0.001436815 | 1 |
| Obesity | mast_cell | *YIPF5* | 4.079603402 | 7.02E-05 | 0.000898663 | 0.34053233 |
| Obesity | mast_cell | *CXCL14* | 2.475667865 | 0.000229264 | 0.000976811 | 1 |
| Obesity | mast_cell | *FBXO38* | 3.905779577 | 0.00018656 | 0.000976811 | 0.905186874 |
| Obesity | mast_cell | *BOD1* | 3.762139442 | 0.000256404 | 0.00098093 | 1 |
| Obesity | mast_cell | *FAM50B* | 5.245206879 | 0.00245555 | 0.003798001 | 1 |
| Obesity | mast_cell | *MYLK4* | -4.33834497 | 0.037203376 | 0.04566425 | 1 |
| Obesity | mast_cell | *GFOD1* | 2.051487139 | 0.000232608 | 0.000976811 | 1 |
| Obesity | mast_cell | *HIGD2A* | 2.157492219 | 0.000540402 | 0.001171639 | 1 |
| Obesity | mast_cell | *FAM193B* | 5.781089896 | 0.003845596 | 0.005633706 | 1 |
| Obesity | mast_cell | *RNF44* | 3.199166642 | 0.000547962 | 0.001175269 | 1 |
| Obesity | mast_cell | *MUT* | 4.946857588 | 0.001902973 | 0.003046263 | 1 |
| Obesity | mast_cell | *DAAM2* | 2.049529627 | 0.000363177 | 0.001037687 | 1 |
| Obesity | mast_cell | *ANO7* | 3.410983207 | 0.002730824 | 0.004150989 | 1 |
| Obesity | mast_cell | *TCTE1* | -3.85030593 | 0.00307556 | 0.004617146 | 1 |
| Obesity | mast_cell | *RPL7L1* | 6.584535567 | 0.000108762 | 0.000976811 | 0.527711986 |
| Obesity | mast_cell | *TPBG* | 1.664829115 | 0.000690606 | 0.001357707 | 1 |
| Obesity | mast_cell | *IRAK1BP1* | 4.047741397 | 1.85E-05 | 0.000495554 | 0.08969519 |
| Obesity | mast_cell | *PHIP* | 3.329665443 | 0.00012855 | 0.000976811 | 0.623723841 |
| Obesity | mast_cell | *PNRC1* | 6.041678723 | 0.00017728 | 0.000976811 | 0.860164515 |
| Obesity | mast_cell | *RNF217* | 2.699572678 | 0.000285134 | 0.000993978 | 1 |
| Obesity | mast_cell | *ARHGAP18* | 2.649422046 | 0.000123531 | 0.000976811 | 0.599371887 |
| Obesity | mast_cell | *SLC18B1* | 1.366528317 | 0.003609673 | 0.005328303 | 1 |
| Obesity | mast_cell | *PNLDC1* | 7.492475132 | 0.003156265 | 0.004723688 | 1 |
| Obesity | mast_cell | *WTAP* | 4.326671863 | 0.000251188 | 0.000976811 | 1 |
| Obesity | mast_cell | *SLC22A3* | 1.680733925 | 0.000511714 | 0.001145411 | 1 |
| Obesity | mast_cell | *GNA12* | 2.582283721 | 0.001258605 | 0.002165515 | 1 |
| Obesity | mast_cell | *SDK1* | 1.003361531 | 0.026853909 | 0.033624559 | 1 |
| Obesity | mast_cell | *PSPH* | 1.824353957 | 0.036731895 | 0.045165524 | 1 |
| Obesity | mast_cell | *GIGYF1* | 4.313057445 | 0.00056925 | 0.001193196 | 1 |
| Obesity | mast_cell | *TMEM209* | 2.917179521 | 0.000306872 | 0.000998908 | 1 |

| | | | | | | |
|---|---|---|---|---|---|---|
| Obesity | mast_cell | *EPHA1* | 6.27472953 | 0.000538652 | 0.001170415 | 1 |
| Obesity | mast_cell | *NCAPG2* | 6.019967003 | 0.001335214 | 0.002278582 | 1 |
| Obesity | mast_cell | *FBXO25* | 2.066553968 | 0.000363349 | 0.001037687 | 1 |
| Obesity | mast_cell | *CSGALNACT1* | 1.788889055 | 0.000196141 | 0.000976811 | 0.951675099 |
| Obesity | mast_cell | *BIN3* | 3.405283987 | 0.000636408 | 0.001284464 | 1 |
| Obesity | mast_cell | *CHMP7* | 2.120452437 | 0.00166812 | 0.002728833 | 1 |
| Obesity | mast_cell | *DOCK5* | 2.35918749 | 0.000296434 | 0.000998908 | 1 |
| Obesity | mast_cell | *TACC1* | 1.973015658 | 0.000465011 | 0.001116682 | 1 |
| Obesity | mast_cell | *RPL7* | 3.523726849 | 0.000402224 | 0.001064698 | 1 |
| Obesity | mast_cell | *ATP6V0D2* | -1.453074963 | 0.002437724 | 0.003778862 | 1 |
| Obesity | mast_cell | *DPYS* | 1.434300217 | 0.000523494 | 0.001154019 | 1 |
| Obesity | mast_cell | *LRP12* | 4.582444722 | 0.000206057 | 0.000976811 | 0.999787984 |
| Obesity | mast_cell | *MAL2* | 1.486832204 | 0.002260222 | 0.003537612 | 1 |
| Obesity | mast_cell | *GSDMC* | -10.66923638 | 0.005426167 | 0.007682452 | 1 |
| Obesity | mast_cell | *AK3* | 2.338720436 | 0.000296455 | 0.000998908 | 1 |
| Obesity | mast_cell | *HAUS6* | 1.837280669 | 0.000486825 | 0.001132608 | 1 |
| Obesity | mast_cell | *C9orf72* | 1.725253905 | 0.002688105 | 0.004101474 | 1 |
| Obesity | mast_cell | *FBXO10* | -1.730589863 | 0.024312874 | 0.030561156 | 1 |
| Obesity | mast_cell | *ZNF462* | 2.278827091 | 0.000150873 | 0.000976811 | 0.732034366 |
| Obesity | mast_cell | *ALAD* | 4.414639703 | 0.000162043 | 0.000976811 | 0.786233132 |
| Obesity | mast_cell | *WDR31* | 2.183665438 | 7.23E-05 | 0.000918732 | 0.350955469 |
| Obesity | mast_cell | *SURF1* | 2.21916102 | 0.000410927 | 0.001075307 | 1 |
| Obesity | mast_cell | *SURF6* | 2.945958545 | 0.000573943 | 0.001200332 | 1 |
| Obesity | mast_cell | *MED22* | -3.619946005 | 0.014501713 | 0.018976834 | 1 |
| Obesity | mast_cell | *REXO4* | 3.95845562 | 0.000227985 | 0.000976811 | 1 |
| Obesity | mast_cell | *SH3GLB2* | 2.522986809 | 0.000403256 | 0.001065105 | 1 |
| Obesity | mast_cell | *INPP5E* | 2.661956544 | 0.000693622 | 0.001362532 | 1 |
| Obesity | mast_cell | *SEC16A* | 2.026263394 | 0.00137465 | 0.002332914 | 1 |
| Obesity | mast_cell | *SLC39A12* | 6.015450093 | 0.000267337 | 0.000985747 | 1 |
| Obesity | mast_cell | *RSU1* | 4.241712341 | 0.000155109 | 0.000976811 | 0.752586848 |
| Obesity | mast_cell | *PARD3* | 1.836810894 | 0.000231189 | 0.000976811 | 1 |
| Obesity | mast_cell | *FRA10AC1* | 4.495327949 | 0.001831376 | 0.002956033 | 1 |
| Obesity | mast_cell | *ADD3* | 3.019528426 | 0.000175084 | 0.000976811 | 0.849508792 |
| Obesity | mast_cell | *LRRC27* | 1.559175051 | 0.004313761 | 0.006249736 | 1 |
| Obesity | mast_cell | *MTG1* | 2.69903182 | 0.012603992 | 0.016690658 | 1 |
| Obesity | mast_cell | *PAOX* | 6.580610322 | 0.000175863 | 0.000976811 | 0.853287825 |
| Obesity | mast_cell | *PDCD11* | 9.542393365 | 2.13E-05 | 0.000524672 | 0.10330739 |
| Obesity | mast_cell | *RGS10* | 4.699316174 | 0.000209853 | 0.000976811 | 1 |
| Obesity | mast_cell | *GAS2* | 8.354870562 | 0.001343244 | 0.002290029 | 1 |
| Obesity | mast_cell | *LIN7C* | 4.691996645 | 0.000228105 | 0.000976811 | 1 |
| Obesity | mast_cell | *HSD17B12* | -1.384209724 | 0.008218163 | 0.011210156 | 1 |
| Obesity | mast_cell | *APIP* | 1.279788118 | 0.012815691 | 0.016935389 | 1 |
| Obesity | mast_cell | *EIF3M* | 2.902677218 | 0.000311951 | 0.000998908 | 1 |
| Obesity | mast_cell | *TNKS1BP1* | 3.791036854 | 1.63E-08 | 2.55E-06 | 7.91E-05 |
| Obesity | mast_cell | *PTPRJ* | 2.135784041 | 0.000342084 | 0.001027099 | 1 |
| Obesity | mast_cell | *ENDOD1* | 3.000979424 | 0.000223933 | 0.000976811 | 1 |
| Obesity | mast_cell | *CCDC82* | 1.351653552 | 0.03451808 | 0.04252964 | 1 |
| Obesity | mast_cell | *CAPN5* | 3.795588483 | 0.000620522 | 0.001263438 | 1 |
| Obesity | mast_cell | *INTS4* | 3.881631356 | 3.77E-05 | 0.000679348 | 0.18305524 |
| Obesity | mast_cell | *RPS3* | 2.346544847 | 0.000431277 | 0.001094432 | 1 |

| | | | | | | |
|---|---|---|---|---|---|---|
| Obesity | mast_cell | *TTC12* | 3.200964631 | 1.03E-06 | 7.48E-05 | 0.004986439 |
| Obesity | mast_cell | *DRD2* | -1.710921665 | 0.001679586 | 0.002742042 | 1 |
| Obesity | mast_cell | *NPAT* | 5.414022373 | 0.000170007 | 0.000976811 | 0.824871907 |
| Obesity | mast_cell | *ATM* | 2.11498543 | 0.000225824 | 0.000976811 | 1 |
| Obesity | mast_cell | *SLX4IP* | 3.391728733 | 4.00E-05 | 0.000698431 | 0.194163803 |
| Obesity | mast_cell | *ST14* | -3.572455647 | 0.01737306 | 0.022430572 | 1 |
| Obesity | mast_cell | *TMC2* | 4.043408255 | 0.001043453 | 0.00185316 | 1 |
| Obesity | mast_cell | *EML3* | 2.212192827 | 0.000868192 | 0.001599875 | 1 |
| Obesity | mast_cell | *INCENP* | -2.912298972 | 0.001753776 | 0.002843074 | 1 |
| Obesity | mast_cell | *CPSF7* | 2.201429426 | 0.000432308 | 0.001094797 | 1 |
| Obesity | mast_cell | *CCDC15* | 1.625977487 | 0.017782303 | 0.022922353 | 1 |
| Obesity | mast_cell | *CHEK1* | 4.163688568 | 0.000513349 | 0.001146236 | 1 |
| Obesity | mast_cell | *CABLES2* | -1.337374915 | 0.000745401 | 0.001438047 | 1 |
| Obesity | mast_cell | *NUDT22* | 1.585900845 | 0.007888801 | 0.010815615 | 1 |
| Obesity | mast_cell | *TBX6* | 4.142385029 | 1.01E-13 | 5.46E-11 | 4.92E-10 |
| Obesity | mast_cell | *ALDOA* | 3.277135697 | 0.000942321 | 0.001709783 | 1 |
| Obesity | mast_cell | *ANXA8L1* | 1.063886687 | 0.018911832 | 0.024275188 | 1 |
| Obesity | mast_cell | *CWC15* | 1.966586116 | 0.000340431 | 0.001025052 | 1 |
| Obesity | mast_cell | *TMEM218* | 2.14487324 | 0.000331247 | 0.001014048 | 1 |
| Obesity | mast_cell | *SAP18* | 5.123544158 | 0.000236169 | 0.000976811 | 1 |
| Obesity | mast_cell | *FAM124A* | 4.801550297 | 0.000285794 | 0.000993978 | 1 |
| Obesity | mast_cell | *WDR17* | 3.688221667 | 0.005562932 | 0.00785317 | 1 |
| Obesity | mast_cell | *CCDC102B* | 1.60429886 | 0.000819742 | 0.001535845 | 1 |
| Obesity | mast_cell | *RGS18* | 3.630937898 | 7.91E-07 | 6.34E-05 | 0.003839583 |
| Obesity | mast_cell | *PRSS23* | 2.465109507 | 0.000519173 | 0.001151636 | 1 |
| Obesity | mast_cell | *PPP1R1C* | 10.19029977 | 0.00020197 | 0.000976811 | 0.979958951 |
| Obesity | mast_cell | *FAM173B* | 3.468354821 | 0.000284629 | 0.000993978 | 1 |
| Obesity | mast_cell | *GPR158* | 7.056455848 | 0.000451564 | 0.001106559 | 1 |
| Obesity | mast_cell | *UBE3B* | 2.975888163 | 2.23E-05 | 0.000536645 | 0.108402311 |
| Obesity | mast_cell | *ANK3* | 1.798909915 | 0.000409774 | 0.001074715 | 1 |
| Obesity | mast_cell | *PLBD2* | 2.919520594 | 0.000233179 | 0.000976811 | 1 |
| Obesity | mast_cell | *DLG5* | 2.277173163 | 0.000949412 | 0.001719712 | 1 |
| Obesity | mast_cell | *GXYLT1* | 3.422809214 | 0.000269488 | 0.000986064 | 1 |
| Obesity | mast_cell | *EXT2* | 3.633692869 | 0.000250854 | 0.000976811 | 1 |
| Obesity | mast_cell | *TMEM18* | 7.072610205 | 0.000169307 | 0.000976811 | 0.821477559 |
| Obesity | mast_cell | *NDUFC2* | 3.528178365 | 0.000402155 | 0.001064698 | 1 |
| Obesity | mast_cell | *ADAMTS12* | 1.650530846 | 0.000577928 | 0.00120507 | 1 |
| Obesity | mast_cell | *NUBPL* | 2.044346545 | 0.000305432 | 0.000998908 | 1 |
| Obesity | mast_cell | *VIPAS39* | 4.485318844 | 0.000314026 | 0.000998908 | 1 |
| Obesity | mast_cell | *ANKRD50* | 3.284977496 | 0.000116456 | 0.000976811 | 0.565046426 |
| Obesity | mast_cell | *UPF2* | 5.268296876 | 0.000873672 | 0.001608751 | 1 |
| Obesity | mast_cell | *C4orf33* | 3.113312541 | 0.000336076 | 0.001017877 | 1 |
| Obesity | mast_cell | *PTPRO* | 2.105262445 | 0.000201235 | 0.000976811 | 0.976392229 |
| Obesity | mast_cell | *EPS8* | 1.978020133 | 0.000254866 | 0.00098093 | 1 |
| Obesity | mast_cell | *QDPR* | 2.771858791 | 0.000545734 | 0.00117408 | 1 |
| Obesity | mast_cell | *FAM160B1* | 2.363701951 | 0.000228604 | 0.000976811 | 1 |
| Obesity | mast_cell | *ZNF827* | 2.379538898 | 0.000388987 | 0.001054382 | 1 |
| Obesity | mast_cell | *AKR1C2* | 1.898302459 | 0.002651355 | 0.004051772 | 1 |
| Obesity | mast_cell | *DPYSL4* | 1.916968935 | 0.000976158 | 0.001753543 | 1 |
| Obesity | mast_cell | *PIGF* | 3.274091359 | 0.000231313 | 0.000976811 | 1 |

| | | | | | | |
|---|---|---|---|---|---|---|
| Obesity | mast_cell | *INPP1* | 2.881529856 | 0.000193368 | 0.000976811 | 0.93822246 |
| Obesity | mast_cell | *MFSD6* | 3.581846007 | 0.000236984 | 0.000976811 | 1 |
| Obesity | mast_cell | *RNF144A* | 2.288836093 | 0.000312924 | 0.000998908 | 1 |
| Obesity | mast_cell | *CENPU* | 4.049558604 | 0.00713987 | 0.009847257 | 1 |
| Obesity | mast_cell | *BICD1* | 3.099070879 | 0.00020058 | 0.000976811 | 0.973215215 |
| Obesity | mast_cell | *GUF1* | 1.859075793 | 0.001736178 | 0.0028202 | 1 |
| Obesity | mast_cell | *GABRA2* | -3.221060485 | 0.03078565 | 0.038183019 | 1 |
| Obesity | mast_cell | *PARP8* | 1.09531573 | 0.001834766 | 0.00296052 | 1 |
| Obesity | mast_cell | *CACUL1* | 2.621064816 | 0.000284774 | 0.000993978 | 1 |
| Obesity | mast_cell | *TIAL1* | 2.893553087 | 0.000147576 | 0.000976811 | 0.716039799 |
| Obesity | mast_cell | *TMEM56* | 4.227736183 | 0.000498852 | 0.001141403 | 1 |
| Obesity | mast_cell | *PTPN14* | 1.351705656 | 0.000423265 | 0.001086605 | 1 |
| Obesity | mast_cell | *MGAT5* | 2.765038105 | 0.000608756 | 0.001246806 | 1 |
| Obesity | mast_cell | *EPG5* | 3.664190405 | 0.000134679 | 0.000976811 | 0.653462754 |
| Obesity | mast_cell | *HAUS1* | 2.198593428 | 0.008185667 | 0.011172111 | 1 |
| Obesity | mast_cell | *SPC25* | 1.603903291 | 0.00269758 | 0.00411205 | 1 |
| Obesity | mast_cell | *KCNK13* | 3.326366101 | 0.000920346 | 0.001674989 | 1 |
| Obesity | mast_cell | *ATG10* | 1.135328184 | 0.030670255 | 0.038059355 | 1 |
| Obesity | mast_cell | *CWF19L2* | 3.38710175 | 2.06E-05 | 0.000522259 | 0.100052033 |
| Obesity | mast_cell | *NMT2* | 2.645361249 | 0.004738201 | 0.006787645 | 1 |
| Obesity | mast_cell | *CCDC50* | 2.515647905 | 0.000198815 | 0.000976811 | 0.964649796 |
| Obesity | mast_cell | *PLEKHH2* | 1.920322619 | 0.000540905 | 0.001171639 | 1 |
| Obesity | mast_cell | *SPARCL1* | 1.768617902 | 0.000295281 | 0.000998908 | 1 |
| Obesity | mast_cell | *MBNL1* | 2.380804362 | 0.000180636 | 0.000976811 | 0.876446853 |
| Obesity | mast_cell | *GPD1L* | 2.064005464 | 0.000125245 | 0.000976811 | 0.607686906 |
| Obesity | mast_cell | *CCNO* | -4.65791882 | 0.001601993 | 0.002643834 | 1 |
| Obesity | mast_cell | *CLEC4F* | 4.685689249 | 0.000145975 | 0.000976811 | 0.708271048 |
| Obesity | mast_cell | *SLC30A6* | 1.847988074 | 0.000613666 | 0.001253586 | 1 |
| Obesity | mast_cell | *RASGRP3* | 1.595681096 | 0.00103498 | 0.001839459 | 1 |
| Obesity | mast_cell | *GPR180* | 1.723871818 | 0.025265521 | 0.031692944 | 1 |
| Obesity | mast_cell | *TCTEX1D1* | -0.666281496 | 0.001738296 | 0.002821751 | 1 |
| Obesity | mast_cell | *IFIT5* | 1.35965994 | 0.035047166 | 0.043130623 | 1 |
| Obesity | mast_cell | *PRDM8* | 2.755724095 | 0.013829829 | 0.018150482 | 1 |
| Obesity | mast_cell | *GGPS1* | 2.473446312 | 0.000267566 | 0.000985747 | 1 |
| Obesity | mast_cell | *ZNF117* | 1.236362783 | 1.08E-05 | 0.000373166 | 0.052335033 |
| Obesity | mast_cell | *MED21* | 4.72438852 | 0.000165829 | 0.000976811 | 0.804602481 |
| Obesity | mast_cell | *JAKMIP1* | 3.078305913 | 0.000389428 | 0.001054382 | 1 |
| Obesity | mast_cell | *SREK1IP1* | 1.9493305 | 0.00040162 | 0.001064698 | 1 |
| Obesity | mast_cell | *MR1* | 2.772785064 | 0.00024898 | 0.000976811 | 1 |
| Obesity | mast_cell | *CENPH* | 4.416137846 | 0.006406755 | 0.008924943 | 1 |
| Obesity | mast_cell | *CARHSP1* | 2.061673454 | 0.000512856 | 0.001145661 | 1 |
| Obesity | mast_cell | *CAST* | 2.731870919 | 0.001665976 | 0.002727165 | 1 |
| Obesity | mast_cell | *SCOC* | 3.4042115 | 0.000777263 | 0.001482613 | 1 |
| Obesity | mast_cell | *CETN3* | 4.21629196 | 2.64E-05 | 0.000562671 | 0.128288986 |
| Obesity | mast_cell | *SYCP2L* | -0.800945275 | 0.002482269 | 0.003833218 | 1 |
| Obesity | mast_cell | *MERTK* | 0.750165631 | 0.002470903 | 0.00381688 | 1 |
| Obesity | mast_cell | *TMEM87B* | 2.716211636 | 0.000278094 | 0.000992498 | 1 |
| Obesity | mast_cell | *PLA2R1* | 2.85630358 | 2.19E-06 | 0.00013132 | 0.010621857 |
| Obesity | mast_cell | *SLC25A27* | 1.810424973 | 0.000478479 | 0.001129371 | 1 |
| Obesity | mast_cell | *FRMD1* | 4.248705631 | 4.50E-05 | 0.000747028 | 0.21813232 |

| | | | | | | |
|---|---|---|---|---|---|---|
| Obesity | mast_cell | *INO80C* | 4.348115807 | 0.000124402 | 0.000976811 | 0.603598141 |
| Obesity | mast_cell | *LPCAT1* | 10.67444077 | 0.000580836 | 0.001209016 | 1 |
| Obesity | mast_cell | *ADPRHL1* | -5.03387412 | 0.011058718 | 0.014777444 | 1 |
| Obesity | mast_cell | *CFDP1* | 2.494165945 | 0.00024235 | 0.000976811 | 1 |
| Obesity | mast_cell | *ZDHHC7* | 6.412132833 | 0.000266624 | 0.000985747 | 1 |
| Obesity | mast_cell | *JAZF1* | 2.921363918 | 2.05E-06 | 0.000129429 | 0.009966054 |
| Obesity | mast_cell | *FBXO36* | 4.619858977 | 0.000236222 | 0.000976811 | 1 |
| Obesity | mast_cell | *MCOLN2* | 2.655014949 | 0.000543573 | 0.00117408 | 1 |
| Obesity | mast_cell | *GDPD1* | 2.052915985 | 0.000580388 | 0.001208894 | 1 |
| Obesity | mast_cell | *SEMA3D* | 1.652802643 | 0.00040663 | 0.001071102 | 1 |
| Obesity | mast_cell | *PPP2R5E* | 3.70572688 | 0.000158526 | 0.000976811 | 0.769167294 |
| Obesity | mast_cell | *ANKRD29* | 1.666820746 | 0.000458516 | 0.001112606 | 1 |
| Obesity | mast_cell | *DNAAF1* | 3.889092857 | 0.019006538 | 0.02438385 | 1 |
| Obesity | mast_cell | *ANKH* | 2.454383351 | 0.000245698 | 0.000976811 | 1 |
| Obesity | mast_cell | *UBASH3B* | 1.702861722 | 0.001429427 | 0.002413215 | 1 |
| Obesity | mast_cell | *ROBO4* | 1.835870131 | 0.000451229 | 0.001106421 | 1 |
| Obesity | mast_cell | *ROBO3* | 1.809638804 | 0.000300481 | 0.000998908 | 1 |
| Obesity | mast_cell | *CC2D1B* | 4.720408808 | 2.60E-05 | 0.00056111 | 0.125974439 |
| Obesity | mast_cell | *LRRK1* | 2.046075503 | 0.000350824 | 0.001031837 | 1 |
| Obesity | mast_cell | *ABCA5* | 2.977891047 | 0.00038459 | 0.001052047 | 1 |
| Obesity | mast_cell | *ENPP3* | -0.781513706 | 0.002175313 | 0.003426824 | 1 |
| Obesity | mast_cell | *MIA3* | 3.138260523 | 0.000136038 | 0.000976811 | 0.660058468 |
| Obesity | mast_cell | *NEIL2* | 3.067188753 | 0.019870549 | 0.025391599 | 1 |
| Obesity | mast_cell | *PGM5* | 1.279091442 | 0.000763445 | 0.001462564 | 1 |
| Obesity | mast_cell | *CCSAP* | 2.545252382 | 0.000494791 | 0.001138685 | 1 |
| Obesity | mast_cell | *GBP5* | 1.530461245 | 0.019306503 | 0.02474905 | 1 |
| Obesity | mast_cell | *ATP5G3* | 2.864616734 | 0.000353817 | 0.001031837 | 1 |
| Obesity | mast_cell | *LY96* | 1.576736927 | 0.002449145 | 0.003789302 | 1 |
| Obesity | mast_cell | *PDE1C* | 1.602065898 | 0.000481406 | 0.001130443 | 1 |
| Obesity | mast_cell | *XPC* | 3.016617921 | 0.00042179 | 0.00108454 | 1 |
| Obesity | mast_cell | *FLCN* | 4.376534783 | 0.002397089 | 0.003725393 | 1 |
| Obesity | mast_cell | *OXNAD1* | 2.082470287 | 0.000364073 | 0.001037687 | 1 |
| Obesity | mast_cell | *APCDD1* | 1.89660966 | 0.004463871 | 0.006442208 | 1 |
| Obesity | mast_cell | *MPPE1* | 2.138770597 | 0.004115778 | 0.005995123 | 1 |
| Obesity | mast_cell | *RAB6B* | 3.170251021 | 0.000548747 | 0.001175269 | 1 |
| Obesity | mast_cell | *ANKRD40* | 2.255594859 | 0.000627917 | 0.001274751 | 1 |
| Obesity | mast_cell | *CYP2U1* | 2.615448664 | 0.001534165 | 0.002562398 | 1 |
| Obesity | mast_cell | *PROM2* | -3.021417187 | 0.028678226 | 0.035742808 | 1 |
| Obesity | mast_cell | *AK9* | 3.369410347 | 4.37E-05 | 0.00074212 | 0.212192933 |
| Obesity | mast_cell | *MARCKS* | 3.292394018 | 0.001000275 | 0.00178826 | 1 |
| Obesity | mast_cell | *MARVELD1* | 2.817005151 | 0.000486018 | 0.001132608 | 1 |
| Obesity | mast_cell | *GOLGA7B* | 13.77367625 | 0.000316846 | 0.000998908 | 1 |
| Obesity | mast_cell | *TRMT44* | 2.703604099 | 0.000275635 | 0.000990086 | 1 |
| Obesity | mast_cell | *MOV10* | 2.616945097 | 0.001297292 | 0.002224192 | 1 |
| Obesity | mast_cell | *HEATR3* | 1.600992403 | 0.004316766 | 0.006250357 | 1 |
| Obesity | mast_cell | *CNOT8* | 2.939648429 | 0.000448955 | 0.001104518 | 1 |
| Obesity | mast_cell | *LRGUK* | 1.339561988 | 0.018690821 | 0.024010554 | 1 |
| Obesity | mast_cell | *NUP205* | 6.097350051 | 0.000249583 | 0.000976811 | 1 |
| Obesity | mast_cell | *PDIA4* | 3.275935072 | 0.000311734 | 0.000998908 | 1 |
| Obesity | mast_cell | *KDM8* | 2.946211613 | 0.000389947 | 0.001054382 | 1 |

| | | | | | | |
|---|---|---|---|---|---|---|
| Obesity | mast_cell | *ALS2CR12* | 3.130551863 | 7.62E-05 | 0.000935479 | 0.369514099 |
| Obesity | mast_cell | *FZD7* | 3.31946948 | 0.006183413 | 0.008640956 | 1 |
| Obesity | mast_cell | *DEPTOR* | 2.903814037 | 0.000207856 | 0.000976811 | 1 |
| Obesity | mast_cell | *RRAGA* | 2.639522184 | 0.000512004 | 0.001145411 | 1 |
| Obesity | mast_cell | *TMBIM4* | 4.191236622 | 8.63E-05 | 0.000971162 | 0.418526221 |
| Obesity | mast_cell | *PSD3* | 2.1887906 | 0.004442601 | 0.006415327 | 1 |
| Obesity | mast_cell | *KCNMA1* | 1.025994897 | 0.003128292 | 0.004684714 | 1 |
| Obesity | mast_cell | *NDUFAF6* | 3.491198102 | 9.55E-05 | 0.000976811 | 0.463533509 |
| Obesity | mast_cell | *DRAM2* | 1.675290599 | 0.000505494 | 0.001144783 | 1 |
| Obesity | mast_cell | *ART3* | 3.47350631 | 0.001547448 | 0.002578372 | 1 |
| Obesity | mast_cell | *WHAMM* | 1.964543026 | 0.002489212 | 0.003841493 | 1 |
| Obesity | mast_cell | *N6AMT1* | 3.250096651 | 0.005557818 | 0.00785052 | 1 |
| Obesity | mast_cell | *NAA11* | 3.371544692 | 0.00327318 | 0.004877601 | 1 |
| Obesity | mast_cell | *SFXN2* | 1.217291856 | 0.0194482 | 0.024903169 | 1 |
| Obesity | mast_cell | *C14orf2* | 2.943740297 | 0.000511379 | 0.001145411 | 1 |
| Obesity | mast_cell | *TDRD9* | 5.255584028 | 0.002203848 | 0.003462166 | 1 |
| Obesity | mast_cell | *RPL30* | 3.472607136 | 0.000115179 | 0.000976811 | 0.558849383 |
| Obesity | mast_cell | *SUPV3L1* | 2.375371531 | 0.000527608 | 0.001157825 | 1 |
| Obesity | mast_cell | *HKDC1* | 9.957409475 | 0.009017306 | 0.012234891 | 1 |
| Obesity | mast_cell | *TYSND1* | 1.880632014 | 0.000623925 | 0.001269302 | 1 |
| Obesity | mast_cell | *NODAL* | 3.511060506 | 0.000544252 | 0.00117408 | 1 |
| Obesity | mast_cell | *ZDHHC5* | 7.441987388 | 0.000224011 | 0.000976811 | 1 |
| Obesity | mast_cell | *MED19* | 1.949091399 | 0.012623439 | 0.016711849 | 1 |
| Obesity | mast_cell | *ATAD2* | 1.825642735 | 0.000338678 | 0.001022567 | 1 |
| Obesity | mast_cell | *FBRS* | 2.093258267 | 0.000371596 | 0.00104107 | 1 |
| Obesity | mast_cell | *SASS6* | 3.165670473 | 0.011184022 | 0.014924334 | 1 |
| Obesity | mast_cell | *ITGAD* | 4.873409096 | 5.94E-06 | 0.000248582 | 0.028835478 |
| Obesity | mast_cell | *VPS8* | 3.122383417 | 0.000153174 | 0.000976811 | 0.743201999 |
| Obesity | mast_cell | *MPV17L* | -3.175864189 | 0.00305306 | 0.00459047 | 1 |
| Obesity | mast_cell | *RPUSD3* | 6.011879387 | 0.00118225 | 0.002060445 | 1 |
| Obesity | mast_cell | *NTAN1* | 7.171100437 | 9.73E-06 | 0.000352485 | 0.047232977 |
| Obesity | mast_cell | *NMNAT2* | 1.832154279 | 0.000558057 | 0.001182832 | 1 |
| Obesity | mast_cell | *SLC6A1* | 7.962444519 | 0.000287343 | 0.00099443 | 1 |
| Obesity | mast_cell | *FCHO2* | 2.538342262 | 7.47E-05 | 0.000934346 | 0.362526356 |
| Obesity | mast_cell | *NECAP2* | 2.787103842 | 0.000303809 | 0.000998908 | 1 |
| Obesity | mast_cell | *STEAP2* | 0.823034732 | 0.037747996 | 0.046250828 | 1 |
| Obesity | mast_cell | *CLDN12* | 2.389692443 | 0.000496639 | 0.001138685 | 1 |
| Obesity | mast_cell | *MMP14* | 4.101528687 | 0.000346963 | 0.001029012 | 1 |
| Obesity | mast_cell | *GATAD1* | 4.125821317 | 0.000363911 | 0.001037687 | 1 |
| Obesity | mast_cell | *CLEC18A* | -10.97252005 | 5.59E-17 | 6.78E-14 | 2.71E-13 |
| Obesity | mast_cell | *DHRS4* | -2.96603835 | 0.018044968 | 0.023242417 | 1 |
| Obesity | mast_cell | *AASDH* | 4.029083364 | 3.07E-05 | 0.000621513 | 0.149169714 |
| Obesity | mast_cell | *ETS2* | 2.720910047 | 1.35E-06 | 9.24E-05 | 0.006561988 |
| Obesity | mast_cell | *LCA5L* | 1.244913091 | 0.018986438 | 0.024364506 | 1 |
| Obesity | mast_cell | *MX1* | 1.807518828 | 0.00051012 | 0.001145411 | 1 |
| Obesity | mast_cell | *SLC38A10* | 2.814553615 | 0.000265096 | 0.000985696 | 1 |
| Obesity | mast_cell | *C9orf43* | -1.449326769 | 0.010736187 | 0.014378134 | 1 |
| Obesity | mast_cell | *DGKI* | 1.481820204 | 0.016874833 | 0.021862935 | 1 |
| Obesity | mast_cell | *SLC34A2* | 33.13192904 | 0.019761099 | 0.025265047 | 1 |
| Obesity | mast_cell | *AP3S2* | 4.439657323 | 0.000380391 | 0.001048903 | 1 |

| | | | | | | |
|---|---|---|---|---|---|---|
| Obesity | mast_cell | *SPPL3* | 3.308832508 | 0.000105642 | 0.000976811 | 0.512575612 |
| Obesity | mast_cell | *C12orf43* | 2.337322 | 0.005808932 | 0.008174286 | 1 |
| Obesity | mast_cell | *PEX10* | 3.289126158 | 0.000201485 | 0.000976811 | 0.977604184 |
| Obesity | mast_cell | *KRTCAP3* | 2.915946365 | 7.00E-05 | 0.000898663 | 0.339735144 |
| Obesity | mast_cell | *EXTL1* | 35.61578303 | 0.00055862 | 0.001183074 | 1 |
| Obesity | mast_cell | *WDR66* | 2.241745049 | 0.000985503 | 0.001766406 | 1 |
| Obesity | mast_cell | *LRRC43* | 6.971510593 | 1.58E-05 | 0.00045715 | 0.076801121 |
| Obesity | mast_cell | *EYA3* | 2.251677116 | 0.000325636 | 0.001005082 | 1 |
| Obesity | mast_cell | *DZIP1L* | 1.964466905 | 0.00109305 | 0.001927075 | 1 |
| Obesity | mast_cell | *FANCC* | 2.247417208 | 0.000467468 | 0.001116682 | 1 |
| Obesity | mast_cell | *ESYT3* | 5.382447548 | 0.000205701 | 0.000976811 | 0.998062922 |
| Obesity | mast_cell | *CLSTN2* | 5.144089818 | 0.003275746 | 0.004879926 | 1 |
| Obesity | mast_cell | *COLEC12* | 1.806729785 | 0.000369174 | 0.00103942 | 1 |
| Obesity | mast_cell | *MITD1* | 2.852322475 | 0.000140576 | 0.000976811 | 0.682076548 |
| Obesity | mast_cell | *TSPAN33* | 3.513621171 | 0.002657826 | 0.004060381 | 1 |
| Obesity | mast_cell | *FAM86C1* | -5.370167943 | 0.007112947 | 0.009815704 | 1 |
| Obesity | mast_cell | *PPP1R9A* | 1.666126564 | 0.00064696 | 0.001300352 | 1 |
| Obesity | mast_cell | *DYNC1I1* | 1.69018296 | 0.000202083 | 0.000976811 | 0.980508009 |
| Obesity | mast_cell | *DUSP23* | 2.290352075 | 0.002439221 | 0.003779355 | 1 |
| Obesity | mast_cell | *RNF166* | 1.879551678 | 0.002915748 | 0.004403115 | 1 |
| Obesity | mast_cell | *F11R* | 2.177575862 | 0.009397004 | 0.012700352 | 1 |
| Obesity | mast_cell | *SPATA2L* | 3.678419695 | 0.013363346 | 0.01758583 | 1 |
| Obesity | mast_cell | *FGF17* | 13.91922017 | 0.00048414 | 0.001131723 | 1 |
| Obesity | mast_cell | *SLC5A11* | -3.363140414 | 1.03E-06 | 7.48E-05 | 0.005013714 |
| Obesity | mast_cell | *CCAR2* | 2.805635349 | 0.000777747 | 0.001482613 | 1 |
| Obesity | mast_cell | *CDC42SE2* | 2.67642172 | 0.000209199 | 0.000976811 | 1 |
| Obesity | mast_cell | *RAPGEF6* | 2.133421903 | 0.022739952 | 0.02871792 | 1 |
| Obesity | mast_cell | *IFNAR2* | 2.295608637 | 0.003782418 | 0.005551066 | 1 |
| Obesity | mast_cell | *CLIC6* | -0.399976161 | 0.001937244 | 0.003092961 | 1 |
| Obesity | mast_cell | *CBR3* | 1.950795472 | 0.000845379 | 0.001572223 | 1 |
| Obesity | mast_cell | *SIM2* | -1.223294047 | 0.018447195 | 0.023729 | 1 |
| Obesity | mast_cell | *HLCS* | 2.849287142 | 0.000278553 | 0.000992498 | 1 |
| Obesity | mast_cell | *ARHGAP27* | 4.351076928 | 0.001842706 | 0.002970369 | 1 |
| Obesity | mast_cell | *IRX6* | 1.644460136 | 0.003118522 | 0.004672967 | 1 |
| Obesity | mast_cell | *HK2* | 1.590409023 | 0.000697334 | 0.001368162 | 1 |
| Obesity | mast_cell | *THEM4* | 1.873254752 | 0.000885699 | 0.001626575 | 1 |
| Obesity | mast_cell | *AMFR* | 3.437650518 | 0.00012732 | 0.000976811 | 0.617755584 |
| Obesity | mast_cell | *TMEM69* | 3.134990628 | 0.00043806 | 0.001098433 | 1 |
| Obesity | mast_cell | *ACE* | 2.70084593 | 0.000464445 | 0.001116682 | 1 |
| Obesity | mast_cell | *TEPP* | -2.163141383 | 0.001565348 | 0.002598381 | 1 |
| Obesity | mast_cell | *CHCHD6* | 2.678461706 | 0.000417997 | 0.00108267 | 1 |
| Obesity | mast_cell | *CTBP1* | 4.43938809 | 0.001093593 | 0.001927075 | 1 |
| Obesity | mast_cell | *LRRC36* | 2.012168729 | 0.000220915 | 0.000976811 | 1 |
| Obesity | mast_cell | *ZFYVE28* | 2.540226457 | 0.000476852 | 0.001128628 | 1 |
| Obesity | mast_cell | *RGS12* | 2.545625403 | 0.000519174 | 0.001151636 | 1 |
| Obesity | mast_cell | *LYPD5* | -1.439586809 | 0.001618204 | 0.002666053 | 1 |
| Obesity | mast_cell | *CCDC107* | 1.838301115 | 0.000900017 | 0.001645397 | 1 |
| Obesity | mast_cell | *ATAD3B* | 3.867979714 | 0.002223418 | 0.00348563 | 1 |
| Obesity | mast_cell | *SSU72* | 6.637381744 | 0.000688113 | 0.001355939 | 1 |
| Obesity | mast_cell | *CPAMD8* | -2.280130725 | 0.008350999 | 0.011381755 | 1 |

| | | | | | | |
|---|---|---|---|---|---|---|
| Obesity | mast_cell | *CCDC58* | 3.530217999 | 0.00031467 | 0.000998908 | 1 |
| Obesity | mast_cell | *CILP2* | -5.934437603 | 0.005102925 | 0.007273618 | 1 |
| Obesity | mast_cell | *SLC37A1* | 2.439453389 | 0.000463345 | 0.001116261 | 1 |
| Obesity | mast_cell | *PDE9A* | 1.899486671 | 0.000250792 | 0.000976811 | 1 |
| Obesity | mast_cell | *NDUFV3* | 1.202964394 | 0.005738374 | 0.008086724 | 1 |
| Obesity | mast_cell | *CSTB* | 3.11523104 | 0.001134727 | 0.001989052 | 1 |
| Obesity | mast_cell | *FTCD* | -1.554630485 | 0.006868513 | 0.009508139 | 1 |
| Obesity | mast_cell | *LSS* | 1.520281976 | 0.000290938 | 0.000997619 | 1 |
| Obesity | mast_cell | *VAV2* | 5.519123025 | 0.000231966 | 0.000976811 | 1 |
| Obesity | mast_cell | *MCM3AP* | 1.457074697 | 0.027146217 | 0.033955515 | 1 |
| Obesity | mast_cell | *PCNT* | 1.87766694 | 0.004266018 | 0.006184261 | 1 |
| Obesity | mast_cell | *DIP2A* | 4.01856176 | 0.000144529 | 0.000976811 | 0.701255287 |
| Obesity | mast_cell | *PRMT2* | 3.211767181 | 8.37E-05 | 0.000964174 | 0.405917221 |
| Obesity | mast_cell | *ZNF208* | 2.849807404 | 0.00011219 | 0.000976811 | 0.544343492 |
| Obesity | mast_cell | *ADAMTS13* | 2.211981698 | 0.00270924 | 0.004127231 | 1 |
| Obesity | mast_cell | *CACFD1* | 5.281558988 | 0.000840417 | 0.001566859 | 1 |
| Obesity | mast_cell | *GPSM1* | 7.500484284 | 3.22E-06 | 0.000162615 | 0.015611076 |
| Obesity | mast_cell | *TAOK1* | 3.144706398 | 0.000142572 | 0.000976811 | 0.691758562 |
| Obesity | mast_cell | *S100A1* | 9.946436454 | 8.71E-05 | 0.000971162 | 0.422455288 |
| Obesity | mast_cell | *ZBTB7B* | 2.827551042 | 0.000602776 | 0.001238217 | 1 |
| Obesity | mast_cell | *VPS11* | 2.890848254 | 0.014932992 | 0.019513837 | 1 |
| Obesity | mast_cell | *FDPS* | 2.346999929 | 0.00050004 | 0.001141738 | 1 |
| Obesity | mast_cell | *CCR5* | 4.293954956 | 0.000817158 | 0.001532014 | 1 |
| Obesity | mast_cell | *NBEAL2* | 5.131483402 | 6.98E-06 | 0.000277505 | 0.033855603 |
| Obesity | mast_cell | *PTH1R* | 2.063397285 | 0.000109596 | 0.000976811 | 0.531759807 |
| Obesity | mast_cell | *FCRL3* | 2.418771838 | 0.01296707 | 0.017120061 | 1 |
| Obesity | mast_cell | *LY6K* | 3.874057634 | 0.001600536 | 0.002642602 | 1 |
| Obesity | mast_cell | *IER2* | 3.019800355 | 0.000191614 | 0.000976811 | 0.929711312 |
| Obesity | mast_cell | *CPSF4* | 2.907239908 | 0.000109865 | 0.000976811 | 0.533066483 |
| Obesity | mast_cell | *VPS28* | 3.669729303 | 0.002137076 | 0.003370966 | 1 |
| Obesity | mast_cell | *MUM1* | 3.541119431 | 0.000197914 | 0.000976811 | 0.960280387 |
| Obesity | mast_cell | *SH2B2* | 2.919117749 | 0.004346312 | 0.006289385 | 1 |
| Obesity | mast_cell | *RPL8* | 2.671069989 | 0.000220874 | 0.000976811 | 1 |
| Obesity | mast_cell | *DVL3* | 4.733980665 | 0.00102354 | 0.001824474 | 1 |
| Obesity | mast_cell | *ABCF3* | 3.92410828 | 2.64E-06 | 0.000144867 | 0.012825442 |
| Obesity | mast_cell | *PCYT1A* | 1.429657876 | 0.008392306 | 0.011432103 | 1 |
| Obesity | mast_cell | *FBXO27* | 1.984766121 | 0.00020621 | 0.000976811 | 1 |
| Obesity | mast_cell | *BDH1* | 2.674127059 | 0.001074896 | 0.001897157 | 1 |
| Obesity | mast_cell | *LRRC56* | -1.674381276 | 0.005039119 | 0.007184779 | 1 |
| Obesity | mast_cell | *PGAP3* | 6.00649596 | 0.000201265 | 0.000976811 | 0.976538162 |
| Obesity | mast_cell | *ZNF577* | 5.248562399 | 5.79E-05 | 0.000833899 | 0.281024057 |
| Obesity | mast_cell | *TMEM143* | 3.030536871 | 0.000370035 | 0.00103942 | 1 |
| Obesity | mast_cell | *ASB16* | 3.11258386 | 0.000631333 | 0.001279531 | 1 |
| Obesity | mast_cell | *IP6K3* | 25.25865097 | 0.003631373 | 0.005355447 | 1 |
| Obesity | mast_cell | *LEMD2* | 3.718119448 | 0.002205776 | 0.003463568 | 1 |
| Obesity | mast_cell | *ASGR2* | 17.45238461 | 0.000208517 | 0.000976811 | 1 |
| Obesity | mast_cell | *POLR3K* | -3.762484539 | 0.002896141 | 0.004376749 | 1 |
| Obesity | mast_cell | *SNRNP25* | 4.409424319 | 0.000230702 | 0.000976811 | 1 |
| Obesity | mast_cell | *WDR90* | 1.729414094 | 0.013780412 | 0.01809052 | 1 |
| Obesity | mast_cell | *JMJD8* | 3.42846888 | 0.000247243 | 0.000976811 | 1 |

| | | | | | | |
|---|---|---|---|---|---|---|
| Obesity | mast_cell | *MEIOB* | 3.341319705 | 0.000233502 | 0.000976811 | 1 |
| Obesity | mast_cell | *SHANK2* | 5.839253908 | 0.000144866 | 0.000976811 | 0.702890423 |
| Obesity | mast_cell | *ASRGL1* | 2.681129358 | 0.000477899 | 0.001129371 | 1 |
| Obesity | mast_cell | *TTC9C* | 2.754749455 | 0.001033589 | 0.001838083 | 1 |
| Obesity | mast_cell | *RPS6KA4* | 8.998900176 | 0.000146905 | 0.000976811 | 0.712784191 |
| Obesity | mast_cell | *TPCN2* | 2.923265023 | 0.00036733 | 0.001037687 | 1 |
| Obesity | mast_cell | *PDZK1IP1* | -2.870504869 | 0.004711327 | 0.006755129 | 1 |
| Obesity | mast_cell | *CMPK1* | 3.605816058 | 0.00553051 | 0.007816496 | 1 |
| Obesity | mast_cell | *COA7* | -4.340171451 | 0.040962131 | 0.049899137 | 1 |
| Obesity | mast_cell | *C1orf123* | 2.963495245 | 0.000445363 | 0.001103405 | 1 |
| Obesity | mast_cell | *USP24* | 4.214600244 | 0.000217269 | 0.000976811 | 1 |
| Obesity | mast_cell | *ZSWIM5* | 3.941452944 | 8.84E-05 | 0.000976811 | 0.429063416 |
| Obesity | mast_cell | *AK4* | 1.703546026 | 0.000602628 | 0.001238217 | 1 |
| Obesity | mast_cell | *RBP7* | 1.780396011 | 0.000582842 | 0.001210077 | 1 |
| Obesity | mast_cell | *SLC25A34* | 3.294542167 | 0.010473387 | 0.01405334 | 1 |
| Obesity | mast_cell | *PDPN* | 2.031247065 | 0.000574938 | 0.001201079 | 1 |
| Obesity | mast_cell | *LAPTM5* | 3.087314324 | 0.000249999 | 0.000976811 | 1 |
| Obesity | mast_cell | *SDC3* | 2.064382871 | 0.000480352 | 0.001129748 | 1 |
| Obesity | mast_cell | *SYNC* | 3.390207278 | 0.003712123 | 0.005457571 | 1 |
| Obesity | mast_cell | *CAMK2N1* | 5.187364637 | 1.01E-06 | 7.48E-05 | 0.004885653 |
| Obesity | mast_cell | *MEGF6* | 3.57468034 | 0.000320709 | 0.000999138 | 1 |
| Obesity | mast_cell | *OMA1* | 2.570628219 | 0.000291391 | 0.000998325 | 1 |
| Obesity | mast_cell | *TM2D1* | 2.649106629 | 0.00024372 | 0.000976811 | 1 |
| Obesity | mast_cell | *NEXN* | 2.52951393 | 0.000251504 | 0.000976811 | 1 |
| Obesity | mast_cell | *LRRIQ3* | 2.395747121 | 0.000442973 | 0.00110084 | 1 |
| Obesity | mast_cell | *TYW3* | 2.137615441 | 0.000835781 | 0.001560296 | 1 |
| Obesity | mast_cell | *HFM1* | 2.001013736 | 0.000423132 | 0.001086605 | 1 |
| Obesity | mast_cell | *ARPC5* | 2.600065662 | 0.001394307 | 0.002360495 | 1 |
| Obesity | mast_cell | *ZNF496* | 4.900019815 | 7.72E-05 | 0.000938986 | 0.374655467 |
| Obesity | mast_cell | *TDRD5* | -4.440019991 | 0.001830691 | 0.002955911 | 1 |
| Obesity | mast_cell | *BROX* | 1.870955342 | 0.029543801 | 0.036736679 | 1 |
| Obesity | mast_cell | *ACP6* | 3.135783647 | 0.000427982 | 0.001092079 | 1 |
| Obesity | mast_cell | *CNST* | 3.499060631 | 0.000159473 | 0.000976811 | 0.773763041 |
| Obesity | mast_cell | *PM20D1* | 5.177988697 | 2.06E-05 | 0.000522259 | 0.099983853 |
| Obesity | mast_cell | *PKDCC* | 1.563803988 | 0.00060267 | 0.001238217 | 1 |
| Obesity | mast_cell | *B3GALNT2* | 2.201986959 | 0.000363851 | 0.001037687 | 1 |
| Obesity | mast_cell | *MRPL55* | 2.410720303 | 0.00015565 | 0.000976811 | 0.755213845 |
| Obesity | mast_cell | *KIAA1841* | 5.235108496 | 0.000147296 | 0.000976811 | 0.714678426 |
| Obesity | mast_cell | *KCNJ3* | 5.430208896 | 0.021175886 | 0.026889661 | 1 |
| Obesity | mast_cell | *CLHC1* | 1.972452018 | 4.99E-06 | 0.000228436 | 0.024214179 |
| Obesity | mast_cell | *DUSP19* | -4.716820061 | 0.0108534 | 0.014531097 | 1 |
| Obesity | mast_cell | *NUP35* | 2.847358792 | 0.000567248 | 0.001191982 | 1 |
| Obesity | mast_cell | *C2orf48* | 23.09556365 | 0.000193752 | 0.000976811 | 0.940083325 |
| Obesity | mast_cell | *CCDC74A* | 3.826036412 | 0.040713923 | 0.049646633 | 1 |
| Obesity | mast_cell | *H3F3A* | 1.791765324 | 0.000624536 | 0.001270012 | 1 |
| Obesity | mast_cell | *NOSTRIN* | 1.829504584 | 0.000549363 | 0.001175269 | 1 |
| Obesity | mast_cell | *INHBB* | 3.920634885 | 0.000651456 | 0.001307763 | 1 |
| Obesity | mast_cell | *RPRD2* | 3.476141159 | 4.25E-05 | 0.000731263 | 0.206216143 |
| Obesity | mast_cell | *CTSS* | 0.706643412 | 0.037362401 | 0.04583625 | 1 |
| Obesity | mast_cell | *PACRGL* | 1.973740764 | 0.00037875 | 0.001048885 | 1 |

| | | | | | | |
|---|---|---|---|---|---|---|
| Obesity | mast_cell | *C1QTNF7* | 2.86345147 | 0.000176275 | 0.000976811 | 0.855287122 |
| Obesity | mast_cell | *VPS72* | 2.672667203 | 0.000383303 | 0.001052047 | 1 |
| Obesity | mast_cell | *ERCC3* | 5.323064998 | 0.000148892 | 0.000976811 | 0.722426376 |
| Obesity | mast_cell | *RNF149* | 2.112226932 | 0.000496272 | 0.001138685 | 1 |
| Obesity | mast_cell | *DHX57* | 2.754678971 | 0.000343425 | 0.001027777 | 1 |
| Obesity | mast_cell | *S100A9* | 1.739403015 | 0.001808999 | 0.002923806 | 1 |
| Obesity | mast_cell | *TDRD10* | 1.665142661 | 0.000497527 | 0.001139218 | 1 |
| Obesity | mast_cell | *DCAF16* | 3.139848239 | 0.001904942 | 0.003048409 | 1 |
| Obesity | mast_cell | *GNPDA2* | 2.887421357 | 0.000128933 | 0.000976811 | 0.625583624 |
| Obesity | mast_cell | *NIPAL1* | 2.17252092 | 0.001193859 | 0.002075005 | 1 |
| Obesity | mast_cell | *ANTXR2* | 5.247995575 | 2.92E-07 | 3.21E-05 | 0.001418124 |
| Obesity | mast_cell | *GPR155* | 1.523505949 | 0.00057986 | 0.001208539 | 1 |
| Obesity | mast_cell | *DAPL1* | -1.806492836 | 0.003782599 | 0.005551066 | 1 |
| Obesity | mast_cell | *PBXIP1* | 6.711895555 | 5.00E-05 | 0.000765648 | 0.242710573 |
| Obesity | mast_cell | *CLDN1* | 1.708428858 | 0.000259429 | 0.00098093 | 1 |
| Obesity | mast_cell | *DCST2* | 4.852168196 | 1.17E-05 | 0.000377668 | 0.056650193 |
| Obesity | mast_cell | *LINC01116* | 4.889867771 | 9.61E-05 | 0.000976811 | 0.466112166 |
| Obesity | mast_cell | *POGLUT1* | 1.908617147 | 0.016207114 | 0.021059699 | 1 |
| Obesity | mast_cell | *LRRC58* | 5.428534055 | 0.040872081 | 0.049814452 | 1 |
| Obesity | mast_cell | *LMOD1* | 1.091790746 | 0.02096322 | 0.026661479 | 1 |
| Obesity | mast_cell | *TMEM169* | 2.565471305 | 0.006877933 | 0.009517717 | 1 |
| Obesity | mast_cell | *ARPC2* | 3.453617432 | 0.000192954 | 0.000976811 | 0.936213816 |
| Obesity | mast_cell | *CCT3* | 2.009896658 | 0.000359313 | 0.001035264 | 1 |
| Obesity | mast_cell | *SERPINI1* | 2.739468995 | 0.000247565 | 0.000976811 | 1 |
| Obesity | mast_cell | *SUCLG1* | 2.551183109 | 0.000185611 | 0.000976811 | 0.900585206 |
| Obesity | mast_cell | *PRKCI* | 1.483275803 | 0.004483734 | 0.00646895 | 1 |
| Obesity | mast_cell | *AIM2* | 3.639872088 | 0.000675216 | 0.001336113 | 1 |
| Obesity | mast_cell | *GTPBP8* | 3.175365075 | 0.000347999 | 0.001029993 | 1 |
| Obesity | mast_cell | *WDFY3* | 2.547179947 | 0.000222956 | 0.000976811 | 1 |
| Obesity | mast_cell | *PTPN13* | 1.458009583 | 0.001535881 | 0.002564381 | 1 |
| Obesity | mast_cell | *C4orf36* | 6.848977446 | 0.000233646 | 0.000976811 | 1 |
| Obesity | mast_cell | *PPM1K* | 4.003175139 | 7.66E-05 | 0.000936854 | 0.371569365 |
| Obesity | mast_cell | *TIPARP* | 1.836615329 | 0.000481418 | 0.001130443 | 1 |
| Obesity | mast_cell | *DCLK3* | 7.85798287 | 0.000907254 | 0.001656362 | 1 |
| Obesity | mast_cell | *RPP14* | 9.682565556 | 0.002441635 | 0.003780982 | 1 |
| Obesity | mast_cell | *DNASE1L3* | 2.46673213 | 0.015592613 | 0.020310164 | 1 |
| Obesity | mast_cell | *IL17RC* | 2.176840775 | 0.001251591 | 0.00215574 | 1 |
| Obesity | mast_cell | *CRELD1* | 1.975657505 | 0.000583471 | 0.001210865 | 1 |
| Obesity | mast_cell | *TTC14* | 2.10013055 | 0.002206791 | 0.003464041 | 1 |
| Obesity | mast_cell | *RCHY1* | 2.625134059 | 0.000782549 | 0.001487241 | 1 |
| Obesity | mast_cell | *CCDC158* | 5.396096143 | 0.010537238 | 0.014131199 | 1 |
| Obesity | mast_cell | *CPA3* | -0.090027302 | 0.001568335 | 0.00260067 | 1 |
| Obesity | mast_cell | *SLC4A1AP* | 2.326589177 | 0.000414629 | 0.001080439 | 1 |
| Obesity | mast_cell | *KIAA1143* | 1.858233188 | 0.002378033 | 0.003702894 | 1 |
| Obesity | mast_cell | *CDCP1* | 1.809729073 | 0.003332328 | 0.004953571 | 1 |
| Obesity | mast_cell | *CLEC3B* | 5.012275022 | 0.000133568 | 0.000976811 | 0.648070526 |
| Obesity | mast_cell | *NMNAT3* | 2.226105656 | 0.000562684 | 0.001188051 | 1 |
| Obesity | mast_cell | *MEAF6* | 1.89470581 | 0.001135187 | 0.00198914 | 1 |
| Obesity | mast_cell | *SNIP1* | 6.477749816 | 0.000175906 | 0.000976811 | 0.853496916 |
| Obesity | mast_cell | *RPN1* | 3.2358792 | 0.000252056 | 0.000976811 | 1 |

| | | | | | | |
|---|---|---|---|---|---|---|
| Obesity | mast_cell | *SENP2* | 2.932304382 | 0.006713841 | 0.009309962 | 1 |
| Obesity | mast_cell | *IFT122* | 2.930556107 | 0.001561072 | 0.002595186 | 1 |
| Obesity | mast_cell | *SFMBT1* | 3.128849599 | 0.000226977 | 0.000976811 | 1 |
| Obesity | mast_cell | *UVSSA* | 2.110109424 | 0.001247255 | 0.002150561 | 1 |
| Obesity | mast_cell | *FAM208A* | 4.051425345 | 0.000191379 | 0.000976811 | 0.92857077 |
| Obesity | mast_cell | *LRPAP1* | 5.667145517 | 0.00035191 | 0.001031837 | 1 |
| Obesity | mast_cell | *PIGX* | 1.483098216 | 0.025897806 | 0.032460902 | 1 |
| Obesity | mast_cell | *ABLIM2* | 2.427627158 | 0.000448954 | 0.001104518 | 1 |
| Obesity | mast_cell | *ERMAP* | 2.007527856 | 0.000664885 | 0.001322683 | 1 |
| Obesity | mast_cell | *METAP1* | 4.974951798 | 0.000963169 | 0.001739949 | 1 |
| Obesity | mast_cell | *BDH2* | 1.925227216 | 0.000667552 | 0.001326001 | 1 |
| Obesity | mast_cell | *CDC25A* | 10.05885939 | 0.000801746 | 0.001513647 | 1 |
| Obesity | mast_cell | *ZNF589* | 3.1887416 | 0.002572404 | 0.003952038 | 1 |
| Obesity | mast_cell | *RNF123* | 9.187773338 | 6.41E-19 | 1.04E-15 | 3.11E-15 |
| Obesity | mast_cell | *MFSD8* | 5.218736076 | 0.000863762 | 0.001594132 | 1 |
| Obesity | mast_cell | *MST1R* | 7.107698504 | 4.22E-19 | 1.02E-15 | 2.05E-15 |
| Obesity | mast_cell | *RAD54L2* | 2.557441292 | 0.000700074 | 0.001372427 | 1 |
| Obesity | mast_cell | *ETNPPL* | 17.65403364 | 0.000511143 | 0.001145411 | 1 |
| Obesity | mast_cell | *PRSS12* | -0.732203638 | 0.00733995 | 0.010103103 | 1 |
| Obesity | mast_cell | *HMGB2* | 4.13083389 | 0.001019024 | 0.001817092 | 1 |
| Obesity | mast_cell | *ANXA5* | 2.093332803 | 0.001310353 | 0.002243413 | 1 |
| Obesity | mast_cell | *MAP9* | 2.934320851 | 0.000360537 | 0.001035657 | 1 |
| Obesity | mast_cell | *CEP44* | 2.218377272 | 0.001807566 | 0.002922462 | 1 |
| Obesity | mast_cell | *HPGD* | -0.126287086 | 0.001583739 | 0.002619946 | 1 |
| Obesity | mast_cell | *C4orf45* | 5.387148198 | 0.003854115 | 0.005644481 | 1 |
| Obesity | mast_cell | *TMEM144* | 2.889492404 | 0.002183439 | 0.003436279 | 1 |
| Obesity | mast_cell | *IL15* | 2.650204233 | 1.00E-04 | 0.000976811 | 0.484990484 |
| Obesity | mast_cell | *ARFIP1* | 3.346235214 | 4.95E-05 | 0.000765648 | 0.240283269 |
| Obesity | mast_cell | *LSM6* | 2.171958318 | 0.002980359 | 0.004490901 | 1 |
| Obesity | mast_cell | *ITGA2* | 2.147737745 | 0.000356563 | 0.001033017 | 1 |
| Obesity | mast_cell | *MOCS2* | 8.401637466 | 0.022814184 | 0.028796676 | 1 |
| Obesity | mast_cell | *NDUFAF2* | 2.494176259 | 0.000161066 | 0.000976811 | 0.781491434 |
| Obesity | mast_cell | *RNF180* | 1.52213689 | 0.000766754 | 0.001466991 | 1 |
| Obesity | mast_cell | *PGGT1B* | 4.681935811 | 0.000460674 | 0.001114279 | 1 |
| Obesity | mast_cell | *F2RL2* | 6.097175596 | 0.000403005 | 0.001065021 | 1 |
| Obesity | mast_cell | *CCDC112* | 3.361769312 | 0.000321681 | 0.00100051 | 1 |
| Obesity | mast_cell | *CMBL* | 2.178950492 | 0.000432323 | 0.001094797 | 1 |
| Obesity | mast_cell | *C5orf63* | 2.136323323 | 0.00038896 | 0.001054382 | 1 |
| Obesity | mast_cell | *WDR41* | 1.908343404 | 0.00026977 | 0.000986064 | 1 |
| Obesity | mast_cell | *NDUFS4* | 2.110015133 | 0.000263045 | 0.000983277 | 1 |
| Obesity | mast_cell | *SERINC5* | 2.238888043 | 0.000373735 | 0.001043962 | 1 |
| Obesity | mast_cell | *PRIMPOL* | 4.861075589 | 0.000318694 | 0.000998908 | 1 |
| Obesity | mast_cell | *ERAP2* | 1.765265838 | 0.000170777 | 0.000976811 | 0.828611391 |
| Obesity | mast_cell | *UTP15* | 3.074400365 | 0.00084977 | 0.001577296 | 1 |
| Obesity | mast_cell | *TLR3* | 6.622622126 | 0.001524724 | 0.002547507 | 1 |
| Obesity | mast_cell | *KLKB1* | -1.234936749 | 0.013433093 | 0.017668031 | 1 |
| Obesity | mast_cell | *NSA2* | 1.594023521 | 0.000297845 | 0.000998908 | 1 |
| Obesity | mast_cell | *CCDC127* | 2.713898019 | 0.001025178 | 0.001826722 | 1 |
| Obesity | mast_cell | *GRIK2* | 1.18427702 | 0.028163231 | 0.035137052 | 1 |
| Obesity | mast_cell | *DCBLD1* | 2.075709578 | 0.000125857 | 0.000976811 | 0.6106595 |

| | | | | | | |
|---|---|---|---|---|---|---|
| Obesity | mast_cell | *PDSS2* | 2.961415087 | 0.000191548 | 0.000976811 | 0.929392365 |
| Obesity | mast_cell | *RAET1E* | 2.478699997 | 0.000576124 | 0.00120282 | 1 |
| Obesity | mast_cell | *DAGLB* | 2.129579607 | 0.002444868 | 0.003783891 | 1 |
| Obesity | mast_cell | *STK17A* | 4.110879254 | 0.000237087 | 0.000976811 | 1 |
| Obesity | mast_cell | *GALNT10* | 2.439043108 | 0.009633871 | 0.012991535 | 1 |
| Obesity | mast_cell | *SAP30L* | 2.791159809 | 0.000354426 | 0.001031837 | 1 |
| Obesity | mast_cell | *RPS14* | 3.274800064 | 0.000154011 | 0.000976811 | 0.747259444 |
| Obesity | mast_cell | *RP9* | 2.21226617 | 0.0004761 | 0.001127398 | 1 |
| Obesity | mast_cell | *CAMLG* | 2.694485439 | 0.000101136 | 0.000976811 | 0.490712 |
| Obesity | mast_cell | *MIOS* | 2.625464948 | 0.000276293 | 0.000990816 | 1 |
| Obesity | mast_cell | *USP49* | 3.697807145 | 0.000225895 | 0.000976811 | 1 |
| Obesity | mast_cell | *CHMP4C* | 7.308268075 | 0.000455702 | 0.00111155 | 1 |
| Obesity | mast_cell | *SLC13A4* | 6.246590843 | 0.000203452 | 0.000976811 | 0.987147663 |
| Obesity | mast_cell | *BRI3* | 2.608465358 | 0.000225531 | 0.000976811 | 1 |
| Obesity | mast_cell | *SOX17* | -2.124324934 | 0.013051657 | 0.017222367 | 1 |
| Obesity | mast_cell | *RAD21* | 3.517624841 | 0.000131802 | 0.000976811 | 0.639502697 |
| Obesity | mast_cell | *TNFRSF11B* | 5.830853076 | 0.000283969 | 0.000993978 | 1 |
| Obesity | mast_cell | *SUN1* | 3.273807889 | 0.000283155 | 0.000993978 | 1 |
| Obesity | mast_cell | *OXR1* | 2.546766192 | 0.000223836 | 0.000976811 | 1 |
| Obesity | mast_cell | *GPER1* | 2.889116487 | 0.000601245 | 0.001237692 | 1 |
| Obesity | mast_cell | *INTS1* | 2.148193822 | 0.000655816 | 0.001312378 | 1 |
| Obesity | mast_cell | *SLC4A2* | 4.864861595 | 0.000115295 | 0.000976811 | 0.559413088 |
| Obesity | mast_cell | *FASTK* | 2.354464676 | 0.000464351 | 0.001116682 | 1 |
| Obesity | mast_cell | *ALDH7A1* | 2.746719096 | 0.00021181 | 0.000976811 | 1 |
| Obesity | mast_cell | *YWHAZ* | 2.099765568 | 0.005229353 | 0.00743417 | 1 |
| Obesity | mast_cell | *FZD6* | 2.137664096 | 0.000122586 | 0.000976811 | 0.594789135 |
| Obesity | mast_cell | *SLC25A32* | 2.429740174 | 0.000949528 | 0.001719712 | 1 |
| Obesity | mast_cell | *DCSTAMP* | 38.59064807 | 0.000205179 | 0.000976811 | 0.995530618 |
| Obesity | mast_cell | *INTS8* | 4.685669945 | 0.000103502 | 0.000976811 | 0.502191825 |
| Obesity | mast_cell | *C9orf24* | 4.733080041 | 8.06E-05 | 0.000955983 | 0.391171628 |
| Obesity | mast_cell | *NUDT2* | 1.244843022 | 0.01006389 | 0.013526315 | 1 |
| Obesity | mast_cell | *TMEM65* | 4.027324309 | 0.000171174 | 0.000976811 | 0.830534621 |
| Obesity | mast_cell | *CCDC171* | 2.058610834 | 0.000557627 | 0.001182832 | 1 |
| Obesity | mast_cell | *ZMAT4* | 4.578405142 | 0.000438674 | 0.00109874 | 1 |
| Obesity | mast_cell | *C8orf34* | 1.152985368 | 0.00602219 | 0.00843037 | 1 |
| Obesity | mast_cell | *ANKS6* | 2.333238924 | 3.28E-05 | 0.000621969 | 0.159224059 |
| Obesity | mast_cell | *AQP7* | 1.441258608 | 0.001026786 | 0.001828778 | 1 |
| Obesity | mast_cell | *NOL6* | 3.198734675 | 0.00015472 | 0.000976811 | 0.750700936 |
| Obesity | mast_cell | *AQP3* | -9.29864799 | 0.019452393 | 0.024903169 | 1 |
| Obesity | mast_cell | *TRMT10B* | 2.721864428 | 0.000350286 | 0.001031837 | 1 |
| Obesity | mast_cell | *VCP* | 3.017415062 | 0.000244706 | 0.000976811 | 1 |
| Obesity | mast_cell | *WRN* | 2.420926616 | 0.000165483 | 0.000976811 | 0.802924524 |
| Obesity | mast_cell | *MARCH8* | 6.900652968 | 0.002502914 | 0.003857731 | 1 |
| Obesity | mast_cell | *TSHR* | 2.168561569 | 0.000619188 | 0.001262311 | 1 |
| Obesity | mast_cell | *CFL2* | 4.043877628 | 0.000207397 | 0.000976811 | 1 |
| Obesity | mast_cell | *SLC16A9* | 3.164033842 | 0.000505325 | 0.001144783 | 1 |
| Obesity | mast_cell | *GJB2* | 13.82536873 | 0.000448577 | 0.001104518 | 1 |
| Obesity | mast_cell | *CRYL1* | 2.105372697 | 0.000781746 | 0.001487241 | 1 |
| Obesity | mast_cell | *REEP3* | 3.683585709 | 0.001936187 | 0.00309229 | 1 |
| Obesity | mast_cell | *MICU2* | 2.425474888 | 0.0002839 | 0.000993978 | 1 |

| | | | | | | |
|---|---|---|---|---|---|---|
| Obesity | mast_cell | *LRR1* | -3.258428979 | 0.012716135 | 0.016820798 | 1 |
| Obesity | mast_cell | *RPL36AL* | 1.975836585 | 0.001537848 | 0.002566783 | 1 |
| Obesity | mast_cell | *TMEM63C* | 10.85441983 | 0.009760995 | 0.013141051 | 1 |
| Obesity | mast_cell | *NOXRED1* | 2.93785859 | 0.00054546 | 0.00117408 | 1 |
| Obesity | mast_cell | *AKR1E2* | 3.583225073 | 0.000303324 | 0.000998908 | 1 |
| Obesity | mast_cell | *ATP5C1* | 4.336321458 | 0.000257963 | 0.00098093 | 1 |
| Obesity | mast_cell | *VDAC2* | 1.901949876 | 0.001471366 | 0.002473796 | 1 |
| Obesity | mast_cell | *PDZD8* | 2.894347871 | 0.00020831 | 0.000976811 | 1 |
| Obesity | mast_cell | *QSOX2* | 2.661783798 | 0.00052439 | 0.001154943 | 1 |
| Obesity | mast_cell | *SNAPC4* | 2.527387101 | 0.034910603 | 0.042989238 | 1 |
| Obesity | mast_cell | *TSC1* | 3.313071074 | 1.25E-05 | 0.000388294 | 0.060479044 |
| Obesity | mast_cell | *STOX1* | 1.894069647 | 0.000159601 | 0.000976811 | 0.77438164 |
| Obesity | mast_cell | *RET* | 3.923139821 | 0.00042033 | 0.00108308 | 1 |
| Obesity | mast_cell | *VWA2* | 12.00671106 | 0.000459449 | 0.001112953 | 1 |
| Obesity | mast_cell | *METTL3* | 4.00083138 | 0.000210114 | 0.000976811 | 1 |
| Obesity | mast_cell | *TRUB1* | 4.38085446 | 0.00061997 | 0.001262844 | 1 |
| Obesity | mast_cell | *HSPA12A* | 1.440388011 | 0.014087419 | 0.018458589 | 1 |
| Obesity | mast_cell | *UBTD1* | 1.929821449 | 0.000437284 | 0.001098433 | 1 |
| Obesity | mast_cell | *OTOGL* | 4.423889801 | 0.000745992 | 0.001438613 | 1 |
| Obesity | mast_cell | *PACSIN3* | -2.828329206 | 0.001095339 | 0.001929068 | 1 |
| Obesity | mast_cell | *TTC7B* | 5.358363444 | 0.004652912 | 0.006679268 | 1 |
| Obesity | mast_cell | *PSMC3* | 2.575638385 | 0.000295878 | 0.000998908 | 1 |
| Obesity | mast_cell | *RAPSN* | -10.5281595 | 5.12E-15 | 3.55E-12 | 2.48E-11 |
| Obesity | mast_cell | *TC2N* | 1.856077871 | 0.000991004 | 0.001774955 | 1 |
| Obesity | mast_cell | *IFI27L1* | 2.043559094 | 0.00043674 | 0.001098433 | 1 |
| Obesity | mast_cell | *PTER* | 5.597005793 | 5.56E-05 | 0.000812749 | 0.269832559 |
| Obesity | mast_cell | *C1QL3* | 3.946456185 | 0.000218203 | 0.000976811 | 1 |
| Obesity | mast_cell | *ARL5B* | 3.181897791 | 0.000231248 | 0.000976811 | 1 |
| Obesity | mast_cell | *LIPC* | -2.031859113 | 0.017036012 | 0.022048207 | 1 |
| Obesity | mast_cell | *CEP57* | 5.051109206 | 0.001444585 | 0.002432036 | 1 |
| Obesity | mast_cell | *TCP11L2* | 3.131641068 | 0.000217637 | 0.000976811 | 1 |
| Obesity | mast_cell | *JAM3* | 2.587601345 | 0.001794752 | 0.002904649 | 1 |
| Obesity | mast_cell | *IKBIP* | 2.404572609 | 0.000170089 | 0.000976811 | 0.825271173 |
| Obesity | mast_cell | *ZFYVE19* | 5.149401825 | 0.000189639 | 0.000976811 | 0.920127541 |
| Obesity | mast_cell | *TRMT61A* | 2.93474075 | 0.000553872 | 0.00118075 | 1 |
| Obesity | mast_cell | *LARP6* | 2.05629117 | 0.000442605 | 0.00110084 | 1 |
| Obesity | mast_cell | *API5* | 5.03562912 | 0.000169026 | 0.000976811 | 0.820114771 |
| Obesity | mast_cell | *ASPG* | 14.41539927 | 7.87E-05 | 0.000944925 | 0.381749509 |
| Obesity | mast_cell | *COX11* | 1.975591964 | 0.000721176 | 0.001403026 | 1 |
| Obesity | mast_cell | *ZNF202* | 2.725010639 | 0.00087528 | 0.001611099 | 1 |
| Obesity | mast_cell | *STXBP4* | 2.120324272 | 0.000466015 | 0.001116682 | 1 |
| Obesity | mast_cell | *WBP1L* | 3.507874242 | 0.000109334 | 0.000976811 | 0.530486645 |
| Obesity | mast_cell | *NUDT13* | 1.513792667 | 0.004242565 | 0.00615762 | 1 |
| Obesity | mast_cell | *WDR88* | 2.319998482 | 0.000351776 | 0.001031837 | 1 |
| Obesity | mast_cell | *XRRA1* | 1.8189918 | 0.000729374 | 0.001413873 | 1 |
| Obesity | mast_cell | *TRIM66* | 1.418433093 | 0.000201478 | 0.000976811 | 0.977572599 |
| Obesity | mast_cell | *RPL27A* | 2.788253431 | 0.00024339 | 0.000976811 | 1 |
| Obesity | mast_cell | *CDYL2* | 1.119163419 | 0.028813853 | 0.035884193 | 1 |
| Obesity | mast_cell | *CENPN* | -3.45807439 | 0.005790222 | 0.008150322 | 1 |
| Obesity | mast_cell | *C16orf46* | -2.392538736 | 0.004315626 | 0.006250357 | 1 |

| | | | | | | |
|---|---|---|---|---|---|---|
| Obesity | mast_cell | *TMEM41B* | 3.926948313 | 0.000219934 | 0.000976811 | 1 |
| Obesity | mast_cell | *TMX3* | 2.164392185 | 0.000504627 | 0.001144783 | 1 |
| Obesity | mast_cell | *PRKCB* | 3.718927298 | 4.45E-05 | 0.00074422 | 0.215787728 |
| Obesity | mast_cell | *NDST2* | -0.827512687 | 0.001584392 | 0.002620133 | 1 |
| Obesity | mast_cell | *CCDC68* | 2.687936531 | 1.20E-05 | 0.000383871 | 0.05816541 |
| Obesity | mast_cell | *RIMKLB* | 1.540247286 | 0.021137496 | 0.02684794 | 1 |
| Obesity | mast_cell | *TMED3* | 1.227976249 | 0.003070584 | 0.004612531 | 1 |
| Obesity | mast_cell | *SEC11C* | 6.725644963 | 0.00014135 | 0.000976811 | 0.685830506 |
| Obesity | mast_cell | *CENPV* | 2.013774758 | 0.001251383 | 0.00215574 | 1 |
| Obesity | mast_cell | *FAM96B* | 2.935768451 | 0.00054881 | 0.001175269 | 1 |
| Obesity | mast_cell | *CHRFAM7A* | 3.327017913 | 0.013312592 | 0.017528547 | 1 |
| Obesity | mast_cell | *SPDYE6* | 2.921499993 | 0.001650442 | 0.002706385 | 1 |
| Obesity | mast_cell | *ZNF592* | 2.59670899 | 0.001157433 | 0.002022278 | 1 |
| Obesity | mast_cell | *ACSM1* | 2.926246684 | 0.0013618 | 0.002315155 | 1 |
| Obesity | mast_cell | *SLFN5* | 3.622892276 | 0.000286513 | 0.000993978 | 1 |
| Obesity | mast_cell | *CATSPER2* | 2.446967498 | 0.000393379 | 0.0010551 | 1 |
| Obesity | mast_cell | *ZNF667-AS1* | 4.884060174 | 0.009121456 | 0.012358924 | 1 |
| Obesity | mast_cell | *SAAL1* | 2.511184326 | 0.000461914 | 0.001114555 | 1 |
| Obesity | mast_cell | *YPEL4* | 4.047750073 | 0.013661262 | 0.017938957 | 1 |
| Obesity | mast_cell | *LDHC* | 1.590715605 | 0.024025698 | 0.030223668 | 1 |
| Obesity | mast_cell | *FAM96A* | 3.47962324 | 0.000398169 | 0.001061493 | 1 |
| Obesity | mast_cell | *TMEM170A* | 2.333718625 | 0.000452101 | 0.001107316 | 1 |
| Obesity | mast_cell | *DCTN5* | 3.070814693 | 0.000121546 | 0.000976811 | 0.589743462 |
| Obesity | mast_cell | *STAT6* | 3.220513335 | 0.000181294 | 0.000976811 | 0.879640312 |
| Obesity | mast_cell | *STX3* | -0.567261891 | 0.00176647 | 0.00286174 | 1 |
| Obesity | mast_cell | *PIP4K2C* | 3.830290698 | 2.45E-05 | 0.000545024 | 0.118924128 |
| Obesity | mast_cell | *MTMR10* | 2.967747968 | 8.08E-05 | 0.000955983 | 0.391953014 |
| Obesity | mast_cell | *MS4A7* | 0.952907686 | 0.0202986 | 0.025878707 | 1 |
| Obesity | mast_cell | *DIS3L* | 1.879426312 | 0.000777975 | 0.001482613 | 1 |
| Obesity | mast_cell | *CCNDBP1* | 3.434663879 | 6.63E-05 | 0.000885378 | 0.321496701 |
| Obesity | mast_cell | *EPB42* | 5.089104394 | 0.000179721 | 0.000976811 | 0.872004317 |
| Obesity | mast_cell | *SMAD3* | 2.532019475 | 0.00048609 | 0.001132608 | 1 |
| Obesity | mast_cell | *RCCD1* | 6.673228468 | 0.007249813 | 0.009990369 | 1 |
| Obesity | mast_cell | *AKTIP* | 2.305873126 | 0.000645695 | 0.001298348 | 1 |
| Obesity | mast_cell | *CNPY4* | 2.795299159 | 0.000150887 | 0.000976811 | 0.732103817 |
| Obesity | mast_cell | *MEI1* | 6.273013855 | 1.13E-05 | 0.000373166 | 0.054855348 |
| Obesity | mast_cell | *ACSF2* | 1.735700205 | 0.000815419 | 0.001531712 | 1 |
| Obesity | mast_cell | *URM1* | 1.98474722 | 0.003796023 | 0.005567806 | 1 |
| Obesity | mast_cell | *ENDOG* | 3.016299995 | 0.007086699 | 0.009785049 | 1 |
| Obesity | mast_cell | *ISLR2* | 6.026480234 | 1.43E-05 | 0.000424191 | 0.069143142 |
| Obesity | mast_cell | *SP2* | 8.667014911 | 0.000416303 | 0.001080612 | 1 |
| Obesity | mast_cell | *COQ7* | 4.497549866 | 7.90E-05 | 0.000946679 | 0.383404887 |
| Obesity | mast_cell | *GPRC5B* | 2.797457674 | 0.000122966 | 0.000976811 | 0.596630021 |
| Obesity | mast_cell | *FBXO22* | 3.229956252 | 0.000285087 | 0.000993978 | 1 |
| Obesity | mast_cell | *TBC1D2B* | 1.694070041 | 0.00150958 | 0.002528299 | 1 |
| Obesity | mast_cell | *LOXHD1* | 1.824213042 | 0.000194735 | 0.000976811 | 0.944854372 |
| Obesity | mast_cell | *KATNAL2* | 2.337360753 | 0.001750727 | 0.00283908 | 1 |
| Obesity | mast_cell | *ZNF91* | 2.95995925 | 0.000151122 | 0.000976811 | 0.733242476 |
| Obesity | mast_cell | *CCL23* | -0.70336474 | 0.021971367 | 0.027819695 | 1 |
| Obesity | mast_cell | *POP5* | 5.425805604 | 0.006057753 | 0.008475264 | 1 |

| | | | | | | |
|---|---|---|---|---|---|---|
| Obesity | mast_cell | *ENGASE* | 2.416962225 | 4.74E-05 | 0.000758703 | 0.229986363 |
| Obesity | mast_cell | *MYO5B* | 3.026667044 | 0.001027086 | 0.001828778 | 1 |
| Obesity | mast_cell | *RRM1* | 2.007926793 | 0.001850558 | 0.002977091 | 1 |
| Obesity | mast_cell | *ZNF23* | 5.065690555 | 0.002035846 | 0.00322808 | 1 |
| Obesity | mast_cell | *VKORC1* | 2.530735249 | 0.000471885 | 0.001122896 | 1 |
| Obesity | mast_cell | *SMG8* | -5.0470723 | 2.36E-05 | 0.000545024 | 0.114679509 |
| Obesity | mast_cell | *RAB8A* | 4.07570353 | 0.001238121 | 0.002136331 | 1 |
| Obesity | mast_cell | *RPL13* | 2.975333866 | 0.000316241 | 0.000998908 | 1 |
| Obesity | mast_cell | *ZNF641* | 3.454310334 | 0.000220083 | 0.000976811 | 1 |
| Obesity | mast_cell | *TP53I13* | 3.059640642 | 0.000153517 | 0.000976811 | 0.744865731 |
| Obesity | mast_cell | *TUBA1C* | 1.70869629 | 0.001980232 | 0.003151226 | 1 |
| Obesity | mast_cell | *ZNF528* | 5.792427912 | 0.000162282 | 0.000976811 | 0.78739313 |
| Obesity | mast_cell | *TMC4* | 2.917270402 | 0.00026726 | 0.000985747 | 1 |
| Obesity | mast_cell | *TTYH1* | 3.107098833 | 0.00051902 | 0.001151636 | 1 |
| Obesity | mast_cell | *LENG8* | 5.254353775 | 0.000234031 | 0.000976811 | 1 |
| Obesity | mast_cell | *CDC42EP5* | 1.658422059 | 0.016714155 | 0.021672122 | 1 |
| Obesity | mast_cell | *FAM57A* | 3.766708428 | 3.16E-05 | 0.000621513 | 0.153083368 |
| Obesity | mast_cell | *KIFC2* | 3.11672753 | 0.000262804 | 0.000983135 | 1 |
| Obesity | mast_cell | *SERPINF2* | 2.913913572 | 0.00267769 | 0.004088153 | 1 |
| Obesity | mast_cell | *SRR* | 3.635740592 | 5.89E-07 | 5.25E-05 | 0.00285896 |
| Obesity | mast_cell | *TRPV3* | 2.967869996 | 0.000481831 | 0.001130485 | 1 |
| Obesity | mast_cell | *CYB5D2* | 2.413745952 | 0.000226309 | 0.000976811 | 1 |
| Obesity | mast_cell | *C19orf48* | 1.628877855 | 0.013448453 | 0.017683441 | 1 |
| Obesity | mast_cell | *KLK5* | 3.38434246 | 0.006001994 | 0.008409006 | 1 |
| Obesity | mast_cell | *KLK6* | 92.85079784 | 0.00070604 | 0.001381207 | 1 |
| Obesity | mast_cell | *ZNF83* | 4.961105007 | 3.50E-05 | 0.000647264 | 0.169834655 |
| Obesity | mast_cell | *SOAT2* | -2.418219925 | 0.020253135 | 0.025832863 | 1 |
| Obesity | mast_cell | *ZNF558* | 3.482392278 | 0.000492641 | 0.001138685 | 1 |
| Obesity | mast_cell | *NDUFV1* | 2.966490806 | 0.000259818 | 0.00098093 | 1 |
| Obesity | mast_cell | *NUDT8* | -3.202478212 | 0.000249456 | 0.000976811 | 1 |
| Obesity | mast_cell | *PRDX2* | 2.356510655 | 0.000475173 | 0.001127398 | 1 |
| Obesity | mast_cell | *ZNF232* | -1.408640344 | 0.00667268 | 0.009266125 | 1 |
| Obesity | mast_cell | *MIS12* | 3.536792632 | 0.000229835 | 0.000976811 | 1 |
| Obesity | mast_cell | *ATP5H* | 4.135811737 | 0.000324362 | 0.001004343 | 1 |
| Obesity | mast_cell | *MGAT5B* | 5.99026953 | 0.000866235 | 0.001596874 | 1 |
| Obesity | mast_cell | *GSDMA* | 2.643816277 | 0.002548203 | 0.003921306 | 1 |
| Obesity | mast_cell | *TMEM99* | 2.365135564 | 0.001600702 | 0.002642602 | 1 |
| Obesity | mast_cell | *ZNF598* | 6.816469037 | 2.47E-05 | 0.000545024 | 0.119905321 |
| Obesity | mast_cell | *ECI1* | 2.638471493 | 0.000134387 | 0.000976811 | 0.652046065 |
| Obesity | mast_cell | *ABCA3* | 2.194020459 | 0.003459819 | 0.005118 | 1 |
| Obesity | mast_cell | *ATG16L2* | 2.272302242 | 0.000349162 | 0.001031121 | 1 |
| Obesity | mast_cell | *RPSA* | 2.559590605 | 0.000275299 | 0.000990086 | 1 |
| Obesity | mast_cell | *ULK4* | 2.390560179 | 0.000191628 | 0.000976811 | 0.929780125 |
| Obesity | mast_cell | *LTBP3* | 3.153642225 | 0.000311127 | 0.000998908 | 1 |
| Obesity | mast_cell | *SCARA5* | 1.631665379 | 0.001093809 | 0.001927075 | 1 |
| Obesity | mast_cell | *FAM83B* | 17.46801185 | 0.010863545 | 0.014536657 | 1 |
| Obesity | mast_cell | *COA6* | 8.197049052 | 0.000143422 | 0.000976811 | 0.695885383 |
| Obesity | mast_cell | *PDHB* | 2.87006815 | 0.000114673 | 0.000976811 | 0.556391298 |
| Obesity | mast_cell | *PXK* | 4.400876213 | 0.002712431 | 0.004130261 | 1 |
| Obesity | mast_cell | *PCMTD1* | 4.65078021 | 0.000205133 | 0.000976811 | 0.995304202 |

| | | | | | | |
|---|---|---|---|---|---|---|
| Obesity | mast_cell | *ACOX2* | 2.66186816 | 0.000902765 | 0.001649799 | 1 |
| Obesity | mast_cell | *FAM107A* | -0.870820136 | 0.002810954 | 0.00425546 | 1 |
| Obesity | mast_cell | *SCN11A* | 1.579783052 | 0.0116877 | 0.015545154 | 1 |
| Obesity | mast_cell | *SEPT2* | 2.668528721 | 0.000166831 | 0.000976811 | 0.809464816 |
| Obesity | mast_cell | *DTYMK* | 4.694214258 | 0.000653719 | 0.001309596 | 1 |
| Obesity | mast_cell | *ING5* | 3.819326163 | 4.75E-05 | 0.000758703 | 0.230645622 |
| Obesity | mast_cell | *MLKL* | 1.600734335 | 0.00166742 | 0.002728608 | 1 |
| Obesity | mast_cell | *RFWD3* | 2.25407129 | 0.000227186 | 0.000976811 | 1 |
| Obesity | mast_cell | *KLHL30* | -2.613246667 | 0.00179885 | 0.00291031 | 1 |
| Obesity | mast_cell | *CCDC110* | -6.223568768 | 0.0222763 | 0.028169041 | 1 |
| Obesity | mast_cell | *FNTA* | 4.7022296 | 8.29E-07 | 6.49E-05 | 0.00402145 |
| Obesity | mast_cell | *SLC20A2* | 2.536880836 | 0.000228815 | 0.000976811 | 1 |
| Obesity | mast_cell | *TMUB2* | 2.423168488 | 0.000796555 | 0.001505604 | 1 |
| Obesity | mast_cell | *ZSWIM1* | 1.413039266 | 0.009478092 | 0.012802812 | 1 |
| Obesity | mast_cell | *NBPF9* | 3.368610273 | 0.000461784 | 0.001114555 | 1 |
| Obesity | mast_cell | *ADAM9* | 2.500400048 | 0.000205333 | 0.000976811 | 0.99627404 |
| Obesity | mast_cell | *NDUFS5* | 3.974076439 | 2.52E-05 | 0.000550066 | 0.122114556 |
| Obesity | mast_cell | *ZNF30* | 3.02124747 | 0.018425216 | 0.023707013 | 1 |
| Obesity | mast_cell | *SLC16A4* | 2.722112078 | 8.03E-05 | 0.000955407 | 0.389806144 |
| Obesity | mast_cell | *DNAJC21* | 4.169368282 | 0.00019464 | 0.000976811 | 0.94439379 |
| Obesity | mast_cell | *NPNT* | 4.27710016 | 3.84E-06 | 0.000187911 | 0.018636427 |
| Obesity | mast_cell | *GSTM4* | 8.826321173 | 3.52E-05 | 0.000647264 | 0.17087767 |
| Obesity | mast_cell | *PPIP5K1* | 1.798745325 | 0.003875272 | 0.005670592 | 1 |
| Obesity | mast_cell | *ADAL* | 1.972813047 | 0.000104392 | 0.000976811 | 0.506508015 |
| Obesity | mast_cell | *SNTB2* | 3.062771787 | 0.000216262 | 0.000976811 | 1 |
| Obesity | mast_cell | *ZNF507* | 2.928552247 | 0.001500827 | 0.002515376 | 1 |
| Obesity | mast_cell | *STX18* | 4.69941485 | 0.000175381 | 0.000976811 | 0.850946584 |
| Obesity | mast_cell | *ZBTB49* | 4.725559219 | 0.000196753 | 0.000976811 | 0.954646493 |
| Obesity | mast_cell | *GFM1* | 3.927234875 | 0.000127399 | 0.000976811 | 0.618138913 |
| Obesity | mast_cell | *ATOH8* | 2.020457136 | 0.000687264 | 0.001355244 | 1 |
| Obesity | mast_cell | *BTNL3* | 3.008691614 | 0.000865248 | 0.001595702 | 1 |
| Obesity | mast_cell | *TMEM129* | 2.504337281 | 0.000409067 | 0.001074603 | 1 |
| Obesity | mast_cell | *CEP120* | 3.877324052 | 0.000234844 | 0.000976811 | 1 |
| Obesity | mast_cell | *MFF* | 2.661203929 | 0.0002451 | 0.000976811 | 1 |
| Obesity | mast_cell | *LGALS9* | 2.483723363 | 0.000509807 | 0.001145411 | 1 |
| Obesity | mast_cell | *CPLX1* | 2.729125229 | 1.23E-05 | 0.000386737 | 0.059513109 |
| Obesity | mast_cell | *MFSD7* | -5.067159766 | 0.031982466 | 0.039576364 | 1 |
| Obesity | mast_cell | *MAP2K1* | 3.983942283 | 0.00017145 | 0.000976811 | 0.831874288 |
| Obesity | mast_cell | *KLK7* | 4.037629186 | 0.000170429 | 0.000976811 | 0.826922548 |
| Obesity | mast_cell | *IRS1* | 2.19097896 | 1.03E-05 | 0.000365431 | 0.049779705 |
| Obesity | mast_cell | *UPF3A* | 2.258119502 | 0.003412125 | 0.005056698 | 1 |
| Obesity | mast_cell | *C8orf46* | 1.764869025 | 2.77E-06 | 0.000149272 | 0.013434506 |
| Obesity | mast_cell | *HSPBAP1* | 1.855964759 | 0.003712993 | 0.005457571 | 1 |
| Obesity | mast_cell | *UBE2V2* | 3.350889598 | 0.000345926 | 0.00102869 | 1 |
| Obesity | mast_cell | *CPT1C* | 2.699033183 | 0.004429252 | 0.006397955 | 1 |
| Obesity | mast_cell | *PCSK9* | 11.52896745 | 0.017092679 | 0.022096856 | 1 |
| Obesity | mast_cell | *XPO6* | 4.42248097 | 0.001435798 | 0.002420602 | 1 |
| Obesity | mast_cell | *RGS14* | 1.773007282 | 0.020517584 | 0.026135815 | 1 |
| Obesity | mast_cell | *RAB24* | -1.820899409 | 0.002113458 | 0.00333588 | 1 |
| Obesity | mast_cell | *THBS3* | 3.316216687 | 0.000110173 | 0.000976811 | 0.534559296 |

| | | | | | | |
|---|---|---|---|---|---|---|
| Obesity | mast_cell | *NPIPB3* | -1.98998222 | 0.000533148 | 0.001162622 | 1 |
| Obesity | mast_cell | *SH3TC2* | 1.897798153 | 0.02059364 | 0.026212052 | 1 |
| Obesity | mast_cell | *P2RY12* | 4.234380001 | 0.001614435 | 0.00266165 | 1 |
| Obesity | mast_cell | *SNUPN* | 2.786746638 | 5.20E-05 | 0.000790977 | 0.252321527 |
| Obesity | mast_cell | *RNASE6* | 4.134461574 | 0.00051553 | 0.001150047 | 1 |
| Obesity | mast_cell | *CD52* | 1.592496509 | 0.001016958 | 0.001814073 | 1 |
| Obesity | mast_cell | *SLC38A11* | 2.140517296 | 0.000861657 | 0.001592064 | 1 |
| Obesity | mast_cell | *CCDC8* | 2.888540512 | 0.003589587 | 0.005300266 | 1 |
| Obesity | mast_cell | *ZEB2* | 1.635925313 | 0.002896484 | 0.004376749 | 1 |
| Obesity | mast_cell | *HINT1* | 2.860656153 | 0.000362321 | 0.001037687 | 1 |
| Obesity | mast_cell | *DTWD2* | 2.052709994 | 0.000775054 | 0.001479954 | 1 |
| Obesity | mast_cell | *CLIC3* | 3.229364673 | 0.000457831 | 0.001112606 | 1 |
| Obesity | mast_cell | *DFFB* | 4.799071924 | 0.000143378 | 0.000976811 | 0.695668442 |
| Obesity | mast_cell | *NFU1* | 3.273147968 | 0.000173944 | 0.000976811 | 0.843974047 |
| Obesity | mast_cell | *C15orf40* | 3.939909568 | 0.001780355 | 0.002882309 | 1 |
| Obesity | mast_cell | *LUZP1* | 4.284199803 | 0.000212398 | 0.000976811 | 1 |
| Obesity | mast_cell | *CHRNA5* | -3.530780618 | 0.004260566 | 0.006180049 | 1 |
| Obesity | mast_cell | *UGP2* | 2.079543319 | 0.000124857 | 0.000976811 | 0.605806323 |
| Obesity | mast_cell | *HNRNPF* | 4.683254721 | 0.000675887 | 0.001336567 | 1 |
| Obesity | mast_cell | *CSGALNACT2* | 2.784314331 | 0.000223497 | 0.000976811 | 1 |
| Obesity | mast_cell | *AVEN* | 2.967938781 | 0.000297503 | 0.000998908 | 1 |
| Obesity | mast_cell | *CALML6* | 34.65809607 | 0.011655298 | 0.015506308 | 1 |
| Obesity | mast_cell | *MUC3A* | 2.928849486 | 0.007630748 | 0.010476624 | 1 |
| Obesity | mast_cell | *TPST1* | 1.562459342 | 0.001144643 | 0.002002815 | 1 |
| Obesity | mast_cell | *TOR1AIP2* | 2.290457884 | 0.000511935 | 0.001145411 | 1 |
| Obesity | mast_cell | *OTUD3* | 2.36548417 | 3.92E-05 | 0.000686693 | 0.190214063 |
| Obesity | mast_cell | *ZFPM2* | 1.408902679 | 0.000287899 | 0.000994669 | 1 |
| Obesity | mast_cell | *MAP3K2* | 2.31598752 | 0.00013229 | 0.000976811 | 0.641869652 |
| Obesity | mast_cell | *MYO7B* | 6.255640233 | 2.47E-05 | 0.000545024 | 0.119716371 |
| Obesity | mast_cell | *UBE2E3* | 1.969074875 | 0.00075353 | 0.001449118 | 1 |
| Obesity | mast_cell | *NIPA1* | 3.002699189 | 0.000301303 | 0.000998908 | 1 |
| Obesity | mast_cell | *HNRNPA3* | 3.679611507 | 0.00023411 | 0.000976811 | 1 |
| Obesity | mast_cell | *RNF150* | 1.76466037 | 0.001046527 | 0.001857261 | 1 |
| Obesity | mast_cell | *PDCD6IP* | 3.368305246 | 0.000185949 | 0.000976811 | 0.902224338 |
| Obesity | mast_cell | *CRTAP* | 2.33491378 | 0.000865271 | 0.001595702 | 1 |
| Obesity | mast_cell | *CNGB3* | 4.898563162 | 1.67E-06 | 0.000109614 | 0.008111461 |
| Obesity | mast_cell | *STX8* | 3.375197032 | 0.00024411 | 0.000976811 | 1 |
| Obesity | mast_cell | *NFRKB* | 2.456108821 | 0.000512662 | 0.001145661 | 1 |
| Obesity | mast_cell | *ADORA2B* | -3.625311522 | 7.99E-09 | 1.75E-06 | 3.88E-05 |
| Obesity | mast_cell | *MGMT* | 3.432924182 | 0.000169381 | 0.000976811 | 0.821837812 |
| Obesity | mast_cell | *NFXL1* | 6.254617247 | 0.00023099 | 0.000976811 | 1 |
| Obesity | mast_cell | *DNAJC18* | 1.877585458 | 0.001443689 | 0.002431371 | 1 |
| Obesity | mast_cell | *ELOVL6* | 1.789437707 | 0.000441184 | 0.001100333 | 1 |
| Obesity | mast_cell | *ARL6IP1* | 3.564760816 | 0.002108932 | 0.00332982 | 1 |
| Obesity | mast_cell | *SERPINB9* | 4.458262727 | 0.000266857 | 0.000985747 | 1 |
| Obesity | mast_cell | *CDH2* | -2.701161905 | 0.017556154 | 0.022642865 | 1 |
| Obesity | mast_cell | *EMB* | 1.35587757 | 0.000436317 | 0.001098433 | 1 |
| Obesity | mast_cell | *STAT2* | 3.4334257 | 0.000257503 | 0.00098093 | 1 |
| Obesity | mast_cell | *SLC26A5* | 5.239723937 | 9.17E-05 | 0.000976811 | 0.445059556 |
| Obesity | mast_cell | *GTSF1* | 5.063497112 | 0.031537874 | 0.039066063 | 1 |

| | | | | | | |
|---|---|---|---|---|---|---|
| Obesity | mast_cell | *ARMC10* | 1.902920981 | 0.000364141 | 0.001037687 | 1 |
| Obesity | mast_cell | *ACYP2* | 2.573331893 | 7.00E-05 | 0.000898663 | 0.339766974 |
| Obesity | mast_cell | *TRABD* | 5.584063021 | 0.000981593 | 0.001762653 | 1 |
| Obesity | mast_cell | *HOXB9* | -2.877557102 | 0.003335938 | 0.004957419 | 1 |
| Obesity | mast_cell | *KCNS3* | 1.698301459 | 0.003319213 | 0.00494013 | 1 |
| Obesity | mast_cell | *FOXN2* | 3.25420767 | 0.000200888 | 0.000976811 | 0.974706621 |
| Obesity | mast_cell | *GPR27* | 2.592758267 | 0.01685804 | 0.021847011 | 1 |
| Obesity | mast_cell | *TRIAP1* | 4.069989323 | 0.00035577 | 0.001033017 | 1 |
| Obesity | mast_cell | *RPS9* | 2.296703601 | 0.000103065 | 0.000976811 | 0.500069066 |
| Obesity | mast_cell | *MSANTD4* | 7.687605709 | 0.00013609 | 0.000976811 | 0.660309129 |
| Obesity | mast_cell | *NDUFA3* | 3.021030456 | 0.000302535 | 0.000998908 | 1 |
| Obesity | mast_cell | *PAQR8* | -3.607377257 | 0.009068341 | 0.012293823 | 1 |
| Obesity | mast_cell | *NUDT6* | 1.681579497 | 0.002787317 | 0.004224948 | 1 |
| Obesity | mast_cell | *ZNF160* | 2.018249001 | 0.000309383 | 0.000998908 | 1 |
| Obesity | mast_cell | *ZNF415* | 2.46716129 | 0.001564874 | 0.002598381 | 1 |
| Obesity | mast_cell | *S1PR1* | 2.456279708 | 0.000459076 | 0.001112606 | 1 |
| Obesity | mast_cell | *PYGO1* | 2.057348311 | 0.000348588 | 0.001030053 | 1 |
| Obesity | mast_cell | *TSNARE1* | 4.137417459 | 7.97E-07 | 6.34E-05 | 0.003865569 |
| Obesity | mast_cell | *FPR1* | 2.197969835 | 0.000704773 | 0.001380207 | 1 |
| Obesity | mast_cell | *FEZ2* | 2.577328593 | 0.00050818 | 0.001144962 | 1 |
| Obesity | mast_cell | *SOX7* | 4.34095388 | 0.000539775 | 0.001171639 | 1 |
| Obesity | mast_cell | *TRMT61B* | 3.481651891 | 2.40E-06 | 0.000137224 | 0.011640846 |
| Obesity | mast_cell | *MFN1* | 4.187561917 | 6.18E-05 | 0.000858912 | 0.299760265 |
| Obesity | mast_cell | *GIMAP8* | 1.801913863 | 0.000277809 | 0.000992498 | 1 |
| Obesity | mast_cell | *ATP6V0E2* | 2.233591761 | 0.015432576 | 0.020112505 | 1 |
| Obesity | mast_cell | *SCAND1* | 2.99952923 | 0.002192203 | 0.003448953 | 1 |
| Obesity | mast_cell | *SOSTDC1* | 5.128139553 | 0.002434994 | 0.003775837 | 1 |
| Obesity | mast_cell | *GAA* | 0.977883038 | 0.040692392 | 0.049632853 | 1 |
| Obesity | mast_cell | *CANT1* | -1.644204158 | 0.037105424 | 0.045555546 | 1 |
| Obesity | mast_cell | *LURAP1* | -2.08127874 | 0.005030811 | 0.007175043 | 1 |
| Obesity | mast_cell | *PDE7B* | 4.371809346 | 0.011403089 | 0.015187424 | 1 |
| Obesity | mast_cell | *MRPL36* | 2.322614194 | 0.000555979 | 0.001182125 | 1 |
| Obesity | mast_cell | *MCC* | 2.436111686 | 0.000248038 | 0.000976811 | 1 |
| Obesity | mast_cell | *ZNF561* | 3.208656928 | 4.62E-05 | 0.000754069 | 0.223958506 |
| Obesity | mast_cell | *HOPX* | 3.096746535 | 0.004201062 | 0.006108347 | 1 |
| Obesity | mast_cell | *PPID* | 5.174226873 | 0.000118358 | 0.000976811 | 0.574274417 |
| Obesity | mast_cell | *ETFDH* | 1.25436958 | 0.019420743 | 0.024875778 | 1 |
| Obesity | mast_cell | *TBCA* | 2.645811696 | 0.000237958 | 0.000976811 | 1 |
| Obesity | mast_cell | *ZNF584* | 1.409649388 | 0.009668841 | 0.01303145 | 1 |
| Obesity | mast_cell | *NMUR1* | 3.728046387 | 0.000387781 | 0.001054069 | 1 |
| Obesity | mast_cell | *PIK3CD* | 2.059374266 | 0.000106195 | 0.000976811 | 0.515256982 |
| Obesity | mast_cell | *RGS19* | 1.872850693 | 0.009690626 | 0.013057183 | 1 |
| Obesity | mast_cell | *ANO5* | 5.786467465 | 0.000370417 | 0.00103942 | 1 |
| Obesity | mast_cell | *VAT1L* | -1.267088613 | 0.002510685 | 0.003868481 | 1 |
| Obesity | mast_cell | *CAMTA1* | 3.909716657 | 2.08E-05 | 0.000524109 | 0.101153055 |
| Obesity | mast_cell | *LGALS4* | 4.28832555 | 0.000106921 | 0.000976811 | 0.518781147 |
| Obesity | mast_cell | *PAH* | 8.515227668 | 0.022797579 | 0.028783205 | 1 |
| Obesity | mast_cell | *SPATA5L1* | 3.098985402 | 0.000784786 | 0.001490475 | 1 |
| Obesity | mast_cell | *RHNO1* | 3.146843152 | 0.03811623 | 0.046678432 | 1 |
| Obesity | mast_cell | *EXOSC10* | 4.307926399 | 0.000577129 | 0.001204399 | 1 |

| | | | | | | |
|---|---|---|---|---|---|---|
| Obesity | mast_cell | *TRAPPC12* | 2.723028756 | 0.00030488 | 0.000998908 | 1 |
| Obesity | mast_cell | *PRNP* | 1.917569187 | 0.000668681 | 0.001326427 | 1 |
| Obesity | mast_cell | *TVP23B* | 2.442416847 | 0.011162872 | 0.014900208 | 1 |
| Obesity | mast_cell | *FBXW10* | 5.748206951 | 0.001230926 | 0.002125429 | 1 |
| Obesity | mast_cell | *SRGAP2C* | 1.592701705 | 0.000415807 | 0.001080612 | 1 |
| Obesity | mast_cell | *ATPAF2* | 1.769756598 | 0.003038469 | 0.004571366 | 1 |
| Obesity | mast_cell | *ZNF554* | 2.497926609 | 0.001329747 | 0.002271806 | 1 |
| Obesity | mast_cell | *ANKRD20A4* | 16.73034665 | 0.0001731 | 0.000976811 | 0.839880576 |
| Obesity | mast_cell | *SERF1A* | 17.87228147 | 0.001183052 | 0.002061101 | 1 |
| Obesity | mast_cell | *EIF2AK3* | 3.384826533 | 0.000198732 | 0.000976811 | 0.964248432 |
| Obesity | mast_cell | *NME6* | 2.919978987 | 0.000609581 | 0.001247969 | 1 |
| Obesity | mast_cell | *MTBP* | -1.830231422 | 0.010484318 | 0.014064117 | 1 |
| Obesity | mast_cell | *TEFM* | 2.420129019 | 0.00039157 | 0.001054889 | 1 |
| Obesity | mast_cell | *MALT1* | 5.863808582 | 0.000209829 | 0.000976811 | 1 |
| Obesity | mast_cell | *MBOAT1* | 2.467258991 | 0.000349938 | 0.001031837 | 1 |
| Obesity | mast_cell | *CXCR6* | 4.499467051 | 0.002304548 | 0.003595391 | 1 |
| Obesity | mast_cell | *PAIP1* | 4.06880088 | 0.000377683 | 0.001048135 | 1 |
| Obesity | mast_cell | *C1QTNF4* | -4.342277674 | 4.74E-11 | 1.77E-08 | 2.30E-07 |
| Obesity | mast_cell | *MACROD2* | 1.67232783 | 8.99E-05 | 0.000976811 | 0.435997063 |
| Obesity | mast_cell | *BSG* | 2.030283008 | 0.000353904 | 0.001031837 | 1 |
| Obesity | mast_cell | *HINFP* | 6.10388201 | 0.00033466 | 0.001017401 | 1 |
| Obesity | mast_cell | *TP53RK* | 3.63889508 | 0.000372088 | 0.00104107 | 1 |
| Obesity | mast_cell | *SUCLG2* | 2.463731047 | 0.000247615 | 0.000976811 | 1 |
| Obesity | mast_cell | *ARNT2* | 3.398961164 | 7.61E-05 | 0.000935479 | 0.36931805 |
| Obesity | mast_cell | *GTPBP2* | 2.949403947 | 0.002006672 | 0.003188072 | 1 |
| Obesity | mast_cell | *FGGY* | 3.026308351 | 0.00010194 | 0.000976811 | 0.494611366 |
| Obesity | mast_cell | *MANEA* | 2.79177896 | 0.000967471 | 0.001744664 | 1 |
| Obesity | mast_cell | *AFF1* | 2.797819099 | 0.000189599 | 0.000976811 | 0.919933849 |
| Obesity | mast_cell | *FIBP* | 3.601633456 | 7.54E-05 | 0.000934364 | 0.365882833 |
| Obesity | mast_cell | *CTSW* | -1.125662509 | 1.88E-06 | 0.000121571 | 0.009117861 |
| Obesity | mast_cell | *SNTG2* | 1.129285882 | 0.002752903 | 0.004179313 | 1 |
| Obesity | mast_cell | *SMPDL3A* | 1.855582187 | 0.000743519 | 0.001436127 | 1 |
| Obesity | mast_cell | *RAD9A* | 3.235971169 | 0.000200939 | 0.000976811 | 0.974957801 |
| Obesity | mast_cell | *EFEMP2* | 2.620726604 | 2.43E-05 | 0.000545024 | 0.118004413 |
| Obesity | mast_cell | *ZFAND4* | 2.546755679 | 0.027076942 | 0.033880087 | 1 |
| Obesity | mast_cell | *ZNF738* | 2.528564222 | 0.000255611 | 0.00098093 | 1 |
| Obesity | mast_cell | *SLFN11* | 1.80506432 | 0.000461813 | 0.001114555 | 1 |
| Obesity | mast_cell | *FAM71D* | -1.571640069 | 0.004793818 | 0.006861241 | 1 |
| Obesity | mast_cell | *ZNF596* | 2.636613627 | 0.000628947 | 0.001275241 | 1 |
| Obesity | mast_cell | *CFL1* | 3.378582436 | 0.000589102 | 0.001219421 | 1 |
| Obesity | mast_cell | *FAM192A* | 1.951492948 | 0.00052811 | 0.001158084 | 1 |
| Obesity | mast_cell | *RAB43* | -3.645420702 | 0.001541296 | 0.002571654 | 1 |
| Obesity | mast_cell | *CBWD1* | 3.295800602 | 0.000325179 | 0.001004947 | 1 |
| Obesity | mast_cell | *DCP2* | 2.061887268 | 0.000476037 | 0.001127398 | 1 |
| Obesity | mast_cell | *SNX32* | 3.016449789 | 2.52E-07 | 3.06E-05 | 0.001223774 |
| Obesity | mast_cell | *ZNF621* | 1.935110813 | 0.000527123 | 0.001157301 | 1 |
| Obesity | mast_cell | *NADSYN1* | 3.838576045 | 4.87E-05 | 0.000765648 | 0.236336425 |
| Obesity | mast_cell | *RNASEH2C* | 3.224090015 | 0.001430627 | 0.0024144 | 1 |
| Obesity | mast_cell | *MYEOV* | 6.826849837 | 0.016943552 | 0.021940249 | 1 |
| Obesity | mast_cell | *ANKRD13D* | 4.686059552 | 0.00127643 | 0.002191522 | 1 |

| | | | | | | |
|---|---|---|---|---|---|---|
| Obesity | mast_cell | *MRGPRF* | 3.91028424 | 1.34E-06 | 9.24E-05 | 0.006500388 |
| Obesity | mast_cell | *LCLAT1* | 1.897955835 | 0.000488794 | 0.001134939 | 1 |
| Obesity | mast_cell | *SH3RF3* | 2.002446199 | 0.000905862 | 0.001654835 | 1 |
| Obesity | mast_cell | *GXYLT2* | 1.376862969 | 0.000343452 | 0.001027777 | 1 |
| Obesity | mast_cell | *DCAKD* | 4.029551987 | 0.001066004 | 0.001883559 | 1 |
| Obesity | mast_cell | *TRMT112* | 1.827668437 | 0.006463305 | 0.008993392 | 1 |
| Obesity | mast_cell | *NOC3L* | 3.245711554 | 0.000197116 | 0.000976811 | 0.956404947 |
| Obesity | mast_cell | *RHOD* | 3.618049284 | 0.000221023 | 0.000976811 | 1 |
| Obesity | mast_cell | *PARP14* | 2.06007339 | 0.001177388 | 0.002054184 | 1 |
| Obesity | mast_cell | *PARP15* | 1.563468813 | 0.001979393 | 0.003151127 | 1 |
| Obesity | mast_cell | *AHSA2* | 2.446152174 | 0.000573925 | 0.001200332 | 1 |
| Obesity | mast_cell | *VANGL1* | 3.317387816 | 0.000218327 | 0.000976811 | 1 |
| Obesity | mast_cell | *IQCB1* | 2.291120315 | 0.005112008 | 0.007282285 | 1 |
| Obesity | mast_cell | *ZNF483* | 2.254563243 | 0.000500555 | 0.001142376 | 1 |
| Obesity | mast_cell | *MMRN2* | 3.275787262 | 3.10E-05 | 0.000621513 | 0.150293205 |
| Obesity | mast_cell | *MAP3K11* | 4.178286397 | 0.010023792 | 0.013479889 | 1 |
| Obesity | mast_cell | *NDNF* | 5.554083899 | 9.87E-05 | 0.000976811 | 0.478910846 |
| Obesity | mast_cell | *ARV1* | 2.345883321 | 0.000593681 | 0.001225761 | 1 |
| Obesity | mast_cell | *CCDC36* | 1.348912974 | 0.002918313 | 0.004405617 | 1 |
| Obesity | mast_cell | *EHBP1L1* | 1.703507537 | 0.00138096 | 0.002341985 | 1 |
| Obesity | mast_cell | *PPP1R14B* | -2.692789313 | 0.002467029 | 0.003812413 | 1 |
| Obesity | mast_cell | *PEAK1* | 2.345439911 | 0.000335145 | 0.001017877 | 1 |
| Obesity | mast_cell | *TNFRSF10D* | 2.375325053 | 0.000359632 | 0.00103557 | 1 |
| Obesity | mast_cell | *GMPPB* | 3.912411938 | 0.00071018 | 0.001387196 | 1 |
| Obesity | mast_cell | *NUDT18* | 6.518013709 | 0.000553803 | 0.00118075 | 1 |
| Obesity | mast_cell | *SULT1B1* | 5.110209697 | 0.003213617 | 0.004794732 | 1 |
| Obesity | mast_cell | *NUDT4* | 2.800735493 | 0.000379501 | 0.001048885 | 1 |
| Obesity | mast_cell | *SCAI* | 2.202595866 | 0.000946801 | 0.001716054 | 1 |
| Obesity | mast_cell | *UQCRH* | 2.35702857 | 0.000253162 | 0.000977535 | 1 |
| Obesity | mast_cell | *C11orf80* | 2.987202745 | 0.013320816 | 0.017534618 | 1 |
| Obesity | mast_cell | *CD7* | 13.703801 | 0.00033583 | 0.001017877 | 1 |
| Obesity | mast_cell | *RNF213* | 2.445633143 | 0.000465722 | 0.001116682 | 1 |
| Obesity | mast_cell | *TUBB8* | 3.763048236 | 0.000504246 | 0.001144783 | 1 |
| Obesity | mast_cell | *GPR160* | 2.731551864 | 0.001952816 | 0.003116798 | 1 |
| Obesity | mast_cell | *GOLIM4* | 2.658920312 | 0.000316811 | 0.000998908 | 1 |
| Obesity | mast_cell | *HOXB2* | 3.972591486 | 5.86E-06 | 0.000247224 | 0.028430741 |
| Obesity | mast_cell | *XXYLT1* | 4.773380444 | 0.000234138 | 0.000976811 | 1 |
| Obesity | mast_cell | *UBXN2A* | 3.830721249 | 6.94E-05 | 0.000898663 | 0.336575368 |
| Obesity | mast_cell | *SLC25A30* | 4.074497538 | 0.000170262 | 0.000976811 | 0.82611329 |
| Obesity | mast_cell | *CTSF* | 4.261452908 | 0.00387545 | 0.005670592 | 1 |
| Obesity | mast_cell | *LEMD3* | 4.538227917 | 0.000224834 | 0.000976811 | 1 |
| Obesity | mast_cell | *GLIS1* | 2.205354284 | 0.000532931 | 0.001162622 | 1 |
| Obesity | mast_cell | *RALGAPA1* | 3.390491319 | 0.000409714 | 0.001074715 | 1 |
| Obesity | mast_cell | *SNAPC5* | -3.58924118 | 0.013584875 | 0.017848311 | 1 |
| Obesity | mast_cell | *ANKRD36C* | 3.747774608 | 0.000243568 | 0.000976811 | 1 |
| Obesity | mast_cell | *MSL2* | 4.302196922 | 0.000173043 | 0.000976811 | 0.839605984 |
| Obesity | mast_cell | *ZNF497* | -8.9652918 | 0.002302082 | 0.003592699 | 1 |
| Obesity | mast_cell | *TRAM1L1* | -1.794136344 | 0.001777066 | 0.002877945 | 1 |
| Obesity | mast_cell | *ANGEL2* | 3.440526308 | 0.000153511 | 0.000976811 | 0.744833651 |
| Obesity | mast_cell | *ZNF266* | 2.125870285 | 0.000365754 | 0.001037687 | 1 |

| | | | | | | |
|---|---|---|---|---|---|---|
| Obesity | mast_cell | *SH3PXD2B* | 2.229619943 | 0.000428824 | 0.001092129 | 1 |
| Obesity | mast_cell | *KIAA1551* | 2.032517774 | 0.000366642 | 0.001037687 | 1 |
| Obesity | mast_cell | *LARP7* | 3.337049662 | 0.000266503 | 0.000985747 | 1 |
| Obesity | mast_cell | *FGFBP3* | 11.26518486 | 0.000152235 | 0.000976811 | 0.738645826 |
| Obesity | mast_cell | *C4orf32* | 2.192512822 | 0.00017032 | 0.000976811 | 0.826391149 |
| Obesity | mast_cell | *HRAS* | 3.905191247 | 0.000108025 | 0.000976811 | 0.524138099 |
| Obesity | mast_cell | *SRP72* | 3.148305853 | 1.07E-05 | 0.000373166 | 0.052144185 |
| Obesity | mast_cell | *CEP135* | 5.986777143 | 0.000113839 | 0.000976811 | 0.552344836 |
| Obesity | mast_cell | *NLRP6* | -16.93886136 | 0.000542069 | 0.001171675 | 1 |
| Obesity | mast_cell | *RSRC1* | 2.640257533 | 0.000992159 | 0.001776335 | 1 |
| Obesity | mast_cell | *PTDSS2* | 5.768028501 | 0.000244357 | 0.000976811 | 1 |
| Obesity | mast_cell | *SEZ6L2* | 7.659520962 | 3.01E-08 | 4.56E-06 | 0.000146075 |
| Obesity | mast_cell | *KCTD13* | 2.517223906 | 2.66E-06 | 0.000144867 | 0.012893155 |
| Obesity | mast_cell | *DHX36* | 2.967346119 | 0.0002209 | 0.000976811 | 1 |
| Obesity | mast_cell | *FBXW8* | 2.78513643 | 0.000158463 | 0.000976811 | 0.768864731 |
| Obesity | mast_cell | *SLC22A1* | -1.471771627 | 0.00250061 | 0.003855405 | 1 |
| Obesity | mast_cell | *CHST2* | -1.859461615 | 0.003162396 | 0.004728488 | 1 |
| Obesity | mast_cell | *TVP23C* | 9.055007247 | 0.000183119 | 0.000976811 | 0.88849341 |
| Obesity | mast_cell | *PACS1* | 2.469080887 | 0.007152394 | 0.009861727 | 1 |
| Obesity | mast_cell | *SH3BP5L* | 8.959894476 | 0.000122959 | 0.000976811 | 0.596597341 |
| Obesity | mast_cell | *CADM2* | 1.98462862 | 7.40E-07 | 6.24E-05 | 0.003591393 |
| Obesity | mast_cell | *PSMD2* | 2.719133195 | 0.000384567 | 0.001052047 | 1 |
| Obesity | mast_cell | *FAM182B* | 2.226850147 | 0.00037054 | 0.00103942 | 1 |
| Obesity | mast_cell | *ARHGAP1* | 4.451375915 | 0.00081659 | 0.001532014 | 1 |
| Obesity | mast_cell | *ATG13* | 2.738165583 | 0.000384992 | 0.001052047 | 1 |
| Obesity | mast_cell | *CCNE2* | 2.813431616 | 0.000493655 | 0.001138685 | 1 |
| Obesity | mast_cell | *ZNF519* | 3.307276277 | 0.013907056 | 0.018237036 | 1 |
| Obesity | mast_cell | *CHRNA7* | 3.326188153 | 0.005749341 | 0.008099826 | 1 |
| Obesity | mast_cell | *TMEM9B* | 2.133225292 | 0.000357912 | 0.001034916 | 1 |
| Obesity | mast_cell | *SCUBE2* | 4.778686067 | 0.001328817 | 0.002271666 | 1 |
| Obesity | mast_cell | *EIF1AD* | 11.50464124 | 1.01E-05 | 0.000363168 | 0.049027634 |
| Obesity | mast_cell | *EIF3F* | 3.46569003 | 0.017005936 | 0.022015155 | 1 |
| Obesity | mast_cell | *ZNF25* | 2.45876421 | 0.031637321 | 0.039179244 | 1 |
| Obesity | mast_cell | *CLTB* | 1.992558697 | 0.000457802 | 0.001112606 | 1 |
| Obesity | mast_cell | *PPP2R2D* | 3.67358916 | 6.94E-05 | 0.000898663 | 0.336614137 |
| Obesity | mast_cell | *MCTP1* | 1.76728506 | 0.000313773 | 0.000998908 | 1 |
| Obesity | mast_cell | *UCP2* | 2.281786501 | 0.005416164 | 0.007670527 | 1 |
| Obesity | mast_cell | *C11orf68* | 3.595354622 | 0.00049181 | 0.001138685 | 1 |
| Obesity | mast_cell | *MRPL48* | 2.779136505 | 0.000782203 | 0.001487241 | 1 |
| Obesity | mast_cell | *RAB6A* | 3.507139702 | 0.000267399 | 0.000985747 | 1 |
| Obesity | mast_cell | *TMEM70* | 2.632768667 | 0.000226088 | 0.000976811 | 1 |
| Obesity | mast_cell | *RPS6KB2* | 5.196571258 | 1.20E-05 | 0.000383871 | 0.058348405 |
| Obesity | mast_cell | *TOM1L2* | 2.945832899 | 0.000104139 | 0.000976811 | 0.505280673 |
| Obesity | mast_cell | *TEX26* | -1.749837654 | 0.001674109 | 0.002735863 | 1 |
| Obesity | mast_cell | *GPR156* | 3.374436347 | 0.014178882 | 0.018568404 | 1 |
| Obesity | mast_cell | *TTLL11* | 4.638957102 | 0.000173202 | 0.000976811 | 0.840376986 |
| Obesity | mast_cell | *HOXD8* | 1.678209685 | 0.037044301 | 0.045503531 | 1 |
| Obesity | mast_cell | *ZDHHC21* | 2.504214633 | 0.000354186 | 0.001031837 | 1 |
| Obesity | mast_cell | *A2M* | 2.639353318 | 0.000196215 | 0.000976811 | 0.952033786 |
| Obesity | mast_cell | *DOK7* | 17.29665044 | 0.000450669 | 0.001106294 | 1 |

| | | | | | | |
|---|---|---|---|---|---|---|
| Obesity | mast_cell | *ORAI3* | 12.76878983 | 1.22E-14 | 7.39E-12 | 5.91E-11 |
| Obesity | mast_cell | *B3GALT6* | -5.597769174 | 0.011934805 | 0.015856427 | 1 |
| Obesity | mast_cell | *ZNF613* | 3.017802592 | 0.000190585 | 0.000976811 | 0.924717029 |
| Obesity | mast_cell | *TCP11L1* | 2.532792975 | 0.00012276 | 0.000976811 | 0.595632538 |
| Obesity | mast_cell | *CCDC57* | 1.131182952 | 0.011235969 | 0.01498541 | 1 |
| Obesity | mast_cell | *MYPOP* | 4.573342847 | 4.41E-05 | 0.00074422 | 0.214108994 |
| Obesity | mast_cell | *CIDEA* | 1.998101534 | 0.00063332 | 0.00128149 | 1 |
| Obesity | mast_cell | *SMIM19* | 2.171162782 | 2.27E-05 | 0.000536798 | 0.109926346 |
| Obesity | mast_cell | *ZNF404* | 1.526058827 | 0.00644324 | 0.008968043 | 1 |
| Obesity | mast_cell | *SLC35G1* | 5.055909577 | 0.000217258 | 0.000976811 | 1 |
| Obesity | mast_cell | *OR4N2* | 7.190925585 | 0.000808384 | 0.001522033 | 1 |
| Obesity | mast_cell | *TAC4* | 2.644126461 | 0.000380845 | 0.001048903 | 1 |
| Obesity | mast_cell | *CDC26* | 1.877004213 | 0.013857966 | 0.01818249 | 1 |
| Obesity | mast_cell | *CRLF3* | 3.04392629 | 0.000261277 | 0.000981964 | 1 |
| Obesity | mast_cell | *VPS37D* | 2.93653513 | 0.001694701 | 0.00275916 | 1 |
| Obesity | mast_cell | *WDR25* | 3.565891776 | 0.000238691 | 0.000976811 | 1 |
| Obesity | mast_cell | *PLA2G16* | 1.800615632 | 0.018480716 | 0.023765819 | 1 |
| Obesity | mast_cell | *RMDN1* | 2.790751506 | 1.29E-05 | 0.000395043 | 0.062416767 |
| Obesity | mast_cell | *RNF152* | 1.604072854 | 0.000728012 | 0.001412361 | 1 |
| Obesity | mast_cell | *LRRC37A* | 2.038154251 | 0.004940064 | 0.007053911 | 1 |
| Obesity | mast_cell | *BDNF* | 7.036812625 | 0.000763535 | 0.001462564 | 1 |
| Obesity | mast_cell | *BOK* | 2.477681919 | 0.000385479 | 0.001052047 | 1 |
| Obesity | mast_cell | *RUFY1* | 3.547861179 | 1.93E-06 | 0.000123138 | 0.00935848 |
| Obesity | mast_cell | *VSIG10* | 3.844857051 | 0.000647286 | 0.001300468 | 1 |
| Obesity | mast_cell | *ANKLE2* | 3.599690172 | 0.000118908 | 0.000976811 | 0.576940896 |
| Obesity | mast_cell | *C8G* | 53.23610522 | 0.00056373 | 0.001189743 | 1 |
| Obesity | mast_cell | *MUC20* | -3.025435729 | 0.003642889 | 0.005367536 | 1 |
| Obesity | mast_cell | *LY6H* | 7.566307973 | 2.55E-05 | 0.000555569 | 0.123891986 |
| Obesity | mast_cell | *SHMT1* | 1.754103041 | 0.000456111 | 0.00111155 | 1 |
| Obesity | mast_cell | *DPP7* | 2.308317367 | 0.000369421 | 0.00103942 | 1 |
| Obesity | mast_cell | *MTHFR* | 2.589832343 | 0.000447632 | 0.001104518 | 1 |
| Obesity | mast_cell | *TMEM80* | 1.809166481 | 0.00440322 | 0.006364142 | 1 |
| Obesity | mast_cell | *WDR73* | 3.290775196 | 0.000270355 | 0.000986064 | 1 |
| Obesity | mast_cell | *DSCAML1* | 2.951007773 | 0.000254843 | 0.00098093 | 1 |
| Obesity | mast_cell | *TALDO1* | 6.940042273 | 0.000206614 | 0.000976811 | 1 |
| Obesity | mast_cell | *AP006621.1* | 5.506974211 | 2.74E-07 | 3.21E-05 | 0.001329793 |
| Obesity | mast_cell | *MAN1B1* | 4.544632272 | 0.000213898 | 0.000976811 | 1 |
| Obesity | mast_cell | *SAMD9L* | 2.621302517 | 0.000116529 | 0.000976811 | 0.565399768 |
| Obesity | mast_cell | *MIEF2* | 3.891747326 | 0.000624822 | 0.001270062 | 1 |
| Obesity | mast_cell | *NAP1L5* | 5.31387167 | 0.000164863 | 0.000976811 | 0.799913556 |
| Obesity | mast_cell | *RBM44* | 5.697811541 | 0.004227594 | 0.006141403 | 1 |
| Obesity | mast_cell | *SLC25A22* | -4.990695478 | 0.021079089 | 0.026780765 | 1 |
| Obesity | mast_cell | *GBA* | 4.019599258 | 6.07E-05 | 0.000855265 | 0.294500168 |
| Obesity | mast_cell | *ACAD9* | 1.294738174 | 0.002572043 | 0.003952038 | 1 |
| Obesity | mast_cell | *IL17RA* | 2.324956611 | 0.000780062 | 0.001486008 | 1 |
| Obesity | mast_cell | *MBOAT4* | 5.599960972 | 0.001636636 | 0.002690938 | 1 |
| Obesity | mast_cell | *AGTRAP* | 2.636099971 | 0.001965441 | 0.003133855 | 1 |
| Obesity | mast_cell | *NAALADL2* | 2.13892337 | 0.000969394 | 0.001746565 | 1 |
| Obesity | mast_cell | *CD151* | 2.46055954 | 6.08E-05 | 0.000855265 | 0.295066346 |
| Obesity | mast_cell | *SLC35G5* | 18.21689274 | 0.00049706 | 0.001138685 | 1 |

| | | | | | | |
|---|---|---|---|---|---|---|
| Obesity | mast_cell | *FAM87B* | 16.9859092 | 0.000474843 | 0.001127398 | 1 |
| Obesity | mast_cell | *PCDHB9* | -3.15649225 | 0.002797543 | 0.004239125 | 1 |
| Obesity | mast_cell | *CENPBD1* | -4.081846578 | 0.006969131 | 0.009633681 | 1 |
| Obesity | mast_cell | *RPS27* | 2.601134903 | 0.00023902 | 0.000976811 | 1 |
| Obesity | mast_cell | *RIC8A* | 2.609539388 | 0.000964112 | 0.001740926 | 1 |
| Obesity | mast_cell | *ASB8* | 6.455086394 | 0.000469838 | 0.001119672 | 1 |
| Obesity | mast_cell | *ODF3B* | 2.225475564 | 0.000527019 | 0.001157301 | 1 |
| Obesity | mast_cell | *DPY19L2* | 2.098670297 | 0.004557031 | 0.006557152 | 1 |
| Obesity | mast_cell | *ADAMTSL1* | 1.57414785 | 0.000352731 | 0.001031837 | 1 |
| Obesity | mast_cell | *IMPDH2* | 2.386309124 | 0.000510605 | 0.001145411 | 1 |
| Obesity | mast_cell | *GRAMD1C* | 1.616824567 | 0.00948211 | 0.0128046 | 1 |
| Obesity | mast_cell | *DALRD3* | 1.866636034 | 0.001116631 | 0.001960694 | 1 |
| Obesity | mast_cell | *ZNF454* | 1.820682822 | 0.00332716 | 0.004947404 | 1 |
| Obesity | mast_cell | *SH2B1* | 4.772209834 | 0.000185672 | 0.000976811 | 0.900879652 |
| Obesity | mast_cell | *KDELC2* | 3.42822963 | 0.000416568 | 0.001080612 | 1 |
| Obesity | mast_cell | *PLEC* | 2.379714116 | 0.019477948 | 0.024929308 | 1 |
| Obesity | mast_cell | *SH2D4B* | 18.51087044 | 0.032506218 | 0.040193723 | 1 |
| Obesity | mast_cell | *RNF212* | 2.051604423 | 0.000758189 | 0.00145577 | 1 |
| Obesity | mast_cell | *PRSS36* | 3.753147242 | 0.002779119 | 0.004216475 | 1 |
| Obesity | mast_cell | *WDR6* | 2.106246922 | 0.000203132 | 0.000976811 | 0.985598613 |
| Obesity | mast_cell | *GEN1* | 1.410232706 | 0.005761799 | 0.008112666 | 1 |
| Obesity | mast_cell | *TMPRSS9* | 4.014763428 | 0.002385881 | 0.003712731 | 1 |
| Obesity | mast_cell | *TMEM11* | 7.086858353 | 0.006045947 | 0.008461187 | 1 |
| Obesity | mast_cell | *ZFAND2A* | 4.100632225 | 0.00075498 | 0.001450502 | 1 |
| Obesity | mast_cell | *FAM220A* | 5.000719742 | 0.005114439 | 0.00728361 | 1 |
| Obesity | mast_cell | *NT5DC1* | 2.241339858 | 0.000276645 | 0.000991132 | 1 |
| Obesity | mast_cell | *GLDC* | 4.248402079 | 0.000429643 | 0.001092129 | 1 |
| Obesity | mast_cell | *P4HTM* | 2.780876784 | 0.000747693 | 0.001440352 | 1 |
| Obesity | mast_cell | *ERN1* | 3.446365685 | 0.000312955 | 0.000998908 | 1 |
| Obesity | mast_cell | *PARP10* | 2.775033179 | 0.00120096 | 0.00208305 | 1 |
| Obesity | mast_cell | *NSUN3* | 2.655698964 | 0.000211382 | 0.000976811 | 1 |
| Obesity | mast_cell | *RPP25* | -2.034439036 | 0.030115099 | 0.037389576 | 1 |
| Obesity | mast_cell | *STX19* | 6.745981837 | 8.30E-09 | 1.75E-06 | 4.03E-05 |
| Obesity | mast_cell | *MPI* | 3.222725636 | 3.16E-06 | 0.000161268 | 0.015320493 |
| Obesity | mast_cell | *TRIM73* | -22.4664619 | 2.81E-15 | 2.27E-12 | 1.36E-11 |
| Obesity | mast_cell | *EFCAB13* | 2.468953347 | 0.024531255 | 0.030819691 | 1 |
| Obesity | mast_cell | *EXOSC4* | -4.323242135 | 0.000520588 | 0.001152325 | 1 |
| Obesity | mast_cell | *C17orf62* | 2.17878626 | 0.000387039 | 0.00105395 | 1 |
| Obesity | mast_cell | *GAK* | 3.842835222 | 0.000186075 | 0.000976811 | 0.902835449 |
| Obesity | mast_cell | *TUFM* | 4.29613158 | 1.00E-08 | 1.87E-06 | 4.87E-05 |
| Obesity | mast_cell | *CTC1* | 3.326438203 | 0.000382041 | 0.00104964 | 1 |
| Obesity | mast_cell | *C3orf38* | 5.729668169 | 1.84E-09 | 5.59E-07 | 8.94E-06 |
| Obesity | mast_cell | *CCDC89* | 8.18273846 | 0.000540063 | 0.001171639 | 1 |
| Obesity | mast_cell | *HTR1F* | 5.891312533 | 0.014502494 | 0.018976834 | 1 |
| Obesity | mast_cell | *SPTY2D1* | 2.133430079 | 0.000449008 | 0.001104518 | 1 |
| Obesity | mast_cell | *ZNF664* | 3.73080259 | 0.00031258 | 0.000998908 | 1 |
| Obesity | mast_cell | *CDH4* | 2.411964179 | 0.001615331 | 0.002662224 | 1 |
| Obesity | mast_cell | *NSUN7* | 1.533742288 | 0.025548154 | 0.032030916 | 1 |
| Obesity | mast_cell | *PACS2* | 2.600708122 | 0.000297036 | 0.000998908 | 1 |
| Obesity | mast_cell | *GEMIN4* | 5.436415033 | 0.036821142 | 0.045252326 | 1 |

| | | | | | | |
|---|---|---|---|---|---|---|
| Obesity | mast_cell | *LBX2* | 2.616242953 | 0.011264212 | 0.015018949 | 1 |
| Obesity | mast_cell | *ZFPM1* | 2.333001481 | 0.000209679 | 0.000976811 | 1 |
| Obesity | mast_cell | *FCER1A* | -1.200135124 | 0.001619609 | 0.002666558 | 1 |
| Obesity | mast_cell | *RPRML* | 21.33312664 | 7.46E-07 | 6.24E-05 | 0.003620457 |
| Obesity | mast_cell | *SERTAD2* | 6.898267038 | 0.000117819 | 0.000976811 | 0.57165907 |
| Obesity | mast_cell | *GIPC3* | 13.31491455 | 0.000407929 | 0.001073348 | 1 |
| Obesity | mast_cell | *PDXDC1* | 4.975619627 | 0.000211612 | 0.000976811 | 1 |
| Obesity | mast_cell | *B3GNT3* | -2.446765524 | 0.00547592 | 0.007745063 | 1 |
| Obesity | mast_cell | *SEPHS2* | 2.814925974 | 0.000477166 | 0.001128819 | 1 |
| Obesity | mast_cell | *SSC5D* | 1.08836032 | 0.038360091 | 0.046965219 | 1 |
| Obesity | mast_cell | *ZADH2* | 3.145118833 | 0.000593641 | 0.001225761 | 1 |
| Obesity | mast_cell | *SEPT1* | 4.03328779 | 0.000400082 | 0.001064254 | 1 |
| Obesity | mast_cell | *EXOC3* | 2.479367284 | 0.000344274 | 0.001027971 | 1 |
| Obesity | mast_cell | *LYNX1* | 2.889751875 | 0.000759107 | 0.001456958 | 1 |
| Obesity | mast_cell | *HCLS1* | 1.468464709 | 0.002178812 | 0.00343011 | 1 |
| Obesity | mast_cell | *MTURN* | 1.859026332 | 0.000589815 | 0.001220377 | 1 |
| Obesity | mast_cell | *CCDC66* | 2.190088395 | 0.000317214 | 0.000998908 | 1 |
| Obesity | mast_cell | *MCFD2* | 2.624052791 | 0.000133845 | 0.000976811 | 0.649417879 |
| Obesity | mast_cell | *GLIPR1L2* | 3.003939517 | 0.004988368 | 0.007118695 | 1 |
| Obesity | mast_cell | *DEFB119* | -0.421124579 | 0.001542054 | 0.002571893 | 1 |
| Obesity | mast_cell | *TSPYL5* | 2.76043025 | 0.004391751 | 0.006349456 | 1 |
| Obesity | mast_cell | *MB21D2* | 2.28234692 | 0.000150575 | 0.000976811 | 0.730591264 |
| Obesity | mast_cell | *SLC47A2* | 2.723119489 | 0.001418451 | 0.002398022 | 1 |
| Obesity | mast_cell | *CHST13* | 19.97916416 | 0.00032667 | 0.001007521 | 1 |
| Obesity | mast_cell | *SLC36A4* | 3.26037043 | 0.000230698 | 0.000976811 | 1 |
| Obesity | mast_cell | *ZDHHC20* | 2.468605818 | 0.000260696 | 0.00098093 | 1 |
| Obesity | mast_cell | *PSMG4* | 2.6325234 | 0.000752452 | 0.001447619 | 1 |
| Obesity | mast_cell | *BHLHE22* | 2.475874462 | 0.040836456 | 0.049783539 | 1 |
| Obesity | mast_cell | *CAPS2* | 6.417317279 | 3.12E-05 | 0.000621513 | 0.151363319 |
| Obesity | mast_cell | *KCTD2* | 1.888840476 | 0.000714039 | 0.001391373 | 1 |
| Obesity | mast_cell | *D2HGDH* | 1.542365717 | 0.001695189 | 0.00275916 | 1 |
| Obesity | mast_cell | *CMTR2* | 5.317898256 | 8.80E-06 | 0.000334041 | 0.042683868 |
| Obesity | mast_cell | *LRRC57* | 2.293041336 | 0.001357574 | 0.002308779 | 1 |
| Obesity | mast_cell | *MRPL14* | 2.508611415 | 0.000167813 | 0.000976811 | 0.814229942 |
| Obesity | mast_cell | *LSMEM1* | 2.190403906 | 0.001374024 | 0.002332667 | 1 |
| Obesity | mast_cell | *NQO1* | 1.47283221 | 0.008749289 | 0.011894522 | 1 |
| Obesity | mast_cell | *FKRP* | 3.038381144 | 1.70E-05 | 0.000474831 | 0.082620508 |
| Obesity | mast_cell | *RPH3AL* | 2.361965566 | 0.000207612 | 0.000976811 | 1 |
| Obesity | mast_cell | *ADIPOQ* | 1.096472099 | 0.002379063 | 0.003703308 | 1 |
| Obesity | mast_cell | *F2R* | 2.021831435 | 0.012341963 | 0.016366003 | 1 |
| Obesity | mast_cell | *PENK* | -4.384979547 | 0.00732397 | 0.010083968 | 1 |
| Obesity | mast_cell | *SLC25A41* | 11.74680376 | 0.000213396 | 0.000976811 | 1 |
| Obesity | mast_cell | *ZNF322* | 1.768875895 | 0.015057539 | 0.019660705 | 1 |
| Obesity | mast_cell | *DDX60L* | 3.372082487 | 0.00019523 | 0.000976811 | 0.947256927 |
| Obesity | mast_cell | *OGFOD3* | 3.656192305 | 0.000265869 | 0.000985747 | 1 |
| Obesity | mast_cell | *TMEM45A* | 3.230173374 | 0.000907777 | 0.001656463 | 1 |
| Obesity | mast_cell | *SGSH* | 4.440753588 | 0.000306872 | 0.000998908 | 1 |
| Obesity | mast_cell | *ZFP41* | 6.619671729 | 1.41E-06 | 9.49E-05 | 0.006832345 |
| Obesity | mast_cell | *ZBTB20* | 2.541619422 | 0.000219164 | 0.000976811 | 1 |
| Obesity | mast_cell | *AMIGO1* | 4.148924202 | 0.000479918 | 0.001129748 | 1 |

| | | | | | | |
|---|---|---|---|---|---|---|
| Obesity | mast_cell | *TMEM252* | 99.15210226 | 0.00041902 | 0.00108267 | 1 |
| Obesity | mast_cell | *RELL1* | 2.630262642 | 8.15E-05 | 0.000957283 | 0.395355351 |
| Obesity | mast_cell | *RFX7* | 3.019845957 | 0.000202656 | 0.000976811 | 0.983285849 |
| Obesity | mast_cell | *CLDN7* | -2.027742965 | 0.000150021 | 0.000976811 | 0.727900787 |
| Obesity | mast_cell | *ZNF329* | 5.487046197 | 0.000204527 | 0.000976811 | 0.992366 |
| Obesity | mast_cell | *MGAT4C* | 1.549365803 | 0.000788756 | 0.001496693 | 1 |
| Obesity | mast_cell | *IDH2* | 2.246261606 | 0.000516442 | 0.001150279 | 1 |
| Obesity | mast_cell | *ZNF708* | 2.514150668 | 0.01175723 | 0.015633346 | 1 |
| Obesity | mast_cell | *ERCC6L2* | 2.081179572 | 0.000340057 | 0.001025052 | 1 |
| Obesity | mast_cell | *MRPL41* | 5.796744403 | 0.000195663 | 0.000976811 | 0.94935653 |
| Obesity | mast_cell | *ARL6IP4* | -2.440454768 | 0.009120892 | 0.012358924 | 1 |
| Obesity | mast_cell | *HHIPL1* | 3.036964861 | 0.000283926 | 0.000993978 | 1 |
| Obesity | mast_cell | *FAM153B* | 2.533414377 | 0.000346732 | 0.001028958 | 1 |
| Obesity | mast_cell | *SYNM* | 6.902870587 | 0.000363115 | 0.001037687 | 1 |
| Obesity | mast_cell | *KCNA4* | 2.85042432 | 0.000507654 | 0.001144962 | 1 |
| Obesity | mast_cell | *FIGN* | 1.625651797 | 0.000345364 | 0.00102869 | 1 |
| Obesity | mast_cell | *IZUMO1* | -1.260404844 | 0.012385171 | 0.016414326 | 1 |
| Obesity | mast_cell | *C8orf33* | 1.789005982 | 0.000854041 | 0.00158376 | 1 |
| Obesity | mast_cell | *KBTBD3* | 2.321716052 | 0.000537914 | 0.001169861 | 1 |
| Obesity | mast_cell | *CLN8* | 2.269355 | 2.04E-05 | 0.000522259 | 0.099215616 |
| Obesity | mast_cell | *NXPH4* | 24.18557953 | 0.000495763 | 0.001138685 | 1 |
| Obesity | mast_cell | *NPLOC4* | 2.45616792 | 0.038531487 | 0.047163162 | 1 |
| Obesity | mast_cell | *CAPN12* | 1.779067563 | 0.020178995 | 0.025751837 | 1 |
| Obesity | mast_cell | *KPNA2* | 3.126512306 | 0.000704895 | 0.001380207 | 1 |
| Obesity | mast_cell | *ORAI1* | 4.960245194 | 2.31E-05 | 0.000544178 | 0.112100612 |
| Obesity | mast_cell | *FES* | 3.089394382 | 9.07E-05 | 0.000976811 | 0.440238462 |
| Obesity | mast_cell | *GLRX5* | 2.243534533 | 0.000485782 | 0.001132608 | 1 |
| Obesity | mast_cell | *MXRA7* | 2.469484383 | 0.000242236 | 0.000976811 | 1 |
| Obesity | mast_cell | *MFSD5* | -2.124110302 | 0.007743433 | 0.010622317 | 1 |
| Obesity | mast_cell | *ADI1* | 1.805501007 | 0.000557428 | 0.001182832 | 1 |
| Obesity | mast_cell | *TRAK1* | 2.373553174 | 0.00293826 | 0.004430217 | 1 |
| Obesity | mast_cell | *SKA2* | 5.898224874 | 0.000185741 | 0.000976811 | 0.901213903 |
| Obesity | mast_cell | *TTC3* | 1.851426981 | 0.000533668 | 0.001163233 | 1 |
| Obesity | mast_cell | *PPP1R27* | -5.601653659 | 7.97E-05 | 0.000949918 | 0.386616426 |
| Obesity | mast_cell | *BRICD5* | 5.010392911 | 0.000484421 | 0.001131723 | 1 |
| Obesity | mast_cell | *GRID1* | 1.325885962 | 0.014188654 | 0.018576188 | 1 |
| Obesity | mast_cell | *ACBD3* | 4.496757594 | 0.000555286 | 0.001181687 | 1 |
| Obesity | mast_cell | *PLCXD3* | 3.735032929 | 0.000191431 | 0.000976811 | 0.928825531 |
| Obesity | mast_cell | *VMO1* | -1.007349063 | 0.008463675 | 0.011522376 | 1 |
| Obesity | mast_cell | *ALG12* | 4.460739562 | 0.000223987 | 0.000976811 | 1 |
| Obesity | mast_cell | *COL18A1* | 4.448757236 | 2.13E-06 | 0.000130935 | 0.010343835 |
| Obesity | mast_cell | *ZNF721* | 2.268993975 | 0.000220058 | 0.000976811 | 1 |
| Obesity | mast_cell | *C11orf54* | 1.28643976 | 0.004574576 | 0.006580446 | 1 |
| Obesity | mast_cell | *CEP63* | 1.769822074 | 0.002433944 | 0.003775414 | 1 |
| Obesity | mast_cell | *HMGN4* | 2.913402896 | 0.010858638 | 0.0145341 | 1 |
| Obesity | mast_cell | *C12orf60* | 2.894932479 | 0.000365294 | 0.001037687 | 1 |
| Obesity | mast_cell | *ABAT* | 3.380723684 | 0.000340769 | 0.001025052 | 1 |
| Obesity | mast_cell | *LYSMD4* | 2.313043275 | 5.00E-05 | 0.000765648 | 0.242505285 |
| Obesity | mast_cell | *WBP2NL* | 1.845263025 | 0.000742577 | 0.001435778 | 1 |
| Obesity | mast_cell | *ARHGEF37* | 2.609076262 | 0.000377337 | 0.001048135 | 1 |

| | | | | | | |
|---|---|---|---|---|---|---|
| Obesity | mast_cell | *CSMD1* | 2.28504441 | 0.000419335 | 0.00108267 | 1 |
| Obesity | mast_cell | *FANCF* | -3.610282942 | 0.012810541 | 0.016935389 | 1 |
| Obesity | mast_cell | *SMDT1* | 1.599894275 | 0.011392316 | 0.015177243 | 1 |
| Obesity | mast_cell | *RUVBL2* | 2.631938247 | 0.000393096 | 0.001054923 | 1 |
| Obesity | mast_cell | *GDPGP1* | 2.745559833 | 0.020579259 | 0.02620062 | 1 |
| Obesity | mast_cell | *PTTG1IP* | 2.276982975 | 0.00582603 | 0.00819597 | 1 |
| Obesity | mast_cell | *PLGLB1* | 4.839038338 | 3.06E-05 | 0.000621513 | 0.148634577 |
| Obesity | mast_cell | *ZNF623* | 4.756080721 | 0.003107454 | 0.004660701 | 1 |
| Obesity | mast_cell | *CCDC125* | 1.718207441 | 0.000468215 | 0.001116901 | 1 |
| Obesity | mast_cell | *C10orf107* | 1.923490042 | 0.000145006 | 0.000976811 | 0.703568906 |
| Obesity | mast_cell | *FHL3* | -1.382752892 | 0.023416899 | 0.029519042 | 1 |
| Obesity | mast_cell | *C19orf71* | 3.506943286 | 0.000548631 | 0.001175269 | 1 |
| Obesity | mast_cell | *LRIT3* | -1.722005949 | 0.001688523 | 0.002752003 | 1 |
| Obesity | mast_cell | *SF3A3* | 3.623323657 | 0.000505069 | 0.001144783 | 1 |
| Obesity | mast_cell | *GPR132* | 1.984708531 | 0.002442209 | 0.003780982 | 1 |
| Obesity | mast_cell | *PSMG1* | 6.518454861 | 3.16E-07 | 3.33E-05 | 0.001533436 |
| Obesity | mast_cell | *PCBP3* | 1.279803395 | 0.001185885 | 0.002063075 | 1 |
| Obesity | mast_cell | *SFXN4* | 5.307640405 | 0.000478842 | 0.001129371 | 1 |
| Obesity | mast_cell | *MRPL54* | 1.75445973 | 0.014694719 | 0.01921282 | 1 |
| Obesity | mast_cell | *HMCES* | 1.611364814 | 0.001476901 | 0.002481276 | 1 |
| Obesity | mast_cell | *NDUFB1* | 2.728716564 | 0.000310402 | 0.000998908 | 1 |
| Obesity | mast_cell | *FAM19A1* | 1.738323595 | 0.001192362 | 0.002073598 | 1 |
| Obesity | mast_cell | *TRMT12* | 3.105482713 | 0.002405462 | 0.003736011 | 1 |
| Obesity | mast_cell | *ALYREF* | 3.26896718 | 0.005215526 | 0.007418861 | 1 |
| Obesity | mast_cell | *MRGPRX2* | -0.485946069 | 0.001612585 | 0.002659504 | 1 |
| Obesity | mast_cell | *UPP1* | 2.903079706 | 3.19E-05 | 0.000621513 | 0.154866117 |
| Obesity | mast_cell | *FIGLA* | -4.454764881 | 0.003783466 | 0.005551066 | 1 |
| Obesity | mast_cell | *ACSM2A* | -5.124996678 | 0.001715334 | 0.002790078 | 1 |
| Obesity | mast_cell | *KREMEN1* | -0.752208762 | 0.008262176 | 0.011267026 | 1 |
| Obesity | mast_cell | *ZNF703* | 4.60145419 | 0.000114481 | 0.000976811 | 0.555460989 |
| Obesity | mast_cell | *EMILIN3* | 9.915731587 | 5.82E-06 | 0.000247224 | 0.028216092 |
| Obesity | mast_cell | *FAM162B* | 1.730369158 | 0.002227485 | 0.00348975 | 1 |
| Obesity | mast_cell | *BTBD9* | 2.686863744 | 0.000260532 | 0.00098093 | 1 |
| Obesity | mast_cell | *NUDT14* | 3.095800691 | 0.003639301 | 0.005365509 | 1 |
| Obesity | mast_cell | *TTC32* | 4.688630809 | 0.000192917 | 0.000976811 | 0.936034778 |
| Obesity | mast_cell | *DENND5A* | 4.279973288 | 0.000151996 | 0.000976811 | 0.737486375 |
| Obesity | mast_cell | *NIPSNAP1* | 3.06260185 | 0.003385671 | 0.005023632 | 1 |
| Obesity | mast_cell | *CNTN2* | -2.19385723 | 0.035442 | 0.043601568 | 1 |
| Obesity | mast_cell | *CRELD2* | 2.662442226 | 0.000186797 | 0.000976811 | 0.906340645 |
| Obesity | mast_cell | *PGP* | 2.390286021 | 0.006831165 | 0.009459136 | 1 |
| Obesity | mast_cell | *ACOT1* | 3.817638933 | 0.000167613 | 0.000976811 | 0.8132601 |
| Obesity | mast_cell | *ALDH1A3* | 1.182435942 | 0.013309365 | 0.017528547 | 1 |
| Obesity | mast_cell | *CLECL1* | 3.184594083 | 0.000603306 | 0.001238782 | 1 |
| Obesity | mast_cell | *MROH7* | -1.348527321 | 0.01726817 | 0.022307018 | 1 |
| Obesity | mast_cell | *A3GALT2* | 2.534292088 | 0.003378286 | 0.005014208 | 1 |
| Obesity | mast_cell | *SS18L1* | 5.53730067 | 0.000136744 | 0.000976811 | 0.663483966 |
| Obesity | mast_cell | *COPB2* | 5.9638883 | 0.00015849 | 0.000976811 | 0.768991946 |
| Obesity | mast_cell | *WDR27* | 3.542268599 | 5.29E-05 | 0.000791601 | 0.256708579 |
| Obesity | mast_cell | *TXNRD2* | 4.417541323 | 0.000188744 | 0.000976811 | 0.915788185 |
| Obesity | mast_cell | *TMEM255B* | 2.640027522 | 0.000503765 | 0.001144783 | 1 |

| | | | | | | |
|---|---|---|---|---|---|---|
| Obesity | mast_cell | *ZFP1* | 5.098174267 | 0.005256205 | 0.00746578 | 1 |
| Obesity | mast_cell | *DUSP8* | 4.443275314 | 0.00200394 | 0.003184774 | 1 |
| Obesity | mast_cell | *SNN* | 7.265252965 | 0.000234975 | 0.000976811 | 1 |
| Obesity | mast_cell | *RNLS* | 1.597064377 | 0.001149916 | 0.002010592 | 1 |
| Obesity | mast_cell | *NDUFA12* | 6.511785351 | 0.000156414 | 0.000976811 | 0.758919902 |
| Obesity | mast_cell | *TCTE3* | 2.240979152 | 0.000600081 | 0.001236345 | 1 |
| Obesity | mast_cell | *UBE2G2* | 2.305712209 | 0.000316273 | 0.000998908 | 1 |
| Obesity | mast_cell | *TMED9* | 2.880976134 | 0.000557326 | 0.001182832 | 1 |
| Obesity | mast_cell | *DRD1* | 56.00822883 | 0.000584993 | 0.001213504 | 1 |
| Obesity | mast_cell | *BTBD6* | 1.917680659 | 0.004503969 | 0.006490424 | 1 |
| Obesity | mast_cell | *NUTM2A* | -5.288646054 | 8.68E-05 | 0.000971162 | 0.420984723 |
| Obesity | mast_cell | *WT1* | 1.2667747 | 0.005452342 | 0.007717259 | 1 |
| Obesity | mast_cell | *ZFP90* | 5.316881079 | 0.000171925 | 0.000976811 | 0.834179268 |
| Obesity | mast_cell | *NOC4L* | 3.469950988 | 0.004254467 | 0.006173049 | 1 |
| Obesity | mast_cell | *NDUFA6* | 2.053419687 | 0.000182882 | 0.000976811 | 0.887344367 |
| Obesity | mast_cell | *BRI3BP* | 2.377406934 | 0.000544853 | 0.00117408 | 1 |
| Obesity | mast_cell | *SEMA4B* | 2.026859471 | 0.000559081 | 0.00118348 | 1 |
| Obesity | mast_cell | *CIB1* | 2.856022304 | 0.002003438 | 0.003184774 | 1 |
| Obesity | mast_cell | *SGCZ* | 1.421549332 | 0.001256965 | 0.002163459 | 1 |
| Obesity | mast_cell | *C6orf120* | 3.74969274 | 0.002204169 | 0.003462166 | 1 |
| Obesity | mast_cell | *SIGIRR* | 5.758283992 | 0.000102733 | 0.000976811 | 0.498459574 |
| Obesity | mast_cell | *IFITM2* | 4.233674173 | 0.00021647 | 0.000976811 | 1 |
| Obesity | mast_cell | *GP1BA* | 6.352296627 | 3.16E-09 | 7.68E-07 | 1.54E-05 |
| Obesity | mast_cell | *CCDC137* | 1.970791607 | 0.00033375 | 0.001016573 | 1 |
| Obesity | mast_cell | *ATP6V0A2* | -1.405622575 | 0.001298766 | 0.002225931 | 1 |
| Obesity | mast_cell | *C14orf80* | 5.093010762 | 0.000811989 | 0.001527043 | 1 |
| Obesity | mast_cell | *SP140L* | 3.485231848 | 0.000298406 | 0.000998908 | 1 |
| Obesity | mast_cell | *TARSL2* | 3.145813869 | 0.000102874 | 0.000976811 | 0.499143886 |
| Obesity | mast_cell | *FAM174B* | 2.027093433 | 0.000411243 | 0.001075307 | 1 |
| Obesity | mast_cell | *SV2B* | 2.053941688 | 0.003797787 | 0.00556871 | 1 |
| Obesity | mast_cell | *AHNAK2* | -1.976431446 | 0.003892202 | 0.005689956 | 1 |
| Obesity | mast_cell | *SP1* | 2.901901777 | 0.000113943 | 0.000976811 | 0.552851434 |
| Obesity | mast_cell | *PCGF3* | 3.030791034 | 5.63E-05 | 0.000817301 | 0.272978638 |
| Obesity | mast_cell | *C6orf201* | 3.473961516 | 0.03801197 | 0.046562504 | 1 |
| Obesity | mast_cell | *MYBL1* | 2.161232432 | 0.004621319 | 0.006637845 | 1 |
| Obesity | mast_cell | *ADARB2* | 10.73059964 | 0.001195775 | 0.002076558 | 1 |
| Obesity | mast_cell | *DMWD* | 4.266904996 | 0.001685079 | 0.002748237 | 1 |
| Obesity | mast_cell | *SLC52A2* | 10.04411412 | 1.10E-05 | 0.000373166 | 0.053323362 |
| Obesity | mast_cell | *ARL17A* | 3.262573985 | 0.000893051 | 0.001638217 | 1 |
| Obesity | mast_cell | *GNB1L* | -2.262403313 | 0.007102087 | 0.009803506 | 1 |
| Obesity | mast_cell | *EVI2B* | 1.53500031 | 0.002648264 | 0.004048322 | 1 |
| Obesity | mast_cell | *TRIM69* | 1.206866359 | 0.037503286 | 0.045997458 | 1 |
| Obesity | mast_cell | *IFITM1* | -1.388022822 | 0.00449297 | 0.006478422 | 1 |
| Obesity | mast_cell | *LINC00839* | 7.281757964 | 0.000512035 | 0.001145411 | 1 |
| Obesity | mast_cell | *RTN4RL1* | 3.063752501 | 0.000899544 | 0.001645151 | 1 |
| Obesity | mast_cell | *RNPC3* | 2.717071251 | 9.07E-05 | 0.000976811 | 0.44019979 |
| Obesity | mast_cell | *RASA3* | 2.034339022 | 0.000359259 | 0.001035264 | 1 |
| Obesity | mast_cell | *ZNF529* | 4.628083124 | 0.0001739 | 0.000976811 | 0.843762174 |
| Obesity | mast_cell | *C15orf41* | 3.473885965 | 3.92E-05 | 0.000686693 | 0.190162153 |
| Obesity | mast_cell | *WWOX* | 2.734907016 | 0.000178176 | 0.000976811 | 0.864510307 |

| | | | | | | |
|---|---|---|---|---|---|---|
| Obesity | mast_cell | *POLR1D* | 3.485226096 | 5.47E-05 | 0.000804876 | 0.26560912 |
| Obesity | mast_cell | *FFAR4* | 2.042114479 | 0.000918648 | 0.001672525 | 1 |
| Obesity | mast_cell | *NBPF12* | 1.664581516 | 0.007375246 | 0.010145929 | 1 |
| Obesity | mast_cell | *TOR3A* | 3.173602316 | 0.000724302 | 0.001406873 | 1 |
| Obesity | mast_cell | *ZNF555* | 2.647049164 | 0.000248706 | 0.000976811 | 1 |
| Obesity | mast_cell | *CCDC30* | 2.005588495 | 0.000297913 | 0.000998908 | 1 |
| Obesity | mast_cell | *GLDN* | 0.73349682 | 0.002823054 | 0.004272445 | 1 |
| Obesity | mast_cell | *ZNF197* | 2.357656468 | 0.000545375 | 0.00117408 | 1 |
| Obesity | mast_cell | *RPS23* | 2.78967836 | 0.000152649 | 0.000976811 | 0.740653441 |
| Obesity | mast_cell | *GNG2* | 3.261539322 | 0.000164418 | 0.000976811 | 0.79775572 |
| Obesity | mast_cell | *BTN3A2* | -2.653013113 | 5.36E-10 | 1.73E-07 | 2.60E-06 |
| Obesity | mast_cell | *TMEM222* | 7.062170903 | 0.00067433 | 0.001335448 | 1 |
| Obesity | mast_cell | *SEPT10* | 1.542363661 | 0.00092817 | 0.001686065 | 1 |
| Obesity | mast_cell | *FAM86B1* | 4.251829812 | 0.023187593 | 0.029245179 | 1 |
| Obesity | mast_cell | *SMYD4* | 3.694065401 | 0.000102626 | 0.000976811 | 0.497939594 |
| Obesity | mast_cell | *MIR22HG* | 2.631809157 | 0.000333866 | 0.001016573 | 1 |
| Obesity | mast_cell | *BCR* | 2.480870481 | 0.000140359 | 0.000976811 | 0.681021662 |
| Obesity | mast_cell | *ZNF732* | 4.292978845 | 0.000536246 | 0.001167804 | 1 |
| Obesity | mast_cell | *VSIG10L* | 5.89841571 | 0.001925153 | 0.003076693 | 1 |
| Obesity | mast_cell | *TRABD2A* | 2.605534035 | 3.09E-05 | 0.000621513 | 0.149882366 |
| Obesity | mast_cell | *QRFPR* | 9.427055137 | 0.025244994 | 0.031675384 | 1 |
| Obesity | mast_cell | *MAPT* | 2.565184688 | 0.000367743 | 0.001037687 | 1 |
| Obesity | mast_cell | *TNFRSF18* | 7.415378922 | 0.002609762 | 0.003999546 | 1 |
| Obesity | mast_cell | *C1QL4* | 28.73307603 | 0.000570555 | 0.001194792 | 1 |
| Obesity | mast_cell | *RTN4RL2* | -3.324349402 | 0.001212126 | 0.002098193 | 1 |
| Obesity | mast_cell | *TMEM232* | 1.768898419 | 0.001146027 | 0.002004514 | 1 |
| Obesity | mast_cell | *EFCAB6* | 1.547845708 | 0.000550805 | 0.001177314 | 1 |
| Obesity | mast_cell | *RHD* | 1.83225561 | 0.001056538 | 0.001870475 | 1 |
| Obesity | mast_cell | *TMPRSS6* | 16.70477728 | 0.000634524 | 0.001281728 | 1 |
| Obesity | mast_cell | *TMEM216* | 4.282445808 | 0.000559575 | 0.001183547 | 1 |
| Obesity | mast_cell | *C3orf70* | 1.758404191 | 0.004609931 | 0.006623449 | 1 |
| Obesity | mast_cell | *ENTPD5* | 2.166528946 | 0.003259076 | 0.004859569 | 1 |
| Obesity | mast_cell | *MRPS21* | 2.032466382 | 0.000163119 | 0.000976811 | 0.791454575 |
| Obesity | mast_cell | *ANGPTL5* | -2.48145703 | 0.000289055 | 0.000995384 | 1 |
| Obesity | mast_cell | *WDR86* | 1.484679132 | 0.01037278 | 0.013926045 | 1 |
| Obesity | mast_cell | *EPOR* | 3.84115904 | 0.002920845 | 0.004406698 | 1 |
| Obesity | mast_cell | *LUZP2* | 4.431686064 | 0.000334234 | 0.001016742 | 1 |
| Obesity | mast_cell | *GJA4* | 4.299528779 | 0.0002511 | 0.000976811 | 1 |
| Obesity | mast_cell | *IFT140* | 2.917659728 | 0.00032939 | 0.001011931 | 1 |
| Obesity | mast_cell | *SBK2* | 22.61714958 | 0.000447451 | 0.001104518 | 1 |
| Obesity | mast_cell | *ZNF385C* | 2.946277729 | 0.006783493 | 0.009395806 | 1 |
| Obesity | mast_cell | *EXD3* | 5.628392844 | 0.000169909 | 0.000976811 | 0.824399768 |
| Obesity | mast_cell | *C17orf97* | 0.664510063 | 0.028189148 | 0.035160345 | 1 |
| Obesity | mast_cell | *DHRS4L2* | 1.538204242 | 0.037645055 | 0.046136349 | 1 |
| Obesity | mast_cell | *SAMD11* | 4.859961596 | 0.038808819 | 0.04749064 | 1 |
| Obesity | mast_cell | *C2orf88* | 2.715404923 | 0.00042059 | 0.001083175 | 1 |
| Obesity | mast_cell | *KBTBD12* | 2.015349508 | 0.000427272 | 0.001091691 | 1 |
| Obesity | mast_cell | *FANCM* | 3.254445267 | 0.003177385 | 0.004746513 | 1 |
| Obesity | mast_cell | *ZFP69* | 3.391238494 | 0.000371717 | 0.00104107 | 1 |
| Obesity | mast_cell | *TMEM220* | 3.472034121 | 0.000439357 | 0.001098845 | 1 |

| | | | | | | |
|---|---|---|---|---|---|---|
| Obesity | mast_cell | *FAM122A* | 2.906764212 | 0.000507553 | 0.001144962 | 1 |
| Obesity | mast_cell | *GFRAL* | 6.291741395 | 0.000166636 | 0.000976811 | 0.808519452 |
| Obesity | mast_cell | *LDLRAD2* | 2.208382942 | 0.006215301 | 0.00867817 | 1 |
| Obesity | mast_cell | *CPSF4L* | 10.90724006 | 0.000436325 | 0.001098433 | 1 |
| Obesity | mast_cell | *KLHL17* | 3.801941792 | 0.008145202 | 0.011123142 | 1 |
| Obesity | mast_cell | *ZSCAN23* | 2.512253167 | 2.20E-05 | 0.000534512 | 0.106902408 |
| Obesity | mast_cell | *TMEM221* | 3.552456744 | 0.035050298 | 0.043130623 | 1 |
| Obesity | mast_cell | *MESP2* | 3.002675524 | 0.030721923 | 0.038113723 | 1 |
| Obesity | mast_cell | *MAPK12* | 4.126467968 | 0.002222719 | 0.00348563 | 1 |
| Obesity | mast_cell | *AGRN* | 2.909645359 | 0.000462915 | 0.001115779 | 1 |
| Obesity | mast_cell | *FAM166A* | -5.336514596 | 0.012795069 | 0.016920598 | 1 |
| Obesity | mast_cell | *ZNF626* | 4.458820885 | 0.000237995 | 0.000976811 | 1 |
| Obesity | mast_cell | *LINC00265* | 3.074630336 | 0.000299328 | 0.000998908 | 1 |
| Obesity | mast_cell | *LAMTOR4* | 3.275848777 | 0.000133256 | 0.000976811 | 0.646560333 |
| Obesity | mast_cell | *PRKAR1B* | 4.149995987 | 0.000786232 | 0.001492486 | 1 |
| Obesity | mast_cell | *NCR3LG1* | -2.970253139 | 0.005369161 | 0.007610624 | 1 |
| Obesity | mast_cell | *ZNF793* | 1.430078273 | 0.00701889 | 0.00969694 | 1 |
| Obesity | mast_cell | *AGAP4* | 2.005455778 | 0.010323347 | 0.013867353 | 1 |
| Obesity | mast_cell | *RUFY4* | 4.915693193 | 0.000484521 | 0.001131723 | 1 |
| Obesity | mast_cell | *ZNF669* | 3.612337714 | 0.002203861 | 0.003462166 | 1 |
| Obesity | mast_cell | *CENPP* | 2.403704762 | 3.85E-07 | 3.89E-05 | 0.00186876 |
| Obesity | mast_cell | *PLSCR1* | 2.574246444 | 0.000207522 | 0.000976811 | 1 |
| Obesity | mast_cell | *C3orf62* | 2.257240581 | 0.000470223 | 0.00112004 | 1 |
| Obesity | mast_cell | *ENO4* | 1.963075772 | 0.000566616 | 0.001191687 | 1 |
| Obesity | mast_cell | *ZP3* | 2.263487919 | 6.65E-05 | 0.000885378 | 0.322457703 |
| Obesity | mast_cell | *H3F3C* | -9.400541813 | 0.001568179 | 0.00260067 | 1 |
| Obesity | mast_cell | *PDCD1* | 17.55217472 | 0.000313053 | 0.000998908 | 1 |
| Obesity | mast_cell | *SELL* | 2.183846687 | 0.021811535 | 0.027631741 | 1 |
| Obesity | mast_cell | *C19orf54* | 3.72185739 | 0.000202692 | 0.000976811 | 0.983462418 |
| Obesity | mast_cell | *C22orf34* | 1.731466162 | 0.000376002 | 0.001047881 | 1 |
| Obesity | mast_cell | *COL25A1* | 1.393469314 | 0.000524112 | 0.001154856 | 1 |
| Obesity | mast_cell | *SRSF10* | 2.39641653 | 0.000777901 | 0.001482613 | 1 |
| Obesity | mast_cell | *NBR1* | 2.539312012 | 0.000276858 | 0.000991132 | 1 |
| Obesity | mast_cell | *NDOR1* | 4.05098615 | 0.000312756 | 0.000998908 | 1 |
| Obesity | mast_cell | *FAM72B* | 5.44619709 | 0.011153975 | 0.014892429 | 1 |
| Obesity | mast_cell | *GOLGA8M* | 2.603260829 | 0.000898668 | 0.001644379 | 1 |
| Obesity | mast_cell | *RHCE* | 8.855974609 | 0.002199689 | 0.003458486 | 1 |
| Obesity | mast_cell | *IDO2* | 6.068467325 | 0.001521015 | 0.002543062 | 1 |
| Obesity | mast_cell | *SLC4A5* | 7.529635715 | 0.000300463 | 0.000998908 | 1 |
| Obesity | mast_cell | *UROS* | 3.197897352 | 0.000275648 | 0.000990086 | 1 |
| Obesity | mast_cell | *ZBED6CL* | -3.565701447 | 0.021075182 | 0.026780765 | 1 |
| Obesity | mast_cell | *OSTN* | 3.958729318 | 4.87E-06 | 0.00022485 | 0.023609211 |
| Obesity | mast_cell | *FSIP2* | 2.254674462 | 0.000282677 | 0.000993978 | 1 |
| Obesity | mast_cell | *NOXA1* | 2.152083444 | 0.000565887 | 0.001190669 | 1 |
| Obesity | mast_cell | *FZD9* | -3.073899068 | 0.001356071 | 0.002307381 | 1 |
| Obesity | mast_cell | *MTF1* | 9.530073413 | 0.000421742 | 0.00108454 | 1 |
| Obesity | mast_cell | *FAM26F* | 2.244378422 | 0.002103077 | 0.003322739 | 1 |
| Obesity | mast_cell | *ENTPD8* | 236.6792114 | 0.001398974 | 0.00236757 | 1 |
| Obesity | mast_cell | *RPL14* | 3.442751356 | 0.000184517 | 0.000976811 | 0.895277597 |
| Obesity | mast_cell | *FBF1* | 3.88426861 | 2.54E-06 | 0.000141479 | 0.012308632 |

| | | | | | | |
|---|---|---|---|---|---|---|
| Obesity | mast_cell | *ASTL* | -6.282361611 | 0.005221927 | 0.007425788 | 1 |
| Obesity | mast_cell | *MSL1* | 3.18851748 | 0.007030952 | 0.009710839 | 1 |
| Obesity | mast_cell | *LRRK2* | 1.912392595 | 0.002305495 | 0.003595713 | 1 |
| Obesity | mast_cell | *FAM196A* | 2.955618217 | 0.000169694 | 0.000976811 | 0.823353394 |
| Obesity | mast_cell | *UTS2B* | 2.447622674 | 0.003726992 | 0.005474831 | 1 |
| Obesity | mast_cell | *C9orf152* | -2.891361955 | 0.007169999 | 0.009883191 | 1 |
| Obesity | mast_cell | *SLC15A5* | 3.037586371 | 0.000790328 | 0.001498503 | 1 |
| Obesity | mast_cell | *ALKBH2* | 3.944971169 | 0.000611752 | 0.001251358 | 1 |
| Obesity | mast_cell | *LITAF* | 3.712472887 | 0.000165804 | 0.000976811 | 0.804482281 |
| Obesity | mast_cell | *SF3B3* | 2.786635954 | 0.000162968 | 0.000976811 | 0.790721461 |
| Obesity | mast_cell | *PLAC9* | 4.777160446 | 9.75E-05 | 0.000976811 | 0.473086247 |
| Obesity | mast_cell | *ZNF573* | 2.793294323 | 0.001367442 | 0.002323933 | 1 |
| Obesity | mast_cell | *FAM47E* | 7.198817346 | 0.002267974 | 0.003547456 | 1 |
| Obesity | mast_cell | *TSPYL1* | 9.400657813 | 0.001312762 | 0.002245952 | 1 |
| Obesity | mast_cell | *PNRC2* | 1.943318325 | 0.000507411 | 0.001144962 | 1 |
| Obesity | mast_cell | *FHIT* | 2.018891338 | 0.000191987 | 0.000976811 | 0.931520932 |
| Obesity | mast_cell | *SLC35E2B* | 2.647235692 | 3.13E-05 | 0.000621513 | 0.152044231 |
| Obesity | mast_cell | *HLA-DRB1* | 1.603028927 | 0.000488376 | 0.001134866 | 1 |
| Obesity | mast_cell | *AKR1C3* | 6.450522874 | 0.000189613 | 0.000976811 | 0.920002203 |
| Obesity | mast_cell | *WDSUB1* | 3.010314182 | 0.000744121 | 0.001436719 | 1 |
| Obesity | mast_cell | *SEMA4A* | 5.701978929 | 2.36E-05 | 0.000545024 | 0.114700627 |
| Obesity | mast_cell | *MPHOSPH8* | 2.095573787 | 0.000156412 | 0.000976811 | 0.758909653 |
| Obesity | mast_cell | *SIRPB2* | 1.160079169 | 0.020977995 | 0.026673279 | 1 |
| Obesity | mast_cell | *SRGAP3* | 4.478884289 | 1.11E-05 | 0.000373166 | 0.054030278 |
| Obesity | mast_cell | *PPIA* | 3.783295714 | 5.42E-05 | 0.000802079 | 0.263081751 |
| Obesity | mast_cell | *ZNF471* | 3.356147672 | 0.000346402 | 0.00102869 | 1 |
| Obesity | mast_cell | *ZNF493* | 2.996465962 | 0.000419911 | 0.00108267 | 1 |
| Obesity | mast_cell | *GTF2IRD2* | 3.140760905 | 0.000231115 | 0.000976811 | 1 |
| Obesity | mast_cell | *SUPT3H* | 3.64798123 | 0.00018237 | 0.000976811 | 0.884859897 |
| Obesity | mast_cell | *GIMAP5* | 2.774742471 | 0.00143288 | 0.002417063 | 1 |
| Obesity | mast_cell | *STK31* | 3.708372052 | 0.000141088 | 0.000976811 | 0.684558436 |
| Obesity | mast_cell | *ZKSCAN7* | 2.8626378 | 6.24E-05 | 0.000864528 | 0.302584845 |
| Obesity | mast_cell | *CD55* | 1.909983046 | 0.000350572 | 0.001031837 | 1 |
| Obesity | mast_cell | *ZNF565* | 4.311246294 | 0.000127774 | 0.000976811 | 0.619958211 |
| Obesity | mast_cell | *LONP1* | 3.469402297 | 5.25E-05 | 0.000791601 | 0.254665207 |
| Obesity | mast_cell | *ZNF34* | 3.785185956 | 0.000402926 | 0.001065021 | 1 |
| Obesity | mast_cell | *EVL* | 2.843893029 | 0.000619888 | 0.001262844 | 1 |
| Obesity | mast_cell | *ZNF124* | 2.791334185 | 0.000435865 | 0.001098433 | 1 |
| Obesity | mast_cell | *PIK3R4* | 3.754852514 | 0.000238667 | 0.000976811 | 1 |
| Obesity | mast_cell | *ZNF605* | 2.33366615 | 1.03E-05 | 0.000365431 | 0.050064059 |
| Obesity | mast_cell | *RFX8* | 1.570721996 | 0.003918202 | 0.005724516 | 1 |
| Obesity | mast_cell | *C20orf96* | 2.535043172 | 0.000425979 | 0.001090111 | 1 |
| Obesity | mast_cell | *SULT1A1* | 2.661638943 | 0.007576614 | 0.010408192 | 1 |
| Obesity | mast_cell | *TPK1* | 2.216084564 | 0.00020365 | 0.000976811 | 0.98811161 |
| Obesity | mast_cell | *MYO18A* | 5.410498511 | 0.001015797 | 0.00181317 | 1 |
| Obesity | mast_cell | *CACNA1H* | 8.885332579 | 5.67E-06 | 0.000243327 | 0.027495927 |
| Obesity | mast_cell | *SULF2* | 1.988937948 | 0.000175792 | 0.000976811 | 0.852942815 |
| Obesity | mast_cell | *XRCC2* | 5.082465156 | 0.000521247 | 0.001152731 | 1 |
| Obesity | mast_cell | *MYO6* | 3.118791804 | 0.001130936 | 0.001983124 | 1 |
| Obesity | mast_cell | *ZNF846* | 1.995243765 | 0.000855864 | 0.001584536 | 1 |

| | | | | | | |
|---|---|---|---|---|---|---|
| Obesity | mast_cell | *ADH1B* | 1.739324156 | 0.000273264 | 0.000987746 | 1 |
| Obesity | mast_cell | *ZKSCAN5* | 4.809655538 | 3.73E-05 | 0.000675733 | 0.18098955 |
| Obesity | mast_cell | *ZNF502* | 4.940253528 | 0.000306923 | 0.000998908 | 1 |
| Obesity | mast_cell | *TRAPPC4* | 2.75523063 | 0.005858735 | 0.008234815 | 1 |
| Obesity | mast_cell | *ZFP62* | 5.418174148 | 1.32E-05 | 0.000401199 | 0.064191796 |
| Obesity | mast_cell | *TOMM7* | 3.048143399 | 0.000227566 | 0.000976811 | 1 |
| Obesity | mast_cell | *ZNF33B* | 3.777984496 | 0.000153403 | 0.000976811 | 0.744311601 |
| Obesity | mast_cell | *AMZ2* | 6.635633197 | 0.000494589 | 0.001138685 | 1 |
| Obesity | mast_cell | *ZNF431* | 3.136645801 | 0.00023212 | 0.000976811 | 1 |
| Obesity | mast_cell | *COL27A1* | 1.681782891 | 0.011438022 | 0.015229771 | 1 |
| Obesity | mast_cell | *GM2A* | 2.161522372 | 0.000234503 | 0.000976811 | 1 |
| Obesity | mast_cell | *ZNF239* | 5.468239409 | 2.27E-05 | 0.000536798 | 0.11004356 |
| Obesity | mast_cell | *ANKRD36B* | 2.905530942 | 0.003256316 | 0.004856945 | 1 |
| Obesity | mast_cell | *FAM3C* | 5.91146571 | 3.09E-05 | 0.000621513 | 0.149956048 |
| Obesity | mast_cell | *SLC39A10* | 3.610093847 | 0.000181293 | 0.000976811 | 0.879636052 |
| Obesity | mast_cell | *AP2A1* | 2.451465294 | 0.00019726 | 0.000976811 | 0.957103193 |
| Obesity | mast_cell | *PCDHB16* | 1.653691804 | 0.034875288 | 0.042958847 | 1 |
| Obesity | mast_cell | *HIST1H3E* | 2.596873594 | 0.002921757 | 0.004406703 | 1 |
| Obesity | mast_cell | *FUT11* | 6.058801151 | 0.000339469 | 0.001024319 | 1 |
| Obesity | mast_cell | *WDR5B* | -2.331341225 | 0.011277312 | 0.015032285 | 1 |
| Obesity | mast_cell | *ZNF138* | 7.627265176 | 6.16E-05 | 0.000858537 | 0.298770901 |
| Obesity | mast_cell | *ZNF429* | 2.810824184 | 0.000295806 | 0.000998908 | 1 |
| Obesity | mast_cell | *ZNF470* | -2.479325943 | 0.029098238 | 0.036210477 | 1 |
| Obesity | mast_cell | *ANXA6* | 3.047809269 | 0.001221605 | 0.002113094 | 1 |
| Obesity | mast_cell | *GMFB* | 5.009570812 | 0.000192219 | 0.000976811 | 0.932646729 |
| Obesity | mast_cell | *ZNF420* | 2.945014275 | 0.000411329 | 0.001075307 | 1 |
| Obesity | mast_cell | *ZMYM1* | 3.775453854 | 0.000209937 | 0.000976811 | 1 |
| Obesity | mast_cell | *IGF2R* | 2.18842341 | 0.000432955 | 0.001095825 | 1 |
| Obesity | mast_cell | *GAL3ST4* | 4.040025933 | 0.0034078 | 0.005051832 | 1 |
| Obesity | mast_cell | *SLC25A29* | 4.358072482 | 3.78E-05 | 0.000679348 | 0.183498087 |
| Obesity | mast_cell | *ZNF257* | 3.317627932 | 0.019213576 | 0.024636436 | 1 |
| Obesity | mast_cell | *ADAM32* | 3.618798232 | 0.000279382 | 0.000993813 | 1 |
| Obesity | mast_cell | *LRRC8B* | 3.584170285 | 8.83E-05 | 0.000976811 | 0.428513716 |
| Obesity | mast_cell | *ABCB8* | 2.258480348 | 0.000270225 | 0.000986064 | 1 |
| Obesity | mast_cell | *SND1* | 2.997357024 | 9.98E-05 | 0.000976811 | 0.484005536 |
| Obesity | mast_cell | *SULT1A2* | 16.34217307 | 1.45E-26 | 7.01E-23 | 7.01E-23 |
| Obesity | mast_cell | *NEK5* | 1.272093968 | 0.016197138 | 0.021052375 | 1 |
| Obesity | mast_cell | *ENTPD4* | 2.231573868 | 0.000385245 | 0.001052047 | 1 |
| Obesity | mast_cell | *TBC1D9B* | 3.079715907 | 0.00091027 | 0.00165895 | 1 |
| Obesity | mast_cell | *FAM110D* | 5.294801393 | 0.000565174 | 0.001190078 | 1 |
| Obesity | mast_cell | *CCL4L2* | 6.772009256 | 0.000408126 | 0.001073348 | 1 |
| Obesity | mast_cell | *ZNF165* | -3.861744169 | 3.36E-05 | 0.000630711 | 0.162948532 |
| Obesity | mast_cell | *RAMP2-AS1* | 1.866578177 | 0.000566968 | 0.001191911 | 1 |
| Obesity | mast_cell | *ZNF720* | 2.681691944 | 0.000419795 | 0.00108267 | 1 |
| Obesity | mast_cell | *ZNF98* | 1.820377055 | 0.036752346 | 0.045179221 | 1 |
| Obesity | mast_cell | *ZNF517* | 10.10352036 | 3.85E-05 | 0.000683409 | 0.186570782 |
| Obesity | mast_cell | *ZNF675* | 4.963040816 | 0.001839874 | 0.002966789 | 1 |
| Obesity | mast_cell | *SLC22A5* | 3.240067178 | 0.000486813 | 0.001132608 | 1 |
| Obesity | mast_cell | *VEPH1* | 2.044106789 | 0.000939851 | 0.001706008 | 1 |
| Obesity | mast_cell | *SHPK* | 2.79503229 | 0.00219949 | 0.003458486 | 1 |

| | | | | | | |
|---|---|---|---|---|---|---|
| Obesity | mast_cell | *MAP3K5* | 1.956421324 | 0.000210744 | 0.000976811 | 1 |
| Obesity | mast_cell | *GSTK1* | 1.766877193 | 0.000528324 | 0.001158084 | 1 |
| Obesity | mast_cell | *GYPE* | 4.008329281 | 0.000677733 | 0.001338364 | 1 |
| Obesity | mast_cell | *SLC2A10* | 4.910159713 | 0.038877816 | 0.047551087 | 1 |
| Obesity | mast_cell | *ZNF665* | 6.801720411 | 0.000162536 | 0.000976811 | 0.78862358 |
| Obesity | mast_cell | *RPF2* | 7.715387971 | 0.000717295 | 0.001396595 | 1 |
| Obesity | mast_cell | *MYO5A* | 1.438482976 | 0.000714031 | 0.001391373 | 1 |
| Obesity | mast_cell | *ATG7* | 1.642143826 | 0.00085703 | 0.001585931 | 1 |
| Obesity | mast_cell | *RAB40C* | 4.072493999 | 0.000214274 | 0.000976811 | 1 |
| Obesity | mast_cell | *PIGN* | 1.818068448 | 0.012654622 | 0.016743994 | 1 |
| Obesity | mast_cell | *HHLA3* | 1.611902598 | 0.016361076 | 0.021237009 | 1 |
| Obesity | mast_cell | *TOPORS* | 8.063458302 | 0.000430878 | 0.001093993 | 1 |
| Obesity | mast_cell | *BCO2* | 1.006422609 | 0.030658602 | 0.038054626 | 1 |
| Obesity | mast_cell | *ENPP1* | 1.828825321 | 0.000765116 | 0.001464435 | 1 |
| Obesity | mast_cell | *MFAP5* | 0.949248081 | 0.026012285 | 0.032587556 | 1 |
| Obesity | mast_cell | *PDCD1LG2* | 1.488584367 | 0.002898825 | 0.004378923 | 1 |
| Obesity | mast_cell | *DNAH10* | 4.091927913 | 0.003405707 | 0.005050272 | 1 |
| Obesity | mast_cell | *NMB* | 4.229938337 | 0.006514465 | 0.009054192 | 1 |
| Obesity | mast_cell | *FAM114A1* | 2.96162526 | 0.00013375 | 0.000976811 | 0.648954368 |
| Obesity | mast_cell | *PSAP* | 1.691428863 | 0.000320398 | 0.000999138 | 1 |
| Obesity | mast_cell | *TXNRD3* | 1.991114225 | 0.000256337 | 0.00098093 | 1 |
| Obesity | mast_cell | *EME2* | -3.485963163 | 0.013970791 | 0.018315665 | 1 |
| Obesity | mast_cell | *AC122129.1* | 21.19243952 | 6.72E-05 | 0.000885378 | 0.326025646 |
| Obesity | mast_cell | *CCDC180* | -2.571988981 | 0.00358365 | 0.005294723 | 1 |
| Obesity | mast_cell | *OCLN* | -1.633095741 | 0.001931149 | 0.003085259 | 1 |
| Obesity | mast_cell | *ZNF181* | 2.071405315 | 0.002022005 | 0.00320928 | 1 |
| Obesity | mast_cell | *SGTB* | 2.077071494 | 0.000443331 | 0.00110084 | 1 |
| Obesity | mast_cell | *ADH5* | 2.941761348 | 0.000276994 | 0.000991132 | 1 |
| Obesity | mast_cell | *HRNR* | 0.780853037 | 0.019912012 | 0.025437884 | 1 |
| Obesity | mast_cell | *ZNF677* | 2.16788923 | 0.000384695 | 0.001052047 | 1 |
| Obesity | mast_cell | *ZNF71* | 2.562231766 | 0.000346224 | 0.00102869 | 1 |
| Obesity | mast_cell | *RPL12* | 2.397294296 | 0.001009425 | 0.001803953 | 1 |
| Obesity | mast_cell | *MPZL1* | 1.673244884 | 0.000356617 | 0.001033017 | 1 |
| Obesity | mast_cell | *VPS13A* | 2.275768604 | 0.00041641 | 0.001080612 | 1 |
| Obesity | mast_cell | *NOL8* | 3.721931397 | 0.004485323 | 0.006469318 | 1 |
| Obesity | mast_cell | *DLGAP2* | 1.660450644 | 0.000139671 | 0.000976811 | 0.677682603 |
| Obesity | mast_cell | *ZNF560* | 4.781060748 | 0.001098193 | 0.001932692 | 1 |
| Obesity | mast_cell | *AGAP9* | 3.164851087 | 0.006770803 | 0.009380906 | 1 |
| Obesity | mast_cell | *MAK16* | 8.983708697 | 0.005377643 | 0.007620421 | 1 |
| Obesity | mast_cell | *ZNF667* | 2.486812022 | 0.002395588 | 0.003725393 | 1 |
| Obesity | mast_cell | *SFI1* | 3.612672179 | 0.000404857 | 0.001068171 | 1 |
| Obesity | mast_cell | *HIBCH* | 1.50616628 | 0.011099143 | 0.01482738 | 1 |
| Obesity | mast_cell | *ZNF544* | 2.955184013 | 0.000372917 | 0.001042278 | 1 |
| Obesity | mast_cell | *SOWAHC* | 5.046943724 | 5.27E-05 | 0.000791601 | 0.255928679 |
| Obesity | mast_cell | *AC135178.1* | -7.533084708 | 0.002049667 | 0.003247872 | 1 |
| Obesity | mast_cell | *MIER1* | 3.302853017 | 0.000112817 | 0.000976811 | 0.547389029 |
| Obesity | mast_cell | *MAN1A2* | 2.91044356 | 0.000206067 | 0.000976811 | 0.999836269 |
| Obesity | mast_cell | *SVIP* | 3.002118484 | 0.000280603 | 0.000993978 | 1 |
| Obesity | mast_cell | *DDRGK1* | 2.44096293 | 0.000379252 | 0.001048885 | 1 |
| Obesity | mast_cell | *ZNF607* | 2.590355284 | 0.014945666 | 0.01952514 | 1 |

| | | | | | | |
|---|---|---|---|---|---|---|
| Obesity | mast_cell | *ZNF334* | 2.9471094 | 0.001371512 | 0.002329219 | 1 |
| Obesity | mast_cell | *HSD17B11* | 4.179208684 | 9.60E-05 | 0.000976811 | 0.465943832 |
| Obesity | mast_cell | *SULT1C2* | 1.626834462 | 0.009402058 | 0.012703643 | 1 |
| Obesity | mast_cell | *RPS6KL1* | 4.648508629 | 0.002784555 | 0.004222081 | 1 |
| Obesity | mast_cell | *DDX42* | 2.462829872 | 0.000140426 | 0.000976811 | 0.681347037 |
| Obesity | mast_cell | *STYX* | 3.995778708 | 0.000102186 | 0.000976811 | 0.495805458 |
| Obesity | mast_cell | *HELZ* | 2.258658832 | 0.000245846 | 0.000976811 | 1 |
| Obesity | mast_cell | *TMEM116* | 1.886222916 | 0.000890378 | 0.001633931 | 1 |
| Obesity | mast_cell | *UCKL1* | 4.369803248 | 0.000236258 | 0.000976811 | 1 |
| Obesity | mast_cell | *ZNF442* | -1.596976571 | 0.023925258 | 0.030120745 | 1 |
| Obesity | mast_cell | *GFPT1* | 3.224029578 | 0.000166112 | 0.000976811 | 0.805976765 |
| Obesity | mast_cell | *ZNF568* | 2.414414172 | 0.000348355 | 0.001029993 | 1 |
| Obesity | mast_cell | *ZNF587* | 3.481909904 | 0.000668574 | 0.001326427 | 1 |
| Obesity | mast_cell | *FLVCR1-AS1* | 1.233846133 | 0.004800754 | 0.006869142 | 1 |
| Obesity | mast_cell | *SH3BGRL2* | 1.95943748 | 2.73E-05 | 0.000577669 | 0.132286098 |
| Obesity | mast_cell | *TMA16* | 2.87550599 | 0.000269956 | 0.000986064 | 1 |
| Obesity | mast_cell | *CNGA1* | 1.749202985 | 0.001624729 | 0.002674079 | 1 |
| Obesity | mast_cell | *ZNF28* | 6.005434407 | 0.000425767 | 0.001090111 | 1 |
| Obesity | mast_cell | *C20orf203* | -1.822281636 | 0.000718119 | 0.001397639 | 1 |
| Obesity | mast_cell | *NCOA6* | 1.986238676 | 0.009204118 | 0.012463963 | 1 |
| Obesity | mast_cell | *FAM19A2* | 3.017544226 | 0.000492244 | 0.001138685 | 1 |
| Obesity | mast_cell | *TTC37* | 3.285198904 | 0.000755146 | 0.001450502 | 1 |
| Obesity | mast_cell | *TUSC1* | 3.523823181 | 0.000628831 | 0.001275241 | 1 |
| Obesity | mast_cell | *PAPSS2* | 1.812597289 | 0.000365538 | 0.001037687 | 1 |
| Obesity | mast_cell | *CEP290* | 3.850994592 | 0.000265114 | 0.000985696 | 1 |
| Obesity | mast_cell | *ANKRD13B* | 4.199814735 | 0.000419713 | 0.00108267 | 1 |
| Obesity | mast_cell | *ECI2* | 5.556382609 | 0.000467859 | 0.001116682 | 1 |
| Obesity | mast_cell | *UNC13B* | 2.005014584 | 0.000657433 | 0.001312467 | 1 |
| Obesity | mast_cell | *PPP1R14C* | 8.613714066 | 0.000508387 | 0.001144962 | 1 |
| Obesity | mast_cell | *RPL10A* | 2.821709716 | 0.000211621 | 0.000976811 | 1 |
| Obesity | mast_cell | *COLGALT2* | 4.246219695 | 0.000390381 | 0.001054382 | 1 |
| Obesity | mast_cell | *RCSD1* | 1.556394919 | 0.000438287 | 0.001098434 | 1 |
| Obesity | mast_cell | *GRIN3A* | -2.994455997 | 0.003513982 | 0.005194954 | 1 |
| Obesity | mast_cell | *TMEM184B* | 1.621069407 | 0.006004101 | 0.008409006 | 1 |
| Obesity | mast_cell | *SFT2D1* | 3.759142688 | 0.000159036 | 0.000976811 | 0.771640938 |
| Obesity | mast_cell | *HMGN2* | 1.904743651 | 0.001638494 | 0.002692168 | 1 |
| Obesity | mast_cell | *TYW1* | 2.441467271 | 0.000595109 | 0.001228188 | 1 |
| Obesity | mast_cell | *ITPRIPL1* | -1.480682593 | 0.027332234 | 0.03417652 | 1 |
| Obesity | mast_cell | *PRMT6* | -2.698152423 | 0.001639602 | 0.002693077 | 1 |
| Obesity | mast_cell | *SHISA4* | -2.558351507 | 0.000191016 | 0.000976811 | 0.926807253 |
| Obesity | mast_cell | *MAP3K3* | 2.688637206 | 0.022205577 | 0.028108912 | 1 |
| Obesity | mast_cell | *C1orf174* | 3.337892644 | 0.000448205 | 0.001104518 | 1 |
| Obesity | mast_cell | *DZIP3* | 1.862899614 | 0.000408671 | 0.001074145 | 1 |
| Obesity | mast_cell | *KIAA0753* | 1.738544764 | 0.000501447 | 0.001143874 | 1 |
| Obesity | mast_cell | *CCDC167* | 8.569010097 | 0.00048918 | 0.001135103 | 1 |
| Obesity | mast_cell | *ZFP2* | 3.400047642 | 0.000509677 | 0.001145411 | 1 |
| Obesity | mast_cell | *MFAP3L* | 3.292459547 | 0.000445903 | 0.001103405 | 1 |
| Obesity | mast_cell | *NAGA* | 2.213584512 | 0.001404812 | 0.002376621 | 1 |
| Obesity | mast_cell | *TGM2* | 1.86364058 | 0.001552857 | 0.002582949 | 1 |
| Obesity | mast_cell | *PJA2* | 2.39774978 | 0.000390909 | 0.001054382 | 1 |

| | | | | | | |
|---|---|---|---|---|---|---|
| Obesity | mast_cell | *AL590822.1* | -1.877184949 | 0.005690558 | 0.008026333 | 1 |
| Obesity | mast_cell | *AP001007.1* | 5.000249547 | 0.000446111 | 0.001103405 | 1 |
| Obesity | mast_cell | *EFCAB2* | 2.362682913 | 0.000265353 | 0.000985747 | 1 |
| Obesity | mast_cell | *TATDN3* | 2.687414789 | 0.000196926 | 0.000976811 | 0.955487107 |
| Obesity | mast_cell | *SAMD5* | 3.10988631 | 0.000484871 | 0.001131723 | 1 |
| Obesity | mast_cell | *ECT2L* | 3.566359629 | 0.00047971 | 0.001129748 | 1 |
| Obesity | mast_cell | *DDO* | 4.005580554 | 0.000688349 | 0.001355939 | 1 |
| Obesity | mast_cell | *RBM20* | 3.788515865 | 0.002460401 | 0.003804291 | 1 |
| Obesity | mast_cell | *PCMTD2* | 4.304728881 | 0.000176989 | 0.000976811 | 0.858752306 |
| Obesity | mast_cell | *SAMD13* | 3.187806974 | 0.00036605 | 0.001037687 | 1 |
| Obesity | mast_cell | *EFCAB7* | 4.633578839 | 0.000152593 | 0.000976811 | 0.740380754 |
| Obesity | mast_cell | *IFIT1B* | 13.83811515 | 0.000656462 | 0.001312378 | 1 |
| Obesity | mast_cell | *LIPN* | 6.321647893 | 0.000475053 | 0.001127398 | 1 |
| Obesity | mast_cell | *NEU4* | 4.119713783 | 0.032575713 | 0.04026939 | 1 |
| Obesity | mast_cell | *C2orf72* | 11.67448785 | 0.039711631 | 0.048509777 | 1 |
| Obesity | mast_cell | *RUFY2* | 2.66891554 | 0.000233278 | 0.000976811 | 1 |
| Obesity | mast_cell | *PHACTR4* | 2.810227782 | 0.001496759 | 0.002511161 | 1 |
| Obesity | mast_cell | *ASAH2B* | 3.591505521 | 0.000482869 | 0.001131281 | 1 |
| Obesity | mast_cell | *AGAP6* | 2.671401874 | 0.000970024 | 0.001747051 | 1 |
| Obesity | mast_cell | *SYT15* | 4.118727185 | 0.032854001 | 0.04060306 | 1 |
| Obesity | mast_cell | *LINC00619* | -2.503338864 | 0.001660059 | 0.002719313 | 1 |
| Obesity | mast_cell | *BMPR2* | 3.892806431 | 0.000112737 | 0.000976811 | 0.54699782 |
| Obesity | mast_cell | *TCEA3* | 2.327609186 | 0.001888024 | 0.003027327 | 1 |
| Obesity | mast_cell | *HSD17B8* | -1.45551125 | 0.002994015 | 0.004508678 | 1 |
| Obesity | mast_cell | *OXLD1* | 1.965973874 | 0.004527557 | 0.006518607 | 1 |
| Obesity | mast_cell | *TAP2* | 1.225925005 | 0.017752723 | 0.022890303 | 1 |
| Obesity | mast_cell | *COL15A1* | 1.38313792 | 0.000443138 | 0.00110084 | 1 |
| Obesity | mast_cell | *NOTCH4* | -0.878486442 | 1.20E-08 | 2.04E-06 | 5.83E-05 |
| Obesity | mast_cell | *LAYN* | 2.698461121 | 0.000568115 | 0.001192771 | 1 |
| Obesity | mast_cell | *C6orf48* | 2.661266673 | 0.000471341 | 0.001122153 | 1 |
| Obesity | mast_cell | *LST1* | 2.069537762 | 0.000142201 | 0.000976811 | 0.689957315 |
| Obesity | mast_cell | *ZNF814* | 4.75176625 | 2.43E-05 | 0.000545024 | 0.117842592 |
| Obesity | mast_cell | *HLA-C* | 2.292633007 | 2.12E-05 | 0.000524672 | 0.102638083 |
| Obesity | mast_cell | *POU5F1* | 1.370205079 | 1.70E-05 | 0.000474831 | 0.082407467 |
| Obesity | mast_cell | *CCHCR1* | 2.007409599 | 2.96E-07 | 3.21E-05 | 0.001438469 |
| Obesity | mast_cell | *DHX16* | 2.909805609 | 0.000499204 | 0.00114144 | 1 |
| Obesity | mast_cell | *ABCF1* | 2.165965588 | 4.29E-05 | 0.00073545 | 0.208132354 |
| Obesity | mast_cell | *LILRB3* | 3.338012194 | 0.000559299 | 0.00118348 | 1 |
| Obesity | mast_cell | *HLA-E* | 2.312731444 | 5.46E-05 | 0.000804558 | 0.26469947 |
| Obesity | mast_cell | *ZNF468* | -5.754251973 | 0.006315247 | 0.008802521 | 1 |
| Obesity | mast_cell | *RNF39* | -5.986942302 | 0.000830914 | 0.001553002 | 1 |
| Obesity | mast_cell | *HCG9* | -1.302588737 | 0.000816634 | 0.001532014 | 1 |
| Obesity | mast_cell | *HLA-G* | 2.399862447 | 0.000555961 | 0.001182125 | 1 |
| Obesity | mast_cell | *ZFP57* | 3.079396714 | 2.37E-09 | 6.78E-07 | 1.15E-05 |
| Obesity | mast_cell | *TRIM27* | 1.953605337 | 0.019380728 | 0.024837637 | 1 |
| Obesity | mast_cell | *MALRD1* | 5.254560727 | 0.000113289 | 0.000976811 | 0.54967701 |
| Obesity | mast_cell | *FAM196B* | 4.053272286 | 0.000296088 | 0.000998908 | 1 |
| Obesity | mast_cell | *DCTN1* | 6.940248318 | 0.039955648 | 0.048795571 | 1 |
| Obesity | mast_cell | *FAM216A* | 3.078436635 | 0.000652253 | 0.001308725 | 1 |
| Obesity | mast_cell | *ZBTB48* | 3.950321125 | 8.70E-05 | 0.000971162 | 0.421993628 |

| | | | | | | |
|---|---|---|---|---|---|---|
| Obesity | mast_cell | *MZT1* | 3.14602839 | 0.000462265 | 0.001114767 | 1 |
| Obesity | mast_cell | *ZNF155* | 2.142429191 | 0.000269817 | 0.000986064 | 1 |
| Obesity | mast_cell | *ZNF783* | 2.910334695 | 0.001063106 | 0.001880492 | 1 |
| Obesity | mast_cell | *C12orf73* | 5.619648555 | 0.008999398 | 0.012214008 | 1 |
| Obesity | mast_cell | *PCDHGA1* | -3.660806788 | 0.003414133 | 0.00505813 | 1 |
| Obesity | mast_cell | *TRIM13* | 3.196580457 | 9.87E-05 | 0.000976811 | 0.478704856 |
| Obesity | mast_cell | *PKHD1L1* | 1.062263959 | 0.000439866 | 0.001099087 | 1 |
| Obesity | mast_cell | *SLFN12L* | 1.242304514 | 0.006506918 | 0.009046294 | 1 |
| Obesity | mast_cell | *SLC35B4* | 4.410340171 | 0.000121847 | 0.000976811 | 0.591200652 |
| Obesity | mast_cell | *TMEM231* | 3.702636654 | 0.000316316 | 0.000998908 | 1 |
| Obesity | mast_cell | *LINC00654* | 3.549823485 | 0.004367785 | 0.006318573 | 1 |
| Obesity | mast_cell | *SCGB2B2* | 3.768635912 | 0.000240223 | 0.000976811 | 1 |
| Obesity | mast_cell | *CCDC144NL* | 5.445931477 | 0.000278808 | 0.000992498 | 1 |
| Obesity | mast_cell | *TMEM170B* | 1.503458432 | 0.008537547 | 0.011619685 | 1 |
| Obesity | mast_cell | *GOLGA6L10* | -4.205546974 | 0.039104571 | 0.047816376 | 1 |
| Obesity | mast_cell | *SAP25* | -4.998189088 | 0.018663629 | 0.023981973 | 1 |
| Obesity | mast_cell | *IPO7* | 4.992071908 | 0.000270259 | 0.000986064 | 1 |
| Obesity | mast_cell | *PRR13* | 2.042261411 | 0.012421157 | 0.016457524 | 1 |
| Obesity | mast_cell | *TECPR1* | 2.299088384 | 0.000323333 | 0.001002436 | 1 |
| Obesity | mast_cell | *MT1A* | -3.76512238 | 0.004038673 | 0.005891653 | 1 |
| Obesity | mast_cell | *CFI* | 1.851962862 | 0.000405468 | 0.0010692 | 1 |
| Obesity | mast_cell | *ATP6AP1L* | 1.232440762 | 0.004408983 | 0.006370574 | 1 |
| Obesity | mast_cell | *RGL3* | 1.72615571 | 0.000675997 | 0.001336567 | 1 |
| Obesity | mast_cell | *NAP1L4* | 3.15326298 | 0.000860382 | 0.001590921 | 1 |
| Obesity | mast_cell | *SMN2* | -2.131915027 | 6.71E-05 | 0.000885378 | 0.325752107 |
| Obesity | mast_cell | *LIN52* | 3.038065619 | 0.000143299 | 0.000976811 | 0.695285277 |
| Obesity | mast_cell | *C17orf107* | 5.825699594 | 4.96E-05 | 0.000765648 | 0.240640957 |
| Obesity | mast_cell | *KLRC2* | 5.322804012 | 0.00013748 | 0.000976811 | 0.667053547 |
| Obesity | mast_cell | *C1QTNF9B* | -1.913003771 | 0.001684072 | 0.002747518 | 1 |
| Obesity | mast_cell | *ZDHHC11B* | 7.483230893 | 0.027576826 | 0.034440865 | 1 |
| Obesity | mast_cell | *GOLGA8O* | -3.748848941 | 0.011025837 | 0.014737565 | 1 |
| Obesity | mast_cell | *HCG27* | 1.18906585 | 0.015028622 | 0.019628229 | 1 |
| Obesity | mast_cell | *COL6A6* | 1.974654749 | 0.000446982 | 0.001104256 | 1 |
| Obesity | mast_cell | *HLA-A* | 2.220814011 | 0.003421644 | 0.005067709 | 1 |
| Obesity | mast_cell | *METTL6* | 2.692441191 | 0.000288638 | 0.00099522 | 1 |
| Obesity | mast_cell | *SACM1L* | 3.916004533 | 0.000203831 | 0.000976811 | 0.988989123 |
| Obesity | mast_cell | *SLC48A1* | 2.191486733 | 0.000406376 | 0.001071013 | 1 |
| Obesity | mast_cell | *TAS2R50* | 37.72097147 | 0.009620232 | 0.012980722 | 1 |
| Obesity | mast_cell | *TAS2R14* | 2.069093955 | 0.008160684 | 0.011141147 | 1 |
| Obesity | mast_cell | *C17orf51* | 9.672738414 | 0.000199096 | 0.000976811 | 0.966011736 |
| Obesity | mast_cell | *ZNF580* | 3.047220249 | 0.001256876 | 0.002163459 | 1 |
| Obesity | mast_cell | *FGFR1OP* | 8.578002002 | 0.000118656 | 0.000976811 | 0.575720294 |
| Obesity | mast_cell | *ZNF254* | 2.088505979 | 0.000994046 | 0.001778433 | 1 |
| Obesity | mast_cell | *TCTEX1D2* | 6.396804589 | 4.72E-05 | 0.000758703 | 0.228858446 |
| Obesity | mast_cell | *ASIC3* | 5.87419166 | 2.92E-05 | 0.000612418 | 0.141468518 |
| Obesity | mast_cell | *GIMAP1* | 2.263923373 | 0.000239648 | 0.000976811 | 1 |
| Obesity | mast_cell | *NOTCH2NL* | 2.448634348 | 0.021047247 | 0.026754321 | 1 |
| Obesity | mast_cell | *SUPT4H1* | 6.462215426 | 1.62E-05 | 0.000463269 | 0.07875574 |
| Obesity | mast_cell | *MXD3* | 3.741981772 | 0.000235606 | 0.000976811 | 1 |
| Obesity | mast_cell | *GSTM2* | 1.864990382 | 0.000108117 | 0.000976811 | 0.524582821 |

| | | | | | | |
|---|---|---|---|---|---|---|
| Obesity | mast_cell | *LCAT* | 2.471759572 | 0.000561811 | 0.001187242 | 1 |
| Obesity | mast_cell | *ERV3-1* | 1.920079473 | 2.96E-06 | 0.000157569 | 0.014338794 |
| Obesity | mast_cell | *SRA1* | 4.189411688 | 1.27E-05 | 0.000393798 | 0.061826364 |
| Obesity | mast_cell | *TMEM110* | 11.51411699 | 1.78E-12 | 8.65E-10 | 8.65E-09 |
| Obesity | mast_cell | *TMX2* | 3.446034075 | 0.002783015 | 0.004221066 | 1 |
| Obesity | mast_cell | *LBH* | 1.936657051 | 0.001640516 | 0.002693592 | 1 |
| Obesity | mast_cell | *PPP1CB* | 1.93797885 | 0.019796151 | 0.025303194 | 1 |
| Obesity | mast_cell | *NCKIPSD* | 2.258273085 | 0.0061326 | 0.008575036 | 1 |
| Obesity | mast_cell | *ATF6B* | 3.093259221 | 0.000825867 | 0.001545951 | 1 |
| Obesity | mast_cell | *SLC35F6* | 3.779578776 | 0.000495466 | 0.001138685 | 1 |
| Obesity | mast_cell | *ATP6V1G2* | -1.837097319 | 0.001682517 | 0.002745904 | 1 |
| Obesity | mast_cell | *EMP2* | 2.792032697 | 0.000173856 | 0.000976811 | 0.843547064 |
| Obesity | mast_cell | *SLC23A3* | 3.60099417 | 1.60E-05 | 0.000459428 | 0.077643249 |
| Obesity | mast_cell | *LTB4R* | 2.777630995 | 0.000774444 | 0.001479447 | 1 |
| Obesity | mast_cell | *IRF9* | 2.061179906 | 0.0003552 | 0.00103227 | 1 |
| Obesity | mast_cell | *GALT* | 10.41847715 | 3.27E-05 | 0.000621513 | 0.158485854 |
| Obesity | mast_cell | *ZNF726* | -5.283429285 | 0.006201094 | 0.008660826 | 1 |
| Obesity | mast_cell | *ZNF90* | 2.41212198 | 0.001181644 | 0.002060128 | 1 |
| Obesity | mast_cell | *REPIN1* | 3.509172845 | 0.000392831 | 0.001054923 | 1 |
| Obesity | mast_cell | *SMIM7* | 3.683553537 | 0.000333969 | 0.001016573 | 1 |
| Obesity | mast_cell | *TSPAN4* | 5.542639739 | 9.51E-06 | 0.000349581 | 0.046144709 |
| Obesity | mast_cell | *PAXIP1-AS2* | 3.52981264 | 0.000138164 | 0.000976811 | 0.670370333 |
| Obesity | mast_cell | *LYRM4* | 2.932912351 | 0.000211392 | 0.000976811 | 1 |
| Obesity | mast_cell | *PRCD* | 2.681441634 | 0.000928026 | 0.001686065 | 1 |
| Obesity | mast_cell | *SH3D21* | 3.445932352 | 0.000569349 | 0.001193196 | 1 |
| Obesity | mast_cell | *ANG* | 2.346325166 | 0.000494916 | 0.001138685 | 1 |
| Obesity | mast_cell | *LCNL1* | 3.624071734 | 0.026378367 | 0.033037645 | 1 |
| Obesity | mast_cell | *GNAT3* | 2.616636093 | 0.000480245 | 0.001129748 | 1 |
| Obesity | mast_cell | *PLIN5* | 1.281313076 | 0.01062712 | 0.014239929 | 1 |
| Obesity | mast_cell | *STARD10* | -1.554024651 | 0.028573309 | 0.035621196 | 1 |
| Obesity | mast_cell | *CPEB1* | 1.931001922 | 0.000653325 | 0.001309349 | 1 |
| Obesity | mast_cell | *EML6* | 6.824014502 | 0.000273386 | 0.000987746 | 1 |
| Obesity | mast_cell | *ARHGEF33* | 1.993447359 | 0.020223675 | 0.02580207 | 1 |
| Obesity | mast_cell | *IFRD2* | 2.226379232 | 0.00978683 | 0.013172177 | 1 |
| Obesity | mast_cell | *AC005082.1* | 7.725578672 | 0.009369705 | 0.012670515 | 1 |
| Obesity | mast_cell | *UBXN2B* | 2.377948832 | 0.000510541 | 0.001145411 | 1 |
| Obesity | mast_cell | *C5orf49* | 26.80241845 | 0.000119892 | 0.000976811 | 0.581716009 |
| Obesity | mast_cell | *GOLGA8B* | 2.560864752 | 3.68E-05 | 0.000670417 | 0.178330853 |
| Obesity | mast_cell | *VPS16* | 2.86184026 | 0.000320743 | 0.000999138 | 1 |
| Obesity | mast_cell | *MYL5* | 2.98957723 | 0.00426452 | 0.006183936 | 1 |
| Obesity | mast_cell | *NPEPL1* | -3.243631015 | 0.025149205 | 0.031563358 | 1 |
| Obesity | mast_cell | *TMEM242* | 2.07840721 | 0.000401289 | 0.001064698 | 1 |
| Obesity | mast_cell | *TSTD1* | 3.530678468 | 0.000707665 | 0.00138284 | 1 |
| Obesity | mast_cell | *AC009403.2* | -2.340905639 | 0.003170623 | 0.00473787 | 1 |
| Obesity | mast_cell | *PAM16* | -2.372765922 | 0.009484693 | 0.0128046 | 1 |
| Obesity | mast_cell | *AC016773.1* | 2.884293197 | 0.001812098 | 0.002927839 | 1 |
| Obesity | mast_cell | *AP000350.4* | 1.027821437 | 0.03926753 | 0.048003541 | 1 |
| Obesity | mast_cell | *RNASEK* | 1.801672418 | 0.000878021 | 0.001614282 | 1 |
| Obesity | mast_cell | *NBPF1* | 4.097211637 | 0.004990163 | 0.007119163 | 1 |
| Obesity | mast_cell | *RNU6ATAC35I* | -5.837478333 | 0.000613849 | 0.001253586 | 1 |

| | | | | | | |
|---|---|---|---|---|---|---|
| Obesity | mast_cell | *PPP3R1* | 2.775749737 | 0.000196334 | 0.000976811 | 0.952610858 |
| Obesity | mast_cell | *FANCG* | -2.843653257 | 0.002425465 | 0.003763466 | 1 |
| Obesity | mast_cell | *FXYD7* | 2.948845077 | 0.003346751 | 0.004971964 | 1 |
| Obesity | mast_cell | *SLC12A8* | 1.640039049 | 0.010617508 | 0.01423098 | 1 |
| Obesity | mast_cell | *APOL6* | 1.982706754 | 0.000351246 | 0.001031837 | 1 |
| Obesity | mast_cell | *OR2A1* | 2.461809916 | 0.033598485 | 0.041459779 | 1 |
| Obesity | mast_cell | *UBA52* | 3.832486489 | 0.00016449 | 0.000976811 | 0.798106312 |
| Obesity | mast_cell | *TIAF1* | 3.493923866 | 0.005972463 | 0.008370419 | 1 |
| Obesity | mast_cell | *RAB6C* | 60.23314498 | 0.002051992 | 0.003250495 | 1 |
| Obesity | mast_cell | *AC062017.1* | 2.42523 | 0.000530117 | 0.00115966 | 1 |
| Obesity | mast_cell | *ZNF844* | 1.718069881 | 0.002291029 | 0.003577752 | 1 |
| Obesity | mast_cell | *LINC00571* | 1.902146854 | 0.006701078 | 0.009297578 | 1 |
| Obesity | mast_cell | *OSER1-AS1* | 2.535768144 | 3.09E-05 | 0.000621513 | 0.149911068 |
| Obesity | mast_cell | *C5orf66* | 3.960380637 | 0.000216424 | 0.000976811 | 1 |
| Obesity | mast_cell | *LINC01133* | 1.640754202 | 0.000923845 | 0.001680726 | 1 |
| Obesity | mast_cell | *C4B* | 7.706334514 | 2.64E-05 | 0.000562671 | 0.128023526 |
| Obesity | mast_cell | *LINC01117* | 2.462368029 | 0.000418898 | 0.00108267 | 1 |
| Obesity | mast_cell | *LINC00240* | 1.880735755 | 3.15E-06 | 0.000161268 | 0.015289931 |
| Obesity | mast_cell | *LINC00184* | 61.13464963 | 0.00056837 | 0.001192791 | 1 |
| Obesity | mast_cell | *AL049840.1* | 2.353117199 | 0.033739097 | 0.041622705 | 1 |
| Obesity | mast_cell | *PLEKHM1* | 1.80916267 | 0.000518776 | 0.001151636 | 1 |
| Obesity | mast_cell | *AP000350.6* | 1.987574827 | 0.001824158 | 0.002946342 | 1 |
| Obesity | mast_cell | *CYP4A22-AS1* | 6.762886309 | 0.000221014 | 0.000976811 | 1 |
| Obesity | mast_cell | *MIAT* | 3.245859146 | 0.000450596 | 0.001106294 | 1 |
| Obesity | mast_cell | *ERCC6* | 3.869905347 | 0.000182315 | 0.000976811 | 0.88459062 |
| Obesity | mast_cell | *LINC00694* | 7.356497953 | 0.000516004 | 0.001150279 | 1 |
| Obesity | mast_cell | *LINC00115* | 9.79517278 | 0.001670402 | 0.002730725 | 1 |
| Obesity | mast_cell | *LINC00937* | -1.74361719 | 0.004334731 | 0.006274498 | 1 |
| Obesity | mast_cell | *FAM138B* | -5.466416498 | 0.001849026 | 0.0029766 | 1 |
| Obesity | mast_cell | *ZNF717* | 1.68646498 | 0.000473108 | 0.001125254 | 1 |
| Obesity | mast_cell | *AC017074.2* | 19.94264056 | 0.019674276 | 0.025167305 | 1 |
| Obesity | mast_cell | *AC009299.3* | 2.479689589 | 0.00051074 | 0.001145411 | 1 |
| Obesity | mast_cell | *SCAMP4* | 2.301176067 | 0.000401705 | 0.001064698 | 1 |
| Obesity | mast_cell | *SLC2A1-AS1* | 4.008017078 | 0.002423495 | 0.003761612 | 1 |
| Obesity | mast_cell | *AC005884.1* | 2.826703863 | 0.028845051 | 0.035913828 | 1 |
| Obesity | mast_cell | *HCG20* | -6.30972227 | 1.66E-05 | 0.000471596 | 0.080642831 |
| Obesity | mast_cell | *POLR2J2* | -10.80323613 | 0.002035292 | 0.00322808 | 1 |
| Obesity | mast_cell | *LINC00578* | 4.037729481 | 0.000284098 | 0.000993978 | 1 |
| Obesity | mast_cell | *C19orf24* | 4.561408604 | 0.000777266 | 0.001482613 | 1 |
| Obesity | mast_cell | *ZNF883* | 2.85330815 | 8.43E-05 | 0.000964176 | 0.408810556 |
| Obesity | mast_cell | *ARL17B* | 3.092623952 | 0.000499523 | 0.001141453 | 1 |
| Obesity | mast_cell | *PATL2* | 2.848870745 | 0.000160561 | 0.000976811 | 0.779040973 |
| Obesity | mast_cell | *CASC20* | -1.42336771 | 0.002749128 | 0.004174889 | 1 |
| Obesity | mast_cell | *PSMG3-AS1* | 2.4779249 | 0.00035472 | 0.001031837 | 1 |
| Obesity | mast_cell | *AC112229.4* | 2.969991085 | 0.008189995 | 0.011174875 | 1 |
| Obesity | mast_cell | *AC092171.4* | 3.331002549 | 0.000441312 | 0.001100333 | 1 |
| Obesity | mast_cell | *RPS18* | 3.795621967 | 4.61E-05 | 0.000754069 | 0.223862366 |
| Obesity | mast_cell | *DIRC3* | 1.922490438 | 0.000248337 | 0.000976811 | 1 |
| Obesity | mast_cell | *TAPBP* | 4.015082241 | 3.33E-06 | 0.000166436 | 0.016144339 |
| Obesity | mast_cell | *AC107021.1* | 2.313532253 | 0.001392972 | 0.002359881 | 1 |

| | | | | | | |
|---|---|---|---|---|---|---|
| Obesity | mast_cell | *TMA7* | 5.274670825 | 6.77E-06 | 0.000271528 | 0.032854858 |
| Obesity | mast_cell | *MTRNR2L4* | 4.509211782 | 0.037092774 | 0.045551541 | 1 |
| Obesity | mast_cell | *LINC00865* | 2.198119914 | 0.023733315 | 0.029902375 | 1 |
| Obesity | mast_cell | *AC009495.3* | 6.765310673 | 0.002526357 | 0.003888922 | 1 |
| Obesity | mast_cell | *HLA-DQB2* | 0.795569366 | 0.005476775 | 0.007745063 | 1 |
| Obesity | mast_cell | *GOLGA8N* | 2.390427383 | 0.001572424 | 0.00260567 | 1 |
| Obesity | mast_cell | *LINC00665* | 2.122390063 | 0.001141249 | 0.001998318 | 1 |
| Obesity | mast_cell | *LYRM9* | 2.190727979 | 0.002617675 | 0.00400862 | 1 |
| Obesity | mast_cell | *LINC00342* | 2.762551666 | 0.00091085 | 0.00165895 | 1 |
| Obesity | mast_cell | *LINC00702* | 4.986561448 | 0.000222656 | 0.000976811 | 1 |
| Obesity | mast_cell | *GPX1* | 5.821391387 | 0.001567098 | 0.002600396 | 1 |
| Obesity | mast_cell | *RPS28* | 3.672412719 | 0.000234728 | 0.000976811 | 1 |
| Obesity | mast_cell | *TRIM26* | 2.645853078 | 0.004037759 | 0.005891653 | 1 |
| Obesity | mast_cell | *ZNF736* | 1.723690268 | 0.000607513 | 0.001245839 | 1 |
| Obesity | mast_cell | *PINLYP* | 2.043701006 | 0.001895956 | 0.003037034 | 1 |
| Obesity | mast_cell | *ANKRD65* | -5.297539088 | 0.005709727 | 0.008051031 | 1 |
| Obesity | mast_cell | *ZSCAN31* | 2.538743575 | 2.52E-09 | 6.80E-07 | 1.22E-05 |
| Obesity | mast_cell | *LINC00957* | 5.968041708 | 0.001619171 | 0.002666558 | 1 |
| Obesity | mast_cell | *KIAA0040* | 6.11289856 | 8.60E-05 | 0.000971162 | 0.417051489 |
| Obesity | mast_cell | *LINC00607* | 2.419315153 | 4.66E-05 | 0.000755533 | 0.225904219 |
| Obesity | mast_cell | *AC019117.2* | -5.863016805 | 0.000657 | 0.001312378 | 1 |
| Obesity | mast_cell | *ZBTB22* | -1.583055624 | 0.025940556 | 0.032506089 | 1 |
| Obesity | mast_cell | *ZBED5* | 3.437290012 | 0.001227562 | 0.002121129 | 1 |
| Obesity | mast_cell | *CDKN2AIPNL* | 2.553111554 | 0.000479498 | 0.001129748 | 1 |
| Obesity | mast_cell | *GS1-124K5.4* | 1.076995108 | 0.031815405 | 0.03937968 | 1 |
| Obesity | mast_cell | *LINC00959* | -2.302537075 | 0.021314848 | 0.027030748 | 1 |
| Obesity | mast_cell | *HLA-DQA2* | 0.874207109 | 0.022542422 | 0.028475874 | 1 |
| Obesity | mast_cell | *LINC01143* | -9.056244961 | 0.006905497 | 0.009551161 | 1 |
| Obesity | mast_cell | *LINC00211* | 15.94800546 | 4.74E-05 | 0.000758703 | 0.229909327 |
| Obesity | mast_cell | *LRRC37A2* | 3.510020719 | 0.001327632 | 0.002270592 | 1 |
| Obesity | mast_cell | *TXNDC5* | 12.58451146 | 0.000454476 | 0.001110332 | 1 |
| Obesity | mast_cell | *LINC01119* | 4.156294958 | 0.000353563 | 0.001031837 | 1 |
| Obesity | mast_cell | *TNFSF12* | 5.210191039 | 0.005337968 | 0.007570834 | 1 |
| Obesity | mast_cell | *CDRT4* | 5.32764817 | 0.006245256 | 0.008712473 | 1 |
| Obesity | mast_cell | *C1orf226* | -4.388236831 | 0.001144448 | 0.002002815 | 1 |
| Obesity | mast_cell | *PSMB9* | 2.929351451 | 9.70E-08 | 1.27E-05 | 0.000470628 |
| Obesity | mast_cell | *C1QTNF9* | -5.681612302 | 0.003001782 | 0.004518971 | 1 |
| Obesity | mast_cell | *PNMA2* | 3.358072083 | 0.002400123 | 0.003728913 | 1 |
| Obesity | mast_cell | *KRBOX1* | 38.59934917 | 0.000585904 | 0.001214874 | 1 |
| Obesity | mast_cell | *ARHGEF25* | 4.14018026 | 9.72E-06 | 0.000352485 | 0.047149938 |
| Obesity | mast_cell | *NSUN6* | 5.590852803 | 0.00043204 | 0.001094797 | 1 |
| Obesity | mast_cell | *LINC00877* | 2.288791686 | 0.016619145 | 0.021554689 | 1 |
| Obesity | mast_cell | *TDGF1* | 2.810008394 | 0.021668218 | 0.02745735 | 1 |
| Obesity | mast_cell | *CD302* | 2.437286755 | 0.000587224 | 0.00121709 | 1 |
| Obesity | mast_cell | *ATP5O* | 2.766252774 | 0.000120035 | 0.000976811 | 0.582408789 |
| Obesity | mast_cell | *C8orf58* | -1.546032772 | 0.001651524 | 0.00270716 | 1 |
| Obesity | mast_cell | *HOGA1* | 4.899400966 | 0.000575042 | 0.001201079 | 1 |
| Obesity | mast_cell | *PI4KA* | 2.41148182 | 0.000148561 | 0.000976811 | 0.720819177 |
| Obesity | mast_cell | *LINC00969* | 2.207310398 | 0.000337749 | 0.00102167 | 1 |
| Obesity | mast_cell | *ARFGAP3* | 7.282239829 | 0.000410545 | 0.001075307 | 1 |

| | | | | | | |
|---|---|---|---|---|---|---|
| Obesity | mast_cell | *MRPL20* | 6.160753174 | 0.000134137 | 0.000976811 | 0.650832179 |
| Obesity | mast_cell | *DECR2* | 3.538088516 | 0.000545698 | 0.00117408 | 1 |
| Obesity | mast_cell | *ERVW-1* | -2.96972981 | 0.003166565 | 0.004733263 | 1 |
| Obesity | mast_cell | *MRPL33* | 3.025280545 | 0.000429789 | 0.001092129 | 1 |
| Obesity | mast_cell | *STON1* | 3.312965721 | 0.000433858 | 0.001096396 | 1 |
| Obesity | mast_cell | *VSIG8* | 4.384397171 | 0.000807206 | 0.001520405 | 1 |
| Obesity | mast_cell | *C7orf73* | 3.704200262 | 0.00034641 | 0.00102869 | 1 |
| Obesity | mast_cell | *FGF14-IT1* | 7.47206151 | 0.000542129 | 0.001171675 | 1 |
| Obesity | mast_cell | *PALM2* | 2.36827238 | 0.00217701 | 0.003428384 | 1 |
| Obesity | mast_cell | *NBPF15* | 2.674643882 | 0.000365472 | 0.001037687 | 1 |
| Obesity | mast_cell | *UPK3B* | 4.143561074 | 0.000227369 | 0.000976811 | 1 |
| Obesity | mast_cell | *IL10RB* | 6.033846634 | 0.000343793 | 0.001027777 | 1 |
| Obesity | mast_cell | *TTC4* | 2.969004792 | 0.000378037 | 0.001048135 | 1 |
| Obesity | mast_cell | *APOBEC3D* | -14.01779924 | 0.018233315 | 0.023472551 | 1 |
| Obesity | mast_cell | *TMEM199* | 2.677032428 | 0.000454914 | 0.001110842 | 1 |
| Obesity | mast_cell | *LILRA6* | 1.505582258 | 0.022229477 | 0.028124491 | 1 |
| Obesity | mast_cell | *SCARF2* | 2.86612674 | 0.000609864 | 0.001248022 | 1 |
| Obesity | mast_cell | *APOBEC3C* | -2.110992088 | 0.002500107 | 0.003855405 | 1 |
| Obesity | mast_cell | *CCDC13* | 2.913859049 | 0.000774173 | 0.001479447 | 1 |
| Obesity | mast_cell | *C4A* | -8.273710499 | 2.10E-06 | 0.000130871 | 0.010207937 |
| Obesity | mast_cell | *NEAT1* | 2.245939146 | 0.000112862 | 0.000976811 | 0.547605555 |
| Obesity | mast_cell | *RAB30-AS1* | 1.976552899 | 0.001422768 | 0.002402809 | 1 |
| Obesity | mast_cell | *UBAP1L* | 2.321799727 | 0.000453056 | 0.001107978 | 1 |
| Obesity | mast_cell | *DNAJC3-AS1* | 2.028324446 | 0.001471429 | 0.002473796 | 1 |
| Obesity | mast_cell | *LINC00926* | 3.846755604 | 0.000488875 | 0.001134939 | 1 |
| Obesity | mast_cell | *DPH6-AS1* | 1.949847272 | 0.000262758 | 0.000983135 | 1 |
| Obesity | mast_cell | *CDK11B* | 2.079883875 | 0.000452922 | 0.001107978 | 1 |
| Obesity | mast_cell | *ABHD14A* | 2.087815368 | 0.000302541 | 0.000998908 | 1 |
| Obesity | mast_cell | *SRP14-AS1* | 1.855142287 | 1.44E-05 | 0.000426352 | 0.069921701 |
| Obesity | mast_cell | *GDNF-AS1* | 4.07244092 | 0.000970574 | 0.001747393 | 1 |
| Obesity | mast_cell | *CCDC153* | 3.981432948 | 0.00025976 | 0.00098093 | 1 |
| Obesity | mast_cell | *AL357673.1* | 3.350681728 | 0.000504297 | 0.001144783 | 1 |
| Obesity | mast_cell | *C5orf17* | 1.706314366 | 0.000525473 | 0.001156279 | 1 |
| Obesity | mast_cell | *TMEM150C* | 2.105806145 | 0.000715869 | 0.001394378 | 1 |
| Obesity | mast_cell | *LINC00939* | 5.011738816 | 0.004624649 | 0.006640661 | 1 |
| Obesity | mast_cell | *LINC01060* | 1.87111858 | 0.001869789 | 0.003003051 | 1 |
| Obesity | mast_cell | *LINC00942* | -7.025643745 | 0.000855737 | 0.001584536 | 1 |
| Obesity | mast_cell | *PDCD6* | 2.874721614 | 0.000285956 | 0.000993978 | 1 |
| Obesity | mast_cell | *TMEM158* | -6.239700346 | 0.020935551 | 0.02663327 | 1 |
| Obesity | mast_cell | *DNAH10OS* | 28.45167456 | 3.56E-08 | 5.23E-06 | 0.000172663 |
| Obesity | mast_cell | *PCDHA10* | 1.83070569 | 0.04005002 | 0.048898514 | 1 |
| Obesity | mast_cell | *ZNF718* | 3.453405141 | 0.000144664 | 0.000976811 | 0.701909227 |
| Obesity | mast_cell | *SMIM20* | 3.543227582 | 0.00031889 | 0.000998908 | 1 |
| Obesity | mast_cell | *LINC00989* | 2.729912405 | 0.001116552 | 0.001960694 | 1 |
| Obesity | mast_cell | *ATP6V1E2* | 1.247270298 | 0.015691609 | 0.020433625 | 1 |
| Obesity | mast_cell | *GLI4* | 2.324324678 | 0.000877746 | 0.001614282 | 1 |
| Obesity | mast_cell | *SNHG18* | 2.071306807 | 0.002758111 | 0.004185909 | 1 |
| Obesity | mast_cell | *ALG1L2* | 3.203440441 | 0.000288173 | 0.000994669 | 1 |
| Obesity | mast_cell | *LINC01094* | 2.826685971 | 0.000518086 | 0.001151636 | 1 |
| Obesity | mast_cell | *TMEM200B* | 3.393653412 | 0.000212932 | 0.000976811 | 1 |

| | | | | | | |
|---|---|---|---|---|---|---|
| Obesity | mast_cell | *PCDHGB6* | 3.800588825 | 0.000139269 | 0.000976811 | 0.675735271 |
| Obesity | mast_cell | *SLC10A5* | -2.548239587 | 0.006986459 | 0.009654884 | 1 |
| Obesity | mast_cell | *PCDHGA6* | 2.424811335 | 0.000743012 | 0.001435778 | 1 |
| Obesity | mast_cell | *UTP14C* | 4.533975954 | 0.001230626 | 0.002125429 | 1 |
| Obesity | mast_cell | *PCDHGB2* | -7.189302482 | 0.013293397 | 0.017512778 | 1 |
| Obesity | mast_cell | *LYN* | 1.986806656 | 0.000836251 | 0.001560573 | 1 |
| Obesity | mast_cell | *DPP3* | 3.074305879 | 4.61E-06 | 0.00021705 | 0.022356196 |
| Obesity | mast_cell | *SNHG9* | 3.628472569 | 3.16E-05 | 0.000621513 | 0.153314632 |
| Obesity | mast_cell | *ZNF432* | 2.753041247 | 0.000621547 | 0.001264995 | 1 |
| Obesity | mast_cell | *HMBS* | 5.12811624 | 3.45E-07 | 3.56E-05 | 0.001674923 |
| Obesity | mast_cell | *AC093668.2* | 2.114096589 | 0.024010554 | 0.03021245 | 1 |
| Obesity | mast_cell | *KIAA1147* | 3.31392178 | 9.42E-05 | 0.000976811 | 0.457252926 |
| Obesity | mast_cell | *ZBED6* | -3.799778207 | 0.004525067 | 0.006518607 | 1 |
| Obesity | mast_cell | *CNPY2* | 5.395524014 | 0.000154328 | 0.000976811 | 0.748801122 |
| Obesity | mast_cell | *LINC00592* | -3.169281754 | 0.001575448 | 0.00260979 | 1 |
| Obesity | mast_cell | *CHURC1* | 2.674504752 | 0.000133281 | 0.000976811 | 0.646677942 |
| Obesity | mast_cell | *PDF* | -11.03140561 | 0.024196289 | 0.030430377 | 1 |
| Obesity | mast_cell | *HIF1A-AS2* | 4.334313913 | 0.000181707 | 0.000976811 | 0.881642243 |
| Obesity | mast_cell | *LINC00638* | 6.751173505 | 0.000161425 | 0.000976811 | 0.783232816 |
| Obesity | mast_cell | *RNASE4* | -6.710757568 | 0.018613935 | 0.023930792 | 1 |
| Obesity | mast_cell | *MC1R* | -1.017958023 | 0.002488149 | 0.003841076 | 1 |
| Obesity | mast_cell | *LINC00639* | 1.829489653 | 0.000467334 | 0.001116682 | 1 |
| Obesity | mast_cell | *MRPL46* | 2.859155804 | 0.00063874 | 0.0012881 | 1 |
| Obesity | mast_cell | *UBE2Q2L* | -3.233434191 | 0.022215739 | 0.028114441 | 1 |
| Obesity | mast_cell | *IQCH-AS1* | 2.915851022 | 0.000832277 | 0.001554951 | 1 |
| Obesity | mast_cell | *AC083843.1* | 46.07147441 | 0.000544958 | 0.00117408 | 1 |
| Obesity | mast_cell | *PWRN1* | 2.759156734 | 0.001498104 | 0.002511679 | 1 |
| Obesity | mast_cell | *LINC01003* | 2.477790417 | 0.000844398 | 0.001572149 | 1 |
| Obesity | mast_cell | *C15orf65* | 2.409118103 | 0.007937298 | 0.010875959 | 1 |
| Obesity | mast_cell | *GOLGA8H* | 1.452522357 | 0.009059081 | 0.012284701 | 1 |
| Obesity | mast_cell | *LINC00662* | 2.111363795 | 0.00079168 | 0.001499309 | 1 |
| Obesity | mast_cell | *C19orf84* | 39.51663841 | 3.86E-05 | 0.000684099 | 0.187443199 |
| Obesity | mast_cell | *LINC00667* | 1.919388673 | 0.001679556 | 0.002742042 | 1 |
| Obesity | mast_cell | *LINC00672* | 5.496751653 | 0.000493946 | 0.001138685 | 1 |
| Obesity | mast_cell | *LINC00909* | 4.249698298 | 0.000549227 | 0.001175269 | 1 |
| Obesity | mast_cell | *DYNLL2* | 2.815078414 | 0.000687398 | 0.001355244 | 1 |
| Obesity | mast_cell | *STRADA* | 2.047865537 | 0.008392653 | 0.011432103 | 1 |
| Obesity | mast_cell | *MYO15B* | 3.871473908 | 0.000135551 | 0.000976811 | 0.657691693 |
| Obesity | mast_cell | *ILF3-AS1* | 2.086962002 | 0.000466566 | 0.001116682 | 1 |
| Obesity | mast_cell | *ZNF582-AS1* | 5.723029193 | 0.000286757 | 0.000993978 | 1 |
| Obesity | mast_cell | *S1PR2* | 3.071167028 | 0.000944309 | 0.001712178 | 1 |
| Obesity | mast_cell | *ZNF224* | 3.258182383 | 0.000248782 | 0.000976811 | 1 |
| Obesity | mast_cell | *LIN37* | -7.89723887 | 0.000145496 | 0.000976811 | 0.705948354 |
| Obesity | mast_cell | *NDUFA7* | 15.87015303 | 0.000352516 | 0.001031837 | 1 |
| Obesity | mast_cell | *AC004466.1* | 3.323757227 | 0.023873331 | 0.030063172 | 1 |
| Obesity | mast_cell | *LINC00664* | -66.23688652 | 0.016887848 | 0.021873955 | 1 |
| Obesity | mast_cell | *ZNF728* | 5.839082512 | 0.000128018 | 0.000976811 | 0.621142788 |
| Obesity | mast_cell | *TRABD2B* | 2.433376129 | 0.00031562 | 0.000998908 | 1 |
| Obesity | mast_cell | *AC093157.1* | 5.132120389 | 0.000460685 | 0.001114279 | 1 |
| Obesity | mast_cell | *FBXO17* | 2.090839268 | 0.000207578 | 0.000976811 | 1 |

| | | | | | | |
|---|---|---|---|---|---|---|
| Obesity | mast_cell | *ZNF587B* | 3.000062601 | 0.000526848 | 0.001157301 | 1 |
| Obesity | mast_cell | *AP001172.2* | 2.94923866 | 1.10E-05 | 0.000373166 | 0.053531476 |
| Obesity | mast_cell | *HCG17* | 2.699581181 | 0.0001554 | 0.000976811 | 0.753999004 |
| Obesity | mast_cell | *YTHDF3-AS1* | 3.678121733 | 0.029647704 | 0.036856433 | 1 |
| Obesity | mast_cell | *MILR1* | 1.275431499 | 0.01099151 | 0.014695731 | 1 |
| Obesity | mast_cell | *GTF2H5* | 2.506418723 | 1.12E-05 | 0.000373166 | 0.054259217 |
| Obesity | mast_cell | *FGF14-AS2* | 3.866695203 | 0.000784865 | 0.001490475 | 1 |
| Obesity | mast_cell | *CASC15* | 2.432459413 | 0.000293192 | 0.000998908 | 1 |
| Obesity | mast_cell | *AP000347.2* | 5.029599763 | 0.004910095 | 0.007013334 | 1 |
| Obesity | mast_cell | *GRIN2B* | 2.40570269 | 2.45E-05 | 0.000545024 | 0.118718163 |
| Obesity | mast_cell | *PAXIP1-AS1* | 2.525607608 | 0.00034298 | 0.001027777 | 1 |
| Obesity | mesothelium | *SEMA3F* | -0.757099554 | 2.17E-12 | 5.54E-10 | 1.05E-08 |
| Obesity | mesothelium | *CYP26B1* | 0.478223819 | 0.002551618 | 0.044856699 | 1 |
| Obesity | mesothelium | *RBM6* | -0.402283403 | 7.67E-26 | 1.86E-22 | 3.72E-22 |
| Obesity | mesothelium | *GGNBP2* | -1.096923776 | 2.62E-05 | 0.001285828 | 0.12729697 |
| Obesity | mesothelium | *CLCN6* | 0.419004461 | 0.001089162 | 0.024399375 | 1 |
| Obesity | mesothelium | *RABEP1* | 0.602769794 | 0.002278856 | 0.041412015 | 1 |
| Obesity | mesothelium | *PIAS1* | 0.73436 | 2.99E-05 | 0.001418929 | 0.145212086 |
| Obesity | mesothelium | *SH3YL1* | -0.659978354 | 1.39E-05 | 0.000739103 | 0.067258391 |
| Obesity | mesothelium | *LSG1* | 0.12168173 | 0.002697279 | 0.046573659 | 1 |
| Obesity | mesothelium | *RCN1* | -0.517775255 | 0.002134379 | 0.039654553 | 1 |
| Obesity | mesothelium | *SDCCAG8* | 0.262358248 | 0.001773789 | 0.034703315 | 1 |
| Obesity | mesothelium | *ITIH4* | 0.148326806 | 8.34E-11 | 1.44E-08 | 4.05E-07 |
| Obesity | mesothelium | *UHRF1BP1* | 0.617213652 | 7.05E-23 | 8.55E-20 | 3.42E-19 |
| Obesity | mesothelium | *TIPIN* | 0.139198321 | 0.001064804 | 0.024142188 | 1 |
| Obesity | mesothelium | *NT5C2* | 0.32046056 | 0.000162108 | 0.005578344 | 0.786546553 |
| Obesity | mesothelium | *PIK3C3* | -0.82823201 | 0.002067243 | 0.038577926 | 1 |
| Obesity | mesothelium | *FDFT1* | 0.77377635 | 1.15E-08 | 1.30E-06 | 5.58E-05 |
| Obesity | mesothelium | *GSTP1* | 0.508272715 | 3.49E-05 | 0.001581021 | 0.169169267 |
| Obesity | mesothelium | *EPS15* | -0.302109423 | 0.002304166 | 0.041560637 | 1 |
| Obesity | mesothelium | *MAST2* | 0.47492741 | 0.001014545 | 0.023552968 | 1 |
| Obesity | mesothelium | *NME8* | 10.60833153 | 0.000481477 | 0.012835868 | 1 |
| Obesity | mesothelium | *TRIP6* | -0.615738501 | 0.002636698 | 0.046071694 | 1 |
| Obesity | mesothelium | *OGFOD1* | -0.62277054 | 8.63E-06 | 0.00050426 | 0.041853594 |
| Obesity | mesothelium | *PPP1R13B* | -0.357978864 | 0.000731394 | 0.018278685 | 1 |
| Obesity | mesothelium | *YPEL3* | 0.570085535 | 6.61E-09 | 7.64E-07 | 3.21E-05 |
| Obesity | mesothelium | *PABPC4* | -0.641694605 | 1.30E-10 | 1.97E-08 | 6.31E-07 |
| Obesity | mesothelium | *RGS17* | -0.623078683 | 2.17E-09 | 2.78E-07 | 1.05E-05 |
| Obesity | mesothelium | *PPP2R3C* | 0.457159541 | 0.000156217 | 0.005452987 | 0.757965136 |
| Obesity | mesothelium | *TBX15* | -0.28018363 | 3.40E-05 | 0.001556632 | 0.165002993 |
| Obesity | mesothelium | *ITPR3* | 0.298644123 | 4.44E-05 | 0.001887762 | 0.215204877 |
| Obesity | mesothelium | *SETD1A* | 0.834049525 | 1.81E-07 | 1.72E-05 | 0.000878112 |
| Obesity | mesothelium | *BCL2L13* | 0.482561184 | 0.000104814 | 0.003973123 | 0.508559695 |
| Obesity | mesothelium | *ANKRD54* | -0.272658967 | 0.000118412 | 0.004419512 | 0.574536575 |
| Obesity | mesothelium | *CENPM* | 0.324986707 | 3.98E-05 | 0.00175566 | 0.19312255 |
| Obesity | mesothelium | *KDELR3* | -0.516781899 | 0.000811584 | 0.0198879 | 1 |
| Obesity | mesothelium | *ABHD12* | 0.521774269 | 2.15E-05 | 0.001087768 | 0.104425766 |
| Obesity | mesothelium | *COG4* | -0.353364435 | 0.00092365 | 0.022194411 | 1 |
| Obesity | mesothelium | *CLUAP1* | 0.151493609 | 0.001793327 | 0.034786281 | 1 |
| Obesity | mesothelium | *RRN3P2* | -0.404029025 | 0.00265987 | 0.046256958 | 1 |

| | | | | | | |
|---|---|---|---|---|---|---|
| Obesity | mesothelium | *PYCARD* | 1.152101233 | 1.01E-09 | 1.40E-07 | 4.89E-06 |
| Obesity | mesothelium | *TMC5* | -0.267631779 | 0.002377481 | 0.042566552 | 1 |
| Obesity | mesothelium | *KNOP1* | -1.684747017 | 5.31E-08 | 5.48E-06 | 0.00025773 |
| Obesity | mesothelium | *RPAP1* | -0.414050076 | 0.002639722 | 0.046071694 | 1 |
| Obesity | mesothelium | *ZDHHC2* | -0.41419466 | 0.001128213 | 0.024995832 | 1 |
| Obesity | mesothelium | *LIN7B* | -1.48335522 | 1.77E-06 | 0.000120869 | 0.008581672 |
| Obesity | mesothelium | *MEST* | -0.218890348 | 0.000134419 | 0.004978618 | 0.652198911 |
| Obesity | mesothelium | *RAD51C* | -0.346483857 | 6.69E-05 | 0.002750823 | 0.324597101 |
| Obesity | mesothelium | *PNPO* | -1.247462474 | 2.76E-07 | 2.57E-05 | 0.001338127 |
| Obesity | mesothelium | *CHRNE* | 0.090909573 | 0.000464066 | 0.012509145 | 1 |
| Obesity | mesothelium | *B3GAT1* | 7.895552266 | 1.93E-06 | 0.000128519 | 0.009381879 |
| Obesity | mesothelium | *KCTD10* | -0.412067874 | 0.001773218 | 0.034703315 | 1 |
| Obesity | mesothelium | *DDX55* | 0.591154427 | 0.001025529 | 0.023694608 | 1 |
| Obesity | mesothelium | *BTN3A3* | 0.402050807 | 3.13E-10 | 4.60E-08 | 1.52E-06 |
| Obesity | mesothelium | *GABRR2* | 0.307478817 | 0.001233647 | 0.027084404 | 1 |
| Obesity | mesothelium | *TULP1* | -0.260134423 | 0.000231487 | 0.00743826 | 1 |
| Obesity | mesothelium | *CUTA* | -0.301549507 | 0.000440052 | 0.011995114 | 1 |
| Obesity | mesothelium | *BTN2A1* | -0.6734403 | 0.000533088 | 0.013831783 | 1 |
| Obesity | mesothelium | *NME5* | 0.161264626 | 0.000438036 | 0.011995114 | 1 |
| Obesity | mesothelium | *TMCO6* | -0.310178647 | 0.000384728 | 0.010852919 | 1 |
| Obesity | mesothelium | *IK* | -0.323118321 | 0.000515945 | 0.013531697 | 1 |
| Obesity | mesothelium | *PCCB* | 0.401107187 | 1.40E-06 | 0.000104388 | 0.0067852 |
| Obesity | mesothelium | *SCAP* | 0.609344084 | 0.001959002 | 0.037174143 | 1 |
| Obesity | mesothelium | *CHMP3* | -0.418253089 | 1.90E-06 | 0.000128339 | 0.009240435 |
| Obesity | mesothelium | *NSUN4* | -0.481907493 | 7.69E-05 | 0.003110431 | 0.373251736 |
| Obesity | mesothelium | *SERPINC1* | 0.833541596 | 9.65E-11 | 1.51E-08 | 4.68E-07 |
| Obesity | mesothelium | *CCDC92* | 0.623349536 | 2.11E-05 | 0.001076122 | 0.102231596 |
| Obesity | mesothelium | *INVS* | 0.466313566 | 2.70E-06 | 0.000172438 | 0.013105316 |
| Obesity | mesothelium | *SNX19* | -0.491301387 | 1.22E-06 | 9.28E-05 | 0.005940036 |
| Obesity | mesothelium | *ADRA1A* | 0.062990814 | 0.001749502 | 0.034647277 | 1 |
| Obesity | mesothelium | *TEX14* | 0.364855106 | 3.98E-05 | 0.00175566 | 0.192869864 |
| Obesity | mesothelium | *CPXM2* | 0.227231075 | 0.001894956 | 0.036198137 | 1 |
| Obesity | mesothelium | *OPN4* | 1.0876728 | 2.89E-06 | 0.000182269 | 0.0140347 |
| Obesity | mesothelium | *SLC25A51* | 0.093983719 | 0.002203871 | 0.040199933 | 1 |
| Obesity | mesothelium | *ITIH5* | 0.161660186 | 0.002952848 | 0.049920618 | 1 |
| Obesity | mesothelium | *BTN2A2* | 0.150113964 | 3.01E-05 | 0.001418929 | 0.146149643 |
| Obesity | mesothelium | *SNRPC* | -0.926425886 | 8.25E-24 | 1.33E-20 | 4.00E-20 |
| Obesity | mesothelium | *GRK4* | 0.082220119 | 0.000147283 | 0.005293455 | 0.714616487 |
| Obesity | mesothelium | *RIBC2* | 0.192558721 | 0.000836959 | 0.020406648 | 1 |
| Obesity | mesothelium | *RPAIN* | -0.43668284 | 3.00E-05 | 0.001418929 | 0.145703411 |
| Obesity | mesothelium | *AP1M2* | -0.189080106 | 0.000230133 | 0.00743826 | 1 |
| Obesity | mesothelium | *ATP8B3* | -0.255787532 | 2.49E-11 | 5.25E-09 | 1.21E-07 |
| Obesity | mesothelium | *TULP4* | 0.412970995 | 0.000860063 | 0.020865137 | 1 |
| Obesity | mesothelium | *PGPEP1* | 0.244621464 | 0.001091233 | 0.024399375 | 1 |
| Obesity | mesothelium | *CYP2E1* | -0.135852243 | 0.001715827 | 0.034647277 | 1 |
| Obesity | mesothelium | *NPAS1* | -1.018877131 | 3.91E-10 | 5.58E-08 | 1.90E-06 |
| Obesity | mesothelium | *C12orf65* | 0.344564111 | 0.001549058 | 0.032821085 | 1 |
| Obesity | mesothelium | *PKDREJ* | -0.185060985 | 0.002163697 | 0.039880157 | 1 |
| Obesity | mesothelium | *ATP6V1E1* | -0.592659577 | 0.002782579 | 0.047876154 | 1 |
| Obesity | mesothelium | *NDUFA2* | 0.382392749 | 0.000164491 | 0.005613052 | 0.798109391 |

| | | | | | | |
|---|---|---|---|---|---|---|
| Obesity | mesothelium | *LRRC41* | -1.043633082 | 0.00145556 | 0.031249458 | 1 |
| Obesity | mesothelium | *MYBBP1A* | 0.23726388 | 0.002474206 | 0.043973806 | 1 |
| Obesity | mesothelium | *FIGNL1* | -0.505550445 | 0.001726082 | 0.034647277 | 1 |
| Obesity | mesothelium | *IGHMBP2* | 0.221153329 | 4.25E-05 | 0.001839589 | 0.20603395 |
| Obesity | mesothelium | *ZBED3* | -0.789820618 | 0.000498387 | 0.01314225 | 1 |
| Obesity | mesothelium | *FBXO44* | -0.664930531 | 0.001878679 | 0.036153596 | 1 |
| Obesity | mesothelium | *LARS* | -0.214748236 | 0.000254977 | 0.007879921 | 1 |
| Obesity | mesothelium | *GSTM5* | 0.162079948 | 0.002965019 | 0.049952341 | 1 |
| Obesity | mesothelium | *ACP2* | 0.480935025 | 0.001742739 | 0.034647277 | 1 |
| Obesity | mesothelium | *TMEM60* | -1.245432303 | 0.000102443 | 0.003913797 | 0.497052159 |
| Obesity | mesothelium | *POM121C* | -1.042091113 | 1.62E-16 | 8.72E-14 | 7.85E-13 |
| Obesity | mesothelium | *MICAL1* | 0.194574667 | 0.001796587 | 0.034786281 | 1 |
| Obesity | mesothelium | *USP37* | 0.454400564 | 0.00214128 | 0.039654553 | 1 |
| Obesity | mesothelium | *SCN2A* | 0.596129635 | 4.38E-05 | 0.001880962 | 0.212548716 |
| Obesity | mesothelium | *DERL1* | -0.734193757 | 0.001031664 | 0.023723377 | 1 |
| Obesity | mesothelium | *TCF19* | 0.224006263 | 0.002865342 | 0.048610628 | 1 |
| Obesity | mesothelium | *SLC37A4* | 0.420746821 | 6.95E-08 | 7.03E-06 | 0.000337375 |
| Obesity | mesothelium | *NUSAP1* | 0.497794704 | 0.000371354 | 0.01053689 | 1 |
| Obesity | mesothelium | *NDUFAF1* | 0.437365894 | 3.24E-05 | 0.001496103 | 0.157090856 |
| Obesity | mesothelium | *RSL24D1* | -0.594762264 | 0.000390547 | 0.010953385 | 1 |
| Obesity | mesothelium | *CENPO* | 1.302421842 | 1.12E-17 | 6.77E-15 | 5.41E-14 |
| Obesity | mesothelium | *KLRG1* | 0.101864152 | 0.001799538 | 0.034786281 | 1 |
| Obesity | mesothelium | *VPS33A* | 0.436883594 | 8.94E-06 | 0.000516334 | 0.043372039 |
| Obesity | mesothelium | *MYO19* | 1.105581868 | 1.53E-09 | 2.01E-07 | 7.45E-06 |
| Obesity | mesothelium | *C18orf8* | -0.99121511 | 6.28E-15 | 2.34E-12 | 3.05E-11 |
| Obesity | mesothelium | *NPC1* | -1.215121503 | 5.39E-07 | 4.59E-05 | 0.002615034 |
| Obesity | mesothelium | *COL6A1* | 0.11631302 | 0.001570933 | 0.03313985 | 1 |
| Obesity | mesothelium | *SAE1* | -1.7286177 | 3.84E-14 | 1.24E-11 | 1.86E-10 |
| Obesity | mesothelium | *EPHA2* | -0.40293669 | 0.002518908 | 0.044442704 | 1 |
| Obesity | mesothelium | *SEMA6C* | 1.228914127 | 1.47E-06 | 0.000108053 | 0.007131483 |
| Obesity | mesothelium | *FAHD2B* | -0.243693029 | 0.000950018 | 0.022485307 | 1 |
| Obesity | mesothelium | *NBEAL1* | 0.572192132 | 0.000928579 | 0.022194411 | 1 |
| Obesity | mesothelium | *LIMD1* | 0.460663184 | 0.000486977 | 0.012911549 | 1 |
| Obesity | mesothelium | *CCDC39* | 0.422167915 | 0.001282398 | 0.027777648 | 1 |
| Obesity | mesothelium | *PCYOX1L* | -1.115577875 | 0.000546162 | 0.014095617 | 1 |
| Obesity | mesothelium | *ABT1* | 0.162277274 | 0.001743272 | 0.034647277 | 1 |
| Obesity | mesothelium | *ADHFE1* | -0.107591807 | 0.000259133 | 0.007957665 | 1 |
| Obesity | mesothelium | *RGR* | -1.205990751 | 0.000274537 | 0.008273617 | 1 |
| Obesity | mesothelium | *MTG1* | 0.188801905 | 0.000440019 | 0.011995114 | 1 |
| Obesity | mesothelium | *HSD17B12* | -0.202570907 | 3.26E-11 | 6.55E-09 | 1.58E-07 |
| Obesity | mesothelium | *TNKS1BP1* | 0.295820164 | 7.57E-07 | 6.02E-05 | 0.003675031 |
| Obesity | mesothelium | *TTC12* | 0.276250807 | 0.000139322 | 0.005114864 | 0.675990826 |
| Obesity | mesothelium | *NUDT22* | -0.391444966 | 0.000971568 | 0.022773168 | 1 |
| Obesity | mesothelium | *TBX6* | 0.169027141 | 3.38E-11 | 6.55E-09 | 1.64E-07 |
| Obesity | mesothelium | *RGS18* | 0.202446326 | 0.000213738 | 0.00700715 | 1 |
| Obesity | mesothelium | *DPYSL4* | 0.080182988 | 0.001728293 | 0.034647277 | 1 |
| Obesity | mesothelium | *POC5* | -0.277479964 | 5.36E-06 | 0.000316967 | 0.025991278 |
| Obesity | mesothelium | *PLA2R1* | 0.171442337 | 0.001731341 | 0.034647277 | 1 |
| Obesity | mesothelium | *JAZF1* | 0.456546026 | 1.02E-05 | 0.000570925 | 0.049670433 |
| Obesity | mesothelium | *NEIL2* | 0.266439727 | 9.59E-05 | 0.003721464 | 0.465183003 |

| | | | | | | |
|---|---|---|---|---|---|---|
| Obesity | mesothelium | *MOV10* | -0.883360171 | 0.001993227 | 0.037630881 | 1 |
| Obesity | mesothelium | *MMP16* | -0.130698586 | 0.000479106 | 0.012835868 | 1 |
| Obesity | mesothelium | *RWDD2B* | -0.216044326 | 0.002490247 | 0.044097367 | 1 |
| Obesity | mesothelium | *ITGAD* | 0.19700526 | 0.002338867 | 0.042030312 | 1 |
| Obesity | mesothelium | *STEAP2* | -0.187209677 | 0.000201301 | 0.006689798 | 0.976710534 |
| Obesity | mesothelium | *TMED6* | 0.864548487 | 0.000140205 | 0.005114864 | 0.68027691 |
| Obesity | mesothelium | *CLEC18A* | -0.470837323 | 2.04E-18 | 1.42E-15 | 9.92E-15 |
| Obesity | mesothelium | *ETS2* | 0.111274526 | 0.001885173 | 0.036153596 | 1 |
| Obesity | mesothelium | *SLC5A11* | -0.159943343 | 1.51E-07 | 1.47E-05 | 0.000734758 |
| Obesity | mesothelium | *CILP2* | -0.401063619 | 4.62E-05 | 0.001949202 | 0.224158274 |
| Obesity | mesothelium | *MCM3AP* | -0.239827984 | 0.001961373 | 0.037174143 | 1 |
| Obesity | mesothelium | *LRWD1* | -0.172661163 | 0.002681453 | 0.046465756 | 1 |
| Obesity | mesothelium | *ABCF3* | 0.212534173 | 0.001603945 | 0.033400605 | 1 |
| Obesity | mesothelium | *CAMK2N1* | 0.262284049 | 0.00053241 | 0.013831783 | 1 |
| Obesity | mesothelium | *DCAF16* | -0.812646821 | 0.001251479 | 0.02735214 | 1 |
| Obesity | mesothelium | *ANTXR2* | 0.45752359 | 2.99E-05 | 0.001418929 | 0.145197365 |
| Obesity | mesothelium | *LMOD1* | -0.581109364 | 6.30E-09 | 7.64E-07 | 3.06E-05 |
| Obesity | mesothelium | *DNALI1* | -0.344259595 | 1.49E-06 | 0.000108115 | 0.007243698 |
| Obesity | mesothelium | *RFT1* | -0.379748038 | 1.41E-07 | 1.39E-05 | 0.000682504 |
| Obesity | mesothelium | *RNF123* | 0.582165862 | 4.80E-19 | 3.88E-16 | 2.33E-15 |
| Obesity | mesothelium | *MST1R* | 0.461377959 | 3.92E-19 | 3.80E-16 | 1.90E-15 |
| Obesity | mesothelium | *TEX264* | -0.330477511 | 0.000370105 | 0.01053689 | 1 |
| Obesity | mesothelium | *ERAP1* | -0.512755525 | 3.21E-05 | 0.001496103 | 0.155651756 |
| Obesity | mesothelium | *GALNT10* | -0.575604395 | 0.000167504 | 0.005643971 | 0.812731831 |
| Obesity | mesothelium | *YWHAZ* | -0.513759458 | 0.000276416 | 0.008278816 | 1 |
| Obesity | mesothelium | *RPP25L* | -0.324110379 | 0.000338986 | 0.009790226 | 1 |
| Obesity | mesothelium | *ANKS6* | 0.431630235 | 0.000573588 | 0.014725138 | 1 |
| Obesity | mesothelium | *VDAC2* | -1.137010459 | 0.000269845 | 0.008183038 | 1 |
| Obesity | mesothelium | *SLC39A13* | 1.234138241 | 9.56E-11 | 1.51E-08 | 4.64E-07 |
| Obesity | mesothelium | *RAPSN* | -0.481627493 | 5.14E-16 | 2.49E-13 | 2.49E-12 |
| Obesity | mesothelium | *CCDC68* | 0.238124012 | 0.001532333 | 0.032609113 | 1 |
| Obesity | mesothelium | *ZSCAN21* | -0.172959167 | 0.001591922 | 0.033293133 | 1 |
| Obesity | mesothelium | *PIP4K2C* | 0.830557771 | 0.000307225 | 0.008979865 | 1 |
| Obesity | mesothelium | *SMG8* | -0.248991613 | 0.002174399 | 0.039880157 | 1 |
| Obesity | mesothelium | *SRR* | 0.138556847 | 2.51E-05 | 0.001254725 | 0.121708316 |
| Obesity | mesothelium | *KIAA1586* | -0.323441284 | 0.001645947 | 0.033983563 | 1 |
| Obesity | mesothelium | *NT5DC2* | -0.261142441 | 3.32E-06 | 0.000203629 | 0.016086706 |
| Obesity | mesothelium | *FNTA* | 0.417467213 | 9.41E-05 | 0.003681603 | 0.456518804 |
| Obesity | mesothelium | *C8orf46* | 0.066191577 | 0.000363402 | 0.010433291 | 1 |
| Obesity | mesothelium | *GPRIN1* | 0.4409456 | 0.001346116 | 0.029028246 | 1 |
| Obesity | mesothelium | *INO80E* | -0.44300346 | 2.74E-15 | 1.11E-12 | 1.33E-11 |
| Obesity | mesothelium | *CALML6* | -8.147012042 | 0.000674611 | 0.01713724 | 1 |
| Obesity | mesothelium | *MYO7B* | 0.216783218 | 0.002863572 | 0.048610628 | 1 |
| Obesity | mesothelium | *ADORA2B* | -0.150948621 | 5.19E-07 | 4.49E-05 | 0.002515925 |
| Obesity | mesothelium | *TTLL6* | -1.138792363 | 4.48E-14 | 1.36E-11 | 2.17E-10 |
| Obesity | mesothelium | *TSNARE1* | 0.438948459 | 1.78E-05 | 0.000927593 | 0.086266118 |
| Obesity | mesothelium | *TRMT61B* | 0.420697258 | 6.07E-05 | 0.002517394 | 0.294535087 |
| Obesity | mesothelium | *RGS19* | 0.103513123 | 0.001634532 | 0.033892091 | 1 |
| Obesity | mesothelium | *C1QTNF4* | -0.241554359 | 1.55E-15 | 6.84E-13 | 7.53E-12 |
| Obesity | mesothelium | *CTSW* | -0.061744496 | 6.58E-09 | 7.64E-07 | 3.19E-05 |

| | | | | | | |
|---|---|---|---|---|---|---|
| Obesity | mesothelium | *EFEMP2* | 0.810559412 | 1.70E-06 | 0.000117778 | 0.008244429 |
| Obesity | mesothelium | *FUT10* | -0.153708546 | 0.000245738 | 0.007692379 | 1 |
| Obesity | mesothelium | *SNX32* | 0.126624051 | 0.00016543 | 0.005613052 | 0.802666379 |
| Obesity | mesothelium | *MRGPRF* | 0.343051745 | 0.000157872 | 0.005471402 | 0.765996245 |
| Obesity | mesothelium | *MST1* | -0.109407204 | 0.000241107 | 0.00761815 | 1 |
| Obesity | mesothelium | *HOXB2* | 0.380053791 | 0.000778551 | 0.019175288 | 1 |
| Obesity | mesothelium | *SEZ6L2* | 0.242002401 | 7.40E-05 | 0.003017898 | 0.35912986 |
| Obesity | mesothelium | *KCTD13* | 0.427190791 | 1.11E-06 | 8.71E-05 | 0.005401991 |
| Obesity | mesothelium | *PACS1* | -0.427297324 | 0.001759003 | 0.034693834 | 1 |
| Obesity | mesothelium | *CADM2* | 0.382769502 | 1.50E-09 | 2.01E-07 | 7.27E-06 |
| Obesity | mesothelium | *BANF1* | -0.817019272 | 4.55E-07 | 4.09E-05 | 0.002209236 |
| Obesity | mesothelium | *ORAI3* | 0.562481158 | 8.20E-12 | 1.81E-09 | 3.98E-08 |
| Obesity | mesothelium | *CCDC57* | -0.252202304 | 0.00061908 | 0.015809345 | 1 |
| Obesity | mesothelium | *SPRYD4* | -0.175603854 | 0.00166852 | 0.03415889 | 1 |
| Obesity | mesothelium | *RMDN1* | 0.277610523 | 0.002852377 | 0.048610628 | 1 |
| Obesity | mesothelium | *RUFY1* | 0.441112339 | 4.19E-05 | 0.001831733 | 0.203322339 |
| Obesity | mesothelium | *AP006621.1* | 0.505316682 | 5.10E-06 | 0.000305785 | 0.024768616 |
| Obesity | mesothelium | *RPP25* | -0.168727958 | 0.000149799 | 0.005305274 | 0.726822577 |
| Obesity | mesothelium | *STX19* | 0.430140015 | 3.07E-06 | 0.000191008 | 0.014898608 |
| Obesity | mesothelium | *MPI* | 0.327637821 | 9.19E-05 | 0.003623751 | 0.44572141 |
| Obesity | mesothelium | *TRIM73* | -1.003008654 | 1.71E-13 | 4.61E-11 | 8.29E-10 |
| Obesity | mesothelium | *ZNF852* | -0.439736053 | 1.82E-05 | 0.000938954 | 0.088261722 |
| Obesity | mesothelium | *TUFM* | 0.597061161 | 4.84E-11 | 9.04E-09 | 2.35E-07 |
| Obesity | mesothelium | *C3orf38* | 0.433302563 | 4.02E-08 | 4.24E-06 | 0.000195073 |
| Obesity | mesothelium | *RPRML* | 0.860302546 | 9.51E-06 | 0.000536301 | 0.046121862 |
| Obesity | mesothelium | *ZNF322* | -0.333256284 | 0.001711333 | 0.034647277 | 1 |
| Obesity | mesothelium | *ZFP41* | 0.308201969 | 0.000970385 | 0.022773168 | 1 |
| Obesity | mesothelium | *CLDN7* | -0.215292955 | 0.002178121 | 0.039880157 | 1 |
| Obesity | mesothelium | *PPP1R27* | -0.974817387 | 0.000205894 | 0.006795919 | 0.999000064 |
| Obesity | mesothelium | *GRID1* | -0.364217195 | 0.000283239 | 0.008379725 | 1 |
| Obesity | mesothelium | *COL18A1* | 0.351241359 | 0.00205538 | 0.038504647 | 1 |
| Obesity | mesothelium | *PLGLB1* | 0.174256381 | 0.001080861 | 0.024392275 | 1 |
| Obesity | mesothelium | *PSMG1* | 0.32719106 | 0.000434436 | 0.011995114 | 1 |
| Obesity | mesothelium | *UPP1* | 0.38417592 | 0.000926156 | 0.022194411 | 1 |
| Obesity | mesothelium | *EMILIN3* | 0.347557479 | 0.000942668 | 0.022420722 | 1 |
| Obesity | mesothelium | *ACOT1* | 0.174204275 | 9.76E-05 | 0.00375751 | 0.473446241 |
| Obesity | mesothelium | *NUTM2A* | -0.256686448 | 1.34E-05 | 0.00072251 | 0.065025897 |
| Obesity | mesothelium | *MAFF* | -0.199765399 | 9.06E-05 | 0.003604362 | 0.439732199 |
| Obesity | mesothelium | *GP1BA* | 0.354533749 | 6.92E-07 | 5.59E-05 | 0.00335627 |
| Obesity | mesothelium | *ATP6V0A2* | -0.626653177 | 0.000290732 | 0.008549268 | 1 |
| Obesity | mesothelium | *BTN3A2* | -0.1240497 | 9.26E-11 | 1.51E-08 | 4.49E-07 |
| Obesity | mesothelium | *ZSCAN23* | 0.090800417 | 0.001064127 | 0.024142188 | 1 |
| Obesity | mesothelium | *CENPP* | 0.216716981 | 1.70E-05 | 0.00089691 | 0.082515705 |
| Obesity | mesothelium | *OSTN* | 0.201976825 | 0.002010497 | 0.037809806 | 1 |
| Obesity | mesothelium | *SKOR1* | 0.395124989 | 0.001040551 | 0.023814875 | 1 |
| Obesity | mesothelium | *FBF1* | 0.288210728 | 0.001471173 | 0.03144552 | 1 |
| Obesity | mesothelium | *ZKSCAN3* | -0.420537229 | 1.81E-08 | 1.95E-06 | 8.79E-05 |
| Obesity | mesothelium | *CPNE4* | 0.332320002 | 2.62E-05 | 0.001285828 | 0.127261947 |
| Obesity | mesothelium | *ZNF605* | 0.285295625 | 0.000266334 | 0.008127387 | 1 |
| Obesity | mesothelium | *SULT1A1* | -0.52066984 | 0.001003141 | 0.023400198 | 1 |

| | | | | | | |
|---|---|---|---|---|---|---|
| Obesity | mesothelium | *SULT1A2* | 0.969690502 | 1.30E-26 | 6.29E-23 | 6.29E-23 |
| Obesity | mesothelium | *ZNF165* | -0.160758136 | 0.000219597 | 0.007150918 | 1 |
| Obesity | mesothelium | *MRPL21* | -0.117279598 | 0.000253146 | 0.007873485 | 1 |
| Obesity | mesothelium | *MFAP5* | -0.238884438 | 0.000241796 | 0.00761815 | 1 |
| Obesity | mesothelium | *DNAH10* | 0.654094683 | 5.13E-12 | 1.19E-09 | 2.49E-08 |
| Obesity | mesothelium | *RPS26* | -0.137492587 | 0.000320005 | 0.009297381 | 1 |
| Obesity | mesothelium | *ZNF780A* | -0.084198811 | 5.26E-05 | 0.002199029 | 0.255087323 |
| Obesity | mesothelium | *NOL8* | -0.866250275 | 0.000282915 | 0.008379725 | 1 |
| Obesity | mesothelium | *ADH4* | -0.139069166 | 0.002845431 | 0.048610628 | 1 |
| Obesity | mesothelium | *TMEM184B* | -0.415802289 | 0.000706294 | 0.017848646 | 1 |
| Obesity | mesothelium | *PRMT6* | -2.603446942 | 0.002423926 | 0.043238569 | 1 |
| Obesity | mesothelium | *SHISA4* | -2.620217817 | 1.23E-14 | 4.26E-12 | 5.96E-11 |
| Obesity | mesothelium | *MAP3K3* | -0.74367435 | 1.29E-05 | 0.000702129 | 0.062489507 |
| Obesity | mesothelium | *NOTCH4* | -0.036143476 | 1.28E-08 | 1.41E-06 | 6.20E-05 |
| Obesity | mesothelium | *SKIV2L* | -0.156253861 | 5.61E-07 | 4.70E-05 | 0.002723653 |
| Obesity | mesothelium | *GPANK1* | -0.575635649 | 3.35E-12 | 8.12E-10 | 1.62E-08 |
| Obesity | mesothelium | *POU5F1* | 0.054942073 | 8.83E-05 | 0.003541861 | 0.428565236 |
| Obesity | mesothelium | *CCHCR1* | 0.112755472 | 4.33E-06 | 0.000262453 | 0.020996239 |
| Obesity | mesothelium | *ABCF1* | 0.273520551 | 0.001588128 | 0.033293133 | 1 |
| Obesity | mesothelium | *RNF39* | -0.362162693 | 2.18E-06 | 0.00014119 | 0.010589233 |
| Obesity | mesothelium | *ZFP57* | 0.136414169 | 5.55E-09 | 6.90E-07 | 2.69E-05 |
| Obesity | mesothelium | *TRIM27* | -0.35992332 | 0.000734613 | 0.018278685 | 1 |
| Obesity | mesothelium | *C17orf107* | 0.225347182 | 0.000400799 | 0.011176302 | 1 |
| Obesity | mesothelium | *KLRC2* | 2.663911745 | 9.47E-06 | 0.000536301 | 0.045928647 |
| Obesity | mesothelium | *AC084018.1* | -0.145960972 | 0.000105805 | 0.003979585 | 0.513366512 |
| Obesity | mesothelium | *SUPT4H1* | 1.191706942 | 0.000200139 | 0.006689798 | 0.971073757 |
| Obesity | mesothelium | *SRA1* | 0.473927357 | 0.002291277 | 0.041482379 | 1 |
| Obesity | mesothelium | *TMEM110* | 0.601161731 | 7.45E-11 | 1.34E-08 | 3.62E-07 |
| Obesity | mesothelium | *PPP1CB* | -0.494798094 | 3.57E-05 | 0.0016029 | 0.173113172 |
| Obesity | mesothelium | *GALT* | 0.607348002 | 0.000142368 | 0.005155011 | 0.690771416 |
| Obesity | mesothelium | *EXOSC6* | -0.931776116 | 9.92E-14 | 2.83E-11 | 4.81E-10 |
| Obesity | mesothelium | *C4B* | 0.335739969 | 0.000148606 | 0.005301724 | 0.721034509 |
| Obesity | mesothelium | *LINC00240* | 0.11784694 | 0.000455071 | 0.01233522 | 1 |
| Obesity | mesothelium | *AL049840.1* | -0.786427435 | 2.02E-06 | 0.000132384 | 0.009796393 |
| Obesity | mesothelium | *HCG20* | -0.309525704 | 6.36E-07 | 5.23E-05 | 0.003084577 |
| Obesity | mesothelium | *TAPBP* | 0.368652888 | 0.001150802 | 0.025380409 | 1 |
| Obesity | mesothelium | *ZSCAN31* | 0.131147266 | 3.45E-07 | 3.16E-05 | 0.001674131 |
| Obesity | mesothelium | *AC019117.2* | -1.338749979 | 0.00110523 | 0.024598971 | 1 |
| Obesity | mesothelium | *PSMB9* | 0.172266894 | 1.18E-06 | 9.11E-05 | 0.005741095 |
| Obesity | mesothelium | *ISY1* | -0.195530799 | 0.000716985 | 0.018024921 | 1 |
| Obesity | mesothelium | *C4A* | -0.348741014 | 4.76E-07 | 4.20E-05 | 0.00230888 |
| Obesity | mesothelium | *DNAH10OS* | 2.468691917 | 1.67E-06 | 0.000117596 | 0.008114111 |
| Obesity | mesothelium | *DPP3* | 0.107248374 | 0.001258144 | 0.02737451 | 1 |
| Obesity | mesothelium | *BRK1* | -0.321876454 | 0.000155055 | 0.005451632 | 0.752325198 |
| Obesity | mesothelium | *PCDHA3* | -0.38169164 | 0.000234983 | 0.007500904 | 1 |
| Obesity | mesothelium | *HMBS* | 0.440290306 | 1.04E-05 | 0.000572841 | 0.05041001 |
| Obesity | mesothelium | *PDF* | -0.691135533 | 0.001659621 | 0.034120677 | 1 |
| Obesity | mesothelium | *C19orf84* | 11.06228209 | 1.62E-06 | 0.000115764 | 0.00787195 |
| Obesity | mesothelium | *AP001172.2* | 0.106841185 | 0.000741407 | 0.018353606 | 1 |
| Obesity | monocyte | *NFYA* | -1.653199775 | 0.010918019 | 0.04214338 | 1 |

| | | | | | | |
|---|---|---|---|---|---|---|
| Obesity | monocyte | *SEMA3F* | -2.104156818 | 3.95E-05 | 0.003504422 | 0.191631637 |
| Obesity | monocyte | *HS3ST1* | -0.818977146 | 0.015245172 | 0.045130917 | 1 |
| Obesity | monocyte | *AOC1* | -3.056793065 | 0.002936431 | 0.035116467 | 1 |
| Obesity | monocyte | *TMEM176A* | 2.882417867 | 0.004927169 | 0.035116467 | 1 |
| Obesity | monocyte | *M6PR* | 3.083994489 | 0.008939564 | 0.041462933 | 1 |
| Obesity | monocyte | *RBM6* | -3.677385108 | 3.29E-15 | 7.99E-12 | 1.60E-11 |
| Obesity | monocyte | *POLR2J* | 4.438214477 | 0.001402635 | 0.03111676 | 1 |
| Obesity | monocyte | *MEOX1* | -2.63413811 | 0.017929469 | 0.048768447 | 1 |
| Obesity | monocyte | *PON1* | -3.653393898 | 0.009260405 | 0.041462933 | 1 |
| Obesity | monocyte | *RHBDD2* | 2.577013654 | 0.000392148 | 0.017455969 | 1 |
| Obesity | monocyte | *GGNBP2* | -1.858828602 | 0.015100952 | 0.044986158 | 1 |
| Obesity | monocyte | *TMEM98* | -1.354747279 | 0.015753608 | 0.045685351 | 1 |
| Obesity | monocyte | *MAP3K9* | -0.973797578 | 0.018612969 | 0.0494579 | 1 |
| Obesity | monocyte | *USH1C* | 1.922324112 | 0.004713663 | 0.035116467 | 1 |
| Obesity | monocyte | *ELAC2* | -1.843571914 | 0.003420308 | 0.035116467 | 1 |
| Obesity | monocyte | *SCIN* | -1.08189225 | 0.003856655 | 0.035116467 | 1 |
| Obesity | monocyte | *NOS2* | -5.634937907 | 0.00885659 | 0.041462933 | 1 |
| Obesity | monocyte | *CD79B* | 0.43679328 | 0.010764734 | 0.041961024 | 1 |
| Obesity | monocyte | *PAX6* | -1.817860386 | 0.006238511 | 0.037094676 | 1 |
| Obesity | monocyte | *BAIAP3* | -3.056925727 | 0.004276661 | 0.035116467 | 1 |
| Obesity | monocyte | *FMO3* | -1.492481245 | 0.000534739 | 0.020269938 | 1 |
| Obesity | monocyte | *NADK* | 1.785558802 | 0.006343377 | 0.037442903 | 1 |
| Obesity | monocyte | *DLEC1* | -1.032253515 | 0.00934923 | 0.041462933 | 1 |
| Obesity | monocyte | *SS18L2* | 5.487934086 | 0.005961317 | 0.036337076 | 1 |
| Obesity | monocyte | *MGST1* | -0.564411278 | 0.006934653 | 0.039124342 | 1 |
| Obesity | monocyte | *IL32* | 6.747115304 | 0.006756734 | 0.038614455 | 1 |
| Obesity | monocyte | *TRAF3IP3* | 0.328272935 | 0.003576274 | 0.035116467 | 1 |
| Obesity | monocyte | *FUZ* | -1.436466742 | 0.016680822 | 0.046940995 | 1 |
| Obesity | monocyte | *SLC6A13* | -2.911302008 | 0.00760475 | 0.040370074 | 1 |
| Obesity | monocyte | *LRRC23* | 3.171454678 | 0.000573198 | 0.021082768 | 1 |
| Obesity | monocyte | *CLCN6* | 4.627461718 | 0.002813889 | 0.035116467 | 1 |
| Obesity | monocyte | *SLC6A7* | 8.21228501 | 0.005425855 | 0.035630311 | 1 |
| Obesity | monocyte | *DPF1* | -4.633746646 | 0.002686824 | 0.035116467 | 1 |
| Obesity | monocyte | *ZBTB32* | -2.479880411 | 0.007713939 | 0.040591959 | 1 |
| Obesity | monocyte | *ALOX5* | 0.372688161 | 0.010847369 | 0.041961024 | 1 |
| Obesity | monocyte | *EHD3* | 3.67621183 | 0.003426373 | 0.035116467 | 1 |
| Obesity | monocyte | *MAN2B2* | 4.075815483 | 0.006712854 | 0.038545286 | 1 |
| Obesity | monocyte | *SLC25A39* | -1.478336383 | 0.01449354 | 0.044883966 | 1 |
| Obesity | monocyte | *DDX11* | -1.388946116 | 0.001684335 | 0.03218121 | 1 |
| Obesity | monocyte | *MTMR11* | 0.899798696 | 0.000685042 | 0.022981804 | 1 |
| Obesity | monocyte | *CCDC88C* | 0.835675077 | 0.003589437 | 0.035116467 | 1 |
| Obesity | monocyte | *CHDH* | -2.201469214 | 0.012125159 | 0.043083715 | 1 |
| Obesity | monocyte | *CLCA4* | -6.310518414 | 0.001262849 | 0.029332492 | 1 |
| Obesity | monocyte | *SLC11A1* | 0.252063797 | 0.004285819 | 0.035116467 | 1 |
| Obesity | monocyte | *VSIG2* | -1.983459934 | 0.0020566 | 0.034173375 | 1 |
| Obesity | monocyte | *MARCO* | -0.893613907 | 0.001911742 | 0.033521987 | 1 |
| Obesity | monocyte | *SLC7A9* | 1.327680402 | 0.008156457 | 0.040894308 | 1 |
| Obesity | monocyte | *CPS1* | -0.994186052 | 0.009516077 | 0.041462933 | 1 |
| Obesity | monocyte | *GRAMD1B* | 1.208136324 | 0.006875647 | 0.03896597 | 1 |
| Obesity | monocyte | *ABCC2* | 3.016953174 | 0.007963671 | 0.040894308 | 1 |

| | | | | | | |
|---|---|---|---|---|---|---|
| Obesity | monocyte | *TYMP* | 1.580917312 | 0.003363046 | 0.035116467 | 1 |
| Obesity | monocyte | *TOMM34* | -1.963549145 | 0.008969848 | 0.041462933 | 1 |
| Obesity | monocyte | *RNASET2* | 1.106175187 | 0.013923614 | 0.044540919 | 1 |
| Obesity | monocyte | *INSRR* | -2.008911139 | 0.004911893 | 0.035116467 | 1 |
| Obesity | monocyte | *B4GALT7* | 2.473899264 | 0.00224819 | 0.035072295 | 1 |
| Obesity | monocyte | *SH2D2A* | 4.251145113 | 0.000868991 | 0.025867134 | 1 |
| Obesity | monocyte | *BAK1* | 1.487102051 | 0.00433595 | 0.035116467 | 1 |
| Obesity | monocyte | *CENPQ* | -1.36512183 | 0.000196523 | 0.011628398 | 0.953528628 |
| Obesity | monocyte | *UBA6* | -2.136848715 | 0.019217483 | 0.049996369 | 1 |
| Obesity | monocyte | *SH3YL1* | -1.391169308 | 0.002537207 | 0.035116467 | 1 |
| Obesity | monocyte | *MYOM2* | -2.055001208 | 0.007377282 | 0.040251891 | 1 |
| Obesity | monocyte | *FLT4* | -1.312682697 | 0.00018976 | 0.011628398 | 0.920716857 |
| Obesity | monocyte | *AGA* | 3.571410632 | 0.011959933 | 0.042874352 | 1 |
| Obesity | monocyte | *PI4K2B* | 11.50197077 | 0.006885073 | 0.03896597 | 1 |
| Obesity | monocyte | *PSMA4* | 3.441574194 | 0.013135782 | 0.044028405 | 1 |
| Obesity | monocyte | *TDP1* | 4.27644069 | 0.002741058 | 0.035116467 | 1 |
| Obesity | monocyte | *SPATA7* | -1.382087763 | 0.011242921 | 0.04231558 | 1 |
| Obesity | monocyte | *MED17* | 4.581055723 | 0.005228835 | 0.035351132 | 1 |
| Obesity | monocyte | *AP2S1* | 3.626509462 | 0.004395123 | 0.035116467 | 1 |
| Obesity | monocyte | *TRIT1* | 2.125713265 | 0.013812487 | 0.044539684 | 1 |
| Obesity | monocyte | *CUL7* | -1.724449667 | 0.014724568 | 0.044883966 | 1 |
| Obesity | monocyte | *ARAP2* | 1.175480691 | 0.00314116 | 0.035116467 | 1 |
| Obesity | monocyte | *ANO2* | -0.736213253 | 0.011612625 | 0.042465454 | 1 |
| Obesity | monocyte | *RCN1* | -2.155538425 | 0.006458611 | 0.037755639 | 1 |
| Obesity | monocyte | *MCUR1* | 2.619628342 | 0.003356664 | 0.035116467 | 1 |
| Obesity | monocyte | *LAMC3* | 1.925707224 | 0.00503825 | 0.035224192 | 1 |
| Obesity | monocyte | *CYBA* | 2.943121184 | 0.00546351 | 0.035630311 | 1 |
| Obesity | monocyte | *THOC3* | 2.96814049 | 0.003048447 | 0.035116467 | 1 |
| Obesity | monocyte | *ALX4* | 2.328020949 | 0.002646307 | 0.035116467 | 1 |
| Obesity | monocyte | *FSTL4* | -3.085805163 | 0.00408172 | 0.035116467 | 1 |
| Obesity | monocyte | *AKR7A2* | -1.152595452 | 0.013033638 | 0.043967382 | 1 |
| Obesity | monocyte | *MRTO4* | -1.125555749 | 0.016781272 | 0.047065162 | 1 |
| Obesity | monocyte | *PLEKHH1* | -1.255561552 | 0.011542692 | 0.042465454 | 1 |
| Obesity | monocyte | *GALC* | -1.852578311 | 0.013476053 | 0.044239385 | 1 |
| Obesity | monocyte | *ITIH4* | 2.543285582 | 1.72E-08 | 6.41E-06 | 8.33E-05 |
| Obesity | monocyte | *NPFFR2* | -1.748919728 | 0.009673915 | 0.041464521 | 1 |
| Obesity | monocyte | *IL17RB* | -2.249663333 | 0.005028325 | 0.035209637 | 1 |
| Obesity | monocyte | *DNAJC25* | 2.976965911 | 0.002909416 | 0.035116467 | 1 |
| Obesity | monocyte | *OGFR* | 3.471627265 | 0.013122412 | 0.044028405 | 1 |
| Obesity | monocyte | *RIMBP2* | -1.750885033 | 0.014491875 | 0.044883966 | 1 |
| Obesity | monocyte | *PRDM6* | -0.999332891 | 0.011259154 | 0.04231558 | 1 |
| Obesity | monocyte | *SPAG4* | -4.80513863 | 0.01060803 | 0.041845661 | 1 |
| Obesity | monocyte | *NCKAP1* | -1.081108609 | 0.017582098 | 0.048196802 | 1 |
| Obesity | monocyte | *LTK* | 23.37098573 | 0.002168471 | 0.035071411 | 1 |
| Obesity | monocyte | *AHRR* | -2.164979106 | 0.005084315 | 0.03528148 | 1 |
| Obesity | monocyte | *CDON* | -0.58141413 | 0.01507571 | 0.044986158 | 1 |
| Obesity | monocyte | *TMEM161A* | -1.77364464 | 0.013555836 | 0.04433272 | 1 |
| Obesity | monocyte | *SLC12A2* | -1.252165739 | 0.018312317 | 0.04903887 | 1 |
| Obesity | monocyte | *ABCA7* | 1.06947514 | 0.014009846 | 0.044610733 | 1 |
| Obesity | monocyte | *BCAS1* | -1.674863263 | 0.009236907 | 0.041462933 | 1 |

| | | | | | | |
|---|---|---|---|---|---|---|
| Obesity | monocyte | *CHI3L2* | 1.679108637 | 0.005203367 | 0.035351132 | 1 |
| Obesity | monocyte | *ANKRD44* | 1.581607055 | 0.001815653 | 0.032842449 | 1 |
| Obesity | monocyte | *TIE1* | -1.985350393 | 0.005512354 | 0.035757287 | 1 |
| Obesity | monocyte | *ZMYND12* | -1.02445424 | 0.017048763 | 0.047509289 | 1 |
| Obesity | monocyte | *NGEF* | 2.997460764 | 0.00931386 | 0.041462933 | 1 |
| Obesity | monocyte | *SPI1* | 0.502858587 | 0.005279903 | 0.035531331 | 1 |
| Obesity | monocyte | *ZNRD1* | -2.859718233 | 0.000745394 | 0.024272824 | 1 |
| Obesity | monocyte | *MTFR1* | -1.597179901 | 0.010531865 | 0.041845661 | 1 |
| Obesity | monocyte | *STOML1* | 6.330277177 | 0.00332551 | 0.035116467 | 1 |
| Obesity | monocyte | *ROGDI* | -1.637287608 | 0.009469178 | 0.041462933 | 1 |
| Obesity | monocyte | *FGFR3* | -2.195058405 | 0.010763544 | 0.041961024 | 1 |
| Obesity | monocyte | *IFI35* | -1.251099752 | 0.014974547 | 0.044986158 | 1 |
| Obesity | monocyte | *POLR1A* | -1.471292706 | 0.010975499 | 0.042169886 | 1 |
| Obesity | monocyte | *IFT80* | -1.028568404 | 0.010791413 | 0.041961024 | 1 |
| Obesity | monocyte | *DRD4* | -1.961770245 | 0.011539081 | 0.042465454 | 1 |
| Obesity | monocyte | *PLA2G10* | -2.84771262 | 0.011443683 | 0.042352977 | 1 |
| Obesity | monocyte | *PIGB* | -0.977939297 | 0.014292991 | 0.044792561 | 1 |
| Obesity | monocyte | *GNB5* | -1.174533606 | 0.015162651 | 0.045042687 | 1 |
| Obesity | monocyte | *DAPP1* | 0.26898947 | 0.015128283 | 0.045004555 | 1 |
| Obesity | monocyte | *GBA2* | -1.581472906 | 0.014336943 | 0.044792561 | 1 |
| Obesity | monocyte | *ASNS* | 1.459209866 | 0.003695948 | 0.035116467 | 1 |
| Obesity | monocyte | *WDR1* | -1.353486214 | 0.018730238 | 0.049532965 | 1 |
| Obesity | monocyte | *TRHDE* | -0.541953343 | 0.004614386 | 0.035116467 | 1 |
| Obesity | monocyte | *CRMP1* | -0.896863825 | 0.010856753 | 0.041961024 | 1 |
| Obesity | monocyte | *NDE1* | 3.975794017 | 0.004914846 | 0.035116467 | 1 |
| Obesity | monocyte | *PVR* | 2.953933572 | 0.000493871 | 0.019613656 | 1 |
| Obesity | monocyte | *SCARB1* | -1.101585163 | 0.011751926 | 0.042569734 | 1 |
| Obesity | monocyte | *MOV10L1* | -1.205107737 | 0.015238774 | 0.045130917 | 1 |
| Obesity | monocyte | *PPP2R2C* | -2.976349515 | 0.017265668 | 0.047761129 | 1 |
| Obesity | monocyte | *TTC38* | -0.776486159 | 0.017978267 | 0.048768447 | 1 |
| Obesity | monocyte | *ADD2* | 2.375726375 | 0.000837103 | 0.025385159 | 1 |
| Obesity | monocyte | *MCM6* | -1.014625029 | 0.01600402 | 0.046138742 | 1 |
| Obesity | monocyte | *FMO4* | -0.701841324 | 0.016755261 | 0.047046601 | 1 |
| Obesity | monocyte | *SPAG5* | -4.80640376 | 0.001254603 | 0.029332492 | 1 |
| Obesity | monocyte | *PAG1* | 0.787299438 | 0.003581242 | 0.035116467 | 1 |
| Obesity | monocyte | *ICAM3* | 3.202669117 | 0.007824935 | 0.040689236 | 1 |
| Obesity | monocyte | *XAB2* | -1.685213191 | 0.01561614 | 0.045652454 | 1 |
| Obesity | monocyte | *LRCH4* | 1.476774978 | 0.00282521 | 0.035116467 | 1 |
| Obesity | monocyte | *SIRT6* | -1.669589831 | 0.016696848 | 0.046940995 | 1 |
| Obesity | monocyte | *NAALAD2* | -0.623152489 | 0.013473776 | 0.044239385 | 1 |
| Obesity | monocyte | *AMPH* | -1.020824401 | 0.013037339 | 0.043967382 | 1 |
| Obesity | monocyte | *FGF20* | 4.122647626 | 0.005329026 | 0.035561878 | 1 |
| Obesity | monocyte | *FKBP7* | -1.074553746 | 0.009006145 | 0.041462933 | 1 |
| Obesity | monocyte | *SP140* | 1.527077109 | 0.003164465 | 0.035116467 | 1 |
| Obesity | monocyte | *DUSP13* | 8.714864882 | 0.014306183 | 0.044792561 | 1 |
| Obesity | monocyte | *SCGN* | -7.022876995 | 0.012537124 | 0.04319079 | 1 |
| Obesity | monocyte | *PGM1* | -1.074041141 | 0.011877907 | 0.042802404 | 1 |
| Obesity | monocyte | *SCTR* | -0.817535619 | 0.009364981 | 0.041462933 | 1 |
| Obesity | monocyte | *KCNN2* | -2.848269675 | 0.005934405 | 0.036288402 | 1 |
| Obesity | monocyte | *PSEN1* | 3.191713472 | 0.004250301 | 0.035116467 | 1 |

| | | | | | | |
|---|---|---|---|---|---|---|
| Obesity | monocyte | *STK17B* | 1.49161551 | 0.004067844 | 0.035116467 | 1 |
| Obesity | monocyte | *PCDHB4* | 6.383000529 | 0.002780133 | 0.035116467 | 1 |
| Obesity | monocyte | *CCNT2* | -1.610608436 | 0.013867222 | 0.044539684 | 1 |
| Obesity | monocyte | *OXCT1* | -1.140760469 | 0.011197548 | 0.042279868 | 1 |
| Obesity | monocyte | *SLC27A5* | -1.128900967 | 0.012363444 | 0.043115778 | 1 |
| Obesity | monocyte | *ZNF264* | -1.164125724 | 0.01494352 | 0.044986158 | 1 |
| Obesity | monocyte | *NOA1* | 2.71372868 | 0.011622861 | 0.042465454 | 1 |
| Obesity | monocyte | *CAD* | 2.316673468 | 0.00532774 | 0.035561878 | 1 |
| Obesity | monocyte | *PILRA* | 0.639702628 | 0.000288898 | 0.014912068 | 1 |
| Obesity | monocyte | *ZNF213* | -1.898505588 | 0.017326423 | 0.047820138 | 1 |
| Obesity | monocyte | *RRN3* | -1.658374306 | 0.013503487 | 0.044299471 | 1 |
| Obesity | monocyte | *DDHD2* | -1.081359644 | 0.00991608 | 0.041669249 | 1 |
| Obesity | monocyte | *EPS15* | -1.922470885 | 0.010341346 | 0.041771331 | 1 |
| Obesity | monocyte | *POMGNT1* | -1.519039472 | 0.014744663 | 0.044883966 | 1 |
| Obesity | monocyte | *NME8* | 2.762388838 | 0.004499865 | 0.035116467 | 1 |
| Obesity | monocyte | *ACOX3* | -1.278105469 | 0.019003912 | 0.049859574 | 1 |
| Obesity | monocyte | *TRIP6* | -1.415842328 | 0.005596211 | 0.035810934 | 1 |
| Obesity | monocyte | *ACHE* | -4.184616135 | 0.00496387 | 0.035116467 | 1 |
| Obesity | monocyte | *ADAMTS2* | -0.848016723 | 0.012137742 | 0.043083715 | 1 |
| Obesity | monocyte | *LPCAT2* | 2.150113483 | 0.002578375 | 0.035116467 | 1 |
| Obesity | monocyte | *PHACTR3* | -2.085994941 | 0.001363228 | 0.030908327 | 1 |
| Obesity | monocyte | *AAMDC* | -1.326942138 | 0.005525765 | 0.035757287 | 1 |
| Obesity | monocyte | *GP6* | -1.817887398 | 0.001312329 | 0.030034993 | 1 |
| Obesity | monocyte | *ASAP3* | -1.003936336 | 0.012002057 | 0.042922686 | 1 |
| Obesity | monocyte | *DOCK3* | -0.824524569 | 0.010442167 | 0.041803129 | 1 |
| Obesity | monocyte | *PPP1R13B* | -2.200552413 | 0.00346273 | 0.035116467 | 1 |
| Obesity | monocyte | *EBF4* | -1.652135265 | 0.011852223 | 0.042787935 | 1 |
| Obesity | monocyte | *CPXM1* | -1.083247362 | 0.008158212 | 0.040894308 | 1 |
| Obesity | monocyte | *RBBP9* | -1.472059228 | 0.0107722 | 0.041961024 | 1 |
| Obesity | monocyte | *TBX5* | 1.614271402 | 0.005707484 | 0.035810934 | 1 |
| Obesity | monocyte | *FXYD5* | 2.633767804 | 0.00153968 | 0.03218121 | 1 |
| Obesity | monocyte | *ZNF302* | -1.241374708 | 0.010188491 | 0.041771331 | 1 |
| Obesity | monocyte | *CDIP1* | -0.93806706 | 0.009574022 | 0.041462933 | 1 |
| Obesity | monocyte | *ANKRD24* | -1.303497431 | 0.009468931 | 0.041462933 | 1 |
| Obesity | monocyte | *MAEA* | -1.202139971 | 0.01335307 | 0.044157522 | 1 |
| Obesity | monocyte | *LYZ* | 0.626521353 | 0.001651258 | 0.03218121 | 1 |
| Obesity | monocyte | *RAB11FIP3* | -0.984833376 | 0.016889915 | 0.047233354 | 1 |
| Obesity | monocyte | *PABPC4* | -4.980436073 | 2.79E-06 | 0.000466582 | 0.013530889 |
| Obesity | monocyte | *CERS4* | -1.227609884 | 0.019011145 | 0.049859574 | 1 |
| Obesity | monocyte | *ABCC6* | -0.898684911 | 0.011273141 | 0.042332379 | 1 |
| Obesity | monocyte | *RGS17* | -1.142085614 | 0.000521331 | 0.019917294 | 1 |
| Obesity | monocyte | *CAPN3* | -2.606823845 | 0.004050899 | 0.035116467 | 1 |
| Obesity | monocyte | *TBX15* | -2.461702014 | 2.31E-05 | 0.002338738 | 0.112259435 |
| Obesity | monocyte | *XYLB* | -1.38171741 | 0.019182015 | 0.049996369 | 1 |
| Obesity | monocyte | *CRAT* | -1.184063339 | 0.009418177 | 0.041462933 | 1 |
| Obesity | monocyte | *NANS* | 4.10095604 | 0.004105982 | 0.035116467 | 1 |
| Obesity | monocyte | *CRTAC1* | -1.394229124 | 0.006668309 | 0.03842593 | 1 |
| Obesity | monocyte | *DSP* | -1.106121233 | 0.009671443 | 0.041464521 | 1 |
| Obesity | monocyte | *ACOT7* | 2.611272868 | 0.010129559 | 0.041757534 | 1 |
| Obesity | monocyte | *SCD* | -0.791672942 | 0.015767911 | 0.045685351 | 1 |

| | | | | | | |
|---|---|---|---|---|---|---|
| Obesity | monocyte | *ERMP1* | -1.363025591 | 0.013316503 | 0.044157522 | 1 |
| Obesity | monocyte | *PRTFDC1* | -0.973774732 | 0.013048852 | 0.043967382 | 1 |
| Obesity | monocyte | *TSPAN15* | -0.795160819 | 0.012004552 | 0.042922686 | 1 |
| Obesity | monocyte | *OCEL1* | -2.57844293 | 0.005903889 | 0.036288402 | 1 |
| Obesity | monocyte | *SETD1A* | 6.994297212 | 0.0004292 | 0.018429008 | 1 |
| Obesity | monocyte | *SERPIND1* | -2.460359168 | 0.002343972 | 0.035116467 | 1 |
| Obesity | monocyte | *LZTR1* | -2.254519416 | 0.013870458 | 0.044539684 | 1 |
| Obesity | monocyte | *CECR2* | -0.975204619 | 0.009736449 | 0.041544165 | 1 |
| Obesity | monocyte | *DDT* | 1.480992459 | 0.006968648 | 0.039147772 | 1 |
| Obesity | monocyte | *SPECC1L* | -2.177032361 | 0.017145023 | 0.047644704 | 1 |
| Obesity | monocyte | *SLC35E4* | 2.222997796 | 0.008189852 | 0.040894308 | 1 |
| Obesity | monocyte | *CYTH4* | 0.609641722 | 0.003880957 | 0.035116467 | 1 |
| Obesity | monocyte | *CARD10* | -1.490107764 | 0.00094765 | 0.026972741 | 1 |
| Obesity | monocyte | *LGALS2* | 1.324008029 | 0.008946442 | 0.041462933 | 1 |
| Obesity | monocyte | *SH3BP1* | 0.554054386 | 0.004386806 | 0.035116467 | 1 |
| Obesity | monocyte | *ANKRD54* | -3.756037047 | 0.002983489 | 0.035116467 | 1 |
| Obesity | monocyte | *EIF3L* | 2.434771982 | 0.003928878 | 0.035116467 | 1 |
| Obesity | monocyte | *CENPM* | 4.025835429 | 0.005611777 | 0.035810934 | 1 |
| Obesity | monocyte | *KDELR3* | -1.441910524 | 0.002312832 | 0.035072295 | 1 |
| Obesity | monocyte | *CYP2D6* | -3.125423332 | 0.001590252 | 0.03218121 | 1 |
| Obesity | monocyte | *CBY1* | -1.885954699 | 0.008366029 | 0.041379919 | 1 |
| Obesity | monocyte | *RAB36* | -2.439807619 | 0.002562017 | 0.035116467 | 1 |
| Obesity | monocyte | *RHBDD3* | -1.046968847 | 0.012209719 | 0.043115778 | 1 |
| Obesity | monocyte | *CHKB* | 5.664043318 | 0.007282082 | 0.039923913 | 1 |
| Obesity | monocyte | *THOC5* | -2.269978246 | 0.016337 | 0.046589332 | 1 |
| Obesity | monocyte | *TTLL12* | -1.245448433 | 0.013616029 | 0.044368686 | 1 |
| Obesity | monocyte | *APOL4* | 2.269423202 | 0.001637033 | 0.03218121 | 1 |
| Obesity | monocyte | *KIAA0930* | 1.083221535 | 0.004923271 | 0.035116467 | 1 |
| Obesity | monocyte | *MLC1* | 0.722652098 | 0.002804069 | 0.035116467 | 1 |
| Obesity | monocyte | *HDAC10* | -1.146486903 | 0.010768219 | 0.041961024 | 1 |
| Obesity | monocyte | *RBM23* | -1.272432256 | 0.015828841 | 0.045769688 | 1 |
| Obesity | monocyte | *PYGL* | -0.823249163 | 0.007029836 | 0.039250591 | 1 |
| Obesity | monocyte | *CNIH1* | -1.560427801 | 0.016995435 | 0.047419121 | 1 |
| Obesity | monocyte | *GSTZ1* | -1.108458788 | 0.013988907 | 0.044596002 | 1 |
| Obesity | monocyte | *LGMN* | -0.81756609 | 0.016506249 | 0.046845773 | 1 |
| Obesity | monocyte | *MTHFD1* | -0.917378979 | 0.015671859 | 0.045669584 | 1 |
| Obesity | monocyte | *EFS* | -1.032550439 | 0.010430117 | 0.041803129 | 1 |
| Obesity | monocyte | *KIAA0391* | 1.143386228 | 0.003763713 | 0.035116467 | 1 |
| Obesity | monocyte | *EMC9* | -1.447792589 | 0.01570104 | 0.045674648 | 1 |
| Obesity | monocyte | *PYGB* | 2.504299351 | 0.017979016 | 0.048768447 | 1 |
| Obesity | monocyte | *PABPC1L* | -1.400535964 | 0.011119384 | 0.042279868 | 1 |
| Obesity | monocyte | *DOK5* | -1.117282665 | 0.009641505 | 0.041464521 | 1 |
| Obesity | monocyte | *PTK6* | -3.334575395 | 0.007679397 | 0.040456498 | 1 |
| Obesity | monocyte | *GMEB2* | -1.751680222 | 0.013594453 | 0.044347765 | 1 |
| Obesity | monocyte | *CDC25B* | -1.174690003 | 0.013076738 | 0.044000232 | 1 |
| Obesity | monocyte | *RASSF2* | 0.535021349 | 0.002254737 | 0.035072295 | 1 |
| Obesity | monocyte | *ANGPT4* | -2.007721767 | 0.013638207 | 0.04438134 | 1 |
| Obesity | monocyte | *SIRPB1* | 0.470201215 | 0.005127473 | 0.03528148 | 1 |
| Obesity | monocyte | *MROH8* | -1.065139394 | 0.01403261 | 0.044610733 | 1 |
| Obesity | monocyte | *ACTR5* | -1.40823939 | 0.011543689 | 0.042465454 | 1 |

| | | | | | | |
|---|---|---|---|---|---|---|
| Obesity | monocyte | *DNTTIP1* | -1.091276792 | 0.011694724 | 0.042490474 | 1 |
| Obesity | monocyte | *MAP1LC3A* | -1.446335149 | 0.017325743 | 0.047820138 | 1 |
| Obesity | monocyte | *MYOM1* | -0.852713668 | 0.011882695 | 0.042802404 | 1 |
| Obesity | monocyte | *LAMA1* | -1.875127834 | 0.002253148 | 0.035072295 | 1 |
| Obesity | monocyte | *NALCN* | -0.986302151 | 0.009371065 | 0.041462933 | 1 |
| Obesity | monocyte | *ACP5* | -1.171927953 | 0.013853405 | 0.044539684 | 1 |
| Obesity | monocyte | *UGGT2* | -1.336765819 | 0.014319121 | 0.044792561 | 1 |
| Obesity | monocyte | *SUPT20H* | -1.57632199 | 0.017960478 | 0.048768447 | 1 |
| Obesity | monocyte | *RGCC* | -0.991448197 | 0.010028793 | 0.041669249 | 1 |
| Obesity | monocyte | *PLLP* | -0.897161046 | 0.005366899 | 0.035574034 | 1 |
| Obesity | monocyte | *CCDC113* | -4.44181774 | 0.001240792 | 0.029258086 | 1 |
| Obesity | monocyte | *MON1B* | 4.968576121 | 0.019095836 | 0.049947706 | 1 |
| Obesity | monocyte | *NPRL3* | -0.979071764 | 0.010025679 | 0.041669249 | 1 |
| Obesity | monocyte | *TAF1C* | -1.540282381 | 0.004973968 | 0.035116467 | 1 |
| Obesity | monocyte | *NAGPA* | -1.438291851 | 0.010862128 | 0.041961024 | 1 |
| Obesity | monocyte | *NME4* | -2.47716533 | 0.009141209 | 0.041462933 | 1 |
| Obesity | monocyte | *CLCN7* | 3.293248625 | 0.000391635 | 0.017455969 | 1 |
| Obesity | monocyte | *RHBDL1* | -4.719013953 | 0.002864881 | 0.035116467 | 1 |
| Obesity | monocyte | *NUBP1* | 5.117304069 | 0.006130493 | 0.036813307 | 1 |
| Obesity | monocyte | *CAPN15* | -1.795597498 | 0.018671366 | 0.049504628 | 1 |
| Obesity | monocyte | *CPPED1* | 0.599140379 | 0.004762048 | 0.035116467 | 1 |
| Obesity | monocyte | *RRN3P2* | -3.92845107 | 0.007677488 | 0.040456498 | 1 |
| Obesity | monocyte | *PYCARD* | 10.37214862 | 6.40E-08 | 1.63E-05 | 0.000310401 |
| Obesity | monocyte | *CDIPT* | -2.002823521 | 0.009329951 | 0.041462933 | 1 |
| Obesity | monocyte | *KAT8* | -1.20394978 | 0.012389432 | 0.043115778 | 1 |
| Obesity | monocyte | *TMC5* | -1.115790759 | 0.002992376 | 0.035116467 | 1 |
| Obesity | monocyte | *KNOP1* | -2.390323453 | 0.006062866 | 0.036679583 | 1 |
| Obesity | monocyte | *FAH* | -0.817283192 | 0.008470199 | 0.041379919 | 1 |
| Obesity | monocyte | *RPAP1* | -2.133533032 | 0.002516948 | 0.035116467 | 1 |
| Obesity | monocyte | *SLC30A4* | -0.932764455 | 0.012405117 | 0.043115778 | 1 |
| Obesity | monocyte | *ZDHHC2* | -2.94892177 | 0.005452351 | 0.035630311 | 1 |
| Obesity | monocyte | *SFRP1* | -1.04140891 | 0.011182356 | 0.042279868 | 1 |
| Obesity | monocyte | *TSTA3* | 2.286733072 | 0.008209204 | 0.040894308 | 1 |
| Obesity | monocyte | *R3HCC1* | 3.07075522 | 0.001039319 | 0.027556163 | 1 |
| Obesity | monocyte | *PEX11G* | -1.392839258 | 0.007832594 | 0.040689236 | 1 |
| Obesity | monocyte | *ERCC2* | -1.308233571 | 0.009108842 | 0.041462933 | 1 |
| Obesity | monocyte | *CD37* | 1.006987902 | 0.006210611 | 0.037019516 | 1 |
| Obesity | monocyte | *AMH* | 2.87079895 | 0.004784075 | 0.035116467 | 1 |
| Obesity | monocyte | *FCER2* | -3.384234854 | 0.009805035 | 0.041585688 | 1 |
| Obesity | monocyte | *CLEC4M* | -3.341914377 | 0.008976115 | 0.041462933 | 1 |
| Obesity | monocyte | *OLFM2* | -0.977416601 | 0.005092325 | 0.03528148 | 1 |
| Obesity | monocyte | *RASAL3* | 0.675874531 | 0.003987046 | 0.035116467 | 1 |
| Obesity | monocyte | *EPHX3* | 0.415308752 | 0.008686094 | 0.041462933 | 1 |
| Obesity | monocyte | *CEACAM4* | 1.492649129 | 0.004778475 | 0.035116467 | 1 |
| Obesity | monocyte | *ICAM5* | 0.879601576 | 0.018391877 | 0.04913953 | 1 |
| Obesity | monocyte | *GRIN2D* | -2.299903603 | 0.007432111 | 0.040291176 | 1 |
| Obesity | monocyte | *CCDC114* | 3.096459128 | 0.008563455 | 0.041462933 | 1 |
| Obesity | monocyte | *LIG1* | -2.011460859 | 0.012226872 | 0.043115778 | 1 |
| Obesity | monocyte | *PLEKHA4* | -0.953090737 | 0.008856574 | 0.041462933 | 1 |
| Obesity | monocyte | *TNPO2* | -1.406094117 | 0.012969001 | 0.043967382 | 1 |

| | | | | | | |
|---|---|---|---|---|---|---|
| Obesity | monocyte | *GCDH* | -3.207177758 | 0.018759071 | 0.049532965 | 1 |
| Obesity | monocyte | *DNASE2* | 3.02413142 | 0.003049687 | 0.035116467 | 1 |
| Obesity | monocyte | *TFPT* | 1.582542808 | 0.003466417 | 0.035116467 | 1 |
| Obesity | monocyte | *RAB3A* | -4.135705454 | 0.009857141 | 0.041637281 | 1 |
| Obesity | monocyte | *MAG* | 2.751983214 | 0.001699455 | 0.03218121 | 1 |
| Obesity | monocyte | *ZNF14* | -1.37502911 | 0.01601639 | 0.046146985 | 1 |
| Obesity | monocyte | *ETHE1* | -1.367545887 | 0.010523453 | 0.041845661 | 1 |
| Obesity | monocyte | *SMG9* | 3.971292394 | 0.014598746 | 0.044883966 | 1 |
| Obesity | monocyte | *RASA4* | -6.862664601 | 0.009272377 | 0.041462933 | 1 |
| Obesity | monocyte | *STEAP1B* | 2.78017915 | 0.003369645 | 0.035116467 | 1 |
| Obesity | monocyte | *PTN* | -1.018486894 | 0.012466767 | 0.043186096 | 1 |
| Obesity | monocyte | *ADAP1* | -3.349540662 | 0.009884848 | 0.041637281 | 1 |
| Obesity | monocyte | *MET* | -1.11047082 | 0.004308681 | 0.035116467 | 1 |
| Obesity | monocyte | *RNF32* | -1.834276987 | 0.01112424 | 0.042279868 | 1 |
| Obesity | monocyte | *IQCE* | -1.329798726 | 0.015175013 | 0.045042687 | 1 |
| Obesity | monocyte | *VIPR2* | -0.679250538 | 0.013280652 | 0.044105218 | 1 |
| Obesity | monocyte | *HIBADH* | -1.280959612 | 0.013222549 | 0.044032814 | 1 |
| Obesity | monocyte | *CPVL* | 2.479098131 | 0.005093347 | 0.03528148 | 1 |
| Obesity | monocyte | *ABHD11* | 1.017505784 | 0.009234998 | 0.041462933 | 1 |
| Obesity | monocyte | *PLEKHA8* | 4.023095132 | 0.017940184 | 0.048768447 | 1 |
| Obesity | monocyte | *CHCHD2* | -1.140287974 | 0.017383114 | 0.047867689 | 1 |
| Obesity | monocyte | *TFR2* | 3.313895903 | 0.011367283 | 0.042352977 | 1 |
| Obesity | monocyte | *MOSPD3* | 3.951008016 | 0.003725212 | 0.035116467 | 1 |
| Obesity | monocyte | *IMPDH1* | 2.302317052 | 0.004329482 | 0.035116467 | 1 |
| Obesity | monocyte | *MEST* | -0.681185 | 0.00119882 | 0.028851404 | 1 |
| Obesity | monocyte | *ACTR3C* | -1.899100947 | 0.01452379 | 0.044883966 | 1 |
| Obesity | monocyte | *POU6F2* | -0.798166821 | 0.008009413 | 0.040894308 | 1 |
| Obesity | monocyte | *TSPAN13* | -1.109309562 | 0.011949577 | 0.042874352 | 1 |
| Obesity | monocyte | *AEBP1* | -1.121916492 | 0.012970911 | 0.043967382 | 1 |
| Obesity | monocyte | *GCK* | -6.30182348 | 0.009000427 | 0.041462933 | 1 |
| Obesity | monocyte | *SPATA6L* | -0.773663763 | 0.011507355 | 0.042465454 | 1 |
| Obesity | monocyte | *FKTN* | -1.116839418 | 0.019112818 | 0.049965191 | 1 |
| Obesity | monocyte | *MEGF9* | 1.151384168 | 0.003069635 | 0.035116467 | 1 |
| Obesity | monocyte | *TRIM14* | 3.334065474 | 0.007055496 | 0.039348581 | 1 |
| Obesity | monocyte | *CORO2A* | 3.644103726 | 0.006700787 | 0.038521587 | 1 |
| Obesity | monocyte | *ECM2* | -0.735315203 | 0.011316216 | 0.042333294 | 1 |
| Obesity | monocyte | *PTGR1* | -0.872371051 | 0.006889561 | 0.03896597 | 1 |
| Obesity | monocyte | *SUSD1* | 1.2147722 | 0.004096671 | 0.035116467 | 1 |
| Obesity | monocyte | *PLGRKT* | -2.242232465 | 0.005355216 | 0.035561878 | 1 |
| Obesity | monocyte | *DOCK8* | 0.859844492 | 0.009069092 | 0.041462933 | 1 |
| Obesity | monocyte | *KANK1* | -1.002964706 | 0.012683465 | 0.043399276 | 1 |
| Obesity | monocyte | *NCS1* | -1.79646784 | 0.002784004 | 0.035116467 | 1 |
| Obesity | monocyte | *PTGDS* | -1.327066578 | 0.015091583 | 0.044986158 | 1 |
| Obesity | monocyte | *HPS1* | -1.853285377 | 0.005220718 | 0.035351132 | 1 |
| Obesity | monocyte | *PKD2L1* | 0.778723446 | 0.001745531 | 0.032449484 | 1 |
| Obesity | monocyte | *CUBN* | -1.800891303 | 0.017262759 | 0.047761129 | 1 |
| Obesity | monocyte | *BMPR1A* | -0.918943258 | 0.01666995 | 0.046940995 | 1 |
| Obesity | monocyte | *SFXN3* | 4.767659367 | 0.013363384 | 0.044157522 | 1 |
| Obesity | monocyte | *PITRM1* | -1.811022426 | 0.014255594 | 0.044792561 | 1 |
| Obesity | monocyte | *DNAJC12* | 1.516380714 | 0.001766997 | 0.032535431 | 1 |

| | | | | | | |
|---|---|---|---|---|---|---|
| Obesity | monocyte | *PBLD* | -0.962131639 | 0.012681359 | 0.043399276 | 1 |
| Obesity | monocyte | *WNT3* | -1.843711046 | 0.007168446 | 0.039704679 | 1 |
| Obesity | monocyte | *ASPA* | -0.636398258 | 0.015006474 | 0.044986158 | 1 |
| Obesity | monocyte | *RAD51C* | -1.203135225 | 0.000890974 | 0.025883824 | 1 |
| Obesity | monocyte | *PNPO* | -2.482697593 | 0.000866565 | 0.025867134 | 1 |
| Obesity | monocyte | *TRIM16L* | 1.491806856 | 0.004294355 | 0.035116467 | 1 |
| Obesity | monocyte | *CDK5RAP3* | -1.34192472 | 0.005096036 | 0.03528148 | 1 |
| Obesity | monocyte | *CHRNE* | 1.530019625 | 0.003261231 | 0.035116467 | 1 |
| Obesity | monocyte | *ALDH3A1* | 4.201243118 | 0.015780843 | 0.045685351 | 1 |
| Obesity | monocyte | *PSMD11* | -1.002732713 | 0.018806256 | 0.049537435 | 1 |
| Obesity | monocyte | *PEX12* | -2.016963865 | 0.014804001 | 0.044883966 | 1 |
| Obesity | monocyte | *DHX58* | 1.552675375 | 0.00293468 | 0.035116467 | 1 |
| Obesity | monocyte | *ABCC3* | 0.579712414 | 0.001591533 | 0.03218121 | 1 |
| Obesity | monocyte | *EFTUD2* | 3.391168877 | 0.004705712 | 0.035116467 | 1 |
| Obesity | monocyte | *HLF* | -1.447272164 | 0.004580814 | 0.035116467 | 1 |
| Obesity | monocyte | *MYH3* | -1.790749335 | 0.011912134 | 0.04284483 | 1 |
| Obesity | monocyte | *TMEM97* | 2.012095505 | 0.00507947 | 0.03528148 | 1 |
| Obesity | monocyte | *KLHL2* | 2.867946087 | 0.009057683 | 0.041462933 | 1 |
| Obesity | monocyte | *CPE* | -1.113526488 | 0.018763724 | 0.049532965 | 1 |
| Obesity | monocyte | *WFS1* | -1.204569848 | 0.011438971 | 0.042352977 | 1 |
| Obesity | monocyte | *AADAT* | -1.495898608 | 0.00889838 | 0.041462933 | 1 |
| Obesity | monocyte | *BST1* | 4.643052611 | 0.004402325 | 0.035116467 | 1 |
| Obesity | monocyte | *FAM149A* | -0.775680123 | 0.008672096 | 0.041462933 | 1 |
| Obesity | monocyte | *TECTA* | -1.772632237 | 0.009349825 | 0.041462933 | 1 |
| Obesity | monocyte | *SC5D* | -2.88285123 | 0.014233127 | 0.044792561 | 1 |
| Obesity | monocyte | *B3GAT1* | 4.132164327 | 0.002861582 | 0.035116467 | 1 |
| Obesity | monocyte | *P2RX3* | 2.927932297 | 0.004627269 | 0.035116467 | 1 |
| Obesity | monocyte | *SNX15* | -4.419198922 | 0.000426124 | 0.018429008 | 1 |
| Obesity | monocyte | *UNC93B1* | 1.490208869 | 0.004611283 | 0.035116467 | 1 |
| Obesity | monocyte | *NRXN2* | -2.010794329 | 0.008896072 | 0.041462933 | 1 |
| Obesity | monocyte | *CCDC86* | 1.294641761 | 0.011584855 | 0.042465454 | 1 |
| Obesity | monocyte | *PANX1* | -1.767163661 | 0.011958437 | 0.042874352 | 1 |
| Obesity | monocyte | *KIAA1549L* | 17.17935486 | 0.004088345 | 0.035116467 | 1 |
| Obesity | monocyte | *SLC22A18* | 1.930163738 | 5.16E-05 | 0.004319877 | 0.250552862 |
| Obesity | monocyte | *C11orf21* | 2.54641189 | 0.01004673 | 0.041669249 | 1 |
| Obesity | monocyte | *NDUFS8* | -1.481559721 | 0.015029071 | 0.044986158 | 1 |
| Obesity | monocyte | *TCIRG1* | 2.629737549 | 0.00901677 | 0.041462933 | 1 |
| Obesity | monocyte | *PRDM4* | 4.172419166 | 0.018201323 | 0.048981042 | 1 |
| Obesity | monocyte | *ASIC1* | -2.586865237 | 0.0105396 | 0.041845661 | 1 |
| Obesity | monocyte | *KCTD10* | -3.101643337 | 0.003458911 | 0.035116467 | 1 |
| Obesity | monocyte | *IL23A* | -6.669817863 | 0.010029148 | 0.041669249 | 1 |
| Obesity | monocyte | *ACSS3* | -0.528173811 | 0.0102228 | 0.041771331 | 1 |
| Obesity | monocyte | *LTA4H* | 1.967447891 | 0.012429933 | 0.043171107 | 1 |
| Obesity | monocyte | *ACAD10* | -0.90655959 | 0.019021037 | 0.049859574 | 1 |
| Obesity | monocyte | *LTBR* | 2.589393373 | 0.00171741 | 0.03218121 | 1 |
| Obesity | monocyte | *ENDOU* | 1.155958697 | 0.005654679 | 0.035810934 | 1 |
| Obesity | monocyte | *VDR* | 0.933115838 | 0.004240693 | 0.035116467 | 1 |
| Obesity | monocyte | *MDM1* | 2.52348111 | 0.003454099 | 0.035116467 | 1 |
| Obesity | monocyte | *CDCA3* | 1.414144442 | 0.006667857 | 0.03842593 | 1 |
| Obesity | monocyte | *ATN1* | -1.066178794 | 0.006987381 | 0.039147772 | 1 |

| | | | | | | |
|---|---|---|---|---|---|---|
| Obesity | monocyte | *C12orf57* | -1.059505402 | 0.009493398 | 0.041462933 | 1 |
| Obesity | monocyte | *SUDS3* | -1.204258217 | 0.017738339 | 0.048500599 | 1 |
| Obesity | monocyte | *BTN3A3* | 4.147594629 | 3.77E-09 | 1.66E-06 | 1.83E-05 |
| Obesity | monocyte | *FRK* | -0.937816013 | 0.009066935 | 0.041462933 | 1 |
| Obesity | monocyte | *RWDD1* | -1.516580128 | 0.014063547 | 0.044657283 | 1 |
| Obesity | monocyte | *GABRR2* | 1.559351217 | 0.000443346 | 0.018547623 | 1 |
| Obesity | monocyte | *HDDC2* | -1.057465355 | 0.013042083 | 0.043967382 | 1 |
| Obesity | monocyte | *MDGA1* | -1.152602311 | 0.007406498 | 0.040251891 | 1 |
| Obesity | monocyte | *CD83* | 1.022980537 | 0.00433083 | 0.035116467 | 1 |
| Obesity | monocyte | *MDN1* | -1.250184004 | 0.012573129 | 0.04320455 | 1 |
| Obesity | monocyte | *GLP1R* | -11.84949336 | 0.000576371 | 0.021082768 | 1 |
| Obesity | monocyte | *BVES* | -1.506443013 | 0.002659598 | 0.035116467 | 1 |
| Obesity | monocyte | *COL9A1* | 1.319632771 | 0.003585598 | 0.035116467 | 1 |
| Obesity | monocyte | *WASF1* | -0.823078972 | 0.014725007 | 0.044883966 | 1 |
| Obesity | monocyte | *VNN1* | -1.50512341 | 0.012338357 | 0.043115778 | 1 |
| Obesity | monocyte | *VNN2* | 0.749742364 | 0.003293627 | 0.035116467 | 1 |
| Obesity | monocyte | *CUTA* | -1.812039607 | 0.000700344 | 0.023274441 | 1 |
| Obesity | monocyte | *PACRG* | -0.950738108 | 0.010255857 | 0.041771331 | 1 |
| Obesity | monocyte | *DNPH1* | 1.416589852 | 0.017992053 | 0.04876952 | 1 |
| Obesity | monocyte | *IMPG1* | -3.647359111 | 0.002026036 | 0.034173375 | 1 |
| Obesity | monocyte | *LY86* | 0.495547919 | 0.004564715 | 0.035116467 | 1 |
| Obesity | monocyte | *PCDHB2* | -4.824137982 | 0.002295869 | 0.035072295 | 1 |
| Obesity | monocyte | *CEP72* | 1.918354495 | 0.005670958 | 0.035810934 | 1 |
| Obesity | monocyte | *KIF20A* | -11.59459143 | 0.004667769 | 0.035116467 | 1 |
| Obesity | monocyte | *TMCO6* | -3.697346364 | 0.000171882 | 0.011119597 | 0.833969759 |
| Obesity | monocyte | *PCDHB15* | -2.261169349 | 0.003882456 | 0.035116467 | 1 |
| Obesity | monocyte | *HAVCR1* | 0.880452153 | 0.009158907 | 0.041462933 | 1 |
| Obesity | monocyte | *ARSB* | 3.659931869 | 0.013131523 | 0.044028405 | 1 |
| Obesity | monocyte | *MSH3* | -1.077588623 | 0.014222175 | 0.044792561 | 1 |
| Obesity | monocyte | *CDH6* | -1.448607214 | 0.019207567 | 0.049996369 | 1 |
| Obesity | monocyte | *LMNB1* | 0.615598693 | 0.004333033 | 0.035116467 | 1 |
| Obesity | monocyte | *TXNDC15* | 4.074502646 | 0.01402594 | 0.044610733 | 1 |
| Obesity | monocyte | *BNIP1* | -2.318467957 | 0.008167835 | 0.040894308 | 1 |
| Obesity | monocyte | *UNC5A* | -4.320919328 | 0.009004638 | 0.041462933 | 1 |
| Obesity | monocyte | *HYAL1* | -1.722931943 | 0.001242202 | 0.029258086 | 1 |
| Obesity | monocyte | *C3orf14* | -3.568268417 | 0.003537486 | 0.035116467 | 1 |
| Obesity | monocyte | *SLC41A3* | -1.173088284 | 0.009167141 | 0.041462933 | 1 |
| Obesity | monocyte | *PLXNA1* | -0.980802281 | 0.015073375 | 0.044986158 | 1 |
| Obesity | monocyte | *KIAA1257* | -2.556912633 | 0.010694033 | 0.041946198 | 1 |
| Obesity | monocyte | *NEK11* | -0.764371163 | 0.012242871 | 0.043115778 | 1 |
| Obesity | monocyte | *HEMK1* | -0.851447555 | 0.015448558 | 0.045400608 | 1 |
| Obesity | monocyte | *ARHGEF26* | -0.911776751 | 0.011565327 | 0.042465454 | 1 |
| Obesity | monocyte | *VIPR1* | 0.955098 | 0.003589098 | 0.035116467 | 1 |
| Obesity | monocyte | *DNAH1* | 3.458604931 | 0.000565619 | 0.021082768 | 1 |
| Obesity | monocyte | *NEK4* | -2.972224771 | 0.009394792 | 0.041462933 | 1 |
| Obesity | monocyte | *ADAM23* | -2.150961935 | 0.016152499 | 0.046359178 | 1 |
| Obesity | monocyte | *LMAN2L* | -1.128228327 | 0.014510058 | 0.044883966 | 1 |
| Obesity | monocyte | *OTOF* | -6.607605872 | 0.003345849 | 0.035116467 | 1 |
| Obesity | monocyte | *INO80B* | -1.638107031 | 0.005155944 | 0.035289693 | 1 |
| Obesity | monocyte | *GRB14* | -0.948586125 | 0.009988584 | 0.041669249 | 1 |

| | | | | | | |
|---|---|---|---|---|---|---|
| Obesity | monocyte | *GALNT3* | 0.794113354 | 0.002277583 | 0.035072295 | 1 |
| Obesity | monocyte | *ACADL* | -0.640062578 | 0.013466322 | 0.044239385 | 1 |
| Obesity | monocyte | *WNT6* | -2.161127431 | 0.001916084 | 0.033521987 | 1 |
| Obesity | monocyte | *IL18R1* | -0.980854181 | 0.00818238 | 0.040894308 | 1 |
| Obesity | monocyte | *MLPH* | -1.295366628 | 0.008202339 | 0.040894308 | 1 |
| Obesity | monocyte | *TPO* | -1.073382121 | 0.011222338 | 0.042279868 | 1 |
| Obesity | monocyte | *PRKD3* | -1.243843105 | 0.018072074 | 0.048795606 | 1 |
| Obesity | monocyte | *WIPF1* | 2.770120747 | 0.006435339 | 0.037710463 | 1 |
| Obesity | monocyte | *MARK1* | -1.070205753 | 0.012790518 | 0.043673183 | 1 |
| Obesity | monocyte | *MORN1* | -1.540838824 | 0.004002058 | 0.035116467 | 1 |
| Obesity | monocyte | *RALGPS2* | -0.948426984 | 0.014636732 | 0.044883966 | 1 |
| Obesity | monocyte | *WRAP73* | -1.687827506 | 0.007390716 | 0.040251891 | 1 |
| Obesity | monocyte | *KCNC4* | -1.195675451 | 0.0143675 | 0.044830297 | 1 |
| Obesity | monocyte | *DNAJC6* | -1.475261084 | 0.003139586 | 0.035116467 | 1 |
| Obesity | monocyte | *PRG4* | -0.954557646 | 0.010516096 | 0.041845661 | 1 |
| Obesity | monocyte | *NCF2* | 0.45612872 | 0.005724831 | 0.035810934 | 1 |
| Obesity | monocyte | *CFHR3* | -3.993009384 | 0.012186564 | 0.043115778 | 1 |
| Obesity | monocyte | *TMEM9* | -2.297848108 | 0.016050956 | 0.046219132 | 1 |
| Obesity | monocyte | *OSCP1* | -0.864713243 | 0.011339934 | 0.042352977 | 1 |
| Obesity | monocyte | *EXOC8* | -1.490155124 | 0.014517773 | 0.044883966 | 1 |
| Obesity | monocyte | *SLAMF1* | 0.595171091 | 0.008154848 | 0.040894308 | 1 |
| Obesity | monocyte | *PADI2* | -3.021206267 | 0.001173064 | 0.028774316 | 1 |
| Obesity | monocyte | *MFAP2* | -2.416668742 | 0.003600362 | 0.035116467 | 1 |
| Obesity | monocyte | *KIF17* | 1.99951407 | 0.018444925 | 0.04918073 | 1 |
| Obesity | monocyte | *EBNA1BP2* | -1.013344882 | 0.01300391 | 0.043967382 | 1 |
| Obesity | monocyte | *CCDC181* | 1.857195135 | 0.007596016 | 0.040367875 | 1 |
| Obesity | monocyte | *NSUN4* | -3.110318369 | 0.000787227 | 0.024802764 | 1 |
| Obesity | monocyte | *IRF6* | 1.583429814 | 0.002791338 | 0.035116467 | 1 |
| Obesity | monocyte | *SERPINC1* | 7.981124207 | 7.09E-09 | 2.87E-06 | 3.44E-05 |
| Obesity | monocyte | *MARC2* | -1.155346755 | 0.014660334 | 0.044883966 | 1 |
| Obesity | monocyte | *CASC1* | -1.05647879 | 0.005104285 | 0.03528148 | 1 |
| Obesity | monocyte | *LRMP* | 1.154988106 | 0.002186082 | 0.035072295 | 1 |
| Obesity | monocyte | *SGIP1* | -0.839757329 | 0.009783156 | 0.041546562 | 1 |
| Obesity | monocyte | *VAMP8* | 3.834833587 | 0.004965808 | 0.035116467 | 1 |
| Obesity | monocyte | *DNAH7* | -1.279642759 | 0.003287659 | 0.035116467 | 1 |
| Obesity | monocyte | *CPSF3* | -1.628168823 | 0.00456999 | 0.035116467 | 1 |
| Obesity | monocyte | *TRIM32* | -1.28996625 | 0.012457138 | 0.043186096 | 1 |
| Obesity | monocyte | *HDHD3* | -1.062297969 | 0.002598546 | 0.035116467 | 1 |
| Obesity | monocyte | *RBM18* | 3.540318024 | 0.003101723 | 0.035116467 | 1 |
| Obesity | monocyte | *HSDL2* | -0.92695963 | 0.015870996 | 0.045836948 | 1 |
| Obesity | monocyte | *TTLL5* | -1.054982306 | 0.017946429 | 0.048768447 | 1 |
| Obesity | monocyte | *FLVCR2* | 1.078887345 | 0.004531129 | 0.035116467 | 1 |
| Obesity | monocyte | *EIF2B2* | -2.07323308 | 0.002327465 | 0.035072295 | 1 |
| Obesity | monocyte | *COQ6* | -1.384330848 | 0.014752607 | 0.044883966 | 1 |
| Obesity | monocyte | *CNRIP1* | -1.667969941 | 0.001382747 | 0.031060598 | 1 |
| Obesity | monocyte | *HELLS* | -1.33428183 | 0.006078713 | 0.036701128 | 1 |
| Obesity | monocyte | *CAAP1* | -1.807695486 | 0.018626389 | 0.04946647 | 1 |
| Obesity | monocyte | *PCDHB10* | 1.689323775 | 0.010969642 | 0.042169886 | 1 |
| Obesity | monocyte | *MASTL* | -1.881699439 | 0.001296434 | 0.029953788 | 1 |
| Obesity | monocyte | *MRC1* | -0.815579625 | 0.013369507 | 0.044157522 | 1 |

| | | | | | | |
|---|---|---|---|---|---|---|
| Obesity | monocyte | *IQSEC3* | 5.080862065 | 0.001418818 | 0.03111676 | 1 |
| Obesity | monocyte | *CCDC77* | 1.625985401 | 0.002521445 | 0.035116467 | 1 |
| Obesity | monocyte | *ARL1* | -1.223097101 | 0.014635765 | 0.044883966 | 1 |
| Obesity | monocyte | *APAF1* | 3.193990135 | 0.003583053 | 0.035116467 | 1 |
| Obesity | monocyte | *AKAP1* | -0.889269251 | 0.014237043 | 0.044792561 | 1 |
| Obesity | monocyte | *TBX2* | -1.617885401 | 0.000891728 | 0.025883824 | 1 |
| Obesity | monocyte | *TRIM6* | -2.763243698 | 0.009762049 | 0.041544165 | 1 |
| Obesity | monocyte | *ECHDC2* | -0.666692826 | 0.006457544 | 0.037755639 | 1 |
| Obesity | monocyte | *PYROXD1* | 2.783706547 | 0.000283803 | 0.014806582 | 1 |
| Obesity | monocyte | *ZNF549* | -1.20337371 | 0.012960821 | 0.043967382 | 1 |
| Obesity | monocyte | *A1BG* | 8.300438198 | 0.001002858 | 0.027556163 | 1 |
| Obesity | monocyte | *ZNF211* | 2.026709108 | 0.00162075 | 0.03218121 | 1 |
| Obesity | monocyte | *PDZRN3* | -0.501640345 | 0.007822363 | 0.040689236 | 1 |
| Obesity | monocyte | *LHX4* | -1.08807658 | 0.009554357 | 0.041462933 | 1 |
| Obesity | monocyte | *CD80* | 0.856340549 | 0.018712335 | 0.04953205 | 1 |
| Obesity | monocyte | *ZNF639* | -1.648766288 | 0.016970988 | 0.047378154 | 1 |
| Obesity | monocyte | *FLT3* | 1.259067092 | 0.004360754 | 0.035116467 | 1 |
| Obesity | monocyte | *CD244* | 0.277883459 | 0.004378995 | 0.035116467 | 1 |
| Obesity | monocyte | *SERAC1* | -1.716171107 | 0.016300137 | 0.046549891 | 1 |
| Obesity | monocyte | *LDB3* | -1.099536613 | 0.014401305 | 0.044843874 | 1 |
| Obesity | monocyte | *NAA60* | -1.584815435 | 0.01297645 | 0.043967382 | 1 |
| Obesity | monocyte | *RWDD3* | -2.154028027 | 0.001005372 | 0.027556163 | 1 |
| Obesity | monocyte | *RBM19* | -1.069055098 | 0.013882337 | 0.044540919 | 1 |
| Obesity | monocyte | *DDX54* | 4.483479129 | 0.001107312 | 0.028083118 | 1 |
| Obesity | monocyte | *ARHGAP9* | 0.746620952 | 0.004559137 | 0.035116467 | 1 |
| Obesity | monocyte | *CDK2* | -1.657603056 | 0.005133703 | 0.03528148 | 1 |
| Obesity | monocyte | *DBH* | -27.94726828 | 0.009088685 | 0.041462933 | 1 |
| Obesity | monocyte | *ACVR1C* | -0.473921148 | 0.017253663 | 0.047761129 | 1 |
| Obesity | monocyte | *SLC36A1* | 1.907704196 | 0.005476078 | 0.035664337 | 1 |
| Obesity | monocyte | *C4BPA* | -1.667869352 | 0.011007906 | 0.042221628 | 1 |
| Obesity | monocyte | *DAW1* | -2.25704144 | 0.0139891 | 0.044596002 | 1 |
| Obesity | monocyte | *OBSL1* | -1.097856574 | 0.014588913 | 0.044883966 | 1 |
| Obesity | monocyte | *FAM210B* | -1.565741523 | 0.015475415 | 0.045417901 | 1 |
| Obesity | monocyte | *MATN4* | -2.924691857 | 0.014741502 | 0.044883966 | 1 |
| Obesity | monocyte | *NEURL2* | 2.610903453 | 0.003115292 | 0.035116467 | 1 |
| Obesity | monocyte | *NAGK* | 2.751032002 | 0.000510136 | 0.01966188 | 1 |
| Obesity | monocyte | *SNRNP27* | -1.216893408 | 0.013534269 | 0.04433272 | 1 |
| Obesity | monocyte | *ZNF576* | 3.063788745 | 0.01122351 | 0.042279868 | 1 |
| Obesity | monocyte | *BTN2A2* | 2.584319308 | 0.000195832 | 0.011628398 | 0.950176652 |
| Obesity | monocyte | *RPS10* | 3.250975065 | 0.003869231 | 0.035116467 | 1 |
| Obesity | monocyte | *TCP11* | 0.684098829 | 0.001201033 | 0.028851404 | 1 |
| Obesity | monocyte | *GNMT* | 0.855515152 | 0.008847814 | 0.041462933 | 1 |
| Obesity | monocyte | *TREM1* | 1.414619728 | 0.000715143 | 0.023604598 | 1 |
| Obesity | monocyte | *KLHL31* | -0.808914793 | 0.004584932 | 0.035116467 | 1 |
| Obesity | monocyte | *SH3TC1* | 1.201919811 | 0.002653853 | 0.035116467 | 1 |
| Obesity | monocyte | *GOT2* | -1.143087196 | 0.015720664 | 0.045674648 | 1 |
| Obesity | monocyte | *NT5C* | 2.198146805 | 0.003221042 | 0.035116467 | 1 |
| Obesity | monocyte | *OPRL1* | 0.502748693 | 0.004986654 | 0.035116467 | 1 |
| Obesity | monocyte | *PPDPF* | 3.07431866 | 0.000466072 | 0.018861406 | 1 |
| Obesity | monocyte | *IL1B* | 0.296208384 | 0.005391275 | 0.035614643 | 1 |

| | | | | | | |
|---|---|---|---|---|---|---|
| Obesity | monocyte | *SH2D3A* | 1.396956056 | 0.012056783 | 0.043014345 | 1 |
| Obesity | monocyte | *SDCBP2* | -2.488617641 | 0.011357535 | 0.042352977 | 1 |
| Obesity | monocyte | *FLRT3* | -0.954037379 | 0.009285511 | 0.041462933 | 1 |
| Obesity | monocyte | *PCSK2* | -4.173557068 | 0.012309094 | 0.043115778 | 1 |
| Obesity | monocyte | *LAMP5* | -23.76193291 | 0.005269528 | 0.035510766 | 1 |
| Obesity | monocyte | *MGME1* | 3.000267038 | 0.003817114 | 0.035116467 | 1 |
| Obesity | monocyte | *UROD* | -1.027980836 | 0.013255322 | 0.04407167 | 1 |
| Obesity | monocyte | *TSKS* | 1.763732684 | 0.001770271 | 0.032535431 | 1 |
| Obesity | monocyte | *ZNF384* | -1.038951355 | 0.013188762 | 0.044032814 | 1 |
| Obesity | monocyte | *L3HYPDH* | -1.575724124 | 0.01663175 | 0.046940995 | 1 |
| Obesity | monocyte | *FGD3* | 0.567972611 | 0.005668092 | 0.035810934 | 1 |
| Obesity | monocyte | *RAB3IP* | 5.637453627 | 0.008831653 | 0.041462933 | 1 |
| Obesity | monocyte | *YEATS4* | 3.506413775 | 0.000345401 | 0.016559891 | 1 |
| Obesity | monocyte | *IDUA* | -1.434797432 | 0.006743538 | 0.038614455 | 1 |
| Obesity | monocyte | *EMC1* | 3.92921977 | 0.013156665 | 0.044028405 | 1 |
| Obesity | monocyte | *WDR24* | -1.78343296 | 0.00554631 | 0.035785501 | 1 |
| Obesity | monocyte | *PEX1* | -0.838063085 | 0.010954647 | 0.042169886 | 1 |
| Obesity | monocyte | *VPREB3* | -6.287483592 | 0.009503944 | 0.041462933 | 1 |
| Obesity | monocyte | *ATF4* | -1.321778534 | 0.01541877 | 0.045365696 | 1 |
| Obesity | monocyte | *A4GALT* | -1.315401938 | 0.014033833 | 0.044610733 | 1 |
| Obesity | monocyte | *CDC42EP1* | -2.183755 | 0.018329967 | 0.04903887 | 1 |
| Obesity | monocyte | *RIBC2* | 2.83443711 | 0.010732814 | 0.041961024 | 1 |
| Obesity | monocyte | *CDHR3* | -0.839820562 | 0.012138384 | 0.043083715 | 1 |
| Obesity | monocyte | *IRF5* | 0.769660608 | 0.006838614 | 0.038883685 | 1 |
| Obesity | monocyte | *OPN1SW* | -2.338881898 | 0.007650827 | 0.040393703 | 1 |
| Obesity | monocyte | *GAD1* | -3.500272528 | 0.00239601 | 0.035116467 | 1 |
| Obesity | monocyte | *PSMG2* | -1.128972655 | 0.009769557 | 0.041544165 | 1 |
| Obesity | monocyte | *KNSTRN* | -0.937051193 | 0.005879523 | 0.036288402 | 1 |
| Obesity | monocyte | *CALML4* | -4.072344643 | 0.007525941 | 0.04034316 | 1 |
| Obesity | monocyte | *ANAPC13* | -1.337616806 | 0.008376401 | 0.041379919 | 1 |
| Obesity | monocyte | *BBOX1* | 15.55264531 | 0.003833919 | 0.035116467 | 1 |
| Obesity | monocyte | *SERGEF* | 3.547214539 | 0.000822132 | 0.025087947 | 1 |
| Obesity | monocyte | *DCTD* | -0.883009959 | 0.012972965 | 0.043967382 | 1 |
| Obesity | monocyte | *RPAIN* | -3.740140236 | 0.001429353 | 0.03111676 | 1 |
| Obesity | monocyte | *USP6* | 0.929281807 | 0.014741563 | 0.044883966 | 1 |
| Obesity | monocyte | *PLD2* | 4.464590787 | 0.00320793 | 0.035116467 | 1 |
| Obesity | monocyte | *AP1M2* | -3.171022885 | 2.12E-05 | 0.002239829 | 0.103032117 |
| Obesity | monocyte | *KLK8* | -7.51064202 | 0.010088094 | 0.041669249 | 1 |
| Obesity | monocyte | *RIPK3* | 1.641626591 | 0.011875279 | 0.042802404 | 1 |
| Obesity | monocyte | *PARP2* | -1.598238222 | 0.018223507 | 0.04900239 | 1 |
| Obesity | monocyte | *RNASE1* | -0.764841945 | 0.018847091 | 0.049618061 | 1 |
| Obesity | monocyte | *GALNT8* | 5.275105603 | 0.018259753 | 0.04900239 | 1 |
| Obesity | monocyte | *CDC16* | -1.765838108 | 0.008620666 | 0.041462933 | 1 |
| Obesity | monocyte | *ATP8B3* | -3.322047642 | 3.22E-06 | 0.000520503 | 0.015615079 |
| Obesity | monocyte | *DDA1* | -1.414482159 | 0.017544874 | 0.048121951 | 1 |
| Obesity | monocyte | *MAP1S* | 5.582275928 | 0.0048463 | 0.035116467 | 1 |
| Obesity | monocyte | *CYP2E1* | -3.104477681 | 8.13E-05 | 0.006360178 | 0.394331011 |
| Obesity | monocyte | *PNPLA7* | -0.722819335 | 0.009888636 | 0.041637281 | 1 |
| Obesity | monocyte | *PAK4* | -1.151171713 | 0.013899431 | 0.044540919 | 1 |
| Obesity | monocyte | *POMT1* | -1.375690005 | 0.006981332 | 0.039147772 | 1 |

| | | | | | | |
|---|---|---|---|---|---|---|
| Obesity | monocyte | *YIPF2* | 4.09400199 | 0.002986318 | 0.035116467 | 1 |
| Obesity | monocyte | *MAP3K10* | -1.669156223 | 0.014831556 | 0.044883966 | 1 |
| Obesity | monocyte | *THEMIS2* | 0.604348956 | 0.002254718 | 0.035072295 | 1 |
| Obesity | monocyte | *ZNF317* | -2.698322593 | 0.009456817 | 0.041462933 | 1 |
| Obesity | monocyte | *C12orf65* | 5.492342613 | 0.000231688 | 0.013071531 | 1 |
| Obesity | monocyte | *PKDREJ* | -4.552589171 | 0.000194882 | 0.011628398 | 0.945568288 |
| Obesity | monocyte | *HSD17B3* | -1.420773256 | 0.010336266 | 0.041771331 | 1 |
| Obesity | monocyte | *TTLL9* | 9.263096605 | 0.004500813 | 0.035116467 | 1 |
| Obesity | monocyte | *COX4I2* | -2.444341662 | 0.007323286 | 0.040059284 | 1 |
| Obesity | monocyte | *IDO1* | 2.506722102 | 0.002793608 | 0.035116467 | 1 |
| Obesity | monocyte | *SLC6A6* | 2.253638271 | 0.002308449 | 0.035072295 | 1 |
| Obesity | monocyte | *KCNC3* | 1.165555368 | 0.010293311 | 0.041771331 | 1 |
| Obesity | monocyte | *NAPSA* | 1.886738504 | 0.003193075 | 0.035116467 | 1 |
| Obesity | monocyte | *PDLIM4* | 2.323644286 | 0.00252855 | 0.035116467 | 1 |
| Obesity | monocyte | *BARX1* | 3.577123637 | 0.004986045 | 0.035116467 | 1 |
| Obesity | monocyte | *MAP1B* | -0.822421992 | 0.010841705 | 0.041961024 | 1 |
| Obesity | monocyte | *CKMT2* | -1.199059223 | 0.006415218 | 0.03766044 | 1 |
| Obesity | monocyte | *CHD1L* | -1.71181188 | 0.010090431 | 0.041669249 | 1 |
| Obesity | monocyte | *GCH1* | 0.445185427 | 0.004851277 | 0.035116467 | 1 |
| Obesity | monocyte | *TRIM21* | -1.417189977 | 0.008667226 | 0.041462933 | 1 |
| Obesity | monocyte | *LRRC41* | -1.878652162 | 0.012859179 | 0.043815124 | 1 |
| Obesity | monocyte | *CNGA4* | -2.26163761 | 0.004283571 | 0.035116467 | 1 |
| Obesity | monocyte | *RRP8* | 3.599422797 | 0.004498349 | 0.035116467 | 1 |
| Obesity | monocyte | *FIGNL1* | -2.598974577 | 0.002809183 | 0.035116467 | 1 |
| Obesity | monocyte | *EIF5A* | 0.841833574 | 0.012348376 | 0.043115778 | 1 |
| Obesity | monocyte | *RGS22* | -0.810845227 | 0.004344192 | 0.035116467 | 1 |
| Obesity | monocyte | *DMGDH* | -2.288225562 | 0.012519072 | 0.04319079 | 1 |
| Obesity | monocyte | *BHMT2* | -0.641958431 | 0.006006896 | 0.036477419 | 1 |
| Obesity | monocyte | *ZBED3* | -1.082403889 | 0.004553772 | 0.035116467 | 1 |
| Obesity | monocyte | *CASP9* | -1.731291449 | 0.015470718 | 0.045417901 | 1 |
| Obesity | monocyte | *WASF3* | -1.080574472 | 0.017397261 | 0.047879473 | 1 |
| Obesity | monocyte | *CNDP2* | -1.491323614 | 0.016668328 | 0.046940995 | 1 |
| Obesity | monocyte | *PDZD2* | -0.696154715 | 0.014407981 | 0.044843874 | 1 |
| Obesity | monocyte | *FAM83F* | -6.434213634 | 0.006230895 | 0.037094676 | 1 |
| Obesity | monocyte | *SFTPD* | 1.426145473 | 0.000753428 | 0.024286258 | 1 |
| Obesity | monocyte | *LARS* | -2.119129047 | 0.000358183 | 0.016872858 | 1 |
| Obesity | monocyte | *SPINK5* | -2.169209681 | 0.011836801 | 0.042787935 | 1 |
| Obesity | monocyte | *RRAS2* | -0.805906338 | 0.019161269 | 0.049996369 | 1 |
| Obesity | monocyte | *CATSPERB* | -0.739876169 | 0.012513791 | 0.04319079 | 1 |
| Obesity | monocyte | *COX16* | 7.917927553 | 0.00234729 | 0.035116467 | 1 |
| Obesity | monocyte | *TTC9* | 3.583474937 | 0.002462926 | 0.035116467 | 1 |
| Obesity | monocyte | *PEBP4* | -1.020947349 | 0.009315258 | 0.041462933 | 1 |
| Obesity | monocyte | *CTIF* | -0.750527048 | 0.009045016 | 0.041462933 | 1 |
| Obesity | monocyte | *MRO* | 7.226695481 | 0.001228391 | 0.029216448 | 1 |
| Obesity | monocyte | *THUMPD3* | -1.905719219 | 0.016523046 | 0.046845773 | 1 |
| Obesity | monocyte | *CNTN6* | -1.457908756 | 0.010173945 | 0.041771331 | 1 |
| Obesity | monocyte | *CHL1* | -0.737673237 | 0.011309364 | 0.042333294 | 1 |
| Obesity | monocyte | *GSTM5* | 3.40500311 | 2.26E-05 | 0.002336103 | 0.10979685 |
| Obesity | monocyte | *GSTM3* | -1.055719947 | 0.013905619 | 0.044540919 | 1 |
| Obesity | monocyte | *TRIM45* | -1.771752839 | 0.008870687 | 0.041462933 | 1 |

| | | | | | | |
|---|---|---|---|---|---|---|
| Obesity | monocyte | *IL15RA* | 4.201479282 | 0.001034938 | 0.027556163 | 1 |
| Obesity | monocyte | *TMEM241* | -0.988318175 | 0.009787246 | 0.041546562 | 1 |
| Obesity | monocyte | *SOX5* | -0.784574408 | 0.016156919 | 0.046359178 | 1 |
| Obesity | monocyte | *HOOK1* | -1.26048989 | 0.002958123 | 0.035116467 | 1 |
| Obesity | monocyte | *TIMM10* | 2.111328005 | 0.004411638 | 0.035116467 | 1 |
| Obesity | monocyte | *FADS2* | 3.785588259 | 0.008376298 | 0.041379919 | 1 |
| Obesity | monocyte | *ERCC5* | -1.766487054 | 0.007250729 | 0.039882912 | 1 |
| Obesity | monocyte | *GOLM1* | -1.21555092 | 0.004070642 | 0.035116467 | 1 |
| Obesity | monocyte | *ADAM19* | -1.155389824 | 0.00380275 | 0.035116467 | 1 |
| Obesity | monocyte | *DTX1* | -0.788311751 | 0.009605928 | 0.041462933 | 1 |
| Obesity | monocyte | *TMEM60* | -2.335573691 | 0.004051756 | 0.035116467 | 1 |
| Obesity | monocyte | *POM121C* | -2.85058224 | 2.44E-05 | 0.002412146 | 0.118195135 |
| Obesity | monocyte | *TES* | 1.441879596 | 0.011964524 | 0.042874352 | 1 |
| Obesity | monocyte | *KHDC1* | -0.73216846 | 0.013600469 | 0.044347765 | 1 |
| Obesity | monocyte | *NT5E* | -1.20064101 | 0.01854177 | 0.049322734 | 1 |
| Obesity | monocyte | *MAP3K7* | 3.334769828 | 0.013866238 | 0.044539684 | 1 |
| Obesity | monocyte | *B4GALNT1* | -4.891803978 | 0.016293869 | 0.046549891 | 1 |
| Obesity | monocyte | *FAIM2* | 1.56349581 | 0.010601851 | 0.041845661 | 1 |
| Obesity | monocyte | *PAN2* | -1.483682728 | 0.013236085 | 0.044047658 | 1 |
| Obesity | monocyte | *MAP7* | -1.291521143 | 0.013847859 | 0.044539684 | 1 |
| Obesity | monocyte | *AHI1* | -1.122113763 | 0.005130921 | 0.03528148 | 1 |
| Obesity | monocyte | *MICAL1* | 2.006923006 | 9.07E-05 | 0.006877787 | 0.440178369 |
| Obesity | monocyte | *SMYD5* | 1.451362767 | 0.00190199 | 0.033521987 | 1 |
| Obesity | monocyte | *KCNK1* | -1.036565782 | 0.013908208 | 0.044540919 | 1 |
| Obesity | monocyte | *CAPN9* | -3.595974767 | 0.006781291 | 0.038663717 | 1 |
| Obesity | monocyte | *NTPCR* | 4.243050242 | 0.01828027 | 0.04903033 | 1 |
| Obesity | monocyte | *MRPL44* | -1.470650702 | 0.013016304 | 0.043967382 | 1 |
| Obesity | monocyte | *SLC19A3* | -0.529456196 | 0.008109421 | 0.040894308 | 1 |
| Obesity | monocyte | *TSGA10* | -0.662404492 | 0.005720756 | 0.035810934 | 1 |
| Obesity | monocyte | *STAB2* | -0.84155384 | 0.009443723 | 0.041462933 | 1 |
| Obesity | monocyte | *RNASEH2B* | -1.191016586 | 0.016535417 | 0.046845773 | 1 |
| Obesity | monocyte | *CKAP2* | -1.132931787 | 0.012464206 | 0.043186096 | 1 |
| Obesity | monocyte | *RCBTB2* | 1.411593176 | 0.002853839 | 0.035116467 | 1 |
| Obesity | monocyte | *AOAH* | 0.281452423 | 0.004142749 | 0.035116467 | 1 |
| Obesity | monocyte | *TBRG4* | 1.917983115 | 0.005027231 | 0.035209637 | 1 |
| Obesity | monocyte | *MTHFS* | 1.35898565 | 0.005899796 | 0.036288402 | 1 |
| Obesity | monocyte | *RTP4* | 1.770734445 | 0.0103906 | 0.041803129 | 1 |
| Obesity | monocyte | *IMP4* | 2.50266349 | 0.004371564 | 0.035116467 | 1 |
| Obesity | monocyte | *NIPSNAP3A* | -1.724932451 | 0.011072629 | 0.042269392 | 1 |
| Obesity | monocyte | *TOR1B* | -1.602998658 | 0.01395699 | 0.044581511 | 1 |
| Obesity | monocyte | *SLC2A8* | 2.984642868 | 0.002080305 | 0.034263531 | 1 |
| Obesity | monocyte | *POLR1E* | 0.950151909 | 0.007520507 | 0.04034316 | 1 |
| Obesity | monocyte | *IL11RA* | -1.892631565 | 0.001039218 | 0.027556163 | 1 |
| Obesity | monocyte | *GMPR* | -0.937113309 | 0.007152362 | 0.039660871 | 1 |
| Obesity | monocyte | *IRF4* | 1.381145029 | 0.010016234 | 0.041669249 | 1 |
| Obesity | monocyte | *LRRC1* | -0.879626153 | 0.017057127 | 0.047509289 | 1 |
| Obesity | monocyte | *TUBB2B* | -2.015274641 | 0.017159091 | 0.047655914 | 1 |
| Obesity | monocyte | *NRM* | 1.069359338 | 0.004503899 | 0.035116467 | 1 |
| Obesity | monocyte | *VARS2* | -1.102171049 | 0.001717835 | 0.03218121 | 1 |
| Obesity | monocyte | *TLR2* | 0.701525071 | 0.003578268 | 0.035116467 | 1 |

| | | | | | | |
|---|---|---|---|---|---|---|
| Obesity | monocyte | *SYTL2* | -0.979933426 | 0.003787774 | 0.035116467 | 1 |
| Obesity | monocyte | *PRCP* | 2.785130304 | 0.012207325 | 0.043115778 | 1 |
| Obesity | monocyte | *GGH* | -1.312807895 | 0.009627218 | 0.041462933 | 1 |
| Obesity | monocyte | *SULF1* | -0.633965607 | 0.009557285 | 0.041462933 | 1 |
| Obesity | monocyte | *NEK1* | -0.934504201 | 0.018344095 | 0.04903887 | 1 |
| Obesity | monocyte | *POU2F3* | 2.085539237 | 0.005461767 | 0.035630311 | 1 |
| Obesity | monocyte | *PPP2R1B* | -0.506134558 | 0.01425444 | 0.044792561 | 1 |
| Obesity | monocyte | *FXYD6* | -1.349763571 | 0.015339363 | 0.045299203 | 1 |
| Obesity | monocyte | *PARP6* | -1.328917056 | 0.016285946 | 0.046549891 | 1 |
| Obesity | monocyte | *SLC44A5* | -0.715533104 | 0.013938308 | 0.044551165 | 1 |
| Obesity | monocyte | *KHK* | -1.748487812 | 0.015868412 | 0.045836948 | 1 |
| Obesity | monocyte | *DYNC2LI1* | -0.768401703 | 0.016454751 | 0.046771209 | 1 |
| Obesity | monocyte | *LHCGR* | -2.022117605 | 0.008119685 | 0.040894308 | 1 |
| Obesity | monocyte | *BTBD16* | -1.569070979 | 0.014669186 | 0.044883966 | 1 |
| Obesity | monocyte | *TACC2* | -0.777428483 | 0.014927653 | 0.044986158 | 1 |
| Obesity | monocyte | *PLCE1* | -0.919219572 | 0.014696262 | 0.044883966 | 1 |
| Obesity | monocyte | *ASCC1* | -1.293985623 | 0.011426806 | 0.042352977 | 1 |
| Obesity | monocyte | *STAT4* | 1.605292698 | 0.001717694 | 0.03218121 | 1 |
| Obesity | monocyte | *FASTKD1* | 3.18658477 | 0.018795828 | 0.049537435 | 1 |
| Obesity | monocyte | *COX17* | 4.444033525 | 0.004929011 | 0.035116467 | 1 |
| Obesity | monocyte | *AP1AR* | -1.561302217 | 0.018920108 | 0.049756294 | 1 |
| Obesity | monocyte | *BMPR1B* | -0.751407748 | 0.011038276 | 0.042237944 | 1 |
| Obesity | monocyte | *MMRN1* | -0.786843619 | 0.001404526 | 0.03111676 | 1 |
| Obesity | monocyte | *PRDM5* | -1.148040776 | 0.018337801 | 0.04903887 | 1 |
| Obesity | monocyte | *TRPC3* | -2.792765763 | 0.002400715 | 0.035116467 | 1 |
| Obesity | monocyte | *NAAA* | 0.630974805 | 0.004150575 | 0.035116467 | 1 |
| Obesity | monocyte | *FBN2* | 0.356585343 | 0.008289188 | 0.041208137 | 1 |
| Obesity | monocyte | *PARVG* | 0.57566557 | 0.005744432 | 0.035871281 | 1 |
| Obesity | monocyte | *B4GALNT3* | 2.308551348 | 0.002003209 | 0.034173375 | 1 |
| Obesity | monocyte | *ERP27* | 2.430024252 | 0.001678807 | 0.03218121 | 1 |
| Obesity | monocyte | *KIF21A* | -0.805776021 | 0.013140058 | 0.044028405 | 1 |
| Obesity | monocyte | *ALG10* | 1.859705434 | 0.006570667 | 0.038158325 | 1 |
| Obesity | monocyte | *CLSTN3* | -0.969311689 | 0.014085767 | 0.044698589 | 1 |
| Obesity | monocyte | *VAMP1* | -1.22979072 | 0.008343522 | 0.041351144 | 1 |
| Obesity | monocyte | *PEX5* | -1.318542403 | 0.017321193 | 0.047820138 | 1 |
| Obesity | monocyte | *PPFIA2* | -1.238468004 | 0.018008301 | 0.048786307 | 1 |
| Obesity | monocyte | *MARCH9* | -1.253259619 | 0.009948213 | 0.041669249 | 1 |
| Obesity | monocyte | *LGR5* | -2.69186718 | 0.007725778 | 0.040591959 | 1 |
| Obesity | monocyte | *POC1B* | 3.506139151 | 0.013132386 | 0.044028405 | 1 |
| Obesity | monocyte | *AMDHD1* | -1.707870088 | 0.002160162 | 0.03505386 | 1 |
| Obesity | monocyte | *RITA1* | -1.996312713 | 0.012054594 | 0.043014345 | 1 |
| Obesity | monocyte | *SDSL* | -1.691244087 | 0.016115414 | 0.046322269 | 1 |
| Obesity | monocyte | *GLTP* | -1.52950684 | 0.015427328 | 0.045365696 | 1 |
| Obesity | monocyte | *BRCA2* | -1.293051603 | 0.008037124 | 0.040894308 | 1 |
| Obesity | monocyte | *LMBR1L* | -1.582230814 | 0.018033725 | 0.048795606 | 1 |
| Obesity | monocyte | *ZNF740* | -1.057807134 | 0.01558443 | 0.045619236 | 1 |
| Obesity | monocyte | *HNRNPA1L2* | 2.900074105 | 0.002622196 | 0.035116467 | 1 |
| Obesity | monocyte | *GRTP1* | -0.910773852 | 0.006156561 | 0.036878563 | 1 |
| Obesity | monocyte | *CBLN3* | -0.707734125 | 0.012640113 | 0.043342634 | 1 |
| Obesity | monocyte | *RTN1* | 1.792679899 | 0.004706156 | 0.035116467 | 1 |

| | | | | | | |
|---|---|---|---|---|---|---|
| Obesity | monocyte | *EFCAB11* | -1.421074372 | 0.005325504 | 0.035561878 | 1 |
| Obesity | monocyte | *SLC24A4* | 0.210184632 | 0.004900963 | 0.035116467 | 1 |
| Obesity | monocyte | *C14orf79* | -1.08865181 | 0.010562716 | 0.041845661 | 1 |
| Obesity | monocyte | *DUOXA1* | -2.939658484 | 0.000589991 | 0.021362967 | 1 |
| Obesity | monocyte | *NEIL1* | -1.673623087 | 0.012245247 | 0.043115778 | 1 |
| Obesity | monocyte | *PIF1* | -1.684122165 | 0.007293979 | 0.039944003 | 1 |
| Obesity | monocyte | *ULK3* | -1.809844257 | 0.01076848 | 0.041961024 | 1 |
| Obesity | monocyte | *PCSK6* | -0.81688029 | 0.008919152 | 0.041462933 | 1 |
| Obesity | monocyte | *CELF6* | 3.014362562 | 0.005727388 | 0.035810934 | 1 |
| Obesity | monocyte | *ITGAX* | 0.199884321 | 0.004869304 | 0.035116467 | 1 |
| Obesity | monocyte | *TGFB1I1* | -3.086574446 | 0.011518677 | 0.042465454 | 1 |
| Obesity | monocyte | *DHX38* | -1.247795823 | 0.015511385 | 0.045468081 | 1 |
| Obesity | monocyte | *GCSH* | -1.246821569 | 0.011696472 | 0.042490474 | 1 |
| Obesity | monocyte | *CMTM3* | 2.109455656 | 0.003723938 | 0.035116467 | 1 |
| Obesity | monocyte | *PRPSAP2* | -0.857964298 | 0.017714738 | 0.048478233 | 1 |
| Obesity | monocyte | *RASL10B* | -3.727225245 | 0.018343394 | 0.04903887 | 1 |
| Obesity | monocyte | *UNC45B* | 1.614561538 | 0.007426331 | 0.040291176 | 1 |
| Obesity | monocyte | *LRRC46* | -2.516628681 | 0.013035916 | 0.043967382 | 1 |
| Obesity | monocyte | *IMPA2* | 1.983293494 | 0.002101652 | 0.034345034 | 1 |
| Obesity | monocyte | *C18orf8* | -6.905485425 | 1.26E-05 | 0.00160399 | 0.060951606 |
| Obesity | monocyte | *SLC14A1* | -1.066106412 | 0.012515256 | 0.04319079 | 1 |
| Obesity | monocyte | *CCDC40* | -0.84607674 | 0.006475077 | 0.037806349 | 1 |
| Obesity | monocyte | *TMC6* | 3.679601939 | 0.019191407 | 0.049996369 | 1 |
| Obesity | monocyte | *ANAPC11* | -1.959094198 | 0.0181768 | 0.048953818 | 1 |
| Obesity | monocyte | *TRIM65* | 2.23994297 | 0.000667394 | 0.02280421 | 1 |
| Obesity | monocyte | *SECTM1* | 1.899496544 | 0.002683666 | 0.035116467 | 1 |
| Obesity | monocyte | *MAPK4* | 1.326661978 | 0.004208628 | 0.035116467 | 1 |
| Obesity | monocyte | *FBXO15* | -1.739596501 | 0.015373614 | 0.045316053 | 1 |
| Obesity | monocyte | *CBLN2* | -1.705052438 | 0.009544654 | 0.041462933 | 1 |
| Obesity | monocyte | *ERBB2* | -1.363026704 | 0.015291819 | 0.045241407 | 1 |
| Obesity | monocyte | *PNMT* | 4.514450486 | 0.008020495 | 0.040894308 | 1 |
| Obesity | monocyte | *CACNA1A* | -1.170799626 | 0.006534357 | 0.038060866 | 1 |
| Obesity | monocyte | *SAMD1* | -2.32830091 | 0.011395118 | 0.042352977 | 1 |
| Obesity | monocyte | *PRDM15* | -1.816397949 | 0.010064987 | 0.041669249 | 1 |
| Obesity | monocyte | *SAE1* | -4.072068009 | 0.001102397 | 0.028083118 | 1 |
| Obesity | monocyte | *GEMIN7* | 1.599221741 | 0.012359822 | 0.043115778 | 1 |
| Obesity | monocyte | *CAPN10* | -1.659491575 | 0.015930043 | 0.045980113 | 1 |
| Obesity | monocyte | *SLC47A1* | 2.401578915 | 0.00384573 | 0.035116467 | 1 |
| Obesity | monocyte | *CTU1* | 3.97414202 | 0.002833832 | 0.035116467 | 1 |
| Obesity | monocyte | *MMEL1* | -2.351872205 | 0.000348126 | 0.016559891 | 1 |
| Obesity | monocyte | *ZNF593* | 2.501173097 | 0.003540624 | 0.035116467 | 1 |
| Obesity | monocyte | *C1orf216* | 2.169576503 | 0.001115015 | 0.028083118 | 1 |
| Obesity | monocyte | *MAEL* | -2.528333721 | 0.009065648 | 0.041462933 | 1 |
| Obesity | monocyte | *GABPB2* | -1.223177325 | 0.013261467 | 0.04407167 | 1 |
| Obesity | monocyte | *SMYD2* | -1.10249446 | 0.013587789 | 0.044347765 | 1 |
| Obesity | monocyte | *CREB3L4* | 1.073060376 | 0.010426783 | 0.041803129 | 1 |
| Obesity | monocyte | *KCNN3* | -1.198382717 | 0.005625643 | 0.035810934 | 1 |
| Obesity | monocyte | *FLG* | -1.26633789 | 0.012311166 | 0.043115778 | 1 |
| Obesity | monocyte | *ACP1* | -1.04754778 | 0.010746853 | 0.041961024 | 1 |
| Obesity | monocyte | *CNIH4* | 2.450432803 | 0.019178354 | 0.049996369 | 1 |

| | | | | | | |
|---|---|---|---|---|---|---|
| Obesity | monocyte | *MBOAT2* | 2.917214487 | 0.002994627 | 0.035116467 | 1 |
| Obesity | monocyte | *PYCR2* | 2.360424722 | 0.011997359 | 0.042922686 | 1 |
| Obesity | monocyte | *ABHD1* | -1.399982556 | 0.009232965 | 0.041462933 | 1 |
| Obesity | monocyte | *TEX261* | -1.178354385 | 0.009531407 | 0.041462933 | 1 |
| Obesity | monocyte | *FBLN7* | -1.28654328 | 0.000848368 | 0.025566965 | 1 |
| Obesity | monocyte | *CNGA3* | -20.55797879 | 0.013204543 | 0.044032814 | 1 |
| Obesity | monocyte | *FAHD2B* | -4.335591033 | 0.000208839 | 0.012208274 | 1 |
| Obesity | monocyte | *POLR2D* | -0.99240115 | 0.010527635 | 0.041845661 | 1 |
| Obesity | monocyte | *GALNT13* | -0.712743761 | 0.009245844 | 0.041462933 | 1 |
| Obesity | monocyte | *PHOSPHO2* | 1.839083211 | 0.018457978 | 0.04918073 | 1 |
| Obesity | monocyte | *ESPNL* | -14.01806527 | 0.008724953 | 0.041462933 | 1 |
| Obesity | monocyte | *STK11IP* | 2.702658559 | 0.012305276 | 0.043115778 | 1 |
| Obesity | monocyte | *EAF1* | -1.392073799 | 0.011223127 | 0.042279868 | 1 |
| Obesity | monocyte | *CNTN4* | -0.989083241 | 0.016883327 | 0.047233354 | 1 |
| Obesity | monocyte | *SLC25A38* | -1.229710953 | 0.009628849 | 0.041462933 | 1 |
| Obesity | monocyte | *PTPRG* | -0.805440331 | 0.017085904 | 0.047562138 | 1 |
| Obesity | monocyte | *SLC25A26* | -1.42254233 | 0.009443135 | 0.041462933 | 1 |
| Obesity | monocyte | *ARL6IP5* | 2.871894702 | 0.003841544 | 0.035116467 | 1 |
| Obesity | monocyte | *LIMD1* | 3.269794372 | 0.019036721 | 0.049873741 | 1 |
| Obesity | monocyte | *PLA1A* | 1.805316235 | 0.003208692 | 0.035116467 | 1 |
| Obesity | monocyte | *BOC* | -0.610845498 | 0.010997487 | 0.042215037 | 1 |
| Obesity | monocyte | *MED12L* | -1.085715049 | 0.013672704 | 0.044440213 | 1 |
| Obesity | monocyte | *EIF2A* | -1.278512697 | 0.003300382 | 0.035116467 | 1 |
| Obesity | monocyte | *AMT* | -1.765222716 | 0.000243258 | 0.013412372 | 1 |
| Obesity | monocyte | *MUC4* | -1.303851151 | 0.010003946 | 0.041669249 | 1 |
| Obesity | monocyte | *SCD5* | -0.872640919 | 0.009544147 | 0.041462933 | 1 |
| Obesity | monocyte | *PYURF* | -1.051433705 | 0.012802983 | 0.043685001 | 1 |
| Obesity | monocyte | *CISD2* | 3.715181132 | 0.001153602 | 0.028703977 | 1 |
| Obesity | monocyte | *BHMT* | -1.950585757 | 0.000471484 | 0.018906113 | 1 |
| Obesity | monocyte | *GTF2H2* | -1.374315719 | 0.010322852 | 0.041771331 | 1 |
| Obesity | monocyte | *SPATA9* | -0.711734172 | 0.009196361 | 0.041462933 | 1 |
| Obesity | monocyte | *TSLP* | -1.405391124 | 0.002327551 | 0.035072295 | 1 |
| Obesity | monocyte | *FEM1C* | -1.558914135 | 0.018775214 | 0.049536344 | 1 |
| Obesity | monocyte | *YIPF5* | 4.486499314 | 0.001050715 | 0.027706905 | 1 |
| Obesity | monocyte | *BOD1* | 3.165054235 | 0.002364176 | 0.035116467 | 1 |
| Obesity | monocyte | *FAM50B* | -2.83414983 | 0.005938374 | 0.036288402 | 1 |
| Obesity | monocyte | *MYLK4* | -1.921320241 | 0.0107124 | 0.041961024 | 1 |
| Obesity | monocyte | *HIGD2A* | 3.623059008 | 0.008430837 | 0.041379919 | 1 |
| Obesity | monocyte | *RNF44* | -2.092825727 | 0.018796125 | 0.049537435 | 1 |
| Obesity | monocyte | *MUT* | -0.991429113 | 0.009160477 | 0.041462933 | 1 |
| Obesity | monocyte | *DOK3* | 0.401315445 | 0.007402823 | 0.040251891 | 1 |
| Obesity | monocyte | *MLIP* | -1.614740073 | 0.004090383 | 0.035116467 | 1 |
| Obesity | monocyte | *ANO7* | 0.606444915 | 0.002839928 | 0.035116467 | 1 |
| Obesity | monocyte | *ABRACL* | 1.427452099 | 0.003114233 | 0.035116467 | 1 |
| Obesity | monocyte | *SLC18B1* | -1.710185483 | 0.00992138 | 0.041669249 | 1 |
| Obesity | monocyte | *PNLDC1* | 1.025466797 | 0.002456011 | 0.035116467 | 1 |
| Obesity | monocyte | *SLC22A3* | -0.732655524 | 0.009308984 | 0.041462933 | 1 |
| Obesity | monocyte | *SDK1* | -1.119449822 | 0.001179159 | 0.028774316 | 1 |
| Obesity | monocyte | *CCZ1B* | -2.533428333 | 0.003684389 | 0.035116467 | 1 |
| Obesity | monocyte | *PSPH* | -1.000488222 | 0.01253707 | 0.04319079 | 1 |

| | | | | | | |
|---|---|---|---|---|---|---|
| Obesity | monocyte | *TMEM209* | -1.163104585 | 0.014221136 | 0.044792561 | 1 |
| Obesity | monocyte | *EPHA1* | -2.754066339 | 0.009257568 | 0.041462933 | 1 |
| Obesity | monocyte | *NCAPG2* | -0.954167875 | 0.010813005 | 0.041961024 | 1 |
| Obesity | monocyte | *ADHFE1* | -1.053436218 | 0.000125561 | 0.008829335 | 0.609224137 |
| Obesity | monocyte | *DPYS* | -0.625929885 | 0.009394105 | 0.041462933 | 1 |
| Obesity | monocyte | *SLC39A4* | 0.550353438 | 0.00545087 | 0.035630311 | 1 |
| Obesity | monocyte | *AK3* | -1.097671073 | 0.019063026 | 0.049915706 | 1 |
| Obesity | monocyte | *C9orf72* | 1.341331411 | 0.018696441 | 0.049516993 | 1 |
| Obesity | monocyte | *POLE3* | -1.251694026 | 0.007342579 | 0.04011959 | 1 |
| Obesity | monocyte | *SURF1* | 2.396644987 | 0.0040722 | 0.035116467 | 1 |
| Obesity | monocyte | *SURF6* | -1.69659215 | 0.016967924 | 0.047378154 | 1 |
| Obesity | monocyte | *REXO4* | -1.421130748 | 0.017981606 | 0.048768447 | 1 |
| Obesity | monocyte | *SH3GLB2* | 4.149591658 | 0.00404239 | 0.035116467 | 1 |
| Obesity | monocyte | *SLC39A12* | 4.257142847 | 0.001617771 | 0.03218121 | 1 |
| Obesity | monocyte | *ST8SIA6* | -0.958593726 | 0.012280627 | 0.043115778 | 1 |
| Obesity | monocyte | *RGR* | -21.14984892 | 4.71E-06 | 0.0007149 | 0.022876813 |
| Obesity | monocyte | *GAS2* | -1.107019599 | 0.012020843 | 0.042949286 | 1 |
| Obesity | monocyte | *HSD17B12* | -2.724337128 | 4.46E-10 | 2.40E-07 | 2.16E-06 |
| Obesity | monocyte | *PTPRJ* | 0.815804167 | 0.003458009 | 0.035116467 | 1 |
| Obesity | monocyte | *CAPN5* | -2.192894549 | 0.011756353 | 0.042569734 | 1 |
| Obesity | monocyte | *DRD2* | -8.693359528 | 0.008329245 | 0.041322595 | 1 |
| Obesity | monocyte | *FADS1* | 1.32443646 | 0.012092315 | 0.043083715 | 1 |
| Obesity | monocyte | *TMC2* | 1.065185557 | 0.005153425 | 0.035289693 | 1 |
| Obesity | monocyte | *EML3* | -1.109868482 | 0.017919134 | 0.048768447 | 1 |
| Obesity | monocyte | *INCENP* | -1.450782302 | 0.004335814 | 0.035116467 | 1 |
| Obesity | monocyte | *SCN2B* | 0.975895521 | 0.01330299 | 0.044149185 | 1 |
| Obesity | monocyte | *CABLES2* | -1.967230997 | 0.001686748 | 0.03218121 | 1 |
| Obesity | monocyte | *TRPT1* | -1.481082246 | 0.000999333 | 0.027556163 | 1 |
| Obesity | monocyte | *NUDT22* | -2.458494651 | 0.001401538 | 0.03111676 | 1 |
| Obesity | monocyte | *TBX6* | 1.797872322 | 1.00E-09 | 4.85E-07 | 4.85E-06 |
| Obesity | monocyte | *ANXA8L1* | -0.645634767 | 0.012337304 | 0.043115778 | 1 |
| Obesity | monocyte | *FAM124A* | -1.76172653 | 0.018754592 | 0.049532965 | 1 |
| Obesity | monocyte | *LYPD6B* | -2.359501031 | 0.001658724 | 0.03218121 | 1 |
| Obesity | monocyte | *RGS18* | 0.302437431 | 0.001785002 | 0.032602913 | 1 |
| Obesity | monocyte | *PRSS23* | -0.737128753 | 0.012290301 | 0.043115778 | 1 |
| Obesity | monocyte | *PPP1R1C* | 1.260824319 | 0.003962445 | 0.035116467 | 1 |
| Obesity | monocyte | *GPR158* | -1.517727814 | 0.012400045 | 0.043115778 | 1 |
| Obesity | monocyte | *PLBD2* | 3.236322635 | 0.002035043 | 0.034173375 | 1 |
| Obesity | monocyte | *DLG5* | -0.879497634 | 0.014178241 | 0.044792561 | 1 |
| Obesity | monocyte | *TMEM18* | 4.445415132 | 0.004610244 | 0.035116467 | 1 |
| Obesity | monocyte | *NDUFC2* | -1.678428794 | 0.015684288 | 0.045674648 | 1 |
| Obesity | monocyte | *ADAMTS12* | -0.706201513 | 0.014823203 | 0.044883966 | 1 |
| Obesity | monocyte | *VIPAS39* | -1.381089022 | 0.015100808 | 0.044986158 | 1 |
| Obesity | monocyte | *ANKRD50* | 4.184679477 | 0.001332824 | 0.030360851 | 1 |
| Obesity | monocyte | *DPYSL4* | 1.863303889 | 0.000269169 | 0.014195746 | 1 |
| Obesity | monocyte | *CCDC122* | -0.994791176 | 0.005836834 | 0.036261612 | 1 |
| Obesity | monocyte | *GUF1* | -1.69071599 | 0.007507124 | 0.04034316 | 1 |
| Obesity | monocyte | *TMEM56* | -1.83213254 | 0.009687476 | 0.041470763 | 1 |
| Obesity | monocyte | *TMEM178A* | -1.878126385 | 0.01083864 | 0.041961024 | 1 |
| Obesity | monocyte | *KCNK13* | 0.527526755 | 0.004912407 | 0.035116467 | 1 |

| | | | | | | |
|---|---|---|---|---|---|---|
| Obesity | monocyte | *POC5* | -3.087607654 | 1.59E-05 | 0.001983951 | 0.077374075 |
| Obesity | monocyte | *NMT2* | -1.643811994 | 0.009880906 | 0.041637281 | 1 |
| Obesity | monocyte | *CCNO* | -12.74830804 | 0.00550331 | 0.035757287 | 1 |
| Obesity | monocyte | *RASGRP3* | -0.799630424 | 0.007069834 | 0.039383277 | 1 |
| Obesity | monocyte | *GPR180* | -0.962050398 | 0.007777127 | 0.04060923 | 1 |
| Obesity | monocyte | *JAKMIP1* | 7.085558408 | 0.011253056 | 0.04231558 | 1 |
| Obesity | monocyte | *CENPH* | -1.940877947 | 0.005156717 | 0.035289693 | 1 |
| Obesity | monocyte | *CLGN* | -5.141983328 | 0.003444147 | 0.035116467 | 1 |
| Obesity | monocyte | *LPCAT1* | 1.988592362 | 0.002652185 | 0.035116467 | 1 |
| Obesity | monocyte | *NMRAL1* | -2.468117522 | 0.014821096 | 0.044883966 | 1 |
| Obesity | monocyte | *ADPRHL1* | -2.113549303 | 0.006745525 | 0.038614455 | 1 |
| Obesity | monocyte | *ZDHHC7* | 3.092053593 | 0.004951723 | 0.035116467 | 1 |
| Obesity | monocyte | *FBXO36* | -1.458357855 | 0.017484902 | 0.048085326 | 1 |
| Obesity | monocyte | *MCOLN2* | 0.72701655 | 0.00355357 | 0.035116467 | 1 |
| Obesity | monocyte | *DNAAF1* | 2.370386951 | 0.018984587 | 0.049857566 | 1 |
| Obesity | monocyte | *UBASH3B* | 0.394155208 | 0.004802896 | 0.035116467 | 1 |
| Obesity | monocyte | *ABCA5* | -0.690067699 | 0.013781279 | 0.044539684 | 1 |
| Obesity | monocyte | *ENPP3* | -1.009886923 | 0.007621912 | 0.040372837 | 1 |
| Obesity | monocyte | *CCSAP* | -1.119144174 | 0.009751981 | 0.041544165 | 1 |
| Obesity | monocyte | *GBP5* | 0.909096013 | 0.013975693 | 0.044596002 | 1 |
| Obesity | monocyte | *FLCN* | -1.210366242 | 0.019142402 | 0.049988663 | 1 |
| Obesity | monocyte | *APCDD1* | -0.657168854 | 0.007013652 | 0.039205347 | 1 |
| Obesity | monocyte | *PROM2* | 1.760482817 | 0.004260851 | 0.035116467 | 1 |
| Obesity | monocyte | *MARVELD1* | -1.396358112 | 0.01176546 | 0.042569734 | 1 |
| Obesity | monocyte | *GOLGA7B* | 1.168934692 | 0.004718552 | 0.035116467 | 1 |
| Obesity | monocyte | *MOV10* | 2.189985731 | 0.010378368 | 0.041803129 | 1 |
| Obesity | monocyte | *SETD9* | -1.164560147 | 0.005699319 | 0.035810934 | 1 |
| Obesity | monocyte | *KDM8* | -1.649762457 | 0.018659907 | 0.049501295 | 1 |
| Obesity | monocyte | *ALS2CR12* | 3.355070483 | 0.000497213 | 0.019613656 | 1 |
| Obesity | monocyte | *RRAGA* | -1.104667895 | 0.009399829 | 0.041462933 | 1 |
| Obesity | monocyte | *TMBIM4* | 3.409030377 | 0.00563209 | 0.035810934 | 1 |
| Obesity | monocyte | *PSD3* | -1.132099851 | 0.015482524 | 0.045417901 | 1 |
| Obesity | monocyte | *KCNMA1* | -0.836786058 | 0.016800126 | 0.04709082 | 1 |
| Obesity | monocyte | *ART3* | 2.091339241 | 0.000651942 | 0.022434186 | 1 |
| Obesity | monocyte | *RWDD2B* | -4.022522326 | 0.000577908 | 0.021082768 | 1 |
| Obesity | monocyte | *CCT8* | -1.658118528 | 0.00383628 | 0.035116467 | 1 |
| Obesity | monocyte | *NAA11* | -1.933331489 | 0.004344205 | 0.035116467 | 1 |
| Obesity | monocyte | *TDRD9* | -4.439804919 | 0.005527198 | 0.035757287 | 1 |
| Obesity | monocyte | *PCDH1* | -1.409374078 | 0.001640361 | 0.03218121 | 1 |
| Obesity | monocyte | *HKDC1* | -6.575889849 | 0.00496887 | 0.035116467 | 1 |
| Obesity | monocyte | *TYSND1* | -0.680577412 | 0.018323895 | 0.04903887 | 1 |
| Obesity | monocyte | *NODAL* | -1.866337049 | 0.010585145 | 0.041845661 | 1 |
| Obesity | monocyte | *MED19* | -2.917148235 | 0.014548399 | 0.044883966 | 1 |
| Obesity | monocyte | *FBRS* | 2.740431275 | 0.002671793 | 0.035116467 | 1 |
| Obesity | monocyte | *MPV17L* | -2.12139821 | 0.00542722 | 0.035630311 | 1 |
| Obesity | monocyte | *GHRL* | -0.984709282 | 0.018650354 | 0.049501295 | 1 |
| Obesity | monocyte | *NMNAT2* | -1.320951573 | 0.016873124 | 0.047233354 | 1 |
| Obesity | monocyte | *SLC6A1* | 1.611549251 | 0.003852136 | 0.035116467 | 1 |
| Obesity | monocyte | *TIMP4* | -1.50774965 | 0.006343025 | 0.037442903 | 1 |
| Obesity | monocyte | *STEAP2* | -1.222155975 | 0.000464676 | 0.018861406 | 1 |

| | | | | | | |
|---|---|---|---|---|---|---|
| Obesity | monocyte | *CLEC18A* | -10.16067091 | 3.78E-18 | 1.83E-14 | 1.83E-14 |
| Obesity | monocyte | *ETS2* | 2.644021274 | 4.67E-05 | 0.004043647 | 0.226444234 |
| Obesity | monocyte | *LCA5L* | -1.123429723 | 0.005722213 | 0.035810934 | 1 |
| Obesity | monocyte | *SLC38A10* | 3.43383835 | 0.012748521 | 0.043560438 | 1 |
| Obesity | monocyte | *DGKI* | -0.829877179 | 0.003066044 | 0.035116467 | 1 |
| Obesity | monocyte | *SLC34A2* | -25.63712066 | 0.005357736 | 0.035561878 | 1 |
| Obesity | monocyte | *AP3S2* | -0.933077956 | 0.00984736 | 0.041637281 | 1 |
| Obesity | monocyte | *C12orf43* | -1.469194087 | 0.00376595 | 0.035116467 | 1 |
| Obesity | monocyte | *PEX10* | 4.314195518 | 0.00391501 | 0.035116467 | 1 |
| Obesity | monocyte | *KRTCAP3* | 2.815126713 | 0.000505677 | 0.01966188 | 1 |
| Obesity | monocyte | *EXTL1* | -15.65885189 | 0.009236889 | 0.041462933 | 1 |
| Obesity | monocyte | *DZIP1L* | -1.357145937 | 0.010103413 | 0.04168517 | 1 |
| Obesity | monocyte | *CLSTN2* | -0.615280436 | 0.008759284 | 0.041462933 | 1 |
| Obesity | monocyte | *TSPAN33* | -1.85951013 | 0.010263254 | 0.041771331 | 1 |
| Obesity | monocyte | *PPP1R9A* | -1.15652274 | 0.017026365 | 0.047478116 | 1 |
| Obesity | monocyte | *DYNC1I1* | -0.64929467 | 0.016942956 | 0.04735439 | 1 |
| Obesity | monocyte | *COPG2* | -0.987182039 | 0.001563561 | 0.03218121 | 1 |
| Obesity | monocyte | *ELK4* | 4.105226211 | 0.007646237 | 0.040393703 | 1 |
| Obesity | monocyte | *DUSP23* | -1.24223328 | 0.006112182 | 0.036748833 | 1 |
| Obesity | monocyte | *RNF166* | 1.004831517 | 0.006588446 | 0.038158325 | 1 |
| Obesity | monocyte | *SPATA2L* | -2.399205571 | 0.003724575 | 0.035116467 | 1 |
| Obesity | monocyte | *CDA* | 0.496263464 | 0.009202226 | 0.041462933 | 1 |
| Obesity | monocyte | *SLC5A11* | -2.238845403 | 6.35E-08 | 1.63E-05 | 0.000308122 |
| Obesity | monocyte | *RAPGEF6* | -4.080085141 | 0.002026081 | 0.034173375 | 1 |
| Obesity | monocyte | *IFNAR2* | 1.150912699 | 0.005663153 | 0.035810934 | 1 |
| Obesity | monocyte | *SV2A* | 1.641344916 | 0.004482883 | 0.035116467 | 1 |
| Obesity | monocyte | *CBR1* | -1.073032084 | 0.003643156 | 0.035116467 | 1 |
| Obesity | monocyte | *IRX6* | -0.850588362 | 0.014892361 | 0.044964366 | 1 |
| Obesity | monocyte | *HK2* | -0.720024935 | 0.008996359 | 0.041462933 | 1 |
| Obesity | monocyte | *ACE* | -1.164833696 | 0.01030564 | 0.041771331 | 1 |
| Obesity | monocyte | *TEPP* | 1.161622121 | 0.00589256 | 0.036288402 | 1 |
| Obesity | monocyte | *ZFYVE28* | -1.095458444 | 0.013428798 | 0.044219637 | 1 |
| Obesity | monocyte | *C16orf86* | -1.41256125 | 0.008043952 | 0.040894308 | 1 |
| Obesity | monocyte | *ATAD3B* | 1.692952868 | 0.00497394 | 0.035116467 | 1 |
| Obesity | monocyte | *CPAMD8* | -1.474471027 | 0.011410295 | 0.042352977 | 1 |
| Obesity | monocyte | *ANKLE1* | -5.55357118 | 0.01422711 | 0.044792561 | 1 |
| Obesity | monocyte | *CILP2* | -8.362272149 | 2.30E-06 | 0.000429324 | 0.01116243 |
| Obesity | monocyte | *C21orf2* | -2.038472152 | 0.003865842 | 0.035116467 | 1 |
| Obesity | monocyte | *VAV2* | 3.640222362 | 0.001378557 | 0.031060598 | 1 |
| Obesity | monocyte | *S100B* | -0.847628367 | 0.004446616 | 0.035116467 | 1 |
| Obesity | monocyte | *ADAMTS13* | -1.172040918 | 0.009653274 | 0.041464521 | 1 |
| Obesity | monocyte | *CACFD1* | 4.841677095 | 0.004533288 | 0.035116467 | 1 |
| Obesity | monocyte | *RDH13* | -1.163450457 | 0.011126565 | 0.042279868 | 1 |
| Obesity | monocyte | *CCR5* | 2.655818177 | 0.001855321 | 0.033300696 | 1 |
| Obesity | monocyte | *NBEAL2* | 1.107071167 | 0.001064575 | 0.02778834 | 1 |
| Obesity | monocyte | *SH2B2* | 0.410513716 | 0.005215587 | 0.035351132 | 1 |
| Obesity | monocyte | *ABCF3* | 2.547747581 | 4.99E-05 | 0.004251349 | 0.24232687 |
| Obesity | monocyte | *PCYT1A* | -2.609659547 | 0.00714016 | 0.039642943 | 1 |
| Obesity | monocyte | *BDH1* | -1.83291028 | 0.014816107 | 0.044883966 | 1 |
| Obesity | monocyte | *PGAP3* | 2.758817446 | 0.002827303 | 0.035116467 | 1 |

| | | | | | | |
|---|---|---|---|---|---|---|
| Obesity | monocyte | *ALDH16A1* | 1.713137021 | 0.002494976 | 0.035116467 | 1 |
| Obesity | monocyte | *PLCD3* | 4.387441209 | 0.003580656 | 0.035116467 | 1 |
| Obesity | monocyte | *ASGR2* | 0.658461341 | 0.004934056 | 0.035116467 | 1 |
| Obesity | monocyte | *POLR3K* | 0.742898478 | 0.005857411 | 0.036288402 | 1 |
| Obesity | monocyte | *WDR90* | -1.306815564 | 0.002050335 | 0.034173375 | 1 |
| Obesity | monocyte | *CCDC78* | 0.809768841 | 0.004419024 | 0.035116467 | 1 |
| Obesity | monocyte | *ASRGL1* | 2.963917915 | 0.002634009 | 0.035116467 | 1 |
| Obesity | monocyte | *RPS6KA4* | 1.294505851 | 0.003474841 | 0.035116467 | 1 |
| Obesity | monocyte | *PDZK1IP1* | -1.716237267 | 0.00918281 | 0.041462933 | 1 |
| Obesity | monocyte | *COA7* | 6.224871709 | 0.003741755 | 0.035116467 | 1 |
| Obesity | monocyte | *SLC1A7* | -1.164356613 | 0.018558378 | 0.049339863 | 1 |
| Obesity | monocyte | *LAPTM5* | 2.351104515 | 0.002678008 | 0.035116467 | 1 |
| Obesity | monocyte | *SYNC* | -0.997894773 | 0.00787422 | 0.040861728 | 1 |
| Obesity | monocyte | *CAMK2N1* | 5.260079048 | 4.91E-06 | 0.000721276 | 0.023802111 |
| Obesity | monocyte | *LRRIQ3* | -0.829323991 | 0.009102223 | 0.041462933 | 1 |
| Obesity | monocyte | *TYW3* | -0.858078225 | 0.007475094 | 0.04034316 | 1 |
| Obesity | monocyte | *HFM1* | -0.96986263 | 0.015588766 | 0.045619236 | 1 |
| Obesity | monocyte | *FCRLB* | 2.033257759 | 0.004303545 | 0.035116467 | 1 |
| Obesity | monocyte | *TDRD5* | -3.660425846 | 0.005336028 | 0.035561878 | 1 |
| Obesity | monocyte | *ACP6* | -1.215448023 | 0.011200975 | 0.042279868 | 1 |
| Obesity | monocyte | *PM20D1* | 3.999776634 | 0.002581098 | 0.035116467 | 1 |
| Obesity | monocyte | *DUSP19* | 2.416025839 | 0.013805494 | 0.044539684 | 1 |
| Obesity | monocyte | *C2orf48* | 4.246309157 | 0.003410863 | 0.035116467 | 1 |
| Obesity | monocyte | *NOSTRIN* | -0.892437784 | 0.015657263 | 0.045669584 | 1 |
| Obesity | monocyte | *C1QTNF7* | -0.893273898 | 0.018034683 | 0.048795606 | 1 |
| Obesity | monocyte | *VPS72* | 4.637092242 | 0.01380236 | 0.044539684 | 1 |
| Obesity | monocyte | *RNF149* | 2.161049646 | 0.009489824 | 0.041462933 | 1 |
| Obesity | monocyte | *S100A9* | -1.413440277 | 0.008937247 | 0.041462933 | 1 |
| Obesity | monocyte | *DCAF16* | -2.275354932 | 0.015303902 | 0.045249563 | 1 |
| Obesity | monocyte | *CGGBP1* | -1.296690829 | 0.00808632 | 0.040894308 | 1 |
| Obesity | monocyte | *DAPL1* | 1.092492366 | 0.004273492 | 0.035116467 | 1 |
| Obesity | monocyte | *PBXIP1* | 5.025318221 | 0.014428162 | 0.044875283 | 1 |
| Obesity | monocyte | *CLDN1* | -0.648369665 | 0.015070954 | 0.044986158 | 1 |
| Obesity | monocyte | *LINC01116* | 3.10110888 | 0.001010478 | 0.027556163 | 1 |
| Obesity | monocyte | *LMOD1* | -2.176204998 | 0.000216042 | 0.012449829 | 1 |
| Obesity | monocyte | *CCT3* | 2.213753493 | 0.001948566 | 0.03376586 | 1 |
| Obesity | monocyte | *GLB1L* | -2.954508166 | 0.004568231 | 0.035116467 | 1 |
| Obesity | monocyte | *SERPINI1* | -1.148304303 | 0.01737718 | 0.047867689 | 1 |
| Obesity | monocyte | *C4orf36* | -0.798716171 | 0.009606494 | 0.041462933 | 1 |
| Obesity | monocyte | *DCLK3* | 6.113208876 | 0.000808421 | 0.025087947 | 1 |
| Obesity | monocyte | *RPP14* | -1.035219752 | 0.010078688 | 0.041669249 | 1 |
| Obesity | monocyte | *DNASE1L3* | 0.892908325 | 0.007518898 | 0.04034316 | 1 |
| Obesity | monocyte | *TTC14* | -1.458540757 | 0.012695615 | 0.043410235 | 1 |
| Obesity | monocyte | *CCDC158* | -1.665105679 | 0.006112177 | 0.036748833 | 1 |
| Obesity | monocyte | *CPA3* | -1.442849305 | 0.008459788 | 0.041379919 | 1 |
| Obesity | monocyte | *SLC4A1AP* | 2.981884851 | 0.003429581 | 0.035116467 | 1 |
| Obesity | monocyte | *CDCP1* | -1.380249539 | 0.00473333 | 0.035116467 | 1 |
| Obesity | monocyte | *FYCO1* | -2.688804127 | 0.016596547 | 0.046940995 | 1 |
| Obesity | monocyte | *TPRA1* | -2.840112652 | 0.008733016 | 0.041462933 | 1 |
| Obesity | monocyte | *DNALI1* | -3.176561794 | 8.66E-06 | 0.001167173 | 0.04201824 |

| | | | | | | |
|---|---|---|---|---|---|---|
| Obesity | monocyte | *RFT1* | -2.64112398 | 1.87E-05 | 0.002156681 | 0.090580591 |
| Obesity | monocyte | *SFMBT1* | 4.619022899 | 0.007494301 | 0.04034316 | 1 |
| Obesity | monocyte | *ABLIM2* | -1.218948587 | 0.015972402 | 0.046074968 | 1 |
| Obesity | monocyte | *ATRIP* | -1.850706942 | 0.006081555 | 0.036701128 | 1 |
| Obesity | monocyte | *RNF123* | 4.23082234 | 2.53E-06 | 0.000438755 | 0.012285153 |
| Obesity | monocyte | *MST1R* | 1.312723831 | 0.013216563 | 0.044032814 | 1 |
| Obesity | monocyte | *ETNPPL* | -7.657988372 | 0.009362715 | 0.041462933 | 1 |
| Obesity | monocyte | *CEP44* | -1.575016257 | 0.010059508 | 0.041669249 | 1 |
| Obesity | monocyte | *HPGD* | -1.463269755 | 0.008166486 | 0.040894308 | 1 |
| Obesity | monocyte | *TMEM144* | -2.197921081 | 0.013340222 | 0.044157522 | 1 |
| Obesity | monocyte | *IL15* | 3.72745598 | 0.014969255 | 0.044986158 | 1 |
| Obesity | monocyte | *MOCS2* | -1.553995222 | 0.012188377 | 0.043115778 | 1 |
| Obesity | monocyte | *RNF180* | -1.395405752 | 0.014857989 | 0.044888519 | 1 |
| Obesity | monocyte | *CCDC112* | 2.389793163 | 0.002830842 | 0.035116467 | 1 |
| Obesity | monocyte | *CMBL* | -1.213449592 | 0.015106087 | 0.044986158 | 1 |
| Obesity | monocyte | *C5orf63* | -0.90845014 | 0.013433583 | 0.044219637 | 1 |
| Obesity | monocyte | *CASP3* | -1.537740403 | 0.015382428 | 0.045316053 | 1 |
| Obesity | monocyte | *PRIMPOL* | -1.043402274 | 0.017980688 | 0.048768447 | 1 |
| Obesity | monocyte | *ERAP1* | -1.715026129 | 0.003041897 | 0.035116467 | 1 |
| Obesity | monocyte | *UTP15* | 1.791514361 | 0.004929734 | 0.035116467 | 1 |
| Obesity | monocyte | *TLR3* | -1.590762372 | 0.009626389 | 0.041462933 | 1 |
| Obesity | monocyte | *KLKB1* | -1.044010081 | 0.013058366 | 0.043968905 | 1 |
| Obesity | monocyte | *CCDC127* | -1.28708245 | 0.008144627 | 0.040894308 | 1 |
| Obesity | monocyte | *RAET1E* | -1.096446623 | 0.009057867 | 0.041462933 | 1 |
| Obesity | monocyte | *GALNT10* | -2.15727726 | 0.007207386 | 0.039861716 | 1 |
| Obesity | monocyte | *RELL2* | 1.16118207 | 0.011399316 | 0.042352977 | 1 |
| Obesity | monocyte | *MIOS* | 3.693783596 | 0.00530094 | 0.035561878 | 1 |
| Obesity | monocyte | *CHMP4C* | -3.144044794 | 0.010029796 | 0.041669249 | 1 |
| Obesity | monocyte | *CTSB* | -1.593613084 | 0.000463575 | 0.018861406 | 1 |
| Obesity | monocyte | *TNFRSF11B* | -1.609729258 | 0.011136715 | 0.042279868 | 1 |
| Obesity | monocyte | *FASTK* | -1.473400386 | 0.01483723 | 0.044883966 | 1 |
| Obesity | monocyte | *YWHAZ* | -2.099082482 | 0.014855442 | 0.044888519 | 1 |
| Obesity | monocyte | *SLC25A32* | -1.698268881 | 0.01537835 | 0.045316053 | 1 |
| Obesity | monocyte | *RPP25L* | -3.659346177 | 9.46E-05 | 0.007064482 | 0.459191346 |
| Obesity | monocyte | *C9orf24* | -0.973346684 | 0.014135706 | 0.044792561 | 1 |
| Obesity | monocyte | *ZMAT4* | -1.95491134 | 0.009832802 | 0.041637281 | 1 |
| Obesity | monocyte | *C8orf34* | -0.745826866 | 0.002770956 | 0.035116467 | 1 |
| Obesity | monocyte | *RASEF* | -1.739665598 | 0.000792411 | 0.02480501 | 1 |
| Obesity | monocyte | *AQP7* | -0.484387413 | 0.014380828 | 0.044843045 | 1 |
| Obesity | monocyte | *NOL6* | 3.707028938 | 0.00143014 | 0.03111676 | 1 |
| Obesity | monocyte | *AQP3* | -3.203688533 | 0.00924282 | 0.041462933 | 1 |
| Obesity | monocyte | *PIGO* | 2.521935423 | 0.005807242 | 0.036124025 | 1 |
| Obesity | monocyte | *TSHR* | -0.720485365 | 0.009543946 | 0.041462933 | 1 |
| Obesity | monocyte | *SLC16A9* | -1.374619189 | 0.009505903 | 0.041462933 | 1 |
| Obesity | monocyte | *GJB2* | -5.930374007 | 0.010295282 | 0.041771331 | 1 |
| Obesity | monocyte | *LRR1* | -2.057628609 | 0.015618935 | 0.045652454 | 1 |
| Obesity | monocyte | *NOXRED1* | -1.181399436 | 0.01507229 | 0.044986158 | 1 |
| Obesity | monocyte | *AKR1E2* | -1.198816194 | 0.014228219 | 0.044792561 | 1 |
| Obesity | monocyte | *VDAC2* | -2.514057601 | 0.011129543 | 0.042279868 | 1 |
| Obesity | monocyte | *CLEC1B* | 0.618970544 | 0.003739582 | 0.035116467 | 1 |

| | | | | | | |
|---|---|---|---|---|---|---|
| Obesity | monocyte | *RET* | -1.656293556 | 0.011175362 | 0.042279868 | 1 |
| Obesity | monocyte | *CASP7* | -2.54026852 | 0.002483271 | 0.035116467 | 1 |
| Obesity | monocyte | *PPP1R36* | -1.489550843 | 0.007590905 | 0.040367875 | 1 |
| Obesity | monocyte | *HSPA12A* | -0.984244614 | 0.004851098 | 0.035116467 | 1 |
| Obesity | monocyte | *OTOGL* | 0.598246403 | 0.003498362 | 0.035116467 | 1 |
| Obesity | monocyte | *PACSIN3* | -2.339691398 | 0.000417692 | 0.018424019 | 1 |
| Obesity | monocyte | *RAPSN* | -2.690161233 | 0.000961932 | 0.027135438 | 1 |
| Obesity | monocyte | *LIPC* | -1.431959112 | 0.016438369 | 0.046771209 | 1 |
| Obesity | monocyte | *JAM3* | -1.207083288 | 0.007277046 | 0.039923913 | 1 |
| Obesity | monocyte | *TRMT61A* | 2.189390468 | 0.003257665 | 0.035116467 | 1 |
| Obesity | monocyte | *GABRB3* | -1.49900461 | 0.002044019 | 0.034173375 | 1 |
| Obesity | monocyte | *COX11* | -1.196355733 | 0.01138351 | 0.042352977 | 1 |
| Obesity | monocyte | *ZNF202* | -1.277330869 | 0.007783715 | 0.04060923 | 1 |
| Obesity | monocyte | *APBB1* | -1.317011804 | 0.006357621 | 0.037481384 | 1 |
| Obesity | monocyte | *WDR88* | 2.587461979 | 0.001859952 | 0.033300696 | 1 |
| Obesity | monocyte | *PLD4* | 0.37865684 | 0.006405562 | 0.03766044 | 1 |
| Obesity | monocyte | *CENPN* | 1.217090026 | 0.004134466 | 0.035116467 | 1 |
| Obesity | monocyte | *PRKCB* | 0.297944411 | 0.003578065 | 0.035116467 | 1 |
| Obesity | monocyte | *NDST2* | -9.810567682 | 0.008116771 | 0.040894308 | 1 |
| Obesity | monocyte | *CLEC4D* | 0.298490157 | 0.005904991 | 0.036288402 | 1 |
| Obesity | monocyte | *ZSCAN21* | -2.079834197 | 0.000186706 | 0.011614101 | 0.905899913 |
| Obesity | monocyte | *RIMKLB* | -2.368998134 | 0.004169182 | 0.035116467 | 1 |
| Obesity | monocyte | *SEC11C* | 2.476836963 | 0.002569963 | 0.035116467 | 1 |
| Obesity | monocyte | *CENPV* | -1.778367308 | 0.010047772 | 0.041669249 | 1 |
| Obesity | monocyte | *CHRFAM7A* | 2.730403341 | 0.000567819 | 0.021082768 | 1 |
| Obesity | monocyte | *SPDYE6* | 2.84479766 | 0.000120149 | 0.008700951 | 0.582963696 |
| Obesity | monocyte | *ACSM1* | -1.368973441 | 0.013559307 | 0.04433272 | 1 |
| Obesity | monocyte | *FAM96A* | 3.2596069 | 0.007140959 | 0.039642943 | 1 |
| Obesity | monocyte | *STX3* | -2.421380607 | 0.01672238 | 0.046981463 | 1 |
| Obesity | monocyte | *DIS3L* | -0.90514643 | 0.012560224 | 0.04319079 | 1 |
| Obesity | monocyte | *MAP1A* | -2.15335046 | 0.004788365 | 0.035116467 | 1 |
| Obesity | monocyte | *RCCD1* | -1.451945527 | 0.013107115 | 0.044028405 | 1 |
| Obesity | monocyte | *SAMD14* | 2.144563143 | 0.001027259 | 0.027556163 | 1 |
| Obesity | monocyte | *URM1* | -1.860645234 | 0.004506576 | 0.035116467 | 1 |
| Obesity | monocyte | *ENDOG* | 2.847980541 | 0.002939272 | 0.035116467 | 1 |
| Obesity | monocyte | *ISLR2* | 2.286820701 | 0.000336473 | 0.016325648 | 1 |
| Obesity | monocyte | *TBC1D2B* | -1.007910563 | 0.007953824 | 0.040894308 | 1 |
| Obesity | monocyte | *LOXHD1* | -0.689701218 | 0.016248342 | 0.046538934 | 1 |
| Obesity | monocyte | *KATNAL2* | -0.784742334 | 0.013345045 | 0.044157522 | 1 |
| Obesity | monocyte | *POP5* | 2.696720384 | 0.015518372 | 0.045468081 | 1 |
| Obesity | monocyte | *MYO5B* | -0.793081225 | 0.007987833 | 0.040894308 | 1 |
| Obesity | monocyte | *ART5* | 1.536210728 | 0.012667217 | 0.043399276 | 1 |
| Obesity | monocyte | *ZNF23* | -2.547963791 | 0.012135361 | 0.043083715 | 1 |
| Obesity | monocyte | *VKORC1* | 2.183647416 | 0.002874962 | 0.035116467 | 1 |
| Obesity | monocyte | *SMG8* | -4.167385372 | 6.32E-05 | 0.005108585 | 0.306515108 |
| Obesity | monocyte | *RAB8A* | 1.396047128 | 0.003496034 | 0.035116467 | 1 |
| Obesity | monocyte | *JSRP1* | 1.354416793 | 0.011360877 | 0.042352977 | 1 |
| Obesity | monocyte | *SPATA33* | -1.442844955 | 0.015242957 | 0.045130917 | 1 |
| Obesity | monocyte | *PROCA1* | 1.937652132 | 0.001547353 | 0.03218121 | 1 |
| Obesity | monocyte | *TP53I13* | -0.832099349 | 0.014136832 | 0.044792561 | 1 |

| | | | | | | |
|---|---|---|---|---|---|---|
| Obesity | monocyte | *SERTAD3* | -1.772846087 | 0.012327194 | 0.043115778 | 1 |
| Obesity | monocyte | *TMC4* | 2.492499228 | 0.001716925 | 0.03218121 | 1 |
| Obesity | monocyte | *TTYH1* | -1.3557284 | 0.009297959 | 0.041462933 | 1 |
| Obesity | monocyte | *CDC42EP5* | -2.31164916 | 0.016312132 | 0.046556743 | 1 |
| Obesity | monocyte | *KIR3DL1* | 5.512939758 | 0.011850284 | 0.042787935 | 1 |
| Obesity | monocyte | *SRR* | 1.402590906 | 0.013681125 | 0.044440213 | 1 |
| Obesity | monocyte | *C19orf48* | 1.382188018 | 0.01806629 | 0.048795606 | 1 |
| Obesity | monocyte | *KLK5* | -1.836622792 | 0.004578852 | 0.035116467 | 1 |
| Obesity | monocyte | *KLK6* | 1.660760188 | 0.004883809 | 0.035116467 | 1 |
| Obesity | monocyte | *SPRYD3* | -1.345771947 | 0.005388924 | 0.035614643 | 1 |
| Obesity | monocyte | *SOAT2* | -2.457212687 | 0.004961602 | 0.035116467 | 1 |
| Obesity | monocyte | *ZNF558* | -0.895545796 | 0.015667991 | 0.045669584 | 1 |
| Obesity | monocyte | *NDUFV1* | -1.120761134 | 0.016688842 | 0.046940995 | 1 |
| Obesity | monocyte | *ZNF232* | -1.160051595 | 0.00863319 | 0.041462933 | 1 |
| Obesity | monocyte | *MIS12* | -1.159743131 | 0.014336324 | 0.044792561 | 1 |
| Obesity | monocyte | *MGAT5B* | -2.664475259 | 0.015398783 | 0.045336709 | 1 |
| Obesity | monocyte | *TMEM99* | -1.194517815 | 0.017763168 | 0.048528654 | 1 |
| Obesity | monocyte | *ZNF598* | 6.243242362 | 0.00091048 | 0.026139948 | 1 |
| Obesity | monocyte | *MLST8* | 0.954731048 | 0.004200139 | 0.035116467 | 1 |
| Obesity | monocyte | *BSCL2* | 0.810600963 | 0.008407208 | 0.041379919 | 1 |
| Obesity | monocyte | *ATG16L2* | 3.675658947 | 0.00799075 | 0.040894308 | 1 |
| Obesity | monocyte | *SCARA5* | -0.783378947 | 0.009722454 | 0.041544165 | 1 |
| Obesity | monocyte | *KIAA1586* | -1.495924371 | 0.001736724 | 0.032409936 | 1 |
| Obesity | monocyte | *FAM83B* | -12.20290893 | 0.003278244 | 0.035116467 | 1 |
| Obesity | monocyte | *POLR2J3* | -0.916265641 | 0.010237853 | 0.041771331 | 1 |
| Obesity | monocyte | *COA6* | 2.893147301 | 0.002600749 | 0.035116467 | 1 |
| Obesity | monocyte | *PXK* | -1.484430999 | 0.016073273 | 0.046231201 | 1 |
| Obesity | monocyte | *FAM107A* | -1.464277354 | 0.016526033 | 0.046845773 | 1 |
| Obesity | monocyte | *SCN11A* | -1.025171522 | 0.016342754 | 0.046589332 | 1 |
| Obesity | monocyte | *DTYMK* | -1.716568457 | 0.011520832 | 0.042465454 | 1 |
| Obesity | monocyte | *KLHL30* | -8.595104587 | 0.012392023 | 0.043115778 | 1 |
| Obesity | monocyte | *CCDC110* | -2.06813627 | 0.009133089 | 0.041462933 | 1 |
| Obesity | monocyte | *FNTA* | 5.950485455 | 8.04E-06 | 0.001114028 | 0.03899097 |
| Obesity | monocyte | *TMUB2* | 2.532020993 | 0.008760713 | 0.041462933 | 1 |
| Obesity | monocyte | *ZSWIM1* | 1.203289536 | 0.002558477 | 0.035116467 | 1 |
| Obesity | monocyte | *NBPF9* | -1.537588554 | 0.011022138 | 0.042237039 | 1 |
| Obesity | monocyte | *TCTN2* | -0.923237205 | 0.002204388 | 0.035072295 | 1 |
| Obesity | monocyte | *PPIP5K1* | -1.609766971 | 0.005205192 | 0.035351132 | 1 |
| Obesity | monocyte | *ZNF507* | -1.981718787 | 0.017140227 | 0.047644704 | 1 |
| Obesity | monocyte | *ATOH8* | 2.428185577 | 0.010438993 | 0.041803129 | 1 |
| Obesity | monocyte | *LGALS9* | 0.964590802 | 0.001869131 | 0.033341993 | 1 |
| Obesity | monocyte | *SIGLEC7* | 0.658078619 | 0.018254705 | 0.04900239 | 1 |
| Obesity | monocyte | *MFSD7* | 2.161022852 | 0.003370662 | 0.035116467 | 1 |
| Obesity | monocyte | *KLK7* | -1.499402365 | 0.017346033 | 0.047847044 | 1 |
| Obesity | monocyte | *UPF3A* | -1.481669899 | 0.018057915 | 0.048795606 | 1 |
| Obesity | monocyte | *CPT1C* | -1.496043145 | 0.012870354 | 0.043822428 | 1 |
| Obesity | monocyte | *PCSK9* | 1.42333282 | 0.006051677 | 0.036657602 | 1 |
| Obesity | monocyte | *RGS14* | 1.029491482 | 0.007213229 | 0.039861716 | 1 |
| Obesity | monocyte | *RAB24* | 5.151436095 | 0.007089283 | 0.039446333 | 1 |
| Obesity | monocyte | *NPIPB3* | -2.897667949 | 0.001122862 | 0.028083118 | 1 |

| | | | | | | |
|---|---|---|---|---|---|---|
| Obesity | monocyte | *SH3TC2* | -1.091374044 | 0.012247195 | 0.043115778 | 1 |
| Obesity | monocyte | *RNASE6* | 2.01742219 | 0.001263498 | 0.029332492 | 1 |
| Obesity | monocyte | *CD52* | 0.452988515 | 0.003420922 | 0.035116467 | 1 |
| Obesity | monocyte | *CCDC8* | -1.583909443 | 0.010283004 | 0.041771331 | 1 |
| Obesity | monocyte | *CLIC3* | -1.384631494 | 0.010042552 | 0.041669249 | 1 |
| Obesity | monocyte | *INO80E* | -4.56006095 | 6.14E-11 | 4.96E-08 | 2.98E-07 |
| Obesity | monocyte | *NFU1* | -0.939584254 | 0.018181117 | 0.048953818 | 1 |
| Obesity | monocyte | *LRRC45* | -1.793311359 | 0.0025405 | 0.035116467 | 1 |
| Obesity | monocyte | *CHRNA5* | 1.544937466 | 0.004597694 | 0.035116467 | 1 |
| Obesity | monocyte | *CSGALNACT2* | 3.016582376 | 0.00297782 | 0.035116467 | 1 |
| Obesity | monocyte | *CTNND2* | -1.241471924 | 0.001514099 | 0.03218121 | 1 |
| Obesity | monocyte | *CALML6* | -25.31005038 | 0.001813938 | 0.032842449 | 1 |
| Obesity | monocyte | *MUC3A* | 1.947828886 | 0.000684499 | 0.022981804 | 1 |
| Obesity | monocyte | *MYO7B* | 4.111739676 | 3.67E-05 | 0.003421783 | 0.177932733 |
| Obesity | monocyte | *NIPA1* | -1.361421587 | 0.019075196 | 0.049920631 | 1 |
| Obesity | monocyte | *RNF150* | -0.717704496 | 0.008415648 | 0.041379919 | 1 |
| Obesity | monocyte | *USP38* | -1.720470861 | 0.005689767 | 0.035810934 | 1 |
| Obesity | monocyte | *ANKK1* | -3.493225817 | 0.007767362 | 0.04060923 | 1 |
| Obesity | monocyte | *ELP5* | -1.386598334 | 0.010831364 | 0.041961024 | 1 |
| Obesity | monocyte | *METTL7B* | -4.061331324 | 0.014242678 | 0.044792561 | 1 |
| Obesity | monocyte | *HSD17B13* | -2.392749376 | 0.011679573 | 0.042490474 | 1 |
| Obesity | monocyte | *ELOVL6* | -1.068131678 | 0.015178234 | 0.045042687 | 1 |
| Obesity | monocyte | *SMAGP* | -1.667603794 | 0.001714715 | 0.03218121 | 1 |
| Obesity | monocyte | *GTSF1* | -4.243148662 | 0.006525457 | 0.038054711 | 1 |
| Obesity | monocyte | *TTLL6* | -16.43547787 | 1.21E-12 | 1.80E-09 | 5.89E-09 |
| Obesity | monocyte | *TRIAP1* | 1.717464685 | 0.003069721 | 0.035116467 | 1 |
| Obesity | monocyte | *TSEN34* | -1.207268531 | 0.011573982 | 0.042465454 | 1 |
| Obesity | monocyte | *PAQR8* | -1.853830869 | 0.002650247 | 0.035116467 | 1 |
| Obesity | monocyte | *NUDT6* | -1.646377023 | 0.008580121 | 0.041462933 | 1 |
| Obesity | monocyte | *ZNF415* | -2.461174609 | 0.018935626 | 0.049770129 | 1 |
| Obesity | monocyte | *S1PR1* | -1.018390349 | 0.014957582 | 0.044986158 | 1 |
| Obesity | monocyte | *PYGO1* | -1.1253799 | 0.017919811 | 0.048768447 | 1 |
| Obesity | monocyte | *FPR1* | 1.731689252 | 0.003906155 | 0.035116467 | 1 |
| Obesity | monocyte | *FEZ2* | -1.071460751 | 0.017224784 | 0.047729671 | 1 |
| Obesity | monocyte | *SOX7* | -1.852512295 | 0.014039693 | 0.044610733 | 1 |
| Obesity | monocyte | *ATP6V0E2* | -2.052831624 | 0.004843295 | 0.035116467 | 1 |
| Obesity | monocyte | *MORN4* | -0.893027027 | 0.006034172 | 0.036597252 | 1 |
| Obesity | monocyte | *SCAND1* | -2.108252568 | 0.008081696 | 0.040894308 | 1 |
| Obesity | monocyte | *LURAP1* | -1.604191058 | 0.000367482 | 0.016908522 | 1 |
| Obesity | monocyte | *HOPX* | 1.686381361 | 0.008081752 | 0.040894308 | 1 |
| Obesity | monocyte | *NMUR1* | 4.695032714 | 0.002141941 | 0.034874831 | 1 |
| Obesity | monocyte | *PIK3CD* | 1.949720081 | 0.000636304 | 0.022052485 | 1 |
| Obesity | monocyte | *RGS19* | 0.607973951 | 0.00044343 | 0.018547623 | 1 |
| Obesity | monocyte | *ANO5* | -0.894924484 | 0.011620232 | 0.042465454 | 1 |
| Obesity | monocyte | *VAT1L* | -1.241854067 | 0.008118325 | 0.040894308 | 1 |
| Obesity | monocyte | *LRRC34* | -0.875796935 | 0.004258366 | 0.035116467 | 1 |
| Obesity | monocyte | *PAH* | -7.071542336 | 0.002474765 | 0.035116467 | 1 |
| Obesity | monocyte | *SPATA5L1* | 2.176471908 | 0.002102324 | 0.034345034 | 1 |
| Obesity | monocyte | *RHNO1* | -2.753858811 | 0.007488485 | 0.04034316 | 1 |
| Obesity | monocyte | *METTL18* | 0.992488244 | 0.006843913 | 0.038883685 | 1 |

| | | | | | | |
|---|---|---|---|---|---|---|
| Obesity | monocyte | *PWWP2B* | 2.570124899 | 0.001880821 | 0.033427634 | 1 |
| Obesity | monocyte | *EXOSC10* | -1.236235487 | 0.013372489 | 0.044157522 | 1 |
| Obesity | monocyte | *FBXW10* | -2.838787803 | 0.006140074 | 0.036825267 | 1 |
| Obesity | monocyte | *ANKRD20A4* | 0.755829809 | 0.004727667 | 0.035116467 | 1 |
| Obesity | monocyte | *SERF1A* | 0.712842604 | 0.005307941 | 0.035561878 | 1 |
| Obesity | monocyte | *NME6* | 1.830336062 | 0.00302631 | 0.035116467 | 1 |
| Obesity | monocyte | *CD8B* | 0.866983749 | 0.003367356 | 0.035116467 | 1 |
| Obesity | monocyte | *TEFM* | -1.052121759 | 0.015826169 | 0.045769688 | 1 |
| Obesity | monocyte | *MBOAT1* | 2.66683836 | 0.004439434 | 0.035116467 | 1 |
| Obesity | monocyte | *CXCR6* | 1.612212845 | 0.00600291 | 0.036477419 | 1 |
| Obesity | monocyte | *PAIP1* | -1.384815439 | 0.01628632 | 0.046549891 | 1 |
| Obesity | monocyte | *C1QTNF4* | -3.645772515 | 2.62E-10 | 1.82E-07 | 1.27E-06 |
| Obesity | monocyte | *TP53RK* | -1.474757478 | 0.011573556 | 0.042465454 | 1 |
| Obesity | monocyte | *CLEC12A* | 0.181342954 | 0.003118726 | 0.035116467 | 1 |
| Obesity | monocyte | *FIBP* | 1.995142095 | 0.000886378 | 0.025883824 | 1 |
| Obesity | monocyte | *CTSW* | -0.765660686 | 8.69E-05 | 0.006691353 | 0.42155524 |
| Obesity | monocyte | *SNTG2* | -0.701810182 | 0.007594638 | 0.040367875 | 1 |
| Obesity | monocyte | *ZFAND4* | -1.441168386 | 0.003247353 | 0.035116467 | 1 |
| Obesity | monocyte | *SLFN11* | -1.011419183 | 0.013870249 | 0.044539684 | 1 |
| Obesity | monocyte | *FAM71D* | -1.457922766 | 0.013703651 | 0.044449752 | 1 |
| Obesity | monocyte | *FUT10* | -1.354720158 | 0.000177901 | 0.011357562 | 0.863174685 |
| Obesity | monocyte | *ZNF596* | -1.774342727 | 0.018063301 | 0.048795606 | 1 |
| Obesity | monocyte | *CFL1* | 2.338350511 | 0.006564661 | 0.038158325 | 1 |
| Obesity | monocyte | *RAB43* | 7.891595022 | 0.0057217 | 0.035810934 | 1 |
| Obesity | monocyte | *SNX32* | 1.361840348 | 1.72E-05 | 0.002038902 | 0.083594995 |
| Obesity | monocyte | *ZNF621* | -1.421388708 | 0.01780667 | 0.048599513 | 1 |
| Obesity | monocyte | *GXYLT2* | -0.676214551 | 0.013925922 | 0.044540919 | 1 |
| Obesity | monocyte | *TRMT112* | -2.106863841 | 0.010383108 | 0.041803129 | 1 |
| Obesity | monocyte | *RHOD* | -1.363843094 | 0.018758608 | 0.049532965 | 1 |
| Obesity | monocyte | *PARP15* | 1.359759873 | 0.018692661 | 0.049516993 | 1 |
| Obesity | monocyte | *VANGL1* | -1.166320383 | 0.016867362 | 0.047233354 | 1 |
| Obesity | monocyte | *SLC2A14* | 1.947042286 | 0.007903048 | 0.040894308 | 1 |
| Obesity | monocyte | *CCDC36* | -0.944802832 | 0.007730208 | 0.040591959 | 1 |
| Obesity | monocyte | *PPP1R14B* | -1.486820018 | 0.00560151 | 0.035810934 | 1 |
| Obesity | monocyte | *ZNF417* | -2.807003077 | 0.007505635 | 0.04034316 | 1 |
| Obesity | monocyte | *TNFRSF10D* | 1.427578025 | 0.002275453 | 0.035072295 | 1 |
| Obesity | monocyte | *MST1* | -1.833144117 | 0.000820049 | 0.025087947 | 1 |
| Obesity | monocyte | *GMPPB* | 1.064321685 | 0.003986749 | 0.035116467 | 1 |
| Obesity | monocyte | *SULT1B1* | 2.224332192 | 0.008472767 | 0.041379919 | 1 |
| Obesity | monocyte | *HSPB7* | -1.227133618 | 0.011613729 | 0.042465454 | 1 |
| Obesity | monocyte | *C11orf80* | -1.031385122 | 0.006109525 | 0.036748833 | 1 |
| Obesity | monocyte | *TUBB8* | 0.492588657 | 0.002909769 | 0.035116467 | 1 |
| Obesity | monocyte | *GPR160* | -1.382629612 | 0.010692399 | 0.041946198 | 1 |
| Obesity | monocyte | *XXYLT1* | 2.934033132 | 0.005084788 | 0.03528148 | 1 |
| Obesity | monocyte | *SLC25A30* | 2.054925393 | 0.002083211 | 0.034263531 | 1 |
| Obesity | monocyte | *GLIS1* | -0.966965013 | 0.009125596 | 0.041462933 | 1 |
| Obesity | monocyte | *SNAPC5* | -1.546511544 | 0.006209341 | 0.037019516 | 1 |
| Obesity | monocyte | *ZNF497* | -8.058965603 | 0.00641904 | 0.03766044 | 1 |
| Obesity | monocyte | *TRAM1L1* | -3.257252384 | 0.017534669 | 0.048121163 | 1 |
| Obesity | monocyte | *FGFBP3* | 9.72698391 | 0.006288903 | 0.037211899 | 1 |

| | | | | | | |
|---|---|---|---|---|---|---|
| Obesity | monocyte | *NLRP6* | -15.2854271 | 0.00625096 | 0.037123203 | 1 |
| Obesity | monocyte | *PTDSS2* | 3.892683846 | 0.004829212 | 0.035116467 | 1 |
| Obesity | monocyte | *KCTD13* | 4.980095097 | 1.85E-07 | 4.28E-05 | 0.000898216 |
| Obesity | monocyte | *SLC22A1* | -3.214465215 | 0.018151598 | 0.048953818 | 1 |
| Obesity | monocyte | *CHST2* | 2.222890799 | 0.003564633 | 0.035116467 | 1 |
| Obesity | monocyte | *TVP23C* | 5.768509725 | 0.002806511 | 0.035116467 | 1 |
| Obesity | monocyte | *PACS1* | -1.756088428 | 0.015660276 | 0.045669584 | 1 |
| Obesity | monocyte | *ZNF519* | -0.945795087 | 0.006636931 | 0.038381871 | 1 |
| Obesity | monocyte | *CHRNA7* | 1.730938423 | 0.007543708 | 0.04034316 | 1 |
| Obesity | monocyte | *TMEM9B* | 4.56732835 | 0.007408273 | 0.040251891 | 1 |
| Obesity | monocyte | *SCUBE2* | -1.760264246 | 0.010171002 | 0.041771331 | 1 |
| Obesity | monocyte | *ZNF25* | -2.949063408 | 0.005574731 | 0.035810934 | 1 |
| Obesity | monocyte | *CLTB* | 3.46457769 | 0.004206306 | 0.035116467 | 1 |
| Obesity | monocyte | *LRRC25* | 0.371455407 | 0.008449152 | 0.041379919 | 1 |
| Obesity | monocyte | *KCNE3* | 0.988389951 | 0.013727701 | 0.044493524 | 1 |
| Obesity | monocyte | *MRPL48* | -1.811655248 | 0.018451074 | 0.04918073 | 1 |
| Obesity | monocyte | *TMEM70* | 5.282972555 | 0.015643646 | 0.045669584 | 1 |
| Obesity | monocyte | *RPS6KB2* | 6.128017906 | 0.000110226 | 0.008103274 | 0.534816099 |
| Obesity | monocyte | *GPR156* | -1.982897335 | 0.004393713 | 0.035116467 | 1 |
| Obesity | monocyte | *PRIMA1* | -1.027041458 | 0.008770854 | 0.041462933 | 1 |
| Obesity | monocyte | *DOK7* | 6.166139749 | 0.003161542 | 0.035116467 | 1 |
| Obesity | monocyte | *ORAI3* | 9.114712385 | 1.48E-12 | 1.80E-09 | 7.19E-09 |
| Obesity | monocyte | *JAKMIP2* | 0.478256463 | 0.009692466 | 0.041470763 | 1 |
| Obesity | monocyte | *CCDC57* | -1.995463312 | 0.00101965 | 0.027556163 | 1 |
| Obesity | monocyte | *CIDEA* | -0.593568258 | 0.008477255 | 0.041379919 | 1 |
| Obesity | monocyte | *ZNF135* | -1.591515382 | 0.015047581 | 0.044986158 | 1 |
| Obesity | monocyte | *CDC26* | -1.445734606 | 0.014331714 | 0.044792561 | 1 |
| Obesity | monocyte | *SPRYD4* | -3.254570935 | 0.000140351 | 0.009591281 | 0.680980975 |
| Obesity | monocyte | *WDR25* | 3.781270359 | 0.01031219 | 0.041771331 | 1 |
| Obesity | monocyte | *PLA2G16* | -0.758614806 | 0.006995284 | 0.039147772 | 1 |
| Obesity | monocyte | *LRRC37A* | 1.41617172 | 0.008976009 | 0.041462933 | 1 |
| Obesity | monocyte | *BDNF* | -1.298878766 | 0.012956799 | 0.043967382 | 1 |
| Obesity | monocyte | *BOK* | -0.797009275 | 0.009360035 | 0.041462933 | 1 |
| Obesity | monocyte | *VSIG10* | -0.727223303 | 0.014757189 | 0.044883966 | 1 |
| Obesity | monocyte | *C8G* | -25.52708245 | 0.009582922 | 0.041462933 | 1 |
| Obesity | monocyte | *MUC20* | -1.759686448 | 0.006682537 | 0.038462243 | 1 |
| Obesity | monocyte | *SHMT1* | -0.940347469 | 0.013171037 | 0.044032814 | 1 |
| Obesity | monocyte | *DPP7* | 2.609878874 | 0.00319942 | 0.035116467 | 1 |
| Obesity | monocyte | *C19orf18* | -1.240465315 | 0.013634391 | 0.04438134 | 1 |
| Obesity | monocyte | *WDR73* | -0.927611925 | 0.01338741 | 0.044157522 | 1 |
| Obesity | monocyte | *DSCAML1* | -0.725810161 | 0.010547633 | 0.041845661 | 1 |
| Obesity | monocyte | *MIEF2* | -1.927967215 | 0.011413775 | 0.042352977 | 1 |
| Obesity | monocyte | *NAP1L5* | 1.062141477 | 0.003231619 | 0.035116467 | 1 |
| Obesity | monocyte | *RBM44* | 3.20902615 | 0.005516755 | 0.035757287 | 1 |
| Obesity | monocyte | *SLC25A22* | -2.541634964 | 0.004796263 | 0.035116467 | 1 |
| Obesity | monocyte | *SAMD12* | -0.910869988 | 0.012279322 | 0.043115778 | 1 |
| Obesity | monocyte | *IL17RA* | 2.137738807 | 0.010431948 | 0.041803129 | 1 |
| Obesity | monocyte | *MBOAT4* | 1.121166459 | 0.003259306 | 0.035116467 | 1 |
| Obesity | monocyte | *THAP5* | -1.252791683 | 0.012128978 | 0.043083715 | 1 |
| Obesity | monocyte | *NAALADL2* | -0.780332262 | 0.01210182 | 0.043083715 | 1 |

| | | | | | | |
|---|---|---|---|---|---|---|
| Obesity | monocyte | *SLC35G5* | 3.752305217 | 0.004362706 | 0.035116467 | 1 |
| Obesity | monocyte | *FAM87B* | 7.906402883 | 0.003448619 | 0.035116467 | 1 |
| Obesity | monocyte | *CENPBD1* | 0.762009184 | 0.004510384 | 0.035116467 | 1 |
| Obesity | monocyte | *ODF3B* | 0.761170047 | 0.00260229 | 0.035116467 | 1 |
| Obesity | monocyte | *ADAMTSL1* | -0.76795028 | 0.016137017 | 0.046356902 | 1 |
| Obesity | monocyte | *DALRD3* | -1.327804462 | 0.009212317 | 0.041462933 | 1 |
| Obesity | monocyte | *ZNF114* | -3.239275128 | 0.004191901 | 0.035116467 | 1 |
| Obesity | monocyte | *SPINK6* | -5.962077462 | 0.00196912 | 0.03400061 | 1 |
| Obesity | monocyte | *SH2B1* | -1.017417217 | 0.014617058 | 0.044883966 | 1 |
| Obesity | monocyte | *VN1R1* | -4.416780971 | 0.001201151 | 0.028851404 | 1 |
| Obesity | monocyte | *SH2D4B* | -3.774608469 | 0.010653923 | 0.041890466 | 1 |
| Obesity | monocyte | *RNF212* | -0.923351365 | 0.010531994 | 0.041845661 | 1 |
| Obesity | monocyte | *PRSS36* | -2.0728948 | 0.00579379 | 0.036086613 | 1 |
| Obesity | monocyte | *GLDC* | -1.758143569 | 0.014408821 | 0.044843874 | 1 |
| Obesity | monocyte | *P4HTM* | -1.588741006 | 0.012190634 | 0.043115778 | 1 |
| Obesity | monocyte | *CA8* | -1.42048183 | 0.010538196 | 0.041845661 | 1 |
| Obesity | monocyte | *RPP25* | -3.489725955 | 5.64E-06 | 0.000804507 | 0.027353229 |
| Obesity | monocyte | *TRIM73* | -9.191784388 | 3.97E-05 | 0.003504422 | 0.192743203 |
| Obesity | monocyte | *EFCAB13* | -1.485132596 | 0.011506406 | 0.042465454 | 1 |
| Obesity | monocyte | *ZNF852* | -4.974639543 | 7.65E-05 | 0.006081176 | 0.370951727 |
| Obesity | monocyte | *C17orf62* | 4.076797359 | 0.002256572 | 0.035072295 | 1 |
| Obesity | monocyte | *TUFM* | 4.787562933 | 0.000169664 | 0.011119597 | 0.82320822 |
| Obesity | monocyte | *C3orf38* | 6.415190328 | 3.57E-08 | 1.15E-05 | 0.00017309 |
| Obesity | monocyte | *CCDC89* | -3.562760887 | 0.009894432 | 0.041637281 | 1 |
| Obesity | monocyte | *ZNF664* | -1.487919568 | 0.014446942 | 0.044883966 | 1 |
| Obesity | monocyte | *NSUN7* | -1.317573436 | 0.018509718 | 0.049291522 | 1 |
| Obesity | monocyte | *LINC00174* | -1.106627674 | 0.002454845 | 0.035116467 | 1 |
| Obesity | monocyte | *LBX2* | 1.9363777 | 0.002006741 | 0.034173375 | 1 |
| Obesity | monocyte | *FCER1A* | -1.75770419 | 0.010977033 | 0.042169886 | 1 |
| Obesity | monocyte | *RPRML* | 12.19041103 | 0.001104677 | 0.028083118 | 1 |
| Obesity | monocyte | *AKAP5* | -1.434086098 | 0.011058183 | 0.042269392 | 1 |
| Obesity | monocyte | *GIPC3* | 24.05910019 | 0.002303015 | 0.035072295 | 1 |
| Obesity | monocyte | *C1orf194* | 0.649725736 | 0.006898331 | 0.03896597 | 1 |
| Obesity | monocyte | *SSC5D* | -1.055041597 | 0.013545653 | 0.04433272 | 1 |
| Obesity | monocyte | *SEPT1* | -1.052486437 | 0.008811102 | 0.041462933 | 1 |
| Obesity | monocyte | *CSNK1A1L* | 1.904681397 | 0.004810638 | 0.035116467 | 1 |
| Obesity | monocyte | *HCLS1* | 0.949686032 | 0.006578201 | 0.038158325 | 1 |
| Obesity | monocyte | *MTURN* | -1.161336966 | 0.014774666 | 0.044883966 | 1 |
| Obesity | monocyte | *GLIPR1L2* | -1.263724043 | 0.007966333 | 0.040894308 | 1 |
| Obesity | monocyte | *DEFB119* | -16.60541301 | 0.014242623 | 0.044792561 | 1 |
| Obesity | monocyte | *SLC47A2* | -1.235125158 | 0.015056353 | 0.044986158 | 1 |
| Obesity | monocyte | *BHLHE22* | 1.600495396 | 0.007782874 | 0.04060923 | 1 |
| Obesity | monocyte | *ZNF443* | 2.260128512 | 0.007258164 | 0.039882912 | 1 |
| Obesity | monocyte | *KCTD2* | 1.932785333 | 0.004240415 | 0.035116467 | 1 |
| Obesity | monocyte | *D2HGDH* | -1.182271812 | 0.010649982 | 0.041890466 | 1 |
| Obesity | monocyte | *NQO1* | -2.774968015 | 0.002263781 | 0.035072295 | 1 |
| Obesity | monocyte | *SLC25A41* | 2.588988318 | 0.003439454 | 0.035116467 | 1 |
| Obesity | monocyte | *TMEM136* | -1.80098954 | 0.012593121 | 0.043212039 | 1 |
| Obesity | monocyte | *DDX60L* | 2.20003753 | 0.002634924 | 0.035116467 | 1 |
| Obesity | monocyte | *TMEM45A* | -0.869135448 | 0.008321813 | 0.041322595 | 1 |

| | | | | | | |
|---|---|---|---|---|---|---|
| Obesity | monocyte | *SGSH* | 5.334978361 | 0.003913519 | 0.035116467 | 1 |
| Obesity | monocyte | *AMIGO1* | 1.618799466 | 0.000686802 | 0.022981804 | 1 |
| Obesity | monocyte | *TMEM252* | -42.54982662 | 0.0106203 | 0.041860027 | 1 |
| Obesity | monocyte | *ODF3L2* | -5.61908168 | 0.001122853 | 0.028083118 | 1 |
| Obesity | monocyte | *CLDN7* | -2.009842237 | 0.00030424 | 0.015460484 | 1 |
| Obesity | monocyte | *ZNF329* | -1.244079109 | 0.015367139 | 0.045316053 | 1 |
| Obesity | monocyte | *ADO* | 1.409508372 | 0.003255592 | 0.035116467 | 1 |
| Obesity | monocyte | *WRB* | -1.700064693 | 0.001920674 | 0.033521987 | 1 |
| Obesity | monocyte | *ZNF708* | -1.396564233 | 0.012556613 | 0.04319079 | 1 |
| Obesity | monocyte | *MRPL41* | -1.27902924 | 0.012404683 | 0.043115778 | 1 |
| Obesity | monocyte | *HHIPL1* | -0.979433049 | 0.014918656 | 0.044986158 | 1 |
| Obesity | monocyte | *FAM153B* | -0.874355444 | 0.011298523 | 0.042332379 | 1 |
| Obesity | monocyte | *SYNM* | -1.250366226 | 0.009586472 | 0.041462933 | 1 |
| Obesity | monocyte | *FIGN* | -0.519904273 | 0.010043231 | 0.041669249 | 1 |
| Obesity | monocyte | *CLN8* | 2.565492223 | 0.002747702 | 0.035116467 | 1 |
| Obesity | monocyte | *NXPH4* | 9.098579473 | 0.003927646 | 0.035116467 | 1 |
| Obesity | monocyte | *NPLOC4* | -1.492542045 | 0.014171071 | 0.044792561 | 1 |
| Obesity | monocyte | *CAPN12* | -2.809347555 | 0.002669343 | 0.035116467 | 1 |
| Obesity | monocyte | *KPNA2* | 0.76862933 | 0.004222534 | 0.035116467 | 1 |
| Obesity | monocyte | *CAV3* | -1.629547986 | 0.001024628 | 0.027556163 | 1 |
| Obesity | monocyte | *CLEC4G* | -6.00054994 | 0.002081641 | 0.034263531 | 1 |
| Obesity | monocyte | *PPP1R27* | -21.76665942 | 2.10E-05 | 0.002239829 | 0.101723238 |
| Obesity | monocyte | *BRICD5* | 1.000502906 | 0.004377083 | 0.035116467 | 1 |
| Obesity | monocyte | *GRID1* | -3.8217933 | 0.000322251 | 0.015954693 | 1 |
| Obesity | monocyte | *PLCXD3* | -0.997158027 | 0.014753347 | 0.044883966 | 1 |
| Obesity | monocyte | *VMO1* | 1.34785165 | 0.001706815 | 0.03218121 | 1 |
| Obesity | monocyte | *ODF3L1* | -6.445616044 | 0.008967816 | 0.041462933 | 1 |
| Obesity | monocyte | *HMGN4* | -1.566314096 | 0.009629733 | 0.041462933 | 1 |
| Obesity | monocyte | *ABAT* | 3.481470259 | 0.011602025 | 0.042465454 | 1 |
| Obesity | monocyte | *ARHGEF37* | -1.319693469 | 0.015056847 | 0.044986158 | 1 |
| Obesity | monocyte | *FANCF* | 5.687720217 | 0.002584503 | 0.035116467 | 1 |
| Obesity | monocyte | *RUVBL2* | -1.509477633 | 0.017205257 | 0.047702803 | 1 |
| Obesity | monocyte | *PTTG1IP* | -1.723587517 | 0.017374638 | 0.047867689 | 1 |
| Obesity | monocyte | *PLGLB1* | 1.537828021 | 0.000130637 | 0.009054986 | 0.633849037 |
| Obesity | monocyte | *C19orf71* | 4.437060666 | 0.005545932 | 0.035785501 | 1 |
| Obesity | monocyte | *LRIT3* | -7.667022789 | 0.008110685 | 0.040894308 | 1 |
| Obesity | monocyte | *NPIPA1* | -1.254982777 | 0.008136211 | 0.040894308 | 1 |
| Obesity | monocyte | *GPR132* | 1.64272908 | 0.015075161 | 0.044986158 | 1 |
| Obesity | monocyte | *AC131971.1* | -5.766090621 | 0.015709133 | 0.045674648 | 1 |
| Obesity | monocyte | *SERHL2* | -1.269426439 | 0.011754083 | 0.042569734 | 1 |
| Obesity | monocyte | *MRPL54* | -1.144785069 | 0.005455026 | 0.035630311 | 1 |
| Obesity | monocyte | *HMCES* | -1.621441055 | 0.008848792 | 0.041462933 | 1 |
| Obesity | monocyte | *TRMT12* | 2.140608956 | 0.011111155 | 0.042279868 | 1 |
| Obesity | monocyte | *FIGLA* | 2.646328631 | 0.003829315 | 0.035116467 | 1 |
| Obesity | monocyte | *ACSM2A* | 13.80687503 | 0.007619067 | 0.040372837 | 1 |
| Obesity | monocyte | *ZNF703* | 1.191362366 | 0.001646672 | 0.03218121 | 1 |
| Obesity | monocyte | *NUDT14* | -1.809311761 | 0.004426743 | 0.035116467 | 1 |
| Obesity | monocyte | *FAM3B* | -0.81644251 | 0.005445664 | 0.035630311 | 1 |
| Obesity | monocyte | *TTC32* | -1.5673258 | 0.018243736 | 0.04900239 | 1 |
| Obesity | monocyte | *PP2D1* | -1.686747063 | 0.015065071 | 0.044986158 | 1 |

| | | | | | | |
|---|---|---|---|---|---|---|
| Obesity | monocyte | *DENND5A* | 3.079323373 | 0.013448876 | 0.044239385 | 1 |
| Obesity | monocyte | *NIPSNAP1* | -1.568020805 | 0.01079485 | 0.041961024 | 1 |
| Obesity | monocyte | *CRELD2* | -0.996796896 | 0.01912348 | 0.049966141 | 1 |
| Obesity | monocyte | *GOLGA6L4* | -2.072448503 | 0.005919402 | 0.036288402 | 1 |
| Obesity | monocyte | *OAF* | -1.682767484 | 0.000620713 | 0.021983207 | 1 |
| Obesity | monocyte | *ALDH1A3* | -0.953593134 | 0.005640859 | 0.035810934 | 1 |
| Obesity | monocyte | *PKP3* | -30.77011431 | 0.000262614 | 0.014002248 | 1 |
| Obesity | monocyte | *A3GALT2* | 1.959586132 | 0.000606352 | 0.021632489 | 1 |
| Obesity | monocyte | *TXNRD2* | -1.103831446 | 0.016277827 | 0.046549891 | 1 |
| Obesity | monocyte | *TMEM255B* | -1.092513822 | 0.010222951 | 0.041771331 | 1 |
| Obesity | monocyte | *ZFP1* | -1.757835025 | 0.013569403 | 0.044335853 | 1 |
| Obesity | monocyte | *DUSP8* | 2.453235227 | 0.003285337 | 0.035116467 | 1 |
| Obesity | monocyte | *SNN* | -1.511285238 | 0.009630817 | 0.041462933 | 1 |
| Obesity | monocyte | *RNLS* | -1.447917546 | 0.014541644 | 0.044883966 | 1 |
| Obesity | monocyte | *TCTE3* | -1.231014295 | 0.010605336 | 0.041845661 | 1 |
| Obesity | monocyte | *DRD1* | -24.81425491 | 0.007966712 | 0.040894308 | 1 |
| Obesity | monocyte | *BTBD6* | -1.14356834 | 0.005120609 | 0.03528148 | 1 |
| Obesity | monocyte | *NUTM2A* | -5.643809043 | 2.43E-06 | 0.000436218 | 0.011777889 |
| Obesity | monocyte | *WT1* | -0.625941625 | 0.004122827 | 0.035116467 | 1 |
| Obesity | monocyte | *NOC4L* | 1.749296338 | 0.00858733 | 0.041462933 | 1 |
| Obesity | monocyte | *SORCS2* | -1.465623975 | 0.006590411 | 0.038158325 | 1 |
| Obesity | monocyte | *TMEM106A* | -1.681558028 | 0.013378894 | 0.044157522 | 1 |
| Obesity | monocyte | *SEMA4B* | 3.114853565 | 0.004596699 | 0.035116467 | 1 |
| Obesity | monocyte | *SGCZ* | -0.652526455 | 0.016538914 | 0.046845773 | 1 |
| Obesity | monocyte | *C6orf120* | 2.550797216 | 0.011957661 | 0.042874352 | 1 |
| Obesity | monocyte | *GP1BA* | 1.601315676 | 3.90E-05 | 0.003504422 | 0.189208652 |
| Obesity | monocyte | *C14orf80* | 3.386153346 | 0.002479896 | 0.035116467 | 1 |
| Obesity | monocyte | *TNFAIP8L1* | -1.909394728 | 0.004401012 | 0.035116467 | 1 |
| Obesity | monocyte | *SH3BGR* | -1.373390372 | 0.010349563 | 0.041771331 | 1 |
| Obesity | monocyte | *SV2B* | -1.165105338 | 0.008048265 | 0.040894308 | 1 |
| Obesity | monocyte | *PSMD13* | -2.858428027 | 0.007238892 | 0.039882912 | 1 |
| Obesity | monocyte | *C6orf201* | 2.884482356 | 0.00820918 | 0.040894308 | 1 |
| Obesity | monocyte | *MYBL1* | 1.438808269 | 0.016445125 | 0.046771209 | 1 |
| Obesity | monocyte | *ADARB2* | 1.801822884 | 0.00182082 | 0.032842449 | 1 |
| Obesity | monocyte | *DMWD* | -1.598800549 | 0.013705035 | 0.044449752 | 1 |
| Obesity | monocyte | *ARL17A* | -0.878053362 | 0.017832727 | 0.048636533 | 1 |
| Obesity | monocyte | *GNB1L* | -1.77349088 | 0.008900136 | 0.041462933 | 1 |
| Obesity | monocyte | *EVI2B* | 0.965827056 | 0.00165821 | 0.03218121 | 1 |
| Obesity | monocyte | *LINC00839* | -3.106348862 | 0.012469852 | 0.043186096 | 1 |
| Obesity | monocyte | *CYP4Z1* | -0.992967611 | 0.00563132 | 0.035810934 | 1 |
| Obesity | monocyte | *FFAR4* | -0.962285771 | 0.00758767 | 0.040367875 | 1 |
| Obesity | monocyte | *ZNF555* | 4.065434282 | 0.001695212 | 0.03218121 | 1 |
| Obesity | monocyte | *GOLGA8R* | -4.814726343 | 0.018454249 | 0.04918073 | 1 |
| Obesity | monocyte | *GNG2* | 2.386369925 | 0.002895519 | 0.035116467 | 1 |
| Obesity | monocyte | *BTN3A2* | -2.442611979 | 2.26E-11 | 2.19E-08 | 1.10E-07 |
| Obesity | monocyte | *TMEM222* | -1.657162984 | 0.014282639 | 0.044792561 | 1 |
| Obesity | monocyte | *CEACAM19* | -1.461206402 | 0.008256731 | 0.041088879 | 1 |
| Obesity | monocyte | *MIR22HG* | -0.85278047 | 0.010356742 | 0.041771331 | 1 |
| Obesity | monocyte | *ZNF732* | 2.384400553 | 0.00334046 | 0.035116467 | 1 |
| Obesity | monocyte | *TRABD2A* | 2.356548576 | 0.001453427 | 0.031482273 | 1 |

| | | | | | | |
|---|---|---|---|---|---|---|
| Obesity | monocyte | *MAPT* | -0.830861976 | 0.009762203 | 0.041544165 | 1 |
| Obesity | monocyte | *C1QL4* | -12.64284743 | 0.008960739 | 0.041462933 | 1 |
| Obesity | monocyte | *RTN4RL2* | -2.716880077 | 0.003836101 | 0.035116467 | 1 |
| Obesity | monocyte | *TMEM232* | -0.66644986 | 0.01022765 | 0.041771331 | 1 |
| Obesity | monocyte | *EFCAB6* | -1.066625679 | 0.014565229 | 0.044883966 | 1 |
| Obesity | monocyte | *TMPRSS6* | -7.486454073 | 0.00855223 | 0.041462933 | 1 |
| Obesity | monocyte | *ANGPTL5* | -2.536810488 | 0.000773833 | 0.024540116 | 1 |
| Obesity | monocyte | *WDR86* | -0.748842439 | 0.005224476 | 0.035351132 | 1 |
| Obesity | monocyte | *LUZP2* | 5.330405033 | 0.003234002 | 0.035116467 | 1 |
| Obesity | monocyte | *GJA4* | -1.42505897 | 0.011699768 | 0.042490474 | 1 |
| Obesity | monocyte | *C17orf97* | 0.559322904 | 0.003082467 | 0.035116467 | 1 |
| Obesity | monocyte | *C2orf88* | -0.812552858 | 0.010249063 | 0.041771331 | 1 |
| Obesity | monocyte | *FANCM* | -1.605901826 | 0.011064088 | 0.042269392 | 1 |
| Obesity | monocyte | *FAM122A* | -1.798923816 | 0.013203854 | 0.044032814 | 1 |
| Obesity | monocyte | *OR7D2* | -7.138927593 | 0.00814513 | 0.040894308 | 1 |
| Obesity | monocyte | *MAPK12* | 1.117732602 | 0.005200028 | 0.035351132 | 1 |
| Obesity | monocyte | *ZNF626* | 4.957404332 | 0.011127438 | 0.042279868 | 1 |
| Obesity | monocyte | *LINC00265* | 5.514288465 | 0.004207946 | 0.035116467 | 1 |
| Obesity | monocyte | *PRKAR1B* | -1.11733879 | 0.016676216 | 0.046940995 | 1 |
| Obesity | monocyte | *NCR3LG1* | -1.724975013 | 0.009668984 | 0.041464521 | 1 |
| Obesity | monocyte | *RUFY4* | 0.426940873 | 0.004891629 | 0.035116467 | 1 |
| Obesity | monocyte | *ENO4* | -1.077326385 | 0.016423595 | 0.046764836 | 1 |
| Obesity | monocyte | *H3F3C* | -35.3973845 | 0.00775165 | 0.04060923 | 1 |
| Obesity | monocyte | *JAKMIP3* | -1.699793018 | 0.000256052 | 0.013868993 | 1 |
| Obesity | monocyte | *PDCD1* | 6.035134451 | 0.0011057 | 0.028083118 | 1 |
| Obesity | monocyte | *SELL* | 0.918175879 | 0.006971813 | 0.039147772 | 1 |
| Obesity | monocyte | *C19orf54* | 7.399131166 | 0.000818202 | 0.025087947 | 1 |
| Obesity | monocyte | *C22orf34* | -1.042975661 | 0.017510579 | 0.048109473 | 1 |
| Obesity | monocyte | *COL25A1* | 2.500684711 | 0.004377321 | 0.035116467 | 1 |
| Obesity | monocyte | *NDOR1* | 1.650568831 | 0.003312652 | 0.035116467 | 1 |
| Obesity | monocyte | *FAM72B* | 0.936182343 | 0.004936248 | 0.035116467 | 1 |
| Obesity | monocyte | *RHCE* | -1.790116123 | 0.01322124 | 0.044032814 | 1 |
| Obesity | monocyte | *SLC4A5* | -2.982868957 | 0.015055106 | 0.044986158 | 1 |
| Obesity | monocyte | *UROS* | -1.339304496 | 0.018520064 | 0.049292019 | 1 |
| Obesity | monocyte | *ZBED6CL* | 2.393328061 | 0.003856485 | 0.035116467 | 1 |
| Obesity | monocyte | *NOXA1* | -1.159811196 | 0.010341678 | 0.041771331 | 1 |
| Obesity | monocyte | *FZD9* | 2.894505978 | 0.006771188 | 0.038651532 | 1 |
| Obesity | monocyte | *MTF1* | 3.068494532 | 0.000950606 | 0.026972741 | 1 |
| Obesity | monocyte | *FAM26F* | 0.310313625 | 0.003900334 | 0.035116467 | 1 |
| Obesity | monocyte | *ENTPD8* | -118.8357982 | 0.00593301 | 0.036288402 | 1 |
| Obesity | monocyte | *ASTL* | 9.812383231 | 0.003418722 | 0.035116467 | 1 |
| Obesity | monocyte | *MSL1* | -1.176581855 | 0.014837981 | 0.044883966 | 1 |
| Obesity | monocyte | *LRRK2* | 1.053928639 | 0.009212638 | 0.041462933 | 1 |
| Obesity | monocyte | *LIPI* | 2.524213515 | 0.009225706 | 0.041462933 | 1 |
| Obesity | monocyte | *ADAT2* | -0.973312102 | 0.00722812 | 0.039882912 | 1 |
| Obesity | monocyte | *VSTM1* | 0.258546898 | 0.004324857 | 0.035116467 | 1 |
| Obesity | monocyte | *FAM47E* | -1.207444447 | 0.013554057 | 0.04433272 | 1 |
| Obesity | monocyte | *ZNF600* | -1.763222624 | 0.001310652 | 0.030034993 | 1 |
| Obesity | monocyte | *TSPYL1* | -1.677828083 | 0.0181682 | 0.048953818 | 1 |
| Obesity | monocyte | *ZKSCAN3* | -3.776896001 | 1.46E-06 | 0.000283473 | 0.007086836 |

| | | | | | | |
|---|---|---|---|---|---|---|
| Obesity | monocyte | *KIAA0408* | -1.515154841 | 0.002220152 | 0.035072295 | 1 |
| Obesity | monocyte | *BLOC1S2* | -1.879999324 | 0.000510593 | 0.01966188 | 1 |
| Obesity | monocyte | *AKR1C3* | -1.299370013 | 0.016074204 | 0.046231201 | 1 |
| Obesity | monocyte | *SEMA4A* | 1.234151653 | 0.001558487 | 0.03218121 | 1 |
| Obesity | monocyte | *HRCT1* | -1.960604782 | 0.002542768 | 0.035116467 | 1 |
| Obesity | monocyte | *SIRPB2* | 0.48591121 | 0.009441306 | 0.041462933 | 1 |
| Obesity | monocyte | *ZNF493* | -1.239176853 | 0.014672646 | 0.044883966 | 1 |
| Obesity | monocyte | *LONP1* | 2.907302935 | 0.009564098 | 0.041462933 | 1 |
| Obesity | monocyte | *ZNF34* | 2.210103687 | 0.003102651 | 0.035116467 | 1 |
| Obesity | monocyte | *ZNF781* | -1.032210105 | 0.012912612 | 0.043935481 | 1 |
| Obesity | monocyte | *RFX8* | -0.770793317 | 0.012585036 | 0.043212039 | 1 |
| Obesity | monocyte | *C20orf96* | -1.073329082 | 0.010563086 | 0.041845661 | 1 |
| Obesity | monocyte | *SULT1A1* | -1.961843851 | 0.004215544 | 0.035116467 | 1 |
| Obesity | monocyte | *SPTSSB* | -4.123496054 | 0.00273138 | 0.035116467 | 1 |
| Obesity | monocyte | *SULF2* | 1.698430107 | 0.001414254 | 0.03111676 | 1 |
| Obesity | monocyte | *XRCC2* | -2.213711389 | 0.009235273 | 0.041462933 | 1 |
| Obesity | monocyte | *MYO6* | -1.189366572 | 0.010844935 | 0.041961024 | 1 |
| Obesity | monocyte | *ADH1B* | -0.429096632 | 0.009879218 | 0.041637281 | 1 |
| Obesity | monocyte | *COL27A1* | -1.019252247 | 0.013795869 | 0.044539684 | 1 |
| Obesity | monocyte | *ANKRD36B* | -0.984972927 | 0.00695428 | 0.039147772 | 1 |
| Obesity | monocyte | *AP2A1* | 2.782205743 | 0.001914021 | 0.033521987 | 1 |
| Obesity | monocyte | *ZNF429* | -0.863622438 | 0.011902251 | 0.04284104 | 1 |
| Obesity | monocyte | *ZNF100* | -1.886402357 | 0.007912394 | 0.040894308 | 1 |
| Obesity | monocyte | *ZNF420* | -1.098587632 | 0.013038939 | 0.043967382 | 1 |
| Obesity | monocyte | *ZMYM1* | -1.113605676 | 0.016213502 | 0.046494039 | 1 |
| Obesity | monocyte | *GAL3ST4* | -2.555378647 | 0.004080124 | 0.035116467 | 1 |
| Obesity | monocyte | *ADAM32* | -1.525428826 | 0.01770315 | 0.048473864 | 1 |
| Obesity | monocyte | *SULT1A2* | 3.334607934 | 0.000426674 | 0.018429008 | 1 |
| Obesity | monocyte | *NEK5* | -1.390113701 | 0.006651853 | 0.038422368 | 1 |
| Obesity | monocyte | *PIWIL2* | -1.288858796 | 0.013820476 | 0.044539684 | 1 |
| Obesity | monocyte | *CCL4L2* | 0.406083782 | 0.004866158 | 0.035116467 | 1 |
| Obesity | monocyte | *ZNF165* | -3.518261088 | 2.11E-05 | 0.002239829 | 0.102452074 |
| Obesity | monocyte | *RAMP2-AS1* | -0.82405121 | 0.009056303 | 0.041462933 | 1 |
| Obesity | monocyte | *ZNF720* | -1.276530665 | 0.017742902 | 0.048500599 | 1 |
| Obesity | monocyte | *MRPL21* | -2.359751475 | 0.000124969 | 0.008829335 | 0.606348106 |
| Obesity | monocyte | *ZNF98* | -1.34970586 | 0.002742711 | 0.035116467 | 1 |
| Obesity | monocyte | *ZNF786* | 1.438271302 | 0.014276807 | 0.044792561 | 1 |
| Obesity | monocyte | *ZNF517* | -1.233340185 | 0.014531119 | 0.044883966 | 1 |
| Obesity | monocyte | *ZNF675* | -1.67001748 | 0.010693163 | 0.041946198 | 1 |
| Obesity | monocyte | *SLC22A5* | -0.849204405 | 0.01129839 | 0.042332379 | 1 |
| Obesity | monocyte | *SLC2A10* | -0.883911365 | 0.014755275 | 0.044883966 | 1 |
| Obesity | monocyte | *PIGN* | -1.744057918 | 0.006274722 | 0.037173321 | 1 |
| Obesity | monocyte | *HHLA3* | -1.274392488 | 0.010189259 | 0.041771331 | 1 |
| Obesity | monocyte | *BCO2* | -0.870299709 | 0.009112859 | 0.041462933 | 1 |
| Obesity | monocyte | *ENTPD6* | -1.079403472 | 0.004741244 | 0.035116467 | 1 |
| Obesity | monocyte | *ENPP1* | -0.82562208 | 0.010034631 | 0.041669249 | 1 |
| Obesity | monocyte | *MFAP5* | -2.658120138 | 0.000218103 | 0.012449829 | 1 |
| Obesity | monocyte | *NMB* | -1.147453368 | 0.011657472 | 0.042490474 | 1 |
| Obesity | monocyte | *EME2* | -1.312171658 | 0.009184435 | 0.041462933 | 1 |
| Obesity | monocyte | *AC122129.1* | 1.423679622 | 0.003892079 | 0.035116467 | 1 |

| | | | | | | |
|---|---|---|---|---|---|---|
| Obesity | monocyte | *OCLN* | -1.70995614 | 0.005913384 | 0.036288402 | 1 |
| Obesity | monocyte | *ADH5* | -1.027331147 | 0.016478243 | 0.04681056 | 1 |
| Obesity | monocyte | *ZNF677* | -1.283969582 | 0.01749188 | 0.048085326 | 1 |
| Obesity | monocyte | *NOL8* | -1.896444664 | 0.008422518 | 0.041379919 | 1 |
| Obesity | monocyte | *MRPL42* | -1.783173337 | 0.004198194 | 0.035116467 | 1 |
| Obesity | monocyte | *AGAP9* | 2.168306877 | 0.016230448 | 0.046515141 | 1 |
| Obesity | monocyte | *ZNF273* | -1.75649306 | 0.011029334 | 0.042237039 | 1 |
| Obesity | monocyte | *ZNF667* | -1.017260024 | 0.00617509 | 0.036931437 | 1 |
| Obesity | monocyte | *ADH4* | -1.11571218 | 0.000635069 | 0.022052485 | 1 |
| Obesity | monocyte | *HIBCH* | -1.259737967 | 0.004315365 | 0.035116467 | 1 |
| Obesity | monocyte | *ZNF544* | -1.624346855 | 0.018125747 | 0.048913305 | 1 |
| Obesity | monocyte | *ZNF334* | -1.137706313 | 0.010090966 | 0.041669249 | 1 |
| Obesity | monocyte | *RPS6KL1* | -1.946235943 | 0.011295632 | 0.042332379 | 1 |
| Obesity | monocyte | *ZNF442* | 4.344849471 | 0.004605073 | 0.035116467 | 1 |
| Obesity | monocyte | *C20orf203* | -6.97264908 | 0.00118015 | 0.028774316 | 1 |
| Obesity | monocyte | *NCOA6* | -1.728220323 | 0.008067903 | 0.040894308 | 1 |
| Obesity | monocyte | *FAM19A2* | -1.11084252 | 0.016586757 | 0.046940995 | 1 |
| Obesity | monocyte | *TUSC1* | -1.578781775 | 0.008432452 | 0.041379919 | 1 |
| Obesity | monocyte | *PAPSS2* | -0.941830698 | 0.017138423 | 0.047644704 | 1 |
| Obesity | monocyte | *ANKRD13B* | 2.84912071 | 0.003077589 | 0.035116467 | 1 |
| Obesity | monocyte | *PPP1R14C* | 11.97128288 | 0.003395529 | 0.035116467 | 1 |
| Obesity | monocyte | *GPATCH3* | 4.840237483 | 0.003423918 | 0.035116467 | 1 |
| Obesity | monocyte | *RCSD1* | 1.603132967 | 0.002606922 | 0.035116467 | 1 |
| Obesity | monocyte | *GRIN3A* | -2.426329943 | 0.00566985 | 0.035810934 | 1 |
| Obesity | monocyte | *TMEM184B* | -2.293551946 | 0.002316562 | 0.035072295 | 1 |
| Obesity | monocyte | *SCAMP5* | -1.842077137 | 0.000760823 | 0.024286258 | 1 |
| Obesity | monocyte | *ITPRIPL1* | -1.295989637 | 0.005349638 | 0.035561878 | 1 |
| Obesity | monocyte | *PRMT6* | -3.304878859 | 0.005111657 | 0.03528148 | 1 |
| Obesity | monocyte | *SHISA4* | -6.119291311 | 3.53E-05 | 0.003355841 | 0.171147882 |
| Obesity | monocyte | *MAP3K3* | -2.407169728 | 0.013822181 | 0.044539684 | 1 |
| Obesity | monocyte | *CCDC167* | -1.67776139 | 0.00899887 | 0.041462933 | 1 |
| Obesity | monocyte | *L3MBTL3* | 3.31381475 | 0.003515678 | 0.035116467 | 1 |
| Obesity | monocyte | *MFAP3L* | -1.386060021 | 0.012504111 | 0.04319079 | 1 |
| Obesity | monocyte | *TGM2* | -0.773672163 | 0.017809137 | 0.048599513 | 1 |
| Obesity | monocyte | *AL590822.1* | -1.203396757 | 0.01118187 | 0.042279868 | 1 |
| Obesity | monocyte | *CHML* | 3.452551689 | 0.003512612 | 0.035116467 | 1 |
| Obesity | monocyte | *TATDN3* | 2.077222383 | 0.001617304 | 0.03218121 | 1 |
| Obesity | monocyte | *SAMD5* | -1.009604973 | 0.01254373 | 0.04319079 | 1 |
| Obesity | monocyte | *FAM229B* | -1.083010212 | 0.00461373 | 0.035116467 | 1 |
| Obesity | monocyte | *RBM20* | -1.476011496 | 0.005225467 | 0.035351132 | 1 |
| Obesity | monocyte | *C6orf163* | -1.984731228 | 0.000305896 | 0.015460484 | 1 |
| Obesity | monocyte | *PCMTD2* | -0.995101707 | 0.015719076 | 0.045674648 | 1 |
| Obesity | monocyte | *SAMD13* | -1.313205956 | 0.012188718 | 0.043115778 | 1 |
| Obesity | monocyte | *IFIT1B* | -6.262321348 | 0.008578874 | 0.041462933 | 1 |
| Obesity | monocyte | *LIPN* | 0.666981856 | 0.004756354 | 0.035116467 | 1 |
| Obesity | monocyte | *INPP5B* | -1.486876447 | 0.004075202 | 0.035116467 | 1 |
| Obesity | monocyte | *NEU4* | 2.454253733 | 0.01057562 | 0.041845661 | 1 |
| Obesity | monocyte | *C2orf72* | 2.474749589 | 0.006898551 | 0.03896597 | 1 |
| Obesity | monocyte | *ASAH2B* | -1.513833041 | 0.013683775 | 0.044440213 | 1 |
| Obesity | monocyte | *LINC00619* | -11.14996952 | 0.008431515 | 0.041379919 | 1 |

| | | | | | | |
|---|---|---|---|---|---|---|
| Obesity | monocyte | *TCEA3* | -0.791826458 | 0.006180611 | 0.036931437 | 1 |
| Obesity | monocyte | *OXLD1* | 1.354647774 | 0.014357056 | 0.044826534 | 1 |
| Obesity | monocyte | *NOTCH4* | -0.725805446 | 2.17E-08 | 7.51E-06 | 0.000105074 |
| Obesity | monocyte | *PRRT1* | -1.131714787 | 0.00755978 | 0.040352091 | 1 |
| Obesity | monocyte | *SKIV2L* | -1.949127924 | 5.45E-07 | 0.000114871 | 0.002642036 |
| Obesity | monocyte | *LAYN* | -1.130446224 | 0.017168701 | 0.047655914 | 1 |
| Obesity | monocyte | *GPANK1* | -2.753214514 | 4.24E-06 | 0.000662893 | 0.020549693 |
| Obesity | monocyte | *LST1* | 0.240082626 | 0.003454081 | 0.035116467 | 1 |
| Obesity | monocyte | *MICA* | -1.88484905 | 0.017196823 | 0.047702803 | 1 |
| Obesity | monocyte | *POU5F1* | 1.039533292 | 0.000369395 | 0.016908522 | 1 |
| Obesity | monocyte | *ABCF1* | 4.358421187 | 0.000735441 | 0.024110545 | 1 |
| Obesity | monocyte | *LILRB3* | 0.310106894 | 0.004771485 | 0.035116467 | 1 |
| Obesity | monocyte | *ZNF468* | -2.070095925 | 0.005991432 | 0.036474814 | 1 |
| Obesity | monocyte | *HCG9* | -2.95892675 | 0.014750064 | 0.044883966 | 1 |
| Obesity | monocyte | *ZFP57* | 2.637008173 | 3.02E-10 | 1.83E-07 | 1.46E-06 |
| Obesity | monocyte | *FAM153C* | -1.14177076 | 0.003067188 | 0.035116467 | 1 |
| Obesity | monocyte | *TRIM27* | -2.411609656 | 0.00203325 | 0.034173375 | 1 |
| Obesity | monocyte | *MALRD1* | -0.680448705 | 0.0104234 | 0.041803129 | 1 |
| Obesity | monocyte | *FAM196B* | 0.999004235 | 0.00342529 | 0.035116467 | 1 |
| Obesity | monocyte | *MROH6* | 0.577529836 | 0.007549792 | 0.04034316 | 1 |
| Obesity | monocyte | *ZBTB48* | 3.686511608 | 0.00152654 | 0.03218121 | 1 |
| Obesity | monocyte | *MZT1* | 4.445343783 | 0.005314412 | 0.035561878 | 1 |
| Obesity | monocyte | *ZNF425* | -1.254866415 | 0.010645828 | 0.041890466 | 1 |
| Obesity | monocyte | *PCDHGA1* | -3.021204117 | 0.010348001 | 0.041771331 | 1 |
| Obesity | monocyte | *PCDHA7* | 4.01892696 | 0.010321396 | 0.041771331 | 1 |
| Obesity | monocyte | *PCDHA4* | -3.694807953 | 0.001226515 | 0.029216448 | 1 |
| Obesity | monocyte | *SPIRE2* | -1.570040732 | 0.0006358 | 0.022052485 | 1 |
| Obesity | monocyte | *TMEM231* | 3.810690672 | 0.002223844 | 0.035072295 | 1 |
| Obesity | monocyte | *LINC00654* | 2.767640961 | 0.011353146 | 0.042352977 | 1 |
| Obesity | monocyte | *SCGB2B2* | -0.937460405 | 0.012513385 | 0.04319079 | 1 |
| Obesity | monocyte | *CCDC144NL* | 1.333660211 | 0.003720136 | 0.035116467 | 1 |
| Obesity | monocyte | *NT5M* | -1.229794079 | 0.008460712 | 0.041379919 | 1 |
| Obesity | monocyte | *PRR13* | -2.794442978 | 0.001630634 | 0.03218121 | 1 |
| Obesity | monocyte | *MT1A* | 1.817096587 | 0.005685585 | 0.035810934 | 1 |
| Obesity | monocyte | *C15orf59* | 4.516885634 | 0.000150247 | 0.010113064 | 0.72899616 |
| Obesity | monocyte | *ATP6AP1L* | -1.179212756 | 0.018448276 | 0.04918073 | 1 |
| Obesity | monocyte | *SMN2* | -2.075339624 | 0.000152155 | 0.010113064 | 0.738253663 |
| Obesity | monocyte | *SERF1B* | -2.697787235 | 0.002728052 | 0.035116467 | 1 |
| Obesity | monocyte | *C17orf107* | 3.783638352 | 0.002849904 | 0.035116467 | 1 |
| Obesity | monocyte | *KLRC2* | 1.714423308 | 0.002516182 | 0.035116467 | 1 |
| Obesity | monocyte | *RFPL3S* | 1.214548841 | 0.009220595 | 0.041462933 | 1 |
| Obesity | monocyte | *C1QTNF9B* | 2.14759535 | 0.004944273 | 0.035116467 | 1 |
| Obesity | monocyte | *ZDHHC11B* | -1.63962199 | 0.016377179 | 0.046660053 | 1 |
| Obesity | monocyte | *HBA1* | -1.773533816 | 0.001065258 | 0.02778834 | 1 |
| Obesity | monocyte | *HCG27* | -1.259559407 | 0.015112827 | 0.044986158 | 1 |
| Obesity | monocyte | *LNP1* | -1.213560043 | 0.011425779 | 0.042352977 | 1 |
| Obesity | monocyte | *TAS2R50* | -23.0260509 | 0.015755314 | 0.045685351 | 1 |
| Obesity | monocyte | *AC084018.1* | -3.070395568 | 5.27E-05 | 0.004336602 | 0.25585949 |
| Obesity | monocyte | *C17orf51* | -1.578226133 | 0.009476156 | 0.041462933 | 1 |
| Obesity | monocyte | *OR2A42* | -2.572834571 | 0.002324286 | 0.035072295 | 1 |

| | | | | | | |
|---|---|---|---|---|---|---|
| Obesity | monocyte | *NUP62* | -1.711642043 | 0.003468446 | 0.035116467 | 1 |
| Obesity | monocyte | *FGFR1OP* | -1.105815242 | 0.015778404 | 0.045685351 | 1 |
| Obesity | monocyte | *NOTCH2NL* | 1.428539966 | 0.000893026 | 0.025883824 | 1 |
| Obesity | monocyte | *SUPT4H1* | 5.242349887 | 0.000237051 | 0.013220357 | 1 |
| Obesity | monocyte | *MXD3* | 3.255010773 | 0.004214317 | 0.035116467 | 1 |
| Obesity | monocyte | *ARHGAP19* | 1.956880812 | 0.016663569 | 0.046940995 | 1 |
| Obesity | monocyte | *GPC2* | -8.911847295 | 0.002038522 | 0.034173375 | 1 |
| Obesity | monocyte | *TMEM110* | 1.818681956 | 2.56E-05 | 0.002487358 | 0.12436789 |
| Obesity | monocyte | *TMX2* | -1.30992147 | 0.006809073 | 0.038776552 | 1 |
| Obesity | monocyte | *NCKIPSD* | -1.769366679 | 0.01921586 | 0.049996369 | 1 |
| Obesity | monocyte | *ATP6V1G2* | -1.561227486 | 0.001787381 | 0.032602913 | 1 |
| Obesity | monocyte | *PPM1N* | 1.050719582 | 0.004421394 | 0.035116467 | 1 |
| Obesity | monocyte | *SLC23A3* | 3.283918867 | 0.000257257 | 0.013868993 | 1 |
| Obesity | monocyte | *ZNF726* | 1.434077771 | 0.010568679 | 0.041845661 | 1 |
| Obesity | monocyte | *ZNF891* | -1.201443389 | 0.015334239 | 0.045299203 | 1 |
| Obesity | monocyte | *CPNE1* | -1.66294173 | 0.000896225 | 0.025883824 | 1 |
| Obesity | monocyte | *ARL16* | -1.199594633 | 0.011692046 | 0.042490474 | 1 |
| Obesity | monocyte | *PAXIP1-AS2* | -1.089716089 | 0.017526361 | 0.048121163 | 1 |
| Obesity | monocyte | *PRCD* | -2.251100936 | 0.01459261 | 0.044883966 | 1 |
| Obesity | monocyte | *SH3D21* | 2.306486271 | 0.002730509 | 0.035116467 | 1 |
| Obesity | monocyte | *ANG* | 5.208938496 | 0.014182369 | 0.044792561 | 1 |
| Obesity | monocyte | *MBLAC1* | -1.595538864 | 0.005908732 | 0.036288402 | 1 |
| Obesity | monocyte | *GNAT3* | -1.131912972 | 0.009328481 | 0.041462933 | 1 |
| Obesity | monocyte | *PLIN5* | -0.60742448 | 0.006975185 | 0.039147772 | 1 |
| Obesity | monocyte | *EML6* | -1.289353854 | 0.012365056 | 0.043115778 | 1 |
| Obesity | monocyte | *ARHGEF33* | -1.889560785 | 0.003315422 | 0.035116467 | 1 |
| Obesity | monocyte | *EVPLL* | -3.362194751 | 0.00245578 | 0.035116467 | 1 |
| Obesity | monocyte | *AC005082.1* | -5.232494736 | 0.003715289 | 0.035116467 | 1 |
| Obesity | monocyte | *C5orf49* | 7.261455804 | 0.002257673 | 0.035072295 | 1 |
| Obesity | monocyte | *MYL5* | -2.948263828 | 0.009088089 | 0.041462933 | 1 |
| Obesity | monocyte | *NPEPL1* | -1.084550149 | 0.009031451 | 0.041462933 | 1 |
| Obesity | monocyte | *SLC35E2* | -2.477383932 | 0.002005997 | 0.034173375 | 1 |
| Obesity | monocyte | *PAM16* | -1.202766271 | 0.010546916 | 0.041845661 | 1 |
| Obesity | monocyte | *RNU6ATAC35P* | -18.03391829 | 0.000369287 | 0.016908522 | 1 |
| Obesity | monocyte | *AP4M1* | -2.43226078 | 0.002234931 | 0.035072295 | 1 |
| Obesity | monocyte | *ZNF880* | -1.89432349 | 0.008677514 | 0.041462933 | 1 |
| Obesity | monocyte | *SLC12A8* | -1.101351478 | 0.004796262 | 0.035116467 | 1 |
| Obesity | monocyte | *APOL6* | -0.854123315 | 0.013403607 | 0.044180912 | 1 |
| Obesity | monocyte | *RAB6C* | -29.89005486 | 0.012837206 | 0.043770995 | 1 |
| Obesity | monocyte | *AC062017.1* | -1.06186717 | 0.009273798 | 0.041462933 | 1 |
| Obesity | monocyte | *ADRA2B* | -6.657112649 | 0.000433869 | 0.018466067 | 1 |
| Obesity | monocyte | *EXOSC6* | -5.723593687 | 3.95E-07 | 8.71E-05 | 0.001917151 |
| Obesity | monocyte | *LINC00571* | -1.613600097 | 0.004555143 | 0.035116467 | 1 |
| Obesity | monocyte | *C5orf66* | 3.600776087 | 0.011763653 | 0.042569734 | 1 |
| Obesity | monocyte | *LINC01133* | -0.869987511 | 0.00813125 | 0.040894308 | 1 |
| Obesity | monocyte | *C4B* | 5.781877876 | 0.001487751 | 0.032082525 | 1 |
| Obesity | monocyte | *LINC01117* | -1.042433917 | 0.010799218 | 0.041961024 | 1 |
| Obesity | monocyte | *LINC00184* | 33.22417132 | 0.003888523 | 0.035116467 | 1 |
| Obesity | monocyte | *AL049840.1* | -2.459489044 | 0.001534932 | 0.03218121 | 1 |
| Obesity | monocyte | *AP000350.6* | 1.199982737 | 0.001170159 | 0.028774316 | 1 |

| | | | | | | |
|---|---|---|---|---|---|---|
| Obesity | monocyte | *SFTA1P* | -4.924832196 | 0.001755693 | 0.032513821 | 1 |
| Obesity | monocyte | *CYP4A22-AS1* | -2.100153775 | 0.016768682 | 0.047057053 | 1 |
| Obesity | monocyte | *LINC00115* | -1.786522575 | 0.009874409 | 0.041637281 | 1 |
| Obesity | monocyte | *HAR1A* | -14.1758761 | 0.007489658 | 0.04034316 | 1 |
| Obesity | monocyte | *LINC00937* | 0.379777158 | 0.006755792 | 0.038614455 | 1 |
| Obesity | monocyte | *FAM138B* | 16.4884885 | 0.003949765 | 0.035116467 | 1 |
| Obesity | monocyte | *AC009299.3* | 3.536859505 | 0.003277893 | 0.035116467 | 1 |
| Obesity | monocyte | *SLC2A1-AS1* | -2.137948911 | 0.00763082 | 0.040375944 | 1 |
| Obesity | monocyte | *AC005884.1* | -3.280051316 | 0.011150117 | 0.042279868 | 1 |
| Obesity | monocyte | *HCG20* | -6.001677297 | 6.30E-08 | 1.63E-05 | 0.000305596 |
| Obesity | monocyte | *C14orf144* | -3.047584367 | 0.010334535 | 0.041771331 | 1 |
| Obesity | monocyte | *POLR2J2* | 3.48389637 | 0.005875557 | 0.036288402 | 1 |
| Obesity | monocyte | *LINC00578* | -1.311167681 | 0.014450597 | 0.044883966 | 1 |
| Obesity | monocyte | *C19orf24* | 0.925527234 | 0.004223398 | 0.035116467 | 1 |
| Obesity | monocyte | *CASC20* | 3.704544771 | 0.002957943 | 0.035116467 | 1 |
| Obesity | monocyte | *AC112229.4* | 0.958345839 | 0.006402887 | 0.03766044 | 1 |
| Obesity | monocyte | *KLHL7-AS1* | 1.744207542 | 0.001936151 | 0.033670989 | 1 |
| Obesity | monocyte | *LINC00332* | -2.51592831 | 0.001557055 | 0.03218121 | 1 |
| Obesity | monocyte | *DIRC3* | -0.653535329 | 0.014827034 | 0.044883966 | 1 |
| Obesity | monocyte | *AC107021.1* | -0.842775586 | 0.008009262 | 0.040894308 | 1 |
| Obesity | monocyte | *TMA7* | 4.693868911 | 0.000184266 | 0.011611124 | 0.894056526 |
| Obesity | monocyte | *AC009495.3* | 0.907670754 | 0.004976987 | 0.035116467 | 1 |
| Obesity | monocyte | *LINC00702* | -1.92534263 | 0.014553906 | 0.044883966 | 1 |
| Obesity | monocyte | *PINLYP* | -1.072880038 | 0.006271362 | 0.037173321 | 1 |
| Obesity | monocyte | *ANKRD65* | 1.348479502 | 0.004434525 | 0.035116467 | 1 |
| Obesity | monocyte | *ZSCAN31* | 1.595975684 | 0.000466482 | 0.018861406 | 1 |
| Obesity | monocyte | *LINC00957* | -2.822761051 | 0.017315988 | 0.047820138 | 1 |
| Obesity | monocyte | *LINC00607* | 4.47187923 | 0.000330314 | 0.016188733 | 1 |
| Obesity | monocyte | *AC019117.2* | -11.03283581 | 0.00096881 | 0.027171493 | 1 |
| Obesity | monocyte | *ZBED5* | -1.441448412 | 0.018989444 | 0.049857566 | 1 |
| Obesity | monocyte | *CDKN2AIPNL* | 2.959088718 | 0.01766934 | 0.048408604 | 1 |
| Obesity | monocyte | *LINC00987* | -1.363956044 | 0.016617563 | 0.046940995 | 1 |
| Obesity | monocyte | *HLA-DQA2* | 0.901733003 | 0.004804269 | 0.035116467 | 1 |
| Obesity | monocyte | *LINC00211* | 2.094131138 | 0.002582193 | 0.035116467 | 1 |
| Obesity | monocyte | *LRRC37A2* | -1.91000096 | 0.015227654 | 0.045130917 | 1 |
| Obesity | monocyte | *LINC00707* | 2.759444945 | 0.00421037 | 0.035116467 | 1 |
| Obesity | monocyte | *TXNDC5* | 1.563762623 | 0.004623073 | 0.035116467 | 1 |
| Obesity | monocyte | *LINC01119* | -1.675394296 | 0.01483091 | 0.044883966 | 1 |
| Obesity | monocyte | *APOBEC3G* | 0.782708684 | 0.007541123 | 0.04034316 | 1 |
| Obesity | monocyte | *C1orf226* | -3.524325804 | 0.00339274 | 0.035116467 | 1 |
| Obesity | monocyte | *LILRA2* | 0.430641371 | 0.004407479 | 0.035116467 | 1 |
| Obesity | monocyte | *PSMB9* | 3.409189878 | 5.06E-08 | 1.53E-05 | 0.000245444 |
| Obesity | monocyte | *PPIL3* | -0.965805751 | 0.01575639 | 0.045685351 | 1 |
| Obesity | monocyte | *PNMA2* | 1.071882939 | 0.00565448 | 0.035810934 | 1 |
| Obesity | monocyte | *KRBOX1* | -17.05879249 | 0.008932193 | 0.041462933 | 1 |
| Obesity | monocyte | *LINC00886* | -0.807676839 | 0.005778166 | 0.036035554 | 1 |
| Obesity | monocyte | *LINC00877* | 0.223273194 | 0.005028912 | 0.035209637 | 1 |
| Obesity | monocyte | *TDGF1* | 2.286946476 | 0.002848799 | 0.035116467 | 1 |
| Obesity | monocyte | *INMT* | -0.999588109 | 0.004491651 | 0.035116467 | 1 |
| Obesity | monocyte | *PLEKHO2* | 3.036917129 | 0.001561281 | 0.03218121 | 1 |

| | | | | | | |
|---|---|---|---|---|---|---|
| Obesity | monocyte | *C8orf58* | -2.188528696 | 0.005010086 | 0.035209637 | 1 |
| Obesity | monocyte | *DECR2* | -1.55403463 | 0.009115882 | 0.041462933 | 1 |
| Obesity | monocyte | *ZNF709* | 5.927752908 | 0.004980276 | 0.035116467 | 1 |
| Obesity | monocyte | *ERVW-1* | -3.469483847 | 0.014265344 | 0.044792561 | 1 |
| Obesity | monocyte | *VSIG8* | -1.957421886 | 0.013157706 | 0.044028405 | 1 |
| Obesity | monocyte | *FGF14-IT1* | -3.26602619 | 0.009488558 | 0.041462933 | 1 |
| Obesity | monocyte | *PALM2* | -1.018096908 | 0.005593086 | 0.035810934 | 1 |
| Obesity | monocyte | *NBPF15* | -1.103322991 | 0.012381672 | 0.043115778 | 1 |
| Obesity | monocyte | *UPK3B* | -1.167123927 | 0.015152867 | 0.045042687 | 1 |
| Obesity | monocyte | *PLA2G4B* | -8.321308331 | 0.001102323 | 0.028083118 | 1 |
| Obesity | monocyte | *LEFTY1* | -22.82684061 | 0.004494138 | 0.035116467 | 1 |
| Obesity | monocyte | *APOBEC3D* | -5.973199713 | 0.011649648 | 0.042490474 | 1 |
| Obesity | monocyte | *LILRA6* | 0.573348542 | 0.004423539 | 0.035116467 | 1 |
| Obesity | monocyte | *CCDC13* | 0.950630225 | 0.002775104 | 0.035116467 | 1 |
| Obesity | monocyte | *C4A* | -7.434793948 | 8.72E-07 | 0.000176203 | 0.004228879 |
| Obesity | monocyte | *RAB30-AS1* | -1.219378559 | 0.0116401 | 0.042490474 | 1 |
| Obesity | monocyte | *LINC00926* | -1.663710798 | 0.010231881 | 0.041771331 | 1 |
| Obesity | monocyte | *DPH6-AS1* | -0.7595513 | 0.016698301 | 0.046940995 | 1 |
| Obesity | monocyte | *CDK11B* | 3.958549492 | 0.005948629 | 0.036305344 | 1 |
| Obesity | monocyte | *GDNF-AS1* | -1.866439111 | 0.011175828 | 0.042279868 | 1 |
| Obesity | monocyte | *CCDC153* | 2.625349001 | 0.001673469 | 0.03218121 | 1 |
| Obesity | monocyte | *C5orf17* | -0.912922416 | 0.010939422 | 0.042169886 | 1 |
| Obesity | monocyte | *LINC00939* | 2.178512609 | 0.004490258 | 0.035116467 | 1 |
| Obesity | monocyte | *LINC01060* | -1.014787562 | 0.007958344 | 0.040894308 | 1 |
| Obesity | monocyte | *PCDHA10* | 0.984594796 | 0.013838079 | 0.044539684 | 1 |
| Obesity | monocyte | *SMIM20* | -0.980728251 | 0.009607297 | 0.041462933 | 1 |
| Obesity | monocyte | *SNHG18* | -1.306602616 | 0.005395046 | 0.035614643 | 1 |
| Obesity | monocyte | *LINC01094* | -1.230148066 | 0.009601036 | 0.041462933 | 1 |
| Obesity | monocyte | *PCDHGB6* | 1.679552746 | 0.000756905 | 0.024286258 | 1 |
| Obesity | monocyte | *SLC10A5* | 2.424095375 | 0.007256826 | 0.039882912 | 1 |
| Obesity | monocyte | *UTP14C* | -1.408347009 | 0.009097083 | 0.041462933 | 1 |
| Obesity | monocyte | *LYN* | 0.334682105 | 0.003699062 | 0.035116467 | 1 |
| Obesity | monocyte | *DPP3* | 2.571066752 | 1.72E-05 | 0.002038902 | 0.083256016 |
| Obesity | monocyte | *BRK1* | -3.938803919 | 0.00060208 | 0.021632489 | 1 |
| Obesity | monocyte | *PCDHA3* | -7.469747638 | 9.69E-06 | 0.00127036 | 0.047003316 |
| Obesity | monocyte | *HMBS* | 7.041145319 | 1.78E-07 | 4.28E-05 | 0.000864232 |
| Obesity | monocyte | *AC093668.2* | -1.806469897 | 0.001519243 | 0.03218121 | 1 |
| Obesity | monocyte | *CLEC12B* | 0.412284044 | 0.003469675 | 0.035116467 | 1 |
| Obesity | monocyte | *ZBED6* | -2.307612352 | 0.008930695 | 0.041462933 | 1 |
| Obesity | monocyte | *RTEL1* | 1.308204431 | 0.010028951 | 0.041669249 | 1 |
| Obesity | monocyte | *PDF* | -15.31879685 | 0.000309666 | 0.015489661 | 1 |
| Obesity | monocyte | *LINC00640* | 0.987739198 | 0.018873779 | 0.049661376 | 1 |
| Obesity | monocyte | *LINC00639* | -0.787417712 | 0.010955748 | 0.042169886 | 1 |
| Obesity | monocyte | *LINC00924* | -0.97127522 | 0.008095347 | 0.040894308 | 1 |
| Obesity | monocyte | *ITGB3* | -2.227684701 | 0.007754182 | 0.04060923 | 1 |
| Obesity | monocyte | *AC083843.1* | 7.376590075 | 0.00467695 | 0.035116467 | 1 |
| Obesity | monocyte | *PWRN1* | -1.387106929 | 0.007825879 | 0.040689236 | 1 |
| Obesity | monocyte | *C15orf65* | -3.205696452 | 0.004067758 | 0.035116467 | 1 |
| Obesity | monocyte | *LINC00675* | -2.640404504 | 0.005411456 | 0.035630311 | 1 |
| Obesity | monocyte | *LINC00667* | -1.23098553 | 0.014495236 | 0.044883966 | 1 |

| | | | | | | |
|---|---|---|---|---|---|---|
| Obesity | monocyte | *LINC00672* | -2.377152418 | 0.009761892 | 0.041544165 | 1 |
| Obesity | monocyte | *DYNLL2* | -2.645830508 | 0.018244988 | 0.04900239 | 1 |
| Obesity | monocyte | *STRADA* | -2.930448136 | 0.007483897 | 0.04034316 | 1 |
| Obesity | monocyte | *LINC00663* | -1.194798342 | 0.003806227 | 0.035116467 | 1 |
| Obesity | monocyte | *LIN37* | -7.824242337 | 0.000384601 | 0.01744005 | 1 |
| Obesity | monocyte | *NDUFA7* | -6.566246124 | 0.01158165 | 0.042465454 | 1 |
| Obesity | monocyte | *AC004466.1* | 2.790238255 | 0.004742449 | 0.035116467 | 1 |
| Obesity | monocyte | *LINC00664* | -63.49786426 | 0.010535951 | 0.041845661 | 1 |
| Obesity | monocyte | *FBXO17* | -0.689968456 | 0.016680618 | 0.046940995 | 1 |
| Obesity | monocyte | *SNHG8* | -1.593180171 | 0.005253279 | 0.035450498 | 1 |
| Obesity | monocyte | *AP001172.2* | 2.562256176 | 2.03E-05 | 0.002239829 | 0.098469611 |
| Obesity | monocyte | *YTHDF3-AS1* | -2.794457701 | 0.014475643 | 0.044883966 | 1 |
| Obesity | monocyte | *MILR1* | 0.747249193 | 0.005231268 | 0.035351132 | 1 |
| Obesity | monocyte | *FGF14-AS2* | -1.763563844 | 0.009311731 | 0.041462933 | 1 |
| Obesity | nk_cell | *NFYA* | -3.116299645 | 2.91E-08 | 1.14E-07 | 0.000141223 |
| Obesity | nk_cell | *ENPP4* | -1.854483523 | 1.29E-05 | 2.19E-05 | 0.062418676 |
| Obesity | nk_cell | *SEMA3F* | -5.115076339 | 1.01E-16 | 5.42E-14 | 4.88E-13 |
| Obesity | nk_cell | *KRIT1* | -2.127896272 | 4.52E-07 | 1.08E-06 | 0.002195253 |
| Obesity | nk_cell | *RAD52* | -2.442800627 | 3.22E-07 | 8.11E-07 | 0.001560701 |
| Obesity | nk_cell | *HS3ST1* | -1.61976274 | 0.007315321 | 0.009094015 | 1 |
| Obesity | nk_cell | *AOC1* | -2.901039152 | 1.22E-09 | 2.08E-08 | 5.94E-06 |
| Obesity | nk_cell | *SNX11* | -1.386873493 | 7.07E-08 | 2.30E-07 | 0.000343102 |
| Obesity | nk_cell | *TMEM176A* | -3.568567961 | 2.32E-08 | 9.60E-08 | 0.000112744 |
| Obesity | nk_cell | *CYP26B1* | -1.330137854 | 2.21E-08 | 9.29E-08 | 0.000107107 |
| Obesity | nk_cell | *DBNDD1* | -1.514949641 | 3.31E-06 | 6.38E-06 | 0.016065498 |
| Obesity | nk_cell | *ALS2* | -2.25841662 | 2.76E-07 | 7.10E-07 | 0.001337759 |
| Obesity | nk_cell | *CD38* | 0.882441515 | 1.76E-09 | 2.16E-08 | 8.55E-06 |
| Obesity | nk_cell | *RBM6* | -3.141172251 | 5.28E-27 | 2.56E-23 | 2.56E-23 |
| Obesity | nk_cell | *RECQL* | -2.739552464 | 0.000794741 | 0.001077419 | 1 |
| Obesity | nk_cell | *SLC22A16* | 1.124441536 | 3.77E-05 | 6.01E-05 | 0.182976823 |
| Obesity | nk_cell | *CDC27* | -1.528052892 | 0.000117944 | 0.000175865 | 0.572263204 |
| Obesity | nk_cell | *POLR2J* | 3.196932898 | 5.56E-09 | 3.68E-08 | 2.70E-05 |
| Obesity | nk_cell | *MEOX1* | -1.853377997 | 1.82E-09 | 2.18E-08 | 8.83E-06 |
| Obesity | nk_cell | *LIG3* | -2.115456019 | 1.90E-07 | 5.18E-07 | 0.000924003 |
| Obesity | nk_cell | *CIAPIN1* | 2.109492258 | 0.041851603 | 0.048661389 | 1 |
| Obesity | nk_cell | *PON1* | -6.457770774 | 3.96E-09 | 3.01E-08 | 1.92E-05 |
| Obesity | nk_cell | *GCFC2* | -2.250047295 | 1.96E-06 | 3.97E-06 | 0.009520745 |
| Obesity | nk_cell | *RHBDD2* | -1.747882606 | 5.52E-07 | 1.28E-06 | 0.002680652 |
| Obesity | nk_cell | *ZNF195* | -1.465526668 | 0.033806856 | 0.039640131 | 1 |
| Obesity | nk_cell | *PDK2* | -1.920473154 | 4.97E-07 | 1.17E-06 | 0.002409556 |
| Obesity | nk_cell | *GGNBP2* | -2.523516675 | 2.87E-08 | 1.13E-07 | 0.000139442 |
| Obesity | nk_cell | *ITGA2B* | -1.423389792 | 9.54E-09 | 5.03E-08 | 4.63E-05 |
| Obesity | nk_cell | *TMEM98* | -1.785938312 | 3.18E-08 | 1.21E-07 | 0.00015437 |
| Obesity | nk_cell | *ABCC8* | -1.819962658 | 5.54E-06 | 1.02E-05 | 0.026857252 |
| Obesity | nk_cell | *ZNF263* | -1.863804335 | 0.035316293 | 0.041320148 | 1 |
| Obesity | nk_cell | *SPATA20* | -1.900501694 | 0.00490978 | 0.006198869 | 1 |
| Obesity | nk_cell | *MAP3K9* | -1.101196651 | 0.000340954 | 0.000480764 | 1 |
| Obesity | nk_cell | *BAIAP2L1* | -1.924029813 | 8.08E-09 | 4.51E-08 | 3.92E-05 |
| Obesity | nk_cell | *AGK* | 2.94707069 | 1.60E-06 | 3.32E-06 | 0.007776624 |
| Obesity | nk_cell | *FARP2* | -1.771213432 | 2.80E-08 | 1.11E-07 | 0.00013604 |

| | | | | | | |
|---|---|---|---|---|---|---|
| Obesity | nk_cell | *USH1C* | -12.5326377 | 6.86E-08 | 2.24E-07 | 0.000332795 |
| Obesity | nk_cell | *IFRD1* | -1.5285147 | 5.67E-07 | 1.31E-06 | 0.002750013 |
| Obesity | nk_cell | *VPS41* | -2.107217496 | 3.25E-07 | 8.17E-07 | 0.001575002 |
| Obesity | nk_cell | *ELAC2* | -2.466963884 | 7.21E-09 | 4.27E-08 | 3.50E-05 |
| Obesity | nk_cell | *SCIN* | -1.894013468 | 6.47E-10 | 1.90E-08 | 3.14E-06 |
| Obesity | nk_cell | *CCDC124* | -1.950796738 | 0.000329039 | 0.000465316 | 1 |
| Obesity | nk_cell | *CEACAM21* | 0.371219775 | 1.07E-09 | 2.05E-08 | 5.18E-06 |
| Obesity | nk_cell | *CD79B* | 1.908608708 | 1.25E-05 | 2.14E-05 | 0.060712508 |
| Obesity | nk_cell | *ST7L* | -1.799377505 | 1.69E-05 | 2.83E-05 | 0.082056582 |
| Obesity | nk_cell | *PIGQ* | -2.369199629 | 3.73E-05 | 5.96E-05 | 0.181048427 |
| Obesity | nk_cell | *FMO3* | -1.562331096 | 3.34E-11 | 3.24E-09 | 1.62E-07 |
| Obesity | nk_cell | *PSMB1* | 2.941054405 | 1.70E-05 | 2.85E-05 | 0.082722126 |
| Obesity | nk_cell | *NADK* | -2.220367728 | 2.78E-09 | 2.51E-08 | 1.35E-05 |
| Obesity | nk_cell | *DLEC1* | -1.877159578 | 1.55E-08 | 7.19E-08 | 7.54E-05 |
| Obesity | nk_cell | *CYB561* | -1.658370323 | 7.16E-07 | 1.62E-06 | 0.003473124 |
| Obesity | nk_cell | *SS18L2* | -2.7179451 | 5.54E-07 | 1.29E-06 | 0.00268803 |
| Obesity | nk_cell | *MGST1* | -1.169765896 | 4.77E-08 | 1.68E-07 | 0.000231587 |
| Obesity | nk_cell | *MMP25* | 1.534031793 | 3.29E-06 | 6.34E-06 | 0.01595566 |
| Obesity | nk_cell | *IL32* | 0.260678241 | 1.00E-09 | 2.05E-08 | 4.86E-06 |
| Obesity | nk_cell | *PKD1* | -1.653607983 | 4.80E-08 | 1.68E-07 | 0.000232776 |
| Obesity | nk_cell | *MAPK8IP2* | 8.206640163 | 2.80E-09 | 2.51E-08 | 1.36E-05 |
| Obesity | nk_cell | *MED24* | -3.252092962 | 2.01E-06 | 4.06E-06 | 0.009751946 |
| Obesity | nk_cell | *TRAF3IP3* | 0.292041066 | 8.08E-10 | 1.98E-08 | 3.92E-06 |
| Obesity | nk_cell | *POMT2* | -1.220469398 | 2.48E-09 | 2.37E-08 | 1.20E-05 |
| Obesity | nk_cell | *MLXIPL* | -0.766304628 | 7.78E-05 | 0.000118314 | 0.37730187 |
| Obesity | nk_cell | *DYRK4* | -1.833198837 | 1.33E-07 | 3.88E-07 | 0.000643939 |
| Obesity | nk_cell | *NCAPD2* | 1.633139021 | 6.93E-06 | 1.24E-05 | 0.033628682 |
| Obesity | nk_cell | *FUZ* | 5.384759119 | 1.80E-06 | 3.68E-06 | 0.008737407 |
| Obesity | nk_cell | *LRRC23* | -1.781035548 | 0.00025868 | 0.00037035 | 1 |
| Obesity | nk_cell | *SLC6A7* | -34.95770151 | 3.63E-09 | 2.86E-08 | 1.76E-05 |
| Obesity | nk_cell | *TSPAN9* | -1.735068757 | 3.27E-06 | 6.31E-06 | 0.015843483 |
| Obesity | nk_cell | *UTP18* | -2.091380542 | 5.62E-05 | 8.74E-05 | 0.2728951 |
| Obesity | nk_cell | *TTC19* | -1.913179882 | 0.000126837 | 0.000188315 | 0.615412425 |
| Obesity | nk_cell | *DPF1* | -2.495494986 | 0.031429567 | 0.037004673 | 1 |
| Obesity | nk_cell | *LARS2* | -2.434393892 | 0.000165579 | 0.00024213 | 0.80338851 |
| Obesity | nk_cell | *DCN* | -1.35455029 | 5.50E-07 | 1.28E-06 | 0.002668097 |
| Obesity | nk_cell | *PPP5C* | 2.222550579 | 0.005685077 | 0.007125806 | 1 |
| Obesity | nk_cell | *CEP68* | -1.808834913 | 2.71E-07 | 7.01E-07 | 0.001315743 |
| Obesity | nk_cell | *ZBTB32* | -4.45759722 | 0.000143373 | 0.000211249 | 0.695644255 |
| Obesity | nk_cell | *ERCC1* | -2.570847484 | 5.13E-06 | 9.48E-06 | 0.024884798 |
| Obesity | nk_cell | *CD22* | 0.915545233 | 8.30E-08 | 2.63E-07 | 0.000402839 |
| Obesity | nk_cell | *MAP4K5* | -2.18250196 | 1.55E-05 | 2.60E-05 | 0.075083648 |
| Obesity | nk_cell | *EHD3* | -3.930102061 | 1.87E-08 | 8.19E-08 | 9.07E-05 |
| Obesity | nk_cell | *MAN2B2* | -2.928909171 | 5.63E-06 | 1.03E-05 | 0.02732302 |
| Obesity | nk_cell | *SLC25A39* | 1.932934297 | 0.00060582 | 0.000831524 | 1 |
| Obesity | nk_cell | *RNF14* | -2.55101623 | 2.69E-06 | 5.29E-06 | 0.013051493 |
| Obesity | nk_cell | *DDX11* | -1.565014403 | 2.31E-10 | 1.23E-08 | 1.12E-06 |
| Obesity | nk_cell | *TACC3* | -3.675734357 | 1.45E-07 | 4.15E-07 | 0.000703155 |
| Obesity | nk_cell | *ZC3H3* | -1.734833291 | 0.000398749 | 0.00055772 | 1 |
| Obesity | nk_cell | *MDH1* | -1.784518439 | 0.000567636 | 0.000780882 | 1 |

| | | | | | | |
|---|---|---|---|---|---|---|
| Obesity | nk_cell | *MTMR11* | -0.990269631 | 0.000169606 | 0.000247647 | 0.82293058 |
| Obesity | nk_cell | *COX15* | -2.290581671 | 5.33E-08 | 1.83E-07 | 0.000258825 |
| Obesity | nk_cell | *CCDC88C* | 0.411963238 | 1.12E-09 | 2.05E-08 | 5.43E-06 |
| Obesity | nk_cell | *NUDCD3* | -1.098092379 | 0.023096301 | 0.027527206 | 1 |
| Obesity | nk_cell | *CHDH* | -4.454059166 | 1.42E-08 | 6.70E-08 | 6.90E-05 |
| Obesity | nk_cell | *CLCA4* | -5.701292826 | 6.02E-10 | 1.84E-08 | 2.92E-06 |
| Obesity | nk_cell | *RUFY3* | -1.991750539 | 2.35E-06 | 4.67E-06 | 0.01140018 |
| Obesity | nk_cell | *SLC11A1* | -0.945678786 | 3.70E-08 | 1.38E-07 | 0.00017938 |
| Obesity | nk_cell | *WWTR1* | -1.739872698 | 6.20E-08 | 2.06E-07 | 0.00030081 |
| Obesity | nk_cell | *AGPS* | -1.922121636 | 0.000189859 | 0.000276056 | 0.921197469 |
| Obesity | nk_cell | *TTC27* | -2.008598467 | 1.25E-06 | 2.65E-06 | 0.006053759 |
| Obesity | nk_cell | *VSIG2* | -1.796606923 | 1.94E-10 | 1.09E-08 | 9.41E-07 |
| Obesity | nk_cell | *MARCO* | -0.910833633 | 0.007766905 | 0.009625804 | 1 |
| Obesity | nk_cell | *HGF* | -3.081297126 | 0.000116023 | 0.000173266 | 0.56294172 |
| Obesity | nk_cell | *SLC7A9* | -0.981756013 | 0.000300628 | 0.000427345 | 1 |
| Obesity | nk_cell | *OSBPL5* | 1.864153785 | 6.39E-09 | 4.02E-08 | 3.10E-05 |
| Obesity | nk_cell | *CPS1* | -2.074684186 | 1.51E-06 | 3.15E-06 | 0.007311283 |
| Obesity | nk_cell | *NLRP2* | 0.977802687 | 4.34E-07 | 1.04E-06 | 0.00210395 |
| Obesity | nk_cell | *SLC45A4* | -1.881739441 | 0.007691345 | 0.009541908 | 1 |
| Obesity | nk_cell | *RNH1* | -1.725200022 | 0.000752958 | 0.001023348 | 1 |
| Obesity | nk_cell | *GLRX2* | -3.327429127 | 1.52E-08 | 7.04E-08 | 7.36E-05 |
| Obesity | nk_cell | *ABCC2* | -3.302018917 | 2.35E-09 | 2.30E-08 | 1.14E-05 |
| Obesity | nk_cell | *GCLM* | -2.202214783 | 0.036710993 | 0.042910561 | 1 |
| Obesity | nk_cell | *UBR2* | -1.947518668 | 5.09E-06 | 9.41E-06 | 0.024672855 |
| Obesity | nk_cell | *RRAGD* | -1.713860791 | 4.36E-09 | 3.19E-08 | 2.12E-05 |
| Obesity | nk_cell | *HSF2* | -2.342151263 | 1.97E-05 | 3.25E-05 | 0.095458289 |
| Obesity | nk_cell | *TYMP* | -1.250242708 | 0.004731285 | 0.005989093 | 1 |
| Obesity | nk_cell | *TOMM34* | 1.689898183 | 5.58E-09 | 3.69E-08 | 2.71E-05 |
| Obesity | nk_cell | *SEC63* | -2.284576385 | 7.64E-06 | 1.36E-05 | 0.037077826 |
| Obesity | nk_cell | *FAS* | -2.671272878 | 0.000296957 | 0.000422532 | 1 |
| Obesity | nk_cell | *RNASET2* | -1.666153113 | 0.000470318 | 0.000652368 | 1 |
| Obesity | nk_cell | *AGPAT4* | -2.596835634 | 8.44E-07 | 1.87E-06 | 0.004093647 |
| Obesity | nk_cell | *BTN3A1* | 1.105867752 | 0.000405857 | 0.000567008 | 1 |
| Obesity | nk_cell | *MIPEP* | -2.389629192 | 6.55E-08 | 2.16E-07 | 0.000318025 |
| Obesity | nk_cell | *INSRR* | 4.869908078 | 6.16E-06 | 1.12E-05 | 0.029872232 |
| Obesity | nk_cell | *B4GALT7* | -1.792543541 | 5.88E-09 | 3.81E-08 | 2.85E-05 |
| Obesity | nk_cell | *SH2D2A* | 0.334950961 | 9.44E-10 | 2.05E-08 | 4.58E-06 |
| Obesity | nk_cell | *VRK2* | -1.89812952 | 8.43E-08 | 2.67E-07 | 0.000408872 |
| Obesity | nk_cell | *VEZT* | -2.167919601 | 1.65E-06 | 3.40E-06 | 0.007997803 |
| Obesity | nk_cell | *BRD9* | -2.491532997 | 7.54E-06 | 1.34E-05 | 0.036583654 |
| Obesity | nk_cell | *ANK1* | 1.70712718 | 1.41E-09 | 2.11E-08 | 6.82E-06 |
| Obesity | nk_cell | *RABEP1* | -1.76003209 | 0.001314683 | 0.001741449 | 1 |
| Obesity | nk_cell | *BAK1* | 1.89001959 | 1.25E-08 | 6.07E-08 | 6.05E-05 |
| Obesity | nk_cell | *IKZF2* | 2.103901564 | 3.56E-08 | 1.33E-07 | 0.000172561 |
| Obesity | nk_cell | *GRN* | -1.843526766 | 0.000199643 | 0.000289413 | 0.968665817 |
| Obesity | nk_cell | *IFT88* | -2.150422832 | 1.00E-06 | 2.19E-06 | 0.00485912 |
| Obesity | nk_cell | *CHPF2* | 1.863712496 | 1.27E-09 | 2.09E-08 | 6.18E-06 |
| Obesity | nk_cell | *UBA6* | -2.021213498 | 0.002013659 | 0.002627124 | 1 |
| Obesity | nk_cell | *ATP6V0A1* | -1.837568067 | 1.89E-06 | 3.85E-06 | 0.009171923 |
| Obesity | nk_cell | *PIAS1* | -1.548335415 | 0.000209991 | 0.000303639 | 1 |

| | | | | | | |
|---|---|---|---|---|---|---|
| Obesity | nk_cell | *ASTE1* | -3.134675229 | 4.45E-08 | 1.59E-07 | 0.000215705 |
| Obesity | nk_cell | *MYOC* | -1.209288583 | 7.25E-09 | 4.28E-08 | 3.52E-05 |
| Obesity | nk_cell | *SH3YL1* | -3.462914264 | 5.19E-07 | 1.22E-06 | 0.002520132 |
| Obesity | nk_cell | *VCL* | -2.415879586 | 2.00E-06 | 4.04E-06 | 0.009703559 |
| Obesity | nk_cell | *TBC1D23* | -2.384973121 | 1.31E-06 | 2.78E-06 | 0.006361388 |
| Obesity | nk_cell | *MYOM2* | 2.693553673 | 2.06E-07 | 5.55E-07 | 0.000999572 |
| Obesity | nk_cell | *TUBG2* | -1.763097625 | 9.31E-10 | 2.05E-08 | 4.52E-06 |
| Obesity | nk_cell | *NSUN2* | -2.409595141 | 0.000217978 | 0.000314396 | 1 |
| Obesity | nk_cell | *MFAP3* | -1.823834643 | 0.001850518 | 0.00242275 | 1 |
| Obesity | nk_cell | *MRI1* | -2.160145601 | 4.23E-06 | 7.94E-06 | 0.020519967 |
| Obesity | nk_cell | *AGA* | -3.939123293 | 2.86E-07 | 7.31E-07 | 0.001389673 |
| Obesity | nk_cell | *PI4K2B* | -8.481058051 | 5.13E-06 | 9.48E-06 | 0.024910859 |
| Obesity | nk_cell | *BOD1L1* | -1.754788426 | 1.86E-06 | 3.79E-06 | 0.009041342 |
| Obesity | nk_cell | *PHLPP2* | -1.679674675 | 5.89E-05 | 9.13E-05 | 0.285837449 |
| Obesity | nk_cell | *SPDL1* | 1.612138171 | 1.95E-09 | 2.20E-08 | 9.45E-06 |
| Obesity | nk_cell | *PQLC2* | -3.301900196 | 4.46E-09 | 3.23E-08 | 2.17E-05 |
| Obesity | nk_cell | *INPP4A* | 1.766611424 | 1.61E-08 | 7.35E-08 | 7.79E-05 |
| Obesity | nk_cell | *PSMA4* | 2.835931832 | 5.63E-07 | 1.31E-06 | 0.002732608 |
| Obesity | nk_cell | *TDP1* | -3.204765558 | 0.000233024 | 0.000335102 | 1 |
| Obesity | nk_cell | *SPATA7* | -2.096081989 | 6.51E-09 | 4.05E-08 | 3.16E-05 |
| Obesity | nk_cell | *MED17* | 3.836804464 | 6.10E-05 | 9.42E-05 | 0.295790073 |
| Obesity | nk_cell | *CAPG* | -1.231849145 | 1.06E-08 | 5.41E-08 | 5.16E-05 |
| Obesity | nk_cell | *AP2S1* | -3.491929327 | 0.00031425 | 0.00044544 | 1 |
| Obesity | nk_cell | *TRIT1* | -1.878425996 | 1.59E-05 | 2.66E-05 | 0.077067169 |
| Obesity | nk_cell | *CUL7* | -2.173257801 | 0.027261741 | 0.032317118 | 1 |
| Obesity | nk_cell | *CTNNA1* | -1.74226479 | 1.07E-07 | 3.24E-07 | 0.00051855 |
| Obesity | nk_cell | *CNTLN* | -1.770444459 | 8.86E-08 | 2.78E-07 | 0.000429696 |
| Obesity | nk_cell | *YTHDC2* | -2.301335126 | 3.48E-05 | 5.58E-05 | 0.1689073 |
| Obesity | nk_cell | *POLR2B* | -1.782374418 | 0.020255989 | 0.024291166 | 1 |
| Obesity | nk_cell | *ARAP2* | 1.304707168 | 6.02E-09 | 3.88E-08 | 2.92E-05 |
| Obesity | nk_cell | *DTNBP1* | -2.208760491 | 0.002848514 | 0.003673841 | 1 |
| Obesity | nk_cell | *ANO2* | -1.345163564 | 6.71E-09 | 4.11E-08 | 3.25E-05 |
| Obesity | nk_cell | *SNX29* | -1.934553929 | 4.79E-06 | 8.91E-06 | 0.02326037 |
| Obesity | nk_cell | *UTS2* | 1.610540404 | 0.025483113 | 0.030282652 | 1 |
| Obesity | nk_cell | *EPN3* | -3.106789428 | 6.62E-05 | 0.000101549 | 0.321301149 |
| Obesity | nk_cell | *RCN1* | -3.486838457 | 1.14E-07 | 3.41E-07 | 0.00055174 |
| Obesity | nk_cell | *JKAMP* | -1.928558955 | 0.000331187 | 0.000467943 | 1 |
| Obesity | nk_cell | *ARHGEF5* | -1.453273306 | 5.17E-09 | 3.54E-08 | 2.51E-05 |
| Obesity | nk_cell | *MCUR1* | -1.946786222 | 8.72E-09 | 4.72E-08 | 4.23E-05 |
| Obesity | nk_cell | *LAMC3* | 1.860000207 | 0.002185779 | 0.002844795 | 1 |
| Obesity | nk_cell | *MAPK9* | -2.681503489 | 0.00171275 | 0.002249665 | 1 |
| Obesity | nk_cell | *FAM160A2* | -2.399619789 | 5.23E-07 | 1.23E-06 | 0.00253692 |
| Obesity | nk_cell | *CYBA* | 1.355056142 | 4.77E-09 | 3.36E-08 | 2.31E-05 |
| Obesity | nk_cell | *THOC3* | 3.392871445 | 2.62E-09 | 2.44E-08 | 1.27E-05 |
| Obesity | nk_cell | *ALX4* | 2.523825284 | 0.002610174 | 0.003377217 | 1 |
| Obesity | nk_cell | *FSTL4* | -1.776324146 | 1.31E-10 | 8.67E-09 | 6.34E-07 |
| Obesity | nk_cell | *MRTO4* | 3.467856082 | 0.001668813 | 0.002192549 | 1 |
| Obesity | nk_cell | *LY75* | 1.353242648 | 7.77E-10 | 1.95E-08 | 3.77E-06 |
| Obesity | nk_cell | *SDCCAG8* | -1.186512905 | 1.54E-05 | 2.59E-05 | 0.074553493 |
| Obesity | nk_cell | *KIF1B* | -1.595997048 | 1.95E-06 | 3.94E-06 | 0.00943794 |

| | | | | | | |
|---|---|---|---|---|---|---|
| Obesity | nk_cell | *TBC1D22A* | -1.769749317 | 0.000888489 | 0.001197486 | 1 |
| Obesity | nk_cell | *GALC* | -2.850548759 | 3.14E-08 | 1.21E-07 | 0.000152329 |
| Obesity | nk_cell | *FAM114A2* | -2.972658634 | 0.000101761 | 0.00015272 | 0.493743332 |
| Obesity | nk_cell | *ITIH4* | 1.058591742 | 1.48E-05 | 2.51E-05 | 0.071963477 |
| Obesity | nk_cell | *NPFFR2* | -1.772201863 | 1.59E-09 | 2.11E-08 | 7.73E-06 |
| Obesity | nk_cell | *PCGF2* | -1.909282437 | 5.19E-07 | 1.22E-06 | 0.002518388 |
| Obesity | nk_cell | *IL17RB* | 2.898976545 | 0.006524113 | 0.008141717 | 1 |
| Obesity | nk_cell | *MSH4* | -0.698224036 | 0.040729533 | 0.047436317 | 1 |
| Obesity | nk_cell | *GDI2* | -2.543555605 | 7.06E-06 | 1.26E-05 | 0.034267763 |
| Obesity | nk_cell | *PITHD1* | -1.465512065 | 4.19E-09 | 3.12E-08 | 2.03E-05 |
| Obesity | nk_cell | *LAMC2* | -1.512576554 | 1.51E-05 | 2.55E-05 | 0.073237659 |
| Obesity | nk_cell | *RASGRF1* | -2.783212824 | 0.000113269 | 0.000169414 | 0.549579356 |
| Obesity | nk_cell | *CROCC* | -0.854211819 | 0.034791933 | 0.040736115 | 1 |
| Obesity | nk_cell | *ALDH18A1* | -2.260253751 | 1.59E-06 | 3.30E-06 | 0.00772596 |
| Obesity | nk_cell | *DNAJC25* | -1.378300432 | 1.76E-09 | 2.16E-08 | 8.52E-06 |
| Obesity | nk_cell | *OGFR* | -2.337245758 | 0.000177566 | 0.000258723 | 0.861548509 |
| Obesity | nk_cell | *GNA15* | -1.353238941 | 6.16E-08 | 2.05E-07 | 0.000298694 |
| Obesity | nk_cell | *PIGV* | -1.642080863 | 0.000650806 | 0.000891002 | 1 |
| Obesity | nk_cell | *RIMBP2* | -1.556764787 | 2.10E-09 | 2.25E-08 | 1.02E-05 |
| Obesity | nk_cell | *PRDM6* | -2.183169904 | 1.06E-08 | 5.41E-08 | 5.16E-05 |
| Obesity | nk_cell | *SPAG4* | -4.308363883 | 1.87E-08 | 8.19E-08 | 9.07E-05 |
| Obesity | nk_cell | *NCKAP1* | -1.787713768 | 2.77E-08 | 1.10E-07 | 0.000134421 |
| Obesity | nk_cell | *LTK* | -8.841123111 | 1.89E-09 | 2.19E-08 | 9.19E-06 |
| Obesity | nk_cell | *EIF4B* | -1.127294489 | 0.042411767 | 0.049269033 | 1 |
| Obesity | nk_cell | *AHRR* | -2.076042437 | 6.56E-10 | 1.90E-08 | 3.18E-06 |
| Obesity | nk_cell | *ADCK1* | -2.169638426 | 6.63E-09 | 4.09E-08 | 3.22E-05 |
| Obesity | nk_cell | *RNF4* | 2.797882496 | 0.000213488 | 0.000308379 | 1 |
| Obesity | nk_cell | *CASP8* | 1.604596042 | 2.59E-08 | 1.05E-07 | 0.00012579 |
| Obesity | nk_cell | *TM7SF3* | -2.275480531 | 0.000458938 | 0.00063713 | 1 |
| Obesity | nk_cell | *CDON* | -0.75778658 | 8.59E-09 | 4.67E-08 | 4.17E-05 |
| Obesity | nk_cell | *TMEM161A* | -4.118104786 | 1.02E-05 | 1.77E-05 | 0.049331165 |
| Obesity | nk_cell | *LPAR2* | 2.468088051 | 1.72E-10 | 1.01E-08 | 8.36E-07 |
| Obesity | nk_cell | *SUGP2* | -2.028075365 | 0.008441176 | 0.010429485 | 1 |
| Obesity | nk_cell | *CNN2* | 1.248485692 | 8.30E-10 | 1.99E-08 | 4.03E-06 |
| Obesity | nk_cell | *ABCA7* | -3.162792916 | 0.000127967 | 0.00018984 | 0.620896634 |
| Obesity | nk_cell | *DDX20* | -1.99572115 | 0.015293334 | 0.018518407 | 1 |
| Obesity | nk_cell | *BTBD1* | 2.869258362 | 3.68E-05 | 5.87E-05 | 0.17847533 |
| Obesity | nk_cell | *FAR2* | -2.418865764 | 9.92E-06 | 1.73E-05 | 0.048115178 |
| Obesity | nk_cell | *BCAS1* | -2.749825525 | 1.25E-08 | 6.07E-08 | 6.06E-05 |
| Obesity | nk_cell | *CHI3L2* | -4.109028803 | 1.94E-08 | 8.43E-08 | 9.42E-05 |
| Obesity | nk_cell | *PMS1* | -2.328092105 | 0.000175227 | 0.000255547 | 0.850203716 |
| Obesity | nk_cell | *WDR18* | 1.08951555 | 0.00393609 | 0.005011259 | 1 |
| Obesity | nk_cell | *ANKRD44* | 1.066582064 | 2.24E-09 | 2.27E-08 | 1.08E-05 |
| Obesity | nk_cell | *ADAT1* | -1.571157952 | 6.53E-05 | 0.000100242 | 0.317064525 |
| Obesity | nk_cell | *MAP2K4* | -1.955680725 | 1.11E-07 | 3.35E-07 | 0.000539223 |
| Obesity | nk_cell | *TMEM206* | -2.057187556 | 2.52E-07 | 6.61E-07 | 0.00122489 |
| Obesity | nk_cell | *SEC61A2* | -2.253444844 | 2.78E-09 | 2.51E-08 | 1.35E-05 |
| Obesity | nk_cell | *TIE1* | -2.706206685 | 4.76E-05 | 7.50E-05 | 0.230949676 |
| Obesity | nk_cell | *ZMYND12* | -1.098619061 | 2.34E-06 | 4.66E-06 | 0.01136145 |
| Obesity | nk_cell | *SLC9A3* | -0.869627629 | 0.003071709 | 0.00394597 | 1 |

| | | | | | | |
|---|---|---|---|---|---|---|
| Obesity | nk_cell | *NGEF* | -3.380975658 | 1.73E-09 | 2.16E-08 | 8.40E-06 |
| Obesity | nk_cell | *SPI1* | -1.969486987 | 5.60E-08 | 1.90E-07 | 0.000271919 |
| Obesity | nk_cell | *ZNRD1* | -2.708750682 | 6.28E-07 | 1.44E-06 | 0.003047479 |
| Obesity | nk_cell | *ATXN3* | -1.418266651 | 7.99E-05 | 0.000121483 | 0.387895089 |
| Obesity | nk_cell | *GOLGA5* | -2.631888144 | 3.33E-07 | 8.36E-07 | 0.001617603 |
| Obesity | nk_cell | *FGFR2* | -1.222001794 | 2.10E-05 | 3.45E-05 | 0.101730404 |
| Obesity | nk_cell | *FECH* | -1.104274589 | 4.10E-07 | 9.96E-07 | 0.001989095 |
| Obesity | nk_cell | *MYO9A* | -1.598110129 | 1.27E-07 | 3.74E-07 | 0.000616396 |
| Obesity | nk_cell | *EVI5* | -1.646208657 | 9.06E-07 | 2.00E-06 | 0.004397464 |
| Obesity | nk_cell | *STOML1* | -5.639868626 | 4.94E-08 | 1.72E-07 | 0.000239845 |
| Obesity | nk_cell | *METTL22* | -1.607651602 | 1.38E-09 | 2.11E-08 | 6.71E-06 |
| Obesity | nk_cell | *TP53BP1* | -2.143399186 | 8.23E-07 | 1.83E-06 | 0.003994854 |
| Obesity | nk_cell | *RRP15* | -1.788355365 | 0.000143586 | 0.0002115 | 0.696681457 |
| Obesity | nk_cell | *DHX8* | -2.775483983 | 2.22E-07 | 5.91E-07 | 0.00107725 |
| Obesity | nk_cell | *ROGDI* | 6.66633167 | 6.29E-05 | 9.70E-05 | 0.305308604 |
| Obesity | nk_cell | *CBFB* | -2.404708249 | 0.000117172 | 0.000174768 | 0.568519431 |
| Obesity | nk_cell | *FGFR3* | -2.104360174 | 0.000770978 | 0.001047253 | 1 |
| Obesity | nk_cell | *IFI35* | -2.996910338 | 3.57E-06 | 6.82E-06 | 0.017340697 |
| Obesity | nk_cell | *HEATR6* | -2.074079563 | 2.11E-08 | 8.99E-08 | 0.000102527 |
| Obesity | nk_cell | *POLR1A* | -2.150221257 | 5.57E-07 | 1.29E-06 | 0.002704264 |
| Obesity | nk_cell | *IP6K2* | -2.56718532 | 1.04E-06 | 2.25E-06 | 0.005038634 |
| Obesity | nk_cell | *STON1-GTF2A* | -0.722636451 | 1.24E-05 | 2.12E-05 | 0.060241153 |
| Obesity | nk_cell | *SRBD1* | -1.448444619 | 0.039291091 | 0.045838032 | 1 |
| Obesity | nk_cell | *CYFIP1* | -2.045397247 | 4.34E-07 | 1.04E-06 | 0.002103846 |
| Obesity | nk_cell | *IFT80* | -3.230431407 | 1.61E-07 | 4.53E-07 | 0.000781525 |
| Obesity | nk_cell | *NUCKS1* | -1.916479798 | 0.003033032 | 0.00389835 | 1 |
| Obesity | nk_cell | *CLEC2D* | 1.432910317 | 7.16E-09 | 4.27E-08 | 3.47E-05 |
| Obesity | nk_cell | *DRD4* | -5.108619716 | 3.68E-08 | 1.37E-07 | 0.000178474 |
| Obesity | nk_cell | *PLA2G10* | 3.339682513 | 1.47E-09 | 2.11E-08 | 7.14E-06 |
| Obesity | nk_cell | *NEDD4* | -2.168413285 | 2.25E-06 | 4.49E-06 | 0.010912673 |
| Obesity | nk_cell | *GNB5* | -2.04511972 | 3.50E-06 | 6.69E-06 | 0.016960399 |
| Obesity | nk_cell | *LRP6* | -1.983188056 | 5.26E-08 | 1.82E-07 | 0.000255016 |
| Obesity | nk_cell | *DAPP1* | -0.820919679 | 0.019811784 | 0.023781983 | 1 |
| Obesity | nk_cell | *CLTCL1* | -2.31222695 | 4.02E-06 | 7.62E-06 | 0.019522076 |
| Obesity | nk_cell | *ZXDC* | -2.378057387 | 1.55E-05 | 2.61E-05 | 0.075368518 |
| Obesity | nk_cell | *WIPI1* | 1.203336791 | 0.039554683 | 0.046123365 | 1 |
| Obesity | nk_cell | *GBA2* | -2.928480087 | 4.06E-08 | 1.49E-07 | 0.000196914 |
| Obesity | nk_cell | *NDST1* | -1.433608235 | 1.77E-08 | 7.90E-08 | 8.60E-05 |
| Obesity | nk_cell | *ASNS* | -2.359331321 | 7.10E-08 | 2.31E-07 | 0.000344621 |
| Obesity | nk_cell | *AP3M2* | -3.354539281 | 1.78E-05 | 2.96E-05 | 0.086254639 |
| Obesity | nk_cell | *ST6GALNAC2* | -1.633382591 | 7.55E-09 | 4.37E-08 | 3.66E-05 |
| Obesity | nk_cell | *EIF2B3* | -2.259739704 | 1.69E-06 | 3.47E-06 | 0.008189295 |
| Obesity | nk_cell | *MAP4K4* | -2.155814216 | 1.99E-05 | 3.28E-05 | 0.096505971 |
| Obesity | nk_cell | *SNX13* | -2.195998885 | 6.00E-06 | 1.09E-05 | 0.02912111 |
| Obesity | nk_cell | *RPS6KA2* | -2.161005575 | 6.97E-07 | 1.59E-06 | 0.003380229 |
| Obesity | nk_cell | *ING3* | -2.370718689 | 1.90E-08 | 8.29E-08 | 9.20E-05 |
| Obesity | nk_cell | *VASH1* | -3.024282332 | 2.73E-06 | 5.36E-06 | 0.013255249 |
| Obesity | nk_cell | *TRIB2* | 2.201556049 | 3.74E-05 | 5.96E-05 | 0.181325997 |
| Obesity | nk_cell | *CPSF1* | -2.697347278 | 1.48E-08 | 6.92E-08 | 7.20E-05 |
| Obesity | nk_cell | *CYBRD1* | -1.59633278 | 6.24E-07 | 1.43E-06 | 0.003027553 |

| | | | | | | |
|---|---|---|---|---|---|---|
| Obesity | nk_cell | *RDH11* | -3.4681195 | 1.82E-06 | 3.72E-06 | 0.008842272 |
| Obesity | nk_cell | *EPN2* | -1.83010867 | 2.56E-07 | 6.68E-07 | 0.001242074 |
| Obesity | nk_cell | *ALDH3A2* | -1.670781577 | 3.82E-07 | 9.40E-07 | 0.001854824 |
| Obesity | nk_cell | *AFF4* | -1.777581608 | 7.10E-06 | 1.27E-05 | 0.034445684 |
| Obesity | nk_cell | *TRHDE* | -1.492010677 | 8.34E-10 | 1.99E-08 | 4.05E-06 |
| Obesity | nk_cell | *FCGR2B* | -2.733532831 | 3.03E-08 | 1.18E-07 | 0.000147182 |
| Obesity | nk_cell | *CRMP1* | -1.769700235 | 6.90E-09 | 4.17E-08 | 3.35E-05 |
| Obesity | nk_cell | *NDE1* | -2.513202661 | 1.78E-06 | 3.64E-06 | 0.008627288 |
| Obesity | nk_cell | *PVR* | -2.2855197 | 0.000363192 | 0.000510784 | 1 |
| Obesity | nk_cell | *SCARB1* | -2.102927901 | 2.84E-08 | 1.12E-07 | 0.000137804 |
| Obesity | nk_cell | *MOV10L1* | -1.277244811 | 3.94E-09 | 3.00E-08 | 1.91E-05 |
| Obesity | nk_cell | *ALPK1* | -1.873703075 | 0.000189256 | 0.000275261 | 0.918271793 |
| Obesity | nk_cell | *LLGL2* | 0.730397715 | 1.50E-09 | 2.11E-08 | 7.28E-06 |
| Obesity | nk_cell | *SDHA* | 1.594300828 | 0.034759031 | 0.040707414 | 1 |
| Obesity | nk_cell | *ADAM11* | -1.621894522 | 1.71E-06 | 3.50E-06 | 0.00827693 |
| Obesity | nk_cell | *PPP2R2C* | -1.845096518 | 1.58E-09 | 2.11E-08 | 7.66E-06 |
| Obesity | nk_cell | *ZNF532* | -2.116431503 | 2.21E-07 | 5.88E-07 | 0.001071019 |
| Obesity | nk_cell | *ZZEF1* | -1.999194477 | 0.000664707 | 0.00090824 | 1 |
| Obesity | nk_cell | *TACR2* | -10.5987793 | 0.000928015 | 0.001247985 | 1 |
| Obesity | nk_cell | *TIPIN* | 1.164044042 | 0.007412319 | 0.009212237 | 1 |
| Obesity | nk_cell | *TTC38* | 1.546632146 | 1.81E-09 | 2.18E-08 | 8.80E-06 |
| Obesity | nk_cell | *GRAMD4* | -2.73644094 | 0.000100284 | 0.000150737 | 0.486579964 |
| Obesity | nk_cell | *ADD2* | -2.729444473 | 0.007562343 | 0.009386794 | 1 |
| Obesity | nk_cell | *VPS9D1* | 2.529730782 | 0.013535434 | 0.016459631 | 1 |
| Obesity | nk_cell | *FNDC3B* | -1.64173195 | 1.89E-06 | 3.85E-06 | 0.009194046 |
| Obesity | nk_cell | *MOCOS* | -1.369634876 | 2.11E-08 | 8.99E-08 | 0.000102603 |
| Obesity | nk_cell | *SEC31B* | -1.571812856 | 0.000936951 | 0.001258955 | 1 |
| Obesity | nk_cell | *KIFAP3* | -2.258007953 | 1.19E-06 | 2.54E-06 | 0.005775592 |
| Obesity | nk_cell | *MKRN2* | -2.114700121 | 3.37E-05 | 5.42E-05 | 0.163743932 |
| Obesity | nk_cell | *MCM6* | -2.659812655 | 4.66E-06 | 8.68E-06 | 0.02260152 |
| Obesity | nk_cell | *MLH1* | -2.319510091 | 0.001391508 | 0.001840676 | 1 |
| Obesity | nk_cell | *FMO4* | -2.111270454 | 2.26E-06 | 4.52E-06 | 0.01098551 |
| Obesity | nk_cell | *RGS11* | -1.37281017 | 0.000482338 | 0.000668658 | 1 |
| Obesity | nk_cell | *SPAG5* | 2.956803407 | 5.38E-08 | 1.85E-07 | 0.000261175 |
| Obesity | nk_cell | *ANKRD13A* | -1.948754597 | 6.82E-09 | 4.14E-08 | 3.31E-05 |
| Obesity | nk_cell | *PAG1* | 2.660182631 | 8.69E-05 | 0.000131288 | 0.421435376 |
| Obesity | nk_cell | *GPATCH1* | -2.938554122 | 4.57E-06 | 8.54E-06 | 0.02219216 |
| Obesity | nk_cell | *ICAM3* | 0.534869558 | 1.29E-09 | 2.09E-08 | 6.28E-06 |
| Obesity | nk_cell | *NT5C2* | -1.111676525 | 0.005659979 | 0.00709618 | 1 |
| Obesity | nk_cell | *XAB2* | -3.904720251 | 8.63E-08 | 2.72E-07 | 0.000418822 |
| Obesity | nk_cell | *IL4R* | -2.357103489 | 2.15E-07 | 5.76E-07 | 0.001042391 |
| Obesity | nk_cell | *DYNC1I2* | -2.113961091 | 2.57E-07 | 6.69E-07 | 0.00124471 |
| Obesity | nk_cell | *LRCH4* | -2.772463587 | 1.70E-06 | 3.50E-06 | 0.008263374 |
| Obesity | nk_cell | *SIRT6* | -3.500935557 | 4.89E-05 | 7.68E-05 | 0.237143126 |
| Obesity | nk_cell | *GPR137B* | -2.018976023 | 1.02E-05 | 1.77E-05 | 0.049542842 |
| Obesity | nk_cell | *NAALAD2* | -1.095759222 | 8.12E-05 | 0.00012335 | 0.393981017 |
| Obesity | nk_cell | *AMPH* | 2.880943979 | 5.25E-09 | 3.56E-08 | 2.54E-05 |
| Obesity | nk_cell | *MCCC1* | -1.429897685 | 1.65E-06 | 3.40E-06 | 0.008007584 |
| Obesity | nk_cell | *ACER3* | -1.284063986 | 6.34E-06 | 1.15E-05 | 0.030754948 |
| Obesity | nk_cell | *PIK3C3* | -2.628746808 | 2.02E-06 | 4.08E-06 | 0.009817172 |

| | | | | | | |
|---|---|---|---|---|---|---|
| Obesity | nk_cell | *RBFOX1* | -1.314987203 | 6.30E-05 | 9.70E-05 | 0.305496066 |
| Obesity | nk_cell | *PCM1* | -1.873672017 | 1.40E-06 | 2.95E-06 | 0.006800698 |
| Obesity | nk_cell | *MYH7B* | -1.628490052 | 2.33E-09 | 2.30E-08 | 1.13E-05 |
| Obesity | nk_cell | *CLUL1* | -1.914380492 | 2.53E-07 | 6.61E-07 | 0.001226476 |
| Obesity | nk_cell | *FKBP7* | -2.168317192 | 9.95E-08 | 3.06E-07 | 0.000482708 |
| Obesity | nk_cell | *XRCC5* | -1.551992043 | 0.019311386 | 0.023210019 | 1 |
| Obesity | nk_cell | *SP140* | 0.79465904 | 1.34E-09 | 2.11E-08 | 6.52E-06 |
| Obesity | nk_cell | *SAR1A* | -2.268796745 | 2.56E-07 | 6.69E-07 | 0.0012444 |
| Obesity | nk_cell | *RAPGEF3* | -0.822959808 | 0.001832978 | 0.002400434 | 1 |
| Obesity | nk_cell | *FDFT1* | 3.314376336 | 1.99E-05 | 3.29E-05 | 0.096628415 |
| Obesity | nk_cell | *SCGN* | 3.030375563 | 1.06E-09 | 2.05E-08 | 5.14E-06 |
| Obesity | nk_cell | *PGM1* | -2.04460624 | 1.15E-08 | 5.68E-08 | 5.59E-05 |
| Obesity | nk_cell | *DDX1* | -2.683636072 | 1.49E-05 | 2.51E-05 | 0.072269126 |
| Obesity | nk_cell | *STX7* | -1.788752302 | 1.60E-07 | 4.51E-07 | 0.000778093 |
| Obesity | nk_cell | *RABL2B* | -3.492845969 | 1.45E-06 | 3.03E-06 | 0.007015995 |
| Obesity | nk_cell | *DCT* | -1.065691924 | 1.57E-05 | 2.64E-05 | 0.076337841 |
| Obesity | nk_cell | *SCTR* | -1.657071764 | 3.95E-09 | 3.00E-08 | 1.92E-05 |
| Obesity | nk_cell | *RIF1* | -1.951020996 | 1.62E-07 | 4.54E-07 | 0.000784868 |
| Obesity | nk_cell | *KCNN2* | 2.16220586 | 5.70E-09 | 3.74E-08 | 2.77E-05 |
| Obesity | nk_cell | *PSEN1* | -2.106727973 | 2.98E-06 | 5.81E-06 | 0.014461433 |
| Obesity | nk_cell | *MOK* | -1.518383872 | 9.14E-07 | 2.01E-06 | 0.00443232 |
| Obesity | nk_cell | *STK17B* | 1.407384337 | 1.08E-08 | 5.45E-08 | 5.26E-05 |
| Obesity | nk_cell | *ZNF506* | -1.896419057 | 0.009177494 | 0.011316188 | 1 |
| Obesity | nk_cell | *PCDHB4* | 2.154713689 | 1.57E-09 | 2.11E-08 | 7.64E-06 |
| Obesity | nk_cell | *CCNT2* | -2.625707893 | 1.16E-05 | 2.00E-05 | 0.056451388 |
| Obesity | nk_cell | *SEMA5B* | -1.61621608 | 7.79E-08 | 2.50E-07 | 0.000378208 |
| Obesity | nk_cell | *DOPEY1* | -1.421078315 | 0.000664696 | 0.00090824 | 1 |
| Obesity | nk_cell | *LYRM2* | -2.038033445 | 5.68E-06 | 1.04E-05 | 0.027571678 |
| Obesity | nk_cell | *ZCCHC6* | -2.073628933 | 0.001400306 | 0.001851304 | 1 |
| Obesity | nk_cell | *ULK2* | -1.547872079 | 3.09E-08 | 1.20E-07 | 0.000150094 |
| Obesity | nk_cell | *PIBF1* | -1.947387576 | 0.000108409 | 0.000162446 | 0.526001335 |
| Obesity | nk_cell | *OXCT1* | -2.498234717 | 4.20E-07 | 1.02E-06 | 0.002036875 |
| Obesity | nk_cell | *SLC27A5* | 3.21955155 | 4.22E-08 | 1.52E-07 | 0.000204544 |
| Obesity | nk_cell | *ZNF586* | -1.6599505 | 0.007279958 | 0.009054693 | 1 |
| Obesity | nk_cell | *ZNF264* | -2.47080309 | 5.49E-08 | 1.87E-07 | 0.000266517 |
| Obesity | nk_cell | *RPS5* | 2.315325153 | 8.89E-05 | 0.000134295 | 0.431355966 |
| Obesity | nk_cell | *CHMP2B* | -2.941311283 | 0.000804921 | 0.001090001 | 1 |
| Obesity | nk_cell | *NOA1* | -3.687552658 | 1.25E-08 | 6.07E-08 | 6.07E-05 |
| Obesity | nk_cell | *WBP11* | 3.297207534 | 2.40E-06 | 4.76E-06 | 0.011661551 |
| Obesity | nk_cell | *NCOA1* | -1.573824951 | 0.003122016 | 0.004005294 | 1 |
| Obesity | nk_cell | *CAD* | -2.516202107 | 0.013137978 | 0.015988329 | 1 |
| Obesity | nk_cell | *CD59* | -1.692872705 | 0.00026678 | 0.000381131 | 1 |
| Obesity | nk_cell | *CD82* | 1.428511576 | 0.014368703 | 0.017442318 | 1 |
| Obesity | nk_cell | *HACE1* | -1.855666522 | 2.40E-08 | 9.87E-08 | 0.000116343 |
| Obesity | nk_cell | *SEH1L* | -2.083068339 | 1.91E-07 | 5.19E-07 | 0.00092545 |
| Obesity | nk_cell | *OVGP1* | -2.040766173 | 1.99E-06 | 4.02E-06 | 0.009661271 |
| Obesity | nk_cell | *SLC25A24* | -2.159277634 | 1.40E-06 | 2.95E-06 | 0.0068117 |
| Obesity | nk_cell | *ZNF213* | -2.49196491 | 7.11E-08 | 2.31E-07 | 0.000344868 |
| Obesity | nk_cell | *CPNE3* | -2.304763133 | 3.49E-06 | 6.68E-06 | 0.016937728 |
| Obesity | nk_cell | *RRN3* | -2.147195464 | 5.06E-09 | 3.50E-08 | 2.46E-05 |

| | | | | | | |
|---|---|---|---|---|---|---|
| Obesity | nk_cell | *DDHD2* | -2.238610334 | 5.83E-08 | 1.96E-07 | 0.000282687 |
| Obesity | nk_cell | *EPS15* | -2.195021283 | 1.22E-07 | 3.61E-07 | 0.000590853 |
| Obesity | nk_cell | *USP40* | -1.757043788 | 4.21E-08 | 1.52E-07 | 0.000204162 |
| Obesity | nk_cell | *POMGNT1* | -1.901013052 | 7.33E-09 | 4.31E-08 | 3.56E-05 |
| Obesity | nk_cell | *MAST2* | -1.847074221 | 0.000651601 | 0.000891588 | 1 |
| Obesity | nk_cell | *EIF2AK1* | -2.379230893 | 1.07E-07 | 3.24E-07 | 0.000519232 |
| Obesity | nk_cell | *NME8* | 4.619944321 | 8.06E-10 | 1.98E-08 | 3.91E-06 |
| Obesity | nk_cell | *SNX10* | 1.074712319 | 0.003668107 | 0.004687294 | 1 |
| Obesity | nk_cell | *ITPKC* | -1.391803242 | 5.59E-08 | 1.90E-07 | 0.000271082 |
| Obesity | nk_cell | *TMED2* | -2.888019811 | 1.94E-06 | 3.94E-06 | 0.009424089 |
| Obesity | nk_cell | *ALG9* | -3.119281212 | 1.09E-08 | 5.48E-08 | 5.31E-05 |
| Obesity | nk_cell | *ACOX3* | -2.68494865 | 4.04E-06 | 7.64E-06 | 0.019615296 |
| Obesity | nk_cell | *HSD17B14* | -1.543010882 | 3.46E-09 | 2.78E-08 | 1.68E-05 |
| Obesity | nk_cell | *TRIP6* | -3.425077428 | 1.11E-08 | 5.56E-08 | 5.40E-05 |
| Obesity | nk_cell | *ACHE* | -3.872626292 | 9.48E-08 | 2.94E-07 | 0.000460126 |
| Obesity | nk_cell | *SRRT* | -2.095903576 | 7.33E-09 | 4.31E-08 | 3.56E-05 |
| Obesity | nk_cell | *ADAMTS2* | -1.770168441 | 1.63E-08 | 7.43E-08 | 7.92E-05 |
| Obesity | nk_cell | *PSMC5* | 2.833219299 | 1.71E-06 | 3.50E-06 | 0.008278441 |
| Obesity | nk_cell | *UIMC1* | -2.197327352 | 0.000495016 | 0.000684864 | 1 |
| Obesity | nk_cell | *LPCAT2* | -2.926541118 | 1.83E-05 | 3.04E-05 | 0.088641465 |
| Obesity | nk_cell | *OGFOD1* | -4.133870306 | 1.83E-08 | 8.08E-08 | 8.86E-05 |
| Obesity | nk_cell | *TXNDC16* | -2.198359919 | 0.000433059 | 0.000602581 | 1 |
| Obesity | nk_cell | *SF3B2* | 1.809307148 | 0.004014626 | 0.005107227 | 1 |
| Obesity | nk_cell | *AAMDC* | -1.985980639 | 4.10E-08 | 1.50E-07 | 0.000199077 |
| Obesity | nk_cell | *GP6* | 0.851249185 | 0.006677882 | 0.008325047 | 1 |
| Obesity | nk_cell | *ASAP3* | -1.285556214 | 2.74E-09 | 2.48E-08 | 1.33E-05 |
| Obesity | nk_cell | *EDEM2* | -2.24652801 | 9.00E-09 | 4.83E-08 | 4.37E-05 |
| Obesity | nk_cell | *ANKRD10* | -1.928805937 | 7.24E-08 | 2.35E-07 | 0.00035141 |
| Obesity | nk_cell | *DOCK3* | -1.963787015 | 1.31E-08 | 6.28E-08 | 6.34E-05 |
| Obesity | nk_cell | *ARHGAP28* | -1.533348775 | 4.83E-08 | 1.69E-07 | 0.000234317 |
| Obesity | nk_cell | *PPP1R13B* | -2.830083926 | 3.21E-06 | 6.20E-06 | 0.015561537 |
| Obesity | nk_cell | *NSFL1C* | -1.850326622 | 7.41E-06 | 1.32E-05 | 0.035959385 |
| Obesity | nk_cell | *ZNF343* | -2.130768811 | 9.30E-07 | 2.04E-06 | 0.004510714 |
| Obesity | nk_cell | *EBF4* | -2.980706907 | 9.03E-09 | 4.84E-08 | 4.38E-05 |
| Obesity | nk_cell | *CPXM1* | -1.849285372 | 5.04E-09 | 3.50E-08 | 2.45E-05 |
| Obesity | nk_cell | *MAVS* | -2.039173254 | 5.26E-05 | 8.21E-05 | 0.255220166 |
| Obesity | nk_cell | *XRN2* | -2.154718279 | 0.000681402 | 0.000929172 | 1 |
| Obesity | nk_cell | *ESF1* | -2.006371018 | 5.25E-08 | 1.82E-07 | 0.000254554 |
| Obesity | nk_cell | *RBBP9* | -2.689805411 | 5.58E-06 | 1.02E-05 | 0.027071504 |
| Obesity | nk_cell | *TMEM230* | -2.350735427 | 1.12E-06 | 2.40E-06 | 0.005429995 |
| Obesity | nk_cell | *KIF16B* | -2.454473207 | 7.72E-06 | 1.37E-05 | 0.037480748 |
| Obesity | nk_cell | *TBX5* | -5.165246443 | 1.68E-07 | 4.69E-07 | 0.000816202 |
| Obesity | nk_cell | *FXYD5* | 2.139507592 | 2.55E-09 | 2.40E-08 | 1.24E-05 |
| Obesity | nk_cell | *ZNF302* | -2.054163547 | 0.000141244 | 0.000208204 | 0.685318154 |
| Obesity | nk_cell | *GRAMD1A* | -2.444164581 | 1.53E-07 | 4.35E-07 | 0.00074308 |
| Obesity | nk_cell | *CDIP1* | -2.990358628 | 4.80E-08 | 1.68E-07 | 0.000233074 |
| Obesity | nk_cell | *GANAB* | -2.433540488 | 4.85E-08 | 1.70E-07 | 0.000235319 |
| Obesity | nk_cell | *MLF2* | -1.590820352 | 2.28E-08 | 9.51E-08 | 0.000110688 |
| Obesity | nk_cell | *ANKRD24* | -2.046124057 | 2.42E-09 | 2.35E-08 | 1.18E-05 |
| Obesity | nk_cell | *DHX32* | -1.720519106 | 1.87E-08 | 8.19E-08 | 9.07E-05 |

| | | | | | | |
|---|---|---|---|---|---|---|
| Obesity | nk_cell | *MRPS33* | -1.795483348 | 3.16E-09 | 2.68E-08 | 1.53E-05 |
| Obesity | nk_cell | *STRN4* | -2.635650009 | 0.001661081 | 0.002184164 | 1 |
| Obesity | nk_cell | *MUL1* | -1.668153733 | 0.000810657 | 0.001097463 | 1 |
| Obesity | nk_cell | *SPG21* | -2.61933286 | 1.27E-07 | 3.73E-07 | 0.000615224 |
| Obesity | nk_cell | *CHRD* | -1.911642733 | 1.12E-08 | 5.57E-08 | 5.44E-05 |
| Obesity | nk_cell | *RAB11FIP3* | -2.421170588 | 4.32E-05 | 6.84E-05 | 0.209825869 |
| Obesity | nk_cell | *GNPTG* | -2.32113499 | 2.27E-08 | 9.47E-08 | 0.000109955 |
| Obesity | nk_cell | *PABPC4* | -4.525003392 | 4.03E-13 | 9.30E-11 | 1.95E-09 |
| Obesity | nk_cell | *USP48* | -2.176911972 | 1.25E-06 | 2.65E-06 | 0.006047932 |
| Obesity | nk_cell | *PDPR* | -2.20352398 | 4.31E-08 | 1.55E-07 | 0.000209109 |
| Obesity | nk_cell | *GLG1* | -2.086981788 | 0.000122355 | 0.000181994 | 0.593664407 |
| Obesity | nk_cell | *DTX2* | -4.017209261 | 4.78E-06 | 8.89E-06 | 0.023194828 |
| Obesity | nk_cell | *LAMB1* | -1.028396537 | 2.97E-05 | 4.82E-05 | 0.144305458 |
| Obesity | nk_cell | *ABCC6* | -1.043450291 | 3.39E-09 | 2.77E-08 | 1.64E-05 |
| Obesity | nk_cell | *RAPGEF4* | -1.145041183 | 1.87E-09 | 2.19E-08 | 9.09E-06 |
| Obesity | nk_cell | *TF* | -3.116429175 | 1.73E-08 | 7.77E-08 | 8.40E-05 |
| Obesity | nk_cell | *RGS17* | -2.913726116 | 2.72E-13 | 6.96E-11 | 1.32E-09 |
| Obesity | nk_cell | *HAUS4* | 0.952768067 | 0.000970024 | 0.001300513 | 1 |
| Obesity | nk_cell | *OSGEP* | -2.449111004 | 0.000940124 | 0.001262519 | 1 |
| Obesity | nk_cell | *SCFD1* | -2.126690713 | 1.58E-05 | 2.65E-05 | 0.076557688 |
| Obesity | nk_cell | *SUPT16H* | -2.820551312 | 5.00E-05 | 7.83E-05 | 0.242612838 |
| Obesity | nk_cell | *CAPN3* | 3.278013734 | 1.74E-07 | 4.81E-07 | 0.000842364 |
| Obesity | nk_cell | *UNC13D* | 1.040927776 | 0.000422368 | 0.000588719 | 1 |
| Obesity | nk_cell | *MFSD11* | -1.944215857 | 6.85E-05 | 0.000104902 | 0.33232977 |
| Obesity | nk_cell | *NUP50* | -1.993044748 | 0.003997177 | 0.005086363 | 1 |
| Obesity | nk_cell | *COMT* | -2.499794767 | 7.84E-07 | 1.76E-06 | 0.003802026 |
| Obesity | nk_cell | *LRRFIP2* | -2.009075339 | 3.56E-06 | 6.79E-06 | 0.017257247 |
| Obesity | nk_cell | *SEC22C* | -2.702487724 | 5.51E-07 | 1.28E-06 | 0.002675041 |
| Obesity | nk_cell | *XYLB* | -1.993853557 | 1.03E-08 | 5.32E-08 | 5.00E-05 |
| Obesity | nk_cell | *GABRP* | 3.045084708 | 0.000944581 | 0.001268154 | 1 |
| Obesity | nk_cell | *CDC6* | -3.660597135 | 8.41E-10 | 1.99E-08 | 4.08E-06 |
| Obesity | nk_cell | *FMO2* | -0.886476303 | 0.000216078 | 0.000311842 | 1 |
| Obesity | nk_cell | *MSH2* | -2.481085748 | 9.92E-08 | 3.05E-07 | 0.000481078 |
| Obesity | nk_cell | *PSMD5* | -2.648089316 | 8.53E-07 | 1.89E-06 | 0.004138038 |
| Obesity | nk_cell | *CRAT* | -2.37753289 | 4.07E-09 | 3.06E-08 | 1.98E-05 |
| Obesity | nk_cell | *NANS* | -2.383887795 | 0.033596284 | 0.039402749 | 1 |
| Obesity | nk_cell | *PDE6C* | -4.701694646 | 1.48E-05 | 2.51E-05 | 0.071974622 |
| Obesity | nk_cell | *CWF19L1* | -1.301420418 | 0.00043633 | 0.000606959 | 1 |
| Obesity | nk_cell | *CRTAC1* | -1.530660085 | 5.80E-06 | 1.06E-05 | 0.028119232 |
| Obesity | nk_cell | *WAC* | -1.979369243 | 1.53E-06 | 3.18E-06 | 0.007405225 |
| Obesity | nk_cell | *IFT74* | -1.530991043 | 1.22E-07 | 3.62E-07 | 0.000593226 |
| Obesity | nk_cell | *IL12RB1* | 0.472840848 | 2.24E-08 | 9.41E-08 | 0.000108927 |
| Obesity | nk_cell | *SCD* | -1.859778623 | 0.000115044 | 0.00017191 | 0.558191141 |
| Obesity | nk_cell | *ERMP1* | -2.850842406 | 4.09E-06 | 7.71E-06 | 0.019833025 |
| Obesity | nk_cell | *RAB18* | -2.537898862 | 6.20E-05 | 9.57E-05 | 0.300887955 |
| Obesity | nk_cell | *PRTFDC1* | -2.095691628 | 0.000657508 | 0.000899163 | 1 |
| Obesity | nk_cell | *TSPAN15* | -2.06594181 | 4.54E-08 | 1.61E-07 | 0.000220112 |
| Obesity | nk_cell | *CATSPERG* | -1.641194073 | 0.000325378 | 0.000460407 | 1 |
| Obesity | nk_cell | *CIRBP* | -1.725693825 | 1.01E-06 | 2.20E-06 | 0.004901532 |
| Obesity | nk_cell | *MARCH2* | -1.501968269 | 0.005304858 | 0.006664726 | 1 |

| | | | | | | |
|---|---|---|---|---|---|---|
| Obesity | nk_cell | *MTAP* | -2.909793571 | 5.79E-08 | 1.95E-07 | 0.000280847 |
| Obesity | nk_cell | *POLR2E* | -1.99492079 | 9.37E-09 | 4.97E-08 | 4.54E-05 |
| Obesity | nk_cell | *MADCAM1* | -1.166457236 | 2.20E-09 | 2.27E-08 | 1.07E-05 |
| Obesity | nk_cell | *MED15* | -1.60991175 | 0.040068371 | 0.046699912 | 1 |
| Obesity | nk_cell | *SERPIND1* | -3.350957469 | 8.71E-10 | 2.00E-08 | 4.23E-06 |
| Obesity | nk_cell | *SNAP29* | -2.324497767 | 1.19E-06 | 2.53E-06 | 0.005750718 |
| Obesity | nk_cell | *LZTR1* | -3.154017621 | 0.000135022 | 0.000199734 | 0.65512804 |
| Obesity | nk_cell | *MMP11* | 2.135831619 | 0.036816962 | 0.043003347 | 1 |
| Obesity | nk_cell | *CECR2* | -1.105729007 | 1.82E-09 | 2.18E-08 | 8.83E-06 |
| Obesity | nk_cell | *BCL2L13* | -1.423519234 | 6.67E-06 | 1.20E-05 | 0.03237384 |
| Obesity | nk_cell | *DDTL* | 1.430775872 | 8.68E-07 | 1.92E-06 | 0.00421084 |
| Obesity | nk_cell | *SPECC1L* | -2.426644913 | 1.16E-07 | 3.47E-07 | 0.000563452 |
| Obesity | nk_cell | *PPIL2* | -2.118729927 | 0.002305508 | 0.00299545 | 1 |
| Obesity | nk_cell | *PES1* | 2.693398399 | 1.90E-05 | 3.14E-05 | 0.092125036 |
| Obesity | nk_cell | *SLC35E4* | -1.484794898 | 1.12E-07 | 3.37E-07 | 0.00054428 |
| Obesity | nk_cell | *CRYBB3* | 3.580244995 | 0.000139935 | 0.000206498 | 0.678965115 |
| Obesity | nk_cell | *CYTH4* | 1.195026546 | 1.79E-09 | 2.17E-08 | 8.68E-06 |
| Obesity | nk_cell | *CARD10* | -2.32451282 | 1.22E-10 | 8.58E-09 | 5.92E-07 |
| Obesity | nk_cell | *LRP5L* | -2.261449221 | 5.18E-06 | 9.55E-06 | 0.025124559 |
| Obesity | nk_cell | *LGALS2* | 1.795326457 | 0.000486927 | 0.000674634 | 1 |
| Obesity | nk_cell | *SH3BP1* | 0.710356893 | 1.15E-09 | 2.05E-08 | 5.59E-06 |
| Obesity | nk_cell | *HPS4* | -1.682558513 | 1.30E-08 | 6.28E-08 | 6.33E-05 |
| Obesity | nk_cell | *TRIOBP* | -2.243654773 | 6.45E-09 | 4.04E-08 | 3.13E-05 |
| Obesity | nk_cell | *ANKRD54* | -1.629294917 | 6.68E-12 | 1.16E-09 | 3.24E-08 |
| Obesity | nk_cell | *EIF3L* | -2.653226389 | 2.14E-07 | 5.75E-07 | 0.001040593 |
| Obesity | nk_cell | *CCDC134* | -2.416481485 | 0.00043704 | 0.000607773 | 1 |
| Obesity | nk_cell | *TTC28* | -1.538797848 | 2.21E-07 | 5.89E-07 | 0.001072719 |
| Obesity | nk_cell | *CENPM* | 2.407157503 | 4.51E-13 | 9.74E-11 | 2.19E-09 |
| Obesity | nk_cell | *SLC5A4* | -0.25424714 | 0.0401043 | 0.046730562 | 1 |
| Obesity | nk_cell | *CYP2D6* | -2.806947536 | 1.09E-10 | 7.86E-09 | 5.27E-07 |
| Obesity | nk_cell | *CBY1* | -2.195120301 | 4.07E-06 | 7.69E-06 | 0.019770282 |
| Obesity | nk_cell | *RTCB* | -2.420469087 | 1.98E-06 | 4.01E-06 | 0.009611576 |
| Obesity | nk_cell | *RAB36* | -2.27813446 | 5.20E-10 | 1.75E-08 | 2.52E-06 |
| Obesity | nk_cell | *TIMP3* | -1.577304903 | 1.82E-07 | 5.01E-07 | 0.000884722 |
| Obesity | nk_cell | *PPP6R2* | -2.116669944 | 4.22E-05 | 6.69E-05 | 0.204925532 |
| Obesity | nk_cell | *CYB5R3* | -2.187425366 | 4.18E-07 | 1.01E-06 | 0.002026654 |
| Obesity | nk_cell | *RHBDD3* | -2.999851055 | 2.15E-06 | 4.31E-06 | 0.010434305 |
| Obesity | nk_cell | *TOM1* | -1.976108495 | 3.84E-05 | 6.12E-05 | 0.186428532 |
| Obesity | nk_cell | *CHKB* | -2.232945145 | 4.54E-08 | 1.61E-07 | 0.000220361 |
| Obesity | nk_cell | *THOC5* | -2.491726999 | 1.09E-05 | 1.88E-05 | 0.052913342 |
| Obesity | nk_cell | *TSPO* | 2.585930462 | 0.002270612 | 0.002952038 | 1 |
| Obesity | nk_cell | *TTLL12* | -3.22773921 | 3.72E-07 | 9.17E-07 | 0.001803164 |
| Obesity | nk_cell | *MTMR3* | -1.969620194 | 1.67E-06 | 3.43E-06 | 0.008083245 |
| Obesity | nk_cell | *APOL4* | -1.455197185 | 3.10E-07 | 7.86E-07 | 0.001505868 |
| Obesity | nk_cell | *SAMM50* | -2.904138123 | 3.59E-08 | 1.34E-07 | 0.000173973 |
| Obesity | nk_cell | *FOXRED2* | -1.276226345 | 8.33E-08 | 2.64E-07 | 0.000404368 |
| Obesity | nk_cell | *IFT27* | -3.875949472 | 6.98E-05 | 0.000106736 | 0.338459955 |
| Obesity | nk_cell | *KIAA0930* | -3.35599899 | 1.84E-05 | 3.06E-05 | 0.089400238 |
| Obesity | nk_cell | *SLC25A17* | -2.326864605 | 3.27E-06 | 6.32E-06 | 0.015866492 |
| Obesity | nk_cell | *FAM118A* | -1.376876567 | 0.000349014 | 0.0004917 | 1 |

| | | | | | | |
|---|---|---|---|---|---|---|
| Obesity | nk_cell | *L3MBTL2* | 3.315836718 | 2.34E-08 | 9.67E-08 | 0.000113659 |
| Obesity | nk_cell | *ZC3H7B* | -2.7183175 | 4.06E-06 | 7.67E-06 | 0.019697175 |
| Obesity | nk_cell | *ACO2* | -1.813094445 | 8.97E-09 | 4.82E-08 | 4.35E-05 |
| Obesity | nk_cell | *DESI1* | -2.615134746 | 9.03E-06 | 1.58E-05 | 0.043811809 |
| Obesity | nk_cell | *MLC1* | 0.526119301 | 6.01E-10 | 1.84E-08 | 2.92E-06 |
| Obesity | nk_cell | *HDAC10* | -2.451696872 | 8.03E-09 | 4.51E-08 | 3.90E-05 |
| Obesity | nk_cell | *KHNYN* | -1.895801051 | 0.000372482 | 0.000522495 | 1 |
| Obesity | nk_cell | *SDR39U1* | -1.355413142 | 1.36E-06 | 2.87E-06 | 0.006595383 |
| Obesity | nk_cell | *RBM23* | -1.462242979 | 0.037694067 | 0.044006645 | 1 |
| Obesity | nk_cell | *AP4S1* | -2.148502015 | 1.12E-08 | 5.56E-08 | 5.43E-05 |
| Obesity | nk_cell | *NIN* | 2.438811903 | 7.86E-07 | 1.76E-06 | 0.003813962 |
| Obesity | nk_cell | *PYGL* | -1.114631936 | 2.83E-07 | 7.23E-07 | 0.001370903 |
| Obesity | nk_cell | *CNIH1* | -2.238487644 | 9.89E-09 | 5.16E-08 | 4.80E-05 |
| Obesity | nk_cell | *ATP6V1D* | -1.934690792 | 3.10E-08 | 1.20E-07 | 0.000150307 |
| Obesity | nk_cell | *PLEK2* | -2.407461122 | 6.01E-05 | 9.30E-05 | 0.291566825 |
| Obesity | nk_cell | *GSTZ1* | -1.174460008 | 6.65E-09 | 4.09E-08 | 3.23E-05 |
| Obesity | nk_cell | *TMED8* | -1.621932221 | 1.72E-07 | 4.77E-07 | 0.000832468 |
| Obesity | nk_cell | *LGMN* | -1.8466277 | 2.04E-06 | 4.10E-06 | 0.00987707 |
| Obesity | nk_cell | *MTHFD1* | -1.517194294 | 1.07E-08 | 5.41E-08 | 5.17E-05 |
| Obesity | nk_cell | *TELO2* | -1.814570944 | 2.96E-09 | 2.58E-08 | 1.44E-05 |
| Obesity | nk_cell | *GSKIP* | -3.134289582 | 1.95E-08 | 8.43E-08 | 9.44E-05 |
| Obesity | nk_cell | *PSMB5* | -1.517003363 | 0.00302638 | 0.003890831 | 1 |
| Obesity | nk_cell | *YY1* | -2.569670374 | 4.90E-05 | 7.69E-05 | 0.237528413 |
| Obesity | nk_cell | *CCNB1IP1* | -2.517827382 | 4.42E-08 | 1.58E-07 | 0.000214493 |
| Obesity | nk_cell | *EFS* | -2.914511021 | 2.89E-08 | 1.13E-07 | 0.000140082 |
| Obesity | nk_cell | *CINP* | -1.665568419 | 6.26E-09 | 3.98E-08 | 3.04E-05 |
| Obesity | nk_cell | *KIAA0391* | 4.952128341 | 4.80E-08 | 1.68E-07 | 0.000232779 |
| Obesity | nk_cell | *EMC9* | -1.875528233 | 5.18E-08 | 1.80E-07 | 0.000251543 |
| Obesity | nk_cell | *TRPC4AP* | -1.837876798 | 1.92E-06 | 3.89E-06 | 0.009301271 |
| Obesity | nk_cell | *PYGB* | -1.418358408 | 0.003932179 | 0.005007594 | 1 |
| Obesity | nk_cell | *CD40* | -1.732147671 | 3.54E-09 | 2.83E-08 | 1.72E-05 |
| Obesity | nk_cell | *UQCC1* | -1.95639708 | 1.59E-06 | 3.30E-06 | 0.007708725 |
| Obesity | nk_cell | *IFT52* | -1.612588541 | 0.011322362 | 0.013841295 | 1 |
| Obesity | nk_cell | *NDRG3* | -2.600653028 | 0.000393844 | 0.000551336 | 1 |
| Obesity | nk_cell | *PABPC1L* | -2.421440392 | 5.76E-09 | 3.76E-08 | 2.80E-05 |
| Obesity | nk_cell | *DOK5* | -1.726625965 | 2.68E-09 | 2.45E-08 | 1.30E-05 |
| Obesity | nk_cell | *CSTF1* | 3.700552767 | 0.007749711 | 0.009606949 | 1 |
| Obesity | nk_cell | *NELFCD* | -2.538607687 | 0.000792216 | 0.001074297 | 1 |
| Obesity | nk_cell | *CTSZ* | -2.333802679 | 1.86E-07 | 5.09E-07 | 0.000903012 |
| Obesity | nk_cell | *MTG2* | -2.195183556 | 3.85E-07 | 9.46E-07 | 0.001870263 |
| Obesity | nk_cell | *TCFL5* | -2.980583273 | 1.12E-07 | 3.37E-07 | 0.000542784 |
| Obesity | nk_cell | *DIDO1* | -2.140183632 | 0.000137147 | 0.000202816 | 0.665438485 |
| Obesity | nk_cell | *NKAIN4* | -1.47273265 | 0.020003712 | 0.024000497 | 1 |
| Obesity | nk_cell | *PTK6* | -3.309447269 | 1.17E-09 | 2.05E-08 | 5.65E-06 |
| Obesity | nk_cell | *GMEB2* | -2.256253395 | 3.35E-09 | 2.76E-08 | 1.63E-05 |
| Obesity | nk_cell | *CDC25B* | 2.523913218 | 2.29E-07 | 6.06E-07 | 0.001109979 |
| Obesity | nk_cell | *RNF24* | -2.320967315 | 4.16E-06 | 7.83E-06 | 0.020189556 |
| Obesity | nk_cell | *NDUFAF5* | -1.383294553 | 0.000280452 | 0.000399634 | 1 |
| Obesity | nk_cell | *TRIB3* | 6.5463199 | 1.51E-07 | 4.30E-07 | 0.000733166 |
| Obesity | nk_cell | *RASSF2* | -1.601942714 | 0.006917854 | 0.008615356 | 1 |

| | | | | | | |
|---|---|---|---|---|---|---|
| Obesity | nk_cell | *ANGPT4* | -2.102236135 | 2.98E-09 | 2.58E-08 | 1.44E-05 |
| Obesity | nk_cell | *RSPO4* | -1.425885128 | 4.76E-08 | 1.68E-07 | 0.000230931 |
| Obesity | nk_cell | *SEC23B* | 1.336388909 | 0.029571877 | 0.034859754 | 1 |
| Obesity | nk_cell | *CRNKL1* | -2.509669606 | 3.57E-05 | 5.71E-05 | 0.173097226 |
| Obesity | nk_cell | *MROH8* | -1.704865043 | 0.000911977 | 0.001227778 | 1 |
| Obesity | nk_cell | *MANBAL* | -1.515599059 | 1.19E-05 | 2.04E-05 | 0.057680184 |
| Obesity | nk_cell | *ACTR5* | 3.61958551 | 1.54E-05 | 2.59E-05 | 0.074499257 |
| Obesity | nk_cell | *MAP1LC3A* | -2.38878634 | 2.13E-08 | 9.05E-08 | 0.000103416 |
| Obesity | nk_cell | *RBFA* | 2.174878464 | 0.019529658 | 0.023454926 | 1 |
| Obesity | nk_cell | *METTL4* | -2.783037188 | 4.36E-07 | 1.05E-06 | 0.002115378 |
| Obesity | nk_cell | *MYOM1* | -1.690864806 | 9.90E-09 | 5.16E-08 | 4.81E-05 |
| Obesity | nk_cell | *LAMA1* | -1.717487529 | 2.65E-10 | 1.29E-08 | 1.29E-06 |
| Obesity | nk_cell | *RNF125* | 4.096346 | 0.000314491 | 0.000445651 | 1 |
| Obesity | nk_cell | *MIB1* | -2.135828228 | 1.47E-06 | 3.08E-06 | 0.007141367 |
| Obesity | nk_cell | *NALCN* | -1.998689709 | 5.61E-08 | 1.90E-07 | 0.00027218 |
| Obesity | nk_cell | *STK24* | -2.245873709 | 2.42E-06 | 4.80E-06 | 0.011760314 |
| Obesity | nk_cell | *DNAJC3* | -2.425534022 | 1.88E-05 | 3.12E-05 | 0.091407986 |
| Obesity | nk_cell | *UGGT2* | -1.925635046 | 1.42E-08 | 6.70E-08 | 6.90E-05 |
| Obesity | nk_cell | *ARHGEF7* | -1.99574815 | 2.87E-07 | 7.31E-07 | 0.001390417 |
| Obesity | nk_cell | *PARP4* | -2.134674671 | 4.50E-05 | 7.10E-05 | 0.218140851 |
| Obesity | nk_cell | *SUPT20H* | -2.477859361 | 8.18E-08 | 2.60E-07 | 0.000396976 |
| Obesity | nk_cell | *MRPS31* | -2.211962114 | 1.87E-07 | 5.12E-07 | 0.000909666 |
| Obesity | nk_cell | *RGCC* | -0.988739116 | 0.026568089 | 0.031525646 | 1 |
| Obesity | nk_cell | *DGKH* | -1.644605028 | 0.000949035 | 0.001273782 | 1 |
| Obesity | nk_cell | *KATNAL1* | -2.176341896 | 2.94E-07 | 7.47E-07 | 0.001425909 |
| Obesity | nk_cell | *INTS6* | -2.389091453 | 3.11E-06 | 6.03E-06 | 0.015094296 |
| Obesity | nk_cell | *CLN5* | -2.803809078 | 8.29E-05 | 0.000125647 | 0.40210134 |
| Obesity | nk_cell | *MGRN1* | -2.306922552 | 1.14E-05 | 1.96E-05 | 0.055265732 |
| Obesity | nk_cell | *ELMO3* | 1.335656229 | 0.006026197 | 0.007533911 | 1 |
| Obesity | nk_cell | *LONP2* | -1.821297498 | 2.27E-06 | 4.53E-06 | 0.011022164 |
| Obesity | nk_cell | *PLLP* | -1.496426163 | 2.90E-09 | 2.56E-08 | 1.41E-05 |
| Obesity | nk_cell | *DHODH* | -1.770216661 | 6.85E-07 | 1.56E-06 | 0.00332429 |
| Obesity | nk_cell | *CCDC113* | 2.149424149 | 1.91E-05 | 3.16E-05 | 0.09281471 |
| Obesity | nk_cell | *PSMD7* | 2.109773935 | 0.014876059 | 0.018035641 | 1 |
| Obesity | nk_cell | *COG4* | -3.23965406 | 1.89E-07 | 5.15E-07 | 0.000917259 |
| Obesity | nk_cell | *MON1B* | -3.94642417 | 1.12E-05 | 1.92E-05 | 0.054137735 |
| Obesity | nk_cell | *NPRL3* | -2.210903848 | 0.000121856 | 0.000181363 | 0.591244493 |
| Obesity | nk_cell | *NAGPA* | -2.132764017 | 1.38E-07 | 3.98E-07 | 0.000668875 |
| Obesity | nk_cell | *ZNF500* | -1.663642005 | 5.28E-06 | 9.72E-06 | 0.025632665 |
| Obesity | nk_cell | *NME4* | -2.759777018 | 1.48E-09 | 2.11E-08 | 7.20E-06 |
| Obesity | nk_cell | *NOMO3* | 1.803816515 | 0.001944099 | 0.002540471 | 1 |
| Obesity | nk_cell | *LMF1* | -2.321500527 | 2.29E-07 | 6.06E-07 | 0.001109875 |
| Obesity | nk_cell | *NARFL* | -2.402769082 | 4.36E-07 | 1.05E-06 | 0.002113107 |
| Obesity | nk_cell | *CLCN7* | 1.850217059 | 0.000379915 | 0.000532605 | 1 |
| Obesity | nk_cell | *HAGHL* | -1.2465327 | 0.007436389 | 0.009239785 | 1 |
| Obesity | nk_cell | *SLC7A5* | 2.892668236 | 2.12E-07 | 5.69E-07 | 0.001026749 |
| Obesity | nk_cell | *RHBDL1* | -4.348709008 | 3.08E-10 | 1.40E-08 | 1.49E-06 |
| Obesity | nk_cell | *NUBP1* | -2.514023382 | 4.08E-06 | 7.70E-06 | 0.019802965 |
| Obesity | nk_cell | *EARS2* | -1.411079244 | 0.021830942 | 0.026096017 | 1 |
| Obesity | nk_cell | *CPPED1* | -2.046257546 | 7.65E-09 | 4.39E-08 | 3.71E-05 |

| | | | | | | |
|---|---|---|---|---|---|---|
| Obesity | nk_cell | *HMOX2* | 2.303280906 | 1.76E-07 | 4.86E-07 | 0.000852667 |
| Obesity | nk_cell | *TOX3* | -3.451110466 | 4.46E-07 | 1.07E-06 | 0.002161893 |
| Obesity | nk_cell | *RRN3P2* | 2.109294219 | 0.000120251 | 0.000179084 | 0.583456819 |
| Obesity | nk_cell | *PYCARD* | 1.667211191 | 2.51E-11 | 2.77E-09 | 1.22E-07 |
| Obesity | nk_cell | *CDIPT* | 2.499979235 | 1.70E-06 | 3.49E-06 | 0.008243747 |
| Obesity | nk_cell | *KAT8* | -2.127141739 | 8.09E-09 | 4.51E-08 | 3.92E-05 |
| Obesity | nk_cell | *TMC5* | -1.634539699 | 2.52E-10 | 1.27E-08 | 1.22E-06 |
| Obesity | nk_cell | *C16orf62* | -1.900393163 | 7.32E-08 | 2.37E-07 | 0.000355167 |
| Obesity | nk_cell | *RNF40* | -3.211518302 | 3.68E-07 | 9.10E-07 | 0.001783641 |
| Obesity | nk_cell | *KNOP1* | -3.267318909 | 4.32E-09 | 3.18E-08 | 2.10E-05 |
| Obesity | nk_cell | *IQCH* | -2.385399503 | 1.14E-07 | 3.41E-07 | 0.000550985 |
| Obesity | nk_cell | *LACTB* | -2.315166865 | 3.73E-08 | 1.39E-07 | 0.000181167 |
| Obesity | nk_cell | *HERC1* | -1.899230118 | 0.000214692 | 0.000310025 | 1 |
| Obesity | nk_cell | *TRIP4* | -2.193606701 | 2.85E-05 | 4.62E-05 | 0.138167725 |
| Obesity | nk_cell | *TTC23* | -1.931868999 | 2.63E-08 | 1.06E-07 | 0.000127454 |
| Obesity | nk_cell | *FAH* | -1.790680463 | 3.51E-09 | 2.82E-08 | 1.70E-05 |
| Obesity | nk_cell | *SLC30A4* | -3.293738783 | 7.89E-06 | 1.39E-05 | 0.038280417 |
| Obesity | nk_cell | *ZDHHC2* | -2.52631348 | 0.002511318 | 0.003253649 | 1 |
| Obesity | nk_cell | *ZFAND1* | -2.714759795 | 1.72E-07 | 4.79E-07 | 0.000835757 |
| Obesity | nk_cell | *EYA1* | -1.053795693 | 1.47E-08 | 6.87E-08 | 7.13E-05 |
| Obesity | nk_cell | *IMPAD1* | -1.731139841 | 3.56E-09 | 2.83E-08 | 1.73E-05 |
| Obesity | nk_cell | *SFRP1* | -3.320913236 | 3.08E-06 | 5.97E-06 | 0.014927563 |
| Obesity | nk_cell | *NIPAL2* | -2.226239832 | 0.002595249 | 0.003359698 | 1 |
| Obesity | nk_cell | *RAB2A* | -2.11949635 | 3.97E-07 | 9.68E-07 | 0.001924615 |
| Obesity | nk_cell | *ARMC1* | -2.239686954 | 6.99E-07 | 1.59E-06 | 0.003392108 |
| Obesity | nk_cell | *SPAG1* | -2.03520447 | 0.00079736 | 0.001080667 | 1 |
| Obesity | nk_cell | *CHRAC1* | -5.088604403 | 0.000201717 | 0.000292159 | 0.978732481 |
| Obesity | nk_cell | *SNX16* | -3.054525139 | 1.80E-05 | 2.99E-05 | 0.087309494 |
| Obesity | nk_cell | *GSDMD* | -2.571907167 | 0.012252027 | 0.014943901 | 1 |
| Obesity | nk_cell | *TSTA3* | -1.491703956 | 8.77E-09 | 4.74E-08 | 4.26E-05 |
| Obesity | nk_cell | *TNFRSF10A* | 1.088669799 | 0.029604851 | 0.034890147 | 1 |
| Obesity | nk_cell | *ERICH1* | -1.986876177 | 2.59E-08 | 1.05E-07 | 0.000125561 |
| Obesity | nk_cell | *TUSC3* | -1.546449156 | 7.23E-09 | 4.27E-08 | 3.51E-05 |
| Obesity | nk_cell | *KLHDC4* | 2.604555947 | 2.56E-08 | 1.04E-07 | 0.000124436 |
| Obesity | nk_cell | *MAN2B1* | -2.698140739 | 5.60E-06 | 1.03E-05 | 0.027165393 |
| Obesity | nk_cell | *TULP2* | 8.357284151 | 7.81E-09 | 4.43E-08 | 3.79E-05 |
| Obesity | nk_cell | *NFKBIB* | 1.583551842 | 2.41E-09 | 2.34E-08 | 1.17E-05 |
| Obesity | nk_cell | *SARS2* | -1.291460623 | 4.46E-06 | 8.34E-06 | 0.021617616 |
| Obesity | nk_cell | *SNRNP70* | -1.912467046 | 5.17E-08 | 1.79E-07 | 0.00025096 |
| Obesity | nk_cell | *LIN7B* | -1.960270245 | 2.20E-10 | 1.19E-08 | 1.07E-06 |
| Obesity | nk_cell | *PPP1R37* | -1.883910932 | 0.015907505 | 0.01923809 | 1 |
| Obesity | nk_cell | *FCGRT* | -1.747051983 | 5.17E-06 | 9.55E-06 | 0.02510507 |
| Obesity | nk_cell | *ARHGEF18* | -4.913778444 | 2.06E-07 | 5.55E-07 | 0.000997737 |
| Obesity | nk_cell | *PEX11G* | -2.169762527 | 1.50E-08 | 6.98E-08 | 7.29E-05 |
| Obesity | nk_cell | *ERCC2* | -4.351400775 | 2.14E-07 | 5.74E-07 | 0.001037919 |
| Obesity | nk_cell | *CD37* | 1.172167112 | 7.09E-09 | 4.25E-08 | 3.44E-05 |
| Obesity | nk_cell | *AMH* | -26.0845209 | 6.57E-09 | 4.07E-08 | 3.19E-05 |
| Obesity | nk_cell | *DKKL1* | -1.414627214 | 0.00300107 | 0.003860337 | 1 |
| Obesity | nk_cell | *FCER2* | -9.80293303 | 2.55E-08 | 1.04E-07 | 0.000123518 |
| Obesity | nk_cell | *CLEC4M* | -3.355674416 | 1.52E-09 | 2.11E-08 | 7.39E-06 |

| | | | | | | |
|---|---|---|---|---|---|---|
| Obesity | nk_cell | *TLE6* | -2.540259262 | 1.16E-07 | 3.46E-07 | 0.000561674 |
| Obesity | nk_cell | *NOVA2* | -0.977188482 | 0.000124026 | 0.000184311 | 0.601773874 |
| Obesity | nk_cell | *SGTA* | -3.686716863 | 0.000655853 | 0.000897153 | 1 |
| Obesity | nk_cell | *KIR3DX1* | 0.244404623 | 1.12E-09 | 2.05E-08 | 5.42E-06 |
| Obesity | nk_cell | *MED25* | -1.9857585 | 5.86E-09 | 3.81E-08 | 2.84E-05 |
| Obesity | nk_cell | *TIMM44* | -2.60658994 | 0.000521977 | 0.000719703 | 1 |
| Obesity | nk_cell | *IL27RA* | 0.628831818 | 1.12E-09 | 2.05E-08 | 5.43E-06 |
| Obesity | nk_cell | *FAM32A* | 4.351921522 | 2.14E-06 | 4.29E-06 | 0.010373836 |
| Obesity | nk_cell | *C19orf44* | -1.243899091 | 6.32E-06 | 1.14E-05 | 0.030672994 |
| Obesity | nk_cell | *OLFM2* | -2.042524289 | 1.77E-08 | 7.90E-08 | 8.61E-05 |
| Obesity | nk_cell | *RASAL3* | 0.314769052 | 1.11E-09 | 2.05E-08 | 5.40E-06 |
| Obesity | nk_cell | *EPHX3* | -2.904763073 | 0.000980124 | 0.001313328 | 1 |
| Obesity | nk_cell | *PDCD5* | -1.437656908 | 3.69E-09 | 2.89E-08 | 1.79E-05 |
| Obesity | nk_cell | *ANKRD27* | -2.311700976 | 0.002305854 | 0.00299545 | 1 |
| Obesity | nk_cell | *POLR2I* | -2.973660599 | 0.000842897 | 0.001138568 | 1 |
| Obesity | nk_cell | *CCDC9* | -3.99900694 | 1.02E-08 | 5.26E-08 | 4.93E-05 |
| Obesity | nk_cell | *TGFB1* | 2.639024105 | 3.03E-05 | 4.89E-05 | 0.146884801 |
| Obesity | nk_cell | *CEACAM4* | -2.411850637 | 0.000892224 | 0.001202185 | 1 |
| Obesity | nk_cell | *ICAM5* | -2.667775137 | 3.21E-08 | 1.22E-07 | 0.000155745 |
| Obesity | nk_cell | *CD33* | -0.83309344 | 0.004258255 | 0.005408653 | 1 |
| Obesity | nk_cell | *RABAC1* | 3.083676733 | 5.94E-07 | 1.37E-06 | 0.002882663 |
| Obesity | nk_cell | *GRIN2D* | -5.173032101 | 1.15E-08 | 5.66E-08 | 5.56E-05 |
| Obesity | nk_cell | *CCDC114* | -5.045700011 | 4.34E-05 | 6.87E-05 | 0.210725269 |
| Obesity | nk_cell | *LIG1* | 2.679136616 | 7.86E-08 | 2.52E-07 | 0.000381402 |
| Obesity | nk_cell | *ZNF175* | -2.206606071 | 2.81E-09 | 2.51E-08 | 1.36E-05 |
| Obesity | nk_cell | *CAPS* | -1.705498468 | 1.95E-09 | 2.20E-08 | 9.47E-06 |
| Obesity | nk_cell | *MIER2* | -2.594909744 | 3.51E-07 | 8.73E-07 | 0.001702795 |
| Obesity | nk_cell | *PLEKHA4* | -1.209337492 | 1.70E-09 | 2.16E-08 | 8.23E-06 |
| Obesity | nk_cell | *TNPO2* | -2.440247615 | 1.84E-08 | 8.11E-08 | 8.92E-05 |
| Obesity | nk_cell | *GCDH* | -3.556296906 | 7.61E-08 | 2.45E-07 | 0.000369365 |
| Obesity | nk_cell | *DNASE2* | -2.282931472 | 0.032998566 | 0.038729812 | 1 |
| Obesity | nk_cell | *TFPT* | -4.780489885 | 6.37E-06 | 1.15E-05 | 0.030915907 |
| Obesity | nk_cell | *RAB3A* | -9.127645088 | 7.30E-09 | 4.30E-08 | 3.54E-05 |
| Obesity | nk_cell | *UPK1A* | -4.420200717 | 4.32E-07 | 1.04E-06 | 0.0020959 |
| Obesity | nk_cell | *COPE* | 2.700230978 | 1.32E-05 | 2.25E-05 | 0.064189673 |
| Obesity | nk_cell | *LSR* | -1.13915566 | 3.34E-08 | 1.26E-07 | 0.000161921 |
| Obesity | nk_cell | *ZNF14* | -3.413283445 | 2.88E-06 | 5.63E-06 | 0.013951005 |
| Obesity | nk_cell | *ZNF85* | 3.883852312 | 6.40E-05 | 9.84E-05 | 0.310514887 |
| Obesity | nk_cell | *SMG9* | -3.693352274 | 6.30E-05 | 9.70E-05 | 0.305584221 |
| Obesity | nk_cell | *GTPBP10* | -1.357699147 | 0.028002245 | 0.033105968 | 1 |
| Obesity | nk_cell | *RASA4* | -6.894443066 | 1.64E-09 | 2.14E-08 | 7.97E-06 |
| Obesity | nk_cell | *BET1* | -1.92696465 | 2.83E-08 | 1.11E-07 | 0.000137223 |
| Obesity | nk_cell | *SP4* | 2.278667606 | 1.10E-06 | 2.37E-06 | 0.005318117 |
| Obesity | nk_cell | *DNAH11* | -3.375553219 | 2.64E-06 | 5.20E-06 | 0.012816625 |
| Obesity | nk_cell | *STEAP1B* | -2.869696036 | 1.99E-06 | 4.02E-06 | 0.00963892 |
| Obesity | nk_cell | *PTN* | -2.394016434 | 6.74E-08 | 2.21E-07 | 0.000327221 |
| Obesity | nk_cell | *ADAP1* | -11.31643083 | 1.04E-07 | 3.15E-07 | 0.000502361 |
| Obesity | nk_cell | *MET* | -1.430279186 | 6.50E-10 | 1.90E-08 | 3.15E-06 |
| Obesity | nk_cell | *RNF32* | -3.00497633 | 7.42E-09 | 4.35E-08 | 3.60E-05 |
| Obesity | nk_cell | *IQCE* | -2.331677358 | 1.45E-08 | 6.81E-08 | 7.04E-05 |

| | | | | | | |
|---|---|---|---|---|---|---|
| Obesity | nk_cell | *VIPR2* | -1.367144463 | 1.38E-08 | 6.58E-08 | 6.70E-05 |
| Obesity | nk_cell | *HIBADH* | -1.888654376 | 2.76E-08 | 1.10E-07 | 0.000133946 |
| Obesity | nk_cell | *CPVL* | -2.218178545 | 2.08E-06 | 4.17E-06 | 0.010071582 |
| Obesity | nk_cell | *CHN2* | -1.283185699 | 0.041463352 | 0.048233082 | 1 |
| Obesity | nk_cell | *GRB10* | -2.065210847 | 5.93E-07 | 1.37E-06 | 0.002876548 |
| Obesity | nk_cell | *ABHD11* | -1.699313367 | 3.46E-07 | 8.63E-07 | 0.001678885 |
| Obesity | nk_cell | *PLEKHA8* | -1.553847696 | 5.47E-09 | 3.65E-08 | 2.65E-05 |
| Obesity | nk_cell | *CRHR2* | 1.775353899 | 1.78E-06 | 3.65E-06 | 0.008653539 |
| Obesity | nk_cell | *CHCHD2* | 1.872951295 | 0.012493929 | 0.015215999 | 1 |
| Obesity | nk_cell | *CCL24* | 2.318088383 | 0.001102434 | 0.001469912 | 1 |
| Obesity | nk_cell | *ZKSCAN1* | -2.283376699 | 2.28E-06 | 4.54E-06 | 0.011053131 |
| Obesity | nk_cell | *EIF3B* | -2.922859772 | 2.19E-06 | 4.37E-06 | 0.010608076 |
| Obesity | nk_cell | *TAF6* | -2.668068487 | 5.28E-10 | 1.75E-08 | 2.56E-06 |
| Obesity | nk_cell | *TFR2* | 0.875763197 | 2.02E-09 | 2.20E-08 | 9.78E-06 |
| Obesity | nk_cell | *MOSPD3* | -1.404065442 | 1.19E-08 | 5.82E-08 | 5.76E-05 |
| Obesity | nk_cell | *RBM28* | -2.25350339 | 8.35E-07 | 1.85E-06 | 0.004049504 |
| Obesity | nk_cell | *IMPDH1* | -2.548592811 | 1.01E-07 | 3.09E-07 | 0.000489721 |
| Obesity | nk_cell | *C1GALT1* | -2.547237376 | 7.35E-06 | 1.31E-05 | 0.035686272 |
| Obesity | nk_cell | *NRF1* | -1.936906089 | 0.000968057 | 0.001298593 | 1 |
| Obesity | nk_cell | *ZNF862* | -2.230779228 | 3.86E-05 | 6.14E-05 | 0.187250071 |
| Obesity | nk_cell | *MEST* | -1.489964826 | 1.96E-11 | 2.32E-09 | 9.53E-08 |
| Obesity | nk_cell | *ACTR3C* | 2.361554754 | 0.00940869 | 0.011577724 | 1 |
| Obesity | nk_cell | *POU6F2* | -1.239319012 | 1.72E-09 | 2.16E-08 | 8.33E-06 |
| Obesity | nk_cell | *TSPAN13* | -1.488301003 | 3.51E-09 | 2.82E-08 | 1.70E-05 |
| Obesity | nk_cell | *RARRES2* | -2.235715236 | 0.000141151 | 0.000208166 | 0.684867069 |
| Obesity | nk_cell | *TMEM176B* | -1.049327313 | 0.006262594 | 0.007821392 | 1 |
| Obesity | nk_cell | *COA1* | -2.001352792 | 0.000220825 | 0.000318251 | 1 |
| Obesity | nk_cell | *BLVRA* | -2.446172398 | 1.51E-05 | 2.55E-05 | 0.073217387 |
| Obesity | nk_cell | *URGCP* | -2.551430808 | 6.56E-06 | 1.18E-05 | 0.031819389 |
| Obesity | nk_cell | *AEBP1* | -3.102302118 | 1.02E-06 | 2.23E-06 | 0.004960055 |
| Obesity | nk_cell | *GCK* | -8.942106461 | 2.04E-09 | 2.21E-08 | 9.89E-06 |
| Obesity | nk_cell | *SPATA6L* | -1.762097459 | 1.84E-08 | 8.11E-08 | 8.91E-05 |
| Obesity | nk_cell | *SLC1A1* | -0.614714044 | 0.005142525 | 0.006477552 | 1 |
| Obesity | nk_cell | *FKTN* | -2.466780949 | 8.84E-07 | 1.95E-06 | 0.004287177 |
| Obesity | nk_cell | *NMRK1* | -2.215035242 | 1.72E-07 | 4.79E-07 | 0.000836347 |
| Obesity | nk_cell | *MEGF9* | -2.573409747 | 6.06E-05 | 9.37E-05 | 0.294163674 |
| Obesity | nk_cell | *TRIM14* | 1.905235737 | 3.70E-10 | 1.52E-08 | 1.80E-06 |
| Obesity | nk_cell | *CORO2A* | 0.958548672 | 1.51E-09 | 2.11E-08 | 7.31E-06 |
| Obesity | nk_cell | *C5* | -1.556320383 | 9.34E-09 | 4.96E-08 | 4.53E-05 |
| Obesity | nk_cell | *OGN* | -1.526813989 | 1.04E-05 | 1.80E-05 | 0.050297194 |
| Obesity | nk_cell | *ECM2* | -2.152800977 | 1.15E-06 | 2.48E-06 | 0.005603023 |
| Obesity | nk_cell | *PTGR1* | -2.07376073 | 3.04E-08 | 1.18E-07 | 0.000147312 |
| Obesity | nk_cell | *SUSD1* | 1.283342965 | 8.35E-08 | 2.64E-07 | 0.000404946 |
| Obesity | nk_cell | *PLGRKT* | -2.208400298 | 0.000455181 | 0.000632276 | 1 |
| Obesity | nk_cell | *KANK1* | -1.845189857 | 1.50E-07 | 4.28E-07 | 0.000727704 |
| Obesity | nk_cell | *NCS1* | -2.733935476 | 2.85E-06 | 5.59E-06 | 0.013851003 |
| Obesity | nk_cell | *FUBP3* | -2.075086973 | 1.29E-07 | 3.79E-07 | 0.000626777 |
| Obesity | nk_cell | *RGP1* | -2.672419477 | 0.011018468 | 0.013483381 | 1 |
| Obesity | nk_cell | *ATRNL1* | -0.432379286 | 0.027438687 | 0.032497728 | 1 |
| Obesity | nk_cell | *HPS1* | -3.432367592 | 3.10E-08 | 1.20E-07 | 0.000150587 |

| | | | | | | |
|---|---|---|---|---|---|---|
| Obesity | nk_cell | *PHYH* | -1.698717552 | 1.23E-05 | 2.10E-05 | 0.059651619 |
| Obesity | nk_cell | *DNMBP* | -1.621269815 | 2.40E-07 | 6.31E-07 | 0.001162287 |
| Obesity | nk_cell | *PKD2L1* | 2.736026455 | 4.33E-11 | 3.96E-09 | 2.10E-07 |
| Obesity | nk_cell | *CUBN* | -3.18523778 | 3.90E-06 | 7.40E-06 | 0.018901674 |
| Obesity | nk_cell | *TRDMT1* | -2.315636773 | 5.46E-08 | 1.86E-07 | 0.000264728 |
| Obesity | nk_cell | *ATE1* | -1.84266769 | 4.25E-06 | 7.98E-06 | 0.020636253 |
| Obesity | nk_cell | *PLEKHA1* | -2.115246494 | 0.001197517 | 0.00159057 | 1 |
| Obesity | nk_cell | *CDH23* | -1.628705991 | 1.19E-08 | 5.82E-08 | 5.76E-05 |
| Obesity | nk_cell | *SPOCK2* | 1.024763564 | 2.79E-07 | 7.15E-07 | 0.00135201 |
| Obesity | nk_cell | *BMPR1A* | -1.642600238 | 3.17E-08 | 1.21E-07 | 0.000153747 |
| Obesity | nk_cell | *LIPA* | -1.250687038 | 0.004363754 | 0.005539752 | 1 |
| Obesity | nk_cell | *SFXN3* | -2.565366079 | 1.83E-08 | 8.10E-08 | 8.90E-05 |
| Obesity | nk_cell | *ANKRD26* | -1.934511793 | 3.69E-07 | 9.13E-07 | 0.001790542 |
| Obesity | nk_cell | *LHPP* | -2.321262827 | 0.000632229 | 0.000867036 | 1 |
| Obesity | nk_cell | *PITRM1* | -2.249742826 | 4.45E-09 | 3.23E-08 | 2.16E-05 |
| Obesity | nk_cell | *RPL28* | 2.102290508 | 2.62E-07 | 6.81E-07 | 0.001271972 |
| Obesity | nk_cell | *DNAJC12* | -2.476433054 | 1.36E-07 | 3.95E-07 | 0.000660523 |
| Obesity | nk_cell | *PBLD* | -2.855130026 | 5.83E-07 | 1.35E-06 | 0.002829558 |
| Obesity | nk_cell | *TSPAN14* | -2.392841805 | 4.75E-06 | 8.85E-06 | 0.023067908 |
| Obesity | nk_cell | *TADA2A* | -1.290175003 | 0.001996889 | 0.002605946 | 1 |
| Obesity | nk_cell | *PSMB3* | 3.059656491 | 0.000104545 | 0.000156801 | 0.507251163 |
| Obesity | nk_cell | *RPL19* | 2.09787014 | 4.65E-05 | 7.33E-05 | 0.225648803 |
| Obesity | nk_cell | *RAPGEFL1* | -1.953509888 | 3.77E-09 | 2.94E-08 | 1.83E-05 |
| Obesity | nk_cell | *WNT3* | -1.833434471 | 1.60E-09 | 2.11E-08 | 7.74E-06 |
| Obesity | nk_cell | *ASPA* | -1.237660085 | 3.07E-08 | 1.19E-07 | 0.000148731 |
| Obesity | nk_cell | *RAD51C* | -3.494556329 | 4.74E-08 | 1.68E-07 | 0.000229892 |
| Obesity | nk_cell | *TRIM37* | -2.615096223 | 0.000157663 | 0.000231251 | 0.764978909 |
| Obesity | nk_cell | *PNPO* | 3.25044791 | 0.004882006 | 0.006168618 | 1 |
| Obesity | nk_cell | *TRIM16L* | -1.367575456 | 1.24E-09 | 2.08E-08 | 6.01E-06 |
| Obesity | nk_cell | *CDK5RAP3* | -4.387437794 | 3.25E-08 | 1.24E-07 | 0.000157743 |
| Obesity | nk_cell | *PIGL* | -2.372006324 | 1.10E-05 | 1.90E-05 | 0.05354991 |
| Obesity | nk_cell | *MED13* | -2.212690504 | 4.16E-07 | 1.01E-06 | 0.002016961 |
| Obesity | nk_cell | *ENO3* | -1.953997394 | 0.011771219 | 0.014379142 | 1 |
| Obesity | nk_cell | *CHRNE* | 1.524620786 | 6.50E-11 | 5.44E-09 | 3.15E-07 |
| Obesity | nk_cell | *RAI1* | 2.997105282 | 0.000231924 | 0.000333916 | 1 |
| Obesity | nk_cell | *GOSR1* | -2.189618215 | 2.09E-07 | 5.63E-07 | 0.001014136 |
| Obesity | nk_cell | *DRG2* | -2.120444164 | 2.84E-08 | 1.12E-07 | 0.000137692 |
| Obesity | nk_cell | *FTSJ3* | -0.936595351 | 0.001105014 | 0.001472947 | 1 |
| Obesity | nk_cell | *ICAM2* | 1.976014804 | 4.08E-09 | 3.06E-08 | 1.98E-05 |
| Obesity | nk_cell | *DDX5* | -1.910068318 | 5.45E-06 | 1.00E-05 | 0.026454357 |
| Obesity | nk_cell | *C17orf75* | -1.792673851 | 2.35E-05 | 3.84E-05 | 0.11393041 |
| Obesity | nk_cell | *PSMD11* | -2.349686077 | 1.96E-05 | 3.24E-05 | 0.095016013 |
| Obesity | nk_cell | *PEX12* | 1.876237209 | 2.16E-09 | 2.26E-08 | 1.05E-05 |
| Obesity | nk_cell | *DHX58* | -1.799295614 | 1.59E-08 | 7.32E-08 | 7.71E-05 |
| Obesity | nk_cell | *RAB5C* | 2.318108718 | 5.80E-05 | 8.99E-05 | 0.28143828 |
| Obesity | nk_cell | *NAGLU* | -3.120258593 | 6.77E-06 | 1.22E-05 | 0.03286927 |
| Obesity | nk_cell | *CNTNAP1* | -1.574792775 | 1.64E-08 | 7.44E-08 | 7.95E-05 |
| Obesity | nk_cell | *EZH1* | -1.774993347 | 2.69E-07 | 6.96E-07 | 0.001304448 |
| Obesity | nk_cell | *EFTUD2* | -2.574786467 | 1.35E-07 | 3.92E-07 | 0.000654523 |
| Obesity | nk_cell | *HLF* | -2.221423226 | 4.16E-09 | 3.11E-08 | 2.02E-05 |

| | | | | | | |
|---|---|---|---|---|---|---|
| Obesity | nk_cell | *EFNB3* | -2.305949114 | 1.97E-07 | 5.35E-07 | 0.000957325 |
| Obesity | nk_cell | *FAM20A* | -1.923605766 | 3.87E-08 | 1.43E-07 | 0.000187645 |
| Obesity | nk_cell | *DPH1* | -1.493115694 | 3.76E-09 | 2.94E-08 | 1.83E-05 |
| Obesity | nk_cell | *DHRS7B* | -2.502037193 | 9.66E-05 | 0.000145501 | 0.468804834 |
| Obesity | nk_cell | *RCVRN* | -1.868834627 | 0.005246646 | 0.006596716 | 1 |
| Obesity | nk_cell | *MYH3* | 5.97650195 | 5.95E-07 | 1.37E-06 | 0.002885933 |
| Obesity | nk_cell | *TMEM97* | 1.813650296 | 0.003110386 | 0.003992485 | 1 |
| Obesity | nk_cell | *OCIAD1* | -2.366095407 | 6.67E-06 | 1.20E-05 | 0.03237447 |
| Obesity | nk_cell | *UBE2D3* | -1.990963298 | 1.52E-06 | 3.16E-06 | 0.007362525 |
| Obesity | nk_cell | *NDUFC1* | 3.041642483 | 1.74E-05 | 2.89E-05 | 0.084198412 |
| Obesity | nk_cell | *KLHL2* | -3.045122092 | 2.84E-06 | 5.57E-06 | 0.013801935 |
| Obesity | nk_cell | *CPE* | -1.63343858 | 2.01E-07 | 5.43E-07 | 0.000973881 |
| Obesity | nk_cell | *WFS1* | -3.916437326 | 3.90E-06 | 7.40E-06 | 0.018905521 |
| Obesity | nk_cell | *GRPEL1* | 3.256811597 | 0.016774675 | 0.020256527 | 1 |
| Obesity | nk_cell | *AADAT* | -1.628995294 | 1.55E-09 | 2.11E-08 | 7.54E-06 |
| Obesity | nk_cell | *TRIM2* | -2.195713694 | 2.51E-07 | 6.57E-07 | 0.001215959 |
| Obesity | nk_cell | *BST1* | -2.151254269 | 1.68E-07 | 4.69E-07 | 0.000816198 |
| Obesity | nk_cell | *KLHL5* | -2.024504165 | 1.01E-07 | 3.09E-07 | 0.000489615 |
| Obesity | nk_cell | *FAM149A* | -2.099147071 | 1.13E-08 | 5.60E-08 | 5.49E-05 |
| Obesity | nk_cell | *DDX25* | -2.081284508 | 7.20E-10 | 1.92E-08 | 3.50E-06 |
| Obesity | nk_cell | *ELP4* | -2.248535495 | 1.47E-06 | 3.08E-06 | 0.007130038 |
| Obesity | nk_cell | *TECTA* | -1.793764412 | 1.48E-09 | 2.11E-08 | 7.20E-06 |
| Obesity | nk_cell | *SC5D* | 2.42903895 | 0.005881055 | 0.007360041 | 1 |
| Obesity | nk_cell | *B3GAT1* | 0.323849009 | 1.12E-09 | 2.05E-08 | 5.44E-06 |
| Obesity | nk_cell | *P2RX3* | 2.102459696 | 1.13E-09 | 2.05E-08 | 5.50E-06 |
| Obesity | nk_cell | *DNAJC4* | 2.209554111 | 1.53E-05 | 2.58E-05 | 0.074383212 |
| Obesity | nk_cell | *SNX15* | -3.173476309 | 1.29E-07 | 3.77E-07 | 0.000623681 |
| Obesity | nk_cell | *UNC93B1* | -2.848258502 | 1.80E-07 | 4.96E-07 | 0.000873317 |
| Obesity | nk_cell | *PUS3* | -1.791041105 | 9.47E-06 | 1.65E-05 | 0.045964267 |
| Obesity | nk_cell | *FOXRED1* | -1.227757162 | 1.90E-07 | 5.18E-07 | 0.000923607 |
| Obesity | nk_cell | *NRXN2* | -6.721133825 | 6.39E-08 | 2.11E-07 | 0.000309832 |
| Obesity | nk_cell | *CCND1* | -2.81734764 | 4.70E-06 | 8.75E-06 | 0.022810386 |
| Obesity | nk_cell | *CCDC86* | -5.638369204 | 1.45E-07 | 4.16E-07 | 0.000704692 |
| Obesity | nk_cell | *PANX1* | -3.383259636 | 4.54E-08 | 1.61E-07 | 0.000220306 |
| Obesity | nk_cell | *CEP164* | -2.178939752 | 1.80E-07 | 4.95E-07 | 0.000872082 |
| Obesity | nk_cell | *GALNT18* | -1.651055772 | 0.01549413 | 0.018747511 | 1 |
| Obesity | nk_cell | *KIAA1549L* | -3.586966057 | 2.13E-09 | 2.26E-08 | 1.03E-05 |
| Obesity | nk_cell | *ACCS* | -1.433060435 | 1.82E-05 | 3.03E-05 | 0.088528896 |
| Obesity | nk_cell | *SLC22A18* | 1.508271354 | 3.11E-07 | 7.86E-07 | 0.001507755 |
| Obesity | nk_cell | *SLC35F2* | -3.070289727 | 5.39E-08 | 1.85E-07 | 0.00026141 |
| Obesity | nk_cell | *C11orf21* | 1.041599196 | 2.01E-09 | 2.20E-08 | 9.74E-06 |
| Obesity | nk_cell | *ELMOD1* | 1.849605207 | 3.00E-08 | 1.17E-07 | 0.000145764 |
| Obesity | nk_cell | *NDUFS8* | -2.791431748 | 0.002907778 | 0.003742766 | 1 |
| Obesity | nk_cell | *TCIRG1* | 2.063348979 | 0.036746948 | 0.042942243 | 1 |
| Obesity | nk_cell | *GTF2H1* | -2.110877096 | 3.16E-06 | 6.13E-06 | 0.015348191 |
| Obesity | nk_cell | *PRDM4* | -2.410942847 | 2.45E-06 | 4.85E-06 | 0.011898168 |
| Obesity | nk_cell | *CORO1C* | -1.635269331 | 1.27E-08 | 6.14E-08 | 6.14E-05 |
| Obesity | nk_cell | *ASIC1* | -2.199580849 | 1.56E-09 | 2.11E-08 | 7.57E-06 |
| Obesity | nk_cell | *KCTD10* | -2.147597091 | 1.68E-10 | 9.97E-09 | 8.15E-07 |
| Obesity | nk_cell | *IL23A* | 16.67236293 | 6.09E-09 | 3.91E-08 | 2.95E-05 |

| | | | | | | |
|---|---|---|---|---|---|---|
| Obesity | nk_cell | *ACSS3* | -1.626537471 | 1.80E-07 | 4.95E-07 | 0.000870941 |
| Obesity | nk_cell | *LTA4H* | -1.786041257 | 1.72E-05 | 2.87E-05 | 0.083500306 |
| Obesity | nk_cell | *PRR4* | -1.793508808 | 0.000266806 | 0.000381131 | 1 |
| Obesity | nk_cell | *PARP11* | -2.557728985 | 7.25E-07 | 1.64E-06 | 0.003516641 |
| Obesity | nk_cell | *GPN3* | -2.365646547 | 4.20E-09 | 3.12E-08 | 2.04E-05 |
| Obesity | nk_cell | *VPS29* | -2.323665709 | 1.02E-05 | 1.78E-05 | 0.049632745 |
| Obesity | nk_cell | *AKAP3* | -1.03520543 | 3.20E-06 | 6.19E-06 | 0.015523695 |
| Obesity | nk_cell | *ACAD10* | -2.295734788 | 0.00015262 | 0.000224398 | 0.740512647 |
| Obesity | nk_cell | *LTBR* | -2.019266426 | 4.69E-07 | 1.11E-06 | 0.002274926 |
| Obesity | nk_cell | *CDK2AP1* | 3.180012554 | 8.42E-10 | 1.99E-08 | 4.09E-06 |
| Obesity | nk_cell | *DDX55* | 2.569099412 | 0.004954527 | 0.006253737 | 1 |
| Obesity | nk_cell | *ENDOU* | -3.143716794 | 8.23E-10 | 1.99E-08 | 3.99E-06 |
| Obesity | nk_cell | *VDR* | -1.888820944 | 1.74E-09 | 2.16E-08 | 8.42E-06 |
| Obesity | nk_cell | *STX2* | -1.745865241 | 1.16E-05 | 1.99E-05 | 0.056307878 |
| Obesity | nk_cell | *CAND1* | -2.475905084 | 5.29E-05 | 8.26E-05 | 0.256772401 |
| Obesity | nk_cell | *MDM1* | -3.19455377 | 3.33E-07 | 8.36E-07 | 0.001616831 |
| Obesity | nk_cell | *NUP107* | -2.196062541 | 8.10E-06 | 1.43E-05 | 0.039290316 |
| Obesity | nk_cell | *KRR1* | -2.802932854 | 9.36E-07 | 2.05E-06 | 0.004540829 |
| Obesity | nk_cell | *MRPL51* | 1.125602707 | 0.014947803 | 0.018118097 | 1 |
| Obesity | nk_cell | *GNB3* | -1.039019172 | 0.00519829 | 0.006539306 | 1 |
| Obesity | nk_cell | *CDCA3* | -3.056745919 | 1.13E-05 | 1.95E-05 | 0.054882453 |
| Obesity | nk_cell | *CHPT1* | -1.838571211 | 3.10E-05 | 5.00E-05 | 0.150195655 |
| Obesity | nk_cell | *GNPTAB* | 2.326122495 | 5.34E-06 | 9.81E-06 | 0.025892713 |
| Obesity | nk_cell | *SPSB2* | -1.607111022 | 1.75E-06 | 3.58E-06 | 0.008471594 |
| Obesity | nk_cell | *ATN1* | -2.137067835 | 2.22E-08 | 9.34E-08 | 0.000107915 |
| Obesity | nk_cell | *C12orf57* | 3.34563359 | 6.68E-07 | 1.53E-06 | 0.003243214 |
| Obesity | nk_cell | *NT5DC3* | -3.131269321 | 1.11E-05 | 1.92E-05 | 0.053940906 |
| Obesity | nk_cell | *SUDS3* | -2.067712491 | 2.31E-08 | 9.58E-08 | 0.000112237 |
| Obesity | nk_cell | *GOLT1B* | 2.882062557 | 0.000329941 | 0.000466456 | 1 |
| Obesity | nk_cell | *C2CD5* | -2.228266641 | 4.10E-06 | 7.73E-06 | 0.019912748 |
| Obesity | nk_cell | *BTN3A3* | 3.995560064 | 2.70E-16 | 1.31E-13 | 1.31E-12 |
| Obesity | nk_cell | *FRK* | -1.981980334 | 4.00E-09 | 3.03E-08 | 1.94E-05 |
| Obesity | nk_cell | *RWDD1* | 1.624287699 | 0.033285149 | 0.039047278 | 1 |
| Obesity | nk_cell | *RSPH4A* | -1.988436372 | 0.00065143 | 0.000891588 | 1 |
| Obesity | nk_cell | *TMEM14C* | -2.901680431 | 3.78E-07 | 9.31E-07 | 0.001834742 |
| Obesity | nk_cell | *SMIM8* | -1.610960738 | 1.40E-06 | 2.95E-06 | 0.006801775 |
| Obesity | nk_cell | *FAM184A* | -1.654043515 | 6.55E-08 | 2.16E-07 | 0.000317766 |
| Obesity | nk_cell | *RNGTT* | -2.304393059 | 1.70E-06 | 3.50E-06 | 0.008248282 |
| Obesity | nk_cell | *GABRR2* | 2.293713973 | 0.021444391 | 0.025665561 | 1 |
| Obesity | nk_cell | *HDDC2* | 1.573483335 | 0.025551226 | 0.030356158 | 1 |
| Obesity | nk_cell | *UST* | -1.217263351 | 1.71E-08 | 7.71E-08 | 8.31E-05 |
| Obesity | nk_cell | *TULP1* | -4.194168674 | 3.56E-10 | 1.52E-08 | 1.73E-06 |
| Obesity | nk_cell | *SLC26A8* | -2.364785398 | 1.39E-06 | 2.94E-06 | 0.006756852 |
| Obesity | nk_cell | *RNF8* | -2.870759949 | 1.91E-06 | 3.88E-06 | 0.009278901 |
| Obesity | nk_cell | *PHACTR1* | -2.097439578 | 4.07E-05 | 6.47E-05 | 0.197616928 |
| Obesity | nk_cell | *MDGA1* | -2.6759967 | 2.65E-09 | 2.45E-08 | 1.28E-05 |
| Obesity | nk_cell | *CD83* | -2.679385916 | 5.15E-09 | 3.53E-08 | 2.50E-05 |
| Obesity | nk_cell | *MDN1* | -1.964834959 | 1.08E-08 | 5.45E-08 | 5.25E-05 |
| Obesity | nk_cell | *GLP1R* | -10.26692709 | 2.16E-09 | 2.26E-08 | 1.05E-05 |
| Obesity | nk_cell | *BVES* | -1.366406296 | 1.85E-08 | 8.15E-08 | 8.98E-05 |

| | | | | | | |
|---|---|---|---|---|---|---|
| Obesity | nk_cell | *COL9A1* | -3.281874168 | 1.00E-08 | 5.22E-08 | 4.87E-05 |
| Obesity | nk_cell | *WASF1* | -1.627626608 | 4.12E-08 | 1.50E-07 | 0.000200027 |
| Obesity | nk_cell | *VNN1* | -1.610593493 | 7.36E-05 | 0.000112443 | 0.357343022 |
| Obesity | nk_cell | *VNN2* | 2.188252907 | 8.59E-10 | 2.00E-08 | 4.17E-06 |
| Obesity | nk_cell | *C6orf62* | -2.699400241 | 1.07E-06 | 2.32E-06 | 0.005196838 |
| Obesity | nk_cell | *HBS1L* | -2.143444572 | 6.69E-07 | 1.53E-06 | 0.003244593 |
| Obesity | nk_cell | *PEX7* | -1.287161485 | 0.007744665 | 0.009603147 | 1 |
| Obesity | nk_cell | *FIG4* | -2.608613432 | 3.64E-05 | 5.81E-05 | 0.176381594 |
| Obesity | nk_cell | *PERP* | -1.983587762 | 3.32E-05 | 5.34E-05 | 0.161022794 |
| Obesity | nk_cell | *PHACTR2* | -2.076714887 | 7.23E-05 | 0.000110488 | 0.350800925 |
| Obesity | nk_cell | *CUTA* | 1.353070147 | 5.41E-06 | 9.94E-06 | 0.026248358 |
| Obesity | nk_cell | *PACRG* | -1.295885645 | 2.35E-09 | 2.30E-08 | 1.14E-05 |
| Obesity | nk_cell | *QKI* | -1.716928311 | 7.41E-07 | 1.67E-06 | 0.003596828 |
| Obesity | nk_cell | *FAM120B* | -2.011683708 | 2.27E-05 | 3.72E-05 | 0.110023177 |
| Obesity | nk_cell | *GUCA1B* | -1.763051375 | 7.35E-10 | 1.92E-08 | 3.57E-06 |
| Obesity | nk_cell | *PRPH2* | 0.39921838 | 0.018461343 | 0.022228268 | 1 |
| Obesity | nk_cell | *TMEM30A* | -2.351453176 | 4.77E-08 | 1.68E-07 | 0.000231624 |
| Obesity | nk_cell | *IMPG1* | -3.297020534 | 1.74E-10 | 1.01E-08 | 8.46E-07 |
| Obesity | nk_cell | *LY86* | -4.078320039 | 0.000375146 | 0.000526072 | 1 |
| Obesity | nk_cell | *PCDHB2* | -2.681117324 | 3.21E-10 | 1.43E-08 | 1.56E-06 |
| Obesity | nk_cell | *NUDT12* | -3.057428888 | 2.81E-07 | 7.20E-07 | 0.001363004 |
| Obesity | nk_cell | *CEP72* | 3.806267202 | 2.15E-05 | 3.53E-05 | 0.104289955 |
| Obesity | nk_cell | *DAP* | 1.539394138 | 0.04122101 | 0.047974176 | 1 |
| Obesity | nk_cell | *NME5* | -0.853560123 | 0.0005694 | 0.000782865 | 1 |
| Obesity | nk_cell | *KIF20A* | 4.54971296 | 1.80E-08 | 8.01E-08 | 8.76E-05 |
| Obesity | nk_cell | *PFDN1* | -1.839922191 | 2.73E-06 | 5.36E-06 | 0.013238435 |
| Obesity | nk_cell | *TMCO6* | -4.059405955 | 2.91E-09 | 2.56E-08 | 1.41E-05 |
| Obesity | nk_cell | *SPARC* | -2.056904226 | 2.09E-06 | 4.20E-06 | 0.010143261 |
| Obesity | nk_cell | *IK* | 1.2761672 | 0.023679745 | 0.028181046 | 1 |
| Obesity | nk_cell | *HAVCR1* | 1.877487016 | 0.027783141 | 0.032870959 | 1 |
| Obesity | nk_cell | *RNF130* | -1.891979754 | 1.25E-05 | 2.14E-05 | 0.060796034 |
| Obesity | nk_cell | *THBS4* | -1.335250298 | 1.65E-08 | 7.49E-08 | 8.02E-05 |
| Obesity | nk_cell | *BTNL8* | -1.019155424 | 6.08E-07 | 1.40E-06 | 0.002949581 |
| Obesity | nk_cell | *MSH3* | -2.021357984 | 1.24E-06 | 2.64E-06 | 0.006022107 |
| Obesity | nk_cell | *RASGRF2* | -1.050383554 | 1.55E-07 | 4.38E-07 | 0.000749932 |
| Obesity | nk_cell | *DROSHA* | -1.975753019 | 7.19E-06 | 1.28E-05 | 0.034862271 |
| Obesity | nk_cell | *CDH6* | -0.960691461 | 2.06E-09 | 2.22E-08 | 9.99E-06 |
| Obesity | nk_cell | *LMNB1* | -2.246594525 | 0.02257528 | 0.026933192 | 1 |
| Obesity | nk_cell | *SLC12A7* | -2.742262831 | 0.006597924 | 0.008231712 | 1 |
| Obesity | nk_cell | *SKP1* | -2.346839538 | 2.20E-07 | 5.86E-07 | 0.001065405 |
| Obesity | nk_cell | *PPWD1* | -2.555585153 | 4.09E-05 | 6.49E-05 | 0.198238108 |
| Obesity | nk_cell | *TXNDC15* | -2.443912336 | 4.63E-08 | 1.64E-07 | 0.000224835 |
| Obesity | nk_cell | *SMAD5* | -2.262266028 | 0.000829664 | 0.00112163 | 1 |
| Obesity | nk_cell | *ERGIC1* | -2.516656239 | 7.65E-05 | 0.000116576 | 0.37140995 |
| Obesity | nk_cell | *ATP6V0E1* | -2.135089113 | 0.000111452 | 0.000166765 | 0.540763686 |
| Obesity | nk_cell | *BNIP1* | 0.905463296 | 0.004400977 | 0.005584085 | 1 |
| Obesity | nk_cell | *UNC5A* | -4.338260007 | 1.48E-09 | 2.11E-08 | 7.17E-06 |
| Obesity | nk_cell | *ACTR8* | -3.560526044 | 4.22E-09 | 3.13E-08 | 2.05E-05 |
| Obesity | nk_cell | *TBCCD1* | -3.608492014 | 5.84E-06 | 1.06E-05 | 0.028318443 |
| Obesity | nk_cell | *CLDN16* | -1.31010714 | 3.31E-09 | 2.74E-08 | 1.61E-05 |

| | | | | | | |
|---|---|---|---|---|---|---|
| Obesity | nk_cell | *NPHP3* | -1.157714034 | 0.007668307 | 0.009515761 | 1 |
| Obesity | nk_cell | *PCCB* | -1.118653216 | 0.014476794 | 0.017569136 | 1 |
| Obesity | nk_cell | *CEP70* | -2.672593314 | 5.21E-05 | 8.14E-05 | 0.25295203 |
| Obesity | nk_cell | *HYAL1* | -2.249687617 | 0.001175494 | 0.001562174 | 1 |
| Obesity | nk_cell | *C3orf14* | -3.0612365 | 2.15E-08 | 9.12E-08 | 0.000104526 |
| Obesity | nk_cell | *IQCG* | -2.458397687 | 5.13E-07 | 1.21E-06 | 0.002489236 |
| Obesity | nk_cell | *NCBP2* | -2.558806898 | 0.008031496 | 0.00993849 | 1 |
| Obesity | nk_cell | *SLC41A3* | -1.722130492 | 4.54E-09 | 3.26E-08 | 2.20E-05 |
| Obesity | nk_cell | *PLXNA1* | -2.444538696 | 1.03E-07 | 3.14E-07 | 0.00050029 |
| Obesity | nk_cell | *SCAP* | -2.226088278 | 0.00013724 | 0.000202829 | 0.665887667 |
| Obesity | nk_cell | *KIAA1257* | -3.017042338 | 2.19E-09 | 2.27E-08 | 1.06E-05 |
| Obesity | nk_cell | *NEK11* | -1.552666922 | 2.26E-08 | 9.45E-08 | 0.000109646 |
| Obesity | nk_cell | *PLSCR4* | -1.565592663 | 3.35E-08 | 1.27E-07 | 0.000162348 |
| Obesity | nk_cell | *HEMK1* | -2.765896203 | 9.18E-06 | 1.61E-05 | 0.04454211 |
| Obesity | nk_cell | *COMMD2* | -1.912910091 | 0.029344754 | 0.034625668 | 1 |
| Obesity | nk_cell | *AADAC* | -0.663098409 | 7.64E-05 | 0.00011634 | 0.370543608 |
| Obesity | nk_cell | *ABHD14B* | -3.410295373 | 9.93E-06 | 1.73E-05 | 0.048174357 |
| Obesity | nk_cell | *ARHGEF26* | -2.174720782 | 2.70E-08 | 1.08E-07 | 0.000131224 |
| Obesity | nk_cell | *VIPR1* | -1.900436016 | 3.30E-09 | 2.73E-08 | 1.60E-05 |
| Obesity | nk_cell | *DNAH1* | -1.779230085 | 1.47E-06 | 3.08E-06 | 0.007140883 |
| Obesity | nk_cell | *NKTR* | -1.778210851 | 4.27E-05 | 6.76E-05 | 0.206976526 |
| Obesity | nk_cell | *NEK4* | -2.308585427 | 5.80E-10 | 1.83E-08 | 2.82E-06 |
| Obesity | nk_cell | *ADAM23* | -3.215829205 | 6.81E-08 | 2.23E-07 | 0.000330577 |
| Obesity | nk_cell | *LMAN2L* | -1.90993273 | 1.47E-08 | 6.89E-08 | 7.16E-05 |
| Obesity | nk_cell | *ACTR1B* | -2.71209248 | 0.003795827 | 0.004844122 | 1 |
| Obesity | nk_cell | *SLC35F5* | -2.012556015 | 6.24E-07 | 1.43E-06 | 0.003025498 |
| Obesity | nk_cell | *OTOF* | 3.51206844 | 0.001167369 | 0.001551801 | 1 |
| Obesity | nk_cell | *CENPA* | -1.556988439 | 5.39E-08 | 1.85E-07 | 0.000261497 |
| Obesity | nk_cell | *NRBP1* | -3.017664622 | 2.12E-05 | 3.48E-05 | 0.10266791 |
| Obesity | nk_cell | *ITGB6* | -1.539701213 | 8.08E-10 | 1.98E-08 | 3.92E-06 |
| Obesity | nk_cell | *PPM1G* | 2.594035465 | 1.70E-07 | 4.72E-07 | 0.000823991 |
| Obesity | nk_cell | *MOGS* | -1.615051229 | 4.99E-05 | 7.82E-05 | 0.242219877 |
| Obesity | nk_cell | *GRB14* | -2.603764693 | 2.02E-08 | 8.68E-08 | 9.82E-05 |
| Obesity | nk_cell | *RTN4* | -1.702728361 | 3.70E-07 | 9.14E-07 | 0.001794419 |
| Obesity | nk_cell | *GALNT3* | 0.743628126 | 4.85E-10 | 1.71E-08 | 2.35E-06 |
| Obesity | nk_cell | *POLE4* | 3.566953334 | 0.000269245 | 0.000384454 | 1 |
| Obesity | nk_cell | *ACADL* | -2.001720841 | 2.82E-05 | 4.59E-05 | 0.137015022 |
| Obesity | nk_cell | *MRPL19* | -2.077780896 | 5.22E-08 | 1.81E-07 | 0.000253264 |
| Obesity | nk_cell | *IGFBP2* | -2.585696504 | 1.88E-05 | 3.11E-05 | 0.091065604 |
| Obesity | nk_cell | *GNLY* | 0.10349995 | 1.13E-09 | 2.05E-08 | 5.47E-06 |
| Obesity | nk_cell | *CHMP3* | -4.036799188 | 9.25E-10 | 2.05E-08 | 4.49E-06 |
| Obesity | nk_cell | *WNT6* | -2.062316809 | 1.32E-10 | 8.67E-09 | 6.42E-07 |
| Obesity | nk_cell | *IL1RL2* | -2.390126807 | 7.15E-08 | 2.32E-07 | 0.000346721 |
| Obesity | nk_cell | *IL18R1* | 1.386037627 | 5.49E-09 | 3.65E-08 | 2.66E-05 |
| Obesity | nk_cell | *MLPH* | -2.193193583 | 7.50E-09 | 4.36E-08 | 3.64E-05 |
| Obesity | nk_cell | *HDLBP* | -2.14494644 | 2.45E-05 | 4.00E-05 | 0.118867765 |
| Obesity | nk_cell | *PASK* | -1.321603635 | 0.000294104 | 0.000418596 | 1 |
| Obesity | nk_cell | *TPO* | -2.574838977 | 1.24E-07 | 3.66E-07 | 0.00060087 |
| Obesity | nk_cell | *TAF1B* | -2.311766526 | 4.27E-07 | 1.03E-06 | 0.002071419 |
| Obesity | nk_cell | *NOL10* | -2.31317541 | 4.08E-06 | 7.70E-06 | 0.019801904 |

| | | | | | | |
|---|---|---|---|---|---|---|
| Obesity | nk_cell | *STRN* | -2.185706495 | 3.26E-07 | 8.20E-07 | 0.001580893 |
| Obesity | nk_cell | *CEBPZ* | -2.743590367 | 1.40E-05 | 2.38E-05 | 0.068049524 |
| Obesity | nk_cell | *PRKD3* | -2.455227842 | 7.28E-06 | 1.30E-05 | 0.035334845 |
| Obesity | nk_cell | *SLC25A12* | -2.277873036 | 2.97E-05 | 4.82E-05 | 0.144315244 |
| Obesity | nk_cell | *DARS* | -2.276180027 | 0.000592035 | 0.000813064 | 1 |
| Obesity | nk_cell | *WIPF1* | 1.914757938 | 6.21E-08 | 2.06E-07 | 0.000301223 |
| Obesity | nk_cell | *PNO1* | -3.879726635 | 1.08E-07 | 3.27E-07 | 0.000524502 |
| Obesity | nk_cell | *ORC4* | -2.001999419 | 2.61E-06 | 5.14E-06 | 0.012655405 |
| Obesity | nk_cell | *CD207* | -2.831051435 | 0.000190364 | 0.000276623 | 0.923645364 |
| Obesity | nk_cell | *GRIN3B* | 1.06549705 | 2.33E-09 | 2.30E-08 | 1.13E-05 |
| Obesity | nk_cell | *NFE2L2* | -1.81208003 | 8.04E-07 | 1.80E-06 | 0.003902599 |
| Obesity | nk_cell | *MSH6* | -2.462970485 | 2.47E-08 | 1.01E-07 | 0.000120021 |
| Obesity | nk_cell | *ALMS1* | -2.25264446 | 2.74E-07 | 7.05E-07 | 0.001327149 |
| Obesity | nk_cell | *DNAJC16* | -1.948159384 | 4.85E-09 | 3.42E-08 | 2.35E-05 |
| Obesity | nk_cell | *MARK1* | -2.0548432 | 3.39E-08 | 1.28E-07 | 0.000164655 |
| Obesity | nk_cell | *MORN1* | -1.842526779 | 1.00E-09 | 2.05E-08 | 4.85E-06 |
| Obesity | nk_cell | *GPX7* | 0.742747895 | 1.58E-08 | 7.29E-08 | 7.65E-05 |
| Obesity | nk_cell | *RALGPS2* | -1.08029092 | 3.97E-09 | 3.01E-08 | 1.93E-05 |
| Obesity | nk_cell | *CEP104* | -2.892640089 | 7.83E-07 | 1.76E-06 | 0.003797616 |
| Obesity | nk_cell | *FAM20B* | -2.903569855 | 4.37E-06 | 8.18E-06 | 0.02118418 |
| Obesity | nk_cell | *WRAP73* | 2.859471984 | 7.39E-07 | 1.67E-06 | 0.003585717 |
| Obesity | nk_cell | *ICMT* | -4.050928407 | 1.62E-07 | 4.55E-07 | 0.000787315 |
| Obesity | nk_cell | *PARK7* | -2.90479332 | 9.02E-06 | 1.58E-05 | 0.043776844 |
| Obesity | nk_cell | *KIAA1324* | 1.195790642 | 1.94E-09 | 2.20E-08 | 9.40E-06 |
| Obesity | nk_cell | *MECR* | -1.790680881 | 1.40E-07 | 4.02E-07 | 0.000677459 |
| Obesity | nk_cell | *KCNC4* | -1.986900376 | 8.10E-07 | 1.81E-06 | 0.003932057 |
| Obesity | nk_cell | *EDEM3* | -2.657006291 | 4.28E-05 | 6.77E-05 | 0.207493017 |
| Obesity | nk_cell | *CAPZA1* | -2.045236104 | 8.29E-05 | 0.000125647 | 0.402194762 |
| Obesity | nk_cell | *ASH1L* | -1.608029243 | 2.32E-05 | 3.79E-05 | 0.112398667 |
| Obesity | nk_cell | *GON4L* | -2.149088899 | 5.59E-08 | 1.90E-07 | 0.000271339 |
| Obesity | nk_cell | *DOCK7* | -1.942293716 | 1.28E-07 | 3.76E-07 | 0.000620855 |
| Obesity | nk_cell | *FBXO2* | 1.345966907 | 2.25E-08 | 9.43E-08 | 0.000109242 |
| Obesity | nk_cell | *DNAJC6* | 1.661435994 | 0.026732362 | 0.031712817 | 1 |
| Obesity | nk_cell | *LEPR* | -2.019491528 | 7.48E-07 | 1.69E-06 | 0.003631465 |
| Obesity | nk_cell | *IVNS1ABP* | 1.621035143 | 0.002367426 | 0.003072967 | 1 |
| Obesity | nk_cell | *PRG4* | -1.803478333 | 5.18E-09 | 3.54E-08 | 2.51E-05 |
| Obesity | nk_cell | *BCAS2* | -2.149728637 | 1.03E-07 | 3.15E-07 | 0.000500789 |
| Obesity | nk_cell | *CFHR3* | -4.049828951 | 1.10E-07 | 3.32E-07 | 0.000534143 |
| Obesity | nk_cell | *CRYZ* | -0.958001817 | 0.002276572 | 0.002958995 | 1 |
| Obesity | nk_cell | *TMEM9* | -2.369077107 | 2.40E-08 | 9.88E-08 | 0.000116526 |
| Obesity | nk_cell | *WARS2* | -1.797508285 | 3.06E-06 | 5.94E-06 | 0.014828247 |
| Obesity | nk_cell | *OSCP1* | -1.734484423 | 3.97E-08 | 1.46E-07 | 0.000192585 |
| Obesity | nk_cell | *MRPS15* | -2.89336606 | 1.19E-07 | 3.53E-07 | 0.00057562 |
| Obesity | nk_cell | *GNPAT* | -2.093047085 | 0.006976203 | 0.008683566 | 1 |
| Obesity | nk_cell | *C1orf109* | -1.686268348 | 8.59E-09 | 4.67E-08 | 4.17E-05 |
| Obesity | nk_cell | *TBCE* | -2.656503079 | 3.88E-07 | 9.49E-07 | 0.001882072 |
| Obesity | nk_cell | *ACADM* | -2.080767043 | 8.14E-07 | 1.81E-06 | 0.003951199 |
| Obesity | nk_cell | *PADI2* | 1.702702021 | 7.74E-05 | 0.000117812 | 0.375584078 |
| Obesity | nk_cell | *SDHB* | -2.834015466 | 2.63E-06 | 5.17E-06 | 0.012748923 |
| Obesity | nk_cell | *MFAP2* | -1.542338832 | 0.000729946 | 0.000993464 | 1 |

| | | | | | | |
|---|---|---|---|---|---|---|
| Obesity | nk_cell | *CTBS* | -1.39222167 | 0.008808075 | 0.010871733 | 1 |
| Obesity | nk_cell | *KLHL12* | -3.353468047 | 9.63E-07 | 2.11E-06 | 0.004671912 |
| Obesity | nk_cell | *ZNHIT6* | -2.517389233 | 1.38E-05 | 2.34E-05 | 0.066867489 |
| Obesity | nk_cell | *KIF17* | -5.800800417 | 3.43E-09 | 2.77E-08 | 1.66E-05 |
| Obesity | nk_cell | *CD46* | -2.065396193 | 1.87E-05 | 3.10E-05 | 0.090565204 |
| Obesity | nk_cell | *EBNA1BP2* | -1.648595569 | 6.24E-09 | 3.98E-08 | 3.03E-05 |
| Obesity | nk_cell | *ERI3* | -2.321933446 | 7.05E-07 | 1.60E-06 | 0.003419538 |
| Obesity | nk_cell | *NSUN4* | -3.539284137 | 3.26E-05 | 5.25E-05 | 0.158176516 |
| Obesity | nk_cell | *DR1* | -1.886041756 | 0.015640203 | 0.018919538 | 1 |
| Obesity | nk_cell | *PRRC2C* | -1.466198441 | 0.010685911 | 0.013112807 | 1 |
| Obesity | nk_cell | *ABCD3* | -2.427226311 | 4.03E-07 | 9.80E-07 | 0.001953036 |
| Obesity | nk_cell | *PTBP2* | -1.613688475 | 4.94E-05 | 7.75E-05 | 0.23962742 |
| Obesity | nk_cell | *DARS2* | -2.317225363 | 0.017518565 | 0.021139039 | 1 |
| Obesity | nk_cell | *MTFR1L* | -2.052069623 | 1.30E-08 | 6.28E-08 | 6.32E-05 |
| Obesity | nk_cell | *RPS6KA1* | 0.906144995 | 2.43E-09 | 2.35E-08 | 1.18E-05 |
| Obesity | nk_cell | *MARC2* | -1.731484322 | 7.17E-09 | 4.27E-08 | 3.48E-05 |
| Obesity | nk_cell | *SLC5A9* | -1.298655687 | 5.80E-09 | 3.77E-08 | 2.81E-05 |
| Obesity | nk_cell | *ESYT2* | -2.254617404 | 1.83E-05 | 3.04E-05 | 0.088644609 |
| Obesity | nk_cell | *CD3EAP* | -1.984009551 | 1.79E-08 | 7.96E-08 | 8.69E-05 |
| Obesity | nk_cell | *RCN2* | -2.60015049 | 3.60E-07 | 8.95E-07 | 0.001748848 |
| Obesity | nk_cell | *A4GNT* | -1.243671882 | 0.006422752 | 0.008017287 | 1 |
| Obesity | nk_cell | *IFT46* | -1.570111745 | 1.12E-06 | 2.40E-06 | 0.005418158 |
| Obesity | nk_cell | *APOA1* | -7.739529172 | 5.46E-06 | 1.00E-05 | 0.026491713 |
| Obesity | nk_cell | *DDX59* | -2.451285435 | 5.10E-06 | 9.43E-06 | 0.02472458 |
| Obesity | nk_cell | *CAMSAP2* | -1.684047648 | 1.23E-07 | 3.63E-07 | 0.000595028 |
| Obesity | nk_cell | *KLF7* | -1.981514805 | 2.89E-06 | 5.65E-06 | 0.014018082 |
| Obesity | nk_cell | *CASC1* | -1.743088593 | 1.43E-06 | 3.01E-06 | 0.006956219 |
| Obesity | nk_cell | *LRMP* | 1.403982583 | 0.000260287 | 0.00037243 | 1 |
| Obesity | nk_cell | *ELOVL4* | -1.880892688 | 0.001057791 | 0.001412718 | 1 |
| Obesity | nk_cell | *CNR1* | -1.316402926 | 7.73E-09 | 4.41E-08 | 3.75E-05 |
| Obesity | nk_cell | *SGIP1* | -2.531872248 | 0.013820852 | 0.016789879 | 1 |
| Obesity | nk_cell | *AKAP7* | -2.381160038 | 2.95E-06 | 5.75E-06 | 0.01431463 |
| Obesity | nk_cell | *ALDH8A1* | -0.64819709 | 0.013096871 | 0.015942303 | 1 |
| Obesity | nk_cell | *MED28* | -2.091945382 | 2.30E-08 | 9.56E-08 | 0.000111627 |
| Obesity | nk_cell | *TMEM5* | -1.788445085 | 0.036771848 | 0.042960994 | 1 |
| Obesity | nk_cell | *VAMP8* | 3.13544372 | 9.14E-06 | 1.60E-05 | 0.044344243 |
| Obesity | nk_cell | *FOXO3* | -1.929557614 | 1.53E-06 | 3.19E-06 | 0.007429655 |
| Obesity | nk_cell | *SPP1* | 2.164543415 | 7.59E-10 | 1.93E-08 | 3.68E-06 |
| Obesity | nk_cell | *MFSD1* | -2.216082989 | 2.32E-07 | 6.15E-07 | 0.001127262 |
| Obesity | nk_cell | *UBN1* | -2.540628272 | 1.88E-05 | 3.12E-05 | 0.091239148 |
| Obesity | nk_cell | *WDR35* | -2.354452847 | 5.19E-07 | 1.22E-06 | 0.002518788 |
| Obesity | nk_cell | *DNAH7* | -2.848738231 | 3.16E-09 | 2.68E-08 | 1.53E-05 |
| Obesity | nk_cell | *C2orf40* | -2.85779137 | 2.69E-05 | 4.37E-05 | 0.130393746 |
| Obesity | nk_cell | *CPSF3* | -3.118172152 | 8.91E-08 | 2.79E-07 | 0.000432123 |
| Obesity | nk_cell | *PIGZ* | -0.83973131 | 0.002225357 | 0.002893977 | 1 |
| Obesity | nk_cell | *SENP5* | -1.903081161 | 7.92E-08 | 2.54E-07 | 0.000384468 |
| Obesity | nk_cell | *CCDC92* | -1.720647366 | 0.000220847 | 0.000318251 | 1 |
| Obesity | nk_cell | *FKBP15* | -1.513756613 | 0.000237843 | 0.000341757 | 1 |
| Obesity | nk_cell | *RAB14* | -2.516771285 | 0.00075158 | 0.001021762 | 1 |
| Obesity | nk_cell | *TRIM32* | 5.472294214 | 4.55E-07 | 1.09E-06 | 0.002206985 |

| | | | | | | |
|---|---|---|---|---|---|---|
| Obesity | nk_cell | *PPP6C* | -2.217614902 | 1.00E-07 | 3.07E-07 | 0.00048568 |
| Obesity | nk_cell | *HDHD3* | -0.975853113 | 0.000259542 | 0.000371475 | 1 |
| Obesity | nk_cell | *RBM18* | -2.103931448 | 0.040145854 | 0.046767751 | 1 |
| Obesity | nk_cell | *HSDL2* | -1.338224189 | 9.31E-08 | 2.90E-07 | 0.000451739 |
| Obesity | nk_cell | *MAPKAP1* | -2.130184653 | 2.70E-05 | 4.40E-05 | 0.131206249 |
| Obesity | nk_cell | *INVS* | -0.955831531 | 0.029112958 | 0.034368874 | 1 |
| Obesity | nk_cell | *DENND1A* | -1.85346252 | 3.28E-06 | 6.33E-06 | 0.015931765 |
| Obesity | nk_cell | *KDSR* | -2.130335482 | 3.50E-08 | 1.32E-07 | 0.000169629 |
| Obesity | nk_cell | *DCAF4* | -2.201739294 | 1.55E-07 | 4.40E-07 | 0.000753665 |
| Obesity | nk_cell | *IFI27L2* | 3.648310612 | 4.01E-06 | 7.60E-06 | 0.019475062 |
| Obesity | nk_cell | *NEK9* | -2.248333323 | 5.12E-08 | 1.78E-07 | 0.00024859 |
| Obesity | nk_cell | *TTLL5* | -2.360808515 | 7.40E-06 | 1.32E-05 | 0.035886116 |
| Obesity | nk_cell | *FLVCR2* | -2.750152096 | 6.89E-08 | 2.25E-07 | 0.000334401 |
| Obesity | nk_cell | *PPP4R4* | -1.528561644 | 1.82E-07 | 5.01E-07 | 0.000884641 |
| Obesity | nk_cell | *ALDH6A1* | -1.827359875 | 2.39E-07 | 6.31E-07 | 0.001161893 |
| Obesity | nk_cell | *COQ6* | -3.207964149 | 0.000846342 | 0.001142903 | 1 |
| Obesity | nk_cell | *DNMT3A* | -2.152276589 | 3.05E-06 | 5.92E-06 | 0.01478298 |
| Obesity | nk_cell | *CNRIP1* | -2.405241309 | 9.41E-05 | 0.000141812 | 0.45661121 |
| Obesity | nk_cell | *CRIPT* | -2.649417605 | 0.006675161 | 0.008323794 | 1 |
| Obesity | nk_cell | *EPCAM* | -2.796350162 | 1.87E-08 | 8.20E-08 | 9.09E-05 |
| Obesity | nk_cell | *TECTB* | -1.933526893 | 0.000686602 | 0.000935522 | 1 |
| Obesity | nk_cell | *ELOVL3* | -3.065295902 | 4.01E-06 | 7.60E-06 | 0.019443611 |
| Obesity | nk_cell | *PPP1R3C* | -1.840808499 | 4.46E-09 | 3.23E-08 | 2.17E-05 |
| Obesity | nk_cell | *PYROXD2* | -1.567743177 | 7.35E-08 | 2.38E-07 | 0.000356593 |
| Obesity | nk_cell | *MXI1* | -1.933860182 | 5.90E-08 | 1.98E-07 | 0.000286098 |
| Obesity | nk_cell | *C10orf88* | -1.797386089 | 0.018233738 | 0.021974689 | 1 |
| Obesity | nk_cell | *HELLS* | 2.312593391 | 3.05E-05 | 4.92E-05 | 0.14786595 |
| Obesity | nk_cell | *TCTN3* | -2.11593193 | 6.90E-09 | 4.17E-08 | 3.35E-05 |
| Obesity | nk_cell | *WDR11* | -2.110513012 | 9.72E-07 | 2.13E-06 | 0.004716567 |
| Obesity | nk_cell | *KANSL1* | -1.741824778 | 0.000247575 | 0.00035529 | 1 |
| Obesity | nk_cell | *HOXB3* | -2.092800656 | 7.45E-08 | 2.41E-07 | 0.000361366 |
| Obesity | nk_cell | *PCDHB10* | -4.278269754 | 2.55E-09 | 2.40E-08 | 1.24E-05 |
| Obesity | nk_cell | *GORAB* | -2.991635717 | 9.35E-08 | 2.91E-07 | 0.000453516 |
| Obesity | nk_cell | *SNX19* | -2.796013384 | 8.46E-12 | 1.32E-09 | 4.11E-08 |
| Obesity | nk_cell | *SLC10A7* | -2.353405349 | 1.02E-06 | 2.22E-06 | 0.004936227 |
| Obesity | nk_cell | *MASTL* | -2.904115872 | 8.59E-09 | 4.67E-08 | 4.17E-05 |
| Obesity | nk_cell | *MRC1* | -2.266920391 | 6.30E-06 | 1.14E-05 | 0.030545816 |
| Obesity | nk_cell | *IQSEC3* | -1.374469009 | 2.63E-09 | 2.45E-08 | 1.28E-05 |
| Obesity | nk_cell | *CCDC77* | -2.976025876 | 0.000306039 | 0.00043469 | 1 |
| Obesity | nk_cell | *DNAJC15* | -2.319448618 | 0.002081089 | 0.002713637 | 1 |
| Obesity | nk_cell | *UFM1* | -2.433558942 | 2.93E-06 | 5.73E-06 | 0.01423495 |
| Obesity | nk_cell | *ALG5* | -2.406012528 | 1.01E-05 | 1.76E-05 | 0.049175566 |
| Obesity | nk_cell | *SIL1* | -1.726771442 | 2.95E-08 | 1.15E-07 | 0.000143274 |
| Obesity | nk_cell | *ARL1* | -3.059858259 | 6.50E-06 | 1.17E-05 | 0.031524774 |
| Obesity | nk_cell | *APAF1* | -2.709022079 | 0.000114369 | 0.000170955 | 0.55491977 |
| Obesity | nk_cell | *EPHX2* | -1.939170894 | 1.64E-06 | 3.39E-06 | 0.007953521 |
| Obesity | nk_cell | *AKAP1* | -1.5266091 | 9.47E-09 | 5.00E-08 | 4.59E-05 |
| Obesity | nk_cell | *SCPEP1* | -1.087925199 | 0.008816422 | 0.010879267 | 1 |
| Obesity | nk_cell | *TBX2* | -1.449524059 | 1.69E-10 | 9.97E-09 | 8.18E-07 |
| Obesity | nk_cell | *TEX14* | -1.444626072 | 3.66E-05 | 5.85E-05 | 0.177518255 |

| | | | | | | |
|---|---|---|---|---|---|---|
| Obesity | nk_cell | *TRIM6* | 8.509204751 | 4.85E-06 | 9.00E-06 | 0.023536145 |
| Obesity | nk_cell | *ADCY7* | 1.179350458 | 8.43E-09 | 4.63E-08 | 4.09E-05 |
| Obesity | nk_cell | *ECHDC2* | -1.844441717 | 3.18E-08 | 1.21E-07 | 0.000154192 |
| Obesity | nk_cell | *TAS2R10* | -1.227063138 | 0.041418101 | 0.048191996 | 1 |
| Obesity | nk_cell | *PYROXD1* | 1.585441297 | 0.023339993 | 0.027797164 | 1 |
| Obesity | nk_cell | *ZNF549* | -3.340929803 | 0.000150768 | 0.000221742 | 0.731527171 |
| Obesity | nk_cell | *A1BG* | 2.954603425 | 3.74E-10 | 1.53E-08 | 1.82E-06 |
| Obesity | nk_cell | *ZSCAN18* | -2.448113085 | 5.44E-07 | 1.27E-06 | 0.002638921 |
| Obesity | nk_cell | *PDZRN3* | -1.467405184 | 8.03E-07 | 1.79E-06 | 0.00389656 |
| Obesity | nk_cell | *LHX4* | -2.383070002 | 5.71E-09 | 3.74E-08 | 2.77E-05 |
| Obesity | nk_cell | *SEC22A* | -2.327442673 | 4.02E-07 | 9.78E-07 | 0.001948552 |
| Obesity | nk_cell | *POPDC2* | -1.76495922 | 1.95E-08 | 8.43E-08 | 9.44E-05 |
| Obesity | nk_cell | *CD80* | -1.879702198 | 5.25E-07 | 1.23E-06 | 0.002545954 |
| Obesity | nk_cell | *MAPK8IP1* | 1.901901605 | 5.69E-10 | 1.82E-08 | 2.76E-06 |
| Obesity | nk_cell | *CAT* | -1.28491097 | 7.40E-07 | 1.67E-06 | 0.003591534 |
| Obesity | nk_cell | *PILRB* | 1.270198462 | 2.02E-06 | 4.07E-06 | 0.009785352 |
| Obesity | nk_cell | *ZCCHC17* | -2.611709654 | 2.73E-07 | 7.03E-07 | 0.001323846 |
| Obesity | nk_cell | *FABP3* | -2.478100099 | 1.06E-09 | 2.05E-08 | 5.15E-06 |
| Obesity | nk_cell | *TNFSF10* | -2.336678078 | 4.76E-07 | 1.13E-06 | 0.002308392 |
| Obesity | nk_cell | *ZNF639* | -3.275363465 | 1.26E-07 | 3.72E-07 | 0.0006115 |
| Obesity | nk_cell | *CPXM2* | -0.975548157 | 2.18E-07 | 5.82E-07 | 0.001055414 |
| Obesity | nk_cell | *FLT3* | 2.344543549 | 1.91E-09 | 2.19E-08 | 9.27E-06 |
| Obesity | nk_cell | *MTIF3* | -2.213693546 | 2.55E-06 | 5.03E-06 | 0.012351626 |
| Obesity | nk_cell | *FMOD* | -1.887423116 | 3.96E-08 | 1.46E-07 | 0.000192046 |
| Obesity | nk_cell | *CD244* | 0.264361567 | 1.01E-09 | 2.05E-08 | 4.88E-06 |
| Obesity | nk_cell | *RBBP6* | -2.332759158 | 9.67E-07 | 2.12E-06 | 0.004693362 |
| Obesity | nk_cell | *SERAC1* | 3.189782185 | 1.64E-06 | 3.39E-06 | 0.007973351 |
| Obesity | nk_cell | *LDB3* | -1.157537673 | 3.53E-09 | 2.82E-08 | 1.71E-05 |
| Obesity | nk_cell | *OPN4* | -3.975432541 | 0.0030004 | 0.003860337 | 1 |
| Obesity | nk_cell | *FAM35A* | -2.19064096 | 1.20E-05 | 2.05E-05 | 0.058118202 |
| Obesity | nk_cell | *ODF2L* | -2.179070449 | 7.90E-07 | 1.77E-06 | 0.003835116 |
| Obesity | nk_cell | *SPATA1* | -2.249721469 | 3.20E-07 | 8.08E-07 | 0.00155203 |
| Obesity | nk_cell | *LRRC39* | -0.924251872 | 0.000797796 | 0.001080957 | 1 |
| Obesity | nk_cell | *RWDD3* | -1.872931213 | 0.00362473 | 0.004634306 | 1 |
| Obesity | nk_cell | *PMS2* | -3.019652452 | 2.27E-06 | 4.53E-06 | 0.011024319 |
| Obesity | nk_cell | *EEPD1* | -1.354876811 | 4.00E-07 | 9.74E-07 | 0.001938726 |
| Obesity | nk_cell | *KLHL7* | -2.191233624 | 1.46E-07 | 4.18E-07 | 0.000709722 |
| Obesity | nk_cell | *NT5C3A* | -2.055928694 | 0.004597257 | 0.005823992 | 1 |
| Obesity | nk_cell | *CCZ1* | -1.501499882 | 0.000252001 | 0.000361427 | 1 |
| Obesity | nk_cell | *POLM* | -1.346968303 | 2.12E-05 | 3.49E-05 | 0.103005236 |
| Obesity | nk_cell | *SMU1* | -2.045730061 | 8.12E-08 | 2.59E-07 | 0.0003942 |
| Obesity | nk_cell | *GLIPR2* | 2.326083113 | 5.71E-07 | 1.32E-06 | 0.002770819 |
| Obesity | nk_cell | *SLC25A51* | 1.455927597 | 8.21E-11 | 6.42E-09 | 3.98E-07 |
| Obesity | nk_cell | *TRIM24* | -2.44178272 | 1.28E-05 | 2.18E-05 | 0.062117603 |
| Obesity | nk_cell | *CISD1* | -1.934206598 | 6.34E-07 | 1.45E-06 | 0.003075728 |
| Obesity | nk_cell | *RBM19* | -2.056440589 | 1.73E-08 | 7.78E-08 | 8.41E-05 |
| Obesity | nk_cell | *DDX54* | 3.437978172 | 0.002148426 | 0.002797681 | 1 |
| Obesity | nk_cell | *SSPN* | -1.430955909 | 8.44E-08 | 2.67E-07 | 0.000409367 |
| Obesity | nk_cell | *CCDC91* | -1.861063985 | 5.98E-05 | 9.27E-05 | 0.290303403 |
| Obesity | nk_cell | *EBPL* | -1.999270117 | 8.67E-06 | 1.53E-05 | 0.042076819 |

| | | | | | | |
|---|---|---|---|---|---|---|
| Obesity | nk_cell | *CENPK* | 0.661436178 | 1.20E-09 | 2.07E-08 | 5.81E-06 |
| Obesity | nk_cell | *ITIH5* | -0.889202083 | 0.011201435 | 0.013696916 | 1 |
| Obesity | nk_cell | *TSFM* | -2.45905365 | 6.72E-08 | 2.20E-07 | 0.000325827 |
| Obesity | nk_cell | *ARHGAP9* | 0.712471238 | 1.49E-09 | 2.11E-08 | 7.23E-06 |
| Obesity | nk_cell | *PFDN5* | 2.589965181 | 2.60E-05 | 4.23E-05 | 0.126046133 |
| Obesity | nk_cell | *PDE1B* | -3.477715198 | 9.24E-08 | 2.88E-07 | 0.000448545 |
| Obesity | nk_cell | *CDK2* | -2.075561432 | 7.20E-09 | 4.27E-08 | 3.49E-05 |
| Obesity | nk_cell | *SARDH* | -1.991777163 | 1.29E-07 | 3.79E-07 | 0.000628012 |
| Obesity | nk_cell | *DBH* | -27.95340094 | 1.74E-09 | 2.16E-08 | 8.45E-06 |
| Obesity | nk_cell | *NDUFAF4* | 2.634059116 | 1.25E-06 | 2.65E-06 | 0.00604143 |
| Obesity | nk_cell | *USP45* | -2.055240208 | 2.03E-07 | 5.48E-07 | 0.000983695 |
| Obesity | nk_cell | *TTC21B* | -2.370186263 | 2.14E-07 | 5.75E-07 | 0.001040547 |
| Obesity | nk_cell | *ACVR1C* | -0.983353105 | 3.80E-07 | 9.35E-07 | 0.001844693 |
| Obesity | nk_cell | *PLA2G12A* | -2.71477622 | 8.92E-06 | 1.56E-05 | 0.043283257 |
| Obesity | nk_cell | *C4BPA* | -1.669358478 | 2.94E-09 | 2.57E-08 | 1.42E-05 |
| Obesity | nk_cell | *C4BPB* | -1.453661278 | 1.36E-07 | 3.95E-07 | 0.000660354 |
| Obesity | nk_cell | *RAB38* | -2.213686396 | 7.75E-08 | 2.49E-07 | 0.000376189 |
| Obesity | nk_cell | *DAW1* | -2.381209466 | 3.77E-09 | 2.94E-08 | 1.83E-05 |
| Obesity | nk_cell | *OBSL1* | -2.821890129 | 6.98E-07 | 1.59E-06 | 0.003388275 |
| Obesity | nk_cell | *SLC12A4* | -3.448487292 | 5.76E-07 | 1.33E-06 | 0.002795148 |
| Obesity | nk_cell | *FAM210B* | -3.117533136 | 1.02E-07 | 3.11E-07 | 0.000493867 |
| Obesity | nk_cell | *WFDC3* | -0.693911402 | 0.000157297 | 0.000230784 | 0.763202908 |
| Obesity | nk_cell | *SLC12A5* | 2.481158203 | 0.001981948 | 0.002587143 | 1 |
| Obesity | nk_cell | *MATN4* | 3.669333978 | 9.33E-10 | 2.05E-08 | 4.53E-06 |
| Obesity | nk_cell | *TOX2* | 1.027605604 | 0.000195003 | 0.000283026 | 0.946154361 |
| Obesity | nk_cell | *SRSF6* | -2.388637795 | 1.69E-07 | 4.70E-07 | 0.000819143 |
| Obesity | nk_cell | *CSE1L* | -2.499708422 | 0.00013767 | 0.000203341 | 0.667976703 |
| Obesity | nk_cell | *PTGIS* | -1.181496679 | 7.82E-06 | 1.38E-05 | 0.037927751 |
| Obesity | nk_cell | *RNF114* | -2.171384696 | 1.77E-07 | 4.90E-07 | 0.000860829 |
| Obesity | nk_cell | *NEURL2* | -1.01125182 | 0.000115629 | 0.000172732 | 0.56103301 |
| Obesity | nk_cell | *MTRR* | -2.142634755 | 5.86E-07 | 1.35E-06 | 0.002842662 |
| Obesity | nk_cell | *PEPD* | -2.079005936 | 2.16E-07 | 5.77E-07 | 0.001046698 |
| Obesity | nk_cell | *CHST8* | -1.739144804 | 2.27E-07 | 6.03E-07 | 0.001101934 |
| Obesity | nk_cell | *STAMBP* | -2.130882218 | 3.38E-08 | 1.28E-07 | 0.000164186 |
| Obesity | nk_cell | *NAGK* | 2.533228933 | 0.001875666 | 0.002454351 | 1 |
| Obesity | nk_cell | *SNRNP27* | -3.407438633 | 0.000178983 | 0.000260711 | 0.868427597 |
| Obesity | nk_cell | *ATP8A1* | 1.13953166 | 1.84E-09 | 2.19E-08 | 8.94E-06 |
| Obesity | nk_cell | *USP22* | -2.45351395 | 7.61E-05 | 0.000115935 | 0.369136419 |
| Obesity | nk_cell | *HIF3A* | -1.442851747 | 2.46E-09 | 2.37E-08 | 1.19E-05 |
| Obesity | nk_cell | *ZNF576* | -1.536016241 | 3.82E-09 | 2.95E-08 | 1.85E-05 |
| Obesity | nk_cell | *BTN2A2* | 0.795806418 | 0.032738037 | 0.03843333 | 1 |
| Obesity | nk_cell | *SERPINB6* | -1.886592605 | 9.20E-08 | 2.87E-07 | 0.00044624 |
| Obesity | nk_cell | *PEX6* | -0.998486009 | 0.000510426 | 0.000704399 | 1 |
| Obesity | nk_cell | *NQO2* | -2.218166864 | 6.14E-08 | 2.05E-07 | 0.000297801 |
| Obesity | nk_cell | *ZNF391* | -0.51084282 | 0.029270887 | 0.03454691 | 1 |
| Obesity | nk_cell | *RPS10* | -2.351609846 | 8.44E-09 | 4.63E-08 | 4.10E-05 |
| Obesity | nk_cell | *TCP11* | 1.775315781 | 0.028460276 | 0.033631091 | 1 |
| Obesity | nk_cell | *GNMT* | 1.643011708 | 2.15E-08 | 9.12E-08 | 0.000104554 |
| Obesity | nk_cell | *TREM1* | -1.718656323 | 2.36E-06 | 4.69E-06 | 0.011460995 |
| Obesity | nk_cell | *MEA1* | 1.672058261 | 5.42E-08 | 1.85E-07 | 0.00026307 |

| | | | | | | |
|---|---|---|---|---|---|---|
| Obesity | nk_cell | *KLHL31* | -1.612979305 | 1.50E-06 | 3.14E-06 | 0.007289795 |
| Obesity | nk_cell | *GLO1* | -2.20115333 | 0.001620339 | 0.002132325 | 1 |
| Obesity | nk_cell | *KCNK17* | -1.965196804 | 0.010265831 | 0.012610079 | 1 |
| Obesity | nk_cell | *RREB1* | -1.581250845 | 3.30E-07 | 8.29E-07 | 0.001601293 |
| Obesity | nk_cell | *RIOK1* | 1.900689819 | 0.014761654 | 0.017901411 | 1 |
| Obesity | nk_cell | *ATXN1* | -1.736136683 | 0.000158265 | 0.000231995 | 0.767903325 |
| Obesity | nk_cell | *RUNX2* | 2.436401406 | 0.00350532 | 0.004484 | 1 |
| Obesity | nk_cell | *CXCL6* | -3.117421949 | 1.20E-09 | 2.08E-08 | 5.84E-06 |
| Obesity | nk_cell | *SH3TC1* | -1.710192667 | 1.97E-06 | 3.99E-06 | 0.009571685 |
| Obesity | nk_cell | *BBS2* | -1.116551394 | 4.36E-05 | 6.90E-05 | 0.21161411 |
| Obesity | nk_cell | *MT1G* | -4.178218272 | 0.003494455 | 0.00447246 | 1 |
| Obesity | nk_cell | *CLYBL* | -2.381963648 | 0.000636918 | 0.000872727 | 1 |
| Obesity | nk_cell | *TMTC4* | -2.669783334 | 5.63E-08 | 1.91E-07 | 0.000272963 |
| Obesity | nk_cell | *ATP5S* | -1.873146552 | 0.001327698 | 0.001757226 | 1 |
| Obesity | nk_cell | *TEKT3* | -0.981785834 | 8.50E-05 | 0.000128669 | 0.4122562 |
| Obesity | nk_cell | *HS3ST3B1* | 0.461309458 | 1.57E-09 | 2.11E-08 | 7.61E-06 |
| Obesity | nk_cell | *GGA3* | -2.595998633 | 0.000153093 | 0.000225025 | 0.742806844 |
| Obesity | nk_cell | *NUP85* | -1.893341791 | 6.32E-09 | 4.00E-08 | 3.07E-05 |
| Obesity | nk_cell | *NT5C* | 3.208798487 | 2.35E-05 | 3.85E-05 | 0.114185092 |
| Obesity | nk_cell | *DDX31* | -2.648179212 | 6.82E-09 | 4.14E-08 | 3.31E-05 |
| Obesity | nk_cell | *OPRL1* | -3.934671913 | 1.86E-09 | 2.19E-08 | 9.03E-06 |
| Obesity | nk_cell | *IL1B* | -5.569717613 | 9.36E-10 | 2.05E-08 | 4.54E-06 |
| Obesity | nk_cell | *PAX8* | -1.426737991 | 9.66E-08 | 2.99E-07 | 0.000468839 |
| Obesity | nk_cell | *POLR1B* | 1.704269316 | 0.000238183 | 0.000342014 | 1 |
| Obesity | nk_cell | *CCDC93* | -2.174951516 | 3.93E-06 | 7.45E-06 | 0.019066867 |
| Obesity | nk_cell | *PSD4* | 0.76417863 | 1.18E-09 | 2.05E-08 | 5.70E-06 |
| Obesity | nk_cell | *GTF2F1* | -3.052321467 | 2.00E-05 | 3.29E-05 | 0.096814643 |
| Obesity | nk_cell | *TNFSF9* | -21.82390313 | 9.85E-08 | 3.04E-07 | 0.000477884 |
| Obesity | nk_cell | *SH2D3A* | -3.540964326 | 3.75E-08 | 1.40E-07 | 0.00018213 |
| Obesity | nk_cell | *GPR108* | -2.220962553 | 0.000154224 | 0.000226618 | 0.748293619 |
| Obesity | nk_cell | *SNRPD2* | 3.571042852 | 3.29E-06 | 6.33E-06 | 0.01594248 |
| Obesity | nk_cell | *SDCBP2* | 2.446789635 | 4.90E-09 | 3.43E-08 | 2.38E-05 |
| Obesity | nk_cell | *PSMF1* | -2.494314297 | 1.50E-06 | 3.13E-06 | 0.007257923 |
| Obesity | nk_cell | *RBCK1* | 1.771309111 | 0.042414829 | 0.049269033 | 1 |
| Obesity | nk_cell | *RRBP1* | -2.088447846 | 2.63E-07 | 6.84E-07 | 0.001277272 |
| Obesity | nk_cell | *FLRT3* | -2.806912834 | 2.05E-08 | 8.74E-08 | 9.93E-05 |
| Obesity | nk_cell | *PCSK2* | -9.235477837 | 0.000333135 | 0.000470422 | 1 |
| Obesity | nk_cell | *MKKS* | -2.981576665 | 1.80E-05 | 2.99E-05 | 0.08709591 |
| Obesity | nk_cell | *LAMP5* | 17.64114631 | 4.72E-09 | 3.34E-08 | 2.29E-05 |
| Obesity | nk_cell | *MGME1* | -2.086622366 | 1.01E-06 | 2.20E-06 | 0.004898624 |
| Obesity | nk_cell | *ITPA* | -2.613696573 | 1.31E-06 | 2.77E-06 | 0.006346908 |
| Obesity | nk_cell | *MCM8* | -1.104834948 | 0.007288007 | 0.009062381 | 1 |
| Obesity | nk_cell | *MRPS26* | -2.144318986 | 0.000202217 | 0.000292708 | 0.98115562 |
| Obesity | nk_cell | *NCLN* | -3.051088438 | 3.03E-06 | 5.89E-06 | 0.014678743 |
| Obesity | nk_cell | *KLC1* | -2.475196687 | 0.000276875 | 0.000395 | 1 |
| Obesity | nk_cell | *XRCC3* | -1.969685269 | 7.71E-09 | 4.40E-08 | 3.74E-05 |
| Obesity | nk_cell | *TUBGCP3* | -2.054082994 | 0.004967293 | 0.00626822 | 1 |
| Obesity | nk_cell | *MCF2L* | -1.438661643 | 4.58E-05 | 7.23E-05 | 0.222413905 |
| Obesity | nk_cell | *PROZ* | -2.623128085 | 0.012467372 | 0.015191283 | 1 |
| Obesity | nk_cell | *PRDX5* | 3.320655894 | 7.20E-05 | 0.000110074 | 0.349373744 |

| | | | | | | |
|---|---|---|---|---|---|---|
| Obesity | nk_cell | *IRF3* | 3.047160113 | 7.96E-06 | 1.40E-05 | 0.038605264 |
| Obesity | nk_cell | *TSKS* | -1.263781326 | 2.26E-08 | 9.45E-08 | 0.000109618 |
| Obesity | nk_cell | *ZNF384* | -2.922088626 | 4.10E-05 | 6.51E-05 | 0.198847284 |
| Obesity | nk_cell | *EMG1* | -2.249177558 | 0.010772881 | 0.013212846 | 1 |
| Obesity | nk_cell | *ATG14* | -1.878872869 | 5.30E-06 | 9.74E-06 | 0.025702586 |
| Obesity | nk_cell | *KTN1* | -2.042375588 | 1.30E-05 | 2.21E-05 | 0.063081111 |
| Obesity | nk_cell | *L3HYPDH* | -1.280004924 | 1.90E-09 | 2.19E-08 | 9.22E-06 |
| Obesity | nk_cell | *ZBTB1* | -2.213530523 | 1.17E-06 | 2.50E-06 | 0.005663605 |
| Obesity | nk_cell | *RHOT1* | -1.661105444 | 3.44E-06 | 6.60E-06 | 0.016695025 |
| Obesity | nk_cell | *WDR60* | -1.919040417 | 2.07E-06 | 4.16E-06 | 0.010023364 |
| Obesity | nk_cell | *AIF1L* | -1.535638657 | 2.47E-09 | 2.37E-08 | 1.20E-05 |
| Obesity | nk_cell | *ZNF484* | -2.509483233 | 0.011439879 | 0.013981435 | 1 |
| Obesity | nk_cell | *FGD3* | 0.692860856 | 3.05E-09 | 2.62E-08 | 1.48E-05 |
| Obesity | nk_cell | *ABHD8* | -1.64132293 | 2.87E-09 | 2.55E-08 | 1.39E-05 |
| Obesity | nk_cell | *ATP13A4* | 3.592880766 | 6.59E-09 | 4.08E-08 | 3.20E-05 |
| Obesity | nk_cell | *HELB* | 3.276211496 | 1.16E-05 | 1.99E-05 | 0.056155571 |
| Obesity | nk_cell | *RAB3IP* | 3.172565857 | 1.36E-08 | 6.51E-08 | 6.59E-05 |
| Obesity | nk_cell | *YEATS4* | -1.048469774 | 0.000488027 | 0.000675772 | 1 |
| Obesity | nk_cell | *TAS2R4* | -1.357574488 | 7.69E-09 | 4.40E-08 | 3.73E-05 |
| Obesity | nk_cell | *LRRC61* | -2.448714576 | 7.49E-08 | 2.42E-07 | 0.000363266 |
| Obesity | nk_cell | *IDUA* | -2.838631357 | 2.65E-09 | 2.45E-08 | 1.28E-05 |
| Obesity | nk_cell | *EMC1* | -2.970646074 | 0.000101596 | 0.00015252 | 0.492943839 |
| Obesity | nk_cell | *PLA2G5* | -1.005087169 | 1.22E-06 | 2.60E-06 | 0.00590788 |
| Obesity | nk_cell | *UBR4* | -2.207353708 | 1.03E-06 | 2.24E-06 | 0.005011258 |
| Obesity | nk_cell | *WFIKKN1* | -2.876667277 | 0.000659102 | 0.000901089 | 1 |
| Obesity | nk_cell | *WDR24* | -1.778742154 | 1.67E-09 | 2.15E-08 | 8.10E-06 |
| Obesity | nk_cell | *SMARCA4* | -1.863248304 | 0.018331356 | 0.022081366 | 1 |
| Obesity | nk_cell | *PNKD* | -2.210253415 | 8.57E-08 | 2.70E-07 | 0.000415882 |
| Obesity | nk_cell | *RNF6* | -1.666966496 | 6.72E-09 | 4.11E-08 | 3.26E-05 |
| Obesity | nk_cell | *HIP1* | -1.933336422 | 0.000317658 | 0.000449877 | 1 |
| Obesity | nk_cell | *STYXL1* | -0.875360449 | 0.008368268 | 0.010342036 | 1 |
| Obesity | nk_cell | *STEAP4* | -1.677207449 | 6.18E-10 | 1.87E-08 | 3.00E-06 |
| Obesity | nk_cell | *PEX1* | -2.775450295 | 1.61E-06 | 3.33E-06 | 0.007799205 |
| Obesity | nk_cell | *SRD5A3* | -1.761376102 | 0.001396236 | 0.001846427 | 1 |
| Obesity | nk_cell | *KDR* | -2.534301404 | 0.003021384 | 0.003885437 | 1 |
| Obesity | nk_cell | *DGCR6L* | -1.625090881 | 6.49E-08 | 2.14E-07 | 0.000314706 |
| Obesity | nk_cell | *VPREB3* | -4.567568306 | 0.000566821 | 0.000779982 | 1 |
| Obesity | nk_cell | *MGAT3* | -2.264865673 | 4.98E-07 | 1.18E-06 | 0.002415838 |
| Obesity | nk_cell | *A4GALT* | -1.807969045 | 2.98E-08 | 1.16E-07 | 0.000144806 |
| Obesity | nk_cell | *CDC42EP1* | -3.82855439 | 2.43E-07 | 6.38E-07 | 0.001176728 |
| Obesity | nk_cell | *RIBC2* | 2.364448481 | 0.000253285 | 0.000363054 | 1 |
| Obesity | nk_cell | *SPECC1* | -2.157866422 | 0.000670648 | 0.000915326 | 1 |
| Obesity | nk_cell | *CDHR3* | -1.987830842 | 3.91E-08 | 1.44E-07 | 0.000189613 |
| Obesity | nk_cell | *PRKRIP1* | -2.505758627 | 3.33E-06 | 6.40E-06 | 0.016149696 |
| Obesity | nk_cell | *IRF5* | -1.011897524 | 3.76E-06 | 7.15E-06 | 0.018227087 |
| Obesity | nk_cell | *OPN1SW* | -1.519264488 | 6.99E-10 | 1.92E-08 | 3.39E-06 |
| Obesity | nk_cell | *HOXD1* | -3.007392077 | 0.000427264 | 0.000595051 | 1 |
| Obesity | nk_cell | *MTX2* | -1.912377764 | 7.65E-08 | 2.46E-07 | 0.000370978 |
| Obesity | nk_cell | *GAD1* | -3.184780527 | 4.87E-10 | 1.71E-08 | 2.36E-06 |
| Obesity | nk_cell | *ORMDL1* | -2.083687699 | 1.00E-05 | 1.74E-05 | 0.048645047 |

| | | | | | | |
|---|---|---|---|---|---|---|
| Obesity | nk_cell | *HOXD9* | 1.103465246 | 6.01E-05 | 9.31E-05 | 0.291809645 |
| Obesity | nk_cell | *HOXD10* | -2.866634144 | 6.09E-08 | 2.03E-07 | 0.000295612 |
| Obesity | nk_cell | *PSMG2* | -2.459568107 | 1.65E-07 | 4.62E-07 | 0.000799944 |
| Obesity | nk_cell | *TWSG1* | -2.891861745 | 1.10E-05 | 1.89E-05 | 0.053176313 |
| Obesity | nk_cell | *WDFY4* | -0.82503615 | 0.024993564 | 0.029715455 | 1 |
| Obesity | nk_cell | *EIF2AK4* | -1.997985133 | 6.36E-05 | 9.79E-05 | 0.308802533 |
| Obesity | nk_cell | *TMOD2* | -2.380705964 | 1.58E-07 | 4.45E-07 | 0.000765294 |
| Obesity | nk_cell | *IVD* | -1.776552035 | 0.000395707 | 0.000553785 | 1 |
| Obesity | nk_cell | *KNSTRN* | -0.922468377 | 1.75E-05 | 2.91E-05 | 0.084843077 |
| Obesity | nk_cell | *ARPP19* | 2.497006214 | 0.003639303 | 0.004651712 | 1 |
| Obesity | nk_cell | *CALML4* | -4.056464204 | 1.53E-09 | 2.11E-08 | 7.41E-06 |
| Obesity | nk_cell | *ANAPC13* | -1.497259973 | 2.52E-07 | 6.60E-07 | 0.001223163 |
| Obesity | nk_cell | *BBOX1* | 2.487846075 | 1.30E-09 | 2.09E-08 | 6.31E-06 |
| Obesity | nk_cell | *SERGEF* | 1.714741877 | 0.027440889 | 0.032497728 | 1 |
| Obesity | nk_cell | *DCTD* | -1.730697613 | 0.028774043 | 0.03397704 | 1 |
| Obesity | nk_cell | *RPAIN* | -2.406831738 | 3.29E-11 | 3.24E-09 | 1.60E-07 |
| Obesity | nk_cell | *USP6* | -1.426834191 | 7.26E-06 | 1.30E-05 | 0.03521572 |
| Obesity | nk_cell | *PLD2* | -3.942814939 | 2.78E-07 | 7.14E-07 | 0.001349977 |
| Obesity | nk_cell | *PHF20L1* | -1.866419041 | 0.005227184 | 0.006573949 | 1 |
| Obesity | nk_cell | *PUS7L* | -2.339703208 | 0.000665853 | 0.000909549 | 1 |
| Obesity | nk_cell | *KRI1* | 1.769576821 | 3.62E-05 | 5.80E-05 | 0.175804149 |
| Obesity | nk_cell | *AP1M2* | -3.019617664 | 2.14E-09 | 2.26E-08 | 1.04E-05 |
| Obesity | nk_cell | *MTUS1* | -1.780100411 | 5.11E-07 | 1.20E-06 | 0.002481227 |
| Obesity | nk_cell | *KLK10* | -1.33849552 | 0.000919581 | 0.001237328 | 1 |
| Obesity | nk_cell | *KLK8* | 3.291595312 | 1.18E-09 | 2.06E-08 | 5.74E-06 |
| Obesity | nk_cell | *RIPK3* | 1.861666777 | 3.99E-05 | 6.34E-05 | 0.193594288 |
| Obesity | nk_cell | *ADCY4* | -2.508292824 | 2.15E-06 | 4.30E-06 | 0.010411649 |
| Obesity | nk_cell | *BCL2L2* | -2.272032396 | 5.04E-05 | 7.89E-05 | 0.244679249 |
| Obesity | nk_cell | *DTD2* | -1.3381758 | 1.16E-07 | 3.46E-07 | 0.000561983 |
| Obesity | nk_cell | *PARP2* | 3.163115901 | 0.000818474 | 0.00110712 | 1 |
| Obesity | nk_cell | *MIS18BP1* | -1.784536359 | 1.51E-06 | 3.14E-06 | 0.007303144 |
| Obesity | nk_cell | *RNASE1* | -1.924645638 | 3.31E-05 | 5.33E-05 | 0.160628937 |
| Obesity | nk_cell | *EPB41L4A* | -1.716032203 | 6.53E-08 | 2.15E-07 | 0.000316781 |
| Obesity | nk_cell | *CDO1* | -1.38082373 | 1.71E-05 | 2.86E-05 | 0.082950322 |
| Obesity | nk_cell | *REEP5* | -2.330449906 | 1.39E-06 | 2.93E-06 | 0.00673187 |
| Obesity | nk_cell | *RHBDF2* | 1.479898553 | 2.50E-05 | 4.08E-05 | 0.121238225 |
| Obesity | nk_cell | *AANAT* | -1.600185922 | 0.029405258 | 0.034680193 | 1 |
| Obesity | nk_cell | *TMEM8A* | -2.973234574 | 0.022575699 | 0.026933192 | 1 |
| Obesity | nk_cell | *MAU2* | -2.47705735 | 4.82E-07 | 1.14E-06 | 0.00233877 |
| Obesity | nk_cell | *SHC2* | -1.906641698 | 1.46E-06 | 3.06E-06 | 0.007080798 |
| Obesity | nk_cell | *SYT5* | -6.206287962 | 0.007035658 | 0.008755326 | 1 |
| Obesity | nk_cell | *ERMARD* | -2.504512221 | 5.29E-08 | 1.83E-07 | 0.000256901 |
| Obesity | nk_cell | *PHF10* | -2.389324191 | 2.17E-08 | 9.15E-08 | 0.000105084 |
| Obesity | nk_cell | *GALNT8* | -1.655156735 | 2.48E-08 | 1.01E-07 | 0.000120319 |
| Obesity | nk_cell | *PRKCSH* | 3.367740417 | 4.54E-07 | 1.08E-06 | 0.00220278 |
| Obesity | nk_cell | *CDC16* | -2.432264642 | 3.03E-05 | 4.90E-05 | 0.14701232 |
| Obesity | nk_cell | *XPO7* | -2.501919282 | 5.61E-05 | 8.73E-05 | 0.272390508 |
| Obesity | nk_cell | *FAM98C* | 2.032199847 | 0.007520259 | 0.009339211 | 1 |
| Obesity | nk_cell | *ATP8B3* | -2.502915178 | 1.73E-18 | 1.40E-15 | 8.38E-15 |
| Obesity | nk_cell | *MRPL34* | -0.971811933 | 0.000593093 | 0.000814286 | 1 |

| | | | | | | |
|---|---|---|---|---|---|---|
| Obesity | nk_cell | *TULP4* | -1.607737711 | 1.47E-05 | 2.48E-05 | 0.071176596 |
| Obesity | nk_cell | *QRSL1* | -1.701087657 | 1.75E-07 | 4.84E-07 | 0.00084773 |
| Obesity | nk_cell | *RSPH3* | -0.845215027 | 0.002602055 | 0.003367611 | 1 |
| Obesity | nk_cell | *STK33* | -1.151020961 | 2.60E-05 | 4.24E-05 | 0.12619121 |
| Obesity | nk_cell | *NDUFA10* | -1.44684399 | 0.000129864 | 0.000192551 | 0.630097848 |
| Obesity | nk_cell | *MAP1S* | -3.687165004 | 2.88E-08 | 1.13E-07 | 0.000139668 |
| Obesity | nk_cell | *SSBP4* | -2.051718224 | 0.00791487 | 0.009799171 | 1 |
| Obesity | nk_cell | *PGPEP1* | 3.238574215 | 8.60E-09 | 4.67E-08 | 4.17E-05 |
| Obesity | nk_cell | *KIAA1683* | -2.784442368 | 8.79E-08 | 2.76E-07 | 0.000426287 |
| Obesity | nk_cell | *LSM4* | 1.78621556 | 3.45E-05 | 5.54E-05 | 0.167400246 |
| Obesity | nk_cell | *TRPM4* | -2.035645843 | 2.94E-10 | 1.36E-08 | 1.43E-06 |
| Obesity | nk_cell | *UBAC1* | 2.220632093 | 0.003800748 | 0.004849126 | 1 |
| Obesity | nk_cell | *ATXN10* | -2.013812283 | 8.70E-07 | 1.92E-06 | 0.004222942 |
| Obesity | nk_cell | *CYP2E1* | -2.28351297 | 3.07E-10 | 1.40E-08 | 1.49E-06 |
| Obesity | nk_cell | *PNPLA7* | -1.824560804 | 1.28E-08 | 6.19E-08 | 6.20E-05 |
| Obesity | nk_cell | *PAK4* | -2.035282946 | 0.039045708 | 0.045562716 | 1 |
| Obesity | nk_cell | *ZNF337* | -1.964782263 | 5.04E-07 | 1.19E-06 | 0.002445524 |
| Obesity | nk_cell | *CEP85* | -1.729865934 | 2.31E-07 | 6.12E-07 | 0.001121515 |
| Obesity | nk_cell | *POMT1* | -2.94270428 | 9.28E-09 | 4.94E-08 | 4.50E-05 |
| Obesity | nk_cell | *CHMP2A* | -1.917949635 | 0.0186778 | 0.022470788 | 1 |
| Obesity | nk_cell | *YIPF2* | 2.387580192 | 0.010826774 | 0.013268882 | 1 |
| Obesity | nk_cell | *MAP3K10* | -2.221839316 | 1.21E-08 | 5.92E-08 | 5.88E-05 |
| Obesity | nk_cell | *LRRC47* | -1.7406538 | 1.39E-07 | 4.00E-07 | 0.000672672 |
| Obesity | nk_cell | *THEMIS2* | 1.518131372 | 6.48E-10 | 1.90E-08 | 3.15E-06 |
| Obesity | nk_cell | *ZNF317* | -3.75513381 | 2.01E-07 | 5.43E-07 | 0.000974587 |
| Obesity | nk_cell | *ANGPTL6* | 1.987392114 | 3.35E-09 | 2.76E-08 | 1.63E-05 |
| Obesity | nk_cell | *DNMT1* | 2.490016673 | 4.00E-08 | 1.47E-07 | 0.000194076 |
| Obesity | nk_cell | *C12orf65* | -1.517429288 | 0.015439648 | 0.018686249 | 1 |
| Obesity | nk_cell | *NOL11* | -2.595473192 | 0.000155826 | 0.000228765 | 0.756067171 |
| Obesity | nk_cell | *PKDREJ* | -3.14761717 | 2.20E-06 | 4.39E-06 | 0.010656008 |
| Obesity | nk_cell | *HSD17B3* | -5.620148899 | 3.60E-05 | 5.76E-05 | 0.174695802 |
| Obesity | nk_cell | *SLC35D2* | -2.744879872 | 2.07E-06 | 4.16E-06 | 0.010030746 |
| Obesity | nk_cell | *ULBP2* | -12.06177655 | 1.21E-07 | 3.60E-07 | 0.000588789 |
| Obesity | nk_cell | *TTLL9* | -2.638481296 | 5.07E-09 | 3.50E-08 | 2.46E-05 |
| Obesity | nk_cell | *ATP6V1E1* | -3.430671242 | 3.37E-06 | 6.48E-06 | 0.016374476 |
| Obesity | nk_cell | *IDO1* | 5.918085983 | 1.15E-08 | 5.66E-08 | 5.57E-05 |
| Obesity | nk_cell | *MRPS25* | -1.948908922 | 1.09E-08 | 5.48E-08 | 5.30E-05 |
| Obesity | nk_cell | *GALNT15* | -1.280132159 | 4.07E-09 | 3.06E-08 | 1.98E-05 |
| Obesity | nk_cell | *KCNC3* | -2.832709426 | 7.39E-10 | 1.92E-08 | 3.59E-06 |
| Obesity | nk_cell | *NAPSA* | 2.885839256 | 0.000308849 | 0.000438297 | 1 |
| Obesity | nk_cell | *KIF3A* | -2.769490845 | 5.03E-05 | 7.88E-05 | 0.244069172 |
| Obesity | nk_cell | *ACLY* | 1.856059327 | 0.019415666 | 0.023329572 | 1 |
| Obesity | nk_cell | *NDUFA2* | -1.563282794 | 0.002132719 | 0.00277872 | 1 |
| Obesity | nk_cell | *EXOC4* | -1.468996461 | 0.000156216 | 0.000229269 | 0.757962037 |
| Obesity | nk_cell | *PPFIA1* | -1.877499496 | 2.40E-07 | 6.33E-07 | 0.001166347 |
| Obesity | nk_cell | *BARX1* | -4.34902698 | 2.01E-08 | 8.63E-08 | 9.75E-05 |
| Obesity | nk_cell | *NINJ1* | -1.253485201 | 1.10E-07 | 3.31E-07 | 0.000531583 |
| Obesity | nk_cell | *NPHP4* | -1.405358567 | 2.17E-09 | 2.26E-08 | 1.05E-05 |
| Obesity | nk_cell | *MAP1B* | -1.814390335 | 7.32E-08 | 2.37E-07 | 0.000355233 |
| Obesity | nk_cell | *CKMT2* | -2.575749308 | 0.000163852 | 0.000239894 | 0.79501014 |

| | | | | | | |
|---|---|---|---|---|---|---|
| Obesity | nk_cell | *KHDRBS3* | -2.377041632 | 3.74E-08 | 1.39E-07 | 0.000181244 |
| Obesity | nk_cell | *CHD1L* | -2.693691958 | 7.06E-05 | 0.000107921 | 0.342432677 |
| Obesity | nk_cell | *ZNF304* | -2.024020375 | 2.74E-06 | 5.38E-06 | 0.013300704 |
| Obesity | nk_cell | *ZNF132* | -1.889010399 | 2.51E-05 | 4.09E-05 | 0.121753766 |
| Obesity | nk_cell | *RHPN2* | -1.053205825 | 0.001292083 | 0.001712424 | 1 |
| Obesity | nk_cell | *GCH1* | 1.76526182 | 0.020127932 | 0.024143567 | 1 |
| Obesity | nk_cell | *MATN3* | -2.47683515 | 4.21E-08 | 1.52E-07 | 0.000204067 |
| Obesity | nk_cell | *TRIM21* | 2.719723802 | 6.86E-09 | 4.16E-08 | 3.33E-05 |
| Obesity | nk_cell | *SPATA6* | -1.422144886 | 9.18E-09 | 4.90E-08 | 4.45E-05 |
| Obesity | nk_cell | *LRRC41* | -2.872452501 | 1.75E-08 | 7.83E-08 | 8.51E-05 |
| Obesity | nk_cell | *GAS2L2* | 4.448766373 | 1.89E-07 | 5.14E-07 | 0.000914995 |
| Obesity | nk_cell | *CCT6B* | -1.886433749 | 0.000280812 | 0.00040003 | 1 |
| Obesity | nk_cell | *CNGA4* | 2.400074243 | 0.000111463 | 0.000166765 | 0.540820001 |
| Obesity | nk_cell | *MYBBP1A* | -1.529572548 | 0.000815361 | 0.001103216 | 1 |
| Obesity | nk_cell | *RPA1* | -1.583655312 | 0.015413534 | 0.018659298 | 1 |
| Obesity | nk_cell | *SERPINF1* | -2.691799522 | 1.65E-05 | 2.76E-05 | 0.080041902 |
| Obesity | nk_cell | *TMEM128* | -2.179510962 | 8.62E-06 | 1.52E-05 | 0.041825933 |
| Obesity | nk_cell | *SEC61G* | 1.773764767 | 0.027066725 | 0.032101626 | 1 |
| Obesity | nk_cell | *FIGNL1* | -2.973400617 | 3.63E-10 | 1.52E-08 | 1.76E-06 |
| Obesity | nk_cell | *DDC* | -5.655086236 | 2.44E-05 | 3.99E-05 | 0.118408138 |
| Obesity | nk_cell | *EIF5A* | 1.575077112 | 7.32E-07 | 1.65E-06 | 0.003550489 |
| Obesity | nk_cell | *RGS22* | -2.057710075 | 4.44E-09 | 3.22E-08 | 2.15E-05 |
| Obesity | nk_cell | *REEP2* | -2.901848169 | 2.53E-06 | 4.99E-06 | 0.012262456 |
| Obesity | nk_cell | *PRMT7* | -2.008993604 | 1.44E-05 | 2.45E-05 | 0.070089459 |
| Obesity | nk_cell | *MUTYH* | 4.028150555 | 4.35E-09 | 3.19E-08 | 2.11E-05 |
| Obesity | nk_cell | *OSER1* | -2.688662139 | 0.001461627 | 0.00193027 | 1 |
| Obesity | nk_cell | *DMGDH* | -2.644671377 | 1.73E-05 | 2.88E-05 | 0.083721463 |
| Obesity | nk_cell | *BHMT2* | -0.928290449 | 1.31E-06 | 2.78E-06 | 0.006358328 |
| Obesity | nk_cell | *ZBED3* | -2.322819917 | 5.72E-10 | 1.82E-08 | 2.77E-06 |
| Obesity | nk_cell | *FBXO44* | -2.147747574 | 4.31E-10 | 1.63E-08 | 2.09E-06 |
| Obesity | nk_cell | *RSG1* | -5.47785141 | 4.97E-05 | 7.79E-05 | 0.241018756 |
| Obesity | nk_cell | *CASP9* | -1.533237595 | 0.000326457 | 0.000461799 | 1 |
| Obesity | nk_cell | *ATP8A2* | -3.063012011 | 0.009506695 | 0.011692391 | 1 |
| Obesity | nk_cell | *ALOX5AP* | 1.113786026 | 1.08E-09 | 2.05E-08 | 5.24E-06 |
| Obesity | nk_cell | *WASF3* | -1.999598149 | 3.98E-08 | 1.46E-07 | 0.000193153 |
| Obesity | nk_cell | *GPR12* | -4.744332268 | 0.004664081 | 0.005905564 | 1 |
| Obesity | nk_cell | *PEMT* | -3.549797388 | 3.24E-07 | 8.17E-07 | 0.001573583 |
| Obesity | nk_cell | *MPRIP* | -2.222613156 | 0.000676375 | 0.000922623 | 1 |
| Obesity | nk_cell | *DSTYK* | -2.698725824 | 1.57E-05 | 2.64E-05 | 0.07614667 |
| Obesity | nk_cell | *CCNA1* | -7.607858473 | 0.001602147 | 0.002109529 | 1 |
| Obesity | nk_cell | *COG6* | -2.279744776 | 3.19E-05 | 5.15E-05 | 0.154963186 |
| Obesity | nk_cell | *EPSTI1* | -2.410372111 | 1.11E-05 | 1.91E-05 | 0.053725907 |
| Obesity | nk_cell | *BTBD2* | -2.590166757 | 8.86E-07 | 1.95E-06 | 0.004298795 |
| Obesity | nk_cell | *CNDP2* | -3.124532636 | 3.04E-07 | 7.70E-07 | 0.001473092 |
| Obesity | nk_cell | *PDZD2* | -1.637939084 | 1.52E-07 | 4.32E-07 | 0.000736364 |
| Obesity | nk_cell | *GSTT2B* | -1.056689679 | 8.20E-06 | 1.45E-05 | 0.039801348 |
| Obesity | nk_cell | *SLC2A11* | 3.013136949 | 7.01E-10 | 1.92E-08 | 3.40E-06 |
| Obesity | nk_cell | *GIMAP4* | 1.072049396 | 3.90E-10 | 1.54E-08 | 1.89E-06 |
| Obesity | nk_cell | *ADCK2* | -2.15959505 | 2.09E-06 | 4.20E-06 | 0.010139412 |
| Obesity | nk_cell | *SFTPD* | -0.6716927 | 5.01E-05 | 7.84E-05 | 0.242967169 |

| | | | | | | |
|---|---|---|---|---|---|---|
| Obesity | nk_cell | *IPO8* | -2.786828061 | 1.47E-05 | 2.49E-05 | 0.07148216 |
| Obesity | nk_cell | *LARS* | -2.159328303 | 1.02E-10 | 7.52E-09 | 4.97E-07 |
| Obesity | nk_cell | *SPINK5* | -1.960122969 | 2.46E-09 | 2.37E-08 | 1.19E-05 |
| Obesity | nk_cell | *IMPA1* | -1.428449518 | 0.009188904 | 0.011327379 | 1 |
| Obesity | nk_cell | *LRRCC1* | 3.041239588 | 0.000300435 | 0.00042723 | 1 |
| Obesity | nk_cell | *CATSPERB* | -1.960299171 | 0.00092916 | 0.001249179 | 1 |
| Obesity | nk_cell | *COX16* | 1.136357017 | 1.10E-09 | 2.05E-08 | 5.32E-06 |
| Obesity | nk_cell | *TTC9* | 5.197638595 | 3.19E-07 | 8.05E-07 | 0.001546258 |
| Obesity | nk_cell | *ELP3* | -2.671010974 | 6.52E-05 | 0.000100049 | 0.316155371 |
| Obesity | nk_cell | *PEBP4* | -1.82220624 | 3.06E-09 | 2.63E-08 | 1.49E-05 |
| Obesity | nk_cell | *CTIF* | -1.464826223 | 6.51E-09 | 4.05E-08 | 3.16E-05 |
| Obesity | nk_cell | *THUMPD3* | -3.598207919 | 3.07E-06 | 5.97E-06 | 0.01491794 |
| Obesity | nk_cell | *CNTN6* | -2.408381718 | 5.54E-09 | 3.68E-08 | 2.69E-05 |
| Obesity | nk_cell | *CHL1* | -1.465676524 | 8.91E-09 | 4.80E-08 | 4.33E-05 |
| Obesity | nk_cell | *GSTM5* | -1.109128941 | 8.36E-05 | 0.000126645 | 0.405518362 |
| Obesity | nk_cell | *GSTM3* | -1.218028874 | 2.78E-07 | 7.14E-07 | 0.001347122 |
| Obesity | nk_cell | *VAV3* | 1.072981535 | 2.98E-08 | 1.16E-07 | 0.000144696 |
| Obesity | nk_cell | *PSRC1* | -2.763445229 | 1.04E-05 | 1.80E-05 | 0.050427914 |
| Obesity | nk_cell | *TRIM45* | -4.905584053 | 5.33E-07 | 1.25E-06 | 0.002583801 |
| Obesity | nk_cell | *CEPT1* | -1.892531571 | 8.93E-08 | 2.80E-07 | 0.000433319 |
| Obesity | nk_cell | *NAPG* | -2.304310716 | 3.93E-07 | 9.61E-07 | 0.001909237 |
| Obesity | nk_cell | *PPHLN1* | -1.963075023 | 2.83E-07 | 7.23E-07 | 0.001370766 |
| Obesity | nk_cell | *FKBP11* | 4.020682953 | 4.84E-07 | 1.15E-06 | 0.002346597 |
| Obesity | nk_cell | *YWHAQ* | -2.191230766 | 4.30E-07 | 1.04E-06 | 0.002084575 |
| Obesity | nk_cell | *KIDINS220* | -2.099344074 | 3.28E-06 | 6.33E-06 | 0.015910214 |
| Obesity | nk_cell | *LPIN1* | -2.055482346 | 0.001101947 | 0.001469666 | 1 |
| Obesity | nk_cell | *NARS* | 2.381088373 | 0.005540729 | 0.006953858 | 1 |
| Obesity | nk_cell | *KIAA1468* | -2.084412882 | 0.001421468 | 0.00187877 | 1 |
| Obesity | nk_cell | *IL15RA* | -2.574572154 | 8.18E-06 | 1.44E-05 | 0.039676076 |
| Obesity | nk_cell | *TMEM241* | -1.415856562 | 7.79E-09 | 4.42E-08 | 3.78E-05 |
| Obesity | nk_cell | *SOX5* | -1.403366751 | 5.82E-08 | 1.96E-07 | 0.000282308 |
| Obesity | nk_cell | *KLRD1* | 0.441855082 | 1.22E-09 | 2.08E-08 | 5.93E-06 |
| Obesity | nk_cell | *ACP2* | 4.758444174 | 7.88E-07 | 1.76E-06 | 0.003823876 |
| Obesity | nk_cell | *HOOK1* | -1.377729358 | 1.96E-07 | 5.30E-07 | 0.000949421 |
| Obesity | nk_cell | *ZCCHC11* | -1.733068787 | 2.36E-05 | 3.86E-05 | 0.114351765 |
| Obesity | nk_cell | *PRPF38A* | 2.497332176 | 0.01863385 | 0.022423472 | 1 |
| Obesity | nk_cell | *ELP2* | -2.389394851 | 4.23E-06 | 7.95E-06 | 0.020536472 |
| Obesity | nk_cell | *TIMM10* | -3.535135807 | 0.00488917 | 0.006176062 | 1 |
| Obesity | nk_cell | *DHX34* | -3.076812669 | 6.78E-06 | 1.22E-05 | 0.032888647 |
| Obesity | nk_cell | *FADS2* | -1.782195556 | 2.81E-09 | 2.51E-08 | 1.36E-05 |
| Obesity | nk_cell | *CLOCK* | -2.267818553 | 7.51E-06 | 1.33E-05 | 0.036456675 |
| Obesity | nk_cell | *GGACT* | -2.173685025 | 4.94E-08 | 1.72E-07 | 0.000239687 |
| Obesity | nk_cell | *CLDN10* | -2.605943741 | 1.29E-08 | 6.21E-08 | 6.24E-05 |
| Obesity | nk_cell | *ARGLU1* | -1.809535435 | 5.08E-05 | 7.94E-05 | 0.246242621 |
| Obesity | nk_cell | *ERCC5* | -3.826489618 | 2.81E-08 | 1.11E-07 | 0.000136187 |
| Obesity | nk_cell | *CARS2* | -2.20697248 | 2.50E-05 | 4.07E-05 | 0.121101099 |
| Obesity | nk_cell | *KLB* | -0.804789463 | 4.02E-06 | 7.61E-06 | 0.019489717 |
| Obesity | nk_cell | *WDR36* | -2.639183968 | 4.76E-07 | 1.13E-06 | 0.002310788 |
| Obesity | nk_cell | *UBQLN1* | -2.473777989 | 7.20E-06 | 1.29E-05 | 0.034943022 |
| Obesity | nk_cell | *GOLM1* | -1.904217532 | 5.14E-05 | 8.04E-05 | 0.249594415 |

| | | | | | | |
|---|---|---|---|---|---|---|
| Obesity | nk_cell | *USP30* | -1.371330638 | 4.83E-06 | 8.96E-06 | 0.023420469 |
| Obesity | nk_cell | *MSI1* | 3.658360825 | 0.000510443 | 0.000704399 | 1 |
| Obesity | nk_cell | *P2RX4* | -2.537531807 | 2.81E-06 | 5.51E-06 | 0.013648679 |
| Obesity | nk_cell | *DTX1* | -1.349874953 | 2.93E-09 | 2.57E-08 | 1.42E-05 |
| Obesity | nk_cell | *DMTF1* | -2.043492345 | 1.38E-06 | 2.92E-06 | 0.006706012 |
| Obesity | nk_cell | *TMEM60* | 3.384312336 | 0.000848107 | 0.001144968 | 1 |
| Obesity | nk_cell | *POM121C* | -2.999315476 | 0.000278402 | 0.000397062 | 1 |
| Obesity | nk_cell | *TES* | 1.750403967 | 2.18E-07 | 5.83E-07 | 0.00105815 |
| Obesity | nk_cell | *MTO1* | -1.968723782 | 4.09E-06 | 7.71E-06 | 0.019862303 |
| Obesity | nk_cell | *KHDC1* | 1.159038937 | 0.002685393 | 0.003469914 | 1 |
| Obesity | nk_cell | *NT5E* | -1.819450316 | 9.34E-07 | 2.05E-06 | 0.004532432 |
| Obesity | nk_cell | *AKIRIN2* | -2.651503422 | 2.15E-06 | 4.32E-06 | 0.010446871 |
| Obesity | nk_cell | *MAP3K7* | -2.003860112 | 6.50E-08 | 2.14E-07 | 0.000315178 |
| Obesity | nk_cell | *PRR5L* | 1.637706368 | 2.80E-08 | 1.11E-07 | 0.000135798 |
| Obesity | nk_cell | *PHF21A* | -1.992356827 | 2.89E-05 | 4.69E-05 | 0.14037591 |
| Obesity | nk_cell | *CAPRIN1* | -2.522228338 | 0.000311043 | 0.000441151 | 1 |
| Obesity | nk_cell | *B4GALNT1* | -5.217927601 | 4.80E-08 | 1.68E-07 | 0.000233122 |
| Obesity | nk_cell | *FAIM2* | -2.135056278 | 3.08E-09 | 2.64E-08 | 1.50E-05 |
| Obesity | nk_cell | *KRT7* | -1.316731333 | 2.15E-05 | 3.53E-05 | 0.104337186 |
| Obesity | nk_cell | *SLC26A10* | -2.585053887 | 1.84E-07 | 5.04E-07 | 0.000892885 |
| Obesity | nk_cell | *MAP7* | -1.859889706 | 0.000489234 | 0.000677251 | 1 |
| Obesity | nk_cell | *NHSL1* | -1.625851566 | 1.27E-07 | 3.73E-07 | 0.000614254 |
| Obesity | nk_cell | *AHI1* | -1.688966647 | 7.57E-08 | 2.44E-07 | 0.000367406 |
| Obesity | nk_cell | *SMPD2* | -1.602361566 | 1.18E-09 | 2.05E-08 | 5.70E-06 |
| Obesity | nk_cell | *MICAL1* | 1.834198093 | 0.005063344 | 0.006379472 | 1 |
| Obesity | nk_cell | *CCT7* | 2.819859265 | 0.000722736 | 0.000983927 | 1 |
| Obesity | nk_cell | *SMYD5* | -2.177657683 | 3.91E-06 | 7.42E-06 | 0.018963004 |
| Obesity | nk_cell | *USP15* | -1.621358603 | 0.006826653 | 0.008503959 | 1 |
| Obesity | nk_cell | *MDM2* | -2.334621249 | 1.08E-05 | 1.87E-05 | 0.052521606 |
| Obesity | nk_cell | *KLHL36* | -1.904106432 | 0.01828509 | 0.022031104 | 1 |
| Obesity | nk_cell | *MPHOSPH6* | 3.185764106 | 5.47E-07 | 1.27E-06 | 0.002652673 |
| Obesity | nk_cell | *CCDC102A* | 2.821015252 | 0.000160544 | 0.000235122 | 0.778958938 |
| Obesity | nk_cell | *AGT* | -1.052801551 | 0.035027743 | 0.041002318 | 1 |
| Obesity | nk_cell | *KCNK1* | -1.124409223 | 1.50E-08 | 6.98E-08 | 7.28E-05 |
| Obesity | nk_cell | *CAPN9* | 4.832704752 | 5.66E-07 | 1.31E-06 | 0.002744273 |
| Obesity | nk_cell | *NTPCR* | -2.871065043 | 2.43E-06 | 4.81E-06 | 0.011805828 |
| Obesity | nk_cell | *PIGC* | -2.685652657 | 3.45E-06 | 6.61E-06 | 0.016737046 |
| Obesity | nk_cell | *SP110* | 2.874599104 | 2.73E-06 | 5.37E-06 | 0.013266711 |
| Obesity | nk_cell | *MRPL44* | 2.751373781 | 5.31E-09 | 3.57E-08 | 2.58E-05 |
| Obesity | nk_cell | *USP37* | -1.893189759 | 8.63E-05 | 0.000130507 | 0.418536353 |
| Obesity | nk_cell | *SLC19A3* | -1.032017551 | 2.29E-09 | 2.28E-08 | 1.11E-05 |
| Obesity | nk_cell | *TMBIM1* | -2.129780781 | 2.30E-08 | 9.55E-08 | 0.000111408 |
| Obesity | nk_cell | *TSGA10* | -1.185956302 | 9.88E-08 | 3.04E-07 | 0.00047926 |
| Obesity | nk_cell | *GCC2* | -2.190229738 | 2.38E-06 | 4.71E-06 | 0.011532371 |
| Obesity | nk_cell | *ANKRD36* | -2.459477014 | 7.51E-06 | 1.33E-05 | 0.036419496 |
| Obesity | nk_cell | *EPC2* | -2.144669384 | 1.65E-06 | 3.40E-06 | 0.007982465 |
| Obesity | nk_cell | *STAB2* | -2.709110276 | 4.77E-08 | 1.68E-07 | 0.000231474 |
| Obesity | nk_cell | *APPL2* | -2.24797115 | 5.59E-07 | 1.30E-06 | 0.002714425 |
| Obesity | nk_cell | *DRAM1* | -2.334675522 | 8.29E-08 | 2.63E-07 | 0.000402082 |
| Obesity | nk_cell | *SLC41A2* | -1.574735304 | 6.88E-08 | 2.25E-07 | 0.000333839 |

| | | | | | | |
|---|---|---|---|---|---|---|
| Obesity | nk_cell | *NEK3* | -1.472412791 | 6.33E-05 | 9.74E-05 | 0.306953782 |
| Obesity | nk_cell | *CKAP2* | -0.993000231 | 0.000205067 | 0.000296745 | 0.994985628 |
| Obesity | nk_cell | *TBC1D4* | -1.508311029 | 4.41E-07 | 1.06E-06 | 0.002140521 |
| Obesity | nk_cell | *SUCLA2* | -1.888413714 | 2.35E-08 | 9.70E-08 | 0.0001142 |
| Obesity | nk_cell | *RCBTB2* | 2.498797243 | 0.001968095 | 0.002570444 | 1 |
| Obesity | nk_cell | *SETDB2* | -2.074938317 | 0.017578348 | 0.021205903 | 1 |
| Obesity | nk_cell | *SCRN1* | -2.926190738 | 1.15E-05 | 1.98E-05 | 0.055837046 |
| Obesity | nk_cell | *C7orf25* | -5.451458645 | 4.42E-07 | 1.06E-06 | 0.002145459 |
| Obesity | nk_cell | *CHST12* | 1.869947882 | 1.47E-08 | 6.87E-08 | 7.13E-05 |
| Obesity | nk_cell | *NUPL2* | -2.630600411 | 2.17E-06 | 4.35E-06 | 0.010532198 |
| Obesity | nk_cell | *ZDHHC4* | 4.278538268 | 0.000263137 | 0.000376397 | 1 |
| Obesity | nk_cell | *AOAH* | 0.312695568 | 1.31E-09 | 2.09E-08 | 6.34E-06 |
| Obesity | nk_cell | *TBRG4* | -3.844230792 | 0.002207605 | 0.002872432 | 1 |
| Obesity | nk_cell | *MTHFS* | -1.963561795 | 0.001476897 | 0.001948845 | 1 |
| Obesity | nk_cell | *MYCBPAP* | 1.084693091 | 0.000833328 | 0.00112627 | 1 |
| Obesity | nk_cell | *BRIP1* | 1.318801568 | 2.17E-08 | 9.16E-08 | 0.000105244 |
| Obesity | nk_cell | *RTP4* | -2.911124263 | 1.41E-08 | 6.67E-08 | 6.86E-05 |
| Obesity | nk_cell | *NDUFB5* | -2.869801982 | 9.38E-06 | 1.64E-05 | 0.045495666 |
| Obesity | nk_cell | *SCN2A* | -1.873614796 | 1.54E-05 | 2.59E-05 | 0.074631842 |
| Obesity | nk_cell | *RPS6KC1* | -2.12820666 | 3.47E-05 | 5.58E-05 | 0.168590579 |
| Obesity | nk_cell | *CCDC115* | 1.762513183 | 2.94E-07 | 7.47E-07 | 0.001426837 |
| Obesity | nk_cell | *IMP4* | -1.665420827 | 1.89E-09 | 2.19E-08 | 9.19E-06 |
| Obesity | nk_cell | *NIPSNAP3A* | -1.922344389 | 2.66E-09 | 2.45E-08 | 1.29E-05 |
| Obesity | nk_cell | *ODF2* | -3.400858331 | 4.12E-07 | 1.00E-06 | 0.001998571 |
| Obesity | nk_cell | *TOR1B* | -3.306827942 | 1.56E-07 | 4.42E-07 | 0.000757648 |
| Obesity | nk_cell | *SLC2A8* | -1.738743565 | 5.10E-09 | 3.51E-08 | 2.47E-05 |
| Obesity | nk_cell | *ZNF189* | -3.043110108 | 0.001525281 | 0.00201105 | 1 |
| Obesity | nk_cell | *STX17* | -2.088346253 | 2.72E-07 | 7.02E-07 | 0.00131989 |
| Obesity | nk_cell | *PRPF4* | -1.033338364 | 0.000332933 | 0.000470274 | 1 |
| Obesity | nk_cell | *ANP32B* | -2.757315612 | 3.86E-07 | 9.46E-07 | 0.001872128 |
| Obesity | nk_cell | *DSCC1* | 1.276191659 | 1.40E-09 | 2.11E-08 | 6.77E-06 |
| Obesity | nk_cell | *DERL1* | -2.844116057 | 0.002798254 | 0.003611899 | 1 |
| Obesity | nk_cell | *NOV* | -2.30115417 | 9.64E-08 | 2.98E-07 | 0.000467847 |
| Obesity | nk_cell | *IL11RA* | -3.55109729 | 1.16E-08 | 5.71E-08 | 5.64E-05 |
| Obesity | nk_cell | *APTX* | -2.578431361 | 2.18E-06 | 4.37E-06 | 0.010598128 |
| Obesity | nk_cell | *TMEM8B* | -2.205857338 | 1.69E-07 | 4.70E-07 | 0.00081784 |
| Obesity | nk_cell | *GRHPR* | -1.839605629 | 6.16E-06 | 1.12E-05 | 0.029872602 |
| Obesity | nk_cell | *DENND4C* | -1.849571382 | 1.81E-06 | 3.70E-06 | 0.008795983 |
| Obesity | nk_cell | *GMPR* | -1.556128637 | 1.93E-09 | 2.20E-08 | 9.37E-06 |
| Obesity | nk_cell | *TMEM14B* | -1.723404958 | 6.09E-09 | 3.91E-08 | 2.96E-05 |
| Obesity | nk_cell | *FRS3* | -3.102126464 | 1.18E-07 | 3.53E-07 | 0.000574128 |
| Obesity | nk_cell | *CAPN11* | -1.771086184 | 3.73E-05 | 5.96E-05 | 0.181092873 |
| Obesity | nk_cell | *IRF4* | 0.834425016 | 0.005190146 | 0.006530754 | 1 |
| Obesity | nk_cell | *LRRC1* | -1.618013192 | 7.99E-08 | 2.55E-07 | 0.000387796 |
| Obesity | nk_cell | *RIPK1* | -2.206867842 | 5.03E-06 | 9.32E-06 | 0.024410337 |
| Obesity | nk_cell | *TUBB2B* | -1.527159642 | 0.00557704 | 0.006997621 | 1 |
| Obesity | nk_cell | *TCF19* | 3.322204881 | 2.17E-10 | 1.19E-08 | 1.05E-06 |
| Obesity | nk_cell | *FLOT1* | -1.621673006 | 0.004215291 | 0.005356886 | 1 |
| Obesity | nk_cell | *NRM* | 10.99611416 | 5.64E-08 | 1.91E-07 | 0.000273838 |
| Obesity | nk_cell | *VARS2* | -1.051903195 | 5.09E-07 | 1.20E-06 | 0.002469198 |

| | | | | | | |
|---|---|---|---|---|---|---|
| Obesity | nk_cell | *TLR2* | 4.61882153 | 7.96E-08 | 2.55E-07 | 0.000386315 |
| Obesity | nk_cell | *NUMA1* | -2.06779274 | 3.46E-07 | 8.63E-07 | 0.001680628 |
| Obesity | nk_cell | *SYTL2* | 1.499835438 | 2.96E-06 | 5.76E-06 | 0.014350347 |
| Obesity | nk_cell | *RAB30* | -1.929557818 | 8.01E-09 | 4.50E-08 | 3.89E-05 |
| Obesity | nk_cell | *PRCP* | -1.928605112 | 5.47E-07 | 1.28E-06 | 0.002654788 |
| Obesity | nk_cell | *NARS2* | -2.065230694 | 6.82E-08 | 2.23E-07 | 0.000331115 |
| Obesity | nk_cell | *GGH* | -1.076966716 | 0.016593422 | 0.020052623 | 1 |
| Obesity | nk_cell | *SULF1* | -1.371700546 | 4.42E-08 | 1.58E-07 | 0.000214672 |
| Obesity | nk_cell | *NEK1* | -2.148203823 | 7.73E-06 | 1.37E-05 | 0.037490609 |
| Obesity | nk_cell | *TRPC6* | -1.40027753 | 5.17E-08 | 1.79E-07 | 0.000250934 |
| Obesity | nk_cell | *MMP27* | -3.219048073 | 0.00833047 | 0.010297947 | 1 |
| Obesity | nk_cell | *POU2F3* | -1.953941794 | 1.89E-05 | 3.12E-05 | 0.091508058 |
| Obesity | nk_cell | *PPP2R1B* | -1.011191733 | 1.19E-07 | 3.55E-07 | 0.000578902 |
| Obesity | nk_cell | *FXYD6* | -1.862818027 | 9.40E-07 | 2.06E-06 | 0.004558643 |
| Obesity | nk_cell | *MAP2K5* | -1.768293066 | 1.26E-06 | 2.67E-06 | 0.006096132 |
| Obesity | nk_cell | *NUSAP1* | 3.343884314 | 1.88E-11 | 2.32E-09 | 9.11E-08 |
| Obesity | nk_cell | *NDUFAF1* | -1.629287263 | 8.68E-05 | 0.000131284 | 0.421290126 |
| Obesity | nk_cell | *PARP6* | -2.83179891 | 8.20E-07 | 1.83E-06 | 0.003980726 |
| Obesity | nk_cell | *ZNF280D* | -1.832157914 | 1.00E-06 | 2.19E-06 | 0.004866035 |
| Obesity | nk_cell | *RSL24D1* | -3.264886117 | 0.000802084 | 0.001086463 | 1 |
| Obesity | nk_cell | *SPTBN5* | 0.562431976 | 0.001739439 | 0.002282866 | 1 |
| Obesity | nk_cell | *SLC44A5* | -1.437755266 | 1.70E-08 | 7.70E-08 | 8.27E-05 |
| Obesity | nk_cell | *DBT* | -2.832696137 | 5.49E-07 | 1.28E-06 | 0.002662506 |
| Obesity | nk_cell | *RTCA* | -2.311385822 | 0.001876194 | 0.002454379 | 1 |
| Obesity | nk_cell | *KHK* | -1.858310056 | 1.41E-08 | 6.67E-08 | 6.85E-05 |
| Obesity | nk_cell | *DYNC2LI1* | -1.977249203 | 5.02E-06 | 9.30E-06 | 0.024354432 |
| Obesity | nk_cell | *LHCGR* | -2.033715017 | 2.17E-09 | 2.26E-08 | 1.05E-05 |
| Obesity | nk_cell | *THUMPD2* | -2.1461215 | 1.35E-07 | 3.92E-07 | 0.000654581 |
| Obesity | nk_cell | *ATRAID* | 3.315053259 | 2.92E-07 | 7.44E-07 | 0.001418017 |
| Obesity | nk_cell | *CENPO* | -1.504279129 | 0.00027918 | 0.000398055 | 1 |
| Obesity | nk_cell | *LRPPRC* | -2.286075219 | 1.21E-06 | 2.58E-06 | 0.005859755 |
| Obesity | nk_cell | *DTNB* | -2.183562782 | 0.000291318 | 0.000414752 | 1 |
| Obesity | nk_cell | *CYP2C8* | -3.095186795 | 0.033276024 | 0.039046014 | 1 |
| Obesity | nk_cell | *MYOF* | -1.444556316 | 4.10E-07 | 9.96E-07 | 0.001989692 |
| Obesity | nk_cell | *BTBD16* | -2.29117977 | 1.66E-08 | 7.52E-08 | 8.05E-05 |
| Obesity | nk_cell | *TACC2* | -1.843303652 | 1.27E-07 | 3.73E-07 | 0.000613818 |
| Obesity | nk_cell | *CALHM2* | -2.563663695 | 3.41E-05 | 5.48E-05 | 0.165445343 |
| Obesity | nk_cell | *PLCE1* | -1.835260774 | 1.54E-06 | 3.21E-06 | 0.007477883 |
| Obesity | nk_cell | *RBP4* | -2.365980248 | 3.35E-07 | 8.39E-07 | 0.001626783 |
| Obesity | nk_cell | *ASCC1* | -1.700283526 | 5.71E-09 | 3.74E-08 | 2.77E-05 |
| Obesity | nk_cell | *DNA2* | -0.776324467 | 0.000421523 | 0.00058771 | 1 |
| Obesity | nk_cell | *ATIC* | -1.748464848 | 1.09E-07 | 3.30E-07 | 0.00052919 |
| Obesity | nk_cell | *SMARCAL1* | -2.559937603 | 0.001016229 | 0.001359455 | 1 |
| Obesity | nk_cell | *BARD1* | 2.567745189 | 3.18E-08 | 1.21E-07 | 0.000154352 |
| Obesity | nk_cell | *STAT4* | 0.392988529 | 9.91E-10 | 2.05E-08 | 4.81E-06 |
| Obesity | nk_cell | *SSB* | -1.741334079 | 6.72E-06 | 1.21E-05 | 0.032620801 |
| Obesity | nk_cell | *FASTKD1* | -2.347767516 | 8.91E-07 | 1.96E-06 | 0.004324527 |
| Obesity | nk_cell | *ITGAV* | -2.104476439 | 2.13E-07 | 5.72E-07 | 0.001033019 |
| Obesity | nk_cell | *SLC35A5* | -2.357151738 | 1.61E-07 | 4.52E-07 | 0.000780387 |
| Obesity | nk_cell | *SENP7* | -2.019429117 | 1.57E-07 | 4.45E-07 | 0.000764111 |

| | | | | | | |
|---|---|---|---|---|---|---|
| Obesity | nk_cell | *COX17* | 3.682206013 | 8.99E-07 | 1.98E-06 | 0.004363071 |
| Obesity | nk_cell | *SPPL2A* | -1.69528831 | 0.010800533 | 0.013243413 | 1 |
| Obesity | nk_cell | *PPCDC* | -2.437665075 | 5.14E-08 | 1.78E-07 | 0.000249329 |
| Obesity | nk_cell | *PCDH10* | 2.386461271 | 9.99E-09 | 5.20E-08 | 4.85E-05 |
| Obesity | nk_cell | *AP1AR* | -3.303686812 | 1.14E-06 | 2.44E-06 | 0.005513736 |
| Obesity | nk_cell | *PRKG2* | 1.721548054 | 3.77E-08 | 1.40E-07 | 0.000182918 |
| Obesity | nk_cell | *KIAA1109* | -2.003325901 | 0.000122937 | 0.000182749 | 0.596492368 |
| Obesity | nk_cell | *BMPR1B* | -1.230037877 | 5.55E-09 | 3.68E-08 | 2.69E-05 |
| Obesity | nk_cell | *PRDM5* | -1.658372316 | 9.48E-09 | 5.00E-08 | 4.60E-05 |
| Obesity | nk_cell | *TRPC3* | 0.405596303 | 2.16E-09 | 2.26E-08 | 1.05E-05 |
| Obesity | nk_cell | *NAAA* | -1.83458038 | 0.002145467 | 0.002794579 | 1 |
| Obesity | nk_cell | *NUP54* | -2.18624955 | 6.12E-05 | 9.46E-05 | 0.296951919 |
| Obesity | nk_cell | *CNOT6L* | 2.030068467 | 0.000486198 | 0.000673817 | 1 |
| Obesity | nk_cell | *ANXA3* | -1.715368151 | 4.17E-06 | 7.85E-06 | 0.020241473 |
| Obesity | nk_cell | *PPA2* | -2.124731897 | 2.36E-06 | 4.69E-06 | 0.01145847 |
| Obesity | nk_cell | *ENPEP* | -1.301772837 | 9.80E-06 | 1.71E-05 | 0.047528611 |
| Obesity | nk_cell | *PAPSS1* | -1.852758859 | 1.73E-08 | 7.77E-08 | 8.40E-05 |
| Obesity | nk_cell | *FBN2* | -0.747921804 | 5.54E-05 | 8.63E-05 | 0.269002624 |
| Obesity | nk_cell | *PARVG* | 1.192775601 | 5.76E-09 | 3.76E-08 | 2.79E-05 |
| Obesity | nk_cell | *B4GALNT3* | 1.744460604 | 7.27E-10 | 1.92E-08 | 3.53E-06 |
| Obesity | nk_cell | *ERP27* | 0.577110345 | 7.14E-10 | 1.92E-08 | 3.46E-06 |
| Obesity | nk_cell | *KIF21A* | -2.114404719 | 0.001129719 | 0.001504225 | 1 |
| Obesity | nk_cell | *ALG10* | -1.99754033 | 1.98E-08 | 8.53E-08 | 9.61E-05 |
| Obesity | nk_cell | *ZCRB1* | -3.001769956 | 0.000114249 | 0.000170828 | 0.554336042 |
| Obesity | nk_cell | *C1RL* | -1.068785059 | 0.003773656 | 0.004818363 | 1 |
| Obesity | nk_cell | *CLSTN3* | 2.436498284 | 3.25E-09 | 2.72E-08 | 1.58E-05 |
| Obesity | nk_cell | *KLRG1* | 0.863913345 | 1.61E-11 | 2.06E-09 | 7.82E-08 |
| Obesity | nk_cell | *VAMP1* | 2.153004846 | 3.78E-07 | 9.31E-07 | 0.001834965 |
| Obesity | nk_cell | *PEX5* | -1.754157375 | 3.12E-08 | 1.20E-07 | 0.000151179 |
| Obesity | nk_cell | *PPFIA2* | -1.532035958 | 4.88E-09 | 3.42E-08 | 2.37E-05 |
| Obesity | nk_cell | *MARCH9* | -3.579103635 | 2.89E-08 | 1.13E-07 | 0.000140354 |
| Obesity | nk_cell | *LGR5* | -3.340682105 | 0.000300692 | 0.000427345 | 1 |
| Obesity | nk_cell | *AMDHD1* | -1.486030735 | 1.52E-05 | 2.57E-05 | 0.07397856 |
| Obesity | nk_cell | *RITA1* | -1.999545507 | 2.01E-09 | 2.20E-08 | 9.75E-06 |
| Obesity | nk_cell | *SDSL* | 2.739028544 | 1.84E-05 | 3.06E-05 | 0.089268583 |
| Obesity | nk_cell | *GLTP* | 2.593206141 | 7.76E-09 | 4.42E-08 | 3.77E-05 |
| Obesity | nk_cell | *FOXN4* | -1.620362944 | 9.19E-09 | 4.90E-08 | 4.46E-05 |
| Obesity | nk_cell | *MTMR6* | 3.015669181 | 4.54E-06 | 8.49E-06 | 0.022041989 |
| Obesity | nk_cell | *CCDC65* | -2.565700039 | 8.82E-07 | 1.95E-06 | 0.004280184 |
| Obesity | nk_cell | *BRCA2* | -4.84290346 | 6.67E-05 | 0.000102184 | 0.323413299 |
| Obesity | nk_cell | *KANSL2* | -2.328113904 | 6.19E-08 | 2.06E-07 | 0.000300312 |
| Obesity | nk_cell | *CERS5* | -2.447016774 | 9.24E-06 | 1.62E-05 | 0.044812854 |
| Obesity | nk_cell | *LMBR1L* | -2.925326706 | 6.96E-05 | 0.000106484 | 0.337554963 |
| Obesity | nk_cell | *ZNF740* | 2.91219402 | 1.00E-05 | 1.74E-05 | 0.048695341 |
| Obesity | nk_cell | *WDFY2* | -2.132841116 | 1.85E-07 | 5.07E-07 | 0.000899145 |
| Obesity | nk_cell | *HNRNPA1L2* | -3.378243231 | 3.10E-08 | 1.20E-07 | 0.000150517 |
| Obesity | nk_cell | *RB1* | -2.06264314 | 7.85E-06 | 1.39E-05 | 0.038067458 |
| Obesity | nk_cell | *VPS33A* | -1.573611522 | 0.001049692 | 0.001402673 | 1 |
| Obesity | nk_cell | *DIAPH3* | 1.643527858 | 0.000513242 | 0.000708061 | 1 |
| Obesity | nk_cell | *GRTP1* | -0.939464472 | 0.028680902 | 0.033883549 | 1 |

| | | | | | | |
|---|---|---|---|---|---|---|
| Obesity | nk_cell | *CBLN3* | -0.670153828 | 7.20E-06 | 1.29E-05 | 0.034913499 |
| Obesity | nk_cell | *NOVA1* | -0.632835649 | 0.001157914 | 0.001540499 | 1 |
| Obesity | nk_cell | *FITM1* | 1.113073559 | 2.42E-10 | 1.25E-08 | 1.17E-06 |
| Obesity | nk_cell | *RTN1* | -1.394431248 | 1.52E-06 | 3.17E-06 | 0.007368976 |
| Obesity | nk_cell | *C14orf37* | -2.221402001 | 4.91E-06 | 9.11E-06 | 0.023831491 |
| Obesity | nk_cell | *RAB15* | -3.970547006 | 0.000364436 | 0.000512238 | 1 |
| Obesity | nk_cell | *EFCAB11* | -2.043709819 | 2.82E-09 | 2.51E-08 | 1.37E-05 |
| Obesity | nk_cell | *SLC24A4* | 2.384942568 | 2.82E-09 | 2.51E-08 | 1.37E-05 |
| Obesity | nk_cell | *C14orf79* | -3.236225012 | 2.42E-06 | 4.79E-06 | 0.011731052 |
| Obesity | nk_cell | *WARS* | -2.602926935 | 3.27E-07 | 8.23E-07 | 0.001588099 |
| Obesity | nk_cell | *DUOXA1* | -2.506490873 | 4.56E-09 | 3.27E-08 | 2.21E-05 |
| Obesity | nk_cell | *SERF2* | 2.229567422 | 0.000878939 | 0.001184944 | 1 |
| Obesity | nk_cell | *DISP2* | -0.721218962 | 0.000532318 | 0.000733543 | 1 |
| Obesity | nk_cell | *UBE2Q2* | -2.470097451 | 0.001116374 | 0.001487273 | 1 |
| Obesity | nk_cell | *HMG20A* | -2.547083306 | 8.96E-05 | 0.000135252 | 0.434835957 |
| Obesity | nk_cell | *SCAPER* | -1.947167122 | 5.01E-07 | 1.18E-06 | 0.002428885 |
| Obesity | nk_cell | *NEIL1* | -2.390920125 | 5.32E-08 | 1.83E-07 | 0.000258007 |
| Obesity | nk_cell | *MAN2C1* | -0.97206887 | 4.03E-06 | 7.63E-06 | 0.019576091 |
| Obesity | nk_cell | *PIF1* | 1.707302591 | 1.05E-08 | 5.37E-08 | 5.09E-05 |
| Obesity | nk_cell | *CYP11A1* | -1.391513198 | 6.17E-06 | 1.12E-05 | 0.029917513 |
| Obesity | nk_cell | *BBS4* | -2.433996312 | 2.88E-05 | 4.67E-05 | 0.13959925 |
| Obesity | nk_cell | *ULK3* | 3.4983049 | 0.003498124 | 0.004475975 | 1 |
| Obesity | nk_cell | *PCSK6* | -2.087935921 | 8.97E-09 | 4.82E-08 | 4.35E-05 |
| Obesity | nk_cell | *CELF6* | 7.396531004 | 2.27E-08 | 9.47E-08 | 0.000110087 |
| Obesity | nk_cell | *HAPLN3* | -1.445481102 | 3.29E-09 | 2.73E-08 | 1.60E-05 |
| Obesity | nk_cell | *ABHD2* | 1.763693806 | 0.042016808 | 0.048841772 | 1 |
| Obesity | nk_cell | *MFGE8* | -2.887718481 | 1.93E-05 | 3.19E-05 | 0.093557966 |
| Obesity | nk_cell | *UNC45A* | -2.093832096 | 2.31E-08 | 9.57E-08 | 0.000111903 |
| Obesity | nk_cell | *CRTC3* | -1.87638902 | 5.44E-07 | 1.27E-06 | 0.002640447 |
| Obesity | nk_cell | *ITGAX* | 0.328864968 | 1.12E-09 | 2.05E-08 | 5.45E-06 |
| Obesity | nk_cell | *TGFB1I1* | -5.585686053 | 1.12E-06 | 2.40E-06 | 0.005416598 |
| Obesity | nk_cell | *DHX38* | 2.253583323 | 0.027342201 | 0.032396669 | 1 |
| Obesity | nk_cell | *NUDT7* | -2.565252903 | 8.83E-09 | 4.76E-08 | 4.28E-05 |
| Obesity | nk_cell | *GCSH* | -1.57259144 | 5.00E-09 | 3.48E-08 | 2.43E-05 |
| Obesity | nk_cell | *CMTM3* | -2.502848659 | 1.61E-08 | 7.36E-08 | 7.81E-05 |
| Obesity | nk_cell | *ZCCHC14* | -2.174106716 | 1.17E-07 | 3.51E-07 | 0.000569979 |
| Obesity | nk_cell | *TLDC1* | -2.157599539 | 8.22E-09 | 4.55E-08 | 3.99E-05 |
| Obesity | nk_cell | *RPL3L* | 5.429162067 | 3.83E-05 | 6.10E-05 | 0.185713527 |
| Obesity | nk_cell | *TIGD7* | -2.0203407 | 4.14E-07 | 1.00E-06 | 0.002007716 |
| Obesity | nk_cell | *DEF8* | -2.841040731 | 5.61E-07 | 1.30E-06 | 0.00272302 |
| Obesity | nk_cell | *TCF25* | -2.102846921 | 0.000137669 | 0.000203341 | 0.667968544 |
| Obesity | nk_cell | *GALNS* | -2.783350573 | 1.10E-07 | 3.33E-07 | 0.000536059 |
| Obesity | nk_cell | *GAS8* | -1.883764022 | 1.31E-07 | 3.84E-07 | 0.000636217 |
| Obesity | nk_cell | *NCOR1* | -1.936624252 | 3.15E-05 | 5.08E-05 | 0.152849134 |
| Obesity | nk_cell | *KSR1* | -2.514836599 | 1.15E-06 | 2.47E-06 | 0.005586052 |
| Obesity | nk_cell | *GFOD2* | -1.656949945 | 0.011884193 | 0.01450984 | 1 |
| Obesity | nk_cell | *PRPSAP2* | -1.964693731 | 0.001055038 | 0.001409428 | 1 |
| Obesity | nk_cell | *RASL10B* | -4.077375251 | 6.39E-09 | 4.02E-08 | 3.10E-05 |
| Obesity | nk_cell | *UNC45B* | -6.36162132 | 1.84E-09 | 2.19E-08 | 8.94E-06 |
| Obesity | nk_cell | *C17orf80* | -1.755000557 | 3.12E-08 | 1.20E-07 | 0.000151298 |

| | | | | | | |
|---|---|---|---|---|---|---|
| Obesity | nk_cell | *VPS53* | -1.815578413 | 9.37E-06 | 1.64E-05 | 0.045469776 |
| Obesity | nk_cell | *SGSM2* | -2.495492571 | 1.49E-07 | 4.25E-07 | 0.000723532 |
| Obesity | nk_cell | *LRRC46* | -2.611081132 | 7.84E-08 | 2.51E-07 | 0.000380476 |
| Obesity | nk_cell | *SSH2* | -1.85507642 | 1.18E-05 | 2.02E-05 | 0.057239249 |
| Obesity | nk_cell | *RHBDL3* | 3.253026065 | 6.13E-09 | 3.92E-08 | 2.98E-05 |
| Obesity | nk_cell | *G6PC3* | 4.036310153 | 0.000103552 | 0.00015536 | 0.502434284 |
| Obesity | nk_cell | *CLTC* | -1.746221708 | 0.000116662 | 0.000174114 | 0.566044193 |
| Obesity | nk_cell | *IMPA2* | -2.756718345 | 1.01E-06 | 2.20E-06 | 0.004908651 |
| Obesity | nk_cell | *GNAL* | -2.94862127 | 8.33E-06 | 1.47E-05 | 0.040394986 |
| Obesity | nk_cell | *RPRD1A* | -2.343694955 | 1.63E-06 | 3.37E-06 | 0.007918623 |
| Obesity | nk_cell | *GALNT1* | -2.257819064 | 9.15E-07 | 2.01E-06 | 0.004440957 |
| Obesity | nk_cell | *ADCYAP1* | -1.421861 | 0.03512873 | 0.041110612 | 1 |
| Obesity | nk_cell | *C18orf8* | -4.038741345 | 2.84E-06 | 5.56E-06 | 0.013762197 |
| Obesity | nk_cell | *NPC1* | 1.484401244 | 0.029563885 | 0.034858802 | 1 |
| Obesity | nk_cell | *SLC14A1* | -1.830766434 | 2.71E-07 | 7.01E-07 | 0.001315209 |
| Obesity | nk_cell | *CCDC40* | -1.543366318 | 4.02E-09 | 3.03E-08 | 1.95E-05 |
| Obesity | nk_cell | *ARHGDIA* | 1.567704146 | 0.02979865 | 0.035110017 | 1 |
| Obesity | nk_cell | *TMC6* | 1.198263665 | 7.86E-10 | 1.96E-08 | 3.81E-06 |
| Obesity | nk_cell | *CARD14* | -1.790328104 | 1.04E-08 | 5.34E-08 | 5.04E-05 |
| Obesity | nk_cell | *ANAPC11* | 2.593881832 | 0.018517263 | 0.022288703 | 1 |
| Obesity | nk_cell | *TBCD* | -2.188122279 | 0.002827189 | 0.003647307 | 1 |
| Obesity | nk_cell | *NARF* | -2.460415941 | 0.000396011 | 0.000554051 | 1 |
| Obesity | nk_cell | *TRIM65* | 2.419741111 | 0.023593607 | 0.028092314 | 1 |
| Obesity | nk_cell | *SECTM1* | -1.190779888 | 5.27E-08 | 1.82E-07 | 0.000255567 |
| Obesity | nk_cell | *RNF157* | 2.291102901 | 5.13E-06 | 9.48E-06 | 0.024909164 |
| Obesity | nk_cell | *ZNF750* | 1.374361867 | 2.65E-09 | 2.45E-08 | 1.29E-05 |
| Obesity | nk_cell | *WDR45B* | -2.743094177 | 1.49E-05 | 2.51E-05 | 0.072114269 |
| Obesity | nk_cell | *RNF165* | 2.742420618 | 4.97E-07 | 1.18E-06 | 0.002412423 |
| Obesity | nk_cell | *MAPK4* | -5.939334302 | 1.02E-08 | 5.26E-08 | 4.93E-05 |
| Obesity | nk_cell | *FBXO15* | -3.204001474 | 3.10E-08 | 1.20E-07 | 0.000150554 |
| Obesity | nk_cell | *CBLN2* | -1.714056293 | 1.56E-09 | 2.11E-08 | 7.55E-06 |
| Obesity | nk_cell | *NT5C3B* | -0.515491501 | 0.033915073 | 0.039757414 | 1 |
| Obesity | nk_cell | *PIP4K2B* | -3.097456978 | 6.41E-07 | 1.47E-06 | 0.003108086 |
| Obesity | nk_cell | *ERBB2* | -1.163791518 | 2.34E-09 | 2.30E-08 | 1.13E-05 |
| Obesity | nk_cell | *MIEN1* | 2.722056443 | 0.029388465 | 0.034668814 | 1 |
| Obesity | nk_cell | *PNMT* | -3.967813809 | 7.00E-06 | 1.26E-05 | 0.033987056 |
| Obesity | nk_cell | *IGFBP4* | -1.579827747 | 2.60E-07 | 6.77E-07 | 0.001263859 |
| Obesity | nk_cell | *TXNL4A* | -1.612325164 | 1.37E-08 | 6.54E-08 | 6.63E-05 |
| Obesity | nk_cell | *CACNA1A* | -1.601820609 | 9.22E-10 | 2.05E-08 | 4.47E-06 |
| Obesity | nk_cell | *SAMD1* | -4.980230378 | 3.62E-07 | 8.97E-07 | 0.001754505 |
| Obesity | nk_cell | *NFIC* | -1.614725854 | 1.67E-07 | 4.67E-07 | 0.000810497 |
| Obesity | nk_cell | *PRDM15* | -2.127822656 | 1.59E-07 | 4.47E-07 | 0.000769224 |
| Obesity | nk_cell | *PFKL* | -2.255882484 | 0.023931979 | 0.028467262 | 1 |
| Obesity | nk_cell | *MVB12A* | -1.866819137 | 0.001941598 | 0.002537886 | 1 |
| Obesity | nk_cell | *DUS3L* | 1.087622138 | 0.006765974 | 0.008432701 | 1 |
| Obesity | nk_cell | *DPP9* | 2.828884595 | 0.001333554 | 0.001764495 | 1 |
| Obesity | nk_cell | *TMEM91* | -1.049146439 | 3.65E-07 | 9.04E-07 | 0.001769458 |
| Obesity | nk_cell | *SIRT3* | -2.224471101 | 0.001242607 | 0.001649557 | 1 |
| Obesity | nk_cell | *HUNK* | -0.775825482 | 0.000445341 | 0.000618961 | 1 |
| Obesity | nk_cell | *COL6A1* | -0.654151946 | 0.005687131 | 0.007126539 | 1 |

| | | | | | | |
|---|---|---|---|---|---|---|
| Obesity | nk_cell | *IFNAR1* | -1.908356256 | 1.35E-07 | 3.92E-07 | 0.00065431 |
| Obesity | nk_cell | *DOPEY2* | -1.845178475 | 0.000216876 | 0.000312899 | 1 |
| Obesity | nk_cell | *URB1* | -2.245663414 | 5.33E-07 | 1.25E-06 | 0.002586177 |
| Obesity | nk_cell | *SAE1* | -3.073803665 | 0.000939007 | 0.001261368 | 1 |
| Obesity | nk_cell | *GEMIN7* | -3.106046533 | 6.13E-08 | 2.05E-07 | 0.000297655 |
| Obesity | nk_cell | *SLC6A3* | -1.430261653 | 0.003891478 | 0.004959667 | 1 |
| Obesity | nk_cell | *CACNG8* | 2.684085069 | 2.00E-08 | 8.60E-08 | 9.70E-05 |
| Obesity | nk_cell | *ZNF787* | -2.727694955 | 0.00025473 | 0.000365017 | 1 |
| Obesity | nk_cell | *FBN3* | -1.512150164 | 8.16E-05 | 0.000123877 | 0.395909548 |
| Obesity | nk_cell | *EVI5L* | 4.039416333 | 1.11E-06 | 2.39E-06 | 0.005382372 |
| Obesity | nk_cell | *SLC47A1* | 3.533310608 | 1.51E-06 | 3.15E-06 | 0.007316688 |
| Obesity | nk_cell | *CTU1* | -1.357865803 | 1.75E-09 | 2.16E-08 | 8.51E-06 |
| Obesity | nk_cell | *RERE* | -1.515389812 | 4.17E-07 | 1.01E-06 | 0.002021274 |
| Obesity | nk_cell | *MMEL1* | -1.85196694 | 6.82E-08 | 2.23E-07 | 0.0003308 |
| Obesity | nk_cell | *EPHA2* | -2.283649309 | 4.06E-10 | 1.59E-08 | 1.97E-06 |
| Obesity | nk_cell | *PEX14* | -1.759951473 | 1.84E-06 | 3.75E-06 | 0.008918769 |
| Obesity | nk_cell | *ZNF593* | -2.910800798 | 3.81E-09 | 2.94E-08 | 1.85E-05 |
| Obesity | nk_cell | *HSPG2* | -1.396302933 | 1.49E-06 | 3.12E-06 | 0.007239892 |
| Obesity | nk_cell | *ITGB3BP* | -1.475953686 | 1.03E-06 | 2.23E-06 | 0.004975431 |
| Obesity | nk_cell | *PIGK* | -2.517865499 | 7.85E-07 | 1.76E-06 | 0.003809353 |
| Obesity | nk_cell | *RPS8* | 1.944734584 | 0.003533854 | 0.00451931 | 1 |
| Obesity | nk_cell | *CYP4B1* | -1.23706142 | 2.80E-08 | 1.11E-07 | 0.000135849 |
| Obesity | nk_cell | *SLC44A3* | -2.991478382 | 1.69E-08 | 7.62E-08 | 8.18E-05 |
| Obesity | nk_cell | *STRIP1* | -2.891830626 | 1.74E-07 | 4.81E-07 | 0.000842467 |
| Obesity | nk_cell | *GPR161* | -2.366765537 | 1.54E-06 | 3.20E-06 | 0.007460559 |
| Obesity | nk_cell | *ALDH9A1* | -2.875574198 | 1.65E-07 | 4.63E-07 | 0.000802195 |
| Obesity | nk_cell | *ATP1B1* | -2.396739622 | 5.73E-05 | 8.90E-05 | 0.278177644 |
| Obesity | nk_cell | *TIPRL* | 2.511913346 | 0.005063217 | 0.006379472 | 1 |
| Obesity | nk_cell | *NME7* | -1.505046821 | 2.05E-09 | 2.21E-08 | 9.93E-06 |
| Obesity | nk_cell | *XCL1* | 1.764172774 | 1.02E-09 | 2.05E-08 | 4.96E-06 |
| Obesity | nk_cell | *XCL2* | 0.514489878 | 2.75E-09 | 2.49E-08 | 1.34E-05 |
| Obesity | nk_cell | *MAEL* | -3.060829275 | 1.67E-09 | 2.15E-08 | 8.08E-06 |
| Obesity | nk_cell | *RFWD2* | -1.859404028 | 0.000408681 | 0.000570624 | 1 |
| Obesity | nk_cell | *RGS16* | 3.015007625 | 5.76E-06 | 1.05E-05 | 0.027964253 |
| Obesity | nk_cell | *TARS2* | -2.97731731 | 1.53E-07 | 4.35E-07 | 0.000744194 |
| Obesity | nk_cell | *SNX27* | -2.083052624 | 1.55E-06 | 3.22E-06 | 0.00751387 |
| Obesity | nk_cell | *SETDB1* | -2.34376844 | 1.90E-06 | 3.86E-06 | 0.009228852 |
| Obesity | nk_cell | *SEMA6C* | -1.198930136 | 1.91E-08 | 8.35E-08 | 9.29E-05 |
| Obesity | nk_cell | *ARNT* | -1.928340593 | 1.10E-07 | 3.31E-07 | 0.000531562 |
| Obesity | nk_cell | *POGZ* | -1.897559763 | 7.23E-07 | 1.64E-06 | 0.003507975 |
| Obesity | nk_cell | *GABPB2* | -1.936379411 | 4.87E-05 | 7.66E-05 | 0.236216483 |
| Obesity | nk_cell | *DYRK3* | -1.776393501 | 4.33E-09 | 3.18E-08 | 2.10E-05 |
| Obesity | nk_cell | *EIF2D* | -2.757915415 | 1.41E-07 | 4.04E-07 | 0.000682005 |
| Obesity | nk_cell | *TAF1A* | -2.41964071 | 1.52E-07 | 4.33E-07 | 0.000739098 |
| Obesity | nk_cell | *RAB13* | 4.316071199 | 3.36E-06 | 6.45E-06 | 0.016280802 |
| Obesity | nk_cell | *S100A8* | -3.283494849 | 5.71E-06 | 1.04E-05 | 0.027723327 |
| Obesity | nk_cell | *CREB3L4* | -3.346303552 | 8.85E-07 | 1.95E-06 | 0.004294576 |
| Obesity | nk_cell | *KCNN3* | -1.728985936 | 7.89E-09 | 4.46E-08 | 3.83E-05 |
| Obesity | nk_cell | *C1orf43* | -2.561510763 | 4.20E-07 | 1.02E-06 | 0.002037441 |
| Obesity | nk_cell | *RIT1* | -2.618580334 | 1.37E-07 | 3.97E-07 | 0.000664437 |

| | | | | | | |
|---|---|---|---|---|---|---|
| Obesity | nk_cell | *FLG* | -1.281249073 | 6.20E-07 | 1.42E-06 | 0.003009468 |
| Obesity | nk_cell | *SCCPDH* | -2.309715823 | 2.76E-08 | 1.10E-07 | 0.000133682 |
| Obesity | nk_cell | *CEP170* | -2.035403582 | 1.44E-06 | 3.02E-06 | 0.006964934 |
| Obesity | nk_cell | *ACP1* | -2.808458726 | 1.22E-07 | 3.61E-07 | 0.000591248 |
| Obesity | nk_cell | *CNIH4* | -2.392647462 | 3.55E-09 | 2.83E-08 | 1.72E-05 |
| Obesity | nk_cell | *MBOAT2* | -1.950545256 | 8.78E-09 | 4.74E-08 | 4.26E-05 |
| Obesity | nk_cell | *PYCR2* | 2.567861802 | 4.51E-07 | 1.08E-06 | 0.002187606 |
| Obesity | nk_cell | *PLEKHA6* | -0.802714338 | 9.40E-09 | 4.99E-08 | 4.56E-05 |
| Obesity | nk_cell | *OSR1* | -1.163197346 | 2.14E-08 | 9.07E-08 | 0.000103892 |
| Obesity | nk_cell | *RHOB* | -2.433511401 | 3.09E-08 | 1.20E-07 | 0.000149981 |
| Obesity | nk_cell | *GALM* | -2.429171413 | 1.51E-07 | 4.30E-07 | 0.000733557 |
| Obesity | nk_cell | *WDPCP* | -1.359419244 | 7.04E-06 | 1.26E-05 | 0.034164843 |
| Obesity | nk_cell | *VPS54* | -1.912168599 | 2.89E-07 | 7.36E-07 | 0.001402544 |
| Obesity | nk_cell | *REG3G* | 4.991674897 | 0.030992739 | 0.036499216 | 1 |
| Obesity | nk_cell | *ABHD1* | -3.709363885 | 4.60E-06 | 8.57E-06 | 0.022305896 |
| Obesity | nk_cell | *CIAO1* | -2.740343657 | 3.70E-07 | 9.14E-07 | 0.001795071 |
| Obesity | nk_cell | *ZNF514* | -3.03094763 | 3.64E-07 | 9.03E-07 | 0.001766675 |
| Obesity | nk_cell | *EXOC6B* | -1.697040776 | 6.35E-07 | 1.45E-06 | 0.00307964 |
| Obesity | nk_cell | *TEX261* | -4.755912425 | 6.85E-05 | 0.000104901 | 0.332220835 |
| Obesity | nk_cell | *RALB* | -3.097563853 | 2.96E-05 | 4.80E-05 | 0.143729391 |
| Obesity | nk_cell | *RABL2A* | -2.417510289 | 2.25E-07 | 5.97E-07 | 0.00109 |
| Obesity | nk_cell | *FBLN7* | -1.804321333 | 1.35E-10 | 8.70E-09 | 6.53E-07 |
| Obesity | nk_cell | *CNGA3* | -21.576886 | 1.16E-08 | 5.70E-08 | 5.62E-05 |
| Obesity | nk_cell | *FAHD2B* | -2.384683311 | 9.87E-12 | 1.45E-09 | 4.79E-08 |
| Obesity | nk_cell | *THSD7B* | -1.064447477 | 9.59E-09 | 5.04E-08 | 4.65E-05 |
| Obesity | nk_cell | *GPR17* | -0.660855397 | 6.89E-06 | 1.24E-05 | 0.033427739 |
| Obesity | nk_cell | *POLR2D* | -3.016468061 | 5.90E-05 | 9.15E-05 | 0.286461154 |
| Obesity | nk_cell | *GALNT13* | -2.172922079 | 2.72E-08 | 1.09E-07 | 0.000132083 |
| Obesity | nk_cell | *PHOSPHO2* | -5.992672951 | 1.72E-05 | 2.87E-05 | 0.08332689 |
| Obesity | nk_cell | *METTL21A* | -2.441208874 | 0.000101267 | 0.000152083 | 0.491349087 |
| Obesity | nk_cell | *NBEAL1* | -1.620777404 | 6.40E-05 | 9.84E-05 | 0.310734524 |
| Obesity | nk_cell | *SUMF1* | -1.866839454 | 5.89E-05 | 9.13E-05 | 0.28581688 |
| Obesity | nk_cell | *ACKR3* | -1.710090319 | 6.02E-06 | 1.09E-05 | 0.029198151 |
| Obesity | nk_cell | *ESPNL* | -13.98878057 | 1.59E-09 | 2.11E-08 | 7.74E-06 |
| Obesity | nk_cell | *ANKMY1* | -2.088484351 | 0.00146368 | 0.001932456 | 1 |
| Obesity | nk_cell | *TAMM41* | -2.32578907 | 4.87E-05 | 7.66E-05 | 0.236449958 |
| Obesity | nk_cell | *STK11IP* | -3.34574851 | 2.32E-08 | 9.59E-08 | 0.000112349 |
| Obesity | nk_cell | *EAF1* | 1.587982166 | 3.19E-09 | 2.69E-08 | 1.55E-05 |
| Obesity | nk_cell | *CNTN4* | -1.321469117 | 5.88E-09 | 3.81E-08 | 2.85E-05 |
| Obesity | nk_cell | *SLC25A38* | 1.605834693 | 2.68E-09 | 2.45E-08 | 1.30E-05 |
| Obesity | nk_cell | *CTDSPL* | -1.865493372 | 1.81E-07 | 4.99E-07 | 0.000879958 |
| Obesity | nk_cell | *PTPRG* | -1.596267807 | 2.77E-07 | 7.14E-07 | 0.001346274 |
| Obesity | nk_cell | *SLC25A26* | -2.57266446 | 1.66E-05 | 2.78E-05 | 0.08052283 |
| Obesity | nk_cell | *LIMD1* | -1.715306775 | 0.000130592 | 0.000193534 | 0.633631194 |
| Obesity | nk_cell | *ZNF660* | -0.687903621 | 0.004771228 | 0.00603808 | 1 |
| Obesity | nk_cell | *PLA1A* | -3.44548875 | 2.60E-08 | 1.05E-07 | 0.00012629 |
| Obesity | nk_cell | *ADPRH* | 2.508127637 | 2.93E-06 | 5.71E-06 | 0.014194272 |
| Obesity | nk_cell | *IGSF11* | -0.942716994 | 6.09E-08 | 2.03E-07 | 0.000295347 |
| Obesity | nk_cell | *BOC* | -1.137902223 | 6.05E-09 | 3.89E-08 | 2.94E-05 |
| Obesity | nk_cell | *MED12L* | -2.490221481 | 4.60E-08 | 1.63E-07 | 0.00022307 |

| | | | | | | |
|---|---|---|---|---|---|---|
| Obesity | nk_cell | *EIF2A* | -2.062073524 | 6.95E-08 | 2.27E-07 | 0.000337292 |
| Obesity | nk_cell | *NCEH1* | -2.06708246 | 4.74E-09 | 3.35E-08 | 2.30E-05 |
| Obesity | nk_cell | *AMT* | -1.418129281 | 4.96E-07 | 1.17E-06 | 0.002405262 |
| Obesity | nk_cell | *CCDC39* | -1.602749719 | 1.90E-05 | 3.14E-05 | 0.092040102 |
| Obesity | nk_cell | *MUC4* | -2.331888624 | 6.70E-09 | 4.11E-08 | 3.25E-05 |
| Obesity | nk_cell | *DGKQ* | 1.565127093 | 8.29E-08 | 2.63E-07 | 0.000402242 |
| Obesity | nk_cell | *CENPC* | -2.115897947 | 1.56E-07 | 4.41E-07 | 0.000756339 |
| Obesity | nk_cell | *SCD5* | -2.485470876 | 1.93E-08 | 8.42E-08 | 9.38E-05 |
| Obesity | nk_cell | *TRMT10A* | -1.460367106 | 6.57E-10 | 1.90E-08 | 3.19E-06 |
| Obesity | nk_cell | *PYURF* | 3.741054626 | 0.000338983 | 0.000478124 | 1 |
| Obesity | nk_cell | *CISD2* | -2.455263001 | 0.010861991 | 0.013308682 | 1 |
| Obesity | nk_cell | *DDIT4L* | -2.973501308 | 1.60E-08 | 7.33E-08 | 7.75E-05 |
| Obesity | nk_cell | *METTL14* | -2.833199271 | 0.000139757 | 0.000206297 | 0.678099547 |
| Obesity | nk_cell | *USP53* | -1.631655347 | 2.29E-06 | 4.55E-06 | 0.011089486 |
| Obesity | nk_cell | *MARCH1* | -2.094810135 | 7.49E-07 | 1.69E-06 | 0.003636361 |
| Obesity | nk_cell | *CBR4* | -2.069223075 | 1.15E-05 | 1.97E-05 | 0.055721339 |
| Obesity | nk_cell | *MARCH6* | -2.083121986 | 2.37E-06 | 4.70E-06 | 0.011506487 |
| Obesity | nk_cell | *NKD2* | 3.269045919 | 1.67E-07 | 4.67E-07 | 0.000810824 |
| Obesity | nk_cell | *MYO10* | -1.074844734 | 3.23E-09 | 2.71E-08 | 1.57E-05 |
| Obesity | nk_cell | *FAM105A* | 0.994262048 | 0.010806813 | 0.013247766 | 1 |
| Obesity | nk_cell | *BHMT* | -1.868015508 | 4.15E-08 | 1.51E-07 | 0.000201184 |
| Obesity | nk_cell | *GTF2H2* | -3.696388861 | 2.63E-08 | 1.06E-07 | 0.000127474 |
| Obesity | nk_cell | *SPATA9* | -1.196583834 | 2.53E-09 | 2.39E-08 | 1.23E-05 |
| Obesity | nk_cell | *TSLP* | -1.220078736 | 1.44E-05 | 2.44E-05 | 0.069779491 |
| Obesity | nk_cell | *FEM1C* | 3.665919786 | 0.000124893 | 0.000185542 | 0.605980775 |
| Obesity | nk_cell | *COMMD10* | -1.942581482 | 3.00E-05 | 4.85E-05 | 0.145424183 |
| Obesity | nk_cell | *YIPF5* | -2.706580352 | 0.001015277 | 0.001358555 | 1 |
| Obesity | nk_cell | *CXCL14* | -1.475476481 | 6.25E-08 | 2.07E-07 | 0.000303016 |
| Obesity | nk_cell | *PCYOX1L* | -2.495665565 | 3.60E-10 | 1.52E-08 | 1.75E-06 |
| Obesity | nk_cell | *BOD1* | 3.807775196 | 6.16E-07 | 1.42E-06 | 0.002989143 |
| Obesity | nk_cell | *TBC1D7* | 2.929247645 | 5.61E-05 | 8.73E-05 | 0.27222515 |
| Obesity | nk_cell | *TRIM7* | -2.148246141 | 6.47E-08 | 2.14E-07 | 0.000313981 |
| Obesity | nk_cell | *HIGD2A* | 2.558760182 | 0.00938426 | 0.011550592 | 1 |
| Obesity | nk_cell | *FAM193B* | -2.402571584 | 1.10E-06 | 2.37E-06 | 0.005319499 |
| Obesity | nk_cell | *RNF44* | 3.112791191 | 3.59E-08 | 1.35E-07 | 0.000174376 |
| Obesity | nk_cell | *MUT* | -2.732119185 | 1.38E-07 | 3.98E-07 | 0.0006674 |
| Obesity | nk_cell | *DOK3* | -1.052841264 | 1.78E-08 | 7.92E-08 | 8.64E-05 |
| Obesity | nk_cell | *ABT1* | 2.060293246 | 1.13E-11 | 1.62E-09 | 5.51E-08 |
| Obesity | nk_cell | *DAAM2* | -2.179813715 | 3.49E-07 | 8.69E-07 | 0.001693665 |
| Obesity | nk_cell | *MLIP* | -1.416749463 | 4.91E-10 | 1.71E-08 | 2.38E-06 |
| Obesity | nk_cell | *ANO7* | -1.550232433 | 3.03E-06 | 5.89E-06 | 0.014684473 |
| Obesity | nk_cell | *TCTE1* | 2.273714211 | 8.44E-08 | 2.67E-07 | 0.00040963 |
| Obesity | nk_cell | *RPL7L1* | -3.004736678 | 1.49E-06 | 3.12E-06 | 0.007248312 |
| Obesity | nk_cell | *TPBG* | -1.814966233 | 7.62E-09 | 4.39E-08 | 3.70E-05 |
| Obesity | nk_cell | *IRAK1BP1* | -1.837067734 | 4.54E-06 | 8.49E-06 | 0.022044113 |
| Obesity | nk_cell | *PHIP* | -1.948486415 | 7.27E-05 | 0.000111134 | 0.352960684 |
| Obesity | nk_cell | *PNRC1* | -2.251289798 | 4.06E-08 | 1.49E-07 | 0.000197088 |
| Obesity | nk_cell | *RNF217* | -1.369307712 | 8.17E-09 | 4.53E-08 | 3.96E-05 |
| Obesity | nk_cell | *ARHGAP18* | -1.665723293 | 0.000100091 | 0.00015054 | 0.485642797 |
| Obesity | nk_cell | *ABRACL* | 1.08250887 | 1.71E-09 | 2.16E-08 | 8.31E-06 |

| | | | | | | |
|---|---|---|---|---|---|---|
| Obesity | nk_cell | *SLC18B1* | -2.083666968 | 3.76E-08 | 1.40E-07 | 0.00018246 |
| Obesity | nk_cell | *PNLDC1* | -2.275007669 | 0.034088001 | 0.039940831 | 1 |
| Obesity | nk_cell | *WTAP* | -1.99249314 | 1.66E-05 | 2.78E-05 | 0.080434287 |
| Obesity | nk_cell | *SLC22A3* | -2.181998312 | 2.11E-08 | 8.99E-08 | 0.000102588 |
| Obesity | nk_cell | *GNA12* | -2.666288658 | 9.50E-07 | 2.08E-06 | 0.00460708 |
| Obesity | nk_cell | *SDK1* | -1.814215997 | 2.21E-09 | 2.27E-08 | 1.07E-05 |
| Obesity | nk_cell | *RBAK* | 3.604661674 | 1.38E-08 | 6.57E-08 | 6.67E-05 |
| Obesity | nk_cell | *POMZP3* | -1.75975962 | 0.002728031 | 0.00352407 | 1 |
| Obesity | nk_cell | *PSPH* | -1.487948883 | 2.70E-07 | 6.98E-07 | 0.001309331 |
| Obesity | nk_cell | *GIGYF1* | -2.397261739 | 4.34E-08 | 1.56E-07 | 0.000210708 |
| Obesity | nk_cell | *TRIM4* | -1.073889066 | 0.002893421 | 0.003726806 | 1 |
| Obesity | nk_cell | *TMEM209* | -3.145991906 | 0.000105497 | 0.000158132 | 0.511872111 |
| Obesity | nk_cell | *EPHA1* | 4.601803314 | 2.96E-09 | 2.58E-08 | 1.44E-05 |
| Obesity | nk_cell | *NCAPG2* | 3.0831931 | 0.000210019 | 0.000303639 | 1 |
| Obesity | nk_cell | *FBXO25* | -2.575242471 | 5.15E-07 | 1.21E-06 | 0.002496817 |
| Obesity | nk_cell | *CSGALNACT1* | -1.259882484 | 9.51E-06 | 1.66E-05 | 0.046143957 |
| Obesity | nk_cell | *CHMP7* | 2.789950808 | 0.000274311 | 0.00039151 | 1 |
| Obesity | nk_cell | *DOCK5* | -2.121652931 | 1.37E-05 | 2.33E-05 | 0.066391829 |
| Obesity | nk_cell | *TACC1* | -1.782236365 | 1.23E-07 | 3.64E-07 | 0.000596947 |
| Obesity | nk_cell | *ADHFE1* | -1.476244285 | 7.26E-10 | 1.92E-08 | 3.52E-06 |
| Obesity | nk_cell | *RPL7* | -2.168559093 | 2.25E-06 | 4.49E-06 | 0.010917109 |
| Obesity | nk_cell | *ATP6V0D2* | -3.885348512 | 1.82E-08 | 8.07E-08 | 8.84E-05 |
| Obesity | nk_cell | *DPYS* | -0.99043529 | 2.48E-09 | 2.37E-08 | 1.20E-05 |
| Obesity | nk_cell | *LRP12* | -2.426781106 | 2.46E-08 | 1.01E-07 | 0.000119595 |
| Obesity | nk_cell | *MAL2* | -0.980546433 | 0.032475755 | 0.038153115 | 1 |
| Obesity | nk_cell | *GSDMC* | -6.603134179 | 0.003139937 | 0.00402722 | 1 |
| Obesity | nk_cell | *SLC39A4* | -1.44857177 | 4.11E-05 | 6.52E-05 | 0.199415739 |
| Obesity | nk_cell | *AK3* | -2.490297865 | 2.03E-06 | 4.09E-06 | 0.009834292 |
| Obesity | nk_cell | *HAUS6* | -2.403398563 | 0.013714914 | 0.016665356 | 1 |
| Obesity | nk_cell | *C9orf72* | -1.087232894 | 1.23E-05 | 2.11E-05 | 0.059882328 |
| Obesity | nk_cell | *FBXO10* | -1.975986571 | 2.30E-06 | 4.58E-06 | 0.01115604 |
| Obesity | nk_cell | *ZNF462* | -1.327920784 | 2.21E-08 | 9.30E-08 | 0.000107378 |
| Obesity | nk_cell | *ALAD* | 1.704245621 | 0.016502932 | 0.019948237 | 1 |
| Obesity | nk_cell | *WDR31* | -1.428132905 | 2.75E-05 | 4.47E-05 | 0.133381415 |
| Obesity | nk_cell | *POLE3* | 1.205804384 | 4.60E-08 | 1.63E-07 | 0.000223326 |
| Obesity | nk_cell | *SURF1* | 1.899359213 | 0.000670004 | 0.000914962 | 1 |
| Obesity | nk_cell | *SURF6* | -1.68497573 | 3.62E-09 | 2.86E-08 | 1.75E-05 |
| Obesity | nk_cell | *MED22* | 1.92981244 | 4.10E-07 | 9.96E-07 | 0.001989006 |
| Obesity | nk_cell | *REXO4* | 3.366384852 | 1.21E-06 | 2.57E-06 | 0.005847005 |
| Obesity | nk_cell | *SH3GLB2* | -3.6220772 | 5.24E-05 | 8.18E-05 | 0.254081981 |
| Obesity | nk_cell | *INPP5E* | -2.474672163 | 5.00E-09 | 3.48E-08 | 2.43E-05 |
| Obesity | nk_cell | *SLC39A12* | -2.646049741 | 6.97E-09 | 4.20E-08 | 3.38E-05 |
| Obesity | nk_cell | *ST8SIA6* | 0.452022779 | 1.01E-09 | 2.05E-08 | 4.88E-06 |
| Obesity | nk_cell | *PARD3* | -1.377855749 | 5.87E-08 | 1.97E-07 | 0.000284612 |
| Obesity | nk_cell | *CDHR1* | 0.779459587 | 2.11E-09 | 2.26E-08 | 1.02E-05 |
| Obesity | nk_cell | *RGR* | -15.75988351 | 8.91E-13 | 1.73E-10 | 4.32E-09 |
| Obesity | nk_cell | *FRA10AC1* | -2.208508405 | 9.97E-07 | 2.18E-06 | 0.004835063 |
| Obesity | nk_cell | *ADD3* | -2.03104255 | 3.32E-05 | 5.34E-05 | 0.160990407 |
| Obesity | nk_cell | *LRRC27* | -1.413911595 | 6.35E-09 | 4.02E-08 | 3.08E-05 |
| Obesity | nk_cell | *PAOX* | 4.561454658 | 4.86E-06 | 9.01E-06 | 0.023566312 |

| | | | | | | |
|---|---|---|---|---|---|---|
| Obesity | nk_cell | *PDCD11* | 3.43914267 | 0.000110836 | 0.00016593 | 0.53777808 |
| Obesity | nk_cell | *GAS2* | -3.077618179 | 2.31E-07 | 6.11E-07 | 0.001119317 |
| Obesity | nk_cell | *LIN7C* | -3.015136928 | 0.001443916 | 0.00190792 | 1 |
| Obesity | nk_cell | *ZNF215* | -0.919196889 | 1.06E-05 | 1.83E-05 | 0.051346504 |
| Obesity | nk_cell | *HSD17B12* | -2.228540734 | 2.91E-17 | 2.02E-14 | 1.41E-13 |
| Obesity | nk_cell | *EIF3M* | -2.014092162 | 0.005270131 | 0.006624528 | 1 |
| Obesity | nk_cell | *PTPRJ* | 1.956809769 | 1.38E-07 | 3.99E-07 | 0.000670501 |
| Obesity | nk_cell | *CCDC82* | -1.456799619 | 0.00064023 | 0.000876771 | 1 |
| Obesity | nk_cell | *CAPN5* | -1.705189439 | 1.58E-09 | 2.11E-08 | 7.68E-06 |
| Obesity | nk_cell | *INTS4* | -2.179410978 | 4.20E-05 | 6.66E-05 | 0.203891113 |
| Obesity | nk_cell | *RPS3* | 2.018719083 | 3.34E-07 | 8.37E-07 | 0.001621515 |
| Obesity | nk_cell | *DRD2* | -8.69384694 | 1.27E-09 | 2.09E-08 | 6.14E-06 |
| Obesity | nk_cell | *ATM* | 1.945973699 | 0.00049634 | 0.0006865 | 1 |
| Obesity | nk_cell | *SLX4IP* | -1.72712372 | 2.46E-05 | 4.01E-05 | 0.119238237 |
| Obesity | nk_cell | *ST14* | -3.120912022 | 9.70E-09 | 5.09E-08 | 4.71E-05 |
| Obesity | nk_cell | *FADS1* | 2.317173142 | 4.61E-06 | 8.58E-06 | 0.022346141 |
| Obesity | nk_cell | *TMC2* | -3.836119264 | 1.89E-09 | 2.19E-08 | 9.15E-06 |
| Obesity | nk_cell | *ROM1* | -1.347922326 | 5.82E-06 | 1.06E-05 | 0.028255235 |
| Obesity | nk_cell | *EML3* | -2.185581226 | 4.04E-07 | 9.83E-07 | 0.001959228 |
| Obesity | nk_cell | *PLCH2* | 0.31095739 | 1.39E-09 | 2.11E-08 | 6.73E-06 |
| Obesity | nk_cell | *CPSF7* | -2.627105882 | 4.03E-05 | 6.40E-05 | 0.195453443 |
| Obesity | nk_cell | *CCDC15* | -1.433780577 | 3.78E-10 | 1.53E-08 | 1.83E-06 |
| Obesity | nk_cell | *CHEK1* | -1.828916275 | 1.70E-09 | 2.16E-08 | 8.25E-06 |
| Obesity | nk_cell | *SCN2B* | -1.285217148 | 0.00453912 | 0.00575485 | 1 |
| Obesity | nk_cell | *DUSP15* | -4.219083568 | 2.25E-05 | 3.69E-05 | 0.109243493 |
| Obesity | nk_cell | *SPATA25* | -1.996447613 | 0.003922438 | 0.004997812 | 1 |
| Obesity | nk_cell | *CABLES2* | -2.667390519 | 2.73E-05 | 4.44E-05 | 0.132639032 |
| Obesity | nk_cell | *TRPT1* | -2.740797798 | 1.75E-09 | 2.16E-08 | 8.51E-06 |
| Obesity | nk_cell | *NUDT22* | -3.925710106 | 1.02E-08 | 5.29E-08 | 4.96E-05 |
| Obesity | nk_cell | *TBX6* | 0.641536555 | 0.009639482 | 0.011849701 | 1 |
| Obesity | nk_cell | *ALDOA* | 2.251983006 | 0.001159462 | 0.001542136 | 1 |
| Obesity | nk_cell | *ANXA8L1* | -0.738715698 | 0.003306741 | 0.00423668 | 1 |
| Obesity | nk_cell | *CWC15* | 2.316067504 | 0.000183633 | 0.000267243 | 0.890988852 |
| Obesity | nk_cell | *FCGR1A* | -2.179095568 | 0.001471239 | 0.001941908 | 1 |
| Obesity | nk_cell | *TMEM218* | -2.889287962 | 2.93E-06 | 5.73E-06 | 0.014240144 |
| Obesity | nk_cell | *FAM124A* | -2.42994694 | 9.89E-09 | 5.16E-08 | 4.80E-05 |
| Obesity | nk_cell | *LYPD6B* | -2.194298085 | 4.77E-10 | 1.70E-08 | 2.31E-06 |
| Obesity | nk_cell | *WDR17* | -2.982755852 | 1.35E-07 | 3.93E-07 | 0.0006556 |
| Obesity | nk_cell | *CCDC102B* | -1.605755287 | 2.94E-08 | 1.15E-07 | 0.000142431 |
| Obesity | nk_cell | *RGS18* | 3.076266671 | 9.55E-10 | 2.05E-08 | 4.64E-06 |
| Obesity | nk_cell | *PRSS23* | -1.559758919 | 5.70E-08 | 1.93E-07 | 0.000276635 |
| Obesity | nk_cell | *PPP1R1C* | -2.23401109 | 2.15E-09 | 2.26E-08 | 1.04E-05 |
| Obesity | nk_cell | *FAM173B* | -2.84081361 | 0.000427461 | 0.000595051 | 1 |
| Obesity | nk_cell | *GPR158* | -2.587839089 | 1.16E-08 | 5.70E-08 | 5.62E-05 |
| Obesity | nk_cell | *ANK3* | 1.572441738 | 7.10E-09 | 4.25E-08 | 3.44E-05 |
| Obesity | nk_cell | *PLBD2* | -2.70275926 | 1.14E-05 | 1.96E-05 | 0.055356589 |
| Obesity | nk_cell | *DLG5* | -1.804146831 | 1.16E-06 | 2.50E-06 | 0.005651235 |
| Obesity | nk_cell | *GXYLT1* | -2.774735795 | 4.90E-05 | 7.69E-05 | 0.237565011 |
| Obesity | nk_cell | *EXT2* | -1.987590961 | 0.000141263 | 0.000208204 | 0.685407287 |
| Obesity | nk_cell | *TMEM18* | -3.933848749 | 6.06E-06 | 1.10E-05 | 0.029386649 |

| | | | | | | |
|---|---|---|---|---|---|---|
| Obesity | nk_cell | *NDUFC2* | -3.313556411 | 0.00327914 | 0.004202426 | 1 |
| Obesity | nk_cell | *ADAMTS12* | -1.501682224 | 3.13E-07 | 7.93E-07 | 0.00152101 |
| Obesity | nk_cell | *NUBPL* | -2.220297266 | 6.74E-06 | 1.21E-05 | 0.032679331 |
| Obesity | nk_cell | *VIPAS39* | -1.973526494 | 7.17E-09 | 4.27E-08 | 3.48E-05 |
| Obesity | nk_cell | *ANKRD50* | -2.461289096 | 8.01E-07 | 1.79E-06 | 0.003888076 |
| Obesity | nk_cell | *UPF2* | -2.20074725 | 3.94E-05 | 6.26E-05 | 0.191001664 |
| Obesity | nk_cell | *C4orf33* | 3.14780449 | 0.000183251 | 0.000266767 | 0.889135481 |
| Obesity | nk_cell | *PTPRO* | -1.798419728 | 2.07E-06 | 4.17E-06 | 0.010061297 |
| Obesity | nk_cell | *EPS8* | -1.732275063 | 4.21E-06 | 7.91E-06 | 0.020414449 |
| Obesity | nk_cell | *QDPR* | -1.557430929 | 9.13E-09 | 4.89E-08 | 4.43E-05 |
| Obesity | nk_cell | *FAM160B1* | -2.090524616 | 2.16E-07 | 5.77E-07 | 0.001045745 |
| Obesity | nk_cell | *ZNF827* | -1.923453992 | 0.008483177 | 0.010478711 | 1 |
| Obesity | nk_cell | *AKR1C2* | -1.453803299 | 1.69E-09 | 2.16E-08 | 8.19E-06 |
| Obesity | nk_cell | *DPYSL4* | 1.405826808 | 4.32E-08 | 1.55E-07 | 0.000209511 |
| Obesity | nk_cell | *PIGF* | -2.429222039 | 2.28E-07 | 6.06E-07 | 0.001108028 |
| Obesity | nk_cell | *INPP1* | -2.416844657 | 1.07E-06 | 2.32E-06 | 0.005200036 |
| Obesity | nk_cell | *MFSD6* | 2.672297336 | 0.000424494 | 0.000591512 | 1 |
| Obesity | nk_cell | *RNF144A* | -1.580510854 | 1.12E-08 | 5.56E-08 | 5.42E-05 |
| Obesity | nk_cell | *CENPU* | -2.015000671 | 0.009299871 | 0.011458347 | 1 |
| Obesity | nk_cell | *BICD1* | -2.268425906 | 1.17E-06 | 2.51E-06 | 0.00568168 |
| Obesity | nk_cell | *CCDC122* | -1.514647111 | 2.54E-05 | 4.13E-05 | 0.123004148 |
| Obesity | nk_cell | *GUF1* | -2.368557079 | 1.97E-09 | 2.20E-08 | 9.55E-06 |
| Obesity | nk_cell | *GABRA2* | 2.887139251 | 5.06E-10 | 1.75E-08 | 2.45E-06 |
| Obesity | nk_cell | *PARP8* | 1.10713343 | 3.86E-07 | 9.46E-07 | 0.001874609 |
| Obesity | nk_cell | *CACUL1* | -2.059458173 | 0.000922442 | 0.001240834 | 1 |
| Obesity | nk_cell | *BEND6* | -1.651183833 | 5.61E-11 | 4.77E-09 | 2.72E-07 |
| Obesity | nk_cell | *TIAL1* | -2.32443854 | 0.000122168 | 0.000181773 | 0.592760452 |
| Obesity | nk_cell | *TMEM56* | -4.244917891 | 8.16E-09 | 4.53E-08 | 3.96E-05 |
| Obesity | nk_cell | *PTPN14* | -1.792216022 | 1.33E-07 | 3.89E-07 | 0.000646542 |
| Obesity | nk_cell | *MGAT5* | -2.27186468 | 5.62E-06 | 1.03E-05 | 0.027269894 |
| Obesity | nk_cell | *TMEM178A* | 2.340884226 | 1.67E-09 | 2.15E-08 | 8.11E-06 |
| Obesity | nk_cell | *EPG5* | -2.393514212 | 0.000200919 | 0.000291177 | 0.974860637 |
| Obesity | nk_cell | *KCNK13* | -4.813461455 | 0.000372344 | 0.000522495 | 1 |
| Obesity | nk_cell | *ATG10* | -1.146447022 | 5.18E-06 | 9.55E-06 | 0.025118765 |
| Obesity | nk_cell | *POC5* | -3.047359905 | 2.01E-08 | 8.63E-08 | 9.76E-05 |
| Obesity | nk_cell | *CWF19L2* | -1.74795941 | 0.000108753 | 0.000162911 | 0.52766763 |
| Obesity | nk_cell | *ZNF547* | -0.970507411 | 0.027141761 | 0.032182753 | 1 |
| Obesity | nk_cell | *NMT2* | -2.671179396 | 7.57E-07 | 1.70E-06 | 0.003675062 |
| Obesity | nk_cell | *ZNF837* | -1.950449374 | 3.21E-07 | 8.09E-07 | 0.001555894 |
| Obesity | nk_cell | *CCDC50* | -1.299023613 | 2.57E-06 | 5.06E-06 | 0.012452828 |
| Obesity | nk_cell | *PLEKHH2* | -1.194814826 | 1.60E-07 | 4.51E-07 | 0.000777071 |
| Obesity | nk_cell | *SPARCL1* | -1.713793427 | 3.40E-07 | 8.50E-07 | 0.001651023 |
| Obesity | nk_cell | *CCNO* | -10.96878246 | 6.85E-10 | 1.91E-08 | 3.33E-06 |
| Obesity | nk_cell | *CLEC4F* | -2.515946505 | 2.43E-07 | 6.39E-07 | 0.001180525 |
| Obesity | nk_cell | *SLC30A6* | -2.182980209 | 4.67E-07 | 1.11E-06 | 0.002264139 |
| Obesity | nk_cell | *RASGRP3* | -2.905284388 | 6.20E-05 | 9.57E-05 | 0.300664403 |
| Obesity | nk_cell | *GPR180* | -1.649737388 | 9.61E-08 | 2.98E-07 | 0.000466289 |
| Obesity | nk_cell | *TCTEX1D1* | -3.081829112 | 1.82E-06 | 3.71E-06 | 0.008831535 |
| Obesity | nk_cell | *IFIT5* | -1.362692318 | 4.39E-05 | 6.94E-05 | 0.213110951 |
| Obesity | nk_cell | *PRDM8* | 1.848433635 | 6.94E-08 | 2.26E-07 | 0.000336726 |

| | | | | | | |
|---|---|---|---|---|---|---|
| Obesity | nk_cell | *ZNF117* | -0.377463602 | 0.000301071 | 0.000427758 | 1 |
| Obesity | nk_cell | *MED21* | -2.689709462 | 8.27E-05 | 0.000125475 | 0.401393531 |
| Obesity | nk_cell | *JAKMIP1* | 0.261864852 | 1.16E-09 | 2.05E-08 | 5.65E-06 |
| Obesity | nk_cell | *SREK1IP1* | -2.641642746 | 1.78E-06 | 3.65E-06 | 0.008656183 |
| Obesity | nk_cell | *MR1* | 2.198888409 | 0.008674107 | 0.010709101 | 1 |
| Obesity | nk_cell | *CENPH* | -3.104002638 | 1.22E-09 | 2.08E-08 | 5.91E-06 |
| Obesity | nk_cell | *CARHSP1* | -2.203833312 | 3.01E-05 | 4.87E-05 | 0.1460829 |
| Obesity | nk_cell | *CAST* | -1.968793616 | 1.42E-06 | 2.99E-06 | 0.006907591 |
| Obesity | nk_cell | *SCOC* | -3.245242292 | 3.10E-06 | 6.01E-06 | 0.015038592 |
| Obesity | nk_cell | *CLGN* | 1.348384219 | 4.96E-09 | 3.46E-08 | 2.41E-05 |
| Obesity | nk_cell | *SYCP2L* | -1.203470969 | 3.93E-09 | 3.00E-08 | 1.91E-05 |
| Obesity | nk_cell | *TMEM87B* | -2.583393518 | 2.72E-06 | 5.34E-06 | 0.013195183 |
| Obesity | nk_cell | *PLA2R1* | -0.996684401 | 5.69E-05 | 8.83E-05 | 0.276152382 |
| Obesity | nk_cell | *SLC25A27* | -1.310803989 | 2.67E-05 | 4.34E-05 | 0.129390961 |
| Obesity | nk_cell | *FRMD1* | -1.528945231 | 4.88E-08 | 1.70E-07 | 0.000236634 |
| Obesity | nk_cell | *INO80C* | -3.261817843 | 1.74E-06 | 3.58E-06 | 0.008458523 |
| Obesity | nk_cell | *LPCAT1* | 0.940762766 | 1.37E-09 | 2.11E-08 | 6.67E-06 |
| Obesity | nk_cell | *NMRAL1* | 1.536803287 | 0.004564851 | 0.00578596 | 1 |
| Obesity | nk_cell | *ADPRHL1* | -2.659889406 | 1.30E-09 | 2.09E-08 | 6.32E-06 |
| Obesity | nk_cell | *CFDP1* | -2.139946743 | 1.78E-06 | 3.64E-06 | 0.008626747 |
| Obesity | nk_cell | *ZDHHC7* | -1.8790216 | 1.40E-08 | 6.64E-08 | 6.79E-05 |
| Obesity | nk_cell | *JAZF1* | 2.023230431 | 0.000366759 | 0.000515204 | 1 |
| Obesity | nk_cell | *FBXO36* | -2.413922939 | 1.82E-08 | 8.07E-08 | 8.83E-05 |
| Obesity | nk_cell | *MCOLN2* | 0.461800323 | 1.04E-09 | 2.05E-08 | 5.02E-06 |
| Obesity | nk_cell | *GDPD1* | 3.452536272 | 4.78E-06 | 8.88E-06 | 0.023173622 |
| Obesity | nk_cell | *SEMA3D* | -1.371827343 | 7.43E-09 | 4.35E-08 | 3.61E-05 |
| Obesity | nk_cell | *PPP2R5E* | -1.682250972 | 0.013065069 | 0.015907582 | 1 |
| Obesity | nk_cell | *ANKRD29* | -2.23716535 | 5.58E-08 | 1.90E-07 | 0.000270668 |
| Obesity | nk_cell | *DNAAF1* | -2.893706764 | 2.58E-09 | 2.41E-08 | 1.25E-05 |
| Obesity | nk_cell | *ANKH* | -2.191766771 | 2.65E-06 | 5.20E-06 | 0.012834013 |
| Obesity | nk_cell | *UBASH3B* | 1.277069854 | 1.39E-08 | 6.60E-08 | 6.74E-05 |
| Obesity | nk_cell | *ROBO4* | -2.5265352 | 1.02E-07 | 3.11E-07 | 0.000493834 |
| Obesity | nk_cell | *ROBO3* | -1.053686943 | 6.99E-09 | 4.20E-08 | 3.39E-05 |
| Obesity | nk_cell | *CC2D1B* | -1.812321657 | 0.00212867 | 0.002774189 | 1 |
| Obesity | nk_cell | *PRKCA* | -2.123040719 | 1.96E-06 | 3.97E-06 | 0.009498836 |
| Obesity | nk_cell | *LRRK1* | -1.991307975 | 6.23E-08 | 2.07E-07 | 0.000302261 |
| Obesity | nk_cell | *ENPP3* | -1.459165941 | 1.58E-09 | 2.11E-08 | 7.67E-06 |
| Obesity | nk_cell | *MIA3* | -2.286779164 | 5.13E-06 | 9.48E-06 | 0.024895128 |
| Obesity | nk_cell | *NEIL2* | -1.232423483 | 0.000675721 | 0.00092199 | 1 |
| Obesity | nk_cell | *PGM5* | -2.268234404 | 2.43E-07 | 6.39E-07 | 0.001179345 |
| Obesity | nk_cell | *CCSAP* | -4.169302387 | 4.70E-07 | 1.12E-06 | 0.002282548 |
| Obesity | nk_cell | *GBP5* | 0.369133919 | 2.24E-09 | 2.27E-08 | 1.09E-05 |
| Obesity | nk_cell | *ATP5G3* | 1.837277267 | 6.52E-05 | 0.000100066 | 0.316310021 |
| Obesity | nk_cell | *LY96* | -2.138209436 | 2.64E-08 | 1.07E-07 | 0.000128221 |
| Obesity | nk_cell | *PDE1C* | -0.986950247 | 2.27E-09 | 2.28E-08 | 1.10E-05 |
| Obesity | nk_cell | *RABGEF1* | -1.23541028 | 7.97E-08 | 2.55E-07 | 0.000386462 |
| Obesity | nk_cell | *MRPL39* | -1.578533645 | 5.17E-05 | 8.07E-05 | 0.250633352 |
| Obesity | nk_cell | *XPC* | -2.685414198 | 4.75E-06 | 8.84E-06 | 0.023044086 |
| Obesity | nk_cell | *FLCN* | -2.238621832 | 5.93E-07 | 1.37E-06 | 0.002877743 |
| Obesity | nk_cell | *OXNAD1* | -2.36856779 | 2.64E-06 | 5.20E-06 | 0.012808221 |

| | | | | | | |
|---|---|---|---|---|---|---|
| Obesity | nk_cell | *APCDD1* | -0.942140906 | 1.92E-09 | 2.19E-08 | 9.30E-06 |
| Obesity | nk_cell | *RAB6B* | 2.461479104 | 0.010079799 | 0.012384701 | 1 |
| Obesity | nk_cell | *ANKRD40* | -1.820717597 | 3.29E-09 | 2.73E-08 | 1.60E-05 |
| Obesity | nk_cell | *CYP2U1* | -1.552741966 | 8.31E-09 | 4.60E-08 | 4.03E-05 |
| Obesity | nk_cell | *MARVELD1* | -1.437949445 | 1.94E-09 | 2.20E-08 | 9.40E-06 |
| Obesity | nk_cell | *GOLGA7B* | 1.905044464 | 1.10E-09 | 2.05E-08 | 5.34E-06 |
| Obesity | nk_cell | *TRMT44* | -2.760875085 | 0.001077658 | 0.001438458 | 1 |
| Obesity | nk_cell | *MOV10* | -3.20037064 | 8.12E-09 | 4.52E-08 | 3.94E-05 |
| Obesity | nk_cell | *CNOT8* | -3.066244715 | 9.45E-06 | 1.65E-05 | 0.045837334 |
| Obesity | nk_cell | *LRGUK* | -0.99817821 | 5.43E-10 | 1.78E-08 | 2.63E-06 |
| Obesity | nk_cell | *SETD9* | -0.95780782 | 6.76E-10 | 1.91E-08 | 3.28E-06 |
| Obesity | nk_cell | *NUP205* | 2.981240296 | 5.93E-06 | 1.08E-05 | 0.028752641 |
| Obesity | nk_cell | *ZKSCAN2* | 2.136704839 | 0.011841877 | 0.014461813 | 1 |
| Obesity | nk_cell | *PDIA4* | 2.889622236 | 3.11E-08 | 1.20E-07 | 0.000150769 |
| Obesity | nk_cell | *KDM8* | 4.784850383 | 7.63E-06 | 1.35E-05 | 0.037039917 |
| Obesity | nk_cell | *ALS2CR12* | -2.635798329 | 0.000459571 | 0.000637825 | 1 |
| Obesity | nk_cell | *FZD7* | -0.994862542 | 2.13E-09 | 2.26E-08 | 1.03E-05 |
| Obesity | nk_cell | *DEPTOR* | -1.265751187 | 9.70E-08 | 3.00E-07 | 0.000470853 |
| Obesity | nk_cell | *RRAGA* | -3.837398751 | 0.001094008 | 0.001459881 | 1 |
| Obesity | nk_cell | *TMBIM4* | -2.015980749 | 5.60E-06 | 1.03E-05 | 0.027178173 |
| Obesity | nk_cell | *PSD3* | -1.664647752 | 2.03E-07 | 5.47E-07 | 0.000982575 |
| Obesity | nk_cell | *UGT2B4* | -0.546114723 | 0.004399459 | 0.005583619 | 1 |
| Obesity | nk_cell | *MMP16* | -1.701713558 | 1.56E-11 | 2.05E-09 | 7.59E-08 |
| Obesity | nk_cell | *KCNMA1* | -1.172322066 | 1.23E-06 | 2.63E-06 | 0.005987561 |
| Obesity | nk_cell | *NDUFAF6* | -1.984542632 | 5.62E-06 | 1.03E-05 | 0.027264825 |
| Obesity | nk_cell | *DRAM2* | -1.242289752 | 0.001181693 | 0.001569982 | 1 |
| Obesity | nk_cell | *C8orf37* | -1.451118036 | 4.08E-08 | 1.49E-07 | 0.000198014 |
| Obesity | nk_cell | *PPEF2* | -3.513012621 | 2.04E-06 | 4.11E-06 | 0.009897387 |
| Obesity | nk_cell | *ART3* | -1.232931159 | 0.005421619 | 0.006807892 | 1 |
| Obesity | nk_cell | *N6AMT1* | -2.286597402 | 0.000560302 | 0.000771449 | 1 |
| Obesity | nk_cell | *RWDD2B* | -3.195840331 | 5.73E-10 | 1.82E-08 | 2.78E-06 |
| Obesity | nk_cell | *CCT8* | -1.084159956 | 0.010556787 | 0.012957635 | 1 |
| Obesity | nk_cell | *NAA11* | -1.844406008 | 4.47E-10 | 1.65E-08 | 2.17E-06 |
| Obesity | nk_cell | *SFXN2* | -1.236773351 | 0.000827807 | 0.001119432 | 1 |
| Obesity | nk_cell | *C14orf2* | -2.588250003 | 1.94E-06 | 3.93E-06 | 0.009396332 |
| Obesity | nk_cell | *TDRD9* | 6.086869139 | 4.68E-07 | 1.11E-06 | 0.002272005 |
| Obesity | nk_cell | *PCDH1* | -0.787938248 | 0.017491678 | 0.021111846 | 1 |
| Obesity | nk_cell | *RPL30* | 2.02109735 | 0.000352509 | 0.00049648 | 1 |
| Obesity | nk_cell | *HKDC1* | -6.353699844 | 7.24E-10 | 1.92E-08 | 3.51E-06 |
| Obesity | nk_cell | *TYSND1* | -1.348545404 | 1.08E-06 | 2.34E-06 | 0.00524871 |
| Obesity | nk_cell | *NODAL* | -2.082286473 | 1.89E-09 | 2.19E-08 | 9.17E-06 |
| Obesity | nk_cell | *MED19* | -2.225359516 | 1.13E-09 | 2.05E-08 | 5.47E-06 |
| Obesity | nk_cell | *WDYHV1* | -1.04195159 | 0.007905581 | 0.009790168 | 1 |
| Obesity | nk_cell | *FBRS* | -2.678630726 | 5.65E-05 | 8.78E-05 | 0.274321654 |
| Obesity | nk_cell | *SASS6* | -1.373408359 | 4.54E-10 | 1.67E-08 | 2.20E-06 |
| Obesity | nk_cell | *ITGAD* | 0.336761859 | 3.26E-10 | 1.44E-08 | 1.58E-06 |
| Obesity | nk_cell | *VPS8* | -1.600794898 | 0.008139833 | 0.010067415 | 1 |
| Obesity | nk_cell | *MPV17L* | -2.073280313 | 1.09E-09 | 2.05E-08 | 5.29E-06 |
| Obesity | nk_cell | *RPUSD3* | -2.762266298 | 2.75E-08 | 1.10E-07 | 0.000133622 |
| Obesity | nk_cell | *GHRL* | -2.617987357 | 3.60E-05 | 5.76E-05 | 0.174735758 |

| | | | | | | |
|---|---|---|---|---|---|---|
| Obesity | nk_cell | *NTAN1* | -1.604611197 | 0.036409086 | 0.042567924 | 1 |
| Obesity | nk_cell | *NMNAT2* | -2.765868262 | 0.000777394 | 0.001055376 | 1 |
| Obesity | nk_cell | *SLC6A1* | -3.318933221 | 2.68E-09 | 2.45E-08 | 1.30E-05 |
| Obesity | nk_cell | *FCHO2* | -1.752760589 | 1.26E-05 | 2.15E-05 | 0.06101529 |
| Obesity | nk_cell | *NECAP2* | -2.926278994 | 1.06E-06 | 2.29E-06 | 0.005125076 |
| Obesity | nk_cell | *STEAP2* | -1.759475606 | 2.01E-11 | 2.32E-09 | 9.75E-08 |
| Obesity | nk_cell | *CLDN12* | -3.860212306 | 3.68E-07 | 9.10E-07 | 0.001784939 |
| Obesity | nk_cell | *MMP14* | -1.481073859 | 2.24E-09 | 2.27E-08 | 1.09E-05 |
| Obesity | nk_cell | *GATAD1* | -2.268133694 | 2.65E-07 | 6.87E-07 | 0.00128398 |
| Obesity | nk_cell | *TMED6* | -4.26212967 | 0.000497777 | 0.000688291 | 1 |
| Obesity | nk_cell | *CLEC18A* | -5.902847562 | 4.78E-20 | 5.80E-17 | 2.32E-16 |
| Obesity | nk_cell | *DHRS4* | -2.557313307 | 7.82E-09 | 4.43E-08 | 3.80E-05 |
| Obesity | nk_cell | *DHRS1* | -1.523796216 | 3.15E-08 | 1.21E-07 | 0.000152798 |
| Obesity | nk_cell | *AASDH* | -2.072317292 | 3.52E-06 | 6.73E-06 | 0.017084946 |
| Obesity | nk_cell | *ETS2* | 1.254819332 | 0.000915886 | 0.001232699 | 1 |
| Obesity | nk_cell | *LCA5L* | -1.603150895 | 1.75E-07 | 4.85E-07 | 0.000850406 |
| Obesity | nk_cell | *MX1* | -1.312510636 | 7.16E-06 | 1.28E-05 | 0.034734699 |
| Obesity | nk_cell | *SLC38A10* | -2.719781817 | 1.70E-05 | 2.85E-05 | 0.082655387 |
| Obesity | nk_cell | *C9orf43* | -1.349476359 | 0.000190701 | 0.000277031 | 0.925282404 |
| Obesity | nk_cell | *DGKI* | -1.123120953 | 3.70E-10 | 1.52E-08 | 1.79E-06 |
| Obesity | nk_cell | *SNX22* | 3.375161792 | 8.33E-09 | 4.60E-08 | 4.04E-05 |
| Obesity | nk_cell | *SLC34A2* | 9.085754536 | 3.17E-09 | 2.68E-08 | 1.54E-05 |
| Obesity | nk_cell | *SPPL3* | -1.946124768 | 5.10E-05 | 7.97E-05 | 0.24729484 |
| Obesity | nk_cell | *C12orf43* | -1.779568266 | 5.41E-10 | 1.78E-08 | 2.63E-06 |
| Obesity | nk_cell | *PEX10* | -2.175679455 | 0.015193611 | 0.018406841 | 1 |
| Obesity | nk_cell | *KRTCAP3* | -1.02067501 | 0.005588106 | 0.007009692 | 1 |
| Obesity | nk_cell | *PAFAH2* | 2.061069813 | 1.42E-05 | 2.42E-05 | 0.069087401 |
| Obesity | nk_cell | *EXTL1* | -15.78685068 | 1.57E-09 | 2.11E-08 | 7.60E-06 |
| Obesity | nk_cell | *WDR66* | -1.42811404 | 3.17E-08 | 1.21E-07 | 0.000153851 |
| Obesity | nk_cell | *RHPN1* | -2.096520661 | 1.64E-08 | 7.44E-08 | 7.95E-05 |
| Obesity | nk_cell | *LRRC43* | 6.739164969 | 1.07E-06 | 2.31E-06 | 0.00518627 |
| Obesity | nk_cell | *DZIP1L* | -1.995263395 | 5.23E-09 | 3.56E-08 | 2.54E-05 |
| Obesity | nk_cell | *FANCC* | -1.685184029 | 1.11E-08 | 5.56E-08 | 5.41E-05 |
| Obesity | nk_cell | *ESYT3* | -2.190815088 | 3.78E-08 | 1.40E-07 | 0.000183278 |
| Obesity | nk_cell | *CLSTN2* | -1.960252696 | 4.42E-08 | 1.58E-07 | 0.000214527 |
| Obesity | nk_cell | *COLEC12* | -1.175488141 | 1.33E-07 | 3.89E-07 | 0.00064551 |
| Obesity | nk_cell | *TSPAN33* | -3.190459524 | 4.31E-08 | 1.55E-07 | 0.00020889 |
| Obesity | nk_cell | *FAM86C1* | -9.568086085 | 7.53E-05 | 0.000114874 | 0.365529763 |
| Obesity | nk_cell | *PPP1R9A* | -2.271884664 | 4.66E-07 | 1.11E-06 | 0.002260114 |
| Obesity | nk_cell | *DYNC1I1* | -1.731706347 | 9.09E-07 | 2.00E-06 | 0.004409276 |
| Obesity | nk_cell | *COPG2* | -1.175291943 | 1.63E-08 | 7.40E-08 | 7.88E-05 |
| Obesity | nk_cell | *ELK4* | 2.680887604 | 0.003212548 | 0.004118172 | 1 |
| Obesity | nk_cell | *DUSP23* | -1.557744963 | 1.04E-09 | 2.05E-08 | 5.07E-06 |
| Obesity | nk_cell | *RNF166* | 0.989675605 | 1.07E-08 | 5.42E-08 | 5.20E-05 |
| Obesity | nk_cell | *SPATA2L* | 2.642808779 | 8.44E-07 | 1.87E-06 | 0.004092877 |
| Obesity | nk_cell | *FGF17* | -5.998450577 | 1.12E-06 | 2.40E-06 | 0.005429199 |
| Obesity | nk_cell | *CDA* | -1.315421379 | 1.60E-06 | 3.31E-06 | 0.007749625 |
| Obesity | nk_cell | *SLC5A11* | -1.589422214 | 4.50E-15 | 1.82E-12 | 2.18E-11 |
| Obesity | nk_cell | *CCAR2* | -3.504275313 | 6.20E-06 | 1.12E-05 | 0.030103573 |
| Obesity | nk_cell | *CDC42SE2* | 1.86148497 | 1.06E-07 | 3.22E-07 | 0.000515491 |

| | | | | | | |
|---|---|---|---|---|---|---|
| Obesity | nk_cell | *IFNAR2* | 1.563721534 | 6.01E-05 | 9.30E-05 | 0.291668786 |
| Obesity | nk_cell | *SV2A* | -4.246079169 | 0.000125138 | 0.000185849 | 0.607167537 |
| Obesity | nk_cell | *CLIC6* | -2.87218668 | 3.32E-06 | 6.40E-06 | 0.016126222 |
| Obesity | nk_cell | *CBR1* | -2.227827314 | 9.15E-06 | 1.60E-05 | 0.044416267 |
| Obesity | nk_cell | *CBR3* | -1.706029352 | 1.37E-07 | 3.98E-07 | 0.000666558 |
| Obesity | nk_cell | *CHAF1B* | -1.75024703 | 0.002051566 | 0.00267586 | 1 |
| Obesity | nk_cell | *SIM2* | 0.912950639 | 1.60E-08 | 7.33E-08 | 7.74E-05 |
| Obesity | nk_cell | *ARHGAP27* | -3.821242371 | 3.83E-07 | 9.40E-07 | 0.001856947 |
| Obesity | nk_cell | *IRX6* | -0.909944248 | 4.63E-09 | 3.29E-08 | 2.25E-05 |
| Obesity | nk_cell | *HK2* | -1.849243696 | 1.87E-08 | 8.20E-08 | 9.09E-05 |
| Obesity | nk_cell | *THEM4* | -1.442758022 | 0.018947779 | 0.022784293 | 1 |
| Obesity | nk_cell | *AMFR* | 3.107650904 | 4.34E-06 | 8.15E-06 | 0.021079276 |
| Obesity | nk_cell | *TMEM69* | -3.436074646 | 1.34E-07 | 3.91E-07 | 0.000650653 |
| Obesity | nk_cell | *ACE* | -1.831331567 | 3.41E-09 | 2.77E-08 | 1.66E-05 |
| Obesity | nk_cell | *TEPP* | 1.606389135 | 2.24E-09 | 2.27E-08 | 1.09E-05 |
| Obesity | nk_cell | *CHCHD6* | -2.340081809 | 3.42E-07 | 8.53E-07 | 0.001657929 |
| Obesity | nk_cell | *CTBP1* | -3.085893917 | 8.51E-05 | 0.00012882 | 0.412998379 |
| Obesity | nk_cell | *LRRC36* | -0.756282233 | 0.000222848 | 0.000321038 | 1 |
| Obesity | nk_cell | *ZFYVE28* | 1.473691915 | 6.42E-09 | 4.03E-08 | 3.11E-05 |
| Obesity | nk_cell | *C16orf86* | -2.057368917 | 7.59E-09 | 4.38E-08 | 3.68E-05 |
| Obesity | nk_cell | *RGS12* | -2.037533255 | 7.68E-08 | 2.47E-07 | 0.000372517 |
| Obesity | nk_cell | *LYPD5* | -2.310713093 | 1.40E-09 | 2.11E-08 | 6.82E-06 |
| Obesity | nk_cell | *CCDC107* | -2.568613725 | 0.007562444 | 0.009386794 | 1 |
| Obesity | nk_cell | *ZNF233* | -3.325980955 | 5.68E-06 | 1.04E-05 | 0.027582968 |
| Obesity | nk_cell | *SSU72* | 2.471577765 | 0.004844953 | 0.006124991 | 1 |
| Obesity | nk_cell | *CPAMD8* | -1.299696501 | 1.53E-09 | 2.11E-08 | 7.40E-06 |
| Obesity | nk_cell | *ANKLE1* | -5.861567667 | 4.93E-05 | 7.74E-05 | 0.239259042 |
| Obesity | nk_cell | *CCDC58* | -2.451873785 | 1.03E-08 | 5.30E-08 | 4.98E-05 |
| Obesity | nk_cell | *SLC37A1* | 3.505444828 | 3.08E-05 | 4.98E-05 | 0.149658761 |
| Obesity | nk_cell | *PDE9A* | -2.110543656 | 4.55E-06 | 8.50E-06 | 0.02207488 |
| Obesity | nk_cell | *HSF2BP* | -1.968662055 | 4.53E-07 | 1.08E-06 | 0.00219891 |
| Obesity | nk_cell | *CSTB* | -1.594755123 | 0.000771742 | 0.001047996 | 1 |
| Obesity | nk_cell | *C21orf33* | 1.697925738 | 0.000577017 | 0.000793112 | 1 |
| Obesity | nk_cell | *C21orf2* | -1.262952456 | 1.53E-10 | 9.43E-09 | 7.45E-07 |
| Obesity | nk_cell | *FTCD* | -1.08068333 | 0.004828351 | 0.006105592 | 1 |
| Obesity | nk_cell | *LSS* | -0.836985899 | 0.001580307 | 0.002081903 | 1 |
| Obesity | nk_cell | *VAV2* | -1.381183616 | 2.23E-07 | 5.94E-07 | 0.001083928 |
| Obesity | nk_cell | *MCM3AP* | -2.241605686 | 7.04E-10 | 1.92E-08 | 3.41E-06 |
| Obesity | nk_cell | *DIP2A* | 1.608620913 | 0.011090499 | 0.013564684 | 1 |
| Obesity | nk_cell | *S100B* | -0.80721926 | 1.53E-05 | 2.58E-05 | 0.074307268 |
| Obesity | nk_cell | *PRMT2* | -1.457256328 | 0.00518198 | 0.006522171 | 1 |
| Obesity | nk_cell | *ZNF208* | -1.672471839 | 4.22E-07 | 1.02E-06 | 0.002046093 |
| Obesity | nk_cell | *ADAMTS13* | -1.389387656 | 3.07E-09 | 2.63E-08 | 1.49E-05 |
| Obesity | nk_cell | *CACFD1* | -6.57481251 | 8.61E-08 | 2.71E-07 | 0.000417821 |
| Obesity | nk_cell | *GPSM1* | 3.880766293 | 3.66E-06 | 6.98E-06 | 0.017774977 |
| Obesity | nk_cell | *RDH13* | 1.996318436 | 7.47E-05 | 0.000113954 | 0.362373409 |
| Obesity | nk_cell | *TAOK1* | -2.015286766 | 2.81E-06 | 5.52E-06 | 0.0136566 |
| Obesity | nk_cell | *TLCD1* | -4.567199231 | 9.70E-05 | 0.000146003 | 0.470568352 |
| Obesity | nk_cell | *S100A1* | -3.644654329 | 3.28E-06 | 6.33E-06 | 0.015914324 |
| Obesity | nk_cell | *ZBTB7B* | -2.119702071 | 3.14E-09 | 2.67E-08 | 1.52E-05 |

| | | | | | | |
|---|---|---|---|---|---|---|
| Obesity | nk_cell | *VPS11* | 2.132038919 | 0.000778062 | 0.001055988 | 1 |
| Obesity | nk_cell | *FDPS* | -2.271498656 | 0.001780308 | 0.002332717 | 1 |
| Obesity | nk_cell | *PAQR6* | 1.233395146 | 0.004491633 | 0.005696133 | 1 |
| Obesity | nk_cell | *CCR5* | 0.655393421 | 6.79E-10 | 1.91E-08 | 3.30E-06 |
| Obesity | nk_cell | *NBEAL2* | 2.108116853 | 1.15E-10 | 8.24E-09 | 5.60E-07 |
| Obesity | nk_cell | *PTH1R* | -1.797171209 | 0.000585056 | 0.000803934 | 1 |
| Obesity | nk_cell | *FCRL3* | 0.272841997 | 6.35E-10 | 1.89E-08 | 3.08E-06 |
| Obesity | nk_cell | *FGFR4* | 4.026010451 | 3.69E-06 | 7.03E-06 | 0.017908361 |
| Obesity | nk_cell | *LY6K* | -2.134835414 | 5.41E-05 | 8.43E-05 | 0.262465225 |
| Obesity | nk_cell | *CPSF4* | 2.61014553 | 2.28E-09 | 2.28E-08 | 1.10E-05 |
| Obesity | nk_cell | *VPS28* | -1.421880379 | 0.041008724 | 0.047747794 | 1 |
| Obesity | nk_cell | *TONSL* | -5.800809236 | 1.09E-09 | 2.05E-08 | 5.29E-06 |
| Obesity | nk_cell | *MUM1* | -1.79602323 | 4.86E-08 | 1.70E-07 | 0.00023567 |
| Obesity | nk_cell | *ZNF333* | 3.136751841 | 5.19E-05 | 8.11E-05 | 0.251939801 |
| Obesity | nk_cell | *SH2B2* | -2.718277775 | 2.47E-06 | 4.89E-06 | 0.012003535 |
| Obesity | nk_cell | *RPL8* | 2.163207198 | 0.000814502 | 0.00110236 | 1 |
| Obesity | nk_cell | *LRWD1* | 1.179579336 | 6.67E-05 | 0.000102275 | 0.323801982 |
| Obesity | nk_cell | *DVL3* | -2.274533337 | 5.17E-07 | 1.21E-06 | 0.002506984 |
| Obesity | nk_cell | *ABCF3* | 3.167137721 | 2.36E-06 | 4.69E-06 | 0.011450793 |
| Obesity | nk_cell | *FBXO27* | -0.875702222 | 6.76E-07 | 1.54E-06 | 0.003281008 |
| Obesity | nk_cell | *DMKN* | -0.399998904 | 0.00489587 | 0.006182915 | 1 |
| Obesity | nk_cell | *U2AF1L4* | -0.636284166 | 0.000487146 | 0.000674746 | 1 |
| Obesity | nk_cell | *BDH1* | -3.48290064 | 4.27E-06 | 8.02E-06 | 0.020728476 |
| Obesity | nk_cell | *LRRC56* | -2.532931191 | 5.69E-07 | 1.32E-06 | 0.002761522 |
| Obesity | nk_cell | *PGAP3* | 4.417536608 | 0.000437287 | 0.000607941 | 1 |
| Obesity | nk_cell | *ZNF577* | -1.627694358 | 3.83E-07 | 9.41E-07 | 0.001859272 |
| Obesity | nk_cell | *TMEM143* | -2.488522764 | 1.59E-08 | 7.32E-08 | 7.69E-05 |
| Obesity | nk_cell | *CCL16* | -2.581633566 | 5.32E-06 | 9.78E-06 | 0.025809698 |
| Obesity | nk_cell | *CCDC155* | -1.91246752 | 0.000382874 | 0.000536503 | 1 |
| Obesity | nk_cell | *DCD* | -6.282483087 | 0.000234534 | 0.000337173 | 1 |
| Obesity | nk_cell | *SIGLEC11* | -1.499576821 | 2.57E-08 | 1.05E-07 | 0.000124848 |
| Obesity | nk_cell | *ASB16* | -2.044756692 | 2.88E-09 | 2.55E-08 | 1.40E-05 |
| Obesity | nk_cell | *PLCD3* | -1.687165701 | 2.36E-09 | 2.31E-08 | 1.15E-05 |
| Obesity | nk_cell | *IP6K3* | -14.23082994 | 6.50E-09 | 4.05E-08 | 3.16E-05 |
| Obesity | nk_cell | *LEMD2* | -2.922751124 | 8.73E-09 | 4.73E-08 | 4.24E-05 |
| Obesity | nk_cell | *ASGR2* | 8.594322836 | 8.43E-10 | 1.99E-08 | 4.09E-06 |
| Obesity | nk_cell | *POLR3K* | 4.456074751 | 0.001245391 | 0.001652801 | 1 |
| Obesity | nk_cell | *SNRNP25* | -2.776304132 | 5.47E-07 | 1.27E-06 | 0.002652423 |
| Obesity | nk_cell | *WDR90* | -2.088958542 | 2.45E-10 | 1.25E-08 | 1.19E-06 |
| Obesity | nk_cell | *JMJD8* | 2.205532009 | 2.48E-07 | 6.49E-07 | 0.001201524 |
| Obesity | nk_cell | *CCDC78* | 2.815784635 | 0.002638398 | 0.003411915 | 1 |
| Obesity | nk_cell | *MEIOB* | -1.376416297 | 3.41E-08 | 1.28E-07 | 0.000165219 |
| Obesity | nk_cell | *HS3ST6* | -1.302862968 | 0.021862342 | 0.026127114 | 1 |
| Obesity | nk_cell | *SHANK2* | -1.223226914 | 3.19E-09 | 2.69E-08 | 1.55E-05 |
| Obesity | nk_cell | *TTC9C* | -2.184899744 | 0.001272098 | 0.001687321 | 1 |
| Obesity | nk_cell | *RPS6KA4* | 3.611387486 | 0.014317789 | 0.017384863 | 1 |
| Obesity | nk_cell | *TPCN2* | -1.638216754 | 1.24E-08 | 6.03E-08 | 6.00E-05 |
| Obesity | nk_cell | *CYP4A22* | -3.184762175 | 0.041016654 | 0.047747794 | 1 |
| Obesity | nk_cell | *PDZK1IP1* | -3.259408104 | 3.61E-09 | 2.86E-08 | 1.75E-05 |
| Obesity | nk_cell | *CMPK1* | -2.214740594 | 3.68E-09 | 2.88E-08 | 1.78E-05 |

| | | | | | | |
|---|---|---|---|---|---|---|
| Obesity | nk_cell | *COA7* | -5.532697791 | 1.89E-06 | 3.85E-06 | 0.009192193 |
| Obesity | nk_cell | *SLC1A7* | -1.293838116 | 1.78E-07 | 4.92E-07 | 0.000864593 |
| Obesity | nk_cell | *C1orf123* | -3.748948766 | 1.30E-05 | 2.21E-05 | 0.062980624 |
| Obesity | nk_cell | *USP24* | -1.708452865 | 0.008504578 | 0.010502472 | 1 |
| Obesity | nk_cell | *ZSWIM5* | -2.035844114 | 1.06E-06 | 2.29E-06 | 0.005133198 |
| Obesity | nk_cell | *AK4* | -1.821943094 | 7.23E-07 | 1.64E-06 | 0.003506537 |
| Obesity | nk_cell | *RBP7* | -2.924923175 | 8.35E-07 | 1.85E-06 | 0.00404986 |
| Obesity | nk_cell | *SLC25A34* | -2.156899113 | 8.75E-10 | 2.00E-08 | 4.24E-06 |
| Obesity | nk_cell | *PDPN* | -2.227943512 | 7.16E-09 | 4.27E-08 | 3.47E-05 |
| Obesity | nk_cell | *LAPTM5* | 3.065821291 | 4.92E-06 | 9.12E-06 | 0.023880238 |
| Obesity | nk_cell | *SDC3* | -2.51245936 | 2.70E-07 | 6.98E-07 | 0.001309465 |
| Obesity | nk_cell | *SYNC* | -2.052589625 | 7.50E-09 | 4.36E-08 | 3.64E-05 |
| Obesity | nk_cell | *CAMK2N1* | -1.360005207 | 2.05E-05 | 3.37E-05 | 0.099223923 |
| Obesity | nk_cell | *MEGF6* | -1.302195686 | 8.79E-09 | 4.74E-08 | 4.27E-05 |
| Obesity | nk_cell | *OMA1* | -1.976399615 | 5.31E-08 | 1.83E-07 | 0.000257534 |
| Obesity | nk_cell | *TM2D1* | -2.347539145 | 5.20E-06 | 9.58E-06 | 0.025225445 |
| Obesity | nk_cell | *NEXN* | -2.265850324 | 3.15E-05 | 5.08E-05 | 0.152740449 |
| Obesity | nk_cell | *LRRIQ3* | -0.834746296 | 1.49E-09 | 2.11E-08 | 7.21E-06 |
| Obesity | nk_cell | *TYW3* | -2.310827241 | 5.60E-09 | 3.70E-08 | 2.72E-05 |
| Obesity | nk_cell | *WDR63* | -1.586043875 | 1.04E-07 | 3.17E-07 | 0.000505737 |
| Obesity | nk_cell | *HFM1* | -2.456362728 | 1.05E-05 | 1.81E-05 | 0.050835995 |
| Obesity | nk_cell | *ARPC5* | -2.476130606 | 0.000238096 | 0.00034199 | 1 |
| Obesity | nk_cell | *ZNF496* | -2.670183377 | 1.46E-05 | 2.47E-05 | 0.070629231 |
| Obesity | nk_cell | *TRIM58* | -1.990892215 | 1.02E-07 | 3.12E-07 | 0.000495633 |
| Obesity | nk_cell | *FCRLB* | -3.507750123 | 3.13E-09 | 2.67E-08 | 1.52E-05 |
| Obesity | nk_cell | *FLVCR1* | -2.947705045 | 2.89E-09 | 2.56E-08 | 1.40E-05 |
| Obesity | nk_cell | *TDRD5* | -2.586336215 | 8.54E-10 | 2.00E-08 | 4.14E-06 |
| Obesity | nk_cell | *BROX* | -1.362796225 | 0.001160813 | 0.001543509 | 1 |
| Obesity | nk_cell | *ACP6* | -2.418444734 | 9.90E-09 | 5.16E-08 | 4.81E-05 |
| Obesity | nk_cell | *CNST* | -2.103312626 | 3.85E-07 | 9.46E-07 | 0.001869649 |
| Obesity | nk_cell | *PM20D1* | 1.622928967 | 0.027488092 | 0.032545686 | 1 |
| Obesity | nk_cell | *PKDCC* | -1.310449627 | 2.65E-08 | 1.07E-07 | 0.000128565 |
| Obesity | nk_cell | *HAAO* | -3.253322228 | 9.97E-06 | 1.74E-05 | 0.04838582 |
| Obesity | nk_cell | *B3GALNT2* | -1.611784156 | 3.52E-05 | 5.64E-05 | 0.170850262 |
| Obesity | nk_cell | *KIAA1841* | -2.722982517 | 3.84E-06 | 7.30E-06 | 0.01862091 |
| Obesity | nk_cell | *KCNJ3* | -4.084619423 | 1.99E-09 | 2.20E-08 | 9.66E-06 |
| Obesity | nk_cell | *DUSP19* | -2.465039442 | 8.58E-10 | 2.00E-08 | 4.16E-06 |
| Obesity | nk_cell | *NUP35* | -1.843711563 | 3.56E-09 | 2.83E-08 | 1.72E-05 |
| Obesity | nk_cell | *C2orf48* | 0.841855225 | 1.07E-09 | 2.05E-08 | 5.19E-06 |
| Obesity | nk_cell | *CCDC74A* | -3.11515228 | 7.31E-06 | 1.30E-05 | 0.035472814 |
| Obesity | nk_cell | *H3F3A* | 1.552140899 | 0.022967058 | 0.027386623 | 1 |
| Obesity | nk_cell | *NOSTRIN* | -1.422637882 | 2.60E-08 | 1.05E-07 | 0.000126019 |
| Obesity | nk_cell | *INHBB* | -1.782198099 | 2.51E-09 | 2.38E-08 | 1.22E-05 |
| Obesity | nk_cell | *RPRD2* | -1.878244488 | 6.33E-05 | 9.74E-05 | 0.307066138 |
| Obesity | nk_cell | *PACRGL* | -2.476458489 | 7.92E-05 | 0.000120389 | 0.384281503 |
| Obesity | nk_cell | *C1QTNF7* | -1.723260247 | 5.75E-08 | 1.94E-07 | 0.000279176 |
| Obesity | nk_cell | *ERCC3* | -2.625336927 | 5.84E-06 | 1.06E-05 | 0.028337834 |
| Obesity | nk_cell | *RNF149* | -2.283652781 | 8.91E-06 | 1.56E-05 | 0.043255551 |
| Obesity | nk_cell | *DHX57* | -2.637097295 | 9.36E-07 | 2.05E-06 | 0.004541353 |
| Obesity | nk_cell | *S100A9* | -2.12359817 | 1.05E-08 | 5.37E-08 | 5.09E-05 |

| | | | | | | |
|---|---|---|---|---|---|---|
| Obesity | nk_cell | *TDRD10* | -2.050430296 | 2.09E-08 | 8.91E-08 | 0.000101268 |
| Obesity | nk_cell | *GNPDA2* | -2.124875103 | 1.81E-05 | 3.02E-05 | 0.088052401 |
| Obesity | nk_cell | *NIPAL1* | -2.094144412 | 6.80E-09 | 4.14E-08 | 3.30E-05 |
| Obesity | nk_cell | *ANTXR2* | -1.34867843 | 3.75E-05 | 5.98E-05 | 0.18197607 |
| Obesity | nk_cell | *CGGBP1* | -1.500652862 | 4.15E-05 | 6.58E-05 | 0.201245343 |
| Obesity | nk_cell | *GPR155* | -2.799630709 | 1.77E-05 | 2.95E-05 | 0.085946149 |
| Obesity | nk_cell | *DAPL1* | -2.447130721 | 1.29E-08 | 6.24E-08 | 6.27E-05 |
| Obesity | nk_cell | *PBXIP1* | -3.240384082 | 0.00021087 | 0.000304688 | 1 |
| Obesity | nk_cell | *CLDN1* | -1.790415659 | 6.97E-07 | 1.59E-06 | 0.003383469 |
| Obesity | nk_cell | *DCST2* | -1.560425703 | 2.50E-07 | 6.54E-07 | 0.001210695 |
| Obesity | nk_cell | *LINC01116* | 6.579537289 | 1.23E-06 | 2.63E-06 | 0.005977773 |
| Obesity | nk_cell | *POGLUT1* | -1.433026069 | 0.000118879 | 0.000177205 | 0.576801627 |
| Obesity | nk_cell | *SLC22A15* | -2.042453568 | 5.09E-07 | 1.20E-06 | 0.002470964 |
| Obesity | nk_cell | *LRRC58* | -2.269464152 | 4.45E-08 | 1.59E-07 | 0.000215863 |
| Obesity | nk_cell | *LMOD1* | -1.190785067 | 1.39E-11 | 1.87E-09 | 6.75E-08 |
| Obesity | nk_cell | *TMEM169* | 1.570930142 | 2.77E-08 | 1.10E-07 | 0.00013438 |
| Obesity | nk_cell | *ARPC2* | 1.983497935 | 1.38E-07 | 3.98E-07 | 0.000668593 |
| Obesity | nk_cell | *CCT3* | -1.578736094 | 0.042588869 | 0.049459356 | 1 |
| Obesity | nk_cell | *GLB1L* | -1.64189408 | 0.03231212 | 0.037979266 | 1 |
| Obesity | nk_cell | *SERPINI1* | -2.098527854 | 2.51E-08 | 1.03E-07 | 0.000121947 |
| Obesity | nk_cell | *SUCLG1* | -2.106704044 | 1.10E-06 | 2.37E-06 | 0.005339807 |
| Obesity | nk_cell | *SPTA1* | -1.150720146 | 0.012074015 | 0.014734185 | 1 |
| Obesity | nk_cell | *PRKCI* | -1.641955518 | 8.37E-09 | 4.61E-08 | 4.06E-05 |
| Obesity | nk_cell | *AIM2* | -2.321492961 | 1.81E-09 | 2.18E-08 | 8.77E-06 |
| Obesity | nk_cell | *EFHB* | -0.51936085 | 0.002613376 | 0.003380459 | 1 |
| Obesity | nk_cell | *GTPBP8* | -3.631894098 | 1.62E-06 | 3.35E-06 | 0.007854786 |
| Obesity | nk_cell | *WDFY3* | -1.817342966 | 7.02E-07 | 1.59E-06 | 0.00340431 |
| Obesity | nk_cell | *PTPN13* | -1.380509056 | 5.40E-07 | 1.26E-06 | 0.002620008 |
| Obesity | nk_cell | *C4orf36* | -1.798318666 | 6.48E-09 | 4.05E-08 | 3.14E-05 |
| Obesity | nk_cell | *TIPARP* | -2.890126904 | 7.15E-07 | 1.62E-06 | 0.003468137 |
| Obesity | nk_cell | *RPP14* | 3.487377784 | 0.002766344 | 0.003571661 | 1 |
| Obesity | nk_cell | *DNASE1L3* | -2.060374321 | 1.60E-08 | 7.34E-08 | 7.77E-05 |
| Obesity | nk_cell | *IL17RC* | -2.943917334 | 4.16E-08 | 1.51E-07 | 0.000201971 |
| Obesity | nk_cell | *CRELD1* | -2.70299804 | 5.40E-08 | 1.85E-07 | 0.000262107 |
| Obesity | nk_cell | *TTC14* | -2.384980011 | 6.56E-07 | 1.50E-06 | 0.003184367 |
| Obesity | nk_cell | *CCDC158* | -4.272936285 | 1.13E-08 | 5.59E-08 | 5.47E-05 |
| Obesity | nk_cell | *CPA3* | -4.083235425 | 1.04E-08 | 5.34E-08 | 5.03E-05 |
| Obesity | nk_cell | *SLC4A1AP* | -2.851907192 | 1.33E-07 | 3.88E-07 | 0.000643112 |
| Obesity | nk_cell | *KIAA1143* | -1.662568234 | 1.54E-08 | 7.14E-08 | 7.48E-05 |
| Obesity | nk_cell | *CDCP1* | -1.950096543 | 0.028092988 | 0.033205158 | 1 |
| Obesity | nk_cell | *CLEC3B* | -2.941701877 | 1.95E-08 | 8.45E-08 | 9.48E-05 |
| Obesity | nk_cell | *NMNAT3* | -1.716182859 | 2.64E-08 | 1.07E-07 | 0.000128237 |
| Obesity | nk_cell | *TPRA1* | -3.319688563 | 6.59E-09 | 4.08E-08 | 3.20E-05 |
| Obesity | nk_cell | *MEAF6* | -3.462650457 | 1.47E-07 | 4.21E-07 | 0.000715277 |
| Obesity | nk_cell | *SNIP1* | -3.162249039 | 1.36E-07 | 3.95E-07 | 0.000660737 |
| Obesity | nk_cell | *DNALI1* | -2.104195193 | 3.55E-13 | 8.62E-11 | 1.72E-09 |
| Obesity | nk_cell | *SENP2* | -2.042171533 | 6.83E-08 | 2.23E-07 | 0.000331188 |
| Obesity | nk_cell | *IFT122* | -2.531181078 | 8.98E-08 | 2.81E-07 | 0.000435918 |
| Obesity | nk_cell | *RFT1* | -4.0938346 | 1.99E-11 | 2.32E-09 | 9.63E-08 |
| Obesity | nk_cell | *SFMBT1* | -2.268697355 | 2.35E-08 | 9.69E-08 | 0.000113979 |

| | | | | | | |
|---|---|---|---|---|---|---|
| Obesity | nk_cell | *UVSSA* | -2.188919577 | 0.000116582 | 0.000174049 | 0.565657939 |
| Obesity | nk_cell | *FAM208A* | -1.650109447 | 0.020488221 | 0.024563589 | 1 |
| Obesity | nk_cell | *LRPAP1* | -2.637287683 | 1.69E-06 | 3.47E-06 | 0.008186523 |
| Obesity | nk_cell | *ABLIM2* | -2.738000175 | 6.24E-05 | 9.63E-05 | 0.302726541 |
| Obesity | nk_cell | *ERMAP* | -1.349955619 | 4.12E-09 | 3.09E-08 | 2.00E-05 |
| Obesity | nk_cell | *ZNF691* | 2.213417283 | 0.001520107 | 0.002004772 | 1 |
| Obesity | nk_cell | *METAP1* | 2.632694747 | 2.62E-05 | 4.27E-05 | 0.127299185 |
| Obesity | nk_cell | *BDH2* | -1.669638976 | 1.05E-08 | 5.38E-08 | 5.11E-05 |
| Obesity | nk_cell | *CDC25A* | -4.695380872 | 8.60E-07 | 1.90E-06 | 0.00417248 |
| Obesity | nk_cell | *ZNF589* | -2.025758153 | 2.65E-05 | 4.31E-05 | 0.128506562 |
| Obesity | nk_cell | *HSPA4L* | -2.878044138 | 3.26E-05 | 5.25E-05 | 0.158327624 |
| Obesity | nk_cell | *MFSD8* | -2.420412247 | 2.38E-07 | 6.29E-07 | 0.001156267 |
| Obesity | nk_cell | *RAD54L2* | -2.685211191 | 3.02E-06 | 5.87E-06 | 0.014642986 |
| Obesity | nk_cell | *TEX264* | -3.616118499 | 6.79E-09 | 4.14E-08 | 3.30E-05 |
| Obesity | nk_cell | *ETNPPL* | 10.78907763 | 2.18E-09 | 2.26E-08 | 1.06E-05 |
| Obesity | nk_cell | *PRSS12* | -1.1503838 | 1.36E-06 | 2.87E-06 | 0.006580356 |
| Obesity | nk_cell | *HMGB2* | -2.578165243 | 9.11E-08 | 2.84E-07 | 0.000441973 |
| Obesity | nk_cell | *ANXA5* | -2.713473987 | 1.79E-06 | 3.66E-06 | 0.008683508 |
| Obesity | nk_cell | *MAP9* | 2.989977828 | 2.98E-08 | 1.16E-07 | 0.000144519 |
| Obesity | nk_cell | *CEP44* | -2.396385133 | 1.06E-08 | 5.41E-08 | 5.15E-05 |
| Obesity | nk_cell | *HPGD* | -2.856616292 | 2.48E-09 | 2.37E-08 | 1.20E-05 |
| Obesity | nk_cell | *C4orf45* | -1.532638566 | 2.74E-08 | 1.10E-07 | 0.000133103 |
| Obesity | nk_cell | *TMEM144* | -3.004762222 | 2.51E-08 | 1.03E-07 | 0.000121973 |
| Obesity | nk_cell | *IL15* | 2.618318176 | 0.000638438 | 0.000874563 | 1 |
| Obesity | nk_cell | *ARFIP1* | -1.987297501 | 1.32E-05 | 2.25E-05 | 0.064030683 |
| Obesity | nk_cell | *LSM6* | -2.273183343 | 8.40E-09 | 4.62E-08 | 4.07E-05 |
| Obesity | nk_cell | *ITGA2* | -1.39524427 | 4.80E-05 | 7.55E-05 | 0.2327928 |
| Obesity | nk_cell | *MOCS2* | -3.898763962 | 1.82E-07 | 5.00E-07 | 0.000882127 |
| Obesity | nk_cell | *NDUFAF2* | -1.443095407 | 0.019683118 | 0.02363338 | 1 |
| Obesity | nk_cell | *LMBRD2* | -3.007608712 | 9.76E-08 | 3.01E-07 | 0.000473532 |
| Obesity | nk_cell | *RNF180* | -2.417194782 | 3.91E-08 | 1.44E-07 | 0.000189633 |
| Obesity | nk_cell | *F2RL2* | 1.181027508 | 1.16E-09 | 2.05E-08 | 5.64E-06 |
| Obesity | nk_cell | *CMBL* | -2.179398537 | 1.98E-08 | 8.52E-08 | 9.59E-05 |
| Obesity | nk_cell | *C5orf63* | -2.589327754 | 0.000357649 | 0.000503135 | 1 |
| Obesity | nk_cell | *WDR41* | -1.970598362 | 4.61E-07 | 1.10E-06 | 0.00223902 |
| Obesity | nk_cell | *NDUFS4* | -1.901693145 | 1.13E-07 | 3.38E-07 | 0.000546524 |
| Obesity | nk_cell | *SERINC5* | -2.03103433 | 3.90E-07 | 9.53E-07 | 0.001890916 |
| Obesity | nk_cell | *PRIMPOL* | -2.38090509 | 3.38E-06 | 6.48E-06 | 0.016386598 |
| Obesity | nk_cell | *ERAP1* | -2.651048193 | 9.58E-06 | 1.67E-05 | 0.046484487 |
| Obesity | nk_cell | *ERAP2* | 1.212334481 | 2.28E-05 | 3.73E-05 | 0.110500468 |
| Obesity | nk_cell | *UTP15* | -2.710559615 | 2.30E-09 | 2.29E-08 | 1.12E-05 |
| Obesity | nk_cell | *TLR3* | -2.91590133 | 7.58E-09 | 4.38E-08 | 3.68E-05 |
| Obesity | nk_cell | *CCDC127* | 3.535534865 | 0.001874194 | 0.002453085 | 1 |
| Obesity | nk_cell | *GRIK2* | -1.022261008 | 1.03E-07 | 3.15E-07 | 0.00050209 |
| Obesity | nk_cell | *DCBLD1* | -1.682152335 | 1.21E-05 | 2.08E-05 | 0.058817058 |
| Obesity | nk_cell | *PDSS2* | -2.04663633 | 0.000158117 | 0.000231848 | 0.767184706 |
| Obesity | nk_cell | *RAET1E* | -1.677242923 | 2.18E-09 | 2.26E-08 | 1.06E-05 |
| Obesity | nk_cell | *DAGLB* | -1.055093912 | 0.000177183 | 0.000258244 | 0.859693422 |
| Obesity | nk_cell | *STK17A* | 0.715717725 | 1.44E-09 | 2.11E-08 | 7.01E-06 |
| Obesity | nk_cell | *GALNT10* | 1.739433366 | 0.000689061 | 0.000938609 | 1 |

| | | | | | | |
|---|---|---|---|---|---|---|
| Obesity | nk_cell | *RPS14* | 2.132306187 | 6.33E-06 | 1.15E-05 | 0.030722942 |
| Obesity | nk_cell | *RP9* | 3.106662165 | 3.59E-08 | 1.34E-07 | 0.000174168 |
| Obesity | nk_cell | *CAMLG* | -2.00585818 | 7.93E-06 | 1.40E-05 | 0.038462799 |
| Obesity | nk_cell | *RELL2* | 0.893149215 | 3.79E-09 | 2.94E-08 | 1.84E-05 |
| Obesity | nk_cell | *MIOS* | -2.328949695 | 8.36E-08 | 2.65E-07 | 0.000405863 |
| Obesity | nk_cell | *USP49* | -2.340959568 | 4.07E-08 | 1.49E-07 | 0.000197443 |
| Obesity | nk_cell | *CHMP4C* | -10.5251068 | 0.004575508 | 0.005797954 | 1 |
| Obesity | nk_cell | *SLC13A4* | -2.468134883 | 1.03E-07 | 3.14E-07 | 0.000499718 |
| Obesity | nk_cell | *BRI3* | -2.451250938 | 8.84E-06 | 1.55E-05 | 0.04289896 |
| Obesity | nk_cell | *CTSB* | -1.608382908 | 1.47E-08 | 6.87E-08 | 7.12E-05 |
| Obesity | nk_cell | *SOX17* | -2.539109585 | 3.51E-07 | 8.73E-07 | 0.001702274 |
| Obesity | nk_cell | *RAD21* | -2.422669923 | 4.88E-05 | 7.67E-05 | 0.236659615 |
| Obesity | nk_cell | *TNFRSF11B* | -1.647984934 | 2.21E-09 | 2.27E-08 | 1.07E-05 |
| Obesity | nk_cell | *DEFB1* | -3.318280097 | 0.000996322 | 0.001334295 | 1 |
| Obesity | nk_cell | *SUN1* | -1.760090615 | 2.98E-07 | 7.55E-07 | 0.001443715 |
| Obesity | nk_cell | *OXR1* | -1.713544303 | 1.22E-07 | 3.61E-07 | 0.000591743 |
| Obesity | nk_cell | *INTS1* | -2.785146603 | 0.017863032 | 0.021538626 | 1 |
| Obesity | nk_cell | *FASTK* | -3.094113472 | 3.96E-08 | 1.46E-07 | 0.000192153 |
| Obesity | nk_cell | *ALDH7A1* | -2.192401364 | 8.75E-07 | 1.94E-06 | 0.004247825 |
| Obesity | nk_cell | *YWHAZ* | -2.568107708 | 8.36E-08 | 2.65E-07 | 0.000405432 |
| Obesity | nk_cell | *FZD6* | -1.494506674 | 1.47E-05 | 2.48E-05 | 0.071255823 |
| Obesity | nk_cell | *SLC25A32* | -2.777411355 | 0.000410903 | 0.000573396 | 1 |
| Obesity | nk_cell | *DCSTAMP* | -15.62086256 | 1.94E-08 | 8.42E-08 | 9.40E-05 |
| Obesity | nk_cell | *INTS8* | -2.511819005 | 3.89E-06 | 7.39E-06 | 0.018871408 |
| Obesity | nk_cell | *RPP25L* | -3.147283413 | 5.48E-12 | 9.85E-10 | 2.66E-08 |
| Obesity | nk_cell | *C9orf24* | 4.382227433 | 5.23E-07 | 1.23E-06 | 0.002537554 |
| Obesity | nk_cell | *CCDC171* | -2.402392367 | 6.45E-06 | 1.16E-05 | 0.031313737 |
| Obesity | nk_cell | *METTL2B* | -0.698550145 | 0.02632796 | 0.031263647 | 1 |
| Obesity | nk_cell | *ZMAT4* | 1.73705477 | 1.39E-09 | 2.11E-08 | 6.74E-06 |
| Obesity | nk_cell | *C8orf34* | -1.661057243 | 4.36E-10 | 1.64E-08 | 2.12E-06 |
| Obesity | nk_cell | *RASEF* | -1.458133166 | 2.23E-05 | 3.66E-05 | 0.108051305 |
| Obesity | nk_cell | *ANKS6* | -1.514423714 | 9.74E-07 | 2.13E-06 | 0.004727956 |
| Obesity | nk_cell | *AQP7* | -0.985212342 | 1.93E-07 | 5.24E-07 | 0.000934682 |
| Obesity | nk_cell | *NOL6* | -1.587491929 | 2.53E-07 | 6.63E-07 | 0.001229651 |
| Obesity | nk_cell | *AQP3* | -3.224855219 | 1.59E-09 | 2.11E-08 | 7.71E-06 |
| Obesity | nk_cell | *TRMT10B* | -1.793383958 | 0.0172721 | 0.02085201 | 1 |
| Obesity | nk_cell | *PIGO* | 1.99985022 | 6.29E-09 | 3.99E-08 | 3.05E-05 |
| Obesity | nk_cell | *WRN* | -2.057091634 | 3.65E-06 | 6.95E-06 | 0.017701211 |
| Obesity | nk_cell | *TSHR* | -1.998893928 | 9.30E-08 | 2.90E-07 | 0.000451312 |
| Obesity | nk_cell | *CFL2* | -3.072582336 | 2.18E-07 | 5.83E-07 | 0.001058911 |
| Obesity | nk_cell | *SLC16A9* | -4.145933574 | 2.91E-08 | 1.14E-07 | 0.000140999 |
| Obesity | nk_cell | *GJB2* | -16.82943997 | 2.75E-08 | 1.10E-07 | 0.000133225 |
| Obesity | nk_cell | *CRYL1* | -2.567371769 | 2.88E-07 | 7.34E-07 | 0.001397187 |
| Obesity | nk_cell | *REEP3* | -2.263935723 | 3.24E-08 | 1.23E-07 | 0.000157163 |
| Obesity | nk_cell | *LRR1* | -2.64363289 | 4.27E-09 | 3.16E-08 | 2.07E-05 |
| Obesity | nk_cell | *C10orf25* | -0.656080019 | 0.014204347 | 0.017251438 | 1 |
| Obesity | nk_cell | *TMEM63C* | 8.155478912 | 1.47E-07 | 4.20E-07 | 0.000712926 |
| Obesity | nk_cell | *NOXRED1* | -2.730521826 | 4.24E-07 | 1.02E-06 | 0.002057764 |
| Obesity | nk_cell | *AKR1E2* | -1.592131699 | 4.63E-09 | 3.29E-08 | 2.25E-05 |
| Obesity | nk_cell | *OXGR1* | 2.577032827 | 3.84E-09 | 2.95E-08 | 1.86E-05 |

| | | | | | | |
|---|---|---|---|---|---|---|
| Obesity | nk_cell | *ATP5C1* | -2.585725078 | 1.17E-05 | 2.02E-05 | 0.056987631 |
| Obesity | nk_cell | *VDAC2* | -2.74284581 | 0.010771857 | 0.013212846 | 1 |
| Obesity | nk_cell | *PDZD8* | -1.438503554 | 0.028763633 | 0.033973016 | 1 |
| Obesity | nk_cell | *QSOX2* | -2.821007978 | 0.032103463 | 0.037761455 | 1 |
| Obesity | nk_cell | *CLEC1B* | -2.30818034 | 0.00016703 | 0.000244106 | 0.810431359 |
| Obesity | nk_cell | *SNAPC4* | -1.479792378 | 5.04E-09 | 3.50E-08 | 2.45E-05 |
| Obesity | nk_cell | *TSC1* | -1.266027436 | 0.00241693 | 0.003134709 | 1 |
| Obesity | nk_cell | *STOX1* | -1.097209414 | 7.54E-05 | 0.000114937 | 0.365845347 |
| Obesity | nk_cell | *RET* | -2.196198126 | 2.91E-09 | 2.56E-08 | 1.41E-05 |
| Obesity | nk_cell | *CASP7* | -2.56772381 | 2.58E-07 | 6.71E-07 | 0.001250059 |
| Obesity | nk_cell | *PPP1R36* | -1.793577074 | 1.60E-09 | 2.11E-08 | 7.76E-06 |
| Obesity | nk_cell | *VWA2* | -2.691190779 | 2.70E-08 | 1.08E-07 | 0.000131185 |
| Obesity | nk_cell | *METTL3* | -2.750974433 | 2.72E-07 | 7.03E-07 | 0.001321969 |
| Obesity | nk_cell | *TRUB1* | -3.056707919 | 5.10E-06 | 9.43E-06 | 0.024747049 |
| Obesity | nk_cell | *HSPA12A* | -1.633456571 | 1.67E-08 | 7.55E-08 | 8.10E-05 |
| Obesity | nk_cell | *UBTD1* | -1.65378967 | 6.22E-09 | 3.97E-08 | 3.02E-05 |
| Obesity | nk_cell | *OTOGL* | 2.889688194 | 0.003939017 | 0.005013671 | 1 |
| Obesity | nk_cell | *TTC7B* | -2.422016045 | 7.30E-07 | 1.65E-06 | 0.003539835 |
| Obesity | nk_cell | *SLC39A13* | 5.267686624 | 6.33E-05 | 9.74E-05 | 0.307115047 |
| Obesity | nk_cell | *PSMC3* | -2.896630179 | 1.76E-07 | 4.86E-07 | 0.000851869 |
| Obesity | nk_cell | *RAPSN* | -4.235916013 | 1.44E-22 | 2.93E-19 | 6.99E-19 |
| Obesity | nk_cell | *TC2N* | 1.537381923 | 6.62E-06 | 1.19E-05 | 0.032124129 |
| Obesity | nk_cell | *CPSF2* | -1.298946641 | 1.46E-06 | 3.07E-06 | 0.007102281 |
| Obesity | nk_cell | *IFI27L1* | -2.073301181 | 1.06E-08 | 5.41E-08 | 5.17E-05 |
| Obesity | nk_cell | *C1QL3* | -1.566781614 | 0.000934866 | 0.001256501 | 1 |
| Obesity | nk_cell | *ARL5B* | 3.565980649 | 5.50E-05 | 8.57E-05 | 0.266864716 |
| Obesity | nk_cell | *LIPC* | -2.333061984 | 7.41E-07 | 1.67E-06 | 0.003595646 |
| Obesity | nk_cell | *CEP57* | -2.308630982 | 0.000315892 | 0.000447506 | 1 |
| Obesity | nk_cell | *TCP11L2* | -1.659841651 | 1.11E-08 | 5.54E-08 | 5.38E-05 |
| Obesity | nk_cell | *JAM3* | -2.273279851 | 9.44E-09 | 5.00E-08 | 4.58E-05 |
| Obesity | nk_cell | *IKBIP* | 3.272620251 | 0.000164778 | 0.000241031 | 0.799500618 |
| Obesity | nk_cell | *ZFYVE19* | -1.630505217 | 0.001009131 | 0.001350705 | 1 |
| Obesity | nk_cell | *TRMT61A* | -1.574207311 | 1.99E-09 | 2.20E-08 | 9.63E-06 |
| Obesity | nk_cell | *LARP6* | -2.158471856 | 5.10E-08 | 1.77E-07 | 0.000247432 |
| Obesity | nk_cell | *API5* | -2.759397551 | 7.02E-06 | 1.26E-05 | 0.034040286 |
| Obesity | nk_cell | *ASPG* | 9.735097004 | 1.72E-09 | 2.16E-08 | 8.34E-06 |
| Obesity | nk_cell | *GABRB3* | -1.882563384 | 0.000399914 | 0.000559027 | 1 |
| Obesity | nk_cell | *COX11* | -2.895345463 | 4.57E-07 | 1.09E-06 | 0.002215682 |
| Obesity | nk_cell | *ZNF202* | -2.689046715 | 3.79E-09 | 2.94E-08 | 1.84E-05 |
| Obesity | nk_cell | *STXBP4* | -2.519793129 | 7.15E-07 | 1.62E-06 | 0.003469278 |
| Obesity | nk_cell | *WBP1L* | -2.240513066 | 1.05E-05 | 1.82E-05 | 0.051059368 |
| Obesity | nk_cell | *APBB1* | -3.309539052 | 2.30E-08 | 9.56E-08 | 0.000111644 |
| Obesity | nk_cell | *NUDT13* | -1.934233717 | 4.43E-05 | 6.99E-05 | 0.214836289 |
| Obesity | nk_cell | *WDR88* | -2.475833651 | 2.59E-08 | 1.05E-07 | 0.000125812 |
| Obesity | nk_cell | *CYB5R2* | -1.648343448 | 2.27E-07 | 6.04E-07 | 0.00110355 |
| Obesity | nk_cell | *SERPINB8* | -1.573694965 | 2.94E-06 | 5.74E-06 | 0.01427213 |
| Obesity | nk_cell | *XRRA1* | -1.861915592 | 2.84E-07 | 7.26E-07 | 0.001378606 |
| Obesity | nk_cell | *CDYL2* | -0.994671491 | 0.021110196 | 0.025271816 | 1 |
| Obesity | nk_cell | *CENPN* | -3.67225079 | 5.57E-09 | 3.68E-08 | 2.70E-05 |
| Obesity | nk_cell | *C16orf46* | -4.333310335 | 0.000232089 | 0.000334053 | 1 |

| | | | | | | |
|---|---|---|---|---|---|---|
| Obesity | nk_cell | *TMEM41B* | -2.651668744 | 1.06E-06 | 2.29E-06 | 0.005124642 |
| Obesity | nk_cell | *TMX3* | -2.580756524 | 1.14E-05 | 1.96E-05 | 0.055354892 |
| Obesity | nk_cell | *PRKCB* | 0.597568164 | 7.51E-10 | 1.93E-08 | 3.64E-06 |
| Obesity | nk_cell | *NDST2* | 4.485596609 | 1.42E-09 | 2.11E-08 | 6.91E-06 |
| Obesity | nk_cell | *CLEC4D* | -5.633202749 | 6.94E-08 | 2.26E-07 | 0.000336774 |
| Obesity | nk_cell | *ZSCAN21* | -2.444338183 | 2.59E-06 | 5.11E-06 | 0.012587672 |
| Obesity | nk_cell | *RIMKLB* | -2.009506987 | 5.11E-10 | 1.75E-08 | 2.48E-06 |
| Obesity | nk_cell | *TMED3* | -1.079293885 | 0.00107535 | 0.001435773 | 1 |
| Obesity | nk_cell | *SEC11C* | 4.056181516 | 3.55E-06 | 6.78E-06 | 0.017212644 |
| Obesity | nk_cell | *GALR1* | -1.07608573 | 0.025582924 | 0.030386377 | 1 |
| Obesity | nk_cell | *CENPV* | -3.069837591 | 1.95E-08 | 8.43E-08 | 9.45E-05 |
| Obesity | nk_cell | *FAM96B* | -1.917646835 | 0.015118926 | 0.018320936 | 1 |
| Obesity | nk_cell | *SPDYE6* | 1.15006268 | 0.011995395 | 0.014641926 | 1 |
| Obesity | nk_cell | *ZNF592* | -2.117611973 | 5.40E-08 | 1.85E-07 | 0.000262069 |
| Obesity | nk_cell | *ACSM1* | -2.065379299 | 1.28E-08 | 6.18E-08 | 6.19E-05 |
| Obesity | nk_cell | *SLFN5* | 2.38196192 | 0.000282084 | 0.000401723 | 1 |
| Obesity | nk_cell | *SAAL1* | 4.199524607 | 6.35E-06 | 1.15E-05 | 0.030824831 |
| Obesity | nk_cell | *LDHC* | 1.322269242 | 5.32E-05 | 8.30E-05 | 0.258274623 |
| Obesity | nk_cell | *FAM96A* | -1.762038904 | 6.81E-09 | 4.14E-08 | 3.30E-05 |
| Obesity | nk_cell | *FAM111A* | -2.332737154 | 1.64E-05 | 2.74E-05 | 0.079333863 |
| Obesity | nk_cell | *TMEM170A* | -2.507338765 | 5.63E-08 | 1.91E-07 | 0.00027326 |
| Obesity | nk_cell | *ANPEP* | -1.022291144 | 0.000175804 | 0.000256311 | 0.853002477 |
| Obesity | nk_cell | *STAT6* | -2.218494422 | 0.001625946 | 0.002139124 | 1 |
| Obesity | nk_cell | *STX3* | 2.006034541 | 0.022292747 | 0.026615258 | 1 |
| Obesity | nk_cell | *PIP4K2C* | 4.344465663 | 6.36E-10 | 1.89E-08 | 3.09E-06 |
| Obesity | nk_cell | *MTMR10* | -1.983326858 | 5.20E-06 | 9.58E-06 | 0.025235595 |
| Obesity | nk_cell | *NYAP1* | -1.319910616 | 4.60E-06 | 8.57E-06 | 0.022295345 |
| Obesity | nk_cell | *MS4A7* | -1.436017471 | 0.000978257 | 0.001311189 | 1 |
| Obesity | nk_cell | *MS4A14* | -1.093668134 | 3.89E-08 | 1.44E-07 | 0.000188546 |
| Obesity | nk_cell | *CCNDBP1* | 2.802902771 | 0.001125261 | 0.0014987 | 1 |
| Obesity | nk_cell | *SMAD3* | -2.249941687 | 0.000129888 | 0.000192551 | 0.630218831 |
| Obesity | nk_cell | *MAP1A* | -2.110801076 | 2.47E-09 | 2.37E-08 | 1.20E-05 |
| Obesity | nk_cell | *RCCD1* | -2.232809545 | 8.47E-09 | 4.63E-08 | 4.11E-05 |
| Obesity | nk_cell | *AKTIP* | 2.07706359 | 1.07E-08 | 5.42E-08 | 5.19E-05 |
| Obesity | nk_cell | *MEI1* | 1.377590099 | 1.66E-10 | 9.97E-09 | 8.03E-07 |
| Obesity | nk_cell | *SAMD14* | 2.256593224 | 2.55E-10 | 1.28E-08 | 1.24E-06 |
| Obesity | nk_cell | *ACSF2* | -2.367381656 | 0.001553348 | 0.002046943 | 1 |
| Obesity | nk_cell | *URM1* | -3.222814234 | 8.93E-05 | 0.000134746 | 0.433073798 |
| Obesity | nk_cell | *ISLR2* | -1.843088002 | 2.32E-05 | 3.79E-05 | 0.112375278 |
| Obesity | nk_cell | *SP2* | -3.001524418 | 1.37E-07 | 3.98E-07 | 0.000665895 |
| Obesity | nk_cell | *COQ7* | -2.376453516 | 3.42E-07 | 8.54E-07 | 0.001661302 |
| Obesity | nk_cell | *GPRC5B* | -1.827749798 | 2.91E-07 | 7.41E-07 | 0.001411893 |
| Obesity | nk_cell | *TBC1D2B* | -1.460716637 | 2.30E-09 | 2.29E-08 | 1.12E-05 |
| Obesity | nk_cell | *LOXHD1* | -2.188988954 | 1.97E-05 | 3.25E-05 | 0.095490633 |
| Obesity | nk_cell | *KATNAL2* | -1.777674989 | 2.89E-06 | 5.64E-06 | 0.01399881 |
| Obesity | nk_cell | *HDHD2* | 1.341083582 | 0.00053377 | 0.000735335 | 1 |
| Obesity | nk_cell | *ZNF91* | -1.964067008 | 1.09E-06 | 2.34E-06 | 0.005268468 |
| Obesity | nk_cell | *ENGASE* | -1.306689999 | 1.21E-05 | 2.07E-05 | 0.058744606 |
| Obesity | nk_cell | *MYO5B* | -1.810835271 | 5.67E-09 | 3.74E-08 | 2.75E-05 |
| Obesity | nk_cell | *ZNF23* | -2.656656037 | 3.38E-09 | 2.77E-08 | 1.64E-05 |

| | | | | | | |
|---|---|---|---|---|---|---|
| Obesity | nk_cell | *SMG8* | -3.263215725 | 2.91E-11 | 3.00E-09 | 1.41E-07 |
| Obesity | nk_cell | *RAB8A* | 1.96669699 | 7.54E-09 | 4.37E-08 | 3.66E-05 |
| Obesity | nk_cell | *JSRP1* | -2.881668891 | 4.84E-05 | 7.62E-05 | 0.234896638 |
| Obesity | nk_cell | *PROCA1* | 1.627337843 | 0.016708977 | 0.02018724 | 1 |
| Obesity | nk_cell | *RPL13* | 1.89919999 | 0.00042746 | 0.000595051 | 1 |
| Obesity | nk_cell | *TP53I13* | -2.655393072 | 9.54E-06 | 1.66E-05 | 0.046268569 |
| Obesity | nk_cell | *TUBA1C* | -1.845563078 | 8.30E-06 | 1.46E-05 | 0.040256846 |
| Obesity | nk_cell | *ZNF528* | -1.905221358 | 7.91E-09 | 4.47E-08 | 3.84E-05 |
| Obesity | nk_cell | *SERTAD3* | -4.508008541 | 5.41E-08 | 1.85E-07 | 0.000262661 |
| Obesity | nk_cell | *TMC4* | -2.486649158 | 3.37E-08 | 1.27E-07 | 0.000163311 |
| Obesity | nk_cell | *TTYH1* | -2.748941903 | 3.91E-09 | 3.00E-08 | 1.90E-05 |
| Obesity | nk_cell | *LENG8* | -1.836500225 | 2.69E-08 | 1.08E-07 | 0.000130745 |
| Obesity | nk_cell | *CDC42EP5* | -2.222424919 | 1.64E-08 | 7.44E-08 | 7.94E-05 |
| Obesity | nk_cell | *KIR3DL1* | 2.987000249 | 2.38E-09 | 2.32E-08 | 1.15E-05 |
| Obesity | nk_cell | *C19orf33* | -1.538364807 | 7.36E-06 | 1.31E-05 | 0.035726195 |
| Obesity | nk_cell | *DNAAF3* | -1.476132865 | 9.38E-06 | 1.64E-05 | 0.045533098 |
| Obesity | nk_cell | *CHAF1A* | -0.781070352 | 0.032324637 | 0.037984776 | 1 |
| Obesity | nk_cell | *FAM57A* | -1.424292139 | 0.002445496 | 0.003170063 | 1 |
| Obesity | nk_cell | *KIFC2* | -1.412743955 | 7.68E-09 | 4.40E-08 | 3.73E-05 |
| Obesity | nk_cell | *SRR* | 1.158626148 | 0.001143628 | 0.001521909 | 1 |
| Obesity | nk_cell | *TRPV3* | -1.420968262 | 0.001602135 | 0.002109529 | 1 |
| Obesity | nk_cell | *CYB5D2* | -2.605300687 | 2.35E-06 | 4.67E-06 | 0.011391537 |
| Obesity | nk_cell | *KLK5* | -1.752175374 | 6.61E-10 | 1.90E-08 | 3.20E-06 |
| Obesity | nk_cell | *KLK6* | -8.76794093 | 1.64E-09 | 2.14E-08 | 7.96E-06 |
| Obesity | nk_cell | *ZNF83* | -1.943374844 | 1.57E-05 | 2.63E-05 | 0.075943942 |
| Obesity | nk_cell | *SPRYD3* | -4.625488028 | 2.13E-08 | 9.04E-08 | 0.000103183 |
| Obesity | nk_cell | *SOAT2* | -2.53790632 | 2.45E-07 | 6.44E-07 | 0.001191106 |
| Obesity | nk_cell | *NDUFV1* | -2.755047245 | 6.49E-05 | 9.97E-05 | 0.314948763 |
| Obesity | nk_cell | *PRDX2* | -2.729657316 | 0.005176697 | 0.006517212 | 1 |
| Obesity | nk_cell | *MIS12* | -3.283882608 | 5.59E-05 | 8.71E-05 | 0.271428828 |
| Obesity | nk_cell | *CD300C* | -0.853096128 | 0.005869258 | 0.007349068 | 1 |
| Obesity | nk_cell | *ATP5H* | 2.686410677 | 0.004486933 | 0.00569166 | 1 |
| Obesity | nk_cell | *MGAT5B* | -2.818358227 | 5.25E-09 | 3.56E-08 | 2.55E-05 |
| Obesity | nk_cell | *GSDMA* | -1.189328187 | 0.00035713 | 0.00050255 | 1 |
| Obesity | nk_cell | *TMEM99* | -2.047505451 | 8.92E-06 | 1.56E-05 | 0.043285904 |
| Obesity | nk_cell | *ZNF598* | -2.021838203 | 2.73E-07 | 7.03E-07 | 0.001323015 |
| Obesity | nk_cell | *MLST8* | -1.617056136 | 1.90E-09 | 2.19E-08 | 9.23E-06 |
| Obesity | nk_cell | *ECI1* | -1.5283405 | 0.009349984 | 0.011517168 | 1 |
| Obesity | nk_cell | *ABCA3* | -2.409214923 | 7.26E-06 | 1.30E-05 | 0.035211568 |
| Obesity | nk_cell | *ATG16L2* | -2.737126173 | 0.0005023 | 0.00069415 | 1 |
| Obesity | nk_cell | *RPSA* | 2.173860289 | 4.41E-05 | 6.96E-05 | 0.213752567 |
| Obesity | nk_cell | *ULK4* | -1.714011501 | 4.02E-06 | 7.62E-06 | 0.019526751 |
| Obesity | nk_cell | *NAALADL1* | 1.792606865 | 6.22E-08 | 2.06E-07 | 0.000301868 |
| Obesity | nk_cell | *SCARA5* | -0.932232421 | 3.26E-09 | 2.72E-08 | 1.58E-05 |
| Obesity | nk_cell | *KIAA1586* | -2.693266613 | 2.93E-10 | 1.36E-08 | 1.42E-06 |
| Obesity | nk_cell | *FAM83B* | -11.39103326 | 4.70E-10 | 1.69E-08 | 2.28E-06 |
| Obesity | nk_cell | *NT5DC2* | -1.660729726 | 0.005814447 | 0.007282317 | 1 |
| Obesity | nk_cell | *COA6* | 2.975598624 | 6.94E-09 | 4.19E-08 | 3.37E-05 |
| Obesity | nk_cell | *PXK* | -2.235046528 | 2.04E-08 | 8.72E-08 | 9.89E-05 |
| Obesity | nk_cell | *PCMTD1* | -2.086815344 | 3.21E-05 | 5.17E-05 | 0.155753135 |

| | | | | | | |
|---|---|---|---|---|---|---|
| Obesity | nk_cell | *ACOX2* | -1.543819905 | 2.05E-09 | 2.21E-08 | 9.93E-06 |
| Obesity | nk_cell | *SCN11A* | -1.313313768 | 1.41E-07 | 4.06E-07 | 0.00068595 |
| Obesity | nk_cell | *SEPT2* | -0.739569037 | 0.030516277 | 0.035946826 | 1 |
| Obesity | nk_cell | *DTYMK* | -4.534316717 | 7.61E-06 | 1.35E-05 | 0.036907357 |
| Obesity | nk_cell | *ING5* | -1.86928499 | 4.30E-07 | 1.04E-06 | 0.002087116 |
| Obesity | nk_cell | *RFWD3* | -2.779327959 | 3.25E-05 | 5.24E-05 | 0.157922595 |
| Obesity | nk_cell | *KLHL30* | -8.922496902 | 2.68E-09 | 2.45E-08 | 1.30E-05 |
| Obesity | nk_cell | *CCDC110* | -2.185330277 | 1.58E-09 | 2.11E-08 | 7.68E-06 |
| Obesity | nk_cell | *FNTA* | -1.518485913 | 2.43E-05 | 3.97E-05 | 0.117856456 |
| Obesity | nk_cell | *SERINC2* | -5.994004438 | 9.05E-11 | 6.97E-09 | 4.39E-07 |
| Obesity | nk_cell | *SLC20A2* | -2.191612453 | 0.000145985 | 0.000214903 | 0.70832155 |
| Obesity | nk_cell | *TMUB2* | -2.174146304 | 6.68E-09 | 4.10E-08 | 3.24E-05 |
| Obesity | nk_cell | *ZSWIM1* | 1.091864798 | 1.61E-06 | 3.34E-06 | 0.007835076 |
| Obesity | nk_cell | *NBPF9* | -1.450071422 | 1.70E-09 | 2.16E-08 | 8.27E-06 |
| Obesity | nk_cell | *ADAM9* | -2.051326969 | 3.55E-07 | 8.83E-07 | 0.001721928 |
| Obesity | nk_cell | *NDUFS5* | 2.063154435 | 0.023251363 | 0.027698407 | 1 |
| Obesity | nk_cell | *ZNF30* | -1.546232663 | 3.99E-07 | 9.72E-07 | 0.001933928 |
| Obesity | nk_cell | *SLC16A4* | -1.183878946 | 0.005059751 | 0.006379472 | 1 |
| Obesity | nk_cell | *NPNT* | -0.988679149 | 0.00287586 | 0.00370714 | 1 |
| Obesity | nk_cell | *GSTM4* | 4.394985688 | 0.000348843 | 0.000491602 | 1 |
| Obesity | nk_cell | *TCTN2* | -0.969411391 | 1.95E-09 | 2.20E-08 | 9.47E-06 |
| Obesity | nk_cell | *PPIP5K1* | -2.904669518 | 5.62E-05 | 8.73E-05 | 0.272593672 |
| Obesity | nk_cell | *ADAL* | -0.59792252 | 0.034328109 | 0.040212454 | 1 |
| Obesity | nk_cell | *SNTB2* | -2.20184963 | 8.32E-07 | 1.85E-06 | 0.004034782 |
| Obesity | nk_cell | *ZNF507* | -3.086534667 | 1.47E-05 | 2.49E-05 | 0.071302966 |
| Obesity | nk_cell | *STX18* | -2.44527116 | 1.26E-06 | 2.67E-06 | 0.006104917 |
| Obesity | nk_cell | *ZBTB49* | -3.23890994 | 8.03E-08 | 2.56E-07 | 0.000389417 |
| Obesity | nk_cell | *GFM1* | -1.589477266 | 8.18E-06 | 1.44E-05 | 0.039703538 |
| Obesity | nk_cell | *TMEM129* | -2.802509407 | 0.000508986 | 0.000702789 | 1 |
| Obesity | nk_cell | *CEP120* | -2.651514176 | 8.79E-07 | 1.94E-06 | 0.004264403 |
| Obesity | nk_cell | *MFF* | -2.362061599 | 4.18E-06 | 7.86E-06 | 0.02026314 |
| Obesity | nk_cell | *LGALS9* | -1.013490887 | 0.019525499 | 0.023454926 | 1 |
| Obesity | nk_cell | *CPLX1* | -1.066292328 | 8.88E-05 | 0.000134191 | 0.430886262 |
| Obesity | nk_cell | *SIGLEC7* | -1.947552316 | 0.000137226 | 0.000202829 | 0.66581997 |
| Obesity | nk_cell | *MFSD7* | -4.117822066 | 9.25E-09 | 4.93E-08 | 4.49E-05 |
| Obesity | nk_cell | *KLK7* | 2.426823724 | 6.00E-10 | 1.84E-08 | 2.91E-06 |
| Obesity | nk_cell | *UPF3A* | -2.152420495 | 5.40E-07 | 1.26E-06 | 0.002619504 |
| Obesity | nk_cell | *HSPBAP1* | -2.577604453 | 3.18E-06 | 6.16E-06 | 0.015453233 |
| Obesity | nk_cell | *UBE2V2* | -2.60729541 | 3.92E-07 | 9.58E-07 | 0.00190256 |
| Obesity | nk_cell | *CPT1C* | 2.223354914 | 0.001097388 | 0.001463988 | 1 |
| Obesity | nk_cell | *PCSK9* | 8.346958342 | 1.15E-07 | 3.44E-07 | 0.000556959 |
| Obesity | nk_cell | *XPO6* | -2.770721405 | 3.78E-07 | 9.31E-07 | 0.001832028 |
| Obesity | nk_cell | *RGS14* | 0.761995659 | 4.36E-09 | 3.19E-08 | 2.12E-05 |
| Obesity | nk_cell | *RAB24* | -2.521148997 | 1.43E-09 | 2.11E-08 | 6.92E-06 |
| Obesity | nk_cell | *THBS3* | -1.94209073 | 6.59E-08 | 2.17E-07 | 0.00031992 |
| Obesity | nk_cell | *SH3TC2* | -1.234659772 | 0.021684858 | 0.02592778 | 1 |
| Obesity | nk_cell | *GPRIN1* | 4.394144907 | 0.004328932 | 0.005496984 | 1 |
| Obesity | nk_cell | *P2RY12* | -1.871784151 | 0.022396284 | 0.026732293 | 1 |
| Obesity | nk_cell | *SNUPN* | 1.957900473 | 0.020751948 | 0.024861346 | 1 |
| Obesity | nk_cell | *RNASE6* | -1.39767576 | 0.013572567 | 0.01650065 | 1 |

| | | | | | | |
|---|---|---|---|---|---|---|
| Obesity | nk_cell | *CD52* | 0.375718536 | 1.08E-09 | 2.05E-08 | 5.24E-06 |
| Obesity | nk_cell | *SLC38A11* | -1.198003493 | 1.01E-08 | 5.26E-08 | 4.91E-05 |
| Obesity | nk_cell | *CCDC8* | -1.715118502 | 4.62E-09 | 3.29E-08 | 2.24E-05 |
| Obesity | nk_cell | *ZEB2* | -1.895083505 | 1.01E-06 | 2.20E-06 | 0.004901248 |
| Obesity | nk_cell | *DTWD2* | -2.321717141 | 1.41E-06 | 2.96E-06 | 0.006823193 |
| Obesity | nk_cell | *CLIC3* | 1.509211292 | 1.44E-09 | 2.11E-08 | 7.00E-06 |
| Obesity | nk_cell | *INO80E* | -4.133109911 | 1.06E-13 | 3.02E-11 | 5.14E-10 |
| Obesity | nk_cell | *NFU1* | -2.311110261 | 9.64E-06 | 1.68E-05 | 0.046762919 |
| Obesity | nk_cell | *C15orf40* | -1.965716307 | 5.82E-06 | 1.06E-05 | 0.028232167 |
| Obesity | nk_cell | *RGPD8* | -0.611562562 | 0.002153822 | 0.002803956 | 1 |
| Obesity | nk_cell | *LUZP1* | -1.851982292 | 0.014665683 | 0.017789474 | 1 |
| Obesity | nk_cell | *LRRC45* | 1.284832224 | 6.74E-06 | 1.21E-05 | 0.032681562 |
| Obesity | nk_cell | *CHRNA5* | 6.956564451 | 3.12E-06 | 6.05E-06 | 0.015151297 |
| Obesity | nk_cell | *UGP2* | -1.643202789 | 1.19E-06 | 2.54E-06 | 0.005775208 |
| Obesity | nk_cell | *CSGALNACT2* | -1.823417187 | 0.032032418 | 0.037687025 | 1 |
| Obesity | nk_cell | *AVEN* | -2.129608076 | 3.61E-08 | 1.35E-07 | 0.000175175 |
| Obesity | nk_cell | *CTNND2* | -1.774679198 | 7.62E-11 | 6.27E-09 | 3.70E-07 |
| Obesity | nk_cell | *CALML6* | -22.76907577 | 1.30E-10 | 8.67E-09 | 6.33E-07 |
| Obesity | nk_cell | *TPST1* | -1.30328152 | 5.19E-09 | 3.54E-08 | 2.52E-05 |
| Obesity | nk_cell | *TOR1AIP2* | -2.275539669 | 1.39E-07 | 4.02E-07 | 0.00067663 |
| Obesity | nk_cell | *OTUD3* | -1.462646042 | 0.003815935 | 0.004867223 | 1 |
| Obesity | nk_cell | *ZFPM2* | -1.105871376 | 1.20E-07 | 3.55E-07 | 0.000580458 |
| Obesity | nk_cell | *MAP3K2* | -1.787396367 | 5.13E-05 | 8.02E-05 | 0.24898809 |
| Obesity | nk_cell | *MYO7B* | -1.352467607 | 3.72E-06 | 7.07E-06 | 0.018031825 |
| Obesity | nk_cell | *UBE2E3* | -2.324441297 | 1.76E-07 | 4.87E-07 | 0.000854746 |
| Obesity | nk_cell | *NIPA1* | 3.044156578 | 0.00053711 | 0.000739727 | 1 |
| Obesity | nk_cell | *HNRNPA3* | -2.224659825 | 3.31E-05 | 5.33E-05 | 0.160570327 |
| Obesity | nk_cell | *RNF150* | -1.70468459 | 3.17E-08 | 1.21E-07 | 0.000153905 |
| Obesity | nk_cell | *HOXD4* | 4.330467308 | 8.32E-07 | 1.85E-06 | 0.004035963 |
| Obesity | nk_cell | *USP38* | -2.163331725 | 0.038095811 | 0.044464969 | 1 |
| Obesity | nk_cell | *ANKK1* | -3.852103768 | 1.06E-07 | 3.21E-07 | 0.000511897 |
| Obesity | nk_cell | *PDCD6IP* | -2.00844888 | 4.01E-07 | 9.77E-07 | 0.001944567 |
| Obesity | nk_cell | *CRTAP* | -2.782657159 | 7.19E-07 | 1.63E-06 | 0.003489801 |
| Obesity | nk_cell | *CNGB3* | -1.621497483 | 0.000834864 | 0.001128031 | 1 |
| Obesity | nk_cell | *ELP5* | -2.793378485 | 8.63E-07 | 1.91E-06 | 0.004186082 |
| Obesity | nk_cell | *STX8* | -1.470566032 | 0.020943602 | 0.025084758 | 1 |
| Obesity | nk_cell | *NFRKB* | -3.353398953 | 4.85E-06 | 9.00E-06 | 0.023520854 |
| Obesity | nk_cell | *SEMA3E* | -1.291957538 | 8.09E-09 | 4.51E-08 | 3.93E-05 |
| Obesity | nk_cell | *ADORA2B* | -2.119798694 | 5.44E-13 | 1.10E-10 | 2.64E-09 |
| Obesity | nk_cell | *MGMT* | -2.06908258 | 6.40E-05 | 9.84E-05 | 0.310402363 |
| Obesity | nk_cell | *METTL7B* | -2.91471397 | 0.008293087 | 0.010254347 | 1 |
| Obesity | nk_cell | *NFXL1* | -2.454457161 | 3.67E-07 | 9.10E-07 | 0.001781312 |
| Obesity | nk_cell | *DNAJC18* | -2.091952089 | 3.08E-07 | 7.81E-07 | 0.001494869 |
| Obesity | nk_cell | *SPATA24* | -1.553728487 | 0.000110408 | 0.00016534 | 0.535700148 |
| Obesity | nk_cell | *HSD17B13* | -7.921921728 | 1.75E-06 | 3.59E-06 | 0.008498017 |
| Obesity | nk_cell | *ELOVL6* | 2.751667368 | 9.91E-07 | 2.17E-06 | 0.004806698 |
| Obesity | nk_cell | *SMAGP* | -1.029896514 | 0.001619729 | 0.002132101 | 1 |
| Obesity | nk_cell | *CDH2* | -1.703369985 | 6.64E-09 | 4.09E-08 | 3.22E-05 |
| Obesity | nk_cell | *EMB* | 1.582694956 | 3.65E-08 | 1.36E-07 | 0.000176856 |
| Obesity | nk_cell | *STAT2* | -2.5757517 | 8.19E-06 | 1.44E-05 | 0.039750074 |

| | | | | | | |
|---|---|---|---|---|---|---|
| Obesity | nk_cell | *SLC26A5* | -2.771491558 | 8.22E-05 | 0.000124754 | 0.398964498 |
| Obesity | nk_cell | *GTSF1* | -4.174429393 | 3.27E-09 | 2.73E-08 | 1.59E-05 |
| Obesity | nk_cell | *ARMC10* | -2.256833084 | 0.00017156 | 0.000250424 | 0.83240843 |
| Obesity | nk_cell | *ACYP2* | -1.639832749 | 1.04E-05 | 1.80E-05 | 0.050500969 |
| Obesity | nk_cell | *TRABD* | 2.625171214 | 2.21E-07 | 5.88E-07 | 0.00107101 |
| Obesity | nk_cell | *TMEM133* | -2.720620193 | 3.52E-06 | 6.73E-06 | 0.017089234 |
| Obesity | nk_cell | *HOXB9* | -4.608516697 | 1.46E-08 | 6.84E-08 | 7.08E-05 |
| Obesity | nk_cell | *TTLL6* | -5.471166257 | 5.71E-17 | 3.46E-14 | 2.77E-13 |
| Obesity | nk_cell | *KCNS3* | -0.971539893 | 1.45E-08 | 6.80E-08 | 7.02E-05 |
| Obesity | nk_cell | *FOXN2* | -2.317445163 | 0.000333326 | 0.000470431 | 1 |
| Obesity | nk_cell | *GPR27* | -1.082911183 | 0.020545766 | 0.024626497 | 1 |
| Obesity | nk_cell | *TRIAP1* | -2.562447526 | 9.43E-09 | 4.99E-08 | 4.57E-05 |
| Obesity | nk_cell | *RPS9* | 1.444680251 | 0.007171706 | 0.008922338 | 1 |
| Obesity | nk_cell | *TSEN34* | -4.050259714 | 4.35E-06 | 8.16E-06 | 0.02112599 |
| Obesity | nk_cell | *MSANTD4* | 3.887135113 | 1.37E-06 | 2.89E-06 | 0.006638967 |
| Obesity | nk_cell | *NDUFA3* | 2.396664964 | 8.37E-09 | 4.61E-08 | 4.06E-05 |
| Obesity | nk_cell | *PAQR8* | -3.004375051 | 2.29E-06 | 4.55E-06 | 0.011095766 |
| Obesity | nk_cell | *NUDT6* | -2.164890933 | 7.93E-09 | 4.47E-08 | 3.85E-05 |
| Obesity | nk_cell | *ZNF160* | -1.639128613 | 1.72E-06 | 3.54E-06 | 0.008361789 |
| Obesity | nk_cell | *ZNF415* | -3.552220037 | 1.23E-07 | 3.64E-07 | 0.000596764 |
| Obesity | nk_cell | *S1PR1* | -2.67832585 | 7.00E-07 | 1.59E-06 | 0.003395398 |
| Obesity | nk_cell | *PYGO1* | -2.585769767 | 4.67E-07 | 1.11E-06 | 0.002264973 |
| Obesity | nk_cell | *TSNARE1* | 3.627675767 | 3.62E-06 | 6.91E-06 | 0.0175729 |
| Obesity | nk_cell | *FPR1* | -2.833692351 | 4.17E-08 | 1.51E-07 | 0.000202367 |
| Obesity | nk_cell | *FEZ2* | -2.151964842 | 5.48E-06 | 1.01E-05 | 0.026599752 |
| Obesity | nk_cell | *SOX7* | -5.181164581 | 8.63E-05 | 0.00013053 | 0.418738965 |
| Obesity | nk_cell | *TRMT61B* | -1.246689393 | 0.023248372 | 0.027698407 | 1 |
| Obesity | nk_cell | *MFN1* | -1.774366712 | 5.90E-07 | 1.36E-06 | 0.002863662 |
| Obesity | nk_cell | *ATP6V0E2* | 1.969187095 | 3.36E-06 | 6.46E-06 | 0.016315922 |
| Obesity | nk_cell | *MORN4* | -1.282392345 | 1.87E-06 | 3.80E-06 | 0.009069867 |
| Obesity | nk_cell | *SCAND1* | 2.863389656 | 6.29E-06 | 1.14E-05 | 0.030512346 |
| Obesity | nk_cell | *SOSTDC1* | -2.572333433 | 1.18E-05 | 2.02E-05 | 0.057016503 |
| Obesity | nk_cell | *GAA* | -0.970396028 | 0.000505228 | 0.000697997 | 1 |
| Obesity | nk_cell | *CANT1* | -1.495663964 | 0.000681559 | 0.000929172 | 1 |
| Obesity | nk_cell | *PGAM1* | -1.953968805 | 4.36E-08 | 1.57E-07 | 0.000211647 |
| Obesity | nk_cell | *LURAP1* | -1.63782408 | 2.23E-08 | 9.34E-08 | 0.000108027 |
| Obesity | nk_cell | *PDE7B* | -1.749056989 | 5.35E-08 | 1.84E-07 | 0.000259605 |
| Obesity | nk_cell | *MRPL36* | 1.703128896 | 1.27E-05 | 2.17E-05 | 0.061687234 |
| Obesity | nk_cell | *MCC* | -2.060643679 | 6.69E-07 | 1.53E-06 | 0.003244956 |
| Obesity | nk_cell | *ZNF561* | -1.038993922 | 0.035569026 | 0.041605813 | 1 |
| Obesity | nk_cell | *HOPX* | 0.358425333 | 1.31E-09 | 2.09E-08 | 6.34E-06 |
| Obesity | nk_cell | *PPID* | -3.238695844 | 3.55E-05 | 5.69E-05 | 0.172292136 |
| Obesity | nk_cell | *ETFDH* | -1.004195781 | 0.000243065 | 0.00034892 | 1 |
| Obesity | nk_cell | *TBCA* | -2.269969628 | 6.33E-06 | 1.14E-05 | 0.03070232 |
| Obesity | nk_cell | *ZNF584* | -1.051908591 | 1.39E-07 | 4.01E-07 | 0.000675928 |
| Obesity | nk_cell | *NMUR1* | 0.458889079 | 1.15E-09 | 2.05E-08 | 5.58E-06 |
| Obesity | nk_cell | *PIK3CD* | 1.279242904 | 5.56E-10 | 1.81E-08 | 2.70E-06 |
| Obesity | nk_cell | *RGS19* | 1.227785404 | 2.36E-11 | 2.66E-09 | 1.14E-07 |
| Obesity | nk_cell | *ANO5* | -1.450371893 | 5.90E-09 | 3.82E-08 | 2.86E-05 |
| Obesity | nk_cell | *VAT1L* | -3.361116925 | 1.14E-08 | 5.62E-08 | 5.52E-05 |

| | | | | | | |
|---|---|---|---|---|---|---|
| Obesity | nk_cell | *CAMTA1* | -1.699965133 | 0.003863134 | 0.004924836 | 1 |
| Obesity | nk_cell | *LGALS4* | -1.515344488 | 0.003076979 | 0.003951695 | 1 |
| Obesity | nk_cell | *LRRC34* | -1.152407007 | 0.000310787 | 0.000440918 | 1 |
| Obesity | nk_cell | *PAH* | -6.489233104 | 3.31E-10 | 1.45E-08 | 1.61E-06 |
| Obesity | nk_cell | *RHNO1* | 0.917245664 | 3.39E-09 | 2.77E-08 | 1.65E-05 |
| Obesity | nk_cell | *METTL18* | 0.923289529 | 2.22E-07 | 5.90E-07 | 0.001074783 |
| Obesity | nk_cell | *PWWP2B* | -1.697581974 | 1.53E-08 | 7.11E-08 | 7.44E-05 |
| Obesity | nk_cell | *EXOSC10* | -2.74223023 | 2.14E-07 | 5.74E-07 | 0.001037986 |
| Obesity | nk_cell | *C3AR1* | -1.158646136 | 0.011489797 | 0.014038905 | 1 |
| Obesity | nk_cell | *PRNP* | -2.239609691 | 2.25E-05 | 3.69E-05 | 0.109203789 |
| Obesity | nk_cell | *TVP23B* | -2.666651736 | 2.14E-05 | 3.52E-05 | 0.103968239 |
| Obesity | nk_cell | *FBXW10* | -2.773181758 | 7.43E-10 | 1.92E-08 | 3.60E-06 |
| Obesity | nk_cell | *SRGAP2C* | -1.261045243 | 8.56E-08 | 2.70E-07 | 0.000415312 |
| Obesity | nk_cell | *ATPAF2* | -3.009338015 | 6.53E-09 | 4.05E-08 | 3.17E-05 |
| Obesity | nk_cell | *ZNF554* | 3.276890572 | 5.49E-07 | 1.28E-06 | 0.002663962 |
| Obesity | nk_cell | *SERF1A* | -8.628234104 | 9.11E-10 | 2.05E-08 | 4.42E-06 |
| Obesity | nk_cell | *NME6* | -2.345963589 | 0.000982808 | 0.001316562 | 1 |
| Obesity | nk_cell | *CD8B* | 0.570374438 | 1.07E-09 | 2.05E-08 | 5.17E-06 |
| Obesity | nk_cell | *SLFN12* | -1.900811334 | 0.006223215 | 0.007774212 | 1 |
| Obesity | nk_cell | *MTBP* | -2.405965144 | 1.06E-07 | 3.21E-07 | 0.000512985 |
| Obesity | nk_cell | *TEFM* | -2.033113201 | 7.75E-08 | 2.49E-07 | 0.000375905 |
| Obesity | nk_cell | *MALT1* | -2.435368039 | 0.00040527 | 0.000566351 | 1 |
| Obesity | nk_cell | *MBOAT1* | -2.20746967 | 5.93E-08 | 1.98E-07 | 0.000287527 |
| Obesity | nk_cell | *CXCR6* | 1.411003305 | 3.02E-09 | 2.61E-08 | 1.47E-05 |
| Obesity | nk_cell | *PAIP1* | -3.072440065 | 1.33E-06 | 2.81E-06 | 0.006437792 |
| Obesity | nk_cell | *C1QTNF4* | -2.84568743 | 1.81E-22 | 2.93E-19 | 8.78E-19 |
| Obesity | nk_cell | *MACROD2* | -1.287885668 | 3.64E-06 | 6.94E-06 | 0.017654158 |
| Obesity | nk_cell | *BSG* | -2.620054634 | 5.35E-06 | 9.82E-06 | 0.025943007 |
| Obesity | nk_cell | *HINFP* | -1.768201432 | 4.39E-09 | 3.20E-08 | 2.13E-05 |
| Obesity | nk_cell | *TP53RK* | 1.406991394 | 1.15E-09 | 2.05E-08 | 5.58E-06 |
| Obesity | nk_cell | *SUCLG2* | -1.885354432 | 0.001768335 | 0.002317656 | 1 |
| Obesity | nk_cell | *ARNT2* | -1.135711887 | 4.48E-06 | 8.38E-06 | 0.021729656 |
| Obesity | nk_cell | *GTPBP2* | -2.491602704 | 2.15E-05 | 3.54E-05 | 0.104544126 |
| Obesity | nk_cell | *FGGY* | -2.129456185 | 7.85E-05 | 0.000119291 | 0.380657657 |
| Obesity | nk_cell | *MANEA* | -3.147601161 | 5.78E-08 | 1.95E-07 | 0.000280384 |
| Obesity | nk_cell | *AFF1* | -1.747324159 | 6.24E-07 | 1.43E-06 | 0.003027459 |
| Obesity | nk_cell | *FIBP* | 1.640362435 | 4.99E-09 | 3.48E-08 | 2.42E-05 |
| Obesity | nk_cell | *CTSW* | 0.168046575 | 1.89E-09 | 2.19E-08 | 9.18E-06 |
| Obesity | nk_cell | *SNTG2* | -1.460496832 | 1.01E-07 | 3.10E-07 | 0.000490898 |
| Obesity | nk_cell | *RAD9A* | -1.254340435 | 0.000188613 | 0.000274407 | 0.915148641 |
| Obesity | nk_cell | *EFEMP2* | -1.315259677 | 7.69E-09 | 4.40E-08 | 3.73E-05 |
| Obesity | nk_cell | *ZFAND4* | -2.87812682 | 3.29E-09 | 2.73E-08 | 1.60E-05 |
| Obesity | nk_cell | *SLFN11* | -2.868186195 | 0.000127981 | 0.00018984 | 0.620965592 |
| Obesity | nk_cell | *FUT10* | -2.050809563 | 2.38E-10 | 1.25E-08 | 1.15E-06 |
| Obesity | nk_cell | *ZNF596* | -2.531829067 | 0.010892935 | 0.013343227 | 1 |
| Obesity | nk_cell | *CFL1* | 1.75817364 | 4.53E-09 | 3.26E-08 | 2.20E-05 |
| Obesity | nk_cell | *FAM192A* | -2.368830569 | 6.53E-05 | 0.000100242 | 0.317024706 |
| Obesity | nk_cell | *CBWD1* | -2.279366378 | 7.51E-09 | 4.36E-08 | 3.64E-05 |
| Obesity | nk_cell | *DCP2* | -2.520834241 | 1.39E-06 | 2.94E-06 | 0.0067609 |
| Obesity | nk_cell | *ZNF621* | -2.287903442 | 0.016772624 | 0.020256527 | 1 |

| | | | | | | |
|---|---|---|---|---|---|---|
| Obesity | nk_cell | *RNASEH2C* | -1.284548654 | 0.008045343 | 0.009953087 | 1 |
| Obesity | nk_cell | *MYEOV* | -4.881161678 | 2.13E-07 | 5.72E-07 | 0.00103328 |
| Obesity | nk_cell | *ANKRD13D* | 2.254291212 | 0.002390566 | 0.003101343 | 1 |
| Obesity | nk_cell | *MRGPRF* | -1.034965236 | 3.81E-07 | 9.37E-07 | 0.001848582 |
| Obesity | nk_cell | *LCLAT1* | -2.42315636 | 0.000144008 | 0.000212057 | 0.69872814 |
| Obesity | nk_cell | *SH3RF3* | -2.18493791 | 8.78E-08 | 2.76E-07 | 0.00042601 |
| Obesity | nk_cell | *GXYLT2* | -1.591533835 | 4.63E-08 | 1.64E-07 | 0.000224728 |
| Obesity | nk_cell | *DCAKD* | -2.916100372 | 9.95E-06 | 1.73E-05 | 0.048298161 |
| Obesity | nk_cell | *TRMT112* | -2.241346741 | 0.000634488 | 0.000869643 | 1 |
| Obesity | nk_cell | *NOC3L* | -2.453979119 | 4.19E-07 | 1.01E-06 | 0.002031663 |
| Obesity | nk_cell | *RHOD* | -1.545162971 | 7.99E-09 | 4.50E-08 | 3.88E-05 |
| Obesity | nk_cell | *PARP15* | 0.391625851 | 1.21E-09 | 2.08E-08 | 5.89E-06 |
| Obesity | nk_cell | *AHSA2* | -1.752082853 | 7.84E-07 | 1.76E-06 | 0.00380526 |
| Obesity | nk_cell | *VANGL1* | -2.076087954 | 4.15E-08 | 1.51E-07 | 0.000201564 |
| Obesity | nk_cell | *IQCB1* | -2.204635789 | 3.45E-09 | 2.78E-08 | 1.67E-05 |
| Obesity | nk_cell | *ZNF483* | -1.737211332 | 1.95E-07 | 5.30E-07 | 0.000948522 |
| Obesity | nk_cell | *SLC2A14* | -1.300598007 | 3.33E-07 | 8.35E-07 | 0.001615316 |
| Obesity | nk_cell | *MMRN2* | -1.926775954 | 0.000732524 | 0.000996693 | 1 |
| Obesity | nk_cell | *MAP3K11* | -2.778149829 | 6.39E-07 | 1.46E-06 | 0.003099244 |
| Obesity | nk_cell | *KCNK7* | 3.843104074 | 5.11E-10 | 1.75E-08 | 2.48E-06 |
| Obesity | nk_cell | *NDNF* | -1.850711325 | 2.71E-08 | 1.08E-07 | 0.00013149 |
| Obesity | nk_cell | *ARV1* | -1.817758046 | 1.28E-07 | 3.76E-07 | 0.000622133 |
| Obesity | nk_cell | *CCDC36* | -1.555987895 | 3.72E-08 | 1.39E-07 | 0.000180354 |
| Obesity | nk_cell | *EHBP1L1* | -1.542403644 | 0.004223825 | 0.005366327 | 1 |
| Obesity | nk_cell | *ZNF417* | -3.579703645 | 6.24E-05 | 9.63E-05 | 0.302818556 |
| Obesity | nk_cell | *PEAK1* | -1.820416078 | 3.18E-07 | 8.04E-07 | 0.001542775 |
| Obesity | nk_cell | *MST1* | -1.506174051 | 1.32E-10 | 8.67E-09 | 6.39E-07 |
| Obesity | nk_cell | *GMPPB* | -3.383286884 | 6.18E-08 | 2.06E-07 | 0.000299764 |
| Obesity | nk_cell | *NUDT18* | -8.026467783 | 5.49E-08 | 1.87E-07 | 0.000266245 |
| Obesity | nk_cell | *SULT1B1* | 5.536145381 | 0.000561745 | 0.000773216 | 1 |
| Obesity | nk_cell | *NUDT4* | -1.783394277 | 7.10E-09 | 4.25E-08 | 3.44E-05 |
| Obesity | nk_cell | *SCAI* | -2.04750574 | 2.37E-06 | 4.70E-06 | 0.011495215 |
| Obesity | nk_cell | *HSPB7* | 1.430933939 | 0.002359044 | 0.003062907 | 1 |
| Obesity | nk_cell | *UQCRH* | -2.531669681 | 1.13E-05 | 1.95E-05 | 0.05496205 |
| Obesity | nk_cell | *C11orf80* | -2.642637329 | 0.000100073 | 0.00015054 | 0.485555458 |
| Obesity | nk_cell | *CD7* | 0.205787328 | 1.03E-09 | 2.05E-08 | 4.98E-06 |
| Obesity | nk_cell | *RNF213* | 1.764690822 | 0.000789746 | 0.001071246 | 1 |
| Obesity | nk_cell | *GPR160* | 2.891184709 | 0.000166518 | 0.00024343 | 0.807944906 |
| Obesity | nk_cell | *GOLIM4* | -2.16546371 | 6.71E-08 | 2.20E-07 | 0.000325352 |
| Obesity | nk_cell | *HOXB2* | -1.792894544 | 6.96E-05 | 0.000106484 | 0.337501422 |
| Obesity | nk_cell | *XXYLT1* | -1.969095051 | 1.40E-08 | 6.64E-08 | 6.81E-05 |
| Obesity | nk_cell | *UBXN2A* | -2.089930332 | 1.79E-05 | 2.98E-05 | 0.086965335 |
| Obesity | nk_cell | *NRROS* | 1.259413995 | 1.87E-05 | 3.10E-05 | 0.090661951 |
| Obesity | nk_cell | *CEP19* | -1.862210835 | 4.55E-05 | 7.18E-05 | 0.220786194 |
| Obesity | nk_cell | *SLC25A30* | -3.690890458 | 7.40E-05 | 0.000113006 | 0.359245621 |
| Obesity | nk_cell | *CTSF* | -2.044617781 | 0.000382917 | 0.000536503 | 1 |
| Obesity | nk_cell | *LEMD3* | -1.751707731 | 0.034007994 | 0.039856712 | 1 |
| Obesity | nk_cell | *CTU2* | 1.966663038 | 1.31E-09 | 2.09E-08 | 6.33E-06 |
| Obesity | nk_cell | *GLIS1* | -1.423672902 | 2.04E-09 | 2.21E-08 | 9.90E-06 |
| Obesity | nk_cell | *RALGAPA1* | -1.828025527 | 1.30E-05 | 2.21E-05 | 0.062876802 |

| | | | | | | |
|---|---|---|---|---|---|---|
| Obesity | nk_cell | *ANKRD36C* | -2.520621208 | 1.56E-06 | 3.23E-06 | 0.007553507 |
| Obesity | nk_cell | *TMEM81* | 2.583299177 | 0.000860623 | 0.001160895 | 1 |
| Obesity | nk_cell | *MSL2* | -2.395730736 | 0.001112396 | 0.001482379 | 1 |
| Obesity | nk_cell | *ZNF497* | 7.408285767 | 1.07E-08 | 5.42E-08 | 5.19E-05 |
| Obesity | nk_cell | *TRAM1L1* | -3.520186369 | 4.57E-08 | 1.62E-07 | 0.000221767 |
| Obesity | nk_cell | *ANGEL2* | -2.548459643 | 4.30E-05 | 6.80E-05 | 0.208429412 |
| Obesity | nk_cell | *SH3PXD2B* | -1.303954511 | 4.26E-09 | 3.16E-08 | 2.07E-05 |
| Obesity | nk_cell | *LARP7* | -2.700714862 | 5.68E-05 | 8.82E-05 | 0.275703013 |
| Obesity | nk_cell | *FGFBP3* | 2.970876347 | 1.87E-09 | 2.19E-08 | 9.08E-06 |
| Obesity | nk_cell | *C4orf32* | -1.603177473 | 9.85E-08 | 3.04E-07 | 0.000477769 |
| Obesity | nk_cell | *HRAS* | 3.0959115 | 2.91E-08 | 1.14E-07 | 0.000141022 |
| Obesity | nk_cell | *PCP2* | 1.880375115 | 0.000492596 | 0.000681711 | 1 |
| Obesity | nk_cell | *CEP135* | -2.393364371 | 0.002224591 | 0.002893758 | 1 |
| Obesity | nk_cell | *NLRP6* | 6.00912878 | 0.004138947 | 0.005262624 | 1 |
| Obesity | nk_cell | *RSRC1* | -2.121608404 | 9.54E-08 | 2.96E-07 | 0.000462719 |
| Obesity | nk_cell | *PTDSS2* | -1.871280903 | 1.04E-06 | 2.25E-06 | 0.005034066 |
| Obesity | nk_cell | *SEZ6L2* | 3.925029279 | 1.48E-10 | 9.24E-09 | 7.20E-07 |
| Obesity | nk_cell | *KCTD13* | 2.375793419 | 0.001504609 | 0.001984873 | 1 |
| Obesity | nk_cell | *DHX36* | -2.237645033 | 0.000266404 | 0.000380846 | 1 |
| Obesity | nk_cell | *FBXW8* | -1.687972974 | 0.026521044 | 0.031477521 | 1 |
| Obesity | nk_cell | *SLC22A1* | -3.672962188 | 4.00E-09 | 3.03E-08 | 1.94E-05 |
| Obesity | nk_cell | *CHST2* | 0.571635499 | 1.02E-09 | 2.05E-08 | 4.94E-06 |
| Obesity | nk_cell | *TVP23C* | 5.09433672 | 2.42E-08 | 9.94E-08 | 0.000117449 |
| Obesity | nk_cell | *PACS1* | -1.660980599 | 0.007864033 | 0.009741202 | 1 |
| Obesity | nk_cell | *PSMD2* | -2.018421138 | 1.35E-08 | 6.49E-08 | 6.57E-05 |
| Obesity | nk_cell | *FAM182B* | 1.009559374 | 0.03154628 | 0.037133079 | 1 |
| Obesity | nk_cell | *ATG13* | -2.916684528 | 1.47E-05 | 2.48E-05 | 0.071207024 |
| Obesity | nk_cell | *CCNE2* | -1.392846274 | 4.23E-08 | 1.53E-07 | 0.000205356 |
| Obesity | nk_cell | *ZNF519* | -1.881103348 | 4.36E-09 | 3.19E-08 | 2.12E-05 |
| Obesity | nk_cell | *CHRNA7* | -2.674896406 | 1.75E-08 | 7.83E-08 | 8.49E-05 |
| Obesity | nk_cell | *TMEM9B* | -2.515184223 | 1.80E-07 | 4.95E-07 | 0.000871996 |
| Obesity | nk_cell | *SCUBE2* | -2.034312767 | 1.90E-09 | 2.19E-08 | 9.24E-06 |
| Obesity | nk_cell | *EIF1AD* | -2.192020074 | 9.19E-09 | 4.90E-08 | 4.46E-05 |
| Obesity | nk_cell | *EIF3F* | 2.04102741 | 0.002643208 | 0.003417225 | 1 |
| Obesity | nk_cell | *ZNF25* | -3.278962054 | 9.95E-08 | 3.06E-07 | 0.000482973 |
| Obesity | nk_cell | *CLTB* | -3.141145085 | 0.000164715 | 0.000241012 | 0.799196786 |
| Obesity | nk_cell | *MCTP1* | -1.848429674 | 2.34E-07 | 6.18E-07 | 0.001133566 |
| Obesity | nk_cell | *LRRC25* | -2.139384732 | 1.81E-06 | 3.69E-06 | 0.008775256 |
| Obesity | nk_cell | *UBQLNL* | -1.995396916 | 0.000257462 | 0.000368714 | 1 |
| Obesity | nk_cell | *KCNE3* | -3.671347261 | 4.07E-06 | 7.68E-06 | 0.019725683 |
| Obesity | nk_cell | *UCP2* | 1.455074411 | 0.01318818 | 0.016045398 | 1 |
| Obesity | nk_cell | *C11orf68* | 4.362336619 | 5.95E-09 | 3.84E-08 | 2.89E-05 |
| Obesity | nk_cell | *MRPL48* | -3.196930593 | 5.00E-07 | 1.18E-06 | 0.002427879 |
| Obesity | nk_cell | *RAB6A* | -2.226381093 | 1.59E-05 | 2.66E-05 | 0.076949653 |
| Obesity | nk_cell | *TMEM70* | -2.537988364 | 1.84E-07 | 5.04E-07 | 0.000892271 |
| Obesity | nk_cell | *RPS6KB2* | 2.099751451 | 2.70E-10 | 1.30E-08 | 1.31E-06 |
| Obesity | nk_cell | *TOM1L2* | -1.766269009 | 3.79E-07 | 9.33E-07 | 0.00183872 |
| Obesity | nk_cell | *TEX26* | -4.687839633 | 8.86E-08 | 2.78E-07 | 0.000429669 |
| Obesity | nk_cell | *GPR156* | -4.462740921 | 9.46E-08 | 2.94E-07 | 0.000458969 |
| Obesity | nk_cell | *TTLL11* | -2.615411943 | 3.58E-06 | 6.83E-06 | 0.017374436 |

| | | | | | | |
|---|---|---|---|---|---|---|
| Obesity | nk_cell | *PRIMA1* | -1.226145196 | 1.60E-09 | 2.11E-08 | 7.75E-06 |
| Obesity | nk_cell | *CTDNEP1* | 3.368112302 | 3.76E-08 | 1.40E-07 | 0.000182536 |
| Obesity | nk_cell | *HOXD8* | -1.009012439 | 0.005943866 | 0.007434813 | 1 |
| Obesity | nk_cell | *ZDHHC21* | -2.158368928 | 3.04E-05 | 4.92E-05 | 0.147625436 |
| Obesity | nk_cell | *A2M* | -1.307812774 | 0.041844677 | 0.048661389 | 1 |
| Obesity | nk_cell | *DOK7* | 5.300109113 | 1.14E-09 | 2.05E-08 | 5.54E-06 |
| Obesity | nk_cell | *ORAI3* | 2.259439115 | 5.13E-15 | 1.91E-12 | 2.49E-11 |
| Obesity | nk_cell | *B3GALT6* | 6.977336306 | 0.015261758 | 0.018484785 | 1 |
| Obesity | nk_cell | *ZNF613* | -1.819579779 | 7.83E-07 | 1.76E-06 | 0.003797943 |
| Obesity | nk_cell | *JAKMIP2* | 0.254877277 | 2.35E-09 | 2.30E-08 | 1.14E-05 |
| Obesity | nk_cell | *TCP11L1* | -1.755787293 | 0.000196564 | 0.000285121 | 0.953730755 |
| Obesity | nk_cell | *CCDC57* | -2.330213957 | 5.22E-10 | 1.75E-08 | 2.53E-06 |
| Obesity | nk_cell | *MYPOP* | -1.86169822 | 3.33E-08 | 1.26E-07 | 0.000161783 |
| Obesity | nk_cell | *CIDEA* | -1.15263019 | 3.67E-09 | 2.88E-08 | 1.78E-05 |
| Obesity | nk_cell | *SLC35G1* | -2.723641558 | 2.04E-08 | 8.72E-08 | 9.87E-05 |
| Obesity | nk_cell | *ZNF135* | -1.686589212 | 0.000314031 | 0.00044526 | 1 |
| Obesity | nk_cell | *OR4N2* | -3.346368175 | 1.30E-08 | 6.26E-08 | 6.29E-05 |
| Obesity | nk_cell | *TAC4* | -3.307006496 | 5.62E-07 | 1.30E-06 | 0.002729127 |
| Obesity | nk_cell | *CDC26* | -1.73801631 | 4.14E-08 | 1.51E-07 | 0.000201067 |
| Obesity | nk_cell | *CRLF3* | -1.693433469 | 0.001933854 | 0.002528446 | 1 |
| Obesity | nk_cell | *DNAJC30* | -4.515772138 | 1.74E-05 | 2.89E-05 | 0.084198498 |
| Obesity | nk_cell | *SPRYD4* | -2.476661363 | 3.41E-11 | 3.25E-09 | 1.66E-07 |
| Obesity | nk_cell | *VPS37D* | -0.998539277 | 0.009035704 | 0.011144188 | 1 |
| Obesity | nk_cell | *ZNF575* | -0.571573972 | 0.020733854 | 0.024845804 | 1 |
| Obesity | nk_cell | *WDR25* | -2.736305232 | 2.58E-07 | 6.71E-07 | 0.001249979 |
| Obesity | nk_cell | *PLA2G16* | -1.389086108 | 1.96E-08 | 8.47E-08 | 9.52E-05 |
| Obesity | nk_cell | *B3GNT5* | -2.075376392 | 2.04E-09 | 2.21E-08 | 9.88E-06 |
| Obesity | nk_cell | *RMDN1* | -1.446742287 | 0.000352969 | 0.000496984 | 1 |
| Obesity | nk_cell | *RNF152* | -1.707398418 | 9.59E-08 | 2.97E-07 | 0.000465354 |
| Obesity | nk_cell | *LRRC37A* | -1.347291749 | 0.000855694 | 0.001154569 | 1 |
| Obesity | nk_cell | *BDNF* | -2.323548416 | 1.62E-08 | 7.38E-08 | 7.85E-05 |
| Obesity | nk_cell | *BOK* | -1.794842533 | 5.47E-09 | 3.65E-08 | 2.66E-05 |
| Obesity | nk_cell | *RUFY1* | -1.430141971 | 0.000146675 | 0.000215853 | 0.711667757 |
| Obesity | nk_cell | *VSIG10* | -1.594228249 | 1.95E-07 | 5.29E-07 | 0.00094545 |
| Obesity | nk_cell | *ANKLE2* | -2.437710537 | 1.29E-06 | 2.75E-06 | 0.006282253 |
| Obesity | nk_cell | *C8G* | -23.50989252 | 1.67E-09 | 2.15E-08 | 8.08E-06 |
| Obesity | nk_cell | *MUC20* | 3.123491376 | 1.86E-07 | 5.09E-07 | 0.000904276 |
| Obesity | nk_cell | *LY6H* | -2.137257442 | 2.96E-05 | 4.79E-05 | 0.143545492 |
| Obesity | nk_cell | *SHMT1* | -2.613847565 | 8.13E-05 | 0.000123448 | 0.394417808 |
| Obesity | nk_cell | *DPP7* | -2.751698769 | 0.000633022 | 0.000867878 | 1 |
| Obesity | nk_cell | *MTHFR* | -2.593619297 | 2.43E-07 | 6.39E-07 | 0.00117854 |
| Obesity | nk_cell | *C19orf18* | -2.144859546 | 7.45E-08 | 2.41E-07 | 0.000361546 |
| Obesity | nk_cell | *TMEM80* | -2.976051626 | 3.38E-09 | 2.77E-08 | 1.64E-05 |
| Obesity | nk_cell | *WDR73* | -2.247031364 | 2.43E-07 | 6.38E-07 | 0.001176757 |
| Obesity | nk_cell | *DSCAML1* | -1.17513442 | 3.41E-09 | 2.77E-08 | 1.65E-05 |
| Obesity | nk_cell | *TALDO1* | -3.140813724 | 1.27E-05 | 2.17E-05 | 0.061810286 |
| Obesity | nk_cell | *AP006621.1* | -1.178133647 | 3.28E-07 | 8.25E-07 | 0.001593811 |
| Obesity | nk_cell | *MAN1B1* | -3.388328264 | 1.16E-06 | 2.48E-06 | 0.005610987 |
| Obesity | nk_cell | *FBXO39* | -1.630302888 | 0.000274347 | 0.00039151 | 1 |
| Obesity | nk_cell | *SAMD9L* | 1.553628585 | 0.004641758 | 0.005878833 | 1 |

| | | | | | | |
|---|---|---|---|---|---|---|
| Obesity | nk_cell | *MIEF2* | -2.607208473 | 3.17E-09 | 2.68E-08 | 1.54E-05 |
| Obesity | nk_cell | *NAP1L5* | -2.106254905 | 6.78E-09 | 4.14E-08 | 3.29E-05 |
| Obesity | nk_cell | *RBM44* | 3.84375905 | 0.00056921 | 0.000782824 | 1 |
| Obesity | nk_cell | *SLC25A22* | 2.14432443 | 3.48E-08 | 1.31E-07 | 0.00016896 |
| Obesity | nk_cell | *GBA* | -1.710750045 | 2.32E-08 | 9.60E-08 | 0.00011274 |
| Obesity | nk_cell | *ACAD9* | -1.267392285 | 3.66E-05 | 5.85E-05 | 0.177600627 |
| Obesity | nk_cell | *IL17RA* | -2.476441231 | 1.20E-06 | 2.57E-06 | 0.005837387 |
| Obesity | nk_cell | *MBOAT4* | -2.803338704 | 1.06E-06 | 2.30E-06 | 0.005153887 |
| Obesity | nk_cell | *AGTRAP* | 1.845162749 | 5.31E-08 | 1.83E-07 | 0.000257471 |
| Obesity | nk_cell | *THAP5* | -3.303645981 | 0.001977745 | 0.002582352 | 1 |
| Obesity | nk_cell | *NAALADL2* | -1.595649087 | 1.26E-07 | 3.73E-07 | 0.000613711 |
| Obesity | nk_cell | *CD151* | -1.947233975 | 0.000499466 | 0.00069043 | 1 |
| Obesity | nk_cell | *FAM87B* | -9.590099861 | 2.51E-09 | 2.38E-08 | 1.22E-05 |
| Obesity | nk_cell | *CENPBD1* | 5.142529815 | 5.35E-09 | 3.59E-08 | 2.59E-05 |
| Obesity | nk_cell | *RPS27* | 1.679167821 | 9.58E-09 | 5.04E-08 | 4.65E-05 |
| Obesity | nk_cell | *RIC8A* | 2.017427295 | 7.48E-09 | 4.36E-08 | 3.63E-05 |
| Obesity | nk_cell | *ODF3B* | -1.150205867 | 5.64E-08 | 1.91E-07 | 0.000273793 |
| Obesity | nk_cell | *DPY19L2* | -0.986946174 | 4.09E-08 | 1.49E-07 | 0.000198554 |
| Obesity | nk_cell | *ADAMTSL1* | -1.233193757 | 1.40E-08 | 6.64E-08 | 6.80E-05 |
| Obesity | nk_cell | *GRAMD1C* | -1.94836605 | 1.74E-08 | 7.78E-08 | 8.42E-05 |
| Obesity | nk_cell | *DALRD3* | -2.607352474 | 2.05E-08 | 8.77E-08 | 9.96E-05 |
| Obesity | nk_cell | *ZNF114* | -5.899190102 | 1.84E-06 | 3.75E-06 | 0.008930233 |
| Obesity | nk_cell | *SPINK6* | -5.446142017 | 1.23E-09 | 2.08E-08 | 5.96E-06 |
| Obesity | nk_cell | *SH2B1* | -2.419193318 | 2.25E-07 | 5.99E-07 | 0.001093556 |
| Obesity | nk_cell | *VN1R1* | -5.964118464 | 8.47E-09 | 4.63E-08 | 4.11E-05 |
| Obesity | nk_cell | *KDELC2* | -3.672934981 | 0.000307554 | 0.000436587 | 1 |
| Obesity | nk_cell | *PLEC* | -2.053750239 | 4.29E-06 | 8.04E-06 | 0.02079637 |
| Obesity | nk_cell | *SH2D4B* | -3.841209292 | 2.25E-09 | 2.27E-08 | 1.09E-05 |
| Obesity | nk_cell | *RNF212* | -2.217379437 | 0.005302482 | 0.006663467 | 1 |
| Obesity | nk_cell | *PRSS36* | -5.726313951 | 1.65E-06 | 3.40E-06 | 0.008007834 |
| Obesity | nk_cell | *WDR6* | -1.362810794 | 0.000409406 | 0.000571473 | 1 |
| Obesity | nk_cell | *TMPRSS9* | -1.315808432 | 5.27E-09 | 3.56E-08 | 2.56E-05 |
| Obesity | nk_cell | *TMEM11* | -1.620372839 | 1.28E-09 | 2.09E-08 | 6.21E-06 |
| Obesity | nk_cell | *ZFAND2A* | -2.033092929 | 4.54E-09 | 3.26E-08 | 2.20E-05 |
| Obesity | nk_cell | *ZNF223* | -2.677638978 | 1.30E-05 | 2.21E-05 | 0.062906887 |
| Obesity | nk_cell | *FAM220A* | -3.483593055 | 1.80E-09 | 2.17E-08 | 8.71E-06 |
| Obesity | nk_cell | *NT5DC1* | -2.251706491 | 7.81E-05 | 0.00011873 | 0.378747652 |
| Obesity | nk_cell | *GLDC* | -1.837453978 | 4.64E-09 | 3.29E-08 | 2.25E-05 |
| Obesity | nk_cell | *P4HTM* | -2.25690054 | 4.20E-09 | 3.12E-08 | 2.04E-05 |
| Obesity | nk_cell | *CA8* | -2.879671094 | 3.51E-08 | 1.32E-07 | 0.000170272 |
| Obesity | nk_cell | *ERN1* | -2.019451888 | 0.018965605 | 0.022800079 | 1 |
| Obesity | nk_cell | *ZNF713* | -1.236379572 | 7.69E-06 | 1.36E-05 | 0.037313228 |
| Obesity | nk_cell | *PARP10* | -2.209972776 | 2.53E-09 | 2.39E-08 | 1.23E-05 |
| Obesity | nk_cell | *NSUN3* | -2.381458507 | 7.04E-07 | 1.60E-06 | 0.003417639 |
| Obesity | nk_cell | *RPP25* | 0.833451807 | 0.002419988 | 0.003137836 | 1 |
| Obesity | nk_cell | *STX19* | -1.321125161 | 7.71E-05 | 0.000117439 | 0.374277793 |
| Obesity | nk_cell | *TRIM73* | -8.967778301 | 2.26E-09 | 2.27E-08 | 1.10E-05 |
| Obesity | nk_cell | *EFCAB13* | -2.001471894 | 1.96E-08 | 8.46E-08 | 9.50E-05 |
| Obesity | nk_cell | *EXOSC4* | -7.307096444 | 1.37E-08 | 6.55E-08 | 6.65E-05 |
| Obesity | nk_cell | *ZNF852* | -4.531289026 | 1.66E-07 | 4.66E-07 | 0.00080768 |

| | | | | | | |
|---|---|---|---|---|---|---|
| Obesity | nk_cell | *HYI* | -1.871090502 | 1.38E-08 | 6.58E-08 | 6.70E-05 |
| Obesity | nk_cell | *C17orf62* | 3.982818875 | 6.29E-06 | 1.14E-05 | 0.030534592 |
| Obesity | nk_cell | *GAK* | -2.620236435 | 0.000194989 | 0.000283026 | 0.946088898 |
| Obesity | nk_cell | *TUFM* | 4.25735883 | 5.82E-08 | 1.96E-07 | 0.000282499 |
| Obesity | nk_cell | *CTC1* | 3.011118319 | 8.92E-05 | 0.000134746 | 0.433003309 |
| Obesity | nk_cell | *C3orf38* | -0.95130206 | 0.02218897 | 0.026497879 | 1 |
| Obesity | nk_cell | *CCDC89* | -3.608906456 | 1.91E-09 | 2.19E-08 | 9.26E-06 |
| Obesity | nk_cell | *HTR1F* | -4.089701976 | 7.68E-10 | 1.94E-08 | 3.73E-06 |
| Obesity | nk_cell | *ZNF664* | -2.921643367 | 2.55E-08 | 1.04E-07 | 0.000123629 |
| Obesity | nk_cell | *CDH4* | -2.005072812 | 8.89E-08 | 2.79E-07 | 0.000431485 |
| Obesity | nk_cell | *NSUN7* | -1.493805998 | 1.63E-07 | 4.58E-07 | 0.000793 |
| Obesity | nk_cell | *PACS2* | -2.269372181 | 8.43E-08 | 2.67E-07 | 0.000409179 |
| Obesity | nk_cell | *LINC00174* | -1.800145849 | 1.42E-05 | 2.42E-05 | 0.069103635 |
| Obesity | nk_cell | *GEMIN4* | -2.975592043 | 4.57E-05 | 7.21E-05 | 0.221765487 |
| Obesity | nk_cell | *LBX2* | 1.65928961 | 2.62E-09 | 2.44E-08 | 1.27E-05 |
| Obesity | nk_cell | *ZFPM1* | 1.47156437 | 1.05E-09 | 2.05E-08 | 5.10E-06 |
| Obesity | nk_cell | *FCER1A* | -1.783501582 | 2.79E-08 | 1.11E-07 | 0.000135571 |
| Obesity | nk_cell | *RPRML* | 1.734661997 | 5.03E-11 | 4.43E-09 | 2.44E-07 |
| Obesity | nk_cell | *SERTAD2* | -2.191459958 | 0.001131429 | 0.001506089 | 1 |
| Obesity | nk_cell | *AKAP5* | 1.020970823 | 1.16E-09 | 2.05E-08 | 5.64E-06 |
| Obesity | nk_cell | *GIPC3* | 6.529779498 | 1.32E-09 | 2.09E-08 | 6.39E-06 |
| Obesity | nk_cell | *PDXDC1* | -1.830325476 | 1.12E-07 | 3.36E-07 | 0.000541303 |
| Obesity | nk_cell | *C1orf194* | -2.982548113 | 0.00599957 | 0.007502555 | 1 |
| Obesity | nk_cell | *B3GNT3* | -2.109554359 | 0.000909578 | 0.001224888 | 1 |
| Obesity | nk_cell | *SEPHS2* | -2.416641012 | 2.18E-08 | 9.19E-08 | 0.000105812 |
| Obesity | nk_cell | *ZADH2* | -2.784018142 | 1.48E-07 | 4.22E-07 | 0.000717272 |
| Obesity | nk_cell | *SEPT1* | 3.257758696 | 1.35E-07 | 3.92E-07 | 0.000653936 |
| Obesity | nk_cell | *TDRD6* | -0.856693744 | 0.000322185 | 0.000456022 | 1 |
| Obesity | nk_cell | *CSNK1A1L* | 8.514257722 | 7.23E-09 | 4.27E-08 | 3.51E-05 |
| Obesity | nk_cell | *LYNX1* | -1.294226444 | 2.17E-06 | 4.35E-06 | 0.010524625 |
| Obesity | nk_cell | *FAHD1* | -0.956177479 | 1.54E-05 | 2.59E-05 | 0.074559669 |
| Obesity | nk_cell | *HCLS1* | 1.195606438 | 4.35E-08 | 1.56E-07 | 0.00021105 |
| Obesity | nk_cell | *MTURN* | -1.762367052 | 8.35E-09 | 4.61E-08 | 4.05E-05 |
| Obesity | nk_cell | *CCDC66* | -1.995262404 | 2.37E-07 | 6.26E-07 | 0.001149586 |
| Obesity | nk_cell | *DEFB124* | -11.32194971 | 2.89E-07 | 7.36E-07 | 0.001402335 |
| Obesity | nk_cell | *GLIPR1L2* | -1.420060325 | 1.98E-09 | 2.20E-08 | 9.60E-06 |
| Obesity | nk_cell | *DEFB119* | -17.48049506 | 6.01E-09 | 3.88E-08 | 2.92E-05 |
| Obesity | nk_cell | *TSPYL5* | -2.035052975 | 4.25E-07 | 1.03E-06 | 0.002064323 |
| Obesity | nk_cell | *MB21D2* | -1.800047116 | 9.65E-08 | 2.98E-07 | 0.000468247 |
| Obesity | nk_cell | *SLC47A2* | -2.49198469 | 1.17E-06 | 2.50E-06 | 0.005656605 |
| Obesity | nk_cell | *CHST13* | -8.53390985 | 7.61E-08 | 2.45E-07 | 0.000369108 |
| Obesity | nk_cell | *SLC36A4* | -2.330558075 | 1.85E-07 | 5.06E-07 | 0.00089666 |
| Obesity | nk_cell | *OR51E1* | -1.752160551 | 1.03E-06 | 2.24E-06 | 0.004998994 |
| Obesity | nk_cell | *PSMG4* | -2.167954092 | 4.88E-08 | 1.71E-07 | 0.00023684 |
| Obesity | nk_cell | *BHLHE22* | 1.439088963 | 0.02648753 | 0.031445436 | 1 |
| Obesity | nk_cell | *ZNF443* | -3.76673415 | 5.64E-10 | 1.82E-08 | 2.74E-06 |
| Obesity | nk_cell | *CAPS2* | -2.176177484 | 5.97E-06 | 1.09E-05 | 0.028963072 |
| Obesity | nk_cell | *KCTD2* | -1.63074915 | 5.43E-09 | 3.63E-08 | 2.64E-05 |
| Obesity | nk_cell | *D2HGDH* | -2.088052526 | 3.77E-07 | 9.29E-07 | 0.001827096 |
| Obesity | nk_cell | *CMTR2* | -2.085058557 | 0.002759043 | 0.003563183 | 1 |

| | | | | | | |
|---|---|---|---|---|---|---|
| Obesity | nk_cell | *ST20* | 0.639084854 | 0.000848455 | 0.001145119 | 1 |
| Obesity | nk_cell | *MRPL14* | -2.537837196 | 0.001737935 | 0.002281509 | 1 |
| Obesity | nk_cell | *NQO1* | -3.397372963 | 5.49E-08 | 1.87E-07 | 0.00026642 |
| Obesity | nk_cell | *FKRP* | -0.68615451 | 0.037373505 | 0.043642899 | 1 |
| Obesity | nk_cell | *RPH3AL* | -2.069412053 | 0.000232576 | 0.000334655 | 1 |
| Obesity | nk_cell | *ADIPOQ* | -0.597954112 | 0.000253175 | 0.000363004 | 1 |
| Obesity | nk_cell | *F2R* | 0.563389368 | 2.01E-09 | 2.20E-08 | 9.77E-06 |
| Obesity | nk_cell | *MUC16* | -2.850879707 | 1.57E-06 | 3.25E-06 | 0.007594866 |
| Obesity | nk_cell | *PENK* | -4.491656759 | 1.04E-05 | 1.81E-05 | 0.050632105 |
| Obesity | nk_cell | *SLC25A41* | 16.50900399 | 9.47E-09 | 5.00E-08 | 4.59E-05 |
| Obesity | nk_cell | *TMEM136* | -2.3754919 | 5.07E-09 | 3.50E-08 | 2.46E-05 |
| Obesity | nk_cell | *ZNF322* | -2.441844 | 0.003112597 | 0.003994267 | 1 |
| Obesity | nk_cell | *DDX60L* | -2.582301073 | 2.27E-05 | 3.72E-05 | 0.110044585 |
| Obesity | nk_cell | *OGFOD3* | -1.597112298 | 0.005062148 | 0.006379472 | 1 |
| Obesity | nk_cell | *TMEM45A* | -2.132726486 | 2.73E-08 | 1.09E-07 | 0.000132293 |
| Obesity | nk_cell | *SGSH* | -3.947464545 | 5.77E-08 | 1.95E-07 | 0.000280132 |
| Obesity | nk_cell | *ZFP41* | 2.694711904 | 8.39E-12 | 1.32E-09 | 4.07E-08 |
| Obesity | nk_cell | *ZBTB20* | -1.505986625 | 4.55E-06 | 8.50E-06 | 0.022092308 |
| Obesity | nk_cell | *C3orf58* | -2.163997273 | 0.022771489 | 0.027160095 | 1 |
| Obesity | nk_cell | *AMIGO1* | -1.565357035 | 0.0001925 | 0.00027956 | 0.934010286 |
| Obesity | nk_cell | *TMEM252* | -42.99563424 | 2.18E-09 | 2.26E-08 | 1.06E-05 |
| Obesity | nk_cell | *ODF3L2* | 2.881047549 | 3.39E-07 | 8.47E-07 | 0.001645651 |
| Obesity | nk_cell | *RELL1* | -2.024098557 | 1.42E-05 | 2.41E-05 | 0.069041403 |
| Obesity | nk_cell | *RFX7* | -2.169215284 | 0.000232924 | 0.000335058 | 1 |
| Obesity | nk_cell | *CLDN7* | -1.662390011 | 2.66E-11 | 2.87E-09 | 1.29E-07 |
| Obesity | nk_cell | *ZNF329* | -1.79866645 | 6.31E-09 | 4.00E-08 | 3.06E-05 |
| Obesity | nk_cell | *ADO* | 2.036602283 | 0.002803672 | 0.00361793 | 1 |
| Obesity | nk_cell | *IDH2* | -3.311494048 | 0.000387864 | 0.000543278 | 1 |
| Obesity | nk_cell | *PTCHD3* | -1.495726697 | 0.010910903 | 0.013361862 | 1 |
| Obesity | nk_cell | *WRB* | -1.897655174 | 2.00E-08 | 8.60E-08 | 9.69E-05 |
| Obesity | nk_cell | *FAM181B* | -3.316042365 | 8.49E-05 | 0.000128669 | 0.412167357 |
| Obesity | nk_cell | *TDRKH* | -2.565209478 | 2.60E-10 | 1.29E-08 | 1.26E-06 |
| Obesity | nk_cell | *ZNF708* | -2.125495195 | 1.59E-06 | 3.30E-06 | 0.007720545 |
| Obesity | nk_cell | *ERCC6L2* | -2.282133843 | 2.44E-06 | 4.82E-06 | 0.011826825 |
| Obesity | nk_cell | *MRPL41* | -3.037846187 | 4.40E-08 | 1.58E-07 | 0.000213577 |
| Obesity | nk_cell | *ARL6IP4* | -2.124363766 | 0.000330553 | 0.000467184 | 1 |
| Obesity | nk_cell | *HHIPL1* | -2.629600053 | 2.78E-07 | 7.14E-07 | 0.001347413 |
| Obesity | nk_cell | *FAM153B* | -2.194633751 | 2.97E-08 | 1.16E-07 | 0.000144103 |
| Obesity | nk_cell | *BACE2* | -1.278682875 | 1.07E-05 | 1.85E-05 | 0.052000504 |
| Obesity | nk_cell | *SYNM* | -3.938149417 | 0.001031806 | 0.001379153 | 1 |
| Obesity | nk_cell | *KCNA4* | -3.916306035 | 0.009370635 | 0.011536747 | 1 |
| Obesity | nk_cell | *FIGN* | -1.944315874 | 3.33E-06 | 6.41E-06 | 0.016173608 |
| Obesity | nk_cell | *IZUMO1* | 2.239821869 | 7.98E-07 | 1.78E-06 | 0.003871522 |
| Obesity | nk_cell | *FBXL6* | 5.273725826 | 9.17E-06 | 1.61E-05 | 0.044489197 |
| Obesity | nk_cell | *NXPH4* | 16.473111 | 2.69E-09 | 2.45E-08 | 1.31E-05 |
| Obesity | nk_cell | *NPLOC4* | -2.224784379 | 3.46E-06 | 6.63E-06 | 0.016789315 |
| Obesity | nk_cell | *CAPN12* | 0.932842833 | 3.09E-08 | 1.20E-07 | 0.000149902 |
| Obesity | nk_cell | *KPNA2* | -3.044491902 | 8.27E-08 | 2.63E-07 | 0.000401071 |
| Obesity | nk_cell | *ORAI1* | 2.222052566 | 1.48E-10 | 9.24E-09 | 7.19E-07 |
| Obesity | nk_cell | *FES* | -2.104455374 | 7.88E-06 | 1.39E-05 | 0.03822933 |

| | | | | | | |
|---|---|---|---|---|---|---|
| Obesity | nk_cell | *GLRX5* | -3.55181082 | 0.001286346 | 0.001705287 | 1 |
| Obesity | nk_cell | *CAV3* | -1.636424554 | 9.71E-11 | 7.34E-09 | 4.71E-07 |
| Obesity | nk_cell | *MFSD5* | 1.717389237 | 1.90E-07 | 5.17E-07 | 0.000921612 |
| Obesity | nk_cell | *ADI1* | -3.118951472 | 0.000132171 | 0.000195696 | 0.641294931 |
| Obesity | nk_cell | *CLEC4G* | -5.524622509 | 9.52E-10 | 2.05E-08 | 4.62E-06 |
| Obesity | nk_cell | *TRAK1* | -2.246746642 | 1.93E-05 | 3.19E-05 | 0.093541367 |
| Obesity | nk_cell | *SKA2* | -2.140899819 | 0.012491758 | 0.015215999 | 1 |
| Obesity | nk_cell | *TTC3* | -0.987690965 | 0.000894234 | 0.00120456 | 1 |
| Obesity | nk_cell | *PPP1R27* | 2.58736397 | 7.76E-08 | 2.49E-07 | 0.000376752 |
| Obesity | nk_cell | *BRICD5* | -2.531410727 | 1.83E-09 | 2.19E-08 | 8.89E-06 |
| Obesity | nk_cell | *GRID1* | -1.469966792 | 3.54E-11 | 3.30E-09 | 1.72E-07 |
| Obesity | nk_cell | *ACBD3* | -2.448138164 | 7.53E-05 | 0.00011487 | 0.365400479 |
| Obesity | nk_cell | *PLCXD3* | -2.71573049 | 5.93E-06 | 1.08E-05 | 0.028763452 |
| Obesity | nk_cell | *VMO1* | 2.567613748 | 1.30E-05 | 2.21E-05 | 0.062994517 |
| Obesity | nk_cell | *ALG12* | 2.755105024 | 1.05E-08 | 5.37E-08 | 5.08E-05 |
| Obesity | nk_cell | *COL18A1* | -1.60761499 | 6.24E-05 | 9.63E-05 | 0.302614934 |
| Obesity | nk_cell | *ZNF721* | -1.952276035 | 0.000420013 | 0.000585773 | 1 |
| Obesity | nk_cell | *CEP63* | -1.591203878 | 7.95E-09 | 4.48E-08 | 3.86E-05 |
| Obesity | nk_cell | *HMGN4* | -2.336391336 | 4.99E-08 | 1.74E-07 | 0.000242182 |
| Obesity | nk_cell | *C12orf60* | -2.879447101 | 4.78E-08 | 1.68E-07 | 0.000231923 |
| Obesity | nk_cell | *ABAT* | -2.909665916 | 0.000265782 | 0.000380069 | 1 |
| Obesity | nk_cell | *LYSMD4* | -1.654160665 | 1.05E-06 | 2.28E-06 | 0.005101853 |
| Obesity | nk_cell | *WBP2NL* | -2.452382202 | 4.00E-08 | 1.47E-07 | 0.000194158 |
| Obesity | nk_cell | *BEGAIN* | -2.32902793 | 5.94E-10 | 1.84E-08 | 2.88E-06 |
| Obesity | nk_cell | *ARHGEF37* | -3.479188098 | 4.22E-06 | 7.92E-06 | 0.020461249 |
| Obesity | nk_cell | *CSMD1* | -2.076254593 | 8.06E-09 | 4.51E-08 | 3.91E-05 |
| Obesity | nk_cell | *RABIF* | 3.266871522 | 4.90E-07 | 1.16E-06 | 0.00237989 |
| Obesity | nk_cell | *FANCF* | 4.44701285 | 1.32E-08 | 6.34E-08 | 6.40E-05 |
| Obesity | nk_cell | *SMDT1* | 1.079805233 | 0.011044895 | 0.013512312 | 1 |
| Obesity | nk_cell | *C2CD4C* | -0.993406789 | 0.001959491 | 0.002559896 | 1 |
| Obesity | nk_cell | *RUVBL2* | 3.165916487 | 1.34E-08 | 6.42E-08 | 6.49E-05 |
| Obesity | nk_cell | *GDPGP1* | -1.685433215 | 0.000416648 | 0.000581246 | 1 |
| Obesity | nk_cell | *PTTG1IP* | -2.382794558 | 4.21E-06 | 7.92E-06 | 0.020442173 |
| Obesity | nk_cell | *ZNF623* | -1.54485511 | 3.50E-08 | 1.32E-07 | 0.000169871 |
| Obesity | nk_cell | *CCDC125* | -1.716350366 | 2.77E-08 | 1.10E-07 | 0.000134459 |
| Obesity | nk_cell | *C10orf107* | -0.821716559 | 0.000195149 | 0.000283153 | 0.946864843 |
| Obesity | nk_cell | *FHL3* | -1.528667888 | 0.0005189 | 0.000715662 | 1 |
| Obesity | nk_cell | *C19orf71* | -1.572841045 | 2.28E-09 | 2.28E-08 | 1.11E-05 |
| Obesity | nk_cell | *LRIT3* | -7.636984826 | 1.25E-09 | 2.09E-08 | 6.06E-06 |
| Obesity | nk_cell | *NPIPA1* | -1.098572618 | 4.29E-09 | 3.17E-08 | 2.08E-05 |
| Obesity | nk_cell | *SF3A3* | -1.80381663 | 0.001787236 | 0.002341164 | 1 |
| Obesity | nk_cell | *GPR132* | 2.10282812 | 0.000506883 | 0.000700085 | 1 |
| Obesity | nk_cell | *PSMG1* | -1.376029515 | 0.027889561 | 0.032988823 | 1 |
| Obesity | nk_cell | *AC131971.1* | -6.128361713 | 4.06E-08 | 1.49E-07 | 0.000197214 |
| Obesity | nk_cell | *SERHL2* | -0.961257819 | 1.54E-09 | 2.11E-08 | 7.46E-06 |
| Obesity | nk_cell | *PCBP3* | -0.975606131 | 1.64E-05 | 2.74E-05 | 0.079355573 |
| Obesity | nk_cell | *MRPL54* | 1.455792188 | 0.000709477 | 0.000966147 | 1 |
| Obesity | nk_cell | *HMCES* | -2.830454235 | 0.000147129 | 0.000216456 | 0.713871739 |
| Obesity | nk_cell | *NDUFB1* | -2.42719613 | 5.66E-07 | 1.31E-06 | 0.002745654 |
| Obesity | nk_cell | *FAM19A1* | 1.000722177 | 4.66E-09 | 3.30E-08 | 2.26E-05 |

| | | | | | | |
|---|---|---|---|---|---|---|
| Obesity | nk_cell | *ALYREF* | 2.727647241 | 3.01E-05 | 4.87E-05 | 0.146147637 |
| Obesity | nk_cell | *MRGPRX2* | 3.297862963 | 1.77E-09 | 2.16E-08 | 8.57E-06 |
| Obesity | nk_cell | *UPP1* | 1.619795402 | 9.83E-11 | 7.34E-09 | 4.77E-07 |
| Obesity | nk_cell | *FIGLA* | -5.265783025 | 5.40E-09 | 3.62E-08 | 2.62E-05 |
| Obesity | nk_cell | *ACSM2A* | -10.11222745 | 7.02E-10 | 1.92E-08 | 3.41E-06 |
| Obesity | nk_cell | *KREMEN1* | -1.214857256 | 3.42E-06 | 6.57E-06 | 0.016596369 |
| Obesity | nk_cell | *ZNF703* | 4.11954937 | 1.93E-08 | 8.41E-08 | 9.36E-05 |
| Obesity | nk_cell | *NPIPA5* | 0.665074258 | 0.003422071 | 0.004382129 | 1 |
| Obesity | nk_cell | *FAM162B* | -2.187468729 | 6.69E-06 | 1.20E-05 | 0.032456533 |
| Obesity | nk_cell | *BTBD9* | -1.930170949 | 6.97E-06 | 1.25E-05 | 0.033817549 |
| Obesity | nk_cell | *NUDT14* | 3.678322663 | 0.00013251 | 0.000196138 | 0.642938904 |
| Obesity | nk_cell | *FAM3B* | -0.684430319 | 3.66E-05 | 5.85E-05 | 0.17743788 |
| Obesity | nk_cell | *PP2D1* | -3.016567024 | 1.59E-08 | 7.32E-08 | 7.71E-05 |
| Obesity | nk_cell | *DENND5A* | -2.163285624 | 3.59E-07 | 8.92E-07 | 0.001742887 |
| Obesity | nk_cell | *CNTN2* | -1.687228625 | 2.00E-08 | 8.60E-08 | 9.71E-05 |
| Obesity | nk_cell | *CRELD2* | 3.397474498 | 1.09E-08 | 5.48E-08 | 5.30E-05 |
| Obesity | nk_cell | *GOLGA6L4* | -2.004816119 | 6.50E-09 | 4.05E-08 | 3.16E-05 |
| Obesity | nk_cell | *PGP* | -3.588040858 | 4.63E-06 | 8.63E-06 | 0.022462011 |
| Obesity | nk_cell | *OAF* | -1.476702818 | 4.66E-10 | 1.69E-08 | 2.26E-06 |
| Obesity | nk_cell | *ALDH1A3* | -0.982454111 | 1.54E-09 | 2.11E-08 | 7.50E-06 |
| Obesity | nk_cell | *CLECL1* | -1.441854611 | 7.97E-08 | 2.55E-07 | 0.000386701 |
| Obesity | nk_cell | *MROH7* | -1.591440061 | 3.47E-05 | 5.57E-05 | 0.168382835 |
| Obesity | nk_cell | *PKP3* | 9.706872724 | 7.75E-06 | 1.37E-05 | 0.037622175 |
| Obesity | nk_cell | *SS18L1* | -1.792776593 | 1.77E-08 | 7.89E-08 | 8.58E-05 |
| Obesity | nk_cell | *TOP1MT* | -1.540238406 | 1.78E-10 | 1.02E-08 | 8.65E-07 |
| Obesity | nk_cell | *COPB2* | -1.766497491 | 0.027988084 | 0.033097291 | 1 |
| Obesity | nk_cell | *WDR27* | -1.808726144 | 2.00E-05 | 3.29E-05 | 0.096853014 |
| Obesity | nk_cell | *TXNRD2* | -1.980950904 | 2.27E-08 | 9.48E-08 | 0.000110206 |
| Obesity | nk_cell | *TMEM255B* | 2.658346052 | 0.036273218 | 0.042419295 | 1 |
| Obesity | nk_cell | *HDDC3* | -2.661118521 | 2.68E-07 | 6.95E-07 | 0.001300519 |
| Obesity | nk_cell | *ZFP1* | 2.742709398 | 9.97E-08 | 3.06E-07 | 0.000483616 |
| Obesity | nk_cell | *DUSP8* | -2.29625222 | 1.34E-07 | 3.91E-07 | 0.000650464 |
| Obesity | nk_cell | *SNN* | 4.840613522 | 3.81E-08 | 1.41E-07 | 0.000184664 |
| Obesity | nk_cell | *KRBA2* | -3.201643213 | 2.08E-10 | 1.16E-08 | 1.01E-06 |
| Obesity | nk_cell | *SEPT5* | -3.769035906 | 0.000173719 | 0.000253422 | 0.842882719 |
| Obesity | nk_cell | *RNLS* | -1.803877893 | 4.64E-09 | 3.29E-08 | 2.25E-05 |
| Obesity | nk_cell | *NDUFA12* | 2.556478742 | 0.002512411 | 0.003254196 | 1 |
| Obesity | nk_cell | *TCTE3* | -2.464457156 | 8.10E-09 | 4.51E-08 | 3.93E-05 |
| Obesity | nk_cell | *UBE2G2* | -2.058973881 | 0.000353705 | 0.000497876 | 1 |
| Obesity | nk_cell | *TMED9* | 2.877748475 | 0.000406329 | 0.000567503 | 1 |
| Obesity | nk_cell | *DRD1* | -24.72092292 | 1.16E-09 | 2.05E-08 | 5.65E-06 |
| Obesity | nk_cell | *BTBD6* | -2.747439421 | 5.36E-05 | 8.35E-05 | 0.259922436 |
| Obesity | nk_cell | *WT1* | -1.202447468 | 7.38E-10 | 1.92E-08 | 3.58E-06 |
| Obesity | nk_cell | *ZFP90* | -2.18684136 | 0.010937197 | 0.013390684 | 1 |
| Obesity | nk_cell | *FAM227A* | -1.349226677 | 0.000248052 | 0.000355869 | 1 |
| Obesity | nk_cell | *NOC4L* | 3.157927833 | 7.70E-07 | 1.73E-06 | 0.003735017 |
| Obesity | nk_cell | *SORCS2* | -1.744658568 | 1.10E-09 | 2.05E-08 | 5.35E-06 |
| Obesity | nk_cell | *TMEM106A* | -1.916492212 | 6.38E-06 | 1.15E-05 | 0.030956929 |
| Obesity | nk_cell | *BRI3BP* | -2.979164486 | 0.014650054 | 0.01777496 | 1 |
| Obesity | nk_cell | *IFNE* | -4.523148426 | 0.021682852 | 0.02592778 | 1 |

| | | | | | | |
|---|---|---|---|---|---|---|
| Obesity | nk_cell | *MAFF* | -1.553927164 | 2.81E-12 | 5.24E-10 | 1.36E-08 |
| Obesity | nk_cell | *SEMA4B* | -3.302630311 | 1.13E-06 | 2.42E-06 | 0.005462307 |
| Obesity | nk_cell | *CIB1* | 1.921593668 | 9.19E-05 | 0.000138536 | 0.445669264 |
| Obesity | nk_cell | *SGCZ* | -1.230214753 | 4.52E-07 | 1.08E-06 | 0.002194828 |
| Obesity | nk_cell | *C6orf120* | -3.201995483 | 0.00175696 | 0.002303992 | 1 |
| Obesity | nk_cell | *SIGIRR* | 1.216434991 | 8.71E-10 | 2.00E-08 | 4.22E-06 |
| Obesity | nk_cell | *IFITM2* | 1.978587972 | 1.38E-08 | 6.58E-08 | 6.70E-05 |
| Obesity | nk_cell | *GP1BA* | -1.10821866 | 0.000748066 | 0.001017555 | 1 |
| Obesity | nk_cell | *ATP6V0A2* | 2.17127216 | 0.000366479 | 0.00051496 | 1 |
| Obesity | nk_cell | *C14orf80* | 2.767947759 | 3.30E-09 | 2.73E-08 | 1.60E-05 |
| Obesity | nk_cell | *TNFAIP8L1* | -5.25225097 | 1.25E-07 | 3.70E-07 | 0.000607974 |
| Obesity | nk_cell | *TARSL2* | -1.927985243 | 1.22E-05 | 2.08E-05 | 0.059032083 |
| Obesity | nk_cell | *SH3BGR* | -1.092609185 | 0.000343096 | 0.000483644 | 1 |
| Obesity | nk_cell | *FAM174B* | -2.051790298 | 3.87E-08 | 1.43E-07 | 0.000187571 |
| Obesity | nk_cell | *SV2B* | -1.639397074 | 7.45E-09 | 4.35E-08 | 3.61E-05 |
| Obesity | nk_cell | *AHNAK2* | -4.312427439 | 1.49E-05 | 2.52E-05 | 0.072435751 |
| Obesity | nk_cell | *SP1* | -2.050075049 | 2.56E-07 | 6.68E-07 | 0.001240706 |
| Obesity | nk_cell | *PCGF3* | -1.974459513 | 0.000167733 | 0.000245059 | 0.813840613 |
| Obesity | nk_cell | *PSMD13* | -3.518966316 | 5.33E-08 | 1.83E-07 | 0.000258408 |
| Obesity | nk_cell | *PMEL* | -1.065061802 | 1.02E-05 | 1.77E-05 | 0.049562571 |
| Obesity | nk_cell | *C6orf201* | -2.222570575 | 3.51E-05 | 5.63E-05 | 0.170435363 |
| Obesity | nk_cell | *MYBL1* | 0.162050279 | 1.51E-09 | 2.11E-08 | 7.33E-06 |
| Obesity | nk_cell | *ADARB2* | 2.639179172 | 4.22E-10 | 1.62E-08 | 2.05E-06 |
| Obesity | nk_cell | *C11orf87* | -4.328023311 | 0.018747918 | 0.022549553 | 1 |
| Obesity | nk_cell | *DMWD* | -2.070308238 | 6.37E-09 | 4.02E-08 | 3.09E-05 |
| Obesity | nk_cell | *SLC52A2* | 3.032953351 | 2.90E-10 | 1.36E-08 | 1.41E-06 |
| Obesity | nk_cell | *PIGP* | -0.71126703 | 0.001758476 | 0.002305357 | 1 |
| Obesity | nk_cell | *ARL17A* | 1.660272139 | 0.010427419 | 0.012802084 | 1 |
| Obesity | nk_cell | *GNB1L* | -1.455200197 | 0.007699265 | 0.009549293 | 1 |
| Obesity | nk_cell | *EVI2B* | 1.134795125 | 1.00E-06 | 2.19E-06 | 0.004864195 |
| Obesity | nk_cell | *NPIPB4* | 2.374019409 | 0.000333335 | 0.000470431 | 1 |
| Obesity | nk_cell | *ZNF829* | -2.37026103 | 1.15E-05 | 1.97E-05 | 0.055598566 |
| Obesity | nk_cell | *IFITM1* | 0.921363452 | 6.74E-10 | 1.91E-08 | 3.27E-06 |
| Obesity | nk_cell | *LINC00839* | -3.211845687 | 3.00E-09 | 2.59E-08 | 1.46E-05 |
| Obesity | nk_cell | *RTN4RL1* | -1.362805013 | 9.89E-09 | 5.16E-08 | 4.80E-05 |
| Obesity | nk_cell | *RNPC3* | -1.342208175 | 7.92E-08 | 2.54E-07 | 0.000384177 |
| Obesity | nk_cell | *RASA3* | 1.935699684 | 7.44E-07 | 1.68E-06 | 0.003611177 |
| Obesity | nk_cell | *ZNF529* | -2.349890344 | 1.36E-07 | 3.94E-07 | 0.000658295 |
| Obesity | nk_cell | *WWOX* | -1.520075807 | 5.78E-07 | 1.34E-06 | 0.002806678 |
| Obesity | nk_cell | *CYP4Z1* | -0.959524938 | 2.45E-08 | 1.01E-07 | 0.000118907 |
| Obesity | nk_cell | *POLR1D* | -2.181138918 | 7.35E-06 | 1.31E-05 | 0.035648034 |
| Obesity | nk_cell | *FFAR4* | -1.25362826 | 1.17E-09 | 2.05E-08 | 5.68E-06 |
| Obesity | nk_cell | *CYP4F12* | -2.065664765 | 1.59E-07 | 4.49E-07 | 0.000773133 |
| Obesity | nk_cell | *ZNF749* | -0.755028202 | 0.00288955 | 0.003722808 | 1 |
| Obesity | nk_cell | *GPAT2* | -3.430443388 | 1.73E-05 | 2.89E-05 | 0.084080406 |
| Obesity | nk_cell | *TOR3A* | -3.382295415 | 3.89E-08 | 1.44E-07 | 0.000188668 |
| Obesity | nk_cell | *ZNF555* | -2.040413082 | 7.31E-08 | 2.37E-07 | 0.000354842 |
| Obesity | nk_cell | *GOLGA8R* | 2.734050842 | 0.002122154 | 0.002766441 | 1 |
| Obesity | nk_cell | *CCDC30* | -1.516848074 | 1.52E-06 | 3.17E-06 | 0.007376625 |
| Obesity | nk_cell | *GLDN* | -0.213876534 | 0.042369042 | 0.049239423 | 1 |

| | | | | | | |
|---|---|---|---|---|---|---|
| Obesity | nk_cell | *ZNF197* | -2.319692239 | 2.05E-07 | 5.54E-07 | 0.000995364 |
| Obesity | nk_cell | *FAM228A* | 1.154877807 | 5.00E-07 | 1.18E-06 | 0.002427405 |
| Obesity | nk_cell | *RPS23* | 2.020800314 | 0.000427524 | 0.000595051 | 1 |
| Obesity | nk_cell | *GNG2* | 0.888510227 | 1.96E-09 | 2.20E-08 | 9.52E-06 |
| Obesity | nk_cell | *BTN3A2* | -0.555654858 | 0.003465251 | 0.004436253 | 1 |
| Obesity | nk_cell | *TMEM222* | -3.783374098 | 7.28E-05 | 0.000111229 | 0.353375999 |
| Obesity | nk_cell | *SEPT10* | -2.238273646 | 2.10E-08 | 8.95E-08 | 0.000101896 |
| Obesity | nk_cell | *FAM86B1* | -3.692180583 | 7.84E-06 | 1.39E-05 | 0.038048875 |
| Obesity | nk_cell | *SMYD4* | -1.797998584 | 1.36E-06 | 2.88E-06 | 0.006621903 |
| Obesity | nk_cell | *CEACAM19* | -1.457774148 | 2.77E-08 | 1.10E-07 | 0.000134209 |
| Obesity | nk_cell | *MIR22HG* | -2.015457345 | 1.03E-08 | 5.33E-08 | 5.02E-05 |
| Obesity | nk_cell | *CYP27C1* | -1.541996886 | 4.77E-08 | 1.68E-07 | 0.000231544 |
| Obesity | nk_cell | *BCR* | -2.128301774 | 0.000621242 | 0.00085245 | 1 |
| Obesity | nk_cell | *FOXI2* | -2.125067978 | 0.003763313 | 0.004806422 | 1 |
| Obesity | nk_cell | *ZNF732* | -1.910977903 | 2.81E-09 | 2.51E-08 | 1.36E-05 |
| Obesity | nk_cell | *HYAL3* | -1.606033246 | 0.003930626 | 0.005006931 | 1 |
| Obesity | nk_cell | *VSIG10L* | -2.896941699 | 7.41E-06 | 1.32E-05 | 0.035949167 |
| Obesity | nk_cell | *TRABD2A* | -0.875836382 | 0.000215215 | 0.000310688 | 1 |
| Obesity | nk_cell | *QRFPR* | -4.738804671 | 4.11E-08 | 1.50E-07 | 0.000199465 |
| Obesity | nk_cell | *MAPT* | -1.668749365 | 4.57E-09 | 3.28E-08 | 2.22E-05 |
| Obesity | nk_cell | *TMEM17* | -2.104166058 | 1.16E-07 | 3.46E-07 | 0.000561351 |
| Obesity | nk_cell | *TNFRSF18* | 0.698859429 | 1.04E-09 | 2.05E-08 | 5.04E-06 |
| Obesity | nk_cell | *C1QL4* | -12.7296639 | 1.46E-09 | 2.11E-08 | 7.09E-06 |
| Obesity | nk_cell | *RTN4RL2* | -2.585896598 | 4.43E-10 | 1.65E-08 | 2.15E-06 |
| Obesity | nk_cell | *TMEM232* | -1.505677061 | 6.71E-08 | 2.20E-07 | 0.000325702 |
| Obesity | nk_cell | *EFCAB6* | -2.55538527 | 4.24E-07 | 1.02E-06 | 0.002056789 |
| Obesity | nk_cell | *RHD* | -2.053050909 | 0.000121642 | 0.0001811 | 0.590205108 |
| Obesity | nk_cell | *TMPRSS6* | -11.07218963 | 1.88E-09 | 2.19E-08 | 9.12E-06 |
| Obesity | nk_cell | *CYP4A11* | 2.803060676 | 2.12E-10 | 1.17E-08 | 1.03E-06 |
| Obesity | nk_cell | *TMEM216* | 2.311203082 | 5.10E-09 | 3.51E-08 | 2.47E-05 |
| Obesity | nk_cell | *C3orf70* | -1.238390976 | 2.49E-09 | 2.37E-08 | 1.21E-05 |
| Obesity | nk_cell | *ENTPD5* | -2.524703369 | 3.39E-07 | 8.46E-07 | 0.001642908 |
| Obesity | nk_cell | *WDR86* | -1.415595294 | 3.43E-09 | 2.77E-08 | 1.66E-05 |
| Obesity | nk_cell | *EPOR* | -1.20845225 | 1.71E-08 | 7.71E-08 | 8.30E-05 |
| Obesity | nk_cell | *LUZP2* | -1.327704219 | 1.75E-09 | 2.16E-08 | 8.47E-06 |
| Obesity | nk_cell | *GJA4* | -4.824986672 | 7.55E-07 | 1.70E-06 | 0.003662709 |
| Obesity | nk_cell | *IFT140* | -1.784515909 | 8.19E-09 | 4.54E-08 | 3.97E-05 |
| Obesity | nk_cell | *SBK2* | 6.228483123 | 1.29E-09 | 2.09E-08 | 6.27E-06 |
| Obesity | nk_cell | *ZNF385C* | 1.556976181 | 7.63E-09 | 4.39E-08 | 3.70E-05 |
| Obesity | nk_cell | *EXD3* | -2.740314161 | 1.46E-06 | 3.06E-06 | 0.00706978 |
| Obesity | nk_cell | *SAMD11* | -1.622128189 | 1.43E-08 | 6.74E-08 | 6.95E-05 |
| Obesity | nk_cell | *TRPV2* | 1.32091392 | 6.52E-09 | 4.05E-08 | 3.16E-05 |
| Obesity | nk_cell | *C2orf88* | -1.635434192 | 6.67E-09 | 4.10E-08 | 3.24E-05 |
| Obesity | nk_cell | *KBTBD12* | -1.761848221 | 4.56E-06 | 8.51E-06 | 0.02212705 |
| Obesity | nk_cell | *FANCA* | 0.886641535 | 5.98E-07 | 1.37E-06 | 0.002899365 |
| Obesity | nk_cell | *DNAH17* | -1.299438964 | 1.02E-09 | 2.05E-08 | 4.94E-06 |
| Obesity | nk_cell | *FANCM* | -2.910162209 | 5.85E-06 | 1.07E-05 | 0.028394779 |
| Obesity | nk_cell | *ZFP69* | 2.101996582 | 2.65E-09 | 2.45E-08 | 1.29E-05 |
| Obesity | nk_cell | *TMEM220* | -1.672670359 | 1.57E-07 | 4.43E-07 | 0.000760634 |
| Obesity | nk_cell | *FAM122A* | -3.604548362 | 1.80E-08 | 7.98E-08 | 8.71E-05 |

| | | | | | | |
|---|---|---|---|---|---|---|
| Obesity | nk_cell | *GFRAL* | -2.49563671 | 1.24E-08 | 6.05E-08 | 6.02E-05 |
| Obesity | nk_cell | *LDLRAD2* | -1.484831968 | 2.33E-09 | 2.30E-08 | 1.13E-05 |
| Obesity | nk_cell | *CPSF4L* | -4.769335647 | 6.53E-08 | 2.15E-07 | 0.000316748 |
| Obesity | nk_cell | *KLHL17* | -1.871065464 | 3.45E-05 | 5.54E-05 | 0.167342136 |
| Obesity | nk_cell | *OR7D2* | 4.91909965 | 0.000182499 | 0.000265751 | 0.885482915 |
| Obesity | nk_cell | *TMEM221* | -2.195892619 | 0.001665169 | 0.002188353 | 1 |
| Obesity | nk_cell | *MESP2* | 2.26719714 | 1.88E-07 | 5.12E-07 | 0.0009111 |
| Obesity | nk_cell | *MAPK12* | -4.299046331 | 0.000372488 | 0.000522495 | 1 |
| Obesity | nk_cell | *FAM166A* | -5.368489808 | 3.94E-05 | 6.27E-05 | 0.191326158 |
| Obesity | nk_cell | *LINC00265* | 3.812033657 | 3.30E-08 | 1.25E-07 | 0.000159879 |
| Obesity | nk_cell | *PRKAR1B* | -2.310190978 | 4.25E-07 | 1.02E-06 | 0.002060282 |
| Obesity | nk_cell | *NCR3LG1* | -2.736350958 | 2.69E-09 | 2.45E-08 | 1.31E-05 |
| Obesity | nk_cell | *ZNF793* | -1.199500806 | 0.003310936 | 0.004240935 | 1 |
| Obesity | nk_cell | *AGAP4* | 1.618013189 | 0.009675854 | 0.011891399 | 1 |
| Obesity | nk_cell | *RUFY4* | -4.676179895 | 4.94E-09 | 3.46E-08 | 2.40E-05 |
| Obesity | nk_cell | *CENPP* | -0.72981193 | 0.009500777 | 0.011688075 | 1 |
| Obesity | nk_cell | *PLSCR1* | -1.62119442 | 0.022151867 | 0.026460084 | 1 |
| Obesity | nk_cell | *C3orf62* | -2.35141282 | 3.49E-06 | 6.68E-06 | 0.01694182 |
| Obesity | nk_cell | *ENO4* | -1.627634366 | 1.39E-08 | 6.60E-08 | 6.73E-05 |
| Obesity | nk_cell | *H3F3C* | -35.20078645 | 2.39E-09 | 2.32E-08 | 1.16E-05 |
| Obesity | nk_cell | *JAKMIP3* | -1.769355933 | 2.65E-08 | 1.07E-07 | 0.000128459 |
| Obesity | nk_cell | *PDCD1* | 1.176733884 | 6.23E-10 | 1.88E-08 | 3.02E-06 |
| Obesity | nk_cell | *SELL* | 0.625957319 | 1.91E-09 | 2.19E-08 | 9.27E-06 |
| Obesity | nk_cell | *C19orf54* | -3.079353555 | 3.18E-08 | 1.21E-07 | 0.000154202 |
| Obesity | nk_cell | *C22orf34* | 2.181487788 | 8.48E-09 | 4.63E-08 | 4.11E-05 |
| Obesity | nk_cell | *COL25A1* | -1.093632778 | 2.89E-09 | 2.56E-08 | 1.40E-05 |
| Obesity | nk_cell | *SRSF10* | -2.141295689 | 1.99E-05 | 3.29E-05 | 0.096759488 |
| Obesity | nk_cell | *NBR1* | -2.142154029 | 0.002487273 | 0.003223358 | 1 |
| Obesity | nk_cell | *NDOR1* | -1.449180518 | 2.07E-09 | 2.22E-08 | 1.00E-05 |
| Obesity | nk_cell | *FAM72B* | -3.607729518 | 1.52E-07 | 4.33E-07 | 0.000739077 |
| Obesity | nk_cell | *GOLGA8M* | -3.147671195 | 6.44E-09 | 4.04E-08 | 3.13E-05 |
| Obesity | nk_cell | *RHCE* | -1.861903504 | 3.66E-09 | 2.88E-08 | 1.78E-05 |
| Obesity | nk_cell | *IDO2* | -3.056917416 | 2.57E-09 | 2.41E-08 | 1.25E-05 |
| Obesity | nk_cell | *SLC4A5* | -3.142813223 | 4.09E-09 | 3.07E-08 | 1.99E-05 |
| Obesity | nk_cell | *UROS* | 2.607285384 | 4.48E-08 | 1.60E-07 | 0.000217203 |
| Obesity | nk_cell | *ZBED6CL* | -3.701626266 | 2.14E-07 | 5.75E-07 | 0.001039542 |
| Obesity | nk_cell | *OSTN* | -1.055602388 | 7.59E-07 | 1.71E-06 | 0.003684481 |
| Obesity | nk_cell | *FSIP2* | -1.614621884 | 4.19E-08 | 1.52E-07 | 0.000203533 |
| Obesity | nk_cell | *NOXA1* | -3.487810616 | 0.001281588 | 0.001699444 | 1 |
| Obesity | nk_cell | *FZD9* | -30.93461076 | 6.37E-05 | 9.79E-05 | 0.308838443 |
| Obesity | nk_cell | *MTF1* | 2.741140246 | 3.79E-09 | 2.94E-08 | 1.84E-05 |
| Obesity | nk_cell | *ZDHHC11* | -2.734024594 | 8.51E-05 | 0.000128807 | 0.412826825 |
| Obesity | nk_cell | *FAM26F* | 1.228767308 | 0.010303855 | 0.012653582 | 1 |
| Obesity | nk_cell | *ENTPD8* | -115.5719704 | 8.20E-10 | 1.99E-08 | 3.98E-06 |
| Obesity | nk_cell | *RPL14* | 2.508809311 | 2.11E-05 | 3.48E-05 | 0.102608957 |
| Obesity | nk_cell | *ASTL* | -3.560465128 | 1.45E-09 | 2.11E-08 | 7.03E-06 |
| Obesity | nk_cell | *MSL1* | -2.170734743 | 6.06E-05 | 9.37E-05 | 0.294087963 |
| Obesity | nk_cell | *LRRK2* | -2.048231514 | 8.68E-08 | 2.73E-07 | 0.00042135 |
| Obesity | nk_cell | *FAM196A* | -2.243874113 | 4.48E-07 | 1.07E-06 | 0.002172938 |
| Obesity | nk_cell | *UTS2B* | -1.803444769 | 1.48E-09 | 2.11E-08 | 7.20E-06 |

| | | | | | | |
|---|---|---|---|---|---|---|
| Obesity | nk_cell | *C9orf152* | -4.186459586 | 1.69E-07 | 4.70E-07 | 0.000819216 |
| Obesity | nk_cell | *NELFB* | 3.558350684 | 1.58E-06 | 3.29E-06 | 0.007682819 |
| Obesity | nk_cell | *SLC15A5* | -1.418797377 | 8.45E-09 | 4.63E-08 | 4.10E-05 |
| Obesity | nk_cell | *LIPI* | -1.21855566 | 6.69E-06 | 1.20E-05 | 0.032444443 |
| Obesity | nk_cell | *ADAT2* | -2.985938472 | 2.31E-08 | 9.58E-08 | 0.000112075 |
| Obesity | nk_cell | *ALKBH2* | 2.268580135 | 0.000749841 | 0.001019683 | 1 |
| Obesity | nk_cell | *LITAF* | 2.505569891 | 0.000458486 | 0.000636684 | 1 |
| Obesity | nk_cell | *SF3B3* | -2.458575506 | 0.00022819 | 0.000328636 | 1 |
| Obesity | nk_cell | *PLAC9* | -2.014750052 | 3.87E-07 | 9.47E-07 | 0.001877153 |
| Obesity | nk_cell | *ZNF573* | -2.664421234 | 7.58E-07 | 1.70E-06 | 0.003676385 |
| Obesity | nk_cell | *FAM47E* | -1.259140324 | 3.62E-09 | 2.86E-08 | 1.76E-05 |
| Obesity | nk_cell | *ZNF600* | 1.158013221 | 0.000525945 | 0.000724967 | 1 |
| Obesity | nk_cell | *TSPYL1* | -3.510406936 | 3.09E-07 | 7.82E-07 | 0.001498398 |
| Obesity | nk_cell | *PNRC2* | -2.638232392 | 4.00E-08 | 1.47E-07 | 0.0001939 |
| Obesity | nk_cell | *FHIT* | -1.491495985 | 1.62E-07 | 4.55E-07 | 0.000787556 |
| Obesity | nk_cell | *ZKSCAN3* | -2.88888213 | 1.59E-14 | 5.51E-12 | 7.72E-11 |
| Obesity | nk_cell | *KIAA0408* | -1.498085531 | 1.12E-08 | 5.57E-08 | 5.45E-05 |
| Obesity | nk_cell | *AKR1C3* | -1.499825356 | 3.53E-09 | 2.82E-08 | 1.71E-05 |
| Obesity | nk_cell | *WDSUB1* | -2.590235463 | 1.51E-06 | 3.15E-06 | 0.007331531 |
| Obesity | nk_cell | *ZNF79* | 0.639677147 | 0.027773344 | 0.032867382 | 1 |
| Obesity | nk_cell | *SEMA4A* | 4.175559862 | 0.00257484 | 0.003334167 | 1 |
| Obesity | nk_cell | *HRCT1* | -2.468499309 | 4.91E-07 | 1.16E-06 | 0.002380383 |
| Obesity | nk_cell | *MPHOSPH8* | -1.821704234 | 1.18E-05 | 2.02E-05 | 0.05705129 |
| Obesity | nk_cell | *SIRPB2* | -1.311654414 | 2.24E-05 | 3.67E-05 | 0.108626592 |
| Obesity | nk_cell | *SRGAP3* | -1.481144027 | 2.26E-06 | 4.50E-06 | 0.010945679 |
| Obesity | nk_cell | *XPNPEP3* | -1.246239284 | 9.91E-08 | 3.05E-07 | 0.000480892 |
| Obesity | nk_cell | *PPIA* | -2.157166431 | 0.000783945 | 0.001063675 | 1 |
| Obesity | nk_cell | *ZNF471* | -1.965299009 | 6.86E-09 | 4.16E-08 | 3.33E-05 |
| Obesity | nk_cell | *ZNF493* | -2.457728013 | 5.48E-08 | 1.87E-07 | 0.000265684 |
| Obesity | nk_cell | *GTF2IRD2* | -1.808262592 | 1.14E-08 | 5.65E-08 | 5.55E-05 |
| Obesity | nk_cell | *SUPT3H* | -2.186703108 | 6.03E-06 | 1.10E-05 | 0.029242981 |
| Obesity | nk_cell | *GIMAP5* | 1.473190883 | 6.09E-09 | 3.91E-08 | 2.96E-05 |
| Obesity | nk_cell | *STK31* | -2.225654351 | 4.91E-08 | 1.71E-07 | 0.000238128 |
| Obesity | nk_cell | *ZKSCAN7* | 4.137569764 | 1.62E-08 | 7.38E-08 | 7.85E-05 |
| Obesity | nk_cell | *CD55* | -2.104977221 | 1.85E-07 | 5.06E-07 | 0.000895768 |
| Obesity | nk_cell | *CPNE4* | -0.897821919 | 2.92E-07 | 7.44E-07 | 0.001418814 |
| Obesity | nk_cell | *ZNF565* | -1.61347296 | 0.00175336 | 0.002299893 | 1 |
| Obesity | nk_cell | *ZNF34* | -3.670522618 | 4.17E-07 | 1.01E-06 | 0.002025497 |
| Obesity | nk_cell | *ZNF781* | -1.129936157 | 8.95E-08 | 2.80E-07 | 0.000434325 |
| Obesity | nk_cell | *EVL* | 1.74347496 | 1.19E-07 | 3.53E-07 | 0.000575919 |
| Obesity | nk_cell | *ZNF124* | -2.903448031 | 4.86E-05 | 7.65E-05 | 0.235985243 |
| Obesity | nk_cell | *PIK3R4* | -2.575138407 | 1.73E-07 | 4.79E-07 | 0.0008378 |
| Obesity | nk_cell | *ZNF605* | -1.352894751 | 0.000196782 | 0.000285352 | 0.954788086 |
| Obesity | nk_cell | *RFX8* | -1.400527547 | 7.33E-07 | 1.66E-06 | 0.003555393 |
| Obesity | nk_cell | *C20orf96* | -3.424742127 | 9.05E-08 | 2.83E-07 | 0.000438918 |
| Obesity | nk_cell | *SULT1A1* | -1.822863576 | 3.49E-10 | 1.51E-08 | 1.69E-06 |
| Obesity | nk_cell | *TPK1* | -1.866317231 | 0.006341395 | 0.007917768 | 1 |
| Obesity | nk_cell | *SPTSSB* | -3.907326455 | 6.01E-10 | 1.84E-08 | 2.91E-06 |
| Obesity | nk_cell | *CACNA1H* | -1.771875758 | 1.49E-08 | 6.95E-08 | 7.24E-05 |
| Obesity | nk_cell | *SULF2* | -1.181463789 | 0.021675627 | 0.02592778 | 1 |

| | | | | | | |
|---|---|---|---|---|---|---|
| Obesity | nk_cell | *XRCC2* | 4.36720818 | 4.18E-09 | 3.12E-08 | 2.03E-05 |
| Obesity | nk_cell | *MYO6* | -2.247877385 | 2.98E-08 | 1.16E-07 | 0.000144806 |
| Obesity | nk_cell | *ZNF846* | -2.474911342 | 1.43E-07 | 4.09E-07 | 0.00069152 |
| Obesity | nk_cell | *ADH1B* | -1.402110828 | 1.11E-07 | 3.35E-07 | 0.000539611 |
| Obesity | nk_cell | *ZKSCAN5* | -1.921003409 | 1.39E-07 | 4.00E-07 | 0.000672875 |
| Obesity | nk_cell | *ZNF502* | -2.764485141 | 6.59E-07 | 1.51E-06 | 0.003195197 |
| Obesity | nk_cell | *TRAPPC4* | -1.437191455 | 0.00286809 | 0.003698106 | 1 |
| Obesity | nk_cell | *ZFP62* | -2.683422766 | 5.51E-05 | 8.59E-05 | 0.267558805 |
| Obesity | nk_cell | *ZNF33B* | -1.636295371 | 0.000101274 | 0.000152083 | 0.491379122 |
| Obesity | nk_cell | *AMZ2* | 1.763486462 | 1.16E-09 | 2.05E-08 | 5.61E-06 |
| Obesity | nk_cell | *ZNF431* | -2.142114049 | 0.000997785 | 0.001335886 | 1 |
| Obesity | nk_cell | *HLA-DQA1* | -1.07407151 | 0.004853928 | 0.006134738 | 1 |
| Obesity | nk_cell | *COL27A1* | -1.171499605 | 0.007973287 | 0.009868977 | 1 |
| Obesity | nk_cell | *SPRR2B* | -11.93610565 | 0.001314701 | 0.001741449 | 1 |
| Obesity | nk_cell | *ANKRD36B* | -2.797208552 | 7.31E-06 | 1.30E-05 | 0.035453472 |
| Obesity | nk_cell | *FAM3C* | 2.957478435 | 2.89E-06 | 5.64E-06 | 0.014006696 |
| Obesity | nk_cell | *SLC39A10* | 2.551550824 | 0.000131032 | 0.000194127 | 0.635766303 |
| Obesity | nk_cell | *AP2A1* | -1.792668673 | 0.006941329 | 0.008642373 | 1 |
| Obesity | nk_cell | *HIST1H3E* | 1.813451904 | 2.03E-07 | 5.48E-07 | 0.000985256 |
| Obesity | nk_cell | *FUT11* | 2.338860471 | 7.50E-09 | 4.36E-08 | 3.64E-05 |
| Obesity | nk_cell | *WDR5B* | -2.564700787 | 5.89E-08 | 1.97E-07 | 0.000285627 |
| Obesity | nk_cell | *NPIPB9* | -3.021549438 | 0.007503564 | 0.009320864 | 1 |
| Obesity | nk_cell | *ZNF138* | -2.074886956 | 1.88E-05 | 3.11E-05 | 0.091040784 |
| Obesity | nk_cell | *ZNF429* | -1.987913984 | 0.012152324 | 0.014826019 | 1 |
| Obesity | nk_cell | *ZNF470* | 3.866954413 | 0.003041994 | 0.003908833 | 1 |
| Obesity | nk_cell | *ZNF100* | -2.312386766 | 6.19E-08 | 2.06E-07 | 0.000300112 |
| Obesity | nk_cell | *ANXA6* | 2.01393876 | 1.34E-06 | 2.83E-06 | 0.006500594 |
| Obesity | nk_cell | *ZNF420* | -2.663637972 | 2.84E-07 | 7.25E-07 | 0.001376017 |
| Obesity | nk_cell | *ZNF763* | -1.464851813 | 3.27E-06 | 6.32E-06 | 0.01588483 |
| Obesity | nk_cell | *ZMYM1* | -2.764917519 | 4.42E-06 | 8.27E-06 | 0.021438085 |
| Obesity | nk_cell | *GAL3ST4* | -4.209867624 | 1.28E-08 | 6.19E-08 | 6.21E-05 |
| Obesity | nk_cell | *SLC25A29* | -1.56999418 | 1.61E-05 | 2.69E-05 | 0.077879943 |
| Obesity | nk_cell | *ZNF772* | -1.833570042 | 2.76E-07 | 7.10E-07 | 0.001337212 |
| Obesity | nk_cell | *ZNF257* | -3.275048629 | 2.09E-06 | 4.20E-06 | 0.010148774 |
| Obesity | nk_cell | *ADAM32* | -1.647242377 | 3.53E-09 | 2.82E-08 | 1.71E-05 |
| Obesity | nk_cell | *LRRC8B* | -1.985735078 | 2.20E-07 | 5.87E-07 | 0.001067231 |
| Obesity | nk_cell | *ABCB8* | -2.520340296 | 3.26E-07 | 8.20E-07 | 0.001581504 |
| Obesity | nk_cell | *SND1* | -1.962051141 | 2.00E-05 | 3.30E-05 | 0.097051016 |
| Obesity | nk_cell | *SULT1A2* | 7.960814766 | 8.67E-19 | 8.41E-16 | 4.21E-15 |
| Obesity | nk_cell | *NEK5* | -1.787836761 | 5.17E-06 | 9.54E-06 | 0.025077542 |
| Obesity | nk_cell | *PIWIL2* | -2.460734598 | 7.42E-06 | 1.32E-05 | 0.035995433 |
| Obesity | nk_cell | *TBC1D9B* | -2.23330435 | 3.18E-06 | 6.15E-06 | 0.015412802 |
| Obesity | nk_cell | *FAM110D* | 2.780051456 | 3.66E-09 | 2.88E-08 | 1.77E-05 |
| Obesity | nk_cell | *CCL4L2* | 0.358016905 | 1.15E-09 | 2.05E-08 | 5.57E-06 |
| Obesity | nk_cell | *RAMP2-AS1* | -1.490821397 | 2.91E-09 | 2.56E-08 | 1.41E-05 |
| Obesity | nk_cell | *MRPL21* | -0.905854996 | 0.008971153 | 0.011067387 | 1 |
| Obesity | nk_cell | *ZNF786* | 1.925926214 | 3.19E-06 | 6.17E-06 | 0.015467374 |
| Obesity | nk_cell | *ZNF517* | -1.690918929 | 5.21E-09 | 3.55E-08 | 2.53E-05 |
| Obesity | nk_cell | *ZNF675* | -3.310695125 | 0.000117066 | 0.000174663 | 0.56800291 |
| Obesity | nk_cell | *SLC22A5* | -2.2502649 | 4.35E-07 | 1.05E-06 | 0.002112608 |

| | | | | | | |
|---|---|---|---|---|---|---|
| Obesity | nk_cell | *VEPH1* | -1.22489325 | 1.28E-09 | 2.09E-08 | 6.22E-06 |
| Obesity | nk_cell | *SHPK* | -1.442763094 | 9.23E-07 | 2.03E-06 | 0.004476909 |
| Obesity | nk_cell | *MAP3K5* | -1.630017308 | 0.005057465 | 0.00637869 | 1 |
| Obesity | nk_cell | *GSTK1* | -1.588917604 | 0.010994989 | 0.013458044 | 1 |
| Obesity | nk_cell | *GYPE* | -2.837456547 | 2.29E-06 | 4.55E-06 | 0.011087009 |
| Obesity | nk_cell | *PCDHB11* | -1.159280511 | 0.000471668 | 0.000654053 | 1 |
| Obesity | nk_cell | *SLC2A10* | -1.379226502 | 1.01E-08 | 5.23E-08 | 4.89E-05 |
| Obesity | nk_cell | *ZNF665* | -3.457404714 | 9.32E-05 | 0.000140581 | 0.452389427 |
| Obesity | nk_cell | *RPF2* | -2.010928685 | 3.25E-08 | 1.24E-07 | 0.000157625 |
| Obesity | nk_cell | *ATG7* | -1.950606242 | 1.63E-07 | 4.58E-07 | 0.00079239 |
| Obesity | nk_cell | *RAB40C* | 2.982795396 | 0.001212422 | 0.001609927 | 1 |
| Obesity | nk_cell | *PIGN* | -1.967092369 | 1.78E-09 | 2.17E-08 | 8.66E-06 |
| Obesity | nk_cell | *HHLA3* | -1.461575815 | 2.85E-08 | 1.12E-07 | 0.000138222 |
| Obesity | nk_cell | *TOPORS* | -3.101546201 | 0.000266839 | 0.000381131 | 1 |
| Obesity | nk_cell | *BCO2* | -1.122396221 | 4.96E-05 | 7.78E-05 | 0.240653785 |
| Obesity | nk_cell | *ENTPD6* | -2.204930919 | 8.47E-10 | 1.99E-08 | 4.11E-06 |
| Obesity | nk_cell | *ENPP1* | -1.902995098 | 2.76E-08 | 1.10E-07 | 0.000134101 |
| Obesity | nk_cell | *CCDC154* | -1.719363551 | 0.000154409 | 0.000226822 | 0.749192072 |
| Obesity | nk_cell | *MFAP5* | -1.703490166 | 9.15E-12 | 1.39E-09 | 4.44E-08 |
| Obesity | nk_cell | *PDCD1LG2* | -0.596527828 | 0.025079084 | 0.029809828 | 1 |
| Obesity | nk_cell | *NMB* | 2.44003504 | 1.82E-07 | 4.99E-07 | 0.000880967 |
| Obesity | nk_cell | *FAM114A1* | -1.859031415 | 8.94E-08 | 2.80E-07 | 0.000433743 |
| Obesity | nk_cell | *PSAP* | -1.843020469 | 8.71E-06 | 1.53E-05 | 0.042284419 |
| Obesity | nk_cell | *TXNRD3* | -1.47713541 | 1.61E-08 | 7.37E-08 | 7.83E-05 |
| Obesity | nk_cell | *EME2* | -1.308104442 | 1.50E-09 | 2.11E-08 | 7.29E-06 |
| Obesity | nk_cell | *ZNF780A* | -1.305614266 | 2.95E-09 | 2.58E-08 | 1.43E-05 |
| Obesity | nk_cell | *AC122129.1* | -3.999522427 | 4.30E-09 | 3.17E-08 | 2.09E-05 |
| Obesity | nk_cell | *CCDC180* | -1.900916326 | 7.54E-09 | 4.37E-08 | 3.66E-05 |
| Obesity | nk_cell | *OCLN* | -1.630824777 | 2.68E-09 | 2.45E-08 | 1.30E-05 |
| Obesity | nk_cell | *SGTB* | 2.562434583 | 0.003719827 | 0.004752133 | 1 |
| Obesity | nk_cell | *ADH5* | -2.479214609 | 6.43E-06 | 1.16E-05 | 0.031174454 |
| Obesity | nk_cell | *ZNF677* | -2.407429799 | 2.67E-08 | 1.07E-07 | 0.000129704 |
| Obesity | nk_cell | *ZNF311* | -1.53118944 | 1.03E-05 | 1.78E-05 | 0.049876579 |
| Obesity | nk_cell | *FCHSD1* | 2.599187318 | 2.46E-05 | 4.01E-05 | 0.119194379 |
| Obesity | nk_cell | *ZNF71* | -1.660701539 | 0.016174003 | 0.01955551 | 1 |
| Obesity | nk_cell | *RPL12* | 1.624724654 | 0.001913766 | 0.002502856 | 1 |
| Obesity | nk_cell | *MPZL1* | -1.653678444 | 3.55E-08 | 1.33E-07 | 0.000172288 |
| Obesity | nk_cell | *VPS13A* | -2.299959781 | 1.89E-05 | 3.13E-05 | 0.091573399 |
| Obesity | nk_cell | *NOL8* | -3.111213805 | 4.79E-08 | 1.68E-07 | 0.000232335 |
| Obesity | nk_cell | *DLGAP2* | -0.944407788 | 1.86E-08 | 8.19E-08 | 9.04E-05 |
| Obesity | nk_cell | *MRPL42* | -2.036878658 | 4.88E-09 | 3.42E-08 | 2.37E-05 |
| Obesity | nk_cell | *AGAP9* | -1.933639323 | 1.14E-09 | 2.05E-08 | 5.56E-06 |
| Obesity | nk_cell | *ZNF273* | -3.078853756 | 1.44E-07 | 4.13E-07 | 0.00069771 |
| Obesity | nk_cell | *MAK16* | -2.739999278 | 5.51E-08 | 1.88E-07 | 0.000267551 |
| Obesity | nk_cell | *ZNF667* | -1.320573909 | 1.29E-09 | 2.09E-08 | 6.24E-06 |
| Obesity | nk_cell | *SFI1* | 1.722552806 | 1.05E-08 | 5.38E-08 | 5.12E-05 |
| Obesity | nk_cell | *ADH4* | -1.294310863 | 3.00E-11 | 3.03E-09 | 1.45E-07 |
| Obesity | nk_cell | *HIBCH* | -1.76811093 | 7.41E-09 | 4.35E-08 | 3.60E-05 |
| Obesity | nk_cell | *ZNF544* | -2.2476339 | 7.64E-09 | 4.39E-08 | 3.71E-05 |
| Obesity | nk_cell | *SOWAHC* | -4.206317959 | 4.58E-05 | 7.23E-05 | 0.222464017 |

| | | | | | | |
|---|---|---|---|---|---|---|
| Obesity | nk_cell | *AC135178.1* | 10.10104775 | 3.70E-07 | 9.14E-07 | 0.001794947 |
| Obesity | nk_cell | *MIER1* | -2.130433161 | 3.52E-05 | 5.64E-05 | 0.170744242 |
| Obesity | nk_cell | *MAN1A2* | -2.066837208 | 0.000761258 | 0.001034338 | 1 |
| Obesity | nk_cell | *SVIP* | -3.036038514 | 3.89E-05 | 6.19E-05 | 0.188828699 |
| Obesity | nk_cell | *DDRGK1* | -2.651567876 | 9.09E-08 | 2.84E-07 | 0.00044123 |
| Obesity | nk_cell | *ZNF607* | -1.887874211 | 0.00166247 | 0.002185399 | 1 |
| Obesity | nk_cell | *ZNF334* | -1.764608738 | 7.23E-09 | 4.27E-08 | 3.51E-05 |
| Obesity | nk_cell | *HSD17B11* | -2.465059567 | 3.84E-06 | 7.30E-06 | 0.018631765 |
| Obesity | nk_cell | *SULT1C2* | 1.29479227 | 6.63E-06 | 1.19E-05 | 0.032149101 |
| Obesity | nk_cell | *RPS6KL1* | -3.912931662 | 6.16E-06 | 1.12E-05 | 0.029882455 |
| Obesity | nk_cell | *DDX42* | -1.97996376 | 2.94E-07 | 7.47E-07 | 0.001425701 |
| Obesity | nk_cell | *HELZ* | -1.950516718 | 6.51E-05 | 1.00E-04 | 0.315837774 |
| Obesity | nk_cell | *TMEM116* | -0.701487097 | 0.017778035 | 0.021441468 | 1 |
| Obesity | nk_cell | *UCKL1* | -2.266977657 | 1.53E-08 | 7.08E-08 | 7.40E-05 |
| Obesity | nk_cell | *GFPT1* | -2.160643297 | 1.28E-06 | 2.71E-06 | 0.006194401 |
| Obesity | nk_cell | *ZNF568* | -2.624280028 | 5.31E-05 | 8.29E-05 | 0.257768513 |
| Obesity | nk_cell | *ZNF587* | -2.728694686 | 9.44E-06 | 1.65E-05 | 0.045822388 |
| Obesity | nk_cell | *SH3BGRL2* | -1.062640175 | 4.69E-05 | 7.40E-05 | 0.227693111 |
| Obesity | nk_cell | *CNGA1* | -1.2324658 | 0.00029729 | 0.000422882 | 1 |
| Obesity | nk_cell | *ZNF28* | 3.17711463 | 1.95E-07 | 5.30E-07 | 0.000948392 |
| Obesity | nk_cell | *C20orf203* | -2.778537072 | 1.28E-10 | 8.67E-09 | 6.19E-07 |
| Obesity | nk_cell | *ZNF534* | -1.409201949 | 9.89E-05 | 0.000148884 | 0.480000611 |
| Obesity | nk_cell | *NCOA6* | -2.075272226 | 0.000450599 | 0.00062609 | 1 |
| Obesity | nk_cell | *FAM19A2* | -1.372207212 | 5.12E-09 | 3.52E-08 | 2.49E-05 |
| Obesity | nk_cell | *TTC37* | -2.165617975 | 0.000255264 | 0.000365675 | 1 |
| Obesity | nk_cell | *TUSC1* | -1.971757503 | 1.58E-09 | 2.11E-08 | 7.67E-06 |
| Obesity | nk_cell | *PAPSS2* | -1.640660504 | 2.17E-08 | 9.16E-08 | 0.000105383 |
| Obesity | nk_cell | *CEP290* | -2.188034991 | 4.04E-06 | 7.64E-06 | 0.019597392 |
| Obesity | nk_cell | *ANKRD13B* | -2.278477545 | 2.53E-09 | 2.39E-08 | 1.23E-05 |
| Obesity | nk_cell | *ECI2* | -2.543977926 | 7.00E-05 | 0.000107059 | 0.339590442 |
| Obesity | nk_cell | *UNC13B* | -2.069267059 | 2.57E-08 | 1.05E-07 | 0.000124842 |
| Obesity | nk_cell | *PPP1R14C* | -10.04092092 | 1.12E-08 | 5.56E-08 | 5.42E-05 |
| Obesity | nk_cell | *GPATCH3* | -5.601161073 | 1.69E-06 | 3.48E-06 | 0.008197848 |
| Obesity | nk_cell | *RPL10A* | 2.008807962 | 0.006812422 | 0.00848841 | 1 |
| Obesity | nk_cell | *COLGALT2* | 1.768521173 | 5.29E-09 | 3.57E-08 | 2.57E-05 |
| Obesity | nk_cell | *RCSD1* | 1.574398409 | 6.13E-09 | 3.92E-08 | 2.97E-05 |
| Obesity | nk_cell | *GRIN3A* | -1.956676801 | 1.64E-09 | 2.14E-08 | 7.95E-06 |
| Obesity | nk_cell | *TMEM184B* | -3.258125297 | 1.84E-08 | 8.13E-08 | 8.95E-05 |
| Obesity | nk_cell | *SCAMP5* | -1.398681084 | 3.14E-10 | 1.41E-08 | 1.52E-06 |
| Obesity | nk_cell | *PNP* | 0.709090003 | 2.04E-05 | 3.37E-05 | 0.099149919 |
| Obesity | nk_cell | *SFT2D1* | -2.713271933 | 2.50E-06 | 4.94E-06 | 0.012124195 |
| Obesity | nk_cell | *HMGN2* | -2.100660424 | 5.34E-09 | 3.59E-08 | 2.59E-05 |
| Obesity | nk_cell | *RYR3* | -1.519475288 | 5.62E-05 | 8.73E-05 | 0.272664881 |
| Obesity | nk_cell | *TYW1* | -2.169235234 | 2.58E-07 | 6.71E-07 | 0.001251367 |
| Obesity | nk_cell | *ITPRIPL1* | -0.801933296 | 0.019916532 | 0.023901809 | 1 |
| Obesity | nk_cell | *PRMT6* | 1.678164687 | 2.21E-09 | 2.27E-08 | 1.07E-05 |
| Obesity | nk_cell | *SHISA4* | -5.113410469 | 2.71E-13 | 6.96E-11 | 1.31E-09 |
| Obesity | nk_cell | *MAP3K3* | -2.639072833 | 8.97E-05 | 0.00013528 | 0.435059952 |
| Obesity | nk_cell | *C1orf174* | -2.23964903 | 6.30E-08 | 2.08E-07 | 0.000305566 |
| Obesity | nk_cell | *DZIP3* | -2.572777515 | 2.94E-06 | 5.73E-06 | 0.014259254 |

| | | | | | | |
|---|---|---|---|---|---|---|
| Obesity | nk_cell | *KIAA0753* | -2.207630393 | 1.45E-07 | 4.15E-07 | 0.000702404 |
| Obesity | nk_cell | *CCDC167* | 4.30200529 | 0.02167089 | 0.02592778 | 1 |
| Obesity | nk_cell | *ZFP2* | -3.180764589 | 1.49E-07 | 4.25E-07 | 0.000721709 |
| Obesity | nk_cell | *L3MBTL3* | 1.840940939 | 0.003774716 | 0.004818448 | 1 |
| Obesity | nk_cell | *MFAP3L* | -3.960409539 | 6.58E-06 | 1.19E-05 | 0.031913464 |
| Obesity | nk_cell | *NAGA* | -2.857581946 | 2.20E-06 | 4.40E-06 | 0.010676848 |
| Obesity | nk_cell | *SMG5* | -2.882038168 | 5.02E-08 | 1.75E-07 | 0.000243342 |
| Obesity | nk_cell | *PJA2* | -2.24634292 | 2.13E-06 | 4.28E-06 | 0.010348765 |
| Obesity | nk_cell | *AL590822.1* | -1.713005546 | 3.43E-09 | 2.77E-08 | 1.66E-05 |
| Obesity | nk_cell | *AP001007.1* | -1.914159644 | 0.000164514 | 0.00024079 | 0.798219723 |
| Obesity | nk_cell | *EFCAB2* | -1.596589929 | 3.94E-07 | 9.61E-07 | 0.001910312 |
| Obesity | nk_cell | *CHML* | -2.295844763 | 1.80E-06 | 3.67E-06 | 0.008711236 |
| Obesity | nk_cell | *TATDN3* | -1.460726625 | 0.000468285 | 0.000649733 | 1 |
| Obesity | nk_cell | *SAMD5* | -2.668017797 | 1.75E-06 | 3.59E-06 | 0.008500107 |
| Obesity | nk_cell | *ECT2L* | -1.825923113 | 2.11E-09 | 2.26E-08 | 1.02E-05 |
| Obesity | nk_cell | *GPR52* | 13.81993089 | 2.18E-09 | 2.26E-08 | 1.06E-05 |
| Obesity | nk_cell | *FAM229B* | -2.741261776 | 1.62E-09 | 2.13E-08 | 7.85E-06 |
| Obesity | nk_cell | *DDO* | -2.115674453 | 1.00E-05 | 1.74E-05 | 0.04861222 |
| Obesity | nk_cell | *RBM20* | -1.418589197 | 6.82E-10 | 1.91E-08 | 3.31E-06 |
| Obesity | nk_cell | *PCMTD2* | -2.554806605 | 1.08E-05 | 1.86E-05 | 0.052159408 |
| Obesity | nk_cell | *SAMD13* | -2.024668146 | 4.49E-09 | 3.24E-08 | 2.18E-05 |
| Obesity | nk_cell | *EFCAB7* | -1.990552785 | 6.00E-08 | 2.01E-07 | 0.000290935 |
| Obesity | nk_cell | *IFIT1B* | 3.481680151 | 1.39E-09 | 2.11E-08 | 6.75E-06 |
| Obesity | nk_cell | *LIPN* | -3.960747084 | 2.24E-09 | 2.27E-08 | 1.09E-05 |
| Obesity | nk_cell | *INPP5B* | -1.301209281 | 0.000127513 | 0.000189261 | 0.618695056 |
| Obesity | nk_cell | *NEU4* | -3.402147485 | 1.01E-07 | 3.10E-07 | 0.000491048 |
| Obesity | nk_cell | *C2orf72* | -10.1806595 | 1.60E-08 | 7.34E-08 | 7.77E-05 |
| Obesity | nk_cell | *RUFY2* | -1.939204801 | 1.20E-07 | 3.55E-07 | 0.000580115 |
| Obesity | nk_cell | *PHACTR4* | -2.01867408 | 2.55E-07 | 6.66E-07 | 0.001236218 |
| Obesity | nk_cell | *ASAH2B* | -2.986526722 | 4.62E-08 | 1.64E-07 | 0.000224029 |
| Obesity | nk_cell | *AGAP6* | -1.782793293 | 5.45E-07 | 1.27E-06 | 0.002643681 |
| Obesity | nk_cell | *SYT15* | -2.436213252 | 4.06E-06 | 7.66E-06 | 0.019683122 |
| Obesity | nk_cell | *LINC00619* | -11.08160794 | 1.55E-09 | 2.11E-08 | 7.52E-06 |
| Obesity | nk_cell | *BMPR2* | -1.738536393 | 2.98E-07 | 7.57E-07 | 0.00144748 |
| Obesity | nk_cell | *TCEA3* | -2.587453342 | 2.61E-08 | 1.06E-07 | 0.000126618 |
| Obesity | nk_cell | *HSD17B8* | -1.765493667 | 1.73E-07 | 4.79E-07 | 0.000838028 |
| Obesity | nk_cell | *OXLD1* | -2.046117373 | 0.00021075 | 0.000304605 | 1 |
| Obesity | nk_cell | *TAP2* | 1.180051484 | 7.79E-06 | 1.38E-05 | 0.037779127 |
| Obesity | nk_cell | *COL15A1* | -1.231629694 | 2.39E-07 | 6.31E-07 | 0.001161085 |
| Obesity | nk_cell | *NOTCH4* | -0.387101976 | 5.28E-08 | 1.82E-07 | 0.000256005 |
| Obesity | nk_cell | *PRRT1* | -0.962883772 | 0.0236544 | 0.028157789 | 1 |
| Obesity | nk_cell | *SMIM5* | -2.914213268 | 6.28E-08 | 2.08E-07 | 0.000304829 |
| Obesity | nk_cell | *STK19* | -1.145686923 | 1.48E-08 | 6.90E-08 | 7.18E-05 |
| Obesity | nk_cell | *SKIV2L* | -1.425167195 | 2.08E-14 | 6.74E-12 | 1.01E-10 |
| Obesity | nk_cell | *LAYN* | -2.158739564 | 0.004114363 | 0.005232737 | 1 |
| Obesity | nk_cell | *LY6G5C* | -0.6917193 | 0.027290634 | 0.032343468 | 1 |
| Obesity | nk_cell | *GPANK1* | -4.850639469 | 2.87E-11 | 3.00E-09 | 1.39E-07 |
| Obesity | nk_cell | *LST1* | 2.5917767 | 8.37E-07 | 1.86E-06 | 0.004059365 |
| Obesity | nk_cell | *ZNF814* | -2.021372993 | 9.61E-05 | 0.000144691 | 0.466050611 |
| Obesity | nk_cell | *MICA* | -2.014915097 | 8.27E-06 | 1.46E-05 | 0.040110734 |

| | | | | | | |
|---|---|---|---|---|---|---|
| Obesity | nk_cell | *HLA-C* | 1.378402138 | 5.25E-10 | 1.75E-08 | 2.55E-06 |
| Obesity | nk_cell | *POU5F1* | 0.440880303 | 0.005613881 | 0.007040205 | 1 |
| Obesity | nk_cell | *CCHCR1* | 1.241419839 | 4.81E-06 | 8.94E-06 | 0.02335248 |
| Obesity | nk_cell | *PSORS1C2* | -1.156557567 | 0.032734487 | 0.03843333 | 1 |
| Obesity | nk_cell | *PSORS1C1* | -0.442589694 | 2.59E-06 | 5.10E-06 | 0.012558307 |
| Obesity | nk_cell | *DHX16* | -2.634695915 | 1.36E-05 | 2.32E-05 | 0.066218766 |
| Obesity | nk_cell | *HLA-E* | 2.250400433 | 1.10E-06 | 2.37E-06 | 0.005346113 |
| Obesity | nk_cell | *ZNF468* | -2.11587488 | 7.43E-10 | 1.92E-08 | 3.60E-06 |
| Obesity | nk_cell | *RNF39* | -2.486101257 | 0.0038354 | 0.004890765 | 1 |
| Obesity | nk_cell | *HLA-G* | 1.776652869 | 5.82E-08 | 1.96E-07 | 0.00028226 |
| Obesity | nk_cell | *ZFP57* | 1.293580443 | 8.25E-07 | 1.84E-06 | 0.004001712 |
| Obesity | nk_cell | *FAM153C* | -1.0717831 | 1.08E-08 | 5.43E-08 | 5.22E-05 |
| Obesity | nk_cell | *TRIM27* | -3.293640284 | 8.57E-07 | 1.90E-06 | 0.004159962 |
| Obesity | nk_cell | *MALRD1* | -2.161236252 | 8.13E-08 | 2.59E-07 | 0.000394334 |
| Obesity | nk_cell | *FAM196B* | 1.99450307 | 1.87E-08 | 8.19E-08 | 9.06E-05 |
| Obesity | nk_cell | *MROH6* | -2.021234283 | 2.08E-07 | 5.61E-07 | 0.001011035 |
| Obesity | nk_cell | *DCTN1* | -3.67224989 | 9.41E-05 | 0.000141812 | 0.45663459 |
| Obesity | nk_cell | *FAM216A* | -3.383891018 | 3.09E-06 | 6.00E-06 | 0.015007081 |
| Obesity | nk_cell | *ZBTB48* | -2.172335335 | 0.002344707 | 0.003045107 | 1 |
| Obesity | nk_cell | *MZT1* | 4.048501062 | 4.49E-07 | 1.07E-06 | 0.0021771 |
| Obesity | nk_cell | *ZNF155* | -2.027523092 | 1.69E-07 | 4.71E-07 | 0.000820506 |
| Obesity | nk_cell | *ZNF783* | -3.48440995 | 3.86E-07 | 9.46E-07 | 0.001874601 |
| Obesity | nk_cell | *ZNF425* | -2.259246444 | 7.03E-08 | 2.29E-07 | 0.000340956 |
| Obesity | nk_cell | *C12orf73* | -2.195503309 | 8.10E-09 | 4.51E-08 | 3.93E-05 |
| Obesity | nk_cell | *PCDHGA1* | -2.516655468 | 4.60E-09 | 3.28E-08 | 2.23E-05 |
| Obesity | nk_cell | *PCDHA7* | -26.5879631 | 2.02E-06 | 4.08E-06 | 0.009814478 |
| Obesity | nk_cell | *PCDHA4* | -3.26629643 | 1.41E-10 | 9.02E-09 | 6.86E-07 |
| Obesity | nk_cell | *TRIM13* | -1.975430606 | 2.45E-05 | 4.00E-05 | 0.118841098 |
| Obesity | nk_cell | *SPIRE2* | -2.06884064 | 4.76E-08 | 1.68E-07 | 0.000230891 |
| Obesity | nk_cell | *PKHD1L1* | -0.90157883 | 0.000369804 | 0.000519181 | 1 |
| Obesity | nk_cell | *SLFN12L* | 0.368694424 | 3.94E-09 | 3.00E-08 | 1.91E-05 |
| Obesity | nk_cell | *SYCE1L* | 0.949102351 | 1.35E-06 | 2.86E-06 | 0.006572597 |
| Obesity | nk_cell | *TMEM231* | -1.652591743 | 1.06E-08 | 5.38E-08 | 5.12E-05 |
| Obesity | nk_cell | *LINC00654* | -1.636144486 | 0.009639237 | 0.011849701 | 1 |
| Obesity | nk_cell | *SCGB2B2* | -1.693273771 | 8.51E-09 | 4.64E-08 | 4.13E-05 |
| Obesity | nk_cell | *CCDC144NL* | 5.126890374 | 0.000969505 | 0.001300176 | 1 |
| Obesity | nk_cell | *TMEM170B* | -1.254490714 | 2.66E-08 | 1.07E-07 | 0.000128867 |
| Obesity | nk_cell | *GOLGA6L10* | -3.213769209 | 1.73E-08 | 7.77E-08 | 8.40E-05 |
| Obesity | nk_cell | *NT5M* | -1.477619282 | 1.47E-09 | 2.11E-08 | 7.12E-06 |
| Obesity | nk_cell | *IPO7* | -2.234369548 | 1.33E-07 | 3.88E-07 | 0.000644169 |
| Obesity | nk_cell | *PRR13* | 2.093048061 | 0.005320293 | 0.006682387 | 1 |
| Obesity | nk_cell | *TECPR1* | -3.00058851 | 5.19E-06 | 9.56E-06 | 0.025179131 |
| Obesity | nk_cell | *MT1M* | 3.295437587 | 1.71E-08 | 7.70E-08 | 8.29E-05 |
| Obesity | nk_cell | *CFI* | -1.578503775 | 1.59E-08 | 7.32E-08 | 7.72E-05 |
| Obesity | nk_cell | *ATP6AP1L* | -1.395352063 | 1.57E-07 | 4.44E-07 | 0.00076207 |
| Obesity | nk_cell | *RGL3* | -1.221571052 | 4.52E-08 | 1.61E-07 | 0.000219249 |
| Obesity | nk_cell | *SERF1B* | -2.517781545 | 6.39E-09 | 4.02E-08 | 3.10E-05 |
| Obesity | nk_cell | *LIN52* | -1.793980666 | 0.000585867 | 0.000804821 | 1 |
| Obesity | nk_cell | *KLRC2* | 0.135951581 | 1.12E-09 | 2.05E-08 | 5.41E-06 |
| Obesity | nk_cell | *LINC00487* | 4.278760917 | 2.34E-06 | 4.66E-06 | 0.011364116 |

| | | | | | | |
|---|---|---|---|---|---|---|
| Obesity | nk_cell | *RFPL3S* | 1.503597452 | 4.36E-07 | 1.05E-06 | 0.002115917 |
| Obesity | nk_cell | *C1QTNF9B* | -9.988499744 | 1.65E-09 | 2.14E-08 | 8.01E-06 |
| Obesity | nk_cell | *ZDHHC11B* | -2.087223172 | 5.46E-09 | 3.65E-08 | 2.65E-05 |
| Obesity | nk_cell | *GOLGA8O* | -2.410007125 | 0.018462473 | 0.022228268 | 1 |
| Obesity | nk_cell | *HBA1* | 0.800811736 | 1.20E-07 | 3.57E-07 | 0.000584093 |
| Obesity | nk_cell | *HCG27* | -1.507904662 | 3.72E-05 | 5.93E-05 | 0.180257406 |
| Obesity | nk_cell | *COL6A6* | -1.793342652 | 2.82E-07 | 7.23E-07 | 0.001369893 |
| Obesity | nk_cell | *HLA-A* | 1.407906715 | 1.11E-06 | 2.39E-06 | 0.005385358 |
| Obesity | nk_cell | *LNP1* | -2.17937157 | 0.001727918 | 0.002268974 | 1 |
| Obesity | nk_cell | *PRSS50* | -5.201717572 | 1.35E-06 | 2.85E-06 | 0.00654524 |
| Obesity | nk_cell | *METTL6* | -2.462647664 | 0.000621993 | 0.000853241 | 1 |
| Obesity | nk_cell | *SACM1L* | 2.421071764 | 0.001327418 | 0.001757226 | 1 |
| Obesity | nk_cell | *SLC48A1* | -2.058407196 | 1.22E-08 | 5.95E-08 | 5.92E-05 |
| Obesity | nk_cell | *TAS2R50* | -24.6923036 | 9.81E-09 | 5.14E-08 | 4.76E-05 |
| Obesity | nk_cell | *TAS2R14* | 1.602151866 | 1.55E-05 | 2.60E-05 | 0.075125998 |
| Obesity | nk_cell | *AC084018.1* | -1.998632762 | 4.62E-13 | 9.74E-11 | 2.24E-09 |
| Obesity | nk_cell | *OR2A42* | -2.2970226 | 8.08E-08 | 2.58E-07 | 0.000392269 |
| Obesity | nk_cell | *ZNF580* | -1.660639576 | 5.67E-07 | 1.31E-06 | 0.00275297 |
| Obesity | nk_cell | *AL021068.1* | -2.304150304 | 0.000168589 | 0.000246235 | 0.81799403 |
| Obesity | nk_cell | *FGFR1OP* | 4.356416445 | 3.51E-06 | 6.72E-06 | 0.017042403 |
| Obesity | nk_cell | *FAM106A* | -2.746122654 | 0.027568603 | 0.032633048 | 1 |
| Obesity | nk_cell | *ZNF254* | -2.471673371 | 3.38E-07 | 8.46E-07 | 0.001641079 |
| Obesity | nk_cell | *TCTEX1D2* | -2.195496032 | 6.97E-06 | 1.25E-05 | 0.033815683 |
| Obesity | nk_cell | *AC022498.1* | 4.350226183 | 2.65E-10 | 1.29E-08 | 1.29E-06 |
| Obesity | nk_cell | *GIMAP1* | 2.444888592 | 1.22E-08 | 5.95E-08 | 5.91E-05 |
| Obesity | nk_cell | *ARHGEF35* | -2.521286071 | 0.000364187 | 0.000512035 | 1 |
| Obesity | nk_cell | *DNLZ* | 0.852985111 | 2.68E-07 | 6.96E-07 | 0.001302441 |
| Obesity | nk_cell | *NOTCH2NL* | 1.421264063 | 4.23E-10 | 1.62E-08 | 2.05E-06 |
| Obesity | nk_cell | *SUPT4H1* | 2.235433253 | 4.14E-10 | 1.61E-08 | 2.01E-06 |
| Obesity | nk_cell | *MXD3* | 2.469673001 | 1.41E-08 | 6.67E-08 | 6.84E-05 |
| Obesity | nk_cell | *GSTM2* | -1.373422192 | 1.26E-05 | 2.15E-05 | 0.061230967 |
| Obesity | nk_cell | *ARHGAP19* | -2.413516802 | 3.65E-10 | 1.52E-08 | 1.77E-06 |
| Obesity | nk_cell | *LCAT* | -3.908700524 | 0.000964594 | 0.001294306 | 1 |
| Obesity | nk_cell | *GPC2* | -6.923140853 | 5.73E-09 | 3.75E-08 | 2.78E-05 |
| Obesity | nk_cell | *ERV3-1* | -0.537771522 | 0.000237863 | 0.000341757 | 1 |
| Obesity | nk_cell | *GBP7* | -1.030199231 | 0.00515417 | 0.006490536 | 1 |
| Obesity | nk_cell | *SRA1* | -1.996302025 | 0.001445544 | 0.001909551 | 1 |
| Obesity | nk_cell | *LBH* | 1.141436408 | 7.44E-09 | 4.35E-08 | 3.61E-05 |
| Obesity | nk_cell | *PPP1CB* | -2.920953125 | 4.50E-08 | 1.61E-07 | 0.000218264 |
| Obesity | nk_cell | *NCKIPSD* | -2.319091862 | 3.44E-06 | 6.59E-06 | 0.01667668 |
| Obesity | nk_cell | *ATF6B* | -2.236136927 | 4.08E-07 | 9.93E-07 | 0.001980855 |
| Obesity | nk_cell | *SLC35F6* | -3.277098126 | 0.00265195 | 0.003427614 | 1 |
| Obesity | nk_cell | *EMP2* | -2.15340527 | 1.29E-06 | 2.74E-06 | 0.006256842 |
| Obesity | nk_cell | *PPM1N* | 1.784105878 | 3.03E-06 | 5.89E-06 | 0.014702168 |
| Obesity | nk_cell | *SLC23A3* | -1.015580774 | 0.00036846 | 0.000517444 | 1 |
| Obesity | nk_cell | *IRF9* | 3.473094285 | 0.000336561 | 0.000474846 | 1 |
| Obesity | nk_cell | *GALT* | 4.367164546 | 1.44E-06 | 3.03E-06 | 0.00701059 |
| Obesity | nk_cell | *ZNF726* | -5.264389976 | 2.80E-07 | 7.17E-07 | 0.001357498 |
| Obesity | nk_cell | *ZNF90* | -1.992717972 | 4.86E-09 | 3.42E-08 | 2.36E-05 |
| Obesity | nk_cell | *REPIN1* | -2.569651481 | 5.84E-09 | 3.80E-08 | 2.83E-05 |

| | | | | | | |
|---|---|---|---|---|---|---|
| Obesity | nk_cell | *ZNF891* | -2.538521572 | 5.60E-08 | 1.90E-07 | 0.000271919 |
| Obesity | nk_cell | *SMIM7* | -2.307667199 | 0.005771734 | 0.007230687 | 1 |
| Obesity | nk_cell | *TSPAN4* | -1.954486556 | 3.78E-05 | 6.02E-05 | 0.183189591 |
| Obesity | nk_cell | *CPNE1* | -1.282309561 | 0.000140199 | 0.000206824 | 0.680244737 |
| Obesity | nk_cell | *ARL16* | -0.778906765 | 0.006158616 | 0.007695494 | 1 |
| Obesity | nk_cell | *PAXIP1-AS2* | -1.891587361 | 1.93E-08 | 8.42E-08 | 9.38E-05 |
| Obesity | nk_cell | *LYRM4* | -2.478534681 | 1.08E-06 | 2.34E-06 | 0.005253997 |
| Obesity | nk_cell | *PRCD* | -2.926640482 | 5.71E-09 | 3.74E-08 | 2.77E-05 |
| Obesity | nk_cell | *SH3D21* | -1.542047145 | 1.10E-08 | 5.53E-08 | 5.35E-05 |
| Obesity | nk_cell | *FIS1* | -1.589785537 | 9.10E-07 | 2.00E-06 | 0.004414551 |
| Obesity | nk_cell | *ANG* | -3.565000493 | 1.39E-07 | 4.00E-07 | 0.000672741 |
| Obesity | nk_cell | *COLCA2* | -1.480811161 | 1.04E-07 | 3.16E-07 | 0.000504437 |
| Obesity | nk_cell | *MBLAC1* | -1.452378211 | 1.96E-07 | 5.30E-07 | 0.000949577 |
| Obesity | nk_cell | *LCNL1* | -2.940318324 | 2.29E-08 | 9.52E-08 | 0.000110964 |
| Obesity | nk_cell | *GNAT3* | -2.370459157 | 4.39E-09 | 3.20E-08 | 2.13E-05 |
| Obesity | nk_cell | *PLIN5* | -0.76759198 | 3.36E-09 | 2.76E-08 | 1.63E-05 |
| Obesity | nk_cell | *STARD10* | -1.402053717 | 1.12E-07 | 3.38E-07 | 0.000545378 |
| Obesity | nk_cell | *NUTM2D* | -13.44212717 | 4.01E-06 | 7.60E-06 | 0.019443989 |
| Obesity | nk_cell | *CPEB1* | -0.873716519 | 4.44E-09 | 3.22E-08 | 2.15E-05 |
| Obesity | nk_cell | *EML6* | -3.769185007 | 1.03E-06 | 2.23E-06 | 0.004984439 |
| Obesity | nk_cell | *ARHGEF33* | -3.221367576 | 4.94E-07 | 1.17E-06 | 0.002397464 |
| Obesity | nk_cell | *IFRD2* | -2.297248484 | 2.53E-08 | 1.03E-07 | 0.000122798 |
| Obesity | nk_cell | *EVPLL* | -3.11040939 | 3.91E-10 | 1.54E-08 | 1.90E-06 |
| Obesity | nk_cell | *AC005082.1* | -4.936145854 | 4.66E-10 | 1.69E-08 | 2.26E-06 |
| Obesity | nk_cell | *SMTNL1* | -0.885341181 | 0.000209982 | 0.000303639 | 1 |
| Obesity | nk_cell | *ACSM4* | -1.062474775 | 0.032258259 | 0.037925144 | 1 |
| Obesity | nk_cell | *C5orf49* | -10.360215 | 1.71E-08 | 7.70E-08 | 8.28E-05 |
| Obesity | nk_cell | *VPS16* | -2.801237554 | 4.31E-08 | 1.55E-07 | 0.00020897 |
| Obesity | nk_cell | *NPEPL1* | -3.155870185 | 2.14E-08 | 9.06E-08 | 0.000103635 |
| Obesity | nk_cell | *SLC35E2* | -3.199712464 | 1.77E-09 | 2.16E-08 | 8.58E-06 |
| Obesity | nk_cell | *LINC01139* | -0.743206254 | 0.000100183 | 0.000150631 | 0.486087003 |
| Obesity | nk_cell | *TSTD1* | 3.438907178 | 3.19E-05 | 5.14E-05 | 0.15459932 |
| Obesity | nk_cell | *AC009403.2* | -2.491107676 | 1.59E-08 | 7.32E-08 | 7.70E-05 |
| Obesity | nk_cell | *AC016773.1* | 2.187128562 | 1.46E-07 | 4.17E-07 | 0.000707307 |
| Obesity | nk_cell | *AP000350.4* | -0.670812733 | 0.00102916 | 0.001375994 | 1 |
| Obesity | nk_cell | *RNASEK* | -2.479972198 | 7.80E-07 | 1.75E-06 | 0.003784149 |
| Obesity | nk_cell | *FAM228B* | -1.472715389 | 0.000321276 | 0.000454868 | 1 |
| Obesity | nk_cell | *RNU6ATAC35I* | -15.06274032 | 4.83E-11 | 4.34E-09 | 2.34E-07 |
| Obesity | nk_cell | *PPP3R1* | -1.995430999 | 7.91E-07 | 1.77E-06 | 0.003840014 |
| Obesity | nk_cell | *FANCG* | -2.507331137 | 2.86E-07 | 7.30E-07 | 0.00138743 |
| Obesity | nk_cell | *AP4M1* | -2.092981386 | 7.86E-10 | 1.96E-08 | 3.81E-06 |
| Obesity | nk_cell | *ZNF880* | -2.002992498 | 2.78E-07 | 7.14E-07 | 0.001349974 |
| Obesity | nk_cell | *FXYD7* | -1.767103784 | 3.09E-09 | 2.64E-08 | 1.50E-05 |
| Obesity | nk_cell | *SLC12A8* | -1.791678561 | 5.26E-09 | 3.56E-08 | 2.55E-05 |
| Obesity | nk_cell | *APOL6* | -2.36640282 | 0.00017247 | 0.000251676 | 0.836823761 |
| Obesity | nk_cell | *OR2A1* | -2.175395509 | 2.79E-06 | 5.46E-06 | 0.013515546 |
| Obesity | nk_cell | *UBA52* | 1.826486743 | 0.00238854 | 0.003099545 | 1 |
| Obesity | nk_cell | *RAB6C* | -31.40512964 | 3.42E-09 | 2.77E-08 | 1.66E-05 |
| Obesity | nk_cell | *AC062017.1* | -1.363275209 | 1.88E-09 | 2.19E-08 | 9.15E-06 |
| Obesity | nk_cell | *ADRA2B* | -6.779035852 | 3.83E-10 | 1.54E-08 | 1.86E-06 |

| | | | | | | |
|---|---|---|---|---|---|---|
| Obesity | nk_cell | *EXOSC6* | -4.644468618 | 5.87E-16 | 2.59E-13 | 2.85E-12 |
| Obesity | nk_cell | *ZNF844* | -2.362242608 | 2.38E-09 | 2.32E-08 | 1.15E-05 |
| Obesity | nk_cell | *LINC00571* | -3.090386291 | 4.50E-08 | 1.61E-07 | 0.000218545 |
| Obesity | nk_cell | *CERS1* | -7.935741681 | 0.000190163 | 0.000276414 | 0.922670415 |
| Obesity | nk_cell | *OSER1-AS1* | -1.300050613 | 0.005902789 | 0.007385336 | 1 |
| Obesity | nk_cell | *C5orf66* | -2.771191154 | 3.33E-06 | 6.40E-06 | 0.016146078 |
| Obesity | nk_cell | *LINC01133* | -1.025303125 | 1.35E-09 | 2.11E-08 | 6.55E-06 |
| Obesity | nk_cell | *C4B* | 2.061299724 | 0.031867479 | 0.037502063 | 1 |
| Obesity | nk_cell | *LINC01117* | -3.820056096 | 1.97E-06 | 4.00E-06 | 0.009582015 |
| Obesity | nk_cell | *RASAL2-AS1* | -2.138481239 | 0.004182435 | 0.005316525 | 1 |
| Obesity | nk_cell | *AC069213.1* | -1.491564006 | 0.021004092 | 0.025151001 | 1 |
| Obesity | nk_cell | *LINC00240* | -0.642954368 | 0.001750607 | 0.002296903 | 1 |
| Obesity | nk_cell | *LINC00184* | -27.13332884 | 1.31E-09 | 2.09E-08 | 6.38E-06 |
| Obesity | nk_cell | *AL049840.1* | -3.876415548 | 9.88E-08 | 3.04E-07 | 0.000479468 |
| Obesity | nk_cell | *PLEKHM1* | -2.629034807 | 4.87E-07 | 1.16E-06 | 0.002365332 |
| Obesity | nk_cell | *LINC00092* | 1.097944528 | 5.79E-05 | 8.97E-05 | 0.280693 |
| Obesity | nk_cell | *AP000350.6* | -0.945144431 | 4.76E-05 | 7.50E-05 | 0.231089408 |
| Obesity | nk_cell | *SFTA1P* | 1.83020159 | 2.35E-07 | 6.21E-07 | 0.001139598 |
| Obesity | nk_cell | *CYP4A22-AS1* | -3.049686249 | 7.78E-09 | 4.42E-08 | 3.77E-05 |
| Obesity | nk_cell | *MIAT* | 0.951553888 | 1.21E-09 | 2.08E-08 | 5.88E-06 |
| Obesity | nk_cell | *ERCC6* | -2.339484383 | 0.001651122 | 0.002171657 | 1 |
| Obesity | nk_cell | *LINC00694* | -3.181559943 | 2.56E-06 | 5.06E-06 | 0.012438934 |
| Obesity | nk_cell | *LINC00115* | 3.085210876 | 1.07E-08 | 5.43E-08 | 5.22E-05 |
| Obesity | nk_cell | *HAR1A* | 18.26175818 | 1.68E-07 | 4.69E-07 | 0.000813821 |
| Obesity | nk_cell | *LINC00937* | -1.326679344 | 7.21E-09 | 4.27E-08 | 3.50E-05 |
| Obesity | nk_cell | *FTCDNL1* | -0.941041455 | 0.005014335 | 0.006325937 | 1 |
| Obesity | nk_cell | *AC010536.1* | -3.887313393 | 0.024193049 | 0.028770753 | 1 |
| Obesity | nk_cell | *FAM138B* | -15.5141656 | 1.39E-09 | 2.11E-08 | 6.77E-06 |
| Obesity | nk_cell | *ZNF717* | -0.523419857 | 0.021934039 | 0.026206343 | 1 |
| Obesity | nk_cell | *AC017074.2* | -9.670534329 | 0.007770241 | 0.009627479 | 1 |
| Obesity | nk_cell | *AC009299.3* | -2.562413814 | 7.00E-09 | 4.20E-08 | 3.39E-05 |
| Obesity | nk_cell | *SCAMP4* | -2.93306903 | 0.001531638 | 0.002018883 | 1 |
| Obesity | nk_cell | *SLC2A1-AS1* | -2.117946998 | 1.74E-09 | 2.16E-08 | 8.46E-06 |
| Obesity | nk_cell | *LINC01068* | -1.913761919 | 0.003193139 | 0.004094374 | 1 |
| Obesity | nk_cell | *AC005884.1* | -3.798725693 | 4.51E-06 | 8.43E-06 | 0.021860496 |
| Obesity | nk_cell | *C14orf144* | -3.091192148 | 1.04E-08 | 5.35E-08 | 5.05E-05 |
| Obesity | nk_cell | *POLR2J2* | 2.975632313 | 1.45E-09 | 2.11E-08 | 7.04E-06 |
| Obesity | nk_cell | *LINC00578* | -1.154298144 | 2.97E-09 | 2.58E-08 | 1.44E-05 |
| Obesity | nk_cell | *C19orf24* | -2.529032449 | 9.13E-09 | 4.89E-08 | 4.43E-05 |
| Obesity | nk_cell | *ARL17B* | -2.088994885 | 1.17E-07 | 3.49E-07 | 0.000567498 |
| Obesity | nk_cell | *DHFR* | -0.934106228 | 5.28E-07 | 1.24E-06 | 0.002560843 |
| Obesity | nk_cell | *PATL2* | 1.466780269 | 1.63E-09 | 2.14E-08 | 7.91E-06 |
| Obesity | nk_cell | *CASC20* | -3.288072209 | 6.25E-09 | 3.98E-08 | 3.03E-05 |
| Obesity | nk_cell | *PSMG3-AS1* | -2.124765476 | 4.53E-08 | 1.61E-07 | 0.000219585 |
| Obesity | nk_cell | *AC112229.4* | -3.04226668 | 0.000119105 | 0.000177487 | 0.577897149 |
| Obesity | nk_cell | *KLHL7-AS1* | -1.965184415 | 9.44E-08 | 2.93E-07 | 0.000458249 |
| Obesity | nk_cell | *LINC00332* | -2.628000542 | 2.48E-08 | 1.01E-07 | 0.000120172 |
| Obesity | nk_cell | *AC092171.4* | -2.456496148 | 3.83E-09 | 2.95E-08 | 1.86E-05 |
| Obesity | nk_cell | *RPS18* | 2.135294564 | 0.000218689 | 0.000315328 | 1 |
| Obesity | nk_cell | *DIRC3* | -1.200684369 | 1.75E-08 | 7.83E-08 | 8.51E-05 |

| | | | | | | |
|---|---|---|---|---|---|---|
| Obesity | nk_cell | *TAPBP* | 2.672847708 | 1.59E-09 | 2.11E-08 | 7.71E-06 |
| Obesity | nk_cell | *AC107021.1* | -1.829277889 | 6.97E-09 | 4.20E-08 | 3.38E-05 |
| Obesity | nk_cell | *TMA7* | 2.644968222 | 1.12E-09 | 2.05E-08 | 5.42E-06 |
| Obesity | nk_cell | *MTRNR2L4* | -3.408646773 | 2.78E-05 | 4.52E-05 | 0.135003237 |
| Obesity | nk_cell | *LINC00865* | -1.676123159 | 1.08E-07 | 3.26E-07 | 0.000522011 |
| Obesity | nk_cell | *AC009495.3* | 0.565340436 | 1.24E-09 | 2.08E-08 | 6.01E-06 |
| Obesity | nk_cell | *GOLGA8N* | -1.230948633 | 0.000670644 | 0.000915326 | 1 |
| Obesity | nk_cell | *LINC00665* | -2.377772347 | 1.42E-05 | 2.41E-05 | 0.068786272 |
| Obesity | nk_cell | *LYRM9* | -2.098095601 | 2.21E-09 | 2.27E-08 | 1.07E-05 |
| Obesity | nk_cell | *LINC01166* | -1.662717413 | 0.023071693 | 0.027504633 | 1 |
| Obesity | nk_cell | *LINC00702* | -2.030607254 | 3.23E-09 | 2.71E-08 | 1.57E-05 |
| Obesity | nk_cell | *GPX1* | -2.391804928 | 0.000158764 | 0.000232655 | 0.770322242 |
| Obesity | nk_cell | *RPS28* | -3.414838333 | 0.000104865 | 0.000157233 | 0.508805075 |
| Obesity | nk_cell | *TRIM26* | -2.499045632 | 4.06E-08 | 1.49E-07 | 0.000196811 |
| Obesity | nk_cell | *ZNF736* | -1.333637536 | 0.000131337 | 0.00019452 | 0.637248248 |
| Obesity | nk_cell | *PINLYP* | -2.039270575 | 5.20E-09 | 3.55E-08 | 2.53E-05 |
| Obesity | nk_cell | *ANKRD65* | -9.038524486 | 3.30E-08 | 1.25E-07 | 0.000160134 |
| Obesity | nk_cell | *ZSCAN31* | 0.611342372 | 0.017874873 | 0.021547549 | 1 |
| Obesity | nk_cell | *AC009495.2* | -30.54285811 | 1.33E-06 | 2.81E-06 | 0.006429448 |
| Obesity | nk_cell | *LINC00957* | -4.28762793 | 1.18E-07 | 3.53E-07 | 0.000573698 |
| Obesity | nk_cell | *LINC00484* | 0.780718917 | 4.17E-05 | 6.62E-05 | 0.202509537 |
| Obesity | nk_cell | *KIAA0040* | 2.325157838 | 5.71E-09 | 3.74E-08 | 2.77E-05 |
| Obesity | nk_cell | *LINC00607* | -1.264309829 | 1.98E-07 | 5.36E-07 | 0.000960257 |
| Obesity | nk_cell | *AP001468.1* | -0.802580967 | 0.032118296 | 0.037769746 | 1 |
| Obesity | nk_cell | *LINC00941* | 1.674713203 | 1.55E-09 | 2.11E-08 | 7.54E-06 |
| Obesity | nk_cell | *AC019117.2* | -9.612998915 | 7.80E-11 | 6.30E-09 | 3.78E-07 |
| Obesity | nk_cell | *ZBED5* | -2.212862603 | 3.27E-08 | 1.24E-07 | 0.00015867 |
| Obesity | nk_cell | *CDKN2AIPNL* | 1.806875711 | 0.013633001 | 0.01656997 | 1 |
| Obesity | nk_cell | *LINC00987* | 1.428533919 | 1.96E-09 | 2.20E-08 | 9.52E-06 |
| Obesity | nk_cell | *GS1-124K5.4* | 1.189552988 | 5.76E-08 | 1.94E-07 | 0.000279516 |
| Obesity | nk_cell | *LINC00959* | -3.16863972 | 4.59E-07 | 1.10E-06 | 0.002228434 |
| Obesity | nk_cell | *HLA-DQA2* | -0.398360397 | 0.009234633 | 0.011380859 | 1 |
| Obesity | nk_cell | *IRGM* | 1.577511821 | 1.44E-07 | 4.14E-07 | 0.000699877 |
| Obesity | nk_cell | *LINC01143* | -6.723933047 | 3.79E-09 | 2.94E-08 | 1.84E-05 |
| Obesity | nk_cell | *LINC00211* | 4.122939921 | 2.45E-10 | 1.25E-08 | 1.19E-06 |
| Obesity | nk_cell | *LRRC37A2* | -2.484118407 | 6.50E-09 | 4.05E-08 | 3.15E-05 |
| Obesity | nk_cell | *TXNDC5* | -5.472463617 | 2.01E-09 | 2.20E-08 | 9.73E-06 |
| Obesity | nk_cell | *LINC01119* | -2.279644181 | 7.58E-09 | 4.38E-08 | 3.68E-05 |
| Obesity | nk_cell | *NME1* | 1.799757153 | 3.04E-09 | 2.62E-08 | 1.48E-05 |
| Obesity | nk_cell | *CDRT4* | -3.417650434 | 2.83E-08 | 1.11E-07 | 0.000137186 |
| Obesity | nk_cell | *APOBEC3G* | 0.343632973 | 1.37E-09 | 2.11E-08 | 6.63E-06 |
| Obesity | nk_cell | *C1orf226* | 3.923259806 | 7.25E-08 | 2.35E-07 | 0.000351939 |
| Obesity | nk_cell | *LILRA2* | -0.846586195 | 0.012276113 | 0.014969514 | 1 |
| Obesity | nk_cell | *LY6G5B* | -0.842328503 | 0.004777271 | 0.006044151 | 1 |
| Obesity | nk_cell | *PSMB9* | 1.777923609 | 8.85E-14 | 2.68E-11 | 4.29E-10 |
| Obesity | nk_cell | *PPIL3* | -0.824978216 | 0.040040985 | 0.046679207 | 1 |
| Obesity | nk_cell | *L1TD1* | -2.617248216 | 2.57E-07 | 6.69E-07 | 0.001244848 |
| Obesity | nk_cell | *C1QTNF9* | 1.061610619 | 1.32E-09 | 2.09E-08 | 6.40E-06 |
| Obesity | nk_cell | *ISY1* | -1.238097174 | 8.02E-12 | 1.32E-09 | 3.89E-08 |
| Obesity | nk_cell | *PNMA2* | 3.469992414 | 6.13E-06 | 1.11E-05 | 0.029765452 |

| | | | | | | |
|---|---|---|---|---|---|---|
| Obesity | nk_cell | *KRBOX1* | -17.17169621 | 1.45E-09 | 2.11E-08 | 7.03E-06 |
| Obesity | nk_cell | *ARHGEF25* | -1.054921453 | 3.00E-06 | 5.85E-06 | 0.014574396 |
| Obesity | nk_cell | *LINC00886* | -1.060742755 | 0.004825662 | 0.006103783 | 1 |
| Obesity | nk_cell | *MIF* | 1.074934183 | 0.005445931 | 0.006836651 | 1 |
| Obesity | nk_cell | *NSUN6* | -2.268737992 | 4.98E-05 | 7.81E-05 | 0.241761686 |
| Obesity | nk_cell | *LINC00877* | -1.672317926 | 3.93E-09 | 3.00E-08 | 1.90E-05 |
| Obesity | nk_cell | *CD302* | -1.779713075 | 4.58E-09 | 3.28E-08 | 2.22E-05 |
| Obesity | nk_cell | *INMT* | -2.718640954 | 3.37E-07 | 8.43E-07 | 0.001635525 |
| Obesity | nk_cell | *ATP5O* | -2.45496002 | 0.000201902 | 0.000292339 | 0.979629312 |
| Obesity | nk_cell | *C8orf58* | -3.967621013 | 9.54E-10 | 2.05E-08 | 4.63E-06 |
| Obesity | nk_cell | *HOGA1* | -2.17357434 | 1.41E-09 | 2.11E-08 | 6.83E-06 |
| Obesity | nk_cell | *LINC00969* | -1.196197973 | 1.05E-06 | 2.28E-06 | 0.00509474 |
| Obesity | nk_cell | *ARFGAP3* | -2.173518032 | 0.012453361 | 0.015178022 | 1 |
| Obesity | nk_cell | *MRPL20* | -2.633179194 | 1.69E-06 | 3.47E-06 | 0.008188008 |
| Obesity | nk_cell | *DECR2* | -1.56427389 | 1.51E-09 | 2.11E-08 | 7.32E-06 |
| Obesity | nk_cell | *ZNF709* | 13.78593628 | 8.44E-09 | 4.63E-08 | 4.10E-05 |
| Obesity | nk_cell | *ERVW-1* | 5.184669154 | 8.06E-09 | 4.51E-08 | 3.91E-05 |
| Obesity | nk_cell | *MRPL33* | -2.491504225 | 8.31E-07 | 1.85E-06 | 0.004033654 |
| Obesity | nk_cell | *STON1* | -2.907633923 | 0.00020118 | 0.000291468 | 0.976126571 |
| Obesity | nk_cell | *VSIG8* | -3.015254946 | 1.94E-08 | 8.43E-08 | 9.41E-05 |
| Obesity | nk_cell | *FGF14-IT1* | -3.29520356 | 1.72E-09 | 2.16E-08 | 8.34E-06 |
| Obesity | nk_cell | *PALM2* | -1.024364592 | 7.58E-10 | 1.93E-08 | 3.68E-06 |
| Obesity | nk_cell | *NBPF15* | -2.391613801 | 2.19E-08 | 9.23E-08 | 0.000106327 |
| Obesity | nk_cell | *UPK3B* | -2.762123064 | 3.22E-07 | 8.12E-07 | 0.001562439 |
| Obesity | nk_cell | *IL10RB* | 1.979324871 | 0.023701053 | 0.028199487 | 1 |
| Obesity | nk_cell | *NME1-NME2* | -4.519155331 | 0.003094813 | 0.003973547 | 1 |
| Obesity | nk_cell | *PLA2G4B* | -6.976008798 | 1.56E-07 | 4.41E-07 | 0.000755229 |
| Obesity | nk_cell | *LEFTY1* | 8.287210084 | 1.45E-08 | 6.81E-08 | 7.04E-05 |
| Obesity | nk_cell | *TTC4* | 2.500770583 | 0.000280069 | 0.000399205 | 1 |
| Obesity | nk_cell | *JMJD7* | -1.067743039 | 0.013371519 | 0.01626438 | 1 |
| Obesity | nk_cell | *APOBEC3D* | 1.100957075 | 1.12E-09 | 2.05E-08 | 5.44E-06 |
| Obesity | nk_cell | *TMEM199* | 1.409579953 | 9.86E-10 | 2.05E-08 | 4.78E-06 |
| Obesity | nk_cell | *SPRR2F* | -2.711143979 | 0.000867725 | 0.00117015 | 1 |
| Obesity | nk_cell | *P2RY11* | 0.84000664 | 3.25E-08 | 1.24E-07 | 0.000157738 |
| Obesity | nk_cell | *DBNDD2* | -6.95915394 | 1.02E-05 | 1.78E-05 | 0.049651814 |
| Obesity | nk_cell | *LILRA6* | -1.066918246 | 2.88E-05 | 4.67E-05 | 0.13962569 |
| Obesity | nk_cell | *SCARF2* | -2.531559261 | 2.39E-08 | 9.84E-08 | 0.00011589 |
| Obesity | nk_cell | *APOBEC3C* | 0.886932616 | 2.25E-09 | 2.27E-08 | 1.09E-05 |
| Obesity | nk_cell | *CCDC13* | 2.101194246 | 0.001320864 | 0.001749135 | 1 |
| Obesity | nk_cell | *C4A* | -4.592372353 | 4.25E-08 | 1.53E-07 | 0.00020627 |
| Obesity | nk_cell | *NEAT1* | -1.232628198 | 3.32E-07 | 8.34E-07 | 0.001611531 |
| Obesity | nk_cell | *RAB30-AS1* | -2.074805808 | 5.97E-08 | 2.00E-07 | 0.000289662 |
| Obesity | nk_cell | *DNAJC3-AS1* | -2.993460703 | 7.86E-09 | 4.45E-08 | 3.81E-05 |
| Obesity | nk_cell | *LINC00926* | -2.544528206 | 3.87E-09 | 2.97E-08 | 1.88E-05 |
| Obesity | nk_cell | *DPH6-AS1* | -1.594974781 | 1.06E-07 | 3.22E-07 | 0.000515609 |
| Obesity | nk_cell | *BCKDHA* | -2.453433121 | 6.16E-05 | 9.51E-05 | 0.298729923 |
| Obesity | nk_cell | *CDK11B* | -3.199661997 | 1.11E-06 | 2.40E-06 | 0.005403119 |
| Obesity | nk_cell | *ABHD14A* | -2.979130707 | 0.001024728 | 0.001370446 | 1 |
| Obesity | nk_cell | *GDNF-AS1* | -1.915869101 | 2.79E-09 | 2.51E-08 | 1.35E-05 |
| Obesity | nk_cell | *CCDC153* | -1.65625163 | 1.18E-08 | 5.77E-08 | 5.71E-05 |

| | | | | | | |
|---|---|---|---|---|---|---|
| Obesity | nk_cell | *AL357673.1* | 1.977832347 | 0.000399146 | 0.000558114 | 1 |
| Obesity | nk_cell | *C5orf17* | -2.643601104 | 1.51E-07 | 4.29E-07 | 0.000731071 |
| Obesity | nk_cell | *TMEM150C* | -1.53408907 | 8.19E-05 | 0.000124332 | 0.397489565 |
| Obesity | nk_cell | *LINC00939* | -2.948410097 | 1.59E-07 | 4.48E-07 | 0.000770157 |
| Obesity | nk_cell | *LINC01060* | -2.137539251 | 1.82E-08 | 8.07E-08 | 8.83E-05 |
| Obesity | nk_cell | *LINC00942* | -6.709554328 | 2.93E-10 | 1.36E-08 | 1.42E-06 |
| Obesity | nk_cell | *PDCD6* | -2.906729613 | 0.000119381 | 0.000177843 | 0.579235911 |
| Obesity | nk_cell | *GOLGA8K* | -2.120115216 | 0.000139565 | 0.000206077 | 0.677167427 |
| Obesity | nk_cell | *TMEM158* | -4.466351898 | 1.11E-08 | 5.55E-08 | 5.39E-05 |
| Obesity | nk_cell | *DNAH10OS* | -6.454722136 | 3.87E-06 | 7.35E-06 | 0.018769784 |
| Obesity | nk_cell | *PCDHA10* | -1.590826204 | 1.90E-08 | 8.31E-08 | 9.23E-05 |
| Obesity | nk_cell | *ZNF718* | 2.737159424 | 2.93E-05 | 4.75E-05 | 0.142059014 |
| Obesity | nk_cell | *SMIM20* | -3.908199941 | 0.000122506 | 0.000182163 | 0.594398518 |
| Obesity | nk_cell | *LINC00989* | -1.3063397 | 1.27E-09 | 2.09E-08 | 6.15E-06 |
| Obesity | nk_cell | *GLI4* | -1.535149844 | 1.09E-09 | 2.05E-08 | 5.29E-06 |
| Obesity | nk_cell | *SNHG18* | -2.69426063 | 1.05E-08 | 5.37E-08 | 5.08E-05 |
| Obesity | nk_cell | *ALG1L2* | -3.330655762 | 2.69E-07 | 6.96E-07 | 0.001304944 |
| Obesity | nk_cell | *ZNF550* | 4.191417624 | 8.77E-06 | 1.54E-05 | 0.042565233 |
| Obesity | nk_cell | *LINC01094* | -1.743382064 | 2.12E-09 | 2.26E-08 | 1.03E-05 |
| Obesity | nk_cell | *TMEM200B* | 3.373733597 | 0.000133107 | 0.000196961 | 0.645835159 |
| Obesity | nk_cell | *SLC10A5* | -2.826345826 | 0.002887545 | 0.003721213 | 1 |
| Obesity | nk_cell | *PCDHGA6* | -1.013949326 | 1.10E-06 | 2.38E-06 | 0.005350939 |
| Obesity | nk_cell | *UTP14C* | 3.555362457 | 1.46E-05 | 2.48E-05 | 0.071006114 |
| Obesity | nk_cell | *PCDHGB2* | -5.444419892 | 7.77E-09 | 4.42E-08 | 3.77E-05 |
| Obesity | nk_cell | *LYN* | 1.503463456 | 2.46E-07 | 6.44E-07 | 0.001191385 |
| Obesity | nk_cell | *SIGLEC14* | -2.118379187 | 0.002900003 | 0.003734292 | 1 |
| Obesity | nk_cell | *FPGT* | -1.743563087 | 2.69E-05 | 4.38E-05 | 0.13055399 |
| Obesity | nk_cell | *MEX3A* | -2.536106589 | 5.46E-07 | 1.27E-06 | 0.002649707 |
| Obesity | nk_cell | *DPP3* | 1.578930149 | 7.11E-06 | 1.27E-05 | 0.034514142 |
| Obesity | nk_cell | *BRK1* | 1.276557517 | 0.009354382 | 0.011519661 | 1 |
| Obesity | nk_cell | *SNHG9* | -1.940665595 | 3.47E-06 | 6.66E-06 | 0.01686053 |
| Obesity | nk_cell | *PCDHA3* | -5.710871195 | 1.33E-11 | 1.85E-09 | 6.48E-08 |
| Obesity | nk_cell | *ZNF432* | -1.91523726 | 3.91E-08 | 1.44E-07 | 0.000189808 |
| Obesity | nk_cell | *HMBS* | -1.306790026 | 1.91E-06 | 3.87E-06 | 0.009249077 |
| Obesity | nk_cell | *TAS2R31* | 1.680712936 | 3.33E-08 | 1.26E-07 | 0.000161767 |
| Obesity | nk_cell | *AC093668.2* | -2.052103807 | 8.05E-11 | 6.40E-09 | 3.90E-07 |
| Obesity | nk_cell | *KIAA1147* | -2.042549185 | 3.67E-07 | 9.10E-07 | 0.001782211 |
| Obesity | nk_cell | *ZBED6* | -2.318924957 | 1.49E-09 | 2.11E-08 | 7.23E-06 |
| Obesity | nk_cell | *CNPY2* | -3.025535388 | 8.74E-06 | 1.54E-05 | 0.042395397 |
| Obesity | nk_cell | *LINC00592* | -4.204544491 | 1.10E-05 | 1.89E-05 | 0.053215346 |
| Obesity | nk_cell | *RTEL1* | 2.18723904 | 0.000154526 | 0.000226925 | 0.749761634 |
| Obesity | nk_cell | *LINC00640* | -1.467685258 | 7.81E-06 | 1.38E-05 | 0.037915334 |
| Obesity | nk_cell | *HIF1A-AS2* | -2.555819481 | 0.000306659 | 0.000435444 | 1 |
| Obesity | nk_cell | *LINC00638* | -2.581750949 | 7.55E-06 | 1.34E-05 | 0.036608377 |
| Obesity | nk_cell | *SLC25A21-AS1* | 4.209652137 | 5.19E-06 | 9.56E-06 | 0.025182424 |
| Obesity | nk_cell | *RNASE4* | -4.72131467 | 9.67E-09 | 5.08E-08 | 4.69E-05 |
| Obesity | nk_cell | *MC1R* | 3.846709798 | 0.000686216 | 0.000935258 | 1 |
| Obesity | nk_cell | *LINC00639* | 2.096062358 | 1.08E-08 | 5.44E-08 | 5.24E-05 |
| Obesity | nk_cell | *SMIM6* | 7.476701538 | 4.21E-05 | 6.67E-05 | 0.20420179 |
| Obesity | nk_cell | *LINC00924* | -1.252801283 | 1.70E-05 | 2.84E-05 | 0.082344347 |

| | | | | | | |
|---|---|---|---|---|---|---|
| Obesity | nk_cell | *ITGB3* | -2.043980237 | 1.61E-06 | 3.33E-06 | 0.007798045 |
| Obesity | nk_cell | *MRPL46* | 3.459799899 | 1.16E-07 | 3.47E-07 | 0.000562403 |
| Obesity | nk_cell | *UBE2Q2L* | -3.813633539 | 7.01E-07 | 1.59E-06 | 0.003399033 |
| Obesity | nk_cell | *IQCH-AS1* | -1.894990783 | 0.005879729 | 0.007360041 | 1 |
| Obesity | nk_cell | *AC083843.1* | -20.32707671 | 1.48E-09 | 2.11E-08 | 7.17E-06 |
| Obesity | nk_cell | *PWRN1* | -1.371777578 | 1.97E-09 | 2.20E-08 | 9.56E-06 |
| Obesity | nk_cell | *LINC01003* | 3.186219483 | 7.04E-07 | 1.60E-06 | 0.003415872 |
| Obesity | nk_cell | *C15orf65* | -3.974898603 | 1.40E-08 | 6.63E-08 | 6.77E-05 |
| Obesity | nk_cell | *GOLGA8H* | -0.62412076 | 0.002908126 | 0.003742766 | 1 |
| Obesity | nk_cell | *LINC00662* | -2.27938795 | 1.83E-07 | 5.04E-07 | 0.000890338 |
| Obesity | nk_cell | *C19orf84* | 0.420173362 | 1.12E-09 | 2.05E-08 | 5.45E-06 |
| Obesity | nk_cell | *LINC00675* | -2.452412958 | 1.86E-07 | 5.08E-07 | 0.000901798 |
| Obesity | nk_cell | *LINC00667* | -2.006071489 | 2.85E-07 | 7.27E-07 | 0.001380706 |
| Obesity | nk_cell | *LINC00672* | -7.593546556 | 0.006674536 | 0.008323794 | 1 |
| Obesity | nk_cell | *LINC00909* | -2.585812865 | 4.14E-05 | 6.57E-05 | 0.201054483 |
| Obesity | nk_cell | *DYNLL2* | 4.454046546 | 6.20E-07 | 1.42E-06 | 0.003009039 |
| Obesity | nk_cell | *STRADA* | -3.379709318 | 9.31E-08 | 2.90E-07 | 0.000451543 |
| Obesity | nk_cell | *MYO15B* | -1.98745059 | 1.20E-06 | 2.56E-06 | 0.005823452 |
| Obesity | nk_cell | *ILF3-AS1* | -2.382746698 | 0.000160372 | 0.000234941 | 0.778125971 |
| Obesity | nk_cell | *ZNF582-AS1* | 5.384158971 | 0.001250652 | 0.001659328 | 1 |
| Obesity | nk_cell | *ZNF224* | -2.874290086 | 1.78E-06 | 3.64E-06 | 0.008641126 |
| Obesity | nk_cell | *LIN37* | -6.618503239 | 5.11E-11 | 4.43E-09 | 2.48E-07 |
| Obesity | nk_cell | *NDUFA7* | 1.460508495 | 1.09E-09 | 2.05E-08 | 5.28E-06 |
| Obesity | nk_cell | *AC004466.1* | 1.611813161 | 0.032737403 | 0.03843333 | 1 |
| Obesity | nk_cell | *LINC00664* | 2.742369692 | 2.00E-09 | 2.20E-08 | 9.69E-06 |
| Obesity | nk_cell | *SCGB1B2P* | -3.967640867 | 1.28E-07 | 3.76E-07 | 0.000621952 |
| Obesity | nk_cell | *AC026740.1* | -3.86499032 | 0.039453756 | 0.046016737 | 1 |
| Obesity | nk_cell | *ZNF728* | -1.610326251 | 5.28E-09 | 3.56E-08 | 2.56E-05 |
| Obesity | nk_cell | *TRABD2B* | -2.997777279 | 8.10E-07 | 1.81E-06 | 0.003930273 |
| Obesity | nk_cell | *AC093157.1* | -1.802171258 | 5.07E-09 | 3.50E-08 | 2.46E-05 |
| Obesity | nk_cell | *FBXO17* | -1.561389062 | 2.67E-07 | 6.93E-07 | 0.001296704 |
| Obesity | nk_cell | *ZNF587B* | -1.389071469 | 1.58E-09 | 2.11E-08 | 7.64E-06 |
| Obesity | nk_cell | *SNHG8* | -2.585725212 | 3.57E-07 | 8.87E-07 | 0.001731162 |
| Obesity | nk_cell | *AP001172.2* | 0.880373883 | 0.012313436 | 0.015011255 | 1 |
| Obesity | nk_cell | *HCG17* | -1.98392196 | 5.62E-06 | 1.03E-05 | 0.027267991 |
| Obesity | nk_cell | *GTF2H5* | -1.22749296 | 0.000388051 | 0.000543383 | 1 |
| Obesity | nk_cell | *FGF14-AS2* | -1.777927188 | 1.73E-09 | 2.16E-08 | 8.37E-06 |
| Obesity | nk_cell | *CASC15* | -1.613173811 | 1.09E-08 | 5.48E-08 | 5.30E-05 |
| Obesity | nk_cell | *AP000347.2* | -2.584063212 | 0.000100903 | 0.00015162 | 0.489581894 |
| Obesity | nk_cell | *DOC2B* | -0.662658748 | 0.006076582 | 0.007594945 | 1 |
| Obesity | nk_cell | *GRIN2B* | -1.025354207 | 5.75E-06 | 1.05E-05 | 0.027875794 |
| Obesity | nk_cell | *PAXIP1-AS1* | -3.331562435 | 5.29E-06 | 9.73E-06 | 0.025653816 |
| Obesity | pericyte | *SEMA3F* | -9.509029838 | 4.59E-09 | 8.82E-07 | 2.23E-05 |
| Obesity | pericyte | *RBM6* | -6.799653206 | 7.46E-23 | 3.62E-19 | 3.62E-19 |
| Obesity | pericyte | *ITIH4* | 2.709620662 | 3.34E-10 | 7.37E-08 | 1.62E-06 |
| Obesity | pericyte | *UHRF1BP1* | 8.822845829 | 3.49E-17 | 3.39E-14 | 1.70E-13 |
| Obesity | pericyte | *NT5C2* | 4.238265072 | 0.001491124 | 0.049019583 | 1 |
| Obesity | pericyte | *FDFT1* | 8.265923314 | 2.25E-05 | 0.001816643 | 0.108998592 |
| Obesity | pericyte | *GSTP1* | 7.291924627 | 0.000143387 | 0.00858905 | 0.695713088 |
| Obesity | pericyte | *OGFOD1* | -7.197828111 | 0.000865835 | 0.033879298 | 1 |

| | | | | | | |
|---|---|---|---|---|---|---|
| Obesity | pericyte | *YPEL3* | 8.558327419 | 1.65E-07 | 2.35E-05 | 0.000799443 |
| Obesity | pericyte | *PABPC4* | -9.170247061 | 2.29E-08 | 3.59E-06 | 0.000111265 |
| Obesity | pericyte | *RGS17* | -8.533685573 | 2.04E-07 | 2.74E-05 | 0.000987757 |
| Obesity | pericyte | *SETD1A* | 12.27502079 | 9.58E-07 | 0.000108056 | 0.004646403 |
| Obesity | pericyte | *ANKRD54* | -4.599850584 | 0.000206985 | 0.011036162 | 1 |
| Obesity | pericyte | *CENPM* | 5.807097369 | 9.03E-05 | 0.005763449 | 0.438022091 |
| Obesity | pericyte | *ABHD12* | 6.705362206 | 0.000355661 | 0.017119827 | 1 |
| Obesity | pericyte | *MEST* | -3.189805133 | 0.000449067 | 0.020363287 | 1 |
| Obesity | pericyte | *RAD51C* | -5.019471476 | 0.000304486 | 0.014922901 | 1 |
| Obesity | pericyte | *PNPO* | -17.31366173 | 1.27E-05 | 0.001143219 | 0.061733841 |
| Obesity | pericyte | *CHRNE* | 1.673650874 | 0.000672201 | 0.027639976 | 1 |
| Obesity | pericyte | *BTN3A3* | 7.548817933 | 9.76E-11 | 2.37E-08 | 4.73E-07 |
| Obesity | pericyte | *TULP1* | -4.912385033 | 0.000186481 | 0.010281898 | 0.904807033 |
| Obesity | pericyte | *CUTA* | -4.468994767 | 0.00122637 | 0.043118451 | 1 |
| Obesity | pericyte | *NME5* | 2.851601035 | 0.000711896 | 0.02878431 | 1 |
| Obesity | pericyte | *TMCO6* | -5.398593483 | 0.000516999 | 0.02301359 | 1 |
| Obesity | pericyte | *IK* | -4.984613272 | 0.001397259 | 0.047079882 | 1 |
| Obesity | pericyte | *PCCB* | 6.101609148 | 2.14E-05 | 0.001816643 | 0.103989296 |
| Obesity | pericyte | *CHMP3* | -6.67210353 | 1.13E-05 | 0.001055855 | 0.054904484 |
| Obesity | pericyte | *NSUN4* | -5.708340965 | 0.000754277 | 0.030075977 | 1 |
| Obesity | pericyte | *SERPINC1* | 12.50821675 | 3.72E-08 | 5.65E-06 | 0.000180732 |
| Obesity | pericyte | *CCDC92* | 6.982219338 | 0.000271589 | 0.013871064 | 1 |
| Obesity | pericyte | *INVS* | 5.535813103 | 0.000145971 | 0.008637209 | 0.708251132 |
| Obesity | pericyte | *SNX19* | -6.680088345 | 2.80E-05 | 0.002153523 | 0.135671922 |
| Obesity | pericyte | *BTN2A2* | 2.816428071 | 3.17E-05 | 0.002327663 | 0.15362577 |
| Obesity | pericyte | *SNRPC* | -12.89551498 | 1.49E-18 | 2.41E-15 | 7.23E-15 |
| Obesity | pericyte | *GRK4* | 1.474195878 | 0.000287036 | 0.014243678 | 1 |
| Obesity | pericyte | *RIBC2* | 3.566858813 | 0.001011354 | 0.03745871 | 1 |
| Obesity | pericyte | *RPAIN* | -5.430836983 | 0.000393434 | 0.018715115 | 1 |
| Obesity | pericyte | *AP1M2* | -3.301334918 | 0.000286189 | 0.014243678 | 1 |
| Obesity | pericyte | *ATP8B3* | -4.732776109 | 2.85E-11 | 8.82E-09 | 1.38E-07 |
| Obesity | pericyte | *PGPEP1* | 4.411380877 | 0.000684713 | 0.027917888 | 1 |
| Obesity | pericyte | *NPAS1* | -12.07742925 | 2.77E-05 | 0.002153523 | 0.134277585 |
| Obesity | pericyte | *PKDREJ* | -3.670552982 | 0.001313447 | 0.045107753 | 1 |
| Obesity | pericyte | *NDUFA2* | 6.325411056 | 0.000287692 | 0.014243678 | 1 |
| Obesity | pericyte | *IGHMBP2* | 3.849930415 | 9.34E-05 | 0.005887755 | 0.45335716 |
| Obesity | pericyte | *LARS* | -3.366108906 | 0.001032614 | 0.037919016 | 1 |
| Obesity | pericyte | *POM121C* | -10.96586132 | 4.28E-10 | 9.03E-08 | 2.08E-06 |
| Obesity | pericyte | *SCN2A* | 7.843137139 | 0.000500498 | 0.022485335 | 1 |
| Obesity | pericyte | *TCF19* | 3.996495861 | 0.001039412 | 0.037919016 | 1 |
| Obesity | pericyte | *NUSAP1* | 6.943062204 | 0.000634298 | 0.026531138 | 1 |
| Obesity | pericyte | *CENPO* | 2.654195657 | 0.000446748 | 0.020363287 | 1 |
| Obesity | pericyte | *VPS33A* | 7.398721135 | 2.76E-05 | 0.002153523 | 0.134092171 |
| Obesity | pericyte | *MYO19* | 8.411274921 | 0.000154107 | 0.008901514 | 0.747727135 |
| Obesity | pericyte | *C18orf8* | -11.86380535 | 3.34E-10 | 7.37E-08 | 1.62E-06 |
| Obesity | pericyte | *COL6A1* | 2.14481619 | 0.001431579 | 0.047575499 | 1 |
| Obesity | pericyte | *SAE1* | -11.11142345 | 2.90E-05 | 0.002197709 | 0.140653382 |
| Obesity | pericyte | *LIMD1* | 6.095124964 | 0.000947061 | 0.035537107 | 1 |
| Obesity | pericyte | *ADHFE1* | -1.91853152 | 0.000202268 | 0.010904492 | 0.981404314 |
| Obesity | pericyte | *RGR* | -23.21515266 | 5.50E-05 | 0.003706691 | 0.26688177 |

| | | | | | | |
|---|---|---|---|---|---|---|
| Obesity | pericyte | *MTG1* | 3.193201842 | 0.001152903 | 0.041436184 | 1 |
| Obesity | pericyte | *HSD17B12* | -3.698318748 | 9.54E-11 | 2.37E-08 | 4.63E-07 |
| Obesity | pericyte | *TNKS1BP1* | 4.694819367 | 6.45E-06 | 0.000625569 | 0.031278426 |
| Obesity | pericyte | *TTC12* | 4.01561403 | 0.000952148 | 0.035537107 | 1 |
| Obesity | pericyte | *TBX6* | 3.276107224 | 1.02E-11 | 3.53E-09 | 4.94E-08 |
| Obesity | pericyte | *POC5* | -4.731851655 | 1.56E-05 | 0.001352058 | 0.075715258 |
| Obesity | pericyte | *JAZF1* | 4.583291073 | 0.001329433 | 0.045107753 | 1 |
| Obesity | pericyte | *MMP16* | -2.442648738 | 0.000411684 | 0.019206646 | 1 |
| Obesity | pericyte | *STEAP2* | -2.861526885 | 0.000667823 | 0.027639976 | 1 |
| Obesity | pericyte | *CLEC18A* | -8.733961973 | 9.48E-18 | 1.15E-14 | 4.60E-14 |
| Obesity | pericyte | *ETS2* | 2.157626593 | 0.001280969 | 0.044394725 | 1 |
| Obesity | pericyte | *SLC5A11* | -2.874799906 | 1.77E-07 | 2.45E-05 | 0.000857041 |
| Obesity | pericyte | *CILP2* | -7.738240512 | 2.23E-05 | 0.001816643 | 0.108243003 |
| Obesity | pericyte | *ABCF3* | 4.048718397 | 0.0012382 | 0.043221202 | 1 |
| Obesity | pericyte | *ANTXR2* | 5.041257108 | 0.001505342 | 0.049019583 | 1 |
| Obesity | pericyte | *DNALI1* | -4.976759087 | 1.25E-05 | 0.001143125 | 0.060585602 |
| Obesity | pericyte | *RFT1* | -6.375465976 | 8.96E-07 | 0.000103482 | 0.00434624 |
| Obesity | pericyte | *RNF123* | 9.363991611 | 5.33E-16 | 4.31E-13 | 2.59E-12 |
| Obesity | pericyte | *MST1R* | 6.428166148 | 6.16E-14 | 3.74E-11 | 2.99E-10 |
| Obesity | pericyte | *RPP25L* | -5.821000875 | 0.000356369 | 0.017119827 | 1 |
| Obesity | pericyte | *SLC39A13* | 10.60477468 | 3.84E-05 | 0.002739738 | 0.186302187 |
| Obesity | pericyte | *RAPSN* | -8.373090168 | 1.46E-15 | 1.01E-12 | 7.06E-12 |
| Obesity | pericyte | *SMG8* | -4.538377436 | 0.000886356 | 0.034404812 | 1 |
| Obesity | pericyte | *SRR* | 2.52408916 | 4.25E-05 | 0.002991124 | 0.206387555 |
| Obesity | pericyte | *FNTA* | 6.828051158 | 0.000134643 | 0.008166097 | 0.653287787 |
| Obesity | pericyte | *INO80E* | -6.786687236 | 2.41E-13 | 1.17E-10 | 1.17E-09 |
| Obesity | pericyte | *ADORA2B* | -2.861299371 | 3.89E-07 | 4.84E-05 | 0.001888553 |
| Obesity | pericyte | *TTLL6* | -14.46573596 | 8.22E-11 | 2.21E-08 | 3.99E-07 |
| Obesity | pericyte | *TSNARE1* | 5.453623769 | 0.000779877 | 0.030763919 | 1 |
| Obesity | pericyte | *C1QTNF4* | -3.877422866 | 1.26E-12 | 5.56E-10 | 6.12E-09 |
| Obesity | pericyte | *CTSW* | -1.173426272 | 4.73E-09 | 8.82E-07 | 2.29E-05 |
| Obesity | pericyte | *EFEMP2* | 6.252345322 | 0.000193005 | 0.010522034 | 0.93646101 |
| Obesity | pericyte | *FUT10* | -2.662267436 | 0.000552211 | 0.024357519 | 1 |
| Obesity | pericyte | *SNX32* | 2.371289908 | 0.000216661 | 0.0114265 | 1 |
| Obesity | pericyte | *MRGPRF* | 6.318858562 | 0.000101482 | 0.006312671 | 0.492388345 |
| Obesity | pericyte | *MST1* | -1.699946907 | 0.001414078 | 0.047317967 | 1 |
| Obesity | pericyte | *HOXB2* | 6.259499806 | 0.000952146 | 0.035537107 | 1 |
| Obesity | pericyte | *SEZ6L2* | 4.661799467 | 2.78E-06 | 0.000286814 | 0.013480269 |
| Obesity | pericyte | *KCTD13* | 6.858013546 | 5.52E-06 | 0.00054699 | 0.026802504 |
| Obesity | pericyte | *CADM2* | 5.578431161 | 2.74E-07 | 3.50E-05 | 0.001328593 |
| Obesity | pericyte | *BANF1* | -8.038018318 | 6.84E-05 | 0.004543217 | 0.331654852 |
| Obesity | pericyte | *ORAI3* | 10.10324414 | 1.01E-11 | 3.53E-09 | 4.90E-08 |
| Obesity | pericyte | *RUFY1* | 5.611571676 | 0.000620261 | 0.026244897 | 1 |
| Obesity | pericyte | *RPP25* | -3.194063153 | 0.000152383 | 0.008901514 | 0.739361726 |
| Obesity | pericyte | *MPI* | 4.912237534 | 0.000756239 | 0.030075977 | 1 |
| Obesity | pericyte | *TRIM73* | -19.05611157 | 1.39E-13 | 7.50E-11 | 6.75E-10 |
| Obesity | pericyte | *ZNF852* | -3.977030665 | 0.000606027 | 0.026244897 | 1 |
| Obesity | pericyte | *TUFM* | 8.633739993 | 1.59E-08 | 2.58E-06 | 7.73E-05 |
| Obesity | pericyte | *C3orf38* | 6.880644237 | 5.91E-07 | 7.00E-05 | 0.002868715 |
| Obesity | pericyte | *RPRML* | 15.15825012 | 3.35E-05 | 0.002422903 | 0.162334486 |

| | | | | | | |
|---|---|---|---|---|---|---|
| Obesity | pericyte | *ZFP41* | 5.721667569 | 0.001086727 | 0.039349256 | 1 |
| Obesity | pericyte | *PPP1R27* | -19.00474156 | 5.29E-05 | 0.003616116 | 0.256744227 |
| Obesity | pericyte | *EMILIN3* | 7.073506862 | 0.000243108 | 0.012548521 | 1 |
| Obesity | pericyte | *ACOT1* | 2.820558329 | 0.000597934 | 0.026136729 | 1 |
| Obesity | pericyte | *NUTM2A* | -4.922432921 | 1.31E-05 | 0.001157674 | 0.063672093 |
| Obesity | pericyte | *MAFF* | -3.383660275 | 0.000175999 | 0.010046413 | 0.853945095 |
| Obesity | pericyte | *GP1BA* | 3.57882319 | 0.000940888 | 0.035537107 | 1 |
| Obesity | pericyte | *BTN3A2* | -2.376162916 | 3.66E-11 | 1.05E-08 | 1.78E-07 |
| Obesity | pericyte | *ZSCAN23* | 1.674833379 | 0.001325246 | 0.045107753 | 1 |
| Obesity | pericyte | *CENPP* | 3.4181727 | 8.62E-05 | 0.005578278 | 0.41837083 |
| Obesity | pericyte | *SKOR1* | 5.460611577 | 0.001205093 | 0.042679645 | 1 |
| Obesity | pericyte | *ZKSCAN3* | -6.258948702 | 1.35E-07 | 1.98E-05 | 0.000654995 |
| Obesity | pericyte | *CPNE4* | 5.53063066 | 0.000107936 | 0.006629156 | 0.523703294 |
| Obesity | pericyte | *SULT1A2* | 14.54238284 | 5.00E-21 | 1.21E-17 | 2.42E-17 |
| Obesity | pericyte | *ZNF165* | -3.078592039 | 0.000181833 | 0.010140838 | 0.882252911 |
| Obesity | pericyte | *MRPL21* | -1.878119823 | 0.001504899 | 0.049019583 | 1 |
| Obesity | pericyte | *MFAP5* | -4.020815701 | 0.000225715 | 0.011775991 | 1 |
| Obesity | pericyte | *DNAH10* | 11.01644591 | 6.04E-12 | 2.44E-09 | 2.93E-08 |
| Obesity | pericyte | *RPS26* | -2.545405459 | 0.000419891 | 0.019402953 | 1 |
| Obesity | pericyte | *ZNF780A* | -1.631833135 | 3.07E-05 | 0.00229255 | 0.149015723 |
| Obesity | pericyte | *SHISA4* | -12.94495364 | 9.15E-06 | 0.000870547 | 0.044397874 |
| Obesity | pericyte | *NOTCH4* | -0.676799606 | 1.30E-08 | 2.18E-06 | 6.32E-05 |
| Obesity | pericyte | *SKIV2L* | -2.965654548 | 4.74E-07 | 5.74E-05 | 0.00229743 |
| Obesity | pericyte | *GPANK1* | -8.429608935 | 3.38E-09 | 6.84E-07 | 1.64E-05 |
| Obesity | pericyte | *POU5F1* | 1.051391711 | 8.03E-05 | 0.005264103 | 0.389543641 |
| Obesity | pericyte | *CCHCR1* | 1.906328349 | 2.21E-05 | 0.001816643 | 0.107151321 |
| Obesity | pericyte | *RNF39* | -7.027409955 | 2.36E-06 | 0.00024842 | 0.011427304 |
| Obesity | pericyte | *ZFP57* | 2.593434414 | 5.43E-09 | 9.76E-07 | 2.64E-05 |
| Obesity | pericyte | *C17orf107* | 4.27625055 | 0.000398125 | 0.018754381 | 1 |
| Obesity | pericyte | *AC084018.1* | -2.696169461 | 5.15E-05 | 0.003569774 | 0.249884164 |
| Obesity | pericyte | *TMEM110* | 9.533989472 | 1.14E-08 | 1.97E-06 | 5.52E-05 |
| Obesity | pericyte | *EXOSC6* | -14.13708418 | 2.91E-11 | 8.82E-09 | 1.41E-07 |
| Obesity | pericyte | *C4B* | 6.202373131 | 0.000180491 | 0.010140838 | 0.87574147 |
| Obesity | pericyte | *LINC00240* | 2.134297166 | 0.000619636 | 0.026244897 | 1 |
| Obesity | pericyte | *HCG20* | -5.972256954 | 2.45E-07 | 3.21E-05 | 0.001186909 |
| Obesity | pericyte | *ZSCAN31* | 2.311137257 | 1.15E-06 | 0.000123491 | 0.005557087 |
| Obesity | pericyte | *PSMB9* | 3.01265953 | 3.63E-06 | 0.000366439 | 0.017589092 |
| Obesity | pericyte | *ISY1* | -3.68561643 | 0.000622045 | 0.026244897 | 1 |
| Obesity | pericyte | *C4A* | -6.959319513 | 1.14E-06 | 0.000123491 | 0.005509029 |
| Obesity | pericyte | *BRK1* | -4.806939818 | 0.001195627 | 0.04265576 | 1 |
| Obesity | pericyte | *PCDHA3* | -6.452712888 | 0.000921445 | 0.035482958 | 1 |
| Obesity | SMC | *SEMA3F* | -4.972721844 | 5.19E-05 | 0.00426575 | 0.251679261 |
| Obesity | SMC | *RBM6* | -6.287458312 | 1.38E-24 | 6.68E-21 | 6.68E-21 |
| Obesity | SMC | *ITIH4* | 2.225504853 | 4.61E-09 | 9.09E-07 | 2.24E-05 |
| Obesity | SMC | *UHRF1BP1* | 7.261640764 | 1.13E-15 | 1.01E-12 | 5.50E-12 |
| Obesity | SMC | *ZNRD1* | -3.008503754 | 0.001188328 | 0.044352069 | 1 |
| Obesity | SMC | *OGFOD1* | -7.980366426 | 1.34E-05 | 0.001295948 | 0.064797401 |
| Obesity | SMC | *YPEL3* | 5.608927039 | 7.75E-05 | 0.005894036 | 0.376246716 |
| Obesity | SMC | *PABPC4* | -8.608190247 | 4.68E-09 | 9.09E-07 | 2.27E-05 |
| Obesity | SMC | *SETD1A* | 6.121807073 | 0.00088806 | 0.036827908 | 1 |

| | | | | | | |
|---|---|---|---|---|---|---|
| Obesity | SMC | *CENPM* | 5.585916834 | 2.13E-05 | 0.001990667 | 0.103514694 |
| Obesity | SMC | *KNOP1* | -7.399637969 | 0.001101349 | 0.042410661 | 1 |
| Obesity | SMC | *RPAP1* | -6.440538417 | 0.000887391 | 0.036827908 | 1 |
| Obesity | SMC | *ZDHHC2* | -6.018617995 | 0.000384571 | 0.020504815 | 1 |
| Obesity | SMC | *LIN7B* | -7.032500701 | 0.000823974 | 0.036017335 | 1 |
| Obesity | SMC | *TAF6* | -3.009914297 | 0.001096881 | 0.042410661 | 1 |
| Obesity | SMC | *RAD51C* | -4.746434509 | 3.43E-05 | 0.00302888 | 0.166588395 |
| Obesity | SMC | *CHRNE* | 1.568438755 | 0.000493759 | 0.025059156 | 1 |
| Obesity | SMC | *BTN3A3* | 4.66029273 | 1.40E-07 | 2.00E-05 | 0.000681023 |
| Obesity | SMC | *TULP1* | -4.65338125 | 0.000103419 | 0.006984933 | 0.50178694 |
| Obesity | SMC | *CUTA* | -5.063196897 | 0.000118074 | 0.007741851 | 0.572896987 |
| Obesity | SMC | *BTN2A1* | -7.401713258 | 0.00066352 | 0.030955756 | 1 |
| Obesity | SMC | *NME5* | 2.034000227 | 0.001026242 | 0.040506679 | 1 |
| Obesity | SMC | *TMCO6* | -4.251578686 | 0.001266505 | 0.046909027 | 1 |
| Obesity | SMC | *IK* | -5.344798037 | 0.000102428 | 0.006984933 | 0.496979895 |
| Obesity | SMC | *PCCB* | 4.7374995 | 0.000223247 | 0.013372779 | 1 |
| Obesity | SMC | *NSUN4* | -5.984839496 | 0.000286385 | 0.015790243 | 1 |
| Obesity | SMC | *SERPINC1* | 11.40225809 | 3.87E-09 | 8.16E-07 | 1.88E-05 |
| Obesity | SMC | *SNX19* | -6.102081536 | 9.89E-06 | 0.000999669 | 0.047984106 |
| Obesity | SMC | *BTN2A2* | 2.278775124 | 5.74E-05 | 0.004639962 | 0.278397728 |
| Obesity | SMC | *SNRPC* | -13.68796519 | 8.66E-22 | 2.10E-18 | 4.20E-18 |
| Obesity | SMC | *GRK4* | 1.32416376 | 0.000381083 | 0.020504815 | 1 |
| Obesity | SMC | *RIBC2* | 3.36524535 | 0.000743352 | 0.03339579 | 1 |
| Obesity | SMC | *ATP8B3* | -4.301118151 | 6.07E-11 | 1.47E-08 | 2.95E-07 |
| Obesity | SMC | *CYP2E1* | -2.198387192 | 0.000795491 | 0.035088397 | 1 |
| Obesity | SMC | *IGHMBP2* | 2.973019107 | 0.000831531 | 0.0360231 | 1 |
| Obesity | SMC | *LARS* | -3.389679719 | 0.000181052 | 0.011119815 | 0.878465423 |
| Obesity | SMC | *POM121C* | -11.17118073 | 1.11E-11 | 3.18E-09 | 5.40E-08 |
| Obesity | SMC | *SLC37A4* | 6.118257467 | 1.94E-06 | 0.000229972 | 0.009428845 |
| Obesity | SMC | *RSL24D1* | -4.814426821 | 0.001071745 | 0.041936345 | 1 |
| Obesity | SMC | *C18orf8* | -10.74463007 | 9.49E-12 | 2.88E-09 | 4.61E-08 |
| Obesity | SMC | *SAE1* | -11.52038853 | 4.86E-08 | 8.43E-06 | 0.000235933 |
| Obesity | SMC | *PCYOX1L* | -12.88336096 | 0.000906393 | 0.037269639 | 1 |
| Obesity | SMC | *ADHFE1* | -1.626481492 | 0.000596466 | 0.028940518 | 1 |
| Obesity | SMC | *HSD17B12* | -3.531734618 | 9.42E-12 | 2.88E-09 | 4.57E-08 |
| Obesity | SMC | *TNKS1BP1* | 3.792072698 | 2.62E-05 | 0.002353572 | 0.127092889 |
| Obesity | SMC | *NUDT22* | -6.046483248 | 0.000419918 | 0.022146104 | 1 |
| Obesity | SMC | *TBX6* | 2.832376333 | 2.73E-11 | 7.35E-09 | 1.32E-07 |
| Obesity | SMC | *POC5* | -4.068142347 | 1.62E-05 | 0.001540378 | 0.078559294 |
| Obesity | SMC | *MMP16* | -2.459821822 | 8.98E-05 | 0.006501962 | 0.435631429 |
| Obesity | SMC | *RWDD2B* | -4.086296182 | 0.000541543 | 0.027088311 | 1 |
| Obesity | SMC | *TMED6* | 9.599608066 | 0.001163872 | 0.043776035 | 1 |
| Obesity | SMC | *CLEC18A* | -4.566867996 | 4.89E-12 | 1.69E-09 | 2.37E-08 |
| Obesity | SMC | *SLC5A11* | -2.799648329 | 6.68E-08 | 1.12E-05 | 0.000323899 |
| Obesity | SMC | *CILP2* | -6.02620081 | 3.77E-05 | 0.003242593 | 0.183008879 |
| Obesity | SMC | *MCM3AP* | -3.850975446 | 0.000767064 | 0.034144906 | 1 |
| Obesity | SMC | *CAMK2N1* | 4.243929762 | 0.000731186 | 0.03315621 | 1 |
| Obesity | SMC | *DNALI1* | -3.537836349 | 0.000638965 | 0.030394692 | 1 |
| Obesity | SMC | *RFT1* | -6.241302739 | 8.73E-08 | 1.41E-05 | 0.00042352 |
| Obesity | SMC | *RNF123* | 8.248231605 | 1.25E-15 | 1.01E-12 | 6.09E-12 |

| | | | | | | |
|---|---|---|---|---|---|---|
| Obesity | SMC | *MST1R* | 6.575306052 | 2.90E-17 | 3.52E-14 | 1.41E-13 |
| Obesity | SMC | *ERAP1* | -5.028037231 | 0.00034165 | 0.018625703 | 1 |
| Obesity | SMC | *YWHAZ* | -5.24143633 | 0.000494136 | 0.025059156 | 1 |
| Obesity | SMC | *RPP25L* | -4.770185742 | 0.000141655 | 0.008926075 | 0.687307787 |
| Obesity | SMC | *RAPSN* | -7.797420123 | 7.02E-15 | 4.19E-12 | 3.41E-11 |
| Obesity | SMC | *ZSCAN21* | -3.076961827 | 0.000710537 | 0.032688831 | 1 |
| Obesity | SMC | *RIMKLB* | -3.59063765 | 0.00091935 | 0.037484749 | 1 |
| Obesity | SMC | *SMG8* | -3.987223738 | 0.000886518 | 0.036827908 | 1 |
| Obesity | SMC | *SRR* | 2.246329516 | 6.75E-05 | 0.005283834 | 0.327597715 |
| Obesity | SMC | *KIAA1586* | -5.09249316 | 0.000260167 | 0.015002612 | 1 |
| Obesity | SMC | *NT5DC2* | -1.659290569 | 0.000951222 | 0.03846106 | 1 |
| Obesity | SMC | *C8orf46* | 1.108350495 | 0.000495812 | 0.025059156 | 1 |
| Obesity | SMC | *GPRIN1* | 7.889660258 | 0.000714142 | 0.032688831 | 1 |
| Obesity | SMC | *INO80E* | -6.630346555 | 7.77E-15 | 4.19E-12 | 3.77E-11 |
| Obesity | SMC | *ADORA2B* | -2.65546987 | 2.42E-07 | 3.26E-05 | 0.001174037 |
| Obesity | SMC | *TTLL6* | -19.16965213 | 4.82E-14 | 2.34E-11 | 2.34E-10 |
| Obesity | SMC | *C1QTNF4* | -4.1352133 | 2.95E-15 | 2.04E-12 | 1.43E-11 |
| Obesity | SMC | *CTSW* | -0.924204376 | 1.14E-07 | 1.73E-05 | 0.000554737 |
| Obesity | SMC | *FUT10* | -2.715792688 | 8.81E-05 | 0.006482134 | 0.427331702 |
| Obesity | SMC | *SNX32* | 1.898123282 | 0.000172516 | 0.010731402 | 0.837049318 |
| Obesity | SMC | *SEZ6L2* | 2.680830534 | 0.000262824 | 0.015002612 | 1 |
| Obesity | SMC | *KCTD13* | 4.608334776 | 0.000117446 | 0.007741851 | 0.569845691 |
| Obesity | SMC | *CADM2* | 3.901678056 | 7.48E-06 | 0.000806448 | 0.036290156 |
| Obesity | SMC | *BANF1* | -8.746458813 | 6.54E-05 | 0.005198719 | 0.317121868 |
| Obesity | SMC | *ORAI3* | 3.860907582 | 5.50E-06 | 0.000635616 | 0.026695868 |
| Obesity | SMC | *CCDC57* | -3.581718145 | 0.001013247 | 0.040506679 | 1 |
| Obesity | SMC | *STX19* | 5.673910139 | 8.82E-05 | 0.006482134 | 0.427820845 |
| Obesity | SMC | *TRIM73* | -17.09716932 | 3.81E-13 | 1.68E-10 | 1.85E-09 |
| Obesity | SMC | *ZNF852* | -7.037469699 | 1.10E-05 | 0.001088411 | 0.053332125 |
| Obesity | SMC | *TUFM* | 7.32168279 | 1.39E-07 | 2.00E-05 | 0.000674243 |
| Obesity | SMC | *C3orf38* | 4.25090735 | 4.77E-05 | 0.003992532 | 0.231566874 |
| Obesity | SMC | *RPRML* | 14.57463135 | 9.51E-06 | 0.000981351 | 0.046123504 |
| Obesity | SMC | *ZNF322* | -5.196235392 | 0.000861348 | 0.036827908 | 1 |
| Obesity | SMC | *PPP1R27* | -16.23513974 | 0.000274721 | 0.015499354 | 1 |
| Obesity | SMC | *PGP* | -6.536199374 | 0.000623773 | 0.029965808 | 1 |
| Obesity | SMC | *ACOT1* | 2.983799486 | 9.22E-05 | 0.006578468 | 0.447335858 |
| Obesity | SMC | *NUTM2A* | -4.30975489 | 2.52E-05 | 0.002305578 | 0.122195611 |
| Obesity | SMC | *MAFF* | -2.891085222 | 0.000553861 | 0.02742179 | 1 |
| Obesity | SMC | *GP1BA* | 5.625041254 | 1.46E-06 | 0.000181987 | 0.007097488 |
| Obesity | SMC | *BTN3A2* | -2.159872239 | 5.56E-11 | 1.42E-08 | 2.70E-07 |
| Obesity | SMC | *CENPP* | 2.482189305 | 0.000653354 | 0.030777409 | 1 |
| Obesity | SMC | *ZKSCAN3* | -5.241495848 | 1.10E-07 | 1.72E-05 | 0.000533045 |
| Obesity | SMC | *CPNE4* | 3.480615871 | 0.001026859 | 0.040506679 | 1 |
| Obesity | SMC | *SULT1A2* | 12.20699624 | 2.71E-20 | 4.39E-17 | 1.32E-16 |
| Obesity | SMC | *ZNF165* | -2.822944038 | 0.000183525 | 0.011130784 | 0.890462706 |
| Obesity | SMC | *MRPL21* | -2.103386484 | 0.000121323 | 0.007848787 | 0.588659011 |
| Obesity | SMC | *DNAH10* | 9.612701943 | 4.28E-10 | 9.90E-08 | 2.08E-06 |
| Obesity | SMC | *ZNF780A* | -1.397278466 | 7.77E-05 | 0.005894036 | 0.377218325 |
| Obesity | SMC | *TMEM184B* | -5.566469098 | 0.000228783 | 0.013537271 | 1 |
| Obesity | SMC | *NOTCH4* | -0.623741997 | 8.73E-09 | 1.57E-06 | 4.24E-05 |

| | | | | | | |
|---|---|---|---|---|---|---|
| Obesity | SMC | *SKIV2L* | -2.678928861 | 2.18E-07 | 3.03E-05 | 0.001059397 |
| Obesity | SMC | *GPANK1* | -9.05845194 | 2.99E-12 | 1.12E-09 | 1.45E-08 |
| Obesity | SMC | *POU5F1* | 0.818447319 | 0.000462325 | 0.024120434 | 1 |
| Obesity | SMC | *CCHCR1* | 1.883308408 | 7.24E-06 | 0.0007983 | 0.035125215 |
| Obesity | SMC | *RNF39* | -5.343659101 | 3.81E-05 | 0.003242593 | 0.184827785 |
| Obesity | SMC | *ZFP57* | 2.350458876 | 5.27E-09 | 9.84E-07 | 2.56E-05 |
| Obesity | SMC | *PRR13* | -5.884286961 | 0.00116235 | 0.043776035 | 1 |
| Obesity | SMC | *AC084018.1* | -2.068216654 | 0.00057222 | 0.028044583 | 1 |
| Obesity | SMC | *TMEM110* | 9.295312566 | 8.47E-10 | 1.87E-07 | 4.11E-06 |
| Obesity | SMC | *PPP1CB* | -5.727964873 | 0.000102906 | 0.006984933 | 0.49930204 |
| Obesity | SMC | *EXOSC6* | -13.4727993 | 5.23E-13 | 2.12E-10 | 2.54E-09 |
| Obesity | SMC | *C4B* | 5.462077441 | 0.000255254 | 0.014921616 | 1 |
| Obesity | SMC | *AL049840.1* | -8.337116024 | 6.24E-06 | 0.000704481 | 0.030292669 |
| Obesity | SMC | *HCG20* | -5.008741966 | 4.33E-07 | 5.68E-05 | 0.002101692 |
| Obesity | SMC | *ZSCAN31* | 2.02010413 | 1.15E-06 | 0.000146749 | 0.005576449 |
| Obesity | SMC | *PSMB9* | 2.536198196 | 7.74E-06 | 0.000816764 | 0.037571135 |
| Obesity | SMC | *ISY1* | -3.498625342 | 0.000283829 | 0.015790243 | 1 |
| Obesity | SMC | *C4A* | -6.277427168 | 1.53E-06 | 0.000186013 | 0.007440519 |
| Obesity | SMC | *DPP3* | 1.735609035 | 0.001154271 | 0.043776035 | 1 |
| Obesity | SMC | *BRK1* | -5.048194533 | 0.000103651 | 0.006984933 | 0.502915206 |
| Obesity | SMC | *HMBS* | 5.569369984 | 0.000126031 | 0.008046066 | 0.611501016 |
| Obesity | SMC | *AP001172.2* | 1.780598763 | 0.000870865 | 0.036827908 | 1 |
| Obesity | t_cell | *SEMA3F* | -5.782423186 | 1.30E-13 | 4.51E-11 | 6.32E-10 |
| Obesity | t_cell | *AOC1* | -3.90251823 | 1.62E-05 | 0.000168143 | 0.07866939 |
| Obesity | t_cell | *TMEM176A* | -6.416665749 | 9.02E-05 | 0.000637091 | 0.437681746 |
| Obesity | t_cell | *M6PR* | 3.011662986 | 0.00456627 | 0.014368057 | 1 |
| Obesity | t_cell | *CYP26B1* | -2.603610492 | 0.002090544 | 0.007678516 | 1 |
| Obesity | t_cell | *ALS2* | -3.445673574 | 0.008971319 | 0.025043856 | 1 |
| Obesity | t_cell | *RBM6* | -4.709639134 | 5.52E-22 | 1.34E-18 | 2.68E-18 |
| Obesity | t_cell | *ARHGAP33* | 3.552983232 | 8.41E-05 | 0.000602603 | 0.407962313 |
| Obesity | t_cell | *MEOX1* | -2.984988426 | 2.52E-06 | 5.90E-05 | 0.012220295 |
| Obesity | t_cell | *PON1* | -7.903983232 | 4.03E-06 | 6.82E-05 | 0.019571067 |
| Obesity | t_cell | *ITGA2B* | -3.177891259 | 0.001287412 | 0.005162414 | 1 |
| Obesity | t_cell | *TMEM98* | -2.250196578 | 0.016809496 | 0.041932995 | 1 |
| Obesity | t_cell | *SYNRG* | 3.349855149 | 9.44E-05 | 0.000657133 | 0.458021585 |
| Obesity | t_cell | *BAIAP2L1* | -4.833684144 | 0.003301399 | 0.010952524 | 1 |
| Obesity | t_cell | *FARP2* | -3.011205271 | 0.018062869 | 0.044533048 | 1 |
| Obesity | t_cell | *USH1C* | -14.85320921 | 0.001534888 | 0.005967367 | 1 |
| Obesity | t_cell | *ELAC2* | -2.985358344 | 0.000896925 | 0.003847817 | 1 |
| Obesity | t_cell | *SCIN* | -3.699427607 | 0.000145623 | 0.000907012 | 0.706562079 |
| Obesity | t_cell | *CEACAM21* | 1.021928028 | 2.94E-06 | 6.01E-05 | 0.014266403 |
| Obesity | t_cell | *CD79B* | -3.676425433 | 0.005408928 | 0.016582323 | 1 |
| Obesity | t_cell | *TSR3* | 8.492310304 | 0.001652557 | 0.006363655 | 1 |
| Obesity | t_cell | *FMO3* | -2.324171779 | 4.08E-08 | 3.97E-06 | 0.00019779 |
| Obesity | t_cell | *NADK* | -5.826794449 | 0.000602604 | 0.002833171 | 1 |
| Obesity | t_cell | *DLEC1* | -2.590168771 | 0.010263959 | 0.028040951 | 1 |
| Obesity | t_cell | *CYB561* | 4.732367811 | 2.36E-06 | 5.65E-05 | 0.011473149 |
| Obesity | t_cell | *SS18L2* | 6.61423734 | 8.54E-06 | 0.000106752 | 0.041443989 |
| Obesity | t_cell | *MMP25* | 1.433532127 | 0.000601444 | 0.002833171 | 1 |
| Obesity | t_cell | *IL32* | 1.280001032 | 9.32E-07 | 3.74E-05 | 0.004521344 |

| | | | | | | |
|---|---|---|---|---|---|---|
| Obesity | t_cell | *PKD1* | -2.96214874 | 0.007399075 | 0.021424767 | 1 |
| Obesity | t_cell | *MAPK8IP2* | -5.245119979 | 0.008975941 | 0.025043856 | 1 |
| Obesity | t_cell | *RPS20* | 3.523828617 | 0.008686609 | 0.024390872 | 1 |
| Obesity | t_cell | *TRAF3IP3* | 0.763072264 | 6.93E-07 | 3.09E-05 | 0.003364333 |
| Obesity | t_cell | *FUZ* | -4.700783934 | 0.003290809 | 0.010941909 | 1 |
| Obesity | t_cell | *SLC6A13* | -2.049831732 | 0.011916238 | 0.03169824 | 1 |
| Obesity | t_cell | *SLC6A7* | -57.17877612 | 2.40E-05 | 0.000225504 | 0.116272963 |
| Obesity | t_cell | *PPP5C* | 3.450882631 | 0.007662352 | 0.022024722 | 1 |
| Obesity | t_cell | *ZBTB32* | -6.781685726 | 7.36E-06 | 9.70E-05 | 0.035708861 |
| Obesity | t_cell | *EHD3* | -6.651663192 | 0.003089834 | 0.010479211 | 1 |
| Obesity | t_cell | *TACC3* | 4.5787591 | 1.25E-05 | 0.000140665 | 0.060485927 |
| Obesity | t_cell | *MDH1* | 2.783703682 | 0.016847154 | 0.042005341 | 1 |
| Obesity | t_cell | *COX15* | -4.093382721 | 0.005224871 | 0.016106146 | 1 |
| Obesity | t_cell | *CCDC88C* | 1.177070524 | 2.85E-06 | 6.01E-05 | 0.013831998 |
| Obesity | t_cell | *CHDH* | -7.969548057 | 8.29E-05 | 0.000595316 | 0.402433468 |
| Obesity | t_cell | *CLCA4* | -7.074763279 | 0.00018874 | 0.001122263 | 0.915766673 |
| Obesity | t_cell | *WWTR1* | -3.204439605 | 0.001999361 | 0.00743364 | 1 |
| Obesity | t_cell | *VSIG2* | -5.714936385 | 9.79E-07 | 3.74E-05 | 0.004750797 |
| Obesity | t_cell | *OSBPL5* | -3.456339721 | 0.003414194 | 0.011253852 | 1 |
| Obesity | t_cell | *CPS1* | -2.110233953 | 0.001108177 | 0.00456441 | 1 |
| Obesity | t_cell | *GRAMD1B* | 1.431680416 | 0.001346703 | 0.005355903 | 1 |
| Obesity | t_cell | *GLRX2* | -6.62167967 | 0.005498626 | 0.01676891 | 1 |
| Obesity | t_cell | *ABCC2* | -5.136453411 | 3.11E-06 | 6.09E-05 | 0.015092532 |
| Obesity | t_cell | *GCLM* | 5.461714432 | 0.000364545 | 0.001881617 | 1 |
| Obesity | t_cell | *TOMM34* | -5.442034018 | 0.007284327 | 0.02118918 | 1 |
| Obesity | t_cell | *AGPAT4* | -4.014953222 | 0.003613611 | 0.011775178 | 1 |
| Obesity | t_cell | *MIPEP* | -4.030219632 | 0.001419617 | 0.005590893 | 1 |
| Obesity | t_cell | *INSRR* | -5.134890769 | 9.02E-07 | 3.71E-05 | 0.004378458 |
| Obesity | t_cell | *SH2D2A* | 1.602498219 | 8.12E-07 | 3.52E-05 | 0.003938127 |
| Obesity | t_cell | *ANK1* | -6.203441466 | 2.77E-05 | 0.00025265 | 0.134189143 |
| Obesity | t_cell | *IKZF2* | 4.367155118 | 6.52E-05 | 0.000487838 | 0.316119121 |
| Obesity | t_cell | *CENPQ* | -1.51512569 | 6.27E-05 | 0.000473432 | 0.304416701 |
| Obesity | t_cell | *PIAS1* | 3.84735475 | 0.001668492 | 0.006413219 | 1 |
| Obesity | t_cell | *MYOC* | -2.929596278 | 0.007097974 | 0.020809287 | 1 |
| Obesity | t_cell | *SH3YL1* | -5.813450753 | 1.13E-05 | 0.000131274 | 0.055003961 |
| Obesity | t_cell | *MYOM2* | -2.944896089 | 0.016356038 | 0.040970313 | 1 |
| Obesity | t_cell | *TUBG2* | -3.707973037 | 0.000302274 | 0.001627385 | 1 |
| Obesity | t_cell | *FLT4* | -1.860694247 | 2.14E-06 | 5.28E-05 | 0.010397721 |
| Obesity | t_cell | *NSUN2* | 4.494009437 | 0.007603695 | 0.021895031 | 1 |
| Obesity | t_cell | *PI4K2B* | -6.720175776 | 7.60E-06 | 9.86E-05 | 0.036879875 |
| Obesity | t_cell | *DNAH5* | -4.90091763 | 0.000118247 | 0.000770853 | 0.573734538 |
| Obesity | t_cell | *PNKP* | 1.931892336 | 0.012448694 | 0.032755456 | 1 |
| Obesity | t_cell | *PQLC2* | -8.166049597 | 4.71E-05 | 0.000377765 | 0.228547743 |
| Obesity | t_cell | *INPP4A* | 2.892196023 | 6.31E-06 | 8.95E-05 | 0.030633068 |
| Obesity | t_cell | *LSG1* | 2.063492929 | 0.000100019 | 0.000683511 | 0.485292512 |
| Obesity | t_cell | *TDP1* | 5.784344582 | 7.36E-06 | 9.70E-05 | 0.035699644 |
| Obesity | t_cell | *SPATA7* | -4.535860614 | 0.000524814 | 0.002529101 | 1 |
| Obesity | t_cell | *CTNNA1* | -2.702777083 | 0.020730056 | 0.04969478 | 1 |
| Obesity | t_cell | *CNTLN* | -2.647827344 | 0.015193826 | 0.038463072 | 1 |
| Obesity | t_cell | *ARAP2* | 2.009949137 | 3.51E-06 | 6.39E-05 | 0.017029044 |

| | | | | | | |
|---|---|---|---|---|---|---|
| Obesity | t_cell | *ANO2* | -1.719194008 | 6.70E-06 | 9.18E-05 | 0.032500293 |
| Obesity | t_cell | *RRM2B* | -5.356510828 | 0.001295654 | 0.005191177 | 1 |
| Obesity | t_cell | *UTS2* | -4.513675283 | 1.79E-05 | 0.000180962 | 0.086861874 |
| Obesity | t_cell | *RCN1* | -5.654528226 | 4.79E-05 | 0.00038313 | 0.2321767 |
| Obesity | t_cell | *ARHGEF5* | -2.046161106 | 0.000338949 | 0.001765048 | 1 |
| Obesity | t_cell | *LAMC3* | -1.967825387 | 0.003329681 | 0.011035253 | 1 |
| Obesity | t_cell | *CYBA* | 4.367187854 | 0.000205429 | 0.001198006 | 0.99674124 |
| Obesity | t_cell | *THOC3* | 3.001099719 | 1.64E-06 | 4.65E-05 | 0.007953451 |
| Obesity | t_cell | *ALX4* | -2.130436536 | 0.006500801 | 0.019362731 | 1 |
| Obesity | t_cell | *AKR7A2* | 4.655614744 | 0.013254855 | 0.034428564 | 1 |
| Obesity | t_cell | *MRTO4* | -3.33117906 | 0.000424468 | 0.002118846 | 1 |
| Obesity | t_cell | *ANAPC4* | 4.501751554 | 0.002802202 | 0.009713933 | 1 |
| Obesity | t_cell | *LY75* | 6.353819126 | 4.73E-07 | 2.34E-05 | 0.002296298 |
| Obesity | t_cell | *PLEKHH1* | -2.245995536 | 0.005801521 | 0.017593113 | 1 |
| Obesity | t_cell | *ITIH4* | 1.798263181 | 5.79E-09 | 8.02E-07 | 2.81E-05 |
| Obesity | t_cell | *NPFFR2* | -3.403451921 | 3.52E-06 | 6.39E-05 | 0.017058879 |
| Obesity | t_cell | *PCGF2* | -1.788052218 | 0.01263127 | 0.033163918 | 1 |
| Obesity | t_cell | *IL17RB* | -4.260972334 | 0.007426465 | 0.021486706 | 1 |
| Obesity | t_cell | *ATP11B* | 4.480651644 | 0.000142198 | 0.000895834 | 0.689942287 |
| Obesity | t_cell | *LAMC2* | -1.451756855 | 0.000625007 | 0.002907512 | 1 |
| Obesity | t_cell | *DNAJC25* | 8.459378966 | 0.002386271 | 0.008519636 | 1 |
| Obesity | t_cell | *RIMBP2* | -3.520495523 | 6.07E-06 | 8.68E-05 | 0.029438125 |
| Obesity | t_cell | *PRDM6* | -4.495735488 | 0.000101206 | 0.000688713 | 0.49105269 |
| Obesity | t_cell | *SPAG4* | -4.787980575 | 0.000309218 | 0.001648863 | 1 |
| Obesity | t_cell | *NCKAP1* | -3.278180713 | 0.006520303 | 0.019380641 | 1 |
| Obesity | t_cell | *LTK* | 1.442200196 | 1.80E-06 | 4.84E-05 | 0.008745071 |
| Obesity | t_cell | *AHRR* | -5.813681086 | 7.26E-07 | 3.20E-05 | 0.003520607 |
| Obesity | t_cell | *ADCK1* | -5.598276242 | 0.001012833 | 0.004243754 | 1 |
| Obesity | t_cell | *CASP8* | 2.359213388 | 1.55E-05 | 0.000163444 | 0.075347895 |
| Obesity | t_cell | *CDON* | -1.1935059 | 0.002202498 | 0.008004883 | 1 |
| Obesity | t_cell | *LPAR2* | 3.976455697 | 1.08E-07 | 8.60E-06 | 0.000524545 |
| Obesity | t_cell | *SLC12A2* | 4.614400317 | 0.00664905 | 0.019683533 | 1 |
| Obesity | t_cell | *CNN2* | 2.98644762 | 3.86E-06 | 6.68E-05 | 0.018705849 |
| Obesity | t_cell | *ABCA7* | 2.121053644 | 0.000600064 | 0.002829459 | 1 |
| Obesity | t_cell | *BTBD1* | 4.326076277 | 0.008168743 | 0.023218946 | 1 |
| Obesity | t_cell | *FAR2* | 3.685205223 | 0.000306776 | 0.001641992 | 1 |
| Obesity | t_cell | *BCAS1* | -3.284531412 | 1.60E-05 | 0.000167796 | 0.077857173 |
| Obesity | t_cell | *CHI3L2* | 5.369852143 | 0.000114543 | 0.00075364 | 0.555761426 |
| Obesity | t_cell | *UHRF1BP1* | 5.909527657 | 5.54E-15 | 4.42E-12 | 2.69E-11 |
| Obesity | t_cell | *ANKRD44* | 1.657761146 | 1.29E-06 | 4.23E-05 | 0.006255841 |
| Obesity | t_cell | *TIE1* | -3.057798693 | 3.95E-06 | 6.77E-05 | 0.019151032 |
| Obesity | t_cell | *NGEF* | -7.464677693 | 0.000217178 | 0.001245357 | 1 |
| Obesity | t_cell | *MTFR1* | 2.613977359 | 0.000660497 | 0.003026188 | 1 |
| Obesity | t_cell | *STOML1* | -8.296597473 | 2.77E-05 | 0.00025265 | 0.134409952 |
| Obesity | t_cell | *METTL22* | -5.606902426 | 0.001632959 | 0.006298184 | 1 |
| Obesity | t_cell | *ROGDI* | -6.478512892 | 1.14E-05 | 0.000131631 | 0.055416745 |
| Obesity | t_cell | *FGFR3* | -2.189792251 | 0.00391475 | 0.012620841 | 1 |
| Obesity | t_cell | *POLR1A* | -2.14684955 | 0.007360441 | 0.021359366 | 1 |
| Obesity | t_cell | *CYFIP1* | -2.950298191 | 0.015059137 | 0.038174992 | 1 |
| Obesity | t_cell | *IFT80* | -5.171460125 | 0.005120965 | 0.015805931 | 1 |

| | | | | | | |
|---|---|---|---|---|---|---|
| Obesity | t_cell | *CLEC2D* | 1.994702914 | 3.14E-06 | 6.09E-05 | 0.015257072 |
| Obesity | t_cell | *DRD4* | -7.31952935 | 3.96E-05 | 0.000331671 | 0.192037281 |
| Obesity | t_cell | *PLA2G10* | 10.9621885 | 3.09E-06 | 6.09E-05 | 0.015015881 |
| Obesity | t_cell | *GNB5* | 1.932819215 | 0.007951742 | 0.022762154 | 1 |
| Obesity | t_cell | *LRP6* | -3.674460225 | 0.000875508 | 0.003775682 | 1 |
| Obesity | t_cell | *WIPI1* | -3.138385192 | 0.003457288 | 0.01135732 | 1 |
| Obesity | t_cell | *GBA2* | -5.62128284 | 0.000353018 | 0.001829961 | 1 |
| Obesity | t_cell | *ASNS* | -2.75953629 | 0.003073902 | 0.010437071 | 1 |
| Obesity | t_cell | *ST6GALNAC2* | -3.273257359 | 0.008263263 | 0.023460125 | 1 |
| Obesity | t_cell | *VASH1* | -3.657926429 | 1.36E-05 | 0.000150238 | 0.065954652 |
| Obesity | t_cell | *CPSF1* | -3.025049925 | 3.37E-05 | 0.000296583 | 0.163714001 |
| Obesity | t_cell | *TRHDE* | -3.688315183 | 7.87E-06 | 0.000100385 | 0.038170393 |
| Obesity | t_cell | *NFATC3* | 3.444053785 | 0.00039035 | 0.001981252 | 1 |
| Obesity | t_cell | *CRMP1* | -3.811594464 | 4.22E-05 | 0.000347849 | 0.204535288 |
| Obesity | t_cell | *NDE1* | 4.886266293 | 0.001580173 | 0.006109162 | 1 |
| Obesity | t_cell | *AP1M1* | -4.415097154 | 0.011086382 | 0.029900569 | 1 |
| Obesity | t_cell | *PVR* | -2.18053764 | 0.003443763 | 0.011343612 | 1 |
| Obesity | t_cell | *SCARB1* | -3.800849262 | 0.0007486 | 0.003344574 | 1 |
| Obesity | t_cell | *MOV10L1* | -3.039911487 | 9.10E-05 | 0.000640611 | 0.441380752 |
| Obesity | t_cell | *LLGL2* | 2.93124167 | 3.39E-06 | 6.34E-05 | 0.016429773 |
| Obesity | t_cell | *SDHA* | 3.781391009 | 0.00121422 | 0.004942444 | 1 |
| Obesity | t_cell | *PPP2R2C* | -3.264097995 | 2.97E-06 | 6.01E-05 | 0.014396978 |
| Obesity | t_cell | *ZNF532* | -3.390010741 | 0.007016916 | 0.02062249 | 1 |
| Obesity | t_cell | *TIPIN* | 1.368413318 | 0.013532363 | 0.03499948 | 1 |
| Obesity | t_cell | *ADD2* | -3.077072926 | 0.004728204 | 0.014810358 | 1 |
| Obesity | t_cell | *VPS9D1* | 6.322508255 | 0.000431757 | 0.002146399 | 1 |
| Obesity | t_cell | *MKRN2* | 3.073857221 | 0.011591996 | 0.03110861 | 1 |
| Obesity | t_cell | *MCM6* | 4.101466031 | 0.000280261 | 0.001535126 | 1 |
| Obesity | t_cell | *SPAG5* | -8.264373266 | 2.21E-07 | 1.34E-05 | 0.001069968 |
| Obesity | t_cell | *PAG1* | 2.2280124 | 3.16E-06 | 6.09E-05 | 0.015339891 |
| Obesity | t_cell | *ICAM3* | 2.346550168 | 5.29E-06 | 7.92E-05 | 0.025667247 |
| Obesity | t_cell | *NT5C2* | 2.310236216 | 0.011700882 | 0.031314219 | 1 |
| Obesity | t_cell | *XAB2* | 9.270459482 | 0.000148295 | 0.000918938 | 0.71952874 |
| Obesity | t_cell | *APBB1IP* | 2.187484131 | 0.000651567 | 0.002993754 | 1 |
| Obesity | t_cell | *LRCH4* | 3.988014256 | 0.000298381 | 0.001612188 | 1 |
| Obesity | t_cell | *FGF20* | 7.386508389 | 0.000482852 | 0.002352209 | 1 |
| Obesity | t_cell | *MYH7B* | -3.220875486 | 0.002838716 | 0.009817141 | 1 |
| Obesity | t_cell | *FKBP7* | -2.362694005 | 0.014768447 | 0.037595229 | 1 |
| Obesity | t_cell | *SP140* | 1.279116585 | 2.05E-06 | 5.24E-05 | 0.009962576 |
| Obesity | t_cell | *FDFT1* | 5.926508726 | 1.48E-05 | 0.000158409 | 0.071757575 |
| Obesity | t_cell | *SCGN* | -13.65467403 | 7.51E-06 | 9.81E-05 | 0.036442119 |
| Obesity | t_cell | *KCNN2* | -4.947290616 | 3.51E-06 | 6.39E-05 | 0.017006385 |
| Obesity | t_cell | *STK17B* | 2.053187855 | 6.86E-06 | 9.38E-05 | 0.033304405 |
| Obesity | t_cell | *PCDHB4* | -4.978389735 | 5.43E-06 | 8.05E-05 | 0.0263225 |
| Obesity | t_cell | *SEMA5B* | -1.788005862 | 0.000156928 | 0.000965036 | 0.761413404 |
| Obesity | t_cell | *DOPEY1* | 2.996653419 | 0.003540285 | 0.011606394 | 1 |
| Obesity | t_cell | *RPS5* | 3.121400976 | 0.010569289 | 0.028777886 | 1 |
| Obesity | t_cell | *PPIE* | 3.375354819 | 0.000287004 | 0.001559398 | 1 |
| Obesity | t_cell | *GSTP1* | 4.891698603 | 0.000241153 | 0.001357395 | 1 |
| Obesity | t_cell | *CD82* | 2.646806071 | 4.49E-07 | 2.31E-05 | 0.002180898 |

| | | | | | | |
|---|---|---|---|---|---|---|
| Obesity | t_cell | *OVGP1* | -1.868147889 | 0.002306453 | 0.008289563 | 1 |
| Obesity | t_cell | *RRN3* | -3.96828628 | 0.020319024 | 0.048854263 | 1 |
| Obesity | t_cell | *POMGNT1* | -3.431697912 | 0.013235218 | 0.034395972 | 1 |
| Obesity | t_cell | *NME8* | -43.22373802 | 8.69E-05 | 0.000620085 | 0.421656849 |
| Obesity | t_cell | *EPDR1* | -1.158759009 | 0.008866812 | 0.024794492 | 1 |
| Obesity | t_cell | *HSD17B14* | -3.384269333 | 4.50E-05 | 0.000363984 | 0.218390413 |
| Obesity | t_cell | *TRIP6* | -6.232605554 | 4.26E-05 | 0.000350029 | 0.206529695 |
| Obesity | t_cell | *ACHE* | -3.815653427 | 0.000837082 | 0.003639357 | 1 |
| Obesity | t_cell | *ADAMTS2* | -3.766463997 | 0.000402654 | 0.002028739 | 1 |
| Obesity | t_cell | *LPCAT2* | -3.862410057 | 0.000110091 | 0.000731725 | 0.534159336 |
| Obesity | t_cell | *OGFOD1* | -4.953904342 | 0.000981214 | 0.004121948 | 1 |
| Obesity | t_cell | *AAMDC* | -2.15904645 | 0.008753096 | 0.024520797 | 1 |
| Obesity | t_cell | *ASAP3* | -3.80959324 | 4.33E-05 | 0.000351724 | 0.209979422 |
| Obesity | t_cell | *DOCK3* | -4.376111954 | 0.00054064 | 0.002592082 | 1 |
| Obesity | t_cell | *ARHGAP28* | -2.536570455 | 0.018598148 | 0.045574855 | 1 |
| Obesity | t_cell | *PPP1R13B* | -4.148193222 | 9.14E-05 | 0.000642301 | 0.443418571 |
| Obesity | t_cell | *ZNF343* | -2.159872631 | 0.018135635 | 0.044667057 | 1 |
| Obesity | t_cell | *EBF4* | -4.502855434 | 8.77E-06 | 0.000108225 | 0.042532317 |
| Obesity | t_cell | *TBX5* | -5.622407125 | 0.002217114 | 0.008039937 | 1 |
| Obesity | t_cell | *FXYD5* | 4.366013599 | 3.87E-06 | 6.69E-05 | 0.018786889 |
| Obesity | t_cell | *GRAMD1A* | 4.450402533 | 0.011891798 | 0.031685341 | 1 |
| Obesity | t_cell | *ANKRD24* | -7.868368528 | 0.001913491 | 0.007169311 | 1 |
| Obesity | t_cell | *YPEL3* | 6.359144422 | 2.40E-08 | 2.53E-06 | 0.000116468 |
| Obesity | t_cell | *STRN4* | -3.758618902 | 0.001514516 | 0.005907099 | 1 |
| Obesity | t_cell | *LYZ* | 4.146402529 | 0.000213507 | 0.001228869 | 1 |
| Obesity | t_cell | *SPG21* | -3.585797969 | 0.003336595 | 0.011043083 | 1 |
| Obesity | t_cell | *PABPC4* | -7.046438388 | 2.73E-09 | 3.90E-07 | 1.33E-05 |
| Obesity | t_cell | *CERS4* | 4.765278545 | 0.002591041 | 0.009090188 | 1 |
| Obesity | t_cell | *GLG1* | 3.237314479 | 0.013910261 | 0.035786101 | 1 |
| Obesity | t_cell | *ABCC6* | -2.373503321 | 0.012821514 | 0.033500261 | 1 |
| Obesity | t_cell | *RAPGEF4* | -3.745405419 | 0.000221815 | 0.001264685 | 1 |
| Obesity | t_cell | *TF* | -5.921431306 | 7.04E-05 | 0.000522497 | 0.341713186 |
| Obesity | t_cell | *RGS17* | -6.31556541 | 5.21E-12 | 1.15E-09 | 2.53E-08 |
| Obesity | t_cell | *PPP2R3C* | 4.740463927 | 7.25E-05 | 0.00053459 | 0.351760406 |
| Obesity | t_cell | *TBX15* | -3.934047225 | 1.26E-10 | 2.26E-08 | 6.10E-07 |
| Obesity | t_cell | *XYLB* | -4.702863012 | 0.001286094 | 0.005162414 | 1 |
| Obesity | t_cell | *GABRP* | -4.11756267 | 2.71E-05 | 0.000248299 | 0.131350366 |
| Obesity | t_cell | *CRAT* | -6.855714841 | 0.000180789 | 0.00108028 | 0.877187363 |
| Obesity | t_cell | *NANS* | 6.300961946 | 0.000844163 | 0.003658911 | 1 |
| Obesity | t_cell | *PDE6C* | -6.494259321 | 9.15E-05 | 0.000642301 | 0.443830092 |
| Obesity | t_cell | *CRTAC1* | -1.310815814 | 0.006160977 | 0.018509634 | 1 |
| Obesity | t_cell | *ITPR3* | 4.267587107 | 3.00E-06 | 6.01E-05 | 0.014537231 |
| Obesity | t_cell | *DSP* | -6.411251923 | 0.005825801 | 0.017633678 | 1 |
| Obesity | t_cell | *IL12RB1* | 0.921109455 | 0.001412341 | 0.005566756 | 1 |
| Obesity | t_cell | *OCEL1* | -2.560289899 | 0.000487067 | 0.002367985 | 1 |
| Obesity | t_cell | *SETD1A* | 9.415386612 | 4.48E-08 | 4.26E-06 | 0.000217434 |
| Obesity | t_cell | *LZTR1* | -3.67632154 | 0.014424423 | 0.036893675 | 1 |
| Obesity | t_cell | *CECR2* | -3.090879109 | 0.013035145 | 0.033957909 | 1 |
| Obesity | t_cell | *OSM* | 1.986958476 | 0.012734066 | 0.03335465 | 1 |
| Obesity | t_cell | *CABIN1* | 3.983763129 | 0.008523879 | 0.024031298 | 1 |

| | | | | | | |
|---|---|---|---|---|---|---|
| Obesity | t_cell | *PES1* | -2.575747208 | 0.007630311 | 0.021958642 | 1 |
| Obesity | t_cell | *CYTH4* | 2.501075313 | 1.75E-06 | 4.79E-05 | 0.008477327 |
| Obesity | t_cell | *CARD10* | -3.841875912 | 2.29E-06 | 5.55E-05 | 0.011094818 |
| Obesity | t_cell | *LGALS2* | -1.908976037 | 0.005423751 | 0.016603178 | 1 |
| Obesity | t_cell | *SH3BP1* | 2.340180546 | 1.41E-06 | 4.46E-05 | 0.006863058 |
| Obesity | t_cell | *TRIOBP* | -4.371841602 | 8.69E-05 | 0.000620085 | 0.421540285 |
| Obesity | t_cell | *ANKRD54* | -4.305943779 | 6.91E-08 | 5.88E-06 | 0.000335129 |
| Obesity | t_cell | *CCDC134* | 3.425463382 | 0.013014425 | 0.033931214 | 1 |
| Obesity | t_cell | *PICK1* | -2.315650965 | 0.006746449 | 0.019935306 | 1 |
| Obesity | t_cell | *CENPM* | 2.811017792 | 0.006649075 | 0.019683533 | 1 |
| Obesity | t_cell | *KDELR3* | -7.238016901 | 1.22E-06 | 4.13E-05 | 0.005913088 |
| Obesity | t_cell | *CYP2D6* | -5.913178475 | 0.017689729 | 0.043746467 | 1 |
| Obesity | t_cell | *RAB36* | -3.535087218 | 3.62E-06 | 6.47E-05 | 0.017587386 |
| Obesity | t_cell | *RHBDD3* | -3.545605571 | 0.006153169 | 0.01849763 | 1 |
| Obesity | t_cell | *CHKB* | -3.227663554 | 0.009292247 | 0.02576342 | 1 |
| Obesity | t_cell | *SAMM50* | -5.166758497 | 0.00074223 | 0.0033253 | 1 |
| Obesity | t_cell | *IFT27* | -5.725230703 | 6.03E-05 | 0.000460343 | 0.292777865 |
| Obesity | t_cell | *KIAA0930* | -4.11717086 | 0.004548811 | 0.014336867 | 1 |
| Obesity | t_cell | *POLR3H* | -4.633689314 | 0.008410946 | 0.023768148 | 1 |
| Obesity | t_cell | *MLC1* | 2.361670321 | 0.010165743 | 0.02783532 | 1 |
| Obesity | t_cell | *AP4S1* | -4.034945857 | 0.000923869 | 0.003932118 | 1 |
| Obesity | t_cell | *NIN* | 3.958936625 | 3.80E-05 | 0.000322564 | 0.184506407 |
| Obesity | t_cell | *CNIH1* | -4.199112987 | 0.019085129 | 0.046416564 | 1 |
| Obesity | t_cell | *GSTZ1* | -2.15114042 | 0.013220948 | 0.034377299 | 1 |
| Obesity | t_cell | *TELO2* | -5.207556632 | 0.000386122 | 0.001969996 | 1 |
| Obesity | t_cell | *GSKIP* | 7.777180905 | 0.000173464 | 0.001048129 | 0.841647875 |
| Obesity | t_cell | *PSMB5* | 3.051737512 | 0.002551174 | 0.009002399 | 1 |
| Obesity | t_cell | *EFS* | -5.433922425 | 0.00028537 | 0.001552261 | 1 |
| Obesity | t_cell | *EMC9* | -1.922019216 | 0.012446215 | 0.032755456 | 1 |
| Obesity | t_cell | *CD40* | -5.280512636 | 0.000281864 | 0.001540095 | 1 |
| Obesity | t_cell | *DOK5* | -4.169054149 | 1.00E-05 | 0.000119547 | 0.048536258 |
| Obesity | t_cell | *CSTF1* | -5.972201074 | 0.001853736 | 0.007003221 | 1 |
| Obesity | t_cell | *PTK6* | 6.779534601 | 3.97E-06 | 6.77E-05 | 0.01924395 |
| Obesity | t_cell | *TRIB3* | 12.50809652 | 0.000153354 | 0.000947865 | 0.744074157 |
| Obesity | t_cell | *RASSF2* | 2.474796313 | 2.18E-07 | 1.34E-05 | 0.001058589 |
| Obesity | t_cell | *ANGPT4* | -6.870262988 | 0.000262764 | 0.001460403 | 1 |
| Obesity | t_cell | *RSPO4* | -1.62314245 | 0.006643476 | 0.019683533 | 1 |
| Obesity | t_cell | *MAP1LC3A* | -4.540164989 | 0.000136199 | 0.000867239 | 0.66083579 |
| Obesity | t_cell | *RBFA* | -3.596981895 | 0.007641381 | 0.021977463 | 1 |
| Obesity | t_cell | *LPIN2* | 3.741216539 | 0.002188508 | 0.007960002 | 1 |
| Obesity | t_cell | *LAMA1* | -4.000835144 | 4.61E-07 | 2.31E-05 | 0.002237479 |
| Obesity | t_cell | *RNF125* | 5.276192117 | 4.71E-06 | 7.44E-05 | 0.022845147 |
| Obesity | t_cell | *NALCN* | -2.547603438 | 0.004609645 | 0.014476374 | 1 |
| Obesity | t_cell | *UGGT2* | -3.762138617 | 0.000645054 | 0.002975097 | 1 |
| Obesity | t_cell | *PARP4* | 3.86180224 | 0.001699294 | 0.006502344 | 1 |
| Obesity | t_cell | *DGKH* | 2.98930499 | 0.002600241 | 0.009115872 | 1 |
| Obesity | t_cell | *ELMO3* | 1.598753027 | 0.007228411 | 0.021064413 | 1 |
| Obesity | t_cell | *PLLP* | -2.38503954 | 0.0010522 | 0.00436348 | 1 |
| Obesity | t_cell | *CCDC113* | -4.737634623 | 0.00040073 | 0.002021148 | 1 |
| Obesity | t_cell | *NME4* | -7.462481221 | 0.000307721 | 0.00164434 | 1 |

| | | | | | | |
|---|---|---|---|---|---|---|
| Obesity | t_cell | *CLCN7* | 2.370952577 | 0.0005075 | 0.002455024 | 1 |
| Obesity | t_cell | *SLC7A5* | 2.883789786 | 0.013675559 | 0.03524995 | 1 |
| Obesity | t_cell | *RHBDL1* | -9.602842053 | 2.61E-07 | 1.48E-05 | 0.001268657 |
| Obesity | t_cell | *CLUAP1* | 1.462438403 | 0.020230438 | 0.048665386 | 1 |
| Obesity | t_cell | *HMOX2* | 3.58945202 | 0.00031254 | 0.001654879 | 1 |
| Obesity | t_cell | *TOX3* | -3.262756733 | 0.007233512 | 0.021066626 | 1 |
| Obesity | t_cell | *RRN3P2* | -5.365933217 | 0.00026392 | 0.001463472 | 1 |
| Obesity | t_cell | *RBL2* | 3.134410791 | 3.82E-05 | 0.000323053 | 0.185357703 |
| Obesity | t_cell | *PYCARD* | 9.240458549 | 4.52E-12 | 1.10E-09 | 2.20E-08 |
| Obesity | t_cell | *TMC5* | -4.337489112 | 1.68E-06 | 4.71E-05 | 0.008163056 |
| Obesity | t_cell | *KNOP1* | -6.856514399 | 1.09E-06 | 3.89E-05 | 0.005275596 |
| Obesity | t_cell | *LACTB* | -3.422904335 | 0.020529353 | 0.049262325 | 1 |
| Obesity | t_cell | *AP3B2* | -0.907452508 | 0.017750638 | 0.043852392 | 1 |
| Obesity | t_cell | *TTC23* | -3.847947472 | 0.003783236 | 0.012245672 | 1 |
| Obesity | t_cell | *FAH* | -4.37474991 | 0.008885644 | 0.024820464 | 1 |
| Obesity | t_cell | *ZFAND1* | -4.232279037 | 0.006512043 | 0.01937414 | 1 |
| Obesity | t_cell | *EYA1* | -1.839823648 | 0.016900961 | 0.042117855 | 1 |
| Obesity | t_cell | *NIPAL2* | -3.392784271 | 0.009443225 | 0.026137208 | 1 |
| Obesity | t_cell | *SPAG1* | 3.229292763 | 0.00957892 | 0.026482577 | 1 |
| Obesity | t_cell | *CHRAC1* | 7.729733154 | 0.017684673 | 0.043746467 | 1 |
| Obesity | t_cell | *TSTA3* | -2.512411819 | 0.008871236 | 0.024794492 | 1 |
| Obesity | t_cell | *TNFRSF10A* | 1.775954579 | 1.71E-06 | 4.72E-05 | 0.008309606 |
| Obesity | t_cell | *TUSC3* | -3.408517066 | 5.76E-05 | 0.000444354 | 0.279498378 |
| Obesity | t_cell | *KLHDC4* | 3.975535086 | 0.001977786 | 0.007359062 | 1 |
| Obesity | t_cell | *MAN2B1* | -2.141157582 | 0.018082239 | 0.044558164 | 1 |
| Obesity | t_cell | *TULP2* | -11.17761018 | 5.07E-06 | 7.73E-05 | 0.024594997 |
| Obesity | t_cell | *NFKBIB* | 3.488938849 | 0.006530781 | 0.019380641 | 1 |
| Obesity | t_cell | *LIN7B* | -8.23972279 | 0.001757159 | 0.006692101 | 1 |
| Obesity | t_cell | *PEX11G* | -2.306029685 | 0.013517391 | 0.034979402 | 1 |
| Obesity | t_cell | *ERCC2* | -6.398694235 | 0.004903459 | 0.015221742 | 1 |
| Obesity | t_cell | *CD37* | 2.293992938 | 4.03E-05 | 0.000335707 | 0.195381517 |
| Obesity | t_cell | *AMH* | -21.49326188 | 3.23E-06 | 6.11E-05 | 0.015649866 |
| Obesity | t_cell | *FCER2* | -7.020921335 | 3.49E-06 | 6.39E-05 | 0.016924086 |
| Obesity | t_cell | *CLEC4M* | -8.531386506 | 3.59E-06 | 6.46E-05 | 0.017427462 |
| Obesity | t_cell | *SGTA* | -5.843271055 | 0.000450428 | 0.002217473 | 1 |
| Obesity | t_cell | *KIR3DX1* | -6.019034938 | 0.016072588 | 0.040343609 | 1 |
| Obesity | t_cell | *IL27RA* | 1.945152482 | 1.36E-06 | 4.35E-05 | 0.006611381 |
| Obesity | t_cell | *FAM32A* | -6.597179637 | 0.005083339 | 0.015719798 | 1 |
| Obesity | t_cell | *OLFM2* | -2.396996735 | 0.004000504 | 0.012837596 | 1 |
| Obesity | t_cell | *RASAL3* | 0.946758258 | 1.50E-06 | 4.50E-05 | 0.007272786 |
| Obesity | t_cell | *POLR2I* | 3.6065745 | 0.007683152 | 0.022058374 | 1 |
| Obesity | t_cell | *CCDC9* | -6.65860914 | 0.001451944 | 0.005699701 | 1 |
| Obesity | t_cell | *TGFB1* | 4.633533754 | 0.000192908 | 0.001141452 | 0.935990776 |
| Obesity | t_cell | *ICAM5* | -2.821838636 | 0.000373408 | 0.001915197 | 1 |
| Obesity | t_cell | *RABAC1* | 3.613053556 | 0.018928684 | 0.04617495 | 1 |
| Obesity | t_cell | *GRIN2D* | -8.180500839 | 0.003451411 | 0.011354552 | 1 |
| Obesity | t_cell | *CARD8* | 3.672967389 | 0.001099049 | 0.004530659 | 1 |
| Obesity | t_cell | *LIG1* | 4.459168351 | 0.000503715 | 0.002441583 | 1 |
| Obesity | t_cell | *MIER2* | -3.221454025 | 0.007037326 | 0.02066895 | 1 |
| Obesity | t_cell | *PLEKHA4* | -3.582680513 | 7.21E-06 | 9.64E-05 | 0.03500599 |

| | | | | | | |
|---|---|---|---|---|---|---|
| Obesity | t_cell | *TNPO2* | -4.477478124 | 0.002224259 | 0.008059823 | 1 |
| Obesity | t_cell | *GCDH* | -3.68824585 | 0.010064508 | 0.027589261 | 1 |
| Obesity | t_cell | *TFPT* | -6.699914622 | 0.004930032 | 0.015294447 | 1 |
| Obesity | t_cell | *UPK1A* | -3.99099516 | 0.00298085 | 0.010188962 | 1 |
| Obesity | t_cell | *ETV2* | -17.2892337 | 0.000648518 | 0.002985399 | 1 |
| Obesity | t_cell | *ZNF85* | -5.596990764 | 0.000378447 | 0.001938993 | 1 |
| Obesity | t_cell | *ETHE1* | -6.027821933 | 0.001836043 | 0.006948893 | 1 |
| Obesity | t_cell | *RASA4* | -23.24644803 | 7.08E-06 | 9.54E-05 | 0.034357028 |
| Obesity | t_cell | *SP4* | 3.145466738 | 0.00026156 | 0.001455378 | 1 |
| Obesity | t_cell | *DNAH11* | -4.940642769 | 0.000261447 | 0.001455378 | 1 |
| Obesity | t_cell | *STEAP1B* | -3.006082249 | 0.002062039 | 0.00758038 | 1 |
| Obesity | t_cell | *PTN* | -4.095706253 | 0.000211594 | 0.001220752 | 1 |
| Obesity | t_cell | *ADAP1* | 12.33069348 | 2.59E-06 | 5.93E-05 | 0.012590178 |
| Obesity | t_cell | *MET* | -2.504102999 | 4.29E-06 | 7.11E-05 | 0.020833413 |
| Obesity | t_cell | *IQCE* | -5.128166085 | 0.00329475 | 0.010941909 | 1 |
| Obesity | t_cell | *VIPR2* | -2.799478475 | 0.000237949 | 0.001342474 | 1 |
| Obesity | t_cell | *HIBADH* | -3.022260911 | 0.004759538 | 0.014898891 | 1 |
| Obesity | t_cell | *GRB10* | -3.185645959 | 0.00103396 | 0.004317665 | 1 |
| Obesity | t_cell | *PLEKHA8* | -4.356293423 | 0.003131831 | 0.010574561 | 1 |
| Obesity | t_cell | *TAF6* | -1.969058928 | 0.007541774 | 0.021742536 | 1 |
| Obesity | t_cell | *TFR2* | 3.631210236 | 2.09E-05 | 0.000200868 | 0.101438429 |
| Obesity | t_cell | *NRF1* | 3.270146015 | 0.010930044 | 0.029577566 | 1 |
| Obesity | t_cell | *MEST* | -3.059738952 | 5.41E-06 | 8.05E-05 | 0.026258364 |
| Obesity | t_cell | *POU6F2* | -4.184785265 | 2.59E-05 | 0.000239304 | 0.125874142 |
| Obesity | t_cell | *TSPAN13* | -4.001090505 | 5.52E-05 | 0.000427962 | 0.267904421 |
| Obesity | t_cell | *COA1* | 3.689650981 | 0.003566749 | 0.011669497 | 1 |
| Obesity | t_cell | *AEBP1* | -4.234435108 | 6.09E-05 | 0.000463406 | 0.29565333 |
| Obesity | t_cell | *GCK* | -12.24885765 | 2.95E-06 | 6.01E-05 | 0.014309399 |
| Obesity | t_cell | *FKTN* | -3.541168801 | 0.012843432 | 0.033539467 | 1 |
| Obesity | t_cell | *TRIM14* | 3.440298761 | 1.17E-06 | 4.10E-05 | 0.005695063 |
| Obesity | t_cell | *CORO2A* | 2.135445374 | 3.25E-06 | 6.11E-05 | 0.015750008 |
| Obesity | t_cell | *C5* | -3.504140377 | 0.001259794 | 0.005085292 | 1 |
| Obesity | t_cell | *ECM2* | -3.419007087 | 0.000618091 | 0.002886408 | 1 |
| Obesity | t_cell | *PTGR1* | -3.204184576 | 0.000633399 | 0.002932492 | 1 |
| Obesity | t_cell | *KANK1* | -2.6496066 | 0.003215501 | 0.010774594 | 1 |
| Obesity | t_cell | *NCS1* | -2.531869777 | 6.63E-06 | 9.16E-05 | 0.03215487 |
| Obesity | t_cell | *PKD2L1* | 2.759780642 | 0.016394617 | 0.04104576 | 1 |
| Obesity | t_cell | *CUBN* | 3.439487206 | 0.003817492 | 0.012348315 | 1 |
| Obesity | t_cell | *CDH23* | -3.208623498 | 0.019652267 | 0.047486453 | 1 |
| Obesity | t_cell | *SPOCK2* | 1.282647827 | 6.69E-07 | 3.06E-05 | 0.003244108 |
| Obesity | t_cell | *SFXN3* | 6.485999798 | 0.002154926 | 0.007867346 | 1 |
| Obesity | t_cell | *TFAM* | 5.043777633 | 0.020714808 | 0.049682772 | 1 |
| Obesity | t_cell | *RPL28* | 3.441209432 | 0.000271818 | 0.001495309 | 1 |
| Obesity | t_cell | *DNAJC12* | -3.196897757 | 0.0025641 | 0.009041435 | 1 |
| Obesity | t_cell | *RPL19* | 3.197501707 | 0.002171495 | 0.007915921 | 1 |
| Obesity | t_cell | *WNT3* | -3.417213478 | 3.14E-05 | 0.000279679 | 0.152424965 |
| Obesity | t_cell | *ASPA* | -1.965147991 | 0.004776016 | 0.014940831 | 1 |
| Obesity | t_cell | *RAD51C* | -2.941931142 | 0.004994374 | 0.015484155 | 1 |
| Obesity | t_cell | *TRIM37* | 4.409917208 | 0.008042348 | 0.022953808 | 1 |
| Obesity | t_cell | *PNPO* | -9.734830129 | 5.88E-10 | 9.20E-08 | 2.85E-06 |

| | | | | | | |
|---|---|---|---|---|---|---|
| Obesity | t_cell | *TRIM16L* | -5.370876334 | 5.03E-06 | 7.73E-05 | 0.024414756 |
| Obesity | t_cell | *CHRNE* | 1.072752767 | 0.002289936 | 0.00824241 | 1 |
| Obesity | t_cell | *DRG2* | 6.451554528 | 5.36E-05 | 0.000418203 | 0.26012206 |
| Obesity | t_cell | *FTSJ3* | -1.154557777 | 0.000976732 | 0.004110238 | 1 |
| Obesity | t_cell | *ICAM2* | 5.915093067 | 0.005034332 | 0.015588115 | 1 |
| Obesity | t_cell | *NAGLU* | -3.758650588 | 0.016687748 | 0.04169359 | 1 |
| Obesity | t_cell | *HLF* | -2.702134841 | 0.011913106 | 0.03169824 | 1 |
| Obesity | t_cell | *EFNB3* | -2.553339397 | 0.001109284 | 0.004565092 | 1 |
| Obesity | t_cell | *FAM20A* | -3.670803805 | 0.003478183 | 0.011410509 | 1 |
| Obesity | t_cell | *WFS1* | -5.883668247 | 0.000910715 | 0.00388636 | 1 |
| Obesity | t_cell | *GAR1* | 6.431905082 | 0.012678669 | 0.033270363 | 1 |
| Obesity | t_cell | *AADAT* | -3.822533219 | 5.11E-06 | 7.78E-05 | 0.024816887 |
| Obesity | t_cell | *TRIM2* | -3.402584779 | 0.001085581 | 0.004486575 | 1 |
| Obesity | t_cell | *STIM2* | 4.376829778 | 0.00021788 | 0.001246645 | 1 |
| Obesity | t_cell | *UFSP2* | -4.002055526 | 0.007196464 | 0.020996539 | 1 |
| Obesity | t_cell | *FAM149A* | -3.935149547 | 5.98E-05 | 0.000457039 | 0.290219912 |
| Obesity | t_cell | *DDX25* | -2.703423127 | 2.61E-06 | 5.93E-05 | 0.012682325 |
| Obesity | t_cell | *TECTA* | -5.530136583 | 9.08E-06 | 0.0001112 | 0.044035223 |
| Obesity | t_cell | *SC5D* | -4.536868989 | 0.012748727 | 0.033363983 | 1 |
| Obesity | t_cell | *B3GAT1* | 3.071575725 | 1.26E-06 | 4.23E-05 | 0.006130082 |
| Obesity | t_cell | *P2RX3* | -22.82779152 | 4.52E-06 | 7.28E-05 | 0.021915209 |
| Obesity | t_cell | *SNX15* | -2.376045651 | 0.016011103 | 0.040247376 | 1 |
| Obesity | t_cell | *DCPS* | 6.52132603 | 3.50E-05 | 0.00030391 | 0.169732978 |
| Obesity | t_cell | *NRXN2* | -5.275920906 | 3.71E-06 | 6.54E-05 | 0.018024393 |
| Obesity | t_cell | *CCDC86* | -4.756042232 | 1.41E-06 | 4.46E-05 | 0.006829553 |
| Obesity | t_cell | *PANX1* | -4.621016143 | 0.014060072 | 0.036094958 | 1 |
| Obesity | t_cell | *GALNT18* | -2.882720276 | 5.28E-05 | 0.000412873 | 0.255981052 |
| Obesity | t_cell | *KIAA1549L* | -7.81325438 | 4.86E-06 | 7.55E-05 | 0.023560149 |
| Obesity | t_cell | *SLC35F2* | 7.201962641 | 0.000316032 | 0.001664915 | 1 |
| Obesity | t_cell | *C11orf21* | 2.828957421 | 9.51E-06 | 0.000114757 | 0.046132265 |
| Obesity | t_cell | *ELMOD1* | -3.101670247 | 0.008100328 | 0.023078562 | 1 |
| Obesity | t_cell | *NDUFS8* | -4.120278446 | 0.013317803 | 0.034536601 | 1 |
| Obesity | t_cell | *CORO1C* | -3.720076414 | 0.000848651 | 0.003673196 | 1 |
| Obesity | t_cell | *KCTD10* | -5.437439225 | 1.10E-06 | 3.89E-05 | 0.005326773 |
| Obesity | t_cell | *IL23A* | -36.12123787 | 0.000126411 | 0.000814521 | 0.613344647 |
| Obesity | t_cell | *ACSS3* | -2.174718105 | 0.006075375 | 0.018297778 | 1 |
| Obesity | t_cell | *PARP11* | 4.805906143 | 0.000800857 | 0.003535722 | 1 |
| Obesity | t_cell | *DDX55* | 4.895459414 | 0.003269268 | 0.010902053 | 1 |
| Obesity | t_cell | *VDR* | -5.969671293 | 9.39E-06 | 0.00011357 | 0.045541482 |
| Obesity | t_cell | *NUP107* | 3.910045541 | 0.009178562 | 0.025506518 | 1 |
| Obesity | t_cell | *KRR1* | 4.259878817 | 0.001281531 | 0.005151607 | 1 |
| Obesity | t_cell | *ATN1* | -2.944493805 | 0.000809064 | 0.00356223 | 1 |
| Obesity | t_cell | *BTN3A3* | 5.488348511 | 1.44E-14 | 8.74E-12 | 6.99E-11 |
| Obesity | t_cell | *FRK* | -4.738667098 | 2.90E-05 | 0.000260354 | 0.140465909 |
| Obesity | t_cell | *RNGTT* | 4.070880064 | 0.000122312 | 0.000794456 | 0.593458999 |
| Obesity | t_cell | *TULP1* | -4.477058041 | 1.22E-06 | 4.13E-05 | 0.005909972 |
| Obesity | t_cell | *SLC26A8* | -2.117358195 | 0.001203159 | 0.004909781 | 1 |
| Obesity | t_cell | *PHACTR1* | -2.884084423 | 0.000156011 | 0.000960618 | 0.756967163 |
| Obesity | t_cell | *MDGA1* | -6.359524818 | 3.78E-05 | 0.000321022 | 0.183303635 |
| Obesity | t_cell | *CD83* | 5.38089186 | 4.61E-05 | 0.000370822 | 0.223605619 |

| | | | | | | |
|---|---|---|---|---|---|---|
| Obesity | t_cell | *GLP1R* | -7.920581225 | 0.00605097 | 0.018246435 | 1 |
| Obesity | t_cell | *BVES* | -1.378387009 | 0.001481367 | 0.005801012 | 1 |
| Obesity | t_cell | *COL9A1* | 9.272885666 | 0.002795911 | 0.00970369 | 1 |
| Obesity | t_cell | *WASF1* | -2.62177328 | 0.001490658 | 0.005818968 | 1 |
| Obesity | t_cell | *VNN1* | -1.324926119 | 0.018012567 | 0.04443161 | 1 |
| Obesity | t_cell | *VNN2* | 3.946917741 | 1.59E-05 | 0.000166634 | 0.076984956 |
| Obesity | t_cell | *PHACTR2* | 3.107450469 | 0.000364922 | 0.001881617 | 1 |
| Obesity | t_cell | *CUTA* | -2.603279889 | 0.014211929 | 0.036407751 | 1 |
| Obesity | t_cell | *PACRG* | -4.420789266 | 5.14E-05 | 0.000403917 | 0.249216737 |
| Obesity | t_cell | *PRPH2* | -0.646854885 | 0.017665422 | 0.043730932 | 1 |
| Obesity | t_cell | *EXOC2* | 4.022963562 | 0.002468058 | 0.008774918 | 1 |
| Obesity | t_cell | *IMPG1* | -9.468758785 | 1.68E-07 | 1.09E-05 | 0.00081756 |
| Obesity | t_cell | *PCDHB2* | -5.96441172 | 1.51E-06 | 4.50E-05 | 0.007308322 |
| Obesity | t_cell | *NUDT12* | -5.024246469 | 0.00071259 | 0.003231294 | 1 |
| Obesity | t_cell | *CEP72* | 4.813243507 | 0.000969093 | 0.004092288 | 1 |
| Obesity | t_cell | *KIF20A* | -11.48848401 | 0.004547791 | 0.014336867 | 1 |
| Obesity | t_cell | *TMCO6* | -3.409179398 | 0.002325865 | 0.008353143 | 1 |
| Obesity | t_cell | *PCDHB15* | -1.976038155 | 0.002918222 | 0.01003488 | 1 |
| Obesity | t_cell | *HAVCR1* | -2.603924761 | 0.017749401 | 0.043852392 | 1 |
| Obesity | t_cell | *BTNL8* | -0.901738233 | 0.002344328 | 0.008411162 | 1 |
| Obesity | t_cell | *CDH6* | -2.381347386 | 0.005027326 | 0.015576365 | 1 |
| Obesity | t_cell | *LMNB1* | 2.749840238 | 9.14E-06 | 0.00011167 | 0.044332834 |
| Obesity | t_cell | *SLC12A7* | -4.818400702 | 0.004535503 | 0.014326992 | 1 |
| Obesity | t_cell | *SKP1* | 3.979914562 | 0.016656734 | 0.041637545 | 1 |
| Obesity | t_cell | *ATP6V0E1* | 3.924690781 | 0.009941893 | 0.02732295 | 1 |
| Obesity | t_cell | *BNIP1* | -2.432981231 | 2.05E-05 | 0.000199705 | 0.099253204 |
| Obesity | t_cell | *UNC5A* | -21.86905308 | 3.58E-05 | 0.00030928 | 0.173505986 |
| Obesity | t_cell | *ACTR8* | -6.147707566 | 0.000387728 | 0.001971965 | 1 |
| Obesity | t_cell | *CLDN16* | -2.330241791 | 2.79E-05 | 0.000253532 | 0.135132372 |
| Obesity | t_cell | *HYAL1* | -2.073488726 | 0.020358447 | 0.048924806 | 1 |
| Obesity | t_cell | *C3orf14* | -2.914590623 | 0.001154991 | 0.004737125 | 1 |
| Obesity | t_cell | *CBLB* | 3.390369987 | 1.81E-05 | 0.000182506 | 0.087785206 |
| Obesity | t_cell | *SLC41A3* | -3.091912002 | 0.0134663 | 0.03486579 | 1 |
| Obesity | t_cell | *KIAA1257* | -11.379696 | 5.98E-05 | 0.000457039 | 0.290087273 |
| Obesity | t_cell | *NEK11* | -2.52300104 | 0.012162888 | 0.032160398 | 1 |
| Obesity | t_cell | *PLSCR4* | -2.841891265 | 0.006022645 | 0.018172806 | 1 |
| Obesity | t_cell | *ABHD14B* | -4.310884717 | 0.000557171 | 0.002658202 | 1 |
| Obesity | t_cell | *ARHGEF26* | -4.395534958 | 0.001096805 | 0.004525252 | 1 |
| Obesity | t_cell | *VIPR1* | 8.881685827 | 0.000775853 | 0.003440987 | 1 |
| Obesity | t_cell | *NEK4* | -4.263944794 | 0.001656288 | 0.006372965 | 1 |
| Obesity | t_cell | *ADAM23* | 2.136700207 | 8.08E-06 | 0.000102066 | 0.039193188 |
| Obesity | t_cell | *LMAN2L* | -3.708010791 | 0.001365555 | 0.005417559 | 1 |
| Obesity | t_cell | *ACTR1B* | 6.907506408 | 7.36E-05 | 0.000541279 | 0.357243969 |
| Obesity | t_cell | *ITGB6* | -3.301763996 | 0.006730167 | 0.019899311 | 1 |
| Obesity | t_cell | *INO80B* | -2.675783523 | 0.006831357 | 0.02011271 | 1 |
| Obesity | t_cell | *GRB14* | -3.232067017 | 9.99E-06 | 0.000119547 | 0.048454773 |
| Obesity | t_cell | *GALNT3* | 2.347651883 | 2.58E-07 | 1.48E-05 | 0.001254076 |
| Obesity | t_cell | *IGFBP2* | -2.826615577 | 0.000328206 | 0.001717861 | 1 |
| Obesity | t_cell | *GNLY* | 2.465530183 | 1.68E-05 | 0.000173104 | 0.081704931 |
| Obesity | t_cell | *CHMP3* | -4.866168009 | 6.36E-06 | 8.95E-05 | 0.030877285 |

| | | | | | | |
|---|---|---|---|---|---|---|
| Obesity | t_cell | *WNT6* | -4.340321569 | 0.000144099 | 0.000898674 | 0.699168231 |
| Obesity | t_cell | *IL1RL2* | -3.272199597 | 0.000699592 | 0.003182653 | 1 |
| Obesity | t_cell | *IL18R1* | 2.419377206 | 2.07E-05 | 0.000200844 | 0.100622704 |
| Obesity | t_cell | *MLPH* | -2.915769215 | 1.12E-05 | 0.000129872 | 0.054286662 |
| Obesity | t_cell | *TPO* | -1.822943589 | 4.13E-06 | 6.91E-05 | 0.020049611 |
| Obesity | t_cell | *WIPF1* | 3.082567631 | 8.05E-06 | 0.000101925 | 0.039037139 |
| Obesity | t_cell | *AAK1* | 3.502028018 | 9.95E-05 | 0.000682141 | 0.482955751 |
| Obesity | t_cell | *DNAJC16* | -3.845344334 | 0.006270335 | 0.018803254 | 1 |
| Obesity | t_cell | *MARK1* | -3.699127107 | 0.000506739 | 0.002453792 | 1 |
| Obesity | t_cell | *MORN1* | -2.920479363 | 0.001319463 | 0.005269165 | 1 |
| Obesity | t_cell | *GPX7* | 0.883787294 | 0.013426639 | 0.034781661 | 1 |
| Obesity | t_cell | *RALGPS2* | -2.597256087 | 0.004837952 | 0.015085953 | 1 |
| Obesity | t_cell | *ICMT* | -6.372430334 | 0.006149724 | 0.01849763 | 1 |
| Obesity | t_cell | *KIAA1324* | 3.486694399 | 1.29E-05 | 0.000143675 | 0.062355139 |
| Obesity | t_cell | *MECR* | -2.608070607 | 0.013957778 | 0.035851316 | 1 |
| Obesity | t_cell | *RAP1A* | 3.516908111 | 0.000707917 | 0.003213108 | 1 |
| Obesity | t_cell | *CAPZA1* | 4.211440838 | 0.000500181 | 0.002426878 | 1 |
| Obesity | t_cell | *FBXO2* | 1.678855158 | 0.008668506 | 0.024368244 | 1 |
| Obesity | t_cell | *PRG4* | -4.48822972 | 8.85E-05 | 0.000627713 | 0.429355657 |
| Obesity | t_cell | *TNNI3K* | -3.69074529 | 0.002145396 | 0.00783845 | 1 |
| Obesity | t_cell | *CFHR3* | -4.266449199 | 0.001082847 | 0.004479093 | 1 |
| Obesity | t_cell | *OSCP1* | -2.24944757 | 0.008184886 | 0.02325121 | 1 |
| Obesity | t_cell | *EXOC8* | 8.423213609 | 0.000306943 | 0.001641992 | 1 |
| Obesity | t_cell | *SLAMF1* | 0.486789359 | 6.47E-07 | 3.01E-05 | 0.003140171 |
| Obesity | t_cell | *PADI2* | -3.37663743 | 2.48E-05 | 0.0002302 | 0.120164175 |
| Obesity | t_cell | *MFAP2* | -1.438473639 | 0.005455528 | 0.016668906 | 1 |
| Obesity | t_cell | *KIF17* | -4.82446529 | 3.79E-07 | 2.04E-05 | 0.001838695 |
| Obesity | t_cell | *EBNA1BP2* | -4.179658315 | 0.000943786 | 0.003999344 | 1 |
| Obesity | t_cell | *NSUN4* | -6.048043655 | 1.33E-06 | 4.32E-05 | 0.006443282 |
| Obesity | t_cell | *PRRC2C* | 3.71260852 | 0.003586297 | 0.011718086 | 1 |
| Obesity | t_cell | *DARS2* | -4.620784748 | 0.001127619 | 0.004636603 | 1 |
| Obesity | t_cell | *SERPINC1* | 6.531195891 | 5.50E-05 | 0.000427306 | 0.266804799 |
| Obesity | t_cell | *MTFR1L* | -3.876913125 | 0.019059935 | 0.046378537 | 1 |
| Obesity | t_cell | *RPS6KA1* | 2.573639255 | 1.22E-05 | 0.000138162 | 0.058994977 |
| Obesity | t_cell | *MARC2* | -4.17384137 | 0.000290011 | 0.001572214 | 1 |
| Obesity | t_cell | *SLC5A9* | -3.288681654 | 0.000836257 | 0.003639029 | 1 |
| Obesity | t_cell | *CD3EAP* | -3.793168521 | 0.001081863 | 0.004478837 | 1 |
| Obesity | t_cell | *RCN2* | -3.554556516 | 0.018901108 | 0.046130872 | 1 |
| Obesity | t_cell | *APOA1* | 8.411924665 | 5.04E-05 | 0.000397951 | 0.244342046 |
| Obesity | t_cell | *CAMSAP2* | -2.54263349 | 0.006548439 | 0.019421164 | 1 |
| Obesity | t_cell | *CASC1* | -1.638821824 | 0.002912303 | 0.010021626 | 1 |
| Obesity | t_cell | *LRMP* | 1.478507383 | 0.003243345 | 0.010830652 | 1 |
| Obesity | t_cell | *ELOVL4* | 3.902342389 | 2.98E-06 | 6.01E-05 | 0.014441525 |
| Obesity | t_cell | *MFSD1* | -3.626910393 | 0.00223061 | 0.008070785 | 1 |
| Obesity | t_cell | *DNAH7* | -4.458282799 | 9.25E-05 | 0.000646813 | 0.448887993 |
| Obesity | t_cell | *CPSF3* | -4.028379426 | 0.000217398 | 0.001245357 | 1 |
| Obesity | t_cell | *CCDC92* | 4.955430959 | 0.000311318 | 0.001653498 | 1 |
| Obesity | t_cell | *INVS* | 3.018142142 | 0.002468624 | 0.008774918 | 1 |
| Obesity | t_cell | *KDSR* | -4.06190446 | 0.003090633 | 0.010479211 | 1 |
| Obesity | t_cell | *DCAF4* | 5.201100279 | 0.000551527 | 0.002636461 | 1 |

| | | | | | | |
|---|---|---|---|---|---|---|
| Obesity | t_cell | *PPP4R4* | -1.737790683 | 0.001932062 | 0.007222161 | 1 |
| Obesity | t_cell | *ALDH6A1* | -1.880698138 | 0.006528317 | 0.019380641 | 1 |
| Obesity | t_cell | *EIF2B2* | -1.812711728 | 0.015635426 | 0.039388933 | 1 |
| Obesity | t_cell | *CNRIP1* | -3.790081192 | 6.49E-06 | 9.10E-05 | 0.031485292 |
| Obesity | t_cell | *PPP1R3C* | -4.899431279 | 0.000106817 | 0.000714864 | 0.518276113 |
| Obesity | t_cell | *HOXB3* | -3.698312907 | 0.004823736 | 0.015060983 | 1 |
| Obesity | t_cell | *PCDHB10* | -6.291937513 | 0.00017513 | 0.001054257 | 0.849731032 |
| Obesity | t_cell | *PCDHB12* | -1.388841728 | 0.013936156 | 0.035827703 | 1 |
| Obesity | t_cell | *SNX19* | -3.798622865 | 0.000970098 | 0.004092969 | 1 |
| Obesity | t_cell | *MASTL* | -3.000493902 | 0.001437678 | 0.005648269 | 1 |
| Obesity | t_cell | *IQSEC3* | 8.964764027 | 0.004797089 | 0.014997084 | 1 |
| Obesity | t_cell | *ARL1* | 3.84337401 | 0.01147461 | 0.03082769 | 1 |
| Obesity | t_cell | *APAF1* | 3.849482875 | 0.018884449 | 0.046115011 | 1 |
| Obesity | t_cell | *TBX2* | -2.301731692 | 7.63E-05 | 0.000556818 | 0.370283639 |
| Obesity | t_cell | *TEX14* | 3.939949339 | 0.000630489 | 0.002924601 | 1 |
| Obesity | t_cell | *ADCY7* | 2.452643987 | 0.000813703 | 0.003572928 | 1 |
| Obesity | t_cell | *ECHDC2* | -2.987857699 | 0.002012139 | 0.007452593 | 1 |
| Obesity | t_cell | *PYROXD1* | 3.027153329 | 0.003243392 | 0.010830652 | 1 |
| Obesity | t_cell | *ZNF549* | 5.767833948 | 0.000737891 | 0.003310639 | 1 |
| Obesity | t_cell | *ZSCAN18* | -3.260016802 | 0.014731876 | 0.037521819 | 1 |
| Obesity | t_cell | *PDZRN3* | -1.581899239 | 0.010146306 | 0.027797785 | 1 |
| Obesity | t_cell | *LHX4* | 7.37481377 | 0.000347978 | 0.001809634 | 1 |
| Obesity | t_cell | *CD80* | -1.546483188 | 0.015614434 | 0.039356486 | 1 |
| Obesity | t_cell | *PILRB* | 1.019693529 | 0.012319568 | 0.032486166 | 1 |
| Obesity | t_cell | *FABP3* | -5.205260605 | 2.06E-06 | 5.24E-05 | 0.009975866 |
| Obesity | t_cell | *CPXM2* | -1.23077462 | 0.017259638 | 0.042857606 | 1 |
| Obesity | t_cell | *FLT3* | -3.753417171 | 5.92E-05 | 0.000454117 | 0.287001937 |
| Obesity | t_cell | *CD244* | 2.108887014 | 6.82E-07 | 3.06E-05 | 0.003310037 |
| Obesity | t_cell | *SERAC1* | -4.177639968 | 0.007106961 | 0.020823052 | 1 |
| Obesity | t_cell | *LDB3* | -3.539898285 | 0.001948029 | 0.00727004 | 1 |
| Obesity | t_cell | *OPN4* | 13.24144943 | 4.27E-11 | 8.64E-09 | 2.07E-07 |
| Obesity | t_cell | *NAA60* | -6.528720287 | 0.010183097 | 0.02786711 | 1 |
| Obesity | t_cell | *CCZ1* | 2.23875488 | 0.010968104 | 0.02966159 | 1 |
| Obesity | t_cell | *GLIPR2* | 2.770988426 | 0.000211165 | 0.001219728 | 1 |
| Obesity | t_cell | *SLC25A51* | 1.060385791 | 0.010683605 | 0.029040252 | 1 |
| Obesity | t_cell | *CCDC91* | 4.063069219 | 0.000212536 | 0.00122473 | 1 |
| Obesity | t_cell | *CENPK* | 1.666935974 | 2.25E-06 | 5.50E-05 | 0.010940315 |
| Obesity | t_cell | *ARHGAP9* | 2.799687183 | 3.16E-06 | 6.09E-05 | 0.015344584 |
| Obesity | t_cell | *PDE1B* | -6.128339224 | 0.001315124 | 0.005256164 | 1 |
| Obesity | t_cell | *CDK2* | -2.680491005 | 0.007181395 | 0.0209778 | 1 |
| Obesity | t_cell | *DBH* | -54.87433668 | 2.72E-06 | 6.01E-05 | 0.013213696 |
| Obesity | t_cell | *C4BPA* | -3.1787172 | 0.000824377 | 0.00361326 | 1 |
| Obesity | t_cell | *RAB38* | -3.019266335 | 0.003985456 | 0.012797773 | 1 |
| Obesity | t_cell | *DAW1* | -4.641828742 | 7.10E-05 | 0.000525885 | 0.344454676 |
| Obesity | t_cell | *OBSL1* | -3.834727865 | 0.004303554 | 0.013646643 | 1 |
| Obesity | t_cell | *SLC12A4* | -4.811503889 | 8.14E-05 | 0.000585391 | 0.395139113 |
| Obesity | t_cell | *FAM210B* | -4.929616156 | 0.002751912 | 0.009571524 | 1 |
| Obesity | t_cell | *MATN4* | -5.609381892 | 1.36E-05 | 0.000150264 | 0.066116115 |
| Obesity | t_cell | *TOX2* | 1.052712925 | 0.002352974 | 0.008431778 | 1 |
| Obesity | t_cell | *CSE1L* | 5.07269562 | 0.000750836 | 0.00334876 | 1 |

| | | | | | | |
|---|---|---|---|---|---|---|
| Obesity | t_cell | *NAGK* | 3.200514989 | 0.014791819 | 0.037634979 | 1 |
| Obesity | t_cell | *ATP8A1* | 3.43448138 | 2.57E-06 | 5.93E-05 | 0.01245214 |
| Obesity | t_cell | *HIF3A* | -3.035002054 | 1.79E-05 | 0.000180962 | 0.086766367 |
| Obesity | t_cell | *BTN2A2* | 2.168164889 | 6.33E-06 | 8.95E-05 | 0.030732497 |
| Obesity | t_cell | *SNRPC* | -7.772184001 | 1.11E-12 | 3.18E-10 | 5.40E-09 |
| Obesity | t_cell | *RPS10* | 4.076468414 | 0.019737237 | 0.047668031 | 1 |
| Obesity | t_cell | *MED20* | 3.100359257 | 0.003622737 | 0.011796993 | 1 |
| Obesity | t_cell | *TCP11* | 2.351720585 | 0.016780225 | 0.041881508 | 1 |
| Obesity | t_cell | *KCNK17* | 5.263955396 | 7.63E-06 | 9.87E-05 | 0.037019492 |
| Obesity | t_cell | *ATXN1* | 3.316168875 | 0.00023044 | 0.001305597 | 1 |
| Obesity | t_cell | *RUNX2* | 2.318559461 | 4.32E-06 | 7.11E-05 | 0.020947521 |
| Obesity | t_cell | *SH3TC1* | -1.679305105 | 0.005493408 | 0.01676891 | 1 |
| Obesity | t_cell | *MT1G* | -4.161452022 | 0.012260912 | 0.032401931 | 1 |
| Obesity | t_cell | *GOT2* | 7.983477918 | 9.98E-06 | 0.000119547 | 0.048405429 |
| Obesity | t_cell | *CLYBL* | 4.523060136 | 0.000407077 | 0.002048899 | 1 |
| Obesity | t_cell | *GRK4* | 0.94228364 | 0.001230543 | 0.005000499 | 1 |
| Obesity | t_cell | *HS3ST3B1* | 0.906991375 | 2.90E-06 | 6.01E-05 | 0.01409444 |
| Obesity | t_cell | *MRPS7* | -4.835892673 | 0.000983586 | 0.004128337 | 1 |
| Obesity | t_cell | *OPRL1* | -6.543773953 | 3.44E-06 | 6.39E-05 | 0.016705371 |
| Obesity | t_cell | *IL1B* | 9.744593194 | 0.001324671 | 0.005285611 | 1 |
| Obesity | t_cell | *PSD4* | 1.892138022 | 1.63E-06 | 4.64E-05 | 0.007889246 |
| Obesity | t_cell | *TNFSF9* | -22.67402266 | 0.001954906 | 0.007285102 | 1 |
| Obesity | t_cell | *SH2D3A* | 1.588537723 | 1.19E-05 | 0.000136377 | 0.057960026 |
| Obesity | t_cell | *GPR108* | 3.26402765 | 0.00733582 | 0.021300656 | 1 |
| Obesity | t_cell | *SDCBP2* | -4.466955261 | 6.34E-05 | 0.000477605 | 0.307577734 |
| Obesity | t_cell | *DTD1* | 3.248554857 | 0.002866358 | 0.009891586 | 1 |
| Obesity | t_cell | *FLRT3* | -4.787282665 | 3.64E-05 | 0.000313041 | 0.176555357 |
| Obesity | t_cell | *PCSK2* | -8.586939703 | 0.019367587 | 0.046915394 | 1 |
| Obesity | t_cell | *MKKS* | -3.620282131 | 0.01195681 | 0.031756032 | 1 |
| Obesity | t_cell | *UROD* | -2.871032189 | 0.004180624 | 0.013301239 | 1 |
| Obesity | t_cell | *TSKS* | -2.073451355 | 0.013213804 | 0.034377145 | 1 |
| Obesity | t_cell | *L3HYPDH* | -1.482578653 | 0.004868829 | 0.015133605 | 1 |
| Obesity | t_cell | *AIF1L* | -3.01627007 | 5.94E-06 | 8.56E-05 | 0.028816298 |
| Obesity | t_cell | *ZNF484* | 5.777340266 | 0.002832676 | 0.009803241 | 1 |
| Obesity | t_cell | *FGD3* | 1.388266798 | 1.10E-05 | 0.000128556 | 0.053487304 |
| Obesity | t_cell | *ABHD8* | -2.412755581 | 0.013038661 | 0.033957909 | 1 |
| Obesity | t_cell | *ATP13A4* | 2.751403097 | 1.88E-06 | 4.99E-05 | 0.009135406 |
| Obesity | t_cell | *HELB* | 1.909312854 | 1.05E-06 | 3.84E-05 | 0.005094591 |
| Obesity | t_cell | *RAB3IP* | 3.850449339 | 1.82E-06 | 4.84E-05 | 0.008815464 |
| Obesity | t_cell | *YEATS4* | 2.674905636 | 6.11E-05 | 0.000463706 | 0.296308161 |
| Obesity | t_cell | *LRRC61* | -2.682880734 | 0.002045827 | 0.007548558 | 1 |
| Obesity | t_cell | *PTPN12* | -2.960733146 | 0.012700549 | 0.033309765 | 1 |
| Obesity | t_cell | *KDR* | -4.27580042 | 0.000578014 | 0.00274415 | 1 |
| Obesity | t_cell | *VPREB3* | -4.174786895 | 0.010781866 | 0.029241818 | 1 |
| Obesity | t_cell | *CDC42EP1* | -4.892852782 | 1.07E-05 | 0.000125619 | 0.051886751 |
| Obesity | t_cell | *RIBC2* | 1.863657294 | 0.015380846 | 0.038868679 | 1 |
| Obesity | t_cell | *CDHR3* | -3.824345664 | 0.002537865 | 0.008961952 | 1 |
| Obesity | t_cell | *GAD1* | -5.637901154 | 1.09E-06 | 3.89E-05 | 0.005310431 |
| Obesity | t_cell | *VPS13C* | 2.862224316 | 0.019049321 | 0.04637597 | 1 |
| Obesity | t_cell | *CALML4* | -8.916024655 | 4.30E-05 | 0.000351528 | 0.208807636 |

| | | | | | | |
|---|---|---|---|---|---|---|
| Obesity | t_cell | *BBOX1* | 25.51018978 | 0.000695872 | 0.003170303 | 1 |
| Obesity | t_cell | *SERGEF* | 4.479446559 | 2.54E-05 | 0.00023497 | 0.123124542 |
| Obesity | t_cell | *RPAIN* | -3.25069808 | 0.005806178 | 0.017596236 | 1 |
| Obesity | t_cell | *PLD2* | -5.108456578 | 3.67E-05 | 0.000314301 | 0.178208619 |
| Obesity | t_cell | *KRI1* | 1.541529276 | 0.013675635 | 0.03524995 | 1 |
| Obesity | t_cell | *AP1M2* | -2.264439805 | 0.001422825 | 0.005598985 | 1 |
| Obesity | t_cell | *MTUS1* | -2.352180536 | 0.018686204 | 0.045744431 | 1 |
| Obesity | t_cell | *KLK8* | -14.70262316 | 3.75E-06 | 6.55E-05 | 0.018198128 |
| Obesity | t_cell | *ADCY4* | -2.626328832 | 0.00063763 | 0.002946456 | 1 |
| Obesity | t_cell | *PARP2* | -4.121688128 | 0.003131555 | 0.010574561 | 1 |
| Obesity | t_cell | *EPB41L4A* | -2.865438717 | 0.015216182 | 0.038492657 | 1 |
| Obesity | t_cell | *ATP8B3* | -3.628355148 | 2.51E-12 | 6.77E-10 | 1.22E-08 |
| Obesity | t_cell | *SSBP4* | 3.931763922 | 6.63E-06 | 9.16E-05 | 0.032151017 |
| Obesity | t_cell | *PGPEP1* | 2.882603917 | 0.002574157 | 0.009054031 | 1 |
| Obesity | t_cell | *LSM4* | 1.857600354 | 0.001691198 | 0.006476475 | 1 |
| Obesity | t_cell | *TRPM4* | -3.922396339 | 1.89E-06 | 4.99E-05 | 0.009175325 |
| Obesity | t_cell | *CYP2E1* | -2.198530785 | 0.000166162 | 0.00101242 | 0.806216609 |
| Obesity | t_cell | *PNPLA7* | -4.121985136 | 0.001560649 | 0.006048137 | 1 |
| Obesity | t_cell | *PAK4* | -4.374183891 | 0.00083498 | 0.003638422 | 1 |
| Obesity | t_cell | *NPAS1* | -12.60567613 | 4.80E-12 | 1.11E-09 | 2.33E-08 |
| Obesity | t_cell | *MAP3K10* | -3.812800225 | 0.001891121 | 0.007118479 | 1 |
| Obesity | t_cell | *THEMIS2* | 2.291980613 | 0.010016379 | 0.027488389 | 1 |
| Obesity | t_cell | *DNMT1* | 5.34265482 | 9.86E-05 | 0.00067849 | 0.47833549 |
| Obesity | t_cell | *C12orf65* | 5.3484257 | 1.37E-07 | 9.95E-06 | 0.000666788 |
| Obesity | t_cell | *PKDREJ* | -2.888737032 | 0.000423399 | 0.002116708 | 1 |
| Obesity | t_cell | *HSD17B3* | -7.026578394 | 9.53E-05 | 0.000661497 | 0.462386391 |
| Obesity | t_cell | *ULBP2* | 13.16887513 | 0.016004114 | 0.040247376 | 1 |
| Obesity | t_cell | *TTLL9* | -7.162038296 | 0.000351535 | 0.00182422 | 1 |
| Obesity | t_cell | *IDO1* | -3.535875284 | 5.04E-06 | 7.73E-05 | 0.024438061 |
| Obesity | t_cell | *GALNT15* | -4.017597179 | 0.000339041 | 0.001765048 | 1 |
| Obesity | t_cell | *SLC6A6* | 5.000421003 | 8.72E-06 | 0.000107983 | 0.042329286 |
| Obesity | t_cell | *PDLIM4* | -4.347749273 | 1.93E-05 | 0.000190853 | 0.093517996 |
| Obesity | t_cell | *KIF3A* | 5.146824084 | 0.006481544 | 0.019329104 | 1 |
| Obesity | t_cell | *NDUFA2* | 5.826868927 | 1.86E-07 | 1.19E-05 | 0.000902207 |
| Obesity | t_cell | *C1orf159* | -5.319290443 | 0.000769711 | 0.003416867 | 1 |
| Obesity | t_cell | *BARX1* | -6.146900719 | 0.000230246 | 0.001305597 | 1 |
| Obesity | t_cell | *MAP1B* | -2.404189439 | 0.011575117 | 0.031080502 | 1 |
| Obesity | t_cell | *CKMT2* | -2.501583433 | 0.020167381 | 0.048537764 | 1 |
| Obesity | t_cell | *KHDRBS3* | -4.573621056 | 0.000621323 | 0.002898709 | 1 |
| Obesity | t_cell | *PRKAB2* | 6.840926945 | 0.000131899 | 0.000844292 | 0.639973022 |
| Obesity | t_cell | *RHPN2* | -1.008700336 | 0.012100924 | 0.032019016 | 1 |
| Obesity | t_cell | *GCH1* | 2.737359818 | 0.000700257 | 0.003182653 | 1 |
| Obesity | t_cell | *MATN3* | -2.923059232 | 0.000651482 | 0.002993754 | 1 |
| Obesity | t_cell | *TRIM21* | 4.405653806 | 6.58E-06 | 9.16E-05 | 0.031949818 |
| Obesity | t_cell | *SPATA6* | -3.357243394 | 0.000612502 | 0.002868592 | 1 |
| Obesity | t_cell | *LRRC41* | -5.845659814 | 9.99E-05 | 0.000683511 | 0.484911352 |
| Obesity | t_cell | *TRIM22* | 4.506831915 | 2.64E-05 | 0.000242456 | 0.128016814 |
| Obesity | t_cell | *MYBBP1A* | 2.893424555 | 0.00464931 | 0.014591494 | 1 |
| Obesity | t_cell | *SERPINF1* | -3.685737672 | 0.01318103 | 0.034310277 | 1 |
| Obesity | t_cell | *FIGNL1* | -5.93537633 | 0.000437118 | 0.002166391 | 1 |

| | | | | | | |
|---|---|---|---|---|---|---|
| Obesity | t_cell | *RGS22* | -3.03344453 | 0.000296189 | 0.001602127 | 1 |
| Obesity | t_cell | *DMGDH* | -3.571170339 | 7.78E-05 | 0.000564129 | 0.377402293 |
| Obesity | t_cell | *BHMT2* | -0.791900193 | 0.00544379 | 0.016654015 | 1 |
| Obesity | t_cell | *ZBED3* | -6.969321121 | 5.75E-06 | 8.42E-05 | 0.027878208 |
| Obesity | t_cell | *FBXO44* | -7.779296291 | 0.000137529 | 0.000871139 | 0.667292369 |
| Obesity | t_cell | *ALOX5AP* | 2.847550676 | 9.56E-07 | 3.74E-05 | 0.004639107 |
| Obesity | t_cell | *WASF3* | -3.597375813 | 0.000134236 | 0.000856989 | 0.651311872 |
| Obesity | t_cell | *CSNK1G2* | 6.035898872 | 0.000104886 | 0.000705833 | 0.508905891 |
| Obesity | t_cell | *FAM83F* | -11.44775247 | 3.23E-06 | 6.11E-05 | 0.015665039 |
| Obesity | t_cell | *GIMAP4* | 2.807406254 | 3.93E-07 | 2.08E-05 | 0.001909026 |
| Obesity | t_cell | *ADCK2* | -2.075361244 | 0.004110876 | 0.013130988 | 1 |
| Obesity | t_cell | *LARS* | -2.903731543 | 4.96E-05 | 0.000392737 | 0.24074801 |
| Obesity | t_cell | *SPINK5* | -6.705618423 | 7.93E-05 | 0.000571487 | 0.384610465 |
| Obesity | t_cell | *CATSPERB* | 3.381175407 | 3.59E-05 | 0.000309505 | 0.174251214 |
| Obesity | t_cell | *COX16* | -5.557722419 | 4.80E-06 | 7.51E-05 | 0.023267201 |
| Obesity | t_cell | *TTC9* | 2.178774441 | 1.58E-06 | 4.58E-05 | 0.007642314 |
| Obesity | t_cell | *PEBP4* | -5.42959318 | 6.26E-05 | 0.000473396 | 0.303920267 |
| Obesity | t_cell | *CTIF* | -2.545299159 | 0.007400641 | 0.021424767 | 1 |
| Obesity | t_cell | *MRO* | -2.981297814 | 0.008862329 | 0.024794492 | 1 |
| Obesity | t_cell | *CNTN6* | -3.44339721 | 8.98E-06 | 0.000110312 | 0.043573328 |
| Obesity | t_cell | *CHL1* | -2.804061663 | 0.000143967 | 0.000898674 | 0.698527111 |
| Obesity | t_cell | *GSTM3* | -1.168645408 | 0.003336043 | 0.011043083 | 1 |
| Obesity | t_cell | *TRIM45* | -5.803712455 | 0.000915255 | 0.003902299 | 1 |
| Obesity | t_cell | *FKBP11* | 7.29580586 | 0.000166302 | 0.00101242 | 0.806899046 |
| Obesity | t_cell | *TMEM241* | -2.001708402 | 0.010767082 | 0.029218056 | 1 |
| Obesity | t_cell | *KLRD1* | 3.988661496 | 6.89E-06 | 9.39E-05 | 0.033411697 |
| Obesity | t_cell | *ACP2* | 4.130450015 | 0.01989948 | 0.048012072 | 1 |
| Obesity | t_cell | *PRPF38A* | 5.365929151 | 0.007488658 | 0.021640838 | 1 |
| Obesity | t_cell | *TIMM10* | 5.205582191 | 0.000472045 | 0.002308834 | 1 |
| Obesity | t_cell | *FADS2* | -4.796547053 | 3.71E-05 | 0.000316028 | 0.180136227 |
| Obesity | t_cell | *OSTF1* | 4.689215133 | 5.18E-05 | 0.00040619 | 0.251431341 |
| Obesity | t_cell | *GOLM1* | -2.165572 | 0.000548925 | 0.00262661 | 1 |
| Obesity | t_cell | *ADAM19* | 1.166978808 | 4.92E-05 | 0.00039032 | 0.238875727 |
| Obesity | t_cell | *MSI1* | -3.303559995 | 0.000275587 | 0.001512609 | 1 |
| Obesity | t_cell | *DTX1* | -3.784405183 | 3.42E-05 | 0.000299177 | 0.165744204 |
| Obesity | t_cell | *TMEM60* | 4.102601983 | 0.012771979 | 0.033406814 | 1 |
| Obesity | t_cell | *POM121C* | -9.38243331 | 5.13E-14 | 2.07E-11 | 2.49E-10 |
| Obesity | t_cell | *NT5E* | -1.771375839 | 0.016440145 | 0.041117312 | 1 |
| Obesity | t_cell | *B4GALNT1* | -5.610886578 | 0.002003859 | 0.007438961 | 1 |
| Obesity | t_cell | *FAIM2* | -3.519113799 | 0.000185682 | 0.001105432 | 0.900927233 |
| Obesity | t_cell | *PAN2* | -3.443100568 | 0.012564394 | 0.033006195 | 1 |
| Obesity | t_cell | *MAP7* | -2.176221424 | 0.002569788 | 0.009054031 | 1 |
| Obesity | t_cell | *NHSL1* | -2.458664861 | 0.007457753 | 0.021564372 | 1 |
| Obesity | t_cell | *AHI1* | -1.664850352 | 0.00986461 | 0.027164069 | 1 |
| Obesity | t_cell | *MICAL1* | 3.417602035 | 7.97E-08 | 6.44E-06 | 0.000386682 |
| Obesity | t_cell | *USP15* | 3.273559813 | 0.005374089 | 0.016492778 | 1 |
| Obesity | t_cell | *CAPN9* | -5.895976509 | 1.01E-05 | 0.000119903 | 0.048800339 |
| Obesity | t_cell | *SP110* | 4.778019364 | 6.67E-06 | 9.16E-05 | 0.032341606 |
| Obesity | t_cell | *MRPL44* | 7.376312384 | 3.28E-05 | 0.000291037 | 0.159197292 |
| Obesity | t_cell | *SLC19A3* | -3.136212901 | 0.005098394 | 0.015746282 | 1 |

| | | | | | | |
|---|---|---|---|---|---|---|
| Obesity | t_cell | *TMBIM1* | -4.321531025 | 0.003546351 | 0.011618431 | 1 |
| Obesity | t_cell | *TSGA10* | -1.1275444 | 0.010697741 | 0.029062396 | 1 |
| Obesity | t_cell | *MRPS9* | 3.140440579 | 0.001214068 | 0.004942444 | 1 |
| Obesity | t_cell | *ANKRD36* | 3.737868469 | 0.015358003 | 0.038831177 | 1 |
| Obesity | t_cell | *STAB2* | -4.905396731 | 0.000263231 | 0.001461326 | 1 |
| Obesity | t_cell | *DRAM1* | -4.078778229 | 0.003456522 | 0.01135732 | 1 |
| Obesity | t_cell | *SETDB2* | 4.695745305 | 0.000738275 | 0.003310639 | 1 |
| Obesity | t_cell | *SCRN1* | -3.704922913 | 0.000170655 | 0.001035022 | 0.828017587 |
| Obesity | t_cell | *CHST12* | 4.3592587 | 0.000236951 | 0.001338402 | 1 |
| Obesity | t_cell | *AOAH* | 1.331169128 | 7.54E-06 | 9.81E-05 | 0.036597874 |
| Obesity | t_cell | *TBRG4* | -5.871054234 | 3.84E-05 | 0.000323656 | 0.18610226 |
| Obesity | t_cell | *MTHFS* | -1.98505753 | 0.006799327 | 0.020042729 | 1 |
| Obesity | t_cell | *NMT1* | 1.72848158 | 0.01923992 | 0.046676046 | 1 |
| Obesity | t_cell | *BRIP1* | 2.175177678 | 0.008386933 | 0.023755633 | 1 |
| Obesity | t_cell | *RTP4* | -3.517463389 | 0.01356111 | 0.035036478 | 1 |
| Obesity | t_cell | *IMP4* | -5.930264899 | 0.001404685 | 0.005550106 | 1 |
| Obesity | t_cell | *ODF2* | -5.545935395 | 0.001799701 | 0.006821991 | 1 |
| Obesity | t_cell | *ZNF189* | -4.685967944 | 0.008993823 | 0.025079326 | 1 |
| Obesity | t_cell | *DERL1* | -5.094247687 | 0.004514959 | 0.014271387 | 1 |
| Obesity | t_cell | *NOV* | -2.90890286 | 0.002443644 | 0.008705259 | 1 |
| Obesity | t_cell | *IL11RA* | -3.13748052 | 0.003967579 | 0.012757252 | 1 |
| Obesity | t_cell | *APTX* | -3.402505209 | 0.017522218 | 0.043420737 | 1 |
| Obesity | t_cell | *GMPR* | -4.05958865 | 0.000306357 | 0.001641992 | 1 |
| Obesity | t_cell | *FRS3* | -3.744663282 | 4.32E-05 | 0.000351724 | 0.209421745 |
| Obesity | t_cell | *CAPN11* | -1.591232533 | 0.005679461 | 0.017244522 | 1 |
| Obesity | t_cell | *IRF4* | 1.179638783 | 0.000722499 | 0.003264029 | 1 |
| Obesity | t_cell | *TUBB2B* | -1.838069705 | 0.004042801 | 0.012947637 | 1 |
| Obesity | t_cell | *NRM* | -7.87765943 | 5.99E-06 | 8.60E-05 | 0.029064758 |
| Obesity | t_cell | *MYO7A* | -2.518992902 | 0.002053432 | 0.007559371 | 1 |
| Obesity | t_cell | *SYTL2* | 1.636180275 | 0.000483433 | 0.002352673 | 1 |
| Obesity | t_cell | *RAB30* | -4.566762159 | 0.000206662 | 0.001203752 | 1 |
| Obesity | t_cell | *GGH* | -1.35995895 | 0.010532425 | 0.028693613 | 1 |
| Obesity | t_cell | *SULF1* | -2.016101853 | 0.000869952 | 0.003755344 | 1 |
| Obesity | t_cell | *SLC37A4* | 2.882952844 | 0.002414466 | 0.008607633 | 1 |
| Obesity | t_cell | *FDX1* | 3.664177127 | 0.008409431 | 0.023768148 | 1 |
| Obesity | t_cell | *FXYD6* | -1.670266779 | 0.012376402 | 0.032618307 | 1 |
| Obesity | t_cell | *NUSAP1* | 5.864984168 | 7.13E-09 | 9.35E-07 | 3.46E-05 |
| Obesity | t_cell | *NDUFAF1* | 5.10847183 | 5.91E-05 | 0.000454117 | 0.286613909 |
| Obesity | t_cell | *ADAM10* | 3.541379024 | 0.011133087 | 0.029976546 | 1 |
| Obesity | t_cell | *KHK* | -2.70977145 | 0.000508414 | 0.002456997 | 1 |
| Obesity | t_cell | *LHCGR* | -3.738599302 | 0.000178974 | 0.001070753 | 0.868380993 |
| Obesity | t_cell | *ACTR2* | 3.967170114 | 0.0022431 | 0.008097859 | 1 |
| Obesity | t_cell | *CENPO* | 3.491704993 | 0.009159866 | 0.025469152 | 1 |
| Obesity | t_cell | *CYP2C8* | 8.918655197 | 1.61E-05 | 0.000168136 | 0.078351557 |
| Obesity | t_cell | *BTBD16* | -3.533463657 | 0.009141658 | 0.025447691 | 1 |
| Obesity | t_cell | *TACC2* | -2.812189305 | 0.012403439 | 0.032671817 | 1 |
| Obesity | t_cell | *RPS24* | 3.72502486 | 0.002510935 | 0.008892741 | 1 |
| Obesity | t_cell | *BARD1* | 4.880236596 | 0.003236989 | 0.01082417 | 1 |
| Obesity | t_cell | *STAT4* | 1.011481961 | 1.05E-06 | 3.84E-05 | 0.005089281 |
| Obesity | t_cell | *COX17* | -5.355273396 | 0.004594501 | 0.014438159 | 1 |

| | | | | | | |
|---|---|---|---|---|---|---|
| Obesity | t_cell | *PPCDC* | -3.123019096 | 0.001204197 | 0.004909886 | 1 |
| Obesity | t_cell | *PCDH10* | -5.178869307 | 0.006490329 | 0.019343414 | 1 |
| Obesity | t_cell | *AP1AR* | -4.639697783 | 0.011692738 | 0.031309693 | 1 |
| Obesity | t_cell | *PRKG2* | 1.947093474 | 0.000330668 | 0.001727017 | 1 |
| Obesity | t_cell | *BMPR1B* | -2.688209432 | 0.001963991 | 0.007313343 | 1 |
| Obesity | t_cell | *MMRN1* | -2.652399362 | 1.47E-06 | 4.50E-05 | 0.007131027 |
| Obesity | t_cell | *PRDM5* | -3.954053707 | 0.000477319 | 0.002329931 | 1 |
| Obesity | t_cell | *TRPC3* | 2.54296818 | 0.000383205 | 0.001957168 | 1 |
| Obesity | t_cell | *CNOT6L* | 2.947334705 | 0.004140218 | 0.013200328 | 1 |
| Obesity | t_cell | *ENPEP* | -1.087604561 | 0.011822932 | 0.03155383 | 1 |
| Obesity | t_cell | *PARVG* | 2.237994518 | 2.20E-05 | 0.000209963 | 0.106661138 |
| Obesity | t_cell | *B4GALNT3* | 2.025117271 | 8.73E-07 | 3.69E-05 | 0.00423802 |
| Obesity | t_cell | *PDE6H* | 5.013563234 | 1.59E-06 | 4.59E-05 | 0.007721221 |
| Obesity | t_cell | *ERP27* | 2.091679064 | 9.22E-07 | 3.73E-05 | 0.004471205 |
| Obesity | t_cell | *KIF21A* | 3.659510114 | 0.008427825 | 0.023788136 | 1 |
| Obesity | t_cell | *ALG10* | -2.951606388 | 0.000939775 | 0.003989317 | 1 |
| Obesity | t_cell | *CLSTN3* | 5.827374619 | 0.000394838 | 0.001999742 | 1 |
| Obesity | t_cell | *KLRG1* | 1.598233095 | 1.84E-09 | 2.70E-07 | 8.92E-06 |
| Obesity | t_cell | *PPFIA2* | -4.273892844 | 0.000433904 | 0.002154866 | 1 |
| Obesity | t_cell | *MARCH9* | 7.552939655 | 2.73E-05 | 0.000249614 | 0.132295606 |
| Obesity | t_cell | *LGR5* | -3.651929282 | 0.000139483 | 0.000881212 | 0.676770654 |
| Obesity | t_cell | *POC1B* | 4.901969095 | 0.000812051 | 0.003568907 | 1 |
| Obesity | t_cell | *AMDHD1* | -1.217510758 | 0.008866684 | 0.024794492 | 1 |
| Obesity | t_cell | *RITA1* | 9.580948351 | 0.009934235 | 0.02732295 | 1 |
| Obesity | t_cell | *GLTP* | 6.448669895 | 0.009952375 | 0.027328197 | 1 |
| Obesity | t_cell | *FOXN4* | -2.757938315 | 0.000265697 | 0.001471647 | 1 |
| Obesity | t_cell | *ACVRL1* | -5.847398383 | 0.000715079 | 0.003233518 | 1 |
| Obesity | t_cell | *HNRNPA1L2* | -5.021009678 | 0.00030477 | 0.001637589 | 1 |
| Obesity | t_cell | *GRTP1* | -1.374154985 | 0.00727259 | 0.021167729 | 1 |
| Obesity | t_cell | *CBLN3* | -0.503175184 | 0.01904461 | 0.04637597 | 1 |
| Obesity | t_cell | *FITM1* | 6.926356753 | 0.000909972 | 0.00388636 | 1 |
| Obesity | t_cell | *RAB15* | 6.107500333 | 0.018316882 | 0.04499479 | 1 |
| Obesity | t_cell | *EFCAB11* | -3.190354374 | 0.00597779 | 0.018059924 | 1 |
| Obesity | t_cell | *WARS* | -3.868165342 | 0.000241671 | 0.001358735 | 1 |
| Obesity | t_cell | *DUOXA1* | -2.802286438 | 2.41E-05 | 0.000225504 | 0.116712347 |
| Obesity | t_cell | *NEIL1* | -3.338974099 | 0.000750917 | 0.00334876 | 1 |
| Obesity | t_cell | *PIF1* | -3.95955908 | 0.000266929 | 0.001474155 | 1 |
| Obesity | t_cell | *ULK3* | 5.799782531 | 0.010376316 | 0.028331955 | 1 |
| Obesity | t_cell | *PCSK6* | -4.092684323 | 4.20E-05 | 0.000347455 | 0.203955792 |
| Obesity | t_cell | *POLG* | 5.002920007 | 0.000199535 | 0.001173506 | 0.968142361 |
| Obesity | t_cell | *MFGE8* | -3.418709722 | 0.010234694 | 0.027976752 | 1 |
| Obesity | t_cell | *ITGAX* | 5.463534973 | 1.69E-06 | 4.71E-05 | 0.008207583 |
| Obesity | t_cell | *TGFB1I1* | -8.153214588 | 1.22E-05 | 0.000138162 | 0.05895718 |
| Obesity | t_cell | *GCSH* | -3.065029767 | 0.001602414 | 0.006190217 | 1 |
| Obesity | t_cell | *CMTM3* | 5.146927333 | 0.000449808 | 0.002217473 | 1 |
| Obesity | t_cell | *TLDC1* | 5.005884215 | 1.07E-05 | 0.000125619 | 0.051926656 |
| Obesity | t_cell | *DEF8* | -2.920722088 | 0.007892459 | 0.022605791 | 1 |
| Obesity | t_cell | *MYO19* | 6.917260347 | 1.61E-05 | 0.000168136 | 0.078230641 |
| Obesity | t_cell | *RASL10B* | -6.350981079 | 0.000207182 | 0.001205331 | 1 |
| Obesity | t_cell | *UNC45B* | 4.418969116 | 1.63E-05 | 0.000168143 | 0.079154087 |

| | | | | | | |
|---|---|---|---|---|---|---|
| Obesity | t_cell | *LRRC46* | -2.633200154 | 0.001345148 | 0.005354108 | 1 |
| Obesity | t_cell | *SSH2* | 3.048671881 | 0.00536667 | 0.016480432 | 1 |
| Obesity | t_cell | *RHBDL3* | -3.125072739 | 0.000142351 | 0.000895834 | 0.690687651 |
| Obesity | t_cell | *G6PC3* | -4.21467154 | 0.007151772 | 0.020916454 | 1 |
| Obesity | t_cell | *IMPA2* | -3.612787689 | 0.009216473 | 0.025582567 | 1 |
| Obesity | t_cell | *GNAL* | -4.00811114 | 0.000103998 | 0.000702186 | 0.504597976 |
| Obesity | t_cell | *C18orf8* | -4.60152906 | 0.00031227 | 0.001654879 | 1 |
| Obesity | t_cell | *SLC14A1* | 1.76405761 | 0.000417476 | 0.00209375 | 1 |
| Obesity | t_cell | *CCDC40* | -2.636200909 | 0.000840284 | 0.003646743 | 1 |
| Obesity | t_cell | *TMC6* | 1.993670428 | 9.08E-07 | 3.71E-05 | 0.004407344 |
| Obesity | t_cell | *TBCD* | 3.43864481 | 0.011228145 | 0.03021573 | 1 |
| Obesity | t_cell | *RNF157* | 2.435732114 | 3.67E-07 | 2.02E-05 | 0.001779251 |
| Obesity | t_cell | *ZNF750* | -6.594112203 | 1.05E-06 | 3.84E-05 | 0.005117532 |
| Obesity | t_cell | *MAPK4* | -4.637480826 | 3.47E-06 | 6.39E-05 | 0.016844814 |
| Obesity | t_cell | *FBXO15* | -3.242318231 | 1.54E-05 | 0.000162814 | 0.074766469 |
| Obesity | t_cell | *PIP4K2B* | -4.538449041 | 0.000243118 | 0.00136529 | 1 |
| Obesity | t_cell | *ERBB2* | -4.09706994 | 0.000746002 | 0.003339117 | 1 |
| Obesity | t_cell | *MIEN1* | 5.19213041 | 0.004071307 | 0.013021741 | 1 |
| Obesity | t_cell | *CACNA1A* | -6.882957923 | 0.001411629 | 0.005566756 | 1 |
| Obesity | t_cell | *SAE1* | -8.195411657 | 3.14E-06 | 6.09E-05 | 0.015223283 |
| Obesity | t_cell | *EVI5L* | -5.935826207 | 0.004857476 | 0.015127389 | 1 |
| Obesity | t_cell | *SLC47A1* | -2.891657192 | 0.00011828 | 0.000770853 | 0.573894145 |
| Obesity | t_cell | *CTU1* | -7.21247589 | 0.006054567 | 0.018246435 | 1 |
| Obesity | t_cell | *MMEL1* | -1.830045265 | 9.55E-05 | 0.000661693 | 0.463185214 |
| Obesity | t_cell | *EPHA2* | -5.662891714 | 6.36E-06 | 8.95E-05 | 0.030842758 |
| Obesity | t_cell | *ZNF593* | -7.05447565 | 0.000147575 | 0.000915644 | 0.716033405 |
| Obesity | t_cell | *RPS8* | 4.149677459 | 0.001355892 | 0.005383623 | 1 |
| Obesity | t_cell | *SLC44A3* | -5.691942092 | 0.000173143 | 0.001047496 | 0.84009146 |
| Obesity | t_cell | *STRIP1* | -4.762522976 | 0.001871717 | 0.007056387 | 1 |
| Obesity | t_cell | *GPR161* | -2.507471531 | 0.012738287 | 0.03335465 | 1 |
| Obesity | t_cell | *TIPRL* | 5.828485948 | 0.000183462 | 0.001093557 | 0.890155562 |
| Obesity | t_cell | *XCL1* | 1.290377598 | 1.45E-06 | 4.50E-05 | 0.007031923 |
| Obesity | t_cell | *XCL2* | 1.863615806 | 0.000447537 | 0.002213508 | 1 |
| Obesity | t_cell | *MAEL* | -8.992966017 | 6.94E-06 | 9.43E-05 | 0.033679395 |
| Obesity | t_cell | *TARS2* | -3.886166596 | 0.003111431 | 0.010535007 | 1 |
| Obesity | t_cell | *DYRK3* | -4.700979523 | 0.000146767 | 0.000911796 | 0.712112476 |
| Obesity | t_cell | *SMYD2* | 5.516846946 | 8.80E-05 | 0.000624816 | 0.42674933 |
| Obesity | t_cell | *CREB3L4* | -3.307817979 | 2.21E-05 | 0.000210404 | 0.107095545 |
| Obesity | t_cell | *KCNN3* | -2.387865576 | 0.000260963 | 0.001455378 | 1 |
| Obesity | t_cell | *RIT1* | 7.632014903 | 0.000259054 | 0.001448076 | 1 |
| Obesity | t_cell | *FLG* | -0.992405622 | 0.010886239 | 0.029491921 | 1 |
| Obesity | t_cell | *SCCPDH* | -4.572576793 | 0.00083089 | 0.003632404 | 1 |
| Obesity | t_cell | *MBOAT2* | -4.7896056 | 0.000693317 | 0.00316163 | 1 |
| Obesity | t_cell | *PLEKHA6* | -1.654951073 | 0.007600875 | 0.021895031 | 1 |
| Obesity | t_cell | *OSR1* | -1.708098297 | 0.006410248 | 0.0191636 | 1 |
| Obesity | t_cell | *RHOB* | -4.807440042 | 0.001896583 | 0.00712798 | 1 |
| Obesity | t_cell | *GALM* | 4.857644036 | 8.69E-05 | 0.000620085 | 0.421657685 |
| Obesity | t_cell | *ABHD1* | -4.173377853 | 0.000525011 | 0.002529101 | 1 |
| Obesity | t_cell | *ZNF514* | -3.49626721 | 0.000832788 | 0.003636983 | 1 |
| Obesity | t_cell | *TEX261* | -5.784431372 | 3.41E-05 | 0.000299177 | 0.165691488 |

| | | | | | | |
|---|---|---|---|---|---|---|
| Obesity | t_cell | *RALB* | -4.484100213 | 0.004509768 | 0.014264271 | 1 |
| Obesity | t_cell | *FBLN7* | -2.119749294 | 0.001902839 | 0.007145861 | 1 |
| Obesity | t_cell | *CNGA3* | -28.1370939 | 0.000231909 | 0.001311449 | 1 |
| Obesity | t_cell | *FAHD2B* | -2.379262154 | 0.014913261 | 0.037904212 | 1 |
| Obesity | t_cell | *GALNT13* | -4.201468693 | 0.000182832 | 0.001091145 | 0.887100899 |
| Obesity | t_cell | *PHOSPHO2* | -5.32279103 | 1.55E-07 | 1.07E-05 | 0.000752275 |
| Obesity | t_cell | *METTL21A* | 5.135446991 | 0.000730924 | 0.003289835 | 1 |
| Obesity | t_cell | *ESPNL* | 37.6053217 | 3.23E-06 | 6.11E-05 | 0.015667481 |
| Obesity | t_cell | *STK11IP* | -4.744593587 | 0.000956407 | 0.004042236 | 1 |
| Obesity | t_cell | *EAF1* | 5.826203735 | 0.000108585 | 0.000723701 | 0.526854532 |
| Obesity | t_cell | *CNTN4* | -3.458096755 | 0.000411482 | 0.002068924 | 1 |
| Obesity | t_cell | *CTDSPL* | -2.754252329 | 0.006406078 | 0.019162942 | 1 |
| Obesity | t_cell | *PTPRG* | -2.379468428 | 0.01206799 | 0.031979184 | 1 |
| Obesity | t_cell | *LIMD1* | 3.132672341 | 0.016183248 | 0.040558429 | 1 |
| Obesity | t_cell | *ADPRH* | -2.1072793 | 0.002579839 | 0.009057439 | 1 |
| Obesity | t_cell | *BOC* | -3.020277786 | 0.000357288 | 0.001850118 | 1 |
| Obesity | t_cell | *MED12L* | 5.075999606 | 0.012311227 | 0.032481824 | 1 |
| Obesity | t_cell | *EIF2A* | -2.218908563 | 0.002953201 | 0.010119303 | 1 |
| Obesity | t_cell | *NCEH1* | -2.625534441 | 0.004723521 | 0.014805248 | 1 |
| Obesity | t_cell | *AMT* | -1.124491446 | 0.003302461 | 0.010952524 | 1 |
| Obesity | t_cell | *MUC4* | -4.058443252 | 3.32E-05 | 0.000293701 | 0.160947976 |
| Obesity | t_cell | *SLC26A1* | 1.360922184 | 0.003012582 | 0.010264782 | 1 |
| Obesity | t_cell | *SCD5* | -5.1109645 | 0.005515627 | 0.016810191 | 1 |
| Obesity | t_cell | *TRMT10A* | -6.112080862 | 5.95E-05 | 0.000456449 | 0.288932374 |
| Obesity | t_cell | *DDIT4L* | -4.546235526 | 0.00020128 | 0.001180907 | 0.976610234 |
| Obesity | t_cell | *MYO10* | -3.391788922 | 0.000104348 | 0.000703188 | 0.506295593 |
| Obesity | t_cell | *BHMT* | -1.939799977 | 1.26E-05 | 0.000142023 | 0.061291591 |
| Obesity | t_cell | *GTF2H2* | -7.608031135 | 0.002484465 | 0.00882476 | 1 |
| Obesity | t_cell | *SPATA9* | -4.181394479 | 0.000145994 | 0.000908158 | 0.708363056 |
| Obesity | t_cell | *TSLP* | -1.236521845 | 0.00045023 | 0.002217473 | 1 |
| Obesity | t_cell | *PCYOX1L* | -11.18357444 | 4.05E-05 | 0.000336662 | 0.196274169 |
| Obesity | t_cell | *FAM50B* | -8.299458903 | 0.014573693 | 0.037197031 | 1 |
| Obesity | t_cell | *GFOD1* | -3.085403635 | 0.000979854 | 0.0041198 | 1 |
| Obesity | t_cell | *HIGD2A* | 4.146713362 | 0.007673591 | 0.02204397 | 1 |
| Obesity | t_cell | *RNF44* | 4.45226342 | 1.26E-05 | 0.000142023 | 0.061353794 |
| Obesity | t_cell | *ABT1* | 2.168569185 | 0.00182901 | 0.006927679 | 1 |
| Obesity | t_cell | *DAAM2* | -2.914594502 | 0.020087644 | 0.048393867 | 1 |
| Obesity | t_cell | *MLIP* | -3.555243626 | 1.29E-06 | 4.23E-05 | 0.006249176 |
| Obesity | t_cell | *TPBG* | -3.266121571 | 2.40E-05 | 0.000225504 | 0.116620387 |
| Obesity | t_cell | *RNF217* | -3.290021338 | 0.002109325 | 0.007735784 | 1 |
| Obesity | t_cell | *ABRACL* | 2.263275042 | 0.000398541 | 0.002014294 | 1 |
| Obesity | t_cell | *SLC18B1* | -2.015131321 | 0.018861325 | 0.046115011 | 1 |
| Obesity | t_cell | *PNLDC1* | 4.728292947 | 0.00262864 | 0.009188876 | 1 |
| Obesity | t_cell | *SLC22A3* | -4.479109021 | 0.00261306 | 0.009141001 | 1 |
| Obesity | t_cell | *GNA12* | -3.545847235 | 0.005329603 | 0.016397739 | 1 |
| Obesity | t_cell | *SDK1* | -2.465556123 | 1.90E-05 | 0.000189814 | 0.092249522 |
| Obesity | t_cell | *EPHA1* | 14.63111472 | 1.14E-05 | 0.000131357 | 0.055169962 |
| Obesity | t_cell | *ADHFE1* | -1.685164957 | 7.88E-06 | 0.000100385 | 0.03824663 |
| Obesity | t_cell | *ATP6V0D2* | -5.956783935 | 0.00339592 | 0.011216475 | 1 |
| Obesity | t_cell | *DPYS* | -3.009299167 | 2.90E-05 | 0.000260354 | 0.140591084 |

| | | | | | | |
|---|---|---|---|---|---|---|
| Obesity | t_cell | *LRP12* | -4.798494296 | 0.005676346 | 0.017244522 | 1 |
| Obesity | t_cell | *MAL2* | -1.467796024 | 0.00403269 | 0.012923784 | 1 |
| Obesity | t_cell | *ZNF462* | -2.587651829 | 0.008136402 | 0.023154147 | 1 |
| Obesity | t_cell | *SURF6* | -4.048367708 | 0.009126712 | 0.02542067 | 1 |
| Obesity | t_cell | *MED22* | -3.227964259 | 0.006941661 | 0.020425071 | 1 |
| Obesity | t_cell | *SLC39A12* | -5.004580758 | 0.000104054 | 0.000702186 | 0.504871733 |
| Obesity | t_cell | *ST8SIA6* | 7.843745633 | 1.78E-05 | 0.000180962 | 0.086601683 |
| Obesity | t_cell | *PARD3* | -2.287687878 | 0.018233895 | 0.04486352 | 1 |
| Obesity | t_cell | *CDHR1* | 2.038464748 | 8.27E-06 | 0.000104261 | 0.040140583 |
| Obesity | t_cell | *RGR* | -21.96955869 | 7.10E-09 | 9.35E-07 | 3.45E-05 |
| Obesity | t_cell | *MTG1* | 2.42064703 | 0.000662561 | 0.003032779 | 1 |
| Obesity | t_cell | *PAOX* | 8.063189792 | 0.000329757 | 0.001724116 | 1 |
| Obesity | t_cell | *RGS10* | 5.319967693 | 0.00010354 | 0.000700662 | 0.502374521 |
| Obesity | t_cell | *GAS2* | -4.862188113 | 0.002534207 | 0.008961952 | 1 |
| Obesity | t_cell | *LIN7C* | -4.93589863 | 0.004410539 | 0.013959515 | 1 |
| Obesity | t_cell | *HSD17B12* | -2.900359554 | 4.26E-13 | 1.38E-10 | 2.07E-09 |
| Obesity | t_cell | *PTPRJ* | 3.630216068 | 9.48E-05 | 0.000659126 | 0.460070283 |
| Obesity | t_cell | *RPS3* | 2.976016325 | 0.015525901 | 0.039174037 | 1 |
| Obesity | t_cell | *TTC12* | 4.225416144 | 2.09E-06 | 5.25E-05 | 0.010137701 |
| Obesity | t_cell | *DRD2* | -34.43650441 | 5.25E-06 | 7.92E-05 | 0.025451198 |
| Obesity | t_cell | *NPAT* | 4.521928636 | 0.007328619 | 0.021292491 | 1 |
| Obesity | t_cell | *ATM* | 3.50223091 | 0.000207726 | 0.001207049 | 1 |
| Obesity | t_cell | *ST14* | -6.140314902 | 0.001156989 | 0.00474131 | 1 |
| Obesity | t_cell | *FADS1* | -5.366238058 | 1.38E-05 | 0.000151925 | 0.067019866 |
| Obesity | t_cell | *TMC2* | -9.516375385 | 1.70E-05 | 0.000174007 | 0.082305129 |
| Obesity | t_cell | *INCENP* | -8.553611087 | 0.000105037 | 0.000705873 | 0.50964024 |
| Obesity | t_cell | *PLCH2* | 1.56197627 | 4.40E-06 | 7.21E-05 | 0.021336633 |
| Obesity | t_cell | *CCDC15* | -2.966596817 | 0.000215634 | 0.001238173 | 1 |
| Obesity | t_cell | *CHEK1* | 6.635689477 | 2.90E-06 | 6.01E-05 | 0.014077155 |
| Obesity | t_cell | *TRPT1* | -2.886104093 | 0.001037475 | 0.004328312 | 1 |
| Obesity | t_cell | *NUDT22* | -4.176316406 | 0.002574781 | 0.009054031 | 1 |
| Obesity | t_cell | *TBX6* | 1.724208103 | 2.59E-07 | 1.48E-05 | 0.001258397 |
| Obesity | t_cell | *FAM124A* | -4.265521245 | 3.82E-05 | 0.000323053 | 0.185432165 |
| Obesity | t_cell | *LYPD6B* | -3.513191722 | 3.57E-06 | 6.46E-05 | 0.017326086 |
| Obesity | t_cell | *WDR17* | -3.70614513 | 1.74E-05 | 0.000176774 | 0.084321073 |
| Obesity | t_cell | *CCDC102B* | -2.452919858 | 0.001750429 | 0.006676951 | 1 |
| Obesity | t_cell | *RGS18* | 3.494611783 | 1.48E-07 | 1.04E-05 | 0.000716421 |
| Obesity | t_cell | *PRSS23* | -2.451190232 | 0.014130042 | 0.036236239 | 1 |
| Obesity | t_cell | *PPP1R1C* | -4.669812715 | 5.50E-06 | 8.10E-05 | 0.026662691 |
| Obesity | t_cell | *GPR158* | -2.859179646 | 1.03E-05 | 0.000122465 | 0.050210561 |
| Obesity | t_cell | *ANK3* | 1.031155857 | 1.51E-06 | 4.50E-05 | 0.007341953 |
| Obesity | t_cell | *TMEM18* | -5.701736186 | 0.005667362 | 0.017229349 | 1 |
| Obesity | t_cell | *ADAMTS12* | -1.643564175 | 0.018971712 | 0.046256656 | 1 |
| Obesity | t_cell | *AKR1C2* | -2.802873355 | 0.000479778 | 0.002339579 | 1 |
| Obesity | t_cell | *DPYSL4* | 1.170714895 | 0.000762954 | 0.003393084 | 1 |
| Obesity | t_cell | *INPP1* | -2.999076434 | 0.001775367 | 0.006745561 | 1 |
| Obesity | t_cell | *MFSD6* | 4.54564755 | 5.14E-05 | 0.000403917 | 0.249195545 |
| Obesity | t_cell | *CCDC122* | -1.324559443 | 0.014481515 | 0.037009683 | 1 |
| Obesity | t_cell | *GUF1* | -4.056874713 | 0.015466076 | 0.039052375 | 1 |
| Obesity | t_cell | *GABRA2* | 5.157427438 | 0.019429815 | 0.047035575 | 1 |

| | | | | | | |
|---|---|---|---|---|---|---|
| Obesity | t_cell | *PARP8* | 1.393567119 | 0.000118361 | 0.000770853 | 0.57428545 |
| Obesity | t_cell | *BEND6* | -3.616185406 | 8.15E-07 | 3.52E-05 | 0.003953725 |
| Obesity | t_cell | *TMEM56* | -9.646144593 | 7.90E-05 | 0.000570515 | 0.383386308 |
| Obesity | t_cell | *PTPN14* | -3.003749981 | 0.002375338 | 0.008488555 | 1 |
| Obesity | t_cell | *MGAT5* | 2.988240996 | 0.003290093 | 0.010941909 | 1 |
| Obesity | t_cell | *TMEM178A* | -10.13028185 | 0.002205766 | 0.008010759 | 1 |
| Obesity | t_cell | *EPG5* | 3.773697362 | 0.016973088 | 0.042254193 | 1 |
| Obesity | t_cell | *POC5* | -3.240141861 | 3.37E-05 | 0.000296383 | 0.163306834 |
| Obesity | t_cell | *CWF19L2* | 3.053057842 | 0.019046969 | 0.04637597 | 1 |
| Obesity | t_cell | *SPARCL1* | -2.50475329 | 0.011928037 | 0.031712239 | 1 |
| Obesity | t_cell | *MBNL1* | 2.869741789 | 0.00011766 | 0.000769388 | 0.570885584 |
| Obesity | t_cell | *CCNO* | 12.08076951 | 5.27E-06 | 7.92E-05 | 0.025547079 |
| Obesity | t_cell | *CLEC4F* | -2.791956739 | 0.000379909 | 0.001943078 | 1 |
| Obesity | t_cell | *RASGRP3* | -4.057470823 | 0.001730175 | 0.006604886 | 1 |
| Obesity | t_cell | *PRDM8* | 2.068176518 | 6.69E-05 | 0.000499969 | 0.324479765 |
| Obesity | t_cell | *GGPS1* | 3.836317418 | 0.005991585 | 0.018090337 | 1 |
| Obesity | t_cell | *JAKMIP1* | 0.892981445 | 1.65E-06 | 4.66E-05 | 0.008006659 |
| Obesity | t_cell | *MR1* | 4.033447054 | 0.002986547 | 0.010197556 | 1 |
| Obesity | t_cell | *CENPH* | -6.373958875 | 0.000306482 | 0.001641992 | 1 |
| Obesity | t_cell | *SCOC* | -3.962123242 | 0.006805118 | 0.020047621 | 1 |
| Obesity | t_cell | *SYCP2L* | -2.319458455 | 0.002881859 | 0.009930953 | 1 |
| Obesity | t_cell | *TMEM87B* | 3.845125847 | 0.018518011 | 0.045401409 | 1 |
| Obesity | t_cell | *LPCAT1* | 5.68399416 | 7.12E-06 | 9.54E-05 | 0.034550047 |
| Obesity | t_cell | *JAZF1* | 4.548605101 | 3.43E-06 | 6.39E-05 | 0.016623511 |
| Obesity | t_cell | *FBXO36* | -4.903969767 | 0.005076448 | 0.015708499 | 1 |
| Obesity | t_cell | *MCOLN2* | 1.526443492 | 1.78E-06 | 4.83E-05 | 0.008644618 |
| Obesity | t_cell | *SEMA3D* | -3.081113166 | 7.57E-05 | 0.000553176 | 0.367308884 |
| Obesity | t_cell | *ANKRD29* | -3.753925187 | 0.000115834 | 0.00076052 | 0.562024623 |
| Obesity | t_cell | *DNAAF1* | -4.279853491 | 0.003002872 | 0.010246085 | 1 |
| Obesity | t_cell | *ANKH* | 3.531551131 | 0.008022854 | 0.022925141 | 1 |
| Obesity | t_cell | *UBASH3B* | 1.889657263 | 7.13E-05 | 0.000526489 | 0.345903303 |
| Obesity | t_cell | *ROBO4* | -4.046996181 | 0.000120121 | 0.000781273 | 0.582829429 |
| Obesity | t_cell | *PRKCA* | 3.368017129 | 0.008596967 | 0.024195178 | 1 |
| Obesity | t_cell | *NEIL2* | 2.696588364 | 0.002526776 | 0.008942317 | 1 |
| Obesity | t_cell | *PGM5* | -3.62025233 | 0.000137392 | 0.000871139 | 0.66662606 |
| Obesity | t_cell | *CCSAP* | 5.051604956 | 4.09E-06 | 6.86E-05 | 0.019827372 |
| Obesity | t_cell | *GBP5* | 0.804198985 | 1.03E-05 | 0.000121742 | 0.049747763 |
| Obesity | t_cell | *PDE1C* | -3.844865996 | 0.000165013 | 0.001007239 | 0.800643262 |
| Obesity | t_cell | *FLCN* | -2.787970604 | 0.002748223 | 0.00956555 | 1 |
| Obesity | t_cell | *OXNAD1* | 3.050150882 | 1.05E-05 | 0.000123607 | 0.050802567 |
| Obesity | t_cell | *APCDD1* | -1.75736304 | 0.016412404 | 0.041069101 | 1 |
| Obesity | t_cell | *ANKRD40* | -4.518038664 | 0.010665629 | 0.029007642 | 1 |
| Obesity | t_cell | *CYP2U1* | -2.654416832 | 0.002279052 | 0.008209326 | 1 |
| Obesity | t_cell | *MARVELD1* | -5.41914836 | 4.59E-05 | 0.000370331 | 0.222568696 |
| Obesity | t_cell | *GOLGA7B* | 4.678352163 | 1.50E-06 | 4.50E-05 | 0.007270273 |
| Obesity | t_cell | *MOV10* | -6.592644391 | 0.000109906 | 0.000731499 | 0.533262539 |
| Obesity | t_cell | *SETD9* | -1.31168468 | 0.007849972 | 0.022497381 | 1 |
| Obesity | t_cell | *KDM8* | 6.28789009 | 0.00541352 | 0.016582323 | 1 |
| Obesity | t_cell | *FZD7* | -2.397994459 | 0.007190585 | 0.020992009 | 1 |
| Obesity | t_cell | *RRAGA* | 5.692220653 | 0.013599733 | 0.035098884 | 1 |

| | | | | | | |
|---|---|---|---|---|---|---|
| Obesity | t_cell | *PSD3* | -1.808434601 | 0.012804635 | 0.033474185 | 1 |
| Obesity | t_cell | *MMP16* | -2.192317026 | 2.82E-06 | 6.01E-05 | 0.013688909 |
| Obesity | t_cell | *PPEF2* | -3.184884983 | 0.011612194 | 0.03112838 | 1 |
| Obesity | t_cell | *RWDD2B* | -3.482432762 | 0.000142608 | 0.000896289 | 0.691935325 |
| Obesity | t_cell | *NAA11* | -5.539803445 | 9.10E-07 | 3.71E-05 | 0.004415134 |
| Obesity | t_cell | *TDRD9* | -12.71785269 | 6.60E-06 | 9.16E-05 | 0.0320361 |
| Obesity | t_cell | *PCDH1* | -1.059401582 | 0.008426251 | 0.023788136 | 1 |
| Obesity | t_cell | *RPL30* | 3.628973729 | 0.001949366 | 0.00727004 | 1 |
| Obesity | t_cell | *SUPV3L1* | -4.574448987 | 0.004864938 | 0.015133605 | 1 |
| Obesity | t_cell | *HKDC1* | 2.292113239 | 3.48E-06 | 6.39E-05 | 0.016891287 |
| Obesity | t_cell | *NODAL* | -4.157781979 | 4.15E-06 | 6.92E-05 | 0.020142495 |
| Obesity | t_cell | *ATAD2* | 2.848182354 | 0.002739489 | 0.009541995 | 1 |
| Obesity | t_cell | *ITGAD* | 1.605128128 | 5.21E-08 | 4.77E-06 | 0.000252869 |
| Obesity | t_cell | *MPV17L* | -4.394183827 | 0.000202019 | 0.001182503 | 0.980198368 |
| Obesity | t_cell | *RPUSD3* | -4.232486989 | 0.014497095 | 0.037021003 | 1 |
| Obesity | t_cell | *NMNAT2* | -4.071280774 | 0.003870724 | 0.012495511 | 1 |
| Obesity | t_cell | *SLC6A1* | -8.472236549 | 2.09E-05 | 0.000200868 | 0.101355306 |
| Obesity | t_cell | *STEAP2* | -2.870410917 | 2.28E-07 | 1.35E-05 | 0.001107671 |
| Obesity | t_cell | *CLDN12* | -6.209656435 | 0.003171554 | 0.010671553 | 1 |
| Obesity | t_cell | *MMP14* | -5.559455177 | 0.00037124 | 0.001908112 | 1 |
| Obesity | t_cell | *CLEC18A* | -5.489816354 | 2.19E-14 | 1.06E-11 | 1.06E-10 |
| Obesity | t_cell | *DHRS4* | -6.486128543 | 0.001912558 | 0.007169311 | 1 |
| Obesity | t_cell | *SLC34A2* | 13.42333037 | 7.24E-06 | 9.65E-05 | 0.035130267 |
| Obesity | t_cell | *AP3S2* | -4.037516724 | 0.014356586 | 0.036758922 | 1 |
| Obesity | t_cell | *C12orf43* | -4.360964129 | 0.002576028 | 0.009054031 | 1 |
| Obesity | t_cell | *LDLRAP1* | 2.631094955 | 0.00782251 | 0.022431927 | 1 |
| Obesity | t_cell | *PAFAH2* | 1.989713795 | 0.012532371 | 0.032939904 | 1 |
| Obesity | t_cell | *EXTL1* | -30.4760685 | 3.13E-06 | 6.09E-05 | 0.015193425 |
| Obesity | t_cell | *RHPN1* | 2.80512965 | 0.000714975 | 0.003233518 | 1 |
| Obesity | t_cell | *DZIP1L* | -3.819535173 | 0.000311334 | 0.001653498 | 1 |
| Obesity | t_cell | *FANCC* | -3.599751795 | 0.002939542 | 0.010086747 | 1 |
| Obesity | t_cell | *CLSTN2* | -3.210869221 | 0.000106573 | 0.000714214 | 0.517090801 |
| Obesity | t_cell | *FAM86C1* | -14.54223139 | 0.000798694 | 0.003529383 | 1 |
| Obesity | t_cell | *PPP1R9A* | -3.629056573 | 0.000793608 | 0.003510105 | 1 |
| Obesity | t_cell | *COPG2* | -1.063196131 | 0.00523891 | 0.016139168 | 1 |
| Obesity | t_cell | *RNF166* | 1.697214597 | 0.000219071 | 0.001251982 | 1 |
| Obesity | t_cell | *SPATA2L* | 3.215695328 | 0.013302161 | 0.034514485 | 1 |
| Obesity | t_cell | *SLC5A11* | -1.988950208 | 1.43E-06 | 4.48E-05 | 0.006950899 |
| Obesity | t_cell | *CDC42SE2* | 1.980301791 | 2.84E-06 | 6.01E-05 | 0.013763868 |
| Obesity | t_cell | *HK2* | -2.632268163 | 0.019185457 | 0.04659051 | 1 |
| Obesity | t_cell | *TMEM69* | -5.741253557 | 0.002132142 | 0.007801773 | 1 |
| Obesity | t_cell | *ACE* | -4.509282284 | 1.92E-05 | 0.000190799 | 0.093300923 |
| Obesity | t_cell | *TEPP* | 8.552649209 | 0.00176108 | 0.006701775 | 1 |
| Obesity | t_cell | *ZFYVE28* | 2.260902331 | 5.47E-06 | 8.10E-05 | 0.026563932 |
| Obesity | t_cell | *C16orf86* | -2.992767136 | 6.92E-05 | 0.000516076 | 0.335965426 |
| Obesity | t_cell | *LYPD5* | -3.90772691 | 7.34E-06 | 9.70E-05 | 0.035613944 |
| Obesity | t_cell | *CCDC107* | 4.034653989 | 0.000835367 | 0.003638422 | 1 |
| Obesity | t_cell | *ZNF233* | -3.567305884 | 0.003165832 | 0.010671553 | 1 |
| Obesity | t_cell | *CPAMD8* | -3.114561682 | 0.001569034 | 0.006075782 | 1 |
| Obesity | t_cell | *ANKLE1* | -4.733648366 | 0.01834752 | 0.045028915 | 1 |

| | | | | | | |
|---|---|---|---|---|---|---|
| Obesity | t_cell | *CCDC58* | -5.716296108 | 0.003171275 | 0.010671553 | 1 |
| Obesity | t_cell | *CILP2* | -6.370363087 | 1.35E-06 | 4.35E-05 | 0.006553955 |
| Obesity | t_cell | *SLC37A1* | 5.519484558 | 0.00057391 | 0.002727336 | 1 |
| Obesity | t_cell | *PDE9A* | -2.199743733 | 0.007979007 | 0.022826734 | 1 |
| Obesity | t_cell | *C21orf2* | -1.32989979 | 0.015686871 | 0.039498028 | 1 |
| Obesity | t_cell | *MCM3AP* | -2.588861816 | 0.004868125 | 0.015133605 | 1 |
| Obesity | t_cell | *CACFD1* | -6.530580624 | 1.24E-05 | 0.000140645 | 0.060336556 |
| Obesity | t_cell | *S100A1* | 7.370289228 | 0.000610087 | 0.002862808 | 1 |
| Obesity | t_cell | *PAQR6* | -1.923267924 | 0.011608507 | 0.03112838 | 1 |
| Obesity | t_cell | *CCR5* | 1.743318325 | 1.22E-06 | 4.13E-05 | 0.005900093 |
| Obesity | t_cell | *NBEAL2* | 4.283294616 | 4.83E-08 | 4.51E-06 | 0.000234533 |
| Obesity | t_cell | *FCRL3* | 1.562621812 | 2.71E-06 | 6.01E-05 | 0.013144493 |
| Obesity | t_cell | *FGFR4* | -9.162032556 | 5.83E-07 | 2.78E-05 | 0.002830651 |
| Obesity | t_cell | *TONSL* | -6.268211089 | 1.39E-07 | 9.95E-06 | 0.000676793 |
| Obesity | t_cell | *MUM1* | -3.02807743 | 0.005662906 | 0.017226595 | 1 |
| Obesity | t_cell | *ZNF333* | -4.337630484 | 0.011771323 | 0.03146802 | 1 |
| Obesity | t_cell | *RPL8* | 3.882757701 | 0.003686945 | 0.011989984 | 1 |
| Obesity | t_cell | *ABCF3* | 2.29463637 | 0.009795225 | 0.02698832 | 1 |
| Obesity | t_cell | *BDH1* | 3.760019986 | 0.001460599 | 0.005729044 | 1 |
| Obesity | t_cell | *TMEM143* | -5.050644085 | 0.002683615 | 0.009367554 | 1 |
| Obesity | t_cell | *CCL16* | -2.15826491 | 0.019123994 | 0.046487784 | 1 |
| Obesity | t_cell | *ALDH16A1* | 1.852567656 | 0.006259484 | 0.018782324 | 1 |
| Obesity | t_cell | *SIGLEC11* | -1.115580402 | 0.014800649 | 0.037637709 | 1 |
| Obesity | t_cell | *PLCD3* | -5.007564683 | 2.01E-05 | 0.000196799 | 0.097593631 |
| Obesity | t_cell | *IP6K3* | -20.98101206 | 8.05E-05 | 0.000579706 | 0.39072218 |
| Obesity | t_cell | *ASGR2* | -13.16558543 | 1.03E-05 | 0.000121742 | 0.049792394 |
| Obesity | t_cell | *POLR3K* | 5.521966745 | 0.002028932 | 0.007503338 | 1 |
| Obesity | t_cell | *WDR90* | -4.105650821 | 1.35E-05 | 0.000149972 | 0.065687808 |
| Obesity | t_cell | *MEIOB* | -2.082064069 | 0.003726589 | 0.012086502 | 1 |
| Obesity | t_cell | *SHANK2* | -3.544744148 | 7.01E-05 | 0.000520823 | 0.340097621 |
| Obesity | t_cell | *ASRGL1* | -6.806703879 | 0.001377232 | 0.005459421 | 1 |
| Obesity | t_cell | *TPCN2* | -3.283585951 | 0.000590538 | 0.002792681 | 1 |
| Obesity | t_cell | *PDZK1IP1* | 9.879862598 | 0.007017257 | 0.02062249 | 1 |
| Obesity | t_cell | *SLC1A7* | -1.303131114 | 0.00942025 | 0.0260885 | 1 |
| Obesity | t_cell | *C1orf123* | -5.456114511 | 0.001432084 | 0.005630853 | 1 |
| Obesity | t_cell | *ZSWIM5* | 4.295024331 | 0.000274531 | 0.001508519 | 1 |
| Obesity | t_cell | *SLC25A34* | -4.010591856 | 3.91E-06 | 6.73E-05 | 0.018988224 |
| Obesity | t_cell | *PDPN* | -3.331271581 | 7.86E-06 | 0.000100385 | 0.038126939 |
| Obesity | t_cell | *LAPTM5* | 5.408342507 | 0.000159198 | 0.000976239 | 0.77243057 |
| Obesity | t_cell | *SDC3* | -3.159892696 | 0.017173629 | 0.042665873 | 1 |
| Obesity | t_cell | *SYNC* | -3.234190296 | 0.007171526 | 0.020961592 | 1 |
| Obesity | t_cell | *CAMK2N1* | 2.550653302 | 0.010759167 | 0.029212914 | 1 |
| Obesity | t_cell | *OMA1* | 3.944217902 | 0.008736985 | 0.024489804 | 1 |
| Obesity | t_cell | *NEXN* | -2.648256166 | 0.02004846 | 0.04832346 | 1 |
| Obesity | t_cell | *LRRIQ3* | -5.150478389 | 0.002044629 | 0.007548558 | 1 |
| Obesity | t_cell | *TYW3* | -4.372313408 | 0.011032439 | 0.029776797 | 1 |
| Obesity | t_cell | *HFM1* | -2.792153861 | 0.000155635 | 0.000959521 | 0.75514332 |
| Obesity | t_cell | *TRIM58* | -1.99459659 | 0.000525419 | 0.002529101 | 1 |
| Obesity | t_cell | *FCRLB* | 8.696201293 | 2.30E-05 | 0.000218314 | 0.111776552 |
| Obesity | t_cell | *TDRD5* | -5.392853802 | 2.39E-06 | 5.69E-05 | 0.011606815 |

| | | | | | | |
|---|---|---|---|---|---|---|
| Obesity | t_cell | *PKDCC* | -1.840657637 | 0.010531999 | 0.028693613 | 1 |
| Obesity | t_cell | *HAAO* | -4.322202665 | 0.001049361 | 0.004362896 | 1 |
| Obesity | t_cell | *KIAA1841* | -3.591493029 | 0.01471708 | 0.03750382 | 1 |
| Obesity | t_cell | *KCNJ3* | -6.876894534 | 0.000726805 | 0.003280427 | 1 |
| Obesity | t_cell | *DUSP19* | -4.156120379 | 1.96E-06 | 5.12E-05 | 0.009529476 |
| Obesity | t_cell | *C2orf48* | -16.54256464 | 3.16E-05 | 0.000281171 | 0.153519266 |
| Obesity | t_cell | *NOSTRIN* | -2.134064741 | 0.000579007 | 0.002746181 | 1 |
| Obesity | t_cell | *INHBB* | -5.04179549 | 0.00012308 | 0.000798374 | 0.597183879 |
| Obesity | t_cell | *C1QTNF7* | -2.902726677 | 0.010924015 | 0.029577566 | 1 |
| Obesity | t_cell | *VPS72* | -4.98967996 | 0.013832839 | 0.035605801 | 1 |
| Obesity | t_cell | *TDRD10* | -3.769814756 | 6.96E-05 | 0.000517693 | 0.337536112 |
| Obesity | t_cell | *DCAF16* | -5.01850143 | 0.004047103 | 0.012952865 | 1 |
| Obesity | t_cell | *NIPAL1* | -3.184012413 | 2.08E-05 | 0.000200868 | 0.100927671 |
| Obesity | t_cell | *ANTXR2* | 4.600591792 | 1.16E-05 | 0.000132737 | 0.056141468 |
| Obesity | t_cell | *PBXIP1* | 3.563052405 | 1.04E-06 | 3.84E-05 | 0.00503964 |
| Obesity | t_cell | *CLDN1* | -2.346597808 | 0.002734447 | 0.00953813 | 1 |
| Obesity | t_cell | *DCST2* | 5.939033667 | 2.83E-05 | 0.000256257 | 0.137353487 |
| Obesity | t_cell | *LINC01116* | -3.529478552 | 0.000732183 | 0.00329245 | 1 |
| Obesity | t_cell | *LRRC58* | 4.726276227 | 0.006432009 | 0.01920499 | 1 |
| Obesity | t_cell | *LMOD1* | -4.552867628 | 1.30E-10 | 2.26E-08 | 6.33E-07 |
| Obesity | t_cell | *ARPC2* | 3.870059188 | 0.000683897 | 0.003124547 | 1 |
| Obesity | t_cell | *GLB1L* | -3.588287667 | 5.48E-05 | 0.000426564 | 0.26574962 |
| Obesity | t_cell | *GTPBP8* | 6.488581075 | 0.000602032 | 0.002833171 | 1 |
| Obesity | t_cell | *PPM1K* | 4.49609251 | 0.00039037 | 0.001981252 | 1 |
| Obesity | t_cell | *DCLK3* | 5.197405541 | 0.000267365 | 0.001474155 | 1 |
| Obesity | t_cell | *RPP14* | -5.140757432 | 0.005497649 | 0.01676891 | 1 |
| Obesity | t_cell | *DNASE1L3* | -2.267468466 | 0.011034353 | 0.029776797 | 1 |
| Obesity | t_cell | *IL17RC* | -4.245062423 | 2.94E-05 | 0.000264091 | 0.142873271 |
| Obesity | t_cell | *CPA3* | -5.255946014 | 4.22E-06 | 7.01E-05 | 0.020480653 |
| Obesity | t_cell | *CDCP1* | -3.824118326 | 7.26E-06 | 9.65E-05 | 0.035214258 |
| Obesity | t_cell | *CLEC3B* | -4.556782083 | 9.09E-05 | 0.000640611 | 0.44105591 |
| Obesity | t_cell | *TPRA1* | -5.620827179 | 0.000110965 | 0.000733516 | 0.538400605 |
| Obesity | t_cell | *DNALI1* | -4.612440912 | 1.09E-10 | 2.11E-08 | 5.28E-07 |
| Obesity | t_cell | *RFT1* | -3.943078816 | 1.96E-05 | 0.000192784 | 0.095042392 |
| Obesity | t_cell | *SFMBT1* | 5.236752767 | 5.86E-06 | 8.51E-05 | 0.028413896 |
| Obesity | t_cell | *METAP1* | 3.445683842 | 0.007071444 | 0.020756591 | 1 |
| Obesity | t_cell | *BDH2* | -2.954383141 | 0.002179092 | 0.007931697 | 1 |
| Obesity | t_cell | *ATRIP* | -1.346251728 | 0.006335439 | 0.018972846 | 1 |
| Obesity | t_cell | *RNF123* | 7.779130049 | 1.63E-22 | 7.92E-19 | 7.92E-19 |
| Obesity | t_cell | *HSPA4L* | -3.312092746 | 0.000568866 | 0.002706018 | 1 |
| Obesity | t_cell | *MST1R* | 3.257352362 | 7.48E-08 | 6.15E-06 | 0.000362869 |
| Obesity | t_cell | *TEX264* | -4.2015797 | 0.000176121 | 0.001057598 | 0.854539022 |
| Obesity | t_cell | *ETNPPL* | -21.54759839 | 4.75E-06 | 7.49E-05 | 0.023058826 |
| Obesity | t_cell | *ANXA5* | -3.274596696 | 0.017619152 | 0.043638655 | 1 |
| Obesity | t_cell | *CEP44* | -3.897429407 | 0.001390016 | 0.005496623 | 1 |
| Obesity | t_cell | *TMEM144* | -5.241217071 | 0.000215553 | 0.001238173 | 1 |
| Obesity | t_cell | *MOCS2* | 6.77387092 | 0.000282568 | 0.001542207 | 1 |
| Obesity | t_cell | *RNF180* | -4.390733355 | 0.000159353 | 0.000976239 | 0.773181124 |
| Obesity | t_cell | *F2RL2* | 2.552572513 | 1.54E-06 | 4.50E-05 | 0.007450413 |
| Obesity | t_cell | *CMBL* | -3.920055464 | 0.000195459 | 0.00115233 | 0.948367291 |

| | | | | | | |
|---|---|---|---|---|---|---|
| Obesity | t_cell | *SERINC5* | 3.142225317 | 0.007379129 | 0.021388013 | 1 |
| Obesity | t_cell | *CASP3* | 3.150490523 | 0.015148815 | 0.038382271 | 1 |
| Obesity | t_cell | *ERAP2* | 1.179803349 | 0.002492573 | 0.008840616 | 1 |
| Obesity | t_cell | *TLR3* | -3.944256373 | 4.51E-06 | 7.28E-05 | 0.021905088 |
| Obesity | t_cell | *GRIK2* | -1.089965401 | 0.004011128 | 0.012863181 | 1 |
| Obesity | t_cell | *RAET1E* | -4.910438377 | 1.94E-05 | 0.000191365 | 0.093960137 |
| Obesity | t_cell | *STK17A* | 1.819946566 | 2.14E-06 | 5.28E-05 | 0.01038634 |
| Obesity | t_cell | *RPS14* | 3.535528848 | 0.002854475 | 0.00986461 | 1 |
| Obesity | t_cell | *CAMLG* | 2.939841147 | 0.01885502 | 0.046115011 | 1 |
| Obesity | t_cell | *MIOS* | -3.712786321 | 0.009944833 | 0.02732295 | 1 |
| Obesity | t_cell | *CHMP4C* | -9.450978708 | 7.50E-06 | 9.81E-05 | 0.036394545 |
| Obesity | t_cell | *CTSB* | -1.48648142 | 0.001128571 | 0.004636603 | 1 |
| Obesity | t_cell | *SOX17* | -2.519968673 | 0.000143419 | 0.000897897 | 0.695869798 |
| Obesity | t_cell | *TNFRSF11B* | -5.899238391 | 4.02E-05 | 0.000335707 | 0.19506056 |
| Obesity | t_cell | *DEFB1* | 3.681774988 | 0.011235285 | 0.030218184 | 1 |
| Obesity | t_cell | *GPER1* | -3.254511212 | 0.017162705 | 0.042660577 | 1 |
| Obesity | t_cell | *ALDH7A1* | -3.156702899 | 0.002802866 | 0.009713933 | 1 |
| Obesity | t_cell | *SLC25A32* | -4.355569519 | 0.004306061 | 0.013646643 | 1 |
| Obesity | t_cell | *DCSTAMP* | -24.27254261 | 0.000312763 | 0.001654879 | 1 |
| Obesity | t_cell | *RPP25L* | -5.437380294 | 5.57E-07 | 2.72E-05 | 0.002702367 |
| Obesity | t_cell | *C9orf24* | -4.052119298 | 0.013680061 | 0.03524995 | 1 |
| Obesity | t_cell | *TMEM65* | 4.546231268 | 0.00059542 | 0.002813028 | 1 |
| Obesity | t_cell | *CCDC171* | -3.009143513 | 0.011803242 | 0.031518619 | 1 |
| Obesity | t_cell | *ZMAT4* | -7.863496828 | 1.63E-05 | 0.000168143 | 0.079098915 |
| Obesity | t_cell | *C8orf34* | -3.000166352 | 2.79E-06 | 6.01E-05 | 0.013523815 |
| Obesity | t_cell | *RASEF* | -1.591382977 | 0.000151434 | 0.000937189 | 0.73475642 |
| Obesity | t_cell | *AQP3* | 7.138691285 | 2.09E-06 | 5.25E-05 | 0.010139757 |
| Obesity | t_cell | *GJB2* | 12.4935191 | 1.47E-06 | 4.50E-05 | 0.007125558 |
| Obesity | t_cell | *CRYL1* | -3.777797346 | 0.001065132 | 0.004413341 | 1 |
| Obesity | t_cell | *NOXRED1* | -2.982787552 | 0.003108061 | 0.010530944 | 1 |
| Obesity | t_cell | *AKR1E2* | -4.07371267 | 0.000825186 | 0.003613542 | 1 |
| Obesity | t_cell | *PDZD8* | 3.16660046 | 0.002869384 | 0.009894989 | 1 |
| Obesity | t_cell | *RET* | -3.319078507 | 5.07E-06 | 7.73E-05 | 0.024596188 |
| Obesity | t_cell | *CASP7* | -2.053563191 | 0.014158893 | 0.036291046 | 1 |
| Obesity | t_cell | *PPP1R36* | -4.738214168 | 5.54E-05 | 0.000428547 | 0.268699199 |
| Obesity | t_cell | *VWA2* | -4.585190144 | 0.001308422 | 0.005233687 | 1 |
| Obesity | t_cell | *HSPA12A* | -1.890000665 | 0.002487505 | 0.008829096 | 1 |
| Obesity | t_cell | *UBTD1* | -4.283211212 | 0.000372691 | 0.001913539 | 1 |
| Obesity | t_cell | *OTOGL* | -2.572666541 | 0.001793064 | 0.006807471 | 1 |
| Obesity | t_cell | *PACSIN3* | -2.048960576 | 0.000135935 | 0.0008667 | 0.659558366 |
| Obesity | t_cell | *TTC7B* | -3.482920053 | 0.010973321 | 0.02966159 | 1 |
| Obesity | t_cell | *SLC39A13* | 6.538781585 | 0.000844596 | 0.003658911 | 1 |
| Obesity | t_cell | *RAPSN* | -6.153042097 | 1.97E-14 | 1.06E-11 | 9.56E-11 |
| Obesity | t_cell | *TC2N* | 1.520542068 | 2.94E-06 | 6.01E-05 | 0.014247882 |
| Obesity | t_cell | *IFI27L1* | -4.520470351 | 0.000349728 | 0.001816789 | 1 |
| Obesity | t_cell | *ARL5B* | -3.470472114 | 0.009604334 | 0.026537717 | 1 |
| Obesity | t_cell | *JAM3* | -3.746798186 | 4.26E-05 | 0.000350029 | 0.206867411 |
| Obesity | t_cell | *ZFYVE19* | 2.738213949 | 0.003123016 | 0.010566856 | 1 |
| Obesity | t_cell | *TRMT61A* | -4.661421563 | 4.90E-05 | 0.000389045 | 0.237706244 |
| Obesity | t_cell | *LARP6* | -3.372754881 | 0.003066892 | 0.010427862 | 1 |

| | | | | | | |
|---|---|---|---|---|---|---|
| Obesity | t_cell | *GABRB3* | -2.164080566 | 0.000646773 | 0.002980193 | 1 |
| Obesity | t_cell | *CYB5R2* | -1.737343005 | 0.003666818 | 0.01193253 | 1 |
| Obesity | t_cell | *RPL27A* | 4.073652517 | 0.008677431 | 0.024379209 | 1 |
| Obesity | t_cell | *CENPN* | -8.240935527 | 9.61E-05 | 0.000664101 | 0.466198775 |
| Obesity | t_cell | *PRKCB* | 1.681538415 | 5.61E-07 | 2.72E-05 | 0.002723202 |
| Obesity | t_cell | *NDST2* | 42.83130231 | 1.98E-05 | 0.000194573 | 0.096118998 |
| Obesity | t_cell | *CLEC4D* | -5.751949356 | 0.000954063 | 0.004039365 | 1 |
| Obesity | t_cell | *ZSCAN21* | -2.819851307 | 4.28E-05 | 0.000350835 | 0.207847044 |
| Obesity | t_cell | *RIMKLB* | -3.449416448 | 2.84E-05 | 0.000256327 | 0.1376477 |
| Obesity | t_cell | *CENPV* | -4.800860399 | 2.53E-05 | 0.000234774 | 0.12278655 |
| Obesity | t_cell | *SLFN5* | 3.560749527 | 3.45E-05 | 0.000301963 | 0.16758947 |
| Obesity | t_cell | *TMEM170A* | 4.958411609 | 0.00087622 | 0.003775682 | 1 |
| Obesity | t_cell | *STX3* | 3.286118412 | 0.00081205 | 0.003568907 | 1 |
| Obesity | t_cell | *PIP4K2C* | 8.756322486 | 1.02E-08 | 1.25E-06 | 4.97E-05 |
| Obesity | t_cell | *MS4A7* | -1.796300655 | 0.001575895 | 0.006097481 | 1 |
| Obesity | t_cell | *MS4A14* | -0.889919326 | 0.003933536 | 0.012664576 | 1 |
| Obesity | t_cell | *SMAD3* | 3.264858812 | 0.001889637 | 0.007118415 | 1 |
| Obesity | t_cell | *AKTIP* | 2.89847203 | 8.65E-06 | 0.000107634 | 0.041977407 |
| Obesity | t_cell | *CNPY4* | 4.270981795 | 0.001298418 | 0.005197958 | 1 |
| Obesity | t_cell | *MEI1* | 2.382780986 | 1.34E-07 | 9.95E-06 | 0.000652336 |
| Obesity | t_cell | *ENDOG* | 1.914380471 | 0.004109594 | 0.013130988 | 1 |
| Obesity | t_cell | *COQ7* | -3.16440023 | 0.020377799 | 0.04894707 | 1 |
| Obesity | t_cell | *GPRC5B* | -2.311816703 | 0.00737411 | 0.021386241 | 1 |
| Obesity | t_cell | *LOXHD1* | -2.391943556 | 0.001331813 | 0.005305383 | 1 |
| Obesity | t_cell | *MYO5B* | -3.399266158 | 0.000426228 | 0.002125447 | 1 |
| Obesity | t_cell | *SMG8* | -4.731112021 | 4.56E-07 | 2.31E-05 | 0.002210296 |
| Obesity | t_cell | *RAB8A* | 2.880616071 | 0.004563866 | 0.014368057 | 1 |
| Obesity | t_cell | *SPATA33* | -5.609778994 | 0.003378893 | 0.011167839 | 1 |
| Obesity | t_cell | *RPL13* | 3.031155381 | 0.008313886 | 0.023576258 | 1 |
| Obesity | t_cell | *TP53I13* | 5.080286599 | 0.001043595 | 0.004346373 | 1 |
| Obesity | t_cell | *TTYH1* | -8.12355444 | 0.000280322 | 0.001535126 | 1 |
| Obesity | t_cell | *CDC42EP5* | -2.142767412 | 2.91E-06 | 6.01E-05 | 0.01412229 |
| Obesity | t_cell | *SRR* | 1.754375156 | 7.54E-05 | 0.000552423 | 0.365704053 |
| Obesity | t_cell | *KLK5* | -5.259266261 | 1.52E-05 | 0.000161776 | 0.073964087 |
| Obesity | t_cell | *KLK6* | -16.85762356 | 3.60E-06 | 6.46E-05 | 0.017452205 |
| Obesity | t_cell | *SPRYD3* | -7.147912892 | 0.000802227 | 0.003538551 | 1 |
| Obesity | t_cell | *NUDT8* | -2.411520171 | 4.31E-07 | 2.25E-05 | 0.002090933 |
| Obesity | t_cell | *MGAT5B* | -5.572441795 | 0.005810736 | 0.017599059 | 1 |
| Obesity | t_cell | *ZNF598* | 5.094572749 | 6.82E-07 | 3.06E-05 | 0.003308818 |
| Obesity | t_cell | *ATG16L2* | 5.09930339 | 0.002007 | 0.007444928 | 1 |
| Obesity | t_cell | *RPSA* | 2.581889416 | 0.002034636 | 0.007518699 | 1 |
| Obesity | t_cell | *NAALADL1* | -2.107347672 | 0.001617115 | 0.006242037 | 1 |
| Obesity | t_cell | *SCARA5* | -2.052996817 | 0.000309247 | 0.001648863 | 1 |
| Obesity | t_cell | *KIAA1586* | -4.459066845 | 0.000117304 | 0.000769136 | 0.569160533 |
| Obesity | t_cell | *FAM83B* | -21.09894103 | 8.82E-07 | 3.69E-05 | 0.00427893 |
| Obesity | t_cell | *PXK* | -3.988779053 | 0.003210318 | 0.010772103 | 1 |
| Obesity | t_cell | *ACOX2* | -3.966944268 | 0.0004624 | 0.002268516 | 1 |
| Obesity | t_cell | *FAM107A* | -3.774986552 | 0.000229684 | 0.001304948 | 1 |
| Obesity | t_cell | *DTYMK* | 5.666071285 | 0.001904286 | 0.007145861 | 1 |
| Obesity | t_cell | *KLHL30* | -16.73661757 | 0.003222068 | 0.010785819 | 1 |

| | | | | | | |
|---|---|---|---|---|---|---|
| Obesity | t_cell | *CCDC110* | -4.029211902 | 3.00E-06 | 6.01E-05 | 0.014537846 |
| Obesity | t_cell | *FNTA* | 3.320616958 | 0.007144501 | 0.020907791 | 1 |
| Obesity | t_cell | *SERINC2* | -8.441278304 | 1.06E-06 | 3.84E-05 | 0.005140411 |
| Obesity | t_cell | *TMUB2* | -4.605033156 | 0.000178206 | 0.001067473 | 0.864653117 |
| Obesity | t_cell | *ZSWIM1* | 0.836948522 | 0.012041166 | 0.031925541 | 1 |
| Obesity | t_cell | *NBPF9* | -7.331050697 | 0.000202039 | 0.001182503 | 0.980295042 |
| Obesity | t_cell | *NDUFS5* | 5.321719039 | 0.000625713 | 0.002908007 | 1 |
| Obesity | t_cell | *TCTN2* | -0.956032068 | 0.003587787 | 0.011718086 | 1 |
| Obesity | t_cell | *MFSD7* | -5.999469964 | 9.38E-06 | 0.00011357 | 0.045505959 |
| Obesity | t_cell | *KLK7* | -4.672604548 | 0.000387416 | 0.001971965 | 1 |
| Obesity | t_cell | *C8orf46* | 0.735303463 | 0.00347463 | 0.011406567 | 1 |
| Obesity | t_cell | *CPT1C* | -2.246785292 | 0.018646717 | 0.045670808 | 1 |
| Obesity | t_cell | *PCSK9* | -12.62517838 | 2.82E-05 | 0.000255776 | 0.136840187 |
| Obesity | t_cell | *XPO6* | 3.56898144 | 0.001537521 | 0.005972819 | 1 |
| Obesity | t_cell | *RGS14* | 1.431611416 | 0.000255478 | 0.001429733 | 1 |
| Obesity | t_cell | *RAB24* | 5.742731542 | 1.51E-05 | 0.000160334 | 0.073112273 |
| Obesity | t_cell | *THBS3* | -3.171204804 | 0.01188408 | 0.031685341 | 1 |
| Obesity | t_cell | *NPIPB3* | -4.558983914 | 0.000623042 | 0.002901153 | 1 |
| Obesity | t_cell | *SH3TC2* | -1.662528345 | 0.007124371 | 0.020861464 | 1 |
| Obesity | t_cell | *GPRIN1* | 8.350210573 | 5.45E-08 | 4.90E-06 | 0.000264485 |
| Obesity | t_cell | *CD52* | 1.266311625 | 2.79E-05 | 0.000253714 | 0.135483013 |
| Obesity | t_cell | *CCDC8* | -2.921335549 | 7.12E-05 | 0.000526304 | 0.345255108 |
| Obesity | t_cell | *ZEB2* | -2.21237677 | 0.015005535 | 0.038064982 | 1 |
| Obesity | t_cell | *HINT1* | 4.00215233 | 0.016156778 | 0.040513016 | 1 |
| Obesity | t_cell | *CLIC3* | 10.27782178 | 0.000134209 | 0.000856989 | 0.651183693 |
| Obesity | t_cell | *INO80E* | -5.76109062 | 2.95E-15 | 2.87E-12 | 1.43E-11 |
| Obesity | t_cell | *RGPD8* | -0.826658749 | 0.000566167 | 0.00269582 | 1 |
| Obesity | t_cell | *LUZP1* | 4.110308097 | 0.003561 | 0.011658552 | 1 |
| Obesity | t_cell | *LRRC45* | -2.229793139 | 0.000393225 | 0.001993654 | 1 |
| Obesity | t_cell | *CSGALNACT2* | 4.763191613 | 3.08E-05 | 0.00027532 | 0.149498882 |
| Obesity | t_cell | *CTNND2* | -3.392274842 | 3.99E-06 | 6.77E-05 | 0.019344732 |
| Obesity | t_cell | *CALML6* | -46.51216609 | 7.23E-08 | 6.05E-06 | 0.000350992 |
| Obesity | t_cell | *TPST1* | -2.054725099 | 0.014935298 | 0.037940349 | 1 |
| Obesity | t_cell | *NIPA1* | 4.990981447 | 0.003073305 | 0.010437071 | 1 |
| Obesity | t_cell | *RNF150* | -2.572760256 | 0.005573263 | 0.016975186 | 1 |
| Obesity | t_cell | *NFRKB* | -5.117641955 | 0.0008978 | 0.003848166 | 1 |
| Obesity | t_cell | *SEMA3E* | -2.531143625 | 9.92E-05 | 0.000681313 | 0.481323261 |
| Obesity | t_cell | *ADORA2B* | -2.290626208 | 1.45E-08 | 1.65E-06 | 7.01E-05 |
| Obesity | t_cell | *METTL7B* | -3.971671392 | 0.001669393 | 0.006413219 | 1 |
| Obesity | t_cell | *DNAJC18* | -2.180311591 | 0.00838043 | 0.023751078 | 1 |
| Obesity | t_cell | *HSD17B13* | -4.657483064 | 5.91E-06 | 8.56E-05 | 0.028671097 |
| Obesity | t_cell | *ELOVL6* | 4.723494776 | 4.86E-05 | 0.000386315 | 0.235608129 |
| Obesity | t_cell | *SERPINB9* | 4.048476578 | 0.0003697 | 0.001902209 | 1 |
| Obesity | t_cell | *SMAGP* | -1.064337703 | 0.008392823 | 0.023758446 | 1 |
| Obesity | t_cell | *CDH2* | -3.612209358 | 0.000429206 | 0.002135907 | 1 |
| Obesity | t_cell | *EMB* | 2.130612097 | 1.94E-05 | 0.00019143 | 0.094183642 |
| Obesity | t_cell | *GTSF1* | 2.068163125 | 7.11E-06 | 9.54E-05 | 0.034473991 |
| Obesity | t_cell | *TRABD* | 4.075357223 | 0.002009537 | 0.007448643 | 1 |
| Obesity | t_cell | *TMEM133* | -2.413490648 | 0.00669333 | 0.019802462 | 1 |
| Obesity | t_cell | *HOXB9* | -6.736283284 | 0.000192089 | 0.001139385 | 0.932016888 |

| | | | | | | |
|---|---|---|---|---|---|---|
| Obesity | t_cell | *TTLL6* | -15.40165925 | 1.64E-17 | 2.65E-14 | 7.95E-14 |
| Obesity | t_cell | *KCNS3* | -1.286581564 | 0.009665148 | 0.026675369 | 1 |
| Obesity | t_cell | *RPS9* | 1.961636697 | 0.017143495 | 0.042634669 | 1 |
| Obesity | t_cell | *NDUFA3* | 6.257470547 | 0.001755907 | 0.006692101 | 1 |
| Obesity | t_cell | *PAQR8* | 1.692699645 | 0.002865753 | 0.009891586 | 1 |
| Obesity | t_cell | *NUDT6* | -3.389945255 | 0.000361958 | 0.001872302 | 1 |
| Obesity | t_cell | *ZNF415* | -4.296478813 | 0.002935519 | 0.01008007 | 1 |
| Obesity | t_cell | *PYGO1* | -3.576583165 | 0.001915874 | 0.007172701 | 1 |
| Obesity | t_cell | *TSNARE1* | 4.966095477 | 4.29E-05 | 0.000350835 | 0.208045198 |
| Obesity | t_cell | *FPR1* | -4.562062151 | 0.000283679 | 0.001546529 | 1 |
| Obesity | t_cell | *SOX7* | -3.748678825 | 1.73E-05 | 0.00017603 | 0.083790483 |
| Obesity | t_cell | *TRMT61B* | 4.498139443 | 0.000143006 | 0.000896466 | 0.693865022 |
| Obesity | t_cell | *MORN4* | -1.130457806 | 0.007509236 | 0.021674004 | 1 |
| Obesity | t_cell | *LURAP1* | -1.651067804 | 0.00013987 | 0.000882507 | 0.67864779 |
| Obesity | t_cell | *PDE7B* | -3.213719845 | 0.001263996 | 0.005098012 | 1 |
| Obesity | t_cell | *HOPX* | 2.087219765 | 1.47E-05 | 0.000158409 | 0.071480256 |
| Obesity | t_cell | *PPID* | 4.882732275 | 0.017306925 | 0.042953044 | 1 |
| Obesity | t_cell | *NMUR1* | 10.08548933 | 8.96E-06 | 0.000110312 | 0.043494967 |
| Obesity | t_cell | *PIK3CD* | 2.82535238 | 1.68E-07 | 1.09E-05 | 0.000812879 |
| Obesity | t_cell | *RGS19* | 1.857782881 | 9.33E-06 | 0.000113501 | 0.045286702 |
| Obesity | t_cell | *ANO5* | -3.20212233 | 0.000205341 | 0.001198006 | 0.996312644 |
| Obesity | t_cell | *VAT1L* | -3.030444509 | 3.08E-06 | 6.09E-05 | 0.014953246 |
| Obesity | t_cell | *LRRC34* | -1.174148024 | 0.004291561 | 0.01362739 | 1 |
| Obesity | t_cell | *PAH* | -15.23970699 | 0.000434415 | 0.002155196 | 1 |
| Obesity | t_cell | *ATPAF2* | -3.925527593 | 0.003693938 | 0.012004681 | 1 |
| Obesity | t_cell | *ANKRD20A4* | -5.959527152 | 3.80E-06 | 6.61E-05 | 0.018449189 |
| Obesity | t_cell | *SERF1A* | -16.92843517 | 1.36E-06 | 4.35E-05 | 0.006590472 |
| Obesity | t_cell | *EIF2AK3* | 3.695058503 | 0.014485035 | 0.037009683 | 1 |
| Obesity | t_cell | *CD8B* | 1.085045263 | 2.79E-06 | 6.01E-05 | 0.013558697 |
| Obesity | t_cell | *SLFN12* | -2.521014217 | 0.009184632 | 0.025508777 | 1 |
| Obesity | t_cell | *MALT1* | 3.930866835 | 0.013336367 | 0.034566267 | 1 |
| Obesity | t_cell | *MBOAT1* | 3.950293604 | 0.01190574 | 0.03169824 | 1 |
| Obesity | t_cell | *CXCR6* | 0.974582752 | 2.63E-06 | 5.93E-05 | 0.012774353 |
| Obesity | t_cell | *C1QTNF4* | -3.438817338 | 5.20E-17 | 6.31E-14 | 2.52E-13 |
| Obesity | t_cell | *HINFP* | -4.4579881 | 0.004192807 | 0.013331259 | 1 |
| Obesity | t_cell | *TP53RK* | 7.399718197 | 1.16E-05 | 0.000132737 | 0.056280352 |
| Obesity | t_cell | *FIBP* | 5.266039988 | 0.000200395 | 0.001177136 | 0.972314345 |
| Obesity | t_cell | *CTSW* | -0.569797438 | 0.000175472 | 0.001055008 | 0.851391511 |
| Obesity | t_cell | *SNTG2* | -1.412710456 | 0.018195414 | 0.044791551 | 1 |
| Obesity | t_cell | *ZFAND4* | -3.811522594 | 0.003698557 | 0.012011647 | 1 |
| Obesity | t_cell | *FAM71D* | -2.775798076 | 0.002975044 | 0.010179769 | 1 |
| Obesity | t_cell | *FUT10* | -2.140888642 | 8.72E-05 | 0.000621546 | 0.423273097 |
| Obesity | t_cell | *ZNF596* | -5.100299404 | 8.98E-05 | 0.000634826 | 0.43549032 |
| Obesity | t_cell | *CFL1* | 4.131117426 | 0.009369983 | 0.02596411 | 1 |
| Obesity | t_cell | *SNX32* | 1.62520996 | 0.000611058 | 0.002864593 | 1 |
| Obesity | t_cell | *ZNF621* | -4.844056968 | 0.001226897 | 0.004989862 | 1 |
| Obesity | t_cell | *MYEOV* | -5.283773979 | 0.002807773 | 0.009723993 | 1 |
| Obesity | t_cell | *LCLAT1* | 3.473225957 | 0.001922024 | 0.007190179 | 1 |
| Obesity | t_cell | *SH3RF3* | -3.520938933 | 0.002233341 | 0.008074644 | 1 |
| Obesity | t_cell | *GXYLT2* | -2.851692697 | 0.008074706 | 0.02301908 | 1 |

| | | | | | | |
|---|---|---|---|---|---|---|
| Obesity | t_cell | *DCAKD* | -3.655115756 | 0.019162511 | 0.046558089 | 1 |
| Obesity | t_cell | *PARP15* | 1.118014759 | 1.24E-05 | 0.000140623 | 0.060186527 |
| Obesity | t_cell | *IQCB1* | -3.608691227 | 0.002467773 | 0.008774918 | 1 |
| Obesity | t_cell | *MAP3K11* | -3.886551443 | 0.003451765 | 0.011354552 | 1 |
| Obesity | t_cell | *NDNF* | -3.052009096 | 0.001896376 | 0.00712798 | 1 |
| Obesity | t_cell | *CCDC36* | -2.033266372 | 0.001243028 | 0.005038575 | 1 |
| Obesity | t_cell | *PPP1R14B* | -7.793558657 | 7.71E-05 | 0.000561203 | 0.374322645 |
| Obesity | t_cell | *ZNF417* | -3.620801548 | 2.07E-05 | 0.000200844 | 0.100582098 |
| Obesity | t_cell | *MST1* | -1.253733544 | 0.000955207 | 0.004040685 | 1 |
| Obesity | t_cell | *GMPPB* | 4.676838088 | 0.006512618 | 0.01937414 | 1 |
| Obesity | t_cell | *NUDT18* | -6.06522877 | 4.78E-06 | 7.51E-05 | 0.023210452 |
| Obesity | t_cell | *SULT1B1* | -5.533228374 | 2.79E-06 | 6.01E-05 | 0.013556691 |
| Obesity | t_cell | *CD7* | 1.934934723 | 8.28E-07 | 3.53E-05 | 0.004019699 |
| Obesity | t_cell | *RNF213* | 2.79667339 | 0.003223297 | 0.010785819 | 1 |
| Obesity | t_cell | *GPR160* | -3.162368937 | 0.000266109 | 0.00147225 | 1 |
| Obesity | t_cell | *GOLIM4* | -3.761182854 | 0.00633862 | 0.018972846 | 1 |
| Obesity | t_cell | *XXYLT1* | -4.365171215 | 0.00075589 | 0.003367839 | 1 |
| Obesity | t_cell | *SLC25A30* | -3.906817936 | 9.74E-05 | 0.000671214 | 0.472534824 |
| Obesity | t_cell | *CTU2* | 2.193445554 | 0.004166726 | 0.013274429 | 1 |
| Obesity | t_cell | *GLIS1* | -5.14092439 | 4.36E-05 | 0.000354062 | 0.211729064 |
| Obesity | t_cell | *ZNF497* | -14.20395099 | 3.70E-06 | 6.54E-05 | 0.01793885 |
| Obesity | t_cell | *TRAM1L1* | -3.267428171 | 0.013541345 | 0.035004051 | 1 |
| Obesity | t_cell | *SH3PXD2B* | -3.414774773 | 0.000125325 | 0.000808611 | 0.608075331 |
| Obesity | t_cell | *KIAA1551* | 2.405613961 | 4.32E-05 | 0.000351724 | 0.209697628 |
| Obesity | t_cell | *FGFBP3* | -10.61803421 | 0.000309683 | 0.001649377 | 1 |
| Obesity | t_cell | *SRP72* | 2.820852321 | 0.002507161 | 0.008885861 | 1 |
| Obesity | t_cell | *NLRP6* | 4.334580255 | 9.69E-05 | 0.000668798 | 0.470165124 |
| Obesity | t_cell | *SEZ6L2* | 3.981615423 | 2.82E-06 | 6.01E-05 | 0.013659099 |
| Obesity | t_cell | *KCTD13* | 6.187567414 | 1.29E-10 | 2.26E-08 | 6.25E-07 |
| Obesity | t_cell | *SLC22A1* | -6.206943074 | 9.83E-06 | 0.000118317 | 0.04768174 |
| Obesity | t_cell | *TVP23C* | -8.796083776 | 0.000110847 | 0.000733516 | 0.537830089 |
| Obesity | t_cell | *PACS1* | 1.934630215 | 0.011647596 | 0.031206039 | 1 |
| Obesity | t_cell | *ZNF519* | -3.495230252 | 0.001254049 | 0.005074766 | 1 |
| Obesity | t_cell | *BANF1* | -6.675498046 | 0.000909004 | 0.00388589 | 1 |
| Obesity | t_cell | *CHRNA7* | -3.198519963 | 2.25E-05 | 0.000213772 | 0.109237664 |
| Obesity | t_cell | *SCUBE2* | -5.544750775 | 1.71E-05 | 0.00017507 | 0.082983188 |
| Obesity | t_cell | *EIF1AD* | 6.273504731 | 0.00815342 | 0.023188975 | 1 |
| Obesity | t_cell | *ZNF25* | -4.487081998 | 1.85E-05 | 0.000186353 | 0.089821906 |
| Obesity | t_cell | *KCNE3* | -4.762737343 | 2.63E-05 | 0.000242394 | 0.127741719 |
| Obesity | t_cell | *C11orf68* | -5.69423135 | 0.001274231 | 0.005130761 | 1 |
| Obesity | t_cell | *TMEM70* | -3.410046985 | 0.014093766 | 0.036162322 | 1 |
| Obesity | t_cell | *RPS6KB2* | 7.028165997 | 4.45E-05 | 0.000360812 | 0.216126526 |
| Obesity | t_cell | *TEX26* | -5.093835958 | 0.002049922 | 0.007557921 | 1 |
| Obesity | t_cell | *PRIMA1* | -3.330047668 | 4.87E-06 | 7.55E-05 | 0.023619688 |
| Obesity | t_cell | *A2M* | 3.251041511 | 1.63E-05 | 0.000168143 | 0.079195124 |
| Obesity | t_cell | *DOK7* | -12.5398202 | 5.50E-05 | 0.000427306 | 0.267066023 |
| Obesity | t_cell | *ORAI3* | 6.040837674 | 1.23E-08 | 1.45E-06 | 5.95E-05 |
| Obesity | t_cell | *B3GALT6* | -9.914052768 | 2.09E-05 | 0.000200868 | 0.101319786 |
| Obesity | t_cell | *ZNF613* | -2.009028452 | 0.011011232 | 0.029747494 | 1 |
| Obesity | t_cell | *JAKMIP2* | 0.750132694 | 1.16E-05 | 0.000132737 | 0.056267393 |

| | | | | | | |
|---|---|---|---|---|---|---|
| Obesity | t_cell | *CCDC57* | -3.147732364 | 0.000317804 | 0.001668815 | 1 |
| Obesity | t_cell | *ZNF135* | -1.92359908 | 0.001528306 | 0.005951315 | 1 |
| Obesity | t_cell | *OR4N2* | -5.164958382 | 0.00011762 | 0.000769388 | 0.57069399 |
| Obesity | t_cell | *DNAJC30* | -5.974839871 | 0.000535868 | 0.002576839 | 1 |
| Obesity | t_cell | *SPRYD4* | -2.618031203 | 0.000423603 | 0.002116708 | 1 |
| Obesity | t_cell | *WDR25* | -4.473688931 | 0.000399032 | 0.002014674 | 1 |
| Obesity | t_cell | *B3GNT5* | -4.346666929 | 8.79E-05 | 0.000624816 | 0.42662711 |
| Obesity | t_cell | *BDNF* | -4.017282933 | 0.001406695 | 0.005553526 | 1 |
| Obesity | t_cell | *BOK* | -4.764422275 | 0.004542535 | 0.014336867 | 1 |
| Obesity | t_cell | *PFN4* | -1.727795257 | 0.001798529 | 0.006821991 | 1 |
| Obesity | t_cell | *RUFY1* | 2.742817584 | 0.013578751 | 0.035063384 | 1 |
| Obesity | t_cell | *C8G* | -45.98815336 | 2.78E-06 | 6.01E-05 | 0.013492373 |
| Obesity | t_cell | *MUC20* | -4.792234337 | 1.54E-05 | 0.000162814 | 0.074894667 |
| Obesity | t_cell | *C19orf18* | -2.641509185 | 0.001195231 | 0.004885644 | 1 |
| Obesity | t_cell | *TMEM80* | -4.713246388 | 0.0001777 | 0.001065758 | 0.862198014 |
| Obesity | t_cell | *DSCAML1* | -3.78088689 | 0.000417714 | 0.00209375 | 1 |
| Obesity | t_cell | *SAMD9L* | 2.253633473 | 0.002762544 | 0.009601622 | 1 |
| Obesity | t_cell | *MIEF2* | -5.441263207 | 1.49E-05 | 0.000158798 | 0.072094074 |
| Obesity | t_cell | *NIM1K* | 4.092025067 | 0.00078214 | 0.003465701 | 1 |
| Obesity | t_cell | *RBM44* | -3.375764045 | 0.002228295 | 0.008068424 | 1 |
| Obesity | t_cell | *SLC25A22* | -4.548043795 | 1.92E-05 | 0.000190799 | 0.093247191 |
| Obesity | t_cell | *MBOAT4* | 2.887669997 | 1.60E-06 | 4.59E-05 | 0.007754596 |
| Obesity | t_cell | *SLC35G5* | 25.23657605 | 2.22E-06 | 5.43E-05 | 0.010751824 |
| Obesity | t_cell | *FAM87B* | 8.918678538 | 1.70E-06 | 4.71E-05 | 0.008240333 |
| Obesity | t_cell | *PCDHB9* | -3.43657565 | 0.000154044 | 0.000950917 | 0.747421137 |
| Obesity | t_cell | *CENPBD1* | -4.143182881 | 3.68E-06 | 6.54E-05 | 0.01785538 |
| Obesity | t_cell | *RPS27* | 4.402588895 | 0.002163101 | 0.007891254 | 1 |
| Obesity | t_cell | *ADAMTSL1* | -2.366161922 | 0.000171344 | 0.001037904 | 0.831361037 |
| Obesity | t_cell | *GRAMD1C* | -2.325832807 | 0.001330543 | 0.005304681 | 1 |
| Obesity | t_cell | *ZNF114* | -5.153869059 | 4.32E-06 | 7.11E-05 | 0.020982505 |
| Obesity | t_cell | *SPINK6* | -7.438148636 | 4.60E-05 | 0.000370822 | 0.223313515 |
| Obesity | t_cell | *SH2B1* | -4.009411306 | 0.002929117 | 0.01006521 | 1 |
| Obesity | t_cell | *VN1R1* | -6.343592033 | 2.33E-06 | 5.61E-05 | 0.011328473 |
| Obesity | t_cell | *SH2D4B* | -7.168480408 | 6.18E-06 | 8.81E-05 | 0.029963746 |
| Obesity | t_cell | *RNF212* | -2.990947637 | 0.002370308 | 0.008481368 | 1 |
| Obesity | t_cell | *TMEM11* | -5.895164846 | 0.005165135 | 0.015932126 | 1 |
| Obesity | t_cell | *ZNF223* | -2.133203031 | 0.01295925 | 0.033805528 | 1 |
| Obesity | t_cell | *GLDC* | -4.830808526 | 0.008568882 | 0.024144145 | 1 |
| Obesity | t_cell | *CA8* | -4.055890465 | 0.000160646 | 0.000982917 | 0.779453098 |
| Obesity | t_cell | *ERN1* | 3.103309209 | 7.75E-06 | 9.97E-05 | 0.0376039 |
| Obesity | t_cell | *NSUN3* | 3.909280597 | 0.019986336 | 0.048197665 | 1 |
| Obesity | t_cell | *RPP25* | -2.94606004 | 1.89E-07 | 1.19E-05 | 0.000919141 |
| Obesity | t_cell | *STX19* | 2.526442215 | 0.017979632 | 0.044372927 | 1 |
| Obesity | t_cell | *MPI* | 3.39934942 | 0.00148378 | 0.005801208 | 1 |
| Obesity | t_cell | *TRIM73* | -14.3376575 | 6.37E-15 | 4.42E-12 | 3.09E-11 |
| Obesity | t_cell | *EFCAB13* | -2.74324965 | 0.0026024 | 0.009116854 | 1 |
| Obesity | t_cell | *EXOSC4* | -8.126703545 | 8.20E-07 | 3.52E-05 | 0.003977841 |
| Obesity | t_cell | *ZNF852* | -6.381906235 | 1.62E-08 | 1.75E-06 | 7.87E-05 |
| Obesity | t_cell | *HYI* | -1.432550521 | 0.019365286 | 0.046915394 | 1 |
| Obesity | t_cell | *C17orf62* | 6.982397137 | 3.50E-05 | 0.00030391 | 0.169885779 |

| | | | | | | |
|---|---|---|---|---|---|---|
| Obesity | t_cell | *GAK* | 3.884142284 | 0.019344434 | 0.046906144 | 1 |
| Obesity | t_cell | *TUFM* | 6.11144707 | 4.09E-08 | 3.97E-06 | 0.000198422 |
| Obesity | t_cell | *CTC1* | 3.945483425 | 0.000165036 | 0.001007239 | 0.800755057 |
| Obesity | t_cell | *C3orf38* | 5.340388568 | 3.87E-08 | 3.91E-06 | 0.000187716 |
| Obesity | t_cell | *CCDC89* | -9.330654479 | 6.65E-06 | 9.16E-05 | 0.03227881 |
| Obesity | t_cell | *HTR1F* | -8.914565596 | 0.000125032 | 0.000808349 | 0.606657484 |
| Obesity | t_cell | *ZNF664* | -5.882281663 | 0.002947563 | 0.010107121 | 1 |
| Obesity | t_cell | *CDH4* | -2.454793491 | 0.000700551 | 0.003182653 | 1 |
| Obesity | t_cell | *LINC00174* | -1.546184303 | 0.011106444 | 0.029921415 | 1 |
| Obesity | t_cell | *GEMIN4* | -2.833849451 | 0.003877554 | 0.012509238 | 1 |
| Obesity | t_cell | *ZFPM1* | 3.071291744 | 0.00031454 | 0.001658857 | 1 |
| Obesity | t_cell | *FCER1A* | -1.983313772 | 0.004137415 | 0.013200328 | 1 |
| Obesity | t_cell | *RPRML* | 8.504101031 | 0.001027496 | 0.004301474 | 1 |
| Obesity | t_cell | *GIPC3* | -19.27820798 | 1.35E-05 | 0.000149972 | 0.065537749 |
| Obesity | t_cell | *SSC5D* | -1.26325621 | 0.000730708 | 0.003289835 | 1 |
| Obesity | t_cell | *SEPT1* | 3.32527459 | 4.48E-06 | 7.28E-05 | 0.021760167 |
| Obesity | t_cell | *HCLS1* | 1.789911116 | 0.000490084 | 0.002380268 | 1 |
| Obesity | t_cell | *MTURN* | -3.660435527 | 0.009611676 | 0.026542888 | 1 |
| Obesity | t_cell | *GLIPR1L2* | -3.238547105 | 0.000930142 | 0.003955346 | 1 |
| Obesity | t_cell | *DEFB119* | -28.87482367 | 3.36E-05 | 0.000296216 | 0.162918725 |
| Obesity | t_cell | *SLC47A2* | -2.636039053 | 0.001234538 | 0.005012533 | 1 |
| Obesity | t_cell | *CHST13* | -10.82652252 | 0.001712812 | 0.006543752 | 1 |
| Obesity | t_cell | *ZDHHC20* | 4.329642671 | 0.000869603 | 0.003755344 | 1 |
| Obesity | t_cell | *PSMG4* | -3.183520478 | 0.005247835 | 0.016156406 | 1 |
| Obesity | t_cell | *ZNF443* | -7.215731488 | 1.02E-06 | 3.84E-05 | 0.004956896 |
| Obesity | t_cell | *KCTD2* | -3.286174855 | 0.006456774 | 0.019267077 | 1 |
| Obesity | t_cell | *NQO1* | -4.527099793 | 2.88E-06 | 6.01E-05 | 0.013956836 |
| Obesity | t_cell | *F2R* | 1.456614509 | 4.83E-05 | 0.000385627 | 0.234461064 |
| Obesity | t_cell | *MUC16* | -3.753742487 | 0.000267279 | 0.001474155 | 1 |
| Obesity | t_cell | *PENK* | -3.604821353 | 0.019446283 | 0.047035575 | 1 |
| Obesity | t_cell | *SLC25A41* | -9.28741008 | 1.50E-05 | 0.000160334 | 0.073005527 |
| Obesity | t_cell | *TMEM136* | -4.496252299 | 0.000271756 | 0.001495309 | 1 |
| Obesity | t_cell | *ZNF322* | -4.683463976 | 7.77E-05 | 0.000564129 | 0.377187168 |
| Obesity | t_cell | *TMEM45A* | -3.406824912 | 0.000189352 | 0.001124527 | 0.918738186 |
| Obesity | t_cell | *SGSH* | -6.966877773 | 0.008408366 | 0.023768148 | 1 |
| Obesity | t_cell | *C3orf58* | -4.493321452 | 0.001530217 | 0.00595398 | 1 |
| Obesity | t_cell | *AMIGO1* | 2.779877122 | 0.000460711 | 0.002262521 | 1 |
| Obesity | t_cell | *TMEM252* | -82.41106165 | 4.87E-06 | 7.55E-05 | 0.023640711 |
| Obesity | t_cell | *ODF3L2* | -7.379726585 | 3.49E-05 | 0.00030391 | 0.169569666 |
| Obesity | t_cell | *RELL1* | 3.232477695 | 0.015995469 | 0.040247376 | 1 |
| Obesity | t_cell | *CLDN7* | -3.935599404 | 4.07E-06 | 6.86E-05 | 0.019760398 |
| Obesity | t_cell | *ZNF329* | -4.730288939 | 0.002120598 | 0.00776539 | 1 |
| Obesity | t_cell | *IDH2* | 4.735594148 | 3.10E-05 | 0.000276247 | 0.15027826 |
| Obesity | t_cell | *WRB* | -1.509189337 | 0.010041347 | 0.027541332 | 1 |
| Obesity | t_cell | *HHIPL1* | -3.127688318 | 0.000317096 | 0.001668632 | 1 |
| Obesity | t_cell | *FAM153B* | -3.932415908 | 0.000367711 | 0.001893986 | 1 |
| Obesity | t_cell | *SYNM* | -6.375205925 | 0.000583671 | 0.002762901 | 1 |
| Obesity | t_cell | *KCNA4* | -4.46757717 | 8.68E-06 | 0.000107764 | 0.042135801 |
| Obesity | t_cell | *FIGN* | -2.928690427 | 0.00211813 | 0.00776221 | 1 |
| Obesity | t_cell | *C8orf33* | -4.54898736 | 0.015553798 | 0.039224027 | 1 |

| | | | | | | |
|---|---|---|---|---|---|---|
| Obesity | t_cell | *NXPH4* | -20.55602197 | 2.98E-06 | 6.01E-05 | 0.014443057 |
| Obesity | t_cell | *CAPN12* | 1.690588172 | 0.003611989 | 0.011775178 | 1 |
| Obesity | t_cell | *KPNA2* | 4.553101725 | 0.001703949 | 0.006515021 | 1 |
| Obesity | t_cell | *ORAI1* | 4.262524118 | 2.26E-07 | 1.35E-05 | 0.00109842 |
| Obesity | t_cell | *CAV3* | -3.25874868 | 2.87E-05 | 0.000258512 | 0.139079635 |
| Obesity | t_cell | *MFSD5* | -2.254877821 | 0.007089019 | 0.020795598 | 1 |
| Obesity | t_cell | *CLEC4G* | -5.309708636 | 0.020162876 | 0.048537764 | 1 |
| Obesity | t_cell | *TRAK1* | -2.702559555 | 0.009910174 | 0.027274057 | 1 |
| Obesity | t_cell | *PPP1R27* | -18.00663708 | 1.59E-08 | 1.75E-06 | 7.70E-05 |
| Obesity | t_cell | *BRICD5* | 12.4821782 | 9.33E-06 | 0.000113501 | 0.045282378 |
| Obesity | t_cell | *GRID1* | -5.566025374 | 1.24E-07 | 9.41E-06 | 0.000601931 |
| Obesity | t_cell | *PLCXD3* | -3.570652542 | 9.40E-05 | 0.000655245 | 0.456050813 |
| Obesity | t_cell | *ODF3L1* | -5.659803977 | 0.002375816 | 0.008488555 | 1 |
| Obesity | t_cell | *C12orf60* | -5.416327623 | 0.002345487 | 0.008411162 | 1 |
| Obesity | t_cell | *BEGAIN* | -3.861742227 | 1.16E-06 | 4.07E-05 | 0.005614774 |
| Obesity | t_cell | *ARHGEF37* | -5.102702098 | 0.000364362 | 0.001881617 | 1 |
| Obesity | t_cell | *CSMD1* | -4.939580394 | 0.010220712 | 0.027954282 | 1 |
| Obesity | t_cell | *RABIF* | 4.350749062 | 0.000450338 | 0.002217473 | 1 |
| Obesity | t_cell | *FANCF* | 10.00898428 | 3.54E-05 | 0.000306727 | 0.171767381 |
| Obesity | t_cell | *RUVBL2* | -4.910141572 | 0.002400836 | 0.008565337 | 1 |
| Obesity | t_cell | *C19orf71* | -5.520683927 | 0.000830991 | 0.003632404 | 1 |
| Obesity | t_cell | *LRIT3* | -14.93965849 | 2.14E-06 | 5.28E-05 | 0.010386983 |
| Obesity | t_cell | *GPR132* | 2.206935788 | 0.000239581 | 0.00135011 | 1 |
| Obesity | t_cell | *SERHL2* | -5.588346148 | 0.007303938 | 0.021233499 | 1 |
| Obesity | t_cell | *HMCES* | -3.639856205 | 0.00544819 | 0.016656974 | 1 |
| Obesity | t_cell | *RP1L1* | -4.23636931 | 0.000380045 | 0.001943078 | 1 |
| Obesity | t_cell | *FAM19A1* | 2.161873637 | 1.86E-05 | 0.000186657 | 0.09034208 |
| Obesity | t_cell | *UPP1* | 6.065497248 | 1.81E-06 | 4.84E-05 | 0.008801864 |
| Obesity | t_cell | *FIGLA* | -8.916825901 | 3.48E-05 | 0.000303778 | 0.168900543 |
| Obesity | t_cell | *ACSM2A* | -52.31992999 | 3.25E-06 | 6.11E-05 | 0.015756826 |
| Obesity | t_cell | *NPIPA5* | 0.643574748 | 0.01919852 | 0.046598909 | 1 |
| Obesity | t_cell | *FAM162B* | -2.410143972 | 0.000596233 | 0.002814128 | 1 |
| Obesity | t_cell | *FAM3B* | -0.624859039 | 0.003945627 | 0.012695081 | 1 |
| Obesity | t_cell | *NPIPA7* | 3.488872856 | 0.000126576 | 0.000814521 | 0.614148693 |
| Obesity | t_cell | *GOLGA6L4* | -2.68281262 | 0.000884542 | 0.00380816 | 1 |
| Obesity | t_cell | *ACOT1* | 2.370897472 | 9.93E-05 | 0.000681313 | 0.481688316 |
| Obesity | t_cell | *OAF* | -1.825732976 | 5.86E-05 | 0.00045141 | 0.284387997 |
| Obesity | t_cell | *ALDH1A3* | -1.515345051 | 0.002062263 | 0.00758038 | 1 |
| Obesity | t_cell | *CLECL1* | 2.558243839 | 7.75E-06 | 9.97E-05 | 0.037603436 |
| Obesity | t_cell | *A3GALT2* | 1.513403024 | 0.004827296 | 0.015062407 | 1 |
| Obesity | t_cell | *TOP1MT* | -1.590522169 | 0.004577775 | 0.014394922 | 1 |
| Obesity | t_cell | *TMEM255B* | -4.64308197 | 0.017102167 | 0.0425537 | 1 |
| Obesity | t_cell | *ZFP1* | -3.748830227 | 0.003406762 | 0.011244633 | 1 |
| Obesity | t_cell | *DUSP8* | -2.648767248 | 0.005089494 | 0.015728805 | 1 |
| Obesity | t_cell | *SNN* | -8.187714609 | 9.16E-05 | 0.000642563 | 0.444653929 |
| Obesity | t_cell | *RNLS* | -3.973274982 | 0.000615258 | 0.002875945 | 1 |
| Obesity | t_cell | *TCTE3* | 5.774061832 | 0.001770029 | 0.00673055 | 1 |
| Obesity | t_cell | *TMED9* | -4.173667352 | 0.006172413 | 0.018532516 | 1 |
| Obesity | t_cell | *DRD1* | -48.18830293 | 2.12E-06 | 5.28E-05 | 0.010274404 |
| Obesity | t_cell | *NUTM2A* | -3.1085225 | 0.000102327 | 0.000693422 | 0.496489849 |

| | | | | | | |
|---|---|---|---|---|---|---|
| Obesity | t_cell | *WT1* | -2.531151209 | 0.000123311 | 0.000798806 | 0.598305782 |
| Obesity | t_cell | *NOC4L* | 3.504252334 | 0.003058839 | 0.010407773 | 1 |
| Obesity | t_cell | *SORCS2* | -4.378642297 | 4.51E-06 | 7.28E-05 | 0.021865057 |
| Obesity | t_cell | *MAFF* | -3.255210869 | 1.59E-07 | 1.09E-05 | 0.000772467 |
| Obesity | t_cell | *SIGIRR* | 3.176193236 | 1.05E-06 | 3.84E-05 | 0.005081468 |
| Obesity | t_cell | *GP1BA* | 5.686035953 | 4.00E-10 | 6.47E-08 | 1.94E-06 |
| Obesity | t_cell | *ATP6V0A2* | -4.131976088 | 0.0139412 | 0.035827703 | 1 |
| Obesity | t_cell | *SP140L* | 3.898046867 | 0.001171056 | 0.004790862 | 1 |
| Obesity | t_cell | *TARSL2* | 3.334735065 | 0.002247807 | 0.008108816 | 1 |
| Obesity | t_cell | *SH3BGR* | -1.248411517 | 0.000614887 | 0.002875945 | 1 |
| Obesity | t_cell | *FAM174B* | -3.775013291 | 0.000471909 | 0.002308834 | 1 |
| Obesity | t_cell | *SV2B* | -2.365215686 | 4.18E-05 | 0.000346306 | 0.202935152 |
| Obesity | t_cell | *AHNAK2* | -4.277270327 | 1.07E-05 | 0.000125619 | 0.052006416 |
| Obesity | t_cell | *PMEL* | 2.198679923 | 6.22E-05 | 0.000470729 | 0.301737385 |
| Obesity | t_cell | *MYBL1* | 0.712281294 | 5.01E-06 | 7.73E-05 | 0.024316839 |
| Obesity | t_cell | *ADARB2* | 5.849110857 | 9.59E-07 | 3.74E-05 | 0.004652206 |
| Obesity | t_cell | *SLC52A2* | 5.994778415 | 0.00021107 | 0.001219728 | 1 |
| Obesity | t_cell | *EVI2B* | 1.108163717 | 0.000807789 | 0.003559847 | 1 |
| Obesity | t_cell | *ZNF829* | -2.162967184 | 0.013298488 | 0.034514485 | 1 |
| Obesity | t_cell | *IFITM1* | -3.072700568 | 0.003171169 | 0.010671553 | 1 |
| Obesity | t_cell | *RTN4RL1* | -2.046738075 | 0.018297079 | 0.044973366 | 1 |
| Obesity | t_cell | *RASA3* | 2.295539987 | 0.012100565 | 0.032019016 | 1 |
| Obesity | t_cell | *CYP4Z1* | -1.043006649 | 0.000627922 | 0.002915481 | 1 |
| Obesity | t_cell | *FFAR4* | -4.764830601 | 0.00859541 | 0.024195178 | 1 |
| Obesity | t_cell | *CYP4F12* | -2.900859572 | 0.010591133 | 0.028821186 | 1 |
| Obesity | t_cell | *TOR3A* | -5.575561884 | 0.001034033 | 0.004317665 | 1 |
| Obesity | t_cell | *ZNF555* | -2.870645025 | 0.018465314 | 0.045295098 | 1 |
| Obesity | t_cell | *RPS23* | 3.215128753 | 0.003284526 | 0.010937899 | 1 |
| Obesity | t_cell | *GNG2* | 3.06602171 | 4.56E-06 | 7.30E-05 | 0.022123326 |
| Obesity | t_cell | *BTN3A2* | -1.042776638 | 2.17E-05 | 0.000207464 | 0.105184284 |
| Obesity | t_cell | *SEPT10* | -4.22755948 | 0.000474484 | 0.002318424 | 1 |
| Obesity | t_cell | *FAM86B1* | -3.355434445 | 0.008028932 | 0.022929004 | 1 |
| Obesity | t_cell | *CEACAM19* | -1.314594401 | 0.004217903 | 0.013402268 | 1 |
| Obesity | t_cell | *MIR22HG* | -3.985721869 | 0.000101117 | 0.000688713 | 0.49061809 |
| Obesity | t_cell | *CYP27C1* | -1.853086829 | 0.00125513 | 0.00507491 | 1 |
| Obesity | t_cell | *HYAL3* | -1.65539208 | 0.01519656 | 0.038463072 | 1 |
| Obesity | t_cell | *TRABD2A* | 1.791096976 | 3.85E-07 | 2.05E-05 | 0.001867051 |
| Obesity | t_cell | *MAPT* | -4.778966368 | 0.002025109 | 0.007494911 | 1 |
| Obesity | t_cell | *TMEM17* | -2.192920383 | 6.86E-05 | 0.000512351 | 0.333028456 |
| Obesity | t_cell | *TNFRSF18* | 1.638741878 | 1.77E-06 | 4.83E-05 | 0.008591891 |
| Obesity | t_cell | *C1QL4* | -78.05401494 | 0.000386897 | 0.001971876 | 1 |
| Obesity | t_cell | *RTN4RL2* | -6.55105421 | 2.01E-05 | 0.000196799 | 0.097612168 |
| Obesity | t_cell | *EFCAB6* | -3.794840844 | 0.000142916 | 0.000896466 | 0.693428042 |
| Obesity | t_cell | *TMPRSS6* | -14.55783062 | 2.54E-06 | 5.90E-05 | 0.012323366 |
| Obesity | t_cell | *C3orf70* | -2.244394696 | 0.000713345 | 0.003231698 | 1 |
| Obesity | t_cell | *ANGPTL5* | -3.083829524 | 0.000225146 | 0.001282171 | 1 |
| Obesity | t_cell | *WDR86* | -2.119629821 | 0.000322081 | 0.001689448 | 1 |
| Obesity | t_cell | *LUZP2* | -5.095358406 | 1.42E-05 | 0.000154036 | 0.068700271 |
| Obesity | t_cell | *GJA4* | -6.34861632 | 0.000220689 | 0.001259746 | 1 |
| Obesity | t_cell | *SBK2* | -18.83683673 | 3.61E-06 | 6.46E-05 | 0.017499056 |

| | | | | | | |
|---|---|---|---|---|---|---|
| Obesity | t_cell | *C17orf97* | 0.525430304 | 0.002176788 | 0.00792926 | 1 |
| Obesity | t_cell | *SAMD11* | -2.509126304 | 0.000157651 | 0.000968257 | 0.76492327 |
| Obesity | t_cell | *TRPV2* | 2.166630454 | 0.00011306 | 0.000745335 | 0.548566689 |
| Obesity | t_cell | *C2orf88* | -4.026495752 | 0.000973801 | 0.004105025 | 1 |
| Obesity | t_cell | *DNAH17* | -2.766044738 | 0.018885104 | 0.046115011 | 1 |
| Obesity | t_cell | *ZFP69* | -6.849686521 | 0.003413957 | 0.011253852 | 1 |
| Obesity | t_cell | *FAM122A* | -5.795266181 | 2.23E-05 | 0.000212381 | 0.108314476 |
| Obesity | t_cell | *GFRAL* | -4.269978201 | 0.000137295 | 0.000871139 | 0.666154598 |
| Obesity | t_cell | *LDLRAD2* | -2.624401743 | 0.000656612 | 0.00301123 | 1 |
| Obesity | t_cell | *CPSF4L* | -6.331556391 | 0.001040823 | 0.004338553 | 1 |
| Obesity | t_cell | *OR7D2* | -7.104137255 | 0.000835021 | 0.003638422 | 1 |
| Obesity | t_cell | *MAPK12* | -3.702975594 | 1.42E-05 | 0.000154414 | 0.069023266 |
| Obesity | t_cell | *AGRN* | -7.127118746 | 0.000455848 | 0.002240905 | 1 |
| Obesity | t_cell | *ZNF626* | -4.809605079 | 4.11E-05 | 0.000340617 | 0.199260724 |
| Obesity | t_cell | *NCR3LG1* | -6.83362798 | 1.40E-05 | 0.000153123 | 0.068139585 |
| Obesity | t_cell | *AGAP4* | -2.735611451 | 0.00125826 | 0.005083329 | 1 |
| Obesity | t_cell | *RUFY4* | -10.60315844 | 3.59E-05 | 0.000309505 | 0.174230363 |
| Obesity | t_cell | *ZNF669* | 3.177929199 | 0.000539532 | 0.002589328 | 1 |
| Obesity | t_cell | *CENPP* | 1.444916677 | 0.015469634 | 0.039052375 | 1 |
| Obesity | t_cell | *H3F3C* | -66.95402623 | 0.00011463 | 0.00075364 | 0.556186101 |
| Obesity | t_cell | *JAKMIP3* | -1.540396822 | 0.000758432 | 0.003376067 | 1 |
| Obesity | t_cell | *PDCD1* | 1.565301982 | 9.71E-07 | 3.74E-05 | 0.004710514 |
| Obesity | t_cell | *SELL* | 1.472933488 | 1.90E-05 | 0.000189814 | 0.092093521 |
| Obesity | t_cell | *C19orf54* | -4.844327911 | 0.000209745 | 0.001217324 | 1 |
| Obesity | t_cell | *C22orf34* | 4.095344755 | 2.52E-06 | 5.90E-05 | 0.012232056 |
| Obesity | t_cell | *NDOR1* | 6.484211626 | 0.011886609 | 0.031685341 | 1 |
| Obesity | t_cell | *GOLGA8M* | -4.354982107 | 6.96E-06 | 9.43E-05 | 0.033769099 |
| Obesity | t_cell | *RHCE* | -3.766423578 | 2.33E-05 | 0.000219931 | 0.113044685 |
| Obesity | t_cell | *SLC4A5* | 13.56542514 | 0.004550448 | 0.014336867 | 1 |
| Obesity | t_cell | *UROS* | -4.095704198 | 0.012861968 | 0.033569807 | 1 |
| Obesity | t_cell | *ZBED6CL* | 4.719389336 | 0.002896532 | 0.009974431 | 1 |
| Obesity | t_cell | *FSIP2* | -2.619772512 | 0.004819331 | 0.01505692 | 1 |
| Obesity | t_cell | *NOXA1* | -5.881438446 | 0.002003262 | 0.007438961 | 1 |
| Obesity | t_cell | *FZD9* | -22.20983836 | 4.61E-07 | 2.31E-05 | 0.002237769 |
| Obesity | t_cell | *MTF1* | -3.320258572 | 0.008724075 | 0.024467754 | 1 |
| Obesity | t_cell | *ZDHHC11* | -2.869525982 | 0.00012905 | 0.000827147 | 0.626150007 |
| Obesity | t_cell | *ENTPD8* | -226.4346937 | 1.20E-06 | 4.13E-05 | 0.005822041 |
| Obesity | t_cell | *RPL14* | 3.589155234 | 0.019836472 | 0.047883862 | 1 |
| Obesity | t_cell | *ASTL* | -22.28495111 | 0.001287065 | 0.005162414 | 1 |
| Obesity | t_cell | *LRRK2* | -2.543337907 | 0.001198288 | 0.004894019 | 1 |
| Obesity | t_cell | *FAM196A* | -3.44015243 | 0.000838304 | 0.003641407 | 1 |
| Obesity | t_cell | *UTS2B* | -3.47632166 | 2.31E-05 | 0.000218805 | 0.112246957 |
| Obesity | t_cell | *C9orf152* | -4.552089001 | 0.000169505 | 0.001029335 | 0.822438427 |
| Obesity | t_cell | *SLC15A5* | -2.369738305 | 0.00017375 | 0.001048548 | 0.843032634 |
| Obesity | t_cell | *LITAF* | 4.545486412 | 0.002210302 | 0.008021229 | 1 |
| Obesity | t_cell | *ZNF573* | -2.932003518 | 0.014678934 | 0.037426267 | 1 |
| Obesity | t_cell | *FAM47E* | -3.646920088 | 0.000888233 | 0.003820661 | 1 |
| Obesity | t_cell | *PNRC2* | -4.655390054 | 0.010876403 | 0.029481735 | 1 |
| Obesity | t_cell | *ZKSCAN3* | -5.217668008 | 5.58E-08 | 4.92E-06 | 0.00027075 |
| Obesity | t_cell | *SLC35E2B* | 2.373385848 | 0.018882446 | 0.046115011 | 1 |

| | | | | | | |
|---|---|---|---|---|---|---|
| Obesity | t_cell | *AKR1C3* | -4.878433391 | 0.001001019 | 0.004197877 | 1 |
| Obesity | t_cell | *COLCA1* | -2.261987774 | 0.003184568 | 0.010700503 | 1 |
| Obesity | t_cell | *HRCT1* | -2.481117104 | 1.59E-05 | 0.000166708 | 0.077185733 |
| Obesity | t_cell | *SIRPB2* | -1.203027736 | 0.006325326 | 0.018956443 | 1 |
| Obesity | t_cell | *ZNF471* | -4.225118356 | 3.65E-05 | 0.000313099 | 0.176900822 |
| Obesity | t_cell | *SUPT3H* | 3.66172952 | 0.003020766 | 0.010285443 | 1 |
| Obesity | t_cell | *GIMAP5* | 2.857464031 | 0.000112403 | 0.000742011 | 0.545377758 |
| Obesity | t_cell | *STK31* | -3.083068601 | 0.000197619 | 0.00116365 | 0.958848003 |
| Obesity | t_cell | *ZNF34* | -4.325622891 | 7.66E-05 | 0.000557988 | 0.37162026 |
| Obesity | t_cell | *EVL* | 2.303078574 | 2.44E-05 | 0.000227928 | 0.118522369 |
| Obesity | t_cell | *RFX8* | -1.348056528 | 0.006524501 | 0.019380641 | 1 |
| Obesity | t_cell | *C20orf96* | -5.755898583 | 0.000654845 | 0.003005969 | 1 |
| Obesity | t_cell | *SULT1A1* | -5.603106104 | 2.48E-06 | 5.88E-05 | 0.012049379 |
| Obesity | t_cell | *TPK1* | 4.337133199 | 0.001052126 | 0.00436348 | 1 |
| Obesity | t_cell | *SPTSSB* | -5.916332662 | 0.00093882 | 0.003988752 | 1 |
| Obesity | t_cell | *CACNA1H* | -3.927216025 | 0.018255974 | 0.044895076 | 1 |
| Obesity | t_cell | *XRCC2* | -7.16363161 | 5.78E-06 | 8.45E-05 | 0.028053657 |
| Obesity | t_cell | *MYO6* | -3.204556445 | 0.014976879 | 0.038026068 | 1 |
| Obesity | t_cell | *ZNF846* | -3.704393622 | 0.00534518 | 0.016435244 | 1 |
| Obesity | t_cell | *ZFP62* | 6.642390385 | 0.00030253 | 0.001627385 | 1 |
| Obesity | t_cell | *TOMM7* | 4.354942471 | 0.004140746 | 0.013200328 | 1 |
| Obesity | t_cell | *ZNF33B* | 2.352965938 | 0.008135425 | 0.023154147 | 1 |
| Obesity | t_cell | *AMZ2* | 4.225557568 | 0.000889675 | 0.003823475 | 1 |
| Obesity | t_cell | *COL27A1* | -1.386567168 | 0.010459766 | 0.028527702 | 1 |
| Obesity | t_cell | *SLC39A10* | 4.467771746 | 9.32E-05 | 0.000650336 | 0.451983339 |
| Obesity | t_cell | *HIST1H3E* | 1.2911517 | 0.016760277 | 0.04185325 | 1 |
| Obesity | t_cell | *ZNF138* | 3.694414697 | 0.006751523 | 0.019938155 | 1 |
| Obesity | t_cell | *ANXA6* | 2.701157277 | 0.007740139 | 0.022208844 | 1 |
| Obesity | t_cell | *GAL3ST4* | 4.199437922 | 7.35E-05 | 0.000540967 | 0.356497108 |
| Obesity | t_cell | *ZNF772* | -1.61655867 | 0.002366224 | 0.008473003 | 1 |
| Obesity | t_cell | *ADAM32* | -4.705161969 | 6.43E-05 | 0.000482501 | 0.312177922 |
| Obesity | t_cell | *LRRC8B* | 3.986848477 | 0.007513536 | 0.021674004 | 1 |
| Obesity | t_cell | *SND1* | 3.686454804 | 0.003174573 | 0.01067431 | 1 |
| Obesity | t_cell | *SULT1A2* | 8.163218928 | 3.70E-14 | 1.63E-11 | 1.80E-10 |
| Obesity | t_cell | *NEK5* | -1.512291141 | 0.012720769 | 0.033344771 | 1 |
| Obesity | t_cell | *PIWIL2* | -2.492846603 | 0.001385579 | 0.005483546 | 1 |
| Obesity | t_cell | *ENTPD4* | 4.753506477 | 0.000516097 | 0.002491644 | 1 |
| Obesity | t_cell | *FAM110D* | -2.426851178 | 0.00105131 | 0.00436348 | 1 |
| Obesity | t_cell | *CCL4L2* | 6.688798333 | 2.53E-06 | 5.90E-05 | 0.012277028 |
| Obesity | t_cell | *ZNF165* | -2.622536924 | 6.61E-06 | 9.16E-05 | 0.032071574 |
| Obesity | t_cell | *RAMP2-AS1* | -4.326718263 | 4.79E-05 | 0.000383239 | 0.232625818 |
| Obesity | t_cell | *MRPL21* | -1.493217943 | 0.000561907 | 0.002678166 | 1 |
| Obesity | t_cell | *ZNF98* | -2.819987606 | 2.47E-05 | 0.000229976 | 0.119817343 |
| Obesity | t_cell | *MAP3K5* | 3.732887185 | 0.000632607 | 0.002931622 | 1 |
| Obesity | t_cell | *PCDHB11* | -1.011621221 | 0.017957577 | 0.044341051 | 1 |
| Obesity | t_cell | *ZNF665* | -4.59011484 | 0.020524524 | 0.049262325 | 1 |
| Obesity | t_cell | *HHLA3* | -1.532298082 | 0.014384054 | 0.036809825 | 1 |
| Obesity | t_cell | *TOPORS* | -4.716575703 | 0.003976014 | 0.012775908 | 1 |
| Obesity | t_cell | *BCO2* | -1.01244753 | 0.008287814 | 0.023516068 | 1 |
| Obesity | t_cell | *ENTPD6* | -4.140330663 | 0.0031273 | 0.01057398 | 1 |

| | | | | | | |
|---|---|---|---|---|---|---|
| Obesity | t_cell | *ENPP1* | -2.909669795 | 0.000892446 | 0.003831988 | 1 |
| Obesity | t_cell | *MFAP5* | -3.684380054 | 3.69E-07 | 2.02E-05 | 0.001789131 |
| Obesity | t_cell | *DNAH10* | 6.1073492 | 2.21E-07 | 1.34E-05 | 0.001070593 |
| Obesity | t_cell | *FAM114A1* | -3.229603575 | 0.002099682 | 0.007706246 | 1 |
| Obesity | t_cell | *RPS26* | -1.767629599 | 0.000622949 | 0.002901153 | 1 |
| Obesity | t_cell | *TXNRD3* | -2.698443235 | 0.004397984 | 0.013928864 | 1 |
| Obesity | t_cell | *EME2* | 8.593481227 | 0.002607968 | 0.009129771 | 1 |
| Obesity | t_cell | *ZNF780A* | -0.950253442 | 0.000736739 | 0.003309868 | 1 |
| Obesity | t_cell | *CCDC180* | -2.434465374 | 0.000299455 | 0.001616193 | 1 |
| Obesity | t_cell | *OCLN* | -2.391211448 | 0.006414503 | 0.019164512 | 1 |
| Obesity | t_cell | *ZNF181* | -2.295316137 | 0.012018454 | 0.031882744 | 1 |
| Obesity | t_cell | *SGTB* | 4.289416885 | 0.000260669 | 0.001455378 | 1 |
| Obesity | t_cell | *ZNF677* | -4.024790034 | 7.83E-05 | 0.000567379 | 0.380143681 |
| Obesity | t_cell | *FCHSD1* | 2.605272612 | 0.005411931 | 0.016582323 | 1 |
| Obesity | t_cell | *MPZL1* | -3.137320266 | 0.004169869 | 0.013275724 | 1 |
| Obesity | t_cell | *DLGAP2* | -1.814590613 | 0.016028278 | 0.040253211 | 1 |
| Obesity | t_cell | *MRPL42* | -2.308469864 | 0.005612362 | 0.01708355 | 1 |
| Obesity | t_cell | *ZNF560* | 2.185385211 | 0.017323268 | 0.042971625 | 1 |
| Obesity | t_cell | *AGAP9* | -4.046528875 | 0.004846116 | 0.015101706 | 1 |
| Obesity | t_cell | *ZNF273* | -4.255739282 | 0.000211089 | 0.001219728 | 1 |
| Obesity | t_cell | *ZNF667* | -3.23618605 | 1.48E-05 | 0.000158409 | 0.071759368 |
| Obesity | t_cell | *SFI1* | 2.934535764 | 8.49E-06 | 0.000106455 | 0.041198232 |
| Obesity | t_cell | *ADH4* | -2.389770851 | 5.08E-05 | 0.00040099 | 0.246609155 |
| Obesity | t_cell | *HIBCH* | -2.30367279 | 0.00386024 | 0.012469963 | 1 |
| Obesity | t_cell | *MIER1* | 3.818772289 | 0.003255307 | 0.010862965 | 1 |
| Obesity | t_cell | *MAN1A2* | 3.775508396 | 0.00321448 | 0.010774594 | 1 |
| Obesity | t_cell | *ZNF334* | -3.182525212 | 0.002292767 | 0.008246484 | 1 |
| Obesity | t_cell | *RPS6KL1* | -4.090302098 | 0.000174603 | 0.00105239 | 0.847174104 |
| Obesity | t_cell | *STYX* | 6.295633113 | 2.42E-05 | 0.000226603 | 0.117607134 |
| Obesity | t_cell | *TMEM116* | 1.309220998 | 0.001280371 | 0.005151211 | 1 |
| Obesity | t_cell | *UCKL1* | -4.640784878 | 0.008070435 | 0.02301908 | 1 |
| Obesity | t_cell | *ZNF442* | -2.139521833 | 0.01229373 | 0.032453307 | 1 |
| Obesity | t_cell | *ZNF587* | -2.824785948 | 0.016017659 | 0.040247376 | 1 |
| Obesity | t_cell | *C20orf203* | -5.844424489 | 1.66E-07 | 1.09E-05 | 0.000805554 |
| Obesity | t_cell | *TUSC1* | -9.542557281 | 8.88E-05 | 0.000629132 | 0.430955555 |
| Obesity | t_cell | *PAPSS2* | -3.414806559 | 0.000975875 | 0.004110194 | 1 |
| Obesity | t_cell | *ANKRD13B* | -4.982015478 | 7.83E-06 | 0.000100385 | 0.03797186 |
| Obesity | t_cell | *UNC13B* | -3.738356371 | 0.001649234 | 0.006355903 | 1 |
| Obesity | t_cell | *PPP1R14C* | -7.28345408 | 3.12E-06 | 6.09E-05 | 0.015144096 |
| Obesity | t_cell | *GPATCH3* | -8.183779374 | 0.000336157 | 0.0017538 | 1 |
| Obesity | t_cell | *RPL10A* | 4.374993038 | 0.001245798 | 0.005045584 | 1 |
| Obesity | t_cell | *COLGALT2* | 3.077167395 | 5.27E-06 | 7.92E-05 | 0.025579868 |
| Obesity | t_cell | *RCSD1* | 2.026678025 | 1.94E-06 | 5.08E-05 | 0.009397685 |
| Obesity | t_cell | *GRIN3A* | -2.91149916 | 0.000918502 | 0.003912707 | 1 |
| Obesity | t_cell | *TMEM184B* | -4.623150598 | 0.000253357 | 0.0014195 | 1 |
| Obesity | t_cell | *SCAMP5* | -1.585987141 | 0.000634569 | 0.002935107 | 1 |
| Obesity | t_cell | *RYR3* | -1.411093435 | 0.013748651 | 0.035407885 | 1 |
| Obesity | t_cell | *PRMT6* | -6.677545906 | 5.76E-07 | 2.77E-05 | 0.002796371 |
| Obesity | t_cell | *SHISA4* | -10.78435697 | 4.15E-12 | 1.06E-09 | 2.01E-08 |
| Obesity | t_cell | *C1orf174* | -2.828009965 | 0.01176875 | 0.03146802 | 1 |

| | | | | | | |
|---|---|---|---|---|---|---|
| Obesity | t_cell | *KIAA0753* | -2.899534096 | 0.018324342 | 0.04499479 | 1 |
| Obesity | t_cell | *CCDC167* | 5.163248242 | 5.83E-06 | 8.49E-05 | 0.028278586 |
| Obesity | t_cell | *ZFP2* | -4.97781463 | 0.00041219 | 0.002070335 | 1 |
| Obesity | t_cell | *L3MBTL3* | 2.785949437 | 0.001094921 | 0.004521324 | 1 |
| Obesity | t_cell | *MFAP3L* | -4.363750901 | 0.000792943 | 0.003510105 | 1 |
| Obesity | t_cell | *SMG5* | -3.488674509 | 0.002053266 | 0.007559371 | 1 |
| Obesity | t_cell | *AL590822.1* | -4.043923701 | 9.60E-05 | 0.000664101 | 0.465614341 |
| Obesity | t_cell | *SAMD5* | -3.087959847 | 0.000101592 | 0.000690372 | 0.492925512 |
| Obesity | t_cell | *GPR52* | 13.34665365 | 2.06E-06 | 5.24E-05 | 0.010000055 |
| Obesity | t_cell | *FAM229B* | -4.822954295 | 1.09E-05 | 0.000127422 | 0.052880263 |
| Obesity | t_cell | *RBM20* | -5.684756812 | 2.63E-06 | 5.93E-05 | 0.01274501 |
| Obesity | t_cell | *C6orf163* | -1.641058699 | 0.003281041 | 0.010933798 | 1 |
| Obesity | t_cell | *SAMD13* | -5.416774913 | 0.009080484 | 0.025306437 | 1 |
| Obesity | t_cell | *IFIT1B* | -35.62067108 | 0.000128954 | 0.000827147 | 0.625683858 |
| Obesity | t_cell | *LIPN* | 20.10192483 | 0.000419792 | 0.002101993 | 1 |
| Obesity | t_cell | *INPP5B* | -1.242565441 | 0.007217985 | 0.021046671 | 1 |
| Obesity | t_cell | *NEU4* | -3.205707586 | 0.015007896 | 0.038064982 | 1 |
| Obesity | t_cell | *C2orf72* | -12.49644734 | 0.000311479 | 0.001653498 | 1 |
| Obesity | t_cell | *ASAH2B* | 4.510579567 | 0.01196179 | 0.031756032 | 1 |
| Obesity | t_cell | *LINC00619* | -26.6195024 | 7.12E-06 | 9.54E-05 | 0.034534987 |
| Obesity | t_cell | *DAXX* | 2.291508792 | 0.003756783 | 0.012168166 | 1 |
| Obesity | t_cell | *TCEA3* | -3.889974656 | 0.001238344 | 0.005023782 | 1 |
| Obesity | t_cell | *COL15A1* | -1.456112124 | 0.012102819 | 0.032019016 | 1 |
| Obesity | t_cell | *NOTCH4* | -0.510218501 | 1.65E-09 | 2.51E-07 | 8.02E-06 |
| Obesity | t_cell | *AGER* | 2.594353789 | 0.000314129 | 0.001658492 | 1 |
| Obesity | t_cell | *PRRT1* | -1.433952704 | 0.0014714 | 0.005766747 | 1 |
| Obesity | t_cell | *SKIV2L* | -2.295673909 | 3.43E-08 | 3.54E-06 | 0.000166534 |
| Obesity | t_cell | *GPANK1* | -6.972040784 | 8.54E-12 | 1.80E-09 | 4.15E-08 |
| Obesity | t_cell | *LST1* | 2.581241293 | 0.003208862 | 0.010772103 | 1 |
| Obesity | t_cell | *HLA-C* | 1.779106651 | 7.55E-05 | 0.000552822 | 0.36652107 |
| Obesity | t_cell | *POU5F1* | 0.670216137 | 0.00046308 | 0.002269562 | 1 |
| Obesity | t_cell | *CCHCR1* | 1.301969566 | 9.20E-05 | 0.000643987 | 0.446283336 |
| Obesity | t_cell | *ABCF1* | 2.920432257 | 0.0053604 | 0.016471602 | 1 |
| Obesity | t_cell | *LILRB3* | -3.707809678 | 4.23E-05 | 0.000348188 | 0.205082541 |
| Obesity | t_cell | *HLA-E* | 3.106704233 | 0.000582202 | 0.002758636 | 1 |
| Obesity | t_cell | *ZNF468* | -9.971697077 | 2.41E-05 | 0.000225504 | 0.116811167 |
| Obesity | t_cell | *RNF39* | -5.766607788 | 8.82E-09 | 1.13E-06 | 4.28E-05 |
| Obesity | t_cell | *HCG9* | -2.935853691 | 0.000143632 | 0.000898067 | 0.69690015 |
| Obesity | t_cell | *HLA-G* | 1.456310701 | 0.011792023 | 0.031506 | 1 |
| Obesity | t_cell | *ZFP57* | 1.81667404 | 1.47E-08 | 1.65E-06 | 7.11E-05 |
| Obesity | t_cell | *FAM153C* | -1.051061497 | 0.001380873 | 0.005469386 | 1 |
| Obesity | t_cell | *MALRD1* | -3.776615807 | 0.000905893 | 0.003876009 | 1 |
| Obesity | t_cell | *FAM196B* | 2.802103906 | 4.69E-06 | 7.43E-05 | 0.022741922 |
| Obesity | t_cell | *MROH6* | -1.992449243 | 0.002778269 | 0.009649365 | 1 |
| Obesity | t_cell | *DCTN1* | -5.457848732 | 0.002654581 | 0.009272878 | 1 |
| Obesity | t_cell | *C12orf73* | -4.464260215 | 0.001544925 | 0.005991986 | 1 |
| Obesity | t_cell | *PCDHGA1* | -2.835366489 | 0.000192445 | 0.001140099 | 0.933740848 |
| Obesity | t_cell | *PCDHA7* | 14.20318272 | 0.009148758 | 0.025452851 | 1 |
| Obesity | t_cell | *PCDHA4* | -5.809359975 | 2.44E-07 | 1.43E-05 | 0.001184015 |
| Obesity | t_cell | *SPIRE2* | -1.777274034 | 0.00298193 | 0.010188962 | 1 |

| | | | | | | |
|---|---|---|---|---|---|---|
| Obesity | t_cell | *SLFN12L* | 0.848493772 | 1.39E-05 | 0.000152151 | 0.067506789 |
| Obesity | t_cell | *TMEM231* | -3.020735089 | 0.005836028 | 0.017653621 | 1 |
| Obesity | t_cell | *SCGB2B2* | -4.109499686 | 0.001680688 | 0.0064464 | 1 |
| Obesity | t_cell | *CCDC144NL* | -8.319021525 | 0.001854728 | 0.007003221 | 1 |
| Obesity | t_cell | *GOLGA6L10* | -4.588067228 | 0.000136936 | 0.000870789 | 0.664411772 |
| Obesity | t_cell | *MT1A* | -4.154503034 | 3.99E-06 | 6.77E-05 | 0.019367658 |
| Obesity | t_cell | *MT1M* | -6.304586259 | 2.14E-05 | 0.0002049 | 0.103679251 |
| Obesity | t_cell | *CFI* | -2.983677082 | 0.000901728 | 0.003861594 | 1 |
| Obesity | t_cell | *SMN2* | -2.192418049 | 3.93E-05 | 0.000330228 | 0.190541632 |
| Obesity | t_cell | *SERF1B* | -2.800013648 | 0.000125118 | 0.000808349 | 0.607070473 |
| Obesity | t_cell | *C17orf107* | 2.641302789 | 0.002141116 | 0.007828708 | 1 |
| Obesity | t_cell | *KLRC2* | 1.088027623 | 1.28E-06 | 4.23E-05 | 0.006203373 |
| Obesity | t_cell | *C1QTNF9B* | 64.85221676 | 0.000747453 | 0.003342527 | 1 |
| Obesity | t_cell | *ZDHHC11B* | -3.642752559 | 1.53E-05 | 0.000161776 | 0.074093222 |
| Obesity | t_cell | *LNP1* | -2.475650688 | 0.000644558 | 0.002975097 | 1 |
| Obesity | t_cell | *SACM1L* | 4.673781449 | 0.00029236 | 0.001583184 | 1 |
| Obesity | t_cell | *SLC48A1* | -4.237069745 | 0.000102204 | 0.000693422 | 0.495892581 |
| Obesity | t_cell | *TAS2R50* | -34.81287881 | 0.000110651 | 0.00073344 | 0.536878054 |
| Obesity | t_cell | *AC084018.1* | -2.560603396 | 1.03E-08 | 1.25E-06 | 4.99E-05 |
| Obesity | t_cell | *C17orf51* | -8.115292084 | 4.70E-05 | 0.000377765 | 0.228182687 |
| Obesity | t_cell | *OR2A42* | -2.029348417 | 0.00186956 | 0.007053735 | 1 |
| Obesity | t_cell | *FGFR1OP* | 5.694085249 | 0.001540364 | 0.005979075 | 1 |
| Obesity | t_cell | *AC022498.1* | -9.735324808 | 0.000685334 | 0.003128165 | 1 |
| Obesity | t_cell | *GIMAP1* | 3.692213886 | 0.000105254 | 0.000706353 | 0.510693019 |
| Obesity | t_cell | *SUPT4H1* | 10.637722 | 2.03E-06 | 5.24E-05 | 0.009869426 |
| Obesity | t_cell | *ARHGAP19* | -6.88262016 | 6.41E-05 | 0.000481223 | 0.310870134 |
| Obesity | t_cell | *LCAT* | -4.395084924 | 1.45E-05 | 0.000156645 | 0.070333803 |
| Obesity | t_cell | *GPC2* | -8.403252371 | 0.000128443 | 0.000825438 | 0.623205632 |
| Obesity | t_cell | *SRA1* | 4.264759337 | 0.004881161 | 0.015162223 | 1 |
| Obesity | t_cell | *TMEM110* | 8.96132959 | 6.30E-14 | 2.35E-11 | 3.06E-10 |
| Obesity | t_cell | *LBH* | 1.792551922 | 1.48E-05 | 0.000158409 | 0.07170902 |
| Obesity | t_cell | *PPP1CB* | -3.317533612 | 0.005717997 | 0.017350671 | 1 |
| Obesity | t_cell | *EMP2* | -2.948675685 | 0.009523559 | 0.026344531 | 1 |
| Obesity | t_cell | *SLC23A3* | 1.47392619 | 0.019444258 | 0.047035575 | 1 |
| Obesity | t_cell | *LTB4R* | 4.044129872 | 3.70E-05 | 0.000315702 | 0.179318971 |
| Obesity | t_cell | *GALT* | 5.956660433 | 0.000428314 | 0.002133652 | 1 |
| Obesity | t_cell | *ZNF726* | 3.484490151 | 0.008654809 | 0.024343844 | 1 |
| Obesity | t_cell | *ZNF90* | -3.576809763 | 0.003010619 | 0.010264782 | 1 |
| Obesity | t_cell | *REPIN1* | -6.993001321 | 0.000537703 | 0.002583102 | 1 |
| Obesity | t_cell | *ZNF891* | -4.210331304 | 0.014585061 | 0.037206475 | 1 |
| Obesity | t_cell | *PRCD* | -4.608448541 | 1.38E-05 | 0.000151925 | 0.067151037 |
| Obesity | t_cell | *SH3D21* | -2.871890465 | 0.001273215 | 0.005130761 | 1 |
| Obesity | t_cell | *ANG* | 7.164622306 | 0.000101159 | 0.000688713 | 0.490823523 |
| Obesity | t_cell | *COLCA2* | -1.599527955 | 0.001152414 | 0.004730553 | 1 |
| Obesity | t_cell | *LCNL1* | -3.756410522 | 0.000193571 | 0.001143981 | 0.939208514 |
| Obesity | t_cell | *GNAT3* | -2.952447309 | 4.60E-06 | 7.34E-05 | 0.022327124 |
| Obesity | t_cell | *CPEB1* | -2.154861362 | 0.001482534 | 0.005801012 | 1 |
| Obesity | t_cell | *ARHGEF33* | -2.625009961 | 9.73E-07 | 3.74E-05 | 0.004718887 |
| Obesity | t_cell | *EVPLL* | -6.052767784 | 5.17E-05 | 0.000405842 | 0.250810497 |
| Obesity | t_cell | *AC005082.1* | -9.140107338 | 9.71E-07 | 3.74E-05 | 0.004710819 |

| | | | | | | |
|---|---|---|---|---|---|---|
| Obesity | t_cell | *C5orf49* | -19.63164361 | 0.011251445 | 0.030244881 | 1 |
| Obesity | t_cell | *GOLGA8B* | 2.671306521 | 1.92E-05 | 0.000190799 | 0.093256578 |
| Obesity | t_cell | *MYL5* | -4.058275367 | 0.008722393 | 0.024467754 | 1 |
| Obesity | t_cell | *NPEPL1* | -6.414333973 | 0.000818015 | 0.003588616 | 1 |
| Obesity | t_cell | *SLC35E2* | -4.707733588 | 3.32E-05 | 0.000293761 | 0.161274799 |
| Obesity | t_cell | *TSTD1* | 4.244978923 | 5.30E-05 | 0.000413934 | 0.25705299 |
| Obesity | t_cell | *AC009403.2* | -2.757811167 | 0.01949092 | 0.047120051 | 1 |
| Obesity | t_cell | *PAM16* | -3.422481417 | 1.29E-05 | 0.000144326 | 0.062781797 |
| Obesity | t_cell | *RNU6ATAC35I* | -25.4120033 | 1.21E-07 | 9.41E-06 | 0.000586628 |
| Obesity | t_cell | *FANCG* | -2.601426949 | 0.00022953 | 0.001304948 | 1 |
| Obesity | t_cell | *AP4M1* | -2.680964741 | 0.000313144 | 0.001655094 | 1 |
| Obesity | t_cell | *FXYD7* | -3.151457164 | 0.000941286 | 0.003992237 | 1 |
| Obesity | t_cell | *SLC12A8* | -2.030438995 | 0.006759246 | 0.019948821 | 1 |
| Obesity | t_cell | *OR2A1* | -1.981331036 | 0.006770135 | 0.019968811 | 1 |
| Obesity | t_cell | *UBA52* | 2.76200421 | 0.003588845 | 0.011718086 | 1 |
| Obesity | t_cell | *RAB6C* | -60.7940393 | 0.000437876 | 0.002167933 | 1 |
| Obesity | t_cell | *AC062017.1* | -5.773185805 | 6.18E-05 | 0.000468839 | 0.300056773 |
| Obesity | t_cell | *ADRA2B* | -8.077694805 | 2.81E-06 | 6.01E-05 | 0.01363251 |
| Obesity | t_cell | *EXOSC6* | -6.077785572 | 1.39E-05 | 0.000152151 | 0.067554843 |
| Obesity | t_cell | *ZNF844* | -4.347730011 | 0.000603386 | 0.002834105 | 1 |
| Obesity | t_cell | *LINC00571* | -3.525254134 | 1.10E-05 | 0.000128556 | 0.053607775 |
| Obesity | t_cell | *LINC01133* | -3.289807494 | 2.55E-05 | 0.000235642 | 0.123712242 |
| Obesity | t_cell | *C4B* | 4.608002976 | 0.000107846 | 0.000720755 | 0.523268144 |
| Obesity | t_cell | *LINC01117* | -4.416248519 | 5.58E-05 | 0.000430758 | 0.270516077 |
| Obesity | t_cell | *RASAL2-AS1* | -2.142376936 | 0.017471984 | 0.043318379 | 1 |
| Obesity | t_cell | *LINC00184* | -71.48629581 | 3.22E-06 | 6.11E-05 | 0.015639493 |
| Obesity | t_cell | *AL049840.1* | -6.611698137 | 8.33E-06 | 0.000104755 | 0.040435272 |
| Obesity | t_cell | *SFTA1P* | -3.910161728 | 0.002736695 | 0.009539111 | 1 |
| Obesity | t_cell | *CYP4A22-AS1* | -6.825549173 | 7.87E-05 | 0.000569315 | 0.382010445 |
| Obesity | t_cell | *MIAT* | 1.535555595 | 1.54E-06 | 4.50E-05 | 0.007471661 |
| Obesity | t_cell | *LINC00115* | 6.466059646 | 0.009694433 | 0.026740984 | 1 |
| Obesity | t_cell | *HAR1A* | 10.13146072 | 5.95E-06 | 8.56E-05 | 0.028851698 |
| Obesity | t_cell | *LINC00937* | -1.646257373 | 0.009776817 | 0.026952908 | 1 |
| Obesity | t_cell | *FAM138B* | -30.15344743 | 2.62E-06 | 5.93E-05 | 0.012705862 |
| Obesity | t_cell | *SCAMP4* | -4.846885163 | 0.002577003 | 0.009054031 | 1 |
| Obesity | t_cell | *AC108463.2* | -5.263871667 | 2.07E-05 | 0.000200844 | 0.100503581 |
| Obesity | t_cell | *HCG20* | -3.670149302 | 1.43E-05 | 0.00015444 | 0.069189235 |
| Obesity | t_cell | *C14orf144* | -4.117011629 | 0.00135391 | 0.005380157 | 1 |
| Obesity | t_cell | *LINC00578* | -3.4073825 | 4.86E-05 | 0.000386315 | 0.235652157 |
| Obesity | t_cell | *PATL2* | 3.104822357 | 2.33E-06 | 5.61E-05 | 0.011314973 |
| Obesity | t_cell | *CASC20* | -4.619546596 | 7.52E-06 | 9.81E-05 | 0.036493854 |
| Obesity | t_cell | *AC112229.4* | -2.888706218 | 3.88E-05 | 0.00032648 | 0.188052238 |
| Obesity | t_cell | *KLHL7-AS1* | -2.485168246 | 0.001690861 | 0.006476475 | 1 |
| Obesity | t_cell | *LINC00332* | -1.943439226 | 0.010417009 | 0.028427069 | 1 |
| Obesity | t_cell | *AC092171.4* | -7.552126885 | 0.002537011 | 0.008961952 | 1 |
| Obesity | t_cell | *RPS18* | 4.193604548 | 0.000108284 | 0.000722689 | 0.525394927 |
| Obesity | t_cell | *TAPBP* | 5.098964017 | 1.23E-06 | 4.13E-05 | 0.005951775 |
| Obesity | t_cell | *AC107021.1* | -3.565965097 | 0.0002451 | 0.001374825 | 1 |
| Obesity | t_cell | *TMA7* | 5.117838895 | 0.000450624 | 0.002217473 | 1 |
| Obesity | t_cell | *AC009495.3* | 2.057592314 | 2.84E-06 | 6.01E-05 | 0.013763687 |

| | | | | | | |
|---|---|---|---|---|---|---|
| Obesity | t_cell | *LINC00665* | -2.570624182 | 0.000859017 | 0.003714751 | 1 |
| Obesity | t_cell | *LYRM9* | -3.823235734 | 0.000168643 | 0.001025385 | 0.818257224 |
| Obesity | t_cell | *LINC01166* | -2.25344011 | 0.006142247 | 0.018487707 | 1 |
| Obesity | t_cell | *LINC00342* | 5.653602351 | 0.001939799 | 0.0072455 | 1 |
| Obesity | t_cell | *LINC00702* | -5.195980412 | 3.71E-05 | 0.000316028 | 0.179864411 |
| Obesity | t_cell | *GPX1* | 3.70430582 | 0.001674703 | 0.006428526 | 1 |
| Obesity | t_cell | *PINLYP* | -2.983313063 | 1.72E-05 | 0.000175502 | 0.083363341 |
| Obesity | t_cell | *ANKRD65* | -6.530471431 | 3.74E-06 | 6.55E-05 | 0.018153248 |
| Obesity | t_cell | *ZSCAN31* | 0.921027776 | 0.003923344 | 0.01264015 | 1 |
| Obesity | t_cell | *LINC00957* | -4.702529341 | 7.53E-05 | 0.000552423 | 0.365372409 |
| Obesity | t_cell | *KIAA0040* | 4.356415432 | 4.44E-06 | 7.25E-05 | 0.021529144 |
| Obesity | t_cell | *AC019117.2* | -18.36924214 | 6.31E-08 | 5.47E-06 | 0.000306353 |
| Obesity | t_cell | *LINC00987* | 2.293970044 | 7.51E-06 | 9.81E-05 | 0.036418658 |
| Obesity | t_cell | *GS1-124K5.4* | 0.903090839 | 0.003722081 | 0.012079957 | 1 |
| Obesity | t_cell | *LINC00959* | -4.484500393 | 0.007513486 | 0.021674004 | 1 |
| Obesity | t_cell | *IRGM* | 1.578947194 | 0.000544077 | 0.002605984 | 1 |
| Obesity | t_cell | *LINC01143* | -10.06637136 | 6.33E-06 | 8.95E-05 | 0.030735931 |
| Obesity | t_cell | *LINC00211* | -6.922538301 | 0.003293536 | 0.010941909 | 1 |
| Obesity | t_cell | *TXNDC5* | -16.63921036 | 8.01E-06 | 0.000101744 | 0.038866071 |
| Obesity | t_cell | *LINC01119* | -5.014278278 | 0.007987351 | 0.022837142 | 1 |
| Obesity | t_cell | *TNFSF12* | 3.272058199 | 0.00465897 | 0.01461236 | 1 |
| Obesity | t_cell | *APOBEC3G* | 0.801183333 | 0.000683689 | 0.003124547 | 1 |
| Obesity | t_cell | *C1orf226* | -6.26467645 | 5.99E-07 | 2.82E-05 | 0.002907495 |
| Obesity | t_cell | *PSMB9* | 2.720860533 | 1.36E-10 | 2.28E-08 | 6.62E-07 |
| Obesity | t_cell | *L1TD1* | -2.512667806 | 0.000767211 | 0.003408892 | 1 |
| Obesity | t_cell | *C1QTNF9* | -9.278400331 | 2.64E-06 | 5.93E-05 | 0.012808707 |
| Obesity | t_cell | *ISY1* | -3.516385366 | 1.37E-07 | 9.95E-06 | 0.000667096 |
| Obesity | t_cell | *PNMA2* | -3.175222662 | 1.86E-05 | 0.000186657 | 0.090314188 |
| Obesity | t_cell | *KRBOX1* | -33.21984019 | 2.83E-06 | 6.01E-05 | 0.013709748 |
| Obesity | t_cell | *LINC00877* | -2.535178193 | 0.002238441 | 0.008087055 | 1 |
| Obesity | t_cell | *CD302* | -3.741360829 | 0.012283079 | 0.032442842 | 1 |
| Obesity | t_cell | *INMT* | -2.97961924 | 0.001490721 | 0.005818968 | 1 |
| Obesity | t_cell | *C8orf58* | -7.388087089 | 2.79E-06 | 6.01E-05 | 0.013547304 |
| Obesity | t_cell | *HOGA1* | -6.847017181 | 4.55E-06 | 7.30E-05 | 0.022092057 |
| Obesity | t_cell | *MRPL20* | 4.547176286 | 0.003748745 | 0.012150241 | 1 |
| Obesity | t_cell | *LINC00960* | -0.771647817 | 0.00337096 | 0.011149214 | 1 |
| Obesity | t_cell | *DECR2* | -7.802948582 | 3.01E-05 | 0.000269805 | 0.146234179 |
| Obesity | t_cell | *ZNF709* | -41.41050907 | 0.000230605 | 0.001305597 | 1 |
| Obesity | t_cell | *ERVW-1* | -6.275753315 | 2.06E-05 | 0.000200566 | 0.099881811 |
| Obesity | t_cell | *FGF14-IT1* | -6.843713177 | 3.72E-06 | 6.54E-05 | 0.018038917 |
| Obesity | t_cell | *PALM2* | -4.260176609 | 0.001046565 | 0.004355004 | 1 |
| Obesity | t_cell | *IL10RB* | 4.117642931 | 0.00297224 | 0.010177354 | 1 |
| Obesity | t_cell | *PLA2G4B* | -6.528667025 | 0.000317425 | 0.001668632 | 1 |
| Obesity | t_cell | *LEFTY1* | -25.19513802 | 0.000193918 | 0.001144633 | 0.940887977 |
| Obesity | t_cell | *APOBEC3D* | 3.854035104 | 1.54E-06 | 4.50E-05 | 0.007460984 |
| Obesity | t_cell | *APOBEC3C* | 4.242970924 | 0.000397564 | 0.002011451 | 1 |
| Obesity | t_cell | *CCDC13* | -1.686267653 | 0.003835548 | 0.012398453 | 1 |
| Obesity | t_cell | *C4A* | -5.208076959 | 3.71E-07 | 2.02E-05 | 0.001799661 |
| Obesity | t_cell | *DNAJC3-AS1* | -5.211852566 | 0.000210063 | 0.001217716 | 1 |
| Obesity | t_cell | *LINC00926* | -6.474354034 | 1.28E-05 | 0.000143164 | 0.061989944 |

| | | | | | | |
|---|---|---|---|---|---|---|
| Obesity | t_cell | *CDK11B* | 4.977428936 | 0.016099454 | 0.040390151 | 1 |
| Obesity | t_cell | *GDNF-AS1* | -4.435807717 | 6.07E-05 | 0.000462216 | 0.29443185 |
| Obesity | t_cell | *CCDC153* | -3.240040168 | 0.001033229 | 0.004317665 | 1 |
| Obesity | t_cell | *LINC01060* | -3.028173652 | 0.000284384 | 0.00154863 | 1 |
| Obesity | t_cell | *LINC00942* | -14.88886015 | 1.23E-07 | 9.41E-06 | 0.000595093 |
| Obesity | t_cell | *DNAH10OS* | 41.85203194 | 9.75E-13 | 2.96E-10 | 4.73E-09 |
| Obesity | t_cell | *PCDHA10* | -1.98649938 | 0.000289153 | 0.001569316 | 1 |
| Obesity | t_cell | *ZNF718* | 4.126765285 | 0.008484701 | 0.023934749 | 1 |
| Obesity | t_cell | *LINC00989* | -3.769568964 | 8.56E-06 | 0.000106752 | 0.041526598 |
| Obesity | t_cell | *GLI4* | -6.464704502 | 1.33E-05 | 0.000147586 | 0.064347349 |
| Obesity | t_cell | *SNHG18* | -3.739741703 | 0.000729402 | 0.003289089 | 1 |
| Obesity | t_cell | *LINC01094* | -6.021408661 | 3.65E-05 | 0.000313219 | 0.177281899 |
| Obesity | t_cell | *UTP14C* | -4.807143138 | 0.001845732 | 0.006980117 | 1 |
| Obesity | t_cell | *PCDHGB2* | -12.79100398 | 0.005852765 | 0.017693218 | 1 |
| Obesity | t_cell | *ARMS2* | -4.114414479 | 0.000326381 | 0.001710152 | 1 |
| Obesity | t_cell | *MEX3A* | -2.422431054 | 0.003158338 | 0.010656644 | 1 |
| Obesity | t_cell | *DPP3* | 1.634355043 | 0.000281051 | 0.001537383 | 1 |
| Obesity | t_cell | *BRK1* | -2.983239777 | 0.004299159 | 0.01364259 | 1 |
| Obesity | t_cell | *SNHG9* | 5.281163748 | 4.06E-05 | 0.000336911 | 0.196756285 |
| Obesity | t_cell | *PCDHA3* | -6.733826265 | 1.64E-07 | 1.09E-05 | 0.000794627 |
| Obesity | t_cell | *HMBS* | 3.238028198 | 0.009258936 | 0.025685739 | 1 |
| Obesity | t_cell | *AC093668.2* | -4.32707927 | 0.002277281 | 0.008209038 | 1 |
| Obesity | t_cell | *ZBED6* | -6.729643177 | 5.32E-06 | 7.94E-05 | 0.025807159 |
| Obesity | t_cell | *CHURC1* | 2.374479303 | 0.011964144 | 0.031756032 | 1 |
| Obesity | t_cell | *RTEL1* | -1.961586406 | 0.016950357 | 0.042219267 | 1 |
| Obesity | t_cell | *PDF* | -11.74811779 | 6.38E-05 | 0.000480282 | 0.309782062 |
| Obesity | t_cell | *MC1R* | -3.225667926 | 0.003595016 | 0.01173034 | 1 |
| Obesity | t_cell | *LINC00639* | -2.286731133 | 1.63E-05 | 0.000168143 | 0.078924035 |
| Obesity | t_cell | *ITGB3* | -1.952863437 | 0.001524255 | 0.005940308 | 1 |
| Obesity | t_cell | *AC083843.1* | -48.38911466 | 3.05E-06 | 6.09E-05 | 0.014790334 |
| Obesity | t_cell | *PWRN1* | -3.478488506 | 0.000110637 | 0.00073344 | 0.536809936 |
| Obesity | t_cell | *LINC01003* | -4.053981192 | 0.000202607 | 0.001184398 | 0.983050268 |
| Obesity | t_cell | *C15orf65* | -3.448234175 | 6.51E-07 | 3.01E-05 | 0.003159625 |
| Obesity | t_cell | *C19orf84* | -25.66701947 | 3.94E-05 | 0.000330476 | 0.19101531 |
| Obesity | t_cell | *LINC00672* | 5.555208207 | 1.97E-06 | 5.12E-05 | 0.009576834 |
| Obesity | t_cell | *STRADA* | -4.095856486 | 0.001168286 | 0.004783566 | 1 |
| Obesity | t_cell | *ZNF582-AS1* | -3.997432978 | 5.21E-06 | 7.90E-05 | 0.025273013 |
| Obesity | t_cell | *LIN37* | -11.28347793 | 1.50E-06 | 4.50E-05 | 0.007301641 |
| Obesity | t_cell | *NDUFA7* | -13.07739025 | 4.62E-06 | 7.35E-05 | 0.022412686 |
| Obesity | t_cell | *SCGB1B2P* | -3.627217144 | 0.012499461 | 0.032871212 | 1 |
| Obesity | t_cell | *ZNF728* | -3.638427085 | 0.000138644 | 0.000877052 | 0.672698629 |
| Obesity | t_cell | *TRABD2B* | -3.450647242 | 4.00E-05 | 0.000334951 | 0.194271331 |
| Obesity | t_cell | *AC093157.1* | -4.145935184 | 0.011101469 | 0.029921415 | 1 |
| Obesity | t_cell | *YTHDF3-AS1* | -4.072240044 | 0.00055678 | 0.002658202 | 1 |
| Obesity | t_cell | *FGF14-AS2* | -3.987698647 | 5.73E-06 | 8.42E-05 | 0.027796165 |
| Obesity | t_cell | *CASC15* | -3.655927249 | 0.000302535 | 0.001627385 | 1 |
| Stroke | ASPC | *CD38* | 0.279008971 | 0.005571108 | 0.039927648 | 1 |
| Stroke | ASPC | *CIAPIN1* | 0.282862357 | 0.004233131 | 0.034901446 | 1 |
| Stroke | ASPC | *GCFC2* | 0.501384671 | 0.002881837 | 0.033849064 | 1 |
| Stroke | ASPC | *GGNBP2* | 0.415024278 | 0.003832008 | 0.034349037 | 1 |

| | | | | | | |
|---|---|---|---|---|---|---|
| Stroke | ASPC | *SYNRG* | 0.464869226 | 0.000409065 | 0.033849064 | 1 |
| Stroke | ASPC | *BAIAP2L1* | 0.673997271 | 0.003615722 | 0.034310859 | 1 |
| Stroke | ASPC | *AGK* | 0.261735001 | 0.000840651 | 0.033849064 | 1 |
| Stroke | ASPC | *VPS41* | 0.451916696 | 0.001032495 | 0.033849064 | 1 |
| Stroke | ASPC | *TSR3* | 0.619984435 | 0.006785562 | 0.044551483 | 1 |
| Stroke | ASPC | *RPS20* | 0.463553006 | 0.001209435 | 0.033849064 | 1 |
| Stroke | ASPC | *CEP68* | 0.237517297 | 0.00107759 | 0.033849064 | 1 |
| Stroke | ASPC | *RNF14* | 0.320969926 | 0.004023011 | 0.034570849 | 1 |
| Stroke | ASPC | *WWTR1* | 0.460000271 | 0.003308696 | 0.033849064 | 1 |
| Stroke | ASPC | *TTC27* | 0.351311459 | 0.007223912 | 0.046240663 | 1 |
| Stroke | ASPC | *ZDHHC6* | 0.500536702 | 0.005248989 | 0.038764222 | 1 |
| Stroke | ASPC | *AGPAT4* | 0.601118165 | 0.002713255 | 0.033849064 | 1 |
| Stroke | ASPC | *VEZT* | 0.487012866 | 0.002755621 | 0.033849064 | 1 |
| Stroke | ASPC | *CENPQ* | -0.060249898 | 0.000675348 | 0.033849064 | 1 |
| Stroke | ASPC | *PIAS1* | 0.331651181 | 0.000878065 | 0.033849064 | 1 |
| Stroke | ASPC | *VCL* | 0.473208563 | 0.003315581 | 0.033849064 | 1 |
| Stroke | ASPC | *ADSS* | 0.618146128 | 0.002594038 | 0.033849064 | 1 |
| Stroke | ASPC | *TBC1D23* | 0.51864569 | 0.003441858 | 0.033962746 | 1 |
| Stroke | ASPC | *PI4K2B* | 1.909375263 | 0.002470587 | 0.033849064 | 1 |
| Stroke | ASPC | *PQLC2* | 1.339285684 | 0.003164593 | 0.033849064 | 1 |
| Stroke | ASPC | *SPATA7* | 0.402185135 | 0.001757749 | 0.033849064 | 1 |
| Stroke | ASPC | *AP2S1* | 0.734894669 | 0.001499892 | 0.033849064 | 1 |
| Stroke | ASPC | *CTNNA1* | 0.452380353 | 0.002665588 | 0.033849064 | 1 |
| Stroke | ASPC | *YTHDC2* | 0.50436896 | 0.002746564 | 0.033849064 | 1 |
| Stroke | ASPC | *MAPK9* | 0.613774132 | 0.003897426 | 0.034349037 | 1 |
| Stroke | ASPC | *CYBA* | 0.527217762 | 0.00249651 | 0.033849064 | 1 |
| Stroke | ASPC | *ANAPC4* | 0.286617346 | 0.004148518 | 0.034824586 | 1 |
| Stroke | ASPC | *TBC1D22A* | 0.443170946 | 0.000369767 | 0.033849064 | 1 |
| Stroke | ASPC | *GALC* | 0.356222186 | 0.001451514 | 0.033849064 | 1 |
| Stroke | ASPC | *DNAJC25* | 0.831875943 | 0.006503725 | 0.043525619 | 1 |
| Stroke | ASPC | *RIMBP2* | 1.009970325 | 0.002956654 | 0.033849064 | 1 |
| Stroke | ASPC | *LTK* | 7.668096719 | 0.00272105 | 0.033849064 | 1 |
| Stroke | ASPC | *ADCK1* | 0.652925672 | 0.00732072 | 0.04640205 | 1 |
| Stroke | ASPC | *RNF4* | 0.62382697 | 0.002644563 | 0.033849064 | 1 |
| Stroke | ASPC | *TMEM161A* | 0.730067976 | 0.000927236 | 0.033849064 | 1 |
| Stroke | ASPC | *SUGP2* | 0.515666671 | 0.000504436 | 0.033849064 | 1 |
| Stroke | ASPC | *SLC12A2* | 0.657942301 | 0.000513812 | 0.033849064 | 1 |
| Stroke | ASPC | *SEC61A2* | 0.247161375 | 0.001541179 | 0.033849064 | 1 |
| Stroke | ASPC | *GOLGA5* | 0.450259975 | 0.006210961 | 0.042169793 | 1 |
| Stroke | ASPC | *STOML1* | 1.486456784 | 0.003223824 | 0.033849064 | 1 |
| Stroke | ASPC | *DHX8* | 0.679421439 | 0.00267143 | 0.033849064 | 1 |
| Stroke | ASPC | *ROGDI* | 1.243995831 | 0.005409416 | 0.039350054 | 1 |
| Stroke | ASPC | *IP6K2* | 0.349601095 | 0.006239783 | 0.042225144 | 1 |
| Stroke | ASPC | *SRBD1* | 0.453369899 | 0.002147271 | 0.033849064 | 1 |
| Stroke | ASPC | *CLEC2D* | 0.385826453 | 0.000565874 | 0.033849064 | 1 |
| Stroke | ASPC | *DRD4* | 0.959173996 | 0.003848247 | 0.034349037 | 1 |
| Stroke | ASPC | *PLA2G10* | 2.036101192 | 0.002985635 | 0.033849064 | 1 |
| Stroke | ASPC | *LRP6* | 0.431594404 | 0.005888559 | 0.041186451 | 1 |
| Stroke | ASPC | *DGCR2* | 0.302963098 | 0.005566042 | 0.039927648 | 1 |
| Stroke | ASPC | *ZXDC* | 0.424670737 | 0.00668412 | 0.044305125 | 1 |

| | | | | | | |
|---|---|---|---|---|---|---|
| Stroke | ASPC | *EIF2B3* | 0.455136857 | 0.004553651 | 0.035984225 | 1 |
| Stroke | ASPC | *WDR1* | 0.564918335 | 0.00078482 | 0.033849064 | 1 |
| Stroke | ASPC | *SNX13* | 0.484936443 | 0.002586435 | 0.033849064 | 1 |
| Stroke | ASPC | *TRNT1* | 0.40998492 | 0.001267391 | 0.033849064 | 1 |
| Stroke | ASPC | *SCARB1* | 0.273365655 | 0.00559733 | 0.039997413 | 1 |
| Stroke | ASPC | *ZNF532* | 0.51639146 | 0.002680128 | 0.033849064 | 1 |
| Stroke | ASPC | *ZZEF1* | 0.362076102 | 0.002121089 | 0.033849064 | 1 |
| Stroke | ASPC | *TIPIN* | 0.067916344 | 0.008097711 | 0.049185335 | 1 |
| Stroke | ASPC | *FNDC3B* | 0.377352145 | 0.002290655 | 0.033849064 | 1 |
| Stroke | ASPC | *KIFAP3* | 0.406927597 | 0.007219445 | 0.046240663 | 1 |
| Stroke | ASPC | *FMO4* | 0.247593327 | 0.006557079 | 0.043761963 | 1 |
| Stroke | ASPC | *ANKRD13A* | 0.727798435 | 8.65E-05 | 0.033849064 | 0.419876566 |
| Stroke | ASPC | *NT5C2* | 0.136852631 | 0.00711088 | 0.046013993 | 1 |
| Stroke | ASPC | *DYNC1I2* | 0.518305529 | 0.002325814 | 0.033849064 | 1 |
| Stroke | ASPC | *PIK3C3* | 0.390916512 | 0.007595928 | 0.047190065 | 1 |
| Stroke | ASPC | *DNM2* | 0.387268288 | 0.003038727 | 0.033849064 | 1 |
| Stroke | ASPC | *SCTR* | 0.573563741 | 0.007095248 | 0.046013993 | 1 |
| Stroke | ASPC | *STK17B* | 0.220137768 | 0.005085456 | 0.03808598 | 1 |
| Stroke | ASPC | *ME2* | 0.585594891 | 0.002901991 | 0.033849064 | 1 |
| Stroke | ASPC | *ZCCHC6* | 0.507203441 | 0.002567547 | 0.033849064 | 1 |
| Stroke | ASPC | *PIBF1* | 0.444272368 | 0.001866109 | 0.033849064 | 1 |
| Stroke | ASPC | *ZNF264* | 0.675331869 | 0.002890815 | 0.033849064 | 1 |
| Stroke | ASPC | *WBP11* | 0.7196278 | 0.002483934 | 0.033849064 | 1 |
| Stroke | ASPC | *CD59* | 0.145895963 | 0.004418552 | 0.03542847 | 1 |
| Stroke | ASPC | *HACE1* | 0.56988737 | 0.001990571 | 0.033849064 | 1 |
| Stroke | ASPC | *SLC25A24* | 0.397536111 | 0.005523153 | 0.03979928 | 1 |
| Stroke | ASPC | *MTIF2* | 0.621052703 | 0.005781902 | 0.040716676 | 1 |
| Stroke | ASPC | *USP40* | 0.488168791 | 0.002878724 | 0.033849064 | 1 |
| Stroke | ASPC | *ALG9* | 1.066427191 | 0.002001882 | 0.033849064 | 1 |
| Stroke | ASPC | *LPCAT2* | 0.67268108 | 0.001873021 | 0.033849064 | 1 |
| Stroke | ASPC | *ANKRD10* | 0.358286453 | 0.007034473 | 0.045883886 | 1 |
| Stroke | ASPC | *NSFL1C* | 0.19679773 | 0.007119955 | 0.046013993 | 1 |
| Stroke | ASPC | *SNX5* | 0.665278426 | 0.002448643 | 0.033849064 | 1 |
| Stroke | ASPC | *KIF16B* | 0.528531839 | 0.003251025 | 0.033849064 | 1 |
| Stroke | ASPC | *TBX5* | -0.291720311 | 0.005720361 | 0.040577764 | 1 |
| Stroke | ASPC | *GRAMD1A* | 0.555566496 | 0.005228356 | 0.038715461 | 1 |
| Stroke | ASPC | *CDIP1* | 0.80957173 | 0.003536446 | 0.033987065 | 1 |
| Stroke | ASPC | *CERS4* | 0.571221726 | 0.004202725 | 0.034901446 | 1 |
| Stroke | ASPC | *GLG1* | 0.418155039 | 0.005864933 | 0.041181845 | 1 |
| Stroke | ASPC | *DTX2* | 0.884780031 | 0.002616529 | 0.033849064 | 1 |
| Stroke | ASPC | *TF* | 0.834383586 | 0.005301627 | 0.038974992 | 1 |
| Stroke | ASPC | *SCFD1* | 0.427801247 | 0.004115305 | 0.034729134 | 1 |
| Stroke | ASPC | *SUPT16H* | 0.617309272 | 0.002796109 | 0.033849064 | 1 |
| Stroke | ASPC | *TOX4* | 0.430660394 | 0.00271463 | 0.033849064 | 1 |
| Stroke | ASPC | *COMT* | 0.56111426 | 0.003159556 | 0.033849064 | 1 |
| Stroke | ASPC | *LRRFIP2* | 0.458892285 | 0.001100803 | 0.033849064 | 1 |
| Stroke | ASPC | *SEC22C* | 0.660799788 | 0.001694099 | 0.033849064 | 1 |
| Stroke | ASPC | *XYLB* | 0.544698816 | 0.008045554 | 0.049165022 | 1 |
| Stroke | ASPC | *MSH2* | 0.652623086 | 0.002583142 | 0.033849064 | 1 |
| Stroke | ASPC | *PSMD5* | 0.578511716 | 0.00327304 | 0.033849064 | 1 |

| | | | | | | |
|---|---|---|---|---|---|---|
| Stroke | ASPC | *CRAT* | 0.979193765 | 0.003897791 | 0.034349037 | 1 |
| Stroke | ASPC | *WAC* | 0.389368721 | 0.003558415 | 0.033987065 | 1 |
| Stroke | ASPC | *RAB18* | 0.556807425 | 0.002796679 | 0.033849064 | 1 |
| Stroke | ASPC | *MARCH2* | 0.143505899 | 0.000531784 | 0.033849064 | 1 |
| Stroke | ASPC | *MED15* | 0.423548788 | 0.004020207 | 0.034570849 | 1 |
| Stroke | ASPC | *SH3BP1* | 1.053513716 | 0.006433654 | 0.043175782 | 1 |
| Stroke | ASPC | *HPS4* | 0.436051295 | 0.006098817 | 0.041914247 | 1 |
| Stroke | ASPC | *TTC28* | 0.365499485 | 0.003194582 | 0.033849064 | 1 |
| Stroke | ASPC | *TIMP3* | 0.386272456 | 0.003028154 | 0.033849064 | 1 |
| Stroke | ASPC | *PPP6R2* | 0.480479576 | 0.001244556 | 0.033849064 | 1 |
| Stroke | ASPC | *TSPO* | 0.610598209 | 0.003084069 | 0.033849064 | 1 |
| Stroke | ASPC | *SAMM50* | 0.39917152 | 0.00639238 | 0.043024 | 1 |
| Stroke | ASPC | *IFT27* | 0.57766493 | 0.002219782 | 0.033849064 | 1 |
| Stroke | ASPC | *ZC3H7B* | 0.35938298 | 0.005422976 | 0.039370903 | 1 |
| Stroke | ASPC | *MLC1* | 0.117413512 | 0.005335717 | 0.03899587 | 1 |
| Stroke | ASPC | *NIN* | 0.406247216 | 0.005095825 | 0.03808598 | 1 |
| Stroke | ASPC | *CNIH1* | 0.719861871 | 1.56E-05 | 0.018927564 | 0.075710256 |
| Stroke | ASPC | *ATP6V1D* | 0.544669002 | 0.00313962 | 0.033849064 | 1 |
| Stroke | ASPC | *MTHFD1* | 0.523738538 | 0.000479264 | 0.033849064 | 1 |
| Stroke | ASPC | *GSKIP* | 0.885226672 | 0.004238757 | 0.034901446 | 1 |
| Stroke | ASPC | *YY1* | 0.537545891 | 0.004100729 | 0.034729134 | 1 |
| Stroke | ASPC | *CCNB1IP1* | 0.696023436 | 0.00284158 | 0.033849064 | 1 |
| Stroke | ASPC | *ABHD12* | 0.1930462 | 0.008119838 | 0.049185335 | 1 |
| Stroke | ASPC | *CD40* | 0.633953627 | 0.004011277 | 0.034570849 | 1 |
| Stroke | ASPC | *NDRG3* | 0.454334808 | 0.004081551 | 0.034729134 | 1 |
| Stroke | ASPC | *CTSZ* | 0.563913216 | 0.002796707 | 0.033849064 | 1 |
| Stroke | ASPC | *DIDO1* | 0.322405706 | 0.006708173 | 0.044319658 | 1 |
| Stroke | ASPC | *METTL4* | 0.548618275 | 0.008211078 | 0.049490872 | 1 |
| Stroke | ASPC | *MYOM1* | 0.53433429 | 0.003916301 | 0.034349037 | 1 |
| Stroke | ASPC | *MIB1* | 0.483140495 | 0.002737833 | 0.033849064 | 1 |
| Stroke | ASPC | *STK24* | 0.510638851 | 0.002245536 | 0.033849064 | 1 |
| Stroke | ASPC | *ARHGEF7* | 0.483259076 | 0.002678829 | 0.033849064 | 1 |
| Stroke | ASPC | *KATNAL1* | 0.476107212 | 0.00463938 | 0.036365543 | 1 |
| Stroke | ASPC | *CLN5* | 0.563763865 | 0.001464798 | 0.033849064 | 1 |
| Stroke | ASPC | *MGRN1* | 0.345092068 | 0.00029576 | 0.033849064 | 1 |
| Stroke | ASPC | *MON1B* | 0.811152154 | 0.003341202 | 0.033849064 | 1 |
| Stroke | ASPC | *LMF1* | 0.56531686 | 0.003234942 | 0.033849064 | 1 |
| Stroke | ASPC | *RBL2* | 0.31933794 | 0.000715042 | 0.033849064 | 1 |
| Stroke | ASPC | *PYCARD* | 0.400662002 | 0.000390236 | 0.033849064 | 1 |
| Stroke | ASPC | *C16orf62* | 0.49386903 | 0.003116684 | 0.033849064 | 1 |
| Stroke | ASPC | *IQCH* | 0.611849359 | 0.003136046 | 0.033849064 | 1 |
| Stroke | ASPC | *TTC23* | 0.573029426 | 0.001753023 | 0.033849064 | 1 |
| Stroke | ASPC | *NIPAL2* | 0.403063283 | 0.005091015 | 0.03808598 | 1 |
| Stroke | ASPC | *CHRAC1* | 1.14107066 | 0.002928605 | 0.033849064 | 1 |
| Stroke | ASPC | *TUSC3* | 0.612789514 | 0.002413956 | 0.033849064 | 1 |
| Stroke | ASPC | *MED25* | 0.718253222 | 0.004828028 | 0.037301895 | 1 |
| Stroke | ASPC | *TGFB1* | 0.558283979 | 0.00273711 | 0.033849064 | 1 |
| Stroke | ASPC | *CAPS* | 1.052018745 | 0.003692947 | 0.034310859 | 1 |
| Stroke | ASPC | *RAB3A* | 3.187855439 | 0.006027857 | 0.041603359 | 1 |
| Stroke | ASPC | *ZNF14* | 0.747332868 | 0.002985435 | 0.033849064 | 1 |

| | | | | | | |
|---|---|---|---|---|---|---|
| Stroke | ASPC | *ZNF85* | 0.752985355 | 0.001078234 | 0.033849064 | 1 |
| Stroke | ASPC | *SP4* | 0.314768553 | 0.007893354 | 0.048742259 | 1 |
| Stroke | ASPC | *DNAH11* | 0.729754965 | 0.003330551 | 0.033849064 | 1 |
| Stroke | ASPC | *IQCE* | 0.736130464 | 0.00303667 | 0.033849064 | 1 |
| Stroke | ASPC | *CHN2* | 0.231769 | 0.001700053 | 0.033849064 | 1 |
| Stroke | ASPC | *GRB10* | 0.429453768 | 0.00798965 | 0.049048657 | 1 |
| Stroke | ASPC | *PLEKHA8* | 0.560513226 | 0.004884276 | 0.037482089 | 1 |
| Stroke | ASPC | *ZKSCAN1* | 0.440772266 | 0.002405715 | 0.033849064 | 1 |
| Stroke | ASPC | *EIF3B* | 0.541342026 | 0.006328041 | 0.042703273 | 1 |
| Stroke | ASPC | *C1GALT1* | 0.555306946 | 0.001769467 | 0.033849064 | 1 |
| Stroke | ASPC | *COA1* | 0.406706004 | 0.002307992 | 0.033849064 | 1 |
| Stroke | ASPC | *TBL2* | 0.168282295 | 0.003508585 | 0.03397935 | 1 |
| Stroke | ASPC | *FKTN* | 0.541112445 | 0.003768483 | 0.034310859 | 1 |
| Stroke | ASPC | *MEGF9* | 0.515808499 | 0.003932887 | 0.034349037 | 1 |
| Stroke | ASPC | *TRIM14* | 0.127128996 | 0.006730247 | 0.044326714 | 1 |
| Stroke | ASPC | *ECM2* | 0.395186601 | 0.007285212 | 0.04640205 | 1 |
| Stroke | ASPC | *CDH23* | 0.46139655 | 0.003890016 | 0.034349037 | 1 |
| Stroke | ASPC | *SFXN3* | 0.852099513 | 0.001594071 | 0.033849064 | 1 |
| Stroke | ASPC | *TNKS2* | 0.532484142 | 0.002681142 | 0.033849064 | 1 |
| Stroke | ASPC | *TFAM* | 0.762892521 | 0.002639148 | 0.033849064 | 1 |
| Stroke | ASPC | *TRIM37* | 0.48453183 | 0.001546138 | 0.033849064 | 1 |
| Stroke | ASPC | *PIGL* | 0.464116987 | 0.001214378 | 0.033849064 | 1 |
| Stroke | ASPC | *MED13* | 0.326953762 | 0.003924687 | 0.034349037 | 1 |
| Stroke | ASPC | *RAI1* | 0.667091902 | 0.00271121 | 0.033849064 | 1 |
| Stroke | ASPC | *GOSR1* | 0.536200702 | 0.002800412 | 0.033849064 | 1 |
| Stroke | ASPC | *ICAM2* | 0.616486777 | 0.005899543 | 0.041186451 | 1 |
| Stroke | ASPC | *RAB5C* | 0.164344679 | 0.007563422 | 0.04713631 | 1 |
| Stroke | ASPC | *EZH1* | 0.215312669 | 0.004372375 | 0.035344127 | 1 |
| Stroke | ASPC | *EFTUD2* | 0.657759197 | 0.002556525 | 0.033849064 | 1 |
| Stroke | ASPC | *DHRS7B* | 0.507845641 | 0.005486044 | 0.039628752 | 1 |
| Stroke | ASPC | *MYH3* | 1.219223356 | 0.001975148 | 0.033849064 | 1 |
| Stroke | ASPC | *UBE2D3* | 0.411918712 | 0.004861999 | 0.037482089 | 1 |
| Stroke | ASPC | *WFS1* | 0.743017558 | 0.005739249 | 0.040593057 | 1 |
| Stroke | ASPC | *STIM2* | 0.495533158 | 0.001381013 | 0.033849064 | 1 |
| Stroke | ASPC | *UFSP2* | 0.574722043 | 0.005155758 | 0.038312767 | 1 |
| Stroke | ASPC | *KLHL5* | 0.507000088 | 0.002748846 | 0.033849064 | 1 |
| Stroke | ASPC | *GALNT18* | 0.213590778 | 0.005329161 | 0.03899587 | 1 |
| Stroke | ASPC | *PRDM4* | 0.345512896 | 0.000715863 | 0.033849064 | 1 |
| Stroke | ASPC | *CORO1C* | 0.545091645 | 0.002643013 | 0.033849064 | 1 |
| Stroke | ASPC | *PARP11* | 0.592076906 | 0.002693839 | 0.033849064 | 1 |
| Stroke | ASPC | *GPN3* | 0.910820732 | 0.001043853 | 0.033849064 | 1 |
| Stroke | ASPC | *VPS29* | 0.449179544 | 0.008068269 | 0.049185335 | 1 |
| Stroke | ASPC | *VDR* | 1.228796217 | 0.004975934 | 0.037664947 | 1 |
| Stroke | ASPC | *CAND1* | 0.586547025 | 0.001069435 | 0.033849064 | 1 |
| Stroke | ASPC | *MDM1* | 0.635532026 | 0.004884397 | 0.037482089 | 1 |
| Stroke | ASPC | *NUP107* | 0.423556428 | 0.001989337 | 0.033849064 | 1 |
| Stroke | ASPC | *GNPTAB* | 0.432307863 | 0.003263482 | 0.033849064 | 1 |
| Stroke | ASPC | *SUDS3* | 0.618780222 | 0.002924661 | 0.033849064 | 1 |
| Stroke | ASPC | *C2CD5* | 0.484852952 | 0.003148287 | 0.033849064 | 1 |
| Stroke | ASPC | *FRK* | 0.740780202 | 0.003013599 | 0.033849064 | 1 |

| | | | | | | |
|---|---|---|---|---|---|---|
| Stroke | ASPC | *TMEM14C* | 0.689639304 | 0.002773085 | 0.033849064 | 1 |
| Stroke | ASPC | *RNGTT* | 0.512370184 | 0.002624315 | 0.033849064 | 1 |
| Stroke | ASPC | *MDN1* | 0.330124979 | 0.00174579 | 0.033849064 | 1 |
| Stroke | ASPC | *ACOT13* | 0.632742303 | 0.002744298 | 0.033849064 | 1 |
| Stroke | ASPC | *PACRG* | 0.675049776 | 0.004550832 | 0.035984225 | 1 |
| Stroke | ASPC | *QKI* | 0.397496902 | 0.002647348 | 0.033849064 | 1 |
| Stroke | ASPC | *TMEM30A* | 0.633596879 | 0.002953904 | 0.033849064 | 1 |
| Stroke | ASPC | *BTN2A1* | 0.32781115 | 0.003985971 | 0.034570849 | 1 |
| Stroke | ASPC | *NUDT12* | 0.719874173 | 0.003195411 | 0.033849064 | 1 |
| Stroke | ASPC | *ARSB* | 0.516951165 | 0.007431049 | 0.046764527 | 1 |
| Stroke | ASPC | *MSH3* | 0.157112127 | 0.00751491 | 0.047048186 | 1 |
| Stroke | ASPC | *RASGRF2* | 0.107058888 | 0.001677343 | 0.033849064 | 1 |
| Stroke | ASPC | *SLC12A7* | 0.732835191 | 0.002432162 | 0.033849064 | 1 |
| Stroke | ASPC | *SKP1* | 0.566507212 | 0.00273526 | 0.033849064 | 1 |
| Stroke | ASPC | *PPWD1* | 0.545857377 | 0.002606167 | 0.033849064 | 1 |
| Stroke | ASPC | *SMAD5* | 0.476167678 | 0.00579737 | 0.04076643 | 1 |
| Stroke | ASPC | *ERGIC1* | 0.50760982 | 0.004426782 | 0.03542847 | 1 |
| Stroke | ASPC | *TBCCD1* | 0.783398884 | 0.002979413 | 0.033849064 | 1 |
| Stroke | ASPC | *IQCG* | 0.502402508 | 0.006742192 | 0.044326714 | 1 |
| Stroke | ASPC | *SCAP* | 0.322768117 | 0.004960736 | 0.037608579 | 1 |
| Stroke | ASPC | *ARHGEF26* | 0.613713876 | 0.003487207 | 0.033975756 | 1 |
| Stroke | ASPC | *NKTR* | 0.311990504 | 0.003359027 | 0.033890905 | 1 |
| Stroke | ASPC | *ACTR1B* | 0.598999331 | 0.001566975 | 0.033849064 | 1 |
| Stroke | ASPC | *PPM1G* | 0.40404177 | 0.004296797 | 0.035191246 | 1 |
| Stroke | ASPC | *RTN4* | 0.276854898 | 0.000868761 | 0.033849064 | 1 |
| Stroke | ASPC | *POLE4* | 0.566952933 | 0.005290994 | 0.038955848 | 1 |
| Stroke | ASPC | *STRN* | 0.506194411 | 0.002801219 | 0.033849064 | 1 |
| Stroke | ASPC | *DARS* | 0.533213886 | 0.002650322 | 0.033849064 | 1 |
| Stroke | ASPC | *WIPF1* | 0.487611245 | 0.000999951 | 0.033849064 | 1 |
| Stroke | ASPC | *PNO1* | 0.823732642 | 0.00639556 | 0.043024 | 1 |
| Stroke | ASPC | *AAK1* | 0.430022803 | 0.002915989 | 0.033849064 | 1 |
| Stroke | ASPC | *ALMS1* | 0.609231123 | 0.000879856 | 0.033849064 | 1 |
| Stroke | ASPC | *EDEM3* | 0.45130947 | 0.003713232 | 0.034310859 | 1 |
| Stroke | ASPC | *DOCK7* | 0.553956679 | 0.000379577 | 0.033849064 | 1 |
| Stroke | ASPC | *PRG4* | 0.604768361 | 0.004103477 | 0.034729134 | 1 |
| Stroke | ASPC | *TBCE* | 0.595519776 | 0.003765626 | 0.034310859 | 1 |
| Stroke | ASPC | *SDHB* | 0.518763107 | 0.003249421 | 0.033849064 | 1 |
| Stroke | ASPC | *ZNHIT6* | 0.370574206 | 0.005234407 | 0.038715461 | 1 |
| Stroke | ASPC | *CD46* | 0.222872609 | 0.001758357 | 0.033849064 | 1 |
| Stroke | ASPC | *ERI3* | 0.520680729 | 0.001090124 | 0.033849064 | 1 |
| Stroke | ASPC | *PRRC2C* | 0.424615234 | 0.000262779 | 0.033849064 | 1 |
| Stroke | ASPC | *CD3EAP* | 0.303242331 | 0.006713716 | 0.044319658 | 1 |
| Stroke | ASPC | *RCN2* | 0.406701409 | 0.004657811 | 0.036392427 | 1 |
| Stroke | ASPC | *KLF7* | 0.351541636 | 0.002789783 | 0.033849064 | 1 |
| Stroke | ASPC | *SGIP1* | 0.667554652 | 0.003663203 | 0.034310859 | 1 |
| Stroke | ASPC | *FOXO3* | 0.375130903 | 0.003361001 | 0.033890905 | 1 |
| Stroke | ASPC | *UBN1* | 0.451113724 | 0.003963732 | 0.034527879 | 1 |
| Stroke | ASPC | *SENP5* | 0.441768243 | 0.008135773 | 0.049220411 | 1 |
| Stroke | ASPC | *RAB14* | 0.528384549 | 0.003413636 | 0.033940496 | 1 |
| Stroke | ASPC | *PPP6C* | 0.551422084 | 0.003216229 | 0.033849064 | 1 |

| | | | | | | |
|---|---|---|---|---|---|---|
| Stroke | ASPC | *DENND1A* | 0.379713924 | 0.004349166 | 0.035282444 | 1 |
| Stroke | ASPC | *KDSR* | 0.479879068 | 0.00734476 | 0.04640205 | 1 |
| Stroke | ASPC | *COQ6* | 0.658692142 | 0.00180161 | 0.033849064 | 1 |
| Stroke | ASPC | *CRIPT* | 0.721133708 | 0.002814038 | 0.033849064 | 1 |
| Stroke | ASPC | *EPCAM* | 0.284982179 | 0.002185299 | 0.033849064 | 1 |
| Stroke | ASPC | *KANSL1* | 0.334423279 | 0.004760625 | 0.036898644 | 1 |
| Stroke | ASPC | *GORAB* | 0.534438866 | 0.001772533 | 0.033849064 | 1 |
| Stroke | ASPC | *SLC10A7* | 0.51551851 | 0.003477565 | 0.033962746 | 1 |
| Stroke | ASPC | *MRC1* | 0.331924242 | 0.006545227 | 0.043743034 | 1 |
| Stroke | ASPC | *IQSEC3* | 0.846666374 | 0.000697354 | 0.033849064 | 1 |
| Stroke | ASPC | *CCDC77* | 0.323855898 | 0.004199445 | 0.034901446 | 1 |
| Stroke | ASPC | *DNAJC15* | 0.446324389 | 0.004385562 | 0.035346754 | 1 |
| Stroke | ASPC | *ALG5* | 0.276432937 | 0.0007428 | 0.033849064 | 1 |
| Stroke | ASPC | *SIL1* | 0.52163099 | 0.000327115 | 0.033849064 | 1 |
| Stroke | ASPC | *A1BG* | 0.28914643 | 0.007896005 | 0.048742259 | 1 |
| Stroke | ASPC | *LHX4* | 0.888502895 | 0.00390822 | 0.034349037 | 1 |
| Stroke | ASPC | *SEC22A* | 0.461417357 | 0.004531303 | 0.035983437 | 1 |
| Stroke | ASPC | *ZCCHC17* | 0.638191679 | 0.002621659 | 0.033849064 | 1 |
| Stroke | ASPC | *ZNF639* | 0.724445759 | 0.000788102 | 0.033849064 | 1 |
| Stroke | ASPC | *MTIF3* | 0.362647275 | 0.001344969 | 0.033849064 | 1 |
| Stroke | ASPC | *RBBP6* | 0.411361478 | 0.006131572 | 0.04206645 | 1 |
| Stroke | ASPC | *NAA60* | 1.006308509 | 0.003008517 | 0.033849064 | 1 |
| Stroke | ASPC | *ODF2L* | 0.520754503 | 0.001430385 | 0.033849064 | 1 |
| Stroke | ASPC | *SPATA1* | 0.542281359 | 0.000715677 | 0.033849064 | 1 |
| Stroke | ASPC | *KLHL7* | 0.556149277 | 0.001943256 | 0.033849064 | 1 |
| Stroke | ASPC | *NT5C3A* | 0.553968199 | 0.000302957 | 0.033849064 | 1 |
| Stroke | ASPC | *RBM19* | 0.63924543 | 0.002914087 | 0.033849064 | 1 |
| Stroke | ASPC | *DDX54* | 0.340947974 | 0.004752561 | 0.036895081 | 1 |
| Stroke | ASPC | *CCDC91* | 0.350996602 | 0.005194631 | 0.038538762 | 1 |
| Stroke | ASPC | *PFDN5* | 0.479489365 | 0.006969591 | 0.045636241 | 1 |
| Stroke | ASPC | *PLA2G12A* | 0.436286747 | 0.004657555 | 0.036392427 | 1 |
| Stroke | ASPC | *DNPEP* | 0.442123494 | 0.002934512 | 0.033849064 | 1 |
| Stroke | ASPC | *STAMBP* | 0.606413532 | 0.002879734 | 0.033849064 | 1 |
| Stroke | ASPC | *NQO2* | 0.594248948 | 0.00285587 | 0.033849064 | 1 |
| Stroke | ASPC | *GLO1* | 0.378938801 | 0.001225792 | 0.033849064 | 1 |
| Stroke | ASPC | *RREB1* | 0.261414949 | 0.002637098 | 0.033849064 | 1 |
| Stroke | ASPC | *RUNX2* | 0.418391919 | 0.002621732 | 0.033849064 | 1 |
| Stroke | ASPC | *CLYBL* | 0.557355213 | 0.001989025 | 0.033849064 | 1 |
| Stroke | ASPC | *SNRPD2* | 0.766398034 | 0.00283414 | 0.033849064 | 1 |
| Stroke | ASPC | *RBCK1* | 0.330266401 | 0.000988218 | 0.033849064 | 1 |
| Stroke | ASPC | *FLRT3* | 0.742754057 | 0.004237254 | 0.034901446 | 1 |
| Stroke | ASPC | *KLC1* | 0.638519683 | 0.00047045 | 0.033849064 | 1 |
| Stroke | ASPC | *TRAP1* | 0.078721179 | 0.001564077 | 0.033849064 | 1 |
| Stroke | ASPC | *KTN1* | 0.446388201 | 0.002723276 | 0.033849064 | 1 |
| Stroke | ASPC | *CANX* | 0.416038654 | 0.003691131 | 0.034310859 | 1 |
| Stroke | ASPC | *HELB* | 0.397563897 | 0.008115058 | 0.049185335 | 1 |
| Stroke | ASPC | *SMARCA4* | 0.333813343 | 0.002380312 | 0.033849064 | 1 |
| Stroke | ASPC | *HIP1* | 0.435115221 | 0.002774663 | 0.033849064 | 1 |
| Stroke | ASPC | *PEX1* | 0.634810135 | 0.001873287 | 0.033849064 | 1 |
| Stroke | ASPC | *CDHR3* | 0.541319569 | 0.003384383 | 0.033898981 | 1 |

| | | | | | | |
|---|---|---|---|---|---|---|
| Stroke | ASPC | *MTX2* | 0.474811439 | 0.006076408 | 0.041819477 | 1 |
| Stroke | ASPC | *TWSG1* | 0.459574241 | 0.007745188 | 0.047994449 | 1 |
| Stroke | ASPC | *ARPP19* | 0.594121747 | 0.004355768 | 0.035282444 | 1 |
| Stroke | ASPC | *BBOX1* | 2.606029018 | 0.006254723 | 0.042267296 | 1 |
| Stroke | ASPC | *PLD2* | 0.867414004 | 0.003648924 | 0.034310859 | 1 |
| Stroke | ASPC | *PHF20L1* | 0.530177075 | 3.81E-05 | 0.024154057 | 0.185086716 |
| Stroke | ASPC | *ADCY4* | 0.164371892 | 0.005937074 | 0.041270322 | 1 |
| Stroke | ASPC | *EPB41L4A* | 0.453789428 | 0.00230176 | 0.033849064 | 1 |
| Stroke | ASPC | *REEP5* | 0.528950841 | 0.002684292 | 0.033849064 | 1 |
| Stroke | ASPC | *TMEM8A* | 0.720820182 | 0.000791866 | 0.033849064 | 1 |
| Stroke | ASPC | *QRSL1* | 0.090845064 | 0.004952909 | 0.037608579 | 1 |
| Stroke | ASPC | *ACTN4* | 0.43988665 | 0.000802542 | 0.033849064 | 1 |
| Stroke | ASPC | *MAP1S* | 1.021509116 | 0.003117367 | 0.033849064 | 1 |
| Stroke | ASPC | *ATXN10* | 0.432282639 | 0.004332883 | 0.03527374 | 1 |
| Stroke | ASPC | *PNPLA7* | 0.534860059 | 0.004889976 | 0.037482089 | 1 |
| Stroke | ASPC | *DNMT1* | 0.660923853 | 0.003854258 | 0.034349037 | 1 |
| Stroke | ASPC | *NOL11* | 0.335876939 | 0.003894566 | 0.034349037 | 1 |
| Stroke | ASPC | *SLC35D2* | 0.640797459 | 0.000827708 | 0.033849064 | 1 |
| Stroke | ASPC | *KIF3A* | 0.556477734 | 0.000407011 | 0.033849064 | 1 |
| Stroke | ASPC | *KHDRBS3* | 0.660840752 | 0.003067599 | 0.033849064 | 1 |
| Stroke | ASPC | *PRKAB2* | 0.495538869 | 0.001929575 | 0.033849064 | 1 |
| Stroke | ASPC | *TRIM22* | 0.43043845 | 0.007319391 | 0.04640205 | 1 |
| Stroke | ASPC | *RGS22* | 0.141171983 | 0.002910794 | 0.033849064 | 1 |
| Stroke | ASPC | *AP3B1* | 0.455668436 | 0.002698525 | 0.033849064 | 1 |
| Stroke | ASPC | *WASF3* | 0.496562799 | 0.005314805 | 0.03899587 | 1 |
| Stroke | ASPC | *PEMT* | 0.806203197 | 0.003556004 | 0.033987065 | 1 |
| Stroke | ASPC | *MPRIP* | 0.518777223 | 0.002885035 | 0.033849064 | 1 |
| Stroke | ASPC | *DSTYK* | 0.583890784 | 0.00282527 | 0.033849064 | 1 |
| Stroke | ASPC | *EPSTI1* | 0.375497028 | 0.006702131 | 0.044319658 | 1 |
| Stroke | ASPC | *CSNK1G2* | 0.711770831 | 0.003725656 | 0.034310859 | 1 |
| Stroke | ASPC | *CNDP2* | 0.386401527 | 0.005336615 | 0.03899587 | 1 |
| Stroke | ASPC | *PDZD2* | 0.408688085 | 0.002630661 | 0.033849064 | 1 |
| Stroke | ASPC | *SLC2A11* | 0.166706218 | 0.000578896 | 0.033849064 | 1 |
| Stroke | ASPC | *FAM83F* | 0.766377068 | 0.006214221 | 0.042169793 | 1 |
| Stroke | ASPC | *ELP3* | 0.550141271 | 0.001295859 | 0.033849064 | 1 |
| Stroke | ASPC | *CEPT1* | 0.389112507 | 0.005407971 | 0.039350054 | 1 |
| Stroke | ASPC | *NAPG* | 0.4752881 | 0.001270453 | 0.033849064 | 1 |
| Stroke | ASPC | *KIDINS220* | 0.451145043 | 0.003266058 | 0.033849064 | 1 |
| Stroke | ASPC | *KIAA1468* | 0.503812253 | 0.002839913 | 0.033849064 | 1 |
| Stroke | ASPC | *KLRD1* | 0.626331076 | 0.001429366 | 0.033849064 | 1 |
| Stroke | ASPC | *ELP2* | 0.497424703 | 0.004111618 | 0.034729134 | 1 |
| Stroke | ASPC | *DHX34* | 0.729588449 | 0.000407707 | 0.033849064 | 1 |
| Stroke | ASPC | *CLOCK* | 0.486109859 | 0.003285066 | 0.033849064 | 1 |
| Stroke | ASPC | *ARGLU1* | 0.38561847 | 0.003497356 | 0.03397935 | 1 |
| Stroke | ASPC | *WDR36* | 0.535095652 | 0.007324427 | 0.04640205 | 1 |
| Stroke | ASPC | *UBQLN1* | 0.455430578 | 0.005536792 | 0.03979928 | 1 |
| Stroke | ASPC | *SRPK2* | 0.388047533 | 0.001937788 | 0.033849064 | 1 |
| Stroke | ASPC | *PHF21A* | 0.429923615 | 0.002954659 | 0.033849064 | 1 |
| Stroke | ASPC | *USP15* | 0.409468769 | 0.003748473 | 0.034310859 | 1 |
| Stroke | ASPC | *KLHL36* | 0.448622181 | 9.88E-06 | 0.015984796 | 0.047954388 |

| | | | | | | |
|---|---|---|---|---|---|---|
| Stroke | ASPC | *MPHOSPH6* | 0.553063655 | 0.003936122 | 0.034349037 | 1 |
| Stroke | ASPC | *NTPCR* | 0.590197994 | 0.00166441 | 0.033849064 | 1 |
| Stroke | ASPC | *SP110* | 0.469264424 | 0.000322433 | 0.033849064 | 1 |
| Stroke | ASPC | *USP37* | 0.261783617 | 0.002613546 | 0.033849064 | 1 |
| Stroke | ASPC | *TMBIM1* | 0.628427688 | 0.002947087 | 0.033849064 | 1 |
| Stroke | ASPC | *EPC2* | 0.374452435 | 0.007921813 | 0.048839439 | 1 |
| Stroke | ASPC | *DRAM1* | 0.519606409 | 0.007337681 | 0.04640205 | 1 |
| Stroke | ASPC | *SUCLA2* | 0.554295716 | 0.003185743 | 0.033849064 | 1 |
| Stroke | ASPC | *RCBTB2* | 0.607497084 | 0.002577638 | 0.033849064 | 1 |
| Stroke | ASPC | *SETDB2* | 0.577556847 | 0.000570787 | 0.033849064 | 1 |
| Stroke | ASPC | *KDELR2* | 0.730620068 | 0.002791306 | 0.033849064 | 1 |
| Stroke | ASPC | *NUPL2* | 0.589349569 | 0.002684078 | 0.033849064 | 1 |
| Stroke | ASPC | *ODF2* | 0.577946285 | 0.003554056 | 0.033987065 | 1 |
| Stroke | ASPC | *ZNF189* | 0.715199174 | 0.004122832 | 0.034729134 | 1 |
| Stroke | ASPC | *STX17* | 0.575969226 | 0.000868362 | 0.033849064 | 1 |
| Stroke | ASPC | *DENND4C* | 0.329369527 | 0.006140545 | 0.04206645 | 1 |
| Stroke | ASPC | *NARS2* | 0.551574461 | 0.002793513 | 0.033849064 | 1 |
| Stroke | ASPC | *FDX1* | 0.237500598 | 0.003796221 | 0.034349037 | 1 |
| Stroke | ASPC | *ACTR2* | 0.493098563 | 0.002791572 | 0.033849064 | 1 |
| Stroke | ASPC | *LRPPRC* | 0.509877059 | 0.002460047 | 0.033849064 | 1 |
| Stroke | ASPC | *RBP4* | 0.405975304 | 0.004204966 | 0.034901446 | 1 |
| Stroke | ASPC | *STAT4* | 0.485947264 | 0.007711096 | 0.047844294 | 1 |
| Stroke | ASPC | *COX17* | 0.821671467 | 0.002471159 | 0.033849064 | 1 |
| Stroke | ASPC | *KIAA1109* | 0.443039334 | 0.00281788 | 0.033849064 | 1 |
| Stroke | ASPC | *PRDM5* | 0.561961289 | 0.003746285 | 0.034310859 | 1 |
| Stroke | ASPC | *NUP54* | 0.158220912 | 0.006201167 | 0.042169793 | 1 |
| Stroke | ASPC | *KIF21A* | 0.472288662 | 0.004010511 | 0.034570849 | 1 |
| Stroke | ASPC | *PPFIA2* | 0.615852584 | 0.003031151 | 0.033849064 | 1 |
| Stroke | ASPC | *MARCH9* | 1.043225867 | 0.002818867 | 0.033849064 | 1 |
| Stroke | ASPC | *RITA1* | 1.224820079 | 0.004121828 | 0.034729134 | 1 |
| Stroke | ASPC | *GLTP* | 0.87388933 | 0.002946338 | 0.033849064 | 1 |
| Stroke | ASPC | *RB1* | 0.368649739 | 0.000498882 | 0.033849064 | 1 |
| Stroke | ASPC | *NOVA1* | 0.057771102 | 0.002928987 | 0.033849064 | 1 |
| Stroke | ASPC | *TMX1* | 0.643663775 | 0.003232708 | 0.033849064 | 1 |
| Stroke | ASPC | *C14orf37* | 0.461931244 | 0.004343615 | 0.035282444 | 1 |
| Stroke | ASPC | *RAB15* | 1.013313548 | 0.000583391 | 0.033849064 | 1 |
| Stroke | ASPC | *WARS* | 0.529064887 | 0.000502631 | 0.033849064 | 1 |
| Stroke | ASPC | *SERF2* | 0.572223614 | 0.000822857 | 0.033849064 | 1 |
| Stroke | ASPC | *HMG20A* | 0.556218876 | 0.002780792 | 0.033849064 | 1 |
| Stroke | ASPC | *DHX38* | 0.720388116 | 0.002625883 | 0.033849064 | 1 |
| Stroke | ASPC | *TLDC1* | 0.749073468 | 0.0039208 | 0.034349037 | 1 |
| Stroke | ASPC | *TCF25* | 0.295407624 | 0.003857541 | 0.034349037 | 1 |
| Stroke | ASPC | *NCOR1* | 0.416272684 | 0.002757758 | 0.033849064 | 1 |
| Stroke | ASPC | *SSH2* | 0.407406592 | 0.0026422 | 0.033849064 | 1 |
| Stroke | ASPC | *IMPA2* | 0.486664006 | 0.000160245 | 0.033849064 | 0.777507089 |
| Stroke | ASPC | *GNAL* | 0.640507116 | 0.002902795 | 0.033849064 | 1 |
| Stroke | ASPC | *ANAPC11* | 0.616390048 | 0.007140506 | 0.046013993 | 1 |
| Stroke | ASPC | *MAPK4* | 1.776821612 | 0.004709409 | 0.036677449 | 1 |
| Stroke | ASPC | *DPP9* | 0.704302517 | 0.001489533 | 0.033849064 | 1 |
| Stroke | ASPC | *DOPEY2* | 0.177706119 | 3.98E-05 | 0.024154057 | 0.193232453 |

| | | | | | | |
|---|---|---|---|---|---|---|
| Stroke | ASPC | *EVI5L* | 0.892767428 | 0.002338756 | 0.033849064 | 1 |
| Stroke | ASPC | *SLC44A3* | 0.522298738 | 0.00320714 | 0.033849064 | 1 |
| Stroke | ASPC | *ALDH9A1* | 0.670486166 | 0.002532107 | 0.033849064 | 1 |
| Stroke | ASPC | *TARS2* | 0.290308709 | 0.001215241 | 0.033849064 | 1 |
| Stroke | ASPC | *SMYD2* | 0.646362226 | 0.003924044 | 0.034349037 | 1 |
| Stroke | ASPC | *RAB13* | 0.979777879 | 0.001269412 | 0.033849064 | 1 |
| Stroke | ASPC | *S100A8* | 0.64515877 | 0.003895143 | 0.034349037 | 1 |
| Stroke | ASPC | *RHOB* | 0.643989258 | 0.003096048 | 0.033849064 | 1 |
| Stroke | ASPC | *GALM* | 0.617259983 | 0.002384173 | 0.033849064 | 1 |
| Stroke | ASPC | *VPS54* | 0.494065775 | 0.001456104 | 0.033849064 | 1 |
| Stroke | ASPC | *EXOC6B* | 0.391578518 | 0.002975733 | 0.033849064 | 1 |
| Stroke | ASPC | *TEX261* | 0.98962066 | 0.003715647 | 0.034310859 | 1 |
| Stroke | ASPC | *RALB* | 0.690071899 | 0.00238851 | 0.033849064 | 1 |
| Stroke | ASPC | *POLR2D* | 0.206642607 | 0.008204902 | 0.049490872 | 1 |
| Stroke | ASPC | *METTL21A* | 0.378416918 | 0.007567845 | 0.04713631 | 1 |
| Stroke | ASPC | *NBEAL1* | 0.469101633 | 1.48E-06 | 0.007164717 | 0.007164717 |
| Stroke | ASPC | *SUMF1* | 0.416607623 | 0.002355644 | 0.033849064 | 1 |
| Stroke | ASPC | *ARL6IP5* | 0.60337901 | 0.000444619 | 0.033849064 | 1 |
| Stroke | ASPC | *MED12L* | 0.560817947 | 0.00623501 | 0.042225144 | 1 |
| Stroke | ASPC | *PYURF* | 0.453233409 | 0.007356787 | 0.046417594 | 1 |
| Stroke | ASPC | *MARCH1* | 0.146369113 | 0.003313861 | 0.033849064 | 1 |
| Stroke | ASPC | *CBR4* | 0.438538551 | 0.000914906 | 0.033849064 | 1 |
| Stroke | ASPC | *MARCH6* | 0.488049039 | 0.001711052 | 0.033849064 | 1 |
| Stroke | ASPC | *GTF2H2* | 1.098003063 | 0.003295729 | 0.033849064 | 1 |
| Stroke | ASPC | *SPATA9* | 0.567490102 | 0.004609106 | 0.036363227 | 1 |
| Stroke | ASPC | *FEM1C* | 0.802698692 | 0.002644472 | 0.033849064 | 1 |
| Stroke | ASPC | *FAM193B* | 0.353970355 | 0.007248166 | 0.046334783 | 1 |
| Stroke | ASPC | *IRAK1BP1* | 0.287114832 | 0.000814781 | 0.033849064 | 1 |
| Stroke | ASPC | *PHIP* | 0.505369339 | 0.000417883 | 0.033849064 | 1 |
| Stroke | ASPC | *PNRC1* | 0.647477388 | 0.002513299 | 0.033849064 | 1 |
| Stroke | ASPC | *RNF217* | 0.456917826 | 0.001576715 | 0.033849064 | 1 |
| Stroke | ASPC | *ARHGAP18* | 0.216745561 | 0.000952521 | 0.033849064 | 1 |
| Stroke | ASPC | *RBAK* | 1.12887684 | 0.002577717 | 0.033849064 | 1 |
| Stroke | ASPC | *GIGYF1* | 0.281307007 | 0.002785358 | 0.033849064 | 1 |
| Stroke | ASPC | *TMEM209* | 0.624755477 | 0.004419828 | 0.03542847 | 1 |
| Stroke | ASPC | *NCAPG2* | 0.226143186 | 0.007513396 | 0.047048186 | 1 |
| Stroke | ASPC | *DOCK5* | 0.338510853 | 0.005920046 | 0.041270206 | 1 |
| Stroke | ASPC | *DPYS* | 0.525568802 | 0.003700264 | 0.034310859 | 1 |
| Stroke | ASPC | *LRP12* | 0.706739525 | 0.003223191 | 0.033849064 | 1 |
| Stroke | ASPC | *HAUS6* | 0.656159669 | 0.003550537 | 0.033987065 | 1 |
| Stroke | ASPC | *REXO4* | 0.743471708 | 0.002822091 | 0.033849064 | 1 |
| Stroke | ASPC | *SH3GLB2* | 0.460781046 | 0.003573375 | 0.034062899 | 1 |
| Stroke | ASPC | *RSU1* | 0.423274036 | 0.00570785 | 0.040548297 | 1 |
| Stroke | ASPC | *ADD3* | 0.386852122 | 0.004169185 | 0.034901446 | 1 |
| Stroke | ASPC | *PAOX* | 0.930809068 | 0.002753073 | 0.033849064 | 1 |
| Stroke | ASPC | *PTPRJ* | 0.520524142 | 0.000154441 | 0.033849064 | 0.749350148 |
| Stroke | ASPC | *NPAT* | 0.537056062 | 0.00502474 | 0.037916075 | 1 |
| Stroke | ASPC | *SAP18* | 0.672795361 | 0.003264602 | 0.033849064 | 1 |
| Stroke | ASPC | *TMEM18* | 0.863013271 | 0.002712484 | 0.033849064 | 1 |
| Stroke | ASPC | *NUBPL* | 0.480521291 | 0.002869751 | 0.033849064 | 1 |

| | | | | | | |
|---|---|---|---|---|---|---|
| Stroke | ASPC | *ANKRD50* | 0.402361292 | 0.001853896 | 0.033849064 | 1 |
| Stroke | ASPC | *UPF2* | 0.457876834 | 0.001797901 | 0.033849064 | 1 |
| Stroke | ASPC | *PIGF* | 0.585324454 | 0.003020469 | 0.033849064 | 1 |
| Stroke | ASPC | *RNF144A* | 0.514539815 | 0.002657615 | 0.033849064 | 1 |
| Stroke | ASPC | *BICD1* | 0.516769178 | 0.002728589 | 0.033849064 | 1 |
| Stroke | ASPC | *CACUL1* | 0.292135126 | 0.008013439 | 0.04909243 | 1 |
| Stroke | ASPC | *TIAL1* | 0.437040973 | 0.008285769 | 0.049879096 | 1 |
| Stroke | ASPC | *PTPN14* | 0.379554193 | 0.002734189 | 0.033849064 | 1 |
| Stroke | ASPC | *TMEM178A* | 1.206304235 | 0.008199867 | 0.049490872 | 1 |
| Stroke | ASPC | *MBNL1* | 0.358709354 | 0.002593842 | 0.033849064 | 1 |
| Stroke | ASPC | *SREK1IP1* | 0.551319174 | 0.003828442 | 0.034349037 | 1 |
| Stroke | ASPC | *TMEM87B* | 0.553856545 | 0.002871003 | 0.033849064 | 1 |
| Stroke | ASPC | *PLA2R1* | 0.101770746 | 0.001843305 | 0.033849064 | 1 |
| Stroke | ASPC | *LPCAT1* | 0.86544635 | 0.002450505 | 0.033849064 | 1 |
| Stroke | ASPC | *CFDP1* | 0.471231032 | 0.00308859 | 0.033849064 | 1 |
| Stroke | ASPC | *ZDHHC7* | 0.582049902 | 0.003474547 | 0.033962746 | 1 |
| Stroke | ASPC | *FBXO36* | 0.626845724 | 0.004177615 | 0.034901446 | 1 |
| Stroke | ASPC | *SEMA3D* | 0.44182059 | 0.006591248 | 0.043929581 | 1 |
| Stroke | ASPC | *PPP2R5E* | 0.501484006 | 0.002213852 | 0.033849064 | 1 |
| Stroke | ASPC | *ANKRD29* | 0.54200152 | 0.00606874 | 0.041819477 | 1 |
| Stroke | ASPC | *ANKH* | 0.387010101 | 0.005376872 | 0.039230953 | 1 |
| Stroke | ASPC | *ROBO4* | 0.635330191 | 0.003336035 | 0.033849064 | 1 |
| Stroke | ASPC | *ROBO3* | 0.192413368 | 0.00619967 | 0.042169793 | 1 |
| Stroke | ASPC | *PRKCA* | 0.385174783 | 0.006602225 | 0.043942382 | 1 |
| Stroke | ASPC | *CCSAP* | 0.970646559 | 0.00254559 | 0.033849064 | 1 |
| Stroke | ASPC | *ATP5G3* | 0.093232133 | 0.003990372 | 0.034570849 | 1 |
| Stroke | ASPC | *OXNAD1* | 0.450927664 | 0.007286066 | 0.04640205 | 1 |
| Stroke | ASPC | *MARCKS* | 0.258498511 | 0.001950224 | 0.033849064 | 1 |
| Stroke | ASPC | *PDIA4* | 0.768525049 | 0.002311641 | 0.033849064 | 1 |
| Stroke | ASPC | *C14orf2* | 0.265756686 | 0.003059689 | 0.033849064 | 1 |
| Stroke | ASPC | *RPL30* | 0.41467659 | 0.001090182 | 0.033849064 | 1 |
| Stroke | ASPC | *SUPV3L1* | 0.598200371 | 0.000205802 | 0.033849064 | 0.998549019 |
| Stroke | ASPC | *NODAL* | 1.175606475 | 0.005470578 | 0.039616784 | 1 |
| Stroke | ASPC | *ZDHHC5* | 0.681177573 | 0.002981938 | 0.033849064 | 1 |
| Stroke | ASPC | *RPUSD3* | 0.352046232 | 0.004300991 | 0.035191246 | 1 |
| Stroke | ASPC | *NECAP2* | 0.667014659 | 0.002624952 | 0.033849064 | 1 |
| Stroke | ASPC | *CLDN12* | 0.852382193 | 0.004212768 | 0.034901446 | 1 |
| Stroke | ASPC | *MMP14* | 0.699254674 | 0.000568939 | 0.033849064 | 1 |
| Stroke | ASPC | *GATAD1* | 0.442131603 | 0.005751991 | 0.040623961 | 1 |
| Stroke | ASPC | *DHRS4* | 0.949879271 | 0.00258334 | 0.033849064 | 1 |
| Stroke | ASPC | *SLC38A10* | 0.594932484 | 0.002665805 | 0.033849064 | 1 |
| Stroke | ASPC | *EXTL1* | 13.6151918 | 0.002571764 | 0.033849064 | 1 |
| Stroke | ASPC | *CCDC58* | 0.803763871 | 0.003476506 | 0.033962746 | 1 |
| Stroke | ASPC | *PCNT* | 0.104129975 | 0.004115184 | 0.034729134 | 1 |
| Stroke | ASPC | *TAOK1* | 0.446570482 | 0.002143257 | 0.033849064 | 1 |
| Stroke | ASPC | *FDPS* | 0.419495166 | 7.02E-05 | 0.033849064 | 0.340791564 |
| Stroke | ASPC | *ZNF333* | 0.682866093 | 0.001601389 | 0.033849064 | 1 |
| Stroke | ASPC | *PGAP3* | 0.893240244 | 0.004543541 | 0.035984225 | 1 |
| Stroke | ASPC | *ZNF577* | 0.232759741 | 0.000271406 | 0.033849064 | 1 |
| Stroke | ASPC | *TMEM143* | 0.516681861 | 0.004201962 | 0.034901446 | 1 |

| | | | | | | |
|---|---|---|---|---|---|---|
| Stroke | ASPC | *PLCD3* | 0.990284983 | 0.002727886 | 0.033849064 | 1 |
| Stroke | ASPC | *RPS6KA4* | 0.962728613 | 0.003643111 | 0.034310859 | 1 |
| Stroke | ASPC | *COA7* | 1.048977209 | 0.00432255 | 0.035248758 | 1 |
| Stroke | ASPC | *PDPN* | 0.640512171 | 0.002163273 | 0.033849064 | 1 |
| Stroke | ASPC | *LAPTM5* | 0.569885376 | 0.004446813 | 0.03542847 | 1 |
| Stroke | ASPC | *TM2D1* | 0.51743789 | 0.002726153 | 0.033849064 | 1 |
| Stroke | ASPC | *CNST* | 0.445473284 | 0.00345953 | 0.033962746 | 1 |
| Stroke | ASPC | *KIAA1841* | 0.439743918 | 0.005954149 | 0.041329802 | 1 |
| Stroke | ASPC | *VPS72* | 0.764424188 | 0.003366739 | 0.033890905 | 1 |
| Stroke | ASPC | *TDRD10* | 0.592573835 | 0.004042186 | 0.034651391 | 1 |
| Stroke | ASPC | *GPR155* | 0.545999 | 0.002356196 | 0.033849064 | 1 |
| Stroke | ASPC | *AIM2* | 0.94417825 | 0.002478682 | 0.033849064 | 1 |
| Stroke | ASPC | *GTPBP8* | 0.824283145 | 0.002253186 | 0.033849064 | 1 |
| Stroke | ASPC | *WDFY3* | 0.379740466 | 0.002127777 | 0.033849064 | 1 |
| Stroke | ASPC | *C4orf36* | 0.689718224 | 0.002787685 | 0.033849064 | 1 |
| Stroke | ASPC | *DNASE1L3* | 0.130721155 | 0.003297927 | 0.033849064 | 1 |
| Stroke | ASPC | *CRELD1* | 0.541048313 | 0.001491499 | 0.033849064 | 1 |
| Stroke | ASPC | *CPA3* | 0.958712286 | 0.006147365 | 0.04206645 | 1 |
| Stroke | ASPC | *SLC4A1AP* | 0.712843761 | 0.002752954 | 0.033849064 | 1 |
| Stroke | ASPC | *SNIP1* | 0.78095156 | 0.003462583 | 0.033962746 | 1 |
| Stroke | ASPC | *RPN1* | 0.679186207 | 0.002366127 | 0.033849064 | 1 |
| Stroke | ASPC | *SFMBT1* | 0.671004604 | 0.005779164 | 0.040716676 | 1 |
| Stroke | ASPC | *HMGB2* | 0.257829606 | 0.003139101 | 0.033849064 | 1 |
| Stroke | ASPC | *MAP9* | 0.618405985 | 0.001549961 | 0.033849064 | 1 |
| Stroke | ASPC | *IL15* | 0.257900672 | 0.007035781 | 0.045883886 | 1 |
| Stroke | ASPC | *RNF180* | 0.351919658 | 0.008025346 | 0.049103376 | 1 |
| Stroke | ASPC | *PDSS2* | 0.471762738 | 0.001996475 | 0.033849064 | 1 |
| Stroke | ASPC | *RP9* | 0.841145031 | 0.002783457 | 0.033849064 | 1 |
| Stroke | ASPC | *USP49* | 0.625423307 | 0.002703789 | 0.033849064 | 1 |
| Stroke | ASPC | *SLC13A4* | 0.632143002 | 0.002858198 | 0.033849064 | 1 |
| Stroke | ASPC | *BRI3* | 0.566628044 | 0.001477184 | 0.033849064 | 1 |
| Stroke | ASPC | *RAD21* | 0.632206948 | 3.48E-05 | 0.024154057 | 0.168829148 |
| Stroke | ASPC | *FASTK* | 0.846408988 | 0.001333465 | 0.033849064 | 1 |
| Stroke | ASPC | *YWHAZ* | 0.290230592 | 0.000451756 | 0.033849064 | 1 |
| Stroke | ASPC | *SLC25A32* | 0.381671474 | 0.002789769 | 0.033849064 | 1 |
| Stroke | ASPC | *TMEM65* | 0.556754655 | 0.002642431 | 0.033849064 | 1 |
| Stroke | ASPC | *WRN* | 0.45670434 | 0.001090027 | 0.033849064 | 1 |
| Stroke | ASPC | *CFL2* | 0.654869633 | 0.003402474 | 0.033898981 | 1 |
| Stroke | ASPC | *REEP3* | 0.383533601 | 0.000942113 | 0.033849064 | 1 |
| Stroke | ASPC | *MICU2* | 0.480482697 | 0.001294213 | 0.033849064 | 1 |
| Stroke | ASPC | *LRR1* | 0.996813244 | 0.004636888 | 0.036365543 | 1 |
| Stroke | ASPC | *QSOX2* | 0.412412109 | 0.007007683 | 0.045823826 | 1 |
| Stroke | ASPC | *HSPA12A* | 0.094880832 | 0.002997079 | 0.033849064 | 1 |
| Stroke | ASPC | *UBTD1* | 0.557438938 | 0.004954263 | 0.037608579 | 1 |
| Stroke | ASPC | *TCP11L2* | 0.591427187 | 0.002276033 | 0.033849064 | 1 |
| Stroke | ASPC | *API5* | 0.596877198 | 0.003046898 | 0.033849064 | 1 |
| Stroke | ASPC | *WBP1L* | 0.39806091 | 0.000621102 | 0.033849064 | 1 |
| Stroke | ASPC | *WDR88* | 0.700307263 | 0.002590174 | 0.033849064 | 1 |
| Stroke | ASPC | *RPL27A* | 0.562506925 | 0.00261725 | 0.033849064 | 1 |
| Stroke | ASPC | *TMEM41B* | 0.610932298 | 0.001363911 | 0.033849064 | 1 |

| | | | | | | |
|---|---|---|---|---|---|---|
| Stroke | ASPC | *SEC11C* | 0.669327633 | 0.007372851 | 0.046458535 | 1 |
| Stroke | ASPC | *CHRFAM7A* | 0.112943958 | 0.001635833 | 0.033849064 | 1 |
| Stroke | ASPC | *SLFN5* | 0.371659115 | 0.007097464 | 0.046013993 | 1 |
| Stroke | ASPC | *SAAL1* | 0.915594156 | 0.002399559 | 0.033849064 | 1 |
| Stroke | ASPC | *MTMR10* | 0.262613052 | 0.001509482 | 0.033849064 | 1 |
| Stroke | ASPC | *EPB42* | 0.527183587 | 0.00278563 | 0.033849064 | 1 |
| Stroke | ASPC | *CNPY4* | 0.23155339 | 0.000682897 | 0.033849064 | 1 |
| Stroke | ASPC | *SP2* | 0.749994593 | 0.002980818 | 0.033849064 | 1 |
| Stroke | ASPC | *TBC1D2B* | 0.218875512 | 0.003275798 | 0.033849064 | 1 |
| Stroke | ASPC | *VKORC1* | 0.841822089 | 0.004958779 | 0.037608579 | 1 |
| Stroke | ASPC | *SPATA33* | 0.763647858 | 0.004720762 | 0.036706947 | 1 |
| Stroke | ASPC | *ZNF641* | 0.570541315 | 0.000579449 | 0.033849064 | 1 |
| Stroke | ASPC | *TP53I13* | 0.363305042 | 0.001556749 | 0.033849064 | 1 |
| Stroke | ASPC | *ZNF528* | 0.589570276 | 0.004913787 | 0.037545977 | 1 |
| Stroke | ASPC | *TTYH1* | 1.166669944 | 0.003455707 | 0.033962746 | 1 |
| Stroke | ASPC | *LENG8* | 0.546721117 | 0.00036549 | 0.033849064 | 1 |
| Stroke | ASPC | *FAM57A* | 0.126058139 | 0.001269108 | 0.033849064 | 1 |
| Stroke | ASPC | *CYB5D2* | 0.55221268 | 0.001066054 | 0.033849064 | 1 |
| Stroke | ASPC | *SPRYD3* | 0.47264408 | 0.000501231 | 0.033849064 | 1 |
| Stroke | ASPC | *NDUFV1* | 0.606627188 | 0.002759362 | 0.033849064 | 1 |
| Stroke | ASPC | *PRDX2* | 0.739814573 | 0.000645277 | 0.033849064 | 1 |
| Stroke | ASPC | *MIS12* | 0.671819253 | 0.003670854 | 0.034310859 | 1 |
| Stroke | ASPC | *ATG16L2* | 0.634998843 | 0.002475102 | 0.033849064 | 1 |
| Stroke | ASPC | *COA6* | 1.040289605 | 0.002328638 | 0.033849064 | 1 |
| Stroke | ASPC | *PCMTD1* | 0.454581886 | 0.002817413 | 0.033849064 | 1 |
| Stroke | ASPC | *RFWD3* | 0.386858865 | 0.005929589 | 0.041270322 | 1 |
| Stroke | ASPC | *SLC20A2* | 0.35564782 | 0.007892689 | 0.048742259 | 1 |
| Stroke | ASPC | *ADAM9* | 0.285239949 | 0.003086375 | 0.033849064 | 1 |
| Stroke | ASPC | *DNAJC21* | 0.670380674 | 0.002705768 | 0.033849064 | 1 |
| Stroke | ASPC | *SNTB2* | 0.468773859 | 0.004136072 | 0.03478028 | 1 |
| Stroke | ASPC | *STX18* | 0.552424744 | 0.002827163 | 0.033849064 | 1 |
| Stroke | ASPC | *CEP120* | 0.515476437 | 0.002651413 | 0.033849064 | 1 |
| Stroke | ASPC | *MFF* | 0.505642025 | 0.003506524 | 0.03397935 | 1 |
| Stroke | ASPC | *MFSD7* | 1.508298269 | 0.00234622 | 0.033849064 | 1 |
| Stroke | ASPC | *MAP2K1* | 0.565098966 | 0.002336338 | 0.033849064 | 1 |
| Stroke | ASPC | *UPF3A* | 0.176693589 | 0.002176364 | 0.033849064 | 1 |
| Stroke | ASPC | *THBS3* | 0.465964577 | 0.000117686 | 0.033849064 | 0.571011525 |
| Stroke | ASPC | *CLIC3* | 0.997674082 | 0.007110891 | 0.046013993 | 1 |
| Stroke | ASPC | *LUZP1* | 0.562747909 | 0.0015785 | 0.033849064 | 1 |
| Stroke | ASPC | *CSGALNACT2* | 0.632594907 | 0.000330883 | 0.033849064 | 1 |
| Stroke | ASPC | *AVEN* | 0.553162458 | 0.001737813 | 0.033849064 | 1 |
| Stroke | ASPC | *MAP3K2* | 0.253927301 | 0.000401785 | 0.033849064 | 1 |
| Stroke | ASPC | *PDCD6IP* | 0.474736434 | 0.00278249 | 0.033849064 | 1 |
| Stroke | ASPC | *CNGB3* | 0.189250405 | 0.002270207 | 0.033849064 | 1 |
| Stroke | ASPC | *STX8* | 0.426321526 | 0.001745579 | 0.033849064 | 1 |
| Stroke | ASPC | *NFRKB* | 0.552553207 | 0.005534009 | 0.03979928 | 1 |
| Stroke | ASPC | *MGMT* | 0.385328443 | 0.002934529 | 0.033849064 | 1 |
| Stroke | ASPC | *HSD17B13* | 0.866871826 | 0.005976669 | 0.041385645 | 1 |
| Stroke | ASPC | *ELOVL6* | 0.536088984 | 0.005678901 | 0.040520629 | 1 |
| Stroke | ASPC | *FOXN2* | 0.519290526 | 0.002748692 | 0.033849064 | 1 |

| | | | | | | |
|---|---|---|---|---|---|---|
| Stroke | ASPC | *PYGO1* | 0.357818852 | 0.00078978 | 0.033849064 | 1 |
| Stroke | ASPC | *FEZ2* | 0.348868127 | 0.002056799 | 0.033849064 | 1 |
| Stroke | ASPC | *PDE7B* | 0.374831718 | 0.001732941 | 0.033849064 | 1 |
| Stroke | ASPC | *MCC* | 0.47373063 | 0.002884311 | 0.033849064 | 1 |
| Stroke | ASPC | *TBCA* | 0.459627903 | 0.004395299 | 0.035366486 | 1 |
| Stroke | ASPC | *NMUR1* | 1.128658936 | 0.007171273 | 0.046147234 | 1 |
| Stroke | ASPC | *EXOSC10* | 0.346134092 | 0.004888376 | 0.037482089 | 1 |
| Stroke | ASPC | *TRAPPC12* | 0.600117274 | 0.000324585 | 0.033849064 | 1 |
| Stroke | ASPC | *TEFM* | 0.453978086 | 8.81E-06 | 0.015984796 | 0.042745977 |
| Stroke | ASPC | *MALT1* | 0.560171222 | 0.002663519 | 0.033849064 | 1 |
| Stroke | ASPC | *SUCLG2* | 0.439402131 | 0.004249619 | 0.034901446 | 1 |
| Stroke | ASPC | *MANEA* | 0.386812707 | 0.001962185 | 0.033849064 | 1 |
| Stroke | ASPC | *AFF1* | 0.405331863 | 0.002602505 | 0.033849064 | 1 |
| Stroke | ASPC | *SLFN11* | 0.407768719 | 0.003720727 | 0.034310859 | 1 |
| Stroke | ASPC | *CFL1* | 0.638076564 | 0.000255951 | 0.033849064 | 1 |
| Stroke | ASPC | *FAM192A* | 0.234620072 | 0.002076302 | 0.033849064 | 1 |
| Stroke | ASPC | *CBWD1* | 0.856078728 | 0.001909342 | 0.033849064 | 1 |
| Stroke | ASPC | *DCP2* | 0.477778422 | 0.001767586 | 0.033849064 | 1 |
| Stroke | ASPC | *ZNF621* | 0.550154625 | 0.00705009 | 0.045915488 | 1 |
| Stroke | ASPC | *RHOD* | 0.292762702 | 0.003999504 | 0.034570849 | 1 |
| Stroke | ASPC | *UQCRH* | 0.41560485 | 0.005699831 | 0.040548297 | 1 |
| Stroke | ASPC | *GOLIM4* | 0.46343045 | 0.004025666 | 0.034570849 | 1 |
| Stroke | ASPC | *XXYLT1* | 0.545753421 | 0.004377951 | 0.035344127 | 1 |
| Stroke | ASPC | *LEMD3* | 0.514023532 | 0.005886329 | 0.041186451 | 1 |
| Stroke | ASPC | *SNAPC5* | 0.236325295 | 0.006618812 | 0.043992433 | 1 |
| Stroke | ASPC | *MSL2* | 0.572323435 | 0.002787174 | 0.033849064 | 1 |
| Stroke | ASPC | *ANGEL2* | 0.522343821 | 0.003656284 | 0.034310859 | 1 |
| Stroke | ASPC | *C4orf32* | 0.256997922 | 0.001237409 | 0.033849064 | 1 |
| Stroke | ASPC | *CEP135* | 0.498112925 | 0.004671713 | 0.036442364 | 1 |
| Stroke | ASPC | *DHX36* | 0.49702655 | 0.003312585 | 0.033849064 | 1 |
| Stroke | ASPC | *TVP23C* | 1.45700671 | 0.002551493 | 0.033849064 | 1 |
| Stroke | ASPC | *SH3BP5L* | 0.914120347 | 0.005695355 | 0.040548297 | 1 |
| Stroke | ASPC | *CLTB* | 0.692786799 | 0.002899028 | 0.033849064 | 1 |
| Stroke | ASPC | *MCTP1* | 0.380762196 | 0.005488566 | 0.039628752 | 1 |
| Stroke | ASPC | *CTDNEP1* | 0.234054895 | 0.000387466 | 0.033849064 | 1 |
| Stroke | ASPC | *A2M* | 0.439567401 | 0.002822186 | 0.033849064 | 1 |
| Stroke | ASPC | *B3GALT6* | 2.068519037 | 0.002215444 | 0.033849064 | 1 |
| Stroke | ASPC | *SLC35G1* | 0.736969845 | 0.005102203 | 0.03808598 | 1 |
| Stroke | ASPC | *TAC4* | 0.750330721 | 0.003070761 | 0.033849064 | 1 |
| Stroke | ASPC | *CRLF3* | 0.279426575 | 3.83E-05 | 0.024154057 | 0.185977083 |
| Stroke | ASPC | *WDR25* | 0.636761881 | 0.00339865 | 0.033898981 | 1 |
| Stroke | ASPC | *DPP7* | 0.535990151 | 0.005056136 | 0.03808598 | 1 |
| Stroke | ASPC | *DSCAML1* | 0.49521681 | 0.004905143 | 0.037539046 | 1 |
| Stroke | ASPC | *TALDO1* | 0.639517564 | 0.003591471 | 0.034168274 | 1 |
| Stroke | ASPC | *MAN1B1* | 0.775898672 | 0.002622719 | 0.033849064 | 1 |
| Stroke | ASPC | *MIEF2* | 0.595736036 | 0.001864417 | 0.033849064 | 1 |
| Stroke | ASPC | *SLC35G5* | 5.788659124 | 0.004439887 | 0.03542847 | 1 |
| Stroke | ASPC | *CENPBD1* | 1.871189437 | 0.002403238 | 0.033849064 | 1 |
| Stroke | ASPC | *RPS27* | 0.561420404 | 0.00184222 | 0.033849064 | 1 |
| Stroke | ASPC | *ASB8* | 0.423236392 | 0.001206104 | 0.033849064 | 1 |

| | | | | | | |
|---|---|---|---|---|---|---|
| Stroke | ASPC | *SH2B1* | 0.578388835 | 0.00305683 | 0.033849064 | 1 |
| Stroke | ASPC | *ERN1* | 0.442244594 | 0.007317239 | 0.04640205 | 1 |
| Stroke | ASPC | *NSUN3* | 0.4920327 | 0.005156273 | 0.038312767 | 1 |
| Stroke | ASPC | *C17orf62* | 0.823329974 | 0.000682107 | 0.033849064 | 1 |
| Stroke | ASPC | *GAK* | 0.447895069 | 0.006844634 | 0.044878603 | 1 |
| Stroke | ASPC | *CTC1* | 0.252933304 | 0.001358298 | 0.033849064 | 1 |
| Stroke | ASPC | *ZNF664* | 0.842291997 | 0.003030176 | 0.033849064 | 1 |
| Stroke | ASPC | *HLA-DQB1* | -0.04592581 | 0.003810729 | 0.034349037 | 1 |
| Stroke | ASPC | *PACS2* | 0.588566938 | 0.002986616 | 0.033849064 | 1 |
| Stroke | ASPC | *PDXDC1* | 0.444050883 | 0.002700188 | 0.033849064 | 1 |
| Stroke | ASPC | *EXOC3* | 0.351889471 | 0.007967004 | 0.049048657 | 1 |
| Stroke | ASPC | *CCDC66* | 0.350551928 | 0.00640217 | 0.043024 | 1 |
| Stroke | ASPC | *ZDHHC20* | 0.518297284 | 0.002877555 | 0.033849064 | 1 |
| Stroke | ASPC | *MRPL14* | 0.289842921 | 0.001032682 | 0.033849064 | 1 |
| Stroke | ASPC | *SGSH* | 1.023363206 | 0.003187394 | 0.033849064 | 1 |
| Stroke | ASPC | *C3orf58* | 0.50073089 | 0.003022478 | 0.033849064 | 1 |
| Stroke | ASPC | *RFX7* | 0.496384414 | 0.002375804 | 0.033849064 | 1 |
| Stroke | ASPC | *ZNF329* | 0.673557437 | 0.002718385 | 0.033849064 | 1 |
| Stroke | ASPC | *MRPL41* | 0.852800631 | 0.00257228 | 0.033849064 | 1 |
| Stroke | ASPC | *FIGN* | 0.414126574 | 0.003266058 | 0.033849064 | 1 |
| Stroke | ASPC | *C8orf33* | 0.537495629 | 0.000988521 | 0.033849064 | 1 |
| Stroke | ASPC | *YBEY* | -0.034296682 | 0.006656479 | 0.044182268 | 1 |
| Stroke | ASPC | *CLN8* | 0.078840332 | 0.003746114 | 0.034310859 | 1 |
| Stroke | ASPC | *GLRX5* | 0.852511366 | 0.002783702 | 0.033849064 | 1 |
| Stroke | ASPC | *TRAK1* | 0.226837326 | 0.000634469 | 0.033849064 | 1 |
| Stroke | ASPC | *TTC3* | 0.066132488 | 0.007502591 | 0.047048186 | 1 |
| Stroke | ASPC | *ALG12* | 0.809287054 | 0.00197723 | 0.033849064 | 1 |
| Stroke | ASPC | *ZNF721* | 0.339185479 | 0.005015035 | 0.037901788 | 1 |
| Stroke | ASPC | *ARHGEF37* | 0.609509734 | 0.007210311 | 0.046240663 | 1 |
| Stroke | ASPC | *CSMD1* | 0.706543507 | 0.004446651 | 0.03542847 | 1 |
| Stroke | ASPC | *FANCF* | 1.182160818 | 0.000246918 | 0.033849064 | 1 |
| Stroke | ASPC | *RUVBL2* | 0.408505254 | 0.006213118 | 0.042169793 | 1 |
| Stroke | ASPC | *SERHL2* | 0.766041919 | 0.004632693 | 0.036365543 | 1 |
| Stroke | ASPC | *BTBD9* | 0.365615568 | 0.003049473 | 0.033849064 | 1 |
| Stroke | ASPC | *TTC32* | 0.78543668 | 0.000625383 | 0.033849064 | 1 |
| Stroke | ASPC | *PP2D1* | 0.914475479 | 0.003389391 | 0.033898981 | 1 |
| Stroke | ASPC | *PGP* | 0.236982931 | 0.008112387 | 0.049185335 | 1 |
| Stroke | ASPC | *TMEM255B* | 0.451819846 | 0.000703874 | 0.033849064 | 1 |
| Stroke | ASPC | *ZFP90* | 0.61017759 | 0.002979266 | 0.033849064 | 1 |
| Stroke | ASPC | *TMEM106A* | 0.089699561 | 0.006155643 | 0.04206645 | 1 |
| Stroke | ASPC | *IFITM2* | 0.549596656 | 0.007328204 | 0.04640205 | 1 |
| Stroke | ASPC | *AHNAK2* | 0.32532239 | 0.007139828 | 0.046013993 | 1 |
| Stroke | ASPC | *ARL17A* | 0.120146468 | 0.007183655 | 0.046165686 | 1 |
| Stroke | ASPC | *WWOX* | 0.346533822 | 0.003180963 | 0.033849064 | 1 |
| Stroke | ASPC | *ZNF749* | -0.044647487 | 0.002438641 | 0.033849064 | 1 |
| Stroke | ASPC | *GNG2* | 0.531251083 | 0.002242103 | 0.033849064 | 1 |
| Stroke | ASPC | *QRFPR* | 0.348403676 | 0.005117409 | 0.038140815 | 1 |
| Stroke | ASPC | *MAPT* | 0.549236733 | 0.007590798 | 0.047190065 | 1 |
| Stroke | ASPC | *EXD3* | 0.612683967 | 0.002838704 | 0.033849064 | 1 |
| Stroke | ASPC | *C2orf88* | 0.460600318 | 0.003102704 | 0.033849064 | 1 |

| | | | | | | |
|---|---|---|---|---|---|---|
| Stroke | ASPC | *ZFP69* | 1.042026353 | 0.002381343 | 0.033849064 | 1 |
| Stroke | ASPC | *FAM122A* | 1.056667896 | 0.003726028 | 0.034310859 | 1 |
| Stroke | ASPC | *ZNF626* | 0.851565772 | 0.002977924 | 0.033849064 | 1 |
| Stroke | ASPC | *C22orf34* | 0.414724073 | 0.007536516 | 0.047079426 | 1 |
| Stroke | ASPC | *COL25A1* | 0.546552646 | 0.000861573 | 0.033849064 | 1 |
| Stroke | ASPC | *NDOR1* | 0.789147041 | 0.005891773 | 0.041186451 | 1 |
| Stroke | ASPC | *RPL14* | 0.537019214 | 0.00272118 | 0.033849064 | 1 |
| Stroke | ASPC | *FBF1* | 0.1555327 | 0.004469974 | 0.035554616 | 1 |
| Stroke | ASPC | *FAM196A* | 0.474257004 | 0.002391205 | 0.033849064 | 1 |
| Stroke | ASPC | *SF3B3* | 0.504898513 | 0.00277566 | 0.033849064 | 1 |
| Stroke | ASPC | *TSPYL1* | 0.562863292 | 0.001920336 | 0.033849064 | 1 |
| Stroke | ASPC | *PNRC2* | 0.612407929 | 0.004633374 | 0.036365543 | 1 |
| Stroke | ASPC | *FHIT* | 0.370465266 | 0.00183905 | 0.033849064 | 1 |
| Stroke | ASPC | *AKR1C3* | 0.697825754 | 0.003017762 | 0.033849064 | 1 |
| Stroke | ASPC | *ZNF471* | 0.633463072 | 0.002948687 | 0.033849064 | 1 |
| Stroke | ASPC | *SUPT3H* | 0.442857265 | 0.001122587 | 0.033849064 | 1 |
| Stroke | ASPC | *STK31* | 0.331606155 | 0.001332811 | 0.033849064 | 1 |
| Stroke | ASPC | *CD55* | 0.53618126 | 0.002322832 | 0.033849064 | 1 |
| Stroke | ASPC | *XRCC2* | 1.596532862 | 0.007539306 | 0.047079426 | 1 |
| Stroke | ASPC | *ADH1B* | 0.342142203 | 0.003557437 | 0.033987065 | 1 |
| Stroke | ASPC | *TOMM7* | 0.56604604 | 0.002823557 | 0.033849064 | 1 |
| Stroke | ASPC | *ZNF431* | 0.303693083 | 0.004251186 | 0.034901446 | 1 |
| Stroke | ASPC | *HLA-DQA1* | -0.063004157 | 0.007141083 | 0.046013993 | 1 |
| Stroke | ASPC | *WDR5B* | 0.294416654 | 0.000138472 | 0.033849064 | 0.671868347 |
| Stroke | ASPC | *ZNF138* | 0.262494482 | 0.004049489 | 0.03465277 | 1 |
| Stroke | ASPC | *GMFB* | 0.757759502 | 0.001092403 | 0.033849064 | 1 |
| Stroke | ASPC | *ZMYM1* | 0.593838163 | 0.003115031 | 0.033849064 | 1 |
| Stroke | ASPC | *ADAM32* | 0.631539575 | 0.005594049 | 0.039997413 | 1 |
| Stroke | ASPC | *ABCB8* | 0.508666445 | 0.001098636 | 0.033849064 | 1 |
| Stroke | ASPC | *RAMP2-AS1* | 0.575215524 | 0.002112444 | 0.033849064 | 1 |
| Stroke | ASPC | *ZNF665* | 0.737279196 | 0.003030271 | 0.033849064 | 1 |
| Stroke | ASPC | *ATG7* | 0.3381406 | 0.000506778 | 0.033849064 | 1 |
| Stroke | ASPC | *RAB40C* | 0.672365608 | 0.003664866 | 0.034310859 | 1 |
| Stroke | ASPC | *TOPORS* | 0.690036072 | 0.003148545 | 0.033849064 | 1 |
| Stroke | ASPC | *PSAP* | 0.289456283 | 0.003341653 | 0.033849064 | 1 |
| Stroke | ASPC | *EME2* | 1.122480425 | 0.003024534 | 0.033849064 | 1 |
| Stroke | ASPC | *ZNF181* | 0.143135624 | 0.003242328 | 0.033849064 | 1 |
| Stroke | ASPC | *ZNF544* | 0.807835942 | 0.001515393 | 0.033849064 | 1 |
| Stroke | ASPC | *MAN1A2* | 0.504932671 | 0.001657502 | 0.033849064 | 1 |
| Stroke | ASPC | *HSD17B11* | 0.469542227 | 0.00383073 | 0.034349037 | 1 |
| Stroke | ASPC | *HELZ* | 0.422100432 | 0.002701719 | 0.033849064 | 1 |
| Stroke | ASPC | *TMEM116* | -0.065790419 | 0.001125895 | 0.033849064 | 1 |
| Stroke | ASPC | *UCKL1* | 0.58564362 | 0.005282307 | 0.03895099 | 1 |
| Stroke | ASPC | *GFPT1* | 0.448862414 | 0.002124485 | 0.033849064 | 1 |
| Stroke | ASPC | *TMA16* | 0.63125993 | 0.002292097 | 0.033849064 | 1 |
| Stroke | ASPC | *C20orf203* | 0.260028759 | 0.006740528 | 0.044326714 | 1 |
| Stroke | ASPC | *TUSC1* | 1.2289595 | 0.003776174 | 0.034310859 | 1 |
| Stroke | ASPC | *PAPSS2* | 0.498996602 | 0.002718442 | 0.033849064 | 1 |
| Stroke | ASPC | *RPL10A* | 0.467741097 | 0.003719062 | 0.034310859 | 1 |
| Stroke | ASPC | *RCSD1* | 0.463980771 | 0.005979253 | 0.041385645 | 1 |

| | | | | | | |
|---|---|---|---|---|---|---|
| Stroke | ASPC | *ZFP2* | 0.407736031 | 0.003436526 | 0.033962746 | 1 |
| Stroke | ASPC | *PCMTD2* | 0.544481735 | 0.003214334 | 0.033849064 | 1 |
| Stroke | ASPC | *RUFY2* | 0.482733164 | 0.003313191 | 0.033849064 | 1 |
| Stroke | ASPC | *BMPR2* | 0.434907466 | 0.002012574 | 0.033849064 | 1 |
| Stroke | ASPC | *COL15A1* | 0.108007838 | 0.002087797 | 0.033849064 | 1 |
| Stroke | ASPC | *LILRB3* | 0.312942166 | 0.002494737 | 0.033849064 | 1 |
| Stroke | ASPC | *DCTN1* | 0.781751521 | 0.002975655 | 0.033849064 | 1 |
| Stroke | ASPC | *MZT1* | 0.852794314 | 0.000107368 | 0.033849064 | 0.520950621 |
| Stroke | ASPC | *SCGB2B2* | 0.582067777 | 0.002536119 | 0.033849064 | 1 |
| Stroke | ASPC | *CCDC144NL* | 1.060730198 | 0.005081908 | 0.03808598 | 1 |
| Stroke | ASPC | *METTL6* | 0.286287741 | 0.007987935 | 0.049048657 | 1 |
| Stroke | ASPC | *SACM1L* | 0.584827535 | 0.002093278 | 0.033849064 | 1 |
| Stroke | ASPC | *SLC48A1* | 0.580758646 | 0.007996185 | 0.049048657 | 1 |
| Stroke | ASPC | *REPIN1* | 0.887598321 | 0.004775579 | 0.036955517 | 1 |
| Stroke | ASPC | *ZNF891* | 0.629156769 | 0.000976676 | 0.033849064 | 1 |
| Stroke | ASPC | *LYRM4* | 0.575958924 | 0.001527665 | 0.033849064 | 1 |
| Stroke | ASPC | *ANG* | 0.842102364 | 0.004121257 | 0.034729134 | 1 |
| Stroke | ASPC | *VPS16* | 0.637458034 | 0.008095209 | 0.049185335 | 1 |
| Stroke | ASPC | *NPEPL1* | 0.874803145 | 0.000384446 | 0.033849064 | 1 |
| Stroke | ASPC | *APOL6* | 0.461223359 | 0.005101576 | 0.03808598 | 1 |
| Stroke | ASPC | *AC062017.1* | 0.776441466 | 0.005988998 | 0.041394044 | 1 |
| Stroke | ASPC | *FAM138B* | 12.67666442 | 0.004229028 | 0.034901446 | 1 |
| Stroke | ASPC | *SCAMP4* | 0.717169559 | 0.002666566 | 0.033849064 | 1 |
| Stroke | ASPC | *AC092171.4* | 1.111130086 | 0.002677464 | 0.033849064 | 1 |
| Stroke | ASPC | *RPS18* | 0.364166022 | 0.003828435 | 0.034349037 | 1 |
| Stroke | ASPC | *DIRC3* | 0.234604975 | 0.00648104 | 0.04343371 | 1 |
| Stroke | ASPC | *AC009495.3* | 0.318956178 | 0.004310269 | 0.035207788 | 1 |
| Stroke | ASPC | *HLA-DQB2* | 0.025900661 | 0.00286119 | 0.033849064 | 1 |
| Stroke | ASPC | *LINC01166* | 0.112293789 | 0.005729074 | 0.040580242 | 1 |
| Stroke | ASPC | *GPX1* | 0.182964 | 0.005428511 | 0.039370903 | 1 |
| Stroke | ASPC | *ANKRD65* | 2.135931591 | 0.004947159 | 0.037608579 | 1 |
| Stroke | ASPC | *TNFSF12* | 0.155401306 | 0.003903509 | 0.034349037 | 1 |
| Stroke | ASPC | *PSMB9* | 0.065926829 | 0.002132798 | 0.033849064 | 1 |
| Stroke | ASPC | *PNMA2* | 0.239790345 | 0.001031657 | 0.033849064 | 1 |
| Stroke | ASPC | *NSUN6* | 0.354208926 | 0.003478872 | 0.033962746 | 1 |
| Stroke | ASPC | *CD302* | 0.284806919 | 0.008102691 | 0.049185335 | 1 |
| Stroke | ASPC | *ARFGAP3* | 0.60770751 | 0.00337374 | 0.033891069 | 1 |
| Stroke | ASPC | *MRPL20* | 0.47033425 | 0.00020142 | 0.033849064 | 0.977289145 |
| Stroke | ASPC | *UPK3B* | 0.589946117 | 0.001053541 | 0.033849064 | 1 |
| Stroke | ASPC | *NEAT1* | 0.306438222 | 0.001693979 | 0.033849064 | 1 |
| Stroke | ASPC | *LINC00926* | 1.333446779 | 0.003556013 | 0.033987065 | 1 |
| Stroke | ASPC | *CDK11B* | 0.722521474 | 0.002839186 | 0.033849064 | 1 |
| Stroke | ASPC | *C5orf17* | 0.643261888 | 0.002130145 | 0.033849064 | 1 |
| Stroke | ASPC | *PDCD6* | 0.64856798 | 0.002673834 | 0.033849064 | 1 |
| Stroke | ASPC | *SMIM20* | 0.875308816 | 0.00261624 | 0.033849064 | 1 |
| Stroke | ASPC | *CNPY2* | 0.649582877 | 0.003280549 | 0.033849064 | 1 |
| Stroke | ASPC | *LINC00672* | 1.683249404 | 0.001476921 | 0.033849064 | 1 |
| Stroke | ASPC | *ZNF582-AS1* | 1.214298663 | 0.003775234 | 0.034310859 | 1 |
| Stroke | ASPC | *ZNF224* | 0.646319276 | 0.002756664 | 0.033849064 | 1 |
| Stroke | ASPC | *LIN37* | 0.337716379 | 0.007504057 | 0.047048186 | 1 |

| | | | | | | |
|---|---|---|---|---|---|---|
| Stroke | ASPC | *ZNF587B* | 1.135879569 | 0.003079064 | 0.033849064 | 1 |
| Stroke | ASPC | *CASC15* | 0.513699486 | 0.003762885 | 0.034310859 | 1 |
| Heart_Failure | adipocyte | *SEMA3F* | -0.459611039 | 0.001629663 | 0.007919528 | 1 |
| Heart_Failure | adipocyte | *KRIT1* | 0.964653728 | 0.000388688 | 0.002782166 | 1 |
| Heart_Failure | adipocyte | *HS3ST1* | 0.253653583 | 0.011797537 | 0.036669858 | 1 |
| Heart_Failure | adipocyte | *ALS2* | 0.984094023 | 0.00107379 | 0.00580828 | 1 |
| Heart_Failure | adipocyte | *CD38* | 0.743967071 | 0.001327538 | 0.006801703 | 1 |
| Heart_Failure | adipocyte | *RBM6* | -0.226272701 | 3.33E-05 | 0.001268787 | 0.161559954 |
| Heart_Failure | adipocyte | *RECQL* | 0.798849614 | 0.007166236 | 0.024943026 | 1 |
| Heart_Failure | adipocyte | *CDC27* | 0.331258548 | 0.014544332 | 0.043029937 | 1 |
| Heart_Failure | adipocyte | *POLR2J* | 1.612831501 | 7.78E-05 | 0.001268787 | 0.377609447 |
| Heart_Failure | adipocyte | *GCFC2* | 1.173264651 | 4.92E-05 | 0.001268787 | 0.238518064 |
| Heart_Failure | adipocyte | *GGNBP2* | 0.795312164 | 0.003145094 | 0.012943168 | 1 |
| Heart_Failure | adipocyte | *ITGA2B* | 0.626445635 | 0.001419217 | 0.007150616 | 1 |
| Heart_Failure | adipocyte | *SYNRG* | 0.873680399 | 0.000551322 | 0.003580247 | 1 |
| Heart_Failure | adipocyte | *ZNF263* | 1.146290419 | 0.000339946 | 0.002557239 | 1 |
| Heart_Failure | adipocyte | *SPATA20* | 0.369507551 | 0.00275603 | 0.011823391 | 1 |
| Heart_Failure | adipocyte | *AGK* | 0.459290251 | 0.009028109 | 0.029637608 | 1 |
| Heart_Failure | adipocyte | *FARP2* | 0.730678713 | 0.002486854 | 0.010831431 | 1 |
| Heart_Failure | adipocyte | *VPS41* | 0.953504404 | 0.000256243 | 0.002143602 | 1 |
| Heart_Failure | adipocyte | *TSR3* | 1.637753136 | 0.000350887 | 0.002611199 | 1 |
| Heart_Failure | adipocyte | *PSMB1* | 1.484056396 | 2.78E-05 | 0.001268787 | 0.135107896 |
| Heart_Failure | adipocyte | *MGST1* | 0.153839333 | 0.008373609 | 0.028097337 | 1 |
| Heart_Failure | adipocyte | *PKD1* | 1.219315438 | 3.55E-05 | 0.001268787 | 0.172245418 |
| Heart_Failure | adipocyte | *MED24* | 1.29953646 | 0.000551942 | 0.003580247 | 1 |
| Heart_Failure | adipocyte | *RPS20* | 0.89896305 | 0.000558203 | 0.003611204 | 1 |
| Heart_Failure | adipocyte | *TSPAN9* | 0.319343381 | 0.006641469 | 0.0234189 | 1 |
| Heart_Failure | adipocyte | *TTC19* | 0.721414298 | 0.000741912 | 0.004438667 | 1 |
| Heart_Failure | adipocyte | *LARS2* | 0.759587236 | 0.001706245 | 0.008199798 | 1 |
| Heart_Failure | adipocyte | *MAP4K5* | 0.879369778 | 0.000698779 | 0.004247059 | 1 |
| Heart_Failure | adipocyte | *RNF14* | 1.017525095 | 2.82E-05 | 0.001268787 | 0.136717694 |
| Heart_Failure | adipocyte | *TACC3* | 0.94770387 | 0.002078109 | 0.009512251 | 1 |
| Heart_Failure | adipocyte | *ZC3H3* | 0.261240197 | 0.006300023 | 0.022426788 | 1 |
| Heart_Failure | adipocyte | *MDH1* | 0.414030484 | 0.001645998 | 0.00797044 | 1 |
| Heart_Failure | adipocyte | *COX15* | 1.226464426 | 0.000212726 | 0.001960237 | 1 |
| Heart_Failure | adipocyte | *RUFY3* | 0.826574251 | 0.000335786 | 0.002541707 | 1 |
| Heart_Failure | adipocyte | *WWTR1* | 1.092928954 | 7.14E-05 | 0.001268787 | 0.346512326 |
| Heart_Failure | adipocyte | *TTC27* | 0.959149308 | 0.00017705 | 0.001756516 | 0.859048231 |
| Heart_Failure | adipocyte | *SLC7A9* | 0.138018829 | 0.005005156 | 0.018781914 | 1 |
| Heart_Failure | adipocyte | *ZDHHC6* | 0.869409986 | 0.008635756 | 0.028679459 | 1 |
| Heart_Failure | adipocyte | *RNH1* | 0.355779822 | 0.015121216 | 0.044492869 | 1 |
| Heart_Failure | adipocyte | *GLRX2* | 1.316587564 | 0.003955973 | 0.015491832 | 1 |
| Heart_Failure | adipocyte | *GCLM* | 0.930590161 | 0.001901285 | 0.008861704 | 1 |
| Heart_Failure | adipocyte | *UBR2* | 0.497044365 | 0.006404285 | 0.022747869 | 1 |
| Heart_Failure | adipocyte | *SEC63* | 0.63530548 | 0.00809101 | 0.027376278 | 1 |
| Heart_Failure | adipocyte | *MIPEP* | 1.030170407 | 3.13E-05 | 0.001268787 | 0.151668432 |
| Heart_Failure | adipocyte | *VRK2* | 0.829553406 | 0.000449288 | 0.003102284 | 1 |
| Heart_Failure | adipocyte | *VEZT* | 1.1454634 | 6.38E-05 | 0.001268787 | 0.309344839 |
| Heart_Failure | adipocyte | *BRD9* | 1.250651187 | 1.81E-05 | 0.001268787 | 0.087990334 |
| Heart_Failure | adipocyte | *SNX1* | 0.74400604 | 0.002234383 | 0.010041405 | 1 |

| | | | | | | |
|---|---|---|---|---|---|---|
| Heart_Failure | adipocyte | *ANK1* | 2.422693437 | 0.000541912 | 0.003553188 | 1 |
| Heart_Failure | adipocyte | *RABEP1* | 0.665187461 | 0.003528116 | 0.014185437 | 1 |
| Heart_Failure | adipocyte | *IKZF2* | 1.005191564 | 0.000814945 | 0.004763495 | 1 |
| Heart_Failure | adipocyte | *CHPF2* | 0.320322487 | 0.016190282 | 0.046786927 | 1 |
| Heart_Failure | adipocyte | *ATP6V0A1* | 0.744390088 | 0.000283956 | 0.002300045 | 1 |
| Heart_Failure | adipocyte | *PIAS1* | 0.660258225 | 0.000716378 | 0.004323218 | 1 |
| Heart_Failure | adipocyte | *VCL* | 1.074353342 | 0.000212375 | 0.001960237 | 1 |
| Heart_Failure | adipocyte | *ADSS* | 1.103026557 | 0.00080638 | 0.00473675 | 1 |
| Heart_Failure | adipocyte | *TBC1D23* | 1.289990287 | 4.50E-05 | 0.001268787 | 0.21853872 |
| Heart_Failure | adipocyte | *NSUN2* | 0.67474037 | 0.005661797 | 0.02077991 | 1 |
| Heart_Failure | adipocyte | *MRI1* | 0.366722355 | 0.001988116 | 0.009160817 | 1 |
| Heart_Failure | adipocyte | *AGA* | 0.996123464 | 0.003138986 | 0.012943168 | 1 |
| Heart_Failure | adipocyte | *BOD1L1* | 0.403590128 | 0.006507984 | 0.023048714 | 1 |
| Heart_Failure | adipocyte | *PHLPP2* | 0.228990499 | 0.013005728 | 0.039500447 | 1 |
| Heart_Failure | adipocyte | *PQLC2* | 2.250333161 | 0.000456074 | 0.003125524 | 1 |
| Heart_Failure | adipocyte | *INPP4A* | 0.435466266 | 0.00659312 | 0.023282254 | 1 |
| Heart_Failure | adipocyte | *PSMA4* | 1.427765994 | 0.00013673 | 0.001561419 | 0.663413946 |
| Heart_Failure | adipocyte | *TDP1* | 1.25838608 | 0.001378814 | 0.007027317 | 1 |
| Heart_Failure | adipocyte | *AP2S1* | 1.53765122 | 0.000263521 | 0.002185645 | 1 |
| Heart_Failure | adipocyte | *CTNNA1* | 1.00889991 | 0.000104267 | 0.001409201 | 0.505903128 |
| Heart_Failure | adipocyte | *YTHDC2* | 1.151186397 | 8.21E-05 | 0.001268787 | 0.398203187 |
| Heart_Failure | adipocyte | *POLR2B* | 1.019914448 | 0.000283789 | 0.002300045 | 1 |
| Heart_Failure | adipocyte | *ARAP2* | 0.4218258 | 0.007829466 | 0.026714885 | 1 |
| Heart_Failure | adipocyte | *DTNBP1* | 1.07169048 | 0.000111719 | 0.001449365 | 0.5420625 |
| Heart_Failure | adipocyte | *SNX29* | 0.632350856 | 0.003110616 | 0.012855801 | 1 |
| Heart_Failure | adipocyte | *JKAMP* | 0.490096963 | 0.005010916 | 0.018788998 | 1 |
| Heart_Failure | adipocyte | *MAPK9* | 1.354975268 | 0.000239586 | 0.002077964 | 1 |
| Heart_Failure | adipocyte | *CYBA* | 0.786272592 | 0.002108253 | 0.009595912 | 1 |
| Heart_Failure | adipocyte | *THOC3* | 4.093725204 | 4.20E-05 | 0.001268787 | 0.203861947 |
| Heart_Failure | adipocyte | *MRTO4* | 0.746242653 | 0.001047328 | 0.005709703 | 1 |
| Heart_Failure | adipocyte | *ANAPC4* | 0.767541286 | 0.00026865 | 0.002210435 | 1 |
| Heart_Failure | adipocyte | *SDCCAG8* | 0.544044902 | 1.56E-06 | 0.000302238 | 0.00755594 |
| Heart_Failure | adipocyte | *KIF1B* | 0.482912698 | 0.001823737 | 0.008582709 | 1 |
| Heart_Failure | adipocyte | *TBC1D22A* | 0.690051974 | 0.001421264 | 0.007153497 | 1 |
| Heart_Failure | adipocyte | *GALC* | 0.881582167 | 0.000268618 | 0.002210435 | 1 |
| Heart_Failure | adipocyte | *FAM114A2* | 0.527081078 | 0.007780905 | 0.026569036 | 1 |
| Heart_Failure | adipocyte | *GINM1* | 0.521253127 | 0.0117727 | 0.036616116 | 1 |
| Heart_Failure | adipocyte | *GDI2* | 0.592409786 | 0.007848809 | 0.026724506 | 1 |
| Heart_Failure | adipocyte | *PITHD1* | 0.668725019 | 0.002893909 | 0.012213225 | 1 |
| Heart_Failure | adipocyte | *ATP11B* | 0.712637724 | 0.000232133 | 0.00204651 | 1 |
| Heart_Failure | adipocyte | *LAMC2* | -0.17860689 | 0.007504223 | 0.025823044 | 1 |
| Heart_Failure | adipocyte | *CROCC* | 0.223074687 | 0.001984112 | 0.009151054 | 1 |
| Heart_Failure | adipocyte | *ALDH18A1* | 0.437496155 | 0.014281054 | 0.042406166 | 1 |
| Heart_Failure | adipocyte | *DNAJC25* | 1.809271992 | 0.000367967 | 0.002684255 | 1 |
| Heart_Failure | adipocyte | *OGFR* | 0.582318954 | 0.002149803 | 0.00975757 | 1 |
| Heart_Failure | adipocyte | *NCKAP1* | 0.761256915 | 0.002687937 | 0.01158784 | 1 |
| Heart_Failure | adipocyte | *SLC6A16* | 0.638064732 | 0.015595807 | 0.045622985 | 1 |
| Heart_Failure | adipocyte | *ADCK1* | 1.924291002 | 2.91E-05 | 0.001268787 | 0.141011533 |
| Heart_Failure | adipocyte | *RNF4* | 1.493958741 | 4.61E-05 | 0.001268787 | 0.22383949 |
| Heart_Failure | adipocyte | *CASP8* | 0.898607359 | 0.00033794 | 0.002550054 | 1 |

| | | | | | | |
|---|---|---|---|---|---|---|
| Heart_Failure | adipocyte | *TM7SF3* | 0.954110251 | 0.000378858 | 0.002739523 | 1 |
| Heart_Failure | adipocyte | *TMEM161A* | 1.367329633 | 0.000467473 | 0.003190128 | 1 |
| Heart_Failure | adipocyte | *SUGP2* | 1.05707744 | 0.00021444 | 0.001960237 | 1 |
| Heart_Failure | adipocyte | *SLC12A2* | 1.331312201 | 7.96E-05 | 0.001268787 | 0.386196975 |
| Heart_Failure | adipocyte | *BTBD1* | 1.033594798 | 9.99E-05 | 0.001368761 | 0.484541493 |
| Heart_Failure | adipocyte | *FAR2* | 0.63499846 | 0.000866562 | 0.004971605 | 1 |
| Heart_Failure | adipocyte | *CHI3L2* | 0.67455836 | 0.007388016 | 0.025586476 | 1 |
| Heart_Failure | adipocyte | *SBNO2* | 1.177074137 | 1.39E-05 | 0.001175323 | 0.067410793 |
| Heart_Failure | adipocyte | *PMS1* | 1.044504577 | 0.000173685 | 0.001756516 | 0.842721516 |
| Heart_Failure | adipocyte | *ANKRD44* | 0.379137557 | 0.012413786 | 0.03807313 | 1 |
| Heart_Failure | adipocyte | *MAP2K4* | 0.902956342 | 0.000867879 | 0.004971605 | 1 |
| Heart_Failure | adipocyte | *MYO9A* | 0.436215006 | 0.010798378 | 0.03425187 | 1 |
| Heart_Failure | adipocyte | *TRAM1* | 0.667516905 | 0.002307313 | 0.010289599 | 1 |
| Heart_Failure | adipocyte | *STOML1* | 2.343021445 | 0.000614096 | 0.003889811 | 1 |
| Heart_Failure | adipocyte | *METTL22* | 0.870535122 | 0.008964648 | 0.029549234 | 1 |
| Heart_Failure | adipocyte | *TP53BP1* | 0.71405673 | 0.006576627 | 0.023257867 | 1 |
| Heart_Failure | adipocyte | *DHX8* | 1.556855277 | 8.22E-05 | 0.001268787 | 0.398809036 |
| Heart_Failure | adipocyte | *ROGDI* | 3.121684533 | 0.000147255 | 0.001616476 | 0.714482332 |
| Heart_Failure | adipocyte | *CBFB* | 0.91444088 | 0.001212332 | 0.006372953 | 1 |
| Heart_Failure | adipocyte | *IP6K2* | 0.873703451 | 0.000776806 | 0.004590958 | 1 |
| Heart_Failure | adipocyte | *SRBD1* | 1.08708936 | 7.02E-05 | 0.001268787 | 0.340658635 |
| Heart_Failure | adipocyte | *IFT80* | 1.068830711 | 0.008398379 | 0.028160978 | 1 |
| Heart_Failure | adipocyte | *NUCKS1* | 0.919829786 | 0.000655326 | 0.004055662 | 1 |
| Heart_Failure | adipocyte | *PLA2G10* | 2.499232936 | 0.001947581 | 0.009016852 | 1 |
| Heart_Failure | adipocyte | *LRP6* | 1.177250037 | 4.98E-05 | 0.001268787 | 0.241548281 |
| Heart_Failure | adipocyte | *CLTCL1* | 0.50182119 | 0.013728911 | 0.041118936 | 1 |
| Heart_Failure | adipocyte | *DGCR2* | 0.632768703 | 0.006146365 | 0.022057812 | 1 |
| Heart_Failure | adipocyte | *ZXDC* | 1.124207352 | 8.07E-05 | 0.001268787 | 0.391446559 |
| Heart_Failure | adipocyte | *GBA2* | 1.805519948 | 2.51E-06 | 0.00039274 | 0.012174927 |
| Heart_Failure | adipocyte | *AP3M2* | 0.969576397 | 0.002368369 | 0.010441952 | 1 |
| Heart_Failure | adipocyte | *EIF2B3* | 0.799205271 | 0.000472776 | 0.003212755 | 1 |
| Heart_Failure | adipocyte | *MAP4K4* | 0.729310193 | 0.001481358 | 0.007371659 | 1 |
| Heart_Failure | adipocyte | *WDR1* | 0.648760654 | 0.016429831 | 0.047338208 | 1 |
| Heart_Failure | adipocyte | *SNX13* | 1.020815703 | 0.000184553 | 0.001798299 | 0.895449962 |
| Heart_Failure | adipocyte | *ING3* | 1.512397821 | 2.85E-05 | 0.001268787 | 0.138214885 |
| Heart_Failure | adipocyte | *CYBRD1* | 0.579241787 | 0.001462965 | 0.007302784 | 1 |
| Heart_Failure | adipocyte | *RDH11* | 0.917546932 | 0.003797209 | 0.015003303 | 1 |
| Heart_Failure | adipocyte | *ALDH3A2* | 0.39571349 | 0.0057141 | 0.020877117 | 1 |
| Heart_Failure | adipocyte | *AFF4* | 0.837856601 | 0.000307636 | 0.002421831 | 1 |
| Heart_Failure | adipocyte | *NFATC3* | 0.586327706 | 0.001184852 | 0.006262421 | 1 |
| Heart_Failure | adipocyte | *TRNT1* | 0.712071367 | 0.005915469 | 0.021451311 | 1 |
| Heart_Failure | adipocyte | *CRMP1* | 1.472964407 | 7.16E-05 | 0.001268787 | 0.347235717 |
| Heart_Failure | adipocyte | *AP1M1* | 1.256171055 | 0.000629973 | 0.003954243 | 1 |
| Heart_Failure | adipocyte | *ALPK1* | 0.459466808 | 0.009414473 | 0.030739586 | 1 |
| Heart_Failure | adipocyte | *LLGL2* | 0.800379093 | 0.013541614 | 0.040708743 | 1 |
| Heart_Failure | adipocyte | *ADAM11* | -0.129732712 | 0.015681656 | 0.045780623 | 1 |
| Heart_Failure | adipocyte | *ZNF532* | 1.181838002 | 8.57E-05 | 0.001299545 | 0.41585425 |
| Heart_Failure | adipocyte | *ZZEF1* | 0.838102523 | 0.000386275 | 0.002772495 | 1 |
| Heart_Failure | adipocyte | *TIPIN* | 0.156442834 | 0.009786803 | 0.031720486 | 1 |
| Heart_Failure | adipocyte | *VPS9D1* | 1.482127088 | 2.03E-06 | 0.000340255 | 0.00986739 |

| | | | | | | |
|---|---|---|---|---|---|---|
| Heart_Failure | adipocyte | *FNDC3B* | 0.912131835 | 3.67E-05 | 0.001268787 | 0.17798597 |
| Heart_Failure | adipocyte | *KIFAP3* | 0.631528241 | 0.007495543 | 0.025823044 | 1 |
| Heart_Failure | adipocyte | *FMO4* | 0.694275711 | 0.000811059 | 0.004751265 | 1 |
| Heart_Failure | adipocyte | *ANKRD13A* | 1.017630401 | 0.002434297 | 0.01064073 | 1 |
| Heart_Failure | adipocyte | *PAG1* | 0.57878321 | 0.006789484 | 0.023836887 | 1 |
| Heart_Failure | adipocyte | *XAB2* | 1.337327975 | 0.005960743 | 0.021582093 | 1 |
| Heart_Failure | adipocyte | *DYNC1I2* | 1.087411364 | 0.000167165 | 0.001734499 | 0.811083061 |
| Heart_Failure | adipocyte | *PIK3C3* | 0.7519368 | 0.005199404 | 0.019361096 | 1 |
| Heart_Failure | adipocyte | *PCM1* | 0.743353714 | 0.000518101 | 0.003453059 | 1 |
| Heart_Failure | adipocyte | *XRCC5* | 0.851842452 | 0.000637283 | 0.003979532 | 1 |
| Heart_Failure | adipocyte | *SAR1A* | 0.607875921 | 0.009939358 | 0.032150508 | 1 |
| Heart_Failure | adipocyte | *FDFT1* | 0.597426213 | 0.00037631 | 0.002725156 | 1 |
| Heart_Failure | adipocyte | *PGM1* | 0.438930644 | 0.010253251 | 0.032972696 | 1 |
| Heart_Failure | adipocyte | *RABL2B* | 1.352948037 | 0.000138815 | 0.001573669 | 0.673530335 |
| Heart_Failure | adipocyte | *SCTR* | 0.631378921 | 0.008144855 | 0.027462708 | 1 |
| Heart_Failure | adipocyte | *RIF1* | 0.864359286 | 0.000524418 | 0.003471315 | 1 |
| Heart_Failure | adipocyte | *PSEN1* | 0.821483213 | 0.000677124 | 0.004158743 | 1 |
| Heart_Failure | adipocyte | *MOK* | 0.34355774 | 0.009535578 | 0.031051425 | 1 |
| Heart_Failure | adipocyte | *STK17B* | 0.39933494 | 0.016965778 | 0.048507927 | 1 |
| Heart_Failure | adipocyte | *PCDHB4* | 3.618663677 | 0.000115283 | 0.001471019 | 0.559350971 |
| Heart_Failure | adipocyte | *ME2* | 1.041224474 | 0.0009082 | 0.005123936 | 1 |
| Heart_Failure | adipocyte | *ZCCHC6* | 0.927547324 | 0.000551506 | 0.003580247 | 1 |
| Heart_Failure | adipocyte | *ULK2* | 0.607384554 | 0.006280793 | 0.022391187 | 1 |
| Heart_Failure | adipocyte | *PIBF1* | 0.962928047 | 9.13E-05 | 0.001325336 | 0.44317448 |
| Heart_Failure | adipocyte | *ZNF264* | 1.448854805 | 0.000174701 | 0.001756516 | 0.84764785 |
| Heart_Failure | adipocyte | *CHMP2B* | 1.224562823 | 6.81E-05 | 0.001268787 | 0.330511678 |
| Heart_Failure | adipocyte | *GSTP1* | 0.585488189 | 0.000369002 | 0.002684255 | 1 |
| Heart_Failure | adipocyte | *WBP11* | 1.687668873 | 5.93E-05 | 0.001268787 | 0.287743225 |
| Heart_Failure | adipocyte | *NCOA1* | 0.893431303 | 0.000108865 | 0.001439756 | 0.528213267 |
| Heart_Failure | adipocyte | *HACE1* | 1.426906822 | 1.48E-05 | 0.001175323 | 0.071694684 |
| Heart_Failure | adipocyte | *SLC25A24* | 1.039504645 | 0.000186231 | 0.001801316 | 0.903594372 |
| Heart_Failure | adipocyte | *RRN3* | 1.027175261 | 6.94E-05 | 0.001268787 | 0.336611212 |
| Heart_Failure | adipocyte | *MTIF2* | 1.419898603 | 0.000124171 | 0.001529136 | 0.602479495 |
| Heart_Failure | adipocyte | *DDHD2* | 0.706760832 | 2.34E-05 | 0.001268787 | 0.113761543 |
| Heart_Failure | adipocyte | *USP40* | 0.99025292 | 0.000128134 | 0.001542589 | 0.621706069 |
| Heart_Failure | adipocyte | *MAST2* | 0.989911173 | 1.17E-07 | 4.71E-05 | 0.000565681 |
| Heart_Failure | adipocyte | *EIF2AK1* | 1.407948108 | 3.18E-05 | 0.001268787 | 0.154485418 |
| Heart_Failure | adipocyte | *TMED2* | 0.471097085 | 0.002870178 | 0.012162534 | 1 |
| Heart_Failure | adipocyte | *ALG9* | 1.203140626 | 0.003345753 | 0.013561901 | 1 |
| Heart_Failure | adipocyte | *ACOX3* | 0.639680774 | 0.002981285 | 0.012502331 | 1 |
| Heart_Failure | adipocyte | *TRIP6* | 0.66665963 | 0.013519925 | 0.040693968 | 1 |
| Heart_Failure | adipocyte | *SRRT* | 0.998719937 | 0.002840135 | 0.012066841 | 1 |
| Heart_Failure | adipocyte | *ADAMTS2* | 1.187753945 | 0.000206067 | 0.00193767 | 0.999837592 |
| Heart_Failure | adipocyte | *PSMC5* | 0.826771934 | 0.000375544 | 0.002723678 | 1 |
| Heart_Failure | adipocyte | *UIMC1* | 0.952583108 | 0.000764121 | 0.004548779 | 1 |
| Heart_Failure | adipocyte | *OGFOD1* | 0.527065236 | 0.005584775 | 0.020559429 | 1 |
| Heart_Failure | adipocyte | *ASAP3* | 1.353013338 | 3.06E-05 | 0.001268787 | 0.148515671 |
| Heart_Failure | adipocyte | *ANKRD10* | 0.652981855 | 0.00890276 | 0.029425198 | 1 |
| Heart_Failure | adipocyte | *DOCK3* | 1.101807788 | 0.000940314 | 0.005292814 | 1 |
| Heart_Failure | adipocyte | *NSFL1C* | 0.593742401 | 0.000390419 | 0.002786946 | 1 |

| | | | | | | |
|---|---|---|---|---|---|---|
| Heart_Failure | adipocyte | *MAVS* | 0.564139812 | 0.000227432 | 0.002021062 | 1 |
| Heart_Failure | adipocyte | *XRN2* | 0.648048734 | 0.009346706 | 0.030559447 | 1 |
| Heart_Failure | adipocyte | *SNX5* | 1.578638536 | 4.93E-05 | 0.001268787 | 0.239099011 |
| Heart_Failure | adipocyte | *RPH3A* | 1.331563245 | 0.004059065 | 0.015806248 | 1 |
| Heart_Failure | adipocyte | *KIF16B* | 1.24172917 | 7.95E-05 | 0.001268787 | 0.385747113 |
| Heart_Failure | adipocyte | *FXYD5* | 0.712683188 | 0.000361251 | 0.002659776 | 1 |
| Heart_Failure | adipocyte | *ZNF302* | 0.323393987 | 0.009012571 | 0.029626691 | 1 |
| Heart_Failure | adipocyte | *GRAMD1A* | 1.312320505 | 0.00016862 | 0.001740728 | 0.818142194 |
| Heart_Failure | adipocyte | *CDIP1* | 1.306843128 | 0.001615426 | 0.00786163 | 1 |
| Heart_Failure | adipocyte | *KCNH4* | 0.149975935 | 0.01176283 | 0.036608884 | 1 |
| Heart_Failure | adipocyte | *DHX32* | 0.470765589 | 0.010800775 | 0.03425187 | 1 |
| Heart_Failure | adipocyte | *SPTLC1* | 0.567105163 | 0.005102922 | 0.019060338 | 1 |
| Heart_Failure | adipocyte | *RAB11FIP3* | 0.713537139 | 0.000633949 | 0.003974055 | 1 |
| Heart_Failure | adipocyte | *GNPTG* | 0.930784536 | 0.003013901 | 0.012587417 | 1 |
| Heart_Failure | adipocyte | *PABPC4* | -0.334856763 | 0.016025616 | 0.046449396 | 1 |
| Heart_Failure | adipocyte | *CERS4* | 1.414188259 | 5.65E-05 | 0.001268787 | 0.274345318 |
| Heart_Failure | adipocyte | *USP48* | 0.588327145 | 0.000243705 | 0.002095769 | 1 |
| Heart_Failure | adipocyte | *PDPR* | 0.678357863 | 0.001457341 | 0.00728971 | 1 |
| Heart_Failure | adipocyte | *GLG1* | 0.91009724 | 0.000455155 | 0.003123635 | 1 |
| Heart_Failure | adipocyte | *DTX2* | 1.573436558 | 0.000746469 | 0.004454944 | 1 |
| Heart_Failure | adipocyte | *RAPGEF4* | 0.345382158 | 0.016411426 | 0.047313274 | 1 |
| Heart_Failure | adipocyte | *PPP2R3C* | 0.544440438 | 0.000793783 | 0.00467407 | 1 |
| Heart_Failure | adipocyte | *SCFD1* | 1.016397649 | 0.000134732 | 0.001561419 | 0.653719041 |
| Heart_Failure | adipocyte | *SUPT16H* | 1.384300256 | 0.00010522 | 0.001416381 | 0.510528727 |
| Heart_Failure | adipocyte | *TOX4* | 0.897634497 | 0.001655082 | 0.008006439 | 1 |
| Heart_Failure | adipocyte | *COMT* | 0.994652113 | 0.000883857 | 0.005027518 | 1 |
| Heart_Failure | adipocyte | *LRRFIP2* | 1.000423885 | 7.04E-05 | 0.001268787 | 0.341370678 |
| Heart_Failure | adipocyte | *SEC22C* | 1.497320284 | 6.88E-05 | 0.001268787 | 0.333722154 |
| Heart_Failure | adipocyte | *XYLB* | 1.579483929 | 7.94E-05 | 0.001268787 | 0.385231154 |
| Heart_Failure | adipocyte | *MSH2* | 1.433156455 | 0.00011618 | 0.001471822 | 0.56370771 |
| Heart_Failure | adipocyte | *PSMD5* | 1.528179015 | 1.05E-05 | 0.001035565 | 0.050742692 |
| Heart_Failure | adipocyte | *CRAT* | 2.078487591 | 0.000139978 | 0.001575809 | 0.679173491 |
| Heart_Failure | adipocyte | *WAC* | 0.914653359 | 0.000129592 | 0.001543804 | 0.628779185 |
| Heart_Failure | adipocyte | *ERMP1* | 0.661505657 | 0.002731307 | 0.011748495 | 1 |
| Heart_Failure | adipocyte | *RAB18* | 1.332968085 | 4.79E-05 | 0.001268787 | 0.232292742 |
| Heart_Failure | adipocyte | *SETD1A* | 1.061940034 | 9.88E-07 | 0.000233889 | 0.004796033 |
| Heart_Failure | adipocyte | *MTAP* | 0.760274722 | 0.000112252 | 0.001452392 | 0.544646981 |
| Heart_Failure | adipocyte | *MED15* | 0.883652483 | 0.00179117 | 0.008487067 | 1 |
| Heart_Failure | adipocyte | *LZTR1* | 0.585760539 | 0.003219179 | 0.013154885 | 1 |
| Heart_Failure | adipocyte | *SMARCB1* | 0.314964182 | 0.000245548 | 0.00210123 | 1 |
| Heart_Failure | adipocyte | *DDT* | 0.513875365 | 0.009947002 | 0.032153799 | 1 |
| Heart_Failure | adipocyte | *CABIN1* | 0.786682171 | 0.000845271 | 0.004894097 | 1 |
| Heart_Failure | adipocyte | *TOP3B* | 0.683975524 | 0.003911563 | 0.015367534 | 1 |
| Heart_Failure | adipocyte | *HPS4* | 0.959521749 | 0.001873109 | 0.008772513 | 1 |
| Heart_Failure | adipocyte | *TRIOBP* | 0.485562694 | 0.01007158 | 0.03244841 | 1 |
| Heart_Failure | adipocyte | *TTC28* | 0.76481209 | 0.000242714 | 0.002095458 | 1 |
| Heart_Failure | adipocyte | *RTCB* | 0.558642885 | 0.006593103 | 0.023282254 | 1 |
| Heart_Failure | adipocyte | *TIMP3* | 0.934289426 | 4.41E-05 | 0.001268787 | 0.213989239 |
| Heart_Failure | adipocyte | *PPP6R2* | 0.971459035 | 0.000258248 | 0.002152669 | 1 |
| Heart_Failure | adipocyte | *TSPO* | 1.527435446 | 4.03E-05 | 0.001268787 | 0.195398638 |

| | | | | | | |
|---|---|---|---|---|---|---|
| Heart_Failure | adipocyte | *MTMR3* | 0.409973882 | 0.003220941 | 0.013154885 | 1 |
| Heart_Failure | adipocyte | *IFT27* | 1.513964737 | 7.80E-05 | 0.001268787 | 0.378439802 |
| Heart_Failure | adipocyte | *ACO2* | 0.683805084 | 0.001630587 | 0.007919528 | 1 |
| Heart_Failure | adipocyte | *POLR3H* | 1.33704049 | 3.67E-05 | 0.001268787 | 0.178304137 |
| Heart_Failure | adipocyte | *DESI1* | 0.992785371 | 0.000108835 | 0.001439756 | 0.528066329 |
| Heart_Failure | adipocyte | *HDAC10* | 1.508669047 | 0.001427943 | 0.00716864 | 1 |
| Heart_Failure | adipocyte | *CNIH1* | 0.719420692 | 0.010989422 | 0.034714531 | 1 |
| Heart_Failure | adipocyte | *ATP6V1D* | 1.288327192 | 5.67E-05 | 0.001268787 | 0.27489457 |
| Heart_Failure | adipocyte | *MTHFD1* | 0.364979433 | 0.016114205 | 0.046650431 | 1 |
| Heart_Failure | adipocyte | *GSKIP* | 1.498405154 | 0.001777901 | 0.008440679 | 1 |
| Heart_Failure | adipocyte | *PSMB5* | 0.351836286 | 0.002328198 | 0.010344706 | 1 |
| Heart_Failure | adipocyte | *YY1* | 1.295830041 | 8.16E-05 | 0.001268787 | 0.395948514 |
| Heart_Failure | adipocyte | *CCNB1IP1* | 1.504165834 | 8.19E-05 | 0.001268787 | 0.397184038 |
| Heart_Failure | adipocyte | *EFS* | 1.53313843 | 0.000134895 | 0.001561419 | 0.654511186 |
| Heart_Failure | adipocyte | *CINP* | 1.352128316 | 0.000461504 | 0.003153825 | 1 |
| Heart_Failure | adipocyte | *TRPC4AP* | 0.749384779 | 0.000107723 | 0.001435913 | 0.522672354 |
| Heart_Failure | adipocyte | *ABHD12* | 0.49556536 | 0.002803609 | 0.011953526 | 1 |
| Heart_Failure | adipocyte | *CD40* | 0.940716603 | 0.014077876 | 0.041956912 | 1 |
| Heart_Failure | adipocyte | *NDRG3* | 0.837395334 | 0.003585524 | 0.014365783 | 1 |
| Heart_Failure | adipocyte | *CSTF1* | 1.927366288 | 7.56E-05 | 0.001268787 | 0.366724468 |
| Heart_Failure | adipocyte | *NELFCD* | 0.988045798 | 0.001004185 | 0.005517898 | 1 |
| Heart_Failure | adipocyte | *CTSZ* | 0.815421463 | 0.003140466 | 0.012943168 | 1 |
| Heart_Failure | adipocyte | *MTG2* | 0.868653269 | 5.21E-05 | 0.001268787 | 0.252563371 |
| Heart_Failure | adipocyte | *DIDO1* | 1.075379341 | 6.98E-06 | 0.00084715 | 0.033885993 |
| Heart_Failure | adipocyte | *GID8* | 0.578472312 | 0.012842413 | 0.03914032 | 1 |
| Heart_Failure | adipocyte | *GMEB2* | 1.309662153 | 0.003623184 | 0.014456981 | 1 |
| Heart_Failure | adipocyte | *TRIB3* | 1.446058763 | 0.009690831 | 0.031514685 | 1 |
| Heart_Failure | adipocyte | *RBFA* | 0.483412517 | 0.012344803 | 0.037909483 | 1 |
| Heart_Failure | adipocyte | *METTL4* | 1.082648825 | 0.001899984 | 0.008861704 | 1 |
| Heart_Failure | adipocyte | *LPIN2* | 0.998210067 | 0.000199889 | 0.001894256 | 0.969859114 |
| Heart_Failure | adipocyte | *RNF125* | 1.563534677 | 0.000322235 | 0.002481351 | 1 |
| Heart_Failure | adipocyte | *MIB1* | 1.137056538 | 6.02E-05 | 0.001268787 | 0.29187259 |
| Heart_Failure | adipocyte | *STK24* | 1.154152158 | 5.60E-05 | 0.001268787 | 0.271819572 |
| Heart_Failure | adipocyte | *DNAJC3* | 0.65036712 | 0.007587767 | 0.026068992 | 1 |
| Heart_Failure | adipocyte | *UGGT2* | 0.604515652 | 0.004348004 | 0.016716732 | 1 |
| Heart_Failure | adipocyte | *ARHGEF7* | 1.156761973 | 4.74E-05 | 0.001268787 | 0.229989446 |
| Heart_Failure | adipocyte | *PARP4* | 0.687376193 | 0.000895767 | 0.005071482 | 1 |
| Heart_Failure | adipocyte | *SUPT20H* | 0.857885975 | 0.000321436 | 0.002479505 | 1 |
| Heart_Failure | adipocyte | *MRPS31* | 1.286165756 | 6.36E-05 | 0.001268787 | 0.308493124 |
| Heart_Failure | adipocyte | *KATNAL1* | 1.229029707 | 6.63E-05 | 0.001268787 | 0.321752711 |
| Heart_Failure | adipocyte | *INTS6* | 0.884394025 | 0.000415662 | 0.002922889 | 1 |
| Heart_Failure | adipocyte | *CLN5* | 1.376931967 | 3.75E-05 | 0.001268787 | 0.181873357 |
| Heart_Failure | adipocyte | *MGRN1* | 0.570043462 | 0.005620411 | 0.020674929 | 1 |
| Heart_Failure | adipocyte | *LONP2* | 0.355181639 | 0.010561105 | 0.033689995 | 1 |
| Heart_Failure | adipocyte | *DHODH* | 0.386376148 | 0.008430726 | 0.028224104 | 1 |
| Heart_Failure | adipocyte | *PSMD7* | 1.230198365 | 0.000173967 | 0.001756516 | 0.844090228 |
| Heart_Failure | adipocyte | *MON1B* | 2.079836133 | 3.72E-05 | 0.001268787 | 0.18026858 |
| Heart_Failure | adipocyte | *NME4* | 0.717769226 | 0.012018977 | 0.037238873 | 1 |
| Heart_Failure | adipocyte | *LMF1* | 1.391483009 | 3.47E-05 | 0.001268787 | 0.168334541 |
| Heart_Failure | adipocyte | *UBE2I* | 1.378941405 | 4.59E-05 | 0.001268787 | 0.22289881 |

| | | | | | | |
|---|---|---|---|---|---|---|
| Heart_Failure | adipocyte | *CPPED1* | 0.800838011 | 0.00155189 | 0.007628947 | 1 |
| Heart_Failure | adipocyte | *TOX3* | 0.477059433 | 0.002279148 | 0.0102015 | 1 |
| Heart_Failure | adipocyte | *RBL2* | 0.801127006 | 5.74E-05 | 0.001268787 | 0.278726462 |
| Heart_Failure | adipocyte | *PYCARD* | 0.685130377 | 0.001803605 | 0.008496203 | 1 |
| Heart_Failure | adipocyte | *KAT8* | 1.231778661 | 0.00170101 | 0.008187796 | 1 |
| Heart_Failure | adipocyte | *C16orf62* | 1.156096025 | 8.56E-05 | 0.001299545 | 0.415276307 |
| Heart_Failure | adipocyte | *RNF40* | 1.450069683 | 0.000417015 | 0.002928157 | 1 |
| Heart_Failure | adipocyte | *KNOP1* | 1.13423216 | 0.000620025 | 0.003912042 | 1 |
| Heart_Failure | adipocyte | *IQCH* | 1.753507114 | 1.79E-06 | 0.000321058 | 0.008668554 |
| Heart_Failure | adipocyte | *HERC1* | 0.933399016 | 0.000109198 | 0.001439756 | 0.529830351 |
| Heart_Failure | adipocyte | *TRIP4* | 0.752098731 | 0.002338771 | 0.010380724 | 1 |
| Heart_Failure | adipocyte | *CTSH* | -0.09318791 | 0.015472073 | 0.045385356 | 1 |
| Heart_Failure | adipocyte | *TTC23* | 1.292866931 | 9.00E-05 | 0.001311406 | 0.436526785 |
| Heart_Failure | adipocyte | *SLC30A4* | 1.516837257 | 0.000145757 | 0.001607298 | 0.707210958 |
| Heart_Failure | adipocyte | *ZFAND1* | 1.084457772 | 0.000847652 | 0.004894234 | 1 |
| Heart_Failure | adipocyte | *IMPAD1* | 0.84821301 | 0.005686119 | 0.020822097 | 1 |
| Heart_Failure | adipocyte | *SFRP1* | 1.121513481 | 0.002300961 | 0.010277198 | 1 |
| Heart_Failure | adipocyte | *NIPAL2* | 0.971364211 | 0.000204036 | 0.001926038 | 0.989983747 |
| Heart_Failure | adipocyte | *RAB2A* | 0.557485449 | 0.006144575 | 0.022057812 | 1 |
| Heart_Failure | adipocyte | *CHRAC1* | 2.665871626 | 7.08E-05 | 0.001268787 | 0.34353027 |
| Heart_Failure | adipocyte | *SNX16* | 0.741801851 | 0.006230672 | 0.022245195 | 1 |
| Heart_Failure | adipocyte | *GSDMD* | 0.751465889 | 0.001951929 | 0.009019772 | 1 |
| Heart_Failure | adipocyte | *ERICH1* | 0.701843014 | 0.002241311 | 0.010041405 | 1 |
| Heart_Failure | adipocyte | *MAN2B1* | 0.263050118 | 0.008884563 | 0.029385072 | 1 |
| Heart_Failure | adipocyte | *SNRNP70* | 1.110757663 | 0.000131726 | 0.001543804 | 0.639135033 |
| Heart_Failure | adipocyte | *LIN7B* | 0.951602699 | 0.01174762 | 0.036585013 | 1 |
| Heart_Failure | adipocyte | *SGTA* | 0.918079681 | 0.003262233 | 0.013301137 | 1 |
| Heart_Failure | adipocyte | *MED25* | 1.750339121 | 9.81E-05 | 0.001356203 | 0.476027421 |
| Heart_Failure | adipocyte | *TIMM44* | 1.211966148 | 2.65E-07 | 9.19E-05 | 0.001286034 |
| Heart_Failure | adipocyte | *FAM32A* | 1.152311145 | 0.008943328 | 0.029519066 | 1 |
| Heart_Failure | adipocyte | *ANKRD27* | 1.176828743 | 0.000343039 | 0.002576511 | 1 |
| Heart_Failure | adipocyte | *TGFB1* | 1.144784498 | 0.000258657 | 0.002152669 | 1 |
| Heart_Failure | adipocyte | *RABAC1* | 0.464178981 | 0.017449833 | 0.049483688 | 1 |
| Heart_Failure | adipocyte | *CARD8* | 0.523940483 | 0.010529351 | 0.033632924 | 1 |
| Heart_Failure | adipocyte | *CAPS* | 1.532906948 | 0.000991798 | 0.005480147 | 1 |
| Heart_Failure | adipocyte | *MIER2* | 0.478872538 | 0.01541263 | 0.045240219 | 1 |
| Heart_Failure | adipocyte | *TNPO2* | 1.010035287 | 0.00018674 | 0.001801316 | 0.906061978 |
| Heart_Failure | adipocyte | *TFPT* | 1.208747676 | 0.005703322 | 0.020853444 | 1 |
| Heart_Failure | adipocyte | *RAB3A* | 5.498537219 | 0.00175192 | 0.008341823 | 1 |
| Heart_Failure | adipocyte | *ZNF14* | 1.675835799 | 6.85E-05 | 0.001268787 | 0.332234778 |
| Heart_Failure | adipocyte | *ZNF85* | 1.377119975 | 0.001796963 | 0.008488751 | 1 |
| Heart_Failure | adipocyte | *ETHE1* | 1.010520071 | 0.006496635 | 0.023025329 | 1 |
| Heart_Failure | adipocyte | *SMG9* | 0.661464566 | 0.016124892 | 0.046653533 | 1 |
| Heart_Failure | adipocyte | *RASA4* | 14.47208208 | 4.01E-05 | 0.001268787 | 0.194397546 |
| Heart_Failure | adipocyte | *BET1* | 1.075801685 | 0.000290621 | 0.00234235 | 1 |
| Heart_Failure | adipocyte | *DNAH11* | 0.88730425 | 0.007497253 | 0.025823044 | 1 |
| Heart_Failure | adipocyte | *ADAP1* | 5.999088444 | 0.000137303 | 0.001563831 | 0.666192097 |
| Heart_Failure | adipocyte | *IQCE* | 1.71374031 | 9.27E-05 | 0.001331689 | 0.449771957 |
| Heart_Failure | adipocyte | *HIBADH* | 0.642244645 | 0.000145007 | 0.001606329 | 0.703571926 |
| Heart_Failure | adipocyte | *CPVL* | 0.451283263 | 0.015891379 | 0.046167205 | 1 |

| | | | | | | |
|---|---|---|---|---|---|---|
| Heart_Failure | adipocyte | *CHN2* | 0.51950038 | 0.000254428 | 0.002132099 | 1 |
| Heart_Failure | adipocyte | *GRB10* | 0.975333071 | 0.000460168 | 0.003149133 | 1 |
| Heart_Failure | adipocyte | *PLEKHA8* | 1.34726726 | 0.000172516 | 0.001756516 | 0.837045244 |
| Heart_Failure | adipocyte | *ZKSCAN1* | 1.025129597 | 0.000204705 | 0.001928603 | 0.993230424 |
| Heart_Failure | adipocyte | *EIF3B* | 1.294214146 | 0.000251385 | 0.002117571 | 1 |
| Heart_Failure | adipocyte | *RBM28* | 0.409327792 | 0.003282737 | 0.0133735 | 1 |
| Heart_Failure | adipocyte | *C1GALT1* | 1.308613332 | 6.28E-05 | 0.001268787 | 0.304556119 |
| Heart_Failure | adipocyte | *NRF1* | 0.555527698 | 0.010996751 | 0.034714531 | 1 |
| Heart_Failure | adipocyte | *ZNF862* | 0.736023096 | 0.00265566 | 0.011514999 | 1 |
| Heart_Failure | adipocyte | *RARRES2* | 0.706371292 | 0.003004523 | 0.012567194 | 1 |
| Heart_Failure | adipocyte | *COA1* | 0.95044035 | 0.00014321 | 0.001590059 | 0.694855634 |
| Heart_Failure | adipocyte | *BLVRA* | 0.557099819 | 0.003732969 | 0.014821902 | 1 |
| Heart_Failure | adipocyte | *GCK* | 7.681315722 | 0.002400584 | 0.010521799 | 1 |
| Heart_Failure | adipocyte | *FKTN* | 1.345634651 | 4.65E-05 | 0.001268787 | 0.22544636 |
| Heart_Failure | adipocyte | *MEGF9* | 1.346526564 | 4.83E-05 | 0.001268787 | 0.234301797 |
| Heart_Failure | adipocyte | *C5* | 0.511727493 | 0.005472203 | 0.020206337 | 1 |
| Heart_Failure | adipocyte | *ECM2* | 0.981916213 | 6.00E-05 | 0.001268787 | 0.291197676 |
| Heart_Failure | adipocyte | *KANK1* | 0.429478245 | 0.002714158 | 0.011685089 | 1 |
| Heart_Failure | adipocyte | *FUBP3* | 0.742618361 | 0.003340417 | 0.013561901 | 1 |
| Heart_Failure | adipocyte | *PTGDS* | 0.802162083 | 0.01418454 | 0.042222938 | 1 |
| Heart_Failure | adipocyte | *PHYH* | 0.234251669 | 0.016765247 | 0.048047832 | 1 |
| Heart_Failure | adipocyte | *DNMBP* | 0.459569289 | 0.007328734 | 0.025421724 | 1 |
| Heart_Failure | adipocyte | *TRDMT1* | 1.322804779 | 0.000131523 | 0.001543804 | 0.63814868 |
| Heart_Failure | adipocyte | *ATE1* | 0.556680041 | 0.000965013 | 0.005381888 | 1 |
| Heart_Failure | adipocyte | *CDH23* | 0.884313 | 0.001284946 | 0.006660852 | 1 |
| Heart_Failure | adipocyte | *BMPR1A* | 0.969182374 | 0.000111356 | 0.001449365 | 0.540296983 |
| Heart_Failure | adipocyte | *SFXN3* | 1.10636261 | 0.002992349 | 0.012537889 | 1 |
| Heart_Failure | adipocyte | *TNKS2* | 1.125211569 | 0.000177877 | 0.001757759 | 0.863059897 |
| Heart_Failure | adipocyte | *ANKRD26* | 0.524097854 | 0.008368219 | 0.028097337 | 1 |
| Heart_Failure | adipocyte | *ACBD5* | 0.77966758 | 0.005990368 | 0.021601642 | 1 |
| Heart_Failure | adipocyte | *LHPP* | 0.482946157 | 0.002191106 | 0.009898739 | 1 |
| Heart_Failure | adipocyte | *PITRM1* | 1.28750619 | 4.46E-05 | 0.001268787 | 0.216400475 |
| Heart_Failure | adipocyte | *TFAM* | 1.646954522 | 0.000142407 | 0.001589239 | 0.690959579 |
| Heart_Failure | adipocyte | *PBLD* | 0.488774139 | 0.008523259 | 0.028383563 | 1 |
| Heart_Failure | adipocyte | *RAPGEFL1* | 0.771830392 | 0.009715339 | 0.03155871 | 1 |
| Heart_Failure | adipocyte | *TRIM37* | 0.996242179 | 0.000847785 | 0.004894234 | 1 |
| Heart_Failure | adipocyte | *PIGL* | 0.95572359 | 0.000312692 | 0.002443124 | 1 |
| Heart_Failure | adipocyte | *MED13* | 0.819194868 | 0.000257554 | 0.002150866 | 1 |
| Heart_Failure | adipocyte | *RAI1* | 1.293980682 | 0.000350567 | 0.002611199 | 1 |
| Heart_Failure | adipocyte | *GOSR1* | 1.286285804 | 4.91E-05 | 0.001268787 | 0.238243994 |
| Heart_Failure | adipocyte | *DRG2* | 0.825247447 | 0.004054633 | 0.015801672 | 1 |
| Heart_Failure | adipocyte | *FTSJ3* | 0.093590514 | 0.014894556 | 0.043932148 | 1 |
| Heart_Failure | adipocyte | *ICAM2* | 0.738054671 | 0.015666995 | 0.045765359 | 1 |
| Heart_Failure | adipocyte | *DDX5* | 0.431349349 | 0.003316078 | 0.013475384 | 1 |
| Heart_Failure | adipocyte | *RAB5C* | 0.501694733 | 0.000431388 | 0.003003006 | 1 |
| Heart_Failure | adipocyte | *NAGLU* | 1.069347218 | 3.48E-05 | 0.001268787 | 0.168941787 |
| Heart_Failure | adipocyte | *EFTUD2* | 1.521775 | 7.18E-05 | 0.001268787 | 0.348496973 |
| Heart_Failure | adipocyte | *FAM20A* | 0.59352661 | 0.010542839 | 0.033653851 | 1 |
| Heart_Failure | adipocyte | *DHRS7B* | 1.222339715 | 7.60E-05 | 0.001268787 | 0.368680454 |
| Heart_Failure | adipocyte | *MYH3* | 1.651718943 | 0.001878871 | 0.008791015 | 1 |

| | | | | | | |
|---|---|---|---|---|---|---|
| Heart_Failure | adipocyte | *SLAIN2* | 1.181614391 | 2.35E-05 | 0.001268787 | 0.114059537 |
| Heart_Failure | adipocyte | *OCIAD1* | 1.030581573 | 0.000209011 | 0.001946488 | 1 |
| Heart_Failure | adipocyte | *UBE2D3* | 0.998186003 | 8.91E-05 | 0.001311406 | 0.432497055 |
| Heart_Failure | adipocyte | *NDUFC1* | 1.222046638 | 0.000659577 | 0.004076773 | 1 |
| Heart_Failure | adipocyte | *KLHL2* | 0.873515599 | 0.001559778 | 0.007652218 | 1 |
| Heart_Failure | adipocyte | *WFS1* | 1.774231142 | 0.000119301 | 0.001495325 | 0.578849071 |
| Heart_Failure | adipocyte | *GRPEL1* | 1.673934775 | 0.000299599 | 0.002383411 | 1 |
| Heart_Failure | adipocyte | *GAR1* | 1.150386016 | 0.010451918 | 0.033407581 | 1 |
| Heart_Failure | adipocyte | *TRIM2* | 0.703494891 | 0.00219862 | 0.009923446 | 1 |
| Heart_Failure | adipocyte | *STIM2* | 0.948973422 | 0.000481122 | 0.003237729 | 1 |
| Heart_Failure | adipocyte | *UFSP2* | 1.288254792 | 0.000217038 | 0.001965494 | 1 |
| Heart_Failure | adipocyte | *KLHL5* | 1.277109205 | 2.59E-05 | 0.001268787 | 0.125821002 |
| Heart_Failure | adipocyte | *FAM149A* | 0.620300045 | 0.011389007 | 0.035582396 | 1 |
| Heart_Failure | adipocyte | *ELP4* | 0.525174848 | 0.007217026 | 0.02508382 | 1 |
| Heart_Failure | adipocyte | *CEP164* | 0.887917937 | 0.000996129 | 0.0054902 | 1 |
| Heart_Failure | adipocyte | *KIAA1549L* | 3.655970833 | 0.004233434 | 0.016327999 | 1 |
| Heart_Failure | adipocyte | *SLC35F2* | 0.754234451 | 0.012266843 | 0.037813673 | 1 |
| Heart_Failure | adipocyte | *RPS13* | 0.404556007 | 0.016568234 | 0.047595662 | 1 |
| Heart_Failure | adipocyte | *PRDM4* | 1.016582146 | 3.72E-06 | 0.000504094 | 0.018025882 |
| Heart_Failure | adipocyte | *CORO1C* | 1.271315989 | 6.61E-05 | 0.001268787 | 0.320715504 |
| Heart_Failure | adipocyte | *IL23A* | 6.953834612 | 0.007651828 | 0.026219401 | 1 |
| Heart_Failure | adipocyte | *ACSS3* | 0.284332053 | 0.009727045 | 0.031568978 | 1 |
| Heart_Failure | adipocyte | *PARP11* | 1.368038185 | 7.61E-05 | 0.001268787 | 0.369041416 |
| Heart_Failure | adipocyte | *GPN3* | 1.951457473 | 7.62E-05 | 0.001268787 | 0.369635219 |
| Heart_Failure | adipocyte | *VPS29* | 1.194000274 | 7.31E-05 | 0.001268787 | 0.354661903 |
| Heart_Failure | adipocyte | *CDK2AP1* | 0.425502364 | 0.00017136 | 0.0017504 | 0.831440125 |
| Heart_Failure | adipocyte | *CAND1* | 1.157094826 | 0.000167301 | 0.001734499 | 0.81174555 |
| Heart_Failure | adipocyte | *MDM1* | 1.741372486 | 3.19E-05 | 0.001268787 | 0.154646716 |
| Heart_Failure | adipocyte | *NUP107* | 0.844498115 | 0.00148284 | 0.007371659 | 1 |
| Heart_Failure | adipocyte | *KRR1* | 0.974501414 | 0.00057843 | 0.003722206 | 1 |
| Heart_Failure | adipocyte | *GNPTAB* | 0.953599282 | 0.000613853 | 0.003889811 | 1 |
| Heart_Failure | adipocyte | *C12orf57* | 1.122099282 | 0.003864491 | 0.015231934 | 1 |
| Heart_Failure | adipocyte | *NT5DC3* | 0.492432378 | 0.009811351 | 0.031757621 | 1 |
| Heart_Failure | adipocyte | *SUDS3* | 1.463864358 | 6.26E-05 | 0.001268787 | 0.303571983 |
| Heart_Failure | adipocyte | *GOLT1B* | 0.814585766 | 0.003178418 | 0.013025072 | 1 |
| Heart_Failure | adipocyte | *C2CD5* | 0.793726003 | 0.00140651 | 0.00713102 | 1 |
| Heart_Failure | adipocyte | *FRK* | 1.237618491 | 0.00210563 | 0.009592976 | 1 |
| Heart_Failure | adipocyte | *TMEM14C* | 1.549093865 | 9.94E-05 | 0.001368761 | 0.482423591 |
| Heart_Failure | adipocyte | *FAM184A* | 0.63594675 | 0.001548264 | 0.00761884 | 1 |
| Heart_Failure | adipocyte | *RNGTT* | 1.013051271 | 0.000363559 | 0.002664638 | 1 |
| Heart_Failure | adipocyte | *RNF8* | 0.77768729 | 0.004029536 | 0.015729129 | 1 |
| Heart_Failure | adipocyte | *CD83* | 1.071928899 | 0.005748795 | 0.020972295 | 1 |
| Heart_Failure | adipocyte | *MDN1* | 0.59915372 | 0.006786471 | 0.023836887 | 1 |
| Heart_Failure | adipocyte | *VNN2* | 0.359857491 | 0.011871574 | 0.036829204 | 1 |
| Heart_Failure | adipocyte | *ACOT13* | 1.327643721 | 9.77E-05 | 0.001354769 | 0.474169179 |
| Heart_Failure | adipocyte | *C6orf62* | 0.605716769 | 0.005253147 | 0.019546217 | 1 |
| Heart_Failure | adipocyte | *FIG4* | 1.049903037 | 0.000343802 | 0.002578252 | 1 |
| Heart_Failure | adipocyte | *PACRG* | 0.930411233 | 0.006178107 | 0.022120675 | 1 |
| Heart_Failure | adipocyte | *QKI* | 0.937231882 | 4.71E-05 | 0.001268787 | 0.228400495 |
| Heart_Failure | adipocyte | *FAM120B* | 0.765098669 | 2.41E-05 | 0.001268787 | 0.116698033 |

| | | | | | | |
|---|---|---|---|---|---|---|
| Heart_Failure | adipocyte | *EXOC2* | 1.149887331 | 5.95E-05 | 0.001268787 | 0.288466352 |
| Heart_Failure | adipocyte | *TMEM30A* | 1.242979288 | 0.000480395 | 0.003237729 | 1 |
| Heart_Failure | adipocyte | *BTN2A1* | 0.613269249 | 0.008991823 | 0.029589143 | 1 |
| Heart_Failure | adipocyte | *PRSS16* | 0.358476953 | 0.011607675 | 0.036172409 | 1 |
| Heart_Failure | adipocyte | *NUDT12* | 1.2262042 | 0.00045507 | 0.003123635 | 1 |
| Heart_Failure | adipocyte | *KIF20A* | -1.747728969 | 0.000130187 | 0.001543804 | 0.631667013 |
| Heart_Failure | adipocyte | *SPARC* | 0.695461832 | 0.001085642 | 0.005852815 | 1 |
| Heart_Failure | adipocyte | *ARSB* | 1.073912408 | 0.001711288 | 0.008204713 | 1 |
| Heart_Failure | adipocyte | *RASGRF2* | 0.21285804 | 0.005399123 | 0.02001264 | 1 |
| Heart_Failure | adipocyte | *DROSHA* | 0.433335872 | 0.005859995 | 0.021313865 | 1 |
| Heart_Failure | adipocyte | *SLC12A7* | 0.754744293 | 0.016992862 | 0.048556752 | 1 |
| Heart_Failure | adipocyte | *SKP1* | 1.253911942 | 0.000173475 | 0.001756516 | 0.841700143 |
| Heart_Failure | adipocyte | *PPWD1* | 1.308562954 | 4.82E-05 | 0.001268787 | 0.233697382 |
| Heart_Failure | adipocyte | *TXNDC15* | 0.983713784 | 0.003169267 | 0.01299855 | 1 |
| Heart_Failure | adipocyte | *SMAD5* | 1.124919943 | 0.000227208 | 0.002021062 | 1 |
| Heart_Failure | adipocyte | *ERGIC1* | 1.03720973 | 0.000492663 | 0.003297101 | 1 |
| Heart_Failure | adipocyte | *ATP6V0E1* | 1.167798259 | 1.83E-05 | 0.001268787 | 0.089024836 |
| Heart_Failure | adipocyte | *BNIP1* | 0.19872152 | 0.002377355 | 0.010467266 | 1 |
| Heart_Failure | adipocyte | *UNC5A* | 4.316259671 | 0.00688984 | 0.024136824 | 1 |
| Heart_Failure | adipocyte | *TBCCD1* | 1.818800733 | 8.36E-05 | 0.001279363 | 0.4055581 |
| Heart_Failure | adipocyte | *PCCB* | 0.351833014 | 0.002099077 | 0.009581113 | 1 |
| Heart_Failure | adipocyte | *CEP70* | 0.710488711 | 0.001573539 | 0.007704151 | 1 |
| Heart_Failure | adipocyte | *HYAL1* | 0.241044676 | 0.012064449 | 0.037355908 | 1 |
| Heart_Failure | adipocyte | *CBLB* | 0.492486022 | 0.008300024 | 0.027902715 | 1 |
| Heart_Failure | adipocyte | *IQCG* | 1.177313397 | 0.000315383 | 0.002455108 | 1 |
| Heart_Failure | adipocyte | *PLXNA1* | 0.661449006 | 0.007781245 | 0.026569036 | 1 |
| Heart_Failure | adipocyte | *PLSCR4* | 0.775332684 | 0.000349683 | 0.002610247 | 1 |
| Heart_Failure | adipocyte | *HEMK1* | 0.612662293 | 0.003528817 | 0.014185437 | 1 |
| Heart_Failure | adipocyte | *COMMD2* | 1.078234321 | 0.000648793 | 0.004030656 | 1 |
| Heart_Failure | adipocyte | *ARHGEF26* | 0.673458934 | 0.003921333 | 0.015382114 | 1 |
| Heart_Failure | adipocyte | *DNAH1* | 0.370392815 | 0.0046289 | 0.017587645 | 1 |
| Heart_Failure | adipocyte | *NKTR* | 0.617180933 | 0.002097086 | 0.009581037 | 1 |
| Heart_Failure | adipocyte | *LMAN2L* | 0.631055411 | 0.003921615 | 0.015382114 | 1 |
| Heart_Failure | adipocyte | *ACTR1B* | 1.169849931 | 0.001115551 | 0.005974229 | 1 |
| Heart_Failure | adipocyte | *NRBP1* | 1.149742884 | 0.00072883 | 0.004382011 | 1 |
| Heart_Failure | adipocyte | *PPM1G* | 0.857237054 | 0.002689181 | 0.01158784 | 1 |
| Heart_Failure | adipocyte | *INO80B* | -0.284196376 | 0.01431506 | 0.042481146 | 1 |
| Heart_Failure | adipocyte | *MOGS* | 0.291667739 | 0.001796994 | 0.008488751 | 1 |
| Heart_Failure | adipocyte | *GRB14* | 0.865492087 | 0.004095438 | 0.015909579 | 1 |
| Heart_Failure | adipocyte | *RTN4* | 0.434294338 | 0.008014557 | 0.027155467 | 1 |
| Heart_Failure | adipocyte | *GALNT3* | 0.628845705 | 0.00155922 | 0.007652218 | 1 |
| Heart_Failure | adipocyte | *POLE4* | 0.920413229 | 0.011187278 | 0.03517866 | 1 |
| Heart_Failure | adipocyte | *MRPL19* | 1.026412212 | 0.000346659 | 0.002591662 | 1 |
| Heart_Failure | adipocyte | *DNAH6* | 0.075416902 | 0.012871881 | 0.039180908 | 1 |
| Heart_Failure | adipocyte | *IL18R1* | 0.933572339 | 2.78E-08 | 1.50E-05 | 0.000134647 |
| Heart_Failure | adipocyte | *HDLBP* | 0.981555577 | 8.71E-06 | 0.000960535 | 0.042263533 |
| Heart_Failure | adipocyte | *TAF1B* | 0.867222804 | 2.39E-05 | 0.001268787 | 0.116010805 |
| Heart_Failure | adipocyte | *NOL10* | 1.136986877 | 0.00016936 | 0.001744662 | 0.821735739 |
| Heart_Failure | adipocyte | *STRN* | 1.09276531 | 0.000148171 | 0.001619166 | 0.71892366 |
| Heart_Failure | adipocyte | *CEBPZ* | 1.224223743 | 1.71E-05 | 0.001268787 | 0.082787075 |

| | | | | | | |
|---|---|---|---|---|---|---|
| Heart_Failure | adipocyte | *PRKD3* | 0.941314042 | 0.000147675 | 0.001617421 | 0.71651739 |
| Heart_Failure | adipocyte | *SLC25A12* | 0.71151123 | 0.002659467 | 0.011521191 | 1 |
| Heart_Failure | adipocyte | *DARS* | 1.27475076 | 4.54E-05 | 0.001268787 | 0.220405659 |
| Heart_Failure | adipocyte | *WIPF1* | 0.7777096 | 0.001011974 | 0.005548136 | 1 |
| Heart_Failure | adipocyte | *PNO1* | 2.325276002 | 4.40E-05 | 0.001268787 | 0.213276764 |
| Heart_Failure | adipocyte | *ORC4* | 0.493528001 | 0.008604153 | 0.02861471 | 1 |
| Heart_Failure | adipocyte | *AAK1* | 1.046354338 | 5.09E-05 | 0.001268787 | 0.247053851 |
| Heart_Failure | adipocyte | *NFE2L2* | 0.60687884 | 0.000882554 | 0.005026 | 1 |
| Heart_Failure | adipocyte | *MSH6* | 1.004284262 | 0.000765707 | 0.004548779 | 1 |
| Heart_Failure | adipocyte | *ALMS1* | 1.36276698 | 2.61E-05 | 0.001268787 | 0.126395964 |
| Heart_Failure | adipocyte | *DNAJC16* | 0.674157547 | 0.003102738 | 0.012839666 | 1 |
| Heart_Failure | adipocyte | *FAM20B* | 0.817354576 | 0.005380645 | 0.019959396 | 1 |
| Heart_Failure | adipocyte | *ICMT* | 1.472355387 | 0.002971762 | 0.012483972 | 1 |
| Heart_Failure | adipocyte | *EDEM3* | 1.150375681 | 8.75E-05 | 0.001310535 | 0.424613409 |
| Heart_Failure | adipocyte | *RAP1A* | 0.767400931 | 0.000666536 | 0.004114545 | 1 |
| Heart_Failure | adipocyte | *CAPZA1* | 0.80293944 | 0.000596111 | 0.003810711 | 1 |
| Heart_Failure | adipocyte | *ASH1L* | 0.459935367 | 0.008604465 | 0.02861471 | 1 |
| Heart_Failure | adipocyte | *DOCK7* | 1.023659242 | 0.000247313 | 0.002108901 | 1 |
| Heart_Failure | adipocyte | *LEPR* | 0.651578225 | 0.001254785 | 0.006540911 | 1 |
| Heart_Failure | adipocyte | *MFN2* | 0.452663093 | 0.013020725 | 0.039510041 | 1 |
| Heart_Failure | adipocyte | *MRPS15* | 0.82423344 | 0.003208939 | 0.013127968 | 1 |
| Heart_Failure | adipocyte | *GNPAT* | 0.972513551 | 0.000164897 | 0.001731774 | 0.800079724 |
| Heart_Failure | adipocyte | *TBCE* | 1.377562021 | 0.000177389 | 0.001756516 | 0.860693006 |
| Heart_Failure | adipocyte | *ACADM* | 0.603291251 | 0.008995051 | 0.029589143 | 1 |
| Heart_Failure | adipocyte | *SDHB* | 0.979857256 | 0.002226607 | 0.010031101 | 1 |
| Heart_Failure | adipocyte | *ZNHIT6* | 0.622134803 | 0.015727073 | 0.045830485 | 1 |
| Heart_Failure | adipocyte | *CD46* | 0.39167481 | 0.012129525 | 0.037533453 | 1 |
| Heart_Failure | adipocyte | *ERI3* | 1.244440652 | 2.81E-05 | 0.001268787 | 0.136223757 |
| Heart_Failure | adipocyte | *PRRC2C* | 0.62868234 | 0.004489325 | 0.017107098 | 1 |
| Heart_Failure | adipocyte | *ABCD3* | 1.331718188 | 7.39E-05 | 0.001268787 | 0.358499565 |
| Heart_Failure | adipocyte | *PTBP2* | 0.439022467 | 0.002315347 | 0.010306481 | 1 |
| Heart_Failure | adipocyte | *MTFR1L* | 0.687746325 | 0.015899711 | 0.046167205 | 1 |
| Heart_Failure | adipocyte | *MARC2* | 0.434270217 | 0.013858388 | 0.041430005 | 1 |
| Heart_Failure | adipocyte | *ESYT2* | 0.699907412 | 0.001428705 | 0.00716864 | 1 |
| Heart_Failure | adipocyte | *CD3EAP* | 0.650155067 | 0.010303696 | 0.033086389 | 1 |
| Heart_Failure | adipocyte | *RCN2* | 0.85333942 | 0.003195523 | 0.013084117 | 1 |
| Heart_Failure | adipocyte | *CAMSAP2* | 0.541564982 | 0.001595332 | 0.007795114 | 1 |
| Heart_Failure | adipocyte | *KLF7* | 0.487351015 | 0.006572782 | 0.023257867 | 1 |
| Heart_Failure | adipocyte | *AKAP7* | 0.819212672 | 0.00012038 | 0.001498571 | 0.584082959 |
| Heart_Failure | adipocyte | *MED28* | 0.819019909 | 2.13E-05 | 0.001268787 | 0.103280385 |
| Heart_Failure | adipocyte | *FOXO3* | 0.912583416 | 8.89E-05 | 0.001311406 | 0.431193419 |
| Heart_Failure | adipocyte | *MFSD1* | 0.747660803 | 0.005131352 | 0.019137062 | 1 |
| Heart_Failure | adipocyte | *UBN1* | 1.079908464 | 0.000297242 | 0.002372071 | 1 |
| Heart_Failure | adipocyte | *WDR35* | 0.716553414 | 0.006396344 | 0.022746107 | 1 |
| Heart_Failure | adipocyte | *C2orf40* | 0.583807242 | 0.014399775 | 0.042656397 | 1 |
| Heart_Failure | adipocyte | *SENP5* | 1.157715847 | 9.15E-05 | 0.001325336 | 0.443987482 |
| Heart_Failure | adipocyte | *FKBP15* | 0.309413596 | 0.002396521 | 0.010513491 | 1 |
| Heart_Failure | adipocyte | *CTNNAL1* | 0.276951487 | 8.87E-05 | 0.001311406 | 0.430242445 |
| Heart_Failure | adipocyte | *RAB14* | 0.868490953 | 0.005642106 | 0.020739014 | 1 |
| Heart_Failure | adipocyte | *TRIM32* | 1.589626916 | 0.000669075 | 0.004119738 | 1 |

| | | | | | | |
|---|---|---|---|---|---|---|
| Heart_Failure | adipocyte | *PPP6C* | 1.240732333 | 7.42E-05 | 0.001268787 | 0.359916049 |
| Heart_Failure | adipocyte | *RBM18* | 0.73494642 | 0.00534557 | 0.019859653 | 1 |
| Heart_Failure | adipocyte | *MAPKAP1* | 1.094917151 | 3.62E-05 | 0.001268787 | 0.175804442 |
| Heart_Failure | adipocyte | *INVS* | 0.337160522 | 0.010671515 | 0.03397519 | 1 |
| Heart_Failure | adipocyte | *DENND1A* | 0.804862924 | 0.000238498 | 0.002073819 | 1 |
| Heart_Failure | adipocyte | *KDSR* | 1.136780043 | 0.00057646 | 0.003714453 | 1 |
| Heart_Failure | adipocyte | *DCAF4* | 0.573815777 | 0.004039494 | 0.015755324 | 1 |
| Heart_Failure | adipocyte | *NEK9* | 0.952688309 | 0.000206554 | 0.00193849 | 1 |
| Heart_Failure | adipocyte | *TTLL5* | 0.857324183 | 0.001302934 | 0.006703751 | 1 |
| Heart_Failure | adipocyte | *FLVCR2* | 0.483114414 | 0.012733903 | 0.038834003 | 1 |
| Heart_Failure | adipocyte | *COQ6* | 1.200864382 | 0.001884865 | 0.008810562 | 1 |
| Heart_Failure | adipocyte | *CRIPT* | 1.531984541 | 8.83E-05 | 0.001311406 | 0.428659843 |
| Heart_Failure | adipocyte | *SLC17A5* | 0.493586803 | 0.016710607 | 0.04794788 | 1 |
| Heart_Failure | adipocyte | *MXI1* | 0.811604993 | 0.002167504 | 0.009828721 | 1 |
| Heart_Failure | adipocyte | *WDR11* | 0.67352578 | 0.004577239 | 0.017404987 | 1 |
| Heart_Failure | adipocyte | *KANSL1* | 0.632722211 | 0.003123193 | 0.012896794 | 1 |
| Heart_Failure | adipocyte | *HOXB3* | 1.2248498 | 0.000139164 | 0.001573949 | 0.67522403 |
| Heart_Failure | adipocyte | *GORAB* | 1.083199798 | 0.001147737 | 0.006112864 | 1 |
| Heart_Failure | adipocyte | *SLC10A7* | 1.134721504 | 0.000142481 | 0.001589239 | 0.691319169 |
| Heart_Failure | adipocyte | *IQSEC3* | 0.848147187 | 0.004458083 | 0.017005205 | 1 |
| Heart_Failure | adipocyte | *CCDC77* | 0.782791596 | 0.001890293 | 0.00882743 | 1 |
| Heart_Failure | adipocyte | *DNAJC15* | 1.04538965 | 0.000366958 | 0.002684255 | 1 |
| Heart_Failure | adipocyte | *SIL1* | 1.040843574 | 5.19E-05 | 0.001268787 | 0.252029972 |
| Heart_Failure | adipocyte | *APAF1* | 0.657249291 | 0.009164141 | 0.030023235 | 1 |
| Heart_Failure | adipocyte | *SCPEP1* | 0.208881612 | 0.008134807 | 0.027462708 | 1 |
| Heart_Failure | adipocyte | *ZNF549* | 1.704914124 | 3.86E-05 | 0.001268787 | 0.187272305 |
| Heart_Failure | adipocyte | *LHX4* | 2.187247608 | 5.30E-05 | 0.001268787 | 0.257299406 |
| Heart_Failure | adipocyte | *SEC22A* | 1.075576493 | 0.000229836 | 0.002032956 | 1 |
| Heart_Failure | adipocyte | *MAPK8IP1* | 0.581239765 | 0.017389824 | 0.049371225 | 1 |
| Heart_Failure | adipocyte | *ZCCHC17* | 1.518490874 | 4.05E-05 | 0.001268787 | 0.196526545 |
| Heart_Failure | adipocyte | *TNFSF10* | 1.062605636 | 0.000315744 | 0.002455108 | 1 |
| Heart_Failure | adipocyte | *ZNF639* | 1.260296723 | 0.002302414 | 0.010277198 | 1 |
| Heart_Failure | adipocyte | *MTIF3* | 0.726991743 | 0.001573148 | 0.007704151 | 1 |
| Heart_Failure | adipocyte | *FMOD* | 0.513306623 | 0.005992541 | 0.021601642 | 1 |
| Heart_Failure | adipocyte | *RBBP6* | 1.005795338 | 0.000325041 | 0.002491471 | 1 |
| Heart_Failure | adipocyte | *NAA60* | 2.361983581 | 8.05E-05 | 0.001268787 | 0.390764591 |
| Heart_Failure | adipocyte | *ODF2L* | 1.068760071 | 0.000171233 | 0.0017504 | 0.830823036 |
| Heart_Failure | adipocyte | *SPATA1* | 1.06816132 | 0.000355968 | 0.002632861 | 1 |
| Heart_Failure | adipocyte | *EEPD1* | 0.277887481 | 0.011014509 | 0.034747982 | 1 |
| Heart_Failure | adipocyte | *KLHL7* | 1.283481065 | 7.37E-05 | 0.001268787 | 0.35763024 |
| Heart_Failure | adipocyte | *NT5C3A* | 0.953803361 | 0.000411619 | 0.002898661 | 1 |
| Heart_Failure | adipocyte | *SMU1* | 1.335349188 | 1.58E-05 | 0.001219853 | 0.076850724 |
| Heart_Failure | adipocyte | *TRIM24* | 0.724708845 | 0.002312094 | 0.010301452 | 1 |
| Heart_Failure | adipocyte | *RBM19* | 1.439781851 | 0.00010999 | 0.00144627 | 0.533673524 |
| Heart_Failure | adipocyte | *CCDC91* | 0.933832308 | 9.00E-05 | 0.001311406 | 0.436698288 |
| Heart_Failure | adipocyte | *TSFM* | 0.356218145 | 0.012256669 | 0.037806331 | 1 |
| Heart_Failure | adipocyte | *PFDN5* | 0.975163388 | 0.001005351 | 0.005518056 | 1 |
| Heart_Failure | adipocyte | *PDE1B* | 0.572054811 | 0.016743509 | 0.048013894 | 1 |
| Heart_Failure | adipocyte | *CDK2* | 0.33071964 | 0.004207115 | 0.01627764 | 1 |
| Heart_Failure | adipocyte | *DBH* | 23.34120528 | 0.0030445 | 0.012647185 | 1 |

| | | | | | | |
|---|---|---|---|---|---|---|
| Heart_Failure | adipocyte | *USP45* | 1.025479567 | 0.000123743 | 0.001527742 | 0.600402566 |
| Heart_Failure | adipocyte | *PLA2G12A* | 1.269747024 | 2.92E-05 | 0.001268787 | 0.141849251 |
| Heart_Failure | adipocyte | *DNPEP* | 1.177592602 | 0.000178709 | 0.001762392 | 0.86709674 |
| Heart_Failure | adipocyte | *SLC12A4* | 1.165561289 | 0.002413522 | 0.01056896 | 1 |
| Heart_Failure | adipocyte | *SRSF6* | 0.838096628 | 0.000984398 | 0.005464874 | 1 |
| Heart_Failure | adipocyte | *CSE1L* | 0.882268295 | 0.000270502 | 0.00221328 | 1 |
| Heart_Failure | adipocyte | *RNF114* | 1.046501469 | 0.0001645 | 0.001731349 | 0.798151838 |
| Heart_Failure | adipocyte | *MTRR* | 0.697712301 | 0.00154367 | 0.007608048 | 1 |
| Heart_Failure | adipocyte | *PEPD* | 1.212155163 | 4.65E-05 | 0.001268787 | 0.225475579 |
| Heart_Failure | adipocyte | *STAMBP* | 1.451409734 | 4.63E-05 | 0.001268787 | 0.224887412 |
| Heart_Failure | adipocyte | *SNRNP27* | 1.734651389 | 3.77E-05 | 0.001268787 | 0.183126384 |
| Heart_Failure | adipocyte | *ATP8A1* | 0.571603446 | 0.007422818 | 0.02567036 | 1 |
| Heart_Failure | adipocyte | *USP22* | 0.96535771 | 0.000735389 | 0.004415975 | 1 |
| Heart_Failure | adipocyte | *MRS2* | 0.296889677 | 0.003228483 | 0.013174599 | 1 |
| Heart_Failure | adipocyte | *SNRPC* | 0.315348401 | 0.015229285 | 0.044783328 | 1 |
| Heart_Failure | adipocyte | *NQO2* | 1.013894051 | 0.000368477 | 0.002684255 | 1 |
| Heart_Failure | adipocyte | *GLO1* | 1.222139981 | 1.01E-06 | 0.000233889 | 0.00491167 |
| Heart_Failure | adipocyte | *RREB1* | 0.477001613 | 0.0077018 | 0.026353409 | 1 |
| Heart_Failure | adipocyte | *ATXN1* | 0.731526866 | 0.000319388 | 0.00247156 | 1 |
| Heart_Failure | adipocyte | *RUNX2* | 1.004759765 | 0.000481959 | 0.003238872 | 1 |
| Heart_Failure | adipocyte | *CLYBL* | 1.195262594 | 0.000233545 | 0.002052827 | 1 |
| Heart_Failure | adipocyte | *TMTC4* | 1.227253638 | 0.000287631 | 0.002322108 | 1 |
| Heart_Failure | adipocyte | *GRK4* | 0.080271414 | 0.009717377 | 0.03155871 | 1 |
| Heart_Failure | adipocyte | *MRPS7* | 1.687094049 | 1.66E-05 | 0.001256213 | 0.080397628 |
| Heart_Failure | adipocyte | *NT5C* | 1.384707056 | 0.000407959 | 0.002889657 | 1 |
| Heart_Failure | adipocyte | *OPRL1* | 6.999805767 | 9.27E-06 | 0.000999937 | 0.044997155 |
| Heart_Failure | adipocyte | *PPDPF* | 0.53852533 | 0.000984315 | 0.005464874 | 1 |
| Heart_Failure | adipocyte | *SNRPD2* | 1.818460703 | 6.36E-05 | 0.001268787 | 0.308351767 |
| Heart_Failure | adipocyte | *RBCK1* | 0.528563408 | 0.016352892 | 0.047200615 | 1 |
| Heart_Failure | adipocyte | *RRBP1* | 0.534833285 | 0.013621565 | 0.040886238 | 1 |
| Heart_Failure | adipocyte | *FLRT3* | 1.069453998 | 0.003303445 | 0.013435302 | 1 |
| Heart_Failure | adipocyte | *MKKS* | 0.555172966 | 0.009981827 | 0.032217911 | 1 |
| Heart_Failure | adipocyte | *NCLN* | 0.577450401 | 0.004177772 | 0.016177615 | 1 |
| Heart_Failure | adipocyte | *KLC1* | 1.247772917 | 8.80E-05 | 0.001311406 | 0.426955151 |
| Heart_Failure | adipocyte | *TUBGCP3* | 1.066780739 | 0.000247297 | 0.002108901 | 1 |
| Heart_Failure | adipocyte | *COX6B1* | 1.134079196 | 0.000317321 | 0.002463429 | 1 |
| Heart_Failure | adipocyte | *PRDX5* | 0.812405874 | 0.001471962 | 0.007332609 | 1 |
| Heart_Failure | adipocyte | *IRF3* | 1.132784868 | 0.000876594 | 0.005013834 | 1 |
| Heart_Failure | adipocyte | *KTN1* | 0.985262887 | 0.000119577 | 0.001495325 | 0.580185972 |
| Heart_Failure | adipocyte | *ZBTB1* | 0.806059272 | 0.000548262 | 0.003570693 | 1 |
| Heart_Failure | adipocyte | *RHOT1* | 0.451818478 | 0.00097221 | 0.005409593 | 1 |
| Heart_Failure | adipocyte | *CANX* | 0.915348697 | 0.000382186 | 0.002754513 | 1 |
| Heart_Failure | adipocyte | *ZNF484* | 0.769714535 | 0.009435835 | 0.030767924 | 1 |
| Heart_Failure | adipocyte | *HELB* | 0.781554801 | 0.008958684 | 0.029549234 | 1 |
| Heart_Failure | adipocyte | *RAB3IP* | 1.265394971 | 0.001266752 | 0.006580599 | 1 |
| Heart_Failure | adipocyte | *EMC1* | 1.120492137 | 5.27E-05 | 0.001268787 | 0.255931587 |
| Heart_Failure | adipocyte | *RNF6* | 1.465904549 | 6.85E-05 | 0.001268787 | 0.332169772 |
| Heart_Failure | adipocyte | *HIP1* | 0.746444269 | 0.000911397 | 0.005136005 | 1 |
| Heart_Failure | adipocyte | *PTPN12* | 0.697590181 | 0.001271886 | 0.006600204 | 1 |
| Heart_Failure | adipocyte | *PEX1* | 1.328992267 | 9.53E-05 | 0.001343879 | 0.46225042 |

| | | | | | | |
|---|---|---|---|---|---|---|
| Heart_Failure | adipocyte | *ATF4* | 1.418600432 | 0.000370615 | 0.002691951 | 1 |
| Heart_Failure | adipocyte | *SPECC1* | 0.857008276 | 0.000962896 | 0.005376259 | 1 |
| Heart_Failure | adipocyte | *CDHR3* | 0.447096094 | 0.010407611 | 0.033314461 | 1 |
| Heart_Failure | adipocyte | *PRKRIP1* | 0.989740232 | 0.000140893 | 0.001582442 | 0.68361503 |
| Heart_Failure | adipocyte | *MTX2* | 1.281349848 | 2.70E-05 | 0.001268787 | 0.131241892 |
| Heart_Failure | adipocyte | *ORMDL1* | 0.437884318 | 0.00844629 | 0.028224104 | 1 |
| Heart_Failure | adipocyte | *PSMG2* | 0.500898143 | 0.003748621 | 0.0148476 | 1 |
| Heart_Failure | adipocyte | *TWSG1* | 1.343986303 | 5.95E-05 | 0.001268787 | 0.288924236 |
| Heart_Failure | adipocyte | *EIF2AK4* | 0.591916702 | 0.007253219 | 0.025191565 | 1 |
| Heart_Failure | adipocyte | *ARPP19* | 1.321135967 | 0.000194999 | 0.001858815 | 0.946136652 |
| Heart_Failure | adipocyte | *VPS13C* | 0.647498461 | 0.002668654 | 0.011540381 | 1 |
| Heart_Failure | adipocyte | *SUMF2* | 1.114499028 | 0.000700257 | 0.004247059 | 1 |
| Heart_Failure | adipocyte | *SERGEF* | 0.508112558 | 0.003296961 | 0.013420179 | 1 |
| Heart_Failure | adipocyte | *PHF20L1* | 0.986915746 | 9.69E-05 | 0.001347241 | 0.470187279 |
| Heart_Failure | adipocyte | *MIS18BP1* | 0.541887825 | 0.001260827 | 0.006556842 | 1 |
| Heart_Failure | adipocyte | *RNASE1* | 0.500559506 | 0.00502842 | 0.018825535 | 1 |
| Heart_Failure | adipocyte | *EPB41L4A* | 1.006489617 | 0.000153457 | 0.001650934 | 0.744571201 |
| Heart_Failure | adipocyte | *REEP5* | 1.215257026 | 7.72E-05 | 0.001268787 | 0.374810843 |
| Heart_Failure | adipocyte | *TMEM8A* | 1.802837123 | 5.78E-05 | 0.001268787 | 0.280483861 |
| Heart_Failure | adipocyte | *MAU2* | 0.78835675 | 0.004491847 | 0.017107098 | 1 |
| Heart_Failure | adipocyte | *PHF10* | 1.210471965 | 0.000519398 | 0.003456954 | 1 |
| Heart_Failure | adipocyte | *XPO7* | 1.035896239 | 0.00052829 | 0.003487436 | 1 |
| Heart_Failure | adipocyte | *TULP4* | 0.521333303 | 0.000726582 | 0.004373918 | 1 |
| Heart_Failure | adipocyte | *ACTN4* | 0.863376259 | 0.000294444 | 0.002356205 | 1 |
| Heart_Failure | adipocyte | *MAP1S* | 2.120939973 | 0.000327187 | 0.00250002 | 1 |
| Heart_Failure | adipocyte | *ATXN10* | 0.95258783 | 0.000229254 | 0.002032956 | 1 |
| Heart_Failure | adipocyte | *PNPLA7* | 0.722132438 | 0.002178771 | 0.009870587 | 1 |
| Heart_Failure | adipocyte | *PAK4* | 0.757900661 | 0.009211508 | 0.030158054 | 1 |
| Heart_Failure | adipocyte | *ZNF337* | 0.480725421 | 0.003159639 | 0.012970022 | 1 |
| Heart_Failure | adipocyte | *MAP3K10* | 0.453964812 | 0.014260613 | 0.042371398 | 1 |
| Heart_Failure | adipocyte | *ANGPTL6* | -0.272472336 | 0.012571429 | 0.038435145 | 1 |
| Heart_Failure | adipocyte | *DNMT1* | 1.024744522 | 0.002185288 | 0.009890875 | 1 |
| Heart_Failure | adipocyte | *C12orf65* | 0.576645755 | 0.000136531 | 0.001561419 | 0.662450387 |
| Heart_Failure | adipocyte | *HSD17B3* | 1.055208676 | 0.012889163 | 0.039208916 | 1 |
| Heart_Failure | adipocyte | *SLC35D2* | 1.485628792 | 1.46E-05 | 0.001175323 | 0.071038261 |
| Heart_Failure | adipocyte | *TTLL9* | 1.270883523 | 0.013993761 | 0.04175752 | 1 |
| Heart_Failure | adipocyte | *ATP6V1E1* | 0.796859615 | 0.001164361 | 0.006187822 | 1 |
| Heart_Failure | adipocyte | *CHMP1A* | 1.54500323 | 1.96E-05 | 0.001268787 | 0.095106864 |
| Heart_Failure | adipocyte | *MRPS25* | 0.769393508 | 0.000368964 | 0.002684255 | 1 |
| Heart_Failure | adipocyte | *PDLIM4* | 1.706154545 | 0.00019646 | 0.001869064 | 0.953222666 |
| Heart_Failure | adipocyte | *KIF3A* | 1.181830129 | 6.34E-05 | 0.001268787 | 0.307654564 |
| Heart_Failure | adipocyte | *ACLY* | 0.490890488 | 0.016591516 | 0.047634341 | 1 |
| Heart_Failure | adipocyte | *NDUFA2* | 0.419099819 | 0.003153429 | 0.012964151 | 1 |
| Heart_Failure | adipocyte | *EXOC4* | 0.721089835 | 8.66E-05 | 0.00130347 | 0.420234713 |
| Heart_Failure | adipocyte | *C1orf159* | 1.416312182 | 0.000425583 | 0.0029754 | 1 |
| Heart_Failure | adipocyte | *PPFIA1* | 0.858119182 | 0.000208353 | 0.001944092 | 1 |
| Heart_Failure | adipocyte | *MAP1B* | 0.525448238 | 0.000161249 | 0.001711991 | 0.782379696 |
| Heart_Failure | adipocyte | *KHDRBS3* | 1.540540655 | 8.24E-05 | 0.001268787 | 0.399607465 |
| Heart_Failure | adipocyte | *PRKAB2* | 0.869478387 | 0.005977672 | 0.021582093 | 1 |
| Heart_Failure | adipocyte | *TRIM21* | 0.816228918 | 0.013387748 | 0.040371258 | 1 |

| | | | | | | |
|---|---|---|---|---|---|---|
| Heart_Failure | adipocyte | *SPATA6* | 0.63212171 | 0.003602962 | 0.014394835 | 1 |
| Heart_Failure | adipocyte | *IMMT* | 0.650145027 | 0.001532737 | 0.007580878 | 1 |
| Heart_Failure | adipocyte | *SERPINF1* | 1.381132905 | 4.38E-05 | 0.001268787 | 0.212394869 |
| Heart_Failure | adipocyte | *MUTYH* | 0.465304415 | 0.004452568 | 0.017005205 | 1 |
| Heart_Failure | adipocyte | *CTNNBL1* | 0.725350915 | 0.006672905 | 0.023495598 | 1 |
| Heart_Failure | adipocyte | *AP3B1* | 0.938163135 | 0.000222596 | 0.00200007 | 1 |
| Heart_Failure | adipocyte | *ALOX5AP* | 0.737907167 | 0.013284401 | 0.040159447 | 1 |
| Heart_Failure | adipocyte | *WASF3* | 1.185827428 | 9.62E-05 | 0.001343879 | 0.466893737 |
| Heart_Failure | adipocyte | *PEMT* | 1.058937501 | 0.001842067 | 0.008652185 | 1 |
| Heart_Failure | adipocyte | *SCO1* | 0.573756027 | 0.00284362 | 0.012071078 | 1 |
| Heart_Failure | adipocyte | *MPRIP* | 1.228965562 | 6.12E-05 | 0.001268787 | 0.297027693 |
| Heart_Failure | adipocyte | *DSTYK* | 0.952993179 | 0.001297892 | 0.006690377 | 1 |
| Heart_Failure | adipocyte | *COG6* | 0.707680834 | 0.000712841 | 0.004308658 | 1 |
| Heart_Failure | adipocyte | *BTBD2* | 1.081582976 | 0.000282932 | 0.002300045 | 1 |
| Heart_Failure | adipocyte | *CSNK1G2* | 1.799991944 | 5.60E-05 | 0.001268787 | 0.271807674 |
| Heart_Failure | adipocyte | *PDZD2* | 0.838488502 | 0.000120454 | 0.001498571 | 0.584442807 |
| Heart_Failure | adipocyte | *FAM83F* | 2.227884959 | 0.001214902 | 0.006379552 | 1 |
| Heart_Failure | adipocyte | *IPO8* | 0.971670317 | 0.000866124 | 0.004971605 | 1 |
| Heart_Failure | adipocyte | *RRAS2* | 0.614277949 | 0.000721744 | 0.004350189 | 1 |
| Heart_Failure | adipocyte | *ELP3* | 1.093379095 | 0.000475157 | 0.00322442 | 1 |
| Heart_Failure | adipocyte | *PEBP4* | 1.867622463 | 0.000174822 | 0.001756516 | 0.848235335 |
| Heart_Failure | adipocyte | *THUMPD3* | 0.903965687 | 0.008081921 | 0.027364607 | 1 |
| Heart_Failure | adipocyte | *CEPT1* | 0.677217676 | 0.001024936 | 0.005606526 | 1 |
| Heart_Failure | adipocyte | *PPHLN1* | 0.919521625 | 0.000268787 | 0.002210435 | 1 |
| Heart_Failure | adipocyte | *FKBP11* | 0.989671655 | 0.008255458 | 0.027797004 | 1 |
| Heart_Failure | adipocyte | *YWHAQ* | 0.835395254 | 0.003024632 | 0.01259701 | 1 |
| Heart_Failure | adipocyte | *KIDINS220* | 1.068338173 | 8.13E-05 | 0.001268787 | 0.394671123 |
| Heart_Failure | adipocyte | *LPIN1* | 0.405679366 | 0.012202215 | 0.037662309 | 1 |
| Heart_Failure | adipocyte | *KIAA1468* | 1.065029857 | 0.000216196 | 0.001964387 | 1 |
| Heart_Failure | adipocyte | *IL15RA* | 0.728789297 | 0.000893863 | 0.005066617 | 1 |
| Heart_Failure | adipocyte | *KLRD1* | 1.292958256 | 0.000135318 | 0.001561419 | 0.656563716 |
| Heart_Failure | adipocyte | *ELP2* | 1.251885235 | 4.92E-05 | 0.001268787 | 0.23868314 |
| Heart_Failure | adipocyte | *TPGS2* | 0.948509252 | 0.000946173 | 0.005319618 | 1 |
| Heart_Failure | adipocyte | *TIMM10* | 0.854694867 | 0.000887352 | 0.005041489 | 1 |
| Heart_Failure | adipocyte | *DHX34* | 1.309921706 | 0.000627684 | 0.003944977 | 1 |
| Heart_Failure | adipocyte | *CLOCK* | 1.164637866 | 6.69E-05 | 0.001268787 | 0.324771895 |
| Heart_Failure | adipocyte | *ARGLU1* | 0.779044393 | 0.000266862 | 0.002209578 | 1 |
| Heart_Failure | adipocyte | *ERCC5* | 0.752152476 | 0.000903238 | 0.005107823 | 1 |
| Heart_Failure | adipocyte | *WDR36* | 1.065853327 | 0.002766822 | 0.011831029 | 1 |
| Heart_Failure | adipocyte | *OSTF1* | 0.744598889 | 0.006131505 | 0.022037084 | 1 |
| Heart_Failure | adipocyte | *UBQLN1* | 1.117943614 | 0.000129648 | 0.001543804 | 0.629054286 |
| Heart_Failure | adipocyte | *ADAM19* | 0.246663802 | 0.001632666 | 0.007921696 | 1 |
| Heart_Failure | adipocyte | *DMTF1* | 0.79872165 | 0.001336711 | 0.006841476 | 1 |
| Heart_Failure | adipocyte | *SRPK2* | 0.708075527 | 0.002296562 | 0.01026997 | 1 |
| Heart_Failure | adipocyte | *AKIRIN2* | 0.84860188 | 0.000878538 | 0.005013834 | 1 |
| Heart_Failure | adipocyte | *MAP3K7* | 1.001823857 | 0.000741648 | 0.004438667 | 1 |
| Heart_Failure | adipocyte | *PHF21A* | 0.899918714 | 0.000219468 | 0.001975619 | 1 |
| Heart_Failure | adipocyte | *CAPRIN1* | 0.797150788 | 0.002764402 | 0.011831029 | 1 |
| Heart_Failure | adipocyte | *ATP5G2* | 0.814544557 | 0.002838564 | 0.012066841 | 1 |
| Heart_Failure | adipocyte | *USP15* | 1.013048094 | 3.95E-05 | 0.001268787 | 0.191673703 |

| | | | | | | |
|---|---|---|---|---|---|---|
| Heart_Failure | adipocyte | *KLHL36* | 0.547871064 | 0.013089208 | 0.039693024 | 1 |
| Heart_Failure | adipocyte | *MPHOSPH6* | 1.041766984 | 0.002770002 | 0.011831029 | 1 |
| Heart_Failure | adipocyte | *NTPCR* | 1.267774587 | 0.000447174 | 0.003095135 | 1 |
| Heart_Failure | adipocyte | *PIGC* | 1.079261621 | 0.000619897 | 0.003912042 | 1 |
| Heart_Failure | adipocyte | *MRPL44* | 2.000103092 | 2.15E-06 | 0.000347512 | 0.010425351 |
| Heart_Failure | adipocyte | *USP37* | 0.769134029 | 5.79E-05 | 0.001268787 | 0.281007071 |
| Heart_Failure | adipocyte | *TMBIM1* | 1.229317862 | 0.000176101 | 0.001756516 | 0.854440817 |
| Heart_Failure | adipocyte | *GCC2* | 0.579303479 | 0.00843656 | 0.028224104 | 1 |
| Heart_Failure | adipocyte | *ANKRD36* | 0.835479763 | 0.002189043 | 0.009898636 | 1 |
| Heart_Failure | adipocyte | *EPC2* | 1.002790254 | 0.00011374 | 0.001456112 | 0.551866528 |
| Heart_Failure | adipocyte | *STAB2* | 1.676399803 | 0.000106132 | 0.001422516 | 0.514950963 |
| Heart_Failure | adipocyte | *APPL2* | 0.521973302 | 0.013399459 | 0.040381475 | 1 |
| Heart_Failure | adipocyte | *TBC1D4* | 0.500641581 | 0.0034124 | 0.013820504 | 1 |
| Heart_Failure | adipocyte | *SUCLA2* | 0.797230759 | 0.001053353 | 0.005723257 | 1 |
| Heart_Failure | adipocyte | *RCBTB2* | 1.200020425 | 0.000339929 | 0.002557239 | 1 |
| Heart_Failure | adipocyte | *SETDB2* | 1.121332491 | 0.000213525 | 0.001960237 | 1 |
| Heart_Failure | adipocyte | *KDELR2* | 1.218943894 | 0.001139788 | 0.006077201 | 1 |
| Heart_Failure | adipocyte | *NUPL2* | 1.371288013 | 6.95E-05 | 0.001268787 | 0.337267602 |
| Heart_Failure | adipocyte | *ZDHHC4* | 1.550009352 | 3.55E-05 | 0.001268787 | 0.172472218 |
| Heart_Failure | adipocyte | *NMT1* | 0.363946861 | 1.42E-05 | 0.001175323 | 0.06868996 |
| Heart_Failure | adipocyte | *NDUFB5* | 0.875319902 | 0.001927041 | 0.008949516 | 1 |
| Heart_Failure | adipocyte | *SCN2A* | 0.43260216 | 0.007616864 | 0.026118039 | 1 |
| Heart_Failure | adipocyte | *RPS6KC1* | 0.726364757 | 0.001723141 | 0.008242495 | 1 |
| Heart_Failure | adipocyte | *ODF2* | 1.084978247 | 0.003892205 | 0.015328716 | 1 |
| Heart_Failure | adipocyte | *ZNF189* | 1.816024227 | 3.67E-05 | 0.001268787 | 0.178000199 |
| Heart_Failure | adipocyte | *STX17* | 1.179819852 | 0.000192457 | 0.001841818 | 0.933801578 |
| Heart_Failure | adipocyte | *ANP32B* | 0.753621217 | 0.007606625 | 0.026101375 | 1 |
| Heart_Failure | adipocyte | *DERL1* | 0.657713204 | 0.017329421 | 0.049302284 | 1 |
| Heart_Failure | adipocyte | *APTX* | 0.963543846 | 0.000450403 | 0.003104195 | 1 |
| Heart_Failure | adipocyte | *DENND4C* | 0.721280274 | 0.001105009 | 0.00594402 | 1 |
| Heart_Failure | adipocyte | *FRS3* | 1.270753408 | 0.002369454 | 0.010441952 | 1 |
| Heart_Failure | adipocyte | *TCF19* | 0.309959507 | 0.002239497 | 0.010041405 | 1 |
| Heart_Failure | adipocyte | *FLOT1* | 0.370039209 | 0.005414534 | 0.020039144 | 1 |
| Heart_Failure | adipocyte | *NRM* | 4.412370772 | 0.000787299 | 0.00464717 | 1 |
| Heart_Failure | adipocyte | *TLR2* | 0.737879656 | 0.007921597 | 0.026884666 | 1 |
| Heart_Failure | adipocyte | *NUMA1* | 1.090550976 | 0.000151518 | 0.001637339 | 0.735165015 |
| Heart_Failure | adipocyte | *RAB30* | 1.236451379 | 0.000605687 | 0.003856684 | 1 |
| Heart_Failure | adipocyte | *NARS2* | 1.317516948 | 5.06E-05 | 0.001268787 | 0.245477204 |
| Heart_Failure | adipocyte | *SULF1* | 0.243606555 | 0.007182202 | 0.024980677 | 1 |
| Heart_Failure | adipocyte | *TGS1* | 1.408727134 | 1.23E-05 | 0.00112616 | 0.059810043 |
| Heart_Failure | adipocyte | *NEK1* | 0.863955731 | 0.000382328 | 0.002754513 | 1 |
| Heart_Failure | adipocyte | *MAP2K5* | 1.154767826 | 7.84E-06 | 0.000905615 | 0.038035826 |
| Heart_Failure | adipocyte | *PARP6* | 1.350272033 | 8.54E-05 | 0.001299545 | 0.414225644 |
| Heart_Failure | adipocyte | *ADAM10* | 0.915874377 | 1.46E-05 | 0.001175323 | 0.071065147 |
| Heart_Failure | adipocyte | *ZNF280D* | 1.042339115 | 2.24E-05 | 0.001268787 | 0.10874924 |
| Heart_Failure | adipocyte | *DBT* | 0.88742932 | 0.001599905 | 0.007801746 | 1 |
| Heart_Failure | adipocyte | *RTCA* | 0.630088681 | 0.006287607 | 0.022399022 | 1 |
| Heart_Failure | adipocyte | *DYNC2LI1* | 0.384198648 | 0.008438195 | 0.028224104 | 1 |
| Heart_Failure | adipocyte | *ACTR2* | 0.95275569 | 0.000357152 | 0.002637595 | 1 |
| Heart_Failure | adipocyte | *LRPPRC* | 1.180169378 | 7.85E-05 | 0.001268787 | 0.380650244 |

| | | | | | | |
|---|---|---|---|---|---|---|
| Heart_Failure | adipocyte | *BTBD16* | 0.488345207 | 0.009423803 | 0.030749354 | 1 |
| Heart_Failure | adipocyte | *TACC2* | 0.46215548 | 0.013384346 | 0.040371258 | 1 |
| Heart_Failure | adipocyte | *ARL3* | 0.326431036 | 0.010752761 | 0.034167284 | 1 |
| Heart_Failure | adipocyte | *RPS24* | 0.558357998 | 0.00354512 | 0.014227396 | 1 |
| Heart_Failure | adipocyte | *SMARCAL1* | 1.281341193 | 0.000249436 | 0.002112148 | 1 |
| Heart_Failure | adipocyte | *STAT4* | 0.948444932 | 0.003631705 | 0.014479073 | 1 |
| Heart_Failure | adipocyte | *FASTKD1* | 0.882723904 | 6.57E-05 | 0.001268787 | 0.318811551 |
| Heart_Failure | adipocyte | *ITGAV* | 0.637651288 | 0.005024583 | 0.018825535 | 1 |
| Heart_Failure | adipocyte | *SLC35A5* | 0.562108675 | 0.000992797 | 0.005480147 | 1 |
| Heart_Failure | adipocyte | *SENP7* | 0.615154818 | 0.006748317 | 0.023742485 | 1 |
| Heart_Failure | adipocyte | *COX17* | 1.245871235 | 0.001762339 | 0.008383206 | 1 |
| Heart_Failure | adipocyte | *AP1AR* | 1.625460512 | 1.16E-05 | 0.001110079 | 0.056336155 |
| Heart_Failure | adipocyte | *KIAA1109* | 0.853557127 | 0.000410186 | 0.002894606 | 1 |
| Heart_Failure | adipocyte | *PRDM5* | 1.162853909 | 0.000351759 | 0.002613679 | 1 |
| Heart_Failure | adipocyte | *G3BP2* | 0.523465872 | 0.000250535 | 0.002114077 | 1 |
| Heart_Failure | adipocyte | *CNOT6L* | 0.536282901 | 0.007116599 | 0.024805846 | 1 |
| Heart_Failure | adipocyte | *PPA2* | 0.775172031 | 0.000745317 | 0.004453547 | 1 |
| Heart_Failure | adipocyte | *PAPSS1* | 0.748085145 | 0.000878133 | 0.005013834 | 1 |
| Heart_Failure | adipocyte | *KIF21A* | 0.810752752 | 0.000543437 | 0.00355358 | 1 |
| Heart_Failure | adipocyte | *ZCRB1* | 0.760384977 | 0.007150393 | 0.024905749 | 1 |
| Heart_Failure | adipocyte | *PPFIA2* | 0.715602853 | 0.002655664 | 0.011514999 | 1 |
| Heart_Failure | adipocyte | *MARCH9* | 1.83101097 | 0.000789428 | 0.004654075 | 1 |
| Heart_Failure | adipocyte | *POC1B* | 1.074266966 | 0.001850054 | 0.008672911 | 1 |
| Heart_Failure | adipocyte | *RITA1* | 3.116815136 | 4.99E-05 | 0.001268787 | 0.242216122 |
| Heart_Failure | adipocyte | *GLTP* | 1.242869567 | 0.003021478 | 0.01259701 | 1 |
| Heart_Failure | adipocyte | *CCDC65* | 0.550506946 | 0.010757808 | 0.034167284 | 1 |
| Heart_Failure | adipocyte | *CERS5* | 0.914248276 | 0.002425662 | 0.010612543 | 1 |
| Heart_Failure | adipocyte | *LMBR1L* | 0.696194405 | 0.00223675 | 0.010041405 | 1 |
| Heart_Failure | adipocyte | *ESYT1* | 1.088257606 | 2.56E-05 | 0.001268787 | 0.12435041 |
| Heart_Failure | adipocyte | *WDFY2* | 0.519397403 | 0.010575866 | 0.033714915 | 1 |
| Heart_Failure | adipocyte | *RB1* | 0.737322429 | 0.000382735 | 0.002754513 | 1 |
| Heart_Failure | adipocyte | *TMX1* | 1.335738389 | 0.000245523 | 0.00210123 | 1 |
| Heart_Failure | adipocyte | *C14orf37* | 1.134912899 | 6.76E-05 | 0.001268787 | 0.327768627 |
| Heart_Failure | adipocyte | *RAB15* | 2.062407097 | 8.15E-05 | 0.001268787 | 0.395258888 |
| Heart_Failure | adipocyte | *C14orf79* | 0.729977043 | 0.000397969 | 0.002827151 | 1 |
| Heart_Failure | adipocyte | *WARS* | 0.592865616 | 0.011516119 | 0.035933253 | 1 |
| Heart_Failure | adipocyte | *SERF2* | 1.068445363 | 0.000185947 | 0.001801316 | 0.902214776 |
| Heart_Failure | adipocyte | *HMG20A* | 1.351670184 | 4.64E-05 | 0.001268787 | 0.225092006 |
| Heart_Failure | adipocyte | *ULK3* | 1.41760746 | 0.003023342 | 0.01259701 | 1 |
| Heart_Failure | adipocyte | *PCSK6* | 1.211101525 | 0.000691747 | 0.004221565 | 1 |
| Heart_Failure | adipocyte | *ABHD2* | 0.847923739 | 0.001398769 | 0.007106627 | 1 |
| Heart_Failure | adipocyte | *UNC45A* | 1.200825777 | 0.000226401 | 0.002021062 | 1 |
| Heart_Failure | adipocyte | *CRTC3* | 0.528434189 | 0.000585487 | 0.003752687 | 1 |
| Heart_Failure | adipocyte | *DHX38* | 1.644128269 | 8.26E-05 | 0.001268787 | 0.400936639 |
| Heart_Failure | adipocyte | *NUDT7* | 0.677106994 | 0.013740841 | 0.041129278 | 1 |
| Heart_Failure | adipocyte | *GCSH* | 0.311229182 | 0.01255075 | 0.03842034 | 1 |
| Heart_Failure | adipocyte | *ZCCHC14* | 0.710137328 | 0.004284235 | 0.016497703 | 1 |
| Heart_Failure | adipocyte | *TLDC1* | 1.689465534 | 0.000163122 | 0.00172058 | 0.791466844 |
| Heart_Failure | adipocyte | *TIGD7* | 0.585584716 | 0.011076826 | 0.034876547 | 1 |
| Heart_Failure | adipocyte | *TCF25* | 1.032187534 | 7.25E-07 | 0.000185069 | 0.003516315 |

| | | | | | | |
|---|---|---|---|---|---|---|
| Heart_Failure | adipocyte | *GALNS* | 0.708943908 | 0.001917842 | 0.008921735 | 1 |
| Heart_Failure | adipocyte | *NCOR1* | 0.989740189 | 5.98E-05 | 0.001268787 | 0.290114882 |
| Heart_Failure | adipocyte | *KSR1* | 1.171338103 | 0.000189476 | 0.001820473 | 0.919338673 |
| Heart_Failure | adipocyte | *MYO19* | 0.885611672 | 0.000125983 | 0.00153782 | 0.61127009 |
| Heart_Failure | adipocyte | *SSH2* | 0.955748792 | 6.01E-05 | 0.001268787 | 0.291697884 |
| Heart_Failure | adipocyte | *IMPA2* | 1.210995363 | 1.01E-05 | 0.001018189 | 0.048873083 |
| Heart_Failure | adipocyte | *GNAL* | 1.537356197 | 4.77E-05 | 0.001268787 | 0.231396643 |
| Heart_Failure | adipocyte | *RPRD1A* | 0.520399271 | 0.014753349 | 0.043595159 | 1 |
| Heart_Failure | adipocyte | *GALNT1* | 0.947280751 | 0.000310018 | 0.002430057 | 1 |
| Heart_Failure | adipocyte | *NPC1* | 0.777638644 | 0.004226958 | 0.016315993 | 1 |
| Heart_Failure | adipocyte | *ANAPC11* | 1.2675688 | 0.002888763 | 0.012213225 | 1 |
| Heart_Failure | adipocyte | *TBCD* | 0.709436679 | 0.002076621 | 0.009512251 | 1 |
| Heart_Failure | adipocyte | *NARF* | 1.101221788 | 2.18E-05 | 0.001268787 | 0.105584014 |
| Heart_Failure | adipocyte | *ZNF750* | 1.220791669 | 0.001323399 | 0.006787664 | 1 |
| Heart_Failure | adipocyte | *MAPK4* | 3.737849173 | 0.000224291 | 0.002007856 | 1 |
| Heart_Failure | adipocyte | *ERBB2* | 1.193271954 | 2.77E-08 | 1.50E-05 | 0.0001345 |
| Heart_Failure | adipocyte | *PNMT* | 1.346069334 | 1.69E-12 | 1.64E-09 | 8.21E-09 |
| Heart_Failure | adipocyte | *NFIC* | 0.888514979 | 7.44E-05 | 0.001268787 | 0.361033138 |
| Heart_Failure | adipocyte | *PFKL* | 0.887784666 | 0.001536591 | 0.007584474 | 1 |
| Heart_Failure | adipocyte | *DPP9* | 1.415646687 | 0.000173429 | 0.001756516 | 0.841479688 |
| Heart_Failure | adipocyte | *SIRT3* | 0.838326928 | 0.000176558 | 0.001756516 | 0.856659164 |
| Heart_Failure | adipocyte | *EVI5L* | 2.162039485 | 3.96E-05 | 0.001268787 | 0.191944567 |
| Heart_Failure | adipocyte | *CTU1* | 1.69672138 | 0.003512072 | 0.014152804 | 1 |
| Heart_Failure | adipocyte | *RERE* | 0.794151337 | 0.000118833 | 0.001495325 | 0.57657844 |
| Heart_Failure | adipocyte | *PEX14* | 0.354390665 | 0.004996081 | 0.018762371 | 1 |
| Heart_Failure | adipocyte | *EVA1B* | 0.442581879 | 1.46E-05 | 0.001175323 | 0.070660488 |
| Heart_Failure | adipocyte | *RPS8* | 0.779769366 | 0.001229475 | 0.006449093 | 1 |
| Heart_Failure | adipocyte | *SLC44A3* | 1.094157526 | 0.002997705 | 0.012549493 | 1 |
| Heart_Failure | adipocyte | *STRIP1* | 1.681878787 | 2.70E-05 | 0.001268787 | 0.130900726 |
| Heart_Failure | adipocyte | *ALDH9A1* | 1.133682313 | 0.000697968 | 0.004247059 | 1 |
| Heart_Failure | adipocyte | *MAEL* | 3.786759604 | 0.000950279 | 0.005324197 | 1 |
| Heart_Failure | adipocyte | *RFWD2* | 0.933606887 | 9.57E-05 | 0.001343879 | 0.464539441 |
| Heart_Failure | adipocyte | *SETDB1* | 0.76228359 | 0.005311335 | 0.019747585 | 1 |
| Heart_Failure | adipocyte | *POGZ* | 0.604210235 | 0.002909484 | 0.012264829 | 1 |
| Heart_Failure | adipocyte | *EIF2D* | 0.71487832 | 0.013964598 | 0.041696142 | 1 |
| Heart_Failure | adipocyte | *SMYD2* | 0.975704257 | 0.00302927 | 0.012605505 | 1 |
| Heart_Failure | adipocyte | *RAB13* | 1.406354235 | 0.002393678 | 0.01051052 | 1 |
| Heart_Failure | adipocyte | *S100A8* | 0.867366237 | 0.007073865 | 0.024727948 | 1 |
| Heart_Failure | adipocyte | *C1orf43* | 1.185344833 | 0.000102795 | 0.001397094 | 0.498762527 |
| Heart_Failure | adipocyte | *FLG* | -0.134087607 | 0.001578403 | 0.007720172 | 1 |
| Heart_Failure | adipocyte | *SCCPDH* | 1.423873929 | 8.97E-05 | 0.001311406 | 0.435260546 |
| Heart_Failure | adipocyte | *CEP170* | 1.305736203 | 1.46E-06 | 0.00029586 | 0.007100639 |
| Heart_Failure | adipocyte | *ACP1* | 1.479201489 | 5.59E-05 | 0.001268787 | 0.271322729 |
| Heart_Failure | adipocyte | *CNIH4* | 1.052049602 | 0.000949806 | 0.005324197 | 1 |
| Heart_Failure | adipocyte | *RHOB* | 1.532442575 | 8.93E-05 | 0.001311406 | 0.433355566 |
| Heart_Failure | adipocyte | *GALM* | 1.41735742 | 7.49E-05 | 0.001268787 | 0.363500017 |
| Heart_Failure | adipocyte | *WDPCP* | 0.415651149 | 5.14E-05 | 0.001268787 | 0.249607539 |
| Heart_Failure | adipocyte | *VPS54* | 1.049452477 | 0.000135566 | 0.001561419 | 0.657767363 |
| Heart_Failure | adipocyte | *EXOC6B* | 0.931641556 | 6.01E-05 | 0.001268787 | 0.291779572 |
| Heart_Failure | adipocyte | *TEX261* | 2.276972058 | 0.00015251 | 0.001644401 | 0.739980587 |

| | | | | | | |
|---|---|---|---|---|---|---|
| Heart_Failure | adipocyte | *RALB* | 0.694576342 | 0.013302963 | 0.040190521 | 1 |
| Heart_Failure | adipocyte | *FBLN7* | 0.184213563 | 0.013134333 | 0.039804987 | 1 |
| Heart_Failure | adipocyte | *GALNT13* | 1.329624619 | 0.000105382 | 0.001416381 | 0.511313496 |
| Heart_Failure | adipocyte | *METTL21A* | 0.831978843 | 0.003041711 | 0.012646429 | 1 |
| Heart_Failure | adipocyte | *NBEAL1* | 0.850314462 | 1.98E-05 | 0.001268787 | 0.095868334 |
| Heart_Failure | adipocyte | *SUMF1* | 0.980192693 | 4.60E-05 | 0.001268787 | 0.222969498 |
| Heart_Failure | adipocyte | *ACKR3* | 0.261328947 | 0.005579136 | 0.020554265 | 1 |
| Heart_Failure | adipocyte | *ESPNL* | 10.40758566 | 0.01248612 | 0.038246626 | 1 |
| Heart_Failure | adipocyte | *TAMM41* | 0.588722727 | 0.01705678 | 0.048682056 | 1 |
| Heart_Failure | adipocyte | *EAF1* | 1.194781958 | 0.002449869 | 0.010699157 | 1 |
| Heart_Failure | adipocyte | *PTPRG* | 0.687745787 | 0.00030797 | 0.002421831 | 1 |
| Heart_Failure | adipocyte | *ARL6IP5* | 1.119848939 | 0.000331092 | 0.002517962 | 1 |
| Heart_Failure | adipocyte | *LIMD1* | 0.440266565 | 0.006615572 | 0.023344549 | 1 |
| Heart_Failure | adipocyte | *BOC* | 0.676125647 | 0.002665284 | 0.01153609 | 1 |
| Heart_Failure | adipocyte | *NCEH1* | 0.274914483 | 0.008445137 | 0.028224104 | 1 |
| Heart_Failure | adipocyte | *CCDC39* | 0.462267032 | 0.005906536 | 0.021434939 | 1 |
| Heart_Failure | adipocyte | *CISD2* | 1.662976899 | 9.77E-06 | 0.001018189 | 0.047387376 |
| Heart_Failure | adipocyte | *METTL14* | 0.968314283 | 0.003837737 | 0.015151098 | 1 |
| Heart_Failure | adipocyte | *USP53* | 0.709039896 | 9.88E-06 | 0.001018189 | 0.047954102 |
| Heart_Failure | adipocyte | *CBR4* | 0.830858209 | 0.000844736 | 0.004894097 | 1 |
| Heart_Failure | adipocyte | *MARCH6* | 1.139725082 | 3.44E-05 | 0.001268787 | 0.166891796 |
| Heart_Failure | adipocyte | *GTF2H2* | 2.659052668 | 3.70E-05 | 0.001268787 | 0.179550926 |
| Heart_Failure | adipocyte | *TSLP* | 0.193246112 | 1.12E-06 | 0.000246771 | 0.005428966 |
| Heart_Failure | adipocyte | *FEM1C* | 1.795025777 | 0.000106463 | 0.001423027 | 0.516558836 |
| Heart_Failure | adipocyte | *FBXO38* | 0.943744183 | 0.000294768 | 0.002356205 | 1 |
| Heart_Failure | adipocyte | *BOD1* | 1.424223341 | 2.75E-06 | 0.00041643 | 0.013325772 |
| Heart_Failure | adipocyte | *MYLK4* | 1.11321928 | 0.000470622 | 0.003207105 | 1 |
| Heart_Failure | adipocyte | *TBC1D7* | 0.476180694 | 0.005882798 | 0.021380775 | 1 |
| Heart_Failure | adipocyte | *GFOD1* | 0.634508316 | 0.008930318 | 0.02949619 | 1 |
| Heart_Failure | adipocyte | *FAM193B* | 0.825054203 | 0.001777272 | 0.008440679 | 1 |
| Heart_Failure | adipocyte | *RNF44* | 0.966691723 | 0.001941473 | 0.00899716 | 1 |
| Heart_Failure | adipocyte | *MUT* | 0.554437395 | 0.011846017 | 0.036796974 | 1 |
| Heart_Failure | adipocyte | *RPL7L1* | 1.264116209 | 0.000210964 | 0.001957164 | 1 |
| Heart_Failure | adipocyte | *IRAK1BP1* | 0.568547417 | 0.002483828 | 0.010827971 | 1 |
| Heart_Failure | adipocyte | *PHIP* | 0.991523375 | 6.54E-05 | 0.001268787 | 0.317466308 |
| Heart_Failure | adipocyte | *PNRC1* | 0.972408653 | 0.001949902 | 0.009018993 | 1 |
| Heart_Failure | adipocyte | *RNF217* | 1.010755575 | 6.50E-05 | 0.001268787 | 0.315189923 |
| Heart_Failure | adipocyte | *GNA12* | 0.580323304 | 0.003742578 | 0.0148476 | 1 |
| Heart_Failure | adipocyte | *RBAK* | 2.494437591 | 0.000115511 | 0.001471019 | 0.56045805 |
| Heart_Failure | adipocyte | *GIGYF1* | 0.653489454 | 0.002056803 | 0.009450388 | 1 |
| Heart_Failure | adipocyte | *TMEM209* | 1.399115189 | 0.000131701 | 0.001543804 | 0.639014238 |
| Heart_Failure | adipocyte | *NCAPG2* | 0.477131867 | 0.013671192 | 0.040996678 | 1 |
| Heart_Failure | adipocyte | *FBXO25* | 1.402168838 | 4.42E-05 | 0.001268787 | 0.214448101 |
| Heart_Failure | adipocyte | *BIN3* | 0.702165431 | 0.007501729 | 0.025823044 | 1 |
| Heart_Failure | adipocyte | *DOCK5* | 0.435986072 | 0.016927476 | 0.048455526 | 1 |
| Heart_Failure | adipocyte | *TACC1* | 0.720300383 | 0.000546246 | 0.003562345 | 1 |
| Heart_Failure | adipocyte | *RPL7* | 0.32017857 | 0.009394508 | 0.030695051 | 1 |
| Heart_Failure | adipocyte | *LRP12* | 1.668323153 | 8.26E-05 | 0.001268787 | 0.400724898 |
| Heart_Failure | adipocyte | *AK3* | 1.076644532 | 0.000713078 | 0.004308658 | 1 |
| Heart_Failure | adipocyte | *HAUS6* | 1.559187958 | 0.000126852 | 0.00153782 | 0.615488157 |

| | | | | | | |
|---|---|---|---|---|---|---|
| Heart_Failure | adipocyte | *ALAD* | 0.311517624 | 0.004255241 | 0.016399071 | 1 |
| Heart_Failure | adipocyte | *REXO4* | 1.729678556 | 7.65E-05 | 0.001268787 | 0.371255677 |
| Heart_Failure | adipocyte | *SH3GLB2* | 0.835241636 | 0.009218501 | 0.030160597 | 1 |
| Heart_Failure | adipocyte | *SEC16A* | 0.771994766 | 0.001389751 | 0.007068208 | 1 |
| Heart_Failure | adipocyte | *SLC39A12* | 1.022879765 | 0.004166788 | 0.016147967 | 1 |
| Heart_Failure | adipocyte | *RSU1* | 0.958895442 | 0.000217938 | 0.001965494 | 1 |
| Heart_Failure | adipocyte | *PARD3* | 0.851439303 | 0.000155278 | 0.001666838 | 0.753410998 |
| Heart_Failure | adipocyte | *FRA10AC1* | 0.56115057 | 0.001534565 | 0.007582189 | 1 |
| Heart_Failure | adipocyte | *ADD3* | 0.8217203 | 0.000544963 | 0.003558765 | 1 |
| Heart_Failure | adipocyte | *PAOX* | 2.317658336 | 4.12E-05 | 0.001268787 | 0.199965306 |
| Heart_Failure | adipocyte | *GAS2* | 1.290497925 | 0.000131608 | 0.001543804 | 0.638561299 |
| Heart_Failure | adipocyte | *LIN7C* | 1.479197298 | 0.000139622 | 0.001575459 | 0.677447368 |
| Heart_Failure | adipocyte | *PTPRJ* | 0.580616587 | 0.006185403 | 0.022120675 | 1 |
| Heart_Failure | adipocyte | *INTS4* | 0.632188162 | 0.010731305 | 0.034143143 | 1 |
| Heart_Failure | adipocyte | *RPS3* | 0.622406985 | 0.00208381 | 0.009529358 | 1 |
| Heart_Failure | adipocyte | *NPAT* | 1.333451101 | 0.000121241 | 0.001504508 | 0.588262762 |
| Heart_Failure | adipocyte | *ATM* | 0.52468509 | 0.006852712 | 0.024024102 | 1 |
| Heart_Failure | adipocyte | *FADS1* | 0.687053087 | 0.000599063 | 0.003824541 | 1 |
| Heart_Failure | adipocyte | *ROM1* | 0.21592209 | 0.000684125 | 0.004191132 | 1 |
| Heart_Failure | adipocyte | *INCENP* | 0.799507865 | 0.014096997 | 0.04198811 | 1 |
| Heart_Failure | adipocyte | *CPSF7* | 0.947298455 | 0.000262352 | 0.002179681 | 1 |
| Heart_Failure | adipocyte | *CHEK1* | 1.841491271 | 0.007682556 | 0.026306112 | 1 |
| Heart_Failure | adipocyte | *ALDOA* | 0.885212677 | 1.87E-05 | 0.001268787 | 0.090614997 |
| Heart_Failure | adipocyte | *TMEM218* | 0.592661346 | 0.007049396 | 0.02466018 | 1 |
| Heart_Failure | adipocyte | *SAP18* | 1.49281789 | 0.000234745 | 0.002059641 | 1 |
| Heart_Failure | adipocyte | *FAM124A* | 0.766536731 | 0.011232752 | 0.035207621 | 1 |
| Heart_Failure | adipocyte | *PRSS23* | 0.449401342 | 0.002456349 | 0.010717811 | 1 |
| Heart_Failure | adipocyte | *FAM173B* | 0.730858965 | 0.003482099 | 0.014067562 | 1 |
| Heart_Failure | adipocyte | *GXYLT1* | 0.680747485 | 0.013596436 | 0.040848241 | 1 |
| Heart_Failure | adipocyte | *TMEM18* | 1.722533949 | 0.00029375 | 0.002355823 | 1 |
| Heart_Failure | adipocyte | *NDUFC2* | 1.700564516 | 0.000213781 | 0.001960237 | 1 |
| Heart_Failure | adipocyte | *NUBPL* | 1.159413351 | 5.21E-05 | 0.001268787 | 0.252901412 |
| Heart_Failure | adipocyte | *VIPAS39* | 1.072624139 | 0.001723593 | 0.008242495 | 1 |
| Heart_Failure | adipocyte | *ANKRD50* | 0.98782545 | 0.000165392 | 0.001733225 | 0.802483149 |
| Heart_Failure | adipocyte | *UPF2* | 1.024342546 | 0.000129484 | 0.001543804 | 0.628255041 |
| Heart_Failure | adipocyte | *FAM160B1* | 0.914784024 | 0.000862999 | 0.004967109 | 1 |
| Heart_Failure | adipocyte | *ZNF827* | 0.45324591 | 0.012333909 | 0.037909483 | 1 |
| Heart_Failure | adipocyte | *PIGF* | 1.410637496 | 5.56E-05 | 0.001268787 | 0.269747711 |
| Heart_Failure | adipocyte | *MFSD6* | 0.982892958 | 0.001066396 | 0.005781179 | 1 |
| Heart_Failure | adipocyte | *RNF144A* | 0.874657863 | 0.000471899 | 0.003211294 | 1 |
| Heart_Failure | adipocyte | *BICD1* | 0.563088489 | 0.010388358 | 0.033314154 | 1 |
| Heart_Failure | adipocyte | *TIAL1* | 1.008110467 | 0.000567648 | 0.003667412 | 1 |
| Heart_Failure | adipocyte | *TMEM56* | 1.97540591 | 0.003904122 | 0.015350731 | 1 |
| Heart_Failure | adipocyte | *PTPN14* | 0.985464583 | 5.51E-05 | 0.001268787 | 0.267581319 |
| Heart_Failure | adipocyte | *MGAT5* | 0.469640454 | 0.016477685 | 0.04744791 | 1 |
| Heart_Failure | adipocyte | *TMEM178A* | 2.459328402 | 0.000570869 | 0.003683318 | 1 |
| Heart_Failure | adipocyte | *EPG5* | 1.078762288 | 0.000301726 | 0.002384324 | 1 |
| Heart_Failure | adipocyte | *CWF19L2* | 0.649732441 | 0.001119467 | 0.005983919 | 1 |
| Heart_Failure | adipocyte | *NMT2* | 0.56020817 | 0.001289138 | 0.00667545 | 1 |
| Heart_Failure | adipocyte | *SPARCL1* | 0.44498984 | 0.007554702 | 0.025978325 | 1 |

| | | | | | | |
|---|---|---|---|---|---|---|
| Heart_Failure | adipocyte | *MBNL1* | 0.822319883 | 7.50E-05 | 0.001268787 | 0.363765705 |
| Heart_Failure | adipocyte | *GGPS1* | 0.626737786 | 0.003962239 | 0.015503857 | 1 |
| Heart_Failure | adipocyte | *ZNF117* | 0.066253637 | 0.005961977 | 0.021582093 | 1 |
| Heart_Failure | adipocyte | *MED21* | 0.818237432 | 0.000536517 | 0.003523108 | 1 |
| Heart_Failure | adipocyte | *JAKMIP1* | 2.60133505 | 2.17E-05 | 0.001268787 | 0.105407055 |
| Heart_Failure | adipocyte | *SREK1IP1* | 1.319265005 | 0.000112727 | 0.001454653 | 0.546949585 |
| Heart_Failure | adipocyte | *CAST* | 0.360525009 | 0.015585348 | 0.045622985 | 1 |
| Heart_Failure | adipocyte | *TMEM87B* | 0.714185143 | 0.005978222 | 0.021582093 | 1 |
| Heart_Failure | adipocyte | *CFDP1* | 1.126402612 | 6.60E-05 | 0.001268787 | 0.320386505 |
| Heart_Failure | adipocyte | *ZDHHC7* | 1.261517334 | 0.000208305 | 0.001944092 | 1 |
| Heart_Failure | adipocyte | *FBXO36* | 1.28710553 | 0.000843369 | 0.004894097 | 1 |
| Heart_Failure | adipocyte | *PPP2R5E* | 1.11371599 | 9.97E-05 | 0.001368761 | 0.483673723 |
| Heart_Failure | adipocyte | *ANKH* | 0.78665463 | 0.000988966 | 0.005471451 | 1 |
| Heart_Failure | adipocyte | *PRKCA* | 0.842708431 | 0.000748007 | 0.004458634 | 1 |
| Heart_Failure | adipocyte | *LRRK1* | 0.435175477 | 0.016555616 | 0.047595662 | 1 |
| Heart_Failure | adipocyte | *ABCA5* | 0.443601744 | 0.00084184 | 0.004891748 | 1 |
| Heart_Failure | adipocyte | *ENPP3* | 0.524312905 | 0.010290738 | 0.033066662 | 1 |
| Heart_Failure | adipocyte | *CCSAP* | 1.848239146 | 0.000517221 | 0.003451936 | 1 |
| Heart_Failure | adipocyte | *XPC* | 0.655577045 | 0.007104964 | 0.024783096 | 1 |
| Heart_Failure | adipocyte | *FLCN* | 0.433727334 | 0.003700325 | 0.014728449 | 1 |
| Heart_Failure | adipocyte | *OXNAD1* | 0.859086494 | 0.00063702 | 0.003979532 | 1 |
| Heart_Failure | adipocyte | *ANKRD40* | 0.771483855 | 0.007848677 | 0.026724506 | 1 |
| Heart_Failure | adipocyte | *TRMT44* | 0.894786364 | 0.004837468 | 0.018279904 | 1 |
| Heart_Failure | adipocyte | *CNOT8* | 1.057873616 | 0.000816123 | 0.004763495 | 1 |
| Heart_Failure | adipocyte | *NUP205* | 0.985221248 | 0.002806535 | 0.011955495 | 1 |
| Heart_Failure | adipocyte | *PDIA4* | 0.931136342 | 0.009811025 | 0.031757621 | 1 |
| Heart_Failure | adipocyte | *FZD7* | 0.42745602 | 0.000186611 | 0.001801316 | 0.905437765 |
| Heart_Failure | adipocyte | *TMBIM4* | 0.604539153 | 0.000252222 | 0.002120939 | 1 |
| Heart_Failure | adipocyte | *NAA11* | 0.700507485 | 0.01613874 | 0.046665772 | 1 |
| Heart_Failure | adipocyte | *C14orf2* | 0.780371938 | 8.62E-05 | 0.001303235 | 0.418338379 |
| Heart_Failure | adipocyte | *RPL30* | 0.937408709 | 2.69E-05 | 0.001268787 | 0.13072143 |
| Heart_Failure | adipocyte | *SUPV3L1* | 0.855294116 | 0.005407712 | 0.020029173 | 1 |
| Heart_Failure | adipocyte | *ZDHHC5* | 1.548231646 | 0.000113095 | 0.00145554 | 0.548738408 |
| Heart_Failure | adipocyte | *FBRS* | 0.426548751 | 0.017090913 | 0.048722157 | 1 |
| Heart_Failure | adipocyte | *VPS8* | 0.652417847 | 0.000605131 | 0.003856684 | 1 |
| Heart_Failure | adipocyte | *RPUSD3* | 0.6532575 | 0.015863148 | 0.046116235 | 1 |
| Heart_Failure | adipocyte | *GHRL* | 0.506259397 | 0.008143289 | 0.027462708 | 1 |
| Heart_Failure | adipocyte | *FCHO2* | 0.559790852 | 0.000682011 | 0.00418346 | 1 |
| Heart_Failure | adipocyte | *NECAP2* | 1.194210445 | 0.00070403 | 0.00426461 | 1 |
| Heart_Failure | adipocyte | *CLDN12* | 1.887809991 | 0.000122999 | 0.001522431 | 0.596792801 |
| Heart_Failure | adipocyte | *MMP14* | 1.978378335 | 1.34E-06 | 0.000283222 | 0.006514104 |
| Heart_Failure | adipocyte | *GATAD1* | 0.931788051 | 0.00107013 | 0.005794944 | 1 |
| Heart_Failure | adipocyte | *DHRS4* | 2.255739129 | 4.90E-05 | 0.001268787 | 0.237659874 |
| Heart_Failure | adipocyte | *LCA5L* | 0.172026685 | 0.005470614 | 0.020206337 | 1 |
| Heart_Failure | adipocyte | *SLC38A10* | 1.421395663 | 4.76E-05 | 0.001268787 | 0.231137899 |
| Heart_Failure | adipocyte | *AP3S2* | 0.63230811 | 0.009154089 | 0.030023235 | 1 |
| Heart_Failure | adipocyte | *SPPL3* | 0.954329822 | 2.82E-05 | 0.001268787 | 0.13702373 |
| Heart_Failure | adipocyte | *EYA3* | 0.526253285 | 0.015856801 | 0.046116235 | 1 |
| Heart_Failure | adipocyte | *DZIP1L* | 0.461787013 | 0.01216965 | 0.037630347 | 1 |
| Heart_Failure | adipocyte | *FANCC* | 0.769508459 | 0.002389446 | 0.010501443 | 1 |

| | | | | | | |
|---|---|---|---|---|---|---|
| Heart_Failure | adipocyte | *MITD1* | 0.922424709 | 0.000109037 | 0.001439756 | 0.529046436 |
| Heart_Failure | adipocyte | *FAM86C1* | 2.758104109 | 0.00061828 | 0.003911204 | 1 |
| Heart_Failure | adipocyte | *PPP1R9A* | 0.687943701 | 0.003485043 | 0.014067745 | 1 |
| Heart_Failure | adipocyte | *ELK4* | 1.239538081 | 0.000235391 | 0.00206158 | 1 |
| Heart_Failure | adipocyte | *ARHGAP27* | 0.827559724 | 0.002035602 | 0.009361838 | 1 |
| Heart_Failure | adipocyte | *AMFR* | 0.810005494 | 0.000879384 | 0.005013834 | 1 |
| Heart_Failure | adipocyte | *TMEM69* | 1.475624042 | 0.000478891 | 0.003236181 | 1 |
| Heart_Failure | adipocyte | *CCDC107* | 0.763379103 | 0.003344498 | 0.013561901 | 1 |
| Heart_Failure | adipocyte | *SSU72* | 1.077565575 | 0.001049038 | 0.005712609 | 1 |
| Heart_Failure | adipocyte | *CCDC58* | 1.100315762 | 0.001544503 | 0.007608048 | 1 |
| Heart_Failure | adipocyte | *TAOK1* | 0.970479636 | 0.000217574 | 0.001965494 | 1 |
| Heart_Failure | adipocyte | *VPS11* | 0.462786303 | 0.001200191 | 0.006322828 | 1 |
| Heart_Failure | adipocyte | *FDPS* | 1.056588935 | 3.10E-06 | 0.000449837 | 0.015036284 |
| Heart_Failure | adipocyte | *FCRL3* | -0.188750369 | 0.004403366 | 0.016889432 | 1 |
| Heart_Failure | adipocyte | *FGFR4* | -0.969186317 | 0.000492531 | 0.003297101 | 1 |
| Heart_Failure | adipocyte | *TONSL* | 0.488753679 | 0.012695325 | 0.038740701 | 1 |
| Heart_Failure | adipocyte | *MUM1* | 0.567488834 | 0.008983009 | 0.029589143 | 1 |
| Heart_Failure | adipocyte | *ZNF333* | 1.628924285 | 2.84E-05 | 0.001268787 | 0.137851255 |
| Heart_Failure | adipocyte | *RPL8* | 0.79045396 | 0.001345893 | 0.006881215 | 1 |
| Heart_Failure | adipocyte | *PGAP3* | 2.066202505 | 0.000230027 | 0.002032956 | 1 |
| Heart_Failure | adipocyte | *TMEM143* | 0.63945171 | 0.010826303 | 0.0343104 | 1 |
| Heart_Failure | adipocyte | *PLCD3* | 2.045285636 | 0.00013329 | 0.00155837 | 0.646723405 |
| Heart_Failure | adipocyte | *LEMD2* | 1.038003922 | 0.000111547 | 0.001449365 | 0.541227348 |
| Heart_Failure | adipocyte | *RPS6KA4* | 1.961505673 | 0.000522308 | 0.003471315 | 1 |
| Heart_Failure | adipocyte | *COA7* | 2.86281068 | 3.28E-05 | 0.001268787 | 0.159170903 |
| Heart_Failure | adipocyte | *C1orf123* | 0.928730544 | 0.016001246 | 0.046406484 | 1 |
| Heart_Failure | adipocyte | *USP24* | 0.734321685 | 0.003730451 | 0.014821902 | 1 |
| Heart_Failure | adipocyte | *ZSWIM5* | 0.716678864 | 0.000312257 | 0.002443124 | 1 |
| Heart_Failure | adipocyte | *RBP7* | 0.889156661 | 0.000425131 | 0.0029754 | 1 |
| Heart_Failure | adipocyte | *LAPTM5* | 0.652366299 | 0.014589248 | 0.04313652 | 1 |
| Heart_Failure | adipocyte | *TM2D1* | 1.212534461 | 6.41E-05 | 0.001268787 | 0.311105075 |
| Heart_Failure | adipocyte | *NEXN* | 0.696869087 | 0.000668257 | 0.004119738 | 1 |
| Heart_Failure | adipocyte | *KLHDC9* | -0.1835947 | 0.002920255 | 0.012288878 | 1 |
| Heart_Failure | adipocyte | *ACP6* | 1.057839503 | 0.00168453 | 0.008132678 | 1 |
| Heart_Failure | adipocyte | *CNST* | 1.07052749 | 0.000148922 | 0.001619166 | 0.72256988 |
| Heart_Failure | adipocyte | *HAAO* | 1.417165402 | 9.58E-05 | 0.001343879 | 0.464702949 |
| Heart_Failure | adipocyte | *KIAA1841* | 0.943022 | 0.001153611 | 0.006137413 | 1 |
| Heart_Failure | adipocyte | *NUP35* | 1.094806522 | 0.00045225 | 0.003112504 | 1 |
| Heart_Failure | adipocyte | *NOSTRIN* | 0.409451671 | 0.000535171 | 0.003523108 | 1 |
| Heart_Failure | adipocyte | *C1QTNF7* | 0.438972792 | 0.00745675 | 0.025750996 | 1 |
| Heart_Failure | adipocyte | *VPS72* | 1.236195705 | 0.000622605 | 0.003923218 | 1 |
| Heart_Failure | adipocyte | *ERCC3* | 0.921494873 | 0.000103834 | 0.001407263 | 0.503800208 |
| Heart_Failure | adipocyte | *RNF149* | 0.755271529 | 0.001304275 | 0.006703751 | 1 |
| Heart_Failure | adipocyte | *DHX57* | 1.041800172 | 0.000324193 | 0.002488896 | 1 |
| Heart_Failure | adipocyte | *DCAF16* | 0.748477484 | 0.010991333 | 0.034714531 | 1 |
| Heart_Failure | adipocyte | *ANTXR2* | 0.431092943 | 0.003103537 | 0.012839666 | 1 |
| Heart_Failure | adipocyte | *CGGBP1* | 0.153968353 | 0.017355379 | 0.049302284 | 1 |
| Heart_Failure | adipocyte | *GPR155* | 1.244903307 | 0.000207213 | 0.001940924 | 1 |
| Heart_Failure | adipocyte | *PBXIP1* | 1.567358956 | 7.27E-05 | 0.001268787 | 0.352843305 |
| Heart_Failure | adipocyte | *LRRC58* | 1.090540738 | 0.000330498 | 0.00251739 | 1 |

| | | | | | | |
|---|---|---|---|---|---|---|
| Heart_Failure | adipocyte | *ARPC2* | 0.676173899 | 0.004338151 | 0.016692077 | 1 |
| Heart_Failure | adipocyte | *SERPINI1* | 0.863662208 | 0.006796709 | 0.023844997 | 1 |
| Heart_Failure | adipocyte | *SUCLG1* | 0.604460306 | 0.000536599 | 0.003523108 | 1 |
| Heart_Failure | adipocyte | *EFHB* | 0.092191141 | 0.002065938 | 0.00948338 | 1 |
| Heart_Failure | adipocyte | *GTPBP8* | 1.885288463 | 7.16E-05 | 0.001268787 | 0.347407768 |
| Heart_Failure | adipocyte | *WDFY3* | 0.702297723 | 0.00123272 | 0.006459133 | 1 |
| Heart_Failure | adipocyte | *C4orf36* | 1.645230773 | 4.22E-05 | 0.001268787 | 0.204738054 |
| Heart_Failure | adipocyte | *TIPARP* | 1.211199925 | 0.000113547 | 0.001456112 | 0.550930483 |
| Heart_Failure | adipocyte | *RPP14* | 0.939757199 | 0.004536584 | 0.017263925 | 1 |
| Heart_Failure | adipocyte | *IL17RC* | 0.604480772 | 0.002835942 | 0.012066841 | 1 |
| Heart_Failure | adipocyte | *CRELD1* | 1.005377062 | 0.000639156 | 0.003986098 | 1 |
| Heart_Failure | adipocyte | *TTC14* | 0.504434357 | 0.004108493 | 0.015947526 | 1 |
| Heart_Failure | adipocyte | *CPA3* | 1.433129949 | 0.003858431 | 0.015220413 | 1 |
| Heart_Failure | adipocyte | *SLC4A1AP* | 1.673077104 | 6.98E-05 | 0.001268787 | 0.338689512 |
| Heart_Failure | adipocyte | *SNIP1* | 1.838375331 | 8.10E-05 | 0.001268787 | 0.393026652 |
| Heart_Failure | adipocyte | *RPN1* | 1.08550042 | 0.001255068 | 0.006540911 | 1 |
| Heart_Failure | adipocyte | *SFMBT1* | 1.682986928 | 2.82E-05 | 0.001268787 | 0.136836528 |
| Heart_Failure | adipocyte | *FAM208A* | 0.998491084 | 0.000170106 | 0.001744938 | 0.825355791 |
| Heart_Failure | adipocyte | *ABLIM2* | 0.574822487 | 0.001243883 | 0.006496576 | 1 |
| Heart_Failure | adipocyte | *BDH2* | 0.369146583 | 0.01559945 | 0.045622985 | 1 |
| Heart_Failure | adipocyte | *RNF123* | 0.481437584 | 2.42E-07 | 9.02E-05 | 0.001172242 |
| Heart_Failure | adipocyte | *HSPA4L* | 0.417480749 | 0.009162645 | 0.030023235 | 1 |
| Heart_Failure | adipocyte | *MFSD8* | 0.693666162 | 0.004210313 | 0.01627764 | 1 |
| Heart_Failure | adipocyte | *MST1R* | 0.411390907 | 5.94E-08 | 2.88E-05 | 0.000288136 |
| Heart_Failure | adipocyte | *ANXA5* | 0.702424628 | 0.000738634 | 0.004429977 | 1 |
| Heart_Failure | adipocyte | *MAP9* | 1.339611184 | 0.00067401 | 0.004144863 | 1 |
| Heart_Failure | adipocyte | *TMEM144* | 0.59752173 | 0.004676785 | 0.017714098 | 1 |
| Heart_Failure | adipocyte | *ARFIP1* | 0.78609685 | 0.000811789 | 0.004751265 | 1 |
| Heart_Failure | adipocyte | *MOCS2* | 1.442186264 | 0.001599139 | 0.007801746 | 1 |
| Heart_Failure | adipocyte | *NDUFAF2* | 0.50547222 | 0.002137162 | 0.009709281 | 1 |
| Heart_Failure | adipocyte | *RNF180* | 0.80986584 | 0.00121201 | 0.006372953 | 1 |
| Heart_Failure | adipocyte | *PGGT1B* | 0.803333276 | 0.00084832 | 0.004894234 | 1 |
| Heart_Failure | adipocyte | *CCDC112* | 0.898115354 | 0.003464193 | 0.014006885 | 1 |
| Heart_Failure | adipocyte | *CMBL* | 1.287037192 | 2.02E-06 | 0.000340255 | 0.0097793 |
| Heart_Failure | adipocyte | *NDUFS4* | 1.22905594 | 2.36E-05 | 0.001268787 | 0.11455438 |
| Heart_Failure | adipocyte | *SERINC5* | 1.106840213 | 5.49E-07 | 0.000156813 | 0.002665826 |
| Heart_Failure | adipocyte | *ERAP1* | 0.510394542 | 0.001371775 | 0.006998794 | 1 |
| Heart_Failure | adipocyte | *UTP15* | 1.442858687 | 0.000867475 | 0.004971605 | 1 |
| Heart_Failure | adipocyte | *CCDC127* | 1.001559939 | 0.000149672 | 0.001621002 | 0.726208735 |
| Heart_Failure | adipocyte | *DCBLD1* | 0.329468092 | 0.00597386 | 0.021582093 | 1 |
| Heart_Failure | adipocyte | *PDSS2* | 1.062887307 | 6.78E-05 | 0.001268787 | 0.32885046 |
| Heart_Failure | adipocyte | *RAET1E* | 1.710198423 | 0.000232405 | 0.00204651 | 1 |
| Heart_Failure | adipocyte | *GALNT10* | 0.430508056 | 0.012955873 | 0.039387153 | 1 |
| Heart_Failure | adipocyte | *SAP30L* | 1.307536153 | 1.75E-06 | 0.000321058 | 0.008503784 |
| Heart_Failure | adipocyte | *RPS14* | 0.80107294 | 0.00044148 | 0.00306009 | 1 |
| Heart_Failure | adipocyte | *RP9* | 1.922184316 | 5.76E-05 | 0.001268787 | 0.279655693 |
| Heart_Failure | adipocyte | *MIOS* | 1.036863652 | 0.00057977 | 0.003725338 | 1 |
| Heart_Failure | adipocyte | *USP49* | 1.113292223 | 0.000192073 | 0.00184177 | 0.931935776 |
| Heart_Failure | adipocyte | *SLC13A4* | 1.170813314 | 0.000217524 | 0.001965494 | 1 |
| Heart_Failure | adipocyte | *BRI3* | 1.1250688 | 0.000292266 | 0.002351702 | 1 |

| | | | | | | |
|---|---|---|---|---|---|---|
| Heart_Failure | adipocyte | *RAD21* | 1.123383925 | 9.58E-05 | 0.001343879 | 0.464835271 |
| Heart_Failure | adipocyte | *TNFRSF11B* | 1.903042211 | 0.000321376 | 0.002479505 | 1 |
| Heart_Failure | adipocyte | *SUN1* | 0.462941554 | 0.004004092 | 0.015654998 | 1 |
| Heart_Failure | adipocyte | *OXR1* | 0.930482144 | 0.000128443 | 0.001542589 | 0.62320611 |
| Heart_Failure | adipocyte | *GPER1* | 0.760499287 | 5.74E-05 | 0.001268787 | 0.278461797 |
| Heart_Failure | adipocyte | *INTS1* | 1.126586517 | 0.00617738 | 0.022120675 | 1 |
| Heart_Failure | adipocyte | *FASTK* | 1.819673864 | 4.33E-05 | 0.001268787 | 0.210007363 |
| Heart_Failure | adipocyte | *SLC25A32* | 0.653213633 | 0.013470976 | 0.040571803 | 1 |
| Heart_Failure | adipocyte | *INTS8* | 1.290962408 | 2.43E-05 | 0.001268787 | 0.11811529 |
| Heart_Failure | adipocyte | *TMEM65* | 0.772851959 | 0.003104066 | 0.012839666 | 1 |
| Heart_Failure | adipocyte | *TRMT10B* | 0.90170253 | 0.001691872 | 0.008151899 | 1 |
| Heart_Failure | adipocyte | *WRN* | 1.0613127 | 5.26E-05 | 0.001268787 | 0.25516112 |
| Heart_Failure | adipocyte | *MARCH8* | 1.305843509 | 6.24E-05 | 0.001268787 | 0.302533676 |
| Heart_Failure | adipocyte | *CFL2* | 1.398303535 | 0.000893526 | 0.005066617 | 1 |
| Heart_Failure | adipocyte | *CRYL1* | 0.587562669 | 0.003604642 | 0.014394835 | 1 |
| Heart_Failure | adipocyte | *REEP3* | 0.612159894 | 0.009499437 | 0.030954514 | 1 |
| Heart_Failure | adipocyte | *MICU2* | 0.875164739 | 0.001079598 | 0.005826704 | 1 |
| Heart_Failure | adipocyte | *LRR1* | 1.262030081 | 0.002894726 | 0.012213225 | 1 |
| Heart_Failure | adipocyte | *ATP5C1* | 0.777577761 | 0.005808243 | 0.02117325 | 1 |
| Heart_Failure | adipocyte | *VDAC2* | 0.82684604 | 0.017012713 | 0.048584863 | 1 |
| Heart_Failure | adipocyte | *PDZD8* | 0.363285295 | 0.015315577 | 0.045009801 | 1 |
| Heart_Failure | adipocyte | *METTL3* | 1.317799768 | 5.64E-05 | 0.001268787 | 0.273737336 |
| Heart_Failure | adipocyte | *HSPA12A* | 0.255294003 | 0.000689757 | 0.004214987 | 1 |
| Heart_Failure | adipocyte | *UBTD1* | 0.833194816 | 0.001418556 | 0.007150616 | 1 |
| Heart_Failure | adipocyte | *TTC7B* | 1.057142079 | 0.000136715 | 0.001561419 | 0.663340434 |
| Heart_Failure | adipocyte | *IFI27L1* | 0.844696681 | 0.003748582 | 0.0148476 | 1 |
| Heart_Failure | adipocyte | *TCP11L2* | 1.236991334 | 0.000134573 | 0.001561419 | 0.652945848 |
| Heart_Failure | adipocyte | *TRMT61A* | 0.933638173 | 0.001106741 | 0.005946739 | 1 |
| Heart_Failure | adipocyte | *API5* | 1.442473071 | 4.94E-05 | 0.001268787 | 0.239842665 |
| Heart_Failure | adipocyte | *COX11* | 0.966527864 | 0.000388769 | 0.002782166 | 1 |
| Heart_Failure | adipocyte | *STXBP4* | 0.837305668 | 0.001182998 | 0.006259761 | 1 |
| Heart_Failure | adipocyte | *WBP1L* | 0.779452682 | 0.001136633 | 0.006067043 | 1 |
| Heart_Failure | adipocyte | *WDR88* | 0.82948314 | 0.003775241 | 0.014940841 | 1 |
| Heart_Failure | adipocyte | *RPL27A* | 0.887330913 | 0.001451499 | 0.007267978 | 1 |
| Heart_Failure | adipocyte | *TMEM41B* | 1.193743482 | 0.000270235 | 0.00221328 | 1 |
| Heart_Failure | adipocyte | *NDST2* | 6.250335796 | 0.010254699 | 0.032972696 | 1 |
| Heart_Failure | adipocyte | *SEC11C* | 0.91244215 | 0.01126361 | 0.035258733 | 1 |
| Heart_Failure | adipocyte | *SLFN5* | 0.875493757 | 0.001290749 | 0.006676669 | 1 |
| Heart_Failure | adipocyte | *SAAL1* | 2.078899395 | 7.35E-05 | 0.001268787 | 0.356747663 |
| Heart_Failure | adipocyte | *FAM96A* | 0.617925537 | 0.012289158 | 0.037836654 | 1 |
| Heart_Failure | adipocyte | *STAT6* | 0.75007573 | 0.005058646 | 0.018924096 | 1 |
| Heart_Failure | adipocyte | *PIP4K2C* | 1.333259431 | 4.70E-06 | 0.000600048 | 0.022801808 |
| Heart_Failure | adipocyte | *MTMR10* | 0.584275743 | 0.000987885 | 0.005471451 | 1 |
| Heart_Failure | adipocyte | *EPB42* | 1.253866403 | 4.20E-05 | 0.001268787 | 0.203720741 |
| Heart_Failure | adipocyte | *SMAD3* | 0.950453617 | 3.18E-05 | 0.001268787 | 0.154500193 |
| Heart_Failure | adipocyte | *AKTIP* | 0.639591777 | 0.000904433 | 0.005108624 | 1 |
| Heart_Failure | adipocyte | *CNPY4* | 0.39315942 | 0.01453964 | 0.043029937 | 1 |
| Heart_Failure | adipocyte | *SP2* | 1.810525036 | 4.76E-05 | 0.001268787 | 0.230983215 |
| Heart_Failure | adipocyte | *FBXO22* | 1.30950796 | 5.47E-05 | 0.001268787 | 0.265622066 |
| Heart_Failure | adipocyte | *HDHD2* | 0.167285654 | 0.002103514 | 0.009592339 | 1 |

| | | | | | | |
|---|---|---|---|---|---|---|
| Heart_Failure | adipocyte | *ZNF91* | 1.034442391 | 1.17E-05 | 0.001110079 | 0.056614054 |
| Heart_Failure | adipocyte | *ZNF23* | 0.887223853 | 0.000145656 | 0.001607298 | 0.7067237 |
| Heart_Failure | adipocyte | *VKORC1* | 0.983761254 | 0.012339737 | 0.037909483 | 1 |
| Heart_Failure | adipocyte | *SPATA33* | 1.966260593 | 3.92E-05 | 0.001268787 | 0.190357999 |
| Heart_Failure | adipocyte | *RPL13* | 0.692379355 | 0.001798524 | 0.008488751 | 1 |
| Heart_Failure | adipocyte | *ZNF641* | 0.914512215 | 0.005826659 | 0.021208515 | 1 |
| Heart_Failure | adipocyte | *ZNF528* | 1.695089371 | 1.58E-05 | 0.001219853 | 0.076531146 |
| Heart_Failure | adipocyte | *SERTAD3* | 2.377651431 | 0.000249256 | 0.002112148 | 1 |
| Heart_Failure | adipocyte | *TTYH1* | 1.123939855 | 0.006454234 | 0.022898445 | 1 |
| Heart_Failure | adipocyte | *LENG8* | 0.830397344 | 0.002914135 | 0.01227377 | 1 |
| Heart_Failure | adipocyte | *FAM57A* | 0.229144845 | 0.013334148 | 0.040259667 | 1 |
| Heart_Failure | adipocyte | *TRPV3* | 0.227198224 | 0.004457199 | 0.017005205 | 1 |
| Heart_Failure | adipocyte | *CYB5D2* | 1.199610096 | 0.000128391 | 0.001542589 | 0.622950798 |
| Heart_Failure | adipocyte | *ZNF83* | 0.575117572 | 0.005119462 | 0.019107406 | 1 |
| Heart_Failure | adipocyte | *NDUFV1* | 1.452535756 | 4.34E-05 | 0.001268787 | 0.210439263 |
| Heart_Failure | adipocyte | *PRDX2* | 1.492498062 | 0.000157924 | 0.001684055 | 0.766244999 |
| Heart_Failure | adipocyte | *MIS12* | 1.279865801 | 0.000253075 | 0.002124432 | 1 |
| Heart_Failure | adipocyte | *ATP5H* | 0.814032916 | 0.008217871 | 0.027689661 | 1 |
| Heart_Failure | adipocyte | *GSDMA* | -0.410261623 | 1.34E-10 | 1.08E-07 | 6.51E-07 |
| Heart_Failure | adipocyte | *MLST8* | 1.218459648 | 0.000765942 | 0.004548779 | 1 |
| Heart_Failure | adipocyte | *ATG16L2* | 0.930860547 | 0.002344564 | 0.010380724 | 1 |
| Heart_Failure | adipocyte | *COA6* | 2.2974685 | 9.60E-05 | 0.001343879 | 0.465708653 |
| Heart_Failure | adipocyte | *PDHB* | 0.724443192 | 0.004956854 | 0.018684219 | 1 |
| Heart_Failure | adipocyte | *PXK* | 0.587008315 | 0.014820108 | 0.043739151 | 1 |
| Heart_Failure | adipocyte | *PCMTD1* | 1.05485122 | 7.67E-05 | 0.001268787 | 0.372024262 |
| Heart_Failure | adipocyte | *ING5* | 0.619041051 | 0.002114398 | 0.009614863 | 1 |
| Heart_Failure | adipocyte | *RFWD3* | 1.171081452 | 6.83E-05 | 0.001268787 | 0.331543547 |
| Heart_Failure | adipocyte | *SLC20A2* | 0.910692164 | 0.000239831 | 0.002077964 | 1 |
| Heart_Failure | adipocyte | *TMUB2* | 0.973335683 | 0.000136051 | 0.001561419 | 0.660118139 |
| Heart_Failure | adipocyte | *NBPF9* | 2.511564145 | 7.72E-05 | 0.001268787 | 0.374578879 |
| Heart_Failure | adipocyte | *DNAJC21* | 1.432286888 | 0.000166117 | 0.001734151 | 0.805999282 |
| Heart_Failure | adipocyte | *SNTB2* | 1.160259034 | 6.32E-05 | 0.001268787 | 0.306427358 |
| Heart_Failure | adipocyte | *STX18* | 1.217754375 | 0.00012624 | 0.00153782 | 0.612516703 |
| Heart_Failure | adipocyte | *ZBTB49* | 1.914828668 | 0.000125738 | 0.00153782 | 0.610078552 |
| Heart_Failure | adipocyte | *ATOH8* | 0.270043475 | 0.011210337 | 0.035205538 | 1 |
| Heart_Failure | adipocyte | *TMEM129* | 0.515763744 | 0.003442266 | 0.013929835 | 1 |
| Heart_Failure | adipocyte | *CEP120* | 1.166703132 | 0.000209692 | 0.00194909 | 1 |
| Heart_Failure | adipocyte | *MFF* | 1.231105758 | 5.87E-05 | 0.001268787 | 0.284826034 |
| Heart_Failure | adipocyte | *MFSD7* | 2.125654069 | 0.002786475 | 0.011890922 | 1 |
| Heart_Failure | adipocyte | *MAP2K1* | 1.214354584 | 0.000129965 | 0.001543804 | 0.630590145 |
| Heart_Failure | adipocyte | *RGS14* | -0.157976294 | 0.010760019 | 0.034167284 | 1 |
| Heart_Failure | adipocyte | *GPRIN1* | 0.508278968 | 0.010340625 | 0.033183011 | 1 |
| Heart_Failure | adipocyte | *SNUPN* | 0.449885894 | 0.002958692 | 0.012439837 | 1 |
| Heart_Failure | adipocyte | *HINT1* | 1.208954923 | 0.000216018 | 0.001964387 | 1 |
| Heart_Failure | adipocyte | *DFFB* | 1.395449296 | 0.000776829 | 0.004590958 | 1 |
| Heart_Failure | adipocyte | *NFU1* | 0.434334689 | 0.009987305 | 0.032217911 | 1 |
| Heart_Failure | adipocyte | *LUZP1* | 1.219148181 | 0.000100551 | 0.001374296 | 0.487874935 |
| Heart_Failure | adipocyte | *UGP2* | 0.574229832 | 0.000300464 | 0.002383417 | 1 |
| Heart_Failure | adipocyte | *HNRNPF* | 0.560524636 | 0.008126292 | 0.027462708 | 1 |
| Heart_Failure | adipocyte | *CSGALNACT2* | 1.10541296 | 0.000162943 | 0.00172058 | 0.790597877 |

| | | | | | | |
|---|---|---|---|---|---|---|
| Heart_Failure | adipocyte | *AVEN* | 1.212690124 | 0.000214527 | 0.001960237 | 1 |
| Heart_Failure | adipocyte | *TOR1AIP2* | 0.860715637 | 0.000523248 | 0.003471315 | 1 |
| Heart_Failure | adipocyte | *MYO7B* | 0.25310355 | 0.015910173 | 0.046169951 | 1 |
| Heart_Failure | adipocyte | *NIPA1* | 0.837027542 | 0.006176284 | 0.022120675 | 1 |
| Heart_Failure | adipocyte | *HNRNPA3* | 0.686398144 | 0.000817917 | 0.004764144 | 1 |
| Heart_Failure | adipocyte | *RNF150* | 0.391581454 | 0.003791015 | 0.01499104 | 1 |
| Heart_Failure | adipocyte | *CHRNB1* | 0.129572273 | 0.000803225 | 0.004723937 | 1 |
| Heart_Failure | adipocyte | *PDCD6IP* | 1.12016752 | 6.04E-05 | 0.001268787 | 0.293066852 |
| Heart_Failure | adipocyte | *STX8* | 1.041725843 | 2.73E-05 | 0.001268787 | 0.13263211 |
| Heart_Failure | adipocyte | *NFRKB* | 1.22177403 | 0.000999075 | 0.005496043 | 1 |
| Heart_Failure | adipocyte | *MGMT* | 0.788557328 | 0.00129571 | 0.006690377 | 1 |
| Heart_Failure | adipocyte | *NFXL1* | 0.616394071 | 0.012615426 | 0.038521111 | 1 |
| Heart_Failure | adipocyte | *ELOVL6* | 1.114834919 | 0.000267871 | 0.002210435 | 1 |
| Heart_Failure | adipocyte | *SERPINB9* | 0.550845044 | 0.005076218 | 0.018975201 | 1 |
| Heart_Failure | adipocyte | *STAT2* | 1.150778981 | 4.54E-05 | 0.001268787 | 0.220104412 |
| Heart_Failure | adipocyte | *SLC26A5* | 1.386306364 | 3.74E-06 | 0.000504094 | 0.018147369 |
| Heart_Failure | adipocyte | *ACYP2* | 0.427958102 | 0.012401264 | 0.03805878 | 1 |
| Heart_Failure | adipocyte | *TRABD* | 0.530423046 | 0.01734818 | 0.049302284 | 1 |
| Heart_Failure | adipocyte | *TMEM133* | 0.288918746 | 0.006752809 | 0.023742485 | 1 |
| Heart_Failure | adipocyte | *FOXN2* | 0.97768723 | 0.000226616 | 0.002021062 | 1 |
| Heart_Failure | adipocyte | *TSEN34* | 0.975920879 | 0.002207647 | 0.009954929 | 1 |
| Heart_Failure | adipocyte | *MSANTD4* | 1.948599742 | 2.10E-05 | 0.001268787 | 0.101980987 |
| Heart_Failure | adipocyte | *NDUFA3* | 1.62814334 | 9.64E-05 | 0.001343879 | 0.467669951 |
| Heart_Failure | adipocyte | *PYGO1* | 0.722726093 | 0.002878155 | 0.012185696 | 1 |
| Heart_Failure | adipocyte | *FEZ2* | 0.799161369 | 0.000177294 | 0.001756516 | 0.860228241 |
| Heart_Failure | adipocyte | *MFN1* | 0.612368051 | 0.000477516 | 0.003235904 | 1 |
| Heart_Failure | adipocyte | *PDE7B* | 0.451254002 | 0.007591815 | 0.026068992 | 1 |
| Heart_Failure | adipocyte | *MCC* | 0.972128579 | 0.000284351 | 0.002300045 | 1 |
| Heart_Failure | adipocyte | *PPID* | 1.76071011 | 6.03E-07 | 0.000162548 | 0.002925861 |
| Heart_Failure | adipocyte | *TBCA* | 1.165176487 | 4.61E-05 | 0.001268787 | 0.223608446 |
| Heart_Failure | adipocyte | *NMUR1* | 1.160553296 | 0.008876293 | 0.029377744 | 1 |
| Heart_Failure | adipocyte | *ANO5* | 0.485479937 | 0.004989714 | 0.018752976 | 1 |
| Heart_Failure | adipocyte | *CAMTA1* | 0.571938651 | 0.007465509 | 0.02576291 | 1 |
| Heart_Failure | adipocyte | *LGALS4* | 0.22913497 | 0.01440049 | 0.042656397 | 1 |
| Heart_Failure | adipocyte | *TRAPPC12* | 1.049624134 | 0.000439714 | 0.003056579 | 1 |
| Heart_Failure | adipocyte | *ATPAF2* | 0.564730571 | 0.000768485 | 0.0045583 | 1 |
| Heart_Failure | adipocyte | *ANKRD20A4* | 1.879425276 | 0.010446853 | 0.033407581 | 1 |
| Heart_Failure | adipocyte | *EIF2AK3* | 0.593715181 | 0.007923552 | 0.026884666 | 1 |
| Heart_Failure | adipocyte | *TEFM* | 0.723002817 | 0.000955155 | 0.005345345 | 1 |
| Heart_Failure | adipocyte | *MALT1* | 1.008394015 | 0.000640208 | 0.003987537 | 1 |
| Heart_Failure | adipocyte | *MBOAT1* | 0.763059479 | 0.008137187 | 0.027462708 | 1 |
| Heart_Failure | adipocyte | *PAIP1* | 0.844737711 | 0.008461507 | 0.028255493 | 1 |
| Heart_Failure | adipocyte | *MACROD2* | 0.423012932 | 0.000345266 | 0.002585235 | 1 |
| Heart_Failure | adipocyte | *BSG* | 1.347671294 | 1.28E-05 | 0.001127656 | 0.062021098 |
| Heart_Failure | adipocyte | *HINFP* | 0.839840553 | 0.004959867 | 0.018684219 | 1 |
| Heart_Failure | adipocyte | *SUCLG2* | 0.997832149 | 0.000148553 | 0.001619166 | 0.720777911 |
| Heart_Failure | adipocyte | *FGGY* | 0.717209657 | 0.000853987 | 0.004921072 | 1 |
| Heart_Failure | adipocyte | *MANEA* | 0.784313816 | 0.003536707 | 0.014205384 | 1 |
| Heart_Failure | adipocyte | *AFF1* | 0.89422538 | 0.000131015 | 0.001543804 | 0.635684882 |
| Heart_Failure | adipocyte | *ZFAND4* | 0.452387528 | 0.006269594 | 0.022367699 | 1 |

| | | | | | | |
|---|---|---|---|---|---|---|
| Heart_Failure | adipocyte | *ZNF596* | 0.959662113 | 0.001844357 | 0.008654567 | 1 |
| Heart_Failure | adipocyte | *CFL1* | 0.860720283 | 0.000961875 | 0.005376259 | 1 |
| Heart_Failure | adipocyte | *FAM192A* | 0.417144342 | 0.014213642 | 0.042257715 | 1 |
| Heart_Failure | adipocyte | *CBWD1* | 1.806372862 | 9.55E-05 | 0.001343879 | 0.463406713 |
| Heart_Failure | adipocyte | *DCP2* | 0.745453131 | 0.006186677 | 0.022120675 | 1 |
| Heart_Failure | adipocyte | *ZNF621* | 1.497055979 | 5.01E-05 | 0.001268787 | 0.243260193 |
| Heart_Failure | adipocyte | *RNASEH2C* | 0.274554834 | 0.004408569 | 0.016896032 | 1 |
| Heart_Failure | adipocyte | *SH3RF3* | 0.418005845 | 0.014363935 | 0.042600131 | 1 |
| Heart_Failure | adipocyte | *GXYLT2* | 1.018921 | 0.000110737 | 0.001449365 | 0.537295587 |
| Heart_Failure | adipocyte | *NOC3L* | 0.727616347 | 0.001499562 | 0.007439543 | 1 |
| Heart_Failure | adipocyte | *PARP15* | 0.13768583 | 0.00869907 | 0.028830525 | 1 |
| Heart_Failure | adipocyte | *MAP3K11* | 0.548743098 | 0.002638751 | 0.011462147 | 1 |
| Heart_Failure | adipocyte | *KCNK7* | 1.358005102 | 0.006011236 | 0.021652946 | 1 |
| Heart_Failure | adipocyte | *PPP1R14B* | 1.012929102 | 0.001297178 | 0.006690377 | 1 |
| Heart_Failure | adipocyte | *PEAK1* | 0.626410911 | 0.001506513 | 0.007466397 | 1 |
| Heart_Failure | adipocyte | *GMPPB* | 0.645420774 | 0.003101131 | 0.012839666 | 1 |
| Heart_Failure | adipocyte | *NUDT4* | 0.800790703 | 0.003503183 | 0.014129212 | 1 |
| Heart_Failure | adipocyte | *SCAI* | 0.34770559 | 0.006908935 | 0.024186258 | 1 |
| Heart_Failure | adipocyte | *UQCRH* | 0.818945316 | 0.001199746 | 0.006322828 | 1 |
| Heart_Failure | adipocyte | *RNF213* | 0.461021762 | 0.009984875 | 0.032217911 | 1 |
| Heart_Failure | adipocyte | *GOLIM4* | 0.811646172 | 0.00224113 | 0.010041405 | 1 |
| Heart_Failure | adipocyte | *XXYLT1* | 1.297512818 | 0.000166553 | 0.001734151 | 0.808114353 |
| Heart_Failure | adipocyte | *UBXN2A* | 0.75484703 | 0.001743732 | 0.008319163 | 1 |
| Heart_Failure | adipocyte | *SLC25A30* | 0.871631355 | 0.005676879 | 0.020819514 | 1 |
| Heart_Failure | adipocyte | *LEMD3* | 1.309955583 | 8.12E-05 | 0.001268787 | 0.393877647 |
| Heart_Failure | adipocyte | *RALGAPA1* | 0.396540073 | 0.008304105 | 0.027902715 | 1 |
| Heart_Failure | adipocyte | *ANKRD36C* | 0.709701218 | 0.003514866 | 0.014152804 | 1 |
| Heart_Failure | adipocyte | *MSL2* | 1.380717243 | 4.48E-05 | 0.001268787 | 0.21755813 |
| Heart_Failure | adipocyte | *ANGEL2* | 1.301814323 | 6.89E-05 | 0.001268787 | 0.334333915 |
| Heart_Failure | adipocyte | *KIAA1551* | 0.286548129 | 0.010234271 | 0.032950686 | 1 |
| Heart_Failure | adipocyte | *LARP7* | 0.956177143 | 0.001242267 | 0.006495128 | 1 |
| Heart_Failure | adipocyte | *FGFBP3* | 2.126832542 | 0.001404204 | 0.007126778 | 1 |
| Heart_Failure | adipocyte | *CEP135* | 0.677247069 | 0.00902312 | 0.029637608 | 1 |
| Heart_Failure | adipocyte | *DHX36* | 1.146677707 | 9.30E-05 | 0.001331689 | 0.451442562 |
| Heart_Failure | adipocyte | *FBXW8* | 0.644503016 | 0.007776573 | 0.026569036 | 1 |
| Heart_Failure | adipocyte | *SLC22A1* | 0.918135261 | 0.015501872 | 0.045419735 | 1 |
| Heart_Failure | adipocyte | *TVP23C* | 3.337003319 | 7.64E-05 | 0.001268787 | 0.370828678 |
| Heart_Failure | adipocyte | *SH3BP5L* | 2.156610311 | 0.000177268 | 0.001756516 | 0.860105217 |
| Heart_Failure | adipocyte | *ATG13* | 1.181495207 | 0.000554256 | 0.003590454 | 1 |
| Heart_Failure | adipocyte | *TMEM9B* | 0.751823852 | 0.002977847 | 0.012498716 | 1 |
| Heart_Failure | adipocyte | *EIF1AD* | 0.740912792 | 0.014797911 | 0.043700221 | 1 |
| Heart_Failure | adipocyte | *CLTB* | 1.43090004 | 0.000249972 | 0.002113007 | 1 |
| Heart_Failure | adipocyte | *PPP2R2D* | 0.453624374 | 0.007867958 | 0.026770921 | 1 |
| Heart_Failure | adipocyte | *MCTP1* | 0.480412656 | 0.012289894 | 0.037836654 | 1 |
| Heart_Failure | adipocyte | *C11orf68* | 0.840054685 | 0.01286662 | 0.039180908 | 1 |
| Heart_Failure | adipocyte | *PAAF1* | -0.10059496 | 0.011243993 | 0.035220047 | 1 |
| Heart_Failure | adipocyte | *MRPL48* | 0.80270987 | 0.005893706 | 0.021404389 | 1 |
| Heart_Failure | adipocyte | *RAB6A* | 0.717742912 | 0.003599087 | 0.014394835 | 1 |
| Heart_Failure | adipocyte | *RPS6KB2* | 0.96415877 | 7.58E-06 | 0.000897533 | 0.03679886 |
| Heart_Failure | adipocyte | *TOM1L2* | 0.668859357 | 0.001934559 | 0.008973689 | 1 |

| | | | | | | |
|---|---|---|---|---|---|---|
| Heart_Failure | adipocyte | *TTLL11* | 1.170472788 | 0.000102554 | 0.001397094 | 0.49759299 |
| Heart_Failure | adipocyte | *PRIMA1* | 1.14776988 | 0.001412693 | 0.007145132 | 1 |
| Heart_Failure | adipocyte | *CTDNEP1* | 0.508810836 | 0.00102076 | 0.005589985 | 1 |
| Heart_Failure | adipocyte | *ZDHHC21* | 0.794941682 | 0.000127095 | 0.00153782 | 0.616665818 |
| Heart_Failure | adipocyte | *A2M* | 0.611795926 | 0.003665628 | 0.014602323 | 1 |
| Heart_Failure | adipocyte | *B3GALT6* | 2.193391226 | 0.011278011 | 0.035281049 | 1 |
| Heart_Failure | adipocyte | *TCP11L1* | 0.333903048 | 0.006116766 | 0.022000407 | 1 |
| Heart_Failure | adipocyte | *MYPOP* | 0.916084247 | 0.001927503 | 0.008949516 | 1 |
| Heart_Failure | adipocyte | *SLC35G1* | 1.081405767 | 0.001075891 | 0.005813164 | 1 |
| Heart_Failure | adipocyte | *OR4N2* | 0.912870909 | 0.001417868 | 0.007150616 | 1 |
| Heart_Failure | adipocyte | *TAC4* | 1.005003579 | 0.001411193 | 0.007145132 | 1 |
| Heart_Failure | adipocyte | *CRLF3* | 0.494260689 | 0.001064572 | 0.005777747 | 1 |
| Heart_Failure | adipocyte | *WDR25* | 1.586546178 | 4.22E-05 | 0.001268787 | 0.204837189 |
| Heart_Failure | adipocyte | *RMDN1* | 0.436651164 | 0.000592392 | 0.003791937 | 1 |
| Heart_Failure | adipocyte | *ANKLE2* | 0.69383947 | 0.01111772 | 0.034982605 | 1 |
| Heart_Failure | adipocyte | *SHMT1* | 0.5002717 | 0.006652773 | 0.023441726 | 1 |
| Heart_Failure | adipocyte | *DPP7* | 1.154777261 | 0.001119827 | 0.005983919 | 1 |
| Heart_Failure | adipocyte | *WDR73* | 0.63805801 | 0.015480784 | 0.045385356 | 1 |
| Heart_Failure | adipocyte | *TALDO1* | 1.240446419 | 0.000249359 | 0.002112148 | 1 |
| Heart_Failure | adipocyte | *MAN1B1* | 1.612914843 | 0.000194176 | 0.001854614 | 0.942144117 |
| Heart_Failure | adipocyte | *IL17RA* | 0.558206611 | 0.002344863 | 0.010380724 | 1 |
| Heart_Failure | adipocyte | *THAP5* | 1.230262991 | 0.00144995 | 0.007267726 | 1 |
| Heart_Failure | adipocyte | *NAALADL2* | 0.495198514 | 0.000235982 | 0.002063037 | 1 |
| Heart_Failure | adipocyte | *CD151* | 0.707555425 | 0.000334364 | 0.002534894 | 1 |
| Heart_Failure | adipocyte | *SLC35G5* | 9.78839528 | 0.001315952 | 0.006756612 | 1 |
| Heart_Failure | adipocyte | *CENPBD1* | 4.617583083 | 2.68E-05 | 0.001268787 | 0.129844848 |
| Heart_Failure | adipocyte | *RPS27* | 1.074741413 | 0.000188387 | 0.001813601 | 0.91405501 |
| Heart_Failure | adipocyte | *DPY19L2* | 0.210946895 | 0.00098717 | 0.005471451 | 1 |
| Heart_Failure | adipocyte | *DALRD3* | 0.515617588 | 0.01632081 | 0.047136053 | 1 |
| Heart_Failure | adipocyte | *ZNF454* | 0.178310071 | 0.001193841 | 0.006303062 | 1 |
| Heart_Failure | adipocyte | *SH2B1* | 0.890690845 | 0.000692573 | 0.004221565 | 1 |
| Heart_Failure | adipocyte | *SH2D4B* | 1.774844562 | 0.006062968 | 0.021823085 | 1 |
| Heart_Failure | adipocyte | *NT5DC1* | 0.997064556 | 0.000353127 | 0.002619838 | 1 |
| Heart_Failure | adipocyte | *LINC00314* | 0.088481255 | 0.015770994 | 0.04593089 | 1 |
| Heart_Failure | adipocyte | *P4HTM* | 1.031335903 | 0.003014546 | 0.012587417 | 1 |
| Heart_Failure | adipocyte | *PARP10* | 0.716069574 | 0.004967594 | 0.018698811 | 1 |
| Heart_Failure | adipocyte | *NSUN3* | 0.914648843 | 0.001520823 | 0.007529627 | 1 |
| Heart_Failure | adipocyte | *TRIM73* | -0.464759366 | 0.017348597 | 0.049302284 | 1 |
| Heart_Failure | adipocyte | *C17orf62* | 1.624512833 | 0.000180005 | 0.001767982 | 0.873383122 |
| Heart_Failure | adipocyte | *GAK* | 1.281048235 | 2.83E-05 | 0.001268787 | 0.13734091 |
| Heart_Failure | adipocyte | *CTC1* | 0.570465588 | 0.001708573 | 0.008199798 | 1 |
| Heart_Failure | adipocyte | *C3orf38* | 0.306509294 | 0.005686166 | 0.020822097 | 1 |
| Heart_Failure | adipocyte | *SPTY2D1* | 1.43093517 | 0.000306446 | 0.002417687 | 1 |
| Heart_Failure | adipocyte | *ZNF664* | 1.082410484 | 0.001663228 | 0.008037829 | 1 |
| Heart_Failure | adipocyte | *HLA-DQB1* | -0.381046936 | 1.55E-23 | 7.51E-20 | 7.51E-20 |
| Heart_Failure | adipocyte | *PACS2* | 1.279571603 | 0.000161654 | 0.001712546 | 0.784346012 |
| Heart_Failure | adipocyte | *LBX2* | 0.254012664 | 0.000393689 | 0.002800847 | 1 |
| Heart_Failure | adipocyte | *GIPC3* | 4.971082324 | 0.014205492 | 0.042257715 | 1 |
| Heart_Failure | adipocyte | *PDXDC1* | 1.034830046 | 0.000136769 | 0.001561419 | 0.663603177 |
| Heart_Failure | adipocyte | *ZADH2* | 0.869660243 | 0.003896479 | 0.015333102 | 1 |

| | | | | | | |
|---|---|---|---|---|---|---|
| Heart_Failure | adipocyte | *SEPT1* | 1.537886607 | 0.00168919 | 0.008147066 | 1 |
| Heart_Failure | adipocyte | *EXOC3* | 0.703353916 | 0.011856242 | 0.036805174 | 1 |
| Heart_Failure | adipocyte | *MTURN* | 0.723376946 | 0.002367442 | 0.010441952 | 1 |
| Heart_Failure | adipocyte | *CCDC66* | 0.641520673 | 0.009547772 | 0.031070281 | 1 |
| Heart_Failure | adipocyte | *SLC36A4* | 0.59328203 | 0.013625937 | 0.040886238 | 1 |
| Heart_Failure | adipocyte | *ZDHHC20* | 1.071154257 | 0.000270328 | 0.00221328 | 1 |
| Heart_Failure | adipocyte | *PSMG4* | 0.45072868 | 0.01107399 | 0.034876547 | 1 |
| Heart_Failure | adipocyte | *CAPS2* | 1.09751288 | 5.44E-05 | 0.001268787 | 0.263861162 |
| Heart_Failure | adipocyte | *D2HGDH* | 0.478277825 | 1.25E-05 | 0.00112616 | 0.060812664 |
| Heart_Failure | adipocyte | *CMTR2* | 0.751755483 | 0.000700204 | 0.004247059 | 1 |
| Heart_Failure | adipocyte | *FKRP* | 0.166951224 | 0.003560085 | 0.014275647 | 1 |
| Heart_Failure | adipocyte | *TMEM136* | 0.473577838 | 0.011229076 | 0.035207621 | 1 |
| Heart_Failure | adipocyte | *SGSH* | 2.328964616 | 0.000156081 | 0.001671758 | 0.757306289 |
| Heart_Failure | adipocyte | *ZBTB20* | 0.643610679 | 0.000530001 | 0.003493974 | 1 |
| Heart_Failure | adipocyte | *C3orf58* | 0.931911927 | 0.004125926 | 0.016002391 | 1 |
| Heart_Failure | adipocyte | *TMEM252* | 45.55001898 | 0.005732092 | 0.020927095 | 1 |
| Heart_Failure | adipocyte | *RELL1* | 0.655425228 | 0.000293132 | 0.00235476 | 1 |
| Heart_Failure | adipocyte | *RFX7* | 1.046868451 | 0.000138121 | 0.001569472 | 0.670164386 |
| Heart_Failure | adipocyte | *ZNF329* | 1.5766419 | 5.85E-05 | 0.001268787 | 0.284017301 |
| Heart_Failure | adipocyte | *IDH2* | 1.041378404 | 0.004881872 | 0.018419006 | 1 |
| Heart_Failure | adipocyte | *WRB* | 0.199296372 | 0.001838756 | 0.008645002 | 1 |
| Heart_Failure | adipocyte | *ERCC6L2* | 0.835541043 | 0.001413711 | 0.007145132 | 1 |
| Heart_Failure | adipocyte | *MRPL41* | 1.818073649 | 7.65E-05 | 0.001268787 | 0.371173076 |
| Heart_Failure | adipocyte | *ARL6IP4* | 0.222177349 | 0.011048477 | 0.034832496 | 1 |
| Heart_Failure | adipocyte | *FIGN* | 0.662950129 | 0.000654612 | 0.004055662 | 1 |
| Heart_Failure | adipocyte | *C8orf33* | 1.193410157 | 0.000274132 | 0.00223921 | 1 |
| Heart_Failure | adipocyte | *KBTBD3* | 1.075468873 | 0.007414247 | 0.025659005 | 1 |
| Heart_Failure | adipocyte | *GLRX5* | 2.069419974 | 4.05E-05 | 0.001268787 | 0.196591295 |
| Heart_Failure | adipocyte | *ADI1* | 0.908975931 | 0.005823638 | 0.021208515 | 1 |
| Heart_Failure | adipocyte | *SKA2* | 1.012845413 | 0.001110237 | 0.00595647 | 1 |
| Heart_Failure | adipocyte | *BRICD5* | 3.668111738 | 0.000170092 | 0.001744938 | 0.825284561 |
| Heart_Failure | adipocyte | *ACBD3* | 0.533654775 | 0.015695547 | 0.045793622 | 1 |
| Heart_Failure | adipocyte | *PLCXD3* | 0.918485632 | 0.001745886 | 0.008321258 | 1 |
| Heart_Failure | adipocyte | *ALG12* | 1.605705728 | 0.001041612 | 0.005684928 | 1 |
| Heart_Failure | adipocyte | *COL18A1* | 0.537585983 | 0.000213452 | 0.001960237 | 1 |
| Heart_Failure | adipocyte | *ZNF721* | 0.980731917 | 3.08E-05 | 0.001268787 | 0.149388672 |
| Heart_Failure | adipocyte | *CEP63* | 0.342706618 | 0.001298915 | 0.006690377 | 1 |
| Heart_Failure | adipocyte | *C12orf60* | 1.337928959 | 0.001102287 | 0.005935958 | 1 |
| Heart_Failure | adipocyte | *ABAT* | 0.855323228 | 0.003155536 | 0.012964151 | 1 |
| Heart_Failure | adipocyte | *ARHGEF37* | 1.817911237 | 5.45E-05 | 0.001268787 | 0.264356261 |
| Heart_Failure | adipocyte | *CSMD1* | 1.528352037 | 0.000525635 | 0.003474636 | 1 |
| Heart_Failure | adipocyte | *FANCF* | 1.784297656 | 0.002759114 | 0.011826165 | 1 |
| Heart_Failure | adipocyte | *CCDC125* | 0.424925368 | 0.00863331 | 0.028679459 | 1 |
| Heart_Failure | adipocyte | *LRIT3* | 3.797461725 | 0.015722488 | 0.045830485 | 1 |
| Heart_Failure | adipocyte | *PSMG1* | -0.315013773 | 0.017077379 | 0.048712195 | 1 |
| Heart_Failure | adipocyte | *SERHL2* | 1.905291229 | 6.19E-05 | 0.001268787 | 0.300110519 |
| Heart_Failure | adipocyte | *SFXN4* | 0.455131105 | 0.010994718 | 0.034714531 | 1 |
| Heart_Failure | adipocyte | *NDUFB1* | 0.521950421 | 0.014949297 | 0.044040067 | 1 |
| Heart_Failure | adipocyte | *BTBD9* | 0.819014962 | 0.000362799 | 0.00266375 | 1 |
| Heart_Failure | adipocyte | *TTC32* | 1.805358251 | 4.17E-05 | 0.001268787 | 0.2021331 |

| | | | | | | |
|---|---|---|---|---|---|---|
| Heart_Failure | adipocyte | *PP2D1* | 1.854611331 | 0.000157061 | 0.001678547 | 0.762060307 |
| Heart_Failure | adipocyte | *DENND5A* | 1.026105153 | 0.00011848 | 0.001495325 | 0.574862946 |
| Heart_Failure | adipocyte | *SS18L1* | 0.742357927 | 0.001468276 | 0.007321764 | 1 |
| Heart_Failure | adipocyte | *COPB2* | 1.390694436 | 2.32E-05 | 0.001268787 | 0.112326012 |
| Heart_Failure | adipocyte | *RNLS* | 0.537112629 | 0.011575343 | 0.036094837 | 1 |
| Heart_Failure | adipocyte | *NDUFA12* | 0.643628816 | 0.004363017 | 0.016761171 | 1 |
| Heart_Failure | adipocyte | *TCTE3* | 1.382735486 | 0.000223565 | 0.002005058 | 1 |
| Heart_Failure | adipocyte | *UBE2G2* | 0.49685853 | 0.005702415 | 0.020853444 | 1 |
| Heart_Failure | adipocyte | *ZFP90* | 1.347432031 | 0.000141566 | 0.001586327 | 0.686879733 |
| Heart_Failure | adipocyte | *UBOX5* | 0.983594444 | 0.004664893 | 0.017682859 | 1 |
| Heart_Failure | adipocyte | *SEMA4B* | 0.726434418 | 0.012195164 | 0.037662309 | 1 |
| Heart_Failure | adipocyte | *SIGIRR* | 0.755667385 | 0.000115917 | 0.001471822 | 0.562428023 |
| Heart_Failure | adipocyte | *IFITM2* | 0.71654018 | 0.010404005 | 0.033314461 | 1 |
| Heart_Failure | adipocyte | *ATP6V0A2* | 0.798630524 | 0.000430596 | 0.003002148 | 1 |
| Heart_Failure | adipocyte | *C14orf80* | 0.358267827 | 0.013250582 | 0.040091869 | 1 |
| Heart_Failure | adipocyte | *FAM174B* | 0.838664323 | 0.001793713 | 0.008488751 | 1 |
| Heart_Failure | adipocyte | *SP1* | 0.819101142 | 0.001030043 | 0.005628118 | 1 |
| Heart_Failure | adipocyte | *RNPC3* | 0.340760026 | 0.013715169 | 0.041103149 | 1 |
| Heart_Failure | adipocyte | *ZNF529* | 1.212325255 | 7.67E-05 | 0.001268787 | 0.372180068 |
| Heart_Failure | adipocyte | *WWOX* | 0.819216482 | 7.68E-05 | 0.001268787 | 0.372876097 |
| Heart_Failure | adipocyte | *POLR1D* | 0.567553154 | 0.016052177 | 0.046498605 | 1 |
| Heart_Failure | adipocyte | *RPS23* | 0.517922239 | 0.002363267 | 0.010441952 | 1 |
| Heart_Failure | adipocyte | *GNG2* | 1.23332828 | 6.16E-05 | 0.001268787 | 0.298895984 |
| Heart_Failure | adipocyte | *TMEM222* | 1.291844422 | 0.001356217 | 0.006926699 | 1 |
| Heart_Failure | adipocyte | *MIR22HG* | 0.631611854 | 0.006456116 | 0.022898445 | 1 |
| Heart_Failure | adipocyte | *BCR* | 0.552427841 | 0.012568962 | 0.038435145 | 1 |
| Heart_Failure | adipocyte | *QRFPR* | 1.049783569 | 0.000430646 | 0.003002148 | 1 |
| Heart_Failure | adipocyte | *TMEM17* | 0.196248325 | 0.017447068 | 0.049483688 | 1 |
| Heart_Failure | adipocyte | *TMEM232* | 0.37286347 | 2.74E-05 | 0.001268787 | 0.132793306 |
| Heart_Failure | adipocyte | *EFCAB6* | 0.959848535 | 0.000609058 | 0.003873066 | 1 |
| Heart_Failure | adipocyte | *MRPS21* | -0.227129825 | 0.016380817 | 0.047253106 | 1 |
| Heart_Failure | adipocyte | *WDR86* | 0.20468956 | 0.007912355 | 0.026884275 | 1 |
| Heart_Failure | adipocyte | *LUZP2* | 1.55011149 | 0.001642241 | 0.007960191 | 1 |
| Heart_Failure | adipocyte | *GJA4* | 1.014230638 | 0.01134453 | 0.035466276 | 1 |
| Heart_Failure | adipocyte | *EXD3* | 1.451222269 | 6.77E-05 | 0.001268787 | 0.32872157 |
| Heart_Failure | adipocyte | *C2orf88* | 0.497025542 | 0.010889241 | 0.034464838 | 1 |
| Heart_Failure | adipocyte | *ZFP69* | 2.447795754 | 4.44E-05 | 0.001268787 | 0.21558329 |
| Heart_Failure | adipocyte | *FAM122A* | 2.620823226 | 6.20E-05 | 0.001268787 | 0.300729824 |
| Heart_Failure | adipocyte | *ZNF626* | 1.66044398 | 0.000166252 | 0.001734151 | 0.806655274 |
| Heart_Failure | adipocyte | *PLSCR1* | 0.533132635 | 0.002323282 | 0.010332325 | 1 |
| Heart_Failure | adipocyte | *C3orf62* | 0.487931316 | 0.00019934 | 0.001892755 | 0.96719786 |
| Heart_Failure | adipocyte | *COL25A1* | 1.210146486 | 7.40E-05 | 0.001268787 | 0.35916185 |
| Heart_Failure | adipocyte | *SRSF10* | 0.454868025 | 0.004082955 | 0.015873798 | 1 |
| Heart_Failure | adipocyte | *NBR1* | 0.873819914 | 0.000409287 | 0.002894606 | 1 |
| Heart_Failure | adipocyte | *NDOR1* | 1.712609782 | 0.000184574 | 0.001798299 | 0.895552749 |
| Heart_Failure | adipocyte | *SLC4A5* | 2.125125655 | 0.000611632 | 0.003884346 | 1 |
| Heart_Failure | adipocyte | *UROS* | 1.064405427 | 0.001257212 | 0.006545055 | 1 |
| Heart_Failure | adipocyte | *NOXA1* | 1.422490416 | 0.000353752 | 0.002620463 | 1 |
| Heart_Failure | adipocyte | *MTF1* | 0.70472891 | 0.013253784 | 0.040091869 | 1 |
| Heart_Failure | adipocyte | *ZDHHC11* | 0.306386346 | 0.009993396 | 0.032217911 | 1 |

| | | | | | | |
|---|---|---|---|---|---|---|
| Heart_Failure | adipocyte | *RPL14* | 0.906234599 | 0.000996881 | 0.0054902 | 1 |
| Heart_Failure | adipocyte | *FAM196A* | 1.034236718 | 0.000322698 | 0.002481351 | 1 |
| Heart_Failure | adipocyte | *NELFB* | 1.667471488 | 7.94E-05 | 0.001268787 | 0.385362952 |
| Heart_Failure | adipocyte | *LITAF* | 1.083206517 | 0.00031365 | 0.00244667 | 1 |
| Heart_Failure | adipocyte | *SF3B3* | 1.138881518 | 0.000284424 | 0.002300045 | 1 |
| Heart_Failure | adipocyte | *PLAC9* | 0.669123703 | 0.000390586 | 0.002786946 | 1 |
| Heart_Failure | adipocyte | *TSPYL1* | 1.174363801 | 0.000542878 | 0.00355358 | 1 |
| Heart_Failure | adipocyte | *PNRC2* | 1.534821603 | 0.000125289 | 0.00153782 | 0.607900743 |
| Heart_Failure | adipocyte | *FHIT* | 0.973649996 | 8.53E-06 | 0.000960535 | 0.041390371 |
| Heart_Failure | adipocyte | *HLA-DRB1* | -0.205783253 | 3.87E-06 | 0.000508137 | 0.018801055 |
| Heart_Failure | adipocyte | *AKR1C3* | 0.497946123 | 0.015635892 | 0.045702017 | 1 |
| Heart_Failure | adipocyte | *MPHOSPH8* | 0.771186152 | 0.000175953 | 0.001756516 | 0.853724748 |
| Heart_Failure | adipocyte | *SRGAP3* | 0.349779587 | 0.004774184 | 0.018055264 | 1 |
| Heart_Failure | adipocyte | *ZNF471* | 1.5015264 | 6.92E-05 | 0.001268787 | 0.335839531 |
| Heart_Failure | adipocyte | *ZNF493* | 1.217797105 | 0.000149169 | 0.001619166 | 0.723767166 |
| Heart_Failure | adipocyte | *SUPT3H* | 0.97857408 | 0.000126613 | 0.00153782 | 0.614326846 |
| Heart_Failure | adipocyte | *STK31* | 0.590980843 | 0.017172782 | 0.048898087 | 1 |
| Heart_Failure | adipocyte | *ZKSCAN7* | 0.428616526 | 0.016566609 | 0.047595662 | 1 |
| Heart_Failure | adipocyte | *CD55* | 0.599986503 | 0.008481885 | 0.028304061 | 1 |
| Heart_Failure | adipocyte | *ZNF565* | 0.321314935 | 0.014904586 | 0.043935025 | 1 |
| Heart_Failure | adipocyte | *LONP1* | 0.235487873 | 0.010888619 | 0.034464838 | 1 |
| Heart_Failure | adipocyte | *EVL* | 0.3321708 | 0.017592347 | 0.049858684 | 1 |
| Heart_Failure | adipocyte | *ZNF124* | 0.942496001 | 0.003100005 | 0.012839666 | 1 |
| Heart_Failure | adipocyte | *PIK3R4* | 1.672317268 | 3.15E-06 | 0.000449837 | 0.015294469 |
| Heart_Failure | adipocyte | *C20orf96* | 1.140094357 | 0.009744225 | 0.031603596 | 1 |
| Heart_Failure | adipocyte | *SULF2* | 0.238958019 | 0.012176349 | 0.037630347 | 1 |
| Heart_Failure | adipocyte | *XRCC2* | 2.035201115 | 0.004383874 | 0.01682797 | 1 |
| Heart_Failure | adipocyte | *ZNF846* | 0.656147697 | 0.003721119 | 0.014799075 | 1 |
| Heart_Failure | adipocyte | *ADH1B* | 0.410515737 | 0.00276884 | 0.011831029 | 1 |
| Heart_Failure | adipocyte | *ZKSCAN5* | 0.827112891 | 6.19E-05 | 0.001268787 | 0.300130055 |
| Heart_Failure | adipocyte | *TOMM7* | 1.116538251 | 0.000328186 | 0.00250371 | 1 |
| Heart_Failure | adipocyte | *AMZ2* | 0.432998253 | 0.007356046 | 0.025493952 | 1 |
| Heart_Failure | adipocyte | *ZNF431* | 0.828985572 | 0.000202231 | 0.001912722 | 0.981226416 |
| Heart_Failure | adipocyte | *HLA-DQA1* | -0.511248784 | 9.79E-20 | 2.38E-16 | 4.75E-16 |
| Heart_Failure | adipocyte | *ZNF239* | 0.571087402 | 0.004424995 | 0.016945602 | 1 |
| Heart_Failure | adipocyte | *SLC39A10* | 0.780545291 | 0.00465334 | 0.017666672 | 1 |
| Heart_Failure | adipocyte | *WDR5B* | 0.629712936 | 0.000478374 | 0.003236181 | 1 |
| Heart_Failure | adipocyte | *ZNF138* | 0.792680791 | 6.68E-05 | 0.001268787 | 0.32403717 |
| Heart_Failure | adipocyte | *ZNF429* | 0.942664368 | 0.000402201 | 0.002853038 | 1 |
| Heart_Failure | adipocyte | *GMFB* | 1.485361237 | 0.000175908 | 0.001756516 | 0.853504733 |
| Heart_Failure | adipocyte | *ZNF420* | 0.793312496 | 0.002073083 | 0.009507184 | 1 |
| Heart_Failure | adipocyte | *ZMYM1* | 1.418303288 | 7.30E-05 | 0.001268787 | 0.35414108 |
| Heart_Failure | adipocyte | *IGF2R* | 0.754160246 | 0.001912399 | 0.00890495 | 1 |
| Heart_Failure | adipocyte | *ADAM32* | 0.843505066 | 0.005456152 | 0.020177782 | 1 |
| Heart_Failure | adipocyte | *ABCB8* | 1.055035949 | 0.000284345 | 0.002300045 | 1 |
| Heart_Failure | adipocyte | *SND1* | 0.816558618 | 0.000337418 | 0.002550054 | 1 |
| Heart_Failure | adipocyte | *RAMP2-AS1* | 0.628348656 | 0.005965523 | 0.021582093 | 1 |
| Heart_Failure | adipocyte | *ZNF720* | 0.739336481 | 0.004663469 | 0.017682859 | 1 |
| Heart_Failure | adipocyte | *MAP3K5* | 0.687717703 | 0.001785994 | 0.008470815 | 1 |
| Heart_Failure | adipocyte | *ZNF665* | 1.605455554 | 9.29E-05 | 0.001331689 | 0.450580845 |

| | | | | | | |
|---|---|---|---|---|---|---|
| Heart_Failure | adipocyte | *RPF2* | 0.925772347 | 0.000948012 | 0.005323787 | 1 |
| Heart_Failure | adipocyte | *ATG7* | 0.549393455 | 0.001993607 | 0.009177402 | 1 |
| Heart_Failure | adipocyte | *RAB40C* | 1.102558519 | 0.000655105 | 0.004055662 | 1 |
| Heart_Failure | adipocyte | *PIGN* | 0.283219428 | 0.012304016 | 0.037856109 | 1 |
| Heart_Failure | adipocyte | *TOPORS* | 1.596639957 | 0.000111006 | 0.001449365 | 0.538602766 |
| Heart_Failure | adipocyte | *PDCD1LG2* | 0.143412678 | 0.000440818 | 0.003059867 | 1 |
| Heart_Failure | adipocyte | *FAM114A1* | 0.849442642 | 0.004014167 | 0.015681753 | 1 |
| Heart_Failure | adipocyte | *RPS26* | 0.205582099 | 0.000182458 | 0.00178514 | 0.885288518 |
| Heart_Failure | adipocyte | *PSAP* | 0.624186238 | 0.000449486 | 0.003102284 | 1 |
| Heart_Failure | adipocyte | *TXNRD3* | 0.513622239 | 0.0047743 | 0.018055264 | 1 |
| Heart_Failure | adipocyte | *EME2* | 1.537008972 | 0.001183059 | 0.006259761 | 1 |
| Heart_Failure | adipocyte | *ADH5* | 0.942798582 | 0.000168547 | 0.001740728 | 0.81778847 |
| Heart_Failure | adipocyte | *ZNF677* | 1.508588555 | 0.000213524 | 0.001960237 | 1 |
| Heart_Failure | adipocyte | *MPZL1* | 0.74636479 | 0.002869908 | 0.012162534 | 1 |
| Heart_Failure | adipocyte | *NOL8* | 0.936970708 | 0.000809963 | 0.004751265 | 1 |
| Heart_Failure | adipocyte | *DLGAP2* | 0.382660931 | 0.005189175 | 0.019337847 | 1 |
| Heart_Failure | adipocyte | *SFI1* | 0.708570327 | 0.010613873 | 0.033813861 | 1 |
| Heart_Failure | adipocyte | *ZNF544* | 1.709356795 | 0.000244046 | 0.002095769 | 1 |
| Heart_Failure | adipocyte | *MIER1* | 0.820701372 | 0.000249029 | 0.002112148 | 1 |
| Heart_Failure | adipocyte | *MAN1A2* | 1.126411081 | 7.22E-05 | 0.001268787 | 0.350072235 |
| Heart_Failure | adipocyte | *SVIP* | 1.540886504 | 4.02E-05 | 0.001268787 | 0.19499357 |
| Heart_Failure | adipocyte | *DDRGK1* | 0.809518267 | 0.008754949 | 0.028995914 | 1 |
| Heart_Failure | adipocyte | *STYX* | 1.178448665 | 0.001173852 | 0.006231432 | 1 |
| Heart_Failure | adipocyte | *HELZ* | 0.973180447 | 4.65E-05 | 0.001268787 | 0.225835818 |
| Heart_Failure | adipocyte | *UCKL1* | 1.358587823 | 0.000383202 | 0.002754513 | 1 |
| Heart_Failure | adipocyte | *GFPT1* | 0.796658207 | 0.000309129 | 0.002427015 | 1 |
| Heart_Failure | adipocyte | *ZNF568* | 0.551452523 | 0.007448328 | 0.025740233 | 1 |
| Heart_Failure | adipocyte | *TMA16* | 1.51324016 | 3.50E-05 | 0.001268787 | 0.169957765 |
| Heart_Failure | adipocyte | *HLA-DRB5* | -0.04622576 | 0.007998919 | 0.027121422 | 1 |
| Heart_Failure | adipocyte | *ZNF28* | 1.364506656 | 0.000159731 | 0.001699592 | 0.77501414 |
| Heart_Failure | adipocyte | *FAM19A2* | 0.498323736 | 0.013529756 | 0.040698312 | 1 |
| Heart_Failure | adipocyte | *TTC37* | 0.704870657 | 0.001051555 | 0.005719893 | 1 |
| Heart_Failure | adipocyte | *TUSC1* | 2.577099591 | 0.000182488 | 0.00178514 | 0.885429525 |
| Heart_Failure | adipocyte | *PAPSS2* | 0.664004649 | 0.004218397 | 0.016295908 | 1 |
| Heart_Failure | adipocyte | *CEP290* | 0.619667961 | 0.001111007 | 0.00595647 | 1 |
| Heart_Failure | adipocyte | *PPP1R14C* | 4.00453742 | 0.005557596 | 0.020504795 | 1 |
| Heart_Failure | adipocyte | *GPATCH3* | 2.198574385 | 9.49E-05 | 0.001343879 | 0.460394206 |
| Heart_Failure | adipocyte | *RPL10A* | 0.752327832 | 0.004983543 | 0.018744302 | 1 |
| Heart_Failure | adipocyte | *SFT2D1* | 1.182515624 | 0.000580453 | 0.003725338 | 1 |
| Heart_Failure | adipocyte | *TYW1* | 0.685301232 | 0.000498238 | 0.003329821 | 1 |
| Heart_Failure | adipocyte | *DZIP3* | 0.952478526 | 0.000362889 | 0.00266375 | 1 |
| Heart_Failure | adipocyte | *ZFP2* | 0.812344008 | 0.006399101 | 0.022746107 | 1 |
| Heart_Failure | adipocyte | *MFAP3L* | 0.589323813 | 0.016952409 | 0.048498284 | 1 |
| Heart_Failure | adipocyte | *PJA2* | 0.570387434 | 0.012467358 | 0.03821328 | 1 |
| Heart_Failure | adipocyte | *EFCAB2* | 0.311621523 | 0.011196209 | 0.035183941 | 1 |
| Heart_Failure | adipocyte | *ECT2L* | 1.731391182 | 0.000143059 | 0.001590059 | 0.694124116 |
| Heart_Failure | adipocyte | *RBM20* | 0.736688324 | 0.013944871 | 0.041662879 | 1 |
| Heart_Failure | adipocyte | *PCMTD2* | 1.114951491 | 0.000300629 | 0.002383417 | 1 |
| Heart_Failure | adipocyte | *RUFY2* | 1.217618316 | 2.83E-05 | 0.001268787 | 0.137232131 |
| Heart_Failure | adipocyte | *BMPR2* | 0.998371998 | 7.52E-05 | 0.001268787 | 0.364717717 |

| | | | | | | |
|---|---|---|---|---|---|---|
| Heart_Failure | adipocyte | *TCEA3* | 0.596221498 | 0.000686157 | 0.004198275 | 1 |
| Heart_Failure | adipocyte | *HSD17B8* | 0.255177532 | 0.001382614 | 0.00703929 | 1 |
| Heart_Failure | adipocyte | *PRRT1* | 0.208685838 | 0.000241871 | 0.0020919 | 1 |
| Heart_Failure | adipocyte | *BAG6* | 0.495507577 | 0.000358865 | 0.002646218 | 1 |
| Heart_Failure | adipocyte | *ZNF814* | 0.711566587 | 0.000825027 | 0.0047998 | 1 |
| Heart_Failure | adipocyte | *MICB* | 0.065152075 | 0.001967091 | 0.009081183 | 1 |
| Heart_Failure | adipocyte | *MICA* | 0.398316277 | 4.05E-07 | 0.000122871 | 0.001965928 |
| Heart_Failure | adipocyte | *LILRB3* | 0.662173367 | 0.002380679 | 0.010472397 | 1 |
| Heart_Failure | adipocyte | *ZNF468* | 1.301199982 | 0.002755312 | 0.011823391 | 1 |
| Heart_Failure | adipocyte | *MALRD1* | 0.936872171 | 0.000627534 | 0.003944977 | 1 |
| Heart_Failure | adipocyte | *DCTN1* | 1.545253375 | 0.000410447 | 0.002894606 | 1 |
| Heart_Failure | adipocyte | *FAM216A* | 1.491583707 | 6.03E-06 | 0.000749815 | 0.029242778 |
| Heart_Failure | adipocyte | *TRIM13* | 0.52682372 | 0.006229704 | 0.022245195 | 1 |
| Heart_Failure | adipocyte | *SLC35B4* | 1.370658939 | 0.00032611 | 0.002495717 | 1 |
| Heart_Failure | adipocyte | *SCGB2B2* | 0.75921306 | 0.001458926 | 0.007290121 | 1 |
| Heart_Failure | adipocyte | *CCDC144NL* | 0.823606397 | 0.016515357 | 0.047528181 | 1 |
| Heart_Failure | adipocyte | *NT5M* | 1.063064316 | 0.007891647 | 0.026832706 | 1 |
| Heart_Failure | adipocyte | *NAP1L4* | 0.818041288 | 0.000237869 | 0.002072062 | 1 |
| Heart_Failure | adipocyte | *LIN52* | 0.501936801 | 8.68E-05 | 0.00130347 | 0.421020729 |
| Heart_Failure | adipocyte | *METTL6* | 0.840314223 | 0.000318643 | 0.002469736 | 1 |
| Heart_Failure | adipocyte | *SACM1L* | 1.221133155 | 0.000299646 | 0.002383411 | 1 |
| Heart_Failure | adipocyte | *C17orf51* | 3.163777885 | 6.85E-05 | 0.001268787 | 0.332515042 |
| Heart_Failure | adipocyte | *ZNF254* | 0.646901325 | 0.003952035 | 0.015488912 | 1 |
| Heart_Failure | adipocyte | *SUPT4H1* | 1.34706156 | 0.00048057 | 0.003237729 | 1 |
| Heart_Failure | adipocyte | *LCAT* | 1.077911351 | 0.00709587 | 0.024769179 | 1 |
| Heart_Failure | adipocyte | *ERV3-1* | 0.09600041 | 0.008665416 | 0.028758276 | 1 |
| Heart_Failure | adipocyte | *SRA1* | 0.515575944 | 0.010409052 | 0.033314461 | 1 |
| Heart_Failure | adipocyte | *PPP1CB* | 0.466172196 | 0.002551048 | 0.011091116 | 1 |
| Heart_Failure | adipocyte | *ATF6B* | 0.803217577 | 1.33E-08 | 9.24E-06 | 6.47E-05 |
| Heart_Failure | adipocyte | *EMP2* | 0.713255569 | 0.002891132 | 0.012213225 | 1 |
| Heart_Failure | adipocyte | *GALT* | 0.70475532 | 0.000967052 | 0.005387066 | 1 |
| Heart_Failure | adipocyte | *REPIN1* | 1.585864139 | 0.000523775 | 0.003471315 | 1 |
| Heart_Failure | adipocyte | *ZNF891* | 1.893891348 | 8.31E-08 | 3.66E-05 | 0.000402967 |
| Heart_Failure | adipocyte | *SMIM7* | 0.885839806 | 0.00123772 | 0.006478336 | 1 |
| Heart_Failure | adipocyte | *CPNE1* | -0.140396217 | 0.003144278 | 0.012943168 | 1 |
| Heart_Failure | adipocyte | *PAXIP1-AS2* | 1.371884337 | 7.77E-05 | 0.001268787 | 0.377105228 |
| Heart_Failure | adipocyte | *LYRM4* | 1.290988072 | 7.38E-05 | 0.001268787 | 0.357936614 |
| Heart_Failure | adipocyte | *PRCD* | 0.659386311 | 0.014076306 | 0.041956912 | 1 |
| Heart_Failure | adipocyte | *SH3D21* | 0.575079159 | 0.000186154 | 0.001801316 | 0.903218859 |
| Heart_Failure | adipocyte | *ANG* | 1.968336014 | 7.64E-05 | 0.001268787 | 0.370873536 |
| Heart_Failure | adipocyte | *GNAT3* | 1.755596226 | 0.000816823 | 0.004763495 | 1 |
| Heart_Failure | adipocyte | *VPS16* | 1.923800731 | 2.12E-05 | 0.001268787 | 0.102648856 |
| Heart_Failure | adipocyte | *NPEPL1* | 1.223060518 | 0.000773204 | 0.004580692 | 1 |
| Heart_Failure | adipocyte | *TSTD1* | 0.94762943 | 0.008291996 | 0.02790067 | 1 |
| Heart_Failure | adipocyte | *PAM16* | 1.019374712 | 0.001724265 | 0.008242495 | 1 |
| Heart_Failure | adipocyte | *RNASEK* | 0.454550823 | 0.00148683 | 0.007383928 | 1 |
| Heart_Failure | adipocyte | *FXYD7* | 0.344508545 | 0.007835447 | 0.026716507 | 1 |
| Heart_Failure | adipocyte | *OR2A1* | 0.30826778 | 0.001179833 | 0.006256339 | 1 |
| Heart_Failure | adipocyte | *UBA52* | 0.366669928 | 0.004064216 | 0.015813614 | 1 |
| Heart_Failure | adipocyte | *C5orf66* | 0.699838156 | 0.005361538 | 0.019903735 | 1 |

| | | | | | | |
|---|---|---|---|---|---|---|
| Heart_Failure | adipocyte | *CYP4A22-AS1* | 1.627699586 | 0.002683026 | 0.011581887 | 1 |
| Heart_Failure | adipocyte | *MIAT* | 0.875750999 | 0.001736314 | 0.008291924 | 1 |
| Heart_Failure | adipocyte | *ERCC6* | 0.696946484 | 0.011901136 | 0.036897323 | 1 |
| Heart_Failure | adipocyte | *FAM138B* | 14.24587013 | 0.003602142 | 0.014394835 | 1 |
| Heart_Failure | adipocyte | *AC009299.3* | 0.920479911 | 0.004849767 | 0.018312115 | 1 |
| Heart_Failure | adipocyte | *SCAMP4* | 1.55327659 | 0.000146329 | 0.001609947 | 0.709986426 |
| Heart_Failure | adipocyte | *AC092171.4* | 1.341227388 | 0.002241154 | 0.010041405 | 1 |
| Heart_Failure | adipocyte | *RPS18* | 0.87471535 | 5.36E-05 | 0.001268787 | 0.260267319 |
| Heart_Failure | adipocyte | *AC107021.1* | 0.336275557 | 0.013768124 | 0.041185534 | 1 |
| Heart_Failure | adipocyte | *HLA-DQB2* | 0.173550787 | 2.58E-17 | 3.13E-14 | 1.25E-13 |
| Heart_Failure | adipocyte | *RPS28* | 0.948331281 | 0.00443814 | 0.016969152 | 1 |
| Heart_Failure | adipocyte | *TRIM26* | 0.38139467 | 0.013009422 | 0.039500447 | 1 |
| Heart_Failure | adipocyte | *ZNF736* | 0.259737357 | 0.000301366 | 0.002384324 | 1 |
| Heart_Failure | adipocyte | *ANKRD65* | 2.095764993 | 0.008677896 | 0.028780009 | 1 |
| Heart_Failure | adipocyte | *ZBTB22* | 0.482636691 | 6.50E-05 | 0.001268787 | 0.315330889 |
| Heart_Failure | adipocyte | *ZBED5* | 0.522579816 | 0.005561482 | 0.020504795 | 1 |
| Heart_Failure | adipocyte | *LINC00959* | 0.951872024 | 0.002549462 | 0.011091116 | 1 |
| Heart_Failure | adipocyte | *HLA-DQA2* | 0.217486578 | 3.05E-18 | 4.93E-15 | 1.48E-14 |
| Heart_Failure | adipocyte | *LINC01119* | 0.817192991 | 0.008522723 | 0.028383563 | 1 |
| Heart_Failure | adipocyte | *TNFSF12* | 0.317397448 | 0.011232769 | 0.035207621 | 1 |
| Heart_Failure | adipocyte | *PSMB9* | 0.247381436 | 3.69E-07 | 0.000119246 | 0.001788683 |
| Heart_Failure | adipocyte | *KRBOX1* | 30.00849069 | 0.00041948 | 0.002941212 | 1 |
| Heart_Failure | adipocyte | *NSUN6* | 0.993183276 | 4.51E-05 | 0.001268787 | 0.219005011 |
| Heart_Failure | adipocyte | *CD302* | 0.594057218 | 0.012600336 | 0.038499264 | 1 |
| Heart_Failure | adipocyte | *PI4KA* | 0.812426673 | 0.000487374 | 0.00327073 | 1 |
| Heart_Failure | adipocyte | *LINC00969* | 0.238354845 | 0.004429961 | 0.01695124 | 1 |
| Heart_Failure | adipocyte | *ARFGAP3* | 1.462797535 | 7.12E-05 | 0.001268787 | 0.345491871 |
| Heart_Failure | adipocyte | *DECR2* | 1.816545069 | 0.001423203 | 0.007155836 | 1 |
| Heart_Failure | adipocyte | *STON1* | 0.498920898 | 0.011499844 | 0.035905561 | 1 |
| Heart_Failure | adipocyte | *C7orf73* | 0.523835451 | 0.013236697 | 0.040090172 | 1 |
| Heart_Failure | adipocyte | *NBPF15* | 1.178064977 | 0.001604451 | 0.007816062 | 1 |
| Heart_Failure | adipocyte | *TTC4* | -0.319168216 | 0.002755991 | 0.011823391 | 1 |
| Heart_Failure | adipocyte | *LILRA6* | 0.149851283 | 0.002343189 | 0.010380724 | 1 |
| Heart_Failure | adipocyte | *C4A* | -0.279435798 | 0.007089741 | 0.024765605 | 1 |
| Heart_Failure | adipocyte | *NEAT1* | 0.734130651 | 1.92E-05 | 0.001268787 | 0.093400397 |
| Heart_Failure | adipocyte | *RAB30-AS1* | 0.446821915 | 0.002364396 | 0.010441952 | 1 |
| Heart_Failure | adipocyte | *UBAP1L* | 1.18833294 | 0.000393116 | 0.002800847 | 1 |
| Heart_Failure | adipocyte | *LINC00926* | 3.618450528 | 1.21E-05 | 0.00112616 | 0.058759107 |
| Heart_Failure | adipocyte | *CDK11B* | 1.735696457 | 5.26E-05 | 0.001268787 | 0.255254152 |
| Heart_Failure | adipocyte | *ABHD14A* | 1.359336029 | 9.18E-05 | 0.001326312 | 0.445640911 |
| Heart_Failure | adipocyte | *C5orf17* | 1.338659884 | 0.000179545 | 0.001767041 | 0.871151391 |
| Heart_Failure | adipocyte | *PDCD6* | 1.519928298 | 6.14E-05 | 0.001268787 | 0.297867593 |
| Heart_Failure | adipocyte | *ZNF718* | 0.960911201 | 0.000635248 | 0.003977063 | 1 |
| Heart_Failure | adipocyte | *SMIM20* | 1.567520387 | 0.00064589 | 0.004017769 | 1 |
| Heart_Failure | adipocyte | *LINC00989* | 0.5666373 | 0.00851012 | 0.028378765 | 1 |
| Heart_Failure | adipocyte | *GLI4* | 0.62261347 | 0.015828981 | 0.046072116 | 1 |
| Heart_Failure | adipocyte | *ZNF550* | 1.74939993 | 0.000237503 | 0.002072062 | 1 |
| Heart_Failure | adipocyte | *TMEM200B* | 0.385597133 | 0.007329965 | 0.025421724 | 1 |
| Heart_Failure | adipocyte | *KIAA1147* | 0.746891579 | 0.000243649 | 0.002095769 | 1 |
| Heart_Failure | adipocyte | *ZBED6* | 1.530293335 | 0.016796208 | 0.048108146 | 1 |

| | | | | | | |
|---|---|---|---|---|---|---|
| Heart_Failure | adipocyte | *CNPY2* | 1.534548826 | 3.74E-05 | 0.001268787 | 0.181682238 |
| Heart_Failure | adipocyte | *HIF1A-AS2* | 1.355138009 | 2.84E-05 | 0.001268787 | 0.137815695 |
| Heart_Failure | adipocyte | *MC1R* | 0.493432533 | 0.002672635 | 0.011547308 | 1 |
| Heart_Failure | adipocyte | *IQCH-AS1* | 0.330824368 | 0.01512134 | 0.044492869 | 1 |
| Heart_Failure | adipocyte | *PWRN1* | 0.320776046 | 0.005647365 | 0.020742629 | 1 |
| Heart_Failure | adipocyte | *DYNLL2* | 2.158058347 | 0.000333625 | 0.00253325 | 1 |
| Heart_Failure | adipocyte | *ZNF582-AS1* | 3.10800179 | 4.09E-05 | 0.001268787 | 0.198514215 |
| Heart_Failure | adipocyte | *ZNF224* | 1.429762937 | 0.000119475 | 0.001495325 | 0.579692595 |
| Heart_Failure | adipocyte | *AC093157.1* | 0.745470409 | 0.017104231 | 0.048731492 | 1 |
| Heart_Failure | adipocyte | *ZNF587B* | 2.516062582 | 8.18E-05 | 0.001268787 | 0.396950518 |
| Heart_Failure | adipocyte | *HCG17* | 0.510086465 | 0.001802617 | 0.008496203 | 1 |
| Heart_Failure | adipocyte | *GTF2H5* | 0.328925483 | 0.000215896 | 0.001964387 | 1 |
| Heart_Failure | adipocyte | *CASC15* | 0.5411693 | 0.015401231 | 0.045234124 | 1 |
| Heart_Failure | adipocyte | *PAXIP1-AS1* | 0.97792712 | 0.001707336 | 0.008199798 | 1 |
| Heart_Failure | ASPC | *SEMA3F* | -0.250541187 | 0.000689783 | 0.010420628 | 1 |
| Heart_Failure | ASPC | *KRIT1* | 0.431255545 | 0.010094149 | 0.043037619 | 1 |
| Heart_Failure | ASPC | *HS3ST1* | 0.140210862 | 0.007840548 | 0.035586845 | 1 |
| Heart_Failure | ASPC | *CD38* | 0.304187718 | 0.008600124 | 0.038212272 | 1 |
| Heart_Failure | ASPC | *RBM6* | -0.122051554 | 4.42E-06 | 0.00112959 | 0.021462213 |
| Heart_Failure | ASPC | *POLR2J* | 0.761527965 | 0.000768904 | 0.010420628 | 1 |
| Heart_Failure | ASPC | *GCFC2* | 0.636626655 | 0.001002471 | 0.010420628 | 1 |
| Heart_Failure | ASPC | *ITGA2B* | 0.390286207 | 0.001488471 | 0.011937289 | 1 |
| Heart_Failure | ASPC | *SYNRG* | 0.381348348 | 0.011714358 | 0.0480457 | 1 |
| Heart_Failure | ASPC | *ZNF263* | 0.516376888 | 0.005201927 | 0.02654022 | 1 |
| Heart_Failure | ASPC | *SPATA20* | 0.160882722 | 0.007477493 | 0.03437319 | 1 |
| Heart_Failure | ASPC | *BAIAP2L1* | 0.892981811 | 0.00080255 | 0.010420628 | 1 |
| Heart_Failure | ASPC | *VPS41* | 0.472798041 | 0.002837009 | 0.017535246 | 1 |
| Heart_Failure | ASPC | *TSR3* | 0.837843495 | 0.001468756 | 0.011818249 | 1 |
| Heart_Failure | ASPC | *PSMB1* | 0.831049321 | 0.000229739 | 0.010420628 | 1 |
| Heart_Failure | ASPC | *PKD1* | 0.507006259 | 0.001463473 | 0.011795301 | 1 |
| Heart_Failure | ASPC | *MED24* | 0.584692582 | 0.010684591 | 0.044806945 | 1 |
| Heart_Failure | ASPC | *RPS20* | 0.486759786 | 0.003127667 | 0.018784468 | 1 |
| Heart_Failure | ASPC | *MAP4K5* | 0.47485573 | 0.005306207 | 0.026976152 | 1 |
| Heart_Failure | ASPC | *RNF14* | 0.454583854 | 0.000397384 | 0.010420628 | 1 |
| Heart_Failure | ASPC | *MDH1* | 0.194591389 | 0.00752339 | 0.03446977 | 1 |
| Heart_Failure | ASPC | *COX15* | 0.581592977 | 0.005062902 | 0.025940024 | 1 |
| Heart_Failure | ASPC | *RUFY3* | 0.392066446 | 0.005018858 | 0.025768782 | 1 |
| Heart_Failure | ASPC | *WWTR1* | 0.594349741 | 0.000967325 | 0.010420628 | 1 |
| Heart_Failure | ASPC | *TTC27* | 0.436597589 | 0.003701054 | 0.021065713 | 1 |
| Heart_Failure | ASPC | *SLC7A9* | 0.064372456 | 0.006746623 | 0.032061327 | 1 |
| Heart_Failure | ASPC | *GCLM* | 0.536957349 | 0.008923966 | 0.039291363 | 1 |
| Heart_Failure | ASPC | *FAS* | 0.536014174 | 0.002787821 | 0.017363935 | 1 |
| Heart_Failure | ASPC | *AGPAT4* | 0.74875159 | 0.001165331 | 0.010788896 | 1 |
| Heart_Failure | ASPC | *MIPEP* | 0.478579791 | 0.000430912 | 0.010420628 | 1 |
| Heart_Failure | ASPC | *VRK2* | 0.360886373 | 0.00749463 | 0.03437319 | 1 |
| Heart_Failure | ASPC | *VEZT* | 0.629956764 | 0.000760376 | 0.010420628 | 1 |
| Heart_Failure | ASPC | *BRD9* | 0.66070161 | 0.000220497 | 0.010420628 | 1 |
| Heart_Failure | ASPC | *IKZF2* | 0.457316493 | 0.009406635 | 0.040862971 | 1 |
| Heart_Failure | ASPC | *ATP6V0A1* | 0.34322305 | 0.002420464 | 0.015913671 | 1 |
| Heart_Failure | ASPC | *PIAS1* | 0.319874341 | 0.005273874 | 0.026850823 | 1 |

| | | | | | | |
|---|---|---|---|---|---|---|
| Heart_Failure | ASPC | *VCL* | 0.521004287 | 0.004934447 | 0.025524454 | 1 |
| Heart_Failure | ASPC | *ADSS* | 0.771204325 | 0.001085797 | 0.010484035 | 1 |
| Heart_Failure | ASPC | *TBC1D23* | 0.644577239 | 0.00157325 | 0.012135784 | 1 |
| Heart_Failure | ASPC | *MRI1* | 0.153449726 | 0.008881901 | 0.039212906 | 1 |
| Heart_Failure | ASPC | *PI4K2B* | 2.379966855 | 0.001036947 | 0.010460014 | 1 |
| Heart_Failure | ASPC | *PQLC2* | 1.707937111 | 0.001067055 | 0.010475827 | 1 |
| Heart_Failure | ASPC | *PSMA4* | 0.778122802 | 0.001013713 | 0.010420628 | 1 |
| Heart_Failure | ASPC | *AP2S1* | 0.844423473 | 0.00151627 | 0.011981993 | 1 |
| Heart_Failure | ASPC | *CTNNA1* | 0.57841679 | 0.000839937 | 0.010420628 | 1 |
| Heart_Failure | ASPC | *YTHDC2* | 0.652550483 | 0.000755236 | 0.010420628 | 1 |
| Heart_Failure | ASPC | *POLR2B* | 0.563513486 | 0.001677062 | 0.012518621 | 1 |
| Heart_Failure | ASPC | *ARAP2* | 0.402583631 | 0.000500605 | 0.010420628 | 1 |
| Heart_Failure | ASPC | *DTNBP1* | 0.693555197 | 0.000160993 | 0.010420628 | 0.781138262 |
| Heart_Failure | ASPC | *ANO2* | 0.51480101 | 0.003203271 | 0.019187986 | 1 |
| Heart_Failure | ASPC | *MAPK9* | 0.801110736 | 0.00105507 | 0.010475827 | 1 |
| Heart_Failure | ASPC | *CYBA* | 0.684386853 | 0.000643315 | 0.010420628 | 1 |
| Heart_Failure | ASPC | *THOC3* | 1.609974794 | 0.002185844 | 0.01485394 | 1 |
| Heart_Failure | ASPC | *MRTO4* | 0.302191693 | 0.007367682 | 0.033948711 | 1 |
| Heart_Failure | ASPC | *ANAPC4* | 0.366465514 | 0.001439182 | 0.01167711 | 1 |
| Heart_Failure | ASPC | *LY75* | 0.770156023 | 0.005062349 | 0.025940024 | 1 |
| Heart_Failure | ASPC | *SDCCAG8* | 0.244419863 | 2.02E-05 | 0.002971698 | 0.098066043 |
| Heart_Failure | ASPC | *TBC1D22A* | 0.406230531 | 0.004542949 | 0.024249053 | 1 |
| Heart_Failure | ASPC | *GALC* | 0.365937765 | 0.004455954 | 0.023969816 | 1 |
| Heart_Failure | ASPC | *ATP11B* | 0.304587091 | 0.003730236 | 0.021138904 | 1 |
| Heart_Failure | ASPC | *LAMC2* | -0.091276782 | 0.004538587 | 0.024249053 | 1 |
| Heart_Failure | ASPC | *CROCC* | 0.100797834 | 0.004164896 | 0.02294767 | 1 |
| Heart_Failure | ASPC | *DNAJC25* | 1.107842161 | 0.001628264 | 0.01232502 | 1 |
| Heart_Failure | ASPC | *OGFR* | 0.255261187 | 0.010228549 | 0.043419878 | 1 |
| Heart_Failure | ASPC | *RIMBP2* | 1.268172736 | 0.00117549 | 0.010788896 | 1 |
| Heart_Failure | ASPC | *LTK* | 10.50077177 | 0.000358387 | 0.010420628 | 1 |
| Heart_Failure | ASPC | *ADCK1* | 0.956321738 | 0.000637359 | 0.010420628 | 1 |
| Heart_Failure | ASPC | *RNF4* | 0.812763892 | 0.000660731 | 0.010420628 | 1 |
| Heart_Failure | ASPC | *CASP8* | 0.427619527 | 0.002075186 | 0.014404581 | 1 |
| Heart_Failure | ASPC | *TM7SF3* | 0.489460202 | 0.002792116 | 0.017368395 | 1 |
| Heart_Failure | ASPC | *TMEM161A* | 0.718296316 | 0.004613195 | 0.024409182 | 1 |
| Heart_Failure | ASPC | *SUGP2* | 0.490993569 | 0.003983038 | 0.022156417 | 1 |
| Heart_Failure | ASPC | *SLC12A2* | 0.806984088 | 0.00021222 | 0.010420628 | 1 |
| Heart_Failure | ASPC | *BTBD1* | 0.61241702 | 0.00020705 | 0.010420628 | 1 |
| Heart_Failure | ASPC | *FAR2* | 0.372617443 | 0.000943299 | 0.010420628 | 1 |
| Heart_Failure | ASPC | *SBNO2* | 0.567569358 | 0.000219082 | 0.010420628 | 1 |
| Heart_Failure | ASPC | *PMS1* | 0.474770337 | 0.003814508 | 0.021421286 | 1 |
| Heart_Failure | ASPC | *MAP2K4* | 0.442238785 | 0.010477598 | 0.044227615 | 1 |
| Heart_Failure | ASPC | *TRAM1* | 0.363500141 | 0.005708062 | 0.028449262 | 1 |
| Heart_Failure | ASPC | *STOML1* | 2.008321651 | 0.000540094 | 0.010420628 | 1 |
| Heart_Failure | ASPC | *DHX8* | 0.881611265 | 0.000703746 | 0.010420628 | 1 |
| Heart_Failure | ASPC | *ROGDI* | 1.715360041 | 0.000853774 | 0.010420628 | 1 |
| Heart_Failure | ASPC | *IP6K2* | 0.374512903 | 0.010857908 | 0.045347233 | 1 |
| Heart_Failure | ASPC | *SRBD1* | 0.572105198 | 0.000759471 | 0.010420628 | 1 |
| Heart_Failure | ASPC | *NUCKS1* | 0.435040623 | 0.010704419 | 0.044812634 | 1 |
| Heart_Failure | ASPC | *PLA2G10* | 2.909870486 | 0.000224827 | 0.010420628 | 1 |

| | | | | | | |
|---|---|---|---|---|---|---|
| Heart_Failure | ASPC | *LRP6* | 0.608609432 | 0.000734516 | 0.010420628 | 1 |
| Heart_Failure | ASPC | *ZXDC* | 0.569274004 | 0.001572374 | 0.012135784 | 1 |
| Heart_Failure | ASPC | *GBA2* | 1.075390977 | 1.73E-05 | 0.002619445 | 0.083822232 |
| Heart_Failure | ASPC | *EIF2B3* | 0.605405614 | 0.001034543 | 0.010457505 | 1 |
| Heart_Failure | ASPC | *MAP4K4* | 0.47454452 | 0.003459686 | 0.020224572 | 1 |
| Heart_Failure | ASPC | *WDR1* | 0.502506663 | 0.009402673 | 0.040862971 | 1 |
| Heart_Failure | ASPC | *SNX13* | 0.618579318 | 0.000833086 | 0.010420628 | 1 |
| Heart_Failure | ASPC | *ING3* | 0.742172457 | 0.000488979 | 0.010420628 | 1 |
| Heart_Failure | ASPC | *AFF4* | 0.399157272 | 0.004142568 | 0.022909416 | 1 |
| Heart_Failure | ASPC | *ADAM11* | -0.065958613 | 0.011650281 | 0.047923908 | 1 |
| Heart_Failure | ASPC | *PPP2R2C* | 0.946976377 | 0.01039468 | 0.043932917 | 1 |
| Heart_Failure | ASPC | *ZNF532* | 0.649656401 | 0.001024036 | 0.010439519 | 1 |
| Heart_Failure | ASPC | *ZZEF1* | 0.389542766 | 0.004050865 | 0.022436983 | 1 |
| Heart_Failure | ASPC | *VPS9D1* | 0.723917575 | 3.58E-05 | 0.004717456 | 0.173653471 |
| Heart_Failure | ASPC | *FNDC3B* | 0.469361584 | 0.000973718 | 0.010420628 | 1 |
| Heart_Failure | ASPC | *FMO4* | 0.267033595 | 0.010798525 | 0.045167624 | 1 |
| Heart_Failure | ASPC | *PAG1* | 0.462672342 | 0.003070023 | 0.018520523 | 1 |
| Heart_Failure | ASPC | *IL4R* | 0.640177157 | 0.001161656 | 0.010788896 | 1 |
| Heart_Failure | ASPC | *DYNC1I2* | 0.647586577 | 0.000940014 | 0.010420628 | 1 |
| Heart_Failure | ASPC | *PCM1* | 0.326766683 | 0.009889847 | 0.042427531 | 1 |
| Heart_Failure | ASPC | *XRCC5* | 0.46970985 | 0.003759156 | 0.021233324 | 1 |
| Heart_Failure | ASPC | *FDFT1* | 0.279955429 | 0.002441233 | 0.015984971 | 1 |
| Heart_Failure | ASPC | *PGM1* | 0.440879818 | 0.004397706 | 0.02386764 | 1 |
| Heart_Failure | ASPC | *RABL2B* | 0.907090193 | 0.000483304 | 0.010420628 | 1 |
| Heart_Failure | ASPC | *SCTR* | 0.76611419 | 0.001768261 | 0.012940575 | 1 |
| Heart_Failure | ASPC | *RIF1* | 0.371494372 | 0.010036057 | 0.042903038 | 1 |
| Heart_Failure | ASPC | *PSEN1* | 0.386939271 | 0.006092338 | 0.029708565 | 1 |
| Heart_Failure | ASPC | *ME2* | 0.758094644 | 0.000802951 | 0.010420628 | 1 |
| Heart_Failure | ASPC | *ZCCHC6* | 0.65448415 | 0.00071751 | 0.010420628 | 1 |
| Heart_Failure | ASPC | *PIBF1* | 0.561666755 | 0.00062798 | 0.010420628 | 1 |
| Heart_Failure | ASPC | *ZNF264* | 0.846147136 | 0.001173429 | 0.010788896 | 1 |
| Heart_Failure | ASPC | *CHMP2B* | 0.570006222 | 0.000821041 | 0.010420628 | 1 |
| Heart_Failure | ASPC | *GSTP1* | 0.264277077 | 0.001651819 | 0.012424316 | 1 |
| Heart_Failure | ASPC | *WBP11* | 0.924395211 | 0.000728271 | 0.010420628 | 1 |
| Heart_Failure | ASPC | *NCOA1* | 0.460153093 | 0.001858532 | 0.013314906 | 1 |
| Heart_Failure | ASPC | *HACE1* | 0.769115834 | 0.000286015 | 0.010420628 | 1 |
| Heart_Failure | ASPC | *SLC25A24* | 0.481301606 | 0.003490747 | 0.020305117 | 1 |
| Heart_Failure | ASPC | *RRN3* | 0.469256037 | 0.000984098 | 0.010420628 | 1 |
| Heart_Failure | ASPC | *MTIF2* | 0.851054605 | 0.001008353 | 0.010420628 | 1 |
| Heart_Failure | ASPC | *DDHD2* | 0.310668396 | 0.00040064 | 0.010420628 | 1 |
| Heart_Failure | ASPC | *USP40* | 0.619718991 | 0.001003571 | 0.010420628 | 1 |
| Heart_Failure | ASPC | *MAST2* | 0.489580454 | 7.74E-07 | 0.000284766 | 0.003755989 |
| Heart_Failure | ASPC | *EIF2AK1* | 0.709372612 | 0.000616574 | 0.010420628 | 1 |
| Heart_Failure | ASPC | *ALG9* | 1.275208159 | 0.001316121 | 0.011242641 | 1 |
| Heart_Failure | ASPC | *PSMC5* | 0.405607457 | 0.000974862 | 0.010420628 | 1 |
| Heart_Failure | ASPC | *UIMC1* | 0.479559568 | 0.00838365 | 0.037522597 | 1 |
| Heart_Failure | ASPC | *LPCAT2* | 0.838923086 | 0.000746832 | 0.010420628 | 1 |
| Heart_Failure | ASPC | *ASAP3* | 0.492600205 | 0.004691959 | 0.024660342 | 1 |
| Heart_Failure | ASPC | *DOCK3* | 0.622268389 | 0.002410245 | 0.015913671 | 1 |
| Heart_Failure | ASPC | *NSFL1C* | 0.253087853 | 0.002620756 | 0.0166221 | 1 |

| | | | | | | |
|---|---|---|---|---|---|---|
| Heart_Failure | ASPC | *MAVS* | 0.244760432 | 0.002047228 | 0.014312895 | 1 |
| Heart_Failure | ASPC | *SNX5* | 0.851964969 | 0.000743049 | 0.010420628 | 1 |
| Heart_Failure | ASPC | *RPH3A* | 0.69237938 | 0.010139908 | 0.043156872 | 1 |
| Heart_Failure | ASPC | *KIF16B* | 0.68272721 | 0.000949222 | 0.010420628 | 1 |
| Heart_Failure | ASPC | *FXYD5* | 0.461274047 | 0.000101405 | 0.008945801 | 0.492019043 |
| Heart_Failure | ASPC | *GRAMD1A* | 0.788828433 | 0.000566003 | 0.010420628 | 1 |
| Heart_Failure | ASPC | *CDIP1* | 1.047751411 | 0.001030612 | 0.010439519 | 1 |
| Heart_Failure | ASPC | *KCNH4* | 0.071880486 | 0.012134111 | 0.049355875 | 1 |
| Heart_Failure | ASPC | *ANKRD24* | 0.790766985 | 0.007132757 | 0.033141513 | 1 |
| Heart_Failure | ASPC | *RAB11FIP3* | 0.374966221 | 0.001619252 | 0.012295165 | 1 |
| Heart_Failure | ASPC | *PABPC4* | -0.201127856 | 0.003454654 | 0.02021952 | 1 |
| Heart_Failure | ASPC | *CERS4* | 0.705914504 | 0.002100732 | 0.014478344 | 1 |
| Heart_Failure | ASPC | *USP48* | 0.249401509 | 0.002769773 | 0.01731822 | 1 |
| Heart_Failure | ASPC | *GLG1* | 0.513791642 | 0.003246773 | 0.019353003 | 1 |
| Heart_Failure | ASPC | *DTX2* | 1.112487607 | 0.001001976 | 0.010420628 | 1 |
| Heart_Failure | ASPC | *TF* | 1.111231982 | 0.001244988 | 0.011023143 | 1 |
| Heart_Failure | ASPC | *PPP2R3C* | 0.225594594 | 0.006737833 | 0.032061327 | 1 |
| Heart_Failure | ASPC | *SCFD1* | 0.519466677 | 0.002453168 | 0.016041468 | 1 |
| Heart_Failure | ASPC | *SUPT16H* | 0.800959962 | 0.000745723 | 0.010420628 | 1 |
| Heart_Failure | ASPC | *COMT* | 0.69465184 | 0.001487503 | 0.011937289 | 1 |
| Heart_Failure | ASPC | *LRRFIP2* | 0.578657154 | 0.0003465 | 0.010420628 | 1 |
| Heart_Failure | ASPC | *SEC22C* | 0.778237999 | 0.001307344 | 0.01123051 | 1 |
| Heart_Failure | ASPC | *XYLB* | 0.639434078 | 0.006833486 | 0.03231586 | 1 |
| Heart_Failure | ASPC | *MSH2* | 0.840340893 | 0.000742044 | 0.010420628 | 1 |
| Heart_Failure | ASPC | *PSMD5* | 0.847526116 | 0.000179789 | 0.010420628 | 0.872337591 |
| Heart_Failure | ASPC | *CRAT* | 1.202179899 | 0.00206372 | 0.014386737 | 1 |
| Heart_Failure | ASPC | *WAC* | 0.504477985 | 0.001025363 | 0.010439519 | 1 |
| Heart_Failure | ASPC | *DSP* | 0.799911386 | 0.00470805 | 0.024660342 | 1 |
| Heart_Failure | ASPC | *RAB18* | 0.709619208 | 0.000925679 | 0.010420628 | 1 |
| Heart_Failure | ASPC | *TSPAN15* | 0.610214314 | 0.003680943 | 0.021030334 | 1 |
| Heart_Failure | ASPC | *SETD1A* | 0.482715674 | 1.07E-05 | 0.001922466 | 0.051906573 |
| Heart_Failure | ASPC | *MTAP* | 0.319846383 | 0.001289841 | 0.01121275 | 1 |
| Heart_Failure | ASPC | *SMARCB1* | 0.142503803 | 0.000652515 | 0.010420628 | 1 |
| Heart_Failure | ASPC | *CABIN1* | 0.347194174 | 0.007495226 | 0.03437319 | 1 |
| Heart_Failure | ASPC | *CYTH4* | 0.690760006 | 0.002488924 | 0.01610168 | 1 |
| Heart_Failure | ASPC | *SH3BP1* | 1.411261132 | 0.001506115 | 0.01196018 | 1 |
| Heart_Failure | ASPC | *TTC28* | 0.461516928 | 0.001208777 | 0.01091745 | 1 |
| Heart_Failure | ASPC | *TIMP3* | 0.487754403 | 0.00113407 | 0.010655995 | 1 |
| Heart_Failure | ASPC | *PPP6R2* | 0.473345379 | 0.00568816 | 0.028398201 | 1 |
| Heart_Failure | ASPC | *TSPO* | 0.818038648 | 0.000567015 | 0.010420628 | 1 |
| Heart_Failure | ASPC | *IFT27* | 0.688567891 | 0.00152273 | 0.011993971 | 1 |
| Heart_Failure | ASPC | *POLR3H* | 1.085995588 | 9.41E-06 | 0.001877595 | 0.045657459 |
| Heart_Failure | ASPC | *DESI1* | 0.437543658 | 0.001176283 | 0.010788896 | 1 |
| Heart_Failure | ASPC | *ATP6V1D* | 0.71033613 | 0.000810582 | 0.010420628 | 1 |
| Heart_Failure | ASPC | *MTHFD1* | 0.541504577 | 0.00169467 | 0.01262006 | 1 |
| Heart_Failure | ASPC | *GSKIP* | 1.188584796 | 0.000842376 | 0.010420628 | 1 |
| Heart_Failure | ASPC | *PSMB5* | 0.150435813 | 0.007932752 | 0.035856645 | 1 |
| Heart_Failure | ASPC | *YY1* | 0.721051614 | 0.000815609 | 0.010420628 | 1 |
| Heart_Failure | ASPC | *CCNB1IP1* | 0.880618836 | 0.001027628 | 0.010439519 | 1 |
| Heart_Failure | ASPC | *CINP* | 0.595888875 | 0.009871505 | 0.042386322 | 1 |

| | | | | | | |
|---|---|---|---|---|---|---|
| Heart_Failure | ASPC | *TRPC4AP* | 0.332961444 | 0.001863633 | 0.013317151 | 1 |
| Heart_Failure | ASPC | *NDRG3* | 0.456473175 | 0.012122408 | 0.049355875 | 1 |
| Heart_Failure | ASPC | *CSTF1* | 0.930875168 | 0.001065773 | 0.010475827 | 1 |
| Heart_Failure | ASPC | *NELFCD* | 0.434231018 | 0.011543046 | 0.047624879 | 1 |
| Heart_Failure | ASPC | *CTSZ* | 0.727083335 | 0.000805376 | 0.010420628 | 1 |
| Heart_Failure | ASPC | *MTG2* | 0.403608966 | 0.000664806 | 0.010420628 | 1 |
| Heart_Failure | ASPC | *DIDO1* | 0.532992607 | 9.76E-05 | 0.008855459 | 0.47338978 |
| Heart_Failure | ASPC | *METTL4* | 0.644224894 | 0.006962826 | 0.032704389 | 1 |
| Heart_Failure | ASPC | *LPIN2* | 0.455167697 | 0.003037825 | 0.018378462 | 1 |
| Heart_Failure | ASPC | *MYOM1* | 0.70839446 | 0.000883604 | 0.010420628 | 1 |
| Heart_Failure | ASPC | *RNF125* | 0.83218161 | 0.002619544 | 0.0166221 | 1 |
| Heart_Failure | ASPC | *MIB1* | 0.618236617 | 0.000859546 | 0.010420628 | 1 |
| Heart_Failure | ASPC | *STK24* | 0.633659948 | 0.000978176 | 0.010420628 | 1 |
| Heart_Failure | ASPC | *ARHGEF7* | 0.624620393 | 0.00074106 | 0.010420628 | 1 |
| Heart_Failure | ASPC | *PARP4* | 0.358161021 | 0.003281206 | 0.019481942 | 1 |
| Heart_Failure | ASPC | *SUPT20H* | 0.399117199 | 0.003309528 | 0.019558867 | 1 |
| Heart_Failure | ASPC | *MRPS31* | 0.65764293 | 0.001311423 | 0.01123051 | 1 |
| Heart_Failure | ASPC | *KATNAL1* | 0.582237863 | 0.002611762 | 0.0166221 | 1 |
| Heart_Failure | ASPC | *INTS6* | 0.414469625 | 0.005710961 | 0.028449262 | 1 |
| Heart_Failure | ASPC | *CLN5* | 0.719819639 | 0.000412672 | 0.010420628 | 1 |
| Heart_Failure | ASPC | *PSMD7* | 0.577258255 | 0.003259712 | 0.019406285 | 1 |
| Heart_Failure | ASPC | *MON1B* | 1.082565064 | 0.000661681 | 0.010420628 | 1 |
| Heart_Failure | ASPC | *LMF1* | 0.750817907 | 0.000673727 | 0.010420628 | 1 |
| Heart_Failure | ASPC | *UBE2I* | 0.712326965 | 0.000729223 | 0.010420628 | 1 |
| Heart_Failure | ASPC | *CPPED1* | 0.354070909 | 0.010976098 | 0.045648552 | 1 |
| Heart_Failure | ASPC | *TOX3* | 0.222120944 | 0.003225639 | 0.019274383 | 1 |
| Heart_Failure | ASPC | *RBL2* | 0.358108194 | 0.000969976 | 0.010420628 | 1 |
| Heart_Failure | ASPC | *PYCARD* | 0.453085194 | 0.000488542 | 0.010420628 | 1 |
| Heart_Failure | ASPC | *C16orf62* | 0.615187325 | 0.001367894 | 0.011518561 | 1 |
| Heart_Failure | ASPC | *RNF40* | 0.628944891 | 0.010569318 | 0.044515913 | 1 |
| Heart_Failure | ASPC | *KNOP1* | 0.527759889 | 0.007856511 | 0.035625974 | 1 |
| Heart_Failure | ASPC | *IQCH* | 1.012455878 | 2.14E-05 | 0.003039348 | 0.103721461 |
| Heart_Failure | ASPC | *HERC1* | 0.51197824 | 0.001125508 | 0.01064017 | 1 |
| Heart_Failure | ASPC | *CTSH* | -0.04664929 | 0.012316443 | 0.049841019 | 1 |
| Heart_Failure | ASPC | *TTC23* | 0.636877494 | 0.002496911 | 0.016131839 | 1 |
| Heart_Failure | ASPC | *SLC30A4* | 0.797225253 | 0.00133109 | 0.011310764 | 1 |
| Heart_Failure | ASPC | *NIPAL2* | 0.579539303 | 0.000462134 | 0.010420628 | 1 |
| Heart_Failure | ASPC | *CHRAC1* | 1.471898802 | 0.000848062 | 0.010420628 | 1 |
| Heart_Failure | ASPC | *GSDMD* | 0.393618956 | 0.003809121 | 0.02141582 | 1 |
| Heart_Failure | ASPC | *TUSC3* | 0.779767324 | 0.000789238 | 0.010420628 | 1 |
| Heart_Failure | ASPC | *SNRNP70* | 0.558356997 | 0.002912492 | 0.017797748 | 1 |
| Heart_Failure | ASPC | *MED25* | 1.006934864 | 0.000592264 | 0.010420628 | 1 |
| Heart_Failure | ASPC | *TIMM44* | 0.559409915 | 6.51E-06 | 0.001475161 | 0.031584006 |
| Heart_Failure | ASPC | *RASAL3* | 0.855912319 | 0.00657999 | 0.031516398 | 1 |
| Heart_Failure | ASPC | *ANKRD27* | 0.543048071 | 0.007531932 | 0.034476354 | 1 |
| Heart_Failure | ASPC | *TGFB1* | 0.727301229 | 0.000690836 | 0.010420628 | 1 |
| Heart_Failure | ASPC | *CAPS* | 1.394252611 | 0.000821859 | 0.010420628 | 1 |
| Heart_Failure | ASPC | *TNPO2* | 0.441381073 | 0.003370234 | 0.019845116 | 1 |
| Heart_Failure | ASPC | *RAB3A* | 4.657078174 | 0.000486433 | 0.010420628 | 1 |
| Heart_Failure | ASPC | *ZNF14* | 0.964277501 | 0.00086555 | 0.010420628 | 1 |

| | | | | | | |
|---|---|---|---|---|---|---|
| Heart_Failure | ASPC | *BET1* | 0.560181091 | 0.004185916 | 0.023001207 | 1 |
| Heart_Failure | ASPC | *DNAH11* | 0.920063069 | 0.001292075 | 0.01121275 | 1 |
| Heart_Failure | ASPC | *ADAP1* | 1.926233499 | 0.010068885 | 0.042967661 | 1 |
| Heart_Failure | ASPC | *IQCE* | 0.976265222 | 0.000631684 | 0.010420628 | 1 |
| Heart_Failure | ASPC | *HIBADH* | 0.26972565 | 0.002822447 | 0.017467492 | 1 |
| Heart_Failure | ASPC | *CPVL* | 0.580082557 | 0.000852856 | 0.010420628 | 1 |
| Heart_Failure | ASPC | *CHN2* | 0.323938113 | 0.000137042 | 0.010198142 | 0.664927776 |
| Heart_Failure | ASPC | *GRB10* | 0.566770136 | 0.002338025 | 0.015531293 | 1 |
| Heart_Failure | ASPC | *PLEKHA8* | 0.738344697 | 0.00126663 | 0.011113358 | 1 |
| Heart_Failure | ASPC | *ZKSCAN1* | 0.475533 | 0.004416588 | 0.023889952 | 1 |
| Heart_Failure | ASPC | *EIF3B* | 0.698948839 | 0.002177513 | 0.01484265 | 1 |
| Heart_Failure | ASPC | *C1GALT1* | 0.695895286 | 0.000658321 | 0.010420628 | 1 |
| Heart_Failure | ASPC | *COA1* | 0.477705614 | 0.00185785 | 0.013314906 | 1 |
| Heart_Failure | ASPC | *FKTN* | 0.733036199 | 0.000644899 | 0.010420628 | 1 |
| Heart_Failure | ASPC | *MEGF9* | 0.696048679 | 0.000716939 | 0.010420628 | 1 |
| Heart_Failure | ASPC | *C5* | 0.354186383 | 0.007589972 | 0.034651536 | 1 |
| Heart_Failure | ASPC | *ECM2* | 0.557042204 | 0.001006508 | 0.010420628 | 1 |
| Heart_Failure | ASPC | *TRDMT1* | 0.661253693 | 0.002084417 | 0.014427377 | 1 |
| Heart_Failure | ASPC | *ATE1* | 0.227939243 | 0.011687487 | 0.048016669 | 1 |
| Heart_Failure | ASPC | *CDH23* | 0.512432711 | 0.005309609 | 0.026976152 | 1 |
| Heart_Failure | ASPC | *BMPR1A* | 0.474865691 | 0.0037464 | 0.021185935 | 1 |
| Heart_Failure | ASPC | *SFXN3* | 1.080873703 | 0.000498239 | 0.010420628 | 1 |
| Heart_Failure | ASPC | *TNKS2* | 0.674150979 | 0.000951325 | 0.010420628 | 1 |
| Heart_Failure | ASPC | *LHPP* | 0.211623761 | 0.008894518 | 0.03923291 | 1 |
| Heart_Failure | ASPC | *PITRM1* | 0.579045466 | 0.000986661 | 0.010420628 | 1 |
| Heart_Failure | ASPC | *TFAM* | 0.971782032 | 0.000868079 | 0.010420628 | 1 |
| Heart_Failure | ASPC | *TRIM37* | 0.449789717 | 0.010614571 | 0.044629026 | 1 |
| Heart_Failure | ASPC | *PIGL* | 0.447580464 | 0.006672066 | 0.031831726 | 1 |
| Heart_Failure | ASPC | *MED13* | 0.368527923 | 0.004707268 | 0.024660342 | 1 |
| Heart_Failure | ASPC | *RAI1* | 0.857436584 | 0.00080496 | 0.010420628 | 1 |
| Heart_Failure | ASPC | *GOSR1* | 0.692190819 | 0.000794936 | 0.010420628 | 1 |
| Heart_Failure | ASPC | *ICAM2* | 0.827522439 | 0.001312386 | 0.01123051 | 1 |
| Heart_Failure | ASPC | *RAB5C* | 0.237169105 | 0.000804532 | 0.010420628 | 1 |
| Heart_Failure | ASPC | *NAGLU* | 0.501942481 | 0.000514575 | 0.010420628 | 1 |
| Heart_Failure | ASPC | *EFTUD2* | 0.840148528 | 0.000808308 | 0.010420628 | 1 |
| Heart_Failure | ASPC | *FAM20A* | 0.46925543 | 0.010056784 | 0.042953799 | 1 |
| Heart_Failure | ASPC | *DHRS7B* | 0.686056482 | 0.001107566 | 0.010614947 | 1 |
| Heart_Failure | ASPC | *MYH3* | 1.4103455 | 0.001860574 | 0.013314906 | 1 |
| Heart_Failure | ASPC | *SLAIN2* | 0.584862792 | 0.000448962 | 0.010420628 | 1 |
| Heart_Failure | ASPC | *OCIAD1* | 0.498547922 | 0.004337315 | 0.023645675 | 1 |
| Heart_Failure | ASPC | *UBE2D3* | 0.564979556 | 0.000784801 | 0.010420628 | 1 |
| Heart_Failure | ASPC | *NDUFC1* | 0.597751913 | 0.006124125 | 0.029803664 | 1 |
| Heart_Failure | ASPC | *KLHL2* | 0.606465499 | 0.001172435 | 0.010788896 | 1 |
| Heart_Failure | ASPC | *WFS1* | 1.005049765 | 0.001159748 | 0.010788896 | 1 |
| Heart_Failure | ASPC | *GRPEL1* | 1.084668081 | 0.001393373 | 0.011637563 | 1 |
| Heart_Failure | ASPC | *STIM2* | 0.562085903 | 0.001608084 | 0.012247119 | 1 |
| Heart_Failure | ASPC | *UFSP2* | 0.781278056 | 0.000946568 | 0.010420628 | 1 |
| Heart_Failure | ASPC | *KLHL5* | 0.690029648 | 0.000395008 | 0.010420628 | 1 |
| Heart_Failure | ASPC | *P2RX3* | 6.492479513 | 0.00692137 | 0.032572731 | 1 |
| Heart_Failure | ASPC | *CEP164* | 0.432687876 | 0.009755172 | 0.042147901 | 1 |

| | | | | | | |
|---|---|---|---|---|---|---|
| Heart_Failure | ASPC | *SLC35F2* | 0.541243992 | 0.008482419 | 0.037827846 | 1 |
| Heart_Failure | ASPC | *PRDM4* | 0.511971124 | 1.31E-05 | 0.002187393 | 0.063434395 |
| Heart_Failure | ASPC | *CORO1C* | 0.703324866 | 0.000744367 | 0.010420628 | 1 |
| Heart_Failure | ASPC | *PARP11* | 0.765721124 | 0.000740846 | 0.010420628 | 1 |
| Heart_Failure | ASPC | *GPN3* | 1.028226568 | 0.001291902 | 0.01121275 | 1 |
| Heart_Failure | ASPC | *VPS29* | 0.631702311 | 0.001198547 | 0.010869813 | 1 |
| Heart_Failure | ASPC | *CDK2AP1* | 0.202841178 | 0.000211579 | 0.010420628 | 1 |
| Heart_Failure | ASPC | *VDR* | 1.596725357 | 0.001508649 | 0.011960724 | 1 |
| Heart_Failure | ASPC | *CAND1* | 0.680190621 | 0.000971927 | 0.010420628 | 1 |
| Heart_Failure | ASPC | *MDM1* | 0.839764197 | 0.001223105 | 0.010929107 | 1 |
| Heart_Failure | ASPC | *KRR1* | 0.444920075 | 0.005769496 | 0.028605723 | 1 |
| Heart_Failure | ASPC | *SUDS3* | 0.789103256 | 0.000969887 | 0.010420628 | 1 |
| Heart_Failure | ASPC | *C2CD5* | 0.615778148 | 0.001111898 | 0.010619941 | 1 |
| Heart_Failure | ASPC | *FRK* | 0.927486145 | 0.001242056 | 0.011023143 | 1 |
| Heart_Failure | ASPC | *TMEM14C* | 0.897692727 | 0.000709399 | 0.010420628 | 1 |
| Heart_Failure | ASPC | *RNGTT* | 0.630035322 | 0.001297668 | 0.011223323 | 1 |
| Heart_Failure | ASPC | *VNN2* | 0.182945184 | 0.009474457 | 0.041044701 | 1 |
| Heart_Failure | ASPC | *ACOT13* | 0.811109808 | 0.00084385 | 0.010420628 | 1 |
| Heart_Failure | ASPC | *FIG4* | 0.59673729 | 0.001400545 | 0.011637563 | 1 |
| Heart_Failure | ASPC | *PACRG* | 0.887682711 | 0.001179631 | 0.010799188 | 1 |
| Heart_Failure | ASPC | *QKI* | 0.509087657 | 0.000816423 | 0.010420628 | 1 |
| Heart_Failure | ASPC | *FAM120B* | 0.354225032 | 0.000285826 | 0.010420628 | 1 |
| Heart_Failure | ASPC | *EXOC2* | 0.594450726 | 0.000676757 | 0.010420628 | 1 |
| Heart_Failure | ASPC | *TMEM30A* | 0.754650497 | 0.002083022 | 0.014427377 | 1 |
| Heart_Failure | ASPC | *PRSS16* | 0.17892805 | 0.009419742 | 0.040880669 | 1 |
| Heart_Failure | ASPC | *NUDT12* | 0.884915802 | 0.001627119 | 0.01232502 | 1 |
| Heart_Failure | ASPC | *KIF20A* | -0.872889186 | 6.88E-05 | 0.007116983 | 0.33373349 |
| Heart_Failure | ASPC | *ARSB* | 0.624458795 | 0.004933186 | 0.025524454 | 1 |
| Heart_Failure | ASPC | *SLC12A7* | 0.789697985 | 0.004506794 | 0.024118544 | 1 |
| Heart_Failure | ASPC | *SKP1* | 0.671074123 | 0.002031229 | 0.014258141 | 1 |
| Heart_Failure | ASPC | *PPWD1* | 0.683600374 | 0.001044888 | 0.010475827 | 1 |
| Heart_Failure | ASPC | *TXNDC15* | 0.695312753 | 0.00459959 | 0.024395914 | 1 |
| Heart_Failure | ASPC | *SMAD5* | 0.581793032 | 0.00338003 | 0.019854605 | 1 |
| Heart_Failure | ASPC | *ERGIC1* | 0.625239689 | 0.002305056 | 0.015383948 | 1 |
| Heart_Failure | ASPC | *ATP6V0E1* | 0.60219888 | 0.000240607 | 0.010420628 | 1 |
| Heart_Failure | ASPC | *BNIP1* | 0.091745843 | 0.003684331 | 0.021030334 | 1 |
| Heart_Failure | ASPC | *UNC5A* | 2.192033943 | 0.010178749 | 0.043284215 | 1 |
| Heart_Failure | ASPC | *TBCCD1* | 0.983625892 | 0.001186855 | 0.01084486 | 1 |
| Heart_Failure | ASPC | *PCCB* | 0.153326864 | 0.007715758 | 0.035086089 | 1 |
| Heart_Failure | ASPC | *IQCG* | 0.585654139 | 0.00603477 | 0.029487112 | 1 |
| Heart_Failure | ASPC | *PLSCR4* | 0.374203738 | 0.004508557 | 0.024118544 | 1 |
| Heart_Failure | ASPC | *COMMD2* | 0.491877976 | 0.012289504 | 0.0498151 | 1 |
| Heart_Failure | ASPC | *ARHGEF26* | 0.75637909 | 0.001747229 | 0.012805971 | 1 |
| Heart_Failure | ASPC | *ACTR1B* | 0.586641199 | 0.007094093 | 0.033091794 | 1 |
| Heart_Failure | ASPC | *INO80B* | -0.159135273 | 0.004982263 | 0.025678324 | 1 |
| Heart_Failure | ASPC | *MOGS* | 0.126783154 | 0.005493173 | 0.027648213 | 1 |
| Heart_Failure | ASPC | *GRB14* | 0.639257969 | 0.005433344 | 0.027403933 | 1 |
| Heart_Failure | ASPC | *GALNT3* | 0.266667322 | 0.005927865 | 0.029081901 | 1 |
| Heart_Failure | ASPC | *MRPL19* | 0.476889362 | 0.007644816 | 0.034796106 | 1 |
| Heart_Failure | ASPC | *IL18R1* | 0.448277316 | 1.00E-07 | 4.42E-05 | 0.000486382 |

| | | | | | | |
|---|---|---|---|---|---|---|
| Heart_Failure | ASPC | *HDLBP* | 0.464877482 | 0.000149126 | 0.010420628 | 0.723559563 |
| Heart_Failure | ASPC | *TAF1B* | 0.412920075 | 0.000177688 | 0.010420628 | 0.862140667 |
| Heart_Failure | ASPC | *NOL10* | 0.554251923 | 0.00351738 | 0.020365545 | 1 |
| Heart_Failure | ASPC | *STRN* | 0.677118501 | 0.000509089 | 0.010420628 | 1 |
| Heart_Failure | ASPC | *CEBPZ* | 0.597716784 | 0.000337569 | 0.010420628 | 1 |
| Heart_Failure | ASPC | *PRKD3* | 0.445629427 | 0.002178064 | 0.01484265 | 1 |
| Heart_Failure | ASPC | *DARS* | 0.685826166 | 0.00077632 | 0.010420628 | 1 |
| Heart_Failure | ASPC | *WIPF1* | 0.564576726 | 0.000924815 | 0.010420628 | 1 |
| Heart_Failure | ASPC | *PNO1* | 1.146898772 | 0.000963997 | 0.010420628 | 1 |
| Heart_Failure | ASPC | *AAK1* | 0.560373576 | 0.000745567 | 0.010420628 | 1 |
| Heart_Failure | ASPC | *NFE2L2* | 0.264130418 | 0.008082249 | 0.036285092 | 1 |
| Heart_Failure | ASPC | *ALMS1* | 0.741919181 | 0.000428447 | 0.010420628 | 1 |
| Heart_Failure | ASPC | *EDEM3* | 0.557798964 | 0.001821247 | 0.013208806 | 1 |
| Heart_Failure | ASPC | *RAP1A* | 0.433288254 | 0.002853432 | 0.017592516 | 1 |
| Heart_Failure | ASPC | *CAPZA1* | 0.365229641 | 0.006892268 | 0.032530433 | 1 |
| Heart_Failure | ASPC | *DOCK7* | 0.511246022 | 0.004349815 | 0.023685181 | 1 |
| Heart_Failure | ASPC | *LEPR* | 0.281901019 | 0.011564629 | 0.047673389 | 1 |
| Heart_Failure | ASPC | *PRG4* | 0.631750483 | 0.009142778 | 0.040072951 | 1 |
| Heart_Failure | ASPC | *GNPAT* | 0.44441787 | 0.001900055 | 0.013537544 | 1 |
| Heart_Failure | ASPC | *TBCE* | 0.711699969 | 0.002610121 | 0.0166221 | 1 |
| Heart_Failure | ASPC | *ERI3* | 0.646130097 | 0.000425278 | 0.010420628 | 1 |
| Heart_Failure | ASPC | *ABCD3* | 0.687120383 | 0.001529158 | 0.012017141 | 1 |
| Heart_Failure | ASPC | *MARC2* | 0.505100324 | 0.007009396 | 0.032859506 | 1 |
| Heart_Failure | ASPC | *KLF7* | 0.422248732 | 0.0017935 | 0.013061689 | 1 |
| Heart_Failure | ASPC | *SGIP1* | 0.876446174 | 0.000910098 | 0.010420628 | 1 |
| Heart_Failure | ASPC | *AKAP7* | 0.422319203 | 0.001000967 | 0.010420628 | 1 |
| Heart_Failure | ASPC | *MED28* | 0.367962461 | 0.000202874 | 0.010420628 | 0.984344281 |
| Heart_Failure | ASPC | *FOXO3* | 0.495128892 | 0.000764533 | 0.010420628 | 1 |
| Heart_Failure | ASPC | *MFSD1* | 0.502226035 | 0.009237185 | 0.040340974 | 1 |
| Heart_Failure | ASPC | *UBN1* | 0.484987191 | 0.007082642 | 0.033091794 | 1 |
| Heart_Failure | ASPC | *SENP5* | 0.601266783 | 0.001738866 | 0.012792849 | 1 |
| Heart_Failure | ASPC | *FKBP15* | 0.142697102 | 0.005258218 | 0.026799239 | 1 |
| Heart_Failure | ASPC | *CTNNAL1* | 0.125008879 | 0.000276153 | 0.010420628 | 1 |
| Heart_Failure | ASPC | *TRIM32* | 0.705814468 | 0.006230757 | 0.030231633 | 1 |
| Heart_Failure | ASPC | *PPP6C* | 0.731580009 | 0.000677602 | 0.010420628 | 1 |
| Heart_Failure | ASPC | *MAPKAP1* | 0.608517347 | 0.000306764 | 0.010420628 | 1 |
| Heart_Failure | ASPC | *DENND1A* | 0.542789629 | 0.000393944 | 0.010420628 | 1 |
| Heart_Failure | ASPC | *NEK9* | 0.418360728 | 0.004725629 | 0.024660342 | 1 |
| Heart_Failure | ASPC | *TTLL5* | 0.511891752 | 0.0048754 | 0.025299939 | 1 |
| Heart_Failure | ASPC | *CRIPT* | 0.928204768 | 0.000828502 | 0.010420628 | 1 |
| Heart_Failure | ASPC | *HOXB3* | 0.564477283 | 0.004706512 | 0.024660342 | 1 |
| Heart_Failure | ASPC | *GORAB* | 0.543305661 | 0.00573005 | 0.028485864 | 1 |
| Heart_Failure | ASPC | *SLC10A7* | 0.695140726 | 0.000613253 | 0.010420628 | 1 |
| Heart_Failure | ASPC | *IQSEC3* | 1.057280402 | 0.000232131 | 0.010420628 | 1 |
| Heart_Failure | ASPC | *DNAJC15* | 0.545153685 | 0.002482309 | 0.016080322 | 1 |
| Heart_Failure | ASPC | *SIL1* | 0.553477431 | 0.000918394 | 0.010420628 | 1 |
| Heart_Failure | ASPC | *ZNF549* | 0.885893528 | 0.00053918 | 0.010420628 | 1 |
| Heart_Failure | ASPC | *LHX4* | 1.171377539 | 0.000941595 | 0.010420628 | 1 |
| Heart_Failure | ASPC | *SEC22A* | 0.550340418 | 0.003244538 | 0.019353003 | 1 |
| Heart_Failure | ASPC | *ZCCHC17* | 0.81839797 | 0.000794374 | 0.010420628 | 1 |

| | | | | | | |
|---|---|---|---|---|---|---|
| Heart_Failure | ASPC | *TNFSF10* | 0.500237666 | 0.006262329 | 0.030293937 | 1 |
| Heart_Failure | ASPC | *RBBP6* | 0.48467303 | 0.004993354 | 0.025684352 | 1 |
| Heart_Failure | ASPC | *NAA60* | 1.309353379 | 0.00079 | 0.010420628 | 1 |
| Heart_Failure | ASPC | *ODF2L* | 0.600953442 | 0.001378832 | 0.011574551 | 1 |
| Heart_Failure | ASPC | *SPATA1* | 0.499042121 | 0.006786488 | 0.03215629 | 1 |
| Heart_Failure | ASPC | *KLHL7* | 0.652781187 | 0.001565441 | 0.012135784 | 1 |
| Heart_Failure | ASPC | *NT5C3A* | 0.559843692 | 0.001501776 | 0.01196018 | 1 |
| Heart_Failure | ASPC | *SMU1* | 0.663983936 | 0.000253382 | 0.010420628 | 1 |
| Heart_Failure | ASPC | *RBM19* | 0.808686871 | 0.001060458 | 0.010475827 | 1 |
| Heart_Failure | ASPC | *CCDC91* | 0.457424355 | 0.001542568 | 0.012034028 | 1 |
| Heart_Failure | ASPC | *ITIH5* | -0.108812683 | 0.003772042 | 0.021256616 | 1 |
| Heart_Failure | ASPC | *PFDN5* | 0.610570432 | 0.002814482 | 0.017446383 | 1 |
| Heart_Failure | ASPC | *PDE1B* | 1.047373109 | 0.000774322 | 0.010420628 | 1 |
| Heart_Failure | ASPC | *CDK2* | 0.157108588 | 0.005689005 | 0.028398201 | 1 |
| Heart_Failure | ASPC | *USP45* | 0.584216107 | 0.001540607 | 0.012034028 | 1 |
| Heart_Failure | ASPC | *PLA2G12A* | 0.60350425 | 0.00066578 | 0.010420628 | 1 |
| Heart_Failure | ASPC | *DNPEP* | 0.486034844 | 0.004469441 | 0.024015204 | 1 |
| Heart_Failure | ASPC | *CSE1L* | 0.42034843 | 0.001591296 | 0.012207591 | 1 |
| Heart_Failure | ASPC | *RNF114* | 0.470626418 | 0.002871576 | 0.017614271 | 1 |
| Heart_Failure | ASPC | *PEPD* | 0.714821412 | 0.000303735 | 0.010420628 | 1 |
| Heart_Failure | ASPC | *STAMBP* | 0.770751673 | 0.000990004 | 0.010420628 | 1 |
| Heart_Failure | ASPC | *SNRNP27* | 0.935240868 | 0.00065828 | 0.010420628 | 1 |
| Heart_Failure | ASPC | *ATP8A1* | 0.580197758 | 0.000605932 | 0.010420628 | 1 |
| Heart_Failure | ASPC | *MRS2* | 0.127488512 | 0.010365985 | 0.043849833 | 1 |
| Heart_Failure | ASPC | *NQO2* | 0.750117103 | 0.001061905 | 0.010475827 | 1 |
| Heart_Failure | ASPC | *GLO1* | 0.582589575 | 1.55E-05 | 0.002428369 | 0.075279428 |
| Heart_Failure | ASPC | *ATXN1* | 0.38718168 | 0.002105568 | 0.014491089 | 1 |
| Heart_Failure | ASPC | *RUNX2* | 0.430165394 | 0.0071526 | 0.033178215 | 1 |
| Heart_Failure | ASPC | *CXCL6* | 0.905083422 | 0.00614749 | 0.029857479 | 1 |
| Heart_Failure | ASPC | *CLYBL* | 0.607493566 | 0.003389935 | 0.019888709 | 1 |
| Heart_Failure | ASPC | *TMTC4* | 0.541299427 | 0.004562484 | 0.024299861 | 1 |
| Heart_Failure | ASPC | *MRPS7* | 1.022016508 | 0.00011799 | 0.009385075 | 0.572489561 |
| Heart_Failure | ASPC | *NT5C* | 0.62193677 | 0.010188741 | 0.043288767 | 1 |
| Heart_Failure | ASPC | *OPRL1* | 1.481908741 | 0.009650812 | 0.041771401 | 1 |
| Heart_Failure | ASPC | *PPDPF* | 0.235136846 | 0.0038942 | 0.02181831 | 1 |
| Heart_Failure | ASPC | *SNRPD2* | 0.993495222 | 0.000766937 | 0.010420628 | 1 |
| Heart_Failure | ASPC | *FLRT3* | 1.044536316 | 0.000470898 | 0.010420628 | 1 |
| Heart_Failure | ASPC | *KLC1* | 0.72459161 | 0.000560029 | 0.010420628 | 1 |
| Heart_Failure | ASPC | *TUBGCP3* | 0.594381373 | 0.001593643 | 0.012207591 | 1 |
| Heart_Failure | ASPC | *COX6B1* | 0.536946357 | 0.003712913 | 0.021094911 | 1 |
| Heart_Failure | ASPC | *PRDX5* | 0.378049966 | 0.006376699 | 0.03072467 | 1 |
| Heart_Failure | ASPC | *KTN1* | 0.578373118 | 0.000734111 | 0.010420628 | 1 |
| Heart_Failure | ASPC | *ZBTB1* | 0.346202954 | 0.008955564 | 0.039349527 | 1 |
| Heart_Failure | ASPC | *RHOT1* | 0.200256953 | 0.006707897 | 0.031971234 | 1 |
| Heart_Failure | ASPC | *CANX* | 0.54048021 | 0.001040416 | 0.010473232 | 1 |
| Heart_Failure | ASPC | *RAB3IP* | 0.588172507 | 0.012057608 | 0.049162618 | 1 |
| Heart_Failure | ASPC | *EMC1* | 0.485800606 | 0.00115551 | 0.010781793 | 1 |
| Heart_Failure | ASPC | *RNF6* | 0.769027639 | 0.000859972 | 0.010420628 | 1 |
| Heart_Failure | ASPC | *HIP1* | 0.580449927 | 0.000520814 | 0.010420628 | 1 |
| Heart_Failure | ASPC | *PTPN12* | 0.359294774 | 0.006902746 | 0.032548092 | 1 |

| | | | | | | |
|---|---|---|---|---|---|---|
| Heart_Failure | ASPC | *PEX1* | 0.767642929 | 0.001078101 | 0.010475827 | 1 |
| Heart_Failure | ASPC | *KDR* | 0.629950607 | 0.002417762 | 0.015913671 | 1 |
| Heart_Failure | ASPC | *ATF4* | 0.789900832 | 0.004605659 | 0.024395914 | 1 |
| Heart_Failure | ASPC | *SPECC1* | 0.460273545 | 0.00722276 | 0.033439726 | 1 |
| Heart_Failure | ASPC | *CDHR3* | 0.67766872 | 0.001424701 | 0.01166347 | 1 |
| Heart_Failure | ASPC | *PRKRIP1* | 0.462428145 | 0.003291907 | 0.01950224 | 1 |
| Heart_Failure | ASPC | *MTX2* | 0.693645558 | 0.00049253 | 0.010420628 | 1 |
| Heart_Failure | ASPC | *TWSG1* | 0.666485266 | 0.000786044 | 0.010420628 | 1 |
| Heart_Failure | ASPC | *ARPP19* | 0.806375701 | 0.000766832 | 0.010420628 | 1 |
| Heart_Failure | ASPC | *BBOX1* | 3.733387764 | 0.00066053 | 0.010420628 | 1 |
| Heart_Failure | ASPC | *PLD2* | 1.064512856 | 0.001923854 | 0.013646985 | 1 |
| Heart_Failure | ASPC | *PHF20L1* | 0.459443485 | 0.001916041 | 0.013611465 | 1 |
| Heart_Failure | ASPC | *MIS18BP1* | 0.223029191 | 0.010926522 | 0.04558511 | 1 |
| Heart_Failure | ASPC | *RNASE1* | 0.27978887 | 0.011342578 | 0.046877503 | 1 |
| Heart_Failure | ASPC | *EPB41L4A* | 0.559835295 | 0.001077115 | 0.010475827 | 1 |
| Heart_Failure | ASPC | *REEP5* | 0.676112018 | 0.000850258 | 0.010420628 | 1 |
| Heart_Failure | ASPC | *TMEM8A* | 0.814979607 | 0.000971777 | 0.010420628 | 1 |
| Heart_Failure | ASPC | *PHF10* | 0.546952555 | 0.009834939 | 0.04230419 | 1 |
| Heart_Failure | ASPC | *XPO7* | 0.576527038 | 0.003379518 | 0.019854605 | 1 |
| Heart_Failure | ASPC | *TULP4* | 0.236559359 | 0.005492597 | 0.027648213 | 1 |
| Heart_Failure | ASPC | *ACTN4* | 0.519997056 | 0.000571101 | 0.010420628 | 1 |
| Heart_Failure | ASPC | *MAP1S* | 1.340923023 | 0.000742031 | 0.010420628 | 1 |
| Heart_Failure | ASPC | *ATXN10* | 0.551050476 | 0.001567891 | 0.012135784 | 1 |
| Heart_Failure | ASPC | *PNPLA7* | 0.645935063 | 0.003128164 | 0.018784468 | 1 |
| Heart_Failure | ASPC | *ANGPTL6* | -0.133438174 | 0.01119724 | 0.046355808 | 1 |
| Heart_Failure | ASPC | *DNMT1* | 0.865122246 | 0.001006217 | 0.010420628 | 1 |
| Heart_Failure | ASPC | *C12orf65* | 0.250056855 | 0.000854836 | 0.010420628 | 1 |
| Heart_Failure | ASPC | *SLC35D2* | 0.854874598 | 0.000105354 | 0.009128154 | 0.511176629 |
| Heart_Failure | ASPC | *CHMP1A* | 0.839753362 | 7.82E-05 | 0.007591573 | 0.379578638 |
| Heart_Failure | ASPC | *IDO1* | 0.998812493 | 0.007983841 | 0.036001484 | 1 |
| Heart_Failure | ASPC | *MRPS25* | 0.350688747 | 0.004723992 | 0.024660342 | 1 |
| Heart_Failure | ASPC | *GALNT15* | 0.596292421 | 0.007936941 | 0.035856645 | 1 |
| Heart_Failure | ASPC | *PDLIM4* | 0.877222345 | 0.0004799 | 0.010420628 | 1 |
| Heart_Failure | ASPC | *KIF3A* | 0.67773682 | 0.000181354 | 0.010420628 | 0.879929178 |
| Heart_Failure | ASPC | *NDUFA2* | 0.174958139 | 0.012216669 | 0.04956127 | 1 |
| Heart_Failure | ASPC | *EXOC4* | 0.349972554 | 0.001132959 | 0.010655995 | 1 |
| Heart_Failure | ASPC | *C1orf159* | 0.655581637 | 0.008831943 | 0.039063435 | 1 |
| Heart_Failure | ASPC | *PPFIA1* | 0.379302695 | 0.00427464 | 0.023435654 | 1 |
| Heart_Failure | ASPC | *MAP1B* | 0.22536417 | 0.001325008 | 0.011278839 | 1 |
| Heart_Failure | ASPC | *KHDRBS3* | 0.862879395 | 0.000775746 | 0.010420628 | 1 |
| Heart_Failure | ASPC | *TRIM21* | 0.661978175 | 0.003302195 | 0.019539329 | 1 |
| Heart_Failure | ASPC | *AP3B1* | 0.57741429 | 0.00094866 | 0.010420628 | 1 |
| Heart_Failure | ASPC | *ALOX5AP* | 0.620746923 | 0.003644265 | 0.020876001 | 1 |
| Heart_Failure | ASPC | *WASF3* | 0.606519595 | 0.003076575 | 0.018520523 | 1 |
| Heart_Failure | ASPC | *PEMT* | 1.068437005 | 0.000782724 | 0.010420628 | 1 |
| Heart_Failure | ASPC | *MPRIP* | 0.658959665 | 0.000999381 | 0.010420628 | 1 |
| Heart_Failure | ASPC | *DSTYK* | 0.762331067 | 0.000699808 | 0.010420628 | 1 |
| Heart_Failure | ASPC | *COG6* | 0.284309522 | 0.010939392 | 0.045599597 | 1 |
| Heart_Failure | ASPC | *EPSTI1* | 0.543317149 | 0.0006477 | 0.010420628 | 1 |
| Heart_Failure | ASPC | *BTBD2* | 0.474437953 | 0.005820003 | 0.028785579 | 1 |

| | | | | | | |
|---|---|---|---|---|---|---|
| Heart_Failure | ASPC | *CSNK1G2* | 0.959935621 | 0.000671352 | 0.010420628 | 1 |
| Heart_Failure | ASPC | *PDZD2* | 0.521411231 | 0.000848561 | 0.010420628 | 1 |
| Heart_Failure | ASPC | *FAM83F* | 0.937968825 | 0.003595459 | 0.020694146 | 1 |
| Heart_Failure | ASPC | *IPO8* | 0.448414179 | 0.009195243 | 0.040230225 | 1 |
| Heart_Failure | ASPC | *RRAS2* | 0.255223681 | 0.009793217 | 0.042206908 | 1 |
| Heart_Failure | ASPC | *TTC9* | 0.8969524 | 0.004446962 | 0.023969816 | 1 |
| Heart_Failure | ASPC | *ELP3* | 0.529649556 | 0.007088139 | 0.033091794 | 1 |
| Heart_Failure | ASPC | *CEPT1* | 0.495559272 | 0.002069551 | 0.01439948 | 1 |
| Heart_Failure | ASPC | *PPHLN1* | 0.450307683 | 0.003653311 | 0.020903145 | 1 |
| Heart_Failure | ASPC | *KIDINS220* | 0.56294956 | 0.001416663 | 0.011650251 | 1 |
| Heart_Failure | ASPC | *KIAA1468* | 0.628298735 | 0.001212106 | 0.01091745 | 1 |
| Heart_Failure | ASPC | *IL15RA* | 0.369549788 | 0.00236035 | 0.015645382 | 1 |
| Heart_Failure | ASPC | *KLRD1* | 0.778394948 | 0.000570257 | 0.010420628 | 1 |
| Heart_Failure | ASPC | *ELP2* | 0.636547158 | 0.001410198 | 0.011637563 | 1 |
| Heart_Failure | ASPC | *TPGS2* | 0.429492143 | 0.00892091 | 0.039291363 | 1 |
| Heart_Failure | ASPC | *TIMM10* | 0.383396948 | 0.002954913 | 0.018011603 | 1 |
| Heart_Failure | ASPC | *DHX34* | 0.596419201 | 0.011989023 | 0.048924086 | 1 |
| Heart_Failure | ASPC | *CLOCK* | 0.630041422 | 0.000924185 | 0.010420628 | 1 |
| Heart_Failure | ASPC | *ARGLU1* | 0.531539864 | 0.000465735 | 0.010420628 | 1 |
| Heart_Failure | ASPC | *ERCC5* | 0.325186928 | 0.003987115 | 0.022156417 | 1 |
| Heart_Failure | ASPC | *OSTF1* | 0.573452813 | 0.005194584 | 0.026532412 | 1 |
| Heart_Failure | ASPC | *UBQLN1* | 0.579238607 | 0.002158865 | 0.014774066 | 1 |
| Heart_Failure | ASPC | *ADAM19* | 0.119902504 | 0.002219074 | 0.01501666 | 1 |
| Heart_Failure | ASPC | *AKIRIN2* | 0.416823847 | 0.007268121 | 0.03361766 | 1 |
| Heart_Failure | ASPC | *MAP3K7* | 0.543168751 | 0.004780989 | 0.02488987 | 1 |
| Heart_Failure | ASPC | *PHF21A* | 0.540975771 | 0.001146587 | 0.010719151 | 1 |
| Heart_Failure | ASPC | *USP15* | 0.6135813 | 0.000159143 | 0.010420628 | 0.772159732 |
| Heart_Failure | ASPC | *MPHOSPH6* | 0.556590931 | 0.011644145 | 0.047923908 | 1 |
| Heart_Failure | ASPC | *NTPCR* | 0.557638418 | 0.009794925 | 0.042206908 | 1 |
| Heart_Failure | ASPC | *PIGC* | 0.614051323 | 0.002477335 | 0.016080322 | 1 |
| Heart_Failure | ASPC | *MRPL44* | 1.185779362 | 6.69E-06 | 0.001475161 | 0.032453541 |
| Heart_Failure | ASPC | *USP37* | 0.343947265 | 0.000585583 | 0.010420628 | 1 |
| Heart_Failure | ASPC | *SLC19A3* | 0.456429303 | 0.000158819 | 0.010420628 | 0.770588537 |
| Heart_Failure | ASPC | *TMBIM1* | 0.815531914 | 0.000794723 | 0.010420628 | 1 |
| Heart_Failure | ASPC | *ANKRD36* | 0.535277291 | 0.006273304 | 0.030316804 | 1 |
| Heart_Failure | ASPC | *EPC2* | 0.52465494 | 0.001217563 | 0.010920212 | 1 |
| Heart_Failure | ASPC | *STAB2* | 0.722816677 | 0.002629051 | 0.016652942 | 1 |
| Heart_Failure | ASPC | *DRAM1* | 0.699295149 | 0.001705972 | 0.012675919 | 1 |
| Heart_Failure | ASPC | *SUCLA2* | 0.722336358 | 0.000833319 | 0.010420628 | 1 |
| Heart_Failure | ASPC | *RCBTB2* | 0.783956267 | 0.000720378 | 0.010420628 | 1 |
| Heart_Failure | ASPC | *SETDB2* | 0.654436592 | 0.000688804 | 0.010420628 | 1 |
| Heart_Failure | ASPC | *KDELR2* | 0.910822007 | 0.00119288 | 0.010859009 | 1 |
| Heart_Failure | ASPC | *NUPL2* | 0.74840732 | 0.000919092 | 0.010420628 | 1 |
| Heart_Failure | ASPC | *ZDHHC4* | 0.70244427 | 0.000636637 | 0.010420628 | 1 |
| Heart_Failure | ASPC | *NMT1* | 0.16101439 | 9.26E-05 | 0.008642034 | 0.449385766 |
| Heart_Failure | ASPC | *ZNF189* | 0.973346074 | 0.000688138 | 0.010420628 | 1 |
| Heart_Failure | ASPC | *STX17* | 0.580723364 | 0.003510281 | 0.020350525 | 1 |
| Heart_Failure | ASPC | *APTX* | 0.430595703 | 0.008961191 | 0.039349527 | 1 |
| Heart_Failure | ASPC | *DENND4C* | 0.349358179 | 0.011505208 | 0.047509166 | 1 |
| Heart_Failure | ASPC | *TCF19* | 0.171237268 | 0.001076586 | 0.010475827 | 1 |

| | | | | | | |
|---|---|---|---|---|---|---|
| Heart_Failure | ASPC | *FLOT1* | 0.18233257 | 0.009407242 | 0.040862971 | 1 |
| Heart_Failure | ASPC | *NUMA1* | 0.566327977 | 0.00278569 | 0.017363935 | 1 |
| Heart_Failure | ASPC | *NARS2* | 0.705345791 | 0.00088779 | 0.010420628 | 1 |
| Heart_Failure | ASPC | *TGS1* | 0.720262006 | 0.000249151 | 0.010420628 | 1 |
| Heart_Failure | ASPC | *NEK1* | 0.373092299 | 0.009436891 | 0.040918496 | 1 |
| Heart_Failure | ASPC | *MAP2K5* | 0.634448321 | 9.86E-05 | 0.008855459 | 0.478194794 |
| Heart_Failure | ASPC | *PARP6* | 0.673083913 | 0.001733695 | 0.012792849 | 1 |
| Heart_Failure | ASPC | *ADAM10* | 0.643018052 | 3.86E-06 | 0.001040634 | 0.018731414 |
| Heart_Failure | ASPC | *ZNF280D* | 0.548689171 | 0.00054228 | 0.010420628 | 1 |
| Heart_Failure | ASPC | *DBT* | 0.495994517 | 0.011987386 | 0.048924086 | 1 |
| Heart_Failure | ASPC | *ACTR2* | 0.666409303 | 0.000442999 | 0.010420628 | 1 |
| Heart_Failure | ASPC | *LRPPRC* | 0.617434823 | 0.001431615 | 0.01166347 | 1 |
| Heart_Failure | ASPC | *SMARCAL1* | 0.549719984 | 0.011215054 | 0.046389974 | 1 |
| Heart_Failure | ASPC | *FASTKD1* | 0.405592825 | 0.000513025 | 0.010420628 | 1 |
| Heart_Failure | ASPC | *SLC35A5* | 0.235441527 | 0.006431475 | 0.030927174 | 1 |
| Heart_Failure | ASPC | *COX17* | 1.044125862 | 0.000825497 | 0.010420628 | 1 |
| Heart_Failure | ASPC | *AP1AR* | 0.858144034 | 0.000148561 | 0.010420628 | 0.720820171 |
| Heart_Failure | ASPC | *KIAA1109* | 0.555639731 | 0.001126175 | 0.01064017 | 1 |
| Heart_Failure | ASPC | *PRDM5* | 0.69394681 | 0.001856999 | 0.013314906 | 1 |
| Heart_Failure | ASPC | *G3BP2* | 0.23505323 | 0.001127174 | 0.01064017 | 1 |
| Heart_Failure | ASPC | *PPA2* | 0.332649902 | 0.010482638 | 0.044227615 | 1 |
| Heart_Failure | ASPC | *PAPSS1* | 0.338228206 | 0.005851175 | 0.028851527 | 1 |
| Heart_Failure | ASPC | *KIF21A* | 0.624119299 | 0.00094686 | 0.010420628 | 1 |
| Heart_Failure | ASPC | *ALG10* | -0.18451521 | 0.005673617 | 0.028379782 | 1 |
| Heart_Failure | ASPC | *PPFIA2* | 0.800317193 | 0.000809584 | 0.010420628 | 1 |
| Heart_Failure | ASPC | *MARCH9* | 1.336033409 | 0.000881965 | 0.010420628 | 1 |
| Heart_Failure | ASPC | *RITA1* | 1.647676264 | 0.000793123 | 0.010420628 | 1 |
| Heart_Failure | ASPC | *GLTP* | 1.095651816 | 0.0011909 | 0.010859009 | 1 |
| Heart_Failure | ASPC | *LMBR1L* | 0.30429067 | 0.012135523 | 0.049355875 | 1 |
| Heart_Failure | ASPC | *ESYT1* | 0.515505064 | 0.000310834 | 0.010420628 | 1 |
| Heart_Failure | ASPC | *RB1* | 0.351825606 | 0.003869037 | 0.021702392 | 1 |
| Heart_Failure | ASPC | *TMX1* | 0.825368873 | 0.001027292 | 0.010439519 | 1 |
| Heart_Failure | ASPC | *C14orf37* | 0.617670267 | 0.000914201 | 0.010420628 | 1 |
| Heart_Failure | ASPC | *RAB15* | 1.066044768 | 0.001656603 | 0.012424316 | 1 |
| Heart_Failure | ASPC | *SLC24A4* | 1.001525441 | 0.001354353 | 0.011448295 | 1 |
| Heart_Failure | ASPC | *C14orf79* | 0.32734024 | 0.001432693 | 0.01166347 | 1 |
| Heart_Failure | ASPC | *WARS* | 0.538106199 | 0.002092823 | 0.014464926 | 1 |
| Heart_Failure | ASPC | *SERF2* | 0.683059991 | 0.000517496 | 0.010420628 | 1 |
| Heart_Failure | ASPC | *HMG20A* | 0.722632766 | 0.000728906 | 0.010420628 | 1 |
| Heart_Failure | ASPC | *ABHD2* | 0.642581462 | 0.000674646 | 0.010420628 | 1 |
| Heart_Failure | ASPC | *UNC45A* | 0.514735178 | 0.005672475 | 0.028379782 | 1 |
| Heart_Failure | ASPC | *CRTC3* | 0.220092955 | 0.00664163 | 0.031780267 | 1 |
| Heart_Failure | ASPC | *ITGAX* | 1.142232953 | 0.000291455 | 0.010420628 | 1 |
| Heart_Failure | ASPC | *DHX38* | 0.928105124 | 0.000752177 | 0.010420628 | 1 |
| Heart_Failure | ASPC | *NUDT7* | 0.830497221 | 0.004726735 | 0.024660342 | 1 |
| Heart_Failure | ASPC | *TLDC1* | 0.953431371 | 0.001412877 | 0.011638841 | 1 |
| Heart_Failure | ASPC | *TCF25* | 0.514137895 | 1.23E-05 | 0.002128616 | 0.05960124 |
| Heart_Failure | ASPC | *GALNS* | 0.298567016 | 0.012316359 | 0.049841019 | 1 |
| Heart_Failure | ASPC | *NCOR1* | 0.52054937 | 0.001135439 | 0.010655995 | 1 |
| Heart_Failure | ASPC | *KSR1* | 0.714241655 | 0.000616627 | 0.010420628 | 1 |

| | | | | | | |
|---|---|---|---|---|---|---|
| Heart_Failure | ASPC | *MYO19* | 0.385218571 | 0.001695853 | 0.01262006 | 1 |
| Heart_Failure | ASPC | *SSH2* | 0.522142918 | 0.000807655 | 0.010420628 | 1 |
| Heart_Failure | ASPC | *IMPA2* | 0.571680929 | 0.000115696 | 0.009385075 | 0.561357 |
| Heart_Failure | ASPC | *GNAL* | 0.8146141 | 0.000991979 | 0.010420628 | 1 |
| Heart_Failure | ASPC | *GALNT1* | 0.587241077 | 0.00066625 | 0.010420628 | 1 |
| Heart_Failure | ASPC | *NARF* | 0.537420916 | 0.000291072 | 0.010420628 | 1 |
| Heart_Failure | ASPC | *ZNF750* | 0.556232717 | 0.003631063 | 0.020837904 | 1 |
| Heart_Failure | ASPC | *MAPK4* | 2.413192576 | 0.000845967 | 0.010420628 | 1 |
| Heart_Failure | ASPC | *ERBB2* | 0.66475422 | 1.67E-08 | 1.16E-05 | 8.10E-05 |
| Heart_Failure | ASPC | *PNMT* | 0.660882377 | 9.25E-13 | 8.97E-10 | 4.49E-09 |
| Heart_Failure | ASPC | *NFIC* | 0.444195914 | 0.001542697 | 0.012034028 | 1 |
| Heart_Failure | ASPC | *DPP9* | 0.861903828 | 0.000724967 | 0.010420628 | 1 |
| Heart_Failure | ASPC | *SIRT3* | 0.365896241 | 0.002315809 | 0.01541331 | 1 |
| Heart_Failure | ASPC | *EVI5L* | 1.146513099 | 0.000678445 | 0.010420628 | 1 |
| Heart_Failure | ASPC | *RERE* | 0.388001003 | 0.002170859 | 0.01483522 | 1 |
| Heart_Failure | ASPC | *EVA1B* | 0.219947276 | 1.49E-05 | 0.002410028 | 0.07230083 |
| Heart_Failure | ASPC | *RPS8* | 0.497309599 | 0.002776318 | 0.017336802 | 1 |
| Heart_Failure | ASPC | *STRIP1* | 1.082743923 | 3.93E-05 | 0.004769707 | 0.190788269 |
| Heart_Failure | ASPC | *ALDH9A1* | 0.766980795 | 0.002673054 | 0.016856236 | 1 |
| Heart_Failure | ASPC | *RFWD2* | 0.53925704 | 0.000628061 | 0.010420628 | 1 |
| Heart_Failure | ASPC | *SMYD2* | 0.862057573 | 0.000824562 | 0.010420628 | 1 |
| Heart_Failure | ASPC | *RAB13* | 1.049351933 | 0.002691265 | 0.016914533 | 1 |
| Heart_Failure | ASPC | *S100A8* | 0.828778876 | 0.00126456 | 0.011113358 | 1 |
| Heart_Failure | ASPC | *C1orf43* | 0.557787487 | 0.002044582 | 0.014312895 | 1 |
| Heart_Failure | ASPC | *FLG* | -0.069437901 | 0.000741679 | 0.010420628 | 1 |
| Heart_Failure | ASPC | *SCCPDH* | 0.682553074 | 0.002708706 | 0.017002123 | 1 |
| Heart_Failure | ASPC | *CEP170* | 0.762995004 | 6.52E-06 | 0.001475161 | 0.031611592 |
| Heart_Failure | ASPC | *ACP1* | 0.732624159 | 0.001349393 | 0.011426276 | 1 |
| Heart_Failure | ASPC | *CNIH4* | 0.471297567 | 0.011840775 | 0.048523177 | 1 |
| Heart_Failure | ASPC | *MBOAT2* | 0.636057381 | 0.008120022 | 0.036412522 | 1 |
| Heart_Failure | ASPC | *RHOB* | 0.792970308 | 0.001541462 | 0.012034028 | 1 |
| Heart_Failure | ASPC | *GALM* | 0.78646515 | 0.000765433 | 0.010420628 | 1 |
| Heart_Failure | ASPC | *WDPCP* | 0.183397708 | 0.000574238 | 0.010420628 | 1 |
| Heart_Failure | ASPC | *VPS54* | 0.538173646 | 0.002572151 | 0.016442786 | 1 |
| Heart_Failure | ASPC | *EXOC6B* | 0.498782575 | 0.001004011 | 0.010420628 | 1 |
| Heart_Failure | ASPC | *TEX261* | 1.192295148 | 0.002372704 | 0.015705813 | 1 |
| Heart_Failure | ASPC | *RALB* | 0.881887532 | 0.000739394 | 0.010420628 | 1 |
| Heart_Failure | ASPC | *NBEAL1* | 0.384285079 | 0.000605999 | 0.010420628 | 1 |
| Heart_Failure | ASPC | *SUMF1* | 0.523322989 | 0.000895343 | 0.010420628 | 1 |
| Heart_Failure | ASPC | *ESPNL* | 13.23864844 | 0.000440044 | 0.010420628 | 1 |
| Heart_Failure | ASPC | *ARL6IP5* | 0.541425133 | 0.006144636 | 0.029857479 | 1 |
| Heart_Failure | ASPC | *MED12L* | 0.593485081 | 0.011851402 | 0.048525742 | 1 |
| Heart_Failure | ASPC | *NCEH1* | 0.146420544 | 0.007994885 | 0.036017813 | 1 |
| Heart_Failure | ASPC | *CISD2* | 0.839586421 | 0.000138722 | 0.010198142 | 0.673077399 |
| Heart_Failure | ASPC | *USP53* | 0.30078066 | 0.000275112 | 0.010420628 | 1 |
| Heart_Failure | ASPC | *MARCH6* | 0.618405846 | 0.000550118 | 0.010420628 | 1 |
| Heart_Failure | ASPC | *GTF2H2* | 1.467994381 | 0.000635578 | 0.010420628 | 1 |
| Heart_Failure | ASPC | *SPATA9* | 0.740614433 | 0.001305893 | 0.01123051 | 1 |
| Heart_Failure | ASPC | *TSLP* | 0.093817189 | 1.40E-06 | 0.000425931 | 0.006814902 |
| Heart_Failure | ASPC | *FEM1C* | 1.037361851 | 0.000730475 | 0.010420628 | 1 |

| | | | | | | |
|---|---|---|---|---|---|---|
| Heart_Failure | ASPC | *FBXO38* | 0.412596699 | 0.004631026 | 0.024476837 | 1 |
| Heart_Failure | ASPC | *BOD1* | 0.641430155 | 5.36E-05 | 0.005916131 | 0.260309758 |
| Heart_Failure | ASPC | *MYLK4* | 0.491854085 | 0.002280916 | 0.015264835 | 1 |
| Heart_Failure | ASPC | *RNF44* | 0.4985878 | 0.003951745 | 0.022013622 | 1 |
| Heart_Failure | ASPC | *RPL7L1* | 0.552565404 | 0.004600041 | 0.024395914 | 1 |
| Heart_Failure | ASPC | *PHIP* | 0.57842919 | 0.000445924 | 0.010420628 | 1 |
| Heart_Failure | ASPC | *PNRC1* | 0.832217642 | 0.000733401 | 0.010420628 | 1 |
| Heart_Failure | ASPC | *RNF217* | 0.546763627 | 0.001009696 | 0.010420628 | 1 |
| Heart_Failure | ASPC | *SLC22A3* | 0.681240812 | 0.001795578 | 0.013061689 | 1 |
| Heart_Failure | ASPC | *RBAK* | 1.429513262 | 0.000905364 | 0.010420628 | 1 |
| Heart_Failure | ASPC | *TMEM209* | 0.833251335 | 0.000964182 | 0.010420628 | 1 |
| Heart_Failure | ASPC | *EPHA1* | 2.400784452 | 0.002117567 | 0.014511912 | 1 |
| Heart_Failure | ASPC | *FBXO25* | 0.732814553 | 0.00065086 | 0.010420628 | 1 |
| Heart_Failure | ASPC | *DOCK5* | 0.394254883 | 0.005322247 | 0.026983846 | 1 |
| Heart_Failure | ASPC | *TACC1* | 0.393076749 | 0.003284466 | 0.019481942 | 1 |
| Heart_Failure | ASPC | *DPYS* | 0.673278749 | 0.001225384 | 0.010929346 | 1 |
| Heart_Failure | ASPC | *LRP12* | 0.879892158 | 0.001430406 | 0.01166347 | 1 |
| Heart_Failure | ASPC | *HAUS6* | 0.810701209 | 0.001736851 | 0.012792849 | 1 |
| Heart_Failure | ASPC | *ALAD* | 0.135391713 | 0.011961691 | 0.048894798 | 1 |
| Heart_Failure | ASPC | *REXO4* | 0.933295732 | 0.001116172 | 0.010639821 | 1 |
| Heart_Failure | ASPC | *SLC39A12* | 0.462408992 | 0.007099868 | 0.033091794 | 1 |
| Heart_Failure | ASPC | *RSU1* | 0.605886166 | 0.000581709 | 0.010420628 | 1 |
| Heart_Failure | ASPC | *PARD3* | 0.359181939 | 0.008821948 | 0.03905483 | 1 |
| Heart_Failure | ASPC | *ADD3* | 0.432445026 | 0.005360296 | 0.027148387 | 1 |
| Heart_Failure | ASPC | *PAOX* | 1.223556486 | 0.000622068 | 0.010420628 | 1 |
| Heart_Failure | ASPC | *GAS2* | 0.636751668 | 0.001588353 | 0.012207591 | 1 |
| Heart_Failure | ASPC | *LIN7C* | 0.857820983 | 0.001428236 | 0.01166347 | 1 |
| Heart_Failure | ASPC | *PTPRJ* | 0.483533329 | 0.002243404 | 0.015097085 | 1 |
| Heart_Failure | ASPC | *RPS3* | 0.316657464 | 0.007137781 | 0.033141513 | 1 |
| Heart_Failure | ASPC | *NPAT* | 0.706375315 | 0.001336036 | 0.011332951 | 1 |
| Heart_Failure | ASPC | *FADS1* | 0.292236947 | 0.003607738 | 0.020740219 | 1 |
| Heart_Failure | ASPC | *ROM1* | 0.10105863 | 0.00097524 | 0.010420628 | 1 |
| Heart_Failure | ASPC | *CPSF7* | 0.417616553 | 0.004485785 | 0.024076357 | 1 |
| Heart_Failure | ASPC | *ALDOA* | 0.412123946 | 0.000150122 | 0.010420628 | 0.728391581 |
| Heart_Failure | ASPC | *SAP18* | 0.794503297 | 0.002525929 | 0.016232856 | 1 |
| Heart_Failure | ASPC | *FAM124A* | 0.481828726 | 0.007974723 | 0.035993818 | 1 |
| Heart_Failure | ASPC | *PRSS23* | 0.281031345 | 0.009373449 | 0.040825828 | 1 |
| Heart_Failure | ASPC | *TMEM18* | 1.122237592 | 0.00069861 | 0.010420628 | 1 |
| Heart_Failure | ASPC | *NDUFC2* | 0.863481566 | 0.002853526 | 0.017592516 | 1 |
| Heart_Failure | ASPC | *NUBPL* | 0.625243189 | 0.000744219 | 0.010420628 | 1 |
| Heart_Failure | ASPC | *ANKRD50* | 0.519910419 | 0.000470712 | 0.010420628 | 1 |
| Heart_Failure | ASPC | *UPF2* | 0.510474948 | 0.00247894 | 0.016080322 | 1 |
| Heart_Failure | ASPC | *PIGF* | 0.731565347 | 0.001270225 | 0.011124786 | 1 |
| Heart_Failure | ASPC | *MFSD6* | 0.641666914 | 0.001559203 | 0.012135784 | 1 |
| Heart_Failure | ASPC | *RNF144A* | 0.643407177 | 0.001087919 | 0.010484035 | 1 |
| Heart_Failure | ASPC | *BICD1* | 0.675131061 | 0.000664281 | 0.010420628 | 1 |
| Heart_Failure | ASPC | *TIAL1* | 0.530633426 | 0.005319282 | 0.026983846 | 1 |
| Heart_Failure | ASPC | *PTPN14* | 0.494427657 | 0.000690501 | 0.010420628 | 1 |
| Heart_Failure | ASPC | *TMEM178A* | 1.364496168 | 0.009323676 | 0.04064553 | 1 |
| Heart_Failure | ASPC | *EPG5* | 0.502318157 | 0.005889163 | 0.028979938 | 1 |

| | | | | | | |
|---|---|---|---|---|---|---|
| Heart_Failure | ASPC | *SPARCL1* | 0.35238291 | 0.004171478 | 0.022947858 | 1 |
| Heart_Failure | ASPC | *MBNL1* | 0.461941218 | 0.000744768 | 0.010420628 | 1 |
| Heart_Failure | ASPC | *ZNF117* | 0.02928372 | 0.009711887 | 0.041998287 | 1 |
| Heart_Failure | ASPC | *MED21* | 0.362107662 | 0.006540243 | 0.031356976 | 1 |
| Heart_Failure | ASPC | *JAKMIP1* | 0.936386209 | 0.001501479 | 0.01196018 | 1 |
| Heart_Failure | ASPC | *SREK1IP1* | 0.673972751 | 0.002114025 | 0.014508134 | 1 |
| Heart_Failure | ASPC | *TMEM87B* | 0.696874449 | 0.001109188 | 0.010614947 | 1 |
| Heart_Failure | ASPC | *LPCAT1* | 1.089219836 | 0.000916924 | 0.010420628 | 1 |
| Heart_Failure | ASPC | *CFDP1* | 0.588200861 | 0.001322813 | 0.011278839 | 1 |
| Heart_Failure | ASPC | *ZDHHC7* | 0.727057216 | 0.001505047 | 0.01196018 | 1 |
| Heart_Failure | ASPC | *SEMA3D* | 0.551728394 | 0.00318111 | 0.019078798 | 1 |
| Heart_Failure | ASPC | *PPP2R5E* | 0.617001482 | 0.001063307 | 0.010475827 | 1 |
| Heart_Failure | ASPC | *ANKRD29* | 0.734003002 | 0.001234049 | 0.010986437 | 1 |
| Heart_Failure | ASPC | *ANKH* | 0.502843841 | 0.001664083 | 0.012440876 | 1 |
| Heart_Failure | ASPC | *ROBO4* | 0.836494425 | 0.000780329 | 0.010420628 | 1 |
| Heart_Failure | ASPC | *PRKCA* | 0.436169479 | 0.007489465 | 0.03437319 | 1 |
| Heart_Failure | ASPC | *LRRK1* | 0.464337317 | 0.002031329 | 0.014258141 | 1 |
| Heart_Failure | ASPC | *ABCA5* | 0.201715954 | 0.00611024 | 0.02976595 | 1 |
| Heart_Failure | ASPC | *CCSAP* | 1.210343849 | 0.001068797 | 0.010475827 | 1 |
| Heart_Failure | ASPC | *OXNAD1* | 0.569342865 | 0.003221527 | 0.019273547 | 1 |
| Heart_Failure | ASPC | *MARVELD1* | 0.766819676 | 0.006648371 | 0.031781178 | 1 |
| Heart_Failure | ASPC | *CNOT8* | 0.457294906 | 0.011099213 | 0.045989223 | 1 |
| Heart_Failure | ASPC | *PDIA4* | 0.845577713 | 0.00356013 | 0.020539536 | 1 |
| Heart_Failure | ASPC | *FZD7* | 0.210424192 | 0.000294711 | 0.010420628 | 1 |
| Heart_Failure | ASPC | *TMBIM4* | 0.265772511 | 0.001714367 | 0.012718822 | 1 |
| Heart_Failure | ASPC | *C14orf2* | 0.340452427 | 0.000971316 | 0.010420628 | 1 |
| Heart_Failure | ASPC | *RPL30* | 0.60235449 | 3.71E-05 | 0.004717456 | 0.179952796 |
| Heart_Failure | ASPC | *NODAL* | 1.576772021 | 0.001197873 | 0.010869813 | 1 |
| Heart_Failure | ASPC | *ZDHHC5* | 0.893911279 | 0.000703611 | 0.010420628 | 1 |
| Heart_Failure | ASPC | *VPS8* | 0.30828763 | 0.003936867 | 0.021980417 | 1 |
| Heart_Failure | ASPC | *FCHO2* | 0.275587582 | 0.002633253 | 0.016657815 | 1 |
| Heart_Failure | ASPC | *NECAP2* | 0.84336482 | 0.000942055 | 0.010420628 | 1 |
| Heart_Failure | ASPC | *CLDN12* | 1.033243816 | 0.002560983 | 0.016393532 | 1 |
| Heart_Failure | ASPC | *MMP14* | 0.934988706 | 6.17E-05 | 0.006655983 | 0.299519237 |
| Heart_Failure | ASPC | *DHRS4* | 1.207142454 | 0.000869966 | 0.010420628 | 1 |
| Heart_Failure | ASPC | *LCA5L* | 0.077396035 | 0.009853719 | 0.042347427 | 1 |
| Heart_Failure | ASPC | *SLC38A10* | 0.764247978 | 0.000793935 | 0.010420628 | 1 |
| Heart_Failure | ASPC | *SPPL3* | 0.479909772 | 0.000375872 | 0.010420628 | 1 |
| Heart_Failure | ASPC | *EXTL1* | 17.93343887 | 0.000555518 | 0.010420628 | 1 |
| Heart_Failure | ASPC | *MITD1* | 0.401865356 | 0.001781853 | 0.013000825 | 1 |
| Heart_Failure | ASPC | *FAM86C1* | 1.531494753 | 0.004656332 | 0.024530428 | 1 |
| Heart_Failure | ASPC | *ELK4* | 0.70162885 | 0.00083769 | 0.010420628 | 1 |
| Heart_Failure | ASPC | *ARHGAP27* | 0.404888337 | 0.005581807 | 0.027992199 | 1 |
| Heart_Failure | ASPC | *AMFR* | 0.347664356 | 0.006909426 | 0.032548092 | 1 |
| Heart_Failure | ASPC | *TMEM69* | 0.62515795 | 0.010860158 | 0.045347233 | 1 |
| Heart_Failure | ASPC | *CCDC58* | 1.047494757 | 0.000929709 | 0.010420628 | 1 |
| Heart_Failure | ASPC | *TAOK1* | 0.470652454 | 0.004912197 | 0.025463656 | 1 |
| Heart_Failure | ASPC | *VPS11* | 0.19753335 | 0.004806484 | 0.024995777 | 1 |
| Heart_Failure | ASPC | *FDPS* | 0.494664558 | 4.59E-05 | 0.005298784 | 0.222548912 |
| Heart_Failure | ASPC | *FCRL3* | -0.089257377 | 0.005405216 | 0.02734735 | 1 |

| | | | | | | |
|---|---|---|---|---|---|---|
| Heart_Failure | ASPC | *FGFR4* | -0.467353729 | 0.000279329 | 0.010420628 | 1 |
| Heart_Failure | ASPC | *ZNF333* | 0.844887654 | 0.000687333 | 0.010420628 | 1 |
| Heart_Failure | ASPC | *RPL8* | 0.565263627 | 0.001407068 | 0.011637563 | 1 |
| Heart_Failure | ASPC | *PGAP3* | 1.207716164 | 0.0008505 | 0.010420628 | 1 |
| Heart_Failure | ASPC | *PLCD3* | 1.209393717 | 0.001462423 | 0.011795301 | 1 |
| Heart_Failure | ASPC | *LEMD2* | 0.488968096 | 0.000900871 | 0.010420628 | 1 |
| Heart_Failure | ASPC | *ASRGL1* | 0.740474945 | 0.008966411 | 0.039349527 | 1 |
| Heart_Failure | ASPC | *RPS6KA4* | 1.253139309 | 0.001000376 | 0.010420628 | 1 |
| Heart_Failure | ASPC | *COA7* | 1.541131234 | 0.000267347 | 0.010420628 | 1 |
| Heart_Failure | ASPC | *ZSWIM5* | 0.312294569 | 0.002859684 | 0.017608104 | 1 |
| Heart_Failure | ASPC | *RBP7* | 0.54246948 | 0.000333041 | 0.010420628 | 1 |
| Heart_Failure | ASPC | *LAPTM5* | 0.757659821 | 0.001008774 | 0.010420628 | 1 |
| Heart_Failure | ASPC | *TM2D1* | 0.656297425 | 0.000949836 | 0.010420628 | 1 |
| Heart_Failure | ASPC | *NEXN* | 0.278857516 | 0.010319556 | 0.04372968 | 1 |
| Heart_Failure | ASPC | *LRRIQ3* | 0.577205097 | 0.005771847 | 0.028605723 | 1 |
| Heart_Failure | ASPC | *KLHDC9* | -0.090161548 | 0.002416075 | 0.015913671 | 1 |
| Heart_Failure | ASPC | *ACP6* | 0.602101385 | 0.009209346 | 0.040255627 | 1 |
| Heart_Failure | ASPC | *CNST* | 0.558554696 | 0.0014368 | 0.01167711 | 1 |
| Heart_Failure | ASPC | *HAAO* | 0.497798225 | 0.009961436 | 0.042621592 | 1 |
| Heart_Failure | ASPC | *NUP35* | 0.502882485 | 0.004604195 | 0.024395914 | 1 |
| Heart_Failure | ASPC | *NOSTRIN* | 0.215025623 | 0.000465751 | 0.010420628 | 1 |
| Heart_Failure | ASPC | *VPS72* | 1.007349015 | 0.000781183 | 0.010420628 | 1 |
| Heart_Failure | ASPC | *ERCC3* | 0.41058861 | 0.001173978 | 0.010788896 | 1 |
| Heart_Failure | ASPC | *RNF149* | 0.385712599 | 0.0085549 | 0.038046174 | 1 |
| Heart_Failure | ASPC | *DHX57* | 0.471624639 | 0.005519026 | 0.027749546 | 1 |
| Heart_Failure | ASPC | *TDRD10* | 0.717509855 | 0.0024744 | 0.016080322 | 1 |
| Heart_Failure | ASPC | *GPR155* | 0.646605049 | 0.001740165 | 0.012792849 | 1 |
| Heart_Failure | ASPC | *PBXIP1* | 0.764549256 | 0.000782729 | 0.010420628 | 1 |
| Heart_Failure | ASPC | *LRRC58* | 0.46481181 | 0.005955671 | 0.029188802 | 1 |
| Heart_Failure | ASPC | *ARPC2* | 0.399677852 | 0.010703821 | 0.044812634 | 1 |
| Heart_Failure | ASPC | *SUCLG1* | 0.281926254 | 0.003523953 | 0.020379285 | 1 |
| Heart_Failure | ASPC | *EFHB* | 0.042831339 | 0.003009656 | 0.018253561 | 1 |
| Heart_Failure | ASPC | *GTPBP8* | 1.022867876 | 0.000981996 | 0.010420628 | 1 |
| Heart_Failure | ASPC | *WDFY3* | 0.382533902 | 0.007137857 | 0.033141513 | 1 |
| Heart_Failure | ASPC | *C4orf36* | 0.896022831 | 0.000731681 | 0.010420628 | 1 |
| Heart_Failure | ASPC | *TIPARP* | 0.545566373 | 0.001903465 | 0.013541955 | 1 |
| Heart_Failure | ASPC | *CRELD1* | 0.568631511 | 0.003703432 | 0.021065713 | 1 |
| Heart_Failure | ASPC | *CPA3* | 1.508000478 | 0.000179051 | 0.010420628 | 0.868755089 |
| Heart_Failure | ASPC | *SLC4A1AP* | 0.881027272 | 0.001294134 | 0.01121275 | 1 |
| Heart_Failure | ASPC | *SNIP1* | 1.012774669 | 0.000980191 | 0.010420628 | 1 |
| Heart_Failure | ASPC | *RPN1* | 0.886835985 | 0.000558338 | 0.010420628 | 1 |
| Heart_Failure | ASPC | *SFMBT1* | 1.046966627 | 0.000180965 | 0.010420628 | 0.87804196 |
| Heart_Failure | ASPC | *FAM208A* | 0.494422968 | 0.002205137 | 0.014955515 | 1 |
| Heart_Failure | ASPC | *ABLIM2* | 0.274628274 | 0.002479172 | 0.016080322 | 1 |
| Heart_Failure | ASPC | *RNF123* | 0.223514182 | 8.22E-07 | 0.000284766 | 0.003986728 |
| Heart_Failure | ASPC | *MST1R* | 0.206973735 | 3.38E-08 | 2.05E-05 | 0.000163767 |
| Heart_Failure | ASPC | *ANXA5* | 0.321190249 | 0.004985363 | 0.025678324 | 1 |
| Heart_Failure | ASPC | *HPGD* | 1.313211829 | 0.000787963 | 0.010420628 | 1 |
| Heart_Failure | ASPC | *ARFIP1* | 0.350606938 | 0.010568751 | 0.044515913 | 1 |
| Heart_Failure | ASPC | *RNF180* | 0.388298692 | 0.010986991 | 0.045648552 | 1 |

| | | | | | | |
|---|---|---|---|---|---|---|
| Heart_Failure | ASPC | *PGGT1B* | 0.35268762 | 0.007787402 | 0.035378721 | 1 |
| Heart_Failure | ASPC | *CMBL* | 0.544638876 | 0.000107809 | 0.009177021 | 0.523090217 |
| Heart_Failure | ASPC | *NDUFS4* | 0.636803915 | 0.000538467 | 0.010420628 | 1 |
| Heart_Failure | ASPC | *SERINC5* | 0.553736782 | 9.67E-06 | 0.001877595 | 0.04693988 |
| Heart_Failure | ASPC | *UTP15* | 0.579706514 | 0.011707581 | 0.0480457 | 1 |
| Heart_Failure | ASPC | *CCDC127* | 0.433866789 | 0.001028749 | 0.010439519 | 1 |
| Heart_Failure | ASPC | *PDSS2* | 0.593682339 | 0.000720513 | 0.010420628 | 1 |
| Heart_Failure | ASPC | *RAET1E* | 0.726303944 | 0.004020032 | 0.022291652 | 1 |
| Heart_Failure | ASPC | *SAP30L* | 0.608146378 | 2.19E-05 | 0.003039348 | 0.106377165 |
| Heart_Failure | ASPC | *RPS14* | 0.519590582 | 0.000835943 | 0.010420628 | 1 |
| Heart_Failure | ASPC | *RP9* | 1.090697629 | 0.000747266 | 0.010420628 | 1 |
| Heart_Failure | ASPC | *MIOS* | 0.486548242 | 0.005584594 | 0.027992199 | 1 |
| Heart_Failure | ASPC | *USP49* | 0.883926889 | 0.000227979 | 0.010420628 | 1 |
| Heart_Failure | ASPC | *SLC13A4* | 0.801382206 | 0.001011193 | 0.010420628 | 1 |
| Heart_Failure | ASPC | *BRI3* | 0.619584832 | 0.002508253 | 0.016183566 | 1 |
| Heart_Failure | ASPC | *RAD21* | 0.594738816 | 0.000709854 | 0.010420628 | 1 |
| Heart_Failure | ASPC | *TNFRSF11B* | 0.624311953 | 0.007160473 | 0.033183015 | 1 |
| Heart_Failure | ASPC | *OXR1* | 0.46347848 | 0.00230914 | 0.01539004 | 1 |
| Heart_Failure | ASPC | *GPER1* | 0.339279332 | 0.000301797 | 0.010420628 | 1 |
| Heart_Failure | ASPC | *FASTK* | 1.110674169 | 0.000251507 | 0.010420628 | 1 |
| Heart_Failure | ASPC | *INTS8* | 0.657533954 | 0.000367916 | 0.010420628 | 1 |
| Heart_Failure | ASPC | *C9orf24* | 0.531741123 | 0.00499712 | 0.025684352 | 1 |
| Heart_Failure | ASPC | *TMEM65* | 0.722180154 | 0.000697293 | 0.010420628 | 1 |
| Heart_Failure | ASPC | *ZMAT4* | 1.392240486 | 0.003941254 | 0.021980417 | 1 |
| Heart_Failure | ASPC | *AQP3* | 2.340003001 | 0.002021971 | 0.014258141 | 1 |
| Heart_Failure | ASPC | *WRN* | 0.534355662 | 0.000892002 | 0.010420628 | 1 |
| Heart_Failure | ASPC | *MARCH8* | 0.735187218 | 0.000396405 | 0.010420628 | 1 |
| Heart_Failure | ASPC | *MICU2* | 0.430440688 | 0.012215941 | 0.04956127 | 1 |
| Heart_Failure | ASPC | *LRR1* | 1.336477538 | 0.00096571 | 0.010420628 | 1 |
| Heart_Failure | ASPC | *METTL3* | 0.607236274 | 0.001071086 | 0.010475827 | 1 |
| Heart_Failure | ASPC | *HSPA12A* | 0.10914766 | 0.002992469 | 0.018172042 | 1 |
| Heart_Failure | ASPC | *UBTD1* | 0.75256671 | 0.000972809 | 0.010420628 | 1 |
| Heart_Failure | ASPC | *TTC7B* | 0.501082321 | 0.003323741 | 0.01961897 | 1 |
| Heart_Failure | ASPC | *TCP11L2* | 0.784129452 | 0.000435393 | 0.010420628 | 1 |
| Heart_Failure | ASPC | *TRMT61A* | 0.442842993 | 0.005417946 | 0.027383201 | 1 |
| Heart_Failure | ASPC | *API5* | 0.79262116 | 0.000624126 | 0.010420628 | 1 |
| Heart_Failure | ASPC | *COX11* | 0.409192444 | 0.00792557 | 0.035856645 | 1 |
| Heart_Failure | ASPC | *WDR88* | 0.847246758 | 0.001530625 | 0.012017141 | 1 |
| Heart_Failure | ASPC | *RPL27A* | 0.718496573 | 0.000831526 | 0.010420628 | 1 |
| Heart_Failure | ASPC | *TMEM41B* | 0.619494232 | 0.0047565 | 0.024788977 | 1 |
| Heart_Failure | ASPC | *NDST2* | 5.39419839 | 0.00628237 | 0.030330408 | 1 |
| Heart_Failure | ASPC | *SEC11C* | 0.917768717 | 0.001401375 | 0.011637563 | 1 |
| Heart_Failure | ASPC | *SAAL1* | 1.190201642 | 0.000601475 | 0.010420628 | 1 |
| Heart_Failure | ASPC | *PIP4K2C* | 0.716228251 | 7.05E-06 | 0.001486412 | 0.034187469 |
| Heart_Failure | ASPC | *MTMR10* | 0.250514632 | 0.008501472 | 0.037863253 | 1 |
| Heart_Failure | ASPC | *EPB42* | 0.674296456 | 0.00088253 | 0.010420628 | 1 |
| Heart_Failure | ASPC | *SMAD3* | 0.418531066 | 0.000973041 | 0.010420628 | 1 |
| Heart_Failure | ASPC | *AKTIP* | 0.271804021 | 0.005194928 | 0.026532412 | 1 |
| Heart_Failure | ASPC | *SP2* | 0.944603242 | 0.001146312 | 0.010719151 | 1 |
| Heart_Failure | ASPC | *FBXO22* | 0.708171842 | 0.000510476 | 0.010420628 | 1 |

| | | | | | | |
|---|---|---|---|---|---|---|
| Heart_Failure | ASPC | *HDHD2* | 0.076473662 | 0.003552808 | 0.020521696 | 1 |
| Heart_Failure | ASPC | *ZNF91* | 0.515166728 | 0.000252001 | 0.010420628 | 1 |
| Heart_Failure | ASPC | *ZNF23* | 0.404005286 | 0.000359514 | 0.010420628 | 1 |
| Heart_Failure | ASPC | *VKORC1* | 1.02555923 | 0.002920293 | 0.017822972 | 1 |
| Heart_Failure | ASPC | *SPATA33* | 1.062082154 | 0.000634093 | 0.010420628 | 1 |
| Heart_Failure | ASPC | *RPL13* | 0.427191083 | 0.004843658 | 0.025162132 | 1 |
| Heart_Failure | ASPC | *ZNF528* | 0.877435038 | 0.000273301 | 0.010420628 | 1 |
| Heart_Failure | ASPC | *SERTAD3* | 1.258492089 | 0.001122461 | 0.01064017 | 1 |
| Heart_Failure | ASPC | *TTYH1* | 1.455575994 | 0.001515306 | 0.011981993 | 1 |
| Heart_Failure | ASPC | *TRPV3* | 0.099256417 | 0.009807706 | 0.042224481 | 1 |
| Heart_Failure | ASPC | *CYB5D2* | 0.588305311 | 0.002436245 | 0.015984971 | 1 |
| Heart_Failure | ASPC | *NDUFV1* | 0.779484011 | 0.000825572 | 0.010420628 | 1 |
| Heart_Failure | ASPC | *PRDX2* | 0.738317789 | 0.00307305 | 0.018520523 | 1 |
| Heart_Failure | ASPC | *MIS12* | 0.91116064 | 0.000613312 | 0.010420628 | 1 |
| Heart_Failure | ASPC | *GSDMA* | -0.198981714 | 9.88E-11 | 7.99E-08 | 4.79E-07 |
| Heart_Failure | ASPC | *MLST8* | 0.534089789 | 0.004411046 | 0.023889952 | 1 |
| Heart_Failure | ASPC | *ATG16L2* | 0.828132993 | 0.000600273 | 0.010420628 | 1 |
| Heart_Failure | ASPC | *COA6* | 1.317126851 | 0.000803273 | 0.010420628 | 1 |
| Heart_Failure | ASPC | *PCMTD1* | 0.572364147 | 0.001076059 | 0.010475827 | 1 |
| Heart_Failure | ASPC | *RFWD3* | 0.519676877 | 0.001310279 | 0.01123051 | 1 |
| Heart_Failure | ASPC | *CCDC110* | 1.952923971 | 0.001561352 | 0.012135784 | 1 |
| Heart_Failure | ASPC | *SLC20A2* | 0.436355823 | 0.004598367 | 0.024395914 | 1 |
| Heart_Failure | ASPC | *TMUB2* | 0.422174469 | 0.002098992 | 0.014478344 | 1 |
| Heart_Failure | ASPC | *DNAJC21* | 0.851843502 | 0.000921138 | 0.010420628 | 1 |
| Heart_Failure | ASPC | *SNTB2* | 0.621536522 | 0.000947621 | 0.010420628 | 1 |
| Heart_Failure | ASPC | *STX18* | 0.705206232 | 0.000919614 | 0.010420628 | 1 |
| Heart_Failure | ASPC | *ZBTB49* | 0.978609889 | 0.002206956 | 0.014955515 | 1 |
| Heart_Failure | ASPC | *CEP120* | 0.63025678 | 0.001398713 | 0.011637563 | 1 |
| Heart_Failure | ASPC | *MFF* | 0.64903734 | 0.001121712 | 0.01064017 | 1 |
| Heart_Failure | ASPC | *MFSD7* | 1.878496318 | 0.000986029 | 0.010420628 | 1 |
| Heart_Failure | ASPC | *MAP2K1* | 0.724042678 | 0.000697226 | 0.010420628 | 1 |
| Heart_Failure | ASPC | *RGS14* | -0.077695336 | 0.009296778 | 0.040564718 | 1 |
| Heart_Failure | ASPC | *GPRIN1* | 0.249937396 | 0.009151826 | 0.040076408 | 1 |
| Heart_Failure | ASPC | *HINT1* | 0.750117322 | 0.000628784 | 0.010420628 | 1 |
| Heart_Failure | ASPC | *CLIC3* | 1.372638241 | 0.001283333 | 0.011199159 | 1 |
| Heart_Failure | ASPC | *LUZP1* | 0.695218518 | 0.00068888 | 0.010420628 | 1 |
| Heart_Failure | ASPC | *UGP2* | 0.308480636 | 0.001817433 | 0.013200877 | 1 |
| Heart_Failure | ASPC | *CSGALNACT2* | 0.774821745 | 0.00013193 | 0.010160708 | 0.640124614 |
| Heart_Failure | ASPC | *AVEN* | 0.579805593 | 0.004317918 | 0.023592949 | 1 |
| Heart_Failure | ASPC | *TOR1AIP2* | 0.411983382 | 0.004161893 | 0.02294767 | 1 |
| Heart_Failure | ASPC | *CHRNB1* | 0.06154445 | 0.000957218 | 0.010420628 | 1 |
| Heart_Failure | ASPC | *PDCD6IP* | 0.61657005 | 0.000732401 | 0.010420628 | 1 |
| Heart_Failure | ASPC | *STX8* | 0.572457825 | 0.00025756 | 0.010420628 | 1 |
| Heart_Failure | ASPC | *NFRKB* | 0.587630041 | 0.010315639 | 0.04372968 | 1 |
| Heart_Failure | ASPC | *ELOVL6* | 0.641814122 | 0.003991078 | 0.022156417 | 1 |
| Heart_Failure | ASPC | *SERPINB9* | 0.309210334 | 0.007066403 | 0.033062861 | 1 |
| Heart_Failure | ASPC | *STAT2* | 0.540152337 | 0.001056684 | 0.010475827 | 1 |
| Heart_Failure | ASPC | *SLC26A5* | 0.678846064 | 7.13E-05 | 0.007116983 | 0.345966052 |
| Heart_Failure | ASPC | *TMEM133* | 0.131296784 | 0.010672336 | 0.04479427 | 1 |
| Heart_Failure | ASPC | *FOXN2* | 0.665261724 | 0.000851851 | 0.010420628 | 1 |

| | | | | | | |
|---|---|---|---|---|---|---|
| Heart_Failure | ASPC | *MSANTD4* | 0.991933595 | 0.000388452 | 0.010420628 | 1 |
| Heart_Failure | ASPC | *NDUFA3* | 0.797457743 | 0.001369788 | 0.011518561 | 1 |
| Heart_Failure | ASPC | *FEZ2* | 0.444171452 | 0.00064612 | 0.010420628 | 1 |
| Heart_Failure | ASPC | *MFN1* | 0.257925001 | 0.004555302 | 0.02428827 | 1 |
| Heart_Failure | ASPC | *PDE7B* | 0.460563925 | 0.000818511 | 0.010420628 | 1 |
| Heart_Failure | ASPC | *MCC* | 0.575456694 | 0.001649308 | 0.012424316 | 1 |
| Heart_Failure | ASPC | *PPID* | 0.887443548 | 1.05E-05 | 0.001922466 | 0.050716683 |
| Heart_Failure | ASPC | *TBCA* | 0.642244781 | 0.000539561 | 0.010420628 | 1 |
| Heart_Failure | ASPC | *NMUR1* | 1.558793196 | 0.001243929 | 0.011023143 | 1 |
| Heart_Failure | ASPC | *TRAPPC12* | 0.527669159 | 0.00599356 | 0.029344856 | 1 |
| Heart_Failure | ASPC | *ATPAF2* | 0.247321431 | 0.002970602 | 0.018061858 | 1 |
| Heart_Failure | ASPC | *ANKRD20A4* | 2.54232583 | 0.001163296 | 0.010788896 | 1 |
| Heart_Failure | ASPC | *MALT1* | 0.733130303 | 0.000630116 | 0.010420628 | 1 |
| Heart_Failure | ASPC | *MACROD2* | 0.176011111 | 0.003721766 | 0.021120478 | 1 |
| Heart_Failure | ASPC | *BSG* | 0.763651747 | 6.96E-05 | 0.007116983 | 0.337524074 |
| Heart_Failure | ASPC | *SUCLG2* | 0.586104396 | 0.00091445 | 0.010420628 | 1 |
| Heart_Failure | ASPC | *FGGY* | 0.310768159 | 0.011023765 | 0.045754755 | 1 |
| Heart_Failure | ASPC | *AFF1* | 0.510995021 | 0.000965287 | 0.010420628 | 1 |
| Heart_Failure | ASPC | *ZNF596* | 0.524995277 | 0.004422201 | 0.023893676 | 1 |
| Heart_Failure | ASPC | *CFL1* | 0.819729316 | 4.43E-05 | 0.005242978 | 0.214962085 |
| Heart_Failure | ASPC | *CBWD1* | 1.049303924 | 0.000940287 | 0.010420628 | 1 |
| Heart_Failure | ASPC | *ZNF621* | 0.819160388 | 0.000486425 | 0.010420628 | 1 |
| Heart_Failure | ASPC | *GXYLT2* | 0.465595654 | 0.002864655 | 0.017608856 | 1 |
| Heart_Failure | ASPC | *NOC3L* | 0.35181376 | 0.005562656 | 0.027939967 | 1 |
| Heart_Failure | ASPC | *PARP15* | 0.0661832 | 0.009777531 | 0.042206908 | 1 |
| Heart_Failure | ASPC | *MAP3K11* | 0.265247333 | 0.005426702 | 0.027398915 | 1 |
| Heart_Failure | ASPC | *KCNK7* | 0.639405594 | 0.00732019 | 0.033794063 | 1 |
| Heart_Failure | ASPC | *PPP1R14B* | 0.481577768 | 0.003469794 | 0.020232214 | 1 |
| Heart_Failure | ASPC | *PEAK1* | 0.339301572 | 0.005796557 | 0.02869887 | 1 |
| Heart_Failure | ASPC | *GMPPB* | 0.282485196 | 0.008466651 | 0.037792265 | 1 |
| Heart_Failure | ASPC | *UQCRH* | 0.45341736 | 0.008734553 | 0.038703242 | 1 |
| Heart_Failure | ASPC | *C11orf80* | -0.209352613 | 0.005851049 | 0.028851527 | 1 |
| Heart_Failure | ASPC | *GOLIM4* | 0.520551125 | 0.004972478 | 0.025666449 | 1 |
| Heart_Failure | ASPC | *XXYLT1* | 0.66438647 | 0.002561067 | 0.016393532 | 1 |
| Heart_Failure | ASPC | *SLC25A30* | 0.482538793 | 0.010109032 | 0.043063233 | 1 |
| Heart_Failure | ASPC | *LEMD3* | 0.701752147 | 0.001079537 | 0.010475827 | 1 |
| Heart_Failure | ASPC | *GLIS1* | 0.807559294 | 0.002962835 | 0.018037236 | 1 |
| Heart_Failure | ASPC | *ANKRD36C* | 0.52685837 | 0.003470831 | 0.020232214 | 1 |
| Heart_Failure | ASPC | *MSL2* | 0.741761196 | 0.000752973 | 0.010420628 | 1 |
| Heart_Failure | ASPC | *ANGEL2* | 0.656022555 | 0.001505714 | 0.01196018 | 1 |
| Heart_Failure | ASPC | *CEP135* | 0.613114573 | 0.002467478 | 0.016080322 | 1 |
| Heart_Failure | ASPC | *DHX36* | 0.648630434 | 0.000860564 | 0.010420628 | 1 |
| Heart_Failure | ASPC | *TVP23C* | 1.868332166 | 0.00076887 | 0.010420628 | 1 |
| Heart_Failure | ASPC | *SH3BP5L* | 1.148621721 | 0.002523817 | 0.016232856 | 1 |
| Heart_Failure | ASPC | *CLTB* | 0.902546138 | 0.000742535 | 0.010420628 | 1 |
| Heart_Failure | ASPC | *MCTP1* | 0.506777075 | 0.001312165 | 0.01123051 | 1 |
| Heart_Failure | ASPC | *PAAF1* | -0.052985233 | 0.005828374 | 0.028797629 | 1 |
| Heart_Failure | ASPC | *RPS6KB2* | 0.433023246 | 4.70E-05 | 0.005304407 | 0.228089502 |
| Heart_Failure | ASPC | *TTLL11* | 0.548758988 | 0.002050472 | 0.01431495 | 1 |
| Heart_Failure | ASPC | *CTDNEP1* | 0.222467099 | 0.003363589 | 0.019830053 | 1 |

| | | | | | | |
|---|---|---|---|---|---|---|
| Heart_Failure | ASPC | *ZDHHC21* | 0.368461263 | 0.001406608 | 0.011637563 | 1 |
| Heart_Failure | ASPC | *A2M* | 0.571830781 | 0.00073068 | 0.010420628 | 1 |
| Heart_Failure | ASPC | *B3GALT6* | 2.584785997 | 0.000886892 | 0.010420628 | 1 |
| Heart_Failure | ASPC | *SLC35G1* | 1.097634958 | 0.000287363 | 0.010420628 | 1 |
| Heart_Failure | ASPC | *OR4N2* | 0.422899377 | 0.002747233 | 0.017221671 | 1 |
| Heart_Failure | ASPC | *TAC4* | 0.996854107 | 0.000626916 | 0.010420628 | 1 |
| Heart_Failure | ASPC | *CRLF3* | 0.201328288 | 0.009903298 | 0.042447707 | 1 |
| Heart_Failure | ASPC | *WDR25* | 0.847438285 | 0.000700713 | 0.010420628 | 1 |
| Heart_Failure | ASPC | *RMDN1* | 0.182502488 | 0.004297142 | 0.023532431 | 1 |
| Heart_Failure | ASPC | *BOK* | 0.676606345 | 0.002675042 | 0.016856236 | 1 |
| Heart_Failure | ASPC | *SHMT1* | 0.430524672 | 0.005753543 | 0.02857338 | 1 |
| Heart_Failure | ASPC | *DSCAML1* | 0.572165184 | 0.004714525 | 0.024660342 | 1 |
| Heart_Failure | ASPC | *TALDO1* | 0.841405197 | 0.000865343 | 0.010420628 | 1 |
| Heart_Failure | ASPC | *MAN1B1* | 0.990018284 | 0.000844291 | 0.010420628 | 1 |
| Heart_Failure | ASPC | *NAALADL2* | 0.194694663 | 0.004942775 | 0.025540302 | 1 |
| Heart_Failure | ASPC | *CD151* | 0.332279253 | 0.001778313 | 0.012994537 | 1 |
| Heart_Failure | ASPC | *SLC35G5* | 8.212407831 | 0.000449257 | 0.010420628 | 1 |
| Heart_Failure | ASPC | *FAM87B* | 5.787228149 | 0.001658413 | 0.012424316 | 1 |
| Heart_Failure | ASPC | *CENPBD1* | 2.492250846 | 0.000440031 | 0.010420628 | 1 |
| Heart_Failure | ASPC | *RPS27* | 0.783705725 | 0.000156809 | 0.010420628 | 0.760839187 |
| Heart_Failure | ASPC | *DPY19L2* | 0.104183312 | 0.001020165 | 0.010439519 | 1 |
| Heart_Failure | ASPC | *ZNF454* | 0.082972938 | 0.00183607 | 0.013251142 | 1 |
| Heart_Failure | ASPC | *SH2B1* | 0.75241393 | 0.000807985 | 0.010420628 | 1 |
| Heart_Failure | ASPC | *NT5DC1* | 0.455738396 | 0.007334953 | 0.03383003 | 1 |
| Heart_Failure | ASPC | *NSUN3* | 0.602383886 | 0.002908052 | 0.017793024 | 1 |
| Heart_Failure | ASPC | *C17orf62* | 0.979463113 | 0.000442767 | 0.010420628 | 1 |
| Heart_Failure | ASPC | *GAK* | 0.733867968 | 0.000116958 | 0.009385075 | 0.567481979 |
| Heart_Failure | ASPC | *CTC1* | 0.242359744 | 0.007615341 | 0.0347271 | 1 |
| Heart_Failure | ASPC | *SPTY2D1* | 0.643477288 | 0.007009335 | 0.032859506 | 1 |
| Heart_Failure | ASPC | *ZNF664* | 1.115690667 | 0.000639678 | 0.010420628 | 1 |
| Heart_Failure | ASPC | *HLA-DQB1* | -0.184275251 | 5.86E-24 | 2.84E-20 | 2.84E-20 |
| Heart_Failure | ASPC | *PACS2* | 0.737397913 | 0.001219859 | 0.010920212 | 1 |
| Heart_Failure | ASPC | *LBX2* | 0.120671991 | 0.000483275 | 0.010420628 | 1 |
| Heart_Failure | ASPC | *GIPC3* | 4.689518969 | 0.002262768 | 0.015185271 | 1 |
| Heart_Failure | ASPC | *PDXDC1* | 0.485428285 | 0.004354324 | 0.023685181 | 1 |
| Heart_Failure | ASPC | *ZDHHC20* | 0.672204267 | 0.000777037 | 0.010420628 | 1 |
| Heart_Failure | ASPC | *CAPS2* | 0.486345706 | 0.002248287 | 0.015108985 | 1 |
| Heart_Failure | ASPC | *D2HGDH* | 0.214025209 | 8.92E-05 | 0.008482803 | 0.432622928 |
| Heart_Failure | ASPC | *CMTR2* | 0.313490598 | 0.006827994 | 0.03231586 | 1 |
| Heart_Failure | ASPC | *FKRP* | 0.075756211 | 0.007044371 | 0.03299159 | 1 |
| Heart_Failure | ASPC | *MUC16* | 0.498108246 | 0.004145603 | 0.022909416 | 1 |
| Heart_Failure | ASPC | *TMEM136* | 0.264077563 | 0.011866582 | 0.048546925 | 1 |
| Heart_Failure | ASPC | *DDX60L* | 0.612992253 | 0.00193282 | 0.013690575 | 1 |
| Heart_Failure | ASPC | *SGSH* | 1.337946096 | 0.000801794 | 0.010420628 | 1 |
| Heart_Failure | ASPC | *ZBTB20* | 0.318724098 | 0.00816965 | 0.036601239 | 1 |
| Heart_Failure | ASPC | *RELL1* | 0.27666987 | 0.002898333 | 0.017755952 | 1 |
| Heart_Failure | ASPC | *RFX7* | 0.653533306 | 0.000504483 | 0.010420628 | 1 |
| Heart_Failure | ASPC | *ZNF329* | 0.836426286 | 0.001209119 | 0.01091745 | 1 |
| Heart_Failure | ASPC | *WRB* | 0.089896439 | 0.003688544 | 0.021030334 | 1 |
| Heart_Failure | ASPC | *MRPL41* | 1.10523558 | 0.000681472 | 0.010420628 | 1 |

| | | | | | | |
|---|---|---|---|---|---|---|
| Heart_Failure | ASPC | *FAM153B* | 0.410063372 | 0.004637278 | 0.024483212 | 1 |
| Heart_Failure | ASPC | *KCNA4* | 0.664286304 | 0.012172697 | 0.0494656 | 1 |
| Heart_Failure | ASPC | *FIGN* | 0.504809311 | 0.001824226 | 0.013210664 | 1 |
| Heart_Failure | ASPC | *C8orf33* | 0.525721879 | 0.005095028 | 0.026077083 | 1 |
| Heart_Failure | ASPC | *GLRX5* | 1.11682738 | 0.000658409 | 0.010420628 | 1 |
| Heart_Failure | ASPC | *BRICD5* | 1.547036199 | 0.004396827 | 0.02386764 | 1 |
| Heart_Failure | ASPC | *COL18A1* | 0.282687807 | 0.000342888 | 0.010420628 | 1 |
| Heart_Failure | ASPC | *ZNF721* | 0.477283207 | 0.000596991 | 0.010420628 | 1 |
| Heart_Failure | ASPC | *CEP63* | 0.144374072 | 0.006241851 | 0.030255205 | 1 |
| Heart_Failure | ASPC | *ARHGEF37* | 0.821402121 | 0.001642394 | 0.01241261 | 1 |
| Heart_Failure | ASPC | *CSMD1* | 0.853602486 | 0.002808626 | 0.017446383 | 1 |
| Heart_Failure | ASPC | *FANCF* | 0.992244557 | 0.007476244 | 0.03437319 | 1 |
| Heart_Failure | ASPC | *PSMG1* | -0.173094261 | 0.006956442 | 0.032704389 | 1 |
| Heart_Failure | ASPC | *SERHL2* | 0.94011022 | 0.00251773 | 0.016223142 | 1 |
| Heart_Failure | ASPC | *BTBD9* | 0.384560653 | 0.006741947 | 0.032061327 | 1 |
| Heart_Failure | ASPC | *TTC32* | 0.862746401 | 0.001089026 | 0.010484035 | 1 |
| Heart_Failure | ASPC | *PP2D1* | 1.213961305 | 0.00072034 | 0.010420628 | 1 |
| Heart_Failure | ASPC | *DENND5A* | 0.585910645 | 0.000572288 | 0.010420628 | 1 |
| Heart_Failure | ASPC | *COPB2* | 0.751908018 | 0.000339428 | 0.010420628 | 1 |
| Heart_Failure | ASPC | *SNN* | 1.287631551 | 0.002815441 | 0.017446383 | 1 |
| Heart_Failure | ASPC | *TCTE3* | 0.898143701 | 0.001012309 | 0.010420628 | 1 |
| Heart_Failure | ASPC | *ZFP90* | 0.764669947 | 0.0012128 | 0.01091745 | 1 |
| Heart_Failure | ASPC | *SIGIRR* | 0.367380768 | 0.000247364 | 0.010420628 | 1 |
| Heart_Failure | ASPC | *IFITM2* | 0.707985255 | 0.002670633 | 0.016856236 | 1 |
| Heart_Failure | ASPC | *ATP6V0A2* | 0.339261769 | 0.003766213 | 0.02124845 | 1 |
| Heart_Failure | ASPC | *FAM174B* | 0.43824767 | 0.00687784 | 0.032493941 | 1 |
| Heart_Failure | ASPC | *SP1* | 0.486214235 | 0.005722844 | 0.028479222 | 1 |
| Heart_Failure | ASPC | *ZNF529* | 0.640484201 | 0.000595474 | 0.010420628 | 1 |
| Heart_Failure | ASPC | *WWOX* | 0.435185436 | 0.001279177 | 0.011183002 | 1 |
| Heart_Failure | ASPC | *RPS23* | 0.252547673 | 0.008505966 | 0.037863253 | 1 |
| Heart_Failure | ASPC | *GNG2* | 0.66955388 | 0.000811562 | 0.010420628 | 1 |
| Heart_Failure | ASPC | *MIR22HG* | 0.44538858 | 0.010344401 | 0.043796714 | 1 |
| Heart_Failure | ASPC | *QRFPR* | 0.515548616 | 0.000316125 | 0.010420628 | 1 |
| Heart_Failure | ASPC | *MAPT* | 0.744241556 | 0.001659307 | 0.012424316 | 1 |
| Heart_Failure | ASPC | *C1QL4* | 7.709214654 | 0.008669916 | 0.038451949 | 1 |
| Heart_Failure | ASPC | *TMEM232* | 0.18467264 | 7.19E-05 | 0.007116983 | 0.348732159 |
| Heart_Failure | ASPC | *EFCAB6* | 0.471724828 | 0.00839077 | 0.037522597 | 1 |
| Heart_Failure | ASPC | *MRPS21* | -0.132126299 | 0.004337193 | 0.023645675 | 1 |
| Heart_Failure | ASPC | *GJA4* | 1.016203143 | 0.000607514 | 0.010420628 | 1 |
| Heart_Failure | ASPC | *EXD3* | 0.802589232 | 0.000675592 | 0.010420628 | 1 |
| Heart_Failure | ASPC | *ZFP69* | 1.311246769 | 0.0008881 | 0.010420628 | 1 |
| Heart_Failure | ASPC | *FAM122A* | 1.437665257 | 0.000601343 | 0.010420628 | 1 |
| Heart_Failure | ASPC | *AGRN* | 0.759173455 | 0.004303336 | 0.023539779 | 1 |
| Heart_Failure | ASPC | *ZNF626* | 1.095416129 | 0.000894908 | 0.010420628 | 1 |
| Heart_Failure | ASPC | *NCR3LG1* | 1.000313894 | 0.009943857 | 0.04258393 | 1 |
| Heart_Failure | ASPC | *RUFY4* | 1.305096604 | 0.008084127 | 0.036285092 | 1 |
| Heart_Failure | ASPC | *PLSCR1* | 0.387459541 | 0.000799695 | 0.010420628 | 1 |
| Heart_Failure | ASPC | *C3orf62* | 0.261323254 | 0.000439069 | 0.010420628 | 1 |
| Heart_Failure | ASPC | *C22orf34* | 0.521344628 | 0.003494388 | 0.020305117 | 1 |
| Heart_Failure | ASPC | *COL25A1* | 0.598812019 | 0.001502864 | 0.01196018 | 1 |

| | | | | | | |
|---|---|---|---|---|---|---|
| Heart_Failure | ASPC | *NBR1* | 0.409641079 | 0.003510591 | 0.020350525 | 1 |
| Heart_Failure | ASPC | *NDOR1* | 1.12268735 | 0.000658955 | 0.010420628 | 1 |
| Heart_Failure | ASPC | *SLC4A5* | 0.942539766 | 0.002235352 | 0.015077872 | 1 |
| Heart_Failure | ASPC | *NOXA1* | 0.714865823 | 0.00363332 | 0.020837904 | 1 |
| Heart_Failure | ASPC | *RPL14* | 0.684149248 | 0.000899441 | 0.010420628 | 1 |
| Heart_Failure | ASPC | *ASTL* | 2.642548252 | 0.003733726 | 0.021138904 | 1 |
| Heart_Failure | ASPC | *MSL1* | -0.158553218 | 0.007591629 | 0.034651536 | 1 |
| Heart_Failure | ASPC | *FAM196A* | 0.54382049 | 0.002465621 | 0.016080322 | 1 |
| Heart_Failure | ASPC | *NELFB* | 0.828368302 | 0.001063871 | 0.010475827 | 1 |
| Heart_Failure | ASPC | *SLC15A5* | 0.169738704 | 0.008030898 | 0.036146491 | 1 |
| Heart_Failure | ASPC | *LITAF* | 0.6117785 | 0.003278479 | 0.019481942 | 1 |
| Heart_Failure | ASPC | *SF3B3* | 0.530745991 | 0.006255427 | 0.030290752 | 1 |
| Heart_Failure | ASPC | *PLAC9* | 0.241755854 | 0.008862428 | 0.039162571 | 1 |
| Heart_Failure | ASPC | *TSPYL1* | 0.722748337 | 0.000533444 | 0.010420628 | 1 |
| Heart_Failure | ASPC | *PNRC2* | 0.793317034 | 0.001427729 | 0.01166347 | 1 |
| Heart_Failure | ASPC | *FHIT* | 0.52419989 | 0.00012699 | 0.009938028 | 0.616157755 |
| Heart_Failure | ASPC | *HLA-DRB1* | -0.106386518 | 1.18E-06 | 0.000381209 | 0.005718132 |
| Heart_Failure | ASPC | *AKR1C3* | 0.906306373 | 0.00081089 | 0.010420628 | 1 |
| Heart_Failure | ASPC | *MPHOSPH8* | 0.335623122 | 0.003902394 | 0.021839006 | 1 |
| Heart_Failure | ASPC | *ZNF471* | 0.758819266 | 0.001959153 | 0.013856867 | 1 |
| Heart_Failure | ASPC | *ZNF493* | 0.634700238 | 0.002420505 | 0.015913671 | 1 |
| Heart_Failure | ASPC | *SUPT3H* | 0.491767702 | 0.00165922 | 0.012424316 | 1 |
| Heart_Failure | ASPC | *CD55* | 0.670673616 | 0.000926228 | 0.010420628 | 1 |
| Heart_Failure | ASPC | *PIK3R4* | 0.935979394 | 3.77E-05 | 0.004717456 | 0.182893567 |
| Heart_Failure | ASPC | *XRCC2* | 2.169677915 | 0.001593272 | 0.012207591 | 1 |
| Heart_Failure | ASPC | *ADH1B* | 0.420264041 | 0.0018519 | 0.013314906 | 1 |
| Heart_Failure | ASPC | *ZKSCAN5* | 0.37671058 | 0.000452044 | 0.010420628 | 1 |
| Heart_Failure | ASPC | *TOMM7* | 0.720207831 | 0.000954807 | 0.010420628 | 1 |
| Heart_Failure | ASPC | *ZNF431* | 0.353648979 | 0.003797986 | 0.021377991 | 1 |
| Heart_Failure | ASPC | *HLA-DQA1* | -0.248336939 | 2.99E-20 | 7.24E-17 | 1.45E-16 |
| Heart_Failure | ASPC | *SLC39A10* | 0.519818093 | 0.008665069 | 0.038451949 | 1 |
| Heart_Failure | ASPC | *WDR5B* | 0.27815233 | 0.001746623 | 0.012805971 | 1 |
| Heart_Failure | ASPC | *ZNF138* | 0.360658467 | 0.000595805 | 0.010420628 | 1 |
| Heart_Failure | ASPC | *ZNF429* | 0.600677757 | 0.002184132 | 0.01485394 | 1 |
| Heart_Failure | ASPC | *GMFB* | 0.873774826 | 0.001060846 | 0.010475827 | 1 |
| Heart_Failure | ASPC | *ZMYM1* | 0.719777156 | 0.001838009 | 0.013251142 | 1 |
| Heart_Failure | ASPC | *ABCB8* | 0.530708027 | 0.003068939 | 0.018520523 | 1 |
| Heart_Failure | ASPC | *SND1* | 0.425780934 | 0.002071483 | 0.01439948 | 1 |
| Heart_Failure | ASPC | *RAMP2-AS1* | 0.58555761 | 0.006515858 | 0.031301925 | 1 |
| Heart_Failure | ASPC | *ZNF665* | 0.902400617 | 0.001605978 | 0.012247119 | 1 |
| Heart_Failure | ASPC | *RPF2* | 0.38960516 | 0.010988769 | 0.045648552 | 1 |
| Heart_Failure | ASPC | *ATG7* | 0.31985577 | 0.004241128 | 0.023278226 | 1 |
| Heart_Failure | ASPC | *RAB40C* | 0.88036662 | 0.000940408 | 0.010420628 | 1 |
| Heart_Failure | ASPC | *TOPORS* | 0.821720933 | 0.002233965 | 0.015077872 | 1 |
| Heart_Failure | ASPC | *PDCD1LG2* | 0.065173484 | 0.001005849 | 0.010420628 | 1 |
| Heart_Failure | ASPC | *RPS26* | 0.096672844 | 0.000269281 | 0.010420628 | 1 |
| Heart_Failure | ASPC | *PSAP* | 0.393810331 | 0.000518421 | 0.010420628 | 1 |
| Heart_Failure | ASPC | *EME2* | 1.476757534 | 0.000694491 | 0.010420628 | 1 |
| Heart_Failure | ASPC | *ADH5* | 0.416584107 | 0.004644659 | 0.024495526 | 1 |
| Heart_Failure | ASPC | *ZNF677* | 0.643035291 | 0.006332346 | 0.030541294 | 1 |

| | | | | | | |
|---|---|---|---|---|---|---|
| Heart_Failure | ASPC | *NOL8* | 0.399111351 | 0.010988708 | 0.045648552 | 1 |
| Heart_Failure | ASPC | *ZNF544* | 0.806285799 | 0.005918106 | 0.029081901 | 1 |
| Heart_Failure | ASPC | *MIER1* | 0.393624701 | 0.002439166 | 0.015984971 | 1 |
| Heart_Failure | ASPC | *MAN1A2* | 0.616567923 | 0.000839243 | 0.010420628 | 1 |
| Heart_Failure | ASPC | *SVIP* | 0.637208687 | 0.00145 | 0.011745243 | 1 |
| Heart_Failure | ASPC | *HELZ* | 0.557156556 | 0.000576174 | 0.010420628 | 1 |
| Heart_Failure | ASPC | *UCKL1* | 0.644165692 | 0.007643579 | 0.034796106 | 1 |
| Heart_Failure | ASPC | *GFPT1* | 0.565298313 | 0.00076823 | 0.010420628 | 1 |
| Heart_Failure | ASPC | *TMA16* | 0.796396153 | 0.000823065 | 0.010420628 | 1 |
| Heart_Failure | ASPC | *HLA-DRB5* | -0.023060593 | 0.006008611 | 0.029388894 | 1 |
| Heart_Failure | ASPC | *ZNF28* | 0.630513395 | 0.001719141 | 0.012721823 | 1 |
| Heart_Failure | ASPC | *TUSC1* | 1.470931953 | 0.002579135 | 0.016465743 | 1 |
| Heart_Failure | ASPC | *PAPSS2* | 0.636570647 | 0.000883975 | 0.010420628 | 1 |
| Heart_Failure | ASPC | *CEP290* | 0.281794149 | 0.006770784 | 0.032113241 | 1 |
| Heart_Failure | ASPC | *PPP1R14C* | 3.59543856 | 0.000940033 | 0.010420628 | 1 |
| Heart_Failure | ASPC | *GPATCH3* | 1.11842641 | 0.000949314 | 0.010420628 | 1 |
| Heart_Failure | ASPC | *RCSD1* | 0.626167217 | 0.001257893 | 0.011106021 | 1 |
| Heart_Failure | ASPC | *TYW1* | 0.288981959 | 0.007543339 | 0.034496023 | 1 |
| Heart_Failure | ASPC | *DZIP3* | 0.447960598 | 0.006762944 | 0.032107443 | 1 |
| Heart_Failure | ASPC | *ECT2L* | 0.781386754 | 0.002686706 | 0.01690778 | 1 |
| Heart_Failure | ASPC | *PCMTD2* | 0.680877469 | 0.001357848 | 0.011457877 | 1 |
| Heart_Failure | ASPC | *RUFY2* | 0.647046273 | 0.000619862 | 0.010420628 | 1 |
| Heart_Failure | ASPC | *BMPR2* | 0.523899997 | 0.001218329 | 0.010920212 | 1 |
| Heart_Failure | ASPC | *TCEA3* | 0.245680741 | 0.005869264 | 0.028911341 | 1 |
| Heart_Failure | ASPC | *HSD17B8* | 0.11705449 | 0.002269591 | 0.015210019 | 1 |
| Heart_Failure | ASPC | *PRRT1* | 0.097003603 | 0.000464846 | 0.010420628 | 1 |
| Heart_Failure | ASPC | *BAG6* | 0.213287618 | 0.001720016 | 0.012721823 | 1 |
| Heart_Failure | ASPC | *ZNF814* | 0.291951353 | 0.011058942 | 0.045861526 | 1 |
| Heart_Failure | ASPC | *MICB* | 0.031465267 | 0.002025397 | 0.014258141 | 1 |
| Heart_Failure | ASPC | *MICA* | 0.181619808 | 1.84E-06 | 0.000524434 | 0.008915386 |
| Heart_Failure | ASPC | *LILRB3* | 0.39344075 | 0.000948314 | 0.010420628 | 1 |
| Heart_Failure | ASPC | *ZNF468* | 0.606031723 | 0.008069221 | 0.036285092 | 1 |
| Heart_Failure | ASPC | *DCTN1* | 0.956933135 | 0.001571747 | 0.012135784 | 1 |
| Heart_Failure | ASPC | *FAM216A* | 0.728347402 | 0.000112138 | 0.009380905 | 0.544092508 |
| Heart_Failure | ASPC | *SLC35B4* | 0.679070818 | 0.003922813 | 0.021927981 | 1 |
| Heart_Failure | ASPC | *SCGB2B2* | 0.739543802 | 0.000853204 | 0.010420628 | 1 |
| Heart_Failure | ASPC | *CCDC144NL* | 1.450980329 | 0.000861719 | 0.010420628 | 1 |
| Heart_Failure | ASPC | *NAP1L4* | 0.356443302 | 0.004499919 | 0.024118544 | 1 |
| Heart_Failure | ASPC | *LIN52* | 0.230418128 | 0.000573745 | 0.010420628 | 1 |
| Heart_Failure | ASPC | *C1QTNF9B* | 10.68124922 | 0.000287915 | 0.010420628 | 1 |
| Heart_Failure | ASPC | *METTL6* | 0.355684026 | 0.004166714 | 0.02294767 | 1 |
| Heart_Failure | ASPC | *SACM1L* | 0.638929569 | 0.003473502 | 0.020232214 | 1 |
| Heart_Failure | ASPC | *SLC48A1* | 0.759992702 | 0.002548018 | 0.01635315 | 1 |
| Heart_Failure | ASPC | *SUPT4H1* | 0.668293325 | 0.002237442 | 0.015077872 | 1 |
| Heart_Failure | ASPC | *PPP1CB* | 0.212665436 | 0.008969616 | 0.039349527 | 1 |
| Heart_Failure | ASPC | *ATF6B* | 0.362877234 | 2.12E-07 | 8.58E-05 | 0.001030126 |
| Heart_Failure | ASPC | *GALT* | 0.29696769 | 0.006522421 | 0.03130246 | 1 |
| Heart_Failure | ASPC | *REPIN1* | 1.228693385 | 0.000683417 | 0.010420628 | 1 |
| Heart_Failure | ASPC | *ZNF891* | 1.172886528 | 9.09E-08 | 4.42E-05 | 0.000440893 |
| Heart_Failure | ASPC | *CPNE1* | -0.073360028 | 0.001410323 | 0.011637563 | 1 |

| | | | | | | |
|---|---|---|---|---|---|---|
| Heart_Failure | ASPC | *PAXIP1-AS2* | 0.641014664 | 0.001395233 | 0.011637563 | 1 |
| Heart_Failure | ASPC | *LYRM4* | 0.691471968 | 0.000939014 | 0.010420628 | 1 |
| Heart_Failure | ASPC | *SH3D21* | 0.250630029 | 0.000679594 | 0.010420628 | 1 |
| Heart_Failure | ASPC | *ANG* | 1.041744599 | 0.002032404 | 0.014258141 | 1 |
| Heart_Failure | ASPC | *GNAT3* | 0.822314307 | 0.00470901 | 0.024660342 | 1 |
| Heart_Failure | ASPC | *VPS16* | 0.979969209 | 0.000401092 | 0.010420628 | 1 |
| Heart_Failure | ASPC | *NPEPL1* | 0.88035502 | 0.001893033 | 0.013507349 | 1 |
| Heart_Failure | ASPC | *PAM16* | 0.51827198 | 0.004456054 | 0.023969816 | 1 |
| Heart_Failure | ASPC | *RNASEK* | 0.193476181 | 0.006058621 | 0.029573872 | 1 |
| Heart_Failure | ASPC | *FXYD7* | 0.17774697 | 0.00711904 | 0.033141513 | 1 |
| Heart_Failure | ASPC | *APOL6* | 0.6191015 | 0.001081791 | 0.01047675 | 1 |
| Heart_Failure | ASPC | *OR2A1* | 0.144396516 | 0.001607136 | 0.012247119 | 1 |
| Heart_Failure | ASPC | *AC062017.1* | 1.079006673 | 0.000897992 | 0.010420628 | 1 |
| Heart_Failure | ASPC | *LINC01117* | 0.684099189 | 0.002033519 | 0.014258141 | 1 |
| Heart_Failure | ASPC | *MIAT* | 0.3640017 | 0.006654989 | 0.031781502 | 1 |
| Heart_Failure | ASPC | *FAM138B* | 17.29312486 | 0.000691935 | 0.010420628 | 1 |
| Heart_Failure | ASPC | *AC009299.3* | 0.893552728 | 0.003411808 | 0.019992868 | 1 |
| Heart_Failure | ASPC | *SCAMP4* | 0.922920501 | 0.000777165 | 0.010420628 | 1 |
| Heart_Failure | ASPC | *SLC2A1-AS1* | 0.267228655 | 0.011655031 | 0.047923908 | 1 |
| Heart_Failure | ASPC | *AC092171.4* | 1.372771689 | 0.001258927 | 0.011106021 | 1 |
| Heart_Failure | ASPC | *RPS18* | 0.596689943 | 3.79E-05 | 0.004717456 | 0.183980777 |
| Heart_Failure | ASPC | *HLA-DQB2* | 0.086060659 | 6.95E-18 | 8.43E-15 | 3.37E-14 |
| Heart_Failure | ASPC | *ZNF736* | 0.120869181 | 0.000566054 | 0.010420628 | 1 |
| Heart_Failure | ASPC | *ANKRD65* | 2.817962832 | 0.001265338 | 0.011113358 | 1 |
| Heart_Failure | ASPC | *ZBTB22* | 0.223203902 | 0.000137929 | 0.010198142 | 0.669232692 |
| Heart_Failure | ASPC | *HLA-DQA2* | 0.105376094 | 2.46E-18 | 3.97E-15 | 1.19E-14 |
| Heart_Failure | ASPC | *PSMB9* | 0.131570438 | 9.89E-08 | 4.42E-05 | 0.000479798 |
| Heart_Failure | ASPC | *NSUN6* | 0.454991769 | 0.001101572 | 0.010583812 | 1 |
| Heart_Failure | ASPC | *HOGA1* | 1.198402875 | 0.010600474 | 0.044608413 | 1 |
| Heart_Failure | ASPC | *PI4KA* | 0.431827803 | 0.00229247 | 0.015321024 | 1 |
| Heart_Failure | ASPC | *ARFGAP3* | 0.761757886 | 0.001399416 | 0.011637563 | 1 |
| Heart_Failure | ASPC | *DECR2* | 1.451243563 | 0.0016104 | 0.012247119 | 1 |
| Heart_Failure | ASPC | *NBPF15* | 0.686989725 | 0.007870749 | 0.035657211 | 1 |
| Heart_Failure | ASPC | *TTC4* | -0.169102069 | 0.001057933 | 0.010475827 | 1 |
| Heart_Failure | ASPC | *LILRA6* | 0.073363731 | 0.002113288 | 0.014508134 | 1 |
| Heart_Failure | ASPC | *C4A* | -0.135309583 | 0.005923411 | 0.029081901 | 1 |
| Heart_Failure | ASPC | *NEAT1* | 0.408105699 | 0.000278151 | 0.010420628 | 1 |
| Heart_Failure | ASPC | *UBAP1L* | 0.507644641 | 0.008437784 | 0.03769809 | 1 |
| Heart_Failure | ASPC | *LINC00926* | 1.880096097 | 0.000352299 | 0.010420628 | 1 |
| Heart_Failure | ASPC | *CDK11B* | 0.947861986 | 0.000663503 | 0.010420628 | 1 |
| Heart_Failure | ASPC | *ABHD14A* | 0.69196169 | 0.001595139 | 0.012207591 | 1 |
| Heart_Failure | ASPC | *C5orf17* | 0.721120142 | 0.002756816 | 0.017259444 | 1 |
| Heart_Failure | ASPC | *PDCD6* | 0.829491047 | 0.000840329 | 0.010420628 | 1 |
| Heart_Failure | ASPC | *ZNF718* | 0.456634894 | 0.006403248 | 0.030821983 | 1 |
| Heart_Failure | ASPC | *SMIM20* | 1.124452693 | 0.000775552 | 0.010420628 | 1 |
| Heart_Failure | ASPC | *GLI4* | 0.696746646 | 0.007308174 | 0.033770726 | 1 |
| Heart_Failure | ASPC | *ZNF550* | 0.831611053 | 0.004436367 | 0.02394355 | 1 |
| Heart_Failure | ASPC | *PCDHGB2* | 1.181671821 | 0.002867064 | 0.017608856 | 1 |
| Heart_Failure | ASPC | *KIAA1147* | 0.317898313 | 0.002619147 | 0.0166221 | 1 |
| Heart_Failure | ASPC | *CNPY2* | 0.875312081 | 0.000572025 | 0.010420628 | 1 |

| | | | | | | |
|---|---|---|---|---|---|---|
| Heart_Failure | ASPC | *HIF1A-AS2* | 0.752222158 | 0.000225361 | 0.010420628 | 1 |
| Heart_Failure | ASPC | *MC1R* | 0.23176892 | 0.004413951 | 0.023889952 | 1 |
| Heart_Failure | ASPC | *LINC00639* | 0.266314753 | 0.010668892 | 0.04479427 | 1 |
| Heart_Failure | ASPC | *PWRN1* | 0.251944684 | 0.001835631 | 0.013251142 | 1 |
| Heart_Failure | ASPC | *LINC00672* | 1.930928819 | 0.001520297 | 0.011993971 | 1 |
| Heart_Failure | ASPC | *DYNLL2* | 1.134820325 | 0.002339937 | 0.015531293 | 1 |
| Heart_Failure | ASPC | *ZNF582-AS1* | 1.534795608 | 0.001458534 | 0.011794677 | 1 |
| Heart_Failure | ASPC | *ZNF224* | 0.820008345 | 0.00095925 | 0.010420628 | 1 |
| Heart_Failure | ASPC | *NDUFA7* | 4.630257103 | 0.003018689 | 0.018285489 | 1 |
| Heart_Failure | ASPC | *ZNF587B* | 1.468012191 | 0.000881955 | 0.010420628 | 1 |
| Heart_Failure | ASPC | *GTF2H5* | 0.154647192 | 0.000415102 | 0.010420628 | 1 |
| Heart_Failure | ASPC | *CASC15* | 0.685954516 | 0.000768858 | 0.010420628 | 1 |
| Heart_Failure | ASPC | *PAXIP1-AS1* | 0.738642465 | 0.003573151 | 0.020590179 | 1 |
| Heart_Failure | b_cell | *TSLP* | 4.14522968 | 1.29E-07 | 0.000124968 | 0.00062484 |
| Heart_Failure | b_cell | *RNF123* | 4.563669709 | 5.75E-06 | 0.004649553 | 0.027897321 |
| Heart_Failure | b_cell | *GSDMA* | -5.028282175 | 1.89E-08 | 3.05E-05 | 9.16E-05 |
| Heart_Failure | b_cell | *HLA-DQA1* | -3.546408856 | 7.93E-08 | 9.62E-05 | 0.000384906 |
| Heart_Failure | b_cell | *HLA-DQB2* | 1.681517016 | 4.04E-11 | 9.81E-08 | 1.96E-07 |
| Heart_Failure | b_cell | *ZBTB22* | 5.265053844 | 3.62E-05 | 0.025101537 | 0.175710756 |
| Heart_Failure | b_cell | *HLA-DQA2* | 3.881494159 | 1.55E-15 | 7.50E-12 | 7.50E-12 |
| Heart_Failure | dendritic_cell | *NFYA* | -4.497342855 | 5.89E-05 | 0.000119295 | 0.285591359 |
| Heart_Failure | dendritic_cell | *ENPP4* | 1.461517248 | 0.024398175 | 0.030557549 | 1 |
| Heart_Failure | dendritic_cell | *SEMA3F* | -2.785680665 | 4.31E-10 | 2.61E-07 | 2.09E-06 |
| Heart_Failure | dendritic_cell | *KRIT1* | -2.615324517 | 1.53E-05 | 3.83E-05 | 0.074224323 |
| Heart_Failure | dendritic_cell | *RAD52* | -2.909790724 | 8.93E-05 | 0.000170708 | 0.43308724 |
| Heart_Failure | dendritic_cell | *HS3ST1* | -1.397233811 | 7.64E-05 | 0.000149589 | 0.370681266 |
| Heart_Failure | dendritic_cell | *AOC1* | -2.407527113 | 0.002168329 | 0.003166035 | 1 |
| Heart_Failure | dendritic_cell | *SNX11* | -1.774584331 | 2.36E-05 | 5.54E-05 | 0.11474075 |
| Heart_Failure | dendritic_cell | *TMEM176A* | -4.176688561 | 2.46E-06 | 9.80E-06 | 0.011911898 |
| Heart_Failure | dendritic_cell | *M6PR* | 2.537753209 | 0.000670684 | 0.001062062 | 1 |
| Heart_Failure | dendritic_cell | *DBNDD1* | 0.921399453 | 5.40E-05 | 0.000111212 | 0.26203379 |
| Heart_Failure | dendritic_cell | *ALS2* | -2.552522601 | 3.02E-06 | 1.14E-05 | 0.01465557 |
| Heart_Failure | dendritic_cell | *CD38* | 4.795420315 | 1.93E-06 | 8.34E-06 | 0.009361018 |
| Heart_Failure | dendritic_cell | *RBM6* | -2.507315695 | 3.42E-11 | 3.32E-08 | 1.66E-07 |
| Heart_Failure | dendritic_cell | *RECQL* | -3.565696759 | 6.41E-05 | 0.000128317 | 0.311041392 |
| Heart_Failure | dendritic_cell | *CDC27* | -1.805101263 | 0.008217588 | 0.01100821 | 1 |
| Heart_Failure | dendritic_cell | *POLR2J* | 2.556103596 | 7.58E-08 | 3.44E-06 | 0.000367802 |
| Heart_Failure | dendritic_cell | *MEOX1* | -2.243384546 | 2.46E-07 | 3.66E-06 | 0.001195987 |
| Heart_Failure | dendritic_cell | *PON1* | -6.388718654 | 3.27E-07 | 3.66E-06 | 0.001586945 |
| Heart_Failure | dendritic_cell | *GCFC2* | -2.381353692 | 6.75E-06 | 2.00E-05 | 0.032738152 |
| Heart_Failure | dendritic_cell | *RHBDD2* | -2.594340473 | 0.000273137 | 0.000467794 | 1 |
| Heart_Failure | dendritic_cell | *ZNF195* | -2.398735871 | 0.002852993 | 0.00405826 | 1 |
| Heart_Failure | dendritic_cell | *PDK2* | -1.841012165 | 9.76E-08 | 3.44E-06 | 0.000473747 |
| Heart_Failure | dendritic_cell | *GGNBP2* | -2.903270609 | 2.48E-05 | 5.74E-05 | 0.120536588 |
| Heart_Failure | dendritic_cell | *ITGA2B* | -2.566673433 | 2.75E-05 | 6.22E-05 | 0.133422726 |
| Heart_Failure | dendritic_cell | *ASB4* | -1.546925553 | 0.00258478 | 0.003702791 | 1 |
| Heart_Failure | dendritic_cell | *TMEM98* | -1.974871021 | 2.06E-06 | 8.74E-06 | 0.010019137 |
| Heart_Failure | dendritic_cell | *ABCC8* | 1.927824613 | 0.011500754 | 0.015114209 | 1 |
| Heart_Failure | dendritic_cell | *SYNRG* | -2.644515146 | 5.99E-05 | 0.000121244 | 0.290742331 |
| Heart_Failure | dendritic_cell | *ZNF263* | -2.845762008 | 6.86E-06 | 2.03E-05 | 0.033296571 |

| | | | | | | |
|---|---|---|---|---|---|---|
| Heart_Failure | dendritic_cell | *SPATA20* | -1.474424466 | 4.62E-05 | 9.67E-05 | 0.224132804 |
| Heart_Failure | dendritic_cell | *BAIAP2L1* | -2.91196406 | 2.09E-06 | 8.79E-06 | 0.010129641 |
| Heart_Failure | dendritic_cell | *AGK* | -2.018187623 | 8.64E-06 | 2.40E-05 | 0.04190848 |
| Heart_Failure | dendritic_cell | *FARP2* | -2.796414377 | 0.001750023 | 0.002595876 | 1 |
| Heart_Failure | dendritic_cell | *USH1C* | -17.09026016 | 2.32E-05 | 5.44E-05 | 0.112505928 |
| Heart_Failure | dendritic_cell | *IFRD1* | -1.775808022 | 1.66E-05 | 4.11E-05 | 0.080616099 |
| Heart_Failure | dendritic_cell | *VPS41* | -2.747827401 | 0.000266121 | 0.000456583 | 1 |
| Heart_Failure | dendritic_cell | *SCIN* | -3.074487686 | 0.001037738 | 0.001596419 | 1 |
| Heart_Failure | dendritic_cell | *CCDC124* | -1.732388847 | 7.99E-06 | 2.25E-05 | 0.038764485 |
| Heart_Failure | dendritic_cell | *CEACAM21* | -1.624859607 | 0.008031339 | 0.010764657 | 1 |
| Heart_Failure | dendritic_cell | *CD79B* | -3.679212659 | 0.001773707 | 0.002626992 | 1 |
| Heart_Failure | dendritic_cell | *ST7L* | -2.210753347 | 8.05E-08 | 3.44E-06 | 0.000390489 |
| Heart_Failure | dendritic_cell | *PAX6* | -1.647252431 | 0.000548108 | 0.000882063 | 1 |
| Heart_Failure | dendritic_cell | *TSR3* | -3.161609708 | 1.80E-06 | 7.98E-06 | 0.008732642 |
| Heart_Failure | dendritic_cell | *PIGQ* | 3.402940404 | 1.13E-06 | 6.03E-06 | 0.005483261 |
| Heart_Failure | dendritic_cell | *FMO3* | -1.582071111 | 9.99E-06 | 2.70E-05 | 0.048494746 |
| Heart_Failure | dendritic_cell | *PSMB1* | -3.235733831 | 4.48E-05 | 9.41E-05 | 0.21754346 |
| Heart_Failure | dendritic_cell | *NADK* | 4.024808195 | 3.42E-05 | 7.51E-05 | 0.165803203 |
| Heart_Failure | dendritic_cell | *CYB561* | -3.440614021 | 8.46E-08 | 3.44E-06 | 0.000410607 |
| Heart_Failure | dendritic_cell | *SS18L2* | -1.821446437 | 4.57E-07 | 3.90E-06 | 0.002215148 |
| Heart_Failure | dendritic_cell | *MGST1* | -0.895803324 | 0.000332914 | 0.000560089 | 1 |
| Heart_Failure | dendritic_cell | *IL32* | -4.932168279 | 0.001401893 | 0.002109143 | 1 |
| Heart_Failure | dendritic_cell | *PKD1* | -2.974909965 | 7.21E-05 | 0.00014231 | 0.349940907 |
| Heart_Failure | dendritic_cell | *MAPK8IP2* | -7.03827372 | 4.17E-05 | 8.88E-05 | 0.202394692 |
| Heart_Failure | dendritic_cell | *MED24* | -3.428879166 | 5.77E-06 | 1.80E-05 | 0.028010525 |
| Heart_Failure | dendritic_cell | *RPS20* | -2.618640693 | 2.76E-05 | 6.23E-05 | 0.13393744 |
| Heart_Failure | dendritic_cell | *TRAF3IP3* | 1.684946945 | 1.00E-07 | 3.44E-06 | 0.000486097 |
| Heart_Failure | dendritic_cell | *POMT2* | -2.248244715 | 9.14E-06 | 2.50E-05 | 0.04433131 |
| Heart_Failure | dendritic_cell | *MLXIPL* | -0.900157805 | 0.000317046 | 0.000535436 | 1 |
| Heart_Failure | dendritic_cell | *DYRK4* | 1.891258141 | 4.79E-06 | 1.57E-05 | 0.023255435 |
| Heart_Failure | dendritic_cell | *FUZ* | -2.556634649 | 1.11E-07 | 3.44E-06 | 0.000538004 |
| Heart_Failure | dendritic_cell | *ZNF200* | -1.907387277 | 0.00218887 | 0.003191226 | 1 |
| Heart_Failure | dendritic_cell | *LRRC23* | -2.402023278 | 0.002479661 | 0.003566948 | 1 |
| Heart_Failure | dendritic_cell | *CLCN6* | -3.142551762 | 0.00011378 | 0.000212168 | 0.55206184 |
| Heart_Failure | dendritic_cell | *SLC6A7* | 40.5952236 | 1.90E-07 | 3.60E-06 | 0.000921586 |
| Heart_Failure | dendritic_cell | *TSPAN9* | -1.648367815 | 0.000139167 | 0.000253278 | 0.675238329 |
| Heart_Failure | dendritic_cell | *UTP18* | -2.264224846 | 2.45E-05 | 5.69E-05 | 0.118682545 |
| Heart_Failure | dendritic_cell | *TTC19* | -2.279455096 | 2.74E-05 | 6.19E-05 | 0.132764616 |
| Heart_Failure | dendritic_cell | *DPF1* | -3.227345033 | 0.00312516 | 0.004416918 | 1 |
| Heart_Failure | dendritic_cell | *LARS2* | -2.697367273 | 2.51E-05 | 5.77E-05 | 0.121615562 |
| Heart_Failure | dendritic_cell | *DCN* | -1.405805658 | 3.33E-06 | 1.22E-05 | 0.016134316 |
| Heart_Failure | dendritic_cell | *PPP5C* | -2.34159806 | 1.53E-06 | 7.21E-06 | 0.007443352 |
| Heart_Failure | dendritic_cell | *CEP68* | -2.019630234 | 3.32E-06 | 1.22E-05 | 0.016090598 |
| Heart_Failure | dendritic_cell | *ZBTB32* | -2.581179789 | 1.00E-07 | 3.44E-06 | 0.000485171 |
| Heart_Failure | dendritic_cell | *ERCC1* | -3.000113646 | 9.49E-05 | 0.000179983 | 0.460216968 |
| Heart_Failure | dendritic_cell | *ALOX5* | 0.96658518 | 6.84E-07 | 4.53E-06 | 0.003320485 |
| Heart_Failure | dendritic_cell | *MAP4K5* | -2.596279297 | 3.01E-05 | 6.70E-05 | 0.146269065 |
| Heart_Failure | dendritic_cell | *EHD3* | -3.454211222 | 2.66E-06 | 1.04E-05 | 0.012922144 |
| Heart_Failure | dendritic_cell | *MAN2B2* | -3.867557143 | 2.24E-05 | 5.30E-05 | 0.108621228 |
| Heart_Failure | dendritic_cell | *SLC25A39* | -1.92646472 | 6.59E-08 | 3.44E-06 | 0.000319661 |

| | | | | | | |
|---|---|---|---|---|---|---|
| Heart_Failure | dendritic_cell | *RNF14* | -2.664251573 | 0.000359195 | 0.000598083 | 1 |
| Heart_Failure | dendritic_cell | *TACC3* | 6.046856942 | 1.31E-05 | 3.37E-05 | 0.063739455 |
| Heart_Failure | dendritic_cell | *MDH1* | -1.957640592 | 0.001835684 | 0.002709687 | 1 |
| Heart_Failure | dendritic_cell | *COX15* | -3.294634264 | 2.50E-05 | 5.77E-05 | 0.121288309 |
| Heart_Failure | dendritic_cell | *CCDC88C* | -1.742417462 | 4.96E-08 | 3.44E-06 | 0.000240775 |
| Heart_Failure | dendritic_cell | *CHDH* | 6.702036146 | 1.27E-07 | 3.44E-06 | 0.000616091 |
| Heart_Failure | dendritic_cell | *CLCA4* | -6.839827522 | 0.000184279 | 0.000326561 | 0.894123466 |
| Heart_Failure | dendritic_cell | *RUFY3* | -2.008942912 | 0.025655134 | 0.031975009 | 1 |
| Heart_Failure | dendritic_cell | *SLC11A1* | -0.956153364 | 0.004315011 | 0.005993826 | 1 |
| Heart_Failure | dendritic_cell | *WWTR1* | -1.996974295 | 3.03E-06 | 1.14E-05 | 0.014701763 |
| Heart_Failure | dendritic_cell | *AGPS* | -2.491610337 | 0.000585587 | 0.000936168 | 1 |
| Heart_Failure | dendritic_cell | *TTC27* | -1.940905066 | 3.01E-06 | 1.14E-05 | 0.014584606 |
| Heart_Failure | dendritic_cell | *VSIG2* | 1.954158677 | 6.10E-08 | 3.44E-06 | 0.000296071 |
| Heart_Failure | dendritic_cell | *HGF* | 3.962869485 | 0.001077735 | 0.001653232 | 1 |
| Heart_Failure | dendritic_cell | *SLC7A9* | 0.91406639 | 0.031054791 | 0.038379482 | 1 |
| Heart_Failure | dendritic_cell | *OSBPL5* | -3.131034756 | 2.23E-05 | 5.27E-05 | 0.107968021 |
| Heart_Failure | dendritic_cell | *CPS1* | -1.512313009 | 3.95E-06 | 1.37E-05 | 0.019147093 |
| Heart_Failure | dendritic_cell | *SLC45A4* | 2.114413745 | 2.68E-05 | 6.08E-05 | 0.129898791 |
| Heart_Failure | dendritic_cell | *ZDHHC6* | 3.477816923 | 0.007946789 | 0.010660166 | 1 |
| Heart_Failure | dendritic_cell | *GRAMD1B* | 0.724920498 | 7.41E-06 | 2.12E-05 | 0.035969463 |
| Heart_Failure | dendritic_cell | *RNH1* | -2.137123786 | 0.000181611 | 0.000322302 | 0.881174886 |
| Heart_Failure | dendritic_cell | *GLRX2* | 5.16558828 | 0.009599374 | 0.012739651 | 1 |
| Heart_Failure | dendritic_cell | *ABCC2* | -2.638534975 | 3.69E-07 | 3.66E-06 | 0.001792427 |
| Heart_Failure | dendritic_cell | *GCLM* | -2.754580526 | 1.72E-06 | 7.75E-06 | 0.008357309 |
| Heart_Failure | dendritic_cell | *UBR2* | -2.172700052 | 0.003926268 | 0.005484804 | 1 |
| Heart_Failure | dendritic_cell | *RRAGD* | -3.127211918 | 4.32E-06 | 1.46E-05 | 0.02097597 |
| Heart_Failure | dendritic_cell | *HSF2* | -2.621708535 | 4.30E-05 | 9.10E-05 | 0.208589811 |
| Heart_Failure | dendritic_cell | *TOMM34* | -5.46060664 | 5.26E-05 | 0.000108608 | 0.255446923 |
| Heart_Failure | dendritic_cell | *SEC63* | -2.781357108 | 3.23E-05 | 7.12E-05 | 0.156666448 |
| Heart_Failure | dendritic_cell | *FAS* | -2.49011892 | 4.60E-06 | 1.53E-05 | 0.022333365 |
| Heart_Failure | dendritic_cell | *RNASET2* | 1.616908237 | 0.004885871 | 0.006748148 | 1 |
| Heart_Failure | dendritic_cell | *AGPAT4* | -2.874493869 | 7.17E-06 | 2.08E-05 | 0.034790792 |
| Heart_Failure | dendritic_cell | *MIPEP* | -2.021137463 | 7.28E-06 | 2.10E-05 | 0.035331525 |
| Heart_Failure | dendritic_cell | *INSRR* | -3.073091031 | 1.18E-06 | 6.14E-06 | 0.005702776 |
| Heart_Failure | dendritic_cell | *B4GALT7* | -4.404969414 | 0.000141877 | 0.000257718 | 0.688387604 |
| Heart_Failure | dendritic_cell | *SH2D2A* | -5.61545837 | 1.05E-06 | 5.80E-06 | 0.005101615 |
| Heart_Failure | dendritic_cell | *VEZT* | -2.642960716 | 4.18E-05 | 8.89E-05 | 0.202702306 |
| Heart_Failure | dendritic_cell | *BRD9* | -2.861311122 | 0.000446075 | 0.000728493 | 1 |
| Heart_Failure | dendritic_cell | *SNX1* | -2.811536917 | 0.000324343 | 0.000546808 | 1 |
| Heart_Failure | dendritic_cell | *ANK1* | -2.594639491 | 3.26E-07 | 3.66E-06 | 0.001582227 |
| Heart_Failure | dendritic_cell | *RABEP1* | -2.592269245 | 4.24E-05 | 9.01E-05 | 0.205830463 |
| Heart_Failure | dendritic_cell | *BAK1* | 1.813619288 | 0.000157089 | 0.000282608 | 0.762195071 |
| Heart_Failure | dendritic_cell | *IKZF2* | -2.691832849 | 6.44E-06 | 1.93E-05 | 0.03124008 |
| Heart_Failure | dendritic_cell | *CENPQ* | -0.795280904 | 0.016837412 | 0.021601038 | 1 |
| Heart_Failure | dendritic_cell | *IFT88* | -2.424650235 | 2.28E-06 | 9.32E-06 | 0.011073893 |
| Heart_Failure | dendritic_cell | *CHPF2* | -2.027585845 | 0.00027659 | 0.000473375 | 1 |
| Heart_Failure | dendritic_cell | *UBA6* | -2.264480152 | 5.62E-05 | 0.000115059 | 0.272919167 |
| Heart_Failure | dendritic_cell | *ATP6V0A1* | -2.116342146 | 0.016267848 | 0.020925662 | 1 |
| Heart_Failure | dendritic_cell | *EFCAB1* | 0.774159357 | 0.020324552 | 0.025727818 | 1 |
| Heart_Failure | dendritic_cell | *ASTE1* | -3.634857865 | 0.000121006 | 0.000223922 | 0.587122478 |

| | | | | | | |
|---|---|---|---|---|---|---|
| Heart_Failure | dendritic_cell | *MYOC* | -2.918559246 | 1.34E-06 | 6.62E-06 | 0.006493139 |
| Heart_Failure | dendritic_cell | *SH3YL1* | -2.543575478 | 6.34E-07 | 4.39E-06 | 0.003076759 |
| Heart_Failure | dendritic_cell | *VCL* | -2.857738557 | 8.30E-05 | 0.000160579 | 0.402893363 |
| Heart_Failure | dendritic_cell | *ADSS* | -2.989478765 | 0.018021791 | 0.022992829 | 1 |
| Heart_Failure | dendritic_cell | *TBC1D23* | -3.029569976 | 0.000122286 | 0.000225773 | 0.593330445 |
| Heart_Failure | dendritic_cell | *TUBG2* | -1.883298366 | 7.56E-06 | 2.16E-05 | 0.036688742 |
| Heart_Failure | dendritic_cell | *FLT4* | -0.901272026 | 0.001153171 | 0.001760599 | 1 |
| Heart_Failure | dendritic_cell | *NSUN2* | -3.207896559 | 0.000756671 | 0.001187376 | 1 |
| Heart_Failure | dendritic_cell | *MRI1* | -1.629646747 | 0.000290961 | 0.000495697 | 1 |
| Heart_Failure | dendritic_cell | *AGA* | 5.586428873 | 1.40E-05 | 3.56E-05 | 0.067958648 |
| Heart_Failure | dendritic_cell | *PI4K2B* | -3.869106749 | 3.16E-07 | 3.66E-06 | 0.001533532 |
| Heart_Failure | dendritic_cell | *BOD1L1* | -1.655197891 | 0.039915921 | 0.048710274 | 1 |
| Heart_Failure | dendritic_cell | *DNAH5* | -2.524566531 | 3.17E-07 | 3.66E-06 | 0.001537068 |
| Heart_Failure | dendritic_cell | *PHLPP2* | -1.376721036 | 0.004112171 | 0.005725181 | 1 |
| Heart_Failure | dendritic_cell | *SPDL1* | -1.964038269 | 0.000554565 | 0.000890976 | 1 |
| Heart_Failure | dendritic_cell | *PQLC2* | -3.362917384 | 4.54E-07 | 3.90E-06 | 0.002203613 |
| Heart_Failure | dendritic_cell | *INPP4A* | 2.483959646 | 0.00870905 | 0.011612067 | 1 |
| Heart_Failure | dendritic_cell | *PSMA4* | -3.109251666 | 6.42E-06 | 1.93E-05 | 0.031135208 |
| Heart_Failure | dendritic_cell | *SPATA7* | -2.432012956 | 7.24E-07 | 4.67E-06 | 0.003513764 |
| Heart_Failure | dendritic_cell | *MED17* | -4.432088052 | 7.58E-06 | 2.16E-05 | 0.036797569 |
| Heart_Failure | dendritic_cell | *CAPG* | 2.18861802 | 0.001377328 | 0.002074758 | 1 |
| Heart_Failure | dendritic_cell | *AP2S1* | 4.189913656 | 7.64E-07 | 4.80E-06 | 0.003709209 |
| Heart_Failure | dendritic_cell | *CUL7* | -3.013671629 | 3.87E-05 | 8.34E-05 | 0.187615529 |
| Heart_Failure | dendritic_cell | *CTNNA1* | -2.447282252 | 4.53E-05 | 9.50E-05 | 0.219955212 |
| Heart_Failure | dendritic_cell | *CNTLN* | -2.52688442 | 0.000194244 | 0.000343217 | 0.942473636 |
| Heart_Failure | dendritic_cell | *YTHDC2* | -2.856057281 | 0.000366071 | 0.000608697 | 1 |
| Heart_Failure | dendritic_cell | *POLR2B* | -2.546896423 | 0.017289163 | 0.022122104 | 1 |
| Heart_Failure | dendritic_cell | *ARAP2* | -2.212095833 | 0.009159429 | 0.012182443 | 1 |
| Heart_Failure | dendritic_cell | *DTNBP1* | 3.452263436 | 0.004041703 | 0.005636776 | 1 |
| Heart_Failure | dendritic_cell | *ANO2* | -1.148676759 | 3.14E-07 | 3.66E-06 | 0.001525768 |
| Heart_Failure | dendritic_cell | *RRM2B* | -3.328582908 | 0.028765369 | 0.035631751 | 1 |
| Heart_Failure | dendritic_cell | *SNX29* | 2.33245837 | 0.01062017 | 0.01400627 | 1 |
| Heart_Failure | dendritic_cell | *UTS2* | 1.305206978 | 1.79E-07 | 3.58E-06 | 0.000867505 |
| Heart_Failure | dendritic_cell | *RCN1* | -3.604418004 | 3.79E-06 | 1.33E-05 | 0.018366042 |
| Heart_Failure | dendritic_cell | *JKAMP* | -2.25813529 | 0.007187749 | 0.009695568 | 1 |
| Heart_Failure | dendritic_cell | *ARHGEF5* | 2.165427221 | 0.000182798 | 0.000324173 | 0.886937767 |
| Heart_Failure | dendritic_cell | *MCUR1* | -4.202739393 | 3.18E-06 | 1.18E-05 | 0.015410122 |
| Heart_Failure | dendritic_cell | *LAMC3* | 1.388795519 | 2.41E-05 | 5.63E-05 | 0.117084853 |
| Heart_Failure | dendritic_cell | *MAPK9* | -3.099529474 | 6.32E-06 | 1.92E-05 | 0.030652614 |
| Heart_Failure | dendritic_cell | *FAM160A2* | -2.073135678 | 9.54E-07 | 5.47E-06 | 0.004626443 |
| Heart_Failure | dendritic_cell | *CYBA* | 3.326259078 | 0.002323812 | 0.003367723 | 1 |
| Heart_Failure | dendritic_cell | *THOC3* | 9.292400827 | 0.01253244 | 0.016359267 | 1 |
| Heart_Failure | dendritic_cell | *FSTL4* | -2.364859523 | 9.17E-07 | 5.37E-06 | 0.004448238 |
| Heart_Failure | dendritic_cell | *AKR7A2* | -2.036712351 | 6.29E-07 | 4.39E-06 | 0.003050813 |
| Heart_Failure | dendritic_cell | *MRTO4* | -2.902049923 | 0.007596374 | 0.010212692 | 1 |
| Heart_Failure | dendritic_cell | *ANAPC4* | -2.168826269 | 2.37E-05 | 5.54E-05 | 0.11497559 |
| Heart_Failure | dendritic_cell | *LY75* | 1.938177572 | 8.26E-08 | 3.44E-06 | 0.000400701 |
| Heart_Failure | dendritic_cell | *OPN3* | -2.188190501 | 1.99E-05 | 4.79E-05 | 0.096542304 |
| Heart_Failure | dendritic_cell | *KIF1B* | -1.879502813 | 7.05E-05 | 0.000139425 | 0.341870567 |
| Heart_Failure | dendritic_cell | *PLEKHH1* | 1.671292247 | 0.00055708 | 0.000894426 | 1 |

| | | | | | | |
|---|---|---|---|---|---|---|
| Heart_Failure | dendritic_cell | *GALC* | 4.097707811 | 0.000507672 | 0.00082382 | 1 |
| Heart_Failure | dendritic_cell | *FAM114A2* | -2.849709418 | 0.000612114 | 0.000975042 | 1 |
| Heart_Failure | dendritic_cell | *GINM1* | -2.831972828 | 0.00070779 | 0.001114999 | 1 |
| Heart_Failure | dendritic_cell | *ITIH4* | 0.713699634 | 1.15E-05 | 3.04E-05 | 0.055958587 |
| Heart_Failure | dendritic_cell | *NPFFR2* | -3.097194007 | 2.80E-07 | 3.66E-06 | 0.001358716 |
| Heart_Failure | dendritic_cell | *PCGF2* | -1.444618725 | 2.95E-05 | 6.57E-05 | 0.142927622 |
| Heart_Failure | dendritic_cell | *IL17RB* | -3.350249254 | 2.60E-05 | 5.94E-05 | 0.126377829 |
| Heart_Failure | dendritic_cell | *MSH4* | -1.349825383 | 2.01E-06 | 8.56E-06 | 0.00975131 |
| Heart_Failure | dendritic_cell | *GDI2* | -2.330689224 | 0.025967586 | 0.03233121 | 1 |
| Heart_Failure | dendritic_cell | *PITHD1* | -2.734182602 | 0.009689463 | 0.012848668 | 1 |
| Heart_Failure | dendritic_cell | *ATP11B* | -2.237088851 | 0.000508594 | 0.000825041 | 1 |
| Heart_Failure | dendritic_cell | *RASGRF1* | 0.821135 | 8.67E-08 | 3.44E-06 | 0.000420784 |
| Heart_Failure | dendritic_cell | *CROCC* | -0.991488486 | 0.003283705 | 0.004627516 | 1 |
| Heart_Failure | dendritic_cell | *FLYWCH1* | -1.483708036 | 4.71E-05 | 9.82E-05 | 0.228438746 |
| Heart_Failure | dendritic_cell | *ALDH18A1* | -2.703001225 | 0.000521438 | 0.000843422 | 1 |
| Heart_Failure | dendritic_cell | *DNAJC25* | -4.300057101 | 2.19E-06 | 9.05E-06 | 0.010617568 |
| Heart_Failure | dendritic_cell | *GNA15* | 0.955975507 | 1.74E-05 | 4.29E-05 | 0.084466074 |
| Heart_Failure | dendritic_cell | *PIGV* | -1.80835494 | 8.09E-05 | 0.000157164 | 0.392596345 |
| Heart_Failure | dendritic_cell | *RIMBP2* | -2.659113285 | 4.99E-07 | 4.02E-06 | 0.002420845 |
| Heart_Failure | dendritic_cell | *PRDM6* | -2.598829189 | 5.89E-07 | 4.30E-06 | 0.002858163 |
| Heart_Failure | dendritic_cell | *SPAG4* | -5.779165636 | 8.11E-07 | 4.93E-06 | 0.003936008 |
| Heart_Failure | dendritic_cell | *NCKAP1* | -1.659538861 | 7.22E-07 | 4.66E-06 | 0.003501598 |
| Heart_Failure | dendritic_cell | *LTK* | 12.36264385 | 1.21E-07 | 3.44E-06 | 0.000588942 |
| Heart_Failure | dendritic_cell | *EIF4B* | -1.670118802 | 0.009166857 | 0.01218898 | 1 |
| Heart_Failure | dendritic_cell | *SLC6A16* | -3.73900472 | 0.000158427 | 0.000284805 | 0.768688385 |
| Heart_Failure | dendritic_cell | *AHRR* | 2.30919318 | 7.54E-07 | 4.79E-06 | 0.003660274 |
| Heart_Failure | dendritic_cell | *ADCK1* | 5.582351578 | 3.88E-05 | 8.35E-05 | 0.18816621 |
| Heart_Failure | dendritic_cell | *RNF4* | 4.045323526 | 7.31E-06 | 2.10E-05 | 0.035488967 |
| Heart_Failure | dendritic_cell | *TM7SF3* | -2.857286214 | 0.001434704 | 0.002153166 | 1 |
| Heart_Failure | dendritic_cell | *CDON* | -0.877128896 | 3.30E-06 | 1.22E-05 | 0.016034857 |
| Heart_Failure | dendritic_cell | *TMEM161A* | 4.438693542 | 8.61E-07 | 5.14E-06 | 0.00417826 |
| Heart_Failure | dendritic_cell | *LPAR2* | 4.534816729 | 6.61E-05 | 0.000132125 | 0.320932413 |
| Heart_Failure | dendritic_cell | *SUGP2* | -2.42671183 | 0.027288964 | 0.033915485 | 1 |
| Heart_Failure | dendritic_cell | *SLC12A2* | -3.128941843 | 2.85E-05 | 6.40E-05 | 0.138346561 |
| Heart_Failure | dendritic_cell | *ABCA7* | 2.419758559 | 4.78E-07 | 3.96E-06 | 0.002318058 |
| Heart_Failure | dendritic_cell | *DDX20* | -2.848340585 | 1.27E-05 | 3.28E-05 | 0.061547514 |
| Heart_Failure | dendritic_cell | *BTBD1* | -2.764509414 | 9.66E-05 | 0.000183199 | 0.468805687 |
| Heart_Failure | dendritic_cell | *BCAS1* | -2.718871405 | 1.14E-06 | 6.05E-06 | 0.005525668 |
| Heart_Failure | dendritic_cell | *CHI3L2* | 1.314221643 | 4.56E-08 | 3.44E-06 | 0.0002212 |
| Heart_Failure | dendritic_cell | *PMS1* | -2.37947826 | 6.44E-06 | 1.93E-05 | 0.031224746 |
| Heart_Failure | dendritic_cell | *UHRF1BP1* | -2.184932884 | 0.000116489 | 0.000216388 | 0.565204703 |
| Heart_Failure | dendritic_cell | *WDR18* | -1.707804662 | 1.23E-05 | 3.20E-05 | 0.059698917 |
| Heart_Failure | dendritic_cell | *ANKRD44* | 1.575770057 | 1.81E-07 | 3.58E-06 | 0.000877575 |
| Heart_Failure | dendritic_cell | *ADAT1* | 1.501679985 | 0.000921729 | 0.001431114 | 1 |
| Heart_Failure | dendritic_cell | *MAP2K4* | -2.470563104 | 0.01261762 | 0.016452753 | 1 |
| Heart_Failure | dendritic_cell | *SEC61A2* | -3.078014666 | 0.00019949 | 0.000351717 | 0.96792573 |
| Heart_Failure | dendritic_cell | *TIE1* | -1.637063769 | 1.95E-08 | 2.99E-06 | 9.48E-05 |
| Heart_Failure | dendritic_cell | *NGEF* | 4.13075797 | 1.55E-06 | 7.25E-06 | 0.007497297 |
| Heart_Failure | dendritic_cell | *SPI1* | 1.516092648 | 2.07E-05 | 4.95E-05 | 0.100332638 |
| Heart_Failure | dendritic_cell | *ZNRD1* | -2.36760228 | 1.88E-08 | 2.99E-06 | 9.12E-05 |

| | | | | | | |
|---|---|---|---|---|---|---|
| Heart_Failure | dendritic_cell | *ATXN3* | -1.930610179 | 4.03E-07 | 3.80E-06 | 0.001955078 |
| Heart_Failure | dendritic_cell | *GOLGA5* | -2.500243816 | 5.85E-07 | 4.29E-06 | 0.002839245 |
| Heart_Failure | dendritic_cell | *FGFR2* | -1.623191247 | 8.35E-08 | 3.44E-06 | 0.000405129 |
| Heart_Failure | dendritic_cell | *MTFR1* | -3.066025186 | 0.001390833 | 0.002094451 | 1 |
| Heart_Failure | dendritic_cell | *MYO9A* | -1.762262216 | 3.98E-06 | 1.38E-05 | 0.019317232 |
| Heart_Failure | dendritic_cell | *EVI5* | -2.760344518 | 0.000348491 | 0.00058279 | 1 |
| Heart_Failure | dendritic_cell | *STOML1* | 4.412710112 | 1.62E-07 | 3.51E-06 | 0.00078492 |
| Heart_Failure | dendritic_cell | *TP53BP1* | -2.893579315 | 1.19E-05 | 3.12E-05 | 0.057929947 |
| Heart_Failure | dendritic_cell | *RRP15* | -2.08483928 | 1.03E-05 | 2.76E-05 | 0.049782426 |
| Heart_Failure | dendritic_cell | *DHX8* | 3.506696213 | 0.020659901 | 0.026111446 | 1 |
| Heart_Failure | dendritic_cell | *ROGDI* | 9.758635281 | 1.76E-05 | 4.33E-05 | 0.085616248 |
| Heart_Failure | dendritic_cell | *CBFB* | -3.071631603 | 0.000694078 | 0.001096246 | 1 |
| Heart_Failure | dendritic_cell | *IFI35* | -3.376992912 | 5.65E-05 | 0.000115343 | 0.274055917 |
| Heart_Failure | dendritic_cell | *HEATR6* | -2.771640759 | 2.88E-05 | 6.44E-05 | 0.139823257 |
| Heart_Failure | dendritic_cell | *POLR1A* | -2.151492467 | 4.47E-07 | 3.90E-06 | 0.002167424 |
| Heart_Failure | dendritic_cell | *IP6K2* | -2.63191751 | 1.82E-05 | 4.44E-05 | 0.088369869 |
| Heart_Failure | dendritic_cell | *STON1-GTF2A* | -0.753375393 | 0.00020828 | 0.000365092 | 1 |
| Heart_Failure | dendritic_cell | *IFT80* | 5.212516342 | 7.52E-05 | 0.00014735 | 0.364984865 |
| Heart_Failure | dendritic_cell | *NUCKS1* | -2.258185622 | 3.96E-06 | 1.37E-05 | 0.019229177 |
| Heart_Failure | dendritic_cell | *CLEC2D* | 2.602561965 | 0.016230266 | 0.020882856 | 1 |
| Heart_Failure | dendritic_cell | *DRD4* | -3.768060537 | 2.62E-07 | 3.66E-06 | 0.001268984 |
| Heart_Failure | dendritic_cell | *PLA2G10* | -4.457000865 | 3.39E-07 | 3.66E-06 | 0.001645098 |
| Heart_Failure | dendritic_cell | *NEDD4* | -2.036201484 | 0.000238752 | 0.000413427 | 1 |
| Heart_Failure | dendritic_cell | *GNB5* | -1.513059191 | 7.96E-07 | 4.90E-06 | 0.003863231 |
| Heart_Failure | dendritic_cell | *LRP6* | -2.223583247 | 4.83E-06 | 1.58E-05 | 0.023421437 |
| Heart_Failure | dendritic_cell | *DAPP1* | 0.39205766 | 2.91E-08 | 3.33E-06 | 0.000140966 |
| Heart_Failure | dendritic_cell | *CLTCL1* | -2.931490019 | 2.86E-05 | 6.42E-05 | 0.138993466 |
| Heart_Failure | dendritic_cell | *DGCR2* | -2.891507222 | 7.86E-05 | 0.000153395 | 0.381493634 |
| Heart_Failure | dendritic_cell | *ZXDC* | -2.884114651 | 0.000725181 | 0.001141284 | 1 |
| Heart_Failure | dendritic_cell | *WIPI1* | -2.086082439 | 0.000106603 | 0.000200158 | 0.517240012 |
| Heart_Failure | dendritic_cell | *GBA2* | -3.226634963 | 1.77E-05 | 4.33E-05 | 0.085774492 |
| Heart_Failure | dendritic_cell | *NDST1* | -1.529091499 | 5.85E-07 | 4.29E-06 | 0.002839237 |
| Heart_Failure | dendritic_cell | *ASNS* | -2.518422499 | 0.001188347 | 0.001811454 | 1 |
| Heart_Failure | dendritic_cell | *AP3M2* | -3.546570865 | 2.41E-05 | 5.62E-05 | 0.116741363 |
| Heart_Failure | dendritic_cell | *ST6GALNAC2* | -3.262191673 | 4.30E-05 | 9.10E-05 | 0.208592647 |
| Heart_Failure | dendritic_cell | *EIF2B3* | -2.377163375 | 8.39E-06 | 2.35E-05 | 0.040722243 |
| Heart_Failure | dendritic_cell | *MAP4K4* | -2.226115131 | 8.93E-06 | 2.46E-05 | 0.043332333 |
| Heart_Failure | dendritic_cell | *WDR1* | 4.348850162 | 6.19E-06 | 1.90E-05 | 0.030057424 |
| Heart_Failure | dendritic_cell | *SNX13* | -2.71647568 | 0.001106972 | 0.001694865 | 1 |
| Heart_Failure | dendritic_cell | *RPS6KA2* | -3.055008509 | 1.96E-05 | 4.71E-05 | 0.094944691 |
| Heart_Failure | dendritic_cell | *ING3* | -3.457223706 | 5.64E-05 | 0.000115339 | 0.273836442 |
| Heart_Failure | dendritic_cell | *VASH1* | 2.508750858 | 8.65E-06 | 2.40E-05 | 0.04194878 |
| Heart_Failure | dendritic_cell | *TRIB2* | -1.942207319 | 0.000245317 | 0.000423889 | 1 |
| Heart_Failure | dendritic_cell | *CPSF1* | -1.76331254 | 0.001272781 | 0.001929855 | 1 |
| Heart_Failure | dendritic_cell | *CYBRD1* | -2.084575059 | 3.95E-05 | 8.48E-05 | 0.191849764 |
| Heart_Failure | dendritic_cell | *RDH11* | 3.974803659 | 3.85E-06 | 1.35E-05 | 0.01867476 |
| Heart_Failure | dendritic_cell | *EPN2* | -1.972403451 | 2.33E-05 | 5.47E-05 | 0.113159313 |
| Heart_Failure | dendritic_cell | *ALDH3A2* | -1.8999598 | 5.79E-05 | 0.000117723 | 0.281049831 |
| Heart_Failure | dendritic_cell | *AFF4* | -2.438178478 | 0.000124482 | 0.000229478 | 0.60398582 |
| Heart_Failure | dendritic_cell | *TRHDE* | -0.888204636 | 1.80E-07 | 3.58E-06 | 0.000871897 |

| | | | | | | |
|---|---|---|---|---|---|---|
| Heart_Failure | dendritic_cell | *FCGR2B* | 0.941122244 | 1.37E-07 | 3.44E-06 | 0.000664904 |
| Heart_Failure | dendritic_cell | *NFATC3* | -2.242789348 | 0.000713955 | 0.001124346 | 1 |
| Heart_Failure | dendritic_cell | *CRMP1* | -1.65240973 | 4.65E-07 | 3.93E-06 | 0.002257719 |
| Heart_Failure | dendritic_cell | *NDE1* | 3.784088596 | 0.000776255 | 0.001215749 | 1 |
| Heart_Failure | dendritic_cell | *AP1M1* | -3.113459264 | 3.75E-06 | 1.32E-05 | 0.018193547 |
| Heart_Failure | dendritic_cell | *PVR* | -2.868683764 | 5.78E-05 | 0.00011751 | 0.280378374 |
| Heart_Failure | dendritic_cell | *SCARB1* | -2.265983853 | 4.65E-06 | 1.54E-05 | 0.022581588 |
| Heart_Failure | dendritic_cell | *MOV10L1* | -3.32448756 | 7.65E-06 | 2.18E-05 | 0.037136845 |
| Heart_Failure | dendritic_cell | *LLGL2* | -4.219726241 | 0.000442748 | 0.000724279 | 1 |
| Heart_Failure | dendritic_cell | *SDHA* | -2.902760109 | 8.36E-06 | 2.34E-05 | 0.040540684 |
| Heart_Failure | dendritic_cell | *SMARCE1* | -2.960021994 | 0.000119958 | 0.000222395 | 0.582037176 |
| Heart_Failure | dendritic_cell | *CD5L* | 2.533356771 | 2.51E-06 | 1.00E-05 | 0.012196229 |
| Heart_Failure | dendritic_cell | *PPP2R2C* | -2.928967966 | 3.34E-07 | 3.66E-06 | 0.001619404 |
| Heart_Failure | dendritic_cell | *ZZEF1* | -2.525539875 | 7.02E-05 | 0.000139092 | 0.340774569 |
| Heart_Failure | dendritic_cell | *TACR2* | 11.19645325 | 0.000108822 | 0.000203548 | 0.528002596 |
| Heart_Failure | dendritic_cell | *TTC38* | -1.923219546 | 3.63E-07 | 3.66E-06 | 0.001761173 |
| Heart_Failure | dendritic_cell | *GRAMD4* | -3.83529845 | 2.83E-05 | 6.37E-05 | 0.137440654 |
| Heart_Failure | dendritic_cell | *ADD2* | 2.95836559 | 0.021350823 | 0.026942573 | 1 |
| Heart_Failure | dendritic_cell | *VPS9D1* | -2.912228355 | 5.61E-05 | 0.000114866 | 0.272347479 |
| Heart_Failure | dendritic_cell | *FNDC3B* | -1.976757734 | 2.80E-05 | 6.32E-05 | 0.135961698 |
| Heart_Failure | dendritic_cell | *SEC31B* | -1.659473852 | 1.15E-07 | 3.44E-06 | 0.000557613 |
| Heart_Failure | dendritic_cell | *KIFAP3* | -1.888352566 | 1.16E-06 | 6.11E-06 | 0.005626737 |
| Heart_Failure | dendritic_cell | *MKRN2* | -2.971424892 | 5.10E-06 | 1.64E-05 | 0.024746156 |
| Heart_Failure | dendritic_cell | *MCM6* | -1.944689499 | 7.91E-08 | 3.44E-06 | 0.000383972 |
| Heart_Failure | dendritic_cell | *MLH1* | -2.730232528 | 2.95E-05 | 6.57E-05 | 0.143044364 |
| Heart_Failure | dendritic_cell | *FMO4* | -2.487558969 | 0.000104394 | 0.000196401 | 0.506517265 |
| Heart_Failure | dendritic_cell | *RGS11* | -1.43606555 | 2.11E-05 | 5.04E-05 | 0.10237398 |
| Heart_Failure | dendritic_cell | *SPAG5* | 4.861870935 | 5.89E-06 | 1.83E-05 | 0.028592908 |
| Heart_Failure | dendritic_cell | *PAG1* | -2.257293577 | 5.66E-06 | 1.77E-05 | 0.027469105 |
| Heart_Failure | dendritic_cell | *GPATCH1* | -2.794445145 | 1.08E-06 | 5.91E-06 | 0.005260068 |
| Heart_Failure | dendritic_cell | *ICAM3* | -2.770601828 | 1.24E-07 | 3.44E-06 | 0.000601324 |
| Heart_Failure | dendritic_cell | *NT5C2* | -1.817677908 | 0.000259854 | 0.000447255 | 1 |
| Heart_Failure | dendritic_cell | *XAB2* | -2.793262879 | 4.45E-07 | 3.90E-06 | 0.002159462 |
| Heart_Failure | dendritic_cell | *DYNC1I2* | -2.766809964 | 3.89E-05 | 8.37E-05 | 0.188685269 |
| Heart_Failure | dendritic_cell | *LRCH4* | -2.528982719 | 0.006325219 | 0.008587006 | 1 |
| Heart_Failure | dendritic_cell | *SIRT6* | -3.332736477 | 0.00161716 | 0.002412811 | 1 |
| Heart_Failure | dendritic_cell | *GPR137B* | 2.239746614 | 0.007748367 | 0.010408382 | 1 |
| Heart_Failure | dendritic_cell | *NAALAD2* | -1.042392377 | 9.81E-05 | 0.000185656 | 0.476021869 |
| Heart_Failure | dendritic_cell | *AMPH* | -1.892344062 | 2.20E-07 | 3.66E-06 | 0.001068908 |
| Heart_Failure | dendritic_cell | *MCCC1* | -1.602405149 | 0.000132323 | 0.000242275 | 0.642029663 |
| Heart_Failure | dendritic_cell | *ACER3* | -1.829329722 | 2.06E-06 | 8.73E-06 | 0.010000971 |
| Heart_Failure | dendritic_cell | *PIK3C3* | -2.952203923 | 3.62E-05 | 7.87E-05 | 0.175693283 |
| Heart_Failure | dendritic_cell | *SYNJ2* | -1.668986371 | 0.000515242 | 0.000835266 | 1 |
| Heart_Failure | dendritic_cell | *RBFOX1* | -1.492761398 | 3.68E-06 | 1.31E-05 | 0.017856997 |
| Heart_Failure | dendritic_cell | *FGF20* | 8.812264404 | 1.30E-06 | 6.52E-06 | 0.006322644 |
| Heart_Failure | dendritic_cell | *PCM1* | -2.236619329 | 5.22E-05 | 0.000107885 | 0.25320662 |
| Heart_Failure | dendritic_cell | *MYH7B* | -3.597502957 | 0.000752643 | 0.00118182 | 1 |
| Heart_Failure | dendritic_cell | *BPIFB2* | -8.626590795 | 0.003493913 | 0.004912334 | 1 |
| Heart_Failure | dendritic_cell | *CLUL1* | -2.736103772 | 0.000287334 | 0.000490205 | 1 |
| Heart_Failure | dendritic_cell | *FKBP7* | -1.859874745 | 1.83E-07 | 3.58E-06 | 0.000886378 |

| | | | | | | |
|---|---|---|---|---|---|---|
| Heart_Failure | dendritic_cell | *XRCC5* | -2.666320872 | 0.000169595 | 0.000302973 | 0.822875701 |
| Heart_Failure | dendritic_cell | *FDFT1* | -2.050026745 | 7.27E-05 | 0.000143229 | 0.352630699 |
| Heart_Failure | dendritic_cell | *SCGN* | -12.61652212 | 4.48E-07 | 3.90E-06 | 0.002172014 |
| Heart_Failure | dendritic_cell | *PGM1* | -2.344171396 | 3.57E-06 | 1.28E-05 | 0.017341545 |
| Heart_Failure | dendritic_cell | *DDX1* | -2.528993065 | 2.12E-06 | 8.84E-06 | 0.010287163 |
| Heart_Failure | dendritic_cell | *DNM2* | -3.116539194 | 0.000175294 | 0.00031212 | 0.85052741 |
| Heart_Failure | dendritic_cell | *STX7* | -2.524483044 | 0.0061799 | 0.008403768 | 1 |
| Heart_Failure | dendritic_cell | *RABL2B* | -2.894994597 | 2.75E-06 | 1.07E-05 | 0.013355593 |
| Heart_Failure | dendritic_cell | *DCT* | -1.005611749 | 0.011520948 | 0.015136647 | 1 |
| Heart_Failure | dendritic_cell | *RIF1* | -2.244036145 | 6.00E-06 | 1.85E-05 | 0.02912228 |
| Heart_Failure | dendritic_cell | *KCNN2* | -4.520488818 | 8.59E-05 | 0.000165208 | 0.416654167 |
| Heart_Failure | dendritic_cell | *MOK* | -1.735646328 | 3.00E-05 | 6.67E-05 | 0.145346273 |
| Heart_Failure | dendritic_cell | *STK17B* | 2.564519432 | 4.84E-07 | 3.96E-06 | 0.002346573 |
| Heart_Failure | dendritic_cell | *PCDHB4* | -3.247885373 | 3.34E-07 | 3.66E-06 | 0.001621945 |
| Heart_Failure | dendritic_cell | *CCNT2* | -2.737689078 | 5.87E-06 | 1.82E-05 | 0.028466832 |
| Heart_Failure | dendritic_cell | *DOPEY1* | -2.519563512 | 7.87E-06 | 2.23E-05 | 0.038203925 |
| Heart_Failure | dendritic_cell | *LYRM2* | -2.392023617 | 1.81E-05 | 4.41E-05 | 0.087853006 |
| Heart_Failure | dendritic_cell | *ULK2* | -2.717481943 | 8.99E-05 | 0.000171866 | 0.436367217 |
| Heart_Failure | dendritic_cell | *PLOD1* | -2.519442963 | 3.74E-07 | 3.66E-06 | 0.001816349 |
| Heart_Failure | dendritic_cell | *PIBF1* | -2.425315256 | 0.00019971 | 0.000351925 | 0.968990919 |
| Heart_Failure | dendritic_cell | *OXCT1* | -1.77230101 | 5.88E-08 | 3.44E-06 | 0.000285321 |
| Heart_Failure | dendritic_cell | *ZNF586* | -3.028037265 | 9.22E-05 | 0.000175794 | 0.4473969 |
| Heart_Failure | dendritic_cell | *ZNF264* | -3.13967423 | 5.20E-06 | 1.66E-05 | 0.02524817 |
| Heart_Failure | dendritic_cell | *RPS5* | -2.740822027 | 3.18E-06 | 1.18E-05 | 0.01543801 |
| Heart_Failure | dendritic_cell | *CHMP2B* | -2.925601067 | 2.25E-05 | 5.32E-05 | 0.109289801 |
| Heart_Failure | dendritic_cell | *NOA1* | -2.798058659 | 3.33E-07 | 3.66E-06 | 0.001614831 |
| Heart_Failure | dendritic_cell | *GSTP1* | -2.019141319 | 0.004575369 | 0.006335529 | 1 |
| Heart_Failure | dendritic_cell | *NCOA1* | -2.13064128 | 2.91E-05 | 6.49E-05 | 0.141065181 |
| Heart_Failure | dendritic_cell | *CAD* | -2.538339929 | 2.07E-06 | 8.74E-06 | 0.010036048 |
| Heart_Failure | dendritic_cell | *CD59* | -1.850521672 | 0.000414289 | 0.000680938 | 1 |
| Heart_Failure | dendritic_cell | *HACE1* | -2.193477166 | 2.38E-06 | 9.58E-06 | 0.011557697 |
| Heart_Failure | dendritic_cell | *SEH1L* | -2.321045615 | 4.89E-06 | 1.59E-05 | 0.023710331 |
| Heart_Failure | dendritic_cell | *OVGP1* | 1.265498466 | 1.88E-06 | 8.18E-06 | 0.009101086 |
| Heart_Failure | dendritic_cell | *SLC25A24* | -2.550364187 | 0.014528366 | 0.018807799 | 1 |
| Heart_Failure | dendritic_cell | *PILRA* | 0.55172939 | 0.00357742 | 0.005022466 | 1 |
| Heart_Failure | dendritic_cell | *ZNF213* | -1.972350579 | 4.02E-06 | 1.38E-05 | 0.019483248 |
| Heart_Failure | dendritic_cell | *CPNE3* | -3.02234799 | 0.000681334 | 0.001077324 | 1 |
| Heart_Failure | dendritic_cell | *RRN3* | -2.483353871 | 1.47E-05 | 3.71E-05 | 0.071522119 |
| Heart_Failure | dendritic_cell | *MTIF2* | -2.557779548 | 1.45E-06 | 6.95E-06 | 0.007031946 |
| Heart_Failure | dendritic_cell | *DDHD2* | -1.939353298 | 0.00073356 | 0.001153348 | 1 |
| Heart_Failure | dendritic_cell | *EPS15* | -1.881998551 | 2.41E-05 | 5.63E-05 | 0.117139395 |
| Heart_Failure | dendritic_cell | *USP40* | -2.131820957 | 3.51E-06 | 1.27E-05 | 0.017012267 |
| Heart_Failure | dendritic_cell | *POMGNT1* | -2.795291894 | 4.02E-06 | 1.38E-05 | 0.019505606 |
| Heart_Failure | dendritic_cell | *EIF2AK1* | -2.856421625 | 0.019589758 | 0.024849544 | 1 |
| Heart_Failure | dendritic_cell | *NME8* | 5.408282978 | 1.43E-07 | 3.45E-06 | 0.000691506 |
| Heart_Failure | dendritic_cell | *SNX10* | 1.57217642 | 4.18E-06 | 1.42E-05 | 0.020270487 |
| Heart_Failure | dendritic_cell | *ITPKC* | -3.517743624 | 2.50E-05 | 5.77E-05 | 0.121309723 |
| Heart_Failure | dendritic_cell | *TMED2* | -2.146988192 | 0.002974228 | 0.004218343 | 1 |
| Heart_Failure | dendritic_cell | *ACOX3* | -2.616750907 | 5.68E-05 | 0.000115795 | 0.275592263 |
| Heart_Failure | dendritic_cell | *HSD17B14* | 5.085336875 | 6.14E-05 | 0.000123724 | 0.297845796 |

| | | | | | | |
|---|---|---|---|---|---|---|
| Heart_Failure | dendritic_cell | *ACHE* | 2.746617141 | 4.00E-05 | 8.56E-05 | 0.194174322 |
| Heart_Failure | dendritic_cell | *SRRT* | -3.134807808 | 0.024501789 | 0.030671486 | 1 |
| Heart_Failure | dendritic_cell | *ADAMTS2* | -2.407540251 | 2.73E-06 | 1.06E-05 | 0.013240986 |
| Heart_Failure | dendritic_cell | *PSMC5* | 5.517168417 | 8.18E-05 | 0.00015857 | 0.396743124 |
| Heart_Failure | dendritic_cell | *UIMC1* | -2.879720605 | 4.71E-05 | 9.82E-05 | 0.228465016 |
| Heart_Failure | dendritic_cell | *LPCAT2* | 2.653735239 | 2.98E-07 | 3.66E-06 | 0.001447231 |
| Heart_Failure | dendritic_cell | *OGFOD1* | -2.036080023 | 6.39E-06 | 1.93E-05 | 0.030995085 |
| Heart_Failure | dendritic_cell | *SF3B2* | -2.28684741 | 7.58E-06 | 2.16E-05 | 0.036802013 |
| Heart_Failure | dendritic_cell | *PHACTR3* | -2.385825831 | 0.00049057 | 0.000797135 | 1 |
| Heart_Failure | dendritic_cell | *AAMDC* | -2.440186064 | 1.18E-05 | 3.10E-05 | 0.05741093 |
| Heart_Failure | dendritic_cell | *ASAP3* | -1.488449808 | 6.47E-07 | 4.45E-06 | 0.003139885 |
| Heart_Failure | dendritic_cell | *EDEM2* | -2.842272031 | 0.001297128 | 0.001964316 | 1 |
| Heart_Failure | dendritic_cell | *ANKRD10* | -2.612369955 | 0.004755729 | 0.006574016 | 1 |
| Heart_Failure | dendritic_cell | *DOCK3* | -1.444145275 | 3.96E-07 | 3.76E-06 | 0.001919372 |
| Heart_Failure | dendritic_cell | *ARHGAP28* | -1.508267332 | 5.23E-07 | 4.09E-06 | 0.002538296 |
| Heart_Failure | dendritic_cell | *PPP1R13B* | -1.797317738 | 1.27E-06 | 6.39E-06 | 0.006140232 |
| Heart_Failure | dendritic_cell | *ZNF343* | -1.642620773 | 6.71E-06 | 1.99E-05 | 0.032551011 |
| Heart_Failure | dendritic_cell | *EBF4* | -3.574087979 | 1.32E-06 | 6.56E-06 | 0.006384731 |
| Heart_Failure | dendritic_cell | *CPXM1* | -2.130614842 | 1.64E-06 | 7.57E-06 | 0.007969689 |
| Heart_Failure | dendritic_cell | *MAVS* | -1.812028069 | 0.000132048 | 0.000242023 | 0.640697816 |
| Heart_Failure | dendritic_cell | *XRN2* | -2.725473416 | 1.17E-05 | 3.08E-05 | 0.056839874 |
| Heart_Failure | dendritic_cell | *SNX5* | 3.624197727 | 0.012352151 | 0.016137555 | 1 |
| Heart_Failure | dendritic_cell | *ESF1* | -2.776781129 | 0.001867258 | 0.002752107 | 1 |
| Heart_Failure | dendritic_cell | *RBBP9* | -2.04954337 | 2.28E-05 | 5.37E-05 | 0.110708962 |
| Heart_Failure | dendritic_cell | *TMEM230* | -3.503644584 | 2.24E-05 | 5.30E-05 | 0.108681065 |
| Heart_Failure | dendritic_cell | *RPH3A* | -4.359675576 | 2.51E-06 | 1.00E-05 | 0.012191912 |
| Heart_Failure | dendritic_cell | *KIF16B* | 3.546568618 | 0.000809033 | 0.001263821 | 1 |
| Heart_Failure | dendritic_cell | *TBX5* | -5.011831802 | 0.001804579 | 0.002668027 | 1 |
| Heart_Failure | dendritic_cell | *FXYD5* | -2.382165814 | 0.019057033 | 0.024205425 | 1 |
| Heart_Failure | dendritic_cell | *ZNF302* | -1.792627565 | 0.002441625 | 0.003521631 | 1 |
| Heart_Failure | dendritic_cell | *GRAMD1A* | -3.516391273 | 0.00016787 | 0.000300223 | 0.814504346 |
| Heart_Failure | dendritic_cell | *CDIP1* | -2.41488628 | 6.50E-07 | 4.45E-06 | 0.003153453 |
| Heart_Failure | dendritic_cell | *GANAB* | -3.001988571 | 4.58E-06 | 1.52E-05 | 0.022233381 |
| Heart_Failure | dendritic_cell | *MLF2* | -3.861545514 | 1.11E-06 | 6.00E-06 | 0.005396886 |
| Heart_Failure | dendritic_cell | *ANKRD24* | -3.504342563 | 8.12E-07 | 4.93E-06 | 0.003938985 |
| Heart_Failure | dendritic_cell | *DHX32* | -2.577227464 | 0.000115358 | 0.000214781 | 0.559719065 |
| Heart_Failure | dendritic_cell | *SPTLC1* | -2.598834834 | 0.002785438 | 0.003970312 | 1 |
| Heart_Failure | dendritic_cell | *YPEL3* | -2.07254222 | 1.78E-05 | 4.35E-05 | 0.08614828 |
| Heart_Failure | dendritic_cell | *MRPS33* | 2.469185362 | 0.006776626 | 0.009179283 | 1 |
| Heart_Failure | dendritic_cell | *STRN4* | -2.32014164 | 1.71E-06 | 7.72E-06 | 0.008303469 |
| Heart_Failure | dendritic_cell | *LYZ* | 2.275888711 | 4.68E-05 | 9.76E-05 | 0.226908277 |
| Heart_Failure | dendritic_cell | *MUL1* | 1.99085515 | 5.02E-06 | 1.62E-05 | 0.0243601 |
| Heart_Failure | dendritic_cell | *SPG21* | 2.762740091 | 0.011610964 | 0.01523017 | 1 |
| Heart_Failure | dendritic_cell | *CHRD* | -1.465750992 | 0.013225488 | 0.017203772 | 1 |
| Heart_Failure | dendritic_cell | *RAB11FIP3* | -1.799726429 | 5.43E-06 | 1.72E-05 | 0.026351197 |
| Heart_Failure | dendritic_cell | *GNPTG* | -3.556323027 | 2.50E-05 | 5.77E-05 | 0.121514173 |
| Heart_Failure | dendritic_cell | *CERS4* | -1.897863887 | 7.79E-07 | 4.85E-06 | 0.003778289 |
| Heart_Failure | dendritic_cell | *USP48* | -1.930655355 | 0.001641321 | 0.0024451 | 1 |
| Heart_Failure | dendritic_cell | *PDPR* | -2.413551042 | 0.013808668 | 0.017943132 | 1 |
| Heart_Failure | dendritic_cell | *GLG1* | -2.668562916 | 0.000911004 | 0.001415821 | 1 |

| | | | | | | |
|---|---|---|---|---|---|---|
| Heart_Failure | dendritic_cell | *DTX2* | -4.997865554 | 0.000755293 | 0.001185598 | 1 |
| Heart_Failure | dendritic_cell | *LAMB1* | -1.269712661 | 9.68E-07 | 5.53E-06 | 0.004698764 |
| Heart_Failure | dendritic_cell | *ABCC6* | -1.970850698 | 4.61E-07 | 3.91E-06 | 0.002238619 |
| Heart_Failure | dendritic_cell | *RAPGEF4* | -1.171401531 | 5.12E-07 | 4.04E-06 | 0.002486486 |
| Heart_Failure | dendritic_cell | *TF* | -2.310329025 | 4.28E-07 | 3.90E-06 | 0.002075301 |
| Heart_Failure | dendritic_cell | *ALKBH5* | -3.957404505 | 7.88E-06 | 2.23E-05 | 0.038248652 |
| Heart_Failure | dendritic_cell | *RGS17* | -1.607458161 | 4.09E-06 | 1.41E-05 | 0.019863285 |
| Heart_Failure | dendritic_cell | *PPP2R3C* | -1.852928791 | 0.016038817 | 0.020642 | 1 |
| Heart_Failure | dendritic_cell | *OSGEP* | -3.262572643 | 3.07E-05 | 6.80E-05 | 0.14875905 |
| Heart_Failure | dendritic_cell | *SCFD1* | -2.645174202 | 0.000443363 | 0.000725041 | 1 |
| Heart_Failure | dendritic_cell | *SUPT16H* | -3.507277049 | 0.000265413 | 0.000455691 | 1 |
| Heart_Failure | dendritic_cell | *TOX4* | -3.22995072 | 0.00099973 | 0.00154235 | 1 |
| Heart_Failure | dendritic_cell | *CAPN3* | -4.783038412 | 1.94E-06 | 8.38E-06 | 0.009435984 |
| Heart_Failure | dendritic_cell | *TBX15* | -2.090244829 | 1.07E-05 | 2.85E-05 | 0.051902021 |
| Heart_Failure | dendritic_cell | *COMT* | -3.033085219 | 2.46E-05 | 5.71E-05 | 0.119276604 |
| Heart_Failure | dendritic_cell | *LRRFIP2* | -2.330119151 | 0.00829681 | 0.011105137 | 1 |
| Heart_Failure | dendritic_cell | *SEC22C* | -3.679737798 | 0.00056938 | 0.000913568 | 1 |
| Heart_Failure | dendritic_cell | *XYLB* | -3.408864272 | 1.16E-05 | 3.05E-05 | 0.056354822 |
| Heart_Failure | dendritic_cell | *GABRP* | 2.259123246 | 1.22E-07 | 3.44E-06 | 0.00059434 |
| Heart_Failure | dendritic_cell | *CDC6* | -4.522255758 | 5.11E-06 | 1.64E-05 | 0.024813878 |
| Heart_Failure | dendritic_cell | *FMO2* | -0.888184851 | 1.18E-05 | 3.10E-05 | 0.057409001 |
| Heart_Failure | dendritic_cell | *MSH2* | -3.55747944 | 0.002521968 | 0.003621365 | 1 |
| Heart_Failure | dendritic_cell | *PSMD5* | -2.360914089 | 2.80E-06 | 1.08E-05 | 0.013568711 |
| Heart_Failure | dendritic_cell | *CRAT* | -4.828595881 | 5.45E-06 | 1.73E-05 | 0.026461274 |
| Heart_Failure | dendritic_cell | *NANS* | 5.133678952 | 3.49E-05 | 7.64E-05 | 0.169572065 |
| Heart_Failure | dendritic_cell | *PDE6C* | -2.440712409 | 8.02E-07 | 4.91E-06 | 0.003891729 |
| Heart_Failure | dendritic_cell | *CRTAC1* | -1.126465825 | 0.002513052 | 0.003609629 | 1 |
| Heart_Failure | dendritic_cell | *WAC* | -2.343427698 | 6.18E-05 | 0.000124373 | 0.299738932 |
| Heart_Failure | dendritic_cell | *ITPR3* | -1.642222408 | 0.000337224 | 0.000565967 | 1 |
| Heart_Failure | dendritic_cell | *DSP* | -1.946945798 | 3.24E-07 | 3.66E-06 | 0.001570822 |
| Heart_Failure | dendritic_cell | *IFT74* | -2.609117764 | 1.57E-06 | 7.34E-06 | 0.007629718 |
| Heart_Failure | dendritic_cell | *IL12RB1* | 1.328639358 | 0.002676703 | 0.003826565 | 1 |
| Heart_Failure | dendritic_cell | *ACOT7* | -1.875341547 | 6.31E-07 | 4.39E-06 | 0.003059364 |
| Heart_Failure | dendritic_cell | *SCD* | -2.432508777 | 6.27E-05 | 0.000125858 | 0.304072683 |
| Heart_Failure | dendritic_cell | *ERMP1* | -2.065435338 | 2.25E-06 | 9.24E-06 | 0.010934888 |
| Heart_Failure | dendritic_cell | *RAB18* | -2.480049898 | 4.20E-06 | 1.43E-05 | 0.020379426 |
| Heart_Failure | dendritic_cell | *TSPAN15* | -2.576763705 | 2.15E-06 | 8.94E-06 | 0.010432682 |
| Heart_Failure | dendritic_cell | *OCEL1* | -1.688011518 | 0.002395635 | 0.003459411 | 1 |
| Heart_Failure | dendritic_cell | *SETD1A* | -1.988130705 | 7.38E-05 | 0.000145008 | 0.358024384 |
| Heart_Failure | dendritic_cell | *CIRBP* | -2.600836313 | 1.88E-05 | 4.56E-05 | 0.091193859 |
| Heart_Failure | dendritic_cell | *MTAP* | -2.082392626 | 4.10E-05 | 8.75E-05 | 0.198918881 |
| Heart_Failure | dendritic_cell | *POLR2E* | -2.828734586 | 2.87E-06 | 1.10E-05 | 0.013932259 |
| Heart_Failure | dendritic_cell | *MADCAM1* | 4.053183126 | 0.000981564 | 0.001516252 | 1 |
| Heart_Failure | dendritic_cell | *MED15* | -2.742580286 | 8.75E-06 | 2.43E-05 | 0.042441266 |
| Heart_Failure | dendritic_cell | *SERPIND1* | -2.292937648 | 1.97E-07 | 3.63E-06 | 0.000955151 |
| Heart_Failure | dendritic_cell | *LZTR1* | -2.661617223 | 0.002761322 | 0.00393941 | 1 |
| Heart_Failure | dendritic_cell | *MMP11* | -3.734495436 | 0.000343296 | 0.000575561 | 1 |
| Heart_Failure | dendritic_cell | *CECR2* | -1.362925834 | 1.37E-07 | 3.44E-06 | 0.00066331 |
| Heart_Failure | dendritic_cell | *SMARCB1* | 1.474652714 | 0.031859552 | 0.039321964 | 1 |
| Heart_Failure | dendritic_cell | *BCL2L13* | -2.629309328 | 1.55E-05 | 3.87E-05 | 0.075147656 |

| | | | | | | |
|---|---|---|---|---|---|---|
| Heart_Failure | dendritic_cell | *DDTL* | 1.308828587 | 1.30E-05 | 3.35E-05 | 0.063221741 |
| Heart_Failure | dendritic_cell | *DDT* | -2.399871452 | 1.04E-05 | 2.80E-05 | 0.050690998 |
| Heart_Failure | dendritic_cell | *CABIN1* | -2.673896235 | 0.004419916 | 0.006132523 | 1 |
| Heart_Failure | dendritic_cell | *SPECC1L* | -2.005448797 | 0.00045751 | 0.000746414 | 1 |
| Heart_Failure | dendritic_cell | *PPIL2* | -2.881417933 | 0.000229546 | 0.000398625 | 1 |
| Heart_Failure | dendritic_cell | *PES1* | -1.851766316 | 4.34E-06 | 1.46E-05 | 0.021046789 |
| Heart_Failure | dendritic_cell | *SLC35E4* | -2.490804786 | 0.000140672 | 0.000255729 | 0.682541352 |
| Heart_Failure | dendritic_cell | *TOP3B* | -2.697676041 | 0.0370368 | 0.045356525 | 1 |
| Heart_Failure | dendritic_cell | *CRYBB3* | -3.431896516 | 0.000344021 | 0.000576576 | 1 |
| Heart_Failure | dendritic_cell | *CYTH4* | 0.875627188 | 1.22E-07 | 3.44E-06 | 0.00059407 |
| Heart_Failure | dendritic_cell | *CARD10* | -1.982306875 | 2.46E-07 | 3.66E-06 | 0.001193477 |
| Heart_Failure | dendritic_cell | *LRP5L* | -3.014023697 | 8.51E-05 | 0.000164166 | 0.413042046 |
| Heart_Failure | dendritic_cell | *LGALS2* | 1.471749289 | 1.32E-06 | 6.56E-06 | 0.006388285 |
| Heart_Failure | dendritic_cell | *SH3BP1* | 3.372739553 | 1.30E-07 | 3.44E-06 | 0.000633049 |
| Heart_Failure | dendritic_cell | *TRIOBP* | -2.551776854 | 9.38E-05 | 0.000178428 | 0.455347292 |
| Heart_Failure | dendritic_cell | *ANKRD54* | -2.451914524 | 0.00035751 | 0.000595481 | 1 |
| Heart_Failure | dendritic_cell | *EIF3L* | 3.363780214 | 0.005751449 | 0.007870725 | 1 |
| Heart_Failure | dendritic_cell | *PICK1* | -2.395966657 | 8.70E-05 | 0.00016708 | 0.422210254 |
| Heart_Failure | dendritic_cell | *TTC28* | -1.493755596 | 2.12E-06 | 8.83E-06 | 0.010264079 |
| Heart_Failure | dendritic_cell | *KDELR3* | 3.884401526 | 0.001990236 | 0.002921823 | 1 |
| Heart_Failure | dendritic_cell | *CYP2D6* | 4.39759121 | 1.38E-06 | 6.73E-06 | 0.006695385 |
| Heart_Failure | dendritic_cell | *CBY1* | -1.379917178 | 0.002366286 | 0.003424164 | 1 |
| Heart_Failure | dendritic_cell | *RAB36* | -2.950867402 | 5.91E-06 | 1.83E-05 | 0.028669147 |
| Heart_Failure | dendritic_cell | *TIMP3* | -1.850402904 | 6.74E-06 | 2.00E-05 | 0.032696017 |
| Heart_Failure | dendritic_cell | *CYB5R3* | -2.908008224 | 3.93E-06 | 1.37E-05 | 0.019048559 |
| Heart_Failure | dendritic_cell | *RHBDD3* | -2.579202076 | 2.10E-06 | 8.81E-06 | 0.010211307 |
| Heart_Failure | dendritic_cell | *TOM1* | -1.816718091 | 0.006181584 | 0.008403768 | 1 |
| Heart_Failure | dendritic_cell | *CHKB* | 3.284428893 | 9.31E-07 | 5.42E-06 | 0.004518704 |
| Heart_Failure | dendritic_cell | *THOC5* | -1.410788558 | 1.72E-05 | 4.25E-05 | 0.083679136 |
| Heart_Failure | dendritic_cell | *ARSA* | -0.963650007 | 0.016910969 | 0.021672483 | 1 |
| Heart_Failure | dendritic_cell | *TSPO* | -2.681030791 | 3.33E-06 | 1.22E-05 | 0.01614197 |
| Heart_Failure | dendritic_cell | *TTLL12* | -2.321181792 | 1.90E-07 | 3.60E-06 | 0.000920354 |
| Heart_Failure | dendritic_cell | *MTMR3* | -1.878799686 | 0.000570372 | 0.000914548 | 1 |
| Heart_Failure | dendritic_cell | *SAMM50* | -2.593901818 | 3.84E-07 | 3.72E-06 | 0.001865099 |
| Heart_Failure | dendritic_cell | *FOXRED2* | -1.963376495 | 0.001855656 | 0.002736669 | 1 |
| Heart_Failure | dendritic_cell | *IFT27* | -2.659175022 | 2.06E-06 | 8.73E-06 | 0.009976667 |
| Heart_Failure | dendritic_cell | *KIAA0930* | 2.231185032 | 5.36E-07 | 4.13E-06 | 0.002598321 |
| Heart_Failure | dendritic_cell | *SLC25A17* | -2.354633846 | 3.72E-05 | 8.05E-05 | 0.180656825 |
| Heart_Failure | dendritic_cell | *L3MBTL2* | -2.473853285 | 3.07E-06 | 1.15E-05 | 0.014901006 |
| Heart_Failure | dendritic_cell | *ZC3H7B* | -3.441695237 | 0.00160301 | 0.002393907 | 1 |
| Heart_Failure | dendritic_cell | *ACO2* | -2.864145223 | 0.000266543 | 0.000457146 | 1 |
| Heart_Failure | dendritic_cell | *POLR3H* | -1.643534953 | 6.93E-07 | 4.58E-06 | 0.003360726 |
| Heart_Failure | dendritic_cell | *DESI1* | 3.151486081 | 5.39E-08 | 3.44E-06 | 0.000261764 |
| Heart_Failure | dendritic_cell | *KHNYN* | -2.780062135 | 0.000655964 | 0.00104045 | 1 |
| Heart_Failure | dendritic_cell | *SDR39U1* | -1.452026516 | 1.20E-05 | 3.13E-05 | 0.058138652 |
| Heart_Failure | dendritic_cell | *RBM23* | -2.69669856 | 0.001078459 | 0.001653819 | 1 |
| Heart_Failure | dendritic_cell | *AP4S1* | -3.5732765 | 0.001506471 | 0.002255291 | 1 |
| Heart_Failure | dendritic_cell | *PYGL* | -0.771811822 | 0.013861088 | 0.018001606 | 1 |
| Heart_Failure | dendritic_cell | *CNIH1* | -3.379608062 | 0.009665194 | 0.01281999 | 1 |
| Heart_Failure | dendritic_cell | *ATP6V1D* | -2.976023145 | 2.67E-05 | 6.07E-05 | 0.12966959 |

| | | | | | | |
|---|---|---|---|---|---|---|
| Heart_Failure | dendritic_cell | *PLEK2* | 1.697505601 | 5.08E-07 | 4.03E-06 | 0.002465173 |
| Heart_Failure | dendritic_cell | *GSTZ1* | -1.734602915 | 1.90E-05 | 4.60E-05 | 0.092149676 |
| Heart_Failure | dendritic_cell | *LGMN* | -2.311396007 | 0.000207291 | 0.000363884 | 1 |
| Heart_Failure | dendritic_cell | *MTHFD1* | -1.929424393 | 1.44E-06 | 6.94E-06 | 0.006990283 |
| Heart_Failure | dendritic_cell | *TELO2* | -4.445829793 | 6.87E-06 | 2.03E-05 | 0.033317535 |
| Heart_Failure | dendritic_cell | *GSKIP* | -2.679736844 | 5.87E-07 | 4.30E-06 | 0.002848223 |
| Heart_Failure | dendritic_cell | *PSMB5* | -1.413043461 | 4.97E-05 | 0.000103173 | 0.24132054 |
| Heart_Failure | dendritic_cell | *YY1* | -2.933595341 | 0.00688056 | 0.009309671 | 1 |
| Heart_Failure | dendritic_cell | *CCNB1IP1* | -3.949284263 | 0.000164292 | 0.000294584 | 0.797145042 |
| Heart_Failure | dendritic_cell | *CINP* | -3.47696017 | 4.91E-06 | 1.60E-05 | 0.023828314 |
| Heart_Failure | dendritic_cell | *KIAA0391* | -3.395703444 | 4.96E-06 | 1.61E-05 | 0.024070521 |
| Heart_Failure | dendritic_cell | *EMC9* | 2.11476179 | 3.88E-05 | 8.35E-05 | 0.188046221 |
| Heart_Failure | dendritic_cell | *TRPC4AP* | -2.026695994 | 4.46E-05 | 9.38E-05 | 0.216491085 |
| Heart_Failure | dendritic_cell | *ABHD12* | 3.4954912 | 0.000182608 | 0.000323955 | 0.886016212 |
| Heart_Failure | dendritic_cell | *CD40* | -3.979457453 | 4.17E-06 | 1.42E-05 | 0.020246052 |
| Heart_Failure | dendritic_cell | *UQCC1* | -2.035441114 | 7.17E-06 | 2.08E-05 | 0.034790791 |
| Heart_Failure | dendritic_cell | *IFT52* | -2.463272747 | 1.22E-06 | 6.27E-06 | 0.005924875 |
| Heart_Failure | dendritic_cell | *NDRG3* | -2.072951638 | 1.06E-06 | 5.83E-06 | 0.005157883 |
| Heart_Failure | dendritic_cell | *PABPC1L* | -3.170639713 | 9.35E-07 | 5.43E-06 | 0.004535088 |
| Heart_Failure | dendritic_cell | *DOK5* | -1.685860712 | 3.15E-07 | 3.66E-06 | 0.001526194 |
| Heart_Failure | dendritic_cell | *CSTF1* | 5.570594424 | 5.31E-07 | 4.12E-06 | 0.002577529 |
| Heart_Failure | dendritic_cell | *NELFCD* | -3.118936892 | 0.007805848 | 0.010482694 | 1 |
| Heart_Failure | dendritic_cell | *MTG2* | -2.464943821 | 0.000436686 | 0.000714845 | 1 |
| Heart_Failure | dendritic_cell | *TCFL5* | -2.564596629 | 1.64E-06 | 7.55E-06 | 0.007937385 |
| Heart_Failure | dendritic_cell | *DIDO1* | -2.413682377 | 0.000436484 | 0.000714756 | 1 |
| Heart_Failure | dendritic_cell | *GID8* | -3.245630254 | 0.000489554 | 0.000795751 | 1 |
| Heart_Failure | dendritic_cell | *NKAIN4* | -2.047627875 | 0.002807344 | 0.003999187 | 1 |
| Heart_Failure | dendritic_cell | *PTK6* | -5.684472214 | 3.50E-07 | 3.66E-06 | 0.001696323 |
| Heart_Failure | dendritic_cell | *GMEB2* | 4.49507832 | 6.36E-06 | 1.92E-05 | 0.030855279 |
| Heart_Failure | dendritic_cell | *C20orf27* | -2.492858221 | 1.29E-06 | 6.51E-06 | 0.006279582 |
| Heart_Failure | dendritic_cell | *CDC25B* | -1.649335421 | 8.41E-07 | 5.05E-06 | 0.004079635 |
| Heart_Failure | dendritic_cell | *RNF24* | -2.909487358 | 1.56E-05 | 3.88E-05 | 0.075453697 |
| Heart_Failure | dendritic_cell | *NDUFAF5* | -1.934972312 | 2.89E-08 | 3.33E-06 | 0.000139981 |
| Heart_Failure | dendritic_cell | *TRIB3* | -6.541051431 | 2.35E-06 | 9.51E-06 | 0.011418366 |
| Heart_Failure | dendritic_cell | *RASSF2* | 0.67313347 | 1.60E-07 | 3.51E-06 | 0.000777641 |
| Heart_Failure | dendritic_cell | *ANGPT4* | 3.445455254 | 4.48E-07 | 3.90E-06 | 0.002174982 |
| Heart_Failure | dendritic_cell | *SEC23B* | -1.663208146 | 0.000146896 | 0.000266245 | 0.712738888 |
| Heart_Failure | dendritic_cell | *MROH8* | -1.514272097 | 0.015587798 | 0.020082846 | 1 |
| Heart_Failure | dendritic_cell | *MANBAL* | 2.991206331 | 5.47E-06 | 1.73E-05 | 0.02653023 |
| Heart_Failure | dendritic_cell | *TTI1* | -3.575731942 | 8.28E-06 | 2.32E-05 | 0.04015859 |
| Heart_Failure | dendritic_cell | *ACTR5* | -3.960572042 | 2.43E-05 | 5.65E-05 | 0.117685088 |
| Heart_Failure | dendritic_cell | *DNTTIP1* | -1.579861814 | 0.024032012 | 0.030145637 | 1 |
| Heart_Failure | dendritic_cell | *MAP1LC3A* | -3.447857185 | 3.54E-06 | 1.28E-05 | 0.01716764 |
| Heart_Failure | dendritic_cell | *RBFA* | 4.360449345 | 8.48E-05 | 0.000163628 | 0.411360746 |
| Heart_Failure | dendritic_cell | *METTL4* | 3.731875713 | 0.008643896 | 0.011531532 | 1 |
| Heart_Failure | dendritic_cell | *LPIN2* | -2.637333727 | 0.010180761 | 0.013445033 | 1 |
| Heart_Failure | dendritic_cell | *MYOM1* | -1.866501533 | 8.61E-07 | 5.14E-06 | 0.00417913 |
| Heart_Failure | dendritic_cell | *CEP192* | -2.192720235 | 0.000228762 | 0.000397548 | 1 |
| Heart_Failure | dendritic_cell | *LAMA1* | -2.798488648 | 0.01692066 | 0.02167842 | 1 |
| Heart_Failure | dendritic_cell | *RNF125* | 4.808007348 | 0.018064702 | 0.023026723 | 1 |

| | | | | | | |
|---|---|---|---|---|---|---|
| Heart_Failure | dendritic_cell | *MIB1* | -2.745463364 | 0.000381259 | 0.000631786 | 1 |
| Heart_Failure | dendritic_cell | *NALCN* | -1.657817221 | 5.56E-07 | 4.19E-06 | 0.002695553 |
| Heart_Failure | dendritic_cell | *STK24* | -2.39498938 | 7.57E-06 | 2.16E-05 | 0.036717281 |
| Heart_Failure | dendritic_cell | *DNAJC3* | -3.001409381 | 0.000168638 | 0.000301374 | 0.818229775 |
| Heart_Failure | dendritic_cell | *UGGT2* | -1.920289307 | 2.30E-06 | 9.37E-06 | 0.011149789 |
| Heart_Failure | dendritic_cell | *ARHGEF7* | -2.692167846 | 9.38E-05 | 0.000178428 | 0.455171946 |
| Heart_Failure | dendritic_cell | *PARP4* | -2.371201669 | 0.004331474 | 0.006014972 | 1 |
| Heart_Failure | dendritic_cell | *SUPT20H* | -2.072170352 | 5.55E-06 | 1.75E-05 | 0.026944227 |
| Heart_Failure | dendritic_cell | *MRPS31* | -2.279850594 | 2.62E-06 | 1.03E-05 | 0.012722978 |
| Heart_Failure | dendritic_cell | *RGCC* | -0.999462453 | 0.038623713 | 0.047216492 | 1 |
| Heart_Failure | dendritic_cell | *DGKH* | -1.979631515 | 0.001237117 | 0.00188107 | 1 |
| Heart_Failure | dendritic_cell | *KATNAL1* | -1.851644516 | 1.19E-06 | 6.17E-06 | 0.005779952 |
| Heart_Failure | dendritic_cell | *INTS6* | -2.075148919 | 3.56E-06 | 1.28E-05 | 0.017255545 |
| Heart_Failure | dendritic_cell | *CLN5* | -2.664256572 | 6.53E-06 | 1.95E-05 | 0.031684026 |
| Heart_Failure | dendritic_cell | *MGRN1* | -2.319338875 | 0.014060475 | 0.018245902 | 1 |
| Heart_Failure | dendritic_cell | *ELMO3* | -1.504602652 | 0.001956445 | 0.002874825 | 1 |
| Heart_Failure | dendritic_cell | *LONP2* | -1.980419883 | 0.000306813 | 0.0005196 | 1 |
| Heart_Failure | dendritic_cell | *PLLP* | -1.828578869 | 2.02E-07 | 3.66E-06 | 0.000979501 |
| Heart_Failure | dendritic_cell | *DHODH* | -2.098679212 | 9.22E-05 | 0.000175779 | 0.447182759 |
| Heart_Failure | dendritic_cell | *PSMD7* | -3.424750405 | 0.000518994 | 0.000840507 | 1 |
| Heart_Failure | dendritic_cell | *MON1B* | 4.233088398 | 5.64E-07 | 4.23E-06 | 0.002737299 |
| Heart_Failure | dendritic_cell | *NPRL3* | -2.541031819 | 2.84E-05 | 6.39E-05 | 0.137890097 |
| Heart_Failure | dendritic_cell | *TAF1C* | -1.261353009 | 4.88E-05 | 0.000101347 | 0.23674641 |
| Heart_Failure | dendritic_cell | *NAGPA* | 2.005646232 | 0.011776494 | 0.01543061 | 1 |
| Heart_Failure | dendritic_cell | *ZNF500* | -2.503445484 | 7.70E-07 | 4.81E-06 | 0.003734419 |
| Heart_Failure | dendritic_cell | *NME4* | -4.161465727 | 1.38E-05 | 3.52E-05 | 0.066947597 |
| Heart_Failure | dendritic_cell | *NOMO3* | -1.871029477 | 0.000114357 | 0.00021308 | 0.554859431 |
| Heart_Failure | dendritic_cell | *LMF1* | -2.334063641 | 2.41E-06 | 9.66E-06 | 0.011680983 |
| Heart_Failure | dendritic_cell | *NARFL* | 2.659501272 | 0.021751777 | 0.027420012 | 1 |
| Heart_Failure | dendritic_cell | *MTHFSD* | 1.471056479 | 0.001691563 | 0.002513771 | 1 |
| Heart_Failure | dendritic_cell | *HAGHL* | -1.325214955 | 0.010676485 | 0.014069067 | 1 |
| Heart_Failure | dendritic_cell | *SLC7A5* | -3.154729005 | 0.000101854 | 0.000192035 | 0.494196943 |
| Heart_Failure | dendritic_cell | *RHBDL1* | 10.28256453 | 0.015187664 | 0.019598549 | 1 |
| Heart_Failure | dendritic_cell | *NUBP1* | 3.038220234 | 6.27E-06 | 1.91E-05 | 0.03042831 |
| Heart_Failure | dendritic_cell | *UBE2I* | -2.199044068 | 1.83E-06 | 8.09E-06 | 0.008887781 |
| Heart_Failure | dendritic_cell | *CAPN15* | -2.755250487 | 1.30E-06 | 6.51E-06 | 0.006301428 |
| Heart_Failure | dendritic_cell | *CLUAP1* | -1.48349996 | 1.65E-05 | 4.07E-05 | 0.079821243 |
| Heart_Failure | dendritic_cell | *EARS2* | -1.830591483 | 9.97E-06 | 2.70E-05 | 0.04838193 |
| Heart_Failure | dendritic_cell | *CPPED1* | -2.986018453 | 7.15E-05 | 0.000141164 | 0.346840403 |
| Heart_Failure | dendritic_cell | *HMOX2* | -3.585827761 | 0.000333268 | 0.00056049 | 1 |
| Heart_Failure | dendritic_cell | *RRN3P2* | -3.714510948 | 0.000295532 | 0.000502249 | 1 |
| Heart_Failure | dendritic_cell | *RBL2* | -2.183302286 | 0.000262539 | 0.000451396 | 1 |
| Heart_Failure | dendritic_cell | *PYCARD* | -2.868765142 | 0.017133621 | 0.021940441 | 1 |
| Heart_Failure | dendritic_cell | *CDIPT* | -2.318375069 | 3.66E-07 | 3.66E-06 | 0.001776571 |
| Heart_Failure | dendritic_cell | *KAT8* | -4.020691382 | 1.38E-05 | 3.52E-05 | 0.067044861 |
| Heart_Failure | dendritic_cell | *TMC5* | -1.635779495 | 7.34E-07 | 4.70E-06 | 0.003559925 |
| Heart_Failure | dendritic_cell | *C16orf62* | -2.62264812 | 2.03E-05 | 4.88E-05 | 0.098593231 |
| Heart_Failure | dendritic_cell | *RNF40* | -3.413709668 | 0.032399067 | 0.039928949 | 1 |
| Heart_Failure | dendritic_cell | *KNOP1* | -3.300506862 | 0.007455977 | 0.01003228 | 1 |
| Heart_Failure | dendritic_cell | *IQCH* | -2.581934396 | 5.30E-06 | 1.69E-05 | 0.025713217 |

| | | | | | | |
|---|---|---|---|---|---|---|
| Heart_Failure | dendritic_cell | *HERC1* | -2.193553752 | 2.21E-05 | 5.23E-05 | 0.107035353 |
| Heart_Failure | dendritic_cell | *TRIP4* | -2.807565337 | 0.004669921 | 0.006462766 | 1 |
| Heart_Failure | dendritic_cell | *AP3B2* | -1.050301187 | 0.000538875 | 0.000868933 | 1 |
| Heart_Failure | dendritic_cell | *TTC23* | -1.982640766 | 1.07E-06 | 5.85E-06 | 0.005192138 |
| Heart_Failure | dendritic_cell | *FAH* | -1.898846574 | 2.07E-07 | 3.66E-06 | 0.001006338 |
| Heart_Failure | dendritic_cell | *RPAP1* | -2.376101034 | 1.38E-07 | 3.44E-06 | 0.000667708 |
| Heart_Failure | dendritic_cell | *SLC30A4* | -2.556152834 | 1.47E-06 | 7.04E-06 | 0.007153866 |
| Heart_Failure | dendritic_cell | *ZFAND1* | -3.628979156 | 0.000197335 | 0.000348297 | 0.957467249 |
| Heart_Failure | dendritic_cell | *EYA1* | -1.490273151 | 4.20E-06 | 1.43E-05 | 0.020392096 |
| Heart_Failure | dendritic_cell | *IMPAD1* | -3.89204674 | 6.13E-05 | 0.000123621 | 0.29735182 |
| Heart_Failure | dendritic_cell | *SFRP1* | -2.145698501 | 4.87E-07 | 3.96E-06 | 0.002362234 |
| Heart_Failure | dendritic_cell | *NIPAL2* | -2.404197433 | 0.027510467 | 0.034173268 | 1 |
| Heart_Failure | dendritic_cell | *RAB2A* | -2.276953882 | 1.23E-05 | 3.20E-05 | 0.059694058 |
| Heart_Failure | dendritic_cell | *ARMC1* | -2.205947536 | 5.38E-05 | 0.00011099 | 0.261271581 |
| Heart_Failure | dendritic_cell | *SPAG1* | -2.545582914 | 5.60E-06 | 1.75E-05 | 0.027149677 |
| Heart_Failure | dendritic_cell | *CHRAC1* | -2.463930612 | 3.66E-07 | 3.66E-06 | 0.001775104 |
| Heart_Failure | dendritic_cell | *SNX16* | -2.718132451 | 2.66E-06 | 1.04E-05 | 0.012902678 |
| Heart_Failure | dendritic_cell | *GSDMD* | -3.20046408 | 0.00088866 | 0.00138198 | 1 |
| Heart_Failure | dendritic_cell | *ERICH1* | -2.169221768 | 0.019545316 | 0.024799653 | 1 |
| Heart_Failure | dendritic_cell | *TUSC3* | -2.871329926 | 6.83E-06 | 2.02E-05 | 0.033152729 |
| Heart_Failure | dendritic_cell | *KLHDC4* | -3.068203322 | 0.00055332 | 0.000889271 | 1 |
| Heart_Failure | dendritic_cell | *MAN2B1* | 2.093726435 | 2.90E-08 | 3.33E-06 | 0.000140607 |
| Heart_Failure | dendritic_cell | *TULP2* | 4.227101614 | 1.85E-07 | 3.58E-06 | 0.000898886 |
| Heart_Failure | dendritic_cell | *NFKBIB* | -2.299432013 | 1.33E-07 | 3.44E-06 | 0.000643472 |
| Heart_Failure | dendritic_cell | *SNRNP70* | -2.804561954 | 0.001947074 | 0.002862789 | 1 |
| Heart_Failure | dendritic_cell | *LIN7B* | -5.026337331 | 0.000156071 | 0.000281194 | 0.757255241 |
| Heart_Failure | dendritic_cell | *PPP1R37* | 2.41591403 | 0.00126724 | 0.001922655 | 1 |
| Heart_Failure | dendritic_cell | *FCGRT* | -1.618915236 | 0.032915922 | 0.04054533 | 1 |
| Heart_Failure | dendritic_cell | *ARHGEF18* | -4.098465834 | 0.001735185 | 0.002576229 | 1 |
| Heart_Failure | dendritic_cell | *ERCC2* | 4.357704479 | 2.22E-06 | 9.16E-06 | 0.010790834 |
| Heart_Failure | dendritic_cell | *CD37* | 2.453443906 | 1.26E-06 | 6.39E-06 | 0.006107067 |
| Heart_Failure | dendritic_cell | *AMH* | -19.29133575 | 3.48E-07 | 3.66E-06 | 0.001687499 |
| Heart_Failure | dendritic_cell | *FCER2* | 3.610898531 | 1.29E-07 | 3.44E-06 | 0.000627071 |
| Heart_Failure | dendritic_cell | *CLEC4M* | 8.23647577 | 1.60E-07 | 3.51E-06 | 0.000777382 |
| Heart_Failure | dendritic_cell | *TLE6* | 3.10264153 | 3.81E-05 | 8.22E-05 | 0.184863967 |
| Heart_Failure | dendritic_cell | *NOVA2* | -1.275791737 | 6.13E-05 | 0.000123621 | 0.297270379 |
| Heart_Failure | dendritic_cell | *SGTA* | -3.033799345 | 4.97E-06 | 1.62E-05 | 0.024106648 |
| Heart_Failure | dendritic_cell | *KIR3DX1* | -8.119427266 | 0.000132756 | 0.000242886 | 0.644133691 |
| Heart_Failure | dendritic_cell | *MED25* | -3.395165071 | 3.68E-06 | 1.31E-05 | 0.017878292 |
| Heart_Failure | dendritic_cell | *TIMM44* | -2.084230697 | 7.10E-05 | 0.000140405 | 0.344554946 |
| Heart_Failure | dendritic_cell | *IL27RA* | 3.401584528 | 3.61E-07 | 3.66E-06 | 0.00175299 |
| Heart_Failure | dendritic_cell | *FAM32A* | -3.460728645 | 1.07E-06 | 5.85E-06 | 0.005176305 |
| Heart_Failure | dendritic_cell | *C19orf44* | -1.08967272 | 0.00518991 | 0.007141837 | 1 |
| Heart_Failure | dendritic_cell | *OLFM2* | -1.727700418 | 0.014871629 | 0.019221403 | 1 |
| Heart_Failure | dendritic_cell | *RASAL3* | 2.923301979 | 1.19E-07 | 3.44E-06 | 0.000579756 |
| Heart_Failure | dendritic_cell | *EPHX3* | -2.261164593 | 0.025365889 | 0.031647028 | 1 |
| Heart_Failure | dendritic_cell | *ZNF419* | -0.850050797 | 0.016895462 | 0.021669781 | 1 |
| Heart_Failure | dendritic_cell | *AURKC* | -1.5722217 | 0.029641561 | 0.036679637 | 1 |
| Heart_Failure | dendritic_cell | *PDCD5* | -2.227089285 | 1.34E-06 | 6.62E-06 | 0.006485136 |
| Heart_Failure | dendritic_cell | *ANKRD27* | -3.04234168 | 1.04E-05 | 2.79E-05 | 0.050529246 |

| | | | | | | |
|---|---|---|---|---|---|---|
| Heart_Failure | dendritic_cell | *CCDC9* | -5.169551299 | 2.74E-06 | 1.06E-05 | 0.013308151 |
| Heart_Failure | dendritic_cell | *CEACAM4* | 2.201432184 | 1.06E-06 | 5.82E-06 | 0.005138618 |
| Heart_Failure | dendritic_cell | *ICAM5* | 1.841736924 | 0.006764677 | 0.009165656 | 1 |
| Heart_Failure | dendritic_cell | *RABAC1* | -2.590047879 | 2.45E-05 | 5.70E-05 | 0.118941995 |
| Heart_Failure | dendritic_cell | *GRIN2D* | 7.392456486 | 9.21E-05 | 0.00017577 | 0.446983438 |
| Heart_Failure | dendritic_cell | *CCDC114* | -5.769504242 | 0.000594172 | 0.000948954 | 1 |
| Heart_Failure | dendritic_cell | *CARD8* | -2.615503782 | 0.002873362 | 0.004084838 | 1 |
| Heart_Failure | dendritic_cell | *LIG1* | 4.092285299 | 7.09E-06 | 2.07E-05 | 0.03440803 |
| Heart_Failure | dendritic_cell | *ZNF175* | -3.38197845 | 0.00010367 | 0.000195116 | 0.50300792 |
| Heart_Failure | dendritic_cell | *PLA2G4C* | -1.030901561 | 2.67E-05 | 6.07E-05 | 0.129451002 |
| Heart_Failure | dendritic_cell | *CAPS* | -3.757436485 | 1.12E-06 | 6.03E-06 | 0.005435343 |
| Heart_Failure | dendritic_cell | *PLEKHA4* | -2.415030355 | 6.18E-07 | 4.35E-06 | 0.002998455 |
| Heart_Failure | dendritic_cell | *TNPO2* | -2.582517243 | 0.022735574 | 0.028593314 | 1 |
| Heart_Failure | dendritic_cell | *GCDH* | -2.389517967 | 7.89E-07 | 4.87E-06 | 0.003828588 |
| Heart_Failure | dendritic_cell | *DNASE2* | -3.375952599 | 0.001280972 | 0.001941061 | 1 |
| Heart_Failure | dendritic_cell | *TFPT* | -2.117631276 | 2.82E-07 | 3.66E-06 | 0.001368701 |
| Heart_Failure | dendritic_cell | *RAB3A* | -7.320665179 | 3.30E-07 | 3.66E-06 | 0.001601967 |
| Heart_Failure | dendritic_cell | *UPK1A* | -3.838422109 | 0.001423521 | 0.002137705 | 1 |
| Heart_Failure | dendritic_cell | *COPE* | -2.415324095 | 1.90E-06 | 8.26E-06 | 0.009239473 |
| Heart_Failure | dendritic_cell | *ETV2* | -15.38672883 | 8.98E-06 | 2.48E-05 | 0.04356491 |
| Heart_Failure | dendritic_cell | *ZNF14* | -3.086625545 | 2.41E-06 | 9.66E-06 | 0.011702496 |
| Heart_Failure | dendritic_cell | *ZNF85* | -4.704645759 | 0.002181042 | 0.003181725 | 1 |
| Heart_Failure | dendritic_cell | *ETHE1* | -4.607343274 | 0.001328756 | 0.002009076 | 1 |
| Heart_Failure | dendritic_cell | *SMG9* | -3.562368918 | 0.008581988 | 0.011458394 | 1 |
| Heart_Failure | dendritic_cell | *BET1* | -3.006223245 | 0.005869133 | 0.0080217 | 1 |
| Heart_Failure | dendritic_cell | *SP4* | -3.197997105 | 1.20E-05 | 3.13E-05 | 0.058136472 |
| Heart_Failure | dendritic_cell | *DNAH11* | -1.481995731 | 3.27E-07 | 3.66E-06 | 0.001584675 |
| Heart_Failure | dendritic_cell | *PTN* | -2.612315933 | 1.17E-06 | 6.11E-06 | 0.005658266 |
| Heart_Failure | dendritic_cell | *ADAP1* | -5.941580573 | 3.13E-07 | 3.66E-06 | 0.001517171 |
| Heart_Failure | dendritic_cell | *MET* | -1.448801443 | 4.82E-06 | 1.58E-05 | 0.02337224 |
| Heart_Failure | dendritic_cell | *RNF32* | -4.892718452 | 0.001243161 | 0.001888484 | 1 |
| Heart_Failure | dendritic_cell | *IQCE* | 4.195619661 | 0.011530047 | 0.015144501 | 1 |
| Heart_Failure | dendritic_cell | *VIPR2* | -1.113792158 | 2.58E-07 | 3.66E-06 | 0.00125386 |
| Heart_Failure | dendritic_cell | *HIBADH* | -1.89146015 | 8.60E-05 | 0.000165413 | 0.417501378 |
| Heart_Failure | dendritic_cell | *CPVL* | 1.003297788 | 1.21E-07 | 3.44E-06 | 0.000586354 |
| Heart_Failure | dendritic_cell | *CHN2* | 2.395312322 | 3.89E-08 | 3.44E-06 | 0.000188813 |
| Heart_Failure | dendritic_cell | *GRB10* | -1.533681891 | 6.56E-07 | 4.46E-06 | 0.003183544 |
| Heart_Failure | dendritic_cell | *ABHD11* | 1.911001415 | 0.000135798 | 0.000248076 | 0.658891075 |
| Heart_Failure | dendritic_cell | *PLEKHA8* | -2.523670521 | 2.02E-06 | 8.61E-06 | 0.00982082 |
| Heart_Failure | dendritic_cell | *EPHB6* | -1.339174115 | 5.67E-06 | 1.77E-05 | 0.027516148 |
| Heart_Failure | dendritic_cell | *CHCHD2* | -2.265063419 | 0.014213747 | 0.018425087 | 1 |
| Heart_Failure | dendritic_cell | *ZKSCAN1* | -2.409967941 | 7.96E-06 | 2.25E-05 | 0.03864437 |
| Heart_Failure | dendritic_cell | *EIF3B* | -2.842030324 | 4.19E-06 | 1.43E-05 | 0.020333176 |
| Heart_Failure | dendritic_cell | *TFR2* | 1.745352266 | 8.84E-08 | 3.44E-06 | 0.00042898 |
| Heart_Failure | dendritic_cell | *MOSPD3* | -2.163579715 | 1.15E-06 | 6.08E-06 | 0.005572798 |
| Heart_Failure | dendritic_cell | *RBM28* | -1.756650663 | 1.95E-05 | 4.70E-05 | 0.094567992 |
| Heart_Failure | dendritic_cell | *IMPDH1* | -2.385307499 | 1.26E-06 | 6.39E-06 | 0.006133436 |
| Heart_Failure | dendritic_cell | *NRF1* | -2.457314097 | 0.007439356 | 0.01001825 | 1 |
| Heart_Failure | dendritic_cell | *ZNF862* | -2.520117355 | 7.00E-06 | 2.05E-05 | 0.033962896 |
| Heart_Failure | dendritic_cell | *MEST* | -1.254708081 | 2.47E-05 | 5.71E-05 | 0.119728132 |

| | | | | | | |
|---|---|---|---|---|---|---|
| Heart_Failure | dendritic_cell | *ACTR3C* | 4.200667229 | 0.00013256 | 0.000242618 | 0.643181434 |
| Heart_Failure | dendritic_cell | *POU6F2* | -1.368802296 | 3.41E-07 | 3.66E-06 | 0.001653473 |
| Heart_Failure | dendritic_cell | *TSPAN13* | -2.996674621 | 9.47E-07 | 5.47E-06 | 0.004596837 |
| Heart_Failure | dendritic_cell | *RARRES2* | -2.550230479 | 1.57E-05 | 3.91E-05 | 0.076207684 |
| Heart_Failure | dendritic_cell | *TMEM176B* | -1.62313896 | 0.000105325 | 0.000197999 | 0.511035571 |
| Heart_Failure | dendritic_cell | *COA1* | -2.207923218 | 1.10E-05 | 2.92E-05 | 0.053244322 |
| Heart_Failure | dendritic_cell | *BLVRA* | -2.418117063 | 8.14E-05 | 0.000158037 | 0.395091916 |
| Heart_Failure | dendritic_cell | *URGCP* | -3.38401312 | 5.52E-06 | 1.74E-05 | 0.026784328 |
| Heart_Failure | dendritic_cell | *AEBP1* | -2.083203098 | 7.34E-07 | 4.70E-06 | 0.003560501 |
| Heart_Failure | dendritic_cell | *GCK* | 2.886328116 | 1.31E-07 | 3.44E-06 | 0.000637948 |
| Heart_Failure | dendritic_cell | *BCL7B* | 2.894538094 | 0.000820924 | 0.001281984 | 1 |
| Heart_Failure | dendritic_cell | *TBL2* | -2.741018668 | 1.06E-05 | 2.83E-05 | 0.051448002 |
| Heart_Failure | dendritic_cell | *SPATA6L* | -1.603218831 | 6.69E-07 | 4.49E-06 | 0.003244031 |
| Heart_Failure | dendritic_cell | *SLC1A1* | -0.640548911 | 0.005421962 | 0.007444074 | 1 |
| Heart_Failure | dendritic_cell | *FKTN* | -3.146541549 | 0.000122064 | 0.000225449 | 0.592255792 |
| Heart_Failure | dendritic_cell | *NMRK1* | -2.286709783 | 7.14E-06 | 2.07E-05 | 0.034647146 |
| Heart_Failure | dendritic_cell | *TRIM14* | 2.539046262 | 1.59E-05 | 3.96E-05 | 0.077151196 |
| Heart_Failure | dendritic_cell | *CORO2A* | -2.309309477 | 4.51E-07 | 3.90E-06 | 0.002188146 |
| Heart_Failure | dendritic_cell | *OGN* | -1.662413228 | 1.45E-05 | 3.66E-05 | 0.070285123 |
| Heart_Failure | dendritic_cell | *ECM2* | -1.465753517 | 9.71E-07 | 5.54E-06 | 0.004712886 |
| Heart_Failure | dendritic_cell | *PTGR1* | -1.623681092 | 6.67E-07 | 4.49E-06 | 0.003237953 |
| Heart_Failure | dendritic_cell | *SUSD1* | -1.029747978 | 0.023648035 | 0.029687003 | 1 |
| Heart_Failure | dendritic_cell | *PLGRKT* | -2.008880039 | 3.96E-05 | 8.49E-05 | 0.192242593 |
| Heart_Failure | dendritic_cell | *KANK1* | -1.316321811 | 2.79E-06 | 1.08E-05 | 0.013523032 |
| Heart_Failure | dendritic_cell | *NCS1* | -2.136752267 | 9.83E-06 | 2.67E-05 | 0.047712246 |
| Heart_Failure | dendritic_cell | *FUBP3* | -2.559684998 | 6.20E-06 | 1.90E-05 | 0.030073681 |
| Heart_Failure | dendritic_cell | *RGP1* | 3.433475116 | 0.018841117 | 0.023949987 | 1 |
| Heart_Failure | dendritic_cell | *PTGDS* | -2.192490166 | 5.70E-07 | 4.26E-06 | 0.002764579 |
| Heart_Failure | dendritic_cell | *ATRNL1* | -0.832068318 | 2.28E-06 | 9.32E-06 | 0.011075545 |
| Heart_Failure | dendritic_cell | *HPS1* | -2.920757504 | 0.000181191 | 0.000321675 | 0.879137494 |
| Heart_Failure | dendritic_cell | *PHYH* | -1.568958414 | 0.002865344 | 0.004074634 | 1 |
| Heart_Failure | dendritic_cell | *DNMBP* | -2.366642803 | 0.000316867 | 0.000535321 | 1 |
| Heart_Failure | dendritic_cell | *PKD2L1* | -2.277392373 | 0.010809886 | 0.014233261 | 1 |
| Heart_Failure | dendritic_cell | *CUBN* | -2.22928106 | 3.00E-07 | 3.66E-06 | 0.001454857 |
| Heart_Failure | dendritic_cell | *TRDMT1* | -2.261921217 | 1.21E-06 | 6.24E-06 | 0.0058912 |
| Heart_Failure | dendritic_cell | *ATE1* | -2.096826561 | 0.001245384 | 0.001891268 | 1 |
| Heart_Failure | dendritic_cell | *PLEKHA1* | -2.236719726 | 9.36E-07 | 5.43E-06 | 0.004540758 |
| Heart_Failure | dendritic_cell | *SPOCK2* | -1.38559968 | 9.29E-06 | 2.54E-05 | 0.045078249 |
| Heart_Failure | dendritic_cell | *BMPR1A* | -2.492725782 | 5.47E-05 | 0.000112328 | 0.265230837 |
| Heart_Failure | dendritic_cell | *SFXN3* | 5.223203236 | 4.22E-06 | 1.43E-05 | 0.020479437 |
| Heart_Failure | dendritic_cell | *TNKS2* | -2.847347511 | 3.52E-05 | 7.69E-05 | 0.170705115 |
| Heart_Failure | dendritic_cell | *ANKRD26* | -2.216526259 | 5.29E-06 | 1.69E-05 | 0.025680958 |
| Heart_Failure | dendritic_cell | *ACBD5* | -1.984748663 | 6.00E-07 | 4.30E-06 | 0.002913213 |
| Heart_Failure | dendritic_cell | *PITRM1* | -2.718307549 | 8.88E-06 | 2.45E-05 | 0.043107786 |
| Heart_Failure | dendritic_cell | *TFAM* | -3.54828875 | 6.36E-06 | 1.92E-05 | 0.030875967 |
| Heart_Failure | dendritic_cell | *RPL28* | -2.280286055 | 0.000302778 | 0.000513665 | 1 |
| Heart_Failure | dendritic_cell | *DNAJC12* | -3.841659775 | 1.72E-05 | 4.24E-05 | 0.083322529 |
| Heart_Failure | dendritic_cell | *PBLD* | -3.10007915 | 0.007817163 | 0.010494984 | 1 |
| Heart_Failure | dendritic_cell | *TSPAN14* | -2.067157327 | 1.72E-05 | 4.25E-05 | 0.083585775 |
| Heart_Failure | dendritic_cell | *KRT23* | 2.773280552 | 0.000532635 | 0.000859821 | 1 |

| | | | | | | |
|---|---|---|---|---|---|---|
| Heart_Failure | dendritic_cell | *TADA2A* | -1.628996086 | 0.000515667 | 0.000835677 | 1 |
| Heart_Failure | dendritic_cell | *PSMB3* | -3.027893049 | 2.17E-07 | 3.66E-06 | 0.001054355 |
| Heart_Failure | dendritic_cell | *RPL19* | -2.553637473 | 1.07E-05 | 2.86E-05 | 0.052042716 |
| Heart_Failure | dendritic_cell | *RAPGEFL1* | -2.399305776 | 7.36E-07 | 4.70E-06 | 0.003572276 |
| Heart_Failure | dendritic_cell | *WNT3* | 2.361734205 | 1.59E-06 | 7.40E-06 | 0.00771792 |
| Heart_Failure | dendritic_cell | *ASPA* | -1.573011552 | 1.41E-06 | 6.81E-06 | 0.006824864 |
| Heart_Failure | dendritic_cell | *TRIM37* | -3.09359245 | 2.51E-05 | 5.78E-05 | 0.122018769 |
| Heart_Failure | dendritic_cell | *PNPO* | 4.383589308 | 2.92E-07 | 3.66E-06 | 0.001414618 |
| Heart_Failure | dendritic_cell | *TRIM16L* | -2.446987306 | 2.82E-07 | 3.66E-06 | 0.001367587 |
| Heart_Failure | dendritic_cell | *CDK5RAP3* | 2.446569199 | 7.06E-07 | 4.63E-06 | 0.003424304 |
| Heart_Failure | dendritic_cell | *PIGL* | -2.267241879 | 4.15E-06 | 1.42E-05 | 0.020116978 |
| Heart_Failure | dendritic_cell | *MED13* | -2.342788662 | 7.00E-05 | 0.000138623 | 0.339487724 |
| Heart_Failure | dendritic_cell | *ENO3* | -2.67419362 | 0.000108109 | 0.000202449 | 0.524545592 |
| Heart_Failure | dendritic_cell | *RAI1* | -3.789556219 | 0.000343039 | 0.000575328 | 1 |
| Heart_Failure | dendritic_cell | *GOSR1* | -2.648261632 | 1.06E-05 | 2.82E-05 | 0.051328371 |
| Heart_Failure | dendritic_cell | *DRG2* | -3.65318237 | 6.42E-05 | 0.000128451 | 0.311513831 |
| Heart_Failure | dendritic_cell | *ALDH3A1* | -4.337428811 | 0.010405312 | 0.013730371 | 1 |
| Heart_Failure | dendritic_cell | *ICAM2* | -2.632077116 | 1.14E-06 | 6.06E-06 | 0.005549804 |
| Heart_Failure | dendritic_cell | *DDX5* | -1.814206985 | 0.002022131 | 0.002964164 | 1 |
| Heart_Failure | dendritic_cell | *PSMD11* | -2.56898019 | 8.99E-07 | 5.31E-06 | 0.004364297 |
| Heart_Failure | dendritic_cell | *PEX12* | 3.535635729 | 0.000666384 | 0.001055597 | 1 |
| Heart_Failure | dendritic_cell | *DHX58* | -2.413187528 | 1.49E-07 | 3.45E-06 | 0.000725325 |
| Heart_Failure | dendritic_cell | *RAB5C* | -1.797557171 | 0.002041305 | 0.00298956 | 1 |
| Heart_Failure | dendritic_cell | *NAGLU* | -2.299823284 | 9.05E-06 | 2.49E-05 | 0.043907596 |
| Heart_Failure | dendritic_cell | *HSD17B1* | -3.464588522 | 0.00083066 | 0.001294293 | 1 |
| Heart_Failure | dendritic_cell | *EZH1* | -2.351695369 | 0.001178087 | 0.001796943 | 1 |
| Heart_Failure | dendritic_cell | *ABCC3* | 1.491627137 | 0.000770267 | 0.00120676 | 1 |
| Heart_Failure | dendritic_cell | *EFTUD2* | -3.019361659 | 5.59E-06 | 1.75E-05 | 0.02711693 |
| Heart_Failure | dendritic_cell | *HLF* | -1.372031433 | 2.71E-07 | 3.66E-06 | 0.001313833 |
| Heart_Failure | dendritic_cell | *EFNB3* | 1.142864074 | 1.53E-06 | 7.21E-06 | 0.007442378 |
| Heart_Failure | dendritic_cell | *FAM20A* | -3.10146319 | 0.000546576 | 0.00087989 | 1 |
| Heart_Failure | dendritic_cell | *DPH1* | -4.603083157 | 4.47E-06 | 1.49E-05 | 0.021685659 |
| Heart_Failure | dendritic_cell | *DHRS7B* | -3.138473661 | 0.000197043 | 0.000347908 | 0.956051127 |
| Heart_Failure | dendritic_cell | *MYH3* | -5.195207751 | 2.08E-06 | 8.77E-06 | 0.010093378 |
| Heart_Failure | dendritic_cell | *GNRHR* | -1.577408381 | 0.000127786 | 0.000234855 | 0.620017064 |
| Heart_Failure | dendritic_cell | *SLAIN2* | -2.059646532 | 4.09E-06 | 1.41E-05 | 0.01984149 |
| Heart_Failure | dendritic_cell | *OCIAD1* | -2.395092172 | 7.98E-06 | 2.25E-05 | 0.038698107 |
| Heart_Failure | dendritic_cell | *UBE2D3* | -2.068283522 | 6.57E-06 | 1.96E-05 | 0.031885164 |
| Heart_Failure | dendritic_cell | *NDUFC1* | -2.480849659 | 1.37E-06 | 6.71E-06 | 0.006649904 |
| Heart_Failure | dendritic_cell | *KLHL2* | -2.918224536 | 0.014606233 | 0.018903559 | 1 |
| Heart_Failure | dendritic_cell | *CPE* | -1.410473832 | 0.000200781 | 0.000353479 | 0.974189158 |
| Heart_Failure | dendritic_cell | *WFS1* | -1.711518869 | 3.21E-07 | 3.66E-06 | 0.001559576 |
| Heart_Failure | dendritic_cell | *GRPEL1* | -5.159131281 | 0.003937194 | 0.005497342 | 1 |
| Heart_Failure | dendritic_cell | *AADAT* | -2.452209776 | 3.27E-07 | 3.66E-06 | 0.001587795 |
| Heart_Failure | dendritic_cell | *TRIM2* | -1.405657242 | 6.56E-07 | 4.46E-06 | 0.003182323 |
| Heart_Failure | dendritic_cell | *STIM2* | -2.870536881 | 0.000363761 | 0.000605063 | 1 |
| Heart_Failure | dendritic_cell | *BST1* | -1.703682122 | 6.16E-07 | 4.35E-06 | 0.002988448 |
| Heart_Failure | dendritic_cell | *UFSP2* | -3.170321705 | 0.008444407 | 0.011284016 | 1 |
| Heart_Failure | dendritic_cell | *KLHL5* | 2.664488185 | 0.02484956 | 0.03102678 | 1 |
| Heart_Failure | dendritic_cell | *FAM149A* | -2.176073573 | 9.19E-07 | 5.38E-06 | 0.004460554 |

| | | | | | | |
|---|---|---|---|---|---|---|
| Heart_Failure | dendritic_cell | *DDX25* | -1.627709877 | 5.84E-05 | 0.000118499 | 0.283449794 |
| Heart_Failure | dendritic_cell | *HTATIP2* | -1.614735389 | 0.005840959 | 0.007985441 | 1 |
| Heart_Failure | dendritic_cell | *ELP4* | -2.300753191 | 1.93E-05 | 4.67E-05 | 0.09362977 |
| Heart_Failure | dendritic_cell | *TECTA* | -3.140652692 | 2.24E-07 | 3.66E-06 | 0.00108473 |
| Heart_Failure | dendritic_cell | *SC5D* | -4.132589622 | 0.000368492 | 0.000612302 | 1 |
| Heart_Failure | dendritic_cell | *B3GAT1* | -13.7952839 | 6.32E-07 | 4.39E-06 | 0.003067887 |
| Heart_Failure | dendritic_cell | *P2RX3* | 37.52683753 | 1.36E-07 | 3.44E-06 | 0.000661856 |
| Heart_Failure | dendritic_cell | *SNX15* | -1.7776185 | 0.036733592 | 0.044996563 | 1 |
| Heart_Failure | dendritic_cell | *PUS3* | 1.65610843 | 0.006889435 | 0.009316482 | 1 |
| Heart_Failure | dendritic_cell | *DCPS* | -2.116962617 | 4.03E-07 | 3.80E-06 | 0.001955955 |
| Heart_Failure | dendritic_cell | *FOXRED1* | -2.228806239 | 0.001394178 | 0.002098836 | 1 |
| Heart_Failure | dendritic_cell | *NRXN2* | 2.502279335 | 1.24E-07 | 3.44E-06 | 0.00060405 |
| Heart_Failure | dendritic_cell | *ST3GAL4* | -3.098735248 | 0.000259744 | 0.000447224 | 1 |
| Heart_Failure | dendritic_cell | *CCND1* | 2.265992875 | 1.79E-06 | 7.95E-06 | 0.008688864 |
| Heart_Failure | dendritic_cell | *CCDC86* | 2.186447797 | 6.50E-07 | 4.45E-06 | 0.003154429 |
| Heart_Failure | dendritic_cell | *PANX1* | -3.002519918 | 3.67E-07 | 3.66E-06 | 0.001781461 |
| Heart_Failure | dendritic_cell | *CEP164* | -2.816504464 | 3.29E-05 | 7.24E-05 | 0.159673887 |
| Heart_Failure | dendritic_cell | *GALNT18* | -1.076179271 | 1.68E-07 | 3.52E-06 | 0.000815547 |
| Heart_Failure | dendritic_cell | *KIAA1549L* | -6.395588962 | 2.55E-07 | 3.66E-06 | 0.001236937 |
| Heart_Failure | dendritic_cell | *SLC22A18* | 0.744716341 | 0.019898864 | 0.025221862 | 1 |
| Heart_Failure | dendritic_cell | *SLC35F2* | -2.814066178 | 2.05E-06 | 8.72E-06 | 0.009953171 |
| Heart_Failure | dendritic_cell | *C11orf21* | 0.83373194 | 7.73E-08 | 3.44E-06 | 0.000374902 |
| Heart_Failure | dendritic_cell | *ELMOD1* | 3.287151278 | 0.00038888 | 0.000642221 | 1 |
| Heart_Failure | dendritic_cell | *RPS13* | -2.234330169 | 3.97E-05 | 8.51E-05 | 0.192705454 |
| Heart_Failure | dendritic_cell | *NDUFS8* | -3.356822119 | 3.98E-06 | 1.38E-05 | 0.019308698 |
| Heart_Failure | dendritic_cell | *TCIRG1* | 4.259548966 | 6.08E-06 | 1.87E-05 | 0.029515897 |
| Heart_Failure | dendritic_cell | *GTF2H1* | -2.974014853 | 0.00031155 | 0.000527255 | 1 |
| Heart_Failure | dendritic_cell | *PRDM4* | -2.330471228 | 0.000634983 | 0.001008489 | 1 |
| Heart_Failure | dendritic_cell | *CORO1C* | -3.087205933 | 0.000219265 | 0.000382826 | 1 |
| Heart_Failure | dendritic_cell | *ASIC1* | -3.833832957 | 3.14E-07 | 3.66E-06 | 0.001524487 |
| Heart_Failure | dendritic_cell | *CAPRIN2* | -1.57653066 | 0.010013348 | 0.01323475 | 1 |
| Heart_Failure | dendritic_cell | *KCTD10* | -2.257839199 | 1.77E-08 | 2.99E-06 | 8.59E-05 |
| Heart_Failure | dendritic_cell | *IL23A* | -32.11609567 | 4.35E-05 | 9.19E-05 | 0.211002941 |
| Heart_Failure | dendritic_cell | *ACSS3* | -1.347112372 | 4.56E-06 | 1.52E-05 | 0.022119917 |
| Heart_Failure | dendritic_cell | *LTA4H* | -1.793943821 | 0.011552649 | 0.015165977 | 1 |
| Heart_Failure | dendritic_cell | *PARP11* | -3.1138399 | 0.005568216 | 0.007634073 | 1 |
| Heart_Failure | dendritic_cell | *VPS29* | -2.86425441 | 7.75E-05 | 0.000151394 | 0.375910505 |
| Heart_Failure | dendritic_cell | *AKAP3* | -1.16139674 | 0.000791663 | 0.001238281 | 1 |
| Heart_Failure | dendritic_cell | *CDK2AP1* | 1.096142355 | 4.80E-09 | 1.79E-06 | 2.33E-05 |
| Heart_Failure | dendritic_cell | *DDX55* | -3.702227337 | 4.29E-06 | 1.45E-05 | 0.020833354 |
| Heart_Failure | dendritic_cell | *ENDOU* | 2.560725162 | 1.12E-07 | 3.44E-06 | 0.000544906 |
| Heart_Failure | dendritic_cell | *VDR* | -8.200742066 | 0.001044272 | 0.001604944 | 1 |
| Heart_Failure | dendritic_cell | *CAND1* | -3.050008414 | 0.000126472 | 0.000232616 | 0.613641836 |
| Heart_Failure | dendritic_cell | *MDM1* | -1.654611143 | 4.47E-07 | 3.90E-06 | 0.002168464 |
| Heart_Failure | dendritic_cell | *NUP107* | -2.956102329 | 0.001562792 | 0.002337444 | 1 |
| Heart_Failure | dendritic_cell | *KRR1* | -3.066066873 | 4.81E-05 | 0.000100093 | 0.233517614 |
| Heart_Failure | dendritic_cell | *MRPL51* | -1.305072398 | 0.004594296 | 0.006359921 | 1 |
| Heart_Failure | dendritic_cell | *GNB3* | 0.872483245 | 0.000949679 | 0.001468401 | 1 |
| Heart_Failure | dendritic_cell | *CDCA3* | -3.600030153 | 0.00023052 | 0.000400078 | 1 |
| Heart_Failure | dendritic_cell | *CHPT1* | -2.566200237 | 9.07E-05 | 0.000173264 | 0.440089843 |

| | | | | | | |
|---|---|---|---|---|---|---|
| Heart_Failure | dendritic_cell | *GNPTAB* | -2.964686923 | 0.000742616 | 0.00116683 | 1 |
| Heart_Failure | dendritic_cell | *SPSB2* | -2.085488832 | 5.89E-06 | 1.83E-05 | 0.028565778 |
| Heart_Failure | dendritic_cell | *ATN1* | -1.618944986 | 3.74E-06 | 1.32E-05 | 0.018167552 |
| Heart_Failure | dendritic_cell | *C12orf57* | -2.679375669 | 8.10E-07 | 4.93E-06 | 0.003932189 |
| Heart_Failure | dendritic_cell | *NT5DC3* | -2.057880952 | 3.52E-06 | 1.27E-05 | 0.017059903 |
| Heart_Failure | dendritic_cell | *SUDS3* | -2.362624402 | 1.63E-06 | 7.52E-06 | 0.007897121 |
| Heart_Failure | dendritic_cell | *GOLT1B* | -2.882478733 | 9.40E-06 | 2.57E-05 | 0.04562821 |
| Heart_Failure | dendritic_cell | *C2CD5* | -2.805461183 | 0.000287871 | 0.000490776 | 1 |
| Heart_Failure | dendritic_cell | *BTN3A3* | 2.093484597 | 0.003508362 | 0.00493122 | 1 |
| Heart_Failure | dendritic_cell | *FRK* | -1.691988887 | 3.47E-07 | 3.66E-06 | 0.001683997 |
| Heart_Failure | dendritic_cell | *RWDD1* | -3.258196207 | 2.77E-05 | 6.25E-05 | 0.134477964 |
| Heart_Failure | dendritic_cell | *RSPH4A* | 2.545225194 | 1.88E-06 | 8.19E-06 | 0.009126581 |
| Heart_Failure | dendritic_cell | *SMIM8* | -2.157258257 | 0.000496073 | 0.000805538 | 1 |
| Heart_Failure | dendritic_cell | *FAM184A* | -2.467462147 | 0.008988662 | 0.011971723 | 1 |
| Heart_Failure | dendritic_cell | *RNGTT* | -2.893094926 | 9.18E-05 | 0.000175273 | 0.445368546 |
| Heart_Failure | dendritic_cell | *GABRR2* | -2.515485479 | 3.64E-05 | 7.91E-05 | 0.17669237 |
| Heart_Failure | dendritic_cell | *HDDC2* | -1.860891623 | 1.94E-05 | 4.69E-05 | 0.094226659 |
| Heart_Failure | dendritic_cell | *UST* | -1.318259663 | 3.40E-06 | 1.24E-05 | 0.016502272 |
| Heart_Failure | dendritic_cell | *MTRF1L* | -2.41141848 | 6.05E-07 | 4.32E-06 | 0.002936243 |
| Heart_Failure | dendritic_cell | *TULP1* | -2.123943935 | 0.010663437 | 0.01405951 | 1 |
| Heart_Failure | dendritic_cell | *SLC26A8* | -1.266909875 | 0.002762217 | 0.003939529 | 1 |
| Heart_Failure | dendritic_cell | *MRPL18* | -2.893655263 | 0.015352507 | 0.019800735 | 1 |
| Heart_Failure | dendritic_cell | *RNF8* | -2.73834039 | 5.96E-06 | 1.84E-05 | 0.028937005 |
| Heart_Failure | dendritic_cell | *PHACTR1* | -2.743064894 | 6.54E-06 | 1.95E-05 | 0.031719732 |
| Heart_Failure | dendritic_cell | *MDGA1* | -1.971449699 | 2.77E-07 | 3.66E-06 | 0.001342549 |
| Heart_Failure | dendritic_cell | *CD83* | 0.631872348 | 1.28E-07 | 3.44E-06 | 0.000620405 |
| Heart_Failure | dendritic_cell | *MDN1* | -2.103585269 | 2.90E-06 | 1.11E-05 | 0.014084997 |
| Heart_Failure | dendritic_cell | *GLP1R* | -6.981725473 | 0.003015491 | 0.004270626 | 1 |
| Heart_Failure | dendritic_cell | *BVES* | -1.42421327 | 0.000136303 | 0.000248811 | 0.66134093 |
| Heart_Failure | dendritic_cell | *COL9A1* | -5.018470941 | 8.86E-06 | 2.45E-05 | 0.043010647 |
| Heart_Failure | dendritic_cell | *WASF1* | -1.556418034 | 5.22E-07 | 4.09E-06 | 0.002534049 |
| Heart_Failure | dendritic_cell | *ALDH5A1* | -1.699205867 | 5.45E-07 | 4.15E-06 | 0.002646589 |
| Heart_Failure | dendritic_cell | *VNN2* | 4.994985804 | 1.62E-06 | 7.49E-06 | 0.007850656 |
| Heart_Failure | dendritic_cell | *ACOT13* | -2.940183819 | 5.94E-06 | 1.84E-05 | 0.02880131 |
| Heart_Failure | dendritic_cell | *HBS1L* | -2.889233194 | 1.02E-06 | 5.71E-06 | 0.004955046 |
| Heart_Failure | dendritic_cell | *FIG4* | -2.884745476 | 2.46E-05 | 5.71E-05 | 0.119421234 |
| Heart_Failure | dendritic_cell | *PERP* | -1.812980652 | 0.000389707 | 0.000643368 | 1 |
| Heart_Failure | dendritic_cell | *CUTA* | 1.978867763 | 0.024601268 | 0.030772197 | 1 |
| Heart_Failure | dendritic_cell | *PACRG* | -1.737077993 | 4.03E-07 | 3.80E-06 | 0.001953473 |
| Heart_Failure | dendritic_cell | *QKI* | -2.206899488 | 8.77E-05 | 0.000168229 | 0.425620262 |
| Heart_Failure | dendritic_cell | *FAM120B* | -1.733916865 | 0.020125411 | 0.025495691 | 1 |
| Heart_Failure | dendritic_cell | *GUCA1B* | -2.278642946 | 8.66E-05 | 0.000166426 | 0.420391683 |
| Heart_Failure | dendritic_cell | *PRPH2* | -0.763874933 | 0.001081697 | 0.001658261 | 1 |
| Heart_Failure | dendritic_cell | *DNPH1* | -1.775070365 | 0.000190386 | 0.000336767 | 0.923751649 |
| Heart_Failure | dendritic_cell | *EXOC2* | -2.450181828 | 0.01806734 | 0.023026723 | 1 |
| Heart_Failure | dendritic_cell | *TMEM30A* | 4.260841886 | 0.000548868 | 0.0008827 | 1 |
| Heart_Failure | dendritic_cell | *IMPG1* | -7.992763961 | 6.31E-08 | 3.44E-06 | 0.000306372 |
| Heart_Failure | dendritic_cell | *LY86* | 0.715409414 | 1.45E-07 | 3.45E-06 | 0.000704877 |
| Heart_Failure | dendritic_cell | *PCDHB2* | -4.240179816 | 2.59E-07 | 3.66E-06 | 0.001257586 |
| Heart_Failure | dendritic_cell | *NUDT12* | -1.456638124 | 3.23E-07 | 3.66E-06 | 0.001567095 |

| | | | | | | |
|---|---|---|---|---|---|---|
| Heart_Failure | dendritic_cell | *CEP72* | -2.460347526 | 1.60E-07 | 3.51E-06 | 0.000776894 |
| Heart_Failure | dendritic_cell | *DAP* | -2.244711106 | 0.001949752 | 0.002865857 | 1 |
| Heart_Failure | dendritic_cell | *NME5* | -1.349345912 | 5.52E-05 | 0.000113105 | 0.267719609 |
| Heart_Failure | dendritic_cell | *KIF20A* | -19.67271706 | 1.16E-10 | 8.06E-08 | 5.64E-07 |
| Heart_Failure | dendritic_cell | *PFDN1* | -2.557167802 | 0.001999486 | 0.002934514 | 1 |
| Heart_Failure | dendritic_cell | *TMCO6* | -2.754053341 | 1.65E-08 | 2.99E-06 | 8.03E-05 |
| Heart_Failure | dendritic_cell | *SPARC* | -1.777979252 | 1.79E-06 | 7.94E-06 | 0.008672107 |
| Heart_Failure | dendritic_cell | *IK* | -2.530006231 | 2.18E-07 | 3.66E-06 | 0.001055819 |
| Heart_Failure | dendritic_cell | *PCDHB15* | 1.122567325 | 0.03016647 | 0.037310149 | 1 |
| Heart_Failure | dendritic_cell | *HAVCR1* | -2.716495513 | 0.00708117 | 0.009562437 | 1 |
| Heart_Failure | dendritic_cell | *ARSB* | -3.194050205 | 0.014327581 | 0.018557774 | 1 |
| Heart_Failure | dendritic_cell | *THBS4* | -2.06494114 | 0.012448172 | 0.016253641 | 1 |
| Heart_Failure | dendritic_cell | *MSH3* | -2.167920618 | 7.42E-05 | 0.000145643 | 0.360029528 |
| Heart_Failure | dendritic_cell | *RASGRF2* | -1.031835148 | 3.46E-05 | 7.57E-05 | 0.167659087 |
| Heart_Failure | dendritic_cell | *DROSHA* | -2.202182199 | 0.000531995 | 0.000859268 | 1 |
| Heart_Failure | dendritic_cell | *CDH6* | -1.864600544 | 1.64E-05 | 4.07E-05 | 0.079682355 |
| Heart_Failure | dendritic_cell | *LMNB1* | -2.490392122 | 1.57E-06 | 7.33E-06 | 0.007605527 |
| Heart_Failure | dendritic_cell | *SLC12A7* | 4.608756386 | 4.91E-06 | 1.60E-05 | 0.023841162 |
| Heart_Failure | dendritic_cell | *SKP1* | -3.194246006 | 4.32E-05 | 9.13E-05 | 0.209368563 |
| Heart_Failure | dendritic_cell | *PPWD1* | -2.663122599 | 8.41E-06 | 2.35E-05 | 0.040819414 |
| Heart_Failure | dendritic_cell | *TXNDC15* | -3.548818141 | 1.48E-05 | 3.71E-05 | 0.071596818 |
| Heart_Failure | dendritic_cell | *SMAD5* | -2.444625385 | 4.48E-06 | 1.49E-05 | 0.021736627 |
| Heart_Failure | dendritic_cell | *ERGIC1* | -2.293064291 | 2.57E-06 | 1.02E-05 | 0.012476334 |
| Heart_Failure | dendritic_cell | *ATP6V0E1* | -2.620125391 | 0.000103628 | 0.000195111 | 0.50280108 |
| Heart_Failure | dendritic_cell | *UNC5A* | 2.907755423 | 1.30E-07 | 3.44E-06 | 0.000631918 |
| Heart_Failure | dendritic_cell | *ACTR8* | -2.55594066 | 1.38E-07 | 3.44E-06 | 0.000668605 |
| Heart_Failure | dendritic_cell | *TBCCD1* | -3.095193709 | 1.82E-06 | 8.06E-06 | 0.008850285 |
| Heart_Failure | dendritic_cell | *NPHP3* | -1.409609594 | 0.003158437 | 0.00446135 | 1 |
| Heart_Failure | dendritic_cell | *PCCB* | -1.299870853 | 1.81E-05 | 4.41E-05 | 0.087663703 |
| Heart_Failure | dendritic_cell | *CEP70* | -1.997181037 | 3.04E-06 | 1.14E-05 | 0.014726076 |
| Heart_Failure | dendritic_cell | *HYAL1* | 2.754260157 | 0.000300907 | 0.000510668 | 1 |
| Heart_Failure | dendritic_cell | *CYB561D2* | 2.945598152 | 0.002024074 | 0.002966115 | 1 |
| Heart_Failure | dendritic_cell | *C3orf14* | -2.288935378 | 2.23E-06 | 9.19E-06 | 0.010829624 |
| Heart_Failure | dendritic_cell | *CBLB* | -2.071674442 | 1.05E-05 | 2.81E-05 | 0.050888819 |
| Heart_Failure | dendritic_cell | *IQCG* | -3.18646643 | 2.01E-05 | 4.83E-05 | 0.097604874 |
| Heart_Failure | dendritic_cell | *NCBP2* | -2.677505962 | 3.66E-06 | 1.30E-05 | 0.017766388 |
| Heart_Failure | dendritic_cell | *SLC41A3* | -2.734858283 | 0.001343916 | 0.002029467 | 1 |
| Heart_Failure | dendritic_cell | *PLXNA1* | 5.459474513 | 0.000379315 | 0.000629208 | 1 |
| Heart_Failure | dendritic_cell | *SCAP* | -2.433235507 | 1.51E-06 | 7.16E-06 | 0.007333022 |
| Heart_Failure | dendritic_cell | *KIAA1257* | -3.983592194 | 3.07E-07 | 3.66E-06 | 0.001490259 |
| Heart_Failure | dendritic_cell | *NEK11* | -2.368130865 | 1.09E-05 | 2.90E-05 | 0.052915652 |
| Heart_Failure | dendritic_cell | *PLSCR4* | -1.69349457 | 2.89E-06 | 1.11E-05 | 0.014034258 |
| Heart_Failure | dendritic_cell | *HEMK1* | -2.07200932 | 2.65E-06 | 1.04E-05 | 0.012860057 |
| Heart_Failure | dendritic_cell | *COMMD2* | -2.209190195 | 1.11E-06 | 6.00E-06 | 0.005396179 |
| Heart_Failure | dendritic_cell | *ABHD14B* | 3.660233444 | 6.68E-05 | 0.000133182 | 0.324031459 |
| Heart_Failure | dendritic_cell | *ARHGEF26* | -2.158156378 | 1.01E-06 | 5.69E-06 | 0.004895171 |
| Heart_Failure | dendritic_cell | *VIPR1* | -2.798244509 | 4.59E-07 | 3.90E-06 | 0.002227216 |
| Heart_Failure | dendritic_cell | *NKTR* | -2.250295286 | 5.49E-05 | 0.000112543 | 0.266164487 |
| Heart_Failure | dendritic_cell | *NEK4* | -2.236866421 | 0.030294975 | 0.037459536 | 1 |
| Heart_Failure | dendritic_cell | *ADAM23* | -1.795358831 | 7.09E-07 | 4.63E-06 | 0.003438504 |

| | | | | | | |
|---|---|---|---|---|---|---|
| Heart_Failure | dendritic_cell | *LMAN2L* | -2.47151976 | 6.93E-06 | 2.04E-05 | 0.033645078 |
| Heart_Failure | dendritic_cell | *ACTR1B* | -3.894995908 | 4.89E-05 | 0.00010147 | 0.237235867 |
| Heart_Failure | dendritic_cell | *SLC35F5* | -2.434728184 | 5.67E-05 | 0.000115681 | 0.275090145 |
| Heart_Failure | dendritic_cell | *CENPA* | -2.320334287 | 1.60E-07 | 3.51E-06 | 0.000778042 |
| Heart_Failure | dendritic_cell | *ITGB6* | -1.870924341 | 5.49E-06 | 1.73E-05 | 0.026641427 |
| Heart_Failure | dendritic_cell | *PPM1G* | -3.489984845 | 0.000161732 | 0.000290316 | 0.784724795 |
| Heart_Failure | dendritic_cell | *INO80B* | -4.288117085 | 1.70E-08 | 2.99E-06 | 8.27E-05 |
| Heart_Failure | dendritic_cell | *MOGS* | -1.279274048 | 0.00143523 | 0.002153194 | 1 |
| Heart_Failure | dendritic_cell | *GRB14* | -4.054140833 | 7.20E-06 | 2.09E-05 | 0.034947712 |
| Heart_Failure | dendritic_cell | *RTN4* | -2.146284763 | 6.14E-05 | 0.000123751 | 0.298115536 |
| Heart_Failure | dendritic_cell | *GALNT3* | -2.444621093 | 4.43E-05 | 9.31E-05 | 0.214755118 |
| Heart_Failure | dendritic_cell | *POLE4* | -4.236970035 | 1.93E-05 | 4.67E-05 | 0.093692568 |
| Heart_Failure | dendritic_cell | *ACADL* | -1.214870274 | 5.10E-07 | 4.04E-06 | 0.002475865 |
| Heart_Failure | dendritic_cell | *MRPL19* | -2.7678421 | 1.27E-05 | 3.27E-05 | 0.061415439 |
| Heart_Failure | dendritic_cell | *IGFBP2* | -1.550978203 | 1.11E-06 | 6.00E-06 | 0.005388291 |
| Heart_Failure | dendritic_cell | *GGCX* | 3.466338082 | 0.000404337 | 0.000665483 | 1 |
| Heart_Failure | dendritic_cell | *WNT6* | -2.922788059 | 1.62E-06 | 7.49E-06 | 0.007854444 |
| Heart_Failure | dendritic_cell | *IL1RL2* | 0.972429433 | 2.06E-07 | 3.66E-06 | 0.001000974 |
| Heart_Failure | dendritic_cell | *IL18R1* | 6.522467463 | 1.37E-09 | 6.06E-07 | 6.67E-06 |
| Heart_Failure | dendritic_cell | *MLPH* | 3.569962137 | 0.004289849 | 0.005962288 | 1 |
| Heart_Failure | dendritic_cell | *HDLBP* | -2.16588908 | 9.19E-05 | 0.000175332 | 0.445694799 |
| Heart_Failure | dendritic_cell | *PASK* | -1.922522999 | 1.26E-06 | 6.39E-06 | 0.006115487 |
| Heart_Failure | dendritic_cell | *TPO* | -1.485632485 | 2.06E-07 | 3.66E-06 | 0.001001167 |
| Heart_Failure | dendritic_cell | *TAF1B* | -2.252681972 | 0.000578862 | 0.000926027 | 1 |
| Heart_Failure | dendritic_cell | *NOL10* | -2.577514821 | 8.06E-06 | 2.27E-05 | 0.03909604 |
| Heart_Failure | dendritic_cell | *STRN* | -2.160346135 | 3.18E-06 | 1.18E-05 | 0.01543251 |
| Heart_Failure | dendritic_cell | *CEBPZ* | -2.716226807 | 6.79E-05 | 0.000135147 | 0.329624353 |
| Heart_Failure | dendritic_cell | *PRKD3* | -2.426813035 | 2.92E-05 | 6.52E-05 | 0.141751764 |
| Heart_Failure | dendritic_cell | *SLC25A12* | -2.31600573 | 6.37E-06 | 1.92E-05 | 0.030905772 |
| Heart_Failure | dendritic_cell | *DARS* | -2.874077747 | 4.11E-05 | 8.77E-05 | 0.199523569 |
| Heart_Failure | dendritic_cell | *PNO1* | -3.889956514 | 2.09E-06 | 8.81E-06 | 0.010161131 |
| Heart_Failure | dendritic_cell | *ORC4* | -2.087660933 | 7.00E-06 | 2.05E-05 | 0.033974185 |
| Heart_Failure | dendritic_cell | *AAK1* | -2.313602472 | 2.73E-05 | 6.18E-05 | 0.132369591 |
| Heart_Failure | dendritic_cell | *NFE2L2* | -2.17052747 | 0.00133486 | 0.002017677 | 1 |
| Heart_Failure | dendritic_cell | *MSH6* | -2.74437955 | 0.026828861 | 0.0333522 | 1 |
| Heart_Failure | dendritic_cell | *ALMS1* | -2.359691322 | 3.75E-06 | 1.32E-05 | 0.018179112 |
| Heart_Failure | dendritic_cell | *DNAJC16* | -2.829752852 | 0.002276582 | 0.003305199 | 1 |
| Heart_Failure | dendritic_cell | *MARK1* | -1.849518913 | 9.96E-07 | 5.63E-06 | 0.004830404 |
| Heart_Failure | dendritic_cell | *GPX7* | -0.8267045 | 0.00735326 | 0.009905058 | 1 |
| Heart_Failure | dendritic_cell | *RALGPS2* | -2.562105248 | 1.85E-06 | 8.10E-06 | 0.008967158 |
| Heart_Failure | dendritic_cell | *CEP104* | -2.982535172 | 0.000451245 | 0.000736442 | 1 |
| Heart_Failure | dendritic_cell | *FAM20B* | -2.887743554 | 4.46E-06 | 1.49E-05 | 0.02162418 |
| Heart_Failure | dendritic_cell | *WRAP73* | -3.596515904 | 1.31E-05 | 3.35E-05 | 0.063353167 |
| Heart_Failure | dendritic_cell | *ICMT* | -5.849765742 | 0.000352985 | 0.000588462 | 1 |
| Heart_Failure | dendritic_cell | *PARK7* | -2.635050527 | 8.80E-07 | 5.23E-06 | 0.004271444 |
| Heart_Failure | dendritic_cell | *KIAA1324* | -4.098772706 | 2.06E-07 | 3.66E-06 | 0.001000654 |
| Heart_Failure | dendritic_cell | *MECR* | -3.288300951 | 6.07E-06 | 1.87E-05 | 0.029461569 |
| Heart_Failure | dendritic_cell | *KCNC4* | -1.545781061 | 1.17E-06 | 6.14E-06 | 0.005695398 |
| Heart_Failure | dendritic_cell | *EDEM3* | -2.740412103 | 2.60E-05 | 5.93E-05 | 0.126133819 |
| Heart_Failure | dendritic_cell | *RAP1A* | -2.523521117 | 0.000322273 | 0.000543821 | 1 |

| | | | | | | |
|---|---|---|---|---|---|---|
| Heart_Failure | dendritic_cell | *CAPZA1* | -2.181104477 | 0.019667908 | 0.024942156 | 1 |
| Heart_Failure | dendritic_cell | *ASH1L* | -2.114154229 | 2.75E-05 | 6.22E-05 | 0.133628229 |
| Heart_Failure | dendritic_cell | *GON4L* | -2.443876669 | 0.002961141 | 0.00420101 | 1 |
| Heart_Failure | dendritic_cell | *DOCK7* | -2.362951645 | 6.76E-06 | 2.00E-05 | 0.032784143 |
| Heart_Failure | dendritic_cell | *FBXO2* | -1.360634653 | 0.016358034 | 0.021025255 | 1 |
| Heart_Failure | dendritic_cell | *DNAJC6* | -2.222143374 | 7.19E-06 | 2.08E-05 | 0.0348871 |
| Heart_Failure | dendritic_cell | *LEPR* | -2.512134575 | 0.000312134 | 0.000528059 | 1 |
| Heart_Failure | dendritic_cell | *MFN2* | -2.327542397 | 1.46E-05 | 3.68E-05 | 0.0706457 |
| Heart_Failure | dendritic_cell | *PRG4* | -1.933144712 | 4.56E-07 | 3.90E-06 | 0.002213907 |
| Heart_Failure | dendritic_cell | *NCF2* | 1.327027771 | 6.88E-05 | 0.000136675 | 0.333898133 |
| Heart_Failure | dendritic_cell | *CFHR3* | 2.874156252 | 0.000148936 | 0.000269541 | 0.722639349 |
| Heart_Failure | dendritic_cell | *WARS2* | -1.761615723 | 6.23E-07 | 4.37E-06 | 0.003024565 |
| Heart_Failure | dendritic_cell | *OSCP1* | -1.811082522 | 2.56E-05 | 5.86E-05 | 0.12432684 |
| Heart_Failure | dendritic_cell | *MRPS15* | -3.89432671 | 0.00017057 | 0.000304378 | 0.82760445 |
| Heart_Failure | dendritic_cell | *EXOC8* | 5.553839958 | 0.009598436 | 0.012739651 | 1 |
| Heart_Failure | dendritic_cell | *GNPAT* | -2.914384555 | 0.001019398 | 0.001569699 | 1 |
| Heart_Failure | dendritic_cell | *C1orf109* | -2.446826186 | 2.40E-07 | 3.66E-06 | 0.001165224 |
| Heart_Failure | dendritic_cell | *ACADM* | -2.680987658 | 1.30E-05 | 3.35E-05 | 0.06322984 |
| Heart_Failure | dendritic_cell | *SLAMF1* | -2.474752592 | 6.97E-06 | 2.04E-05 | 0.033797104 |
| Heart_Failure | dendritic_cell | *PADI2* | 2.139616862 | 4.65E-06 | 1.54E-05 | 0.022544488 |
| Heart_Failure | dendritic_cell | *SDHB* | -3.407090821 | 2.09E-05 | 5.00E-05 | 0.101542146 |
| Heart_Failure | dendritic_cell | *CTBS* | -1.591242671 | 0.00365529 | 0.005128823 | 1 |
| Heart_Failure | dendritic_cell | *KLHL12* | -2.644633634 | 7.62E-06 | 2.17E-05 | 0.036985452 |
| Heart_Failure | dendritic_cell | *ZNHIT6* | -3.181860864 | 0.000130273 | 0.000239245 | 0.632084059 |
| Heart_Failure | dendritic_cell | *KIF17* | -4.224150083 | 3.72E-06 | 1.32E-05 | 0.018039257 |
| Heart_Failure | dendritic_cell | *CD46* | -2.147289466 | 8.83E-05 | 0.000169128 | 0.42856972 |
| Heart_Failure | dendritic_cell | *EBNA1BP2* | -1.551062449 | 4.79E-07 | 3.96E-06 | 0.002322478 |
| Heart_Failure | dendritic_cell | *ERI3* | -2.642106228 | 2.28E-05 | 5.37E-05 | 0.110588421 |
| Heart_Failure | dendritic_cell | *PRDX1* | -2.151997373 | 4.42E-07 | 3.90E-06 | 0.002144149 |
| Heart_Failure | dendritic_cell | *CCDC181* | -2.534831035 | 4.33E-07 | 3.90E-06 | 0.00209865 |
| Heart_Failure | dendritic_cell | *NSUN4* | -3.073221575 | 5.93E-07 | 4.30E-06 | 0.002878655 |
| Heart_Failure | dendritic_cell | *PRRC2C* | -2.382477293 | 1.75E-05 | 4.30E-05 | 0.084804453 |
| Heart_Failure | dendritic_cell | *ABCD3* | -2.680593277 | 5.57E-06 | 1.75E-05 | 0.02700403 |
| Heart_Failure | dendritic_cell | *PTBP2* | -1.784630285 | 5.96E-05 | 0.000120731 | 0.289391035 |
| Heart_Failure | dendritic_cell | *DARS2* | -3.034592284 | 2.90E-06 | 1.11E-05 | 0.014077363 |
| Heart_Failure | dendritic_cell | *SERPINC1* | 3.09093839 | 6.10E-06 | 1.87E-05 | 0.029582786 |
| Heart_Failure | dendritic_cell | *MTFR1L* | -2.895295116 | 2.37E-06 | 9.55E-06 | 0.011480651 |
| Heart_Failure | dendritic_cell | *RPS6KA1* | 1.296573423 | 4.84E-07 | 3.96E-06 | 0.002349829 |
| Heart_Failure | dendritic_cell | *RPA2* | -2.83059492 | 3.05E-06 | 1.14E-05 | 0.014793078 |
| Heart_Failure | dendritic_cell | *MARC2* | -1.605196904 | 5.81E-07 | 4.28E-06 | 0.002820268 |
| Heart_Failure | dendritic_cell | *SLC5A9* | -2.6092477 | 5.02E-06 | 1.62E-05 | 0.024376847 |
| Heart_Failure | dendritic_cell | *ESYT2* | -2.315610447 | 1.92E-05 | 4.65E-05 | 0.093042856 |
| Heart_Failure | dendritic_cell | *CD3EAP* | 5.037603952 | 9.78E-06 | 2.65E-05 | 0.047433524 |
| Heart_Failure | dendritic_cell | *RCN2* | -2.737212694 | 3.88E-06 | 1.36E-05 | 0.01882462 |
| Heart_Failure | dendritic_cell | *APOA1* | -5.62382586 | 0.026161446 | 0.03255587 | 1 |
| Heart_Failure | dendritic_cell | *CAMSAP2* | -1.758410731 | 9.79E-06 | 2.66E-05 | 0.047502658 |
| Heart_Failure | dendritic_cell | *KLF7* | -2.20918094 | 1.93E-05 | 4.67E-05 | 0.093645448 |
| Heart_Failure | dendritic_cell | *CASC1* | -1.525419668 | 9.26E-07 | 5.40E-06 | 0.004494886 |
| Heart_Failure | dendritic_cell | *LRMP* | 1.171204838 | 0.009101134 | 0.01211487 | 1 |
| Heart_Failure | dendritic_cell | *ELOVL4* | -2.11987004 | 0.011565128 | 0.015174149 | 1 |

| | | | | | | |
|---|---|---|---|---|---|---|
| Heart_Failure | dendritic_cell | *CNR1* | -2.500719236 | 8.81E-05 | 0.000168796 | 0.427560377 |
| Heart_Failure | dendritic_cell | *SGIP1* | -1.481331169 | 3.27E-07 | 3.66E-06 | 0.001585884 |
| Heart_Failure | dendritic_cell | *ALDH8A1* | -1.290275655 | 1.93E-06 | 8.34E-06 | 0.009374072 |
| Heart_Failure | dendritic_cell | *TMEM5* | -2.651149553 | 6.64E-07 | 4.49E-06 | 0.003221599 |
| Heart_Failure | dendritic_cell | *VAMP8* | -3.397082891 | 0.000193641 | 0.000342276 | 0.939547533 |
| Heart_Failure | dendritic_cell | *FOXO3* | -2.148615839 | 2.76E-05 | 6.23E-05 | 0.134040527 |
| Heart_Failure | dendritic_cell | *SPP1* | -2.584484389 | 4.51E-07 | 3.90E-06 | 0.002189184 |
| Heart_Failure | dendritic_cell | *MFSD1* | -2.51545949 | 0.026692206 | 0.033190821 | 1 |
| Heart_Failure | dendritic_cell | *UBN1* | -2.632152368 | 6.63E-06 | 1.97E-05 | 0.03216616 |
| Heart_Failure | dendritic_cell | *WDR35* | -1.624401516 | 5.90E-07 | 4.30E-06 | 0.002863123 |
| Heart_Failure | dendritic_cell | *DNAH7* | -1.626659331 | 1.78E-07 | 3.58E-06 | 0.000864729 |
| Heart_Failure | dendritic_cell | *CPSF3* | -2.701431369 | 0.000634775 | 0.001008489 | 1 |
| Heart_Failure | dendritic_cell | *PIGZ* | -1.168212323 | 0.001116423 | 0.001707718 | 1 |
| Heart_Failure | dendritic_cell | *SENP5* | -2.781401212 | 4.18E-05 | 8.90E-05 | 0.203041324 |
| Heart_Failure | dendritic_cell | *CCDC92* | -2.343347387 | 2.06E-06 | 8.73E-06 | 0.009995684 |
| Heart_Failure | dendritic_cell | *CTNNAL1* | -0.857068209 | 0.001848823 | 0.002727421 | 1 |
| Heart_Failure | dendritic_cell | *TRIM32* | -2.295107735 | 5.97E-07 | 4.30E-06 | 0.002895103 |
| Heart_Failure | dendritic_cell | *RBM18* | -3.579363226 | 0.000627458 | 0.000997191 | 1 |
| Heart_Failure | dendritic_cell | *HSDL2* | -1.940009199 | 5.21E-07 | 4.08E-06 | 0.002525745 |
| Heart_Failure | dendritic_cell | *MAPKAP1* | -2.335758397 | 0.009831425 | 0.013033353 | 1 |
| Heart_Failure | dendritic_cell | *INVS* | -1.861838478 | 0.001226138 | 0.001865545 | 1 |
| Heart_Failure | dendritic_cell | *KDSR* | -1.977549042 | 8.00E-07 | 4.91E-06 | 0.003880123 |
| Heart_Failure | dendritic_cell | *DCAF4* | 5.082312438 | 2.38E-05 | 5.56E-05 | 0.115407895 |
| Heart_Failure | dendritic_cell | *IFI27L2* | 5.150039834 | 9.75E-05 | 0.000184837 | 0.473181468 |
| Heart_Failure | dendritic_cell | *NEK9* | -2.832269339 | 0.000133103 | 0.000243428 | 0.645815332 |
| Heart_Failure | dendritic_cell | *TTLL5* | -2.704273212 | 2.26E-05 | 5.32E-05 | 0.109575853 |
| Heart_Failure | dendritic_cell | *FLVCR2* | 2.521485855 | 1.43E-07 | 3.45E-06 | 0.000695649 |
| Heart_Failure | dendritic_cell | *PPP4R4* | -1.505907511 | 0.000107196 | 0.00020105 | 0.520115169 |
| Heart_Failure | dendritic_cell | *ALDH6A1* | -1.544169377 | 6.50E-06 | 1.95E-05 | 0.031532521 |
| Heart_Failure | dendritic_cell | *ESRRB* | -2.289539754 | 1.92E-06 | 8.30E-06 | 0.009305217 |
| Heart_Failure | dendritic_cell | *EIF2B2* | -1.247098624 | 0.020236521 | 0.025629757 | 1 |
| Heart_Failure | dendritic_cell | *COQ6* | -4.147630545 | 1.84E-05 | 4.47E-05 | 0.089055018 |
| Heart_Failure | dendritic_cell | *DNMT3A* | -2.767740147 | 2.86E-05 | 6.41E-05 | 0.138609026 |
| Heart_Failure | dendritic_cell | *CNRIP1* | -1.749640721 | 0.00275292 | 0.003928578 | 1 |
| Heart_Failure | dendritic_cell | *CRIPT* | -2.336514146 | 8.73E-07 | 5.19E-06 | 0.004235831 |
| Heart_Failure | dendritic_cell | *EPCAM* | -4.055446445 | 1.41E-05 | 3.59E-05 | 0.068590351 |
| Heart_Failure | dendritic_cell | *SLC17A5* | -2.596134064 | 1.31E-05 | 3.35E-05 | 0.063327861 |
| Heart_Failure | dendritic_cell | *TECTB* | 1.353979201 | 3.13E-05 | 6.91E-05 | 0.151738694 |
| Heart_Failure | dendritic_cell | *ELOVL3* | -2.757524813 | 0.004250994 | 0.005911027 | 1 |
| Heart_Failure | dendritic_cell | *PPP1R3C* | -5.392142897 | 0.013002144 | 0.016940495 | 1 |
| Heart_Failure | dendritic_cell | *MXI1* | -2.985649158 | 5.20E-05 | 0.000107634 | 0.252401231 |
| Heart_Failure | dendritic_cell | *HELLS* | -2.695495156 | 9.41E-05 | 0.000178791 | 0.456453979 |
| Heart_Failure | dendritic_cell | *TCTN3* | -2.925873769 | 0.000466733 | 0.000760439 | 1 |
| Heart_Failure | dendritic_cell | *WDR11* | -2.690661471 | 2.49E-05 | 5.75E-05 | 0.120660792 |
| Heart_Failure | dendritic_cell | *KANSL1* | -2.255008644 | 3.47E-05 | 7.60E-05 | 0.168423399 |
| Heart_Failure | dendritic_cell | *HOXB3* | -1.97525727 | 1.17E-06 | 6.11E-06 | 0.00565952 |
| Heart_Failure | dendritic_cell | *CAAP1* | -2.450456805 | 0.000151524 | 0.000273714 | 0.735195122 |
| Heart_Failure | dendritic_cell | *PCDHB10* | -4.345067668 | 0.000374182 | 0.000621544 | 1 |
| Heart_Failure | dendritic_cell | *PCDHB12* | -1.387831913 | 0.00607859 | 0.008277664 | 1 |
| Heart_Failure | dendritic_cell | *GORAB* | -2.118407793 | 9.92E-07 | 5.62E-06 | 0.004812648 |

| | | | | | | |
|---|---|---|---|---|---|---|
| Heart_Failure | dendritic_cell | *ACAT2* | -2.077613156 | 0.018029219 | 0.02299626 | 1 |
| Heart_Failure | dendritic_cell | *MASTL* | -2.382135035 | 1.74E-05 | 4.29E-05 | 0.084620548 |
| Heart_Failure | dendritic_cell | *MRC1* | 1.747496062 | 5.09E-07 | 4.03E-06 | 0.002469112 |
| Heart_Failure | dendritic_cell | *DNAJC15* | -2.911320677 | 0.00838746 | 0.011214096 | 1 |
| Heart_Failure | dendritic_cell | *ALG5* | -2.503821703 | 4.52E-07 | 3.90E-06 | 0.002190857 |
| Heart_Failure | dendritic_cell | *SIL1* | -2.38700606 | 2.16E-05 | 5.15E-05 | 0.104801004 |
| Heart_Failure | dendritic_cell | *ARL1* | 3.744261894 | 1.03E-05 | 2.77E-05 | 0.049959417 |
| Heart_Failure | dendritic_cell | *ADRA1A* | -0.628937661 | 0.000116027 | 0.000215777 | 0.562961612 |
| Heart_Failure | dendritic_cell | *EPHX2* | -1.544275217 | 1.32E-07 | 3.44E-06 | 0.000641771 |
| Heart_Failure | dendritic_cell | *AKAP1* | -2.011529659 | 7.32E-07 | 4.70E-06 | 0.003552379 |
| Heart_Failure | dendritic_cell | *SCPEP1* | -1.069311956 | 0.018525735 | 0.023579976 | 1 |
| Heart_Failure | dendritic_cell | *TBX2* | -1.96986768 | 4.05E-05 | 8.65E-05 | 0.196338333 |
| Heart_Failure | dendritic_cell | *TEX14* | 3.160209136 | 1.35E-06 | 6.63E-06 | 0.00653083 |
| Heart_Failure | dendritic_cell | *TRIM6* | -9.354623351 | 3.70E-05 | 8.02E-05 | 0.179566159 |
| Heart_Failure | dendritic_cell | *ADCY7* | 2.162421353 | 7.12E-08 | 3.44E-06 | 0.000345289 |
| Heart_Failure | dendritic_cell | *ECHDC2* | -1.450666715 | 4.86E-07 | 3.96E-06 | 0.002356784 |
| Heart_Failure | dendritic_cell | *PYROXD1* | -1.733291646 | 0.011411685 | 0.015005284 | 1 |
| Heart_Failure | dendritic_cell | *ZNF549* | -3.949751144 | 0.002389204 | 0.003453207 | 1 |
| Heart_Failure | dendritic_cell | *A1BG* | 5.325073442 | 5.20E-07 | 4.08E-06 | 0.002524344 |
| Heart_Failure | dendritic_cell | *ZSCAN18* | -2.587653935 | 3.11E-06 | 1.16E-05 | 0.015105431 |
| Heart_Failure | dendritic_cell | *PDZRN3* | -1.015276815 | 1.85E-06 | 8.10E-06 | 0.008954036 |
| Heart_Failure | dendritic_cell | *LHX4* | -5.071529203 | 0.002910996 | 0.004135916 | 1 |
| Heart_Failure | dendritic_cell | *SEC22A* | -2.917072565 | 0.001300795 | 0.001969254 | 1 |
| Heart_Failure | dendritic_cell | *POPDC2* | 1.78096298 | 0.000177623 | 0.000316035 | 0.861826528 |
| Heart_Failure | dendritic_cell | *CD80* | 1.375805536 | 5.18E-08 | 3.44E-06 | 0.000251462 |
| Heart_Failure | dendritic_cell | *MAPK8IP1* | -3.533363896 | 4.21E-05 | 8.95E-05 | 0.204340709 |
| Heart_Failure | dendritic_cell | *CAT* | -1.383524594 | 3.44E-05 | 7.54E-05 | 0.166815921 |
| Heart_Failure | dendritic_cell | *PILRB* | -0.902904786 | 0.006170257 | 0.008393072 | 1 |
| Heart_Failure | dendritic_cell | *ZCCHC17* | -3.609077696 | 0.00033109 | 0.000557214 | 1 |
| Heart_Failure | dendritic_cell | *FABP3* | -4.631871527 | 1.07E-06 | 5.85E-06 | 0.005197558 |
| Heart_Failure | dendritic_cell | *TNFSF10* | -2.315642322 | 2.76E-06 | 1.07E-05 | 0.013400414 |
| Heart_Failure | dendritic_cell | *ZNF639* | -2.928353202 | 8.99E-07 | 5.31E-06 | 0.00436173 |
| Heart_Failure | dendritic_cell | *CPXM2* | -2.521146949 | 2.53E-05 | 5.80E-05 | 0.122698414 |
| Heart_Failure | dendritic_cell | *FLT3* | 0.131951825 | 1.39E-07 | 3.44E-06 | 0.000672927 |
| Heart_Failure | dendritic_cell | *MTIF3* | -2.330339734 | 0.022161412 | 0.027900148 | 1 |
| Heart_Failure | dendritic_cell | *FMOD* | -1.585616496 | 2.33E-06 | 9.45E-06 | 0.011297774 |
| Heart_Failure | dendritic_cell | *RBBP6* | -2.13414671 | 2.76E-06 | 1.07E-05 | 0.013394227 |
| Heart_Failure | dendritic_cell | *LDB3* | -3.571851096 | 0.000506486 | 0.000822172 | 1 |
| Heart_Failure | dendritic_cell | *OPN4* | -4.618698396 | 0.032299535 | 0.039816399 | 1 |
| Heart_Failure | dendritic_cell | *FAM35A* | -2.816340596 | 0.001915129 | 0.002819988 | 1 |
| Heart_Failure | dendritic_cell | *NAA60* | -4.027259357 | 1.93E-06 | 8.34E-06 | 0.009357188 |
| Heart_Failure | dendritic_cell | *ODF2L* | -2.32292855 | 5.68E-06 | 1.77E-05 | 0.027574618 |
| Heart_Failure | dendritic_cell | *LRRC39* | -0.864966672 | 0.005833587 | 0.007979861 | 1 |
| Heart_Failure | dendritic_cell | *RWDD3* | -1.803152269 | 0.00062201 | 0.000990155 | 1 |
| Heart_Failure | dendritic_cell | *PMS2* | -3.408753123 | 0.000324718 | 0.000547249 | 1 |
| Heart_Failure | dendritic_cell | *EEPD1* | -1.247305649 | 3.22E-06 | 1.19E-05 | 0.015611999 |
| Heart_Failure | dendritic_cell | *KLHL7* | -2.597194002 | 6.41E-06 | 1.93E-05 | 0.031088263 |
| Heart_Failure | dendritic_cell | *NT5C3A* | -2.680946386 | 2.37E-05 | 5.54E-05 | 0.1149097 |
| Heart_Failure | dendritic_cell | *SMU1* | -2.928352807 | 5.73E-05 | 0.000116556 | 0.277868397 |
| Heart_Failure | dendritic_cell | *SLC25A51* | -0.661624213 | 0.03209165 | 0.039570187 | 1 |

| | | | | | | |
|---|---|---|---|---|---|---|
| Heart_Failure | dendritic_cell | *TRIM24* | -2.345947726 | 5.34E-06 | 1.70E-05 | 0.025904441 |
| Heart_Failure | dendritic_cell | *C7orf49* | -2.023734821 | 6.96E-06 | 2.04E-05 | 0.033785847 |
| Heart_Failure | dendritic_cell | *CISD1* | -2.177825524 | 3.26E-05 | 7.19E-05 | 0.158415847 |
| Heart_Failure | dendritic_cell | *RBM19* | -3.35834625 | 2.19E-05 | 5.19E-05 | 0.106051531 |
| Heart_Failure | dendritic_cell | *DDX54* | -3.325394382 | 0.02972351 | 0.036771665 | 1 |
| Heart_Failure | dendritic_cell | *SSPN* | -1.477709516 | 4.31E-06 | 1.45E-05 | 0.020902654 |
| Heart_Failure | dendritic_cell | *EBPL* | -1.974027094 | 0.000224441 | 0.000390598 | 1 |
| Heart_Failure | dendritic_cell | *ATP7B* | -0.599857107 | 0.001615539 | 0.002411134 | 1 |
| Heart_Failure | dendritic_cell | *CENPK* | -2.440682418 | 1.53E-06 | 7.21E-06 | 0.007418265 |
| Heart_Failure | dendritic_cell | *ITIH5* | -1.629297752 | 2.56E-09 | 1.04E-06 | 1.24E-05 |
| Heart_Failure | dendritic_cell | *TSFM* | -1.484601925 | 4.40E-06 | 1.48E-05 | 0.021347645 |
| Heart_Failure | dendritic_cell | *ARHGAP9* | 2.269000191 | 3.74E-07 | 3.66E-06 | 0.001813758 |
| Heart_Failure | dendritic_cell | *PFDN5* | -3.209045242 | 0.000664962 | 0.001054033 | 1 |
| Heart_Failure | dendritic_cell | *PDE1B* | -2.683976535 | 8.84E-07 | 5.24E-06 | 0.004289496 |
| Heart_Failure | dendritic_cell | *CDK2* | 3.501958913 | 1.54E-08 | 2.99E-06 | 7.48E-05 |
| Heart_Failure | dendritic_cell | *SARDH* | 2.205675017 | 4.96E-08 | 3.44E-06 | 0.000240759 |
| Heart_Failure | dendritic_cell | *DBH* | 11.53830277 | 1.35E-07 | 3.44E-06 | 0.00065456 |
| Heart_Failure | dendritic_cell | *NDUFAF4* | -2.036415549 | 0.002982531 | 0.004226414 | 1 |
| Heart_Failure | dendritic_cell | *USP45* | -2.478423004 | 1.11E-05 | 2.94E-05 | 0.053970604 |
| Heart_Failure | dendritic_cell | *TTC21B* | -2.991250009 | 6.54E-06 | 1.95E-05 | 0.031755669 |
| Heart_Failure | dendritic_cell | *ACVR1C* | -0.936335144 | 1.23E-07 | 3.44E-06 | 0.000596521 |
| Heart_Failure | dendritic_cell | *SLC36A1* | -1.037529418 | 0.003328403 | 0.004689144 | 1 |
| Heart_Failure | dendritic_cell | *PLA2G12A* | -2.248904852 | 3.56E-06 | 1.28E-05 | 0.017267748 |
| Heart_Failure | dendritic_cell | *C4BPA* | -2.78265359 | 6.94E-05 | 0.000137578 | 0.336654165 |
| Heart_Failure | dendritic_cell | *C4BPB* | -1.963387225 | 4.73E-05 | 9.87E-05 | 0.22959904 |
| Heart_Failure | dendritic_cell | *RAB38* | -1.650358173 | 4.41E-06 | 1.48E-05 | 0.021375801 |
| Heart_Failure | dendritic_cell | *DAW1* | -3.861994946 | 1.05E-06 | 5.81E-06 | 0.005118422 |
| Heart_Failure | dendritic_cell | *DNPEP* | -3.522355156 | 0.000485965 | 0.000790447 | 1 |
| Heart_Failure | dendritic_cell | *OBSL1* | -1.900245212 | 4.71E-07 | 3.95E-06 | 0.002285725 |
| Heart_Failure | dendritic_cell | *SLC12A4* | -3.527716242 | 0.031794054 | 0.039253117 | 1 |
| Heart_Failure | dendritic_cell | *FAM210B* | -3.272785359 | 5.70E-07 | 4.26E-06 | 0.002764112 |
| Heart_Failure | dendritic_cell | *WFDC3* | -0.682658417 | 0.003399561 | 0.00478245 | 1 |
| Heart_Failure | dendritic_cell | *MATN4* | 5.802861511 | 5.49E-07 | 4.16E-06 | 0.002664158 |
| Heart_Failure | dendritic_cell | *VAPB* | -1.656987303 | 0.00021974 | 0.000383241 | 1 |
| Heart_Failure | dendritic_cell | *TOX2* | -0.949657789 | 0.003815043 | 0.005336002 | 1 |
| Heart_Failure | dendritic_cell | *SRSF6* | -2.899072312 | 7.87E-05 | 0.000153449 | 0.381779896 |
| Heart_Failure | dendritic_cell | *CSE1L* | -2.803198943 | 0.000279592 | 0.000478343 | 1 |
| Heart_Failure | dendritic_cell | *PTGIS* | -1.417368945 | 3.58E-05 | 7.80E-05 | 0.173909372 |
| Heart_Failure | dendritic_cell | *RNF114* | -2.836282079 | 5.25E-05 | 0.000108496 | 0.254857287 |
| Heart_Failure | dendritic_cell | *PEPD* | -2.541729622 | 6.40E-06 | 1.93E-05 | 0.031069065 |
| Heart_Failure | dendritic_cell | *CHST8* | 3.68716479 | 6.13E-07 | 4.34E-06 | 0.002975033 |
| Heart_Failure | dendritic_cell | *STAMBP* | -3.242203307 | 3.04E-05 | 6.75E-05 | 0.147611442 |
| Heart_Failure | dendritic_cell | *ATP8A1* | -2.814234728 | 7.14E-05 | 0.000141028 | 0.346364287 |
| Heart_Failure | dendritic_cell | *USP22* | -2.870542008 | 0.003743744 | 0.005242322 | 1 |
| Heart_Failure | dendritic_cell | *HIF3A* | -2.641260303 | 6.91E-05 | 0.000137144 | 0.335179685 |
| Heart_Failure | dendritic_cell | *ZNF576* | 3.025047399 | 1.05E-05 | 2.80E-05 | 0.050766564 |
| Heart_Failure | dendritic_cell | *BTN2A2* | 0.97488962 | 1.74E-07 | 3.54E-06 | 0.000844908 |
| Heart_Failure | dendritic_cell | *MRS2* | -1.447563448 | 0.00245049 | 0.003532317 | 1 |
| Heart_Failure | dendritic_cell | *SNRPC* | -1.865026155 | 0.009899756 | 0.013106034 | 1 |
| Heart_Failure | dendritic_cell | *SERPINB6* | -2.453473694 | 2.12E-06 | 8.83E-06 | 0.01027005 |

| | | | | | | |
|---|---|---|---|---|---|---|
| Heart_Failure | dendritic_cell | *PEX6* | -0.858771931 | 0.025956134 | 0.032325247 | 1 |
| Heart_Failure | dendritic_cell | *NQO2* | -2.580269231 | 3.42E-06 | 1.24E-05 | 0.016598271 |
| Heart_Failure | dendritic_cell | *MED20* | -2.000471348 | 0.035598356 | 0.043727398 | 1 |
| Heart_Failure | dendritic_cell | *TREM1* | 1.892136633 | 4.53E-07 | 3.90E-06 | 0.002196537 |
| Heart_Failure | dendritic_cell | *MEA1* | 1.829806773 | 0.000158138 | 0.000284391 | 0.767285602 |
| Heart_Failure | dendritic_cell | *KLHL31* | -1.087533262 | 5.96E-06 | 1.84E-05 | 0.028923671 |
| Heart_Failure | dendritic_cell | *GLO1* | -2.143593418 | 0.034879181 | 0.042887427 | 1 |
| Heart_Failure | dendritic_cell | *KCNK17* | -3.161356197 | 0.000978817 | 0.001512972 | 1 |
| Heart_Failure | dendritic_cell | *RREB1* | -2.320218183 | 8.42E-05 | 0.000162606 | 0.408466381 |
| Heart_Failure | dendritic_cell | *ATXN1* | -2.136331857 | 0.000119246 | 0.00022117 | 0.578579943 |
| Heart_Failure | dendritic_cell | *RUNX2* | 1.164163359 | 1.08E-07 | 3.44E-06 | 0.000524199 |
| Heart_Failure | dendritic_cell | *CXCL6* | 5.545384436 | 7.52E-08 | 3.44E-06 | 0.000364687 |
| Heart_Failure | dendritic_cell | *BBS2* | -1.580461769 | 0.000114303 | 0.000213061 | 0.554597031 |
| Heart_Failure | dendritic_cell | *MT1G* | -4.889388635 | 0.000842713 | 0.00131221 | 1 |
| Heart_Failure | dendritic_cell | *TMTC4* | -3.472850042 | 2.43E-05 | 5.66E-05 | 0.11797274 |
| Heart_Failure | dendritic_cell | *ABCC4* | 2.132103422 | 2.27E-07 | 3.66E-06 | 0.001099842 |
| Heart_Failure | dendritic_cell | *TEKT3* | 1.121244722 | 0.014905408 | 0.019259144 | 1 |
| Heart_Failure | dendritic_cell | *HS3ST3B1* | 5.57054818 | 3.15E-07 | 3.66E-06 | 0.001525964 |
| Heart_Failure | dendritic_cell | *MRPS7* | -2.980269191 | 4.00E-06 | 1.38E-05 | 0.019428035 |
| Heart_Failure | dendritic_cell | *NT5C* | -3.887673162 | 4.27E-05 | 9.06E-05 | 0.207413196 |
| Heart_Failure | dendritic_cell | *DDX31* | -3.088021732 | 1.86E-06 | 8.14E-06 | 0.009030637 |
| Heart_Failure | dendritic_cell | *OPRL1* | 0.800710847 | 1.34E-07 | 3.44E-06 | 0.000652581 |
| Heart_Failure | dendritic_cell | *PPDPF* | -2.100870137 | 0.008412457 | 0.011244419 | 1 |
| Heart_Failure | dendritic_cell | *IL1B* | -7.210317584 | 2.18E-05 | 5.18E-05 | 0.105723204 |
| Heart_Failure | dendritic_cell | *POLR1B* | -1.541864932 | 0.001240973 | 0.001886341 | 1 |
| Heart_Failure | dendritic_cell | *CCDC93* | -2.62709599 | 4.42E-06 | 1.48E-05 | 0.021468542 |
| Heart_Failure | dendritic_cell | *TNFSF9* | -27.28531136 | 2.13E-05 | 5.08E-05 | 0.103341855 |
| Heart_Failure | dendritic_cell | *SH2D3A* | -3.061100892 | 7.36E-05 | 0.000144768 | 0.357176782 |
| Heart_Failure | dendritic_cell | *GPR108* | -2.958241532 | 0.003161334 | 0.004464143 | 1 |
| Heart_Failure | dendritic_cell | *SNRPD2* | -3.787098434 | 9.84E-06 | 2.67E-05 | 0.047766858 |
| Heart_Failure | dendritic_cell | *SDCBP2* | 5.893128114 | 0.000123228 | 0.000227339 | 0.597902293 |
| Heart_Failure | dendritic_cell | *PSMF1* | -2.222589694 | 0.006298991 | 0.008553794 | 1 |
| Heart_Failure | dendritic_cell | *DTD1* | -2.797349127 | 4.09E-05 | 8.74E-05 | 0.198464666 |
| Heart_Failure | dendritic_cell | *RBCK1* | -3.00375971 | 7.25E-05 | 0.00014299 | 0.351754412 |
| Heart_Failure | dendritic_cell | *RRBP1* | -2.453372579 | 7.44E-06 | 2.13E-05 | 0.036114371 |
| Heart_Failure | dendritic_cell | *FLRT3* | -1.858345306 | 4.78E-07 | 3.96E-06 | 0.002317558 |
| Heart_Failure | dendritic_cell | *PCSK2* | -6.680136842 | 3.67E-06 | 1.30E-05 | 0.017798135 |
| Heart_Failure | dendritic_cell | *MKKS* | -2.497362398 | 8.56E-06 | 2.39E-05 | 0.041514126 |
| Heart_Failure | dendritic_cell | *LAMP5* | -34.90676442 | 2.89E-06 | 1.11E-05 | 0.014042107 |
| Heart_Failure | dendritic_cell | *ITPA* | -3.49569442 | 0.000940932 | 0.001458596 | 1 |
| Heart_Failure | dendritic_cell | *MCM8* | -1.662245044 | 1.89E-07 | 3.60E-06 | 0.000919444 |
| Heart_Failure | dendritic_cell | *MRPS26* | -1.582977053 | 0.035905984 | 0.044059951 | 1 |
| Heart_Failure | dendritic_cell | *NCLN* | -2.439730666 | 0.027788205 | 0.034500607 | 1 |
| Heart_Failure | dendritic_cell | *KLC1* | -3.130462461 | 0.000137836 | 0.000251138 | 0.668781659 |
| Heart_Failure | dendritic_cell | *XRCC3* | -2.223170145 | 0.007568178 | 0.010177605 | 1 |
| Heart_Failure | dendritic_cell | *MCF2L* | -1.377952976 | 3.35E-07 | 3.66E-06 | 0.001623603 |
| Heart_Failure | dendritic_cell | *IGFLR1* | 1.549588193 | 2.54E-07 | 3.66E-06 | 0.001232604 |
| Heart_Failure | dendritic_cell | *COX6B1* | -2.630608176 | 4.97E-06 | 1.62E-05 | 0.024119276 |
| Heart_Failure | dendritic_cell | *PRDX5* | -2.977617868 | 3.49E-05 | 7.64E-05 | 0.169509303 |
| Heart_Failure | dendritic_cell | *IRF3* | -3.049459908 | 6.31E-06 | 1.92E-05 | 0.030601362 |

| | | | | | | |
|---|---|---|---|---|---|---|
| Heart_Failure | dendritic_cell | *TSKS* | -2.020000668 | 4.45E-06 | 1.49E-05 | 0.021576596 |
| Heart_Failure | dendritic_cell | *TRAP1* | -1.217685805 | 0.00057531 | 0.000921256 | 1 |
| Heart_Failure | dendritic_cell | *ZNF384* | -4.729363441 | 2.83E-05 | 6.36E-05 | 0.137076959 |
| Heart_Failure | dendritic_cell | *EMG1* | -3.829224297 | 1.33E-05 | 3.40E-05 | 0.064388109 |
| Heart_Failure | dendritic_cell | *ATG14* | -2.34248046 | 0.000393956 | 0.00064994 | 1 |
| Heart_Failure | dendritic_cell | *KTN1* | -2.426295154 | 4.65E-05 | 9.72E-05 | 0.225514476 |
| Heart_Failure | dendritic_cell | *L3HYPDH* | -1.004641209 | 0.024277042 | 0.030443742 | 1 |
| Heart_Failure | dendritic_cell | *ZBTB1* | -2.488438888 | 0.00382033 | 0.005341857 | 1 |
| Heart_Failure | dendritic_cell | *RHOT1* | -1.800841737 | 0.005193759 | 0.007144916 | 1 |
| Heart_Failure | dendritic_cell | *WDR60* | -2.261154949 | 5.70E-06 | 1.78E-05 | 0.027645048 |
| Heart_Failure | dendritic_cell | *AIF1L* | 6.437359745 | 4.66E-06 | 1.54E-05 | 0.022615919 |
| Heart_Failure | dendritic_cell | *CANX* | -2.330600835 | 7.72E-06 | 2.19E-05 | 0.037467505 |
| Heart_Failure | dendritic_cell | *ZNF484* | -3.178876114 | 5.13E-06 | 1.64E-05 | 0.024908917 |
| Heart_Failure | dendritic_cell | *FGD3* | 1.829962936 | 1.02E-06 | 5.71E-06 | 0.00494553 |
| Heart_Failure | dendritic_cell | *ABHD8* | -2.677352491 | 0.000764111 | 0.001198277 | 1 |
| Heart_Failure | dendritic_cell | *ATP13A4* | -3.588424987 | 1.84E-07 | 3.58E-06 | 0.000893626 |
| Heart_Failure | dendritic_cell | *HELB* | -3.74308724 | 0.000521016 | 0.000843422 | 1 |
| Heart_Failure | dendritic_cell | *RAB3IP* | -1.842010158 | 3.65E-07 | 3.66E-06 | 0.001772656 |
| Heart_Failure | dendritic_cell | *YEATS4* | -1.667870179 | 0.003554534 | 0.004993225 | 1 |
| Heart_Failure | dendritic_cell | *TAS2R4* | 3.893008107 | 6.51E-07 | 4.45E-06 | 0.003156401 |
| Heart_Failure | dendritic_cell | *LRRC61* | -2.223792785 | 1.14E-05 | 3.01E-05 | 0.055478951 |
| Heart_Failure | dendritic_cell | *IDUA* | 4.36151652 | 0.002734485 | 0.003904568 | 1 |
| Heart_Failure | dendritic_cell | *EMC1* | -1.906339869 | 2.38E-06 | 9.57E-06 | 0.011527595 |
| Heart_Failure | dendritic_cell | *PLA2G5* | -0.928484276 | 0.001688274 | 0.002509653 | 1 |
| Heart_Failure | dendritic_cell | *UBR4* | -2.076390644 | 4.39E-05 | 9.25E-05 | 0.212794422 |
| Heart_Failure | dendritic_cell | *WFIKKN1* | 2.339454016 | 0.024545379 | 0.030718127 | 1 |
| Heart_Failure | dendritic_cell | *WDR24* | -2.710521639 | 5.53E-06 | 1.74E-05 | 0.026807504 |
| Heart_Failure | dendritic_cell | *SMARCA4* | -2.783297556 | 0.013748225 | 0.017874166 | 1 |
| Heart_Failure | dendritic_cell | *PNKD* | -3.607893302 | 0.000193181 | 0.000341587 | 0.937315493 |
| Heart_Failure | dendritic_cell | *RNF6* | -2.428925425 | 1.45E-06 | 6.95E-06 | 0.007017297 |
| Heart_Failure | dendritic_cell | *HIP1* | -2.508192839 | 0.000336863 | 0.000565722 | 1 |
| Heart_Failure | dendritic_cell | *PTPN12* | -2.452984364 | 0.000124859 | 0.000230086 | 0.605816266 |
| Heart_Failure | dendritic_cell | *STYXL1* | -1.324150908 | 0.005926119 | 0.008088194 | 1 |
| Heart_Failure | dendritic_cell | *PEX1* | -3.534602018 | 0.000144692 | 0.000262349 | 0.70204549 |
| Heart_Failure | dendritic_cell | *SRD5A3* | -1.780804332 | 4.49E-07 | 3.90E-06 | 0.002179531 |
| Heart_Failure | dendritic_cell | *KDR* | -1.292038751 | 3.25E-07 | 3.66E-06 | 0.001577053 |
| Heart_Failure | dendritic_cell | *VPREB3* | -3.350383939 | 0.024968762 | 0.031167593 | 1 |
| Heart_Failure | dendritic_cell | *MGAT3* | -2.439629725 | 1.02E-06 | 5.71E-06 | 0.004949064 |
| Heart_Failure | dendritic_cell | *ATF4* | -2.489758693 | 7.55E-07 | 4.79E-06 | 0.003662726 |
| Heart_Failure | dendritic_cell | *A4GALT* | -1.748643951 | 0.00309483 | 0.004375325 | 1 |
| Heart_Failure | dendritic_cell | *CDC42EP1* | -2.111597243 | 2.32E-07 | 3.66E-06 | 0.001126764 |
| Heart_Failure | dendritic_cell | *SPECC1* | 3.0027418 | 0.005541834 | 0.007600051 | 1 |
| Heart_Failure | dendritic_cell | *CDHR3* | -2.698023504 | 7.91E-06 | 2.24E-05 | 0.038387679 |
| Heart_Failure | dendritic_cell | *PRKRIP1* | -2.73682821 | 0.001042083 | 0.001602595 | 1 |
| Heart_Failure | dendritic_cell | *IRF5* | 0.945796742 | 0.001886103 | 0.002779039 | 1 |
| Heart_Failure | dendritic_cell | *HOXD3* | -0.798336188 | 0.002995857 | 0.004244057 | 1 |
| Heart_Failure | dendritic_cell | *MTX2* | -2.596221226 | 1.42E-05 | 3.60E-05 | 0.068832214 |
| Heart_Failure | dendritic_cell | *PDE11A* | -1.063056944 | 0.02430278 | 0.030454899 | 1 |
| Heart_Failure | dendritic_cell | *GAD1* | 3.457005623 | 8.81E-07 | 5.23E-06 | 0.004274124 |
| Heart_Failure | dendritic_cell | *ORMDL1* | -2.320159885 | 0.000914791 | 0.001420796 | 1 |

| | | | | | | |
|---|---|---|---|---|---|---|
| Heart_Failure | dendritic_cell | *HOXD10* | -4.083589249 | 1.88E-05 | 4.57E-05 | 0.091416537 |
| Heart_Failure | dendritic_cell | *PSMG2* | -1.999498159 | 0.024672856 | 0.030837892 | 1 |
| Heart_Failure | dendritic_cell | *TWSG1* | -2.900987221 | 1.03E-05 | 2.77E-05 | 0.049948006 |
| Heart_Failure | dendritic_cell | *WDFY4* | 0.472962771 | 5.48E-08 | 3.44E-06 | 0.000265756 |
| Heart_Failure | dendritic_cell | *EIF2AK4* | -2.471036135 | 0.012587908 | 0.016418422 | 1 |
| Heart_Failure | dendritic_cell | *TMOD2* | -2.49041749 | 3.22E-06 | 1.19E-05 | 0.015629157 |
| Heart_Failure | dendritic_cell | *IVD* | -1.712876831 | 0.006877802 | 0.009308534 | 1 |
| Heart_Failure | dendritic_cell | *KNSTRN* | -0.850969018 | 0.001111105 | 0.00170012 | 1 |
| Heart_Failure | dendritic_cell | *ARPP19* | -3.6116758 | 0.000361998 | 0.000602544 | 1 |
| Heart_Failure | dendritic_cell | *VPS13C* | -2.097802386 | 0.015029293 | 0.01940967 | 1 |
| Heart_Failure | dendritic_cell | *CALML4* | 3.852125555 | 1.75E-06 | 7.84E-06 | 0.008477708 |
| Heart_Failure | dendritic_cell | *ANAPC13* | 1.645139938 | 0.000136408 | 0.000248909 | 0.661849402 |
| Heart_Failure | dendritic_cell | *SUMF2* | -3.494732507 | 0.000197713 | 0.000348838 | 0.959304408 |
| Heart_Failure | dendritic_cell | *BBOX1* | 5.180134943 | 1.22E-07 | 3.44E-06 | 0.0005913 |
| Heart_Failure | dendritic_cell | *SERGEF* | -2.312373426 | 0.001832528 | 0.002705851 | 1 |
| Heart_Failure | dendritic_cell | *DCTD* | -3.054396786 | 0.000353841 | 0.000589573 | 1 |
| Heart_Failure | dendritic_cell | *USP6* | -1.333959311 | 0.001964874 | 0.002886337 | 1 |
| Heart_Failure | dendritic_cell | *PLD2* | -5.452197943 | 0.00115233 | 0.001759869 | 1 |
| Heart_Failure | dendritic_cell | *PHF20L1* | -2.614656517 | 0.000122006 | 0.000225427 | 0.59197196 |
| Heart_Failure | dendritic_cell | *PUS7L* | -2.976031373 | 6.40E-06 | 1.93E-05 | 0.031033818 |
| Heart_Failure | dendritic_cell | *KRI1* | -1.991430525 | 0.000430085 | 0.000705229 | 1 |
| Heart_Failure | dendritic_cell | *AP1M2* | -1.670704241 | 0.001804712 | 0.002668027 | 1 |
| Heart_Failure | dendritic_cell | *MTUS1* | -1.759119519 | 4.12E-07 | 3.86E-06 | 0.002001384 |
| Heart_Failure | dendritic_cell | *KLK10* | -1.496347961 | 1.23E-05 | 3.20E-05 | 0.059577913 |
| Heart_Failure | dendritic_cell | *KLK8* | -13.22780297 | 3.61E-07 | 3.66E-06 | 0.0017519 |
| Heart_Failure | dendritic_cell | *ADCY4* | -2.310198387 | 2.84E-06 | 1.09E-05 | 0.013795954 |
| Heart_Failure | dendritic_cell | *BCL2L2* | -2.237507774 | 0.004083264 | 0.005691468 | 1 |
| Heart_Failure | dendritic_cell | *DTD2* | -1.584607579 | 2.51E-05 | 5.77E-05 | 0.12160042 |
| Heart_Failure | dendritic_cell | *PARP2* | -2.611956426 | 1.52E-06 | 7.19E-06 | 0.00738094 |
| Heart_Failure | dendritic_cell | *RNASE1* | -2.219781078 | 7.64E-05 | 0.000149593 | 0.370841593 |
| Heart_Failure | dendritic_cell | *EPB41L4A* | -1.87803955 | 2.11E-06 | 8.82E-06 | 0.010240657 |
| Heart_Failure | dendritic_cell | *CDO1* | -1.168789967 | 4.47E-05 | 9.39E-05 | 0.216917842 |
| Heart_Failure | dendritic_cell | *REEP5* | -2.946553618 | 0.001361821 | 0.002052674 | 1 |
| Heart_Failure | dendritic_cell | *TMEM8A* | -3.984222492 | 1.28E-05 | 3.30E-05 | 0.06208888 |
| Heart_Failure | dendritic_cell | *MAU2* | -2.879616343 | 9.13E-06 | 2.50E-05 | 0.044286207 |
| Heart_Failure | dendritic_cell | *SHC2* | -1.825301067 | 1.23E-06 | 6.30E-06 | 0.005974164 |
| Heart_Failure | dendritic_cell | *SYT5* | 4.368984261 | 2.65E-06 | 1.04E-05 | 0.012867816 |
| Heart_Failure | dendritic_cell | *ERMARD* | -3.036662175 | 6.29E-06 | 1.91E-05 | 0.030526311 |
| Heart_Failure | dendritic_cell | *PHF10* | -3.518550561 | 1.80E-05 | 4.39E-05 | 0.087100304 |
| Heart_Failure | dendritic_cell | *GALNT8* | -2.453836543 | 2.29E-07 | 3.66E-06 | 0.001112681 |
| Heart_Failure | dendritic_cell | *PRKCSH* | -2.710631261 | 3.96E-06 | 1.37E-05 | 0.019214541 |
| Heart_Failure | dendritic_cell | *CDC16* | -2.362734169 | 8.03E-05 | 0.000156196 | 0.389551626 |
| Heart_Failure | dendritic_cell | *XPO7* | -3.150832039 | 0.000199753 | 0.000351925 | 0.969201235 |
| Heart_Failure | dendritic_cell | *FAM98C* | 3.456997559 | 3.99E-06 | 1.38E-05 | 0.019358416 |
| Heart_Failure | dendritic_cell | *DDA1* | 3.607661433 | 0.002793497 | 0.003980631 | 1 |
| Heart_Failure | dendritic_cell | *MRPL34* | -1.096054088 | 0.003927084 | 0.005484804 | 1 |
| Heart_Failure | dendritic_cell | *LSM7* | -1.960183799 | 0.001366733 | 0.002059437 | 1 |
| Heart_Failure | dendritic_cell | *TULP4* | -1.683425892 | 1.31E-05 | 3.35E-05 | 0.063373087 |
| Heart_Failure | dendritic_cell | *RSPH3* | -0.815816646 | 0.028151042 | 0.034924279 | 1 |
| Heart_Failure | dendritic_cell | *ACTN4* | -2.573558299 | 0.000126185 | 0.000232265 | 0.612250707 |

| | | | | | | |
|---|---|---|---|---|---|---|
| Heart_Failure | dendritic_cell | *STK33* | -1.174660927 | 1.83E-06 | 8.09E-06 | 0.008902993 |
| Heart_Failure | dendritic_cell | *NDUFA10* | -1.415473434 | 0.000724605 | 0.001140747 | 1 |
| Heart_Failure | dendritic_cell | *MAP1S* | 4.132255813 | 2.51E-07 | 3.66E-06 | 0.001219015 |
| Heart_Failure | dendritic_cell | *SSBP4* | -3.233811695 | 2.61E-06 | 1.02E-05 | 0.012674423 |
| Heart_Failure | dendritic_cell | *PGPEP1* | 1.859587068 | 0.002712455 | 0.003874251 | 1 |
| Heart_Failure | dendritic_cell | *KIAA1683* | -2.821470192 | 0.000169841 | 0.000303296 | 0.824069612 |
| Heart_Failure | dendritic_cell | *LSM4* | 1.858098371 | 0.00256365 | 0.003676864 | 1 |
| Heart_Failure | dendritic_cell | *TRPM4* | -1.801972161 | 1.03E-05 | 2.76E-05 | 0.049868835 |
| Heart_Failure | dendritic_cell | *ATXN10* | -2.588724278 | 0.001279437 | 0.00193934 | 1 |
| Heart_Failure | dendritic_cell | *PNPLA7* | -2.934437028 | 9.92E-06 | 2.69E-05 | 0.048116229 |
| Heart_Failure | dendritic_cell | *PAK4* | -1.932932774 | 6.76E-07 | 4.52E-06 | 0.003278949 |
| Heart_Failure | dendritic_cell | *ZNF337* | -1.874326154 | 0.025415454 | 0.031692568 | 1 |
| Heart_Failure | dendritic_cell | *CEP85* | 2.169260994 | 3.63E-06 | 1.29E-05 | 0.017633056 |
| Heart_Failure | dendritic_cell | *POMT1* | 3.826770305 | 7.76E-05 | 0.000151537 | 0.376417203 |
| Heart_Failure | dendritic_cell | *CHMP2A* | 2.871312238 | 0.003247166 | 0.004578684 | 1 |
| Heart_Failure | dendritic_cell | *YIPF2* | -2.8122996 | 0.004246008 | 0.00590643 | 1 |
| Heart_Failure | dendritic_cell | *NPAS1* | -2.570864746 | 2.57E-07 | 3.66E-06 | 0.001248743 |
| Heart_Failure | dendritic_cell | *LRRC47* | -2.43295039 | 1.25E-06 | 6.36E-06 | 0.006046388 |
| Heart_Failure | dendritic_cell | *THEMIS2* | 1.414302292 | 2.59E-06 | 1.02E-05 | 0.012573389 |
| Heart_Failure | dendritic_cell | *ZNF317* | -3.115649328 | 6.00E-05 | 0.000121383 | 0.291198038 |
| Heart_Failure | dendritic_cell | *ANGPTL6* | -3.792257784 | 1.48E-05 | 3.73E-05 | 0.072029315 |
| Heart_Failure | dendritic_cell | *DNMT1* | -3.003268154 | 0.037943247 | 0.046431433 | 1 |
| Heart_Failure | dendritic_cell | *C12orf65* | -1.664952194 | 7.43E-05 | 0.00014572 | 0.360365319 |
| Heart_Failure | dendritic_cell | *NOL11* | -3.180144221 | 0.014169631 | 0.018377719 | 1 |
| Heart_Failure | dendritic_cell | *SLC35D2* | -3.281795137 | 0.000404045 | 0.000665235 | 1 |
| Heart_Failure | dendritic_cell | *ULBP2* | -10.72007459 | 0.005449245 | 0.007479417 | 1 |
| Heart_Failure | dendritic_cell | *TTLL9* | 1.744445226 | 1.18E-07 | 3.44E-06 | 0.000571374 |
| Heart_Failure | dendritic_cell | *COX4I2* | -3.331843648 | 8.56E-06 | 2.39E-05 | 0.041528961 |
| Heart_Failure | dendritic_cell | *ATP6V1E1* | -2.719137569 | 1.99E-05 | 4.80E-05 | 0.096790622 |
| Heart_Failure | dendritic_cell | *CHMP1A* | -1.960818432 | 1.30E-06 | 6.51E-06 | 0.006289366 |
| Heart_Failure | dendritic_cell | *IDO1* | 1.469187739 | 1.26E-07 | 3.44E-06 | 0.000612496 |
| Heart_Failure | dendritic_cell | *MRPS25* | -2.348849112 | 2.19E-05 | 5.19E-05 | 0.106071657 |
| Heart_Failure | dendritic_cell | *GALNT15* | -3.823605106 | 0.000117448 | 0.000218085 | 0.569856363 |
| Heart_Failure | dendritic_cell | *SLC6A6* | 1.561793961 | 2.17E-07 | 3.66E-06 | 0.00105517 |
| Heart_Failure | dendritic_cell | *KCNC3* | 3.69639994 | 3.88E-08 | 3.44E-06 | 0.000188088 |
| Heart_Failure | dendritic_cell | *NAPSA* | -2.818756472 | 0.001621288 | 0.00241674 | 1 |
| Heart_Failure | dendritic_cell | *PDLIM4* | -3.030890009 | 2.16E-06 | 8.95E-06 | 0.010463204 |
| Heart_Failure | dendritic_cell | *KIF3A* | -1.614745589 | 1.10E-06 | 5.95E-06 | 0.005317926 |
| Heart_Failure | dendritic_cell | *ACLY* | -2.950118032 | 0.001189532 | 0.001812691 | 1 |
| Heart_Failure | dendritic_cell | *NDUFA2* | -1.748594128 | 4.24E-05 | 9.01E-05 | 0.205869617 |
| Heart_Failure | dendritic_cell | *EXOC4* | -1.85268371 | 8.06E-05 | 0.00015661 | 0.390899342 |
| Heart_Failure | dendritic_cell | *PPFIA1* | -2.285589455 | 2.43E-05 | 5.66E-05 | 0.117889294 |
| Heart_Failure | dendritic_cell | *BARX1* | -6.169093741 | 1.36E-05 | 3.47E-05 | 0.065901931 |
| Heart_Failure | dendritic_cell | *MAP1B* | -1.397111155 | 2.50E-05 | 5.77E-05 | 0.121510403 |
| Heart_Failure | dendritic_cell | *CKMT2* | -2.436929093 | 2.34E-06 | 9.48E-06 | 0.011368167 |
| Heart_Failure | dendritic_cell | *KHDRBS3* | -3.444140355 | 1.75E-05 | 4.29E-05 | 0.084670613 |
| Heart_Failure | dendritic_cell | *PRKAB2* | -2.746660517 | 1.82E-06 | 8.06E-06 | 0.008853339 |
| Heart_Failure | dendritic_cell | *ZNF304* | -1.74075724 | 9.79E-05 | 0.000185373 | 0.475110029 |
| Heart_Failure | dendritic_cell | *ZNF132* | -1.130748435 | 0.026496741 | 0.032964663 | 1 |
| Heart_Failure | dendritic_cell | *RHPN2* | -0.993752382 | 0.000204206 | 0.000359248 | 0.990807136 |

| | | | | | | |
|---|---|---|---|---|---|---|
| Heart_Failure | dendritic_cell | *MATN3* | 1.83986011 | 0.001824165 | 0.002694323 | 1 |
| Heart_Failure | dendritic_cell | *TRIM21* | 3.50199325 | 6.29E-08 | 3.44E-06 | 0.000305399 |
| Heart_Failure | dendritic_cell | *SPATA6* | -1.612667643 | 1.16E-06 | 6.11E-06 | 0.005610638 |
| Heart_Failure | dendritic_cell | *LRRC41* | -3.64476309 | 1.55E-06 | 7.25E-06 | 0.00750756 |
| Heart_Failure | dendritic_cell | *CCT6B* | 2.088488477 | 4.98E-06 | 1.62E-05 | 0.024186393 |
| Heart_Failure | dendritic_cell | *CNGA4* | -2.87597498 | 1.99E-06 | 8.52E-06 | 0.009659679 |
| Heart_Failure | dendritic_cell | *RRP8* | -3.514528371 | 0.00093682 | 0.001453149 | 1 |
| Heart_Failure | dendritic_cell | *TIMM10B* | -2.018756072 | 8.78E-05 | 0.000168339 | 0.426235166 |
| Heart_Failure | dendritic_cell | *IMMT* | -2.576139513 | 0.001158195 | 0.001767714 | 1 |
| Heart_Failure | dendritic_cell | *CARD6* | -1.517165823 | 9.24E-06 | 2.53E-05 | 0.044855649 |
| Heart_Failure | dendritic_cell | *MYBBP1A* | -2.432938397 | 5.11E-06 | 1.64E-05 | 0.024800704 |
| Heart_Failure | dendritic_cell | *SERPINF1* | -2.806193081 | 8.69E-06 | 2.41E-05 | 0.042161878 |
| Heart_Failure | dendritic_cell | *TMEM128* | -1.822520978 | 2.25E-05 | 5.31E-05 | 0.108936817 |
| Heart_Failure | dendritic_cell | *FIGNL1* | -2.301652106 | 2.62E-08 | 3.33E-06 | 0.000126978 |
| Heart_Failure | dendritic_cell | *DDC* | 3.938146457 | 2.10E-06 | 8.81E-06 | 0.010179617 |
| Heart_Failure | dendritic_cell | *RGS22* | -1.194916715 | 1.66E-06 | 7.60E-06 | 0.00803536 |
| Heart_Failure | dendritic_cell | *REEP2* | -4.316638138 | 0.000479366 | 0.000780498 | 1 |
| Heart_Failure | dendritic_cell | *PRMT7* | -1.940385008 | 0.000180385 | 0.000320362 | 0.875229602 |
| Heart_Failure | dendritic_cell | *MUTYH* | 3.563894122 | 7.05E-08 | 3.44E-06 | 0.000341931 |
| Heart_Failure | dendritic_cell | *CTNNBL1* | -3.267812077 | 0.000700869 | 0.001105173 | 1 |
| Heart_Failure | dendritic_cell | *OSER1* | -2.899996039 | 0.035960071 | 0.044104718 | 1 |
| Heart_Failure | dendritic_cell | *SERINC3* | -2.178379055 | 0.002629126 | 0.003760767 | 1 |
| Heart_Failure | dendritic_cell | *DMGDH* | -2.123449196 | 4.19E-08 | 3.44E-06 | 0.000203203 |
| Heart_Failure | dendritic_cell | *BHMT2* | -0.914894442 | 7.19E-07 | 4.65E-06 | 0.003486169 |
| Heart_Failure | dendritic_cell | *ZBED3* | -2.527141515 | 1.15E-06 | 6.09E-06 | 0.005594118 |
| Heart_Failure | dendritic_cell | *RSG1* | 0.932761926 | 2.06E-07 | 3.66E-06 | 0.000998047 |
| Heart_Failure | dendritic_cell | *ATP8A2* | -2.639960828 | 5.98E-07 | 4.30E-06 | 0.002901244 |
| Heart_Failure | dendritic_cell | *ALOX5AP* | 3.383874647 | 1.82E-07 | 3.58E-06 | 0.00088515 |
| Heart_Failure | dendritic_cell | *WASF3* | -1.607627911 | 6.29E-07 | 4.39E-06 | 0.003050125 |
| Heart_Failure | dendritic_cell | *GPR12* | -5.753523886 | 0.007062226 | 0.00953951 | 1 |
| Heart_Failure | dendritic_cell | *PEMT* | -3.569976047 | 2.86E-06 | 1.10E-05 | 0.013853148 |
| Heart_Failure | dendritic_cell | *SCO1* | -2.637953364 | 0.002265802 | 0.003291519 | 1 |
| Heart_Failure | dendritic_cell | *MPRIP* | -2.536885199 | 9.08E-06 | 2.49E-05 | 0.044059986 |
| Heart_Failure | dendritic_cell | *COG6* | -2.430813053 | 0.003689962 | 0.005172984 | 1 |
| Heart_Failure | dendritic_cell | *EPSTI1* | 3.302097961 | 9.90E-07 | 5.62E-06 | 0.00480517 |
| Heart_Failure | dendritic_cell | *BTBD2* | -2.890065998 | 1.05E-05 | 2.81E-05 | 0.051072833 |
| Heart_Failure | dendritic_cell | *CNDP2* | 3.898703985 | 7.26E-06 | 2.10E-05 | 0.035233596 |
| Heart_Failure | dendritic_cell | *PDZD2* | -1.536786238 | 1.51E-06 | 7.15E-06 | 0.007316438 |
| Heart_Failure | dendritic_cell | *GSTT2B* | -0.694884054 | 0.028314921 | 0.035109633 | 1 |
| Heart_Failure | dendritic_cell | *SLC2A11* | -1.769281256 | 7.83E-06 | 2.22E-05 | 0.037967468 |
| Heart_Failure | dendritic_cell | *FAM83F* | -8.328121128 | 4.41E-05 | 9.30E-05 | 0.214123185 |
| Heart_Failure | dendritic_cell | *GIMAP4* | -2.281100425 | 0.002465339 | 0.003550481 | 1 |
| Heart_Failure | dendritic_cell | *ADCK2* | 2.357351872 | 0.000645901 | 0.001024824 | 1 |
| Heart_Failure | dendritic_cell | *LARS* | -1.566335829 | 0.003807366 | 0.0053268 | 1 |
| Heart_Failure | dendritic_cell | *SPINK5* | -7.362573657 | 0.003130218 | 0.004422777 | 1 |
| Heart_Failure | dendritic_cell | *IMPA1* | -1.777012022 | 0.001321804 | 0.001999187 | 1 |
| Heart_Failure | dendritic_cell | *LRRCC1* | -3.943275559 | 4.49E-05 | 9.42E-05 | 0.217793696 |
| Heart_Failure | dendritic_cell | *RRAS2* | -1.583079446 | 3.25E-06 | 1.20E-05 | 0.015774668 |
| Heart_Failure | dendritic_cell | *CATSPERB* | -1.266844411 | 9.07E-07 | 5.33E-06 | 0.004400627 |
| Heart_Failure | dendritic_cell | *COX16* | -6.314051177 | 4.86E-07 | 3.96E-06 | 0.002359627 |

| | | | | | | |
|---|---|---|---|---|---|---|
| Heart_Failure | dendritic_cell | *TTC9* | -2.623169428 | 3.95E-07 | 3.76E-06 | 0.001918303 |
| Heart_Failure | dendritic_cell | *ELP3* | -2.431452206 | 2.70E-06 | 1.05E-05 | 0.013086867 |
| Heart_Failure | dendritic_cell | *PEBP4* | -1.78902405 | 3.35E-07 | 3.66E-06 | 0.001625303 |
| Heart_Failure | dendritic_cell | *CTIF* | -1.952730027 | 1.16E-06 | 6.11E-06 | 0.00564316 |
| Heart_Failure | dendritic_cell | *MRO* | -3.94534378 | 1.71E-06 | 7.72E-06 | 0.00828611 |
| Heart_Failure | dendritic_cell | *THUMPD3* | -3.205501031 | 2.05E-06 | 8.71E-06 | 0.009935851 |
| Heart_Failure | dendritic_cell | *CNTN6* | -2.703457176 | 8.04E-08 | 3.44E-06 | 0.00039031 |
| Heart_Failure | dendritic_cell | *CHL1* | -1.411074652 | 8.02E-07 | 4.91E-06 | 0.003889586 |
| Heart_Failure | dendritic_cell | *GSTM5* | -1.554057104 | 0.001293592 | 0.001959572 | 1 |
| Heart_Failure | dendritic_cell | *VAV3* | 1.32569972 | 5.28E-06 | 1.68E-05 | 0.025610778 |
| Heart_Failure | dendritic_cell | *PSRC1* | -3.236199697 | 0.000600327 | 0.000957452 | 1 |
| Heart_Failure | dendritic_cell | *TRIM45* | -2.91767 | 9.59E-07 | 5.49E-06 | 0.004651551 |
| Heart_Failure | dendritic_cell | *CEPT1* | -1.76977957 | 1.56E-06 | 7.32E-06 | 0.007588804 |
| Heart_Failure | dendritic_cell | *NAPG* | -2.833399403 | 0.021372127 | 0.026962444 | 1 |
| Heart_Failure | dendritic_cell | *PPHLN1* | -2.03555162 | 0.038035266 | 0.046532302 | 1 |
| Heart_Failure | dendritic_cell | *FKBP11* | -2.471484472 | 6.52E-07 | 4.45E-06 | 0.003162444 |
| Heart_Failure | dendritic_cell | *YWHAQ* | -2.8554261 | 1.01E-05 | 2.72E-05 | 0.048991984 |
| Heart_Failure | dendritic_cell | *KIDINS220* | -2.498155475 | 3.63E-05 | 7.89E-05 | 0.176216515 |
| Heart_Failure | dendritic_cell | *LPIN1* | -2.439000005 | 0.000116321 | 0.000216159 | 0.564391348 |
| Heart_Failure | dendritic_cell | *NARS* | -2.924414624 | 3.36E-06 | 1.23E-05 | 0.016289023 |
| Heart_Failure | dendritic_cell | *KIAA1468* | -2.664611924 | 0.006070305 | 0.008268703 | 1 |
| Heart_Failure | dendritic_cell | *IL15RA* | -2.606498724 | 0.011406797 | 0.015002921 | 1 |
| Heart_Failure | dendritic_cell | *TMEM241* | -1.86376612 | 4.50E-06 | 1.50E-05 | 0.021844271 |
| Heart_Failure | dendritic_cell | *SOX5* | -1.252664894 | 2.40E-07 | 3.66E-06 | 0.001162126 |
| Heart_Failure | dendritic_cell | *KLRD1* | -2.610637519 | 4.20E-06 | 1.43E-05 | 0.020379244 |
| Heart_Failure | dendritic_cell | *ACP2* | -3.565689336 | 0.001341629 | 0.002026646 | 1 |
| Heart_Failure | dendritic_cell | *HOOK1* | -0.876149495 | 0.00531027 | 0.007296922 | 1 |
| Heart_Failure | dendritic_cell | *ZCCHC11* | -2.224831766 | 2.20E-05 | 5.23E-05 | 0.106808696 |
| Heart_Failure | dendritic_cell | *PRPF38A* | -4.224581062 | 2.63E-05 | 5.98E-05 | 0.127460558 |
| Heart_Failure | dendritic_cell | *ELP2* | -2.833087819 | 0.003557739 | 0.00499628 | 1 |
| Heart_Failure | dendritic_cell | *TPGS2* | -3.342654508 | 0.000983454 | 0.001518688 | 1 |
| Heart_Failure | dendritic_cell | *TIMM10* | 4.498359252 | 0.014705943 | 0.019022457 | 1 |
| Heart_Failure | dendritic_cell | *DHX34* | -3.656655811 | 0.00652248 | 0.008842434 | 1 |
| Heart_Failure | dendritic_cell | *FADS2* | 2.748002222 | 7.66E-07 | 4.80E-06 | 0.003714893 |
| Heart_Failure | dendritic_cell | *TMEM258* | -2.612057763 | 1.03E-05 | 2.76E-05 | 0.049740311 |
| Heart_Failure | dendritic_cell | *CLOCK* | -2.33717465 | 6.42E-06 | 1.93E-05 | 0.031130404 |
| Heart_Failure | dendritic_cell | *GGACT* | -2.969544566 | 0.001438297 | 0.002156556 | 1 |
| Heart_Failure | dendritic_cell | *CLDN10* | -3.100421306 | 8.57E-06 | 2.39E-05 | 0.04156502 |
| Heart_Failure | dendritic_cell | *ARGLU1* | -2.206378857 | 0.000117884 | 0.000218727 | 0.571971419 |
| Heart_Failure | dendritic_cell | *CARS2* | -1.888933876 | 5.17E-06 | 1.65E-05 | 0.025091014 |
| Heart_Failure | dendritic_cell | *KLB* | -0.953855489 | 2.20E-06 | 9.09E-06 | 0.010682462 |
| Heart_Failure | dendritic_cell | *WDR36* | -3.439932647 | 9.83E-06 | 2.66E-05 | 0.047672446 |
| Heart_Failure | dendritic_cell | *OSTF1* | 3.119415474 | 0.012880723 | 0.016786803 | 1 |
| Heart_Failure | dendritic_cell | *UBQLN1* | -2.372688752 | 4.05E-06 | 1.39E-05 | 0.019649649 |
| Heart_Failure | dendritic_cell | *GOLM1* | -1.761921804 | 2.45E-06 | 9.80E-06 | 0.011891849 |
| Heart_Failure | dendritic_cell | *ADAM19* | 3.070446407 | 2.00E-08 | 2.99E-06 | 9.68E-05 |
| Heart_Failure | dendritic_cell | *MSI1* | -3.549683044 | 3.99E-06 | 1.38E-05 | 0.019338647 |
| Heart_Failure | dendritic_cell | *DTX1* | -1.39623059 | 2.78E-07 | 3.66E-06 | 0.001350857 |
| Heart_Failure | dendritic_cell | *DMTF1* | -2.341870642 | 8.31E-06 | 2.33E-05 | 0.040336472 |
| Heart_Failure | dendritic_cell | *TMEM60* | 3.695632934 | 5.95E-07 | 4.30E-06 | 0.002887536 |

| | | | | | | |
|---|---|---|---|---|---|---|
| Heart_Failure | dendritic_cell | *POM121C* | -4.233440327 | 2.02E-07 | 3.66E-06 | 0.000978519 |
| Heart_Failure | dendritic_cell | *SRPK2* | -2.323240483 | 0.005211819 | 0.007163668 | 1 |
| Heart_Failure | dendritic_cell | *TES* | -3.66297882 | 0.000729686 | 0.001148001 | 1 |
| Heart_Failure | dendritic_cell | *KHDC1* | -1.609490832 | 6.23E-05 | 0.000125349 | 0.302467516 |
| Heart_Failure | dendritic_cell | *NT5E* | -1.61431711 | 2.82E-05 | 6.35E-05 | 0.136860968 |
| Heart_Failure | dendritic_cell | *MAP3K7* | -2.071961784 | 1.36E-06 | 6.68E-06 | 0.006602306 |
| Heart_Failure | dendritic_cell | *PRR5L* | -2.469364467 | 7.40E-07 | 4.72E-06 | 0.003592314 |
| Heart_Failure | dendritic_cell | *PHF21A* | -2.456749514 | 0.000161336 | 0.000289713 | 0.782803248 |
| Heart_Failure | dendritic_cell | *CAPRIN1* | -2.872911496 | 1.83E-05 | 4.46E-05 | 0.088881816 |
| Heart_Failure | dendritic_cell | *ATP5G2* | -3.181961183 | 0.000784043 | 0.00122755 | 1 |
| Heart_Failure | dendritic_cell | *B4GALNT1* | -5.097692886 | 0.000830674 | 0.001294293 | 1 |
| Heart_Failure | dendritic_cell | *FAIM2* | -2.812905209 | 2.25E-05 | 5.32E-05 | 0.109390619 |
| Heart_Failure | dendritic_cell | *PAN2* | -2.035500883 | 1.49E-06 | 7.08E-06 | 0.007224539 |
| Heart_Failure | dendritic_cell | *SLC26A10* | -3.9123701 | 1.00E-05 | 2.71E-05 | 0.048614967 |
| Heart_Failure | dendritic_cell | *MAP7* | -1.58266766 | 0.000382713 | 0.00063333 | 1 |
| Heart_Failure | dendritic_cell | *NHSL1* | -2.279300437 | 0.000304996 | 0.000517065 | 1 |
| Heart_Failure | dendritic_cell | *AHI1* | -1.983711405 | 2.56E-06 | 1.01E-05 | 0.012422679 |
| Heart_Failure | dendritic_cell | *CCT7* | -3.685639618 | 0.000163728 | 0.000293681 | 0.794408004 |
| Heart_Failure | dendritic_cell | *SMYD5* | -2.016168289 | 1.61E-06 | 7.46E-06 | 0.007802998 |
| Heart_Failure | dendritic_cell | *MDM2* | -2.909484455 | 0.000732697 | 0.001152365 | 1 |
| Heart_Failure | dendritic_cell | *MPHOSPH6* | -3.621260411 | 1.61E-05 | 4.00E-05 | 0.078245131 |
| Heart_Failure | dendritic_cell | *CCDC102A* | -2.16501088 | 1.44E-05 | 3.65E-05 | 0.069959685 |
| Heart_Failure | dendritic_cell | *ZNF670* | 3.897448305 | 0.000349366 | 0.000583721 | 1 |
| Heart_Failure | dendritic_cell | *KCNK1* | -1.418871819 | 2.88E-05 | 6.44E-05 | 0.139577248 |
| Heart_Failure | dendritic_cell | *CAPN9* | -5.084027737 | 6.03E-06 | 1.86E-05 | 0.029255759 |
| Heart_Failure | dendritic_cell | *NTPCR* | -2.383399573 | 1.19E-06 | 6.17E-06 | 0.005781529 |
| Heart_Failure | dendritic_cell | *MRPL44* | -2.313863822 | 1.46E-06 | 6.97E-06 | 0.007061828 |
| Heart_Failure | dendritic_cell | *SLC19A3* | -0.89030287 | 5.95E-07 | 4.30E-06 | 0.002884654 |
| Heart_Failure | dendritic_cell | *TMBIM1* | -3.278476639 | 1.64E-05 | 4.06E-05 | 0.079553969 |
| Heart_Failure | dendritic_cell | *TSGA10* | -1.505732156 | 3.64E-07 | 3.66E-06 | 0.001764495 |
| Heart_Failure | dendritic_cell | *GCC2* | -2.557232844 | 0.003249654 | 0.00458086 | 1 |
| Heart_Failure | dendritic_cell | *MRPS9* | -1.821352256 | 0.000206165 | 0.000362038 | 1 |
| Heart_Failure | dendritic_cell | *ANKRD36* | -2.244476516 | 2.19E-06 | 9.07E-06 | 0.010649781 |
| Heart_Failure | dendritic_cell | *EPC2* | -2.318949153 | 1.34E-05 | 3.43E-05 | 0.065092596 |
| Heart_Failure | dendritic_cell | *STAB2* | -1.479456356 | 3.17E-07 | 3.66E-06 | 0.00153578 |
| Heart_Failure | dendritic_cell | *APPL2* | -2.713093889 | 0.000298534 | 0.000506995 | 1 |
| Heart_Failure | dendritic_cell | *SLC41A2* | 2.577352534 | 0.005380737 | 0.007389566 | 1 |
| Heart_Failure | dendritic_cell | *NEK3* | -1.946589937 | 0.000120102 | 0.000222502 | 0.582733179 |
| Heart_Failure | dendritic_cell | *TBC1D4* | -1.902913752 | 8.71E-06 | 2.42E-05 | 0.042272633 |
| Heart_Failure | dendritic_cell | *SUCLA2* | -2.322961084 | 2.37E-06 | 9.56E-06 | 0.011498761 |
| Heart_Failure | dendritic_cell | *RCBTB1* | -2.206513636 | 3.63E-05 | 7.89E-05 | 0.176330207 |
| Heart_Failure | dendritic_cell | *RCBTB2* | 3.159682136 | 0.020688256 | 0.026133668 | 1 |
| Heart_Failure | dendritic_cell | *SETDB2* | 4.439885377 | 0.000123016 | 0.000227035 | 0.59687501 |
| Heart_Failure | dendritic_cell | *C7orf25* | 3.67842499 | 0.000423777 | 0.00069559 | 1 |
| Heart_Failure | dendritic_cell | *CHST12* | -3.045735963 | 4.57E-06 | 1.52E-05 | 0.02215668 |
| Heart_Failure | dendritic_cell | *KDELR2* | 3.748423336 | 0.023094858 | 0.029015083 | 1 |
| Heart_Failure | dendritic_cell | *NUPL2* | -3.330093818 | 0.000213397 | 0.000373118 | 1 |
| Heart_Failure | dendritic_cell | *ZDHHC4* | -3.672366092 | 2.47E-05 | 5.72E-05 | 0.119871243 |
| Heart_Failure | dendritic_cell | *AOAH* | 0.815028647 | 1.73E-07 | 3.52E-06 | 0.000837458 |
| Heart_Failure | dendritic_cell | *TBRG4* | 4.861462893 | 0.001776507 | 0.002629535 | 1 |

| | | | | | | |
|---|---|---|---|---|---|---|
| Heart_Failure | dendritic_cell | *MYCBPAP* | -0.940419526 | 0.024646612 | 0.030820969 | 1 |
| Heart_Failure | dendritic_cell | *BRIP1* | -2.182268796 | 1.05E-06 | 5.79E-06 | 0.005070808 |
| Heart_Failure | dendritic_cell | *RTP4* | -3.520425864 | 1.13E-06 | 6.03E-06 | 0.00546684 |
| Heart_Failure | dendritic_cell | *NDUFB5* | -2.893589786 | 6.25E-06 | 1.91E-05 | 0.030317727 |
| Heart_Failure | dendritic_cell | *SCN2A* | -1.563084397 | 1.71E-06 | 7.72E-06 | 0.008306275 |
| Heart_Failure | dendritic_cell | *RPS6KC1* | -2.612161426 | 0.000150498 | 0.000272165 | 0.730218551 |
| Heart_Failure | dendritic_cell | *SMPD4* | -2.999326717 | 1.53E-05 | 3.83E-05 | 0.074107736 |
| Heart_Failure | dendritic_cell | *IMP4* | -4.673355138 | 8.39E-07 | 5.05E-06 | 0.004071644 |
| Heart_Failure | dendritic_cell | *NIPSNAP3A* | -2.974788795 | 1.17E-05 | 3.06E-05 | 0.056545977 |
| Heart_Failure | dendritic_cell | *ODF2* | -2.829827789 | 1.10E-06 | 5.95E-06 | 0.005328813 |
| Heart_Failure | dendritic_cell | *TOR1B* | 3.891068239 | 1.14E-05 | 3.01E-05 | 0.055470534 |
| Heart_Failure | dendritic_cell | *SLC2A8* | -4.521341908 | 0.000829989 | 0.001294293 | 1 |
| Heart_Failure | dendritic_cell | *ZNF189* | -4.004616319 | 3.55E-05 | 7.75E-05 | 0.172300157 |
| Heart_Failure | dendritic_cell | *STX17* | -3.223912911 | 0.000417181 | 0.000685228 | 1 |
| Heart_Failure | dendritic_cell | *PRPF4* | -1.016682486 | 0.009868093 | 0.01307839 | 1 |
| Heart_Failure | dendritic_cell | *ANP32B* | -3.002384014 | 6.10E-06 | 1.87E-05 | 0.029579623 |
| Heart_Failure | dendritic_cell | *DSCC1* | -1.019212006 | 1.84E-05 | 4.48E-05 | 0.089390901 |
| Heart_Failure | dendritic_cell | *DERL1* | 3.902180316 | 2.57E-05 | 5.86E-05 | 0.124601313 |
| Heart_Failure | dendritic_cell | *NOV* | -2.665485631 | 1.78E-06 | 7.93E-06 | 0.008651989 |
| Heart_Failure | dendritic_cell | *POLR1E* | -0.984111231 | 0.013951773 | 0.018114531 | 1 |
| Heart_Failure | dendritic_cell | *IL11RA* | -2.948344325 | 4.72E-07 | 3.95E-06 | 0.002292047 |
| Heart_Failure | dendritic_cell | *APTX* | -2.805091187 | 2.62E-05 | 5.96E-05 | 0.126887649 |
| Heart_Failure | dendritic_cell | *TMEM8B* | -2.2796996 | 1.97E-07 | 3.63E-06 | 0.00095579 |
| Heart_Failure | dendritic_cell | *GRHPR* | -1.780311803 | 0.006520858 | 0.008842434 | 1 |
| Heart_Failure | dendritic_cell | *DENND4C* | -2.417873682 | 0.000336708 | 0.000565687 | 1 |
| Heart_Failure | dendritic_cell | *GMPR* | -3.101603612 | 4.13E-06 | 1.41E-05 | 0.020020044 |
| Heart_Failure | dendritic_cell | *TMEM14B* | -3.525973537 | 3.90E-05 | 8.40E-05 | 0.189431214 |
| Heart_Failure | dendritic_cell | *FRS3* | -4.053000533 | 4.21E-06 | 1.43E-05 | 0.020423268 |
| Heart_Failure | dendritic_cell | *CAPN11* | -1.745222997 | 0.000793914 | 0.001241003 | 1 |
| Heart_Failure | dendritic_cell | *IRF4* | 0.815208905 | 2.98E-08 | 3.33E-06 | 0.000144832 |
| Heart_Failure | dendritic_cell | *LRRC1* | -1.151998956 | 4.48E-07 | 3.90E-06 | 0.002172265 |
| Heart_Failure | dendritic_cell | *RIPK1* | -2.463117583 | 2.08E-05 | 4.98E-05 | 0.100900418 |
| Heart_Failure | dendritic_cell | *TCF19* | -1.348262973 | 0.031445288 | 0.038842296 | 1 |
| Heart_Failure | dendritic_cell | *FLOT1* | -1.815341954 | 0.000259137 | 0.000446497 | 1 |
| Heart_Failure | dendritic_cell | *NRM* | -5.608174596 | 3.28E-07 | 3.66E-06 | 0.00159073 |
| Heart_Failure | dendritic_cell | *VARS2* | -0.946166462 | 0.000183063 | 0.000324523 | 0.888219831 |
| Heart_Failure | dendritic_cell | *TLR2* | 2.301748081 | 5.36E-08 | 3.44E-06 | 0.000260229 |
| Heart_Failure | dendritic_cell | *MYO7A* | -2.151605579 | 0.000379073 | 0.000629023 | 1 |
| Heart_Failure | dendritic_cell | *NUMA1* | -2.889397168 | 5.26E-05 | 0.000108606 | 0.255332697 |
| Heart_Failure | dendritic_cell | *SYTL2* | -2.41343242 | 4.32E-05 | 9.13E-05 | 0.209589474 |
| Heart_Failure | dendritic_cell | *RAB30* | -3.399633508 | 0.008239362 | 0.011031286 | 1 |
| Heart_Failure | dendritic_cell | *PRCP* | -2.43465487 | 0.002369802 | 0.003428229 | 1 |
| Heart_Failure | dendritic_cell | *NARS2* | -3.141934533 | 0.000411103 | 0.00067593 | 1 |
| Heart_Failure | dendritic_cell | *GGH* | -1.795832125 | 7.35E-05 | 0.000144537 | 0.356427851 |
| Heart_Failure | dendritic_cell | *SULF1* | -1.123921962 | 1.54E-05 | 3.85E-05 | 0.074746907 |
| Heart_Failure | dendritic_cell | *TGS1* | -2.991462522 | 7.17E-05 | 0.000141476 | 0.347748468 |
| Heart_Failure | dendritic_cell | *NEK1* | -2.203077334 | 8.54E-06 | 2.38E-05 | 0.041439794 |
| Heart_Failure | dendritic_cell | *TRPC6* | -3.505306353 | 7.95E-06 | 2.25E-05 | 0.038573809 |
| Heart_Failure | dendritic_cell | *MMP27* | -3.451886382 | 0.0370585 | 0.045371648 | 1 |
| Heart_Failure | dendritic_cell | *SLC37A4* | 2.509179204 | 9.67E-06 | 2.63E-05 | 0.046935114 |

| | | | | | | |
|---|---|---|---|---|---|---|
| Heart_Failure | dendritic_cell | *POU2F3* | -1.995324009 | 0.002700025 | 0.003858769 | 1 |
| Heart_Failure | dendritic_cell | *PPP2R1B* | -1.121846944 | 2.09E-06 | 8.80E-06 | 0.010142326 |
| Heart_Failure | dendritic_cell | *FDX1* | -2.660483416 | 4.29E-06 | 1.45E-05 | 0.02083865 |
| Heart_Failure | dendritic_cell | *FXYD6* | -1.153144166 | 0.034742873 | 0.042730651 | 1 |
| Heart_Failure | dendritic_cell | *FXYD2* | 2.139362793 | 6.33E-06 | 1.92E-05 | 0.030720571 |
| Heart_Failure | dendritic_cell | *NUSAP1* | -2.605095359 | 1.56E-05 | 3.88E-05 | 0.075492337 |
| Heart_Failure | dendritic_cell | *NDUFAF1* | -1.624204708 | 1.96E-06 | 8.41E-06 | 0.009506048 |
| Heart_Failure | dendritic_cell | *PARP6* | -3.029827018 | 1.46E-05 | 3.68E-05 | 0.070732403 |
| Heart_Failure | dendritic_cell | *ADAM10* | -2.195373779 | 0.002280972 | 0.003310582 | 1 |
| Heart_Failure | dendritic_cell | *ZNF280D* | -2.056044879 | 1.26E-05 | 3.26E-05 | 0.061107711 |
| Heart_Failure | dendritic_cell | *RSL24D1* | -3.265178615 | 0.000166071 | 0.000297445 | 0.805778611 |
| Heart_Failure | dendritic_cell | *SLC44A5* | -1.605043295 | 5.71E-07 | 4.26E-06 | 0.002770813 |
| Heart_Failure | dendritic_cell | *DBT* | -2.249187862 | 1.18E-06 | 6.16E-06 | 0.005745144 |
| Heart_Failure | dendritic_cell | *RTCA* | 3.050385425 | 0.012778152 | 0.016657602 | 1 |
| Heart_Failure | dendritic_cell | *KHK* | -2.037740563 | 0.012352613 | 0.016137555 | 1 |
| Heart_Failure | dendritic_cell | *DYNC2LI1* | -1.552706715 | 5.08E-06 | 1.63E-05 | 0.02463112 |
| Heart_Failure | dendritic_cell | *LHCGR* | -2.752410087 | 1.58E-05 | 3.92E-05 | 0.076451361 |
| Heart_Failure | dendritic_cell | *THUMPD2* | -2.972412133 | 4.50E-06 | 1.50E-05 | 0.021855382 |
| Heart_Failure | dendritic_cell | *ATRAID* | -3.030685345 | 0.000120734 | 0.000223589 | 0.585803479 |
| Heart_Failure | dendritic_cell | *CENPO* | -3.199553708 | 3.21E-05 | 7.08E-05 | 0.155844466 |
| Heart_Failure | dendritic_cell | *LRPPRC* | -2.335493479 | 4.79E-06 | 1.57E-05 | 0.023225844 |
| Heart_Failure | dendritic_cell | *DTNB* | -2.55761832 | 7.24E-06 | 2.09E-05 | 0.035122046 |
| Heart_Failure | dendritic_cell | *MYOF* | -1.377686743 | 0.006559351 | 0.008889936 | 1 |
| Heart_Failure | dendritic_cell | *BTBD16* | -2.348550668 | 2.55E-05 | 5.84E-05 | 0.123507386 |
| Heart_Failure | dendritic_cell | *TACC2* | -1.120350125 | 3.34E-07 | 3.66E-06 | 0.001622948 |
| Heart_Failure | dendritic_cell | *CALHM2* | -2.19250156 | 0.016740005 | 0.021487435 | 1 |
| Heart_Failure | dendritic_cell | *PLCE1* | -1.441640175 | 8.34E-06 | 2.34E-05 | 0.040484874 |
| Heart_Failure | dendritic_cell | *RBP4* | -2.195100893 | 7.93E-07 | 4.88E-06 | 0.003848634 |
| Heart_Failure | dendritic_cell | *ASCC1* | -2.126020407 | 1.14E-06 | 6.06E-06 | 0.005549288 |
| Heart_Failure | dendritic_cell | *RPS24* | -2.353502298 | 8.29E-05 | 0.000160435 | 0.402371785 |
| Heart_Failure | dendritic_cell | *ATIC* | -1.512393728 | 0.024767332 | 0.030932602 | 1 |
| Heart_Failure | dendritic_cell | *SMARCAL1* | -3.388949672 | 3.09E-05 | 6.84E-05 | 0.149962161 |
| Heart_Failure | dendritic_cell | *BARD1* | 3.506872668 | 0.007556567 | 0.010164809 | 1 |
| Heart_Failure | dendritic_cell | *STAT4* | -2.891481252 | 3.11E-06 | 1.16E-05 | 0.015104702 |
| Heart_Failure | dendritic_cell | *SSB* | -1.963600819 | 2.56E-05 | 5.86E-05 | 0.124428545 |
| Heart_Failure | dendritic_cell | *FASTKD1* | -2.388117855 | 0.005338709 | 0.007333923 | 1 |
| Heart_Failure | dendritic_cell | *ITGAV* | -2.638784648 | 2.47E-05 | 5.71E-05 | 0.11978064 |
| Heart_Failure | dendritic_cell | *SENP7* | -2.458800846 | 1.02E-05 | 2.74E-05 | 0.049376919 |
| Heart_Failure | dendritic_cell | *SPPL2A* | -2.789448064 | 0.000679105 | 0.001074345 | 1 |
| Heart_Failure | dendritic_cell | *PPCDC* | -2.666003766 | 1.61E-05 | 4.00E-05 | 0.078230487 |
| Heart_Failure | dendritic_cell | *SEMA7A* | -2.085638518 | 0.011219244 | 0.014764245 | 1 |
| Heart_Failure | dendritic_cell | *PCDH10* | -4.92037428 | 0.001620372 | 0.00241674 | 1 |
| Heart_Failure | dendritic_cell | *AP1AR* | -3.784647189 | 0.000583102 | 0.000932502 | 1 |
| Heart_Failure | dendritic_cell | *KIAA1109* | -2.353132003 | 2.63E-05 | 6.00E-05 | 0.127796091 |
| Heart_Failure | dendritic_cell | *BMPR1B* | -1.312400093 | 5.17E-07 | 4.07E-06 | 0.002508353 |
| Heart_Failure | dendritic_cell | *MMRN1* | -0.935001311 | 1.15E-05 | 3.02E-05 | 0.055633394 |
| Heart_Failure | dendritic_cell | *PRDM5* | -3.181553507 | 3.44E-05 | 7.54E-05 | 0.167041288 |
| Heart_Failure | dendritic_cell | *NAAA* | 2.758392242 | 0.000431816 | 0.000707827 | 1 |
| Heart_Failure | dendritic_cell | *NUP54* | -2.247951796 | 1.97E-05 | 4.73E-05 | 0.095426154 |
| Heart_Failure | dendritic_cell | *ANXA3* | -1.497636094 | 3.07E-07 | 3.66E-06 | 0.001490965 |

| | | | | | | |
|---|---|---|---|---|---|---|
| Heart_Failure | dendritic_cell | *PPA2* | -2.597662111 | 0.000232084 | 0.0004026 | 1 |
| Heart_Failure | dendritic_cell | *ENPEP* | -0.934045485 | 0.000943998 | 0.00146288 | 1 |
| Heart_Failure | dendritic_cell | *PAPSS1* | -2.756276133 | 0.000684671 | 0.00108174 | 1 |
| Heart_Failure | dendritic_cell | *FBN2* | -1.026400238 | 3.95E-05 | 8.47E-05 | 0.191552636 |
| Heart_Failure | dendritic_cell | *PARVG* | 1.172196031 | 1.09E-07 | 3.44E-06 | 0.000528135 |
| Heart_Failure | dendritic_cell | *B4GALNT3* | 4.178214736 | 8.38E-08 | 3.44E-06 | 0.000406385 |
| Heart_Failure | dendritic_cell | *PDE6H* | -4.453455736 | 0.034925781 | 0.042933846 | 1 |
| Heart_Failure | dendritic_cell | *ERP27* | 1.811926819 | 2.13E-05 | 5.08E-05 | 0.103316215 |
| Heart_Failure | dendritic_cell | *KIF21A* | -1.285580906 | 4.81E-07 | 3.96E-06 | 0.002335056 |
| Heart_Failure | dendritic_cell | *ALG10* | -4.91640083 | 7.82E-08 | 3.44E-06 | 0.000379491 |
| Heart_Failure | dendritic_cell | *ZCRB1* | -3.467402224 | 2.36E-05 | 5.52E-05 | 0.114316069 |
| Heart_Failure | dendritic_cell | *C1RL* | -1.52362388 | 1.77E-05 | 4.33E-05 | 0.085692059 |
| Heart_Failure | dendritic_cell | *CLSTN3* | -1.974875682 | 6.67E-07 | 4.49E-06 | 0.003233973 |
| Heart_Failure | dendritic_cell | *KLRG1* | -0.837310016 | 0.016358272 | 0.021025255 | 1 |
| Heart_Failure | dendritic_cell | *VAMP1* | -1.964929098 | 9.85E-07 | 5.60E-06 | 0.004780694 |
| Heart_Failure | dendritic_cell | *PEX5* | -2.204775244 | 3.10E-05 | 6.85E-05 | 0.150220072 |
| Heart_Failure | dendritic_cell | *PPFIA2* | -1.702857343 | 5.54E-07 | 4.18E-06 | 0.002685878 |
| Heart_Failure | dendritic_cell | *LGR5* | -3.812247687 | 0.000479676 | 0.00078074 | 1 |
| Heart_Failure | dendritic_cell | *POC1B* | -2.276420767 | 8.12E-07 | 4.93E-06 | 0.003941913 |
| Heart_Failure | dendritic_cell | *RITA1* | -7.525857376 | 0.001423316 | 0.002137705 | 1 |
| Heart_Failure | dendritic_cell | *SDSL* | 1.565093496 | 2.84E-06 | 1.09E-05 | 0.013777846 |
| Heart_Failure | dendritic_cell | *GLTP* | 3.035572562 | 2.01E-07 | 3.66E-06 | 0.000976349 |
| Heart_Failure | dendritic_cell | *FOXN4* | -3.049512323 | 1.47E-08 | 2.99E-06 | 7.15E-05 |
| Heart_Failure | dendritic_cell | *MTMR6* | -2.789643402 | 4.01E-07 | 3.80E-06 | 0.001946282 |
| Heart_Failure | dendritic_cell | *ACVRL1* | -2.848490622 | 7.91E-07 | 4.87E-06 | 0.003835702 |
| Heart_Failure | dendritic_cell | *BRCA2* | 2.625704348 | 2.51E-07 | 3.66E-06 | 0.001216437 |
| Heart_Failure | dendritic_cell | *KANSL2* | -2.681956104 | 9.29E-05 | 0.000176879 | 0.45086512 |
| Heart_Failure | dendritic_cell | *CERS5* | -3.290773319 | 0.000185794 | 0.000329124 | 0.901470771 |
| Heart_Failure | dendritic_cell | *ESYT1* | 4.282756734 | 0.001215383 | 0.001850342 | 1 |
| Heart_Failure | dendritic_cell | *ZNF740* | -2.071228302 | 7.45E-07 | 4.74E-06 | 0.003612601 |
| Heart_Failure | dendritic_cell | *HNRNPA1L2* | -4.968993963 | 2.21E-05 | 5.24E-05 | 0.107392911 |
| Heart_Failure | dendritic_cell | *RB1* | -2.275588815 | 0.000204286 | 0.000359259 | 0.991194484 |
| Heart_Failure | dendritic_cell | *DIAPH3* | -1.022998868 | 0.000264197 | 0.000453764 | 1 |
| Heart_Failure | dendritic_cell | *GRTP1* | -1.253647398 | 4.98E-06 | 1.62E-05 | 0.024186135 |
| Heart_Failure | dendritic_cell | *NOVA1* | -0.684116233 | 0.000374773 | 0.000622312 | 1 |
| Heart_Failure | dendritic_cell | *FITM1* | -4.136458563 | 0.016924538 | 0.02167842 | 1 |
| Heart_Failure | dendritic_cell | *TMX1* | -3.734183484 | 0.000291912 | 0.000496967 | 1 |
| Heart_Failure | dendritic_cell | *RTN1* | 0.762597383 | 2.69E-07 | 3.66E-06 | 0.001306516 |
| Heart_Failure | dendritic_cell | *C14orf37* | -2.463588751 | 0.010198599 | 0.013464926 | 1 |
| Heart_Failure | dendritic_cell | *RAB15* | 4.815287535 | 1.01E-06 | 5.68E-06 | 0.004877801 |
| Heart_Failure | dendritic_cell | *EFCAB11* | 3.160452376 | 9.44E-06 | 2.58E-05 | 0.045812025 |
| Heart_Failure | dendritic_cell | *PTGR2* | -0.713132496 | 0.013754242 | 0.017877199 | 1 |
| Heart_Failure | dendritic_cell | *SLC24A4* | 1.090789409 | 1.14E-07 | 3.44E-06 | 0.000552657 |
| Heart_Failure | dendritic_cell | *C14orf79* | 3.679349426 | 0.013209315 | 0.017187342 | 1 |
| Heart_Failure | dendritic_cell | *WARS* | 4.388478875 | 1.18E-05 | 3.10E-05 | 0.057333239 |
| Heart_Failure | dendritic_cell | *DUOXA1* | 1.233330788 | 0.022047106 | 0.027770654 | 1 |
| Heart_Failure | dendritic_cell | *SERF2* | -2.901116541 | 0.000157035 | 0.000282608 | 0.761935019 |
| Heart_Failure | dendritic_cell | *SRP14* | -2.705129616 | 5.23E-06 | 1.67E-05 | 0.025359667 |
| Heart_Failure | dendritic_cell | *UBE2Q2* | -3.182659547 | 0.001764207 | 0.002614518 | 1 |
| Heart_Failure | dendritic_cell | *HMG20A* | -3.098082929 | 5.68E-05 | 0.000115756 | 0.275383888 |

| | | | | | | |
|---|---|---|---|---|---|---|
| Heart_Failure | dendritic_cell | *SCAPER* | -2.138723525 | 3.55E-05 | 7.75E-05 | 0.172436184 |
| Heart_Failure | dendritic_cell | *NEIL1* | -2.435109946 | 5.58E-06 | 1.75E-05 | 0.027053682 |
| Heart_Failure | dendritic_cell | *MAN2C1* | -0.849389186 | 0.003338738 | 0.004700975 | 1 |
| Heart_Failure | dendritic_cell | *PIF1* | -3.185054392 | 1.16E-07 | 3.44E-06 | 0.0005645 |
| Heart_Failure | dendritic_cell | *BBS4* | -3.050225905 | 0.000283788 | 0.000484497 | 1 |
| Heart_Failure | dendritic_cell | *ULK3* | -5.142372944 | 2.55E-05 | 5.84E-05 | 0.123631989 |
| Heart_Failure | dendritic_cell | *PCSK6* | -2.140713159 | 7.09E-07 | 4.63E-06 | 0.003441953 |
| Heart_Failure | dendritic_cell | *CELF6* | 2.448975964 | 2.08E-07 | 3.66E-06 | 0.001009025 |
| Heart_Failure | dendritic_cell | *HAPLN3* | -2.591962037 | 0.001601152 | 0.002391868 | 1 |
| Heart_Failure | dendritic_cell | *POLG* | -2.582081857 | 2.63E-06 | 1.03E-05 | 0.012772842 |
| Heart_Failure | dendritic_cell | *MFGE8* | -2.105606374 | 9.05E-07 | 5.32E-06 | 0.004388769 |
| Heart_Failure | dendritic_cell | *UNC45A* | -3.57684667 | 0.000121227 | 0.000224245 | 0.58819427 |
| Heart_Failure | dendritic_cell | *IQGAP1* | -2.601589708 | 6.97E-05 | 0.000138095 | 0.338057017 |
| Heart_Failure | dendritic_cell | *CRTC3* | -1.70299331 | 2.75E-05 | 6.22E-05 | 0.133498394 |
| Heart_Failure | dendritic_cell | *ITGAX* | 0.223352515 | 1.35E-07 | 3.44E-06 | 0.000654716 |
| Heart_Failure | dendritic_cell | *TGFB1I1* | -2.217412893 | 4.83E-07 | 3.96E-06 | 0.002345453 |
| Heart_Failure | dendritic_cell | *DHX38* | -4.046346473 | 0.000136052 | 0.000248448 | 0.660125672 |
| Heart_Failure | dendritic_cell | *NUDT7* | -1.94790033 | 3.55E-07 | 3.66E-06 | 0.001721987 |
| Heart_Failure | dendritic_cell | *GCSH* | 4.078318579 | 0.000279955 | 0.00047863 | 1 |
| Heart_Failure | dendritic_cell | *ZCCHC14* | -2.31663013 | 3.20E-06 | 1.18E-05 | 0.015504328 |
| Heart_Failure | dendritic_cell | *TLDC1* | -4.455880802 | 0.000260498 | 0.000448205 | 1 |
| Heart_Failure | dendritic_cell | *ADAD2* | -1.384547754 | 0.038456835 | 0.047024335 | 1 |
| Heart_Failure | dendritic_cell | *RPL3L* | 5.925216684 | 8.44E-06 | 2.36E-05 | 0.040969269 |
| Heart_Failure | dendritic_cell | *TIGD7* | 3.784783858 | 0.004411386 | 0.006122438 | 1 |
| Heart_Failure | dendritic_cell | *DEF8* | 2.819603753 | 7.06E-06 | 2.06E-05 | 0.034274281 |
| Heart_Failure | dendritic_cell | *TCF25* | -1.846277402 | 0.012255516 | 0.01602365 | 1 |
| Heart_Failure | dendritic_cell | *GALNS* | -2.97613963 | 0.001622477 | 0.00241777 | 1 |
| Heart_Failure | dendritic_cell | *NCOR1* | -2.381234533 | 0.000156449 | 0.000281753 | 0.759091766 |
| Heart_Failure | dendritic_cell | *KSR1* | -3.172928732 | 0.001927235 | 0.002835338 | 1 |
| Heart_Failure | dendritic_cell | *GFOD2* | -1.971207567 | 1.62E-06 | 7.49E-06 | 0.00785008 |
| Heart_Failure | dendritic_cell | *PRPSAP2* | -2.516878291 | 0.001414466 | 0.002125422 | 1 |
| Heart_Failure | dendritic_cell | *MYO19* | -2.569936841 | 0.005199822 | 0.007149203 | 1 |
| Heart_Failure | dendritic_cell | *RASL10B* | -5.524790525 | 3.29E-05 | 7.24E-05 | 0.159585638 |
| Heart_Failure | dendritic_cell | *UNC45B* | -8.578815974 | 7.11E-06 | 2.07E-05 | 0.03448056 |
| Heart_Failure | dendritic_cell | *C17orf80* | -4.088741731 | 6.90E-06 | 2.03E-05 | 0.033459951 |
| Heart_Failure | dendritic_cell | *VPS53* | -2.47463877 | 0.000218431 | 0.000381507 | 1 |
| Heart_Failure | dendritic_cell | *SGSM2* | -3.16245622 | 0.007733706 | 0.010391564 | 1 |
| Heart_Failure | dendritic_cell | *LRRC46* | -1.912361979 | 0.008165324 | 0.010941219 | 1 |
| Heart_Failure | dendritic_cell | *SCRN2* | -0.658664012 | 0.002583484 | 0.003702027 | 1 |
| Heart_Failure | dendritic_cell | *SSH2* | -2.242593489 | 7.46E-05 | 0.000146193 | 0.361828645 |
| Heart_Failure | dendritic_cell | *RHBDL3* | -3.410868702 | 0.0002167 | 0.000378756 | 1 |
| Heart_Failure | dendritic_cell | *G6PC3* | -2.288394407 | 5.11E-07 | 4.04E-06 | 0.00247834 |
| Heart_Failure | dendritic_cell | *CLTC* | -2.336233369 | 1.44E-05 | 3.65E-05 | 0.069869885 |
| Heart_Failure | dendritic_cell | *IMPA2* | 5.285135251 | 8.04E-05 | 0.000156427 | 0.390285896 |
| Heart_Failure | dendritic_cell | *GNAL* | -2.531382684 | 1.95E-06 | 8.38E-06 | 0.009440059 |
| Heart_Failure | dendritic_cell | *RPRD1A* | -2.357845311 | 5.19E-06 | 1.66E-05 | 0.025195829 |
| Heart_Failure | dendritic_cell | *GALNT1* | -2.277729483 | 6.21E-06 | 1.90E-05 | 0.03013669 |
| Heart_Failure | dendritic_cell | *C18orf8* | -2.446638841 | 7.61E-07 | 4.79E-06 | 0.003693863 |
| Heart_Failure | dendritic_cell | *NPC1* | -2.796664898 | 7.64E-06 | 2.17E-05 | 0.037052236 |
| Heart_Failure | dendritic_cell | *SLC14A1* | -1.73805675 | 9.48E-05 | 0.000179886 | 0.459789302 |

| | | | | | | |
|---|---|---|---|---|---|---|
| Heart_Failure | dendritic_cell | *ARHGDIA* | -1.871411513 | 1.07E-05 | 2.85E-05 | 0.051836794 |
| Heart_Failure | dendritic_cell | *TMC6* | -2.675941616 | 6.09E-05 | 0.000122943 | 0.295308994 |
| Heart_Failure | dendritic_cell | *CARD14* | -2.403600115 | 2.54E-06 | 1.01E-05 | 0.012332704 |
| Heart_Failure | dendritic_cell | *ANAPC11* | -2.350457048 | 4.53E-07 | 3.90E-06 | 0.00219714 |
| Heart_Failure | dendritic_cell | *TBCD* | -2.528478657 | 2.05E-05 | 4.92E-05 | 0.099672454 |
| Heart_Failure | dendritic_cell | *NARF* | -2.627364035 | 6.51E-05 | 0.000130118 | 0.315796596 |
| Heart_Failure | dendritic_cell | *TRIM65* | -2.890087165 | 0.017199963 | 0.022013775 | 1 |
| Heart_Failure | dendritic_cell | *CBX8* | 2.436880265 | 1.54E-05 | 3.85E-05 | 0.074694743 |
| Heart_Failure | dendritic_cell | *SECTM1* | 2.035634489 | 0.002781393 | 0.003965713 | 1 |
| Heart_Failure | dendritic_cell | *RNF157* | -1.346411525 | 2.94E-06 | 1.12E-05 | 0.014242107 |
| Heart_Failure | dendritic_cell | *ZNF750* | -5.316876753 | 0.00076961 | 0.00120612 | 1 |
| Heart_Failure | dendritic_cell | *RNF165* | -2.49959325 | 6.11E-06 | 1.88E-05 | 0.029660798 |
| Heart_Failure | dendritic_cell | *MAPK4* | -4.362485082 | 3.52E-07 | 3.66E-06 | 0.001707757 |
| Heart_Failure | dendritic_cell | *FBXO15* | -2.937685424 | 1.48E-06 | 7.06E-06 | 0.007189604 |
| Heart_Failure | dendritic_cell | *CBLN2* | -2.77531719 | 3.36E-07 | 3.66E-06 | 0.001629874 |
| Heart_Failure | dendritic_cell | *ERBB2* | -2.004242972 | 0.000399854 | 0.000658998 | 1 |
| Heart_Failure | dendritic_cell | *MIEN1* | 2.551055413 | 2.75E-05 | 6.21E-05 | 0.13324193 |
| Heart_Failure | dendritic_cell | *PNMT* | 3.357282934 | 0.012545429 | 0.016371819 | 1 |
| Heart_Failure | dendritic_cell | *IGFBP4* | -2.028598233 | 3.43E-07 | 3.66E-06 | 0.001662368 |
| Heart_Failure | dendritic_cell | *TXNL4A* | -3.308091952 | 5.01E-05 | 0.000103899 | 0.243122639 |
| Heart_Failure | dendritic_cell | *CACNA1A* | 4.740695377 | 0.003531211 | 0.004961899 | 1 |
| Heart_Failure | dendritic_cell | *SAMD1* | -6.249748978 | 2.24E-06 | 9.20E-06 | 0.010857386 |
| Heart_Failure | dendritic_cell | *NFIC* | -2.170932315 | 2.49E-05 | 5.76E-05 | 0.120937527 |
| Heart_Failure | dendritic_cell | *PRDM15* | -1.912586538 | 0.001817123 | 0.002685074 | 1 |
| Heart_Failure | dendritic_cell | *PFKL* | -3.460694661 | 0.001171044 | 0.001786763 | 1 |
| Heart_Failure | dendritic_cell | *DUS3L* | -1.620840828 | 1.89E-06 | 8.21E-06 | 0.009158955 |
| Heart_Failure | dendritic_cell | *DPP9* | 4.312486899 | 0.002001657 | 0.002936813 | 1 |
| Heart_Failure | dendritic_cell | *SIRT3* | -2.725837266 | 0.000697557 | 0.001101024 | 1 |
| Heart_Failure | dendritic_cell | *HUNK* | -0.653065222 | 0.00869239 | 0.01159304 | 1 |
| Heart_Failure | dendritic_cell | *COL6A1* | -1.037317754 | 0.000327511 | 0.000551765 | 1 |
| Heart_Failure | dendritic_cell | *TMEM50B* | -1.350392243 | 0.031927799 | 0.039388172 | 1 |
| Heart_Failure | dendritic_cell | *SAE1* | -3.275368287 | 1.04E-06 | 5.77E-06 | 0.005042824 |
| Heart_Failure | dendritic_cell | *CACNG8* | -2.039101833 | 0.034117739 | 0.041993727 | 1 |
| Heart_Failure | dendritic_cell | *ZNF787* | 3.612399817 | 3.56E-05 | 7.76E-05 | 0.172717781 |
| Heart_Failure | dendritic_cell | *EVI5L* | -4.27770467 | 8.02E-06 | 2.26E-05 | 0.038899968 |
| Heart_Failure | dendritic_cell | *CTU1* | 6.435216928 | 3.14E-06 | 1.17E-05 | 0.015241895 |
| Heart_Failure | dendritic_cell | *RERE* | -1.929117735 | 2.32E-05 | 5.44E-05 | 0.112482387 |
| Heart_Failure | dendritic_cell | *MMEL1* | 1.130402089 | 0.003416051 | 0.004804254 | 1 |
| Heart_Failure | dendritic_cell | *EPHA2* | -2.137233138 | 3.64E-07 | 3.66E-06 | 0.001764995 |
| Heart_Failure | dendritic_cell | *NBPF3* | -0.809581692 | 0.005682842 | 0.007789025 | 1 |
| Heart_Failure | dendritic_cell | *HSPG2* | -1.862034729 | 1.00E-05 | 2.71E-05 | 0.048585408 |
| Heart_Failure | dendritic_cell | *ITGB3BP* | -1.838430433 | 0.000134973 | 0.000246756 | 0.654889809 |
| Heart_Failure | dendritic_cell | *PIGK* | -3.296951577 | 1.11E-05 | 2.94E-05 | 0.053957024 |
| Heart_Failure | dendritic_cell | *RPS8* | -2.195615402 | 5.40E-06 | 1.71E-05 | 0.026203337 |
| Heart_Failure | dendritic_cell | *CYP4B1* | -1.993898846 | 0.000106955 | 0.000200676 | 0.518947944 |
| Heart_Failure | dendritic_cell | *SLC44A3* | -2.706282814 | 1.22E-06 | 6.27E-06 | 0.00593965 |
| Heart_Failure | dendritic_cell | *STRIP1* | -4.010224368 | 0.00076882 | 0.001205271 | 1 |
| Heart_Failure | dendritic_cell | *ALDH9A1* | -1.489483832 | 3.40E-07 | 3.66E-06 | 0.001650647 |
| Heart_Failure | dendritic_cell | *ATP1B1* | 2.244879143 | 0.000247227 | 0.000427036 | 1 |
| Heart_Failure | dendritic_cell | *TIPRL* | -2.778184409 | 0.028434597 | 0.035249021 | 1 |

| | | | | | | |
|---|---|---|---|---|---|---|
| Heart_Failure | dendritic_cell | *NME7* | -3.012580055 | 8.62E-07 | 5.14E-06 | 0.004183563 |
| Heart_Failure | dendritic_cell | *XCL1* | -5.226562863 | 2.93E-05 | 6.53E-05 | 0.142144078 |
| Heart_Failure | dendritic_cell | *XCL2* | -2.291681328 | 0.004565828 | 0.006325927 | 1 |
| Heart_Failure | dendritic_cell | *RFWD2* | -2.347741344 | 0.001411924 | 0.00212226 | 1 |
| Heart_Failure | dendritic_cell | *RGS16* | -2.001760324 | 1.86E-05 | 4.53E-05 | 0.090426891 |
| Heart_Failure | dendritic_cell | *TARS2* | 4.031563973 | 4.80E-06 | 1.57E-05 | 0.023271803 |
| Heart_Failure | dendritic_cell | *SNX27* | -2.967058577 | 0.001405666 | 0.002114164 | 1 |
| Heart_Failure | dendritic_cell | *SETDB1* | -2.748165181 | 0.014703636 | 0.019022457 | 1 |
| Heart_Failure | dendritic_cell | *ARNT* | -2.684569367 | 1.44E-05 | 3.64E-05 | 0.069635072 |
| Heart_Failure | dendritic_cell | *POGZ* | -2.316823628 | 4.36E-05 | 9.20E-05 | 0.21139599 |
| Heart_Failure | dendritic_cell | *GABPB2* | -1.511971144 | 2.96E-06 | 1.12E-05 | 0.014379126 |
| Heart_Failure | dendritic_cell | *DYRK3* | -5.46332801 | 0.000305644 | 0.000517982 | 1 |
| Heart_Failure | dendritic_cell | *EIF2D* | -3.811936209 | 6.93E-05 | 0.000137385 | 0.336042536 |
| Heart_Failure | dendritic_cell | *TAF1A* | -2.041106157 | 1.56E-06 | 7.29E-06 | 0.007549286 |
| Heart_Failure | dendritic_cell | *SMYD2* | -2.062358439 | 6.77E-07 | 4.52E-06 | 0.003284943 |
| Heart_Failure | dendritic_cell | *ADAM15* | -1.249592396 | 0.013033557 | 0.016976864 | 1 |
| Heart_Failure | dendritic_cell | *RAB13* | -2.683206163 | 5.79E-07 | 4.27E-06 | 0.00281125 |
| Heart_Failure | dendritic_cell | *S100A8* | -3.957107817 | 0.002568231 | 0.003682345 | 1 |
| Heart_Failure | dendritic_cell | *CREB3L4* | -2.06952647 | 1.39E-05 | 3.53E-05 | 0.067360125 |
| Heart_Failure | dendritic_cell | *KCNN3* | -2.15475702 | 7.90E-07 | 4.87E-06 | 0.003832777 |
| Heart_Failure | dendritic_cell | *C1orf43* | -2.153851847 | 1.97E-06 | 8.43E-06 | 0.009546664 |
| Heart_Failure | dendritic_cell | *RIT1* | -2.205251775 | 1.18E-07 | 3.44E-06 | 0.000573489 |
| Heart_Failure | dendritic_cell | *FLG* | -1.262668087 | 0.000625614 | 0.000995019 | 1 |
| Heart_Failure | dendritic_cell | *SCCPDH* | -3.594135161 | 6.36E-05 | 0.000127519 | 0.308595101 |
| Heart_Failure | dendritic_cell | *ACP1* | -3.491953251 | 5.18E-05 | 0.000107262 | 0.251315494 |
| Heart_Failure | dendritic_cell | *CNIH4* | -3.578135487 | 0.000198318 | 0.000349778 | 0.962240307 |
| Heart_Failure | dendritic_cell | *PYCR2* | -2.702944378 | 0.000481049 | 0.000782713 | 1 |
| Heart_Failure | dendritic_cell | *PLEKHA6* | -1.360096722 | 1.02E-05 | 2.74E-05 | 0.049356034 |
| Heart_Failure | dendritic_cell | *OSR1* | -1.468918381 | 2.87E-05 | 6.42E-05 | 0.139255995 |
| Heart_Failure | dendritic_cell | *RHOB* | -3.950732711 | 0.000734755 | 0.001154853 | 1 |
| Heart_Failure | dendritic_cell | *GALM* | 4.144349497 | 2.43E-05 | 5.66E-05 | 0.118029999 |
| Heart_Failure | dendritic_cell | *WDPCP* | -1.266939938 | 0.001357786 | 0.002047228 | 1 |
| Heart_Failure | dendritic_cell | *VPS54* | -2.668588118 | 8.85E-05 | 0.000169427 | 0.429498269 |
| Heart_Failure | dendritic_cell | *ABHD1* | -2.329577453 | 6.99E-07 | 4.61E-06 | 0.003389785 |
| Heart_Failure | dendritic_cell | *CIAO1* | -3.695703495 | 0.000242913 | 0.000420333 | 1 |
| Heart_Failure | dendritic_cell | *ZNF514* | -1.998976735 | 1.26E-05 | 3.26E-05 | 0.061208017 |
| Heart_Failure | dendritic_cell | *EXOC6B* | -1.789279179 | 4.74E-06 | 1.56E-05 | 0.022981617 |
| Heart_Failure | dendritic_cell | *TEX261* | -2.231436686 | 3.52E-07 | 3.66E-06 | 0.001709066 |
| Heart_Failure | dendritic_cell | *THNSL2* | -1.082286839 | 0.004064754 | 0.005667294 | 1 |
| Heart_Failure | dendritic_cell | *RALB* | -3.359667688 | 1.04E-05 | 2.78E-05 | 0.050248448 |
| Heart_Failure | dendritic_cell | *RABL2A* | -2.434070905 | 3.89E-06 | 1.36E-05 | 0.018880053 |
| Heart_Failure | dendritic_cell | *FBLN7* | -1.094514676 | 0.0023359 | 0.00338423 | 1 |
| Heart_Failure | dendritic_cell | *LIPT1* | -1.51769407 | 0.00448812 | 0.006221817 | 1 |
| Heart_Failure | dendritic_cell | *CNGA3* | -23.28890512 | 0.000404049 | 0.000665235 | 1 |
| Heart_Failure | dendritic_cell | *FAHD2B* | -2.396178577 | 3.13E-07 | 3.66E-06 | 0.001519221 |
| Heart_Failure | dendritic_cell | *THSD7B* | -1.948651996 | 4.43E-07 | 3.90E-06 | 0.002151508 |
| Heart_Failure | dendritic_cell | *GPR17* | -0.950877376 | 0.000207549 | 0.000364074 | 1 |
| Heart_Failure | dendritic_cell | *POLR2D* | -2.527340304 | 0.014712356 | 0.01902568 | 1 |
| Heart_Failure | dendritic_cell | *GALNT13* | -1.563803015 | 4.57E-07 | 3.90E-06 | 0.002218497 |
| Heart_Failure | dendritic_cell | *PHOSPHO2* | 4.520357361 | 0.000138937 | 0.000252954 | 0.674122856 |

| | | | | | | |
|---|---|---|---|---|---|---|
| Heart_Failure | dendritic_cell | *METTL21A* | -3.244261024 | 0.000121608 | 0.000224801 | 0.590040995 |
| Heart_Failure | dendritic_cell | *NBEAL1* | -1.767874237 | 1.80E-05 | 4.40E-05 | 0.08741518 |
| Heart_Failure | dendritic_cell | *SUMF1* | -2.312471582 | 0.000946582 | 0.001464793 | 1 |
| Heart_Failure | dendritic_cell | *ACKR3* | -1.284219399 | 0.000131892 | 0.000241852 | 0.639939549 |
| Heart_Failure | dendritic_cell | *ESPNL* | 3.589255019 | 1.34E-07 | 3.44E-06 | 0.000651213 |
| Heart_Failure | dendritic_cell | *TAMM41* | -2.615056969 | 3.68E-06 | 1.31E-05 | 0.017842357 |
| Heart_Failure | dendritic_cell | *STK11IP* | -3.309322979 | 0.002233702 | 0.003251702 | 1 |
| Heart_Failure | dendritic_cell | *EAF1* | -2.860009279 | 9.68E-07 | 5.53E-06 | 0.004694712 |
| Heart_Failure | dendritic_cell | *CNTN4* | -1.309669756 | 5.08E-07 | 4.03E-06 | 0.002466594 |
| Heart_Failure | dendritic_cell | *SLC25A38* | -4.142999484 | 0.000529032 | 0.000854766 | 1 |
| Heart_Failure | dendritic_cell | *CTDSPL* | -1.844515241 | 1.10E-06 | 5.97E-06 | 0.005351255 |
| Heart_Failure | dendritic_cell | *PTPRG* | -1.247143459 | 1.08E-06 | 5.91E-06 | 0.005263007 |
| Heart_Failure | dendritic_cell | *SLC25A26* | -2.628646203 | 1.44E-06 | 6.94E-06 | 0.006994121 |
| Heart_Failure | dendritic_cell | *ARL6IP5* | -3.184995153 | 0.0002171 | 0.00037932 | 1 |
| Heart_Failure | dendritic_cell | *LIMD1* | -2.058476003 | 0.012132387 | 0.015875497 | 1 |
| Heart_Failure | dendritic_cell | *PLA1A* | -5.696004525 | 1.33E-06 | 6.61E-06 | 0.006455561 |
| Heart_Failure | dendritic_cell | *ADPRH* | 2.372941047 | 0.000109117 | 0.000204022 | 0.529436773 |
| Heart_Failure | dendritic_cell | *BOC* | -1.165828324 | 4.56E-07 | 3.90E-06 | 0.002213044 |
| Heart_Failure | dendritic_cell | *MED12L* | 1.396548905 | 1.52E-07 | 3.48E-06 | 0.000737017 |
| Heart_Failure | dendritic_cell | *EIF2A* | -1.645341381 | 8.15E-05 | 0.000158158 | 0.395554028 |
| Heart_Failure | dendritic_cell | *NCEH1* | 3.063905881 | 2.50E-06 | 9.97E-06 | 0.012129187 |
| Heart_Failure | dendritic_cell | *AMT* | -1.194399464 | 8.29E-05 | 0.000160435 | 0.40224687 |
| Heart_Failure | dendritic_cell | *CCDC39* | -1.753851043 | 5.33E-06 | 1.69E-05 | 0.025839603 |
| Heart_Failure | dendritic_cell | *MUC4* | -2.260180852 | 4.28E-07 | 3.90E-06 | 0.002076775 |
| Heart_Failure | dendritic_cell | *DGKQ* | 2.582622945 | 3.65E-07 | 3.66E-06 | 0.001771353 |
| Heart_Failure | dendritic_cell | *CENPC* | -2.196861348 | 1.69E-06 | 7.68E-06 | 0.008184808 |
| Heart_Failure | dendritic_cell | *SCD5* | -1.536142217 | 3.21E-07 | 3.66E-06 | 0.001559198 |
| Heart_Failure | dendritic_cell | *TRMT10A* | -4.19571355 | 3.41E-06 | 1.24E-05 | 0.016528929 |
| Heart_Failure | dendritic_cell | *PYURF* | 4.504134365 | 0.002159322 | 0.003153832 | 1 |
| Heart_Failure | dendritic_cell | *CISD2* | -2.567647283 | 3.63E-06 | 1.29E-05 | 0.01763169 |
| Heart_Failure | dendritic_cell | *DDIT4L* | -4.641219819 | 5.98E-06 | 1.85E-05 | 0.029031783 |
| Heart_Failure | dendritic_cell | *METTL14* | -3.05454014 | 3.11E-06 | 1.16E-05 | 0.01507347 |
| Heart_Failure | dendritic_cell | *USP53* | -1.754998098 | 0.003054516 | 0.004324632 | 1 |
| Heart_Failure | dendritic_cell | *MARCH1* | 2.018254103 | 6.10E-06 | 1.87E-05 | 0.029613392 |
| Heart_Failure | dendritic_cell | *CBR4* | -2.029227541 | 2.91E-06 | 1.11E-05 | 0.014096258 |
| Heart_Failure | dendritic_cell | *MARCH6* | -2.490655514 | 3.72E-05 | 8.04E-05 | 0.180277801 |
| Heart_Failure | dendritic_cell | *NKD2* | 1.860139352 | 0.031467954 | 0.0388604 | 1 |
| Heart_Failure | dendritic_cell | *MYO10* | -3.048728705 | 7.85E-05 | 0.000153133 | 0.380687987 |
| Heart_Failure | dendritic_cell | *FAM105A* | 0.634850224 | 1.23E-06 | 6.31E-06 | 0.005990447 |
| Heart_Failure | dendritic_cell | *BHMT* | -1.831138423 | 4.74E-06 | 1.56E-05 | 0.022999933 |
| Heart_Failure | dendritic_cell | *SPATA9* | -1.246885604 | 3.17E-07 | 3.66E-06 | 0.001536535 |
| Heart_Failure | dendritic_cell | *TSLP* | 1.975816381 | 1.47E-11 | 1.78E-08 | 7.12E-08 |
| Heart_Failure | dendritic_cell | *FEM1C* | 4.190192965 | 0.018928054 | 0.024054196 | 1 |
| Heart_Failure | dendritic_cell | *COMMD10* | -2.020031434 | 2.46E-05 | 5.71E-05 | 0.119369801 |
| Heart_Failure | dendritic_cell | *YIPF5* | -2.866146233 | 3.09E-07 | 3.66E-06 | 0.001501465 |
| Heart_Failure | dendritic_cell | *CXCL14* | -2.093026577 | 2.21E-06 | 9.12E-06 | 0.010722081 |
| Heart_Failure | dendritic_cell | *FBXO38* | -2.533611963 | 1.46E-05 | 3.69E-05 | 0.071040697 |
| Heart_Failure | dendritic_cell | *PCYOX1L* | 6.824331269 | 1.51E-05 | 3.78E-05 | 0.073183899 |
| Heart_Failure | dendritic_cell | *BOD1* | 6.855582646 | 7.40E-06 | 2.12E-05 | 0.035889845 |
| Heart_Failure | dendritic_cell | *FAM50B* | -4.51516486 | 6.09E-07 | 4.33E-06 | 0.002957145 |

| | | | | | | |
|---|---|---|---|---|---|---|
| Heart_Failure | dendritic_cell | *MYLK4* | 6.996291275 | 1.15E-07 | 3.44E-06 | 0.000560181 |
| Heart_Failure | dendritic_cell | *TBC1D7* | 4.287329415 | 1.73E-06 | 7.78E-06 | 0.008406069 |
| Heart_Failure | dendritic_cell | *GFOD1* | -1.901695266 | 9.48E-07 | 5.47E-06 | 0.004601298 |
| Heart_Failure | dendritic_cell | *TRIM7* | 3.223880571 | 5.36E-07 | 4.13E-06 | 0.002599925 |
| Heart_Failure | dendritic_cell | *HIGD2A* | -3.519090367 | 0.00472053 | 0.00652908 | 1 |
| Heart_Failure | dendritic_cell | *FAM193B* | -2.381660028 | 4.10E-06 | 1.41E-05 | 0.019916517 |
| Heart_Failure | dendritic_cell | *RNF44* | 4.134842241 | 1.10E-07 | 3.44E-06 | 0.000533314 |
| Heart_Failure | dendritic_cell | *MUT* | -3.192769898 | 0.000352321 | 0.000587848 | 1 |
| Heart_Failure | dendritic_cell | *ABT1* | -1.200416741 | 0.008721773 | 0.011625835 | 1 |
| Heart_Failure | dendritic_cell | *DAAM2* | -2.195728753 | 7.64E-07 | 4.80E-06 | 0.003705453 |
| Heart_Failure | dendritic_cell | *MLIP* | -3.003106609 | 1.91E-06 | 8.28E-06 | 0.009269715 |
| Heart_Failure | dendritic_cell | *ANO7* | -1.660846553 | 0.002421742 | 0.00349503 | 1 |
| Heart_Failure | dendritic_cell | *RPL7L1* | -3.037265857 | 7.37E-06 | 2.12E-05 | 0.035765482 |
| Heart_Failure | dendritic_cell | *TPBG* | -1.500109813 | 4.66E-07 | 3.93E-06 | 0.00226225 |
| Heart_Failure | dendritic_cell | *IRAK1BP1* | -2.212967407 | 3.02E-05 | 6.71E-05 | 0.146633223 |
| Heart_Failure | dendritic_cell | *PHIP* | -2.420577934 | 0.000179204 | 0.00031838 | 0.869495852 |
| Heart_Failure | dendritic_cell | *PNRC1* | -3.052586459 | 7.16E-06 | 2.08E-05 | 0.034736317 |
| Heart_Failure | dendritic_cell | *RNF217* | -2.054278287 | 4.75E-06 | 1.56E-05 | 0.023036627 |
| Heart_Failure | dendritic_cell | *ABRACL* | 1.176983776 | 3.64E-06 | 1.30E-05 | 0.01765977 |
| Heart_Failure | dendritic_cell | *SLC18B1* | -2.979759005 | 1.13E-05 | 2.98E-05 | 0.054782208 |
| Heart_Failure | dendritic_cell | *WTAP* | -2.245868555 | 4.03E-05 | 8.62E-05 | 0.1955229 |
| Heart_Failure | dendritic_cell | *SLC22A3* | -1.285180525 | 3.20E-07 | 3.66E-06 | 0.001551573 |
| Heart_Failure | dendritic_cell | *GNA12* | -2.397488407 | 2.92E-05 | 6.52E-05 | 0.141808104 |
| Heart_Failure | dendritic_cell | *SDK1* | -1.553405377 | 2.64E-05 | 6.02E-05 | 0.128328886 |
| Heart_Failure | dendritic_cell | *RBAK* | -4.889007716 | 3.74E-06 | 1.32E-05 | 0.018161839 |
| Heart_Failure | dendritic_cell | *POMZP3* | -1.862445425 | 0.017345742 | 0.022188648 | 1 |
| Heart_Failure | dendritic_cell | *PSPH* | -1.43246693 | 0.000349362 | 0.000583721 | 1 |
| Heart_Failure | dendritic_cell | *GIGYF1* | -2.222101014 | 0.040034082 | 0.048842184 | 1 |
| Heart_Failure | dendritic_cell | *TMEM209* | -3.152392723 | 4.78E-06 | 1.57E-05 | 0.023198948 |
| Heart_Failure | dendritic_cell | *EPHA1* | -4.805256655 | 3.53E-07 | 3.66E-06 | 0.001711136 |
| Heart_Failure | dendritic_cell | *FBXO25* | -3.403822408 | 0.000514879 | 0.000834957 | 1 |
| Heart_Failure | dendritic_cell | *CSGALNACT1* | -1.06437625 | 4.25E-05 | 9.02E-05 | 0.206293949 |
| Heart_Failure | dendritic_cell | *BIN3* | -3.375445812 | 0.000388573 | 0.000641932 | 1 |
| Heart_Failure | dendritic_cell | *CHMP7* | -4.047608609 | 9.63E-06 | 2.62E-05 | 0.046704241 |
| Heart_Failure | dendritic_cell | *TACC1* | -2.009611189 | 1.40E-05 | 3.56E-05 | 0.06796903 |
| Heart_Failure | dendritic_cell | *ADHFE1* | -1.076765565 | 0.001091942 | 0.001672908 | 1 |
| Heart_Failure | dendritic_cell | *RPL7* | 2.261632746 | 0.025374232 | 0.031649299 | 1 |
| Heart_Failure | dendritic_cell | *ATP6V0D2* | 4.845152107 | 3.16E-07 | 3.66E-06 | 0.001531595 |
| Heart_Failure | dendritic_cell | *DPYS* | -1.099071675 | 3.24E-07 | 3.66E-06 | 0.001573507 |
| Heart_Failure | dendritic_cell | *LRP12* | -3.282751401 | 4.74E-06 | 1.56E-05 | 0.023011338 |
| Heart_Failure | dendritic_cell | *MAL2* | -1.011882922 | 6.83E-05 | 0.000135773 | 0.331421131 |
| Heart_Failure | dendritic_cell | *AK3* | -2.664184513 | 3.83E-06 | 1.34E-05 | 0.018568157 |
| Heart_Failure | dendritic_cell | *ZNF462* | -2.194566847 | 4.03E-06 | 1.39E-05 | 0.019532039 |
| Heart_Failure | dendritic_cell | *ALAD* | 3.081195917 | 1.70E-07 | 3.52E-06 | 0.000826084 |
| Heart_Failure | dendritic_cell | *WDR31* | -1.379210423 | 1.69E-06 | 7.68E-06 | 0.008191063 |
| Heart_Failure | dendritic_cell | *POLE3* | -1.604722455 | 5.47E-05 | 0.000112328 | 0.265432143 |
| Heart_Failure | dendritic_cell | *SURF1* | -1.513640274 | 0.020457873 | 0.025883077 | 1 |
| Heart_Failure | dendritic_cell | *MED22* | -2.148186839 | 1.58E-06 | 7.39E-06 | 0.007690123 |
| Heart_Failure | dendritic_cell | *REXO4* | -4.247877136 | 0.000396101 | 0.000653256 | 1 |
| Heart_Failure | dendritic_cell | *SH3GLB2* | -1.688096943 | 4.24E-07 | 3.90E-06 | 0.002057722 |

| | | | | | | |
|---|---|---|---|---|---|---|
| Heart_Failure | dendritic_cell | *INPP5E* | -5.127259975 | 6.27E-06 | 1.91E-05 | 0.030407179 |
| Heart_Failure | dendritic_cell | *SEC16A* | -2.80606653 | 5.47E-05 | 0.000112328 | 0.26539455 |
| Heart_Failure | dendritic_cell | *SLC39A12* | -3.756711395 | 1.25E-05 | 3.24E-05 | 0.060634371 |
| Heart_Failure | dendritic_cell | *TMEM236* | 1.163011823 | 2.10E-06 | 8.81E-06 | 0.010171313 |
| Heart_Failure | dendritic_cell | *RSU1* | -2.336964357 | 0.017629319 | 0.022515782 | 1 |
| Heart_Failure | dendritic_cell | *ST8SIA6* | -1.896130204 | 3.27E-07 | 3.66E-06 | 0.001586961 |
| Heart_Failure | dendritic_cell | *PARD3* | -1.444048627 | 1.28E-06 | 6.42E-06 | 0.006186966 |
| Heart_Failure | dendritic_cell | *CDHR1* | 3.232997461 | 0.007603044 | 0.010218827 | 1 |
| Heart_Failure | dendritic_cell | *RGR* | 3.427636123 | 9.09E-08 | 3.44E-06 | 0.000441029 |
| Heart_Failure | dendritic_cell | *FRA10AC1* | -2.141362219 | 5.56E-05 | 0.000113879 | 0.269778558 |
| Heart_Failure | dendritic_cell | *ADD3* | -2.229272955 | 1.25E-05 | 3.23E-05 | 0.060409615 |
| Heart_Failure | dendritic_cell | *LRRC27* | -1.985241045 | 8.44E-06 | 2.36E-05 | 0.040956237 |
| Heart_Failure | dendritic_cell | *MTG1* | -1.161200099 | 0.011647589 | 0.015269954 | 1 |
| Heart_Failure | dendritic_cell | *PAOX* | -4.116010071 | 3.55E-06 | 1.28E-05 | 0.017204053 |
| Heart_Failure | dendritic_cell | *PDCD11* | -3.268830417 | 2.85E-05 | 6.40E-05 | 0.138481864 |
| Heart_Failure | dendritic_cell | *RGS10* | 1.207274417 | 1.85E-07 | 3.58E-06 | 0.000899771 |
| Heart_Failure | dendritic_cell | *GAS2* | -2.089105628 | 1.02E-06 | 5.71E-06 | 0.00495412 |
| Heart_Failure | dendritic_cell | *LIN7C* | -4.138212709 | 0.000666024 | 0.001055372 | 1 |
| Heart_Failure | dendritic_cell | *HSD17B12* | -1.432393883 | 2.85E-07 | 3.66E-06 | 0.001383091 |
| Heart_Failure | dendritic_cell | *EIF3M* | -2.945743507 | 7.43E-05 | 0.000145782 | 0.360663947 |
| Heart_Failure | dendritic_cell | *TNKS1BP1* | -1.589624871 | 3.16E-05 | 6.98E-05 | 0.153516084 |
| Heart_Failure | dendritic_cell | *PTPRJ* | 2.491479939 | 6.02E-07 | 4.30E-06 | 0.002918501 |
| Heart_Failure | dendritic_cell | *ENDOD1* | -2.503468553 | 1.19E-06 | 6.17E-06 | 0.005776224 |
| Heart_Failure | dendritic_cell | *CCDC82* | -1.260823334 | 0.002750545 | 0.003926344 | 1 |
| Heart_Failure | dendritic_cell | *CAPN5* | -2.924364749 | 4.43E-07 | 3.90E-06 | 0.002148697 |
| Heart_Failure | dendritic_cell | *INTS4* | -2.605812696 | 4.67E-06 | 1.54E-05 | 0.022675352 |
| Heart_Failure | dendritic_cell | *RPS3* | -2.425725785 | 0.000105962 | 0.000199043 | 0.514129299 |
| Heart_Failure | dendritic_cell | *TTC12* | -1.459864806 | 0.029233986 | 0.036193749 | 1 |
| Heart_Failure | dendritic_cell | *DRD2* | -15.02840249 | 3.31E-07 | 3.66E-06 | 0.001604059 |
| Heart_Failure | dendritic_cell | *NPAT* | -3.420354625 | 8.57E-05 | 0.000164923 | 0.415605229 |
| Heart_Failure | dendritic_cell | *ST14* | 1.448974674 | 2.22E-07 | 3.66E-06 | 0.001079352 |
| Heart_Failure | dendritic_cell | *FADS1* | -2.251445421 | 0.028287802 | 0.035084974 | 1 |
| Heart_Failure | dendritic_cell | *TMC2* | -3.24673659 | 4.17E-07 | 3.88E-06 | 0.002022602 |
| Heart_Failure | dendritic_cell | *ROM1* | 1.306942562 | 0.01597032 | 0.020559297 | 1 |
| Heart_Failure | dendritic_cell | *PLCH2* | -5.688362481 | 0.00118202 | 0.001802376 | 1 |
| Heart_Failure | dendritic_cell | *CPSF7* | -2.752478031 | 5.25E-05 | 0.000108496 | 0.254792839 |
| Heart_Failure | dendritic_cell | *CCDC15* | -1.91413001 | 0.008463936 | 0.011306998 | 1 |
| Heart_Failure | dendritic_cell | *CHEK1* | -3.377278095 | 3.35E-07 | 3.66E-06 | 0.001626415 |
| Heart_Failure | dendritic_cell | *DUSP15* | -3.272322525 | 0.021632974 | 0.027277336 | 1 |
| Heart_Failure | dendritic_cell | *CABLES2* | -2.404930937 | 1.48E-06 | 7.06E-06 | 0.007190873 |
| Heart_Failure | dendritic_cell | *TRPT1* | -2.144829986 | 1.64E-08 | 2.99E-06 | 7.95E-05 |
| Heart_Failure | dendritic_cell | *NUDT22* | -3.033139788 | 3.91E-06 | 1.36E-05 | 0.018966852 |
| Heart_Failure | dendritic_cell | *ALDOA* | -2.017814641 | 4.88E-05 | 0.000101414 | 0.237004533 |
| Heart_Failure | dendritic_cell | *ANXA8L1* | -0.852947122 | 1.37E-05 | 3.49E-05 | 0.066431964 |
| Heart_Failure | dendritic_cell | *CWC15* | -2.380768269 | 3.19E-06 | 1.18E-05 | 0.015472727 |
| Heart_Failure | dendritic_cell | *TMEM218* | -3.137851198 | 6.21E-05 | 0.000124975 | 0.301315018 |
| Heart_Failure | dendritic_cell | *SAP18* | -2.618898611 | 1.23E-06 | 6.31E-06 | 0.005991724 |
| Heart_Failure | dendritic_cell | *FAM124A* | -2.071524451 | 7.16E-07 | 4.65E-06 | 0.003474876 |
| Heart_Failure | dendritic_cell | *LYPD6B* | -2.132625074 | 0.000400747 | 0.000660245 | 1 |
| Heart_Failure | dendritic_cell | *WDR17* | -1.791951156 | 6.20E-07 | 4.36E-06 | 0.003009173 |

| | | | | | | |
|---|---|---|---|---|---|---|
| Heart_Failure | dendritic_cell | *CCDC102B* | -1.759523088 | 7.68E-06 | 2.18E-05 | 0.037248147 |
| Heart_Failure | dendritic_cell | *RGS18* | 0.541461952 | 1.34E-07 | 3.44E-06 | 0.000650451 |
| Heart_Failure | dendritic_cell | *PRSS23* | -1.693655326 | 5.12E-06 | 1.64E-05 | 0.024854932 |
| Heart_Failure | dendritic_cell | *PPP1R1C* | -3.70310251 | 5.00E-07 | 4.02E-06 | 0.002423622 |
| Heart_Failure | dendritic_cell | *GPR158* | 4.783920082 | 3.13E-05 | 6.92E-05 | 0.152016144 |
| Heart_Failure | dendritic_cell | *ANK3* | -1.784653746 | 9.05E-07 | 5.32E-06 | 0.004388901 |
| Heart_Failure | dendritic_cell | *PLBD2* | -3.536772628 | 0.001774395 | 0.002627209 | 1 |
| Heart_Failure | dendritic_cell | *DLG5* | -1.656290478 | 6.31E-06 | 1.92E-05 | 0.030627516 |
| Heart_Failure | dendritic_cell | *GXYLT1* | -2.990671011 | 5.80E-06 | 1.80E-05 | 0.028134457 |
| Heart_Failure | dendritic_cell | *EXT2* | -2.516333841 | 0.000444922 | 0.000727345 | 1 |
| Heart_Failure | dendritic_cell | *TMEM18* | -3.78830355 | 3.81E-06 | 1.34E-05 | 0.018488677 |
| Heart_Failure | dendritic_cell | *NDUFC2* | -2.222173788 | 5.55E-07 | 4.19E-06 | 0.002694215 |
| Heart_Failure | dendritic_cell | *ADAMTS12* | -1.551419479 | 2.93E-07 | 3.66E-06 | 0.001421051 |
| Heart_Failure | dendritic_cell | *NUBPL* | -2.633268444 | 4.26E-05 | 9.03E-05 | 0.206512439 |
| Heart_Failure | dendritic_cell | *VIPAS39* | -3.331327188 | 5.74E-06 | 1.79E-05 | 0.027840415 |
| Heart_Failure | dendritic_cell | *ANKRD50* | -2.485833168 | 1.43E-05 | 3.62E-05 | 0.069206843 |
| Heart_Failure | dendritic_cell | *UPF2* | -2.680336745 | 0.00060048 | 0.000957452 | 1 |
| Heart_Failure | dendritic_cell | *PTPRO* | 1.702221777 | 6.12E-06 | 1.88E-05 | 0.029698906 |
| Heart_Failure | dendritic_cell | *EPS8* | -2.320021167 | 0.000169901 | 0.000303296 | 0.824357354 |
| Heart_Failure | dendritic_cell | *QDPR* | -2.800677383 | 0.000447077 | 0.000729885 | 1 |
| Heart_Failure | dendritic_cell | *FAM160B1* | -2.387013293 | 3.91E-06 | 1.36E-05 | 0.018960738 |
| Heart_Failure | dendritic_cell | *ZNF827* | -1.483704776 | 6.06E-07 | 4.32E-06 | 0.002939912 |
| Heart_Failure | dendritic_cell | *AKR1C2* | -1.530534702 | 6.12E-07 | 4.34E-06 | 0.002967541 |
| Heart_Failure | dendritic_cell | *DPYSL4* | 0.723145687 | 0.023486449 | 0.029499417 | 1 |
| Heart_Failure | dendritic_cell | *PIGF* | -2.633349121 | 4.01E-06 | 1.38E-05 | 0.019436782 |
| Heart_Failure | dendritic_cell | *INPP1* | -2.402064329 | 5.04E-07 | 4.03E-06 | 0.002445612 |
| Heart_Failure | dendritic_cell | *MFSD6* | -2.32658173 | 1.95E-06 | 8.38E-06 | 0.009455197 |
| Heart_Failure | dendritic_cell | *RNF144A* | -2.088516753 | 1.85E-06 | 8.10E-06 | 0.008956733 |
| Heart_Failure | dendritic_cell | *CENPU* | -2.267588156 | 2.30E-05 | 5.41E-05 | 0.111721343 |
| Heart_Failure | dendritic_cell | *BICD1* | -2.444506008 | 6.28E-06 | 1.91E-05 | 0.030457827 |
| Heart_Failure | dendritic_cell | *CCDC122* | -1.06400059 | 0.0023639 | 0.003421731 | 1 |
| Heart_Failure | dendritic_cell | *GUF1* | -3.573213584 | 9.45E-05 | 0.000179455 | 0.458508474 |
| Heart_Failure | dendritic_cell | *GABRA2* | -5.074006693 | 4.23E-07 | 3.90E-06 | 0.002052516 |
| Heart_Failure | dendritic_cell | *BEND6* | -2.10083986 | 0.001049066 | 0.001611801 | 1 |
| Heart_Failure | dendritic_cell | *TIAL1* | -2.568184375 | 6.38E-06 | 1.93E-05 | 0.030949383 |
| Heart_Failure | dendritic_cell | *TMEM56* | -3.263623942 | 2.61E-07 | 3.66E-06 | 0.001264874 |
| Heart_Failure | dendritic_cell | *PTPN14* | -1.625757162 | 1.94E-06 | 8.35E-06 | 0.009393162 |
| Heart_Failure | dendritic_cell | *MGAT5* | -2.518308829 | 4.56E-05 | 9.54E-05 | 0.221068507 |
| Heart_Failure | dendritic_cell | *TMEM178A* | -3.04861515 | 3.24E-07 | 3.66E-06 | 0.001573825 |
| Heart_Failure | dendritic_cell | *EPG5* | -2.941005982 | 3.48E-05 | 7.62E-05 | 0.168988152 |
| Heart_Failure | dendritic_cell | *SPC25* | -1.463620354 | 4.56E-06 | 1.52E-05 | 0.022145359 |
| Heart_Failure | dendritic_cell | *KCNK13* | 0.423286839 | 1.29E-07 | 3.44E-06 | 0.000625009 |
| Heart_Failure | dendritic_cell | *POC5* | -2.582808982 | 4.98E-06 | 1.62E-05 | 0.024181178 |
| Heart_Failure | dendritic_cell | *CWF19L2* | -2.260095408 | 5.41E-05 | 0.000111254 | 0.262336644 |
| Heart_Failure | dendritic_cell | *ZNF773* | -1.581576618 | 1.27E-05 | 3.28E-05 | 0.061696152 |
| Heart_Failure | dendritic_cell | *NMT2* | -2.085298806 | 4.14E-05 | 8.82E-05 | 0.200839976 |
| Heart_Failure | dendritic_cell | *ZNF837* | -2.661166669 | 0.000233499 | 0.000404765 | 1 |
| Heart_Failure | dendritic_cell | *CCDC50* | -1.819390627 | 0.000590383 | 0.000943524 | 1 |
| Heart_Failure | dendritic_cell | *PLEKHH2* | -1.178089558 | 4.42E-06 | 1.48E-05 | 0.021426719 |
| Heart_Failure | dendritic_cell | *SPARCL1* | -2.128852647 | 4.30E-05 | 9.11E-05 | 0.208692662 |

| | | | | | | |
|---|---|---|---|---|---|---|
| Heart_Failure | dendritic_cell | *GPD1L* | -2.515352461 | 0.000178713 | 0.000317624 | 0.86711314 |
| Heart_Failure | dendritic_cell | *CCNO* | -17.59937799 | 1.14E-06 | 6.05E-06 | 0.00552974 |
| Heart_Failure | dendritic_cell | *CLEC4F* | 0.541282982 | 1.07E-07 | 3.44E-06 | 0.000518143 |
| Heart_Failure | dendritic_cell | *SLC30A6* | -2.354809867 | 2.33E-05 | 5.47E-05 | 0.113213539 |
| Heart_Failure | dendritic_cell | *RASGRP3* | -1.692761528 | 1.06E-06 | 5.82E-06 | 0.0051423 |
| Heart_Failure | dendritic_cell | *GPR180* | -1.622437958 | 1.52E-06 | 7.19E-06 | 0.007391635 |
| Heart_Failure | dendritic_cell | *TCTEX1D1* | -2.751719458 | 0.00409372 | 0.005703654 | 1 |
| Heart_Failure | dendritic_cell | *PRDM8* | -2.975015121 | 3.76E-07 | 3.66E-06 | 0.001823282 |
| Heart_Failure | dendritic_cell | *GGPS1* | -2.832740695 | 0.000294918 | 0.000501556 | 1 |
| Heart_Failure | dendritic_cell | *JAKMIP1* | 8.344041063 | 2.38E-05 | 5.56E-05 | 0.115363806 |
| Heart_Failure | dendritic_cell | *SREK1IP1* | -3.319378762 | 9.24E-05 | 0.000176054 | 0.448410501 |
| Heart_Failure | dendritic_cell | *MR1* | 3.33919881 | 0.003695914 | 0.005179831 | 1 |
| Heart_Failure | dendritic_cell | *CENPH* | -2.728144102 | 2.66E-07 | 3.66E-06 | 0.001291699 |
| Heart_Failure | dendritic_cell | *CARHSP1* | -2.521218631 | 4.24E-06 | 1.44E-05 | 0.020562619 |
| Heart_Failure | dendritic_cell | *CAST* | -2.058438196 | 0.000123329 | 0.000227439 | 0.598391541 |
| Heart_Failure | dendritic_cell | *SCOC* | -3.542844967 | 0.005043789 | 0.006952405 | 1 |
| Heart_Failure | dendritic_cell | *CLGN* | -4.620612804 | 0.000542494 | 0.000873609 | 1 |
| Heart_Failure | dendritic_cell | *CETN3* | -2.181266583 | 5.74E-07 | 4.27E-06 | 0.002785896 |
| Heart_Failure | dendritic_cell | *MERTK* | -0.340937395 | 0.028046665 | 0.034803688 | 1 |
| Heart_Failure | dendritic_cell | *TMEM87B* | -3.272900442 | 0.000245243 | 0.000423889 | 1 |
| Heart_Failure | dendritic_cell | *PLA2R1* | -1.324024585 | 4.59E-06 | 1.52E-05 | 0.022284228 |
| Heart_Failure | dendritic_cell | *SLC25A27* | -1.594458533 | 1.69E-06 | 7.68E-06 | 0.008186036 |
| Heart_Failure | dendritic_cell | *FRMD1* | -2.489038848 | 5.15E-06 | 1.65E-05 | 0.024976136 |
| Heart_Failure | dendritic_cell | *INO80C* | -2.293191994 | 1.41E-06 | 6.81E-06 | 0.00682784 |
| Heart_Failure | dendritic_cell | *LPCAT1* | -4.11649371 | 0.016288959 | 0.020947265 | 1 |
| Heart_Failure | dendritic_cell | *NMRAL1* | 1.879010203 | 0.018657342 | 0.023728813 | 1 |
| Heart_Failure | dendritic_cell | *ADPRHL1* | -3.494831504 | 3.00E-07 | 3.66E-06 | 0.001455566 |
| Heart_Failure | dendritic_cell | *CFDP1* | -2.566352127 | 3.14E-05 | 6.94E-05 | 0.15254191 |
| Heart_Failure | dendritic_cell | *ZDHHC7* | -3.145277808 | 2.08E-05 | 4.98E-05 | 0.100917686 |
| Heart_Failure | dendritic_cell | *JAZF1* | -2.569134435 | 1.47E-05 | 3.69E-05 | 0.071187354 |
| Heart_Failure | dendritic_cell | *FBXO36* | -3.436655718 | 3.59E-06 | 1.28E-05 | 0.017425316 |
| Heart_Failure | dendritic_cell | *MCOLN2* | 0.576590389 | 4.30E-07 | 3.90E-06 | 0.002085547 |
| Heart_Failure | dendritic_cell | *GDPD1* | 4.765314258 | 1.79E-05 | 4.37E-05 | 0.086774863 |
| Heart_Failure | dendritic_cell | *SEMA3D* | -1.01444683 | 3.26E-07 | 3.66E-06 | 0.001583232 |
| Heart_Failure | dendritic_cell | *PPP2R5E* | -2.804875646 | 0.000353053 | 0.000588462 | 1 |
| Heart_Failure | dendritic_cell | *ANKRD29* | -1.464884068 | 3.87E-07 | 3.72E-06 | 0.001877617 |
| Heart_Failure | dendritic_cell | *DNAAF1* | -3.979580198 | 5.40E-06 | 1.71E-05 | 0.026214407 |
| Heart_Failure | dendritic_cell | *UBASH3B* | 1.438044324 | 5.49E-05 | 0.000112562 | 0.266321879 |
| Heart_Failure | dendritic_cell | *ROBO4* | -2.468348597 | 1.40E-06 | 6.80E-06 | 0.006793267 |
| Heart_Failure | dendritic_cell | *ROBO3* | -1.738850499 | 1.02E-06 | 5.71E-06 | 0.004930749 |
| Heart_Failure | dendritic_cell | *CC2D1B* | -3.119722178 | 2.69E-05 | 6.11E-05 | 0.130681683 |
| Heart_Failure | dendritic_cell | *LRRK1* | 2.631262935 | 2.27E-07 | 3.66E-06 | 0.001101155 |
| Heart_Failure | dendritic_cell | *ABCA5* | -1.711894782 | 8.40E-05 | 0.000162224 | 0.4073434 |
| Heart_Failure | dendritic_cell | *ENPP3* | -2.041602012 | 1.33E-06 | 6.61E-06 | 0.006467502 |
| Heart_Failure | dendritic_cell | *MIA3* | -3.058563307 | 0.002395246 | 0.003459411 | 1 |
| Heart_Failure | dendritic_cell | *NEIL2* | -1.582296731 | 6.09E-05 | 0.000122961 | 0.295474641 |
| Heart_Failure | dendritic_cell | *PGM5* | -1.506376436 | 6.76E-07 | 4.52E-06 | 0.00328031 |
| Heart_Failure | dendritic_cell | *CCSAP* | -4.27268121 | 0.036703666 | 0.044971259 | 1 |
| Heart_Failure | dendritic_cell | *GBP5* | -1.384327671 | 6.40E-05 | 0.000128137 | 0.310475234 |
| Heart_Failure | dendritic_cell | *LY96* | 2.182530135 | 0.002417091 | 0.003489357 | 1 |

| | | | | | | |
|---|---|---|---|---|---|---|
| Heart_Failure | dendritic_cell | *RABGEF1* | -1.029409962 | 0.004547892 | 0.006302878 | 1 |
| Heart_Failure | dendritic_cell | *MRPL39* | -1.345568272 | 6.85E-05 | 0.000136035 | 0.332196665 |
| Heart_Failure | dendritic_cell | *XPC* | -3.065760287 | 0.000294061 | 0.000500275 | 1 |
| Heart_Failure | dendritic_cell | *FLCN* | -2.124405116 | 0.00084427 | 0.001314211 | 1 |
| Heart_Failure | dendritic_cell | *OXNAD1* | -2.162942429 | 2.16E-06 | 8.95E-06 | 0.010475011 |
| Heart_Failure | dendritic_cell | *APCDD1* | -1.560690615 | 9.52E-07 | 5.47E-06 | 0.004619884 |
| Heart_Failure | dendritic_cell | *RAB6B* | -2.123526416 | 1.81E-05 | 4.41E-05 | 0.087689772 |
| Heart_Failure | dendritic_cell | *ANKRD40* | 4.058709674 | 0.003657725 | 0.005130755 | 1 |
| Heart_Failure | dendritic_cell | *CYP2U1* | -2.467152046 | 0.000228911 | 0.000397665 | 1 |
| Heart_Failure | dendritic_cell | *AK9* | -2.255388979 | 3.46E-07 | 3.66E-06 | 0.001677694 |
| Heart_Failure | dendritic_cell | *MARCKS* | -3.15510897 | 0.00027996 | 0.00047863 | 1 |
| Heart_Failure | dendritic_cell | *MARVELD1* | -1.964370175 | 4.20E-07 | 3.88E-06 | 0.002036483 |
| Heart_Failure | dendritic_cell | *GOLGA7B* | -10.10581001 | 3.63E-07 | 3.66E-06 | 0.001760399 |
| Heart_Failure | dendritic_cell | *TRMT44* | -1.975669931 | 4.78E-07 | 3.96E-06 | 0.002318836 |
| Heart_Failure | dendritic_cell | *MOV10* | -4.604733358 | 0.000446008 | 0.000728493 | 1 |
| Heart_Failure | dendritic_cell | *LRGUK* | -1.204159797 | 0.004294068 | 0.005966442 | 1 |
| Heart_Failure | dendritic_cell | *NUP205* | -2.954049247 | 3.33E-06 | 1.22E-05 | 0.016151132 |
| Heart_Failure | dendritic_cell | *ZKSCAN2* | -2.17879657 | 3.78E-05 | 8.17E-05 | 0.183558784 |
| Heart_Failure | dendritic_cell | *PDIA4* | -3.376832753 | 2.37E-06 | 9.56E-06 | 0.011510531 |
| Heart_Failure | dendritic_cell | *ALS2CR12* | -2.271055354 | 3.68E-07 | 3.66E-06 | 0.001785837 |
| Heart_Failure | dendritic_cell | *FZD7* | -1.387856833 | 0.000458506 | 0.000747789 | 1 |
| Heart_Failure | dendritic_cell | *DEPTOR* | -1.872810256 | 0.001773333 | 0.002626992 | 1 |
| Heart_Failure | dendritic_cell | *RRAGA* | -1.968277501 | 2.05E-07 | 3.66E-06 | 0.000992363 |
| Heart_Failure | dendritic_cell | *TMBIM4* | -1.978088597 | 0.005053245 | 0.00696346 | 1 |
| Heart_Failure | dendritic_cell | *PSD3* | -1.472444881 | 0.019039274 | 0.024189201 | 1 |
| Heart_Failure | dendritic_cell | *MMP16* | -1.194075034 | 0.000205018 | 0.000360415 | 0.994745022 |
| Heart_Failure | dendritic_cell | *NDUFAF6* | -2.781379747 | 1.47E-06 | 7.03E-06 | 0.007136428 |
| Heart_Failure | dendritic_cell | *C8orf37* | 1.962094365 | 0.001341137 | 0.002026533 | 1 |
| Heart_Failure | dendritic_cell | *PPEF2* | -2.786625809 | 0.012406252 | 0.016203266 | 1 |
| Heart_Failure | dendritic_cell | *WHAMM* | -2.754945475 | 2.77E-07 | 3.66E-06 | 0.001342143 |
| Heart_Failure | dendritic_cell | *N6AMT1* | -1.882832771 | 1.70E-07 | 3.52E-06 | 0.000823434 |
| Heart_Failure | dendritic_cell | *CCT8* | -1.168984407 | 0.014204637 | 0.018418198 | 1 |
| Heart_Failure | dendritic_cell | *NAA11* | -2.807866085 | 2.64E-06 | 1.03E-05 | 0.012810087 |
| Heart_Failure | dendritic_cell | *SFXN2* | -0.892469102 | 0.005943695 | 0.008108905 | 1 |
| Heart_Failure | dendritic_cell | *C14orf2* | -2.32286155 | 0.003084971 | 0.00436393 | 1 |
| Heart_Failure | dendritic_cell | *TDRD9* | -4.335878034 | 1.25E-06 | 6.37E-06 | 0.006073032 |
| Heart_Failure | dendritic_cell | *RPL30* | -2.139772452 | 5.83E-05 | 0.000118247 | 0.282727815 |
| Heart_Failure | dendritic_cell | *SUPV3L1* | -3.441467978 | 0.009917431 | 0.013125853 | 1 |
| Heart_Failure | dendritic_cell | *HKDC1* | -9.169966561 | 4.98E-06 | 1.62E-05 | 0.024151499 |
| Heart_Failure | dendritic_cell | *TYSND1* | -1.241443184 | 1.25E-06 | 6.37E-06 | 0.006061301 |
| Heart_Failure | dendritic_cell | *NODAL* | -4.081158412 | 6.84E-07 | 4.53E-06 | 0.003317677 |
| Heart_Failure | dendritic_cell | *ZDHHC5* | -3.815255586 | 0.002447661 | 0.003529287 | 1 |
| Heart_Failure | dendritic_cell | *WDYHV1* | -0.955964234 | 0.016906009 | 0.021672114 | 1 |
| Heart_Failure | dendritic_cell | *ATAD2* | -1.743116362 | 0.02591248 | 0.032279167 | 1 |
| Heart_Failure | dendritic_cell | *FBRS* | 3.817296476 | 3.20E-06 | 1.18E-05 | 0.015516994 |
| Heart_Failure | dendritic_cell | *SASS6* | -3.160298795 | 8.31E-06 | 2.33E-05 | 0.040301011 |
| Heart_Failure | dendritic_cell | *ITGAD* | -2.179887912 | 0.000141755 | 0.000257601 | 0.687794598 |
| Heart_Failure | dendritic_cell | *VPS8* | -2.062169479 | 0.009028639 | 0.012021667 | 1 |
| Heart_Failure | dendritic_cell | *MPV17L* | -3.08700121 | 1.73E-06 | 7.77E-06 | 0.008383057 |
| Heart_Failure | dendritic_cell | *NTAN1* | -2.162964818 | 1.37E-06 | 6.71E-06 | 0.006659617 |

| | | | | | | |
|---|---|---|---|---|---|---|
| Heart_Failure | dendritic_cell | *NMNAT2* | -3.947058781 | 0.000189115 | 0.000334885 | 0.917583834 |
| Heart_Failure | dendritic_cell | *SLC6A1* | -5.853866364 | 3.02E-07 | 3.66E-06 | 0.001467006 |
| Heart_Failure | dendritic_cell | *FCHO2* | -1.890980497 | 0.007930663 | 0.010641476 | 1 |
| Heart_Failure | dendritic_cell | *TIMP4* | -1.559880923 | 0.001561979 | 0.002336948 | 1 |
| Heart_Failure | dendritic_cell | *NECAP2* | 4.13687641 | 3.60E-06 | 1.29E-05 | 0.017452509 |
| Heart_Failure | dendritic_cell | *STEAP2* | -1.107148944 | 4.28E-06 | 1.45E-05 | 0.020745711 |
| Heart_Failure | dendritic_cell | *CLDN12* | -1.824584892 | 3.23E-07 | 3.66E-06 | 0.001565048 |
| Heart_Failure | dendritic_cell | *MMP14* | -3.112096754 | 1.14E-05 | 3.01E-05 | 0.055274333 |
| Heart_Failure | dendritic_cell | *GATAD1* | -2.647134558 | 5.57E-06 | 1.75E-05 | 0.027002299 |
| Heart_Failure | dendritic_cell | *TMED6* | 3.904309367 | 8.11E-07 | 4.93E-06 | 0.003936024 |
| Heart_Failure | dendritic_cell | *DHRS4* | 6.215292392 | 0.000946682 | 0.001464793 | 1 |
| Heart_Failure | dendritic_cell | *DHRS1* | -1.749620132 | 0.002508114 | 0.003604671 | 1 |
| Heart_Failure | dendritic_cell | *AASDH* | -2.510991488 | 2.75E-06 | 1.07E-05 | 0.013335919 |
| Heart_Failure | dendritic_cell | *ETS2* | 0.909801439 | 0.00292811 | 0.004157796 | 1 |
| Heart_Failure | dendritic_cell | *MX1* | -1.801819437 | 0.00034775 | 0.000582224 | 1 |
| Heart_Failure | dendritic_cell | *SLC38A10* | -2.597410953 | 3.78E-06 | 1.33E-05 | 0.018326267 |
| Heart_Failure | dendritic_cell | *C9orf43* | -1.196276368 | 0.00587452 | 0.008026802 | 1 |
| Heart_Failure | dendritic_cell | *DGKI* | -1.430711133 | 3.94E-05 | 8.46E-05 | 0.191128138 |
| Heart_Failure | dendritic_cell | *SNX22* | 2.831308569 | 3.46E-07 | 3.66E-06 | 0.001680931 |
| Heart_Failure | dendritic_cell | *SLC34A2* | -33.86794884 | 6.52E-06 | 1.95E-05 | 0.031628566 |
| Heart_Failure | dendritic_cell | *AP3S2* | -2.859539989 | 3.90E-06 | 1.36E-05 | 0.018934622 |
| Heart_Failure | dendritic_cell | *SPPL3* | -2.31016021 | 0.001043694 | 0.001604564 | 1 |
| Heart_Failure | dendritic_cell | *C12orf43* | -2.262208121 | 3.21E-07 | 3.66E-06 | 0.001555924 |
| Heart_Failure | dendritic_cell | *PEX10* | -2.782982192 | 5.02E-06 | 1.62E-05 | 0.024333432 |
| Heart_Failure | dendritic_cell | *LDLRAP1* | -2.272856598 | 0.013199568 | 0.017183875 | 1 |
| Heart_Failure | dendritic_cell | *KRTCAP3* | -1.448975061 | 0.00020812 | 0.000364943 | 1 |
| Heart_Failure | dendritic_cell | *PAFAH2* | -1.773620684 | 0.011741669 | 0.015389135 | 1 |
| Heart_Failure | dendritic_cell | *EXTL1* | 12.37280772 | 1.27E-07 | 3.44E-06 | 0.000614453 |
| Heart_Failure | dendritic_cell | *WDR66* | -1.622340112 | 9.30E-06 | 2.54E-05 | 0.045147844 |
| Heart_Failure | dendritic_cell | *RHPN1* | -2.39246297 | 0.000297491 | 0.000505402 | 1 |
| Heart_Failure | dendritic_cell | *LRRC43* | -4.016642008 | 2.62E-06 | 1.03E-05 | 0.012732898 |
| Heart_Failure | dendritic_cell | *EYA3* | -2.566911339 | 0.005918309 | 0.008079808 | 1 |
| Heart_Failure | dendritic_cell | *FANCC* | -2.143455146 | 1.99E-06 | 8.52E-06 | 0.009659849 |
| Heart_Failure | dendritic_cell | *ESYT3* | -3.518025962 | 3.96E-06 | 1.37E-05 | 0.019203588 |
| Heart_Failure | dendritic_cell | *CLSTN2* | -1.096366393 | 1.54E-07 | 3.51E-06 | 0.000747888 |
| Heart_Failure | dendritic_cell | *COLEC12* | -1.603548235 | 3.79E-06 | 1.33E-05 | 0.018396256 |
| Heart_Failure | dendritic_cell | *MITD1* | -2.588942857 | 0.004871299 | 0.006731854 | 1 |
| Heart_Failure | dendritic_cell | *TSPAN33* | 0.392776033 | 1.64E-07 | 3.52E-06 | 0.000796899 |
| Heart_Failure | dendritic_cell | *FAM86C1* | -4.114040408 | 4.83E-07 | 3.96E-06 | 0.002345802 |
| Heart_Failure | dendritic_cell | *PPP1R9A* | -1.983865768 | 1.77E-06 | 7.90E-06 | 0.008593039 |
| Heart_Failure | dendritic_cell | *DYNC1I1* | -2.816926797 | 0.000230548 | 0.000400078 | 1 |
| Heart_Failure | dendritic_cell | *COPG2* | -1.019691524 | 0.000489374 | 0.000795725 | 1 |
| Heart_Failure | dendritic_cell | *ELK4* | -3.063119865 | 0.01804219 | 0.023006756 | 1 |
| Heart_Failure | dendritic_cell | *DUSP23* | 2.510495207 | 1.75E-05 | 4.30E-05 | 0.085009472 |
| Heart_Failure | dendritic_cell | *F11R* | -1.934640826 | 2.64E-05 | 6.01E-05 | 0.128065024 |
| Heart_Failure | dendritic_cell | *SPATA2L* | -2.412803717 | 1.00E-07 | 3.44E-06 | 0.000486357 |
| Heart_Failure | dendritic_cell | *FGF17* | 4.188586701 | 6.16E-08 | 3.44E-06 | 0.000298723 |
| Heart_Failure | dendritic_cell | *CDA* | -0.946906004 | 0.015106475 | 0.019498966 | 1 |
| Heart_Failure | dendritic_cell | *SLC5A11* | -1.249305375 | 3.44E-05 | 7.54E-05 | 0.166860957 |
| Heart_Failure | dendritic_cell | *CCAR2* | -2.530567407 | 1.13E-06 | 6.03E-06 | 0.005462899 |

| | | | | | | |
|---|---|---|---|---|---|---|
| Heart_Failure | dendritic_cell | *SV2A* | 2.353514737 | 1.35E-07 | 3.44E-06 | 0.000655069 |
| Heart_Failure | dendritic_cell | *CLIC6* | 2.350294 | 3.05E-07 | 3.66E-06 | 0.001479529 |
| Heart_Failure | dendritic_cell | *CBR1* | -1.778405032 | 3.60E-05 | 7.84E-05 | 0.174886839 |
| Heart_Failure | dendritic_cell | *CHAF1B* | -1.359165226 | 0.002493451 | 0.003585721 | 1 |
| Heart_Failure | dendritic_cell | *SIM2* | -1.391142741 | 0.00181735 | 0.002685074 | 1 |
| Heart_Failure | dendritic_cell | *HLCS* | -2.464569692 | 2.21E-05 | 5.23E-05 | 0.107081573 |
| Heart_Failure | dendritic_cell | *ARHGAP27* | -3.126887498 | 2.68E-05 | 6.08E-05 | 0.130002206 |
| Heart_Failure | dendritic_cell | *IRX6* | -1.153772136 | 8.13E-05 | 0.000157769 | 0.394264248 |
| Heart_Failure | dendritic_cell | *HK2* | -1.265059645 | 2.41E-07 | 3.66E-06 | 0.001169232 |
| Heart_Failure | dendritic_cell | *AMFR* | -2.740832183 | 0.009962104 | 0.013181382 | 1 |
| Heart_Failure | dendritic_cell | *TMEM69* | -3.718702979 | 5.56E-06 | 1.75E-05 | 0.026981666 |
| Heart_Failure | dendritic_cell | *ACE* | -3.673264695 | 9.53E-07 | 5.47E-06 | 0.004625979 |
| Heart_Failure | dendritic_cell | *TEPP* | -10.28032204 | 2.96E-08 | 3.33E-06 | 0.000143513 |
| Heart_Failure | dendritic_cell | *CHCHD6* | -1.663151186 | 1.50E-06 | 7.11E-06 | 0.00727009 |
| Heart_Failure | dendritic_cell | *CTBP1* | -3.265915717 | 0.009134507 | 0.012155959 | 1 |
| Heart_Failure | dendritic_cell | *LRRC36* | -0.897949428 | 0.002057831 | 0.003012854 | 1 |
| Heart_Failure | dendritic_cell | *ZFYVE28* | -2.57526286 | 8.17E-06 | 2.30E-05 | 0.039617419 |
| Heart_Failure | dendritic_cell | *C16orf86* | -3.03335605 | 0.001483714 | 0.002221908 | 1 |
| Heart_Failure | dendritic_cell | *RGS12* | -3.024572694 | 0.000176092 | 0.000313426 | 0.854399126 |
| Heart_Failure | dendritic_cell | *LYPD5* | -3.331893019 | 5.08E-06 | 1.63E-05 | 0.024629089 |
| Heart_Failure | dendritic_cell | *CCDC107* | -3.240528085 | 5.20E-05 | 0.000107616 | 0.25225256 |
| Heart_Failure | dendritic_cell | *ZNF233* | -4.47675646 | 0.000258155 | 0.000445121 | 1 |
| Heart_Failure | dendritic_cell | *SSU72* | -3.578553844 | 5.63E-05 | 0.000115192 | 0.273350402 |
| Heart_Failure | dendritic_cell | *ANKLE1* | 3.565616339 | 7.03E-05 | 0.000139171 | 0.341109099 |
| Heart_Failure | dendritic_cell | *CCDC58* | -1.668168367 | 3.24E-07 | 3.66E-06 | 0.001574394 |
| Heart_Failure | dendritic_cell | *SLC37A1* | -4.227189676 | 0.000829551 | 0.001294206 | 1 |
| Heart_Failure | dendritic_cell | *PDE9A* | -1.701314476 | 6.65E-06 | 1.98E-05 | 0.032284067 |
| Heart_Failure | dendritic_cell | *HSF2BP* | -2.418285535 | 8.25E-05 | 0.000159941 | 0.400492323 |
| Heart_Failure | dendritic_cell | *CSTB* | -1.656686226 | 1.69E-05 | 4.18E-05 | 0.082021836 |
| Heart_Failure | dendritic_cell | *C21orf33* | 1.274044144 | 0.000306792 | 0.0005196 | 1 |
| Heart_Failure | dendritic_cell | *C21orf2* | -1.344895985 | 6.74E-05 | 0.000134207 | 0.326928527 |
| Heart_Failure | dendritic_cell | *LSS* | -0.99306564 | 0.000348569 | 0.00058279 | 1 |
| Heart_Failure | dendritic_cell | *MCM3AP* | -1.917474324 | 8.78E-05 | 0.000168339 | 0.426168557 |
| Heart_Failure | dendritic_cell | *DIP2A* | -2.550239588 | 0.00066426 | 0.001053265 | 1 |
| Heart_Failure | dendritic_cell | *PRMT2* | -1.817493355 | 0.020502299 | 0.025925763 | 1 |
| Heart_Failure | dendritic_cell | *ZNF208* | -1.624802068 | 7.31E-07 | 4.70E-06 | 0.003547179 |
| Heart_Failure | dendritic_cell | *ADAMTS13* | -1.74506538 | 7.74E-06 | 2.19E-05 | 0.037533727 |
| Heart_Failure | dendritic_cell | *CACFD1* | -7.999385505 | 0.000280056 | 0.00047863 | 1 |
| Heart_Failure | dendritic_cell | *GPSM1* | -2.17758822 | 0.000338801 | 0.000568417 | 1 |
| Heart_Failure | dendritic_cell | *RDH13* | -1.601552474 | 0.022294029 | 0.028059825 | 1 |
| Heart_Failure | dendritic_cell | *TAOK1* | -2.668490032 | 0.000295508 | 0.000502249 | 1 |
| Heart_Failure | dendritic_cell | *TLCD1* | 2.028512519 | 2.60E-07 | 3.66E-06 | 0.001262171 |
| Heart_Failure | dendritic_cell | *S100A1* | -6.773194088 | 5.42E-07 | 4.13E-06 | 0.002628333 |
| Heart_Failure | dendritic_cell | *FDPS* | -2.185580094 | 9.23E-05 | 0.000175931 | 0.447920411 |
| Heart_Failure | dendritic_cell | *PAQR6* | -1.351194214 | 0.03892866 | 0.047553338 | 1 |
| Heart_Failure | dendritic_cell | *CCR5* | 1.292667187 | 3.99E-08 | 3.44E-06 | 0.00019348 |
| Heart_Failure | dendritic_cell | *PTH1R* | -1.522813105 | 6.58E-06 | 1.96E-05 | 0.031902745 |
| Heart_Failure | dendritic_cell | *FCRL3* | -2.430832679 | 4.48E-06 | 1.49E-05 | 0.021731453 |
| Heart_Failure | dendritic_cell | *FGFR4* | 3.050784606 | 0.022787257 | 0.028650887 | 1 |
| Heart_Failure | dendritic_cell | *LY6K* | -2.108883358 | 0.004714829 | 0.006523054 | 1 |

| | | | | | | |
|---|---|---|---|---|---|---|
| Heart_Failure | dendritic_cell | *IER2* | -3.691761717 | 1.40E-05 | 3.56E-05 | 0.06801544 |
| Heart_Failure | dendritic_cell | *CPSF4* | -2.011117527 | 3.23E-07 | 3.66E-06 | 0.001565804 |
| Heart_Failure | dendritic_cell | *VPS28* | -2.332963824 | 0.00897566 | 0.011957689 | 1 |
| Heart_Failure | dendritic_cell | *TONSL* | 4.363113954 | 4.88E-08 | 3.44E-06 | 0.000236611 |
| Heart_Failure | dendritic_cell | *MUM1* | -2.553942607 | 1.12E-05 | 2.96E-05 | 0.054278562 |
| Heart_Failure | dendritic_cell | *ZNF333* | -3.598784059 | 4.65E-05 | 9.72E-05 | 0.225801125 |
| Heart_Failure | dendritic_cell | *SH2B2* | 1.384292269 | 2.69E-07 | 3.66E-06 | 0.001306532 |
| Heart_Failure | dendritic_cell | *RPL8* | -2.311837604 | 6.57E-06 | 1.96E-05 | 0.031870177 |
| Heart_Failure | dendritic_cell | *ABCF3* | -2.074686451 | 0.00178642 | 0.002643401 | 1 |
| Heart_Failure | dendritic_cell | *FBXO27* | -1.258294225 | 8.29E-06 | 2.33E-05 | 0.040205314 |
| Heart_Failure | dendritic_cell | *DMKN* | -0.541489227 | 0.000619438 | 0.000986384 | 1 |
| Heart_Failure | dendritic_cell | *BDH1* | -2.374053039 | 2.29E-05 | 5.38E-05 | 0.110919412 |
| Heart_Failure | dendritic_cell | *LRRC56* | -2.41362126 | 0.002849101 | 0.004053911 | 1 |
| Heart_Failure | dendritic_cell | *PGAP3* | -5.695523163 | 0.000426418 | 0.000699452 | 1 |
| Heart_Failure | dendritic_cell | *ZNF577* | 2.759523556 | 2.68E-06 | 1.04E-05 | 0.012999411 |
| Heart_Failure | dendritic_cell | *TMEM143* | -2.508111073 | 9.57E-07 | 5.48E-06 | 0.00464329 |
| Heart_Failure | dendritic_cell | *SIGLEC11* | -0.960784979 | 0.010000749 | 0.013225302 | 1 |
| Heart_Failure | dendritic_cell | *DBF4B* | -1.142026181 | 3.47E-05 | 7.60E-05 | 0.168561401 |
| Heart_Failure | dendritic_cell | *PLCD3* | -1.973164671 | 3.21E-07 | 3.66E-06 | 0.001558486 |
| Heart_Failure | dendritic_cell | *IP6K3* | -15.78258228 | 0.000460618 | 0.00075098 | 1 |
| Heart_Failure | dendritic_cell | *LEMD2* | 5.275967068 | 1.22E-05 | 3.18E-05 | 0.059144017 |
| Heart_Failure | dendritic_cell | *CXCL16* | 0.691048371 | 5.85E-06 | 1.82E-05 | 0.028373968 |
| Heart_Failure | dendritic_cell | *ASGR2* | 1.889517972 | 1.28E-07 | 3.44E-06 | 0.000621727 |
| Heart_Failure | dendritic_cell | *POLR3K* | -6.487015777 | 6.69E-06 | 1.99E-05 | 0.032468098 |
| Heart_Failure | dendritic_cell | *SNRNP25* | -2.460099585 | 4.48E-06 | 1.49E-05 | 0.021741507 |
| Heart_Failure | dendritic_cell | *WDR90* | 2.02245703 | 1.69E-06 | 7.68E-06 | 0.008184443 |
| Heart_Failure | dendritic_cell | *MEIOB* | -1.987825455 | 3.14E-05 | 6.94E-05 | 0.152533167 |
| Heart_Failure | dendritic_cell | *SHANK2* | -2.830606567 | 2.33E-06 | 9.46E-06 | 0.011318046 |
| Heart_Failure | dendritic_cell | *ASRGL1* | -2.053439127 | 3.10E-07 | 3.66E-06 | 0.001502644 |
| Heart_Failure | dendritic_cell | *TTC9C* | -2.755816326 | 2.87E-05 | 6.42E-05 | 0.139086993 |
| Heart_Failure | dendritic_cell | *RPS6KA4* | -2.89298844 | 5.79E-07 | 4.27E-06 | 0.002807379 |
| Heart_Failure | dendritic_cell | *TPCN2* | -2.93254329 | 0.000946742 | 0.001464793 | 1 |
| Heart_Failure | dendritic_cell | *PDZK1IP1* | -3.669040479 | 2.81E-07 | 3.66E-06 | 0.001361984 |
| Heart_Failure | dendritic_cell | *CMPK1* | -1.700886532 | 3.60E-07 | 3.66E-06 | 0.00174591 |
| Heart_Failure | dendritic_cell | *COA7* | 4.47820251 | 1.58E-07 | 3.51E-06 | 0.00076486 |
| Heart_Failure | dendritic_cell | *C1orf123* | -3.989977926 | 3.52E-06 | 1.27E-05 | 0.01706062 |
| Heart_Failure | dendritic_cell | *USP24* | -2.863069165 | 0.000166521 | 0.000298031 | 0.807961629 |
| Heart_Failure | dendritic_cell | *ZSWIM5* | -2.296473777 | 0.000132125 | 0.000242023 | 0.641068707 |
| Heart_Failure | dendritic_cell | *AK4* | -1.427613524 | 3.56E-06 | 1.28E-05 | 0.017275806 |
| Heart_Failure | dendritic_cell | *RBP7* | -2.756279619 | 3.58E-05 | 7.80E-05 | 0.173924345 |
| Heart_Failure | dendritic_cell | *SLC25A34* | -3.316757566 | 9.41E-07 | 5.45E-06 | 0.004563489 |
| Heart_Failure | dendritic_cell | *PDPN* | -1.583258151 | 2.42E-07 | 3.66E-06 | 0.001175322 |
| Heart_Failure | dendritic_cell | *LAPTM5* | 3.210221183 | 5.47E-07 | 4.15E-06 | 0.002652029 |
| Heart_Failure | dendritic_cell | *SDC3* | -2.250587405 | 1.67E-06 | 7.64E-06 | 0.008087509 |
| Heart_Failure | dendritic_cell | *SYNC* | -3.03552411 | 1.51E-05 | 3.79E-05 | 0.073389248 |
| Heart_Failure | dendritic_cell | *CAMK2N1* | 2.203527744 | 6.67E-05 | 0.000133078 | 0.323645107 |
| Heart_Failure | dendritic_cell | *MEGF6* | -1.844598575 | 4.99E-06 | 1.62E-05 | 0.024214863 |
| Heart_Failure | dendritic_cell | *TM2D1* | -2.801304639 | 4.35E-05 | 9.19E-05 | 0.211166366 |
| Heart_Failure | dendritic_cell | *NEXN* | -2.26706814 | 3.37E-05 | 7.41E-05 | 0.163516702 |
| Heart_Failure | dendritic_cell | *LRRIQ3* | -1.451274733 | 3.15E-07 | 3.66E-06 | 0.001528126 |

| | | | | | | |
|---|---|---|---|---|---|---|
| Heart_Failure | dendritic_cell | *TYW3* | -2.95278864 | 5.72E-07 | 4.26E-06 | 0.002774287 |
| Heart_Failure | dendritic_cell | *HFM1* | -1.752305932 | 4.70E-07 | 3.95E-06 | 0.002280625 |
| Heart_Failure | dendritic_cell | *ARPC5* | -2.312431906 | 0.020496525 | 0.025925219 | 1 |
| Heart_Failure | dendritic_cell | *ZNF496* | -3.296500807 | 0.001967937 | 0.002889961 | 1 |
| Heart_Failure | dendritic_cell | *FCRLB* | 2.165323501 | 2.87E-07 | 3.66E-06 | 0.001391306 |
| Heart_Failure | dendritic_cell | *KLHDC9* | -2.264752424 | 9.63E-06 | 2.62E-05 | 0.04672739 |
| Heart_Failure | dendritic_cell | *TDRD5* | -4.278449288 | 2.37E-07 | 3.66E-06 | 0.001151781 |
| Heart_Failure | dendritic_cell | *BROX* | 1.319641455 | 0.002523541 | 0.00362255 | 1 |
| Heart_Failure | dendritic_cell | *ACP6* | -2.865309758 | 1.37E-06 | 6.71E-06 | 0.006637726 |
| Heart_Failure | dendritic_cell | *CNST* | -2.057060559 | 2.41E-06 | 9.66E-06 | 0.011697512 |
| Heart_Failure | dendritic_cell | *PM20D1* | -1.93286066 | 0.005965407 | 0.008134175 | 1 |
| Heart_Failure | dendritic_cell | *PKDCC* | -1.453927076 | 5.10E-06 | 1.64E-05 | 0.024755246 |
| Heart_Failure | dendritic_cell | *HAAO* | -3.635694391 | 0.006889407 | 0.009316482 | 1 |
| Heart_Failure | dendritic_cell | *B3GALNT2* | -1.959998337 | 0.001017511 | 0.00156729 | 1 |
| Heart_Failure | dendritic_cell | *MRPL55* | -2.863557484 | 1.22E-05 | 3.17E-05 | 0.058991561 |
| Heart_Failure | dendritic_cell | *KIAA1841* | -2.176851374 | 1.11E-06 | 6.00E-06 | 0.005397263 |
| Heart_Failure | dendritic_cell | *KCNJ3* | -6.329889831 | 3.43E-07 | 3.66E-06 | 0.001663086 |
| Heart_Failure | dendritic_cell | *DUSP19* | -2.296355067 | 1.19E-06 | 6.17E-06 | 0.005789871 |
| Heart_Failure | dendritic_cell | *NUP35* | -3.476510919 | 0.000125409 | 0.000231012 | 0.608486866 |
| Heart_Failure | dendritic_cell | *C2orf48* | -16.40607239 | 3.79E-07 | 3.68E-06 | 0.001836548 |
| Heart_Failure | dendritic_cell | *CCDC74A* | 3.291943979 | 0.0002053 | 0.000360781 | 0.996115417 |
| Heart_Failure | dendritic_cell | *H3F3A* | -1.984488728 | 0.014908768 | 0.019259144 | 1 |
| Heart_Failure | dendritic_cell | *NOSTRIN* | -1.521505054 | 0.005136664 | 0.007072388 | 1 |
| Heart_Failure | dendritic_cell | *INHBB* | -2.89174475 | 1.71E-06 | 7.72E-06 | 0.008294479 |
| Heart_Failure | dendritic_cell | *RPRD2* | -2.813599497 | 9.95E-06 | 2.69E-05 | 0.048261584 |
| Heart_Failure | dendritic_cell | *PACRGL* | -2.346918439 | 3.00E-06 | 1.14E-05 | 0.01457342 |
| Heart_Failure | dendritic_cell | *C1QTNF7* | -1.601481748 | 1.02E-06 | 5.71E-06 | 0.004958756 |
| Heart_Failure | dendritic_cell | *LYSMD1* | -1.539226171 | 0.000144249 | 0.000261643 | 0.699893942 |
| Heart_Failure | dendritic_cell | *VPS72* | -4.235899325 | 2.85E-05 | 6.40E-05 | 0.138489799 |
| Heart_Failure | dendritic_cell | *ERCC3* | -2.370972635 | 0.022012049 | 0.027733695 | 1 |
| Heart_Failure | dendritic_cell | *DHX57* | -2.109273464 | 1.85E-06 | 8.10E-06 | 0.008971055 |
| Heart_Failure | dendritic_cell | *S100A9* | -2.183591471 | 6.58E-07 | 4.46E-06 | 0.003191108 |
| Heart_Failure | dendritic_cell | *TDRD10* | -2.609257613 | 2.33E-06 | 9.45E-06 | 0.011289773 |
| Heart_Failure | dendritic_cell | *DCAF16* | -3.671838043 | 0.0005702 | 0.000914548 | 1 |
| Heart_Failure | dendritic_cell | *GNPDA2* | -2.530904402 | 0.001456787 | 0.002182931 | 1 |
| Heart_Failure | dendritic_cell | *NIPAL1* | -1.722655219 | 8.24E-07 | 4.99E-06 | 0.003998019 |
| Heart_Failure | dendritic_cell | *ANTXR2* | -1.9053602 | 0.002241705 | 0.003259441 | 1 |
| Heart_Failure | dendritic_cell | *GPR155* | 2.804319135 | 0.03591449 | 0.044059951 | 1 |
| Heart_Failure | dendritic_cell | *DAPL1* | 5.213928277 | 1.45E-06 | 6.95E-06 | 0.007030211 |
| Heart_Failure | dendritic_cell | *PBXIP1* | -3.860516873 | 0.005190049 | 0.007141837 | 1 |
| Heart_Failure | dendritic_cell | *CLDN1* | -1.095703958 | 7.31E-07 | 4.70E-06 | 0.003548374 |
| Heart_Failure | dendritic_cell | *LINC01116* | -3.283361686 | 6.47E-07 | 4.45E-06 | 0.003138938 |
| Heart_Failure | dendritic_cell | *SLC22A15* | -2.343845971 | 2.90E-06 | 1.11E-05 | 0.014067183 |
| Heart_Failure | dendritic_cell | *LRRC58* | -1.941992447 | 1.17E-06 | 6.11E-06 | 0.005660307 |
| Heart_Failure | dendritic_cell | *LMOD1* | -2.083192578 | 1.56E-07 | 3.51E-06 | 0.000756102 |
| Heart_Failure | dendritic_cell | *ARPC2* | -2.753596385 | 0.002084992 | 0.003051698 | 1 |
| Heart_Failure | dendritic_cell | *CCT3* | -2.273012668 | 6.37E-06 | 1.92E-05 | 0.030901089 |
| Heart_Failure | dendritic_cell | *GLB1L* | -2.514423299 | 5.21E-05 | 0.000107816 | 0.252935277 |
| Heart_Failure | dendritic_cell | *SERPINI1* | -3.090110388 | 2.42E-06 | 9.71E-06 | 0.011762902 |
| Heart_Failure | dendritic_cell | *SUCLG1* | -2.142819821 | 6.68E-05 | 0.000133218 | 0.324253432 |

| | | | | | | |
|---|---|---|---|---|---|---|
| Heart_Failure | dendritic_cell | *PRKCI* | -1.598616863 | 3.91E-05 | 8.40E-05 | 0.189568051 |
| Heart_Failure | dendritic_cell | *AIM2* | -5.008904078 | 5.37E-07 | 4.13E-06 | 0.002603668 |
| Heart_Failure | dendritic_cell | *GTPBP8* | -3.518848266 | 3.50E-06 | 1.27E-05 | 0.016979569 |
| Heart_Failure | dendritic_cell | *WDFY3* | -2.147085466 | 1.26E-05 | 3.27E-05 | 0.061309953 |
| Heart_Failure | dendritic_cell | *PTPN13* | -1.186002155 | 0.00032813 | 0.000552617 | 1 |
| Heart_Failure | dendritic_cell | *PPM1K* | -2.729940211 | 3.59E-06 | 1.28E-05 | 0.017416672 |
| Heart_Failure | dendritic_cell | *TIPARP* | -3.105624113 | 3.71E-05 | 8.04E-05 | 0.180246625 |
| Heart_Failure | dendritic_cell | *RPP14* | -4.339001777 | 3.06E-05 | 6.79E-05 | 0.148632538 |
| Heart_Failure | dendritic_cell | *DNASE1L3* | -2.176778802 | 0.001228481 | 0.001868524 | 1 |
| Heart_Failure | dendritic_cell | *IL17RC* | 3.89114517 | 1.92E-07 | 3.61E-06 | 0.000934001 |
| Heart_Failure | dendritic_cell | *CRELD1* | -3.531997776 | 0.003073679 | 0.004350494 | 1 |
| Heart_Failure | dendritic_cell | *TTC14* | -2.193758486 | 6.22E-05 | 0.000125126 | 0.301803253 |
| Heart_Failure | dendritic_cell | *RCHY1* | -3.163552846 | 1.23E-05 | 3.21E-05 | 0.059794272 |
| Heart_Failure | dendritic_cell | *CCDC158* | -2.519932869 | 2.30E-06 | 9.38E-06 | 0.011181698 |
| Heart_Failure | dendritic_cell | *CPA3* | -2.576189832 | 4.78E-07 | 3.96E-06 | 0.002318149 |
| Heart_Failure | dendritic_cell | *SLC4A1AP* | -3.541828079 | 0.012568206 | 0.016397132 | 1 |
| Heart_Failure | dendritic_cell | *KIAA1143* | -2.010774147 | 0.00057867 | 0.000926025 | 1 |
| Heart_Failure | dendritic_cell | *CDCP1* | 0.671660624 | 1.27E-07 | 3.44E-06 | 0.000616364 |
| Heart_Failure | dendritic_cell | *CLEC3B* | 10.27494269 | 0.000202526 | 0.000356422 | 0.982655979 |
| Heart_Failure | dendritic_cell | *FYCO1* | -2.50240114 | 0.003783229 | 0.005294556 | 1 |
| Heart_Failure | dendritic_cell | *NMNAT3* | -2.642053026 | 5.65E-05 | 0.000115339 | 0.273930458 |
| Heart_Failure | dendritic_cell | *TPRA1* | -2.78896268 | 1.18E-06 | 6.14E-06 | 0.005710182 |
| Heart_Failure | dendritic_cell | *SNIP1* | 5.444052771 | 0.000315223 | 0.000532913 | 1 |
| Heart_Failure | dendritic_cell | *DNALI1* | -2.944996351 | 1.73E-08 | 2.99E-06 | 8.40E-05 |
| Heart_Failure | dendritic_cell | *RPN1* | 4.498403099 | 1.50E-05 | 3.76E-05 | 0.072664157 |
| Heart_Failure | dendritic_cell | *SENP2* | -2.315330985 | 7.41E-06 | 2.12E-05 | 0.035945709 |
| Heart_Failure | dendritic_cell | *IFT122* | -1.620451039 | 7.20E-07 | 4.65E-06 | 0.003494775 |
| Heart_Failure | dendritic_cell | *RFT1* | -2.356618646 | 1.95E-05 | 4.70E-05 | 0.094667001 |
| Heart_Failure | dendritic_cell | *SFMBT1* | -4.010030153 | 0.002269315 | 0.003295636 | 1 |
| Heart_Failure | dendritic_cell | *UVSSA* | -1.961334812 | 4.36E-05 | 9.21E-05 | 0.211785449 |
| Heart_Failure | dendritic_cell | *FAM208A* | -2.775477844 | 0.000381058 | 0.000631669 | 1 |
| Heart_Failure | dendritic_cell | *LRPAP1* | -3.676945343 | 0.000131671 | 0.000241538 | 0.638868877 |
| Heart_Failure | dendritic_cell | *ABLIM2* | -2.296004899 | 0.000239358 | 0.00041433 | 1 |
| Heart_Failure | dendritic_cell | *ERMAP* | 2.905988601 | 2.24E-06 | 9.22E-06 | 0.010892173 |
| Heart_Failure | dendritic_cell | *ZNF691* | -2.183451049 | 0.002109319 | 0.003084514 | 1 |
| Heart_Failure | dendritic_cell | *METAP1* | -3.12534581 | 0.001251602 | 0.001900116 | 1 |
| Heart_Failure | dendritic_cell | *BDH2* | -1.855705135 | 8.57E-06 | 2.39E-05 | 0.041575137 |
| Heart_Failure | dendritic_cell | *CDC25A* | 3.02564165 | 1.34E-06 | 6.63E-06 | 0.006510717 |
| Heart_Failure | dendritic_cell | *ZNF589* | -1.769780526 | 1.35E-06 | 6.63E-06 | 0.006534604 |
| Heart_Failure | dendritic_cell | *HSPA4L* | -2.268314199 | 7.90E-05 | 0.000153996 | 0.3833777 |
| Heart_Failure | dendritic_cell | *MFSD8* | -2.629761883 | 1.08E-05 | 2.87E-05 | 0.052301012 |
| Heart_Failure | dendritic_cell | *RAD54L2* | -2.250602567 | 1.49E-07 | 3.45E-06 | 0.000723954 |
| Heart_Failure | dendritic_cell | *TEX264* | -2.881424014 | 0.000147905 | 0.000267875 | 0.717636102 |
| Heart_Failure | dendritic_cell | *ETNPPL* | -13.39167875 | 3.48E-07 | 3.66E-06 | 0.001687533 |
| Heart_Failure | dendritic_cell | *PRSS12* | -1.498838565 | 0.000944518 | 0.00146322 | 1 |
| Heart_Failure | dendritic_cell | *ANXA5* | -2.47913192 | 0.0016209 | 0.00241674 | 1 |
| Heart_Failure | dendritic_cell | *MAP9* | -1.679680452 | 3.76E-07 | 3.66E-06 | 0.001824136 |
| Heart_Failure | dendritic_cell | *CEP44* | -2.277112646 | 7.56E-06 | 2.16E-05 | 0.036687375 |
| Heart_Failure | dendritic_cell | *HPGD* | 2.013617696 | 1.23E-07 | 3.44E-06 | 0.000599056 |
| Heart_Failure | dendritic_cell | *C4orf45* | 2.721581557 | 0.009152535 | 0.01217661 | 1 |

| | | | | | | |
|---|---|---|---|---|---|---|
| Heart_Failure | dendritic_cell | *TMEM144* | -3.067925725 | 0.001349137 | 0.002035452 | 1 |
| Heart_Failure | dendritic_cell | *IL15* | 3.084645906 | 3.11E-06 | 1.16E-05 | 0.015095334 |
| Heart_Failure | dendritic_cell | *ARFIP1* | -2.629465865 | 0.000170781 | 0.000304644 | 0.828630924 |
| Heart_Failure | dendritic_cell | *ITGA2* | 1.756437539 | 0.003194512 | 0.004508368 | 1 |
| Heart_Failure | dendritic_cell | *MOCS2* | -1.897353072 | 3.14E-07 | 3.66E-06 | 0.001524768 |
| Heart_Failure | dendritic_cell | *NDUFAF2* | -2.141287478 | 0.000259692 | 0.000447224 | 1 |
| Heart_Failure | dendritic_cell | *LMBRD2* | -3.182967169 | 0.002139237 | 0.003126379 | 1 |
| Heart_Failure | dendritic_cell | *RNF180* | -2.218810092 | 2.24E-06 | 9.21E-06 | 0.010880338 |
| Heart_Failure | dendritic_cell | *PGGT1B* | 3.008195345 | 0.013206311 | 0.017187342 | 1 |
| Heart_Failure | dendritic_cell | *F2RL2* | 1.122918614 | 1.30E-07 | 3.44E-06 | 0.000628433 |
| Heart_Failure | dendritic_cell | *CCDC112* | 2.987622403 | 2.49E-07 | 3.66E-06 | 0.001207003 |
| Heart_Failure | dendritic_cell | *CMBL* | -1.936773816 | 5.82E-06 | 1.81E-05 | 0.028223009 |
| Heart_Failure | dendritic_cell | *C5orf63* | -1.930545341 | 9.51E-07 | 5.47E-06 | 0.004612742 |
| Heart_Failure | dendritic_cell | *F2RL1* | -3.829211446 | 0.011560094 | 0.015171646 | 1 |
| Heart_Failure | dendritic_cell | *WDR41* | -2.755519641 | 6.51E-06 | 1.95E-05 | 0.031570431 |
| Heart_Failure | dendritic_cell | *NDUFS4* | -2.404549356 | 1.11E-05 | 2.94E-05 | 0.05400104 |
| Heart_Failure | dendritic_cell | *CASP3* | -3.562262443 | 0.002175643 | 0.003174803 | 1 |
| Heart_Failure | dendritic_cell | *PRIMPOL* | -3.219246007 | 8.00E-05 | 0.000155622 | 0.387966154 |
| Heart_Failure | dendritic_cell | *ERAP1* | -1.871563908 | 0.01219672 | 0.015955374 | 1 |
| Heart_Failure | dendritic_cell | *UTP15* | 7.11562812 | 1.68E-06 | 7.68E-06 | 0.008164615 |
| Heart_Failure | dendritic_cell | *TLR3* | -3.709585189 | 0.023832216 | 0.02991048 | 1 |
| Heart_Failure | dendritic_cell | *KLKB1* | 0.964039511 | 0.00192863 | 0.00283653 | 1 |
| Heart_Failure | dendritic_cell | *NSA2* | -0.949795681 | 0.016567274 | 0.021288245 | 1 |
| Heart_Failure | dendritic_cell | *CCDC127* | -2.604559739 | 2.12E-05 | 5.05E-05 | 0.10265882 |
| Heart_Failure | dendritic_cell | *GRIK2* | -0.834421497 | 0.016589867 | 0.021311633 | 1 |
| Heart_Failure | dendritic_cell | *DCBLD1* | -1.738102358 | 0.002240946 | 0.003259314 | 1 |
| Heart_Failure | dendritic_cell | *PDSS2* | -2.553376047 | 0.00011068 | 0.000206863 | 0.537017247 |
| Heart_Failure | dendritic_cell | *RAET1E* | -1.895770191 | 4.19E-07 | 3.88E-06 | 0.002034257 |
| Heart_Failure | dendritic_cell | *STK17A* | -3.301879168 | 8.09E-05 | 0.000157164 | 0.392592868 |
| Heart_Failure | dendritic_cell | *GALNT10* | -1.965247499 | 0.035462259 | 0.04358229 | 1 |
| Heart_Failure | dendritic_cell | *SAP30L* | -2.84266876 | 0.0016055 | 0.002396888 | 1 |
| Heart_Failure | dendritic_cell | *RPS14* | -2.214463967 | 1.76E-05 | 4.32E-05 | 0.085431173 |
| Heart_Failure | dendritic_cell | *RP9* | -3.335781084 | 1.95E-06 | 8.39E-06 | 0.009476102 |
| Heart_Failure | dendritic_cell | *CAMLG* | -1.448387862 | 2.74E-07 | 3.66E-06 | 0.001328521 |
| Heart_Failure | dendritic_cell | *RELL2* | 1.382229995 | 7.32E-08 | 3.44E-06 | 0.000355053 |
| Heart_Failure | dendritic_cell | *MIOS* | -3.317116579 | 9.36E-05 | 0.00017801 | 0.453925908 |
| Heart_Failure | dendritic_cell | *USP49* | -2.928592509 | 7.13E-06 | 2.07E-05 | 0.034593379 |
| Heart_Failure | dendritic_cell | *CHMP4C* | 6.968560771 | 1.44E-07 | 3.45E-06 | 0.000700812 |
| Heart_Failure | dendritic_cell | *SLC13A4* | -3.419904741 | 0.003768953 | 0.005276099 | 1 |
| Heart_Failure | dendritic_cell | *BRI3* | -3.101444808 | 0.000106638 | 0.000200158 | 0.517409555 |
| Heart_Failure | dendritic_cell | *RAD21* | -2.384084912 | 6.88E-06 | 2.03E-05 | 0.033391083 |
| Heart_Failure | dendritic_cell | *TNFRSF11B* | -2.792790092 | 7.60E-07 | 4.79E-06 | 0.003685494 |
| Heart_Failure | dendritic_cell | *DEFB1* | -2.782005435 | 0.037230558 | 0.045570804 | 1 |
| Heart_Failure | dendritic_cell | *SUN1* | -2.05937097 | 6.28E-05 | 0.000126054 | 0.304673248 |
| Heart_Failure | dendritic_cell | *OXR1* | -2.042735735 | 7.17E-06 | 2.08E-05 | 0.034811093 |
| Heart_Failure | dendritic_cell | *GPER1* | -1.838599926 | 2.93E-05 | 6.53E-05 | 0.14206647 |
| Heart_Failure | dendritic_cell | *INTS1* | -2.835637685 | 7.19E-07 | 4.65E-06 | 0.003490483 |
| Heart_Failure | dendritic_cell | *SLC4A2* | -1.744940208 | 2.35E-07 | 3.66E-06 | 0.001141642 |
| Heart_Failure | dendritic_cell | *FASTK* | -2.045962833 | 5.48E-07 | 4.15E-06 | 0.002657756 |
| Heart_Failure | dendritic_cell | *ALDH7A1* | -2.443139936 | 2.61E-06 | 1.02E-05 | 0.012645537 |

| | | | | | | |
|---|---|---|---|---|---|---|
| Heart_Failure | dendritic_cell | *YWHAZ* | -2.783067434 | 5.42E-05 | 0.000111487 | 0.262997635 |
| Heart_Failure | dendritic_cell | *FZD6* | -1.337128501 | 3.97E-06 | 1.38E-05 | 0.019254784 |
| Heart_Failure | dendritic_cell | *SLC25A32* | -1.952894564 | 6.06E-07 | 4.32E-06 | 0.00294019 |
| Heart_Failure | dendritic_cell | *DCSTAMP* | -25.78444628 | 1.84E-06 | 8.10E-06 | 0.008941733 |
| Heart_Failure | dendritic_cell | *INTS8* | -2.387074142 | 6.17E-06 | 1.89E-05 | 0.029953215 |
| Heart_Failure | dendritic_cell | *RPP25L* | 1.847775906 | 0.019541595 | 0.024799653 | 1 |
| Heart_Failure | dendritic_cell | *C9orf24* | -1.763907525 | 5.98E-07 | 4.30E-06 | 0.002901158 |
| Heart_Failure | dendritic_cell | *CCDC171* | -1.694578321 | 6.73E-07 | 4.51E-06 | 0.003264236 |
| Heart_Failure | dendritic_cell | *METTL2B* | -1.423726454 | 9.84E-05 | 0.000186156 | 0.47748937 |
| Heart_Failure | dendritic_cell | *ZMAT4* | 3.597713599 | 1.35E-07 | 3.44E-06 | 0.000653024 |
| Heart_Failure | dendritic_cell | *C8orf34* | -1.11490149 | 3.59E-07 | 3.66E-06 | 0.001741442 |
| Heart_Failure | dendritic_cell | *RASEF* | -1.089396447 | 0.002293671 | 0.003327022 | 1 |
| Heart_Failure | dendritic_cell | *ANKS6* | -2.924763479 | 5.52E-06 | 1.74E-05 | 0.026779177 |
| Heart_Failure | dendritic_cell | *AQP7* | -0.846326475 | 3.02E-05 | 6.71E-05 | 0.146568844 |
| Heart_Failure | dendritic_cell | *NOL6* | -2.222932919 | 0.002174146 | 0.003173574 | 1 |
| Heart_Failure | dendritic_cell | *AQP3* | 1.510554884 | 1.30E-07 | 3.44E-06 | 0.000632669 |
| Heart_Failure | dendritic_cell | *TRMT10B* | -2.409218604 | 0.031866027 | 0.039321964 | 1 |
| Heart_Failure | dendritic_cell | *VCP* | -2.774693466 | 2.84E-05 | 6.39E-05 | 0.137987111 |
| Heart_Failure | dendritic_cell | *PIGO* | 3.133480815 | 9.15E-07 | 5.37E-06 | 0.004438551 |
| Heart_Failure | dendritic_cell | *WRN* | -2.28410902 | 1.32E-05 | 3.38E-05 | 0.0638557 |
| Heart_Failure | dendritic_cell | *MARCH8* | -3.261436028 | 0.001011017 | 0.001557781 | 1 |
| Heart_Failure | dendritic_cell | *TSHR* | -2.464998539 | 0.009319119 | 0.012384652 | 1 |
| Heart_Failure | dendritic_cell | *SLC16A9* | -2.421399116 | 3.21E-07 | 3.66E-06 | 0.001555492 |
| Heart_Failure | dendritic_cell | *GJB2* | -10.52492744 | 3.29E-07 | 3.66E-06 | 0.001597596 |
| Heart_Failure | dendritic_cell | *CRYL1* | -2.321984743 | 0.017184805 | 0.022000178 | 1 |
| Heart_Failure | dendritic_cell | *REEP3* | -2.158679325 | 1.70E-06 | 7.72E-06 | 0.008265433 |
| Heart_Failure | dendritic_cell | *MICU2* | -2.799987106 | 7.36E-05 | 0.000144768 | 0.357287883 |
| Heart_Failure | dendritic_cell | *LRR1* | -6.329470994 | 0.000219411 | 0.000382944 | 1 |
| Heart_Failure | dendritic_cell | *RPL36AL* | -2.25331029 | 2.89E-05 | 6.47E-05 | 0.140410123 |
| Heart_Failure | dendritic_cell | *C10orf25* | -0.781776794 | 0.013987248 | 0.018155733 | 1 |
| Heart_Failure | dendritic_cell | *TMEM63C* | -7.866569074 | 0.000526077 | 0.000850275 | 1 |
| Heart_Failure | dendritic_cell | *AKR1E2* | -1.982805951 | 1.47E-07 | 3.45E-06 | 0.000712988 |
| Heart_Failure | dendritic_cell | *OXGR1* | 4.376801598 | 0.000153313 | 0.000276431 | 0.743876957 |
| Heart_Failure | dendritic_cell | *ATP5C1* | -3.277744628 | 0.000939609 | 0.001457009 | 1 |
| Heart_Failure | dendritic_cell | *VDAC2* | -4.185425741 | 1.62E-05 | 4.03E-05 | 0.078747801 |
| Heart_Failure | dendritic_cell | *PDZD8* | -1.982360425 | 2.25E-05 | 5.31E-05 | 0.109042606 |
| Heart_Failure | dendritic_cell | *QSOX2* | -4.314868792 | 2.58E-05 | 5.89E-05 | 0.125237819 |
| Heart_Failure | dendritic_cell | *CLEC1B* | 1.474998513 | 6.38E-05 | 0.000127774 | 0.309468434 |
| Heart_Failure | dendritic_cell | *SNAPC4* | -2.180685412 | 0.000255667 | 0.000441145 | 1 |
| Heart_Failure | dendritic_cell | *TSC1* | -1.561830509 | 0.001243146 | 0.001888484 | 1 |
| Heart_Failure | dendritic_cell | *STOX1* | -1.074519403 | 4.83E-05 | 0.000100426 | 0.23439388 |
| Heart_Failure | dendritic_cell | *RET* | -2.650441608 | 2.91E-07 | 3.66E-06 | 0.001414076 |
| Heart_Failure | dendritic_cell | *CASP7* | -1.519504173 | 0.018621468 | 0.023695611 | 1 |
| Heart_Failure | dendritic_cell | *VWA2* | -4.127137479 | 1.29E-05 | 3.32E-05 | 0.062478951 |
| Heart_Failure | dendritic_cell | *METTL3* | -2.399216569 | 2.89E-06 | 1.11E-05 | 0.014037713 |
| Heart_Failure | dendritic_cell | *TRUB1* | -2.590746755 | 3.62E-07 | 3.66E-06 | 0.001754723 |
| Heart_Failure | dendritic_cell | *HSPA12A* | -1.003912446 | 0.001076487 | 0.001651838 | 1 |
| Heart_Failure | dendritic_cell | *UBTD1* | -2.435809746 | 1.78E-06 | 7.93E-06 | 0.008647864 |
| Heart_Failure | dendritic_cell | *OTOGL* | -3.486920914 | 4.72E-07 | 3.95E-06 | 0.002292253 |
| Heart_Failure | dendritic_cell | *PACSIN3* | -1.034958559 | 0.018630596 | 0.02370101 | 1 |

| | | | | | | |
|---|---|---|---|---|---|---|
| Heart_Failure | dendritic_cell | *TTC7B* | -2.504097502 | 1.11E-05 | 2.94E-05 | 0.053892928 |
| Heart_Failure | dendritic_cell | *SLC39A13* | -3.204962785 | 1.07E-05 | 2.86E-05 | 0.052110066 |
| Heart_Failure | dendritic_cell | *RAPSN* | 1.482940722 | 0.006046393 | 0.008240758 | 1 |
| Heart_Failure | dendritic_cell | *TC2N* | -1.205102246 | 3.88E-06 | 1.36E-05 | 0.018843891 |
| Heart_Failure | dendritic_cell | *CPSF2* | -1.100787296 | 0.00583766 | 0.007983181 | 1 |
| Heart_Failure | dendritic_cell | *IFI27L1* | -2.386261725 | 9.02E-07 | 5.32E-06 | 0.004377767 |
| Heart_Failure | dendritic_cell | *PTER* | -2.016897715 | 6.01E-07 | 4.30E-06 | 0.002917957 |
| Heart_Failure | dendritic_cell | *C1QL3* | -1.7731252 | 0.019149113 | 0.024316016 | 1 |
| Heart_Failure | dendritic_cell | *ARL5B* | -2.32864636 | 1.39E-06 | 6.77E-06 | 0.006741769 |
| Heart_Failure | dendritic_cell | *LIPC* | 2.696882165 | 3.23E-05 | 7.13E-05 | 0.156929065 |
| Heart_Failure | dendritic_cell | *CEP57* | -2.982238867 | 0.000135325 | 0.000247306 | 0.656598147 |
| Heart_Failure | dendritic_cell | *TCP11L2* | -2.714105587 | 7.26E-06 | 2.10E-05 | 0.035231159 |
| Heart_Failure | dendritic_cell | *JAM3* | -2.933202174 | 2.85E-06 | 1.09E-05 | 0.01382959 |
| Heart_Failure | dendritic_cell | *ZFYVE19* | -2.177040351 | 0.010168616 | 0.013432651 | 1 |
| Heart_Failure | dendritic_cell | *TRMT61A* | 6.106178336 | 2.39E-07 | 3.66E-06 | 0.001159398 |
| Heart_Failure | dendritic_cell | *LARP6* | -2.091882316 | 1.38E-06 | 6.73E-06 | 0.006689005 |
| Heart_Failure | dendritic_cell | *API5* | -3.378331903 | 0.000115582 | 0.000215115 | 0.560804191 |
| Heart_Failure | dendritic_cell | *ASPG* | -7.368807759 | 0.00014212 | 0.000257974 | 0.689565268 |
| Heart_Failure | dendritic_cell | *COX11* | -2.87468045 | 2.55E-05 | 5.85E-05 | 0.123866473 |
| Heart_Failure | dendritic_cell | *ZNF202* | -2.156287755 | 4.47E-07 | 3.90E-06 | 0.002170042 |
| Heart_Failure | dendritic_cell | *STXBP4* | -2.031666793 | 2.18E-06 | 9.03E-06 | 0.01058134 |
| Heart_Failure | dendritic_cell | *WBP1L* | -2.598517399 | 2.82E-05 | 6.35E-05 | 0.136793086 |
| Heart_Failure | dendritic_cell | *NUDT13* | -1.762409971 | 3.98E-05 | 8.52E-05 | 0.193020714 |
| Heart_Failure | dendritic_cell | *WDR88* | -4.616450293 | 0.000308951 | 0.000523038 | 1 |
| Heart_Failure | dendritic_cell | *SERPINB8* | 1.032902869 | 0.002282596 | 0.003311949 | 1 |
| Heart_Failure | dendritic_cell | *PLD4* | 0.804457629 | 1.82E-07 | 3.58E-06 | 0.000880772 |
| Heart_Failure | dendritic_cell | *XRRA1* | -2.269815604 | 3.07E-06 | 1.15E-05 | 0.014915668 |
| Heart_Failure | dendritic_cell | *TRIM66* | -0.82018125 | 0.002209152 | 0.003219828 | 1 |
| Heart_Failure | dendritic_cell | *RPL27A* | -3.141173299 | 0.001122469 | 0.001716426 | 1 |
| Heart_Failure | dendritic_cell | *CDYL2* | -1.184128984 | 0.007181114 | 0.009689312 | 1 |
| Heart_Failure | dendritic_cell | *CENPN* | 6.616761669 | 7.87E-07 | 4.87E-06 | 0.003820209 |
| Heart_Failure | dendritic_cell | *C16orf46* | -6.833442776 | 2.29E-05 | 5.38E-05 | 0.110890911 |
| Heart_Failure | dendritic_cell | *TMEM41B* | -3.442459007 | 0.000156497 | 0.000281753 | 0.759324126 |
| Heart_Failure | dendritic_cell | *TMX3* | -2.998665479 | 2.17E-05 | 5.17E-05 | 0.105351349 |
| Heart_Failure | dendritic_cell | *PRKCB* | 2.438598749 | 3.14E-07 | 3.66E-06 | 0.001525271 |
| Heart_Failure | dendritic_cell | *NDST2* | 10.29283826 | 1.15E-07 | 3.44E-06 | 0.000560216 |
| Heart_Failure | dendritic_cell | *CCDC68* | -1.569727405 | 1.26E-06 | 6.39E-06 | 0.006113708 |
| Heart_Failure | dendritic_cell | *CLEC4D* | 4.019998174 | 0.000388036 | 0.000641264 | 1 |
| Heart_Failure | dendritic_cell | *ZSCAN21* | -1.684657292 | 5.55E-05 | 0.000113621 | 0.269055348 |
| Heart_Failure | dendritic_cell | *RIMKLB* | -1.799856564 | 2.21E-07 | 3.66E-06 | 0.001072582 |
| Heart_Failure | dendritic_cell | *TMED3* | -1.145232228 | 0.000321879 | 0.000543408 | 1 |
| Heart_Failure | dendritic_cell | *SEC11C* | -2.130412776 | 4.58E-07 | 3.90E-06 | 0.002224363 |
| Heart_Failure | dendritic_cell | *CENPV* | -3.550771109 | 6.93E-05 | 0.000137385 | 0.336033542 |
| Heart_Failure | dendritic_cell | *FAM96B* | -2.848874673 | 3.95E-05 | 8.47E-05 | 0.191557832 |
| Heart_Failure | dendritic_cell | *ZNF592* | -1.976908814 | 0.015577741 | 0.020075219 | 1 |
| Heart_Failure | dendritic_cell | *ACSM1* | -2.5380908 | 5.79E-07 | 4.27E-06 | 0.002810689 |
| Heart_Failure | dendritic_cell | *SLFN5* | -2.528031855 | 4.76E-06 | 1.57E-05 | 0.023110579 |
| Heart_Failure | dendritic_cell | *CATSPER2* | -3.591299346 | 1.67E-06 | 7.64E-06 | 0.008079926 |
| Heart_Failure | dendritic_cell | *ZNF667-AS1* | -2.416980004 | 1.24E-05 | 3.23E-05 | 0.060164956 |
| Heart_Failure | dendritic_cell | *SAAL1* | 3.096557619 | 2.04E-07 | 3.66E-06 | 0.000988359 |

| | | | | | | |
|---|---|---|---|---|---|---|
| Heart_Failure | dendritic_cell | *YPEL4* | -3.009570825 | 4.77E-05 | 9.93E-05 | 0.231284946 |
| Heart_Failure | dendritic_cell | *LDHC* | -0.853566056 | 0.024303662 | 0.030454899 | 1 |
| Heart_Failure | dendritic_cell | *FAM96A* | 3.816073655 | 5.03E-06 | 1.62E-05 | 0.024421853 |
| Heart_Failure | dendritic_cell | *TMEM170A* | -1.937370703 | 2.42E-08 | 3.27E-06 | 0.000117452 |
| Heart_Failure | dendritic_cell | *ANPEP* | 0.767838787 | 3.26E-06 | 1.20E-05 | 0.015808135 |
| Heart_Failure | dendritic_cell | *DCTN5* | -1.726227484 | 3.15E-06 | 1.17E-05 | 0.015293853 |
| Heart_Failure | dendritic_cell | *STAT6* | -3.123272121 | 8.50E-05 | 0.000164062 | 0.41261711 |
| Heart_Failure | dendritic_cell | *STX3* | 3.379505211 | 7.67E-07 | 4.80E-06 | 0.00371922 |
| Heart_Failure | dendritic_cell | *PIP4K2C* | -2.184158706 | 8.83E-06 | 2.44E-05 | 0.042851449 |
| Heart_Failure | dendritic_cell | *MTMR10* | -2.318777976 | 0.000784865 | 0.00122844 | 1 |
| Heart_Failure | dendritic_cell | *MS4A7* | 1.254185803 | 0.001449178 | 0.002172201 | 1 |
| Heart_Failure | dendritic_cell | *DIS3L* | -1.786660026 | 1.25E-05 | 3.25E-05 | 0.060748268 |
| Heart_Failure | dendritic_cell | *CCNDBP1* | -2.786483175 | 1.90E-06 | 8.23E-06 | 0.009196895 |
| Heart_Failure | dendritic_cell | *EPB42* | -2.855595978 | 4.23E-05 | 8.98E-05 | 0.205119693 |
| Heart_Failure | dendritic_cell | *SMAD3* | -2.30095002 | 0.000137345 | 0.000250337 | 0.666396886 |
| Heart_Failure | dendritic_cell | *MAP1A* | 2.044502181 | 0.012236767 | 0.016003449 | 1 |
| Heart_Failure | dendritic_cell | *RCCD1* | -2.772439201 | 5.32E-08 | 3.44E-06 | 0.000258277 |
| Heart_Failure | dendritic_cell | *AKTIP* | -2.465394594 | 0.00011263 | 0.000210185 | 0.546479713 |
| Heart_Failure | dendritic_cell | *CNPY4* | -2.41514771 | 0.000405217 | 0.000666478 | 1 |
| Heart_Failure | dendritic_cell | *MEI1* | -2.172942502 | 7.50E-06 | 2.14E-05 | 0.036376755 |
| Heart_Failure | dendritic_cell | *SAMD14* | -2.594401655 | 0.002900338 | 0.004121981 | 1 |
| Heart_Failure | dendritic_cell | *ACSF2* | -3.293960584 | 1.08E-06 | 5.91E-06 | 0.005251908 |
| Heart_Failure | dendritic_cell | *URM1* | -3.028910717 | 2.08E-05 | 4.97E-05 | 0.100752499 |
| Heart_Failure | dendritic_cell | *ISLR2* | -3.004124226 | 4.86E-05 | 0.000100989 | 0.235808309 |
| Heart_Failure | dendritic_cell | *SP2* | -3.241793519 | 3.14E-06 | 1.17E-05 | 0.015216263 |
| Heart_Failure | dendritic_cell | *COQ7* | -2.80504224 | 1.60E-06 | 7.42E-06 | 0.00775882 |
| Heart_Failure | dendritic_cell | *GPRC5B* | -2.337970096 | 1.04E-05 | 2.80E-05 | 0.050615199 |
| Heart_Failure | dendritic_cell | *FBXO22* | -3.128379136 | 0.002093829 | 0.003063709 | 1 |
| Heart_Failure | dendritic_cell | *TBC1D2B* | -2.353658285 | 1.26E-06 | 6.39E-06 | 0.006111281 |
| Heart_Failure | dendritic_cell | *LOXHD1* | -1.287331829 | 4.35E-07 | 3.90E-06 | 0.00211177 |
| Heart_Failure | dendritic_cell | *KATNAL2* | -2.839256048 | 1.69E-06 | 7.68E-06 | 0.008185029 |
| Heart_Failure | dendritic_cell | *ZNF91* | -2.278989879 | 6.72E-05 | 0.000133807 | 0.325818978 |
| Heart_Failure | dendritic_cell | *CCL23* | -0.754522183 | 0.007131349 | 0.009624841 | 1 |
| Heart_Failure | dendritic_cell | *POP5* | -3.781633898 | 2.33E-05 | 5.47E-05 | 0.113289311 |
| Heart_Failure | dendritic_cell | *MYO5B* | -1.306814552 | 2.70E-07 | 3.66E-06 | 0.001307755 |
| Heart_Failure | dendritic_cell | *RRM1* | -1.032834008 | 5.82E-05 | 0.000118218 | 0.282540489 |
| Heart_Failure | dendritic_cell | *ZNF23* | -2.838607759 | 0.001005657 | 0.001550507 | 1 |
| Heart_Failure | dendritic_cell | *ZNF229* | -1.35052552 | 0.036509925 | 0.044745177 | 1 |
| Heart_Failure | dendritic_cell | *VKORC1* | 4.911164977 | 0.021403024 | 0.026994404 | 1 |
| Heart_Failure | dendritic_cell | *SMG8* | 2.091912538 | 7.61E-06 | 2.17E-05 | 0.036942202 |
| Heart_Failure | dendritic_cell | *RAB8A* | -1.926159711 | 0.033299942 | 0.041007949 | 1 |
| Heart_Failure | dendritic_cell | *SPATA33* | -2.921161165 | 1.27E-06 | 6.41E-06 | 0.006166424 |
| Heart_Failure | dendritic_cell | *PROCA1* | -1.699508038 | 4.08E-05 | 8.72E-05 | 0.197957794 |
| Heart_Failure | dendritic_cell | *RPL13* | -2.393017371 | 3.73E-05 | 8.06E-05 | 0.180953935 |
| Heart_Failure | dendritic_cell | *ZNF641* | 4.08608529 | 9.31E-06 | 2.54E-05 | 0.045163771 |
| Heart_Failure | dendritic_cell | *TP53I13* | -3.361487122 | 4.81E-06 | 1.57E-05 | 0.023320042 |
| Heart_Failure | dendritic_cell | *TUBA1C* | -1.713570749 | 0.000701522 | 0.001105842 | 1 |
| Heart_Failure | dendritic_cell | *ZNF528* | -2.977858193 | 5.72E-06 | 1.78E-05 | 0.027760052 |
| Heart_Failure | dendritic_cell | *SERTAD3* | 3.370694293 | 1.18E-07 | 3.44E-06 | 0.000572299 |
| Heart_Failure | dendritic_cell | *TMC4* | -3.885247363 | 3.61E-06 | 1.29E-05 | 0.017511012 |

| | | | | | | |
|---|---|---|---|---|---|---|
| Heart_Failure | dendritic_cell | *TTYH1* | -2.378573766 | 3.19E-07 | 3.66E-06 | 0.001547966 |
| Heart_Failure | dendritic_cell | *LENG8* | -3.213999826 | 8.27E-05 | 0.000160117 | 0.401093551 |
| Heart_Failure | dendritic_cell | *CDC42EP5* | -1.973466192 | 2.17E-05 | 5.16E-05 | 0.105200603 |
| Heart_Failure | dendritic_cell | *KIR3DL1* | -9.512459738 | 0.035765172 | 0.043910074 | 1 |
| Heart_Failure | dendritic_cell | *C19orf33* | 1.502348147 | 0.001667039 | 0.002480966 | 1 |
| Heart_Failure | dendritic_cell | *DNAAF3* | -1.786965012 | 0.000137097 | 0.00024998 | 0.665196764 |
| Heart_Failure | dendritic_cell | *CHAF1A* | -1.084805509 | 0.005088732 | 0.007010371 | 1 |
| Heart_Failure | dendritic_cell | *FAM57A* | -1.393738342 | 0.002391131 | 0.003454963 | 1 |
| Heart_Failure | dendritic_cell | *KIFC2* | 4.783534685 | 0.000104779 | 0.00019705 | 0.508390094 |
| Heart_Failure | dendritic_cell | *SERPINF2* | -1.695077179 | 0.000835313 | 0.001301104 | 1 |
| Heart_Failure | dendritic_cell | *TRPV3* | -1.007347726 | 0.0257738 | 0.032114658 | 1 |
| Heart_Failure | dendritic_cell | *CYB5D2* | -2.902737917 | 1.19E-05 | 3.12E-05 | 0.057875174 |
| Heart_Failure | dendritic_cell | *C19orf48* | -0.654255728 | 0.039398979 | 0.048103636 | 1 |
| Heart_Failure | dendritic_cell | *KLK5* | -2.948609068 | 1.62E-07 | 3.51E-06 | 0.000785403 |
| Heart_Failure | dendritic_cell | *KLK6* | -15.17472843 | 3.56E-07 | 3.66E-06 | 0.001726762 |
| Heart_Failure | dendritic_cell | *ZNF83* | -2.583938365 | 9.96E-05 | 0.000188167 | 0.483401442 |
| Heart_Failure | dendritic_cell | *SPRYD3* | -3.962060164 | 0.030949618 | 0.038259247 | 1 |
| Heart_Failure | dendritic_cell | *NDUFV1* | -3.37013739 | 8.27E-05 | 0.000160199 | 0.401457712 |
| Heart_Failure | dendritic_cell | *NUDT8* | -2.186890282 | 3.26E-08 | 3.33E-06 | 0.000158364 |
| Heart_Failure | dendritic_cell | *ZNF232* | 0.735621212 | 0.023883235 | 0.029966758 | 1 |
| Heart_Failure | dendritic_cell | *CD300C* | 0.773749938 | 0.017627521 | 0.022515782 | 1 |
| Heart_Failure | dendritic_cell | *ATP5H* | -3.507991274 | 1.53E-05 | 3.83E-05 | 0.074240857 |
| Heart_Failure | dendritic_cell | *MGAT5B* | -5.799102194 | 8.30E-07 | 5.01E-06 | 0.004026785 |
| Heart_Failure | dendritic_cell | *GSDMA* | -4.078714601 | 1.25E-15 | 6.06E-12 | 6.06E-12 |
| Heart_Failure | dendritic_cell | *TMEM99* | 1.84482947 | 0.007274084 | 0.009806573 | 1 |
| Heart_Failure | dendritic_cell | *ZNF598* | -2.96601913 | 4.12E-06 | 1.41E-05 | 0.019998334 |
| Heart_Failure | dendritic_cell | *MLST8* | 2.787112271 | 4.78E-08 | 3.44E-06 | 0.000231998 |
| Heart_Failure | dendritic_cell | *ABCA3* | -2.546192323 | 3.43E-05 | 7.54E-05 | 0.166649802 |
| Heart_Failure | dendritic_cell | *ATG16L2* | 3.911374936 | 3.07E-06 | 1.15E-05 | 0.014918196 |
| Heart_Failure | dendritic_cell | *RPSA* | -2.28382734 | 0.00059602 | 0.000951279 | 1 |
| Heart_Failure | dendritic_cell | *ULK4* | -1.81891135 | 7.38E-06 | 2.12E-05 | 0.035795922 |
| Heart_Failure | dendritic_cell | *LTBP3* | -1.688123634 | 3.14E-07 | 3.66E-06 | 0.001522261 |
| Heart_Failure | dendritic_cell | *SCARA5* | -1.316043858 | 5.78E-07 | 4.27E-06 | 0.002805841 |
| Heart_Failure | dendritic_cell | *KIAA1586* | -1.822277259 | 0.039465699 | 0.048172974 | 1 |
| Heart_Failure | dendritic_cell | *FAM83B* | -15.9437154 | 8.27E-06 | 2.32E-05 | 0.040124843 |
| Heart_Failure | dendritic_cell | *COA6* | -2.944225766 | 6.36E-07 | 4.39E-06 | 0.003084345 |
| Heart_Failure | dendritic_cell | *PXK* | -2.974872164 | 0.00229967 | 0.003334727 | 1 |
| Heart_Failure | dendritic_cell | *PCMTD1* | -2.220778951 | 9.02E-06 | 2.48E-05 | 0.04374794 |
| Heart_Failure | dendritic_cell | *ACOX2* | -2.683929928 | 4.77E-06 | 1.57E-05 | 0.023120319 |
| Heart_Failure | dendritic_cell | *FAM107A* | -6.089037653 | 0.000127599 | 0.0002346 | 0.619108143 |
| Heart_Failure | dendritic_cell | *SEPT2* | -1.370748752 | 0.000168336 | 0.000300945 | 0.816765176 |
| Heart_Failure | dendritic_cell | *DTYMK* | -4.740268341 | 7.91E-05 | 0.000153996 | 0.383571972 |
| Heart_Failure | dendritic_cell | *ING5* | -2.529693879 | 0.005906921 | 0.0080688 | 1 |
| Heart_Failure | dendritic_cell | *MLKL* | -1.498584054 | 5.44E-05 | 0.0001118 | 0.263847753 |
| Heart_Failure | dendritic_cell | *RFWD3* | 4.802951475 | 0.000125795 | 0.000231635 | 0.610358469 |
| Heart_Failure | dendritic_cell | *KLHL30* | -7.306750053 | 3.85E-07 | 3.72E-06 | 0.001869126 |
| Heart_Failure | dendritic_cell | *CCDC110* | -3.62342278 | 3.23E-07 | 3.66E-06 | 0.001567019 |
| Heart_Failure | dendritic_cell | *FNTA* | -2.439187373 | 0.000382059 | 0.000632679 | 1 |
| Heart_Failure | dendritic_cell | *SERINC2* | -5.878251437 | 8.08E-07 | 4.93E-06 | 0.003920704 |
| Heart_Failure | dendritic_cell | *SLC20A2* | -2.165127487 | 5.34E-06 | 1.70E-05 | 0.025915223 |

| | | | | | | |
|---|---|---|---|---|---|---|
| Heart_Failure | dendritic_cell | *NBPF9* | -2.354747309 | 3.50E-07 | 3.66E-06 | 0.001699291 |
| Heart_Failure | dendritic_cell | *ADAM9* | -2.878363369 | 1.34E-05 | 3.44E-05 | 0.065227932 |
| Heart_Failure | dendritic_cell | *NDUFS5* | -2.690671262 | 5.09E-06 | 1.64E-05 | 0.02471704 |
| Heart_Failure | dendritic_cell | *ZNF30* | 2.431696608 | 0.000275756 | 0.000472112 | 1 |
| Heart_Failure | dendritic_cell | *SLC16A4* | -1.439712481 | 0.00131755 | 0.001993997 | 1 |
| Heart_Failure | dendritic_cell | *DNAJC21* | -2.261526118 | 1.03E-06 | 5.74E-06 | 0.005005212 |
| Heart_Failure | dendritic_cell | *NPNT* | -1.628371884 | 2.79E-06 | 1.08E-05 | 0.013540091 |
| Heart_Failure | dendritic_cell | *GSTM4* | -2.220481078 | 3.41E-07 | 3.66E-06 | 0.001654776 |
| Heart_Failure | dendritic_cell | *ADAL* | -0.781523093 | 0.040104744 | 0.048916093 | 1 |
| Heart_Failure | dendritic_cell | *SNTB2* | -2.188892642 | 4.12E-06 | 1.41E-05 | 0.019970321 |
| Heart_Failure | dendritic_cell | *ZNF507* | -1.789119394 | 1.94E-07 | 3.62E-06 | 0.000940264 |
| Heart_Failure | dendritic_cell | *STX18* | -3.094180339 | 0.000105695 | 0.000198618 | 0.512830934 |
| Heart_Failure | dendritic_cell | *ZBTB49* | -2.838975426 | 7.88E-07 | 4.87E-06 | 0.003823879 |
| Heart_Failure | dendritic_cell | *GFM1* | -1.702004542 | 0.008524867 | 0.011385261 | 1 |
| Heart_Failure | dendritic_cell | *ATOH8* | -1.322836693 | 2.25E-05 | 5.32E-05 | 0.109403147 |
| Heart_Failure | dendritic_cell | *BTNL3* | -1.153646238 | 0.018350162 | 0.023362631 | 1 |
| Heart_Failure | dendritic_cell | *TMEM129* | -2.316860935 | 8.55E-05 | 0.000164712 | 0.414908883 |
| Heart_Failure | dendritic_cell | *CEP120* | -2.621148183 | 5.56E-06 | 1.75E-05 | 0.026965226 |
| Heart_Failure | dendritic_cell | *MFF* | -2.638263168 | 1.46E-05 | 3.68E-05 | 0.070870542 |
| Heart_Failure | dendritic_cell | *CPLX1* | -1.467608112 | 0.000154582 | 0.000278614 | 0.750029932 |
| Heart_Failure | dendritic_cell | *SIGLEC7* | 1.369974372 | 2.17E-05 | 5.16E-05 | 0.105056714 |
| Heart_Failure | dendritic_cell | *MFSD7* | -2.93897637 | 3.25E-07 | 3.66E-06 | 0.001574735 |
| Heart_Failure | dendritic_cell | *MAP2K1* | 2.923047894 | 0.017903844 | 0.022848356 | 1 |
| Heart_Failure | dendritic_cell | *KLK7* | -4.825889222 | 1.18E-06 | 6.14E-06 | 0.005706517 |
| Heart_Failure | dendritic_cell | *IRS1* | -0.850723129 | 4.50E-05 | 9.43E-05 | 0.218132669 |
| Heart_Failure | dendritic_cell | *UPF3A* | -1.95417244 | 1.53E-06 | 7.21E-06 | 0.007441078 |
| Heart_Failure | dendritic_cell | *UBE2V2* | -3.64118268 | 0.000143017 | 0.000259506 | 0.693918894 |
| Heart_Failure | dendritic_cell | *CPT1C* | 2.328907266 | 0.005966514 | 0.008134175 | 1 |
| Heart_Failure | dendritic_cell | *PCSK9* | -9.101653304 | 0.000381585 | 0.00063211 | 1 |
| Heart_Failure | dendritic_cell | *XPO6* | -3.401168927 | 0.000139921 | 0.000254554 | 0.678896719 |
| Heart_Failure | dendritic_cell | *RGS14* | 0.845430487 | 0.004935052 | 0.006812197 | 1 |
| Heart_Failure | dendritic_cell | *RAB24* | -4.101272394 | 1.20E-06 | 6.22E-06 | 0.005842686 |
| Heart_Failure | dendritic_cell | *THBS3* | -2.083501305 | 4.20E-07 | 3.88E-06 | 0.002038087 |
| Heart_Failure | dendritic_cell | *NPIPB3* | -3.653040419 | 5.06E-07 | 4.03E-06 | 0.002453791 |
| Heart_Failure | dendritic_cell | *SH3TC2* | -1.597703309 | 8.10E-06 | 2.28E-05 | 0.039294269 |
| Heart_Failure | dendritic_cell | *PGM2* | 4.510197532 | 0.000281727 | 0.000481148 | 1 |
| Heart_Failure | dendritic_cell | *P2RY12* | 1.282335889 | 3.04E-07 | 3.66E-06 | 0.001476681 |
| Heart_Failure | dendritic_cell | *RNASE6* | 2.151560833 | 1.10E-05 | 2.93E-05 | 0.053610227 |
| Heart_Failure | dendritic_cell | *SLC38A11* | -1.796857397 | 3.02E-06 | 1.14E-05 | 0.014651992 |
| Heart_Failure | dendritic_cell | *CCDC8* | -1.904346067 | 0.00026972 | 0.000462268 | 1 |
| Heart_Failure | dendritic_cell | *ZEB2* | -1.897854407 | 0.003880397 | 0.005423154 | 1 |
| Heart_Failure | dendritic_cell | *HINT1* | -3.022321451 | 1.62E-05 | 4.03E-05 | 0.078830512 |
| Heart_Failure | dendritic_cell | *DTWD2* | -2.667585616 | 0.006792576 | 0.00919832 | 1 |
| Heart_Failure | dendritic_cell | *CLIC3* | -2.448056616 | 3.35E-07 | 3.66E-06 | 0.001626474 |
| Heart_Failure | dendritic_cell | *INO80E* | -1.671833407 | 0.000299843 | 0.00050904 | 1 |
| Heart_Failure | dendritic_cell | *DFFB* | -2.64959013 | 8.49E-07 | 5.09E-06 | 0.004119878 |
| Heart_Failure | dendritic_cell | *NFU1* | -2.189184889 | 9.45E-06 | 2.58E-05 | 0.045832858 |
| Heart_Failure | dendritic_cell | *C15orf40* | -2.178427768 | 0.000132135 | 0.000242023 | 0.641119078 |
| Heart_Failure | dendritic_cell | *LUZP1* | -2.706746793 | 1.24E-05 | 3.23E-05 | 0.06028025 |
| Heart_Failure | dendritic_cell | *LRRC45* | -1.828039846 | 6.07E-05 | 0.000122698 | 0.294597553 |

| | | | | | | |
|---|---|---|---|---|---|---|
| Heart_Failure | dendritic_cell | *CHRNA5* | -6.514110308 | 0.001925865 | 0.002834181 | 1 |
| Heart_Failure | dendritic_cell | *UGP2* | -1.92328143 | 5.40E-05 | 0.000111166 | 0.261796553 |
| Heart_Failure | dendritic_cell | *HNRNPF* | -2.494587733 | 1.68E-05 | 4.16E-05 | 0.081639881 |
| Heart_Failure | dendritic_cell | *CSGALNACT2* | 4.389220514 | 0.000159842 | 0.000287136 | 0.775555599 |
| Heart_Failure | dendritic_cell | *AVEN* | -2.294869273 | 1.71E-06 | 7.72E-06 | 0.008315163 |
| Heart_Failure | dendritic_cell | *CTNND2* | -1.705995773 | 3.68E-07 | 3.66E-06 | 0.001787684 |
| Heart_Failure | dendritic_cell | *CALML6* | 26.85233528 | 1.40E-07 | 3.44E-06 | 0.000679677 |
| Heart_Failure | dendritic_cell | *MUC3A* | -1.369236805 | 0.022554218 | 0.028379944 | 1 |
| Heart_Failure | dendritic_cell | *TPST1* | -2.048881915 | 2.87E-06 | 1.10E-05 | 0.013943395 |
| Heart_Failure | dendritic_cell | *TOR1AIP2* | -2.546224355 | 2.56E-05 | 5.86E-05 | 0.124443981 |
| Heart_Failure | dendritic_cell | *OTUD3* | -1.936793737 | 0.000435686 | 0.000713689 | 1 |
| Heart_Failure | dendritic_cell | *ZFPM2* | -1.137961716 | 1.81E-06 | 8.01E-06 | 0.00877765 |
| Heart_Failure | dendritic_cell | *MAP3K2* | -2.071579999 | 0.011495398 | 0.015111263 | 1 |
| Heart_Failure | dendritic_cell | *MYO7B* | -1.588845386 | 0.00259426 | 0.003715274 | 1 |
| Heart_Failure | dendritic_cell | *UBE2E3* | -1.807019781 | 6.81E-07 | 4.53E-06 | 0.003303098 |
| Heart_Failure | dendritic_cell | *NIPA1* | -2.631142037 | 1.52E-06 | 7.19E-06 | 0.007376093 |
| Heart_Failure | dendritic_cell | *HNRNPA3* | -2.158924415 | 0.017496393 | 0.022357782 | 1 |
| Heart_Failure | dendritic_cell | *RNF150* | -1.793775349 | 3.66E-05 | 7.93E-05 | 0.177446672 |
| Heart_Failure | dendritic_cell | *USP38* | -3.721128926 | 5.92E-05 | 0.000119864 | 0.287074058 |
| Heart_Failure | dendritic_cell | *ANKK1* | -3.431754104 | 0.00034793 | 0.000582237 | 1 |
| Heart_Failure | dendritic_cell | *PDCD6IP* | -2.602273251 | 5.28E-05 | 0.000108805 | 0.256018218 |
| Heart_Failure | dendritic_cell | *CRTAP* | -3.062902589 | 0.001401828 | 0.002109143 | 1 |
| Heart_Failure | dendritic_cell | *CNGB3* | 3.02171561 | 2.80E-06 | 1.08E-05 | 0.013588848 |
| Heart_Failure | dendritic_cell | *ELP5* | -2.085678275 | 1.19E-06 | 6.17E-06 | 0.005786536 |
| Heart_Failure | dendritic_cell | *STX8* | -2.432170053 | 0.000535598 | 0.000864224 | 1 |
| Heart_Failure | dendritic_cell | *NFRKB* | -2.804755817 | 1.77E-06 | 7.90E-06 | 0.008584635 |
| Heart_Failure | dendritic_cell | *ADORA2B* | 0.692764663 | 0.002014369 | 0.002953678 | 1 |
| Heart_Failure | dendritic_cell | *MGMT* | -2.39677177 | 1.25E-05 | 3.25E-05 | 0.060733156 |
| Heart_Failure | dendritic_cell | *NFXL1* | -1.430186542 | 3.83E-07 | 3.72E-06 | 0.001860274 |
| Heart_Failure | dendritic_cell | *DNAJC18* | -2.437792972 | 3.24E-08 | 3.33E-06 | 0.000157249 |
| Heart_Failure | dendritic_cell | *HSD17B13* | -4.460692006 | 1.38E-07 | 3.44E-06 | 0.000669102 |
| Heart_Failure | dendritic_cell | *ELOVL6* | -1.774315391 | 6.35E-07 | 4.39E-06 | 0.003080854 |
| Heart_Failure | dendritic_cell | *ARL6IP1* | -2.514862081 | 0.000102945 | 0.000193976 | 0.49948833 |
| Heart_Failure | dendritic_cell | *SERPINB9* | -2.707463483 | 0.000396426 | 0.000653571 | 1 |
| Heart_Failure | dendritic_cell | *CDH2* | 2.669372149 | 1.95E-07 | 3.62E-06 | 0.000944282 |
| Heart_Failure | dendritic_cell | *EMB* | 1.99889799 | 0.000334559 | 0.000562272 | 1 |
| Heart_Failure | dendritic_cell | *STAT2* | -2.756828563 | 5.71E-05 | 0.00011638 | 0.277246904 |
| Heart_Failure | dendritic_cell | *SLC26A5* | -1.797921113 | 2.13E-06 | 8.88E-06 | 0.01035151 |
| Heart_Failure | dendritic_cell | *GTSF1* | -4.514835971 | 0.000323896 | 0.000546243 | 1 |
| Heart_Failure | dendritic_cell | *ARMC10* | -2.275921314 | 2.52E-05 | 5.79E-05 | 0.122383017 |
| Heart_Failure | dendritic_cell | *ACYP2* | -2.057373014 | 9.28E-06 | 2.54E-05 | 0.045008921 |
| Heart_Failure | dendritic_cell | *TRABD* | 3.297277484 | 0.011255645 | 0.014808131 | 1 |
| Heart_Failure | dendritic_cell | *HOXB9* | -6.642069494 | 1.30E-05 | 3.34E-05 | 0.063023925 |
| Heart_Failure | dendritic_cell | *TTLL6* | -5.423819526 | 2.57E-06 | 1.02E-05 | 0.01249263 |
| Heart_Failure | dendritic_cell | *FOXN2* | -2.610666941 | 0.013168063 | 0.017147461 | 1 |
| Heart_Failure | dendritic_cell | *TRIAP1* | -4.012713592 | 2.65E-07 | 3.66E-06 | 0.001285057 |
| Heart_Failure | dendritic_cell | *RPS9* | -2.334971367 | 0.000175002 | 0.000311829 | 0.849110479 |
| Heart_Failure | dendritic_cell | *TSEN34* | -2.146995418 | 1.03E-06 | 5.74E-06 | 0.004996274 |
| Heart_Failure | dendritic_cell | *MSANTD4* | -2.469160803 | 9.84E-07 | 5.60E-06 | 0.004776588 |
| Heart_Failure | dendritic_cell | *NDUFA3* | -2.463307425 | 1.13E-06 | 6.03E-06 | 0.005459774 |

| | | | | | | |
|---|---|---|---|---|---|---|
| Heart_Failure | dendritic_cell | *PAQR8* | 2.482532571 | 0.000248123 | 0.000428432 | 1 |
| Heart_Failure | dendritic_cell | *NUDT6* | -2.084742233 | 2.19E-07 | 3.66E-06 | 0.001064311 |
| Heart_Failure | dendritic_cell | *ZNF160* | -1.78389858 | 0.000165365 | 0.000296289 | 0.802351636 |
| Heart_Failure | dendritic_cell | *ZNF415* | -3.532466028 | 6.34E-07 | 4.39E-06 | 0.003076834 |
| Heart_Failure | dendritic_cell | *S1PR1* | -1.699044293 | 3.87E-06 | 1.35E-05 | 0.018784958 |
| Heart_Failure | dendritic_cell | *PYGO1* | -2.173642656 | 2.41E-06 | 9.66E-06 | 0.011677069 |
| Heart_Failure | dendritic_cell | *TSNARE1* | 2.534387822 | 0.012006973 | 0.015724111 | 1 |
| Heart_Failure | dendritic_cell | *FPR1* | 0.415283679 | 1.40E-07 | 3.44E-06 | 0.00067926 |
| Heart_Failure | dendritic_cell | *FEZ2* | -2.228539357 | 9.92E-05 | 0.000187526 | 0.481379171 |
| Heart_Failure | dendritic_cell | *SOX7* | -4.675555357 | 0.000382214 | 0.00063272 | 1 |
| Heart_Failure | dendritic_cell | *TRMT61B* | -2.121439738 | 9.84E-07 | 5.60E-06 | 0.004773178 |
| Heart_Failure | dendritic_cell | *MFN1* | -2.115626846 | 0.003197048 | 0.004510635 | 1 |
| Heart_Failure | dendritic_cell | *GIMAP8* | -2.228581033 | 1.16E-06 | 6.11E-06 | 0.005631317 |
| Heart_Failure | dendritic_cell | *ATP6V0E2* | -2.56164637 | 1.89E-06 | 8.21E-06 | 0.009149429 |
| Heart_Failure | dendritic_cell | *MORN4* | -1.43680653 | 2.50E-05 | 5.76E-05 | 0.121096743 |
| Heart_Failure | dendritic_cell | *SCAND1* | -2.023474907 | 3.76E-07 | 3.66E-06 | 0.001824979 |
| Heart_Failure | dendritic_cell | *GAA* | -1.037091834 | 0.001794461 | 0.002654489 | 1 |
| Heart_Failure | dendritic_cell | *PGAM1* | -1.935313518 | 0.000702632 | 0.001107233 | 1 |
| Heart_Failure | dendritic_cell | *LURAP1* | -1.133106574 | 0.000637324 | 0.001011546 | 1 |
| Heart_Failure | dendritic_cell | *PDE7B* | -1.654207306 | 3.01E-06 | 1.14E-05 | 0.014596425 |
| Heart_Failure | dendritic_cell | *MRPL36* | -1.747854149 | 2.96E-06 | 1.12E-05 | 0.014342656 |
| Heart_Failure | dendritic_cell | *MCC* | -2.587982751 | 3.37E-05 | 7.41E-05 | 0.163524103 |
| Heart_Failure | dendritic_cell | *HOPX* | -2.703523388 | 0.000947826 | 0.001466004 | 1 |
| Heart_Failure | dendritic_cell | *PPID* | -3.231954841 | 9.77E-05 | 0.000185133 | 0.474125708 |
| Heart_Failure | dendritic_cell | *ETFDH* | -1.0577582 | 1.11E-05 | 2.94E-05 | 0.053923009 |
| Heart_Failure | dendritic_cell | *TBCA* | -2.767118979 | 0.000347998 | 0.000582237 | 1 |
| Heart_Failure | dendritic_cell | *NMUR1* | -2.770138156 | 3.64E-07 | 3.66E-06 | 0.001765723 |
| Heart_Failure | dendritic_cell | *PIK3CD* | 3.157566284 | 0.000827879 | 0.001292014 | 1 |
| Heart_Failure | dendritic_cell | *ANO5* | -1.484692875 | 1.41E-06 | 6.82E-06 | 0.006838748 |
| Heart_Failure | dendritic_cell | *VAT1L* | -2.215039553 | 1.10E-07 | 3.44E-06 | 0.000532639 |
| Heart_Failure | dendritic_cell | *CAMTA1* | -2.706639299 | 0.000349638 | 0.000583974 | 1 |
| Heart_Failure | dendritic_cell | *LGALS4* | 2.301544428 | 1.38E-08 | 2.99E-06 | 6.71E-05 |
| Heart_Failure | dendritic_cell | *LRRC34* | -0.739010611 | 0.006391144 | 0.008671652 | 1 |
| Heart_Failure | dendritic_cell | *PAH* | 1.691598114 | 2.12E-07 | 3.66E-06 | 0.001029957 |
| Heart_Failure | dendritic_cell | *PWWP2B* | -2.286643122 | 8.13E-07 | 4.93E-06 | 0.003942603 |
| Heart_Failure | dendritic_cell | *EXOSC10* | -3.59605126 | 2.51E-05 | 5.78E-05 | 0.121962108 |
| Heart_Failure | dendritic_cell | *TRAPPC12* | -2.336154512 | 0.035871572 | 0.044029564 | 1 |
| Heart_Failure | dendritic_cell | *C3AR1* | 1.481413605 | 1.30E-06 | 6.51E-06 | 0.006304989 |
| Heart_Failure | dendritic_cell | *TVP23B* | -2.030732771 | 7.71E-05 | 0.00015076 | 0.374036252 |
| Heart_Failure | dendritic_cell | *FBXW10* | -4.625220227 | 5.26E-07 | 4.10E-06 | 0.002552726 |
| Heart_Failure | dendritic_cell | *SRGAP2C* | 1.958537574 | 0.002435318 | 0.003513578 | 1 |
| Heart_Failure | dendritic_cell | *ATPAF2* | 3.179327839 | 2.42E-08 | 3.27E-06 | 0.000117642 |
| Heart_Failure | dendritic_cell | *ZNF554* | -2.106712822 | 1.92E-07 | 3.60E-06 | 0.000929181 |
| Heart_Failure | dendritic_cell | *ANKRD20A4* | 3.487875192 | 1.21E-07 | 3.44E-06 | 0.000586438 |
| Heart_Failure | dendritic_cell | *SERF1A* | -26.08990849 | 2.78E-06 | 1.07E-05 | 0.013476089 |
| Heart_Failure | dendritic_cell | *EIF2AK3* | -2.992129051 | 0.0014117 | 0.00212226 | 1 |
| Heart_Failure | dendritic_cell | *MTBP* | -3.366440479 | 3.87E-05 | 8.34E-05 | 0.187683418 |
| Heart_Failure | dendritic_cell | *TEFM* | -2.467517111 | 0.006998999 | 0.009462007 | 1 |
| Heart_Failure | dendritic_cell | *MALT1* | -3.148893479 | 0.001318656 | 0.001995048 | 1 |
| Heart_Failure | dendritic_cell | *MBOAT1* | -2.980643797 | 6.97E-06 | 2.04E-05 | 0.03383411 |

| | | | | | | |
|---|---|---|---|---|---|---|
| Heart_Failure | dendritic_cell | *CXCR6* | -3.790389024 | 8.18E-07 | 4.95E-06 | 0.003968325 |
| Heart_Failure | dendritic_cell | *PAIP1* | -2.175535144 | 5.09E-07 | 4.03E-06 | 0.002468781 |
| Heart_Failure | dendritic_cell | *C1QTNF4* | -1.021022975 | 0.004572623 | 0.006333533 | 1 |
| Heart_Failure | dendritic_cell | *MACROD2* | -1.416740553 | 0.000101875 | 0.000192035 | 0.494297022 |
| Heart_Failure | dendritic_cell | *BSG* | -2.262363321 | 5.07E-06 | 1.63E-05 | 0.024611486 |
| Heart_Failure | dendritic_cell | *HINFP* | 4.418076067 | 0.001001979 | 0.001545328 | 1 |
| Heart_Failure | dendritic_cell | *TP53RK* | -5.39285967 | 1.42E-06 | 6.87E-06 | 0.006896788 |
| Heart_Failure | dendritic_cell | *SUCLG2* | -2.567930246 | 8.60E-05 | 0.000165349 | 0.417174888 |
| Heart_Failure | dendritic_cell | *ARNT2* | -1.446544518 | 9.30E-07 | 5.42E-06 | 0.004514315 |
| Heart_Failure | dendritic_cell | *GTPBP2* | -2.634312524 | 2.47E-05 | 5.71E-05 | 0.119641773 |
| Heart_Failure | dendritic_cell | *FGGY* | -2.149689085 | 7.01E-06 | 2.05E-05 | 0.03402407 |
| Heart_Failure | dendritic_cell | *MANEA* | -3.482302958 | 0.000140403 | 0.000255336 | 0.681236143 |
| Heart_Failure | dendritic_cell | *AFF1* | -1.879689371 | 0.02428221 | 0.030443742 | 1 |
| Heart_Failure | dendritic_cell | *FIBP* | -3.262739191 | 0.017453622 | 0.022309002 | 1 |
| Heart_Failure | dendritic_cell | *SNTG2* | -1.005639131 | 3.95E-07 | 3.76E-06 | 0.001917762 |
| Heart_Failure | dendritic_cell | *SMPDL3A* | -1.094534818 | 0.000159704 | 0.000286993 | 0.774882343 |
| Heart_Failure | dendritic_cell | *RAD9A* | -1.225407589 | 0.013824174 | 0.017958472 | 1 |
| Heart_Failure | dendritic_cell | *EFEMP2* | -3.019068372 | 8.21E-05 | 0.000159173 | 0.398411044 |
| Heart_Failure | dendritic_cell | *ZNF738* | -1.211307649 | 0.024767757 | 0.030932602 | 1 |
| Heart_Failure | dendritic_cell | *SLFN11* | -3.971645518 | 3.95E-05 | 8.47E-05 | 0.191509991 |
| Heart_Failure | dendritic_cell | *FAM71D* | -2.22580396 | 1.70E-06 | 7.71E-06 | 0.008246685 |
| Heart_Failure | dendritic_cell | *FUT10* | -0.878133744 | 0.026080558 | 0.032463537 | 1 |
| Heart_Failure | dendritic_cell | *ZNF596* | -1.53406925 | 5.07E-07 | 4.03E-06 | 0.002461834 |
| Heart_Failure | dendritic_cell | *CFL1* | -3.438281331 | 0.000600823 | 0.000957686 | 1 |
| Heart_Failure | dendritic_cell | *FAM192A* | -2.45199454 | 0.00103086 | 0.001586844 | 1 |
| Heart_Failure | dendritic_cell | *DCP2* | -3.205633921 | 0.000346004 | 0.0005797 | 1 |
| Heart_Failure | dendritic_cell | *ZNF621* | -2.44022917 | 2.00E-06 | 8.53E-06 | 0.009687194 |
| Heart_Failure | dendritic_cell | *RNASEH2C* | -1.291644991 | 3.44E-05 | 7.54E-05 | 0.166878546 |
| Heart_Failure | dendritic_cell | *ANKRD13D* | 3.377052372 | 0.002382594 | 0.00344468 | 1 |
| Heart_Failure | dendritic_cell | *MRGPRF* | 2.361328239 | 6.50E-06 | 1.95E-05 | 0.031542844 |
| Heart_Failure | dendritic_cell | *LCLAT1* | -2.740773042 | 1.32E-07 | 3.44E-06 | 0.000640046 |
| Heart_Failure | dendritic_cell | *SH3RF3* | -2.035851018 | 8.81E-06 | 2.44E-05 | 0.042728182 |
| Heart_Failure | dendritic_cell | *GXYLT2* | -1.758578092 | 1.65E-06 | 7.59E-06 | 0.008010443 |
| Heart_Failure | dendritic_cell | *DCAKD* | -2.472501923 | 2.26E-07 | 3.66E-06 | 0.001094778 |
| Heart_Failure | dendritic_cell | *TRMT112* | -2.912340281 | 5.86E-07 | 4.29E-06 | 0.00284185 |
| Heart_Failure | dendritic_cell | *NOC3L* | -2.275477058 | 8.72E-06 | 2.42E-05 | 0.042301875 |
| Heart_Failure | dendritic_cell | *RHOD* | -2.537741619 | 7.17E-07 | 4.65E-06 | 0.003478255 |
| Heart_Failure | dendritic_cell | *AHSA2* | -2.251732313 | 0.001036907 | 0.001595646 | 1 |
| Heart_Failure | dendritic_cell | *VANGL1* | -1.889179067 | 6.18E-07 | 4.35E-06 | 0.003000325 |
| Heart_Failure | dendritic_cell | *IQCB1* | -2.501195433 | 3.12E-05 | 6.89E-05 | 0.151146308 |
| Heart_Failure | dendritic_cell | *ZNF483* | -1.47488675 | 5.54E-06 | 1.75E-05 | 0.026897497 |
| Heart_Failure | dendritic_cell | *SLC2A14* | -1.960197195 | 0.000383216 | 0.000633731 | 1 |
| Heart_Failure | dendritic_cell | *MMRN2* | -2.149049495 | 2.56E-06 | 1.01E-05 | 0.012397711 |
| Heart_Failure | dendritic_cell | *MZT2A* | -1.893148572 | 0.002940139 | 0.004172434 | 1 |
| Heart_Failure | dendritic_cell | *MAP3K11* | -2.291646937 | 2.87E-05 | 6.42E-05 | 0.139081519 |
| Heart_Failure | dendritic_cell | *NDNF* | -2.306321103 | 1.27E-05 | 3.28E-05 | 0.061708124 |
| Heart_Failure | dendritic_cell | *ARV1* | -1.979761559 | 0.021315013 | 0.026904382 | 1 |
| Heart_Failure | dendritic_cell | *CCDC36* | -1.401191233 | 1.64E-05 | 4.06E-05 | 0.079376573 |
| Heart_Failure | dendritic_cell | *EHBP1L1* | 2.091808693 | 0.000700261 | 0.001104933 | 1 |
| Heart_Failure | dendritic_cell | *PPP1R14B* | 4.786961493 | 4.17E-08 | 3.44E-06 | 0.00020245 |

| | | | | | | |
|---|---|---|---|---|---|---|
| Heart_Failure | dendritic_cell | *PEAK1* | -2.264846426 | 0.000207546 | 0.000364074 | 1 |
| Heart_Failure | dendritic_cell | *TNFRSF10D* | -1.73169208 | 5.66E-05 | 0.000115443 | 0.274407773 |
| Heart_Failure | dendritic_cell | *MST1* | -1.044921304 | 4.50E-05 | 9.43E-05 | 0.218121041 |
| Heart_Failure | dendritic_cell | *GMPPB* | 1.958802627 | 4.17E-08 | 3.44E-06 | 0.000202107 |
| Heart_Failure | dendritic_cell | *NUDT18* | 3.132440379 | 1.24E-07 | 3.44E-06 | 0.000601412 |
| Heart_Failure | dendritic_cell | *SULT1B1* | 1.439586628 | 3.20E-07 | 3.66E-06 | 0.001554888 |
| Heart_Failure | dendritic_cell | *NUDT4* | -3.399276302 | 6.03E-05 | 0.000121844 | 0.292424519 |
| Heart_Failure | dendritic_cell | *SCAI* | -1.655822997 | 0.018310889 | 0.023318748 | 1 |
| Heart_Failure | dendritic_cell | *UQCRH* | -2.681639816 | 7.23E-06 | 2.09E-05 | 0.035080593 |
| Heart_Failure | dendritic_cell | *C11orf80* | -2.871736947 | 7.98E-09 | 2.58E-06 | 3.87E-05 |
| Heart_Failure | dendritic_cell | *RNF213* | -2.30881319 | 0.001144289 | 0.001748139 | 1 |
| Heart_Failure | dendritic_cell | *TUBB8* | 1.379734618 | 0.006430356 | 0.008722418 | 1 |
| Heart_Failure | dendritic_cell | *GPR160* | 2.008507534 | 0.002706972 | 0.003867559 | 1 |
| Heart_Failure | dendritic_cell | *GOLIM4* | -3.100140095 | 8.77E-05 | 0.000168229 | 0.425615636 |
| Heart_Failure | dendritic_cell | *HOXB2* | -2.437549132 | 7.23E-06 | 2.09E-05 | 0.035098814 |
| Heart_Failure | dendritic_cell | *XXYLT1* | -2.587247379 | 3.02E-06 | 1.14E-05 | 0.014651382 |
| Heart_Failure | dendritic_cell | *UBXN2A* | -2.366350329 | 6.59E-06 | 1.96E-05 | 0.031961962 |
| Heart_Failure | dendritic_cell | *NRROS* | 1.329870571 | 0.002462296 | 0.003547226 | 1 |
| Heart_Failure | dendritic_cell | *CEP19* | -1.487603056 | 0.002557687 | 0.003670481 | 1 |
| Heart_Failure | dendritic_cell | *SLC25A30* | 5.979306868 | 9.37E-06 | 2.56E-05 | 0.045484959 |
| Heart_Failure | dendritic_cell | *CTSF* | -2.160985608 | 0.001667953 | 0.002480966 | 1 |
| Heart_Failure | dendritic_cell | *LEMD3* | -3.254690366 | 0.000625681 | 0.000995019 | 1 |
| Heart_Failure | dendritic_cell | *CTU2* | 1.382312085 | 1.39E-05 | 3.55E-05 | 0.06766125 |
| Heart_Failure | dendritic_cell | *SNX31* | -0.957959342 | 0.008839756 | 0.011779867 | 1 |
| Heart_Failure | dendritic_cell | *GLIS1* | -1.692583989 | 3.15E-07 | 3.66E-06 | 0.001528098 |
| Heart_Failure | dendritic_cell | *RALGAPA1* | -1.965837374 | 8.52E-05 | 0.000164248 | 0.413511102 |
| Heart_Failure | dendritic_cell | *SNAPC5* | -3.441451719 | 0.000404785 | 0.000665994 | 1 |
| Heart_Failure | dendritic_cell | *ANKRD36C* | -2.751595219 | 1.95E-05 | 4.70E-05 | 0.094697394 |
| Heart_Failure | dendritic_cell | *MSL2* | -2.723354963 | 7.01E-06 | 2.05E-05 | 0.034024809 |
| Heart_Failure | dendritic_cell | *ZNF497* | -11.93328544 | 2.97E-06 | 1.12E-05 | 0.014418907 |
| Heart_Failure | dendritic_cell | *TRAM1L1* | 3.31911954 | 5.48E-05 | 0.00011242 | 0.265760652 |
| Heart_Failure | dendritic_cell | *ANGEL2* | -2.628583024 | 0.024588609 | 0.030764294 | 1 |
| Heart_Failure | dendritic_cell | *ZNF266* | -1.123226819 | 0.007340445 | 0.009890542 | 1 |
| Heart_Failure | dendritic_cell | *SH3PXD2B* | -2.219331106 | 5.03E-06 | 1.62E-05 | 0.024413017 |
| Heart_Failure | dendritic_cell | *LARP7* | -3.265945146 | 8.59E-05 | 0.000165208 | 0.416589206 |
| Heart_Failure | dendritic_cell | *FGFBP3* | -3.293897293 | 3.18E-07 | 3.66E-06 | 0.00154417 |
| Heart_Failure | dendritic_cell | *C4orf32* | -1.883949311 | 8.25E-07 | 4.99E-06 | 0.004004735 |
| Heart_Failure | dendritic_cell | *HRAS* | -2.28790022 | 7.16E-07 | 4.65E-06 | 0.00347414 |
| Heart_Failure | dendritic_cell | *SRP72* | -1.559977189 | 1.55E-05 | 3.86E-05 | 0.074979776 |
| Heart_Failure | dendritic_cell | *CEP135* | -3.012595949 | 0.010941763 | 0.014402993 | 1 |
| Heart_Failure | dendritic_cell | *NLRP6* | 2.110476381 | 3.28E-07 | 3.66E-06 | 0.001593291 |
| Heart_Failure | dendritic_cell | *RSRC1* | -2.527017766 | 0.006164569 | 0.008387687 | 1 |
| Heart_Failure | dendritic_cell | *PTDSS2* | -2.577932647 | 0.000210121 | 0.000367921 | 1 |
| Heart_Failure | dendritic_cell | *SEZ6L2* | -1.570410355 | 6.13E-05 | 0.000123621 | 0.297431347 |
| Heart_Failure | dendritic_cell | *KCTD13* | -2.293947531 | 0.000793154 | 0.001240214 | 1 |
| Heart_Failure | dendritic_cell | *DHX36* | -2.785763467 | 6.37E-05 | 0.000127597 | 0.308911316 |
| Heart_Failure | dendritic_cell | *FBXW8* | -2.416222211 | 3.56E-06 | 1.28E-05 | 0.01728195 |
| Heart_Failure | dendritic_cell | *SLC22A1* | -3.82080338 | 1.05E-06 | 5.79E-06 | 0.00508745 |
| Heart_Failure | dendritic_cell | *CHST2* | -5.207248247 | 1.44E-05 | 3.65E-05 | 0.069947374 |
| Heart_Failure | dendritic_cell | *TVP23C* | 10.0057011 | 9.86E-05 | 0.000186378 | 0.478246688 |

| | | | | | | |
|---|---|---|---|---|---|---|
| Heart_Failure | dendritic_cell | *SH3BP5L* | 4.180682232 | 3.28E-07 | 3.66E-06 | 0.001593137 |
| Heart_Failure | dendritic_cell | *CADM2* | -1.483334717 | 0.010005784 | 0.013228356 | 1 |
| Heart_Failure | dendritic_cell | *PSMD2* | -3.847574865 | 4.80E-05 | 9.99E-05 | 0.232942296 |
| Heart_Failure | dendritic_cell | *FAM182B* | -1.274588991 | 7.28E-06 | 2.10E-05 | 0.03532729 |
| Heart_Failure | dendritic_cell | *ARHGAP1* | -2.568167455 | 7.47E-05 | 0.000146414 | 0.36252147 |
| Heart_Failure | dendritic_cell | *ATG13* | -3.590050365 | 0.000521255 | 0.000843422 | 1 |
| Heart_Failure | dendritic_cell | *ZNF519* | -2.099364398 | 9.43E-07 | 5.45E-06 | 0.004573543 |
| Heart_Failure | dendritic_cell | *BANF1* | -2.759223618 | 2.30E-06 | 9.37E-06 | 0.011157054 |
| Heart_Failure | dendritic_cell | *CHRNA7* | -3.389998751 | 5.26E-05 | 0.000108517 | 0.255014918 |
| Heart_Failure | dendritic_cell | *TMEM9B* | 3.822801969 | 4.11E-07 | 3.85E-06 | 0.001992995 |
| Heart_Failure | dendritic_cell | *SCUBE2* | -2.932202113 | 2.95E-07 | 3.66E-06 | 0.001432506 |
| Heart_Failure | dendritic_cell | *CLTB* | -2.677868877 | 1.74E-06 | 7.81E-06 | 0.008439474 |
| Heart_Failure | dendritic_cell | *PPP2R2D* | 2.069690172 | 0.038825317 | 0.047439042 | 1 |
| Heart_Failure | dendritic_cell | *MCTP1* | -2.300534765 | 0.01163363 | 0.015255776 | 1 |
| Heart_Failure | dendritic_cell | *LRRC25* | 1.222431434 | 3.41E-06 | 1.24E-05 | 0.016544384 |
| Heart_Failure | dendritic_cell | *KCNE3* | -2.842460784 | 0.002251375 | 0.00327154 | 1 |
| Heart_Failure | dendritic_cell | *UCP2* | -1.690448038 | 0.002215826 | 0.003228584 | 1 |
| Heart_Failure | dendritic_cell | *C11orf68* | -5.167105258 | 0.00053913 | 0.000869057 | 1 |
| Heart_Failure | dendritic_cell | *PAAF1* | -1.453043825 | 3.43E-06 | 1.24E-05 | 0.016648809 |
| Heart_Failure | dendritic_cell | *MRPL48* | -3.312713065 | 1.59E-05 | 3.95E-05 | 0.076970022 |
| Heart_Failure | dendritic_cell | *RAB6A* | -2.532311765 | 1.14E-05 | 3.01E-05 | 0.055256195 |
| Heart_Failure | dendritic_cell | *TMEM70* | 1.991787846 | 2.66E-07 | 3.66E-06 | 0.001292191 |
| Heart_Failure | dendritic_cell | *RPS6KB2* | -2.381069808 | 0.001681738 | 0.002500703 | 1 |
| Heart_Failure | dendritic_cell | *RMI2* | -2.303907776 | 3.42E-07 | 3.66E-06 | 0.001659295 |
| Heart_Failure | dendritic_cell | *TOM1L2* | -2.57177725 | 0.000315427 | 0.000533072 | 1 |
| Heart_Failure | dendritic_cell | *TEX26* | -5.41176446 | 0.000112464 | 0.000210038 | 0.54567751 |
| Heart_Failure | dendritic_cell | *GPR156* | 3.247680149 | 6.14E-05 | 0.000123724 | 0.297927899 |
| Heart_Failure | dendritic_cell | *TTLL11* | -2.022269685 | 1.85E-06 | 8.11E-06 | 0.008983001 |
| Heart_Failure | dendritic_cell | *PRIMA1* | -1.783027207 | 3.51E-07 | 3.66E-06 | 0.001703781 |
| Heart_Failure | dendritic_cell | *CTDNEP1* | -2.062790652 | 0.001903678 | 0.002804083 | 1 |
| Heart_Failure | dendritic_cell | *HOXD8* | -1.146477699 | 0.007835997 | 0.01051736 | 1 |
| Heart_Failure | dendritic_cell | *ZDHHC21* | -1.896010907 | 1.09E-05 | 2.90E-05 | 0.052966732 |
| Heart_Failure | dendritic_cell | *A2M* | -2.49854605 | 7.27E-05 | 0.000143229 | 0.352572146 |
| Heart_Failure | dendritic_cell | *DOK7* | -12.51319826 | 1.30E-06 | 6.52E-06 | 0.006329547 |
| Heart_Failure | dendritic_cell | *ORAI3* | 3.083694234 | 6.31E-05 | 0.000126683 | 0.306320699 |
| Heart_Failure | dendritic_cell | *B3GALT6* | -5.258801617 | 4.87E-07 | 3.96E-06 | 0.002360998 |
| Heart_Failure | dendritic_cell | *JAKMIP2* | -2.2087962 | 0.007059709 | 0.009538765 | 1 |
| Heart_Failure | dendritic_cell | *TCP11L1* | 1.951839428 | 0.0277566 | 0.034470188 | 1 |
| Heart_Failure | dendritic_cell | *CCDC57* | -2.129456506 | 3.38E-06 | 1.23E-05 | 0.016395273 |
| Heart_Failure | dendritic_cell | *MYPOP* | -3.880550125 | 0.001351842 | 0.002038899 | 1 |
| Heart_Failure | dendritic_cell | *CIDEA* | -1.301645095 | 9.54E-07 | 5.47E-06 | 0.004631159 |
| Heart_Failure | dendritic_cell | *SMIM19* | -1.339456816 | 0.002846528 | 0.004052627 | 1 |
| Heart_Failure | dendritic_cell | *ZNF404* | -0.934590548 | 0.018183128 | 0.023162126 | 1 |
| Heart_Failure | dendritic_cell | *SLC35G1* | -1.651789667 | 3.70E-07 | 3.66E-06 | 0.001794548 |
| Heart_Failure | dendritic_cell | *ZNF135* | -1.381425881 | 1.18E-05 | 3.10E-05 | 0.057424759 |
| Heart_Failure | dendritic_cell | *OR4N2* | -3.878147236 | 0.000570744 | 0.000914548 | 1 |
| Heart_Failure | dendritic_cell | *CDC26* | -2.061040209 | 0.000676755 | 0.001070977 | 1 |
| Heart_Failure | dendritic_cell | *DNAJC30* | -2.022135475 | 8.50E-07 | 5.09E-06 | 0.004123186 |
| Heart_Failure | dendritic_cell | *SPRYD4* | -2.691230291 | 9.60E-08 | 3.44E-06 | 0.000465564 |
| Heart_Failure | dendritic_cell | *VPS37D* | 1.594747627 | 0.000103343 | 0.00019465 | 0.501418503 |

| | | | | | | |
|---|---|---|---|---|---|---|
| Heart_Failure | dendritic_cell | *WDR25* | -2.76515524 | 2.77E-06 | 1.07E-05 | 0.013454399 |
| Heart_Failure | dendritic_cell | *PLA2G16* | -1.186420814 | 3.33E-06 | 1.22E-05 | 0.016175614 |
| Heart_Failure | dendritic_cell | *B3GNT5* | -3.034569441 | 4.80E-05 | 9.99E-05 | 0.232869879 |
| Heart_Failure | dendritic_cell | *RMDN1* | -1.514726677 | 0.015405811 | 0.019858926 | 1 |
| Heart_Failure | dendritic_cell | *RNF152* | -2.121424253 | 8.52E-05 | 0.000164248 | 0.413577517 |
| Heart_Failure | dendritic_cell | *BDNF* | -3.455836421 | 2.83E-05 | 6.37E-05 | 0.137329892 |
| Heart_Failure | dendritic_cell | *BOK* | -1.455552613 | 3.35E-07 | 3.66E-06 | 0.001625564 |
| Heart_Failure | dendritic_cell | *PFN4* | 1.291075849 | 0.009489271 | 0.012603872 | 1 |
| Heart_Failure | dendritic_cell | *RUFY1* | -2.0601571 | 0.003619535 | 0.005080123 | 1 |
| Heart_Failure | dendritic_cell | *VSIG10* | -2.309005149 | 5.29E-07 | 4.12E-06 | 0.002564932 |
| Heart_Failure | dendritic_cell | *ANKLE2* | -3.356334442 | 7.35E-05 | 0.000144537 | 0.356386876 |
| Heart_Failure | dendritic_cell | *C8G* | 5.76746294 | 1.37E-07 | 3.44E-06 | 0.000665653 |
| Heart_Failure | dendritic_cell | *MUC20* | -3.762550569 | 9.54E-05 | 0.000180888 | 0.462711126 |
| Heart_Failure | dendritic_cell | *LY6H* | -2.665566644 | 0.00176408 | 0.002614518 | 1 |
| Heart_Failure | dendritic_cell | *SHMT1* | -2.324236609 | 3.57E-06 | 1.28E-05 | 0.017341786 |
| Heart_Failure | dendritic_cell | *DPP7* | -3.431613105 | 1.43E-05 | 3.62E-05 | 0.069308253 |
| Heart_Failure | dendritic_cell | *MTHFR* | -3.460649327 | 0.000312638 | 0.000528728 | 1 |
| Heart_Failure | dendritic_cell | *C19orf18* | -2.086957091 | 3.51E-07 | 3.66E-06 | 0.001700669 |
| Heart_Failure | dendritic_cell | *TMEM80* | -2.775086794 | 0.003740861 | 0.005239798 | 1 |
| Heart_Failure | dendritic_cell | *WDR73* | -3.008688364 | 9.04E-06 | 2.48E-05 | 0.043859744 |
| Heart_Failure | dendritic_cell | *DSCAML1* | -2.25541485 | 2.06E-06 | 8.73E-06 | 0.009994348 |
| Heart_Failure | dendritic_cell | *TALDO1* | -3.422333556 | 1.25E-05 | 3.25E-05 | 0.060873609 |
| Heart_Failure | dendritic_cell | *AP006621.1* | -1.895495985 | 1.69E-07 | 3.52E-06 | 0.000819748 |
| Heart_Failure | dendritic_cell | *MIEF2* | -2.423442982 | 1.92E-07 | 3.60E-06 | 0.000929187 |
| Heart_Failure | dendritic_cell | *NAP1L5* | -4.385620375 | 3.77E-06 | 1.33E-05 | 0.018276621 |
| Heart_Failure | dendritic_cell | *NIM1K* | -3.364209206 | 1.94E-05 | 4.68E-05 | 0.093906071 |
| Heart_Failure | dendritic_cell | *RBM44* | -3.252542765 | 0.00184358 | 0.002720514 | 1 |
| Heart_Failure | dendritic_cell | *SLC25A22* | -3.741788463 | 7.03E-07 | 4.63E-06 | 0.003410743 |
| Heart_Failure | dendritic_cell | *SAMD12* | -0.631133532 | 0.002502851 | 0.003598173 | 1 |
| Heart_Failure | dendritic_cell | *C18orf32* | -1.840010295 | 1.33E-07 | 3.44E-06 | 0.000644014 |
| Heart_Failure | dendritic_cell | *GBA* | -4.886742625 | 0.001647606 | 0.002453709 | 1 |
| Heart_Failure | dendritic_cell | *ACAD9* | -1.414188402 | 0.000535821 | 0.000864296 | 1 |
| Heart_Failure | dendritic_cell | *IL17RA* | 3.219102583 | 1.59E-06 | 7.40E-06 | 0.00771704 |
| Heart_Failure | dendritic_cell | *MBOAT4* | -3.902301281 | 3.72E-05 | 8.05E-05 | 0.18069882 |
| Heart_Failure | dendritic_cell | *AGTRAP* | 2.621599959 | 7.40E-06 | 2.12E-05 | 0.03591407 |
| Heart_Failure | dendritic_cell | *THAP5* | 5.323716836 | 2.56E-05 | 5.86E-05 | 0.124263061 |
| Heart_Failure | dendritic_cell | *NAALADL2* | -1.464554584 | 4.44E-05 | 9.33E-05 | 0.215417734 |
| Heart_Failure | dendritic_cell | *CD151* | -2.283535477 | 9.28E-05 | 0.000176794 | 0.450472173 |
| Heart_Failure | dendritic_cell | *SLC35G5* | 1.310824133 | 1.37E-07 | 3.44E-06 | 0.000665931 |
| Heart_Failure | dendritic_cell | *FAM87B* | -13.07503386 | 3.17E-07 | 3.66E-06 | 0.001537421 |
| Heart_Failure | dendritic_cell | *PCDHB9* | -2.476360618 | 0.000555314 | 0.000891885 | 1 |
| Heart_Failure | dendritic_cell | *CENPBD1* | 12.69054005 | 0.000635579 | 0.001009106 | 1 |
| Heart_Failure | dendritic_cell | *RPS27* | -2.658714875 | 1.71E-05 | 4.21E-05 | 0.082760699 |
| Heart_Failure | dendritic_cell | *RIC8A* | -3.698016373 | 3.54E-06 | 1.28E-05 | 0.017157088 |
| Heart_Failure | dendritic_cell | *ASB8* | -1.9914049 | 4.18E-07 | 3.88E-06 | 0.00202657 |
| Heart_Failure | dendritic_cell | *ODF3B* | 1.284806396 | 0.015275665 | 0.019706867 | 1 |
| Heart_Failure | dendritic_cell | *ADAMTSL1* | -1.035269515 | 6.14E-07 | 4.35E-06 | 0.002981165 |
| Heart_Failure | dendritic_cell | *IMPDH2* | -2.548894709 | 0.002452924 | 0.003534776 | 1 |
| Heart_Failure | dendritic_cell | *GRAMD1C* | -1.53458501 | 2.00E-06 | 8.54E-06 | 0.009723435 |
| Heart_Failure | dendritic_cell | *DALRD3* | 4.201768858 | 7.74E-05 | 0.000151257 | 0.375418719 |

| | | | | | | |
|---|---|---|---|---|---|---|
| Heart_Failure | dendritic_cell | *ZNF114* | -4.789433237 | 0.005697729 | 0.00780502 | 1 |
| Heart_Failure | dendritic_cell | *SPINK6* | 3.635489973 | 1.04E-07 | 3.44E-06 | 0.000504577 |
| Heart_Failure | dendritic_cell | *ZNF454* | 1.724814739 | 0.000224558 | 0.000390661 | 1 |
| Heart_Failure | dendritic_cell | *SH2B1* | -2.639323615 | 4.45E-06 | 1.49E-05 | 0.021601393 |
| Heart_Failure | dendritic_cell | *VN1R1* | 4.035087087 | 0.000263713 | 0.000453094 | 1 |
| Heart_Failure | dendritic_cell | *KDELC2* | -2.462362735 | 5.21E-07 | 4.08E-06 | 0.002525719 |
| Heart_Failure | dendritic_cell | *PLEC* | -1.81857695 | 0.001052695 | 0.001616864 | 1 |
| Heart_Failure | dendritic_cell | *SH2D4B* | 8.34312837 | 6.13E-08 | 3.44E-06 | 0.000297526 |
| Heart_Failure | dendritic_cell | *RNF212* | -1.580460019 | 6.57E-07 | 4.46E-06 | 0.003189944 |
| Heart_Failure | dendritic_cell | *PRSS36* | -3.767185036 | 1.14E-06 | 6.05E-06 | 0.005515865 |
| Heart_Failure | dendritic_cell | *WDR6* | -1.726221931 | 0.00059404 | 0.000948954 | 1 |
| Heart_Failure | dendritic_cell | *GEN1* | -1.604379623 | 3.05E-05 | 6.78E-05 | 0.148188668 |
| Heart_Failure | dendritic_cell | *TMEM11* | -5.040611257 | 0.001693538 | 0.002515936 | 1 |
| Heart_Failure | dendritic_cell | *ZFAND2A* | 2.506302075 | 7.38E-08 | 3.44E-06 | 0.000358105 |
| Heart_Failure | dendritic_cell | *ZNF223* | -1.710421082 | 0.027489246 | 0.034155652 | 1 |
| Heart_Failure | dendritic_cell | *FAM220A* | -4.740782813 | 0.012294563 | 0.016070372 | 1 |
| Heart_Failure | dendritic_cell | *NT5DC1* | -2.346411532 | 4.40E-06 | 1.48E-05 | 0.021351598 |
| Heart_Failure | dendritic_cell | *GLDC* | -3.053934883 | 1.44E-06 | 6.94E-06 | 0.006994835 |
| Heart_Failure | dendritic_cell | *P4HTM* | -3.556706038 | 9.01E-06 | 2.48E-05 | 0.043699224 |
| Heart_Failure | dendritic_cell | *CA8* | -2.276238109 | 2.90E-06 | 1.11E-05 | 0.014055722 |
| Heart_Failure | dendritic_cell | *GPR35* | -0.852930415 | 0.040727739 | 0.049651003 | 1 |
| Heart_Failure | dendritic_cell | *ZNF713* | -1.131164884 | 0.003974124 | 0.00554731 | 1 |
| Heart_Failure | dendritic_cell | *PARP10* | -1.620337118 | 5.99E-07 | 4.30E-06 | 0.002905149 |
| Heart_Failure | dendritic_cell | *NSUN3* | -3.272135647 | 0.000375367 | 0.000623085 | 1 |
| Heart_Failure | dendritic_cell | *RPP25* | -1.78476222 | 0.00046975 | 0.000765098 | 1 |
| Heart_Failure | dendritic_cell | *STX19* | -1.842567454 | 3.66E-05 | 7.93E-05 | 0.177536336 |
| Heart_Failure | dendritic_cell | *MPI* | -1.950132608 | 0.001737264 | 0.002578527 | 1 |
| Heart_Failure | dendritic_cell | *TRIM73* | -5.436134588 | 0.001221172 | 0.001858571 | 1 |
| Heart_Failure | dendritic_cell | *EFCAB13* | -2.195530478 | 0.000119998 | 0.000222395 | 0.582230239 |
| Heart_Failure | dendritic_cell | *EXOSC4* | -4.223835741 | 9.09E-06 | 2.49E-05 | 0.044110542 |
| Heart_Failure | dendritic_cell | *HYI* | -1.68068736 | 2.02E-05 | 4.85E-05 | 0.098014765 |
| Heart_Failure | dendritic_cell | *GAK* | -2.66080399 | 0.018137803 | 0.023110457 | 1 |
| Heart_Failure | dendritic_cell | *TUFM* | -2.293727265 | 2.21E-05 | 5.23E-05 | 0.107043901 |
| Heart_Failure | dendritic_cell | *CTC1* | -2.439328601 | 0.000441488 | 0.000722463 | 1 |
| Heart_Failure | dendritic_cell | *CCDC89* | -6.121418637 | 6.66E-07 | 4.49E-06 | 0.003231458 |
| Heart_Failure | dendritic_cell | *HTR1F* | -6.742506723 | 9.91E-08 | 3.44E-06 | 0.00048079 |
| Heart_Failure | dendritic_cell | *SPTY2D1* | -4.069216224 | 6.82E-05 | 0.000135629 | 0.330935537 |
| Heart_Failure | dendritic_cell | *ZNF664* | 5.178698136 | 2.59E-06 | 1.02E-05 | 0.012553683 |
| Heart_Failure | dendritic_cell | *CDH4* | 1.420801775 | 4.53E-07 | 3.90E-06 | 0.002198598 |
| Heart_Failure | dendritic_cell | *NSUN7* | -1.159534213 | 0.000681432 | 0.001077324 | 1 |
| Heart_Failure | dendritic_cell | *PACS2* | -2.566454014 | 3.34E-06 | 1.22E-05 | 0.016183492 |
| Heart_Failure | dendritic_cell | *LINC00174* | -2.055410333 | 2.62E-07 | 3.66E-06 | 0.001270936 |
| Heart_Failure | dendritic_cell | *GEMIN4* | -3.235037501 | 0.00411038 | 0.00572433 | 1 |
| Heart_Failure | dendritic_cell | *ZFPM1* | -1.745471796 | 1.71E-06 | 7.72E-06 | 0.008307995 |
| Heart_Failure | dendritic_cell | *FCER1A* | 0.1646022 | 1.47E-07 | 3.45E-06 | 0.000714557 |
| Heart_Failure | dendritic_cell | *AKAP5* | 2.119660215 | 2.39E-07 | 3.66E-06 | 0.001161333 |
| Heart_Failure | dendritic_cell | *GIPC3* | -10.07150712 | 3.31E-07 | 3.66E-06 | 0.001603778 |
| Heart_Failure | dendritic_cell | *PDXDC1* | -2.518813885 | 2.36E-05 | 5.54E-05 | 0.11473018 |
| Heart_Failure | dendritic_cell | *C1orf194* | -4.266933044 | 0.000270091 | 0.000462741 | 1 |
| Heart_Failure | dendritic_cell | *SEPHS2* | -3.615278722 | 5.40E-05 | 0.000111212 | 0.262127642 |

| | | | | | | |
|---|---|---|---|---|---|---|
| Heart_Failure | dendritic_cell | *SSC5D* | -0.911421003 | 0.000594483 | 0.000949139 | 1 |
| Heart_Failure | dendritic_cell | *ZADH2* | -3.057551746 | 8.56E-06 | 2.39E-05 | 0.041530242 |
| Heart_Failure | dendritic_cell | *SEPT1* | -2.264369766 | 3.17E-07 | 3.66E-06 | 0.001536722 |
| Heart_Failure | dendritic_cell | *LYNX1* | -1.690606821 | 0.000200753 | 0.000353479 | 0.974053352 |
| Heart_Failure | dendritic_cell | *FAHD1* | -0.958066007 | 0.000107479 | 0.000201424 | 0.521487934 |
| Heart_Failure | dendritic_cell | *HCLS1* | 1.230129247 | 5.02E-06 | 1.62E-05 | 0.024365653 |
| Heart_Failure | dendritic_cell | *MTURN* | -1.927918045 | 1.01E-06 | 5.71E-06 | 0.004914741 |
| Heart_Failure | dendritic_cell | *CCDC66* | -2.805807606 | 2.04E-05 | 4.89E-05 | 0.099017413 |
| Heart_Failure | dendritic_cell | *DEFB124* | 2.927984092 | 9.65E-08 | 3.44E-06 | 0.000468185 |
| Heart_Failure | dendritic_cell | *MCFD2* | -2.626458295 | 5.06E-06 | 1.63E-05 | 0.024544543 |
| Heart_Failure | dendritic_cell | *GLIPR1L2* | -2.881923608 | 0.000830228 | 0.001294293 | 1 |
| Heart_Failure | dendritic_cell | *DEFB119* | -24.23992471 | 2.71E-05 | 6.15E-05 | 0.131569544 |
| Heart_Failure | dendritic_cell | *TSPYL5* | 3.089557964 | 9.63E-06 | 2.62E-05 | 0.046706976 |
| Heart_Failure | dendritic_cell | *MB21D2* | 3.480534474 | 0.000165228 | 0.000296152 | 0.801684387 |
| Heart_Failure | dendritic_cell | *SLC47A2* | -2.796475459 | 3.08E-05 | 6.83E-05 | 0.14956365 |
| Heart_Failure | dendritic_cell | *CHST13* | -13.80877964 | 2.34E-06 | 9.48E-06 | 0.011365193 |
| Heart_Failure | dendritic_cell | *SLC36A4* | -3.26513557 | 0.000151259 | 0.000273439 | 0.733909736 |
| Heart_Failure | dendritic_cell | *ZDHHC20* | -2.857319645 | 4.13E-05 | 8.80E-05 | 0.200149446 |
| Heart_Failure | dendritic_cell | *OR51E1* | -1.612484397 | 0.00284778 | 0.00405322 | 1 |
| Heart_Failure | dendritic_cell | *PSMG4* | -2.299490004 | 2.14E-05 | 5.11E-05 | 0.104033252 |
| Heart_Failure | dendritic_cell | *BHLHE22* | 1.66294704 | 2.28E-07 | 3.66E-06 | 0.001107416 |
| Heart_Failure | dendritic_cell | *ZNF443* | -6.266216913 | 2.95E-07 | 3.66E-06 | 0.001431784 |
| Heart_Failure | dendritic_cell | *CAPS2* | -2.269803471 | 6.99E-06 | 2.04E-05 | 0.03391155 |
| Heart_Failure | dendritic_cell | *KCTD2* | -2.94374888 | 2.53E-06 | 1.01E-05 | 0.012293504 |
| Heart_Failure | dendritic_cell | *D2HGDH* | -1.132055472 | 0.034698572 | 0.042686985 | 1 |
| Heart_Failure | dendritic_cell | *LRRC57* | -2.727062375 | 0.004094358 | 0.005703654 | 1 |
| Heart_Failure | dendritic_cell | *MRPL14* | -3.045591557 | 4.65E-05 | 9.72E-05 | 0.225852549 |
| Heart_Failure | dendritic_cell | *NQO1* | -1.961354122 | 6.34E-06 | 1.92E-05 | 0.030752999 |
| Heart_Failure | dendritic_cell | *RPH3AL* | -1.787181888 | 1.77E-06 | 7.90E-06 | 0.008571568 |
| Heart_Failure | dendritic_cell | *OR52N4* | 2.866042962 | 0.005035162 | 0.006944459 | 1 |
| Heart_Failure | dendritic_cell | *ADIPOQ* | -0.866953714 | 6.05E-06 | 1.86E-05 | 0.029344713 |
| Heart_Failure | dendritic_cell | *F2R* | -1.925525769 | 7.99E-06 | 2.25E-05 | 0.038771394 |
| Heart_Failure | dendritic_cell | *MUC16* | -1.415241511 | 4.83E-07 | 3.96E-06 | 0.002344907 |
| Heart_Failure | dendritic_cell | *PENK* | 1.650519086 | 1.60E-07 | 3.51E-06 | 0.000776316 |
| Heart_Failure | dendritic_cell | *SLC25A41* | 4.418515098 | 1.66E-07 | 3.52E-06 | 0.000803796 |
| Heart_Failure | dendritic_cell | *TMEM136* | -2.73757477 | 0.000166223 | 0.000297606 | 0.80651316 |
| Heart_Failure | dendritic_cell | *TMEM132E* | 0.804461031 | 1.51E-06 | 7.17E-06 | 0.007348013 |
| Heart_Failure | dendritic_cell | *ZNF322* | 2.198462566 | 0.027994719 | 0.034748114 | 1 |
| Heart_Failure | dendritic_cell | *DDX60L* | -3.070629209 | 7.81E-05 | 0.000152585 | 0.379174446 |
| Heart_Failure | dendritic_cell | *OGFOD3* | -1.397898407 | 2.50E-05 | 5.77E-05 | 0.121396586 |
| Heart_Failure | dendritic_cell | *TMEM45A* | -2.457649455 | 3.98E-05 | 8.52E-05 | 0.193113165 |
| Heart_Failure | dendritic_cell | *SGSH* | -3.370042064 | 7.86E-07 | 4.87E-06 | 0.003815341 |
| Heart_Failure | dendritic_cell | *ZBTB20* | -1.701743995 | 1.16E-05 | 3.05E-05 | 0.056221961 |
| Heart_Failure | dendritic_cell | *C3orf58* | 4.358800386 | 5.86E-05 | 0.000118914 | 0.284562033 |
| Heart_Failure | dendritic_cell | *AMIGO1* | -1.355367874 | 0.019818262 | 0.025126263 | 1 |
| Heart_Failure | dendritic_cell | *TMEM252* | -76.07232557 | 2.98E-07 | 3.66E-06 | 0.001444691 |
| Heart_Failure | dendritic_cell | *ODF3L2* | -4.728476663 | 0.000385238 | 0.000636857 | 1 |
| Heart_Failure | dendritic_cell | *RFX7* | -2.75947378 | 0.000350048 | 0.000584458 | 1 |
| Heart_Failure | dendritic_cell | *ZNF329* | -3.682972394 | 3.59E-05 | 7.82E-05 | 0.174406057 |
| Heart_Failure | dendritic_cell | *MGAT4C* | -0.957127307 | 6.78E-05 | 0.00013495 | 0.329009049 |

| | | | | | | |
|---|---|---|---|---|---|---|
| Heart_Failure | dendritic_cell | *PTCHD3* | -1.554997071 | 0.022889262 | 0.028764232 | 1 |
| Heart_Failure | dendritic_cell | *WRB* | -0.804030601 | 0.017879233 | 0.022822951 | 1 |
| Heart_Failure | dendritic_cell | *TDRKH* | -2.848934203 | 1.51E-08 | 2.99E-06 | 7.33E-05 |
| Heart_Failure | dendritic_cell | *ZNF708* | -1.83424174 | 0.000521489 | 0.000843422 | 1 |
| Heart_Failure | dendritic_cell | *ERCC6L2* | -2.656131023 | 1.71E-05 | 4.23E-05 | 0.083191718 |
| Heart_Failure | dendritic_cell | *MRPL41* | 5.168862072 | 3.06E-06 | 1.15E-05 | 0.01486179 |
| Heart_Failure | dendritic_cell | *HHIPL1* | -1.767347789 | 2.52E-06 | 1.00E-05 | 0.012225008 |
| Heart_Failure | dendritic_cell | *FAM153B* | -1.374600749 | 5.92E-07 | 4.30E-06 | 0.002874709 |
| Heart_Failure | dendritic_cell | *BACE2* | -1.07426593 | 0.000141926 | 0.000257718 | 0.688623011 |
| Heart_Failure | dendritic_cell | *SYNM* | -5.104141072 | 9.42E-05 | 0.000178977 | 0.45710823 |
| Heart_Failure | dendritic_cell | *KCNA4* | -3.072782074 | 6.31E-07 | 4.39E-06 | 0.003059613 |
| Heart_Failure | dendritic_cell | *FIGN* | -1.159193213 | 5.45E-07 | 4.15E-06 | 0.002642277 |
| Heart_Failure | dendritic_cell | *C8orf33* | -3.640496787 | 0.000153122 | 0.000276188 | 0.74294652 |
| Heart_Failure | dendritic_cell | *FBXL6* | -2.779951295 | 7.52E-07 | 4.78E-06 | 0.00364684 |
| Heart_Failure | dendritic_cell | *NXPH4* | -18.36679064 | 3.31E-07 | 3.66E-06 | 0.001607533 |
| Heart_Failure | dendritic_cell | *NPLOC4* | -2.160694879 | 2.14E-07 | 3.66E-06 | 0.00103619 |
| Heart_Failure | dendritic_cell | *KPNA2* | -3.326542143 | 6.34E-05 | 0.000127247 | 0.307809572 |
| Heart_Failure | dendritic_cell | *ORAI1* | -2.707590728 | 1.12E-06 | 6.03E-06 | 0.005445819 |
| Heart_Failure | dendritic_cell | *FES* | 2.308628186 | 0.000195194 | 0.000344769 | 0.947079275 |
| Heart_Failure | dendritic_cell | *GLRX5* | -4.888203159 | 0.000212455 | 0.000371739 | 1 |
| Heart_Failure | dendritic_cell | *CAV3* | 1.861174944 | 0.003393374 | 0.00477513 | 1 |
| Heart_Failure | dendritic_cell | *MXRA7* | -1.855785572 | 3.34E-05 | 7.35E-05 | 0.162117304 |
| Heart_Failure | dendritic_cell | *MFSD5* | -2.005146763 | 0.000137096 | 0.00024998 | 0.665187801 |
| Heart_Failure | dendritic_cell | *ADI1* | -4.094604469 | 0.000166597 | 0.000298056 | 0.808327011 |
| Heart_Failure | dendritic_cell | *CLEC4G* | 4.622828448 | 0.000126361 | 0.0002325 | 0.61310164 |
| Heart_Failure | dendritic_cell | *TRAK1* | -2.268152915 | 0.002977282 | 0.004220209 | 1 |
| Heart_Failure | dendritic_cell | *SKA2* | -3.037417192 | 1.01E-05 | 2.72E-05 | 0.048883921 |
| Heart_Failure | dendritic_cell | *TTC3* | -1.325909878 | 0.00076035 | 0.001192764 | 1 |
| Heart_Failure | dendritic_cell | *PPP1R27* | -8.242408446 | 0.003671637 | 0.005148781 | 1 |
| Heart_Failure | dendritic_cell | *BRICD5* | -10.76677655 | 0.0001178 | 0.000218656 | 0.57156607 |
| Heart_Failure | dendritic_cell | *GRID1* | -1.780874534 | 1.16E-07 | 3.44E-06 | 0.000565126 |
| Heart_Failure | dendritic_cell | *ACBD3* | -2.911449398 | 0.000352719 | 0.00058831 | 1 |
| Heart_Failure | dendritic_cell | *PLCXD3* | -1.423423069 | 4.37E-07 | 3.90E-06 | 0.002122112 |
| Heart_Failure | dendritic_cell | *VMO1* | 0.477594862 | 1.10E-07 | 3.44E-06 | 0.000536021 |
| Heart_Failure | dendritic_cell | *ALG12* | -3.790809711 | 1.88E-06 | 8.19E-06 | 0.009124751 |
| Heart_Failure | dendritic_cell | *COL18A1* | -1.545859033 | 4.50E-05 | 9.44E-05 | 0.218458439 |
| Heart_Failure | dendritic_cell | *ZNF721* | -1.997084663 | 1.10E-05 | 2.92E-05 | 0.053291263 |
| Heart_Failure | dendritic_cell | *ODF3L1* | -4.281124957 | 0.009893311 | 0.013101077 | 1 |
| Heart_Failure | dendritic_cell | *HMGN4* | -1.830084017 | 5.63E-06 | 1.76E-05 | 0.027298006 |
| Heart_Failure | dendritic_cell | *ZNF320* | 0.676111652 | 0.032073446 | 0.039557793 | 1 |
| Heart_Failure | dendritic_cell | *C12orf60* | -3.197224442 | 2.10E-06 | 8.81E-06 | 0.010201991 |
| Heart_Failure | dendritic_cell | *LYSMD4* | -2.37281313 | 2.60E-06 | 1.02E-05 | 0.012624884 |
| Heart_Failure | dendritic_cell | *ARHGEF37* | -1.660787482 | 3.62E-07 | 3.66E-06 | 0.001756831 |
| Heart_Failure | dendritic_cell | *CSMD1* | -1.966484966 | 4.08E-07 | 3.83E-06 | 0.001978823 |
| Heart_Failure | dendritic_cell | *RABIF* | -3.811881521 | 2.31E-05 | 5.42E-05 | 0.111935602 |
| Heart_Failure | dendritic_cell | *FANCF* | -5.133959884 | 2.61E-06 | 1.02E-05 | 0.012664454 |
| Heart_Failure | dendritic_cell | *C2CD4C* | -1.500724014 | 0.00011593 | 0.00021568 | 0.562494437 |
| Heart_Failure | dendritic_cell | *RUVBL2* | -1.966400475 | 3.59E-07 | 3.66E-06 | 0.001744183 |
| Heart_Failure | dendritic_cell | *GDPGP1* | -1.466353815 | 0.007965812 | 0.010682731 | 1 |
| Heart_Failure | dendritic_cell | *PTTG1IP* | -1.67042479 | 0.028473655 | 0.035288422 | 1 |

| | | | | | | |
|---|---|---|---|---|---|---|
| Heart_Failure | dendritic_cell | *ZNF623* | -2.106414917 | 3.25E-07 | 3.66E-06 | 0.001575325 |
| Heart_Failure | dendritic_cell | *CCDC125* | -2.494557186 | 0.000417031 | 0.000685213 | 1 |
| Heart_Failure | dendritic_cell | *C10orf107* | -0.879417404 | 0.001091939 | 0.001672908 | 1 |
| Heart_Failure | dendritic_cell | *FHL3* | 1.324372758 | 1.81E-05 | 4.41E-05 | 0.087755634 |
| Heart_Failure | dendritic_cell | *C19orf71* | -2.747365689 | 2.19E-07 | 3.66E-06 | 0.001062397 |
| Heart_Failure | dendritic_cell | *LRIT3* | 4.158847435 | 1.21E-07 | 3.44E-06 | 0.000585835 |
| Heart_Failure | dendritic_cell | *NPIPA1* | -0.850583221 | 0.015706011 | 0.020229776 | 1 |
| Heart_Failure | dendritic_cell | *SF3A3* | -2.570138861 | 0.000151519 | 0.000273714 | 0.735171799 |
| Heart_Failure | dendritic_cell | *GPR132* | 0.848745984 | 9.39E-07 | 5.45E-06 | 0.004557947 |
| Heart_Failure | dendritic_cell | *PSMG1* | -4.515016146 | 6.41E-08 | 3.44E-06 | 0.000311056 |
| Heart_Failure | dendritic_cell | *AC131971.1* | -5.743074585 | 0.001346578 | 0.002032223 | 1 |
| Heart_Failure | dendritic_cell | *MRPL54* | 1.491588625 | 0.021955762 | 0.027669963 | 1 |
| Heart_Failure | dendritic_cell | *HMCES* | -1.580294964 | 4.53E-07 | 3.90E-06 | 0.002198117 |
| Heart_Failure | dendritic_cell | *RP1L1* | 3.036400111 | 0.00605044 | 0.008243959 | 1 |
| Heart_Failure | dendritic_cell | *NDUFB1* | -2.459356111 | 6.26E-06 | 1.91E-05 | 0.030382902 |
| Heart_Failure | dendritic_cell | *FAM19A1* | -3.115087597 | 1.33E-07 | 3.44E-06 | 0.0006461 |
| Heart_Failure | dendritic_cell | *TRMT12* | 4.475372828 | 0.009888411 | 0.013098163 | 1 |
| Heart_Failure | dendritic_cell | *ALYREF* | -2.445240857 | 0.000826653 | 0.001290515 | 1 |
| Heart_Failure | dendritic_cell | *MRGPRX2* | -11.42040003 | 8.47E-05 | 0.00016347 | 0.410799376 |
| Heart_Failure | dendritic_cell | *UPP1* | 3.035498959 | 0.010470814 | 0.013813047 | 1 |
| Heart_Failure | dendritic_cell | *FIGLA* | 9.647653725 | 1.55E-05 | 3.87E-05 | 0.075253834 |
| Heart_Failure | dendritic_cell | *ACSM2A* | -24.24545723 | 1.85E-06 | 8.10E-06 | 0.008957308 |
| Heart_Failure | dendritic_cell | *KREMEN1* | -1.374020064 | 7.67E-05 | 0.000149966 | 0.371914942 |
| Heart_Failure | dendritic_cell | *ZNF703* | -3.225691208 | 1.50E-08 | 2.99E-06 | 7.26E-05 |
| Heart_Failure | dendritic_cell | *FAM162B* | -1.660803293 | 6.24E-05 | 0.000125398 | 0.302711532 |
| Heart_Failure | dendritic_cell | *BTBD9* | -2.228565383 | 2.54E-05 | 5.83E-05 | 0.123260572 |
| Heart_Failure | dendritic_cell | *NUDT14* | -2.766832442 | 6.82E-07 | 4.53E-06 | 0.00330904 |
| Heart_Failure | dendritic_cell | *TTC32* | -3.333844559 | 3.85E-06 | 1.35E-05 | 0.018666553 |
| Heart_Failure | dendritic_cell | *PP2D1* | -5.280278274 | 0.002154139 | 0.003147209 | 1 |
| Heart_Failure | dendritic_cell | *NIPSNAP1* | -2.421852376 | 1.19E-06 | 6.17E-06 | 0.00577986 |
| Heart_Failure | dendritic_cell | *CNTN2* | -2.182918494 | 6.42E-05 | 0.000128451 | 0.31162207 |
| Heart_Failure | dendritic_cell | *CRELD2* | 2.255571218 | 1.05E-06 | 5.79E-06 | 0.00508235 |
| Heart_Failure | dendritic_cell | *GOLGA6L4* | 1.938053852 | 0.020552994 | 0.025983098 | 1 |
| Heart_Failure | dendritic_cell | *PGP* | 2.908244255 | 0.000575647 | 0.000921491 | 1 |
| Heart_Failure | dendritic_cell | *ACOT1* | 1.504235351 | 0.000291231 | 0.000495983 | 1 |
| Heart_Failure | dendritic_cell | *OAF* | -1.447160318 | 3.59E-06 | 1.28E-05 | 0.017413296 |
| Heart_Failure | dendritic_cell | *ALDH1A3* | -0.928286403 | 0.000224069 | 0.000390231 | 1 |
| Heart_Failure | dendritic_cell | *CLECL1* | 0.649972455 | 1.64E-07 | 3.52E-06 | 0.000798093 |
| Heart_Failure | dendritic_cell | *A3GALT2* | -1.92243006 | 4.64E-05 | 9.70E-05 | 0.225130685 |
| Heart_Failure | dendritic_cell | *SS18L1* | -3.007927436 | 0.000266694 | 0.000457244 | 1 |
| Heart_Failure | dendritic_cell | *TOP1MT* | 0.970090658 | 0.032784068 | 0.040393168 | 1 |
| Heart_Failure | dendritic_cell | *COPB2* | -3.151932048 | 0.000294015 | 0.000500275 | 1 |
| Heart_Failure | dendritic_cell | *WDR27* | -2.238683197 | 4.44E-06 | 1.49E-05 | 0.021559258 |
| Heart_Failure | dendritic_cell | *TXNRD2* | -3.339928376 | 0.002115438 | 0.003092529 | 1 |
| Heart_Failure | dendritic_cell | *TMEM255B* | -1.970692705 | 1.88E-07 | 3.60E-06 | 0.000910685 |
| Heart_Failure | dendritic_cell | *HDDC3* | -1.803771675 | 4.64E-07 | 3.92E-06 | 0.002252594 |
| Heart_Failure | dendritic_cell | *ZFP1* | -2.048443385 | 2.97E-06 | 1.12E-05 | 0.014409418 |
| Heart_Failure | dendritic_cell | *SNN* | -5.697170104 | 3.05E-06 | 1.14E-05 | 0.014786422 |
| Heart_Failure | dendritic_cell | *KRBA2* | -4.316665937 | 5.07E-07 | 4.03E-06 | 0.002461119 |
| Heart_Failure | dendritic_cell | *SEPT5* | 3.138644879 | 7.28E-06 | 2.10E-05 | 0.035328246 |

| | | | | | | |
|---|---|---|---|---|---|---|
| Heart_Failure | dendritic_cell | *RNLS* | -1.917943906 | 1.36E-06 | 6.68E-06 | 0.006596345 |
| Heart_Failure | dendritic_cell | *NDUFA12* | -2.964297209 | 9.95E-05 | 0.000187935 | 0.482615971 |
| Heart_Failure | dendritic_cell | *TCTE3* | 4.646445197 | 1.33E-06 | 6.61E-06 | 0.006450569 |
| Heart_Failure | dendritic_cell | *UBE2G2* | -2.393933174 | 0.001561767 | 0.002336948 | 1 |
| Heart_Failure | dendritic_cell | *TMED9* | -3.97249747 | 0.000261521 | 0.000449805 | 1 |
| Heart_Failure | dendritic_cell | *DRD1* | 2.425166948 | 1.39E-07 | 3.44E-06 | 0.00067555 |
| Heart_Failure | dendritic_cell | *WT1* | -1.126079991 | 5.35E-07 | 4.13E-06 | 0.002594137 |
| Heart_Failure | dendritic_cell | *ZFP90* | -3.525907514 | 0.000696487 | 0.001099692 | 1 |
| Heart_Failure | dendritic_cell | *FAM227A* | -1.408668831 | 3.12E-05 | 6.90E-05 | 0.151446666 |
| Heart_Failure | dendritic_cell | *NOC4L* | -2.684317231 | 2.55E-05 | 5.84E-05 | 0.123664043 |
| Heart_Failure | dendritic_cell | *SORCS2* | -2.04275816 | 1.16E-07 | 3.44E-06 | 0.000560416 |
| Heart_Failure | dendritic_cell | *BRI3BP* | 4.040210528 | 7.40E-06 | 2.12E-05 | 0.035926249 |
| Heart_Failure | dendritic_cell | *UBOX5* | -3.181909498 | 1.79E-06 | 7.95E-06 | 0.008695116 |
| Heart_Failure | dendritic_cell | *MAFF* | -1.552133275 | 1.35E-06 | 6.63E-06 | 0.006528776 |
| Heart_Failure | dendritic_cell | *SEMA4B* | -2.126887317 | 7.56E-07 | 4.79E-06 | 0.003668475 |
| Heart_Failure | dendritic_cell | *CIB1* | -1.839166541 | 4.80E-06 | 1.57E-05 | 0.023290831 |
| Heart_Failure | dendritic_cell | *SGCZ* | -1.295672481 | 2.23E-08 | 3.18E-06 | 0.000107997 |
| Heart_Failure | dendritic_cell | *C6orf120* | -3.604783687 | 0.000684291 | 0.001081492 | 1 |
| Heart_Failure | dendritic_cell | *SIGIRR* | -2.225391036 | 0.000163559 | 0.000293487 | 0.793588871 |
| Heart_Failure | dendritic_cell | *IFITM2* | -3.530205243 | 0.002342496 | 0.003391761 | 1 |
| Heart_Failure | dendritic_cell | *CDK10* | -1.853939529 | 4.16E-05 | 8.86E-05 | 0.201864327 |
| Heart_Failure | dendritic_cell | *TNFAIP8L1* | -2.11184543 | 1.71E-07 | 3.52E-06 | 0.000831308 |
| Heart_Failure | dendritic_cell | *SP140L* | 3.409440939 | 0.000107727 | 0.000201811 | 0.522689227 |
| Heart_Failure | dendritic_cell | *TARSL2* | -2.112542495 | 1.95E-05 | 4.71E-05 | 0.094790538 |
| Heart_Failure | dendritic_cell | *FAM174B* | -3.033471667 | 5.58E-05 | 0.000114332 | 0.270966745 |
| Heart_Failure | dendritic_cell | *SV2B* | -1.68751739 | 6.64E-06 | 1.97E-05 | 0.032202987 |
| Heart_Failure | dendritic_cell | *AHNAK2* | -1.992203853 | 9.25E-07 | 5.40E-06 | 0.004487982 |
| Heart_Failure | dendritic_cell | *SP1* | -3.020683542 | 4.32E-05 | 9.13E-05 | 0.209512128 |
| Heart_Failure | dendritic_cell | *PCGF3* | -2.732904659 | 8.95E-05 | 0.000171091 | 0.434228333 |
| Heart_Failure | dendritic_cell | *PMEL* | -1.75043656 | 1.21E-05 | 3.16E-05 | 0.058683823 |
| Heart_Failure | dendritic_cell | *MYBL1* | -2.441498425 | 1.36E-07 | 3.44E-06 | 0.000660604 |
| Heart_Failure | dendritic_cell | *ADARB2* | -3.663751755 | 1.39E-06 | 6.77E-06 | 0.006753643 |
| Heart_Failure | dendritic_cell | *DMWD* | -1.982350436 | 7.60E-07 | 4.79E-06 | 0.003685856 |
| Heart_Failure | dendritic_cell | *SLC52A2* | 2.629783524 | 2.35E-06 | 9.49E-06 | 0.01138302 |
| Heart_Failure | dendritic_cell | *ARL17A* | -2.423187502 | 0.000108444 | 0.00020292 | 0.526170377 |
| Heart_Failure | dendritic_cell | *NPIPB4* | -2.310789308 | 0.000859545 | 0.001337559 | 1 |
| Heart_Failure | dendritic_cell | *IFITM1* | -3.75262969 | 2.41E-05 | 5.63E-05 | 0.117108014 |
| Heart_Failure | dendritic_cell | *LINC00839* | -5.271985878 | 1.45E-06 | 6.95E-06 | 0.007032663 |
| Heart_Failure | dendritic_cell | *RTN4RL1* | -2.008301117 | 1.48E-06 | 7.06E-06 | 0.007195508 |
| Heart_Failure | dendritic_cell | *RNPC3* | -1.984251992 | 1.60E-05 | 3.99E-05 | 0.077847965 |
| Heart_Failure | dendritic_cell | *RASA3* | -2.083893843 | 1.87E-06 | 8.16E-06 | 0.009058237 |
| Heart_Failure | dendritic_cell | *ZNF529* | -2.853339826 | 2.71E-05 | 6.15E-05 | 0.131627995 |
| Heart_Failure | dendritic_cell | *C15orf41* | -2.15210488 | 2.53E-06 | 1.01E-05 | 0.012281219 |
| Heart_Failure | dendritic_cell | *WWOX* | -1.747689334 | 1.04E-05 | 2.78E-05 | 0.05030377 |
| Heart_Failure | dendritic_cell | *POLR1D* | -2.093447186 | 1.21E-06 | 6.22E-06 | 0.005859106 |
| Heart_Failure | dendritic_cell | *FFAR4* | -2.610977978 | 5.27E-08 | 3.44E-06 | 0.000255527 |
| Heart_Failure | dendritic_cell | *CYP4F12* | -1.721662299 | 7.81E-07 | 4.86E-06 | 0.003791347 |
| Heart_Failure | dendritic_cell | *NBPF12* | 1.000646867 | 0.035598336 | 0.043727398 | 1 |
| Heart_Failure | dendritic_cell | *GPAT2* | -2.101747828 | 0.038755164 | 0.047365254 | 1 |
| Heart_Failure | dendritic_cell | *TOR3A* | 1.550869276 | 5.70E-07 | 4.26E-06 | 0.002767188 |

| | | | | | | |
|---|---|---|---|---|---|---|
| Heart_Failure | dendritic_cell | *ZNF555* | -3.78772403 | 3.37E-06 | 1.23E-05 | 0.01634983 |
| Heart_Failure | dendritic_cell | *GOLGA8R* | -4.505639071 | 0.000550843 | 0.000885583 | 1 |
| Heart_Failure | dendritic_cell | *CCDC30* | -1.739278306 | 0.00440914 | 0.006121072 | 1 |
| Heart_Failure | dendritic_cell | *ZNF197* | -2.502662784 | 3.73E-06 | 1.32E-05 | 0.018085942 |
| Heart_Failure | dendritic_cell | *RPS23* | -2.181223069 | 0.00020867 | 0.000365644 | 1 |
| Heart_Failure | dendritic_cell | *GNG2* | -2.729770196 | 0.006117681 | 0.008328561 | 1 |
| Heart_Failure | dendritic_cell | *TMEM222* | -3.582064113 | 3.85E-06 | 1.35E-05 | 0.018700192 |
| Heart_Failure | dendritic_cell | *CLCNKA* | 1.524457997 | 0.00701871 | 0.009486011 | 1 |
| Heart_Failure | dendritic_cell | *SEPT10* | -2.787905504 | 3.61E-06 | 1.29E-05 | 0.0175088 |
| Heart_Failure | dendritic_cell | *FAM86B1* | -3.503893578 | 0.000460975 | 0.000751311 | 1 |
| Heart_Failure | dendritic_cell | *CEACAM19* | -1.303405451 | 1.51E-05 | 3.78E-05 | 0.073136562 |
| Heart_Failure | dendritic_cell | *MIR22HG* | -2.515436945 | 1.92E-06 | 8.30E-06 | 0.009300812 |
| Heart_Failure | dendritic_cell | *CYP27C1* | -1.34883225 | 0.005040919 | 0.006950423 | 1 |
| Heart_Failure | dendritic_cell | *BCR* | -2.474768508 | 6.36E-06 | 1.92E-05 | 0.030853998 |
| Heart_Failure | dendritic_cell | *ZNF732* | 2.131318668 | 2.98E-07 | 3.66E-06 | 0.00144408 |
| Heart_Failure | dendritic_cell | *HYAL3* | 1.410179792 | 0.015099569 | 0.019495239 | 1 |
| Heart_Failure | dendritic_cell | *VSIG10L* | -3.8960005 | 0.000129794 | 0.000238455 | 0.629760194 |
| Heart_Failure | dendritic_cell | *TRABD2A* | 0.929376094 | 4.42E-07 | 3.90E-06 | 0.002142778 |
| Heart_Failure | dendritic_cell | *QRFPR* | 5.604886644 | 1.14E-09 | 5.54E-07 | 5.54E-06 |
| Heart_Failure | dendritic_cell | *MAPT* | -1.366587037 | 3.20E-07 | 3.66E-06 | 0.001551328 |
| Heart_Failure | dendritic_cell | *TMEM17* | -1.234881012 | 0.005944637 | 0.008108905 | 1 |
| Heart_Failure | dendritic_cell | *TNFRSF18* | 5.230046066 | 0.014090155 | 0.018279527 | 1 |
| Heart_Failure | dendritic_cell | *C1QL4* | -22.03633568 | 3.40E-07 | 3.66E-06 | 0.001647286 |
| Heart_Failure | dendritic_cell | *RTN4RL2* | 3.496404723 | 0.006389512 | 0.008671652 | 1 |
| Heart_Failure | dendritic_cell | *TMEM232* | -1.086422765 | 0.002008051 | 0.002945304 | 1 |
| Heart_Failure | dendritic_cell | *EFCAB6* | -2.04240299 | 1.21E-06 | 6.22E-06 | 0.005862851 |
| Heart_Failure | dendritic_cell | *TMPRSS6* | 2.01340135 | 1.40E-07 | 3.44E-06 | 0.000679122 |
| Heart_Failure | dendritic_cell | *CYP4A11* | -5.202644469 | 3.93E-05 | 8.45E-05 | 0.190869946 |
| Heart_Failure | dendritic_cell | *TMEM216* | -2.231021608 | 5.47E-06 | 1.73E-05 | 0.026551394 |
| Heart_Failure | dendritic_cell | *C3orf70* | -1.467455566 | 6.95E-06 | 2.04E-05 | 0.033715685 |
| Heart_Failure | dendritic_cell | *ENTPD5* | -2.491915801 | 0.000570681 | 0.000914548 | 1 |
| Heart_Failure | dendritic_cell | *MRPS21* | -3.080435772 | 1.16E-08 | 2.99E-06 | 5.63E-05 |
| Heart_Failure | dendritic_cell | *ANGPTL5* | -1.639962364 | 0.004722749 | 0.006530287 | 1 |
| Heart_Failure | dendritic_cell | *WDR86* | -1.013263425 | 3.57E-05 | 7.78E-05 | 0.173168833 |
| Heart_Failure | dendritic_cell | *EPOR* | 1.972669426 | 0.01423971 | 0.018448884 | 1 |
| Heart_Failure | dendritic_cell | *LUZP2* | 5.661592777 | 7.10E-07 | 4.63E-06 | 0.00344331 |
| Heart_Failure | dendritic_cell | *GJA4* | 4.735866057 | 1.00E-07 | 3.44E-06 | 0.000485407 |
| Heart_Failure | dendritic_cell | *IFT140* | -3.217942426 | 2.27E-06 | 9.28E-06 | 0.011007764 |
| Heart_Failure | dendritic_cell | *SBK2* | -21.95764123 | 5.57E-07 | 4.19E-06 | 0.002704953 |
| Heart_Failure | dendritic_cell | *ZNF385C* | -2.407723839 | 0.002339419 | 0.003388316 | 1 |
| Heart_Failure | dendritic_cell | *EXD3* | -2.963492644 | 7.13E-06 | 2.07E-05 | 0.034615612 |
| Heart_Failure | dendritic_cell | *SAMD11* | -3.101056361 | 0.000189485 | 0.000335296 | 0.919381244 |
| Heart_Failure | dendritic_cell | *TRPV2* | -2.050913593 | 0.009882122 | 0.013093407 | 1 |
| Heart_Failure | dendritic_cell | *C2orf88* | -2.51380482 | 3.38E-06 | 1.23E-05 | 0.016387234 |
| Heart_Failure | dendritic_cell | *KBTBD12* | -1.592382309 | 0.004209871 | 0.005859523 | 1 |
| Heart_Failure | dendritic_cell | *DNAH17* | 2.341491261 | 0.011548793 | 0.015165019 | 1 |
| Heart_Failure | dendritic_cell | *FANCM* | -2.136254743 | 1.40E-06 | 6.80E-06 | 0.006784072 |
| Heart_Failure | dendritic_cell | *ZFP69* | -4.654301939 | 4.65E-06 | 1.54E-05 | 0.022566303 |
| Heart_Failure | dendritic_cell | *TMEM220* | -2.228629266 | 0.000178336 | 0.000317074 | 0.865284943 |
| Heart_Failure | dendritic_cell | *FAM122A* | -5.745269661 | 1.93E-05 | 4.66E-05 | 0.093483669 |

| | | | | | | |
|---|---|---|---|---|---|---|
| Heart_Failure | dendritic_cell | *GFRAL* | -4.526842798 | 2.48E-07 | 3.66E-06 | 0.001203852 |
| Heart_Failure | dendritic_cell | *LDLRAD2* | -3.061589818 | 7.06E-05 | 0.000139693 | 0.342666704 |
| Heart_Failure | dendritic_cell | *CPSF4L* | 3.458569753 | 4.34E-07 | 3.90E-06 | 0.002104254 |
| Heart_Failure | dendritic_cell | *KLHL17* | -2.059042802 | 0.001344494 | 0.002029709 | 1 |
| Heart_Failure | dendritic_cell | *OR7D2* | -5.631367437 | 0.002916355 | 0.004142318 | 1 |
| Heart_Failure | dendritic_cell | *S100A3* | 2.579559762 | 0.020028171 | 0.02537913 | 1 |
| Heart_Failure | dendritic_cell | *TMEM221* | 1.717077959 | 6.12E-06 | 1.88E-05 | 0.029704668 |
| Heart_Failure | dendritic_cell | *MESP2* | -3.083169572 | 3.02E-05 | 6.71E-05 | 0.146454041 |
| Heart_Failure | dendritic_cell | *MAPK12* | -3.65734282 | 2.78E-07 | 3.66E-06 | 0.001350821 |
| Heart_Failure | dendritic_cell | *AGRN* | -6.003525201 | 0.000322347 | 0.000543821 | 1 |
| Heart_Failure | dendritic_cell | *FAM166A* | -4.553151859 | 0.010697471 | 0.014092894 | 1 |
| Heart_Failure | dendritic_cell | *ZNF626* | -3.102585346 | 1.34E-06 | 6.62E-06 | 0.006484443 |
| Heart_Failure | dendritic_cell | *LINC00265* | 4.056185289 | 0.017854486 | 0.022797359 | 1 |
| Heart_Failure | dendritic_cell | *PRKAR1B* | -1.914672626 | 2.78E-07 | 3.66E-06 | 0.001348851 |
| Heart_Failure | dendritic_cell | *NCR3LG1* | -5.258254489 | 1.13E-06 | 6.04E-06 | 0.005498579 |
| Heart_Failure | dendritic_cell | *ZNF793* | -1.498928362 | 5.76E-05 | 0.000117094 | 0.279269248 |
| Heart_Failure | dendritic_cell | *AGAP4* | -1.995986268 | 0.001915639 | 0.002819988 | 1 |
| Heart_Failure | dendritic_cell | *RUFY4* | -3.725152048 | 3.43E-07 | 3.66E-06 | 0.00166287 |
| Heart_Failure | dendritic_cell | *ZNF669* | -3.153698343 | 0.001254181 | 0.001903436 | 1 |
| Heart_Failure | dendritic_cell | *CENPP* | -1.539649936 | 0.002467482 | 0.003550482 | 1 |
| Heart_Failure | dendritic_cell | *PLSCR1* | -2.360886007 | 0.002242411 | 0.00325949 | 1 |
| Heart_Failure | dendritic_cell | *C3orf62* | -1.76860438 | 0.002309364 | 0.003347784 | 1 |
| Heart_Failure | dendritic_cell | *ENO4* | 1.441502481 | 6.08E-07 | 4.33E-06 | 0.002950905 |
| Heart_Failure | dendritic_cell | *ZNF559* | -0.673622435 | 0.040399823 | 0.04926362 | 1 |
| Heart_Failure | dendritic_cell | *ZP3* | -1.77012524 | 3.52E-07 | 3.66E-06 | 0.001706958 |
| Heart_Failure | dendritic_cell | *H3F3C* | -45.88176655 | 4.39E-05 | 9.25E-05 | 0.212931728 |
| Heart_Failure | dendritic_cell | *JAKMIP3* | -1.310925185 | 0.000113047 | 0.000210881 | 0.548502354 |
| Heart_Failure | dendritic_cell | *PDCD1* | -7.283113103 | 0.005195286 | 0.007144991 | 1 |
| Heart_Failure | dendritic_cell | *SELL* | 1.091475268 | 3.54E-06 | 1.28E-05 | 0.01718971 |
| Heart_Failure | dendritic_cell | *C19orf54* | 6.705526909 | 2.72E-05 | 6.17E-05 | 0.132113352 |
| Heart_Failure | dendritic_cell | *C22orf34* | 3.797996686 | 1.49E-06 | 7.07E-06 | 0.007213027 |
| Heart_Failure | dendritic_cell | *COL25A1* | -1.341772225 | 4.59E-07 | 3.90E-06 | 0.002225607 |
| Heart_Failure | dendritic_cell | *SRSF10* | -1.993860875 | 7.41E-05 | 0.000145581 | 0.359729557 |
| Heart_Failure | dendritic_cell | *NBR1* | -2.749896501 | 0.000171206 | 0.000305288 | 0.830689881 |
| Heart_Failure | dendritic_cell | *NDOR1* | -4.525860265 | 0.014357981 | 0.018592186 | 1 |
| Heart_Failure | dendritic_cell | *FAM72B* | 4.844900358 | 8.84E-06 | 2.45E-05 | 0.04291451 |
| Heart_Failure | dendritic_cell | *GOLGA8M* | -2.073627456 | 3.87E-07 | 3.72E-06 | 0.001877084 |
| Heart_Failure | dendritic_cell | *RHCE* | 3.560304148 | 1.64E-06 | 7.57E-06 | 0.007968408 |
| Heart_Failure | dendritic_cell | *IDO2* | 0.708353328 | 1.45E-07 | 3.45E-06 | 0.000701473 |
| Heart_Failure | dendritic_cell | *SLC4A5* | 1.169647366 | 8.94E-08 | 3.44E-06 | 0.000433592 |
| Heart_Failure | dendritic_cell | *UROS* | -3.769893574 | 0.000946495 | 0.001464793 | 1 |
| Heart_Failure | dendritic_cell | *ZBED6CL* | -4.358851965 | 0.000263261 | 0.000452477 | 1 |
| Heart_Failure | dendritic_cell | *OSTN* | -1.756557481 | 1.18E-05 | 3.10E-05 | 0.057432836 |
| Heart_Failure | dendritic_cell | *FSIP2* | -3.089038053 | 0.000420704 | 0.00069078 | 1 |
| Heart_Failure | dendritic_cell | *NOXA1* | -2.191999898 | 5.39E-07 | 4.13E-06 | 0.002616191 |
| Heart_Failure | dendritic_cell | *FZD9* | -18.16197497 | 6.66E-07 | 4.49E-06 | 0.003230199 |
| Heart_Failure | dendritic_cell | *SKOR1* | 3.300251963 | 4.25E-07 | 3.90E-06 | 0.002060041 |
| Heart_Failure | dendritic_cell | *MTF1* | -3.583881777 | 0.004473011 | 0.006204416 | 1 |
| Heart_Failure | dendritic_cell | *ZDHHC11* | -1.797572046 | 0.001666771 | 0.002480966 | 1 |
| Heart_Failure | dendritic_cell | *FAM26F* | 0.999261807 | 7.04E-06 | 2.05E-05 | 0.034180938 |

| | | | | | | |
|---|---|---|---|---|---|---|
| Heart_Failure | dendritic_cell | *ENTPD8* | -191.0995287 | 6.71E-07 | 4.50E-06 | 0.003255712 |
| Heart_Failure | dendritic_cell | *RPL14* | -2.498197282 | 6.25E-06 | 1.91E-05 | 0.030343944 |
| Heart_Failure | dendritic_cell | *FBF1* | -1.496531952 | 5.42E-06 | 1.72E-05 | 0.026317481 |
| Heart_Failure | dendritic_cell | *ASTL* | -6.18173801 | 3.25E-07 | 3.66E-06 | 0.001574626 |
| Heart_Failure | dendritic_cell | *MSL1* | -2.226214054 | 5.94E-09 | 2.06E-06 | 2.88E-05 |
| Heart_Failure | dendritic_cell | *FAM196A* | -2.997455864 | 6.66E-05 | 0.000132906 | 0.323093672 |
| Heart_Failure | dendritic_cell | *UTS2B* | -2.479830851 | 1.95E-08 | 2.99E-06 | 9.46E-05 |
| Heart_Failure | dendritic_cell | *NELFB* | -3.8870623 | 0.00997264 | 0.013191726 | 1 |
| Heart_Failure | dendritic_cell | *SLC15A5* | 3.500423073 | 1.89E-08 | 2.99E-06 | 9.18E-05 |
| Heart_Failure | dendritic_cell | *ADAT2* | -3.514513678 | 0.010796593 | 0.014219617 | 1 |
| Heart_Failure | dendritic_cell | *ALKBH2* | -2.313729604 | 0.000900374 | 0.001399748 | 1 |
| Heart_Failure | dendritic_cell | *LITAF* | -3.071747789 | 0.004481248 | 0.006214065 | 1 |
| Heart_Failure | dendritic_cell | *SF3B3* | -3.255968063 | 0.000913947 | 0.001419939 | 1 |
| Heart_Failure | dendritic_cell | *PLAC9* | -2.156894241 | 7.30E-05 | 0.000143842 | 0.354425612 |
| Heart_Failure | dendritic_cell | *ZNF573* | -2.73939295 | 0.000700864 | 0.001105173 | 1 |
| Heart_Failure | dendritic_cell | *FAM47E* | -1.982268698 | 4.16E-06 | 1.42E-05 | 0.020180058 |
| Heart_Failure | dendritic_cell | *PNRC2* | -1.511110137 | 3.88E-07 | 3.72E-06 | 0.001880267 |
| Heart_Failure | dendritic_cell | *FHIT* | -2.070503598 | 0.000287216 | 0.000490177 | 1 |
| Heart_Failure | dendritic_cell | *ZKSCAN3* | 2.193088677 | 0.006255919 | 0.008502443 | 1 |
| Heart_Failure | dendritic_cell | *RRP7A* | -3.524628562 | 1.55E-05 | 3.88E-05 | 0.07544042 |
| Heart_Failure | dendritic_cell | *KIAA0408* | -0.971880472 | 0.00864268 | 0.011531532 | 1 |
| Heart_Failure | dendritic_cell | *BLOC1S2* | -1.256724625 | 0.004908942 | 0.006778083 | 1 |
| Heart_Failure | dendritic_cell | *HLA-DRB1* | 0.628716773 | 0.000934242 | 0.001449613 | 1 |
| Heart_Failure | dendritic_cell | *AKR1C3* | -2.242820683 | 7.82E-07 | 4.86E-06 | 0.003792728 |
| Heart_Failure | dendritic_cell | *WDSUB1* | -2.53231777 | 4.88E-06 | 1.59E-05 | 0.02368216 |
| Heart_Failure | dendritic_cell | *COLCA1* | -2.675034588 | 3.32E-06 | 1.22E-05 | 0.016097259 |
| Heart_Failure | dendritic_cell | *SEMA4A* | 1.891830762 | 1.49E-07 | 3.45E-06 | 0.00072422 |
| Heart_Failure | dendritic_cell | *HRCT1* | 1.696598784 | 7.30E-05 | 0.000143842 | 0.354366392 |
| Heart_Failure | dendritic_cell | *MPHOSPH8* | -2.257836709 | 0.000994006 | 0.001534006 | 1 |
| Heart_Failure | dendritic_cell | *SIRPB2* | 0.973918667 | 0.00054866 | 0.000882658 | 1 |
| Heart_Failure | dendritic_cell | *SRGAP3* | -1.55291854 | 0.02207036 | 0.02779273 | 1 |
| Heart_Failure | dendritic_cell | *XPNPEP3* | -1.226647795 | 0.005734891 | 0.007851493 | 1 |
| Heart_Failure | dendritic_cell | *PPIA* | -2.744408229 | 7.94E-05 | 0.00015467 | 0.385438856 |
| Heart_Failure | dendritic_cell | *ZNF493* | -1.746738192 | 6.51E-07 | 4.45E-06 | 0.003160157 |
| Heart_Failure | dendritic_cell | *GTF2IRD2* | -2.257244606 | 4.50E-07 | 3.90E-06 | 0.00218117 |
| Heart_Failure | dendritic_cell | *SUPT3H* | -2.410268962 | 2.47E-05 | 5.71E-05 | 0.119616172 |
| Heart_Failure | dendritic_cell | *GIMAP5* | -2.418569807 | 1.40E-07 | 3.44E-06 | 0.000680379 |
| Heart_Failure | dendritic_cell | *STK31* | -2.579158253 | 2.58E-06 | 1.02E-05 | 0.012520057 |
| Heart_Failure | dendritic_cell | *ZKSCAN7* | -2.199227933 | 7.85E-06 | 2.22E-05 | 0.038090136 |
| Heart_Failure | dendritic_cell | *CD55* | -2.990217307 | 0.000245228 | 0.000423889 | 1 |
| Heart_Failure | dendritic_cell | *ZNF565* | -1.892768666 | 0.005538501 | 0.007597627 | 1 |
| Heart_Failure | dendritic_cell | *ZNF34* | -4.087177327 | 4.23E-06 | 1.44E-05 | 0.020543807 |
| Heart_Failure | dendritic_cell | *ZNF781* | -0.862550352 | 0.02652426 | 0.03299044 | 1 |
| Heart_Failure | dendritic_cell | *EVL* | -1.773056301 | 0.020669018 | 0.026116166 | 1 |
| Heart_Failure | dendritic_cell | *ZNF124* | -3.500943205 | 4.43E-05 | 9.31E-05 | 0.214718093 |
| Heart_Failure | dendritic_cell | *PIK3R4* | -2.20983403 | 2.27E-06 | 9.29E-06 | 0.011030882 |
| Heart_Failure | dendritic_cell | *ZNF605* | -2.187493873 | 0.000107472 | 0.000201424 | 0.521451933 |
| Heart_Failure | dendritic_cell | *RFX8* | -0.960222815 | 0.000221881 | 0.00038656 | 1 |
| Heart_Failure | dendritic_cell | *C20orf96* | -5.639182507 | 0.000379805 | 0.000629807 | 1 |
| Heart_Failure | dendritic_cell | *SULT1A1* | -2.714554972 | 6.99E-06 | 2.04E-05 | 0.033907838 |

| | | | | | | |
|---|---|---|---|---|---|---|
| Heart_Failure | dendritic_cell | *SPTSSB* | 5.285025616 | 0.001583465 | 0.002366176 | 1 |
| Heart_Failure | dendritic_cell | *CACNA1H* | -7.092312241 | 0.000608318 | 0.000969314 | 1 |
| Heart_Failure | dendritic_cell | *SULF2* | 2.729883516 | 2.58E-06 | 1.02E-05 | 0.012503377 |
| Heart_Failure | dendritic_cell | *XRCC2* | -3.870850678 | 3.33E-07 | 3.66E-06 | 0.001617995 |
| Heart_Failure | dendritic_cell | *MYO6* | -2.340181632 | 4.14E-07 | 3.87E-06 | 0.002010847 |
| Heart_Failure | dendritic_cell | *ADH1B* | -1.304261292 | 1.35E-06 | 6.67E-06 | 0.006572826 |
| Heart_Failure | dendritic_cell | *ZKSCAN5* | -1.688472151 | 5.46E-06 | 1.73E-05 | 0.026507131 |
| Heart_Failure | dendritic_cell | *ZNF502* | -1.967923337 | 4.30E-07 | 3.90E-06 | 0.002086181 |
| Heart_Failure | dendritic_cell | *TOMM7* | -2.892675227 | 1.59E-05 | 3.97E-05 | 0.077385006 |
| Heart_Failure | dendritic_cell | *ZNF33B* | -1.741971747 | 0.022794555 | 0.028652638 | 1 |
| Heart_Failure | dendritic_cell | *AMZ2* | -2.369159117 | 0.000148559 | 0.000268959 | 0.720809125 |
| Heart_Failure | dendritic_cell | *ZNF431* | -2.56049022 | 0.001107728 | 0.001695488 | 1 |
| Heart_Failure | dendritic_cell | *HLA-DQA1* | 0.362595087 | 0.000178338 | 0.000317074 | 0.865295559 |
| Heart_Failure | dendritic_cell | *COL27A1* | -1.314545967 | 8.61E-07 | 5.14E-06 | 0.004177134 |
| Heart_Failure | dendritic_cell | *ZNF239* | -2.264414474 | 1.46E-05 | 3.68E-05 | 0.070820231 |
| Heart_Failure | dendritic_cell | *SPRR2B* | -13.4786951 | 0.002582568 | 0.003701809 | 1 |
| Heart_Failure | dendritic_cell | *ANKRD36B* | -2.414260777 | 0.003199494 | 0.004512775 | 1 |
| Heart_Failure | dendritic_cell | *FAM3C* | -2.709936733 | 5.94E-06 | 1.84E-05 | 0.0288284 |
| Heart_Failure | dendritic_cell | *AP2A1* | 2.45993966 | 0.014810178 | 0.019147078 | 1 |
| Heart_Failure | dendritic_cell | *HIST1H3E* | 1.623217207 | 8.89E-06 | 2.45E-05 | 0.043112203 |
| Heart_Failure | dendritic_cell | *FUT11* | -2.971622678 | 1.12E-05 | 2.97E-05 | 0.054554144 |
| Heart_Failure | dendritic_cell | *WDR5B* | -2.050225338 | 5.11E-05 | 0.000105799 | 0.247781602 |
| Heart_Failure | dendritic_cell | *ZNF138* | -2.24562277 | 0.001435611 | 0.002153194 | 1 |
| Heart_Failure | dendritic_cell | *ZNF429* | -2.234179542 | 2.16E-06 | 8.95E-06 | 0.010465818 |
| Heart_Failure | dendritic_cell | *ZNF470* | -3.601633657 | 2.71E-05 | 6.14E-05 | 0.131277345 |
| Heart_Failure | dendritic_cell | *ZNF100* | -2.26634228 | 0.003337276 | 0.004700279 | 1 |
| Heart_Failure | dendritic_cell | *ANXA6* | -2.023778395 | 4.51E-07 | 3.90E-06 | 0.002187877 |
| Heart_Failure | dendritic_cell | *GMFB* | 4.763238712 | 2.18E-05 | 5.18E-05 | 0.105727692 |
| Heart_Failure | dendritic_cell | *ZNF420* | -2.838686685 | 1.77E-05 | 4.33E-05 | 0.085655939 |
| Heart_Failure | dendritic_cell | *ZNF763* | 1.309474045 | 0.003880707 | 0.005423154 | 1 |
| Heart_Failure | dendritic_cell | *ZMYM1* | -2.41607304 | 1.96E-06 | 8.42E-06 | 0.009520277 |
| Heart_Failure | dendritic_cell | *IGF2R* | -2.559184947 | 1.79E-05 | 4.37E-05 | 0.086641449 |
| Heart_Failure | dendritic_cell | *GAL3ST4* | -3.355050818 | 1.68E-06 | 7.68E-06 | 0.008151482 |
| Heart_Failure | dendritic_cell | *ZNF772* | -1.111283341 | 0.008609363 | 0.011491782 | 1 |
| Heart_Failure | dendritic_cell | *ZNF257* | 2.15809962 | 1.25E-05 | 3.25E-05 | 0.060752065 |
| Heart_Failure | dendritic_cell | *ADAM32* | -1.945545648 | 3.73E-07 | 3.66E-06 | 0.001811274 |
| Heart_Failure | dendritic_cell | *LRRC8B* | -3.083127752 | 5.82E-05 | 0.000118136 | 0.282227848 |
| Heart_Failure | dendritic_cell | *ABCB8* | -2.165277038 | 1.68E-06 | 7.67E-06 | 0.008133774 |
| Heart_Failure | dendritic_cell | *NEK5* | -2.001546565 | 4.13E-05 | 8.81E-05 | 0.200518389 |
| Heart_Failure | dendritic_cell | *PIWIL2* | -2.476584024 | 1.26E-05 | 3.27E-05 | 0.06135243 |
| Heart_Failure | dendritic_cell | *ENTPD4* | -3.591071744 | 0.000209577 | 0.0003671 | 1 |
| Heart_Failure | dendritic_cell | *TBC1D9B* | -1.700375425 | 0.028672174 | 0.03552538 | 1 |
| Heart_Failure | dendritic_cell | *FAM110D* | -2.724716712 | 9.02E-06 | 2.48E-05 | 0.0437796 |
| Heart_Failure | dendritic_cell | *CCL4L2* | -5.990471912 | 2.26E-06 | 9.26E-06 | 0.01097031 |
| Heart_Failure | dendritic_cell | *RAMP2-AS1* | -2.442700495 | 1.40E-06 | 6.81E-06 | 0.006813187 |
| Heart_Failure | dendritic_cell | *ZNF720* | -2.320922606 | 1.75E-06 | 7.84E-06 | 0.008496907 |
| Heart_Failure | dendritic_cell | *MRPL21* | -0.807410744 | 0.008347107 | 0.011166298 | 1 |
| Heart_Failure | dendritic_cell | *ZNF98* | -1.615364396 | 5.96E-08 | 3.44E-06 | 0.000289212 |
| Heart_Failure | dendritic_cell | *ZNF786* | -1.918523141 | 1.36E-05 | 3.47E-05 | 0.065935669 |
| Heart_Failure | dendritic_cell | *ZNF517* | -2.291355106 | 3.32E-07 | 3.66E-06 | 0.001610003 |

| | | | | | | |
|---|---|---|---|---|---|---|
| Heart_Failure | dendritic_cell | *ZNF675* | 3.208651062 | 1.47E-06 | 7.03E-06 | 0.007128242 |
| Heart_Failure | dendritic_cell | *SLC22A5* | -1.814792028 | 9.88E-08 | 3.44E-06 | 0.000479401 |
| Heart_Failure | dendritic_cell | *SHPK* | 1.473541998 | 0.008222766 | 0.011012106 | 1 |
| Heart_Failure | dendritic_cell | *MAP3K5* | -2.539065866 | 0.00062639 | 0.000995821 | 1 |
| Heart_Failure | dendritic_cell | *GYPE* | 2.297541041 | 2.46E-05 | 5.71E-05 | 0.119449599 |
| Heart_Failure | dendritic_cell | *PCDHB11* | -1.027439209 | 0.005133004 | 0.007069355 | 1 |
| Heart_Failure | dendritic_cell | *SLC2A10* | -1.717170394 | 1.32E-07 | 3.44E-06 | 0.000641328 |
| Heart_Failure | dendritic_cell | *ZNF665* | 3.967907162 | 0.020436983 | 0.025863391 | 1 |
| Heart_Failure | dendritic_cell | *RAB40C* | -1.895734781 | 5.30E-07 | 4.12E-06 | 0.002572322 |
| Heart_Failure | dendritic_cell | *PIGN* | -1.482086891 | 0.016777589 | 0.021529981 | 1 |
| Heart_Failure | dendritic_cell | *TOPORS* | -3.969127966 | 8.38E-05 | 0.000162024 | 0.406679839 |
| Heart_Failure | dendritic_cell | *BCO2* | -0.916868617 | 0.001866581 | 0.002751945 | 1 |
| Heart_Failure | dendritic_cell | *ENTPD6* | -3.723861119 | 0.000112611 | 0.000210185 | 0.546387155 |
| Heart_Failure | dendritic_cell | *ENPP1* | -2.56794337 | 0.000131426 | 0.000241179 | 0.637677943 |
| Heart_Failure | dendritic_cell | *CCDC154* | 1.218589506 | 0.005014168 | 0.00691747 | 1 |
| Heart_Failure | dendritic_cell | *MFAP5* | -1.998846565 | 9.72E-08 | 3.44E-06 | 0.000471378 |
| Heart_Failure | dendritic_cell | *DNAH10* | -3.907888713 | 2.02E-07 | 3.66E-06 | 0.000982307 |
| Heart_Failure | dendritic_cell | *FAM114A1* | -2.223082498 | 1.16E-06 | 6.11E-06 | 0.005644541 |
| Heart_Failure | dendritic_cell | *RPS26* | 1.885056684 | 5.72E-05 | 0.00011638 | 0.277332367 |
| Heart_Failure | dendritic_cell | *TXNRD3* | -2.586464372 | 0.000336961 | 0.000565722 | 1 |
| Heart_Failure | dendritic_cell | *EME2* | 6.494334907 | 1.65E-06 | 7.58E-06 | 0.007999414 |
| Heart_Failure | dendritic_cell | *ZNF780A* | -0.711521107 | 0.001667679 | 0.002480966 | 1 |
| Heart_Failure | dendritic_cell | *ATAD3A* | 2.80422206 | 0.019465988 | 0.024711925 | 1 |
| Heart_Failure | dendritic_cell | *AC122129.1* | -5.511810213 | 4.57E-07 | 3.90E-06 | 0.002218448 |
| Heart_Failure | dendritic_cell | *CCDC180* | 2.513534628 | 1.68E-07 | 3.52E-06 | 0.000816843 |
| Heart_Failure | dendritic_cell | *ZNF181* | -2.028624814 | 0.000517766 | 0.000838797 | 1 |
| Heart_Failure | dendritic_cell | *SGTB* | -3.35225698 | 6.87E-06 | 2.03E-05 | 0.033335358 |
| Heart_Failure | dendritic_cell | *ADH5* | -2.222720742 | 5.79E-06 | 1.80E-05 | 0.028085925 |
| Heart_Failure | dendritic_cell | *ZNF677* | 5.51579359 | 0.000152715 | 0.000275557 | 0.740972349 |
| Heart_Failure | dendritic_cell | *ZNF311* | 1.654674897 | 0.0022314 | 0.003249325 | 1 |
| Heart_Failure | dendritic_cell | *ZNF71* | -1.691657851 | 6.50E-06 | 1.95E-05 | 0.031533289 |
| Heart_Failure | dendritic_cell | *RPL12* | -1.79713819 | 4.79E-05 | 9.97E-05 | 0.232233262 |
| Heart_Failure | dendritic_cell | *MPZL1* | -1.686300284 | 8.38E-07 | 5.05E-06 | 0.004068215 |
| Heart_Failure | dendritic_cell | *VPS13A* | -2.014404135 | 1.09E-06 | 5.94E-06 | 0.0053044 |
| Heart_Failure | dendritic_cell | *NOL8* | -3.188517239 | 0.00013805 | 0.000251434 | 0.669818849 |
| Heart_Failure | dendritic_cell | *DLGAP2* | -1.747211395 | 4.43E-05 | 9.31E-05 | 0.214709601 |
| Heart_Failure | dendritic_cell | *MRPL42* | -1.59088597 | 0.000328779 | 0.000553517 | 1 |
| Heart_Failure | dendritic_cell | *ZNF560* | -2.29532184 | 0.004037164 | 0.005632064 | 1 |
| Heart_Failure | dendritic_cell | *AGAP9* | -3.036279086 | 4.44E-05 | 9.33E-05 | 0.215251212 |
| Heart_Failure | dendritic_cell | *ZNF273* | 3.173101094 | 0.004958515 | 0.006842638 | 1 |
| Heart_Failure | dendritic_cell | *MAK16* | -3.072619336 | 1.12E-06 | 6.01E-06 | 0.005414631 |
| Heart_Failure | dendritic_cell | *ZNF667* | -1.619823208 | 1.28E-06 | 6.45E-06 | 0.006216308 |
| Heart_Failure | dendritic_cell | *ZNF649* | 1.888311019 | 0.004878684 | 0.006740141 | 1 |
| Heart_Failure | dendritic_cell | *ADH4* | -1.354722692 | 7.35E-07 | 4.70E-06 | 0.003566204 |
| Heart_Failure | dendritic_cell | *HIBCH* | -1.540237472 | 4.70E-06 | 1.55E-05 | 0.022784918 |
| Heart_Failure | dendritic_cell | *ZNF544* | -1.994874628 | 4.03E-07 | 3.80E-06 | 0.001953145 |
| Heart_Failure | dendritic_cell | *SOWAHC* | -3.191039806 | 1.04E-06 | 5.77E-06 | 0.005035983 |
| Heart_Failure | dendritic_cell | *AC135178.1* | 5.257784855 | 2.25E-06 | 9.24E-06 | 0.010925054 |
| Heart_Failure | dendritic_cell | *MIER1* | -2.506080904 | 0.000390517 | 0.000644486 | 1 |
| Heart_Failure | dendritic_cell | *MAN1A2* | -2.351278274 | 8.82E-06 | 2.44E-05 | 0.042797124 |

| | | | | | | |
|---|---|---|---|---|---|---|
| Heart_Failure | dendritic_cell | *SVIP* | -1.507487287 | 4.60E-07 | 3.90E-06 | 0.002232295 |
| Heart_Failure | dendritic_cell | *DDRGK1* | -3.409486786 | 7.68E-06 | 2.18E-05 | 0.037261954 |
| Heart_Failure | dendritic_cell | *ZNF607* | 2.536221497 | 2.45E-05 | 5.70E-05 | 0.119079464 |
| Heart_Failure | dendritic_cell | *ZNF334* | -1.920890409 | 6.96E-06 | 2.04E-05 | 0.03375994 |
| Heart_Failure | dendritic_cell | *HSD17B11* | -3.308520115 | 0.002617733 | 0.003747784 | 1 |
| Heart_Failure | dendritic_cell | *SULT1C2* | -1.178763794 | 0.000424532 | 0.000696595 | 1 |
| Heart_Failure | dendritic_cell | *RPS6KL1* | -2.262661102 | 6.18E-06 | 1.89E-05 | 0.029966809 |
| Heart_Failure | dendritic_cell | *DDX42* | -3.164749375 | 3.20E-06 | 1.18E-05 | 0.015515241 |
| Heart_Failure | dendritic_cell | *STYX* | -3.714051993 | 0.006126516 | 0.008338249 | 1 |
| Heart_Failure | dendritic_cell | *HELZ* | -2.277511141 | 3.84E-05 | 8.29E-05 | 0.186498995 |
| Heart_Failure | dendritic_cell | *TMEM116* | -1.419259094 | 3.83E-06 | 1.34E-05 | 0.018590429 |
| Heart_Failure | dendritic_cell | *UCKL1* | -3.19012711 | 0.033340549 | 0.041047537 | 1 |
| Heart_Failure | dendritic_cell | *ZNF442* | -3.015116528 | 0.00098777 | 0.001524867 | 1 |
| Heart_Failure | dendritic_cell | *GFPT1* | -1.979485374 | 3.33E-06 | 1.22E-05 | 0.016133187 |
| Heart_Failure | dendritic_cell | *ZNF568* | -2.302909392 | 8.60E-06 | 2.39E-05 | 0.041708101 |
| Heart_Failure | dendritic_cell | *ZNF587* | -2.478772881 | 0.00552662 | 0.007583473 | 1 |
| Heart_Failure | dendritic_cell | *SH3BGRL2* | -1.400223486 | 3.76E-05 | 8.13E-05 | 0.182670447 |
| Heart_Failure | dendritic_cell | *TMA16* | -2.911957875 | 6.96E-06 | 2.04E-05 | 0.033793788 |
| Heart_Failure | dendritic_cell | *HLA-DRB5* | 0.425797396 | 0.000596752 | 0.000952134 | 1 |
| Heart_Failure | dendritic_cell | *CNGA1* | -1.488841681 | 0.001137105 | 0.001737711 | 1 |
| Heart_Failure | dendritic_cell | *C20orf203* | 4.134450238 | 1.04E-07 | 3.44E-06 | 0.000504753 |
| Heart_Failure | dendritic_cell | *ZNF534* | 1.353407401 | 0.000149851 | 0.000271096 | 0.727078582 |
| Heart_Failure | dendritic_cell | *NCOA6* | -2.191287374 | 0.001064431 | 0.001633855 | 1 |
| Heart_Failure | dendritic_cell | *FAM19A2* | -2.795050542 | 7.91E-05 | 0.000153996 | 0.383604567 |
| Heart_Failure | dendritic_cell | *TTC37* | -2.472847724 | 0.000225984 | 0.000393002 | 1 |
| Heart_Failure | dendritic_cell | *TUSC1* | 7.420114415 | 1.02E-06 | 5.71E-06 | 0.00494059 |
| Heart_Failure | dendritic_cell | *PAPSS2* | -2.580997526 | 1.87E-05 | 4.55E-05 | 0.090855951 |
| Heart_Failure | dendritic_cell | *CEP290* | -1.815745025 | 6.84E-06 | 2.02E-05 | 0.033183147 |
| Heart_Failure | dendritic_cell | *ANKRD13B* | -2.837571081 | 3.71E-07 | 3.66E-06 | 0.001800743 |
| Heart_Failure | dendritic_cell | *ECI2* | -2.563339251 | 4.42E-06 | 1.48E-05 | 0.021452934 |
| Heart_Failure | dendritic_cell | *UNC13B* | -2.171956042 | 1.38E-06 | 6.73E-06 | 0.006683136 |
| Heart_Failure | dendritic_cell | *PPP1R14C* | -6.555786315 | 3.38E-07 | 3.66E-06 | 0.001641326 |
| Heart_Failure | dendritic_cell | *GPATCH3* | 2.834745892 | 9.90E-08 | 3.44E-06 | 0.000480339 |
| Heart_Failure | dendritic_cell | *RPL10A* | -2.730409793 | 8.10E-06 | 2.28E-05 | 0.039285486 |
| Heart_Failure | dendritic_cell | *COLGALT2* | -2.443262517 | 3.82E-06 | 1.34E-05 | 0.018515493 |
| Heart_Failure | dendritic_cell | *RCSD1* | 2.659221642 | 4.36E-07 | 3.90E-06 | 0.002116172 |
| Heart_Failure | dendritic_cell | *GRIN3A* | -2.311640353 | 4.63E-05 | 9.68E-05 | 0.224530825 |
| Heart_Failure | dendritic_cell | *TMEM184B* | -1.663100771 | 1.26E-06 | 6.39E-06 | 0.006122579 |
| Heart_Failure | dendritic_cell | *SCAMP5* | -1.35758403 | 4.44E-06 | 1.49E-05 | 0.021523877 |
| Heart_Failure | dendritic_cell | *SFT2D1* | -3.272982438 | 1.32E-05 | 3.38E-05 | 0.064007477 |
| Heart_Failure | dendritic_cell | *HMGN2* | -2.875963088 | 1.87E-05 | 4.55E-05 | 0.090884228 |
| Heart_Failure | dendritic_cell | *RYR3* | -1.679928452 | 1.74E-05 | 4.28E-05 | 0.084186998 |
| Heart_Failure | dendritic_cell | *TYW1* | -2.379729641 | 0.000333665 | 0.000560965 | 1 |
| Heart_Failure | dendritic_cell | *PRMT6* | -5.535715705 | 1.46E-07 | 3.45E-06 | 0.000710109 |
| Heart_Failure | dendritic_cell | *SHISA4* | 5.908170817 | 7.61E-06 | 2.17E-05 | 0.036923281 |
| Heart_Failure | dendritic_cell | *MAP3K3* | -2.576112776 | 2.52E-05 | 5.79E-05 | 0.122321921 |
| Heart_Failure | dendritic_cell | *C1orf174* | 3.130983973 | 0.002026477 | 0.00296874 | 1 |
| Heart_Failure | dendritic_cell | *DZIP3* | -2.544917749 | 6.96E-06 | 2.04E-05 | 0.033771722 |
| Heart_Failure | dendritic_cell | *KIAA0753* | -2.902453862 | 8.92E-05 | 0.000170674 | 0.432829301 |
| Heart_Failure | dendritic_cell | *CCDC167* | -2.842519112 | 6.79E-07 | 4.52E-06 | 0.003293249 |

| | | | | | | |
|---|---|---|---|---|---|---|
| Heart_Failure | dendritic_cell | *ZFP2* | -1.853706726 | 6.22E-07 | 4.37E-06 | 0.003017669 |
| Heart_Failure | dendritic_cell | *MFAP3L* | -2.312158674 | 2.31E-06 | 9.39E-06 | 0.011198314 |
| Heart_Failure | dendritic_cell | *NAGA* | 3.841337068 | 2.97E-05 | 6.61E-05 | 0.143945794 |
| Heart_Failure | dendritic_cell | *TGM2* | -1.008573057 | 0.03573187 | 0.043880292 | 1 |
| Heart_Failure | dendritic_cell | *PJA2* | -2.365471881 | 4.83E-06 | 1.58E-05 | 0.023441044 |
| Heart_Failure | dendritic_cell | *AL590822.1* | -3.481480995 | 5.71E-06 | 1.78E-05 | 0.027707571 |
| Heart_Failure | dendritic_cell | *AP001007.1* | -1.653295247 | 0.031299227 | 0.038671722 | 1 |
| Heart_Failure | dendritic_cell | *EFCAB2* | -1.797758576 | 0.000351492 | 0.000586666 | 1 |
| Heart_Failure | dendritic_cell | *CHML* | -1.931648021 | 1.88E-06 | 8.18E-06 | 0.009100162 |
| Heart_Failure | dendritic_cell | *CR1* | 0.471553121 | 0.000116171 | 0.000215962 | 0.563662057 |
| Heart_Failure | dendritic_cell | *SAMD5* | -1.632378829 | 4.44E-07 | 3.90E-06 | 0.002154846 |
| Heart_Failure | dendritic_cell | *ECT2L* | 4.144947443 | 1.52E-07 | 3.48E-06 | 0.000737057 |
| Heart_Failure | dendritic_cell | *GPR52* | 19.50960666 | 4.25E-06 | 1.44E-05 | 0.020633438 |
| Heart_Failure | dendritic_cell | *FAM229B* | -1.66428671 | 8.37E-07 | 5.05E-06 | 0.004063042 |
| Heart_Failure | dendritic_cell | *DDO* | -3.099244632 | 7.71E-07 | 4.81E-06 | 0.003739352 |
| Heart_Failure | dendritic_cell | *RBM20* | -2.307782386 | 9.96E-07 | 5.63E-06 | 0.004832897 |
| Heart_Failure | dendritic_cell | *C6orf163* | 1.223483469 | 0.003079006 | 0.004356761 | 1 |
| Heart_Failure | dendritic_cell | *PCMTD2* | -2.391775537 | 3.20E-06 | 1.18E-05 | 0.015521762 |
| Heart_Failure | dendritic_cell | *EFCAB7* | -2.485094873 | 1.99E-06 | 8.53E-06 | 0.009677137 |
| Heart_Failure | dendritic_cell | *IFIT1B* | -10.84993399 | 3.15E-07 | 3.66E-06 | 0.001527405 |
| Heart_Failure | dendritic_cell | *LIPN* | 6.095786153 | 1.39E-07 | 3.44E-06 | 0.000676229 |
| Heart_Failure | dendritic_cell | *NEU4* | -4.037350375 | 8.63E-05 | 0.000165823 | 0.418702297 |
| Heart_Failure | dendritic_cell | *C2orf72* | -11.80606645 | 8.71E-05 | 0.000167117 | 0.422472746 |
| Heart_Failure | dendritic_cell | *RUFY2* | -2.141957231 | 4.01E-06 | 1.38E-05 | 0.019440987 |
| Heart_Failure | dendritic_cell | *PHACTR4* | -2.302453987 | 7.41E-05 | 0.000145477 | 0.359327974 |
| Heart_Failure | dendritic_cell | *ASAH2B* | -2.544885093 | 2.18E-06 | 9.02E-06 | 0.010567284 |
| Heart_Failure | dendritic_cell | *AGAP6* | -1.805217739 | 0.000796203 | 0.00124418 | 1 |
| Heart_Failure | dendritic_cell | *SYT15* | 2.561534475 | 0.000623491 | 0.000992188 | 1 |
| Heart_Failure | dendritic_cell | *LINC00619* | -18.67002416 | 5.41E-07 | 4.13E-06 | 0.002622746 |
| Heart_Failure | dendritic_cell | *DAXX* | 1.929661889 | 0.002619665 | 0.003749444 | 1 |
| Heart_Failure | dendritic_cell | *BMPR2* | -2.17303006 | 1.17E-05 | 3.08E-05 | 0.056886248 |
| Heart_Failure | dendritic_cell | *TCEA3* | -1.609541217 | 5.60E-06 | 1.75E-05 | 0.027160005 |
| Heart_Failure | dendritic_cell | *HSD17B8* | 2.834297749 | 1.17E-08 | 2.99E-06 | 5.66E-05 |
| Heart_Failure | dendritic_cell | *OXLD1* | -1.905561384 | 3.66E-05 | 7.93E-05 | 0.177463979 |
| Heart_Failure | dendritic_cell | *TAP2* | -1.049383906 | 0.007453976 | 0.01003228 | 1 |
| Heart_Failure | dendritic_cell | *COL15A1* | -1.13274161 | 1.13E-06 | 6.03E-06 | 0.005464138 |
| Heart_Failure | dendritic_cell | *AGER* | 1.470771833 | 1.84E-07 | 3.58E-06 | 0.000892704 |
| Heart_Failure | dendritic_cell | *PRRT1* | 1.773951918 | 0.000151714 | 0.000273955 | 0.736117242 |
| Heart_Failure | dendritic_cell | *SMIM5* | 5.33030267 | 4.14E-06 | 1.42E-05 | 0.020081823 |
| Heart_Failure | dendritic_cell | *STK19* | -3.189186831 | 0.000673681 | 0.001066461 | 1 |
| Heart_Failure | dendritic_cell | *SKIV2L* | -1.302016305 | 0.000303292 | 0.000514356 | 1 |
| Heart_Failure | dendritic_cell | *LAYN* | -2.408521887 | 2.88E-07 | 3.66E-06 | 0.001395132 |
| Heart_Failure | dendritic_cell | *C6orf48* | -1.915137188 | 0.017448157 | 0.022307894 | 1 |
| Heart_Failure | dendritic_cell | *LST1* | 1.245272144 | 1.61E-07 | 3.51E-06 | 0.000780996 |
| Heart_Failure | dendritic_cell | *ZNF814* | -2.520867592 | 0.000131331 | 0.000241096 | 0.637216587 |
| Heart_Failure | dendritic_cell | *MICB* | 0.873163907 | 2.03E-08 | 2.99E-06 | 9.87E-05 |
| Heart_Failure | dendritic_cell | *MICA* | 3.44442008 | 8.63E-11 | 6.98E-08 | 4.19E-07 |
| Heart_Failure | dendritic_cell | *HLA-C* | -1.332195462 | 0.000224209 | 0.000390335 | 1 |
| Heart_Failure | dendritic_cell | *PSORS1C1* | -0.373815759 | 0.002560884 | 0.003673982 | 1 |
| Heart_Failure | dendritic_cell | *DHX16* | -2.073679067 | 2.67E-06 | 1.04E-05 | 0.012975926 |

| | | | | | | |
|---|---|---|---|---|---|---|
| Heart_Failure | dendritic_cell | *ABCF1* | -1.929776462 | 0.028834404 | 0.035708149 | 1 |
| Heart_Failure | dendritic_cell | *LILRB3* | 0.84697985 | 5.30E-08 | 3.44E-06 | 0.000257106 |
| Heart_Failure | dendritic_cell | *HLA-E* | -2.364675588 | 6.89E-06 | 2.03E-05 | 0.033430326 |
| Heart_Failure | dendritic_cell | *ZNF468* | 4.717690673 | 4.71E-08 | 3.44E-06 | 0.000228519 |
| Heart_Failure | dendritic_cell | *RNF39* | -2.030288756 | 0.029544827 | 0.03656926 | 1 |
| Heart_Failure | dendritic_cell | *HCG9* | -2.10298387 | 0.000532692 | 0.000859821 | 1 |
| Heart_Failure | dendritic_cell | *HLA-G* | -1.472069004 | 0.006279702 | 0.008529987 | 1 |
| Heart_Failure | dendritic_cell | *FAM153C* | -0.935853362 | 0.000574793 | 0.000920732 | 1 |
| Heart_Failure | dendritic_cell | *TRIM27* | -2.309145466 | 6.71E-06 | 1.99E-05 | 0.032560275 |
| Heart_Failure | dendritic_cell | *MALRD1* | -1.731335634 | 7.07E-07 | 4.63E-06 | 0.003431286 |
| Heart_Failure | dendritic_cell | *FAM196B* | 1.379865841 | 1.64E-07 | 3.52E-06 | 0.000797869 |
| Heart_Failure | dendritic_cell | *MROH6* | 1.173072262 | 9.49E-06 | 2.59E-05 | 0.046067162 |
| Heart_Failure | dendritic_cell | *DCTN1* | -1.673504357 | 3.52E-07 | 3.66E-06 | 0.001706472 |
| Heart_Failure | dendritic_cell | *FAM216A* | 4.77854952 | 0.000930011 | 0.001443511 | 1 |
| Heart_Failure | dendritic_cell | *MZT1* | -4.767660256 | 2.49E-06 | 9.95E-06 | 0.012094867 |
| Heart_Failure | dendritic_cell | *ZNF155* | -2.869036731 | 6.67E-08 | 3.44E-06 | 0.000323431 |
| Heart_Failure | dendritic_cell | *ZNF783* | -3.237181831 | 0.024714076 | 0.030881457 | 1 |
| Heart_Failure | dendritic_cell | *ZNF425* | -2.218410334 | 0.000445231 | 0.000727605 | 1 |
| Heart_Failure | dendritic_cell | *C12orf73* | -2.955360538 | 4.18E-06 | 1.43E-05 | 0.020300136 |
| Heart_Failure | dendritic_cell | *PCDHGA1* | -2.174064359 | 0.000747634 | 0.001174335 | 1 |
| Heart_Failure | dendritic_cell | *PCDHA7* | -22.56974141 | 0.001463696 | 0.002192607 | 1 |
| Heart_Failure | dendritic_cell | *SPIRE2* | -1.222759566 | 0.009205077 | 0.012236448 | 1 |
| Heart_Failure | dendritic_cell | *PKHD1L1* | -0.777175686 | 2.39E-06 | 9.61E-06 | 0.011599162 |
| Heart_Failure | dendritic_cell | *SLFN12L* | -2.346072656 | 3.64E-08 | 3.44E-06 | 0.000176677 |
| Heart_Failure | dendritic_cell | *SLC35B4* | -3.296518051 | 6.50E-06 | 1.95E-05 | 0.0315561 |
| Heart_Failure | dendritic_cell | *TMEM231* | 1.550227675 | 1.43E-07 | 3.45E-06 | 0.00069396 |
| Heart_Failure | dendritic_cell | *LINC00654* | 2.005410631 | 0.007327672 | 0.009876074 | 1 |
| Heart_Failure | dendritic_cell | *SCGB2B2* | -2.090775775 | 1.30E-06 | 6.52E-06 | 0.006326878 |
| Heart_Failure | dendritic_cell | *CCDC144NL* | -6.626616493 | 0.002466018 | 0.003550481 | 1 |
| Heart_Failure | dendritic_cell | *TMEM170B* | -1.337108505 | 0.001097935 | 0.001681559 | 1 |
| Heart_Failure | dendritic_cell | *GOLGA6L10* | -8.152389605 | 1.27E-05 | 3.28E-05 | 0.061778101 |
| Heart_Failure | dendritic_cell | *SAP25* | -3.748349527 | 0.017372356 | 0.022216835 | 1 |
| Heart_Failure | dendritic_cell | *NT5M* | -2.139059928 | 2.91E-07 | 3.66E-06 | 0.001412212 |
| Heart_Failure | dendritic_cell | *IPO7* | 2.849227351 | 0.011998234 | 0.015716909 | 1 |
| Heart_Failure | dendritic_cell | *PRR13* | -2.993823784 | 1.57E-06 | 7.33E-06 | 0.007610773 |
| Heart_Failure | dendritic_cell | *TECPR1* | 3.345235002 | 0.016661475 | 0.021392293 | 1 |
| Heart_Failure | dendritic_cell | *MT1A* | -3.576033918 | 1.69E-06 | 7.69E-06 | 0.008207606 |
| Heart_Failure | dendritic_cell | *MT1M* | -4.706793132 | 1.20E-06 | 6.18E-06 | 0.005798401 |
| Heart_Failure | dendritic_cell | *CFI* | -2.240208046 | 4.39E-06 | 1.48E-05 | 0.021303508 |
| Heart_Failure | dendritic_cell | *ATP6AP1L* | -1.484614882 | 5.68E-05 | 0.000115844 | 0.275824721 |
| Heart_Failure | dendritic_cell | *RGL3* | -1.693566334 | 0.000287523 | 0.000490355 | 1 |
| Heart_Failure | dendritic_cell | *NAP1L4* | -2.446717926 | 0.000219563 | 0.000383071 | 1 |
| Heart_Failure | dendritic_cell | *SMN2* | -1.469383188 | 0.000265536 | 0.000455742 | 1 |
| Heart_Failure | dendritic_cell | *SERF1B* | 1.804874662 | 0.000280225 | 0.000478751 | 1 |
| Heart_Failure | dendritic_cell | *LIN52* | -1.540621971 | 0.009649276 | 0.012802376 | 1 |
| Heart_Failure | dendritic_cell | *KLRC2* | -3.612556569 | 4.74E-07 | 3.96E-06 | 0.002301847 |
| Heart_Failure | dendritic_cell | *C1QTNF9B* | 17.59375643 | 1.21E-07 | 3.44E-06 | 0.000585667 |
| Heart_Failure | dendritic_cell | *ZDHHC11B* | -2.550815021 | 8.27E-08 | 3.44E-06 | 0.000401285 |
| Heart_Failure | dendritic_cell | *HBA1* | 1.343194659 | 0.00745036 | 0.010030286 | 1 |
| Heart_Failure | dendritic_cell | *COL6A6* | -2.004373191 | 1.06E-05 | 2.82E-05 | 0.051329108 |

| | | | | | | |
|---|---|---|---|---|---|---|
| Heart_Failure | dendritic_cell | *HLA-A* | -2.028369765 | 0.000244534 | 0.000422986 | 1 |
| Heart_Failure | dendritic_cell | *LNP1* | 2.690154164 | 1.55E-07 | 3.51E-06 | 0.000751334 |
| Heart_Failure | dendritic_cell | *PRSS50* | -4.315609082 | 0.006273907 | 0.008524502 | 1 |
| Heart_Failure | dendritic_cell | *SACM1L* | -2.891320292 | 0.014232755 | 0.018444799 | 1 |
| Heart_Failure | dendritic_cell | *SLC48A1* | -2.915472526 | 2.07E-06 | 8.74E-06 | 0.010045882 |
| Heart_Failure | dendritic_cell | *TAS2R50* | -23.22995008 | 0.002512967 | 0.003609629 | 1 |
| Heart_Failure | dendritic_cell | *TAS2R14* | -0.97898318 | 0.016594349 | 0.021311747 | 1 |
| Heart_Failure | dendritic_cell | *AC084018.1* | -1.619972453 | 4.80E-05 | 9.99E-05 | 0.232890987 |
| Heart_Failure | dendritic_cell | *C17orf51* | -5.315677762 | 1.60E-06 | 7.42E-06 | 0.007758956 |
| Heart_Failure | dendritic_cell | *NUP62* | 1.713973241 | 0.002222341 | 0.003237105 | 1 |
| Heart_Failure | dendritic_cell | *AL021068.1* | 1.757704607 | 0.009380879 | 0.012463314 | 1 |
| Heart_Failure | dendritic_cell | *FGFR1OP* | 4.05517386 | 0.007107721 | 0.009595621 | 1 |
| Heart_Failure | dendritic_cell | *ZNF254* | -2.444657349 | 8.86E-06 | 2.45E-05 | 0.042989427 |
| Heart_Failure | dendritic_cell | *TCTEX1D2* | -2.979223906 | 0.000362458 | 0.000603103 | 1 |
| Heart_Failure | dendritic_cell | *AC022498.1* | 7.579046112 | 3.29E-08 | 3.33E-06 | 0.0001597 |
| Heart_Failure | dendritic_cell | *ASIC3* | 1.444661965 | 6.65E-05 | 0.000132782 | 0.322659682 |
| Heart_Failure | dendritic_cell | *GIMAP1* | -1.823955563 | 6.32E-07 | 4.39E-06 | 0.003066062 |
| Heart_Failure | dendritic_cell | *NOTCH2NL* | 2.061243389 | 1.46E-05 | 3.68E-05 | 0.070759007 |
| Heart_Failure | dendritic_cell | *SUPT4H1* | 6.819530458 | 3.40E-06 | 1.24E-05 | 0.016482465 |
| Heart_Failure | dendritic_cell | *MXD3* | -1.93693158 | 1.81E-05 | 4.42E-05 | 0.087988861 |
| Heart_Failure | dendritic_cell | *GSTM2* | -1.332314622 | 1.18E-06 | 6.15E-06 | 0.005728265 |
| Heart_Failure | dendritic_cell | *ARHGAP19* | 4.253512338 | 7.12E-05 | 0.00014063 | 0.345247263 |
| Heart_Failure | dendritic_cell | *LCAT* | 6.018572574 | 0.000189301 | 0.000335093 | 0.918490668 |
| Heart_Failure | dendritic_cell | *GPC2* | -7.297258498 | 9.97E-05 | 0.000188302 | 0.483936775 |
| Heart_Failure | dendritic_cell | *ERV3-1* | -0.486597749 | 0.013688026 | 0.01780067 | 1 |
| Heart_Failure | dendritic_cell | *GBP7* | 1.198340323 | 0.000112432 | 0.000210038 | 0.545519533 |
| Heart_Failure | dendritic_cell | *SRA1* | -2.872588164 | 0.000257716 | 0.000444521 | 1 |
| Heart_Failure | dendritic_cell | *TMX2* | -2.850586235 | 2.56E-07 | 3.66E-06 | 0.001239874 |
| Heart_Failure | dendritic_cell | *LBH* | 3.623057327 | 0.001566196 | 0.002341813 | 1 |
| Heart_Failure | dendritic_cell | *PPP1CB* | 2.313599048 | 0.015939988 | 0.020525696 | 1 |
| Heart_Failure | dendritic_cell | *SLC35F6* | -2.127291846 | 6.53E-07 | 4.46E-06 | 0.003169606 |
| Heart_Failure | dendritic_cell | *EMP2* | -2.020816167 | 2.10E-06 | 8.81E-06 | 0.01019098 |
| Heart_Failure | dendritic_cell | *SLC23A3* | -1.343124779 | 0.002622798 | 0.003752822 | 1 |
| Heart_Failure | dendritic_cell | *LTB4R* | -2.97177214 | 0.023630396 | 0.029672536 | 1 |
| Heart_Failure | dendritic_cell | *IRF9* | 3.823790525 | 3.39E-06 | 1.23E-05 | 0.016438214 |
| Heart_Failure | dendritic_cell | *ZNF726* | -3.052690884 | 3.03E-07 | 3.66E-06 | 0.001469041 |
| Heart_Failure | dendritic_cell | *ZNF90* | -1.70293498 | 2.95E-06 | 1.12E-05 | 0.014330141 |
| Heart_Failure | dendritic_cell | *REPIN1* | 3.567583437 | 2.13E-07 | 3.66E-06 | 0.001032532 |
| Heart_Failure | dendritic_cell | *ZNF891* | -2.563978385 | 1.46E-05 | 3.68E-05 | 0.07085231 |
| Heart_Failure | dendritic_cell | *SMIM7* | -2.725149637 | 0.022732624 | 0.028593314 | 1 |
| Heart_Failure | dendritic_cell | *TSPAN4* | -2.416961672 | 1.34E-05 | 3.43E-05 | 0.065070806 |
| Heart_Failure | dendritic_cell | *CPNE1* | -1.896342887 | 8.78E-08 | 3.44E-06 | 0.000425789 |
| Heart_Failure | dendritic_cell | *ARL16* | -0.778603519 | 0.005913912 | 0.008076076 | 1 |
| Heart_Failure | dendritic_cell | *PAXIP1-AS2* | -2.642018049 | 3.83E-06 | 1.34E-05 | 0.018565811 |
| Heart_Failure | dendritic_cell | *LYRM4* | -2.542586007 | 5.01E-06 | 1.62E-05 | 0.0242883 |
| Heart_Failure | dendritic_cell | *PRCD* | -2.076789327 | 8.14E-07 | 4.94E-06 | 0.003950537 |
| Heart_Failure | dendritic_cell | *SH3D21* | -1.909184467 | 0.003368805 | 0.004741932 | 1 |
| Heart_Failure | dendritic_cell | *FIS1* | -1.623077876 | 0.000790745 | 0.001237244 | 1 |
| Heart_Failure | dendritic_cell | *ANG* | -2.417328471 | 5.39E-07 | 4.13E-06 | 0.002616849 |
| Heart_Failure | dendritic_cell | *COLCA2* | -1.59952078 | 0.000221392 | 0.000385846 | 1 |

| | | | | | | |
|---|---|---|---|---|---|---|
| Heart_Failure | dendritic_cell | *MBLAC1* | -0.868584856 | 0.020724751 | 0.026172955 | 1 |
| Heart_Failure | dendritic_cell | *SOGA3* | -1.563532303 | 1.85E-05 | 4.50E-05 | 0.089853659 |
| Heart_Failure | dendritic_cell | *LCNL1* | -3.108375483 | 0.000347333 | 0.000581726 | 1 |
| Heart_Failure | dendritic_cell | *GNAT3* | -1.989090305 | 3.10E-07 | 3.66E-06 | 0.001501793 |
| Heart_Failure | dendritic_cell | *PLIN5* | -0.998070808 | 1.72E-07 | 3.52E-06 | 0.000834597 |
| Heart_Failure | dendritic_cell | *SEC14L6* | -0.676869066 | 0.000108183 | 0.000202509 | 0.524903968 |
| Heart_Failure | dendritic_cell | *NUTM2D* | 6.486294344 | 6.35E-07 | 4.39E-06 | 0.003082364 |
| Heart_Failure | dendritic_cell | *CPEB1* | -1.613747098 | 3.09E-06 | 1.16E-05 | 0.015008555 |
| Heart_Failure | dendritic_cell | *EML6* | 5.302598112 | 0.000234331 | 0.000406062 | 1 |
| Heart_Failure | dendritic_cell | *ARHGEF33* | -2.106417435 | 2.96E-06 | 1.12E-05 | 0.014384671 |
| Heart_Failure | dendritic_cell | *IFRD2* | -3.627906948 | 4.12E-06 | 1.41E-05 | 0.019967345 |
| Heart_Failure | dendritic_cell | *EVPLL* | -3.378314421 | 0.000205611 | 0.000361196 | 0.997623023 |
| Heart_Failure | dendritic_cell | *AC005082.1* | 5.821197218 | 3.60E-07 | 3.66E-06 | 0.001745029 |
| Heart_Failure | dendritic_cell | *ACSM4* | 0.874483097 | 2.98E-08 | 3.33E-06 | 0.00014475 |
| Heart_Failure | dendritic_cell | *UBXN2B* | -2.000821011 | 0.00218219 | 0.003182442 | 1 |
| Heart_Failure | dendritic_cell | *C5orf49* | -19.08367717 | 1.67E-07 | 3.52E-06 | 0.000811219 |
| Heart_Failure | dendritic_cell | *GOLGA8B* | -1.434304035 | 2.10E-05 | 5.03E-05 | 0.102074234 |
| Heart_Failure | dendritic_cell | *VPS16* | 3.966109512 | 0.036135133 | 0.044297035 | 1 |
| Heart_Failure | dendritic_cell | *MYL5* | -2.645424479 | 1.37E-06 | 6.71E-06 | 0.00664487 |
| Heart_Failure | dendritic_cell | *NPEPL1* | -2.452565529 | 7.58E-07 | 4.79E-06 | 0.003678575 |
| Heart_Failure | dendritic_cell | *TMEM242* | -2.011571795 | 2.37E-07 | 3.66E-06 | 0.001149087 |
| Heart_Failure | dendritic_cell | *SLC35E2* | 2.369461368 | 6.79E-06 | 2.01E-05 | 0.032936257 |
| Heart_Failure | dendritic_cell | *TSTD1* | -3.606543421 | 3.59E-06 | 1.28E-05 | 0.017414976 |
| Heart_Failure | dendritic_cell | *AC009403.2* | -3.236774016 | 2.56E-05 | 5.86E-05 | 0.124191869 |
| Heart_Failure | dendritic_cell | *PAM16* | -2.293775078 | 1.31E-06 | 6.56E-06 | 0.006374142 |
| Heart_Failure | dendritic_cell | *RNASEK* | 3.85677935 | 2.01E-05 | 4.84E-05 | 0.097750197 |
| Heart_Failure | dendritic_cell | *NBPF1* | -1.968216183 | 0.001272294 | 0.001929718 | 1 |
| Heart_Failure | dendritic_cell | *RNU6ATAC35I* | -20.55100345 | 4.93E-07 | 3.99E-06 | 0.002390586 |
| Heart_Failure | dendritic_cell | *FANCG* | 1.398063144 | 0.001210126 | 0.001843495 | 1 |
| Heart_Failure | dendritic_cell | *ZNF880* | -1.722600116 | 9.21E-07 | 5.38E-06 | 0.004466824 |
| Heart_Failure | dendritic_cell | *TIGD1* | 1.913391578 | 0.008304835 | 0.011112813 | 1 |
| Heart_Failure | dendritic_cell | *FXYD7* | -1.882194809 | 0.000383162 | 0.000633731 | 1 |
| Heart_Failure | dendritic_cell | *XKR9* | -0.779746014 | 0.000433224 | 0.000709897 | 1 |
| Heart_Failure | dendritic_cell | *APOL6* | -1.976697126 | 1.54E-06 | 7.23E-06 | 0.007464479 |
| Heart_Failure | dendritic_cell | *UBA52* | -1.795874757 | 0.00143161 | 0.002149186 | 1 |
| Heart_Failure | dendritic_cell | *TIAF1* | 1.945189399 | 1.21E-06 | 6.23E-06 | 0.005877637 |
| Heart_Failure | dendritic_cell | *RAB6C* | -51.3306389 | 5.34E-07 | 4.13E-06 | 0.002592248 |
| Heart_Failure | dendritic_cell | *AC062017.1* | 2.908489231 | 1.88E-07 | 3.60E-06 | 0.000909939 |
| Heart_Failure | dendritic_cell | *ADRA2B* | 4.560015014 | 1.22E-06 | 6.27E-06 | 0.005927409 |
| Heart_Failure | dendritic_cell | *EXOSC6* | -2.244071993 | 2.61E-06 | 1.02E-05 | 0.012659867 |
| Heart_Failure | dendritic_cell | *ZNF844* | -2.386839687 | 6.26E-06 | 1.91E-05 | 0.030391537 |
| Heart_Failure | dendritic_cell | *LINC00571* | -2.757368856 | 3.23E-06 | 1.19E-05 | 0.015686839 |
| Heart_Failure | dendritic_cell | *OSER1-AS1* | -1.885520238 | 0.000366772 | 0.000609654 | 1 |
| Heart_Failure | dendritic_cell | *C5orf66* | 3.916456951 | 0.000539458 | 0.000869295 | 1 |
| Heart_Failure | dendritic_cell | *LINC01133* | -1.268387313 | 5.91E-07 | 4.30E-06 | 0.002867777 |
| Heart_Failure | dendritic_cell | *C4B* | 2.40440801 | 0.018789247 | 0.023890311 | 1 |
| Heart_Failure | dendritic_cell | *LINC01117* | -1.832173521 | 4.95E-07 | 4.00E-06 | 0.002401626 |
| Heart_Failure | dendritic_cell | *RASAL2-AS1* | -1.947455491 | 0.015386052 | 0.019838725 | 1 |
| Heart_Failure | dendritic_cell | *LINC00240* | -1.006450658 | 0.002371523 | 0.003429696 | 1 |
| Heart_Failure | dendritic_cell | *LINC00184* | -46.87189724 | 3.28E-07 | 3.66E-06 | 0.001591975 |

| | | | | | | |
|---|---|---|---|---|---|---|
| Heart_Failure | dendritic_cell | *AL049840.1* | -3.515816917 | 6.93E-06 | 2.04E-05 | 0.033613533 |
| Heart_Failure | dendritic_cell | *PLEKHM1* | -2.500769372 | 1.05E-06 | 5.79E-06 | 0.005087277 |
| Heart_Failure | dendritic_cell | *LINC00092* | 1.216142781 | 4.40E-05 | 9.27E-05 | 0.213432856 |
| Heart_Failure | dendritic_cell | *AP000350.6* | 1.417947704 | 0.000542097 | 0.000873258 | 1 |
| Heart_Failure | dendritic_cell | *CYP4A22-AS1* | -2.216081192 | 3.22E-07 | 3.66E-06 | 0.001562871 |
| Heart_Failure | dendritic_cell | *MIAT* | -2.924884408 | 1.05E-05 | 2.81E-05 | 0.050936894 |
| Heart_Failure | dendritic_cell | *ERCC6* | -3.451447838 | 0.000101608 | 0.000191681 | 0.493002421 |
| Heart_Failure | dendritic_cell | *LINC00694* | 3.4469801 | 4.50E-07 | 3.90E-06 | 0.002183494 |
| Heart_Failure | dendritic_cell | *LINC00115* | -2.623943248 | 5.05E-07 | 4.03E-06 | 0.002452109 |
| Heart_Failure | dendritic_cell | *HAR1A* | 9.327130124 | 1.33E-06 | 6.61E-06 | 0.006442551 |
| Heart_Failure | dendritic_cell | *LINC00937* | 1.643002991 | 7.31E-06 | 2.10E-05 | 0.0354903 |
| Heart_Failure | dendritic_cell | *FAM138B* | -26.89336375 | 3.33E-07 | 3.66E-06 | 0.001617981 |
| Heart_Failure | dendritic_cell | *AC017074.2* | -10.47420397 | 0.024655403 | 0.030824019 | 1 |
| Heart_Failure | dendritic_cell | *AC009299.3* | 3.048247686 | 1.84E-07 | 3.58E-06 | 0.000894671 |
| Heart_Failure | dendritic_cell | *SCAMP4* | -3.1020468 | 0.038421289 | 0.046992714 | 1 |
| Heart_Failure | dendritic_cell | *SLC2A1-AS1* | -2.838465358 | 4.26E-05 | 9.04E-05 | 0.206843571 |
| Heart_Failure | dendritic_cell | *AC005884.1* | -2.801884628 | 9.02E-06 | 2.48E-05 | 0.043753707 |
| Heart_Failure | dendritic_cell | *AC108463.2* | 2.551713111 | 0.024336102 | 0.030487676 | 1 |
| Heart_Failure | dendritic_cell | *HCG20* | 1.181861393 | 1.12E-07 | 3.44E-06 | 0.000544442 |
| Heart_Failure | dendritic_cell | *C14orf144* | -4.216477981 | 7.48E-06 | 2.14E-05 | 0.036309413 |
| Heart_Failure | dendritic_cell | *C19orf24* | -4.201212911 | 1.65E-06 | 7.57E-06 | 0.007982981 |
| Heart_Failure | dendritic_cell | *ZNF883* | -1.878614711 | 3.96E-06 | 1.37E-05 | 0.0192227 |
| Heart_Failure | dendritic_cell | *ARL17B* | -2.427287255 | 7.73E-07 | 4.82E-06 | 0.003751617 |
| Heart_Failure | dendritic_cell | *PATL2* | -2.720817712 | 6.25E-05 | 0.000125557 | 0.303219613 |
| Heart_Failure | dendritic_cell | *CASC20* | -2.793604249 | 5.64E-07 | 4.23E-06 | 0.002736881 |
| Heart_Failure | dendritic_cell | *AC112229.4* | -2.827415221 | 0.001059105 | 0.001626196 | 1 |
| Heart_Failure | dendritic_cell | *KLHL7-AS1* | 1.293933917 | 1.69E-07 | 3.52E-06 | 0.000820014 |
| Heart_Failure | dendritic_cell | *LINC00332* | -1.320551331 | 0.021004283 | 0.026519068 | 1 |
| Heart_Failure | dendritic_cell | *AC092171.4* | -3.132518746 | 5.92E-07 | 4.30E-06 | 0.002874224 |
| Heart_Failure | dendritic_cell | *RPS18* | -2.212241275 | 0.000173062 | 0.000308485 | 0.839695143 |
| Heart_Failure | dendritic_cell | *DIRC3* | -1.349572422 | 7.06E-07 | 4.63E-06 | 0.003425496 |
| Heart_Failure | dendritic_cell | *AC107021.1* | -1.528057848 | 1.78E-06 | 7.92E-06 | 0.008629641 |
| Heart_Failure | dendritic_cell | *TMA7* | -2.776558302 | 1.07E-06 | 5.85E-06 | 0.005190528 |
| Heart_Failure | dendritic_cell | *MTRNR2L4* | 2.946075577 | 2.48E-05 | 5.73E-05 | 0.1203148 |
| Heart_Failure | dendritic_cell | *AC009495.3* | -5.069668767 | 1.78E-05 | 4.36E-05 | 0.086353856 |
| Heart_Failure | dendritic_cell | *HLA-DQB2* | 0.52550605 | 1.61E-13 | 2.60E-10 | 7.79E-10 |
| Heart_Failure | dendritic_cell | *GOLGA8N* | -1.034731023 | 0.039110114 | 0.047762968 | 1 |
| Heart_Failure | dendritic_cell | *LINC00665* | -1.385868017 | 3.02E-06 | 1.14E-05 | 0.014662449 |
| Heart_Failure | dendritic_cell | *LYRM9* | -3.022830246 | 0.002466862 | 0.003550482 | 1 |
| Heart_Failure | dendritic_cell | *LINC01166* | -2.239679319 | 0.002240198 | 0.003259203 | 1 |
| Heart_Failure | dendritic_cell | *LINC00342* | -2.373011999 | 4.85E-07 | 3.96E-06 | 0.002352745 |
| Heart_Failure | dendritic_cell | *GPX1* | 3.600637501 | 6.95E-06 | 2.04E-05 | 0.033709158 |
| Heart_Failure | dendritic_cell | *RPS28* | -4.006666912 | 5.10E-05 | 0.000105779 | 0.247628876 |
| Heart_Failure | dendritic_cell | *ZNF736* | 2.227609697 | 0.000100913 | 0.000190443 | 0.48962897 |
| Heart_Failure | dendritic_cell | *PINLYP* | -2.27911218 | 0.006823133 | 0.00923712 | 1 |
| Heart_Failure | dendritic_cell | *ANKRD65* | -9.883843509 | 2.11E-06 | 8.82E-06 | 0.010243619 |
| Heart_Failure | dendritic_cell | *ZSCAN31* | -0.919995242 | 0.00014756 | 0.000267349 | 0.715960254 |
| Heart_Failure | dendritic_cell | *AC009495.2* | -23.41872718 | 0.010672569 | 0.014067728 | 1 |
| Heart_Failure | dendritic_cell | *LINC00957* | -4.143056226 | 3.57E-05 | 7.77E-05 | 0.173048426 |
| Heart_Failure | dendritic_cell | *LINC00484* | -0.800544668 | 0.002838254 | 0.004042034 | 1 |

| | | | | | | |
|---|---|---|---|---|---|---|
| Heart_Failure | dendritic_cell | *KIAA0040* | -3.334745587 | 0.000120794 | 0.000223615 | 0.586094759 |
| Heart_Failure | dendritic_cell | *LINC00607* | -2.163745423 | 0.00022048 | 0.000384394 | 1 |
| Heart_Failure | dendritic_cell | *AP001468.1* | -1.213232081 | 0.005752183 | 0.007870725 | 1 |
| Heart_Failure | dendritic_cell | *AC019117.2* | -12.60177514 | 9.52E-06 | 2.59E-05 | 0.046210577 |
| Heart_Failure | dendritic_cell | *ZBTB22* | -1.4524078 | 0.012098537 | 0.015835474 | 1 |
| Heart_Failure | dendritic_cell | *ZBED5* | -2.303823838 | 2.26E-05 | 5.32E-05 | 0.1095562 |
| Heart_Failure | dendritic_cell | *CDKN2AIPNL* | -2.424937878 | 1.94E-05 | 4.69E-05 | 0.094179042 |
| Heart_Failure | dendritic_cell | *LINC00959* | -3.413985368 | 5.58E-06 | 1.75E-05 | 0.027090427 |
| Heart_Failure | dendritic_cell | *HLA-DQA2* | 0.800105351 | 6.73E-15 | 1.63E-11 | 3.26E-11 |
| Heart_Failure | dendritic_cell | *IRGM* | -0.975818191 | 0.010337734 | 0.013644909 | 1 |
| Heart_Failure | dendritic_cell | *LINC01143* | 3.788522196 | 1.30E-07 | 3.44E-06 | 0.000628832 |
| Heart_Failure | dendritic_cell | *LINC00211* | 2.40788845 | 6.31E-08 | 3.44E-06 | 0.000306125 |
| Heart_Failure | dendritic_cell | *LRRC37A2* | -3.833455237 | 0.000258594 | 0.000445719 | 1 |
| Heart_Failure | dendritic_cell | *TXNDC5* | 3.710448729 | 1.28E-07 | 3.44E-06 | 0.000619717 |
| Heart_Failure | dendritic_cell | *LINC01119* | 6.427790871 | 0.002936715 | 0.004168795 | 1 |
| Heart_Failure | dendritic_cell | *CDRT4* | -3.180334555 | 0.00835496 | 0.011173722 | 1 |
| Heart_Failure | dendritic_cell | *C1orf226* | -4.538142948 | 1.35E-05 | 3.45E-05 | 0.065415998 |
| Heart_Failure | dendritic_cell | *LILRA2* | -0.88112658 | 0.025543492 | 0.031844045 | 1 |
| Heart_Failure | dendritic_cell | *PSMB9* | 1.803704361 | 3.00E-05 | 6.67E-05 | 0.145413873 |
| Heart_Failure | dendritic_cell | *PPIL3* | -0.927449785 | 0.034552057 | 0.04251752 | 1 |
| Heart_Failure | dendritic_cell | *L1TD1* | 1.901363268 | 0.000210594 | 0.000368616 | 1 |
| Heart_Failure | dendritic_cell | *C1QTNF9* | -7.562469538 | 3.51E-07 | 3.66E-06 | 0.001701019 |
| Heart_Failure | dendritic_cell | *ISY1* | -1.390053089 | 0.007254859 | 0.009783373 | 1 |
| Heart_Failure | dendritic_cell | *PNMA2* | -2.905112967 | 5.41E-07 | 4.13E-06 | 0.002625996 |
| Heart_Failure | dendritic_cell | *KRBOX1* | -29.83829409 | 3.09E-07 | 3.66E-06 | 0.001500201 |
| Heart_Failure | dendritic_cell | *ARHGEF25* | -1.832769556 | 6.53E-06 | 1.95E-05 | 0.031680351 |
| Heart_Failure | dendritic_cell | *NSUN6* | -2.290827423 | 2.21E-05 | 5.23E-05 | 0.106988374 |
| Heart_Failure | dendritic_cell | *HLA-DOB* | 1.241133709 | 0.00097981 | 0.001514024 | 1 |
| Heart_Failure | dendritic_cell | *LINC00877* | 1.930528185 | 4.32E-08 | 3.44E-06 | 0.000209615 |
| Heart_Failure | dendritic_cell | *TDGF1* | -1.400486813 | 0.036013204 | 0.044158723 | 1 |
| Heart_Failure | dendritic_cell | *INMT* | -1.588456643 | 2.54E-06 | 1.01E-05 | 0.012343873 |
| Heart_Failure | dendritic_cell | *ATP5O* | -3.050336193 | 4.89E-06 | 1.59E-05 | 0.023714011 |
| Heart_Failure | dendritic_cell | *PLEKHO2* | -1.858615952 | 0.004251748 | 0.005911027 | 1 |
| Heart_Failure | dendritic_cell | *C8orf58* | -5.202079276 | 0.024500933 | 0.030671486 | 1 |
| Heart_Failure | dendritic_cell | *HOGA1* | -3.766627717 | 3.31E-07 | 3.66E-06 | 0.001605243 |
| Heart_Failure | dendritic_cell | *PI4KA* | -2.545960766 | 0.002664515 | 0.003810264 | 1 |
| Heart_Failure | dendritic_cell | *LINC00969* | -1.074327279 | 0.016906214 | 0.021672114 | 1 |
| Heart_Failure | dendritic_cell | *ARFGAP3* | -3.5663345 | 0.000213151 | 0.000372823 | 1 |
| Heart_Failure | dendritic_cell | *MRPL20* | -3.449000959 | 0.002975828 | 0.00421938 | 1 |
| Heart_Failure | dendritic_cell | *DECR2* | -6.931770552 | 1.35E-05 | 3.46E-05 | 0.065692556 |
| Heart_Failure | dendritic_cell | *ZNF709* | -14.86392241 | 3.39E-07 | 3.66E-06 | 0.00164272 |
| Heart_Failure | dendritic_cell | *ERVW-1* | -8.648561638 | 1.24E-05 | 3.23E-05 | 0.060335638 |
| Heart_Failure | dendritic_cell | *MRPL33* | -2.808389947 | 1.11E-05 | 2.94E-05 | 0.053696442 |
| Heart_Failure | dendritic_cell | *STON1* | -2.558608315 | 1.36E-05 | 3.47E-05 | 0.06602391 |
| Heart_Failure | dendritic_cell | *VSIG8* | 2.738360146 | 5.94E-08 | 3.44E-06 | 0.000288105 |
| Heart_Failure | dendritic_cell | *C7orf73* | -2.884385157 | 9.79E-05 | 0.000185339 | 0.474839442 |
| Heart_Failure | dendritic_cell | *FGF14-IT1* | -5.743462528 | 3.35E-07 | 3.66E-06 | 0.001626784 |
| Heart_Failure | dendritic_cell | *PALM2* | -1.737385783 | 3.70E-07 | 3.66E-06 | 0.001797598 |
| Heart_Failure | dendritic_cell | *NBPF15* | -2.320229161 | 7.57E-07 | 4.79E-06 | 0.003672999 |
| Heart_Failure | dendritic_cell | *UPK3B* | -2.089295112 | 4.88E-07 | 3.96E-06 | 0.002365765 |

| | | | | | | |
|---|---|---|---|---|---|---|
| Heart_Failure | dendritic_cell | *IL10RB* | -3.104358096 | 2.88E-06 | 1.10E-05 | 0.013983047 |
| Heart_Failure | dendritic_cell | *PLA2G4B* | 2.336979763 | 3.05E-08 | 3.33E-06 | 0.000147772 |
| Heart_Failure | dendritic_cell | *LEFTY1* | -20.46733926 | 0.000525828 | 0.000850155 | 1 |
| Heart_Failure | dendritic_cell | *TTC4* | -2.71421301 | 6.22E-10 | 3.35E-07 | 3.02E-06 |
| Heart_Failure | dendritic_cell | *JMJD7* | 1.463431573 | 3.21E-08 | 3.33E-06 | 0.000155513 |
| Heart_Failure | dendritic_cell | *TMEM199* | 1.552207278 | 3.91E-05 | 8.40E-05 | 0.189670002 |
| Heart_Failure | dendritic_cell | *DBNDD2* | 8.555040327 | 6.56E-05 | 0.000131099 | 0.318308339 |
| Heart_Failure | dendritic_cell | *LILRA6* | 1.916850833 | 1.01E-07 | 3.44E-06 | 0.000491224 |
| Heart_Failure | dendritic_cell | *SCARF2* | -1.912613462 | 5.33E-07 | 4.13E-06 | 0.002584441 |
| Heart_Failure | dendritic_cell | *APOBEC3C* | 4.510629557 | 0.000882298 | 0.001372526 | 1 |
| Heart_Failure | dendritic_cell | *C4A* | -2.727391732 | 0.003170024 | 0.004475111 | 1 |
| Heart_Failure | dendritic_cell | *CRYBB2* | 2.669785426 | 0.000115132 | 0.000214442 | 0.558622228 |
| Heart_Failure | dendritic_cell | *NEAT1* | -1.547708004 | 2.80E-05 | 6.32E-05 | 0.136039512 |
| Heart_Failure | dendritic_cell | *RAB30-AS1* | -1.865006594 | 0.010161096 | 0.013426372 | 1 |
| Heart_Failure | dendritic_cell | *UBAP1L* | -2.540814128 | 2.08E-06 | 8.77E-06 | 0.010097664 |
| Heart_Failure | dendritic_cell | *DNAJC3-AS1* | -3.00268607 | 4.06E-07 | 3.82E-06 | 0.001971773 |
| Heart_Failure | dendritic_cell | *LINC00926* | -3.64816943 | 3.91E-07 | 3.75E-06 | 0.00189836 |
| Heart_Failure | dendritic_cell | *DPH6-AS1* | -2.456140183 | 6.54E-07 | 4.46E-06 | 0.003175157 |
| Heart_Failure | dendritic_cell | *CDK11B* | -4.129764166 | 0.00022857 | 0.000397356 | 1 |
| Heart_Failure | dendritic_cell | *ABHD14A* | -3.961963246 | 0.000290399 | 0.000494913 | 1 |
| Heart_Failure | dendritic_cell | *SRP14-AS1* | -0.771663297 | 0.004222652 | 0.005875625 | 1 |
| Heart_Failure | dendritic_cell | *GDNF-AS1* | -3.068998786 | 1.77E-06 | 7.90E-06 | 0.008586829 |
| Heart_Failure | dendritic_cell | *CCDC153* | -2.790792555 | 1.05E-06 | 5.81E-06 | 0.005116426 |
| Heart_Failure | dendritic_cell | *AL357673.1* | -1.368574953 | 0.007991247 | 0.01071388 | 1 |
| Heart_Failure | dendritic_cell | *C5orf17* | -1.646231216 | 4.41E-07 | 3.90E-06 | 0.002137648 |
| Heart_Failure | dendritic_cell | *TMEM150C* | -1.351401604 | 0.002237638 | 0.003256454 | 1 |
| Heart_Failure | dendritic_cell | *LINC00939* | 2.790174618 | 0.000254238 | 0.000438833 | 1 |
| Heart_Failure | dendritic_cell | *LINC01060* | -1.621282661 | 2.22E-06 | 9.13E-06 | 0.010751513 |
| Heart_Failure | dendritic_cell | *LINC00942* | -11.16317875 | 2.37E-07 | 3.66E-06 | 0.001150604 |
| Heart_Failure | dendritic_cell | *GOLGA8K* | -1.703394105 | 0.012044622 | 0.015769159 | 1 |
| Heart_Failure | dendritic_cell | *TMEM158* | 4.470487699 | 2.75E-07 | 3.66E-06 | 0.001334911 |
| Heart_Failure | dendritic_cell | *DNAH10OS* | -14.71748635 | 4.95E-08 | 3.44E-06 | 0.000239976 |
| Heart_Failure | dendritic_cell | *PCDHA10* | -1.54813967 | 0.001123633 | 0.001717665 | 1 |
| Heart_Failure | dendritic_cell | *ZNF718* | -2.927675615 | 3.54E-05 | 7.73E-05 | 0.17175512 |
| Heart_Failure | dendritic_cell | *LINC00989* | -2.019848593 | 3.58E-06 | 1.28E-05 | 0.017379898 |
| Heart_Failure | dendritic_cell | *GLI4* | -3.321085277 | 1.75E-06 | 7.84E-06 | 0.008503578 |
| Heart_Failure | dendritic_cell | *SNHG18* | -1.908769075 | 1.27E-07 | 3.44E-06 | 0.000614867 |
| Heart_Failure | dendritic_cell | *ALG1L2* | 4.431185613 | 2.13E-06 | 8.88E-06 | 0.010353249 |
| Heart_Failure | dendritic_cell | *ZNF550* | -2.323434672 | 4.94E-07 | 4.00E-06 | 0.002397333 |
| Heart_Failure | dendritic_cell | *LINC01094* | 6.355790122 | 2.00E-06 | 8.53E-06 | 0.009695122 |
| Heart_Failure | dendritic_cell | *TMEM200B* | -1.924505985 | 5.95E-05 | 0.000120439 | 0.288571347 |
| Heart_Failure | dendritic_cell | *PCDHGB6* | 1.352899376 | 3.60E-07 | 3.66E-06 | 0.001744995 |
| Heart_Failure | dendritic_cell | *SLC10A5* | 3.041230618 | 1.02E-06 | 5.71E-06 | 0.004937035 |
| Heart_Failure | dendritic_cell | *PCDHGA6* | -1.120139515 | 0.005714777 | 0.007826164 | 1 |
| Heart_Failure | dendritic_cell | *PCDHGB2* | -8.156011041 | 1.75E-05 | 4.30E-05 | 0.084970167 |
| Heart_Failure | dendritic_cell | *LYN* | 1.439643169 | 0.000175218 | 0.000312099 | 0.850157907 |
| Heart_Failure | dendritic_cell | *SIGLEC14* | 1.048581962 | 1.70E-06 | 7.72E-06 | 0.008255111 |
| Heart_Failure | dendritic_cell | *ARMS2* | -4.913645544 | 2.48E-07 | 3.66E-06 | 0.001201229 |
| Heart_Failure | dendritic_cell | *MEX3A* | -3.323201419 | 7.14E-07 | 4.65E-06 | 0.003462707 |
| Heart_Failure | dendritic_cell | *DPP3* | 1.624937789 | 6.83E-07 | 4.53E-06 | 0.003312326 |

| | | | | | | |
|---|---|---|---|---|---|---|
| Heart_Failure | dendritic_cell | *BRK1* | -2.218287088 | 0.003086013 | 0.004364131 | 1 |
| Heart_Failure | dendritic_cell | *SNHG9* | -2.879688616 | 0.001006012 | 0.001550563 | 1 |
| Heart_Failure | dendritic_cell | *ZNF432* | -2.358223361 | 0.002105259 | 0.003079505 | 1 |
| Heart_Failure | dendritic_cell | *HMBS* | -2.416949441 | 4.76E-06 | 1.57E-05 | 0.023072349 |
| Heart_Failure | dendritic_cell | *TAS2R31* | -2.144662621 | 0.001724344 | 0.002560917 | 1 |
| Heart_Failure | dendritic_cell | *KIAA1147* | -2.11415353 | 0.025091967 | 0.03131333 | 1 |
| Heart_Failure | dendritic_cell | *ZBED6* | 1.457246831 | 1.33E-07 | 3.44E-06 | 0.000645998 |
| Heart_Failure | dendritic_cell | *CHURC1* | -1.873761317 | 9.77E-06 | 2.65E-05 | 0.047393322 |
| Heart_Failure | dendritic_cell | *SLC25A21-AS1* | -2.441665656 | 1.84E-06 | 8.09E-06 | 0.008904754 |
| Heart_Failure | dendritic_cell | *RNASE4* | -7.807999871 | 9.00E-07 | 5.31E-06 | 0.004366948 |
| Heart_Failure | dendritic_cell | *LINC00639* | -1.598062038 | 1.44E-06 | 6.94E-06 | 0.006987542 |
| Heart_Failure | dendritic_cell | *LINC00924* | -1.25549211 | 1.45E-05 | 3.66E-05 | 0.070170066 |
| Heart_Failure | dendritic_cell | *MRPL46* | -3.998547014 | 0.000238133 | 0.000412502 | 1 |
| Heart_Failure | dendritic_cell | *UBE2Q2L* | 1.944296824 | 8.63E-07 | 5.14E-06 | 0.004188724 |
| Heart_Failure | dendritic_cell | *AC083843.1* | -35.39036463 | 3.30E-07 | 3.66E-06 | 0.001599743 |
| Heart_Failure | dendritic_cell | *PWRN1* | -2.21763955 | 0.001747317 | 0.002592656 | 1 |
| Heart_Failure | dendritic_cell | *GOLGA8H* | -0.916430824 | 0.001213031 | 0.001847341 | 1 |
| Heart_Failure | dendritic_cell | *LINC00662* | -1.73141512 | 5.76E-07 | 4.27E-06 | 0.002793349 |
| Heart_Failure | dendritic_cell | *C19orf84* | -24.07165847 | 7.03E-07 | 4.63E-06 | 0.003410115 |
| Heart_Failure | dendritic_cell | *LINC00675* | 1.983001236 | 2.69E-07 | 3.66E-06 | 0.001306018 |
| Heart_Failure | dendritic_cell | *LINC00667* | -2.51748558 | 1.13E-06 | 6.03E-06 | 0.005473015 |
| Heart_Failure | dendritic_cell | *LINC00672* | 4.482545859 | 1.09E-07 | 3.44E-06 | 0.000530228 |
| Heart_Failure | dendritic_cell | *LINC00909* | -2.339565131 | 2.31E-06 | 9.39E-06 | 0.011212527 |
| Heart_Failure | dendritic_cell | *DYNLL2* | 4.206532095 | 2.73E-07 | 3.66E-06 | 0.001322799 |
| Heart_Failure | dendritic_cell | *STRADA* | 2.529027989 | 0.001569148 | 0.002345503 | 1 |
| Heart_Failure | dendritic_cell | *MYO15B* | -2.602791617 | 0.005693821 | 0.00780187 | 1 |
| Heart_Failure | dendritic_cell | *LINC00663* | -1.034116179 | 0.000557483 | 0.000894776 | 1 |
| Heart_Failure | dendritic_cell | *ILF3-AS1* | -2.573992743 | 7.94E-06 | 2.24E-05 | 0.038501471 |
| Heart_Failure | dendritic_cell | *ZNF582-AS1* | 7.234412092 | 1.26E-06 | 6.39E-06 | 0.006135572 |
| Heart_Failure | dendritic_cell | *S1PR2* | -2.068306795 | 3.64E-05 | 7.91E-05 | 0.176833589 |
| Heart_Failure | dendritic_cell | *ZNF224* | -3.644134244 | 0.000152458 | 0.000275196 | 0.739725724 |
| Heart_Failure | dendritic_cell | *LIN37* | 2.663427537 | 4.57E-07 | 3.90E-06 | 0.002217033 |
| Heart_Failure | dendritic_cell | *NDUFA7* | -11.89072466 | 3.35E-07 | 3.66E-06 | 0.001623419 |
| Heart_Failure | dendritic_cell | *AC004466.1* | 1.92558027 | 0.020270702 | 0.025666348 | 1 |
| Heart_Failure | dendritic_cell | *SCGB1B2P* | -3.035854062 | 0.009584836 | 0.012727319 | 1 |
| Heart_Failure | dendritic_cell | *AC026740.1* | -6.524509088 | 0.003726794 | 0.005221601 | 1 |
| Heart_Failure | dendritic_cell | *ZNF728* | -5.093918261 | 1.14E-06 | 6.05E-06 | 0.005533519 |
| Heart_Failure | dendritic_cell | *TRABD2B* | -2.987532757 | 2.44E-06 | 9.75E-06 | 0.011821116 |
| Heart_Failure | dendritic_cell | *AC093157.1* | -2.462152558 | 7.04E-07 | 4.63E-06 | 0.003416625 |
| Heart_Failure | dendritic_cell | *FBXO17* | -1.319571427 | 3.93E-07 | 3.76E-06 | 0.001906023 |
| Heart_Failure | dendritic_cell | *ZNF587B* | -6.373921954 | 5.79E-05 | 0.000117723 | 0.281121793 |
| Heart_Failure | dendritic_cell | *SNHG8* | -2.911375088 | 0.000490854 | 0.00079733 | 1 |
| Heart_Failure | dendritic_cell | *HCG17* | -2.008675662 | 0.000232725 | 0.000403568 | 1 |
| Heart_Failure | dendritic_cell | *YTHDF3-AS1* | 1.441127873 | 9.74E-08 | 3.44E-06 | 0.000472645 |
| Heart_Failure | dendritic_cell | *MILR1* | 0.822731783 | 0.00176363 | 0.002614518 | 1 |
| Heart_Failure | dendritic_cell | *FGF14-AS2* | -2.933314652 | 1.03E-06 | 5.74E-06 | 0.004992928 |
| Heart_Failure | dendritic_cell | *CASC15* | -3.098918311 | 0.00012162 | 0.000224801 | 0.590102212 |
| Heart_Failure | dendritic_cell | *AP000347.2* | 1.420519837 | 1.09E-06 | 5.94E-06 | 0.005305958 |
| Heart_Failure | dendritic_cell | *GRIN2B* | -1.154453915 | 6.76E-05 | 0.000134605 | 0.328032719 |
| Heart_Failure | dendritic_cell | *PAXIP1-AS1* | -4.001522874 | 0.003979943 | 0.005553835 | 1 |

| | | | | | | |
|---|---|---|---|---|---|---|
| Heart_Failure | endometrium | *KRIT1* | 4.079485201 | 0.000412411 | 0.01886646 | 1 |
| Heart_Failure | endometrium | *RBM6* | -1.668269246 | 0.003252964 | 0.040889589 | 1 |
| Heart_Failure | endometrium | *RHBDD2* | 4.580057244 | 0.000860446 | 0.02198015 | 1 |
| Heart_Failure | endometrium | *AGK* | 4.514947558 | 0.001305091 | 0.025636846 | 1 |
| Heart_Failure | endometrium | *LARS2* | 4.785590024 | 0.000302317 | 0.018633666 | 1 |
| Heart_Failure | endometrium | *DCN* | 3.130482792 | 0.000842141 | 0.02198015 | 1 |
| Heart_Failure | endometrium | *COX15* | 5.518373777 | 0.000815356 | 0.021978364 | 1 |
| Heart_Failure | endometrium | *VSIG2* | 7.454129467 | 0.00051799 | 0.019813539 | 1 |
| Heart_Failure | endometrium | *MIPEP* | 5.59150036 | 0.000136042 | 0.018633666 | 0.660075122 |
| Heart_Failure | endometrium | *CHPF2* | 5.782270079 | 0.000736133 | 0.02152328 | 1 |
| Heart_Failure | endometrium | *ASTE1* | 4.149074509 | 0.001898596 | 0.030706624 | 1 |
| Heart_Failure | endometrium | *VCL* | 4.272813328 | 0.000539033 | 0.019813539 | 1 |
| Heart_Failure | endometrium | *TUBG2* | 2.63335399 | 0.002028995 | 0.031859817 | 1 |
| Heart_Failure | endometrium | *MRI1* | 4.821455099 | 6.15E-05 | 0.014913561 | 0.29827122 |
| Heart_Failure | endometrium | *PQLC2* | 4.392886371 | 0.002460934 | 0.035537059 | 1 |
| Heart_Failure | endometrium | *SPATA7* | 4.044431748 | 0.002892576 | 0.037903629 | 1 |
| Heart_Failure | endometrium | *DTNBP1* | 4.163378965 | 0.003965303 | 0.047741071 | 1 |
| Heart_Failure | endometrium | *RRM2B* | 5.066498186 | 0.002138107 | 0.032622947 | 1 |
| Heart_Failure | endometrium | *GALC* | 3.906743556 | 0.000220236 | 0.018633666 | 1 |
| Heart_Failure | endometrium | *ALDH18A1* | 2.985459713 | 0.000936791 | 0.022739652 | 1 |
| Heart_Failure | endometrium | *OGFR* | 5.415450842 | 0.000124478 | 0.018633666 | 0.603967763 |
| Heart_Failure | endometrium | *SLC12A2* | 2.51657532 | 0.001238886 | 0.025131936 | 1 |
| Heart_Failure | endometrium | *CHI3L2* | 6.050418424 | 0.000780826 | 0.021856201 | 1 |
| Heart_Failure | endometrium | *SEC61A2* | 4.415996291 | 0.002159999 | 0.032853655 | 1 |
| Heart_Failure | endometrium | *ZMYND12* | 1.468526505 | 0.002763742 | 0.03726176 | 1 |
| Heart_Failure | endometrium | *STOML1* | 7.058237127 | 0.001002004 | 0.023373667 | 1 |
| Heart_Failure | endometrium | *ROGDI* | 12.66555415 | 0.000610261 | 0.020408542 | 1 |
| Heart_Failure | endometrium | *CRMP1* | 5.048006437 | 0.000325764 | 0.018633666 | 1 |
| Heart_Failure | endometrium | *PPP2R2C* | 8.463424265 | 0.000601227 | 0.020383381 | 1 |
| Heart_Failure | endometrium | *MLH1* | 4.465346083 | 0.000813335 | 0.021978364 | 1 |
| Heart_Failure | endometrium | *ANKRD13A* | 5.702503573 | 0.000908523 | 0.022490591 | 1 |
| Heart_Failure | endometrium | *GSTP1* | 5.348711696 | 0.000280748 | 0.018633666 | 1 |
| Heart_Failure | endometrium | *ZNF213* | 3.863491265 | 0.001885033 | 0.030706624 | 1 |
| Heart_Failure | endometrium | *EIF2AK1* | 5.395127614 | 0.000224965 | 0.018633666 | 1 |
| Heart_Failure | endometrium | *ADAMTS2* | 4.507428801 | 0.0021197 | 0.032622947 | 1 |
| Heart_Failure | endometrium | *ANKRD24* | 3.244775939 | 0.00323202 | 0.040731845 | 1 |
| Heart_Failure | endometrium | *YPEL3* | 4.020887077 | 0.002593219 | 0.036633871 | 1 |
| Heart_Failure | endometrium | *RAB11FIP3* | 2.046727984 | 0.001215696 | 0.025100245 | 1 |
| Heart_Failure | endometrium | *CERS4* | 4.993087821 | 0.00031483 | 0.018633666 | 1 |
| Heart_Failure | endometrium | *PDPR* | 2.520286238 | 0.000920018 | 0.022659519 | 1 |
| Heart_Failure | endometrium | *RAPGEF4* | 2.682637267 | 0.00079207 | 0.021856201 | 1 |
| Heart_Failure | endometrium | *SEC22C* | 5.613793307 | 0.000712646 | 0.02134419 | 1 |
| Heart_Failure | endometrium | *PSMD5* | 4.798455026 | 0.000210265 | 0.018633666 | 1 |
| Heart_Failure | endometrium | *ERMP1* | 2.58164147 | 0.001102228 | 0.024090147 | 1 |
| Heart_Failure | endometrium | *PRTFDC1* | 3.348047247 | 0.001722411 | 0.029843291 | 1 |
| Heart_Failure | endometrium | *MTAP* | 2.391267174 | 0.000799018 | 0.021856201 | 1 |
| Heart_Failure | endometrium | *DDT* | 2.100031177 | 0.0027954 | 0.03726176 | 1 |
| Heart_Failure | endometrium | *TOP3B* | 7.439579528 | 0.000174776 | 0.018633666 | 0.84801159 |
| Heart_Failure | endometrium | *L3MBTL2* | 5.699067825 | 0.002128832 | 0.032622947 | 1 |
| Heart_Failure | endometrium | *EFS* | 7.510315273 | 0.000154828 | 0.018633666 | 0.751225131 |

| | | | | | | |
|---|---|---|---|---|---|---|
| Heart_Failure | endometrium | *MTG2* | 5.166321684 | 6.96E-05 | 0.015964789 | 0.337468937 |
| Heart_Failure | endometrium | *SEC23B* | 3.623077303 | 0.000489035 | 0.019813539 | 1 |
| Heart_Failure | endometrium | *NAGPA* | 2.869621275 | 0.00313171 | 0.039882044 | 1 |
| Heart_Failure | endometrium | *NME4* | 4.437732678 | 0.001019118 | 0.02365915 | 1 |
| Heart_Failure | endometrium | *LMF1* | 4.829771859 | 0.00139059 | 0.026563563 | 1 |
| Heart_Failure | endometrium | *UBE2I* | 4.161866735 | 0.000308044 | 0.018633666 | 1 |
| Heart_Failure | endometrium | *TOX3* | 4.430560158 | 0.00041337 | 0.01886646 | 1 |
| Heart_Failure | endometrium | *KAT8* | 3.378782836 | 0.001424956 | 0.026902274 | 1 |
| Heart_Failure | endometrium | *ZFAND1* | 3.209528115 | 0.000965292 | 0.023020211 | 1 |
| Heart_Failure | endometrium | *NIPAL2* | 5.222708004 | 0.000472891 | 0.019813539 | 1 |
| Heart_Failure | endometrium | *GSDMD* | 4.022241671 | 0.000321345 | 0.018633666 | 1 |
| Heart_Failure | endometrium | *CLEC4M* | 22.08919122 | 0.004208915 | 0.049687727 | 1 |
| Heart_Failure | endometrium | *MED25* | 6.151841578 | 0.0027467 | 0.03726176 | 1 |
| Heart_Failure | endometrium | *PDCD5* | 3.592717801 | 0.001860818 | 0.030706624 | 1 |
| Heart_Failure | endometrium | *POLR2I* | 5.320397773 | 0.002223258 | 0.033279425 | 1 |
| Heart_Failure | endometrium | *DNASE2* | 4.554875783 | 0.000685053 | 0.021171187 | 1 |
| Heart_Failure | endometrium | *SP4* | 4.962541337 | 0.00226263 | 0.033675705 | 1 |
| Heart_Failure | endometrium | *EIF3B* | 4.953709902 | 0.002780077 | 0.03726176 | 1 |
| Heart_Failure | endometrium | *AEBP1* | 4.772669077 | 0.000357158 | 0.018633666 | 1 |
| Heart_Failure | endometrium | *C5* | 5.023253645 | 0.003167069 | 0.040121723 | 1 |
| Heart_Failure | endometrium | *PTGDS* | 5.426724908 | 0.000400732 | 0.01886646 | 1 |
| Heart_Failure | endometrium | *LHPP* | 4.736329192 | 0.002451814 | 0.035511047 | 1 |
| Heart_Failure | endometrium | *ASPA* | 3.127455969 | 0.003959895 | 0.047741071 | 1 |
| Heart_Failure | endometrium | *FTSJ3* | 1.801147923 | 0.001181371 | 0.024600092 | 1 |
| Heart_Failure | endometrium | *CNTNAP1* | 4.851741022 | 0.001366169 | 0.026514613 | 1 |
| Heart_Failure | endometrium | *FAM20A* | 2.32818025 | 0.001245497 | 0.025131936 | 1 |
| Heart_Failure | endometrium | *NDUFC1* | 4.164038852 | 0.000530644 | 0.019813539 | 1 |
| Heart_Failure | endometrium | *GRPEL1* | 6.812263152 | 0.000337743 | 0.018633666 | 1 |
| Heart_Failure | endometrium | *FOXRED1* | 2.51698329 | 0.00351241 | 0.043586223 | 1 |
| Heart_Failure | endometrium | *SLC35F2* | 5.271848384 | 0.000372939 | 0.01886646 | 1 |
| Heart_Failure | endometrium | *TCIRG1* | 3.146600544 | 0.001522543 | 0.028088894 | 1 |
| Heart_Failure | endometrium | *PRDM4* | 5.706513429 | 0.000558007 | 0.020253481 | 1 |
| Heart_Failure | endometrium | *ACSS3* | 2.842511339 | 0.000287736 | 0.018633666 | 1 |
| Heart_Failure | endometrium | *CDK2AP1* | 6.385751354 | 5.74E-06 | 0.003977612 | 0.027843284 |
| Heart_Failure | endometrium | *MDM1* | 3.687718944 | 0.000739882 | 0.02152328 | 1 |
| Heart_Failure | endometrium | *KRR1* | 4.827712914 | 0.002738881 | 0.03726176 | 1 |
| Heart_Failure | endometrium | *ATN1* | 3.358802422 | 0.001086026 | 0.023843425 | 1 |
| Heart_Failure | endometrium | *SUDS3* | 4.532247372 | 0.000329742 | 0.018633666 | 1 |
| Heart_Failure | endometrium | *FRK* | 3.379496081 | 0.001379333 | 0.026563563 | 1 |
| Heart_Failure | endometrium | *TMEM14C* | 4.148940626 | 0.00049179 | 0.019813539 | 1 |
| Heart_Failure | endometrium | *CUTA* | 3.618710954 | 0.003582756 | 0.044120636 | 1 |
| Heart_Failure | endometrium | *TMEM30A* | 5.449474145 | 0.001049543 | 0.023843425 | 1 |
| Heart_Failure | endometrium | *DAP* | 3.187547517 | 0.000596172 | 0.020383381 | 1 |
| Heart_Failure | endometrium | *DROSHA* | 4.358925785 | 0.000450813 | 0.01947507 | 1 |
| Heart_Failure | endometrium | *ERGIC1* | 4.631512621 | 0.000311494 | 0.018633666 | 1 |
| Heart_Failure | endometrium | *HYAL1* | 4.271913899 | 0.000299133 | 0.018633666 | 1 |
| Heart_Failure | endometrium | *NEK11* | 2.219417676 | 0.002074031 | 0.032369836 | 1 |
| Heart_Failure | endometrium | *DNAH1* | 1.995492282 | 0.001557701 | 0.02830699 | 1 |
| Heart_Failure | endometrium | *MOGS* | 4.160636717 | 0.000186286 | 0.018633666 | 0.903858386 |
| Heart_Failure | endometrium | *GGCX* | 5.056282417 | 0.001060045 | 0.023843425 | 1 |

| | | | | | | |
|---|---|---|---|---|---|---|
| Heart_Failure | endometrium | *FAM20B* | 5.248514652 | 0.000852517 | 0.02198015 | 1 |
| Heart_Failure | endometrium | *MTFR1L* | 6.609552316 | 0.001598827 | 0.02886811 | 1 |
| Heart_Failure | endometrium | *MARC2* | 4.735744655 | 0.000336908 | 0.018633666 | 1 |
| Heart_Failure | endometrium | *SGIP1* | 5.752579984 | 0.000291823 | 0.018633666 | 1 |
| Heart_Failure | endometrium | *MED28* | 4.801365808 | 0.001081072 | 0.023843425 | 1 |
| Heart_Failure | endometrium | *WDR35* | 2.783470439 | 0.00114171 | 0.024296385 | 1 |
| Heart_Failure | endometrium | *NEK9* | 4.474921135 | 0.001897419 | 0.030706624 | 1 |
| Heart_Failure | endometrium | *PPP1R3C* | 5.714150799 | 0.001116983 | 0.024158762 | 1 |
| Heart_Failure | endometrium | *IQSEC3* | 9.982811491 | 0.001226882 | 0.025117436 | 1 |
| Heart_Failure | endometrium | *CCDC77* | 5.117311939 | 0.000224053 | 0.018633666 | 1 |
| Heart_Failure | endometrium | *RBBP6* | 3.995271576 | 0.00033806 | 0.018633666 | 1 |
| Heart_Failure | endometrium | *NAA60* | 5.423413206 | 0.000704711 | 0.021281234 | 1 |
| Heart_Failure | endometrium | *PMS2* | 3.506144846 | 0.001600478 | 0.02886811 | 1 |
| Heart_Failure | endometrium | *SSPN* | 3.68503364 | 0.001062032 | 0.023843425 | 1 |
| Heart_Failure | endometrium | *EBPL* | 4.272934762 | 0.0017244 | 0.029843291 | 1 |
| Heart_Failure | endometrium | *MTRR* | 3.790920569 | 0.000302229 | 0.018633666 | 1 |
| Heart_Failure | endometrium | *CHST8* | 3.304613857 | 0.004110152 | 0.049119357 | 1 |
| Heart_Failure | endometrium | *TMTC4* | 6.268551865 | 0.000257083 | 0.018633666 | 1 |
| Heart_Failure | endometrium | *MRPS7* | 4.037841333 | 0.000511899 | 0.019813539 | 1 |
| Heart_Failure | endometrium | *PPDPF* | 2.613348476 | 0.000573125 | 0.020383381 | 1 |
| Heart_Failure | endometrium | *COX6B1* | 5.530784299 | 0.000353434 | 0.018633666 | 1 |
| Heart_Failure | endometrium | *ZBTB1* | 4.302783312 | 0.000636784 | 0.020597844 | 1 |
| Heart_Failure | endometrium | *ZNF484* | 6.521080253 | 0.00051707 | 0.019813539 | 1 |
| Heart_Failure | endometrium | *ANAPC13* | 1.515052776 | 0.001130323 | 0.024266934 | 1 |
| Heart_Failure | endometrium | *BBOX1* | 11.4209512 | 0.001710852 | 0.029843291 | 1 |
| Heart_Failure | endometrium | *PLD2* | 4.185516951 | 0.001253807 | 0.025131936 | 1 |
| Heart_Failure | endometrium | *PRKCSH* | 5.673350668 | 0.002022389 | 0.031859817 | 1 |
| Heart_Failure | endometrium | *LSM7* | 4.275389511 | 0.000264146 | 0.018633666 | 1 |
| Heart_Failure | endometrium | *LSM4* | 2.228255028 | 0.000518526 | 0.019813539 | 1 |
| Heart_Failure | endometrium | *YIPF2* | 2.993879996 | 0.002402343 | 0.035003515 | 1 |
| Heart_Failure | endometrium | *ZNF317* | 2.137414207 | 0.003539838 | 0.043703041 | 1 |
| Heart_Failure | endometrium | *ANGPTL6* | -4.492967658 | 0.002755439 | 0.03726176 | 1 |
| Heart_Failure | endometrium | *C12orf65* | 5.918961704 | 1.10E-05 | 0.004849318 | 0.053342495 |
| Heart_Failure | endometrium | *NOL11* | 2.354486645 | 0.00332628 | 0.041603855 | 1 |
| Heart_Failure | endometrium | *NDUFA2* | 1.422951552 | 0.003037413 | 0.0388853 | 1 |
| Heart_Failure | endometrium | *MAP1B* | 4.602722263 | 2.97E-05 | 0.009619633 | 0.144294501 |
| Heart_Failure | endometrium | *SPATA6* | 4.068404165 | 0.00069704 | 0.021281234 | 1 |
| Heart_Failure | endometrium | *LRRC41* | 2.928577629 | 0.002885636 | 0.037903629 | 1 |
| Heart_Failure | endometrium | *TRIM22* | 5.060290021 | 0.000518343 | 0.019813539 | 1 |
| Heart_Failure | endometrium | *SERPINF1* | 4.732407774 | 0.000297327 | 0.018633666 | 1 |
| Heart_Failure | endometrium | *TMEM128* | 3.59339261 | 0.001120305 | 0.024158762 | 1 |
| Heart_Failure | endometrium | *RGS22* | 1.995426793 | 0.002134956 | 0.032622947 | 1 |
| Heart_Failure | endometrium | *WASF3* | 4.354625171 | 0.002984389 | 0.03840917 | 1 |
| Heart_Failure | endometrium | *PEMT* | 5.177065619 | 0.000484769 | 0.019813539 | 1 |
| Heart_Failure | endometrium | *SLC2A11* | 3.152102214 | 0.002806362 | 0.037279279 | 1 |
| Heart_Failure | endometrium | *COX16* | 19.22381399 | 0.000937331 | 0.022739652 | 1 |
| Heart_Failure | endometrium | *TMEM258* | 3.835625326 | 0.001693846 | 0.029843291 | 1 |
| Heart_Failure | endometrium | *ERCC5* | 2.483850824 | 0.002698922 | 0.03726176 | 1 |
| Heart_Failure | endometrium | *CARS2* | 4.015802177 | 0.001055831 | 0.023843425 | 1 |
| Heart_Failure | endometrium | *MTO1* | 2.561804948 | 0.002097519 | 0.032507759 | 1 |

| | | | | | | |
|---|---|---|---|---|---|---|
| Heart_Failure | endometrium | *PAN2* | 2.398411674 | 0.002301122 | 0.034110273 | 1 |
| Heart_Failure | endometrium | *STAB2* | 6.343988614 | 0.000387429 | 0.01886646 | 1 |
| Heart_Failure | endometrium | *NMT1* | 2.682811728 | 0.001786482 | 0.029889995 | 1 |
| Heart_Failure | endometrium | *NIPSNAP3A* | 3.971198254 | 0.00175713 | 0.029889995 | 1 |
| Heart_Failure | endometrium | *TCF19* | 5.021462625 | 0.000635083 | 0.020597844 | 1 |
| Heart_Failure | endometrium | *NARS2* | 4.365997972 | 0.00259729 | 0.036633871 | 1 |
| Heart_Failure | endometrium | *TGS1* | 4.116817501 | 0.000238178 | 0.018633666 | 1 |
| Heart_Failure | endometrium | *RSL24D1* | 5.070568269 | 0.000427723 | 0.01886646 | 1 |
| Heart_Failure | endometrium | *RBP4* | 4.032881929 | 0.000717501 | 0.021357772 | 1 |
| Heart_Failure | endometrium | *AP1AR* | 6.087172998 | 0.001500312 | 0.027890857 | 1 |
| Heart_Failure | endometrium | *BMPR1B* | 4.040864817 | 0.002742449 | 0.03726176 | 1 |
| Heart_Failure | endometrium | *G3BP2* | 4.567149945 | 2.73E-05 | 0.009447784 | 0.132268976 |
| Heart_Failure | endometrium | *B4GALNT3* | 5.058997495 | 0.001650778 | 0.029555621 | 1 |
| Heart_Failure | endometrium | *ZCRB1* | 4.70739984 | 0.000517157 | 0.019813539 | 1 |
| Heart_Failure | endometrium | *PPFIA2* | 4.813586726 | 0.000307101 | 0.018633666 | 1 |
| Heart_Failure | endometrium | *MARCH9* | 9.043565922 | 0.000423092 | 0.01886646 | 1 |
| Heart_Failure | endometrium | *RITA1* | 11.66862437 | 0.000667322 | 0.020755427 | 1 |
| Heart_Failure | endometrium | *SRP14* | 4.504710113 | 0.000972618 | 0.023020211 | 1 |
| Heart_Failure | endometrium | *PCSK6* | 3.252827747 | 0.001118142 | 0.024158762 | 1 |
| Heart_Failure | endometrium | *ABHD2* | 5.556935374 | 0.000276834 | 0.018633666 | 1 |
| Heart_Failure | endometrium | *G6PC3* | 3.544766534 | 0.003676795 | 0.044955066 | 1 |
| Heart_Failure | endometrium | *ARHGDIA* | 3.061687108 | 0.001730894 | 0.029843291 | 1 |
| Heart_Failure | endometrium | *ANAPC11* | 4.528517483 | 0.000840547 | 0.02198015 | 1 |
| Heart_Failure | endometrium | *NARF* | 5.707084906 | 0.000172858 | 0.018633666 | 0.838707072 |
| Heart_Failure | endometrium | *TRIM65* | 6.250382899 | 0.000830312 | 0.02198015 | 1 |
| Heart_Failure | endometrium | *NT5C3B* | 0.881499961 | 0.003326936 | 0.041603855 | 1 |
| Heart_Failure | endometrium | *PNMT* | 9.361185901 | 1.82E-05 | 0.007343299 | 0.088119583 |
| Heart_Failure | endometrium | *EVA1B* | 2.034494815 | 0.00021808 | 0.018633666 | 1 |
| Heart_Failure | endometrium | *ALDH9A1* | 3.919826502 | 0.000706158 | 0.021281234 | 1 |
| Heart_Failure | endometrium | *EIF2D* | 4.722254417 | 0.000800789 | 0.021856201 | 1 |
| Heart_Failure | endometrium | *CNIH4* | 5.119663966 | 0.002685241 | 0.03726176 | 1 |
| Heart_Failure | endometrium | *PYCR2* | 2.996121362 | 0.002006429 | 0.031859817 | 1 |
| Heart_Failure | endometrium | *THSD7B* | 2.299585976 | 0.004093053 | 0.049119357 | 1 |
| Heart_Failure | endometrium | *POLR2D* | 2.430158624 | 0.002414196 | 0.035070895 | 1 |
| Heart_Failure | endometrium | *CNTN4* | 4.05067838 | 0.000355793 | 0.018633666 | 1 |
| Heart_Failure | endometrium | *SLC25A26* | 3.183929814 | 0.004233573 | 0.049857509 | 1 |
| Heart_Failure | endometrium | *DGKQ* | 1.979130784 | 0.002783744 | 0.03726176 | 1 |
| Heart_Failure | endometrium | *TSLP* | 3.017776808 | 9.53E-07 | 0.000770255 | 0.00462153 |
| Heart_Failure | endometrium | *PCYOX1L* | 6.489820072 | 0.001268845 | 0.025154252 | 1 |
| Heart_Failure | endometrium | *RNF44* | 8.501930542 | 9.24E-05 | 0.018633666 | 0.44847843 |
| Heart_Failure | endometrium | *GNA12* | 4.507022429 | 0.000740805 | 0.02152328 | 1 |
| Heart_Failure | endometrium | *SURF6* | 2.626701912 | 0.003524815 | 0.043628579 | 1 |
| Heart_Failure | endometrium | *SLC39A12* | 12.72743544 | 0.002019327 | 0.031859817 | 1 |
| Heart_Failure | endometrium | *GAS2* | 4.959069165 | 0.00025338 | 0.018633666 | 1 |
| Heart_Failure | endometrium | *INTS4* | 4.295825916 | 0.000863523 | 0.02198015 | 1 |
| Heart_Failure | endometrium | *ROM1* | 3.483887614 | 0.000282573 | 0.018633666 | 1 |
| Heart_Failure | endometrium | *ALDOA* | 4.592478382 | 0.00171789 | 0.029843291 | 1 |
| Heart_Failure | endometrium | *FAM160B1* | 4.397672619 | 0.002721794 | 0.03726176 | 1 |
| Heart_Failure | endometrium | *RNF144A* | 4.55048264 | 0.000978041 | 0.023036192 | 1 |
| Heart_Failure | endometrium | *GUF1* | 4.131342346 | 0.001258669 | 0.025131936 | 1 |

| | | | | | | |
|---|---|---|---|---|---|---|
| Heart_Failure | endometrium | *TMEM56* | 13.46114579 | 0.001081502 | 0.023843425 | 1 |
| Heart_Failure | endometrium | *EPG5* | 4.440843403 | 0.000340624 | 0.018633666 | 1 |
| Heart_Failure | endometrium | *PLEKHH2* | 2.46978308 | 0.002397855 | 0.035003515 | 1 |
| Heart_Failure | endometrium | *INO80C* | 7.381461736 | 0.000374563 | 0.01886646 | 1 |
| Heart_Failure | endometrium | *ENPP3* | 6.722401468 | 0.000226387 | 0.018633666 | 1 |
| Heart_Failure | endometrium | *MARVELD1* | 7.804138395 | 0.000526222 | 0.019813539 | 1 |
| Heart_Failure | endometrium | *PDIA4* | 6.299391595 | 0.000376084 | 0.01886646 | 1 |
| Heart_Failure | endometrium | *NAA11* | 3.817056879 | 0.00230691 | 0.034110273 | 1 |
| Heart_Failure | endometrium | *C14orf2* | 5.184581557 | 4.46E-05 | 0.01272617 | 0.216344883 |
| Heart_Failure | endometrium | *NODAL* | 6.297412175 | 0.001031029 | 0.023708792 | 1 |
| Heart_Failure | endometrium | *GHRL* | 5.036103973 | 0.004148707 | 0.04933708 | 1 |
| Heart_Failure | endometrium | *NTAN1* | 4.648109378 | 0.001784564 | 0.029889995 | 1 |
| Heart_Failure | endometrium | *LCA5L* | 1.086171938 | 0.001874107 | 0.030706624 | 1 |
| Heart_Failure | endometrium | *SLC38A10* | 4.585192488 | 0.003221186 | 0.040701029 | 1 |
| Heart_Failure | endometrium | *PEX10* | 3.542540217 | 0.0022194 | 0.033279425 | 1 |
| Heart_Failure | endometrium | *FANCC* | 3.051622657 | 0.000755939 | 0.021607062 | 1 |
| Heart_Failure | endometrium | *ARHGAP27* | 6.544617248 | 0.000925046 | 0.022668294 | 1 |
| Heart_Failure | endometrium | *VAV2* | 3.254034468 | 0.003114013 | 0.03976103 | 1 |
| Heart_Failure | endometrium | *MCM3AP* | 3.065887509 | 0.002952104 | 0.038362182 | 1 |
| Heart_Failure | endometrium | *VPS11* | 5.951991381 | 2.72E-05 | 0.009447784 | 0.132036579 |
| Heart_Failure | endometrium | *PTH1R* | 1.815875243 | 0.002182679 | 0.033094874 | 1 |
| Heart_Failure | endometrium | *FCRL3* | -2.879319689 | 0.00145517 | 0.027366225 | 1 |
| Heart_Failure | endometrium | *FGFR4* | -7.99450658 | 0.001224508 | 0.025117436 | 1 |
| Heart_Failure | endometrium | *RPL8* | 4.286860617 | 0.001136722 | 0.024296385 | 1 |
| Heart_Failure | endometrium | *BDH1* | 4.771133326 | 0.000266368 | 0.018633666 | 1 |
| Heart_Failure | endometrium | *PGAP3* | 4.335273002 | 0.001186402 | 0.024600092 | 1 |
| Heart_Failure | endometrium | *LEMD2* | 5.791709532 | 5.97E-05 | 0.014913561 | 0.289705195 |
| Heart_Failure | endometrium | *OMA1* | 5.303812455 | 0.001826098 | 0.030239687 | 1 |
| Heart_Failure | endometrium | *KLHDC9* | -2.710218635 | 0.000881966 | 0.022288015 | 1 |
| Heart_Failure | endometrium | *FLVCR1* | 4.634482177 | 0.002312918 | 0.034110273 | 1 |
| Heart_Failure | endometrium | *ACP6* | 5.838376899 | 0.000425712 | 0.01886646 | 1 |
| Heart_Failure | endometrium | *HAAO* | 5.594145255 | 0.000211328 | 0.018633666 | 1 |
| Heart_Failure | endometrium | *KIAA1841* | 3.765025954 | 0.000595163 | 0.020383381 | 1 |
| Heart_Failure | endometrium | *DUSP19* | 3.180684086 | 0.00275269 | 0.03726176 | 1 |
| Heart_Failure | endometrium | *NUP35* | 3.268606467 | 0.000753907 | 0.021607062 | 1 |
| Heart_Failure | endometrium | *CCT3* | 3.981041373 | 0.000853786 | 0.02198015 | 1 |
| Heart_Failure | endometrium | *AIM2* | 7.263773816 | 0.001495336 | 0.027890857 | 1 |
| Heart_Failure | endometrium | *C4orf36* | 5.999898268 | 0.000662888 | 0.020755427 | 1 |
| Heart_Failure | endometrium | *TIPARP* | 1.939229676 | 0.002537435 | 0.036317511 | 1 |
| Heart_Failure | endometrium | *CPA3* | 4.238335211 | 0.0017865 | 0.029889995 | 1 |
| Heart_Failure | endometrium | *SLC4A1AP* | 5.590643563 | 0.002898237 | 0.037903629 | 1 |
| Heart_Failure | endometrium | *TPRA1* | 4.407466838 | 0.002791189 | 0.03726176 | 1 |
| Heart_Failure | endometrium | *MEAF6* | 5.324668607 | 0.001181622 | 0.024600092 | 1 |
| Heart_Failure | endometrium | *ERMAP* | 4.013559885 | 0.000958755 | 0.023020211 | 1 |
| Heart_Failure | endometrium | *RNF123* | 2.693716778 | 0.00367831 | 0.044955066 | 1 |
| Heart_Failure | endometrium | *HSPA4L* | 2.530885553 | 0.001164846 | 0.024466818 | 1 |
| Heart_Failure | endometrium | *CEP44* | 2.788807346 | 0.001270155 | 0.025154252 | 1 |
| Heart_Failure | endometrium | *CMBL* | 3.108345907 | 0.000446883 | 0.01947507 | 1 |
| Heart_Failure | endometrium | *NDUFS4* | 4.530263649 | 0.000403281 | 0.01886646 | 1 |
| Heart_Failure | endometrium | *DCBLD1* | 3.825364632 | 0.000138341 | 0.018633666 | 0.671228477 |

| | | | | | | |
|---|---|---|---|---|---|---|
| Heart_Failure | endometrium | *USP49* | 4.27095055 | 0.000329025 | 0.018633666 | 1 |
| Heart_Failure | endometrium | *BRI3* | 4.379626218 | 0.002229145 | 0.033279425 | 1 |
| Heart_Failure | endometrium | *RAD21* | 3.525632661 | 0.00034317 | 0.018633666 | 1 |
| Heart_Failure | endometrium | *OXR1* | 3.571035511 | 0.001804405 | 0.029982784 | 1 |
| Heart_Failure | endometrium | *SLC4A2* | 2.623334708 | 0.004131407 | 0.049252062 | 1 |
| Heart_Failure | endometrium | *FASTK* | 7.907883732 | 0.000207641 | 0.018633666 | 1 |
| Heart_Failure | endometrium | *FZD6* | 3.195217804 | 0.001966139 | 0.031484187 | 1 |
| Heart_Failure | endometrium | *INTS8* | 3.557668043 | 0.000330074 | 0.018633666 | 1 |
| Heart_Failure | endometrium | *SLC39A13* | 2.974459409 | 0.002497772 | 0.035961992 | 1 |
| Heart_Failure | endometrium | *IFI27L1* | 2.036315383 | 0.003791213 | 0.045987413 | 1 |
| Heart_Failure | endometrium | *PTER* | 3.855980708 | 0.002578604 | 0.036633871 | 1 |
| Heart_Failure | endometrium | *ZNF202* | 3.238874055 | 0.002977481 | 0.03840917 | 1 |
| Heart_Failure | endometrium | *WDR88* | 3.863624868 | 0.000951429 | 0.022966833 | 1 |
| Heart_Failure | endometrium | *CENPV* | 5.362092792 | 0.000618772 | 0.020423691 | 1 |
| Heart_Failure | endometrium | *SAAL1* | 7.739104579 | 0.000328156 | 0.018633666 | 1 |
| Heart_Failure | endometrium | *FAM96A* | 5.295827993 | 0.002677567 | 0.03726176 | 1 |
| Heart_Failure | endometrium | *MTMR10* | 3.509035439 | 0.000215185 | 0.018633666 | 1 |
| Heart_Failure | endometrium | *AKTIP* | 6.517710891 | 3.74E-05 | 0.011339054 | 0.181424868 |
| Heart_Failure | endometrium | *COQ7* | 4.671802413 | 0.001290849 | 0.025460156 | 1 |
| Heart_Failure | endometrium | *HDHD2* | 2.935846135 | 0.00030665 | 0.018633666 | 1 |
| Heart_Failure | endometrium | *VKORC1* | 7.430194471 | 0.000287411 | 0.018633666 | 1 |
| Heart_Failure | endometrium | *ZNF641* | 4.745253385 | 0.000600626 | 0.020383381 | 1 |
| Heart_Failure | endometrium | *ZNF528* | 4.23448111 | 0.000390419 | 0.01886646 | 1 |
| Heart_Failure | endometrium | *SERTAD3* | 10.08176051 | 0.000210361 | 0.018633666 | 1 |
| Heart_Failure | endometrium | *KIFC2* | 4.868172045 | 0.00055935 | 0.020253481 | 1 |
| Heart_Failure | endometrium | *ATP5H* | 5.424895041 | 0.000489431 | 0.019813539 | 1 |
| Heart_Failure | endometrium | *GSDMA* | -4.821765018 | 1.17E-09 | 1.89E-06 | 5.67E-06 |
| Heart_Failure | endometrium | *ULK4* | 3.262016596 | 0.001782539 | 0.029889995 | 1 |
| Heart_Failure | endometrium | *DTYMK* | 3.372360979 | 0.002567118 | 0.036633871 | 1 |
| Heart_Failure | endometrium | *RFWD3* | 3.96700566 | 0.000304865 | 0.018633666 | 1 |
| Heart_Failure | endometrium | *NDUFS5* | 4.903688796 | 0.001040915 | 0.023823209 | 1 |
| Heart_Failure | endometrium | *ZNF30* | 3.191873811 | 0.000736165 | 0.02152328 | 1 |
| Heart_Failure | endometrium | *STX18* | 4.657378092 | 0.000323722 | 0.018633666 | 1 |
| Heart_Failure | endometrium | *MFSD7* | 12.13846178 | 0.000313855 | 0.018633666 | 1 |
| Heart_Failure | endometrium | *CLIC3* | 4.569700039 | 0.00174643 | 0.029889995 | 1 |
| Heart_Failure | endometrium | *AVEN* | 4.994780363 | 0.000402555 | 0.01886646 | 1 |
| Heart_Failure | endometrium | *OTUD3* | 3.878878587 | 0.001155316 | 0.024372141 | 1 |
| Heart_Failure | endometrium | *RNF150* | 3.144973467 | 0.003016501 | 0.038719741 | 1 |
| Heart_Failure | endometrium | *S1PR1* | 6.284002267 | 9.43E-05 | 0.018633666 | 0.457756588 |
| Heart_Failure | endometrium | *SOX7* | 3.506486409 | 0.002880481 | 0.037903629 | 1 |
| Heart_Failure | endometrium | *TVP23B* | 2.902360803 | 0.002024875 | 0.031859817 | 1 |
| Heart_Failure | endometrium | *SERF1A* | 51.77945687 | 0.00082159 | 0.02198015 | 1 |
| Heart_Failure | endometrium | *MTBP* | 2.482483072 | 0.003631912 | 0.04461275 | 1 |
| Heart_Failure | endometrium | *TEFM* | 4.563032137 | 0.001072335 | 0.023843425 | 1 |
| Heart_Failure | endometrium | *BSG* | 5.211078183 | 0.000136396 | 0.018633666 | 0.661792976 |
| Heart_Failure | endometrium | *HINFP* | 5.998887851 | 0.000604948 | 0.020383381 | 1 |
| Heart_Failure | endometrium | *FGGY* | 4.541747648 | 0.000415672 | 0.01886646 | 1 |
| Heart_Failure | endometrium | *FUT10* | 1.773367046 | 0.001878393 | 0.030706624 | 1 |
| Heart_Failure | endometrium | *FAM192A* | 4.102016886 | 0.003350168 | 0.041786669 | 1 |
| Heart_Failure | endometrium | *DCP2* | 4.172508038 | 0.000586031 | 0.020383381 | 1 |

| | | | | | | |
|---|---|---|---|---|---|---|
| Heart_Failure | endometrium | *RNASEH2C* | 2.494333832 | 0.000427403 | 0.01886646 | 1 |
| Heart_Failure | endometrium | *VANGL1* | 4.391197905 | 0.001734503 | 0.029843291 | 1 |
| Heart_Failure | endometrium | *MMRN2* | 3.881268367 | 0.002582953 | 0.036633871 | 1 |
| Heart_Failure | endometrium | *NDNF* | 7.013782187 | 0.000152603 | 0.018633666 | 0.740428012 |
| Heart_Failure | endometrium | *NUDT4* | 2.954282241 | 0.001233371 | 0.025131936 | 1 |
| Heart_Failure | endometrium | *XXYLT1* | 3.415831506 | 0.000614107 | 0.020408542 | 1 |
| Heart_Failure | endometrium | *ANKRD36C* | 4.803853476 | 0.001000544 | 0.023373667 | 1 |
| Heart_Failure | endometrium | *CEP135* | 4.360149784 | 0.000341127 | 0.018633666 | 1 |
| Heart_Failure | endometrium | *CLTB* | 3.978179145 | 0.000582333 | 0.020383381 | 1 |
| Heart_Failure | endometrium | *PRIMA1* | 2.913942536 | 0.00235909 | 0.034685769 | 1 |
| Heart_Failure | endometrium | *MYPOP* | 5.212686812 | 0.000328376 | 0.018633666 | 1 |
| Heart_Failure | endometrium | *SLC35G1* | 4.329130931 | 0.000654063 | 0.020755427 | 1 |
| Heart_Failure | endometrium | *WDR25* | 1.709424644 | 0.003854685 | 0.04664073 | 1 |
| Heart_Failure | endometrium | *B3GNT5* | 2.961398881 | 0.002074818 | 0.032369836 | 1 |
| Heart_Failure | endometrium | *ANKLE2* | 4.172435202 | 0.000603765 | 0.020383381 | 1 |
| Heart_Failure | endometrium | *MTHFR* | 4.675487925 | 0.004188342 | 0.049565455 | 1 |
| Heart_Failure | endometrium | *WDR73* | 4.26338088 | 0.002700189 | 0.03726176 | 1 |
| Heart_Failure | endometrium | *RPS27* | 4.734860272 | 0.000972294 | 0.023020211 | 1 |
| Heart_Failure | endometrium | *ASB8* | 3.231830981 | 0.001887474 | 0.030706624 | 1 |
| Heart_Failure | endometrium | *TMEM11* | 4.305762099 | 0.002720307 | 0.03726176 | 1 |
| Heart_Failure | endometrium | *CA8* | 7.204796011 | 0.000846196 | 0.02198015 | 1 |
| Heart_Failure | endometrium | *CTC1* | 2.811069852 | 0.000657104 | 0.020755427 | 1 |
| Heart_Failure | endometrium | *ZNF664* | 7.462870658 | 0.000418814 | 0.01886646 | 1 |
| Heart_Failure | endometrium | *HLA-DQB1* | -2.908499961 | 1.04E-14 | 5.06E-11 | 5.06E-11 |
| Heart_Failure | endometrium | *PACS2* | 3.009819632 | 0.000796042 | 0.021856201 | 1 |
| Heart_Failure | endometrium | *SEPHS2* | 6.828569904 | 0.001719078 | 0.029843291 | 1 |
| Heart_Failure | endometrium | *ZADH2* | 4.249780761 | 0.000526727 | 0.019813539 | 1 |
| Heart_Failure | endometrium | *EXOC3* | 5.346253998 | 0.001721669 | 0.029843291 | 1 |
| Heart_Failure | endometrium | *MCFD2* | 3.054626262 | 0.001380053 | 0.026563563 | 1 |
| Heart_Failure | endometrium | *CMTR2* | 5.621937097 | 0.000121574 | 0.018633666 | 0.589874779 |
| Heart_Failure | endometrium | *MRPL14* | 6.030373897 | 0.001553271 | 0.02830699 | 1 |
| Heart_Failure | endometrium | *OGFOD3* | 4.345248623 | 0.000865253 | 0.02198015 | 1 |
| Heart_Failure | endometrium | *ZNF329* | 2.542227655 | 0.001756283 | 0.029889995 | 1 |
| Heart_Failure | endometrium | *WRB* | 2.747018188 | 0.001703673 | 0.029843291 | 1 |
| Heart_Failure | endometrium | *KPNA2* | 3.273511437 | 0.002605044 | 0.036636734 | 1 |
| Heart_Failure | endometrium | *CEP63* | 3.56727762 | 0.001153103 | 0.024372141 | 1 |
| Heart_Failure | endometrium | *LYSMD4* | 2.55663759 | 0.004102955 | 0.049119357 | 1 |
| Heart_Failure | endometrium | *CSMD1* | 2.40153351 | 0.002957019 | 0.038362182 | 1 |
| Heart_Failure | endometrium | *HMCES* | 4.117177415 | 0.002097223 | 0.032507759 | 1 |
| Heart_Failure | endometrium | *UPP1* | 4.946341577 | 0.001251636 | 0.025131936 | 1 |
| Heart_Failure | endometrium | *TXNRD2* | 4.241179996 | 0.000769641 | 0.021838011 | 1 |
| Heart_Failure | endometrium | *HDDC3* | 5.560982636 | 0.003469504 | 0.043164194 | 1 |
| Heart_Failure | endometrium | *BRI3BP* | 6.191361105 | 0.004177483 | 0.049557817 | 1 |
| Heart_Failure | endometrium | *SH3BGR* | 1.576411341 | 0.00133301 | 0.026079689 | 1 |
| Heart_Failure | endometrium | *POLR1D* | 3.821310087 | 0.000700976 | 0.021281234 | 1 |
| Heart_Failure | endometrium | *MIR22HG* | 2.552684425 | 0.001947014 | 0.03128117 | 1 |
| Heart_Failure | endometrium | *TMEM17* | 2.419871866 | 0.001083272 | 0.023843425 | 1 |
| Heart_Failure | endometrium | *KBTBD12* | 3.109497421 | 0.000892294 | 0.022316553 | 1 |
| Heart_Failure | endometrium | *AGRN* | 9.526799441 | 0.001028841 | 0.023708792 | 1 |
| Heart_Failure | endometrium | *C3orf62* | 5.57264592 | 8.71E-06 | 0.004226721 | 0.042267206 |

| | | | | | | |
|---|---|---|---|---|---|---|
| Heart_Failure | endometrium | *COL25A1* | 2.800593549 | 0.000757049 | 0.021607062 | 1 |
| Heart_Failure | endometrium | *FBF1* | 4.423033131 | 0.000276219 | 0.018633666 | 1 |
| Heart_Failure | endometrium | *ASTL* | 16.89690564 | 0.000663678 | 0.020755427 | 1 |
| Heart_Failure | endometrium | *LRRK2* | 3.015782647 | 0.002533435 | 0.036317511 | 1 |
| Heart_Failure | endometrium | *TSPYL1* | 4.578490231 | 0.000385802 | 0.01886646 | 1 |
| Heart_Failure | endometrium | *PNRC2* | 2.655386714 | 0.001769892 | 0.029889995 | 1 |
| Heart_Failure | endometrium | *ZKSCAN3* | 3.003106234 | 0.003765927 | 0.045795177 | 1 |
| Heart_Failure | endometrium | *RRP7A* | 4.230885532 | 0.003720897 | 0.045361292 | 1 |
| Heart_Failure | endometrium | *HLA-DRB1* | -0.906666435 | 0.001539841 | 0.028193609 | 1 |
| Heart_Failure | endometrium | *SRGAP3* | 4.245288738 | 0.000160561 | 0.018633666 | 0.779043758 |
| Heart_Failure | endometrium | *LONP1* | 3.967147111 | 0.000309601 | 0.018633666 | 1 |
| Heart_Failure | endometrium | *EVL* | 3.778026764 | 0.002103759 | 0.032507759 | 1 |
| Heart_Failure | endometrium | *ZNF124* | 4.441042491 | 0.000593425 | 0.020383381 | 1 |
| Heart_Failure | endometrium | *PIK3R4* | 5.028233684 | 0.002395362 | 0.035003515 | 1 |
| Heart_Failure | endometrium | *SULT1A1* | 4.752422645 | 0.000892184 | 0.022316553 | 1 |
| Heart_Failure | endometrium | *ZKSCAN5* | 4.960423162 | 5.12E-05 | 0.013797913 | 0.248362434 |
| Heart_Failure | endometrium | *ZFP62* | 3.599017158 | 0.001797176 | 0.029965278 | 1 |
| Heart_Failure | endometrium | *TOMM7* | 4.571812527 | 0.000294686 | 0.018633666 | 1 |
| Heart_Failure | endometrium | *HLA-DQA1* | -2.091098272 | 4.28E-08 | 4.15E-05 | 0.000207562 |
| Heart_Failure | endometrium | *GM2A* | 1.170849121 | 0.002956563 | 0.038362182 | 1 |
| Heart_Failure | endometrium | *ANKRD36B* | 4.183124497 | 0.000627618 | 0.020575683 | 1 |
| Heart_Failure | endometrium | *ABCB8* | 5.837874268 | 0.000499445 | 0.019813539 | 1 |
| Heart_Failure | endometrium | *SULT1A2* | 4.377220871 | 0.001387525 | 0.026563563 | 1 |
| Heart_Failure | endometrium | *EME2* | 4.329888866 | 0.001630498 | 0.029300651 | 1 |
| Heart_Failure | endometrium | *RPL12* | 3.27125931 | 0.002716556 | 0.03726176 | 1 |
| Heart_Failure | endometrium | *MRPL42* | 3.342184303 | 0.00053523 | 0.019813539 | 1 |
| Heart_Failure | endometrium | *ZNF667* | 4.642656463 | 0.001697906 | 0.029843291 | 1 |
| Heart_Failure | endometrium | *ZNF544* | 2.432739612 | 0.003147698 | 0.039980707 | 1 |
| Heart_Failure | endometrium | *MIER1* | 3.964429982 | 0.002213278 | 0.033279425 | 1 |
| Heart_Failure | endometrium | *SVIP* | 3.726483659 | 0.000537894 | 0.019813539 | 1 |
| Heart_Failure | endometrium | *STYX* | 3.728367274 | 0.000899112 | 0.022371753 | 1 |
| Heart_Failure | endometrium | *UCKL1* | 6.05731805 | 0.000498015 | 0.019813539 | 1 |
| Heart_Failure | endometrium | *HLA-DRB5* | -1.010780665 | 0.000351022 | 0.018633666 | 1 |
| Heart_Failure | endometrium | *CCDC167* | 8.468619596 | 0.000343729 | 0.018633666 | 1 |
| Heart_Failure | endometrium | *PRRT1* | 3.157682574 | 8.30E-06 | 0.004226721 | 0.040259242 |
| Heart_Failure | endometrium | *TRIM27* | 3.741617722 | 0.001462634 | 0.027400394 | 1 |
| Heart_Failure | endometrium | *FAM216A* | 5.88679981 | 7.24E-05 | 0.015964789 | 0.351225356 |
| Heart_Failure | endometrium | *ZNF783* | 4.342762125 | 0.001924561 | 0.031023158 | 1 |
| Heart_Failure | endometrium | *ATF6B* | 3.733016557 | 0.001341954 | 0.026149237 | 1 |
| Heart_Failure | endometrium | *ZNF891* | 6.885220519 | 0.000246187 | 0.018633666 | 1 |
| Heart_Failure | endometrium | *PRCD* | 7.52109741 | 0.001416558 | 0.026848203 | 1 |
| Heart_Failure | endometrium | *VPS16* | 7.302836035 | 0.000797581 | 0.021856201 | 1 |
| Heart_Failure | endometrium | *TSTD1* | 6.592848254 | 0.001534608 | 0.028193609 | 1 |
| Heart_Failure | endometrium | *PPP3R1* | 2.869764608 | 0.002190082 | 0.033103662 | 1 |
| Heart_Failure | endometrium | *EXOSC6* | 5.215485722 | 0.002815161 | 0.037279279 | 1 |
| Heart_Failure | endometrium | *HLA-DQB2* | 1.747910708 | 7.64E-11 | 1.85E-07 | 3.71E-07 |
| Heart_Failure | endometrium | *LINC00665* | 4.308189432 | 0.000801814 | 0.021856201 | 1 |
| Heart_Failure | endometrium | *LINC00342* | 8.35777863 | 0.000844836 | 0.02198015 | 1 |
| Heart_Failure | endometrium | *RPS28* | 6.167621091 | 0.002656353 | 0.03725036 | 1 |
| Heart_Failure | endometrium | *ZNF736* | 4.081392149 | 7.84E-06 | 0.004226721 | 0.038056823 |

| | | | | | | |
|---|---|---|---|---|---|---|
| Heart_Failure | endometrium | *HLA-DQA2* | 1.69674835 | 1.12E-08 | 1.36E-05 | 5.44E-05 |
| Heart_Failure | endometrium | *TNFSF12* | 5.518319608 | 0.000165726 | 0.018633666 | 0.8041048 |
| Heart_Failure | endometrium | *PSMB9* | 2.025866107 | 0.000284442 | 0.018633666 | 1 |
| Heart_Failure | endometrium | *ARFGAP3* | 3.914908978 | 0.000453562 | 0.01947507 | 1 |
| Heart_Failure | endometrium | *DECR2* | 4.171551149 | 0.002795376 | 0.03726176 | 1 |
| Heart_Failure | endometrium | *CDK11B* | 3.960956022 | 0.000643254 | 0.020669341 | 1 |
| Heart_Failure | endometrium | *C5orf17* | 2.724375758 | 0.001396705 | 0.026575731 | 1 |
| Heart_Failure | endometrium | *SMIM20* | 3.412434463 | 0.001516197 | 0.02807858 | 1 |
| Heart_Failure | endometrium | *TMEM200B* | 2.342086798 | 0.001985504 | 0.031689693 | 1 |
| Heart_Failure | endometrium | *CHURC1* | 2.34745213 | 0.002819764 | 0.037279279 | 1 |
| Heart_Failure | endometrium | *AC083843.1* | 120.3171423 | 0.000341542 | 0.018633666 | 1 |
| Heart_Failure | endometrium | *ZNF582-AS1* | 3.742598153 | 0.002967049 | 0.038389657 | 1 |
| Heart_Failure | endothelial | *TSLP* | 0.369843658 | 4.97E-06 | 0.003014915 | 0.024119323 |
| Heart_Failure | endothelial | *MST1R* | 0.786194601 | 2.66E-07 | 0.000214767 | 0.001288601 |
| Heart_Failure | endothelial | *GSDMA* | -0.740082087 | 1.18E-09 | 1.14E-06 | 5.70E-06 |
| Heart_Failure | endothelial | *HLA-DQB1* | -0.693796111 | 4.69E-21 | 2.28E-17 | 2.28E-17 |
| Heart_Failure | endothelial | *HLA-DQA1* | -0.927520333 | 4.83E-18 | 7.81E-15 | 2.34E-14 |
| Heart_Failure | endothelial | *HLA-DQB2* | 0.381305929 | 1.13E-18 | 2.74E-15 | 5.47E-15 |
| Heart_Failure | endothelial | *HLA-DQA2* | 0.424207701 | 2.94E-17 | 3.56E-14 | 1.42E-13 |
| Heart_Failure | endothelial | *PSMB9* | 0.407302094 | 4.43E-06 | 0.003014915 | 0.021493939 |
| Heart_Failure | LEC | *RBM6* | -0.774676635 | 3.68E-05 | 0.012770338 | 0.178784737 |
| Heart_Failure | LEC | *PNMT* | 3.552307805 | 5.46E-10 | 4.41E-07 | 2.65E-06 |
| Heart_Failure | LEC | *TSLP* | 0.669172824 | 3.80E-06 | 0.001845754 | 0.018457545 |
| Heart_Failure | LEC | *RNF123* | 1.545865029 | 2.05E-06 | 0.00124291 | 0.00994328 |
| Heart_Failure | LEC | *MST1R* | 1.546154268 | 3.74E-08 | 2.59E-05 | 0.000181313 |
| Heart_Failure | LEC | *GSDMA* | -1.492204335 | 1.29E-10 | 1.26E-07 | 6.28E-07 |
| Heart_Failure | LEC | *HLA-DQB1* | -1.3796659 | 1.88E-23 | 9.14E-20 | 9.14E-20 |
| Heart_Failure | LEC | *HLA-DRB1* | -0.717280667 | 4.86E-06 | 0.002142094 | 0.023563034 |
| Heart_Failure | LEC | *HLA-DQA1* | -1.782500865 | 3.40E-19 | 8.25E-16 | 1.65E-15 |
| Heart_Failure | LEC | *MICA* | 1.216078032 | 6.84E-06 | 0.002551296 | 0.033166848 |
| Heart_Failure | LEC | *ATF6B* | 2.037356043 | 5.76E-06 | 0.002329289 | 0.02795147 |
| Heart_Failure | LEC | *HLA-DQB2* | 0.61633857 | 9.94E-17 | 1.61E-13 | 4.82E-13 |
| Heart_Failure | LEC | *HLA-DQA2* | 0.714812235 | 3.39E-16 | 4.11E-13 | 1.65E-12 |
| Heart_Failure | LEC | *PSMB9* | 0.794343961 | 2.85E-06 | 0.001538257 | 0.013844309 |
| Heart_Failure | macrophage | *KRIT1* | 0.786896367 | 0.004690186 | 0.025598182 | 1 |
| Heart_Failure | macrophage | *HS3ST1* | 0.451355317 | 0.006805808 | 0.031912157 | 1 |
| Heart_Failure | macrophage | *ALS2* | 0.905053152 | 0.005465222 | 0.027888457 | 1 |
| Heart_Failure | macrophage | *RBM6* | -0.328073181 | 0.000134583 | 0.010532241 | 0.652998926 |
| Heart_Failure | macrophage | *CDC27* | 0.490181023 | 0.011046037 | 0.043397061 | 1 |
| Heart_Failure | macrophage | *POLR2J* | 1.869879927 | 0.000304829 | 0.013632389 | 1 |
| Heart_Failure | macrophage | *GCFC2* | 0.721739927 | 0.004634993 | 0.025382605 | 1 |
| Heart_Failure | macrophage | *GGNBP2* | 0.828658952 | 0.00934121 | 0.038704997 | 1 |
| Heart_Failure | macrophage | *ITGA2B* | 0.68076455 | 0.00516974 | 0.027066078 | 1 |
| Heart_Failure | macrophage | *SYNRG* | 0.951907624 | 0.002574843 | 0.018426459 | 1 |
| Heart_Failure | macrophage | *ZNF263* | 1.340909516 | 0.000978123 | 0.013998911 | 1 |
| Heart_Failure | macrophage | *SPATA20* | 0.565145899 | 0.003829818 | 0.023041698 | 1 |
| Heart_Failure | macrophage | *BAIAP2L1* | 1.372671033 | 0.001640747 | 0.015548043 | 1 |
| Heart_Failure | macrophage | *AGK* | 0.644075174 | 0.008311408 | 0.036038382 | 1 |
| Heart_Failure | macrophage | *FARP2* | 0.908076813 | 0.004218121 | 0.024078028 | 1 |
| Heart_Failure | macrophage | *VPS41* | 1.022249922 | 0.001990991 | 0.016627005 | 1 |

| | | | | | | |
|---|---|---|---|---|---|---|
| Heart_Failure | macrophage | *TSR3* | 1.495561419 | 0.007679416 | 0.034278311 | 1 |
| Heart_Failure | macrophage | *PSMB1* | 1.593292018 | 0.000283487 | 0.013225747 | 1 |
| Heart_Failure | macrophage | *MGST1* | 0.208805955 | 0.010104073 | 0.040718406 | 1 |
| Heart_Failure | macrophage | *PKD1* | 0.877597405 | 0.001956809 | 0.016534361 | 1 |
| Heart_Failure | macrophage | *MED24* | 1.386008654 | 0.00255615 | 0.018426459 | 1 |
| Heart_Failure | macrophage | *RPS20* | 1.065724077 | 0.001051382 | 0.014193744 | 1 |
| Heart_Failure | macrophage | *TSPAN9* | 0.398243492 | 0.007656639 | 0.034217225 | 1 |
| Heart_Failure | macrophage | *TTC19* | 0.631584918 | 0.005361338 | 0.027699259 | 1 |
| Heart_Failure | macrophage | *LARS2* | 0.966585369 | 0.002496601 | 0.018350936 | 1 |
| Heart_Failure | macrophage | *MAP4K5* | 0.63830558 | 0.007336751 | 0.033622873 | 1 |
| Heart_Failure | macrophage | *EHD3* | 1.444835819 | 0.00863128 | 0.036962903 | 1 |
| Heart_Failure | macrophage | *RNF14* | 1.135547112 | 0.000540515 | 0.013744636 | 1 |
| Heart_Failure | macrophage | *ZC3H3* | 0.428036878 | 0.004833545 | 0.026058178 | 1 |
| Heart_Failure | macrophage | *MDH1* | 0.669664231 | 0.001289472 | 0.014557357 | 1 |
| Heart_Failure | macrophage | *COX15* | 1.304362963 | 0.001345796 | 0.014806416 | 1 |
| Heart_Failure | macrophage | *RUFY3* | 1.017228182 | 0.000791433 | 0.013744636 | 1 |
| Heart_Failure | macrophage | *TTC27* | 0.953002081 | 0.001128131 | 0.014193744 | 1 |
| Heart_Failure | macrophage | *SLC7A9* | 0.225767116 | 0.004663833 | 0.025511745 | 1 |
| Heart_Failure | macrophage | *ZDHHC6* | 1.082422817 | 0.011980394 | 0.045484252 | 1 |
| Heart_Failure | macrophage | *RNH1* | 0.543920759 | 0.011445552 | 0.044320684 | 1 |
| Heart_Failure | macrophage | *GCLM* | 1.211968245 | 0.003145083 | 0.020604359 | 1 |
| Heart_Failure | macrophage | *UBR2* | 0.593717135 | 0.010063328 | 0.040621685 | 1 |
| Heart_Failure | macrophage | *SEC63* | 0.776363186 | 0.009360328 | 0.03875112 | 1 |
| Heart_Failure | macrophage | *FAS* | 0.982233078 | 0.00185371 | 0.016293846 | 1 |
| Heart_Failure | macrophage | *MIPEP* | 1.233215154 | 0.000119955 | 0.010034832 | 0.582020257 |
| Heart_Failure | macrophage | *VRK2* | 0.988819854 | 0.001505568 | 0.015102525 | 1 |
| Heart_Failure | macrophage | *VEZT* | 0.804045718 | 0.002754567 | 0.019289709 | 1 |
| Heart_Failure | macrophage | *BRD9* | 0.880172942 | 0.001413284 | 0.015044204 | 1 |
| Heart_Failure | macrophage | *SNX1* | 0.824206874 | 0.008739464 | 0.037294531 | 1 |
| Heart_Failure | macrophage | *RABEP1* | 0.791895482 | 0.008579475 | 0.036806023 | 1 |
| Heart_Failure | macrophage | *IKZF2* | 1.151101621 | 0.002228555 | 0.017328441 | 1 |
| Heart_Failure | macrophage | *PIAS1* | 0.702455457 | 0.005088653 | 0.026837114 | 1 |
| Heart_Failure | macrophage | *VCL* | 1.010766511 | 0.001848455 | 0.016277137 | 1 |
| Heart_Failure | macrophage | *ADSS* | 1.40909694 | 0.000908607 | 0.013744636 | 1 |
| Heart_Failure | macrophage | *TBC1D23* | 0.969042122 | 0.0019798 | 0.016578327 | 1 |
| Heart_Failure | macrophage | *NSUN2* | 0.90771754 | 0.008680032 | 0.037138904 | 1 |
| Heart_Failure | macrophage | *MRI1* | 0.541273694 | 0.001898195 | 0.016377125 | 1 |
| Heart_Failure | macrophage | *BOD1L1* | 0.638117831 | 0.004458733 | 0.024908187 | 1 |
| Heart_Failure | macrophage | *PHLPP2* | 0.368135794 | 0.007441489 | 0.033712515 | 1 |
| Heart_Failure | macrophage | *PSMA4* | 1.320451219 | 0.003312593 | 0.021120503 | 1 |
| Heart_Failure | macrophage | *TDP1* | 1.416021569 | 0.005007188 | 0.026720217 | 1 |
| Heart_Failure | macrophage | *CTNNA1* | 1.031675407 | 0.000760277 | 0.013744636 | 1 |
| Heart_Failure | macrophage | *CNTLN* | 0.67095731 | 0.01320274 | 0.048714595 | 1 |
| Heart_Failure | macrophage | *YTHDC2* | 1.164206985 | 0.001086627 | 0.014193744 | 1 |
| Heart_Failure | macrophage | *POLR2B* | 1.15930281 | 0.000741502 | 0.013744636 | 1 |
| Heart_Failure | macrophage | *DTNBP1* | 0.912672129 | 0.004820773 | 0.026058178 | 1 |
| Heart_Failure | macrophage | *JKAMP* | 0.694895318 | 0.007189354 | 0.033158505 | 1 |
| Heart_Failure | macrophage | *MAPK9* | 1.522510853 | 0.001063809 | 0.014193744 | 1 |
| Heart_Failure | macrophage | *MRTO4* | 1.13247691 | 0.001194651 | 0.014405948 | 1 |
| Heart_Failure | macrophage | *ANAPC4* | 1.102269088 | 0.000232078 | 0.012453892 | 1 |

| | | | | | | |
|---|---|---|---|---|---|---|
| Heart_Failure | macrophage | *SDCCAG8* | 0.771015527 | 4.50E-06 | 0.001213821 | 0.021848773 |
| Heart_Failure | macrophage | *KIF1B* | 0.665712533 | 0.002861336 | 0.019714284 | 1 |
| Heart_Failure | macrophage | *TBC1D22A* | 0.684283408 | 0.010169337 | 0.040879557 | 1 |
| Heart_Failure | macrophage | *GALC* | 1.041651089 | 0.001722964 | 0.015726742 | 1 |
| Heart_Failure | macrophage | *FAM114A2* | 0.797129822 | 0.007595903 | 0.034083874 | 1 |
| Heart_Failure | macrophage | *GINM1* | 0.652826737 | 0.012195471 | 0.04604858 | 1 |
| Heart_Failure | macrophage | *PITHD1* | 0.995102606 | 0.002438536 | 0.018242378 | 1 |
| Heart_Failure | macrophage | *ATP11B* | 0.813864874 | 0.000868979 | 0.013744636 | 1 |
| Heart_Failure | macrophage | *CROCC* | 0.356119808 | 0.001437165 | 0.015100028 | 1 |
| Heart_Failure | macrophage | *ALDH18A1* | 0.63479653 | 0.013431508 | 0.049228836 | 1 |
| Heart_Failure | macrophage | *DNAJC25* | 1.26377834 | 0.008850757 | 0.03747284 | 1 |
| Heart_Failure | macrophage | *OGFR* | 0.786602928 | 0.004151565 | 0.023980231 | 1 |
| Heart_Failure | macrophage | *PRDM6* | 1.397946737 | 0.005736146 | 0.028811366 | 1 |
| Heart_Failure | macrophage | *RNF4* | 1.468541922 | 0.000665261 | 0.013744636 | 1 |
| Heart_Failure | macrophage | *CASP8* | 1.17257921 | 0.000524081 | 0.013744636 | 1 |
| Heart_Failure | macrophage | *TM7SF3* | 1.14892959 | 0.000929617 | 0.013744636 | 1 |
| Heart_Failure | macrophage | *TMEM161A* | 0.887436038 | 0.012158048 | 0.045963902 | 1 |
| Heart_Failure | macrophage | *SUGP2* | 1.119567807 | 0.001252007 | 0.014557357 | 1 |
| Heart_Failure | macrophage | *SLC12A2* | 0.996546068 | 0.001962267 | 0.016534361 | 1 |
| Heart_Failure | macrophage | *BTBD1* | 1.344656396 | 0.000262795 | 0.012750837 | 1 |
| Heart_Failure | macrophage | *FAR2* | 0.844763634 | 0.001948913 | 0.01650319 | 1 |
| Heart_Failure | macrophage | *SBNO2* | 1.432545658 | 0.000126749 | 0.01024973 | 0.614983796 |
| Heart_Failure | macrophage | *PMS1* | 1.135531628 | 0.000917621 | 0.013744636 | 1 |
| Heart_Failure | macrophage | *MAP2K4* | 0.907876963 | 0.003215224 | 0.0208381 | 1 |
| Heart_Failure | macrophage | *MYO9A* | 0.472950664 | 0.013306757 | 0.049023831 | 1 |
| Heart_Failure | macrophage | *TRAM1* | 1.014908622 | 0.001811208 | 0.016138719 | 1 |
| Heart_Failure | macrophage | *STOML1* | 2.463693194 | 0.002918731 | 0.019890006 | 1 |
| Heart_Failure | macrophage | *DHX8* | 1.558970428 | 0.00116779 | 0.01427233 | 1 |
| Heart_Failure | macrophage | *CBFB* | 0.968388072 | 0.006378483 | 0.030886628 | 1 |
| Heart_Failure | macrophage | *IP6K2* | 0.868484101 | 0.003235995 | 0.0208381 | 1 |
| Heart_Failure | macrophage | *SRBD1* | 1.01492858 | 0.002028992 | 0.01674837 | 1 |
| Heart_Failure | macrophage | *NUCKS1* | 0.889447314 | 0.00323484 | 0.0208381 | 1 |
| Heart_Failure | macrophage | *LRP6* | 0.498340867 | 0.012256941 | 0.046208761 | 1 |
| Heart_Failure | macrophage | *DGCR2* | 0.703461453 | 0.010506261 | 0.041990427 | 1 |
| Heart_Failure | macrophage | *ZXDC* | 1.106573886 | 0.000924371 | 0.013744636 | 1 |
| Heart_Failure | macrophage | *GBA2* | 1.61754251 | 0.000182927 | 0.012453892 | 0.887563055 |
| Heart_Failure | macrophage | *AP3M2* | 0.982930781 | 0.006489804 | 0.031084432 | 1 |
| Heart_Failure | macrophage | *EIF2B3* | 0.862304953 | 0.002205271 | 0.017230233 | 1 |
| Heart_Failure | macrophage | *MAP4K4* | 0.532400246 | 0.009587033 | 0.039329411 | 1 |
| Heart_Failure | macrophage | *ING3* | 1.626271625 | 0.000397321 | 0.013744636 | 1 |
| Heart_Failure | macrophage | *CYBRD1* | 0.632423595 | 0.003440518 | 0.021527597 | 1 |
| Heart_Failure | macrophage | *RDH11* | 1.153220218 | 0.005432139 | 0.027827314 | 1 |
| Heart_Failure | macrophage | *ALDH3A2* | 0.502203125 | 0.007397558 | 0.033664185 | 1 |
| Heart_Failure | macrophage | *AFF4* | 0.92908742 | 0.001148163 | 0.014193744 | 1 |
| Heart_Failure | macrophage | *NFATC3* | 0.753494262 | 0.003062919 | 0.020448353 | 1 |
| Heart_Failure | macrophage | *TRNT1* | 0.909678933 | 0.007413353 | 0.033664185 | 1 |
| Heart_Failure | macrophage | *AP1M1* | 1.385311957 | 0.002498121 | 0.018350936 | 1 |
| Heart_Failure | macrophage | *ZNF532* | 1.198665095 | 0.000744811 | 0.013744636 | 1 |
| Heart_Failure | macrophage | *ZZEF1* | 0.995166694 | 0.001525662 | 0.01520515 | 1 |
| Heart_Failure | macrophage | *TIPIN* | 0.256281685 | 0.007348557 | 0.033622873 | 1 |

| | | | | | | |
|---|---|---|---|---|---|---|
| Heart_Failure | macrophage | *VPS9D1* | 1.721792082 | 2.77E-05 | 0.003955379 | 0.134482873 |
| Heart_Failure | macrophage | *FNDC3B* | 0.766583639 | 0.001069811 | 0.014193744 | 1 |
| Heart_Failure | macrophage | *KIFAP3* | 0.802523241 | 0.004755867 | 0.025840386 | 1 |
| Heart_Failure | macrophage | *FMO4* | 0.997977741 | 0.001023543 | 0.014193744 | 1 |
| Heart_Failure | macrophage | *XAB2* | 1.730080978 | 0.009086183 | 0.038236046 | 1 |
| Heart_Failure | macrophage | *IL4R* | 0.867758874 | 0.011330862 | 0.044087683 | 1 |
| Heart_Failure | macrophage | *DYNC1I2* | 1.026328213 | 0.001196591 | 0.014405948 | 1 |
| Heart_Failure | macrophage | *PIK3C3* | 0.897198593 | 0.007454088 | 0.033738094 | 1 |
| Heart_Failure | macrophage | *PCM1* | 0.732378804 | 0.002821673 | 0.01956279 | 1 |
| Heart_Failure | macrophage | *XRCC5* | 1.040029712 | 0.001351427 | 0.014806416 | 1 |
| Heart_Failure | macrophage | *FDFT1* | 0.725828561 | 0.00110954 | 0.014193744 | 1 |
| Heart_Failure | macrophage | *PGM1* | 0.635525116 | 0.00927719 | 0.038538463 | 1 |
| Heart_Failure | macrophage | *RABL2B* | 1.627358404 | 0.000561411 | 0.013744636 | 1 |
| Heart_Failure | macrophage | *RIF1* | 0.982156563 | 0.001610638 | 0.015528127 | 1 |
| Heart_Failure | macrophage | *MOK* | 0.401519592 | 0.01322986 | 0.04877757 | 1 |
| Heart_Failure | macrophage | *ULK2* | 0.783505061 | 0.009122154 | 0.038340073 | 1 |
| Heart_Failure | macrophage | *PIBF1* | 0.86900684 | 0.001043382 | 0.014193744 | 1 |
| Heart_Failure | macrophage | *ZNF264* | 1.606622672 | 0.000867271 | 0.013744636 | 1 |
| Heart_Failure | macrophage | *CHMP2B* | 1.439648524 | 0.000537767 | 0.013744636 | 1 |
| Heart_Failure | macrophage | *GSTP1* | 0.939988805 | 0.000193086 | 0.012453892 | 0.936852021 |
| Heart_Failure | macrophage | *WBP11* | 1.670320079 | 0.000994738 | 0.013998911 | 1 |
| Heart_Failure | macrophage | *NCOA1* | 0.904411871 | 0.000794105 | 0.013744636 | 1 |
| Heart_Failure | macrophage | *HACE1* | 0.917627509 | 0.002156795 | 0.017199722 | 1 |
| Heart_Failure | macrophage | *SLC25A24* | 1.120394784 | 0.001628289 | 0.015548043 | 1 |
| Heart_Failure | macrophage | *RRN3* | 1.232458914 | 0.000304365 | 0.013632389 | 1 |
| Heart_Failure | macrophage | *MTIF2* | 1.581492772 | 0.001293121 | 0.014557357 | 1 |
| Heart_Failure | macrophage | *DDHD2* | 0.721265509 | 0.000309061 | 0.013632389 | 1 |
| Heart_Failure | macrophage | *USP40* | 0.789333478 | 0.003093724 | 0.020493053 | 1 |
| Heart_Failure | macrophage | *MAST2* | 1.39425937 | 4.04E-07 | 0.000196049 | 0.001960494 |
| Heart_Failure | macrophage | *EIF2AK1* | 1.43815383 | 0.000451792 | 0.013744636 | 1 |
| Heart_Failure | macrophage | *TMED2* | 0.724167048 | 0.002534033 | 0.018350936 | 1 |
| Heart_Failure | macrophage | *ALG9* | 1.139129004 | 0.013138333 | 0.048550794 | 1 |
| Heart_Failure | macrophage | *ACOX3* | 0.691962356 | 0.006977536 | 0.032490409 | 1 |
| Heart_Failure | macrophage | *TRIP6* | 0.925974988 | 0.011514566 | 0.044481427 | 1 |
| Heart_Failure | macrophage | *SRRT* | 0.951516997 | 0.009484029 | 0.039129686 | 1 |
| Heart_Failure | macrophage | *ADAMTS2* | 0.87396373 | 0.004449428 | 0.024908187 | 1 |
| Heart_Failure | macrophage | *PSMC5* | 1.335437645 | 0.000209309 | 0.012453892 | 1 |
| Heart_Failure | macrophage | *UIMC1* | 0.917939159 | 0.0033521 | 0.021316371 | 1 |
| Heart_Failure | macrophage | *OGFOD1* | 0.739002581 | 0.005222438 | 0.027217261 | 1 |
| Heart_Failure | macrophage | *DOCK3* | 0.831413445 | 0.00671661 | 0.031761858 | 1 |
| Heart_Failure | macrophage | *NSFL1C* | 0.666227782 | 0.003932169 | 0.023296977 | 1 |
| Heart_Failure | macrophage | *MAVS* | 0.788088088 | 0.000357648 | 0.013744636 | 1 |
| Heart_Failure | macrophage | *SNX5* | 1.005654999 | 0.012355328 | 0.046438004 | 1 |
| Heart_Failure | macrophage | *RPH3A* | 1.600097958 | 0.007012187 | 0.03262045 | 1 |
| Heart_Failure | macrophage | *KIF16B* | 0.895910002 | 0.008540634 | 0.036731333 | 1 |
| Heart_Failure | macrophage | *FXYD5* | 1.432015306 | 2.97E-05 | 0.004112525 | 0.14393836 |
| Heart_Failure | macrophage | *ZNF302* | 0.458095563 | 0.008121354 | 0.035571937 | 1 |
| Heart_Failure | macrophage | *GRAMD1A* | 1.105372711 | 0.005851476 | 0.029038751 | 1 |
| Heart_Failure | macrophage | *CDIP1* | 1.516045809 | 0.001894136 | 0.016377125 | 1 |
| Heart_Failure | macrophage | *KCNH4* | 0.245948641 | 0.010737848 | 0.042635055 | 1 |

| | | | | | | |
|---|---|---|---|---|---|---|
| Heart_Failure | macrophage | *ANKRD24* | 1.408423923 | 0.007541347 | 0.034000632 | 1 |
| Heart_Failure | macrophage | *DHX32* | 0.664446214 | 0.01368803 | 0.049898063 | 1 |
| Heart_Failure | macrophage | *SPTLC1* | 0.786163054 | 0.004966097 | 0.026566153 | 1 |
| Heart_Failure | macrophage | *RAB11FIP3* | 0.700216912 | 0.002282001 | 0.017547177 | 1 |
| Heart_Failure | macrophage | *CERS4* | 0.724693499 | 0.008884849 | 0.037584382 | 1 |
| Heart_Failure | macrophage | *USP48* | 0.755557909 | 0.000474824 | 0.013744636 | 1 |
| Heart_Failure | macrophage | *PDPR* | 0.940483596 | 0.002088463 | 0.016973572 | 1 |
| Heart_Failure | macrophage | *GLG1* | 1.033506033 | 0.001269544 | 0.014557357 | 1 |
| Heart_Failure | macrophage | *PPP2R3C* | 0.743923982 | 0.001686129 | 0.015642632 | 1 |
| Heart_Failure | macrophage | *SCFD1* | 0.966992495 | 0.001231694 | 0.014505287 | 1 |
| Heart_Failure | macrophage | *SUPT16H* | 1.472087644 | 0.00074813 | 0.013744636 | 1 |
| Heart_Failure | macrophage | *TOX4* | 0.981684661 | 0.004548265 | 0.025106007 | 1 |
| Heart_Failure | macrophage | *LRRFIP2* | 1.161323371 | 0.000351735 | 0.013744636 | 1 |
| Heart_Failure | macrophage | *SEC22C* | 0.904294712 | 0.005182677 | 0.027077595 | 1 |
| Heart_Failure | macrophage | *XYLB* | 1.529375149 | 0.000880038 | 0.013744636 | 1 |
| Heart_Failure | macrophage | *MSH2* | 1.04064562 | 0.003001868 | 0.020257391 | 1 |
| Heart_Failure | macrophage | *PSMD5* | 1.067433127 | 0.001325061 | 0.014712122 | 1 |
| Heart_Failure | macrophage | *WAC* | 1.005983617 | 0.000600009 | 0.013744636 | 1 |
| Heart_Failure | macrophage | *DSP* | 1.196607879 | 0.008171698 | 0.035687739 | 1 |
| Heart_Failure | macrophage | *ERMP1* | 0.728485223 | 0.006757548 | 0.031863578 | 1 |
| Heart_Failure | macrophage | *RAB18* | 1.147237327 | 0.001149596 | 0.014193744 | 1 |
| Heart_Failure | macrophage | *TSPAN15* | 0.853938991 | 0.006587248 | 0.031396194 | 1 |
| Heart_Failure | macrophage | *SETD1A* | 1.572980788 | 2.08E-06 | 0.000719805 | 0.010077268 |
| Heart_Failure | macrophage | *MTAP* | 1.090067137 | 0.000260611 | 0.012750837 | 1 |
| Heart_Failure | macrophage | *MED15* | 0.965604395 | 0.006907802 | 0.032258573 | 1 |
| Heart_Failure | macrophage | *LZTR1* | 0.68916776 | 0.00485069 | 0.026121583 | 1 |
| Heart_Failure | macrophage | *SMARCB1* | 0.50359367 | 0.00022671 | 0.012453892 | 1 |
| Heart_Failure | macrophage | *DDT* | 0.795855155 | 0.008573708 | 0.036806023 | 1 |
| Heart_Failure | macrophage | *CABIN1* | 1.013100385 | 0.001279677 | 0.014557357 | 1 |
| Heart_Failure | macrophage | *TOP3B* | 1.040209381 | 0.003832758 | 0.023041698 | 1 |
| Heart_Failure | macrophage | *HPS4* | 0.949923237 | 0.007104667 | 0.032868649 | 1 |
| Heart_Failure | macrophage | *TRIOBP* | 0.710513124 | 0.005366303 | 0.027699259 | 1 |
| Heart_Failure | macrophage | *TTC28* | 0.529424469 | 0.004813359 | 0.026058178 | 1 |
| Heart_Failure | macrophage | *PPP6R2* | 0.938613863 | 0.002195229 | 0.017199722 | 1 |
| Heart_Failure | macrophage | *MTMR3* | 0.58734367 | 0.002862321 | 0.019714284 | 1 |
| Heart_Failure | macrophage | *IFT27* | 1.48315438 | 0.000809209 | 0.013744636 | 1 |
| Heart_Failure | macrophage | *ACO2* | 0.860777614 | 0.003145753 | 0.020604359 | 1 |
| Heart_Failure | macrophage | *POLR3H* | 1.193014409 | 0.000911956 | 0.013744636 | 1 |
| Heart_Failure | macrophage | *DESI1* | 1.320072693 | 0.000244139 | 0.012453892 | 1 |
| Heart_Failure | macrophage | *HDAC10* | 1.868656029 | 0.002521337 | 0.018350936 | 1 |
| Heart_Failure | macrophage | *CNIH1* | 1.235966647 | 0.002531458 | 0.018350936 | 1 |
| Heart_Failure | macrophage | *ATP6V1D* | 1.267514226 | 0.000839754 | 0.013744636 | 1 |
| Heart_Failure | macrophage | *MTHFD1* | 0.798025991 | 0.003945464 | 0.023296977 | 1 |
| Heart_Failure | macrophage | *GSKIP* | 2.009506429 | 0.002503716 | 0.018350936 | 1 |
| Heart_Failure | macrophage | *PSMB5* | 0.559154738 | 0.002050674 | 0.016835648 | 1 |
| Heart_Failure | macrophage | *YY1* | 1.324614316 | 0.000921591 | 0.013744636 | 1 |
| Heart_Failure | macrophage | *CCNB1IP1* | 1.014652164 | 0.004437522 | 0.024891165 | 1 |
| Heart_Failure | macrophage | *CINP* | 1.131188527 | 0.005151173 | 0.027049231 | 1 |
| Heart_Failure | macrophage | *TRPC4AP* | 0.893797719 | 0.000437514 | 0.013744636 | 1 |
| Heart_Failure | macrophage | *NDRG3* | 1.285446947 | 0.001682073 | 0.015642632 | 1 |

| | | | | | | |
|---|---|---|---|---|---|---|
| Heart_Failure | macrophage | *CSTF1* | 2.232364549 | 0.000500185 | 0.013744636 | 1 |
| Heart_Failure | macrophage | *NELFCD* | 1.133057326 | 0.002385404 | 0.018030253 | 1 |
| Heart_Failure | macrophage | *MTG2* | 1.035002042 | 0.000250629 | 0.012497964 | 1 |
| Heart_Failure | macrophage | *DIDO1* | 1.165298572 | 7.55E-05 | 0.007965661 | 0.366420401 |
| Heart_Failure | macrophage | *GID8* | 0.842965507 | 0.011152695 | 0.043604253 | 1 |
| Heart_Failure | macrophage | *GMEB2* | 1.202722159 | 0.011839572 | 0.045268405 | 1 |
| Heart_Failure | macrophage | *RBFA* | 0.629468631 | 0.012028634 | 0.04559604 | 1 |
| Heart_Failure | macrophage | *METTL4* | 1.168678818 | 0.003453049 | 0.021562667 | 1 |
| Heart_Failure | macrophage | *RNF125* | 1.820902832 | 0.003146709 | 0.020604359 | 1 |
| Heart_Failure | macrophage | *MIB1* | 0.841268332 | 0.002193973 | 0.017199722 | 1 |
| Heart_Failure | macrophage | *STK24* | 0.98822564 | 0.001401444 | 0.015035788 | 1 |
| Heart_Failure | macrophage | *UGGT2* | 0.625663274 | 0.010075197 | 0.04063579 | 1 |
| Heart_Failure | macrophage | *ARHGEF7* | 0.908664004 | 0.001606036 | 0.015528127 | 1 |
| Heart_Failure | macrophage | *PARP4* | 0.882877335 | 0.002132187 | 0.017128099 | 1 |
| Heart_Failure | macrophage | *SUPT20H* | 1.0010665 | 0.000809238 | 0.013744636 | 1 |
| Heart_Failure | macrophage | *MRPS31* | 0.987153721 | 0.001981746 | 0.016578327 | 1 |
| Heart_Failure | macrophage | *INTS6* | 0.867214767 | 0.002073009 | 0.016973572 | 1 |
| Heart_Failure | macrophage | *CLN5* | 1.306484166 | 0.000511903 | 0.013744636 | 1 |
| Heart_Failure | macrophage | *MGRN1* | 0.73360814 | 0.007986034 | 0.035193674 | 1 |
| Heart_Failure | macrophage | *LONP2* | 0.505422481 | 0.010541607 | 0.042097017 | 1 |
| Heart_Failure | macrophage | *DHODH* | 0.504317536 | 0.00921819 | 0.038467142 | 1 |
| Heart_Failure | macrophage | *PSMD7* | 1.331536591 | 0.001036168 | 0.014193744 | 1 |
| Heart_Failure | macrophage | *NME4* | 1.160902419 | 0.009229628 | 0.038472643 | 1 |
| Heart_Failure | macrophage | *LMF1* | 1.228893144 | 0.000897234 | 0.013744636 | 1 |
| Heart_Failure | macrophage | *UBE2I* | 1.415811818 | 0.000450736 | 0.013744636 | 1 |
| Heart_Failure | macrophage | *TOX3* | 0.672305287 | 0.002502447 | 0.018350936 | 1 |
| Heart_Failure | macrophage | *RBL2* | 0.844879229 | 0.000456274 | 0.013744636 | 1 |
| Heart_Failure | macrophage | *KAT8* | 1.264700242 | 0.010796972 | 0.042778057 | 1 |
| Heart_Failure | macrophage | *C16orf62* | 1.192746036 | 0.000766796 | 0.013744636 | 1 |
| Heart_Failure | macrophage | *RNF40* | 1.596063133 | 0.001911295 | 0.016395644 | 1 |
| Heart_Failure | macrophage | *KNOP1* | 1.300230101 | 0.001610525 | 0.015528127 | 1 |
| Heart_Failure | macrophage | *IQCH* | 0.945527263 | 0.001920113 | 0.016431018 | 1 |
| Heart_Failure | macrophage | *HERC1* | 0.955250306 | 0.001272378 | 0.014557357 | 1 |
| Heart_Failure | macrophage | *TRIP4* | 0.897560521 | 0.007352405 | 0.033622873 | 1 |
| Heart_Failure | macrophage | *TTC23* | 1.235624714 | 0.001152584 | 0.014193744 | 1 |
| Heart_Failure | macrophage | *SLC30A4* | 1.299936085 | 0.001722805 | 0.015726742 | 1 |
| Heart_Failure | macrophage | *ZFAND1* | 0.755557 | 0.011267617 | 0.043927413 | 1 |
| Heart_Failure | macrophage | *IMPAD1* | 1.075656134 | 0.009201897 | 0.038456162 | 1 |
| Heart_Failure | macrophage | *NIPAL2* | 1.313992381 | 0.000354467 | 0.013744636 | 1 |
| Heart_Failure | macrophage | *RAB2A* | 0.670312575 | 0.009695037 | 0.039496489 | 1 |
| Heart_Failure | macrophage | *CHRAC1* | 2.153341176 | 0.004516279 | 0.024957841 | 1 |
| Heart_Failure | macrophage | *SNX16* | 0.963358108 | 0.008243504 | 0.035840035 | 1 |
| Heart_Failure | macrophage | *GSDMD* | 1.211619354 | 0.001222376 | 0.014488552 | 1 |
| Heart_Failure | macrophage | *ERICH1* | 0.846255317 | 0.003442996 | 0.021527597 | 1 |
| Heart_Failure | macrophage | *SNRNP70* | 0.85432406 | 0.003076595 | 0.02044882 | 1 |
| Heart_Failure | macrophage | *AMH* | 17.11172096 | 0.002268395 | 0.017479059 | 1 |
| Heart_Failure | macrophage | *SGTA* | 1.261746838 | 0.003544365 | 0.021879464 | 1 |
| Heart_Failure | macrophage | *MED25* | 1.474904045 | 0.004257587 | 0.024189473 | 1 |
| Heart_Failure | macrophage | *TIMM44* | 1.193111696 | 1.91E-05 | 0.003189731 | 0.092502199 |
| Heart_Failure | macrophage | *FAM32A* | 1.568919264 | 0.010843601 | 0.042879503 | 1 |

| | | | | | | |
|---|---|---|---|---|---|---|
| Heart_Failure | macrophage | *RASAL3* | 3.098554552 | 0.000408635 | 0.013744636 | 1 |
| Heart_Failure | macrophage | *ANKRD27* | 1.212914215 | 0.002101201 | 0.017048544 | 1 |
| Heart_Failure | macrophage | *RABAC1* | 0.843927552 | 0.005193598 | 0.027096064 | 1 |
| Heart_Failure | macrophage | *CAPS* | 1.606269907 | 0.004129247 | 0.023913414 | 1 |
| Heart_Failure | macrophage | *MIER2* | 0.757372247 | 0.011758027 | 0.045098774 | 1 |
| Heart_Failure | macrophage | *TNPO2* | 0.930648206 | 0.00187675 | 0.016371728 | 1 |
| Heart_Failure | macrophage | *TFPT* | 1.610400419 | 0.007430044 | 0.033692127 | 1 |
| Heart_Failure | macrophage | *RAB3A* | 7.811618014 | 0.0019126 | 0.016395644 | 1 |
| Heart_Failure | macrophage | *ZNF14* | 0.885624571 | 0.009011603 | 0.038021129 | 1 |
| Heart_Failure | macrophage | *ZNF85* | 1.585125543 | 0.005016892 | 0.026720217 | 1 |
| Heart_Failure | macrophage | *BET1* | 0.739770396 | 0.007605167 | 0.034083874 | 1 |
| Heart_Failure | macrophage | *DNAH11* | 0.918671954 | 0.008123188 | 0.035571937 | 1 |
| Heart_Failure | macrophage | *IQCE* | 1.738690452 | 0.000733923 | 0.013744636 | 1 |
| Heart_Failure | macrophage | *HIBADH* | 0.721240927 | 0.000634539 | 0.013744636 | 1 |
| Heart_Failure | macrophage | *PLEKHA8* | 1.55093147 | 0.00084115 | 0.013744636 | 1 |
| Heart_Failure | macrophage | *ZKSCAN1* | 0.925092083 | 0.002213658 | 0.017241117 | 1 |
| Heart_Failure | macrophage | *EIF3B* | 1.32650088 | 0.001292193 | 0.014557357 | 1 |
| Heart_Failure | macrophage | *TFR2* | 1.369844774 | 0.012359415 | 0.046438004 | 1 |
| Heart_Failure | macrophage | *RBM28* | 0.529531446 | 0.004947969 | 0.026527676 | 1 |
| Heart_Failure | macrophage | *C1GALT1* | 0.85564715 | 0.011940402 | 0.045484252 | 1 |
| Heart_Failure | macrophage | *COA1* | 1.017680301 | 0.000764032 | 0.013744636 | 1 |
| Heart_Failure | macrophage | *FKTN* | 1.09290104 | 0.00125055 | 0.014557357 | 1 |
| Heart_Failure | macrophage | *MEGF9* | 1.269252679 | 0.001331374 | 0.014716944 | 1 |
| Heart_Failure | macrophage | *C5* | 0.831655999 | 0.003550613 | 0.021890185 | 1 |
| Heart_Failure | macrophage | *ECM2* | 0.482518116 | 0.011035004 | 0.043388847 | 1 |
| Heart_Failure | macrophage | *FUBP3* | 0.806227536 | 0.007344945 | 0.033622873 | 1 |
| Heart_Failure | macrophage | *DNMBP* | 0.758154737 | 0.003183795 | 0.020758363 | 1 |
| Heart_Failure | macrophage | *TRDMT1* | 0.90372224 | 0.005045959 | 0.02672816 | 1 |
| Heart_Failure | macrophage | *ATE1* | 0.710940577 | 0.001775282 | 0.016040353 | 1 |
| Heart_Failure | macrophage | *CDH23* | 0.91568734 | 0.003105386 | 0.020493053 | 1 |
| Heart_Failure | macrophage | *BMPR1A* | 0.728833753 | 0.003390051 | 0.021417354 | 1 |
| Heart_Failure | macrophage | *TNKS2* | 1.252376452 | 0.000884142 | 0.013744636 | 1 |
| Heart_Failure | macrophage | *LHPP* | 0.706000234 | 0.004484088 | 0.024925587 | 1 |
| Heart_Failure | macrophage | *PITRM1* | 1.286541009 | 0.000481646 | 0.013744636 | 1 |
| Heart_Failure | macrophage | *TFAM* | 1.800614888 | 0.000805614 | 0.013744636 | 1 |
| Heart_Failure | macrophage | *PBLD* | 1.083439758 | 0.002818704 | 0.01956279 | 1 |
| Heart_Failure | macrophage | *TRIM37* | 1.206413459 | 0.001815722 | 0.016138719 | 1 |
| Heart_Failure | macrophage | *PIGL* | 0.701068248 | 0.00607264 | 0.029716691 | 1 |
| Heart_Failure | macrophage | *MED13* | 0.900466095 | 0.001151626 | 0.014193744 | 1 |
| Heart_Failure | macrophage | *RAI1* | 0.92290386 | 0.005038953 | 0.026720217 | 1 |
| Heart_Failure | macrophage | *GOSR1* | 1.174312713 | 0.000899652 | 0.013744636 | 1 |
| Heart_Failure | macrophage | *DRG2* | 1.151313877 | 0.004614842 | 0.025300804 | 1 |
| Heart_Failure | macrophage | *FTSJ3* | 0.155441616 | 0.011952223 | 0.045484252 | 1 |
| Heart_Failure | macrophage | *ICAM2* | 1.451179128 | 0.004031545 | 0.023595969 | 1 |
| Heart_Failure | macrophage | *DDX5* | 0.538958253 | 0.004605435 | 0.025277795 | 1 |
| Heart_Failure | macrophage | *NAGLU* | 1.434734845 | 0.000185694 | 0.012453892 | 0.9009863 |
| Heart_Failure | macrophage | *EFTUD2* | 1.145986806 | 0.006586438 | 0.031396194 | 1 |
| Heart_Failure | macrophage | *DHRS7B* | 1.240362597 | 0.000878117 | 0.013744636 | 1 |
| Heart_Failure | macrophage | *MYH3* | 2.403706724 | 0.001931444 | 0.016469888 | 1 |
| Heart_Failure | macrophage | *SLAIN2* | 1.303657231 | 0.00021254 | 0.012453892 | 1 |

| | | | | | | |
|---|---|---|---|---|---|---|
| Heart_Failure | macrophage | *OCIAD1* | 0.937427895 | 0.002034066 | 0.016756007 | 1 |
| Heart_Failure | macrophage | *UBE2D3* | 1.053544572 | 0.000635354 | 0.013744636 | 1 |
| Heart_Failure | macrophage | *NDUFC1* | 1.419452354 | 0.001724367 | 0.015726742 | 1 |
| Heart_Failure | macrophage | *GRPEL1* | 2.122959565 | 0.000856021 | 0.013744636 | 1 |
| Heart_Failure | macrophage | *STIM2* | 1.150900371 | 0.001066261 | 0.014193744 | 1 |
| Heart_Failure | macrophage | *UFSP2* | 1.121409254 | 0.001974604 | 0.016575738 | 1 |
| Heart_Failure | macrophage | *KLHL5* | 1.23960767 | 0.000540002 | 0.013744636 | 1 |
| Heart_Failure | macrophage | *ELP4* | 0.702868354 | 0.007084883 | 0.032849883 | 1 |
| Heart_Failure | macrophage | *B3GAT1* | 12.86500684 | 0.001541034 | 0.01520515 | 1 |
| Heart_Failure | macrophage | *CEP164* | 0.711277282 | 0.006567165 | 0.031362089 | 1 |
| Heart_Failure | macrophage | *KIAA1549L* | 4.671942934 | 0.007658678 | 0.034217225 | 1 |
| Heart_Failure | macrophage | *SLC35F2* | 0.952551539 | 0.006792122 | 0.031902592 | 1 |
| Heart_Failure | macrophage | *RPS13* | 0.63580541 | 0.009570462 | 0.039329411 | 1 |
| Heart_Failure | macrophage | *PRDM4* | 1.438169832 | 9.88E-06 | 0.002397166 | 0.047943318 |
| Heart_Failure | macrophage | *CORO1C* | 1.033879336 | 0.004201252 | 0.024038296 | 1 |
| Heart_Failure | macrophage | *ASIC1* | 2.148206331 | 0.011494338 | 0.044438666 | 1 |
| Heart_Failure | macrophage | *PARP11* | 1.319587769 | 0.001494295 | 0.015102525 | 1 |
| Heart_Failure | macrophage | *VPS29* | 1.183113781 | 0.000842968 | 0.013744636 | 1 |
| Heart_Failure | macrophage | *CDK2AP1* | 0.689877484 | 9.84E-05 | 0.00918566 | 0.477654312 |
| Heart_Failure | macrophage | *CAND1* | 0.80889557 | 0.003936269 | 0.023296977 | 1 |
| Heart_Failure | macrophage | *MDM1* | 0.973970494 | 0.005098133 | 0.026857917 | 1 |
| Heart_Failure | macrophage | *NUP107* | 0.763327959 | 0.007533466 | 0.034000632 | 1 |
| Heart_Failure | macrophage | *KRR1* | 1.219101819 | 0.001387109 | 0.014989426 | 1 |
| Heart_Failure | macrophage | *GNPTAB* | 1.031011856 | 0.003032941 | 0.020353843 | 1 |
| Heart_Failure | macrophage | *NT5DC3* | 0.695734164 | 0.0110961 | 0.043448188 | 1 |
| Heart_Failure | macrophage | *SUDS3* | 1.419139653 | 0.000793988 | 0.013744636 | 1 |
| Heart_Failure | macrophage | *GOLT1B* | 1.120738235 | 0.003544266 | 0.021879464 | 1 |
| Heart_Failure | macrophage | *C2CD5* | 0.753535347 | 0.012527541 | 0.046937166 | 1 |
| Heart_Failure | macrophage | *TMEM14C* | 1.490224405 | 0.000900631 | 0.013744636 | 1 |
| Heart_Failure | macrophage | *RNGTT* | 1.199075629 | 0.000983889 | 0.013998911 | 1 |
| Heart_Failure | macrophage | *RNF8* | 0.779169271 | 0.009248137 | 0.038485348 | 1 |
| Heart_Failure | macrophage | *MDN1* | 0.704631397 | 0.009528448 | 0.039279548 | 1 |
| Heart_Failure | macrophage | *ACOT13* | 1.393597882 | 0.000884769 | 0.013744636 | 1 |
| Heart_Failure | macrophage | *PHACTR2* | 0.678004906 | 0.01303596 | 0.048319694 | 1 |
| Heart_Failure | macrophage | *PACRG* | 0.987887505 | 0.006644345 | 0.03157528 | 1 |
| Heart_Failure | macrophage | *QKI* | 0.763809181 | 0.003948887 | 0.023296977 | 1 |
| Heart_Failure | macrophage | *FAM120B* | 0.958102113 | 7.06E-05 | 0.007789864 | 0.342754035 |
| Heart_Failure | macrophage | *EXOC2* | 1.253863246 | 0.000505582 | 0.013744636 | 1 |
| Heart_Failure | macrophage | *TMEM30A* | 1.398145494 | 0.002014555 | 0.016703608 | 1 |
| Heart_Failure | macrophage | *PRSS16* | 0.603655961 | 0.007412844 | 0.033664185 | 1 |
| Heart_Failure | macrophage | *NUDT12* | 1.239517639 | 0.002755105 | 0.019289709 | 1 |
| Heart_Failure | macrophage | *KIF20A* | -2.323830995 | 0.000873738 | 0.013744636 | 1 |
| Heart_Failure | macrophage | *RASGRF2* | 0.271384294 | 0.008005667 | 0.035248181 | 1 |
| Heart_Failure | macrophage | *DROSHA* | 0.613369895 | 0.004432992 | 0.024891165 | 1 |
| Heart_Failure | macrophage | *SKP1* | 1.075895546 | 0.00210795 | 0.017074746 | 1 |
| Heart_Failure | macrophage | *PPWD1* | 1.034754023 | 0.001670431 | 0.015616438 | 1 |
| Heart_Failure | macrophage | *TXNDC15* | 1.356195688 | 0.00173584 | 0.015772092 | 1 |
| Heart_Failure | macrophage | *SMAD5* | 0.99480382 | 0.001962859 | 0.016534361 | 1 |
| Heart_Failure | macrophage | *ERGIC1* | 0.948429114 | 0.002385431 | 0.018030253 | 1 |
| Heart_Failure | macrophage | *ATP6V0E1* | 1.189326425 | 0.000491594 | 0.013744636 | 1 |

| | | | | | | |
|---|---|---|---|---|---|---|
| Heart_Failure | macrophage | *BNIP1* | 0.326960873 | 0.00189881 | 0.016377125 | 1 |
| Heart_Failure | macrophage | *TBCCD1* | 1.531898056 | 0.003893493 | 0.023236443 | 1 |
| Heart_Failure | macrophage | *PCCB* | 0.57402842 | 0.001794229 | 0.016131335 | 1 |
| Heart_Failure | macrophage | *HYAL1* | 0.400232431 | 0.00955669 | 0.039329411 | 1 |
| Heart_Failure | macrophage | *CBLB* | 0.558312006 | 0.009851102 | 0.039997947 | 1 |
| Heart_Failure | macrophage | *IQCG* | 1.140563051 | 0.002480439 | 0.018350936 | 1 |
| Heart_Failure | macrophage | *HEMK1* | 0.865109573 | 0.003702723 | 0.022513298 | 1 |
| Heart_Failure | macrophage | *COMMD2* | 1.0190453 | 0.004196764 | 0.024038296 | 1 |
| Heart_Failure | macrophage | *DNAH1* | 0.570776214 | 0.006958399 | 0.032432421 | 1 |
| Heart_Failure | macrophage | *NKTR* | 0.647169576 | 0.00449955 | 0.024925587 | 1 |
| Heart_Failure | macrophage | *LMAN2L* | 0.646562157 | 0.010887812 | 0.043019272 | 1 |
| Heart_Failure | macrophage | *ACTR1B* | 1.190668948 | 0.003130435 | 0.020604359 | 1 |
| Heart_Failure | macrophage | *NRBP1* | 1.0916148 | 0.004197008 | 0.024038296 | 1 |
| Heart_Failure | macrophage | *PPM1G* | 1.123741639 | 0.004501237 | 0.024925587 | 1 |
| Heart_Failure | macrophage | *MOGS* | 0.461345846 | 0.001227287 | 0.014488552 | 1 |
| Heart_Failure | macrophage | *RTN4* | 0.544472021 | 0.011972331 | 0.045484252 | 1 |
| Heart_Failure | macrophage | *GALNT3* | 0.991929862 | 0.001453121 | 0.015102525 | 1 |
| Heart_Failure | macrophage | *MRPL19* | 0.887814302 | 0.003846772 | 0.023071121 | 1 |
| Heart_Failure | macrophage | *DNAH6* | 0.124648164 | 0.009772656 | 0.039712671 | 1 |
| Heart_Failure | macrophage | *IL18R1* | 0.937724174 | 2.60E-06 | 0.000842515 | 0.012637727 |
| Heart_Failure | macrophage | *HDLBP* | 1.176095158 | 6.52E-05 | 0.007535902 | 0.3165079 |
| Heart_Failure | macrophage | *TAF1B* | 1.207090228 | 4.31E-05 | 0.005222304 | 0.208892176 |
| Heart_Failure | macrophage | *NOL10* | 1.069446473 | 0.001638002 | 0.015548043 | 1 |
| Heart_Failure | macrophage | *STRN* | 1.181818668 | 0.000625279 | 0.013744636 | 1 |
| Heart_Failure | macrophage | *CEBPZ* | 1.095151711 | 0.000450022 | 0.013744636 | 1 |
| Heart_Failure | macrophage | *PRKD3* | 0.998403933 | 0.000681478 | 0.013744636 | 1 |
| Heart_Failure | macrophage | *SLC25A12* | 0.606387298 | 0.011354378 | 0.044108439 | 1 |
| Heart_Failure | macrophage | *DARS* | 1.261318156 | 0.000788506 | 0.013744636 | 1 |
| Heart_Failure | macrophage | *PNO1* | 2.328432859 | 0.000869019 | 0.013744636 | 1 |
| Heart_Failure | macrophage | *ORC4* | 0.590657569 | 0.01099744 | 0.043347494 | 1 |
| Heart_Failure | macrophage | *AAK1* | 0.951264253 | 0.001826083 | 0.016138719 | 1 |
| Heart_Failure | macrophage | *NFE2L2* | 0.76720875 | 0.002427086 | 0.018201271 | 1 |
| Heart_Failure | macrophage | *MSH6* | 0.971483639 | 0.004158258 | 0.023990328 | 1 |
| Heart_Failure | macrophage | *ALMS1* | 1.155769959 | 0.000834274 | 0.013744636 | 1 |
| Heart_Failure | macrophage | *DNAJC16* | 0.912142372 | 0.004505305 | 0.024925587 | 1 |
| Heart_Failure | macrophage | *FAM20B* | 1.090844547 | 0.006310859 | 0.03065094 | 1 |
| Heart_Failure | macrophage | *EDEM3* | 1.185051062 | 0.000745572 | 0.013744636 | 1 |
| Heart_Failure | macrophage | *RAP1A* | 0.839987907 | 0.004000875 | 0.023473089 | 1 |
| Heart_Failure | macrophage | *CAPZA1* | 0.821200548 | 0.00539426 | 0.027717297 | 1 |
| Heart_Failure | macrophage | *DOCK7* | 0.954749407 | 0.00413014 | 0.023913414 | 1 |
| Heart_Failure | macrophage | *LEPR* | 0.769962915 | 0.00178602 | 0.01610738 | 1 |
| Heart_Failure | macrophage | *MFN2* | 0.692966197 | 0.012163572 | 0.045963902 | 1 |
| Heart_Failure | macrophage | *MRPS15* | 1.228057087 | 0.003607093 | 0.022174968 | 1 |
| Heart_Failure | macrophage | *GNPAT* | 1.256738476 | 0.000334364 | 0.013744636 | 1 |
| Heart_Failure | macrophage | *TBCE* | 1.422938275 | 0.001451806 | 0.015102525 | 1 |
| Heart_Failure | macrophage | *SDHB* | 1.065951929 | 0.009647716 | 0.039451323 | 1 |
| Heart_Failure | macrophage | *ERI3* | 1.168351779 | 0.000475754 | 0.013744636 | 1 |
| Heart_Failure | macrophage | *PRRC2C* | 0.793560816 | 0.006022042 | 0.029550899 | 1 |
| Heart_Failure | macrophage | *ABCD3* | 1.141975848 | 0.001473256 | 0.015102525 | 1 |
| Heart_Failure | macrophage | *PTBP2* | 0.517369728 | 0.00429476 | 0.02433649 | 1 |

| | | | | | | |
|---|---|---|---|---|---|---|
| Heart_Failure | macrophage | *ESYT2* | 0.622766983 | 0.007639324 | 0.034193728 | 1 |
| Heart_Failure | macrophage | *CD3EAP* | 0.951038549 | 0.007374609 | 0.033654566 | 1 |
| Heart_Failure | macrophage | *CAMSAP2* | 0.659280228 | 0.002530924 | 0.018350936 | 1 |
| Heart_Failure | macrophage | *KLF7* | 0.88617857 | 0.002246578 | 0.017385005 | 1 |
| Heart_Failure | macrophage | *AKAP7* | 1.117468923 | 0.00040055 | 0.013744636 | 1 |
| Heart_Failure | macrophage | *MED28* | 1.170982348 | 3.23E-05 | 0.004351462 | 0.156652636 |
| Heart_Failure | macrophage | *UBN1* | 1.107026043 | 0.001900314 | 0.016377125 | 1 |
| Heart_Failure | macrophage | *C2orf40* | 0.70532858 | 0.011346944 | 0.044108439 | 1 |
| Heart_Failure | macrophage | *SENP5* | 1.125809254 | 0.001101103 | 0.014193744 | 1 |
| Heart_Failure | macrophage | *CTNNAL1* | 0.390580953 | 0.000114844 | 0.009786465 | 0.557224124 |
| Heart_Failure | macrophage | *RAB14* | 1.043931601 | 0.01078886 | 0.042778057 | 1 |
| Heart_Failure | macrophage | *PPP6C* | 1.176292858 | 0.00111991 | 0.014193744 | 1 |
| Heart_Failure | macrophage | *RBM18* | 1.012489333 | 0.008237139 | 0.035840035 | 1 |
| Heart_Failure | macrophage | *MAPKAP1* | 1.046079674 | 0.001106633 | 0.014193744 | 1 |
| Heart_Failure | macrophage | *INVS* | 0.472813564 | 0.008977055 | 0.037908331 | 1 |
| Heart_Failure | macrophage | *KDSR* | 1.142422464 | 0.002330856 | 0.017754027 | 1 |
| Heart_Failure | macrophage | *DCAF4* | 0.901047316 | 0.003632369 | 0.022224786 | 1 |
| Heart_Failure | macrophage | *NEK9* | 0.999594135 | 0.001314746 | 0.014698493 | 1 |
| Heart_Failure | macrophage | *TTLL5* | 0.962319006 | 0.00230083 | 0.017636064 | 1 |
| Heart_Failure | macrophage | *COQ6* | 1.167336299 | 0.007903652 | 0.034925794 | 1 |
| Heart_Failure | macrophage | *CRIPT* | 0.76067398 | 0.012275387 | 0.046242374 | 1 |
| Heart_Failure | macrophage | *MXI1* | 0.954204137 | 0.003030318 | 0.020353843 | 1 |
| Heart_Failure | macrophage | *WDR11* | 0.838333505 | 0.008326248 | 0.036070497 | 1 |
| Heart_Failure | macrophage | *KANSL1* | 0.628255917 | 0.008236737 | 0.035840035 | 1 |
| Heart_Failure | macrophage | *HOXB3* | 0.641927041 | 0.01031741 | 0.041371962 | 1 |
| Heart_Failure | macrophage | *GORAB* | 1.601741371 | 0.000924908 | 0.013744636 | 1 |
| Heart_Failure | macrophage | *SLC10A7* | 1.280194498 | 0.000571757 | 0.013744636 | 1 |
| Heart_Failure | macrophage | *IQSEC3* | 2.724182546 | 9.70E-05 | 0.00918566 | 0.470636085 |
| Heart_Failure | macrophage | *CCDC77* | 1.065339474 | 0.003702021 | 0.022513298 | 1 |
| Heart_Failure | macrophage | *DNAJC15* | 1.239117943 | 0.001416983 | 0.015044204 | 1 |
| Heart_Failure | macrophage | *SIL1* | 0.94121622 | 0.001058991 | 0.014193744 | 1 |
| Heart_Failure | macrophage | *SCPEP1* | 0.325304344 | 0.009591939 | 0.039329411 | 1 |
| Heart_Failure | macrophage | *ZNF549* | 1.48261329 | 0.000868745 | 0.013744636 | 1 |
| Heart_Failure | macrophage | *LHX4* | 1.742902738 | 0.004600672 | 0.025277795 | 1 |
| Heart_Failure | macrophage | *SEC22A* | 0.955336207 | 0.002151157 | 0.017198249 | 1 |
| Heart_Failure | macrophage | *ZCCHC17* | 1.422756564 | 0.000789228 | 0.013744636 | 1 |
| Heart_Failure | macrophage | *TNFSF10* | 0.669378758 | 0.009274279 | 0.038538463 | 1 |
| Heart_Failure | macrophage | *MTIF3* | 0.81442181 | 0.004093397 | 0.023785825 | 1 |
| Heart_Failure | macrophage | *FMOD* | 0.599748167 | 0.007547131 | 0.034000632 | 1 |
| Heart_Failure | macrophage | *RBBP6* | 0.870406921 | 0.002890453 | 0.019780646 | 1 |
| Heart_Failure | macrophage | *NAA60* | 1.747773864 | 0.00758879 | 0.034083874 | 1 |
| Heart_Failure | macrophage | *ODF2L* | 0.665870024 | 0.00607563 | 0.029716691 | 1 |
| Heart_Failure | macrophage | *SPATA1* | 0.992683248 | 0.005771279 | 0.028868296 | 1 |
| Heart_Failure | macrophage | *EEPD1* | 0.425610191 | 0.008489138 | 0.036612709 | 1 |
| Heart_Failure | macrophage | *KLHL7* | 0.880054961 | 0.003227616 | 0.0208381 | 1 |
| Heart_Failure | macrophage | *NT5C3A* | 1.034964494 | 0.001103355 | 0.014193744 | 1 |
| Heart_Failure | macrophage | *SMU1* | 1.495534351 | 0.000145659 | 0.011106235 | 0.706738163 |
| Heart_Failure | macrophage | *TRIM24* | 0.908840076 | 0.003939691 | 0.023296977 | 1 |
| Heart_Failure | macrophage | *RBM19* | 1.513118101 | 0.000902421 | 0.013744636 | 1 |
| Heart_Failure | macrophage | *CCDC91* | 0.716371637 | 0.007570974 | 0.034076407 | 1 |

| | | | | | | |
|---|---|---|---|---|---|---|
| Heart_Failure | macrophage | *TSFM* | 0.573571517 | 0.009439016 | 0.038977112 | 1 |
| Heart_Failure | macrophage | *PFDN5* | 1.007590291 | 0.006434046 | 0.030965061 | 1 |
| Heart_Failure | macrophage | *PDE1B* | 1.975851972 | 0.001101532 | 0.014193744 | 1 |
| Heart_Failure | macrophage | *CDK2* | 0.565177952 | 0.002734236 | 0.019199008 | 1 |
| Heart_Failure | macrophage | *USP45* | 0.696872551 | 0.00530085 | 0.027419747 | 1 |
| Heart_Failure | macrophage | *PLA2G12A* | 1.125194175 | 0.000708851 | 0.013744636 | 1 |
| Heart_Failure | macrophage | *DNPEP* | 1.310085238 | 0.000703537 | 0.013744636 | 1 |
| Heart_Failure | macrophage | *SRSF6* | 0.813996124 | 0.004359893 | 0.024569337 | 1 |
| Heart_Failure | macrophage | *CSE1L* | 1.148518512 | 0.000793263 | 0.013744636 | 1 |
| Heart_Failure | macrophage | *RNF114* | 0.845490183 | 0.009153117 | 0.038340073 | 1 |
| Heart_Failure | macrophage | *MTRR* | 0.99633188 | 0.001936596 | 0.016484846 | 1 |
| Heart_Failure | macrophage | *STAMBP* | 1.30136062 | 0.000993837 | 0.013998911 | 1 |
| Heart_Failure | macrophage | *SNRNP27* | 1.390165361 | 0.001497811 | 0.015102525 | 1 |
| Heart_Failure | macrophage | *ATP8A1* | 1.173551145 | 0.00050569 | 0.013744636 | 1 |
| Heart_Failure | macrophage | *USP22* | 0.959919212 | 0.003381792 | 0.021393032 | 1 |
| Heart_Failure | macrophage | *MRS2* | 0.473192191 | 0.002656358 | 0.018762541 | 1 |
| Heart_Failure | macrophage | *NQO2* | 1.370019286 | 0.00079162 | 0.013744636 | 1 |
| Heart_Failure | macrophage | *GLO1* | 1.192144934 | 3.89E-05 | 0.004844418 | 0.188932297 |
| Heart_Failure | macrophage | *ATXN1* | 0.878920371 | 0.00093206 | 0.013744636 | 1 |
| Heart_Failure | macrophage | *RUNX2* | 0.839148601 | 0.004142322 | 0.02395536 | 1 |
| Heart_Failure | macrophage | *CLYBL* | 1.140285843 | 0.003369194 | 0.021366443 | 1 |
| Heart_Failure | macrophage | *TMTC4* | 1.179066264 | 0.002304557 | 0.017636769 | 1 |
| Heart_Failure | macrophage | *ABCC4* | 0.903876157 | 0.003523872 | 0.021836305 | 1 |
| Heart_Failure | macrophage | *GRK4* | 0.133090716 | 0.007892111 | 0.034925794 | 1 |
| Heart_Failure | macrophage | *MRPS7* | 1.782378512 | 0.000245173 | 0.012453892 | 1 |
| Heart_Failure | macrophage | *NT5C* | 1.349483936 | 0.002472763 | 0.018350936 | 1 |
| Heart_Failure | macrophage | *PPDPF* | 0.862895536 | 0.000596442 | 0.013744636 | 1 |
| Heart_Failure | macrophage | *SNRPD2* | 1.71724587 | 0.000847766 | 0.013744636 | 1 |
| Heart_Failure | macrophage | *RBCK1* | 0.789040173 | 0.011812185 | 0.045234982 | 1 |
| Heart_Failure | macrophage | *RRBP1* | 0.912466075 | 0.004899938 | 0.026328349 | 1 |
| Heart_Failure | macrophage | *NCLN* | 0.813973942 | 0.007845412 | 0.034844376 | 1 |
| Heart_Failure | macrophage | *KLC1* | 1.248265569 | 0.00064282 | 0.013744636 | 1 |
| Heart_Failure | macrophage | *TUBGCP3* | 1.183612607 | 0.00150963 | 0.015102525 | 1 |
| Heart_Failure | macrophage | *COX6B1* | 1.325709062 | 0.001221539 | 0.014488552 | 1 |
| Heart_Failure | macrophage | *PRDX5* | 1.23734899 | 0.001120019 | 0.014193744 | 1 |
| Heart_Failure | macrophage | *IRF3* | 1.155808469 | 0.003276854 | 0.020982667 | 1 |
| Heart_Failure | macrophage | *KTN1* | 0.88276973 | 0.001310768 | 0.014687862 | 1 |
| Heart_Failure | macrophage | *ZBTB1* | 0.964571414 | 0.001623641 | 0.015548043 | 1 |
| Heart_Failure | macrophage | *RHOT1* | 0.689909172 | 0.001035736 | 0.014193744 | 1 |
| Heart_Failure | macrophage | *CANX* | 0.976067873 | 0.003090911 | 0.020493053 | 1 |
| Heart_Failure | macrophage | *ZNF484* | 1.146723031 | 0.005502517 | 0.027936396 | 1 |
| Heart_Failure | macrophage | *HELB* | 1.094461711 | 0.008056576 | 0.035408066 | 1 |
| Heart_Failure | macrophage | *TAS2R4* | 0.534413069 | 0.0115773 | 0.044511141 | 1 |
| Heart_Failure | macrophage | *EMC1* | 1.237481628 | 0.0003574 | 0.013744636 | 1 |
| Heart_Failure | macrophage | *RNF6* | 1.228979989 | 0.00561887 | 0.028339666 | 1 |
| Heart_Failure | macrophage | *HIP1* | 0.963975035 | 0.000795544 | 0.013744636 | 1 |
| Heart_Failure | macrophage | *PTPN12* | 0.941961713 | 0.001538455 | 0.01520515 | 1 |
| Heart_Failure | macrophage | *PEX1* | 1.030507246 | 0.002874615 | 0.019714284 | 1 |
| Heart_Failure | macrophage | *ATF4* | 1.427160578 | 0.002482138 | 0.018350936 | 1 |
| Heart_Failure | macrophage | *SPECC1* | 0.608497941 | 0.008090566 | 0.035493153 | 1 |

| | | | | | | |
|---|---|---|---|---|---|---|
| Heart_Failure | macrophage | *PRKRIP1* | 0.827701518 | 0.002560834 | 0.018426459 | 1 |
| Heart_Failure | macrophage | *MTX2* | 0.938749009 | 0.00165264 | 0.015548043 | 1 |
| Heart_Failure | macrophage | *ORMDL1* | 0.606251074 | 0.009159152 | 0.038340073 | 1 |
| Heart_Failure | macrophage | *PSMG2* | 0.721004979 | 0.005106173 | 0.026871095 | 1 |
| Heart_Failure | macrophage | *TWSG1* | 1.21526636 | 0.000863024 | 0.013744636 | 1 |
| Heart_Failure | macrophage | *TMOD2* | 0.73670325 | 0.009968658 | 0.040306608 | 1 |
| Heart_Failure | macrophage | *IVD* | 0.459992588 | 0.010800313 | 0.042778057 | 1 |
| Heart_Failure | macrophage | *ARPP19* | 1.377006924 | 0.001816858 | 0.016138719 | 1 |
| Heart_Failure | macrophage | *SUMF2* | 1.205643696 | 0.002565127 | 0.018426459 | 1 |
| Heart_Failure | macrophage | *SERGEF* | 0.699597336 | 0.005859456 | 0.029039917 | 1 |
| Heart_Failure | macrophage | *PHF20L1* | 1.073475598 | 0.000624366 | 0.013744636 | 1 |
| Heart_Failure | macrophage | *MIS18BP1* | 0.630201602 | 0.006399501 | 0.030957506 | 1 |
| Heart_Failure | macrophage | *REEP5* | 1.224554364 | 0.001042352 | 0.014193744 | 1 |
| Heart_Failure | macrophage | *MAU2* | 0.751497837 | 0.013454617 | 0.049269285 | 1 |
| Heart_Failure | macrophage | *PHF10* | 0.978555645 | 0.005967269 | 0.029364287 | 1 |
| Heart_Failure | macrophage | *XPO7* | 1.141837259 | 0.002647474 | 0.018762541 | 1 |
| Heart_Failure | macrophage | *TULP4* | 0.747055208 | 0.000994156 | 0.013998911 | 1 |
| Heart_Failure | macrophage | *ACTN4* | 0.825908281 | 0.001414852 | 0.015044204 | 1 |
| Heart_Failure | macrophage | *ATXN10* | 0.93274323 | 0.001263839 | 0.014557357 | 1 |
| Heart_Failure | macrophage | *PNPLA7* | 0.76510917 | 0.006946256 | 0.032406957 | 1 |
| Heart_Failure | macrophage | *PAK4* | 1.124175718 | 0.005293399 | 0.027410427 | 1 |
| Heart_Failure | macrophage | *ZNF337* | 0.657645455 | 0.003195898 | 0.020758363 | 1 |
| Heart_Failure | macrophage | *MAP3K10* | 0.659590396 | 0.013477065 | 0.049314269 | 1 |
| Heart_Failure | macrophage | *C12orf65* | 0.78479808 | 0.000232911 | 0.012453892 | 1 |
| Heart_Failure | macrophage | *SLC35D2* | 1.571242122 | 0.000207265 | 0.012453892 | 1 |
| Heart_Failure | macrophage | *TTLL9* | 1.631534262 | 0.010824456 | 0.042838713 | 1 |
| Heart_Failure | macrophage | *ATP6V1E1* | 0.975847341 | 0.004496181 | 0.024925587 | 1 |
| Heart_Failure | macrophage | *CHMP1A* | 1.913526364 | 0.000104572 | 0.009328149 | 0.507384014 |
| Heart_Failure | macrophage | *IDO1* | 2.592615227 | 0.002502555 | 0.018350936 | 1 |
| Heart_Failure | macrophage | *MRPS25* | 0.872771748 | 0.001368016 | 0.014915987 | 1 |
| Heart_Failure | macrophage | *PDLIM4* | 0.865984872 | 0.012016724 | 0.045586508 | 1 |
| Heart_Failure | macrophage | *KIF3A* | 0.826281875 | 0.002160711 | 0.017199722 | 1 |
| Heart_Failure | macrophage | *NDUFA2* | 0.604845986 | 0.005236067 | 0.02725901 | 1 |
| Heart_Failure | macrophage | *EXOC4* | 0.828298043 | 0.00039039 | 0.013744636 | 1 |
| Heart_Failure | macrophage | *C1orf159* | 1.453098355 | 0.002452991 | 0.018310635 | 1 |
| Heart_Failure | macrophage | *PPFIA1* | 1.006471803 | 0.000851637 | 0.013744636 | 1 |
| Heart_Failure | macrophage | *MAP1B* | 0.440201989 | 0.002417037 | 0.018153971 | 1 |
| Heart_Failure | macrophage | *PRKAB2* | 1.277236123 | 0.005029691 | 0.026720217 | 1 |
| Heart_Failure | macrophage | *SPATA6* | 0.603426405 | 0.006675778 | 0.031693613 | 1 |
| Heart_Failure | macrophage | *IMMT* | 0.881668399 | 0.002587615 | 0.018490589 | 1 |
| Heart_Failure | macrophage | *SERPINF1* | 0.682962648 | 0.008804642 | 0.037452304 | 1 |
| Heart_Failure | macrophage | *MUTYH* | 0.700863692 | 0.003414134 | 0.02142726 | 1 |
| Heart_Failure | macrophage | *CTNNBL1* | 0.954346498 | 0.009084406 | 0.038236046 | 1 |
| Heart_Failure | macrophage | *AP3B1* | 0.930263707 | 0.002706894 | 0.019034563 | 1 |
| Heart_Failure | macrophage | *PEMT* | 0.865949446 | 0.013505156 | 0.049358108 | 1 |
| Heart_Failure | macrophage | *SCO1* | 0.847594394 | 0.003136852 | 0.020604359 | 1 |
| Heart_Failure | macrophage | *MPRIP* | 0.690785627 | 0.006023462 | 0.029550899 | 1 |
| Heart_Failure | macrophage | *DSTYK* | 1.376557262 | 0.000711002 | 0.013744636 | 1 |
| Heart_Failure | macrophage | *COG6* | 0.795665901 | 0.00197137 | 0.016575738 | 1 |
| Heart_Failure | macrophage | *BTBD2* | 1.075501465 | 0.002029687 | 0.01674837 | 1 |

| | | | | | | |
|---|---|---|---|---|---|---|
| Heart_Failure | macrophage | *CSNK1G2* | 1.816331017 | 0.000706574 | 0.013744636 | 1 |
| Heart_Failure | macrophage | *IPO8* | 1.168709358 | 0.0023439 | 0.017825399 | 1 |
| Heart_Failure | macrophage | *RRAS2* | 0.444824161 | 0.009758162 | 0.039687011 | 1 |
| Heart_Failure | macrophage | *ELP3* | 1.189741021 | 0.002121637 | 0.017128099 | 1 |
| Heart_Failure | macrophage | *THUMPD3* | 1.105061391 | 0.012591803 | 0.047068896 | 1 |
| Heart_Failure | macrophage | *CEPT1* | 0.852814021 | 0.005957112 | 0.029344068 | 1 |
| Heart_Failure | macrophage | *PPHLN1* | 0.924072711 | 0.001472212 | 0.015102525 | 1 |
| Heart_Failure | macrophage | *FKBP11* | 1.483988913 | 0.003531835 | 0.021857733 | 1 |
| Heart_Failure | macrophage | *YWHAQ* | 0.995025224 | 0.003780597 | 0.022860676 | 1 |
| Heart_Failure | macrophage | *KIDINS220* | 0.978756933 | 0.001036806 | 0.014193744 | 1 |
| Heart_Failure | macrophage | *KIAA1468* | 0.945319905 | 0.005002768 | 0.026720217 | 1 |
| Heart_Failure | macrophage | *IL15RA* | 1.126884813 | 0.000810728 | 0.013744636 | 1 |
| Heart_Failure | macrophage | *ELP2* | 0.888102033 | 0.002529084 | 0.018350936 | 1 |
| Heart_Failure | macrophage | *TPGS2* | 1.123382403 | 0.001546935 | 0.015218954 | 1 |
| Heart_Failure | macrophage | *TIMM10* | 1.288505451 | 0.001094497 | 0.014193744 | 1 |
| Heart_Failure | macrophage | *CLOCK* | 1.1550637 | 0.000804676 | 0.013744636 | 1 |
| Heart_Failure | macrophage | *ARGLU1* | 0.661082987 | 0.002196774 | 0.017199722 | 1 |
| Heart_Failure | macrophage | *ERCC5* | 1.202641885 | 0.00082049 | 0.013744636 | 1 |
| Heart_Failure | macrophage | *UBQLN1* | 1.096574431 | 0.001354969 | 0.014807007 | 1 |
| Heart_Failure | macrophage | *ADAM19* | 0.401543791 | 0.000665835 | 0.013744636 | 1 |
| Heart_Failure | macrophage | *DMTF1* | 0.708937891 | 0.00637553 | 0.030886628 | 1 |
| Heart_Failure | macrophage | *SRPK2* | 0.786527883 | 0.006807313 | 0.031912157 | 1 |
| Heart_Failure | macrophage | *AKIRIN2* | 1.100757533 | 0.003611888 | 0.022174968 | 1 |
| Heart_Failure | macrophage | *MAP3K7* | 1.201548654 | 0.001592281 | 0.015513549 | 1 |
| Heart_Failure | macrophage | *PHF21A* | 0.980654666 | 0.000799161 | 0.013744636 | 1 |
| Heart_Failure | macrophage | *CAPRIN1* | 0.945981294 | 0.00594938 | 0.029335765 | 1 |
| Heart_Failure | macrophage | *ATP5G2* | 1.007141854 | 0.004464013 | 0.024908187 | 1 |
| Heart_Failure | macrophage | *MICAL1* | 0.363040252 | 0.01177098 | 0.045112791 | 1 |
| Heart_Failure | macrophage | *USP15* | 0.953400155 | 0.001277499 | 0.014557357 | 1 |
| Heart_Failure | macrophage | *MPHOSPH6* | 1.396166536 | 0.003418116 | 0.02142726 | 1 |
| Heart_Failure | macrophage | *NTPCR* | 1.326953743 | 0.001809539 | 0.016138719 | 1 |
| Heart_Failure | macrophage | *PIGC* | 0.923415649 | 0.003783414 | 0.022860676 | 1 |
| Heart_Failure | macrophage | *MRPL44* | 2.628775625 | 2.36E-05 | 0.003694574 | 0.114531794 |
| Heart_Failure | macrophage | *USP37* | 1.086844461 | 8.23E-05 | 0.008480527 | 0.399536876 |
| Heart_Failure | macrophage | *SLC19A3* | 0.355676131 | 0.012555894 | 0.046970854 | 1 |
| Heart_Failure | macrophage | *TMBIM1* | 1.307011305 | 0.002822332 | 0.01956279 | 1 |
| Heart_Failure | macrophage | *GCC2* | 0.739149036 | 0.00924854 | 0.038485348 | 1 |
| Heart_Failure | macrophage | *ANKRD36* | 0.808099213 | 0.004741867 | 0.025802467 | 1 |
| Heart_Failure | macrophage | *EPC2* | 1.135951717 | 0.000483426 | 0.013744636 | 1 |
| Heart_Failure | macrophage | *TBC1D4* | 0.695360635 | 0.005033312 | 0.026720217 | 1 |
| Heart_Failure | macrophage | *SUCLA2* | 1.022050711 | 0.002017377 | 0.016703608 | 1 |
| Heart_Failure | macrophage | *SETDB2* | 0.943471429 | 0.007277211 | 0.033436578 | 1 |
| Heart_Failure | macrophage | *KDELR2* | 1.693810828 | 0.00115957 | 0.014216721 | 1 |
| Heart_Failure | macrophage | *NUPL2* | 1.197897339 | 0.002812279 | 0.01956279 | 1 |
| Heart_Failure | macrophage | *ZDHHC4* | 1.947740566 | 0.000239125 | 0.012453892 | 1 |
| Heart_Failure | macrophage | *NMT1* | 0.561955392 | 1.49E-05 | 0.002842801 | 0.072487422 |
| Heart_Failure | macrophage | *NDUFB5* | 1.025575166 | 0.004234834 | 0.024116686 | 1 |
| Heart_Failure | macrophage | *RPS6KC1* | 0.773189144 | 0.008334884 | 0.036075696 | 1 |
| Heart_Failure | macrophage | *ZNF189* | 1.764876753 | 0.000916304 | 0.013744636 | 1 |
| Heart_Failure | macrophage | *STX17* | 0.99641439 | 0.002480265 | 0.018350936 | 1 |

| | | | | | | |
|---|---|---|---|---|---|---|
| Heart_Failure | macrophage | *ANP32B* | 0.944229 | 0.012740892 | 0.047370736 | 1 |
| Heart_Failure | macrophage | *APTX* | 0.943166215 | 0.002876693 | 0.019714284 | 1 |
| Heart_Failure | macrophage | *GRHPR* | 0.454623626 | 0.013429518 | 0.049228836 | 1 |
| Heart_Failure | macrophage | *DENND4C* | 0.862705718 | 0.002163995 | 0.017199722 | 1 |
| Heart_Failure | macrophage | *FRS3* | 1.293274043 | 0.007391422 | 0.033664185 | 1 |
| Heart_Failure | macrophage | *TCF19* | 0.54914577 | 0.001284567 | 0.014557357 | 1 |
| Heart_Failure | macrophage | *FLOT1* | 0.667748156 | 0.001702702 | 0.015706292 | 1 |
| Heart_Failure | macrophage | *NUMA1* | 0.966558399 | 0.002169268 | 0.017199722 | 1 |
| Heart_Failure | macrophage | *NARS2* | 1.202444874 | 0.000894242 | 0.013744636 | 1 |
| Heart_Failure | macrophage | *SULF1* | 0.299142275 | 0.006408806 | 0.030961419 | 1 |
| Heart_Failure | macrophage | *TGS1* | 1.282448657 | 0.000355418 | 0.013744636 | 1 |
| Heart_Failure | macrophage | *NEK1* | 0.760750526 | 0.003305202 | 0.021101106 | 1 |
| Heart_Failure | macrophage | *MAP2K5* | 1.108915118 | 0.000216571 | 0.012453892 | 1 |
| Heart_Failure | macrophage | *PARP6* | 1.257677578 | 0.001062746 | 0.014193744 | 1 |
| Heart_Failure | macrophage | *ADAM10* | 1.348950068 | 1.09E-05 | 0.002404114 | 0.052890506 |
| Heart_Failure | macrophage | *ZNF280D* | 0.874024908 | 0.000840554 | 0.013744636 | 1 |
| Heart_Failure | macrophage | *DBT* | 0.863845722 | 0.007241968 | 0.033337789 | 1 |
| Heart_Failure | macrophage | *RTCA* | 0.883275895 | 0.007106186 | 0.032868649 | 1 |
| Heart_Failure | macrophage | *DYNC2LI1* | 0.43091141 | 0.012243344 | 0.046193395 | 1 |
| Heart_Failure | macrophage | *LRPPRC* | 1.102909801 | 0.001083873 | 0.014193744 | 1 |
| Heart_Failure | macrophage | *CALHM2* | 0.734236527 | 0.003933041 | 0.023296977 | 1 |
| Heart_Failure | macrophage | *ARL3* | 0.535250027 | 0.007599381 | 0.034083874 | 1 |
| Heart_Failure | macrophage | *RPS24* | 0.835667634 | 0.002000007 | 0.016673602 | 1 |
| Heart_Failure | macrophage | *SMARCAL1* | 1.108212775 | 0.002999792 | 0.020257391 | 1 |
| Heart_Failure | macrophage | *STAT4* | 0.927721071 | 0.00707625 | 0.032849883 | 1 |
| Heart_Failure | macrophage | *FASTKD1* | 0.96281087 | 0.000965307 | 0.013998911 | 1 |
| Heart_Failure | macrophage | *ITGAV* | 0.922291793 | 0.003750482 | 0.022718277 | 1 |
| Heart_Failure | macrophage | *SLC35A5* | 0.814490739 | 0.000901133 | 0.013744636 | 1 |
| Heart_Failure | macrophage | *SENP7* | 0.807048661 | 0.006086909 | 0.029741874 | 1 |
| Heart_Failure | macrophage | *COX17* | 1.549142829 | 0.004049844 | 0.023665742 | 1 |
| Heart_Failure | macrophage | *AP1AR* | 1.810290561 | 0.000212123 | 0.012453892 | 1 |
| Heart_Failure | macrophage | *KIAA1109* | 0.991716725 | 0.000831976 | 0.013744636 | 1 |
| Heart_Failure | macrophage | *G3BP2* | 0.795270839 | 0.000307145 | 0.013632389 | 1 |
| Heart_Failure | macrophage | *CNOT6L* | 0.83460193 | 0.00407467 | 0.02376238 | 1 |
| Heart_Failure | macrophage | *PPA2* | 0.937542761 | 0.001868286 | 0.016362677 | 1 |
| Heart_Failure | macrophage | *PAPSS1* | 0.983138775 | 0.001898401 | 0.016377125 | 1 |
| Heart_Failure | macrophage | *ZCRB1* | 0.838703702 | 0.012550844 | 0.046970854 | 1 |
| Heart_Failure | macrophage | *POC1B* | 1.103749513 | 0.005723867 | 0.028779485 | 1 |
| Heart_Failure | macrophage | *GLTP* | 2.024357746 | 0.001267384 | 0.014557357 | 1 |
| Heart_Failure | macrophage | *CCDC65* | 0.767088001 | 0.011384224 | 0.044189004 | 1 |
| Heart_Failure | macrophage | *ACVRL1* | 1.848803211 | 0.001504 | 0.015102525 | 1 |
| Heart_Failure | macrophage | *LMBR1L* | 0.955082526 | 0.001927447 | 0.016464742 | 1 |
| Heart_Failure | macrophage | *ESYT1* | 1.340268857 | 0.00012562 | 0.01024973 | 0.609508372 |
| Heart_Failure | macrophage | *RB1* | 0.834276477 | 0.002087055 | 0.016973572 | 1 |
| Heart_Failure | macrophage | *NOVA1* | 0.165610972 | 0.012405094 | 0.046550285 | 1 |
| Heart_Failure | macrophage | *TMX1* | 1.146813245 | 0.00655792 | 0.031348795 | 1 |
| Heart_Failure | macrophage | *C14orf37* | 0.779005399 | 0.002969026 | 0.020092244 | 1 |
| Heart_Failure | macrophage | *RAB15* | 1.041110746 | 0.010379223 | 0.041551148 | 1 |
| Heart_Failure | macrophage | *C14orf79* | 1.04843292 | 0.000378307 | 0.013744636 | 1 |
| Heart_Failure | macrophage | *SERF2* | 0.99787598 | 0.00357899 | 0.022037131 | 1 |

| | | | | | | |
|---|---|---|---|---|---|---|
| Heart_Failure | macrophage | *HMG20A* | 1.347033759 | 0.000668599 | 0.013744636 | 1 |
| Heart_Failure | macrophage | *ABHD2* | 0.955025363 | 0.006432772 | 0.030965061 | 1 |
| Heart_Failure | macrophage | *UNC45A* | 1.044903871 | 0.002874383 | 0.019714284 | 1 |
| Heart_Failure | macrophage | *CRTC3* | 0.587487347 | 0.002121609 | 0.017128099 | 1 |
| Heart_Failure | macrophage | *ITGAX* | 1.507510193 | 0.011826289 | 0.045253276 | 1 |
| Heart_Failure | macrophage | *DHX38* | 1.488640758 | 0.001084737 | 0.014193744 | 1 |
| Heart_Failure | macrophage | *NUDT7* | 0.996771908 | 0.011756638 | 0.045098774 | 1 |
| Heart_Failure | macrophage | *GCSH* | 0.503944275 | 0.009926664 | 0.040184219 | 1 |
| Heart_Failure | macrophage | *ZCCHC14* | 0.870757078 | 0.005165115 | 0.027066078 | 1 |
| Heart_Failure | macrophage | *TLDC1* | 0.804866992 | 0.013727979 | 0.049968608 | 1 |
| Heart_Failure | macrophage | *TIGD7* | 0.788331319 | 0.011576193 | 0.044511141 | 1 |
| Heart_Failure | macrophage | *TCF25* | 1.114875479 | 1.77E-05 | 0.003064185 | 0.085797187 |
| Heart_Failure | macrophage | *GALNS* | 1.04416393 | 0.001941558 | 0.016498145 | 1 |
| Heart_Failure | macrophage | *NCOR1* | 0.773203768 | 0.001866642 | 0.016362677 | 1 |
| Heart_Failure | macrophage | *KSR1* | 1.077862468 | 0.001374254 | 0.014947577 | 1 |
| Heart_Failure | macrophage | *MYO19* | 1.169567037 | 0.000243473 | 0.012453892 | 1 |
| Heart_Failure | macrophage | *SSH2* | 0.898190556 | 0.001559689 | 0.015288099 | 1 |
| Heart_Failure | macrophage | *IMPA2* | 1.627245666 | 2.77E-05 | 0.003955379 | 0.134196068 |
| Heart_Failure | macrophage | *GALNT1* | 0.901547073 | 0.004779383 | 0.02591013 | 1 |
| Heart_Failure | macrophage | *NPC1* | 0.905304076 | 0.007704679 | 0.03432792 | 1 |
| Heart_Failure | macrophage | *ANAPC11* | 1.507472808 | 0.005251796 | 0.027282348 | 1 |
| Heart_Failure | macrophage | *TBCD* | 0.948314156 | 0.002260318 | 0.01746348 | 1 |
| Heart_Failure | macrophage | *NARF* | 1.134840466 | 0.000290158 | 0.013375324 | 1 |
| Heart_Failure | macrophage | *ZNF750* | 1.582569863 | 0.005784155 | 0.028902903 | 1 |
| Heart_Failure | macrophage | *MAPK4* | 4.245366103 | 0.002652073 | 0.018762541 | 1 |
| Heart_Failure | macrophage | *ERBB2* | 1.051680218 | 7.09E-06 | 0.00180937 | 0.034378039 |
| Heart_Failure | macrophage | *PNMT* | 2.157687465 | 7.69E-13 | 9.33E-10 | 3.73E-09 |
| Heart_Failure | macrophage | *NFIC* | 0.920666149 | 0.000703467 | 0.013744636 | 1 |
| Heart_Failure | macrophage | *DPP9* | 1.293189681 | 0.003837117 | 0.023041698 | 1 |
| Heart_Failure | macrophage | *SIRT3* | 0.824630794 | 0.001394435 | 0.015035108 | 1 |
| Heart_Failure | macrophage | *EVI5L* | 1.992733218 | 0.000805345 | 0.013744636 | 1 |
| Heart_Failure | macrophage | *RERE* | 0.802099398 | 0.001030766 | 0.014193744 | 1 |
| Heart_Failure | macrophage | *PEX14* | 0.529936687 | 0.004826307 | 0.026058178 | 1 |
| Heart_Failure | macrophage | *EVA1B* | 0.788720525 | 2.89E-06 | 0.000877135 | 0.014034155 |
| Heart_Failure | macrophage | *RPS8* | 0.950990829 | 0.001437801 | 0.015100028 | 1 |
| Heart_Failure | macrophage | *STRIP1* | 1.922164059 | 0.000230736 | 0.012453892 | 1 |
| Heart_Failure | macrophage | *ALDH9A1* | 1.577993187 | 0.00153434 | 0.01520515 | 1 |
| Heart_Failure | macrophage | *RFWD2* | 0.912065526 | 0.001698622 | 0.015698506 | 1 |
| Heart_Failure | macrophage | *SETDB1* | 0.936669324 | 0.009159748 | 0.038340073 | 1 |
| Heart_Failure | macrophage | *POGZ* | 0.675545971 | 0.005455752 | 0.027888457 | 1 |
| Heart_Failure | macrophage | *SMYD2* | 1.600053903 | 0.00110806 | 0.014193744 | 1 |
| Heart_Failure | macrophage | *RAB13* | 2.110610239 | 0.001199506 | 0.014405948 | 1 |
| Heart_Failure | macrophage | *C1orf43* | 1.306823064 | 0.000846567 | 0.013744636 | 1 |
| Heart_Failure | macrophage | *FLG* | -0.214851286 | 0.001681162 | 0.015642632 | 1 |
| Heart_Failure | macrophage | *SCCPDH* | 1.154857909 | 0.002129076 | 0.017128099 | 1 |
| Heart_Failure | macrophage | *CEP170* | 0.704363493 | 0.007297413 | 0.033497681 | 1 |
| Heart_Failure | macrophage | *ACP1* | 1.391948895 | 0.000854223 | 0.013744636 | 1 |
| Heart_Failure | macrophage | *CNIH4* | 1.24417216 | 0.002129165 | 0.017128099 | 1 |
| Heart_Failure | macrophage | *RHOB* | 1.580463675 | 0.00122204 | 0.014488552 | 1 |
| Heart_Failure | macrophage | *WDPCP* | 0.509768948 | 0.000210071 | 0.012453892 | 1 |

| | | | | | | |
|---|---|---|---|---|---|---|
| Heart_Failure | macrophage | *VPS54* | 1.013425024 | 0.001320448 | 0.014712122 | 1 |
| Heart_Failure | macrophage | *EXOC6B* | 0.92142859 | 0.000783833 | 0.013744636 | 1 |
| Heart_Failure | macrophage | *FBLN7* | 0.304610411 | 0.006774174 | 0.031902592 | 1 |
| Heart_Failure | macrophage | *NBEAL1* | 0.604552847 | 0.001565825 | 0.015317305 | 1 |
| Heart_Failure | macrophage | *SUMF1* | 0.968789728 | 0.000754349 | 0.013744636 | 1 |
| Heart_Failure | macrophage | *ACKR3* | 0.36116765 | 0.005853235 | 0.029038751 | 1 |
| Heart_Failure | macrophage | *EAF1* | 1.183784473 | 0.008020411 | 0.035281081 | 1 |
| Heart_Failure | macrophage | *ARL6IP5* | 1.148152736 | 0.002653998 | 0.018762541 | 1 |
| Heart_Failure | macrophage | *SCD5* | 0.93260536 | 0.008754013 | 0.037323787 | 1 |
| Heart_Failure | macrophage | *METTL14* | 0.988682033 | 0.011103824 | 0.043448188 | 1 |
| Heart_Failure | macrophage | *USP53* | 0.593927432 | 0.000459707 | 0.013744636 | 1 |
| Heart_Failure | macrophage | *CBR4* | 0.621512993 | 0.008765221 | 0.037338765 | 1 |
| Heart_Failure | macrophage | *MARCH6* | 0.979942909 | 0.000894495 | 0.013744636 | 1 |
| Heart_Failure | macrophage | *MYO10* | 0.569323354 | 0.011577067 | 0.044511141 | 1 |
| Heart_Failure | macrophage | *GTF2H2* | 2.673985154 | 0.000656168 | 0.013744636 | 1 |
| Heart_Failure | macrophage | *TSLP* | 0.315119326 | 1.39E-06 | 0.000532321 | 0.00676228 |
| Heart_Failure | macrophage | *FEM1C* | 1.870761861 | 0.000719552 | 0.013744636 | 1 |
| Heart_Failure | macrophage | *FBXO38* | 1.153304174 | 0.000978385 | 0.013998911 | 1 |
| Heart_Failure | macrophage | *BOD1* | 1.831808889 | 1.16E-05 | 0.002448568 | 0.056317053 |
| Heart_Failure | macrophage | *MYLK4* | 1.332181329 | 0.0009256 | 0.013744636 | 1 |
| Heart_Failure | macrophage | *TBC1D7* | 0.67898413 | 0.01 | 0.040399666 | 1 |
| Heart_Failure | macrophage | *GFOD1* | 0.718504152 | 0.009159605 | 0.038340073 | 1 |
| Heart_Failure | macrophage | *FAM193B* | 0.789780886 | 0.006788186 | 0.031902592 | 1 |
| Heart_Failure | macrophage | *RPL7L1* | 1.310965551 | 0.001210484 | 0.014466177 | 1 |
| Heart_Failure | macrophage | *IRAK1BP1* | 0.592485237 | 0.005847542 | 0.029038751 | 1 |
| Heart_Failure | macrophage | *PHIP* | 1.066630088 | 0.000472474 | 0.013744636 | 1 |
| Heart_Failure | macrophage | *PNRC1* | 1.509164164 | 0.000917852 | 0.013744636 | 1 |
| Heart_Failure | macrophage | *RNF217* | 0.869326374 | 0.001480399 | 0.015102525 | 1 |
| Heart_Failure | macrophage | *GNA12* | 0.740061185 | 0.005436994 | 0.027827314 | 1 |
| Heart_Failure | macrophage | *GIGYF1* | 0.92548118 | 0.0032207 | 0.0208381 | 1 |
| Heart_Failure | macrophage | *TMEM209* | 1.409966361 | 0.001243894 | 0.014557357 | 1 |
| Heart_Failure | macrophage | *EPHA1* | 5.179852218 | 0.001665432 | 0.015616438 | 1 |
| Heart_Failure | macrophage | *FBXO25* | 0.860194122 | 0.003951651 | 0.023296977 | 1 |
| Heart_Failure | macrophage | *CSGALNACT1* | 0.312246229 | 0.009659557 | 0.039451323 | 1 |
| Heart_Failure | macrophage | *BIN3* | 0.790126031 | 0.010577973 | 0.042172822 | 1 |
| Heart_Failure | macrophage | *TACC1* | 0.617719732 | 0.003503435 | 0.021783227 | 1 |
| Heart_Failure | macrophage | *RPL7* | 0.504023593 | 0.006717223 | 0.031761858 | 1 |
| Heart_Failure | macrophage | *LRP12* | 1.40769454 | 0.001474772 | 0.015102525 | 1 |
| Heart_Failure | macrophage | *AK3* | 0.673343565 | 0.0135937 | 0.049628769 | 1 |
| Heart_Failure | macrophage | *HAUS6* | 1.445383428 | 0.001400679 | 0.015035788 | 1 |
| Heart_Failure | macrophage | *ALAD* | 0.479073846 | 0.003361309 | 0.021346951 | 1 |
| Heart_Failure | macrophage | *REXO4* | 1.506647905 | 0.001242211 | 0.014557357 | 1 |
| Heart_Failure | macrophage | *SH3GLB2* | 1.082094528 | 0.007155364 | 0.033033135 | 1 |
| Heart_Failure | macrophage | *SEC16A* | 1.034964864 | 0.002782346 | 0.01942438 | 1 |
| Heart_Failure | macrophage | *RSU1* | 1.220427632 | 0.000376598 | 0.013744636 | 1 |
| Heart_Failure | macrophage | *FRA10AC1* | 0.639835619 | 0.003252862 | 0.020904487 | 1 |
| Heart_Failure | macrophage | *ADD3* | 0.609309215 | 0.00542184 | 0.02780842 | 1 |
| Heart_Failure | macrophage | *PAOX* | 1.435299063 | 0.013064276 | 0.048387685 | 1 |
| Heart_Failure | macrophage | *GAS2* | 0.843030289 | 0.006031201 | 0.029558978 | 1 |
| Heart_Failure | macrophage | *LIN7C* | 1.592899963 | 0.000995388 | 0.013998911 | 1 |

| | | | | | | |
|---|---|---|---|---|---|---|
| Heart_Failure | macrophage | *ENDOD1* | 0.89457649 | 0.009863641 | 0.040015373 | 1 |
| Heart_Failure | macrophage | *RPS3* | 0.892540987 | 0.001289846 | 0.014557357 | 1 |
| Heart_Failure | macrophage | *NPAT* | 1.399281649 | 0.001146534 | 0.014193744 | 1 |
| Heart_Failure | macrophage | *FADS1* | 1.030743383 | 0.00075061 | 0.013744636 | 1 |
| Heart_Failure | macrophage | *ROM1* | 0.35134025 | 0.000485388 | 0.013744636 | 1 |
| Heart_Failure | macrophage | *CPSF7* | 1.046042843 | 0.001182196 | 0.014375975 | 1 |
| Heart_Failure | macrophage | *ALDOA* | 1.240130813 | 3.62E-05 | 0.004748912 | 0.175709752 |
| Heart_Failure | macrophage | *TMEM218* | 0.94893424 | 0.007478673 | 0.03381782 | 1 |
| Heart_Failure | macrophage | *SAP18* | 1.522804852 | 0.002151553 | 0.017198249 | 1 |
| Heart_Failure | macrophage | *FAM173B* | 0.878708174 | 0.007032024 | 0.032681399 | 1 |
| Heart_Failure | macrophage | *TMEM18* | 2.007702861 | 0.000718605 | 0.013744636 | 1 |
| Heart_Failure | macrophage | *NDUFC2* | 1.875339554 | 0.000979175 | 0.013998911 | 1 |
| Heart_Failure | macrophage | *NUBPL* | 1.092397449 | 0.000815964 | 0.013744636 | 1 |
| Heart_Failure | macrophage | *VIPAS39* | 1.175835876 | 0.008730933 | 0.037290923 | 1 |
| Heart_Failure | macrophage | *ANKRD50* | 1.453955889 | 0.000163982 | 0.011875236 | 0.795640836 |
| Heart_Failure | macrophage | *UPF2* | 0.941158512 | 0.001472521 | 0.015102525 | 1 |
| Heart_Failure | macrophage | *FAM160B1* | 0.726206694 | 0.007849321 | 0.034844376 | 1 |
| Heart_Failure | macrophage | *PIGF* | 1.405433499 | 0.000806955 | 0.013744636 | 1 |
| Heart_Failure | macrophage | *MFSD6* | 1.044362695 | 0.002151389 | 0.017198249 | 1 |
| Heart_Failure | macrophage | *BICD1* | 1.225977155 | 0.000709137 | 0.013744636 | 1 |
| Heart_Failure | macrophage | *TIAL1* | 1.146252119 | 0.001451845 | 0.015102525 | 1 |
| Heart_Failure | macrophage | *EPG5* | 1.165730235 | 0.001541825 | 0.01520515 | 1 |
| Heart_Failure | macrophage | *CWF19L2* | 0.720407218 | 0.003108592 | 0.020493053 | 1 |
| Heart_Failure | macrophage | *NMT2* | 0.523940873 | 0.006413072 | 0.030961419 | 1 |
| Heart_Failure | macrophage | *SPARCL1* | 0.456468132 | 0.008849743 | 0.03747284 | 1 |
| Heart_Failure | macrophage | *MBNL1* | 0.822344628 | 0.000712849 | 0.013744636 | 1 |
| Heart_Failure | macrophage | *GGPS1* | 0.778158658 | 0.005667354 | 0.0285249 | 1 |
| Heart_Failure | macrophage | *ZNF117* | 0.097122425 | 0.007380144 | 0.033654566 | 1 |
| Heart_Failure | macrophage | *MED21* | 1.075498993 | 0.001192945 | 0.014405948 | 1 |
| Heart_Failure | macrophage | *JAKMIP1* | 1.292236544 | 0.005768513 | 0.028868296 | 1 |
| Heart_Failure | macrophage | *SREK1IP1* | 1.248588935 | 0.00232343 | 0.017739426 | 1 |
| Heart_Failure | macrophage | *CAST* | 0.521685385 | 0.01155359 | 0.044511141 | 1 |
| Heart_Failure | macrophage | *TMEM87B* | 1.278458247 | 0.00093217 | 0.013744636 | 1 |
| Heart_Failure | macrophage | *CFDP1* | 0.962961434 | 0.0012782 | 0.014557357 | 1 |
| Heart_Failure | macrophage | *PPP2R5E* | 1.152291232 | 0.000934817 | 0.013744636 | 1 |
| Heart_Failure | macrophage | *PRKCA* | 0.858649395 | 0.002087738 | 0.016973572 | 1 |
| Heart_Failure | macrophage | *ABCA5* | 0.412524756 | 0.005504368 | 0.027936396 | 1 |
| Heart_Failure | macrophage | *PGM5* | 0.719630109 | 0.007151029 | 0.033033135 | 1 |
| Heart_Failure | macrophage | *XPC* | 0.903701669 | 0.006273083 | 0.030522744 | 1 |
| Heart_Failure | macrophage | *FLCN* | 0.673844828 | 0.002962153 | 0.020092244 | 1 |
| Heart_Failure | macrophage | *OXNAD1* | 0.63790368 | 0.0095691 | 0.039329411 | 1 |
| Heart_Failure | macrophage | *CNOT8* | 1.094086305 | 0.006439354 | 0.030965061 | 1 |
| Heart_Failure | macrophage | *NUP205* | 1.209262611 | 0.005534402 | 0.027942684 | 1 |
| Heart_Failure | macrophage | *FZD7* | 0.575161378 | 0.000202397 | 0.012453892 | 0.982032344 |
| Heart_Failure | macrophage | *TMBIM4* | 0.899118546 | 0.000281211 | 0.013225747 | 1 |
| Heart_Failure | macrophage | *C14orf2* | 1.091914082 | 0.000146496 | 0.011106235 | 0.710799046 |
| Heart_Failure | macrophage | *RPL30* | 1.061315525 | 0.00010574 | 0.009328149 | 0.513048195 |
| Heart_Failure | macrophage | *ZDHHC5* | 1.65145537 | 0.000712747 | 0.013744636 | 1 |
| Heart_Failure | macrophage | *FBRS* | 0.675522915 | 0.013029398 | 0.048319694 | 1 |
| Heart_Failure | macrophage | *GHRL* | 0.697179055 | 0.009039663 | 0.038106383 | 1 |

| | | | | | | |
|---|---|---|---|---|---|---|
| Heart_Failure | macrophage | *MMP14* | 1.693653313 | 0.000417025 | 0.013744636 | 1 |
| Heart_Failure | macrophage | *GATAD1* | 0.927465292 | 0.005171116 | 0.027066078 | 1 |
| Heart_Failure | macrophage | *LCA5L* | 0.271559291 | 0.005017364 | 0.026720217 | 1 |
| Heart_Failure | macrophage | *SLC38A10* | 0.902101971 | 0.012634267 | 0.04715497 | 1 |
| Heart_Failure | macrophage | *AP3S2* | 0.887981062 | 0.009912574 | 0.040180291 | 1 |
| Heart_Failure | macrophage | *SPPL3* | 1.071428936 | 0.000273123 | 0.012992079 | 1 |
| Heart_Failure | macrophage | *DZIP1L* | 0.689476051 | 0.010997685 | 0.043347494 | 1 |
| Heart_Failure | macrophage | *MITD1* | 1.176716141 | 0.000337042 | 0.013744636 | 1 |
| Heart_Failure | macrophage | *ELK4* | 1.395007326 | 0.001403795 | 0.015035788 | 1 |
| Heart_Failure | macrophage | *AMFR* | 1.101840348 | 0.001472944 | 0.015102525 | 1 |
| Heart_Failure | macrophage | *TMEM69* | 1.695140707 | 0.001667621 | 0.015616438 | 1 |
| Heart_Failure | macrophage | *CHCHD6* | 0.691339095 | 0.010330318 | 0.041389515 | 1 |
| Heart_Failure | macrophage | *CCDC107* | 1.00768997 | 0.004504634 | 0.024925587 | 1 |
| Heart_Failure | macrophage | *SSU72* | 1.242569945 | 0.003721264 | 0.022597713 | 1 |
| Heart_Failure | macrophage | *CCDC58* | 1.758301373 | 0.001150815 | 0.014193744 | 1 |
| Heart_Failure | macrophage | *TAOK1* | 0.990203074 | 0.00204805 | 0.016835648 | 1 |
| Heart_Failure | macrophage | *VPS11* | 0.728981465 | 0.001150542 | 0.014193744 | 1 |
| Heart_Failure | macrophage | *FDPS* | 1.034149165 | 9.10E-05 | 0.009013271 | 0.441650256 |
| Heart_Failure | macrophage | *FCRL3* | -0.279328551 | 0.008807312 | 0.037452304 | 1 |
| Heart_Failure | macrophage | *FGFR4* | -1.23415746 | 0.003396367 | 0.02142726 | 1 |
| Heart_Failure | macrophage | *TONSL* | 0.772298341 | 0.009930107 | 0.040184219 | 1 |
| Heart_Failure | macrophage | *ZNF333* | 1.608670622 | 0.000501616 | 0.013744636 | 1 |
| Heart_Failure | macrophage | *RPL8* | 1.156494453 | 0.000784866 | 0.013744636 | 1 |
| Heart_Failure | macrophage | *TMEM143* | 1.358254181 | 0.003058492 | 0.020448353 | 1 |
| Heart_Failure | macrophage | *LEMD2* | 1.429181091 | 0.000232021 | 0.012453892 | 1 |
| Heart_Failure | macrophage | *COA7* | 2.401309053 | 0.002560649 | 0.018426459 | 1 |
| Heart_Failure | macrophage | *USP24* | 0.832769326 | 0.008145951 | 0.035607345 | 1 |
| Heart_Failure | macrophage | *ZSWIM5* | 0.485478935 | 0.007088588 | 0.032849883 | 1 |
| Heart_Failure | macrophage | *RBP7* | 1.366999678 | 0.00015407 | 0.011326447 | 0.747545509 |
| Heart_Failure | macrophage | *TM2D1* | 1.146987925 | 0.00084544 | 0.013744636 | 1 |
| Heart_Failure | macrophage | *NEXN* | 0.507721226 | 0.00836189 | 0.036128131 | 1 |
| Heart_Failure | macrophage | *WDR63* | 0.283451024 | 0.012679379 | 0.047248444 | 1 |
| Heart_Failure | macrophage | *KLHDC9* | -0.285522337 | 0.003975149 | 0.02337213 | 1 |
| Heart_Failure | macrophage | *ACP6* | 0.952869249 | 0.009410333 | 0.038891769 | 1 |
| Heart_Failure | macrophage | *CNST* | 1.124805797 | 0.000847702 | 0.013744636 | 1 |
| Heart_Failure | macrophage | *HAAO* | 1.448810906 | 0.000837027 | 0.013744636 | 1 |
| Heart_Failure | macrophage | *KIAA1841* | 1.096983935 | 0.003373185 | 0.021366443 | 1 |
| Heart_Failure | macrophage | *NUP35* | 1.137188326 | 0.001713781 | 0.015726742 | 1 |
| Heart_Failure | macrophage | *NOSTRIN* | 0.578819528 | 0.000374302 | 0.013744636 | 1 |
| Heart_Failure | macrophage | *PACRGL* | 0.748989761 | 0.013337392 | 0.04909272 | 1 |
| Heart_Failure | macrophage | *VPS72* | 1.178162014 | 0.004206914 | 0.024042342 | 1 |
| Heart_Failure | macrophage | *ERCC3* | 1.222386998 | 0.000213313 | 0.012453892 | 1 |
| Heart_Failure | macrophage | *DHX57* | 1.200117813 | 0.001549498 | 0.015218954 | 1 |
| Heart_Failure | macrophage | *ANTXR2* | 0.629531014 | 0.003187889 | 0.020758363 | 1 |
| Heart_Failure | macrophage | *CGGBP1* | 0.246277077 | 0.013385041 | 0.049162922 | 1 |
| Heart_Failure | macrophage | *PBXIP1* | 1.735320252 | 0.000399901 | 0.013744636 | 1 |
| Heart_Failure | macrophage | *LRRC58* | 1.239497001 | 0.002184741 | 0.017199722 | 1 |
| Heart_Failure | macrophage | *SUCLG1* | 0.865690833 | 0.000658436 | 0.013744636 | 1 |
| Heart_Failure | macrophage | *EFHB* | 0.151694911 | 0.001723155 | 0.015726742 | 1 |
| Heart_Failure | macrophage | *GTPBP8* | 1.690449659 | 0.001112029 | 0.014193744 | 1 |

| | | | | | | |
|---|---|---|---|---|---|---|
| Heart_Failure | macrophage | *C4orf36* | 1.028157383 | 0.003971311 | 0.02337213 | 1 |
| Heart_Failure | macrophage | *TIPARP* | 1.307427957 | 0.001324293 | 0.014712122 | 1 |
| Heart_Failure | macrophage | *RPP14* | 1.131851644 | 0.008900204 | 0.03761654 | 1 |
| Heart_Failure | macrophage | *IL17RC* | 0.708881715 | 0.005531498 | 0.027942684 | 1 |
| Heart_Failure | macrophage | *CRELD1* | 1.019127385 | 0.003238237 | 0.0208381 | 1 |
| Heart_Failure | macrophage | *TTC14* | 0.620119934 | 0.006710987 | 0.031761858 | 1 |
| Heart_Failure | macrophage | *CPA3* | 1.151476065 | 0.010619933 | 0.042305347 | 1 |
| Heart_Failure | macrophage | *SLC4A1AP* | 1.665413592 | 0.00088965 | 0.013744636 | 1 |
| Heart_Failure | macrophage | *CLEC3B* | 1.471739082 | 0.011030579 | 0.043388847 | 1 |
| Heart_Failure | macrophage | *SNIP1* | 1.720477337 | 0.000947655 | 0.0138913 | 1 |
| Heart_Failure | macrophage | *SFMBT1* | 1.437971177 | 0.002969114 | 0.020092244 | 1 |
| Heart_Failure | macrophage | *FAM208A* | 1.04835397 | 0.000887821 | 0.013744636 | 1 |
| Heart_Failure | macrophage | *ABLIM2* | 0.653256265 | 0.002189815 | 0.017199722 | 1 |
| Heart_Failure | macrophage | *RNF123* | 0.768261935 | 2.44E-07 | 0.0001318 | 0.0011862 |
| Heart_Failure | macrophage | *HSPA4L* | 0.527889584 | 0.011657543 | 0.044784162 | 1 |
| Heart_Failure | macrophage | *MFSD8* | 0.900600206 | 0.006709461 | 0.031761858 | 1 |
| Heart_Failure | macrophage | *MST1R* | 0.620390923 | 6.10E-08 | 3.70E-05 | 0.000295925 |
| Heart_Failure | macrophage | *HPGD* | 2.897744854 | 0.000881265 | 0.013744636 | 1 |
| Heart_Failure | macrophage | *ARFIP1* | 0.981005379 | 0.001498862 | 0.015102525 | 1 |
| Heart_Failure | macrophage | *MOCS2* | 1.629832967 | 0.00397885 | 0.02337213 | 1 |
| Heart_Failure | macrophage | *NDUFAF2* | 0.676931415 | 0.002967225 | 0.020092244 | 1 |
| Heart_Failure | macrophage | *PGGT1B* | 0.957535767 | 0.003413729 | 0.02142726 | 1 |
| Heart_Failure | macrophage | *CMBL* | 0.988231973 | 0.000351332 | 0.013744636 | 1 |
| Heart_Failure | macrophage | *NDUFS4* | 1.050303179 | 0.000739326 | 0.013744636 | 1 |
| Heart_Failure | macrophage | *SERINC5* | 1.292348551 | 1.06E-05 | 0.002404114 | 0.051669451 |
| Heart_Failure | macrophage | *ERAP1* | 0.628226847 | 0.002691007 | 0.018950311 | 1 |
| Heart_Failure | macrophage | *UTP15* | 1.522486501 | 0.003187605 | 0.020758363 | 1 |
| Heart_Failure | macrophage | *TLR3* | 1.252540069 | 0.009395465 | 0.038863422 | 1 |
| Heart_Failure | macrophage | *CCDC127* | 1.533298743 | 0.00013057 | 0.010385631 | 0.633523517 |
| Heart_Failure | macrophage | *DCBLD1* | 0.524138793 | 0.002940438 | 0.020009827 | 1 |
| Heart_Failure | macrophage | *PDSS2* | 1.092239483 | 0.000657508 | 0.013744636 | 1 |
| Heart_Failure | macrophage | *SAP30L* | 1.625371867 | 2.22E-05 | 0.00359475 | 0.107842493 |
| Heart_Failure | macrophage | *RPS14* | 0.979423495 | 0.00064945 | 0.013744636 | 1 |
| Heart_Failure | macrophage | *RP9* | 1.835423654 | 0.000927298 | 0.013744636 | 1 |
| Heart_Failure | macrophage | *USP49* | 1.575345946 | 0.000340411 | 0.013744636 | 1 |
| Heart_Failure | macrophage | *SLC13A4* | 0.738393544 | 0.009155755 | 0.038340073 | 1 |
| Heart_Failure | macrophage | *BRI3* | 0.842191308 | 0.013433425 | 0.049228836 | 1 |
| Heart_Failure | macrophage | *RAD21* | 1.095781418 | 0.000773211 | 0.013744636 | 1 |
| Heart_Failure | macrophage | *SUN1* | 0.467861285 | 0.009651978 | 0.039451323 | 1 |
| Heart_Failure | macrophage | *OXR1* | 0.823220872 | 0.001612979 | 0.015528127 | 1 |
| Heart_Failure | macrophage | *GPER1* | 0.852312698 | 0.000323593 | 0.013744636 | 1 |
| Heart_Failure | macrophage | *FASTK* | 1.581945256 | 0.001188711 | 0.014405948 | 1 |
| Heart_Failure | macrophage | *INTS8* | 1.39997358 | 0.000244163 | 0.012453892 | 1 |
| Heart_Failure | macrophage | *TMEM65* | 1.159893469 | 0.002325283 | 0.017739426 | 1 |
| Heart_Failure | macrophage | *TRMT10B* | 0.890447444 | 0.006187416 | 0.030172205 | 1 |
| Heart_Failure | macrophage | *WRN* | 0.748006011 | 0.002402958 | 0.018076208 | 1 |
| Heart_Failure | macrophage | *MARCH8* | 1.462534782 | 0.000384177 | 0.013744636 | 1 |
| Heart_Failure | macrophage | *CFL2* | 1.083456003 | 0.008223098 | 0.035840035 | 1 |
| Heart_Failure | macrophage | *GJB2* | 8.325973399 | 0.003301038 | 0.021101106 | 1 |
| Heart_Failure | macrophage | *MICU2* | 0.998800649 | 0.002804061 | 0.019547851 | 1 |

| | | | | | | |
|---|---|---|---|---|---|---|
| Heart_Failure | macrophage | *ATP5C1* | 0.890702229 | 0.011935865 | 0.045484252 | 1 |
| Heart_Failure | macrophage | *PDZD8* | 0.586640265 | 0.009735123 | 0.039626526 | 1 |
| Heart_Failure | macrophage | *QSOX2* | 1.176532374 | 0.009597219 | 0.039329411 | 1 |
| Heart_Failure | macrophage | *METTL3* | 1.411625084 | 0.000390468 | 0.013744636 | 1 |
| Heart_Failure | macrophage | *HSPA12A* | 0.340233284 | 0.001038809 | 0.014193744 | 1 |
| Heart_Failure | macrophage | *UBTD1* | 0.659203932 | 0.012685521 | 0.047248444 | 1 |
| Heart_Failure | macrophage | *TTC7B* | 0.983723377 | 0.001636348 | 0.015548043 | 1 |
| Heart_Failure | macrophage | *SLC39A13* | 0.838981661 | 0.013714412 | 0.049956703 | 1 |
| Heart_Failure | macrophage | *TCP11L2* | 0.778535222 | 0.00507528 | 0.026824898 | 1 |
| Heart_Failure | macrophage | *TRMT61A* | 1.292803551 | 0.001531042 | 0.01520515 | 1 |
| Heart_Failure | macrophage | *API5* | 1.419646739 | 0.000645845 | 0.013744636 | 1 |
| Heart_Failure | macrophage | *COX11* | 0.759494939 | 0.005382207 | 0.027717297 | 1 |
| Heart_Failure | macrophage | *STXBP4* | 0.856496404 | 0.003105815 | 0.020493053 | 1 |
| Heart_Failure | macrophage | *WBP1L* | 0.921365052 | 0.00429851 | 0.02433649 | 1 |
| Heart_Failure | macrophage | *WDR88* | 0.851825226 | 0.011530938 | 0.044509236 | 1 |
| Heart_Failure | macrophage | *RPL27A* | 1.33038666 | 0.000692467 | 0.013744636 | 1 |
| Heart_Failure | macrophage | *TMEM41B* | 1.136695384 | 0.002440087 | 0.018242378 | 1 |
| Heart_Failure | macrophage | *SLFN5* | 0.716771333 | 0.009599497 | 0.039329411 | 1 |
| Heart_Failure | macrophage | *SAAL1* | 2.168992837 | 0.000683613 | 0.013744636 | 1 |
| Heart_Failure | macrophage | *PIP4K2C* | 1.970416423 | 3.26E-06 | 0.000931215 | 0.015830648 |
| Heart_Failure | macrophage | *MTMR10* | 0.753341601 | 0.00386668 | 0.023161891 | 1 |
| Heart_Failure | macrophage | *EPB42* | 0.702625374 | 0.005841846 | 0.029038751 | 1 |
| Heart_Failure | macrophage | *SMAD3* | 0.551575436 | 0.003666962 | 0.022397116 | 1 |
| Heart_Failure | macrophage | *AKTIP* | 0.792126096 | 0.001892105 | 0.016377125 | 1 |
| Heart_Failure | macrophage | *CNPY4* | 0.57359392 | 0.011011166 | 0.043365403 | 1 |
| Heart_Failure | macrophage | *SP2* | 1.559591849 | 0.001160309 | 0.014216721 | 1 |
| Heart_Failure | macrophage | *FBXO22* | 1.114916075 | 0.001048885 | 0.014193744 | 1 |
| Heart_Failure | macrophage | *HDHD2* | 0.266956706 | 0.002397666 | 0.018064401 | 1 |
| Heart_Failure | macrophage | *ZNF91* | 0.733494975 | 0.001309869 | 0.014687862 | 1 |
| Heart_Failure | macrophage | *ZNF23* | 1.196482441 | 0.000729068 | 0.013744636 | 1 |
| Heart_Failure | macrophage | *VKORC1* | 2.283910237 | 0.000696388 | 0.013744636 | 1 |
| Heart_Failure | macrophage | *SPATA33* | 1.053173694 | 0.006826623 | 0.031971788 | 1 |
| Heart_Failure | macrophage | *RPL13* | 0.905003568 | 0.001582815 | 0.015452349 | 1 |
| Heart_Failure | macrophage | *ZNF528* | 1.156478098 | 0.001637423 | 0.015548043 | 1 |
| Heart_Failure | macrophage | *FAM57A* | 0.38452257 | 0.008358177 | 0.036128131 | 1 |
| Heart_Failure | macrophage | *TRPV3* | 0.340634774 | 0.004091454 | 0.023785825 | 1 |
| Heart_Failure | macrophage | *CYB5D2* | 0.938495543 | 0.003164226 | 0.02069114 | 1 |
| Heart_Failure | macrophage | *ZNF83* | 0.563560892 | 0.011404147 | 0.044195623 | 1 |
| Heart_Failure | macrophage | *NDUFV1* | 1.021843506 | 0.002507086 | 0.018350936 | 1 |
| Heart_Failure | macrophage | *PRDX2* | 1.10332296 | 0.004086335 | 0.023785825 | 1 |
| Heart_Failure | macrophage | *MIS12* | 1.032505497 | 0.004393906 | 0.024703628 | 1 |
| Heart_Failure | macrophage | *ATP5H* | 1.0807781 | 0.011065786 | 0.04340436 | 1 |
| Heart_Failure | macrophage | *GSDMA* | -0.338030383 | 1.43E-06 | 0.000532321 | 0.006920167 |
| Heart_Failure | macrophage | *MLST8* | 1.751125963 | 0.00168585 | 0.015642632 | 1 |
| Heart_Failure | macrophage | *COA6* | 1.930070272 | 0.004290978 | 0.02433649 | 1 |
| Heart_Failure | macrophage | *PDHB* | 0.975336067 | 0.005835982 | 0.029038751 | 1 |
| Heart_Failure | macrophage | *PCMTD1* | 1.001847564 | 0.00088666 | 0.013744636 | 1 |
| Heart_Failure | macrophage | *ING5* | 0.606463961 | 0.007944965 | 0.035058047 | 1 |
| Heart_Failure | macrophage | *RFWD3* | 1.117408077 | 0.002599702 | 0.018546041 | 1 |
| Heart_Failure | macrophage | *SLC20A2* | 0.754511308 | 0.003508455 | 0.021783227 | 1 |

| | | | | | | |
|---|---|---|---|---|---|---|
| Heart_Failure | macrophage | *TMUB2* | 1.08778336 | 0.001756521 | 0.015930167 | 1 |
| Heart_Failure | macrophage | *DNAJC21* | 1.541240128 | 0.000774802 | 0.013744636 | 1 |
| Heart_Failure | macrophage | *SNTB2* | 0.918175347 | 0.001602308 | 0.015528127 | 1 |
| Heart_Failure | macrophage | *STX18* | 1.301769553 | 0.000726306 | 0.013744636 | 1 |
| Heart_Failure | macrophage | *ZBTB49* | 1.915132501 | 0.002005382 | 0.016682333 | 1 |
| Heart_Failure | macrophage | *ATOH8* | 0.5158024 | 0.003343708 | 0.021290908 | 1 |
| Heart_Failure | macrophage | *TMEM129* | 0.759311307 | 0.005495085 | 0.027936396 | 1 |
| Heart_Failure | macrophage | *CEP120* | 1.366443912 | 0.000658582 | 0.013744636 | 1 |
| Heart_Failure | macrophage | *MFF* | 1.097780698 | 0.001040849 | 0.014193744 | 1 |
| Heart_Failure | macrophage | *RGS14* | -0.250724035 | 0.012115518 | 0.045853738 | 1 |
| Heart_Failure | macrophage | *PGM2* | 0.806898893 | 0.012733981 | 0.047370736 | 1 |
| Heart_Failure | macrophage | *SNUPN* | 0.686104676 | 0.002910593 | 0.019883862 | 1 |
| Heart_Failure | macrophage | *CCDC8* | 0.429705139 | 0.011557509 | 0.044511141 | 1 |
| Heart_Failure | macrophage | *HINT1* | 1.400747945 | 0.00055521 | 0.013744636 | 1 |
| Heart_Failure | macrophage | *DFFB* | 1.546758736 | 0.002385701 | 0.018030253 | 1 |
| Heart_Failure | macrophage | *NFU1* | 0.671120512 | 0.007861814 | 0.034867934 | 1 |
| Heart_Failure | macrophage | *LUZP1* | 1.21001207 | 0.001488399 | 0.015102525 | 1 |
| Heart_Failure | macrophage | *UGP2* | 0.611081034 | 0.001653502 | 0.015548043 | 1 |
| Heart_Failure | macrophage | *HNRNPF* | 0.826235072 | 0.007262271 | 0.033399563 | 1 |
| Heart_Failure | macrophage | *CSGALNACT2* | 1.295213751 | 0.000897944 | 0.013744636 | 1 |
| Heart_Failure | macrophage | *AVEN* | 0.887866985 | 0.004669612 | 0.02551459 | 1 |
| Heart_Failure | macrophage | *TOR1AIP2* | 0.964901714 | 0.001431795 | 0.015100028 | 1 |
| Heart_Failure | macrophage | *NIPA1* | 1.066786058 | 0.010561088 | 0.042140132 | 1 |
| Heart_Failure | macrophage | *HNRNPA3* | 0.834664712 | 0.001814737 | 0.016138719 | 1 |
| Heart_Failure | macrophage | *RNF150* | 0.511909262 | 0.011271564 | 0.043927413 | 1 |
| Heart_Failure | macrophage | *CHRNB1* | 0.214903732 | 0.000500169 | 0.013744636 | 1 |
| Heart_Failure | macrophage | *PDCD6IP* | 1.12160024 | 0.000904611 | 0.013744636 | 1 |
| Heart_Failure | macrophage | *STX8* | 1.119020623 | 0.000252432 | 0.012497964 | 1 |
| Heart_Failure | macrophage | *NFRKB* | 1.296412143 | 0.002913732 | 0.019883862 | 1 |
| Heart_Failure | macrophage | *MGMT* | 0.767886664 | 0.004957401 | 0.026548909 | 1 |
| Heart_Failure | macrophage | *SERPINB9* | 0.996819465 | 0.001481212 | 0.015102525 | 1 |
| Heart_Failure | macrophage | *STAT2* | 1.256918436 | 0.000571269 | 0.013744636 | 1 |
| Heart_Failure | macrophage | *SLC26A5* | 1.064350604 | 0.000479809 | 0.013744636 | 1 |
| Heart_Failure | macrophage | *TMEM133* | 0.466812492 | 0.005529806 | 0.027942684 | 1 |
| Heart_Failure | macrophage | *FOXN2* | 1.101231886 | 0.002503311 | 0.018350936 | 1 |
| Heart_Failure | macrophage | *MSANTD4* | 2.051699409 | 0.000339958 | 0.013744636 | 1 |
| Heart_Failure | macrophage | *NDUFA3* | 1.779339483 | 0.000585101 | 0.013744636 | 1 |
| Heart_Failure | macrophage | *FEZ2* | 0.881454545 | 0.00058007 | 0.013744636 | 1 |
| Heart_Failure | macrophage | *SOX7* | 1.024011213 | 0.011952229 | 0.045484252 | 1 |
| Heart_Failure | macrophage | *MFN1* | 0.688935989 | 0.00141322 | 0.015044204 | 1 |
| Heart_Failure | macrophage | *PDE7B* | 0.780444599 | 0.001123514 | 0.014193744 | 1 |
| Heart_Failure | macrophage | *MCC* | 0.693498569 | 0.00474357 | 0.025802467 | 1 |
| Heart_Failure | macrophage | *PPID* | 1.952515497 | 2.53E-05 | 0.003841802 | 0.122937679 |
| Heart_Failure | macrophage | *TBCA* | 1.154345328 | 0.000535344 | 0.013744636 | 1 |
| Heart_Failure | macrophage | *ANO5* | 0.77625938 | 0.004053222 | 0.023665742 | 1 |
| Heart_Failure | macrophage | *CAMTA1* | 0.686883961 | 0.008252129 | 0.035845418 | 1 |
| Heart_Failure | macrophage | *LGALS4* | 0.377791358 | 0.011471964 | 0.044387535 | 1 |
| Heart_Failure | macrophage | *TRAPPC12* | 1.055918477 | 0.002080646 | 0.016973572 | 1 |
| Heart_Failure | macrophage | *ATPAF2* | 0.880701064 | 0.000697242 | 0.013744636 | 1 |
| Heart_Failure | macrophage | *EIF2AK3* | 0.904952167 | 0.004179403 | 0.024038296 | 1 |

| | | | | | | |
|---|---|---|---|---|---|---|
| Heart_Failure | macrophage | *TEFM* | 0.883049616 | 0.00180027 | 0.016138719 | 1 |
| Heart_Failure | macrophage | *MALT1* | 1.323204339 | 0.000912545 | 0.013744636 | 1 |
| Heart_Failure | macrophage | *MACROD2* | 0.400799565 | 0.002566405 | 0.018426459 | 1 |
| Heart_Failure | macrophage | *BSG* | 1.266840126 | 0.000242418 | 0.012453892 | 1 |
| Heart_Failure | macrophage | *HINFP* | 1.059273101 | 0.007365948 | 0.03365309 | 1 |
| Heart_Failure | macrophage | *SUCLG2* | 1.088801509 | 0.000909714 | 0.013744636 | 1 |
| Heart_Failure | macrophage | *FGGY* | 0.735472875 | 0.003674383 | 0.022397116 | 1 |
| Heart_Failure | macrophage | *MANEA* | 0.966659077 | 0.005653975 | 0.028487108 | 1 |
| Heart_Failure | macrophage | *AFF1* | 0.914016469 | 0.000874802 | 0.013744636 | 1 |
| Heart_Failure | macrophage | *ZFAND4* | 0.70741841 | 0.004723356 | 0.025750249 | 1 |
| Heart_Failure | macrophage | *FAM192A* | 0.65351927 | 0.010132193 | 0.040797844 | 1 |
| Heart_Failure | macrophage | *CBWD1* | 1.834076532 | 0.00099491 | 0.013998911 | 1 |
| Heart_Failure | macrophage | *ZNF621* | 1.58200074 | 0.000331466 | 0.013744636 | 1 |
| Heart_Failure | macrophage | *RNASEH2C* | 0.435593708 | 0.003407573 | 0.02142726 | 1 |
| Heart_Failure | macrophage | *SH3RF3* | 0.763110778 | 0.005471931 | 0.027888457 | 1 |
| Heart_Failure | macrophage | *NOC3L* | 1.080729528 | 0.00084279 | 0.013744636 | 1 |
| Heart_Failure | macrophage | *KCNK7* | 2.08270035 | 0.006424475 | 0.030965061 | 1 |
| Heart_Failure | macrophage | *NDNF* | 0.719772261 | 0.009300919 | 0.03857099 | 1 |
| Heart_Failure | macrophage | *GMPPB* | 1.054549645 | 0.002393577 | 0.01806164 | 1 |
| Heart_Failure | macrophage | *SCAI* | 0.436928663 | 0.011248555 | 0.043927413 | 1 |
| Heart_Failure | macrophage | *UQCRH* | 0.943218033 | 0.003471052 | 0.02164723 | 1 |
| Heart_Failure | macrophage | *XXYLT1* | 1.292538044 | 0.000992048 | 0.013998911 | 1 |
| Heart_Failure | macrophage | *LEMD3* | 1.322760733 | 0.000748876 | 0.013744636 | 1 |
| Heart_Failure | macrophage | *RALGAPA1* | 0.479697848 | 0.009691459 | 0.039496489 | 1 |
| Heart_Failure | macrophage | *ANKRD36C* | 0.965215458 | 0.001652286 | 0.015548043 | 1 |
| Heart_Failure | macrophage | *MSL2* | 1.362005262 | 0.000844897 | 0.013744636 | 1 |
| Heart_Failure | macrophage | *ANGEL2* | 1.228319055 | 0.000932847 | 0.013744636 | 1 |
| Heart_Failure | macrophage | *KIAA1551* | 0.457618007 | 0.008844456 | 0.03747284 | 1 |
| Heart_Failure | macrophage | *LARP7* | 1.062223105 | 0.005184478 | 0.027077595 | 1 |
| Heart_Failure | macrophage | *FGFBP3* | 1.938042458 | 0.008503143 | 0.036640542 | 1 |
| Heart_Failure | macrophage | *CEP135* | 1.265012143 | 0.002213771 | 0.017241117 | 1 |
| Heart_Failure | macrophage | *DHX36* | 1.020671311 | 0.001209712 | 0.014466177 | 1 |
| Heart_Failure | macrophage | *FBXW8* | 0.734621425 | 0.01097777 | 0.043339415 | 1 |
| Heart_Failure | macrophage | *SH3BP5L* | 1.993991738 | 0.004935497 | 0.026490077 | 1 |
| Heart_Failure | macrophage | *ATG13* | 1.238291411 | 0.003108573 | 0.020493053 | 1 |
| Heart_Failure | macrophage | *EIF1AD* | 1.058439904 | 0.011093338 | 0.043448188 | 1 |
| Heart_Failure | macrophage | *CLTB* | 1.275096096 | 0.001879442 | 0.016371728 | 1 |
| Heart_Failure | macrophage | *PPP2R2D* | 0.643472362 | 0.007607757 | 0.034083874 | 1 |
| Heart_Failure | macrophage | *C11orf68* | 1.298132109 | 0.011956275 | 0.045484252 | 1 |
| Heart_Failure | macrophage | *MRPL48* | 1.005811155 | 0.008591075 | 0.036823229 | 1 |
| Heart_Failure | macrophage | *RAB6A* | 0.861982911 | 0.005034189 | 0.026720217 | 1 |
| Heart_Failure | macrophage | *RPS6KB2* | 1.421420149 | 1.52E-05 | 0.002842801 | 0.073912825 |
| Heart_Failure | macrophage | *TOM1L2* | 0.608625975 | 0.008297189 | 0.036008911 | 1 |
| Heart_Failure | macrophage | *TTLL11* | 1.00514919 | 0.001421295 | 0.015057034 | 1 |
| Heart_Failure | macrophage | *CTDNEP1* | 0.707683476 | 0.00260302 | 0.018546041 | 1 |
| Heart_Failure | macrophage | *ZDHHC21* | 0.956669577 | 0.000376382 | 0.013744636 | 1 |
| Heart_Failure | macrophage | *TCP11L1* | 0.543396107 | 0.00435856 | 0.024569337 | 1 |
| Heart_Failure | macrophage | *MYPOP* | 1.241549933 | 0.004229471 | 0.024114447 | 1 |
| Heart_Failure | macrophage | *SLC35G1* | 1.169057566 | 0.003906731 | 0.023286805 | 1 |
| Heart_Failure | macrophage | *OR4N2* | 0.834152498 | 0.008069256 | 0.035431702 | 1 |

| | | | | | | |
|---|---|---|---|---|---|---|
| Heart_Failure | macrophage | *CRLF3* | 0.688659858 | 0.00126105 | 0.014557357 | 1 |
| Heart_Failure | macrophage | *WDR25* | 1.088733546 | 0.002775103 | 0.019401728 | 1 |
| Heart_Failure | macrophage | *RMDN1* | 0.643162168 | 0.000834664 | 0.013744636 | 1 |
| Heart_Failure | macrophage | *ANKLE2* | 0.967461524 | 0.008830724 | 0.03747284 | 1 |
| Heart_Failure | macrophage | *DSCAML1* | 0.862819248 | 0.012825088 | 0.047647264 | 1 |
| Heart_Failure | macrophage | *TALDO1* | 1.119954667 | 0.009166217 | 0.038340073 | 1 |
| Heart_Failure | macrophage | *MAN1B1* | 1.837918769 | 0.000764245 | 0.013744636 | 1 |
| Heart_Failure | macrophage | *THAP5* | 1.273742513 | 0.005283975 | 0.027404157 | 1 |
| Heart_Failure | macrophage | *NAALADL2* | 0.532612003 | 0.001130963 | 0.014193744 | 1 |
| Heart_Failure | macrophage | *CD151* | 0.942863315 | 0.000417107 | 0.013744636 | 1 |
| Heart_Failure | macrophage | *SLC35G5* | 16.97081673 | 0.000399232 | 0.013744636 | 1 |
| Heart_Failure | macrophage | *CENPBD1* | 3.646855446 | 0.003493471 | 0.02175908 | 1 |
| Heart_Failure | macrophage | *RPS27* | 1.171765508 | 0.000539168 | 0.013744636 | 1 |
| Heart_Failure | macrophage | *DPY19L2* | 0.363591475 | 0.000338992 | 0.013744636 | 1 |
| Heart_Failure | macrophage | *ZNF454* | 0.275922322 | 0.00087688 | 0.013744636 | 1 |
| Heart_Failure | macrophage | *SH2B1* | 0.765109835 | 0.006452301 | 0.030989333 | 1 |
| Heart_Failure | macrophage | *SH2D4B* | 2.490794561 | 0.006735956 | 0.031792666 | 1 |
| Heart_Failure | macrophage | *NT5DC1* | 0.787830777 | 0.013509392 | 0.049358108 | 1 |
| Heart_Failure | macrophage | *ERN1* | 0.938694125 | 0.010718561 | 0.042635055 | 1 |
| Heart_Failure | macrophage | *PARP10* | 0.985138094 | 0.010724025 | 0.042635055 | 1 |
| Heart_Failure | macrophage | *NSUN3* | 1.025865106 | 0.005991691 | 0.029454593 | 1 |
| Heart_Failure | macrophage | *STX19* | 0.49384917 | 0.013345692 | 0.04909272 | 1 |
| Heart_Failure | macrophage | *GAK* | 1.164023422 | 0.001506674 | 0.015102525 | 1 |
| Heart_Failure | macrophage | *CTC1* | 0.880049315 | 0.001635379 | 0.015548043 | 1 |
| Heart_Failure | macrophage | *C3orf38* | 0.464910155 | 0.006901671 | 0.032258573 | 1 |
| Heart_Failure | macrophage | *SPTY2D1* | 1.391661786 | 0.004254545 | 0.024189473 | 1 |
| Heart_Failure | macrophage | *ZNF664* | 1.162855027 | 0.005468972 | 0.027888457 | 1 |
| Heart_Failure | macrophage | *HLA-DQB1* | -0.456170691 | 3.26E-18 | 1.58E-14 | 1.58E-14 |
| Heart_Failure | macrophage | *PACS2* | 1.278528544 | 0.002007931 | 0.016682333 | 1 |
| Heart_Failure | macrophage | *LBX2* | 0.431915496 | 0.000203981 | 0.012453892 | 0.98971721 |
| Heart_Failure | macrophage | *PDXDC1* | 0.827158789 | 0.002672281 | 0.018845793 | 1 |
| Heart_Failure | macrophage | *ZADH2* | 1.102597409 | 0.00587282 | 0.029076452 | 1 |
| Heart_Failure | macrophage | *SLC36A4* | 0.909714423 | 0.01072917 | 0.042635055 | 1 |
| Heart_Failure | macrophage | *PSMG4* | 0.65533034 | 0.007835661 | 0.034844376 | 1 |
| Heart_Failure | macrophage | *CAPS2* | 1.062843056 | 0.000719943 | 0.013744636 | 1 |
| Heart_Failure | macrophage | *D2HGDH* | 0.715606868 | 1.67E-05 | 0.002997833 | 0.080941489 |
| Heart_Failure | macrophage | *CMTR2* | 1.05433422 | 0.001115024 | 0.014193744 | 1 |
| Heart_Failure | macrophage | *FKRP* | 0.27238978 | 0.00362564 | 0.022211622 | 1 |
| Heart_Failure | macrophage | *TMEM136* | 0.508874783 | 0.013094582 | 0.048462936 | 1 |
| Heart_Failure | macrophage | *ZBTB20* | 0.510991612 | 0.005083145 | 0.026837114 | 1 |
| Heart_Failure | macrophage | *C3orf58* | 1.191645208 | 0.006278174 | 0.030522744 | 1 |
| Heart_Failure | macrophage | *RELL1* | 0.685651883 | 0.00435494 | 0.024569337 | 1 |
| Heart_Failure | macrophage | *RFX7* | 1.185182564 | 0.000647368 | 0.013744636 | 1 |
| Heart_Failure | macrophage | *ZNF329* | 1.04769842 | 0.003733462 | 0.022643449 | 1 |
| Heart_Failure | macrophage | *WRB* | 0.319487074 | 0.001821291 | 0.016138719 | 1 |
| Heart_Failure | macrophage | *ERCC6L2* | 0.969771479 | 0.003195274 | 0.020758363 | 1 |
| Heart_Failure | macrophage | *MRPL41* | 2.009963296 | 0.000810459 | 0.013744636 | 1 |
| Heart_Failure | macrophage | *ARL6IP4* | 0.359686003 | 0.011061409 | 0.04340436 | 1 |
| Heart_Failure | macrophage | *C8orf33* | 0.822076805 | 0.006722883 | 0.031761858 | 1 |
| Heart_Failure | macrophage | *GLRX5* | 1.90093779 | 0.000842234 | 0.013744636 | 1 |

| | | | | | | |
|---|---|---|---|---|---|---|
| Heart_Failure | macrophage | *SKA2* | 0.827719175 | 0.007495037 | 0.03386026 | 1 |
| Heart_Failure | macrophage | *ALG12* | 1.511248967 | 0.005069839 | 0.026824898 | 1 |
| Heart_Failure | macrophage | *COL18A1* | 0.590233963 | 0.000608021 | 0.013744636 | 1 |
| Heart_Failure | macrophage | *ZNF721* | 0.812530042 | 0.000975784 | 0.013998911 | 1 |
| Heart_Failure | macrophage | *CEP63* | 0.532505911 | 0.001141902 | 0.014193744 | 1 |
| Heart_Failure | macrophage | *CSMD1* | 1.381451503 | 0.003069178 | 0.02044882 | 1 |
| Heart_Failure | macrophage | *FANCF* | 2.439596699 | 0.00184657 | 0.016277137 | 1 |
| Heart_Failure | macrophage | *CCDC125* | 0.67023212 | 0.007720263 | 0.034365795 | 1 |
| Heart_Failure | macrophage | *LRIT3* | 9.381902654 | 0.00226954 | 0.017479059 | 1 |
| Heart_Failure | macrophage | *SERHL2* | 0.982645843 | 0.009298717 | 0.03857099 | 1 |
| Heart_Failure | macrophage | *SFXN4* | 0.762420513 | 0.008456872 | 0.036505999 | 1 |
| Heart_Failure | macrophage | *BTBD9* | 0.749112289 | 0.00287278 | 0.019714284 | 1 |
| Heart_Failure | macrophage | *TTC32* | 1.890579064 | 0.000429405 | 0.013744636 | 1 |
| Heart_Failure | macrophage | *PP2D1* | 2.168514221 | 0.000787865 | 0.013744636 | 1 |
| Heart_Failure | macrophage | *DENND5A* | 0.943856736 | 0.003258712 | 0.020914377 | 1 |
| Heart_Failure | macrophage | *SS18L1* | 0.997159127 | 0.002197821 | 0.017199722 | 1 |
| Heart_Failure | macrophage | *COPB2* | 1.226407203 | 0.000601489 | 0.013744636 | 1 |
| Heart_Failure | macrophage | *NDUFA12* | 0.929320804 | 0.004171217 | 0.024036513 | 1 |
| Heart_Failure | macrophage | *TCTE3* | 0.921833232 | 0.007898469 | 0.034925794 | 1 |
| Heart_Failure | macrophage | *UBE2G2* | 0.641608215 | 0.007692259 | 0.034304081 | 1 |
| Heart_Failure | macrophage | *ZFP90* | 1.077717014 | 0.00218256 | 0.017199722 | 1 |
| Heart_Failure | macrophage | *UBOX5* | 1.297733804 | 0.007871849 | 0.034880558 | 1 |
| Heart_Failure | macrophage | *SIGIRR* | 1.106416555 | 0.000214051 | 0.012453892 | 1 |
| Heart_Failure | macrophage | *IFITM2* | 0.844123785 | 0.007213577 | 0.03323863 | 1 |
| Heart_Failure | macrophage | *ATP6V0A2* | 1.089802501 | 0.00059112 | 0.013744636 | 1 |
| Heart_Failure | macrophage | *FAM174B* | 0.953656153 | 0.002875766 | 0.019714284 | 1 |
| Heart_Failure | macrophage | *SP1* | 1.139912879 | 0.00152851 | 0.01520515 | 1 |
| Heart_Failure | macrophage | *ZNF529* | 1.187350002 | 0.000654139 | 0.013744636 | 1 |
| Heart_Failure | macrophage | *WWOX* | 0.641945628 | 0.001912106 | 0.016395644 | 1 |
| Heart_Failure | macrophage | *RPS23* | 0.806703509 | 0.001132433 | 0.014193744 | 1 |
| Heart_Failure | macrophage | *TMEM222* | 1.474789997 | 0.004578062 | 0.02521312 | 1 |
| Heart_Failure | macrophage | *MIR22HG* | 0.816938 | 0.008546924 | 0.036731333 | 1 |
| Heart_Failure | macrophage | *QRFPR* | 1.79403006 | 0.000246408 | 0.012453892 | 1 |
| Heart_Failure | macrophage | *TMEM17* | 0.308813775 | 0.012467534 | 0.046748435 | 1 |
| Heart_Failure | macrophage | *TMEM232* | 0.539408917 | 3.78E-05 | 0.004831528 | 0.183598073 |
| Heart_Failure | macrophage | *WDR86* | 0.279770098 | 0.008847136 | 0.03747284 | 1 |
| Heart_Failure | macrophage | *GJA4* | 1.741975898 | 0.001731512 | 0.01576228 | 1 |
| Heart_Failure | macrophage | *EXD3* | 1.252281407 | 0.001227016 | 0.014488552 | 1 |
| Heart_Failure | macrophage | *ZFP69* | 2.051433369 | 0.001286827 | 0.014557357 | 1 |
| Heart_Failure | macrophage | *AGRN* | 1.036212585 | 0.012879478 | 0.047812722 | 1 |
| Heart_Failure | macrophage | *NCR3LG1* | 3.37156178 | 0.001380749 | 0.014954006 | 1 |
| Heart_Failure | macrophage | *PLSCR1* | 1.117452955 | 0.00018886 | 0.012453892 | 0.916347718 |
| Heart_Failure | macrophage | *C3orf62* | 0.913208461 | 7.06E-05 | 0.007789864 | 0.342655408 |
| Heart_Failure | macrophage | *SRSF10* | 0.552509946 | 0.005248554 | 0.027282348 | 1 |
| Heart_Failure | macrophage | *NBR1* | 1.091978719 | 0.001174184 | 0.014314427 | 1 |
| Heart_Failure | macrophage | *NDOR1* | 1.76083059 | 0.00403081 | 0.023595969 | 1 |
| Heart_Failure | macrophage | *SLC4A5* | 1.328152968 | 0.012611874 | 0.047107631 | 1 |
| Heart_Failure | macrophage | *UROS* | 1.307572492 | 0.00251679 | 0.018350936 | 1 |
| Heart_Failure | macrophage | *NOXA1* | 1.883799478 | 0.000933594 | 0.013744636 | 1 |
| Heart_Failure | macrophage | *ZDHHC11* | 0.39596953 | 0.01196607 | 0.045484252 | 1 |

| | | | | | | |
|---|---|---|---|---|---|---|
| Heart_Failure | macrophage | *RPL14* | 1.205172233 | 0.000797747 | 0.013744636 | 1 |
| Heart_Failure | macrophage | *FAM196A* | 0.659174417 | 0.007416944 | 0.033664185 | 1 |
| Heart_Failure | macrophage | *NELFB* | 1.777302711 | 0.000921607 | 0.013744636 | 1 |
| Heart_Failure | macrophage | *SLC15A5* | 0.464276367 | 0.005841599 | 0.029038751 | 1 |
| Heart_Failure | macrophage | *SF3B3* | 1.100582079 | 0.002185363 | 0.017199722 | 1 |
| Heart_Failure | macrophage | *PLAC9* | 0.659555947 | 0.001948955 | 0.01650319 | 1 |
| Heart_Failure | macrophage | *FAM47E* | 0.653598588 | 0.00960539 | 0.039329411 | 1 |
| Heart_Failure | macrophage | *TSPYL1* | 1.58222671 | 0.001146532 | 0.014193744 | 1 |
| Heart_Failure | macrophage | *PNRC2* | 1.686985039 | 0.000863757 | 0.013744636 | 1 |
| Heart_Failure | macrophage | *FHIT* | 0.771779302 | 0.001872289 | 0.016368189 | 1 |
| Heart_Failure | macrophage | *HLA-DRB1* | -0.226868692 | 7.39E-05 | 0.007964209 | 0.358389408 |
| Heart_Failure | macrophage | *MPHOSPH8* | 0.704034841 | 0.001795326 | 0.016131335 | 1 |
| Heart_Failure | macrophage | *ZNF471* | 0.689780124 | 0.013606968 | 0.049639856 | 1 |
| Heart_Failure | macrophage | *ZNF493* | 0.781000257 | 0.006791809 | 0.031902592 | 1 |
| Heart_Failure | macrophage | *SUPT3H* | 0.930571852 | 0.001081565 | 0.014193744 | 1 |
| Heart_Failure | macrophage | *LONP1* | 0.374180311 | 0.010237327 | 0.041118799 | 1 |
| Heart_Failure | macrophage | *PIK3R4* | 1.564544082 | 0.000153411 | 0.011326447 | 0.744350774 |
| Heart_Failure | macrophage | *RFX8* | 0.230496578 | 0.012688525 | 0.047248444 | 1 |
| Heart_Failure | macrophage | *MYO18A* | 0.760029301 | 0.011400551 | 0.044195623 | 1 |
| Heart_Failure | macrophage | *SULF2* | 0.445281074 | 0.004568034 | 0.02518648 | 1 |
| Heart_Failure | macrophage | *ZKSCAN5* | 1.215334925 | 8.39E-05 | 0.008480527 | 0.407065303 |
| Heart_Failure | macrophage | *TOMM7* | 1.11803008 | 0.001332989 | 0.014716944 | 1 |
| Heart_Failure | macrophage | *AMZ2* | 0.690271855 | 0.004764331 | 0.025857422 | 1 |
| Heart_Failure | macrophage | *ZNF431* | 1.024172054 | 0.000662555 | 0.013744636 | 1 |
| Heart_Failure | macrophage | *HLA-DQA1* | -0.385308664 | 1.77E-10 | 1.72E-07 | 8.60E-07 |
| Heart_Failure | macrophage | *ZNF239* | 0.590512951 | 0.010140987 | 0.040799393 | 1 |
| Heart_Failure | macrophage | *SLC39A10* | 1.163253282 | 0.002626583 | 0.018686485 | 1 |
| Heart_Failure | macrophage | *WDR5B* | 1.018019139 | 0.000440131 | 0.013744636 | 1 |
| Heart_Failure | macrophage | *ZNF138* | 0.851096927 | 0.000465766 | 0.013744636 | 1 |
| Heart_Failure | macrophage | *ZNF429* | 0.753655762 | 0.005910995 | 0.029216655 | 1 |
| Heart_Failure | macrophage | *GMFB* | 1.667703677 | 0.000723409 | 0.013744636 | 1 |
| Heart_Failure | macrophage | *ZNF420* | 0.725796244 | 0.008141306 | 0.035607345 | 1 |
| Heart_Failure | macrophage | *ZMYM1* | 1.312425546 | 0.001052393 | 0.014193744 | 1 |
| Heart_Failure | macrophage | *IGF2R* | 0.857517667 | 0.003671149 | 0.022397116 | 1 |
| Heart_Failure | macrophage | *ABCB8* | 1.157029014 | 0.001489967 | 0.015102525 | 1 |
| Heart_Failure | macrophage | *SND1* | 0.830650009 | 0.003615086 | 0.022174968 | 1 |
| Heart_Failure | macrophage | *ZNF720* | 0.791968854 | 0.01024598 | 0.041119515 | 1 |
| Heart_Failure | macrophage | *MAP3K5* | 0.751741533 | 0.00538847 | 0.027717297 | 1 |
| Heart_Failure | macrophage | *RPF2* | 1.134822142 | 0.00301859 | 0.020313729 | 1 |
| Heart_Failure | macrophage | *RAB40C* | 1.015855079 | 0.004880916 | 0.026255218 | 1 |
| Heart_Failure | macrophage | *PIGN* | 0.444946615 | 0.009682772 | 0.039496489 | 1 |
| Heart_Failure | macrophage | *TOPORS* | 1.582292541 | 0.001149195 | 0.014193744 | 1 |
| Heart_Failure | macrophage | *PDCD1LG2* | 0.223626609 | 0.000695388 | 0.013744636 | 1 |
| Heart_Failure | macrophage | *RPS26* | 0.332327912 | 0.00018418 | 0.012453892 | 0.893641713 |
| Heart_Failure | macrophage | *TXNRD3* | 0.59229405 | 0.0086958 | 0.037173589 | 1 |
| Heart_Failure | macrophage | *EME2* | 2.520827871 | 0.001506823 | 0.015102525 | 1 |
| Heart_Failure | macrophage | *ADH5* | 0.66554905 | 0.005397586 | 0.027717297 | 1 |
| Heart_Failure | macrophage | *ZNF677* | 0.928650679 | 0.007948032 | 0.035058047 | 1 |
| Heart_Failure | macrophage | *MPZL1* | 0.645636409 | 0.00922038 | 0.038467142 | 1 |
| Heart_Failure | macrophage | *NOL8* | 1.163098657 | 0.001823605 | 0.016138719 | 1 |

| | | | | | | |
|---|---|---|---|---|---|---|
| Heart_Failure | macrophage | *DLGAP2* | 0.457828862 | 0.005926251 | 0.029251445 | 1 |
| Heart_Failure | macrophage | *SFI1* | 0.919703277 | 0.00851908 | 0.036676642 | 1 |
| Heart_Failure | macrophage | *ZNF544* | 1.186596536 | 0.005913181 | 0.029216655 | 1 |
| Heart_Failure | macrophage | *MIER1* | 1.050023637 | 0.000423524 | 0.013744636 | 1 |
| Heart_Failure | macrophage | *MAN1A2* | 1.065295641 | 0.000882711 | 0.013744636 | 1 |
| Heart_Failure | macrophage | *DDRGK1* | 1.050931094 | 0.010423364 | 0.041693454 | 1 |
| Heart_Failure | macrophage | *STYX* | 1.312589949 | 0.003883573 | 0.023205782 | 1 |
| Heart_Failure | macrophage | *HELZ* | 0.978874789 | 0.000667003 | 0.013744636 | 1 |
| Heart_Failure | macrophage | *UCKL1* | 1.066051276 | 0.005478861 | 0.027894472 | 1 |
| Heart_Failure | macrophage | *GFPT1* | 0.687381926 | 0.003928681 | 0.023296977 | 1 |
| Heart_Failure | macrophage | *ZNF568* | 0.720109325 | 0.005827302 | 0.029038751 | 1 |
| Heart_Failure | macrophage | *TMA16* | 1.025462154 | 0.002570152 | 0.018426459 | 1 |
| Heart_Failure | macrophage | *HLA-DRB5* | -0.071585598 | 0.006865966 | 0.032125038 | 1 |
| Heart_Failure | macrophage | *ZNF28* | 1.661358572 | 0.000675777 | 0.013744636 | 1 |
| Heart_Failure | macrophage | *TTC37* | 0.780631519 | 0.002515653 | 0.018350936 | 1 |
| Heart_Failure | macrophage | *TUSC1* | 2.658565157 | 0.001428658 | 0.015100028 | 1 |
| Heart_Failure | macrophage | *CEP290* | 0.680948004 | 0.002531578 | 0.018350936 | 1 |
| Heart_Failure | macrophage | *GPATCH3* | 2.399022926 | 0.001706718 | 0.015713468 | 1 |
| Heart_Failure | macrophage | *RPL10A* | 0.978660745 | 0.003811606 | 0.023002379 | 1 |
| Heart_Failure | macrophage | *SFT2D1* | 1.222707883 | 0.004108704 | 0.023846211 | 1 |
| Heart_Failure | macrophage | *TYW1* | 0.714059347 | 0.002176768 | 0.017199722 | 1 |
| Heart_Failure | macrophage | *DZIP3* | 0.747406452 | 0.005398361 | 0.027717297 | 1 |
| Heart_Failure | macrophage | *PCMTD2* | 0.793032693 | 0.004830019 | 0.026058178 | 1 |
| Heart_Failure | macrophage | *RUFY2* | 0.721940731 | 0.003836717 | 0.023041698 | 1 |
| Heart_Failure | macrophage | *BMPR2* | 0.771782549 | 0.002082535 | 0.016973572 | 1 |
| Heart_Failure | macrophage | *TCEA3* | 0.492753054 | 0.006512553 | 0.031162629 | 1 |
| Heart_Failure | macrophage | *HSD17B8* | 0.402250935 | 0.001774511 | 0.016040353 | 1 |
| Heart_Failure | macrophage | *PRRT1* | 0.308512146 | 0.000292206 | 0.013375324 | 1 |
| Heart_Failure | macrophage | *BAG6* | 0.762810419 | 0.00041459 | 0.013744636 | 1 |
| Heart_Failure | macrophage | *ZNF814* | 0.668369936 | 0.004376739 | 0.024635656 | 1 |
| Heart_Failure | macrophage | *MICB* | 0.095999814 | 0.004197008 | 0.024038296 | 1 |
| Heart_Failure | macrophage | *MICA* | 0.61903269 | 4.97E-07 | 0.000219225 | 0.002411478 |
| Heart_Failure | macrophage | *DCTN1* | 1.713848326 | 0.000961842 | 0.013998911 | 1 |
| Heart_Failure | macrophage | *FAM216A* | 1.251929747 | 0.000329594 | 0.013744636 | 1 |
| Heart_Failure | macrophage | *TRIM13* | 0.753351479 | 0.006101766 | 0.029784478 | 1 |
| Heart_Failure | macrophage | *SLC35B4* | 1.61064715 | 0.001123714 | 0.014193744 | 1 |
| Heart_Failure | macrophage | *SCGB2B2* | 0.749291039 | 0.00662446 | 0.031511646 | 1 |
| Heart_Failure | macrophage | *NAP1L4* | 0.837179625 | 0.001351864 | 0.014806416 | 1 |
| Heart_Failure | macrophage | *LIN52* | 0.754576466 | 9.84E-05 | 0.00918566 | 0.477238119 |
| Heart_Failure | macrophage | *METTL6* | 0.858503834 | 0.001652985 | 0.015548043 | 1 |
| Heart_Failure | macrophage | *SACM1L* | 1.239585736 | 0.002571646 | 0.018426459 | 1 |
| Heart_Failure | macrophage | *SLC48A1* | 1.01953782 | 0.005770204 | 0.028868296 | 1 |
| Heart_Failure | macrophage | *ZNF254* | 0.73274675 | 0.00821558 | 0.035840035 | 1 |
| Heart_Failure | macrophage | *AC022498.1* | 2.511660612 | 0.012095649 | 0.045814277 | 1 |
| Heart_Failure | macrophage | *SUPT4H1* | 1.739077068 | 0.001377075 | 0.014947577 | 1 |
| Heart_Failure | macrophage | *ERV3-1* | 0.131248584 | 0.013364124 | 0.049123282 | 1 |
| Heart_Failure | macrophage | *SRA1* | 0.737999743 | 0.012351993 | 0.046438004 | 1 |
| Heart_Failure | macrophage | *PPP1CB* | 0.734169692 | 0.001605988 | 0.015528127 | 1 |
| Heart_Failure | macrophage | *ATF6B* | 1.187031569 | 3.99E-08 | 2.76E-05 | 0.000193541 |
| Heart_Failure | macrophage | *EMP2* | 0.558286548 | 0.013155853 | 0.048578537 | 1 |

| | | | | | | |
|---|---|---|---|---|---|---|
| Heart_Failure | macrophage | *GALT* | 1.002786148 | 0.001148577 | 0.014193744 | 1 |
| Heart_Failure | macrophage | *ZNF90* | 0.653052486 | 0.011860902 | 0.045314251 | 1 |
| Heart_Failure | macrophage | *REPIN1* | 2.45053999 | 0.0006235 | 0.013744636 | 1 |
| Heart_Failure | macrophage | *ZNF891* | 1.703878562 | 1.33E-05 | 0.002688904 | 0.064533694 |
| Heart_Failure | macrophage | *SMIM7* | 1.070319716 | 0.002656609 | 0.018762541 | 1 |
| Heart_Failure | macrophage | *CPNE1* | -0.213648264 | 0.005124885 | 0.02694035 | 1 |
| Heart_Failure | macrophage | *PAXIP1-AS2* | 1.622265401 | 0.000315659 | 0.013744636 | 1 |
| Heart_Failure | macrophage | *LYRM4* | 1.290888862 | 0.000649176 | 0.013744636 | 1 |
| Heart_Failure | macrophage | *SH3D21* | 0.772989354 | 0.000267215 | 0.012836909 | 1 |
| Heart_Failure | macrophage | *VPS16* | 2.027541143 | 0.000241888 | 0.012453892 | 1 |
| Heart_Failure | macrophage | *NPEPL1* | 1.950690408 | 0.000680461 | 0.013744636 | 1 |
| Heart_Failure | macrophage | *RNASEK* | 0.6048553 | 0.004456343 | 0.024908187 | 1 |
| Heart_Failure | macrophage | *FXYD7* | 0.631956086 | 0.002240712 | 0.017367309 | 1 |
| Heart_Failure | macrophage | *APOL6* | 0.823108814 | 0.003510817 | 0.021783227 | 1 |
| Heart_Failure | macrophage | *OR2A1* | 0.467068719 | 0.00102166 | 0.014193744 | 1 |
| Heart_Failure | macrophage | *UBA52* | 0.568008254 | 0.003924581 | 0.023296977 | 1 |
| Heart_Failure | macrophage | *AC062017.1* | 1.603129545 | 0.002297468 | 0.017636064 | 1 |
| Heart_Failure | macrophage | *MIAT* | 0.807613746 | 0.006597502 | 0.031414212 | 1 |
| Heart_Failure | macrophage | *AC009299.3* | 1.275225272 | 0.005516483 | 0.027942684 | 1 |
| Heart_Failure | macrophage | *SCAMP4* | 1.11745827 | 0.01132844 | 0.044087683 | 1 |
| Heart_Failure | macrophage | *AC092171.4* | 2.092350224 | 0.004590937 | 0.02525536 | 1 |
| Heart_Failure | macrophage | *RPS18* | 1.067226361 | 0.000102216 | 0.009328149 | 0.495953695 |
| Heart_Failure | macrophage | *DIRC3* | 0.618313245 | 0.012365602 | 0.046438004 | 1 |
| Heart_Failure | macrophage | *HLA-DQB2* | 0.288112047 | 1.32E-17 | 2.14E-14 | 6.42E-14 |
| Heart_Failure | macrophage | *RPS28* | 1.0958605 | 0.006479144 | 0.031064036 | 1 |
| Heart_Failure | macrophage | *TRIM26* | 0.602297533 | 0.011262429 | 0.043927413 | 1 |
| Heart_Failure | macrophage | *ZNF736* | 0.423959384 | 0.000226078 | 0.012453892 | 1 |
| Heart_Failure | macrophage | *ZBTB22* | 0.790231585 | 5.13E-05 | 0.006068035 | 0.248789443 |
| Heart_Failure | macrophage | *ZBED5* | 0.660383264 | 0.006202324 | 0.030214534 | 1 |
| Heart_Failure | macrophage | *LINC00959* | 1.218612662 | 0.004466225 | 0.024908187 | 1 |
| Heart_Failure | macrophage | *HLA-DQA2* | 0.344318759 | 1.04E-17 | 2.14E-14 | 5.02E-14 |
| Heart_Failure | macrophage | *PSMB9* | 0.449747815 | 3.60E-08 | 2.76E-05 | 0.000174521 |
| Heart_Failure | macrophage | *NSUN6* | 0.886267174 | 0.000849526 | 0.013744636 | 1 |
| Heart_Failure | macrophage | *PI4KA* | 0.845498189 | 0.0041968 | 0.024038296 | 1 |
| Heart_Failure | macrophage | *LINC00969* | 0.361217564 | 0.003278001 | 0.020982667 | 1 |
| Heart_Failure | macrophage | *ARFGAP3* | 1.334088917 | 0.000983496 | 0.013998911 | 1 |
| Heart_Failure | macrophage | *DECR2* | 2.224045867 | 0.003076213 | 0.02044882 | 1 |
| Heart_Failure | macrophage | *NBPF15* | 1.2347021 | 0.003012027 | 0.020297718 | 1 |
| Heart_Failure | macrophage | *TTC4* | -0.485142368 | 0.004488989 | 0.024925587 | 1 |
| Heart_Failure | macrophage | *C4A* | -0.44311663 | 0.006457175 | 0.030989333 | 1 |
| Heart_Failure | macrophage | *NEAT1* | 0.756395921 | 0.00044963 | 0.013744636 | 1 |
| Heart_Failure | macrophage | *RAB30-AS1* | 0.584385979 | 0.003414907 | 0.02142726 | 1 |
| Heart_Failure | macrophage | *UBAP1L* | 0.985824995 | 0.004313418 | 0.02439243 | 1 |
| Heart_Failure | macrophage | *LINC00926* | 2.842322318 | 0.002860915 | 0.019714284 | 1 |
| Heart_Failure | macrophage | *CDK11B* | 1.680648065 | 0.000722659 | 0.013744636 | 1 |
| Heart_Failure | macrophage | *ABHD14A* | 0.875892699 | 0.005524626 | 0.027942684 | 1 |
| Heart_Failure | macrophage | *C5orf17* | 1.339170274 | 0.001623145 | 0.015548043 | 1 |
| Heart_Failure | macrophage | *PDCD6* | 1.529807241 | 0.000717655 | 0.013744636 | 1 |
| Heart_Failure | macrophage | *ZNF718* | 0.957201619 | 0.002473118 | 0.018350936 | 1 |
| Heart_Failure | macrophage | *SMIM20* | 1.533620686 | 0.006355979 | 0.030839209 | 1 |

| | | | | | | |
|---|---|---|---|---|---|---|
| Heart_Failure | macrophage | *LINC00989* | 1.144343137 | 0.001448806 | 0.015102525 | 1 |
| Heart_Failure | macrophage | *GLI4* | 1.821723719 | 0.001485944 | 0.015102525 | 1 |
| Heart_Failure | macrophage | *ZNF550* | 1.275415253 | 0.00528654 | 0.027404157 | 1 |
| Heart_Failure | macrophage | *PCDHGB2* | 2.572376952 | 0.003043012 | 0.020393221 | 1 |
| Heart_Failure | macrophage | *KIAA1147* | 0.814489945 | 0.003063881 | 0.020448353 | 1 |
| Heart_Failure | macrophage | *CNPY2* | 1.015354251 | 0.003229529 | 0.0208381 | 1 |
| Heart_Failure | macrophage | *HIF1A-AS2* | 1.243784565 | 0.001277988 | 0.014557357 | 1 |
| Heart_Failure | macrophage | *MC1R* | 0.770644867 | 0.002366589 | 0.017969783 | 1 |
| Heart_Failure | macrophage | *IQCH-AS1* | 0.522730073 | 0.01312318 | 0.048531761 | 1 |
| Heart_Failure | macrophage | *PWRN1* | 0.674120007 | 0.001479204 | 0.015102525 | 1 |
| Heart_Failure | macrophage | *DYNLL2* | 2.273334726 | 0.001334595 | 0.014716944 | 1 |
| Heart_Failure | macrophage | *ZNF224* | 1.429959869 | 0.000854656 | 0.013744636 | 1 |
| Heart_Failure | macrophage | *NDUFA7* | 12.03184292 | 0.002232787 | 0.017333572 | 1 |
| Heart_Failure | macrophage | *ZNF587B* | 2.186515119 | 0.001697147 | 0.015698506 | 1 |
| Heart_Failure | macrophage | *HCG17* | 0.53210078 | 0.005765974 | 0.028868296 | 1 |
| Heart_Failure | macrophage | *GTF2H5* | 0.515680831 | 0.000114969 | 0.009786465 | 0.557828523 |
| Heart_Failure | macrophage | *PAXIP1-AS1* | 1.162174027 | 0.003872447 | 0.023167836 | 1 |
| Heart_Failure | mast_cell | *PNMT* | 8.382805279 | 1.45E-07 | 0.000116977 | 0.000701863 |
| Heart_Failure | mast_cell | *EVA1B* | 4.161594376 | 4.39E-05 | 0.026608369 | 0.212866949 |
| Heart_Failure | mast_cell | *TSLP* | 1.784061515 | 1.78E-06 | 0.001235481 | 0.008648367 |
| Heart_Failure | mast_cell | *GSDMA* | -3.432010383 | 1.09E-09 | 1.06E-06 | 5.30E-06 |
| Heart_Failure | mast_cell | *HLA-DQB1* | -4.199516816 | 2.00E-26 | 9.71E-23 | 9.71E-23 |
| Heart_Failure | mast_cell | *HLA-DQA1* | -5.135658416 | 9.13E-20 | 2.21E-16 | 4.43E-16 |
| Heart_Failure | mast_cell | *HLA-DQB2* | 1.301953884 | 1.51E-12 | 1.83E-09 | 7.33E-09 |
| Heart_Failure | mast_cell | *HLA-DQA2* | 2.016275036 | 2.67E-16 | 4.32E-13 | 1.30E-12 |
| Heart_Failure | mesothelium | *NFYA* | -0.5652093 | 0.000334867 | 0.003626728 | 1 |
| Heart_Failure | mesothelium | *SEMA3F* | -0.334566194 | 1.36E-06 | 0.000508439 | 0.006609707 |
| Heart_Failure | mesothelium | *KRIT1* | -0.569106304 | 0.000893813 | 0.007017442 | 1 |
| Heart_Failure | mesothelium | *TMEM176A* | -0.603687992 | 0.004910505 | 0.024386665 | 1 |
| Heart_Failure | mesothelium | *ALS2* | -0.55048766 | 0.002263656 | 0.013809292 | 1 |
| Heart_Failure | mesothelium | *RBM6* | -0.140508105 | 1.11E-08 | 7.71E-06 | 5.39E-05 |
| Heart_Failure | mesothelium | *RECQL* | -0.498224799 | 0.004800072 | 0.024059866 | 1 |
| Heart_Failure | mesothelium | *GCFC2* | -0.787546484 | 0.000140943 | 0.002614833 | 0.683854435 |
| Heart_Failure | mesothelium | *GGNBP2* | -0.667149092 | 6.93E-05 | 0.002273164 | 0.336192384 |
| Heart_Failure | mesothelium | *SYNRG* | -0.435164679 | 0.003877678 | 0.020406174 | 1 |
| Heart_Failure | mesothelium | *BAIAP2L1* | -0.844991067 | 0.000188238 | 0.002744551 | 0.913331999 |
| Heart_Failure | mesothelium | *VPS41* | -0.422555295 | 0.008472816 | 0.036745705 | 1 |
| Heart_Failure | mesothelium | *SCIN* | -0.15668469 | 0.006650118 | 0.030965811 | 1 |
| Heart_Failure | mesothelium | *ST7L* | -0.130549766 | 0.001199568 | 0.008635463 | 1 |
| Heart_Failure | mesothelium | *PSMB1* | -0.618683562 | 0.008211688 | 0.035991971 | 1 |
| Heart_Failure | mesothelium | *NADK* | -0.630242838 | 0.000564175 | 0.005004347 | 1 |
| Heart_Failure | mesothelium | *CYB561* | -0.205124387 | 0.002206823 | 0.013588206 | 1 |
| Heart_Failure | mesothelium | *MED24* | -0.768784719 | 0.001008129 | 0.007702226 | 1 |
| Heart_Failure | mesothelium | *RPS20* | -0.462030719 | 0.005736474 | 0.027530535 | 1 |
| Heart_Failure | mesothelium | *FUZ* | -0.450279118 | 0.002453052 | 0.014607614 | 1 |
| Heart_Failure | mesothelium | *ZBTB32* | -0.537670795 | 0.000275193 | 0.003180204 | 1 |
| Heart_Failure | mesothelium | *MAP4K5* | -0.666799046 | 0.000188775 | 0.002744551 | 0.915934096 |
| Heart_Failure | mesothelium | *MAN2B2* | -0.359491469 | 0.010638812 | 0.043025098 | 1 |
| Heart_Failure | mesothelium | *COX15* | -0.607249776 | 0.003958354 | 0.02070168 | 1 |
| Heart_Failure | mesothelium | *WWTR1* | -0.685108419 | 0.000408302 | 0.004059597 | 1 |

| | | | | | | |
|---|---|---|---|---|---|---|
| Heart_Failure | mesothelium | *HGF* | -0.438550213 | 0.004503733 | 0.022929813 | 1 |
| Heart_Failure | mesothelium | *SLC7A9* | 0.055989242 | 0.010882954 | 0.043762318 | 1 |
| Heart_Failure | mesothelium | *ZDHHC6* | -0.842336162 | 5.70E-05 | 0.002273164 | 0.276772643 |
| Heart_Failure | mesothelium | *GLRX2* | -0.699191358 | 0.007911048 | 0.03521505 | 1 |
| Heart_Failure | mesothelium | *SEC63* | -0.456072814 | 0.001108544 | 0.00817425 | 1 |
| Heart_Failure | mesothelium | *AGPAT4* | -0.975066213 | 6.01E-05 | 0.002273164 | 0.291609271 |
| Heart_Failure | mesothelium | *VEZT* | -0.765737933 | 0.00013263 | 0.002542967 | 0.643521096 |
| Heart_Failure | mesothelium | *SNX1* | -0.381676334 | 0.006049652 | 0.028692974 | 1 |
| Heart_Failure | mesothelium | *RABEP1* | -0.326632708 | 0.010080389 | 0.041414098 | 1 |
| Heart_Failure | mesothelium | *IKZF2* | -0.547212551 | 0.001948948 | 0.012475322 | 1 |
| Heart_Failure | mesothelium | *IFT88* | -0.344921943 | 0.001080009 | 0.00804947 | 1 |
| Heart_Failure | mesothelium | *SH3YL1* | -0.300256625 | 0.00208904 | 0.013095637 | 1 |
| Heart_Failure | mesothelium | *VCL* | -0.737183681 | 0.000153332 | 0.002672801 | 0.74396873 |
| Heart_Failure | mesothelium | *ADSS* | -1.014215725 | 5.55E-05 | 0.002273164 | 0.269234466 |
| Heart_Failure | mesothelium | *TBC1D23* | -0.824139878 | 0.000190196 | 0.002744551 | 0.922832672 |
| Heart_Failure | mesothelium | *PI4K2B* | -1.687230381 | 0.01122411 | 0.044785675 | 1 |
| Heart_Failure | mesothelium | *PQLC2* | -1.918331517 | 0.000431309 | 0.004185424 | 1 |
| Heart_Failure | mesothelium | *PSMA4* | -0.630847062 | 0.009198961 | 0.038410807 | 1 |
| Heart_Failure | mesothelium | *TDP1* | -0.829014207 | 0.000663285 | 0.005616504 | 1 |
| Heart_Failure | mesothelium | *SPATA7* | -0.603299644 | 3.24E-05 | 0.002273164 | 0.157312675 |
| Heart_Failure | mesothelium | *MED17* | -0.610677949 | 0.001361493 | 0.009464134 | 1 |
| Heart_Failure | mesothelium | *AP2S1* | -0.723327677 | 0.007266009 | 0.033102983 | 1 |
| Heart_Failure | mesothelium | *CTNNA1* | -0.689415407 | 0.000183159 | 0.002737794 | 0.888685428 |
| Heart_Failure | mesothelium | *YTHDC2* | -0.745502971 | 0.000287652 | 0.003276266 | 1 |
| Heart_Failure | mesothelium | *POLR2B* | -0.46661934 | 0.010561024 | 0.04288041 | 1 |
| Heart_Failure | mesothelium | *RRM2B* | -0.636222187 | 0.00017135 | 0.002708119 | 0.831392489 |
| Heart_Failure | mesothelium | *SNX29* | -0.320206858 | 0.00916834 | 0.038366745 | 1 |
| Heart_Failure | mesothelium | *RCN1* | -0.344508891 | 0.001472769 | 0.010036338 | 1 |
| Heart_Failure | mesothelium | *MCUR1* | -0.370964977 | 0.001901784 | 0.01227047 | 1 |
| Heart_Failure | mesothelium | *MAPK9* | -1.035245633 | 6.78E-05 | 0.002273164 | 0.329041051 |
| Heart_Failure | mesothelium | *CYBA* | -0.646114192 | 0.001505812 | 0.010189958 | 1 |
| Heart_Failure | mesothelium | *TBC1D22A* | -0.51048931 | 0.000559965 | 0.004976097 | 1 |
| Heart_Failure | mesothelium | *LAMC2* | -0.098128055 | 0.001013481 | 0.007702226 | 1 |
| Heart_Failure | mesothelium | *DNAJC25* | -1.394967523 | 0.000150453 | 0.002656078 | 0.729998221 |
| Heart_Failure | mesothelium | *RIMBP2* | -0.405376643 | 0.009402011 | 0.039056985 | 1 |
| Heart_Failure | mesothelium | *PRDM6* | -0.7653491 | 0.000927835 | 0.00722659 | 1 |
| Heart_Failure | mesothelium | *SPAG4* | -0.19627963 | 0.00908377 | 0.038225892 | 1 |
| Heart_Failure | mesothelium | *NCKAP1* | -0.583712538 | 0.000188165 | 0.002744551 | 0.912976824 |
| Heart_Failure | mesothelium | *AHRR* | -0.902038523 | 6.20E-05 | 0.002273164 | 0.300890824 |
| Heart_Failure | mesothelium | *ADCK1* | -1.176684069 | 0.000200353 | 0.002777753 | 0.972112999 |
| Heart_Failure | mesothelium | *RNF4* | -0.92343611 | 0.000277585 | 0.003196348 | 1 |
| Heart_Failure | mesothelium | *TM7SF3* | -0.418600889 | 0.011220953 | 0.044785675 | 1 |
| Heart_Failure | mesothelium | *TMEM161A* | -0.698688987 | 0.00537333 | 0.026123643 | 1 |
| Heart_Failure | mesothelium | *SUGP2* | -0.449288173 | 0.008844183 | 0.037769181 | 1 |
| Heart_Failure | mesothelium | *SLC12A2* | -0.573737771 | 0.012578762 | 0.049179816 | 1 |
| Heart_Failure | mesothelium | *MAP2K4* | -0.707677759 | 7.40E-05 | 0.002273164 | 0.359288991 |
| Heart_Failure | mesothelium | *TIE1* | -0.161945783 | 0.003200759 | 0.017607802 | 1 |
| Heart_Failure | mesothelium | *ZNRD1* | -0.104438231 | 0.005926718 | 0.028275749 | 1 |
| Heart_Failure | mesothelium | *ATXN3* | -0.13746118 | 0.004877525 | 0.024322456 | 1 |
| Heart_Failure | mesothelium | *GOLGA5* | -0.782614846 | 3.47E-05 | 0.002273164 | 0.16856333 |

| | | | | | | |
|---|---|---|---|---|---|---|
| Heart_Failure | mesothelium | *MTFR1* | -0.347216491 | 0.001848163 | 0.012036629 | 1 |
| Heart_Failure | mesothelium | *EVI5* | -0.364603336 | 1.82E-05 | 0.002003718 | 0.088163613 |
| Heart_Failure | mesothelium | *STOML1* | -1.948047996 | 0.001312433 | 0.009228876 | 1 |
| Heart_Failure | mesothelium | *METTL22* | -0.535135418 | 0.00359023 | 0.019269683 | 1 |
| Heart_Failure | mesothelium | *TP53BP1* | -0.500952107 | 0.000829101 | 0.006671304 | 1 |
| Heart_Failure | mesothelium | *DHX8* | -1.043305995 | 0.000180099 | 0.00272738 | 0.873838201 |
| Heart_Failure | mesothelium | *ROGDI* | -2.199049968 | 7.22E-05 | 0.002273164 | 0.350289189 |
| Heart_Failure | mesothelium | *CBFB* | -0.565870348 | 0.001234105 | 0.008824657 | 1 |
| Heart_Failure | mesothelium | *CYFIP1* | -0.802292737 | 3.36E-06 | 0.000811728 | 0.016301005 |
| Heart_Failure | mesothelium | *IFT80* | -0.800846169 | 0.000419348 | 0.004098206 | 1 |
| Heart_Failure | mesothelium | *NUCKS1* | -0.705333922 | 7.03E-05 | 0.002273164 | 0.341009715 |
| Heart_Failure | mesothelium | *CLEC2D* | -0.324994533 | 0.010233197 | 0.041864646 | 1 |
| Heart_Failure | mesothelium | *DRD4* | -1.115276368 | 0.001348016 | 0.009397378 | 1 |
| Heart_Failure | mesothelium | *LRP6* | -0.521753067 | 0.009042658 | 0.038152152 | 1 |
| Heart_Failure | mesothelium | *ZXDC* | -0.545457014 | 0.003101455 | 0.017237409 | 1 |
| Heart_Failure | mesothelium | *EIF2B3* | -0.591012576 | 0.001684533 | 0.011150548 | 1 |
| Heart_Failure | mesothelium | *MAP4K4* | -0.626957052 | 0.000238925 | 0.002946978 | 1 |
| Heart_Failure | mesothelium | *WDR1* | -0.520513738 | 0.007162656 | 0.032724299 | 1 |
| Heart_Failure | mesothelium | *SNX13* | -0.725075717 | 0.000223328 | 0.002905056 | 1 |
| Heart_Failure | mesothelium | *RPS6KA2* | -0.422386292 | 0.000110273 | 0.002412406 | 0.535046961 |
| Heart_Failure | mesothelium | *AFF4* | -0.368236743 | 0.008417541 | 0.036596693 | 1 |
| Heart_Failure | mesothelium | *CRMP1* | -1.047219561 | 4.22E-05 | 0.002273164 | 0.204538187 |
| Heart_Failure | mesothelium | *AP1M1* | -0.66602097 | 0.002579056 | 0.015094791 | 1 |
| Heart_Failure | mesothelium | *LLGL2* | -0.419896969 | 0.012005435 | 0.0472428 | 1 |
| Heart_Failure | mesothelium | *ADAM11* | -0.064600493 | 0.007606575 | 0.034141629 | 1 |
| Heart_Failure | mesothelium | *ZNF532* | -0.794463956 | 0.000165687 | 0.002704817 | 0.803914718 |
| Heart_Failure | mesothelium | *GRAMD4* | -0.350596912 | 0.003536348 | 0.019064847 | 1 |
| Heart_Failure | mesothelium | *ADD2* | -0.363899818 | 0.000899066 | 0.007035916 | 1 |
| Heart_Failure | mesothelium | *FNDC3B* | -0.534550658 | 0.00044318 | 0.00425804 | 1 |
| Heart_Failure | mesothelium | *KIFAP3* | -0.761187423 | 4.21E-05 | 0.002273164 | 0.204055502 |
| Heart_Failure | mesothelium | *MCM6* | -0.197287757 | 0.009099903 | 0.038226758 | 1 |
| Heart_Failure | mesothelium | *ANKRD13A* | -0.648646155 | 0.002359901 | 0.01418865 | 1 |
| Heart_Failure | mesothelium | *GPATCH1* | -0.364094729 | 0.010381038 | 0.042291181 | 1 |
| Heart_Failure | mesothelium | *IL4R* | -0.54887564 | 0.007523878 | 0.033854871 | 1 |
| Heart_Failure | mesothelium | *DYNC1I2* | -0.729910136 | 0.00045022 | 0.004301299 | 1 |
| Heart_Failure | mesothelium | *PIK3C3* | -0.718545606 | 3.20E-05 | 0.002273164 | 0.155332389 |
| Heart_Failure | mesothelium | *PCM1* | -0.356331155 | 0.005003171 | 0.024745551 | 1 |
| Heart_Failure | mesothelium | *MYH7B* | -0.168702885 | 0.010197766 | 0.041754902 | 1 |
| Heart_Failure | mesothelium | *XRCC5* | -0.546695359 | 0.001052977 | 0.007933296 | 1 |
| Heart_Failure | mesothelium | *SAR1A* | -0.409426537 | 0.006771935 | 0.031292789 | 1 |
| Heart_Failure | mesothelium | *DNM2* | -0.574612245 | 0.000139046 | 0.002608902 | 0.674650966 |
| Heart_Failure | mesothelium | *STX7* | -0.413958251 | 0.000726595 | 0.006036714 | 1 |
| Heart_Failure | mesothelium | *RABL2B* | -0.75895879 | 0.004857564 | 0.024272814 | 1 |
| Heart_Failure | mesothelium | *SCTR* | -1.175813062 | 7.58E-05 | 0.002273164 | 0.367781945 |
| Heart_Failure | mesothelium | *ME2* | -0.834525384 | 0.000535927 | 0.004815408 | 1 |
| Heart_Failure | mesothelium | *CCNT2* | -0.350524486 | 0.0019043 | 0.01227047 | 1 |
| Heart_Failure | mesothelium | *DOPEY1* | -0.162666899 | 0.007744522 | 0.034632644 | 1 |
| Heart_Failure | mesothelium | *ZCCHC6* | -0.806769794 | 9.60E-05 | 0.002294209 | 0.465724339 |
| Heart_Failure | mesothelium | *ULK2* | -0.318657976 | 0.007118253 | 0.032552088 | 1 |
| Heart_Failure | mesothelium | *PIBF1* | -0.630981119 | 0.000312754 | 0.003472497 | 1 |

| | | | | | | |
|---|---|---|---|---|---|---|
| Heart_Failure | mesothelium | *OXCT1* | -0.499552123 | 2.25E-06 | 0.000682274 | 0.010916386 |
| Heart_Failure | mesothelium | *ZNF586* | -0.28721164 | 0.003166789 | 0.017480389 | 1 |
| Heart_Failure | mesothelium | *ZNF264* | -1.062754487 | 0.000137444 | 0.002596678 | 0.666879973 |
| Heart_Failure | mesothelium | *RPS5* | -0.308936074 | 0.001454012 | 0.009922453 | 1 |
| Heart_Failure | mesothelium | *WBP11* | -1.105696615 | 0.000161105 | 0.002704817 | 0.781681844 |
| Heart_Failure | mesothelium | *NCOA1* | -0.504850562 | 0.001158863 | 0.008417372 | 1 |
| Heart_Failure | mesothelium | *HACE1* | -0.678346442 | 0.002337498 | 0.014106394 | 1 |
| Heart_Failure | mesothelium | *MTIF2* | -0.981273966 | 0.000381644 | 0.003893896 | 1 |
| Heart_Failure | mesothelium | *USP40* | -0.759757524 | 0.000155682 | 0.002678614 | 0.755369269 |
| Heart_Failure | mesothelium | *EIF2AK1* | -0.546960856 | 0.009981878 | 0.041183736 | 1 |
| Heart_Failure | mesothelium | *ITPKC* | -0.193070404 | 0.007946699 | 0.035314193 | 1 |
| Heart_Failure | mesothelium | *ALG9* | -1.485914014 | 0.000482887 | 0.004523107 | 1 |
| Heart_Failure | mesothelium | *SRRT* | -0.550717952 | 0.00369226 | 0.019724749 | 1 |
| Heart_Failure | mesothelium | *ADAMTS2* | -0.832850552 | 0.000100804 | 0.002318018 | 0.489101814 |
| Heart_Failure | mesothelium | *UIMC1* | -0.751742575 | 7.33E-05 | 0.002273164 | 0.355714938 |
| Heart_Failure | mesothelium | *LPCAT2* | -0.691114873 | 0.003579373 | 0.019232689 | 1 |
| Heart_Failure | mesothelium | *AAMDC* | -0.132512864 | 0.004306201 | 0.022067281 | 1 |
| Heart_Failure | mesothelium | *ASAP3* | -0.525647605 | 0.006209304 | 0.029335486 | 1 |
| Heart_Failure | mesothelium | *ANKRD10* | -0.694815024 | 8.40E-06 | 0.001164647 | 0.040762653 |
| Heart_Failure | mesothelium | *DOCK3* | -0.291784124 | 0.006555101 | 0.03062164 | 1 |
| Heart_Failure | mesothelium | *ARHGAP28* | -0.319928927 | 0.001907447 | 0.012274447 | 1 |
| Heart_Failure | mesothelium | *EBF4* | -0.467296403 | 0.004687723 | 0.023667881 | 1 |
| Heart_Failure | mesothelium | *XRN2* | -0.473532881 | 0.001395107 | 0.009615141 | 1 |
| Heart_Failure | mesothelium | *SNX5* | -0.916406721 | 0.000527778 | 0.004768677 | 1 |
| Heart_Failure | mesothelium | *ESF1* | -0.274863162 | 0.012458499 | 0.048867127 | 1 |
| Heart_Failure | mesothelium | *TMEM230* | -0.419907105 | 0.000225974 | 0.002908664 | 1 |
| Heart_Failure | mesothelium | *KIF16B* | -0.677619633 | 0.001100424 | 0.008139111 | 1 |
| Heart_Failure | mesothelium | *GRAMD1A* | -0.933246776 | 0.000245036 | 0.002972283 | 1 |
| Heart_Failure | mesothelium | *CDIP1* | -1.309585538 | 5.37E-05 | 0.002273164 | 0.260377278 |
| Heart_Failure | mesothelium | *GANAB* | -0.346931404 | 0.005694578 | 0.02738364 | 1 |
| Heart_Failure | mesothelium | *MLF2* | -0.236486477 | 0.000844119 | 0.006758528 | 1 |
| Heart_Failure | mesothelium | *ANKRD24* | -1.5752017 | 0.000290127 | 0.00328901 | 1 |
| Heart_Failure | mesothelium | *MRPS33* | -0.250261636 | 0.002661692 | 0.015485049 | 1 |
| Heart_Failure | mesothelium | *GNPTG* | -0.471365804 | 0.008142626 | 0.035832979 | 1 |
| Heart_Failure | mesothelium | *PABPC4* | -0.297635804 | 3.51E-06 | 0.000811728 | 0.017046279 |
| Heart_Failure | mesothelium | *CERS4* | -0.995597735 | 7.23E-05 | 0.002273164 | 0.350908347 |
| Heart_Failure | mesothelium | *GLG1* | -0.71674079 | 0.000106274 | 0.002398327 | 0.515640403 |
| Heart_Failure | mesothelium | *DTX2* | -1.273943542 | 5.89E-05 | 0.002273164 | 0.285543402 |
| Heart_Failure | mesothelium | *TF* | -0.861603367 | 0.009156524 | 0.038365675 | 1 |
| Heart_Failure | mesothelium | *ALKBH5* | -0.35271892 | 0.001021581 | 0.007720732 | 1 |
| Heart_Failure | mesothelium | *SCFD1* | -0.671688471 | 0.000235899 | 0.002946978 | 1 |
| Heart_Failure | mesothelium | *SUPT16H* | -0.99200317 | 8.82E-05 | 0.002275786 | 0.427847769 |
| Heart_Failure | mesothelium | *TOX4* | -0.458234134 | 0.005221372 | 0.025538402 | 1 |
| Heart_Failure | mesothelium | *COMT* | -0.847498343 | 0.000290118 | 0.00328901 | 1 |
| Heart_Failure | mesothelium | *SEC22C* | -0.866407764 | 0.000592906 | 0.005174062 | 1 |
| Heart_Failure | mesothelium | *XYLB* | -1.042914365 | 0.000183949 | 0.002737794 | 0.892520992 |
| Heart_Failure | mesothelium | *MSH2* | -1.026907083 | 0.000110589 | 0.002412406 | 0.536576031 |
| Heart_Failure | mesothelium | *PSMD5* | -0.739579765 | 0.00204161 | 0.012881523 | 1 |
| Heart_Failure | mesothelium | *CRAT* | -1.585827545 | 0.000190625 | 0.002744551 | 0.92491369 |
| Heart_Failure | mesothelium | *NANS* | -0.656988196 | 0.004136462 | 0.021442428 | 1 |

| | | | | | | |
|---|---|---|---|---|---|---|
| Heart_Failure | mesothelium | *WAC* | -0.470971355 | 0.003069066 | 0.017175442 | 1 |
| Heart_Failure | mesothelium | *DSP* | -0.647950761 | 0.002075062 | 0.013058627 | 1 |
| Heart_Failure | mesothelium | *IFT74* | -0.269425255 | 0.0021469 | 0.013389149 | 1 |
| Heart_Failure | mesothelium | *RAB18* | -0.84441915 | 0.000208197 | 0.002796296 | 1 |
| Heart_Failure | mesothelium | *CIRBP* | -0.289156521 | 0.00254615 | 0.01499262 | 1 |
| Heart_Failure | mesothelium | *MED15* | -0.623232071 | 0.000301887 | 0.00338281 | 1 |
| Heart_Failure | mesothelium | *BCL2L13* | -0.207427932 | 0.009469236 | 0.039269001 | 1 |
| Heart_Failure | mesothelium | *HPS4* | -0.613372672 | 0.000852247 | 0.006789985 | 1 |
| Heart_Failure | mesothelium | *TTC28* | -0.596187611 | 7.77E-05 | 0.002273164 | 0.377069632 |
| Heart_Failure | mesothelium | *TIMP3* | -0.514867111 | 0.000840425 | 0.006740067 | 1 |
| Heart_Failure | mesothelium | *PPP6R2* | -0.733030584 | 4.87E-05 | 0.002273164 | 0.236350921 |
| Heart_Failure | mesothelium | *TSPO* | -0.693082953 | 0.003525028 | 0.019024955 | 1 |
| Heart_Failure | mesothelium | *SAMM50* | -0.526893883 | 0.001144686 | 0.008364486 | 1 |
| Heart_Failure | mesothelium | *ZC3H7B* | -0.400233652 | 0.005414186 | 0.026269629 | 1 |
| Heart_Failure | mesothelium | *NIN* | -0.527624723 | 0.001391723 | 0.009607077 | 1 |
| Heart_Failure | mesothelium | *CNIH1* | -0.481289564 | 0.007321524 | 0.033231087 | 1 |
| Heart_Failure | mesothelium | *ATP6V1D* | -0.907128519 | 7.96E-05 | 0.002273164 | 0.386388244 |
| Heart_Failure | mesothelium | *GSKIP* | -1.54931724 | 5.23E-05 | 0.002273164 | 0.253907948 |
| Heart_Failure | mesothelium | *YY1* | -0.874117281 | 0.000147136 | 0.002637401 | 0.7139048 |
| Heart_Failure | mesothelium | *CCNB1IP1* | -1.067831418 | 0.000189545 | 0.002744551 | 0.919670726 |
| Heart_Failure | mesothelium | *CD40* | -0.790581947 | 0.001277525 | 0.009009523 | 1 |
| Heart_Failure | mesothelium | *PABPC1L* | -0.571416073 | 0.002032093 | 0.012854907 | 1 |
| Heart_Failure | mesothelium | *DOK5* | -1.054055032 | 0.000143896 | 0.002637401 | 0.698182809 |
| Heart_Failure | mesothelium | *CTSZ* | -0.638341289 | 0.002977189 | 0.016777377 | 1 |
| Heart_Failure | mesothelium | *GMEB2* | -1.357010748 | 3.09E-06 | 0.000811728 | 0.014978254 |
| Heart_Failure | mesothelium | *RNF24* | -0.369113587 | 0.001195969 | 0.008622349 | 1 |
| Heart_Failure | mesothelium | *NDUFAF5* | -0.095362563 | 0.001952495 | 0.012481563 | 1 |
| Heart_Failure | mesothelium | *TRIB3* | -1.192475353 | 0.00640309 | 0.030104452 | 1 |
| Heart_Failure | mesothelium | *TTI1* | -0.525601754 | 0.001610422 | 0.01077761 | 1 |
| Heart_Failure | mesothelium | *MAP1LC3A* | -0.580760933 | 0.001557701 | 0.010453614 | 1 |
| Heart_Failure | mesothelium | *METTL4* | -0.935660834 | 0.000156763 | 0.002683421 | 0.760612692 |
| Heart_Failure | mesothelium | *MIB1* | -0.771564128 | 9.51E-05 | 0.002294209 | 0.461340258 |
| Heart_Failure | mesothelium | *STK24* | -0.713627752 | 0.000396936 | 0.003987441 | 1 |
| Heart_Failure | mesothelium | *DNAJC3* | -0.373935818 | 0.007387937 | 0.033376414 | 1 |
| Heart_Failure | mesothelium | *UGGT2* | -0.313175461 | 0.007833641 | 0.034902503 | 1 |
| Heart_Failure | mesothelium | *ARHGEF7* | -0.638031017 | 0.000810522 | 0.006576342 | 1 |
| Heart_Failure | mesothelium | *MRPS31* | -0.724829914 | 0.000769746 | 0.00629816 | 1 |
| Heart_Failure | mesothelium | *KATNAL1* | -0.845490177 | 8.34E-05 | 0.002273164 | 0.404623109 |
| Heart_Failure | mesothelium | *INTS6* | -0.411604593 | 0.006095918 | 0.028884175 | 1 |
| Heart_Failure | mesothelium | *PSMD7* | -0.536425721 | 0.007296505 | 0.033165623 | 1 |
| Heart_Failure | mesothelium | *MON1B* | -1.221543713 | 0.000450342 | 0.004301299 | 1 |
| Heart_Failure | mesothelium | *LMF1* | -0.803970526 | 0.000531335 | 0.004783002 | 1 |
| Heart_Failure | mesothelium | *NUBP1* | -0.404942448 | 0.002268344 | 0.013809292 | 1 |
| Heart_Failure | mesothelium | *UBE2I* | -0.621254549 | 0.004418684 | 0.02256776 | 1 |
| Heart_Failure | mesothelium | *TOX3* | 0.182760732 | 0.008843141 | 0.037769181 | 1 |
| Heart_Failure | mesothelium | *KAT8* | -1.141935505 | 1.28E-05 | 0.001557659 | 0.062245193 |
| Heart_Failure | mesothelium | *C16orf62* | -0.778687004 | 0.000166543 | 0.002704817 | 0.808065362 |
| Heart_Failure | mesothelium | *RNF40* | -0.761449924 | 0.002340752 | 0.014108482 | 1 |
| Heart_Failure | mesothelium | *KNOP1* | -0.537303844 | 0.006937728 | 0.031906972 | 1 |
| Heart_Failure | mesothelium | *IQCH* | -0.679860212 | 0.005975014 | 0.028407099 | 1 |

| | | | | | | |
|---|---|---|---|---|---|---|
| Heart_Failure | mesothelium | *HERC1* | -0.485313061 | 0.00221232 | 0.013604784 | 1 |
| Heart_Failure | mesothelium | *CTSH* | -0.050664588 | 0.003306592 | 0.018101389 | 1 |
| Heart_Failure | mesothelium | *TTC23* | -0.817109832 | 0.00024052 | 0.002946978 | 1 |
| Heart_Failure | mesothelium | *FAH* | -0.552431371 | 0.000625624 | 0.005389776 | 1 |
| Heart_Failure | mesothelium | *RPAP1* | -0.232757327 | 0.008520718 | 0.036853388 | 1 |
| Heart_Failure | mesothelium | *SLC30A4* | -0.650376106 | 0.01061226 | 0.04300974 | 1 |
| Heart_Failure | mesothelium | *ZFAND1* | -0.544751071 | 0.00366407 | 0.019600954 | 1 |
| Heart_Failure | mesothelium | *IMPAD1* | -0.672330237 | 0.000717442 | 0.005974896 | 1 |
| Heart_Failure | mesothelium | *SFRP1* | -0.516749026 | 0.010788347 | 0.043475961 | 1 |
| Heart_Failure | mesothelium | *RAB2A* | -0.329117409 | 0.003986081 | 0.020818584 | 1 |
| Heart_Failure | mesothelium | *CHRAC1* | -1.524624524 | 0.000758778 | 0.006220115 | 1 |
| Heart_Failure | mesothelium | *ERICH1* | -0.423311637 | 0.002638302 | 0.015367398 | 1 |
| Heart_Failure | mesothelium | *TUSC3* | -0.908613542 | 5.86E-05 | 0.002273164 | 0.284367621 |
| Heart_Failure | mesothelium | *SNRNP70* | -0.803920661 | 5.17E-05 | 0.002273164 | 0.250671631 |
| Heart_Failure | mesothelium | *ERCC2* | -0.609889888 | 0.001544556 | 0.010393583 | 1 |
| Heart_Failure | mesothelium | *MED25* | -1.171568405 | 8.12E-05 | 0.002273164 | 0.393838884 |
| Heart_Failure | mesothelium | *ANKRD27* | -0.691959127 | 0.000854322 | 0.006795359 | 1 |
| Heart_Failure | mesothelium | *CCDC9* | -0.458856059 | 0.002380384 | 0.014276417 | 1 |
| Heart_Failure | mesothelium | *TGFB1* | -0.722821232 | 0.00107101 | 0.008019351 | 1 |
| Heart_Failure | mesothelium | *CD33* | -0.044884696 | 0.011604495 | 0.046105142 | 1 |
| Heart_Failure | mesothelium | *CAPS* | -1.354484444 | 0.001317913 | 0.009240625 | 1 |
| Heart_Failure | mesothelium | *PLEKHA4* | -0.487365103 | 0.003084371 | 0.017231146 | 1 |
| Heart_Failure | mesothelium | *TFPT* | -0.950386451 | 0.001566286 | 0.010496716 | 1 |
| Heart_Failure | mesothelium | *RAB3A* | -3.573523606 | 0.000631198 | 0.005418262 | 1 |
| Heart_Failure | mesothelium | *ZNF14* | -1.209687497 | 9.58E-05 | 0.002294209 | 0.465017538 |
| Heart_Failure | mesothelium | *ZNF85* | -0.870162136 | 0.001168624 | 0.00846293 | 1 |
| Heart_Failure | mesothelium | *ETHE1* | -0.632227583 | 0.005217215 | 0.025538402 | 1 |
| Heart_Failure | mesothelium | *RASA4* | -5.954820318 | 0.00669851 | 0.031101598 | 1 |
| Heart_Failure | mesothelium | *BET1* | -0.661257545 | 0.001049934 | 0.007922674 | 1 |
| Heart_Failure | mesothelium | *DNAH11* | -0.631794767 | 0.000758926 | 0.006220115 | 1 |
| Heart_Failure | mesothelium | *PTN* | -0.524003633 | 0.002864997 | 0.016296558 | 1 |
| Heart_Failure | mesothelium | *IQCE* | -1.133597581 | 6.91E-05 | 0.002273164 | 0.335224363 |
| Heart_Failure | mesothelium | *VIPR2* | -0.244187227 | 0.004464756 | 0.022755248 | 1 |
| Heart_Failure | mesothelium | *CPVL* | -0.550036143 | 0.002496904 | 0.014792403 | 1 |
| Heart_Failure | mesothelium | *GRB10* | -0.74386431 | 0.000135487 | 0.00257797 | 0.657382239 |
| Heart_Failure | mesothelium | *PLEKHA8* | -0.680723648 | 0.004088124 | 0.021259998 | 1 |
| Heart_Failure | mesothelium | *ZKSCAN1* | -0.500372941 | 0.003312868 | 0.018101389 | 1 |
| Heart_Failure | mesothelium | *EIF3B* | -0.90906976 | 0.000121305 | 0.002465411 | 0.588570914 |
| Heart_Failure | mesothelium | *C1GALT1* | -0.828181487 | 0.000170183 | 0.002704817 | 0.825729792 |
| Heart_Failure | mesothelium | *NRF1* | -0.320684535 | 0.006724581 | 0.03113327 | 1 |
| Heart_Failure | mesothelium | *ZNF862* | -0.378169046 | 0.006907922 | 0.031854415 | 1 |
| Heart_Failure | mesothelium | *POU6F2* | -0.41407285 | 0.008960189 | 0.037928602 | 1 |
| Heart_Failure | mesothelium | *RARRES2* | -0.407917533 | 0.00226693 | 0.013809292 | 1 |
| Heart_Failure | mesothelium | *COA1* | -0.443042157 | 0.004743424 | 0.023849837 | 1 |
| Heart_Failure | mesothelium | *URGCP* | -0.591941948 | 6.49E-05 | 0.002273164 | 0.314830979 |
| Heart_Failure | mesothelium | *AEBP1* | -0.913528085 | 0.000212282 | 0.002837451 | 1 |
| Heart_Failure | mesothelium | *GCK* | -5.078038911 | 0.001243183 | 0.00883272 | 1 |
| Heart_Failure | mesothelium | *FKTN* | -0.829292273 | 0.000266742 | 0.003131364 | 1 |
| Heart_Failure | mesothelium | *MEGF9* | -0.87467493 | 8.97E-05 | 0.002278979 | 0.435284987 |
| Heart_Failure | mesothelium | *ECM2* | -0.632617144 | 0.000572438 | 0.005049943 | 1 |

| | | | | | | |
|---|---|---|---|---|---|---|
| Heart_Failure | mesothelium | *FUBP3* | -0.362674012 | 0.010065373 | 0.041414098 | 1 |
| Heart_Failure | mesothelium | *RGP1* | -0.772904372 | 2.52E-06 | 0.000719772 | 0.012236131 |
| Heart_Failure | mesothelium | *PTGDS* | -0.780495928 | 0.003787381 | 0.020083465 | 1 |
| Heart_Failure | mesothelium | *CUBN* | -0.277732868 | 0.01108609 | 0.044380947 | 1 |
| Heart_Failure | mesothelium | *PLEKHA1* | -0.292604767 | 0.002704242 | 0.015676205 | 1 |
| Heart_Failure | mesothelium | *CDH23* | -0.73421043 | 9.81E-05 | 0.0022988 | 0.475851681 |
| Heart_Failure | mesothelium | *BMPR1A* | -0.677979793 | 0.0001299 | 0.00251464 | 0.630277029 |
| Heart_Failure | mesothelium | *SFXN3* | -0.92474093 | 0.003192718 | 0.017603484 | 1 |
| Heart_Failure | mesothelium | *TNKS2* | -0.819962231 | 0.000163452 | 0.002704817 | 0.793070396 |
| Heart_Failure | mesothelium | *TFAM* | -1.137807872 | 0.000236128 | 0.002946978 | 1 |
| Heart_Failure | mesothelium | *PSMB3* | -0.49958117 | 0.000234541 | 0.002946978 | 1 |
| Heart_Failure | mesothelium | *RPL19* | -0.361359733 | 0.002557565 | 0.015024293 | 1 |
| Heart_Failure | mesothelium | *TRIM37* | -0.444292964 | 0.011258857 | 0.044887407 | 1 |
| Heart_Failure | mesothelium | *TRIM16L* | -1.093423009 | 0.000417003 | 0.004098206 | 1 |
| Heart_Failure | mesothelium | *PIGL* | -0.511373482 | 0.002290188 | 0.013924802 | 1 |
| Heart_Failure | mesothelium | *RAI1* | -1.060652181 | 6.82E-05 | 0.002273164 | 0.330750465 |
| Heart_Failure | mesothelium | *GOSR1* | -0.844995348 | 0.000130085 | 0.00251464 | 0.631174534 |
| Heart_Failure | mesothelium | *PSMD11* | -0.220604215 | 0.010787931 | 0.043475961 | 1 |
| Heart_Failure | mesothelium | *EFTUD2* | -1.027975565 | 0.000127814 | 0.00251464 | 0.620153498 |
| Heart_Failure | mesothelium | *FAM20A* | -0.750209422 | 0.000527635 | 0.004768677 | 1 |
| Heart_Failure | mesothelium | *DPH1* | -0.509224624 | 0.008013368 | 0.035507635 | 1 |
| Heart_Failure | mesothelium | *DHRS7B* | -0.749262184 | 0.00056944 | 0.005032649 | 1 |
| Heart_Failure | mesothelium | *MYH3* | -1.442293436 | 0.00245367 | 0.014607614 | 1 |
| Heart_Failure | mesothelium | *OCIAD1* | -0.641419374 | 0.000411004 | 0.004069779 | 1 |
| Heart_Failure | mesothelium | *UBE2D3* | -0.647213256 | 0.000297219 | 0.003338215 | 1 |
| Heart_Failure | mesothelium | *NDUFC1* | -0.659254973 | 0.002932256 | 0.016562637 | 1 |
| Heart_Failure | mesothelium | *WFS1* | -1.149262715 | 0.000713513 | 0.005958635 | 1 |
| Heart_Failure | mesothelium | *GRPEL1* | -0.877596091 | 0.00866772 | 0.037349714 | 1 |
| Heart_Failure | mesothelium | *GAR1* | -1.330197425 | 5.22E-05 | 0.002273164 | 0.253337282 |
| Heart_Failure | mesothelium | *STIM2* | -0.708897901 | 0.000240093 | 0.002946978 | 1 |
| Heart_Failure | mesothelium | *UFSP2* | -0.954857561 | 6.03E-05 | 0.002273164 | 0.292567508 |
| Heart_Failure | mesothelium | *P2RX3* | -11.6832911 | 0.003742307 | 0.019887925 | 1 |
| Heart_Failure | mesothelium | *CEP164* | -0.534816386 | 0.001675131 | 0.011103464 | 1 |
| Heart_Failure | mesothelium | *GALNT18* | -0.252443174 | 0.001855583 | 0.012068748 | 1 |
| Heart_Failure | mesothelium | *NDUFS8* | -0.370845711 | 0.01105196 | 0.04428085 | 1 |
| Heart_Failure | mesothelium | *CORO1C* | -0.828010717 | 0.00019629 | 0.002776681 | 0.952401446 |
| Heart_Failure | mesothelium | *ASIC1* | -3.100809334 | 3.45E-05 | 0.002273164 | 0.167191678 |
| Heart_Failure | mesothelium | *KCTD10* | -0.347849729 | 4.01E-05 | 0.002273164 | 0.194610169 |
| Heart_Failure | mesothelium | *IL23A* | -9.646739172 | 4.50E-05 | 0.002273164 | 0.218226225 |
| Heart_Failure | mesothelium | *PARP11* | -0.897594608 | 0.000208627 | 0.002796296 | 1 |
| Heart_Failure | mesothelium | *GPN3* | -1.033057025 | 0.001546613 | 0.010393583 | 1 |
| Heart_Failure | mesothelium | *VPS29* | -0.734258774 | 0.000343327 | 0.003685453 | 1 |
| Heart_Failure | mesothelium | *ACAD10* | -0.274701641 | 0.00911549 | 0.038226758 | 1 |
| Heart_Failure | mesothelium | *CDK2AP1* | 0.175027337 | 0.000548307 | 0.004899424 | 1 |
| Heart_Failure | mesothelium | *DDX55* | -0.396009494 | 0.000620486 | 0.005366489 | 1 |
| Heart_Failure | mesothelium | *VDR* | -2.078841216 | 6.25E-05 | 0.002273164 | 0.303414704 |
| Heart_Failure | mesothelium | *CAND1* | -0.808926821 | 0.000118505 | 0.002465411 | 0.574984844 |
| Heart_Failure | mesothelium | *MDM1* | -0.829529197 | 0.002571379 | 0.01506803 | 1 |
| Heart_Failure | mesothelium | *NUP107* | -0.481358543 | 0.002194012 | 0.01352649 | 1 |
| Heart_Failure | mesothelium | *CHPT1* | -0.158722756 | 0.003328206 | 0.018123968 | 1 |

| | | | | | | |
|---|---|---|---|---|---|---|
| Heart_Failure | mesothelium | *GNPTAB* | -0.49412091 | 0.002774344 | 0.015949191 | 1 |
| Heart_Failure | mesothelium | *C12orf57* | -0.929453572 | 0.000404952 | 0.00404066 | 1 |
| Heart_Failure | mesothelium | *SUDS3* | -0.985548426 | 0.000114678 | 0.002462027 | 0.556418068 |
| Heart_Failure | mesothelium | *C2CD5* | -0.796761077 | 7.17E-05 | 0.002273164 | 0.347939838 |
| Heart_Failure | mesothelium | *FRK* | -1.144238976 | 0.000106042 | 0.002398327 | 0.514516997 |
| Heart_Failure | mesothelium | *TMEM14C* | -1.065520095 | 0.000167751 | 0.002704817 | 0.813926571 |
| Heart_Failure | mesothelium | *RNGTT* | -0.789642725 | 0.00014572 | 0.002637401 | 0.707033401 |
| Heart_Failure | mesothelium | *PHACTR1* | -0.217877004 | 0.000493366 | 0.004577076 | 1 |
| Heart_Failure | mesothelium | *MDGA1* | -0.454239898 | 0.009058789 | 0.03815386 | 1 |
| Heart_Failure | mesothelium | *ACOT13* | -0.857239026 | 0.000646318 | 0.005521008 | 1 |
| Heart_Failure | mesothelium | *HBS1L* | -0.397072979 | 8.92E-05 | 0.002278979 | 0.432762438 |
| Heart_Failure | mesothelium | *PACRG* | -0.371230763 | 0.005047441 | 0.024913715 | 1 |
| Heart_Failure | mesothelium | *QKI* | -0.503878898 | 0.001084434 | 0.008057695 | 1 |
| Heart_Failure | mesothelium | *TMEM30A* | -1.021523548 | 8.39E-05 | 0.002274545 | 0.40714351 |
| Heart_Failure | mesothelium | *NUDT12* | -1.192849977 | 7.74E-05 | 0.002273164 | 0.37530609 |
| Heart_Failure | mesothelium | *KIF20A* | -0.838065814 | 3.71E-05 | 0.002273164 | 0.179901538 |
| Heart_Failure | mesothelium | *TMCO6* | -0.173387693 | 0.002011947 | 0.012744082 | 1 |
| Heart_Failure | mesothelium | *SPARC* | -0.44057121 | 0.001902895 | 0.01227047 | 1 |
| Heart_Failure | mesothelium | *IK* | -0.150649124 | 0.01174704 | 0.046424408 | 1 |
| Heart_Failure | mesothelium | *ARSB* | -0.848854784 | 0.000421249 | 0.004104215 | 1 |
| Heart_Failure | mesothelium | *SLC12A7* | -0.650176663 | 0.008160588 | 0.035865193 | 1 |
| Heart_Failure | mesothelium | *SKP1* | -0.894501681 | 0.000101291 | 0.00231823 | 0.491464859 |
| Heart_Failure | mesothelium | *PPWD1* | -0.769446715 | 0.000455757 | 0.004344469 | 1 |
| Heart_Failure | mesothelium | *TXNDC15* | -0.96795622 | 0.000161623 | 0.002704817 | 0.784195462 |
| Heart_Failure | mesothelium | *SMAD5* | -0.812322955 | 0.000183902 | 0.002737794 | 0.892293823 |
| Heart_Failure | mesothelium | *ERGIC1* | -0.881411584 | 0.00010977 | 0.002412406 | 0.532604609 |
| Heart_Failure | mesothelium | *ACTR8* | -0.326113009 | 0.005509215 | 0.026571282 | 1 |
| Heart_Failure | mesothelium | *TBCCD1* | -1.074576372 | 0.000650585 | 0.005547691 | 1 |
| Heart_Failure | mesothelium | *CBLB* | -0.270374118 | 0.011699809 | 0.046340795 | 1 |
| Heart_Failure | mesothelium | *IQCG* | -0.870146836 | 9.68E-05 | 0.0022988 | 0.46951795 |
| Heart_Failure | mesothelium | *PLSCR4* | -0.377898648 | 0.004798792 | 0.024059866 | 1 |
| Heart_Failure | mesothelium | *COMMD2* | -0.569268158 | 0.003544798 | 0.019089189 | 1 |
| Heart_Failure | mesothelium | *ARHGEF26* | -0.903283706 | 0.000398027 | 0.003989732 | 1 |
| Heart_Failure | mesothelium | *NKTR* | -0.32391848 | 0.007792219 | 0.03474986 | 1 |
| Heart_Failure | mesothelium | *NRBP1* | -0.642592966 | 0.001768426 | 0.011595141 | 1 |
| Heart_Failure | mesothelium | *INO80B* | -0.190622773 | 0.000283322 | 0.003242169 | 1 |
| Heart_Failure | mesothelium | *POLE4* | -0.784907964 | 0.000867282 | 0.006875903 | 1 |
| Heart_Failure | mesothelium | *MRPL19* | -0.553753968 | 0.002232453 | 0.013705318 | 1 |
| Heart_Failure | mesothelium | *IL18R1* | 0.292066041 | 0.000196251 | 0.002776681 | 0.952210255 |
| Heart_Failure | mesothelium | *NOL10* | -0.687284114 | 0.000499366 | 0.004597575 | 1 |
| Heart_Failure | mesothelium | *STRN* | -0.734060333 | 0.000338579 | 0.003650637 | 1 |
| Heart_Failure | mesothelium | *SLC25A12* | -0.335842578 | 0.011174423 | 0.044697692 | 1 |
| Heart_Failure | mesothelium | *DARS* | -0.796345285 | 0.000238281 | 0.002946978 | 1 |
| Heart_Failure | mesothelium | *PNO1* | -1.57930019 | 7.04E-05 | 0.002273164 | 0.341571636 |
| Heart_Failure | mesothelium | *AAK1* | -0.584376336 | 0.000739138 | 0.006119107 | 1 |
| Heart_Failure | mesothelium | *ALMS1* | -0.74044203 | 0.000849855 | 0.006789985 | 1 |
| Heart_Failure | mesothelium | *RALGPS2* | -0.227135126 | 0.001060043 | 0.007974156 | 1 |
| Heart_Failure | mesothelium | *FAM20B* | -0.514152268 | 0.0025257 | 0.014926549 | 1 |
| Heart_Failure | mesothelium | *ICMT* | -0.710571345 | 0.010942156 | 0.043913435 | 1 |
| Heart_Failure | mesothelium | *PARK7* | -0.56991103 | 0.00033823 | 0.003650637 | 1 |

| | | | | | | |
|---|---|---|---|---|---|---|
| Heart_Failure | mesothelium | *MECR* | -0.308202298 | 0.008387825 | 0.036532968 | 1 |
| Heart_Failure | mesothelium | *EDEM3* | -0.464371529 | 0.010164553 | 0.041654065 | 1 |
| Heart_Failure | mesothelium | *ASH1L* | -0.242417828 | 0.010304393 | 0.042079547 | 1 |
| Heart_Failure | mesothelium | *DOCK7* | -0.733371846 | 0.000128558 | 0.00251464 | 0.623763605 |
| Heart_Failure | mesothelium | *PRG4* | -0.331584714 | 0.004546377 | 0.023056011 | 1 |
| Heart_Failure | mesothelium | *TBCE* | -0.984521965 | 8.34E-05 | 0.002273164 | 0.404544191 |
| Heart_Failure | mesothelium | *ACADM* | -0.400001822 | 0.002303871 | 0.013990463 | 1 |
| Heart_Failure | mesothelium | *SDHB* | -0.835025272 | 5.94E-05 | 0.002273164 | 0.288277285 |
| Heart_Failure | mesothelium | *ZNHIT6* | -0.528724482 | 0.000485734 | 0.004528961 | 1 |
| Heart_Failure | mesothelium | *ERI3* | -0.491315617 | 0.008780328 | 0.037667685 | 1 |
| Heart_Failure | mesothelium | *NSUN4* | -0.206950412 | 0.008243815 | 0.036067621 | 1 |
| Heart_Failure | mesothelium | *PRRC2C* | -0.442064709 | 0.000996142 | 0.007635516 | 1 |
| Heart_Failure | mesothelium | *ABCD3* | -0.862431801 | 0.000173722 | 0.002725564 | 0.842897974 |
| Heart_Failure | mesothelium | *RPS6KA1* | -0.400689225 | 0.001243353 | 0.00883272 | 1 |
| Heart_Failure | mesothelium | *ESYT2* | -0.32735019 | 0.008913475 | 0.037879761 | 1 |
| Heart_Failure | mesothelium | *RCN2* | -0.476377677 | 0.003324198 | 0.018122484 | 1 |
| Heart_Failure | mesothelium | *ALDH8A1* | -0.043328948 | 0.009290426 | 0.03869283 | 1 |
| Heart_Failure | mesothelium | *TMEM5* | -0.30009516 | 0.005154848 | 0.025315106 | 1 |
| Heart_Failure | mesothelium | *FOXO3* | -0.446642851 | 0.003506184 | 0.018944326 | 1 |
| Heart_Failure | mesothelium | *MFSD1* | -0.783787342 | 5.02E-05 | 0.002273164 | 0.24333067 |
| Heart_Failure | mesothelium | *UBN1* | -0.503763831 | 0.005231984 | 0.025564537 | 1 |
| Heart_Failure | mesothelium | *WDR35* | -0.383572063 | 0.00691317 | 0.031854415 | 1 |
| Heart_Failure | mesothelium | *SENP5* | -0.762364745 | 0.000163401 | 0.002704817 | 0.792823688 |
| Heart_Failure | mesothelium | *RAB14* | -0.892202432 | 3.04E-05 | 0.002273164 | 0.147346092 |
| Heart_Failure | mesothelium | *PPP6C* | -0.862859167 | 0.000231079 | 0.002942773 | 1 |
| Heart_Failure | mesothelium | *HSDL2* | -0.101341072 | 0.006440489 | 0.030221714 | 1 |
| Heart_Failure | mesothelium | *MAPKAP1* | -0.465692049 | 0.00752206 | 0.033854871 | 1 |
| Heart_Failure | mesothelium | *DENND1A* | -0.429452603 | 0.007115466 | 0.032552088 | 1 |
| Heart_Failure | mesothelium | *KDSR* | -0.942070325 | 3.61E-05 | 0.002273164 | 0.17523491 |
| Heart_Failure | mesothelium | *TTLL5* | -0.745908097 | 4.76E-05 | 0.002273164 | 0.230839247 |
| Heart_Failure | mesothelium | *COQ6* | -0.917565088 | 0.000206099 | 0.002777753 | 0.999991123 |
| Heart_Failure | mesothelium | *DNMT3A* | -0.208096722 | 0.011695256 | 0.046340795 | 1 |
| Heart_Failure | mesothelium | *CRIPT* | -1.135838831 | 0.000126815 | 0.00251464 | 0.615305918 |
| Heart_Failure | mesothelium | *PPP1R3C* | -0.66172725 | 0.012373164 | 0.048611004 | 1 |
| Heart_Failure | mesothelium | *MXI1* | -0.456734199 | 0.004096926 | 0.021282959 | 1 |
| Heart_Failure | mesothelium | *WDR11* | -0.352663173 | 0.008079084 | 0.03566853 | 1 |
| Heart_Failure | mesothelium | *KANSL1* | -0.633560896 | 7.47E-06 | 0.001067457 | 0.036261857 |
| Heart_Failure | mesothelium | *HOXB3* | -0.811814064 | 0.000228434 | 0.002923871 | 1 |
| Heart_Failure | mesothelium | *SLC10A7* | -0.748730935 | 0.000486313 | 0.004528961 | 1 |
| Heart_Failure | mesothelium | *MRC1* | -0.411499098 | 0.002262902 | 0.013809292 | 1 |
| Heart_Failure | mesothelium | *UFM1* | -0.629871508 | 2.77E-05 | 0.002273164 | 0.134632292 |
| Heart_Failure | mesothelium | *ALG5* | -0.254549659 | 0.004219982 | 0.021736042 | 1 |
| Heart_Failure | mesothelium | *SIL1* | -0.571397195 | 0.001010512 | 0.007702226 | 1 |
| Heart_Failure | mesothelium | *AKAP1* | -0.55594493 | 0.001441601 | 0.00986395 | 1 |
| Heart_Failure | mesothelium | *ECHDC2* | -0.230912979 | 0.003134775 | 0.017391229 | 1 |
| Heart_Failure | mesothelium | *ZNF549* | -0.787578996 | 0.003429113 | 0.018610803 | 1 |
| Heart_Failure | mesothelium | *LHX4* | -1.361003599 | 0.000270036 | 0.003149564 | 1 |
| Heart_Failure | mesothelium | *SEC22A* | -0.727108569 | 0.000205939 | 0.002777753 | 0.999215195 |
| Heart_Failure | mesothelium | *ZCCHC17* | -0.872449428 | 0.00057578 | 0.005053432 | 1 |
| Heart_Failure | mesothelium | *TNFSF10* | -0.723431907 | 0.000179195 | 0.002725564 | 0.869454873 |

| | | | | | | |
|---|---|---|---|---|---|---|
| Heart_Failure | mesothelium | *ZNF639* | -0.964816665 | 0.000140978 | 0.002614833 | 0.684022898 |
| Heart_Failure | mesothelium | *CD244* | -0.435941281 | 0.002784732 | 0.015971064 | 1 |
| Heart_Failure | mesothelium | *RBBP6* | -0.581988575 | 0.000961296 | 0.007427083 | 1 |
| Heart_Failure | mesothelium | *FAM35A* | -0.283458298 | 0.005080527 | 0.02505154 | 1 |
| Heart_Failure | mesothelium | *NAA60* | -1.639474856 | 9.77E-05 | 0.0022988 | 0.473869443 |
| Heart_Failure | mesothelium | *ODF2L* | -0.718276768 | 0.000333481 | 0.003626728 | 1 |
| Heart_Failure | mesothelium | *SPATA1* | -0.560401235 | 0.002190508 | 0.01352649 | 1 |
| Heart_Failure | mesothelium | *KLHL7* | -0.812006738 | 0.000205214 | 0.002777753 | 0.995700653 |
| Heart_Failure | mesothelium | *NT5C3A* | -0.480092698 | 0.007542242 | 0.033884219 | 1 |
| Heart_Failure | mesothelium | *C7orf49* | -0.078699424 | 0.011297659 | 0.045005124 | 1 |
| Heart_Failure | mesothelium | *RBM19* | -1.047120692 | 6.25E-05 | 0.002273164 | 0.303408653 |
| Heart_Failure | mesothelium | *CCDC91* | -0.404419805 | 0.006543163 | 0.030614686 | 1 |
| Heart_Failure | mesothelium | *ITIH5* | -0.139630309 | 6.46E-05 | 0.002273164 | 0.313226599 |
| Heart_Failure | mesothelium | *PFDN5* | -0.771331666 | 0.000294497 | 0.003330765 | 1 |
| Heart_Failure | mesothelium | *USP45* | -0.636647484 | 0.000867205 | 0.006875903 | 1 |
| Heart_Failure | mesothelium | *TTC21B* | -0.611647609 | 5.42E-05 | 0.002273164 | 0.262819285 |
| Heart_Failure | mesothelium | *OBSL1* | -0.32308868 | 0.009109253 | 0.038226758 | 1 |
| Heart_Failure | mesothelium | *SLC12A4* | -0.792598743 | 0.002268103 | 0.013809292 | 1 |
| Heart_Failure | mesothelium | *FAM210B* | -0.473201467 | 0.003295734 | 0.018068813 | 1 |
| Heart_Failure | mesothelium | *STAMBP* | -0.945860613 | 0.000153692 | 0.002672801 | 0.745711375 |
| Heart_Failure | mesothelium | *SNRNP27* | -0.775455345 | 0.005446952 | 0.026305984 | 1 |
| Heart_Failure | mesothelium | *USP22* | -0.627992293 | 0.000440427 | 0.004248409 | 1 |
| Heart_Failure | mesothelium | *SERPINB6* | -0.36915282 | 0.001992793 | 0.012655801 | 1 |
| Heart_Failure | mesothelium | *NQO2* | -0.672872851 | 0.002544688 | 0.01499262 | 1 |
| Heart_Failure | mesothelium | *ATXN1* | -0.336297952 | 0.008108234 | 0.035764685 | 1 |
| Heart_Failure | mesothelium | *CLYBL* | -0.905642801 | 4.78E-05 | 0.002273164 | 0.231964951 |
| Heart_Failure | mesothelium | *GGA3* | -0.427145965 | 0.001486867 | 0.010118204 | 1 |
| Heart_Failure | mesothelium | *NUP85* | -0.294973615 | 0.002798621 | 0.016031769 | 1 |
| Heart_Failure | mesothelium | *NT5C* | -0.950180148 | 0.000155124 | 0.002678614 | 0.752662755 |
| Heart_Failure | mesothelium | *DDX31* | -0.17791043 | 0.008072668 | 0.03566853 | 1 |
| Heart_Failure | mesothelium | *CCDC93* | -0.279004511 | 0.005915038 | 0.0282478 | 1 |
| Heart_Failure | mesothelium | *GTF2F1* | -0.25593474 | 0.008522156 | 0.036853388 | 1 |
| Heart_Failure | mesothelium | *SNRPD2* | -1.205143492 | 0.000147177 | 0.002637401 | 0.714104877 |
| Heart_Failure | mesothelium | *KLC1* | -0.67685622 | 0.001982233 | 0.012621777 | 1 |
| Heart_Failure | mesothelium | *TUBGCP3* | -0.562441649 | 0.003723175 | 0.019807947 | 1 |
| Heart_Failure | mesothelium | *IRF3* | -0.758681038 | 0.000410356 | 0.004069779 | 1 |
| Heart_Failure | mesothelium | *ZNF384* | -0.347953184 | 0.000363993 | 0.003806239 | 1 |
| Heart_Failure | mesothelium | *EMG1* | -0.243809446 | 0.010320244 | 0.042079547 | 1 |
| Heart_Failure | mesothelium | *KTN1* | -0.703781646 | 0.000122402 | 0.002465411 | 0.593896326 |
| Heart_Failure | mesothelium | *HELB* | -0.47037288 | 0.006888809 | 0.031802569 | 1 |
| Heart_Failure | mesothelium | *RAB3IP* | -0.686461716 | 0.006741638 | 0.031182488 | 1 |
| Heart_Failure | mesothelium | *SMARCA4* | -0.377749549 | 0.001864363 | 0.012093436 | 1 |
| Heart_Failure | mesothelium | *PNKD* | -0.515972838 | 0.000754898 | 0.006208073 | 1 |
| Heart_Failure | mesothelium | *HIP1* | -0.633545027 | 0.000393287 | 0.003975479 | 1 |
| Heart_Failure | mesothelium | *PEX1* | -0.932153703 | 0.000195326 | 0.002776681 | 0.947722915 |
| Heart_Failure | mesothelium | *KDR* | -0.60061373 | 0.000514883 | 0.004695887 | 1 |
| Heart_Failure | mesothelium | *ATF4* | -1.085410447 | 0.00016942 | 0.002704817 | 0.822027881 |
| Heart_Failure | mesothelium | *CDC42EP1* | -0.682702966 | 0.000217497 | 0.00288242 | 1 |
| Heart_Failure | mesothelium | *SPECC1* | -0.714327778 | 0.000462767 | 0.00438544 | 1 |
| Heart_Failure | mesothelium | *CDHR3* | -0.515503069 | 0.007780987 | 0.034731691 | 1 |

| | | | | | | |
|---|---|---|---|---|---|---|
| Heart_Failure | mesothelium | *PRKRIP1* | -0.399176178 | 0.011984117 | 0.047197189 | 1 |
| Heart_Failure | mesothelium | *MTX2* | -0.74661313 | 0.000417755 | 0.004098206 | 1 |
| Heart_Failure | mesothelium | *ARPP19* | -0.910142152 | 0.000340954 | 0.003668093 | 1 |
| Heart_Failure | mesothelium | *VPS13C* | -0.40297293 | 0.00314538 | 0.017421672 | 1 |
| Heart_Failure | mesothelium | *SUMF2* | -0.726942378 | 0.000429487 | 0.004176097 | 1 |
| Heart_Failure | mesothelium | *BBOX1* | -2.652444561 | 0.001157814 | 0.008417372 | 1 |
| Heart_Failure | mesothelium | *PLD2* | -1.299010459 | 0.000174871 | 0.002725564 | 0.848476346 |
| Heart_Failure | mesothelium | *PUS7L* | -0.249266852 | 0.00921684 | 0.038419335 | 1 |
| Heart_Failure | mesothelium | *MTUS1* | -0.336104588 | 0.000999393 | 0.007648352 | 1 |
| Heart_Failure | mesothelium | *EPB41L4A* | -0.723554327 | 9.31E-05 | 0.002294209 | 0.451624488 |
| Heart_Failure | mesothelium | *REEP5* | -0.75376929 | 0.000395202 | 0.003978253 | 1 |
| Heart_Failure | mesothelium | *MAU2* | -0.596311524 | 0.000297143 | 0.003338215 | 1 |
| Heart_Failure | mesothelium | *PHF10* | -0.658603789 | 0.002267178 | 0.013809292 | 1 |
| Heart_Failure | mesothelium | *XPO7* | -0.727587219 | 0.000381161 | 0.003893896 | 1 |
| Heart_Failure | mesothelium | *DDA1* | -0.514501993 | 0.00017916 | 0.002725564 | 0.869284534 |
| Heart_Failure | mesothelium | *MAP1S* | -1.524089279 | 0.000442891 | 0.00425804 | 1 |
| Heart_Failure | mesothelium | *SSBP4* | -0.295526518 | 0.003234271 | 0.017756544 | 1 |
| Heart_Failure | mesothelium | *ATXN10* | -0.734413602 | 8.17E-05 | 0.002273164 | 0.396344953 |
| Heart_Failure | mesothelium | *PNPLA7* | -0.740739922 | 0.001237094 | 0.008827031 | 1 |
| Heart_Failure | mesothelium | *ANGPTL6* | -0.129760787 | 0.007641286 | 0.034234092 | 1 |
| Heart_Failure | mesothelium | *DNMT1* | -0.810597323 | 0.002086845 | 0.013095637 | 1 |
| Heart_Failure | mesothelium | *GALNT15* | -0.948018424 | 5.04E-05 | 0.002273164 | 0.244492129 |
| Heart_Failure | mesothelium | *SLC6A6* | -0.812656955 | 3.51E-05 | 0.002273164 | 0.170127298 |
| Heart_Failure | mesothelium | *C1orf159* | -1.048545064 | 5.83E-05 | 0.002273164 | 0.28310241 |
| Heart_Failure | mesothelium | *KHDRBS3* | -1.015749636 | 6.23E-05 | 0.002273164 | 0.302150646 |
| Heart_Failure | mesothelium | *SPATA6* | -0.627842226 | 0.00020231 | 0.002777753 | 0.981606219 |
| Heart_Failure | mesothelium | *LRRC41* | -1.161304058 | 3.55E-08 | 1.91E-05 | 0.000172044 |
| Heart_Failure | mesothelium | *TRIM22* | -0.774361492 | 0.000385376 | 0.003911807 | 1 |
| Heart_Failure | mesothelium | *SERPINF1* | -0.595846223 | 0.004933544 | 0.024476027 | 1 |
| Heart_Failure | mesothelium | *FIGNL1* | -0.411470728 | 7.21E-05 | 0.002273164 | 0.349605648 |
| Heart_Failure | mesothelium | *CTNNBL1* | -0.414269651 | 0.007218674 | 0.032918237 | 1 |
| Heart_Failure | mesothelium | *DMGDH* | -0.180986063 | 0.001768015 | 0.011595141 | 1 |
| Heart_Failure | mesothelium | *AP3B1* | -0.652596126 | 0.000383512 | 0.00390105 | 1 |
| Heart_Failure | mesothelium | *WASF3* | -0.908045274 | 4.84E-05 | 0.002273164 | 0.234932272 |
| Heart_Failure | mesothelium | *PEMT* | -1.069881636 | 0.001180181 | 0.008533885 | 1 |
| Heart_Failure | mesothelium | *MPRIP* | -0.738265159 | 0.000437356 | 0.004235632 | 1 |
| Heart_Failure | mesothelium | *DSTYK* | -0.864458961 | 7.08E-05 | 0.002273164 | 0.343601213 |
| Heart_Failure | mesothelium | *CSNK1G2* | -1.199604624 | 0.000121598 | 0.002465411 | 0.589994718 |
| Heart_Failure | mesothelium | *PDZD2* | -0.665684295 | 4.90E-05 | 0.002273164 | 0.237506545 |
| Heart_Failure | mesothelium | *ELP3* | -0.747411482 | 0.000218263 | 0.00288242 | 1 |
| Heart_Failure | mesothelium | *PEBP4* | -1.122524291 | 0.000492012 | 0.004573259 | 1 |
| Heart_Failure | mesothelium | *CTIF* | -0.186929599 | 0.008446445 | 0.036689482 | 1 |
| Heart_Failure | mesothelium | *THUMPD3* | -0.569285341 | 0.002825382 | 0.016127943 | 1 |
| Heart_Failure | mesothelium | *CNTN6* | -0.362993561 | 0.002515323 | 0.014883351 | 1 |
| Heart_Failure | mesothelium | *NAPG* | -0.643055707 | 0.000133123 | 0.002542967 | 0.645913741 |
| Heart_Failure | mesothelium | *PPHLN1* | -0.456260893 | 0.00379327 | 0.020092737 | 1 |
| Heart_Failure | mesothelium | *FKBP11* | -0.600474225 | 0.005189191 | 0.025457994 | 1 |
| Heart_Failure | mesothelium | *YWHAQ* | -0.725241483 | 0.000144119 | 0.002637401 | 0.699264135 |
| Heart_Failure | mesothelium | *KIDINS220* | -0.711270687 | 0.000185717 | 0.002744551 | 0.901099992 |
| Heart_Failure | mesothelium | *KIAA1468* | -0.773135785 | 0.000169107 | 0.002704817 | 0.820508978 |

| | | | | | | |
|---|---|---|---|---|---|---|
| Heart_Failure | mesothelium | *KLRD1* | -0.885799283 | 0.000248656 | 0.003001194 | 1 |
| Heart_Failure | mesothelium | *PRPF38A* | -0.557825775 | 0.001897328 | 0.01227047 | 1 |
| Heart_Failure | mesothelium | *ELP2* | -0.795268203 | 0.0002028 | 0.002777753 | 0.983983552 |
| Heart_Failure | mesothelium | *DHX34* | -0.962825243 | 9.87E-05 | 0.002301699 | 0.478947305 |
| Heart_Failure | mesothelium | *FADS2* | -0.767647368 | 0.000100789 | 0.002318018 | 0.489029612 |
| Heart_Failure | mesothelium | *CLOCK* | -0.773382706 | 0.000141197 | 0.002614833 | 0.685086159 |
| Heart_Failure | mesothelium | *ARGLU1* | -0.560598071 | 0.000543627 | 0.004875557 | 1 |
| Heart_Failure | mesothelium | *WDR36* | -1.0500606 | 7.48E-06 | 0.001067457 | 0.036293522 |
| Heart_Failure | mesothelium | *OSTF1* | -0.539593754 | 0.004721636 | 0.023789594 | 1 |
| Heart_Failure | mesothelium | *UBQLN1* | -0.675528017 | 0.000655702 | 0.005571748 | 1 |
| Heart_Failure | mesothelium | *DMTF1* | -0.62579618 | 9.77E-05 | 0.0022988 | 0.473976459 |
| Heart_Failure | mesothelium | *POM121C* | -0.369554497 | 5.34E-06 | 0.000997284 | 0.025929381 |
| Heart_Failure | mesothelium | *SRPK2* | -0.554650103 | 0.00013754 | 0.002596678 | 0.667346314 |
| Heart_Failure | mesothelium | *MAP3K7* | -0.709942909 | 0.000356409 | 0.003759027 | 1 |
| Heart_Failure | mesothelium | *PHF21A* | -0.686385951 | 0.000122241 | 0.002465411 | 0.593111692 |
| Heart_Failure | mesothelium | *CAPRIN1* | -0.434486232 | 0.004190193 | 0.021651562 | 1 |
| Heart_Failure | mesothelium | *ATP5G2* | -0.407723928 | 0.008183507 | 0.035900884 | 1 |
| Heart_Failure | mesothelium | *AHI1* | -0.10614932 | 0.002728514 | 0.015779202 | 1 |
| Heart_Failure | mesothelium | *USP15* | -0.469360295 | 0.005564397 | 0.02681078 | 1 |
| Heart_Failure | mesothelium | *MPHOSPH6* | -0.647176739 | 0.001365201 | 0.009471532 | 1 |
| Heart_Failure | mesothelium | *NTPCR* | -0.664517757 | 0.002809058 | 0.016053653 | 1 |
| Heart_Failure | mesothelium | *PIGC* | -0.601228927 | 0.003695341 | 0.019724749 | 1 |
| Heart_Failure | mesothelium | *SP110* | -0.474348814 | 0.001315355 | 0.009236039 | 1 |
| Heart_Failure | mesothelium | *TMBIM1* | -0.766423283 | 0.001703025 | 0.01124228 | 1 |
| Heart_Failure | mesothelium | *TSGA10* | -0.061268331 | 0.00871582 | 0.037457183 | 1 |
| Heart_Failure | mesothelium | *GCC2* | -0.360787407 | 0.00392457 | 0.02056373 | 1 |
| Heart_Failure | mesothelium | *ANKRD36* | -0.906387333 | 4.19E-05 | 0.002273164 | 0.203335229 |
| Heart_Failure | mesothelium | *EPC2* | -0.433131235 | 0.00949907 | 0.039325499 | 1 |
| Heart_Failure | mesothelium | *STAB2* | -0.818220032 | 0.002781268 | 0.015970076 | 1 |
| Heart_Failure | mesothelium | *DRAM1* | -0.698276542 | 0.002723833 | 0.015770926 | 1 |
| Heart_Failure | mesothelium | *RNASEH2B* | -0.270382785 | 0.002600783 | 0.015203613 | 1 |
| Heart_Failure | mesothelium | *SUCLA2* | -0.711784022 | 0.001168464 | 0.00846293 | 1 |
| Heart_Failure | mesothelium | *RCBTB2* | -0.791285531 | 0.000958887 | 0.007420284 | 1 |
| Heart_Failure | mesothelium | *KDELR2* | -1.105189823 | 0.000236447 | 0.002946978 | 1 |
| Heart_Failure | mesothelium | *NUPL2* | -0.910453503 | 0.000155188 | 0.002678614 | 0.752971028 |
| Heart_Failure | mesothelium | *NMT1* | 0.099889124 | 0.009055589 | 0.03815386 | 1 |
| Heart_Failure | mesothelium | *RPS6KC1* | -0.397148682 | 0.004225995 | 0.02174393 | 1 |
| Heart_Failure | mesothelium | *IMP4* | -0.517218221 | 0.001391957 | 0.009607077 | 1 |
| Heart_Failure | mesothelium | *ODF2* | -0.924028453 | 7.10E-05 | 0.002273164 | 0.34442548 |
| Heart_Failure | mesothelium | *TOR1B* | -0.484087309 | 0.002634363 | 0.015362897 | 1 |
| Heart_Failure | mesothelium | *ZNF189* | -1.059453112 | 0.000461908 | 0.00438544 | 1 |
| Heart_Failure | mesothelium | *STX17* | -0.716407055 | 0.000527568 | 0.004768677 | 1 |
| Heart_Failure | mesothelium | *ANP32B* | -0.390689141 | 0.01061947 | 0.04300974 | 1 |
| Heart_Failure | mesothelium | *APTX* | -0.457820278 | 0.005632965 | 0.027114234 | 1 |
| Heart_Failure | mesothelium | *DENND4C* | -0.484075476 | 0.000614589 | 0.005334503 | 1 |
| Heart_Failure | mesothelium | *FRS3* | -0.681429438 | 0.005131332 | 0.025225148 | 1 |
| Heart_Failure | mesothelium | *TCF19* | 0.131258612 | 0.006538964 | 0.030614686 | 1 |
| Heart_Failure | mesothelium | *NUMA1* | -0.723108339 | 0.00026274 | 0.003102849 | 1 |
| Heart_Failure | mesothelium | *RAB30* | -1.053713992 | 4.30E-05 | 0.002273164 | 0.20869112 |
| Heart_Failure | mesothelium | *NARS2* | -0.87350707 | 0.000115766 | 0.002463815 | 0.561694216 |

| | | | | | | |
|---|---|---|---|---|---|---|
| Heart_Failure | mesothelium | *NEK1* | -0.397291508 | 0.006311338 | 0.029759583 | 1 |
| Heart_Failure | mesothelium | *PARP6* | -0.74292269 | 0.00085116 | 0.006789985 | 1 |
| Heart_Failure | mesothelium | *ZNF280D* | -0.502923927 | 0.002318189 | 0.014027963 | 1 |
| Heart_Failure | mesothelium | *DBT* | -0.552942344 | 0.004790318 | 0.024059866 | 1 |
| Heart_Failure | mesothelium | *THUMPD2* | -0.775415467 | 6.09E-06 | 0.001001679 | 0.029530263 |
| Heart_Failure | mesothelium | *ACTR2* | -0.62568378 | 0.001412639 | 0.009722159 | 1 |
| Heart_Failure | mesothelium | *LRPPRC* | -0.772314656 | 0.000166868 | 0.002704817 | 0.809644767 |
| Heart_Failure | mesothelium | *DTNB* | -0.275563316 | 0.009730756 | 0.040216039 | 1 |
| Heart_Failure | mesothelium | *TACC2* | -0.30266112 | 0.011385697 | 0.045318623 | 1 |
| Heart_Failure | mesothelium | *RBP4* | -0.466345759 | 0.003338585 | 0.018160108 | 1 |
| Heart_Failure | mesothelium | *SMARCAL1* | -0.785358027 | 0.000498534 | 0.004597575 | 1 |
| Heart_Failure | mesothelium | *BARD1* | -0.37535113 | 0.007205541 | 0.032889261 | 1 |
| Heart_Failure | mesothelium | *SENP7* | -0.412889463 | 0.002617456 | 0.015282669 | 1 |
| Heart_Failure | mesothelium | *COX17* | -1.178773035 | 0.000373289 | 0.003853609 | 1 |
| Heart_Failure | mesothelium | *KIAA1109* | -0.684651652 | 0.000178887 | 0.002725564 | 0.867959653 |
| Heart_Failure | mesothelium | *PRDM5* | -0.946789972 | 3.54E-05 | 0.002273164 | 0.171702653 |
| Heart_Failure | mesothelium | *KIF21A* | -0.748897956 | 5.60E-05 | 0.002273164 | 0.271494897 |
| Heart_Failure | mesothelium | *ALG10* | -0.225789033 | 0.000258206 | 0.003085755 | 1 |
| Heart_Failure | mesothelium | *MARCH9* | -1.209261942 | 0.002178754 | 0.013506926 | 1 |
| Heart_Failure | mesothelium | *POC1B* | -1.04586523 | 4.72E-06 | 0.000952371 | 0.022887738 |
| Heart_Failure | mesothelium | *RITA1* | -2.106554798 | 5.45E-05 | 0.002273164 | 0.264507275 |
| Heart_Failure | mesothelium | *GLTP* | -1.205782069 | 0.000712169 | 0.005957666 | 1 |
| Heart_Failure | mesothelium | *MTMR6* | -0.844171071 | 9.65E-06 | 0.001300796 | 0.046828651 |
| Heart_Failure | mesothelium | *CERS5* | -0.814918066 | 2.47E-05 | 0.002273164 | 0.119820973 |
| Heart_Failure | mesothelium | *TMX1* | -0.948086 | 0.000355945 | 0.003759027 | 1 |
| Heart_Failure | mesothelium | *C14orf37* | -0.731379035 | 0.000246375 | 0.002981076 | 1 |
| Heart_Failure | mesothelium | *RAB15* | -1.389311919 | 6.89E-05 | 0.002273164 | 0.334227024 |
| Heart_Failure | mesothelium | *SERF2* | -0.628433643 | 0.001958448 | 0.012503145 | 1 |
| Heart_Failure | mesothelium | *SRP14* | -0.321411214 | 0.012690182 | 0.04949579 | 1 |
| Heart_Failure | mesothelium | *UBE2Q2* | -0.397414117 | 0.003199233 | 0.017607802 | 1 |
| Heart_Failure | mesothelium | *HMG20A* | -0.808648899 | 0.000357154 | 0.003759027 | 1 |
| Heart_Failure | mesothelium | *ULK3* | -0.865584198 | 0.003024367 | 0.0169975 | 1 |
| Heart_Failure | mesothelium | *IQGAP1* | -0.23302248 | 0.00082651 | 0.006661506 | 1 |
| Heart_Failure | mesothelium | *DHX38* | -1.11533778 | 0.000150441 | 0.002656078 | 0.729939035 |
| Heart_Failure | mesothelium | *NUDT7* | -1.18165497 | 0.000239972 | 0.002946978 | 1 |
| Heart_Failure | mesothelium | *TLDC1* | -1.259593487 | 3.60E-05 | 0.002273164 | 0.174611786 |
| Heart_Failure | mesothelium | *NCOR1* | -0.620874868 | 0.000243377 | 0.002972283 | 1 |
| Heart_Failure | mesothelium | *KSR1* | -0.702804364 | 0.001260576 | 0.008902929 | 1 |
| Heart_Failure | mesothelium | *C17orf80* | -0.265288946 | 0.010097527 | 0.041449409 | 1 |
| Heart_Failure | mesothelium | *SSH2* | -0.534225682 | 0.000825673 | 0.006661506 | 1 |
| Heart_Failure | mesothelium | *GNAL* | -1.016028101 | 5.11E-05 | 0.002273164 | 0.247905617 |
| Heart_Failure | mesothelium | *GALNT1* | -0.48593657 | 0.00641327 | 0.030123124 | 1 |
| Heart_Failure | mesothelium | *NPC1* | -0.479826258 | 0.002072675 | 0.013058627 | 1 |
| Heart_Failure | mesothelium | *ANAPC11* | -1.030738671 | 0.000128937 | 0.00251464 | 0.625601624 |
| Heart_Failure | mesothelium | *FN3KRP* | -0.061489595 | 0.00499438 | 0.024727277 | 1 |
| Heart_Failure | mesothelium | *PIP4K2B* | -0.414317327 | 0.000529512 | 0.004775448 | 1 |
| Heart_Failure | mesothelium | *ERBB2* | 0.262657486 | 0.010460114 | 0.04257758 | 1 |
| Heart_Failure | mesothelium | *MIEN1* | -0.374606842 | 0.000317012 | 0.003507027 | 1 |
| Heart_Failure | mesothelium | *PNMT* | 0.596039437 | 4.11E-12 | 3.99E-09 | 1.99E-08 |
| Heart_Failure | mesothelium | *IGFBP4* | -0.241048563 | 0.002132771 | 0.013318154 | 1 |

| | | | | | | |
|---|---|---|---|---|---|---|
| Heart_Failure | mesothelium | *CACNA1A* | -0.536659756 | 0.006151968 | 0.029121317 | 1 |
| Heart_Failure | mesothelium | *SAMD1* | -0.429046693 | 0.004197038 | 0.02166386 | 1 |
| Heart_Failure | mesothelium | *DPP9* | -0.84580354 | 0.001256772 | 0.008897449 | 1 |
| Heart_Failure | mesothelium | *SAE1* | -0.615500779 | 2.76E-05 | 0.002273164 | 0.133972139 |
| Heart_Failure | mesothelium | *EVI5L* | -1.435070306 | 8.69E-05 | 0.002275786 | 0.421576045 |
| Heart_Failure | mesothelium | *CTU1* | -1.036530583 | 0.002193945 | 0.01352649 | 1 |
| Heart_Failure | mesothelium | *RERE* | -0.371405609 | 0.004064922 | 0.02116202 | 1 |
| Heart_Failure | mesothelium | *EVA1B* | 0.172349117 | 0.000262793 | 0.003102849 | 1 |
| Heart_Failure | mesothelium | *PIGK* | -0.199897249 | 0.008861231 | 0.037780927 | 1 |
| Heart_Failure | mesothelium | *RPS8* | -0.568122686 | 0.000941495 | 0.007320729 | 1 |
| Heart_Failure | mesothelium | *ALDH9A1* | -0.806171306 | 0.001660497 | 0.011021523 | 1 |
| Heart_Failure | mesothelium | *ATP1B1* | -0.191181208 | 0.00405432 | 0.021129497 | 1 |
| Heart_Failure | mesothelium | *TIPRL* | -0.362076538 | 0.005977669 | 0.028407099 | 1 |
| Heart_Failure | mesothelium | *NME7* | -0.358880366 | 2.14E-05 | 0.002254546 | 0.103709124 |
| Heart_Failure | mesothelium | *RFWD2* | -0.503573695 | 0.002036953 | 0.012868876 | 1 |
| Heart_Failure | mesothelium | *SETDB1* | -0.460353466 | 0.002990471 | 0.016832674 | 1 |
| Heart_Failure | mesothelium | *ARNT* | -0.224051334 | 0.010704653 | 0.043210462 | 1 |
| Heart_Failure | mesothelium | *EIF2D* | -0.431837089 | 0.006557272 | 0.03062164 | 1 |
| Heart_Failure | mesothelium | *SMYD2* | -0.952654142 | 0.000558466 | 0.004971881 | 1 |
| Heart_Failure | mesothelium | *RAB13* | -1.494247256 | 5.73E-05 | 0.002273164 | 0.277824212 |
| Heart_Failure | mesothelium | *S100A8* | -0.729502743 | 0.004625206 | 0.023425363 | 1 |
| Heart_Failure | mesothelium | *RIT1* | -0.909546474 | 6.27E-06 | 0.001001679 | 0.030412833 |
| Heart_Failure | mesothelium | *FLG* | -0.067252718 | 0.000419787 | 0.004098206 | 1 |
| Heart_Failure | mesothelium | *SCCPDH* | -0.781115503 | 0.00094597 | 0.007332021 | 1 |
| Heart_Failure | mesothelium | *ACP1* | -0.722431358 | 0.002318719 | 0.014027963 | 1 |
| Heart_Failure | mesothelium | *MBOAT2* | -1.141087938 | 6.30E-05 | 0.002273164 | 0.305726543 |
| Heart_Failure | mesothelium | *RHOB* | -0.927350261 | 0.000523476 | 0.004756379 | 1 |
| Heart_Failure | mesothelium | *GALM* | -0.870002078 | 0.000405565 | 0.00404066 | 1 |
| Heart_Failure | mesothelium | *VPS54* | -0.745861435 | 8.14E-05 | 0.002273164 | 0.394900752 |
| Heart_Failure | mesothelium | *CIAO1* | -0.356624414 | 0.00486925 | 0.024306172 | 1 |
| Heart_Failure | mesothelium | *EXOC6B* | -0.593892488 | 0.00021638 | 0.00287637 | 1 |
| Heart_Failure | mesothelium | *TEX261* | -1.675863777 | 9.20E-05 | 0.002294209 | 0.446410625 |
| Heart_Failure | mesothelium | *RALB* | -0.866532693 | 0.001062574 | 0.007980821 | 1 |
| Heart_Failure | mesothelium | *GALNT13* | -1.028081865 | 5.68E-05 | 0.002273164 | 0.275631255 |
| Heart_Failure | mesothelium | *METTL21A* | -0.451356904 | 0.005895603 | 0.028182723 | 1 |
| Heart_Failure | mesothelium | *SUMF1* | -0.617050432 | 0.000224358 | 0.002908664 | 1 |
| Heart_Failure | mesothelium | *TAMM41* | -0.37348124 | 0.004964373 | 0.024603819 | 1 |
| Heart_Failure | mesothelium | *CNTN4* | -0.636251025 | 8.78E-05 | 0.002275786 | 0.42608722 |
| Heart_Failure | mesothelium | *CTDSPL* | -0.285370442 | 0.004806592 | 0.024067685 | 1 |
| Heart_Failure | mesothelium | *PTPRG* | -0.304917627 | 0.007361183 | 0.0333487 | 1 |
| Heart_Failure | mesothelium | *SLC25A26* | -0.341065806 | 0.000501837 | 0.004604045 | 1 |
| Heart_Failure | mesothelium | *ARL6IP5* | -0.715674424 | 0.000376923 | 0.003874637 | 1 |
| Heart_Failure | mesothelium | *BOC* | -0.641608851 | 8.47E-05 | 0.002275786 | 0.41117551 |
| Heart_Failure | mesothelium | *CENPC* | -0.373446398 | 0.000419107 | 0.004098206 | 1 |
| Heart_Failure | mesothelium | *SCD5* | -0.965335486 | 0.000111326 | 0.002412406 | 0.540154899 |
| Heart_Failure | mesothelium | *PYURF* | -0.598562468 | 0.001532927 | 0.010344592 | 1 |
| Heart_Failure | mesothelium | *METTL14* | -0.891300129 | 2.33E-05 | 0.002273164 | 0.113126213 |
| Heart_Failure | mesothelium | *CBR4* | -0.592058631 | 0.000188424 | 0.002744551 | 0.9142346 |
| Heart_Failure | mesothelium | *MARCH6* | -0.661465628 | 0.00049713 | 0.004597575 | 1 |
| Heart_Failure | mesothelium | *GTF2H2* | -1.727909251 | 0.000147307 | 0.002637401 | 0.714735598 |

| | | | | | | |
|---|---|---|---|---|---|---|
| Heart_Failure | mesothelium | *TSLP* | 0.08373754 | 3.20E-06 | 0.000811728 | 0.01554887 |
| Heart_Failure | mesothelium | *FEM1C* | -1.262276538 | 0.000127509 | 0.00251464 | 0.618671451 |
| Heart_Failure | mesothelium | *YIPF5* | -0.699725906 | 3.35E-05 | 0.002273164 | 0.162642444 |
| Heart_Failure | mesothelium | *GFOD1* | -0.412528517 | 0.006246543 | 0.029482709 | 1 |
| Heart_Failure | mesothelium | *FAM193B* | -0.408040195 | 0.006575022 | 0.030675007 | 1 |
| Heart_Failure | mesothelium | *DAAM2* | -0.336474381 | 0.004358684 | 0.022308373 | 1 |
| Heart_Failure | mesothelium | *PHIP* | -0.584506616 | 0.000801615 | 0.006514969 | 1 |
| Heart_Failure | mesothelium | *PNRC1* | -1.035052307 | 6.91E-05 | 0.002273164 | 0.33516314 |
| Heart_Failure | mesothelium | *RNF217* | -0.565874056 | 0.001070693 | 0.008019351 | 1 |
| Heart_Failure | mesothelium | *SLC18B1* | -0.152527621 | 0.000814454 | 0.006597217 | 1 |
| Heart_Failure | mesothelium | *RBAK* | -1.759467352 | 5.06E-05 | 0.002273164 | 0.24553829 |
| Heart_Failure | mesothelium | *TMEM209* | -1.130971945 | 7.31E-05 | 0.002273164 | 0.354503443 |
| Heart_Failure | mesothelium | *EPHA1* | -1.868325923 | 0.001882487 | 0.012194698 | 1 |
| Heart_Failure | mesothelium | *FBXO25* | -0.661337296 | 0.002940916 | 0.01659224 | 1 |
| Heart_Failure | mesothelium | *DOCK5* | -0.444096075 | 0.001655797 | 0.011005382 | 1 |
| Heart_Failure | mesothelium | *LRP12* | -1.162913181 | 9.36E-05 | 0.002294209 | 0.454299488 |
| Heart_Failure | mesothelium | *AK3* | -0.887712238 | 1.98E-05 | 0.002138716 | 0.096242203 |
| Heart_Failure | mesothelium | *HAUS6* | -1.080206384 | 6.61E-05 | 0.002273164 | 0.320914411 |
| Heart_Failure | mesothelium | *ZNF462* | -0.320124201 | 0.00580782 | 0.02781791 | 1 |
| Heart_Failure | mesothelium | *REXO4* | -1.154553459 | 0.000162674 | 0.002704817 | 0.78929599 |
| Heart_Failure | mesothelium | *INPP5E* | -0.442789216 | 0.007333542 | 0.033254527 | 1 |
| Heart_Failure | mesothelium | *PARD3* | -0.642041445 | 3.37E-05 | 0.002273164 | 0.163544678 |
| Heart_Failure | mesothelium | *ADD3* | -0.665000753 | 0.000228992 | 0.002923871 | 1 |
| Heart_Failure | mesothelium | *PAOX* | -1.155658305 | 0.001976225 | 0.012600054 | 1 |
| Heart_Failure | mesothelium | *LIN7C* | -1.040790998 | 0.000257923 | 0.003085755 | 1 |
| Heart_Failure | mesothelium | *HSD17B12* | -0.051616698 | 0.008518317 | 0.036853388 | 1 |
| Heart_Failure | mesothelium | *PTPRJ* | -0.466133438 | 0.004897781 | 0.02437337 | 1 |
| Heart_Failure | mesothelium | *INTS4* | -0.374776829 | 0.005390109 | 0.026178986 | 1 |
| Heart_Failure | mesothelium | *DRD2* | -3.963524415 | 0.005448787 | 0.026305984 | 1 |
| Heart_Failure | mesothelium | *NPAT* | -0.724189563 | 0.00150418 | 0.010189958 | 1 |
| Heart_Failure | mesothelium | *ROM1* | 0.082635765 | 0.003492329 | 0.018890503 | 1 |
| Heart_Failure | mesothelium | *CHEK1* | -2.510343395 | 9.57E-05 | 0.002294209 | 0.464439215 |
| Heart_Failure | mesothelium | *TRPT1* | -0.116838338 | 0.007427376 | 0.033523376 | 1 |
| Heart_Failure | mesothelium | *SAP18* | -1.042757373 | 0.000160529 | 0.002704817 | 0.77888825 |
| Heart_Failure | mesothelium | *PPP1R1C* | -1.146332561 | 0.000969051 | 0.007475093 | 1 |
| Heart_Failure | mesothelium | *GXYLT1* | -0.448586243 | 0.003714101 | 0.019795771 | 1 |
| Heart_Failure | mesothelium | *TMEM18* | -1.352744249 | 5.95E-05 | 0.002273164 | 0.288898919 |
| Heart_Failure | mesothelium | *NDUFC2* | -0.959365537 | 0.001354203 | 0.009426959 | 1 |
| Heart_Failure | mesothelium | *NUBPL* | -0.787856498 | 6.38E-05 | 0.002273164 | 0.309469899 |
| Heart_Failure | mesothelium | *VIPAS39* | -0.490609167 | 0.010649864 | 0.043025098 | 1 |
| Heart_Failure | mesothelium | *UPF2* | -0.58696533 | 0.000786158 | 0.006410824 | 1 |
| Heart_Failure | mesothelium | *EPS8* | -0.203732891 | 0.008474535 | 0.036745705 | 1 |
| Heart_Failure | mesothelium | *FAM160B1* | -0.448409579 | 0.005093385 | 0.025077928 | 1 |
| Heart_Failure | mesothelium | *PIGF* | -0.865638368 | 0.000317309 | 0.003507027 | 1 |
| Heart_Failure | mesothelium | *MFSD6* | -0.672515921 | 0.001634724 | 0.010895164 | 1 |
| Heart_Failure | mesothelium | *RNF144A* | -0.444853448 | 0.012492871 | 0.048962366 | 1 |
| Heart_Failure | mesothelium | *BICD1* | -0.831516278 | 5.88E-05 | 0.002273164 | 0.285399397 |
| Heart_Failure | mesothelium | *CACUL1* | -0.546742608 | 1.28E-05 | 0.001557659 | 0.062306378 |
| Heart_Failure | mesothelium | *TIAL1* | -0.725686029 | 0.000286846 | 0.003274767 | 1 |
| Heart_Failure | mesothelium | *TMEM56* | -2.21515345 | 3.57E-05 | 0.002273164 | 0.17300183 |

| | | | | | | |
|---|---|---|---|---|---|---|
| Heart_Failure | mesothelium | *PTPN14* | -0.381644064 | 0.009310314 | 0.038742403 | 1 |
| Heart_Failure | mesothelium | *TMEM178A* | -1.538902349 | 0.004636513 | 0.023458146 | 1 |
| Heart_Failure | mesothelium | *EPG5* | -0.583502536 | 0.001735993 | 0.011428817 | 1 |
| Heart_Failure | mesothelium | *POC5* | -0.111566325 | 0.004420171 | 0.02256776 | 1 |
| Heart_Failure | mesothelium | *MBNL1* | -0.465834615 | 0.000903871 | 0.007062128 | 1 |
| Heart_Failure | mesothelium | *SREK1IP1* | -0.8658007 | 0.000178515 | 0.002725564 | 0.866152997 |
| Heart_Failure | mesothelium | *CARHSP1* | -0.204292923 | 0.010078052 | 0.041414098 | 1 |
| Heart_Failure | mesothelium | *TMEM87B* | -0.923874375 | 4.13E-05 | 0.002273164 | 0.200464299 |
| Heart_Failure | mesothelium | *LPCAT1* | -1.312198272 | 0.00018288 | 0.002737794 | 0.88733161 |
| Heart_Failure | mesothelium | *CFDP1* | -0.723178864 | 0.000226602 | 0.002908664 | 1 |
| Heart_Failure | mesothelium | *ZDHHC7* | -0.850195008 | 0.000478772 | 0.004500858 | 1 |
| Heart_Failure | mesothelium | *JAZF1* | -0.232194277 | 0.000479584 | 0.004500858 | 1 |
| Heart_Failure | mesothelium | *FBXO36* | -0.995065607 | 5.81E-05 | 0.002273164 | 0.281812606 |
| Heart_Failure | mesothelium | *SEMA3D* | -0.398995566 | 0.001110846 | 0.00817879 | 1 |
| Heart_Failure | mesothelium | *PPP2R5E* | -0.747542218 | 0.000185083 | 0.002744551 | 0.898021406 |
| Heart_Failure | mesothelium | *ANKRD29* | -0.961692998 | 0.000178896 | 0.002725564 | 0.868001761 |
| Heart_Failure | mesothelium | *ANKH* | -0.408121578 | 0.011619488 | 0.046105142 | 1 |
| Heart_Failure | mesothelium | *ROBO4* | -1.007491802 | 5.70E-05 | 0.002273164 | 0.276538629 |
| Heart_Failure | mesothelium | *PRKCA* | -0.678326449 | 0.000278001 | 0.003196348 | 1 |
| Heart_Failure | mesothelium | *MIA3* | -0.54182447 | 0.000205995 | 0.002777753 | 0.999486491 |
| Heart_Failure | mesothelium | *PDE1C* | -1.048341278 | 6.63E-06 | 0.001004666 | 0.032149312 |
| Heart_Failure | mesothelium | *OXNAD1* | -0.664353405 | 0.000576999 | 0.005053432 | 1 |
| Heart_Failure | mesothelium | *AK9* | -0.180654171 | 0.008906532 | 0.037879761 | 1 |
| Heart_Failure | mesothelium | *MARVELD1* | -0.999460748 | 0.000750608 | 0.006183274 | 1 |
| Heart_Failure | mesothelium | *TRMT44* | -0.538011567 | 0.002808652 | 0.016053653 | 1 |
| Heart_Failure | mesothelium | *MOV10* | -0.668843596 | 0.000270037 | 0.003149564 | 1 |
| Heart_Failure | mesothelium | *NUP205* | -0.798938661 | 0.000311842 | 0.003470312 | 1 |
| Heart_Failure | mesothelium | *PDIA4* | -1.178163543 | 0.000119001 | 0.002465411 | 0.577391435 |
| Heart_Failure | mesothelium | *RRAGA* | -0.600855695 | 0.002683881 | 0.015595437 | 1 |
| Heart_Failure | mesothelium | *NDUFAF6* | -0.410022322 | 3.38E-05 | 0.002273164 | 0.164097119 |
| Heart_Failure | mesothelium | *WHAMM* | -0.143732285 | 0.001346578 | 0.009397378 | 1 |
| Heart_Failure | mesothelium | *SUPV3L1* | -0.545498003 | 0.002739294 | 0.015803873 | 1 |
| Heart_Failure | mesothelium | *NODAL* | -1.505600749 | 0.002759362 | 0.015900742 | 1 |
| Heart_Failure | mesothelium | *ZDHHC5* | -1.115203729 | 9.36E-05 | 0.002294209 | 0.454134601 |
| Heart_Failure | mesothelium | *NMNAT2* | -0.418610988 | 0.003997874 | 0.020857724 | 1 |
| Heart_Failure | mesothelium | *NECAP2* | -0.958444687 | 0.000347504 | 0.003697569 | 1 |
| Heart_Failure | mesothelium | *CLDN12* | -1.507286554 | 6.25E-05 | 0.002273164 | 0.303337177 |
| Heart_Failure | mesothelium | *GATAD1* | -0.787289208 | 3.57E-05 | 0.002273164 | 0.173336286 |
| Heart_Failure | mesothelium | *DHRS4* | -1.278794012 | 0.000680276 | 0.005730383 | 1 |
| Heart_Failure | mesothelium | *DHRS1* | -0.080479288 | 0.01149206 | 0.04570449 | 1 |
| Heart_Failure | mesothelium | *AASDH* | -0.335870929 | 0.007286091 | 0.033163334 | 1 |
| Heart_Failure | mesothelium | *SLC38A10* | -0.842099776 | 0.000418165 | 0.004098206 | 1 |
| Heart_Failure | mesothelium | *FANCC* | -0.508877511 | 0.000654488 | 0.005571187 | 1 |
| Heart_Failure | mesothelium | *CLSTN2* | -0.270669167 | 0.006951601 | 0.031940499 | 1 |
| Heart_Failure | mesothelium | *PPP1R9A* | -0.430259988 | 0.000976492 | 0.007520535 | 1 |
| Heart_Failure | mesothelium | *RNF166* | -0.102457143 | 0.003093225 | 0.017231146 | 1 |
| Heart_Failure | mesothelium | *SPATA2L* | -0.27866588 | 0.008177675 | 0.035900884 | 1 |
| Heart_Failure | mesothelium | *CDC42SE2* | -0.516881848 | 1.87E-06 | 0.000604662 | 0.009069924 |
| Heart_Failure | mesothelium | *TMEM69* | -0.618488191 | 0.011893416 | 0.04691614 | 1 |
| Heart_Failure | mesothelium | *TEPP* | -0.588535928 | 0.0020827 | 0.013089713 | 1 |

| | | | | | | |
|---|---|---|---|---|---|---|
| Heart_Failure | mesothelium | *CTBP1* | -0.34734427 | 0.004216054 | 0.021736042 | 1 |
| Heart_Failure | mesothelium | *SSU72* | -0.485616565 | 0.009998228 | 0.041216143 | 1 |
| Heart_Failure | mesothelium | *CCDC58* | -1.272892956 | 0.000202444 | 0.002777753 | 0.982256527 |
| Heart_Failure | mesothelium | *TAOK1* | -0.620059419 | 0.000305106 | 0.003403159 | 1 |
| Heart_Failure | mesothelium | *ZBTB7B* | -0.707970362 | 0.000499318 | 0.004597575 | 1 |
| Heart_Failure | mesothelium | *FCRL3* | -0.086738739 | 0.003481829 | 0.018854728 | 1 |
| Heart_Failure | mesothelium | *FGFR4* | -0.488934178 | 8.69E-05 | 0.002275786 | 0.42152925 |
| Heart_Failure | mesothelium | *IER2* | -0.372352229 | 0.002122922 | 0.013273736 | 1 |
| Heart_Failure | mesothelium | *ZNF333* | -0.754275531 | 0.00331286 | 0.018101389 | 1 |
| Heart_Failure | mesothelium | *RPL8* | -0.582021834 | 0.001518478 | 0.010261356 | 1 |
| Heart_Failure | mesothelium | *PGAP3* | -1.609117259 | 9.44E-05 | 0.002294209 | 0.45816159 |
| Heart_Failure | mesothelium | *ASB16* | -0.733109991 | 0.000225387 | 0.002908664 | 1 |
| Heart_Failure | mesothelium | *PLCD3* | -1.218630018 | 0.000116285 | 0.002463815 | 0.56421371 |
| Heart_Failure | mesothelium | *ASRGL1* | -0.903072565 | 0.007615854 | 0.034151686 | 1 |
| Heart_Failure | mesothelium | *RPS6KA4* | -1.103565084 | 0.003861854 | 0.020344968 | 1 |
| Heart_Failure | mesothelium | *PDZK1IP1* | -0.494512167 | 0.006541945 | 0.030614686 | 1 |
| Heart_Failure | mesothelium | *CMPK1* | -0.29244049 | 0.008116234 | 0.035767456 | 1 |
| Heart_Failure | mesothelium | *COA7* | -1.105429042 | 0.003912869 | 0.020524584 | 1 |
| Heart_Failure | mesothelium | *C1orf123* | -0.708444073 | 0.001013743 | 0.007702226 | 1 |
| Heart_Failure | mesothelium | *USP24* | -0.566571012 | 0.000204204 | 0.002777753 | 0.990798876 |
| Heart_Failure | mesothelium | *PDPN* | -0.408052982 | 0.001784257 | 0.011667408 | 1 |
| Heart_Failure | mesothelium | *SYNC* | -0.21645779 | 0.008710111 | 0.037457183 | 1 |
| Heart_Failure | mesothelium | *TM2D1* | -0.788704811 | 0.000191284 | 0.002745889 | 0.928110391 |
| Heart_Failure | mesothelium | *TYW3* | -0.515377438 | 0.000458367 | 0.004360779 | 1 |
| Heart_Failure | mesothelium | *KLHDC9* | -0.090247717 | 0.00101437 | 0.007702226 | 1 |
| Heart_Failure | mesothelium | *FLVCR1* | -0.492643399 | 0.001384786 | 0.009584855 | 1 |
| Heart_Failure | mesothelium | *ACP6* | -0.981912806 | 0.000157068 | 0.002683421 | 0.762091529 |
| Heart_Failure | mesothelium | *CNST* | -0.599695201 | 0.001203038 | 0.008647615 | 1 |
| Heart_Failure | mesothelium | *RPRD2* | -0.596132181 | 3.69E-06 | 0.000814006 | 0.017908143 |
| Heart_Failure | mesothelium | *VPS72* | -1.17458121 | 0.000304993 | 0.003403159 | 1 |
| Heart_Failure | mesothelium | *RNF149* | -0.396362663 | 0.006637675 | 0.030937558 | 1 |
| Heart_Failure | mesothelium | *TDRD10* | -0.895959437 | 5.87E-05 | 0.002273164 | 0.284715734 |
| Heart_Failure | mesothelium | *DCAF16* | -0.484191365 | 0.002767871 | 0.015930857 | 1 |
| Heart_Failure | mesothelium | *GPR155* | -0.629087522 | 0.0031363 | 0.017391229 | 1 |
| Heart_Failure | mesothelium | *LMOD1* | -0.190370331 | 0.003066717 | 0.017175442 | 1 |
| Heart_Failure | mesothelium | *ARPC2* | -0.487559463 | 0.002179704 | 0.013506926 | 1 |
| Heart_Failure | mesothelium | *EFHB* | 0.036409818 | 0.006386729 | 0.030085835 | 1 |
| Heart_Failure | mesothelium | *GTPBP8* | -1.296302853 | 8.96E-05 | 0.002278979 | 0.434960036 |
| Heart_Failure | mesothelium | *WDFY3* | -0.458686093 | 0.001443406 | 0.00986395 | 1 |
| Heart_Failure | mesothelium | *C4orf36* | -1.106097007 | 6.72E-05 | 0.002273164 | 0.325905901 |
| Heart_Failure | mesothelium | *RCHY1* | -0.171036336 | 0.001243308 | 0.00883272 | 1 |
| Heart_Failure | mesothelium | *SLC4A1AP* | -1.120143277 | 0.000122966 | 0.002465411 | 0.596629376 |
| Heart_Failure | mesothelium | *MEAF6* | -0.634181047 | 0.000664953 | 0.005620818 | 1 |
| Heart_Failure | mesothelium | *SNIP1* | -1.200383349 | 0.000236623 | 0.002946978 | 1 |
| Heart_Failure | mesothelium | *DNALI1* | -0.139797768 | 0.002359205 | 0.01418865 | 1 |
| Heart_Failure | mesothelium | *RPN1* | -1.040842371 | 0.000157885 | 0.002687922 | 0.766057715 |
| Heart_Failure | mesothelium | *SFMBT1* | -0.816225855 | 0.004742237 | 0.023849837 | 1 |
| Heart_Failure | mesothelium | *FAM208A* | -0.428832691 | 0.008939999 | 0.037928602 | 1 |
| Heart_Failure | mesothelium | *LRPAP1* | -0.433259854 | 0.001311228 | 0.009228876 | 1 |
| Heart_Failure | mesothelium | *RNF123* | 0.167290503 | 6.68E-05 | 0.002273164 | 0.323883941 |

| | | | | | | |
|---|---|---|---|---|---|---|
| Heart_Failure | mesothelium | *MST1R* | 0.151537711 | 4.91E-06 | 0.000952371 | 0.023809274 |
| Heart_Failure | mesothelium | *RAD54L2* | -0.530452073 | 6.40E-06 | 0.001001679 | 0.031052061 |
| Heart_Failure | mesothelium | *MAP9* | -0.503198288 | 0.010886463 | 0.043762318 | 1 |
| Heart_Failure | mesothelium | *MOCS2* | -0.720792475 | 0.005714511 | 0.027452284 | 1 |
| Heart_Failure | mesothelium | *WDR41* | -0.346510356 | 0.001841642 | 0.012010275 | 1 |
| Heart_Failure | mesothelium | *NDUFS4* | -0.583964425 | 0.002529923 | 0.014933315 | 1 |
| Heart_Failure | mesothelium | *CASP3* | -0.238586089 | 0.008008477 | 0.035507635 | 1 |
| Heart_Failure | mesothelium | *PRIMPOL* | -0.482581695 | 0.000350003 | 0.003716004 | 1 |
| Heart_Failure | mesothelium | *PDSS2* | -0.691686938 | 0.000218617 | 0.00288242 | 1 |
| Heart_Failure | mesothelium | *STK17A* | -0.245234691 | 0.010056375 | 0.041414098 | 1 |
| Heart_Failure | mesothelium | *RPS14* | -0.45374726 | 0.004695253 | 0.023681255 | 1 |
| Heart_Failure | mesothelium | *RP9* | -1.216982185 | 0.000378196 | 0.003879506 | 1 |
| Heart_Failure | mesothelium | *CHMP4C* | -2.890711147 | 0.000616019 | 0.005337363 | 1 |
| Heart_Failure | mesothelium | *SLC13A4* | -0.873488998 | 0.00057411 | 0.005053432 | 1 |
| Heart_Failure | mesothelium | *BRI3* | -0.78814998 | 0.000220681 | 0.002900583 | 1 |
| Heart_Failure | mesothelium | *RAD21* | -0.458440065 | 0.008971248 | 0.037928602 | 1 |
| Heart_Failure | mesothelium | *OXR1* | -0.528859723 | 0.00079629 | 0.006482545 | 1 |
| Heart_Failure | mesothelium | *INTS1* | -0.806250114 | 0.000883182 | 0.006956491 | 1 |
| Heart_Failure | mesothelium | *SLC4A2* | -0.380259446 | 0.009017099 | 0.038077427 | 1 |
| Heart_Failure | mesothelium | *FASTK* | -0.826454633 | 0.008803024 | 0.0377036 | 1 |
| Heart_Failure | mesothelium | *ALDH7A1* | -0.341429565 | 0.005822844 | 0.027862367 | 1 |
| Heart_Failure | mesothelium | *YWHAZ* | -0.281620256 | 0.001924581 | 0.01235194 | 1 |
| Heart_Failure | mesothelium | *SLC25A32* | -0.372006998 | 0.008804241 | 0.0377036 | 1 |
| Heart_Failure | mesothelium | *TMEM65* | -0.851863484 | 0.000180439 | 0.00272738 | 0.875488825 |
| Heart_Failure | mesothelium | *ZMAT4* | -2.422361016 | 0.000566937 | 0.005019669 | 1 |
| Heart_Failure | mesothelium | *TRMT10B* | -0.651247511 | 0.000239915 | 0.002946978 | 1 |
| Heart_Failure | mesothelium | *WRN* | -0.436460513 | 0.007947877 | 0.035314193 | 1 |
| Heart_Failure | mesothelium | *TSHR* | -0.227936541 | 0.005956223 | 0.028385176 | 1 |
| Heart_Failure | mesothelium | *CFL2* | -0.945750821 | 0.000323389 | 0.003549967 | 1 |
| Heart_Failure | mesothelium | *SLC16A9* | -1.455730022 | 0.000166448 | 0.002704817 | 0.807607093 |
| Heart_Failure | mesothelium | *GJB2* | -2.915575427 | 0.003852598 | 0.020340377 | 1 |
| Heart_Failure | mesothelium | *REEP3* | -0.344014594 | 0.007069232 | 0.032388963 | 1 |
| Heart_Failure | mesothelium | *MICU2* | -0.717636187 | 6.18E-05 | 0.002273164 | 0.299638133 |
| Heart_Failure | mesothelium | *LRR1* | -1.452371932 | 0.000896512 | 0.007027264 | 1 |
| Heart_Failure | mesothelium | *ATP5C1* | -0.605185569 | 0.000266127 | 0.003131364 | 1 |
| Heart_Failure | mesothelium | *VDAC2* | -0.715426164 | 0.000361862 | 0.003792122 | 1 |
| Heart_Failure | mesothelium | *PPP1R36* | -0.272181365 | 0.00543477 | 0.026302863 | 1 |
| Heart_Failure | mesothelium | *TRUB1* | -0.274488845 | 0.008554352 | 0.036926795 | 1 |
| Heart_Failure | mesothelium | *UBTD1* | -1.027225482 | 6.46E-05 | 0.002273164 | 0.31332672 |
| Heart_Failure | mesothelium | *TTC7B* | -0.429818405 | 0.010852381 | 0.043697719 | 1 |
| Heart_Failure | mesothelium | *PSMC3* | -0.414480317 | 0.003350407 | 0.018204003 | 1 |
| Heart_Failure | mesothelium | *IFI27L1* | -0.497079724 | 0.012399698 | 0.048675837 | 1 |
| Heart_Failure | mesothelium | *CEP57* | -0.300808323 | 0.008077818 | 0.03566853 | 1 |
| Heart_Failure | mesothelium | *TCP11L2* | -0.868274424 | 0.000193169 | 0.002764768 | 0.937256486 |
| Heart_Failure | mesothelium | *ZFYVE19* | -0.120899163 | 0.00921558 | 0.038419335 | 1 |
| Heart_Failure | mesothelium | *API5* | -0.928831303 | 0.000178681 | 0.002725564 | 0.866958771 |
| Heart_Failure | mesothelium | *RPL27A* | -0.8431956 | 0.000223292 | 0.002905056 | 1 |
| Heart_Failure | mesothelium | *TMEM41B* | -0.911347327 | 7.88E-05 | 0.002273164 | 0.382211038 |
| Heart_Failure | mesothelium | *TMX3* | -0.465331993 | 0.001155966 | 0.008417372 | 1 |
| Heart_Failure | mesothelium | *SEC11C* | -1.364127091 | 5.61E-05 | 0.002273164 | 0.272127461 |

| | | | | | | |
|---|---|---|---|---|---|---|
| Heart_Failure | mesothelium | *SLFN5* | -0.425184742 | 0.006687248 | 0.031079048 | 1 |
| Heart_Failure | mesothelium | *CATSPER2* | -0.68507682 | 0.00015054 | 0.002656078 | 0.730421397 |
| Heart_Failure | mesothelium | *SAAL1* | -1.376234439 | 0.000203393 | 0.002777753 | 0.986863405 |
| Heart_Failure | mesothelium | *TMEM170A* | -0.475913504 | 1.15E-05 | 0.001470495 | 0.055878818 |
| Heart_Failure | mesothelium | *STAT6* | -0.495279478 | 0.002234317 | 0.013705318 | 1 |
| Heart_Failure | mesothelium | *EPB42* | -0.754599764 | 0.000398809 | 0.003989732 | 1 |
| Heart_Failure | mesothelium | *RCCD1* | -0.328576265 | 0.001539351 | 0.010373518 | 1 |
| Heart_Failure | mesothelium | *ACSF2* | -0.224388372 | 0.003816829 | 0.020185725 | 1 |
| Heart_Failure | mesothelium | *SP2* | -1.202517286 | 0.000112143 | 0.002418295 | 0.544116406 |
| Heart_Failure | mesothelium | *COQ7* | -0.341028818 | 0.012782702 | 0.04969685 | 1 |
| Heart_Failure | mesothelium | *FBXO22* | -0.657878268 | 0.00185926 | 0.012076478 | 1 |
| Heart_Failure | mesothelium | *TBC1D2B* | -0.289379452 | 0.000323235 | 0.003549967 | 1 |
| Heart_Failure | mesothelium | *KATNAL2* | -0.146879131 | 0.000510233 | 0.004671037 | 1 |
| Heart_Failure | mesothelium | *HDHD2* | 0.062661015 | 0.009828725 | 0.040586361 | 1 |
| Heart_Failure | mesothelium | *ZNF23* | 0.284124984 | 0.00667054 | 0.03103112 | 1 |
| Heart_Failure | mesothelium | *VKORC1* | -1.452010094 | 0.000201104 | 0.002777753 | 0.975755929 |
| Heart_Failure | mesothelium | *SPATA33* | -1.142097443 | 0.000238667 | 0.002946978 | 1 |
| Heart_Failure | mesothelium | *RPL13* | -0.532338854 | 0.000660652 | 0.005603992 | 1 |
| Heart_Failure | mesothelium | *ZNF641* | -0.600867746 | 0.001339847 | 0.009367344 | 1 |
| Heart_Failure | mesothelium | *TTYH1* | -1.995416011 | 4.33E-05 | 0.002273164 | 0.210101552 |
| Heart_Failure | mesothelium | *LENG8* | -0.704728607 | 8.32E-05 | 0.002273164 | 0.403625329 |
| Heart_Failure | mesothelium | *CYB5D2* | -0.513487781 | 0.00891564 | 0.037879761 | 1 |
| Heart_Failure | mesothelium | *NDUFV1* | -0.94599696 | 0.000143089 | 0.002637401 | 0.69426551 |
| Heart_Failure | mesothelium | *NUDT8* | -0.079606047 | 0.005436254 | 0.026302863 | 1 |
| Heart_Failure | mesothelium | *PRDX2* | -0.892019553 | 0.000576981 | 0.005053432 | 1 |
| Heart_Failure | mesothelium | *MIS12* | -1.037806019 | 0.000345695 | 0.003689721 | 1 |
| Heart_Failure | mesothelium | *GSDMA* | -0.191145315 | 1.93E-11 | 1.56E-08 | 9.36E-08 |
| Heart_Failure | mesothelium | *ATG16L2* | -0.730500633 | 0.00242251 | 0.014475394 | 1 |
| Heart_Failure | mesothelium | *LTBP3* | -0.246199523 | 0.009100915 | 0.038226758 | 1 |
| Heart_Failure | mesothelium | *COA6* | -1.462567614 | 0.000388184 | 0.003932081 | 1 |
| Heart_Failure | mesothelium | *PXK* | -0.361289948 | 0.005786362 | 0.027742517 | 1 |
| Heart_Failure | mesothelium | *PCMTD1* | -0.677664546 | 0.00026182 | 0.003102849 | 1 |
| Heart_Failure | mesothelium | *SLC20A2* | -0.407556548 | 0.008614686 | 0.037154182 | 1 |
| Heart_Failure | mesothelium | *NBPF9* | -1.397940612 | 0.0008809 | 0.006949802 | 1 |
| Heart_Failure | mesothelium | *DNAJC21* | -1.076154588 | 8.74E-05 | 0.002275786 | 0.424260055 |
| Heart_Failure | mesothelium | *SNTB2* | -0.643722084 | 0.001155045 | 0.008417372 | 1 |
| Heart_Failure | mesothelium | *ZNF507* | -0.517380101 | 8.80E-05 | 0.002275786 | 0.426925021 |
| Heart_Failure | mesothelium | *STX18* | -0.869397965 | 0.000131797 | 0.002537611 | 0.63947791 |
| Heart_Failure | mesothelium | *ZBTB49* | -1.389685331 | 8.03E-05 | 0.002273164 | 0.389687897 |
| Heart_Failure | mesothelium | *CEP120* | -0.528986924 | 0.008288686 | 0.0361661 | 1 |
| Heart_Failure | mesothelium | *MFF* | -0.8025425 | 0.000164565 | 0.002704817 | 0.798471467 |
| Heart_Failure | mesothelium | *MAP2K1* | -0.647295241 | 0.002157471 | 0.013437804 | 1 |
| Heart_Failure | mesothelium | *UBE2V2* | -0.455917284 | 0.001366462 | 0.009471532 | 1 |
| Heart_Failure | mesothelium | *XPO6* | -0.677445967 | 0.000148052 | 0.002640979 | 0.718346231 |
| Heart_Failure | mesothelium | *RGS14* | -0.081996183 | 0.003027552 | 0.0169975 | 1 |
| Heart_Failure | mesothelium | *ZEB2* | -0.199896151 | 0.002484113 | 0.01477073 | 1 |
| Heart_Failure | mesothelium | *HINT1* | -0.736552981 | 0.001336038 | 0.009354194 | 1 |
| Heart_Failure | mesothelium | *DTWD2* | -0.229370711 | 0.000579571 | 0.005066807 | 1 |
| Heart_Failure | mesothelium | *CLIC3* | -0.524432136 | 0.008242888 | 0.036067621 | 1 |
| Heart_Failure | mesothelium | *LUZP1* | -0.658006663 | 0.001619454 | 0.010823129 | 1 |

| | | | | | | |
|---|---|---|---|---|---|---|
| Heart_Failure | mesothelium | *AVEN* | -0.724845805 | 0.000551789 | 0.004921467 | 1 |
| Heart_Failure | mesothelium | *UBE2E3* | -0.403678938 | 0.001690883 | 0.011177333 | 1 |
| Heart_Failure | mesothelium | *CHRNB1* | 0.052988758 | 0.002111763 | 0.013221 | 1 |
| Heart_Failure | mesothelium | *PDCD6IP* | -0.68585453 | 0.000366724 | 0.003818332 | 1 |
| Heart_Failure | mesothelium | *STX8* | -0.416105631 | 0.010320416 | 0.042079547 | 1 |
| Heart_Failure | mesothelium | *NFRKB* | -0.825726778 | 0.000322406 | 0.003549967 | 1 |
| Heart_Failure | mesothelium | *MGMT* | -0.527783146 | 0.000513411 | 0.004691283 | 1 |
| Heart_Failure | mesothelium | *DNAJC18* | -0.143948396 | 0.003091887 | 0.017231146 | 1 |
| Heart_Failure | mesothelium | *HSD17B13* | -1.03574496 | 0.002391741 | 0.014326825 | 1 |
| Heart_Failure | mesothelium | *ELOVL6* | -0.887609157 | 0.000345027 | 0.003689721 | 1 |
| Heart_Failure | mesothelium | *FOXN2* | -0.509269657 | 0.006401569 | 0.030104452 | 1 |
| Heart_Failure | mesothelium | *RPS9* | -0.128144311 | 0.005263767 | 0.025693961 | 1 |
| Heart_Failure | mesothelium | *MCC* | -0.789075984 | 5.60E-05 | 0.002273164 | 0.271497806 |
| Heart_Failure | mesothelium | *TBCA* | -0.648212285 | 0.000943141 | 0.007321795 | 1 |
| Heart_Failure | mesothelium | *NMUR1* | -1.495447753 | 0.003883383 | 0.020414056 | 1 |
| Heart_Failure | mesothelium | *PIK3CD* | -0.384110234 | 0.001081902 | 0.008051207 | 1 |
| Heart_Failure | mesothelium | *VAT1L* | -0.513621197 | 0.000359834 | 0.003779033 | 1 |
| Heart_Failure | mesothelium | *EXOSC10* | -0.394144262 | 0.003857124 | 0.020342137 | 1 |
| Heart_Failure | mesothelium | *TRAPPC12* | -0.79021464 | 8.67E-05 | 0.002275786 | 0.42048645 |
| Heart_Failure | mesothelium | *ZNF554* | -0.508583182 | 0.00466764 | 0.023591033 | 1 |
| Heart_Failure | mesothelium | *ANKRD20A4* | -3.669861883 | 0.000237072 | 0.002946978 | 1 |
| Heart_Failure | mesothelium | *MALT1* | -0.891335064 | 0.00010862 | 0.002412406 | 0.527025612 |
| Heart_Failure | mesothelium | *MBOAT1* | -0.627938934 | 0.000872513 | 0.006906093 | 1 |
| Heart_Failure | mesothelium | *PAIP1* | -0.676416068 | 0.000346006 | 0.003689721 | 1 |
| Heart_Failure | mesothelium | *SUCLG2* | -0.707578022 | 0.000172509 | 0.00271758 | 0.837014498 |
| Heart_Failure | mesothelium | *AFF1* | -0.638261092 | 0.000110149 | 0.002412406 | 0.534441988 |
| Heart_Failure | mesothelium | *FIBP* | -0.218601496 | 0.008074043 | 0.03566853 | 1 |
| Heart_Failure | mesothelium | *SLFN11* | -0.548194598 | 0.000226557 | 0.002908664 | 1 |
| Heart_Failure | mesothelium | *CBWD1* | -1.319992715 | 0.000122834 | 0.002465411 | 0.595991554 |
| Heart_Failure | mesothelium | *DCP2* | -0.646973843 | 0.000272373 | 0.003169192 | 1 |
| Heart_Failure | mesothelium | *ZNF621* | -0.619854148 | 0.011886889 | 0.04691614 | 1 |
| Heart_Failure | mesothelium | *LCLAT1* | -0.320276051 | 0.000176537 | 0.002725564 | 0.856558685 |
| Heart_Failure | mesothelium | *GXYLT2* | -0.323201937 | 0.008281196 | 0.0361661 | 1 |
| Heart_Failure | mesothelium | *DCAKD* | -0.722652443 | 8.97E-07 | 0.000362718 | 0.004352618 |
| Heart_Failure | mesothelium | *UQCRH* | -0.466264564 | 0.007378792 | 0.033366167 | 1 |
| Heart_Failure | mesothelium | *C11orf80* | -0.362652063 | 1.49E-07 | 7.25E-05 | 0.000725086 |
| Heart_Failure | mesothelium | *GPR160* | -0.190596336 | 0.003030263 | 0.0169975 | 1 |
| Heart_Failure | mesothelium | *GOLIM4* | -0.596846048 | 0.003160153 | 0.017463626 | 1 |
| Heart_Failure | mesothelium | *XXYLT1* | -0.837706031 | 0.00054581 | 0.004886105 | 1 |
| Heart_Failure | mesothelium | *UBXN2A* | -0.40167642 | 0.003911798 | 0.020524584 | 1 |
| Heart_Failure | mesothelium | *LEMD3* | -0.772573188 | 0.000606849 | 0.005286235 | 1 |
| Heart_Failure | mesothelium | *ANKRD36C* | -0.606356043 | 0.003438052 | 0.018638465 | 1 |
| Heart_Failure | mesothelium | *MSL2* | -0.838099383 | 0.000326608 | 0.003577202 | 1 |
| Heart_Failure | mesothelium | *ANGEL2* | -0.683636189 | 0.001437739 | 0.009852977 | 1 |
| Heart_Failure | mesothelium | *LARP7* | -0.498235366 | 0.003959431 | 0.02070168 | 1 |
| Heart_Failure | mesothelium | *CEP135* | -0.66232871 | 0.000632056 | 0.005418262 | 1 |
| Heart_Failure | mesothelium | *RSRC1* | -0.61753326 | 6.21E-06 | 0.001001679 | 0.030128817 |
| Heart_Failure | mesothelium | *DHX36* | -0.800027451 | 0.000118356 | 0.002465411 | 0.574264175 |
| Heart_Failure | mesothelium | *TVP23C* | -2.164874749 | 6.67E-05 | 0.002273164 | 0.32375547 |
| Heart_Failure | mesothelium | *PACS1* | -0.232246244 | 0.008145873 | 0.035832979 | 1 |

| | | | | | | |
|---|---|---|---|---|---|---|
| Heart_Failure | mesothelium | *SH3BP5L* | -1.478127677 | 0.000267186 | 0.003131364 | 1 |
| Heart_Failure | mesothelium | *ATG13* | -0.778444766 | 0.000331332 | 0.00361264 | 1 |
| Heart_Failure | mesothelium | *CCNE2* | -0.140816733 | 0.002857566 | 0.016273367 | 1 |
| Heart_Failure | mesothelium | *SCUBE2* | -0.494733688 | 0.002427331 | 0.014486359 | 1 |
| Heart_Failure | mesothelium | *CLTB* | -0.924690677 | 0.000111105 | 0.002412406 | 0.539080592 |
| Heart_Failure | mesothelium | *PAAF1* | -0.058034125 | 0.001106596 | 0.008172303 | 1 |
| Heart_Failure | mesothelium | *RAB6A* | -0.602504964 | 0.000152222 | 0.002666362 | 0.738582204 |
| Heart_Failure | mesothelium | *PRIMA1* | -1.060376404 | 0.001809767 | 0.011818292 | 1 |
| Heart_Failure | mesothelium | *A2M* | -0.420797467 | 0.007751728 | 0.034632949 | 1 |
| Heart_Failure | mesothelium | *B3GALT6* | -2.00785524 | 0.005437298 | 0.026302863 | 1 |
| Heart_Failure | mesothelium | *SLC35G1* | -1.089385283 | 0.000984 | 0.007557727 | 1 |
| Heart_Failure | mesothelium | *TAC4* | -1.000993761 | 0.000984436 | 0.007557727 | 1 |
| Heart_Failure | mesothelium | *SPRYD4* | -0.104467404 | 0.003609746 | 0.019331666 | 1 |
| Heart_Failure | mesothelium | *WDR25* | -1.050190836 | 5.87E-05 | 0.002273164 | 0.284890729 |
| Heart_Failure | mesothelium | *BOK* | -0.662788853 | 0.007009369 | 0.032145047 | 1 |
| Heart_Failure | mesothelium | *ANKLE2* | -0.477752858 | 0.002842676 | 0.016207596 | 1 |
| Heart_Failure | mesothelium | *DPP7* | -0.715362499 | 0.000887291 | 0.006977527 | 1 |
| Heart_Failure | mesothelium | *WDR73* | -0.545545313 | 0.000278668 | 0.003196446 | 1 |
| Heart_Failure | mesothelium | *DSCAML1* | -0.605853821 | 0.000222515 | 0.002905056 | 1 |
| Heart_Failure | mesothelium | *TALDO1* | -0.821186443 | 0.001647659 | 0.010966313 | 1 |
| Heart_Failure | mesothelium | *MAN1B1* | -1.23597512 | 6.97E-05 | 0.002273164 | 0.33823204 |
| Heart_Failure | mesothelium | *MIEF2* | -0.60190135 | 0.004547527 | 0.023056011 | 1 |
| Heart_Failure | mesothelium | *GBA* | -0.307047298 | 0.00840728 | 0.036584861 | 1 |
| Heart_Failure | mesothelium | *CENPBD1* | -2.721542357 | 0.000374322 | 0.003856074 | 1 |
| Heart_Failure | mesothelium | *RPS27* | -0.672813776 | 0.002188194 | 0.01352649 | 1 |
| Heart_Failure | mesothelium | *ASB8* | -0.361513131 | 0.012595448 | 0.049205404 | 1 |
| Heart_Failure | mesothelium | *DPY19L2* | 0.074404625 | 0.011207109 | 0.044785675 | 1 |
| Heart_Failure | mesothelium | *ZNF454* | 0.065933956 | 0.007368304 | 0.033349826 | 1 |
| Heart_Failure | mesothelium | *SH2B1* | -0.907041706 | 0.000166309 | 0.002704817 | 0.806933007 |
| Heart_Failure | mesothelium | *KDELC2* | -0.514451819 | 0.009397973 | 0.039056985 | 1 |
| Heart_Failure | mesothelium | *TMEM11* | -0.627297093 | 0.000690641 | 0.005807608 | 1 |
| Heart_Failure | mesothelium | *NT5DC1* | -0.744392634 | 7.12E-05 | 0.002273164 | 0.345500353 |
| Heart_Failure | mesothelium | *P4HTM* | -0.569590283 | 0.002326342 | 0.014056551 | 1 |
| Heart_Failure | mesothelium | *ERN1* | -0.794638266 | 1.50E-05 | 0.001697586 | 0.072996203 |
| Heart_Failure | mesothelium | *NSUN3* | -0.717724245 | 0.000623465 | 0.005382656 | 1 |
| Heart_Failure | mesothelium | *TRIM73* | -0.219777198 | 0.011977583 | 0.047197189 | 1 |
| Heart_Failure | mesothelium | *C17orf62* | -0.755576956 | 0.010114174 | 0.041482648 | 1 |
| Heart_Failure | mesothelium | *SPTY2D1* | -0.734462907 | 0.002493742 | 0.01479173 | 1 |
| Heart_Failure | mesothelium | *ZNF664* | -1.299360806 | 0.00022119 | 0.002900583 | 1 |
| Heart_Failure | mesothelium | *HLA-DQB1* | -0.180007887 | 3.19E-26 | 1.55E-22 | 1.55E-22 |
| Heart_Failure | mesothelium | *PACS2* | -0.892655607 | 0.000232307 | 0.002946978 | 1 |
| Heart_Failure | mesothelium | *LINC00174* | -0.091923313 | 0.003819146 | 0.020185725 | 1 |
| Heart_Failure | mesothelium | *LBX2* | 0.109180437 | 0.000672797 | 0.005677237 | 1 |
| Heart_Failure | mesothelium | *SERTAD2* | -0.404107446 | 0.002918927 | 0.016546936 | 1 |
| Heart_Failure | mesothelium | *AKAP5* | -0.773610621 | 0.003235116 | 0.017756544 | 1 |
| Heart_Failure | mesothelium | *PDXDC1* | -0.633578156 | 0.00041403 | 0.004091391 | 1 |
| Heart_Failure | mesothelium | *ZADH2* | -0.423092308 | 0.012745619 | 0.049592418 | 1 |
| Heart_Failure | mesothelium | *SEPT1* | -1.6237588 | 4.41E-07 | 0.000194349 | 0.002137836 |
| Heart_Failure | mesothelium | *MTURN* | -0.543381216 | 0.001757606 | 0.011555425 | 1 |
| Heart_Failure | mesothelium | *CCDC66* | -0.597581499 | 5.74E-05 | 0.002273164 | 0.278731922 |

| | | | | | | |
|---|---|---|---|---|---|---|
| Heart_Failure | mesothelium | *MB21D2* | -0.5000756 | 0.000708395 | 0.005936326 | 1 |
| Heart_Failure | mesothelium | *ZDHHC20* | -0.706853668 | 0.000717923 | 0.005974896 | 1 |
| Heart_Failure | mesothelium | *LRRC57* | -0.143586357 | 0.012705979 | 0.049517597 | 1 |
| Heart_Failure | mesothelium | *DDX60L* | -0.522140966 | 0.008902062 | 0.037879761 | 1 |
| Heart_Failure | mesothelium | *SGSH* | -1.740888044 | 8.63E-05 | 0.002275786 | 0.418710493 |
| Heart_Failure | mesothelium | *ZBTB20* | -0.503721578 | 7.64E-05 | 0.002273164 | 0.370787263 |
| Heart_Failure | mesothelium | *C3orf58* | -0.570617828 | 0.00315946 | 0.017463626 | 1 |
| Heart_Failure | mesothelium | *RFX7* | -0.724262227 | 0.000296944 | 0.003338215 | 1 |
| Heart_Failure | mesothelium | *ZNF329* | -1.031372728 | 0.000170584 | 0.002704817 | 0.827673971 |
| Heart_Failure | mesothelium | *IDH2* | -0.656666897 | 0.001996752 | 0.012664364 | 1 |
| Heart_Failure | mesothelium | *TDRKH* | -0.127093417 | 0.005285002 | 0.02577169 | 1 |
| Heart_Failure | mesothelium | *ERCC6L2* | -0.501858964 | 0.001626484 | 0.010855156 | 1 |
| Heart_Failure | mesothelium | *MRPL41* | -1.082034632 | 0.001094951 | 0.008119798 | 1 |
| Heart_Failure | mesothelium | *FIGN* | -0.696312344 | 7.17E-05 | 0.002273164 | 0.347894391 |
| Heart_Failure | mesothelium | *KBTBD3* | -0.613032677 | 0.005365689 | 0.026112663 | 1 |
| Heart_Failure | mesothelium | *NPLOC4* | -0.160366591 | 0.003321626 | 0.018122484 | 1 |
| Heart_Failure | mesothelium | *GLRX5* | -1.285581546 | 0.000254271 | 0.003061341 | 1 |
| Heart_Failure | mesothelium | *ADI1* | -0.735527618 | 0.000262083 | 0.003102849 | 1 |
| Heart_Failure | mesothelium | *SKA2* | -0.490113195 | 0.006182237 | 0.029236076 | 1 |
| Heart_Failure | mesothelium | *ALG12* | -1.276614345 | 3.92E-05 | 0.002273164 | 0.189973195 |
| Heart_Failure | mesothelium | *C12orf60* | -1.107432223 | 0.000467996 | 0.004417736 | 1 |
| Heart_Failure | mesothelium | *ARHGEF37* | -1.232599629 | 9.51E-05 | 0.002294209 | 0.461598408 |
| Heart_Failure | mesothelium | *CSMD1* | -0.955737871 | 0.001018431 | 0.007708938 | 1 |
| Heart_Failure | mesothelium | *LRIT3* | -4.826352719 | 0.011621309 | 0.046105142 | 1 |
| Heart_Failure | mesothelium | *PSMG1* | -0.233248918 | 9.50E-05 | 0.002294209 | 0.460908448 |
| Heart_Failure | mesothelium | *SERHL2* | -1.365069722 | 6.31E-05 | 0.002273164 | 0.305960666 |
| Heart_Failure | mesothelium | *FAM19A1* | -0.395116401 | 0.000121325 | 0.002465411 | 0.588668255 |
| Heart_Failure | mesothelium | *ZNF703* | -0.205256 | 0.002738977 | 0.015803873 | 1 |
| Heart_Failure | mesothelium | *BTBD9* | -0.549475703 | 0.000197143 | 0.002777753 | 0.956538643 |
| Heart_Failure | mesothelium | *PP2D1* | -1.351590066 | 0.000444919 | 0.0042663 | 1 |
| Heart_Failure | mesothelium | *PGP* | -0.258996104 | 0.006722233 | 0.03113327 | 1 |
| Heart_Failure | mesothelium | *A3GALT2* | -0.066014937 | 0.010477629 | 0.042613124 | 1 |
| Heart_Failure | mesothelium | *COPB2* | -0.699295112 | 0.001491464 | 0.010135273 | 1 |
| Heart_Failure | mesothelium | *RNLS* | -0.345812097 | 0.002919798 | 0.016546936 | 1 |
| Heart_Failure | mesothelium | *TCTE3* | -0.802403074 | 0.004423318 | 0.02256776 | 1 |
| Heart_Failure | mesothelium | *ZFP90* | -0.963630095 | 0.000168918 | 0.002704817 | 0.819588028 |
| Heart_Failure | mesothelium | *SORCS2* | -0.301286015 | 0.00530257 | 0.025831393 | 1 |
| Heart_Failure | mesothelium | *BRI3BP* | -0.721317723 | 0.000262834 | 0.003102849 | 1 |
| Heart_Failure | mesothelium | *UBOX5* | -0.595443974 | 0.004838305 | 0.024201501 | 1 |
| Heart_Failure | mesothelium | *SGCZ* | -0.077677833 | 0.004300459 | 0.022067281 | 1 |
| Heart_Failure | mesothelium | *IFITM2* | -1.006166825 | 3.28E-05 | 0.002273164 | 0.15938113 |
| Heart_Failure | mesothelium | *CDK10* | -0.07656467 | 0.002922934 | 0.016546936 | 1 |
| Heart_Failure | mesothelium | *PSMD13* | -0.205399165 | 0.008974039 | 0.037928602 | 1 |
| Heart_Failure | mesothelium | *WWOX* | -0.559744025 | 0.000121215 | 0.002465411 | 0.588135338 |
| Heart_Failure | mesothelium | *FFAR4* | -0.448640033 | 0.00037203 | 0.0038488 | 1 |
| Heart_Failure | mesothelium | *TOR3A* | -0.542169309 | 0.000244468 | 0.002972283 | 1 |
| Heart_Failure | mesothelium | *TMEM222* | -0.712875311 | 0.002568294 | 0.01506803 | 1 |
| Heart_Failure | mesothelium | *SEPT10* | -0.422853328 | 0.001129155 | 0.008288443 | 1 |
| Heart_Failure | mesothelium | *SMYD4* | -0.178752167 | 0.004527559 | 0.023026958 | 1 |
| Heart_Failure | mesothelium | *MAPT* | -0.712838924 | 0.004130727 | 0.021435601 | 1 |

| | | | | | | |
|---|---|---|---|---|---|---|
| Heart_Failure | mesothelium | *RTN4RL2* | -0.268441774 | 0.008352665 | 0.036412518 | 1 |
| Heart_Failure | mesothelium | *TMEM232* | 0.107542238 | 0.012652675 | 0.049389203 | 1 |
| Heart_Failure | mesothelium | *EFCAB6* | -0.478896409 | 0.00268924 | 0.015607888 | 1 |
| Heart_Failure | mesothelium | *MRPS21* | -0.179069268 | 3.22E-05 | 0.002273164 | 0.156340994 |
| Heart_Failure | mesothelium | *IFT140* | -0.804548454 | 1.01E-05 | 0.001322916 | 0.048947889 |
| Heart_Failure | mesothelium | *ZNF385C* | -0.108308114 | 0.008701485 | 0.037457183 | 1 |
| Heart_Failure | mesothelium | *EXD3* | -0.966937335 | 0.000151378 | 0.002661178 | 0.734485064 |
| Heart_Failure | mesothelium | *C2orf88* | -0.628435576 | 0.000517097 | 0.004707232 | 1 |
| Heart_Failure | mesothelium | *ZFP69* | -1.623026645 | 0.000115788 | 0.002463815 | 0.561802818 |
| Heart_Failure | mesothelium | *FAM122A* | -1.759688146 | 0.000122377 | 0.002465411 | 0.593773444 |
| Heart_Failure | mesothelium | *ZNF626* | -1.389007007 | 7.90E-05 | 0.002273164 | 0.383151362 |
| Heart_Failure | mesothelium | *ZP3* | -0.063489094 | 0.009172594 | 0.038366745 | 1 |
| Heart_Failure | mesothelium | *COL25A1* | -0.451201564 | 0.011749623 | 0.046424408 | 1 |
| Heart_Failure | mesothelium | *NDOR1* | -1.133447787 | 0.001722997 | 0.01135867 | 1 |
| Heart_Failure | mesothelium | *NOXA1* | -0.961694868 | 0.000394435 | 0.003978253 | 1 |
| Heart_Failure | mesothelium | *RPL14* | -0.842905684 | 0.000127724 | 0.00251464 | 0.619717803 |
| Heart_Failure | mesothelium | *MSL1* | -0.241070277 | 1.35E-05 | 0.001602322 | 0.065722027 |
| Heart_Failure | mesothelium | *UTS2B* | -0.179380895 | 0.006716011 | 0.03113327 | 1 |
| Heart_Failure | mesothelium | *ADAT2* | -0.46832731 | 0.003759723 | 0.019958618 | 1 |
| Heart_Failure | mesothelium | *LITAF* | -0.564896844 | 0.00359892 | 0.019294982 | 1 |
| Heart_Failure | mesothelium | *SF3B3* | -0.645094359 | 0.001144002 | 0.008364486 | 1 |
| Heart_Failure | mesothelium | *PNRC2* | -0.759210209 | 0.002311223 | 0.014017567 | 1 |
| Heart_Failure | mesothelium | *FHIT* | -0.387700291 | 0.006966828 | 0.031980179 | 1 |
| Heart_Failure | mesothelium | *HLA-DRB1* | -0.11216609 | 3.04E-08 | 1.85E-05 | 0.000147696 |
| Heart_Failure | mesothelium | *ZNF471* | -0.878455074 | 0.000706817 | 0.005933349 | 1 |
| Heart_Failure | mesothelium | *ZNF493* | -0.714080484 | 0.000927899 | 0.00722659 | 1 |
| Heart_Failure | mesothelium | *GTF2IRD2* | -0.507462075 | 0.00216917 | 0.013493348 | 1 |
| Heart_Failure | mesothelium | *CD55* | -0.795889739 | 0.000203992 | 0.002777753 | 0.989769127 |
| Heart_Failure | mesothelium | *ZNF124* | -0.717104491 | 0.000175931 | 0.002725564 | 0.853616257 |
| Heart_Failure | mesothelium | *C20orf96* | -0.939214917 | 0.000194217 | 0.002771595 | 0.942342173 |
| Heart_Failure | mesothelium | *TPK1* | -0.420210147 | 0.001503171 | 0.010189958 | 1 |
| Heart_Failure | mesothelium | *XRCC2* | -2.43349591 | 0.001205536 | 0.008652756 | 1 |
| Heart_Failure | mesothelium | *MYO6* | -0.348405171 | 0.000779222 | 0.006364962 | 1 |
| Heart_Failure | mesothelium | *ADH1B* | -0.515203446 | 0.000334413 | 0.003626728 | 1 |
| Heart_Failure | mesothelium | *TOMM7* | -0.876180553 | 0.000167382 | 0.002704817 | 0.812135891 |
| Heart_Failure | mesothelium | *HLA-DQA1* | -0.252267961 | 1.42E-23 | 3.46E-20 | 6.91E-20 |
| Heart_Failure | mesothelium | *SLC39A10* | -0.837189356 | 3.72E-05 | 0.002273164 | 0.180311827 |
| Heart_Failure | mesothelium | *ZNF429* | -0.598854339 | 0.002900332 | 0.016478232 | 1 |
| Heart_Failure | mesothelium | *GMFB* | -1.090655826 | 8.28E-05 | 0.002273164 | 0.401507784 |
| Heart_Failure | mesothelium | *ZMYM1* | -0.940827187 | 0.000146501 | 0.002637401 | 0.71082361 |
| Heart_Failure | mesothelium | *IGF2R* | -0.355377026 | 0.011009395 | 0.044146766 | 1 |
| Heart_Failure | mesothelium | *SND1* | -0.352876747 | 0.010903395 | 0.043794101 | 1 |
| Heart_Failure | mesothelium | *NEK5* | -0.076227112 | 0.012560792 | 0.049149163 | 1 |
| Heart_Failure | mesothelium | *ZNF98* | -0.11325814 | 0.008850692 | 0.037769181 | 1 |
| Heart_Failure | mesothelium | *SLC22A5* | -0.170829245 | 0.009438182 | 0.039173702 | 1 |
| Heart_Failure | mesothelium | *MAP3K5* | -0.384297867 | 0.003096998 | 0.017232381 | 1 |
| Heart_Failure | mesothelium | *SLC2A10* | -0.250598722 | 0.00774291 | 0.034632644 | 1 |
| Heart_Failure | mesothelium | *ZNF665* | -1.096102325 | 0.000355087 | 0.003759027 | 1 |
| Heart_Failure | mesothelium | *RAB40C* | -1.136022705 | 0.000107809 | 0.002412406 | 0.523087175 |
| Heart_Failure | mesothelium | *TOPORS* | -1.12761348 | 7.98E-05 | 0.002273164 | 0.387212066 |

| | | | | | | |
|---|---|---|---|---|---|---|
| Heart_Failure | mesothelium | *PDCD1LG2* | 0.051324183 | 0.005096215 | 0.025077928 | 1 |
| Heart_Failure | mesothelium | *DNAH10* | -0.183721248 | 0.002557722 | 0.015024293 | 1 |
| Heart_Failure | mesothelium | *FAM114A1* | -0.87447428 | 5.69E-06 | 0.001001679 | 0.027592817 |
| Heart_Failure | mesothelium | *RPS26* | 0.079254975 | 0.001246826 | 0.008844442 | 1 |
| Heart_Failure | mesothelium | *EME2* | -1.164823372 | 0.004887458 | 0.024346965 | 1 |
| Heart_Failure | mesothelium | *MPZL1* | -0.666282505 | 0.000111372 | 0.002412406 | 0.540378896 |
| Heart_Failure | mesothelium | *VPS13A* | -0.483240357 | 0.00020538 | 0.002777753 | 0.996502283 |
| Heart_Failure | mesothelium | *ZNF544* | -1.166305336 | 7.91E-05 | 0.002273164 | 0.383793729 |
| Heart_Failure | mesothelium | *MAN1A2* | -0.713770262 | 0.000273065 | 0.003169645 | 1 |
| Heart_Failure | mesothelium | *SVIP* | -0.779508716 | 0.001138877 | 0.00834718 | 1 |
| Heart_Failure | mesothelium | *HSD17B11* | -0.703621813 | 0.000176351 | 0.002725564 | 0.855654923 |
| Heart_Failure | mesothelium | *DDX42* | -0.59025891 | 3.29E-05 | 0.002273164 | 0.159422652 |
| Heart_Failure | mesothelium | *STYX* | -0.634366257 | 0.003088184 | 0.017231146 | 1 |
| Heart_Failure | mesothelium | *HELZ* | -0.60631477 | 0.000439432 | 0.004247262 | 1 |
| Heart_Failure | mesothelium | *UCKL1* | -0.779995067 | 0.001416052 | 0.009731847 | 1 |
| Heart_Failure | mesothelium | *TMA16* | -0.954460104 | 0.000183746 | 0.002737794 | 0.891536949 |
| Heart_Failure | mesothelium | *HLA-DRB5* | -0.022631731 | 0.003568651 | 0.019196333 | 1 |
| Heart_Failure | mesothelium | *TUSC1* | -1.84354897 | 0.000469875 | 0.00442686 | 1 |
| Heart_Failure | mesothelium | *PAPSS2* | -0.68100891 | 8.20E-05 | 0.002273164 | 0.39790471 |
| Heart_Failure | mesothelium | *ANKRD13B* | -1.049889092 | 0.000880864 | 0.006949802 | 1 |
| Heart_Failure | mesothelium | *RPL10A* | -0.753972788 | 7.28E-05 | 0.002273164 | 0.353070544 |
| Heart_Failure | mesothelium | *RCSD1* | -0.487423892 | 0.012331618 | 0.048487044 | 1 |
| Heart_Failure | mesothelium | *SFT2D1* | -0.822058047 | 0.000189904 | 0.002744551 | 0.921415353 |
| Heart_Failure | mesothelium | *PRMT6* | -1.924734242 | 0.000484533 | 0.004528961 | 1 |
| Heart_Failure | mesothelium | *PJA2* | -0.41216011 | 0.001431685 | 0.009825372 | 1 |
| Heart_Failure | mesothelium | *TATDN3* | -0.117842507 | 0.002926066 | 0.016546936 | 1 |
| Heart_Failure | mesothelium | *PCMTD2* | -0.852904409 | 5.34E-05 | 0.002273164 | 0.259063066 |
| Heart_Failure | mesothelium | *SAMD13* | -0.431375438 | 0.010522493 | 0.042759744 | 1 |
| Heart_Failure | mesothelium | *EFCAB7* | -0.291609691 | 0.01064404 | 0.043025098 | 1 |
| Heart_Failure | mesothelium | *IFIT1B* | -3.495746293 | 0.010589491 | 0.042960043 | 1 |
| Heart_Failure | mesothelium | *RUFY2* | -0.743250958 | 0.000236542 | 0.002946978 | 1 |
| Heart_Failure | mesothelium | *BMPR2* | -0.536631345 | 0.001234943 | 0.008824657 | 1 |
| Heart_Failure | mesothelium | *HSD17B8* | 0.095512447 | 0.007300265 | 0.033165623 | 1 |
| Heart_Failure | mesothelium | *PRRT1* | 0.079777813 | 0.001938908 | 0.012427454 | 1 |
| Heart_Failure | mesothelium | *STK19* | -0.216865735 | 0.000129883 | 0.00251464 | 0.630193715 |
| Heart_Failure | mesothelium | *MICB* | 0.028566043 | 0.002405356 | 0.014390614 | 1 |
| Heart_Failure | mesothelium | *MICA* | 0.141569365 | 6.02E-05 | 0.002273164 | 0.291891015 |
| Heart_Failure | mesothelium | *HLA-C* | -0.061876254 | 0.009192385 | 0.038410807 | 1 |
| Heart_Failure | mesothelium | *HLA-G* | -0.075480446 | 0.00692114 | 0.031860884 | 1 |
| Heart_Failure | mesothelium | *MALRD1* | -0.60933386 | 0.001774401 | 0.011618615 | 1 |
| Heart_Failure | mesothelium | *DCTN1* | -1.205454627 | 0.000255147 | 0.003064285 | 1 |
| Heart_Failure | mesothelium | *MZT1* | -0.924422368 | 0.000245004 | 0.002972283 | 1 |
| Heart_Failure | mesothelium | *ZNF155* | -0.295460888 | 0.00036827 | 0.003826225 | 1 |
| Heart_Failure | mesothelium | *ZNF783* | -0.551974512 | 0.000501966 | 0.004604045 | 1 |
| Heart_Failure | mesothelium | *SLFN12L* | -0.167081902 | 0.001096139 | 0.008119798 | 1 |
| Heart_Failure | mesothelium | *SLC35B4* | -0.630906069 | 0.008538444 | 0.036890945 | 1 |
| Heart_Failure | mesothelium | *SCGB2B2* | -0.835310093 | 0.000365972 | 0.003818332 | 1 |
| Heart_Failure | mesothelium | *CCDC144NL* | -1.871765926 | 7.11E-05 | 0.002273164 | 0.34478247 |
| Heart_Failure | mesothelium | *NT5M* | -0.970851395 | 0.000736502 | 0.006108559 | 1 |
| Heart_Failure | mesothelium | *IPO7* | -0.545640275 | 0.000199922 | 0.002777753 | 0.970022067 |

| | | | | | | |
|---|---|---|---|---|---|---|
| Heart_Failure | mesothelium | *TECPR1* | -0.72245831 | 0.000186626 | 0.002744551 | 0.905510804 |
| Heart_Failure | mesothelium | *MT1M* | -0.923391994 | 0.000467593 | 0.004417736 | 1 |
| Heart_Failure | mesothelium | *ZDHHC11B* | -0.699839592 | 1.39E-05 | 0.001602322 | 0.067297524 |
| Heart_Failure | mesothelium | *SACM1L* | -0.953679391 | 4.55E-05 | 0.002273164 | 0.220652459 |
| Heart_Failure | mesothelium | *SLC48A1* | -0.888801352 | 0.000120476 | 0.002465411 | 0.584547636 |
| Heart_Failure | mesothelium | *C17orf51* | -2.257481577 | 8.07E-05 | 0.002273164 | 0.391374852 |
| Heart_Failure | mesothelium | *ATF6B* | 0.22361194 | 0.000641578 | 0.005490187 | 1 |
| Heart_Failure | mesothelium | *IRF9* | -0.727828902 | 0.000215292 | 0.002869773 | 1 |
| Heart_Failure | mesothelium | *REPIN1* | -1.574505227 | 0.000170154 | 0.002704817 | 0.825589522 |
| Heart_Failure | mesothelium | *CPNE1* | -0.080118787 | 0.000166485 | 0.002704817 | 0.807783008 |
| Heart_Failure | mesothelium | *LYRM4* | -0.676218475 | 0.001913957 | 0.012300027 | 1 |
| Heart_Failure | mesothelium | *SH3D21* | 0.184064252 | 0.008011793 | 0.035507635 | 1 |
| Heart_Failure | mesothelium | *ANG* | -1.224348938 | 0.000626511 | 0.005389776 | 1 |
| Heart_Failure | mesothelium | *IFRD2* | -0.16731155 | 0.000817167 | 0.006608154 | 1 |
| Heart_Failure | mesothelium | *TMEM242* | -0.30493868 | 0.004147424 | 0.02147631 | 1 |
| Heart_Failure | mesothelium | *TSTD1* | -0.485032807 | 0.004907646 | 0.024386665 | 1 |
| Heart_Failure | mesothelium | *APOL6* | -0.54163195 | 0.004542142 | 0.023056011 | 1 |
| Heart_Failure | mesothelium | *OR2A1* | 0.122276847 | 0.003716807 | 0.019795771 | 1 |
| Heart_Failure | mesothelium | *AC062017.1* | -0.863341211 | 0.008972002 | 0.037928602 | 1 |
| Heart_Failure | mesothelium | *LINC00184* | -36.89253354 | 0.000145644 | 0.002637401 | 0.706665639 |
| Heart_Failure | mesothelium | *AL049840.1* | -0.323231594 | 0.002371539 | 0.014240973 | 1 |
| Heart_Failure | mesothelium | *CYP4A22-AS1* | -1.094651828 | 0.000382006 | 0.003893896 | 1 |
| Heart_Failure | mesothelium | *ERCC6* | -0.524014028 | 0.000742636 | 0.006128011 | 1 |
| Heart_Failure | mesothelium | *FAM138B* | -9.506566604 | 0.002172764 | 0.013498402 | 1 |
| Heart_Failure | mesothelium | *AC009299.3* | -0.781444638 | 0.010276221 | 0.042005243 | 1 |
| Heart_Failure | mesothelium | *SCAMP4* | -1.158875793 | 7.47E-05 | 0.002273164 | 0.362445881 |
| Heart_Failure | mesothelium | *LINC00578* | -0.278770135 | 0.00875946 | 0.037611416 | 1 |
| Heart_Failure | mesothelium | *ARL17B* | -0.312075338 | 0.001257966 | 0.008897449 | 1 |
| Heart_Failure | mesothelium | *TAPBP* | -0.188794261 | 0.009583371 | 0.03964068 | 1 |
| Heart_Failure | mesothelium | *HLA-DQB2* | 0.077514279 | 2.84E-17 | 3.44E-14 | 1.38E-13 |
| Heart_Failure | mesothelium | *ZNF736* | 0.081086515 | 0.012518163 | 0.049021896 | 1 |
| Heart_Failure | mesothelium | *ANKRD65* | -2.385675565 | 0.007435417 | 0.03352848 | 1 |
| Heart_Failure | mesothelium | *ZBTB22* | 0.176740543 | 0.001115837 | 0.008203092 | 1 |
| Heart_Failure | mesothelium | *HLA-DQA2* | 0.094779511 | 1.86E-17 | 3.00E-14 | 9.00E-14 |
| Heart_Failure | mesothelium | *LRRC37A2* | -0.35370956 | 0.008834011 | 0.037769181 | 1 |
| Heart_Failure | mesothelium | *PSMB9* | 0.105192838 | 3.91E-06 | 0.000823873 | 0.018949083 |
| Heart_Failure | mesothelium | *KRBOX1* | -21.83233763 | 0.000203722 | 0.002777753 | 0.988456845 |
| Heart_Failure | mesothelium | *HOGA1* | -1.478035194 | 0.001220303 | 0.008745808 | 1 |
| Heart_Failure | mesothelium | *ARFGAP3* | -0.812030788 | 0.001018317 | 0.007708938 | 1 |
| Heart_Failure | mesothelium | *MRPL20* | -0.385312624 | 0.006002248 | 0.028495994 | 1 |
| Heart_Failure | mesothelium | *DECR2* | -1.564737976 | 0.000202499 | 0.002777753 | 0.98252589 |
| Heart_Failure | mesothelium | *MRPL33* | -0.30418942 | 0.007528732 | 0.033854871 | 1 |
| Heart_Failure | mesothelium | *FGF14-IT1* | -2.135702377 | 0.005961355 | 0.028385176 | 1 |
| Heart_Failure | mesothelium | *NBPF15* | -1.209902019 | 4.13E-05 | 0.002273164 | 0.20043723 |
| Heart_Failure | mesothelium | *UPK3B* | -0.378099008 | 0.00118238 | 0.008537063 | 1 |
| Heart_Failure | mesothelium | *TTC4* | -0.190946991 | 6.50E-05 | 0.002273164 | 0.315509905 |
| Heart_Failure | mesothelium | *LILRA6* | 0.063035644 | 0.004307031 | 0.022067281 | 1 |
| Heart_Failure | mesothelium | *C4A* | -0.124889864 | 0.005011636 | 0.024762179 | 1 |
| Heart_Failure | mesothelium | *NEAT1* | -0.294537201 | 0.00948575 | 0.039303895 | 1 |
| Heart_Failure | mesothelium | *DNAJC3-AS1* | -0.312490657 | 0.004266648 | 0.021929847 | 1 |

| | | | | | | |
|---|---|---|---|---|---|---|
| Heart_Failure | mesothelium | *DPH6-AS1* | -0.454014428 | 1.61E-06 | 0.000557209 | 0.007800931 |
| Heart_Failure | mesothelium | *CDK11B* | -1.162878205 | 9.91E-05 | 0.002301699 | 0.481055098 |
| Heart_Failure | mesothelium | *C5orf17* | -1.042710182 | 4.25E-05 | 0.002273164 | 0.20600651 |
| Heart_Failure | mesothelium | *PDCD6* | -0.941208758 | 0.000327506 | 0.003578957 | 1 |
| Heart_Failure | mesothelium | *DNAH10OS* | -0.874572495 | 0.008283031 | 0.0361661 | 1 |
| Heart_Failure | mesothelium | *SMIM20* | -1.382862807 | 0.000105799 | 0.002398327 | 0.513334391 |
| Heart_Failure | mesothelium | *ZNF550* | -0.988125912 | 0.001075585 | 0.008028846 | 1 |
| Heart_Failure | mesothelium | *LINC01094* | -1.181918873 | 0.002492359 | 0.01479173 | 1 |
| Heart_Failure | mesothelium | *ARMS2* | -0.179258451 | 0.004189081 | 0.021651562 | 1 |
| Heart_Failure | mesothelium | *MEX3A* | -0.114211261 | 0.011716612 | 0.046369494 | 1 |
| Heart_Failure | mesothelium | *CNPY2* | -0.985599407 | 0.000275286 | 0.003180204 | 1 |
| Heart_Failure | mesothelium | *HIF1A-AS2* | -0.556005491 | 0.008962297 | 0.037928602 | 1 |
| Heart_Failure | mesothelium | *AC083843.1* | -15.74573574 | 0.012717668 | 0.049523375 | 1 |
| Heart_Failure | mesothelium | *DYNLL2* | -1.41012886 | 0.000371353 | 0.0038488 | 1 |
| Heart_Failure | mesothelium | *ZNF582-AS1* | -2.037699242 | 9.52E-05 | 0.002294209 | 0.461915592 |
| Heart_Failure | mesothelium | *ZNF224* | -1.01057326 | 0.000139263 | 0.002608902 | 0.675705561 |
| Heart_Failure | mesothelium | *ZNF728* | -0.422750055 | 0.000740296 | 0.006119107 | 1 |
| Heart_Failure | mesothelium | *TRABD2B* | -0.638303828 | 0.005219295 | 0.025538402 | 1 |
| Heart_Failure | mesothelium | *AC093157.1* | -0.602939044 | 0.000613125 | 0.005331328 | 1 |
| Heart_Failure | mesothelium | *ZNF587B* | -1.610974647 | 7.77E-05 | 0.002273164 | 0.377005445 |
| Heart_Failure | mesothelium | *SNHG8* | -0.169168401 | 0.001990343 | 0.012655801 | 1 |
| Heart_Failure | mesothelium | *CASC15* | -0.469606227 | 0.000837484 | 0.006727603 | 1 |
| Heart_Failure | mesothelium | *PAXIP1-AS1* | -0.850724371 | 0.001075587 | 0.008028846 | 1 |
| Heart_Failure | monocyte | *PNMT* | 5.981793833 | 4.59E-08 | 4.45E-05 | 0.000222615 |
| Heart_Failure | monocyte | *GSDMA* | -2.44838491 | 2.10E-08 | 2.55E-05 | 0.000101817 |
| Heart_Failure | monocyte | *HLA-DQB1* | -3.110761002 | 4.85E-22 | 2.35E-18 | 2.35E-18 |
| Heart_Failure | monocyte | *ATF6B* | 3.211615349 | 2.05E-05 | 0.014197888 | 0.099385219 |
| Heart_Failure | monocyte | *HLA-DQB2* | 1.406151579 | 3.58E-15 | 5.78E-12 | 1.73E-11 |
| Heart_Failure | monocyte | *HLA-DQA2* | 1.78888177 | 3.16E-18 | 7.68E-15 | 1.54E-14 |
| Heart_Failure | monocyte | *PSMB9* | 2.083574353 | 2.20E-07 | 0.000177721 | 0.001066325 |
| Heart_Failure | nk_cell | *EVA1B* | 1.901407344 | 5.98E-05 | 0.036247797 | 0.289982379 |
| Heart_Failure | nk_cell | *TSLP* | 0.726828238 | 5.78E-05 | 0.036247797 | 0.280489683 |
| Heart_Failure | nk_cell | *GSDMA* | -0.987609278 | 3.97E-06 | 0.003852781 | 0.019263906 |
| Heart_Failure | nk_cell | *HLA-DQB1* | -1.737145196 | 1.69E-21 | 8.22E-18 | 8.22E-18 |
| Heart_Failure | nk_cell | *HLA-DRB1* | -0.914158035 | 1.04E-05 | 0.008413148 | 0.050478885 |
| Heart_Failure | nk_cell | *HLA-DQA1* | -1.985505437 | 5.35E-16 | 1.30E-12 | 2.59E-12 |
| Heart_Failure | nk_cell | *HLA-DQB2* | 0.640606567 | 9.36E-13 | 1.51E-09 | 4.54E-09 |
| Heart_Failure | nk_cell | *HLA-DQA2* | 0.64972164 | 3.94E-11 | 4.77E-08 | 1.91E-07 |
| Heart_Failure | pericyte | *RBM6* | -2.010062152 | 6.00E-06 | 0.00290882 | 0.029088195 |
| Heart_Failure | pericyte | *IL18R1* | 4.822062749 | 1.05E-05 | 0.004247935 | 0.050975215 |
| Heart_Failure | pericyte | *ERBB2* | 6.68404156 | 4.54E-05 | 0.015721879 | 0.22010631 |
| Heart_Failure | pericyte | *PNMT* | 7.56863559 | 1.12E-09 | 1.09E-06 | 5.45E-06 |
| Heart_Failure | pericyte | *EVA1B* | 2.547980494 | 0.000102618 | 0.031118935 | 0.497902959 |
| Heart_Failure | pericyte | *TSLP* | 1.574841723 | 3.83E-06 | 0.00206733 | 0.018605974 |
| Heart_Failure | pericyte | *FGFR4* | -8.277595088 | 0.000152491 | 0.041104792 | 0.739886262 |
| Heart_Failure | pericyte | *RNF123* | 3.130109541 | 2.50E-05 | 0.009313907 | 0.121080794 |
| Heart_Failure | pericyte | *MST1R* | 2.186203454 | 7.14E-05 | 0.023099695 | 0.346495424 |
| Heart_Failure | pericyte | *GSDMA* | -2.574990364 | 1.08E-07 | 8.76E-05 | 0.000525796 |
| Heart_Failure | pericyte | *HLA-DQB1* | -3.179859961 | 1.75E-23 | 8.51E-20 | 8.51E-20 |
| Heart_Failure | pericyte | *HLA-DRB1* | -1.847295986 | 7.51E-07 | 0.000520739 | 0.003645173 |

| | | | | | | |
|---|---|---|---|---|---|---|
| Heart_Failure | pericyte | *HLA-DQA1* | -4.192604219 | 1.66E-19 | 4.02E-16 | 8.04E-16 |
| Heart_Failure | pericyte | *ATF6B* | 4.854967488 | 7.69E-06 | 0.003390278 | 0.037293059 |
| Heart_Failure | pericyte | *HLA-DQB2* | 1.42339281 | 2.15E-16 | 2.61E-13 | 1.04E-12 |
| Heart_Failure | pericyte | *ZBTB22* | 3.854869125 | 0.000130653 | 0.037289923 | 0.633928698 |
| Heart_Failure | pericyte | *HLA-DQA2* | 1.803115714 | 1.32E-17 | 2.13E-14 | 6.40E-14 |
| Heart_Failure | pericyte | *PSMB9* | 1.982812854 | 2.10E-06 | 0.001274573 | 0.010196581 |
| Heart_Failure | SMC | *SEMA3F* | -3.31693606 | 2.65E-05 | 0.01287348 | 0.128734797 |
| Heart_Failure | SMC | *RBM6* | -2.10266067 | 1.00E-07 | 6.95E-05 | 0.00048664 |
| Heart_Failure | SMC | *PABPC4* | -3.941201265 | 3.01E-05 | 0.013280185 | 0.146082037 |
| Heart_Failure | SMC | *KIF20A* | -12.52779853 | 9.54E-06 | 0.005140753 | 0.046266773 |
| Heart_Failure | SMC | *PNMT* | 5.951827518 | 1.67E-06 | 0.001013529 | 0.008108235 |
| Heart_Failure | SMC | *FLG* | -1.16513908 | 0.000147544 | 0.044742723 | 0.715883569 |
| Heart_Failure | SMC | *FGFR4* | -5.976789705 | 6.71E-05 | 0.02324442 | 0.325421874 |
| Heart_Failure | SMC | *GSDMA* | -2.897930297 | 2.52E-11 | 2.45E-08 | 1.22E-07 |
| Heart_Failure | SMC | *HLA-DQB1* | -3.001372207 | 8.30E-26 | 4.03E-22 | 4.03E-22 |
| Heart_Failure | SMC | *MRPS21* | -2.589409545 | 0.000126415 | 0.040891034 | 0.613365508 |
| Heart_Failure | SMC | *MSL1* | -3.158646529 | 5.39E-05 | 0.020103621 | 0.261347067 |
| Heart_Failure | SMC | *HLA-DRB1* | -1.836252112 | 3.18E-08 | 2.58E-05 | 0.000154518 |
| Heart_Failure | SMC | *HLA-DQA1* | -3.634710807 | 2.83E-21 | 6.87E-18 | 1.37E-17 |
| Heart_Failure | SMC | *HLA-DQB2* | 1.349844226 | 2.84E-17 | 4.59E-14 | 1.38E-13 |
| Heart_Failure | SMC | *HLA-DQA2* | 1.419982482 | 2.47E-14 | 3.00E-11 | 1.20E-10 |
| Heart_Failure | SMC | *TTC4* | -2.757493219 | 3.55E-05 | 0.014371569 | 0.172458833 |
| Heart_Failure | t_cell | *RBM6* | -1.171333251 | 0.000191585 | 0.026559156 | 0.929570462 |
| Heart_Failure | t_cell | *ANAPC4* | 3.310218929 | 0.000628473 | 0.048842924 | 1 |
| Heart_Failure | t_cell | *SDCCAG8* | 2.190338588 | 8.70E-05 | 0.017590841 | 0.422180185 |
| Heart_Failure | t_cell | *BTBD1* | 3.666471523 | 0.000488095 | 0.044683692 | 1 |
| Heart_Failure | t_cell | *VPS9D1* | 4.371885756 | 0.000152798 | 0.023697317 | 0.74137799 |
| Heart_Failure | t_cell | *GSTP1* | 2.975250979 | 0.000511373 | 0.045213444 | 1 |
| Heart_Failure | t_cell | *MAST2* | 3.59911296 | 3.19E-05 | 0.008592586 | 0.154666555 |
| Heart_Failure | t_cell | *PSMC5* | 3.899983621 | 0.000310793 | 0.036779733 | 1 |
| Heart_Failure | t_cell | *SETD1A* | 5.120950483 | 3.67E-06 | 0.001772091 | 0.017826243 |
| Heart_Failure | t_cell | *MTAP* | 3.496855155 | 0.000549552 | 0.045665588 | 1 |
| Heart_Failure | t_cell | *DIDO1* | 3.296156006 | 0.000560565 | 0.045665588 | 1 |
| Heart_Failure | t_cell | *TIMM44* | 4.322634977 | 5.56E-05 | 0.01348031 | 0.269606197 |
| Heart_Failure | t_cell | *CHN2* | 2.78780105 | 9.09E-05 | 0.017642299 | 0.441057484 |
| Heart_Failure | t_cell | *RAB5C* | 2.840862775 | 0.000467586 | 0.043629399 | 1 |
| Heart_Failure | t_cell | *PRDM4* | 4.576338758 | 2.25E-05 | 0.00681922 | 0.109107524 |
| Heart_Failure | t_cell | *FAM120B* | 2.783689148 | 0.000464766 | 0.043629399 | 1 |
| Heart_Failure | t_cell | *KIF20A* | -10.45662202 | 5.85E-05 | 0.013512174 | 0.283755653 |
| Heart_Failure | t_cell | *CEBPZ* | 3.539682627 | 0.000699241 | 0.048842924 | 1 |
| Heart_Failure | t_cell | *MED28* | 3.256413501 | 0.000453072 | 0.043629399 | 1 |
| Heart_Failure | t_cell | *IQSEC3* | 7.346588592 | 0.000323579 | 0.037079109 | 1 |
| Heart_Failure | t_cell | *CSE1L* | 3.319019763 | 0.000600947 | 0.047799925 | 1 |
| Heart_Failure | t_cell | *C12orf65* | 2.385067642 | 0.000255896 | 0.034134894 | 1 |
| Heart_Failure | t_cell | *KIF3A* | 4.421658136 | 0.000273287 | 0.034206301 | 1 |
| Heart_Failure | t_cell | *ERCC5* | 4.441281838 | 0.000686254 | 0.048842924 | 1 |
| Heart_Failure | t_cell | *NMT1* | 2.01692005 | 2.14E-05 | 0.00681922 | 0.103616707 |
| Heart_Failure | t_cell | *ADAM10* | 3.451098249 | 0.000118924 | 0.020607752 | 0.577017067 |
| Heart_Failure | t_cell | *FASTKD1* | 3.423174376 | 0.000543583 | 0.045665588 | 1 |
| Heart_Failure | t_cell | *G3BP2* | 2.787802088 | 0.000260303 | 0.034134894 | 1 |

| | | | | | | |
|---|---|---|---|---|---|---|
| Heart_Failure | t_cell | *ESYT1* | 4.074355173 | 0.000347579 | 0.037079109 | 1 |
| Heart_Failure | t_cell | *TCF25* | 3.17147894 | 0.000112608 | 0.020236058 | 0.546373561 |
| Heart_Failure | t_cell | *PNMT* | 6.358105691 | 2.02E-09 | 1.64E-06 | 9.81E-06 |
| Heart_Failure | t_cell | *EVA1B* | 2.782909688 | 7.16E-06 | 0.002894826 | 0.03473791 |
| Heart_Failure | t_cell | *FLG* | -0.851072423 | 0.000677459 | 0.048842924 | 1 |
| Heart_Failure | t_cell | *WDPCP* | 1.75979007 | 0.000704659 | 0.048842924 | 1 |
| Heart_Failure | t_cell | *CISD2* | 4.769988132 | 0.000673048 | 0.048842924 | 1 |
| Heart_Failure | t_cell | *TSLP* | 0.919654157 | 4.88E-05 | 0.012473135 | 0.236989561 |
| Heart_Failure | t_cell | *MYLK4* | 5.798155703 | 0.000359391 | 0.037101367 | 1 |
| Heart_Failure | t_cell | *ALDOA* | 3.661679662 | 0.000167079 | 0.024565676 | 0.810667295 |
| Heart_Failure | t_cell | *TMBIM4* | 2.931980361 | 0.000342428 | 0.037079109 | 1 |
| Heart_Failure | t_cell | *RPL30* | 2.615979472 | 0.000512518 | 0.045213444 | 1 |
| Heart_Failure | t_cell | *VPS11* | 2.724894995 | 0.000564702 | 0.045665588 | 1 |
| Heart_Failure | t_cell | *FDPS* | 3.736885122 | 0.000173056 | 0.024696123 | 0.839668167 |
| Heart_Failure | t_cell | *LEMD2* | 4.26063094 | 0.000351533 | 0.037079109 | 1 |
| Heart_Failure | t_cell | *RNF123* | 2.678248923 | 1.69E-07 | 0.000102655 | 0.000821236 |
| Heart_Failure | t_cell | *MST1R* | 1.554566609 | 6.47E-05 | 0.014270897 | 0.313959745 |
| Heart_Failure | t_cell | *SERINC5* | 2.856907697 | 0.000150826 | 0.023697317 | 0.731806043 |
| Heart_Failure | t_cell | *SAP30L* | 5.178069835 | 1.53E-05 | 0.005286303 | 0.074008236 |
| Heart_Failure | t_cell | *PIP4K2C* | 3.715202666 | 0.000156289 | 0.023697317 | 0.758314134 |
| Heart_Failure | t_cell | *ZNF23* | 4.970497153 | 0.000279275 | 0.034206301 | 1 |
| Heart_Failure | t_cell | *GSDMA* | -2.406948824 | 1.34E-10 | 1.30E-07 | 6.48E-07 |
| Heart_Failure | t_cell | *PPID* | 5.146543866 | 9.50E-05 | 0.017721767 | 0.460765946 |
| Heart_Failure | t_cell | *CFL1* | 3.526470562 | 0.000559002 | 0.045665588 | 1 |
| Heart_Failure | t_cell | *RPS6KB2* | 4.947478492 | 7.76E-06 | 0.002896836 | 0.037658865 |
| Heart_Failure | t_cell | *GAK* | 3.638294178 | 0.000654119 | 0.048842924 | 1 |
| Heart_Failure | t_cell | *HLA-DQB1* | -2.199488305 | 4.33E-22 | 2.10E-18 | 2.10E-18 |
| Heart_Failure | t_cell | *LBX2* | 1.550408378 | 0.000379104 | 0.038321062 | 1 |
| Heart_Failure | t_cell | *D2HGDH* | 2.360640008 | 8.47E-05 | 0.017590841 | 0.411188815 |
| Heart_Failure | t_cell | *SLC4A5* | 10.81052055 | 0.000434783 | 0.043052372 | 1 |
| Heart_Failure | t_cell | *HLA-DRB1* | -1.108914526 | 2.59E-05 | 0.00738513 | 0.125547217 |
| Heart_Failure | t_cell | *HLA-DQA1* | -2.84754405 | 7.88E-18 | 1.91E-14 | 3.82E-14 |
| Heart_Failure | t_cell | *ZNF138* | 3.142352898 | 0.000337836 | 0.037079109 | 1 |
| Heart_Failure | t_cell | *RPS26* | 1.273938853 | 0.00012444 | 0.020820029 | 0.603780853 |
| Heart_Failure | t_cell | *MICA* | 2.098388422 | 4.02E-06 | 0.001772091 | 0.019492997 |
| Heart_Failure | t_cell | *LIN52* | 2.543485863 | 0.000281998 | 0.034206301 | 1 |
| Heart_Failure | t_cell | *ATF6B* | 3.831396591 | 3.00E-07 | 0.000161933 | 0.0014574 |
| Heart_Failure | t_cell | *RPS18* | 2.375974915 | 0.000643713 | 0.048842924 | 1 |
| Heart_Failure | t_cell | *HLA-DQB2* | 1.028957828 | 1.24E-16 | 1.50E-13 | 6.01E-13 |
| Heart_Failure | t_cell | *HLA-DQA2* | 1.215615956 | 1.01E-16 | 1.50E-13 | 4.89E-13 |
| Heart_Failure | t_cell | *PSMB9* | 1.464306929 | 7.60E-08 | 5.27E-05 | 0.000368913 |
| Heart_Failure | t_cell | *KIAA1147* | 3.209508671 | 0.000655761 | 0.048842924 | 1 |

Table S11 Comparison of genes identified from causal inference and CTS-TWAS analysis

a. Comparison of genes identified from causal inference and CTS-TWAS analysis with adjustment for multiple testing correction(FDR<=0.05)

| Tissue | Disorder | Cell type | No. of causal genes | No. of TWAS genes | No. of novel genes from causal inference | No. of novel genes from TWAS | No. of common genes |
|---|---|---|---|---|---|---|---|
| Frontal Cortex | Anxiety | Astros | 12 | 0 | 12 | 0 | 0 |
| Frontal Cortex | Anxiety | Ex | 60 | 0 | 60 | 0 | 0 |
| Frontal Cortex | Anxiety | Inhib | 53 | 0 | 53 | 0 | 0 |
| Frontal Cortex | Anxiety | Micro_Macro | 4 | 0 | 4 | 0 | 0 |
| Frontal Cortex | Anxiety | Oligos | 19 | 0 | 19 | 0 | 0 |
| Frontal Cortex | Anxiety | OPCs | 13 | 0 | 13 | 0 | 0 |
| Frontal Cortex | Bipolar | Astros | 7 | 0 | 7 | 0 | 0 |
| Frontal Cortex | Bipolar | Ex | 63 | 185 | 44 | 166 | 19 |
| Frontal Cortex | Bipolar | Inhib | 42 | 1 | 41 | 0 | 1 |
| Frontal Cortex | Bipolar | Micro_Macro | 5 | 0 | 5 | 0 | 0 |
| Frontal Cortex | Bipolar | Oligos | 13 | 0 | 13 | 0 | 0 |
| Frontal Cortex | Bipolar | OPCs | 10 | 0 | 10 | 0 | 0 |
| Frontal Cortex | Depression | Astros | 13 | 2 | 12 | 1 | 1 |
| Frontal Cortex | Depression | Ex | 78 | 2 | 77 | 1 | 1 |
| Frontal Cortex | Depression | Inhib | 72 | 2 | 71 | 1 | 1 |
| Frontal Cortex | Depression | Micro_Macro | 9 | 2 | 7 | 0 | 2 |
| Frontal Cortex | Depression | Oligos | 21 | 1 | 20 | 0 | 1 |
| Frontal Cortex | Depression | OPCs | 16 | 1 | 15 | 0 | 1 |
| Frontal Cortex | Psychosis | Astros | 6 | 0 | 6 | 0 | 0 |
| Frontal Cortex | Psychosis | Ex | 53 | 0 | 53 | 0 | 0 |
| Frontal Cortex | Psychosis | Inhib | 63 | 0 | 63 | 0 | 0 |
| Frontal Cortex | Psychosis | Micro_Macro | 6 | 0 | 6 | 0 | 0 |
| Frontal Cortex | Psychosis | Oligos | 21 | 0 | 21 | 0 | 0 |
| Frontal Cortex | Psychosis | OPCs | 9 | 0 | 9 | 0 | 0 |
| Pancreas | Prediabetes | acinar_cell | 16 | 0 | 16 | 0 | 0 |
| Pancreas | Prediabetes | alpha_cell | 27 | 0 | 27 | 0 | 0 |
| Pancreas | Prediabetes | beta_cell | 63 | 0 | 63 | 0 | 0 |
| Pancreas | Prediabetes | delta_cell | 10 | 0 | 10 | 0 | 0 |
| Pancreas | Prediabetes | duct_epithelial_cell | 12 | 1275 | 2 | 1265 | 10 |
| Pancreas | Prediabetes | endothelial_cell | 13 | 0 | 13 | 0 | 0 |
| Pancreas | Prediabetes | epsilon_cell | 4 | 0 | 4 | 0 | 0 |
| Pancreas | Prediabetes | leukocyte | 5 | 0 | 5 | 0 | 0 |
| Pancreas | Prediabetes | mesenchymal_cell | 10 | 0 | 10 | 0 | 0 |
| Pancreas | Prediabetes | PP_cell | 8 | 1636 | 0 | 1628 | 8 |
| Pancreas | T1DM | acinar_cell | 24 | 34 | 18 | 28 | 6 |
| Pancreas | T1DM | alpha_cell | 50 | 35 | 45 | 30 | 5 |
| Pancreas | T1DM | beta_cell | 114 | 36 | 107 | 29 | 7 |
| Pancreas | T1DM | delta_cell | 12 | 29 | 8 | 25 | 4 |
| Pancreas | T1DM | duct_epithelial_cell | 15 | 32 | 12 | 29 | 3 |
| Pancreas | T1DM | endothelial_cell | 10 | 70 | 7 | 67 | 3 |
| Pancreas | T1DM | epsilon_cell | 6 | 16 | 3 | 13 | 3 |
| Pancreas | T1DM | leukocyte | 5 | 3632 | 2 | 3629 | 3 |
| Pancreas | T1DM | mesenchymal_cell | 11 | 37 | 7 | 33 | 4 |
| Pancreas | T1DM | PP_cell | 7 | 29 | 3 | 25 | 4 |
| Pancreas | T2DM | acinar_cell | 44 | 54 | 17 | 27 | 27 |
| Pancreas | T2DM | alpha_cell | 81 | 72 | 39 | 30 | 42 |
| Pancreas | T2DM | beta_cell | 163 | 91 | 102 | 30 | 61 |
| Pancreas | T2DM | delta_cell | 19 | 33 | 9 | 23 | 10 |
| Pancreas | T2DM | duct_epithelial_cell | 31 | 48 | 14 | 31 | 17 |
| Pancreas | T2DM | endothelial_cell | 27 | 167 | 6 | 146 | 21 |
| Pancreas | T2DM | epsilon_cell | 6 | 5 | 4 | 3 | 2 |
| Pancreas | T2DM | leukocyte | 12 | 3171 | 0 | 3159 | 12 |
| Pancreas | T2DM | mesenchymal_cell | 23 | 88 | 8 | 73 | 15 |
| Pancreas | T2DM | PP_cell | 17 | 26 | 6 | 15 | 11 |
| Pancreas | T2DM_admission | acinar_cell | 51 | 69 | 15 | 33 | 36 |
| Pancreas | T2DM_admission | alpha_cell | 82 | 86 | 34 | 38 | 48 |
| Pancreas | T2DM_admission | beta_cell | 185 | 116 | 105 | 36 | 80 |
| Pancreas | T2DM_admission | delta_cell | 22 | 44 | 8 | 30 | 14 |
| Pancreas | T2DM_admission | duct_epithelial_cell | 34 | 120 | 8 | 94 | 26 |
| Pancreas | T2DM_admission | endothelial_cell | 29 | 113 | 9 | 93 | 20 |
| Pancreas | T2DM_admission | epsilon_cell | 7 | 10 | 3 | 6 | 4 |
| Pancreas | T2DM_admission | leukocyte | 11 | 3313 | 0 | 3302 | 11 |
| Pancreas | T2DM_admission | mesenchymal_cell | 24 | 112 | 6 | 94 | 18 |
| Pancreas | T2DM_admission | PP_cell | 18 | 62 | 5 | 49 | 13 |
| Pancreas | T2DM_insulin | acinar_cell | 37 | 11 | 33 | 7 | 4 |
| Pancreas | T2DM_insulin | alpha_cell | 57 | 12 | 52 | 7 | 5 |
| Pancreas | T2DM_insulin | beta_cell | 102 | 15 | 95 | 8 | 7 |
| Pancreas | T2DM_insulin | delta_cell | 13 | 11 | 10 | 8 | 3 |
| Pancreas | T2DM_insulin | duct_epithelial_cell | 23 | 11 | 20 | 8 | 3 |
| Pancreas | T2DM_insulin | endothelial_cell | 22 | 15 | 17 | 10 | 5 |
| Pancreas | T2DM_insulin | epsilon_cell | 9 | 6 | 8 | 5 | 1 |
| Pancreas | T2DM_insulin | leukocyte | 7 | 3136 | 0 | 3129 | 7 |
| Pancreas | T2DM_insulin | mesenchymal_cell | 17 | 13 | 13 | 9 | 4 |
| Pancreas | T2DM_insulin | PP_cell | 12 | 11 | 9 | 8 | 3 |
| SAT | Obesity | adipocyte | 123 | 2398 | 5 | 2280 | 118 |
| SAT | Obesity | ASPC | 331 | 422 | 117 | 208 | 214 |
| SAT | Obesity | b_cell | 14 | 645 | 0 | 631 | 14 |
| SAT | Obesity | dendritic_cell | 22 | 113 | 2 | 93 | 20 |
| SAT | Obesity | endothelial | 25 | 4730 | 0 | 4705 | 25 |
| SAT | Obesity | LEC | 16 | 5102 | 0 | 5086 | 16 |
| SAT | Obesity | macrophage | 88 | 2189 | 4 | 2105 | 84 |
| SAT | Obesity | mast_cell | 28 | 5036 | 0 | 5008 | 28 |

| | | | | | | | |
|---|---|---|---|---|---|---|---|
| SAT | Obesity | monocyte | 16 | 251 | 5 | 240 | 11 |
| SAT | Obesity | neutrophil | 11 | 5195 | 0 | 5184 | 11 |
| SAT | Obesity | nk_cell | 26 | 2495 | 0 | 2469 | 26 |
| SAT | Obesity | pericyte | 41 | 4814 | 2 | 4775 | 39 |
| SAT | Obesity | SMC | 51 | 252 | 6 | 207 | 45 |
| SAT | Obesity | t_cell | 33 | 3948 | 0 | 3915 | 33 |
| SAT | CAD | adipocyte | 97 | 42 | 70 | 15 | 27 |
| SAT | CAD | ASPC | 193 | 51 | 155 | 13 | 38 |
| SAT | CAD | b_cell | 8 | 0 | 8 | 0 | 0 |
| SAT | CAD | dendritic_cell | 11 | 10 | 9 | 8 | 2 |
| SAT | CAD | endothelial | 14 | 1 | 13 | 0 | 1 |
| SAT | CAD | LEC | 13 | 0 | 13 | 0 | 0 |
| SAT | CAD | macrophage | 72 | 37 | 49 | 14 | 23 |
| SAT | CAD | mast_cell | 16 | 6 | 12 | 2 | 4 |
| SAT | CAD | monocyte | 11 | 4 | 8 | 1 | 3 |
| SAT | CAD | neutrophil | 7 | 1 | 6 | 0 | 1 |
| SAT | CAD | nk_cell | 16 | 4 | 14 | 2 | 2 |
| SAT | CAD | pericyte | 29 | 625 | 7 | 603 | 22 |
| SAT | CAD | SMC | 35 | 18 | 25 | 8 | 10 |
| SAT | CAD | t_cell | 18 | 13 | 13 | 8 | 5 |
| SAT | T2DM | adipocyte | 104 | 1367 | 11 | 1274 | 93 |
| SAT | T2DM | ASPC | 226 | 194 | 115 | 83 | 111 |
| SAT | T2DM | b_cell | 10 | 5037 | 1 | 5028 | 9 |
| SAT | T2DM | dendritic_cell | 16 | 4976 | 0 | 4960 | 16 |
| SAT | T2DM | endothelial | 21 | 3956 | 1 | 3936 | 20 |
| SAT | T2DM | LEC | 15 | 4698 | 2 | 4685 | 13 |
| SAT | T2DM | macrophage | 68 | 1686 | 7 | 1625 | 61 |
| SAT | T2DM | mast_cell | 28 | 4943 | 1 | 4916 | 27 |
| SAT | T2DM | monocyte | 13 | 4953 | 1 | 4941 | 12 |
| SAT | T2DM | neutrophil | 8 | 11 | 4 | 7 | 4 |
| SAT | T2DM | nk_cell | 20 | 50 | 4 | 34 | 16 |
| SAT | T2DM | pericyte | 29 | 3829 | 2 | 3802 | 27 |
| SAT | T2DM | SMC | 44 | 83 | 20 | 59 | 24 |
| SAT | T2DM | t_cell | 27 | 394 | 6 | 373 | 21 |
| SAT | HTN | adipocyte | 134 | 381 | 20 | 267 | 114 |
| SAT | HTN | ASPC | 329 | 446 | 92 | 209 | 237 |
| SAT | HTN | b_cell | 10 | 5114 | 0 | 5104 | 10 |
| SAT | HTN | dendritic_cell | 27 | 115 | 0 | 88 | 27 |
| SAT | HTN | endothelial | 26 | 242 | 8 | 224 | 18 |
| SAT | HTN | LEC | 9 | 168 | 0 | 159 | 9 |
| SAT | HTN | macrophage | 87 | 271 | 28 | 212 | 59 |
| SAT | HTN | mast_cell | 26 | 148 | 8 | 130 | 18 |
| SAT | HTN | monocyte | 16 | 49 | 4 | 37 | 12 |
| SAT | HTN | neutrophil | 8 | 14 | 1 | 7 | 7 |
| SAT | HTN | nk_cell | 43 | 3014 | 0 | 2971 | 43 |
| SAT | HTN | pericyte | 60 | 299 | 6 | 245 | 54 |
| SAT | HTN | SMC | 75 | 330 | 4 | 259 | 71 |
| SAT | HTN | t_cell | 38 | 2001 | 6 | 1969 | 32 |
| SAT | Stroke | adipocyte | 44 | 0 | 44 | 0 | 0 |
| SAT | Stroke | ASPC | 127 | 2 | 125 | 0 | 2 |
| SAT | Stroke | b_cell | 4 | 0 | 4 | 0 | 0 |
| SAT | Stroke | dendritic_cell | 7 | 0 | 7 | 0 | 0 |
| SAT | Stroke | endothelial | 10 | 0 | 10 | 0 | 0 |
| SAT | Stroke | LEC | 8 | 0 | 8 | 0 | 0 |
| SAT | Stroke | macrophage | 31 | 0 | 31 | 0 | 0 |
| SAT | Stroke | mast_cell | 6 | 0 | 6 | 0 | 0 |
| SAT | Stroke | monocyte | 6 | 0 | 6 | 0 | 0 |
| SAT | Stroke | neutrophil | 3 | 0 | 3 | 0 | 0 |
| SAT | Stroke | nk_cell | 10 | 0 | 10 | 0 | 0 |
| SAT | Stroke | pericyte | 10 | 0 | 10 | 0 | 0 |
| SAT | Stroke | SMC | 15 | 4 | 14 | 3 | 1 |
| SAT | Stroke | t_cell | 15 | 0 | 15 | 0 | 0 |
| SAT | Heart_Failure | adipocyte | 76 | 36 | 59 | 19 | 17 |
| SAT | Heart_Failure | ASPC | 154 | 36 | 131 | 13 | 23 |
| SAT | Heart_Failure | b_cell | 6 | 3834 | 1 | 3829 | 5 |
| SAT | Heart_Failure | dendritic_cell | 24 | 5680 | 0 | 5656 | 24 |
| SAT | Heart_Failure | endothelial | 18 | 14 | 13 | 9 | 5 |
| SAT | Heart_Failure | LEC | 12 | 9 | 11 | 8 | 1 |
| SAT | Heart_Failure | macrophage | 56 | 22 | 47 | 13 | 9 |
| SAT | Heart_Failure | mast_cell | 17 | 25 | 12 | 20 | 5 |
| SAT | Heart_Failure | monocyte | 9 | 1139 | 1 | 1131 | 8 |
| SAT | Heart_Failure | neutrophil | 4 | 1 | 4 | 1 | 0 |
| SAT | Heart_Failure | nk_cell | 10 | 23 | 4 | 17 | 6 |
| SAT | Heart_Failure | pericyte | 31 | 1159 | 5 | 1133 | 26 |
| SAT | Heart_Failure | SMC | 40 | 20 | 32 | 12 | 8 |
| SAT | Heart_Failure | t_cell | 18 | 23 | 12 | 17 | 6 |
| VAT | Obesity | adipocyte | 121 | 1146 | 4 | 1029 | 117 |
| VAT | Obesity | ASPC | 172 | 305 | 42 | 175 | 130 |
| VAT | Obesity | b_cell | 12 | 28 | 4 | 20 | 8 |
| VAT | Obesity | dendritic_cell | 20 | 161 | 3 | 144 | 17 |
| VAT | Obesity | endometrium | 19 | 4232 | 0 | 4213 | 19 |
| VAT | Obesity | endothelial | 22 | 3424 | 0 | 3402 | 22 |
| VAT | Obesity | LEC | 52 | 175 | 9 | 132 | 43 |
| VAT | Obesity | macrophage | 69 | 167 | 17 | 115 | 52 |
| VAT | Obesity | mast_cell | 24 | 3983 | 0 | 3959 | 24 |
| VAT | Obesity | mesothelium | 173 | 288 | 48 | 163 | 125 |
| VAT | Obesity | monocyte | 15 | 1865 | 0 | 1850 | 15 |
| VAT | Obesity | nk_cell | 14 | 4178 | 0 | 4164 | 14 |
| VAT | Obesity | pericyte | 42 | 149 | 8 | 115 | 34 |
| VAT | Obesity | SMC | 36 | 131 | 9 | 104 | 27 |
| VAT | Obesity | t_cell | 39 | 2024 | 1 | 1986 | 38 |

| | | | | | | | |
|---|---|---|---|---|---|---|---|
| VAT | CAD | adipocyte | 66 | 1220 | 15 | 1169 | 51 |
| VAT | CAD | ASPC | 114 | 56 | 86 | 28 | 28 |
| VAT | CAD | b_cell | 6 | 0 | 6 | 0 | 0 |
| VAT | CAD | dendritic_cell | 16 | 0 | 16 | 0 | 0 |
| VAT | CAD | endometrium | 7 | 0 | 7 | 0 | 0 |
| VAT | CAD | endothelial | 21 | 0 | 21 | 0 | 0 |
| VAT | CAD | LEC | 23 | 3 | 21 | 1 | 2 |
| VAT | CAD | macrophage | 34 | 1169 | 6 | 1141 | 28 |
| VAT | CAD | mast_cell | 15 | 0 | 15 | 0 | 0 |
| VAT | CAD | mesothelium | 106 | 143 | 61 | 98 | 45 |
| VAT | CAD | monocyte | 10 | 0 | 10 | 0 | 0 |
| VAT | CAD | nk_cell | 9 | 0 | 9 | 0 | 0 |
| VAT | CAD | pericyte | 33 | 0 | 33 | 0 | 0 |
| VAT | CAD | SMC | 19 | 0 | 19 | 0 | 0 |
| VAT | CAD | t_cell | 20 | 78 | 9 | 67 | 11 |
| VAT | T2DM | adipocyte | 74 | 183 | 34 | 143 | 40 |
| VAT | T2DM | ASPC | 102 | 178 | 48 | 124 | 54 |
| VAT | T2DM | b_cell | 8 | 4211 | 2 | 4205 | 6 |
| VAT | T2DM | dendritic_cell | 21 | 99 | 6 | 84 | 15 |
| VAT | T2DM | endometrium | 13 | 4284 | 2 | 4273 | 11 |
| VAT | T2DM | endothelial | 22 | 3323 | 1 | 3302 | 21 |
| VAT | T2DM | LEC | 34 | 59 | 16 | 41 | 18 |
| VAT | T2DM | macrophage | 52 | 64 | 24 | 36 | 28 |
| VAT | T2DM | mast_cell | 21 | 4140 | 3 | 4122 | 18 |
| VAT | T2DM | mesothelium | 119 | 115 | 63 | 59 | 56 |
| VAT | T2DM | monocyte | 13 | 19 | 9 | 15 | 4 |
| VAT | T2DM | nk_cell | 11 | 3964 | 1 | 3954 | 10 |
| VAT | T2DM | pericyte | 39 | 55 | 16 | 32 | 23 |
| VAT | T2DM | SMC | 26 | 68 | 11 | 53 | 15 |
| VAT | T2DM | t_cell | 34 | 1353 | 2 | 1321 | 32 |
| VAT | HTN | adipocyte | 134 | 214 | 27 | 107 | 107 |
| VAT | HTN | ASPC | 199 | 1139 | 18 | 958 | 181 |
| VAT | HTN | b_cell | 13 | 18 | 7 | 12 | 6 |
| VAT | HTN | dendritic_cell | 30 | 58 | 9 | 37 | 21 |
| VAT | HTN | endometrium | 17 | 3714 | 0 | 3697 | 17 |
| VAT | HTN | endothelial | 42 | 65 | 10 | 33 | 32 |
| VAT | HTN | LEC | 56 | 946 | 4 | 894 | 52 |
| VAT | HTN | macrophage | 89 | 187 | 19 | 117 | 70 |
| VAT | HTN | mast_cell | 27 | 34 | 7 | 14 | 20 |
| VAT | HTN | mesothelium | 193 | 361 | 45 | 213 | 148 |
| VAT | HTN | monocyte | 24 | 2828 | 1 | 2805 | 23 |
| VAT | HTN | nk_cell | 14 | 4001 | 0 | 3987 | 14 |
| VAT | HTN | pericyte | 60 | 1265 | 2 | 1207 | 58 |
| VAT | HTN | SMC | 42 | 2859 | 2 | 2819 | 40 |
| VAT | HTN | t_cell | 67 | 911 | 1 | 845 | 66 |
| VAT | Stroke | adipocyte | 51 | 0 | 51 | 0 | 0 |
| VAT | Stroke | ASPC | 70 | 806 | 29 | 765 | 41 |

b. Comparison of genes identified from causal inference and CTS-TWAS analysis with adjustment for multiple testing correction(Bonferroni<=0.05)

| Tissue | Disorder | Cell type | No. of causal genes | No. of TWAS genes | No. of novel genes from causal inference | No. of novel genes from TWAS | No. of common genes |
|---|---|---|---|---|---|---|---|
| Frontal Cortex | Anxiety | Astros | 12 | 0 | 12 | 0 | 0 |
| Frontal Cortex | Anxiety | Ex | 60 | 0 | 60 | 0 | 0 |
| Frontal Cortex | Anxiety | Inhib | 53 | 0 | 53 | 0 | 0 |
| Frontal Cortex | Anxiety | Micro_Macro | 4 | 0 | 4 | 0 | 0 |
| Frontal Cortex | Anxiety | Oligos | 19 | 0 | 19 | 0 | 0 |
| Frontal Cortex | Anxiety | OPCs | 13 | 0 | 13 | 0 | 0 |
| Frontal Cortex | Bipolar | Astros | 7 | 0 | 7 | 0 | 0 |
| Frontal Cortex | Bipolar | Ex | 63 | 0 | 63 | 0 | 0 |
| Frontal Cortex | Bipolar | Inhib | 42 | 1 | 41 | 0 | 1 |
| Frontal Cortex | Bipolar | Micro_Macro | 5 | 0 | 5 | 0 | 0 |
| Frontal Cortex | Bipolar | Oligos | 13 | 0 | 13 | 0 | 0 |
| Frontal Cortex | Bipolar | OPCs | 10 | 0 | 10 | 0 | 0 |
| Frontal Cortex | Depression | Astros | 13 | 2 | 12 | 1 | 1 |
| Frontal Cortex | Depression | Ex | 78 | 2 | 77 | 1 | 1 |
| Frontal Cortex | Depression | Inhib | 72 | 1 | 71 | 0 | 1 |
| Frontal Cortex | Depression | Micro_Macro | 9 | 1 | 8 | 0 | 1 |
| Frontal Cortex | Depression | Oligos | 21 | 1 | 20 | 0 | 1 |
| Frontal Cortex | Depression | OPCs | 16 | 1 | 15 | 0 | 1 |
| Frontal Cortex | Psychosis | Astros | 6 | 0 | 6 | 0 | 0 |
| Frontal Cortex | Psychosis | Ex | 53 | 0 | 53 | 0 | 0 |
| Frontal Cortex | Psychosis | Inhib | 63 | 0 | 63 | 0 | 0 |
| Frontal Cortex | Psychosis | Micro_Macro | 6 | 0 | 6 | 0 | 0 |
| Frontal Cortex | Psychosis | Oligos | 21 | 0 | 21 | 0 | 0 |
| Frontal Cortex | Psychosis | OPCs | 9 | 0 | 9 | 0 | 0 |
| Pancreas | Prediabetes | acinar_cell | 16 | 0 | 16 | 0 | 0 |
| Pancreas | Prediabetes | alpha_cell | 27 | 0 | 27 | 0 | 0 |
| Pancreas | Prediabetes | beta_cell | 63 | 0 | 63 | 0 | 0 |
| Pancreas | Prediabetes | delta_cell | 10 | 0 | 10 | 0 | 0 |
| Pancreas | Prediabetes | duct_epithelial_cell | 12 | 3 | 11 | 2 | 1 |
| Pancreas | Prediabetes | endothelial_cell | 13 | 0 | 13 | 0 | 0 |
| Pancreas | Prediabetes | epsilon_cell | 4 | 0 | 4 | 0 | 0 |
| Pancreas | Prediabetes | leukocyte | 5 | 0 | 5 | 0 | 0 |
| Pancreas | Prediabetes | mesenchymal_cell | 10 | 0 | 10 | 0 | 0 |
| Pancreas | Prediabetes | PP_cell | 8 | 2 | 7 | 1 | 1 |
| Pancreas | T1DM | acinar_cell | 24 | 27 | 19 | 22 | 5 |
| Pancreas | T1DM | alpha_cell | 50 | 27 | 45 | 22 | 5 |
| Pancreas | T1DM | beta_cell | 114 | 27 | 108 | 21 | 6 |
| Pancreas | T1DM | delta_cell | 12 | 26 | 8 | 22 | 4 |
| Pancreas | T1DM | duct_epithelial_cell | 15 | 27 | 12 | 24 | 3 |
| Pancreas | T1DM | endothelial_cell | 10 | 23 | 8 | 21 | 2 |
| Pancreas | T1DM | epsilon_cell | 6 | 15 | 3 | 12 | 3 |
| Pancreas | T1DM | leukocyte | 5 | 3168 | 2 | 3165 | 3 |
| Pancreas | T1DM | mesenchymal_cell | 11 | 28 | 8 | 25 | 3 |
| Pancreas | T1DM | PP_cell | 7 | 21 | 3 | 17 | 4 |
| Pancreas | T2DM | acinar_cell | 44 | 20 | 33 | 9 | 11 |
| Pancreas | T2DM | alpha_cell | 81 | 22 | 67 | 8 | 14 |
| Pancreas | T2DM | beta_cell | 163 | 24 | 148 | 9 | 15 |
| Pancreas | T2DM | delta_cell | 19 | 12 | 12 | 5 | 7 |
| Pancreas | T2DM | duct_epithelial_cell | 31 | 17 | 24 | 10 | 7 |
| Pancreas | T2DM | endothelial_cell | 27 | 20 | 16 | 9 | 11 |
| Pancreas | T2DM | epsilon_cell | 6 | 5 | 4 | 3 | 2 |
| Pancreas | T2DM | leukocyte | 12 | 21 | 7 | 16 | 5 |
| Pancreas | T2DM | mesenchymal_cell | 23 | 20 | 16 | 13 | 7 |
| Pancreas | T2DM | PP_cell | 17 | 11 | 13 | 7 | 4 |
| Pancreas | T2DM_admission | acinar_cell | 51 | 27 | 35 | 11 | 16 |
| Pancreas | T2DM_admission | alpha_cell | 82 | 32 | 65 | 15 | 17 |
| Pancreas | T2DM_admission | beta_cell | 185 | 34 | 164 | 13 | 21 |
| Pancreas | T2DM_admission | delta_cell | 22 | 20 | 12 | 10 | 10 |
| Pancreas | T2DM_admission | duct_epithelial_cell | 34 | 30 | 23 | 19 | 11 |
| Pancreas | T2DM_admission | endothelial_cell | 29 | 25 | 17 | 13 | 12 |
| Pancreas | T2DM_admission | epsilon_cell | 7 | 6 | 4 | 3 | 3 |
| Pancreas | T2DM_admission | leukocyte | 11 | 62 | 5 | 56 | 6 |
| Pancreas | T2DM_admission | mesenchymal_cell | 24 | 26 | 12 | 14 | 12 |
| Pancreas | T2DM_admission | PP_cell | 18 | 17 | 12 | 11 | 6 |
| Pancreas | T2DM_insulin | acinar_cell | 37 | 7 | 35 | 5 | 2 |
| Pancreas | T2DM_insulin | alpha_cell | 57 | 8 | 55 | 6 | 2 |
| Pancreas | T2DM_insulin | beta_cell | 102 | 8 | 100 | 6 | 2 |
| Pancreas | T2DM_insulin | delta_cell | 13 | 7 | 10 | 4 | 3 |
| Pancreas | T2DM_insulin | duct_epithelial_cell | 23 | 7 | 22 | 6 | 1 |
| Pancreas | T2DM_insulin | endothelial_cell | 22 | 4 | 22 | 4 | 0 |
| Pancreas | T2DM_insulin | epsilon_cell | 9 | 3 | 8 | 2 | 1 |
| Pancreas | T2DM_insulin | leukocyte | 7 | 4 | 5 | 2 | 2 |
| Pancreas | T2DM_insulin | mesenchymal_cell | 17 | 7 | 15 | 5 | 2 |
| Pancreas | T2DM_insulin | PP_cell | 12 | 6 | 11 | 5 | 1 |
| SAT | Obesity | adipocyte | 123 | 365 | 85 | 327 | 38 |
| SAT | Obesity | ASPC | 331 | 112 | 271 | 52 | 60 |
| SAT | Obesity | b_cell | 14 | 32 | 4 | 22 | 10 |
| SAT | Obesity | dendritic_cell | 22 | 45 | 10 | 33 | 12 |
| SAT | Obesity | endothelial | 25 | 1634 | 7 | 1616 | 18 |
| SAT | Obesity | LEC | 16 | 464 | 5 | 453 | 11 |
| SAT | Obesity | macrophage | 88 | 186 | 49 | 147 | 39 |
| SAT | Obesity | mast_cell | 28 | 2777 | 10 | 2759 | 18 |
| SAT | Obesity | monocyte | 16 | 47 | 6 | 37 | 10 |
| SAT | Obesity | neutrophil | 11 | 25 | 6 | 20 | 5 |

| | | | | | | | |
|---|---|---|---|---|---|---|---|
| SAT | Obesity | nk_cell | 26 | 106 | 10 | 90 | 16 |
| SAT | Obesity | pericyte | 41 | 2417 | 17 | 2393 | 24 |
| SAT | Obesity | SMC | 51 | 69 | 29 | 47 | 22 |
| SAT | Obesity | t_cell | 33 | 378 | 8 | 353 | 25 |
| SAT | CAD | adipocyte | 97 | 8 | 92 | 3 | 5 |
| SAT | CAD | ASPC | 193 | 12 | 185 | 4 | 8 |
| SAT | CAD | b_cell | 8 | 0 | 8 | 0 | 0 |
| SAT | CAD | dendritic_cell | 11 | 6 | 9 | 4 | 2 |
| SAT | CAD | endothelial | 14 | 1 | 13 | 0 | 1 |
| SAT | CAD | LEC | 13 | 0 | 13 | 0 | 0 |
| SAT | CAD | macrophage | 72 | 9 | 65 | 2 | 7 |
| SAT | CAD | mast_cell | 16 | 3 | 14 | 1 | 2 |
| SAT | CAD | monocyte | 11 | 2 | 9 | 0 | 2 |
| SAT | CAD | neutrophil | 7 | 1 | 6 | 0 | 1 |
| SAT | CAD | nk_cell | 16 | 2 | 15 | 1 | 1 |
| SAT | CAD | pericyte | 29 | 8 | 24 | 3 | 5 |
| SAT | CAD | SMC | 35 | 4 | 32 | 1 | 3 |
| SAT | CAD | t_cell | 18 | 3 | 16 | 1 | 2 |
| SAT | T2DM | adipocyte | 104 | 87 | 82 | 65 | 22 |
| SAT | T2DM | ASPC | 226 | 55 | 193 | 22 | 33 |
| SAT | T2DM | b_cell | 10 | 33 | 6 | 29 | 4 |
| SAT | T2DM | dendritic_cell | 16 | 361 | 8 | 353 | 8 |
| SAT | T2DM | endothelial | 21 | 1004 | 10 | 993 | 11 |
| SAT | T2DM | LEC | 15 | 75 | 7 | 67 | 8 |
| SAT | T2DM | macrophage | 68 | 277 | 52 | 261 | 16 |
| SAT | T2DM | mast_cell | 28 | 2628 | 14 | 2614 | 14 |
| SAT | T2DM | monocyte | 13 | 138 | 6 | 131 | 7 |
| SAT | T2DM | neutrophil | 8 | 8 | 5 | 5 | 3 |
| SAT | T2DM | nk_cell | 20 | 23 | 11 | 14 | 9 |
| SAT | T2DM | pericyte | 29 | 135 | 15 | 121 | 14 |
| SAT | T2DM | SMC | 44 | 29 | 32 | 17 | 12 |
| SAT | T2DM | t_cell | 27 | 32 | 18 | 23 | 9 |
| SAT | HTN | adipocyte | 134 | 88 | 82 | 36 | 52 |
| SAT | HTN | ASPC | 329 | 112 | 257 | 40 | 72 |
| SAT | HTN | b_cell | 10 | 92 | 4 | 86 | 6 |
| SAT | HTN | dendritic_cell | 27 | 30 | 12 | 15 | 15 |
| SAT | HTN | endothelial | 26 | 37 | 18 | 29 | 8 |
| SAT | HTN | LEC | 9 | 18 | 3 | 12 | 6 |
| SAT | HTN | macrophage | 87 | 73 | 59 | 45 | 28 |
| SAT | HTN | mast_cell | 26 | 36 | 16 | 26 | 10 |
| SAT | HTN | monocyte | 16 | 18 | 11 | 13 | 5 |
| SAT | HTN | neutrophil | 8 | 5 | 5 | 2 | 3 |
| SAT | HTN | nk_cell | 43 | 85 | 21 | 63 | 22 |
| SAT | HTN | pericyte | 60 | 55 | 41 | 36 | 19 |
| SAT | HTN | SMC | 75 | 70 | 41 | 36 | 34 |
| SAT | HTN | t_cell | 38 | 71 | 26 | 59 | 12 |
| SAT | Stroke | adipocyte | 44 | 0 | 44 | 0 | 0 |
| SAT | Stroke | ASPC | 127 | 2 | 125 | 0 | 2 |
| SAT | Stroke | b_cell | 4 | 0 | 4 | 0 | 0 |
| SAT | Stroke | dendritic_cell | 7 | 0 | 7 | 0 | 0 |
| SAT | Stroke | endothelial | 10 | 0 | 10 | 0 | 0 |
| SAT | Stroke | LEC | 8 | 0 | 8 | 0 | 0 |
| SAT | Stroke | macrophage | 31 | 0 | 31 | 0 | 0 |
| SAT | Stroke | mast_cell | 6 | 0 | 6 | 0 | 0 |
| SAT | Stroke | monocyte | 6 | 0 | 6 | 0 | 0 |
| SAT | Stroke | neutrophil | 3 | 0 | 3 | 0 | 0 |
| SAT | Stroke | nk_cell | 10 | 0 | 10 | 0 | 0 |
| SAT | Stroke | pericyte | 10 | 0 | 10 | 0 | 0 |
| SAT | Stroke | SMC | 15 | 3 | 14 | 2 | 1 |
| SAT | Stroke | t_cell | 15 | 0 | 15 | 0 | 0 |
| SAT | Heart_Failure | adipocyte | 76 | 15 | 73 | 12 | 3 |
| SAT | Heart_Failure | ASPC | 154 | 17 | 147 | 10 | 7 |
| SAT | Heart_Failure | b_cell | 6 | 23 | 3 | 20 | 3 |
| SAT | Heart_Failure | dendritic_cell | 24 | 4757 | 9 | 4742 | 15 |
| SAT | Heart_Failure | endothelial | 18 | 6 | 16 | 4 | 2 |
| SAT | Heart_Failure | LEC | 12 | 7 | 12 | 7 | 0 |
| SAT | Heart_Failure | macrophage | 56 | 14 | 53 | 11 | 3 |
| SAT | Heart_Failure | mast_cell | 17 | 9 | 15 | 7 | 2 |
| SAT | Heart_Failure | monocyte | 9 | 10 | 7 | 8 | 2 |
| SAT | Heart_Failure | neutrophil | 4 | 1 | 4 | 1 | 0 |
| SAT | Heart_Failure | nk_cell | 10 | 9 | 7 | 6 | 3 |
| SAT | Heart_Failure | pericyte | 31 | 31 | 25 | 25 | 6 |
| SAT | Heart_Failure | SMC | 40 | 11 | 38 | 9 | 2 |
| SAT | Heart_Failure | t_cell | 18 | 13 | 16 | 11 | 2 |
| VAT | Obesity | adipocyte | 121 | 112 | 80 | 71 | 41 |
| VAT | Obesity | ASPC | 172 | 91 | 123 | 42 | 49 |
| VAT | Obesity | b_cell | 12 | 18 | 4 | 10 | 8 |
| VAT | Obesity | dendritic_cell | 20 | 42 | 5 | 27 | 15 |
| VAT | Obesity | endometrium | 19 | 2868 | 8 | 2857 | 11 |
| VAT | Obesity | endothelial | 22 | 739 | 6 | 723 | 16 |
| VAT | Obesity | LEC | 52 | 55 | 30 | 33 | 22 |
| VAT | Obesity | macrophage | 69 | 62 | 44 | 37 | 25 |
| VAT | Obesity | mast_cell | 24 | 136 | 11 | 123 | 13 |
| VAT | Obesity | mesothelium | 173 | 87 | 127 | 41 | 46 |
| VAT | Obesity | monocyte | 15 | 37 | 4 | 26 | 11 |
| VAT | Obesity | nk_cell | 14 | 2798 | 4 | 2788 | 10 |
| VAT | Obesity | pericyte | 42 | 51 | 23 | 32 | 19 |
| VAT | Obesity | SMC | 36 | 48 | 19 | 31 | 17 |
| VAT | Obesity | t_cell | 39 | 409 | 17 | 387 | 22 |
| VAT | CAD | adipocyte | 66 | 17 | 58 | 9 | 8 |
| VAT | CAD | ASPC | 114 | 2 | 112 | 0 | 2 |

| | | | | | | | |
|---|---|---|---|---|---|---|---|
| VAT | CAD | b_cell | 6 | 0 | 6 | 0 | 0 |
| VAT | CAD | dendritic_cell | 16 | 0 | 16 | 0 | 0 |
| VAT | CAD | endometrium | 7 | 0 | 7 | 0 | 0 |
| VAT | CAD | endothelial | 21 | 0 | 21 | 0 | 0 |
| VAT | CAD | LEC | 23 | 1 | 22 | 0 | 1 |
| VAT | CAD | macrophage | 34 | 12 | 29 | 7 | 5 |
| VAT | CAD | mast_cell | 15 | 0 | 15 | 0 | 0 |
| VAT | CAD | mesothelium | 106 | 12 | 97 | 3 | 9 |
| VAT | CAD | monocyte | 10 | 0 | 10 | 0 | 0 |
| VAT | CAD | nk_cell | 9 | 0 | 9 | 0 | 0 |
| VAT | CAD | pericyte | 33 | 0 | 33 | 0 | 0 |
| VAT | CAD | SMC | 19 | 0 | 19 | 0 | 0 |
| VAT | CAD | t_cell | 20 | 6 | 17 | 3 | 3 |
| VAT | T2DM | adipocyte | 74 | 33 | 58 | 17 | 16 |
| VAT | T2DM | ASPC | 102 | 34 | 83 | 15 | 19 |
| VAT | T2DM | b_cell | 8 | 21 | 6 | 19 | 2 |
| VAT | T2DM | dendritic_cell | 21 | 17 | 14 | 10 | 7 |
| VAT | T2DM | endometrium | 13 | 3007 | 7 | 3001 | 6 |
| VAT | T2DM | endothelial | 22 | 758 | 10 | 746 | 12 |
| VAT | T2DM | LEC | 34 | 22 | 25 | 13 | 9 |
| VAT | T2DM | macrophage | 52 | 29 | 34 | 11 | 18 |
| VAT | T2DM | mast_cell | 21 | 257 | 14 | 250 | 7 |
| VAT | T2DM | mesothelium | 119 | 41 | 99 | 21 | 20 |
| VAT | T2DM | monocyte | 13 | 12 | 10 | 9 | 3 |
| VAT | T2DM | nk_cell | 11 | 296 | 4 | 289 | 7 |
| VAT | T2DM | pericyte | 39 | 24 | 26 | 11 | 13 |
| VAT | T2DM | SMC | 26 | 23 | 17 | 14 | 9 |
| VAT | T2DM | t_cell | 34 | 52 | 24 | 42 | 10 |
| VAT | HTN | adipocyte | 134 | 60 | 83 | 9 | 51 |
| VAT | HTN | ASPC | 199 | 216 | 126 | 143 | 73 |
| VAT | HTN | b_cell | 13 | 6 | 10 | 3 | 3 |
| VAT | HTN | dendritic_cell | 30 | 13 | 21 | 4 | 9 |
| VAT | HTN | endometrium | 17 | 56 | 11 | 50 | 6 |
| VAT | HTN | endothelial | 42 | 27 | 23 | 8 | 19 |
| VAT | HTN | LEC | 56 | 60 | 30 | 34 | 26 |
| VAT | HTN | macrophage | 89 | 49 | 53 | 13 | 36 |
| VAT | HTN | mast_cell | 27 | 13 | 16 | 2 | 11 |
| VAT | HTN | mesothelium | 193 | 81 | 133 | 21 | 60 |
| VAT | HTN | monocyte | 24 | 38 | 11 | 25 | 13 |
| VAT | HTN | nk_cell | 14 | 1681 | 5 | 1672 | 9 |
| VAT | HTN | pericyte | 60 | 72 | 30 | 42 | 30 |
| VAT | HTN | SMC | 42 | 150 | 19 | 127 | 23 |
| VAT | HTN | t_cell | 67 | 59 | 36 | 28 | 31 |
| VAT | Stroke | adipocyte | 51 | 0 | 51 | 0 | 0 |
| VAT | Stroke | ASPC | 70 | 3 | 68 | 1 | 2 |
